# Metabolic and proteomic signatures of type 2 diabetes subtypes in an Arab population

**DOI:** 10.1101/2022.01.13.22269204

**Authors:** Shaza B. Zaghlool, Anna Halama, Nisha Stephan, Manonanthini Thangam, Emma Ahlqvist, Omar M. E. Albagha, Abdul Badi Abou⍰Samra, Karsten Suhre

## Abstract

**Background:** Type 2 diabetes (T2D) has a heterogeneous etiology which is increasingly recognized to influence the risk of complications and choice of treatment. A data driven cluster analysis in four separate European populations of patients with type 2 diabetes identified four subtypes of severe insulin dependent (SIDD), severe insulin resistant (SIRD), mild obesity-related (MOD), and mild age-related (MARD) (Ahlqvist *et al*., Lancet Diabetes Endocrinology, 2018). Our aim was to extend this classification to the Arab population of Qatar and characterize the biological processes that differentiate these subtypes in relation to metabolomic and proteomic signatures.

**Methods:** The Ahlqvist *et al*. subtype clustering approach was applied to 631 individuals with T2D from the Qatar Biobank (QBB) and validated in an independent set of 420 participants from the same population. The association between blood metabolites (n=1,159) and protein levels (n=1,305) with each cluster were established.

**Findings:** The four subtypes of T2D were reproduced and validated in the population of Qatar. Cluster-specific metabolomic and proteomic associations revealed subtype-specific molecular processes. Activation of the complement system with many features of autoimmune diabetes and reduced 1,5-anhydroglucitol (1,5-AG) characterized SIDD, with evidence of impaired insulin signaling in SIRD, elevated leptin and fatty acid binding protein in MOD, whilst MARD appeared to be the healthiest subgroup.

**Interpretation:** We have replicated the four T2D clusters in an Arab population and identified distinct metabolic and proteomic signatures, providing insights into underlying etiology with the potential to deploy subtype-specific treatment options.

## INTRODUCTION

Type 2 diabetes (T2D) is a complex metabolic disorder defined by dysregulated glucose homeostasis, driven by imbalanced energy intake and expenditure, dysfunction of insulin signaling and chronic inflammation ^1-3^. Multiple therapies are now available to improve glycemic control ^4^ and provide additional benefits in relation to complications ^5-7^. Indeed, individualized therapies targeting the underlying pathophysiology and complications should be a major goal in the treatment of patients with T2D^1^.

Ahlqvist et al. ^8^ used data on age at diagnosis, BMI, HbA_1c_, homeostasis model assessment (HOMA) estimates of β-cell function (HOMA2-B), insulin resistance (HOMA2-IR), and presence or absence of glutamic acid decarboxylase antibodies (GADA) to stratify subjects into four clusters representing T2D subtypes and one cluster with severe autoimmune diabetes (SAID) ^8^. The four T2D clusters were named in reference to their characterizing phenotypic signatures as *Severe Insulin Dependent Diabetes* (SIDD), *Severe Insulin Resistant Diabetes* (SIRD), *Mild Obesity-related Diabetes* (MOD) and *Mild Age-related Diabetes* (MARD). Since its publication in 2018, the paper has been cited over 600 times and discussed in multiple reviews ^1,9-13^. The clusters have been replicated in British ^14^, German ^15,16^, Mexican American and Chinese^17-19^, Japanese ^20^, Asian Indian ^21^, Mexican ^22^ and Icelandic ^23^ cohorts, suggesting a generalizability to other ethnicities. In the original analysis predisposition to retinopathy and nephropathy were identified in different clusters, and more recently a German cohort cluster analysis has revealed predisposition to non-alcoholic fatty liver disease and diabetic neuropathy ^15^. A study clustering genetic risk loci for T2D associated traits observed some overlap with the clusters of Ahlqvist ^24^. More recently, evidence for distinct genetic backgrounds of the subtypes has been found ^25^. Schüssler-Fiorenza Rose et al. ^26^ used multi-omics measurements in a longitudinal study to develop prediction models for insulin resistance and the German Diabetes Study (GDS) showed differences in protein biomarkers of inflammation between subgroups ^16^. These studies suggest that deep molecular phenotyping may provide key insights into the underlying pathophysiology of glucose dysregulation and development and progression of comorbidities in patients with T2D.

Whilst a mechanistic link has been suggested between increased fat storage and compromised glucose homeostasis ^27^, body mass index (BMI) alone does not explain the difference between normal and dysfunctional glucose metabolism. Specific metabolic and proteomic processes may help to characterize the broader spectrum of physiological perturbations associated with impaired glucose metabolism to enable subtype-specific individualization of therapies. We hypothesized that there are distinctive alterations in metabolic and proteomic components of signaling pathways underlying the different T2D clusters.

We have analyzed data from the Qatar Biobank (QBB) population and translated the Ahlqvist et al. clustering approach to an Arab population. Further, applying broad non-targeted metabolomics and affinity proteomics profiling we have identified cluster specific physiological and biochemical processes in relation to their predominant treatment regimens (**Figure 1**).

**Figure 1.**
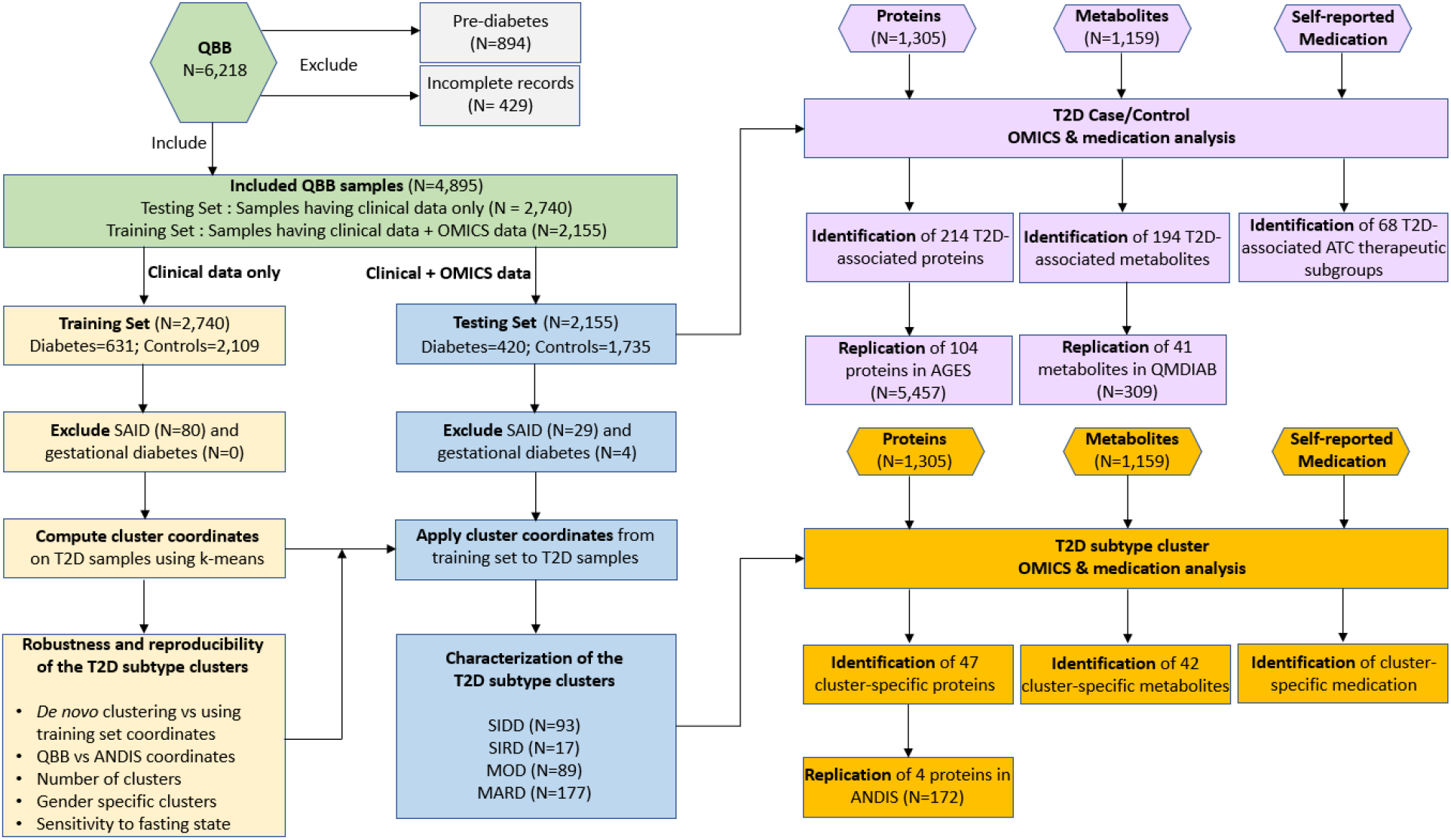
Study flow.

## METHODS

### Study population

Qatar Biobank (QBB) includes a population of Qatar nationals or long-term residents (≥15 years living in Qatar), aged 18 years and older in the State of Qatar ^28^. Extensive baseline socio-demographic data, clinical and behavioral phenotypic data and serum concentrations of HbA_1c_, triglycerides, glucose, C-peptide, creatinine, total cholesterol, LDL-C and HDL-C, and multiple other clinical biochemistry parameters ^29^ have been measured at the central laboratory of Hamad Medical Corporation (HMC), accredited by the College of American Pathologists.

All QBB participants signed an informed consent form prior to their participation. The study was approved by HMC ethics committee and the QBB institutional review board. At the time of analysis, QBB data was available for 6,218 participants. Over 96% of the participants reported having grandparents that were Qatari nationals. For 2,155 individuals, metabolomics and proteomics data had been collected in parallel. 429 participants with incomplete records and 894 individuals with HbA_1c_ ranging between 5.7 and 6.4 who did not match our diabetes definition (see below) were excluded, leaving 4,895 samples for analysis. Blood samples were collected more than 2 hours after their last meal or calorie-containing drink in 77% of participants. 50.7% of the participants (52.8% of the T2D cases and 40.6% of the controls) had been fasting for over 8 hours. This dataset was split into two groups, using the samples without omics data as a training set for the clustering (N=2,740), and the samples with omics data as a testing set for validation, and to further evaluate the associations of the metabolite and protein levels with T2D in a case-control setting and with T2D subtypes (N=2,155).

The group without omics data contained 631 individuals with T2D which were used to define the cluster coordinates. The group with omics data contained 420 individuals with T2D and was used for cluster validation, and then further for metabolomics and proteomics associations analyses (**Figure 1**). The study demographics for the two groups together are shown in **Table 1**. Both groups of data were similar, ie. clinical variables had comparable mean values and percentages in both the T2D cases and controls (**Supplementary Table 1**).

**Table 1.**
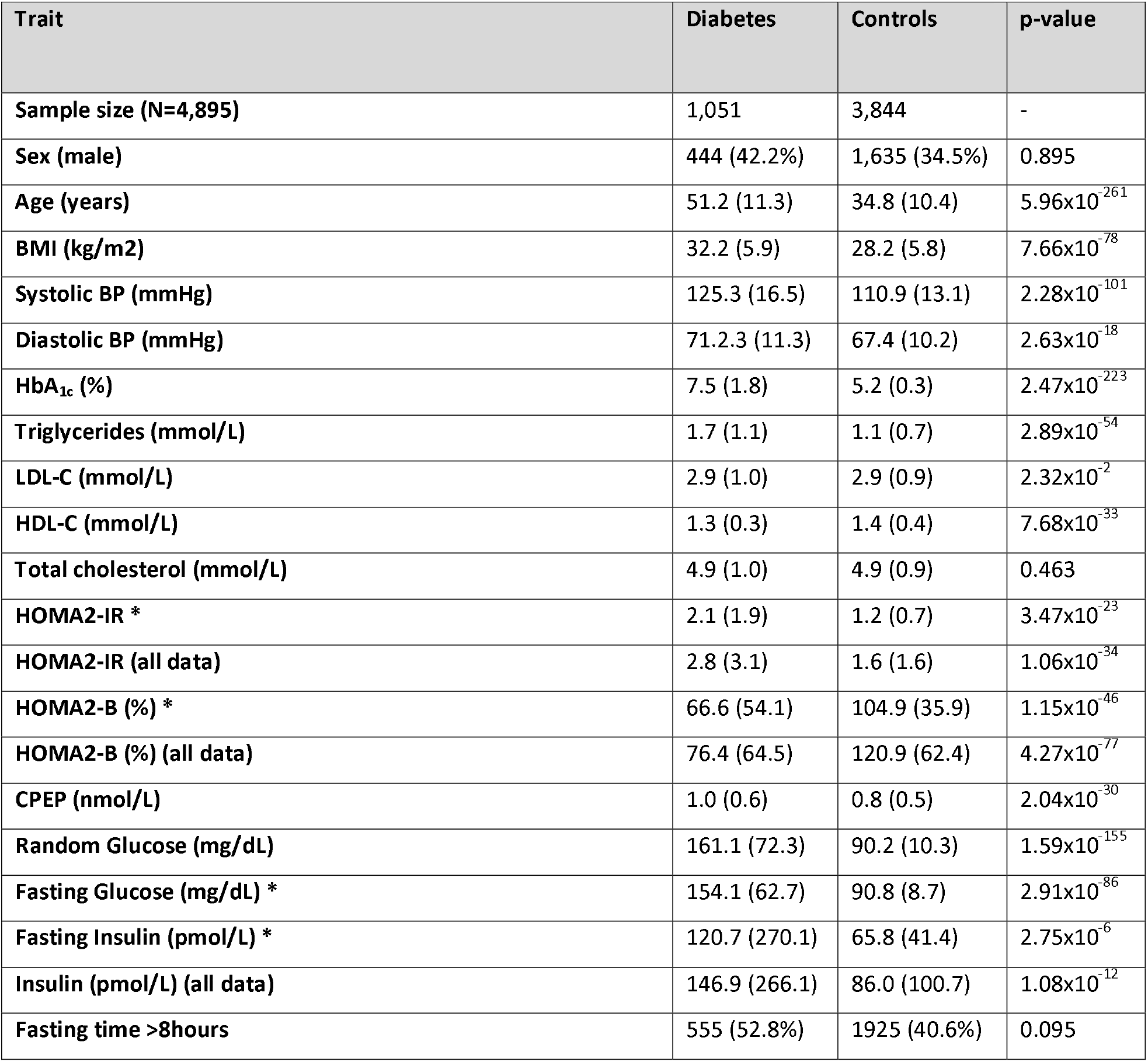
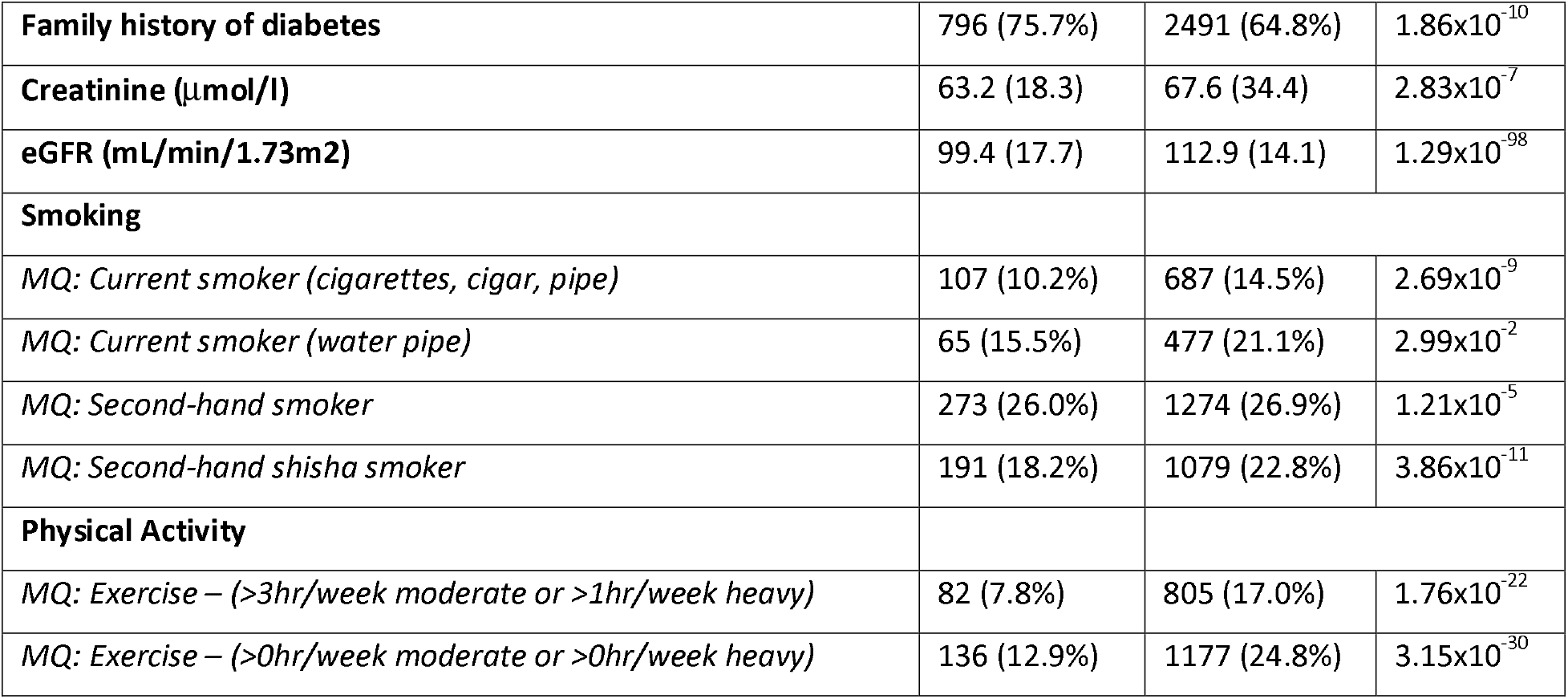
Demographics of the QBB Diabetes and control groups. The clinical traits that are marked with an asterisk were computed for individuals who fasted for eight or more hours at the time of blood drawing. HOMA2-IR: homeostasis model of assessment of insulin resistance. HOMA2-B: homeostasis model assessment of beta cell function. Family history is defined as either parent having a history record of diabetes. The data are number (%) or means (SD), as appropriate, p-values are from Fisher or student t-tests for categorical or continuous variables, respectively.

| Trait | Diabetes | Controls | p-value |
| --- | --- | --- | --- |
| Sample size (N=4,895) | 1,051 | 3,844 | - |
| Sex (male) | 444 (42.2%) | 1,635 (34.5%) | 0.895 |
| Age (years) | 51.2 (11.3) | 34.8 (10.4) | $5.96 \times 10^{-261}$ |
| BMI (kg/m<sup>2</sup>) | 32.2 (5.9) | 28.2 (5.8) | $7.66 \times 10^{-78}$ |
| Systolic BP (mmHg) | 125.3 (16.5) | 110.9 (13.1) | $2.28 \times 10^{-101}$ |
| Diastolic BP (mmHg) | 71.2.3 (11.3) | 67.4 (10.2) | $2.63 \times 10^{-18}$ |
| HbA<sub>1c</sub> (%) | 7.5 (1.8) | 5.2 (0.3) | $2.47 \times 10^{-223}$ |
| Triglycerides (mmol/L) | 1.7 (1.1) | 1.1 (0.7) | $2.89 \times 10^{-54}$ |
| LDL-C (mmol/L) | 2.9 (1.0) | 2.9 (0.9) | $2.32 \times 10^{-2}$ |
| HDL-C (mmol/L) | 1.3 (0.3) | 1.4 (0.4) | $7.68 \times 10^{-33}$ |
| Total cholesterol (mmol/L) | 4.9 (1.0) | 4.9 (0.9) | 0.463 |
| HOMA2-IR * | 2.1 (1.9) | 1.2 (0.7) | $3.47 \times 10^{-23}$ |
| HOMA2-IR (all data) | 2.8 (3.1) | 1.6 (1.6) | $1.06 \times 10^{-34}$ |
| HOMA2-B (%) * | 66.6 (54.1) | 104.9 (35.9) | $1.15 \times 10^{-46}$ |
| HOMA2-B (%) (all data) | 76.4 (64.5) | 120.9 (62.4) | $4.27 \times 10^{-77}$ |
| CPEP (nmol/L) | 1.0 (0.6) | 0.8 (0.5) | $2.04 \times 10^{-30}$ |
| Random Glucose (mg/dL) | 161.1 (72.3) | 90.2 (10.3) | $1.59 \times 10^{-155}$ |
| Fasting Glucose (mg/dL) * | 154.1 (62.7) | 90.8 (8.7) | $2.91 \times 10^{-86}$ |
| Fasting Insulin (pmol/L) * | 120.7 (270.1) | 65.8 (41.4) | $2.75 \times 10^{-6}$ |
| Insulin (pmol/L) (all data) | 146.9 (266.1) | 86.0 (100.7) | $1.08 \times 10^{-12}$ |
| Fasting time >8hours | 555 (52.8%) | 1925 (40.6%) | 0.095 |
| **Family history of diabetes** | 796 (75.7%) | 2491 (64.8%) | $1.86 \times 10^{-10}$ |
| **Creatinine ($\mu\text{mol/l}$)** | 63.2 (18.3) | 67.6 (34.4) | $2.83 \times 10^{-7}$ |
| **eGFR ($\text{mL/min/1.73m}^2$)** | 99.4 (17.7) | 112.9 (14.1) | $1.29 \times 10^{-98}$ |
| **Smoking** |  |  |  |
| *MQ: Current smoker (cigarettes, cigar, pipe)* | 107 (10.2%) | 687 (14.5%) | $2.69 \times 10^{-9}$ |
| *MQ: Current smoker (water pipe)* | 65 (15.5%) | 477 (21.1%) | $2.99 \times 10^{-2}$ |
| *MQ: Second-hand smoker* | 273 (26.0%) | 1274 (26.9%) | $1.21 \times 10^{-5}$ |
| *MQ: Second-hand shisha smoker* | 191 (18.2%) | 1079 (22.8%) | $3.86 \times 10^{-11}$ |
| **Physical Activity** |  |  |  |
| *MQ: Exercise – (>3hr/week moderate or >1hr/week heavy)* | 82 (7.8%) | 805 (17.0%) | $1.76 \times 10^{-22}$ |
| *MQ: Exercise – (>0hr/week moderate or >0hr/week heavy)* | 136 (12.9%) | 1177 (24.8%) | $3.15 \times 10^{-30}$ |

### Definition of T2D and controls

Subjects were defined as controls if all the following four conditions were met: first, no self-reported physician diagnosis of diabetes; second, no self-reported treatment with any diabetes specific medication; third, HbA_1c_ < 5.7%; and fourth, random glucose level < 200 mg/dL. T2D was defined if any one of the following four conditions was met: first, having a physician diagnosis of diabetes based on the questionnaire (13.3% of all participants), second, being under diabetes treatment based on the QBB questionnaire (11.6%), third, having an HbA_1c_ > 6.5% (10.5%), or fourth, having a random glucose level > 200 mg/dL (3.3%). Based on this definition, 15.4% of individuals were defined as T2D cases. Most individuals with a physician diagnosis of diabetes were on oral anti-diabetic treatment (73.7%), insulin treatment (23.9%), diet treatment (38.3%) and/or physical activity treatment (16.2%) (see Supplementary Figure 1 for a Venn diagram). Individuals with HbA_1c_ between 5.7% and 6.4% (N=894) or self-reported gestational diabetes (N=4) were excluded. Ahlqvist *et al*. used glutamic acid decarboxylase antibodies (GADA) to define an additional subtype of severe autoimmune diabetes (SAID). As GADA measurements were not available in QBB, individuals with self-reported type 1 diabetes (T1D) or C-peptide concentrations below 0.5 nmol/L and on insulin treatment were classified as SAID (N=109). These individuals were excluded from the statistical analysis, but the proteomic and metabolic levels of this subgroup are shown where appropriate.

### Training and testing sets

The cohort was split into two sets, a training set (N=2,740) and a testing set (N=2,155). The latter was chosen to overlap with available proteomics, metabolomics, and medication usage data. There were no substantial differences in demographics between the training and the testing set (**Supplementary Table 1**). The training set included 631 individuals with T2D, and the testing set included 420 individuals with T2D.

### Medication

QBB study participants provided information on their regular usage of over the counter and prescription medication as free text, which required annotation and homogenization. The questionnaire included the following question: “Are you taking any over-the-counter medication or prescription medicines regularly? For example, daily, weekly, monthly or every few months - such as depot injections?” and allowed participants to provide up to 30 free text entries. Anatomical Therapeutic Chemical (ATC)^30^ annotation was retrieved from the DrugBank annotation file. We annotated all entries from the questionnaire with a unique active molecule from the DrugBank repository, molecular class, indication, and its corresponding ATC code where available. The medication data covered 394 unique molecules, 529 ATC codes, 218 molecular classes, and 117 indications.

### Proteomics

Levels of 1,305 blood circulating proteins (**Supplementary Table 2**) were measured for 2,935 samples using the aptamer-based SOMAscan platform (kit version 1.3, Somalogic, Boulder, CO) ^31^ implemented at Weill Cornell Medicine – Qatar, as previously described ^32^. A detailed description of the platform can be found in the “SOMAmer Reagent Specificity Technical White Paper SM-500–102015”, which was originally available on Somalogic’s web-site http://info.somalogic.com/hubfs/January_2016/SSM-002-Rev-3-SOMAscan-Technical-White-Paper.pdf (accessed November 26, 2016) and is now archived and available at https://studyres.com/doc/7837606/technical-white-paper. Briefly, EDTA-plasma was incubated with bead-coupled epitope-specific aptamers (SOMAmers). Bead-bound proteins were then biotinylated and complexes comprising biotinylated target proteins and fluorescence-labelled SOMAmers were photocleaved and recaptured on streptavidin beads. SOMAmers were then eluted and quantified by hybridization to custom arrays of SOMAmer-complementary oligonucleotides. The resulting raw intensities were processed using different standards as a reference, including hybridization normalization, median signal normalization and signal calibration to control for inter-plate differences. No samples or data points were excluded. Overlapping phenotype data was obtained for 2,155 of the 2,935 samples and proteomics data for these samples were used. Quality control was performed using repeated measures of two QC samples. The median coefficient of variance (CV) was 0.073 for both QC samples, based on 51 and 54 repeated measures, respectively. 95% of the aptamers had a CV below 0.172 and 0.176, respectively, and 5% had a CV below 0.046 and 0.041, respectively.

**Table 2.** Numbers and percentages for drugs taken by the T2D cases versus controls according to ATC anatomical main groups.

| ID | Anatomical main group | T2D<br>(N=420) | No T2D<br>(N=1,735) | p-value |
| --- | --- | --- | --- | --- |
| A | ALIMENTARY TRACT AND METABOLISM | 273 (65.0%) | 240 (13.8%) | $1.53 \times 10^{-107}$ |
| B | BLOOD AND BLOOD FORMING ORGANS | 98 (23.3%) | 104 (6.0%) | $2.07 \times 10^{-27}$ |
| C | CARDIOVASCULAR SYSTEM | 151 (36.0%) | 125 (7.2%) | $8.33 \times 10^{-56}$ |
| D | DERMATOLOGICALS | 4 (1.0%) | 33 (1.9%) | 0.256 |
| G | GENITO URINARY SYSTEM AND SEX HORMONES | 61 (14.5%) | 95 (5.5%) | $2.68 \times 10^{-10}$ |
| H | SYSTEMIC HORMONAL PREPARATIONS, EXCL. SEX HORMONES AND INSULINS | 46 (11.0%) | 112 (6.5%) | $2.15 \times 10^{-3}$ |
| J | ANTIINFECTIVES FOR SYSTEMIC USE | 3 (0.7%) | 4 (0.2%) | 0.278 |
| L | ANTINEOPLASTIC AND IMMUNOMODULATING AGENTS | 6 (1.4%) | 31 (1.8%) | 0.766 |
| M | MUSCULO-SKELETAL SYSTEM | 104 (24.8%) | 120 (6.9%) | $1.51 \times 10^{-26}$ |
| N | NERVOUS SYSTEM | 79 (18.8%) | 71 (4.1%) | $6.43 \times 10^{-26}$ |
| P | ANTIPARASITIC PRODUCTS, INSECTICIDES AND REPELLENTS | 1 (0.2%) | 2 (0.1%) | 0.962 |
| R | RESPIRATORY SYSTEM | 13 (3.1%) | 48 (2.8%) | 0.841 |
| S | SENSORY ORGANS | 16 (3.8%) | 36 (2.1%) | 0.057 |
| V | VARIOUS | 2 (0.5%) | 4 (0.2%) | 0.733 |

### Metabolomics

1,159 metabolites (937 named compounds and 222 compounds of unknown structural identity) were quantified using Metabolon HD4 technology (Metabolon Inc., Durham, NC) (**Supplementary Table 3**) for 3,000 samples as previously described ^33,34^. All measurements were performed on a Metabolon HD4 platform implemented at the Anti-Doping Laboratory in Qatar (ADLQ) under a joint laboratory agreement with Metabolon and support from Weill Cornell Medicine – Qatar, the Qatar Biomedical Research Institute, and the interim Translational Research Institute (iTRI) of HMC. For 2,155 of the 3,000 samples, we obtained overlapping phenotype data for this study. Metabolomics data for these samples were used. Instrument variability, based on measurement of internal standards, was 12% and total process variability, based on endogenous biochemicals measured in repeated reference samples, was 16%.

### Statistical analysis

Statistical analysis was conducted using R (version 4.0.5) and RStudio (version 1.4.1106). T-tests, Fisher exact tests, and linear and logistic regression models with co-variates as indicated were conducted as appropriate based on the respective variable types, using subroutines implemented in base R. Multiple testing was accounted for using conservative Bonferroni correction at a significance level of 0.05 divided by the number of tests conducted in the specific cases (number of metabolites, proteins, traits taken forward for replication, etc. as indicated).

### Cluster Analysis

Model parameters were selected based on five commonly measured variables as in Ahlqvist *et al*. ^8^. We used BMI, age at onset of diabetes, and homeostasis model assessment HOMA2 estimates of ß-cell function (HOMA2-B) and insulin resistance (HOMA2-IR) based on C-peptide concentrations calculated with the HOMA calculator (University of Oxford, Oxford, UK) ^35^. Patients defined as SAID (see above) were excluded from the clustering and assigned to their own subtype, and clustering was carried out on the T2D patients only. Cluster analysis was carried out on standardized values centered to mean=0 and s.d.=1. The optimal number of clusters was determined from the training set using the Mclust function in the “mclust” R library v.5.4.5. Clusters coordinates were identified in the training set of QBB individuals (N=631) and applied to a separate QBB set which was deeply phenotyped (N=420). We determined the optimal number of clusters using the Bayesian Information Criterion (BIC) for expectation-maximization, initialized by hierarchical clustering for parameterized Gaussian mixture models. We computed the BIC for various cluster sizes (two to 15). Using *k*-means cluster analysis on standardized variables, we derived cluster center coordinates for different values of *k* (**Supplementary Figure 2**). Using a voting scheme^36^, we determined the optimal number of clusters to be *k*=4 (**Supplementary Figure 3**). This finding was consistent with that observed by Ahlqvist *et al*. Cluster stability was assessed by Jaccard similarity ^37^ using 2000 re-runs of the *k*-means procedure. The Jaccard similarity to the original clusters was greater than 0.75, which is generally considered an acceptable threshold for cluster stability ^37^. The cluster variables in QBB followed a similar trend to ANDIS. We therefore assigned cluster labels based on the variable averages that were characteristic of each T2D subtype following Ahlqvist *et al*.

### Robustness and reproducibility analysis

To ensure robustness and reproducibility of our results, we undertook a number of sensitivity tests on the cluster analysis. First, we replicated the clustering in the testing set and compared that to using cluster coordinates from the training set. In the case of clustering in the testing set, the assigned clusters from the *k*-means algorithm were used directly. However, when using the training set coordinates, cluster membership was determined by assigning every individual in the testing set to the cluster with the minimum Euclidean distance to the training coordinates. We then repeated the clustering with varying numbers of clusters. We also repeated the cluster analysis separately for females and males. As QBB participants were not all in a fully fasting state, we further tested the sensitivity of the cluster assignments to self-reported time-since-last-meal. Finally, we tested whether ANDIS cluster centers could be used directly to classify QBB participants into T2D subtype clusters.

### Identification of T2D-associated proteins and metabolites

For proteins, logistic regression models (glm) were used to test for association with the T2D state, including age, sex, and technical covariates into the model. Technical covariates include log(HSP90) which is a measure of cell lysis, week of sample collection, fasting minutes, and SomaLogic tube number. For metabolites, linear regression models (lm) were used to test for association with T2D using age, sex, BMI, and technical covariates into the model. Different types of models (glm for proteins and lm for metabolites) were used for consistency with previously published work^23,38^. Protein and metabolite levels were log-scaled, z-scored, and outliers were winsorized to 5 s.d. before computing the association.

### Replication of association of T2D with proteomics

Association data for the replication of the T2D protein association was obtained from the published AGES study ^23^. In that study, serum levels of 4,137 proteins, targeted by 4,782 SOMAmers, were measured at SomaLogic (Boulder, CO) in samples from 5,457 AGES-Reykjavik participants, as previously described ^39^. The AGES-Reykjavik cohort included 654 individuals with T2D and 4,784 controls. After applying a Box-Cox transformation on the protein data, associations between serum protein levels and prevalent or incident type 2 diabetes were determined using a logistic regression adjusted for age and sex. After following the same preprocessing steps and statistical methods, we replicated the associations for proteins that were shared between QBB and AGES (N=107).

### Replication of association of T2D with metabolomics

Metabolomics associations with T2D have been previously reported for the QMDiab study using the Metabolon HD2 platform ^38,40^. However, here we are using data that has been recently remeasured on the more recent Metabolon HD4 platform in Durham (NC), which is compatible with the QBB metabolomics data annotation. Data for 1,104 metabolites from 309 samples of QMDiab were used. From 194 metabolites that associated with T2D in QBB at a Bonferroni level (p<0.05/1,159), data was also available for 175 metabolites in QMDiab. For the replication, the same processing of the metabolomics data was performed, including log-scaling, z-scoring, and outlier winsorization to 5 s.d., before computing the association.

### T2D subtype cluster omics and medication analysis

Cluster specific proteins and metabolites were identified using linear models without covariates and following two criteria. First, the omics levels for a given cluster were compared to all others combined, requiring Bonferroni significance levels (p<0.05/Nmetabolites and p<0.05/Nproteins). Second, the omics levels for a given cluster were compared to all other clusters individually, requiring nominal significance (p<0.05). Cluster specific drug usage was identified using a Fisher test comparing usage of a given drug in a given cluster to all other clusters combined, requiring nominal significance (p<0.05).

### Replication of associations of T2D subtypes with proteomics

Replication of the subgroup specific proteins was attempted in the ANDIS study^8^ using a linear regression model for each cluster versus all other clusters, while adjusting for sex. Protein levels were measured in the ANDIS study for N=176 individuals (44 individuals per subtype) using the Olink platform (Olink Proteomics, Uppsala, Sweden). Equal numbers of men and women were selected based on Euclidean distance to the cluster centers to be representative of their subtype. All selected individuals were between age 38.1 and 75.2 years, of European decent, and had their blood samples taken within 3 months of diabetes diagnosis. The following 13 Olink panels were used: Olink CARDIOMETABOLIC, Olink CARDIOVASCULAR II, Olink CARDIOVASCULAR III, Olink CELL REGULATION, Olink DEVELOPMENT, Olink IMMUNE RESPONSE, Olink INFLAMMATION, Olink METABOLISM, Olink NEURO EXPLORATORY, Olink NEUROLOGY, Olink ONCOLOGY II, Olink ONCOLOGY III, and Olink ORGAN DAMAGE, covering a total of 1,161 distinct protein assays. The biomarker expression was measured using logarithm of the relative biomarker/protein concentration in each panel, expressed as normalized protein expression (NPX) values. Outlier analysis was performed using an unsupervised clustering algorithm using a One Class Support Vector Machine. Four samples, one from each subtype, were identified as outliers and excluded from the analysis, leaving N=172 individuals (43 individuals per subtype). Data was available for 30 of the 47 subtype specific proteins (matched by Uniprot identifiers) that were shared between the Olink data in ANDIS and the Somalogic data in QBB.

## RESULTS

### The T2D subtype clustering scheme defined for Caucasians can be translated to an Arab population

Following the approach of Ahlqvist *et al*. (**Figure 1**) we used *k*-means clustering of age at diagnosis, BMI, HbA_1c_, HOMA2-B, and HOMA2-IR and identified four clusters with clinical properties similar to those in the ANDIS study (**Figure 2A**). The SIDD cluster was characterized by young age at onset, low BMI, low insulin secretion (HOMA2-B) and poor glycemic control (high HbA_1c_); the SIRD cluster had the highest level of insulin resistance (HOMA2-IR) and high BMI; the MOD cluster had a high BMI with low insulin resistance; and the MARD cluster, like the MOD clusters, had low insulin resistance, but a much lower age of onset of T2D. The relative cluster sizes in QBB were comparable to those found in the ANDIS study, except for SIRD, which made up only 4% of the T2D cases in QBB compared to 15% in ANDIS.

**Figure 2.**
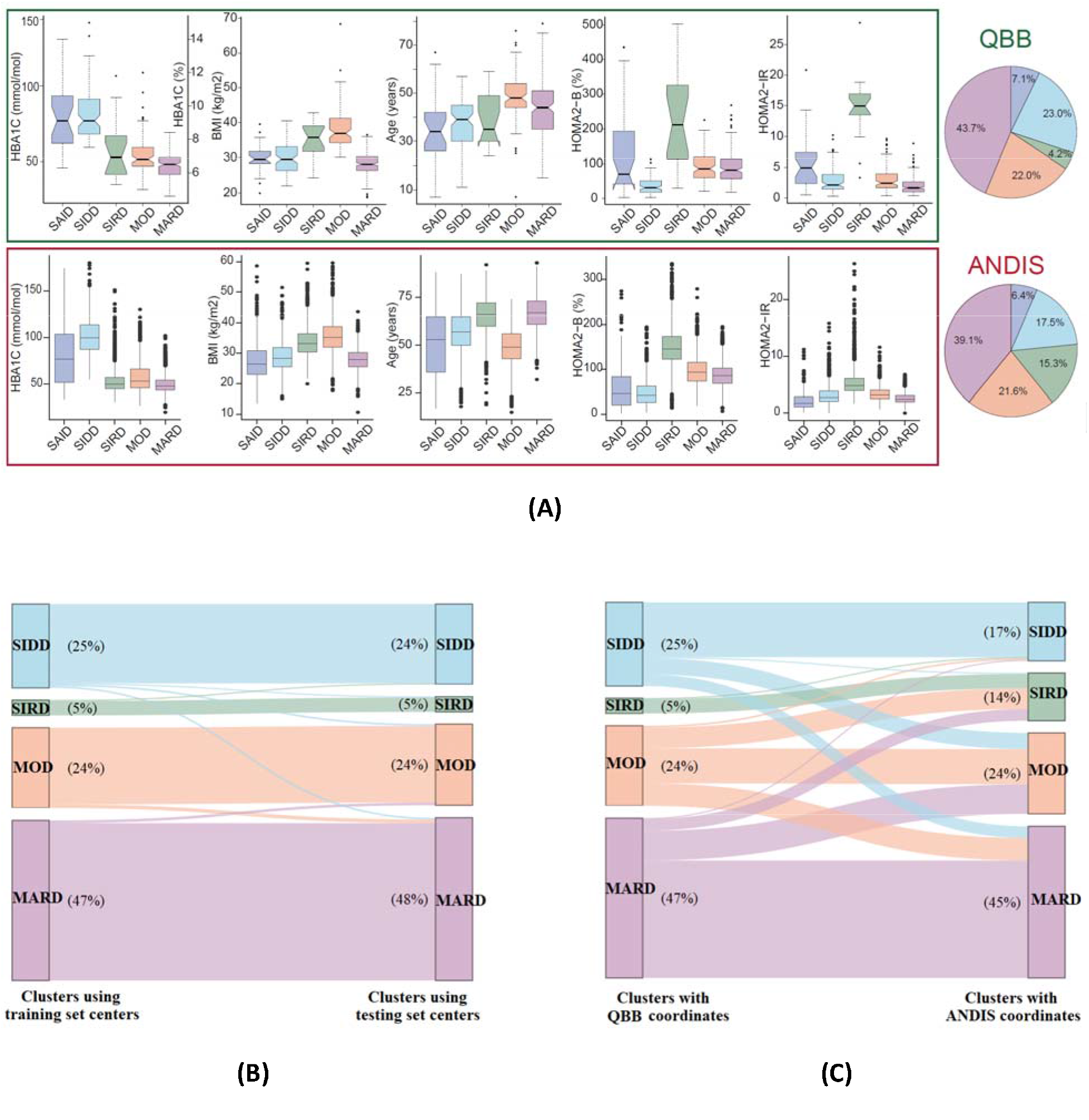
Cluster characteristics and cluster distribution in QBB. K-means clusters were derived using the QBB training set and classification was applied to the testing set using the training set cluster coordinates. **A)** Distributions of HbA_1c_, BMI, age, HOMA2-B, and HOMA2-IR are shown for each cluster in QBB and ANDIS. HbA_1c_, BMI, HOMA2-B, and HOMA2-IR all followed the same trend in QBB and ANDIS, but individuals in the MOD cluster were younger than the other clusters in QBB. **B)** The testing set clusters were similar to the training set clusters, regardless of whether they were assigned based on the training set coordinate centers or derived de novo for the testing set using K-means clustering. Minor changes in the cluster assignments (2%) were observed when clustering the data using the training set coordinates versus the testing set coordinates. **C)** After using the ANDIS coordinates instead of the QBB coordinates to classify QBB patients, 35% changes in the cluster assignments were observed.

We performed several sensitivity tests on the way the clusters were derived. First, we replicated the clustering in an independent testing set within QBB. We obtained very similar results compared to using cluster coordinates from the training set (**Figure 2B**) and found that cluster assignments were identical for 98% of the study participants. We then repeated the clustering allowing for a varying number of cluster (**Supplementary Figure 4A**). Consistent with the observations of Ahlqvist *et al*., four clusters were identified in QBB (**Supplementary Figure 3**). Allowing for a fifth cluster led to a split of the MARD cluster into one cluster with a lower and one with a higher age of T2D onset (**Supplementary Figure 4A**). Repeating the cluster analysis separately for females and males showed that most individuals (93%) were assigned to the same cluster as in the initial analysis (**Supplementary Figure 4B**).

HOMA2-IR and HOMA2-B estimates are based on plasma glucose and C-peptide (Cpep) levels and are sensitive to the fasting state. Other studies have reported on clustering using non-fasting values by using HDL-cholesterol and C-peptide, which is a proxy for insulin resistance ^41^. As QBB participants were not all in a fully fasting state (77% of the individuals had fasted for over two hours at the time of enrolment and 50.7% had fasted for over 8 hours), we tested the sensitivity of the cluster assignments to self-reported time-since-last-meal. Using linear regression, we estimated fasting HOMA2-IR and HOMA2-B and used the corrected values for clustering. 98% of the cluster assignments remained unchanged, indicating that the clustering is robust to fasting state (**Supplementary Figure 4C**).

Finally, we tested whether ANDIS-derived cluster centers could be used directly to classify QBB participants into T2D subtype clusters. We observed consistent cluster assignments for 65% of the individuals when using the ANDIS-derived cluster centers instead of the QBB-derived cluster centers for classification (**Figure 2C**). In addition, we noted a gender bias in the misclassification with 23% of misclassified males compared to 46% of females. A comparison of the cohort-specific cluster centers (standardized values centered to mean=0 and SD=1) is presented in **Supplementary Table 4**. Apart, from the age variable, gender-specific variables for the different clusters were directionally consistent between ANDIS and QBB (**Supplementary Figure 5**). Although the trends of four out of five cluster variables were consistent across the T2D subtypes in both cohorts, the limited overall agreement between T2D subtype classifications obtained using ANDIS versus QBB coordinates suggested that population specific coordinates should be used. We therefore used sex-independent coordinates derived from the QBB training set to classify the QBB training set in the following metabolomics and proteomics analyses.

### Metabolomics and proteomics associations with T2D replicate other populations

Diabetes-specific alterations of protein and metabolite levels have previously been described in different populations ^42-45^. To validate the metabolomic and proteomic data in QBB and its ability to characterize T2D participants, we investigated omics-associations with T2D. Deep molecular phenotyping data was available for 420 QBB participants with T2D and 1,735 controls and covered semi-quantitative measures of 1,159 blood circulating metabolites and 1,305 plasma proteins. We compared the protein levels of the T2D cases (N=420) to those of the controls (N=1,735) in a linear model with covariates as described in the methods. We identified 214 proteins associated with T2D at a Bonferroni level of significance (P < 0.05/1,305 = 3.83×10^−5^) (**Figure 3A & Supplementary Table 5**). We checked the associations for replication in the independent European AGES population (N=5,457). Of 214 proteins associated with T2D in QBB, 107 were also measured on the SOMAscan platform used in AGES. One hundred and four (97.2%) of these proteins replicated at a Bonferroni-corrected significance level (P < 0.05/107). All replicated associations were directionally concordant (**Figure 3B**).

**Figure 3.**
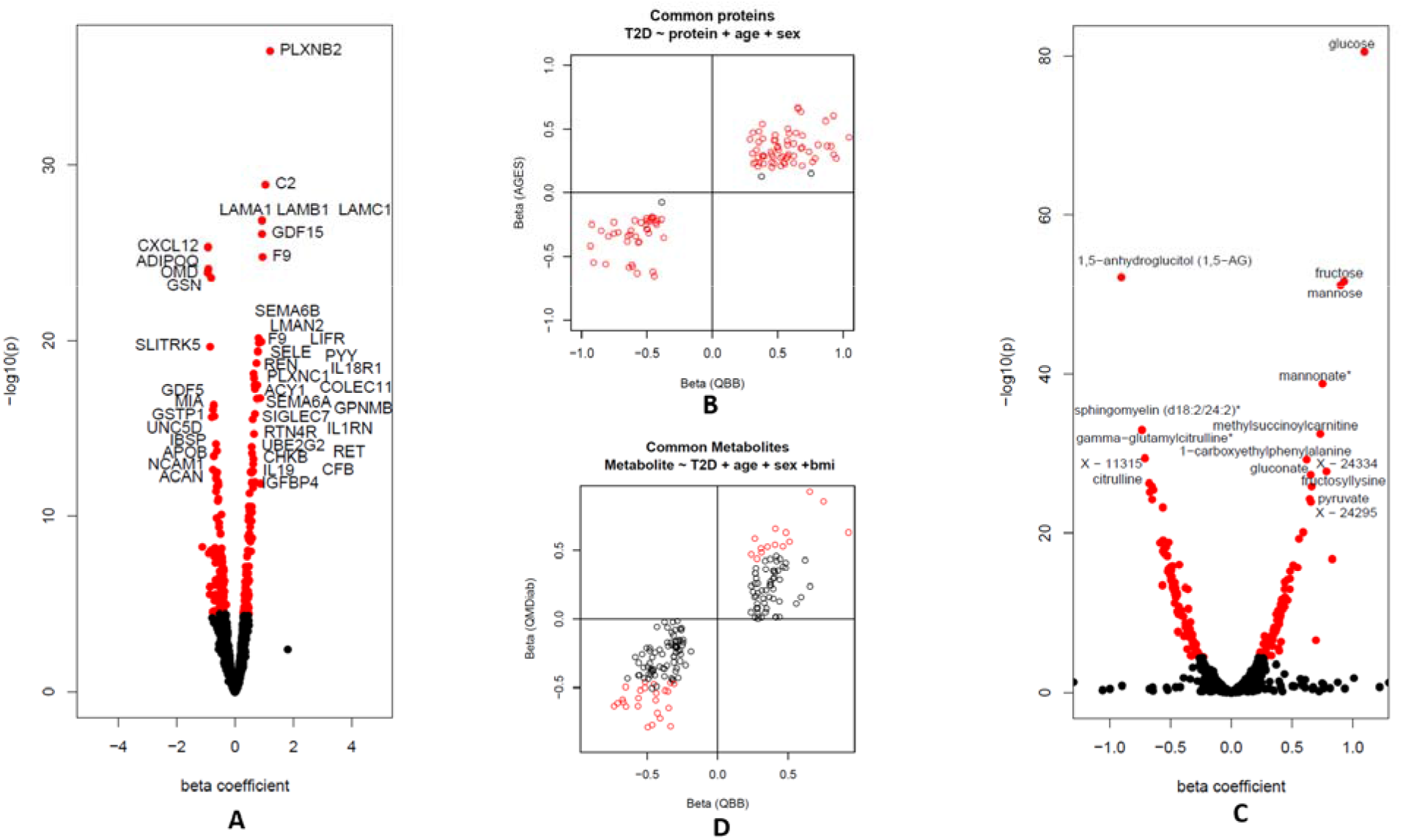
Associations of proteomics and metabolomics levels with T2D. **A)** 214 proteins were significantly associated (p<0.05/1,305) with T2D in QBB after adjusting for age and sex. **B)** Comparison of effect sizes between QBB and AGES. The replication status of 107 common proteins is shown red: Bonferroni significant (p<0.05/107) in both studies, black: significant only in QBB. **C)** 194 metabolites were significantly associated (p<0.05/1,159) with T2D after adjusting for age, sex, and bmi. **D)** Comparison between QBB and QMDiab. The replication status of the 175 common metabolites is shown in red/black – Bonferroni significant (p<0.05/175) or significant only in QBB.

We further identified 194 metabolites that were associated with T2D (P < 0.05/1,159 = 4.31×10^−5^) (**Figure 3C & Supplementary Table 6**). We confirmed previous T2D associations with sugars (glucose, mannose, 1,5-AG, etc.), with branched chain amino acids (BCAAs) (incl. isoleucine, leucine, valine), various lipids and markers of kidney function. We attempted replication of these T2D-metabolite associations in the multi-ethnic Qatar Metabolomics Study of Diabetes (QMDiab) cohort (**Supplementary Table 7**). From the 194 T2D associated metabolites identified in QBB, 175 were also measured in QMDiab. All associations were directionally concordant, and 41 (23%) of these associations were statistically significant at a Bonferroni level (p<0.05/175) (**Figure 3D**).

### Cluster-specific metabolomics and proteomics associations reveal diabetes subtype-specific processes

We identified all proteins and metabolites that were differentially expressed in one of the four T2D subtype clusters. We required (1) that their means were different from those of all other clusters combined at a Bonferroni significance level and (2) that their means were different from all other clusters in a pair-wise comparison at a nominal level of significance (see methods). Based on this criterion, 47 proteins and 42 metabolites were specific to a given T2D subtype. **Figures 4 and 5** represent an overview of the central findings (data in **Supplementary Tables 8 and 9, Figure 6** and **Supplementary Figure 7 and 8** have detailed boxplots). In the following we report highlights of these associations and possible rationalizations for the observed subtype specificities. We start with proteins, followed by metabolites, and address which are – in our view – the most interesting findings, always following the same order, that is, SIDD, SIRD, MOD, and then MARD.

**Figure 4.**
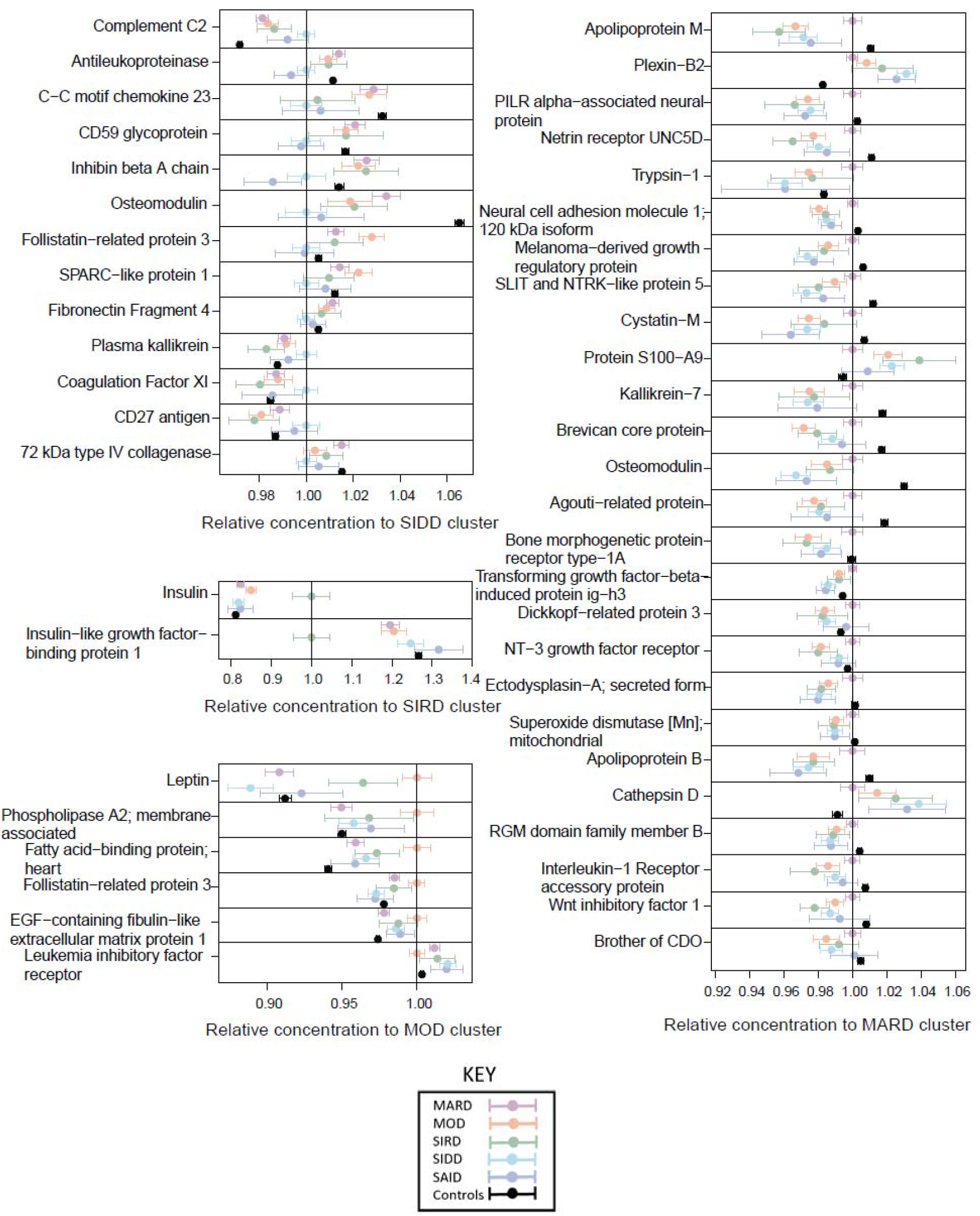
Proteins that distinguish individual diabetes subtypes. The dots and bars represent the mean protein values and the 95% confidence intervals of the means for proteins that are different in one of the four T2D subtypes compared to all others. Values are normalized by the mean of the respective reference subtype. In addition, data for SAID and the control group are shown for reference.

**Figure 5.**
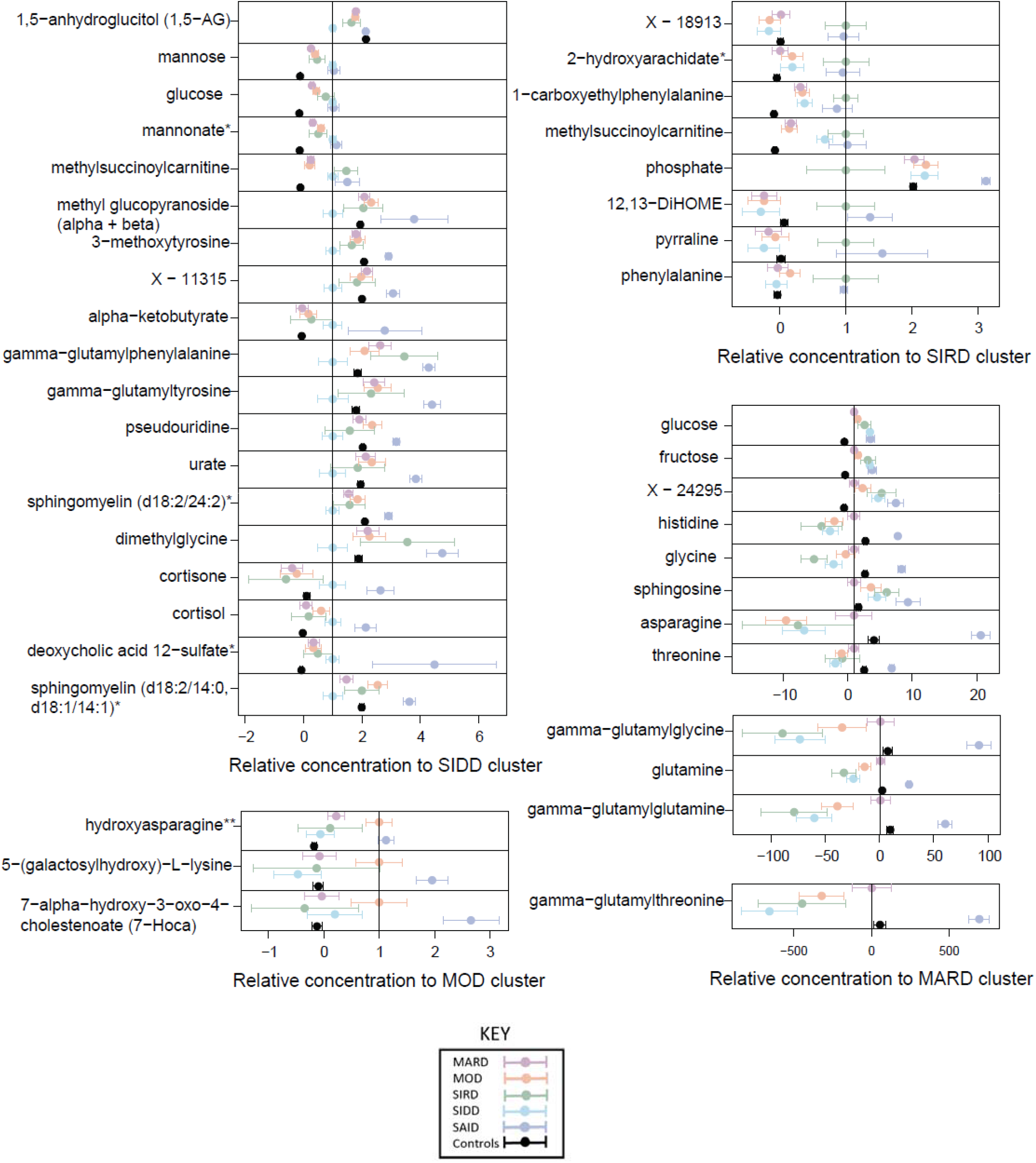
Metabolites that distinguish individual diabetes subtypes. The dots and bar represent the mean metabolite values and the 95% confidence intervals of the means for metabolites that are different in one of the four T2D subtypes compared to all others. Values are normalized by the mean of the respective reference cluster. In addition, SAID and the control group are shown for reference.

**Figure 6.**
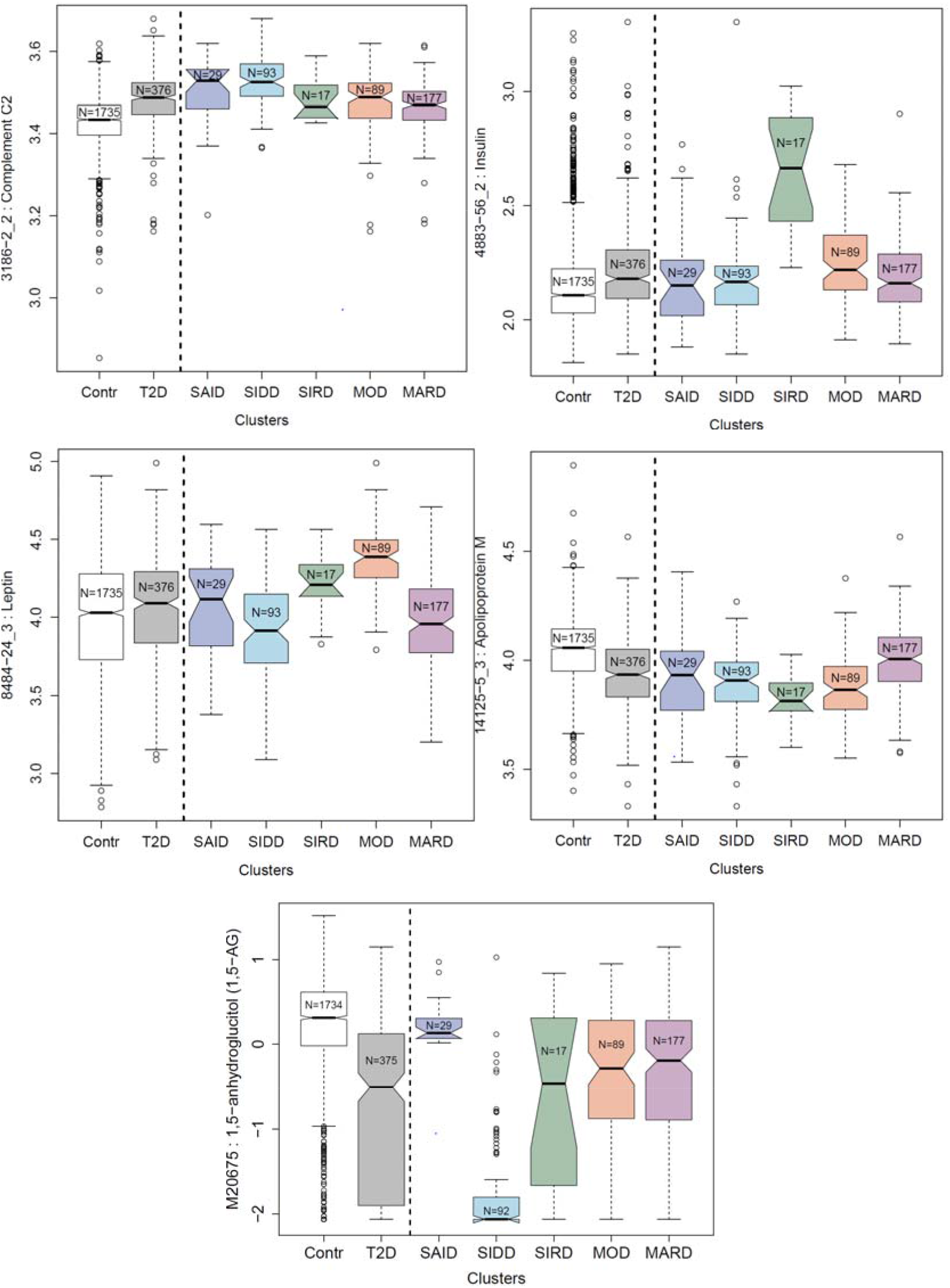
Boxplots of the significantly altered proteins or metabolites in specific diabetes subtypes. Complement C2 (C2) insulin (INS), leptin (LEP), and Apolipoprotein M (APOM) are significantly higher in SIDD, SIRD, MOD, and MARD, respectively, while 1,5 AG is significantly lower in SIDD. The complete set of box plots for all protein and metabolite levels for each cluster are in **Supplementary Figure 7 and 8**.

We observed subtype-specific elevated levels of Complement C2 (C2) in the SIDD cluster. Type 1 diabetes has a well-established association with HLA antigens ^46^ and C2 has been linked to HLA in T1DM ^47^. SIDD is most similar to SAID in terms of being the most severely insulin deficient, but is not auto-antibody positive, which is the primary diagnostic feature in SAID. Complement activation extends beyond microbial defense and can be involved in obesity, insulin resistance, diabetes, and dyslipidemia, indicating an inflammatory component ^48-52^. Studies have shown that metabolic inflammatory signaling can affect pathways that lead to insulin resistance^53,54^. Accumulating evidence supports activation of the complement system with the development of insulin resistance^55^. The SIRD cluster had the highest levels of insulin (INS) and the lowest levels of insulin-like growth factor-binding protein 1 (IGFBP1). Individuals could develop insulin resistance due to low IGFBP1 which directly affects insulin sensitivity through its RGD domain^56^. Proteins (C59 glycoprotein, inhibin beta A chain, osteomodulin, Follistatin-related protein 3, C27 antigen) specifically dysregulated in SIDD were often also dysregulated in SAID, possibly reflecting shared underlying processes. The MOD cluster had the highest leptin (LEP) levels and enzymes involved in lipid metabolism, such as phospholipase A2 (PLA2G2A) and fatty acid-binding protein (FABP3). The MARD cluster had the highest levels of APOM, APOB, UNC5D, NCAM1, Cystatin-M, and the lowest levels of Plexin-B2 (PLXNB2). All protein levels specific to MARD were closer (or comparable) to those of the controls when compared to the other subtypes, suggesting that MARD individuals were the “healthiest” among the T2D subtypes, and that the proteins associated with MARD are more strongly dysregulated in all other subtypes.

In relation to plasma metabolites, individuals in the SIDD cluster had the lowest 1,5-anhydroglucitol (1,5-AG) levels of all groups. 1,5-AG is a marker of short-term glycemic control and is implemented as a clinical test in the GlycoMark™ assay ^57^. The blood sugars mannose, glucose, fructose, mannonate, and gluconate were considerably higher in the SIDD cluster, indicating a greater level of hyperglycemia. Individuals in the SIDD cluster also exhibited elevated levels of cortisone and cortisol, which are stress markers associated with dysregulated glucose metabolism^58^ and a number of chronic complications of T2D^59^. We also observed a decrease in the level of dimethylglycine (DMG) in SIDD, a product of betaine catabolism and low betaine and DMG levels, which has been associated with higher glucose levels and the development of T2D^60^. Furthermore, the SIDD cluster had decreased levels of gammaglutamyl amino acids (gamma-glutamylphenylalanine and gamma-glutamyltyrosine), indicating perturbed glutathione metabolism. The levels of two sphingomyelin species (sphingomyelin (d18:2/14:0, d18:1/14:1) and sphingomyelin (d18:2/24:2)) were also lower in the SIDD cluster. Downregulated sphingolipid metabolism can affect insulin sensitivity and lead to β cell dysfunction^61^.

The SIRD cluster had elevated levels of lipokine-related metabolites, including 12,13-DiHOME, a linoleic acid metabolite, and 2-hydroxyarachidate, an arachidic acid metabolite. 12,13-DiHOME was previously recognized as an important lipid mediator stimulating fatty acid uptake by skeletal muscles^62^, which could serve as an alternative energy source for SIRD subjects given their potentially limited access to glucose, due to insulin resistance. The plasma level of phosphate was lower in the SIRD cluster compared to other clusters, and could potentially be linked to hypophosphatemia which is frequently observed in conditions of diabetic ketoacidosis driven by hyperglycemia-induced osmotic diuresis ^63^. Although diabetic ketoacidosis is rare in SIRD, it has been previously reported ^64^.

The MOD cluster had high levels of hydroxyasparagine, 5−(galactosylhydroxy)−L−lysine, and 7-alpha-hydroxy-3-oxo-4-cholestenoate (7−Hoca), which play a role in lipid metabolism and obesity^65^. The elevated levels of 5−(galactosylhydroxy)−L−lysine, an important post-translationally modified amino acid present in collagen-like proteins, such as adiponectin ^66^, may be the result of adipogenic collagen turnover^67,68^. Metalloproteinase, MT1-MMP, a pericellular collagenase and a member of the matrix metalloproteinase (MMP) gene family, directs interactions that control adipogenesis ^67^ and is critical to white adipose tissue development by remodeling of the 3-D type I collagen scaffolding that dominates primordial white fat deposits. Hence, its absence leads to disruption of transcription factor cascades required for adipocyte maturation and would broadly occur in individuals in the MOD cluster with high BMI. Adiponectin is an important target in obesity treatment, is a key regulator of fatty acid oxidation and lipid synthesis and is well known to decrease triglyceride concentrations and increase insulin sensitivity ^69^. Oxysterols play a signaling role in lipid and glucose metabolism which may be implicated in obesity through the control of lipogenesis ^70,71^. They also play an important role in cholesterol uptake, transport, excretion, and gene regulation ^72-74^. The elevated levels of the oxysterol, 7-Hoca, may be a result of dysregulated fatty acid metabolism and lipid homeostasis.

As in the case of proteins, the metabolic profiles of individuals in the MARD cluster were closest to the controls. Blood carbohydrates levels (glucose and fructose) were higher than normal but were the lowest among the T2D clusters. The levels of glycine, glutamine, histidine, and gamma-glutamyl amino acids (gamma-glutamylglycine, gamma-glutamylglutamine, and gamma-glutamylthreonine) were the lowest in MARD and comparable to individuals without diabetes. Glycine and glutamine are both implicated in insulin secretion^75,76^. Glycine acts on the pancreas through glycine receptors and as a co-ligand for N-methyl-d-aspartate glutamate receptors to control insulin secretion and glutamine regulates beta-cell gene expression, signaling, and insulin secretion. In addition, histidine and gamma-glutamyl amino acids play a role in anti-inflammatory and antioxidative responses^77,78^. Histidine supplementation has been shown to improve insulin resistance by suppressing pro-inflammatory cytokine expression, possibly through the nuclear factor kappa-B (NF-κB) pathway^77^. Serum gamma-glutamyltransferase (GGT) is strongly linked to obesity and nonalcoholic fatty liver disease, which may lead to systemic and hepatic insulin resistance, respectively^79^. Overall, the MARD patients had the least metabolic dysregulation among the T2D subtypes.

### Replication of T2D subtype specific protein associations in a different population using a different proteomics platform

We attempted replication of 30 out of 47 subtype specific protein associations (**Supplementary Table 10**) using proteomics measurements from the Olink platform in ANDIS. Data was available for 43 individuals in each of the four T2D subgroups (N=172). Four protein associations were replicated after accounting for multiple testing (p<0.00167; 0.05/30). These were Follistatin-related protein 3 (FSTL3) with the SIDD cluster, Plexin-B2 (PLXNB2) and Cathepsin D (CTSD) with the MARD cluster, and leptin (LEP) with the MOD cluster. Five additional proteins showed concordant directionality at a nominal significance level (p<0.05), that is, CD59 glycoprotein (CD59), Complement C2 (C2) and 72 kDa type IV collagenase (MMP-2) with SIDD, Leukemia inhibitory factor receptor (LIF-4) with MOD, and NT-3 growth factor receptor (NTRK-3) with MARD.

### Medication patterns are cluster specific

Self-reported drug usage was annotated using unique active molecule identifiers obtained from the Drugbank repository ^80^ and their corresponding ATC codes. We compared difference in medication usage from all ATC anatomical main and subgroup groups between T2D cases and controls (**Table 2**). The most common drugs used by patients with T2D were in the ATC anatomical main groups A (Alimentary tract and metabolism) and C (Cardiovascular system). In descending order, the most frequently administered drug subgroups by T2D subjects were A10: Drugs used in diabetes (p=8.70×10^−220^), C10: lipid modifying agents (p=1.04×10^−64^), B01: anti-thrombotic agents (p=2.43×10^−52^), C07: beta blocking agents (p=1.74×10^−51^), A01: stomatological preparations (p=7.51×10^−41^), N02: analgesics (p=1.17×10^−39^), M01: anti-inflammatory and anti-rheumatic products (p=9.40×10^−32^), and C09: agents acting on the renin-angiotensin system (p=3.40×10^−30^) (**Supplementary Table 11**).

The medication patterns were compared across the diabetes subtype clusters in QBB (**Supplementary Table 12**). Patients in the SIDD cluster were more frequently using insulin (Fisher test p-value=2.62×10^−4^), or Metformin (p=1.12×10^−3^) and Pioglitazone (3.56×10^−3^). SIDD patients were also more frequently prescribed sulfonylureas (p=5.34×10^−4^) which increase insulin release, and Sitagliptin (p=6.02×10^−3^), a DPP-4 inhibitor that increases glucose dependent insulin release. Patients in the SIRD cluster most frequently took anti-depressant medications (p=5.63×10^−3^), medication for diabetic kidney disease e.g. Losartan (p=2.60×10^−2^), and medication for rheumatoid arthritis (4.52×10^−2^). In the MOD cluster, medications used to treat high blood pressure and heart failure, such as Lisinopril (8.71×10^−4^) and esomeprazole (p=3.31×10^−3^) were used. The MARD cluster also had the lowest percentage of individuals on insulin treatment (p=2.62×10^−4^), whilst perindopril usage was more frequent (4.82×10^−2^).

## DISCUSSION

The T2D subtype classification scheme proposed by Ahlqvist el al.^8^ has been replicated in many populations^14-17,19-22,81^ (**Supplementary Figure 6**), but it may not be generalized to all as shown in an Asian Indian population ^21^. Type 2 diabetes has a huge prevalence in the MENA region, with some of the highest rates and predicted increases over the next decade, especially in Qatar (IDF Diabetes Atlas 2021^82^). No previous study has examined the generalizability of the diabetes cluster classification scheme to Arab populations or characterized it using the latest high-throughput proteomics and metabolomics platforms. All protein and metabolite associations with T2D subtypes are made available as a resource in the Figures and Supplementary files.

Our study shows that the T2D subtypes identified in the Scandinavian population are present in individuals of Arab/Middle Eastern descent. However, the age of diabetes onset in the QBB population was lower compared to ANDIS, especially in the MOD cluster, which could be attributed to the high incidence of obesity in younger Arab individuals. Leptin (LEP) and Follistatin-related protein 3 (FLSTL3) were significantly higher in the MOD cluster and of course increased leptin levels in obesity reflects resistance to leptin action. Also, increased FLSTL3 levels are associated with insulin resistance and have been shown to regulate body composition and glucose homeostasis in human population studies ^83,84^. Interestingly, an FLSTL3 knockout mouse has been shown to improve glucose metabolism and increase beta cell mass^84^.

Among the diabetes subtypes, the MARD subtype appeared to be the “healthiest” group and their HbA_1c_, BMI, HOMA2-B and HOMA-IR were closest to the control group. In contrast, proteins and metabolites that were specific to SIDD were often most similar to those found in the autoimmune (SAID) group. Plexin-B2 (PLXNB2) was the most differentially regulated protein in T2D and this association was further validated in the AGES study ^23^. Previous Mendelian randomization studies have suggested that PLXNB2 may have a causal effect on the development of T2D ^23^. In the present study, PLXNB2 was lowest in the “healthy” MARD subtype and closest to levels observed in controls. This could make PLXNB2 a potential drug target in individuals with subtypes other than MARD.

Cluster membership has been shown to be associated with individuals who may be more or less prone to the development of long-term complications ^15,21^ such as nephropathy ^8,15,20^ and fatty liver in SIRD ^8,15^, and neuropathy ^20,22^ and retinopathy ^8,20^ in SIDD. Alterations in protein biomarkers of retinopathy such as antileukoproteinase have been previously reported in the SIDD cluster ^85^. We have observed an association between complement factors like Complement C2 (C2), immune-regulatory proteins like C-C motif chemokine 23 (CCL23) and other proteins of the immune system (CD59 glycoprotein (CD59), CD27 antigen (CD27), and Inhibin beta A chain (INHBA), with the SIDD subtype, suggesting that these proteins may play an important feedback role in the resolution of inflammation. A recent review highlighted a paradigm where targeting complement factors may be a possible therapeutic avenue in slowing down diabetic complications ^55^. Chemokines also link obesity to inflammation and the subsequent development of insulin resistance ^86^. Interestingly, 72 kDa type IV collagenase (MMP-2) was significantly lower in the SIDD subtype. MMP-2 activity has been shown to be lower in rat mesangial cells cultured in high glucose and is believed to contribute to matrix accumulation leading to the development of diabetic nephropathy ^87^. Therefore, MMP-2 could potentially be developed into a protein biomarker for nephropathy in SIDD. Plexins are also receptors for semaphorins, a large family of proteins involved in various physiological processes ^88^. Semaphorins are involved in a number of diabetic complications including diabetic retinopathy, nephropathy, and neuropathy, osteoporosis, and wound healing ^89^. Interestingly, we observed significantly higher levels of CD72 in the SIDD subtype and as CD72 appears to mediate the function of semaphorins in some immune cells ^90^, it may provide a potential functional link to PLXNB2. Cathepsin D (CTSD) was significantly lower in the MARD subtype and is an aspartic endopeptidase implicated in cell growth, apoptosis and collagen biosynthesis in wounded skin of rats with diabetes and has been correlated with retinopathy and foot ulcers^17^, suggesting that MARD individuals are less likely to develop such complications compared to other subtypes. CTSD also correlates significantly with HOMA-IR and the Tei index, a measure of myocardial performance ^91^ and may paradoxically constitute a marker for cardiac dysfunction in the more “severe” subtypes. However, a biomarker does not implicate causality and there may be a bidirectional relationship which requires further analysis.

Metabolic associations observed with T2D were consistent in magnitude and directionality to previously reported T2D and pre-diabetes associations^92^. Although elevated blood glucose levels are the defining feature of diabetes mellitus, multiple biochemicals alter the metabolism of fats and amino acids and are associated with impaired insulin action, obesity, and BCAA catabolic enzymatic activity^93^. BCAAs have been consistently linked to T2D development^94^ and we observed alterations in BCAAs and major glucogenic amino acids. Previous studies have shown that cortisol and its metabolite cortisone, were both higher in individuals with diabetes compared to controls and altered cortisol metabolism is specifically characteristic of T2D patients requiring insulin^95^. Consistent with this observation, we observed higher cortisol in the SIDD cluster. Many amino acids have been associated with insulin resistance and decreased insulin secretion, including phenylalanine^96^ and here, in SIRD, we observed significantly higher phenylalanine and 1-carboxyethylphenylalanine. Previous studies have shown that certain lipokines can affect glucose metabolism in adipose, liver, and skeletal muscle tissue ^97^. For instance, a lipokine, 12,13-diHOME increases fatty acid uptake and oxidation^62^ and here we show significantly higher 12,13 diHOME in the SIRD cluster. Furthermore, a statistically significant association has been shown between 7-HOCA and asparagine and body mass index^98^. We have observed significantly higher levels of 7-HOCA and hydroxyasparagine in MOD.

The majority of proteins and metabolites which were subtype-specific were also discriminative in previous protein ^23^ and metabolite ^38^ T2D case-control studies. However, some molecules only distinguished specific subtypes but not cases from controls. This was especially true for the MARD subtype which showed levels similar to the controls. Inhibin beta A chain (INHBA), SPARC-like protein-1 (SPARCL1), and Fibronectin Fragment 4 (FN1) were significantly lower in SIDD compared to other subtypes but were not significantly different between T2D cases and controls. Also, proteins including Leptin (LEP), Phospholipase A2; membrane associated (PLA2G2A), Follistatin-related protein 3 (FSTL3), and EGF-containing fibulin-like extracellular matrix protein 1 (EFEMP1) were significantly higher in the MOD subtype as they all associate strongly with obesity but did not differentiate between T2D cases and controls when adjusted for BMI.

Studies showing differences in protein biomarkers amongst T2D clusters are limited with the Chinese REACTION study showing that the Angiopoietin-related protein 8 (ANGPTL8) levels were significantly higher in the MARD, SIRD, and SIDD clusters compared to the MOD cluster ^99^. We have found a similar trend for both ANGPTL4 and the cell-surface receptor for ANGPTL4 (TEK), two proteins which interact closely with ANGPTL8^100^. The GDS study ^16^, examined 77 protein biomarkers from the Olink inflammation panel and reported lower levels of Protein S100-A12 (EN-RAGE) and IL6 in SIDD compared to the other subtypes. We have also observed lower mean EN-RAGE, Interleukin-6 (IL6), and Interleukin-6 receptor subunit alpha (IL6R) levels in SIDD.

The efficacy of different diabetes drugs would be expected to differ between different subtypes according to the underlying pathophysiology. Hence, we assessed whether ongoing treatment may reflect the clustering of individuals with T2D. Dennis et al. ^14^, reported differences in glycemic response among clusters in ADOPT and suggested a benefit in using thiazolidinediones for the SIRD individuals and sulfonylureas for the MARD individuals. However, in this cohort we found that sulfonylurea use was highest in the SIDD and MOD clusters (**Supplementary Table 12**) whilst thiazolidinedione utilization was infrequent, reflecting reduced prescription of this class in clinical practice, and being paradoxically higher in the SIDD cluster. A current ongoing phase 2 clinical trial is investigating whether the effect of Semaglutide and Dapagliflozin differ between SIDD and SIRD individuals (ClinicalTrials.gov number, NCT04451837). The outcome of such trials may provide insight towards tailored treatment plans and pave the way for personalized medicine in specific subgroups.

We acknowledge some limitations of our current study. The cohort size was relatively small compared to other population-based studies and phenotyping of diabetes complications was limited to population-study-level questionnaires and biochemical measurements. The deep molecular phenotyping using the SOMAscan and Metabolon technologies provided only relative abundances of protein and metabolite levels respectively, not absolute concentrations. However, this is not a concern for this kind of study. The subtype specific proteins reported here are limited to the specific protein set targeted by the SOMAscan panel, and to protein associations that are detectable in blood. Therefore, the list of subtype specific proteins we report here is not comprehensive, and future studies using other technologies and other biological sample types may reveal further associations. Another limitation is that since GADA was not measured, the SIDD subtype could include some individuals with autoimmune diabetes and may explain the observation of autoimmune features in this group.

In summary, we have identified a wealth of diabetes subtype-specific metabolite and protein signatures which have the potential to identify novel pathways involved in the development and progression of T2D complications, improve risk prediction, and enable more personalized treatment approaches. Our study adds further support to the medical relevance and clinical applicability of the Ahlvist et al. diabetes subtyping approach.

## Data Availability

The informed consent given by the study participants does not cover posting of participant level phenotype data in public databases. However, data are available upon request from Qatar Biobank (QBB) (https://www.qatarbiobank.org.qa/research/how-to-apply). Requests are submitted online and are subject to approval by the QBB board.

## MANUSCRIPT INFORMATION

### Funding

This study is supported by the Biomedical Research Program at Weill Cornell Medicine in Qatar, a program funded by the Qatar Foundation, and by QNRF grant NPRP11C-0115-180010. Qatar Biobank is supported by Qatar Foundation. The statements made herein are solely the responsibility of the authors. EA was funded by grants from the Swedish Research Council (2020-02191) and the Novo Nordisk foundation (NNF18OC0034408). Olink measurements in ANDIS were sponsored by Olink Proteomics (Uppsala, Sweden).

## Acknowledgements

We are grateful to all study participants of Qatar Biobank for their invaluable contributions to this study. We thank Olle Melander and Rayaz Malek for their suggestions and comments on the present manuscript.

## Author contributions

Conceived and designed the study: S.Z., K.S.

Analyzed data: S.Z.

Contributed reagents/materials/analysis tools: N.S., M.T.

Wrote the paper: S.Z., A.H, E.A., O.A., A.B.A., K.S.

All authors discussed the results and reviewed the final manuscript.

### Ethics approval and consent to participate

Use of the Qatar Biobank data was approved by the QBB IRB under reference Ex -2019-RES-ACC-0160-0083. All study participants provided written informed consent.

### Statistical analysis

Statistical analysis was conducted using R and Rstudio.

## SUPPLEMENTARY FIGURES

**Supplementary Figure 1.**
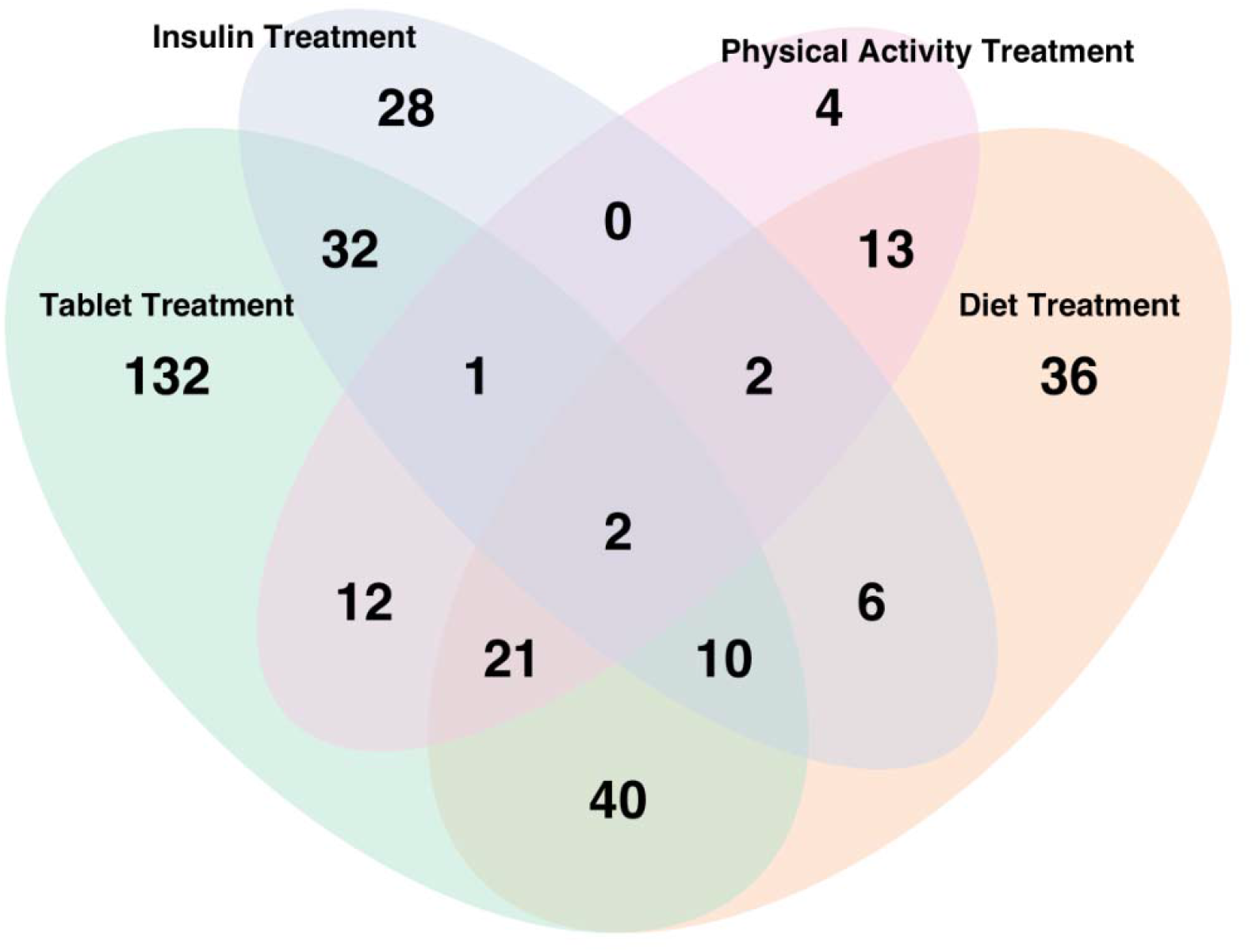
Counts of treatment types for individuals with diabetes. Most individuals with a doctor diagnosis of diabetes were exclusively on tablet treatment, while others were on some combination of insulin, diet and/or physical activity.

**Supplementary Figure 2.**
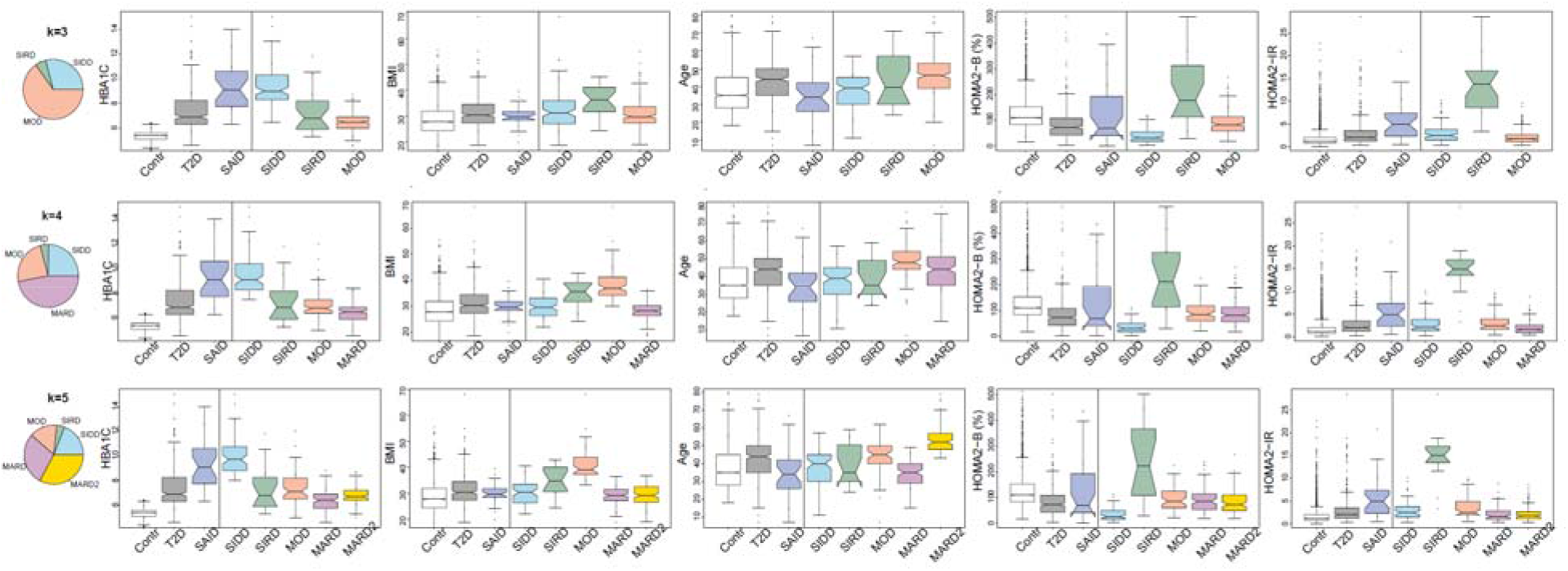
Changes in cluster distribution for different k. Some changes can be observed in the cluster assignments when clustering QBB data using different k (3,4, and 5). The most prominent change is seen in the split of the MOD cluster to discriminate between older and younger individuals. SAID and control data were not included in the clustering but are visualized for comparison with T2D and the individual clusters.

**Supplementary Figure 3.**
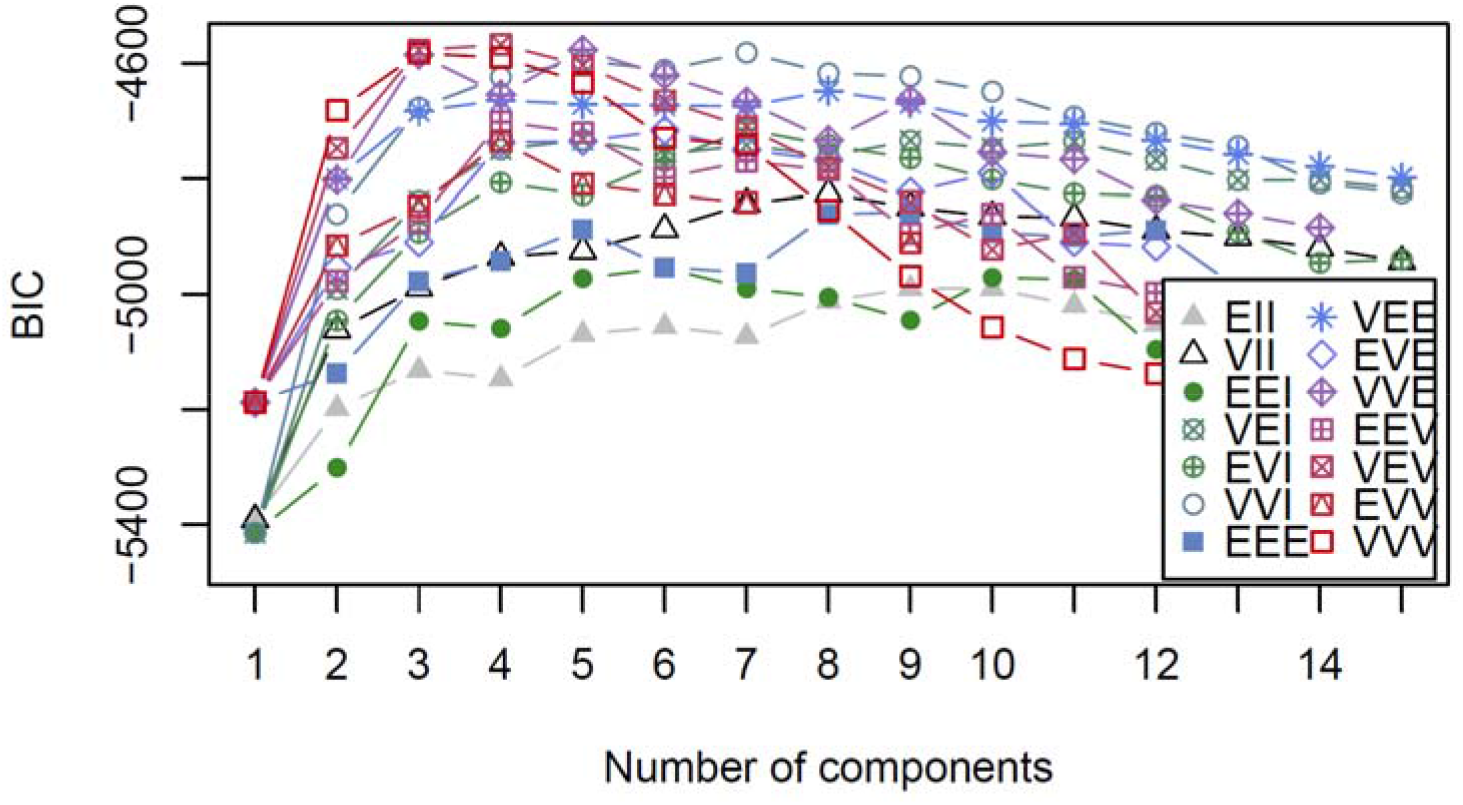
Data-driven clustering based on Gaussian mixture models. Models are estimated by the expectation-maximization (EM) algorithm initialized by hierarchical model-based agglomerative clustering. The optimal model and number of clusters was determined using the Bayesian Information Criterion (BIC) for EM. The top 3 models based on the BIC criterion, were k=3, k=4, and k=5. The spherical and diagonal models fitted in the EM phase of clustering are shown above. The three-character code represents the volume, shape, and orientation. The codes represent the following multivariate mixture models, “EII”: spherical, equal volume, “VII”: spherical, unequal volume, “EEI”: diagonal, equal volume and shape, “VEI”: diagonal, varying volume, equal shape, “EVI”: diagonal, equal volume, varying shape, “VVI”: diagonal, varying volume and shape, “EEE”: ellipsoidal, equal volume, shape, and orientation, “VEE”: ellipsoidal, equal shape and orientation, “EVE”: ellipsoidal, equal volume and orientation, “VVE”: ellipsoidal, equal orientation, “EEV”: ellipsoidal, equal volume and equal shape, “VEV”: ellipsoidal, equal shape, “EVV”: ellipsoidal, equal volume, “VVV”: ellipsoidal, varying volume, shape, and orientation.

**Supplementary Figure 4.**
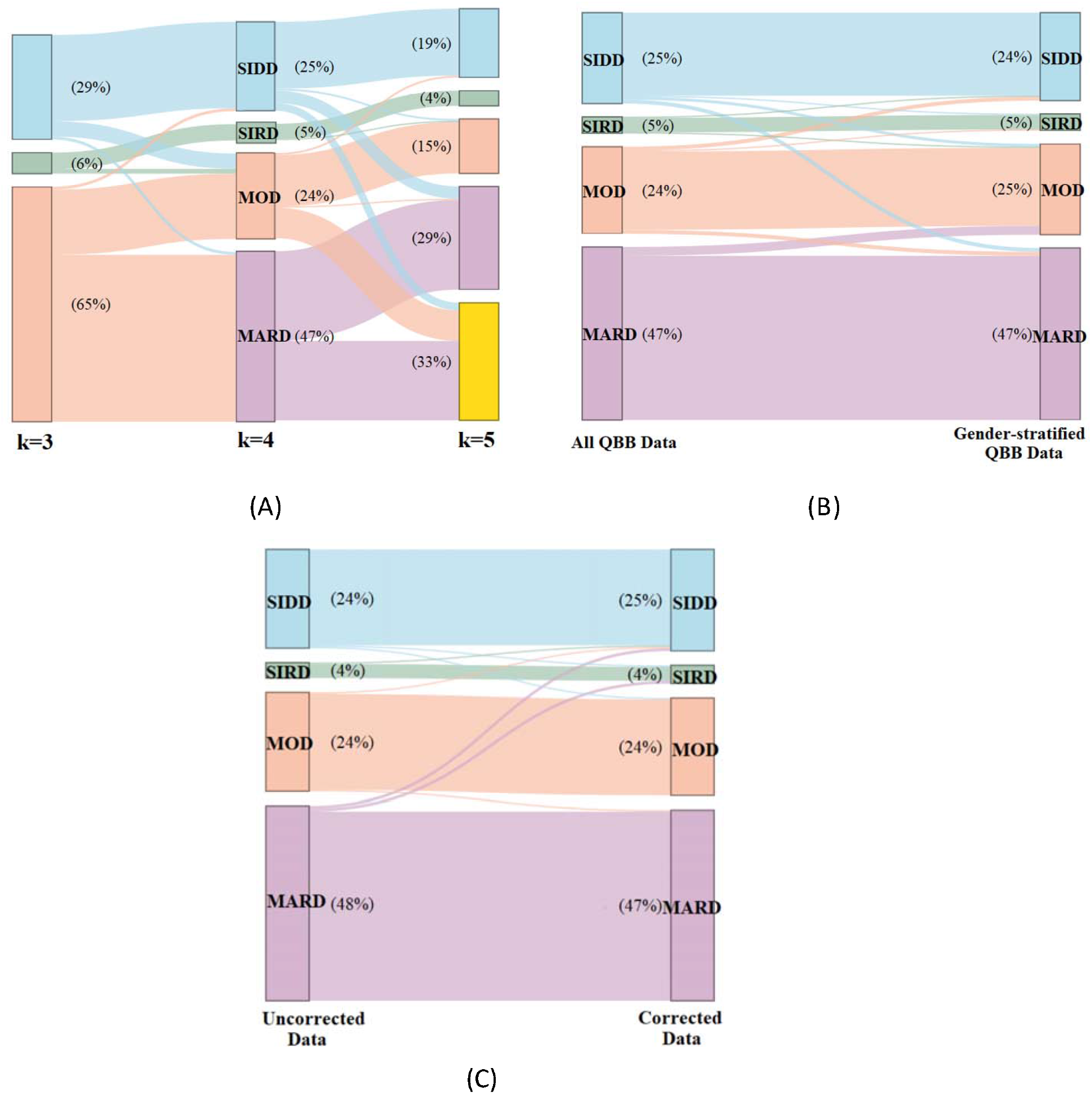
Sankey diagrams showing changes in cluster assignment for various scenarios. **A)** The cluster assignments slightly changed when clustering QBB data using different k (3,4, and 5). To a large extent, clusters 1 and 2 kept the same cluster label using different k, while cluster 3 split into clusters 4 and 5, at k=4 and k=5 respectively. **B)** Males and females were pooled, then clustered separately. Minor changes in the cluster assignments (7%) were observed when clustering the full data set compared to clustering the gender specific datasets. **C)** When clustering using uncorrected vs. corrected HOMA variables, most individuals retained the same cluster assignment using the fasting time - corrected data.

**Supplementary Figure 5.**
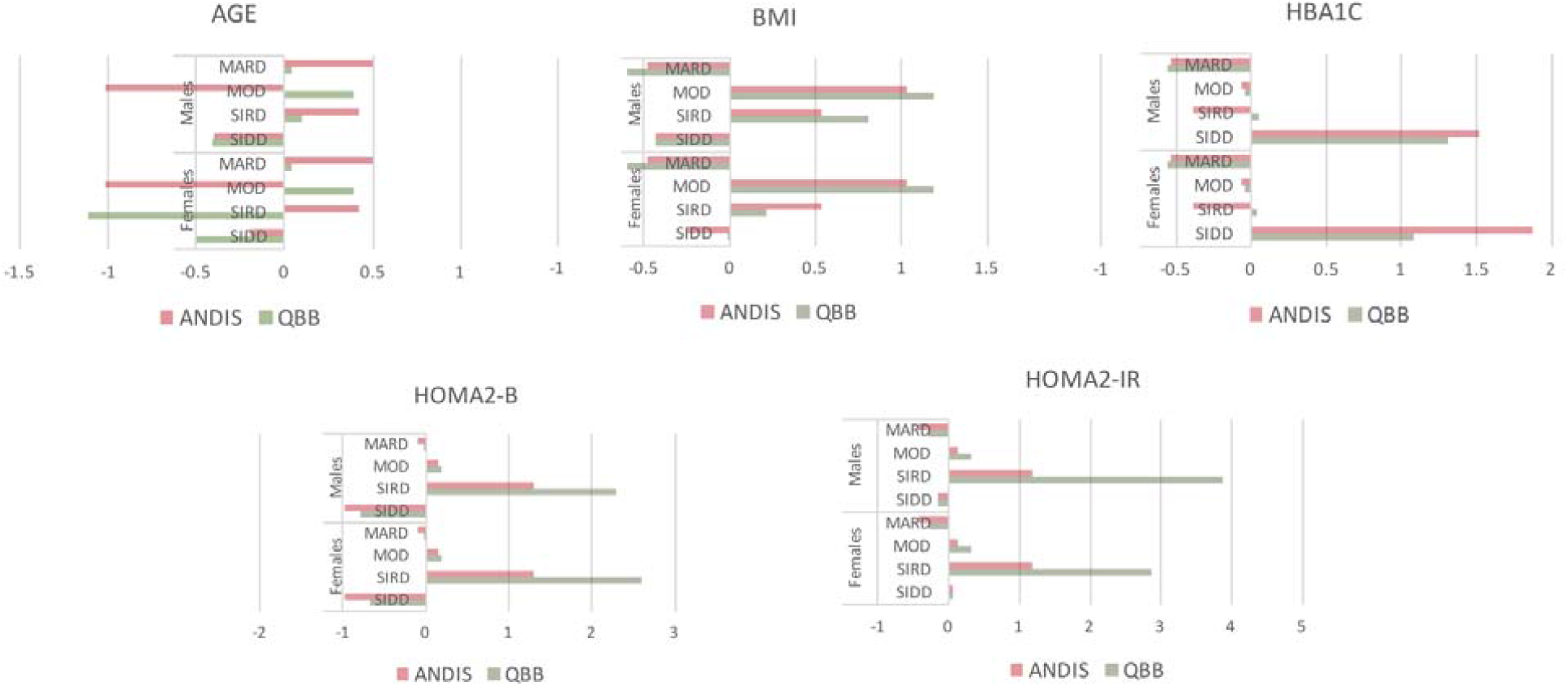
Comparison of cluster coordinates for cluster allocations between QBB and ANDIS. Gender-specific variables including age, BMI, HbA_1c_, HOMA2-B, and HOMA2-IR are shown for the SIDD, SIRD, MOD, and MARD diabetes subtypes. Differences between QBB and ANDIS were mainly observed in the age variable.

**Supplementary Figure 6.**
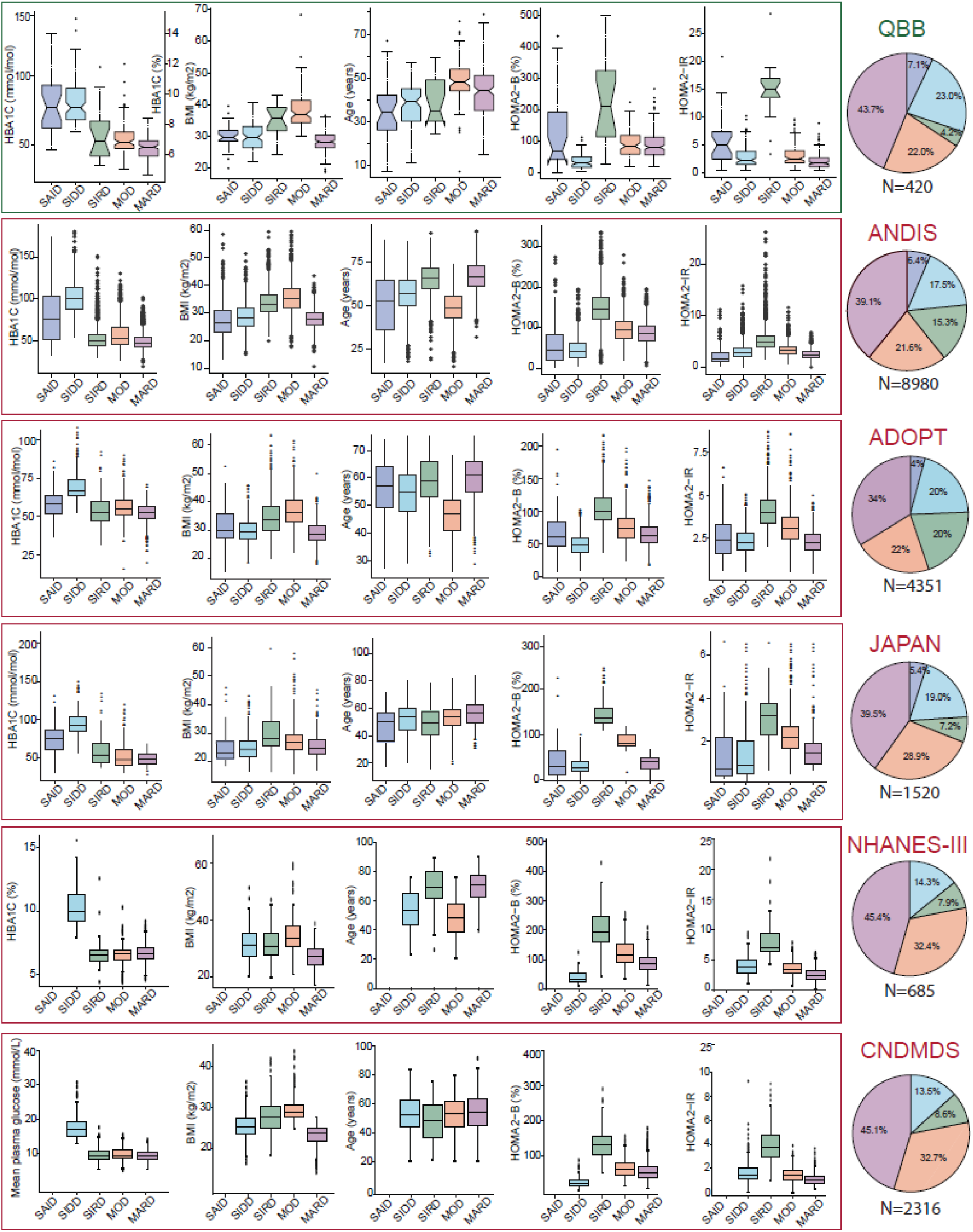
Comparison of cluster variables across studies. The cluster variables were similar across many ethnicities (Arab, Scandinavian, British, Japanese, American, and Chinese).

[Figure provided as multi-page PDF file]

**Supplementary Figure 7 and 8**. Box plots for all protein and metabolite levels associated with a T2D specific cluster (see Figure 6 for an example).

