## Supplementary figures and images for "Metabolic and proteomic signatures of type 2 diabetes subtypes in an Arab population"

### Supplementary Figure 1

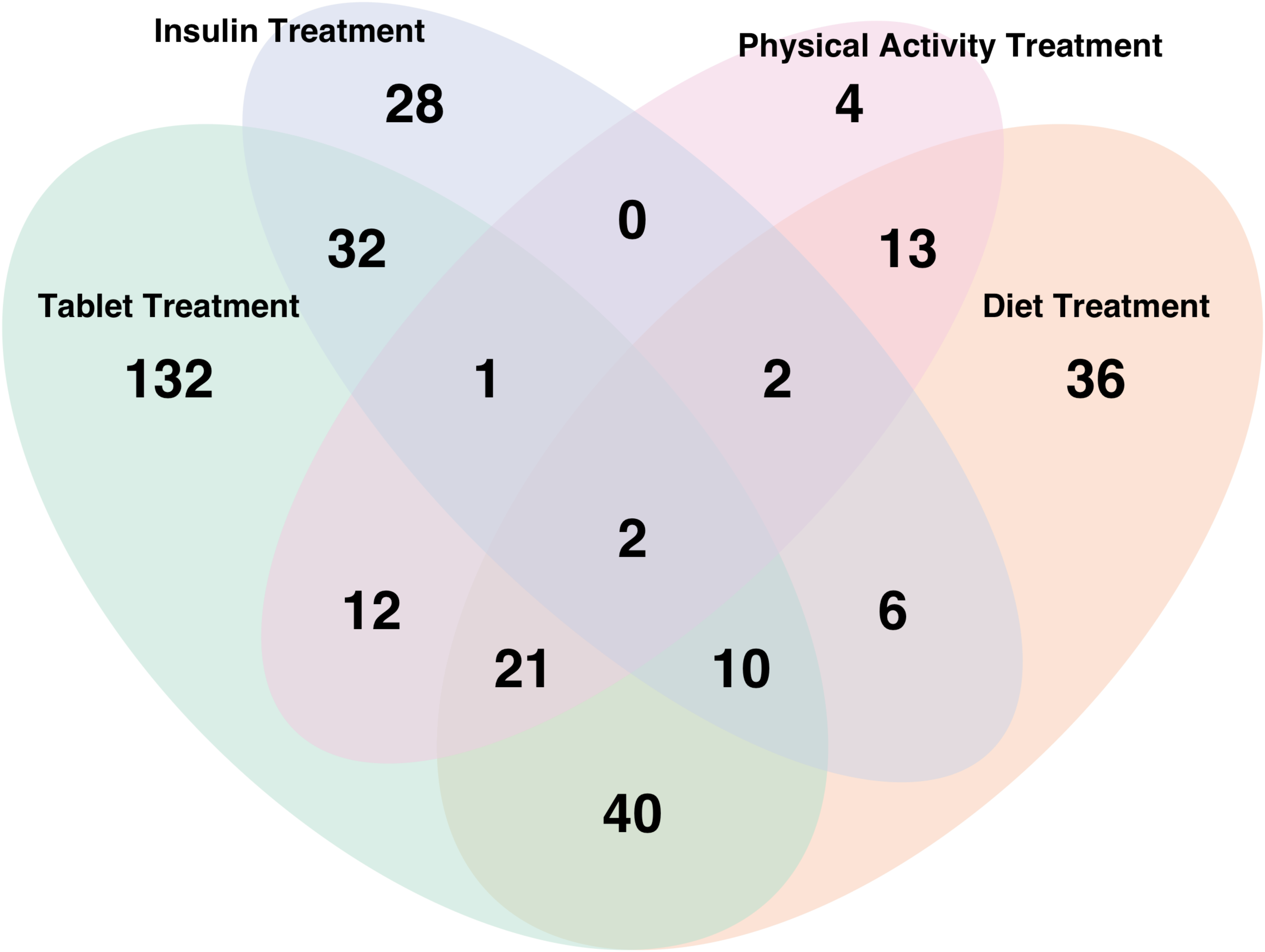

### Supplementary Figure 2

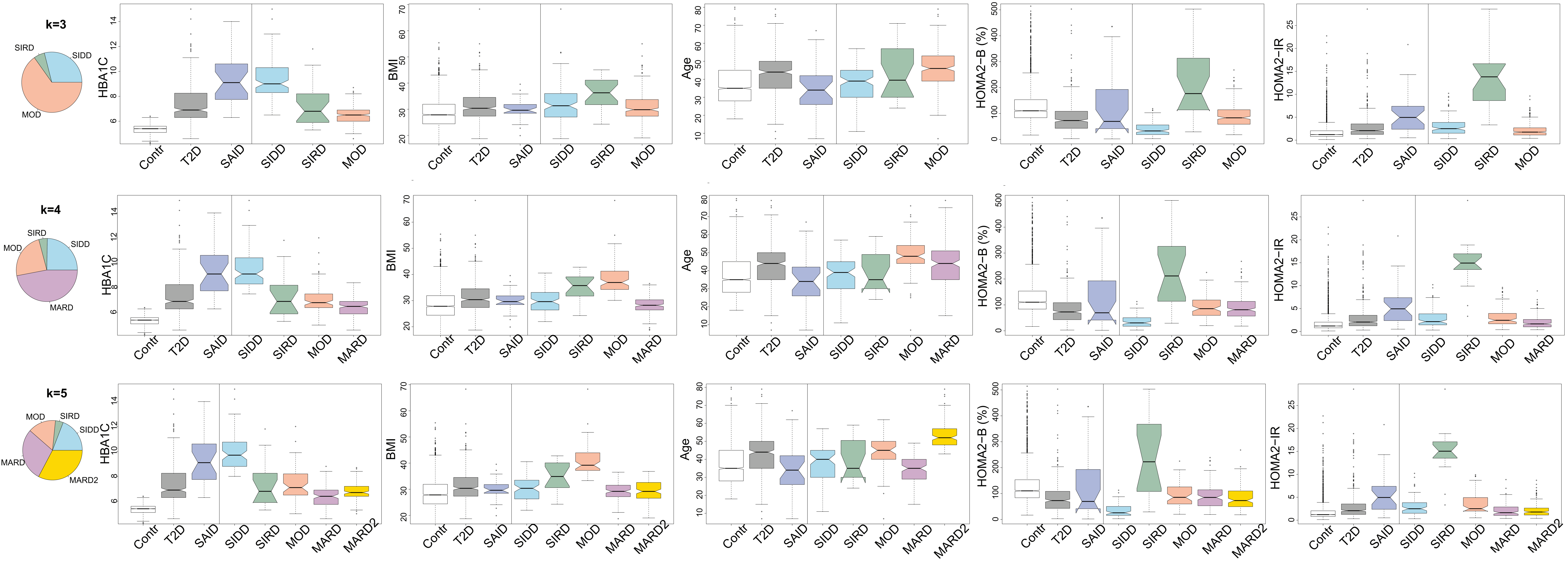

### Supplementary Figure 3

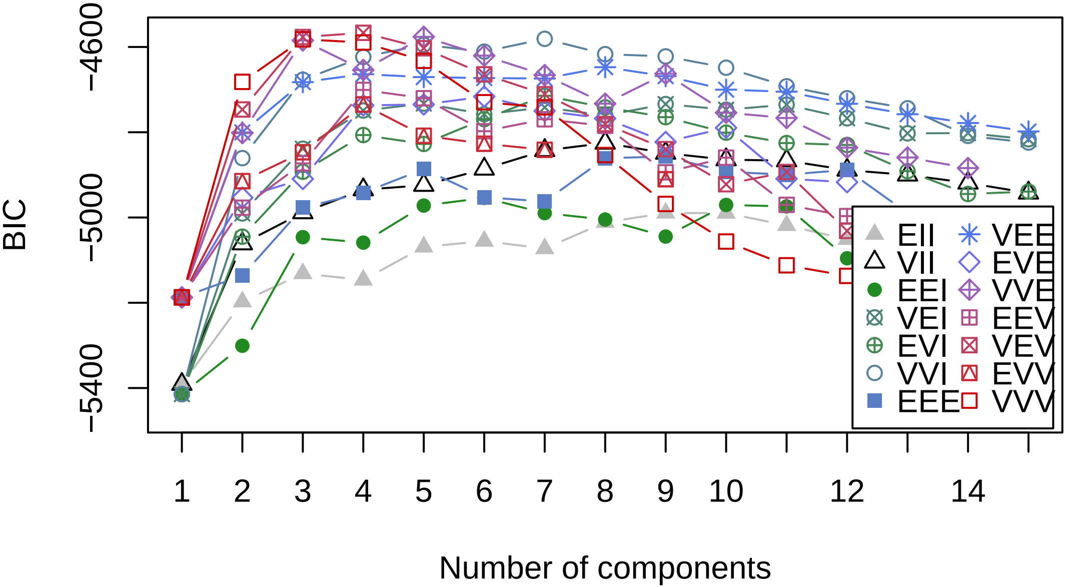

### Supplementary Figure 4a

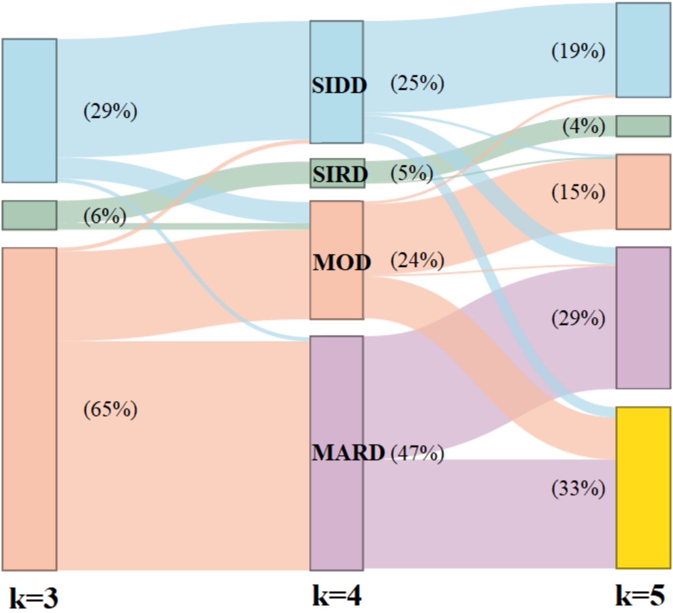

### Supplementary Figure 4b

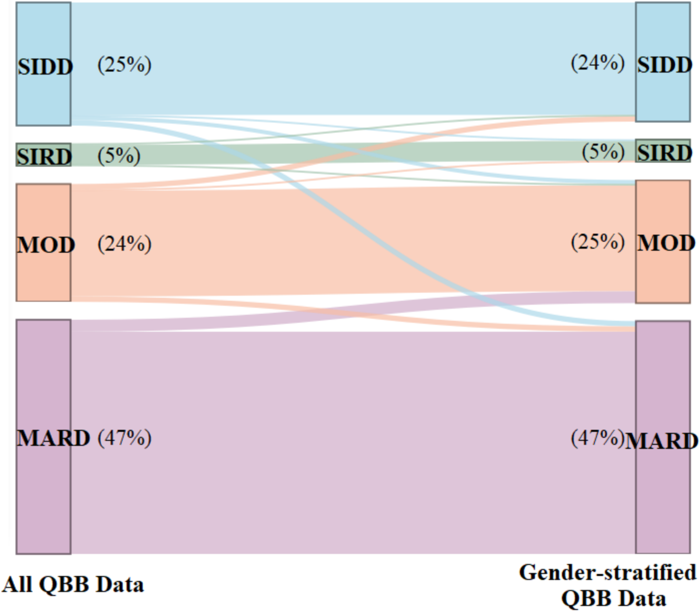

### Supplementary Figure 4c

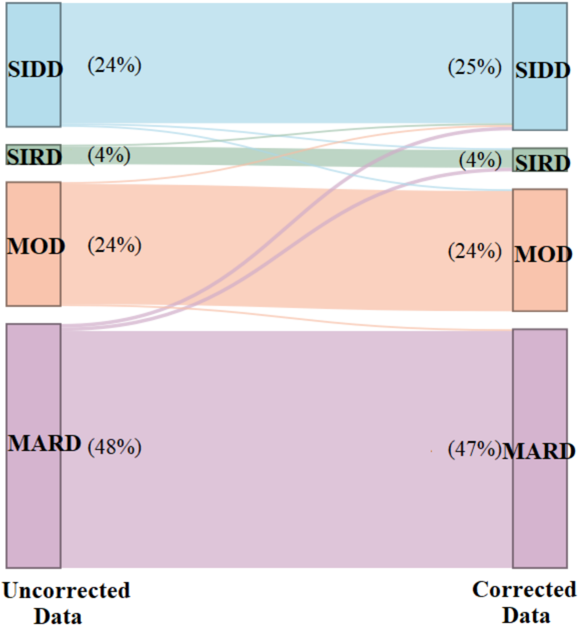

### Supplementary Figure 5a

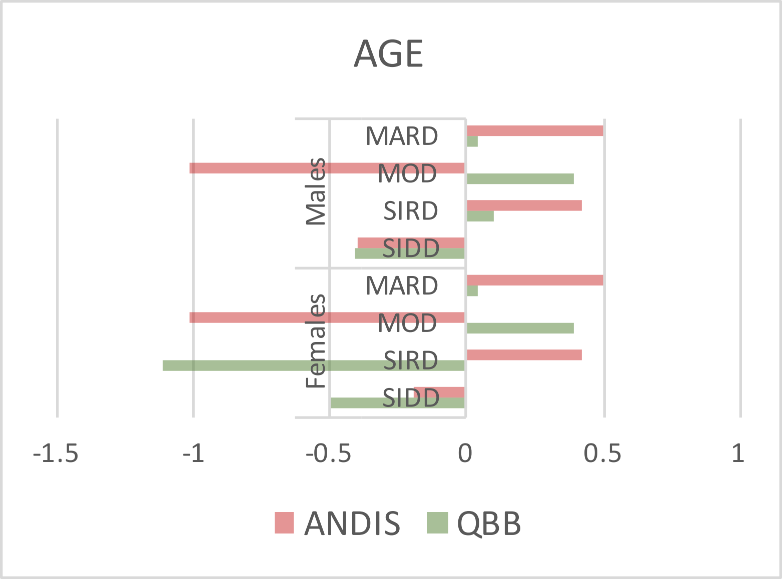

### Supplementary Figure 5b

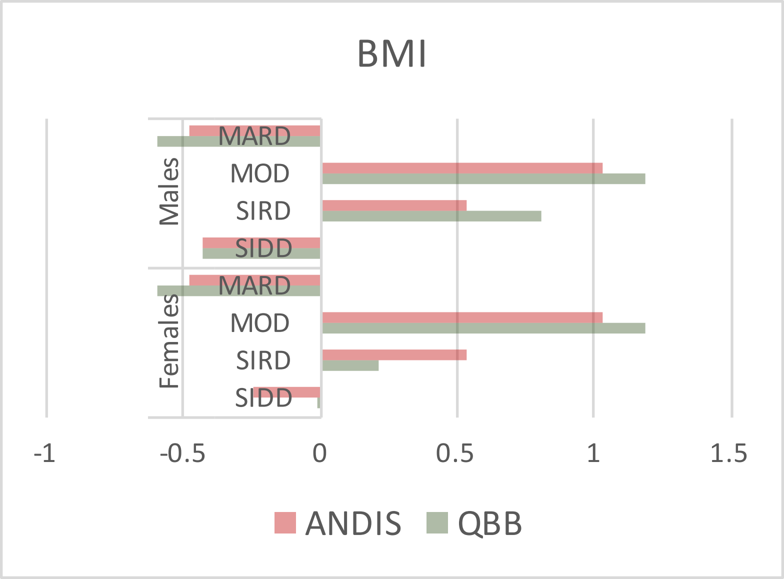

### Supplementary Figure 5c

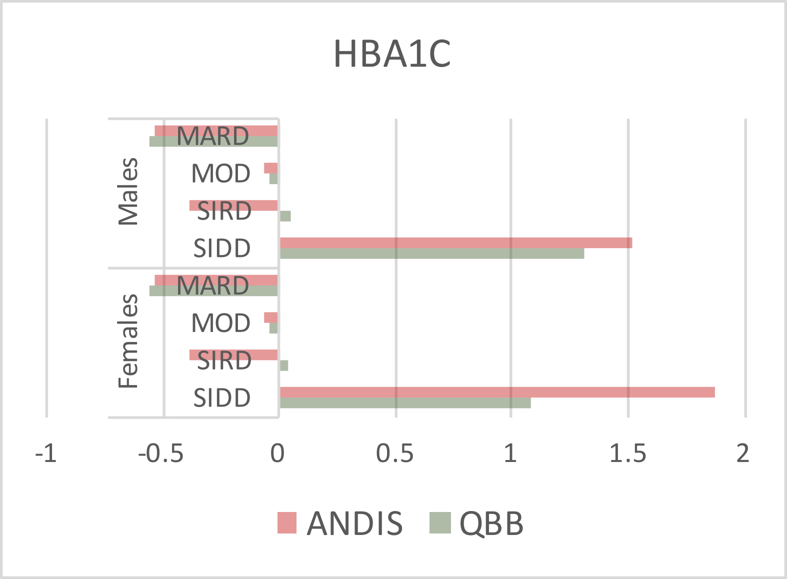

### Supplementary Figure 5d

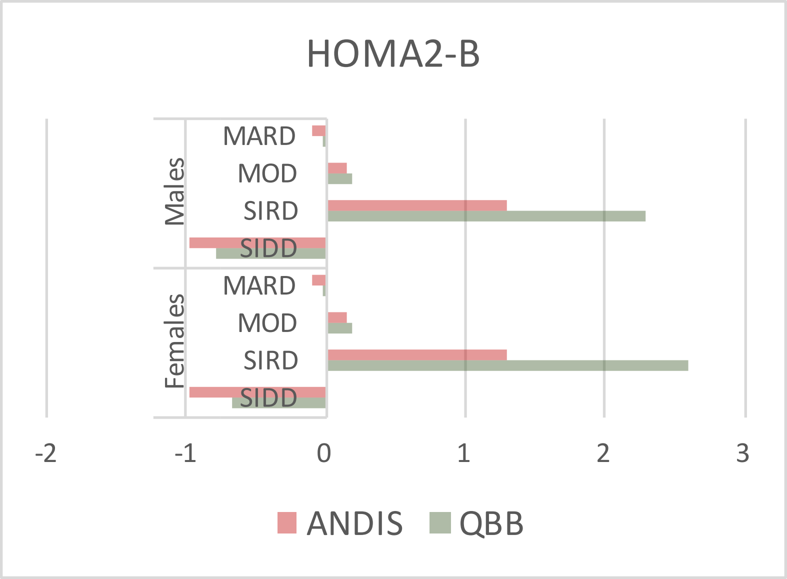

### Supplementary Figure 5e

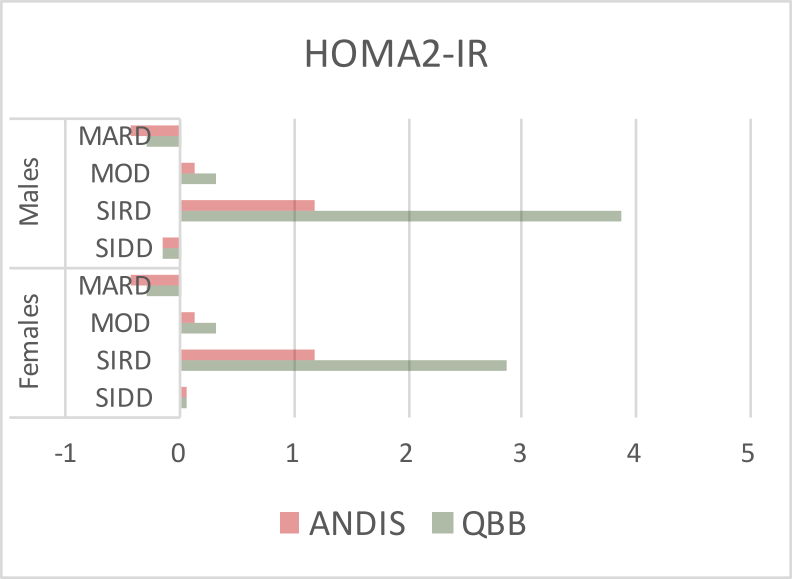

### Supplementary Figure 6

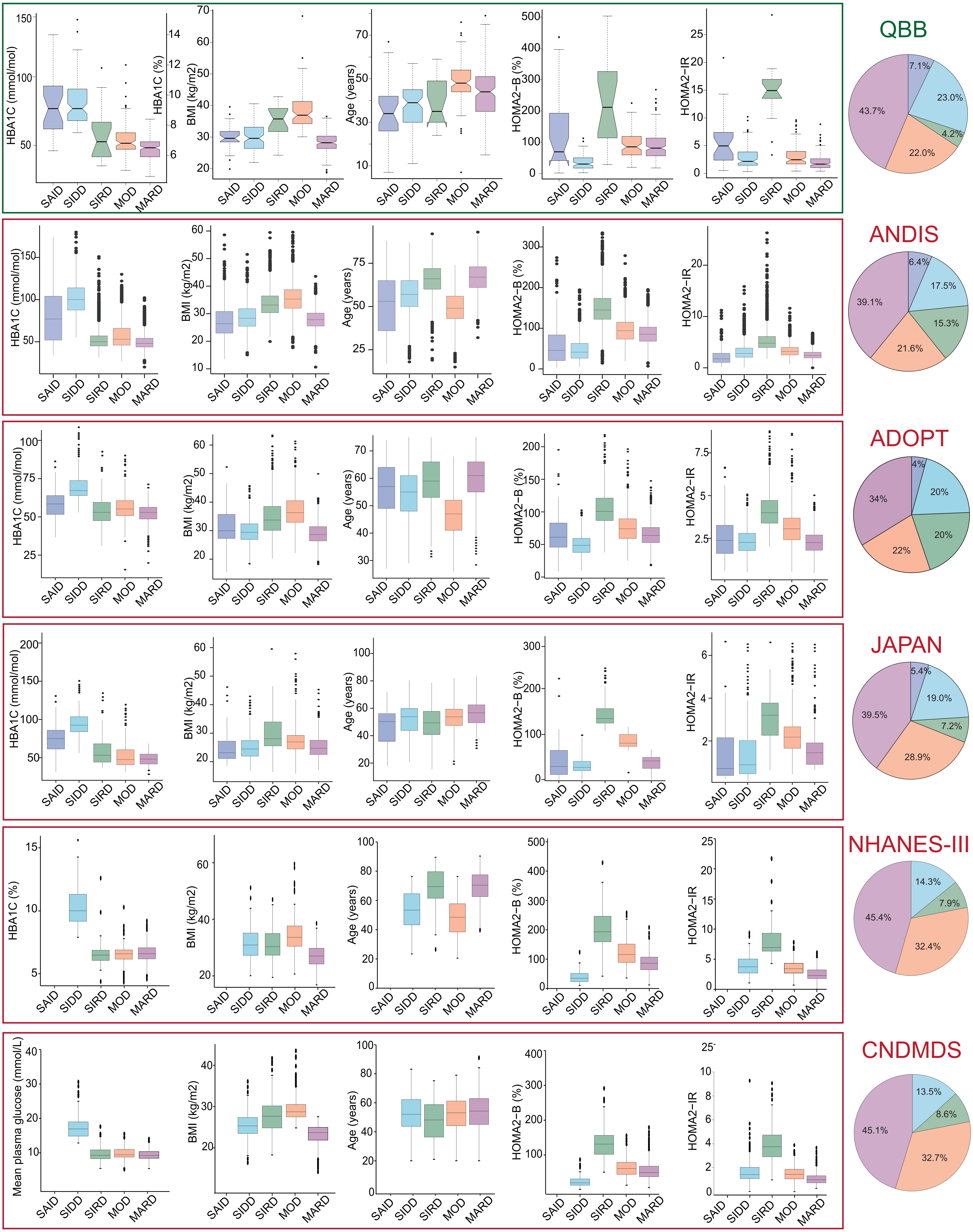
