## Supplementary Figure 7 for "Metabolic and proteomic signatures of type 2 diabetes subtypes in an Arab population"

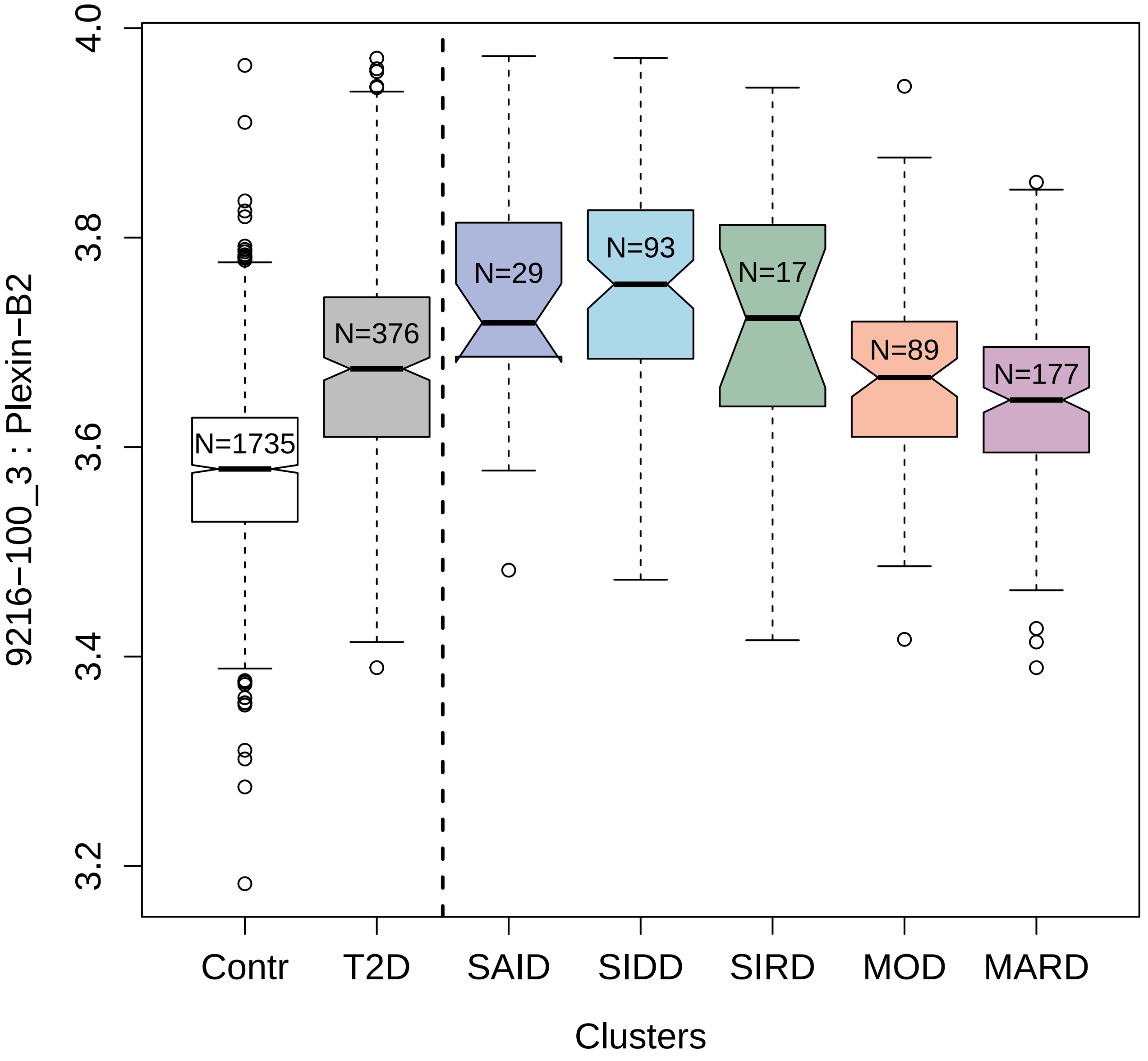

9216−100_3 : Plexin−B2
4.0
3.8
3.6
3.4
3.2
N=1735
N=376
N=29
N=93
N=17
N=89
N=177
Contr
T2D
SAID
SIDD
SIRD
MOD
MARD
Clusters

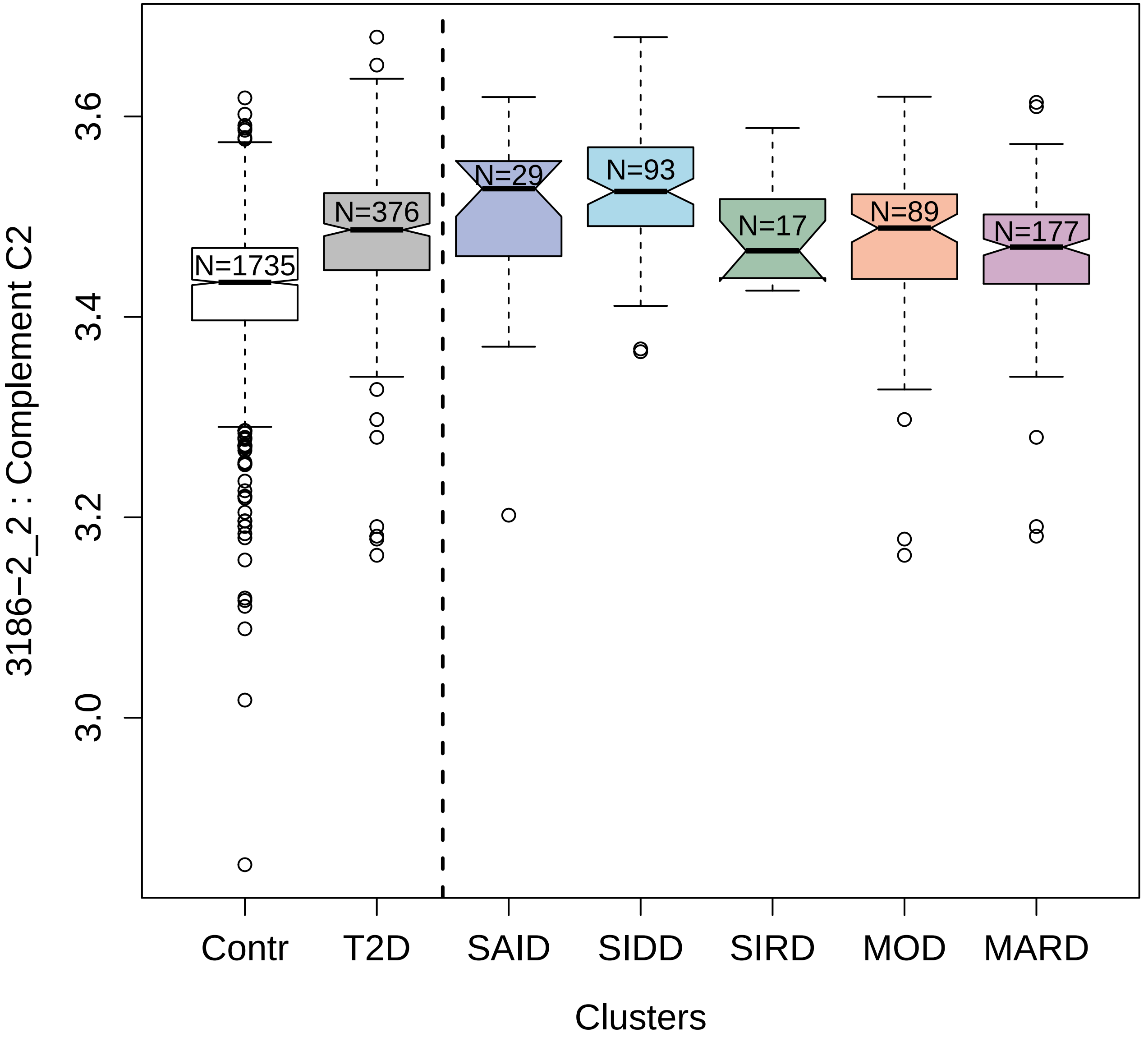

3186−2_2 : Complement C2
3.6
3.4
3.2
3.0
N=1735
N=376
N=29
N=93
N=17
N=89
N=177
Contr
T2D
SAID
SIDD
SIRD
MOD
MARD
Clusters

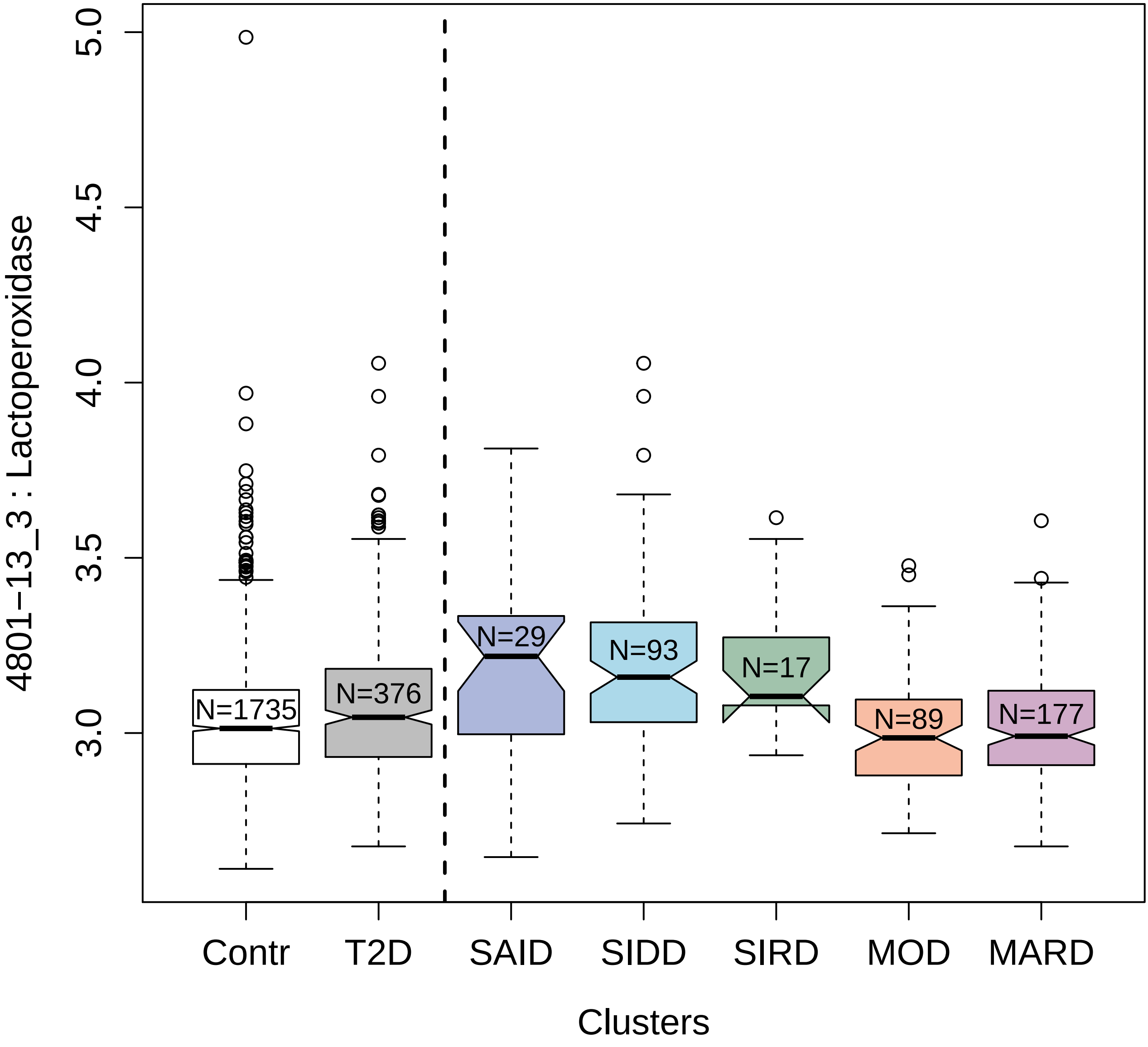

4801−13_3 : Lactoperoxidase
5.0
4.5
4.0
3.5
3.0
N=1735
N=376
N=29
N=93
N=17
N=89
N=177
Contr
T2D
SAID
SIDD
SIRD
MOD
MARD
Clusters

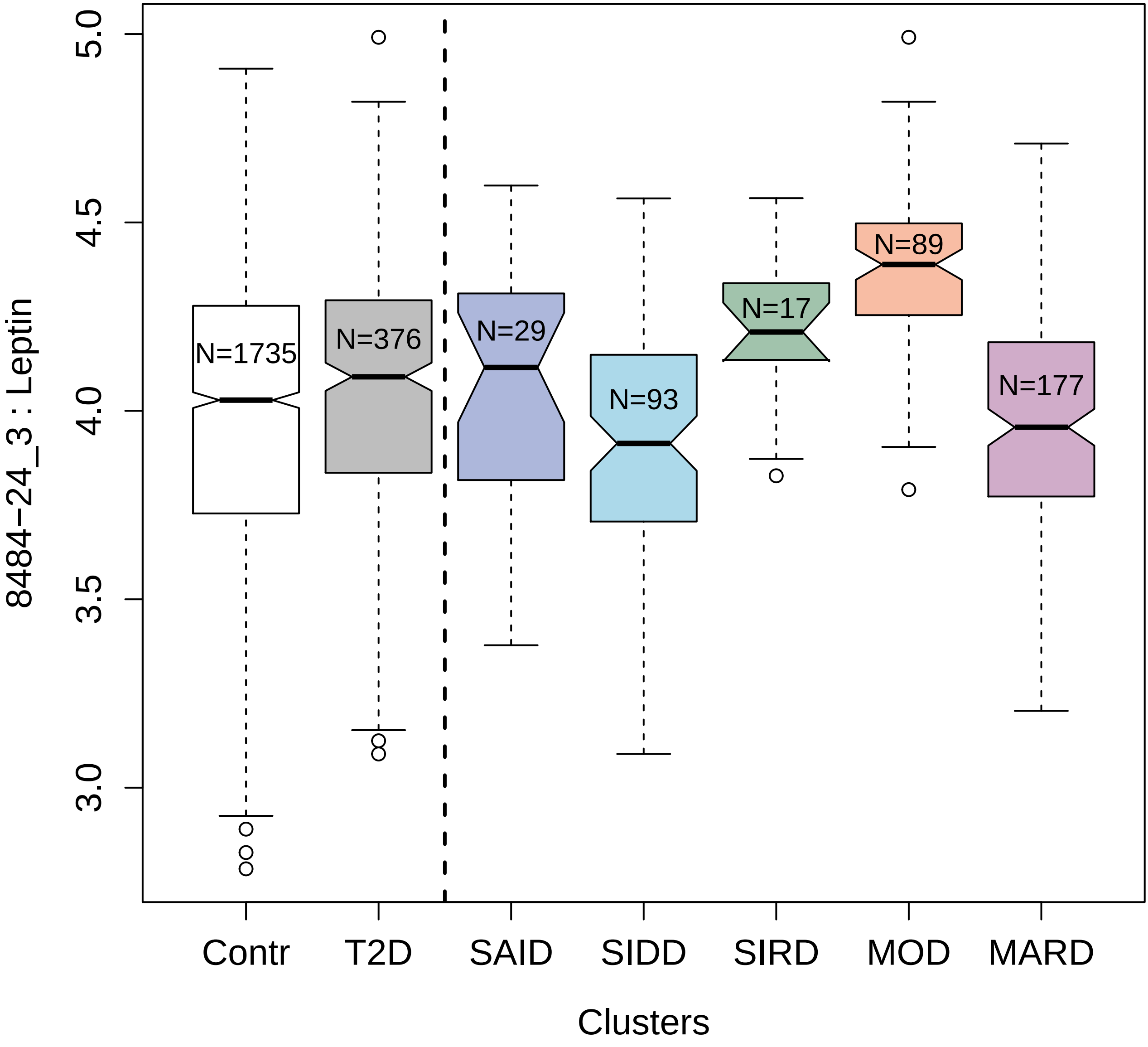

8484−24_3 : Leptin
5.0
4.5
4.0
3.5
3.0
N=1735
N=376
N=29
N=93
N=17
N=89
N=177
Contr
T2D
SAID
SIDD
SIRD
MOD
MARD
Clusters

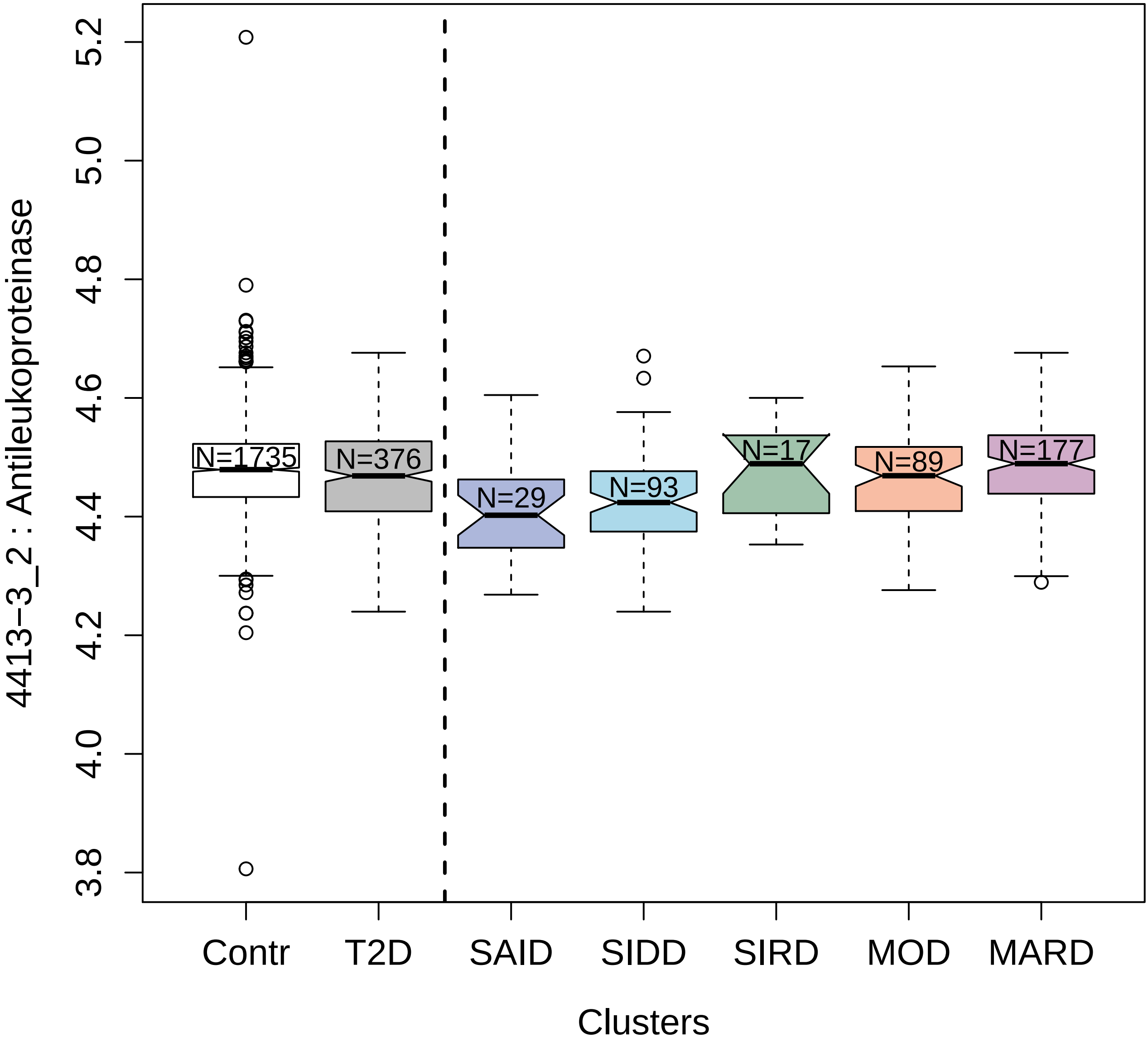
4413−3_2 : Antileukoproteinase
5.2
5.0
4.8
4.6
4.4
4.2
4.0
3.8
N=1735
N=376
N=29
N=93
N=17
N=89
N=177
Contr
T2D
SAID
SIDD
SIRD
MOD
MARD
Clusters

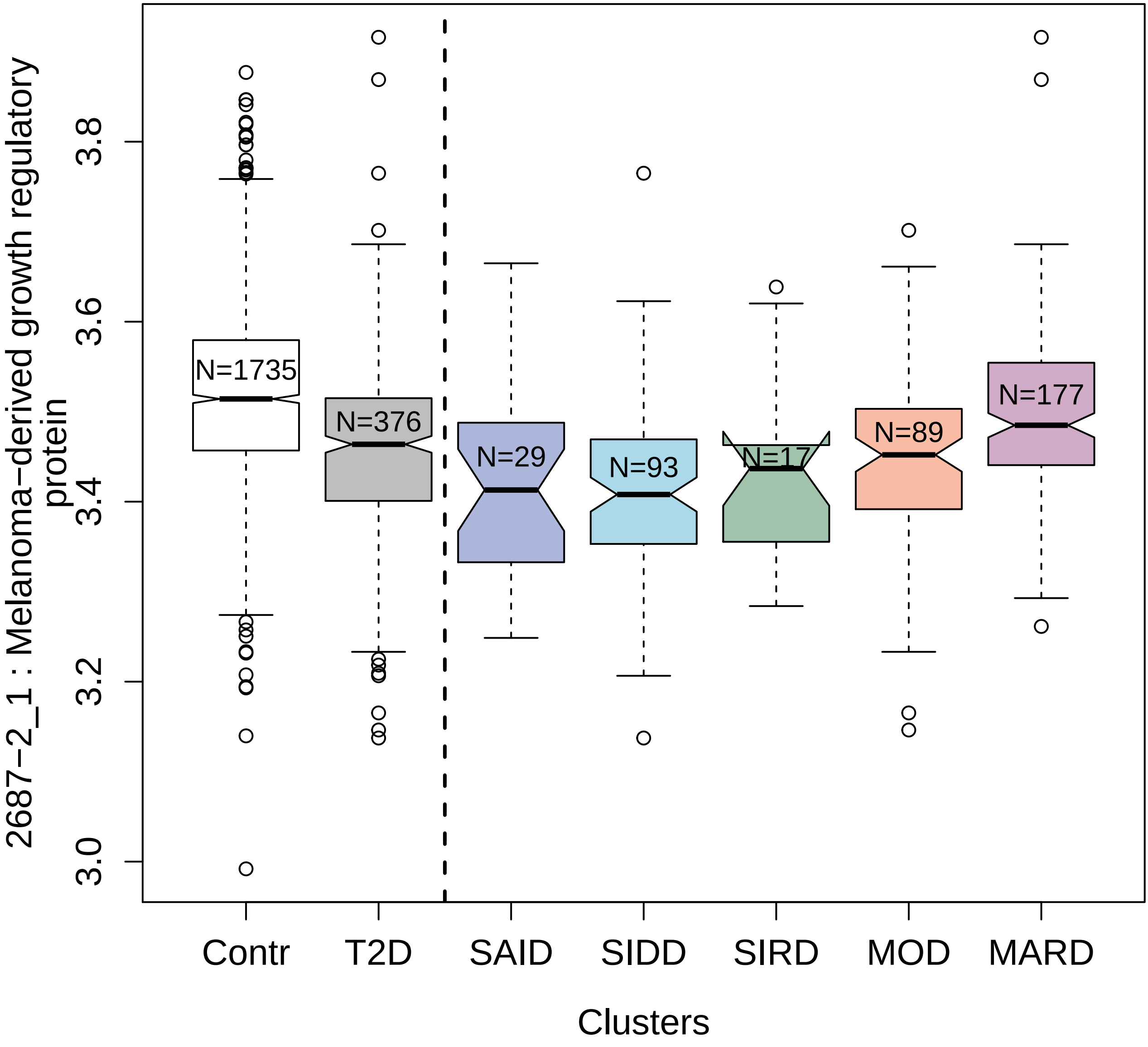
2687−2_1 : Melanoma−derived growth regulatory protein
3.0
3.2
3.4
3.6
3.8
N=1735
N=376
N=29
N=93
N=17
N=89
N=177
Contr
T2D
SAID
SIDD
SIRD
MOD
MARD
Clusters

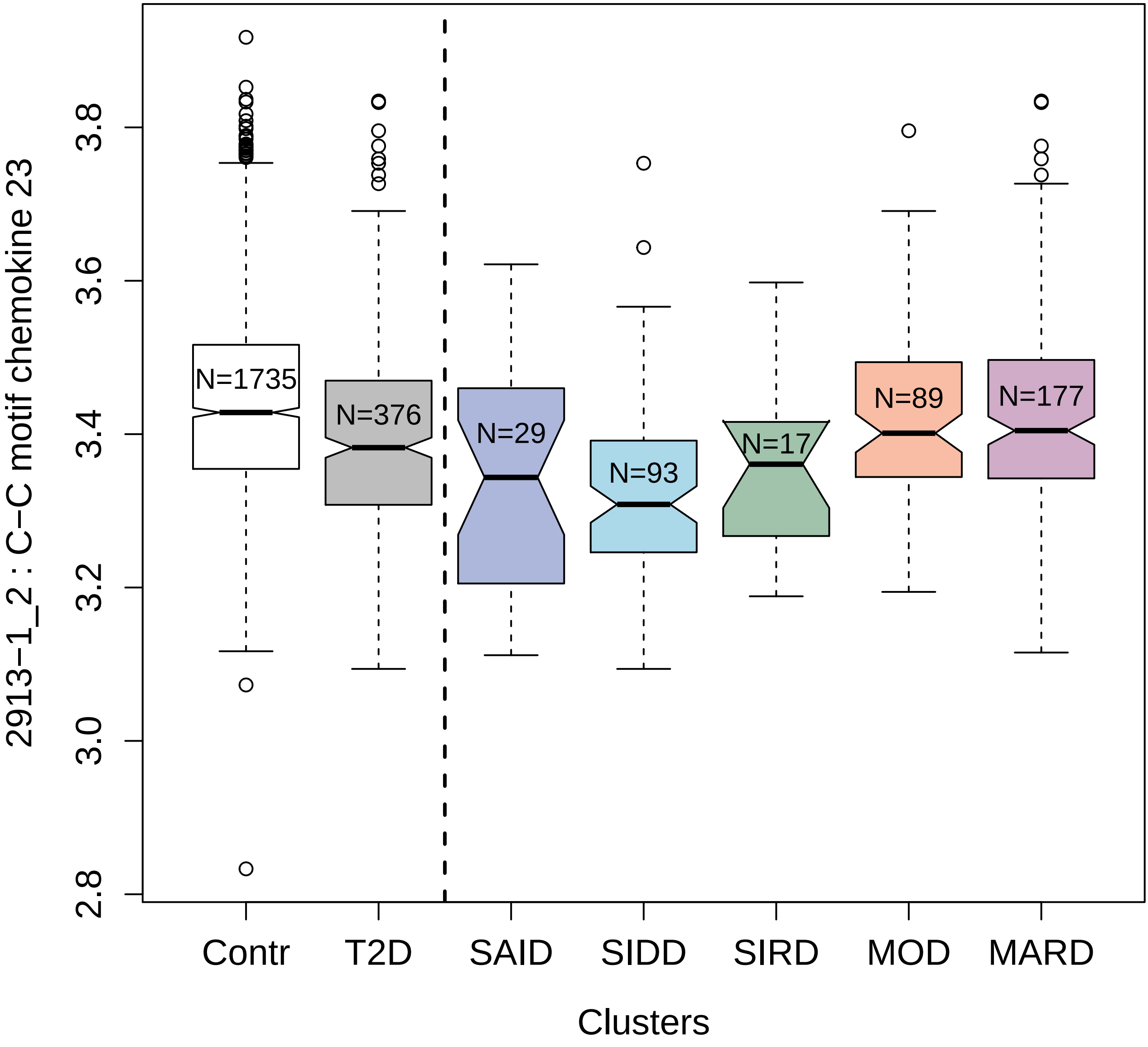

2913−1_2 : C−C motif chemokine 23
2.8
3.0
3.2
3.4
3.6
3.8
N=1735
N=376
N=29
N=93
N=17
N=89
N=177
Contr
T2D
SAID
SIDD
SIRD
MOD
MARD
Clusters

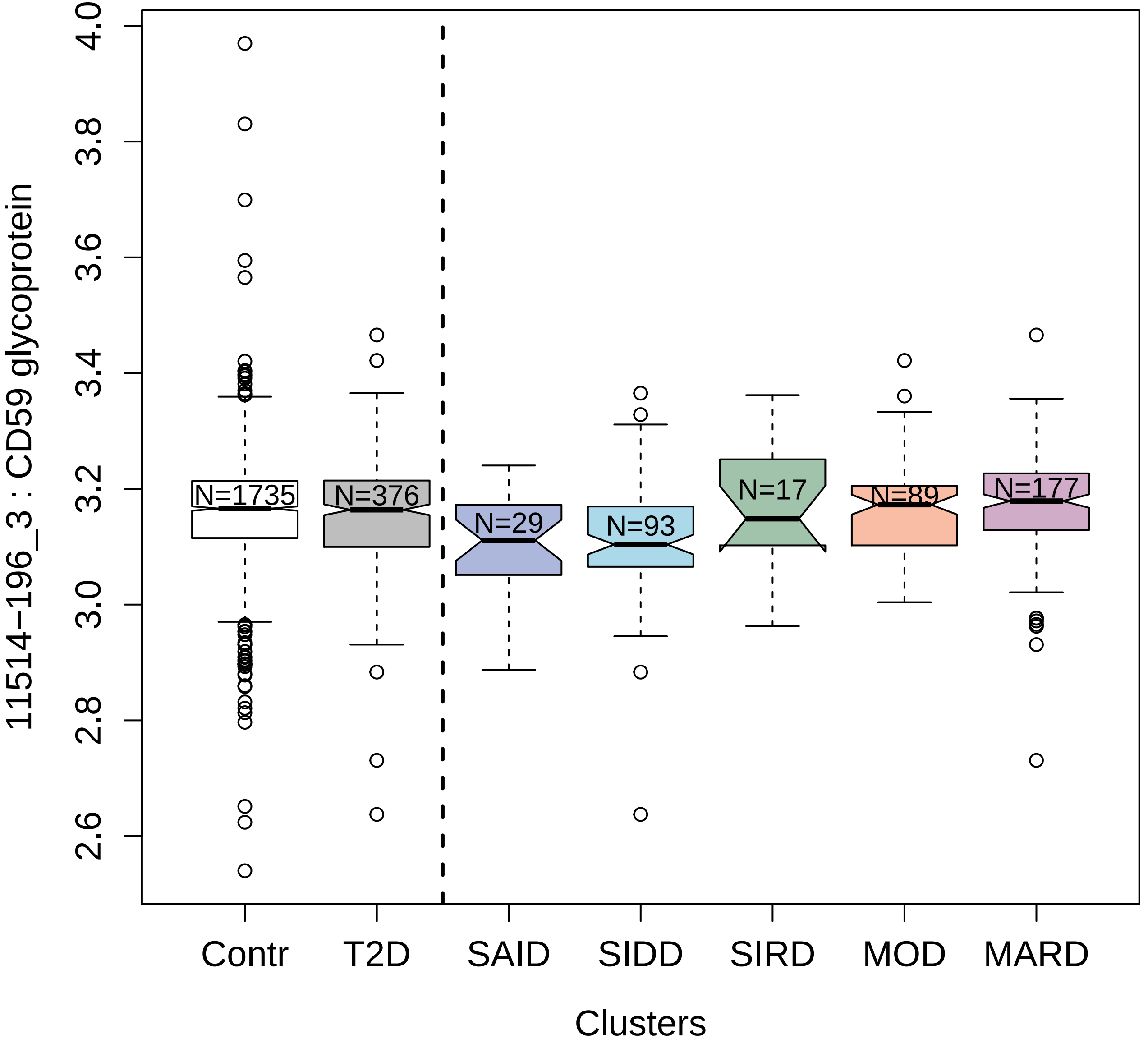

11514−196_3 : CD59 glycoprotein
4.0
3.8
3.6
3.4
3.2
3.0
2.8
2.6
N=1735
N=376
N=29
N=93
N=17
N=89
N=177
Contr
T2D
SAID
SIDD
SIRD
MOD
MARD
Clusters

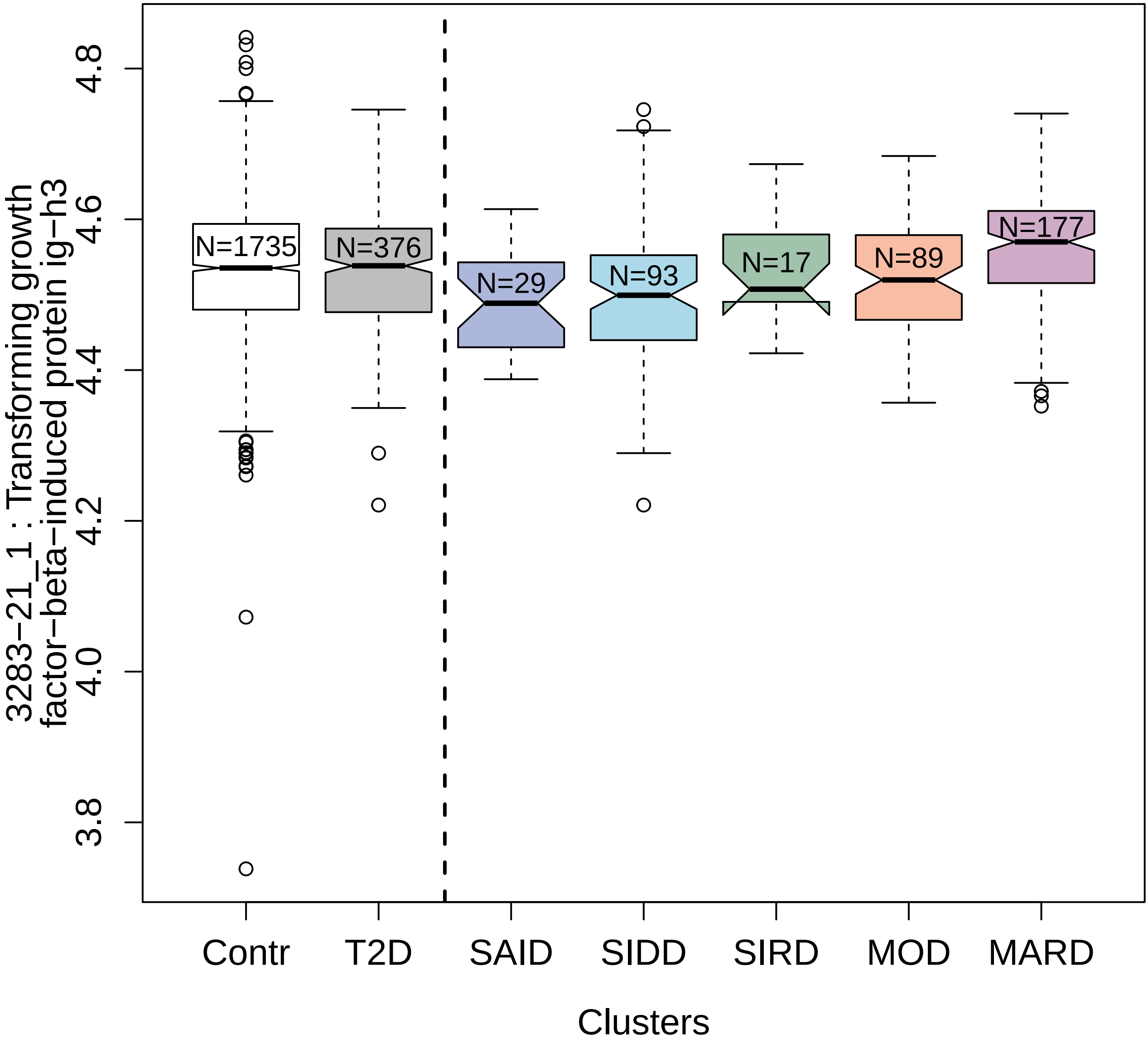

3283−21_1 : Transforming growth
factor−beta−induced protein ig−h3
4.8
4.6
4.4
4.2
4.0
3.8
N=1735
N=376
N=29
N=93
N=17
N=89
N=177
Contr
T2D
SAID
SIDD
SIRD
MOD
MARD
Clusters

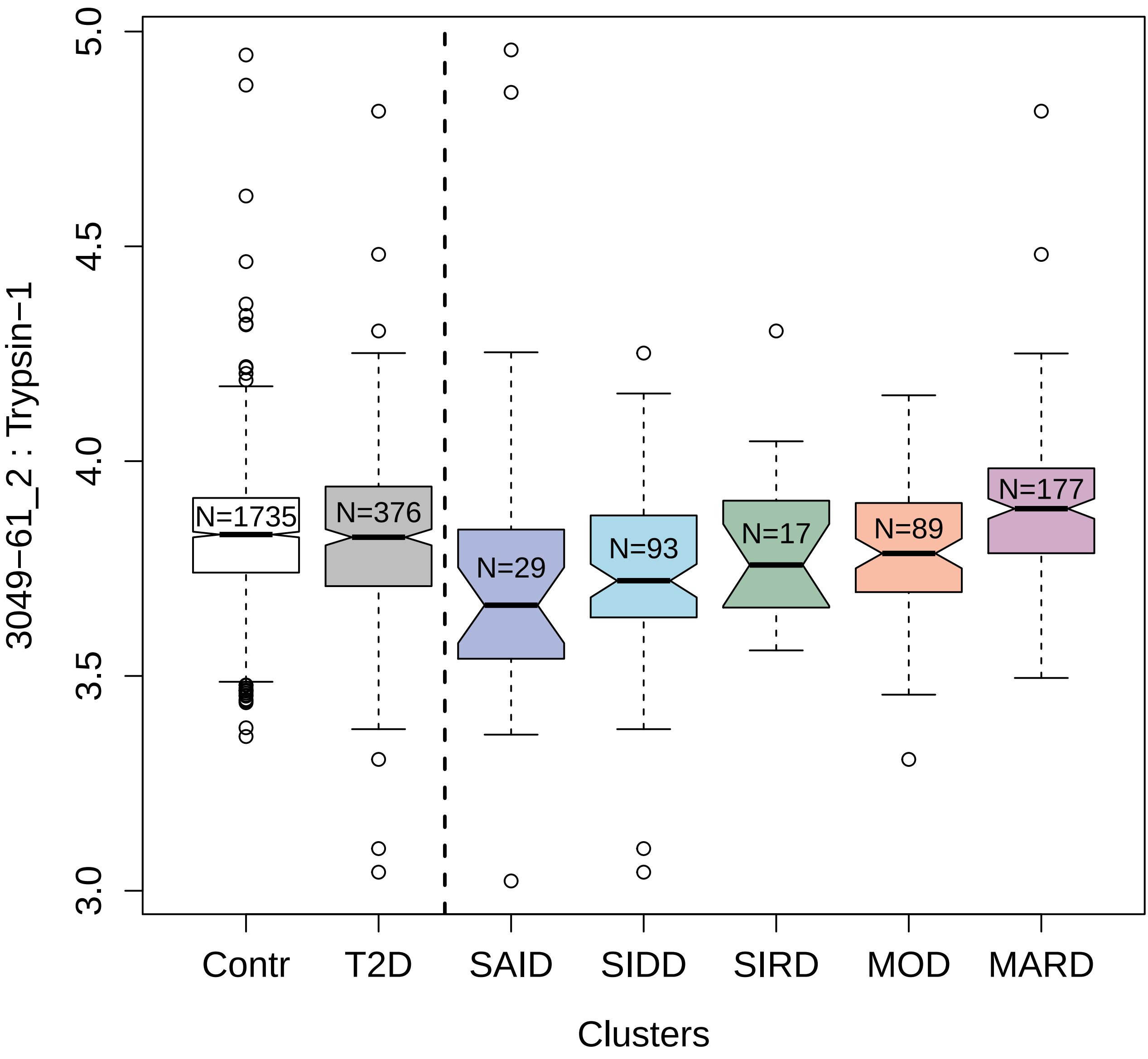

3049−61_2 : Trypsin−1
5.0
4.5
4.0
3.5
3.0
N=1735
N=376
N=29
N=93
N=17
N=89
N=177
Contr
T2D
SAID
SIDD
SIRD
MOD
MARD
Clusters

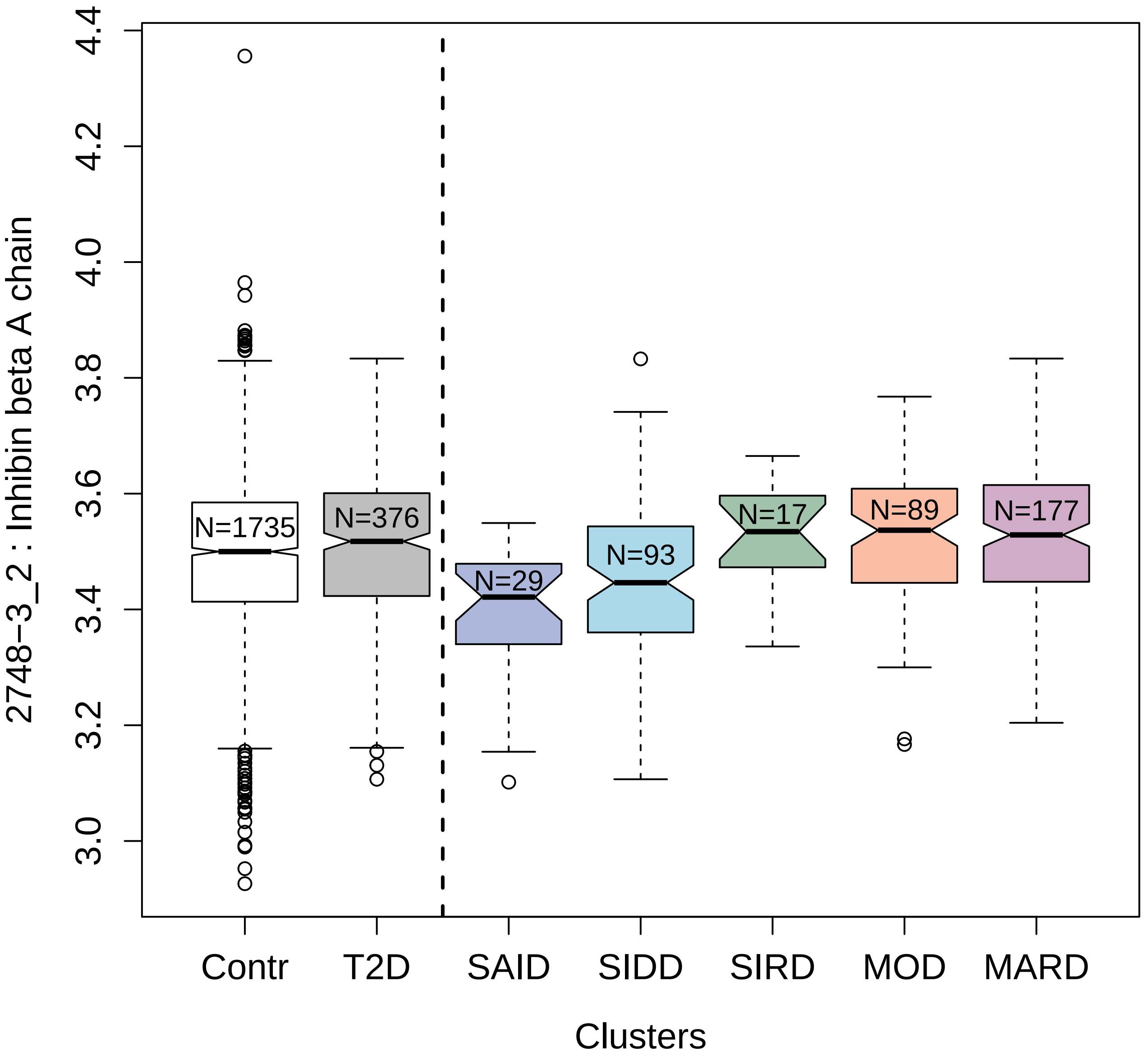

2748−3_2 : Inhibin beta A chain
4.4
4.2
4.0
3.8
3.6
3.4
3.2
3.0
N=1735
N=376
N=29
N=93
N=17
N=89
N=177
Contr
T2D
SAID
SIDD
SIRD
MOD
MARD
Clusters

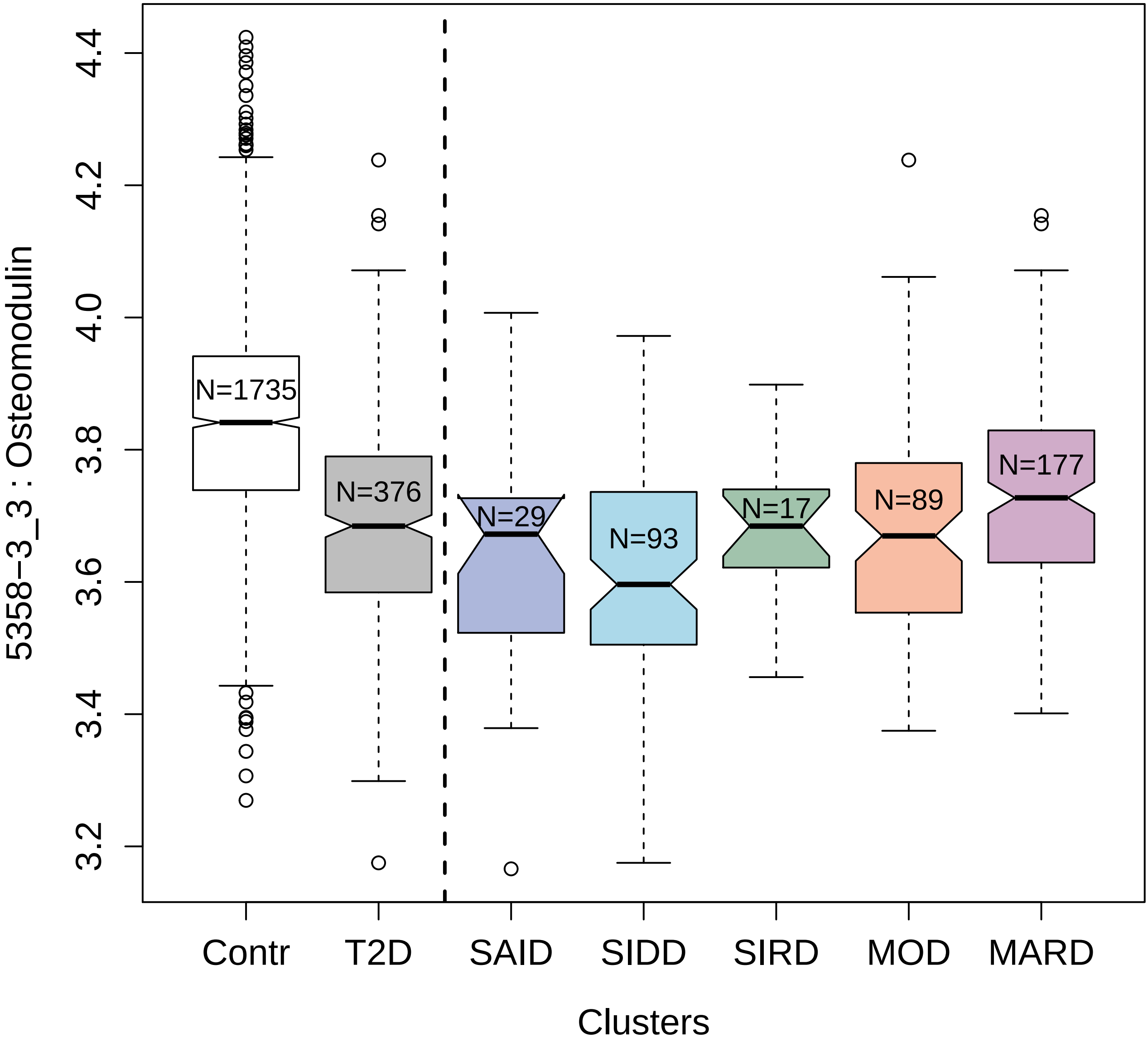

5358−3_3 : Osteomodulin
4.4
4.2
4.0
3.8
3.6
3.4
3.2
N=1735
N=376
N=29
N=93
N=17
N=89
N=177
Contr
T2D
SAID
SIDD
SIRD
MOD
MARD
Clusters

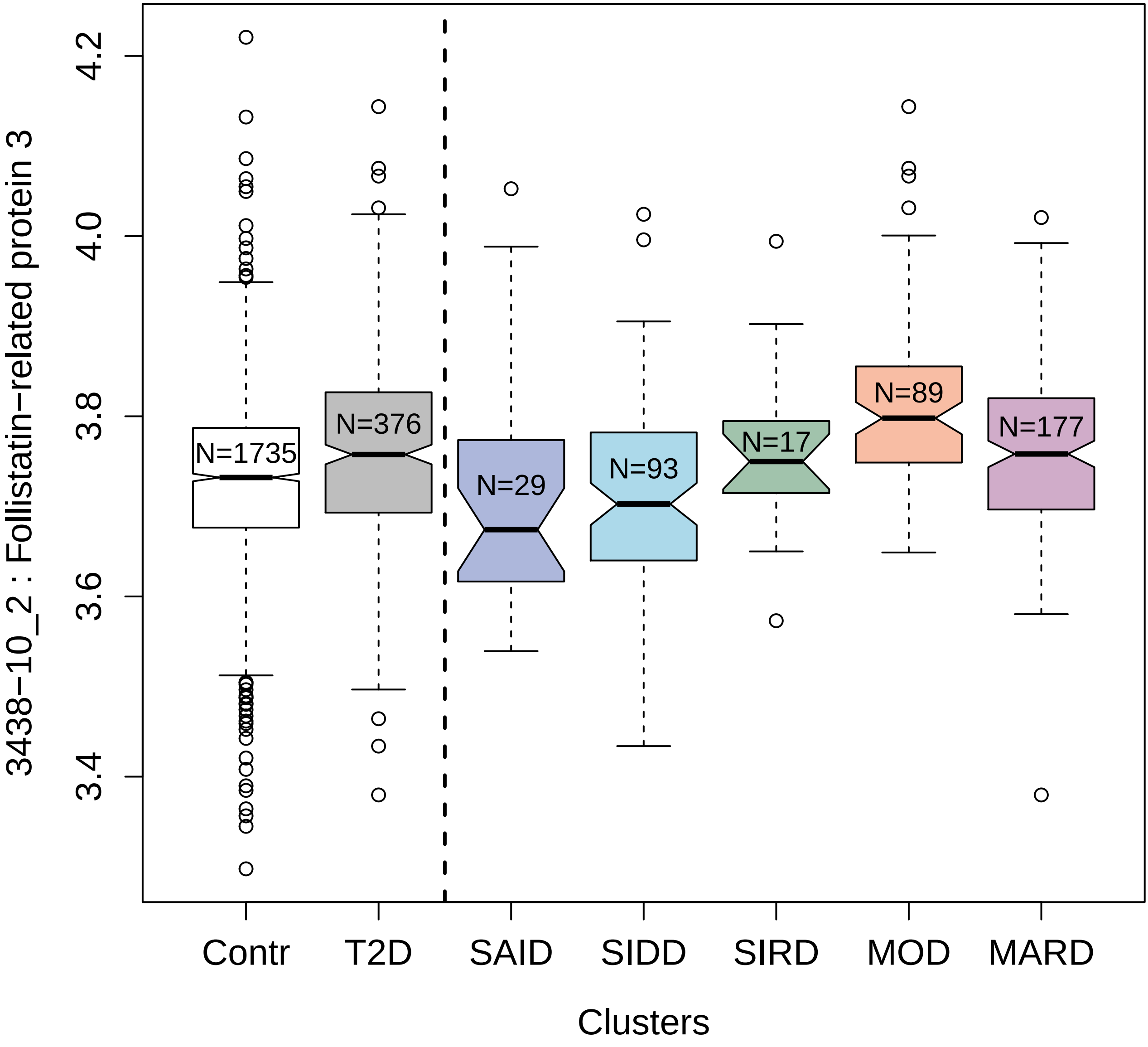

3438−10_2 : Follistatin−related protein 3
4.2
4.0
3.8
3.6
3.4
N=1735
N=376
N=29
N=93
N=17
N=89
N=177
Contr
T2D
SAID
SIDD
SIRD
MOD
MARD
Clusters

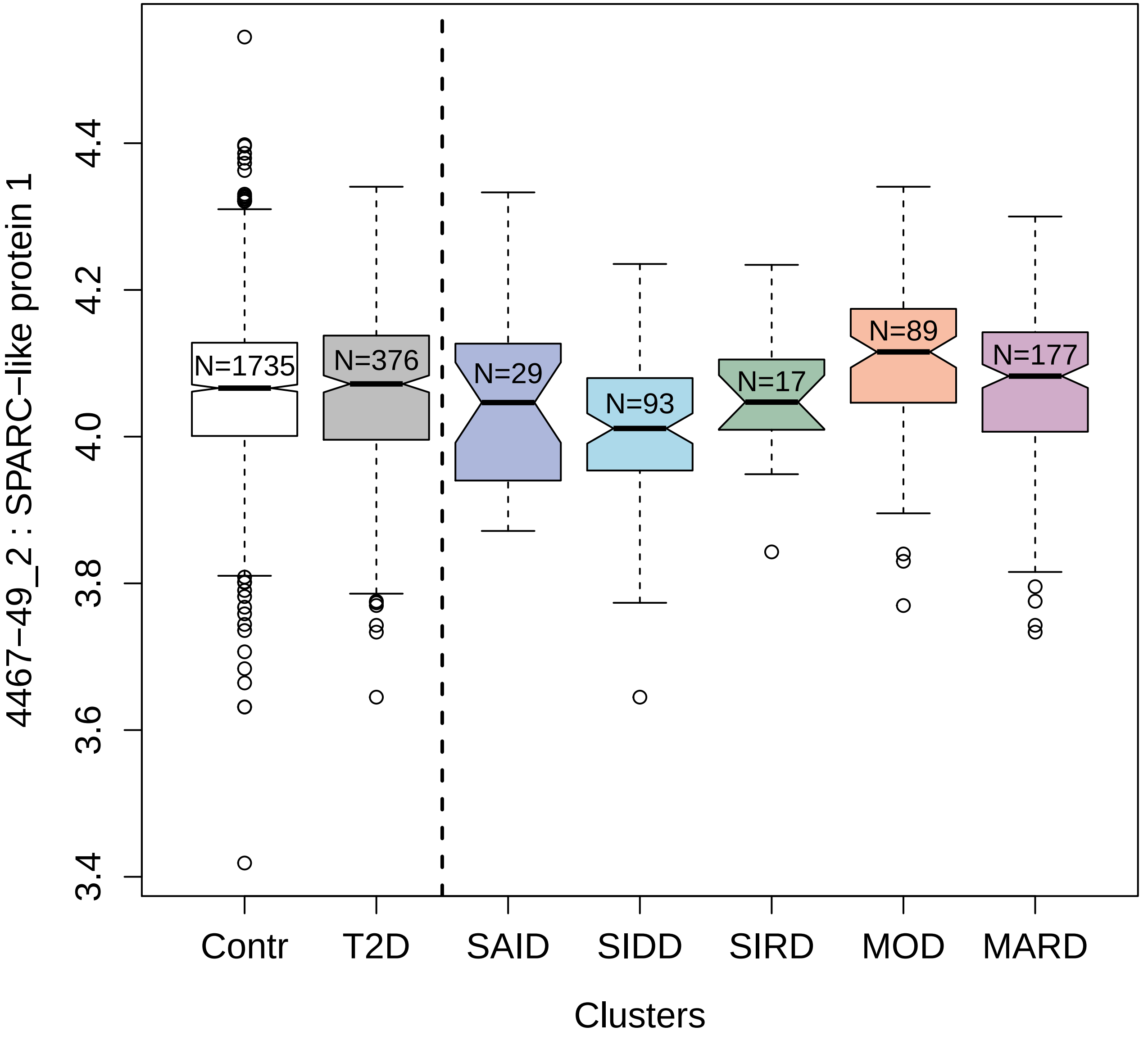
4467−49_2 : SPARC−like protein 1
3.4
3.6
3.8
4.0
4.2
4.4
N=1735
N=376
N=29
N=93
N=17
N=89
N=177
Contr
T2D
SAID
SIDD
SIRD
MOD
MARD
Clusters

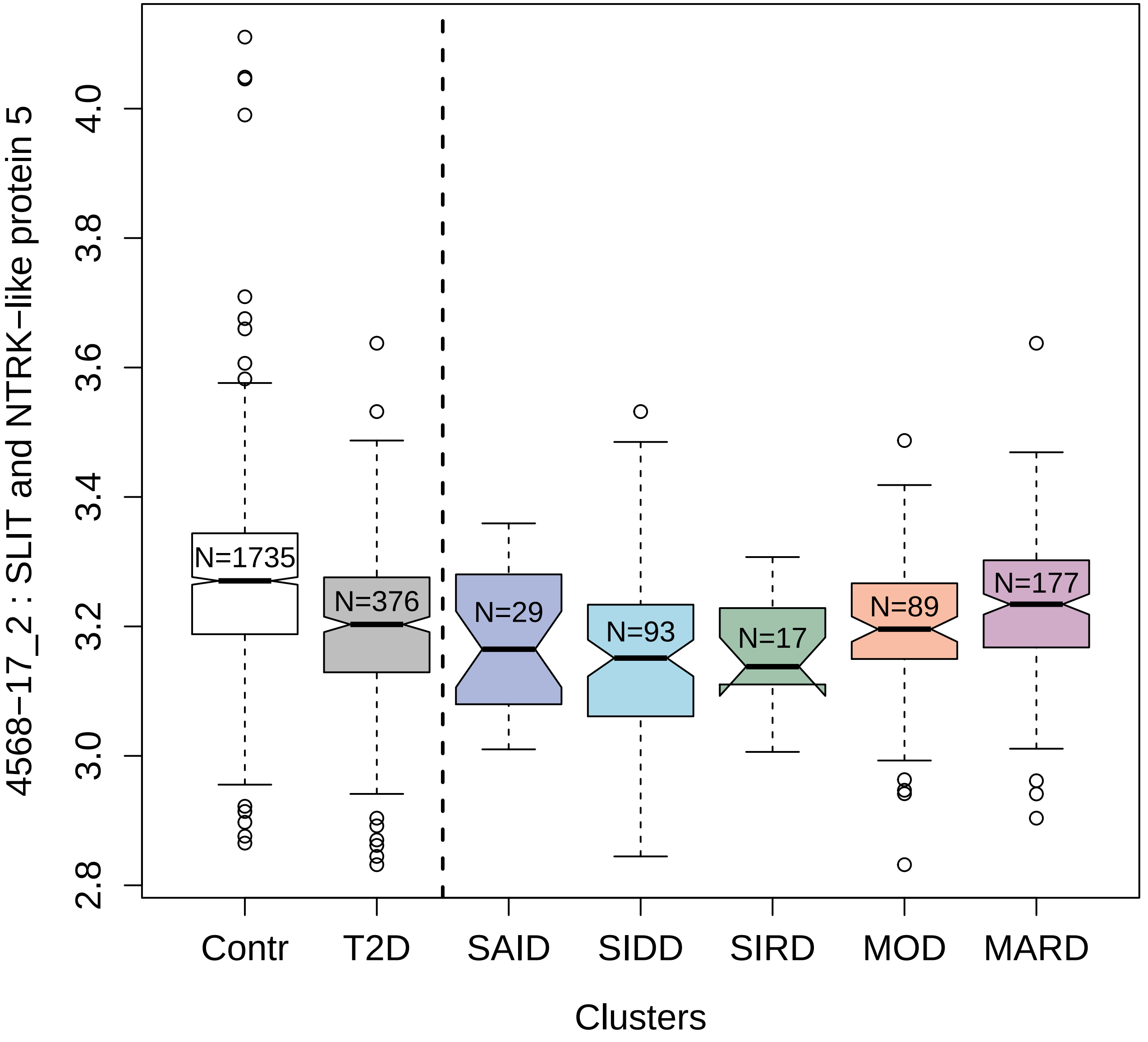

4568−17_2 : SLIT and NTRK−like protein 5
2.8
3.0
3.2
3.4
3.6
3.8
4.0
N=1735
N=376
N=29
N=93
N=17
N=89
N=177
Contr
T2D
SAID
SIDD
SIRD
MOD
MARD
Clusters

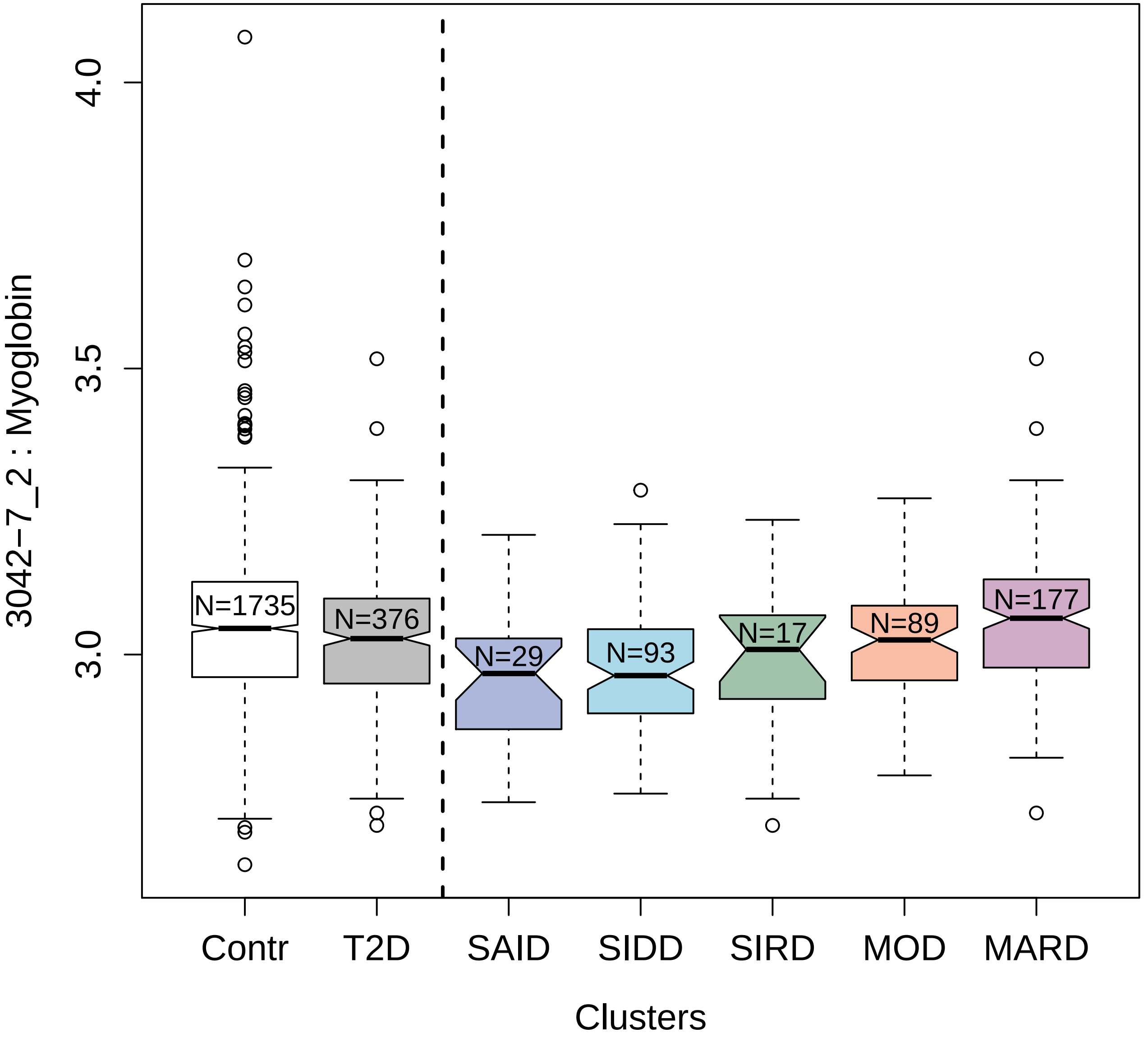

3042−7_2 : Myoglobin
4.0
3.5
3.0
N=1735
N=376
N=29
N=93
N=17
N=89
N=177
Contr
T2D
SAID
SIDD
SIRD
MOD
MARD
Clusters

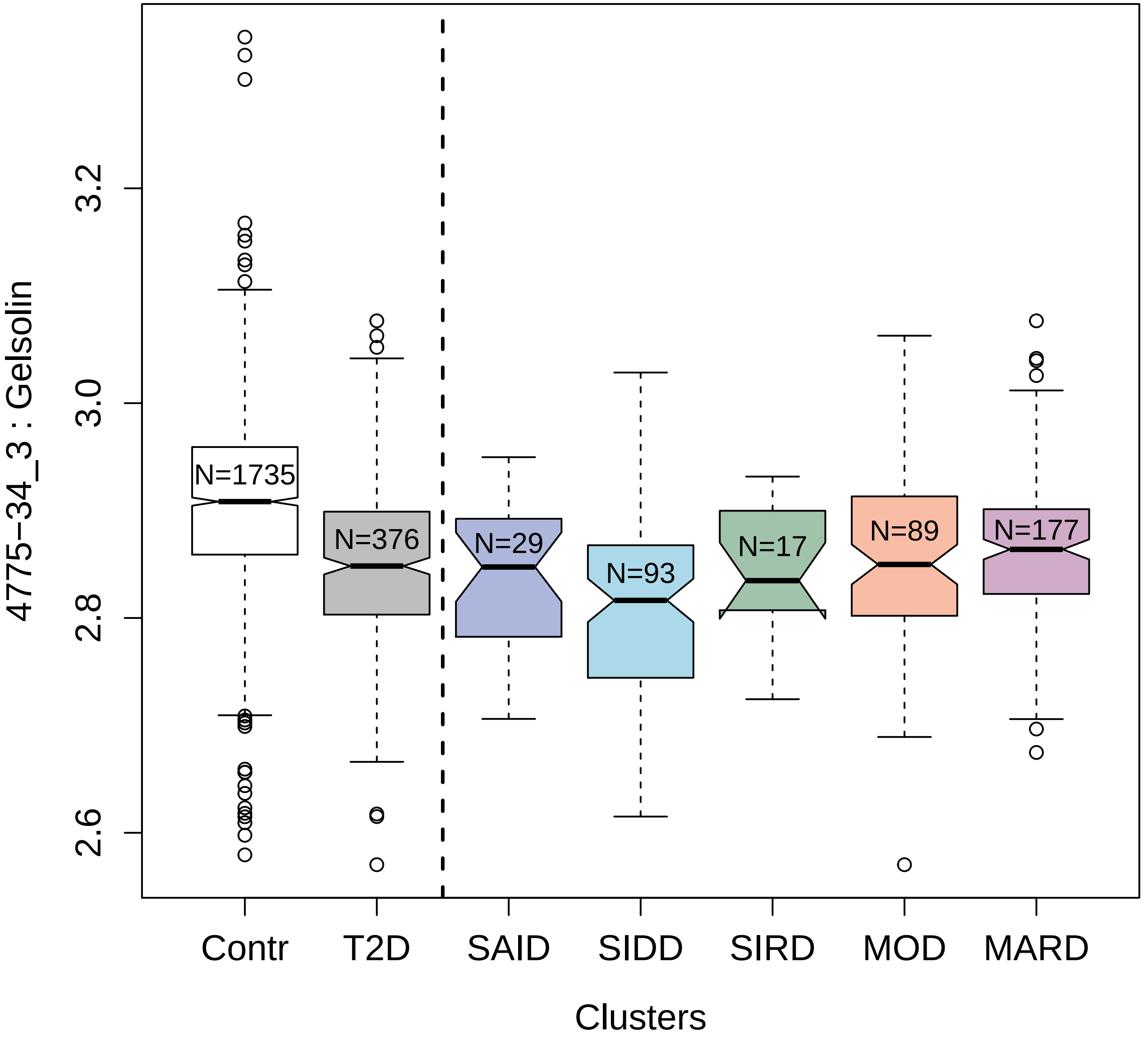

4775−34_3 : Gelsolin
3.2
3.0
2.8
2.6
N=1735
N=376
N=29
N=93
N=17
N=89
N=177
Contr
T2D
SAID
SIDD
SIRD
MOD
MARD
Clusters

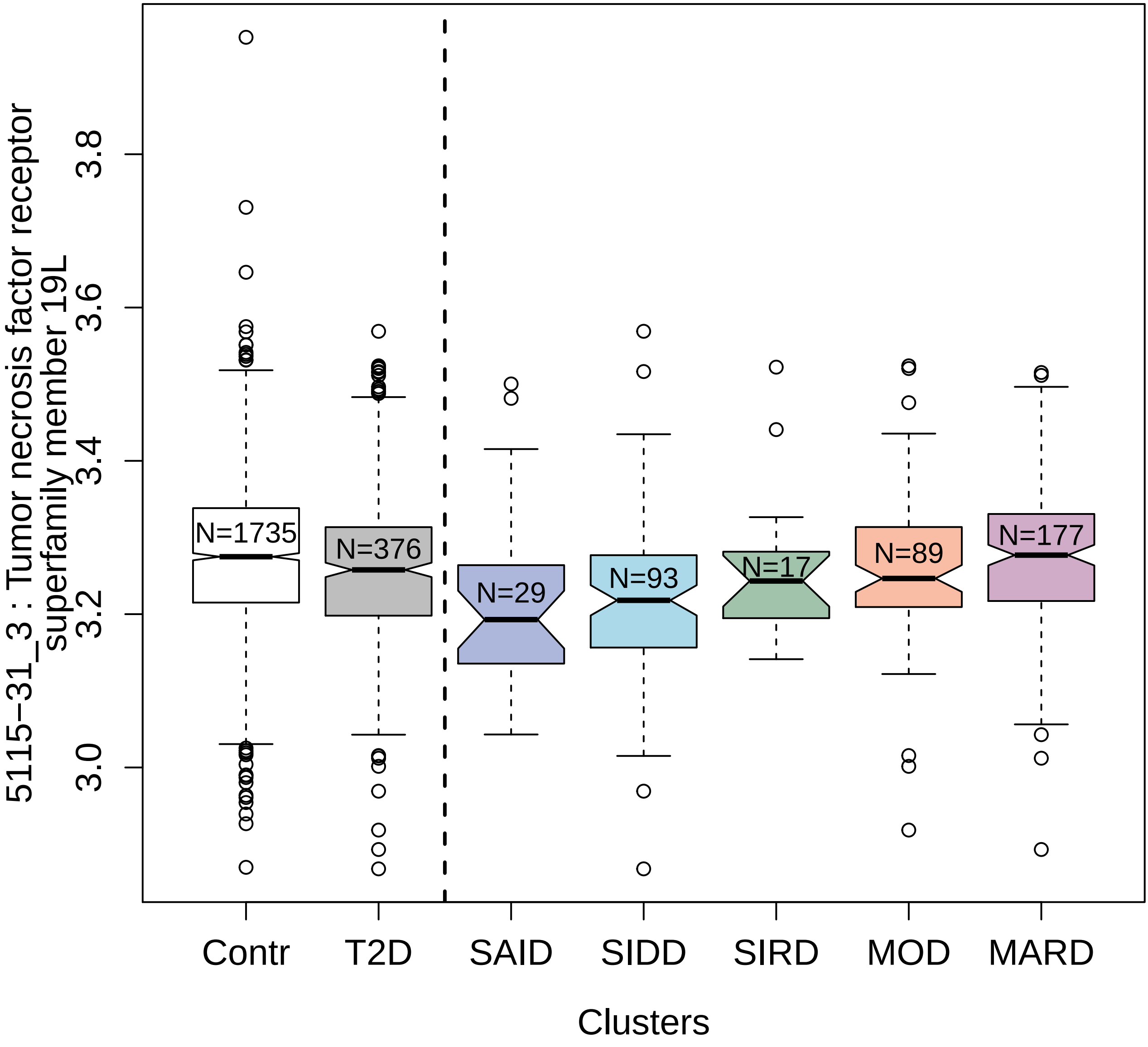

5115−31_3 : Tumor necrosis factor receptor
superfamily member 19L
3.0
3.2
3.4
3.6
3.8
N=1735
N=376
N=29
N=93
N=17
N=89
N=177
Contr
T2D
SAID
SIDD
SIRD
MOD
MARD
Clusters

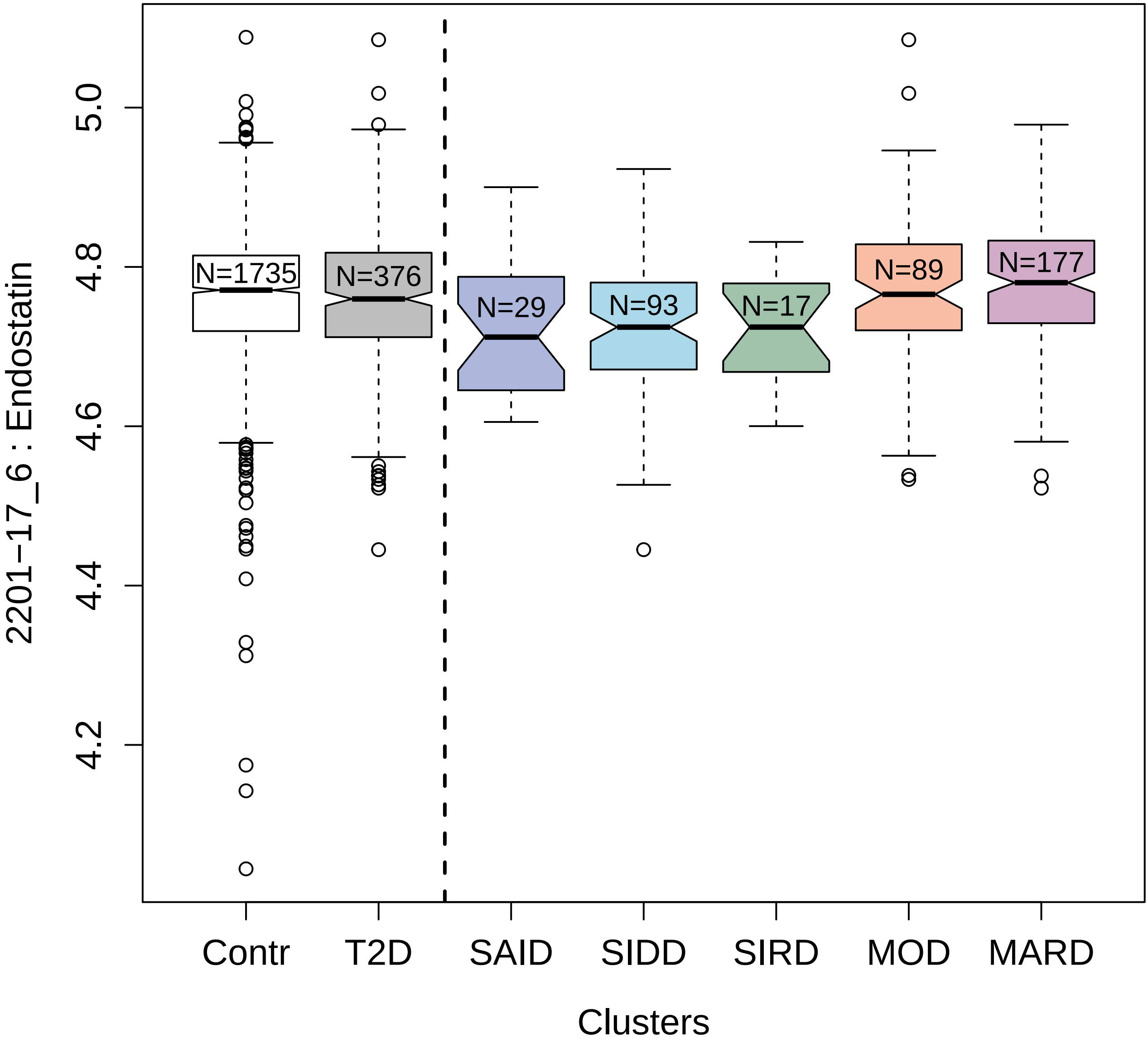

2201−17_6 : Endostatin
5.0
4.8
4.6
4.4
4.2
N=1735
N=376
N=29
N=93
N=17
N=89
N=177
Contr
T2D
SAID
SIDD
SIRD
MOD
MARD
Clusters

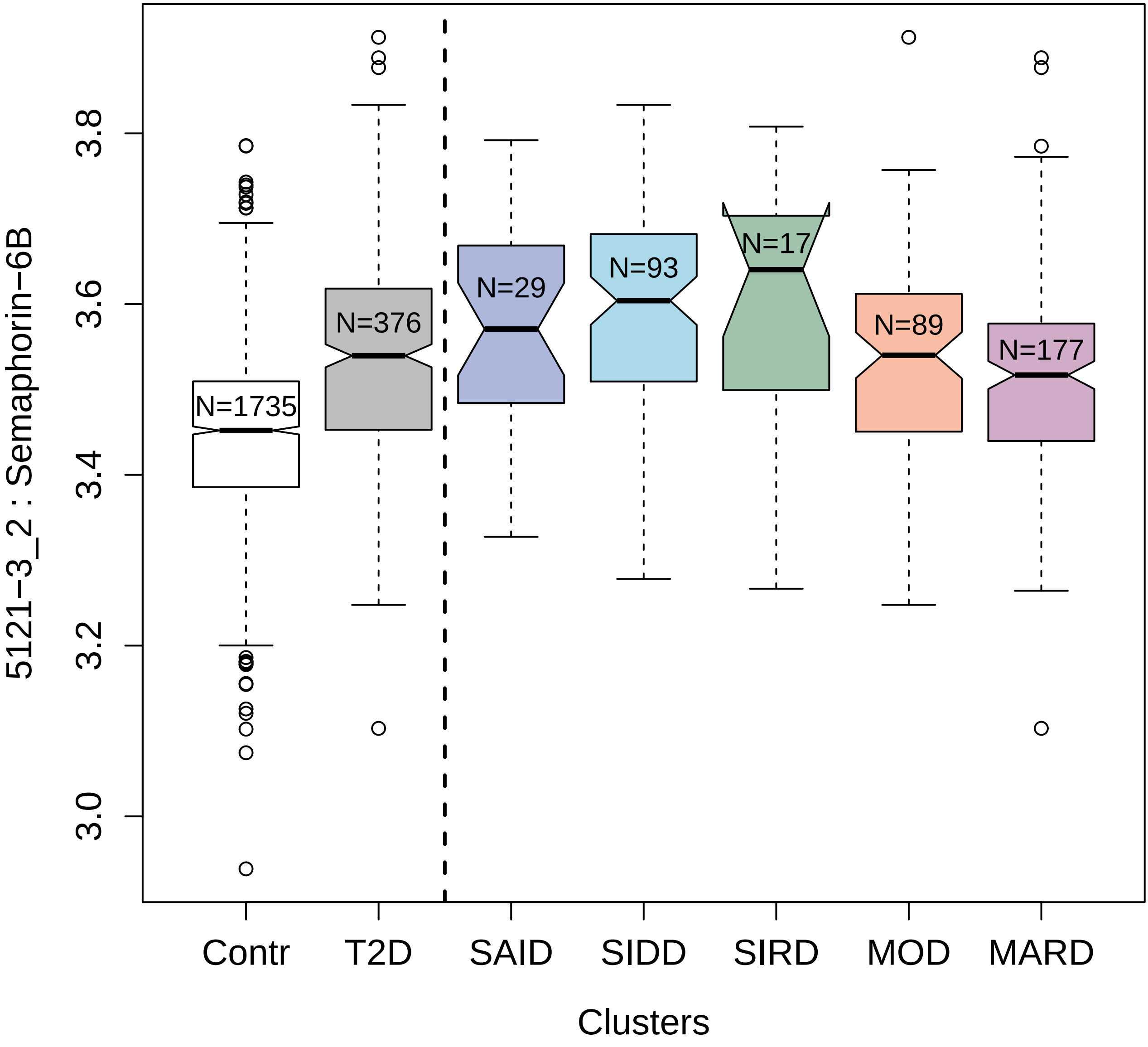

5121−3_2 : Semaphorin−6B
3.0
3.2
3.4
3.6
3.8
N=1735
N=376
N=29
N=93
N=17
N=89
N=177
Contr
T2D
SAID
SIDD
SIRD
MOD
MARD
Clusters

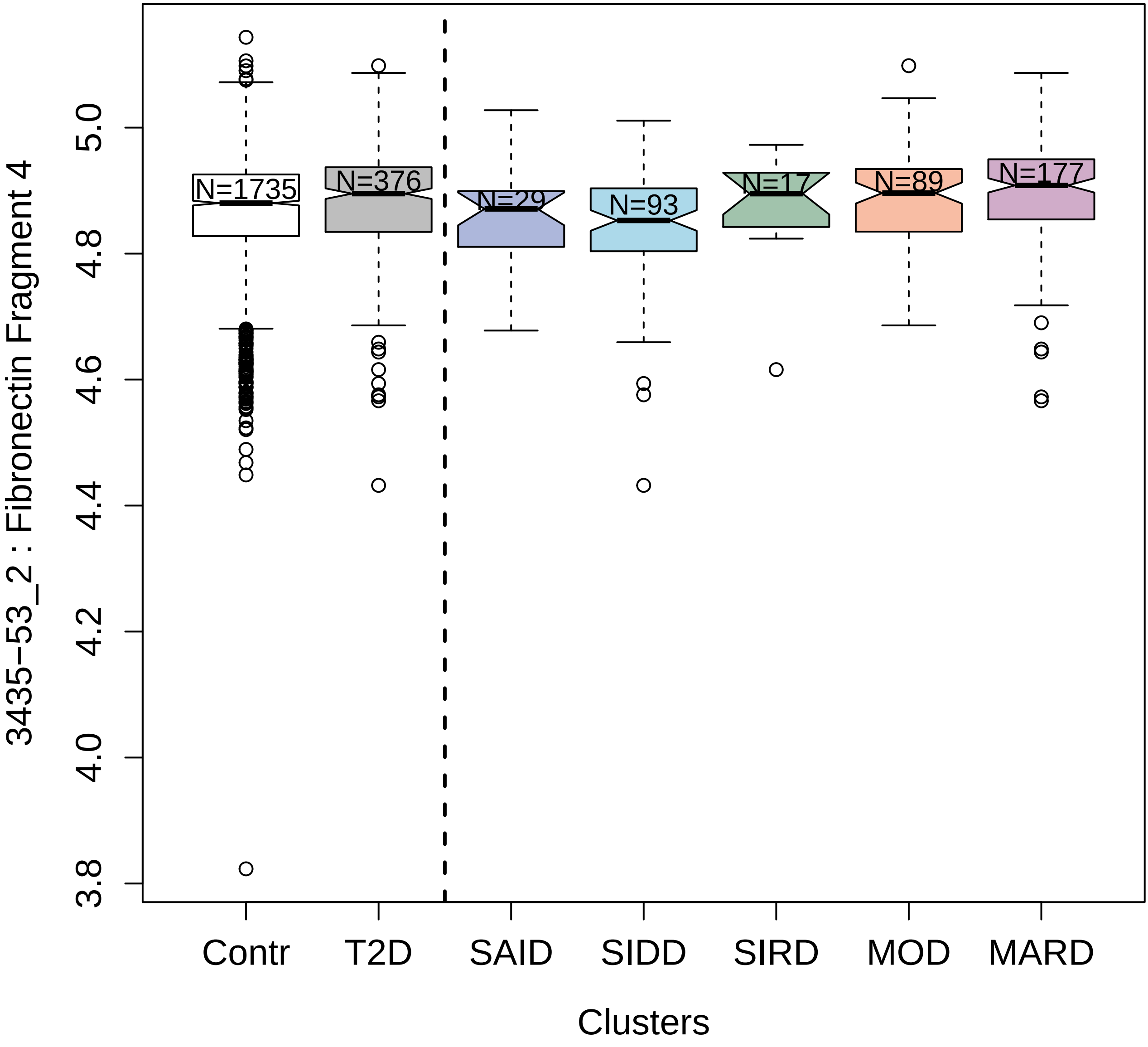

N=1735
N=376
N=29
N=93
N=17
N=89
N=177
5.0
4.8
4.6
4.4
4.2
4.0
3.8
3435−53_2 : Fibronectin Fragment 4
Contr
T2D
SAID
SIDD
SIRD
MOD
MARD
Clusters

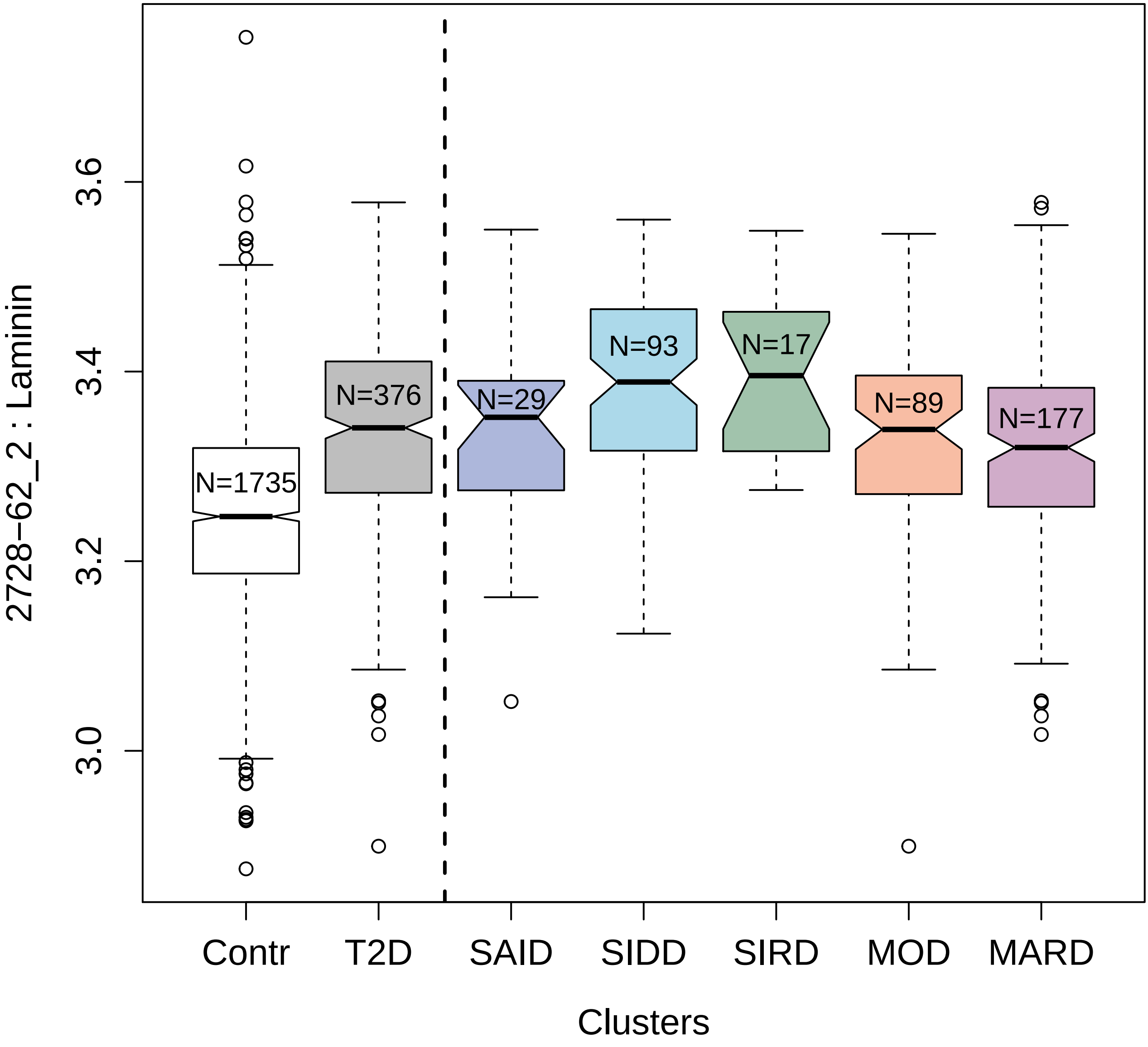

2728−62_2 : Laminin
3.0
3.2
3.4
3.6
N=1735
N=376
N=29
N=93
N=17
N=89
N=177
Contr
T2D
SAID
SIDD
SIRD
MOD
MARD
Clusters

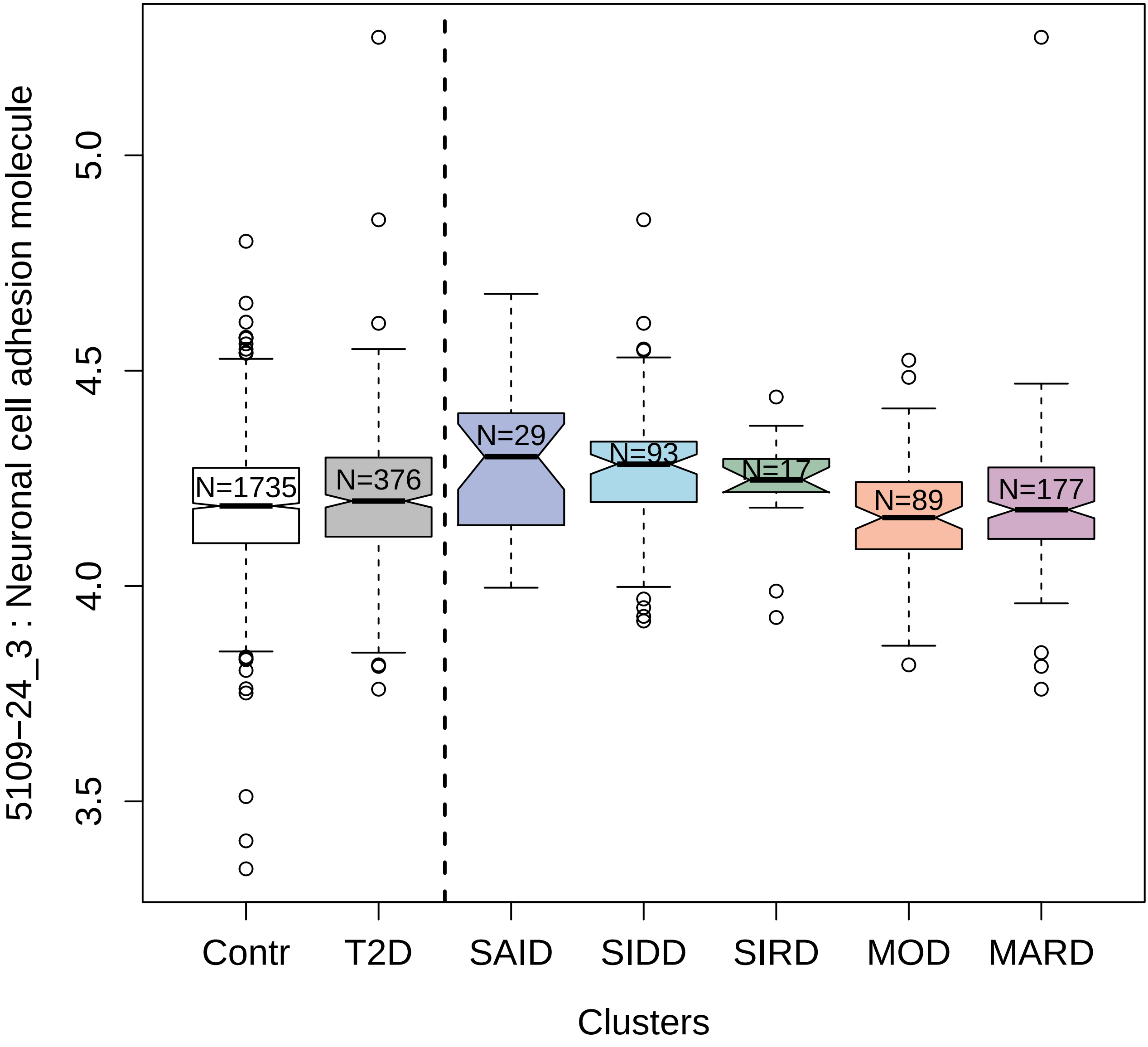

5109−24_3 : Neuronal cell adhesion molecule
3.5
4.0
4.5
5.0
N=1735
N=376
N=29
N=93
N=17
N=89
N=177
Contr
T2D
SAID
SIDD
SIRD
MOD
MARD
Clusters

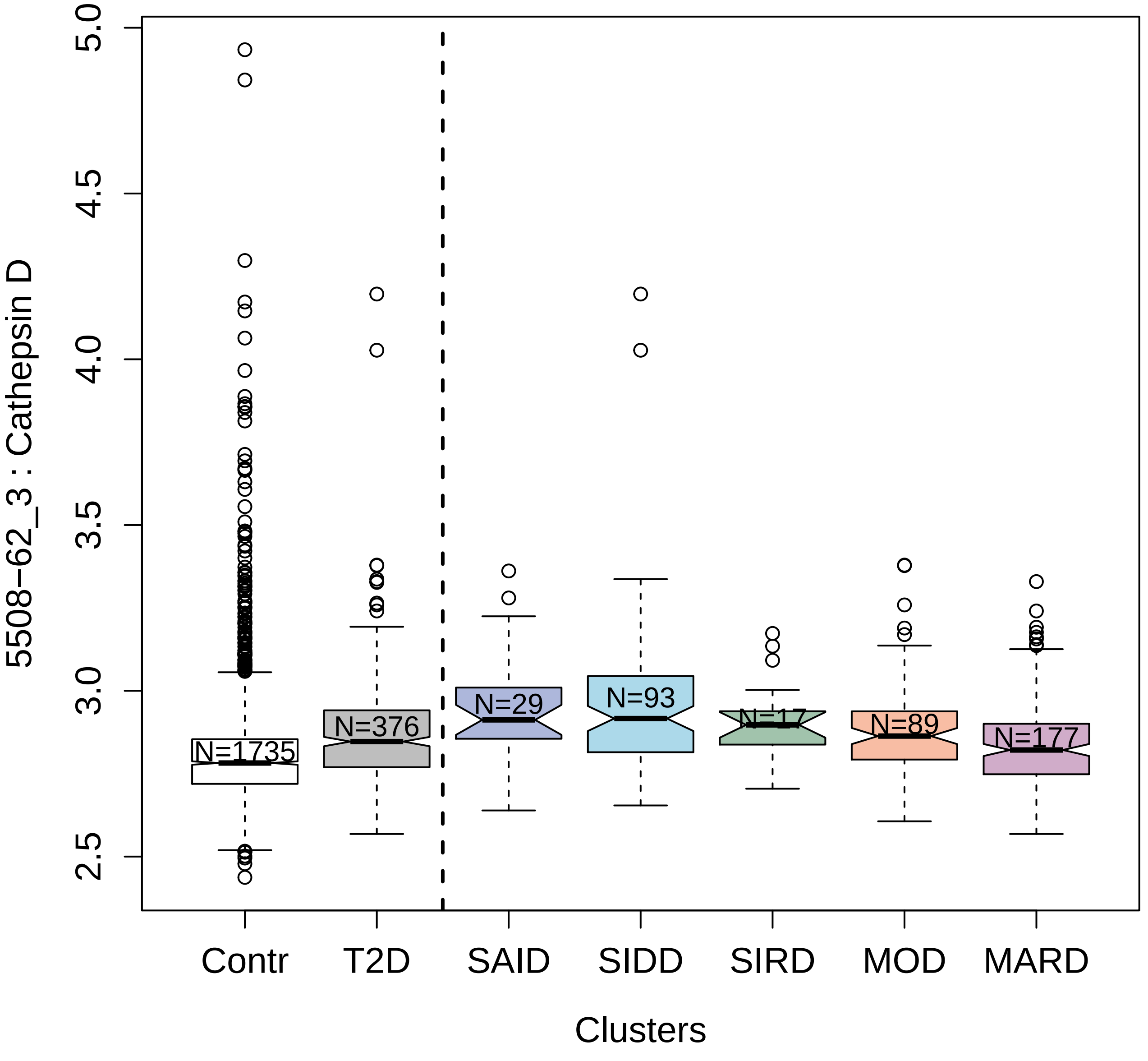

5508−62_3 : Cathepsin D
5.0
4.5
4.0
3.5
3.0
2.5
N=1735
N=376
N=29
N=93
N=17
N=89
N=177
Contr
T2D
SAID
SIDD
SIRD
MOD
MARD
Clusters

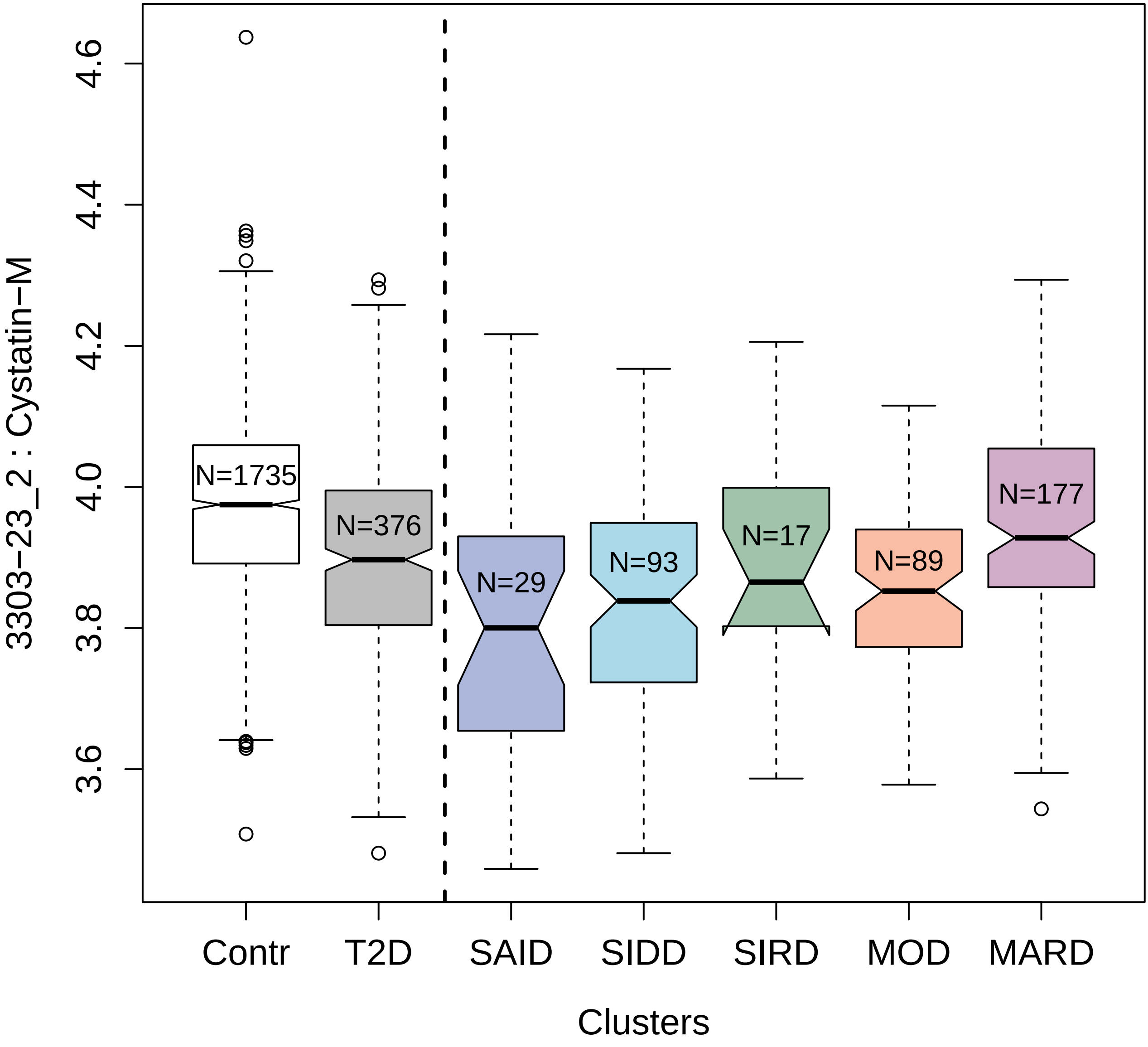
3303−23_2 : Cystatin−M
4.6
4.4
4.2
4.0
3.8
3.6
N=1735
N=376
N=29
N=93
N=17
N=89
N=177
Contr
T2D
SAID
SIDD
SIRD
MOD
MARD
Clusters

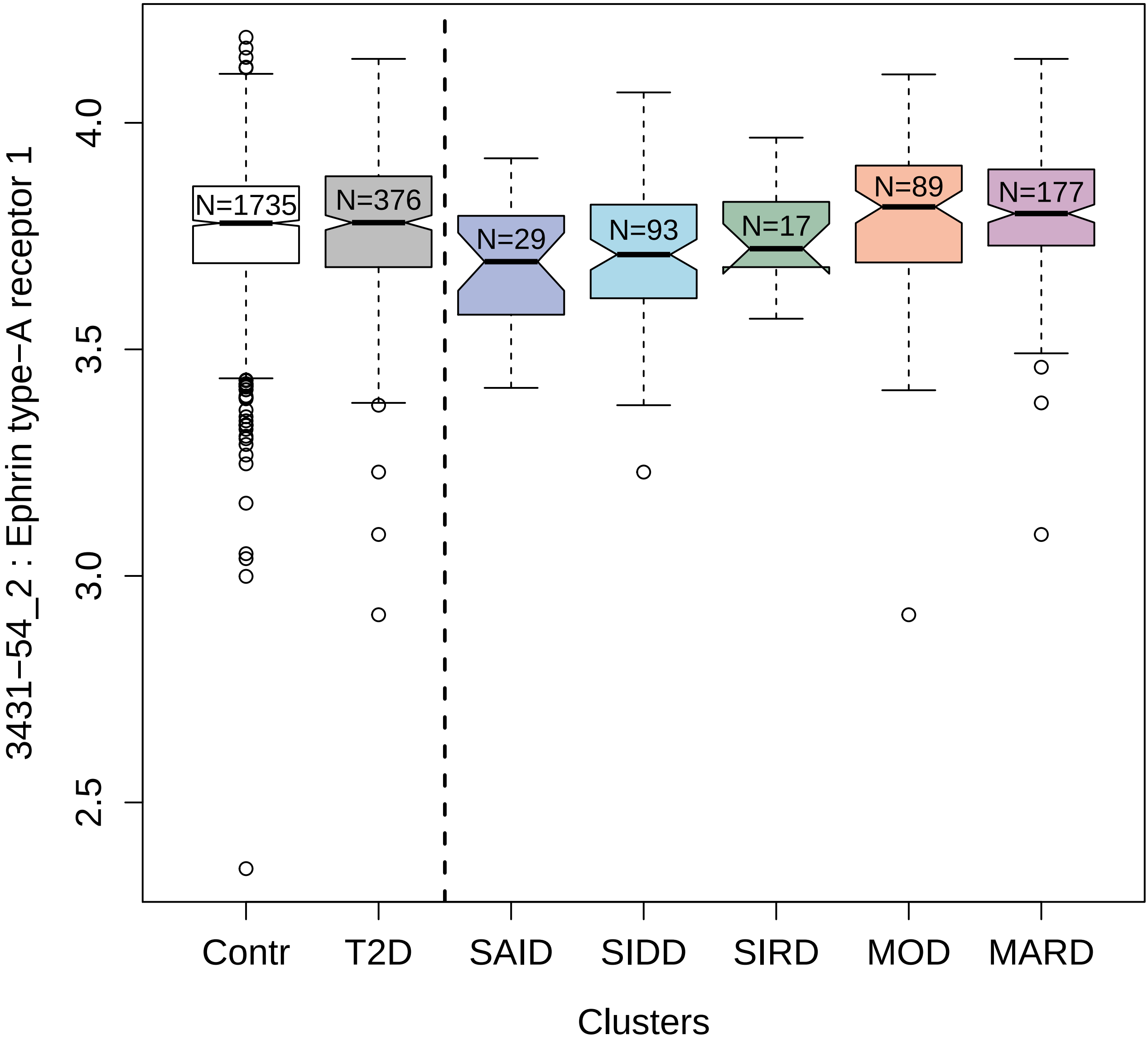

N=1735
N=376
N=29
N=93
N=17
N=89
N=177
4.0
3.5
3.0
2.5
3431−54_2 : Ephrin type−A receptor 1
Contr
T2D
SAID
SIDD
SIRD
MOD
MARD
Clusters

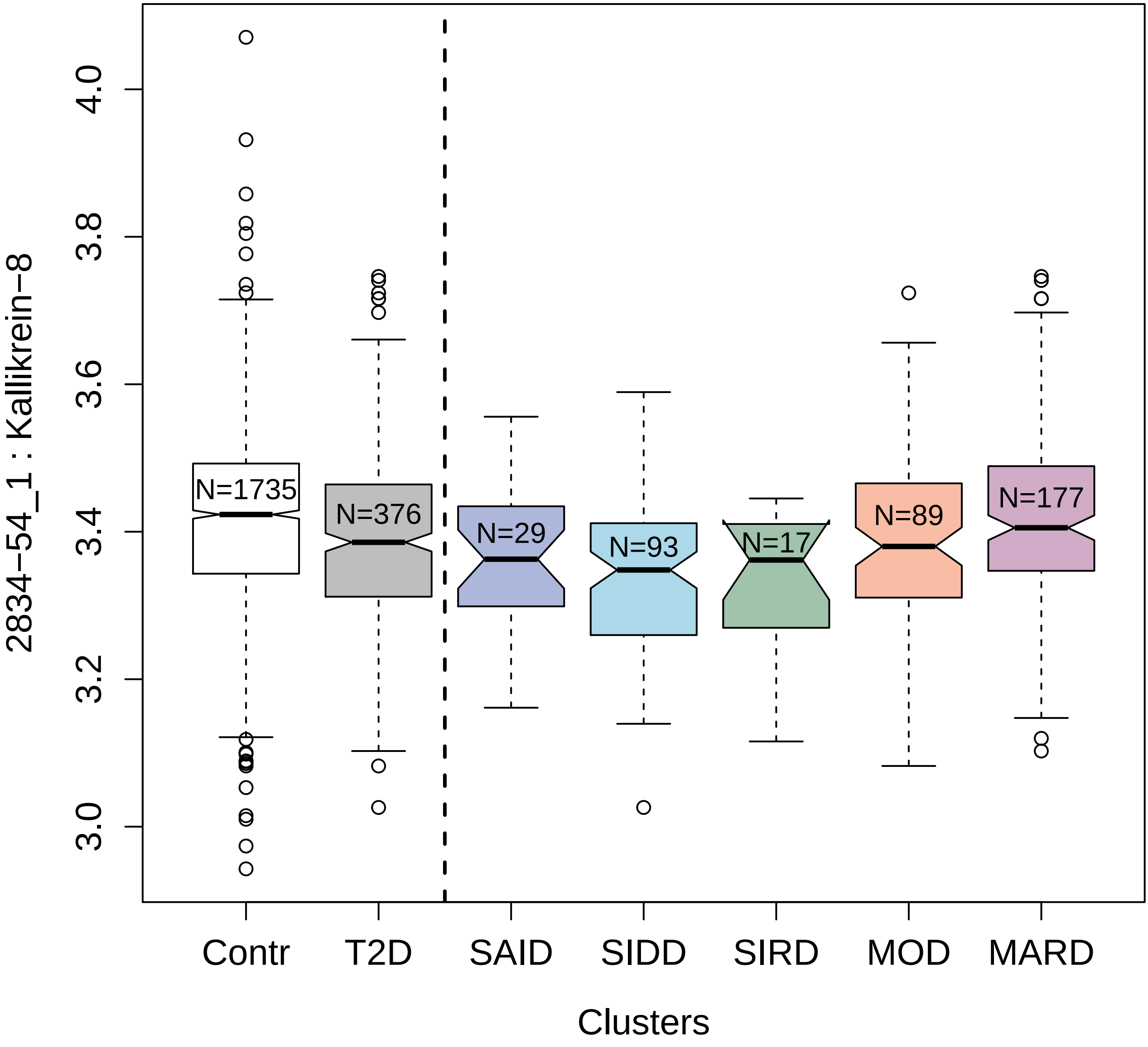
2834−54_1 : Kallikrein−8
4.0
3.8
3.6
3.4
3.2
3.0
N=1735
N=376
N=29
N=93
N=17
N=89
N=177
Contr
T2D
SAID
SIDD
SIRD
MOD
MARD
Clusters

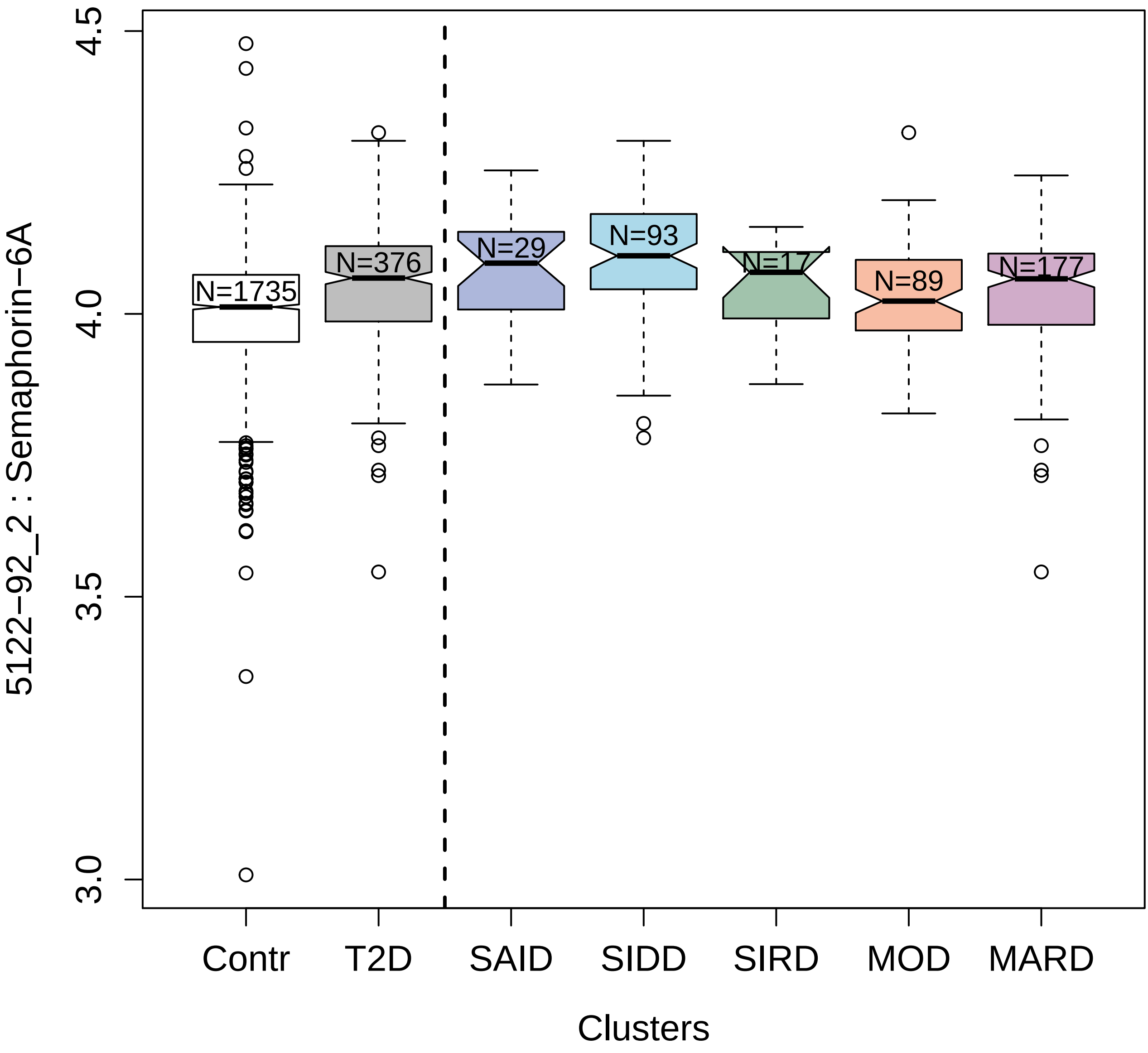

5122−92_2 : Semaphorin−6A
4.5
4.0
3.5
3.0
N=1735
N=376
N=29
N=93
N=17
N=89
N=177
Contr
T2D
SAID
SIDD
SIRD
MOD
MARD
Clusters

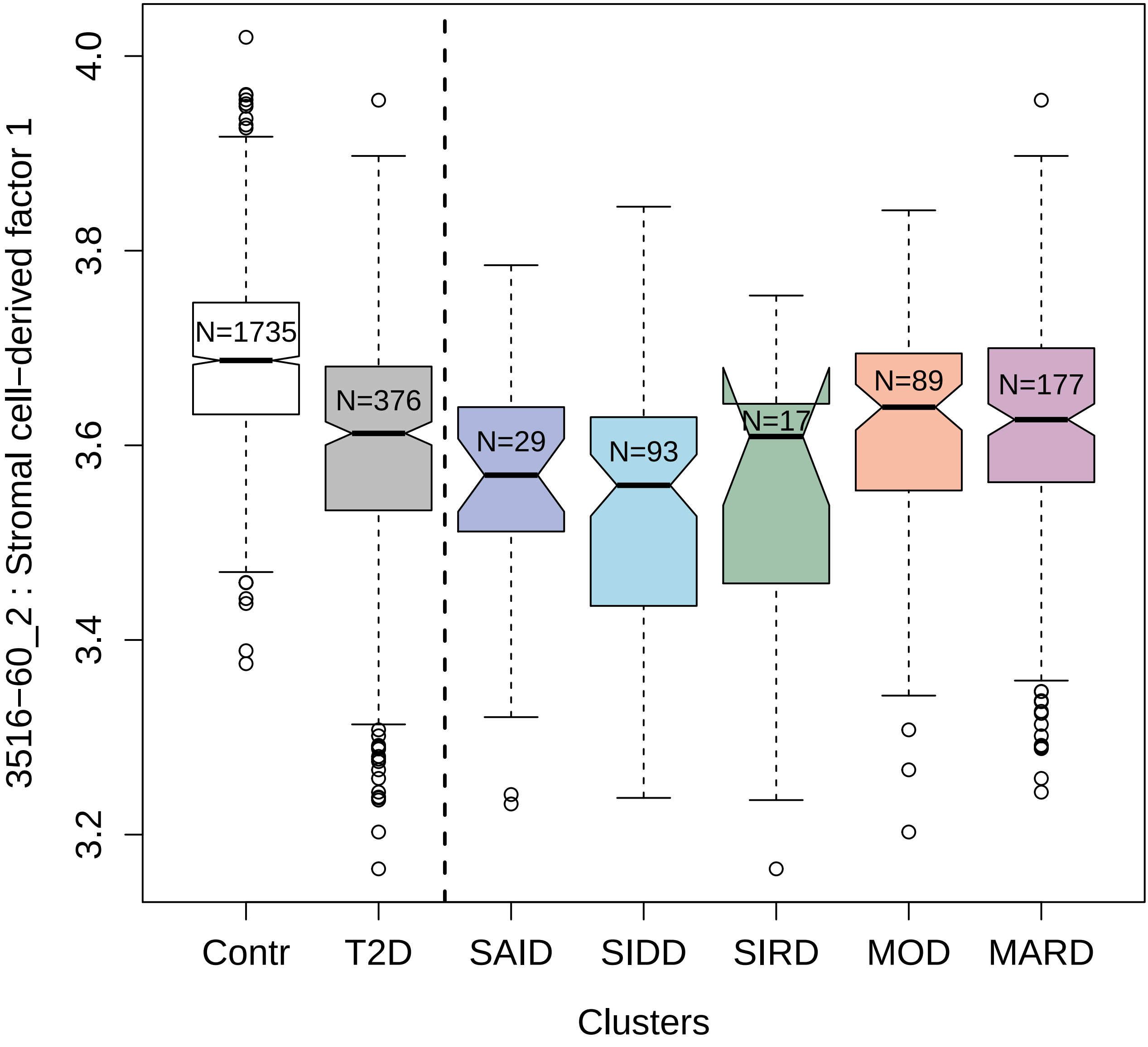
3516−60_2 : Stromal cell−derived factor 1
4.0
3.8
3.6
3.4
3.2
N=1735
N=376
N=29
N=93
N=17
N=89
N=177
Contr
T2D
SAID
SIDD
SIRD
MOD
MARD
Clusters

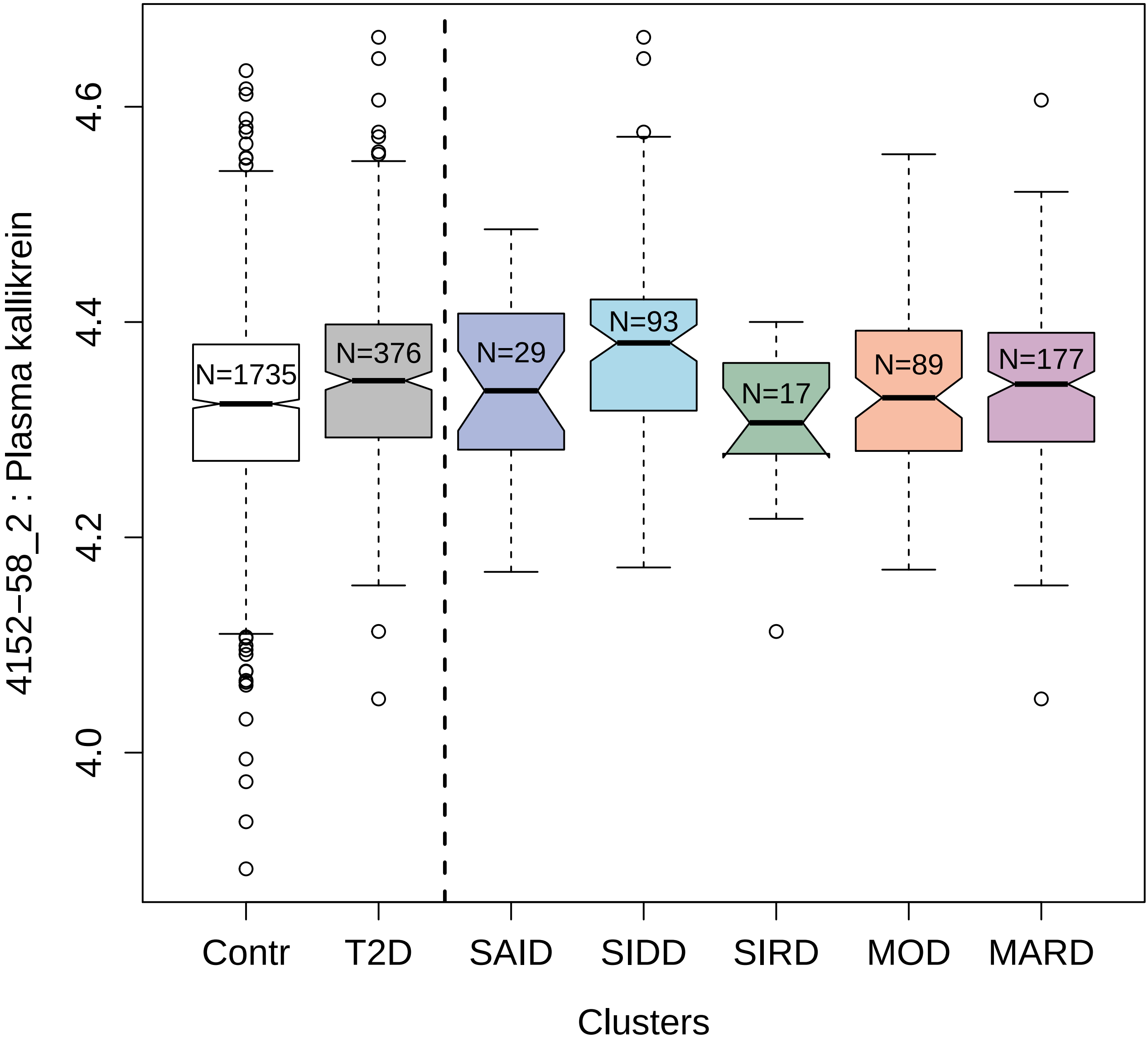
4152−58_2 : Plasma kallikrein
4.0
4.2
4.4
4.6
N=1735
N=376
N=29
N=93
N=17
N=89
N=177
Contr
T2D
SAID
SIDD
SIRD
MOD
MARD
Clusters

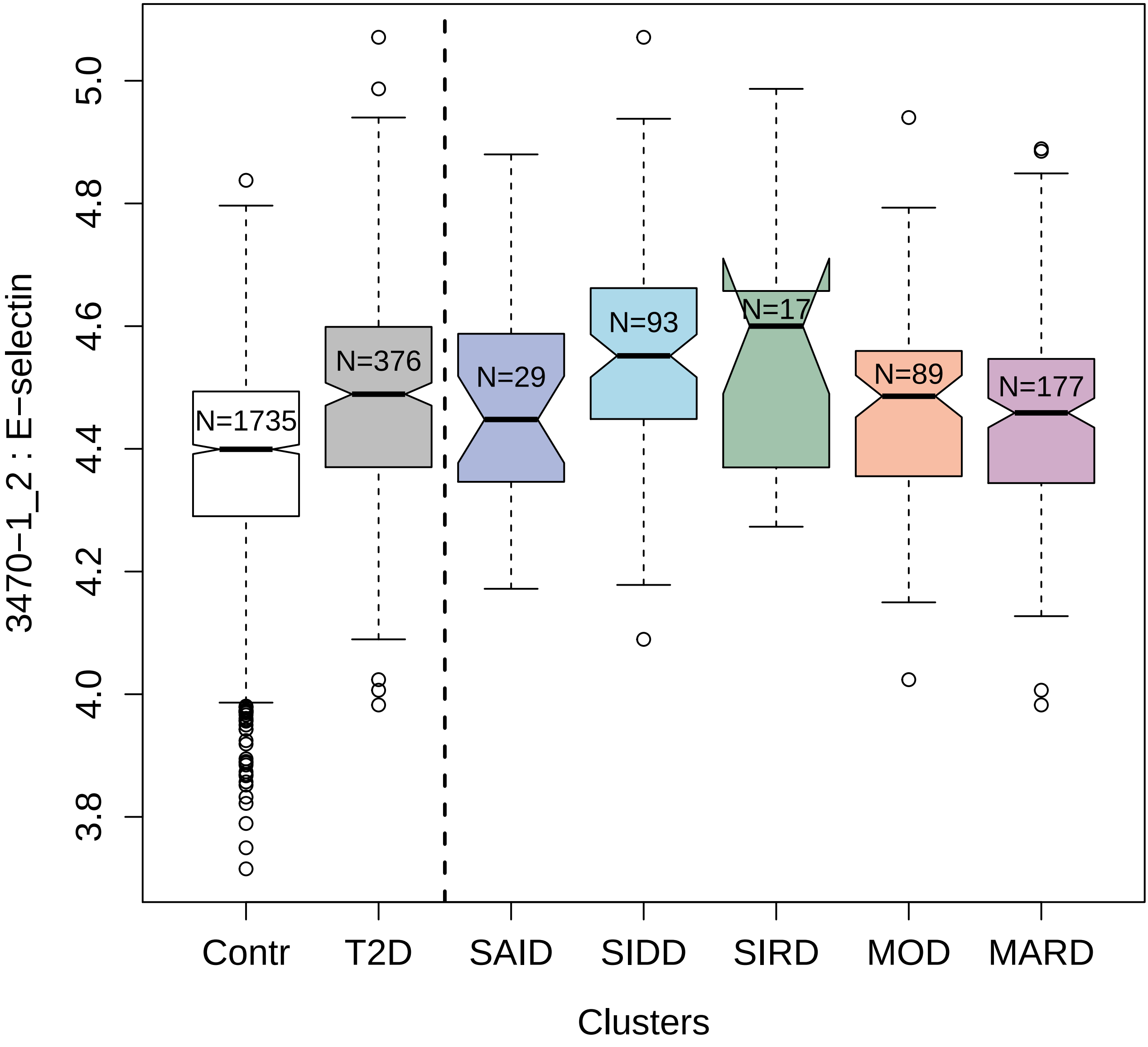
3470−1_2 : E−selectin
5.0
4.8
4.6
4.4
4.2
4.0
3.8
N=1735
N=376
N=29
N=93
N=17
N=89
N=177
Contr
T2D
SAID
SIDD
SIRD
MOD
MARD
Clusters

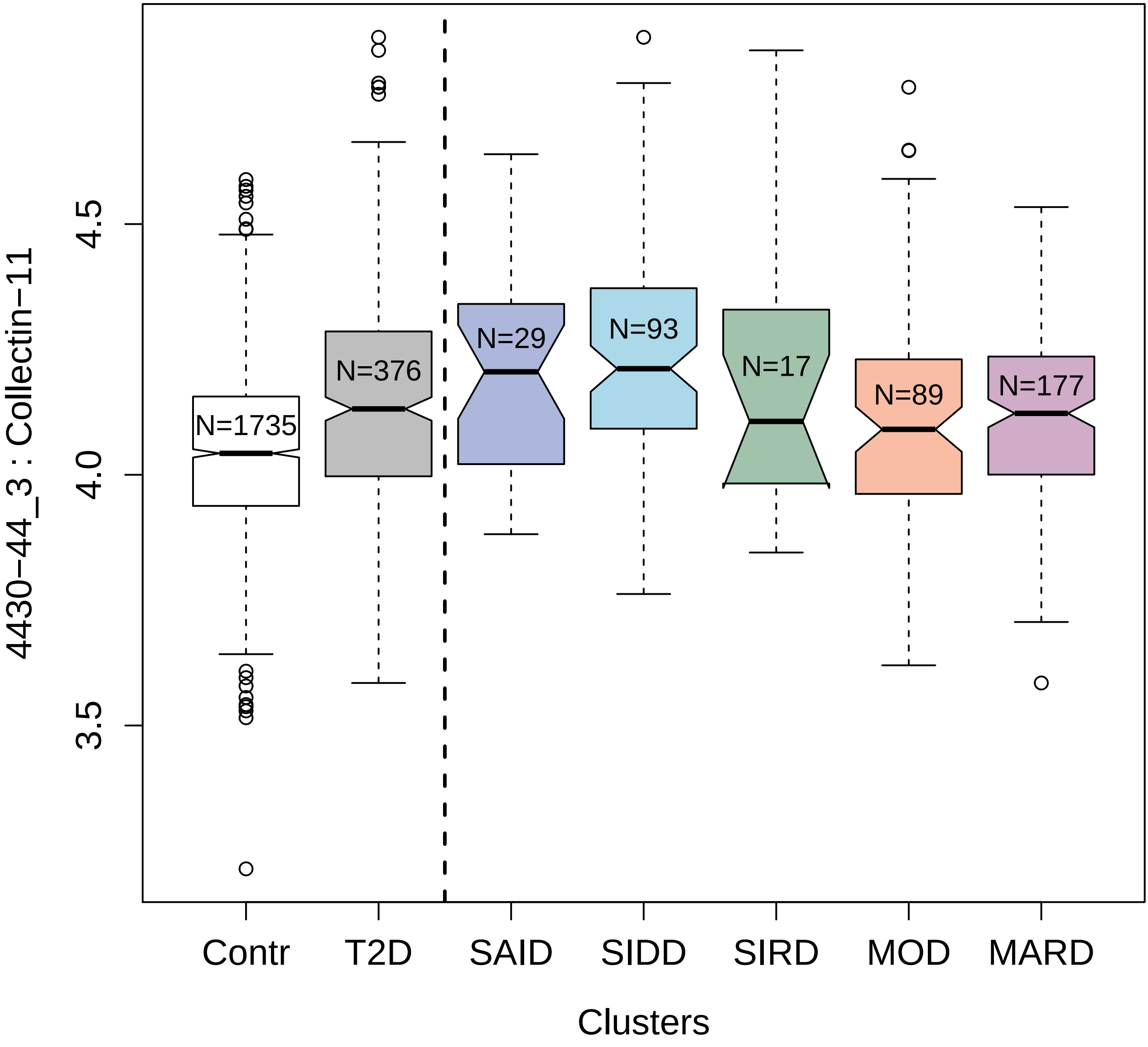
4430−44_3 : Collectin−11
3.5
4.0
4.5
N=1735
N=376
N=29
N=93
N=17
N=89
N=177
Contr
T2D
SAID
SIDD
SIRD
MOD
MARD
Clusters

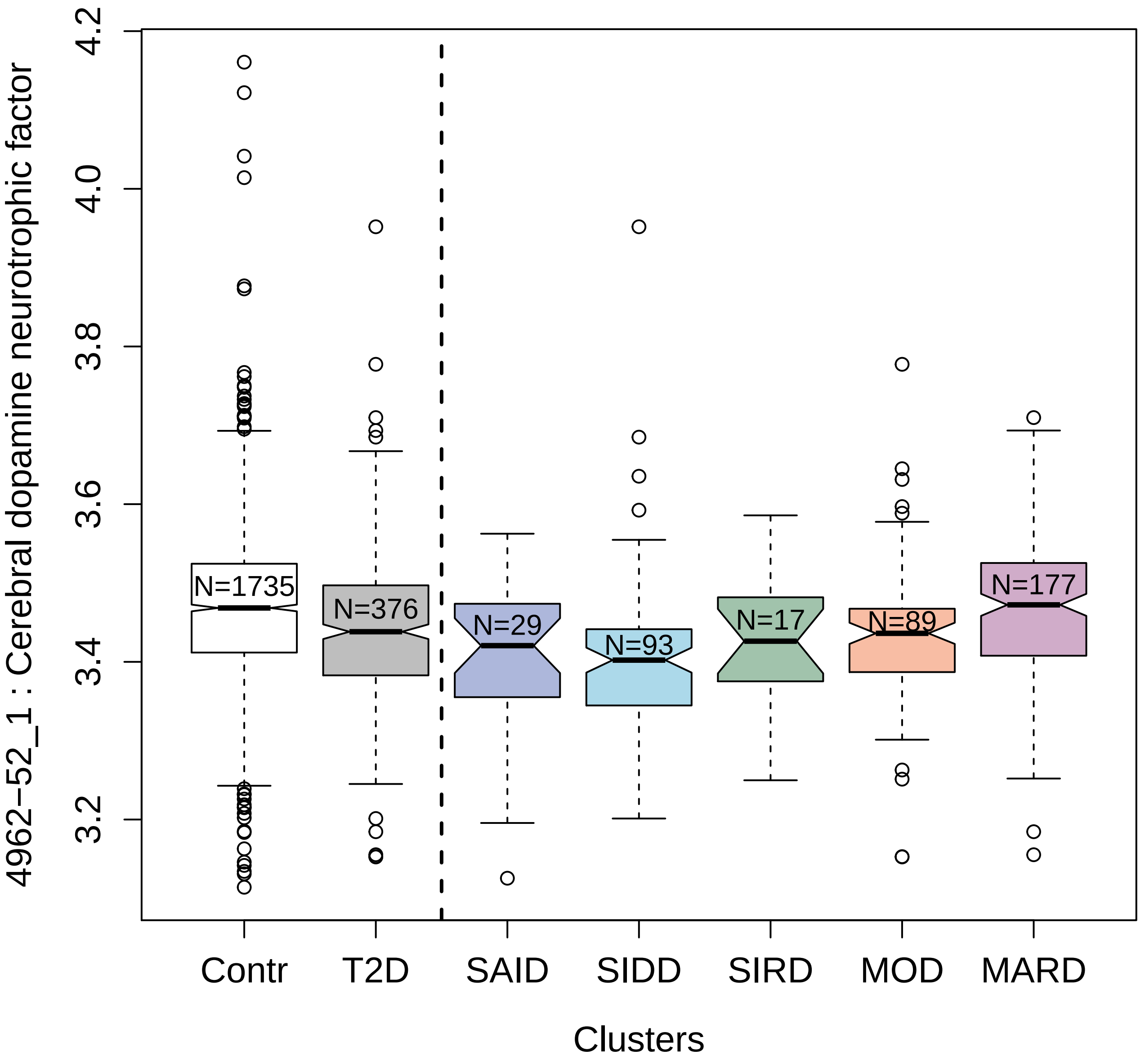
4962−52_1 : Cerebral dopamine neurotrophic factor
4.2
4.0
3.8
3.6
3.4
3.2
N=1735
N=376
N=29
N=93
N=17
N=89
N=177
Contr
T2D
SAID
SIDD
SIRD
MOD
MARD
Clusters

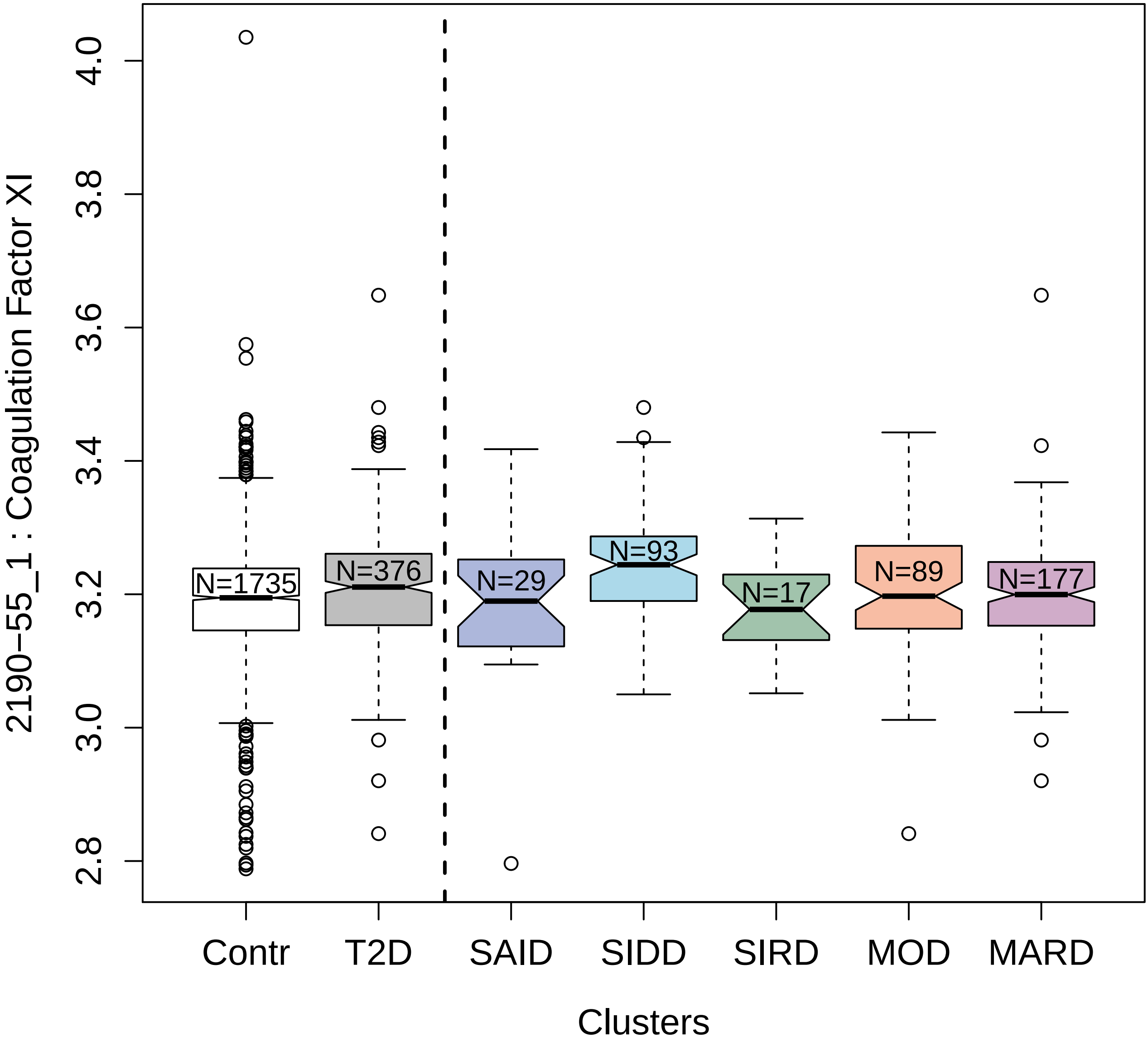
2190−55_1 : Coagulation Factor XI
4.0
3.8
3.6
3.4
3.2
3.0
2.8
N=1735
N=376
N=29
N=93
N=17
N=89
N=177
Contr
T2D
SAID
SIDD
SIRD
MOD
MARD
Clusters

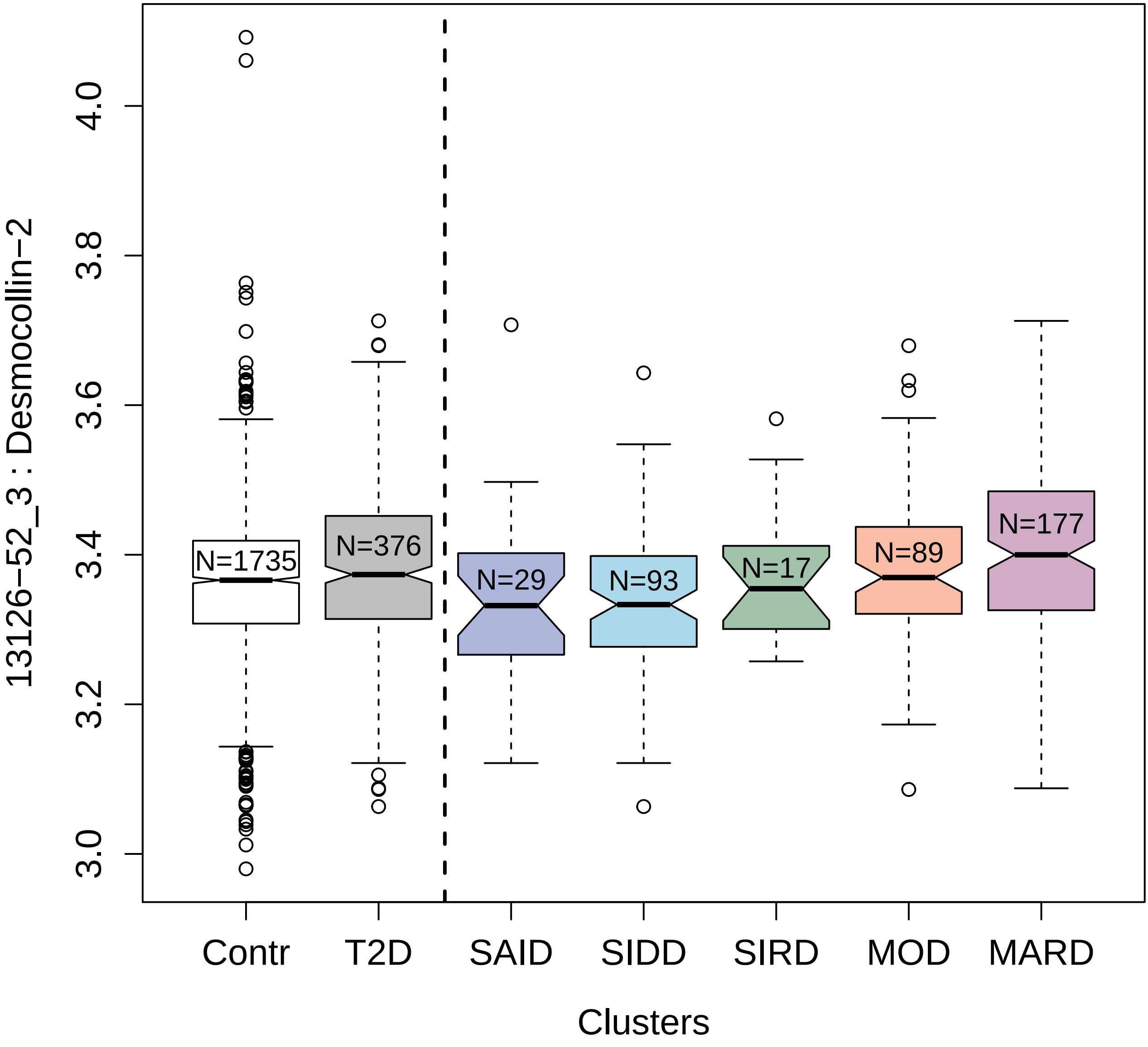
13126−52_3 : Desmocollin−2
4.0
3.8
3.6
3.4
3.2
3.0
N=1735
N=376
N=29
N=93
N=17
N=89
N=177
Contr
T2D
SAID
SIDD
SIRD
MOD
MARD
Clusters

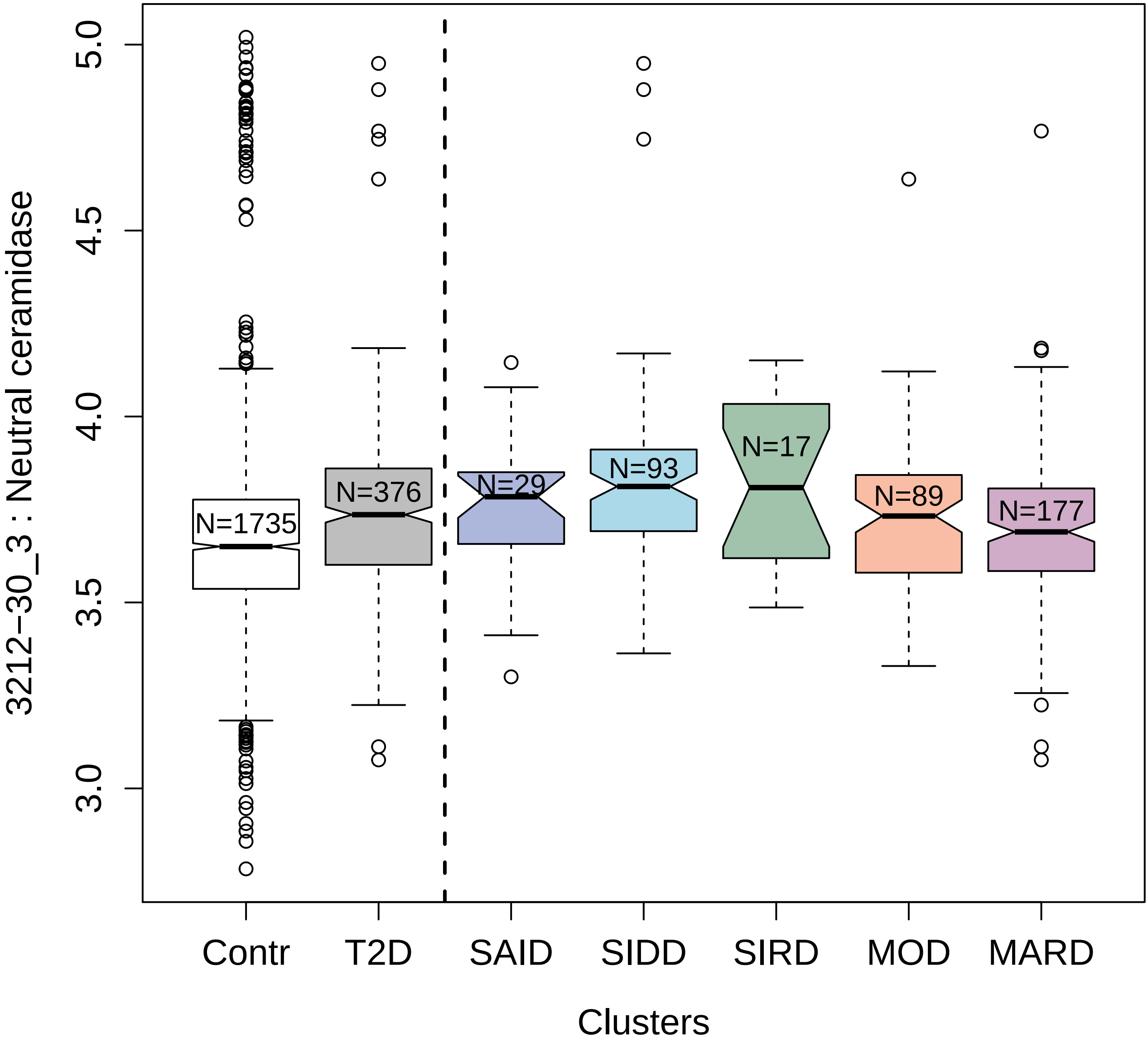

3212−30_3 : Neutral ceramidase
5.0
4.5
4.0
3.5
3.0
N=1735
N=376
N=29
N=93
N=17
N=89
N=177
Contr
T2D
SAID
SIDD
SIRD
MOD
MARD
Clusters

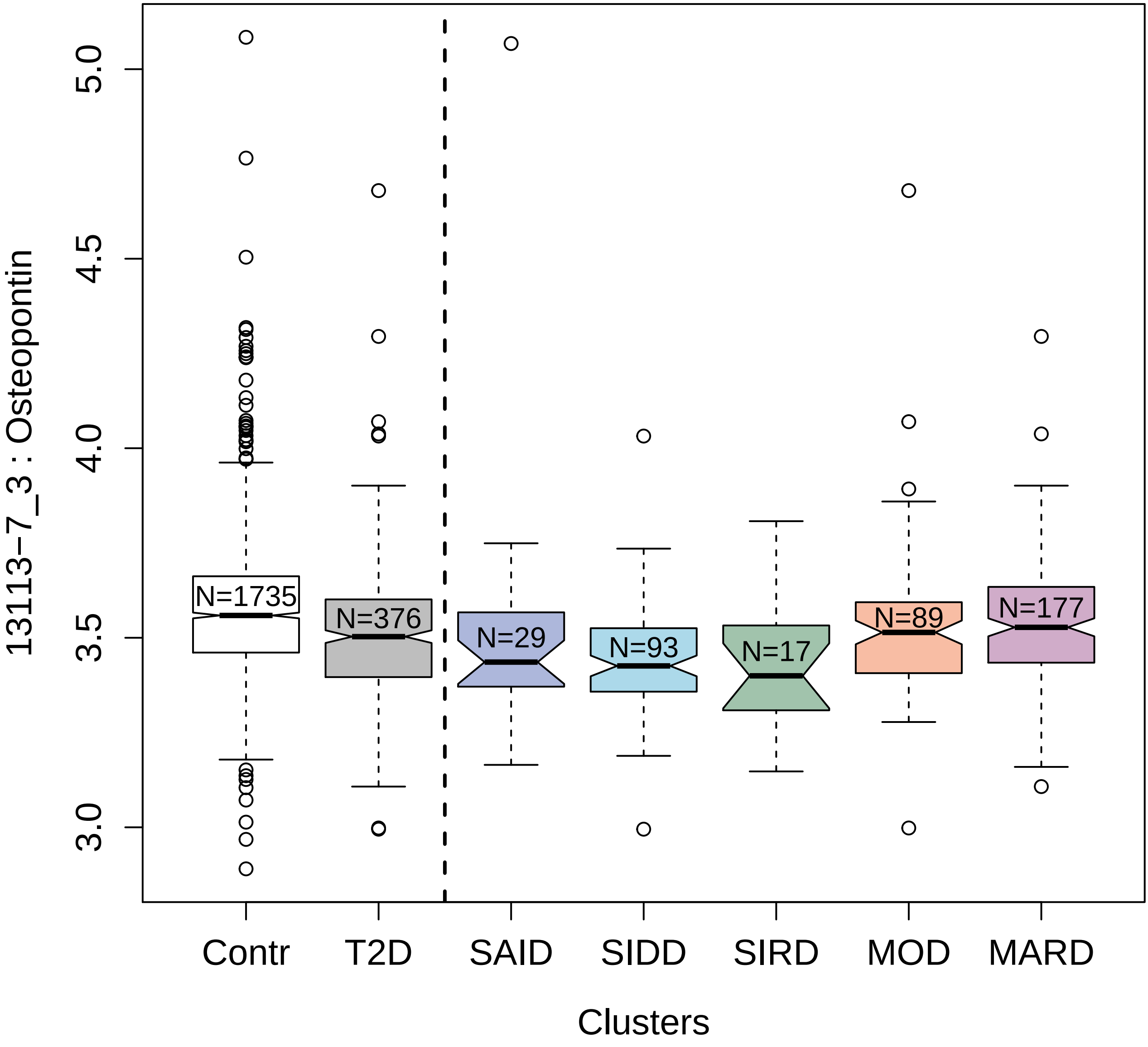

13113−7_3 : Osteopontin
5.0
4.5
4.0
3.5
3.0
N=1735
N=376
N=29
N=93
N=17
N=89
N=177
Contr
T2D
SAID
SIDD
SIRD
MOD
MARD
Clusters

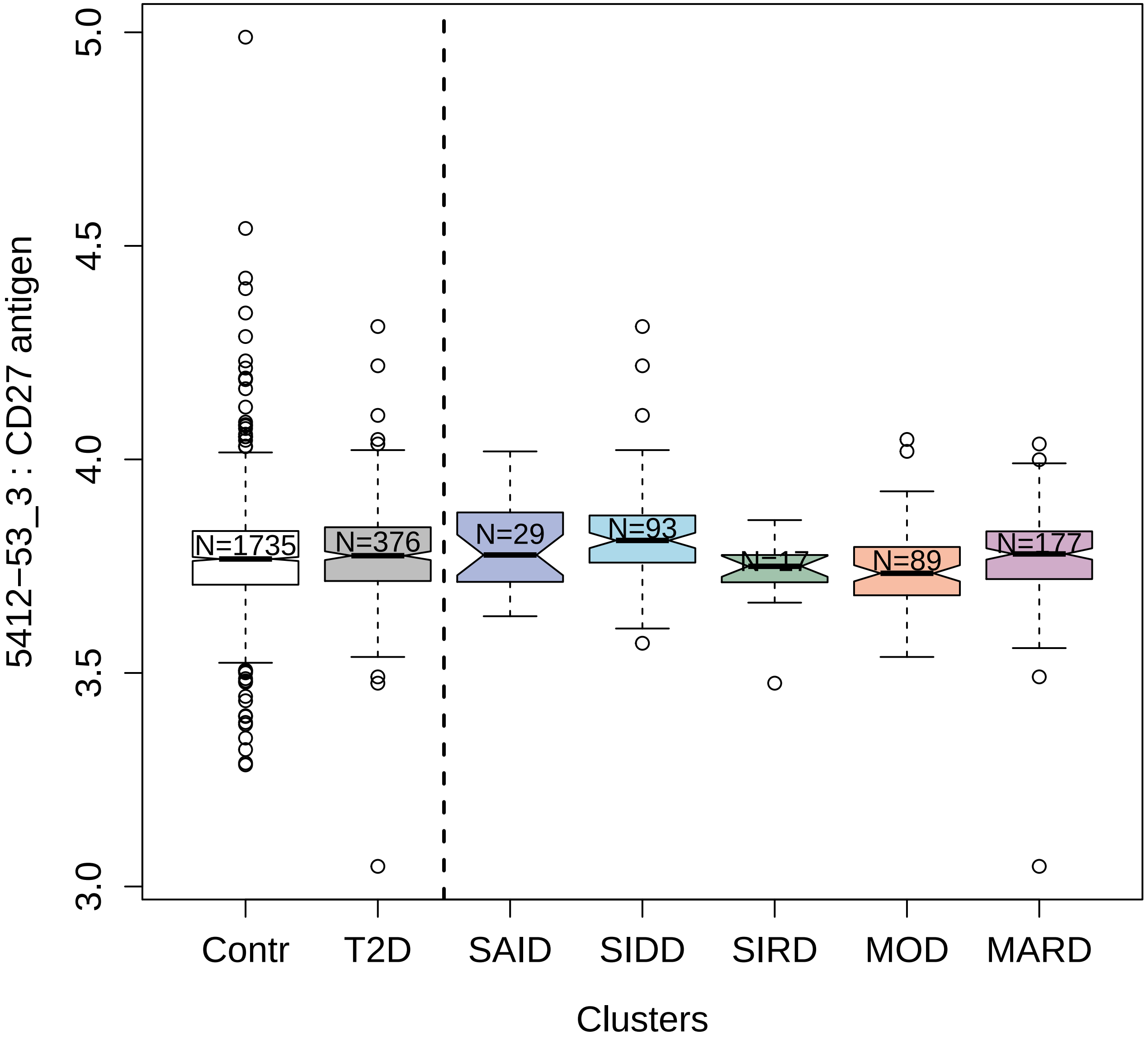
5412−53_3 : CD27 antigen
5.0
4.5
4.0
3.5
3.0
N=1735
N=376
N=29
N=93
N=17
N=89
N=177
Contr
T2D
SAID
SIDD
SIRD
MOD
MARD
Clusters

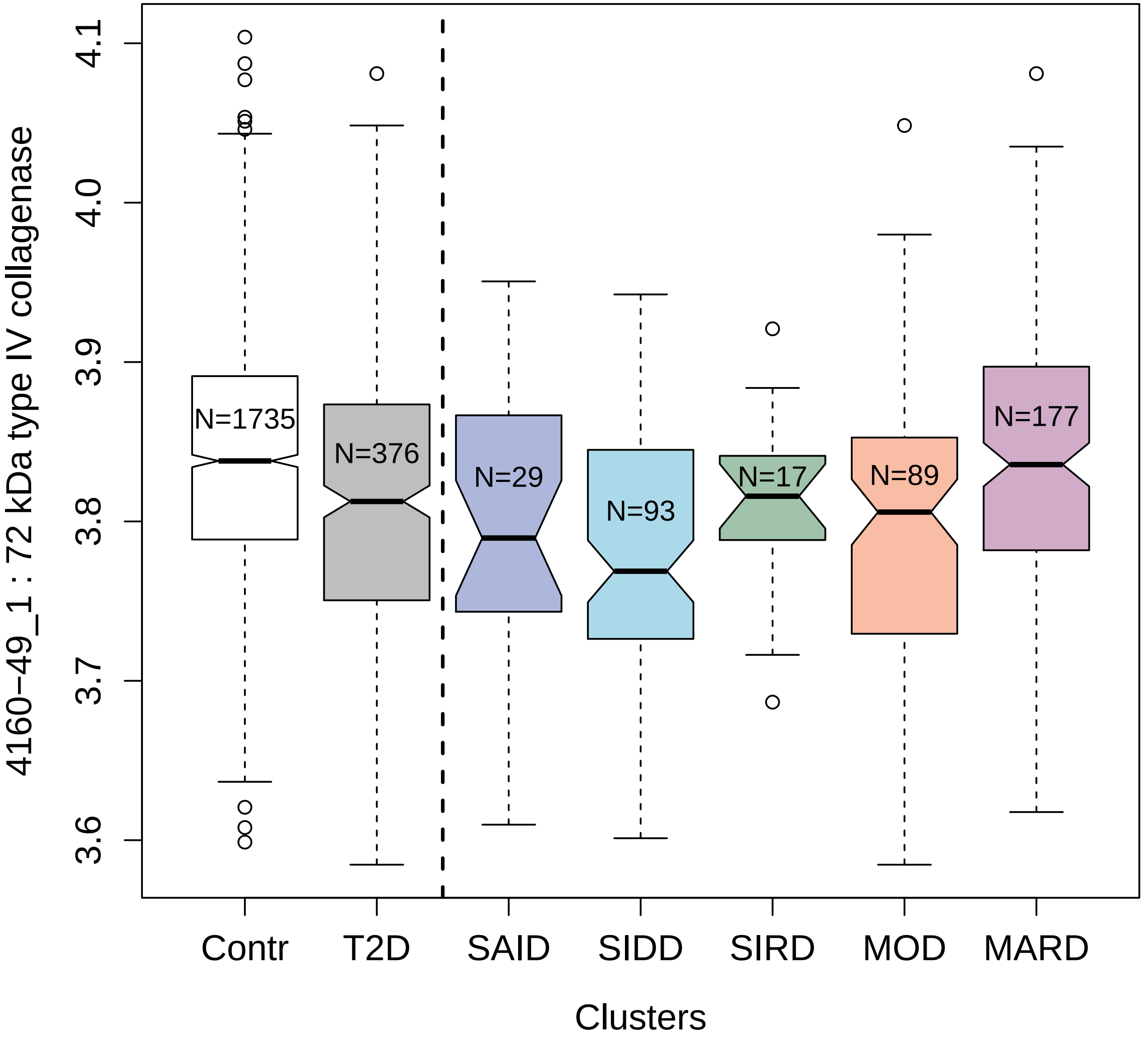
4160−49_1 : 72 kDa type IV collagenase
4.1
4.0
3.9
3.8
3.7
3.6
N=1735
N=376
N=29
N=93
N=17
N=89
N=177
Contr
T2D
SAID
SIDD
SIRD
MOD
MARD
Clusters

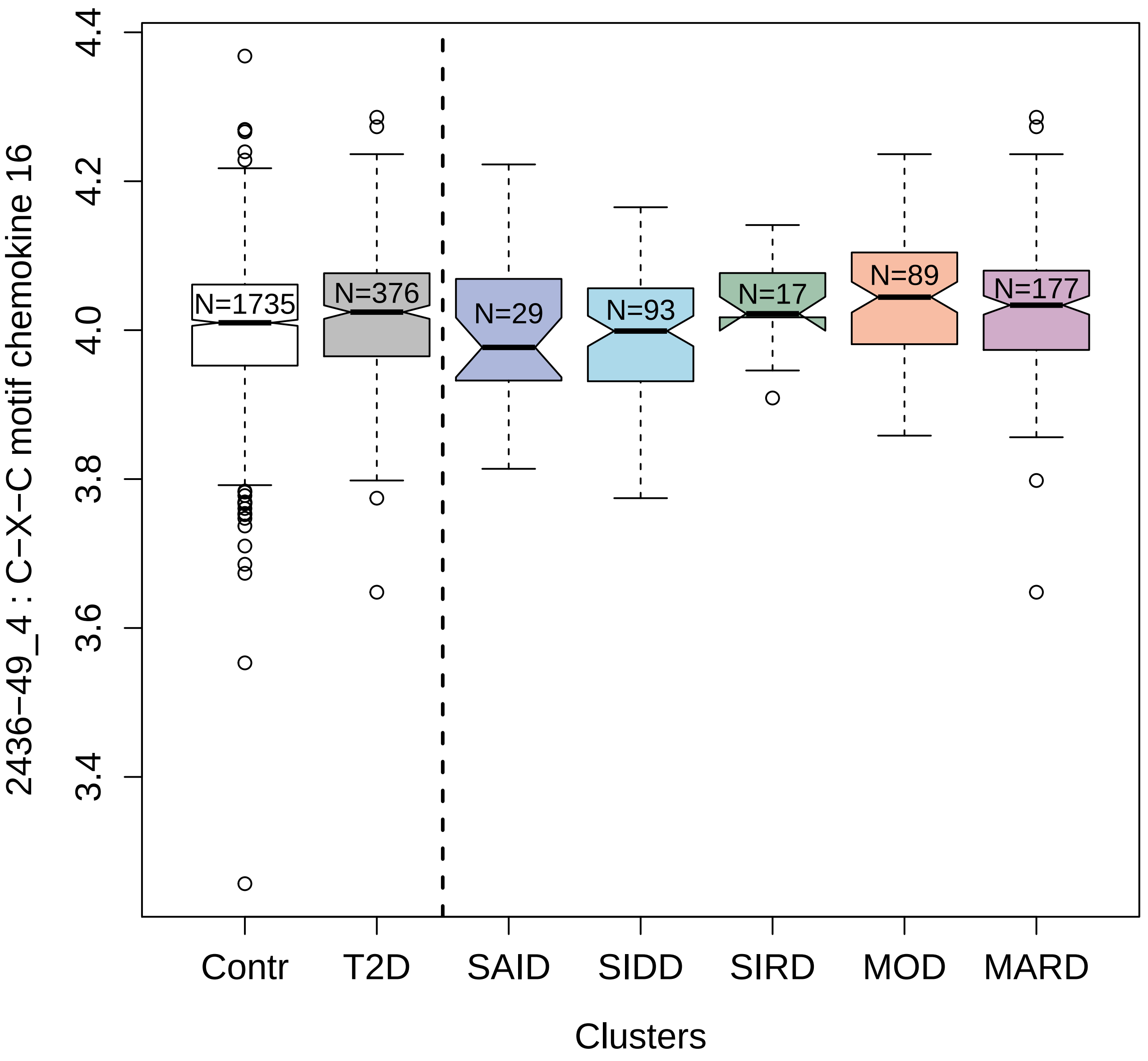

2436−49_4 : C−X−C motif chemokine 16
4.4
4.2
4.0
3.8
3.6
3.4
N=1735
N=376
N=29
N=93
N=17
N=89
N=177
Contr
T2D
SAID
SIDD
SIRD
MOD
MARD
Clusters

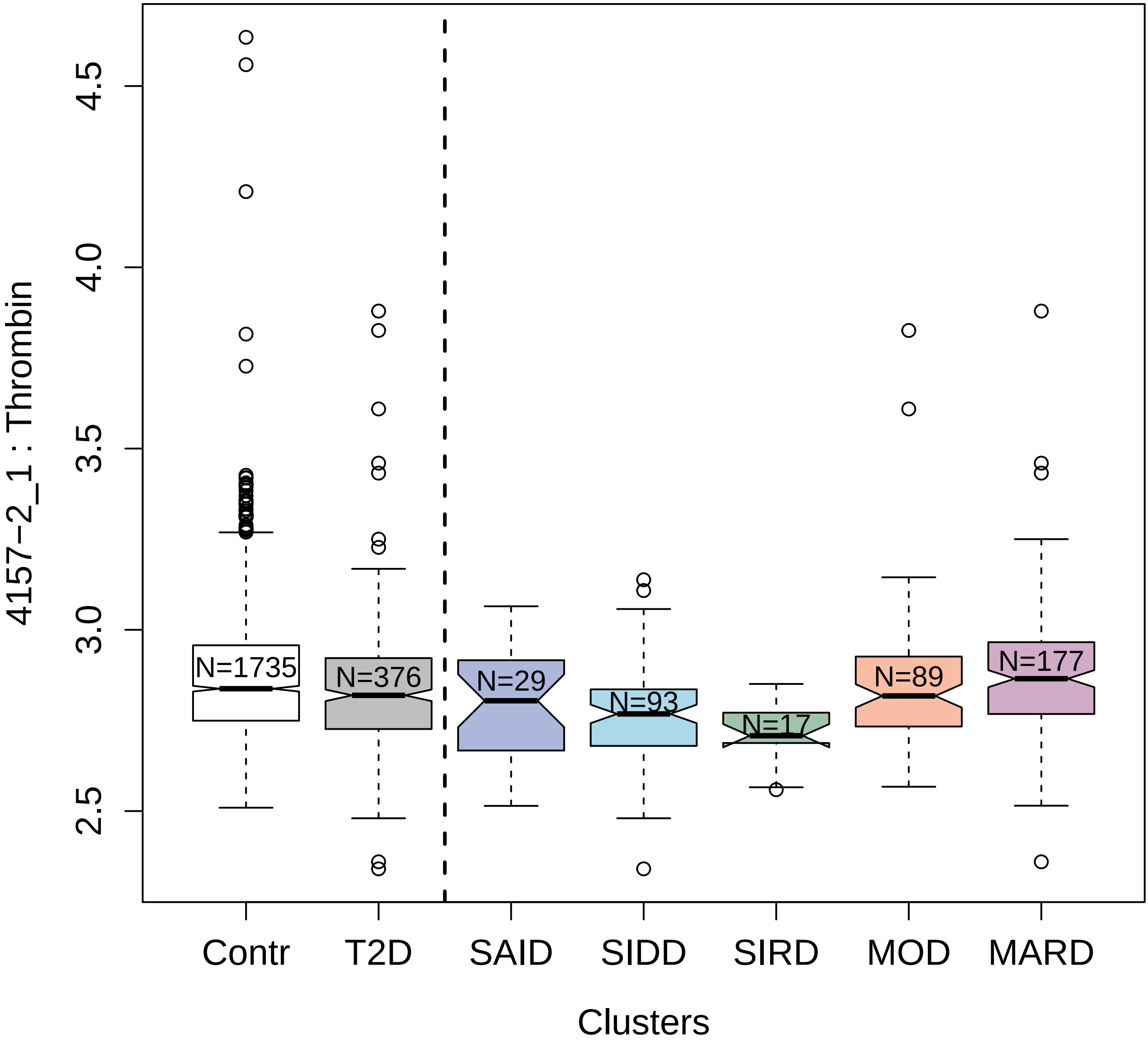
4157−2_1 : Thrombin
2.5
3.0
3.5
4.0
4.5
N=1735
N=376
N=29
N=93
N=17
N=89
N=177
Contr
T2D
SAID
SIDD
SIRD
MOD
MARD
Clusters

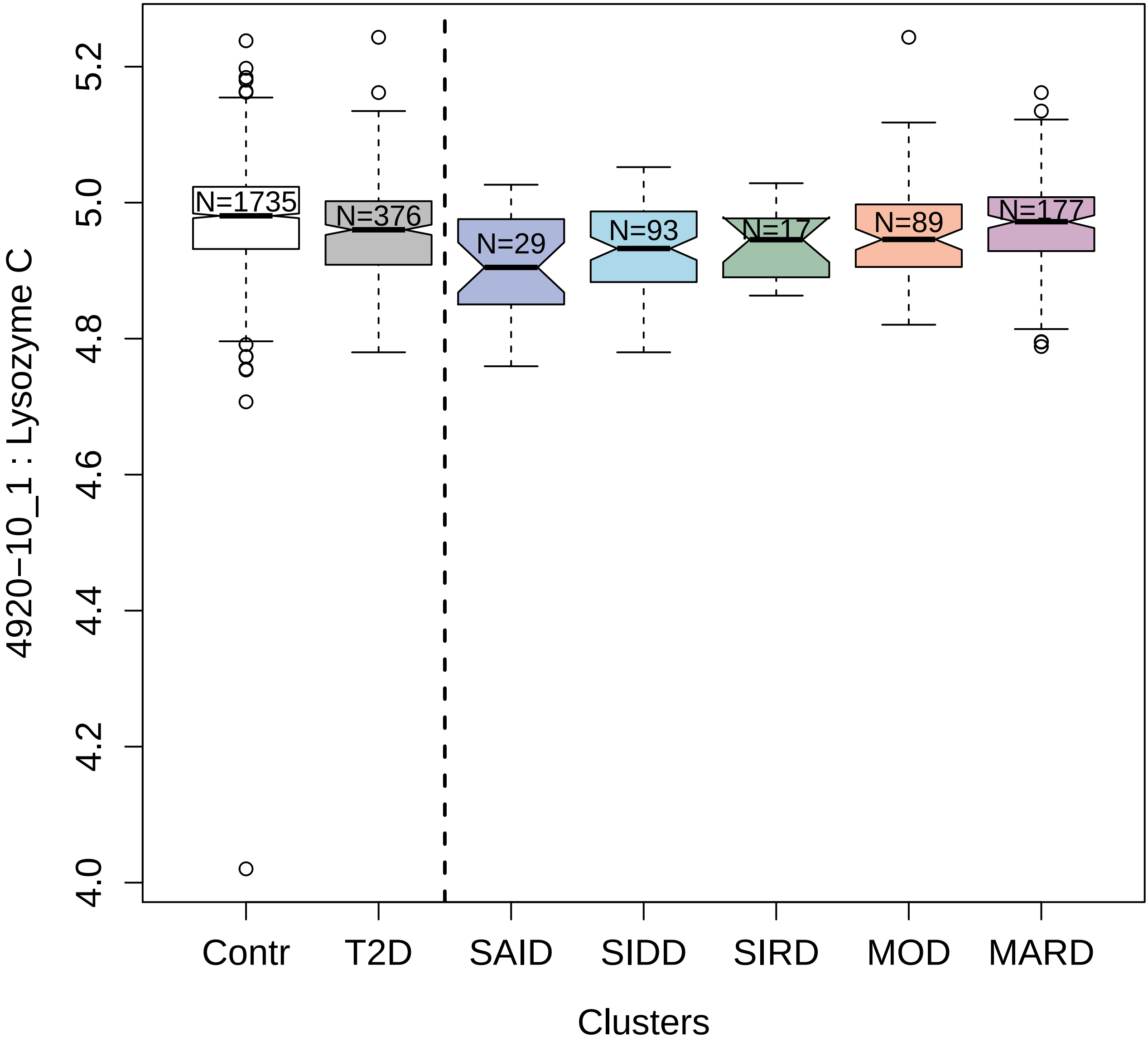

4920−10_1 : Lysozyme C
5.2
5.0
4.8
4.6
4.4
4.2
4.0
N=1735
N=376
N=29
N=93
N=17
N=89
N=177
Contr
T2D
SAID
SIDD
SIRD
MOD
MARD
Clusters

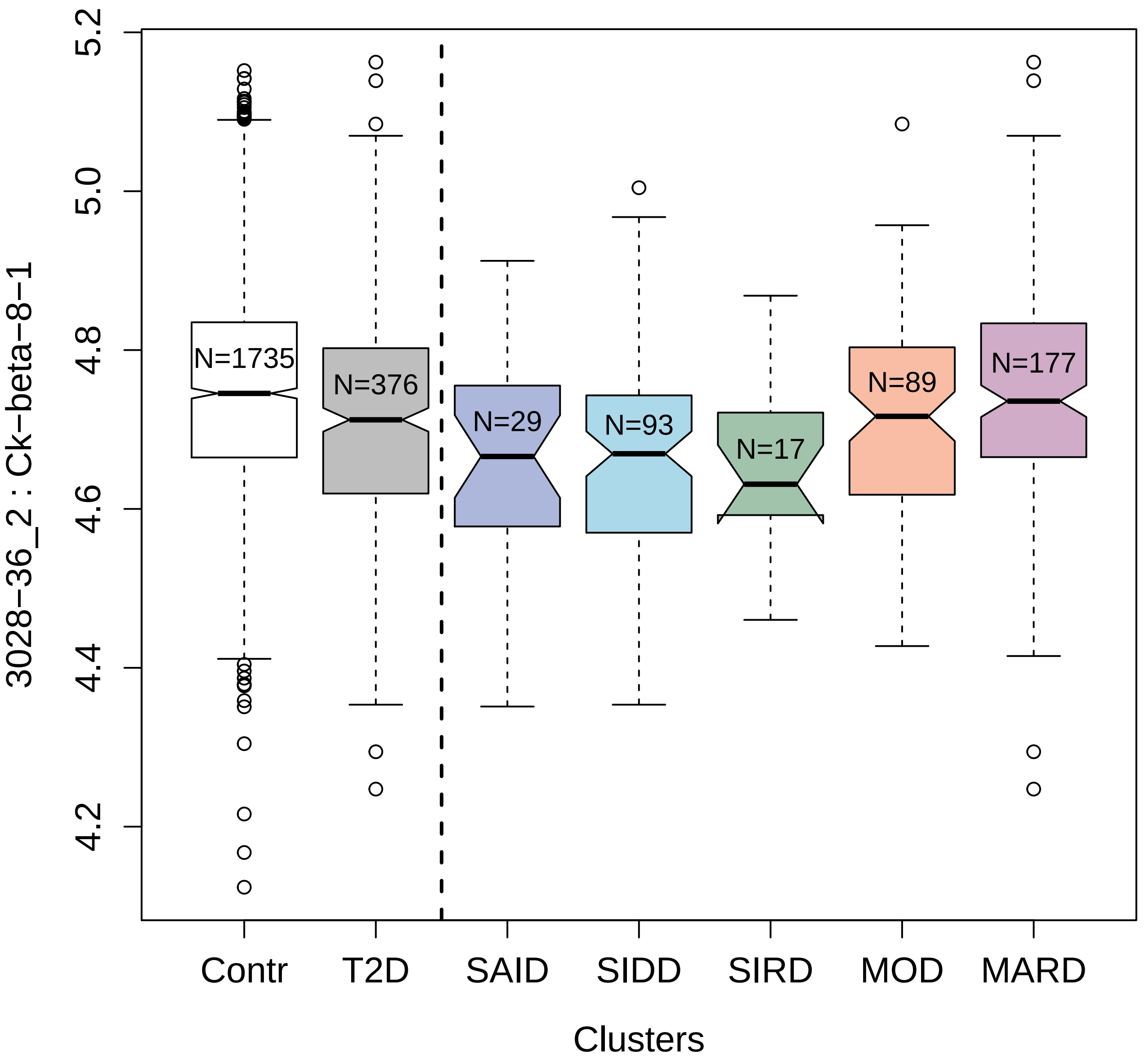

5.2
5.0
4.8
4.6
4.4
4.2
3028−36_2 : Ck−beta−8−1
N=1735
N=376
N=29
N=93
N=17
N=89
N=177
Contr
T2D
SAID
SIDD
SIRD
MOD
MARD
Clusters

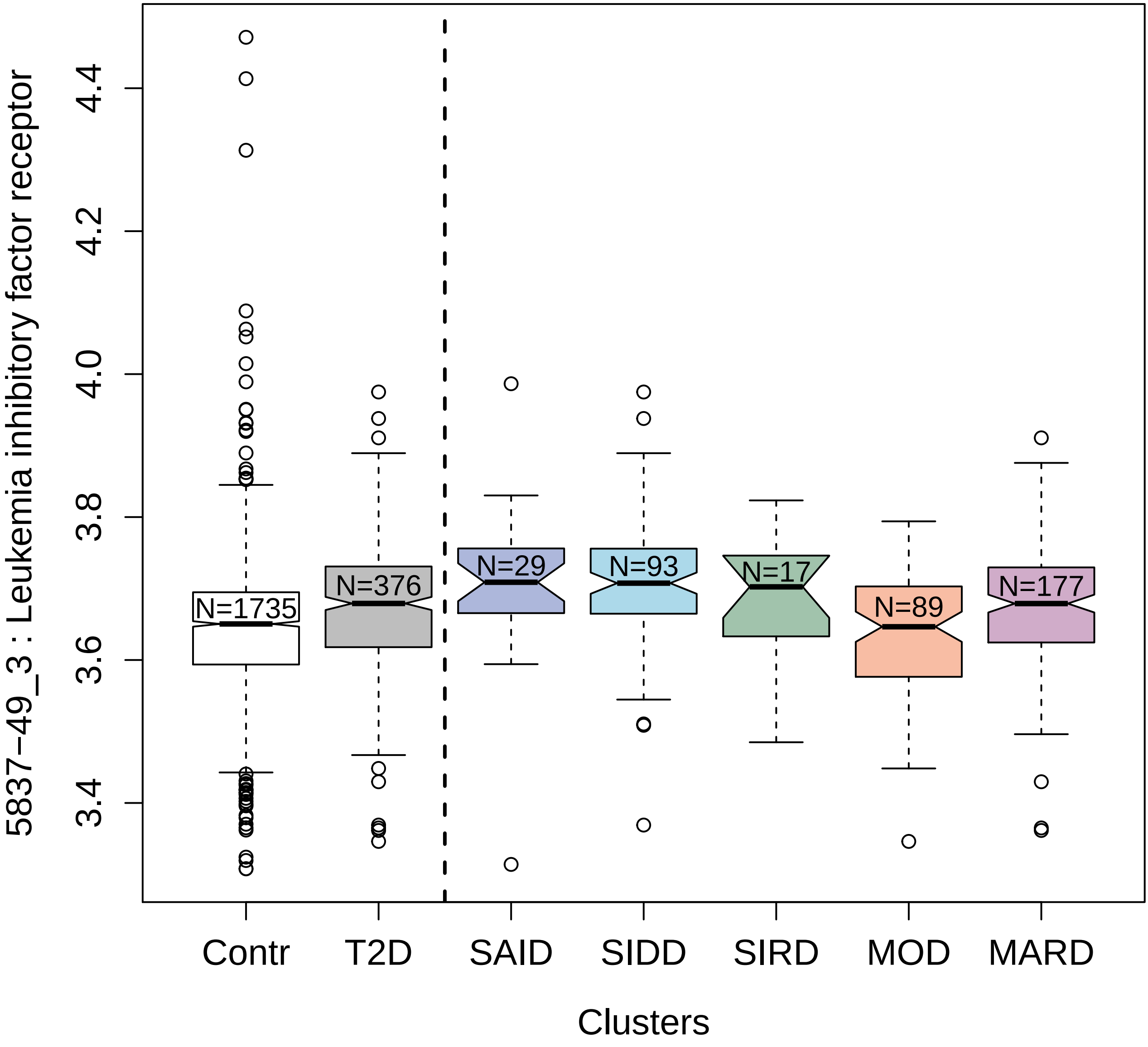

5837−49_3 : Leukemia inhibitory factor receptor
3.4
3.6
3.8
4.0
4.2
4.4
N=1735
N=376
N=29
N=93
N=17
N=89
N=177
Contr
T2D
SAID
SIDD
SIRD
MOD
MARD
Clusters

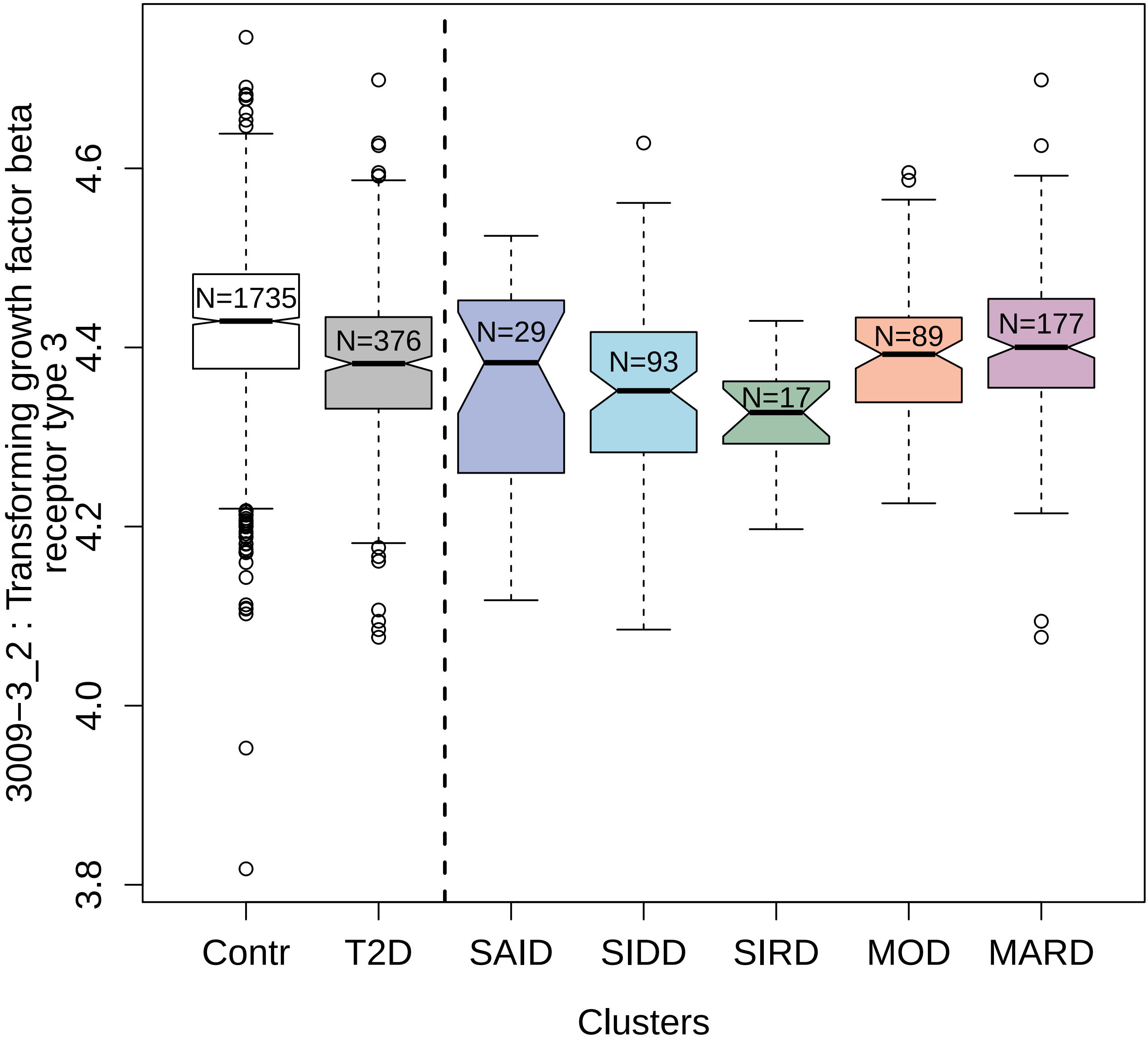

3009−3_2 : Transforming growth factor beta
receptor type 3
3.8
4.0
4.2
4.4
4.6
N=1735
N=376
N=29
N=93
N=17
N=89
N=177
Contr
T2D
SAID
SIDD
SIRD
MOD
MARD
Clusters

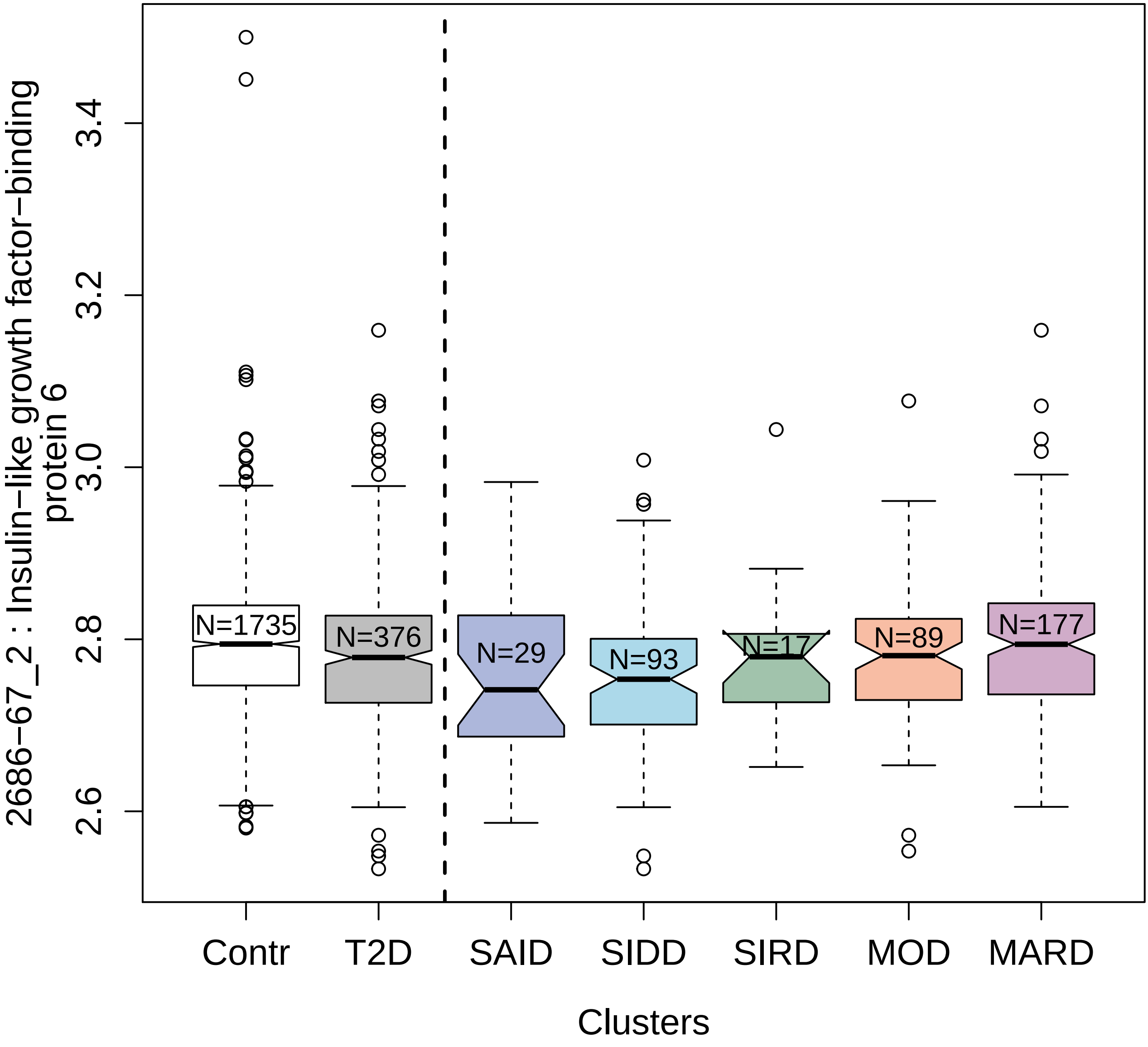

2686−67_2 : Insulin−like growth factor−binding protein 6
3.4
3.2
3.0
2.8
2.6
N=1735
N=376
N=29
N=93
N=17
N=89
N=177
Contr
T2D
SAID
SIDD
SIRD
MOD
MARD
Clusters

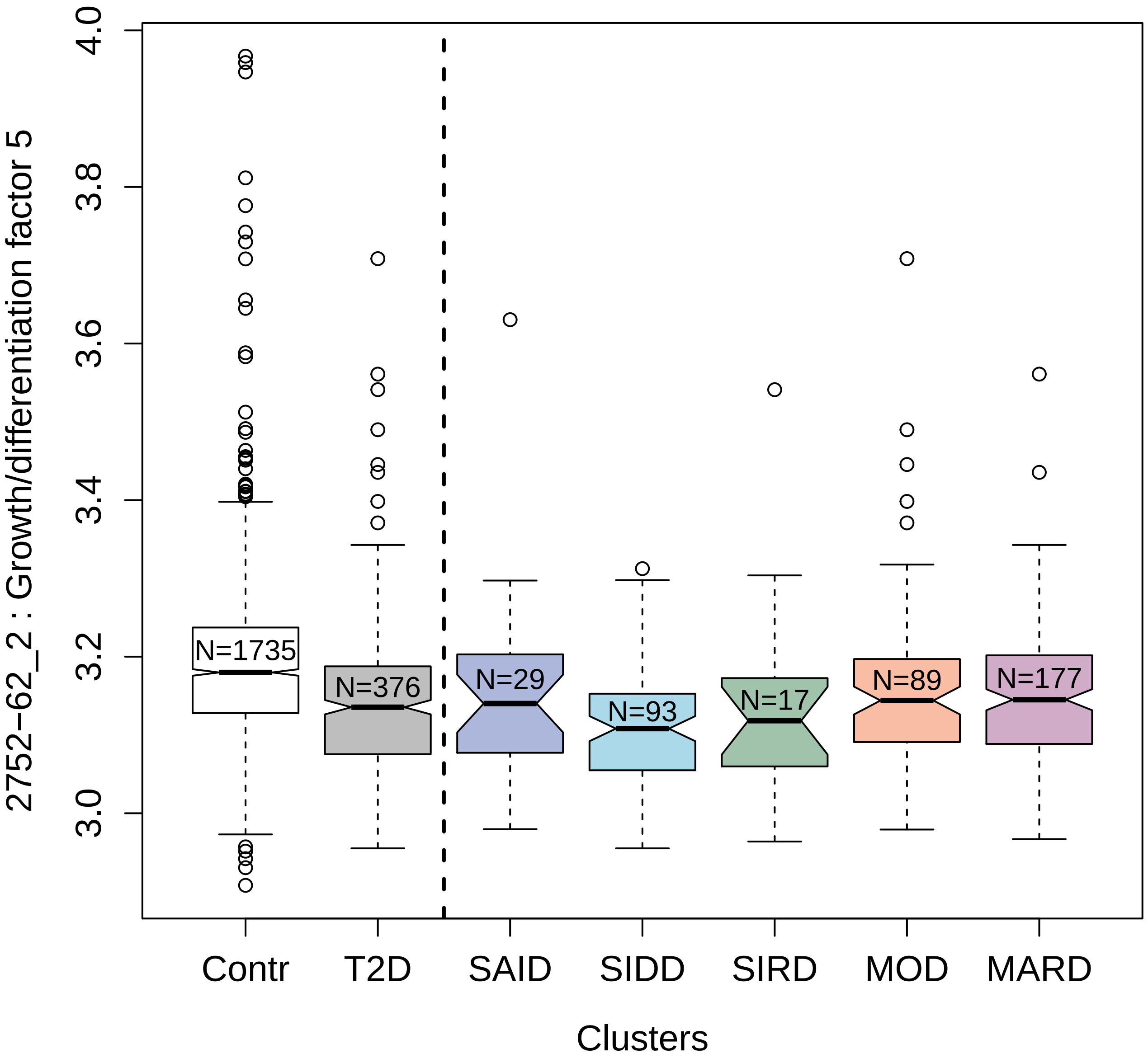
2752−62_2 : Growth/differentiation factor 5
4.0
3.8
3.6
3.4
3.2
3.0
N=1735
N=376
N=29
N=93
N=17
N=89
N=177
Contr
T2D
SAID
SIDD
SIRD
MOD
MARD
Clusters

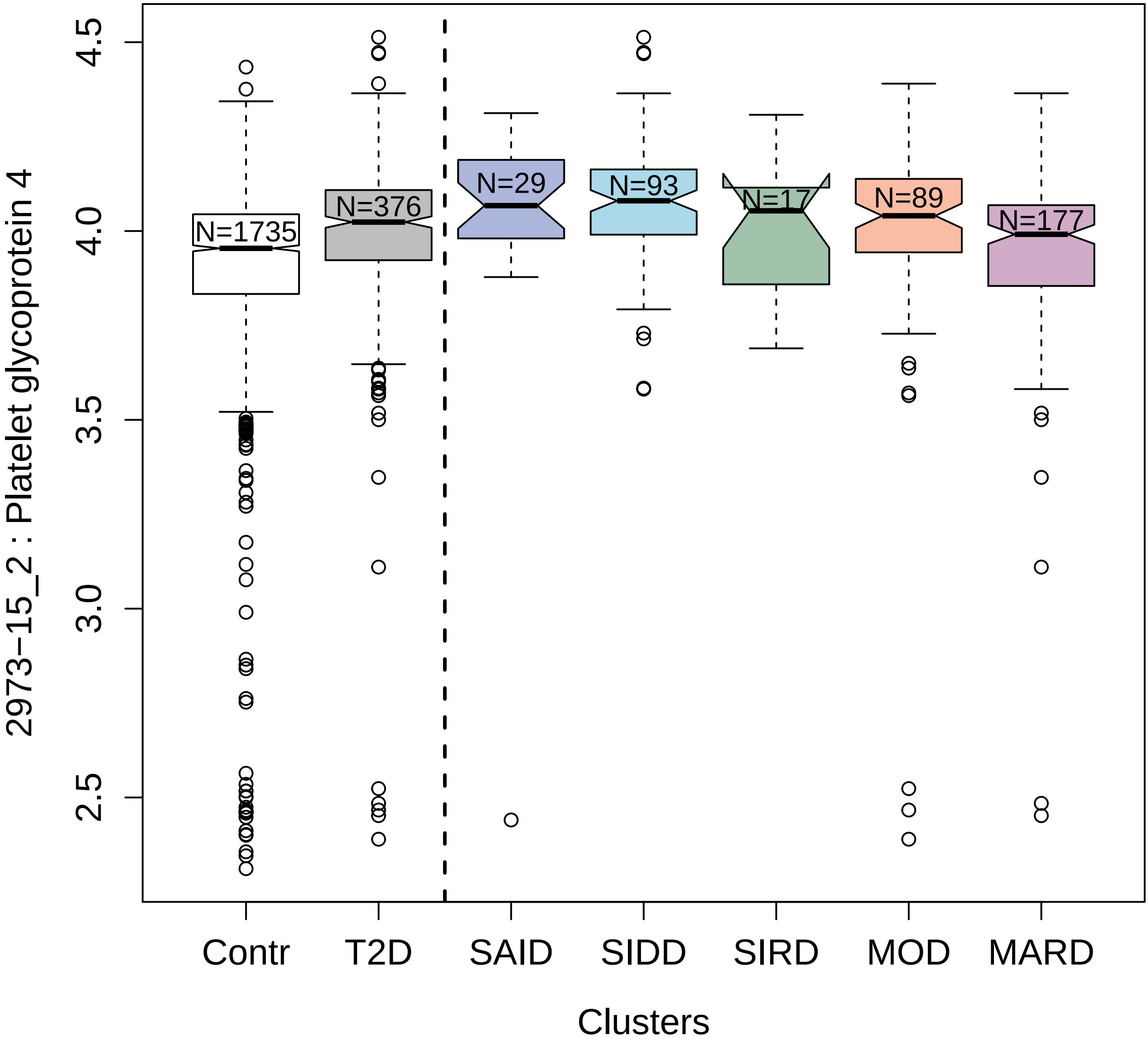

2973−15_2 : Platelet glycoprotein 4
4.5
4.0
3.5
3.0
2.5
N=1735
N=376
N=29
N=93
N=17
N=89
N=177
Contr
T2D
SAID
SIDD
SIRD
MOD
MARD
Clusters

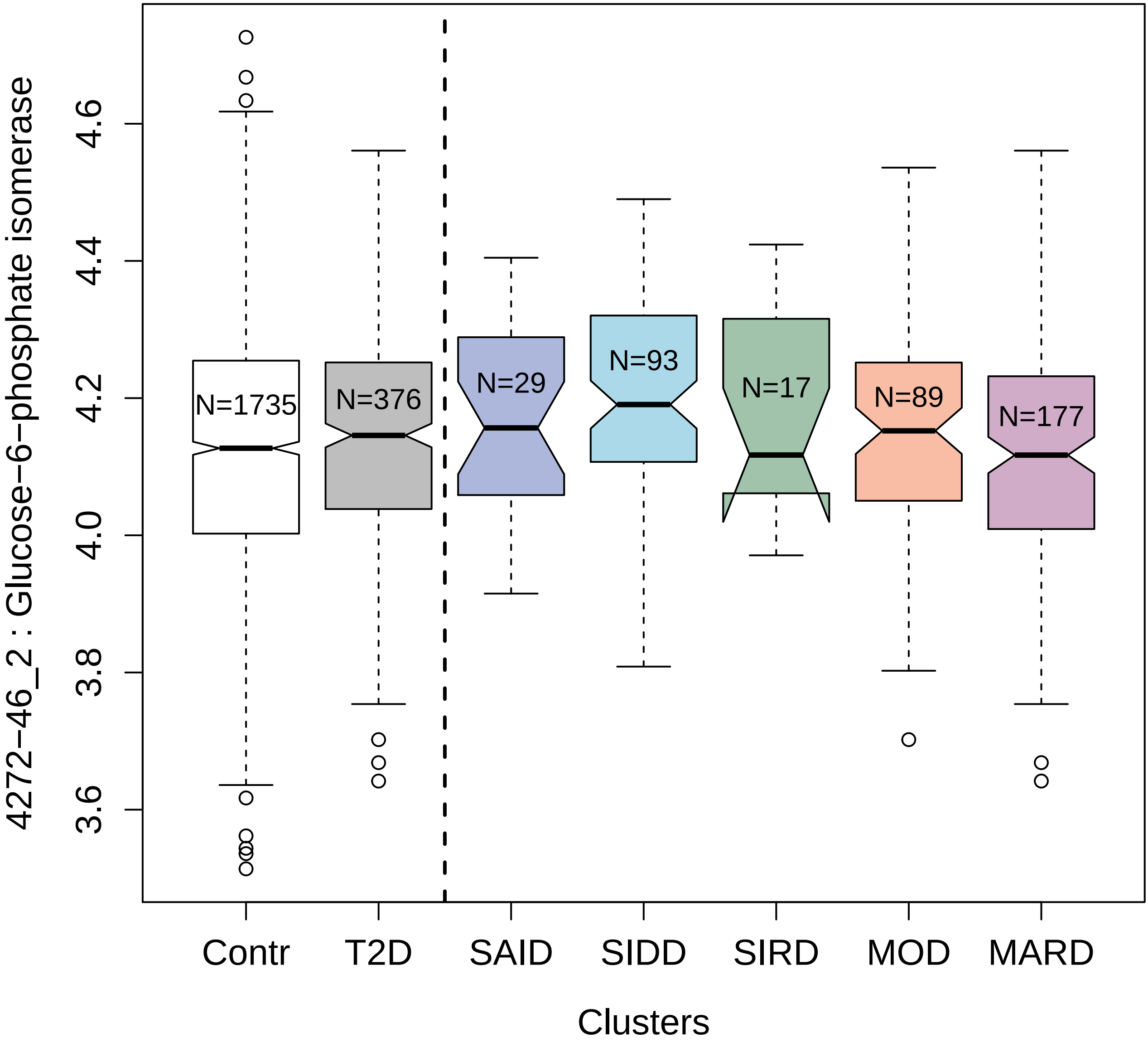

4272−46_2 : Glucose−6−phosphate isomerase
3.6
3.8
4.0
4.2
4.4
4.6
N=1735
N=376
N=29
N=93
N=17
N=89
N=177
Contr
T2D
SAID
SIDD
SIRD
MOD
MARD
Clusters

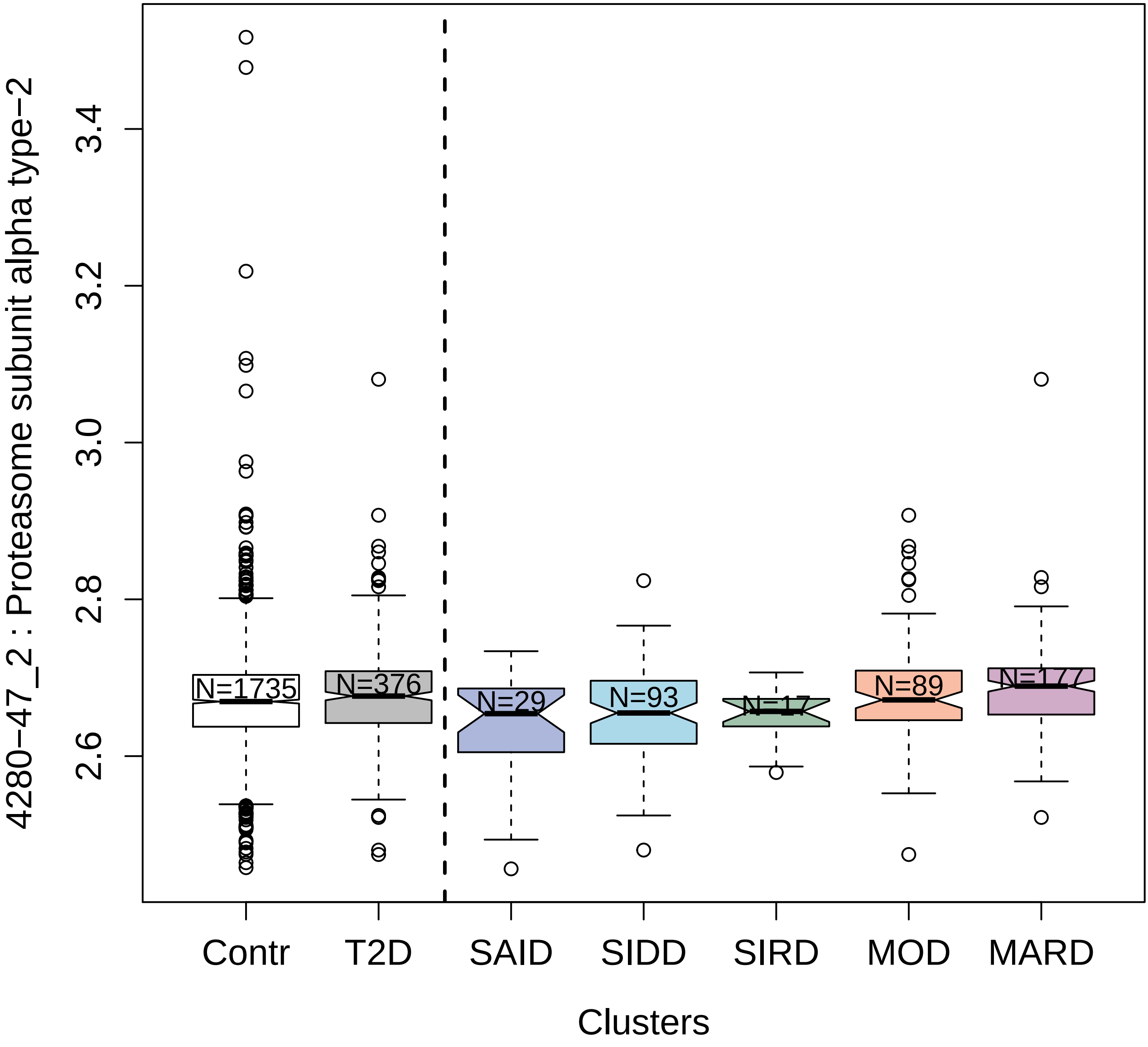
4280−47_2 : Proteasome subunit alpha type−2
3.4
3.2
3.0
2.8
2.6
N=1735
N=376
N=29
N=93
N=17
N=89
N=177
Contr
T2D
SAID
SIDD
SIRD
MOD
MARD
Clusters

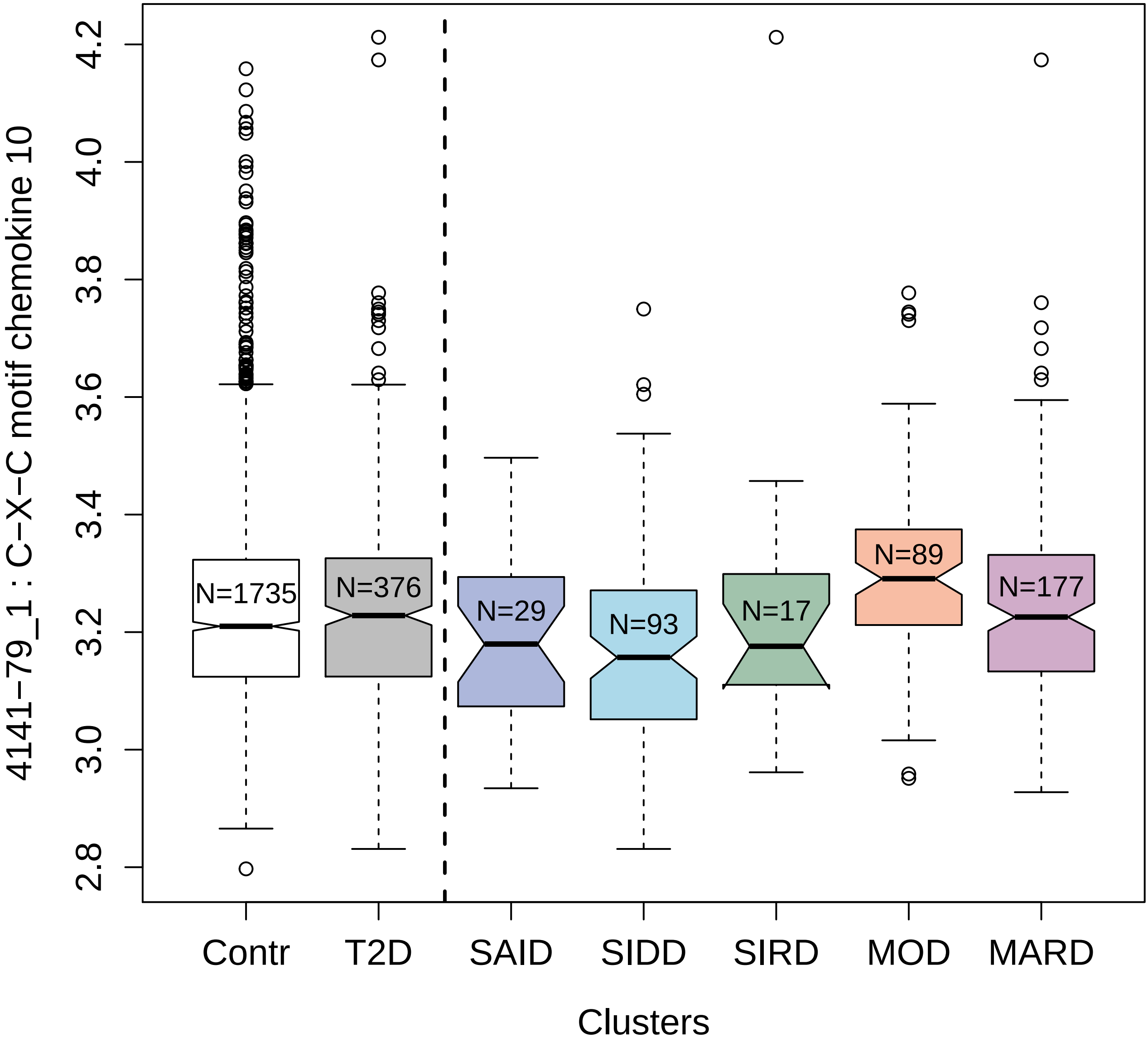

4141−79_1 : C−X−C motif chemokine 10
2.8
3.0
3.2
3.4
3.6
3.8
4.0
4.2
N=1735
N=376
N=29
N=93
N=17
N=89
N=177
Contr
T2D
SAID
SIDD
SIRD
MOD
MARD
Clusters

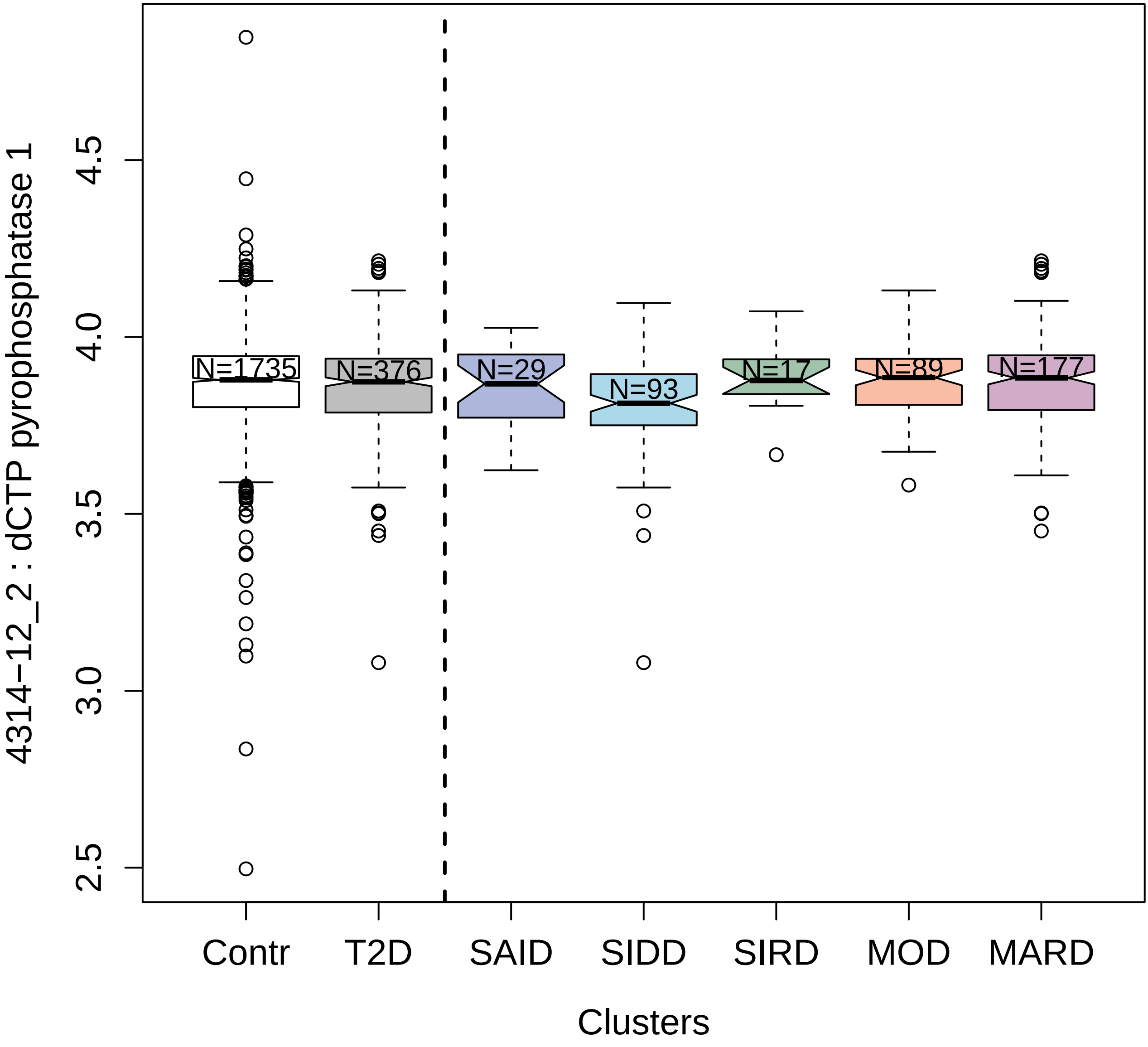

4314−12_2 : dCTP pyrophosphatase 1
2.5
3.0
3.5
4.0
4.5
N=1735
N=376
N=29
N=93
N=17
N=89
N=177
Contr
T2D
SAID
SIDD
SIRD
MOD
MARD
Clusters

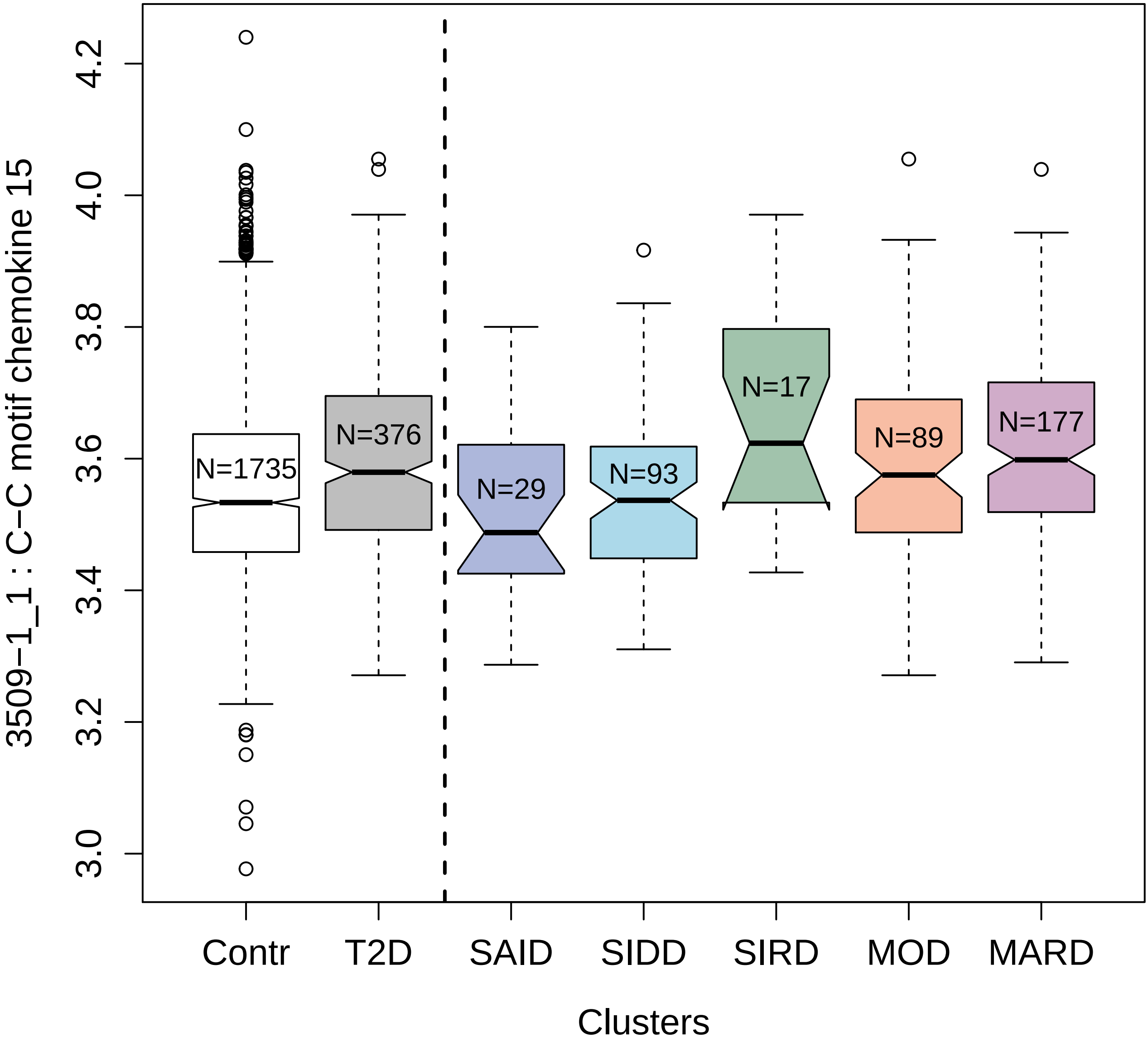
3509−1_1 : C−C motif chemokine 15
4.2
4.0
3.8
3.6
3.4
3.2
3.0
N=1735
N=376
N=29
N=93
N=17
N=89
N=177
Contr
T2D
SAID
SIDD
SIRD
MOD
MARD
Clusters

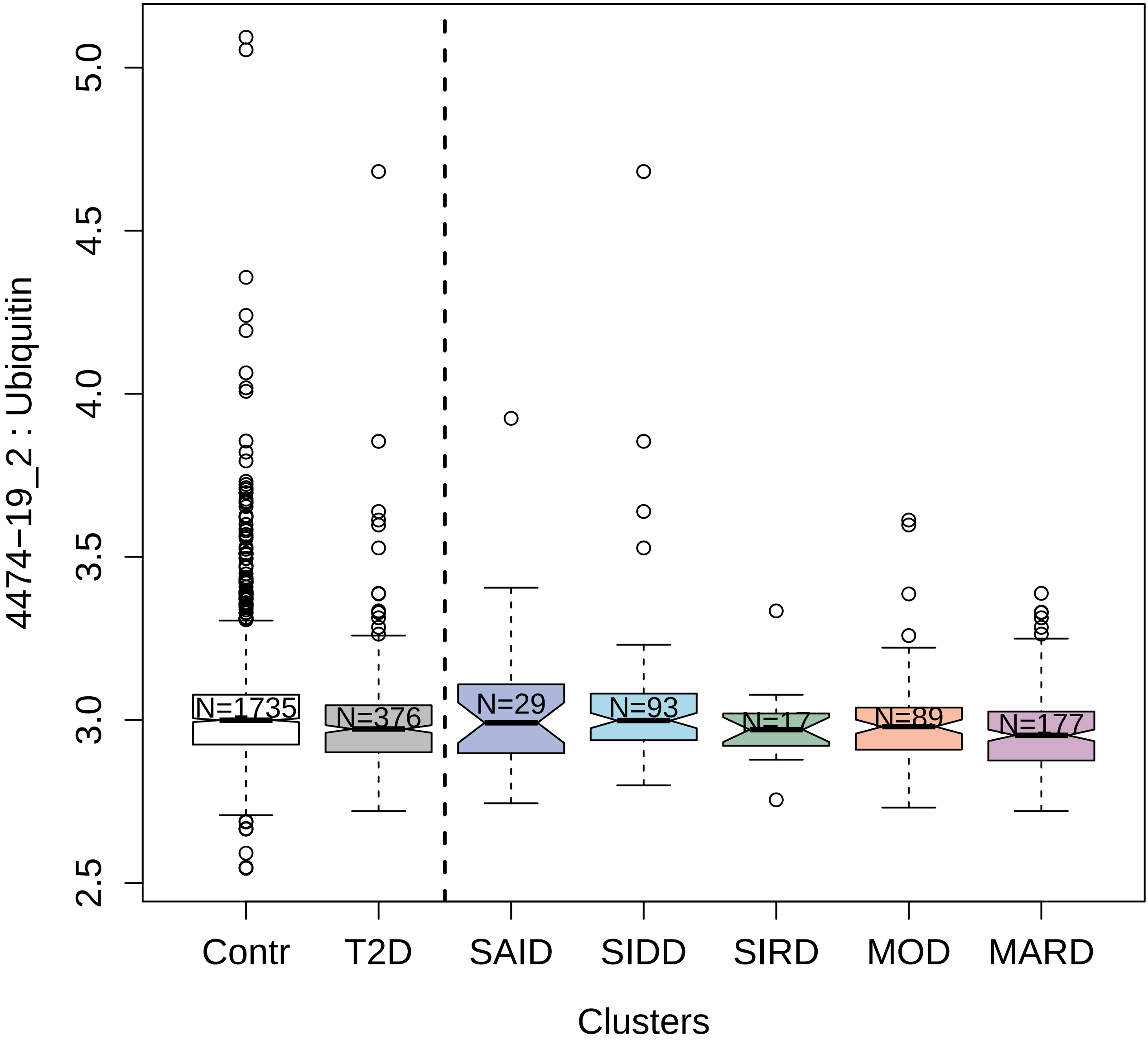
4474−19_2 : Ubiquitin
5.0
4.5
4.0
3.5
3.0
2.5
N=1735
N=376
N=29
N=93
N=17
N=89
N=177
Contr
T2D
SAID
SIDD
SIRD
MOD
MARD
Clusters

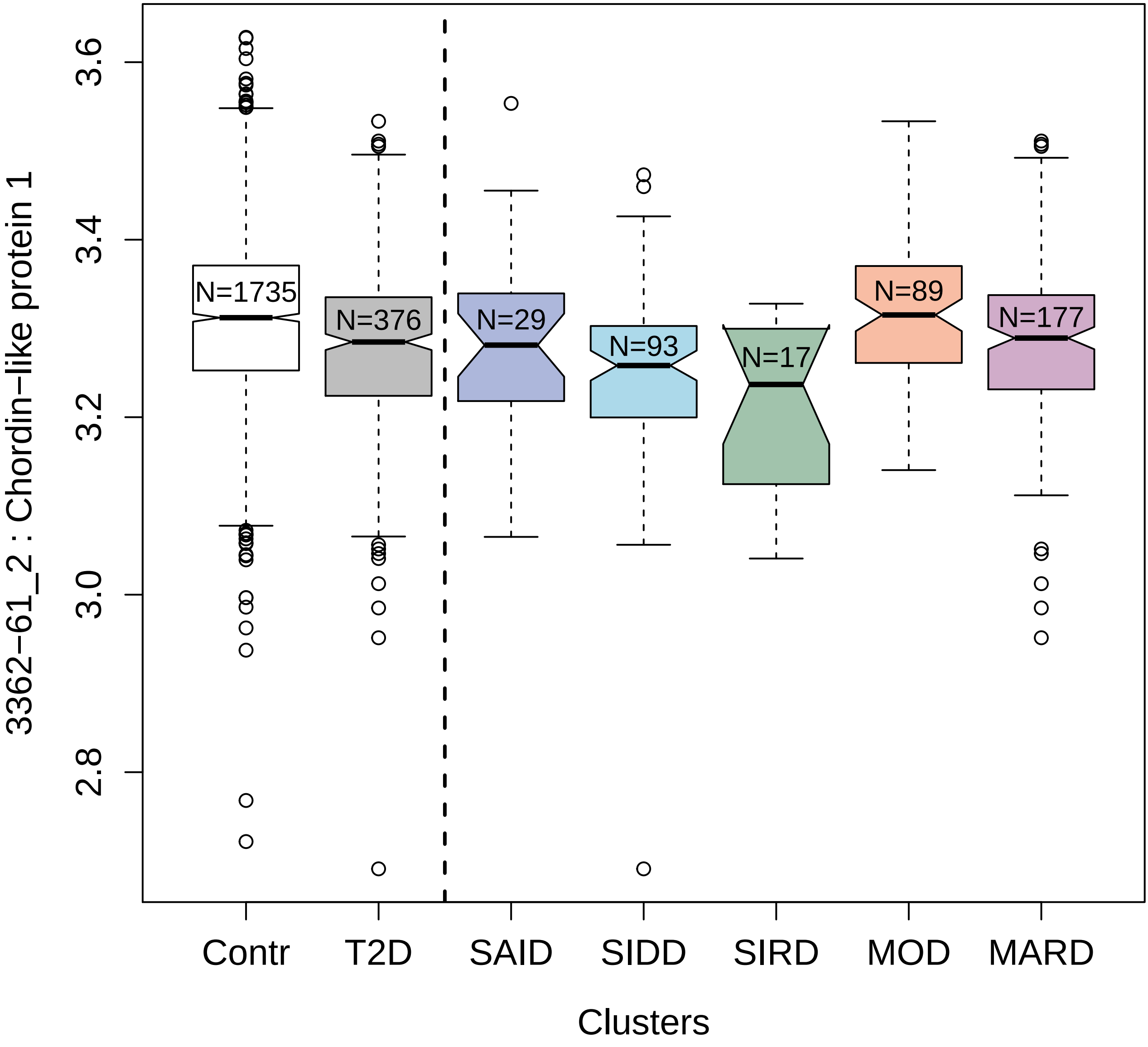

3362−61_2 : Chordin−like protein 1
3.6
3.4
3.2
3.0
2.8
N=1735
N=376
N=29
N=93
N=17
N=89
N=177
Contr
T2D
SAID
SIDD
SIRD
MOD
MARD
Clusters

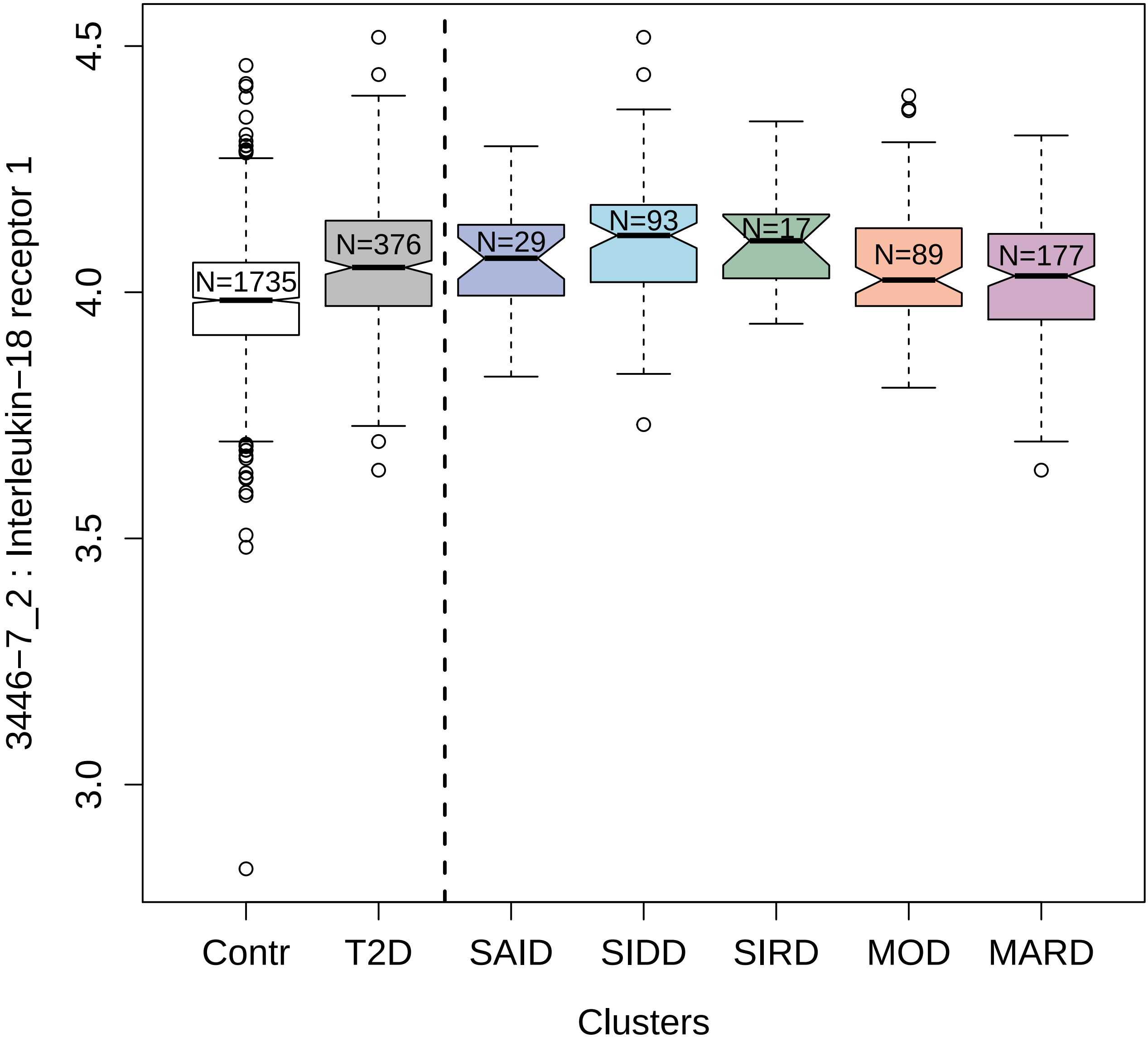

3446−7_2 : Interleukin−18 receptor 1
4.5
4.0
3.5
3.0
N=1735
N=376
N=29
N=93
N=17
N=89
N=177
Contr
T2D
SAID
SIDD
SIRD
MOD
MARD
Clusters

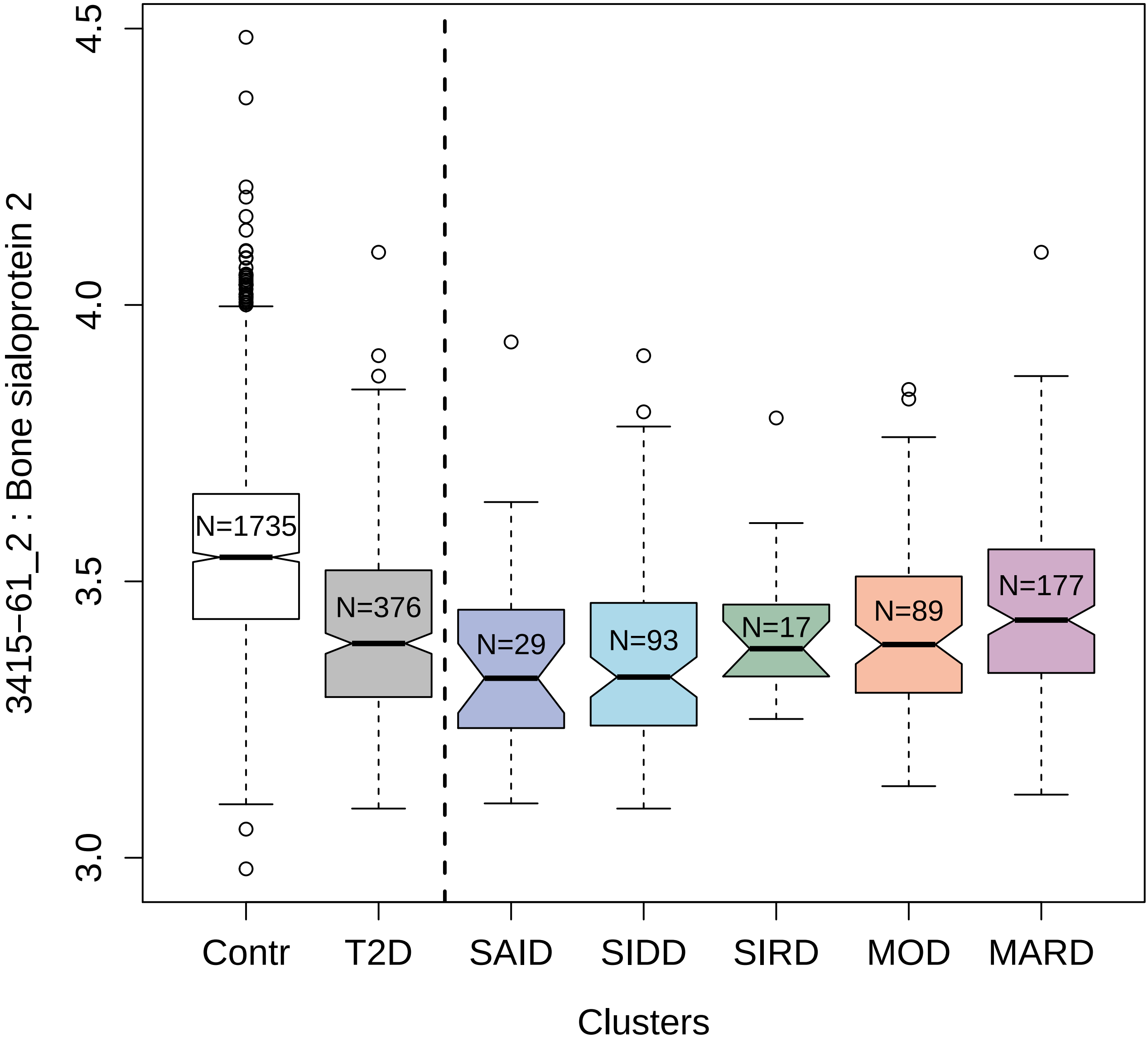

3415−61_2 : Bone sialoprotein 2
4.5
4.0
3.5
3.0
N=1735
N=376
N=29
N=93
N=17
N=89
N=177
Contr
T2D
SAID
SIDD
SIRD
MOD
MARD
Clusters

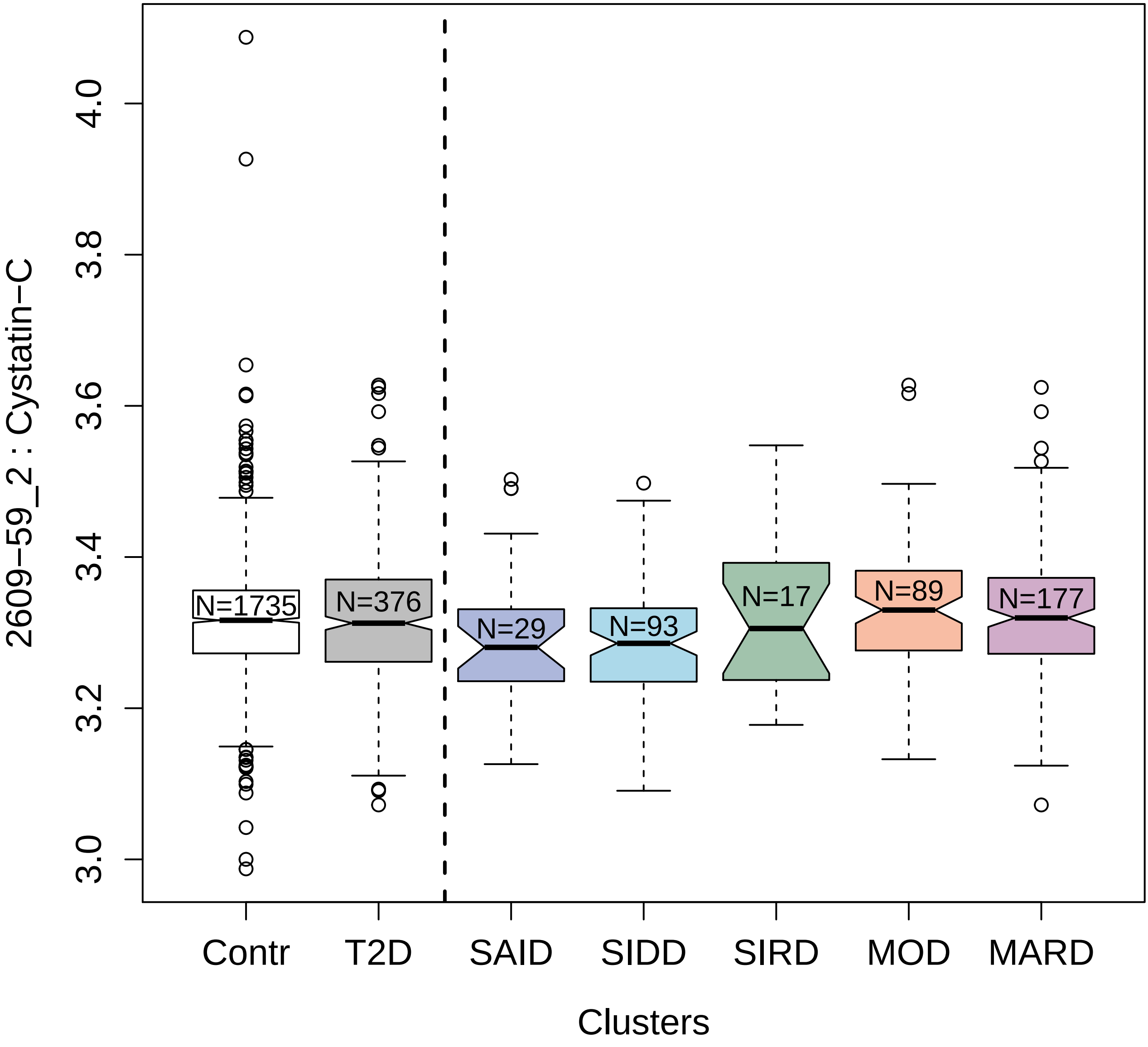
2609−59_2 : Cystatin−C
3.0
3.2
3.4
3.6
3.8
4.0
N=1735
N=376
N=29
N=93
N=17
N=89
N=177
Contr
T2D
SAID
SIDD
SIRD
MOD
MARD
Clusters

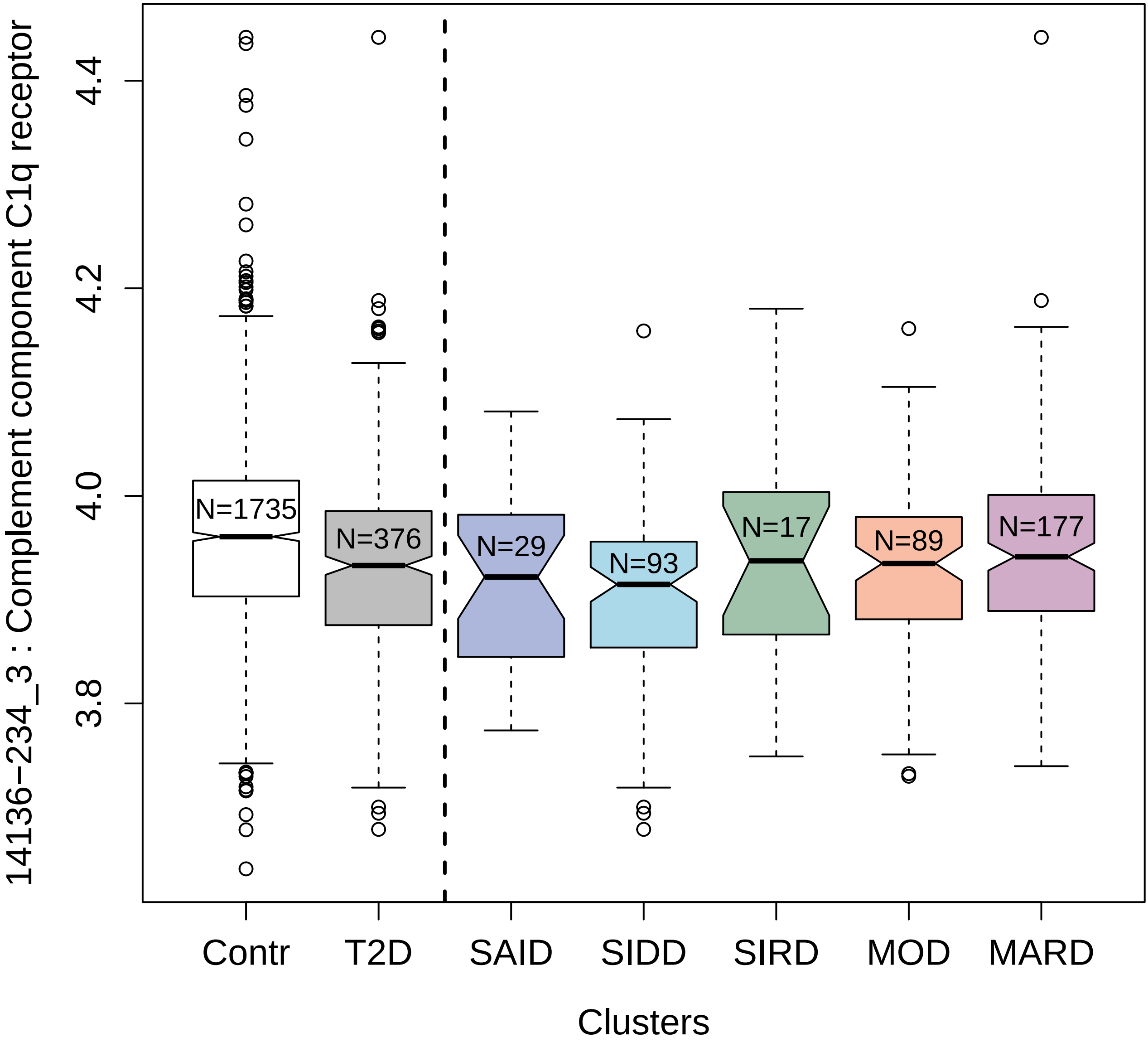
14136−234_3 : Complement component C1q receptor
4.4
4.2
4.0
3.8
N=1735
N=376
N=29
N=93
N=17
N=89
N=177
Contr
T2D
SAID
SIDD
SIRD
MOD
MARD
Clusters

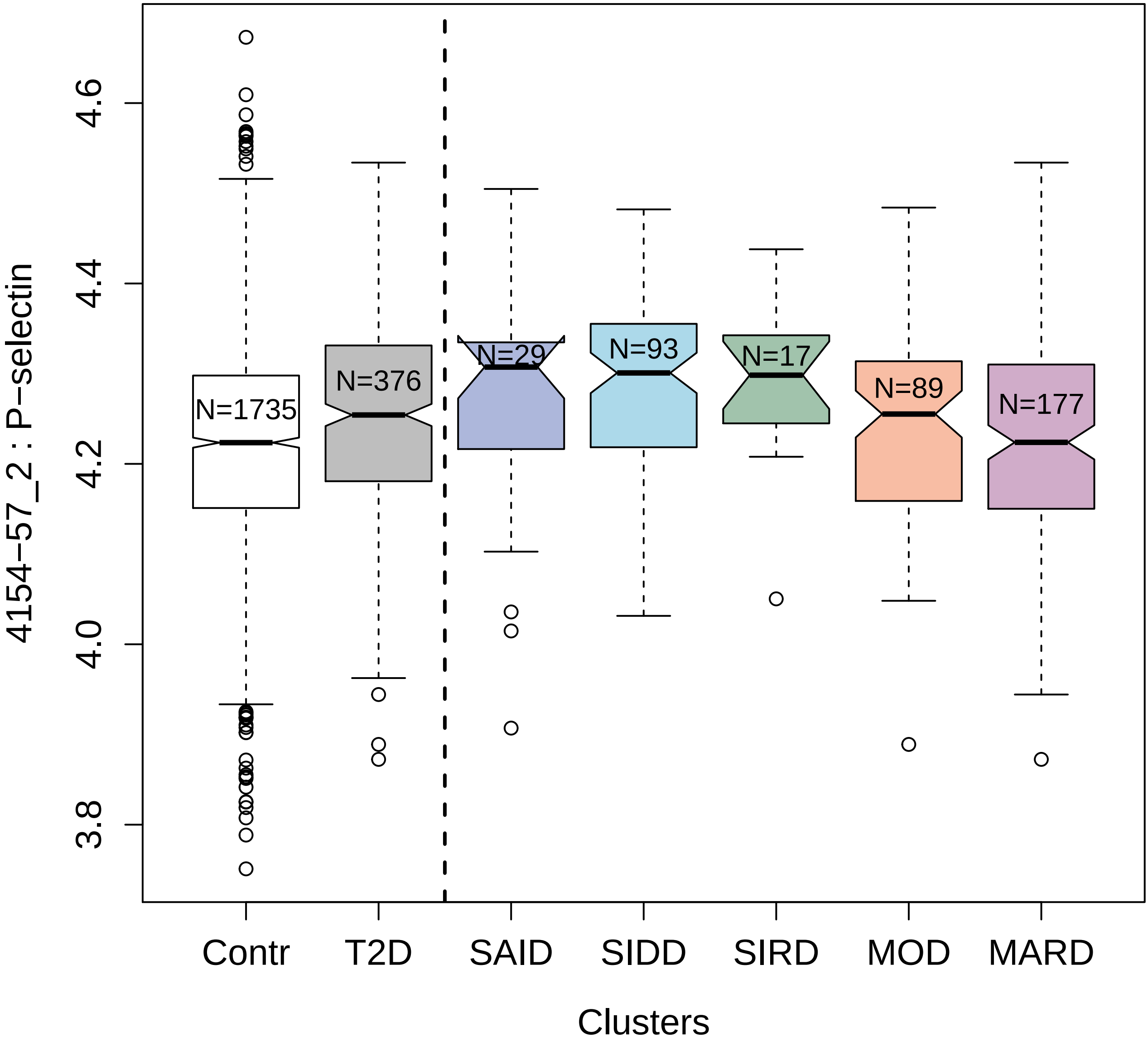
4154−57_2 : P−selectin
4.6
4.4
4.2
4.0
3.8
N=1735
N=376
N=29
N=93
N=17
N=89
N=177
Contr
T2D
SAID
SIDD
SIRD
MOD
MARD
Clusters

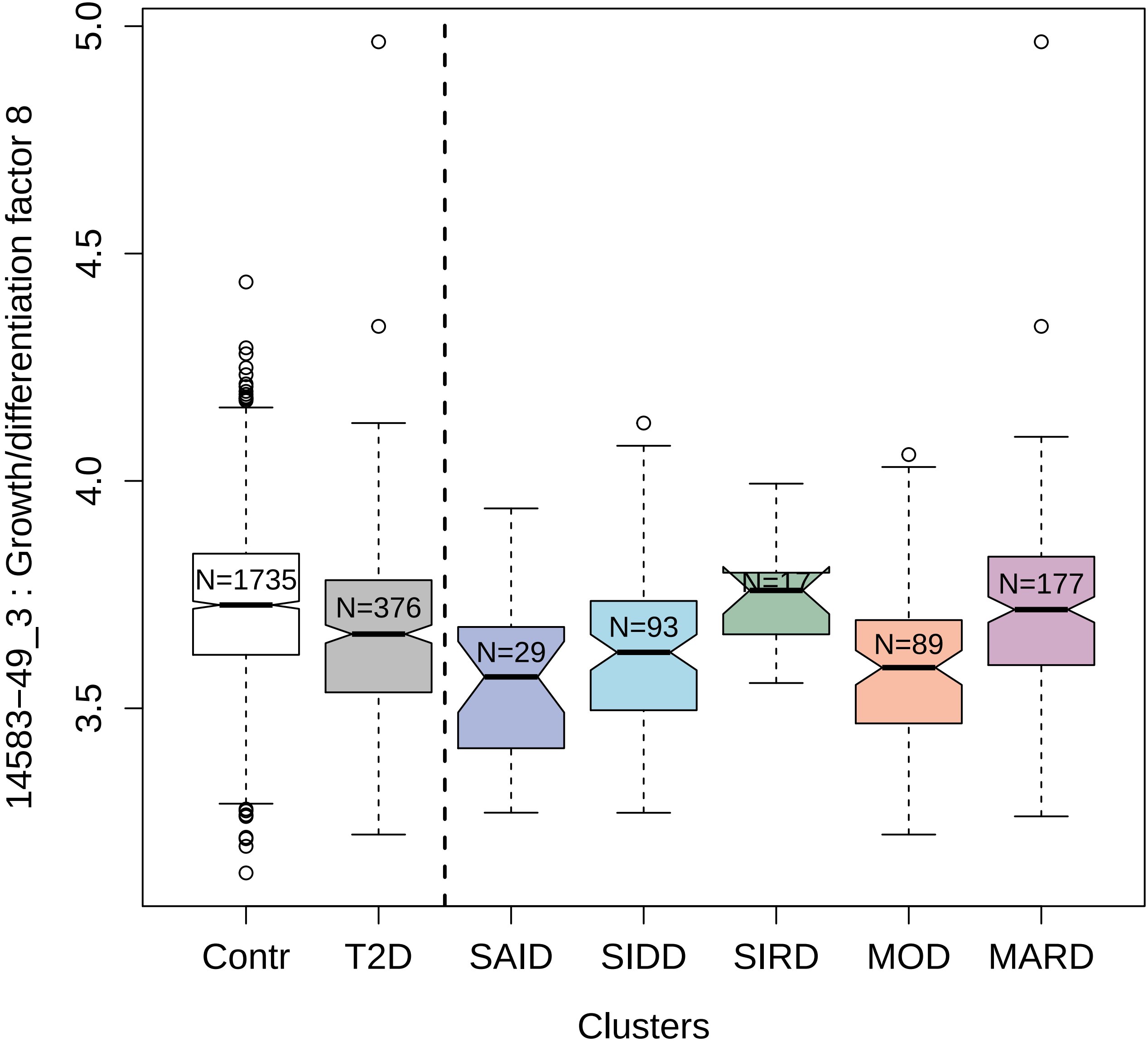
14583−49_3 : Growth/differentiation factor 8
5.0
4.5
4.0
3.5
N=1735
N=376
N=29
N=93
N=17
N=89
N=177
Contr
T2D
SAID
SIDD
SIRD
MOD
MARD
Clusters

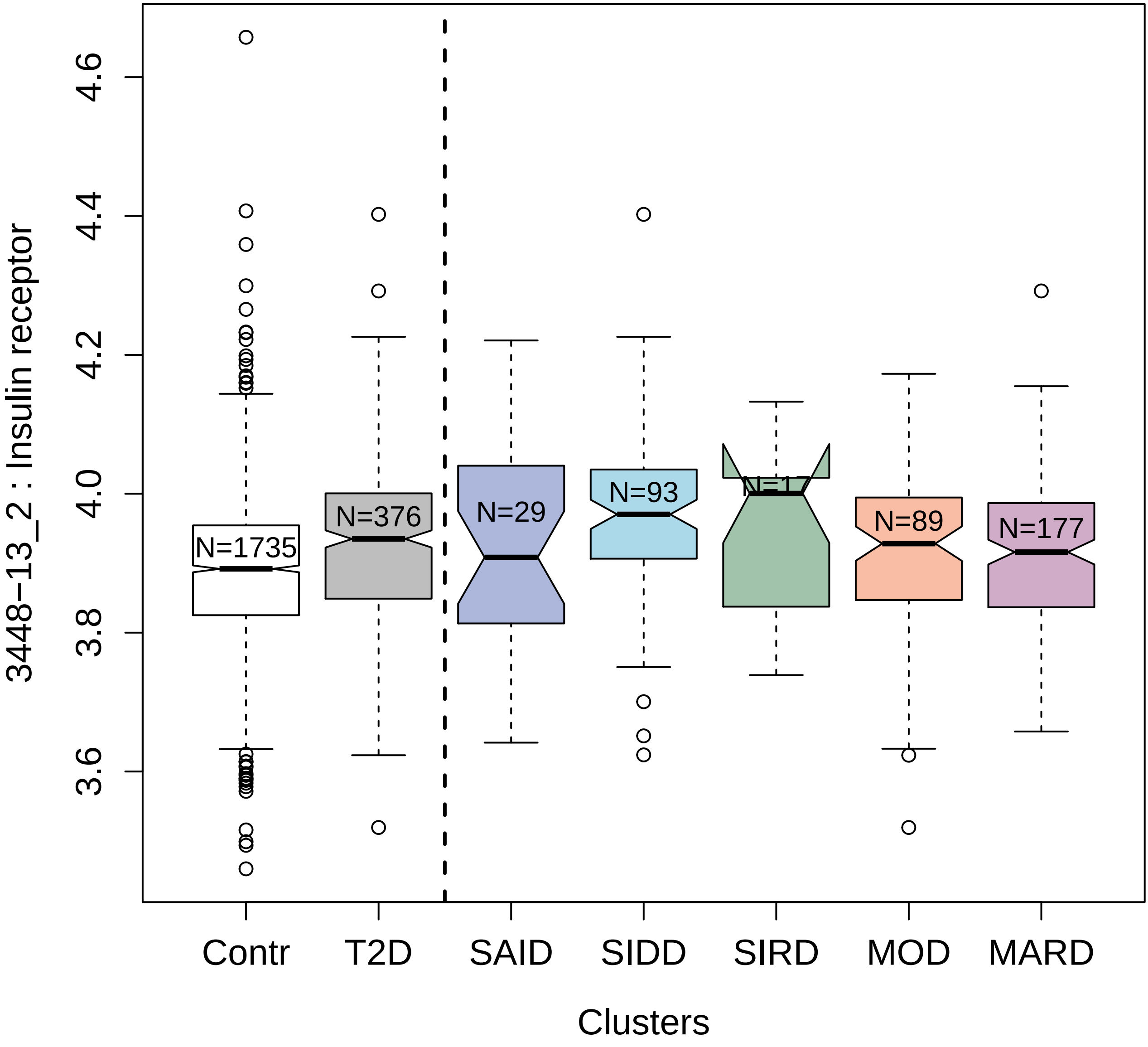
3448−13_2 : Insulin receptor
4.6
4.4
4.2
4.0
3.8
3.6
N=1735
N=376
N=29
N=93
N=17
N=89
N=177
Contr
T2D
SAID
SIDD
SIRD
MOD
MARD
Clusters

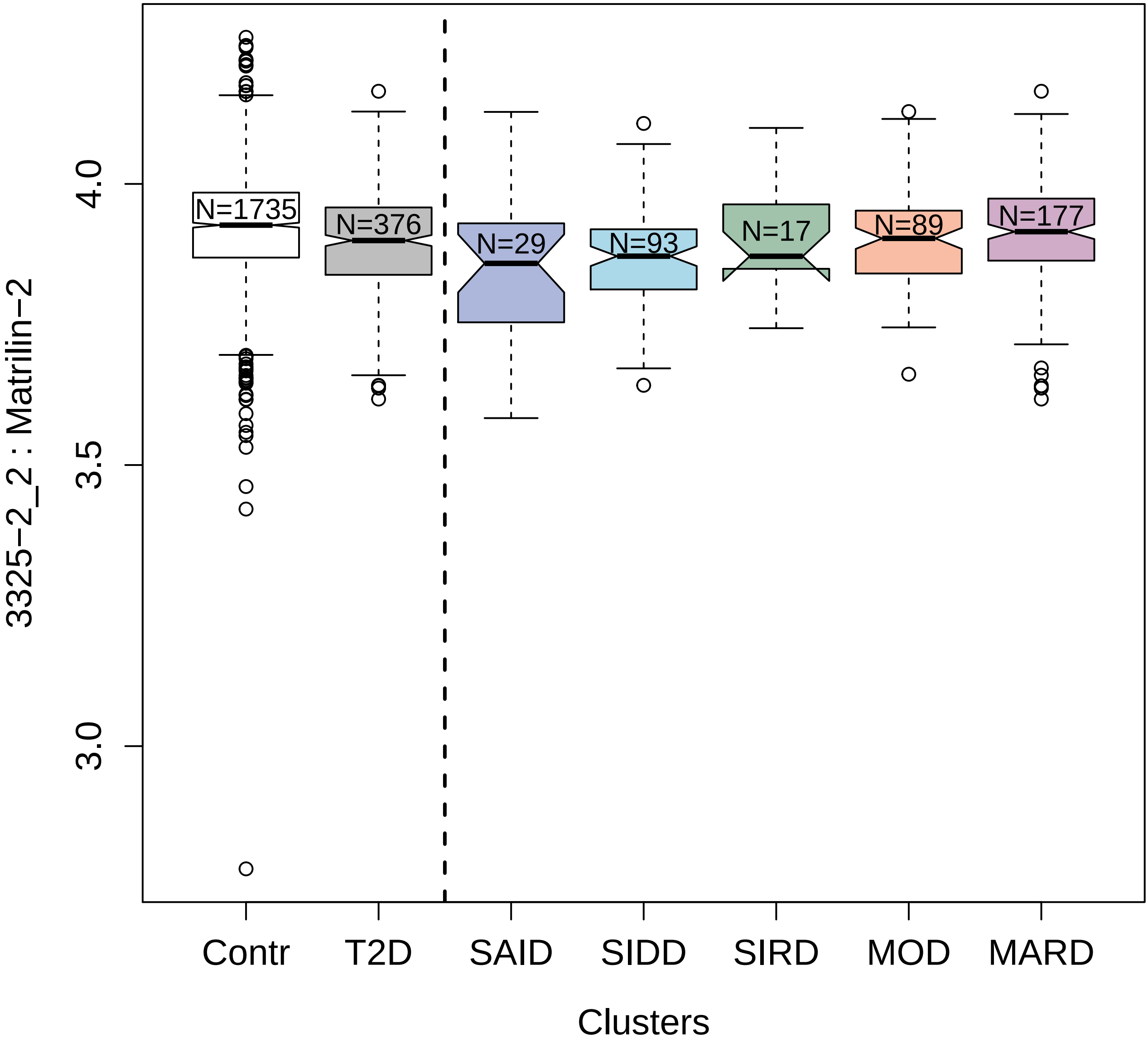
3325−2_2 : Matrilin−2
4.0
3.5
3.0
N=1735
N=376
N=29
N=93
N=17
N=89
N=177
Contr
T2D
SAID
SIDD
SIRD
MOD
MARD
Clusters

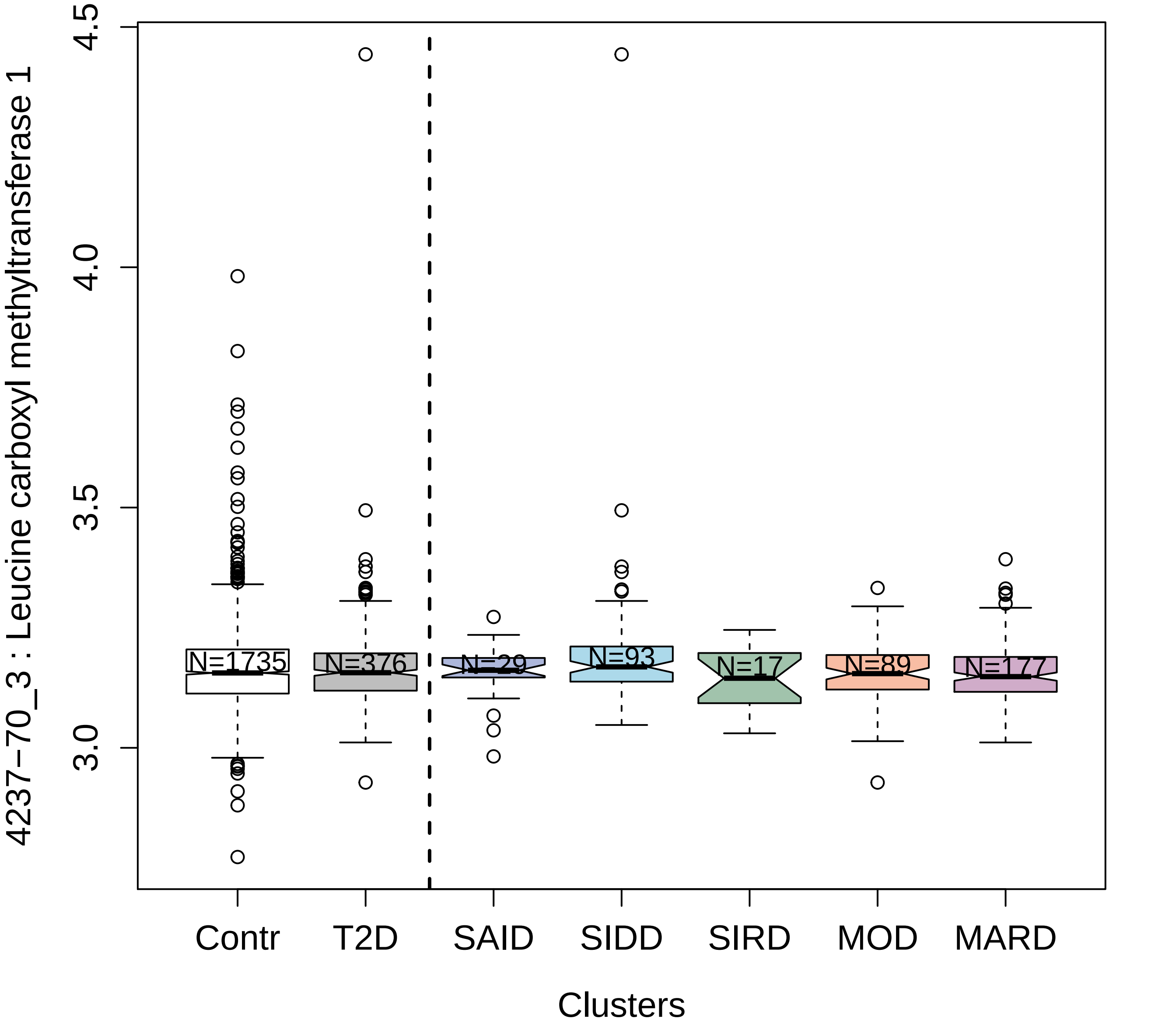

4237−70_3 : Leucine carboxyl methyltransferase 1
4.5
4.0
3.5
3.0
N=1735
N=376
N=29
N=93
N=17
N=89
N=177
Contr
T2D
SAID
SIDD
SIRD
MOD
MARD
Clusters

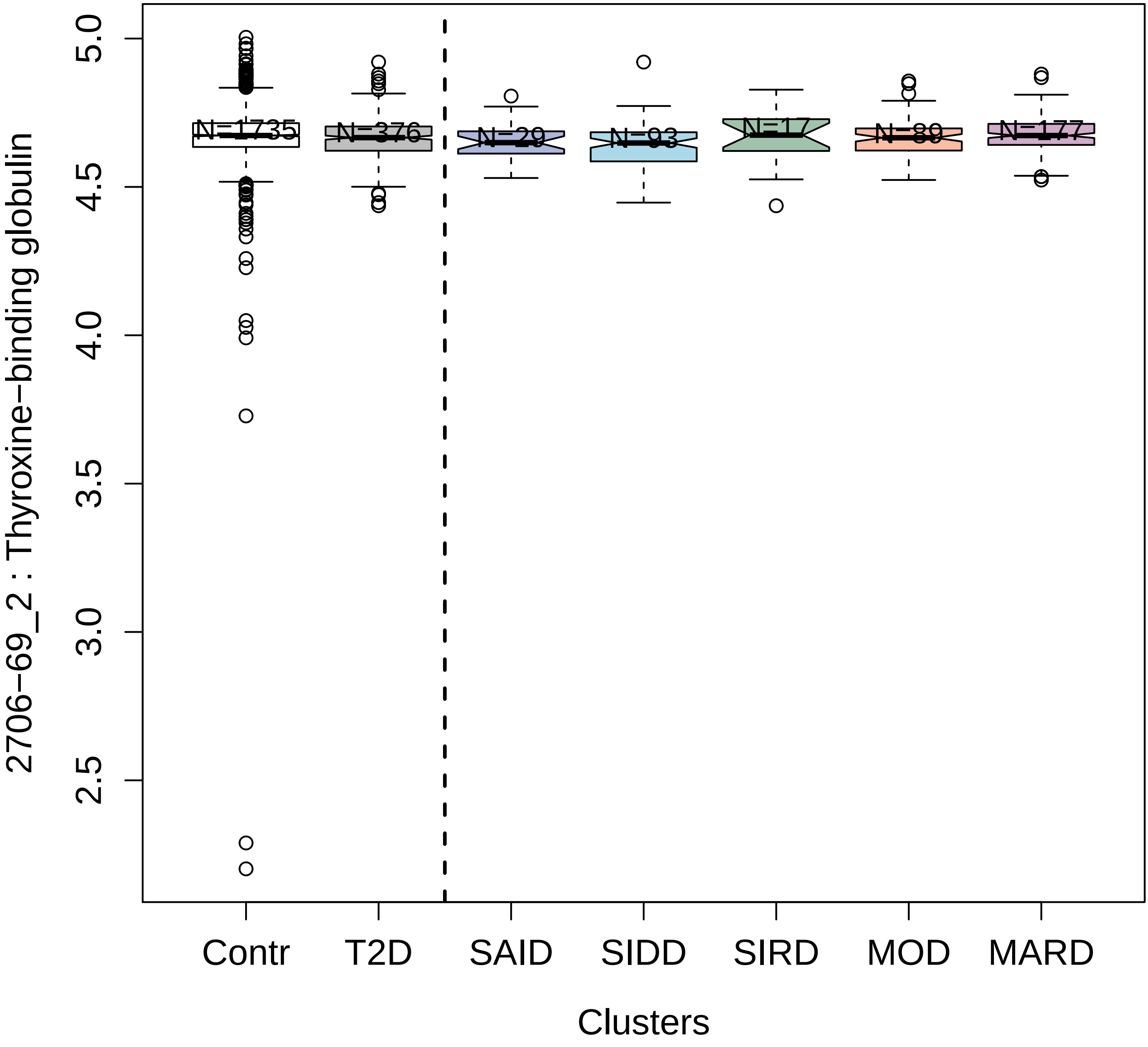
2706−69_2 : Thyroxine−binding globulin
5.0
4.5
4.0
3.5
3.0
2.5
N=1735
N=376
N=29
N=93
N=17
N=89
N=177
Contr
T2D
SAID
SIDD
SIRD
MOD
MARD
Clusters

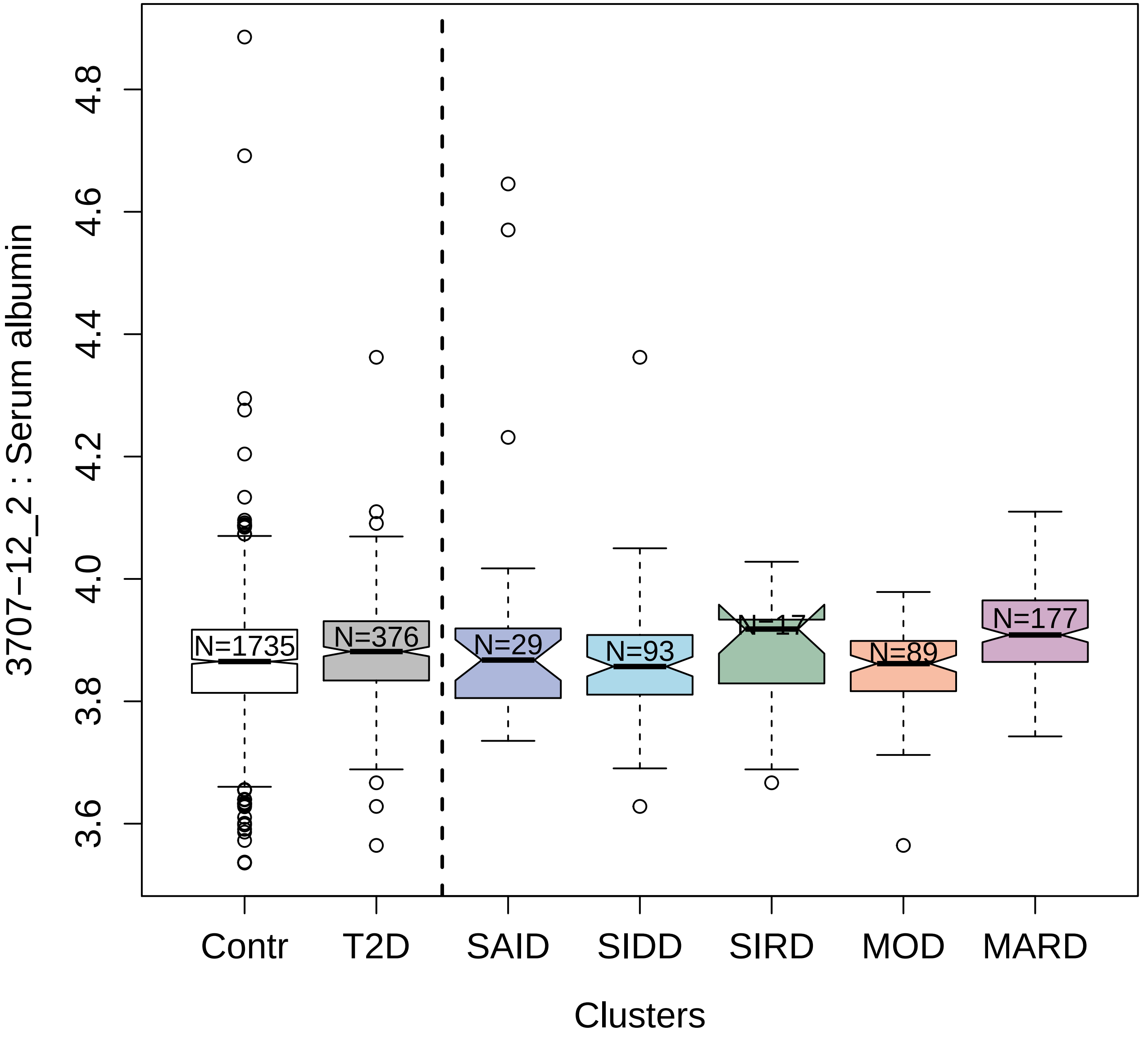
3707−12_2 : Serum albumin
4.8
4.6
4.4
4.2
4.0
3.8
3.6
N=1735
N=376
N=29
N=93
N=89
N=177
Contr
T2D
SAID
SIDD
SIRD
MOD
MARD
Clusters

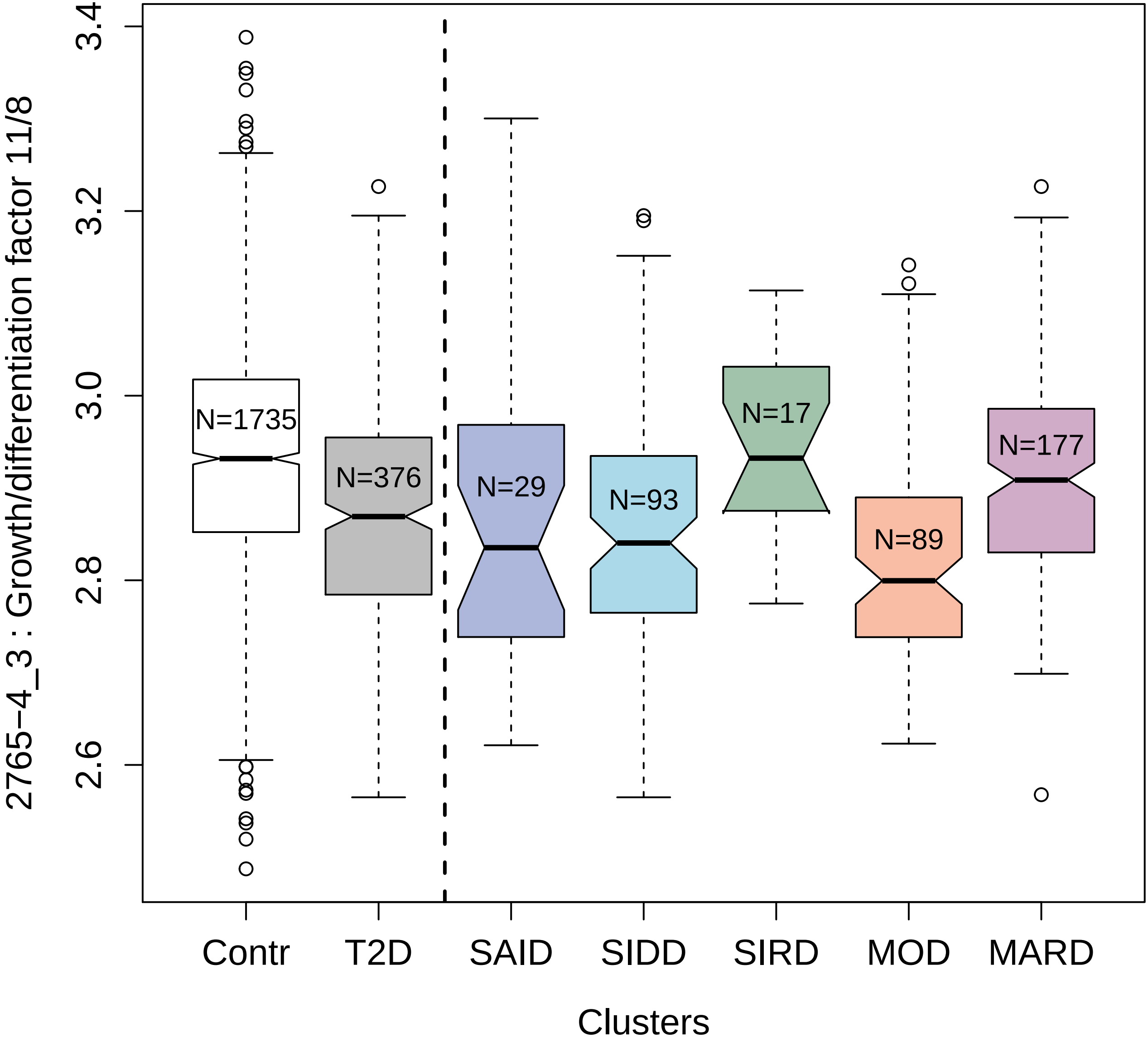
2765−4_3 : Growth/differentiation factor 11/8
3.4
3.2
3.0
2.8
2.6
N=1735
N=376
N=29
N=93
N=17
N=89
N=177
Contr
T2D
SAID
SIDD
SIRD
MOD
MARD
Clusters

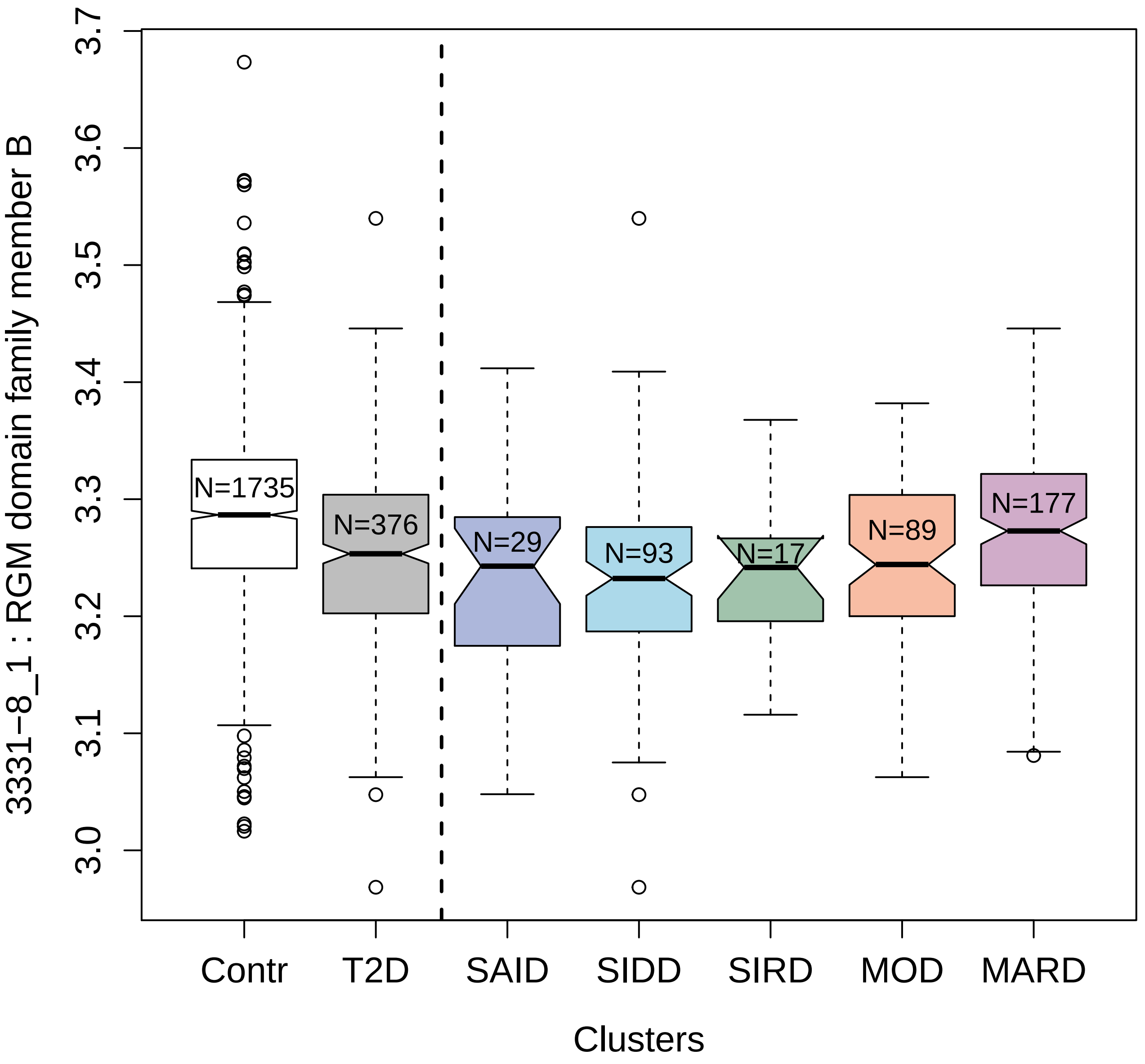
3331−8_1 : RGM domain family member B
3.0
3.1
3.2
3.3
3.4
3.5
3.6
3.7
N=1735
N=376
N=29
N=93
N=17
N=89
N=177
Contr
T2D
SAID
SIDD
SIRD
MOD
MARD
Clusters

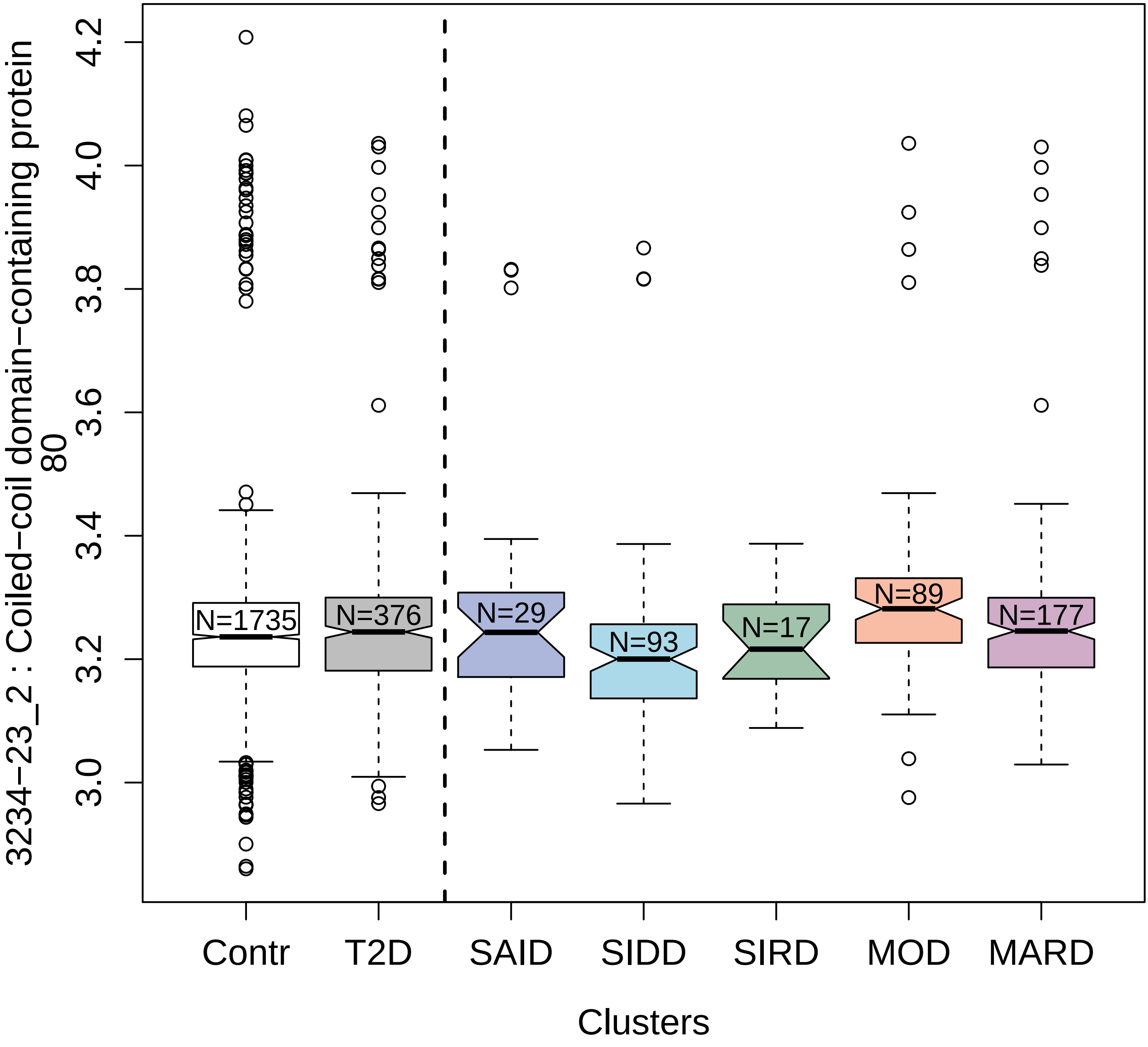

3234−23_2 : Coiled−coil domain−containing protein 80
4.2
4.0
3.8
3.6
3.4
3.2
3.0
N=1735
N=376
N=29
N=93
N=17
N=89
N=177
Contr
T2D
SAID
SIDD
SIRD
MOD
MARD
Clusters

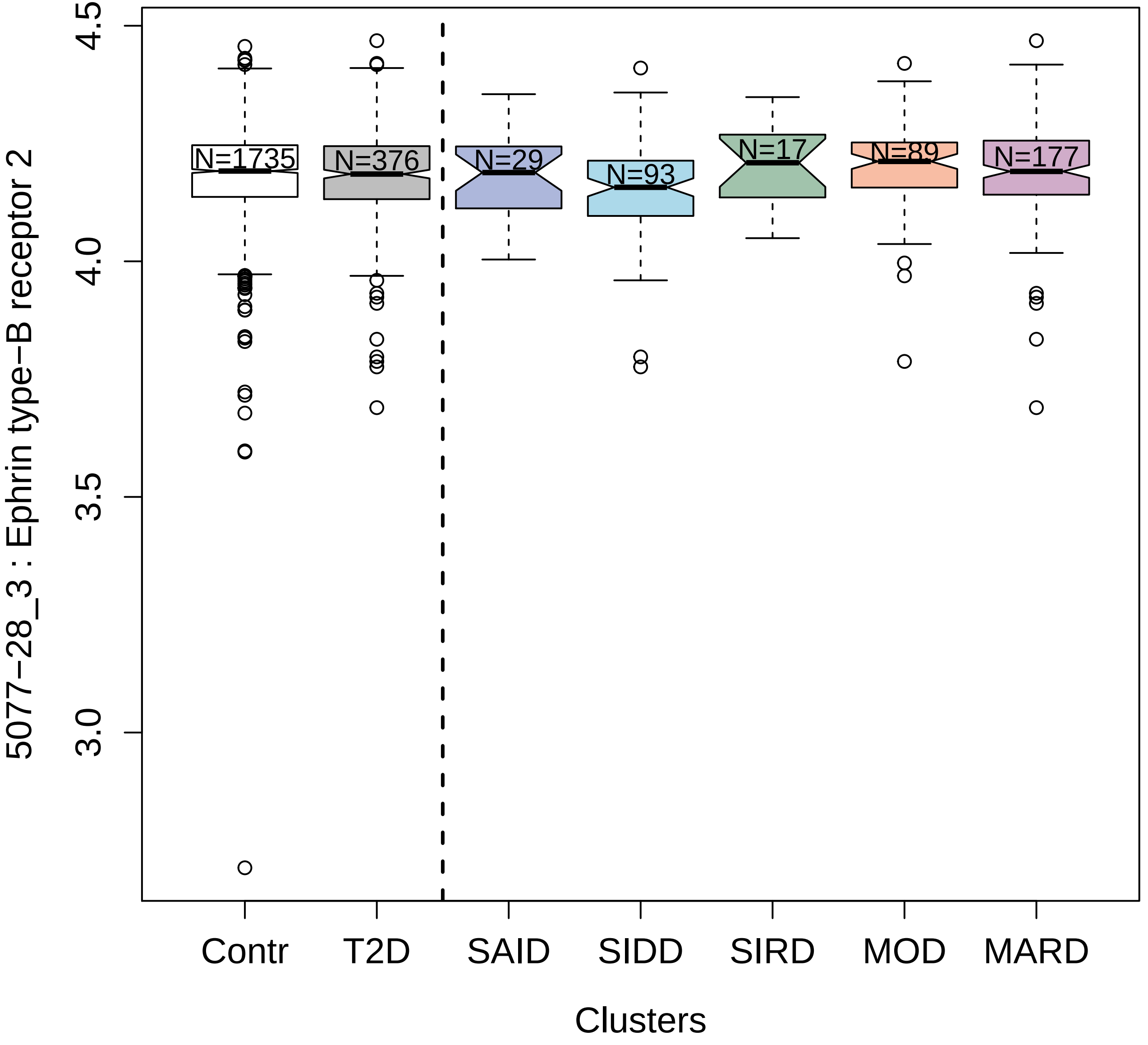

5077−28_3 : Ephrin type−B receptor 2
4.5
4.0
3.5
3.0
N=1735
N=376
N=29
N=93
N=17
N=89
N=177
Contr
T2D
SAID
SIDD
SIRD
MOD
MARD
Clusters

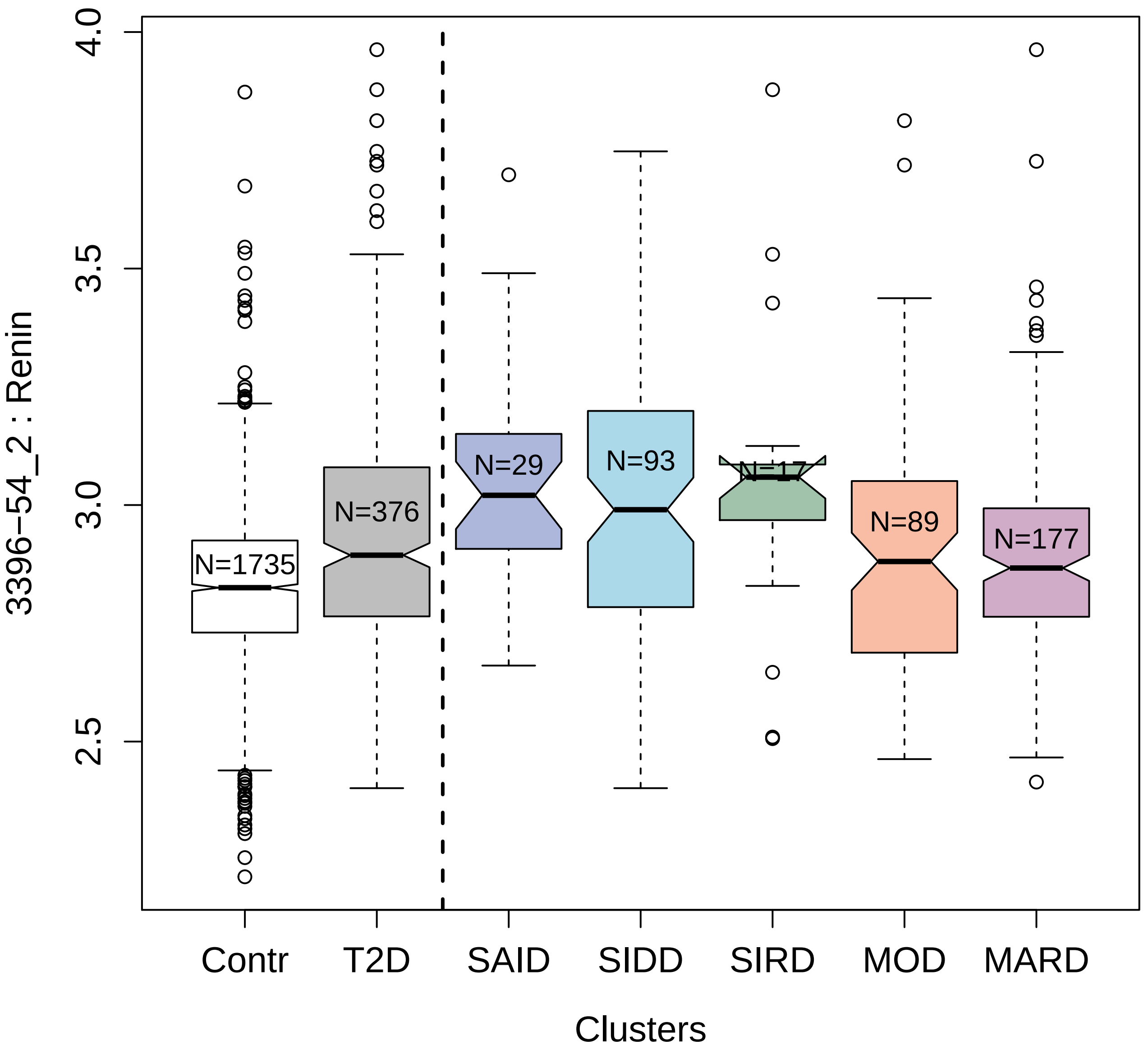
3396−54_2 : Renin
4.0
3.5
3.0
2.5
N=1735
N=376
N=29
N=93
N=89
N=177
Contr
T2D
SAID
SIDD
SIRD
MOD
MARD
Clusters

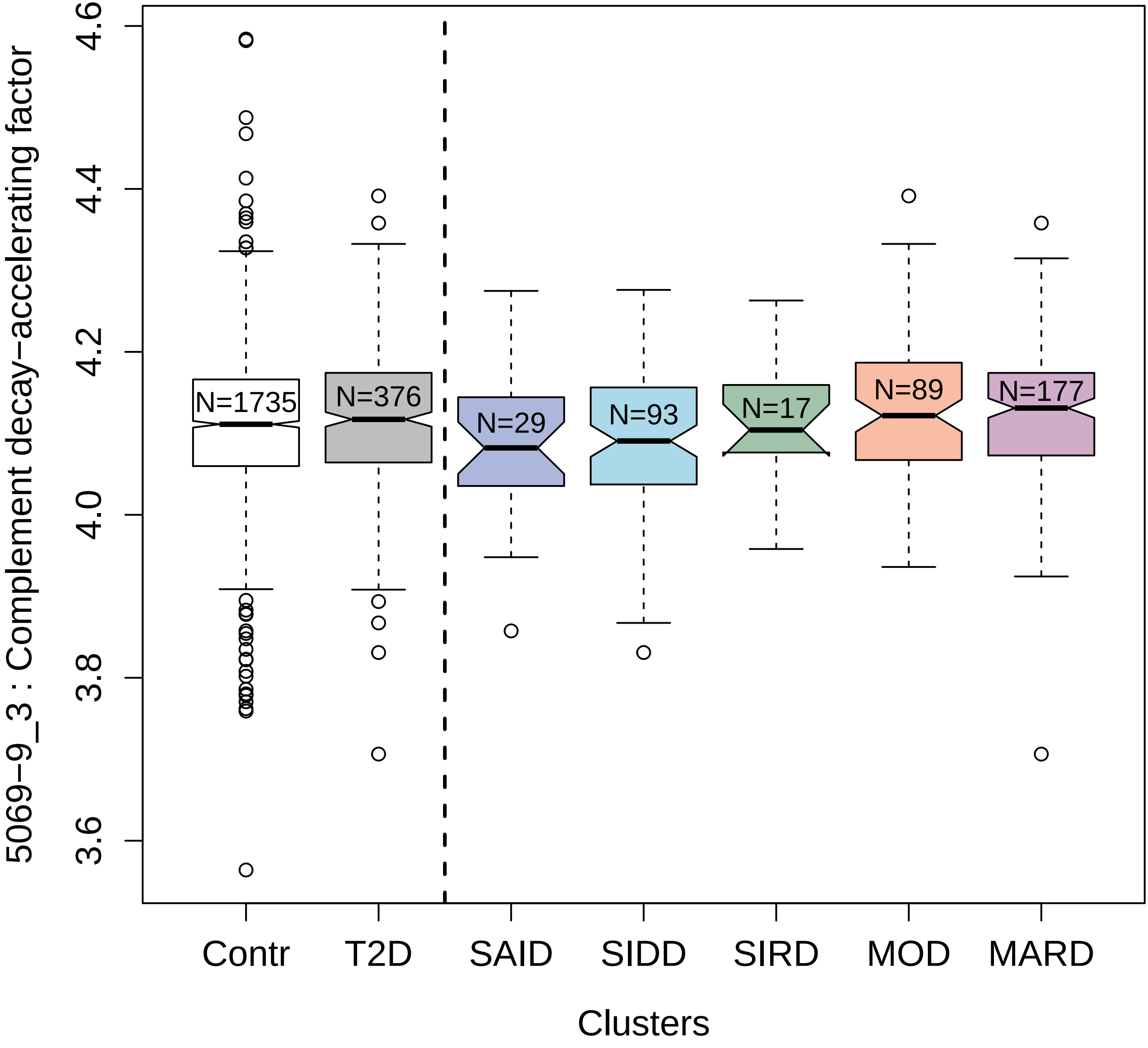
5069−9_3 : Complement decay−accelerating factor
4.6
4.4
4.2
4.0
3.8
3.6
N=1735
N=376
N=29
N=93
N=17
N=89
N=177
Contr
T2D
SAID
SIDD
SIRD
MOD
MARD
Clusters

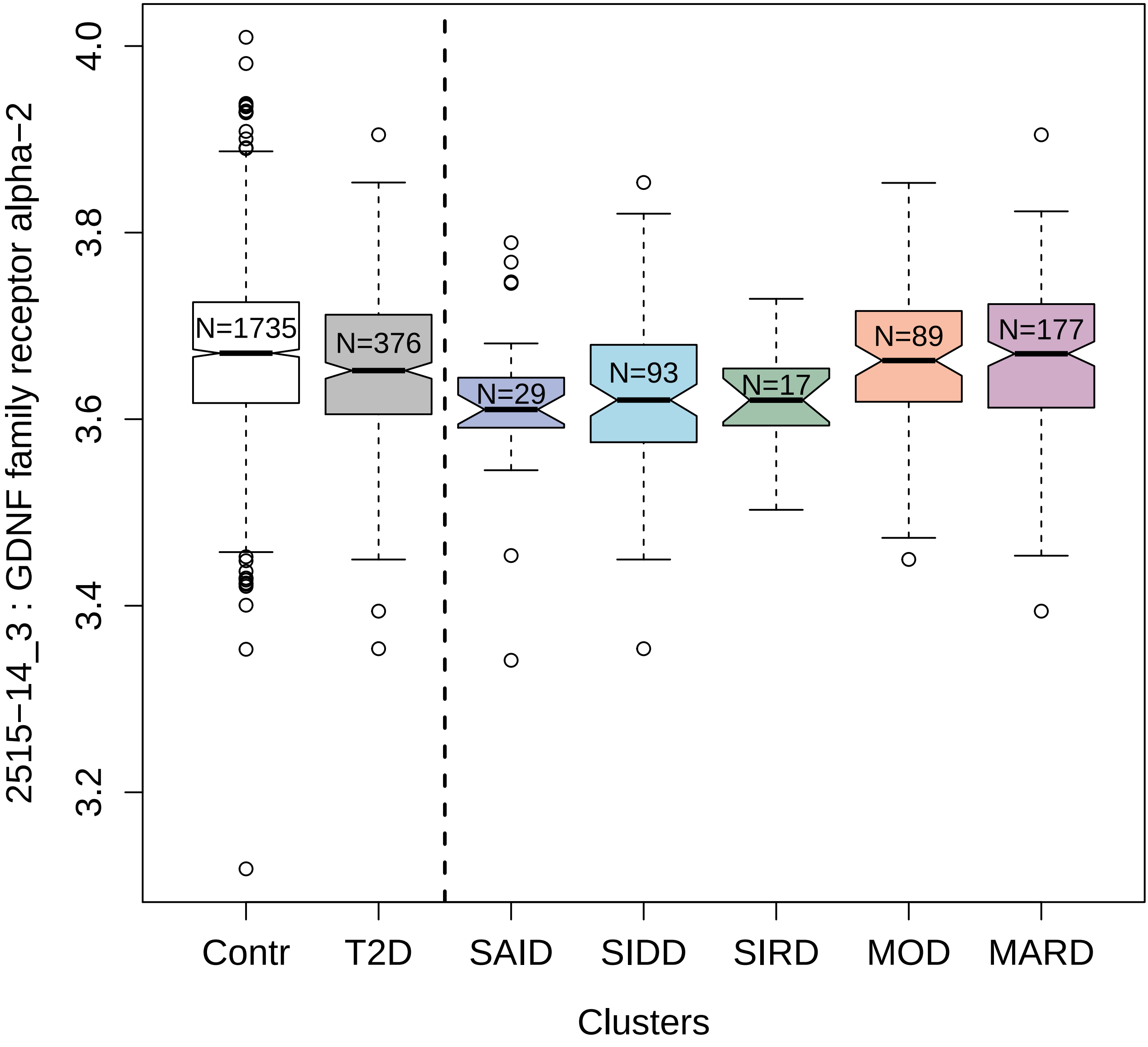

2515−14_3 : GDNF family receptor alpha−2
3.2
3.4
3.6
3.8
4.0
N=1735
N=376
N=29
N=93
N=17
N=89
N=177
Contr
T2D
SAID
SIDD
SIRD
MOD
MARD
Clusters

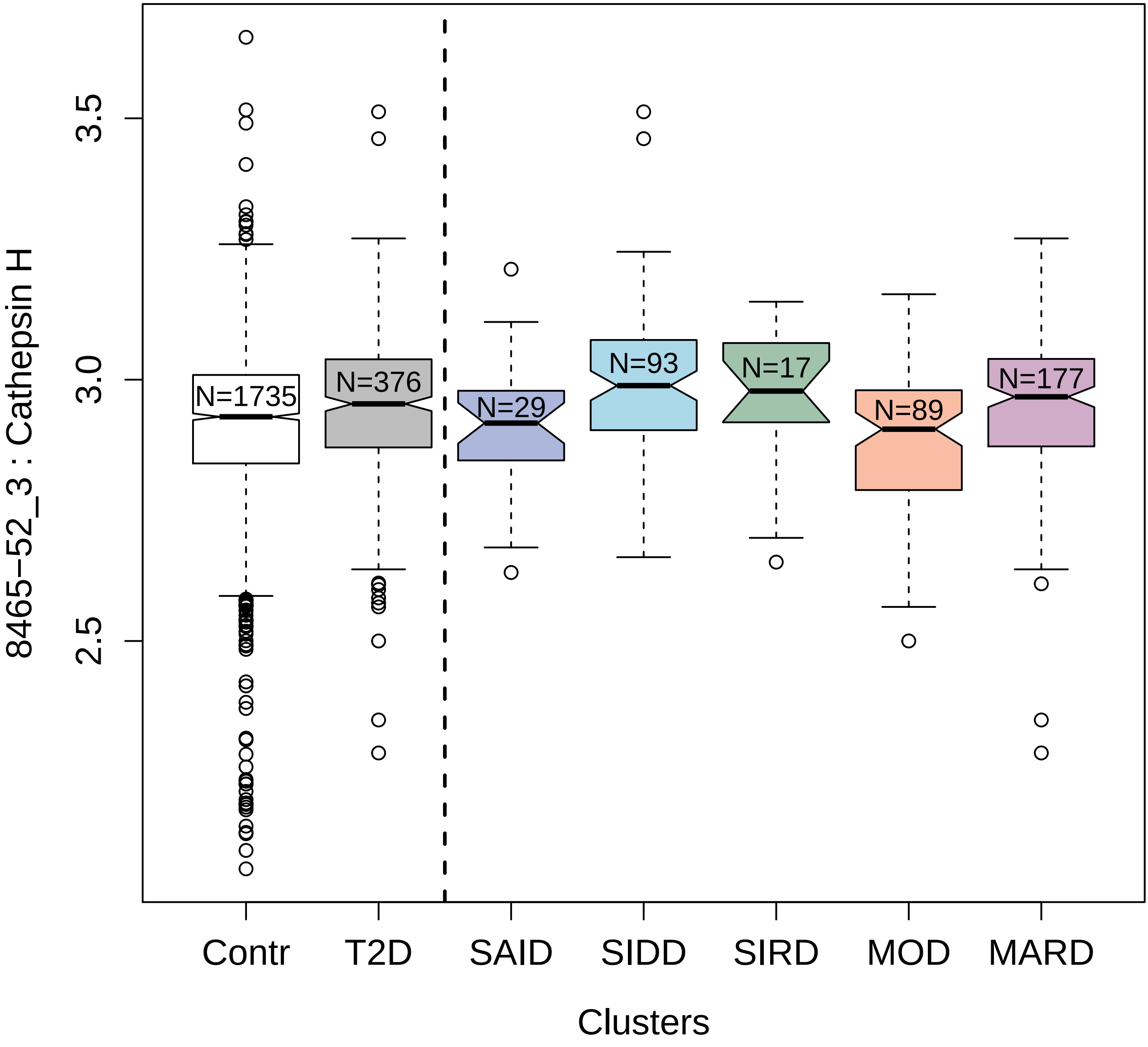

8465−52_3 : Cathepsin H
3.5
3.0
2.5
N=1735
N=376
N=29
N=93
N=17
N=89
N=177
Contr
T2D
SAID
SIDD
SIRD
MOD
MARD
Clusters

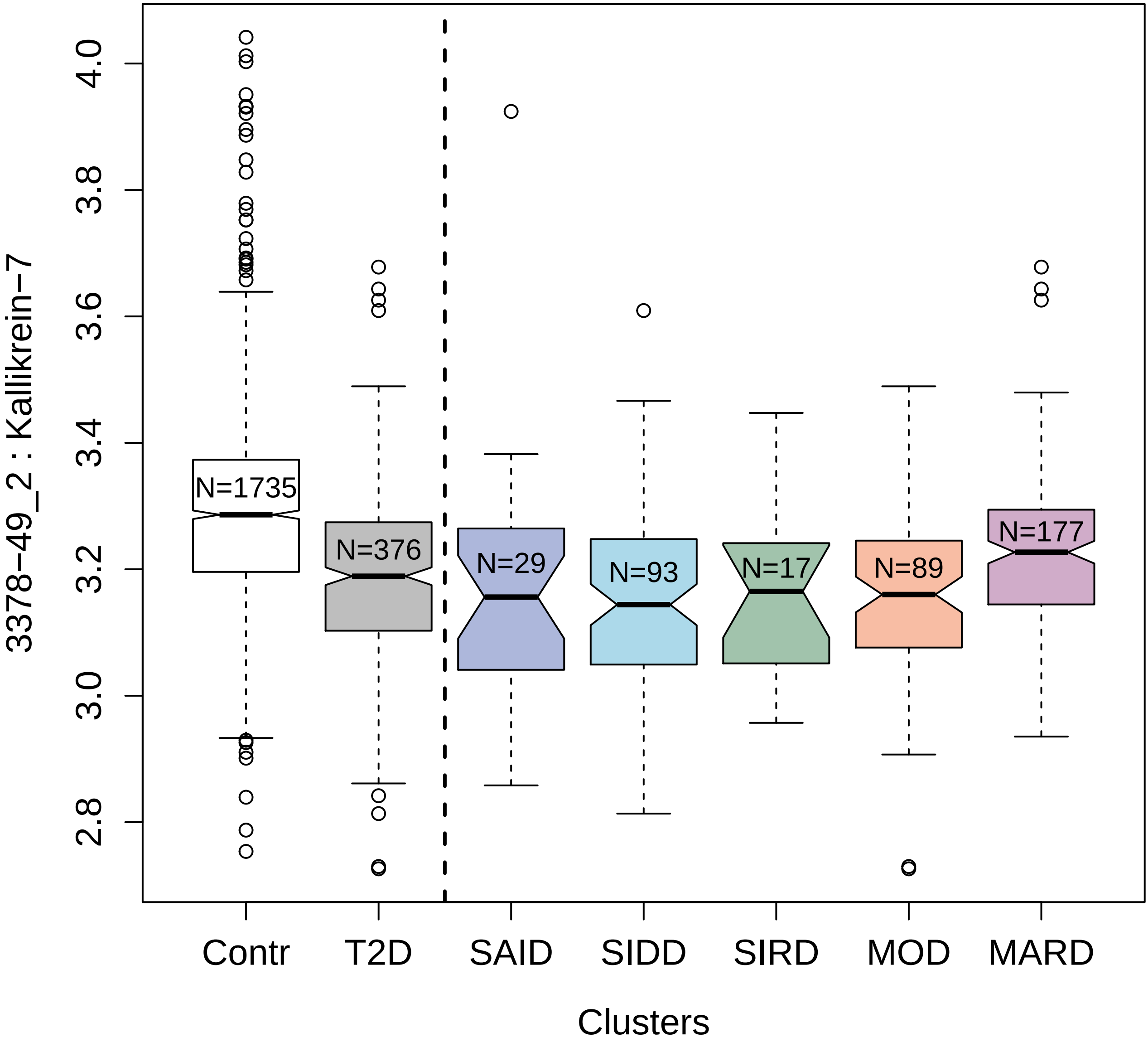

3378−49_2 : Kallikrein−7
4.0
3.8
3.6
3.4
3.2
3.0
2.8
N=1735
N=376
N=29
N=93
N=17
N=89
N=177
Contr
T2D
SAID
SIDD
SIRD
MOD
MARD
Clusters

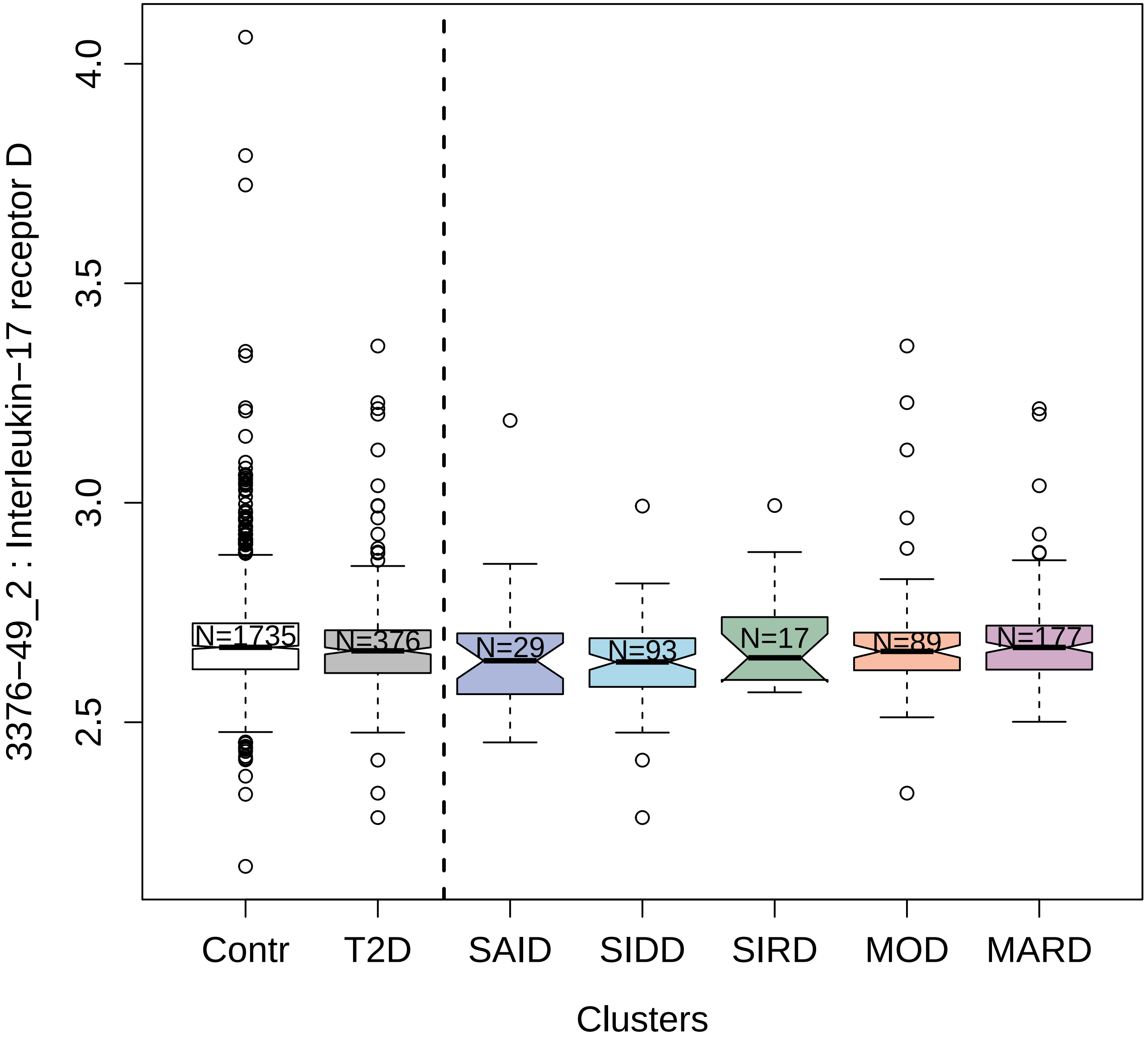

3376−49_2 : Interleukin−17 receptor D
4.0
3.5
3.0
2.5
N=1735
N=376
N=29
N=93
N=17
N=89
N=177
Contr
T2D
SAID
SIDD
SIRD
MOD
MARD
Clusters

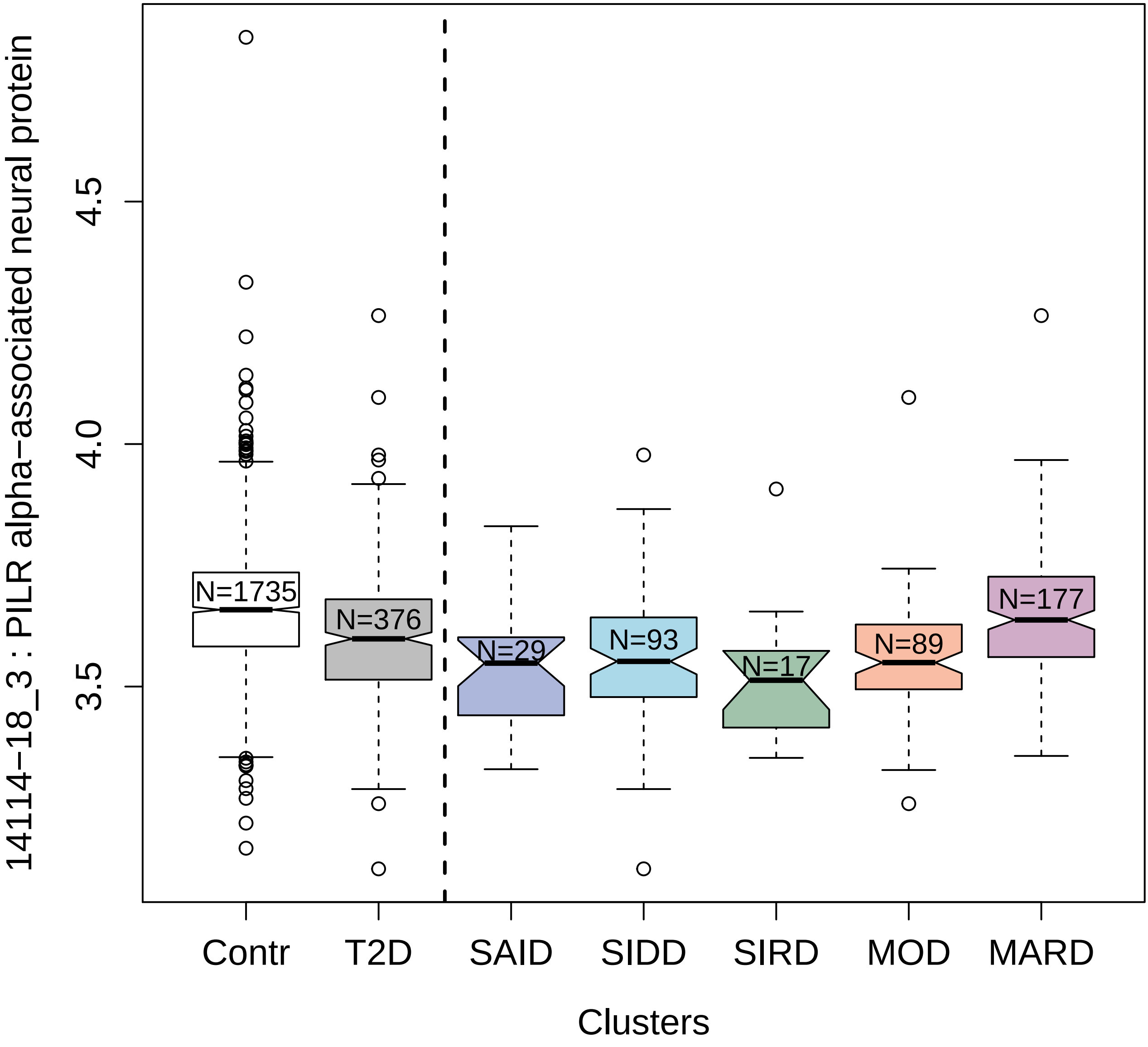

14114−18_3 : PILR alpha−associated neural protein
4.5
4.0
3.5
N=1735
N=376
N=29
N=93
N=17
N=89
N=177
Contr
T2D
SAID
SIDD
SIRD
MOD
MARD
Clusters

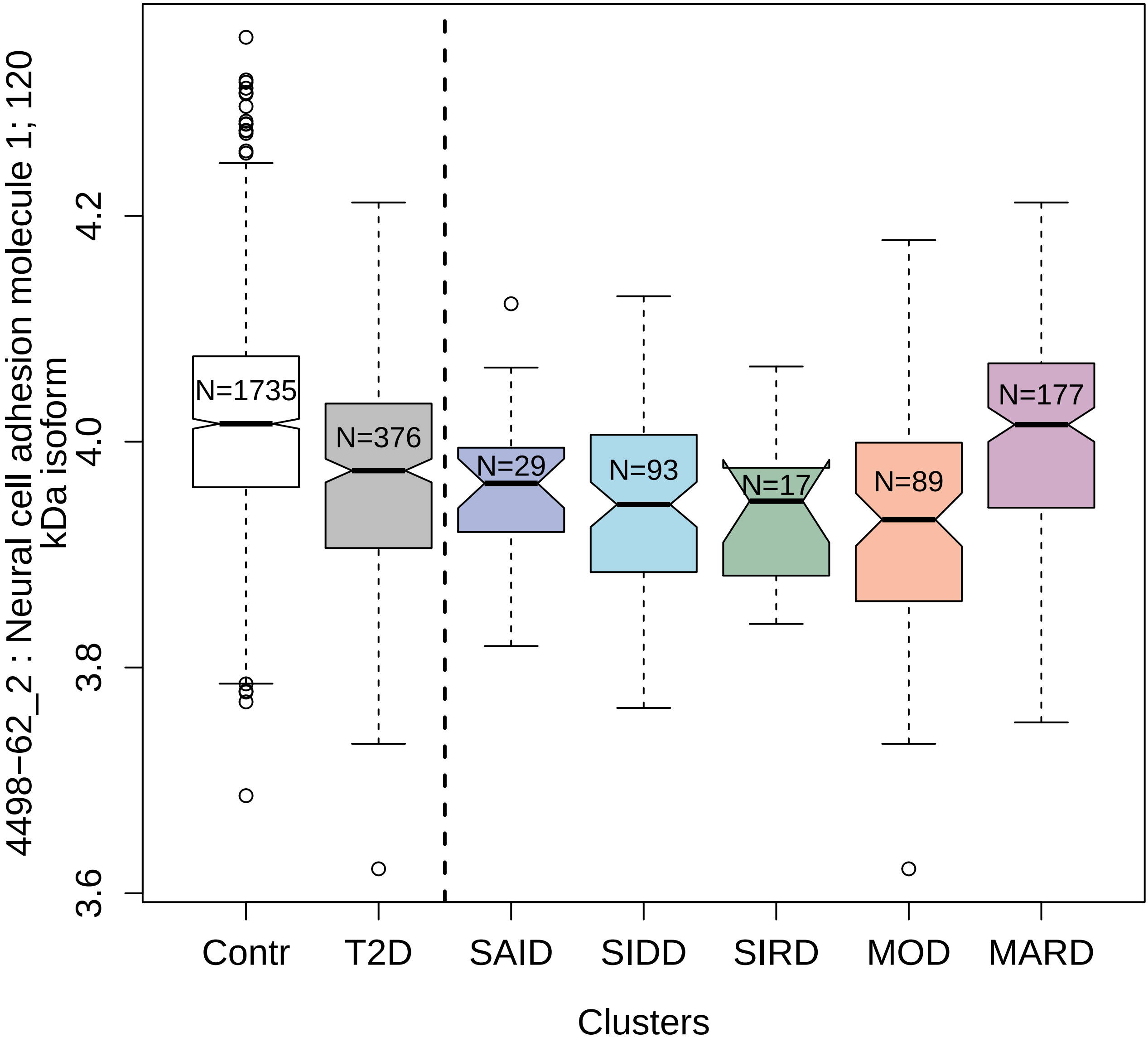
4498−62_2 : Neural cell adhesion molecule 1; 120 kDa isoform
4.2
4.0
3.8
3.6
N=1735
N=376
N=29
N=93
N=17
N=89
N=177
Contr
T2D
SAID
SIDD
SIRD
MOD
MARD
Clusters

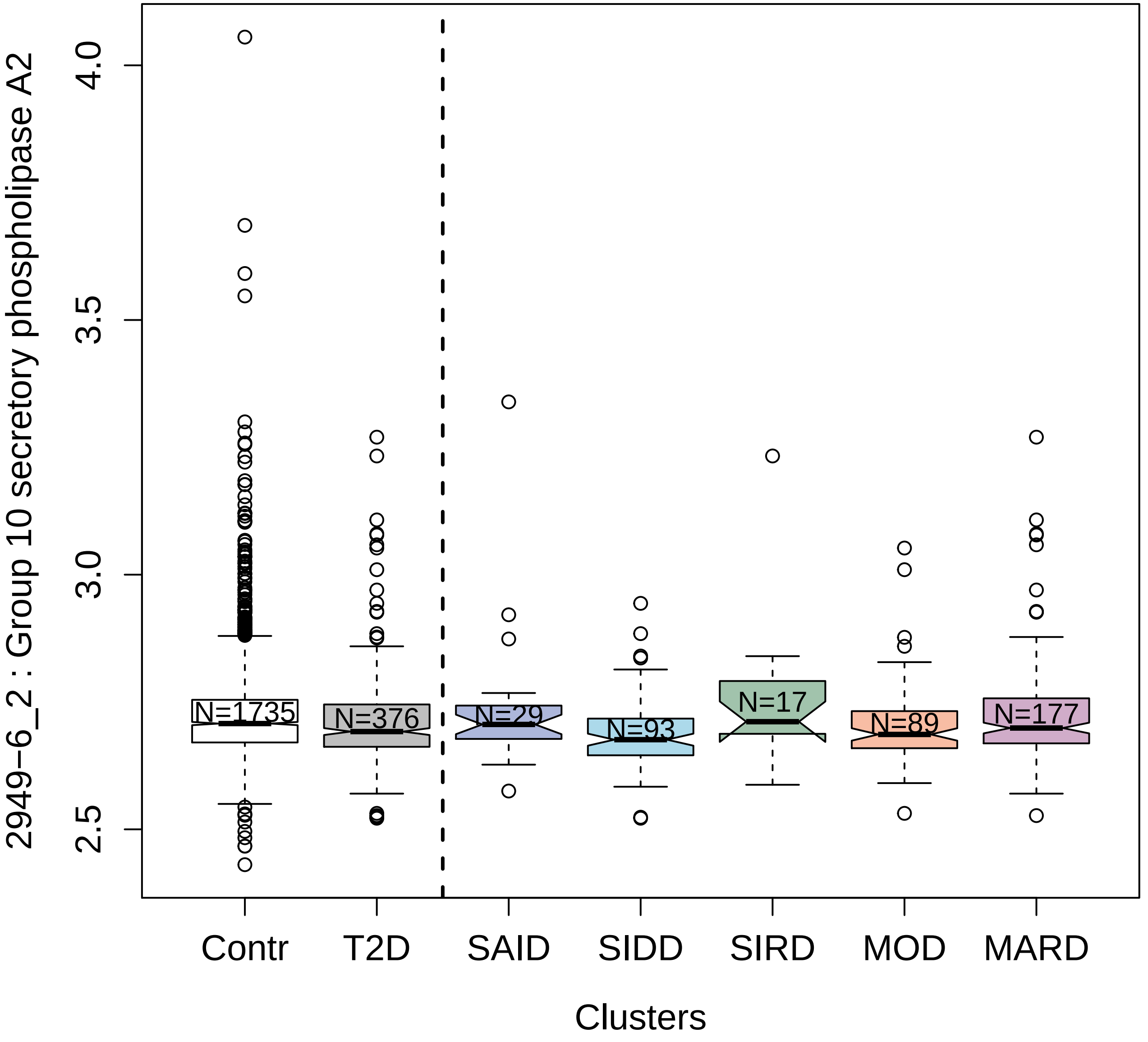
2949−6_2 : Group 10 secretory phospholipase A2
4.0
3.5
3.0
2.5
N=1735
N=376
N=29
N=93
N=17
N=89
N=177
Contr
T2D
SAID
SIDD
SIRD
MOD
MARD
Clusters

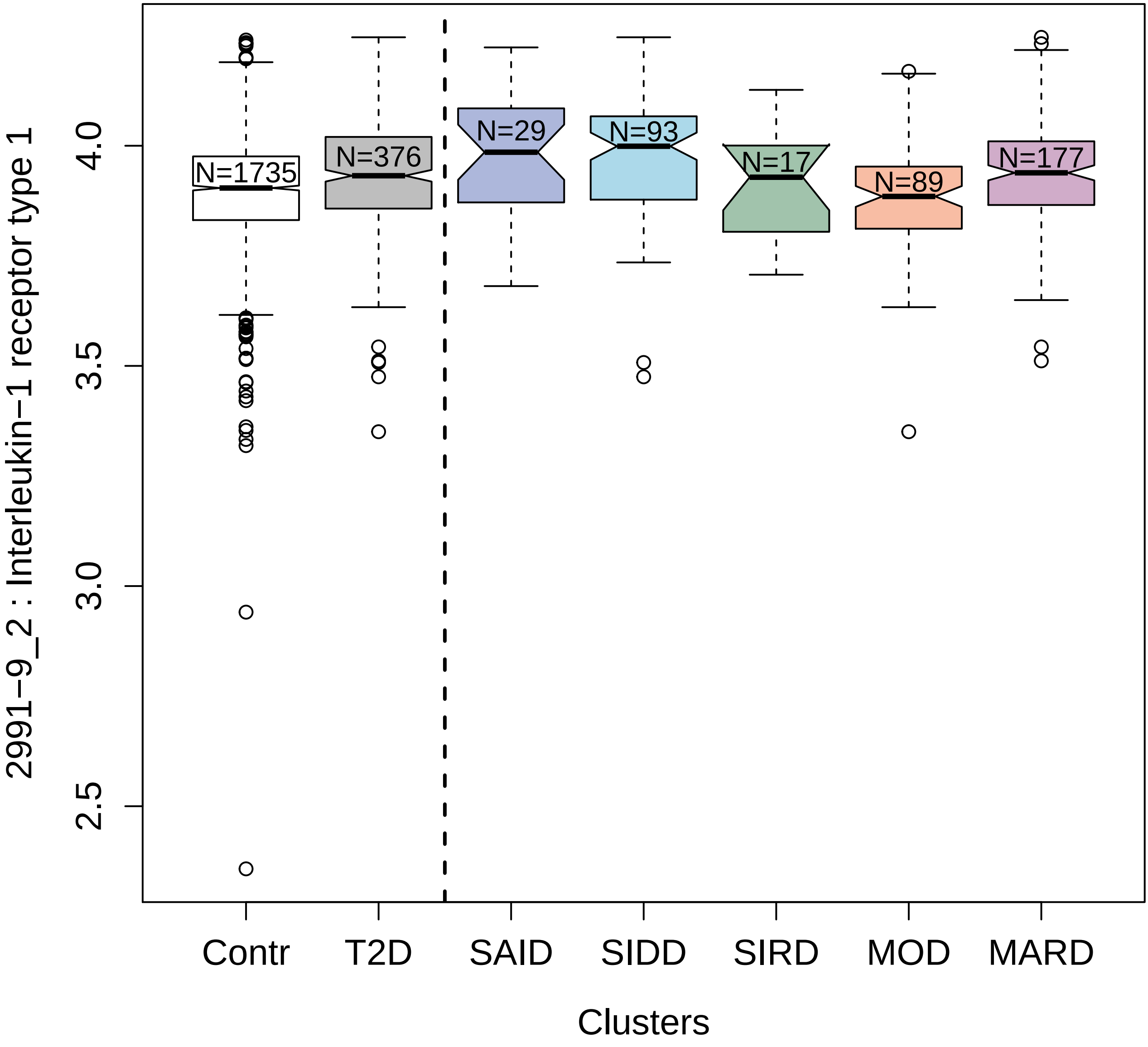

2991−9_2 : Interleukin−1 receptor type 1
2.5
3.0
3.5
4.0
N=1735
N=376
N=29
N=93
N=17
N=89
N=177
Contr
T2D
SAID
SIDD
SIRD
MOD
MARD
Clusters

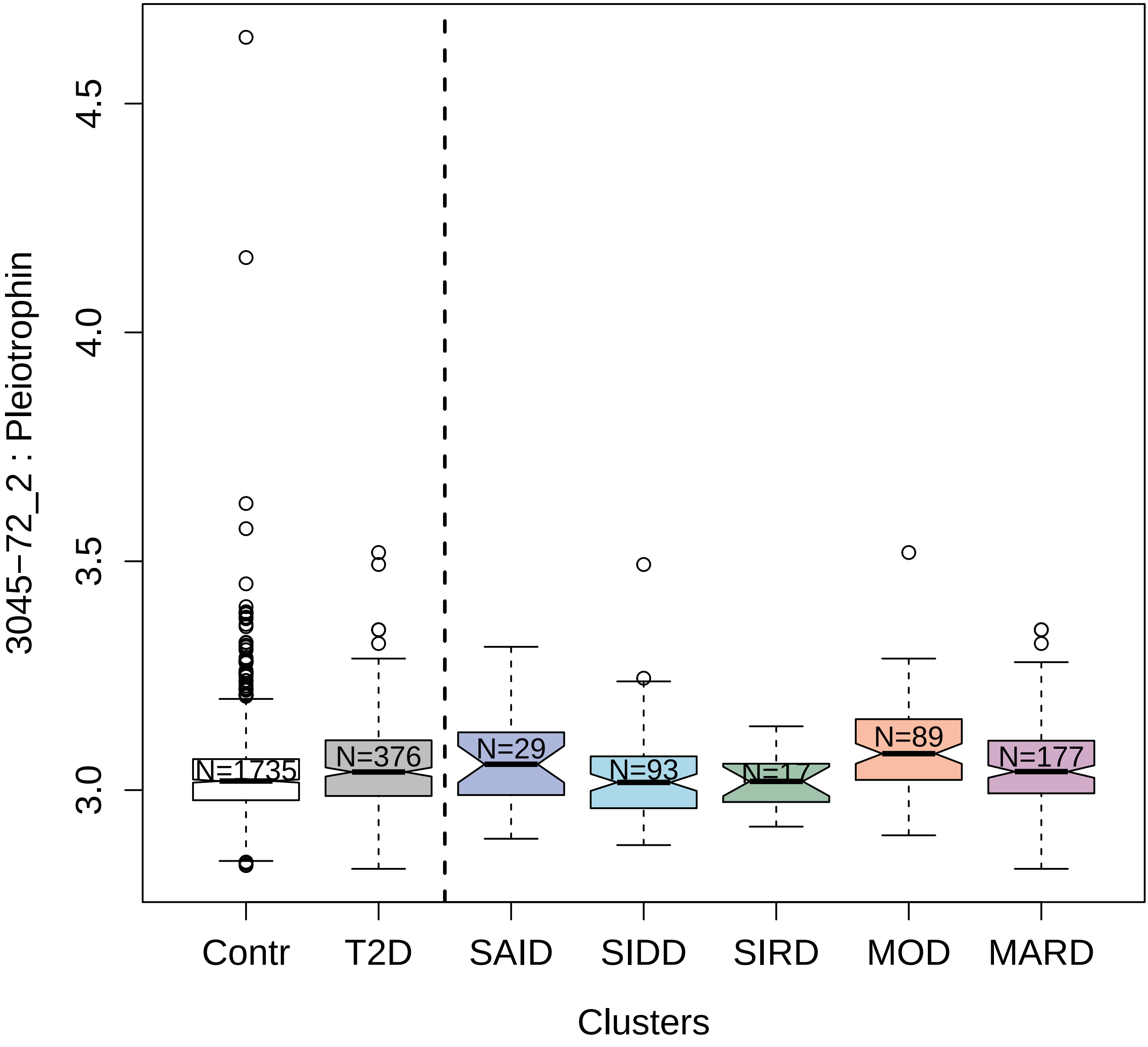

3045−72_2 : Pleiotrophin
4.5
4.0
3.5
3.0
N=1735
N=376
N=29
N=93
N=17
N=89
N=177
Contr
T2D
SAID
SIDD
SIRD
MOD
MARD
Clusters

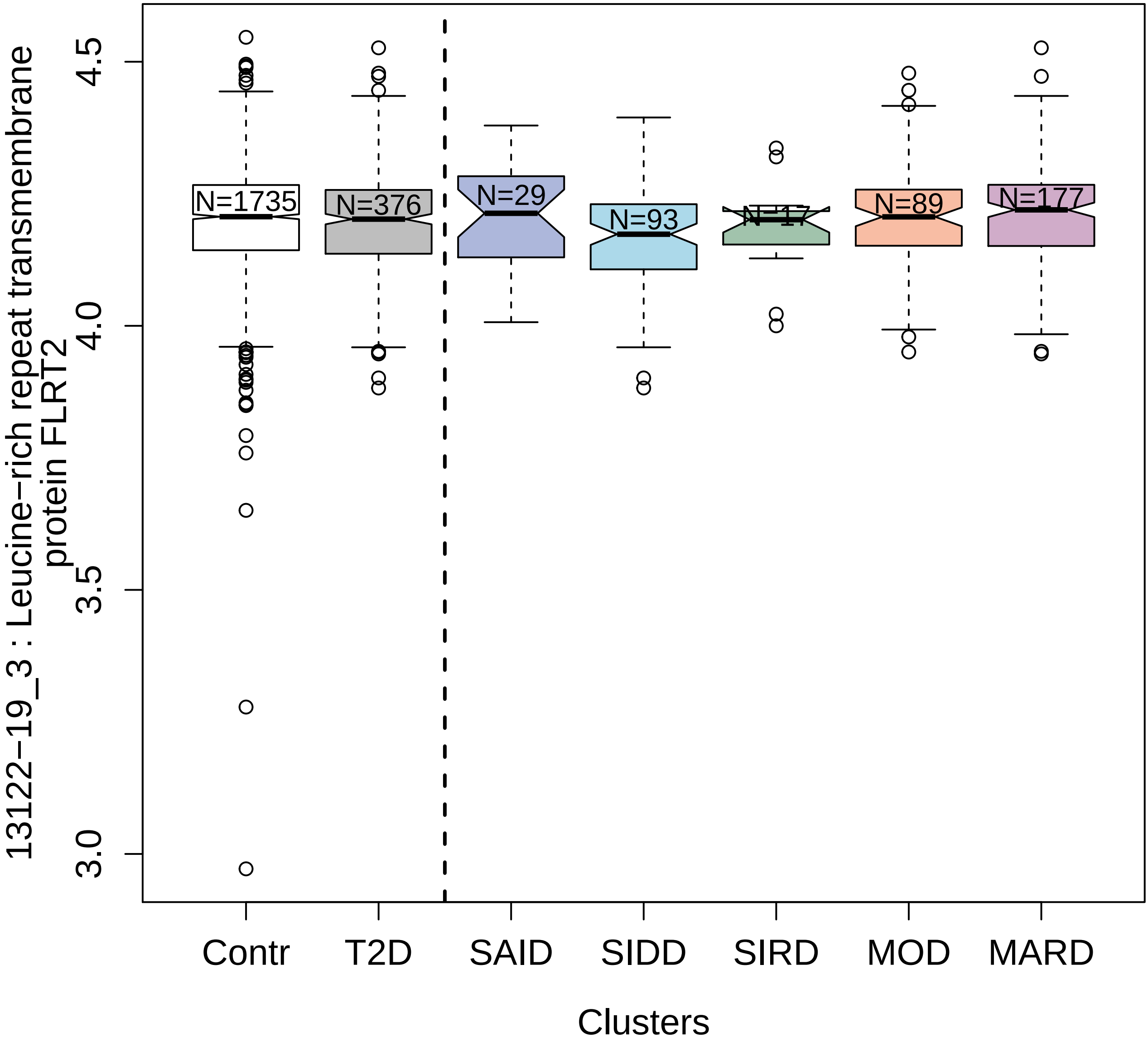
13122−19_3 : Leucine−rich repeat transmembrane protein FLRT2
3.0
3.5
4.0
4.5
N=1735
N=376
N=29
N=93
N=17
N=89
N=177
Contr
T2D
SAID
SIDD
SIRD
MOD
MARD
Clusters

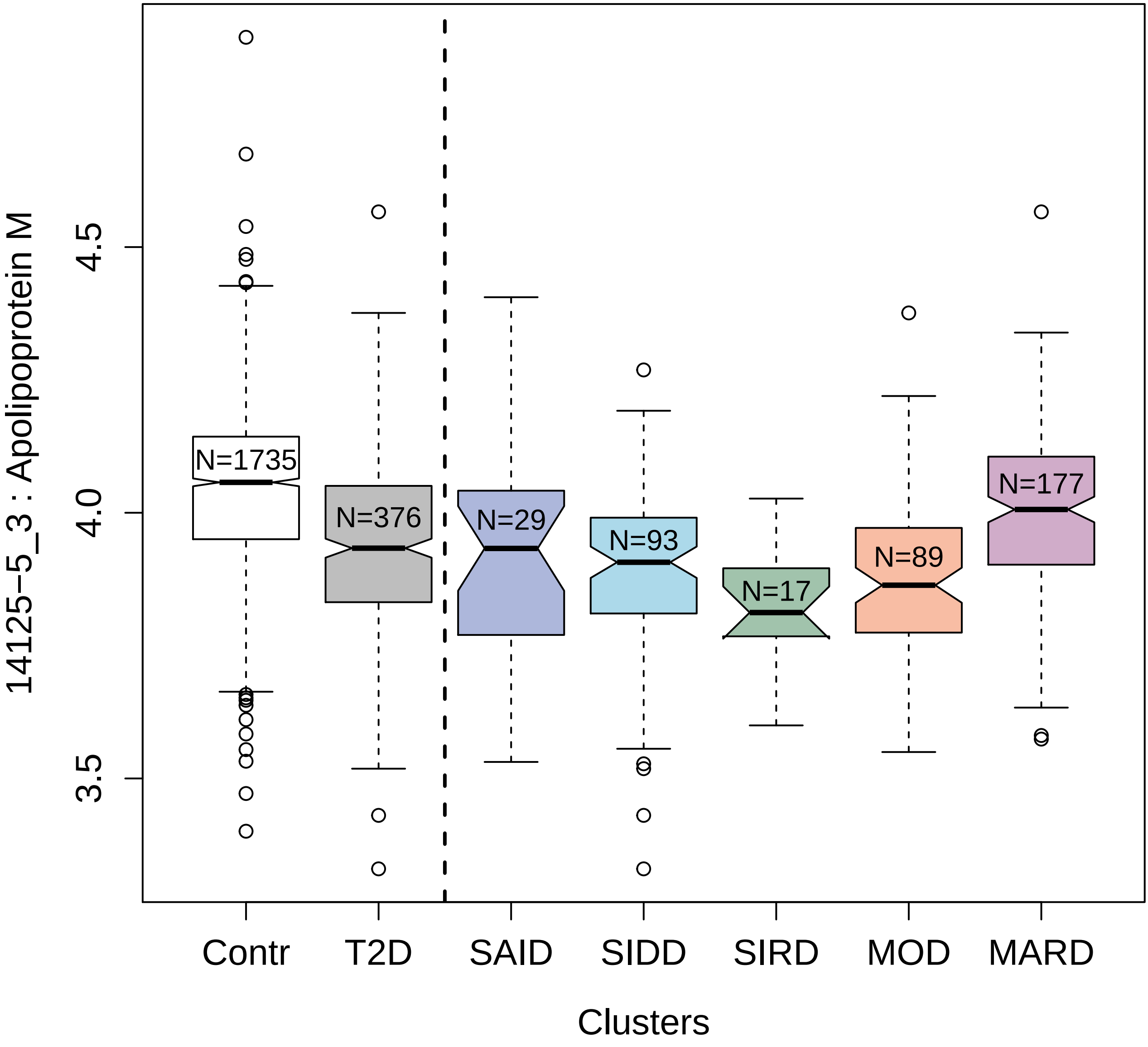

14125−5_3 : Apolipoprotein M
3.5
4.0
4.5
N=1735
N=376
N=29
N=93
N=17
N=89
N=177
Contr
T2D
SAID
SIDD
SIRD
MOD
MARD
Clusters

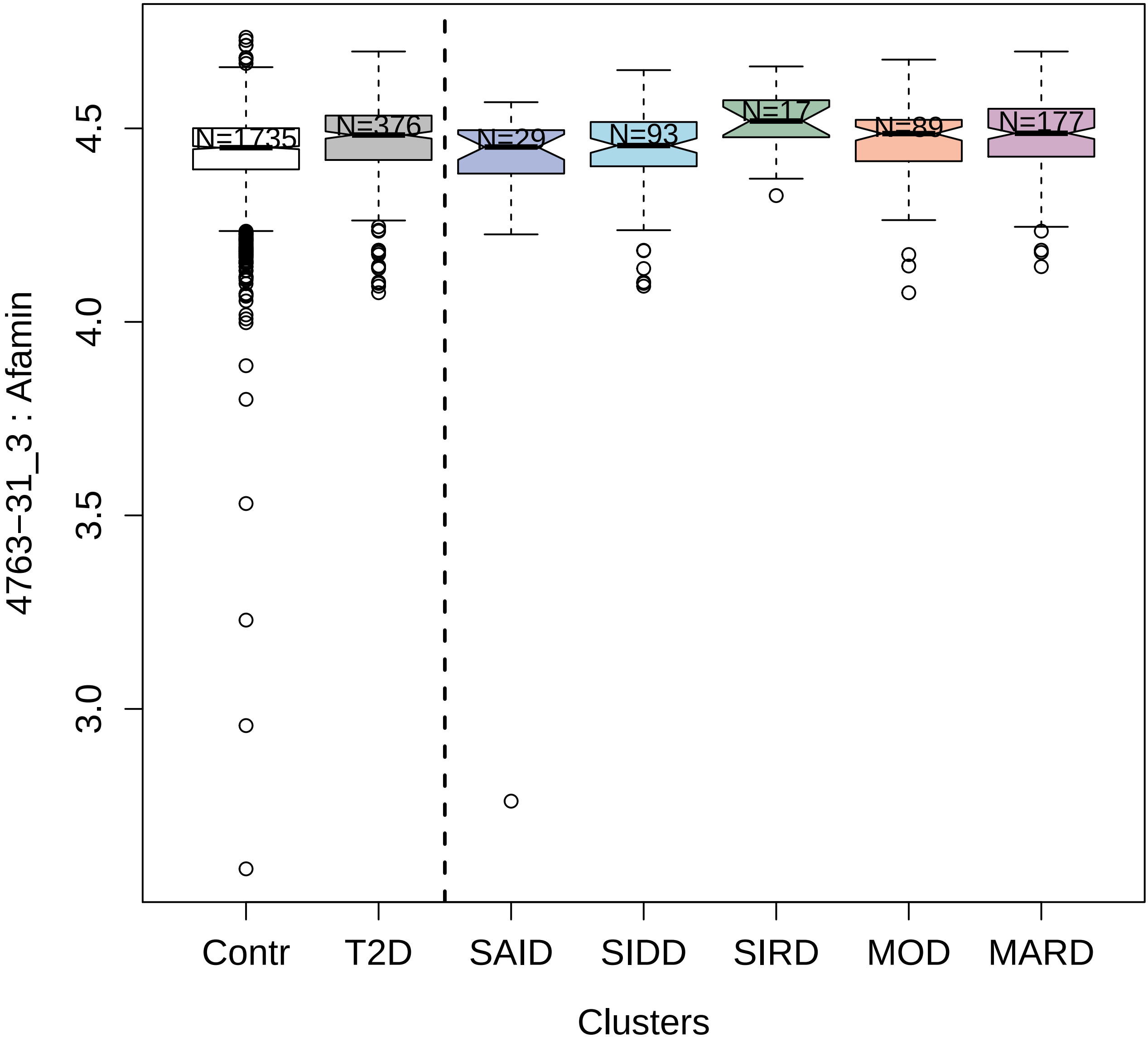
4763−31_3 : Afamin
4.5
4.0
3.5
3.0
N=1735
N=376
N=29
N=93
N=17
N=89
N=177
Contr
T2D
SAID
SIDD
SIRD
MOD
MARD
Clusters

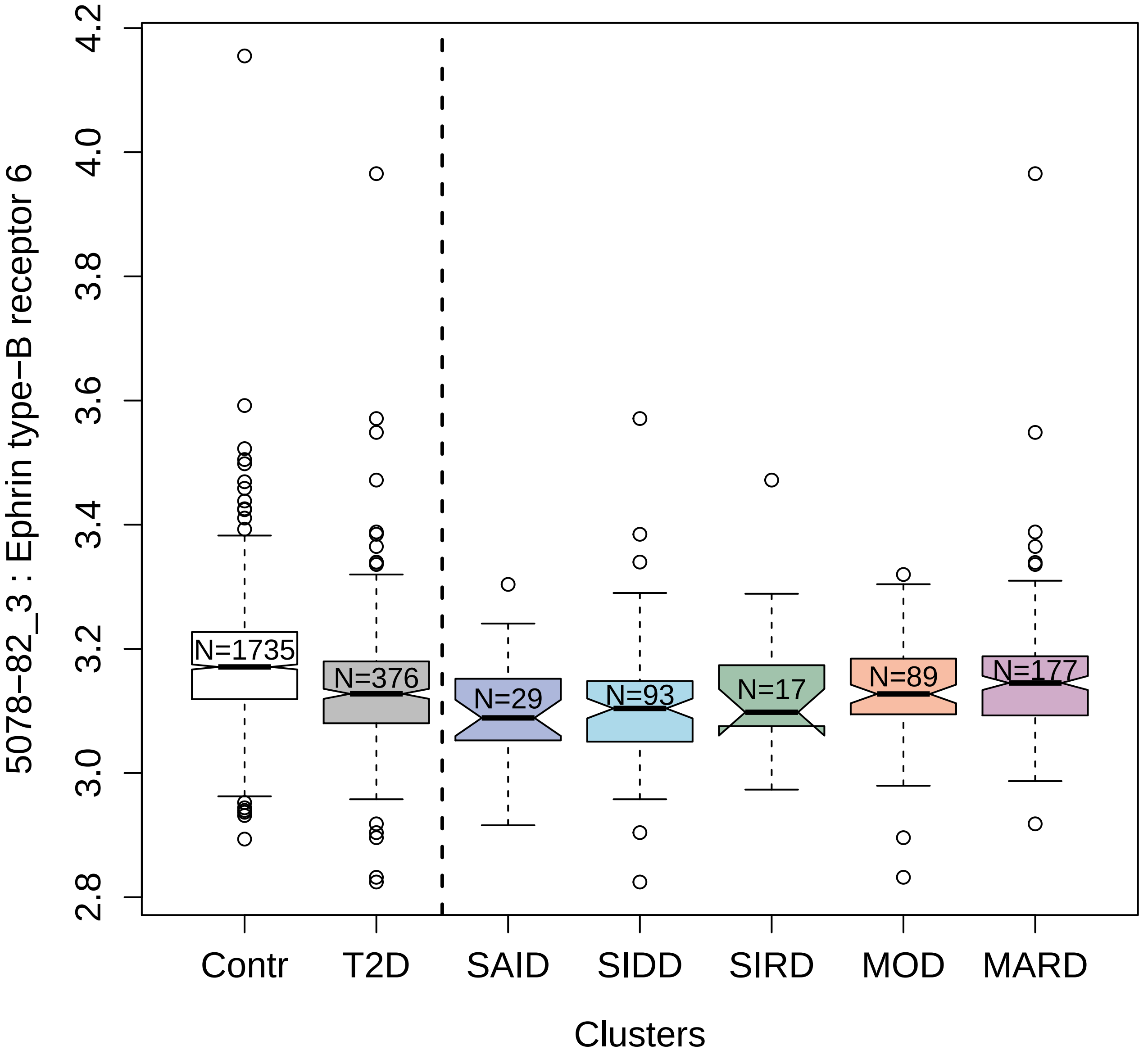

5078−82_3 : Ephrin type−B receptor 6
4.2
4.0
3.8
3.6
3.4
3.2
3.0
2.8
N=1735
N=376
N=29
N=93
N=17
N=89
N=177
Contr
T2D
SAID
SIDD
SIRD
MOD
MARD
Clusters

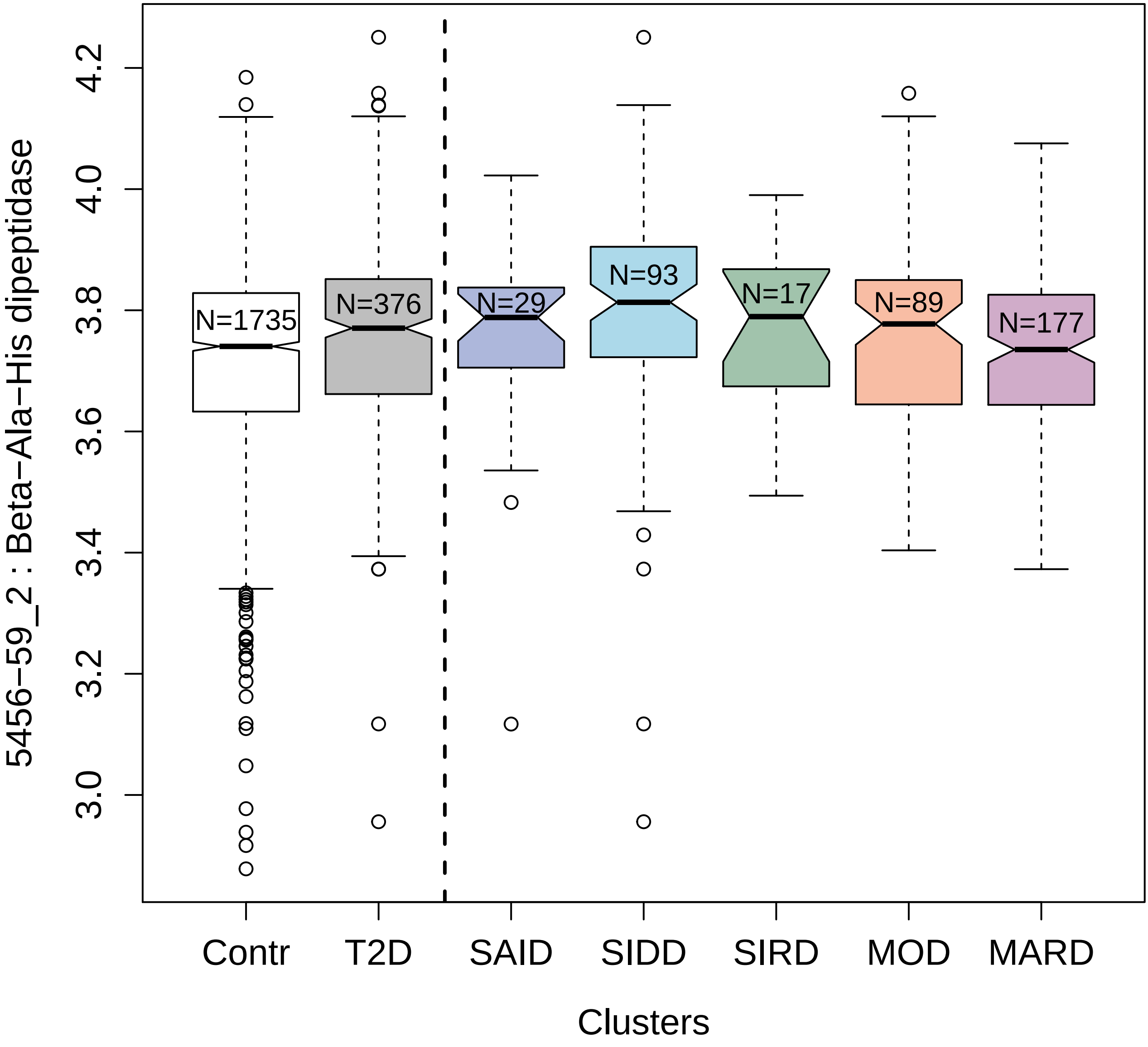

5456−59_2 : Beta−Ala−His dipeptidase
3.0
3.2
3.4
3.6
3.8
4.0
4.2
N=1735
N=376
N=29
N=93
N=17
N=89
N=177
Contr
T2D
SAID
SIDD
SIRD
MOD
MARD
Clusters

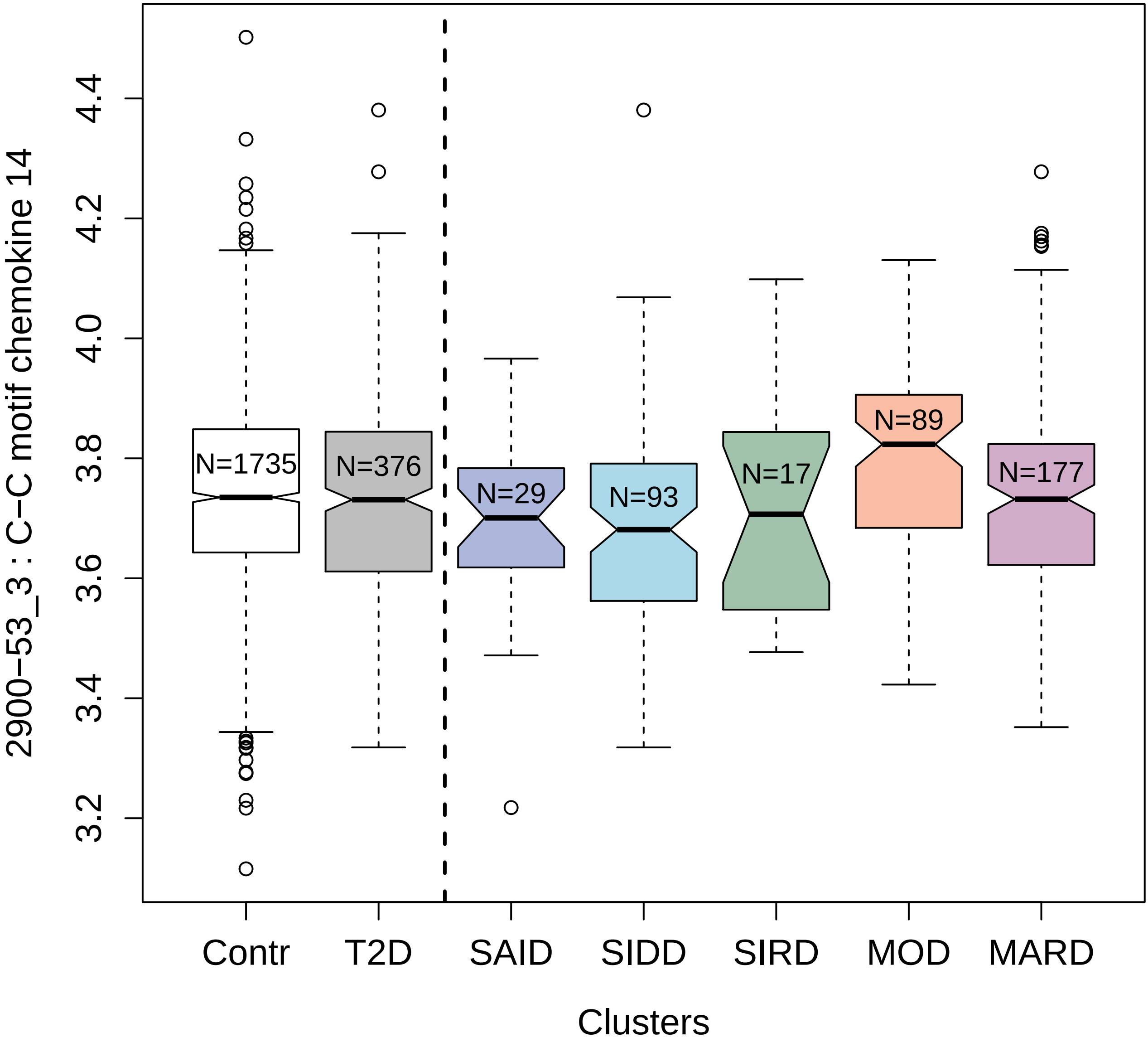
2900−53_3 : C−C motif chemokine 14
4.4
4.2
4.0
3.8
3.6
3.4
3.2
N=1735
N=376
N=29
N=93
N=17
N=89
N=177
Contr
T2D
SAID
SIDD
SIRD
MOD
MARD
Clusters

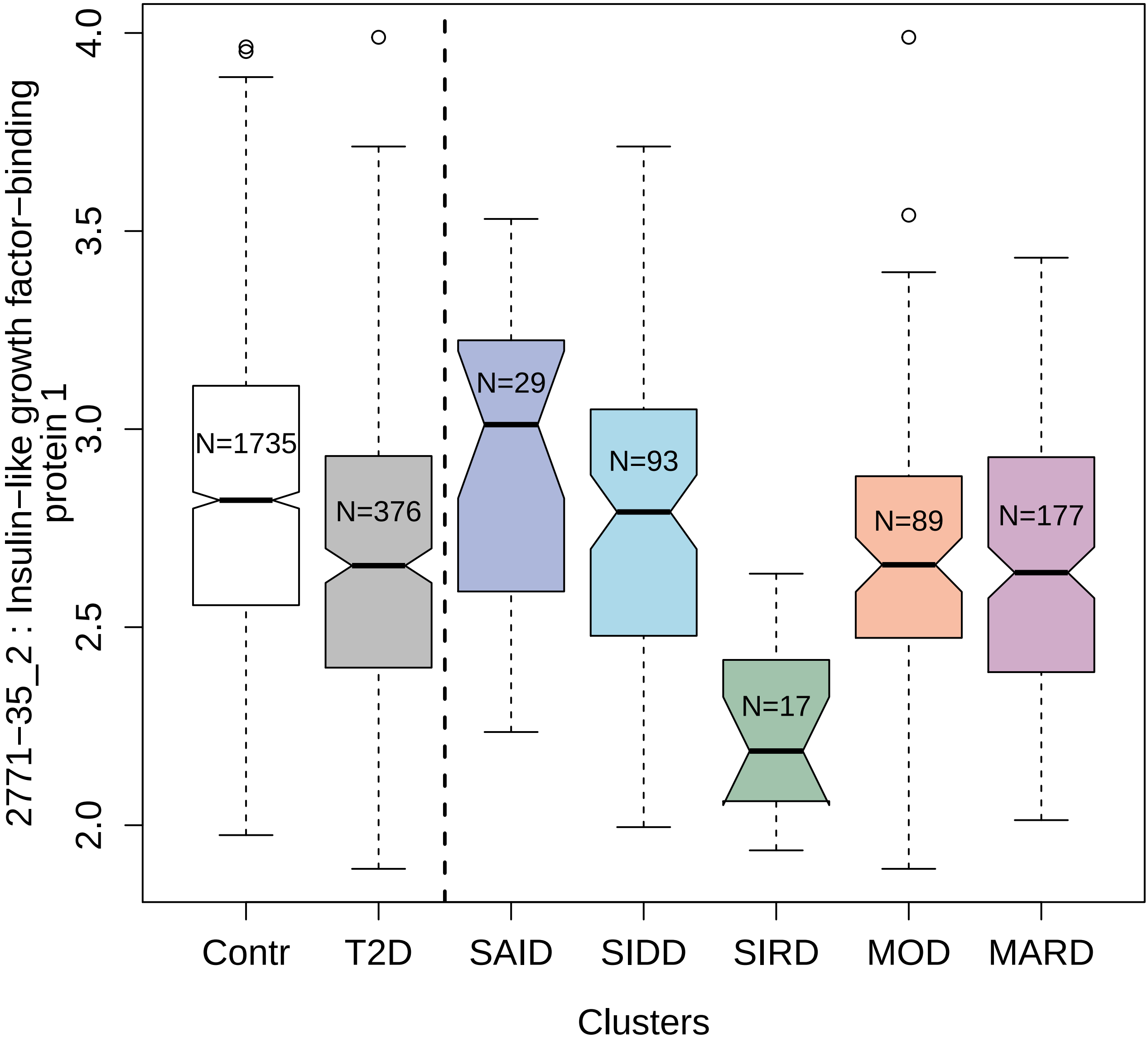

2771−35_2 : Insulin−like growth factor−binding protein 1
4.0
3.5
3.0
2.5
2.0
N=1735
N=376
N=29
N=93
N=17
N=89
N=177
Contr
T2D
SAID
SIDD
SIRD
MOD
MARD
Clusters

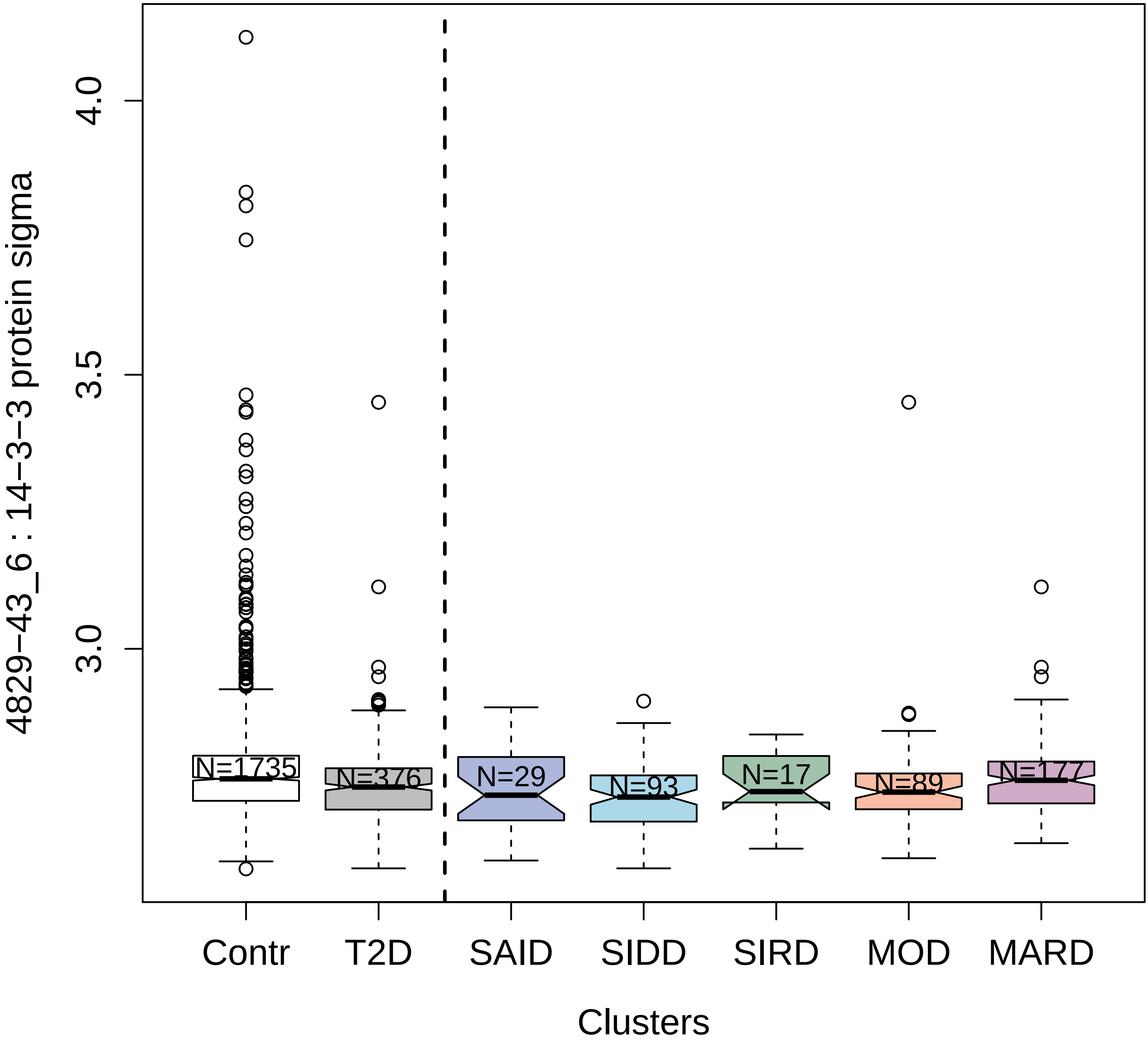
4829−43_6 : 14−3−3 protein sigma
4.0
3.5
3.0
N=1735
N=376
N=29
N=93
N=17
N=89
N=177
Contr
T2D
SAID
SIDD
SIRD
MOD
MARD
Clusters

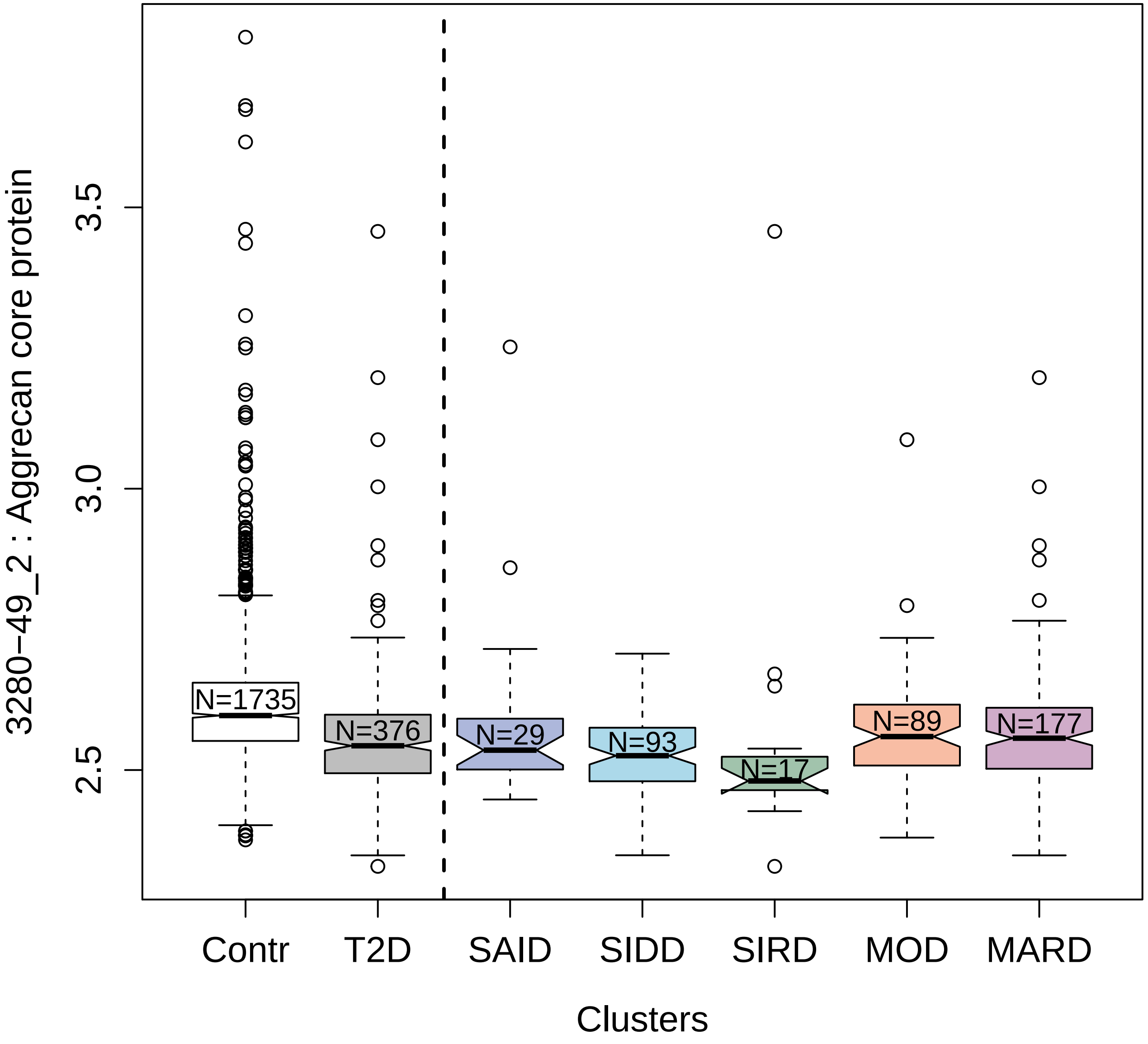

3280−49_2 : Aggrecan core protein
3.5
3.0
2.5
N=1735
N=376
N=29
N=93
N=17
N=89
N=177
Contr
T2D
SAID
SIDD
SIRD
MOD
MARD
Clusters

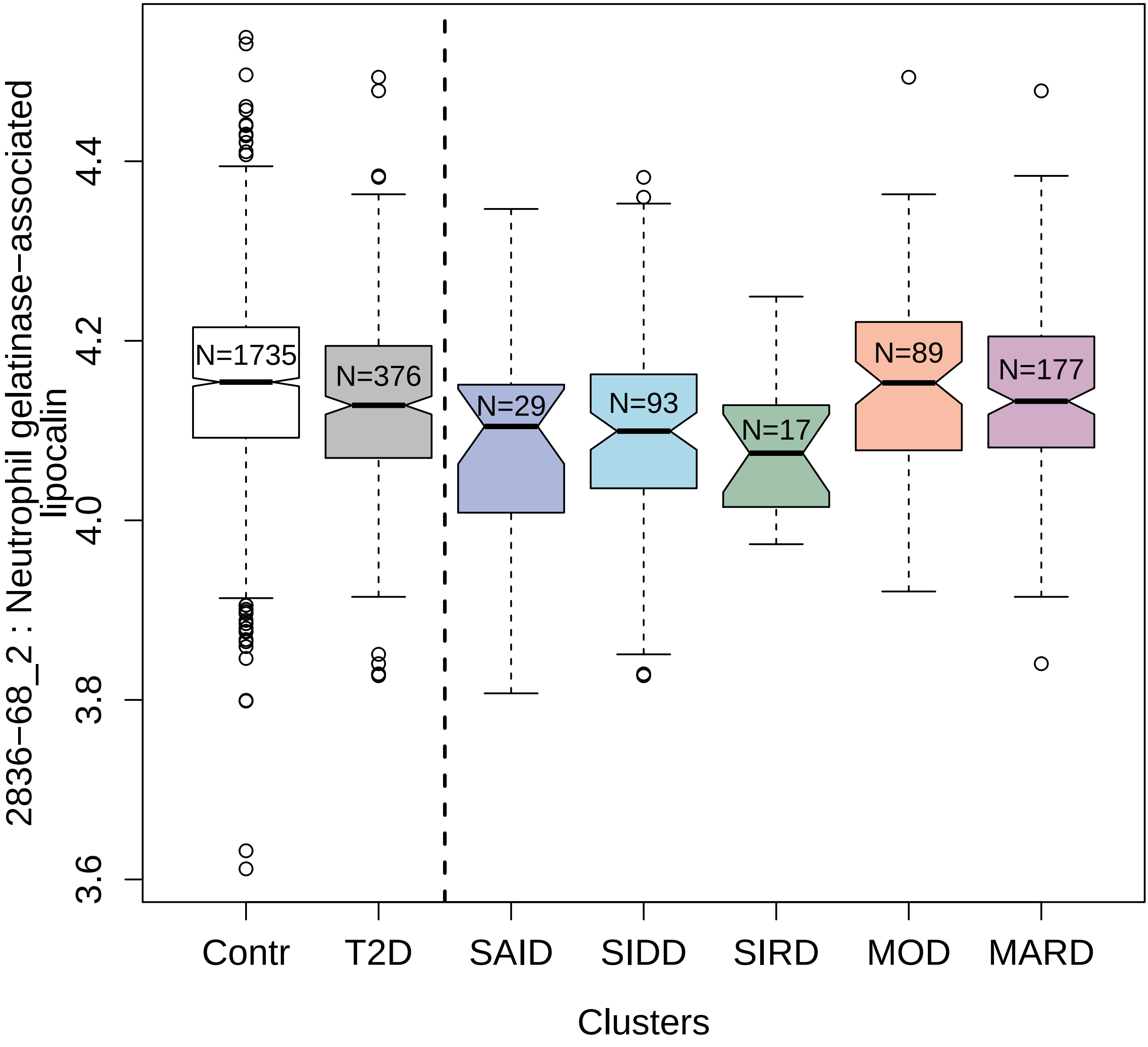
2836−68_2 : Neutrophil gelatinase−associated lipocalin
3.6
3.8
4.0
4.2
4.4
N=1735
N=376
N=29
N=93
N=17
N=89
N=177
Contr
T2D
SAID
SIDD
SIRD
MOD
MARD
Clusters

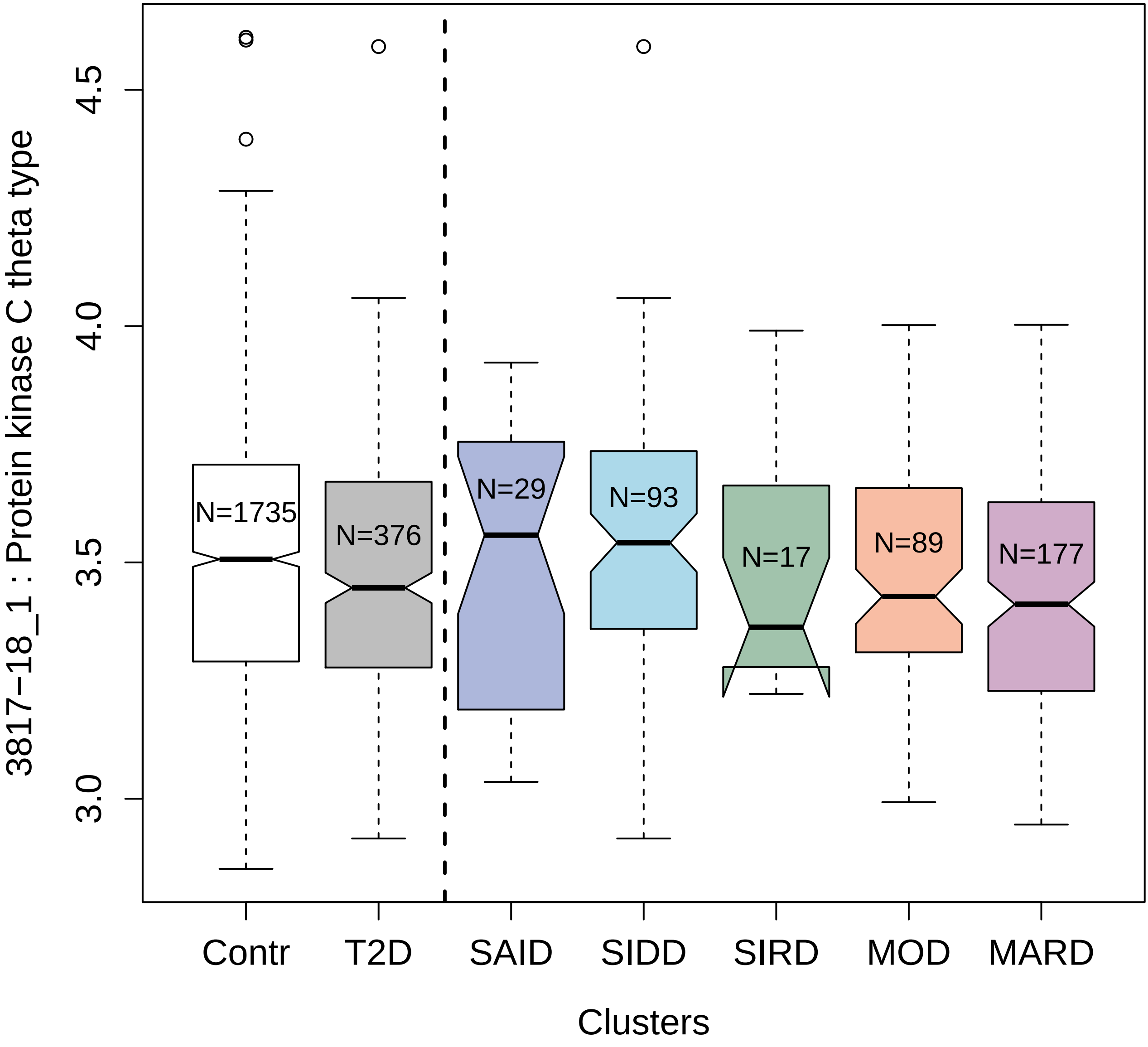
3817−18_1 : Protein kinase C theta type
3.0
3.5
4.0
4.5
N=1735
N=376
N=29
N=93
N=17
N=89
N=177
Contr
T2D
SAID
SIDD
SIRD
MOD
MARD
Clusters

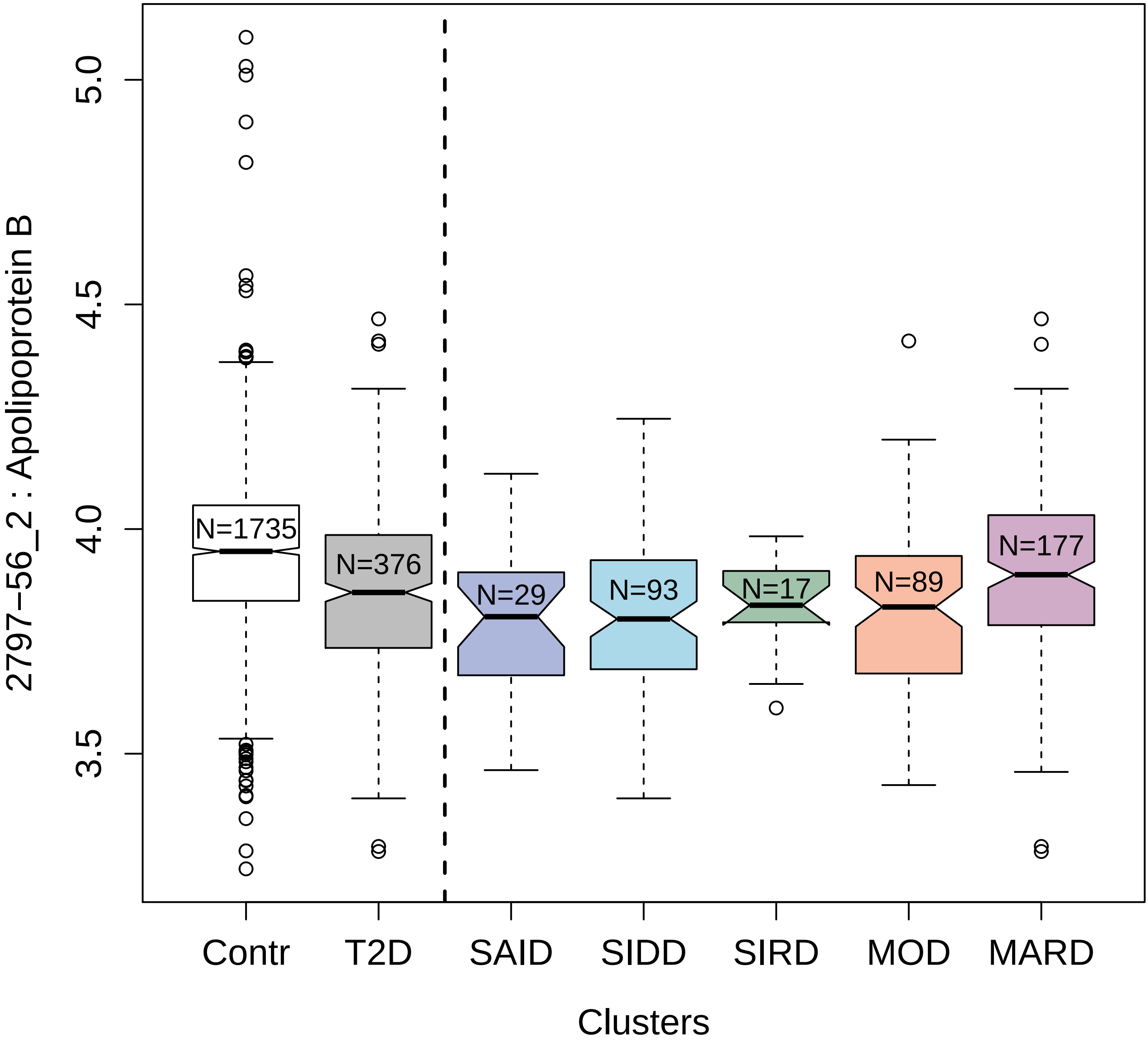

2797−56_2 : Apolipoprotein B
5.0
4.5
4.0
3.5
N=1735
N=376
N=29
N=93
N=17
N=89
N=177
Contr
T2D
SAID
SIDD
SIRD
MOD
MARD
Clusters

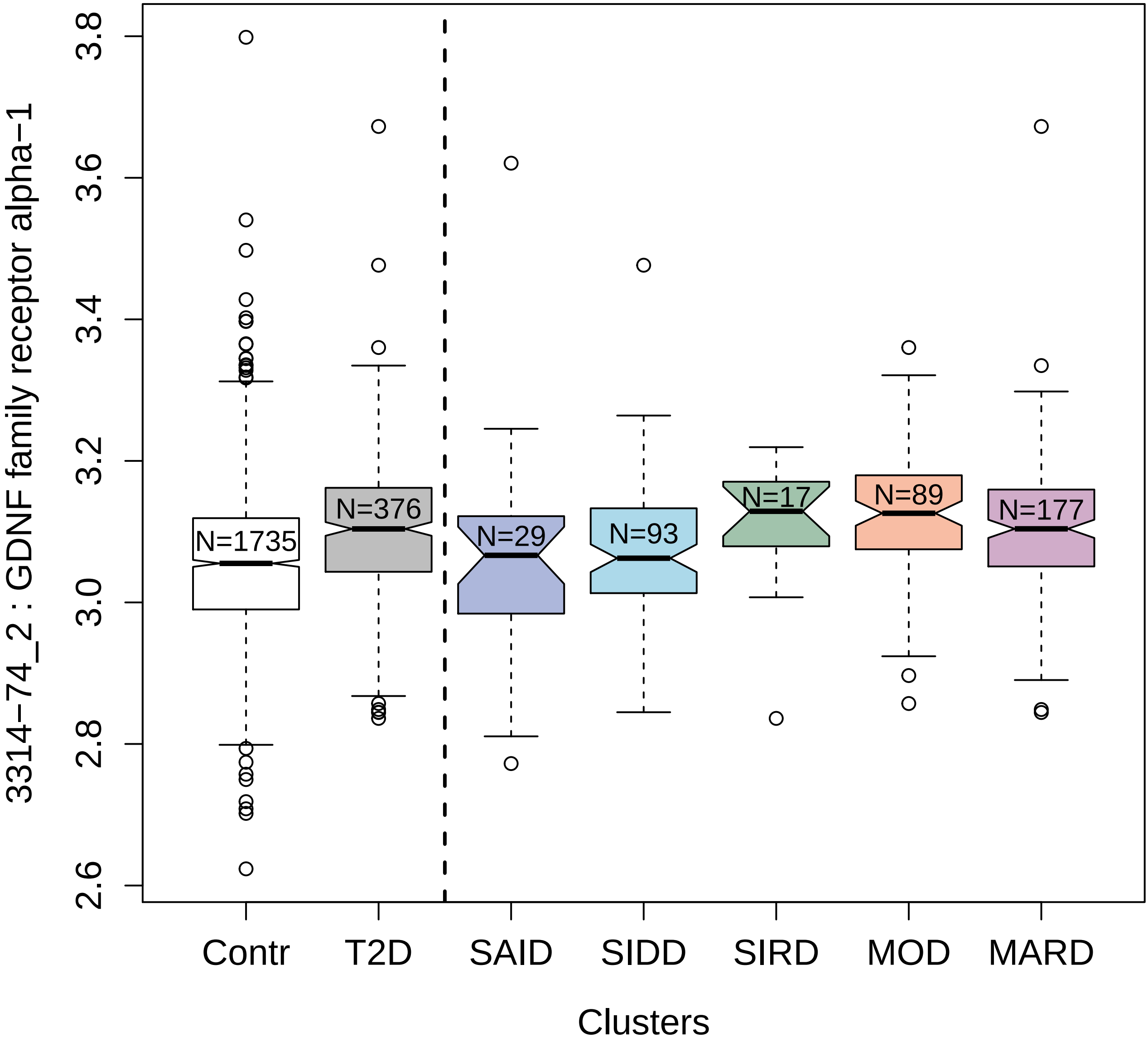
3314−74_2 : GDNF family receptor alpha−1
3.8
3.6
3.4
3.2
3.0
2.8
2.6
N=1735
N=376
N=29
N=93
N=17
N=89
N=177
Contr
T2D
SAID
SIDD
SIRD
MOD
MARD
Clusters

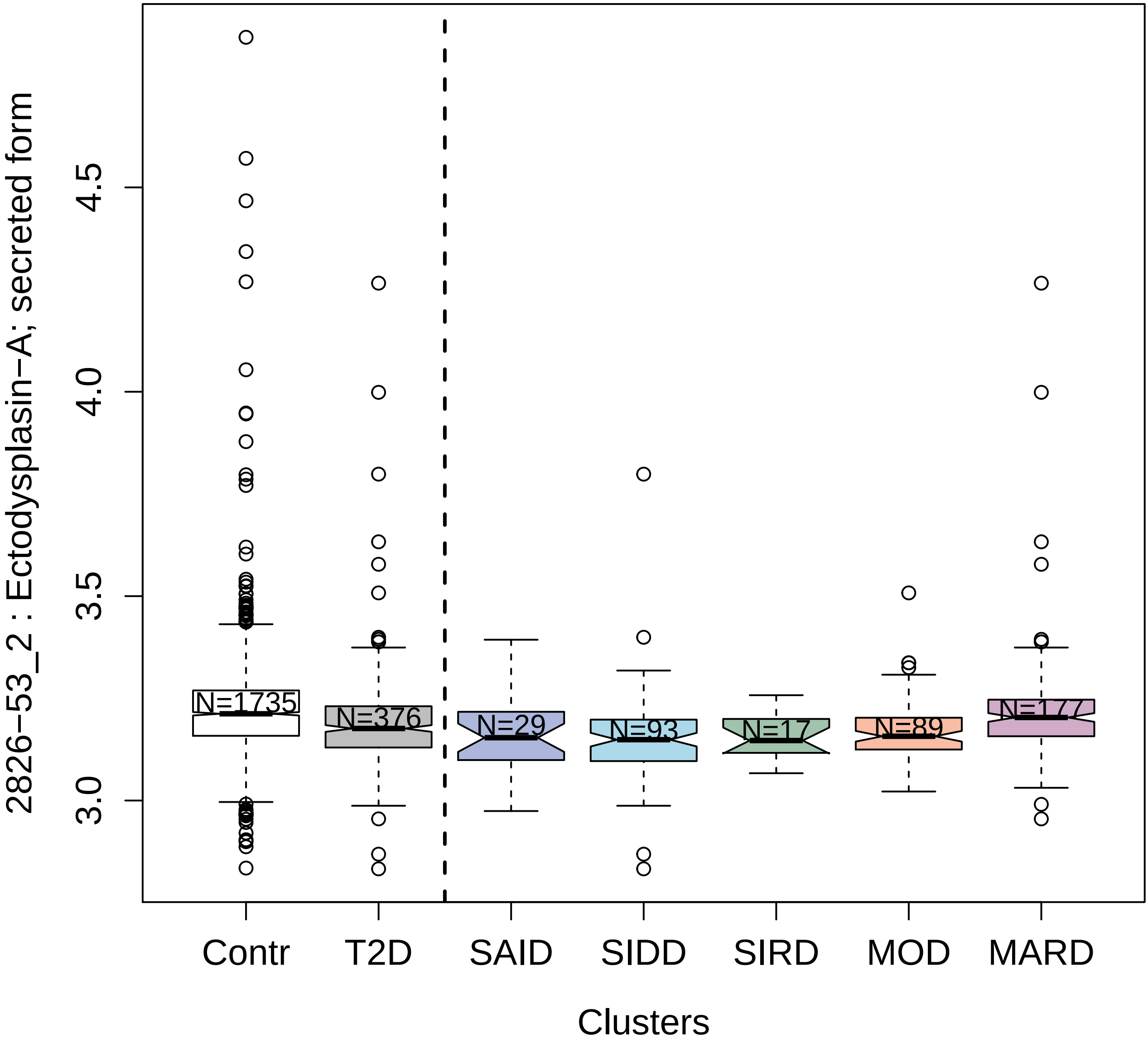
2826−53_2 : Ectodysplasin−A; secreted form
3.0
3.5
4.0
4.5
N=1735
N=376
N=29
N=93
N=17
N=89
N=177
Contr
T2D
SAID
SIDD
SIRD
MOD
MARD
Clusters

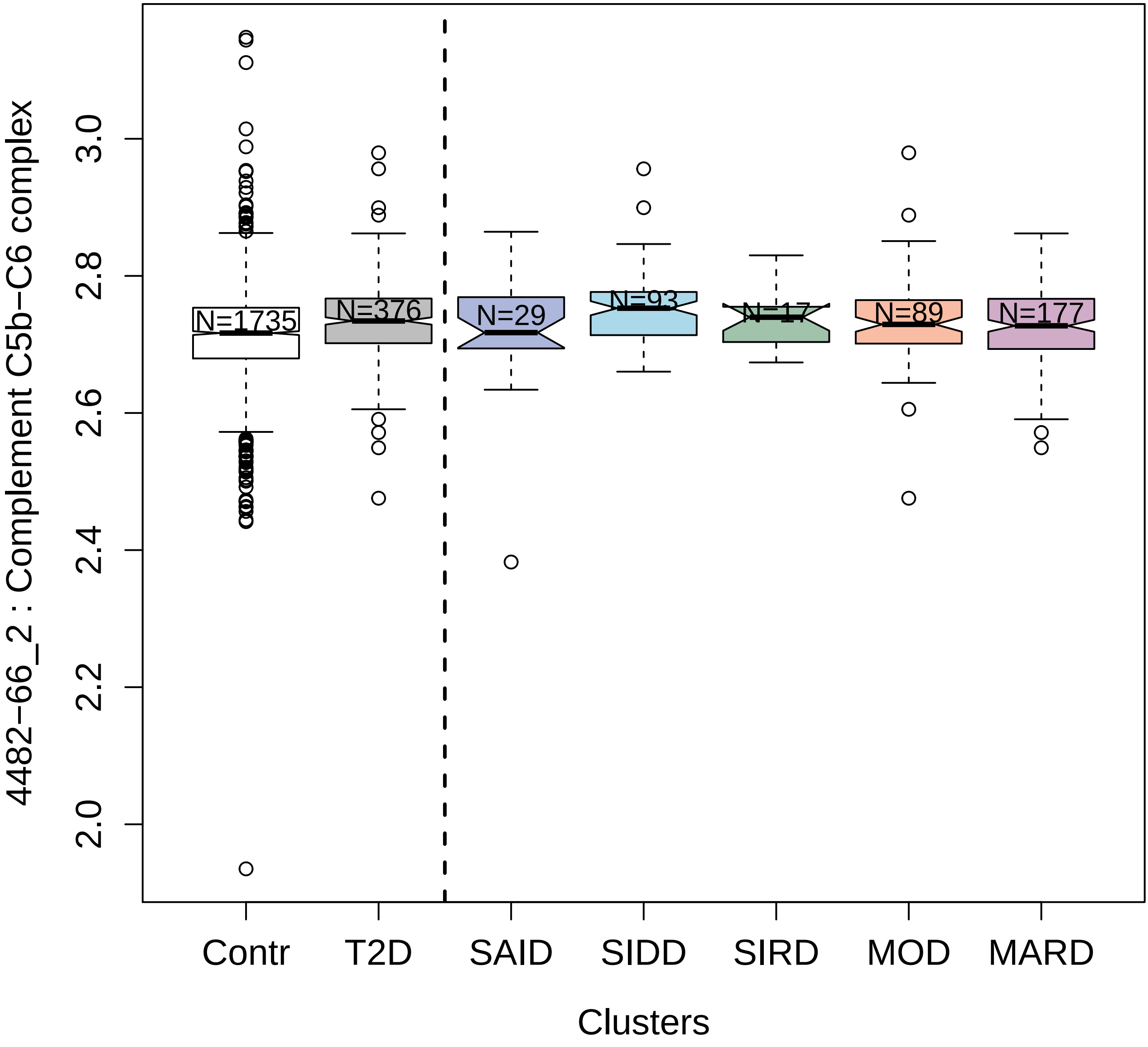

4482−66_2 : Complement C5b−C6 complex
2.0
2.2
2.4
2.6
2.8
3.0
N=1735
N=376
N=29
N=93
N=17
N=89
N=177
Contr
T2D
SAID
SIDD
SIRD
MOD
MARD
Clusters

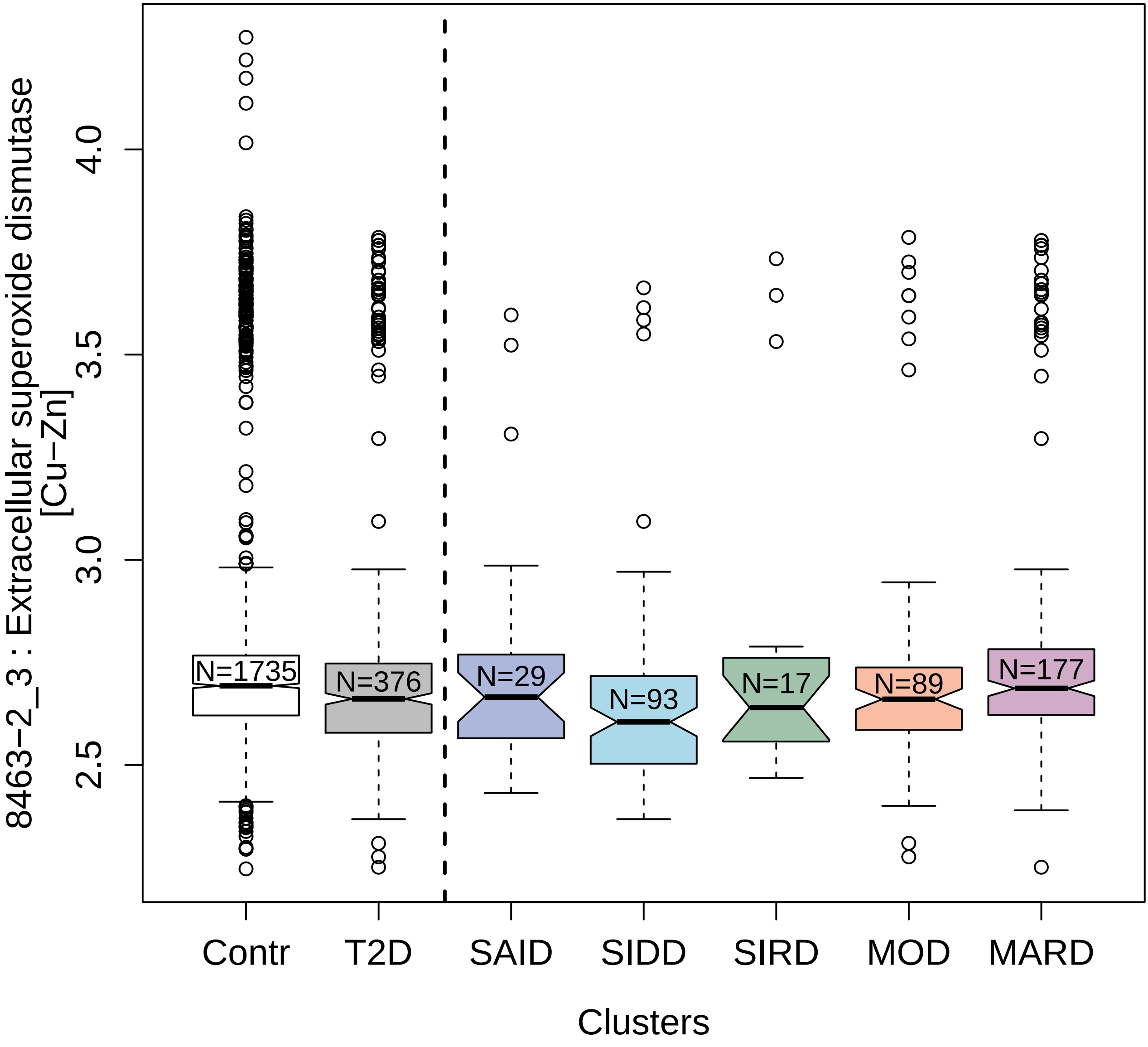
8463−2_3 : Extracellular superoxide dismutase [Cu−Zn]
4.0
3.5
3.0
2.5
N=1735
N=376
N=29
N=93
N=17
N=89
N=177
Contr
T2D
SAID
SIDD
SIRD
MOD
MARD
Clusters

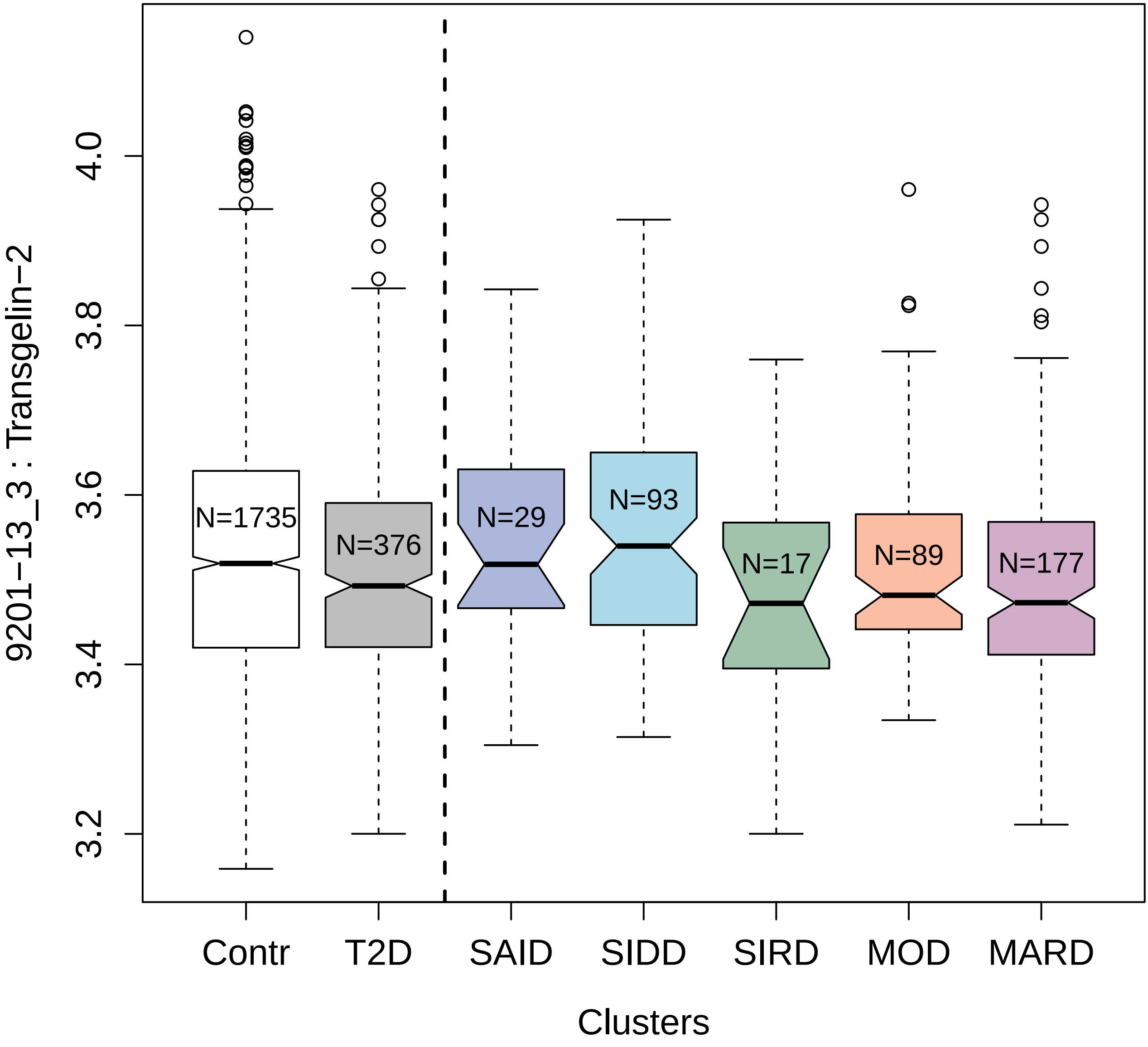
9201−13_3 : Transgelin−2
3.2
3.4
3.6
3.8
4.0
N=1735
N=376
N=29
N=93
N=17
N=89
N=177
Contr
T2D
SAID
SIDD
SIRD
MOD
MARD
Clusters

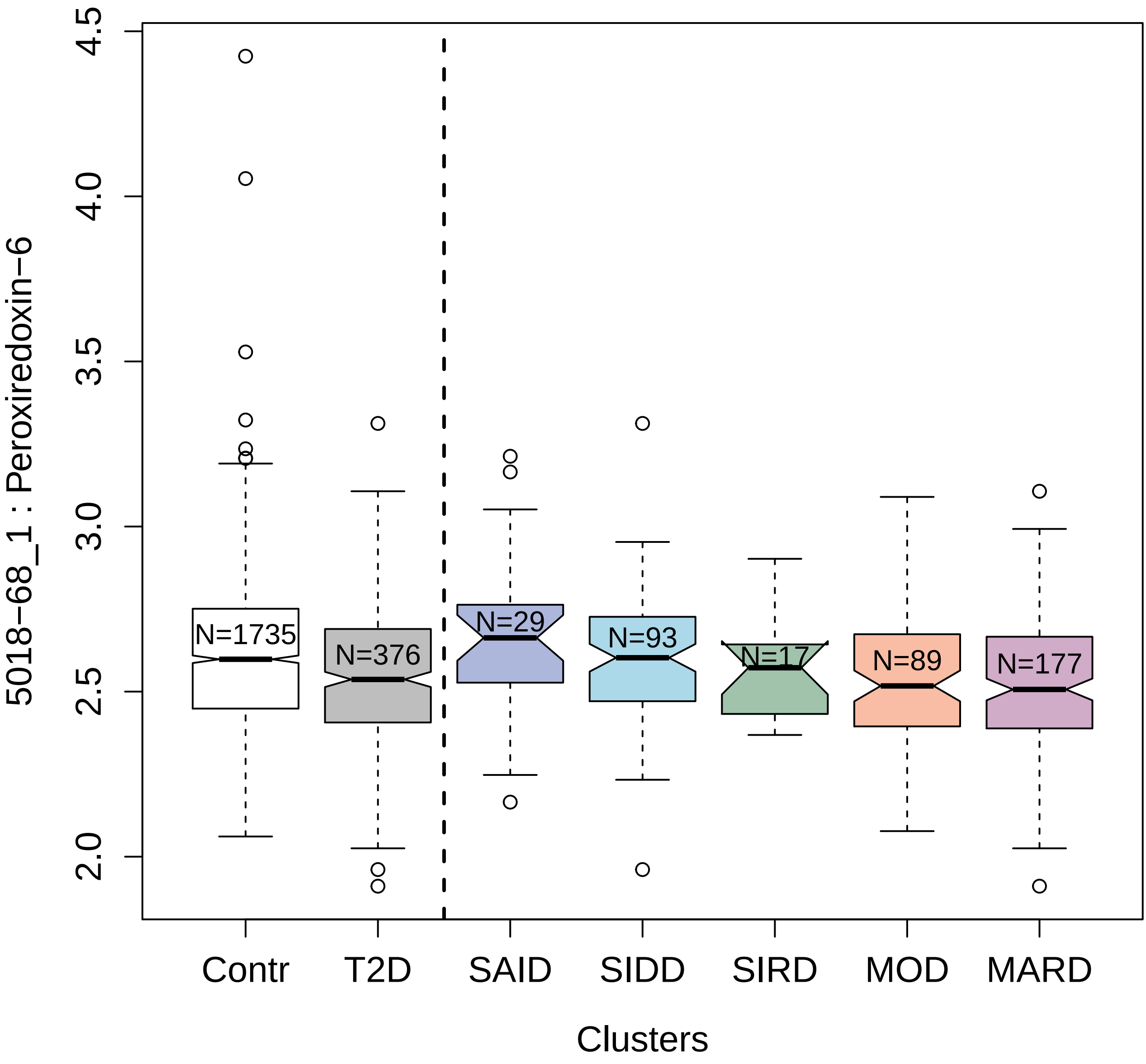

5018−68_1 : Peroxiredoxin−6
4.5
4.0
3.5
3.0
2.5
2.0
N=1735
N=376
N=29
N=93
N=17
N=89
N=177
Contr
T2D
SAID
SIDD
SIRD
MOD
MARD
Clusters

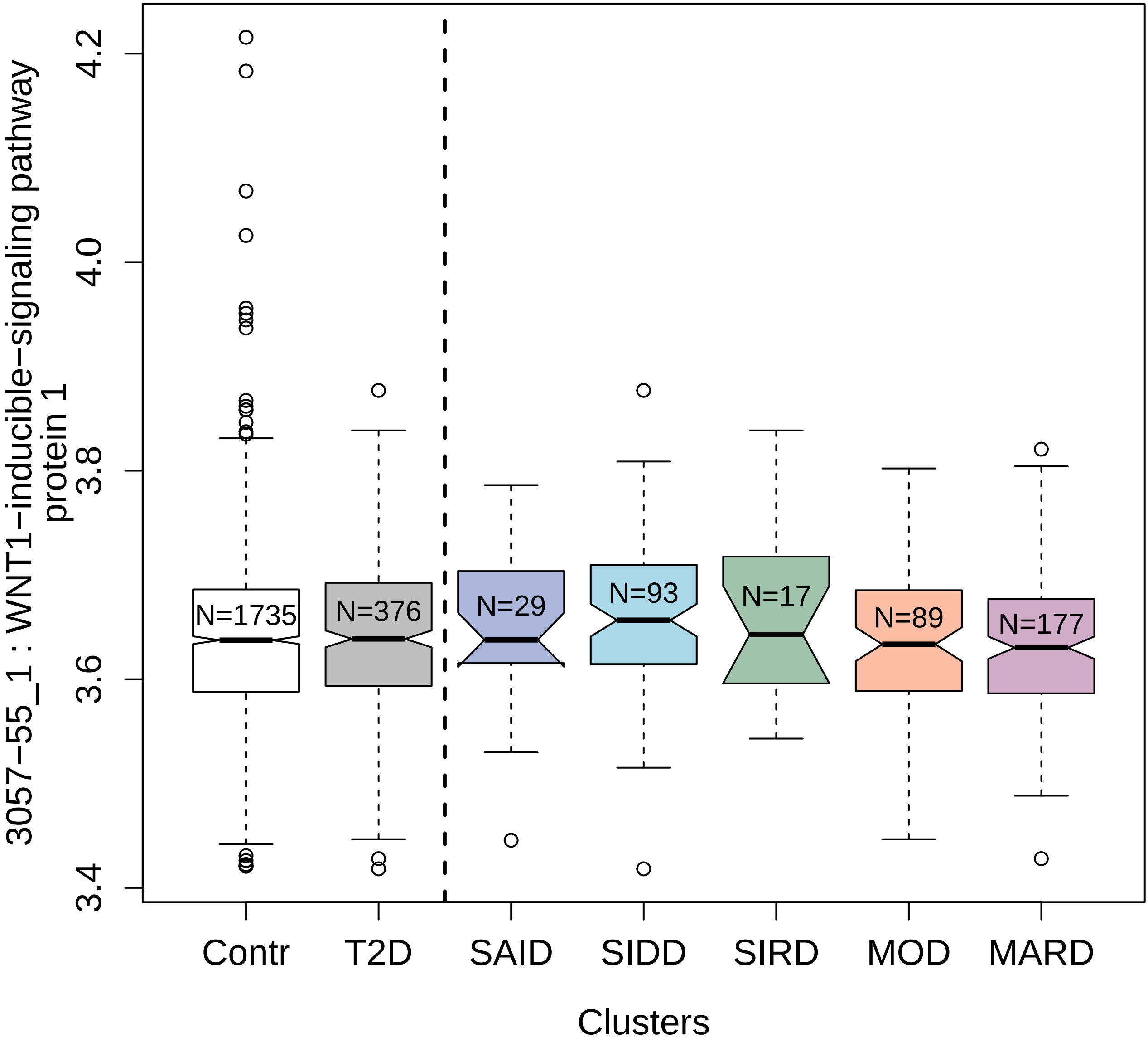
3057−55_1 : WNT1−inducible−signaling pathway protein 1
4.2
4.0
3.8
3.6
3.4
N=1735
N=376
N=29
N=93
N=17
N=89
N=177
Contr
T2D
SAID
SIDD
SIRD
MOD
MARD
Clusters

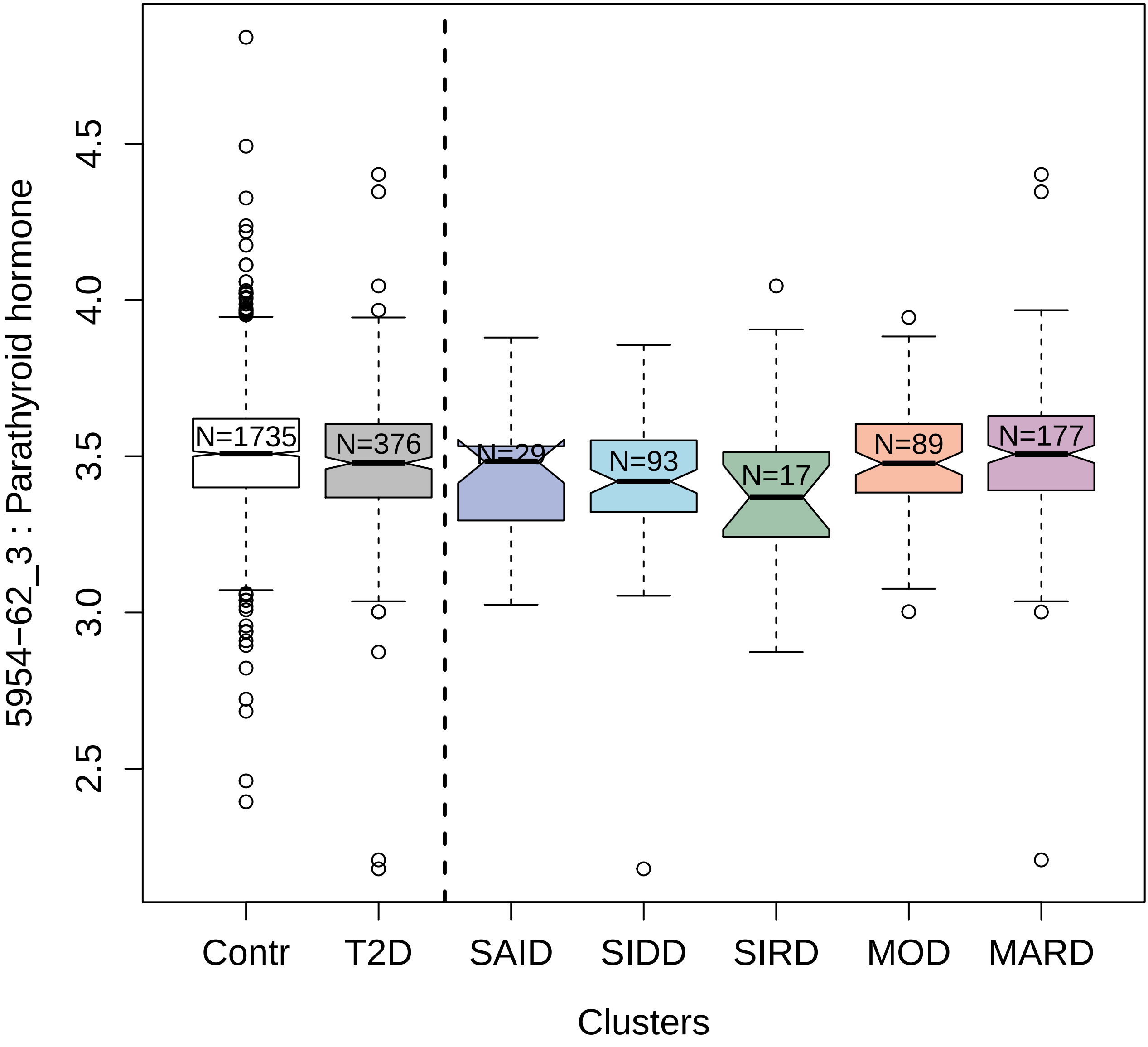

5954−62_3 : Parathyroid hormone
4.5
4.0
3.5
3.0
2.5
N=1735
N=376
N=29
N=93
N=17
N=89
N=177
Contr
T2D
SAID
SIDD
SIRD
MOD
MARD
Clusters

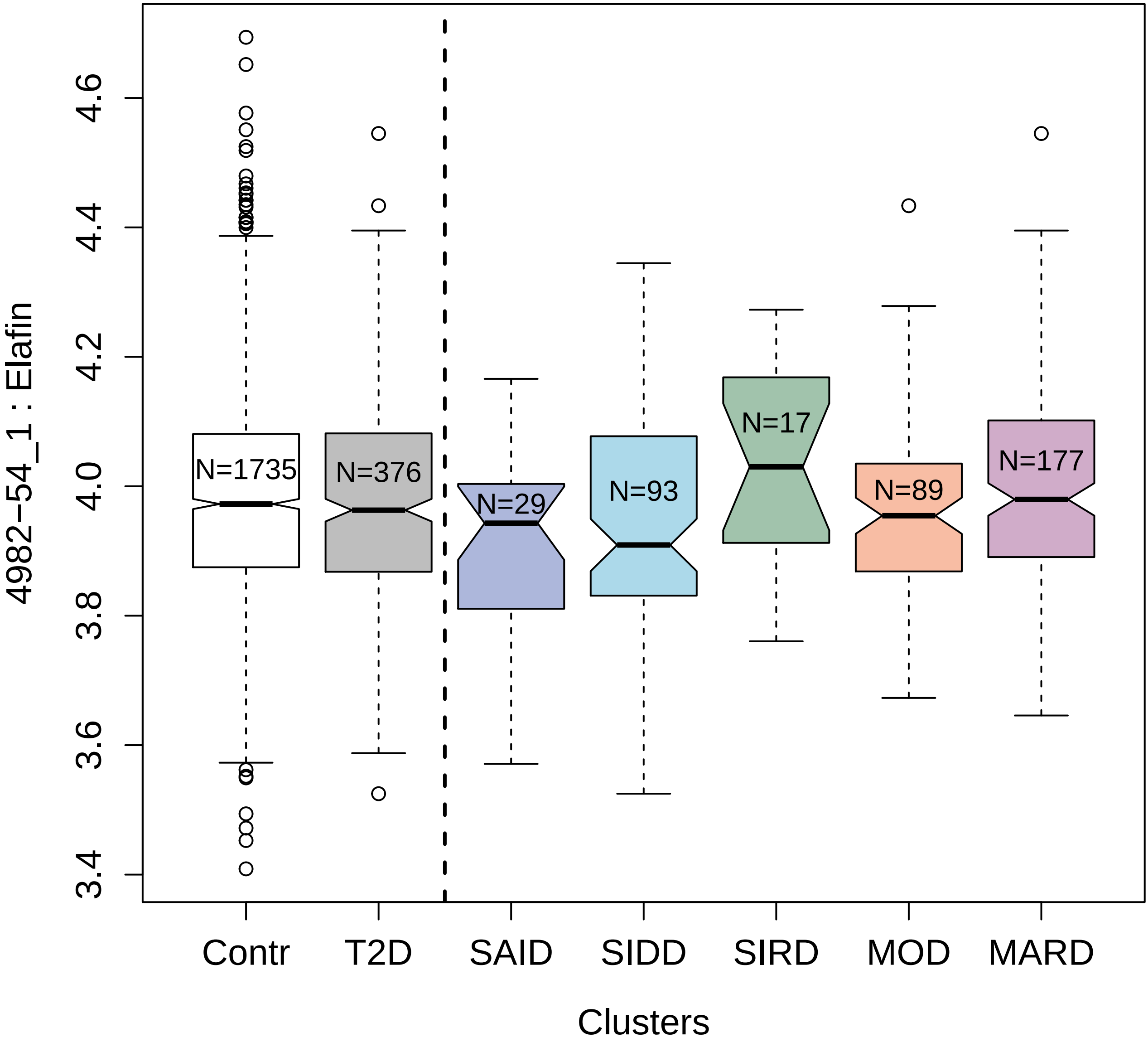

4982−54_1 : Elafin
3.4
3.6
3.8
4.0
4.2
4.4
4.6
N=1735
N=376
N=29
N=93
N=17
N=89
N=177
Contr
T2D
SAID
SIDD
SIRD
MOD
MARD
Clusters

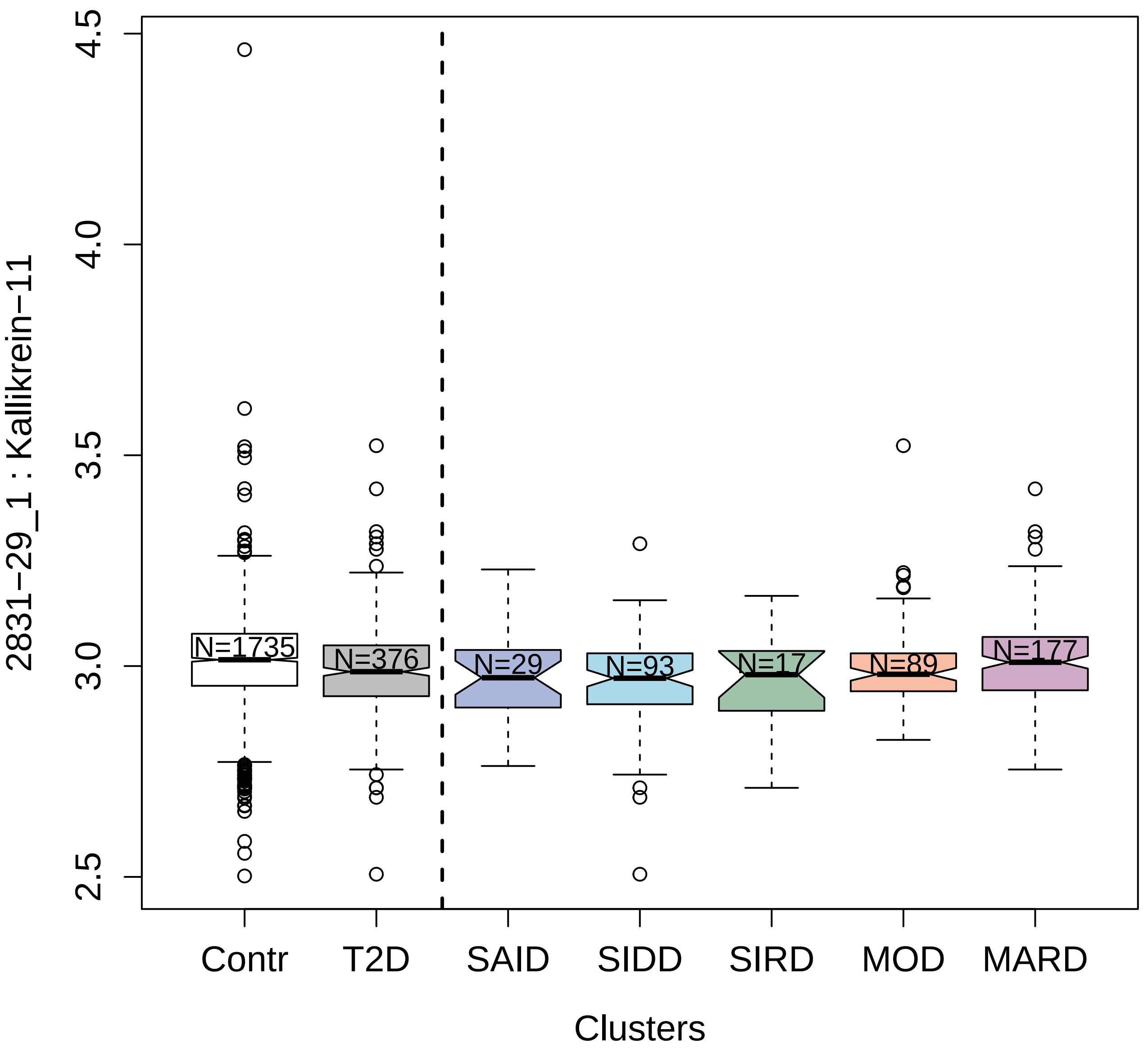
2831−29_1 : Kallikrein−11
4.5
4.0
3.5
3.0
2.5
N=1735
N=376
N=29
N=93
N=17
N=89
N=177
Contr
T2D
SAID
SIDD
SIRD
MOD
MARD
Clusters

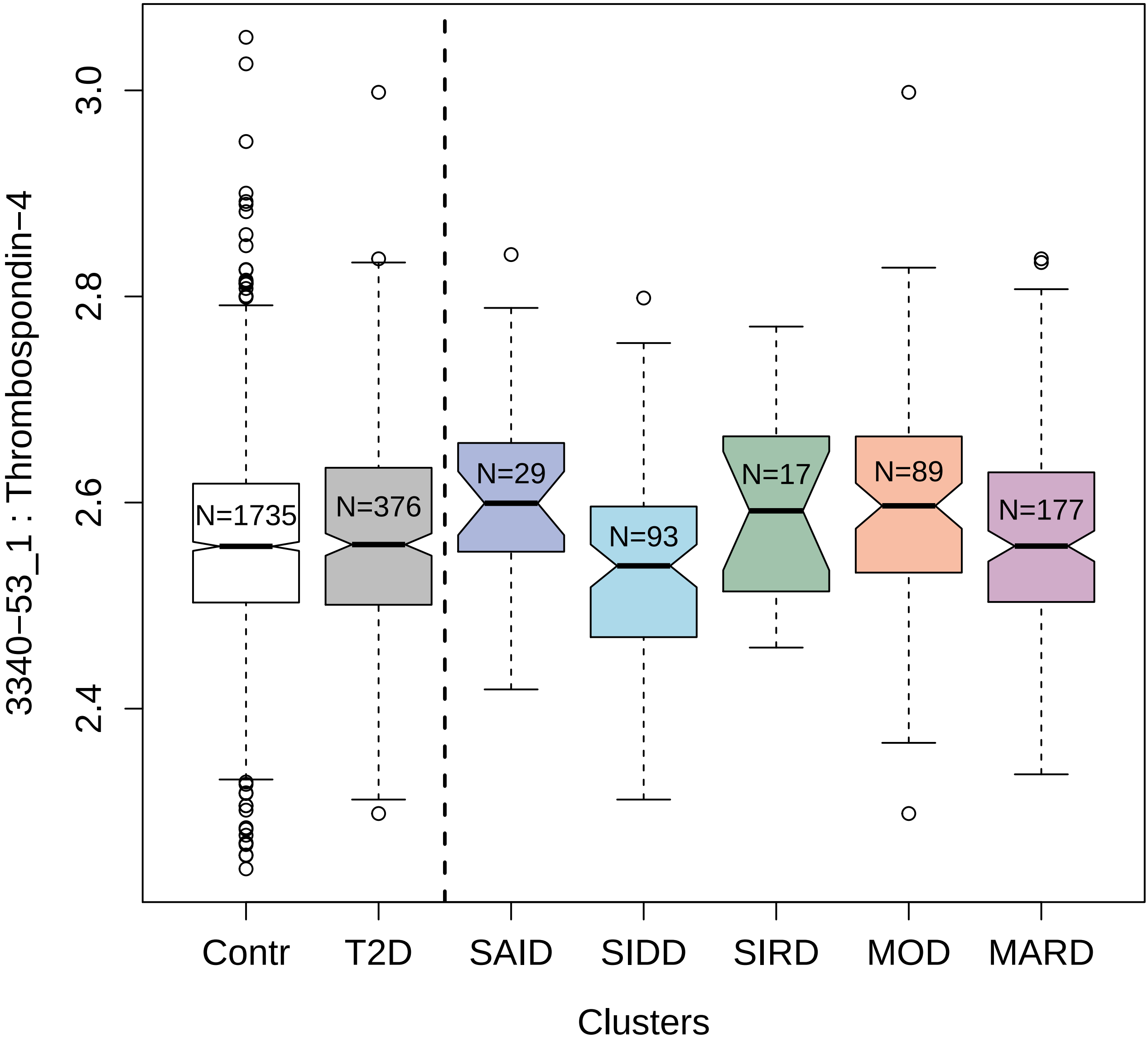
3340−53_1 : Thrombospondin−4
3.0
2.8
2.6
2.4
N=1735
N=376
N=29
N=93
N=17
N=89
N=177
Contr
T2D
SAID
SIDD
SIRD
MOD
MARD
Clusters

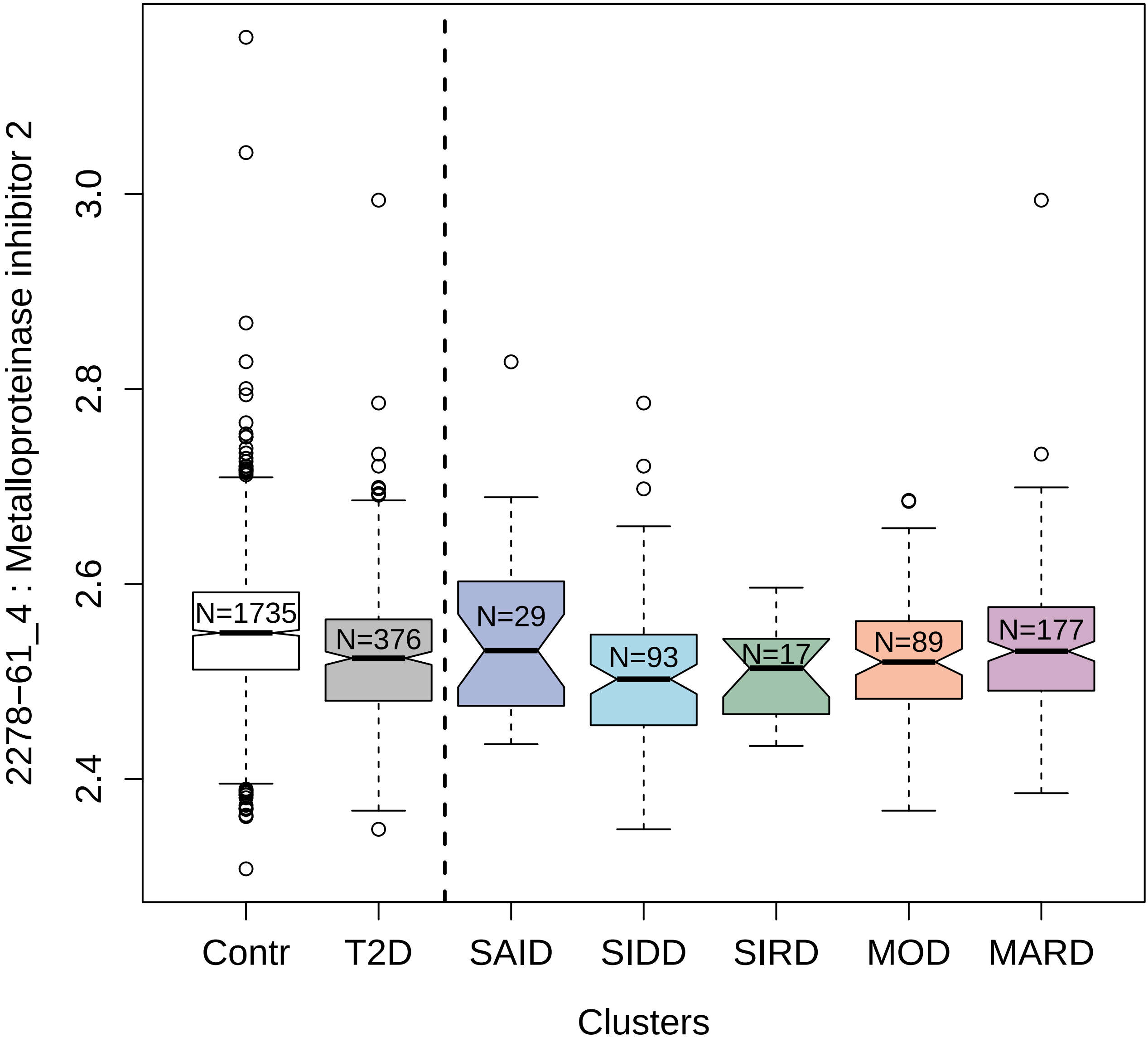
2278−61_4 : Metalloproteinase inhibitor 2
Clusters
Contr
T2D
SAID
SIDD
SIRD
MOD
MARD
N=1735
N=376
N=29
N=93
N=17
N=89
N=177
2.4
2.6
2.8
3.0

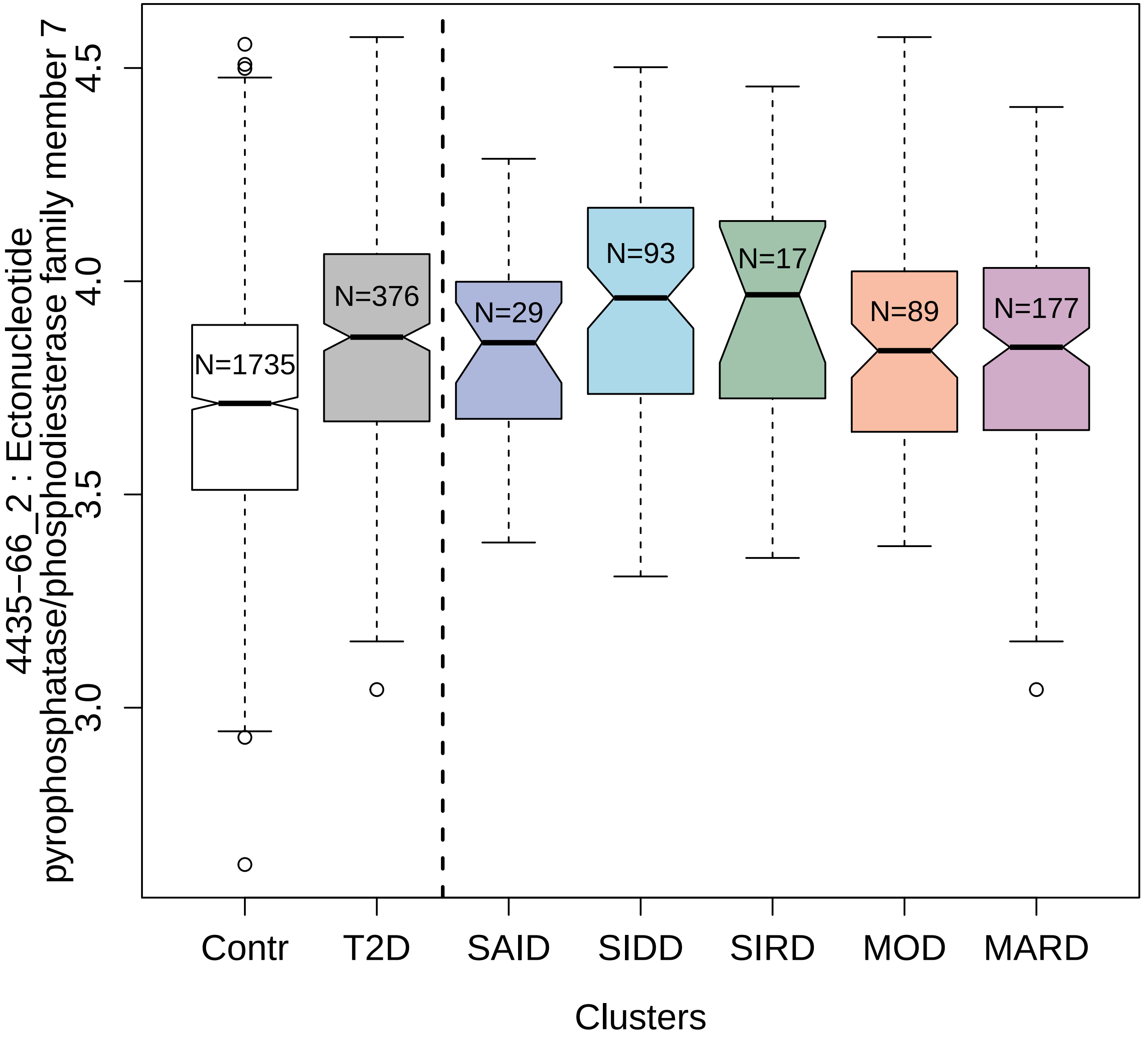

4435−66_2 : Ectonucleotide
pyrophosphatase/phosphodiesterase family member 7
4.5
4.0
3.5
3.0
N=1735
N=376
N=29
N=93
N=17
N=89
N=177
Contr
T2D
SAID
SIDD
SIRD
MOD
MARD
Clusters

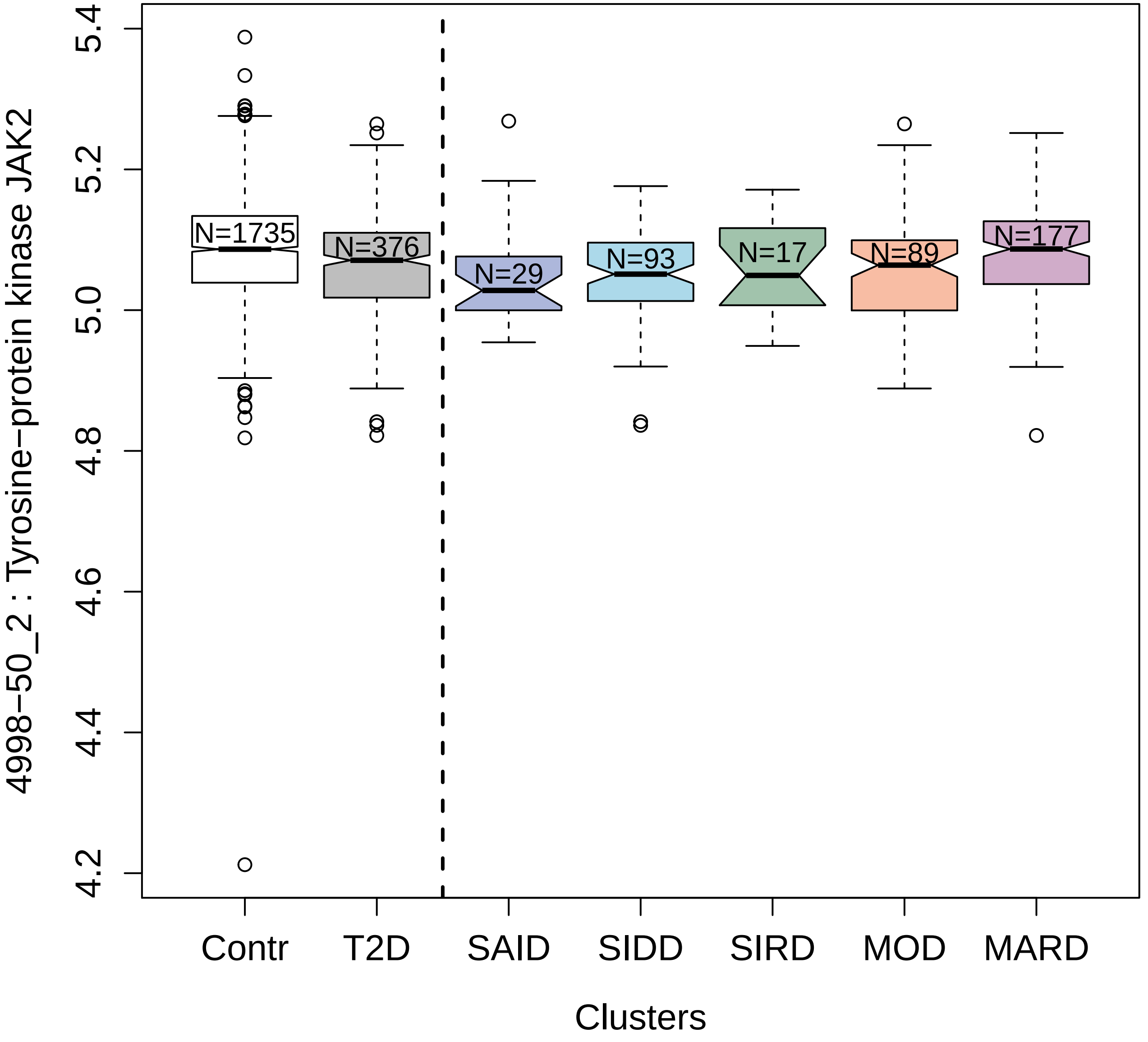
4998−50_2 : Tyrosine−protein kinase JAK2
5.4
5.2
5.0
4.8
4.6
4.4
4.2
N=1735
N=376
N=29
N=93
N=17
N=89
N=177
Contr
T2D
SAID
SIDD
SIRD
MOD
MARD
Clusters

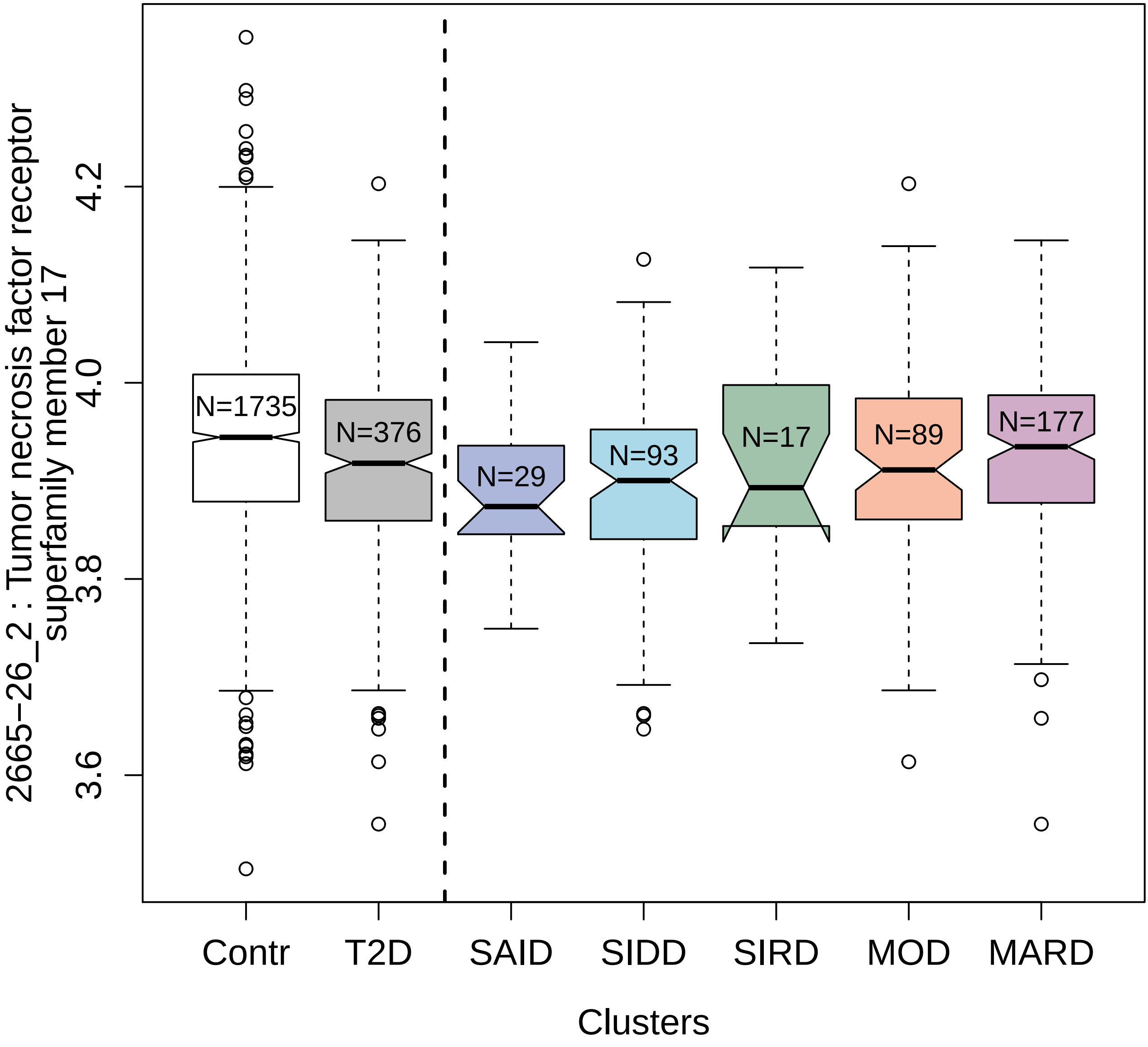

2665−26_2 : Tumor necrosis factor receptor superfamily member 17
3.6
3.8
4.0
4.2
N=1735
N=376
N=29
N=93
N=17
N=89
N=177
Contr
T2D
SAID
SIDD
SIRD
MOD
MARD
Clusters

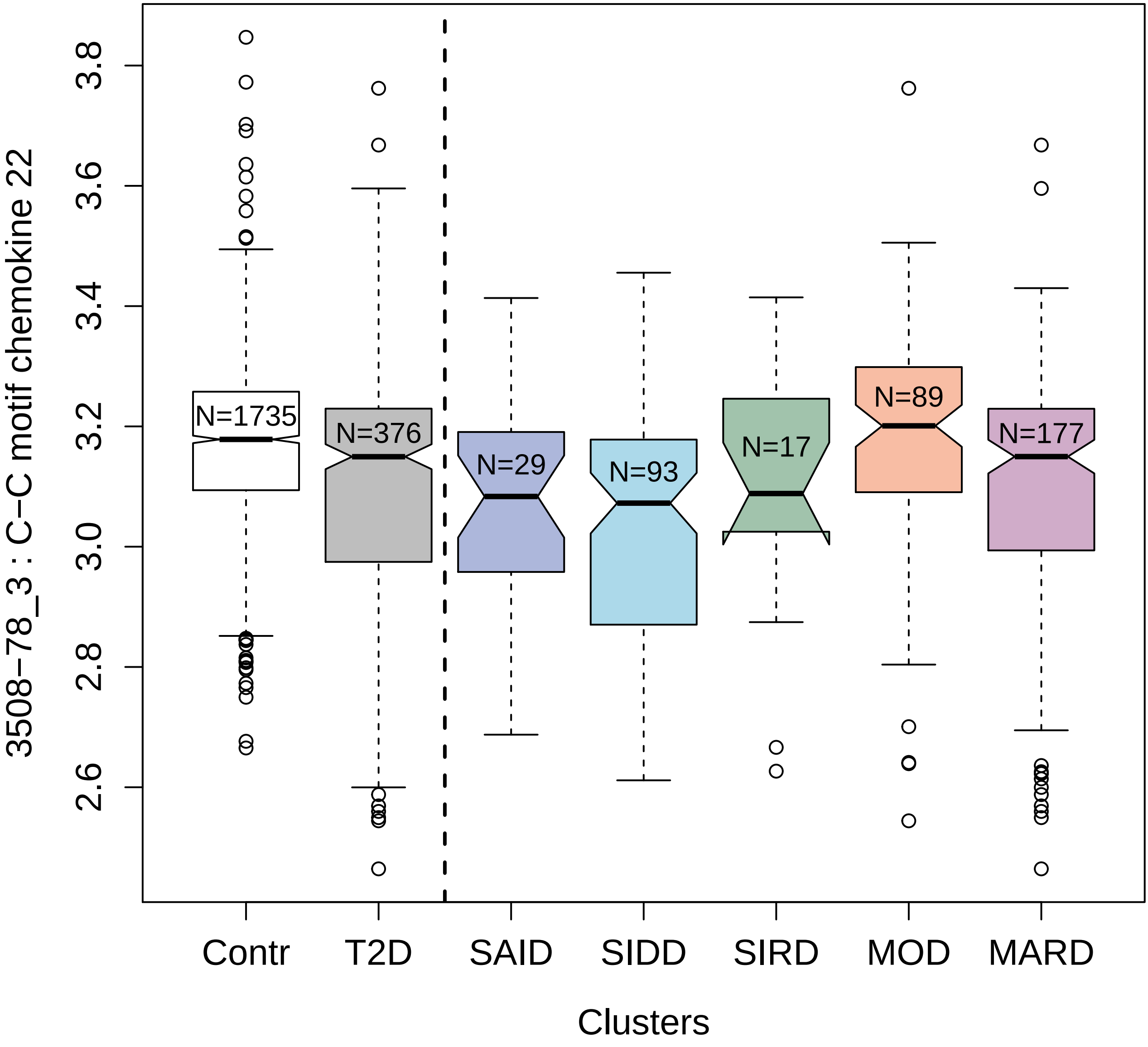

3508−78_3 : C−C motif chemokine 22
3.8
3.6
3.4
3.2
3.0
2.8
2.6
N=1735
N=376
N=29
N=93
N=17
N=89
N=177
Contr
T2D
SAID
SIDD
SIRD
MOD
MARD
Clusters

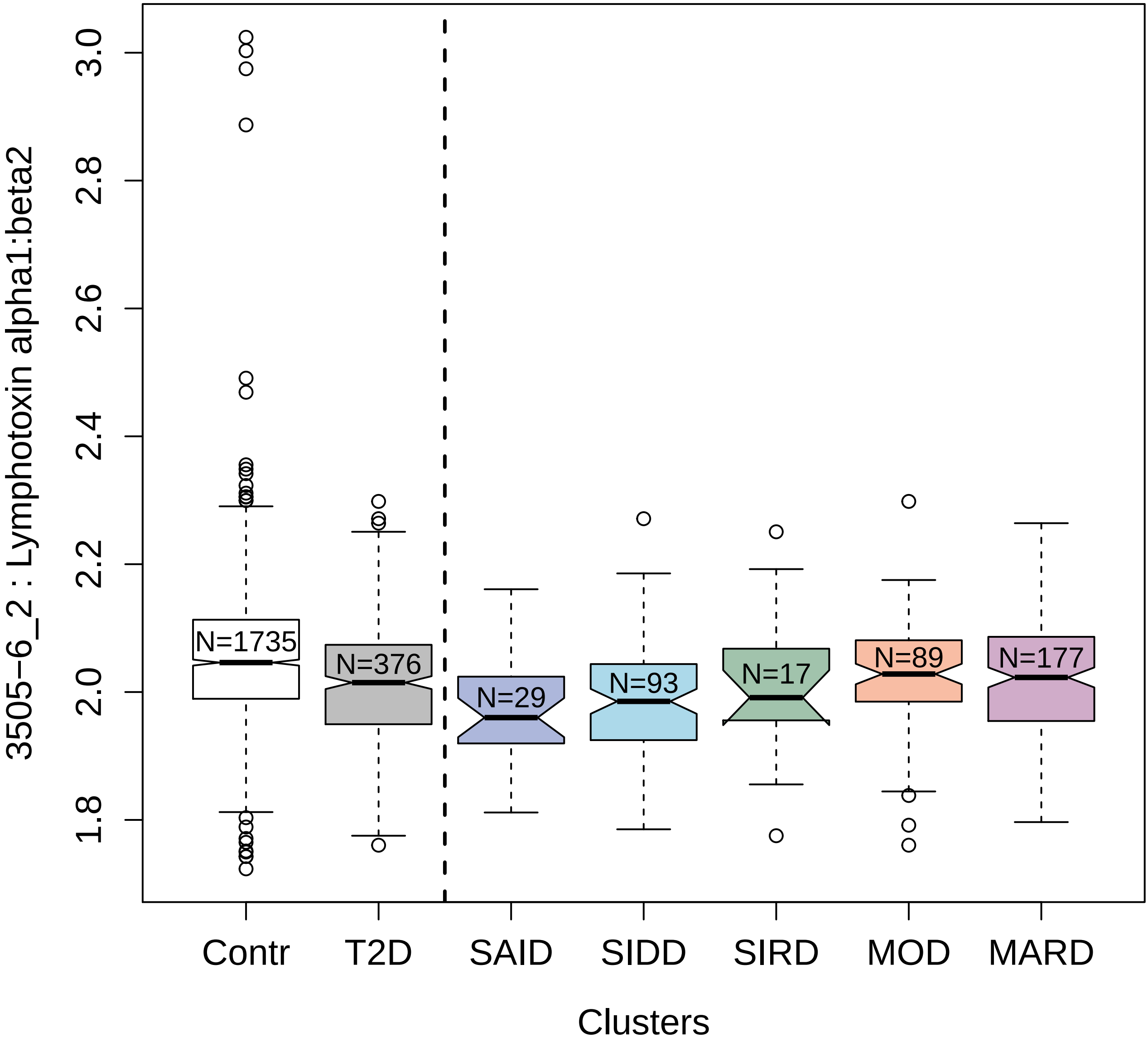
3505−6_2 : Lymphotoxin alpha1:beta2
3.0
2.8
2.6
2.4
2.2
2.0
1.8
N=1735
N=376
N=29
N=93
N=17
N=89
N=177
Contr
T2D
SAID
SIDD
SIRD
MOD
MARD
Clusters

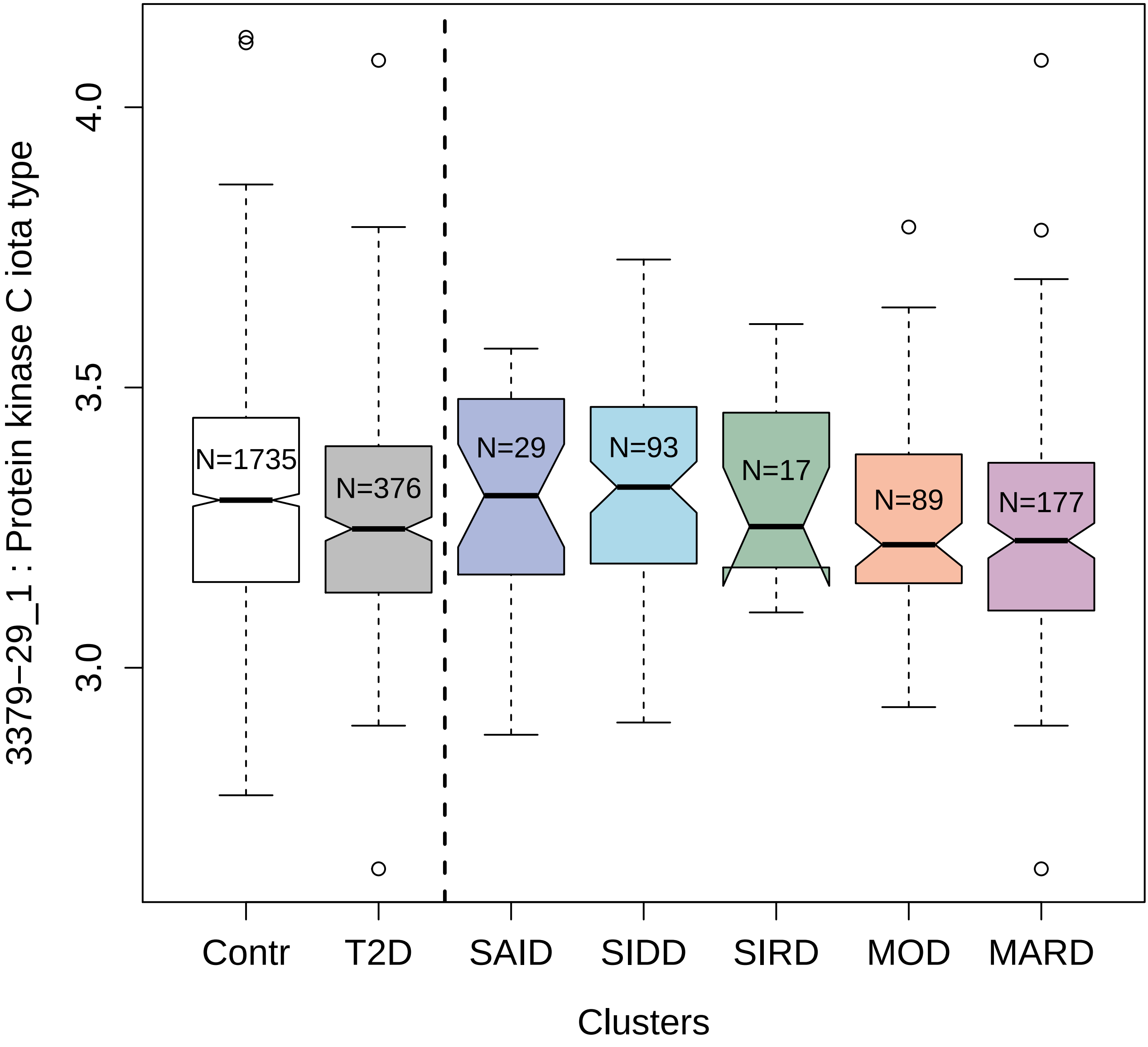
3379−29_1 : Protein kinase C iota type
4.0
3.5
3.0
N=1735
N=376
N=29
N=93
N=17
N=89
N=177
Contr
T2D
SAID
SIDD
SIRD
MOD
MARD
Clusters

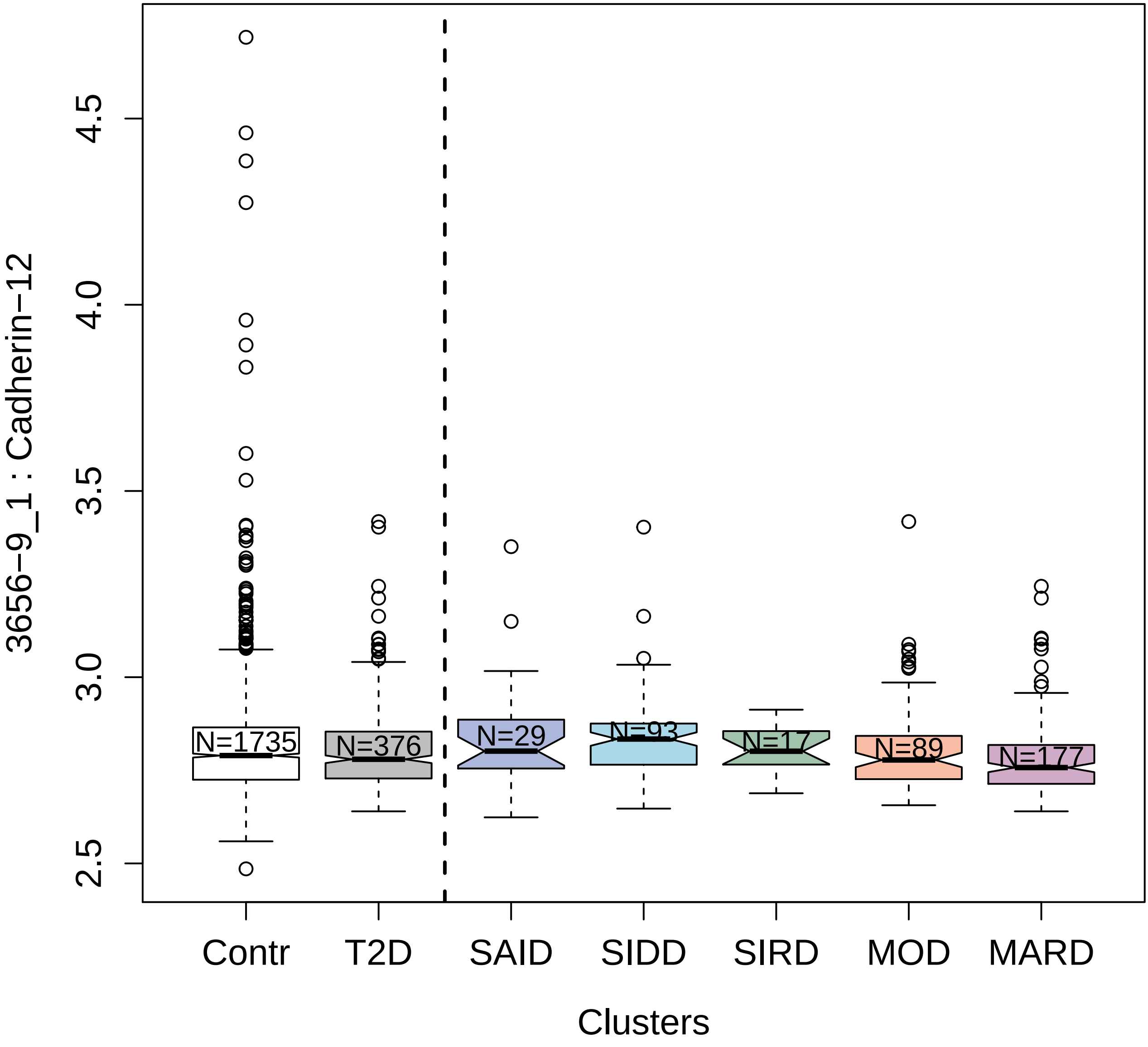

3656−9_1 : Cadherin−12
2.5
3.0
3.5
4.0
4.5
N=1735
N=376
N=29
N=93
N=17
N=89
N=177
Contr
T2D
SAID
SIDD
SIRD
MOD
MARD
Clusters

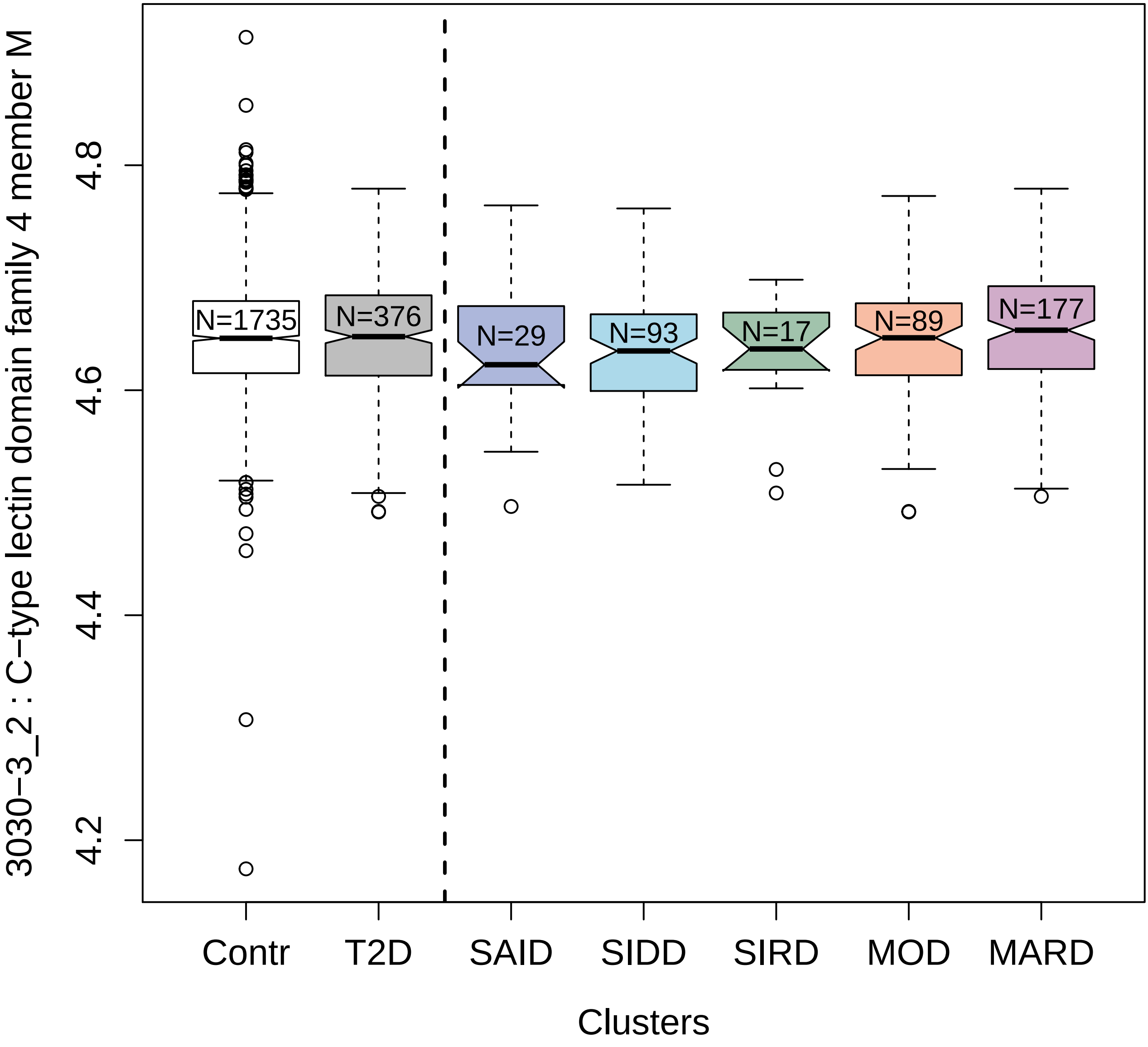

3030−3_2 : C−type lectin domain family 4 member M
4.2
4.4
4.6
4.8
N=1735
N=376
N=29
N=93
N=17
N=89
N=177
Contr
T2D
SAID
SIDD
SIRD
MOD
MARD
Clusters

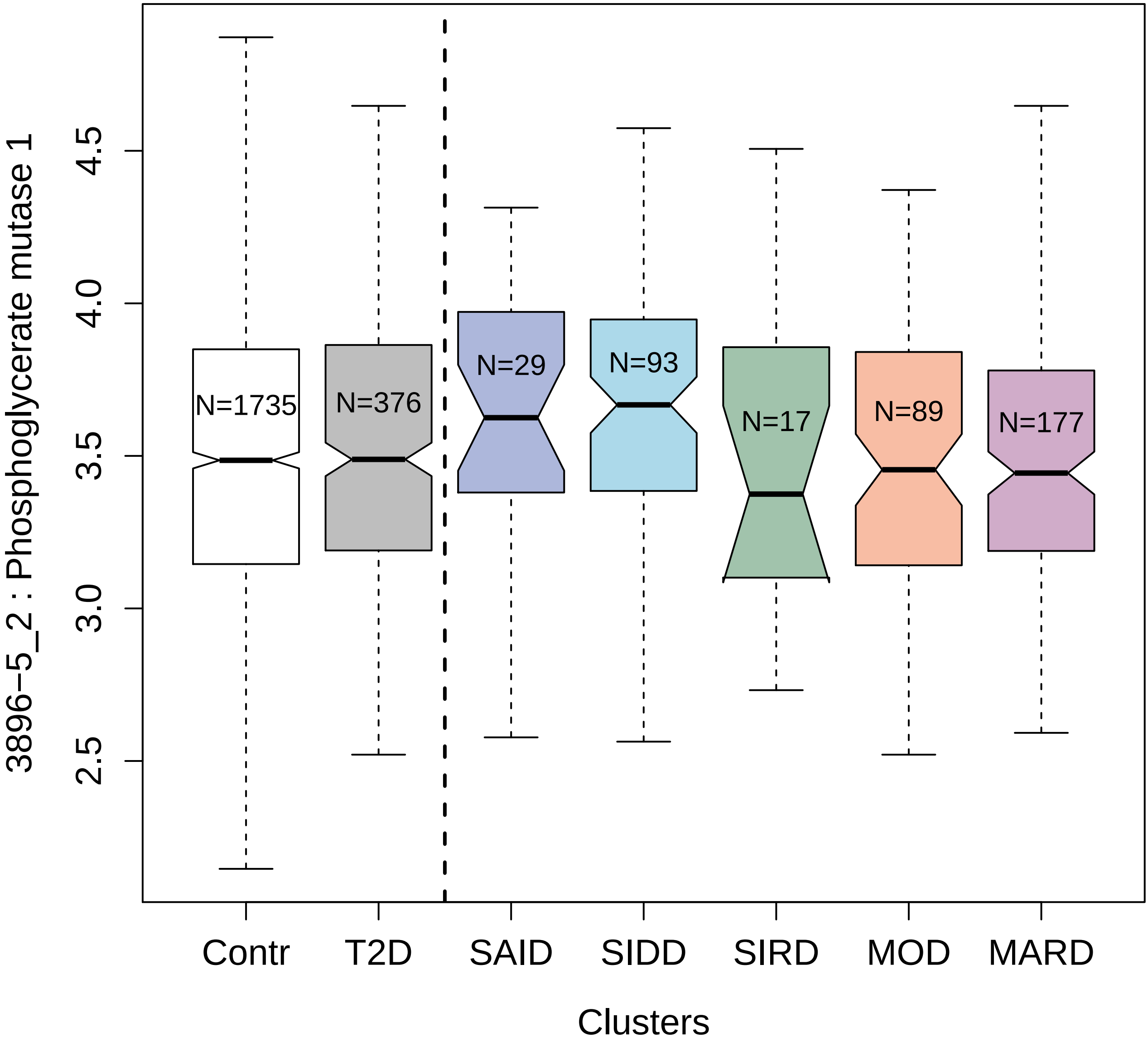

3896–5_2 : Phosphoglycerate mutase 1
2.5
3.0
3.5
4.0
4.5
N=1735
N=376
N=29
N=93
N=17
N=89
N=177
Contr
T2D
SAID
SIDD
SIRD
MOD
MARD
Clusters

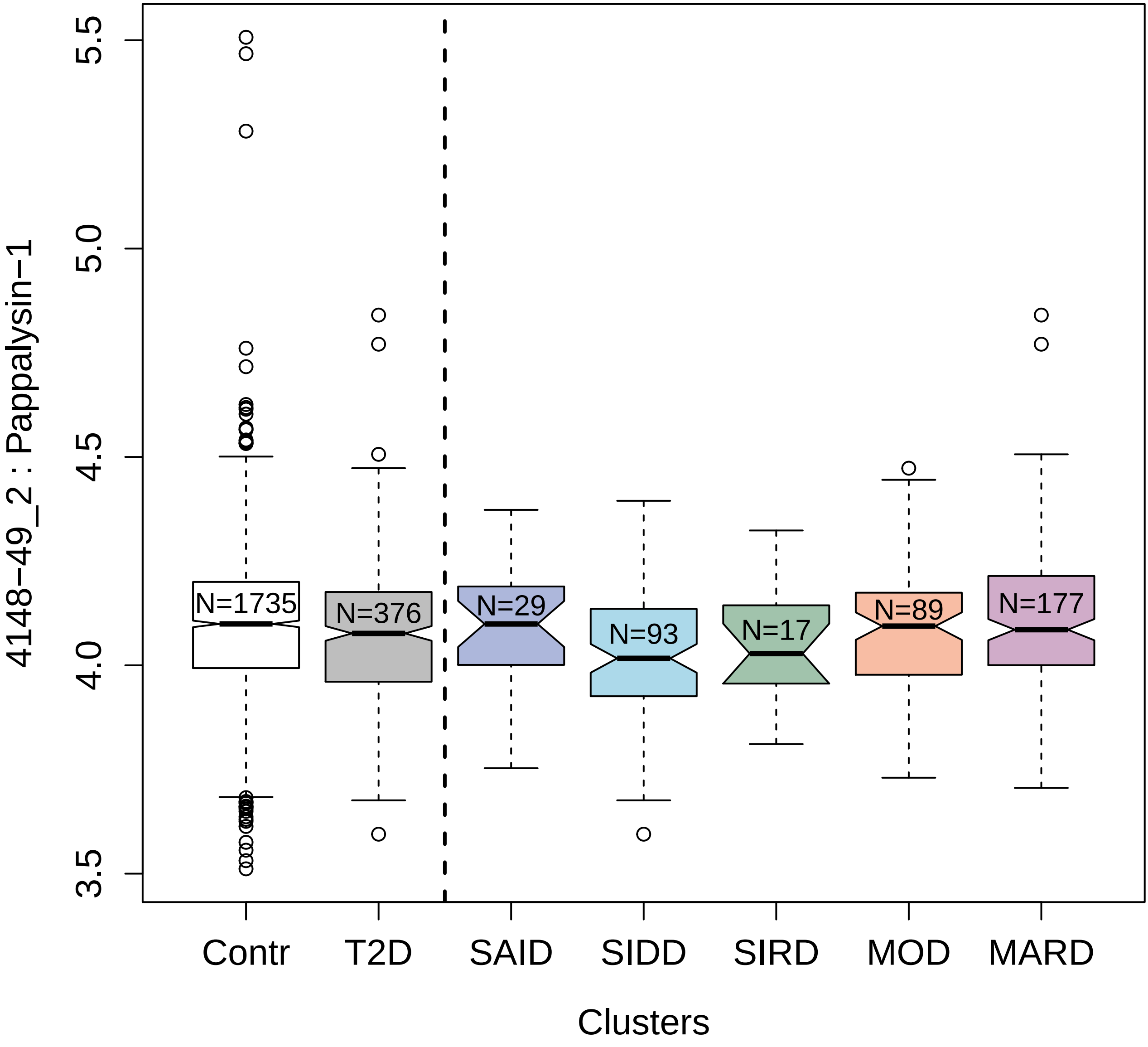

4148−49_2 : Pappalysin−1
5.5
5.0
4.5
4.0
3.5
N=1735
N=376
N=29
N=93
N=17
N=89
N=177
Contr
T2D
SAID
SIDD
SIRD
MOD
MARD
Clusters

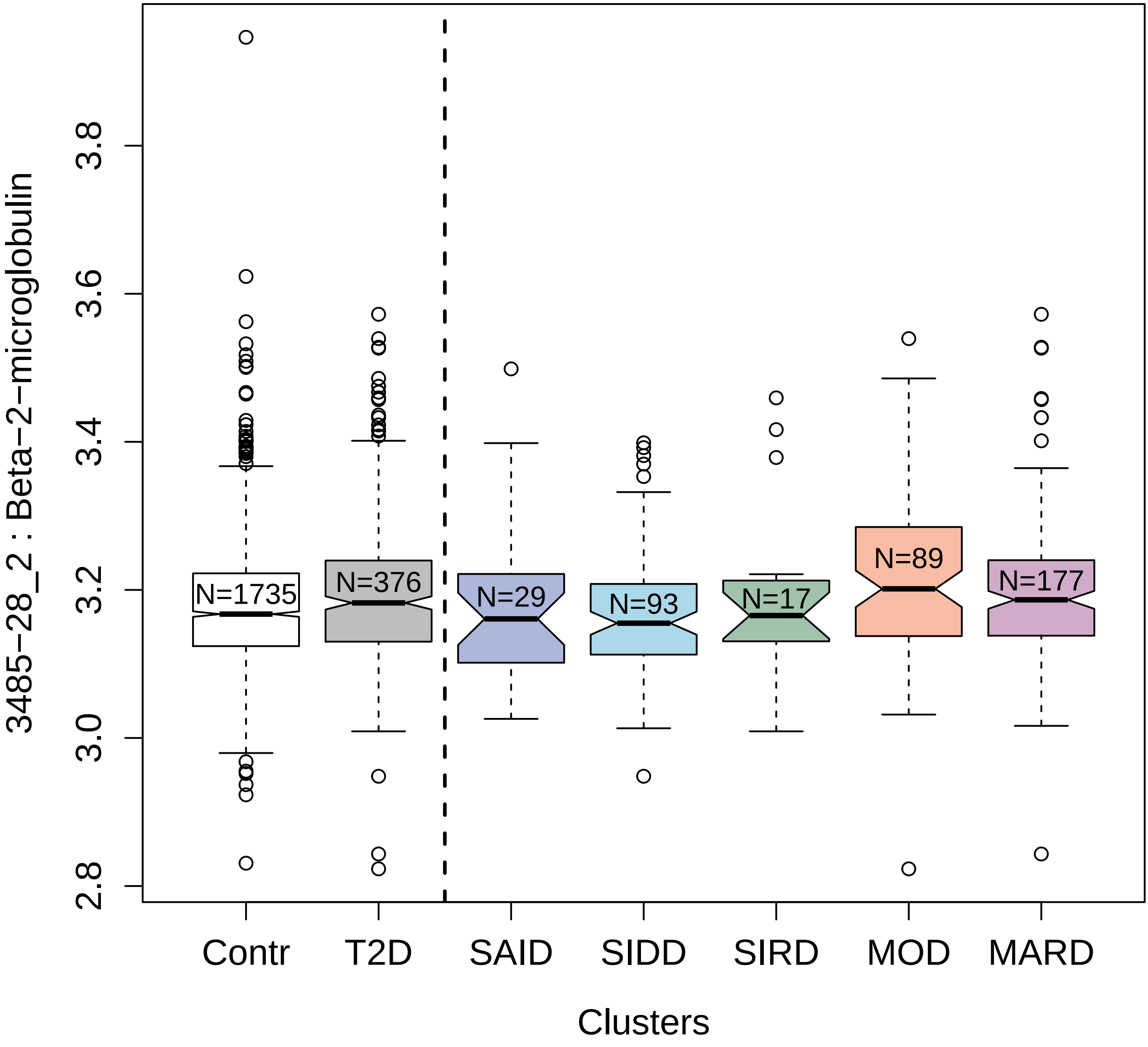

3485−28_2 : Beta−2−microglobulin
Clusters
Contr
T2D
SAID
SIDD
SIRD
MOD
MARD
N=1735
N=376
N=29
N=93
N=17
N=89
N=177
2.8
3.0
3.2
3.4
3.6
3.8

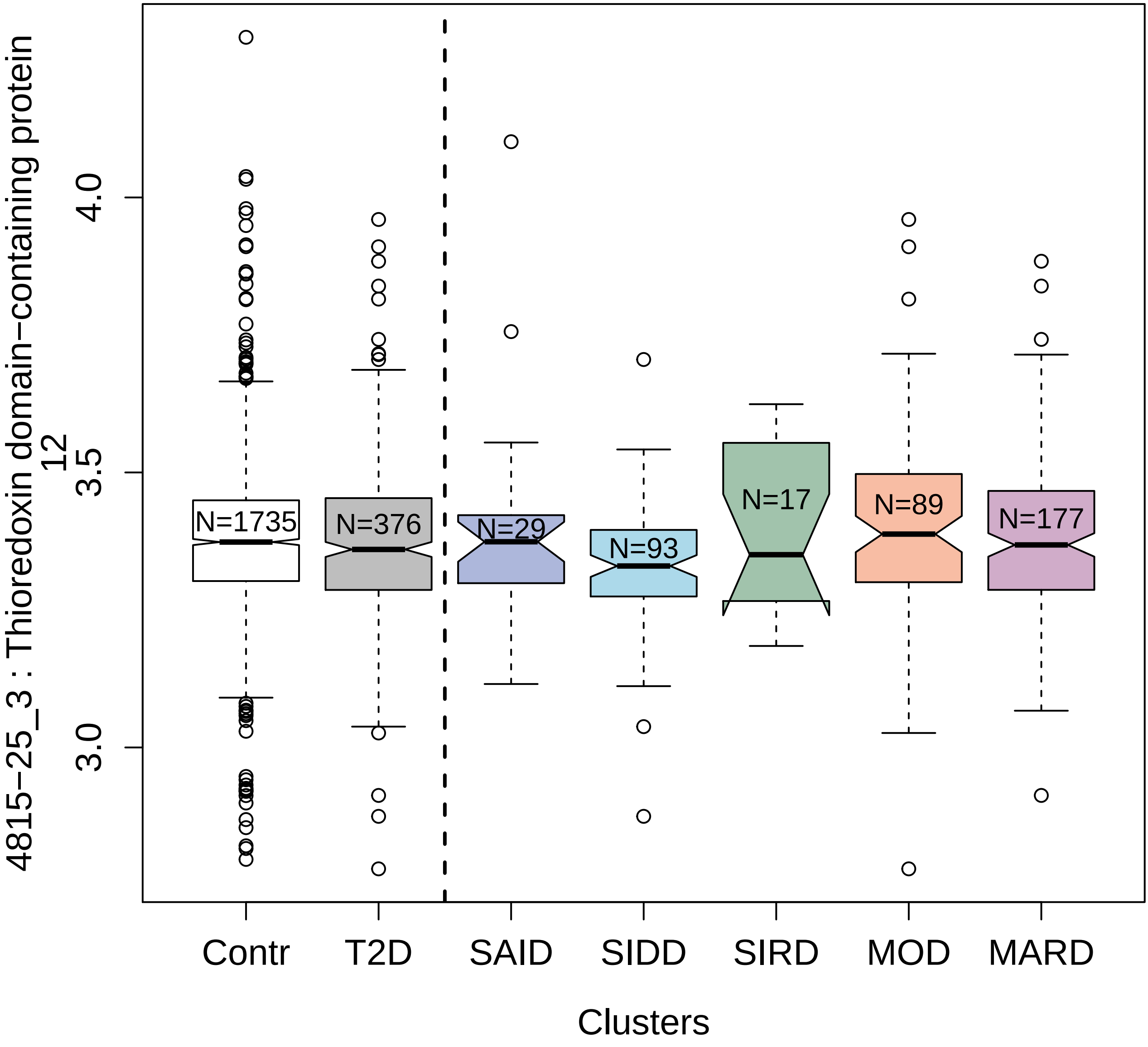
4815−25_3 : Thioredoxin domain−containing protein 12
3.0
3.5
4.0
N=1735
N=376
N=29
N=93
N=17
N=89
N=177
Contr
T2D
SAID
SIDD
SIRD
MOD
MARD
Clusters

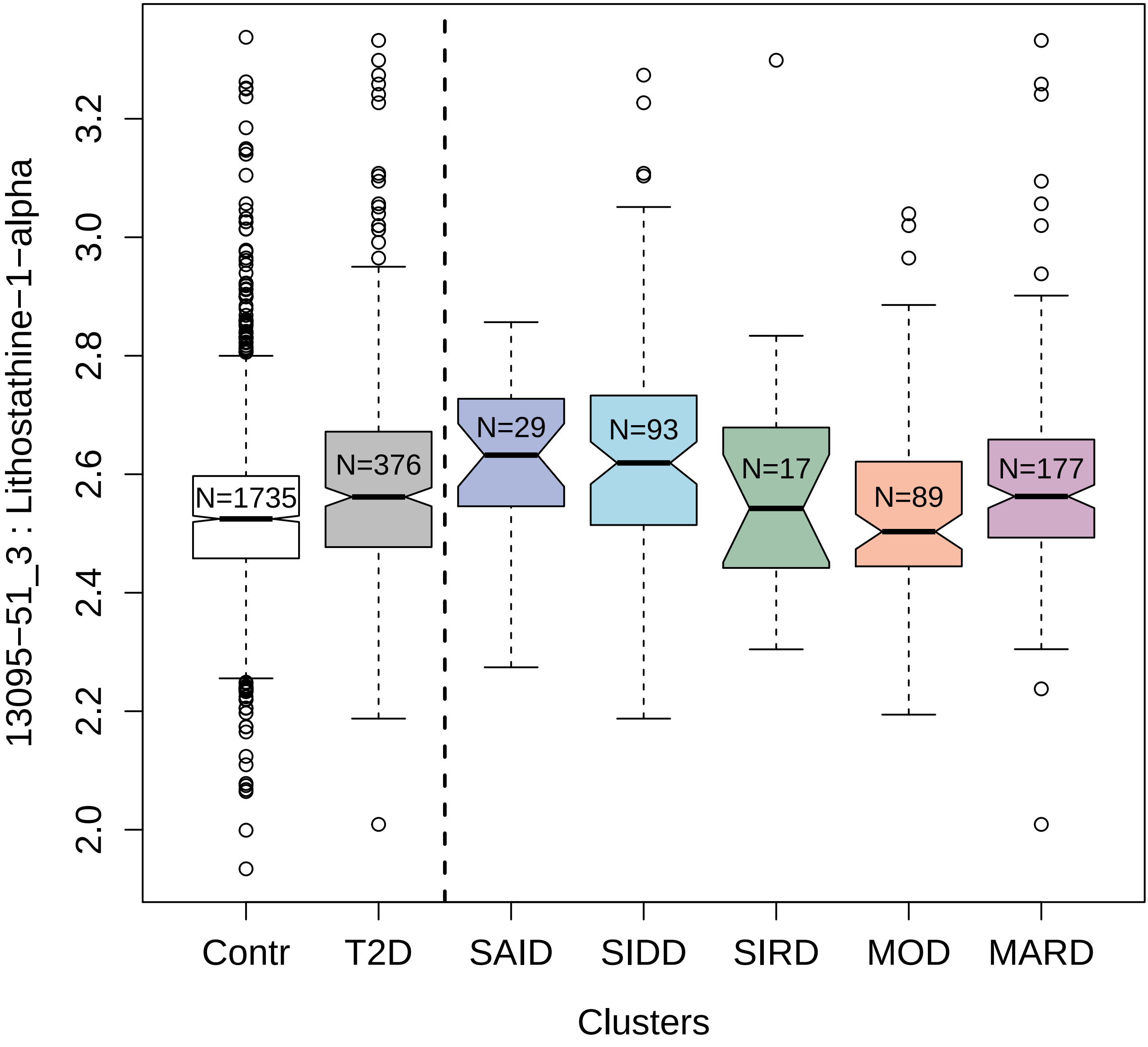

13095−51_3 : Lithostathine−1−alpha
2.0
2.2
2.4
2.6
2.8
3.0
3.2
N=1735
N=376
N=29
N=93
N=17
N=89
N=177
Contr
T2D
SAID
SIDD
SIRD
MOD
MARD
Clusters

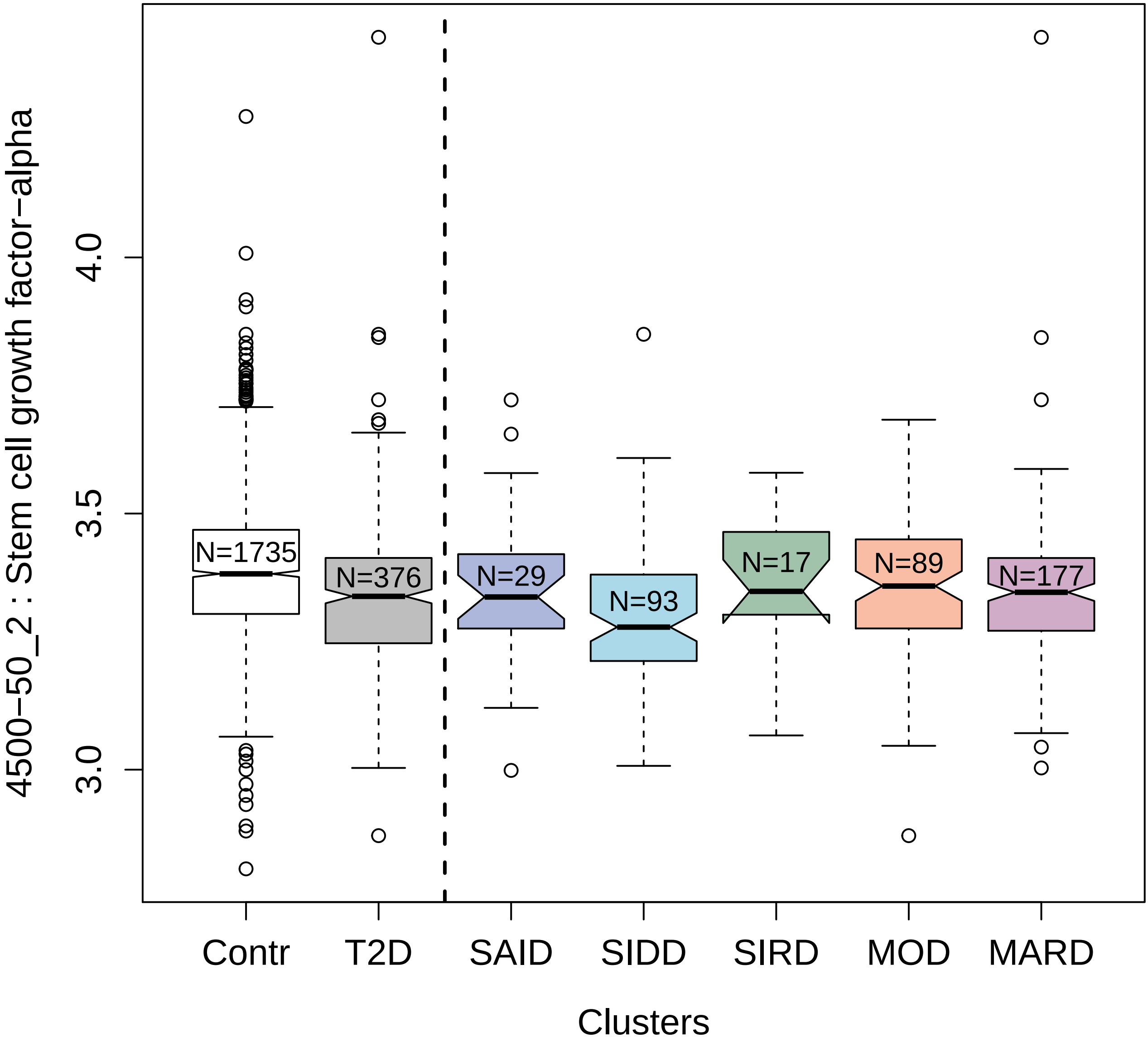

4500−50_2 : Stem cell growth factor−alpha
4.0
3.5
3.0
N=1735
N=376
N=29
N=93
N=17
N=89
N=177
Contr
T2D
SAID
SIDD
SIRD
MOD
MARD
Clusters

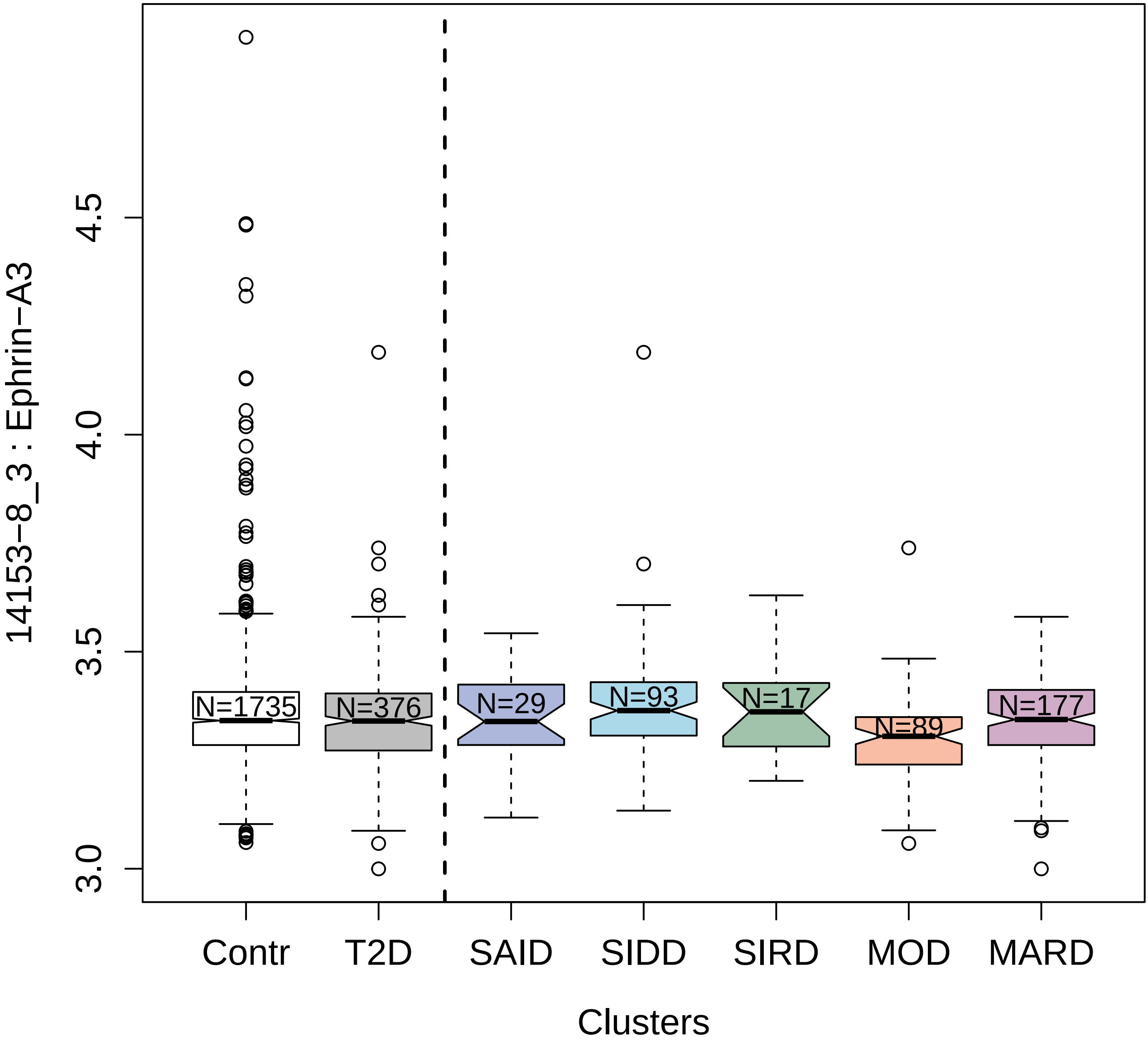

14153−8_3 : Ephrin−A3
3.0
3.5
4.0
4.5
N=1735
N=376
N=29
N=93
N=17
N=89
N=177
Contr
T2D
SAID
SIDD
SIRD
MOD
MARD
Clusters

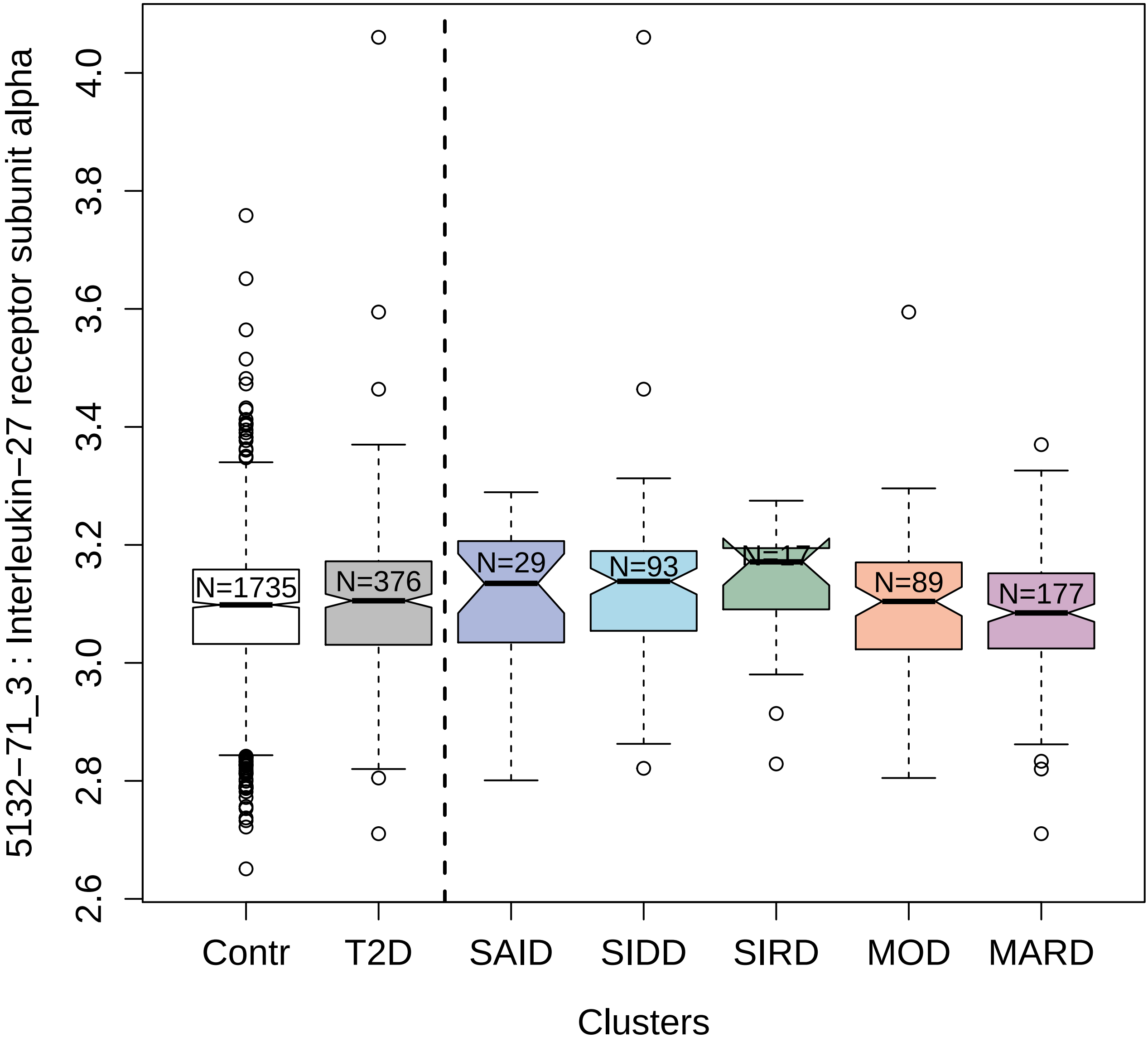

5132−71_3 : Interleukin−27 receptor subunit alpha
4.0
3.8
3.6
3.4
3.2
3.0
2.8
2.6
N=1735
N=376
N=29
N=93
N=17
N=89
N=177
Contr
T2D
SAID
SIDD
SIRD
MOD
MARD
Clusters

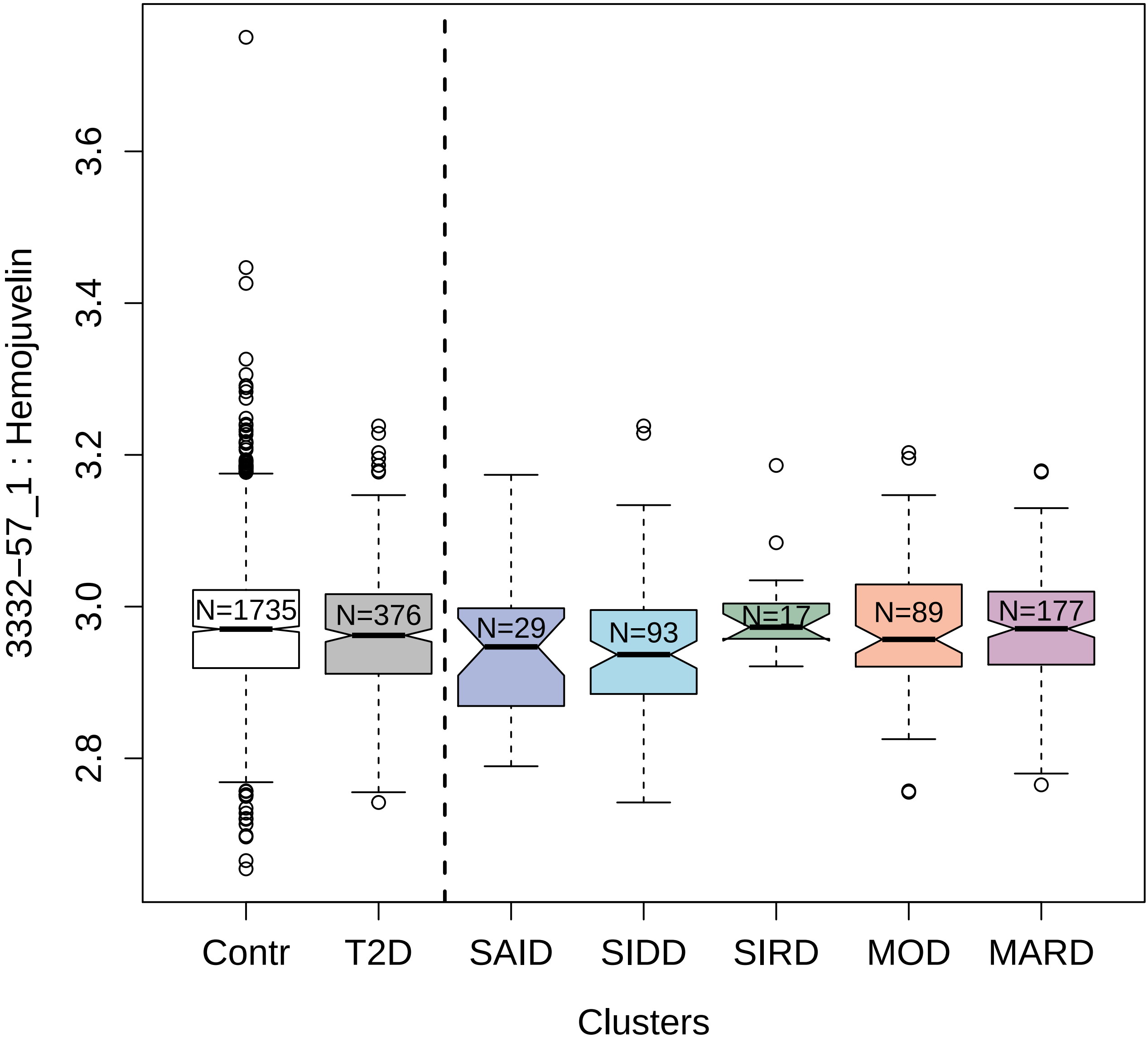
3332−57_1 : Hemojuvelin
3.6
3.4
3.2
3.0
2.8
N=1735
N=376
N=29
N=93
N=17
N=89
N=177
Contr
T2D
SAID
SIDD
SIRD
MOD
MARD
Clusters

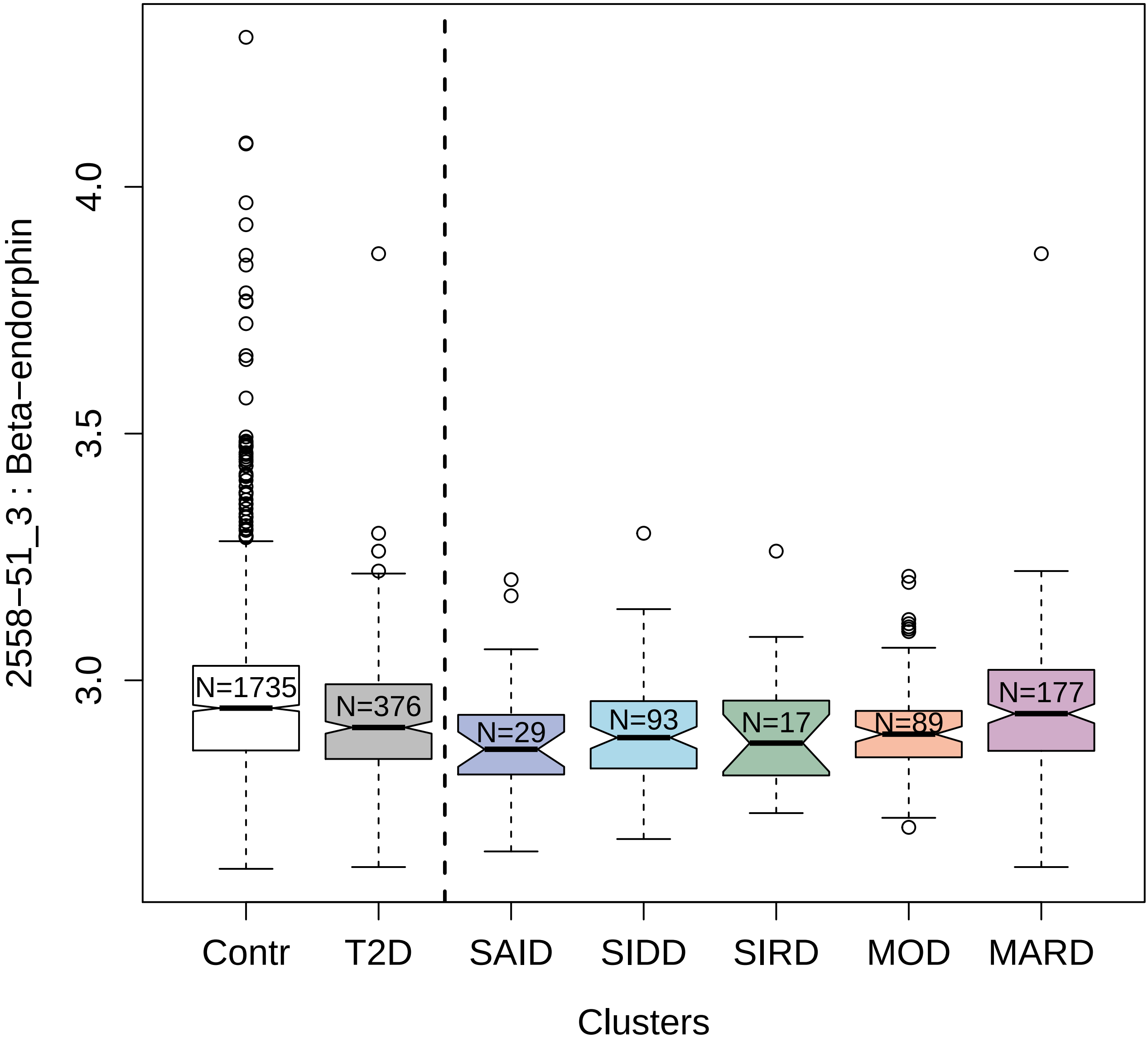
2558−51_3 : Beta−endorphin
4.0
3.5
3.0
N=1735
N=376
N=29
N=93
N=17
N=89
N=177
Contr
T2D
SAID
SIDD
SIRD
MOD
MARD
Clusters

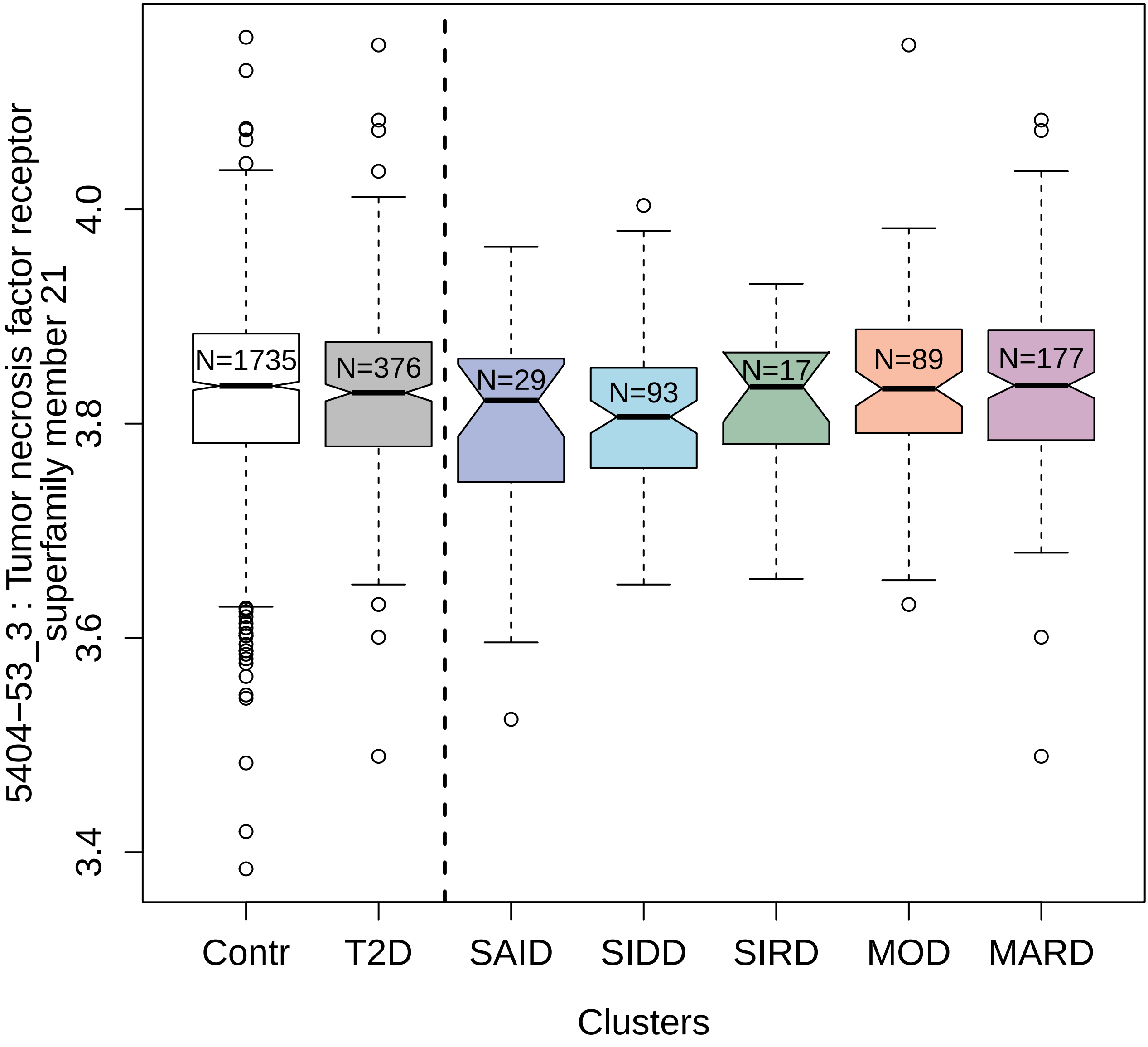

5404−53_3 : Tumor necrosis factor receptor superfamily member 21
3.4
3.6
3.8
4.0
N=1735
N=376
N=29
N=93
N=17
N=89
N=177
Contr
T2D
SAID
SIDD
SIRD
MOD
MARD
Clusters

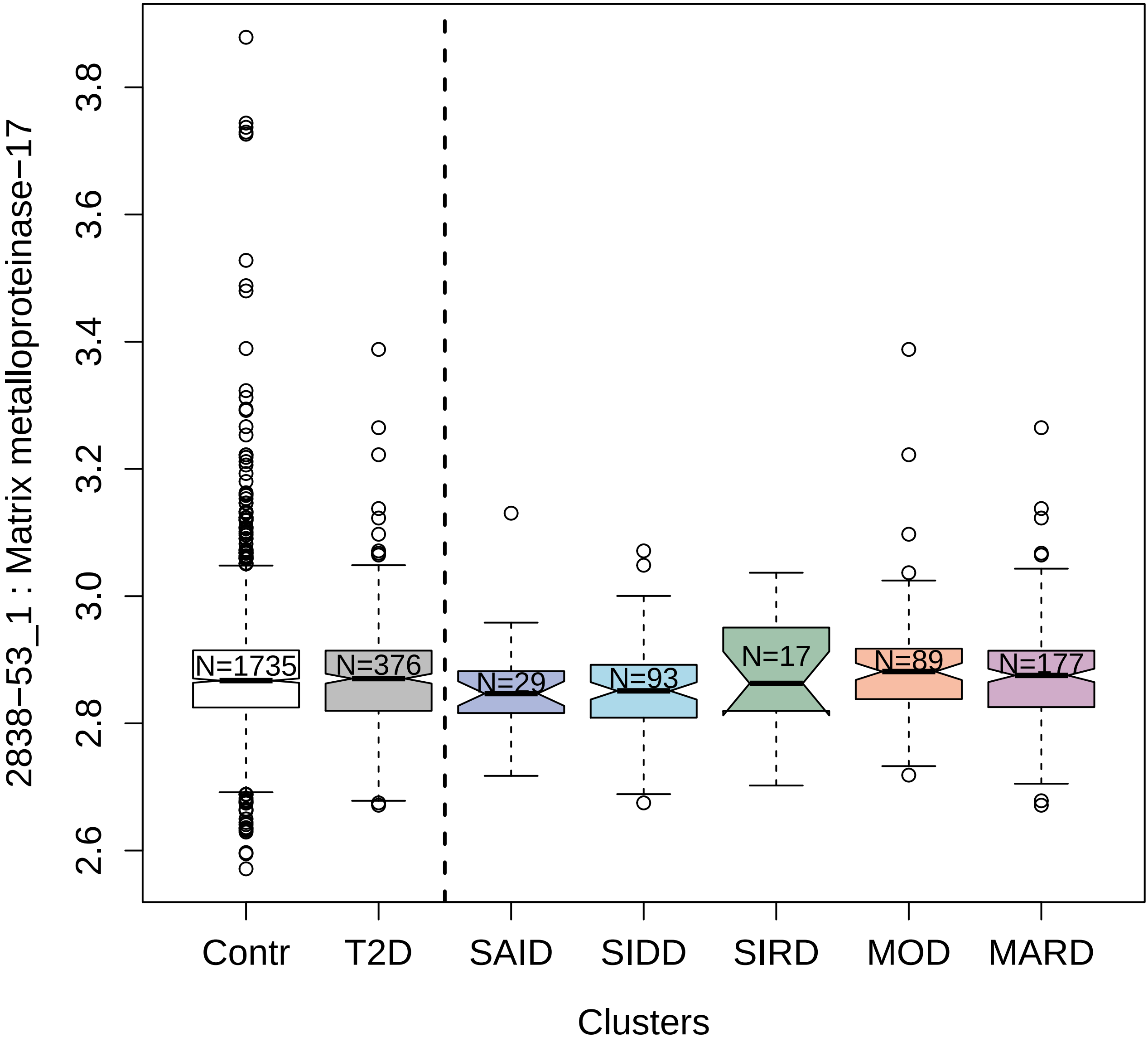

2838−53_1 : Matrix metalloproteinase−17
2.6
2.8
3.0
3.2
3.4
3.6
3.8
N=1735
N=376
N=29
N=93
N=17
N=89
N=177
Contr
T2D
SAID
SIDD
SIRD
MOD
MARD
Clusters

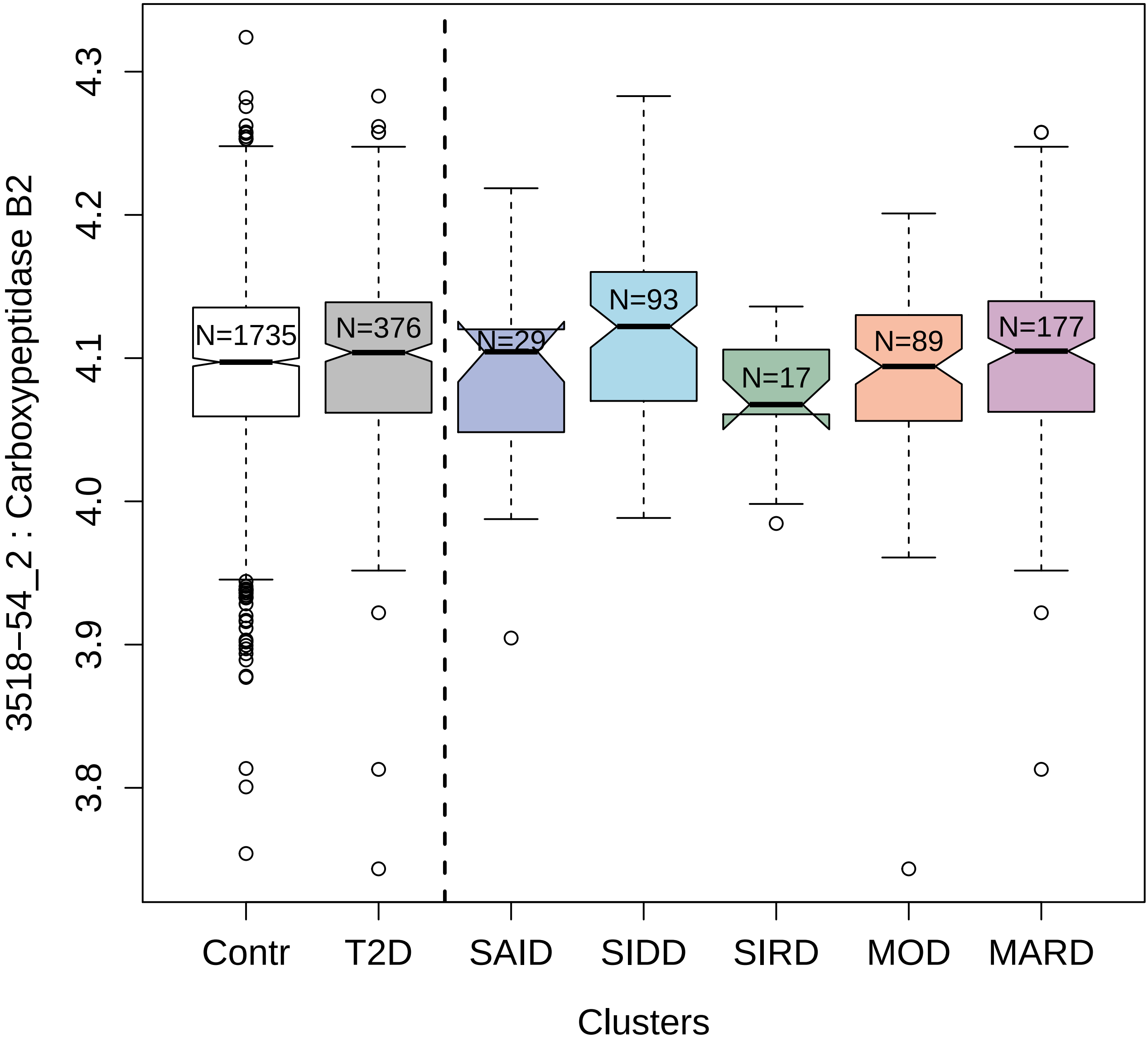

3518−54_2 : Carboxypeptidase B2
4.3
4.2
4.1
4.0
3.9
3.8
N=1735
N=376
N=29
N=93
N=17
N=89
N=177
Contr
T2D
SAID
SIDD
SIRD
MOD
MARD
Clusters

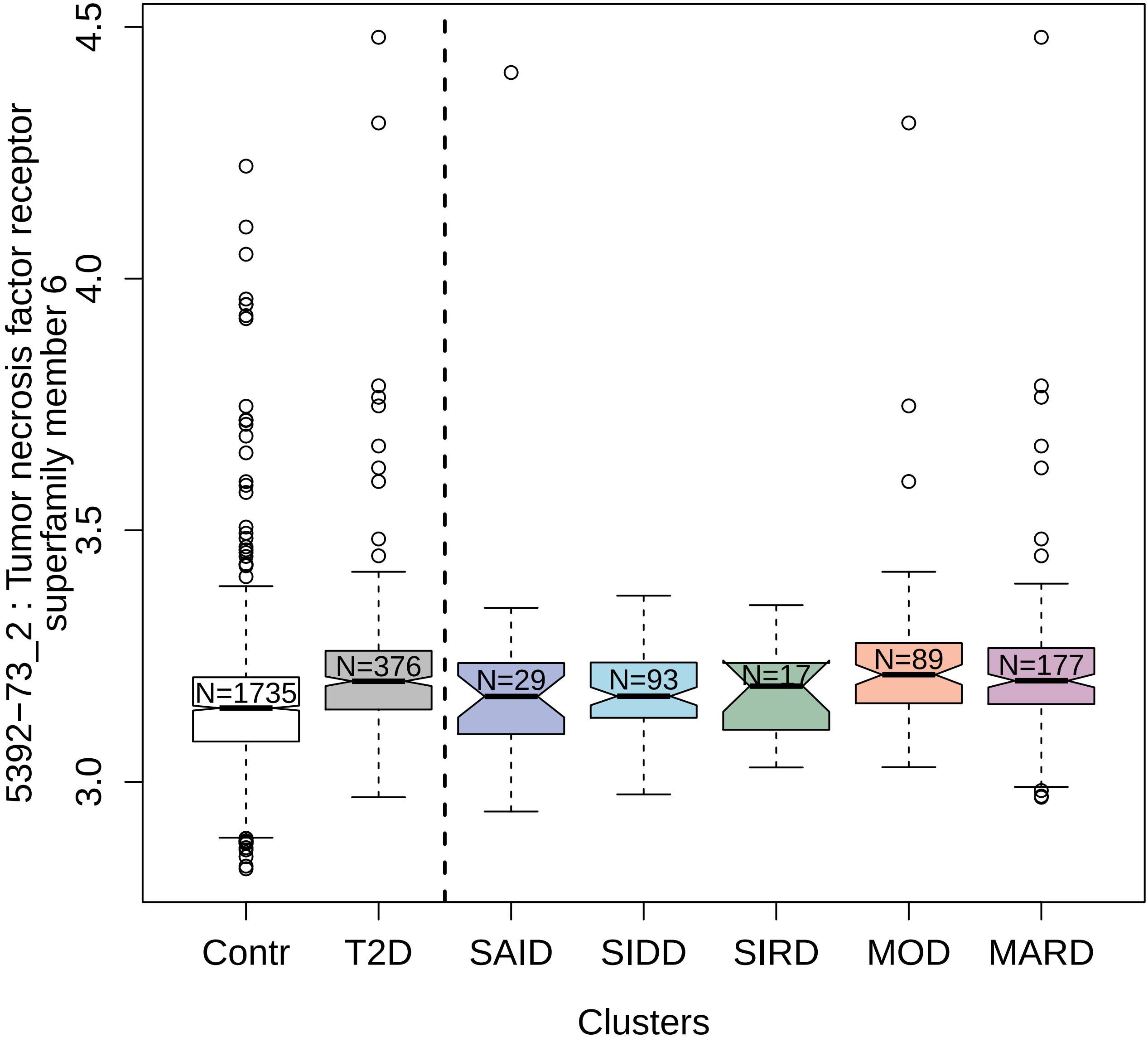

5392−73_2 : Tumor necrosis factor receptor superfamily member 6
4.5
4.0
3.5
3.0
N=1735
N=376
N=29
N=93
N=17
N=89
N=177
Contr
T2D
SAID
SIDD
SIRD
MOD
MARD
Clusters

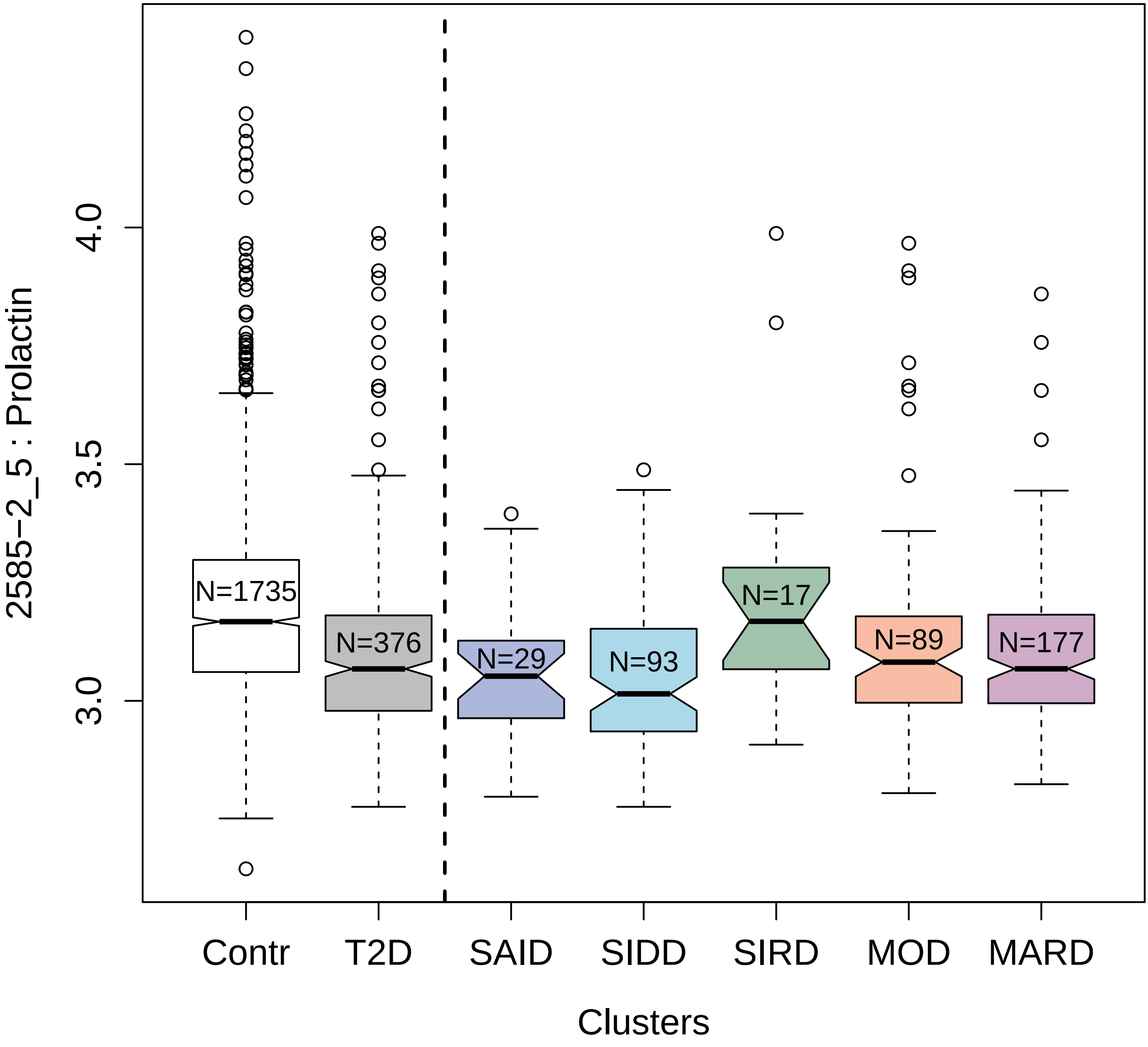
2585−2_5 : Prolactin
4.0
3.5
3.0
N=1735
N=376
N=29
N=93
N=17
N=89
N=177
Contr
T2D
SAID
SIDD
SIRD
MOD
MARD
Clusters

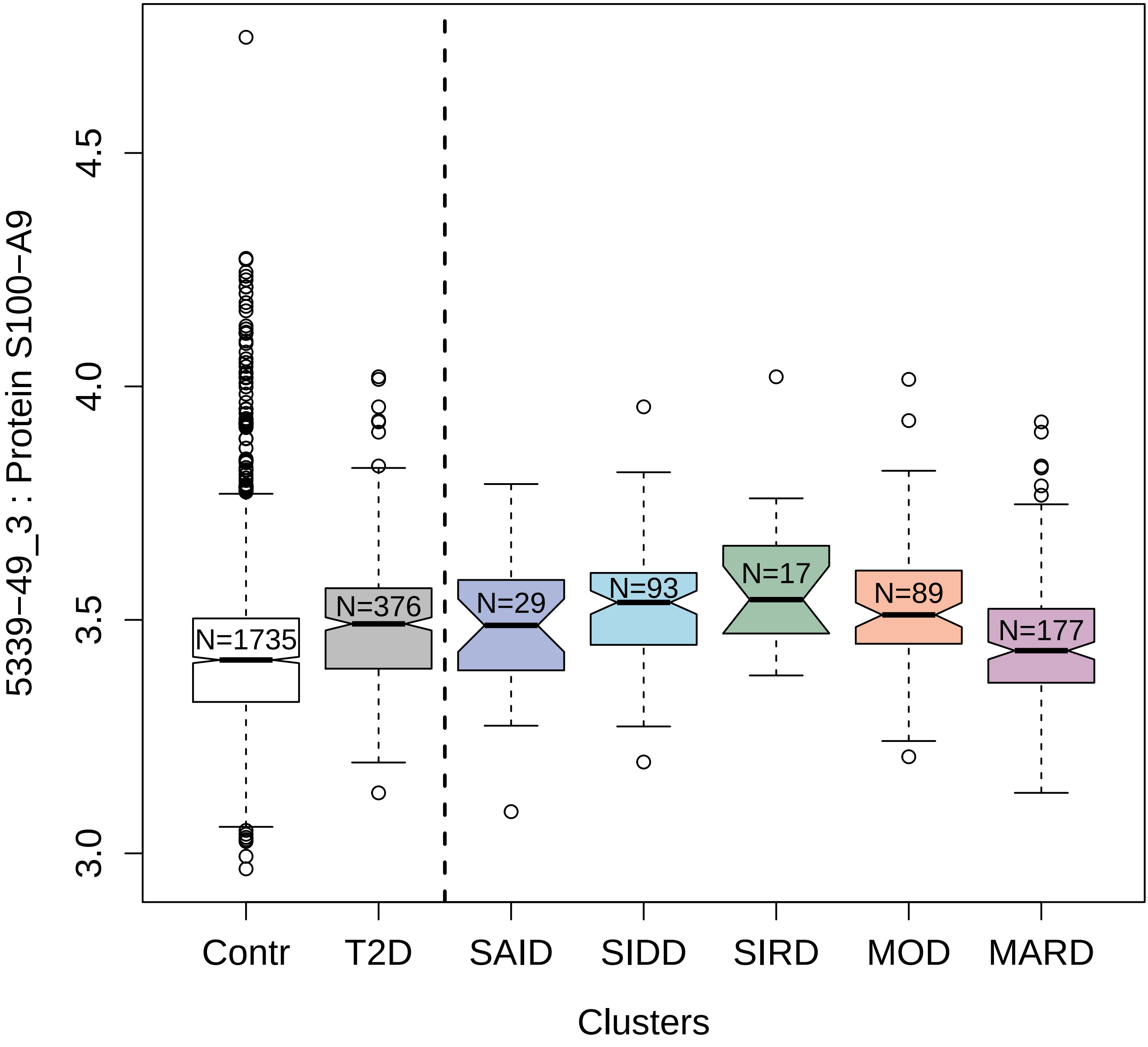

5339−49_3 : Protein S100−A9
4.5
4.0
3.5
3.0
N=1735
N=376
N=29
N=93
N=17
N=89
N=177
Contr
T2D
SAID
SIDD
SIRD
MOD
MARD
Clusters

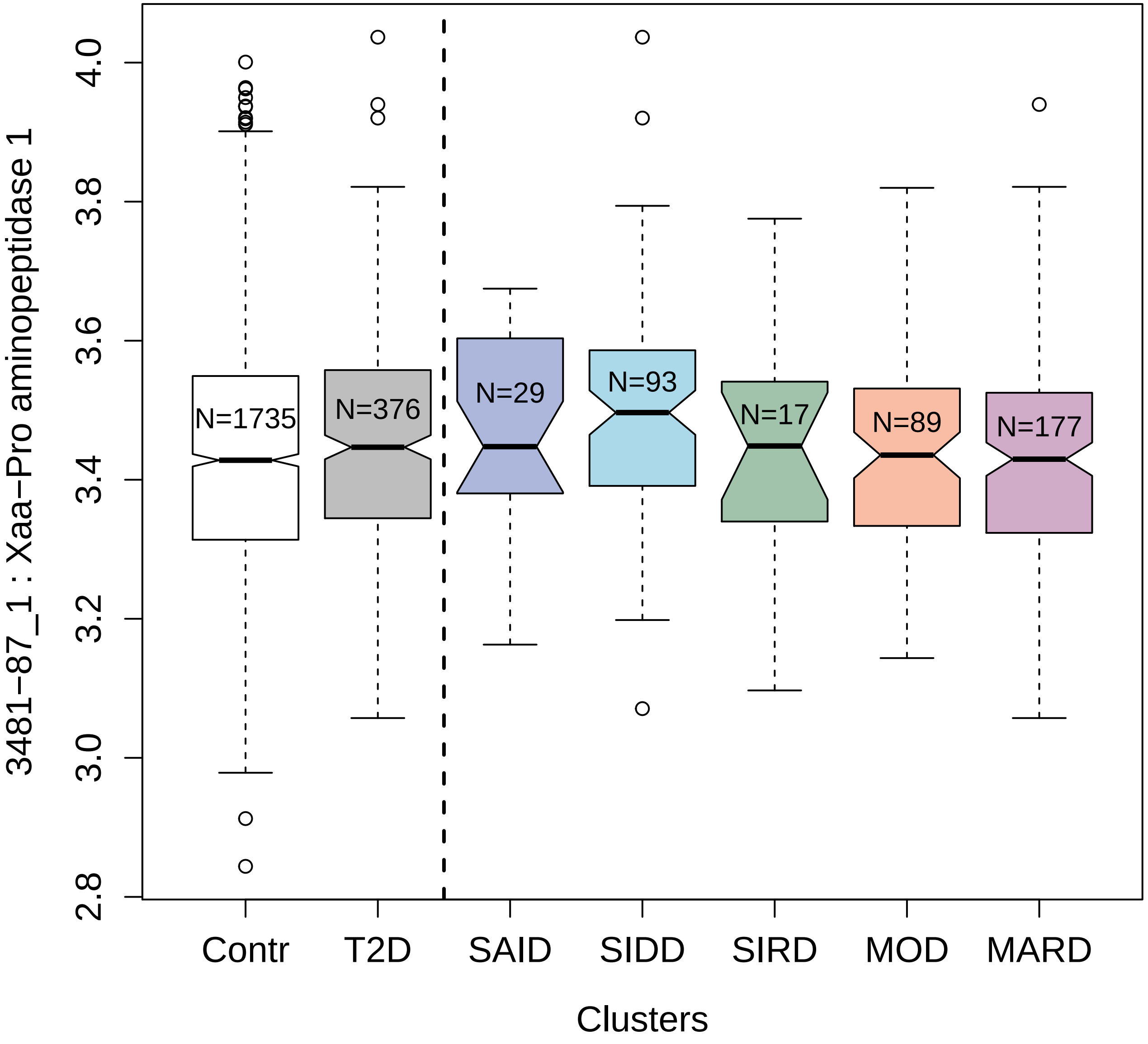
3481−87_1 : Xaa−Pro aminopeptidase 1
2.8
3.0
3.2
3.4
3.6
3.8
4.0
N=1735
N=376
N=29
N=93
N=17
N=89
N=177
Contr
T2D
SAID
SIDD
SIRD
MOD
MARD
Clusters

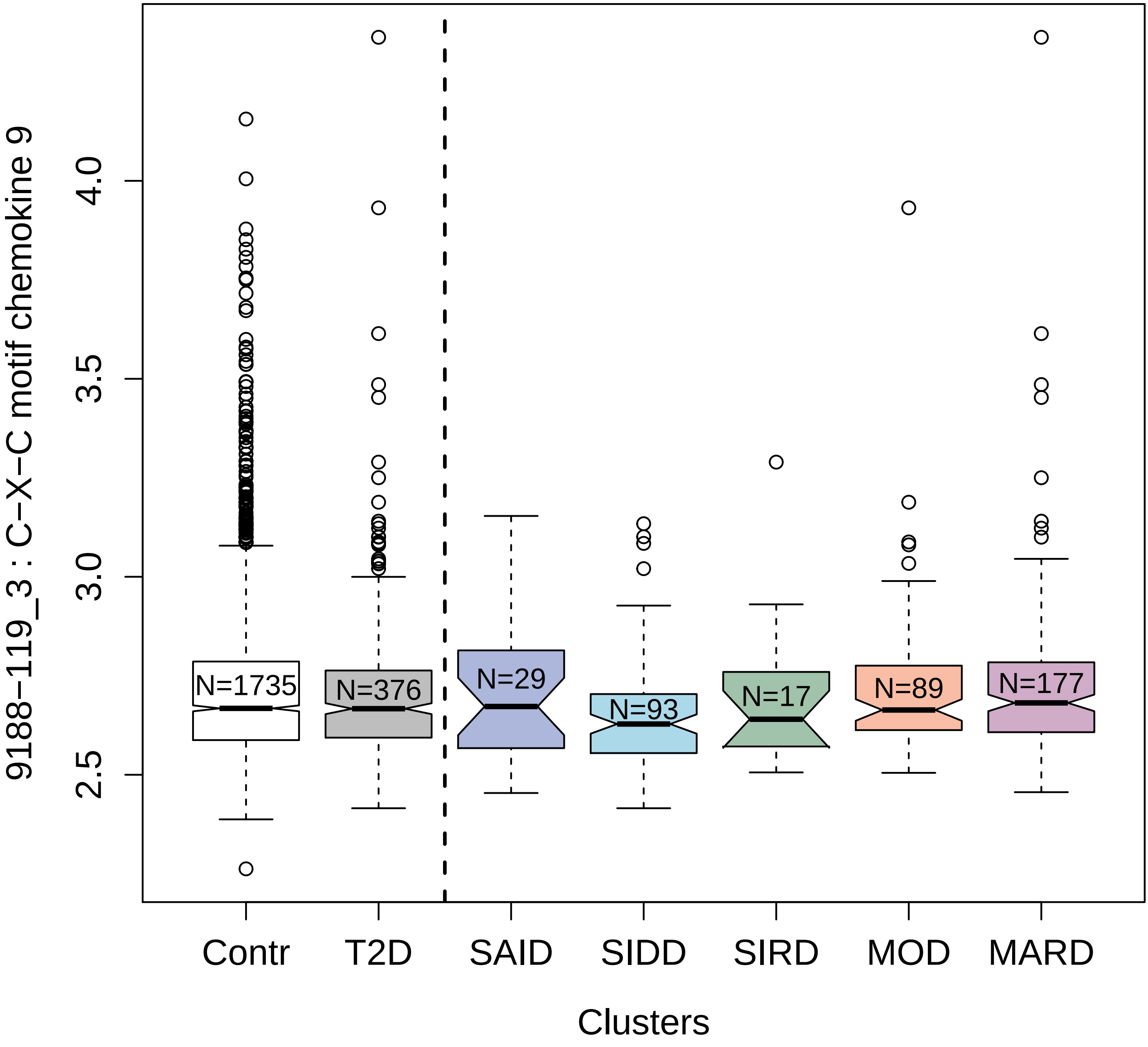
9188−119_3 : C−X−C motif chemokine 9
4.0
3.5
3.0
2.5
N=1735
N=376
N=29
N=93
N=17
N=89
N=177
Contr
T2D
SAID
SIDD
SIRD
MOD
MARD
Clusters

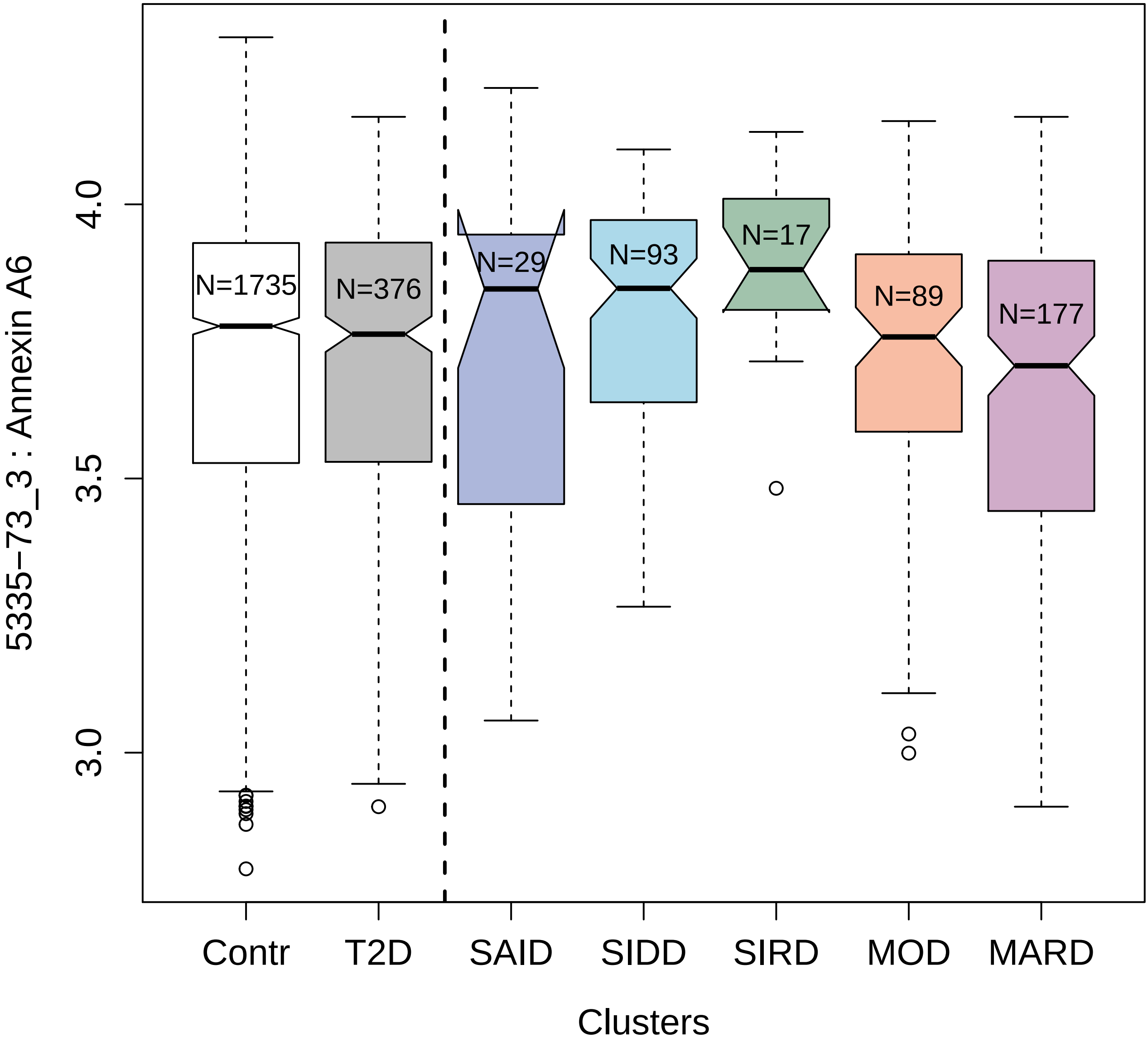
5335−73_3 : Annexin A6
3.0
3.5
4.0
N=1735
N=376
N=29
N=93
N=17
N=89
N=177
Contr
T2D
SAID
SIDD
SIRD
MOD
MARD
Clusters

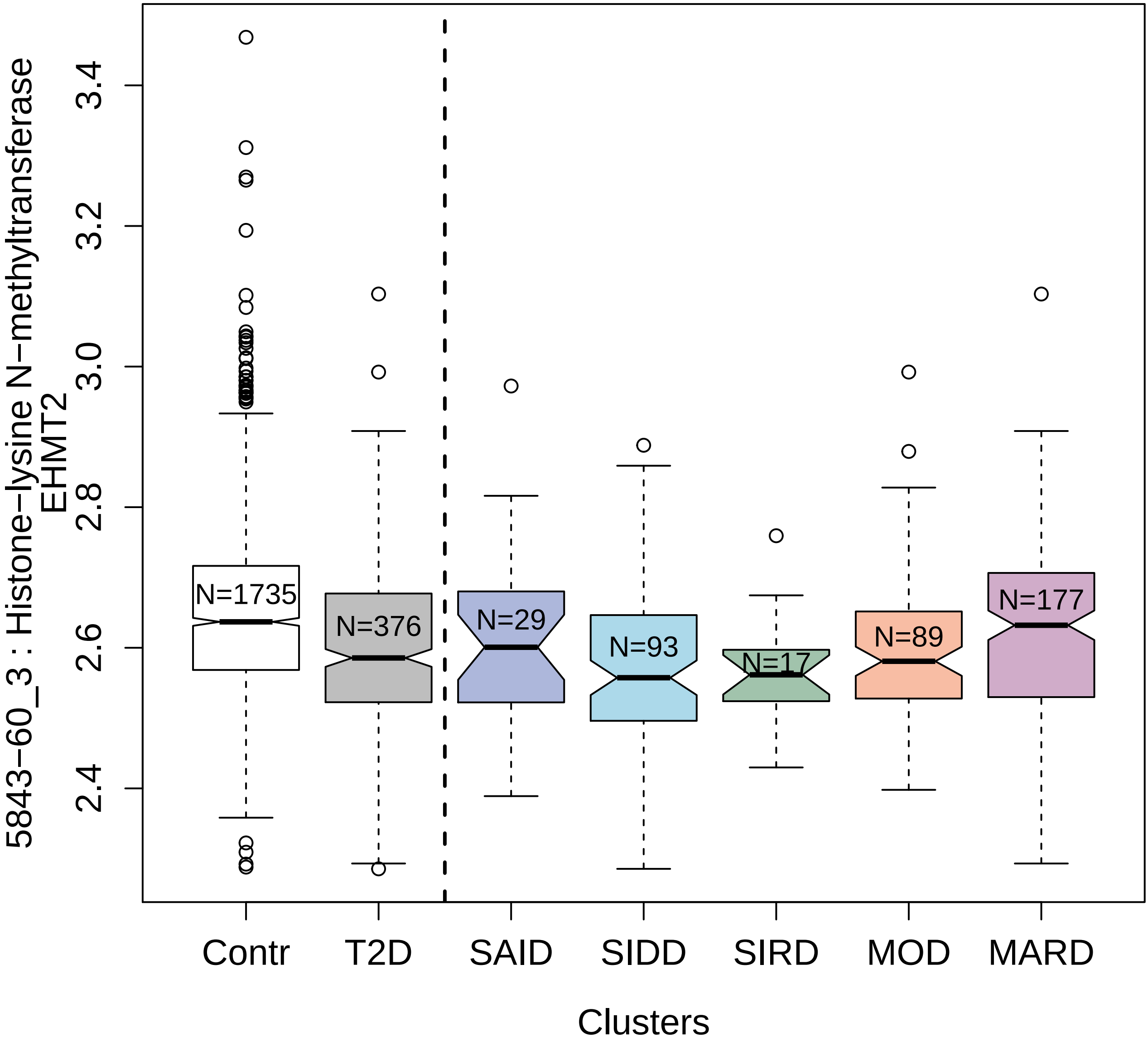

5843−60_3 : Histone−lysine N−methyltransferase
EHMT2
3.4
3.2
3.0
2.8
2.6
2.4
N=1735
N=376
N=29
N=93
N=17
N=89
N=177
Contr
T2D
SAID
SIDD
SIRD
MOD
MARD
Clusters

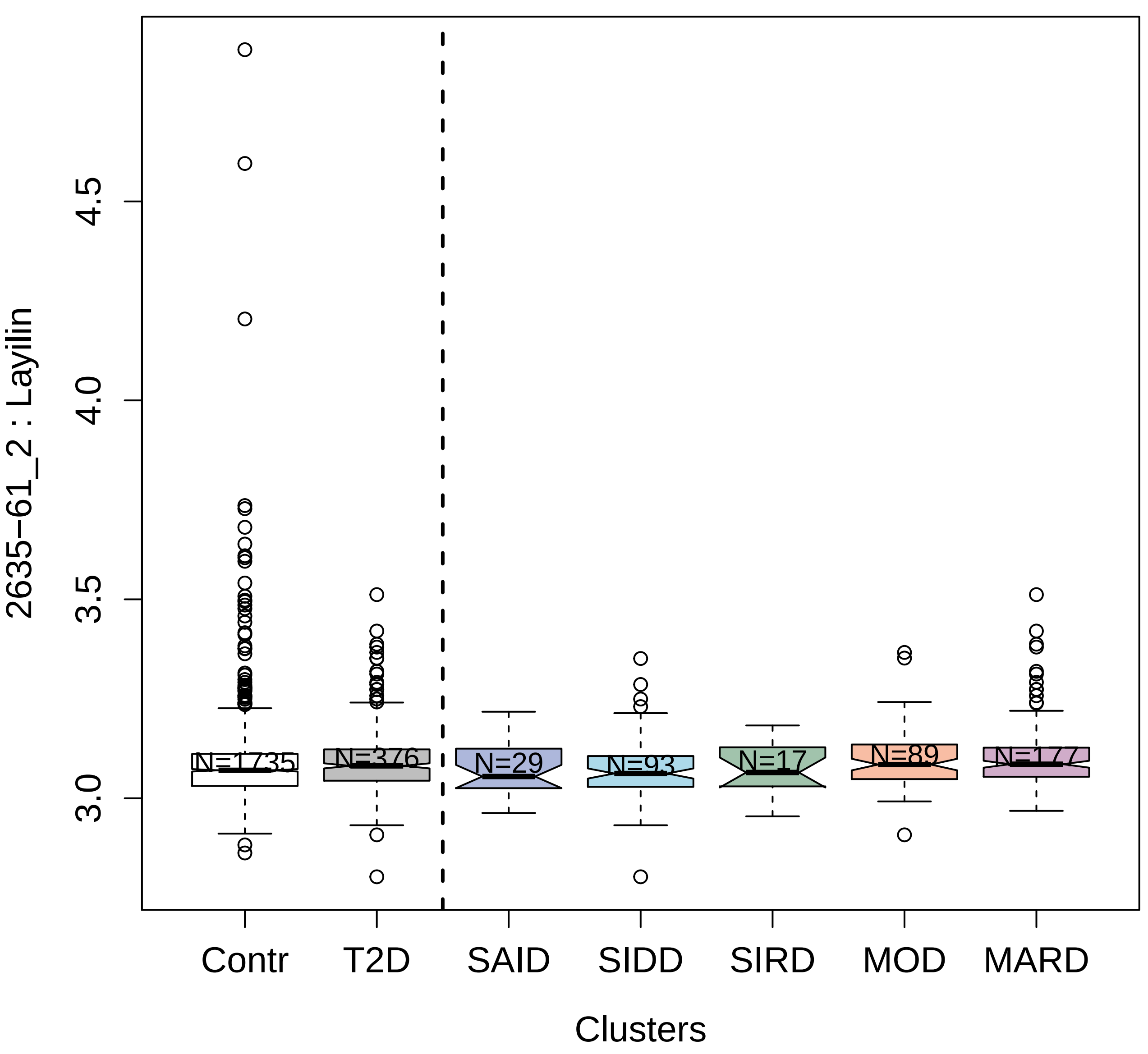
2635−61_2 : Layilin
3.0
3.5
4.0
4.5
N=1735
N=376
N=29
N=93
N=17
N=89
N=177
Contr
T2D
SAID
SIDD
SIRD
MOD
MARD
Clusters

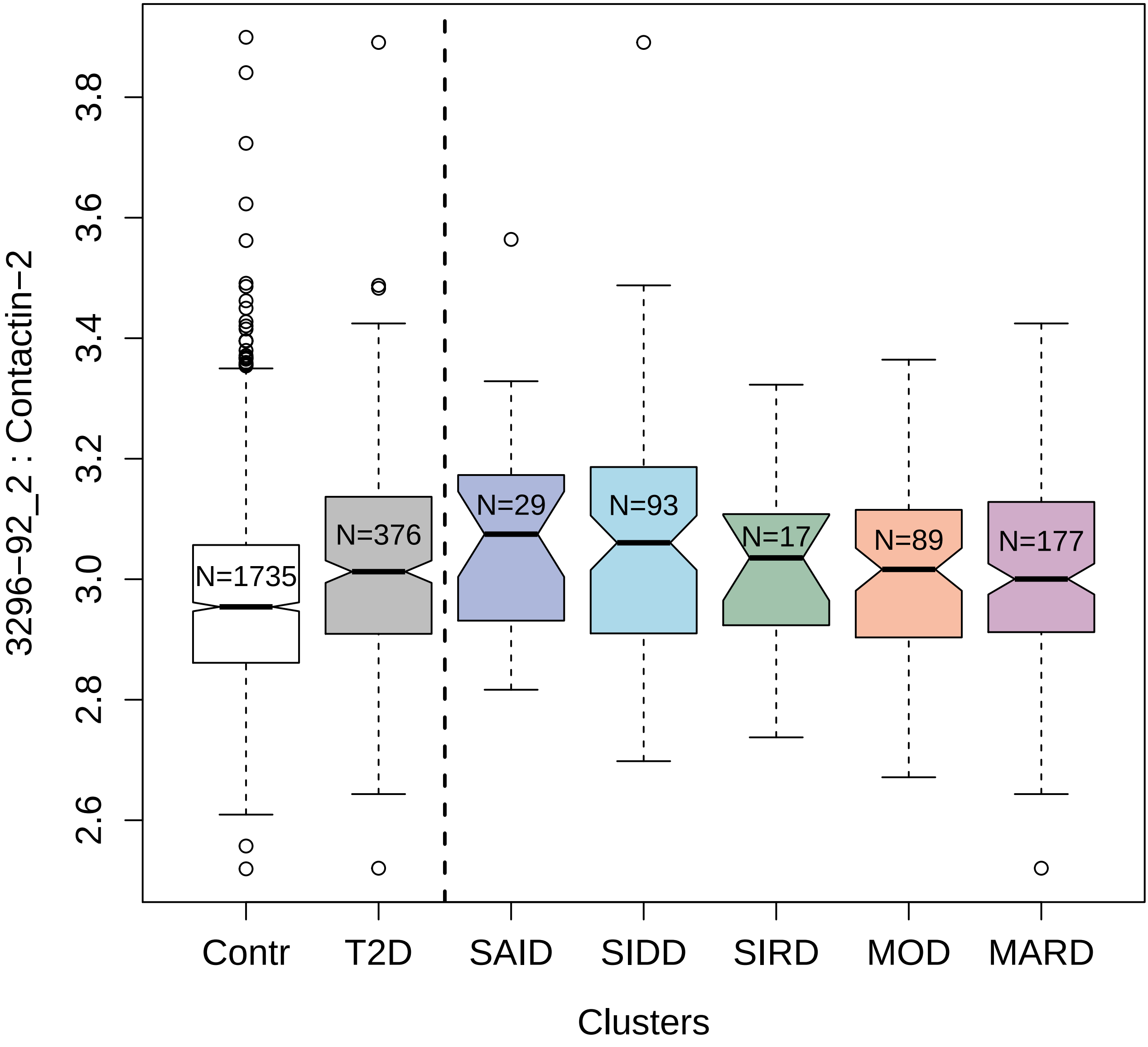

3296−92_2 : Contactin−2
3.8
3.6
3.4
3.2
3.0
2.8
2.6
N=1735
N=376
N=29
N=93
N=17
N=89
N=177
Contr
T2D
SAID
SIDD
SIRD
MOD
MARD
Clusters

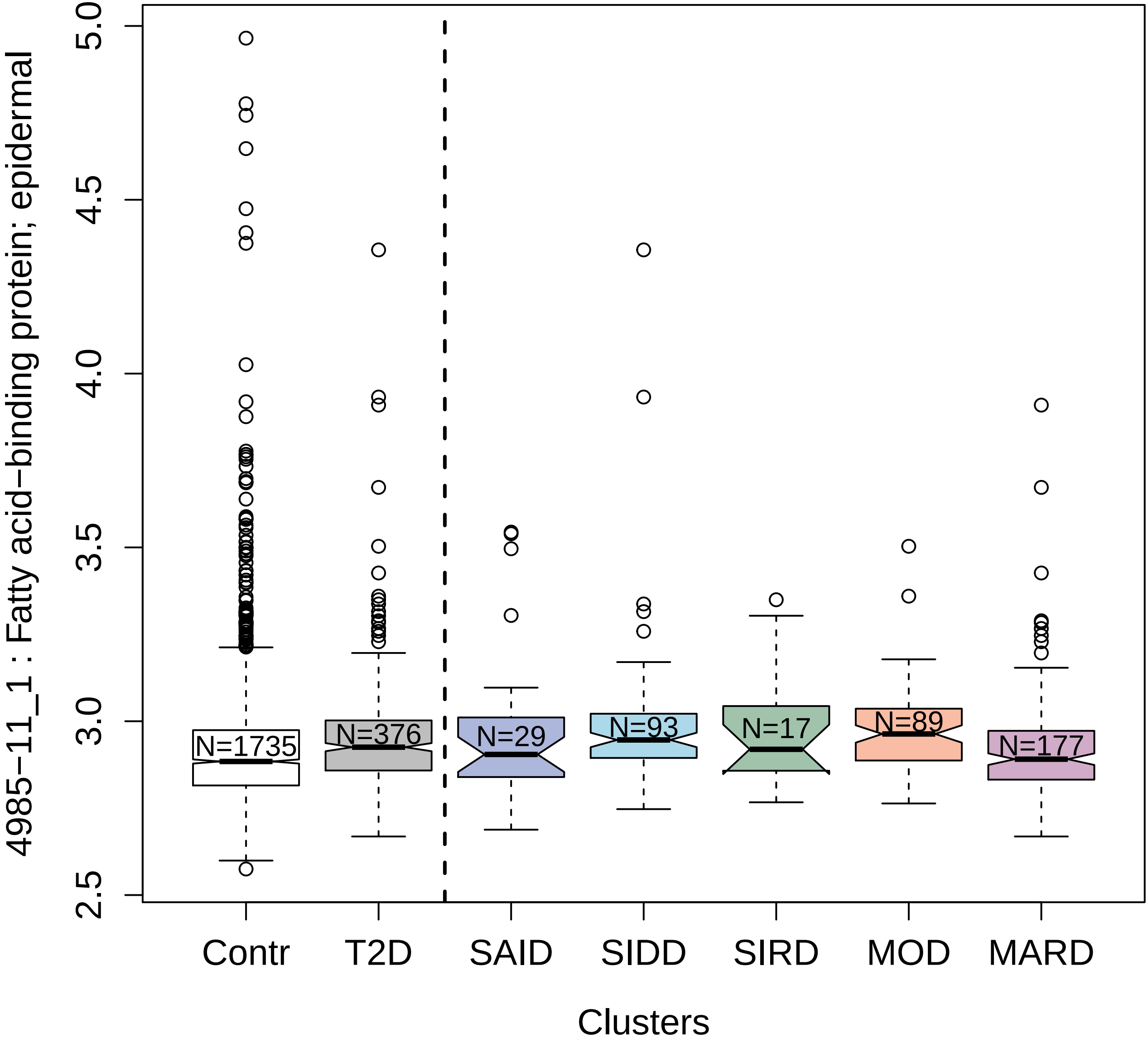
4985−11_1 : Fatty acid−binding protein; epidermal
2.5
3.0
3.5
4.0
4.5
5.0
N=1735
N=376
N=29
N=93
N=17
N=89
N=177
Contr
T2D
SAID
SIDD
SIRD
MOD
MARD
Clusters

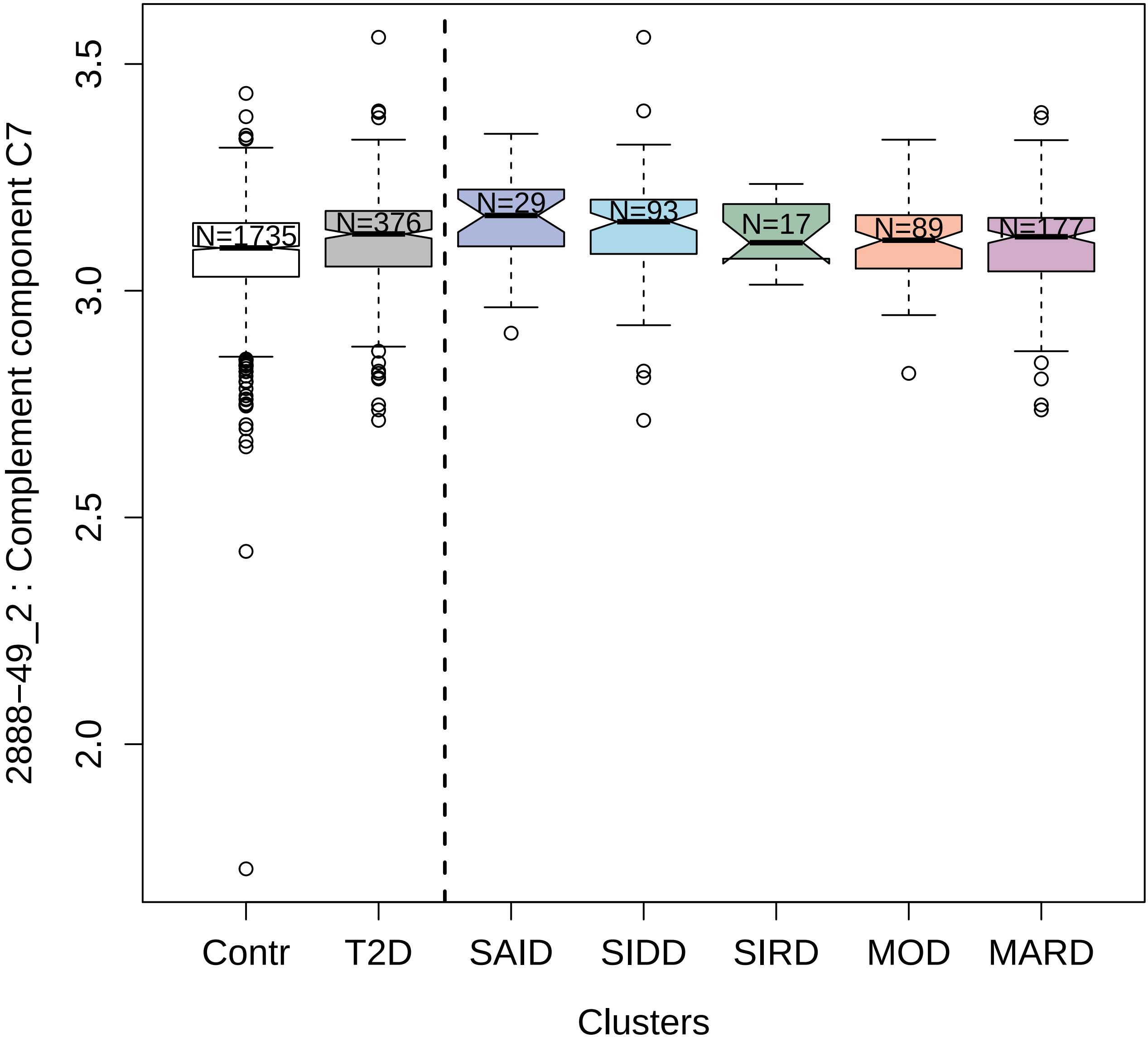
2888−49_2 : Complement component C7
3.5
3.0
2.5
2.0
N=1735
N=376
N=29
N=93
N=17
N=89
N=177
Contr
T2D
SAID
SIDD
SIRD
MOD
MARD
Clusters

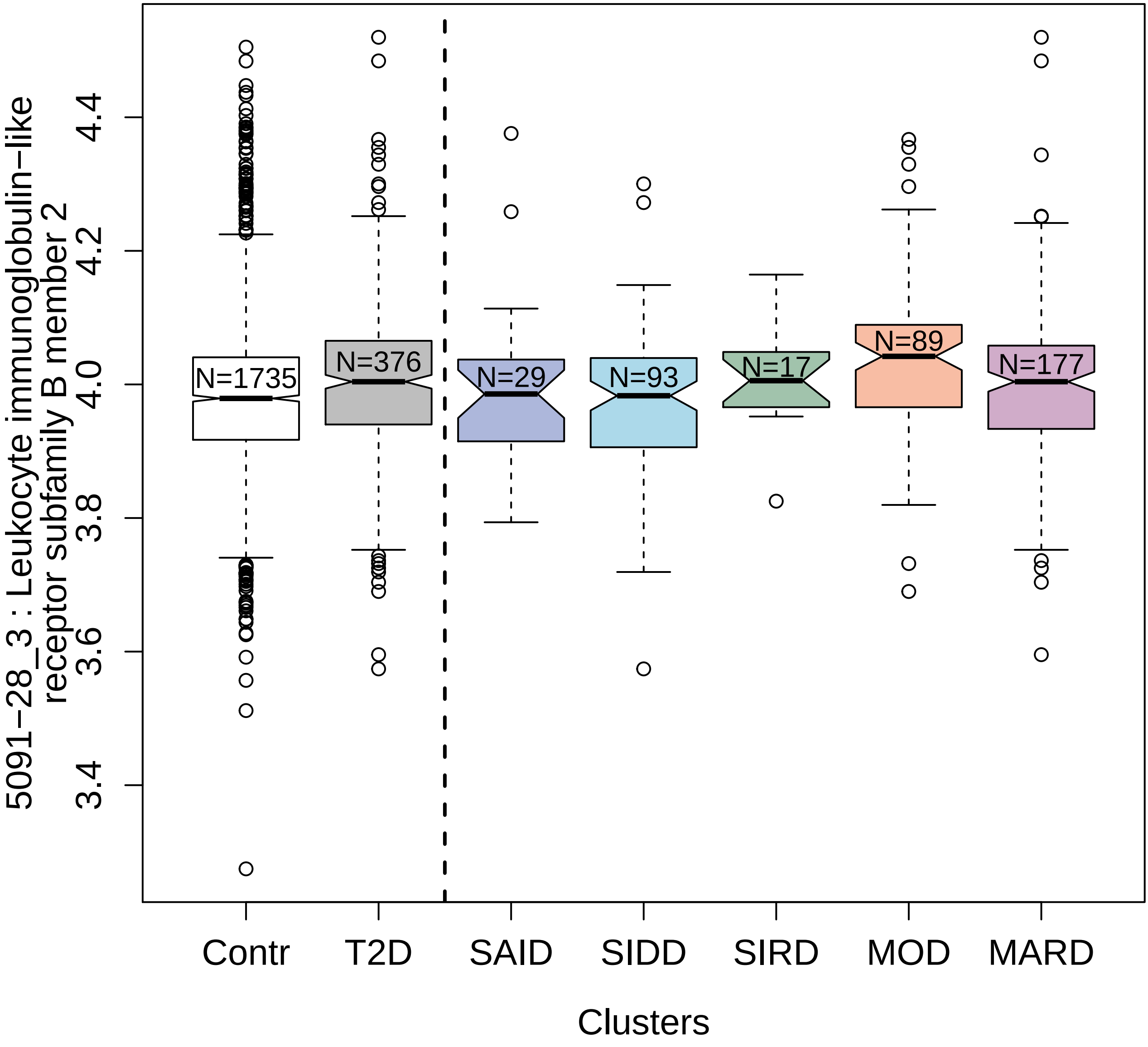

5091−28_3 : Leukocyte immunoglobulin−like receptor subfamily B member 2
3.4
3.6
3.8
4.0
4.2
4.4
N=1735
N=376
N=29
N=93
N=17
N=89
N=177
Contr
T2D
SAID
SIDD
SIRD
MOD
MARD
Clusters

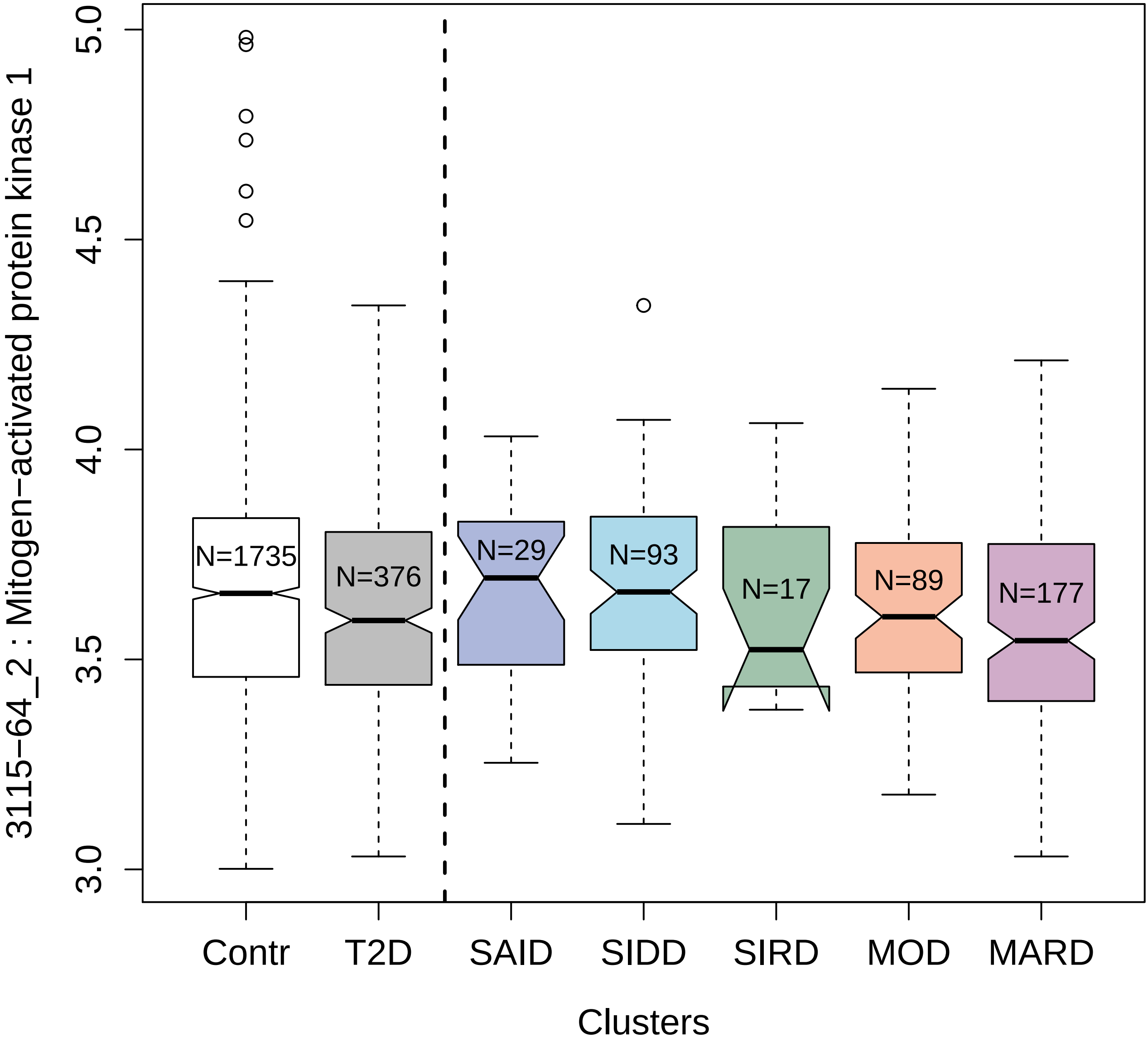

3115−64_2 : Mitogen−activated protein kinase 1
5.0
4.5
4.0
3.5
3.0
N=1735
N=376
N=29
N=93
N=17
N=89
N=177
Contr
T2D
SAID
SIDD
SIRD
MOD
MARD
Clusters

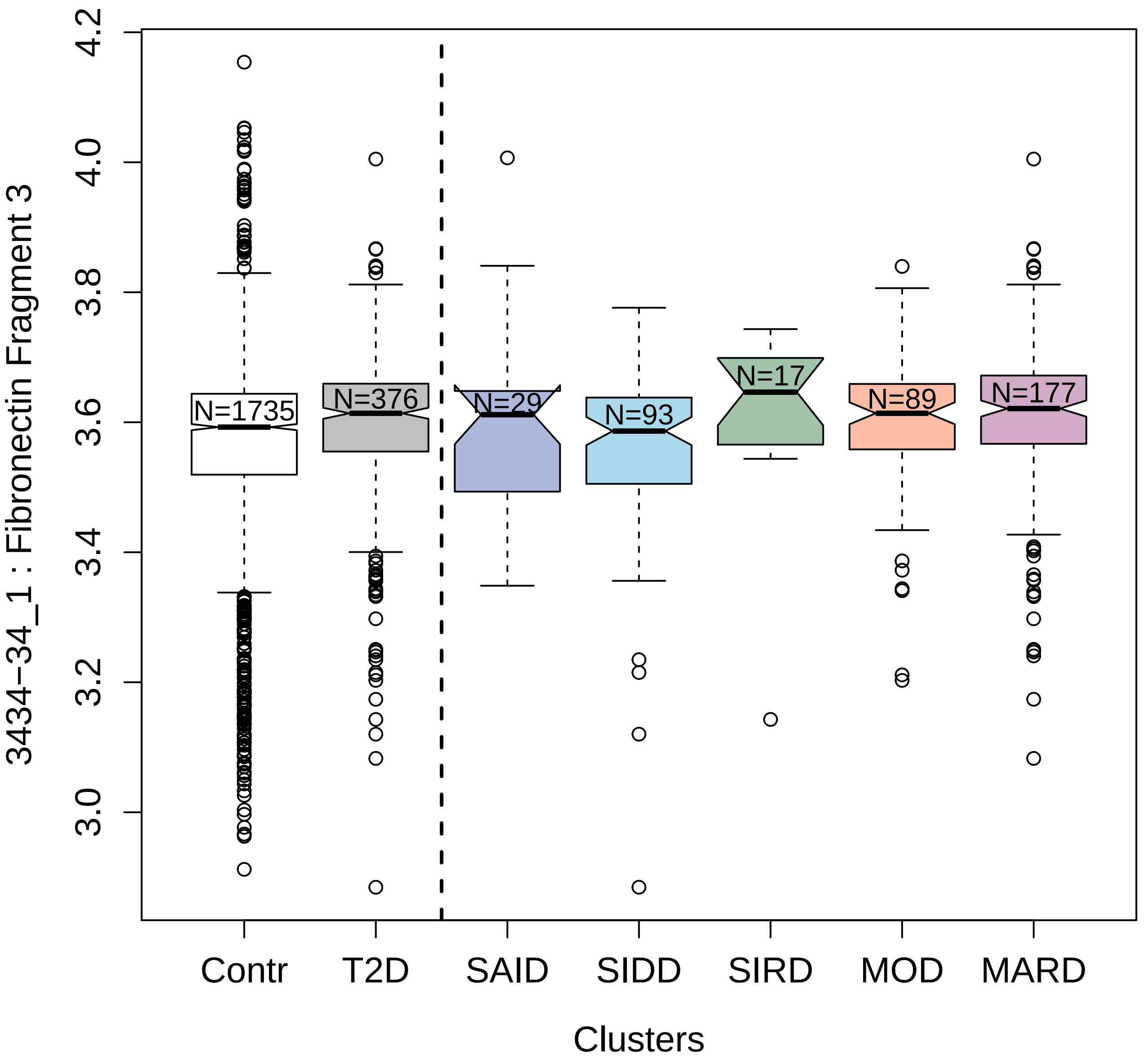

3434−34_1 : Fibronectin Fragment 3
4.2
4.0
3.8
3.6
3.4
3.2
3.0
N=1735
N=376
N=29
N=93
N=17
N=89
N=177
Contr
T2D
SAID
SIDD
SIRD
MOD
MARD
Clusters

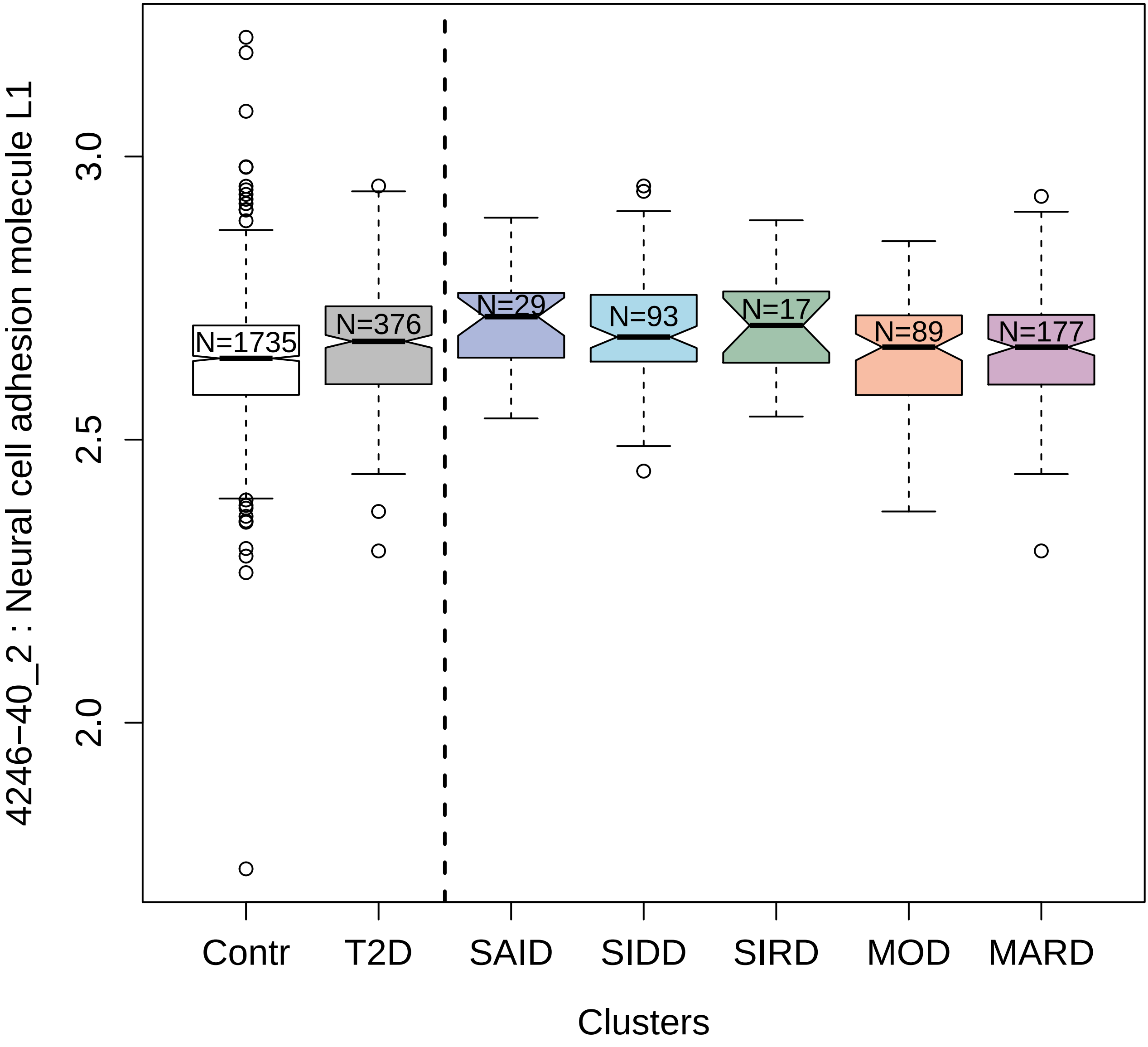
4246−40_2 : Neural cell adhesion molecule L1
3.0
2.5
2.0
N=1735
N=376
N=29
N=93
N=17
N=89
N=177
Contr
T2D
SAID
SIDD
SIRD
MOD
MARD
Clusters

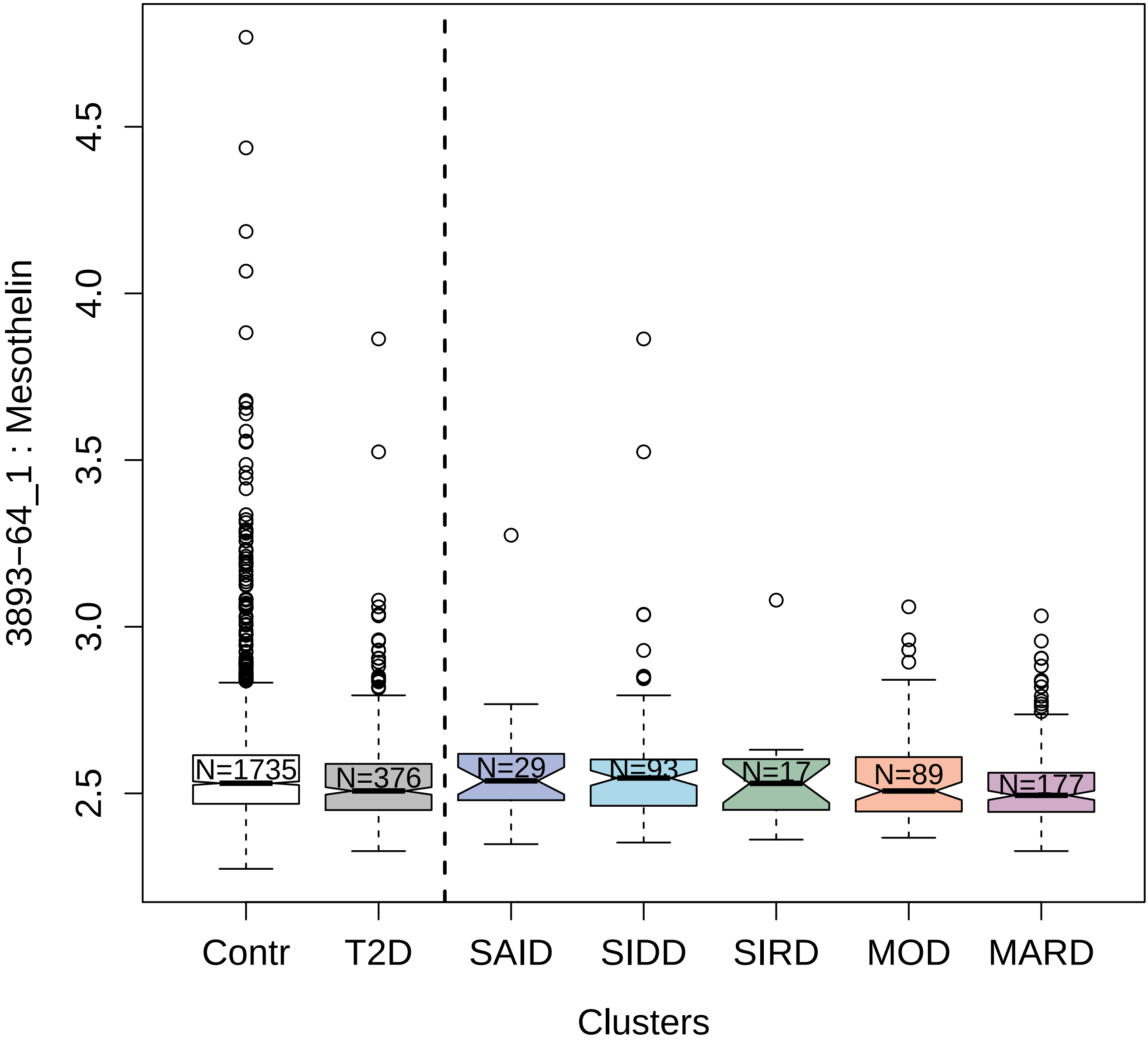

3893−64_1 : Mesothelin
4.5
4.0
3.5
3.0
2.5
N=1735
N=376
N=29
N=93
N=17
N=89
N=177
Contr
T2D
SAID
SIDD
SIRD
MOD
MARD
Clusters

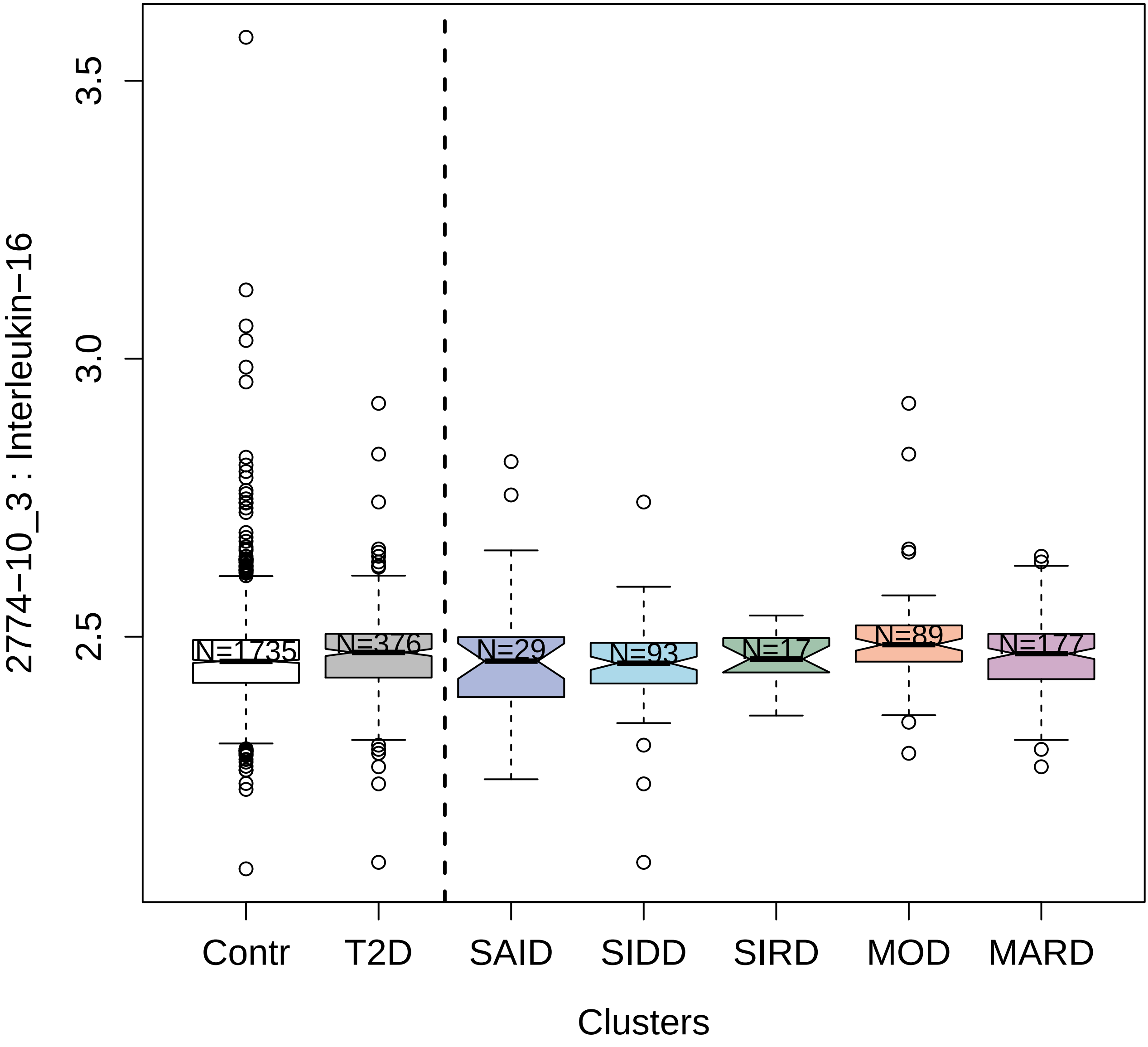
2774−10_3 : Interleukin−16
3.5
3.0
2.5
N=1735
N=376
N=29
N=93
N=17
N=89
N=177
Contr
T2D
SAID
SIDD
SIRD
MOD
MARD
Clusters

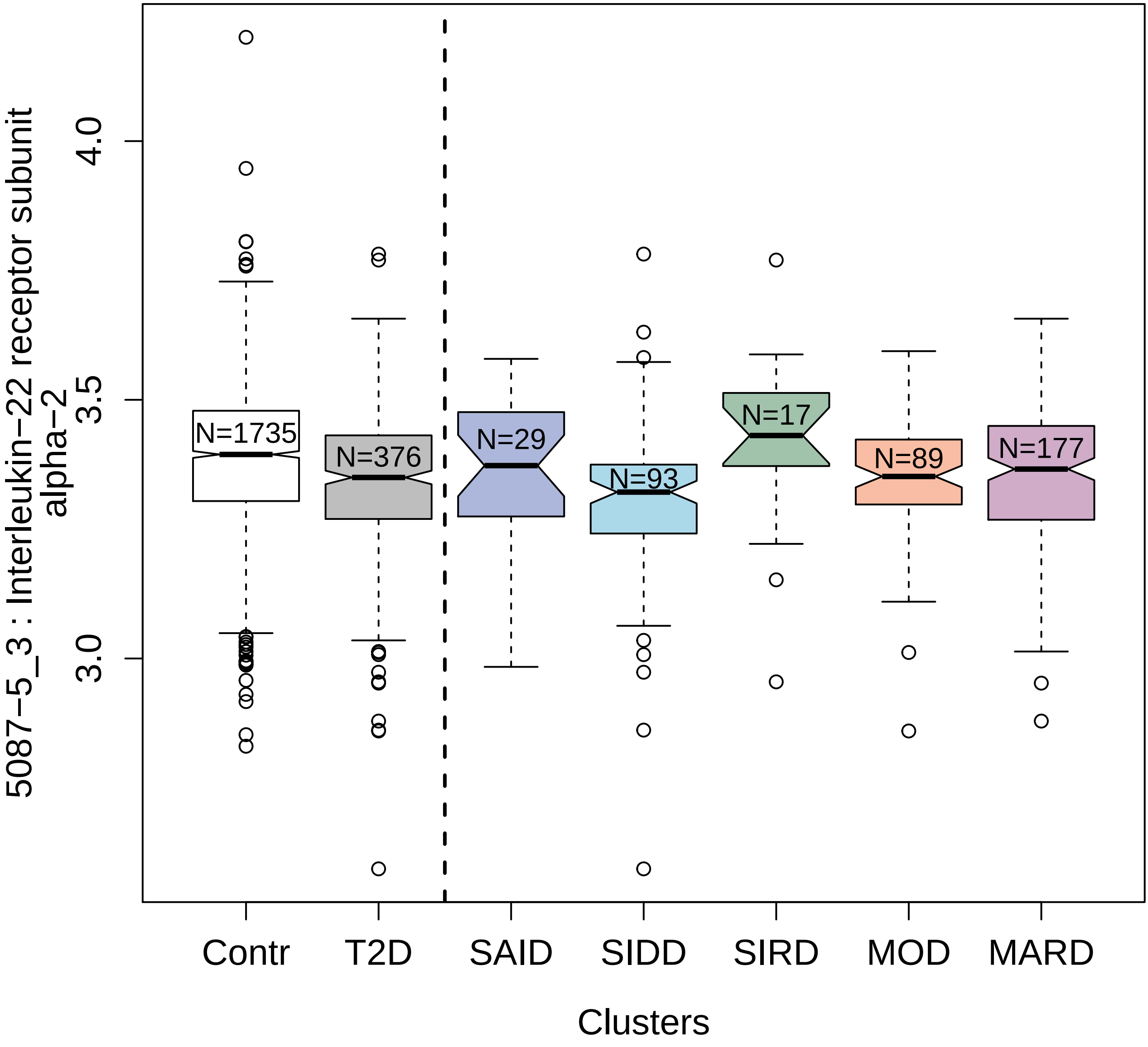
5087−5_3 : Interleukin−22 receptor subunit alpha−2
4.0
3.5
3.0
N=1735
N=376
N=29
N=93
N=17
N=89
N=177
Contr
T2D
SAID
SIDD
SIRD
MOD
MARD
Clusters

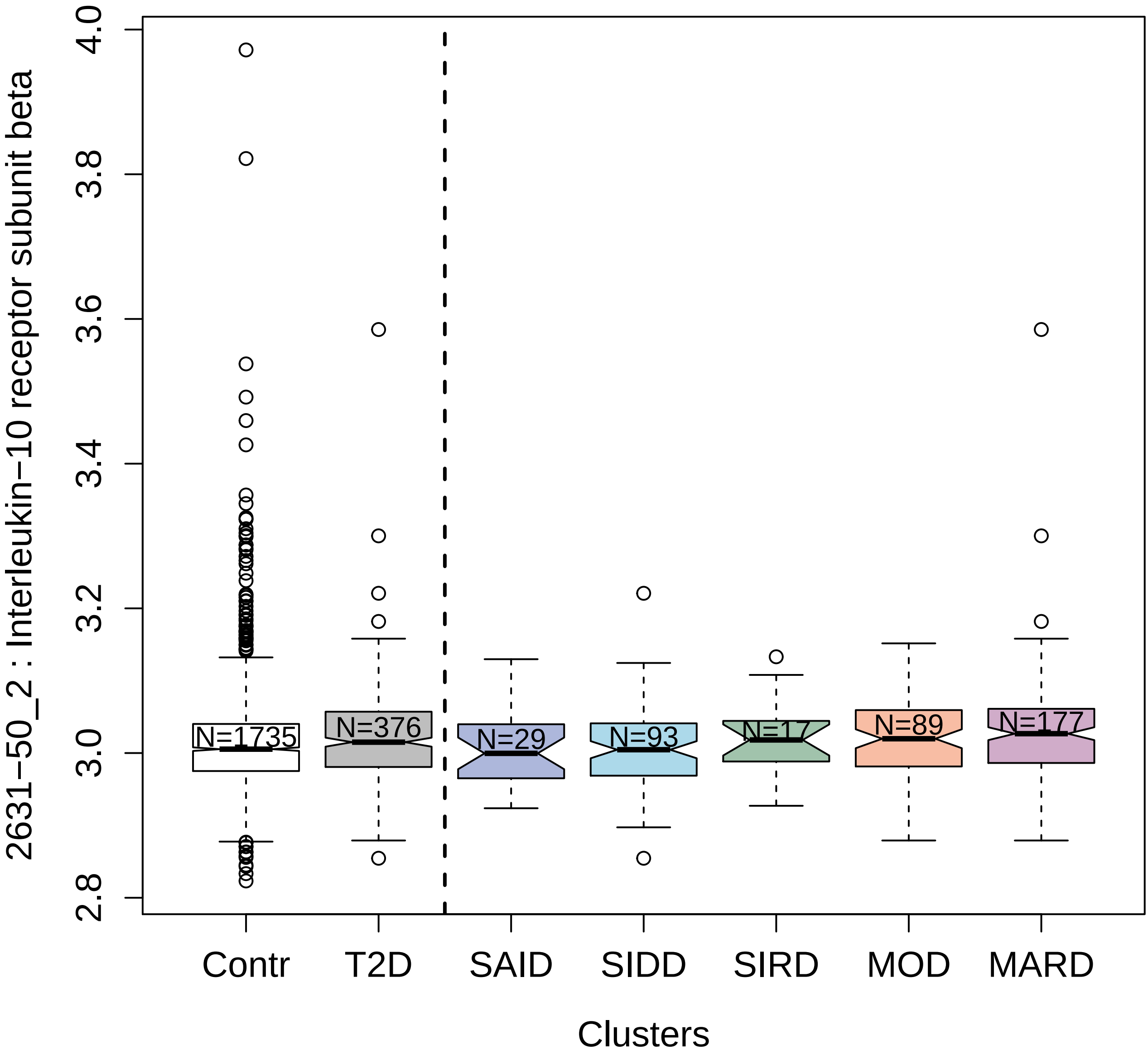
2631−50_2 : Interleukin−10 receptor subunit beta
4.0
3.8
3.6
3.4
3.2
3.0
2.8
N=1735
N=376
N=29
N=93
N=17
N=89
N=177
Contr
T2D
SAID
SIDD
SIRD
MOD
MARD
Clusters

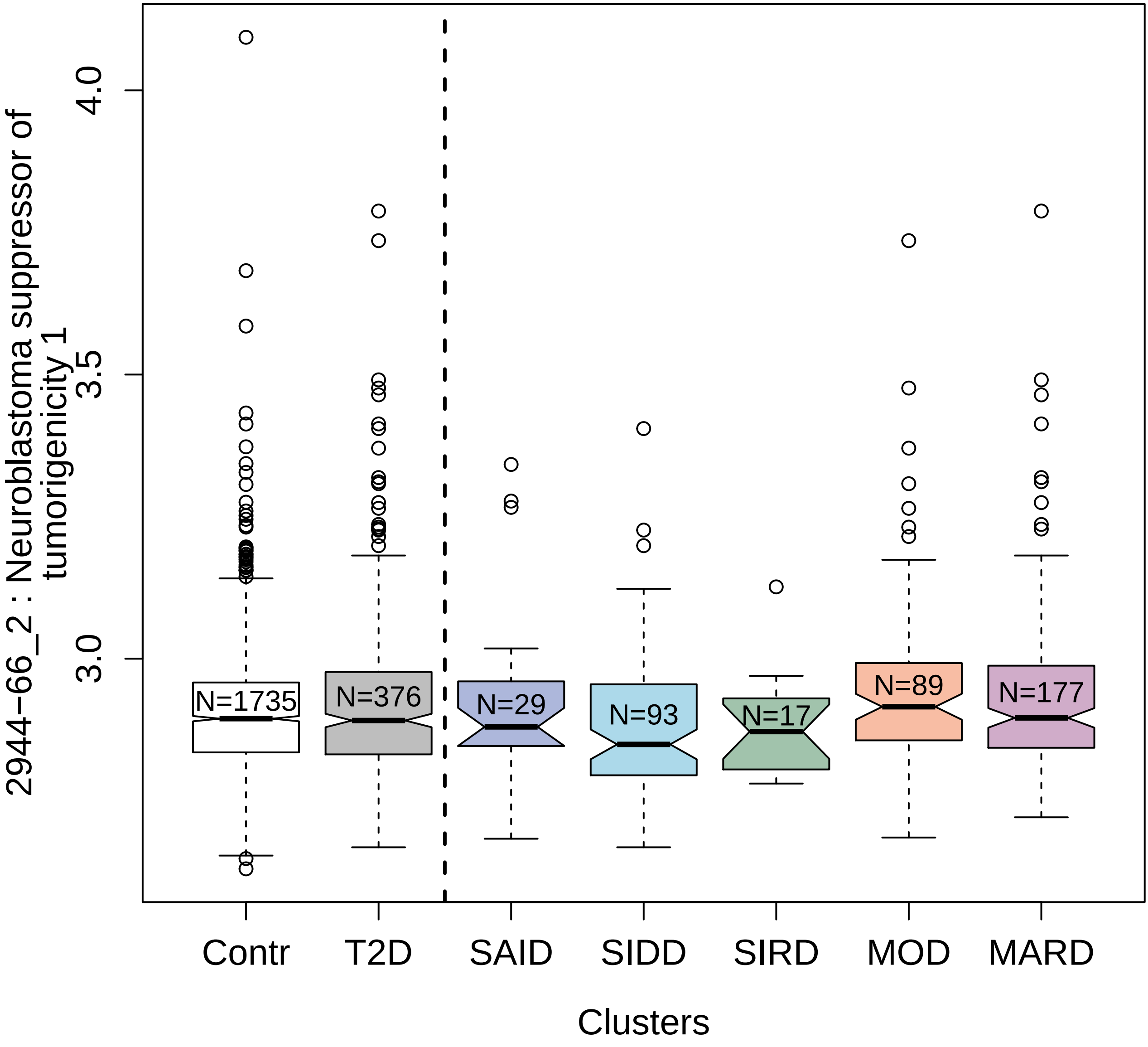

2944−66_2 : Neuroblastoma suppressor of tumorigenicity 1
4.0
3.5
3.0
N=1735
N=376
N=29
N=93
N=17
N=89
N=177
Contr
T2D
SAID
SIDD
SIRD
MOD
MARD
Clusters

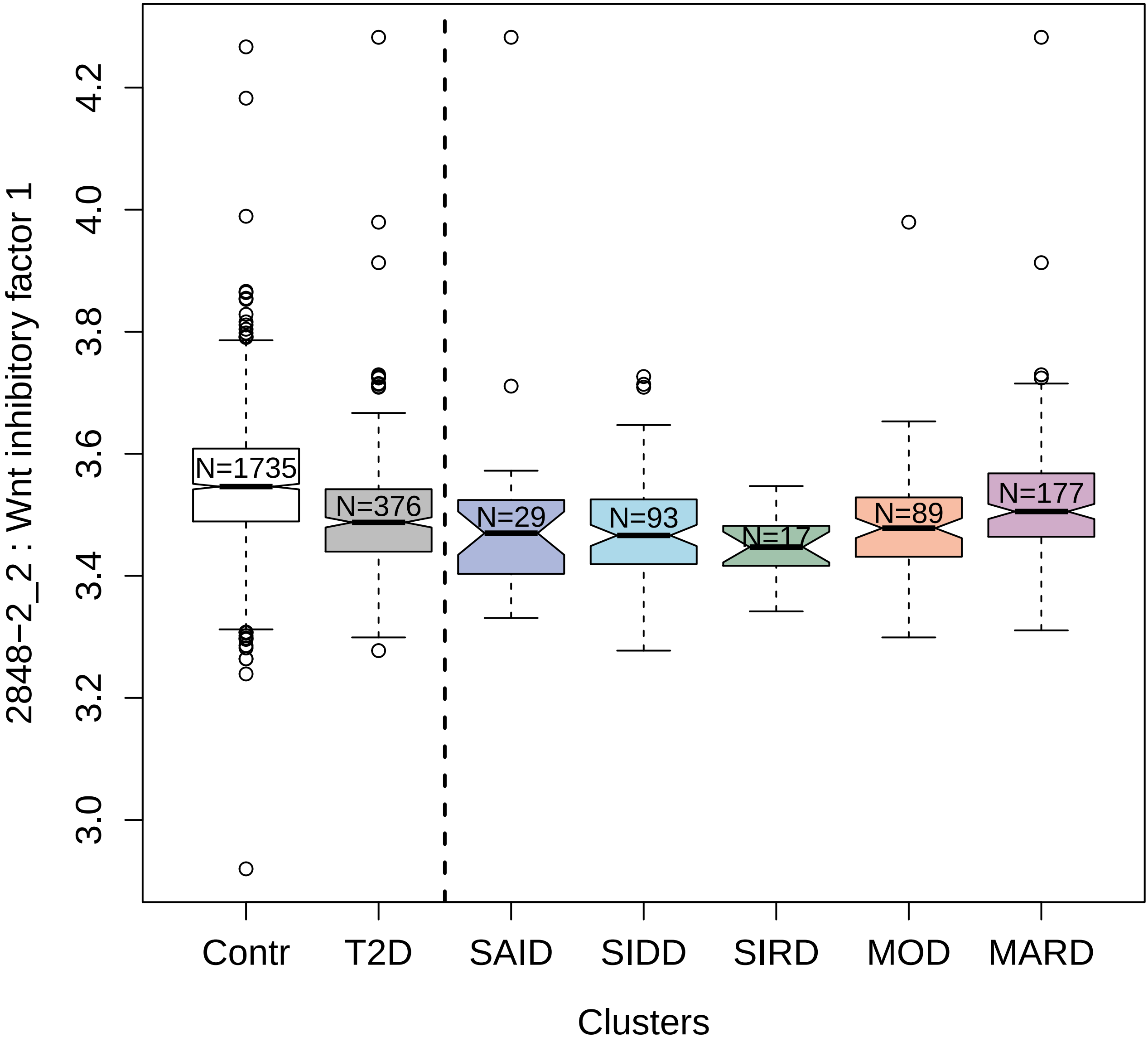

2848−2_2 : Wnt inhibitory factor 1
4.2
4.0
3.8
3.6
3.4
3.2
3.0
N=1735
N=376
N=29
N=93
N=17
N=89
N=177
Contr
T2D
SAID
SIDD
SIRD
MOD
MARD
Clusters

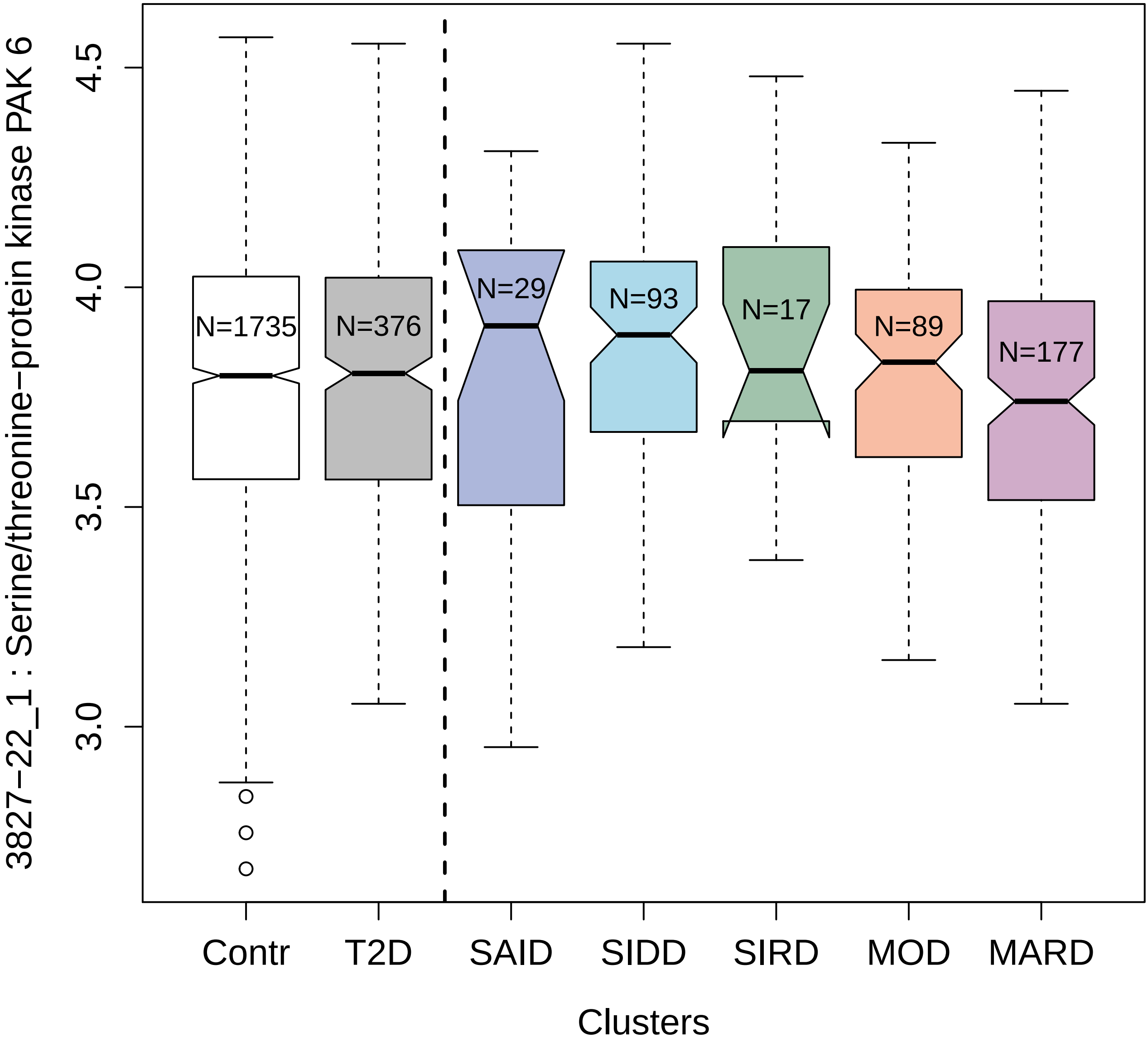

3827−22_1 : Serine/threonine−protein kinase PAK 6
4.5
4.0
3.5
3.0
N=1735
N=376
N=29
N=93
N=17
N=89
N=177
Contr
T2D
SAID
SIDD
SIRD
MOD
MARD
Clusters

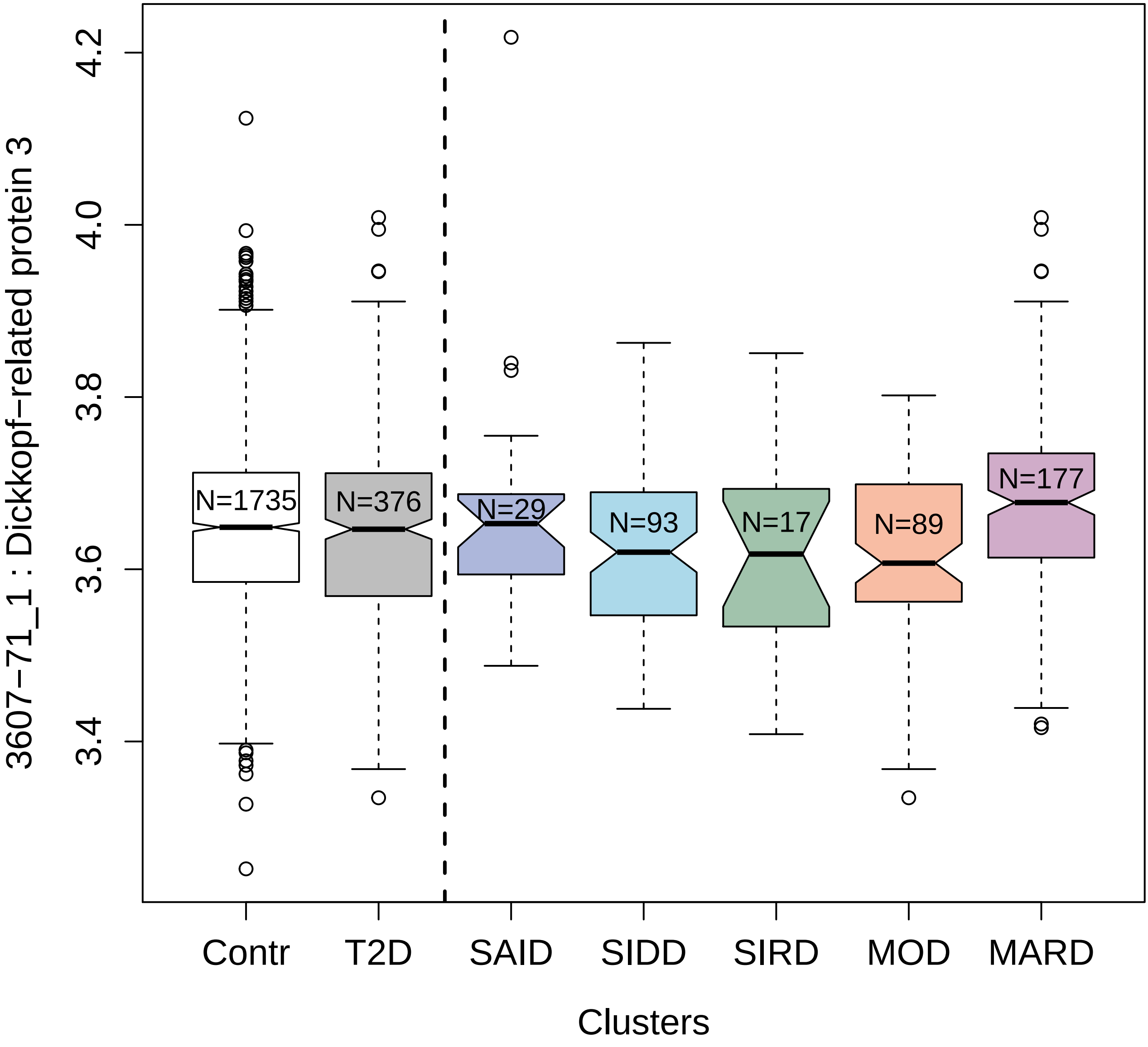

3607−71_1 : Dickkopf−related protein 3
4.2
4.0
3.8
3.6
3.4
N=1735
N=376
N=29
N=93
N=17
N=89
N=177
Contr
T2D
SAID
SIDD
SIRD
MOD
MARD
Clusters

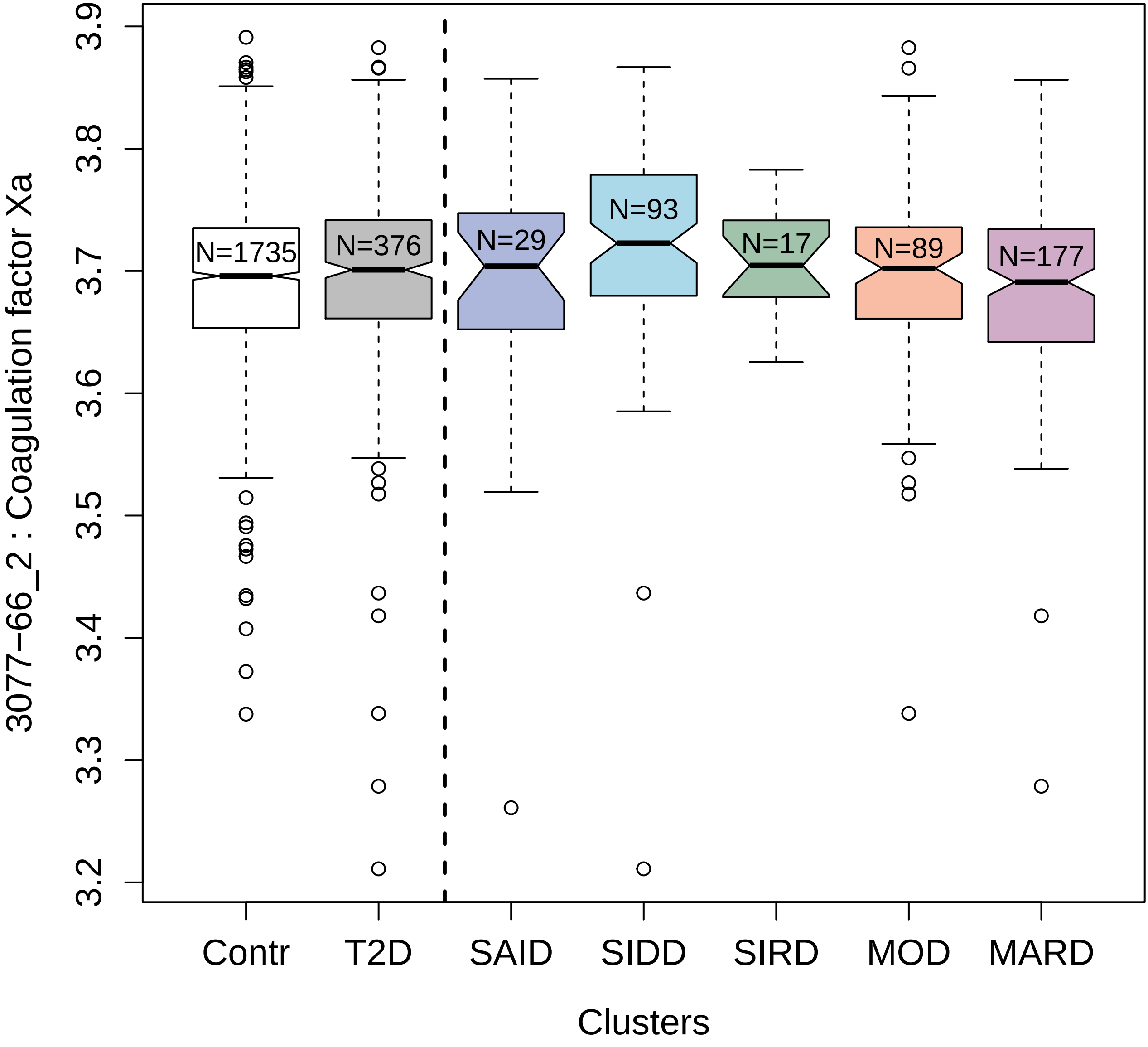
3077−66_2 : Coagulation factor Xa
3.9
3.8
3.7
3.6
3.5
3.4
3.3
3.2
N=1735
N=376
N=29
N=93
N=17
N=89
N=177
Contr
T2D
SAID
SIDD
SIRD
MOD
MARD
Clusters

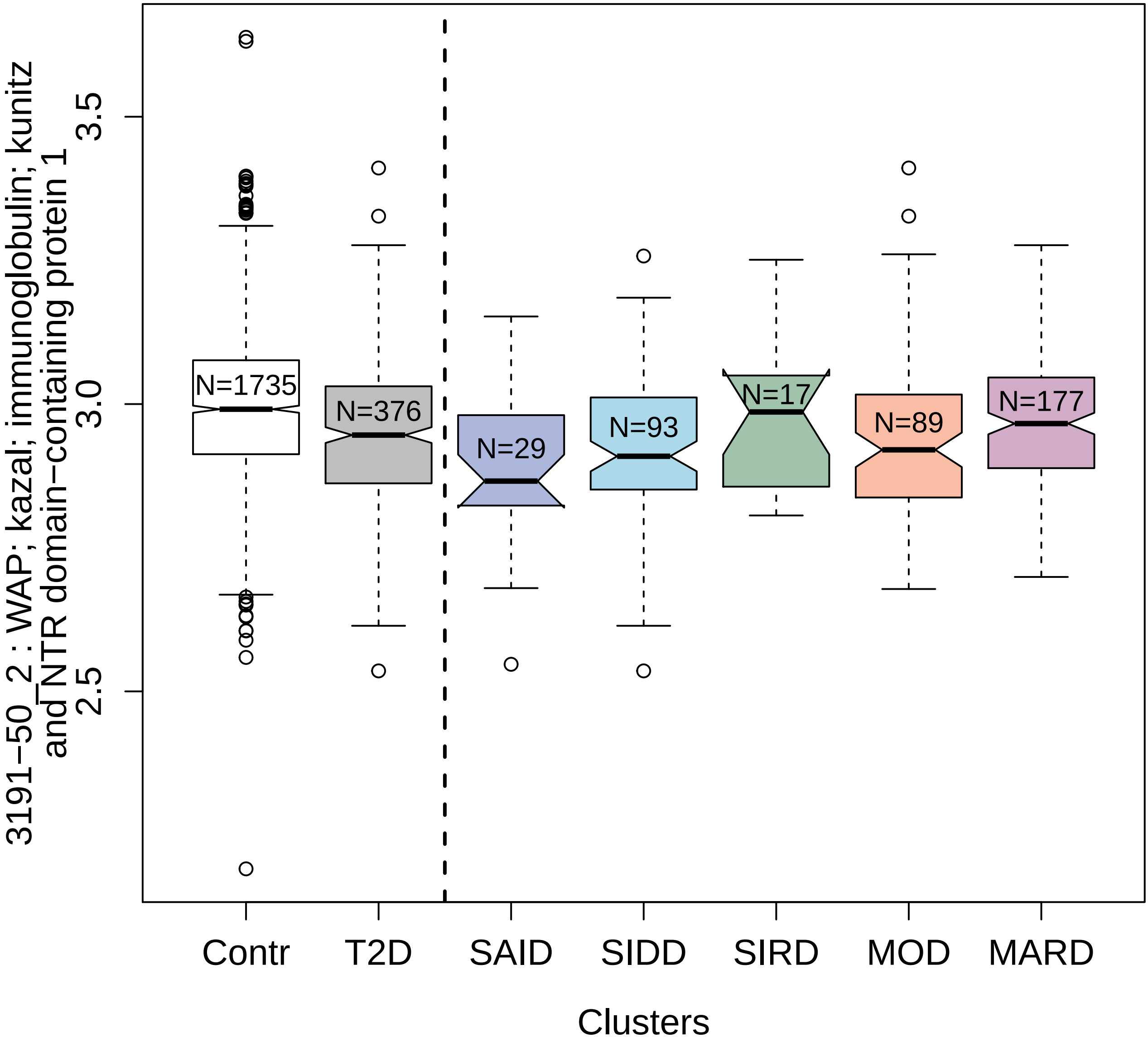

3191−50_2 : WAP; kazal; immunoglobulin; kunitz
and NTR domain−containing protein 1
3.5
3.0
2.5
N=1735
N=376
N=29
N=93
N=17
N=89
N=177
Contr
T2D
SAID
SIDD
SIRD
MOD
MARD
Clusters

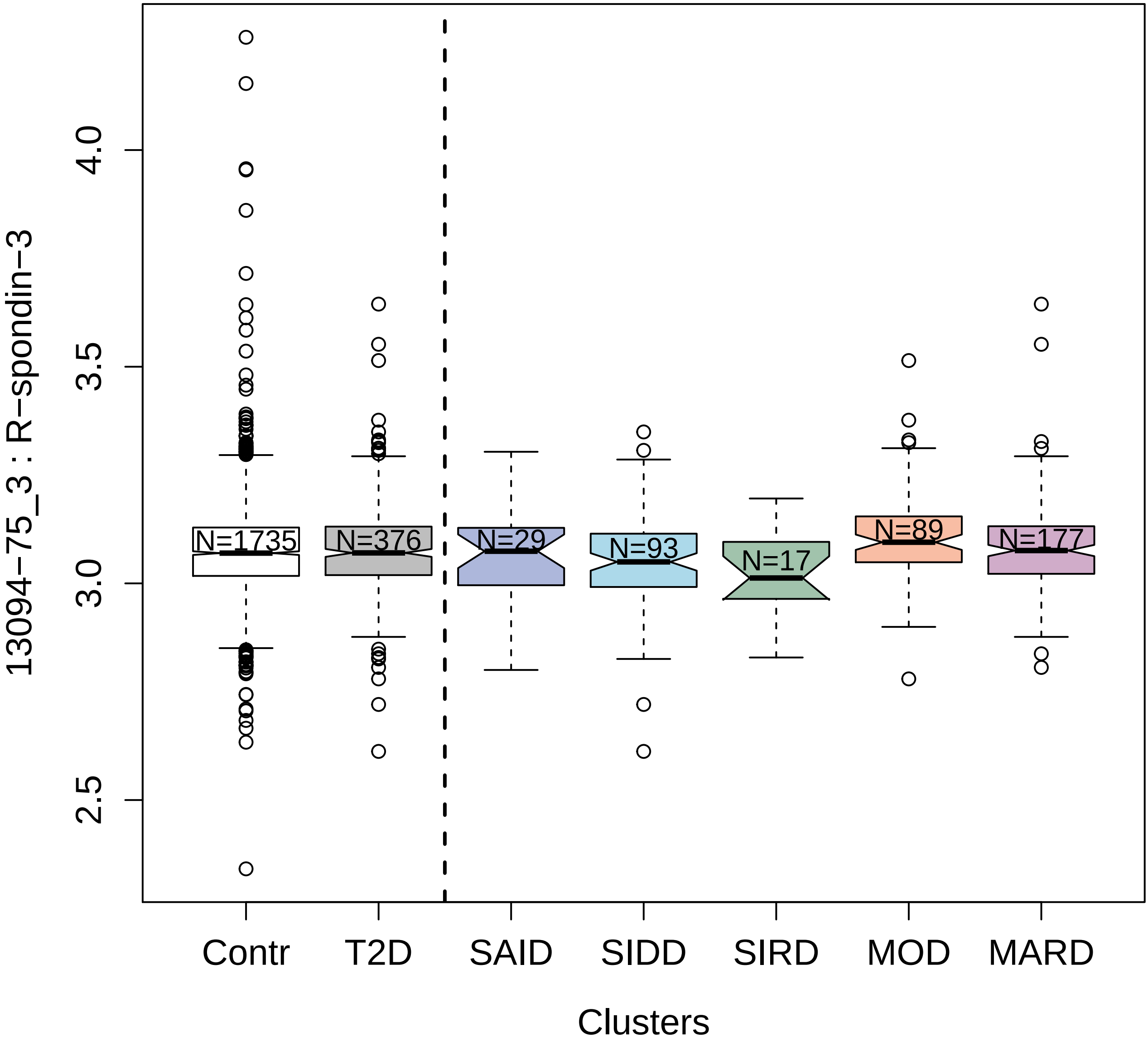
13094−75_3 : R−spondin−3
4.0
3.5
3.0
2.5
N=1735
N=376
N=29
N=93
N=17
N=89
N=177
Contr
T2D
SAID
SIDD
SIRD
MOD
MARD
Clusters

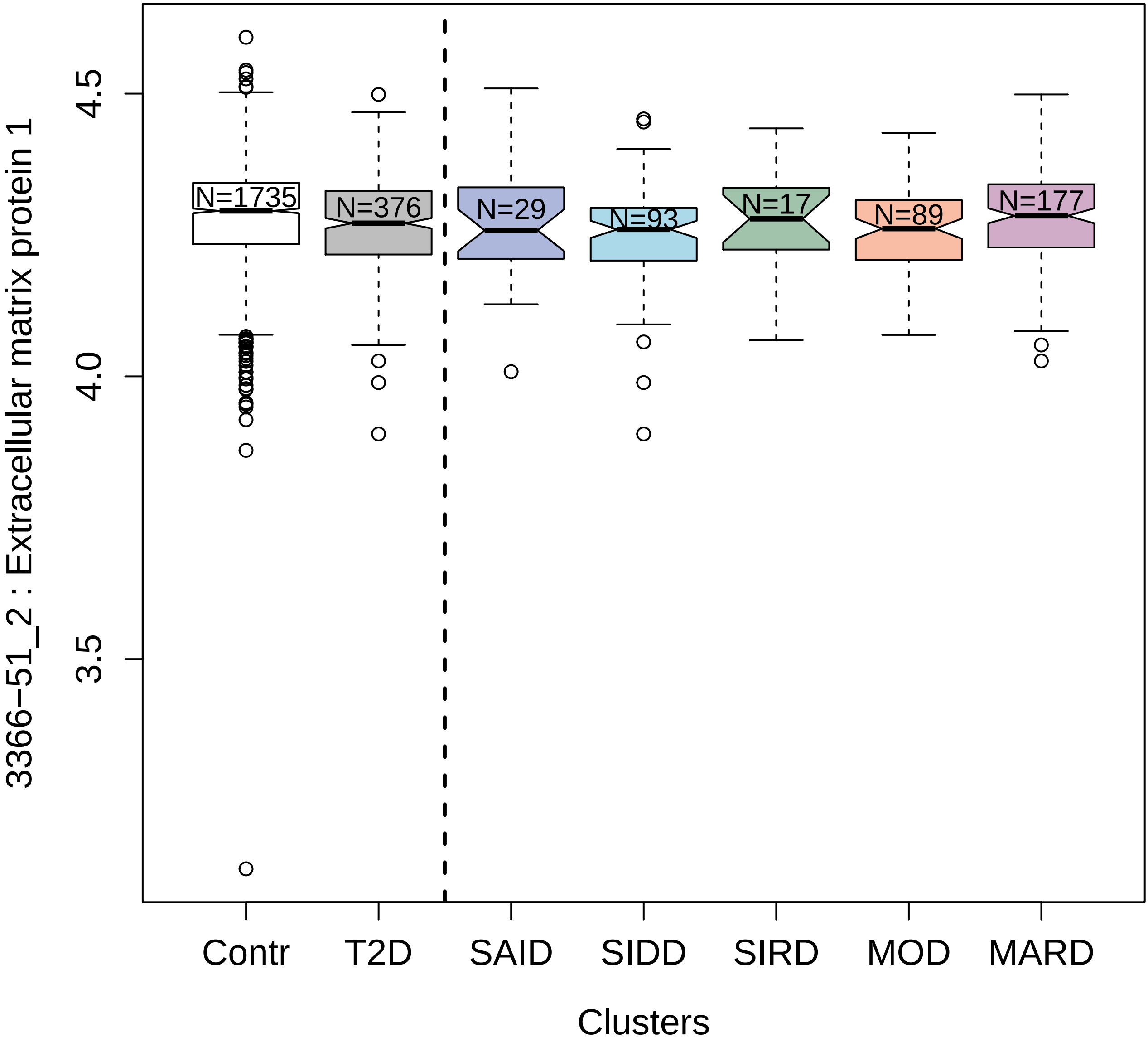
3366−51_2 : Extracellular matrix protein 1
4.5
4.0
3.5
N=1735
N=376
N=29
N=93
N=17
N=89
N=177
Contr
T2D
SAID
SIDD
SIRD
MOD
MARD
Clusters

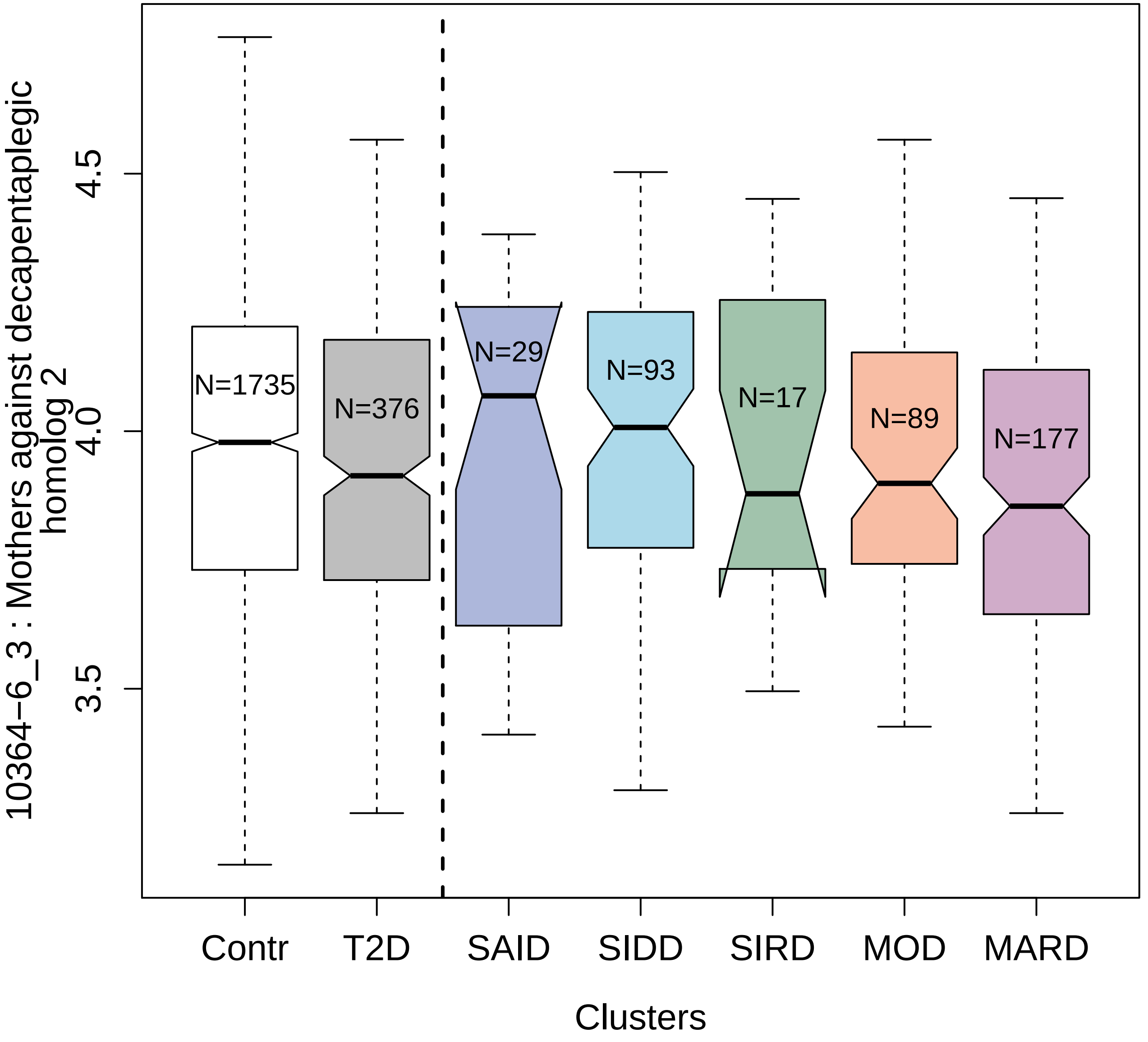

10364−6_3 : Mothers against decapentaplegic homolog 2
4.5
4.0
3.5
N=1735
N=376
N=29
N=93
N=17
N=89
N=177
Contr
T2D
SAID
SIDD
SIRD
MOD
MARD
Clusters

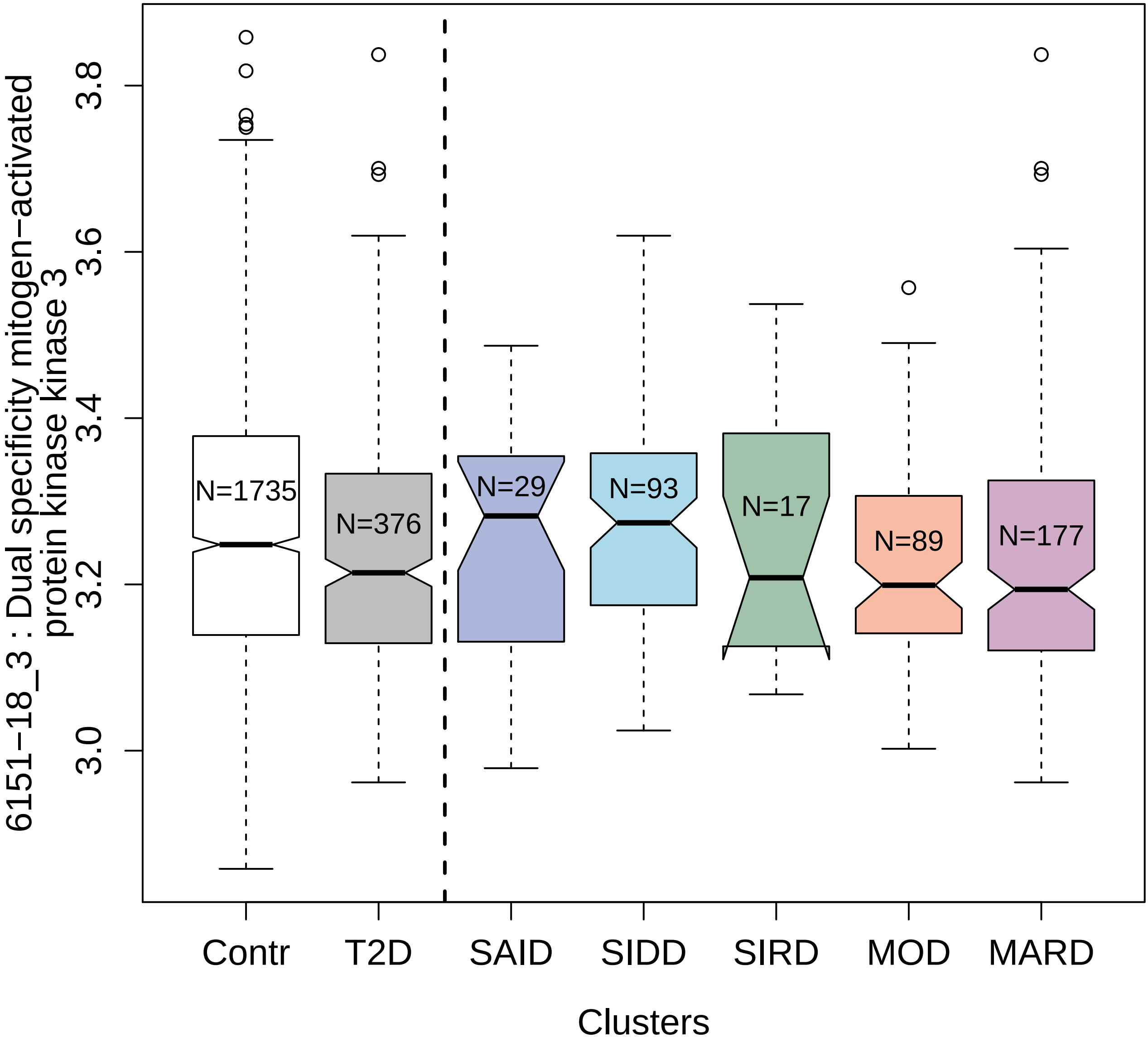
6151−18_3 : Dual specificity mitogen−activated protein kinase kinase 3
3.0
3.2
3.4
3.6
3.8
N=1735
N=376
N=29
N=93
N=17
N=89
N=177
Contr
T2D
SAID
SIDD
SIRD
MOD
MARD
Clusters

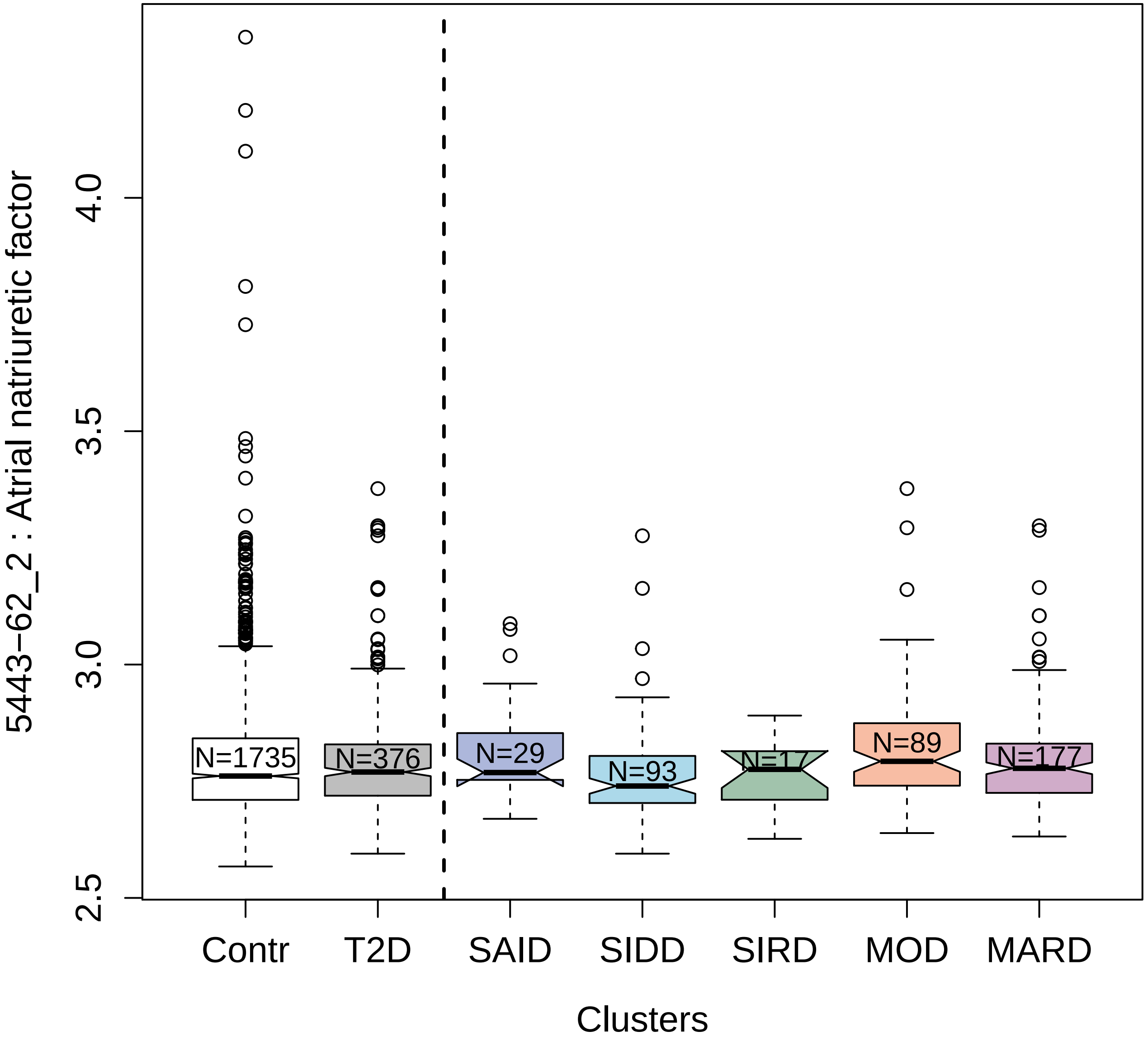
5443−62_2 : Atrial natriuretic factor
2.5
3.0
3.5
4.0
N=1735
N=376
N=29
N=93
N=17
N=89
N=177
Contr
T2D
SAID
SIDD
SIRD
MOD
MARD
Clusters

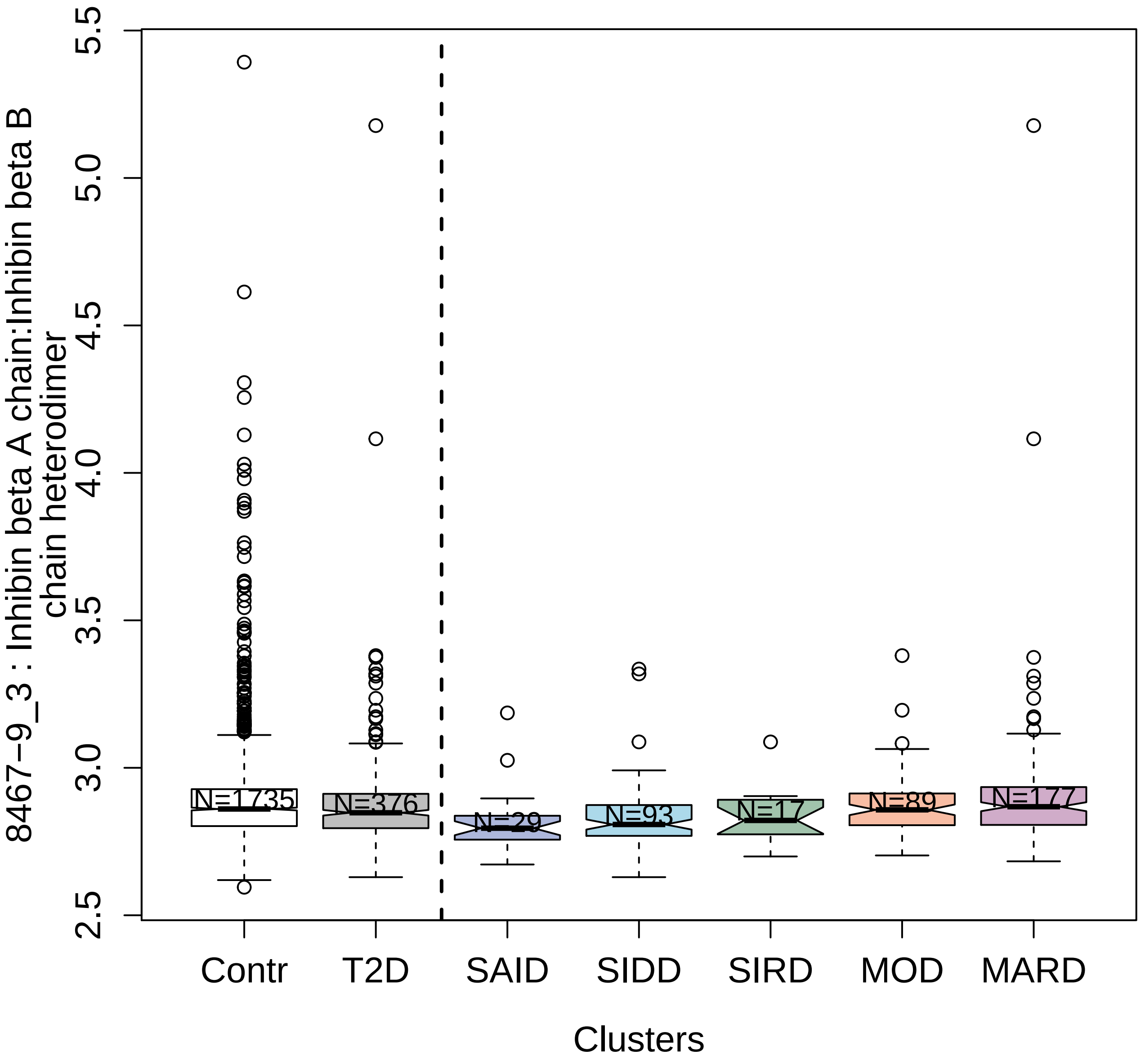

8467−9_3 : Inhibin beta A chain:Inhibin beta B chain heterodimer
5.5
5.0
4.5
4.0
3.5
3.0
2.5
N=1735
N=376
N=29
N=93
N=17
N=89
N=177
Contr
T2D
SAID
SIDD
SIRD
MOD
MARD
Clusters

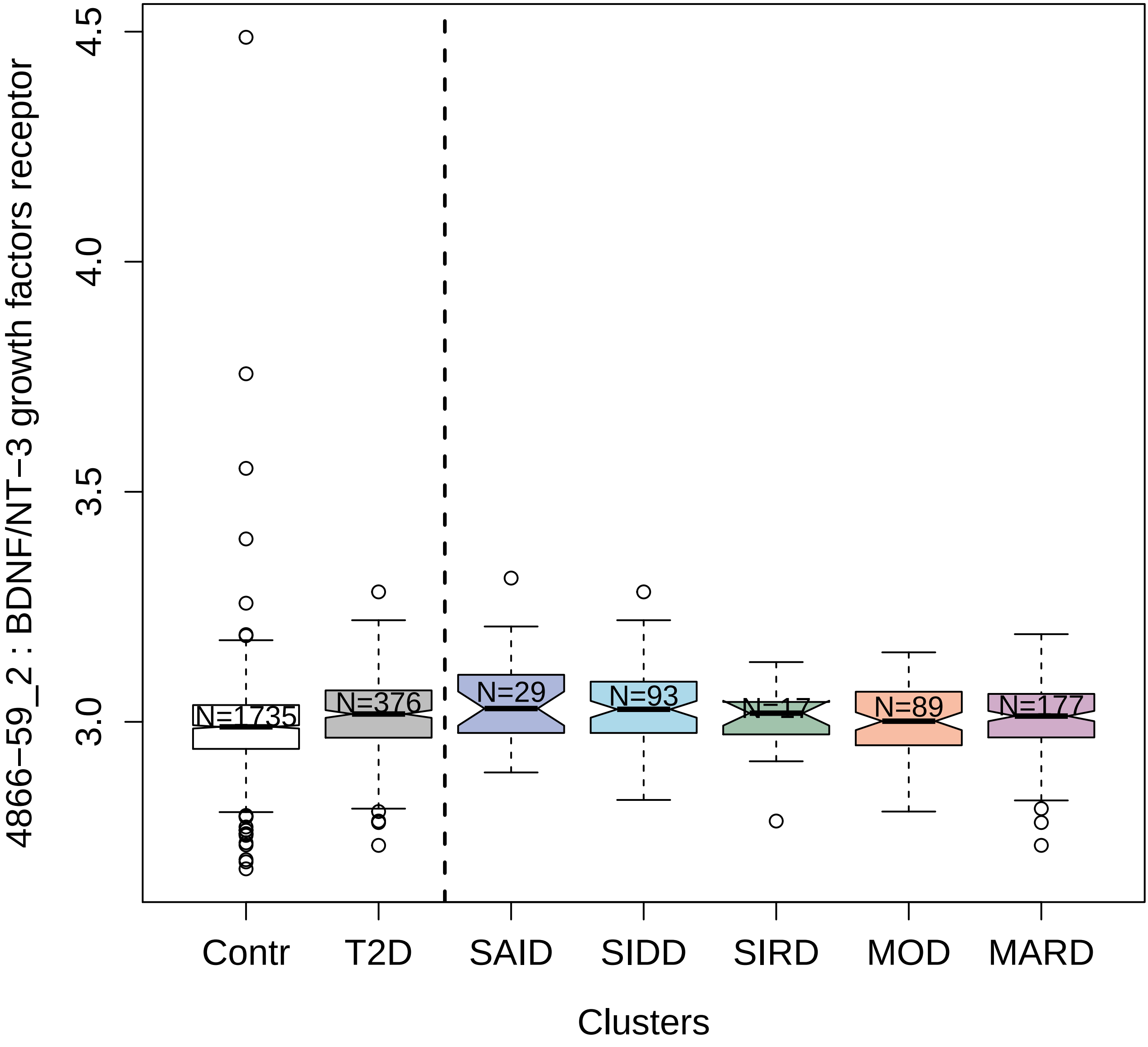

4866−59_2 : BDNF/NT−3 growth factors receptor
4.5
4.0
3.5
3.0
N=1735
N=376
N=29
N=93
N=17
N=89
N=177
Contr
T2D
SAID
SIDD
SIRD
MOD
MARD
Clusters

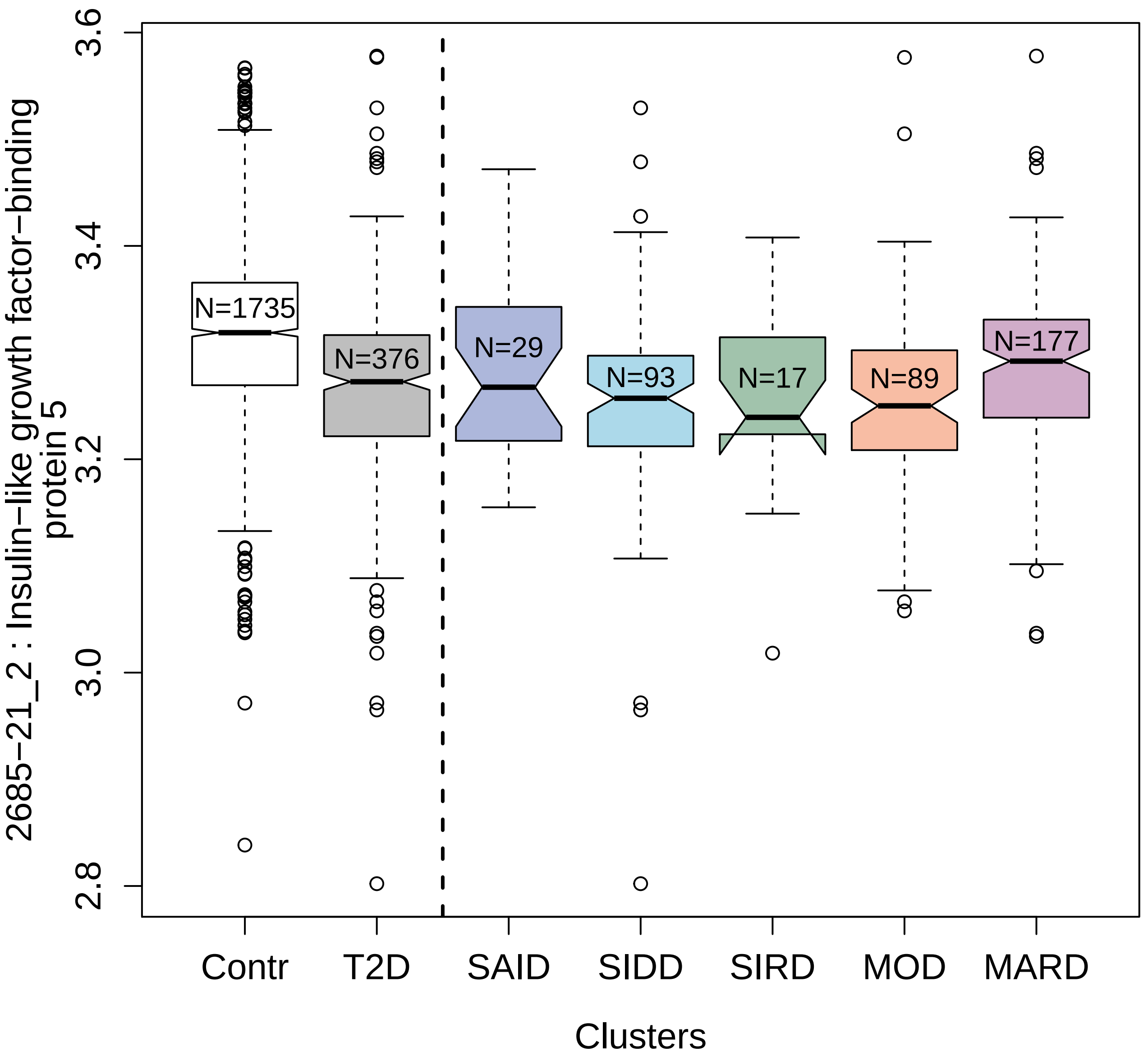

2685−21_2 : Insulin−like growth factor−binding protein 5
3.6
3.4
3.2
3.0
2.8
N=1735
N=376
N=29
N=93
N=17
N=89
N=177
Contr
T2D
SAID
SIDD
SIRD
MOD
MARD
Clusters

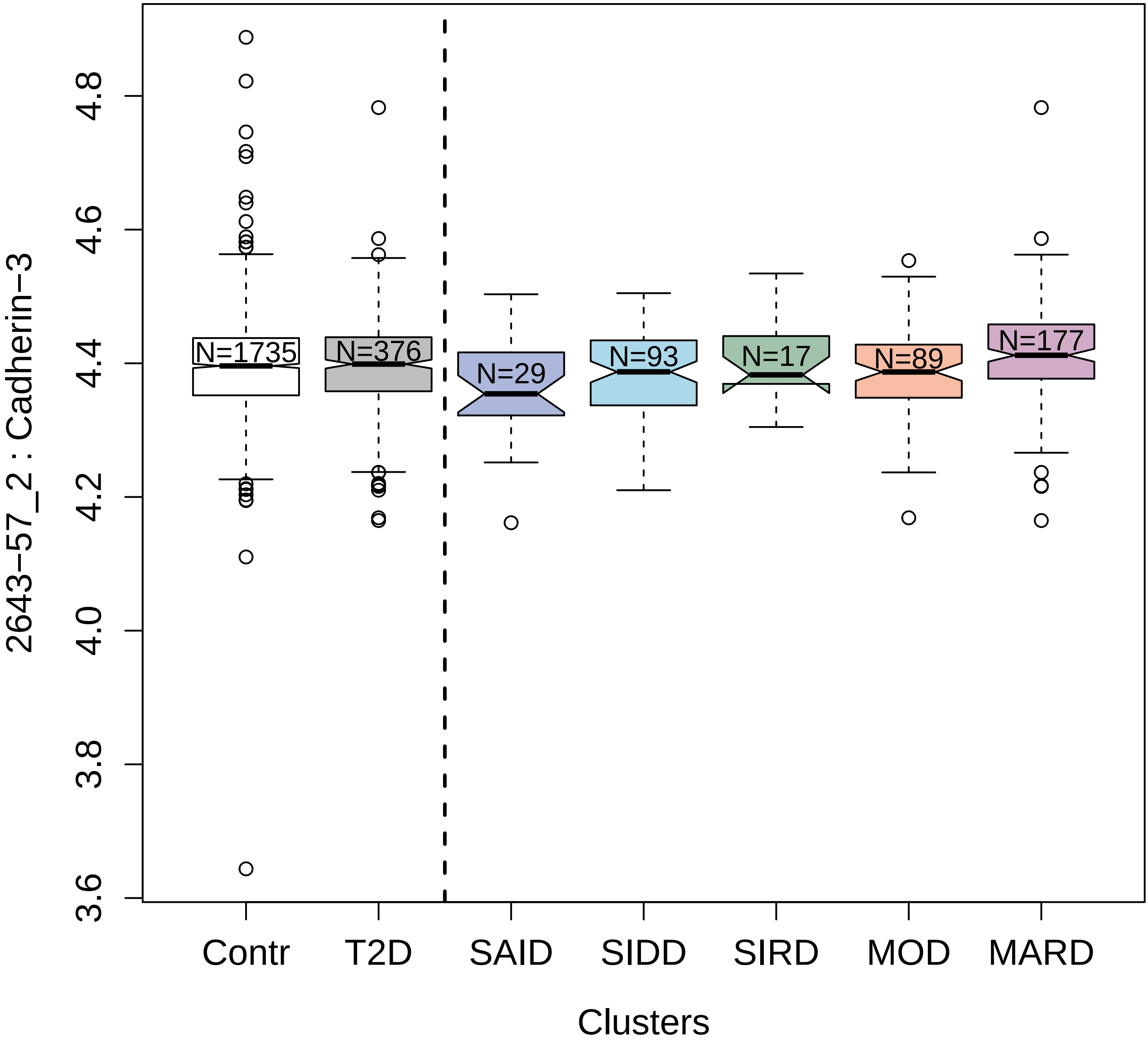

2643−57_2 : Cadherin−3
3.6
3.8
4.0
4.2
4.4
4.6
4.8
N=1735
N=376
N=29
N=93
N=17
N=89
N=177
Contr
T2D
SAID
SIDD
SIRD
MOD
MARD
Clusters

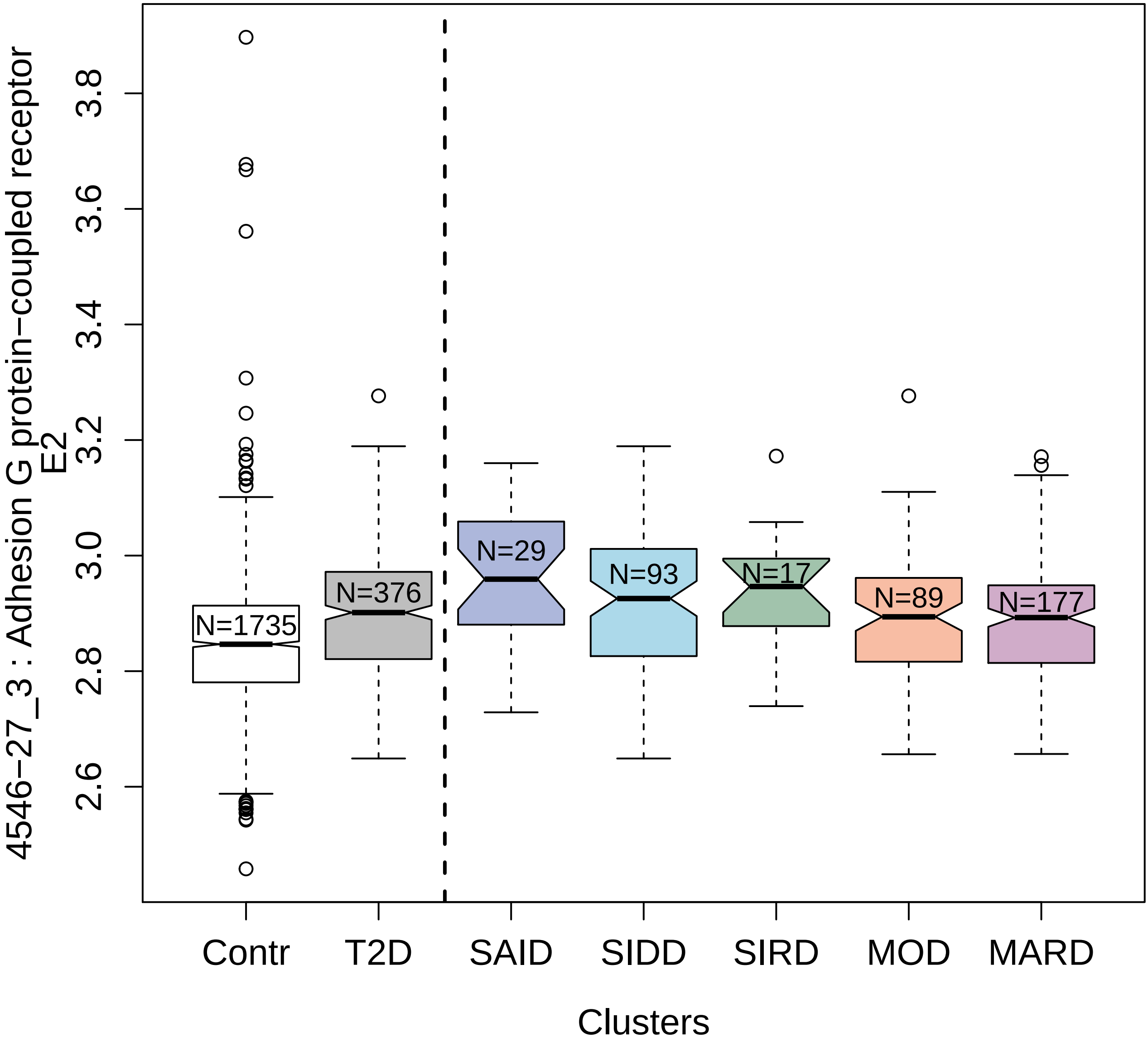
4546−27_3 : Adhesion G protein−coupled receptor E2
3.8
3.6
3.4
3.2
3.0
2.8
2.6
N=1735
N=376
N=29
N=93
N=17
N=89
N=177
Contr
T2D
SAID
SIDD
SIRD
MOD
MARD
Clusters

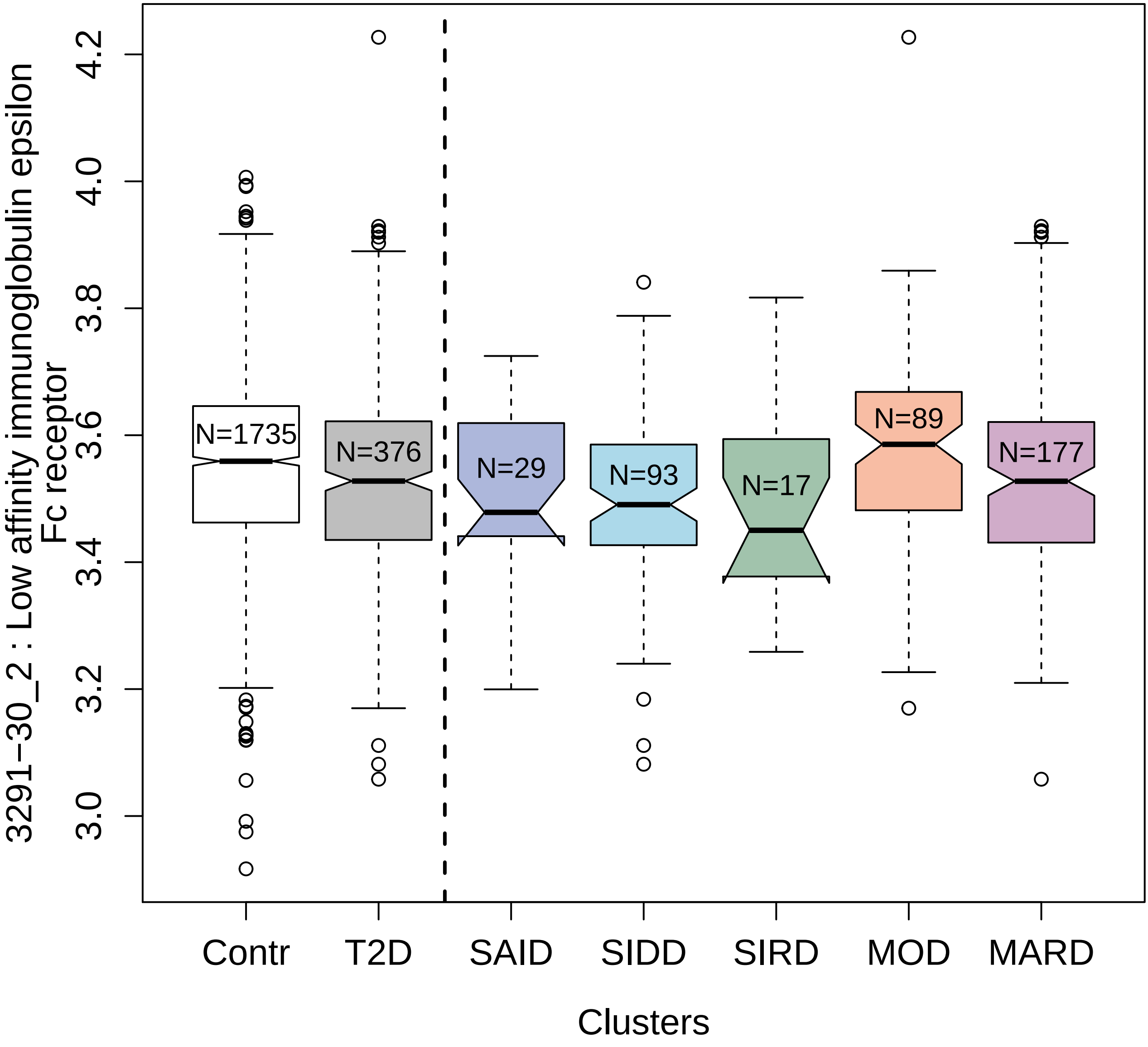
3291−30_2 : Low affinity immunoglobulin epsilon
Fc receptor
4.2
4.0
3.8
3.6
3.4
3.2
3.0
N=1735
N=376
N=29
N=93
N=17
N=89
N=177
Contr
T2D
SAID
SIDD
SIRD
MOD
MARD
Clusters

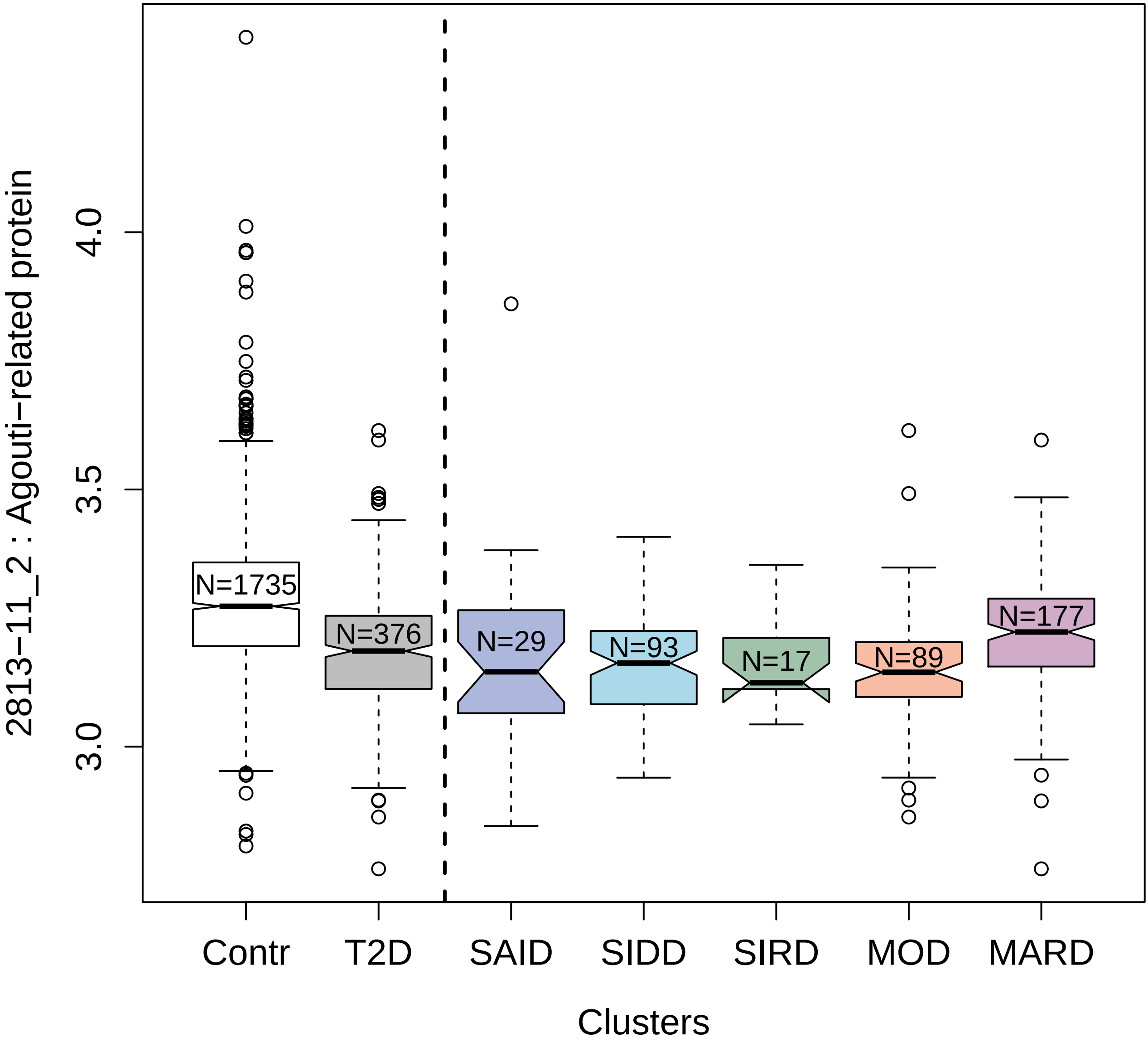

2813−11_2 : Agouti−related protein
4.0
3.5
3.0
N=1735
N=376
N=29
N=93
N=17
N=89
N=177
Contr
T2D
SAID
SIDD
SIRD
MOD
MARD
Clusters

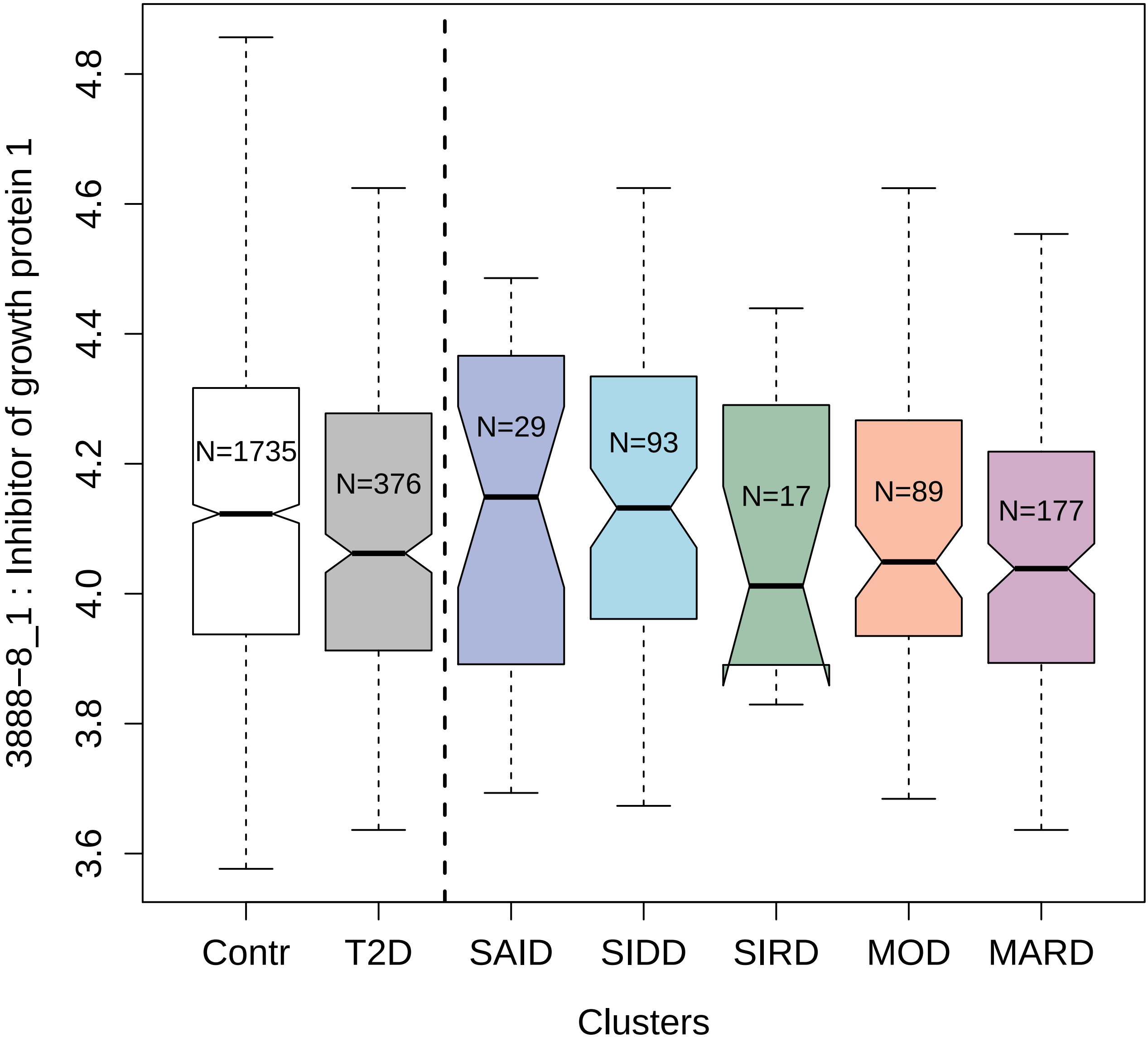
3888−8_1 : Inhibitor of growth protein 1
4.8
4.6
4.4
4.2
4.0
3.8
3.6
N=1735
N=376
N=29
N=93
N=17
N=89
N=177
Contr
T2D
SAID
SIDD
SIRD
MOD
MARD
Clusters

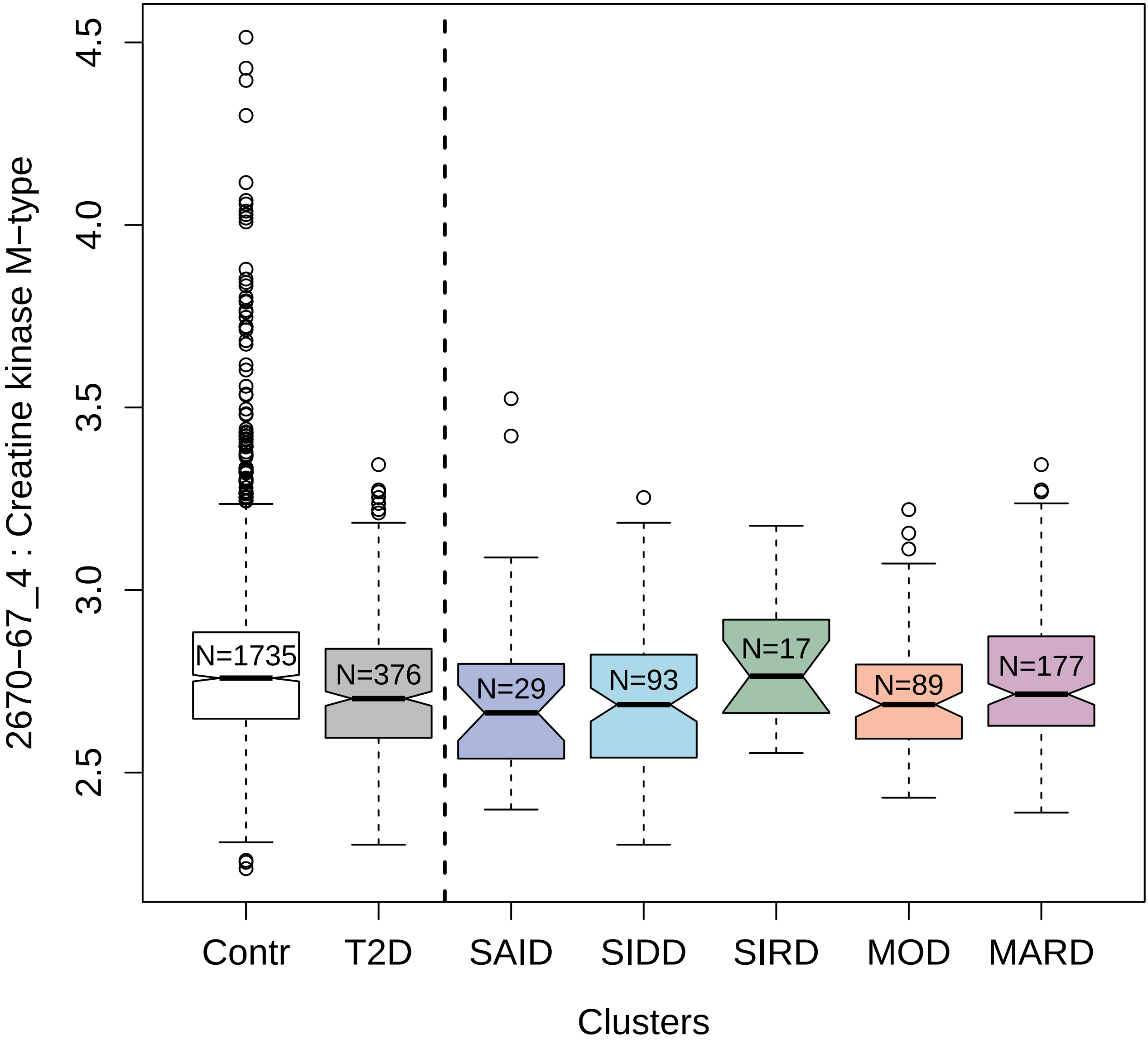
2670−67_4 : Creatine kinase M−type
4.5
4.0
3.5
3.0
2.5
N=1735
N=376
N=29
N=93
N=17
N=89
N=177
Contr
T2D
SAID
SIDD
SIRD
MOD
MARD
Clusters

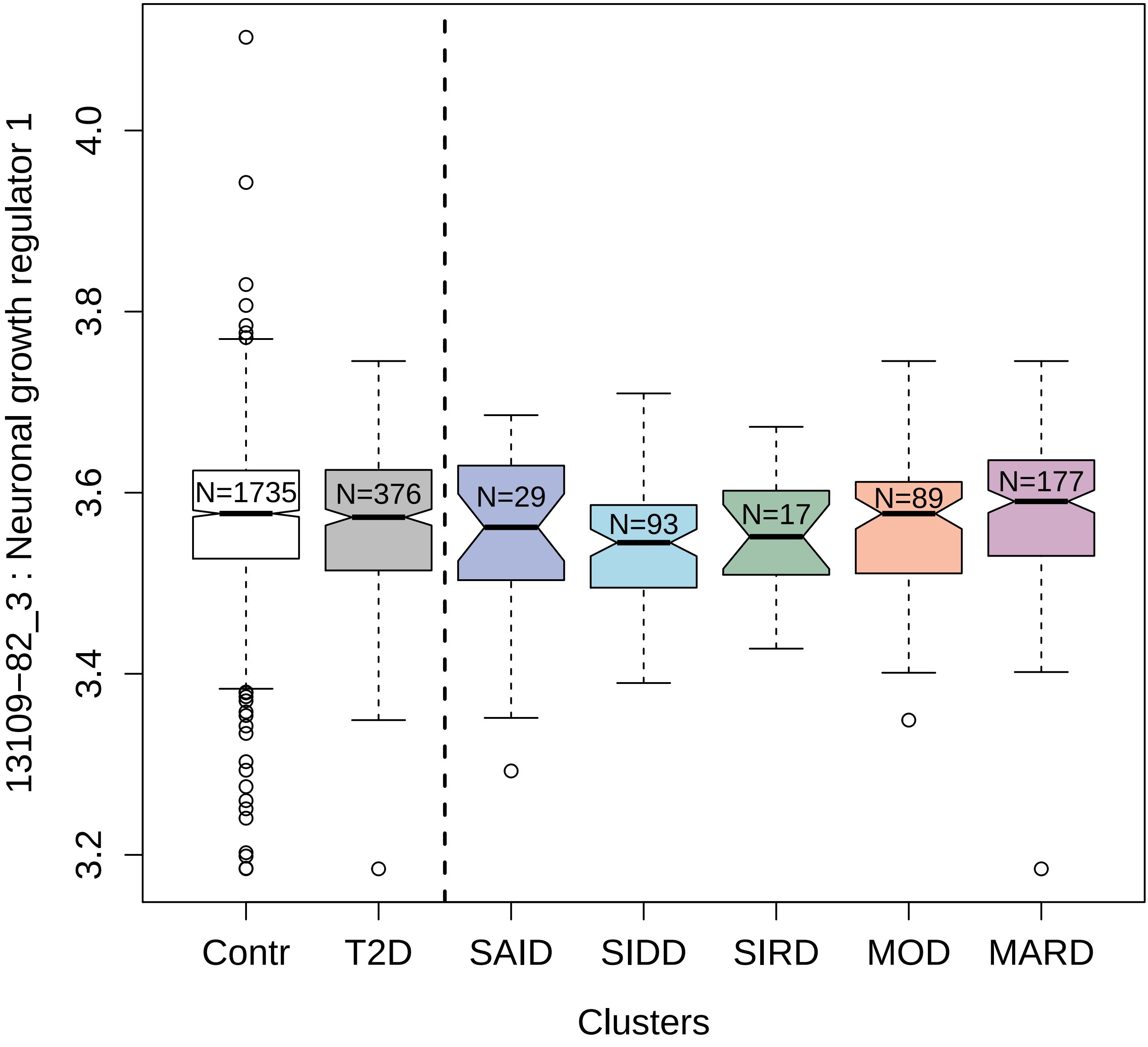
13109−82_3 : Neuronal growth regulator 1
Clusters
Contr
T2D
SAID
SIDD
SIRD
MOD
MARD
N=1735
N=376
N=29
N=93
N=17
N=89
N=177
3.2
3.4
3.6
3.8
4.0

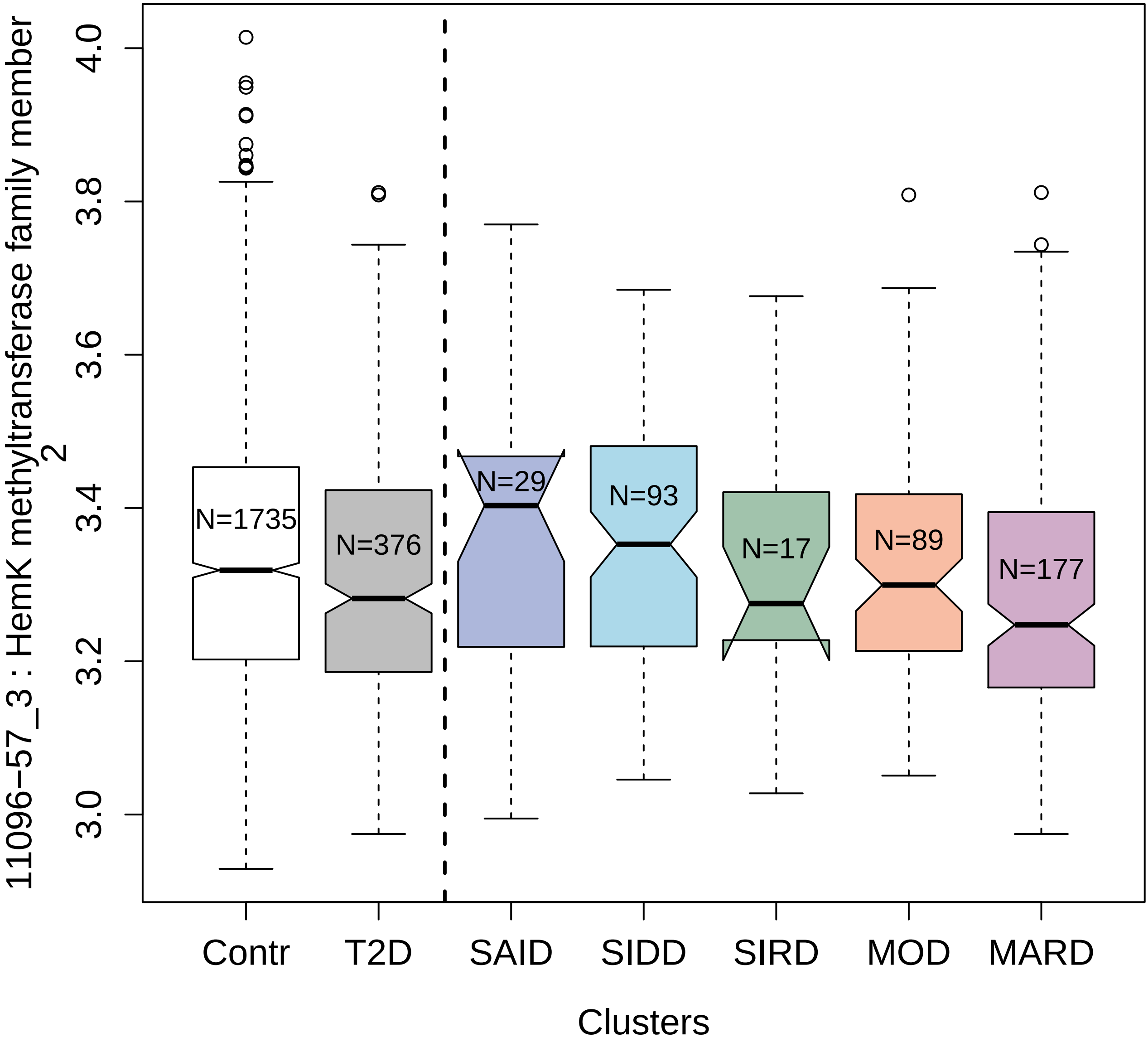
11096−57_3 : HemK methyltransferase family member 2
4.0
3.8
3.6
3.4
3.2
3.0
N=1735
N=376
N=29
N=93
N=17
N=89
N=177
Contr
T2D
SAID
SIDD
SIRD
MOD
MARD
Clusters

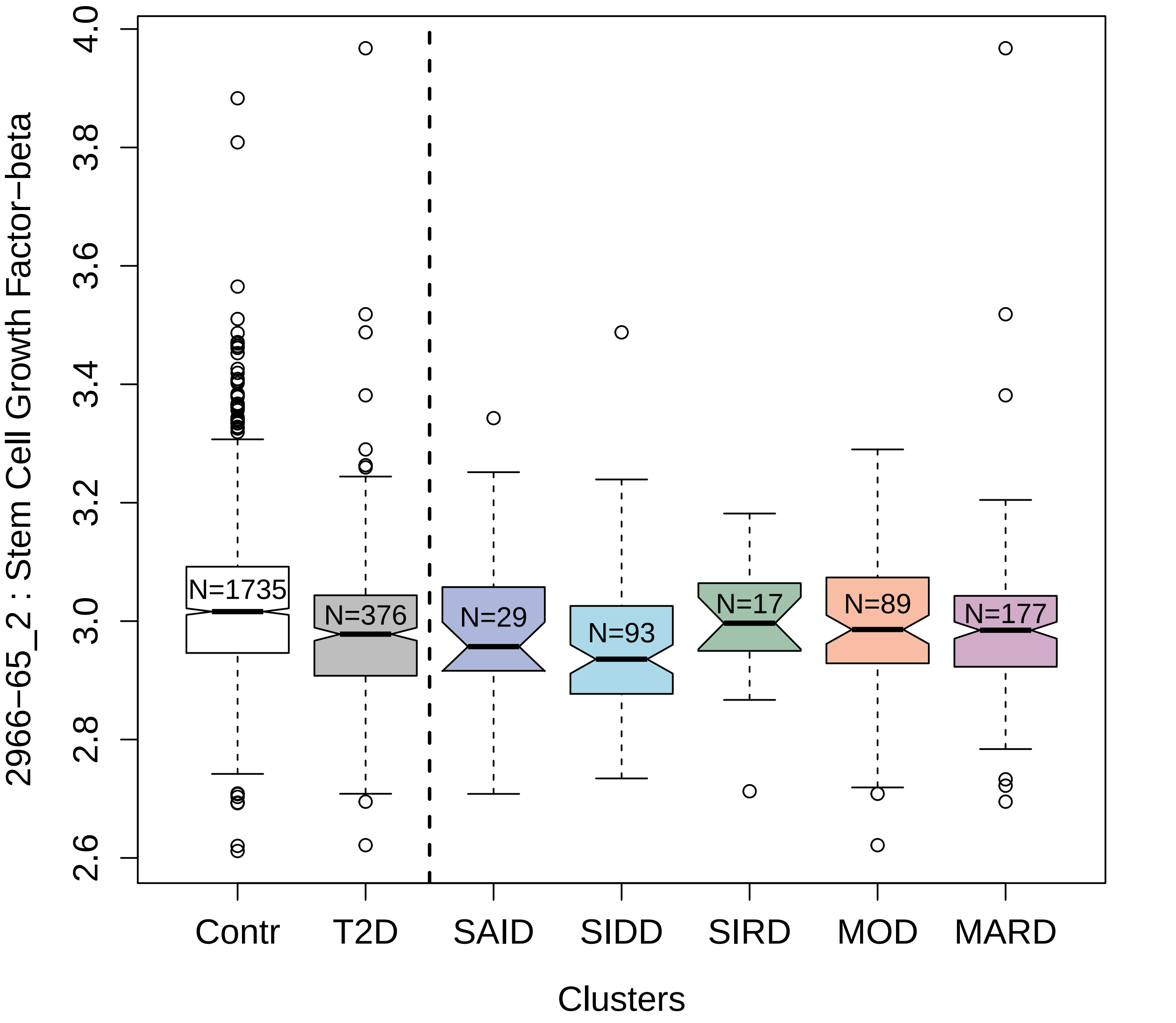
2966−65_2 : Stem Cell Growth Factor−beta
4.0
3.8
3.6
3.4
3.2
3.0
2.8
2.6
N=1735
N=376
N=29
N=93
N=17
N=89
N=177
Contr
T2D
SAID
SIDD
SIRD
MOD
MARD
Clusters

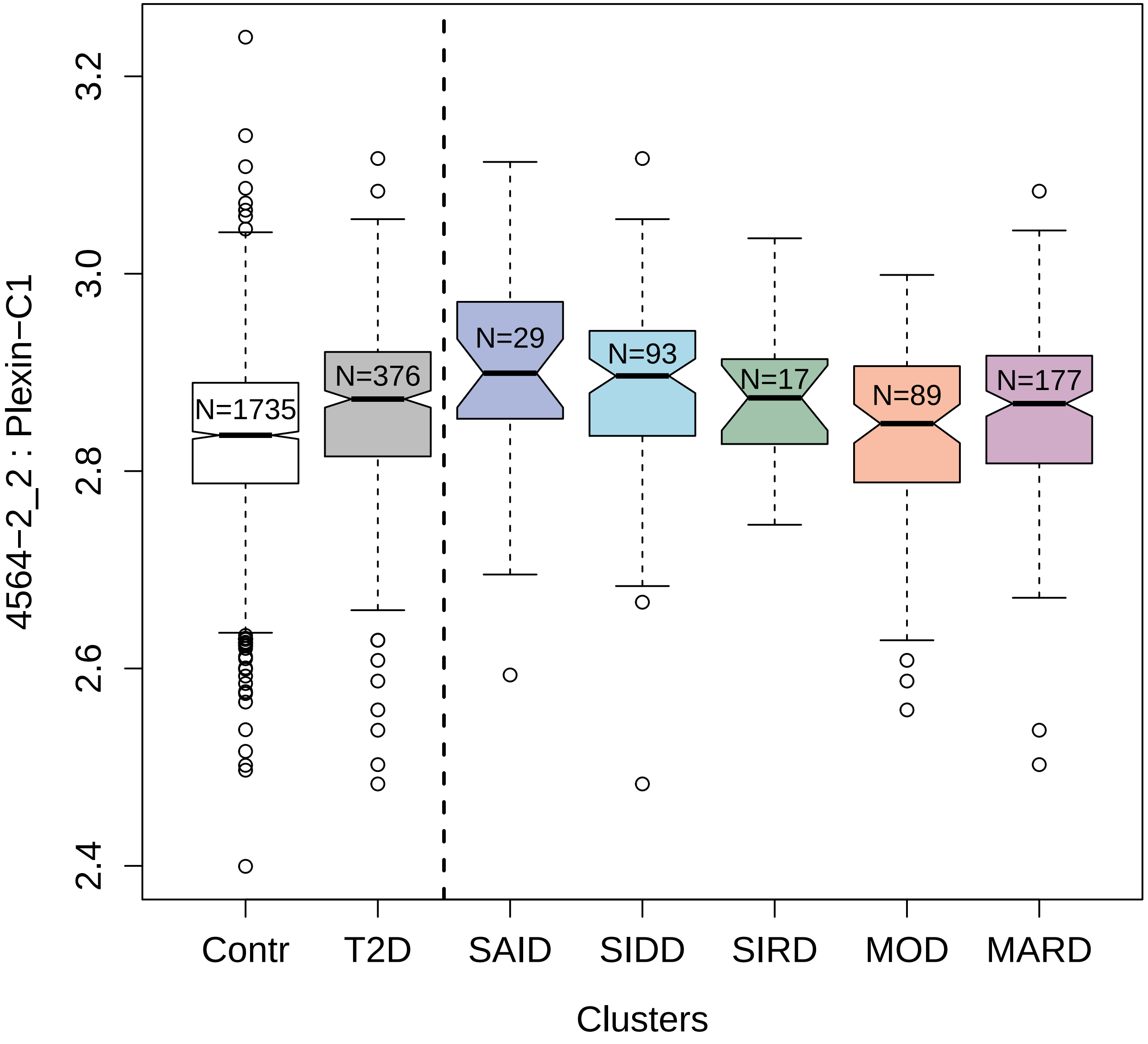

4564−2_2 : Plexin−C1
3.2
3.0
2.8
2.6
2.4
N=1735
N=376
N=29
N=93
N=17
N=89
N=177
Contr
T2D
SAID
SIDD
SIRD
MOD
MARD
Clusters

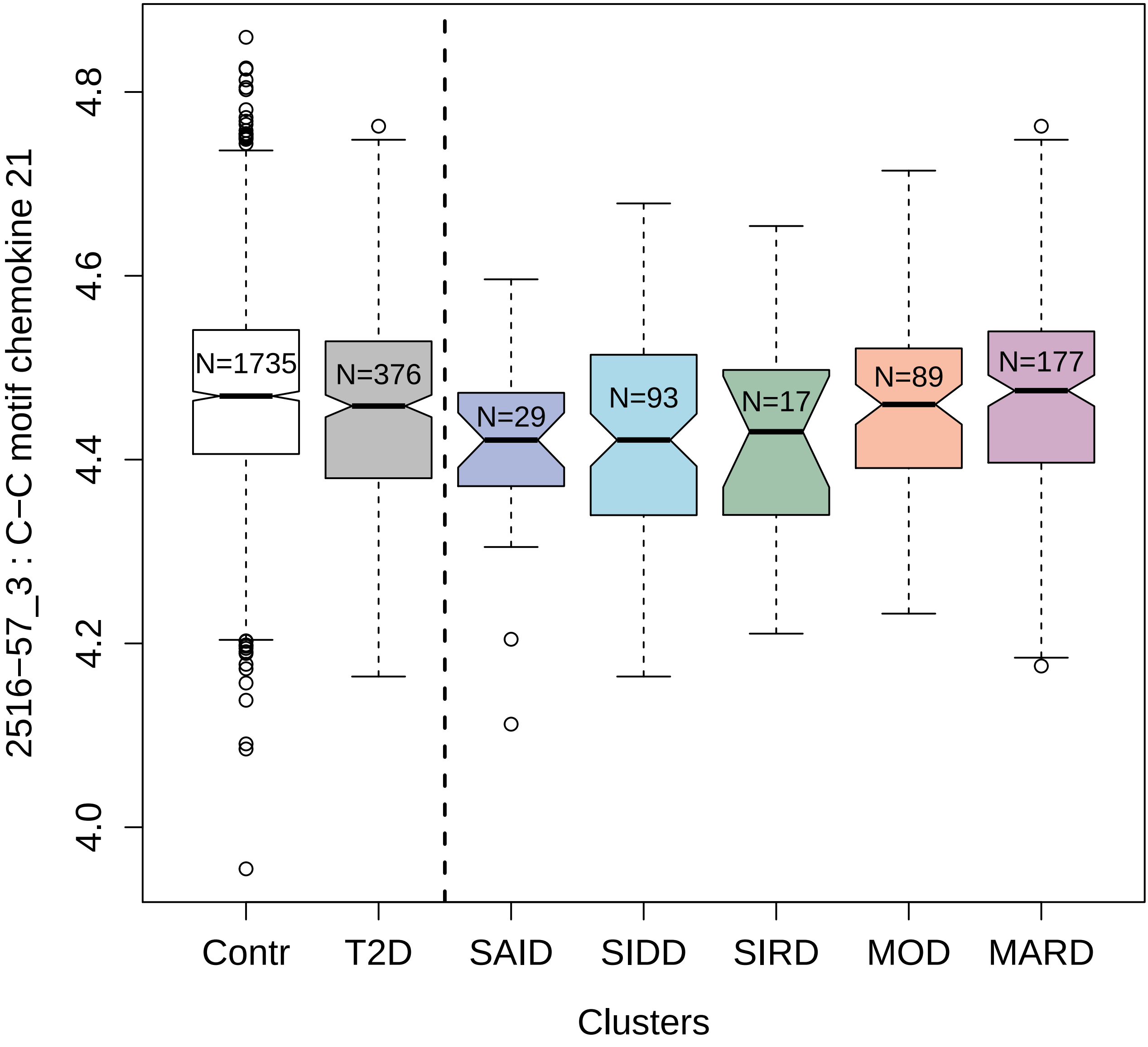

2516−57_3 : C−C motif chemokine 21
4.0
4.2
4.4
4.6
4.8
N=1735
N=376
N=29
N=93
N=17
N=89
N=177
Contr
T2D
SAID
SIDD
SIRD
MOD
MARD
Clusters

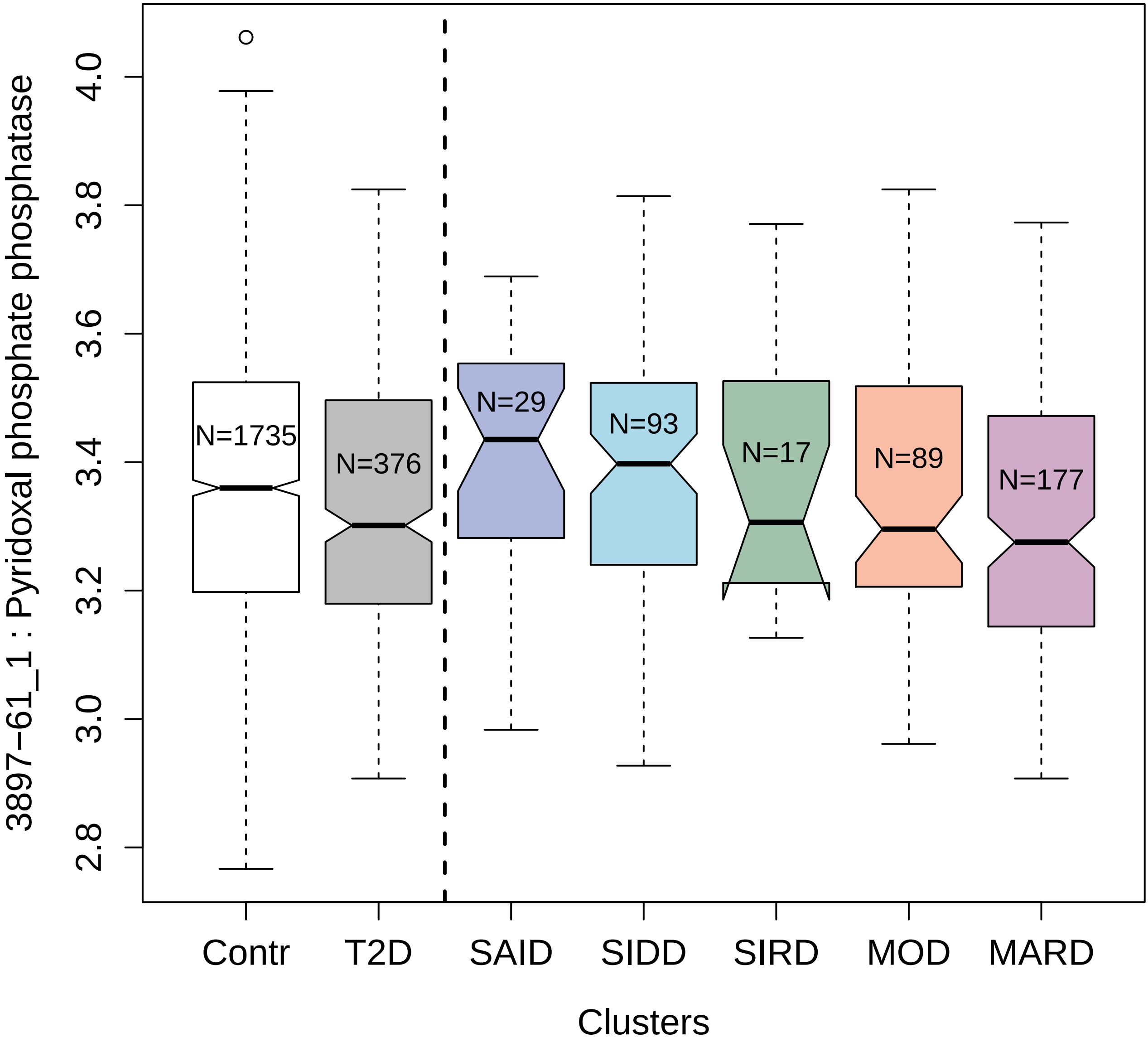
3897−61_1 : Pyridoxal phosphate phosphatase
2.8
3.0
3.2
3.4
3.6
3.8
4.0
N=1735
N=376
N=29
N=93
N=17
N=89
N=177
Contr
T2D
SAID
SIDD
SIRD
MOD
MARD
Clusters

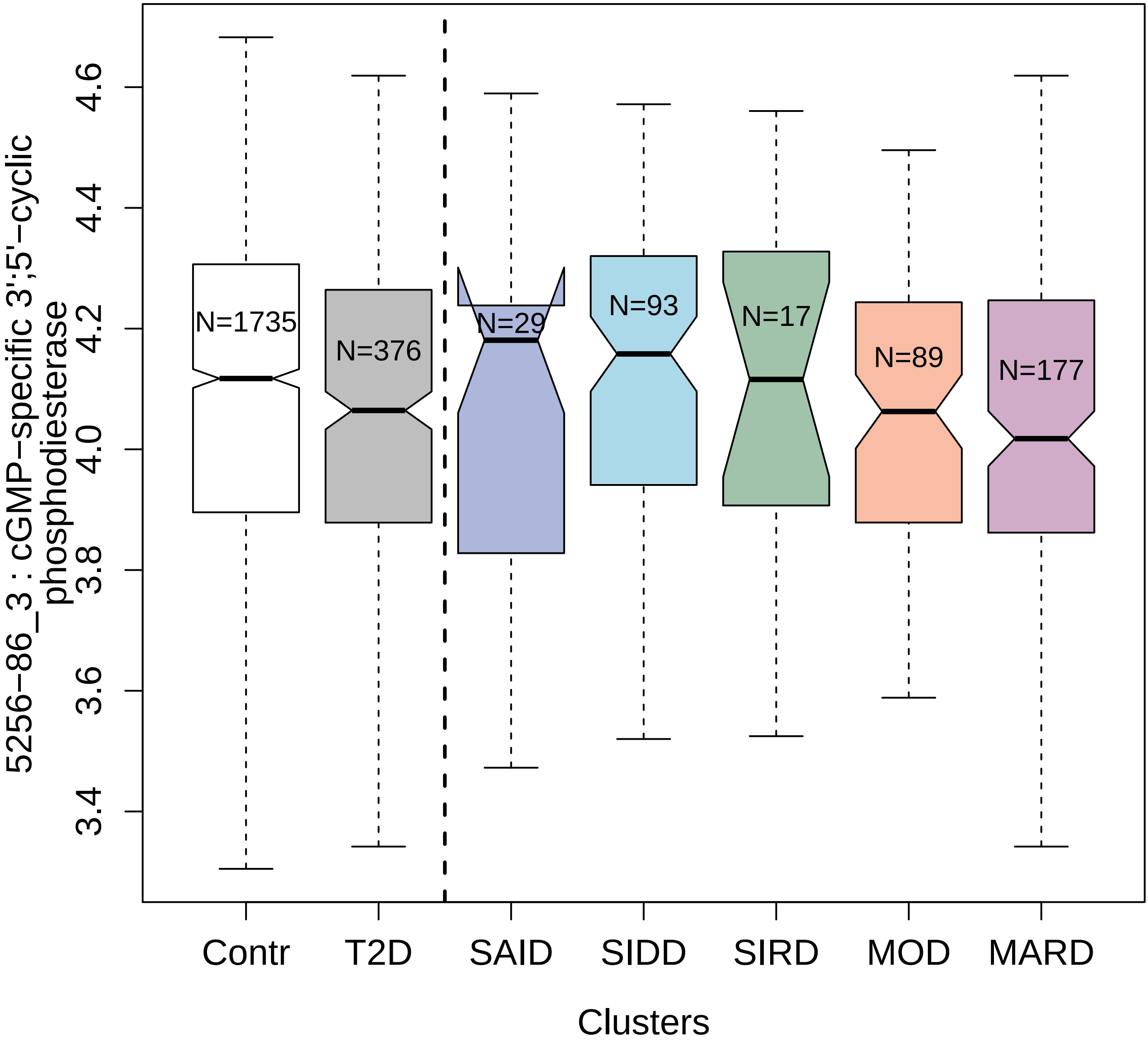

5256−86_3 : cGMP−specific 3',5'−cyclic phosphodiesterase
4.6
4.4
4.2
4.0
3.8
3.6
3.4
N=1735
N=376
N=29
N=93
N=17
N=89
N=177
Contr
T2D
SAID
SIDD
SIRD
MOD
MARD
Clusters

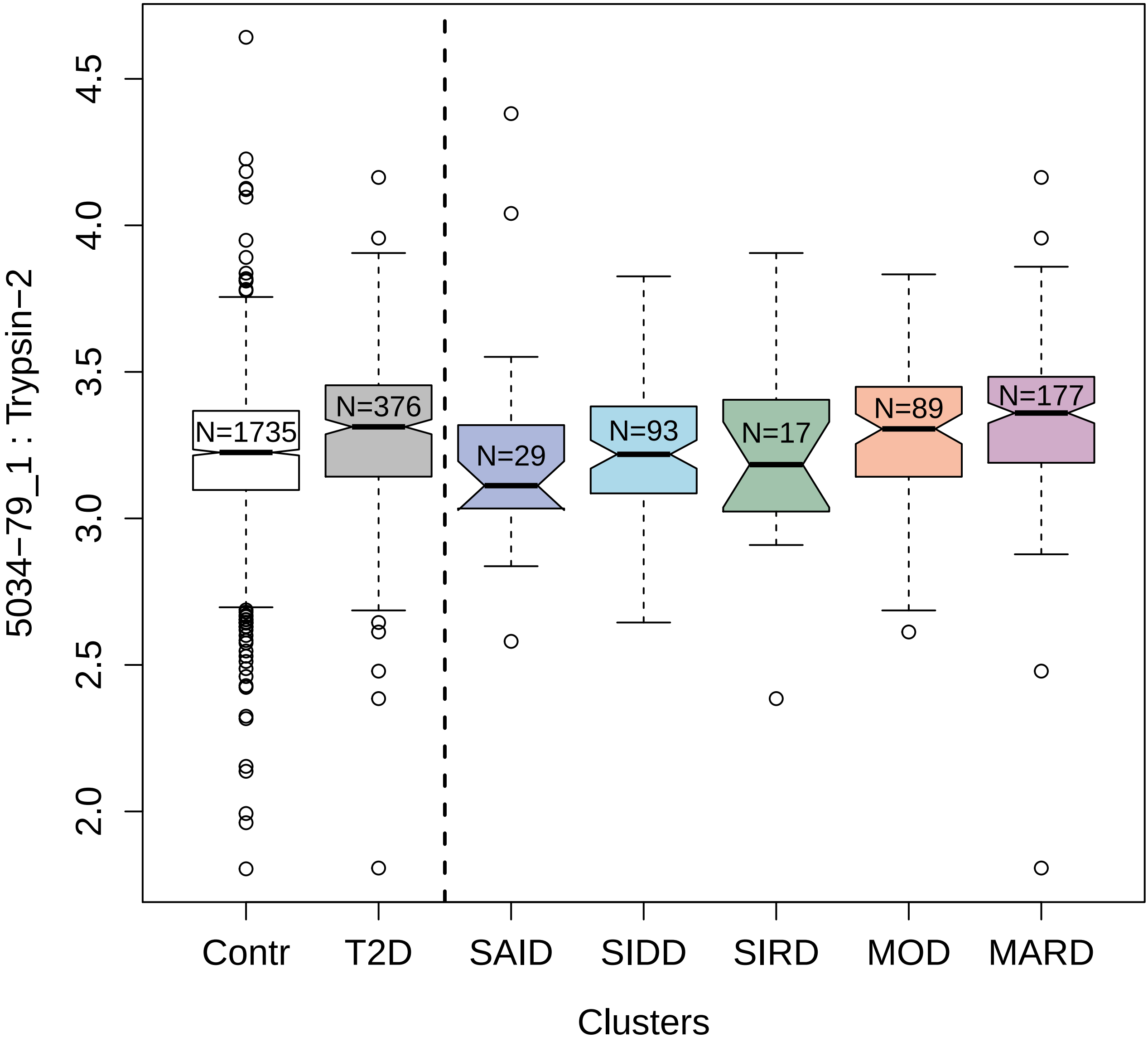

5034−79_1 : Trypsin−2
4.5
4.0
3.5
3.0
2.5
2.0
N=1735
N=376
N=29
N=93
N=17
N=89
N=177
Contr
T2D
SAID
SIDD
SIRD
MOD
MARD
Clusters

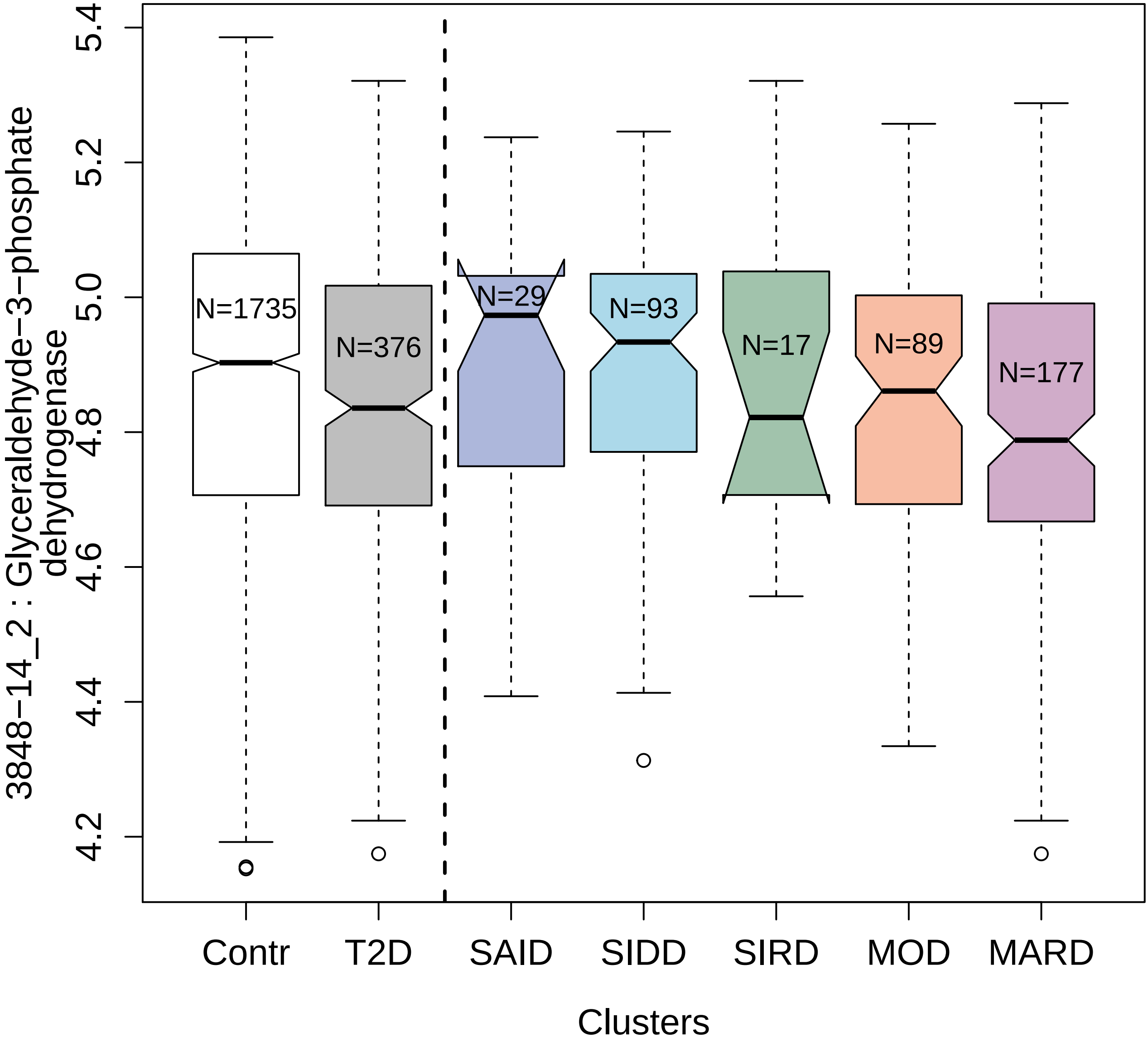
3848−14_2 : Glyceraldehyde−3−phosphate dehydrogenase
5.4
5.2
5.0
4.8
4.6
4.4
4.2
N=1735
N=376
N=29
N=93
N=17
N=89
N=177
Contr
T2D
SAID
SIDD
SIRD
MOD
MARD
Clusters

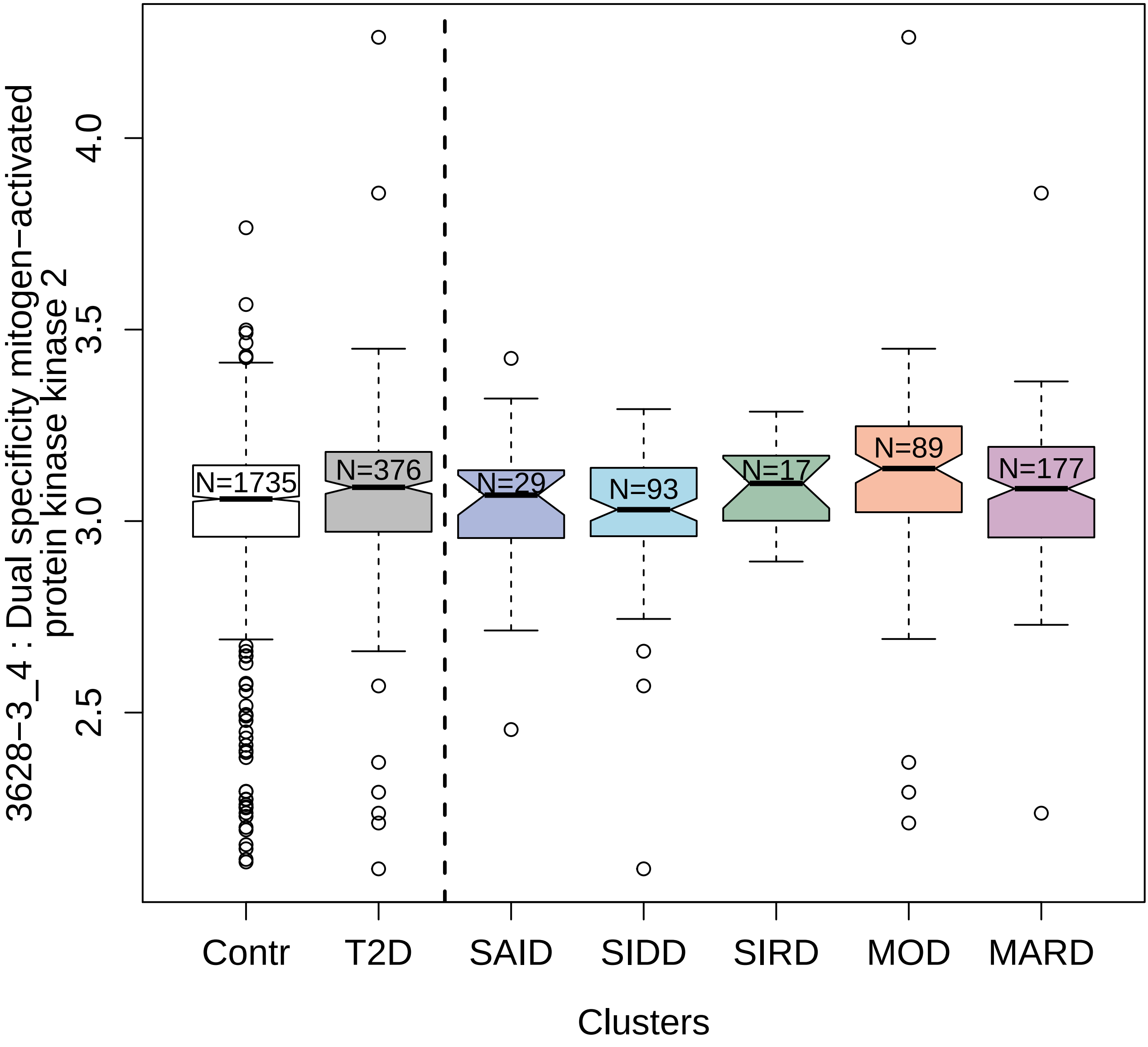

3628−3_4 : Dual specificity mitogen−activated
protein kinase kinase 2
4.0
3.5
3.0
2.5
N=1735
N=376
N=29
N=93
N=17
N=89
N=177
Contr
T2D
SAID
SIDD
SIRD
MOD
MARD
Clusters

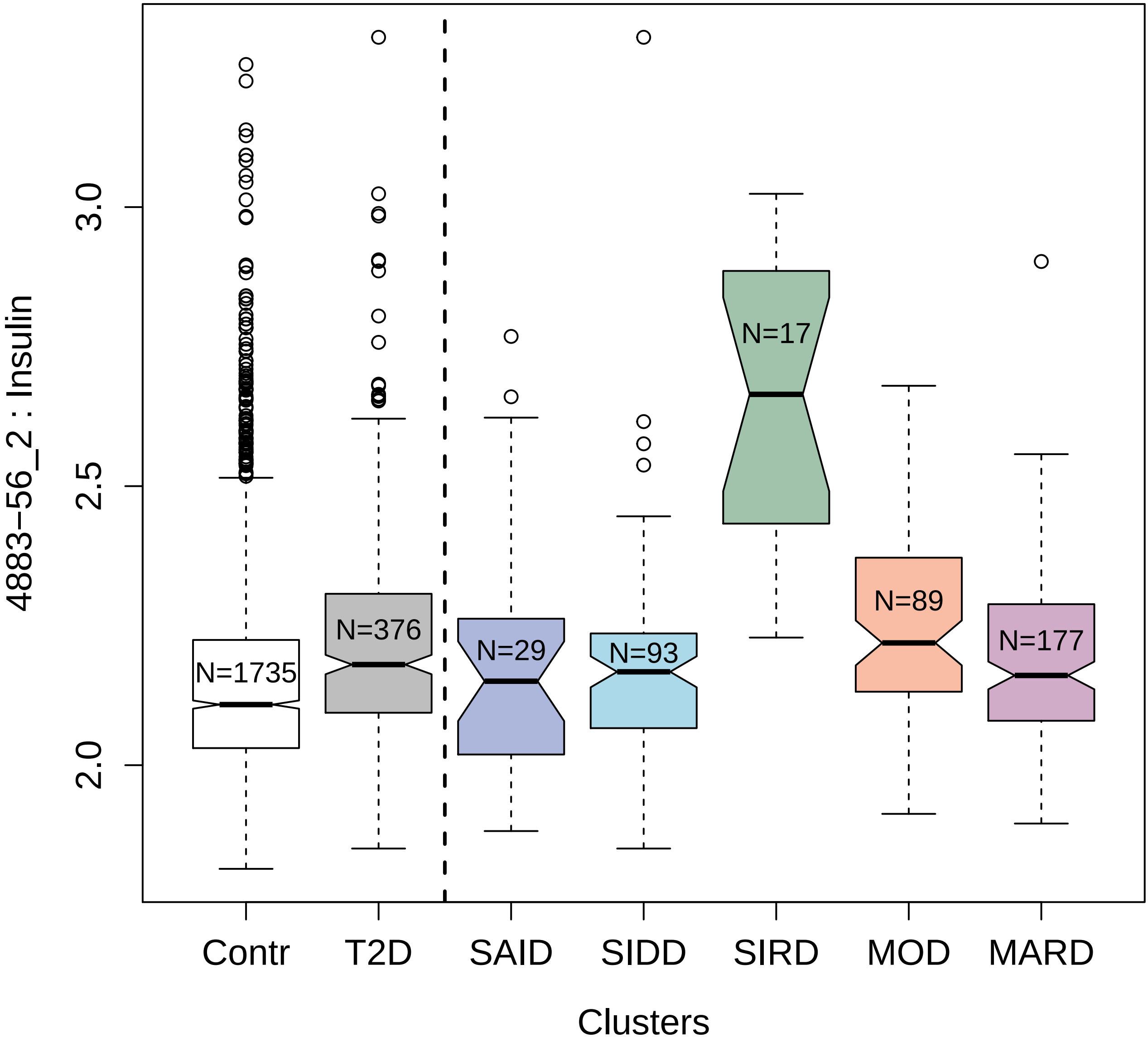
4883−56_2 : Insulin
3.0
2.5
2.0
N=1735
N=376
N=29
N=93
N=17
N=89
N=177
Contr
T2D
SAID
SIDD
SIRD
MOD
MARD
Clusters

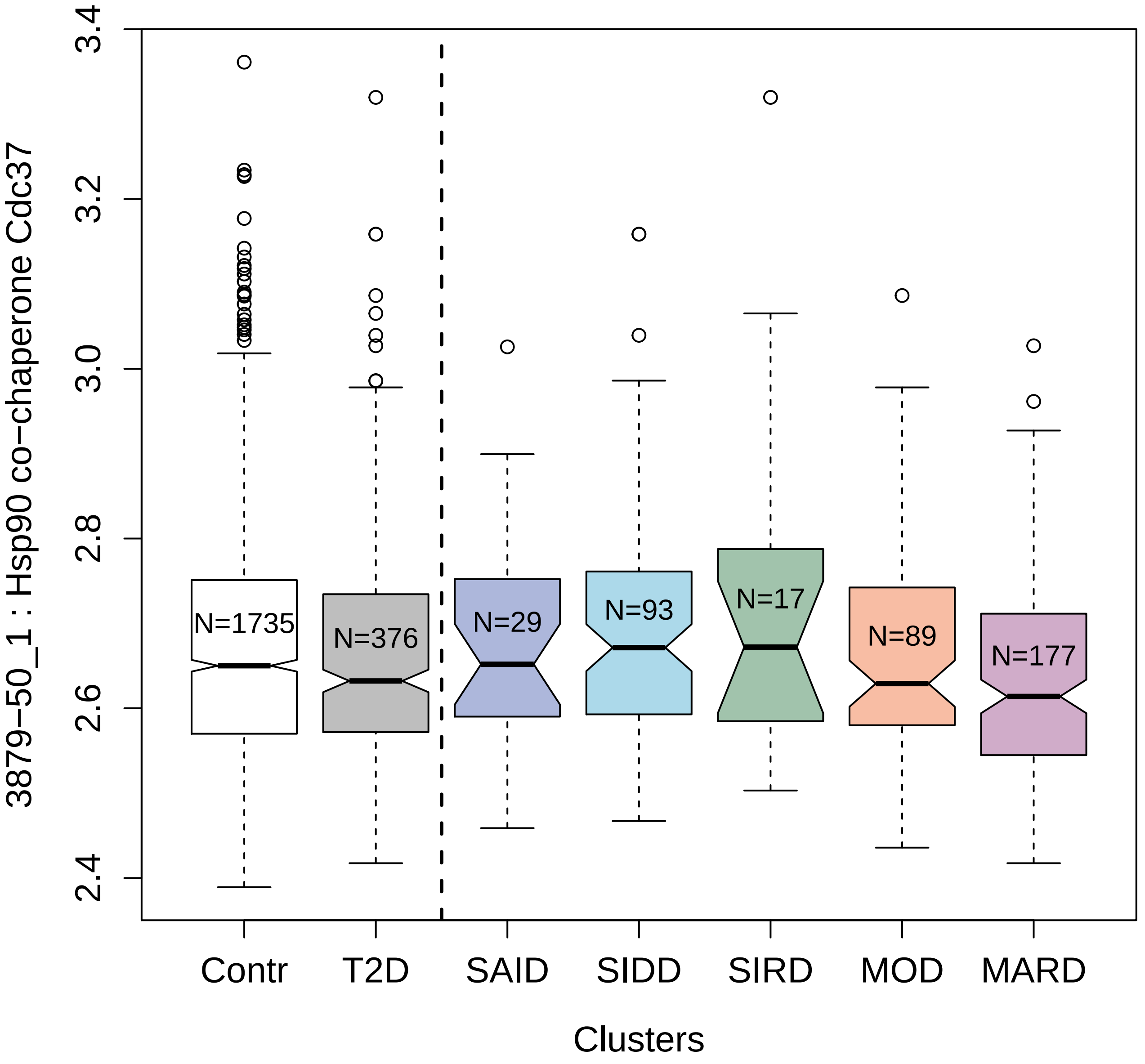

3879−50_1 : Hsp90 co−chaperone Cdc37
3.4
3.2
3.0
2.8
2.6
2.4
N=1735
N=376
N=29
N=93
N=17
N=89
N=177
Contr
T2D
SAID
SIDD
SIRD
MOD
MARD
Clusters

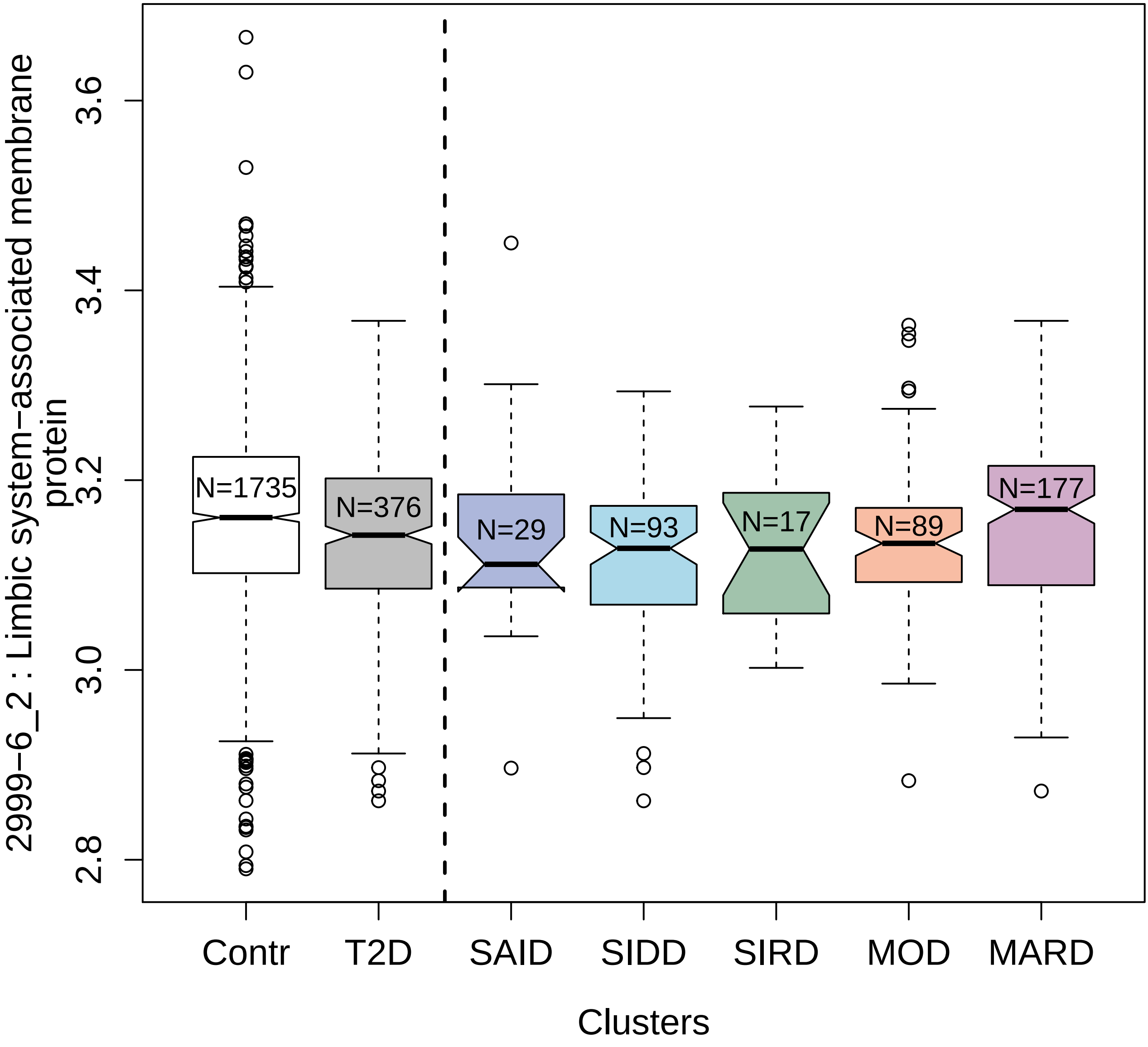
2999−6_2 : Limbic system−associated membrane protein
3.6
3.4
3.2
3.0
2.8
N=1735
N=376
N=29
N=93
N=17
N=89
N=177
Contr
T2D
SAID
SIDD
SIRD
MOD
MARD
Clusters

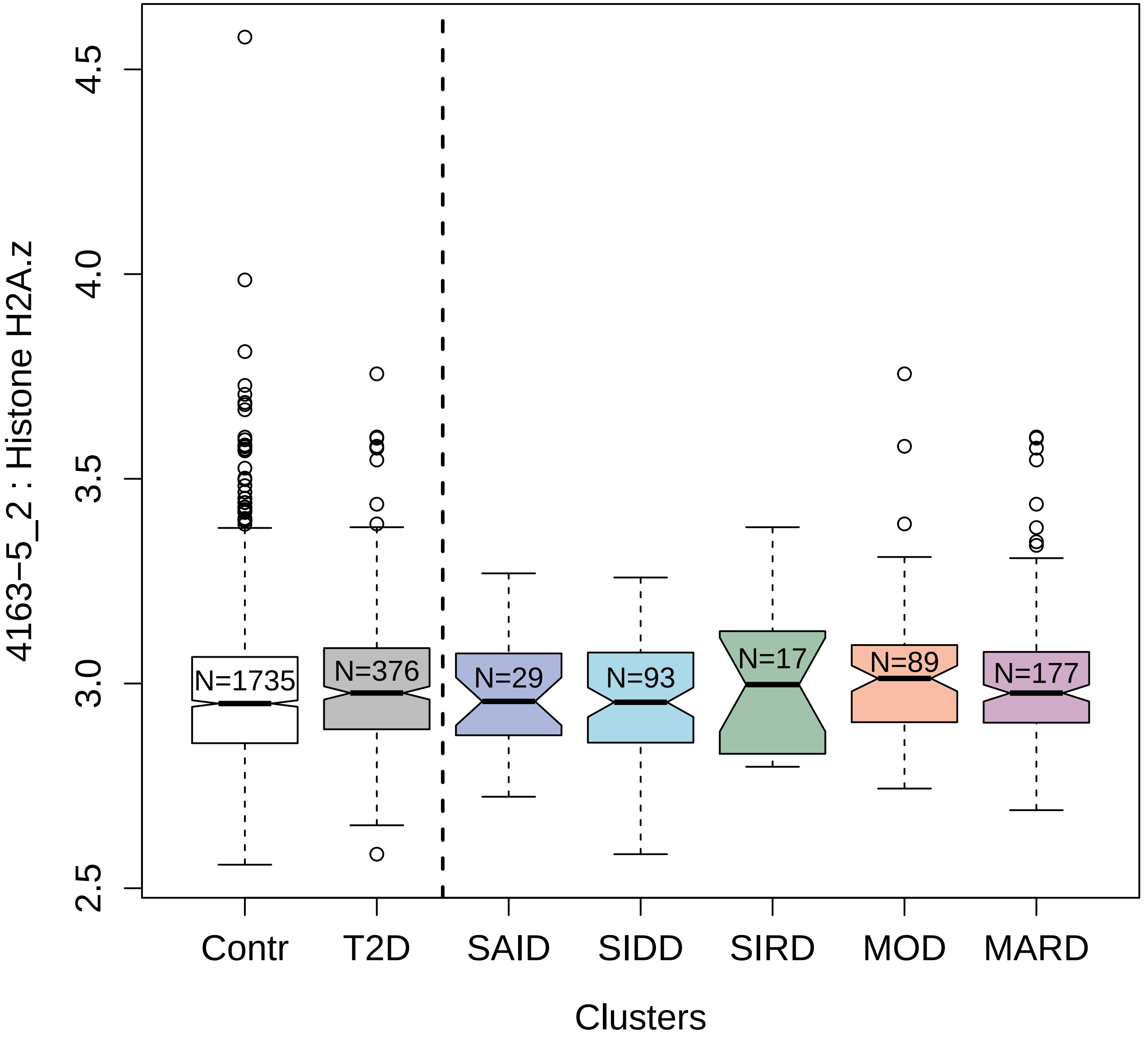
4163−5_2 : Histone H2A.z
4.5
4.0
3.5
3.0
2.5
N=1735
N=376
N=29
N=93
N=17
N=89
N=177
Contr
T2D
SAID
SIDD
SIRD
MOD
MARD
Clusters

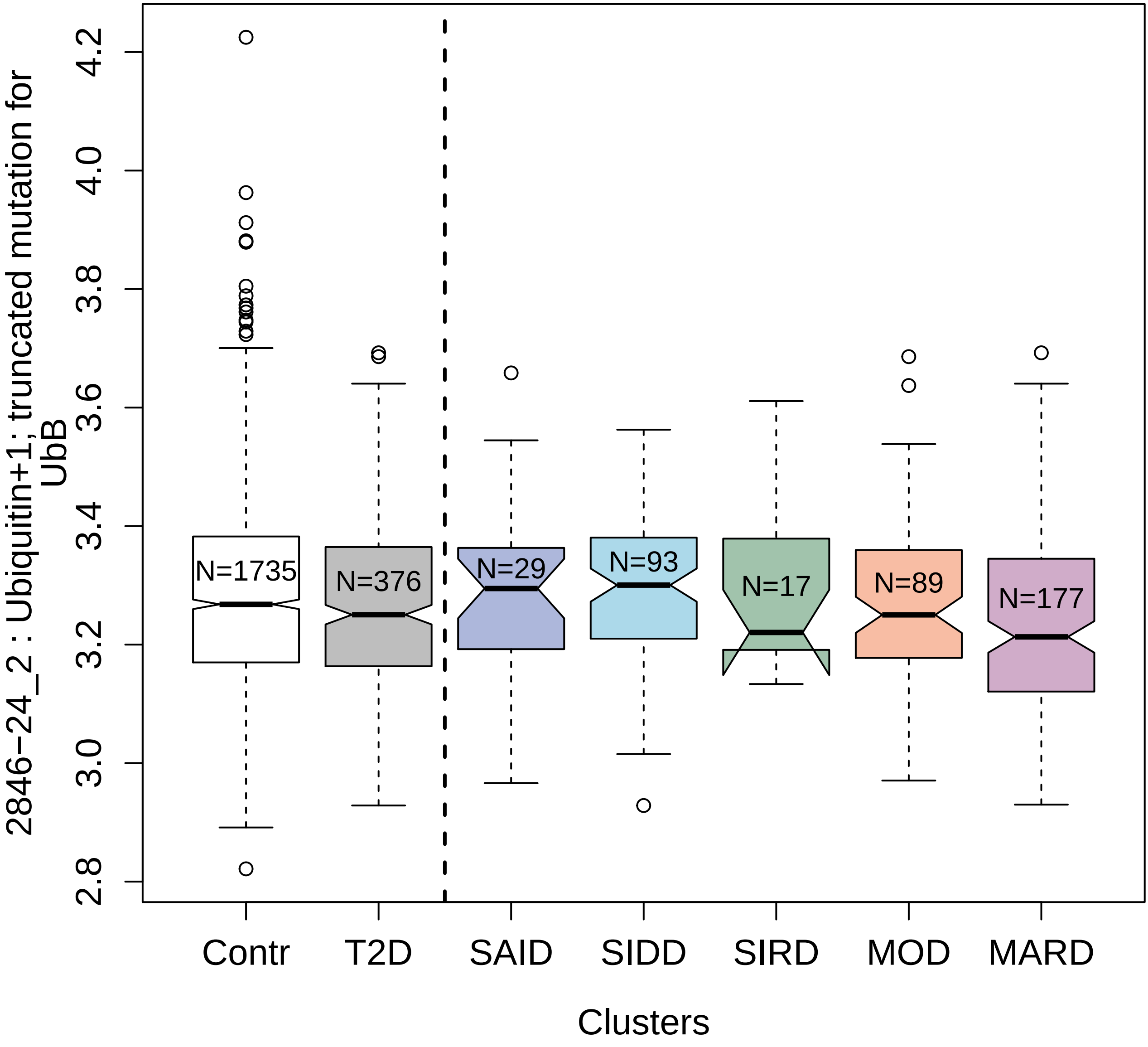
2846−24_2 : Ubiquitin+1; truncated mutation for UbB
4.2
4.0
3.8
3.6
3.4
3.2
3.0
2.8
N=1735
N=376
N=29
N=93
N=17
N=89
N=177
Contr
T2D
SAID
SIDD
SIRD
MOD
MARD
Clusters

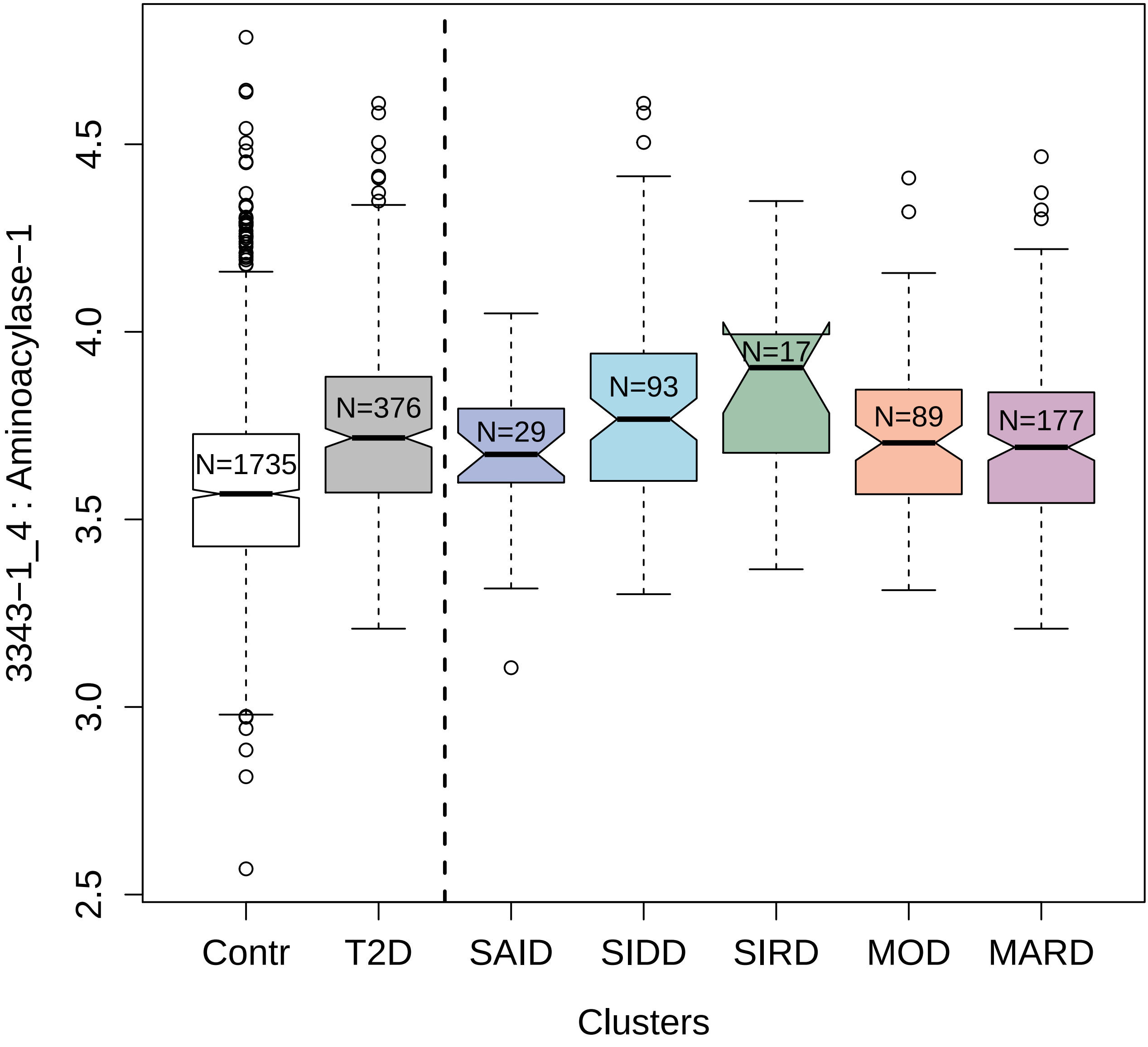

3343−1_4 : Aminoacylase−1
2.5
3.0
3.5
4.0
4.5
N=1735
N=376
N=29
N=93
N=17
N=89
N=177
Contr
T2D
SAID
SIDD
SIRD
MOD
MARD
Clusters

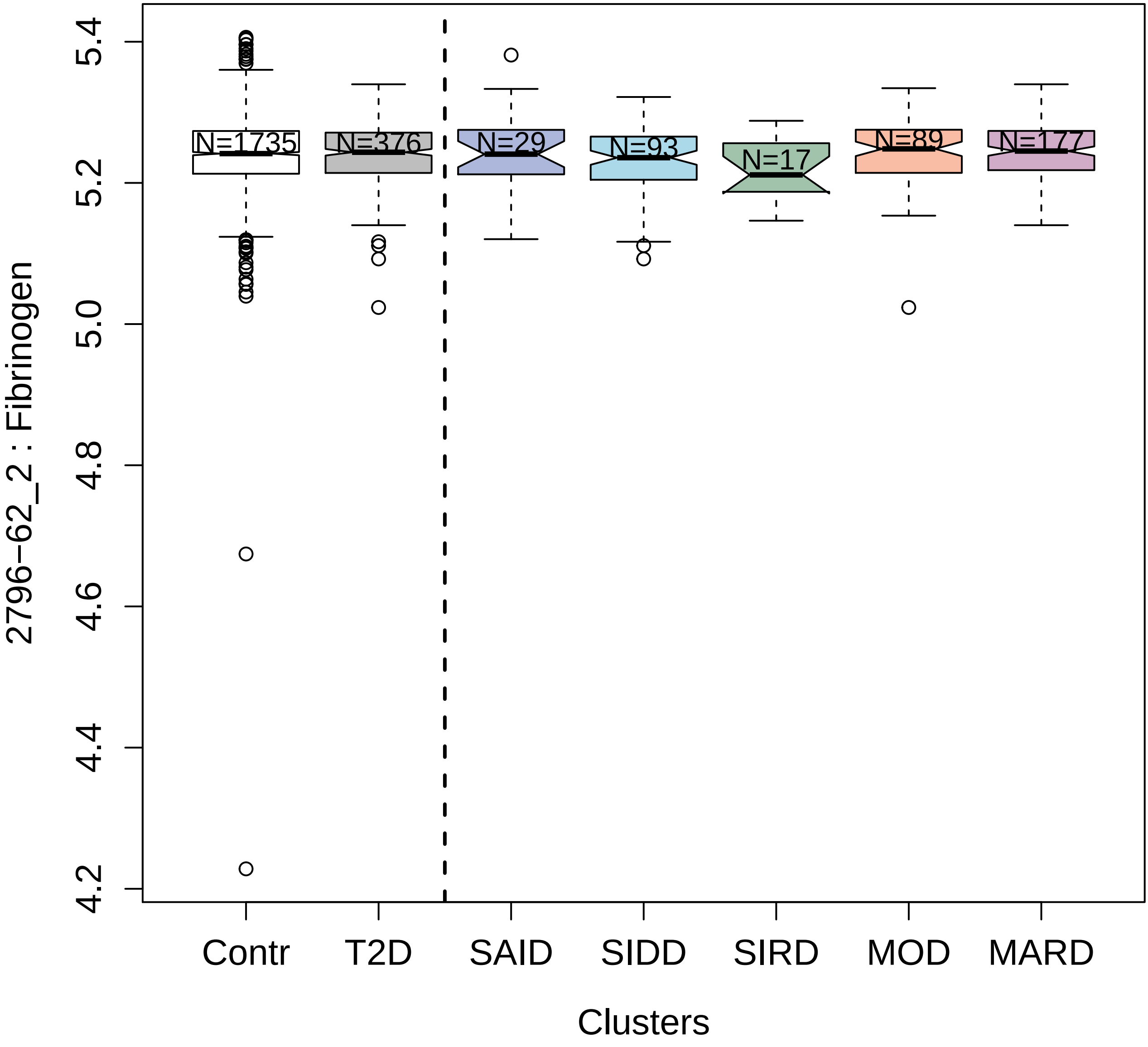

N=1735
N=376
N=29
N=93
N=17
N=89
N=177
5.4
5.2
5.0
4.8
4.6
4.4
4.2
2796−62_2 : Fibrinogen
Contr
T2D
SAID
SIDD
SIRD
MOD
MARD
Clusters

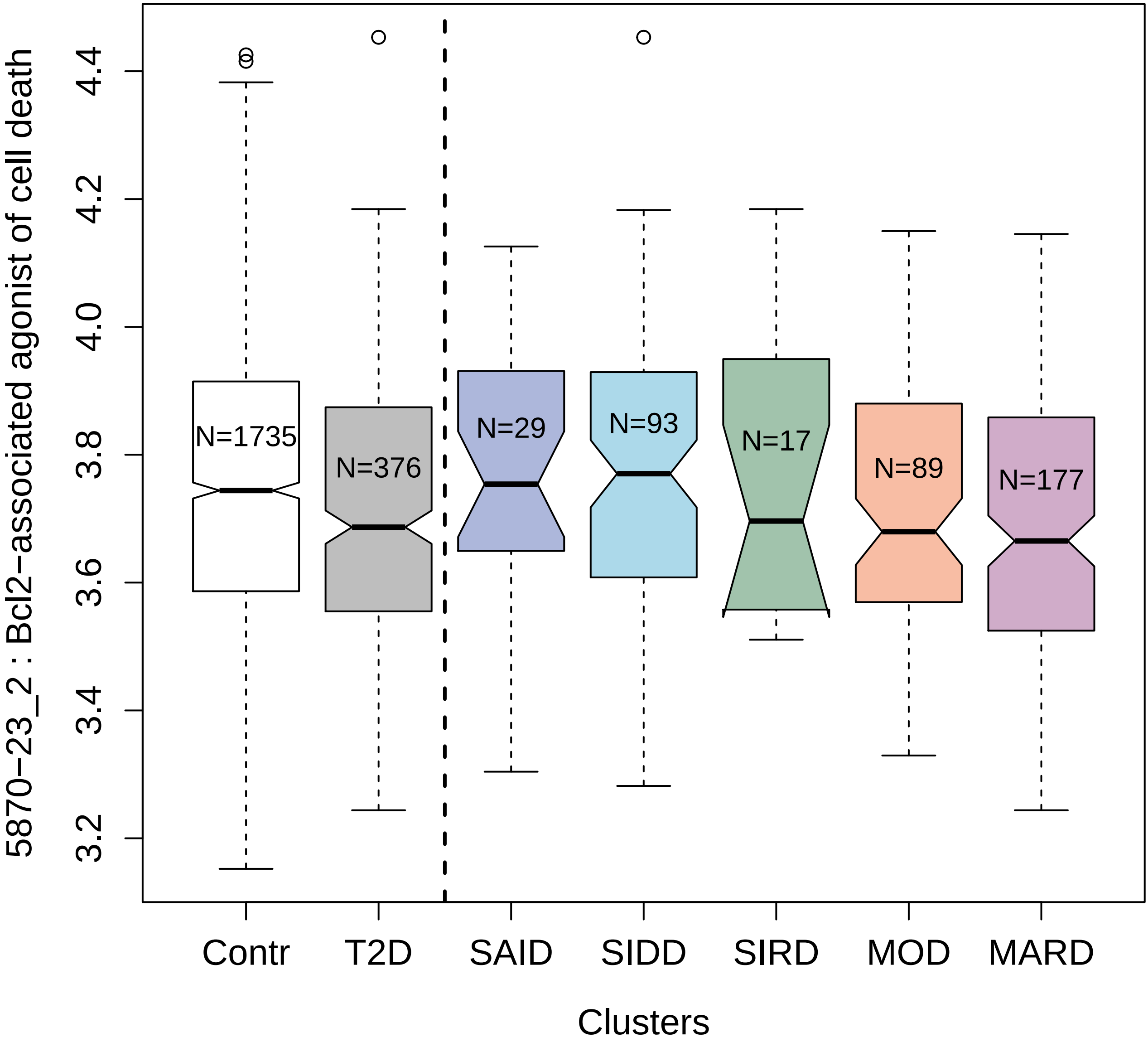
5870−23_2 : Bcl2−associated agonist of cell death
3.2
3.4
3.6
3.8
4.0
4.2
4.4
N=1735
N=376
N=29
N=93
N=17
N=89
N=177
Contr
T2D
SAID
SIDD
SIRD
MOD
MARD
Clusters

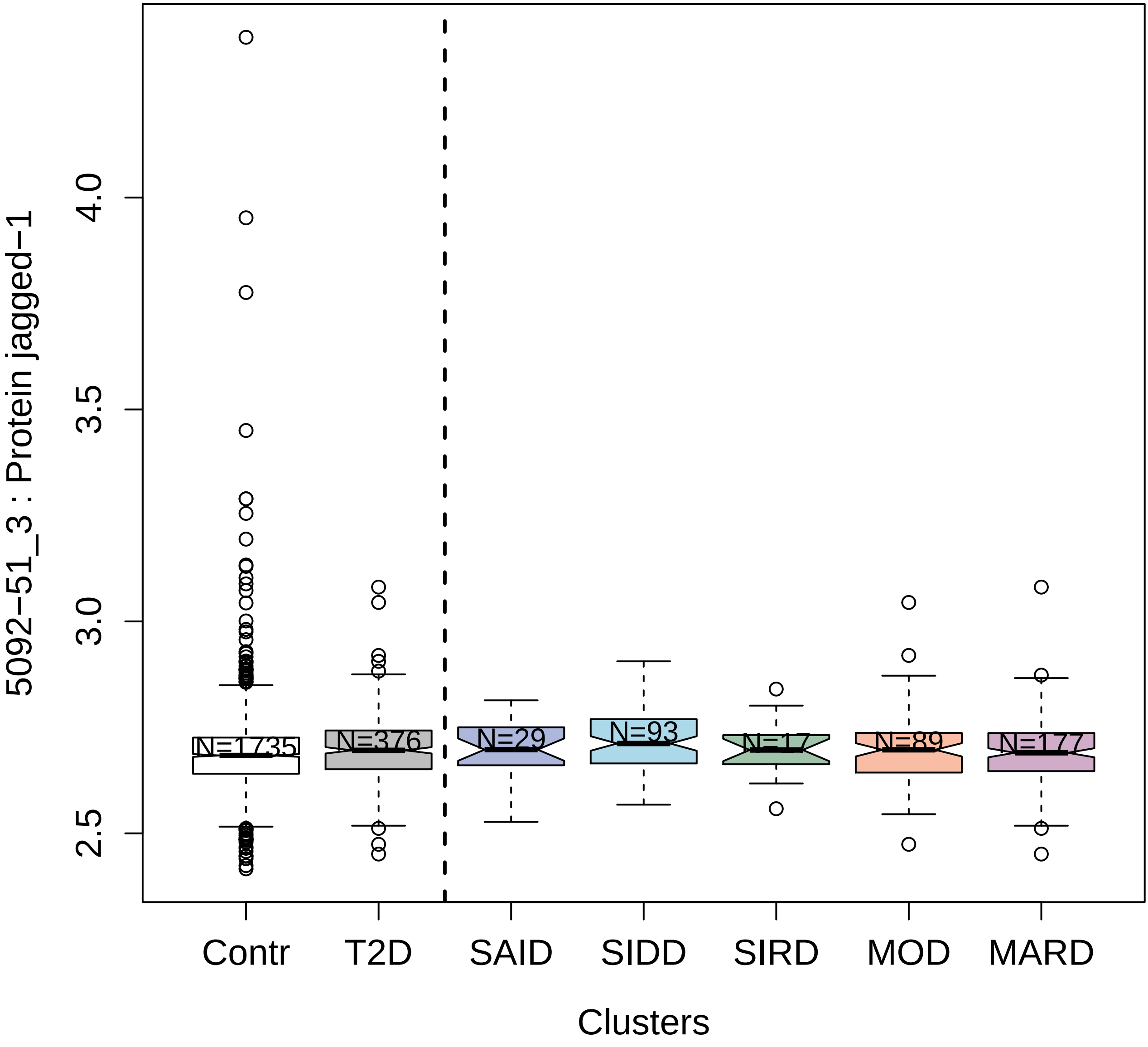

5092−51_3 : Protein jagged−1
4.0
3.5
3.0
2.5
N=1735
N=376
N=29
N=93
N=17
N=89
N=177
Contr
T2D
SAID
SIDD
SIRD
MOD
MARD
Clusters

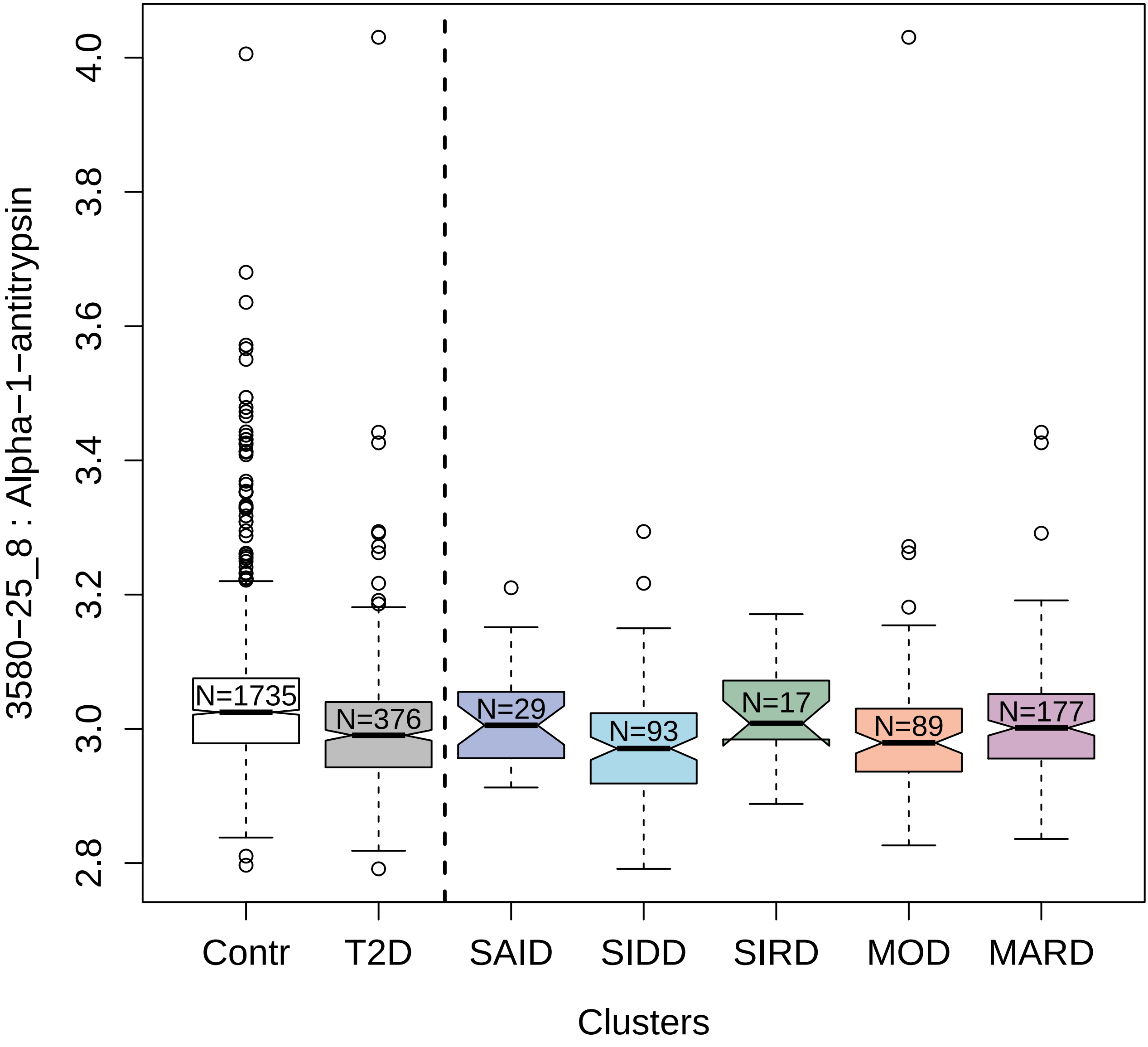
3580−25_8 : Alpha−1−antitrypsin
4.0
3.8
3.6
3.4
3.2
3.0
2.8
N=1735
N=376
N=29
N=93
N=17
N=89
N=177
Contr
T2D
SAID
SIDD
SIRD
MOD
MARD
Clusters

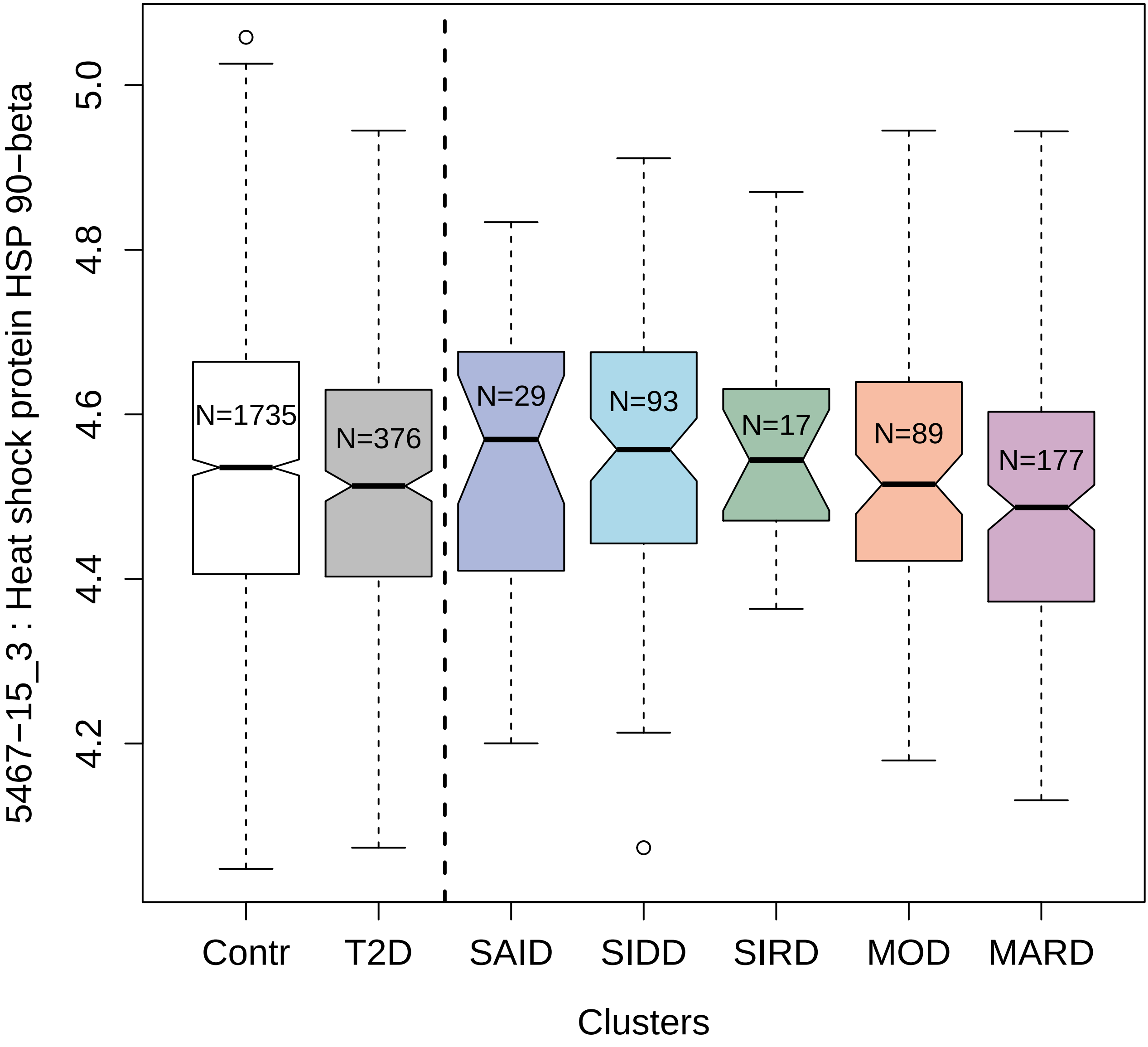
5467−15_3 : Heat shock protein HSP 90−beta
5.0
4.8
4.6
4.4
4.2
N=1735
N=376
N=29
N=93
N=17
N=89
N=177
Contr
T2D
SAID
SIDD
SIRD
MOD
MARD
Clusters

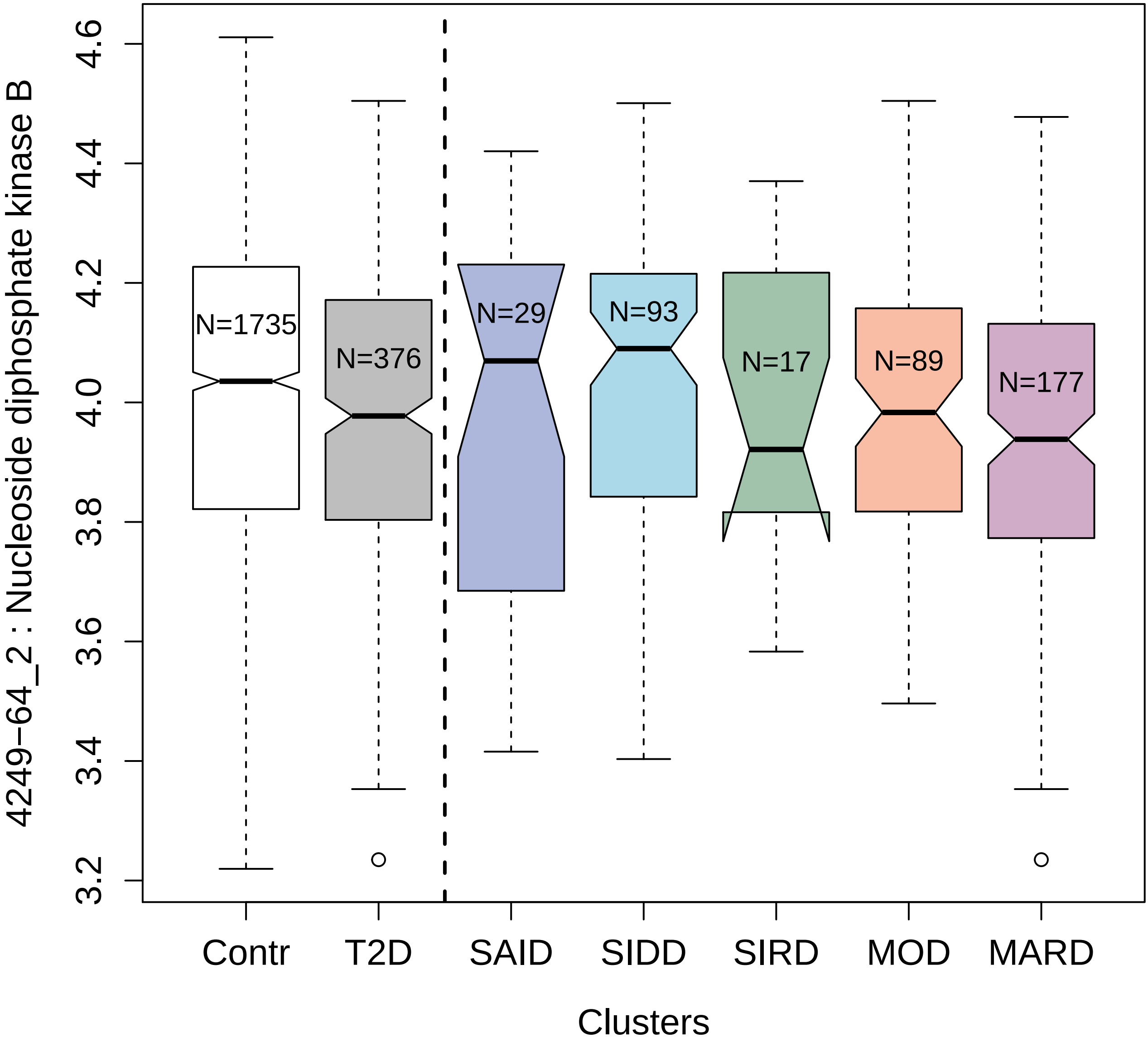
4249−64_2 : Nucleoside diphosphate kinase B
3.2
3.4
3.6
3.8
4.0
4.2
4.4
4.6
N=1735
N=376
N=29
N=93
N=17
N=89
N=177
Contr
T2D
SAID
SIDD
SIRD
MOD
MARD
Clusters

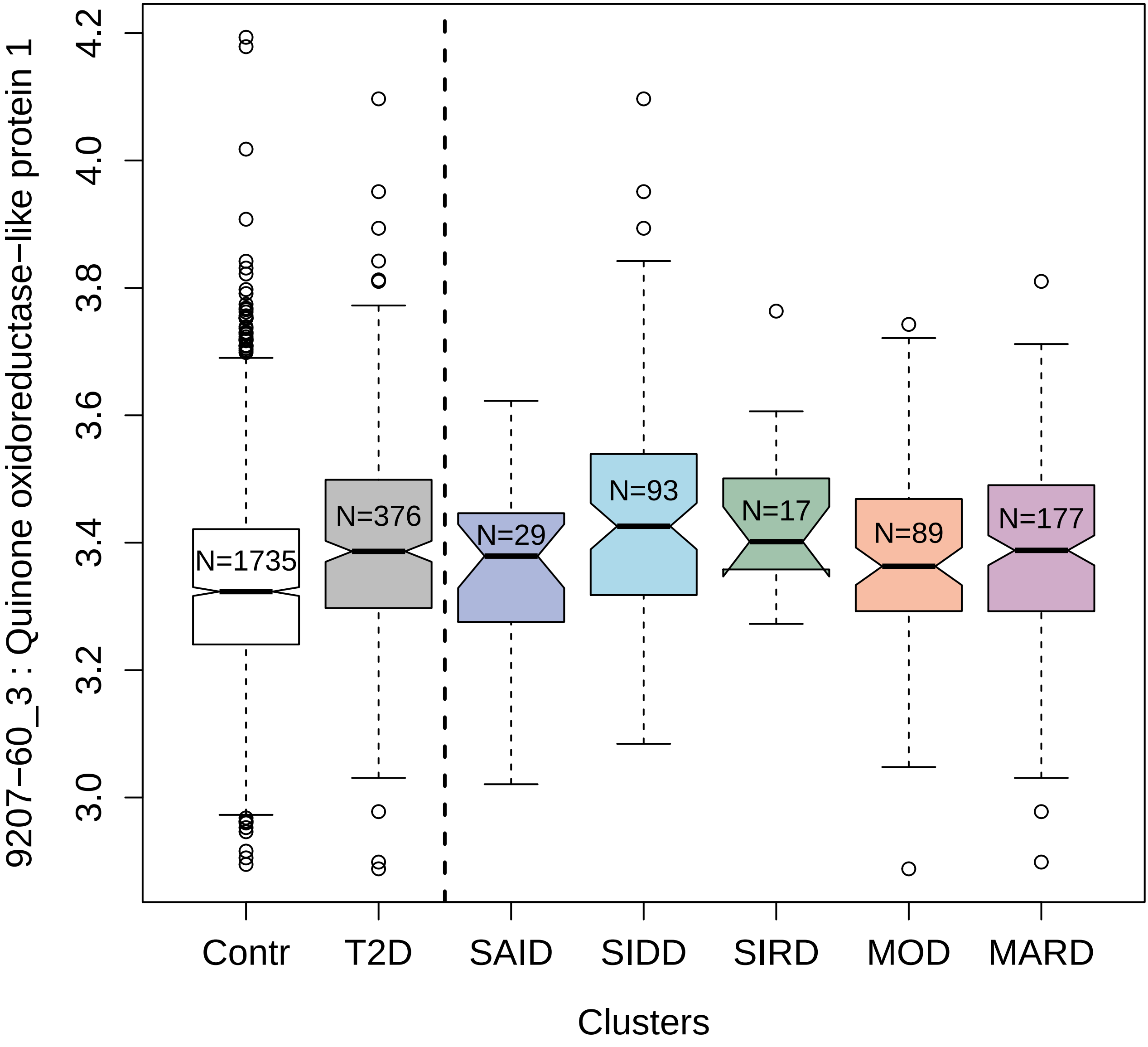

9207−60_3 : Quinone oxidoreductase−like protein 1
4.2
4.0
3.8
3.6
3.4
3.2
3.0
N=1735
N=376
N=29
N=93
N=17
N=89
N=177
Contr
T2D
SAID
SIDD
SIRD
MOD
MARD
Clusters

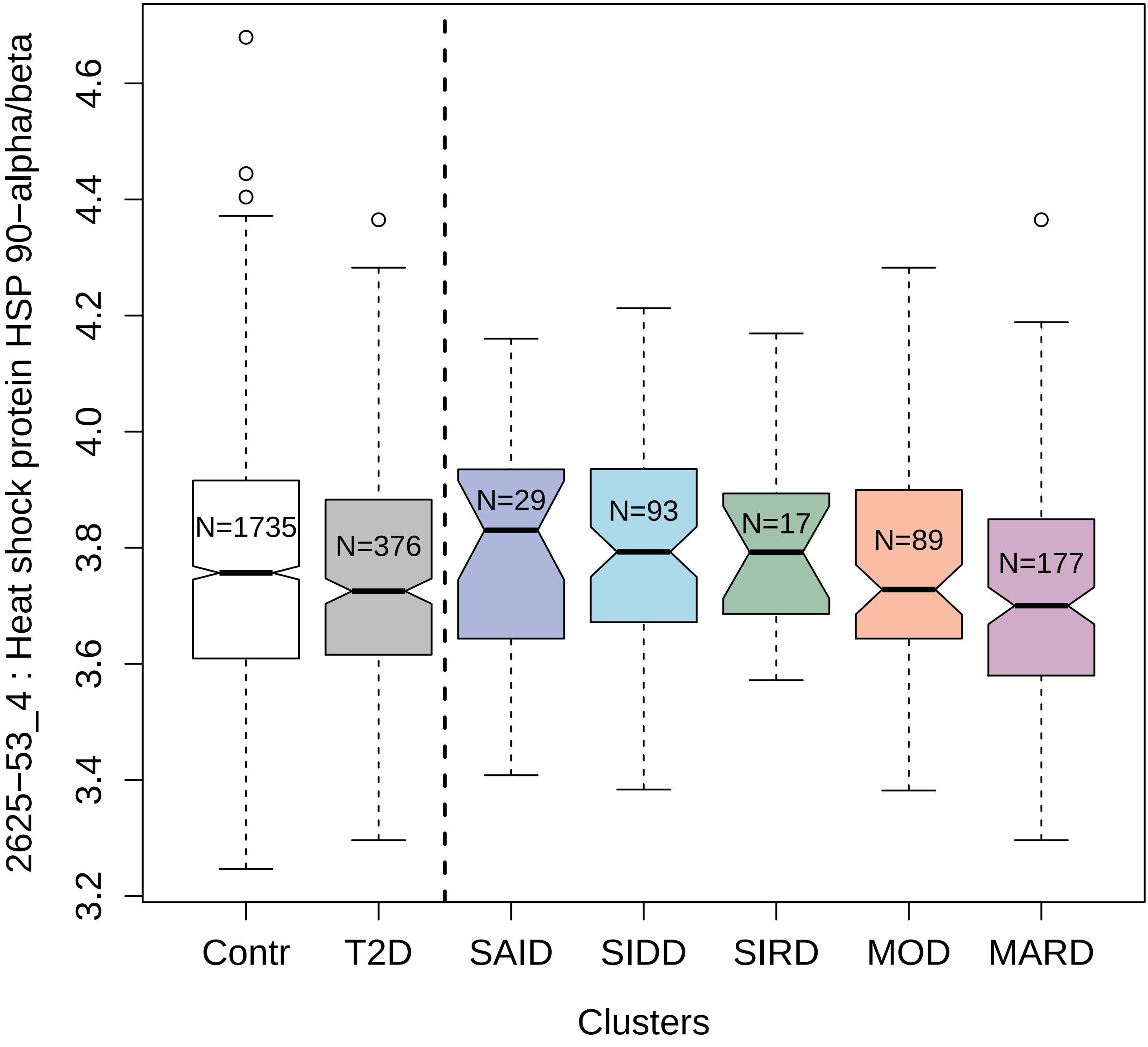
2625−53_4 : Heat shock protein HSP 90−alpha/beta
3.2
3.4
3.6
3.8
4.0
4.2
4.4
4.6
N=1735
N=376
N=29
N=93
N=17
N=89
N=177
Contr
T2D
SAID
SIDD
SIRD
MOD
MARD
Clusters

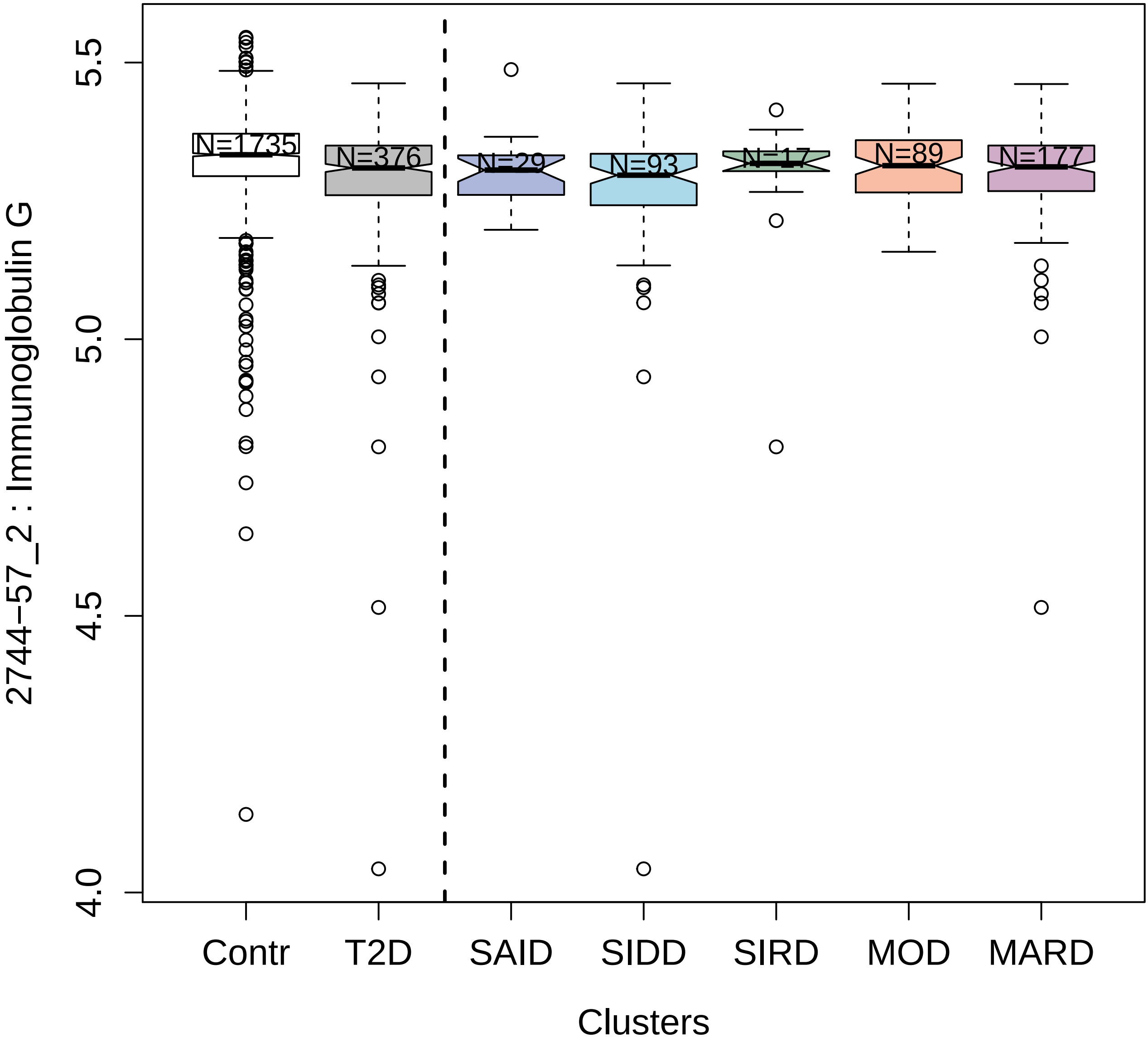
2744−57_2 : Immunoglobulin G
5.5
5.0
4.5
4.0
N=1735
N=376
N=29
N=93
N=17
N=89
N=177
Contr
T2D
SAID
SIDD
SIRD
MOD
MARD
Clusters

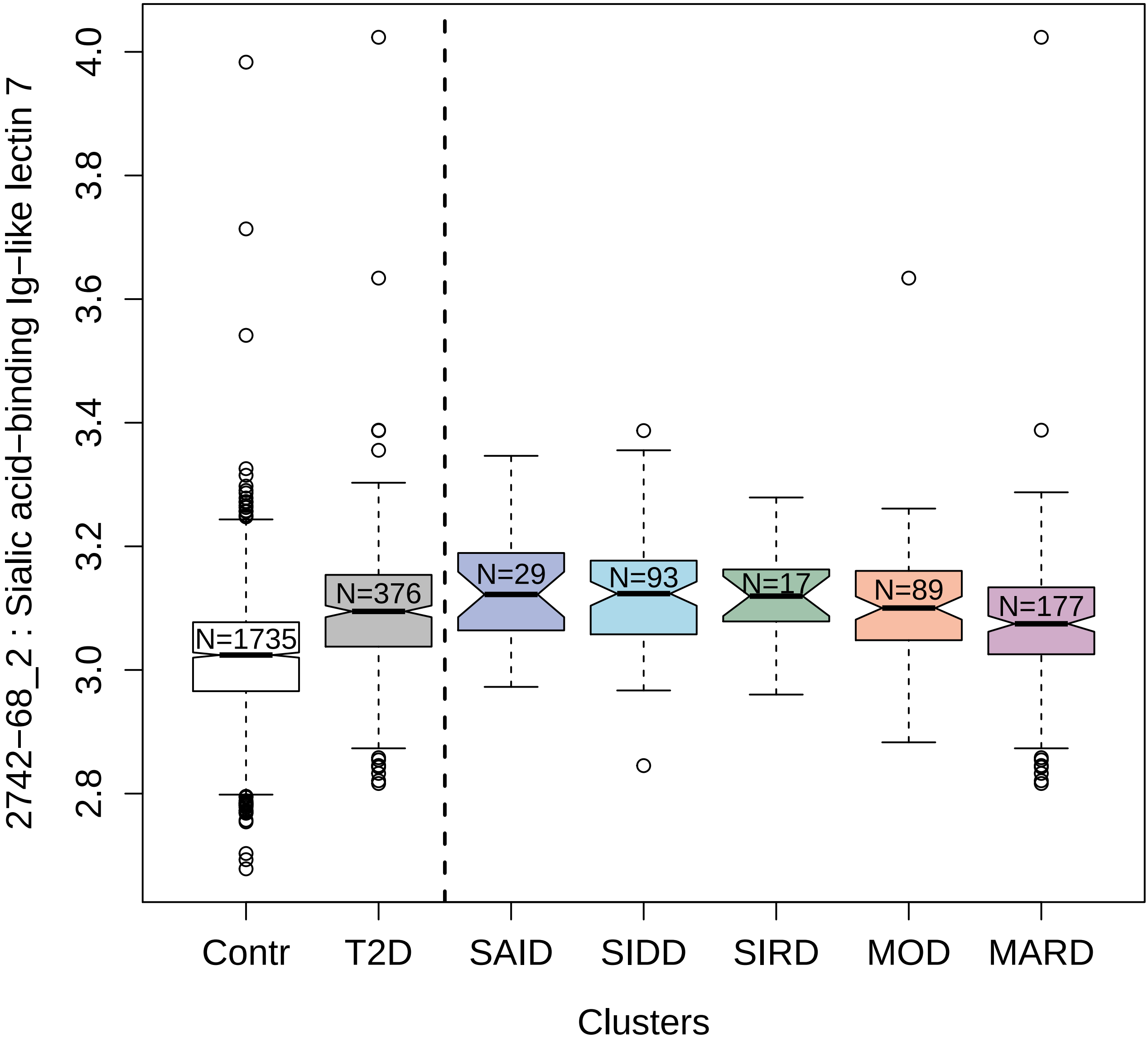

2742−68_2 : Sialic acid−binding Ig−like lectin 7
4.0
3.8
3.6
3.4
3.2
3.0
2.8
N=1735
N=376
N=29
N=93
N=17
N=89
N=177
Contr
T2D
SAID
SIDD
SIRD
MOD
MARD
Clusters

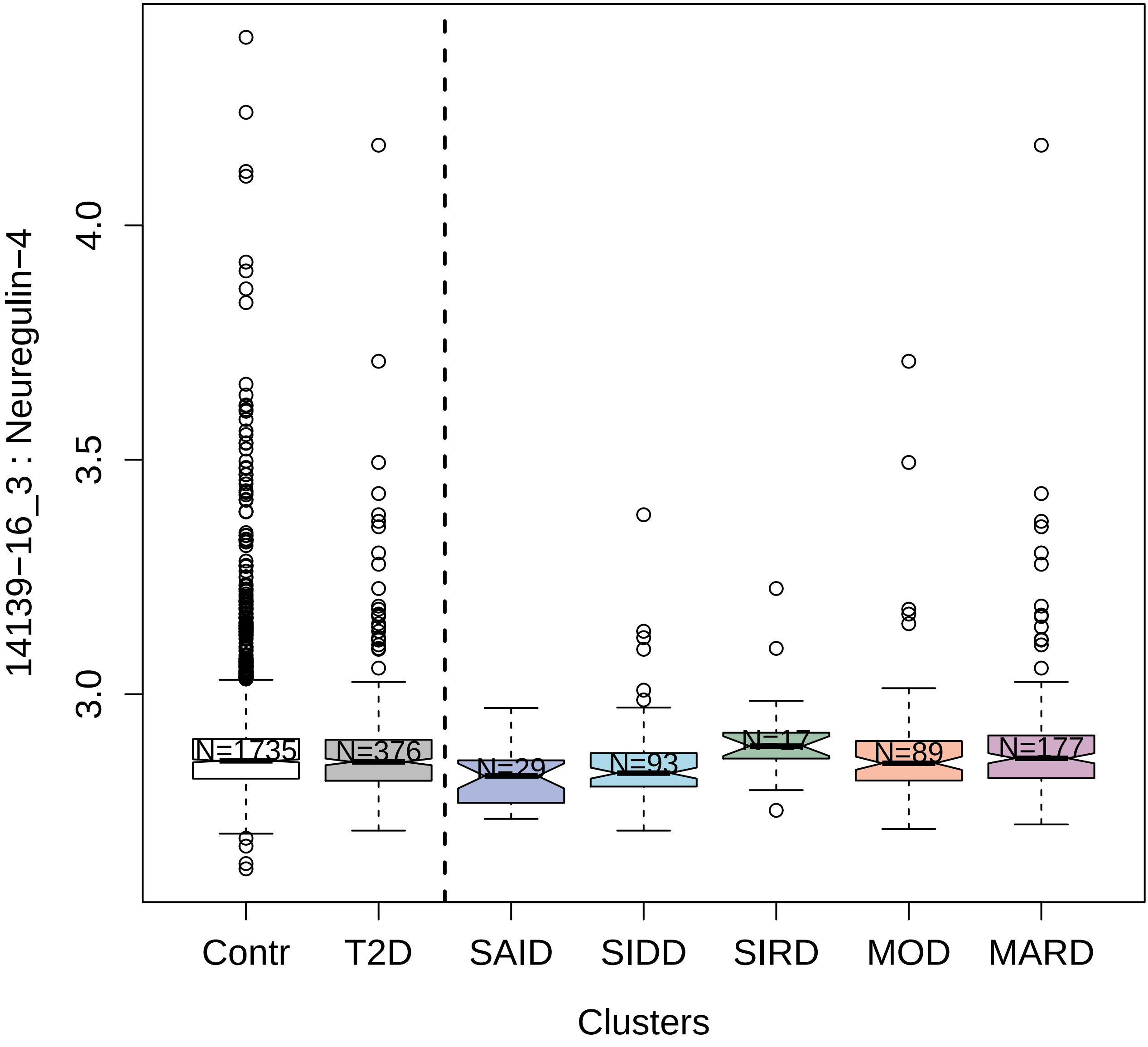

14139−16_3 : Neuregulin−4
4.0
3.5
3.0
N=1735
N=376
N=29
N=93
N=17
N=89
N=177
Contr
T2D
SAID
SIDD
SIRD
MOD
MARD
Clusters

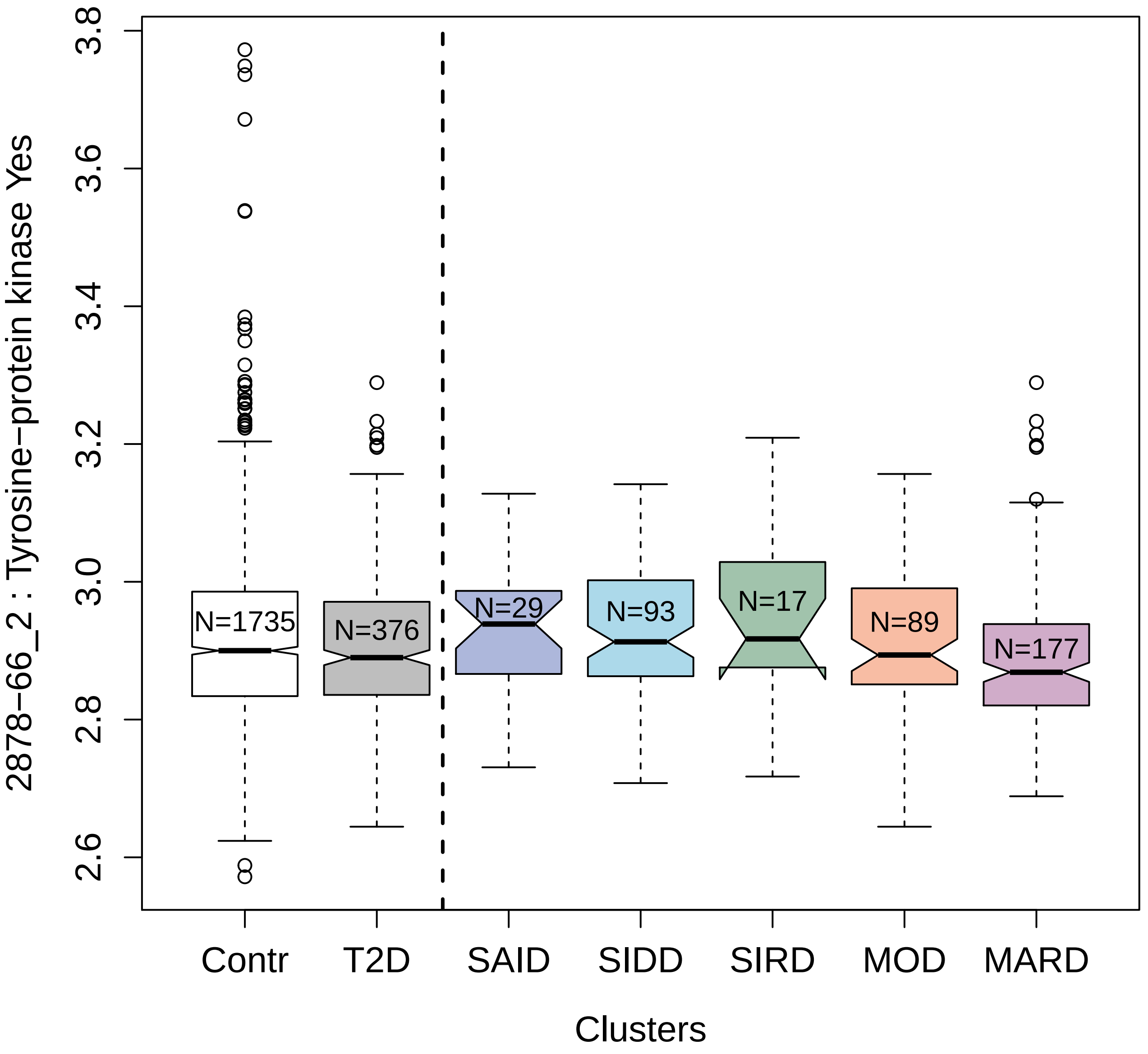
2878−66_2 : Tyrosine−protein kinase Yes
3.8
3.6
3.4
3.2
3.0
2.8
2.6
N=1735
N=376
N=29
N=93
N=17
N=89
N=177
Contr
T2D
SAID
SIDD
SIRD
MOD
MARD
Clusters

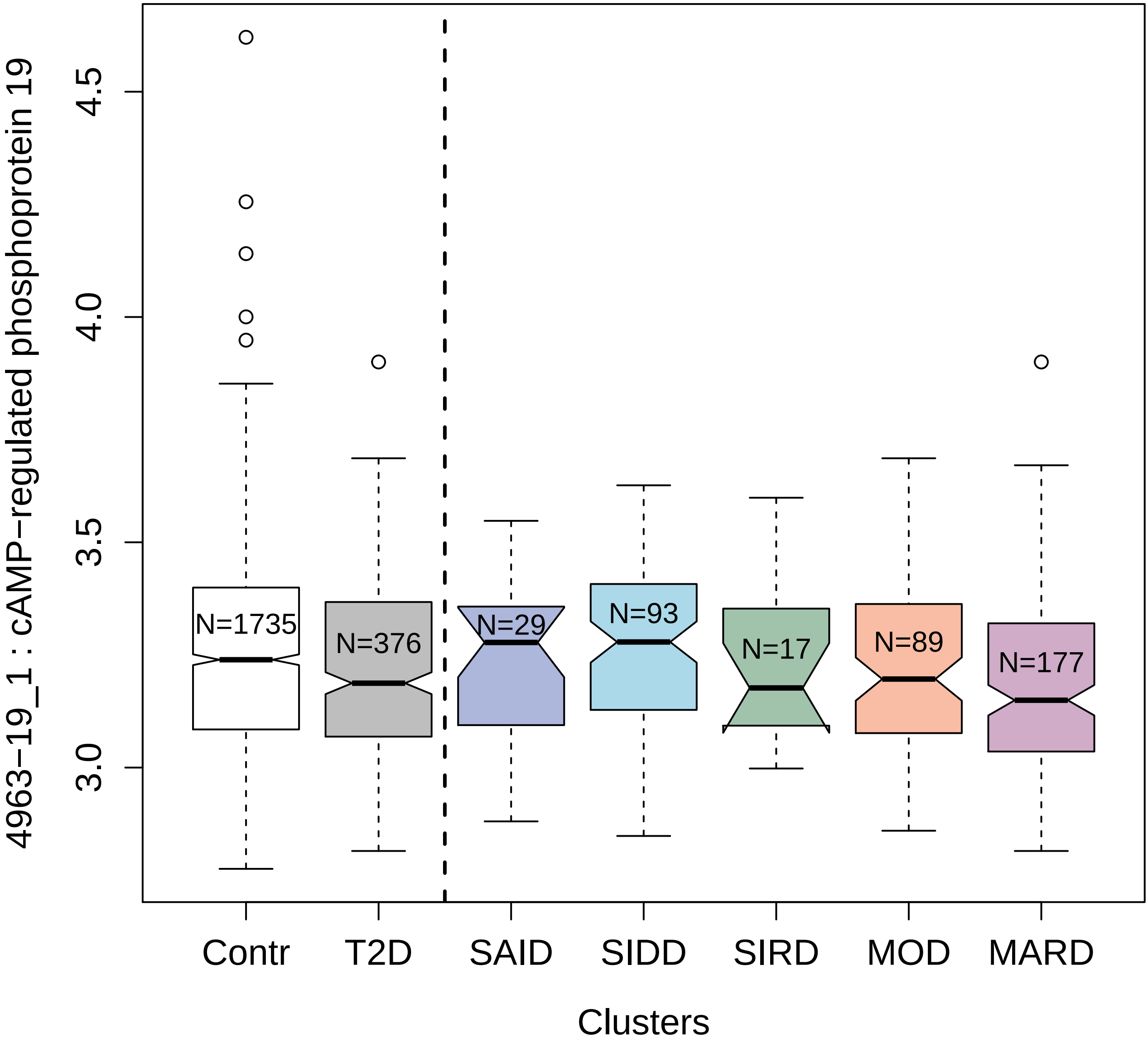
4963−19_1 : cAMP−regulated phosphoprotein 19
4.5
4.0
3.5
3.0
N=1735
N=376
N=29
N=93
N=17
N=89
N=177
Contr
T2D
SAID
SIDD
SIRD
MOD
MARD
Clusters

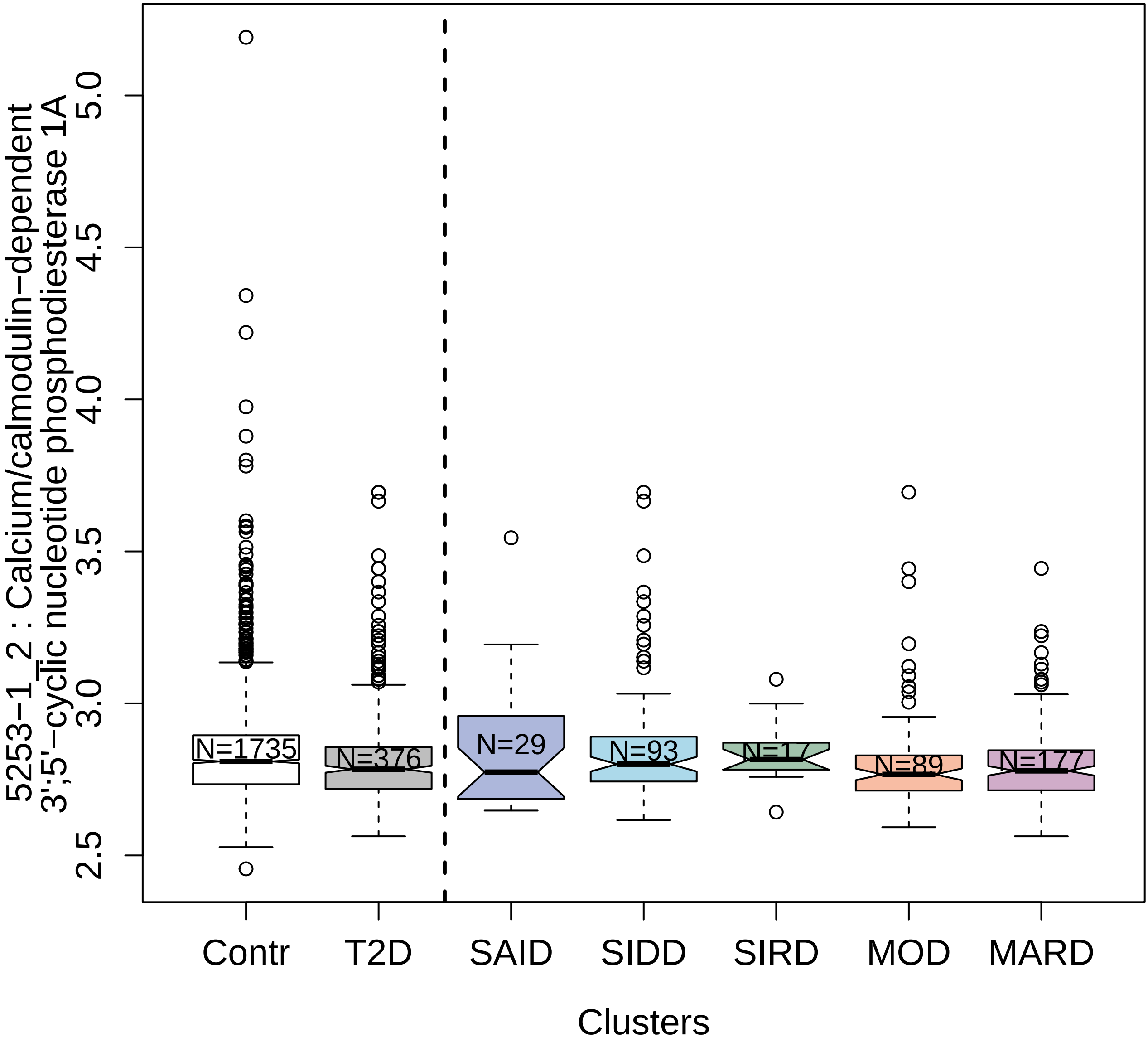

5253−1_2 : Calcium/calmodulin−dependent
3',5'−cyclic nucleotide phosphodiesterase 1A
2.5
3.0
3.5
4.0
4.5
5.0
N=1735
N=376
N=29
N=93
N=17
N=89
N=177
Contr
T2D
SAID
SIDD
SIRD
MOD
MARD
Clusters

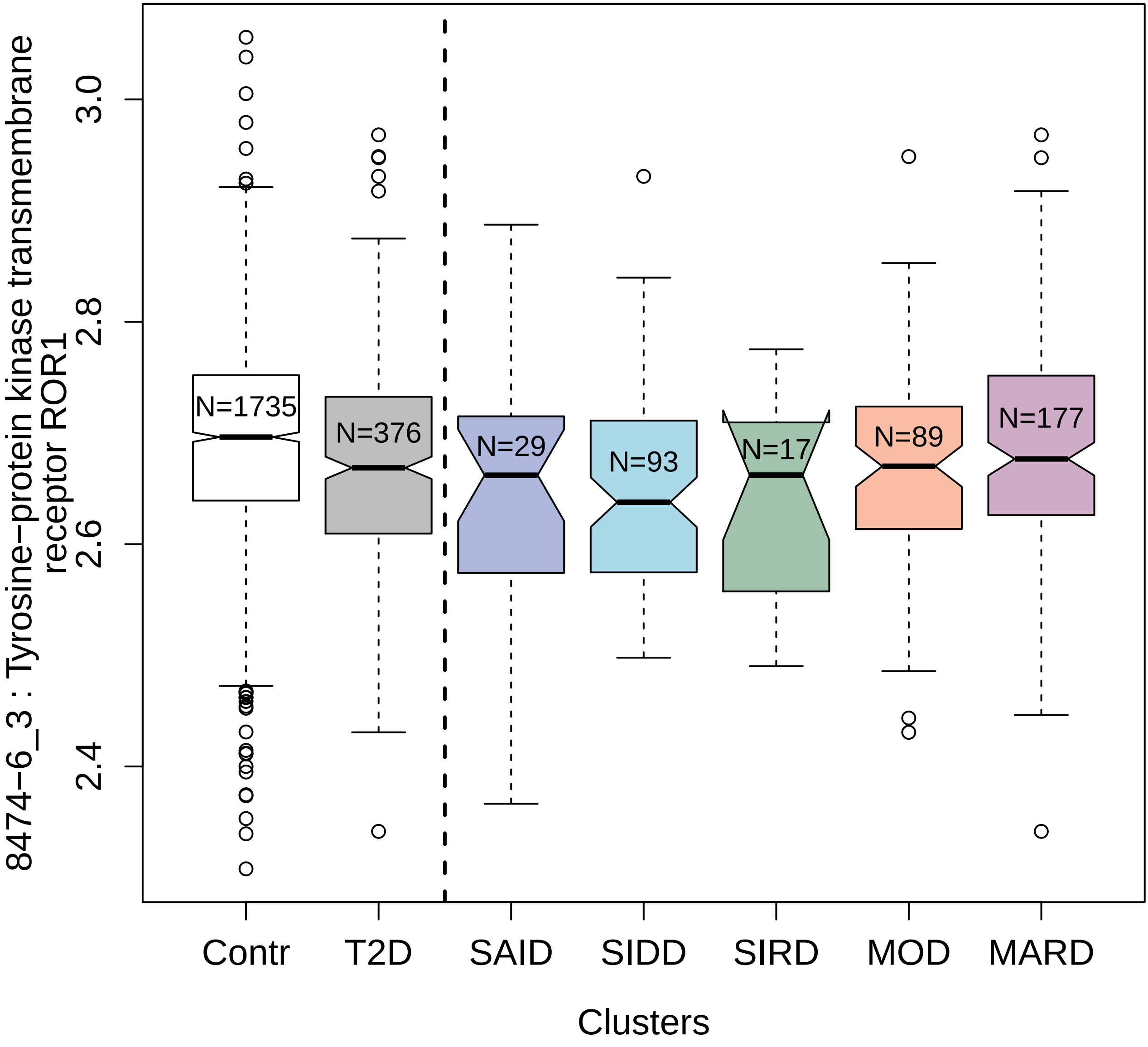

8474−6_3 : Tyrosine−protein kinase transmembrane receptor ROR1
3.0
2.8
2.6
2.4
N=1735
N=376
N=29
N=93
N=17
N=89
N=177
Contr
T2D
SAID
SIDD
SIRD
MOD
MARD
Clusters

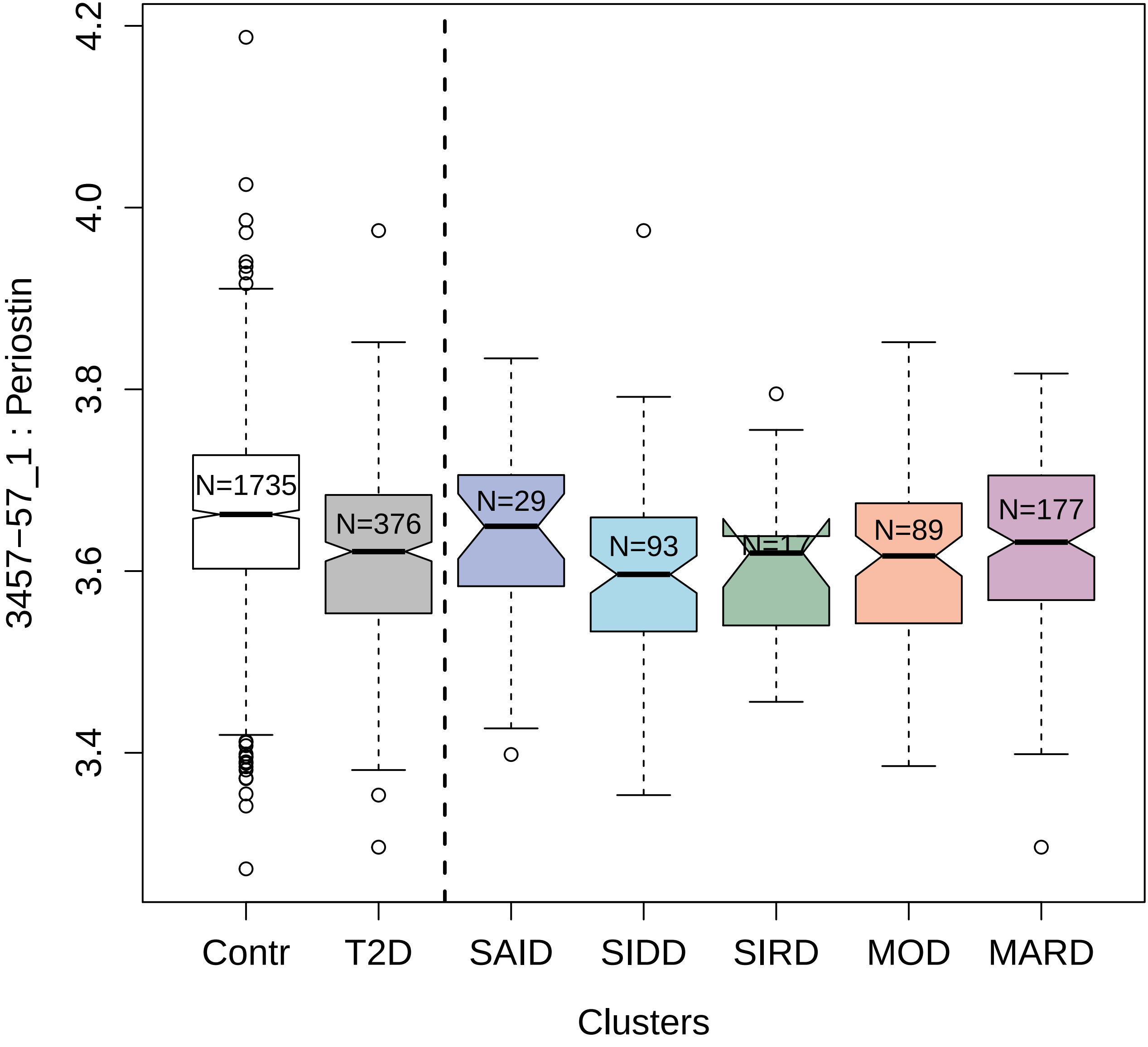

3457−57_1 : Periostin
4.2
4.0
3.8
3.6
3.4
N=1735
N=376
N=29
N=93
N=89
N=177
Contr
T2D
SAID
SIDD
SIRD
MOD
MARD
Clusters

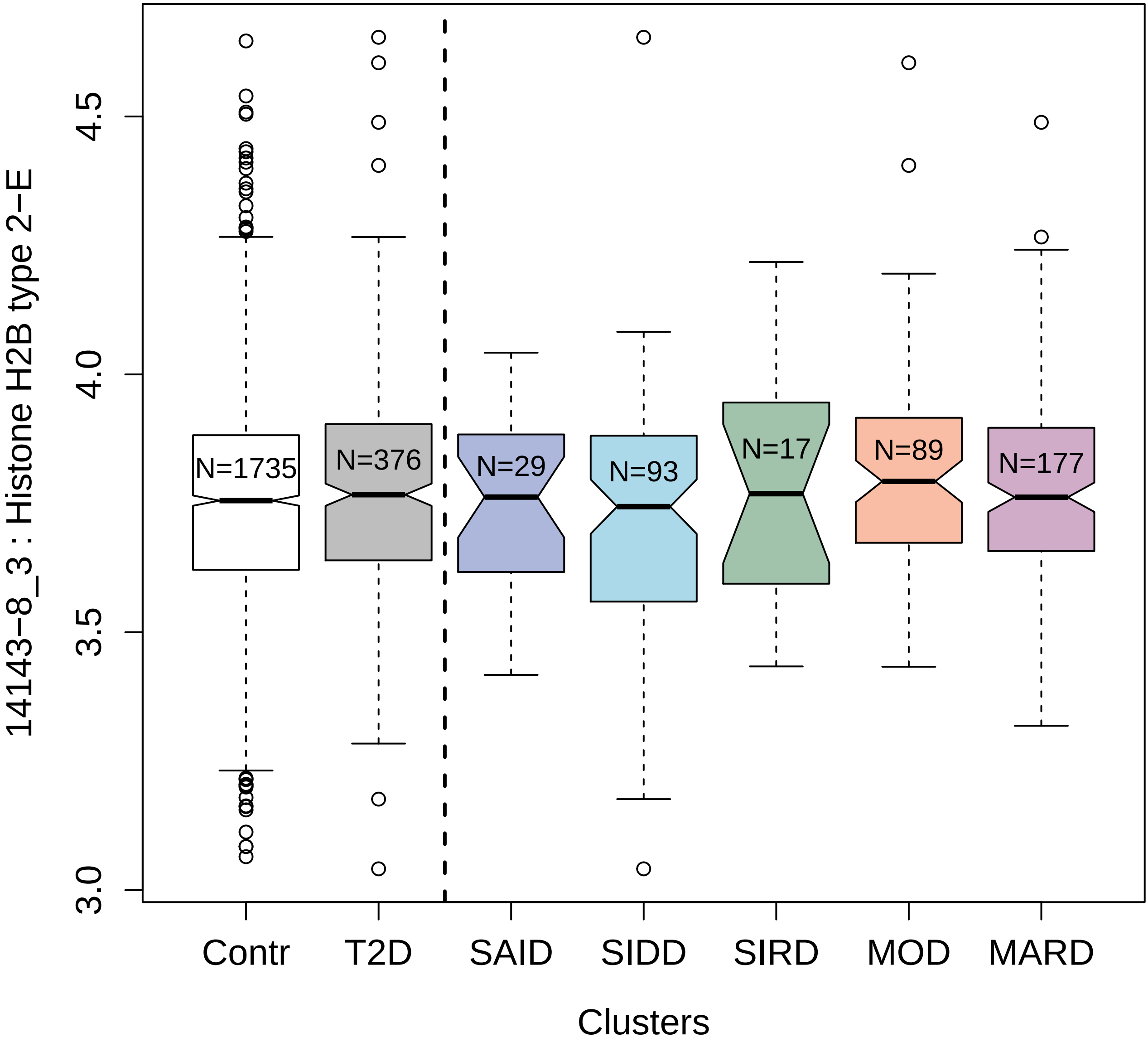

14143−8_3 : Histone H2B type 2−E
3.0
3.5
4.0
4.5
N=1735
N=376
N=29
N=93
N=17
N=89
N=177
Contr
T2D
SAID
SIDD
SIRD
MOD
MARD
Clusters

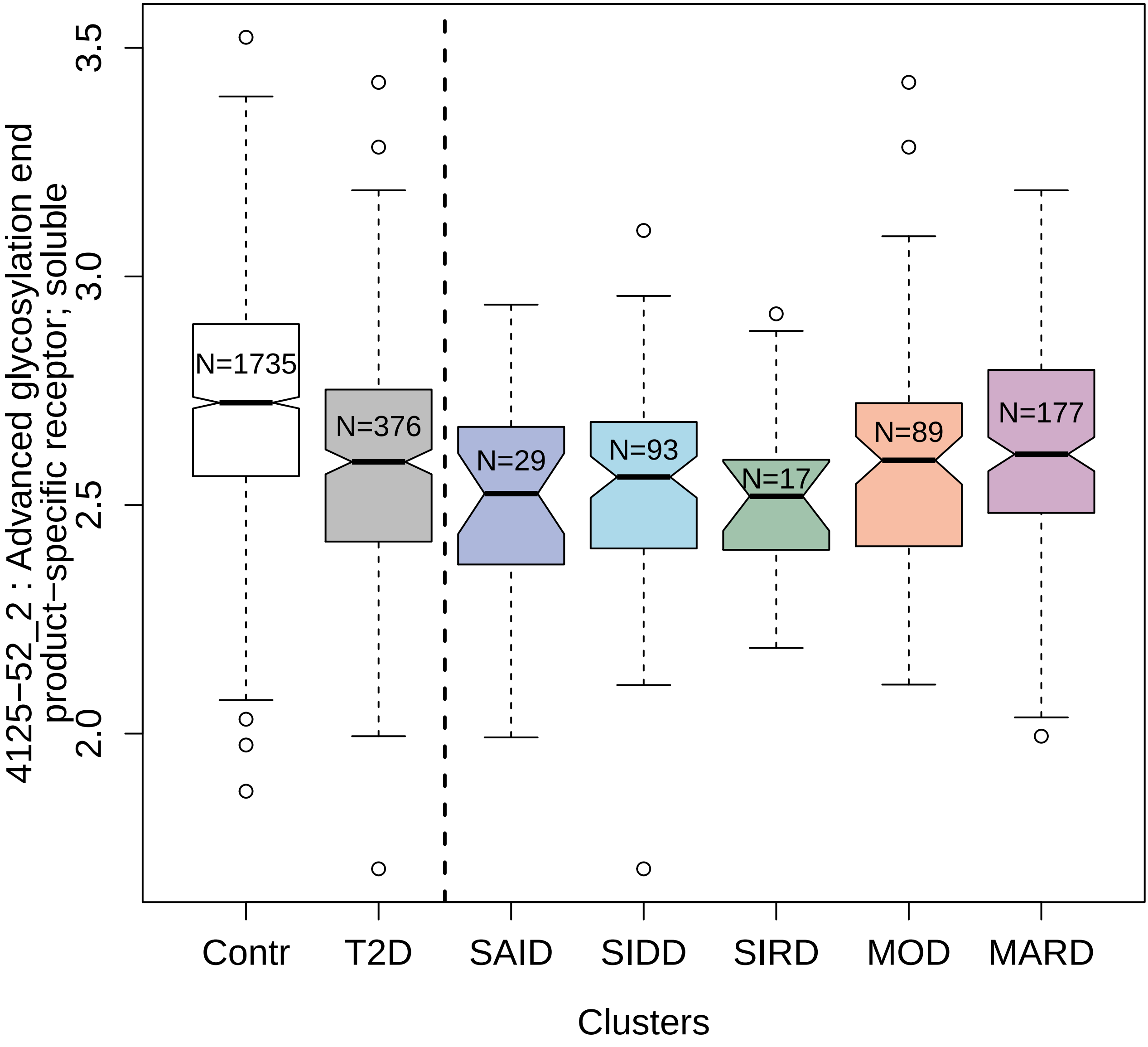
4125−52_2 : Advanced glycosylation end
product−specific receptor; soluble
3.5
3.0
2.5
2.0
N=1735
N=376
N=29
N=93
N=17
N=89
N=177
Contr
T2D
SAID
SIDD
SIRD
MOD
MARD
Clusters

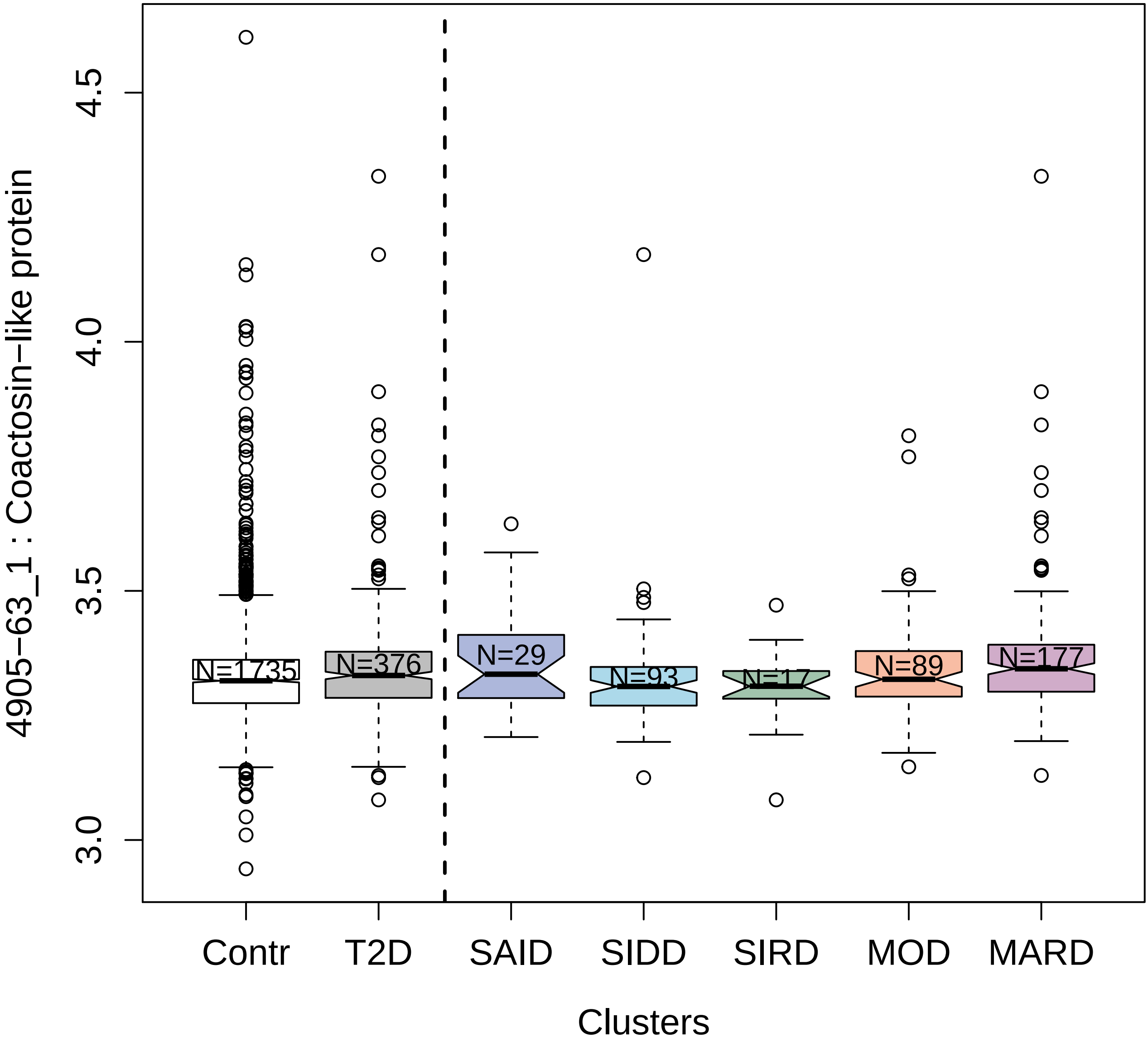
4905−63_1 : Coactosin−like protein
4.5
4.0
3.5
3.0
N=1735
N=376
N=29
N=93
N=17
N=89
N=177
Contr
T2D
SAID
SIDD
SIRD
MOD
MARD
Clusters

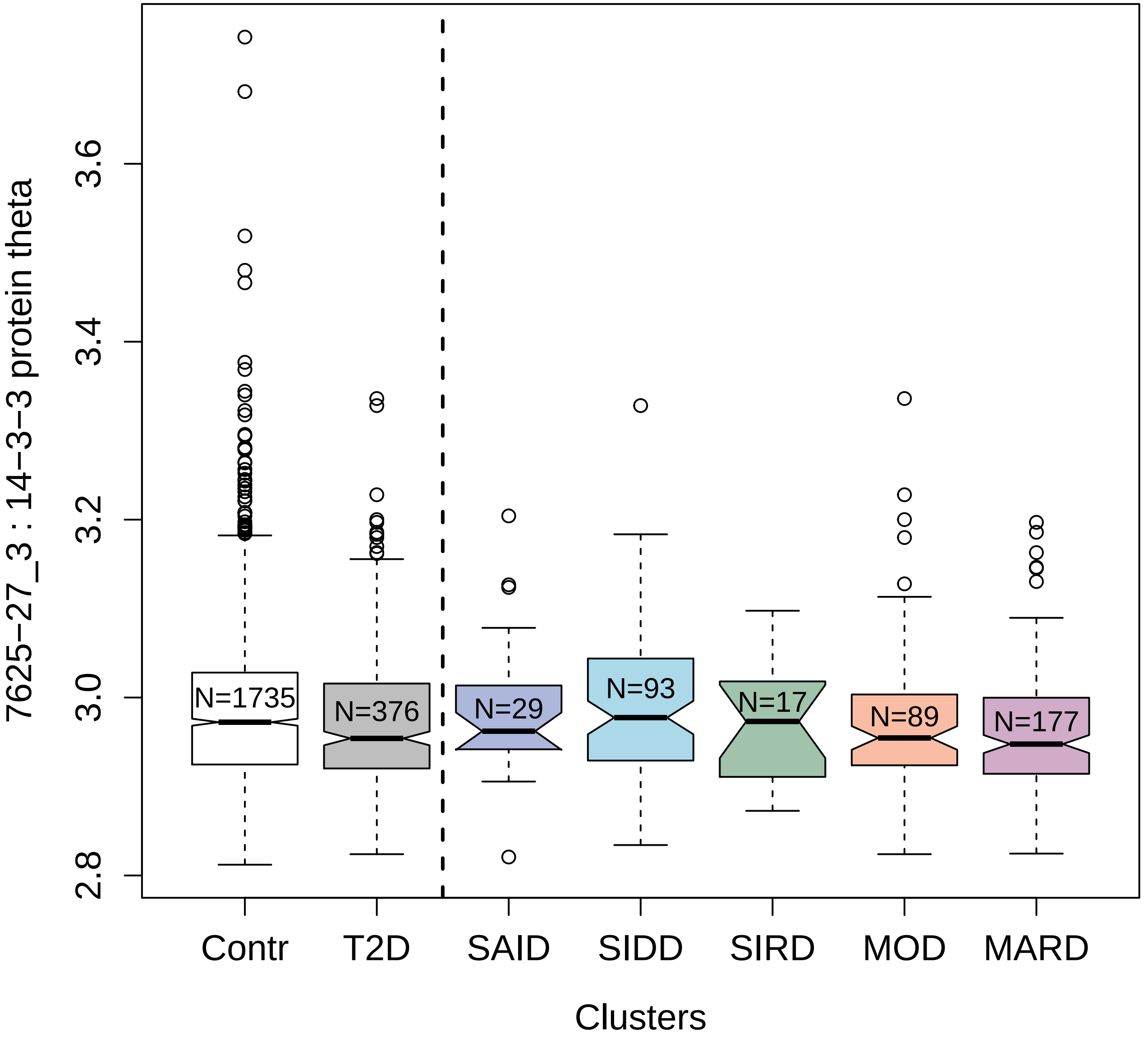
7625−27_3 : 14−3−3 protein theta
3.6
3.4
3.2
3.0
2.8
N=1735
N=376
N=29
N=93
N=17
N=89
N=177
Contr
T2D
SAID
SIDD
SIRD
MOD
MARD
Clusters

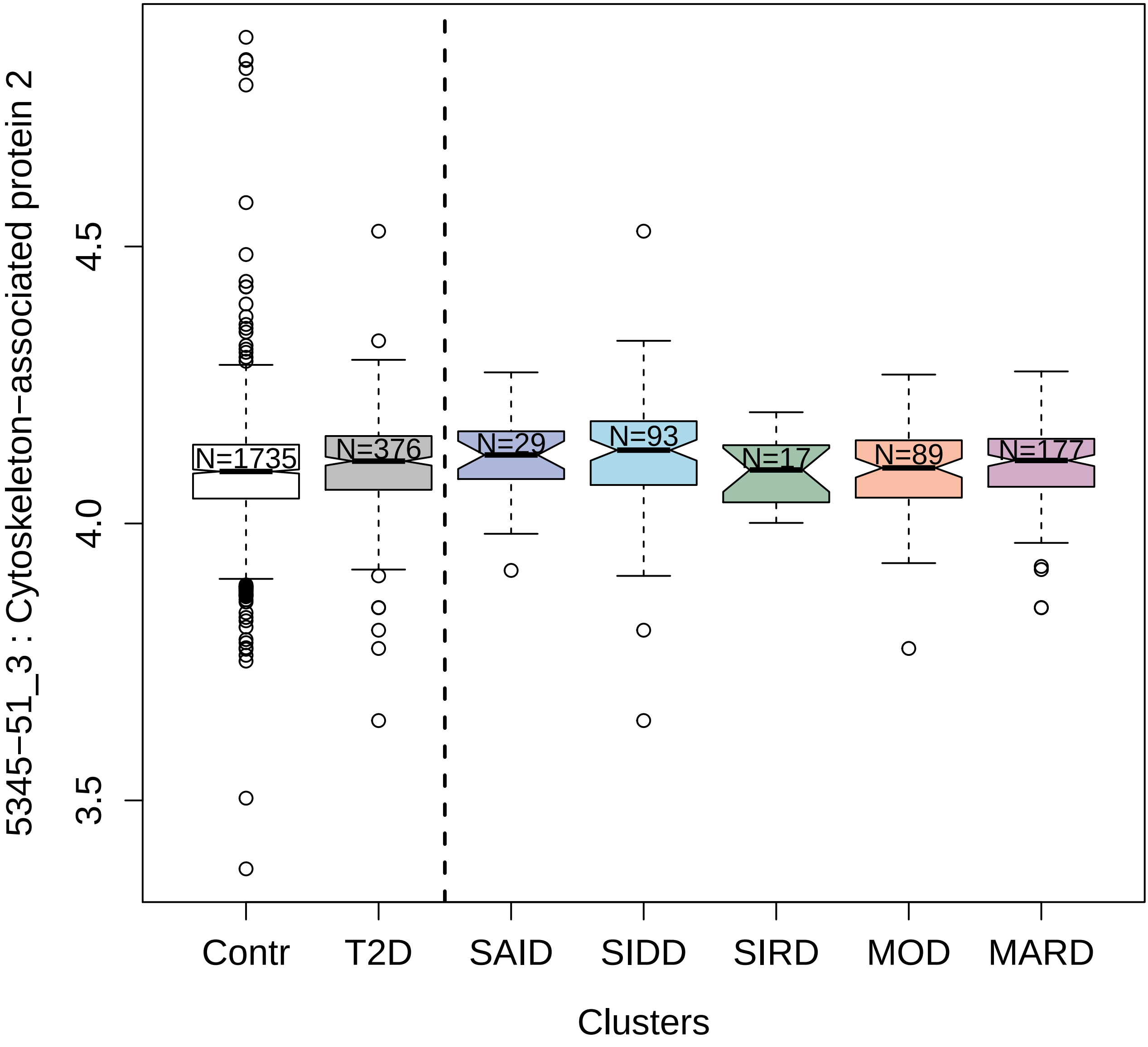
5345−51_3 : Cytoskeleton−associated protein 2
3.5
4.0
4.5
N=1735
N=376
N=29
N=93
N=17
N=89
N=177
Contr
T2D
SAID
SIDD
SIRD
MOD
MARD
Clusters

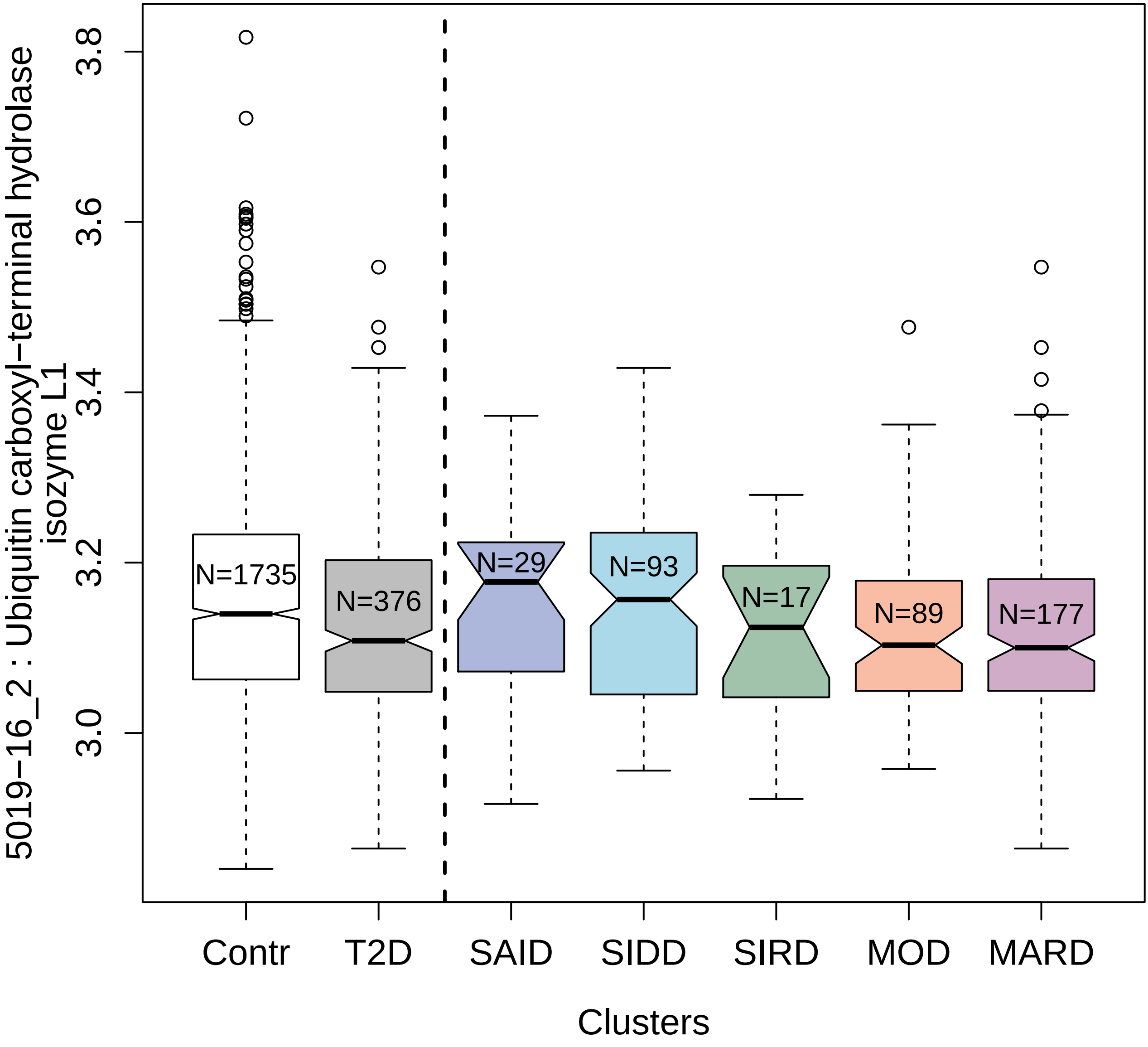

5019−16_2 : Ubiquitin carboxyl−terminal hydrolase isozyme L1
3.0
3.2
3.4
3.6
3.8
N=1735
N=376
N=29
N=93
N=17
N=89
N=177
Contr
T2D
SAID
SIDD
SIRD
MOD
MARD
Clusters

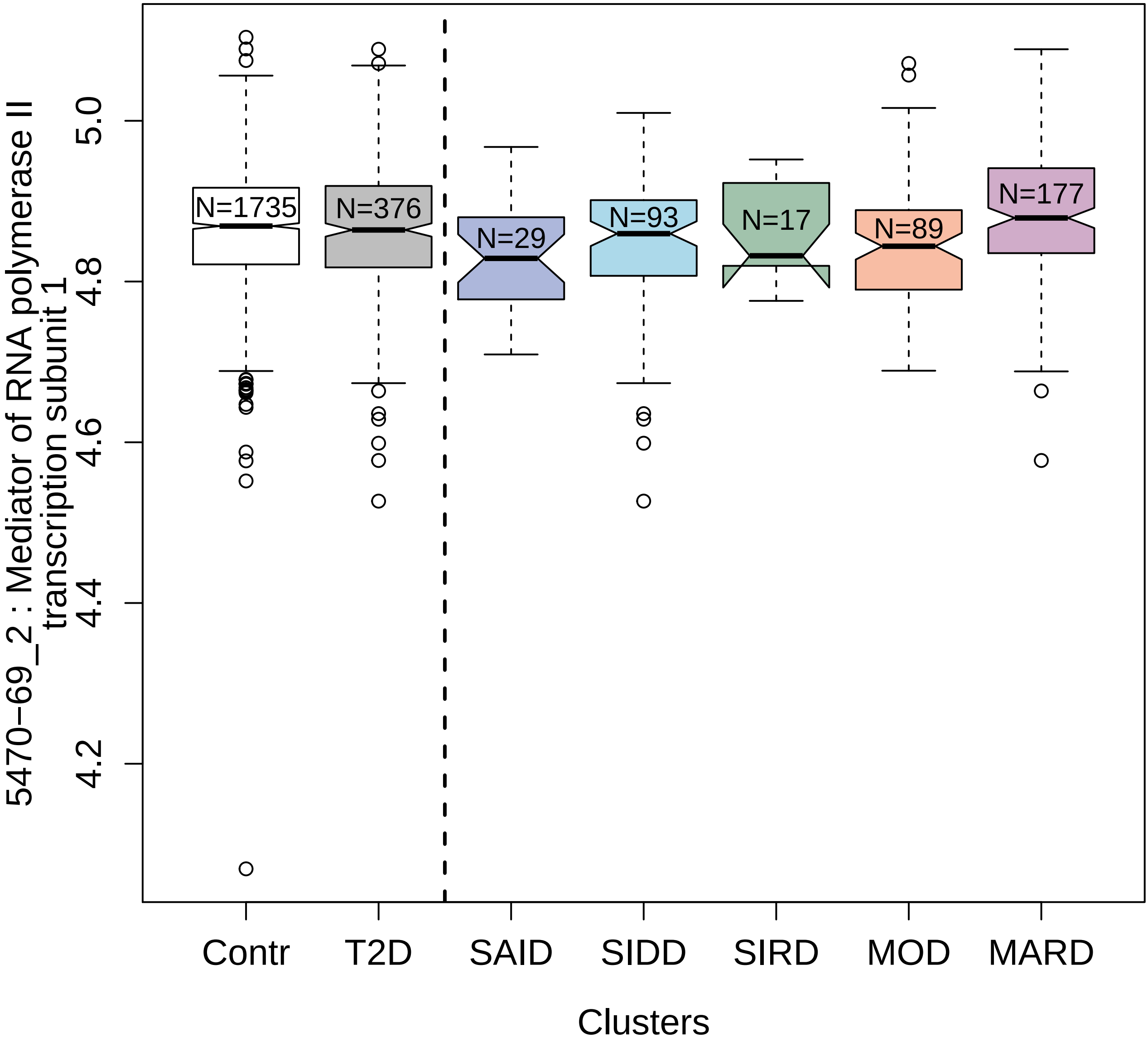

5470−69_2 : Mediator of RNA polymerase II transcription subunit 1
5.0
4.8
4.6
4.4
4.2
N=1735
N=376
N=29
N=93
N=17
N=89
N=177
Contr
T2D
SAID
SIDD
SIRD
MOD
MARD
Clusters

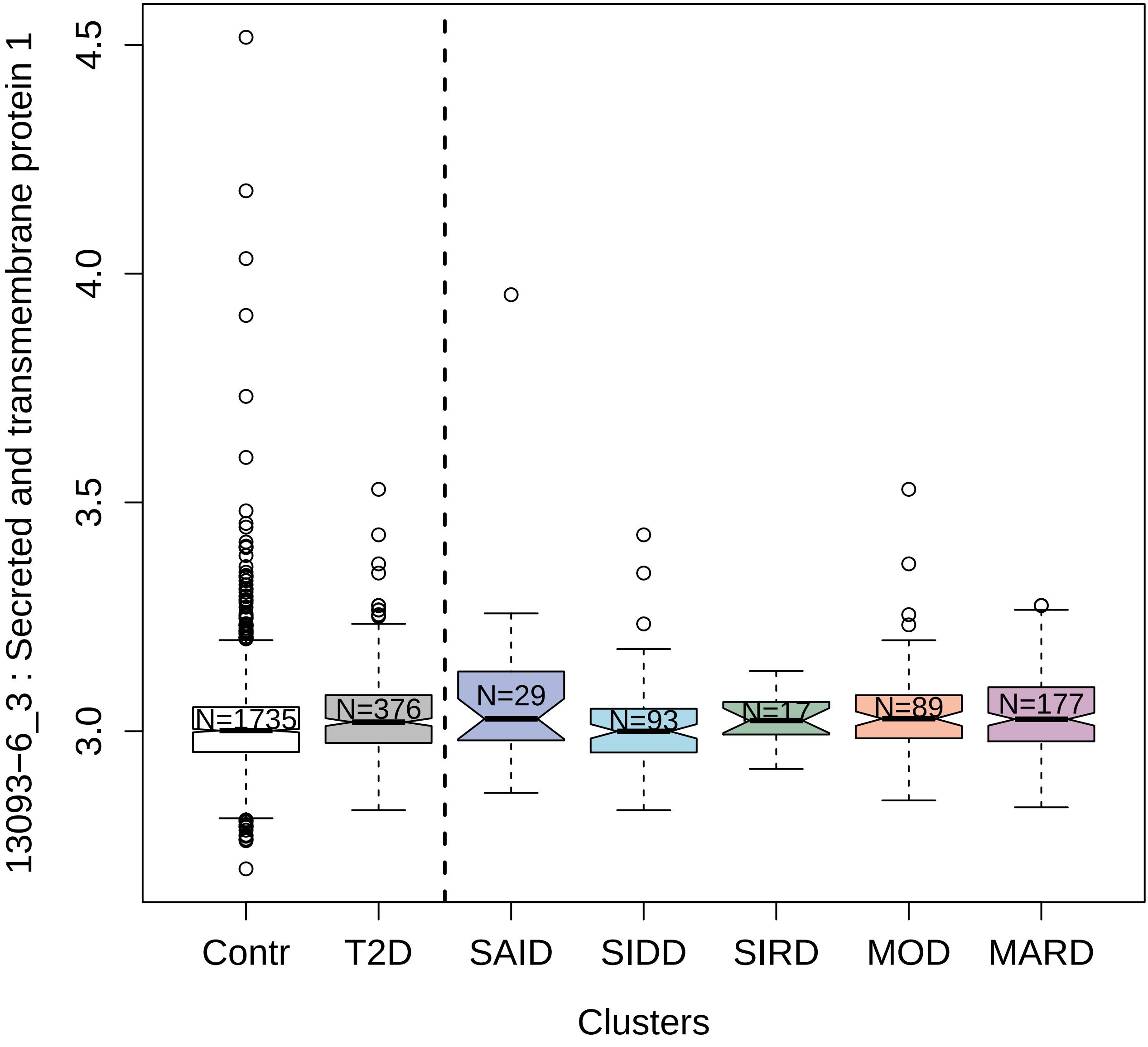

13093−6_3 : Secreted and transmembrane protein 1
4.5
4.0
3.5
3.0
N=1735
N=376
N=29
N=93
N=17
N=89
N=177
Contr
T2D
SAID
SIDD
SIRD
MOD
MARD
Clusters

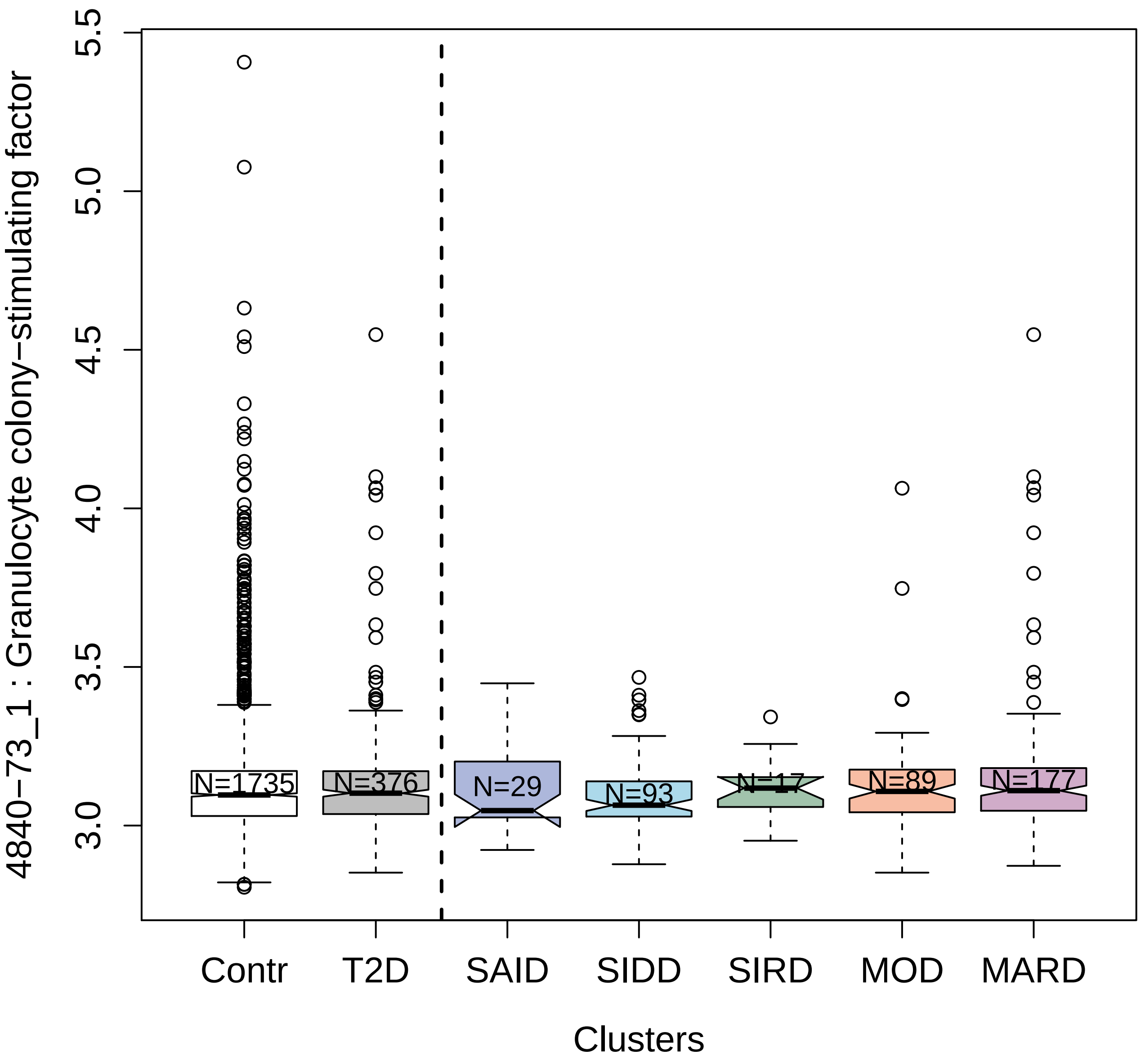

4840−73_1 : Granulocyte colony−stimulating factor
5.5
5.0
4.5
4.0
3.5
3.0
N=1735
N=376
N=29
N=93
N=17
N=89
N=177
Contr
T2D
SAID
SIDD
SIRD
MOD
MARD
Clusters

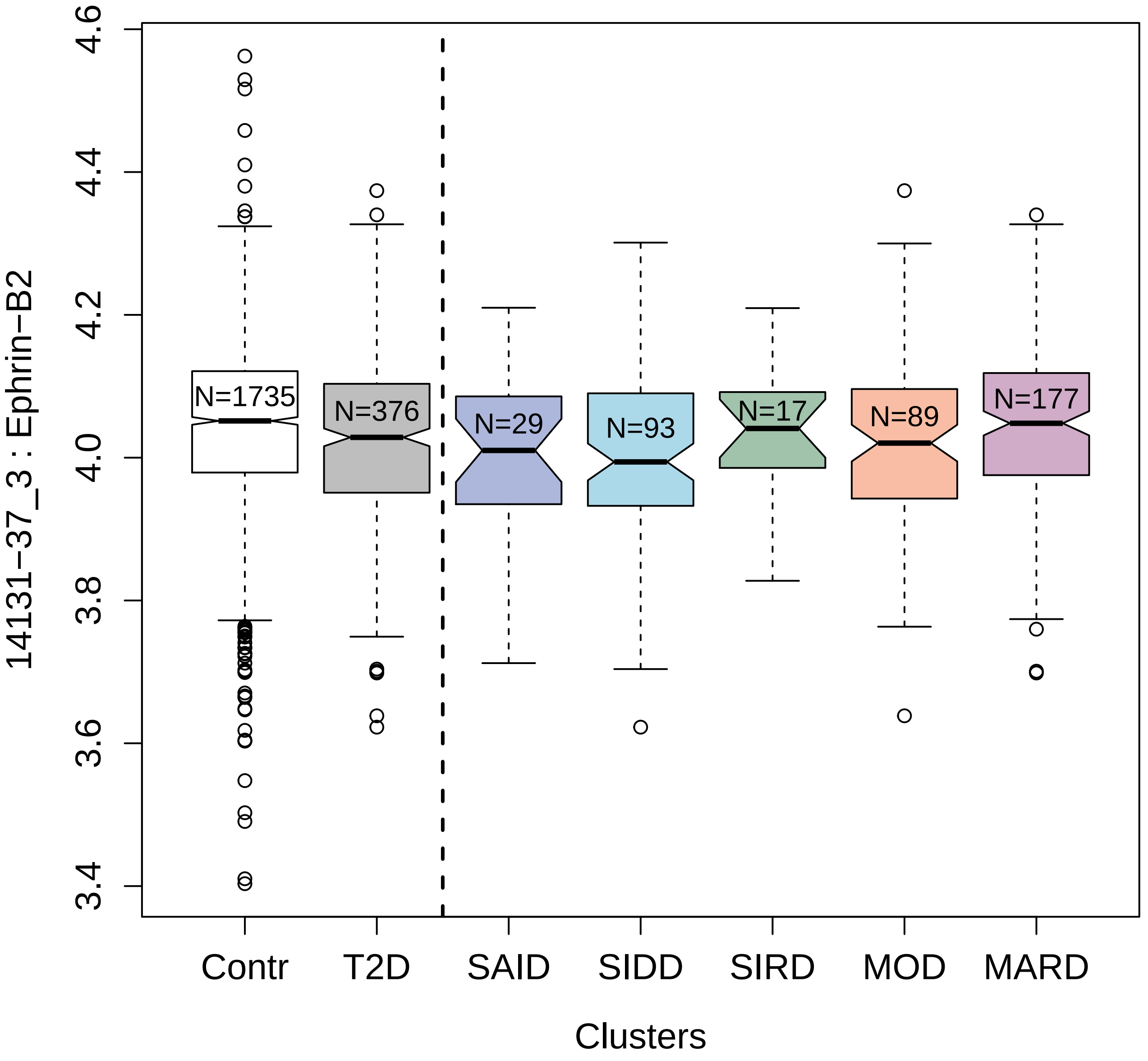
14131−37_3 : Ephrin−B2
4.6
4.4
4.2
4.0
3.8
3.6
3.4
N=1735
N=376
N=29
N=93
N=17
N=89
N=177
Contr
T2D
SAID
SIDD
SIRD
MOD
MARD
Clusters

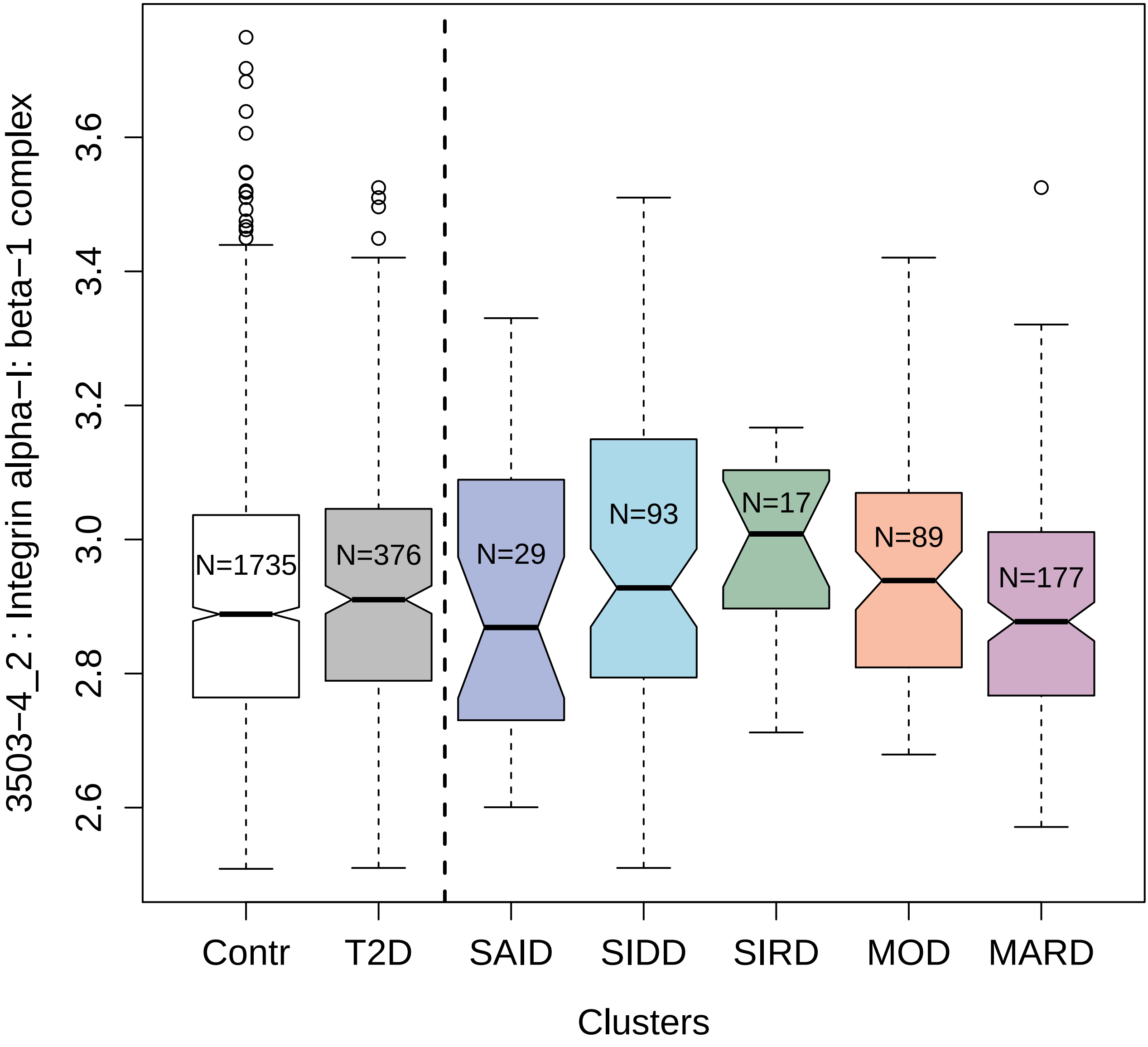
3503−4_2 : Integrin alpha−I: beta−1 complex
3.6
3.4
3.2
3.0
2.8
2.6
N=1735
N=376
N=29
N=93
N=17
N=89
N=177
Contr
T2D
SAID
SIDD
SIRD
MOD
MARD
Clusters

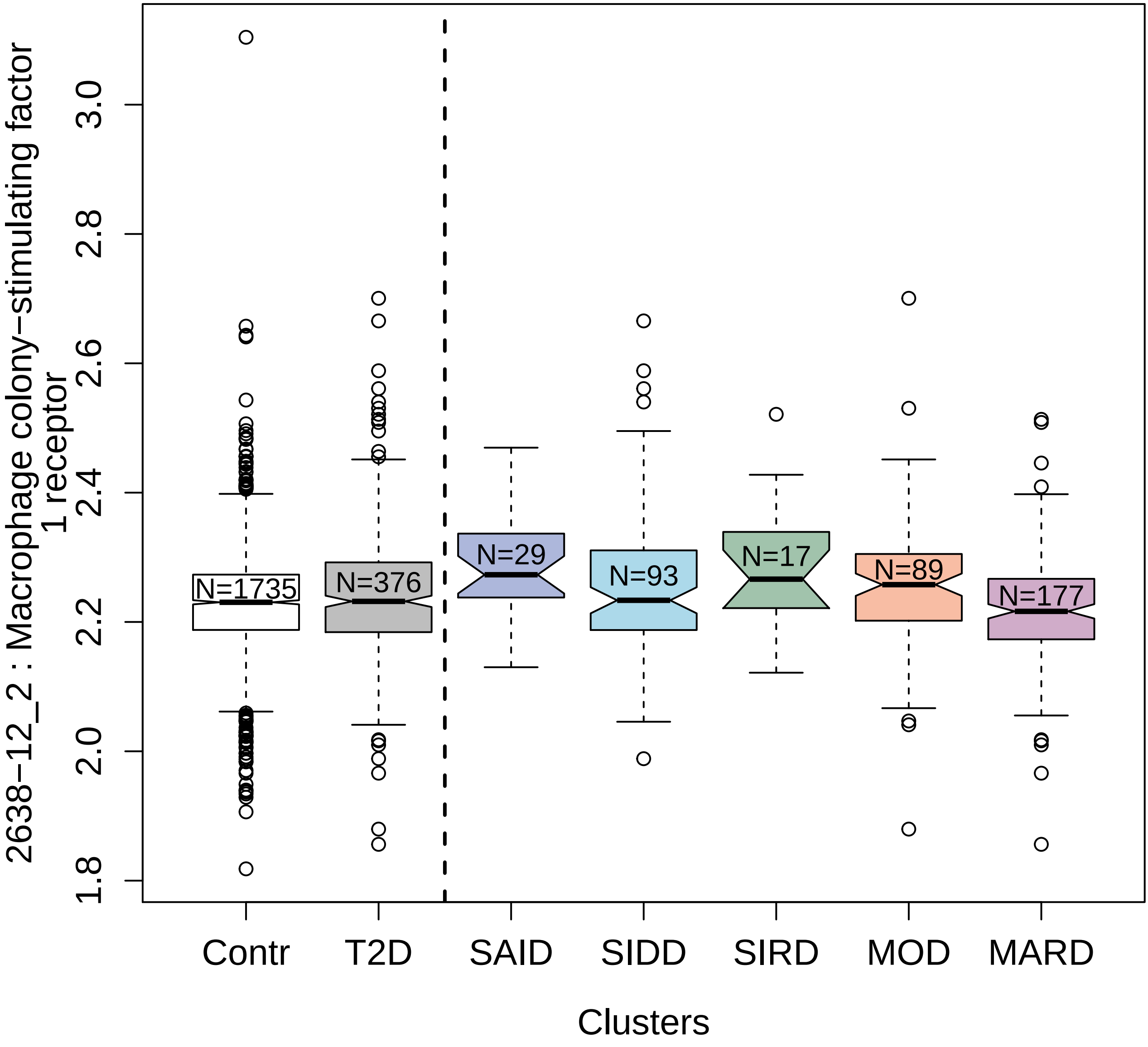

2638−12_2 : Macrophage colony−stimulating factor 1 receptor
1.8
2.0
2.2
2.4
2.6
2.8
3.0
N=1735
N=376
N=29
N=93
N=17
N=89
N=177
Contr
T2D
SAID
SIDD
SIRD
MOD
MARD
Clusters

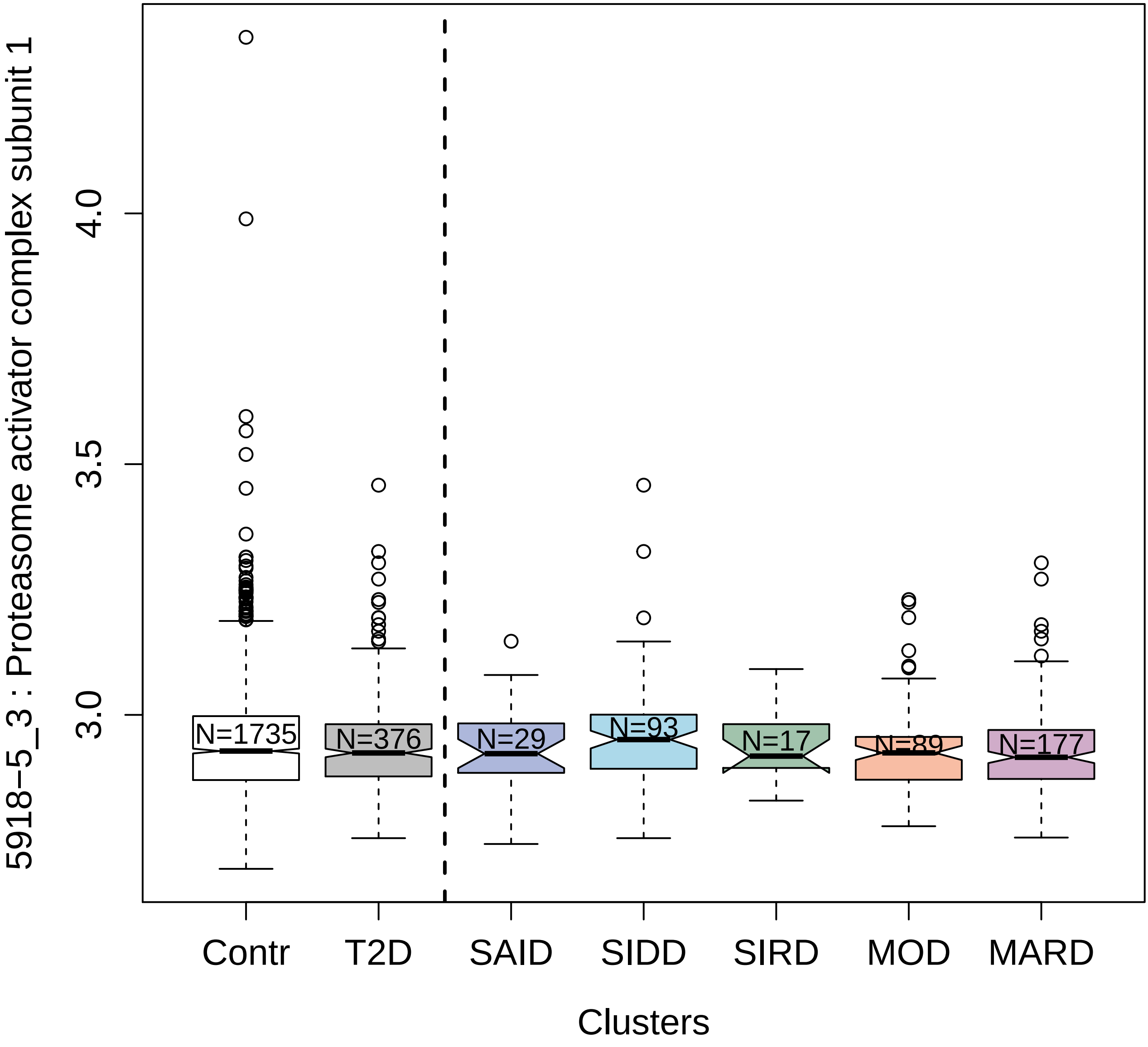

5918−5_3 : Proteasome activator complex subunit 1
4.0
3.5
3.0
N=1735
N=376
N=29
N=93
N=17
N=89
N=177
Contr
T2D
SAID
SIDD
SIRD
MOD
MARD
Clusters

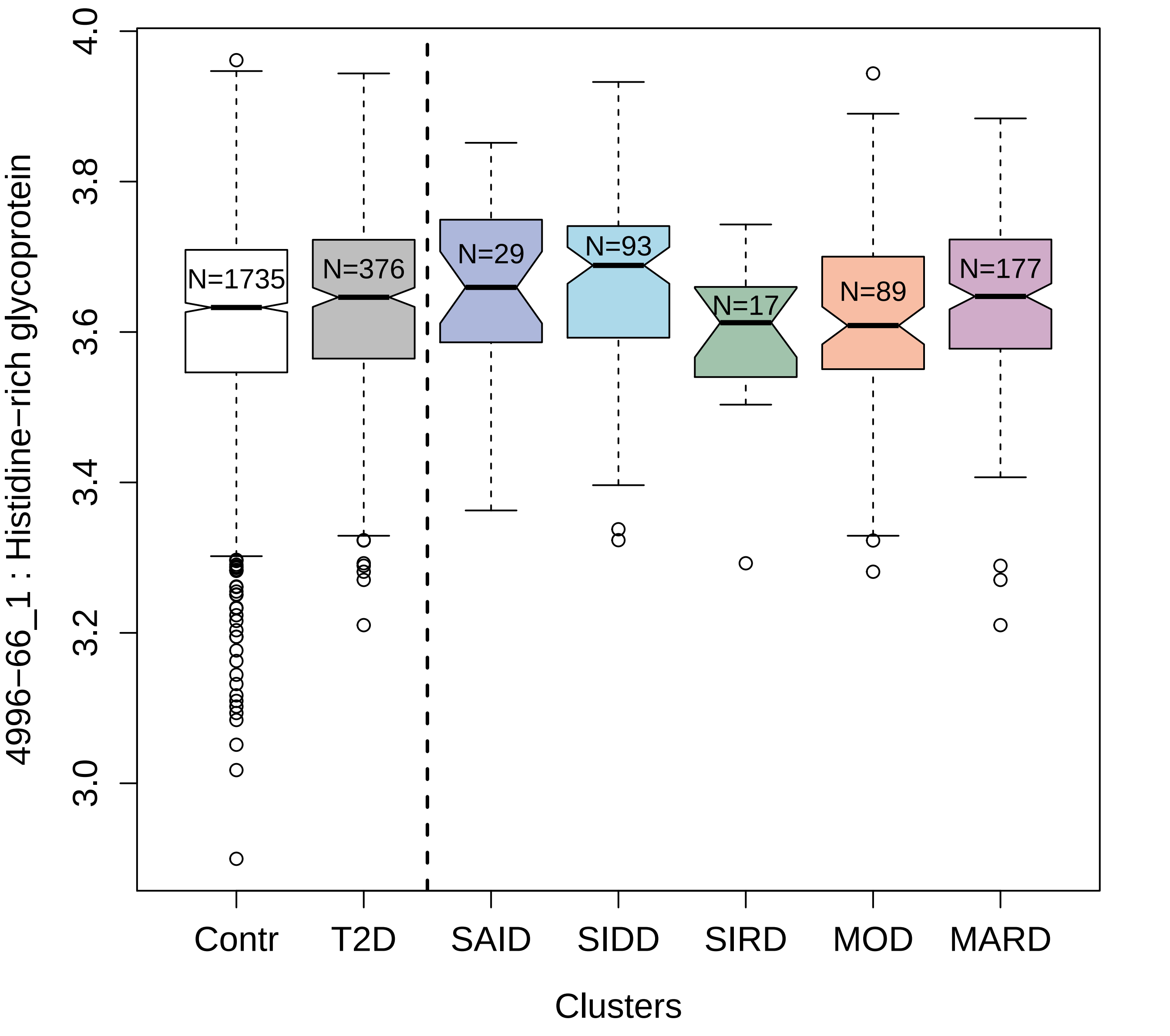

4996−66_1 : Histidine−rich glycoprotein
4.0
3.8
3.6
3.4
3.2
3.0
N=1735
N=376
N=29
N=93
N=17
N=89
N=177
Contr
T2D
SAID
SIDD
SIRD
MOD
MARD
Clusters

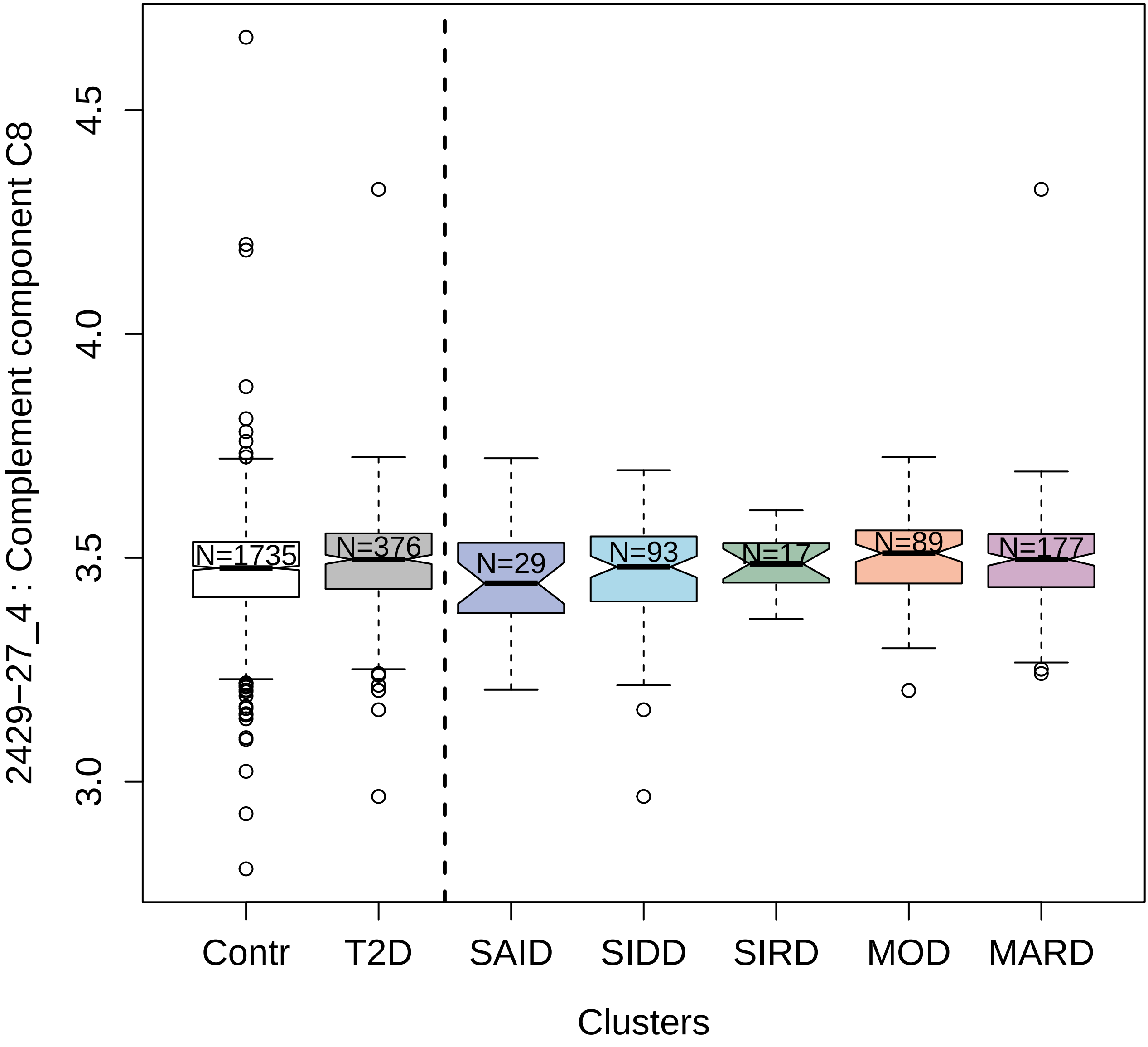

2429−27_4 : Complement component C8
4.5
4.0
3.5
3.0
N=1735
N=376
N=29
N=93
N=17
N=89
N=177
Contr
T2D
SAID
SIDD
SIRD
MOD
MARD
Clusters

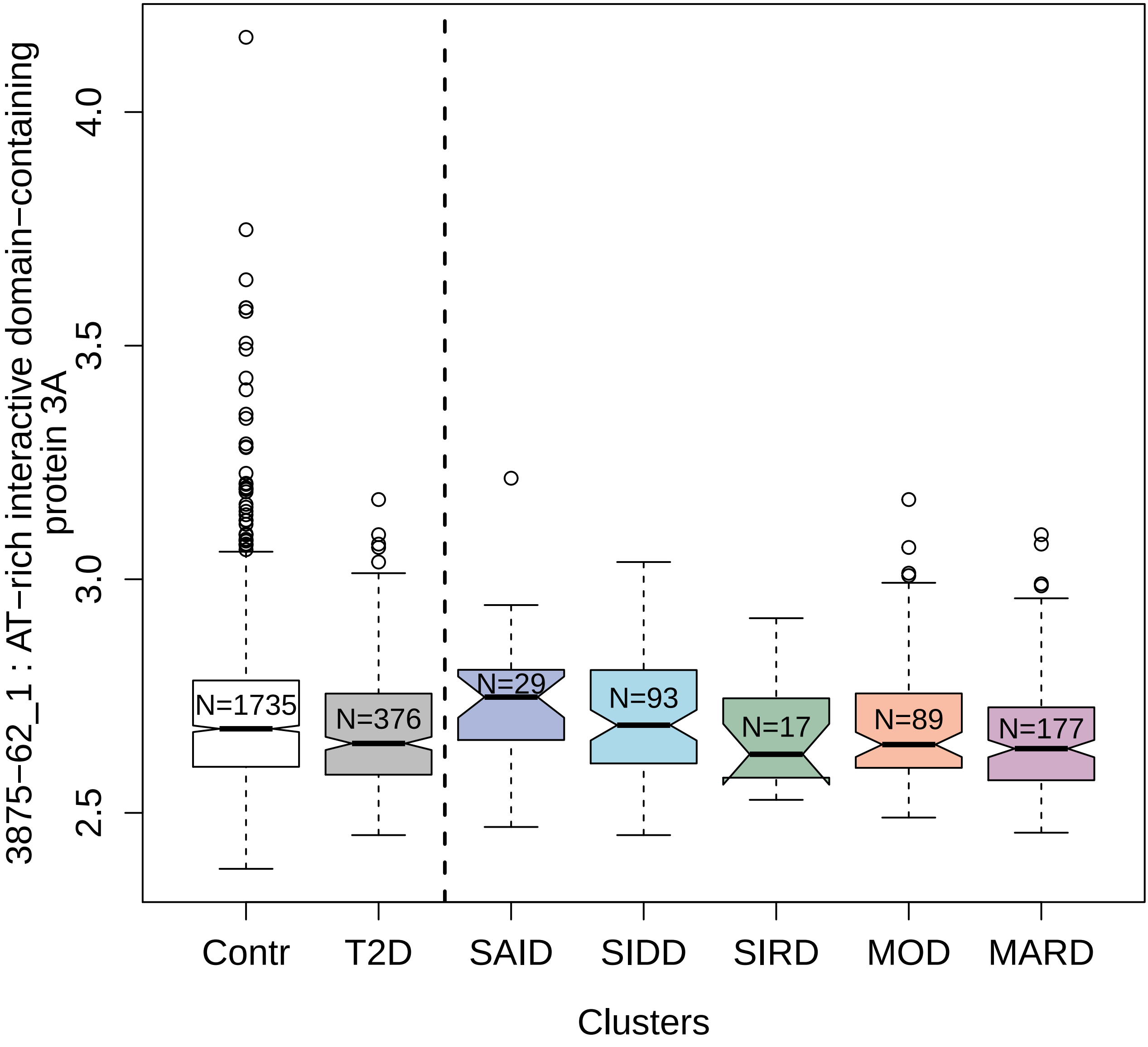
3875−62_1 : AT−rich interactive domain−containing protein 3A
4.0
3.5
3.0
2.5
N=1735
N=376
N=29
N=93
N=17
N=89
N=177
Contr
T2D
SAID
SIDD
SIRD
MOD
MARD
Clusters

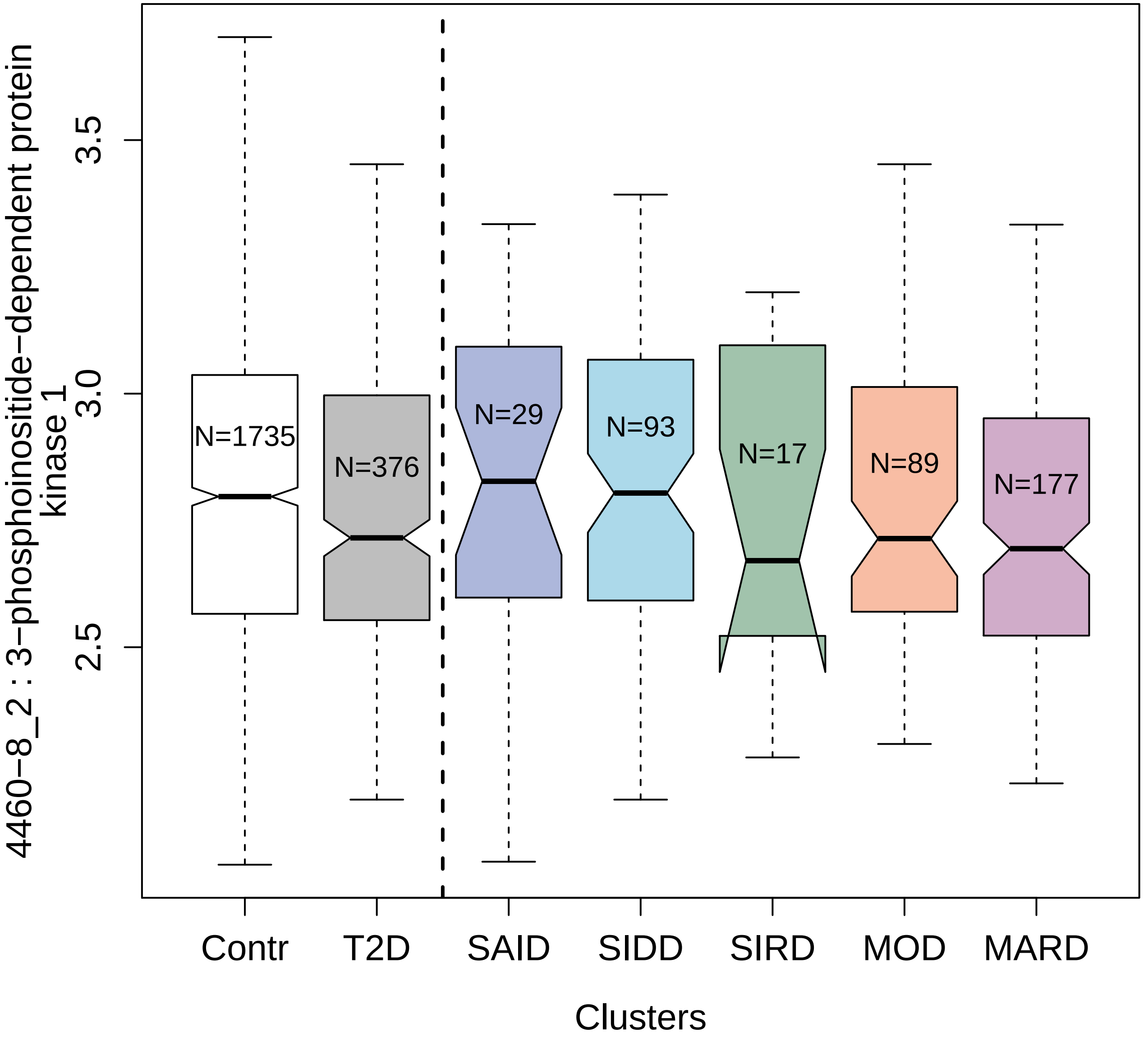

4460−8_2 : 3−phosphoinositide−dependent protein
kinase 1
3.5
3.0
2.5
N=1735
N=376
N=29
N=93
N=17
N=89
N=177
Contr
T2D
SAID
SIDD
SIRD
MOD
MARD
Clusters

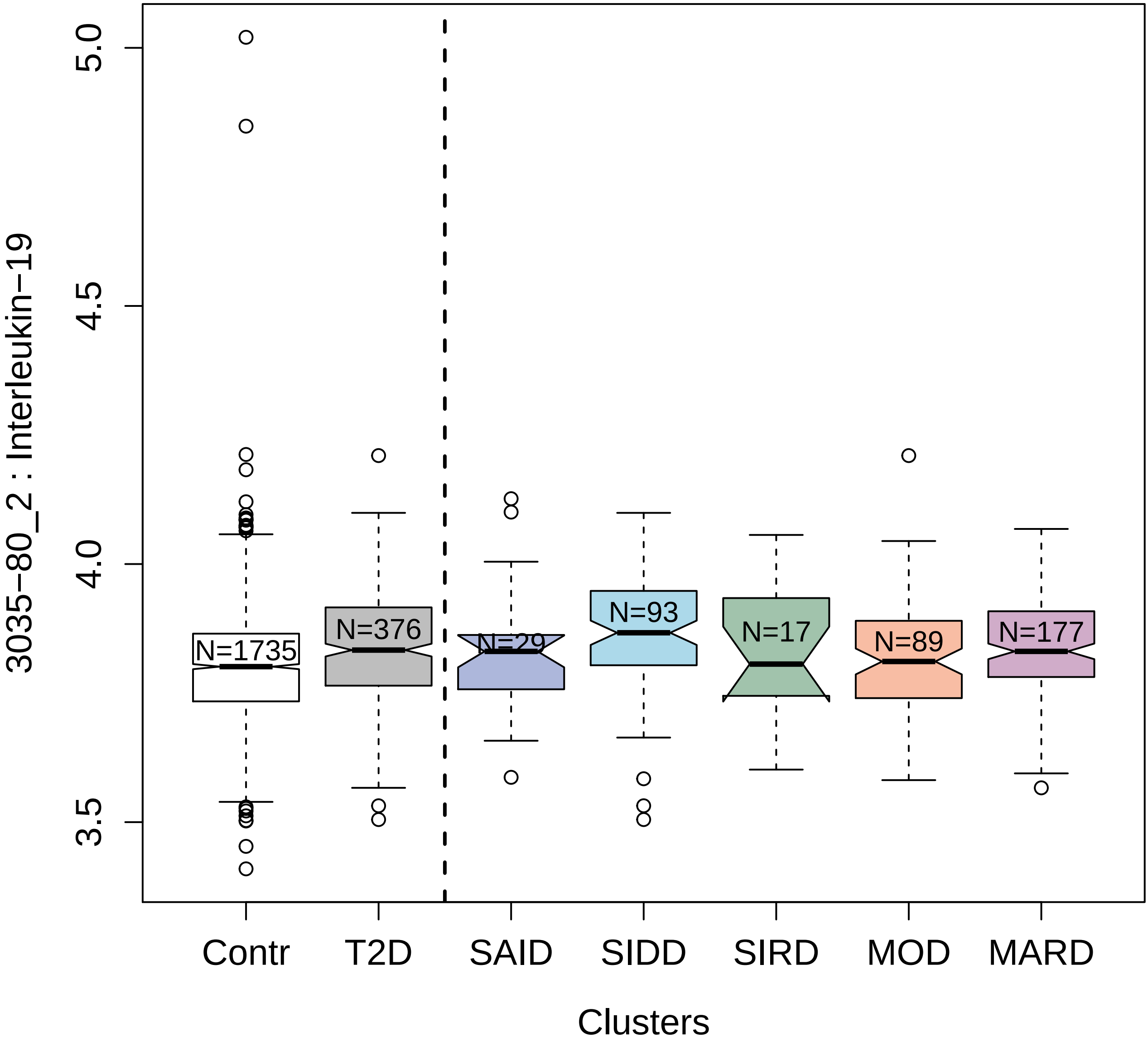
3035−80_2 : Interleukin−19
5.0
4.5
4.0
3.5
N=1735
N=376
N=29
N=93
N=17
N=89
N=177
Contr
T2D
SAID
SIDD
SIRD
MOD
MARD
Clusters

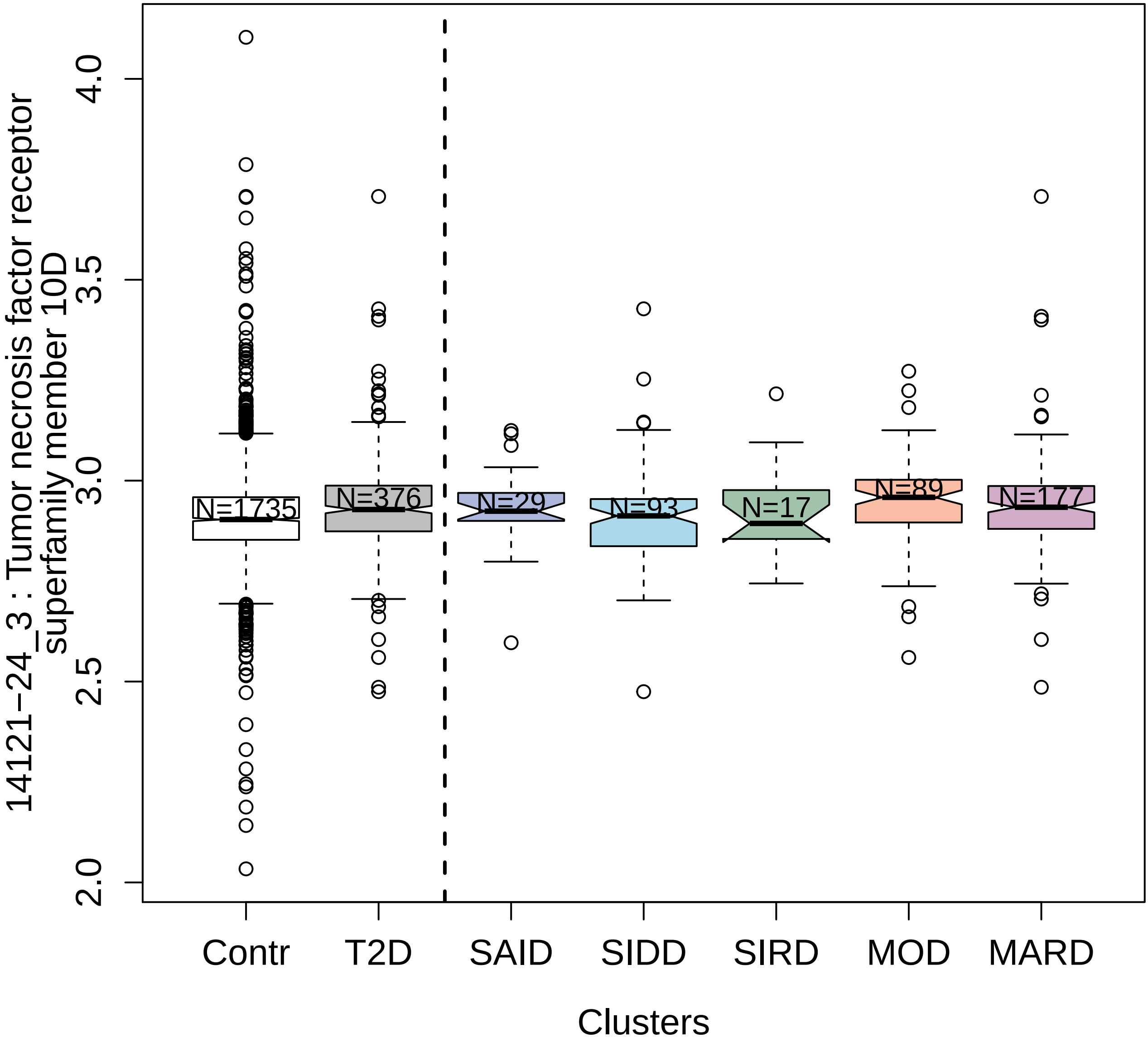
14121−24_3 : Tumor necrosis factor receptor superfamily member 10D
2.0
2.5
3.0
3.5
4.0
N=1735
N=376
N=29
N=93
N=17
N=89
N=177
Contr
T2D
SAID
SIDD
SIRD
MOD
MARD
Clusters

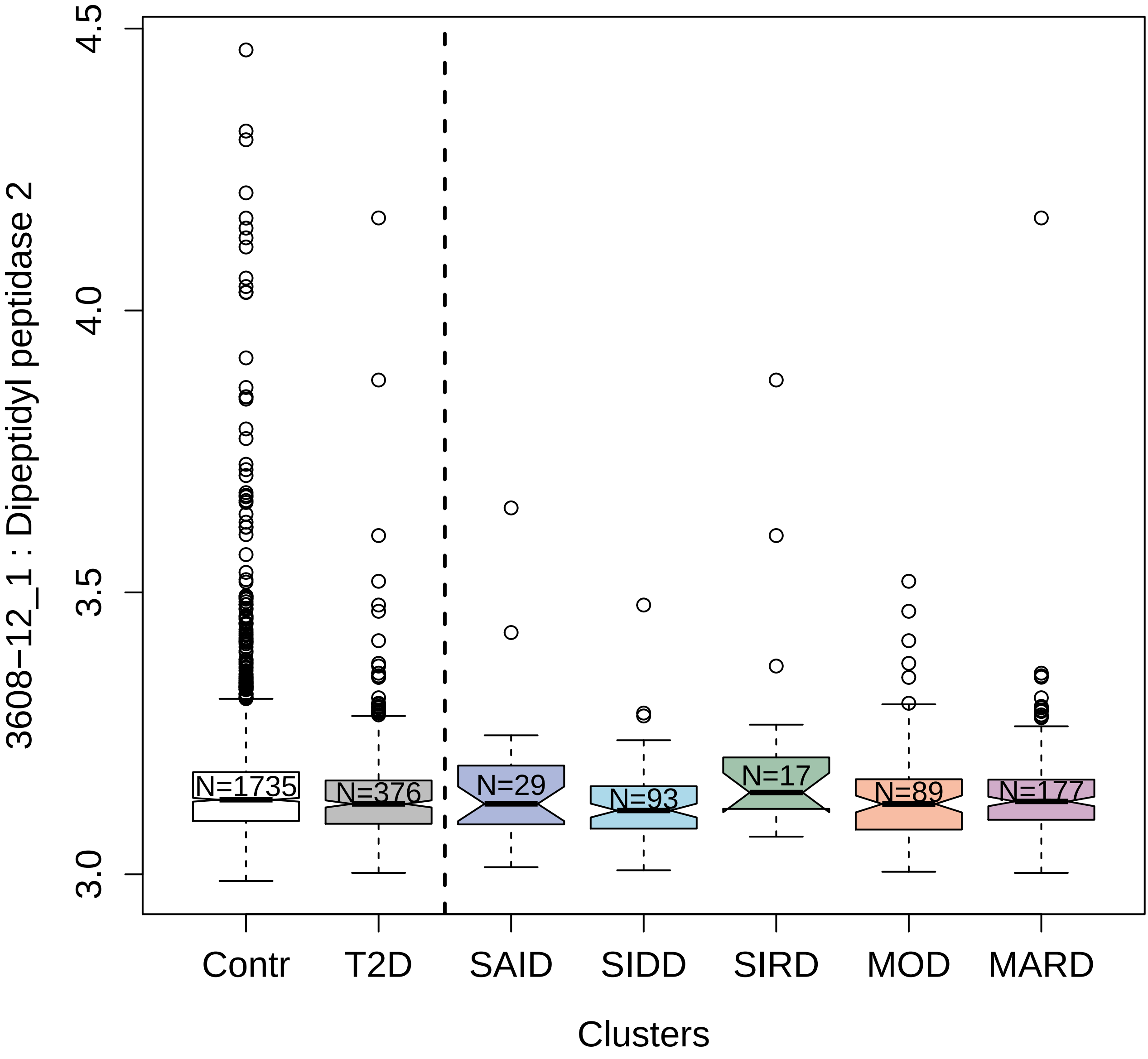
3608−12_1 : Dipeptidyl peptidase 2
4.5
4.0
3.5
3.0
N=1735
N=376
N=29
N=93
N=17
N=89
N=177
Contr
T2D
SAID
SIDD
SIRD
MOD
MARD
Clusters

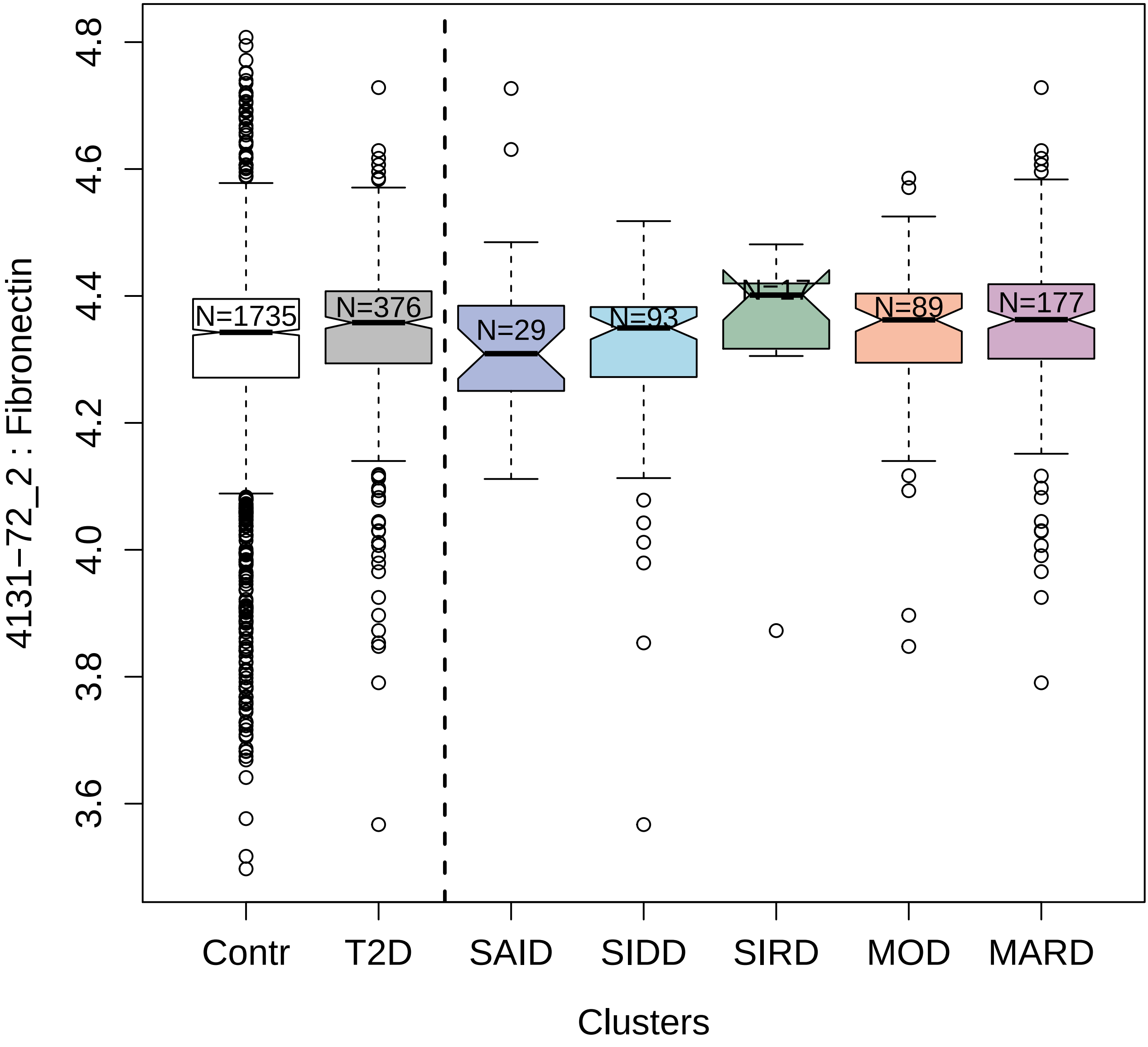

4131−72_2 : Fibronectin
4.8
4.6
4.4
4.2
4.0
3.8
3.6
N=1735
N=376
N=29
N=93
N=17
N=89
N=177
Contr
T2D
SAID
SIDD
SIRD
MOD
MARD
Clusters

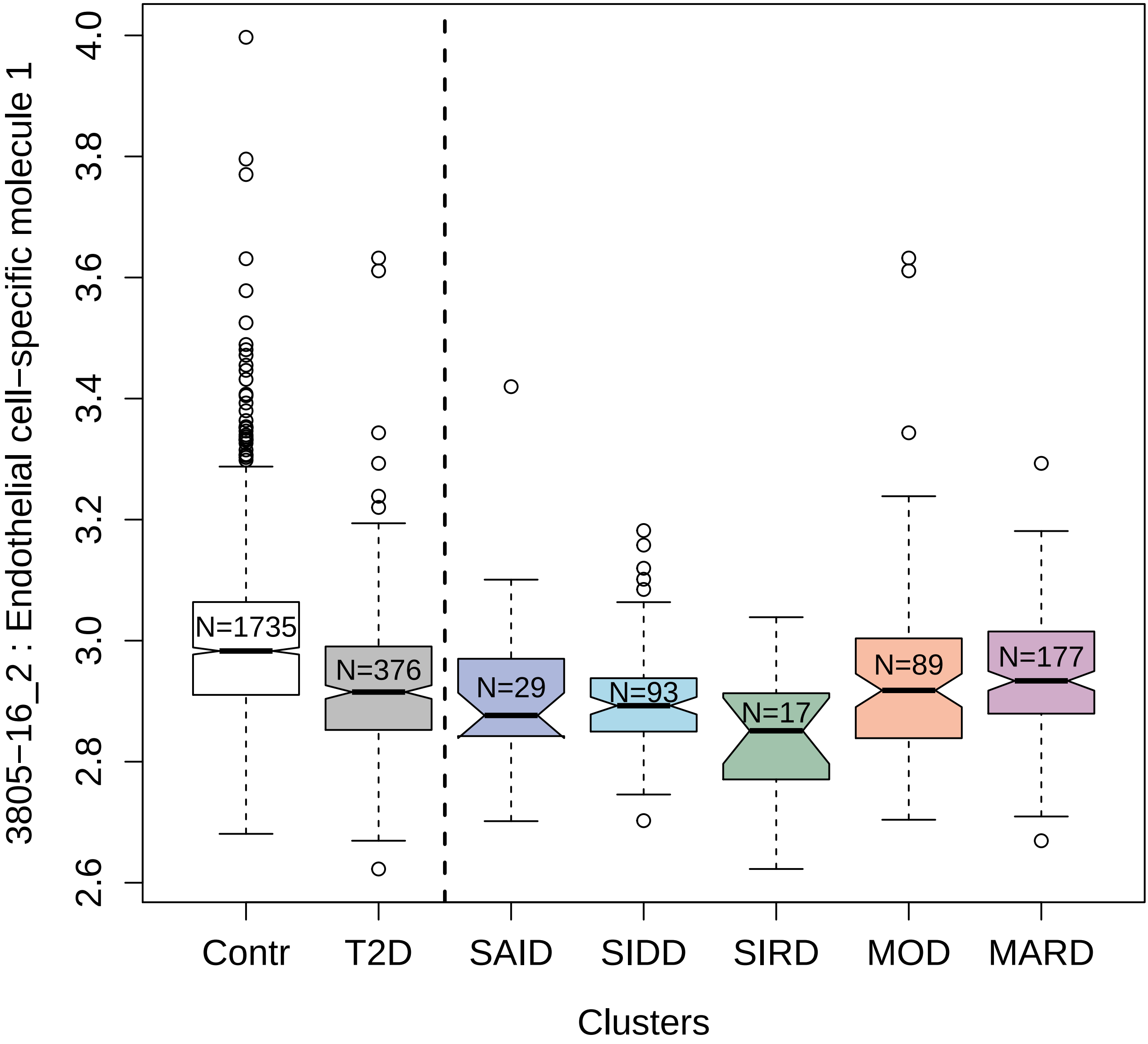
3805−16_2 : Endothelial cell−specific molecule 1
4.0
3.8
3.6
3.4
3.2
3.0
2.8
2.6
N=1735
N=376
N=29
N=93
N=17
N=89
N=177
Contr
T2D
SAID
SIDD
SIRD
MOD
MARD
Clusters

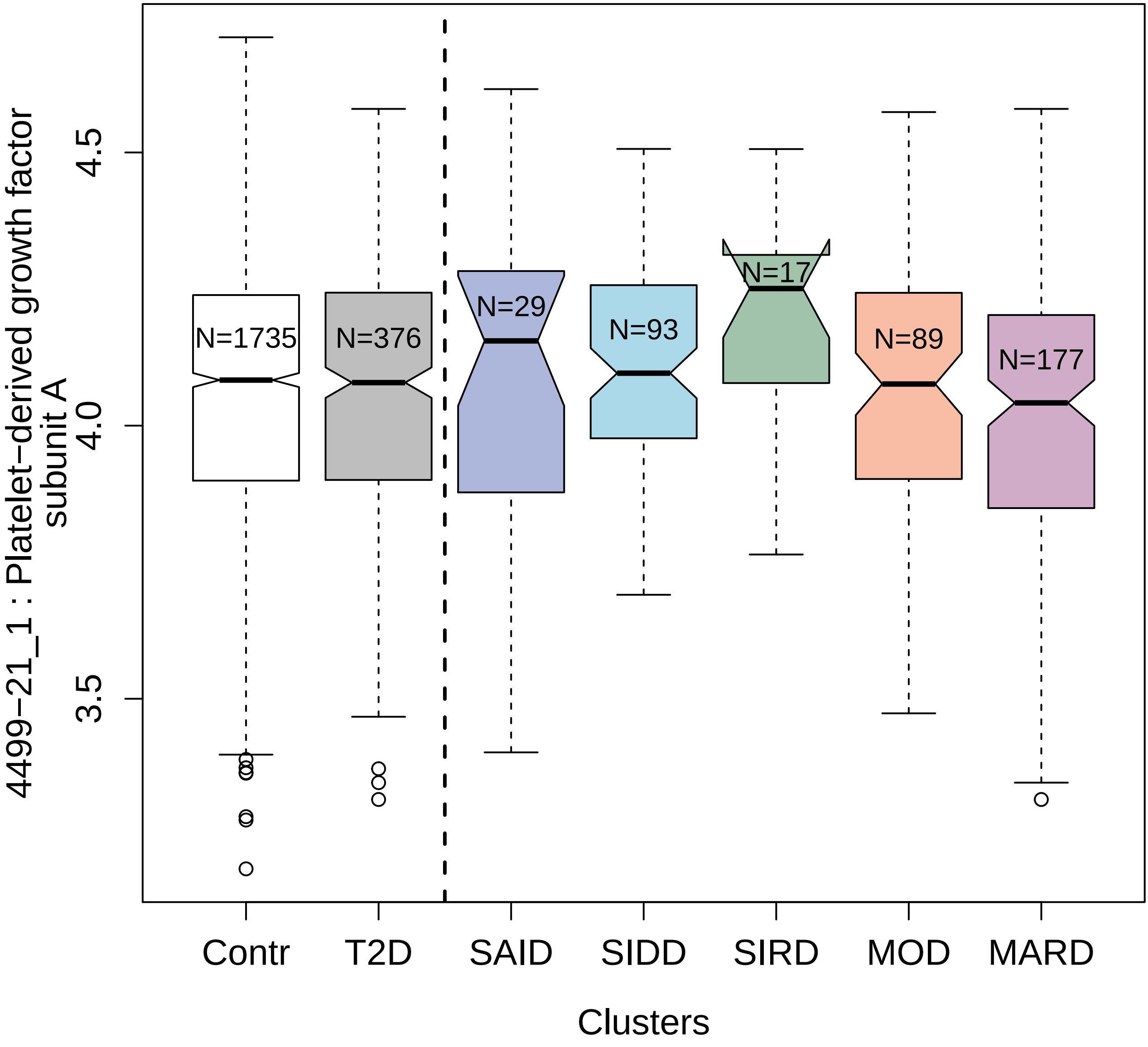

4499−21_1 : Platelet−derived growth factor
subunit A
4.5
4.0
3.5
N=1735
N=376
N=29
N=93
N=17
N=89
N=177
Contr
T2D
SAID
SIDD
SIRD
MOD
MARD
Clusters

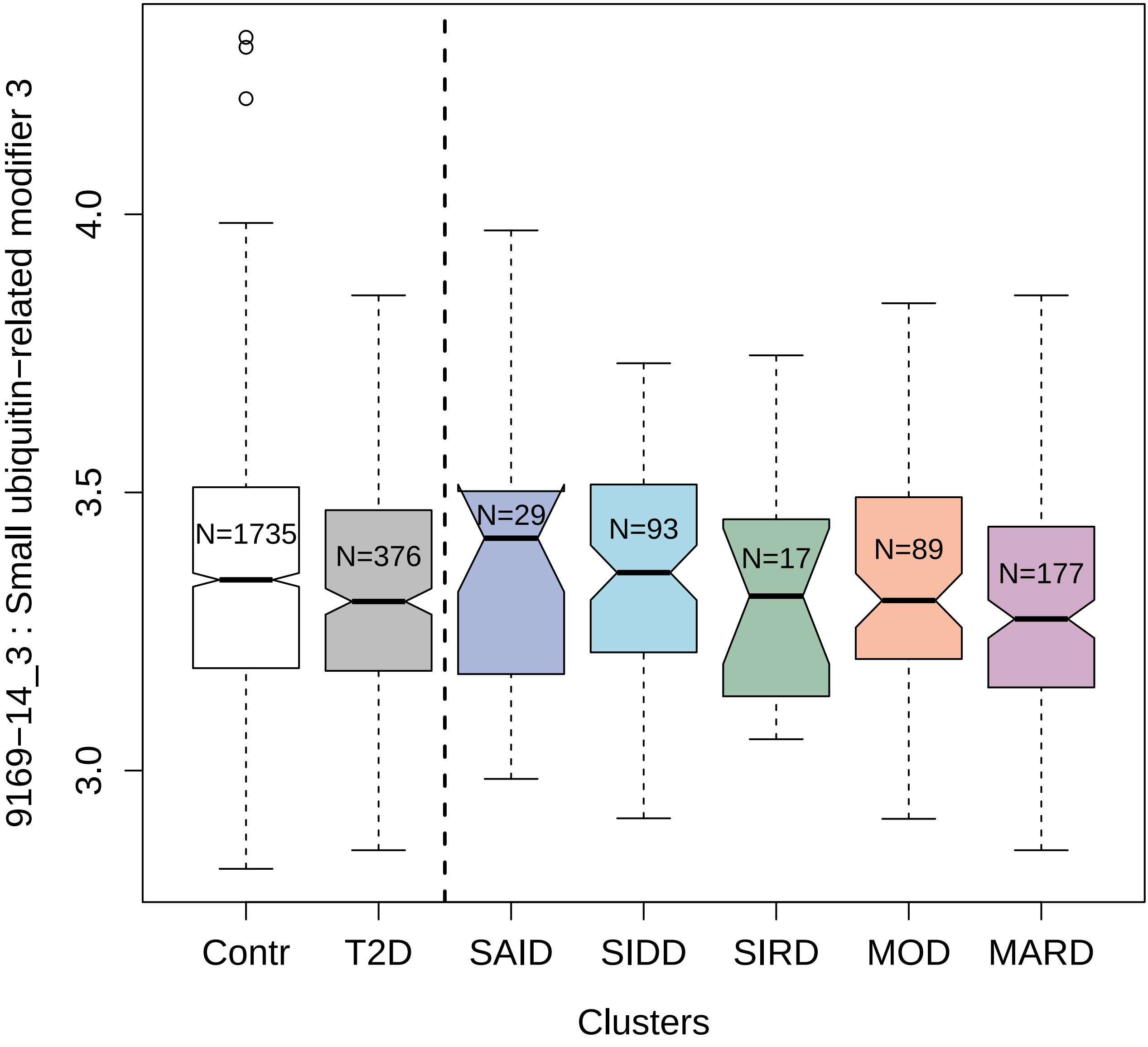
9169−14_3 : Small ubiquitin−related modifier 3
3.0
3.5
4.0
N=1735
N=376
N=29
N=93
N=17
N=89
N=177
Contr
T2D
SAID
SIDD
SIRD
MOD
MARD
Clusters

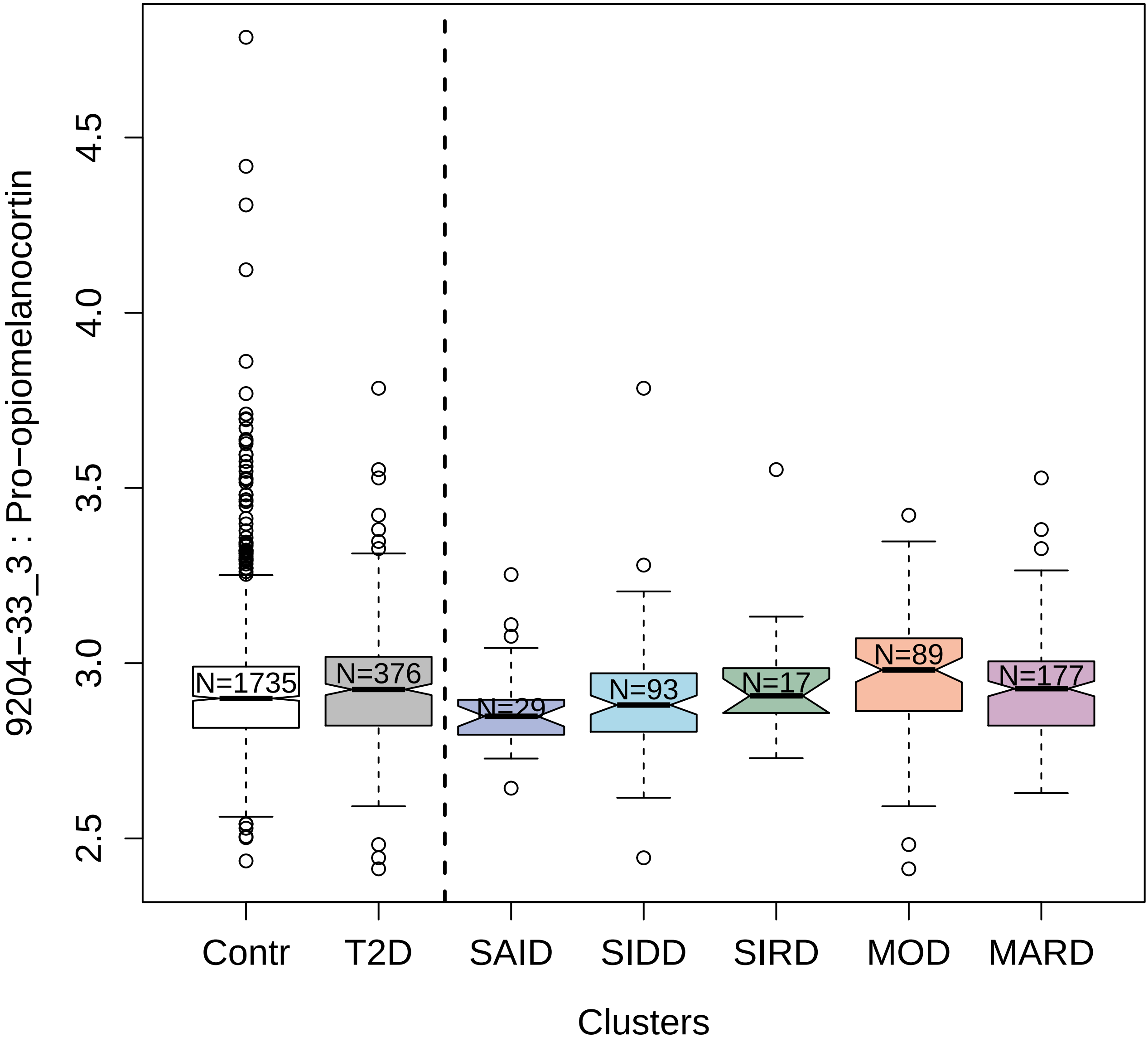

9204−33_3 : Pro−opiomelanocortin
4.5
4.0
3.5
3.0
2.5
N=1735
N=376
N=29
N=93
N=17
N=89
N=177
Contr
T2D
SAID
SIDD
SIRD
MOD
MARD
Clusters

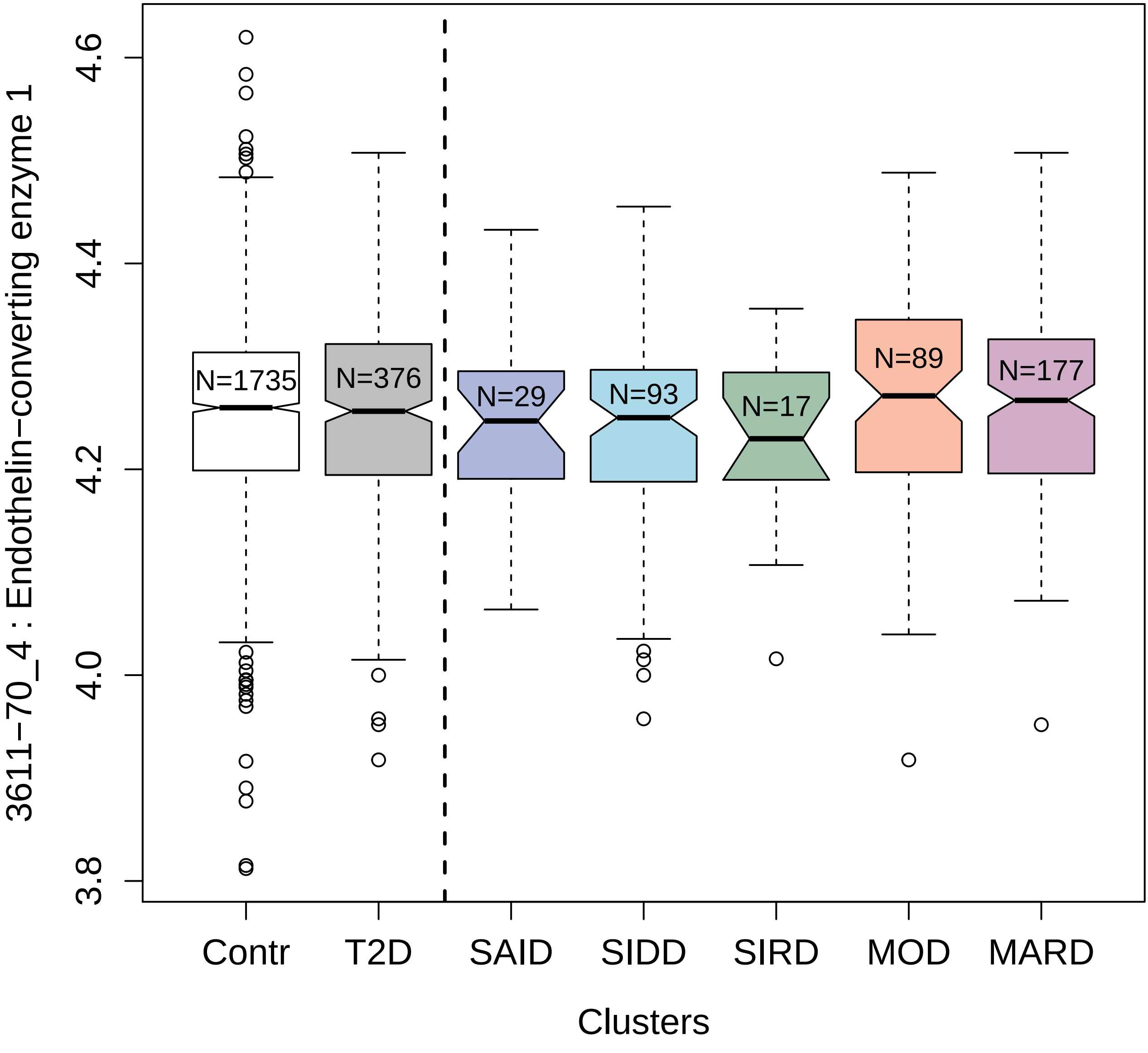

3611−70_4 : Endothelin−converting enzyme 1
4.6
4.4
4.2
4.0
3.8
N=1735
N=376
N=29
N=93
N=17
N=89
N=177
Contr
T2D
SAID
SIDD
SIRD
MOD
MARD
Clusters

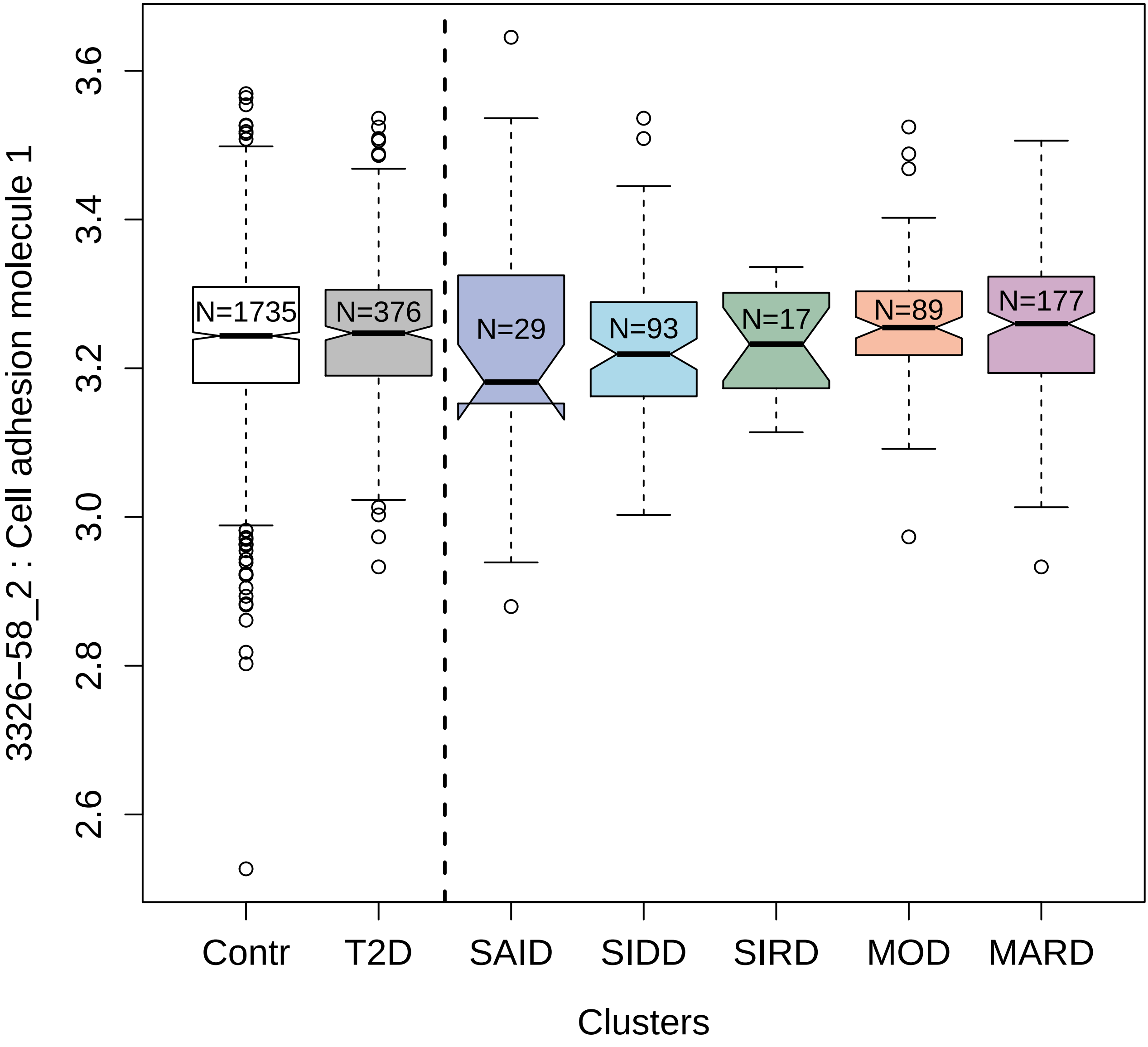

3326−58_2 : Cell adhesion molecule 1
3.6
3.4
3.2
3.0
2.8
2.6
N=1735
N=376
N=29
N=93
N=17
N=89
N=177
Contr
T2D
SAID
SIDD
SIRD
MOD
MARD
Clusters

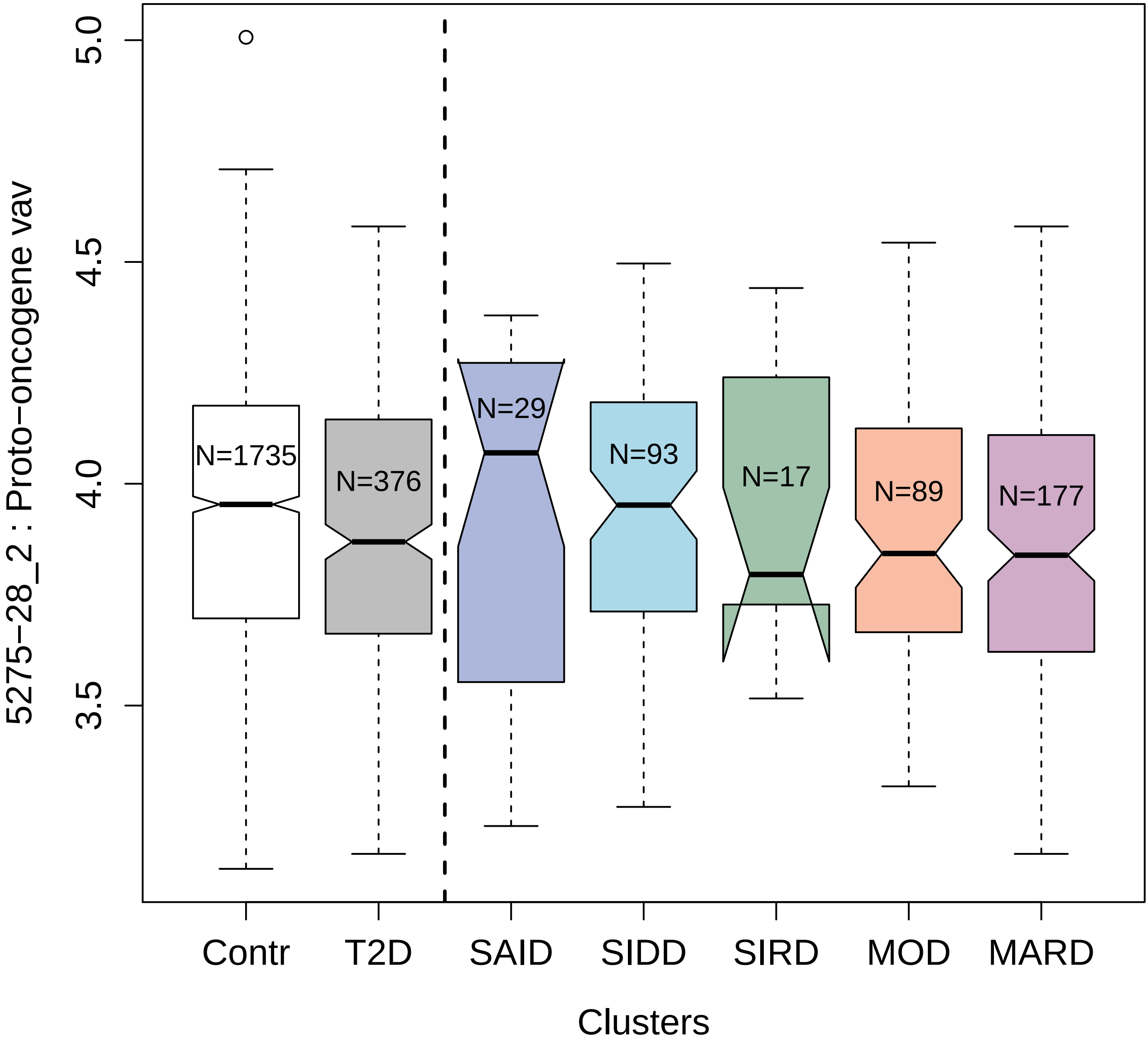
5275−28_2 : Proto−oncogene vav
5.0
4.5
4.0
3.5
N=1735
N=376
N=29
N=93
N=17
N=89
N=177
Contr
T2D
SAID
SIDD
SIRD
MOD
MARD
Clusters

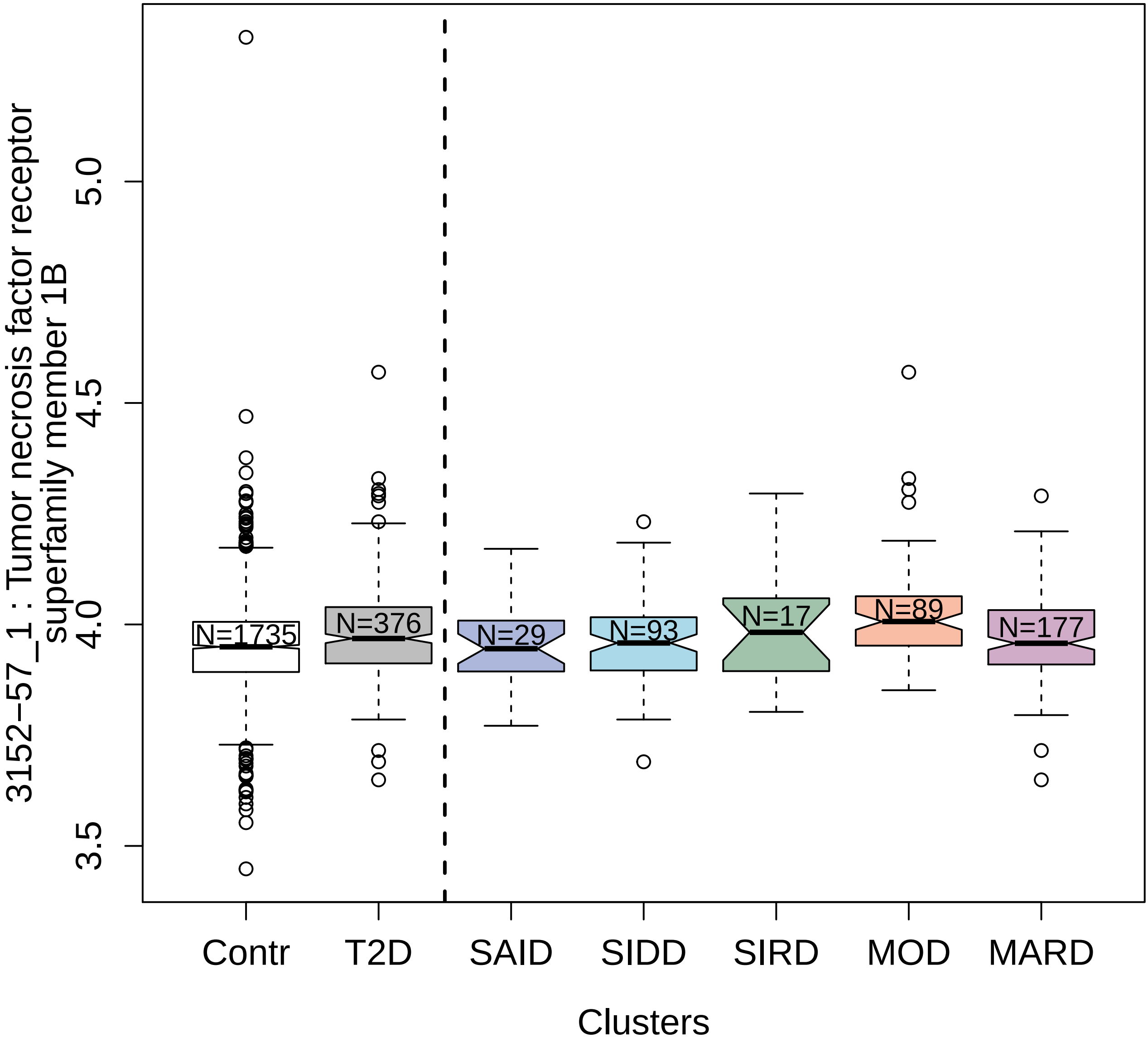

3152−57_1 : Tumor necrosis factor receptor superfamily member 1B
3.5
4.0
4.5
5.0
N=1735
N=376
N=29
N=93
N=17
N=89
N=177
Contr
T2D
SAID
SIDD
SIRD
MOD
MARD
Clusters

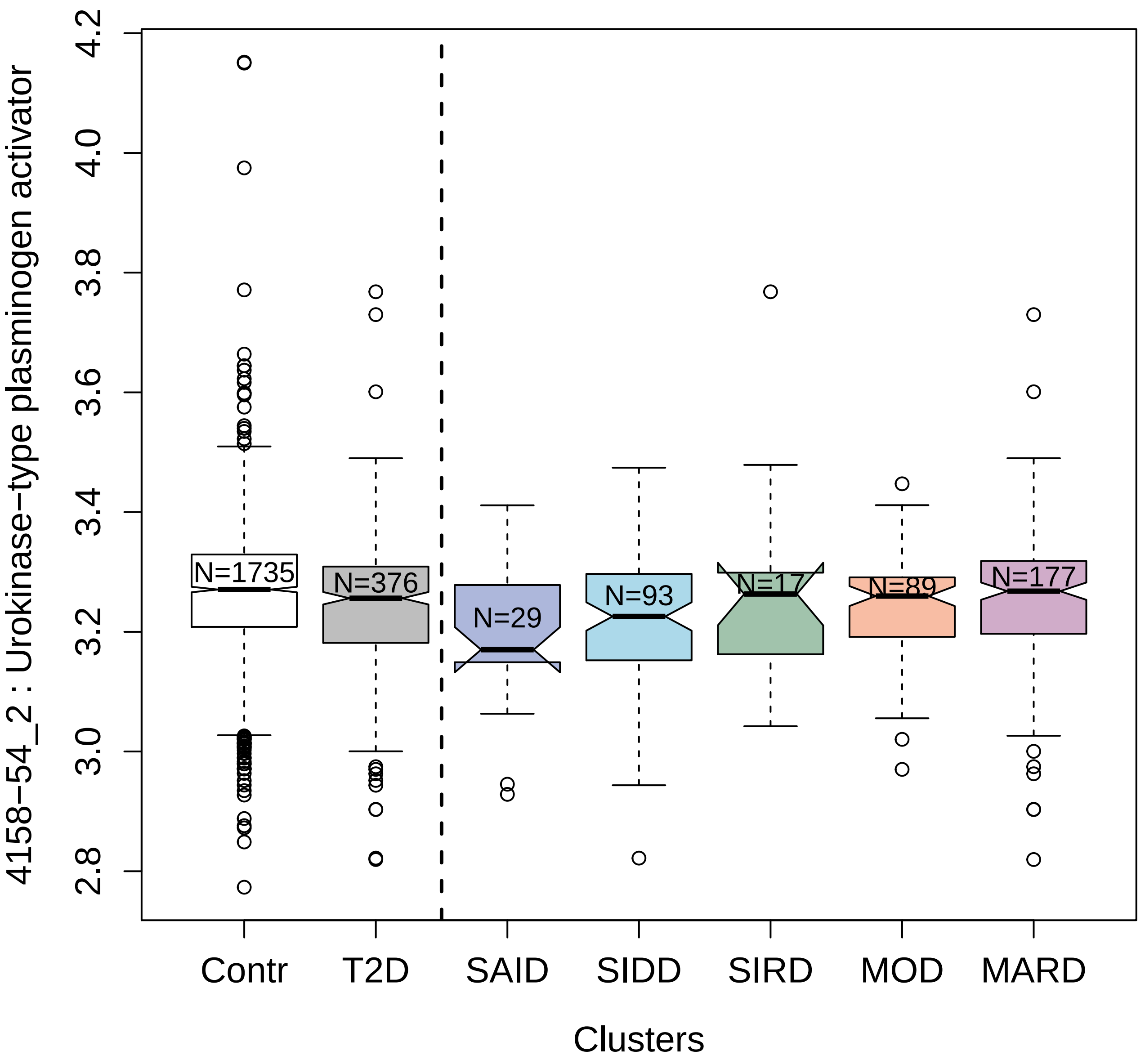

4158−54_2 : Urokinase−type plasminogen activator
4.2
4.0
3.8
3.6
3.4
3.2
3.0
2.8
N=1735
N=376
N=29
N=93
N=17
N=89
N=177
Contr
T2D
SAID
SIDD
SIRD
MOD
MARD
Clusters

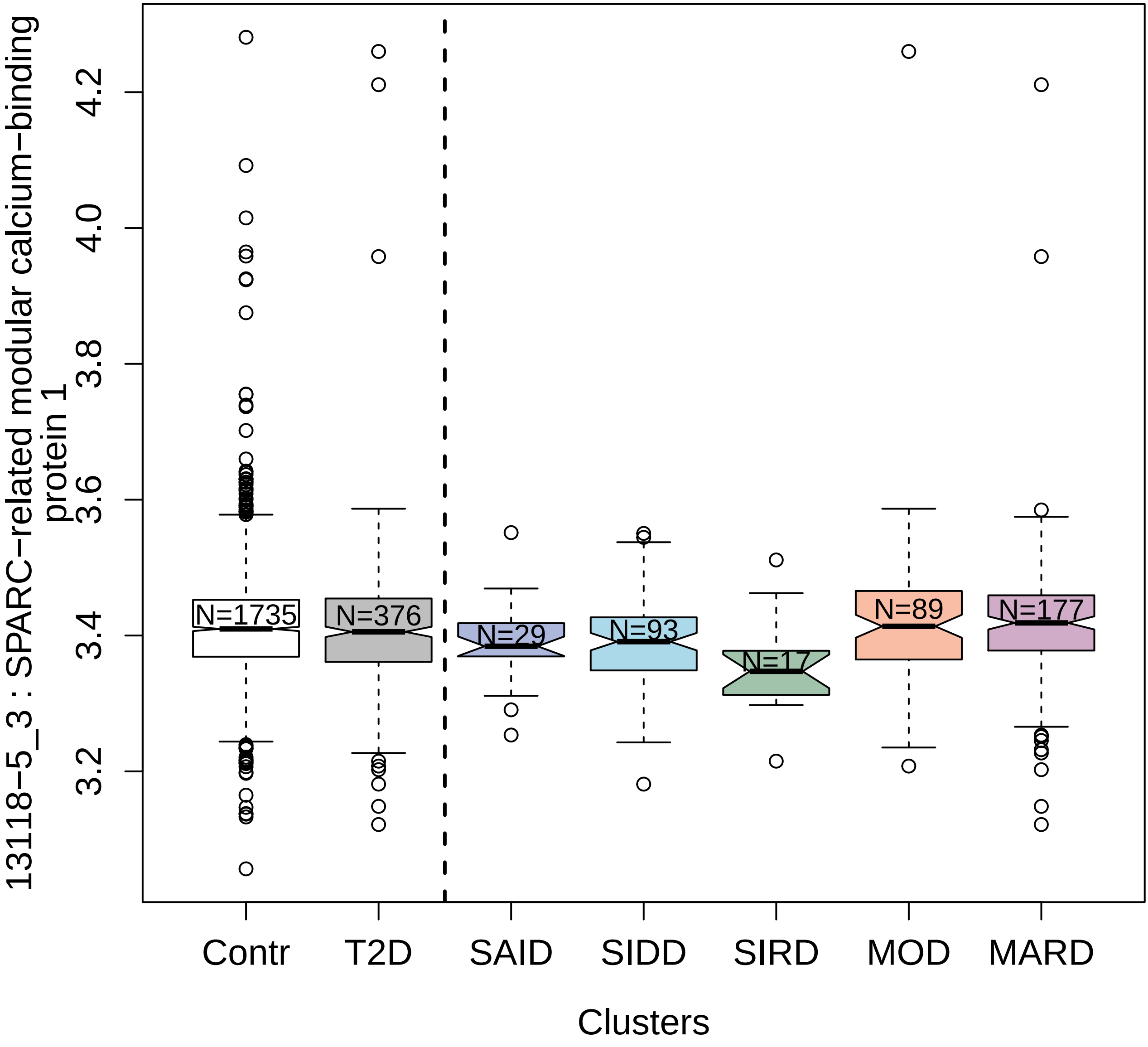

13118−5_3 : SPARC−related modular calcium−binding protein 1
3.2
3.4
3.6
3.8
4.0
4.2
N=1735
N=376
N=29
N=93
N=17
N=89
N=177
Contr
T2D
SAID
SIDD
SIRD
MOD
MARD
Clusters

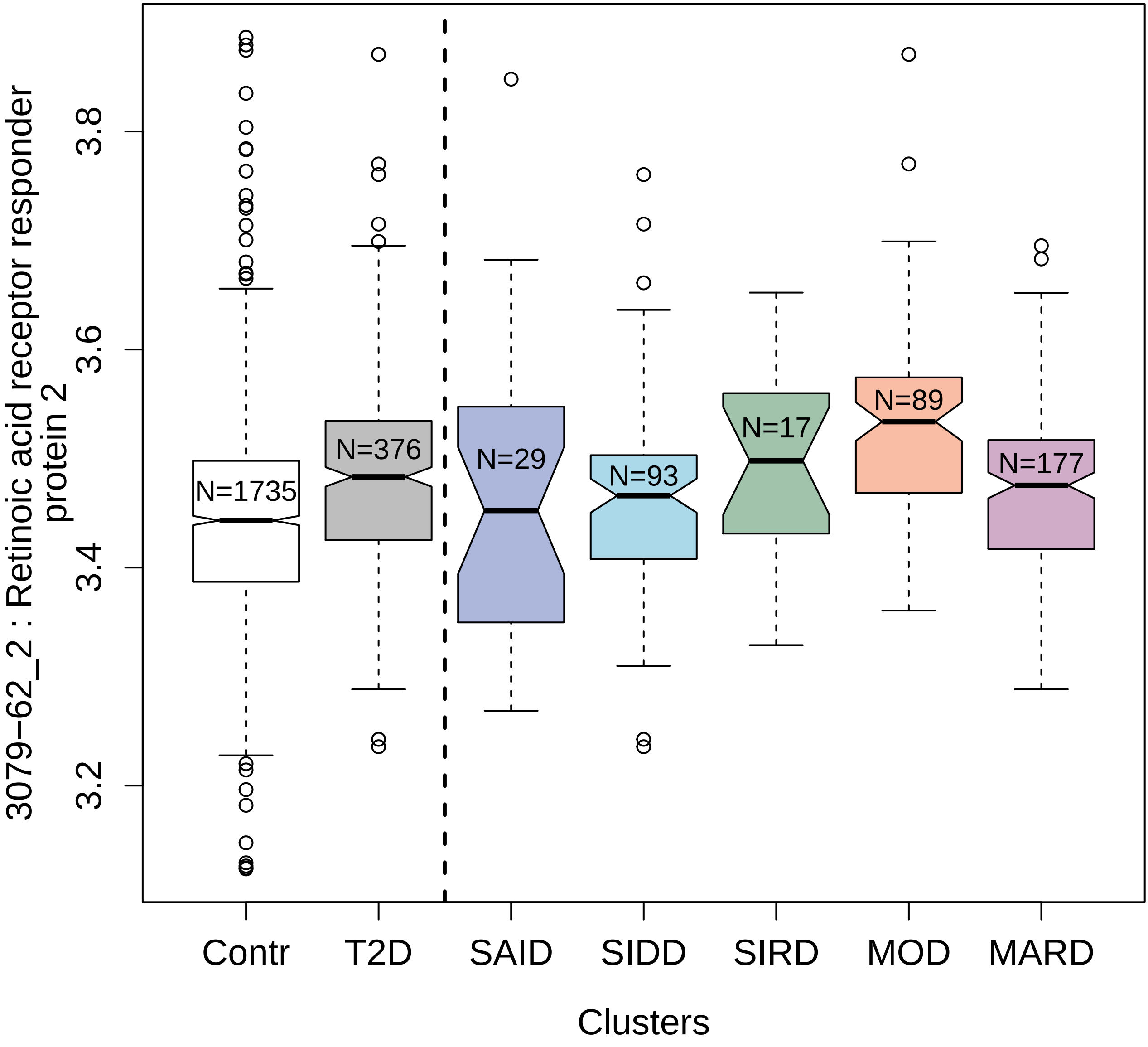

3079−62_2 : Retinoic acid receptor responder protein 2
3.2
3.4
3.6
3.8
N=1735
N=376
N=29
N=93
N=17
N=89
N=177
Contr
T2D
SAID
SIDD
SIRD
MOD
MARD
Clusters

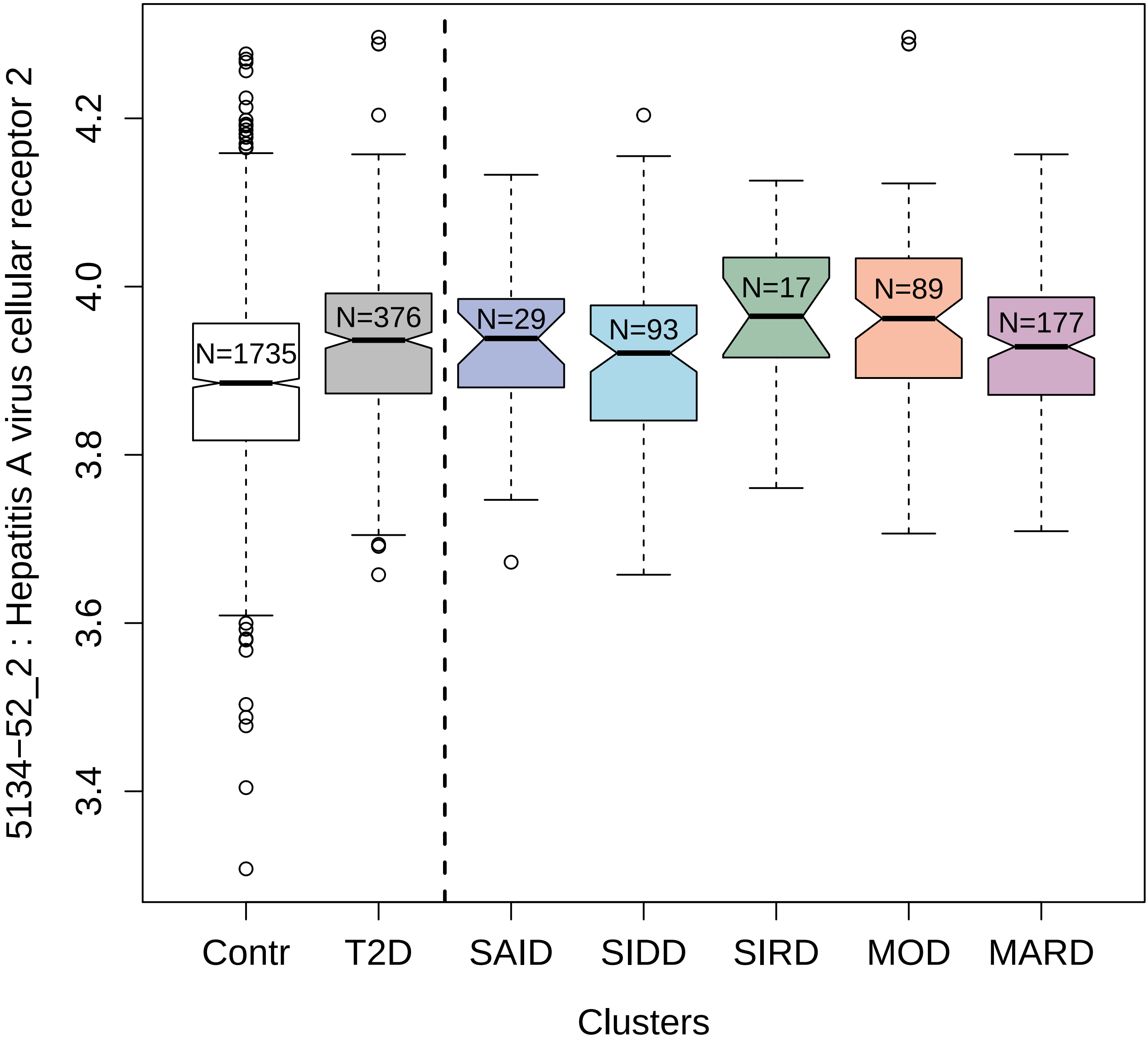

5134−52_2 : Hepatitis A virus cellular receptor 2
3.4
3.6
3.8
4.0
4.2
N=1735
N=376
N=29
N=93
N=17
N=89
N=177
Contr
T2D
SAID
SIDD
SIRD
MOD
MARD
Clusters

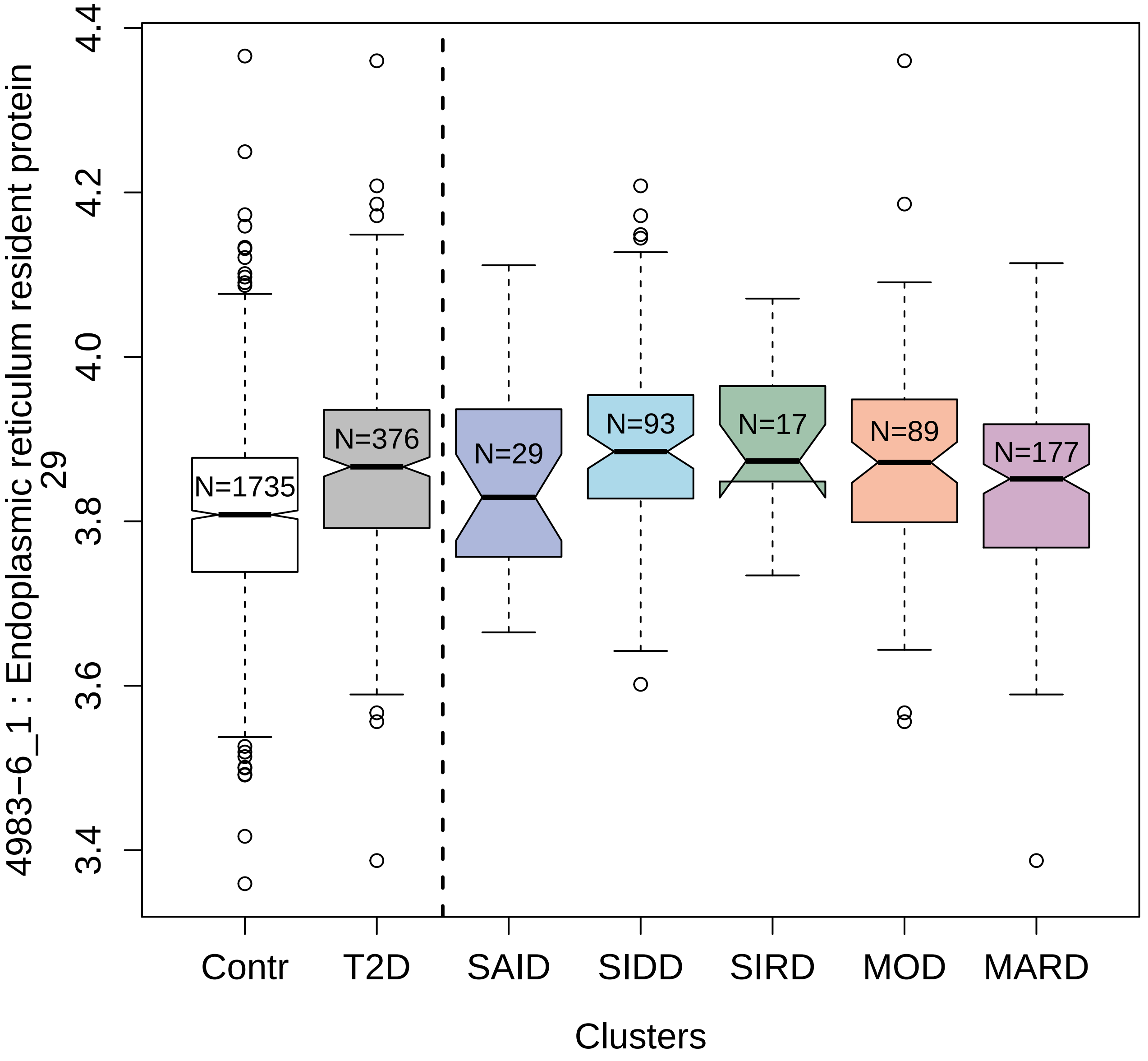

4983−6_1 : Endoplasmic reticulum resident protein 29
3.4
3.6
3.8
4.0
4.2
4.4
N=1735
N=376
N=29
N=93
N=17
N=89
N=177
Contr
T2D
SAID
SIDD
SIRD
MOD
MARD
Clusters

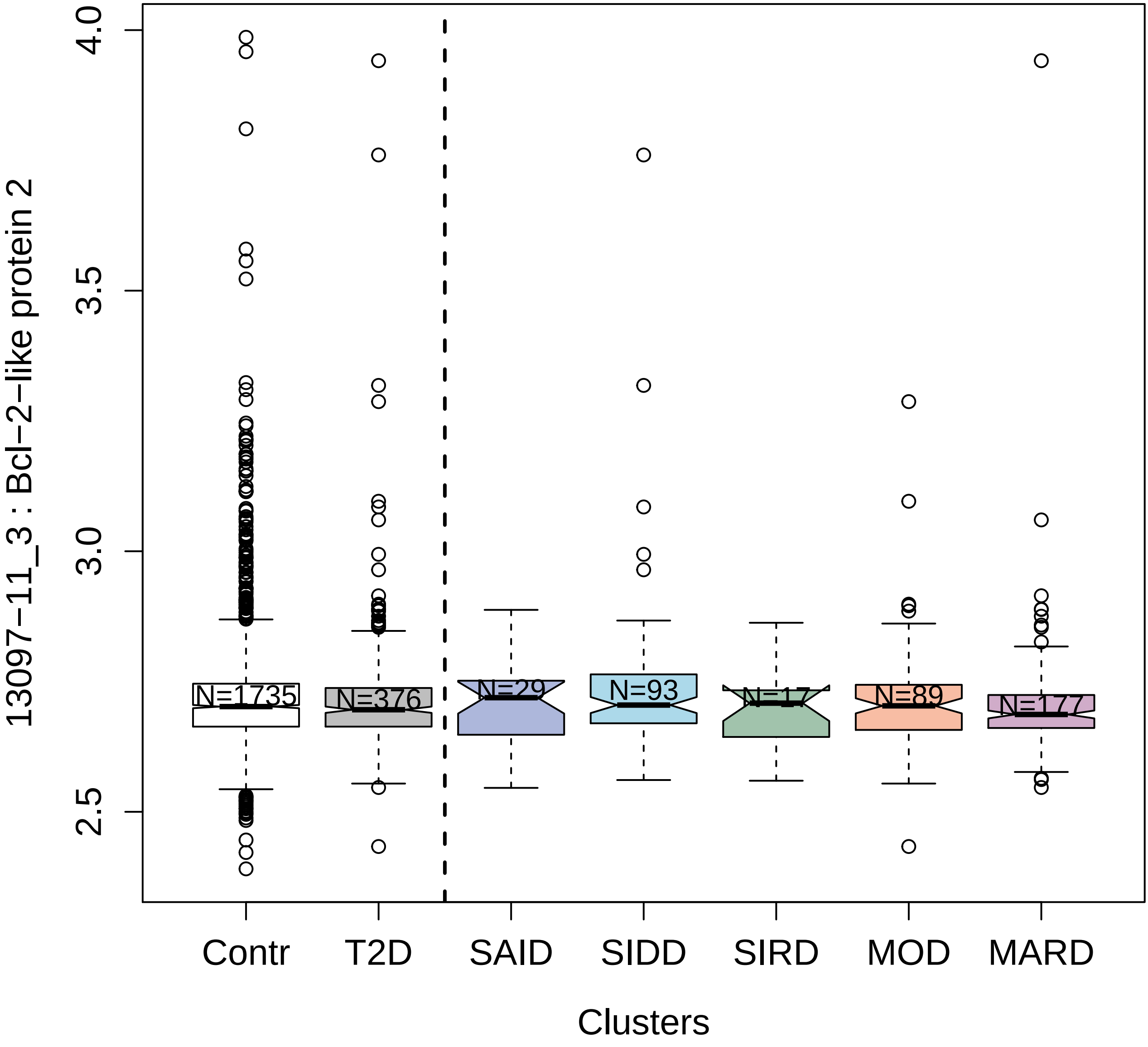

13097−11_3 : Bcl−2−like protein 2
4.0
3.5
3.0
2.5
N=1735
N=376
N=29
N=93
N=17
N=89
N=177
Contr
T2D
SAID
SIDD
SIRD
MOD
MARD
Clusters

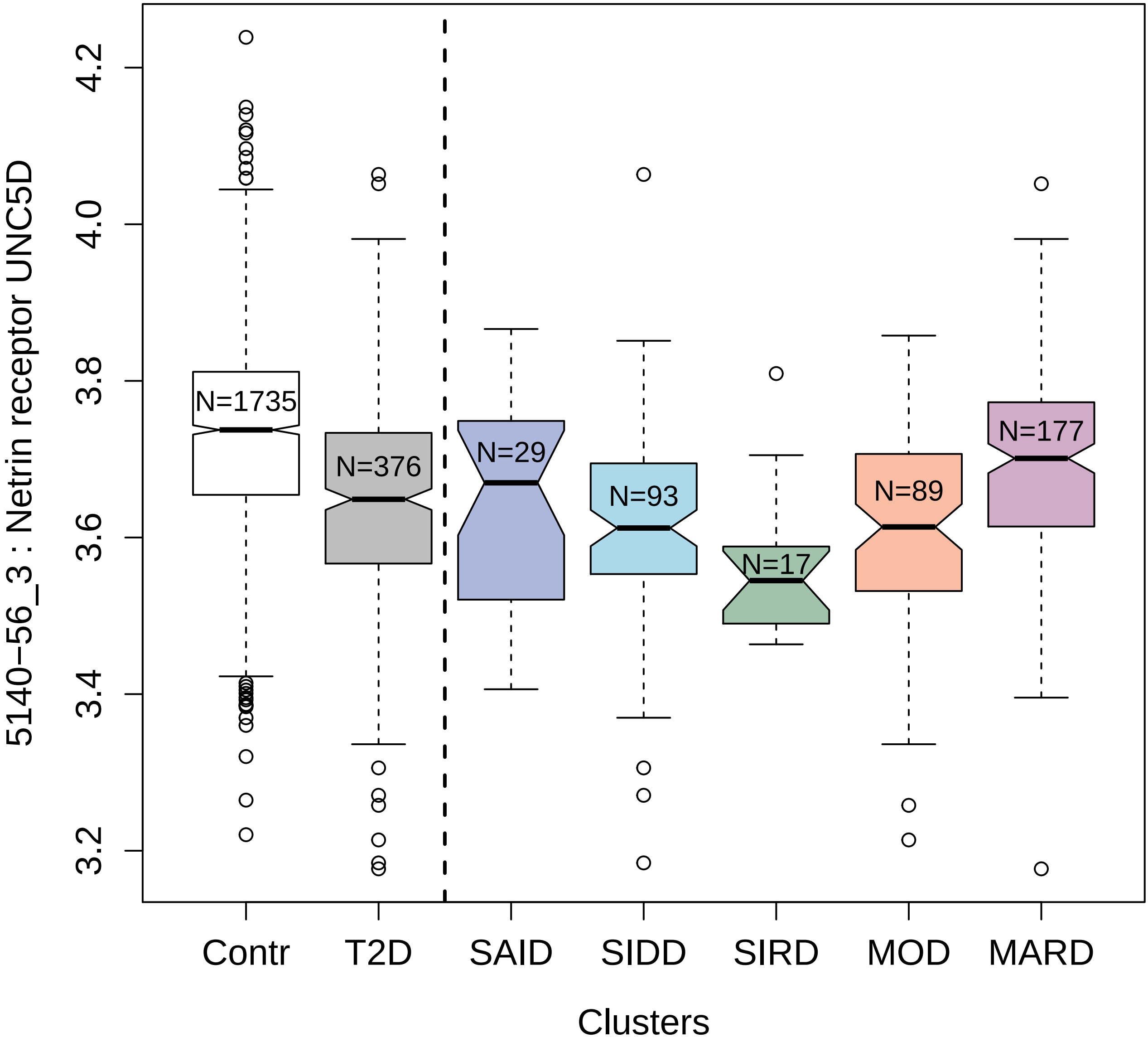

5140−56_3 : Netrin receptor UNC5D
3.2
3.4
3.6
3.8
4.0
4.2
N=1735
N=376
N=29
N=93
N=17
N=89
N=177
Contr
T2D
SAID
SIDD
SIRD
MOD
MARD
Clusters

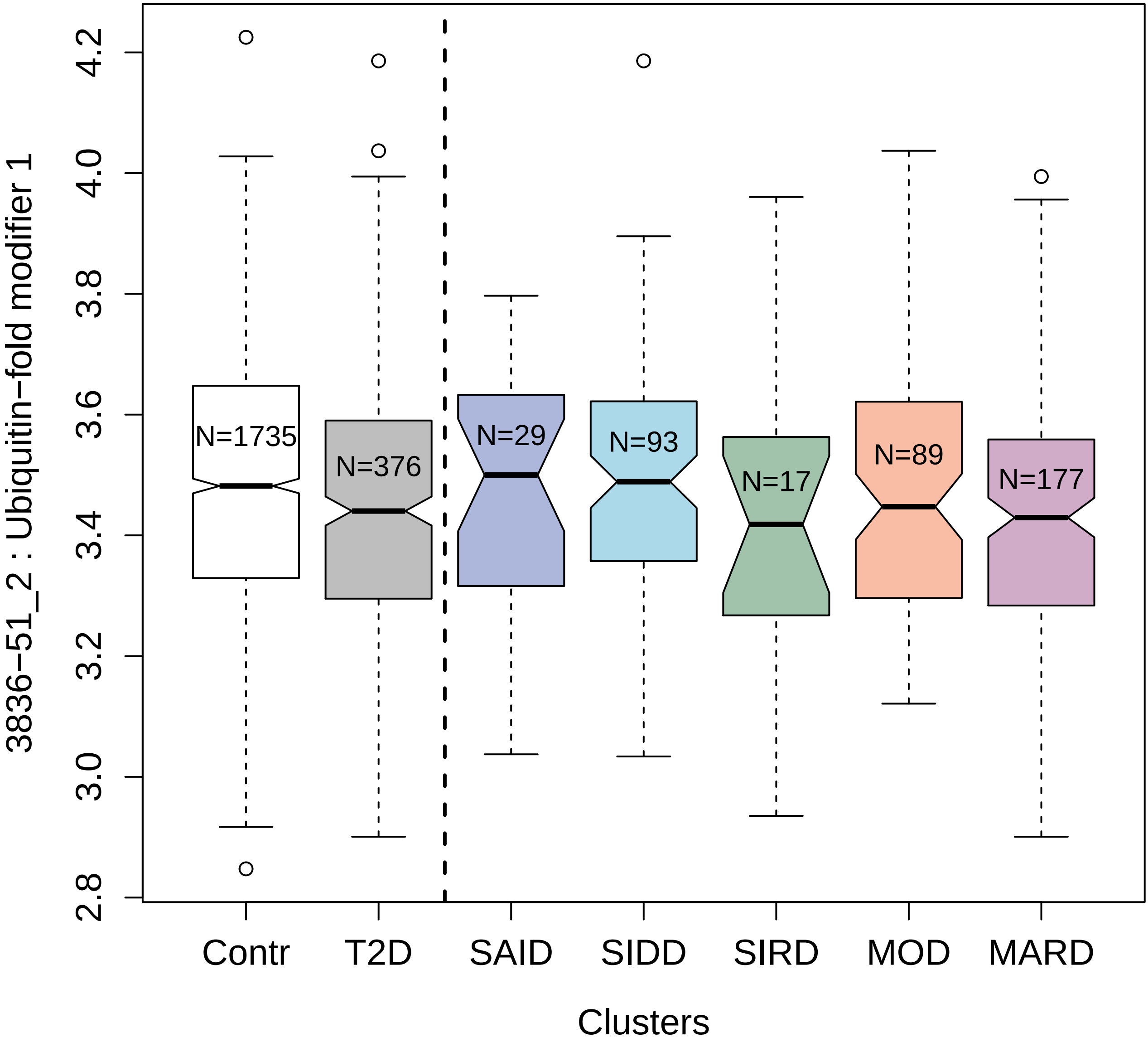
3836−51_2 : Ubiquitin−fold modifier 1
2.8
3.0
3.2
3.4
3.6
3.8
4.0
4.2
N=1735
N=376
N=29
N=93
N=17
N=89
N=177
Contr
T2D
SAID
SIDD
SIRD
MOD
MARD
Clusters

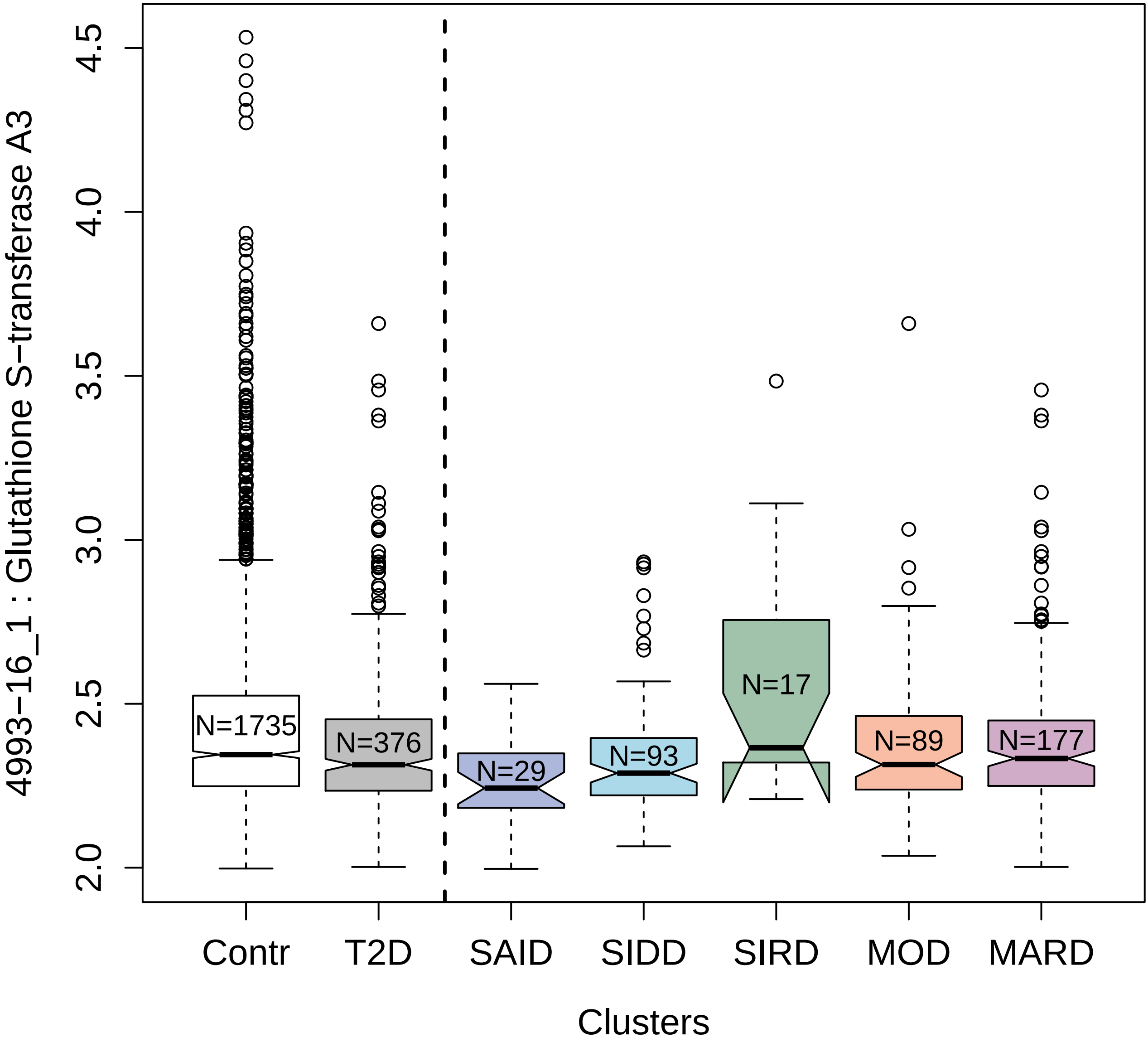

4993−16_1 : Glutathione S−transferase A3
4.5
4.0
3.5
3.0
2.5
2.0
N=1735
N=376
N=29
N=93
N=17
N=89
N=177
Contr
T2D
SAID
SIDD
SIRD
MOD
MARD
Clusters

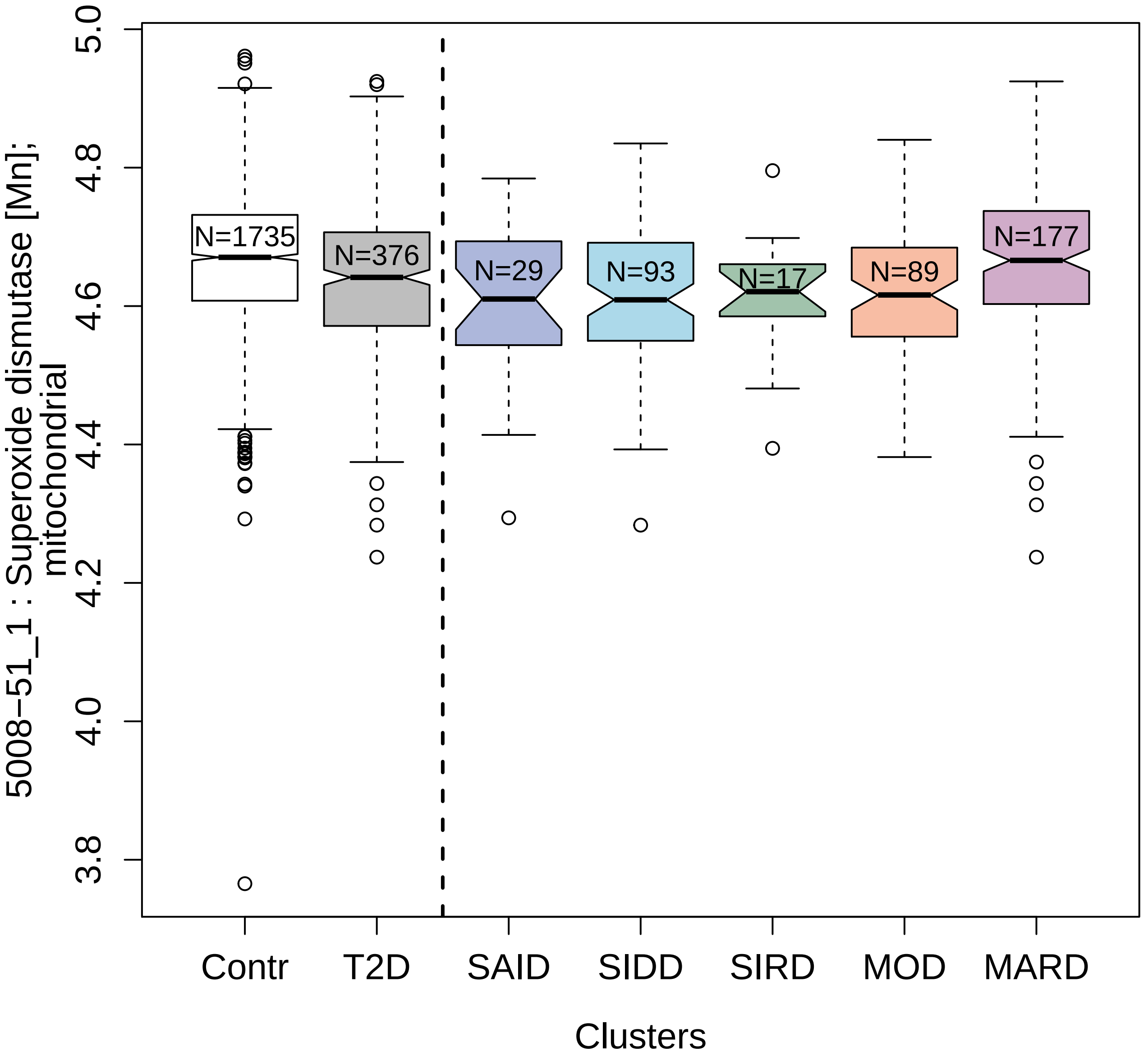
5008−51_1 : Superoxide dismutase [Mn];
mitochondrial
5.0
4.8
4.6
4.4
4.2
4.0
3.8
N=1735
N=376
N=29
N=93
N=17
N=89
N=177
Contr
T2D
SAID
SIDD
SIRD
MOD
MARD
Clusters

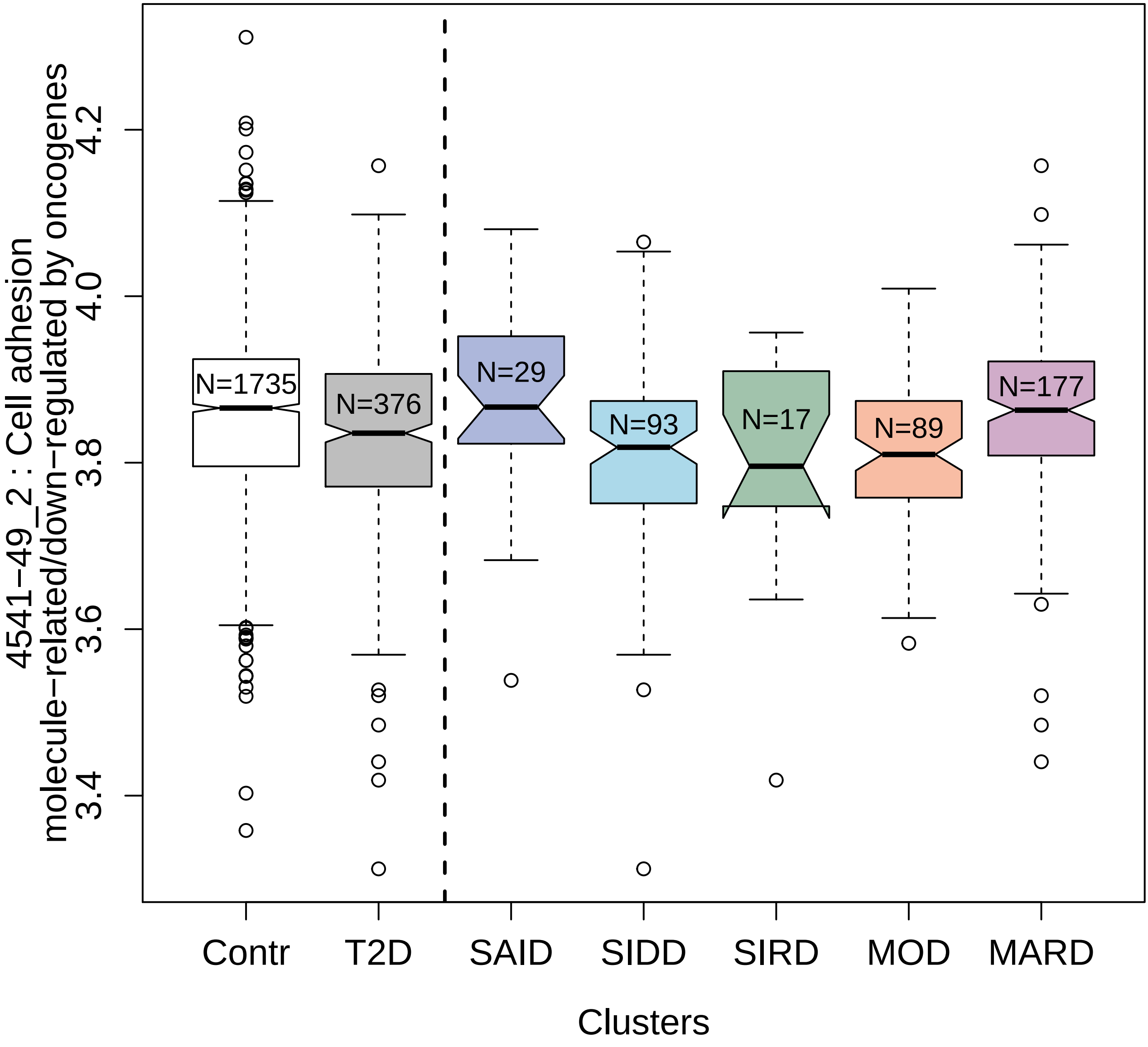
4541−49_2 : Cell adhesion
molecule−related/down−regulated by oncogenes
4.2
4.0
3.8
3.6
3.4
N=1735
N=376
N=29
N=93
N=17
N=89
N=177
Contr
T2D
SAID
SIDD
SIRD
MOD
MARD
Clusters

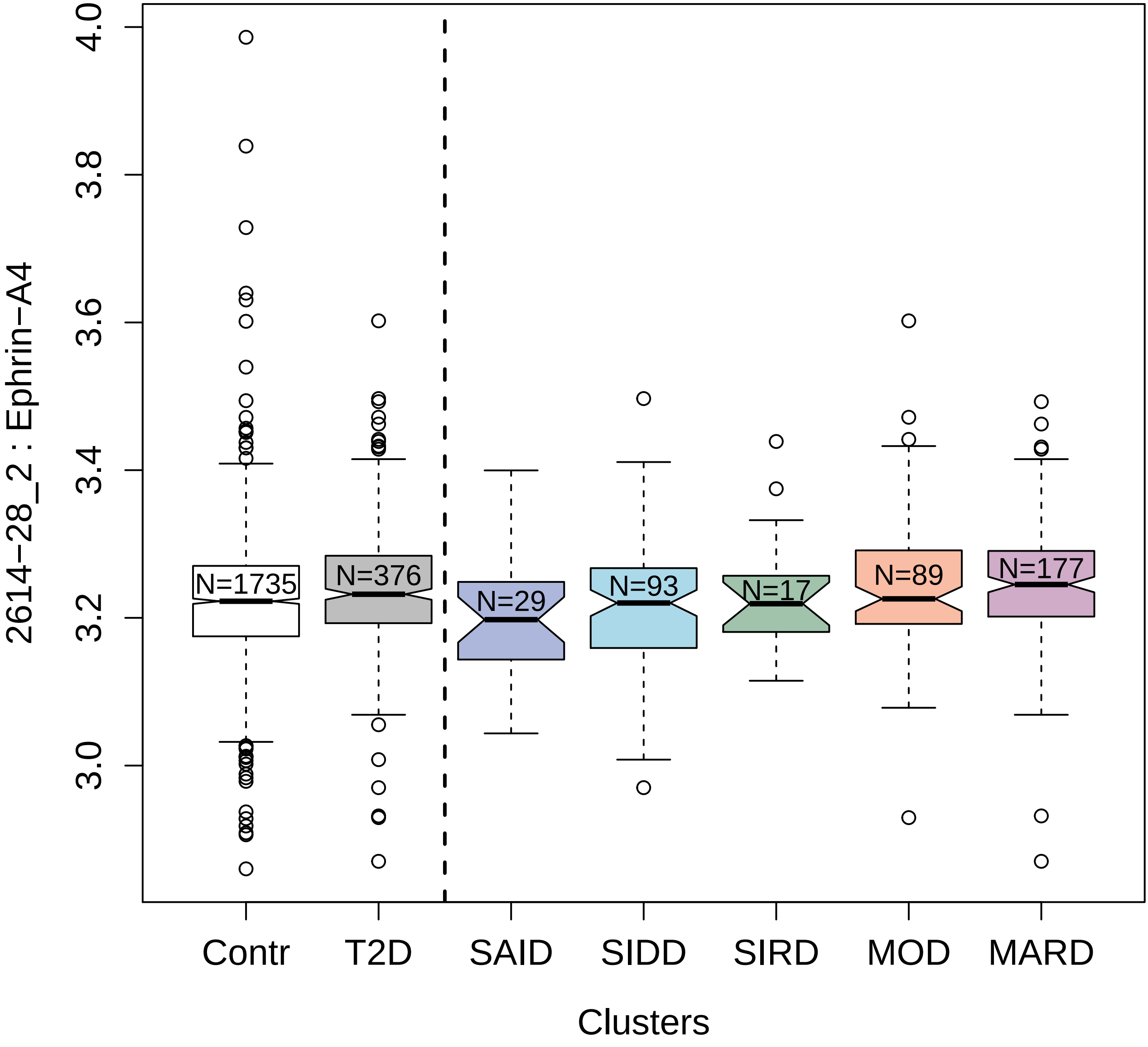

2614−28_2 : Ephrin−A4
4.0
3.8
3.6
3.4
3.2
3.0
N=1735
N=376
N=29
N=93
N=17
N=89
N=177
Contr
T2D
SAID
SIDD
SIRD
MOD
MARD
Clusters

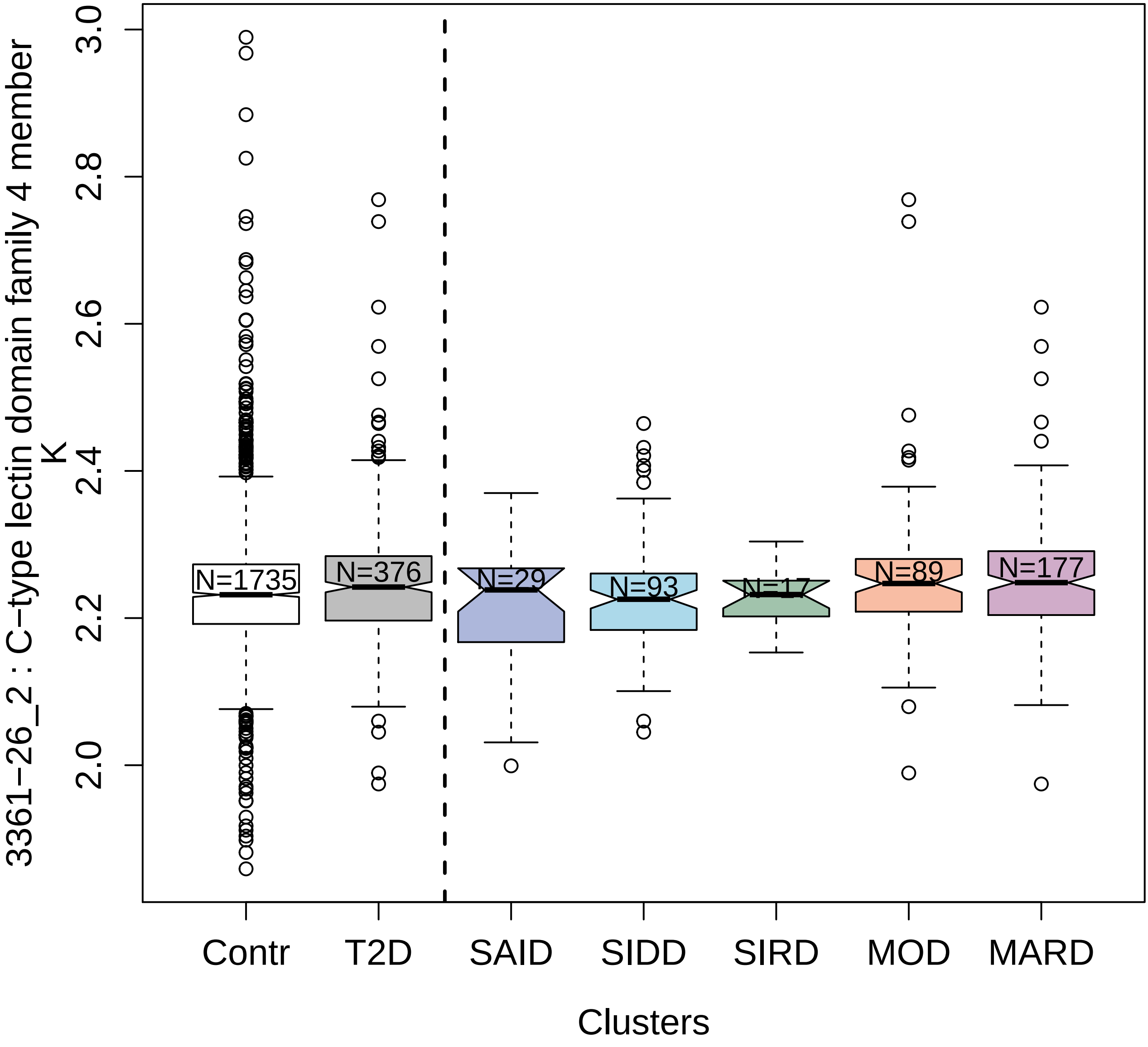

3361−26_2 : C−type lectin domain family 4 member K
3.0
2.8
2.6
2.4
2.2
2.0
N=1735
N=376
N=29
N=93
N=17
N=89
N=177
Contr
T2D
SAID
SIDD
SIRD
MOD
MARD
Clusters

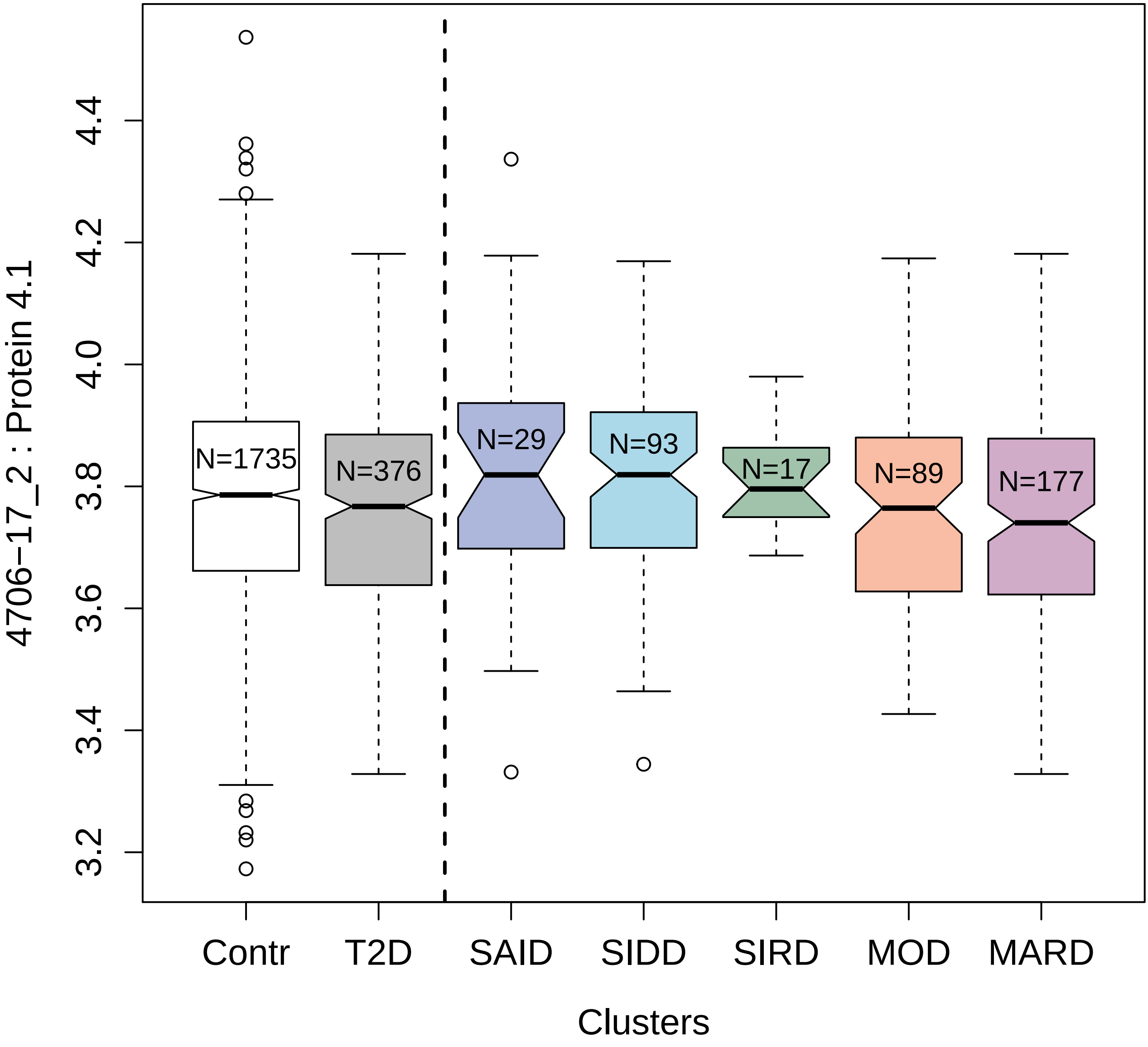
4706−17_2 : Protein 4.1
3.2
3.4
3.6
3.8
4.0
4.2
4.4
N=1735
N=376
N=29
N=93
N=17
N=89
N=177
Contr
T2D
SAID
SIDD
SIRD
MOD
MARD
Clusters

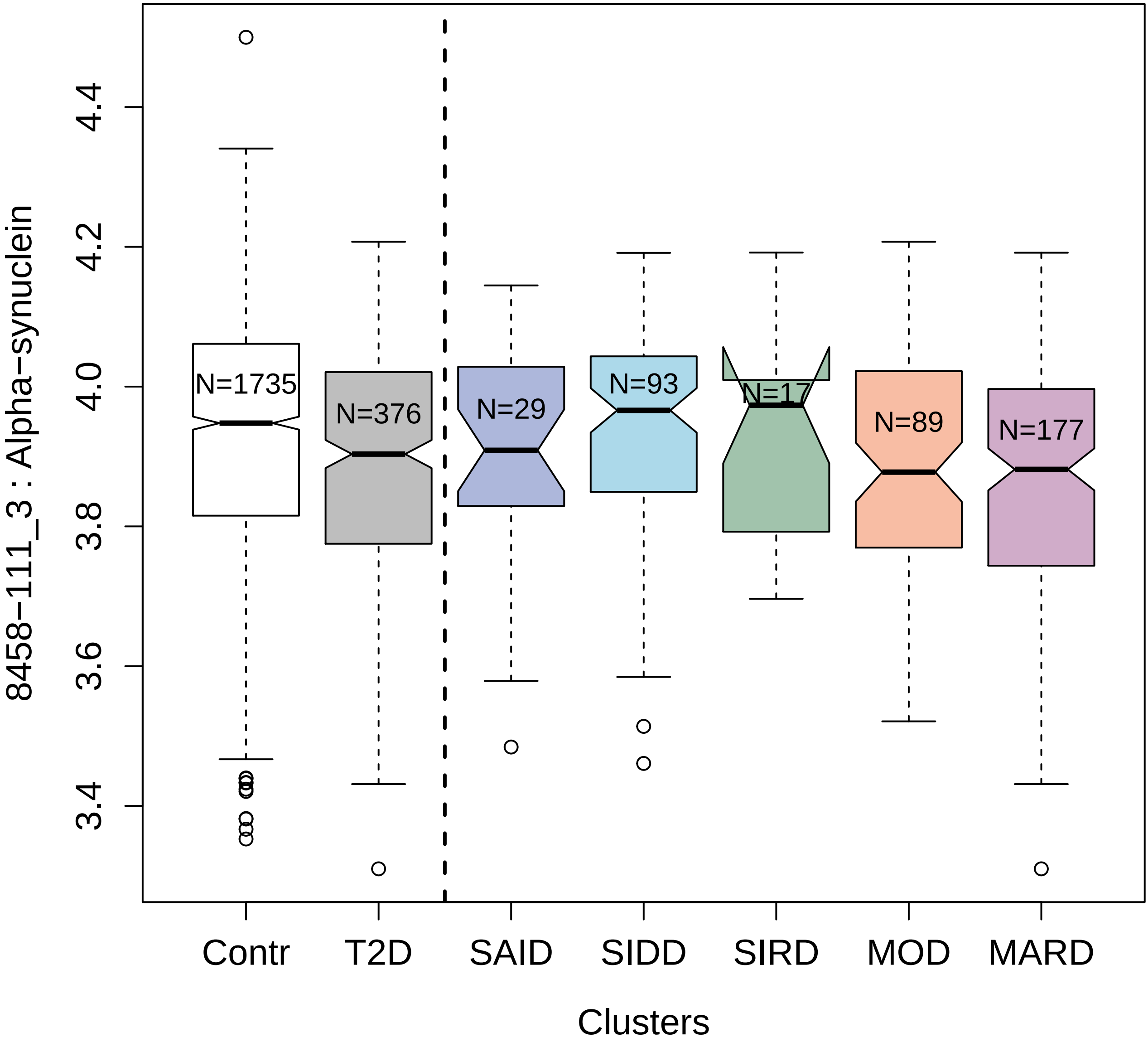

8458−111_3 : Alpha−synuclein
4.4
4.2
4.0
3.8
3.6
3.4
N=1735
N=376
N=29
N=93
N=17
N=89
N=177
Contr
T2D
SAID
SIDD
SIRD
MOD
MARD
Clusters

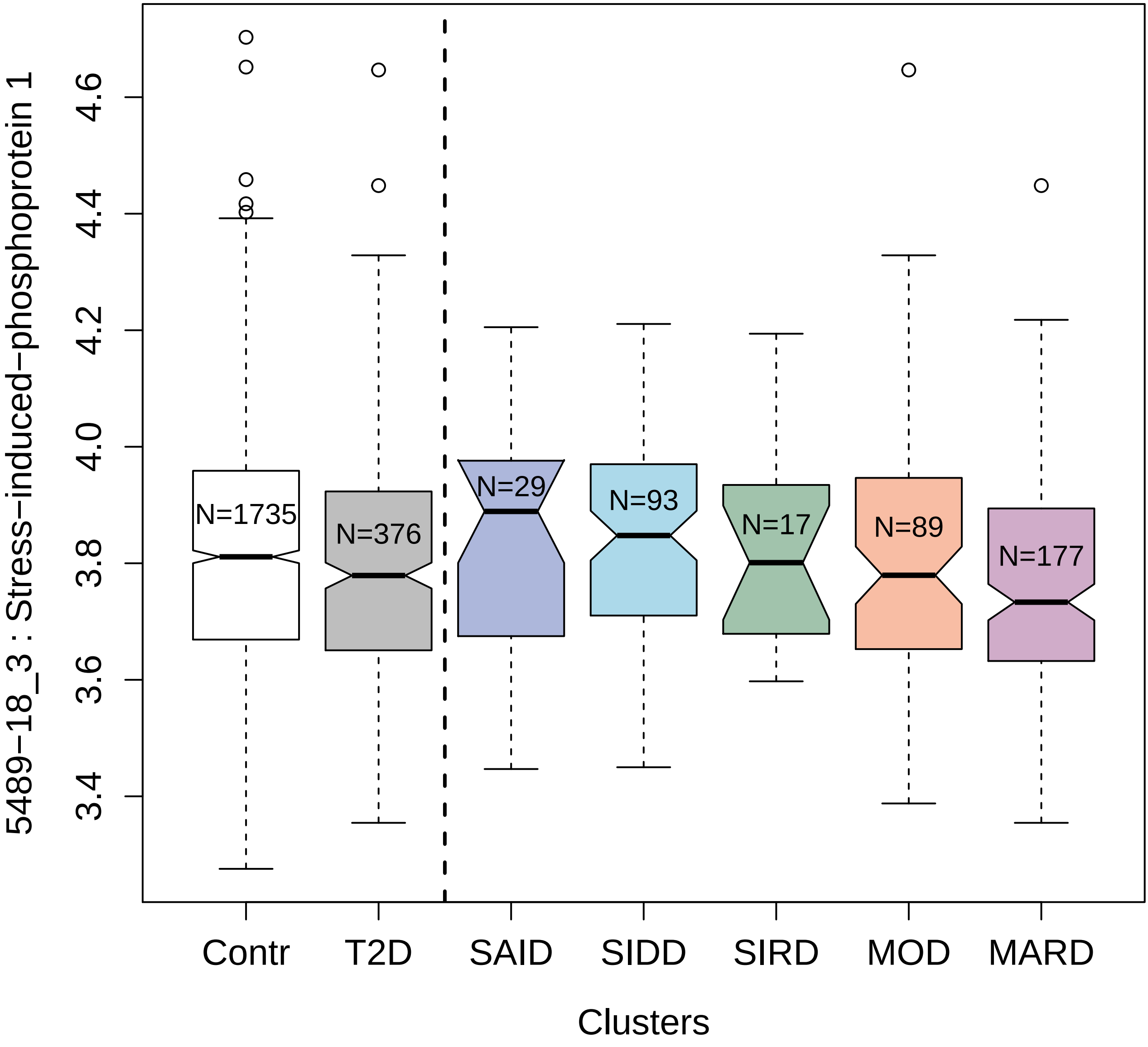

5489−18_3 : Stress−induced−phosphoprotein 1
3.4
3.6
3.8
4.0
4.2
4.4
4.6
N=1735
N=376
N=29
N=93
N=17
N=89
N=177
Contr
T2D
SAID
SIDD
SIRD
MOD
MARD
Clusters

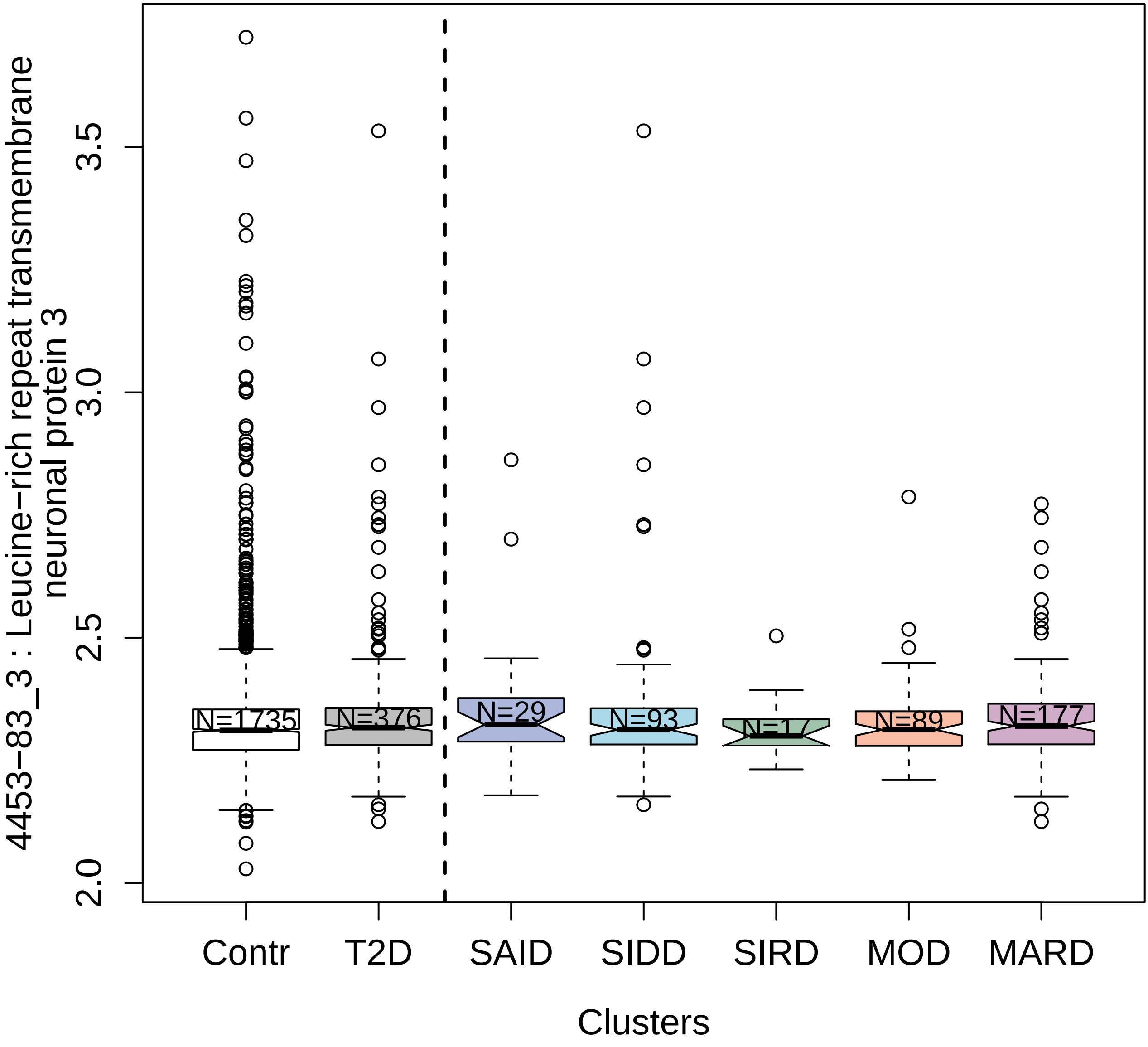
4453−83_3 : Leucine−rich repeat transmembrane neuronal protein 3
2.0
2.5
3.0
3.5
N=1735
N=376
N=29
N=93
N=17
N=89
N=177
Contr
T2D
SAID
SIDD
SIRD
MOD
MARD
Clusters

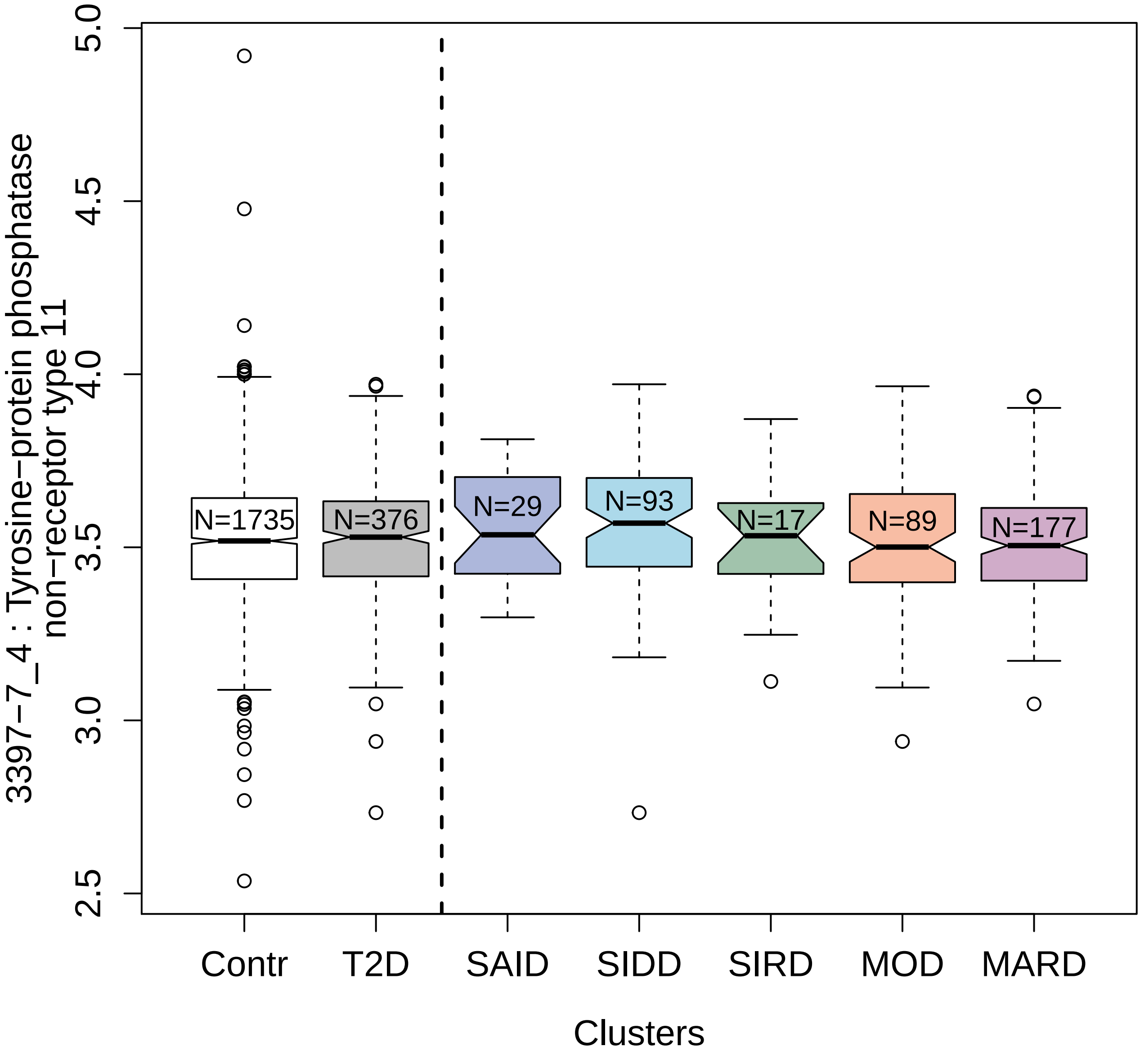
3397−7_4 : Tyrosine−protein phosphatase
non−receptor type 11
5.0
4.5
4.0
3.5
3.0
2.5
N=1735
N=376
N=29
N=93
N=17
N=89
N=177
Contr
T2D
SAID
SIDD
SIRD
MOD
MARD
Clusters

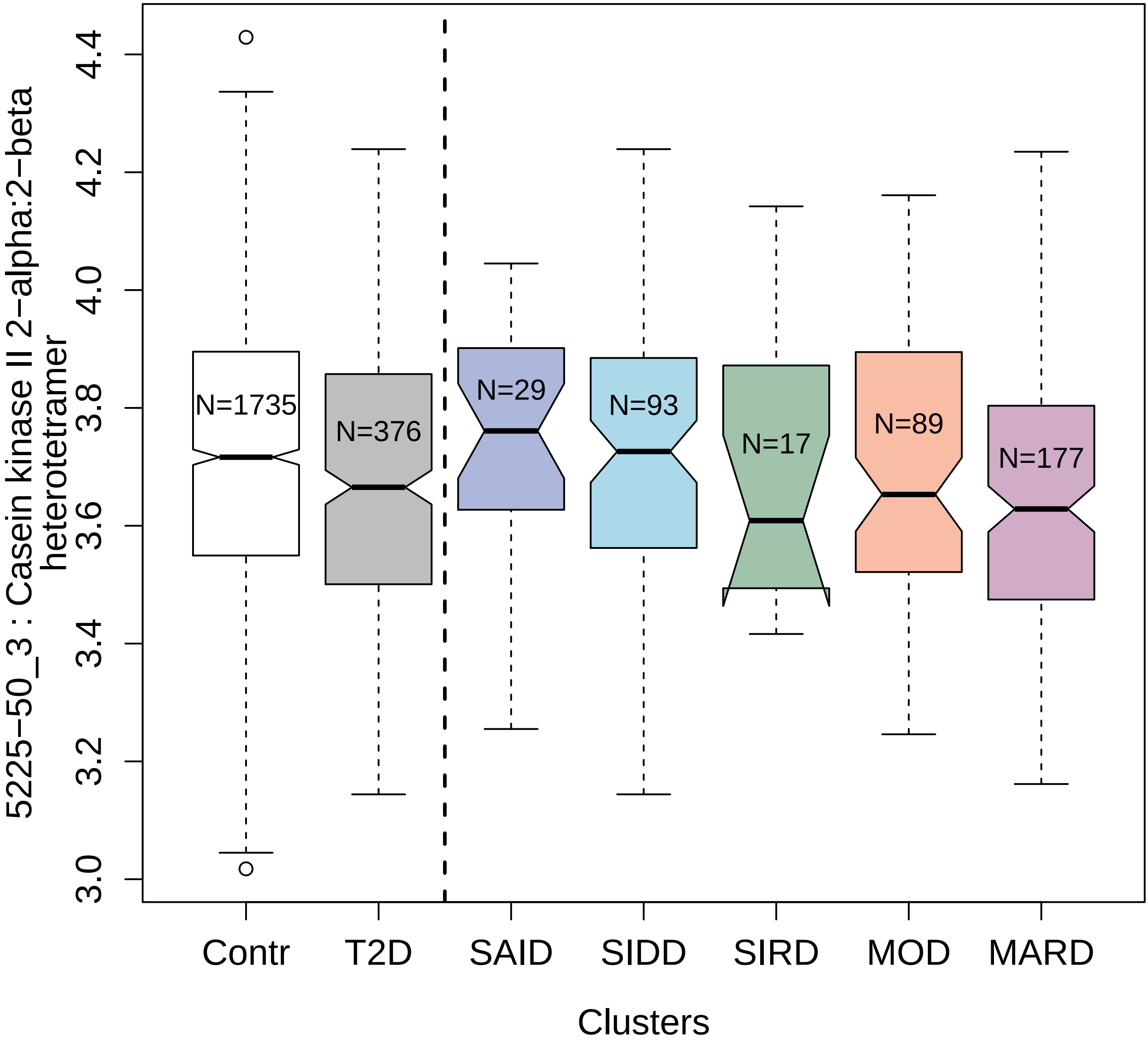
5225−50_3 : Casein kinase II 2−alpha:2−beta heterotetramer
4.4
4.2
4.0
3.8
3.6
3.4
3.2
3.0
N=1735
N=376
N=29
N=93
N=17
N=89
N=177
Contr
T2D
SAID
SIDD
SIRD
MOD
MARD
Clusters

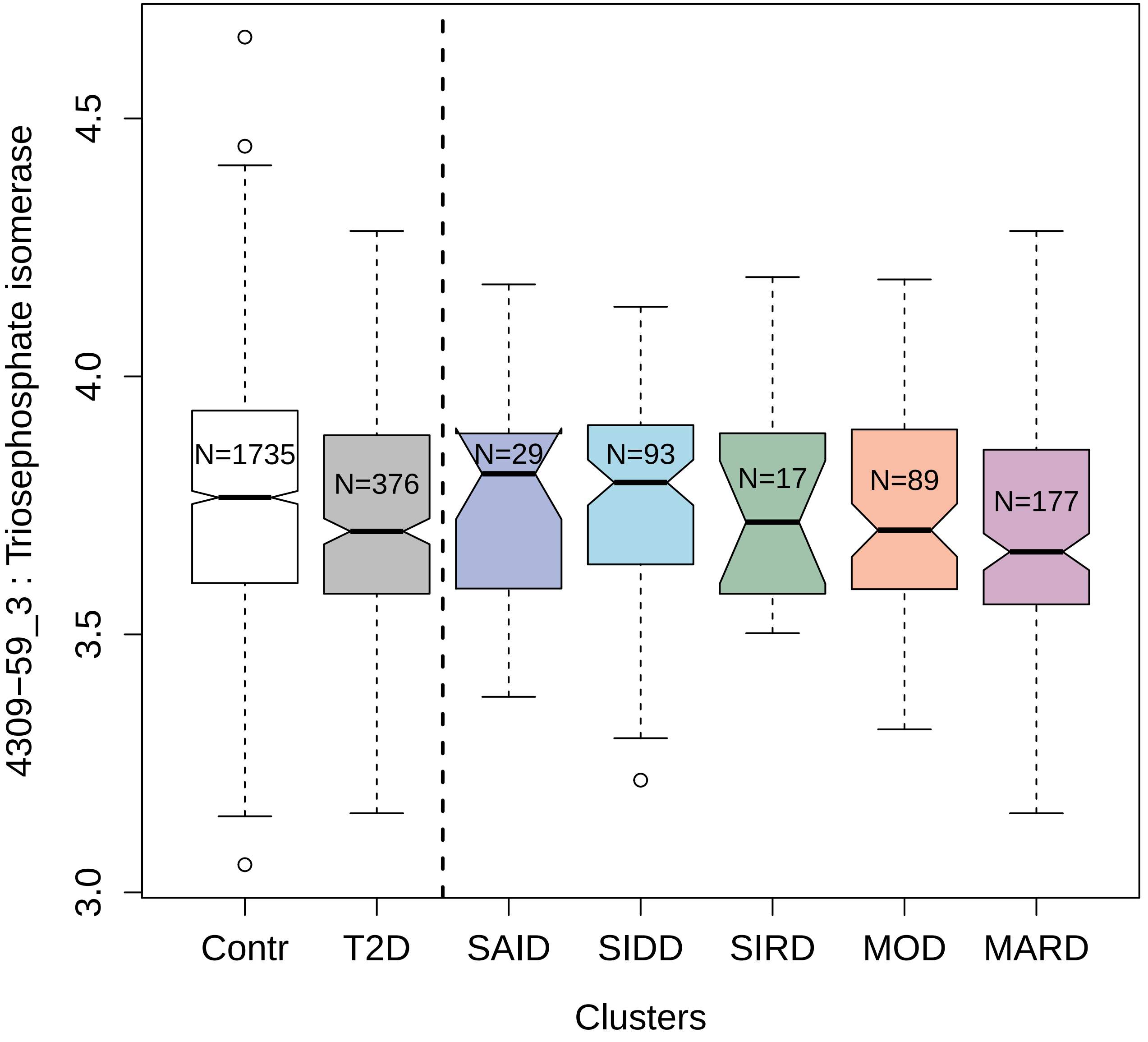

4309−59_3 : Triosephosphate isomerase
3.0
3.5
4.0
4.5
N=1735
N=376
N=29
N=93
N=17
N=89
N=177
Contr
T2D
SAID
SIDD
SIRD
MOD
MARD
Clusters

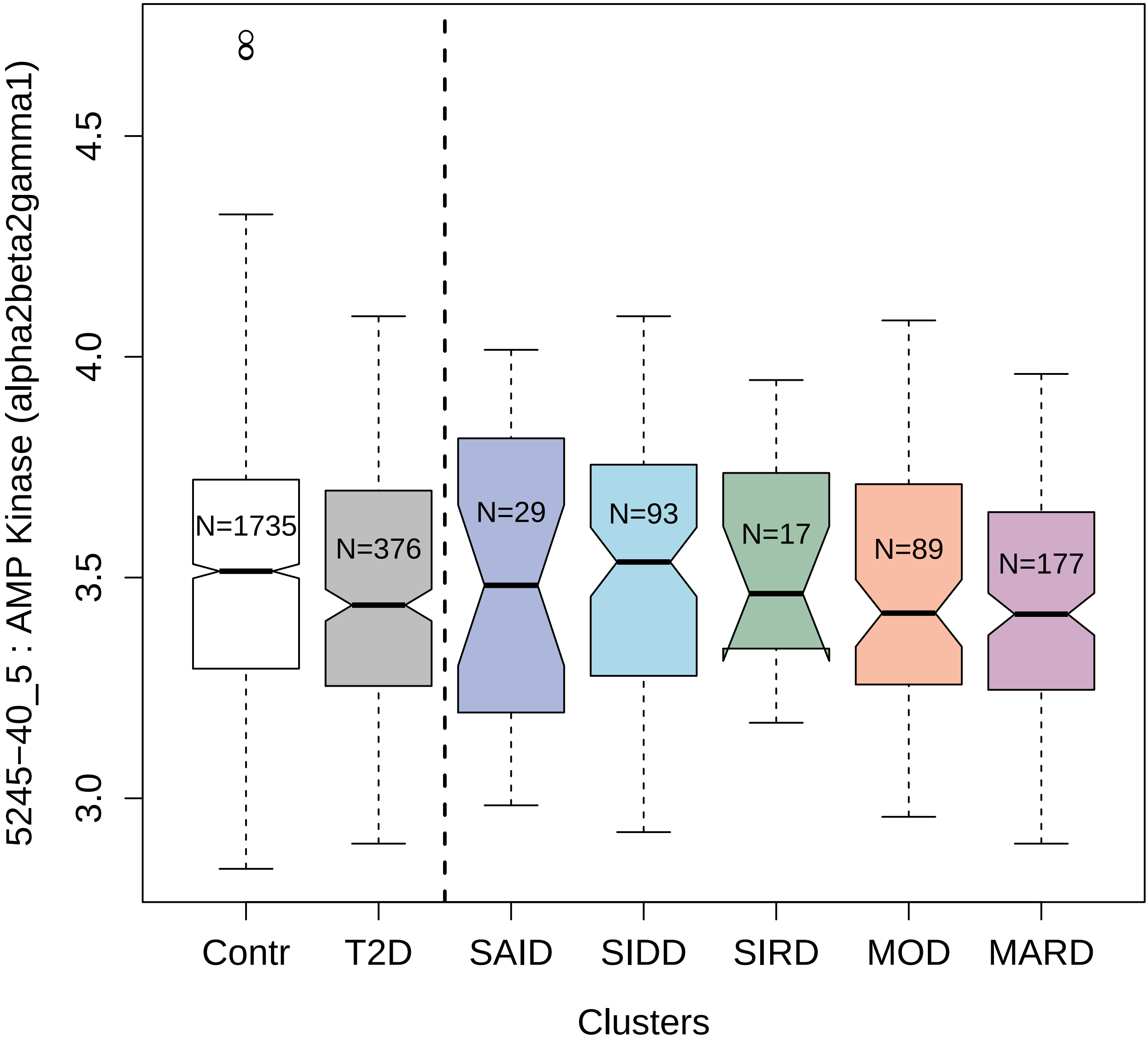

5245−40_5 : AMP Kinase (alpha2beta2gamma1)
4.5
4.0
3.5
3.0
N=1735
N=376
N=29
N=93
N=17
N=89
N=177
Contr
T2D
SAID
SIDD
SIRD
MOD
MARD
Clusters

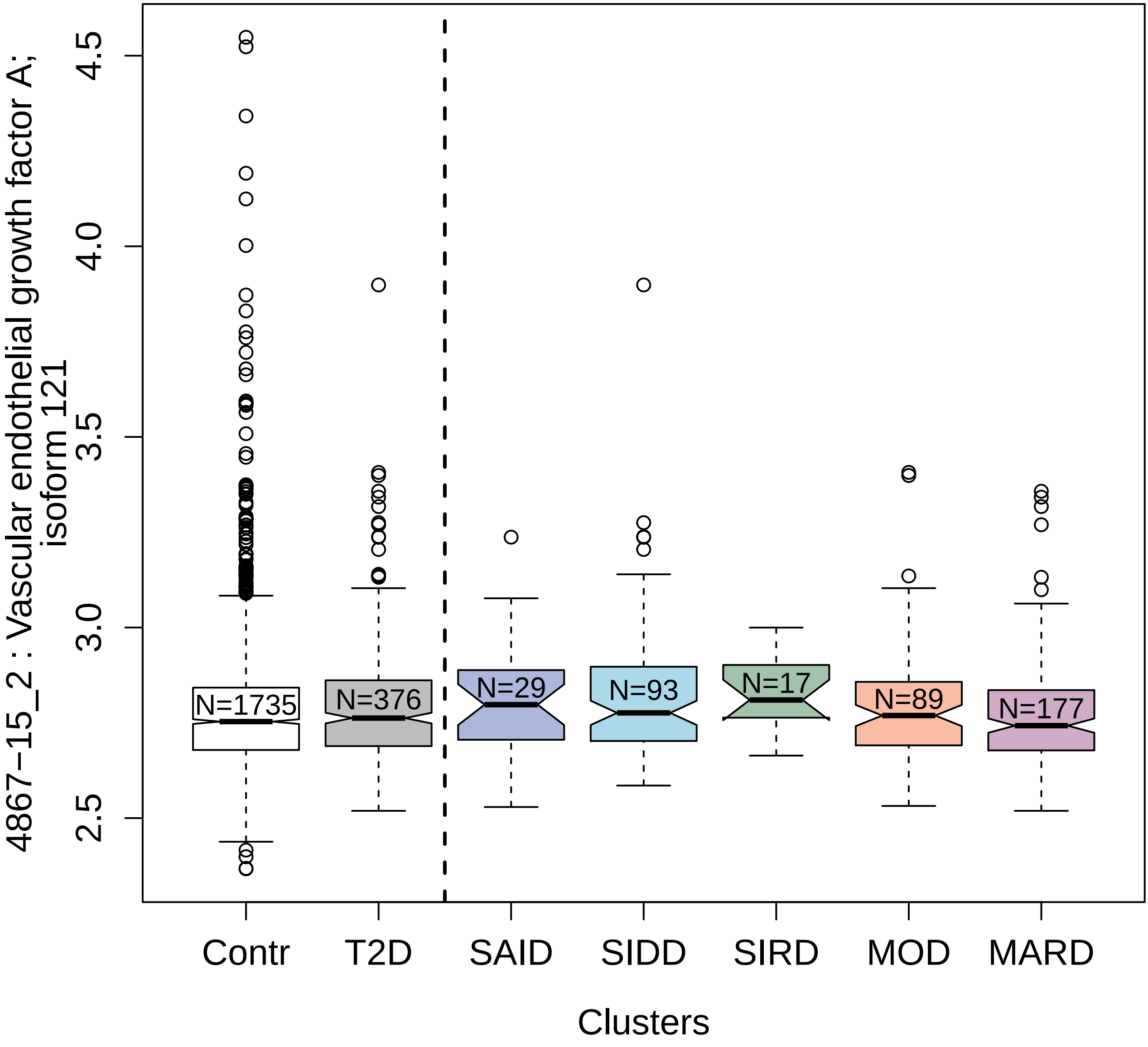
4867−15_2 : Vascular endothelial growth factor A;
isoform 121
2.5
3.0
3.5
4.0
4.5
N=1735
N=376
N=29
N=93
N=17
N=89
N=177
Contr
T2D
SAID
SIDD
SIRD
MOD
MARD
Clusters

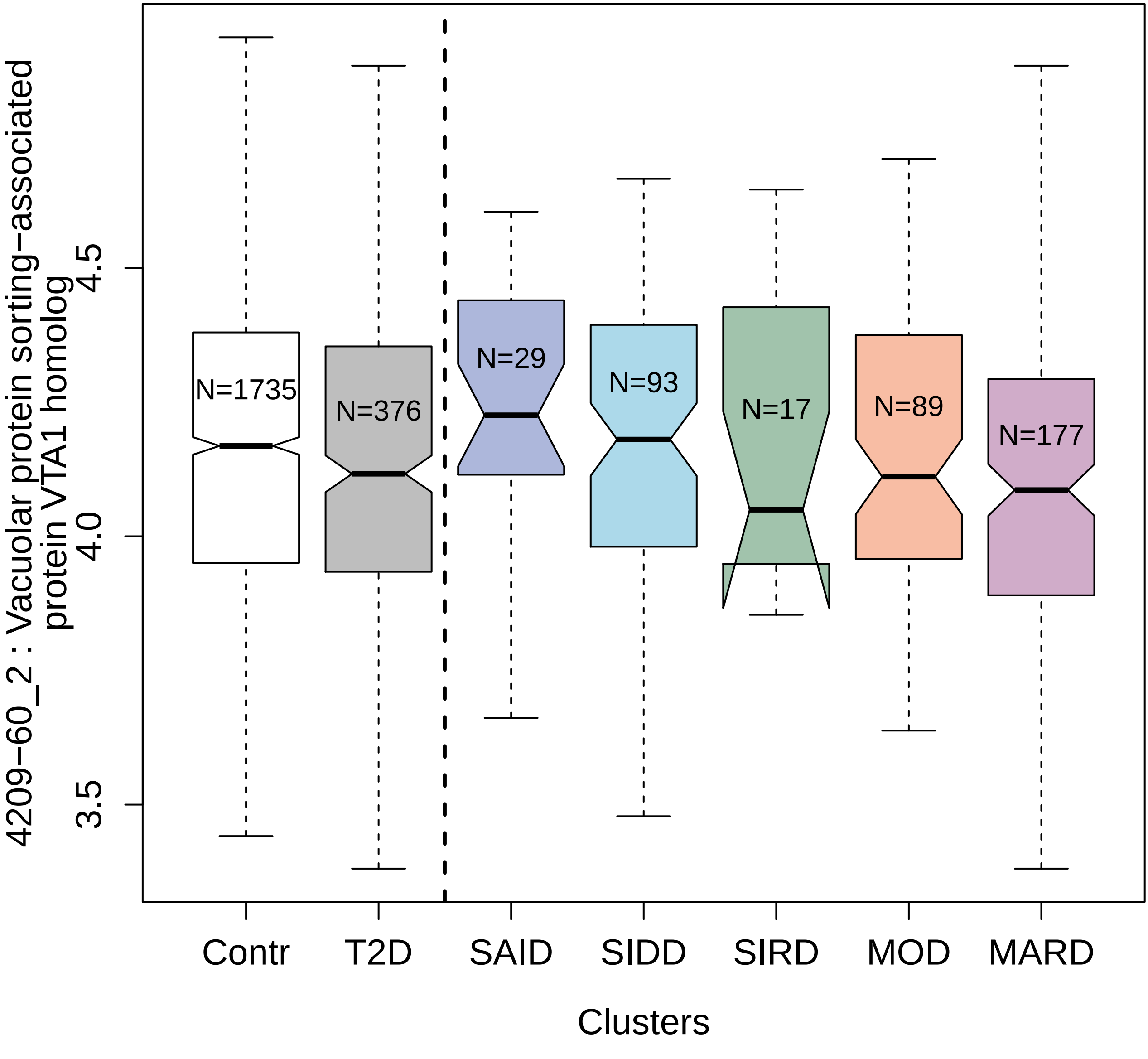

4209−60_2 : Vacuolar protein sorting−associated protein VTA1 homolog
3.5
4.0
4.5
N=1735
N=376
N=29
N=93
N=17
N=89
N=177
Contr
T2D
SAID
SIDD
SIRD
MOD
MARD
Clusters

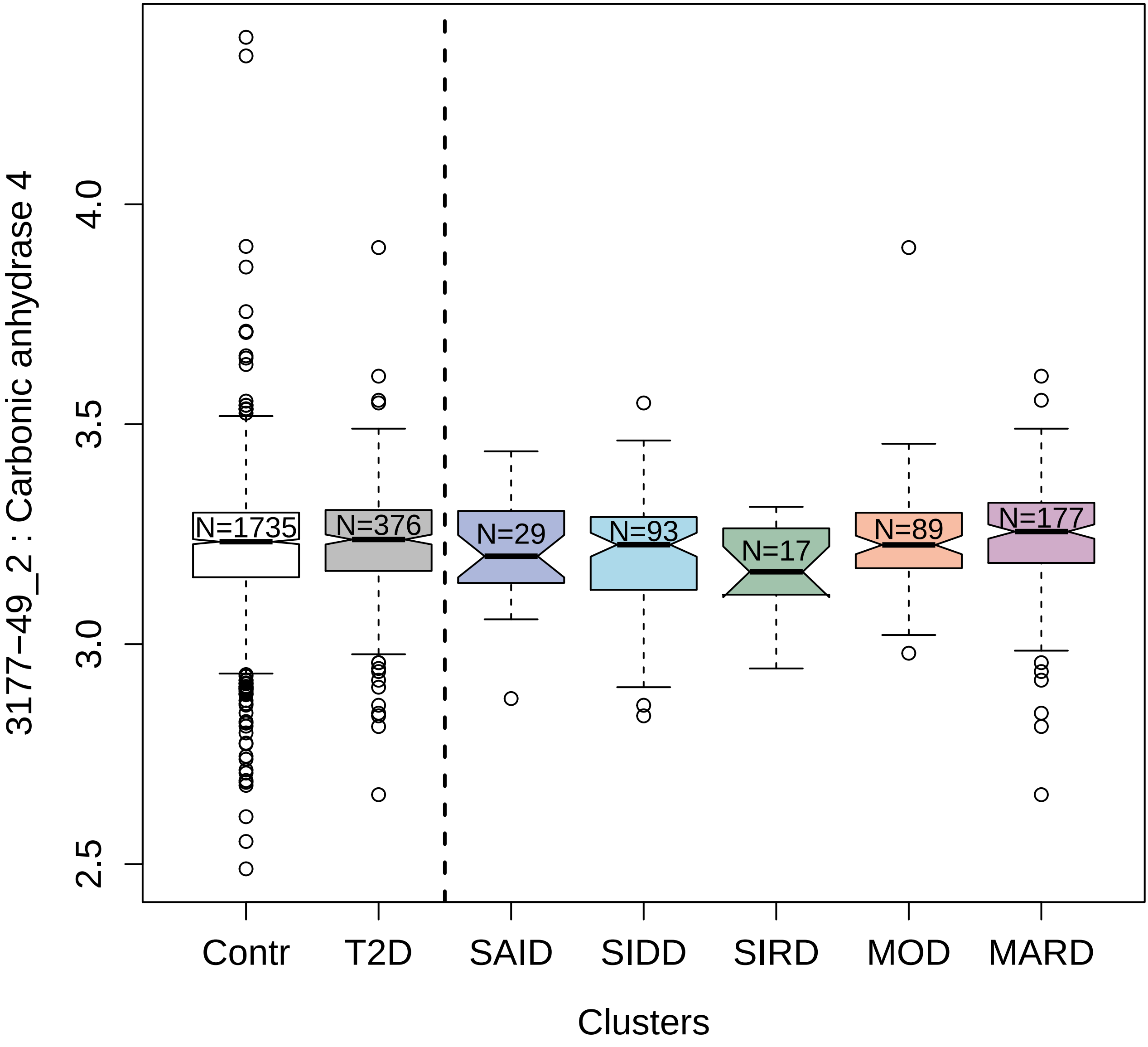
3177−49_2 : Carbonic anhydrase 4
Clusters
Contr
T2D
SAID
SIDD
SIRD
MOD
MARD
N=1735
N=376
N=29
N=93
N=17
N=89
N=177
2.5
3.0
3.5
4.0

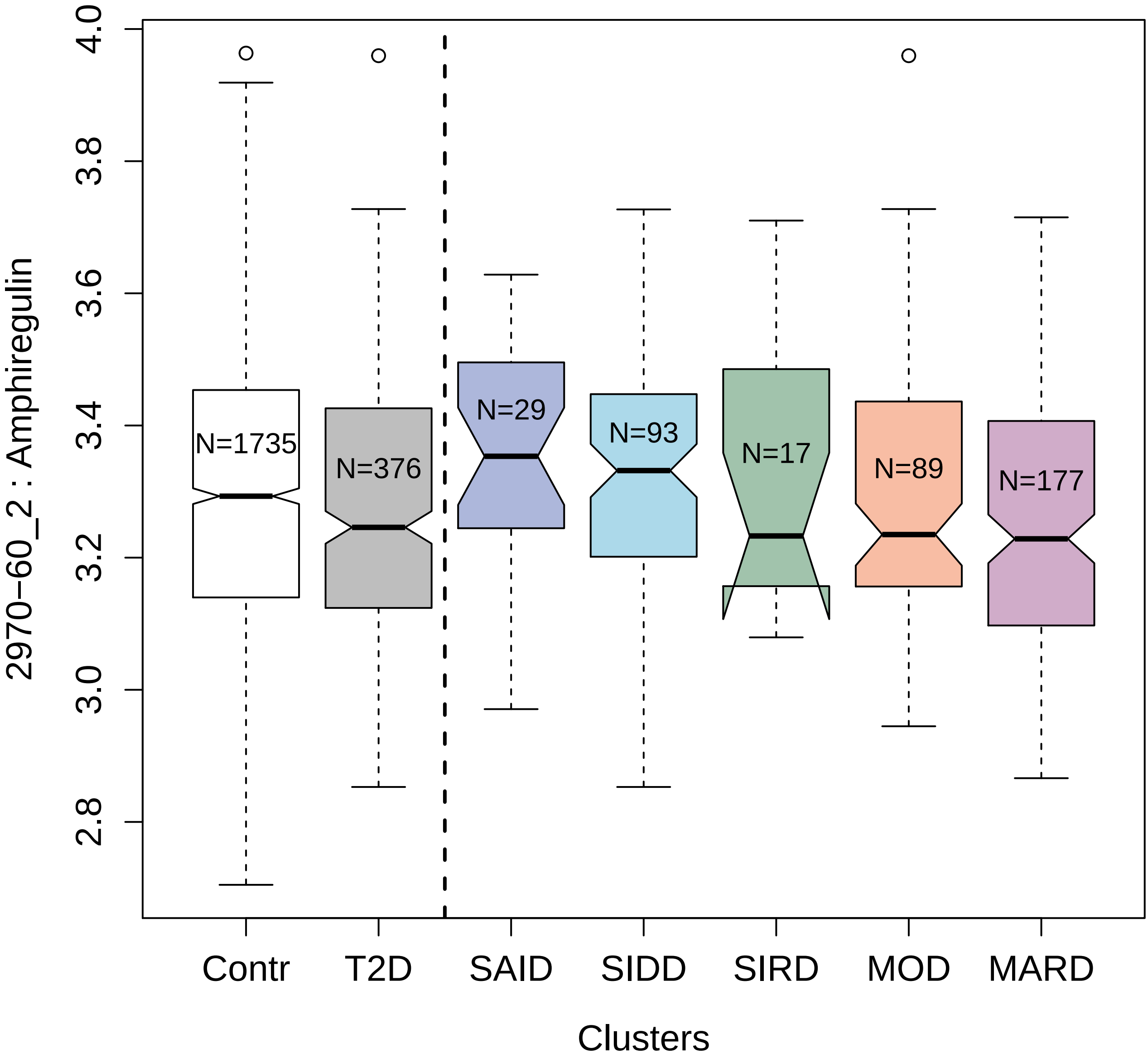

2970−60_2 : Amphiregulin
4.0
3.8
3.6
3.4
3.2
3.0
2.8
N=1735
N=376
N=29
N=93
N=17
N=89
N=177
Contr
T2D
SAID
SIDD
SIRD
MOD
MARD
Clusters

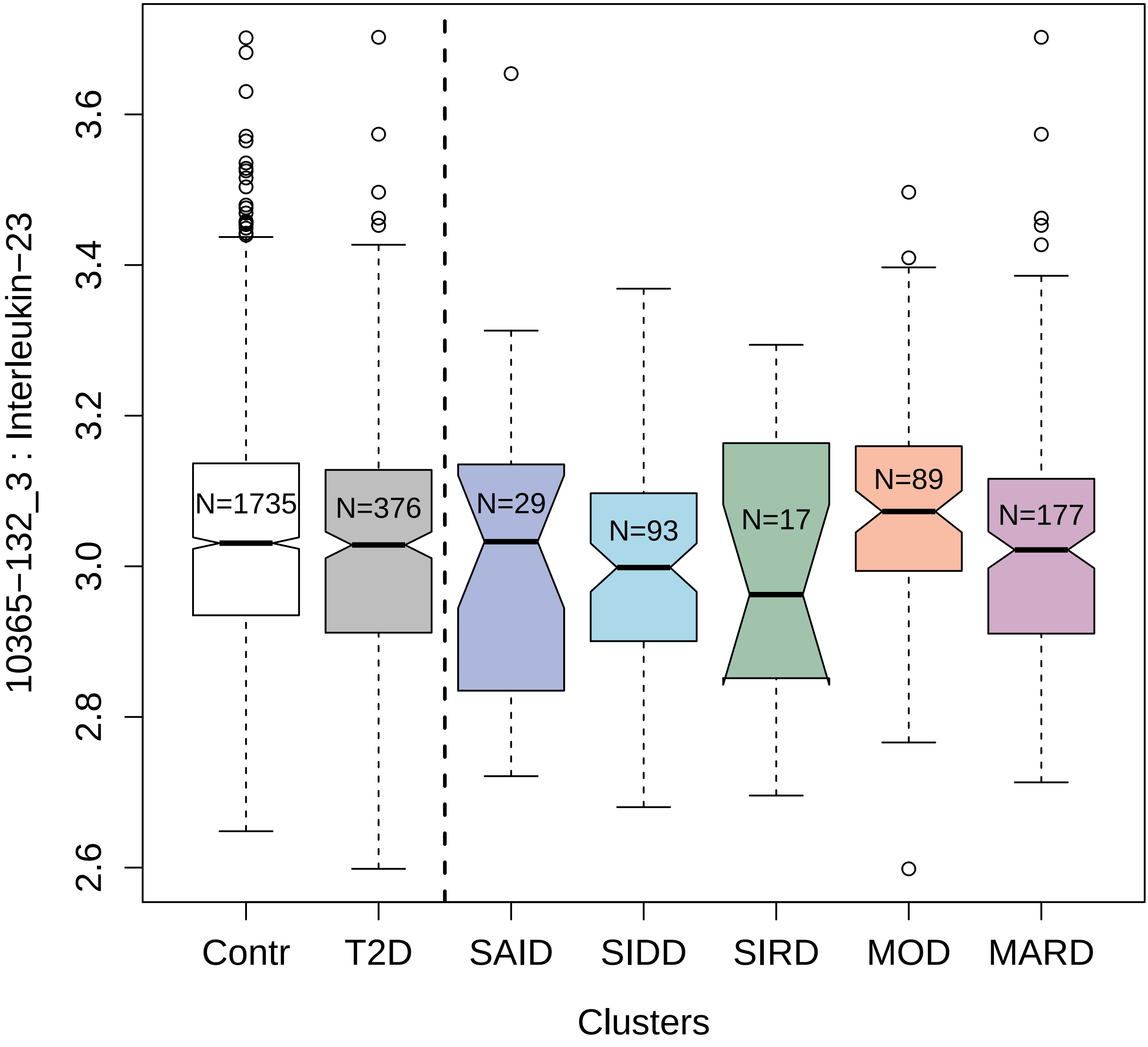

10365−132_3 : Interleukin−23
3.6
3.4
3.2
3.0
2.8
2.6
N=1735
N=376
N=29
N=93
N=17
N=89
N=177
Contr
T2D
SAID
SIDD
SIRD
MOD
MARD
Clusters

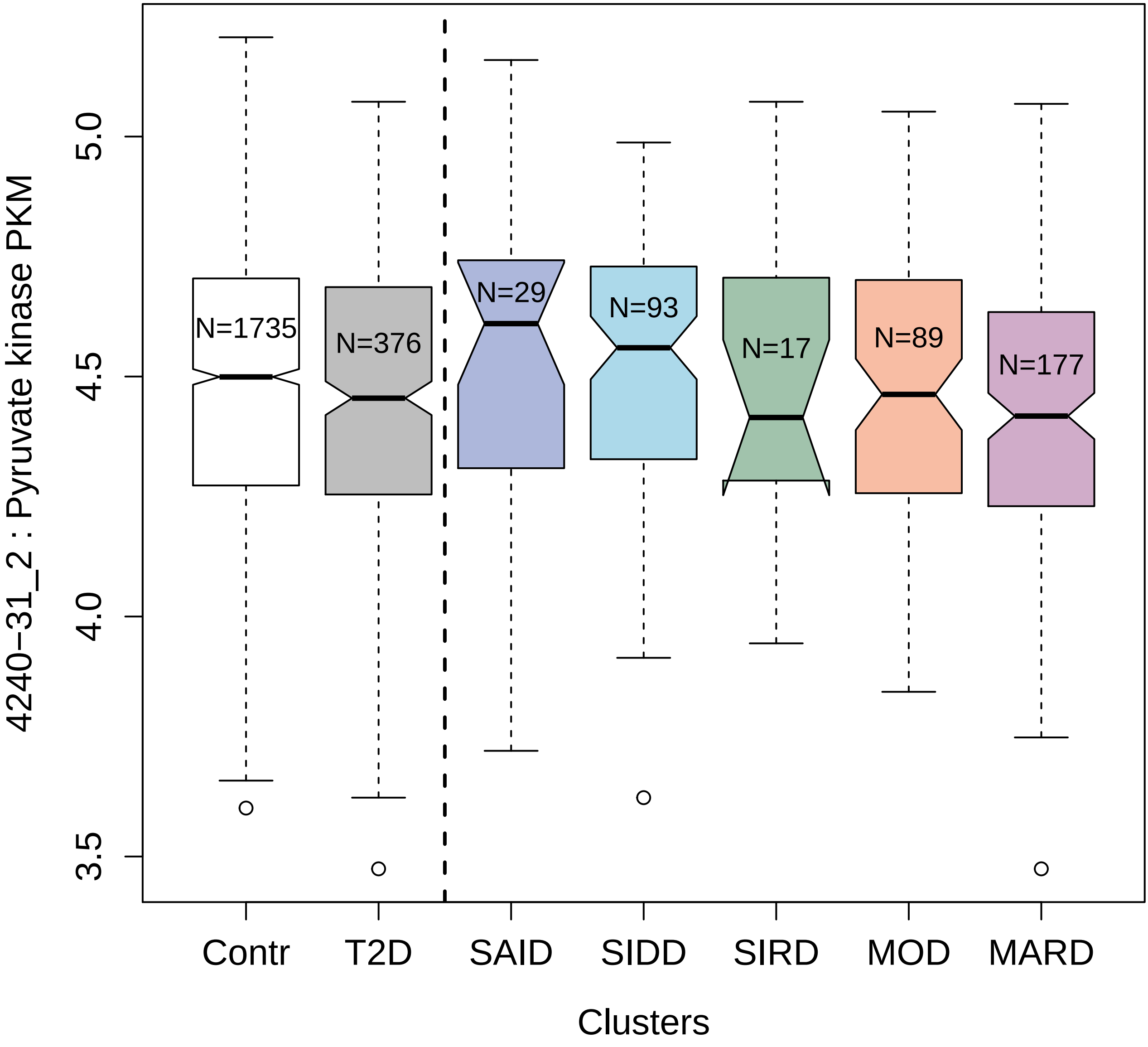

4240−31_2 : Pyruvate kinase PKM
5.0
4.5
4.0
3.5
N=1735
N=376
N=29
N=93
N=17
N=89
N=177
Contr
T2D
SAID
SIDD
SIRD
MOD
MARD
Clusters

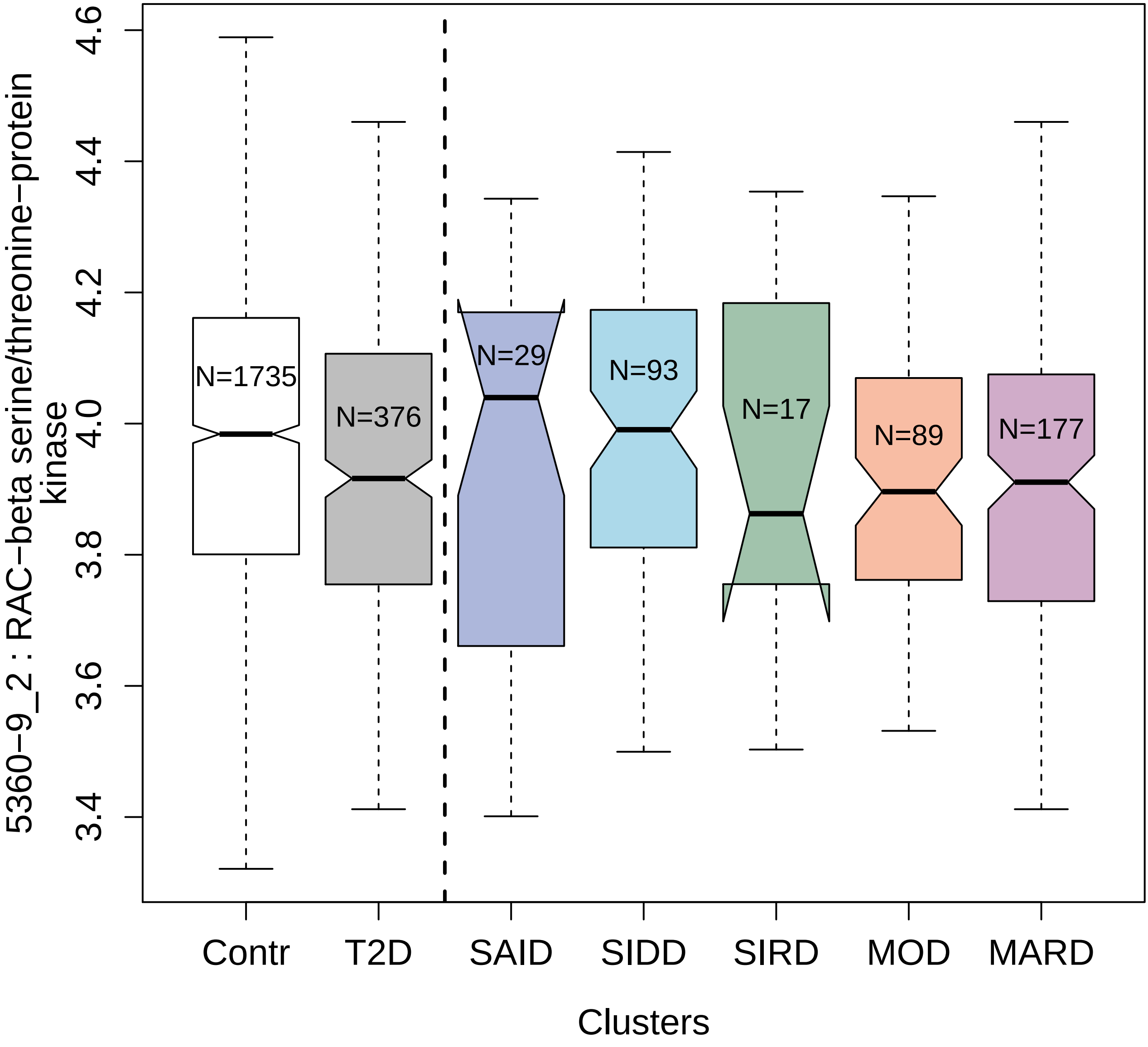
5360−9_2 : RAC−beta serine/threonine−protein kinase
4.6
4.4
4.2
4.0
3.8
3.6
3.4
N=1735
N=376
N=29
N=93
N=17
N=89
N=177
Contr
T2D
SAID
SIDD
SIRD
MOD
MARD
Clusters

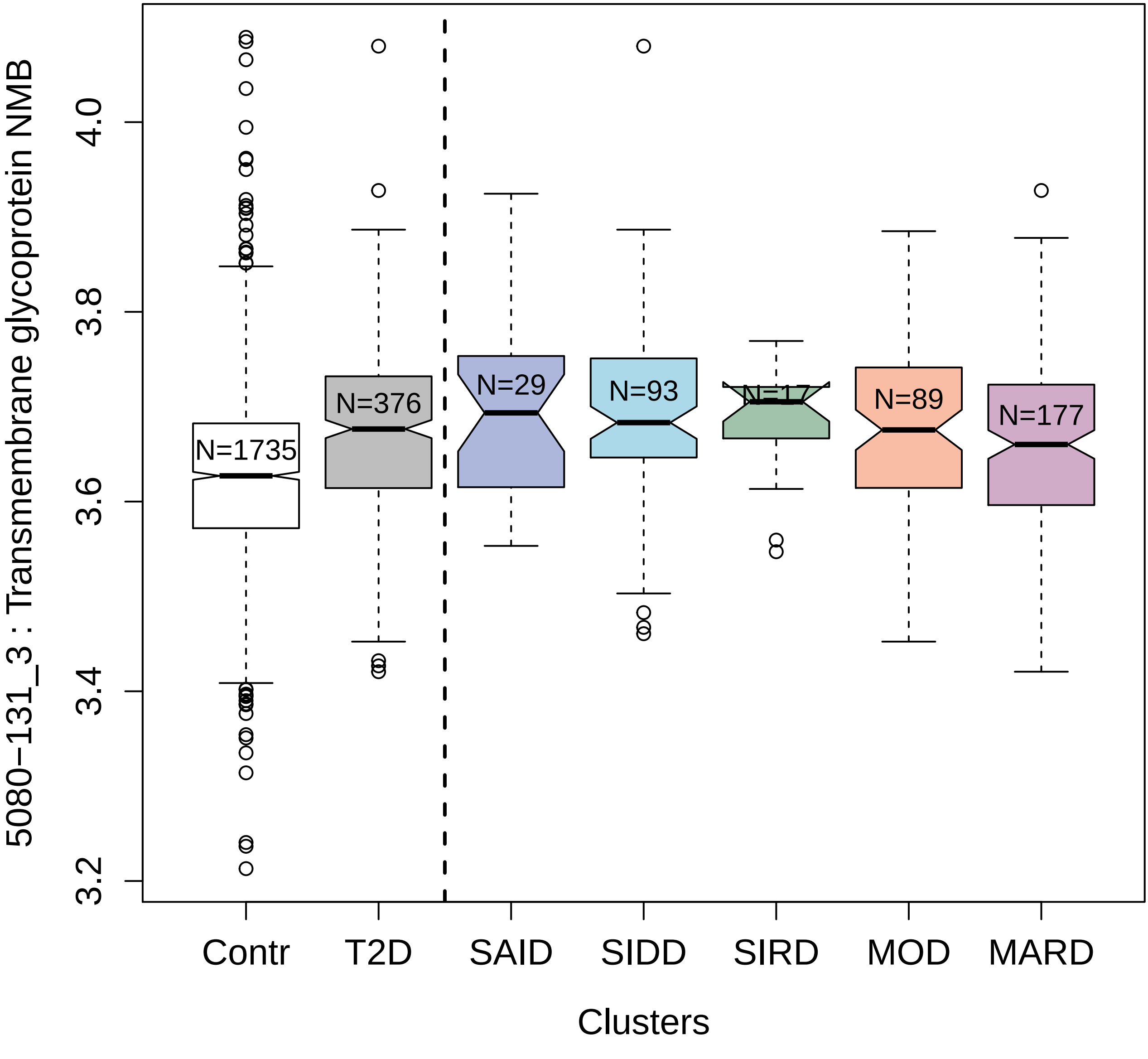

5080−131_3 : Transmembrane glycoprotein NMB
4.0
3.8
3.6
3.4
3.2
N=1735
N=376
N=29
N=93
N=17
N=89
N=177
Contr
T2D
SAID
SIDD
SIRD
MOD
MARD
Clusters

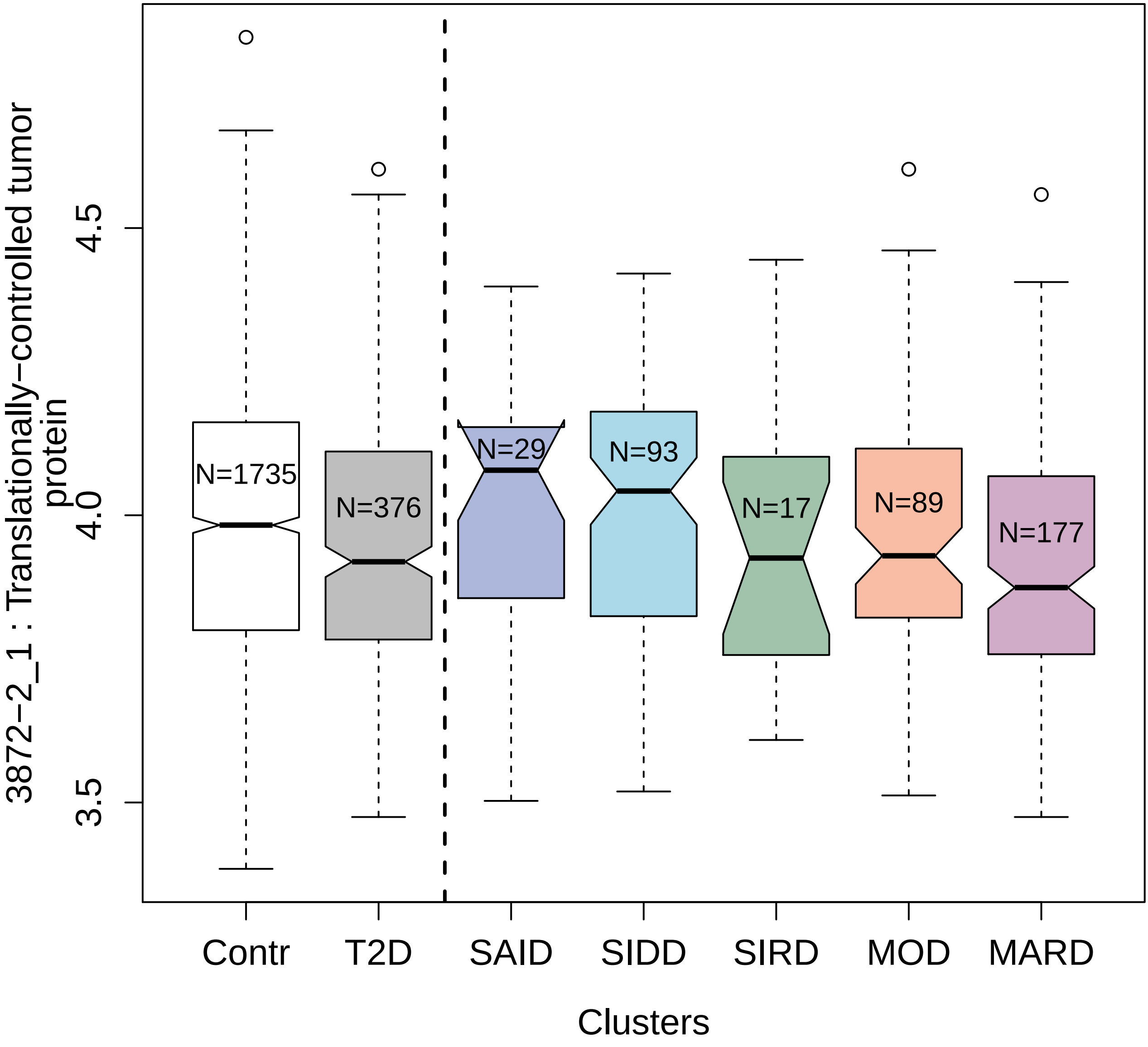
3872−2_1 : Translationally−controlled tumor protein
3.5
4.0
4.5
N=1735
N=376
N=29
N=93
N=17
N=89
N=177
Contr
T2D
SAID
SIDD
SIRD
MOD
MARD
Clusters

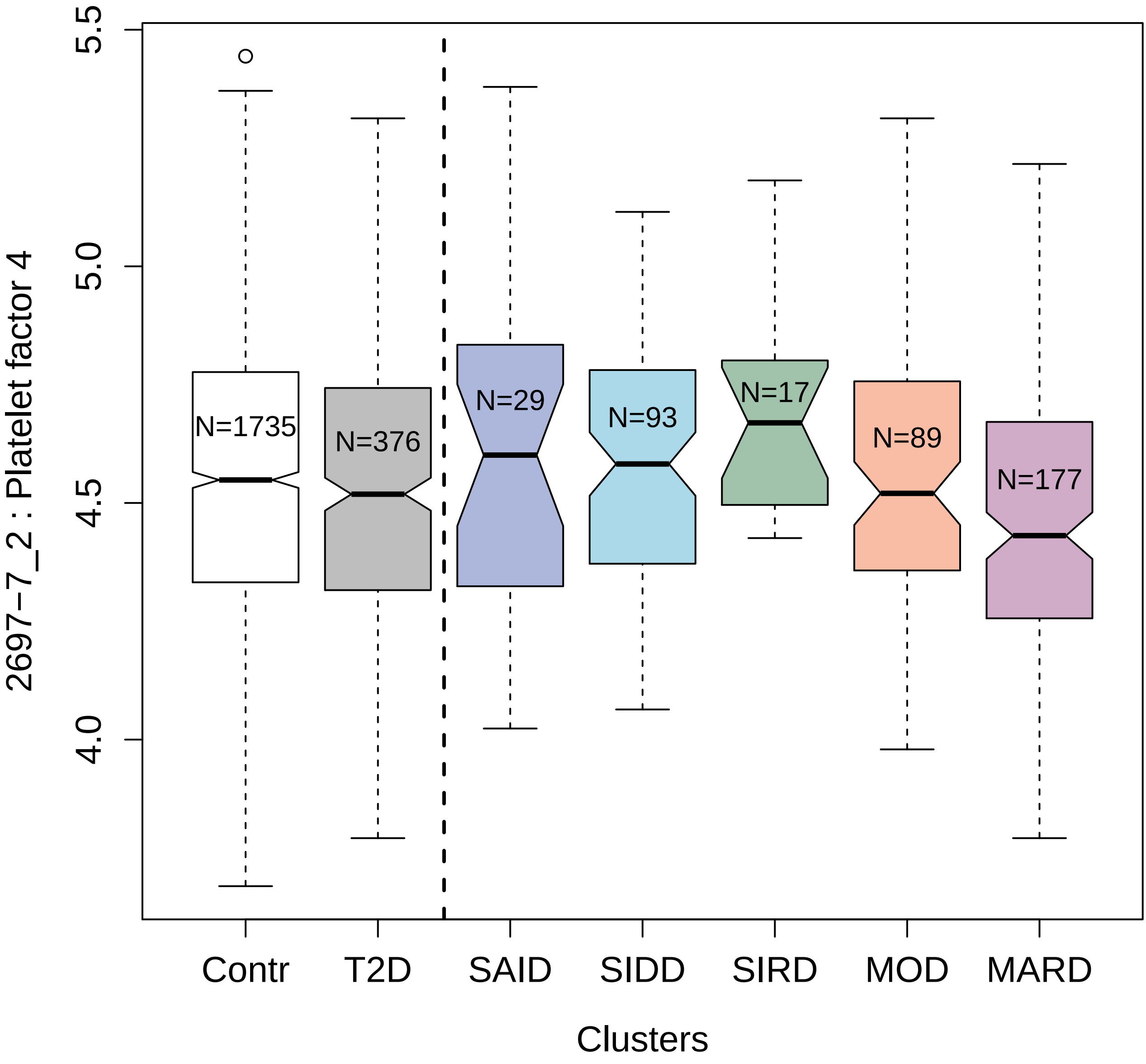
2697−7_2 : Platelet factor 4
5.5
5.0
4.5
4.0
N=1735
N=376
N=29
N=93
N=17
N=89
N=177
Contr
T2D
SAID
SIDD
SIRD
MOD
MARD
Clusters

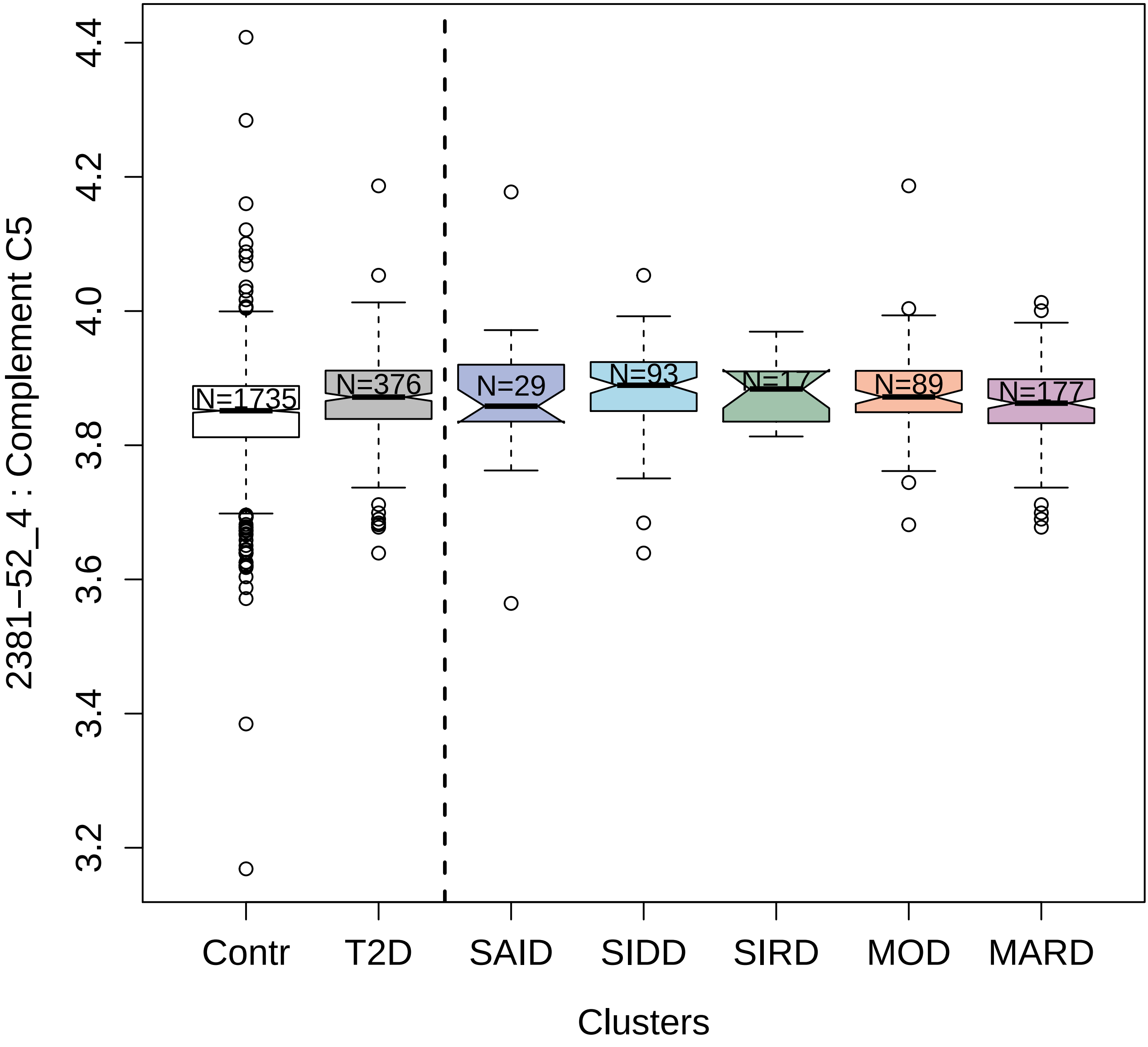

2381−52_4 : Complement C5
4.4
4.2
4.0
3.8
3.6
3.4
3.2
N=1735
N=376
N=29
N=93
N=17
N=89
N=177
Contr
T2D
SAID
SIDD
SIRD
MOD
MARD
Clusters

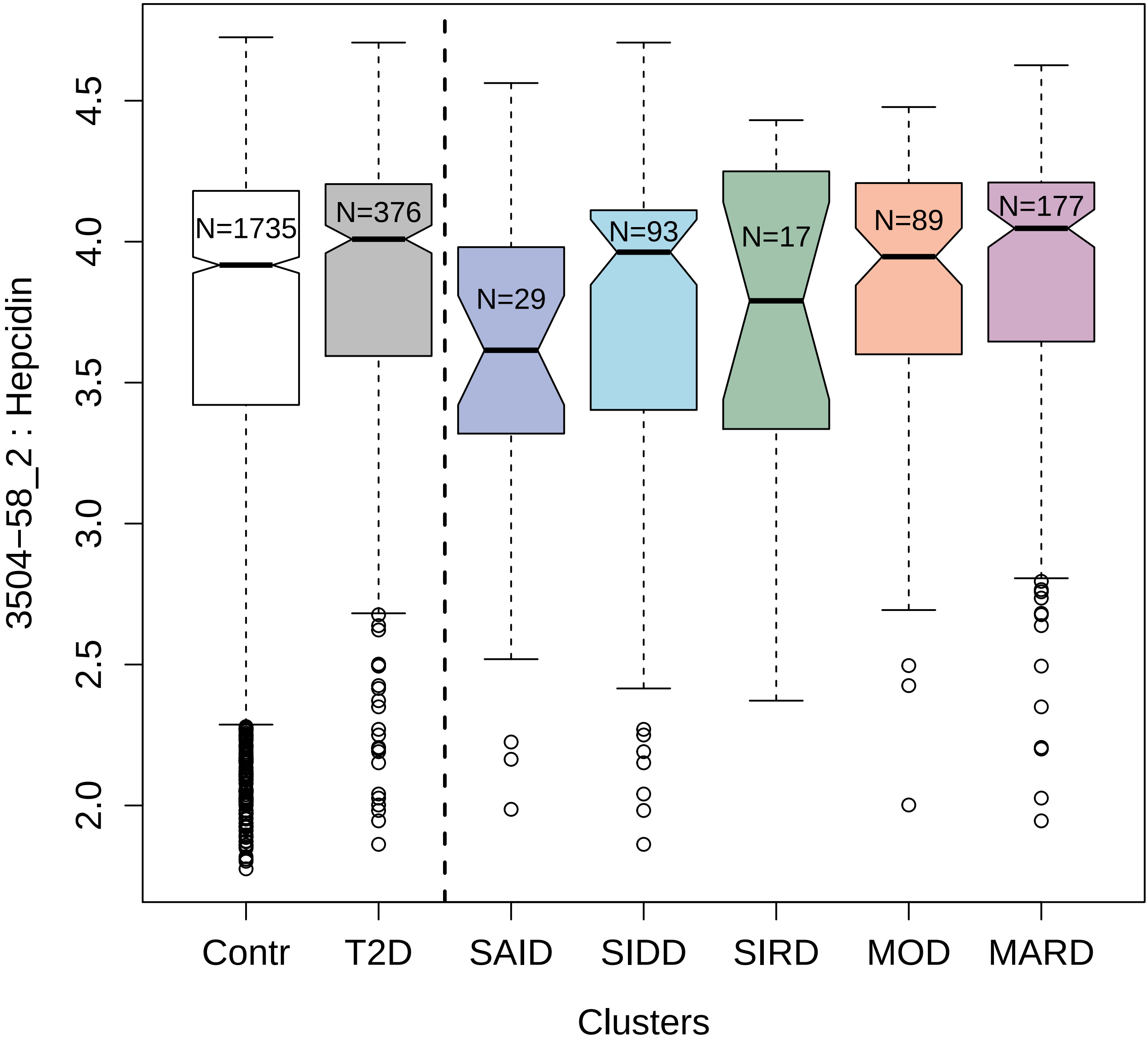

3504−58_2 : Hepcidin
4.5
4.0
3.5
3.0
2.5
2.0
N=1735
N=376
N=29
N=93
N=17
N=89
N=177
Contr
T2D
SAID
SIDD
SIRD
MOD
MARD
Clusters

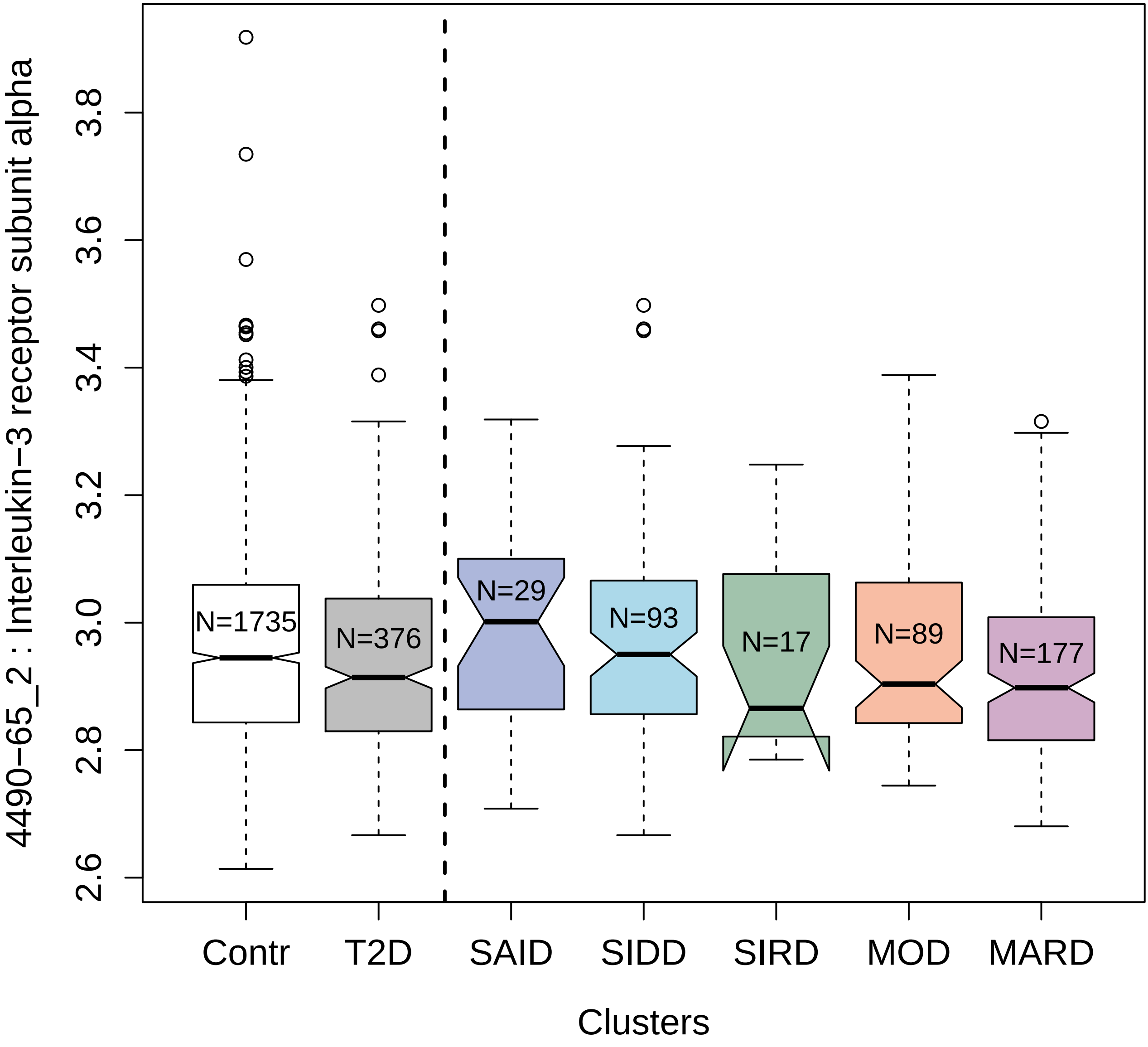

4490−65_2 : Interleukin−3 receptor subunit alpha
2.6
2.8
3.0
3.2
3.4
3.6
3.8
N=1735
N=376
N=29
N=93
N=17
N=89
N=177
Contr
T2D
SAID
SIDD
SIRD
MOD
MARD
Clusters

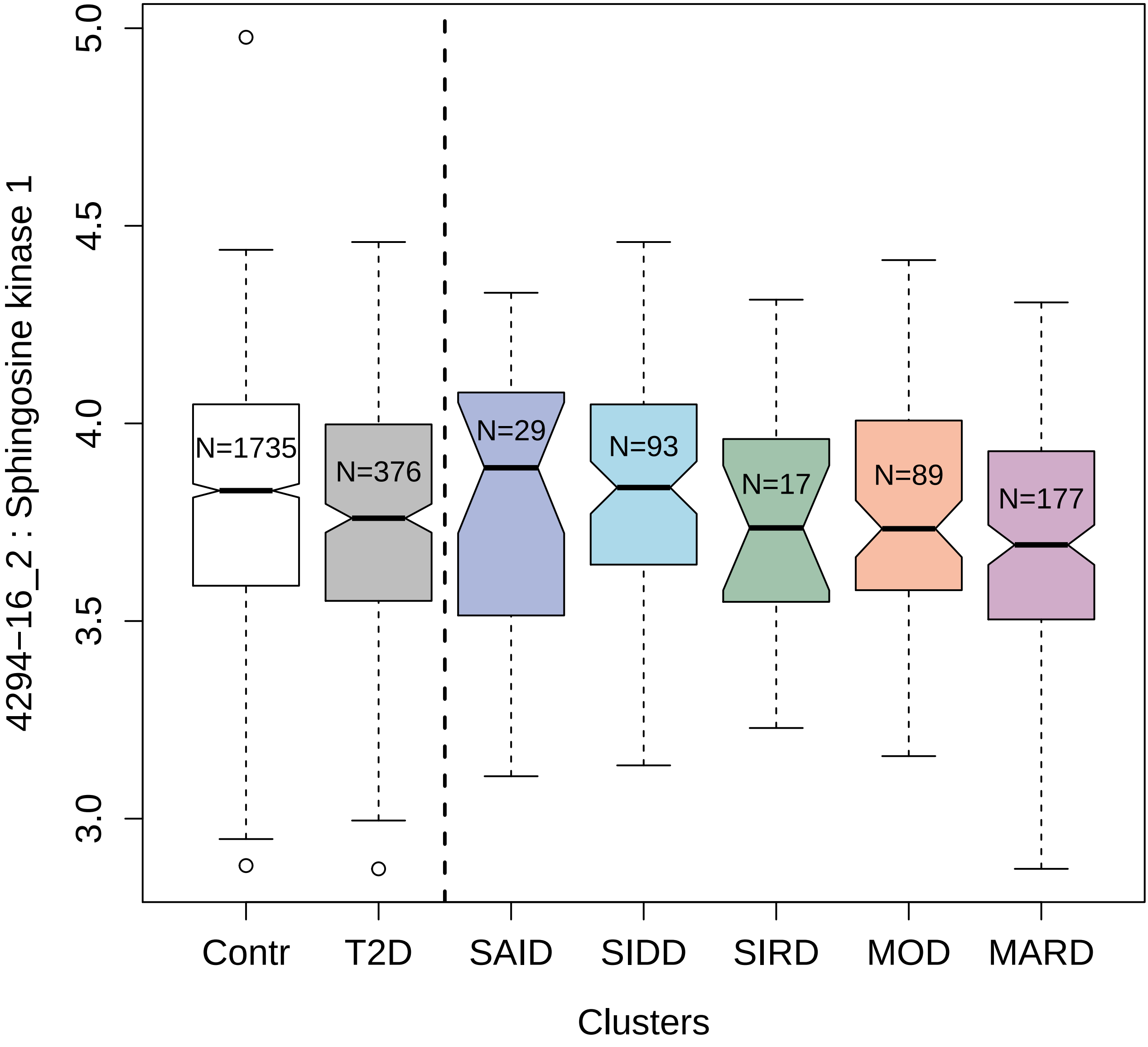

4294−16_2 : Sphingosine kinase 1
5.0
4.5
4.0
3.5
3.0
N=1735
N=376
N=29
N=93
N=17
N=89
N=177
Contr
T2D
SAID
SIDD
SIRD
MOD
MARD
Clusters

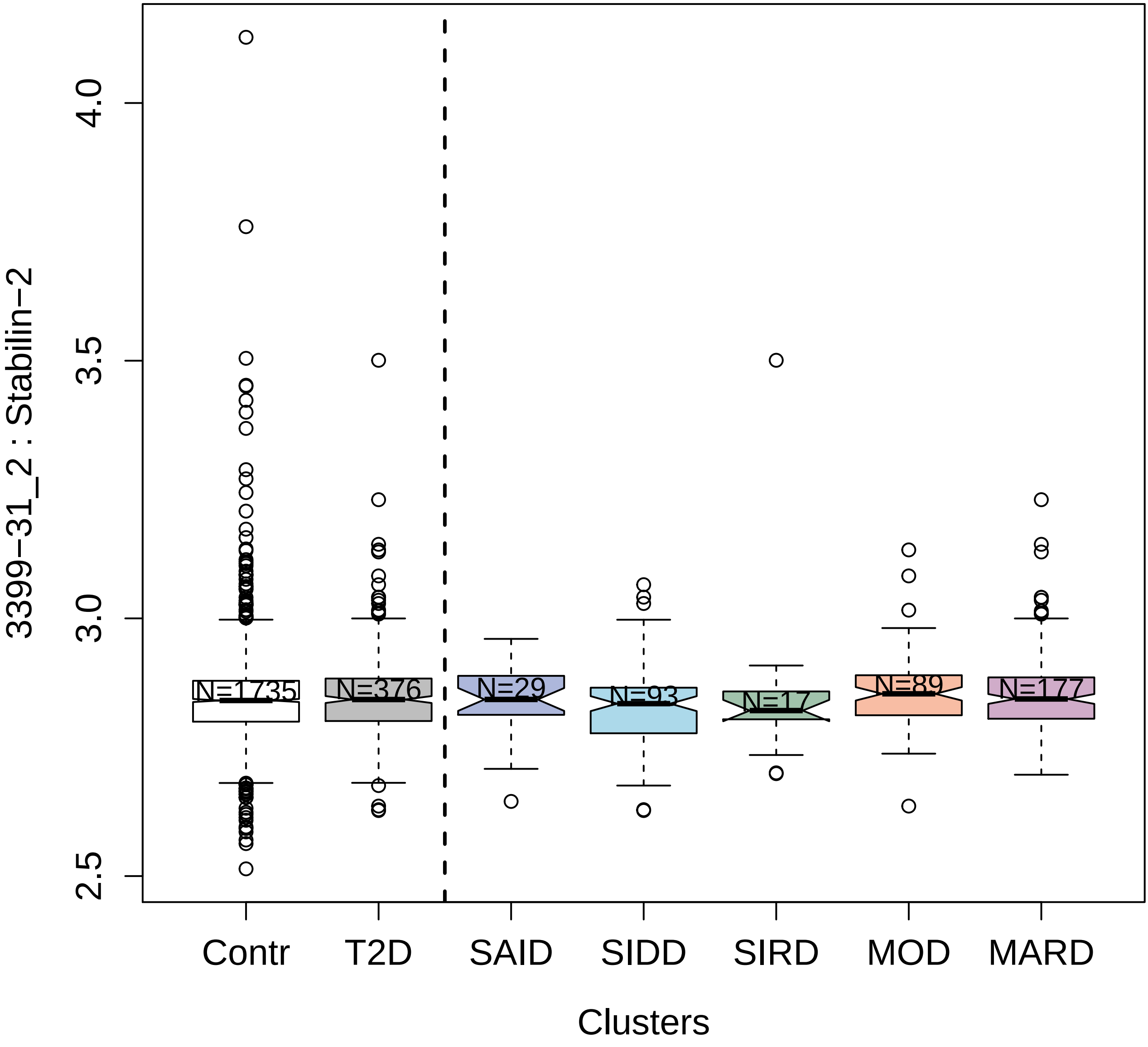
3399−31_2 : Stabilin−2
4.0
3.5
3.0
2.5
N=1735
N=376
N=29
N=93
N=17
N=89
N=177
Contr
T2D
SAID
SIDD
SIRD
MOD
MARD
Clusters

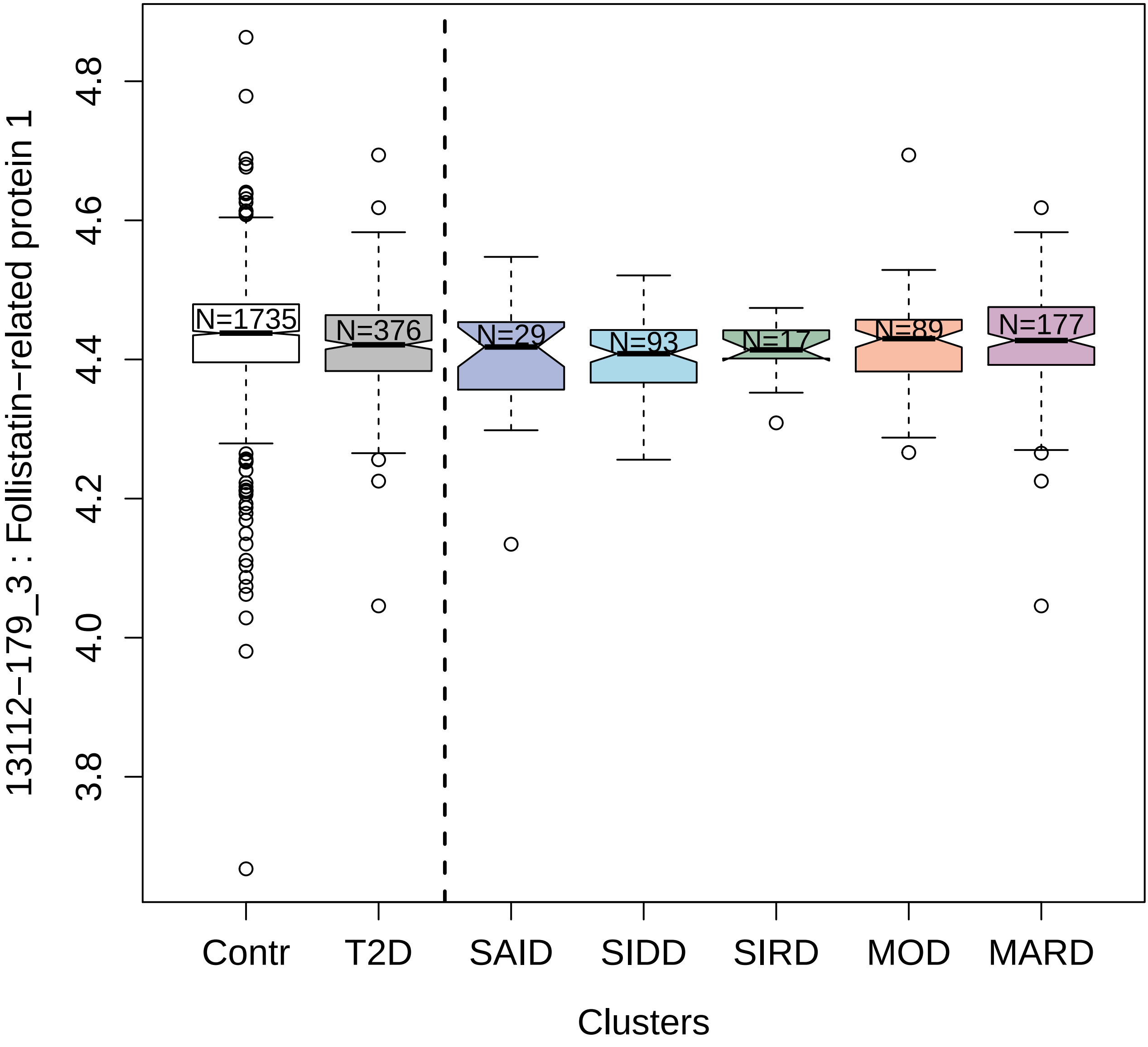

13112−179_3 : Follistatin−related protein 1
3.8
4.0
4.2
4.4
4.6
4.8
N=1735
N=376
N=29
N=93
N=17
N=89
N=177
Contr
T2D
SAID
SIDD
SIRD
MOD
MARD
Clusters

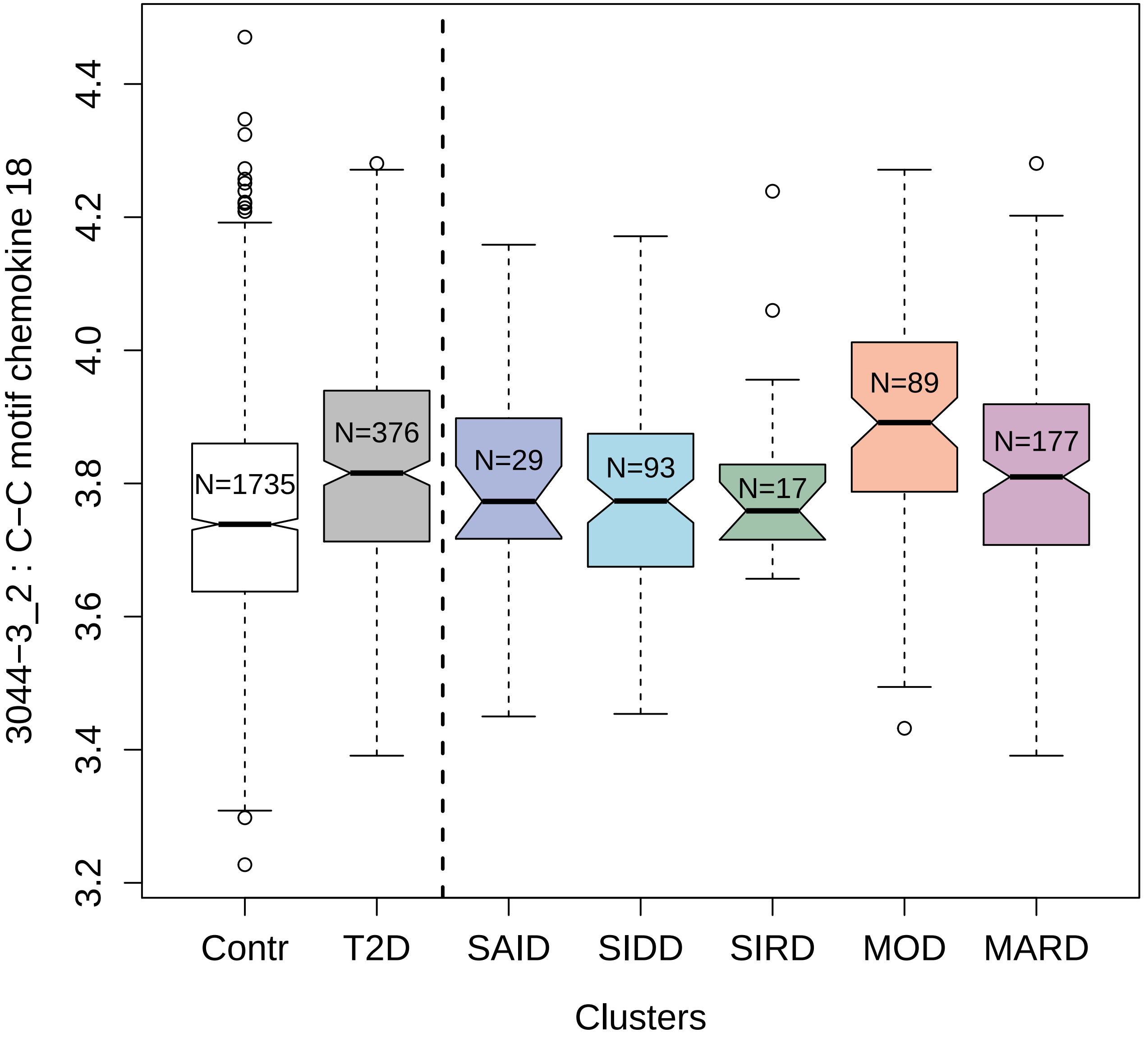

3044−3_2 : C−C motif chemokine 18
3.2
3.4
3.6
3.8
4.0
4.2
4.4
N=1735
N=376
N=29
N=93
N=17
N=89
N=177
Contr
T2D
SAID
SIDD
SIRD
MOD
MARD
Clusters

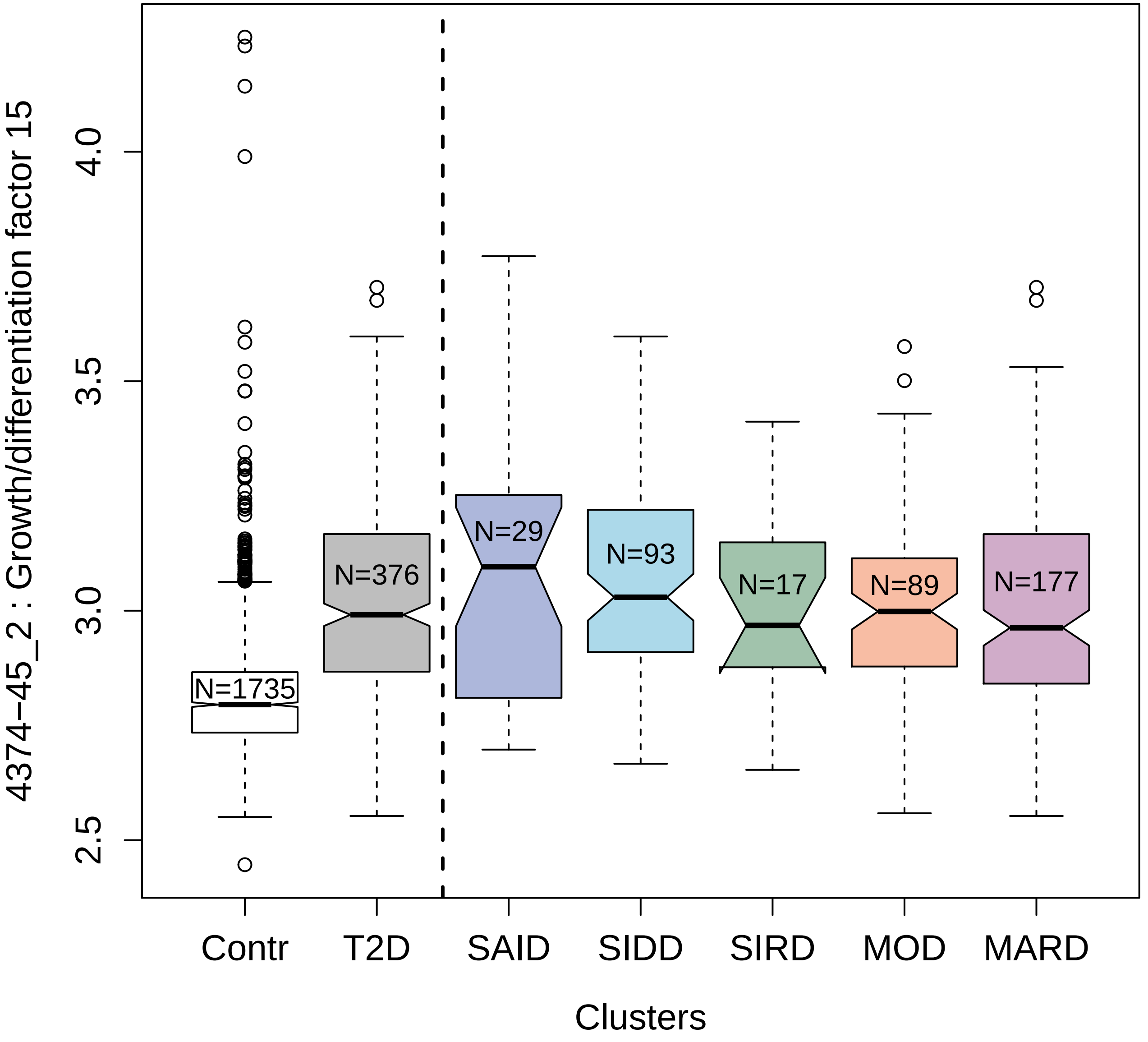

4374−45_2 : Growth/differentiation factor 15
2.5
3.0
3.5
4.0
N=1735
N=376
N=29
N=93
N=17
N=89
N=177
Contr
T2D
SAID
SIDD
SIRD
MOD
MARD
Clusters

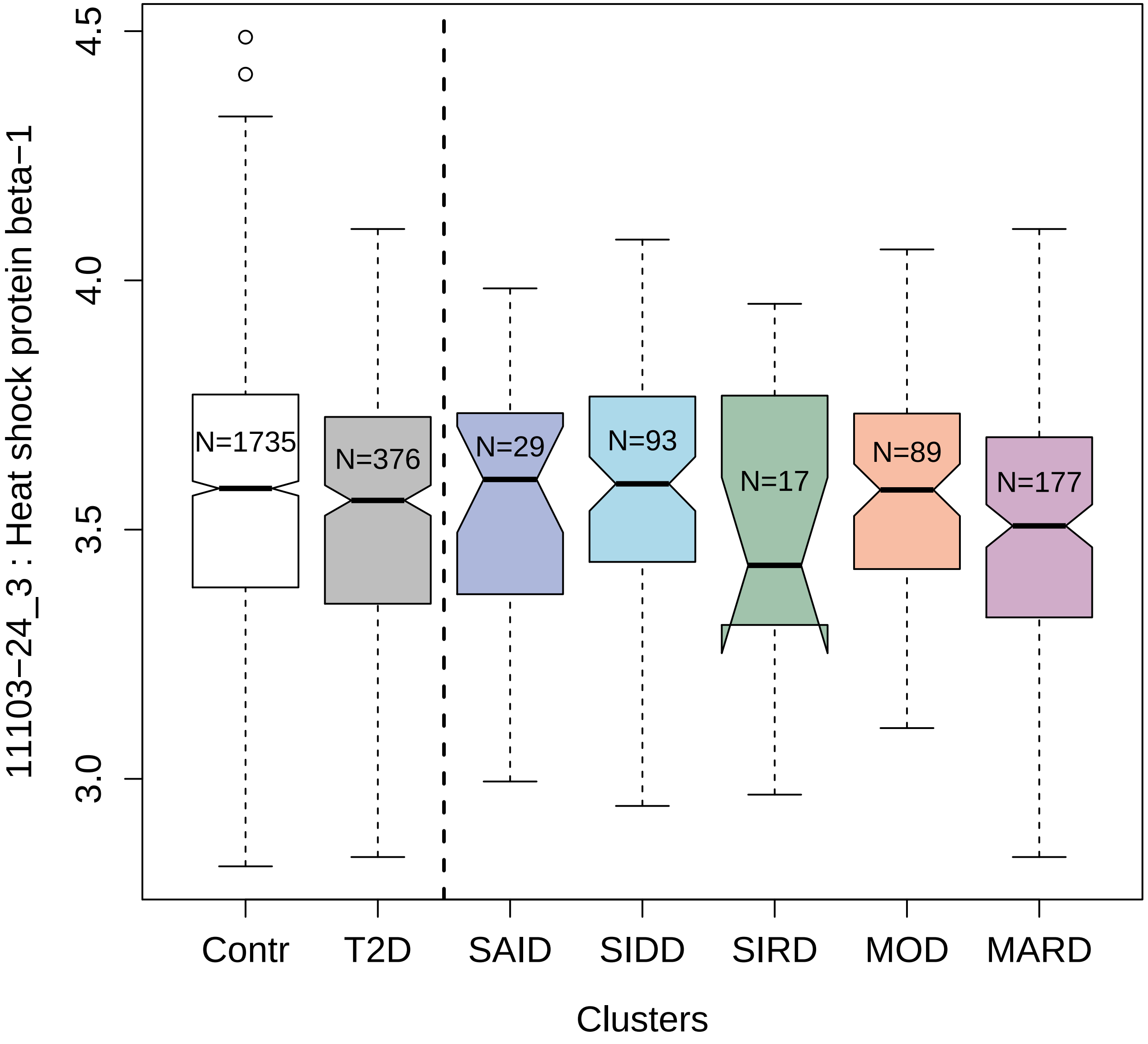

11103−24_3 : Heat shock protein beta−1
4.5
4.0
3.5
3.0
N=1735
N=376
N=29
N=93
N=17
N=89
N=177
Contr
T2D
SAID
SIDD
SIRD
MOD
MARD
Clusters

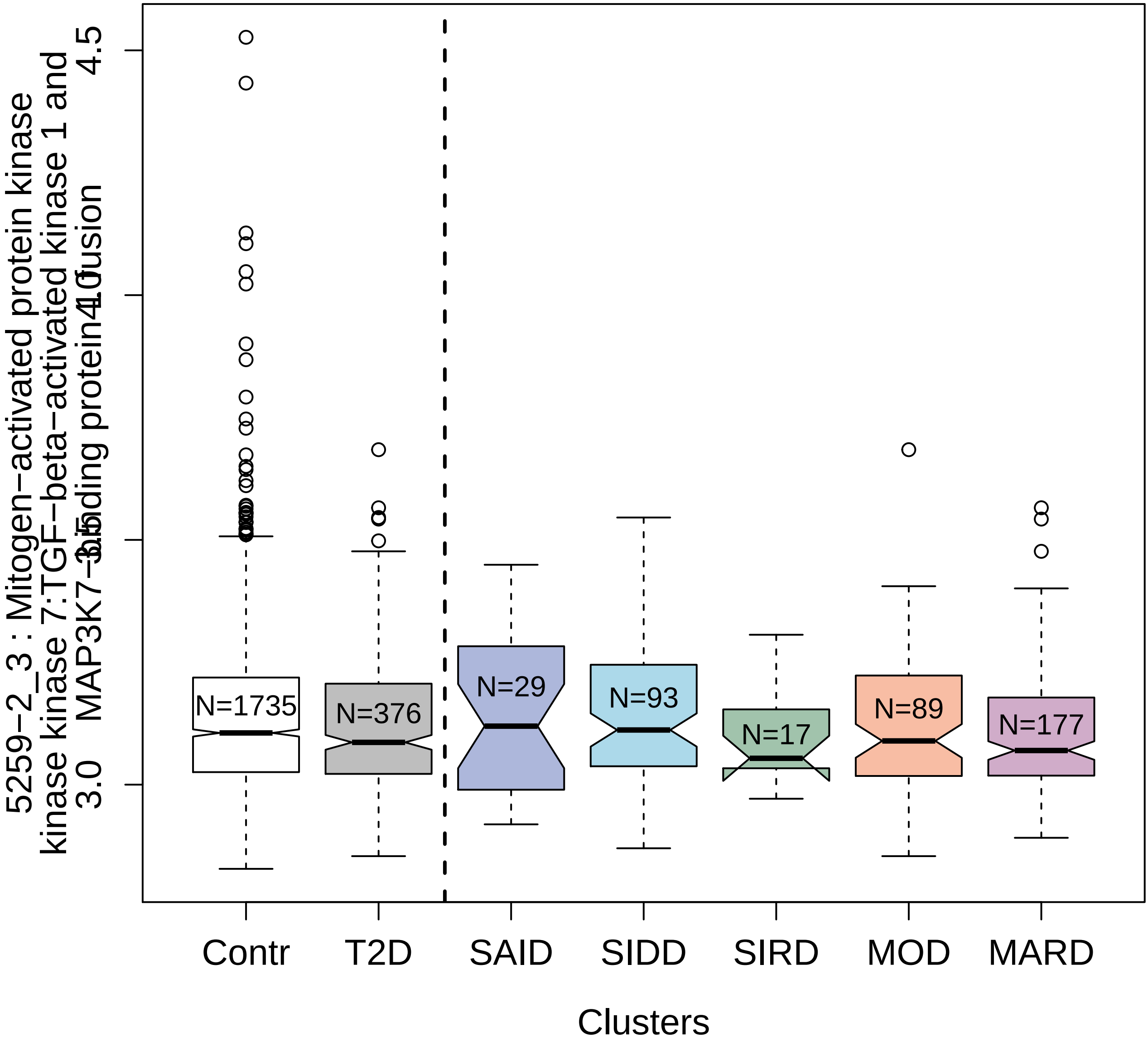

5259−2_3 : Mitogen−activated protein kinase kinase kinase 7:TGF−beta−activated kinase 1 and MAP3K7−binding protein 1 fusion
4.5
4.0
3.5
3.0
N=1735
N=376
N=29
N=93
N=17
N=89
N=177
Contr
T2D
SAID
SIDD
SIRD
MOD
MARD
Clusters

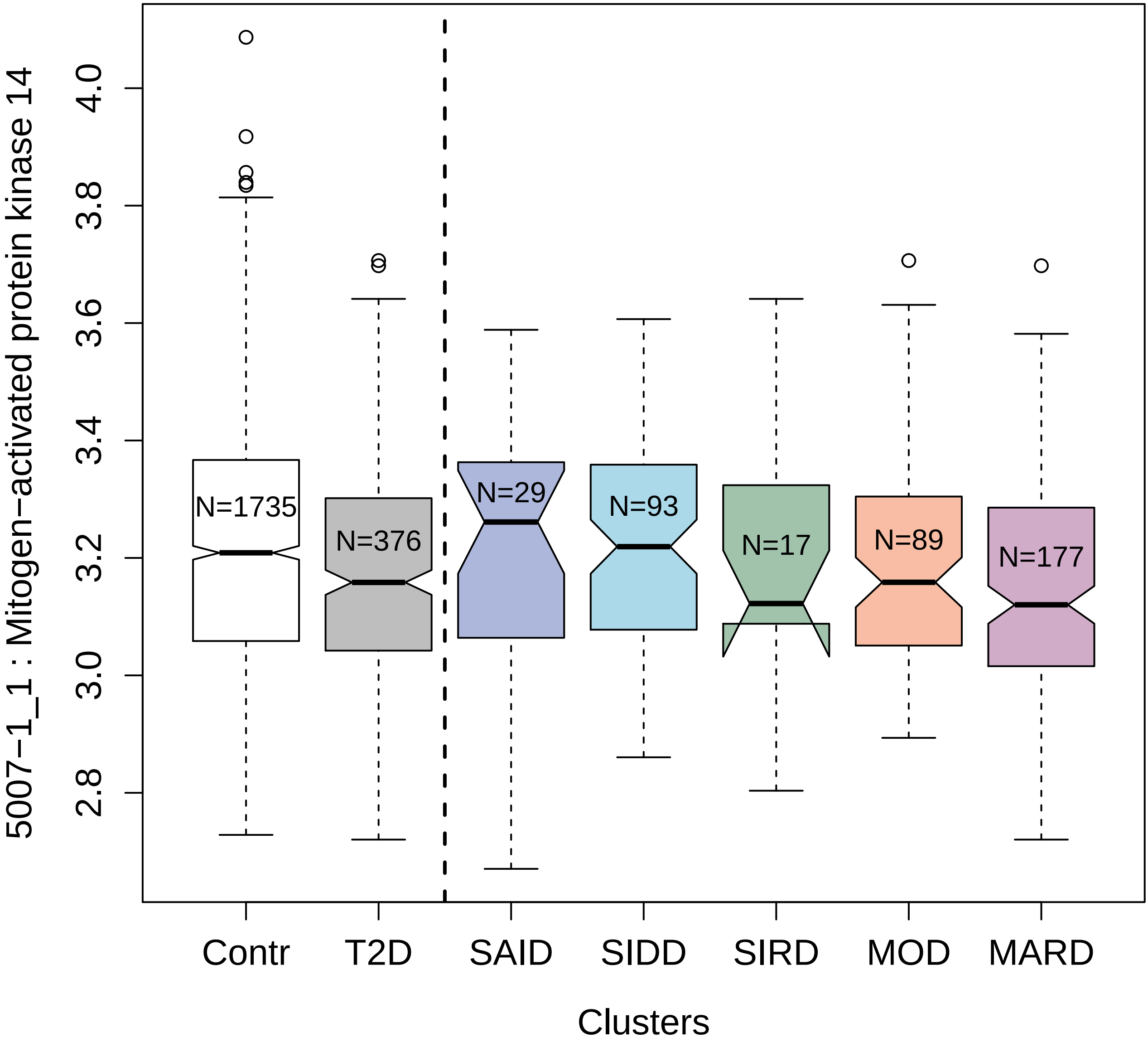

5007−1_1 : Mitogen−activated protein kinase 14
2.8
3.0
3.2
3.4
3.6
3.8
4.0
N=1735
N=376
N=29
N=93
N=17
N=89
N=177
Contr
T2D
SAID
SIDD
SIRD
MOD
MARD
Clusters

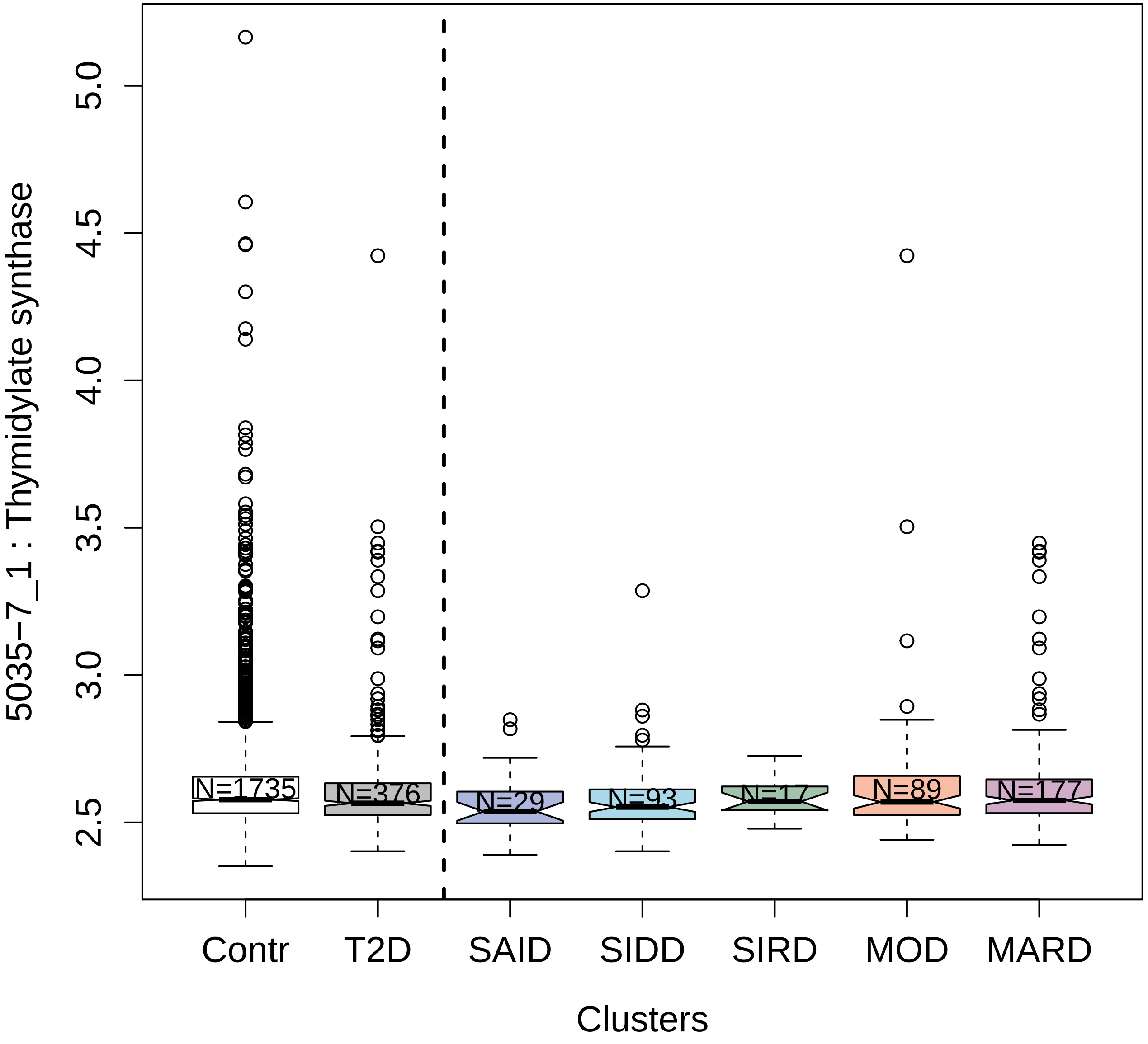
5035−7_1 : Thymidylate synthase
5.0
4.5
4.0
3.5
3.0
2.5
N=1735
N=376
N=29
N=93
N=17
N=89
N=177
Contr
T2D
SAID
SIDD
SIRD
MOD
MARD
Clusters

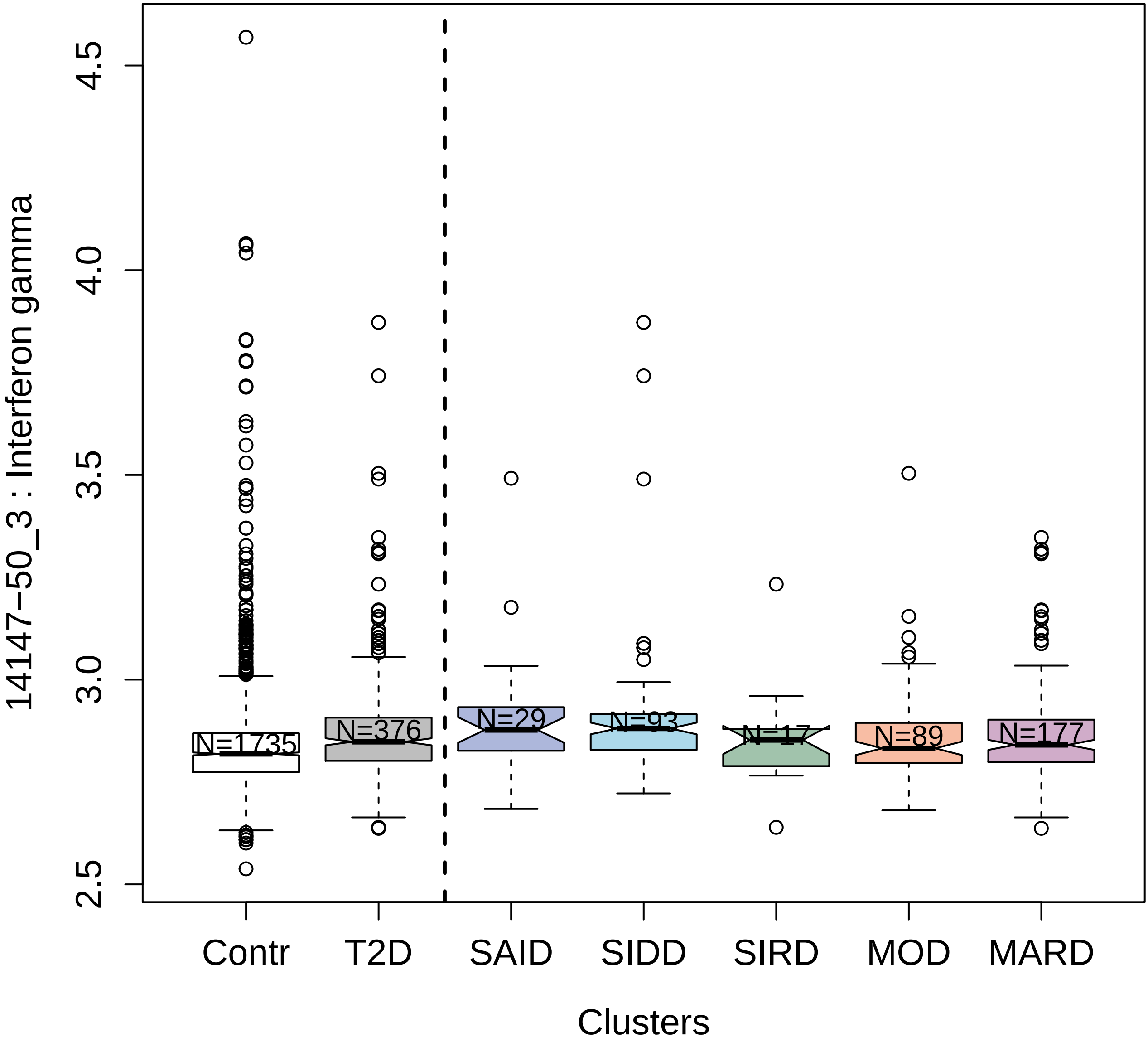

14147−50_3 : Interferon gamma
2.5
3.0
3.5
4.0
4.5
N=1735
N=376
N=29
N=93
N=17
N=89
N=177
Contr
T2D
SAID
SIDD
SIRD
MOD
MARD
Clusters

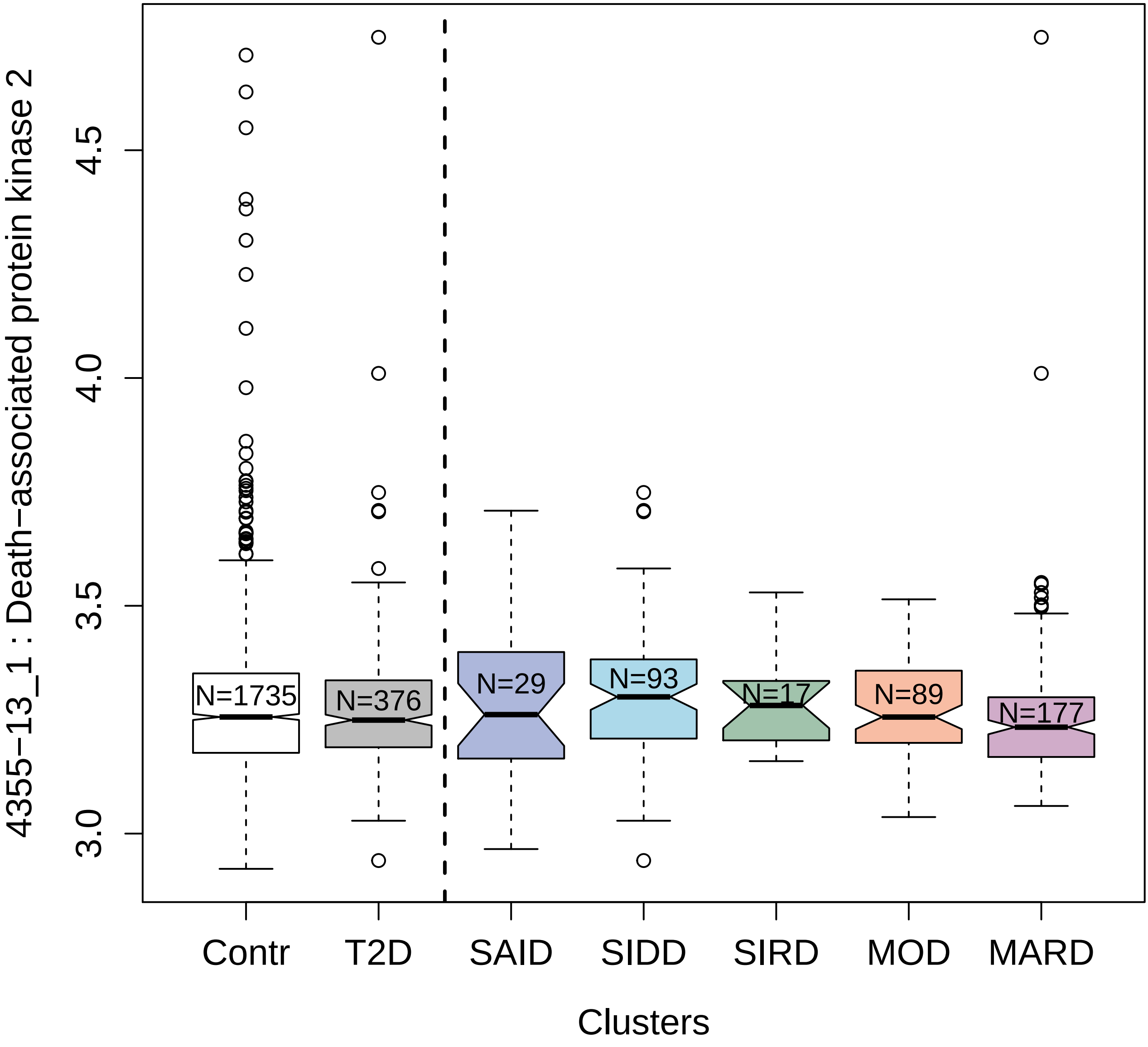

4355−13_1 : Death−associated protein kinase 2
3.0
3.5
4.0
4.5
N=1735
N=376
N=29
N=93
N=17
N=89
N=177
Contr
T2D
SAID
SIDD
SIRD
MOD
MARD
Clusters

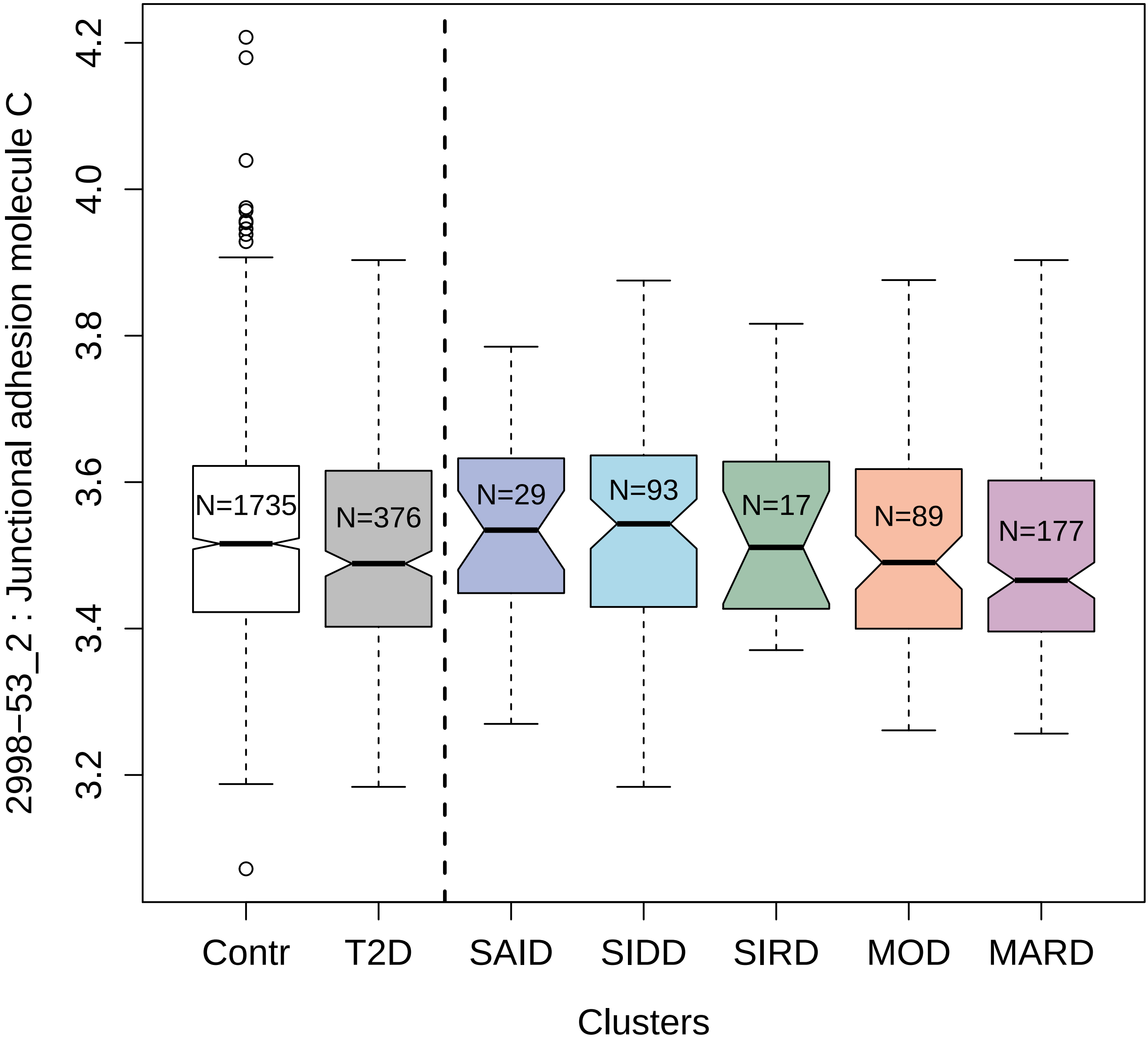

2998−53_2 : Junctional adhesion molecule C
3.2
3.4
3.6
3.8
4.0
4.2
N=1735
N=376
N=29
N=93
N=17
N=89
N=177
Contr
T2D
SAID
SIDD
SIRD
MOD
MARD
Clusters

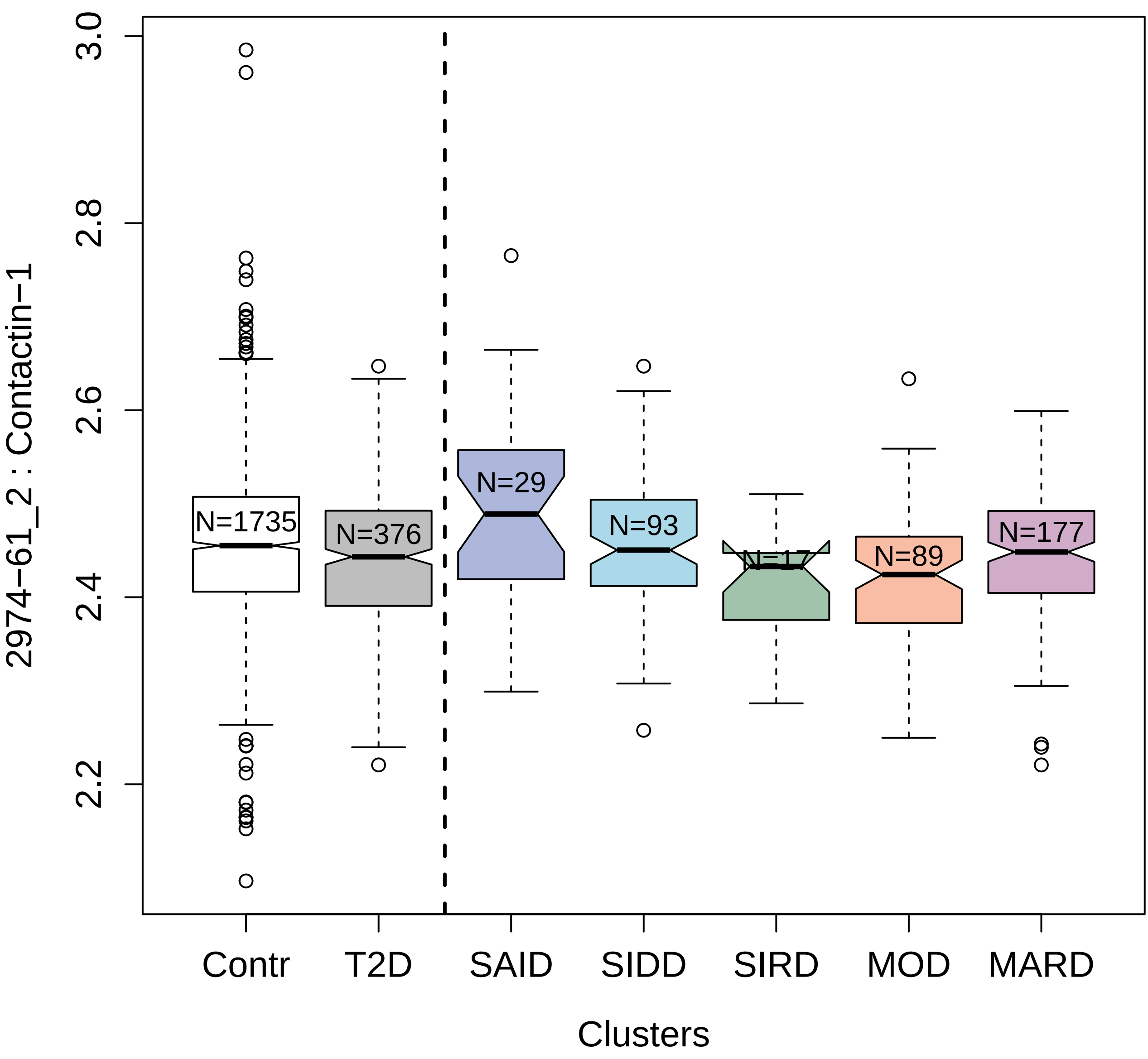

2974−61_2 : Contactin−1
3.0
2.8
2.6
2.4
2.2
N=1735
N=376
N=29
N=93
N=17
N=89
N=177
Contr
T2D
SAID
SIDD
SIRD
MOD
MARD
Clusters

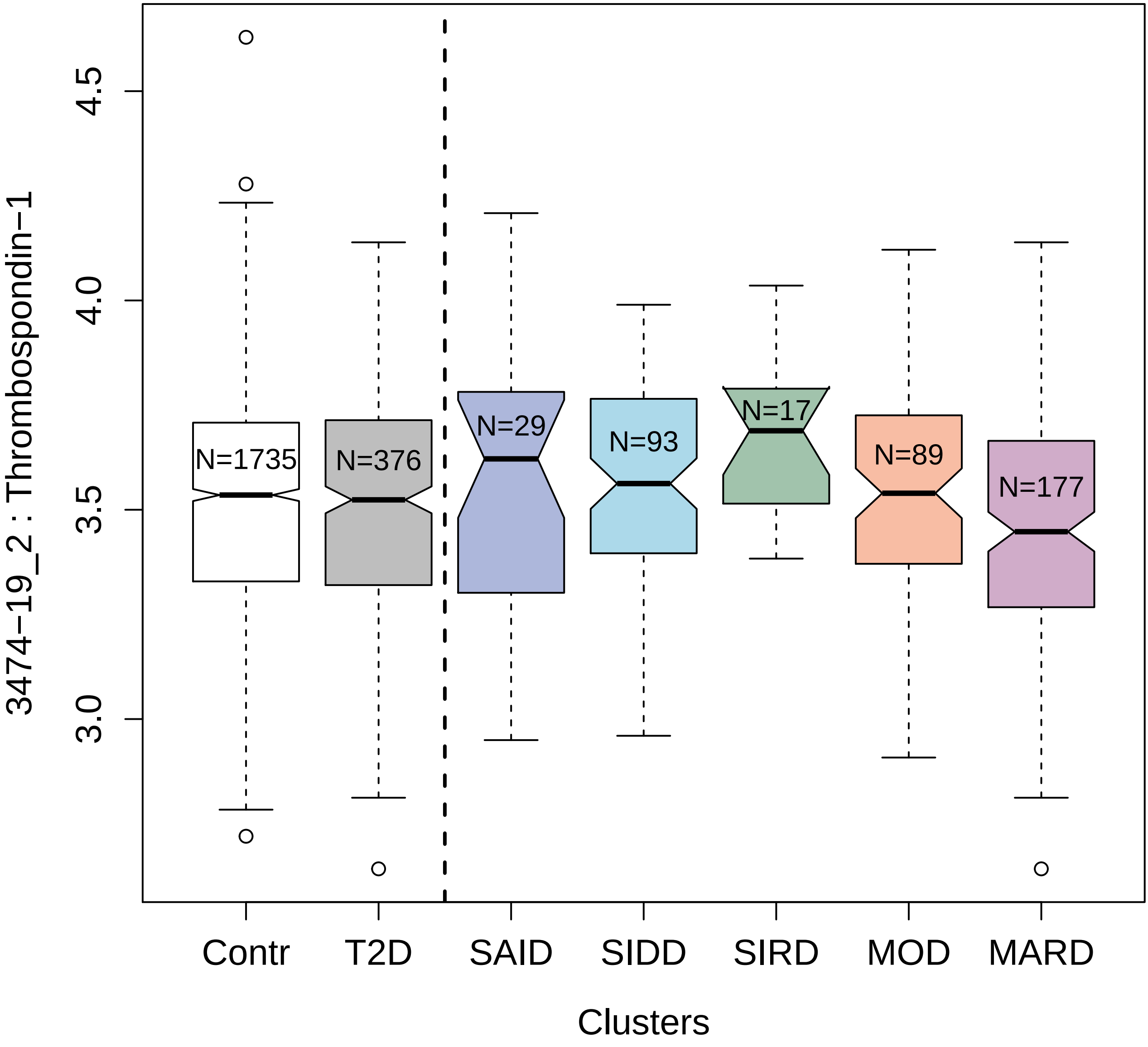
3474−19_2 : Thrombospondin−1
4.5
4.0
3.5
3.0
N=1735
N=376
N=29
N=93
N=17
N=89
N=177
Contr
T2D
SAID
SIDD
SIRD
MOD
MARD
Clusters

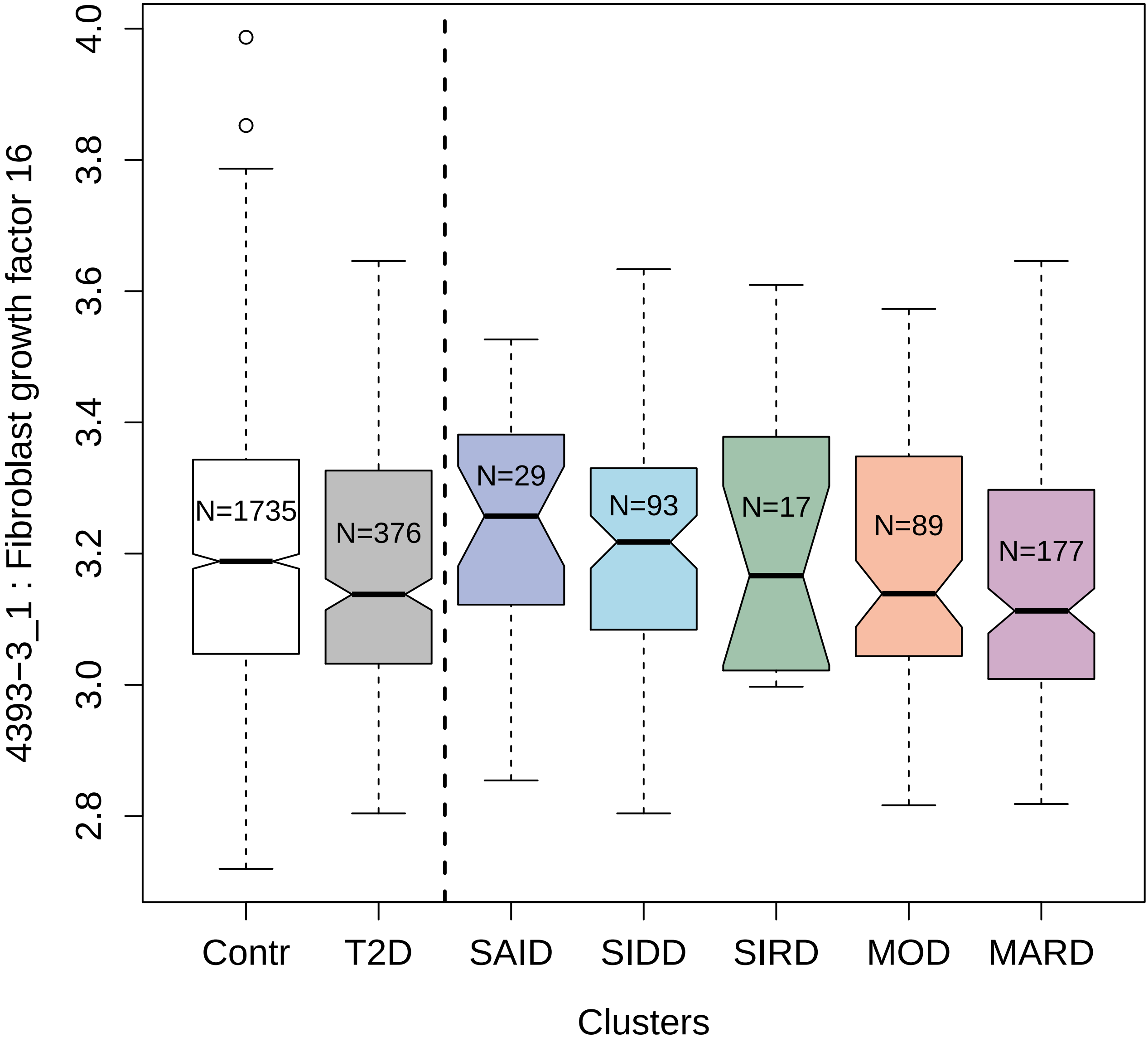
4393−3_1 : Fibroblast growth factor 16
4.0
3.8
3.6
3.4
3.2
3.0
2.8
N=1735
N=376
N=29
N=93
N=17
N=89
N=177
Contr
T2D
SAID
SIDD
SIRD
MOD
MARD
Clusters

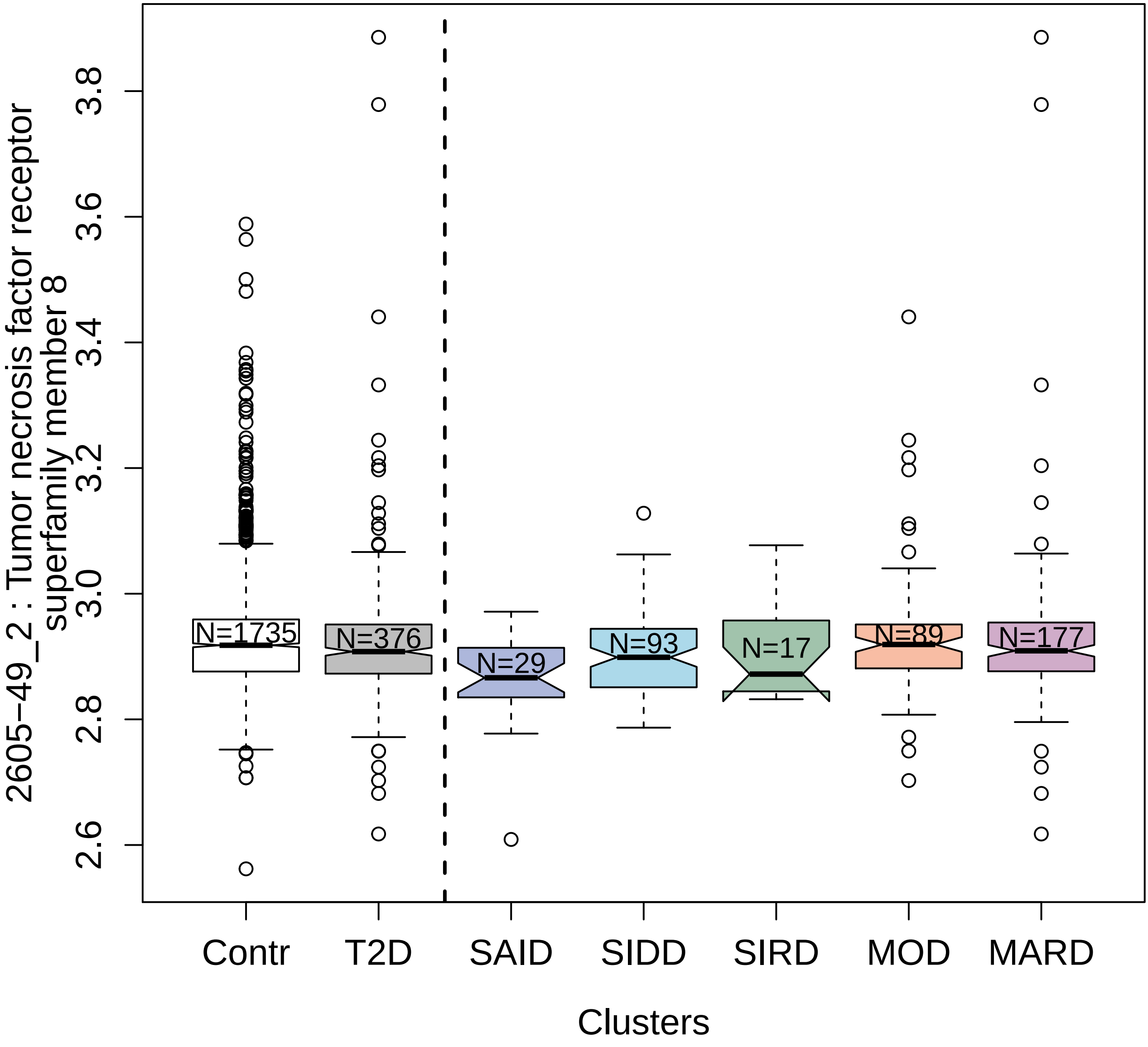
2605−49_2 : Tumor necrosis factor receptor
superfamily member 8
2.6
2.8
3.0
3.2
3.4
3.6
3.8
N=1735
N=376
N=29
N=93
N=17
N=89
N=177
Contr
T2D
SAID
SIDD
SIRD
MOD
MARD
Clusters

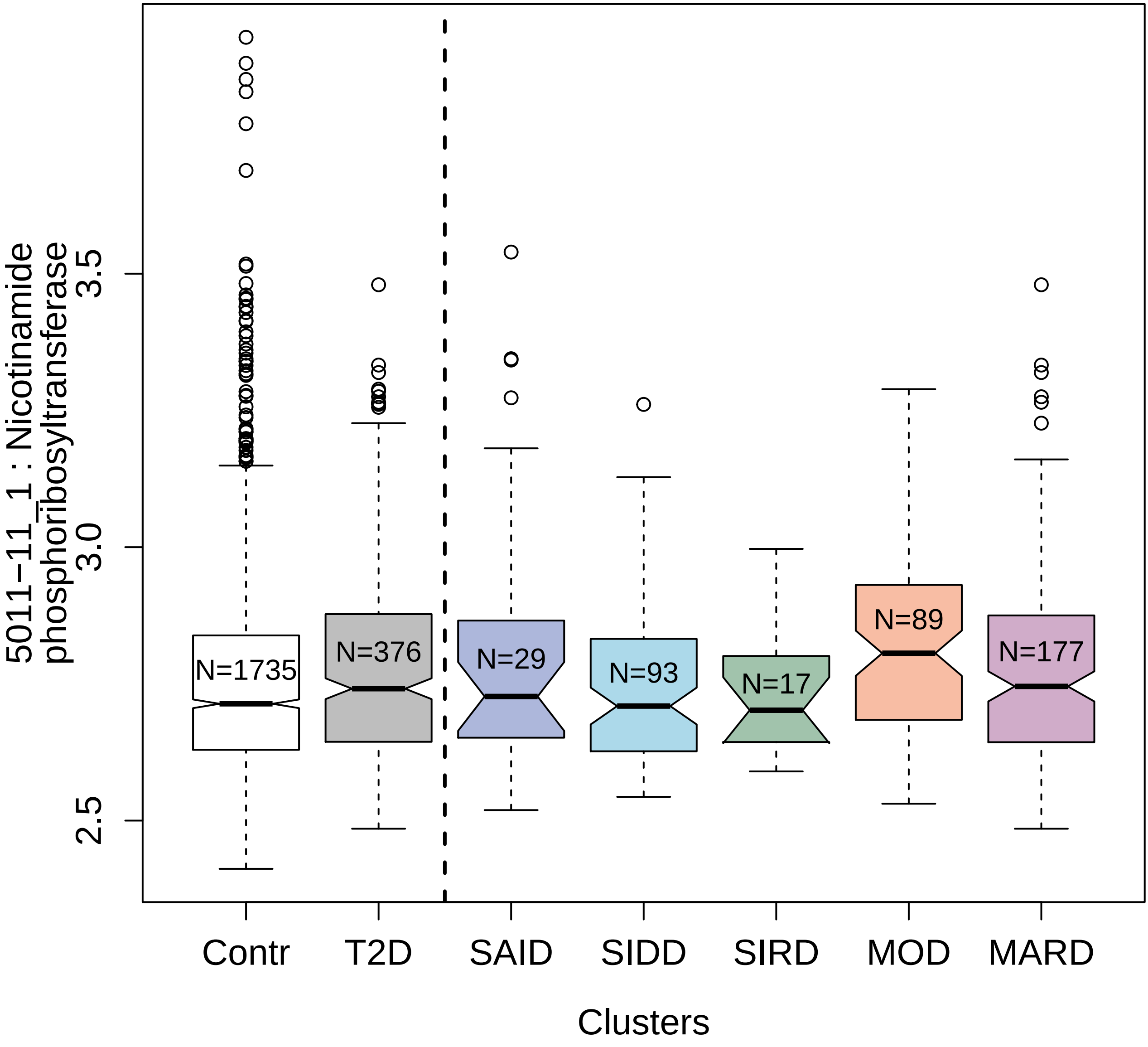
5011−11_1 : Nicotinamide phosphoribosyltransferase
3.5
3.0
2.5
N=1735
N=376
N=29
N=93
N=17
N=89
N=177
Contr
T2D
SAID
SIDD
SIRD
MOD
MARD
Clusters

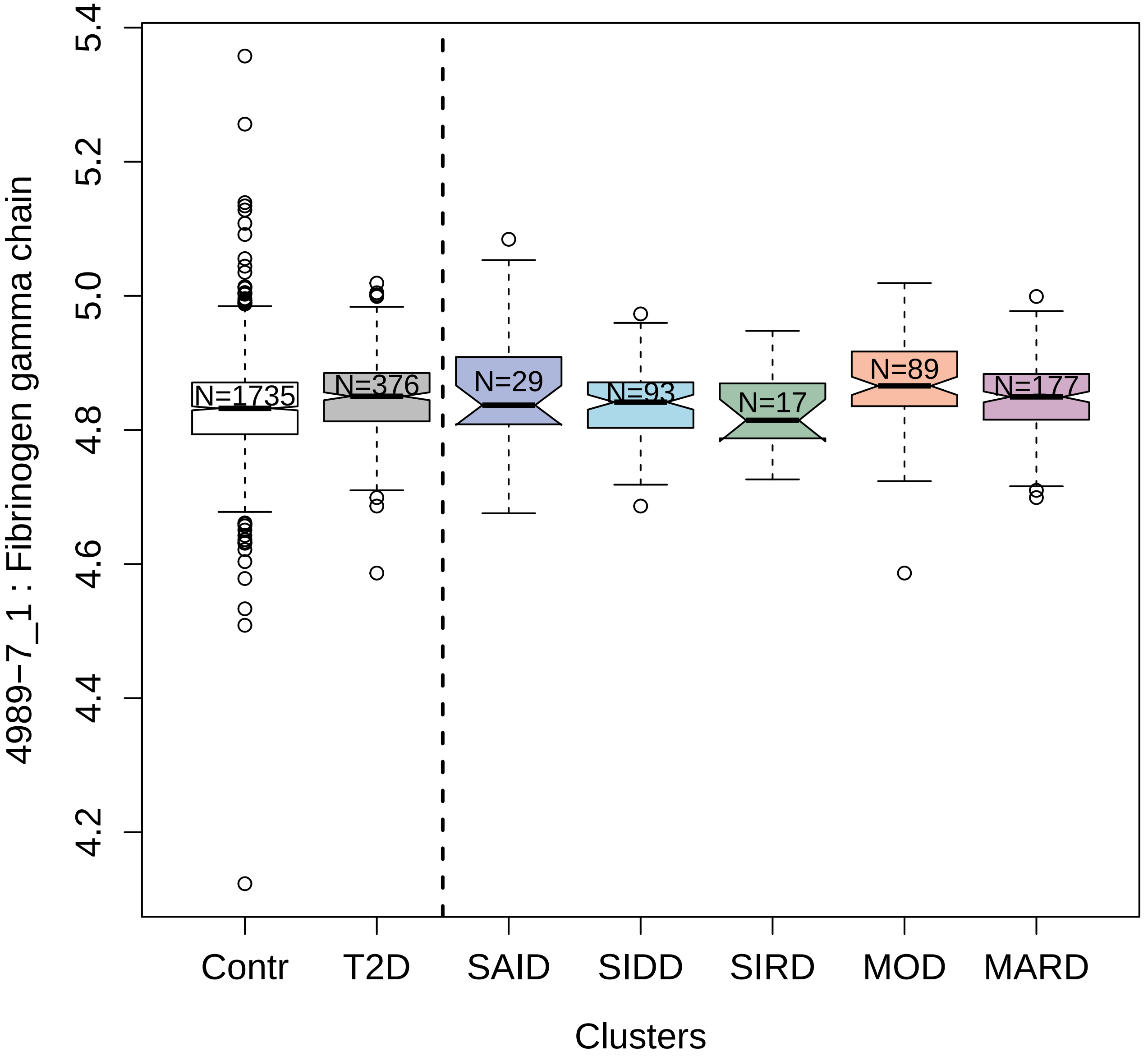
4989−7_1 : Fibrinogen gamma chain
5.4
5.2
5.0
4.8
4.6
4.4
4.2
N=1735
N=376
N=29
N=93
N=17
N=89
N=177
Contr
T2D
SAID
SIDD
SIRD
MOD
MARD
Clusters

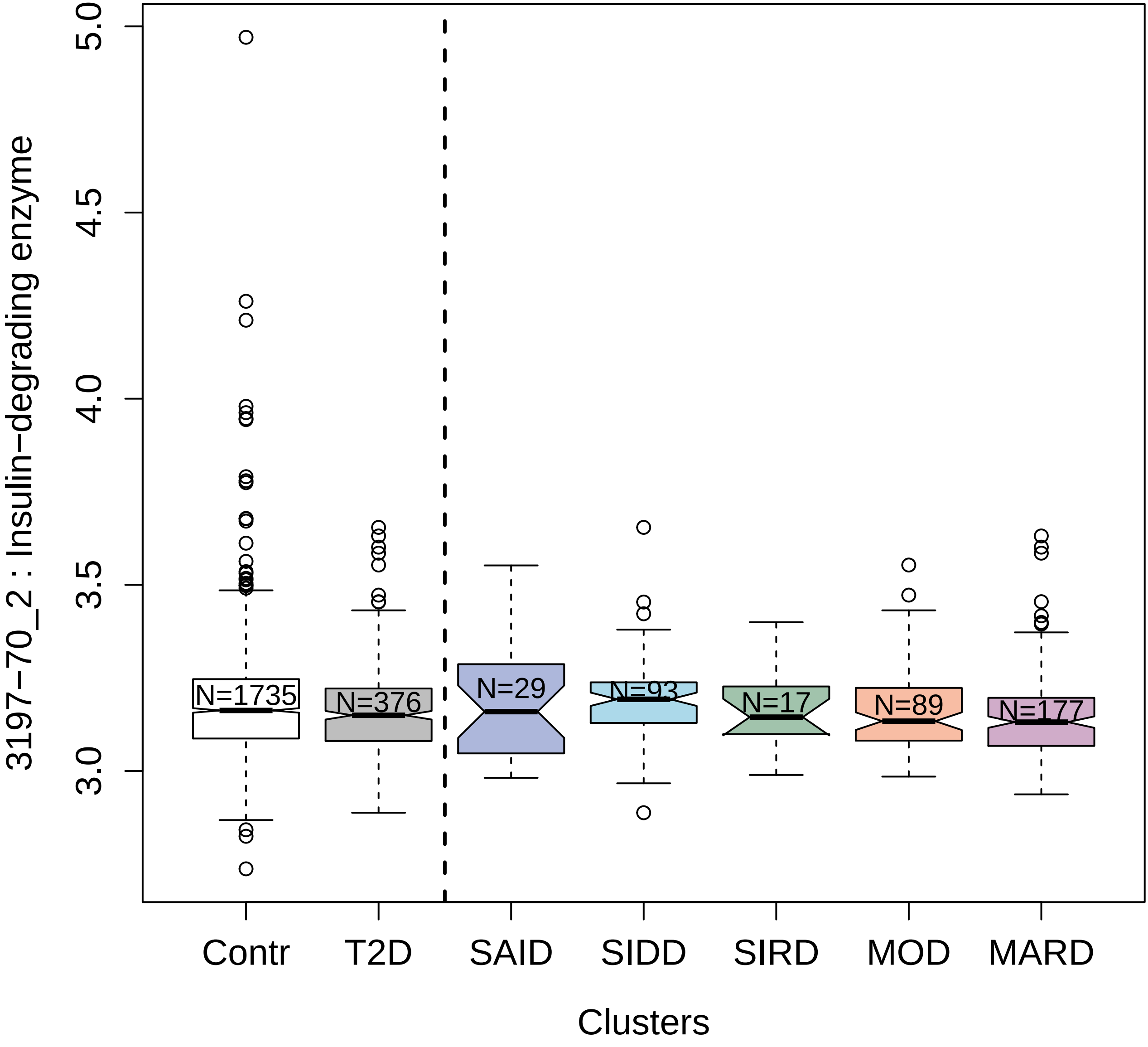
3197−70_2 : Insulin−degrading enzyme
5.0
4.5
4.0
3.5
3.0
N=1735
N=376
N=29
N=93
N=17
N=89
N=177
Contr
T2D
SAID
SIDD
SIRD
MOD
MARD
Clusters

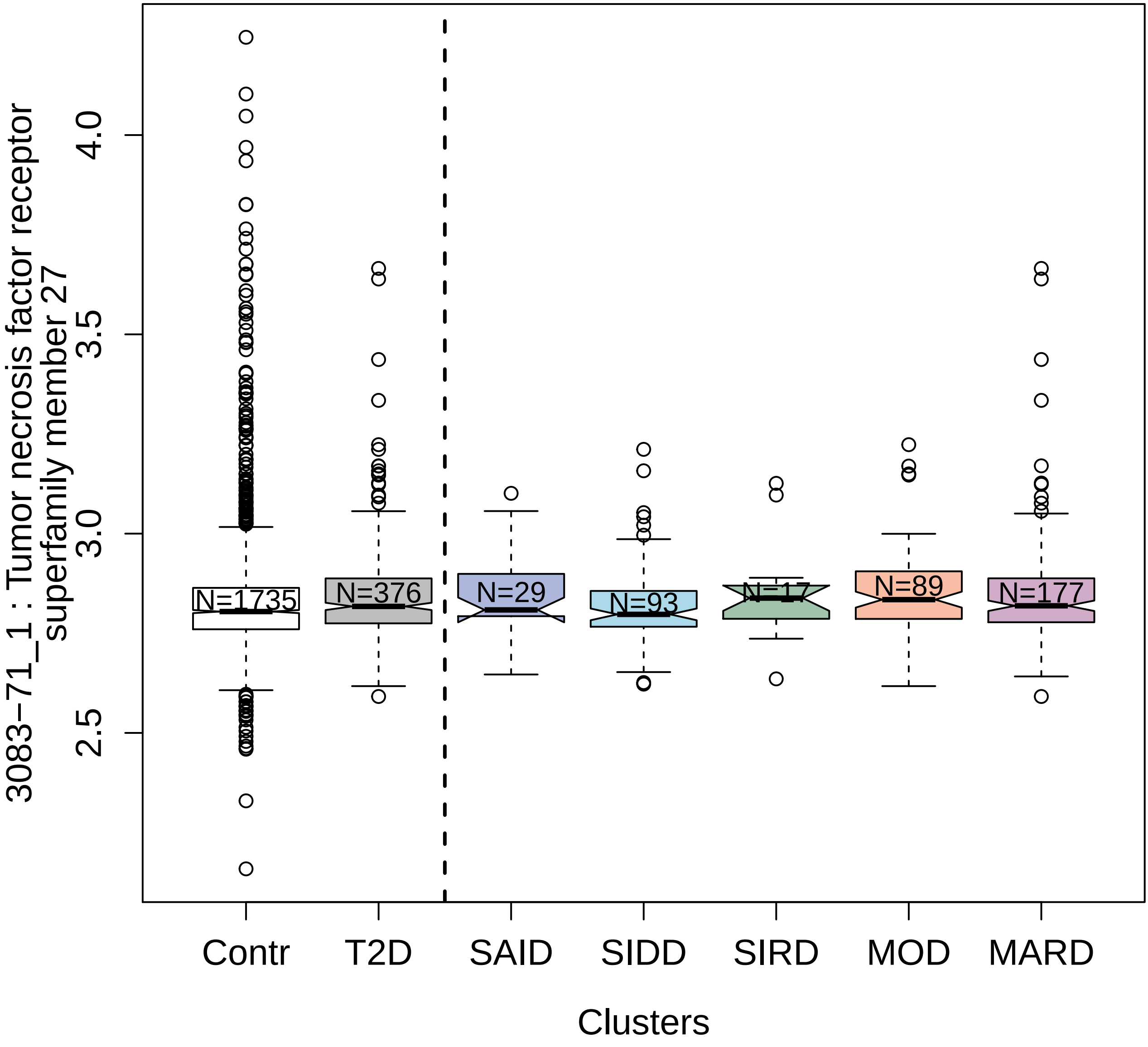

3083−71_1 : Tumor necrosis factor receptor superfamily member 27
4.0
3.5
3.0
2.5
N=1735
N=376
N=29
N=93
N=17
N=89
N=177
Contr
T2D
SAID
SIDD
SIRD
MOD
MARD
Clusters

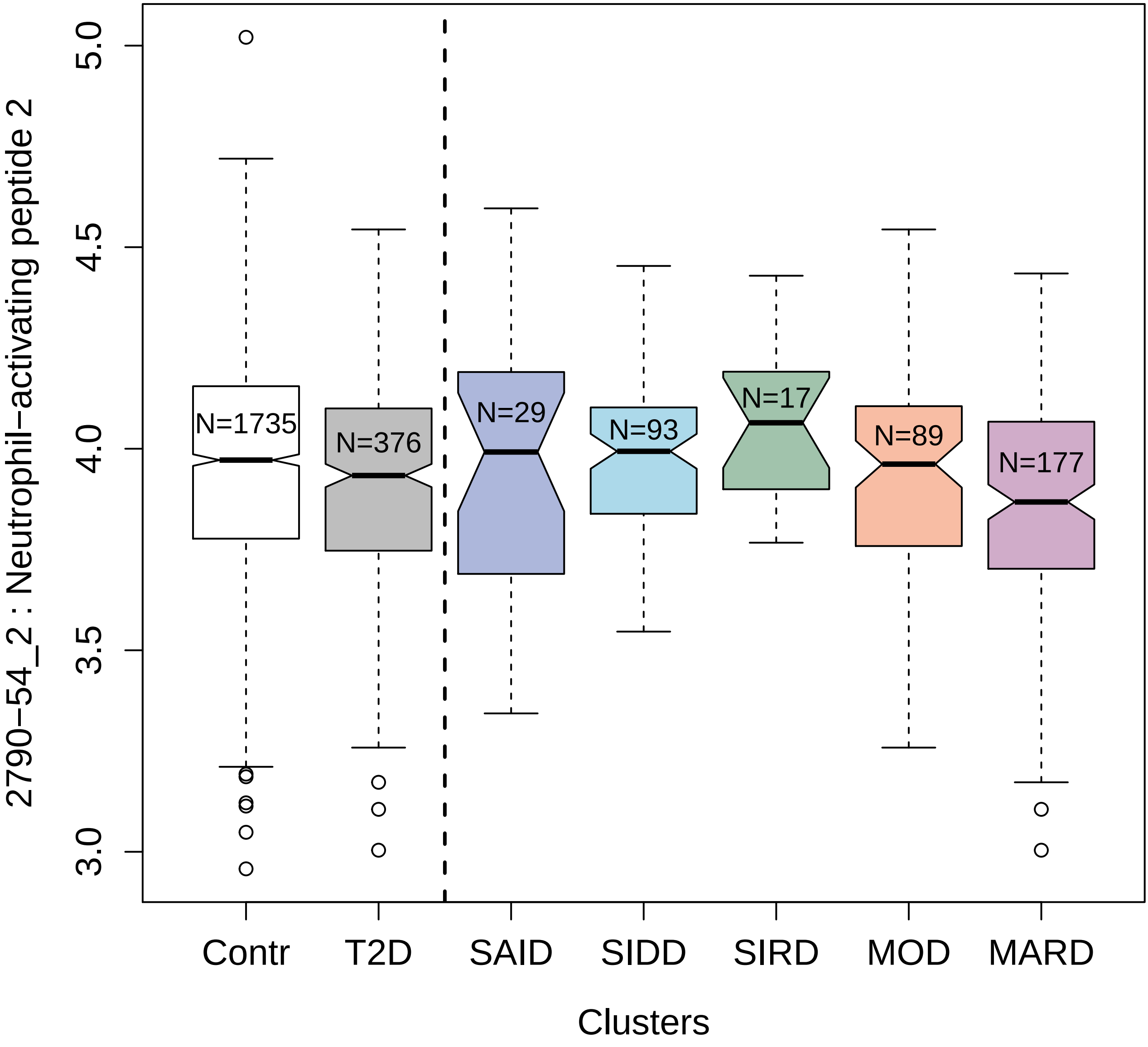
2790−54_2 : Neutrophil−activating peptide 2
5.0
4.5
4.0
3.5
3.0
N=1735
N=376
N=29
N=93
N=17
N=89
N=177
Contr
T2D
SAID
SIDD
SIRD
MOD
MARD
Clusters

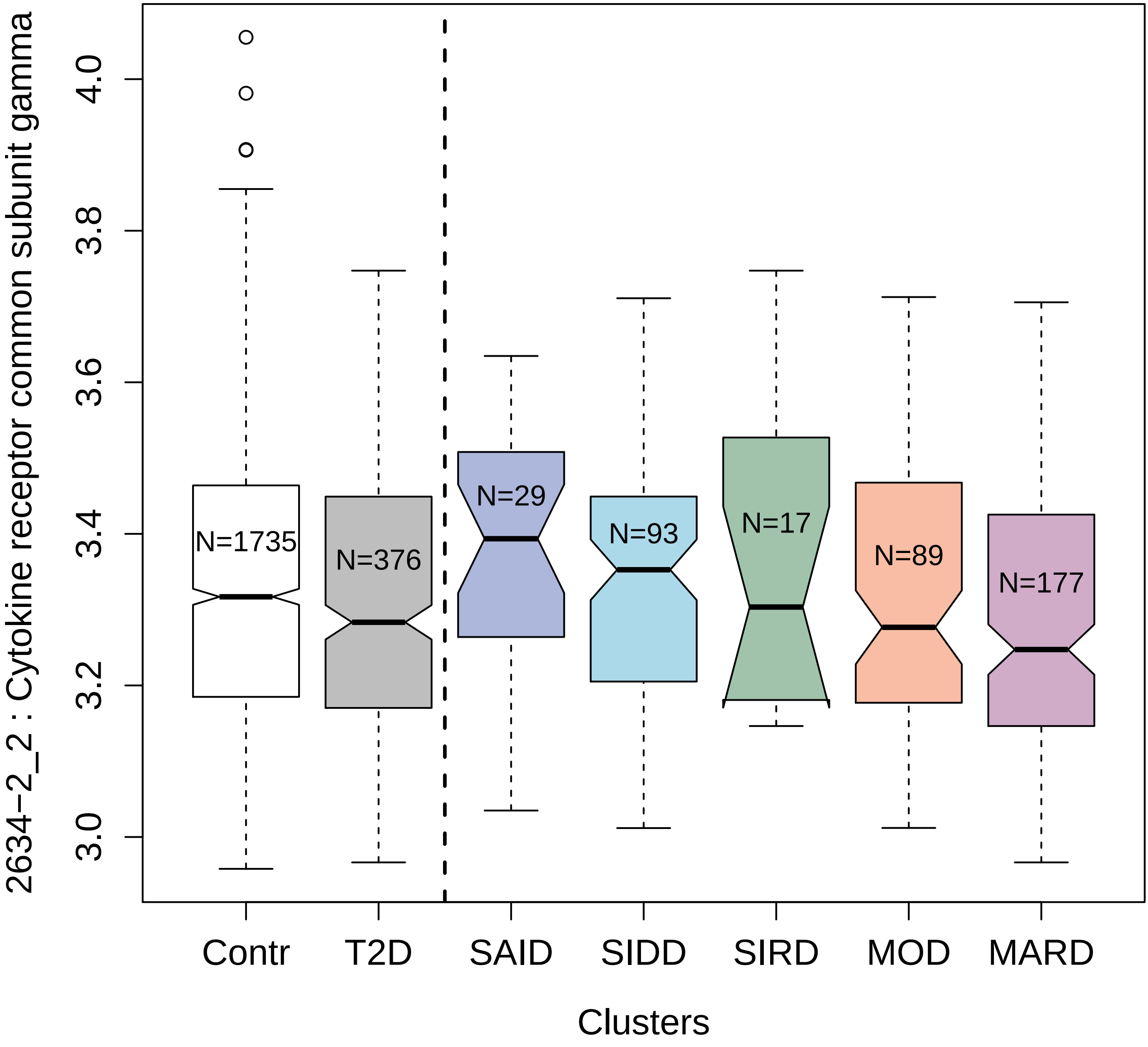

2634−2_2 : Cytokine receptor common subunit gamma
4.0
3.8
3.6
3.4
3.2
3.0
N=1735
N=376
N=29
N=93
N=17
N=89
N=177
Contr
T2D
SAID
SIDD
SIRD
MOD
MARD
Clusters

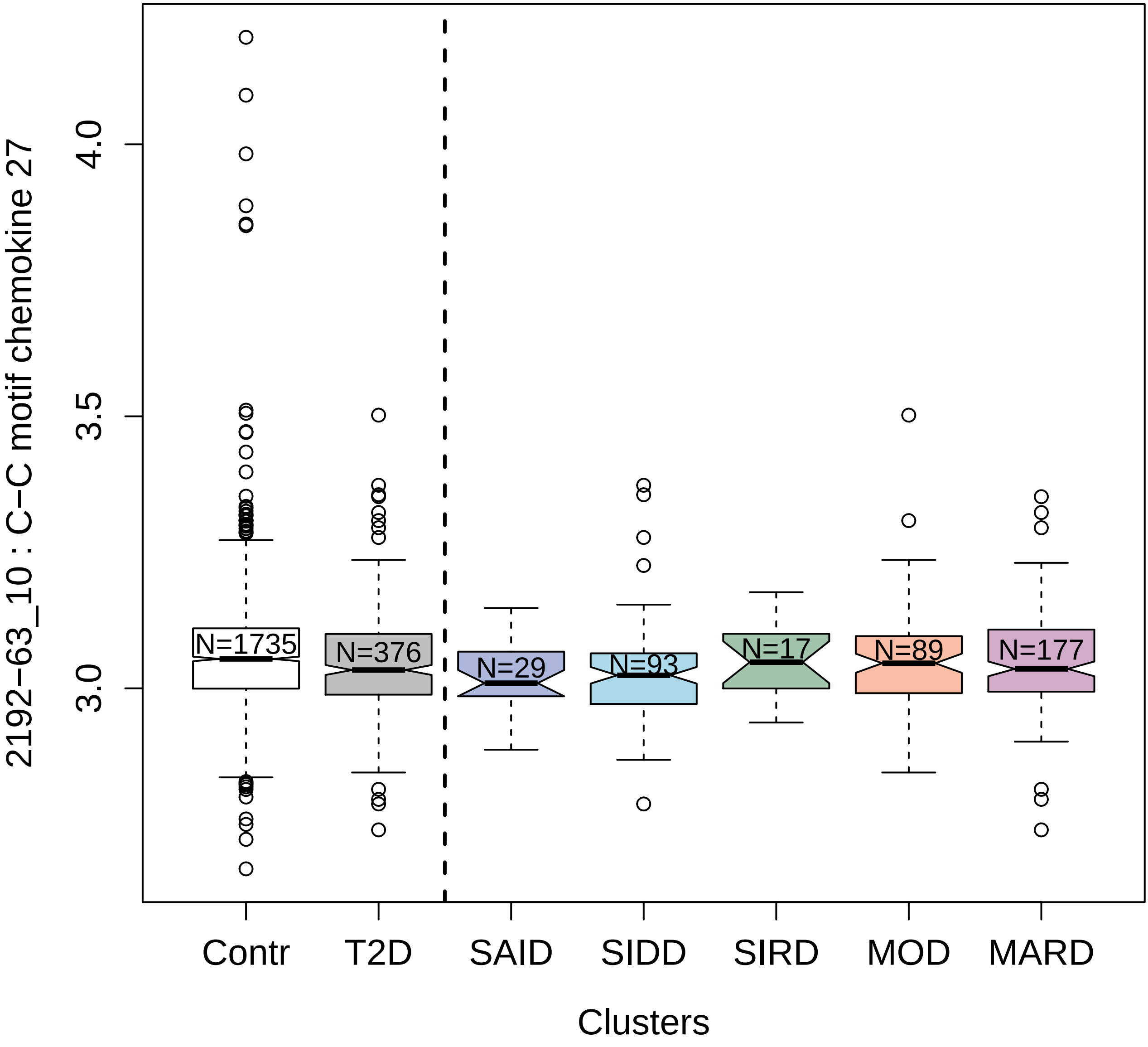
2192−63_10 : C−C motif chemokine 27
4.0
3.5
3.0
N=1735
N=376
N=29
N=93
N=17
N=89
N=177
Contr
T2D
SAID
SIDD
SIRD
MOD
MARD
Clusters

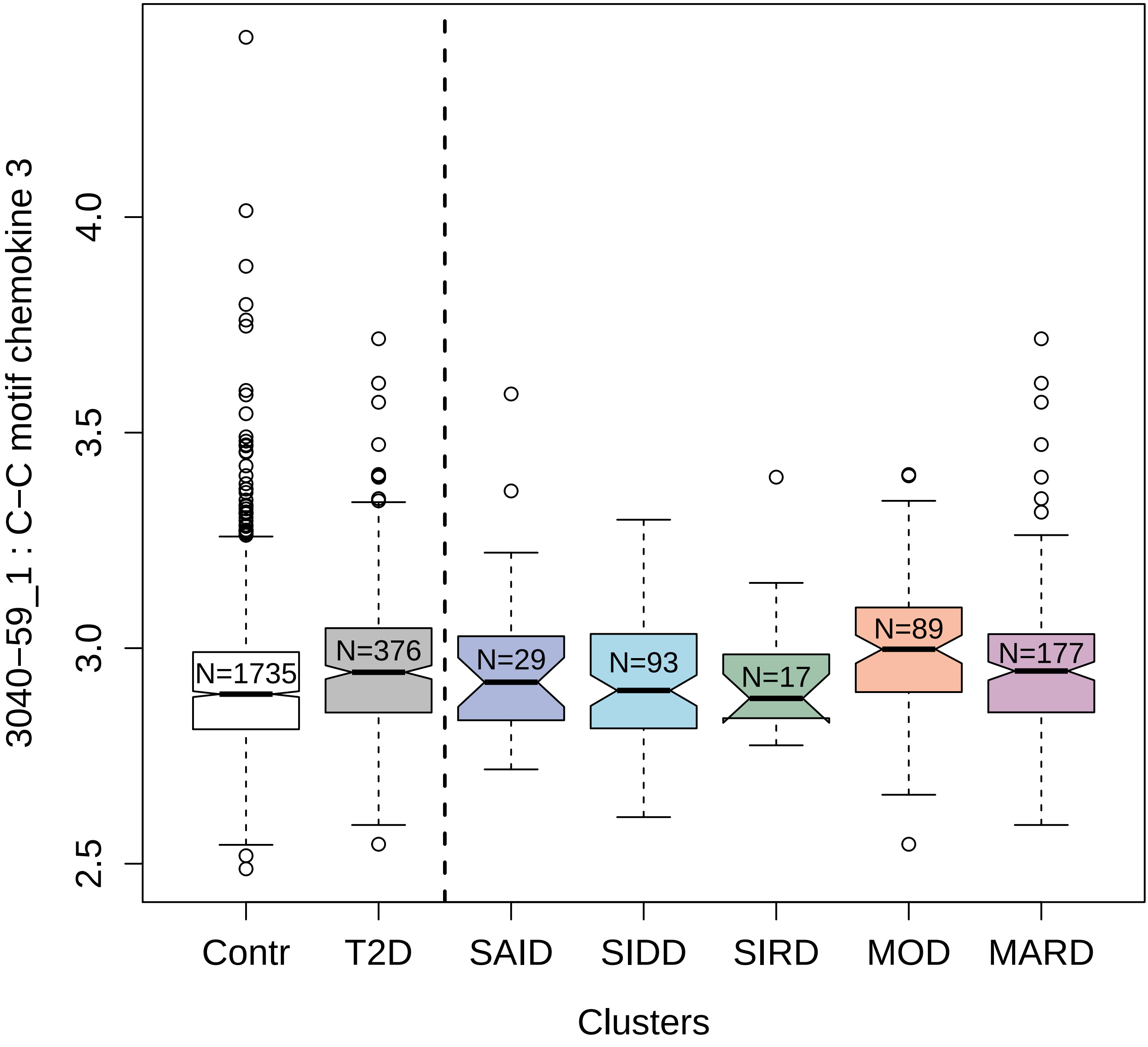

3040−59_1 : C−C motif chemokine 3
2.5
3.0
3.5
4.0
N=1735
N=376
N=29
N=93
N=17
N=89
N=177
Contr
T2D
SAID
SIDD
SIRD
MOD
MARD
Clusters

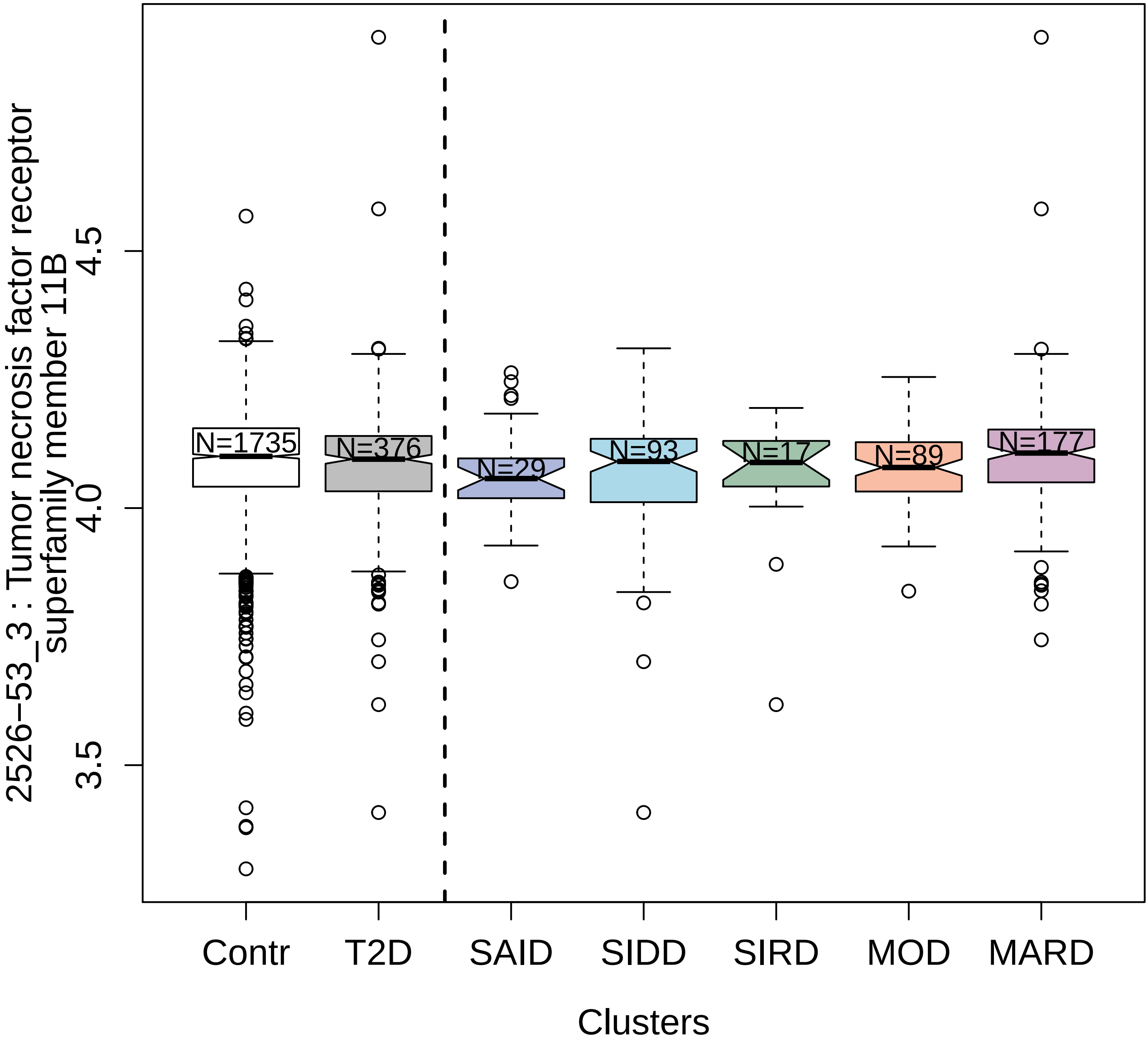

2526−53_3 : Tumor necrosis factor receptor superfamily member 11B
3.5
4.0
4.5
N=1735
N=376
N=29
N=93
N=17
N=89
N=177
Contr
T2D
SAID
SIDD
SIRD
MOD
MARD
Clusters

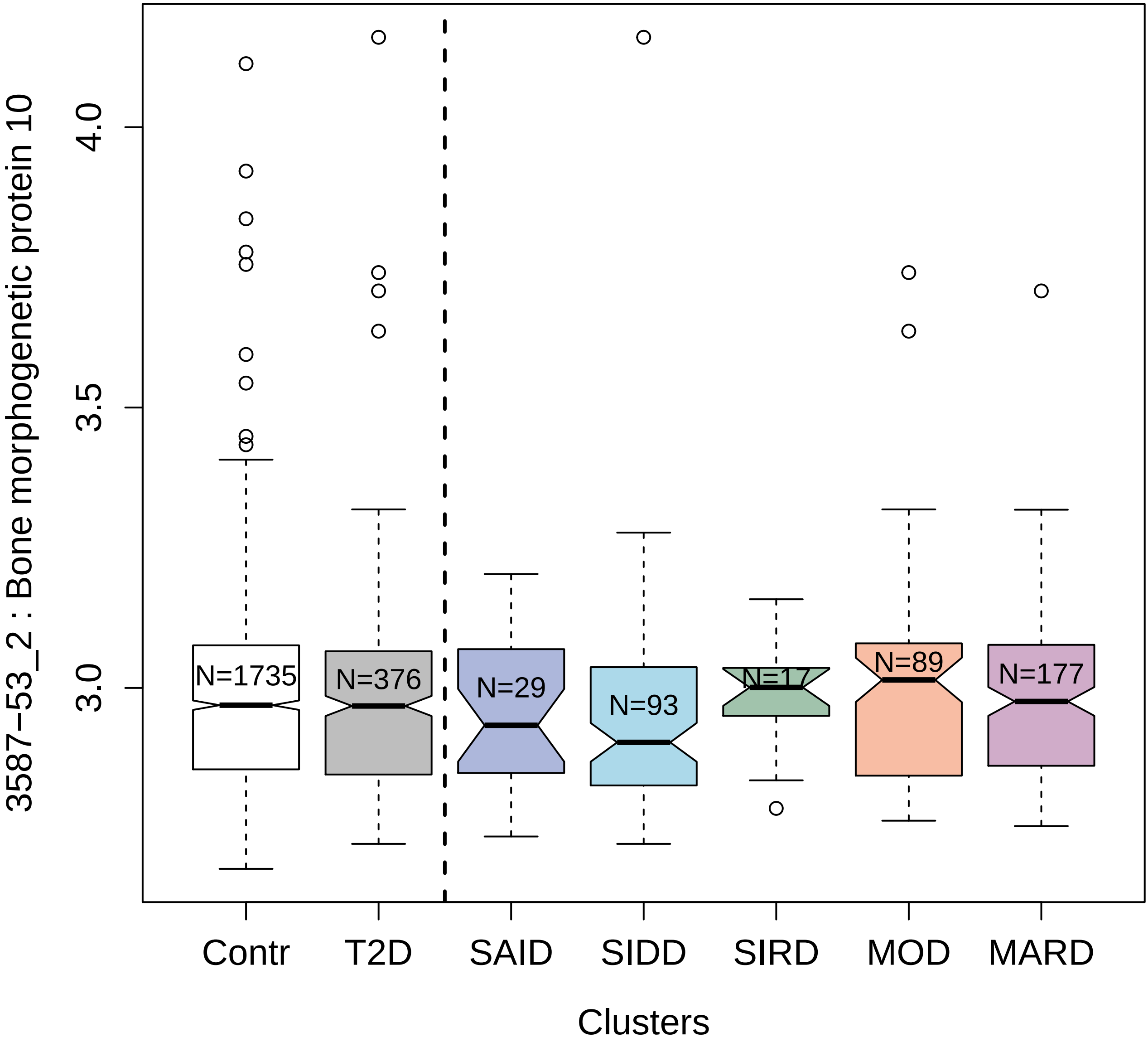

3587−53_2 : Bone morphogenetic protein 10
3.0
3.5
4.0
N=1735
N=376
N=29
N=93
N=17
N=89
N=177
Contr
T2D
SAID
SIDD
SIRD
MOD
MARD
Clusters

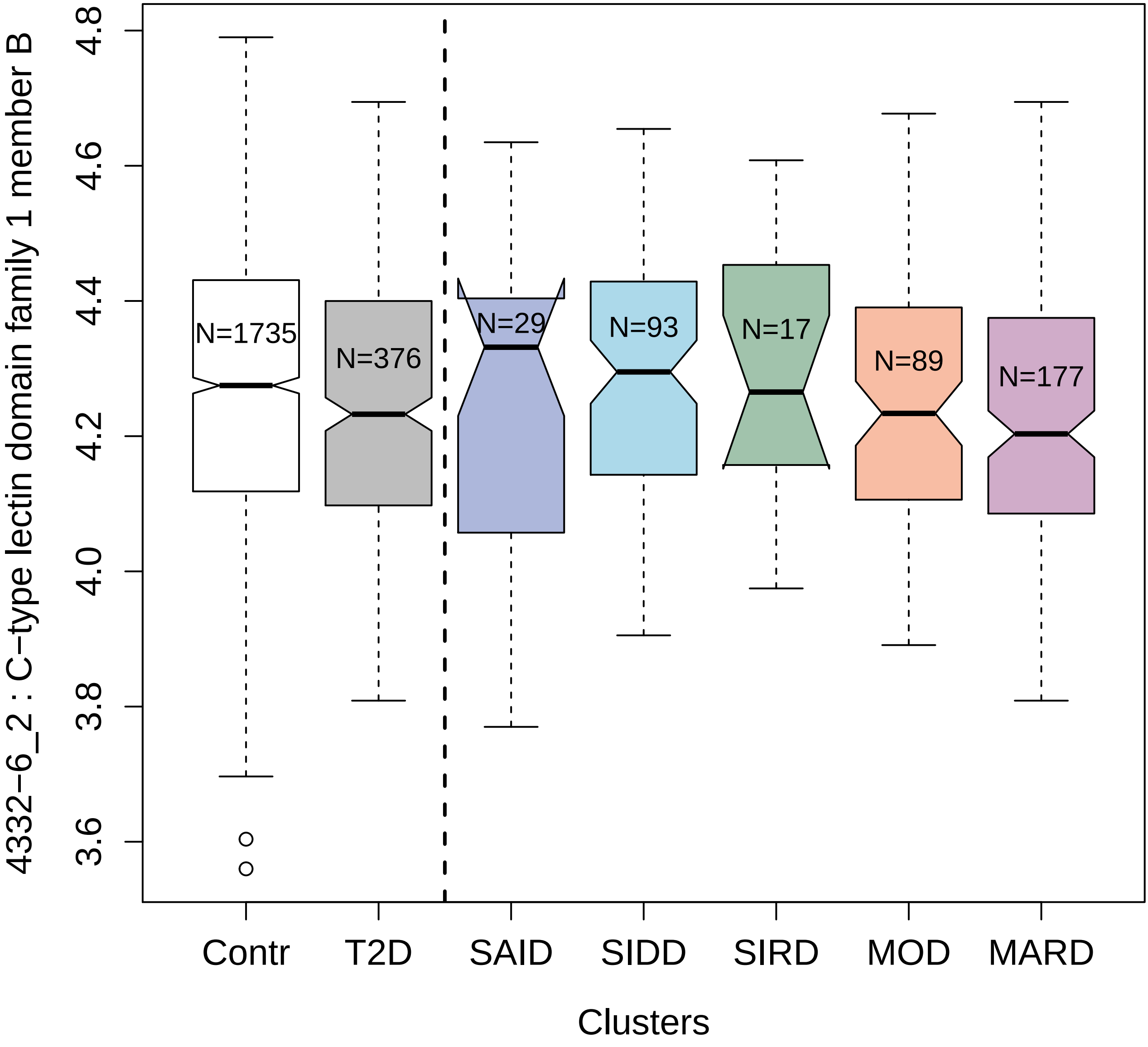

4332−6_2 : C−type lectin domain family 1 member B
4.8
4.6
4.4
4.2
4.0
3.8
3.6
N=1735
N=376
N=29
N=93
N=17
N=89
N=177
Contr
T2D
SAID
SIDD
SIRD
MOD
MARD
Clusters

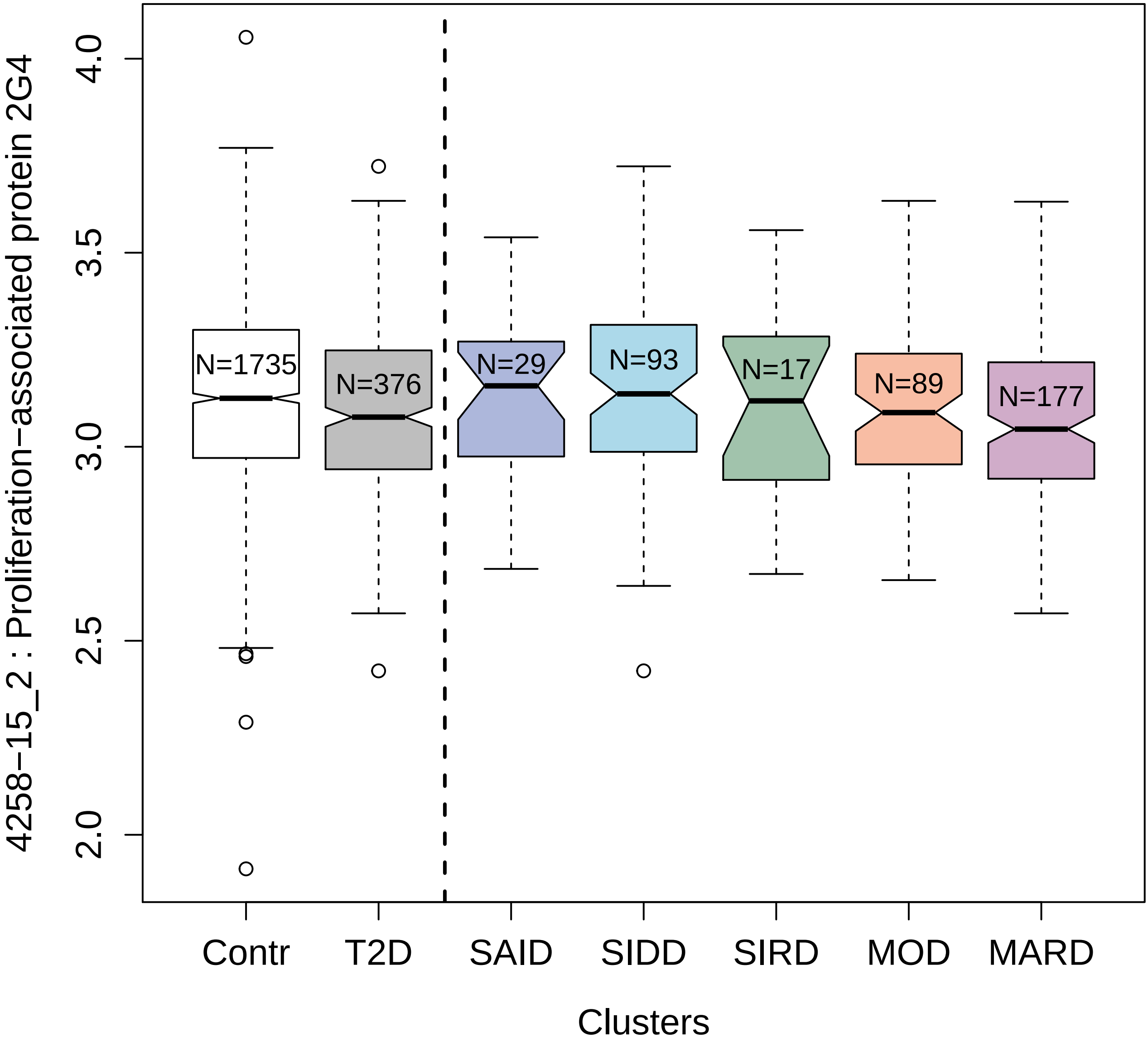

4258−15_2 : Proliferation−associated protein 2G4
4.0
3.5
3.0
2.5
2.0
N=1735
N=376
N=29
N=93
N=17
N=89
N=177
Contr
T2D
SAID
SIDD
SIRD
MOD
MARD
Clusters

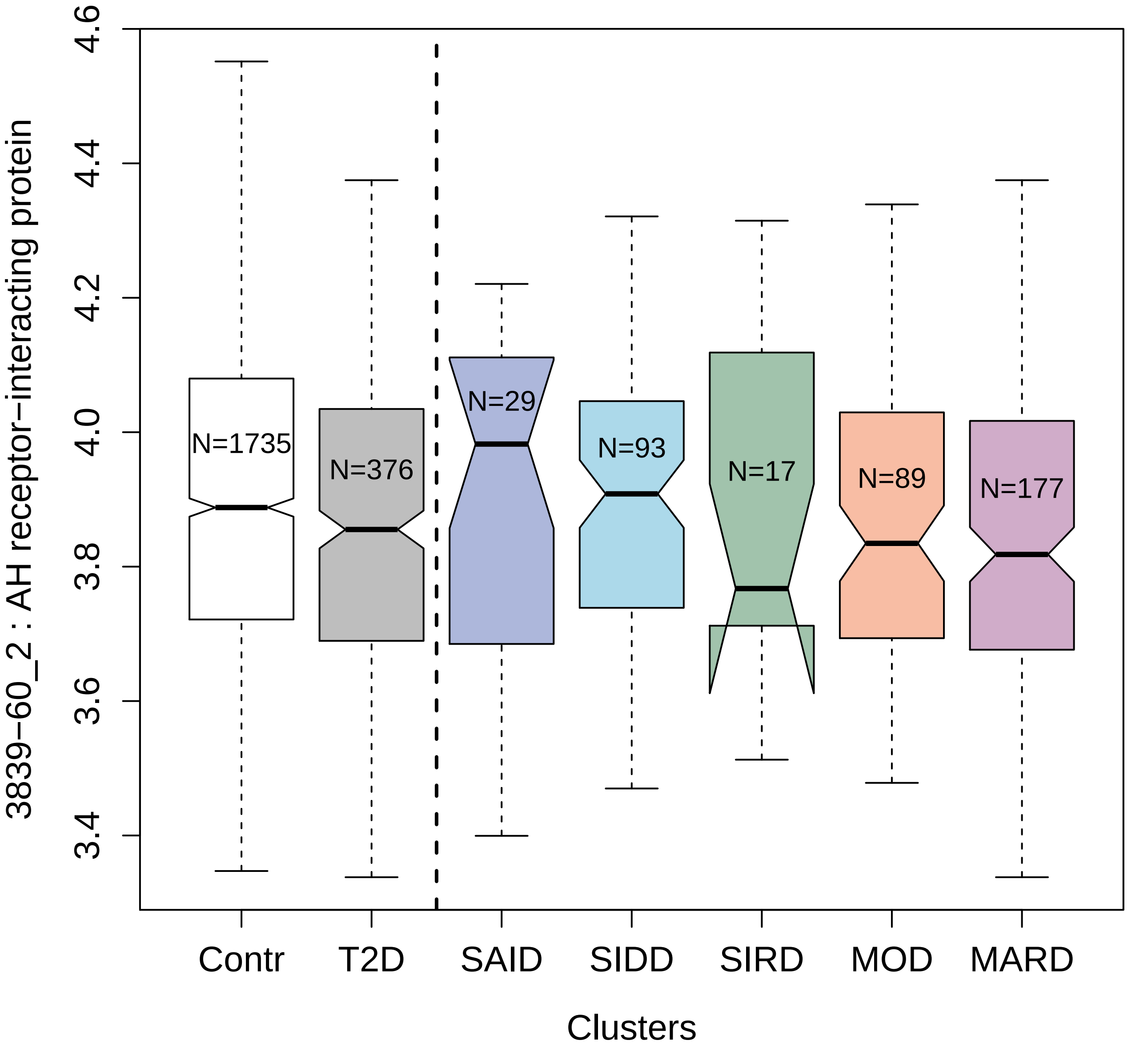
3839−60_2 : AH receptor−interacting protein
4.6
4.4
4.2
4.0
3.8
3.6
3.4
N=1735
N=376
N=29
N=93
N=17
N=89
N=177
Contr
T2D
SAID
SIDD
SIRD
MOD
MARD
Clusters

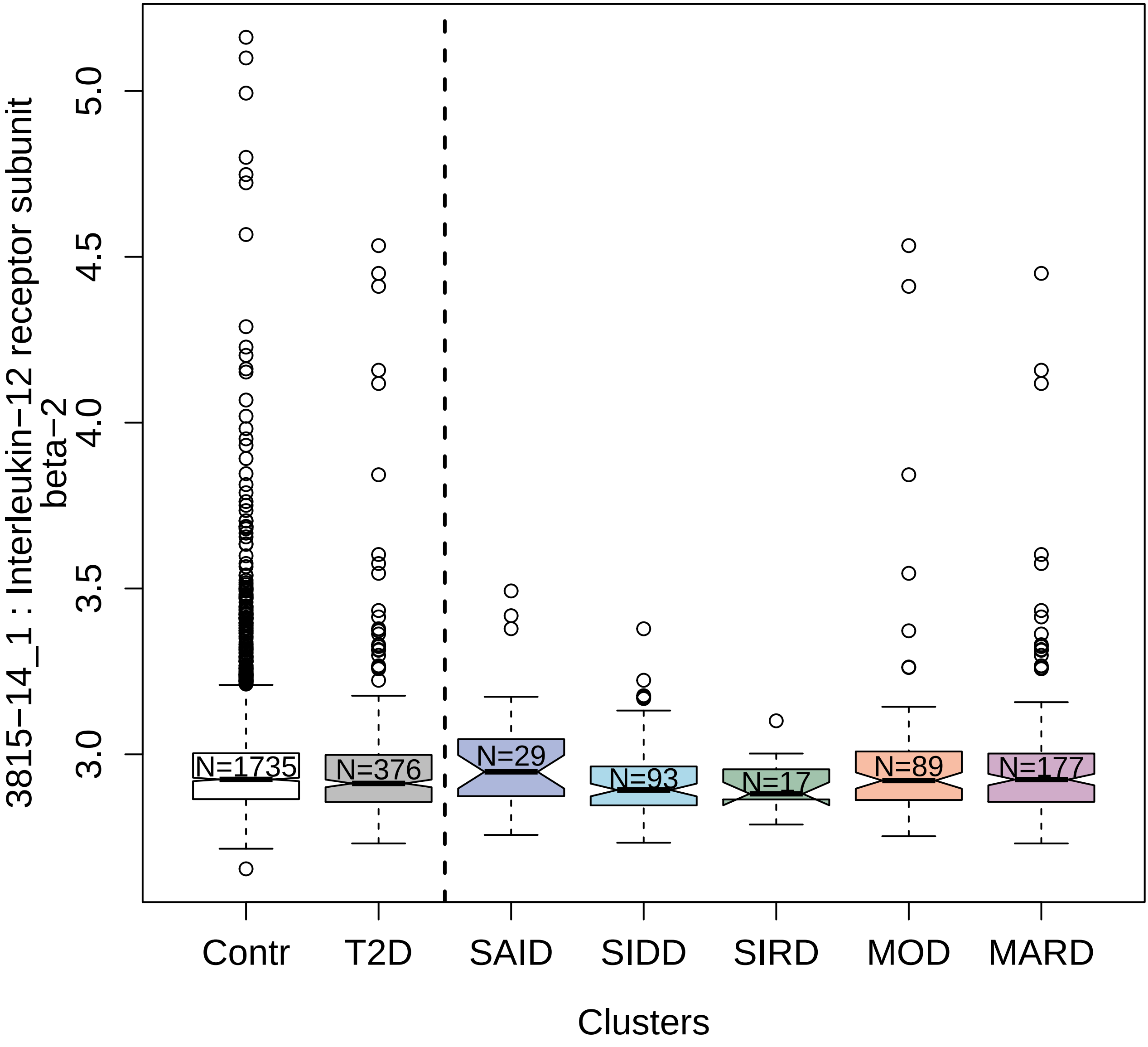

3815−14_1 : Interleukin−12 receptor subunit beta−2
5.0
4.5
4.0
3.5
3.0
N=1735
N=376
N=29
N=93
N=17
N=89
N=177
Contr
T2D
SAID
SIDD
SIRD
MOD
MARD
Clusters

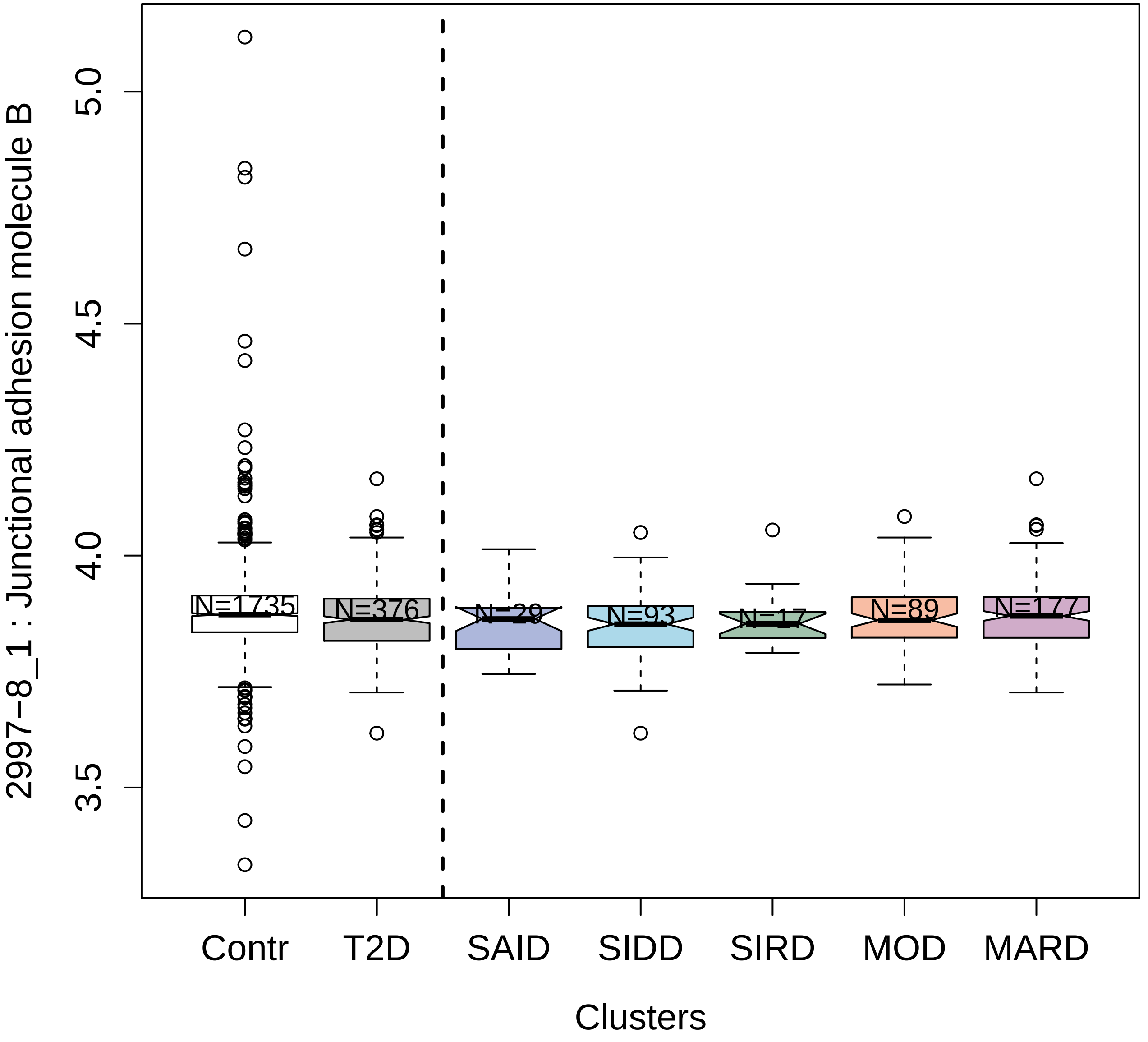

2997−8_1 : Junctional adhesion molecule B
5.0
4.5
4.0
3.5
N=1735
N=376
N=29
N=93
N=17
N=89
N=177
Contr
T2D
SAID
SIDD
SIRD
MOD
MARD
Clusters

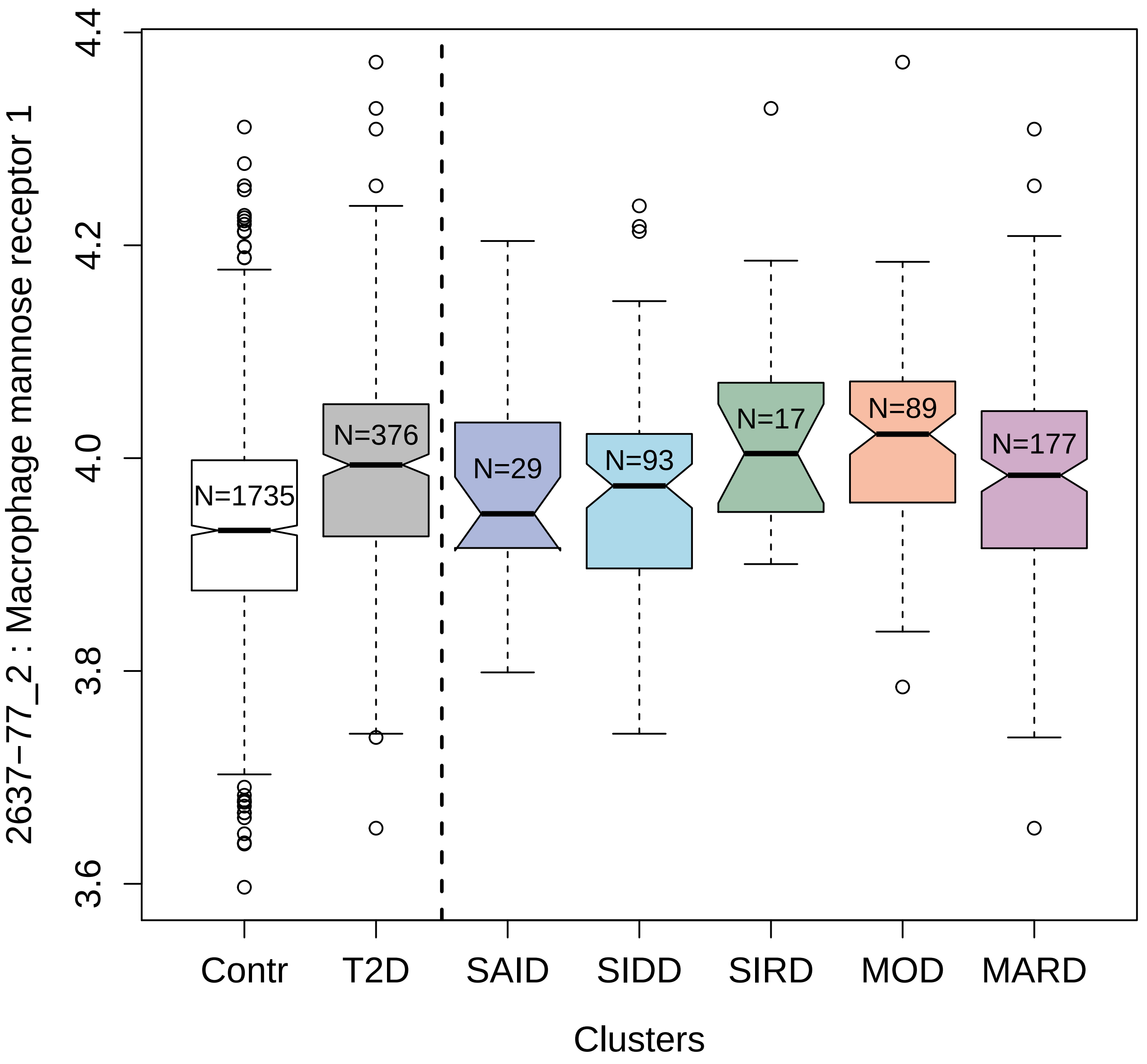

2637−77_2 : Macrophage mannose receptor 1
4.4
4.2
4.0
3.8
3.6
N=1735
N=376
N=29
N=93
N=17
N=89
N=177
Contr
T2D
SAID
SIDD
SIRD
MOD
MARD
Clusters

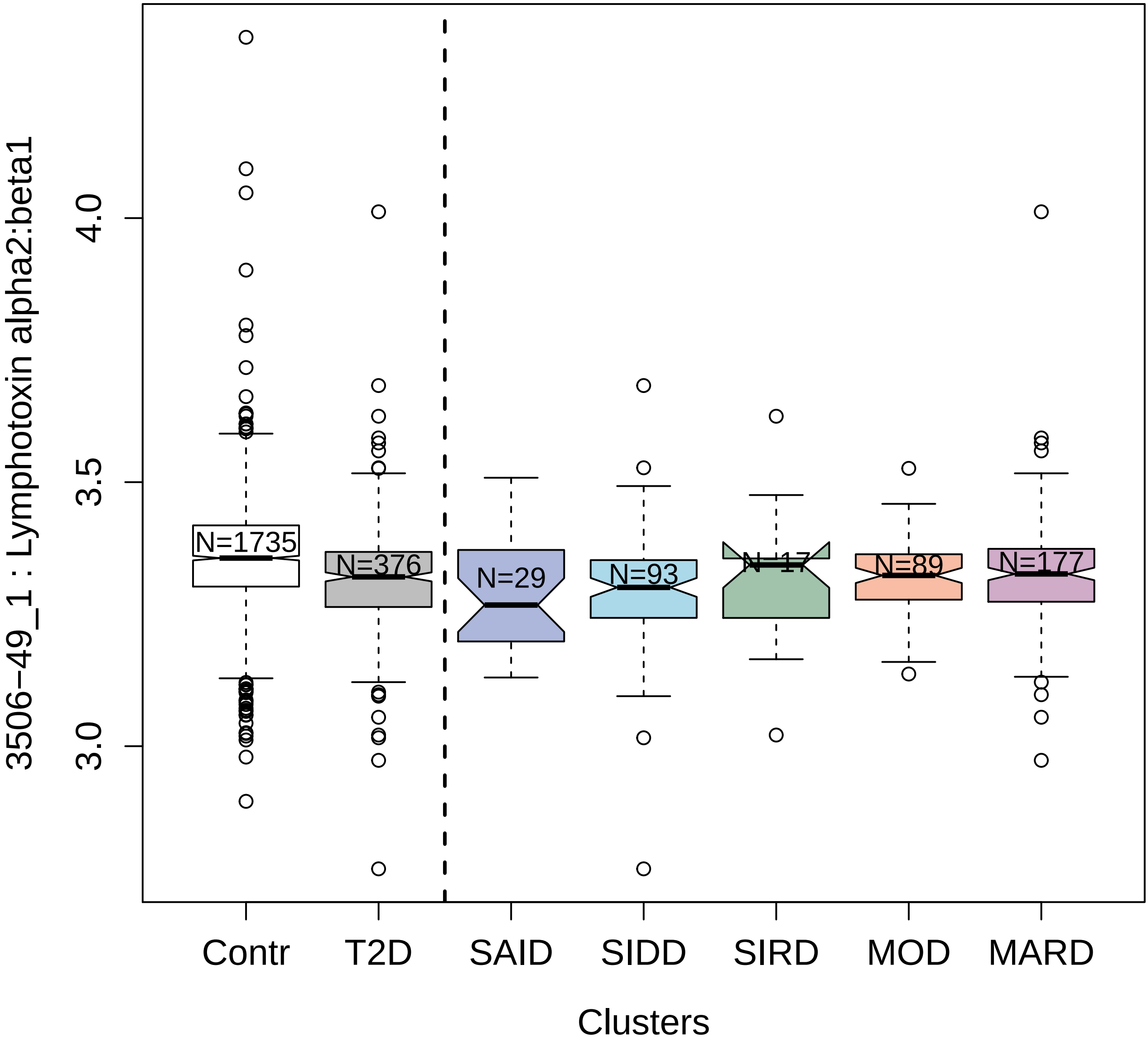

3506−49_1 : Lymphotoxin alpha2:beta1
4.0
3.5
3.0
N=1735
N=376
N=29
N=93
N=17
N=89
N=177
Contr
T2D
SAID
SIDD
SIRD
MOD
MARD
Clusters

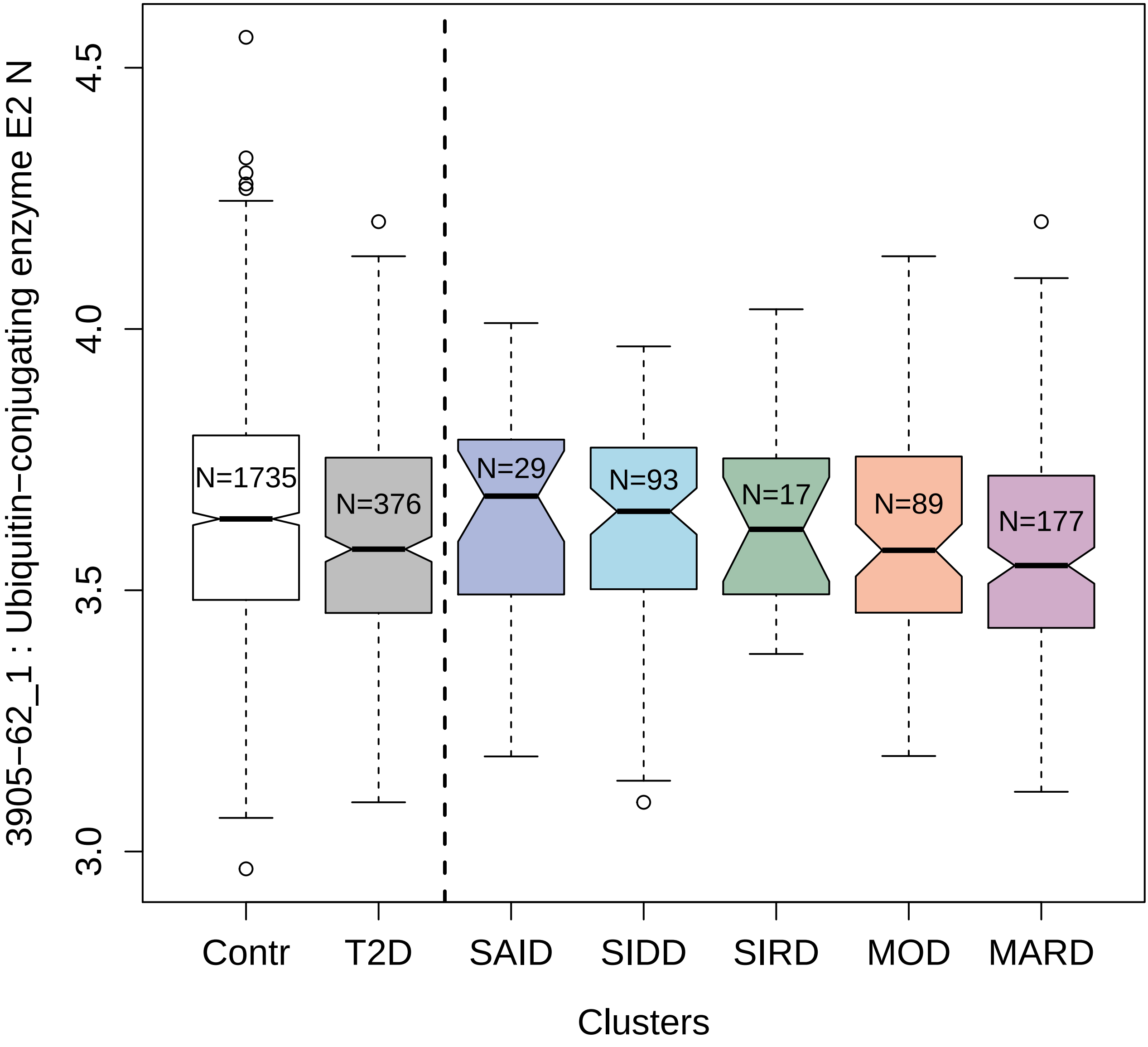
3905−62_1 : Ubiquitin−conjugating enzyme E2 N
4.5
4.0
3.5
3.0
N=1735
N=376
N=29
N=93
N=17
N=89
N=177
Contr
T2D
SAID
SIDD
SIRD
MOD
MARD
Clusters

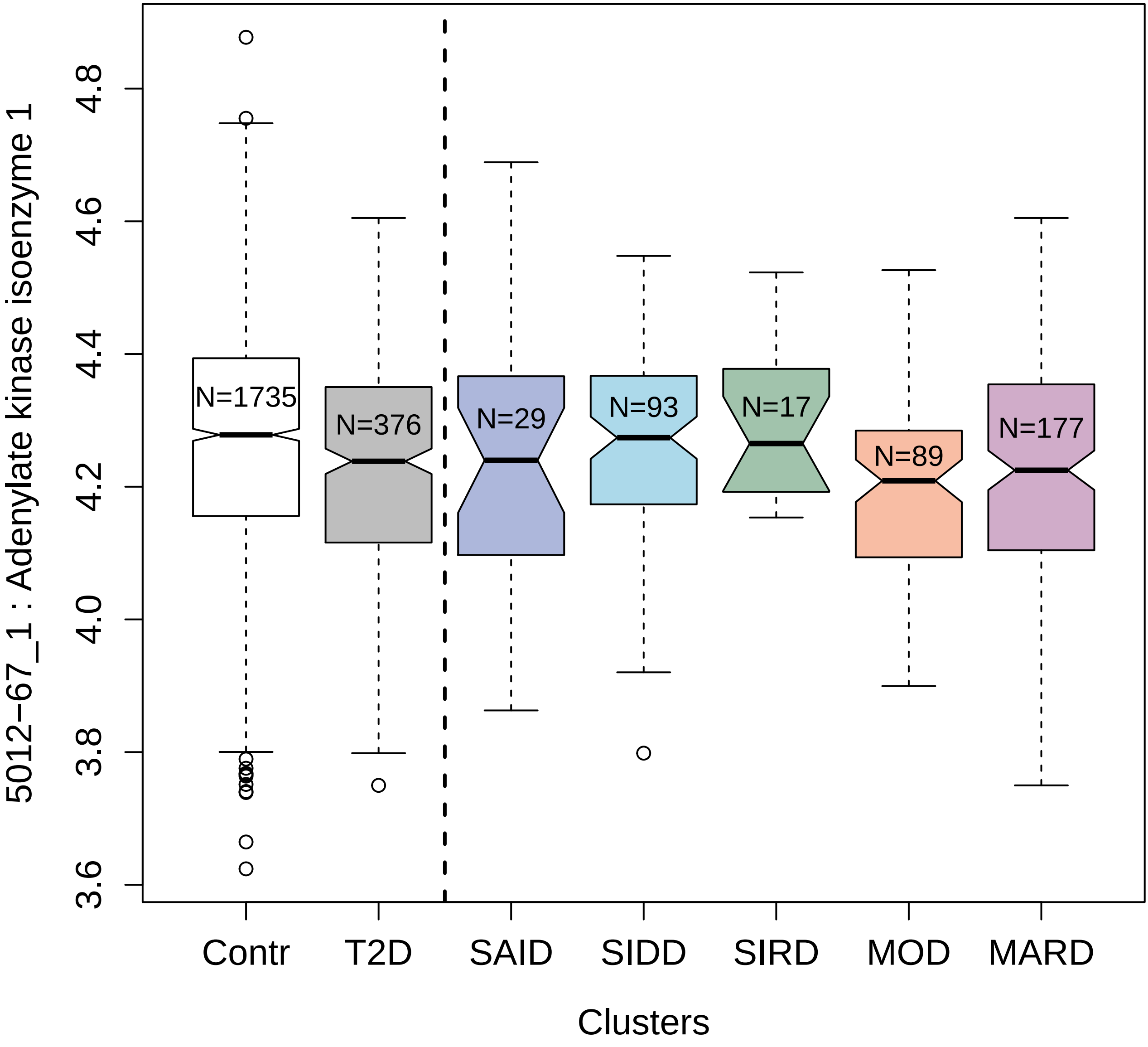

5012−67_1 : Adenylate kinase isoenzyme 1
3.6
3.8
4.0
4.2
4.4
4.6
4.8
N=1735
N=376
N=29
N=93
N=17
N=89
N=177
Contr
T2D
SAID
SIDD
SIRD
MOD
MARD
Clusters

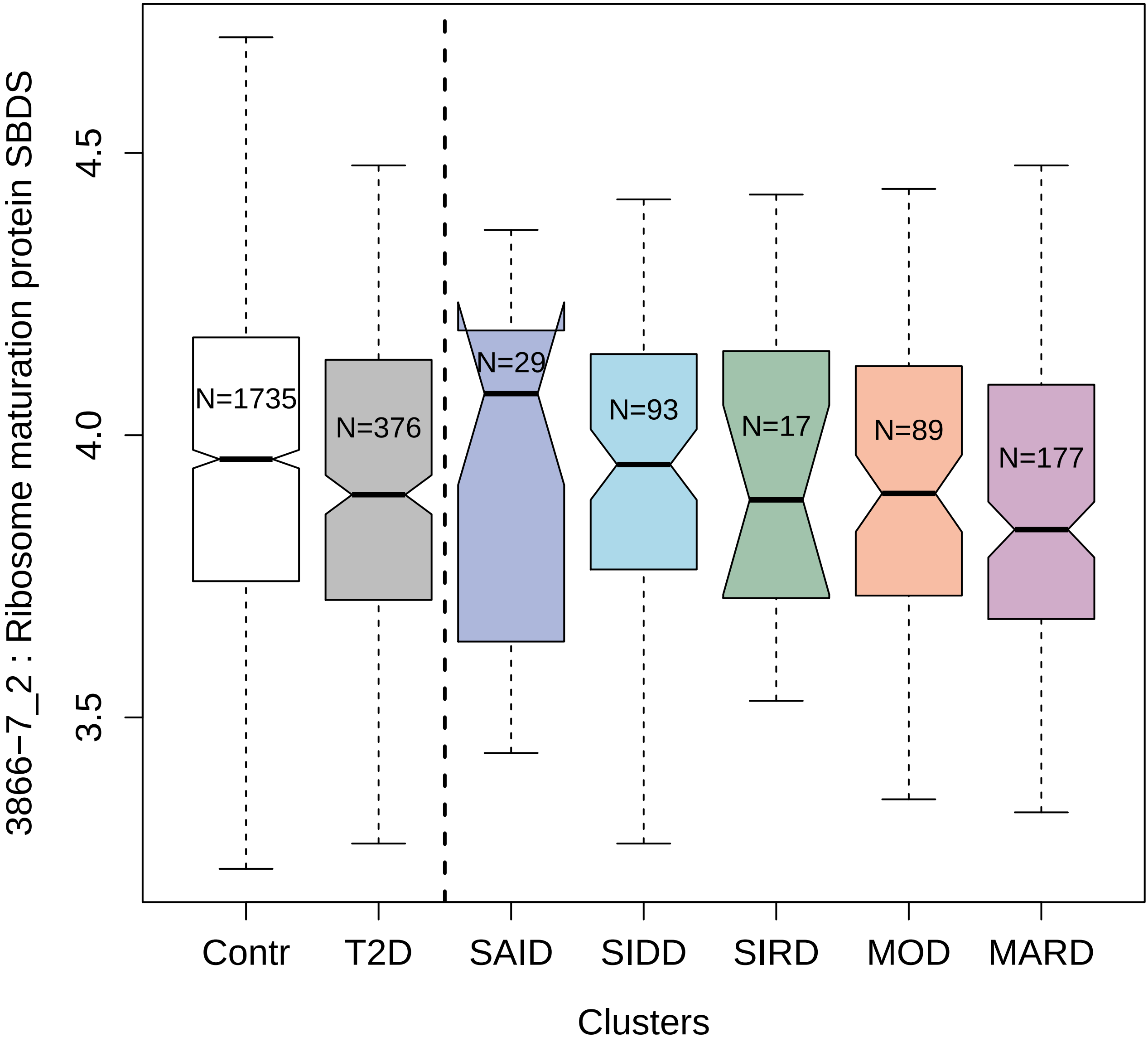
3866−7_2 : Ribosome maturation protein SBDS
3.5
4.0
4.5
N=1735
N=376
N=29
N=93
N=17
N=89
N=177
Contr
T2D
SAID
SIDD
SIRD
MOD
MARD
Clusters

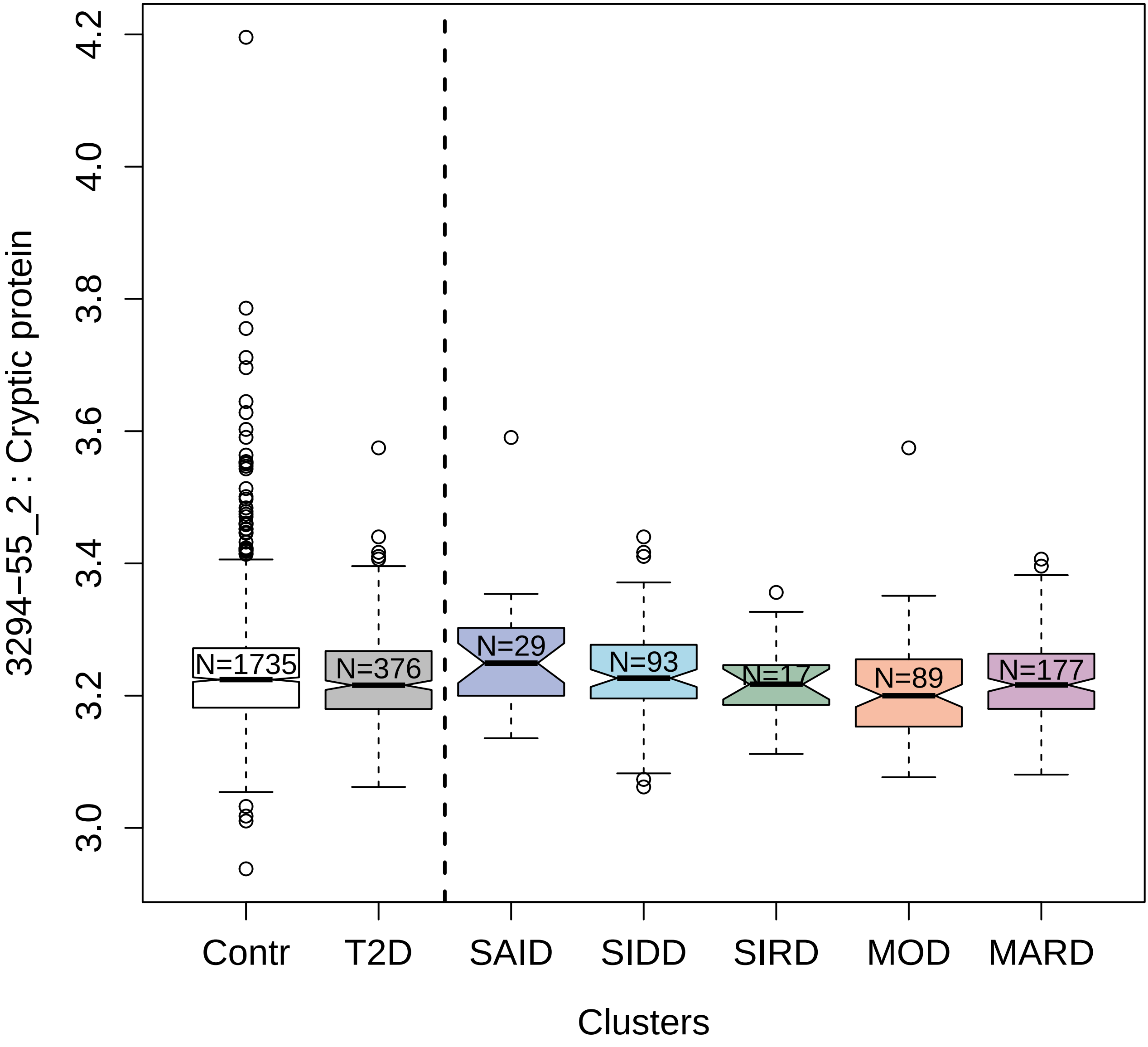
3294−55_2 : Cryptic protein
4.2
4.0
3.8
3.6
3.4
3.2
3.0
N=1735
N=376
N=29
N=93
N=17
N=89
N=177
Contr
T2D
SAID
SIDD
SIRD
MOD
MARD
Clusters

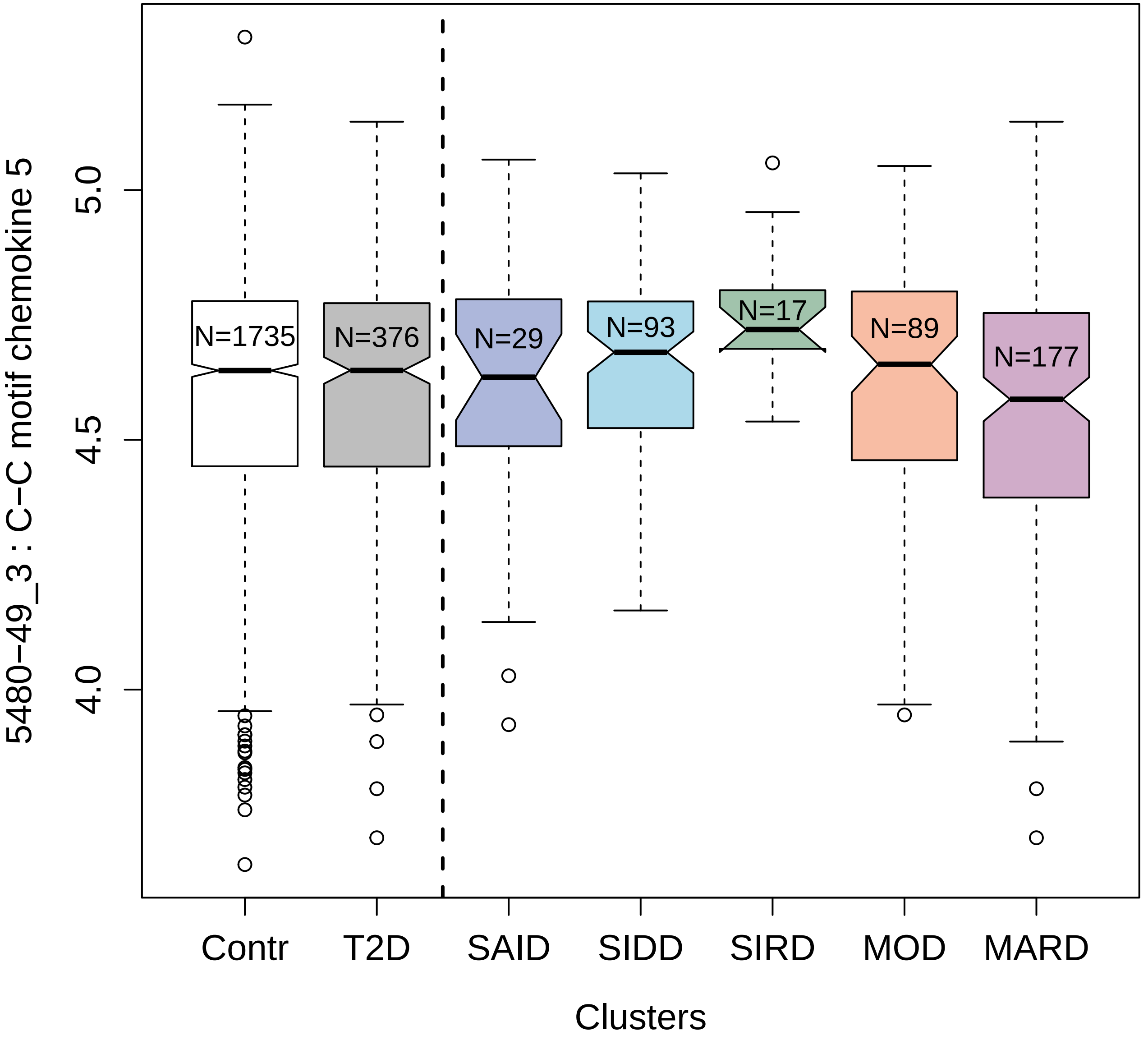

5480−49_3 : C−C motif chemokine 5
5.0
4.5
4.0
N=1735
N=376
N=29
N=93
N=17
N=89
N=177
Contr
T2D
SAID
SIDD
SIRD
MOD
MARD
Clusters

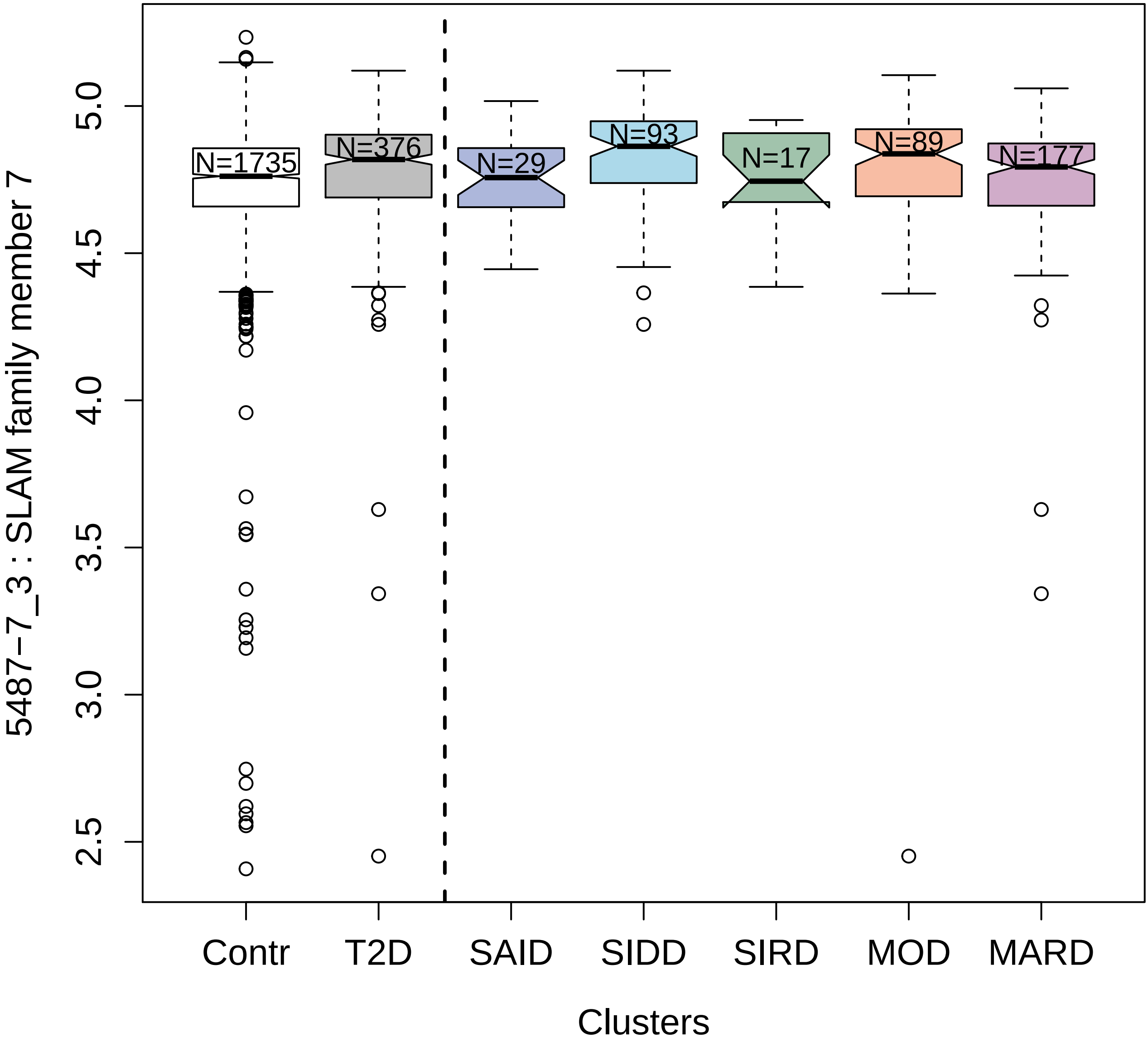
5487−7_3 : SLAM family member 7
5.0
4.5
4.0
3.5
3.0
2.5
N=1735
N=376
N=29
N=93
N=17
N=89
N=177
Contr
T2D
SAID
SIDD
SIRD
MOD
MARD
Clusters

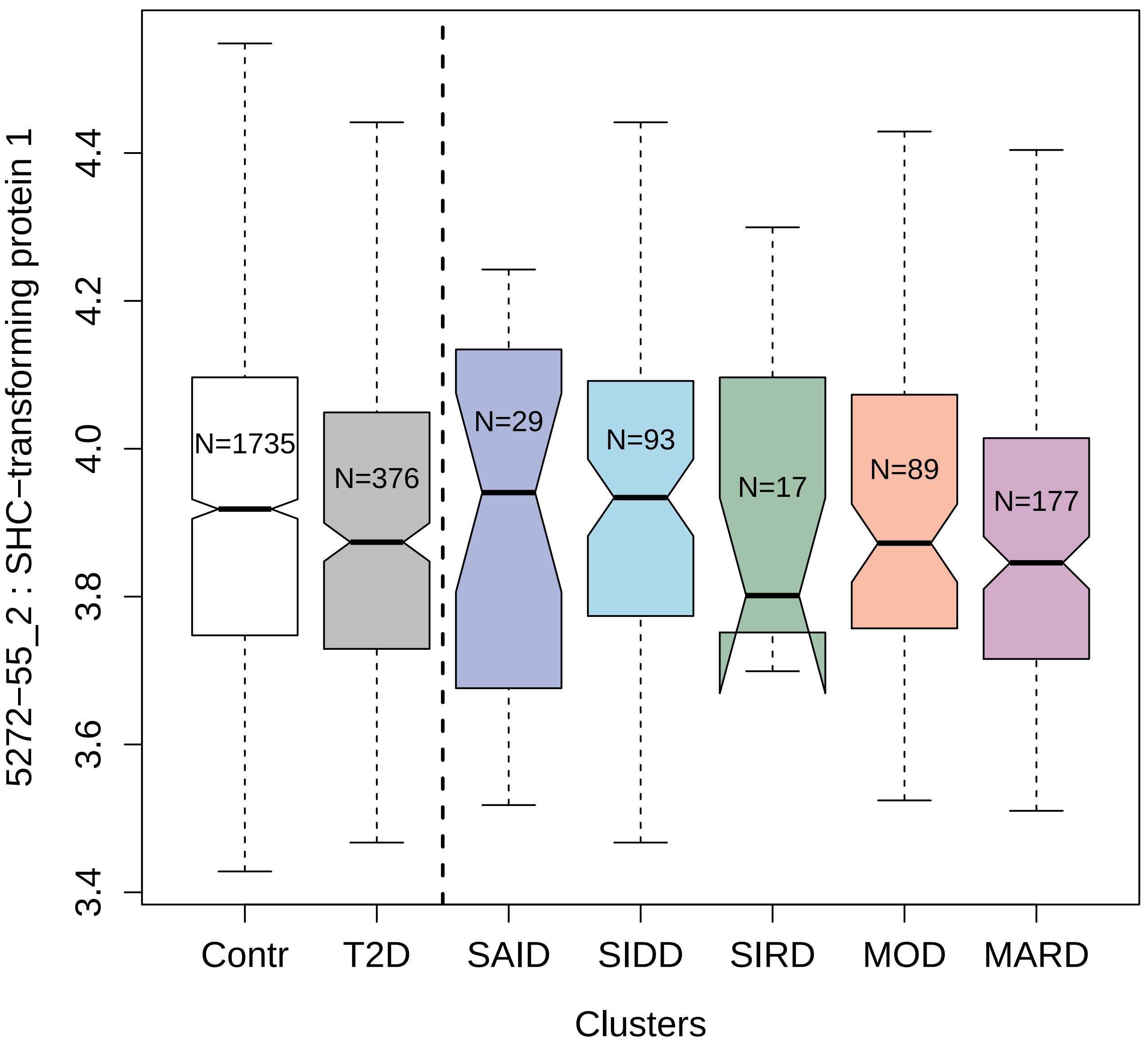
5272−55_2 : SHC−transforming protein 1
3.4
3.6
3.8
4.0
4.2
4.4
N=1735
N=376
N=29
N=93
N=17
N=89
N=177
Contr
T2D
SAID
SIDD
SIRD
MOD
MARD
Clusters

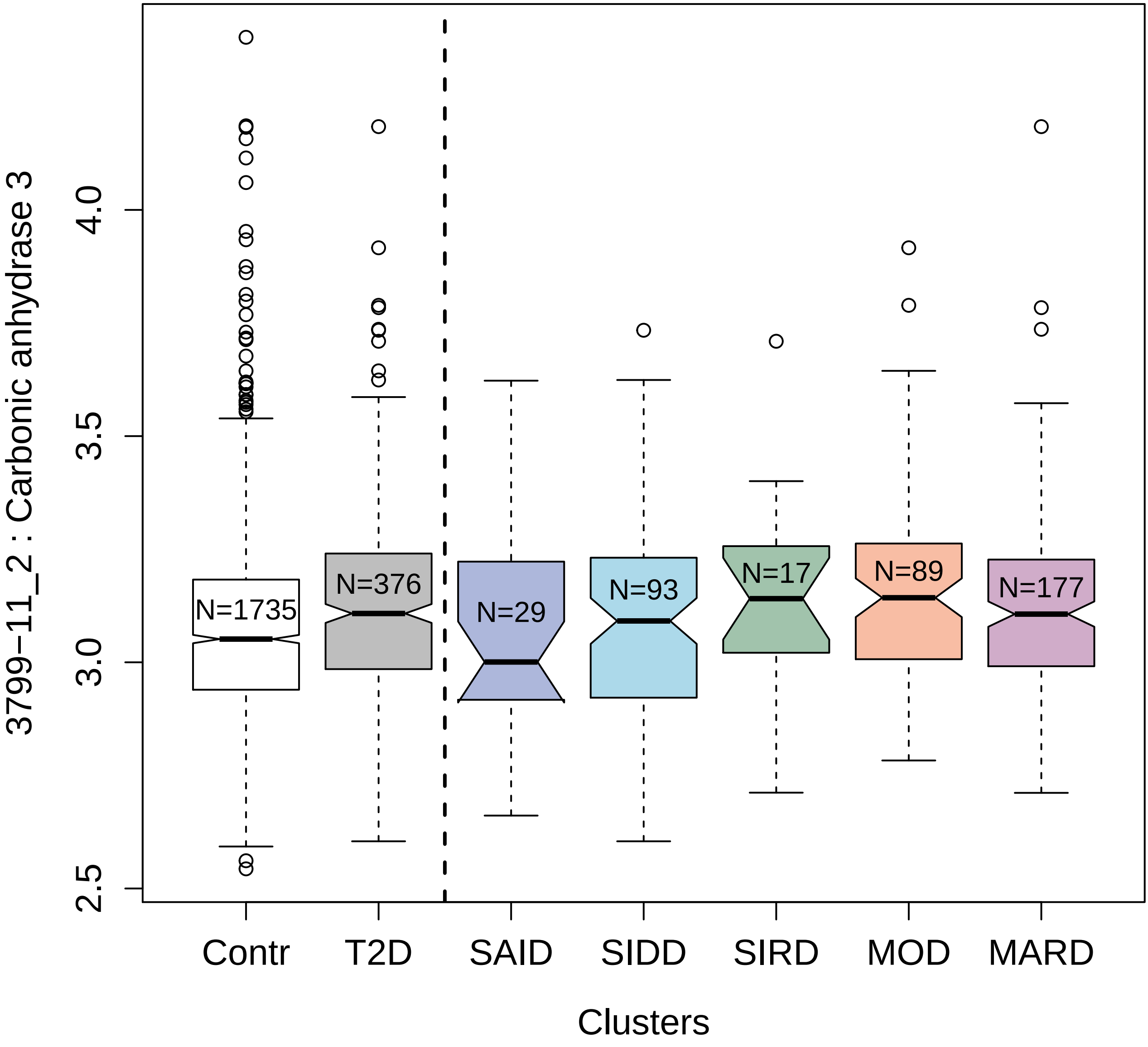

3799−11_2 : Carbonic anhydrase 3
2.5
3.0
3.5
4.0
N=1735
N=376
N=29
N=93
N=17
N=89
N=177
Contr
T2D
SAID
SIDD
SIRD
MOD
MARD
Clusters

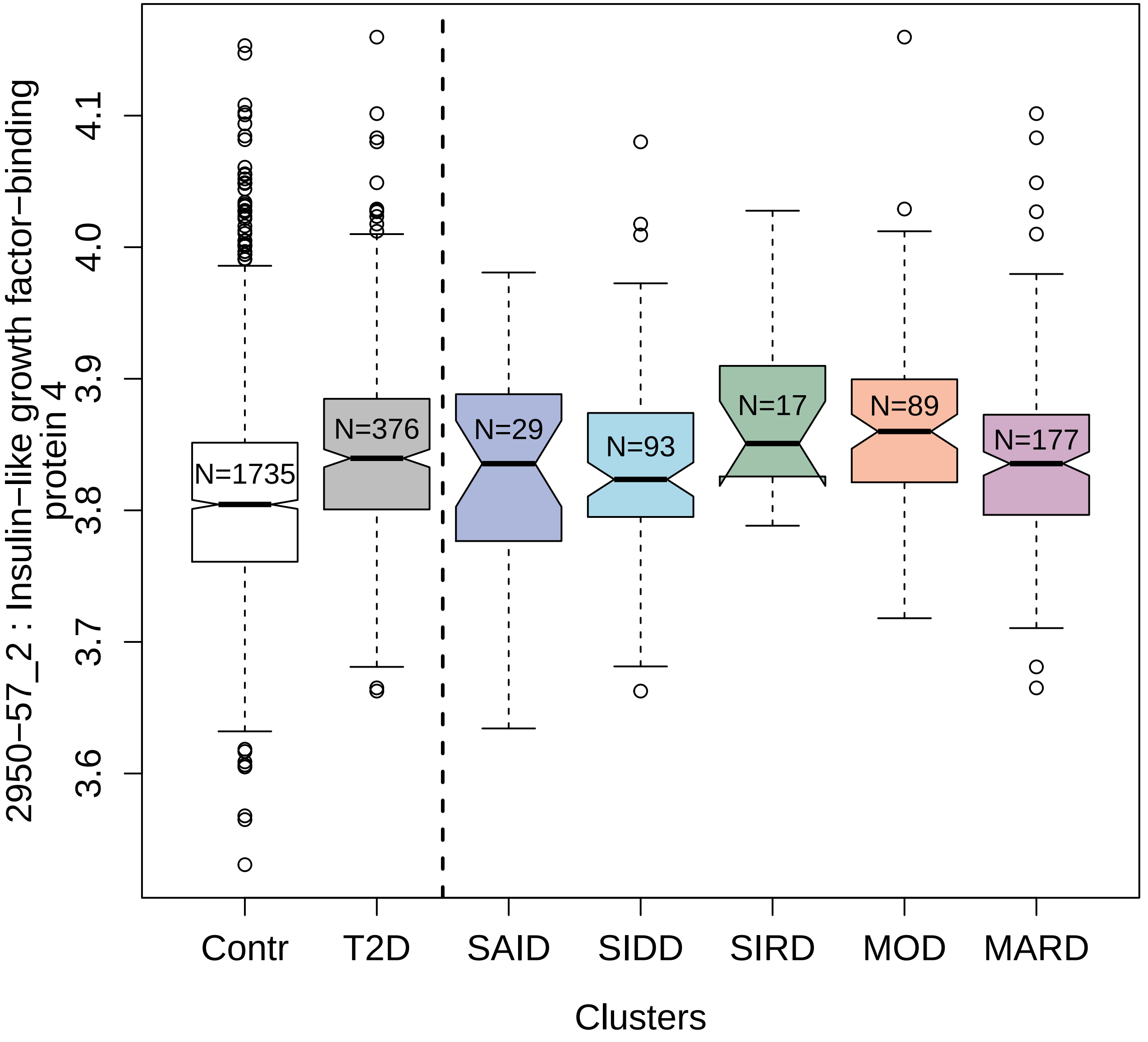

2950−57_2 : Insulin−like growth factor−binding protein 4
3.6
3.7
3.8
3.9
4.0
4.1
N=1735
N=376
N=29
N=93
N=17
N=89
N=177
Contr
T2D
SAID
SIDD
SIRD
MOD
MARD
Clusters

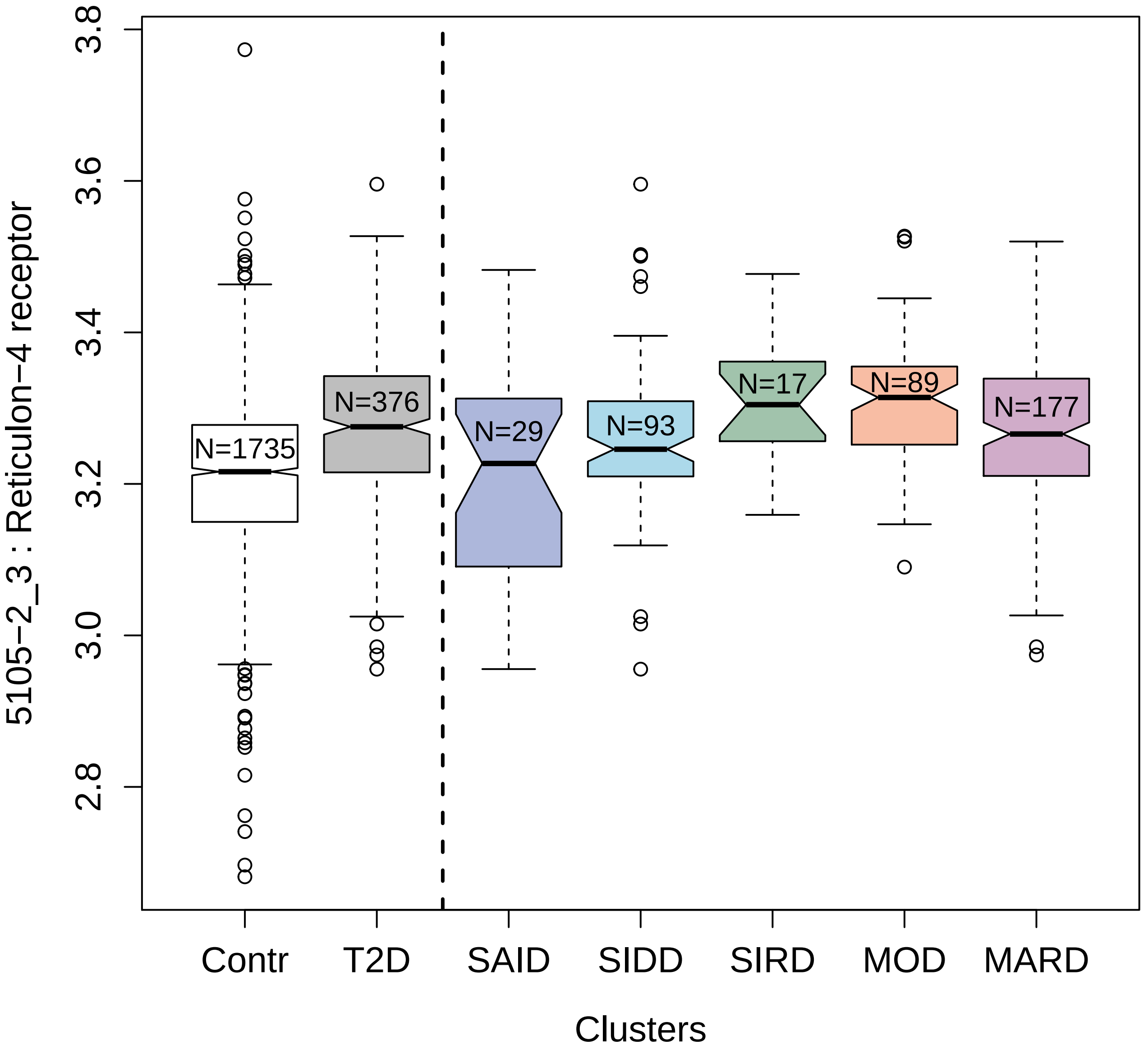

5105−2_3 : Reticulon−4 receptor
3.8
3.6
3.4
3.2
3.0
2.8
N=1735
N=376
N=29
N=93
N=17
N=89
N=177
Contr
T2D
SAID
SIDD
SIRD
MOD
MARD
Clusters

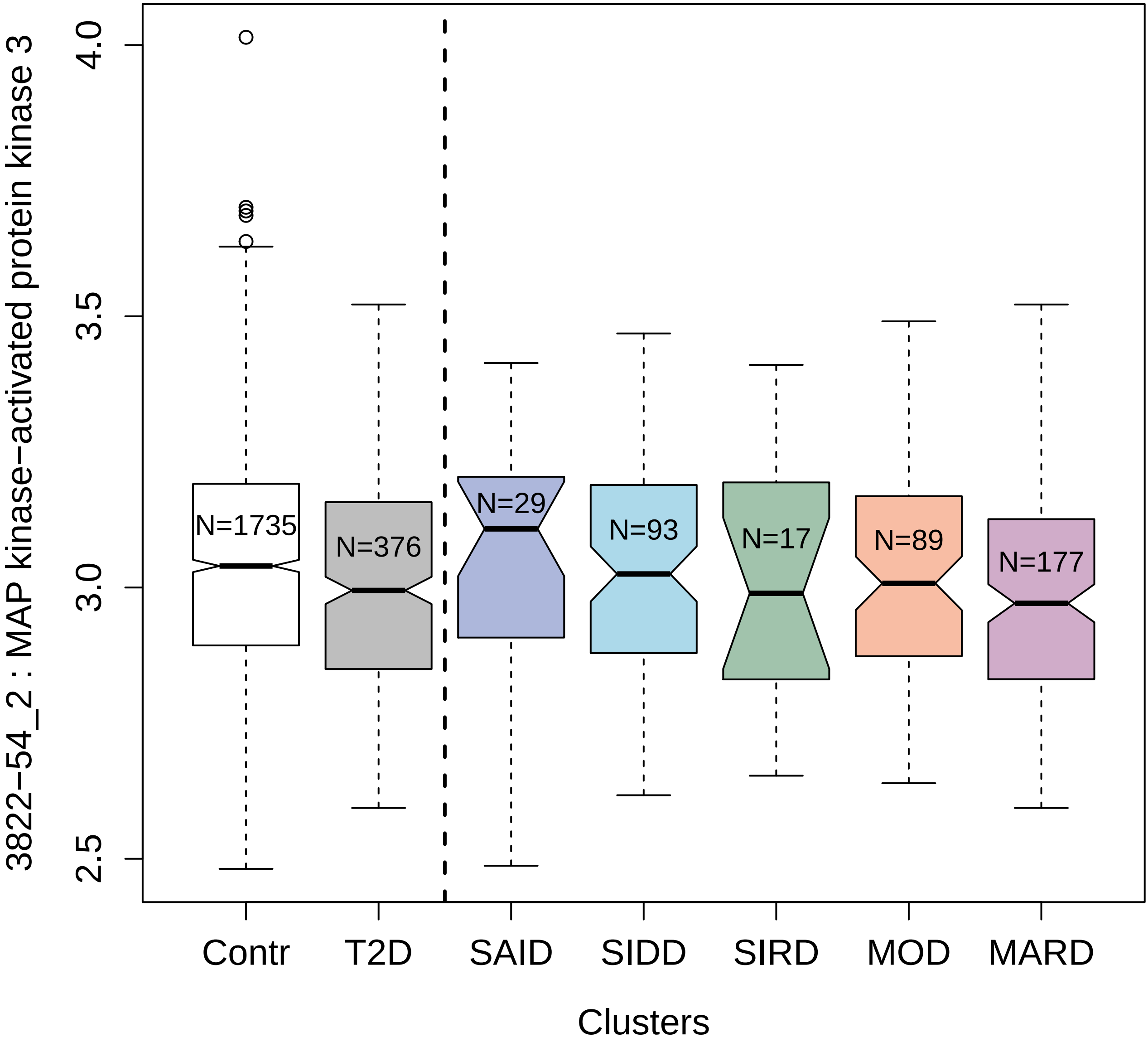

3822−54_2 : MAP kinase−activated protein kinase 3
4.0
3.5
3.0
2.5
N=1735
N=376
N=29
N=93
N=17
N=89
N=177
Contr
T2D
SAID
SIDD
SIRD
MOD
MARD
Clusters

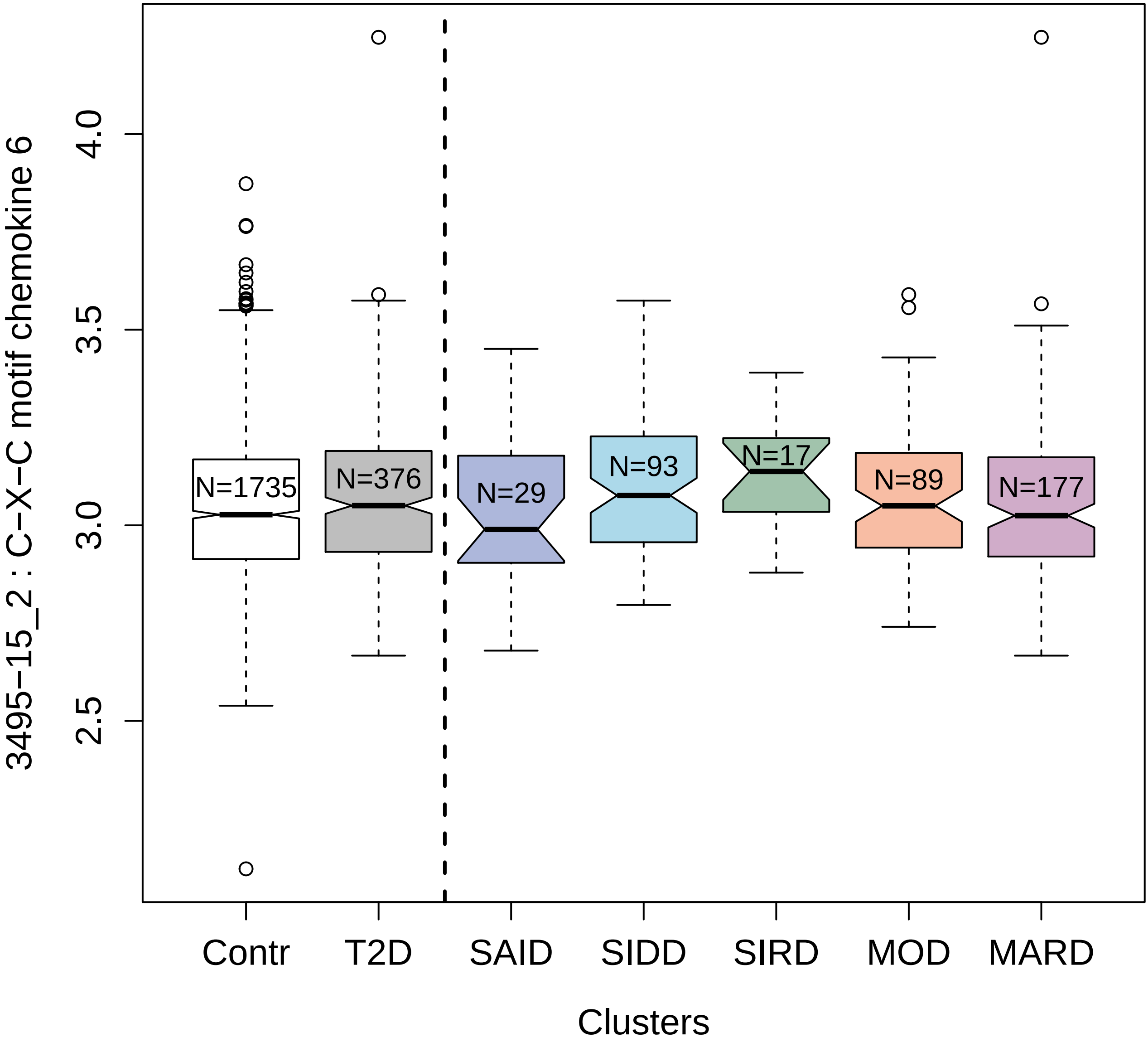

3495−15_2 : C−X−C motif chemokine 6
2.5
3.0
3.5
4.0
N=1735
N=376
N=29
N=93
N=17
N=89
N=177
Contr
T2D
SAID
SIDD
SIRD
MOD
MARD
Clusters

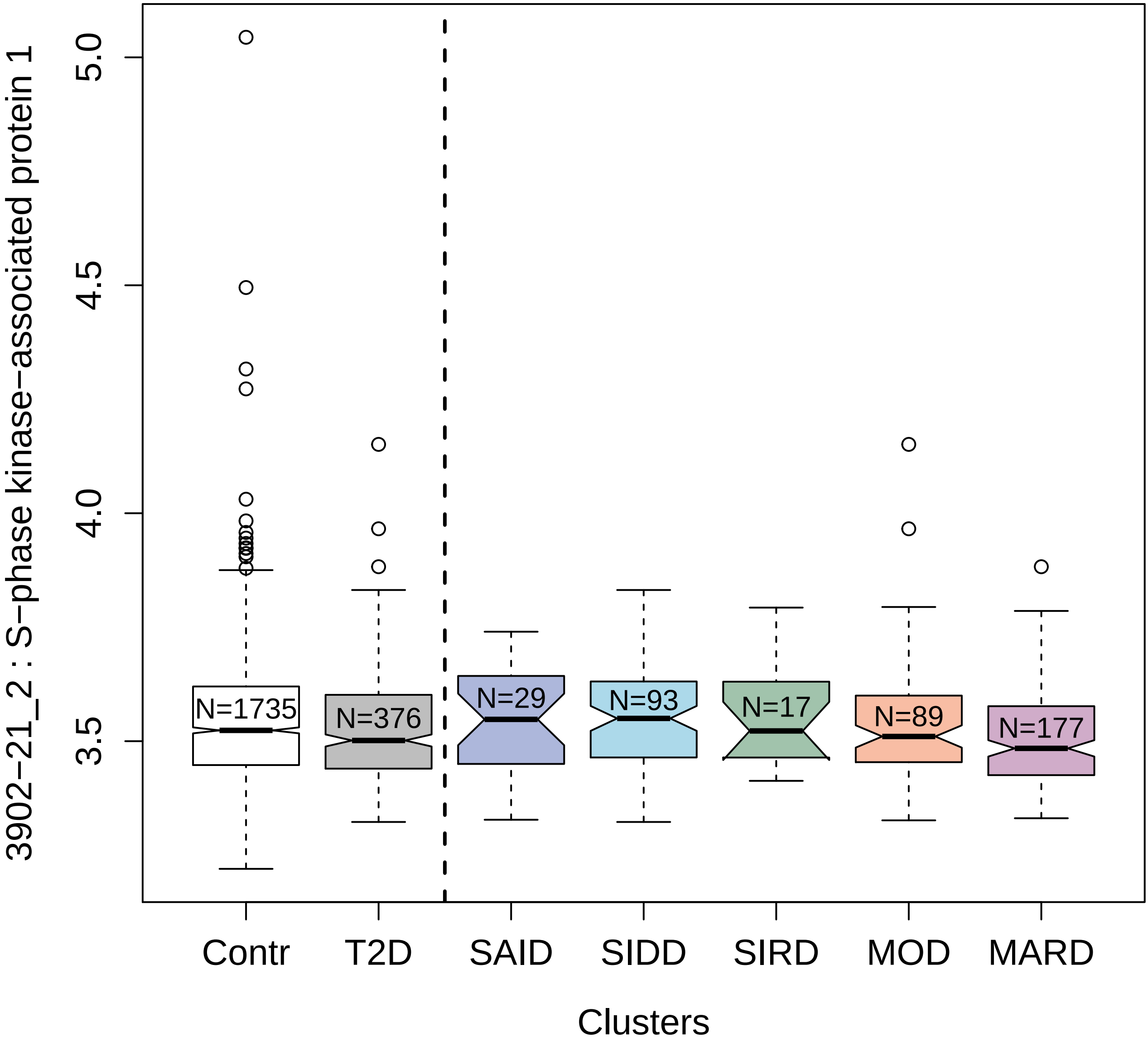
3902−21_2 : S−phase kinase−associated protein 1
5.0
4.5
4.0
3.5
N=1735
N=376
N=29
N=93
N=17
N=89
N=177
Contr
T2D
SAID
SIDD
SIRD
MOD
MARD
Clusters

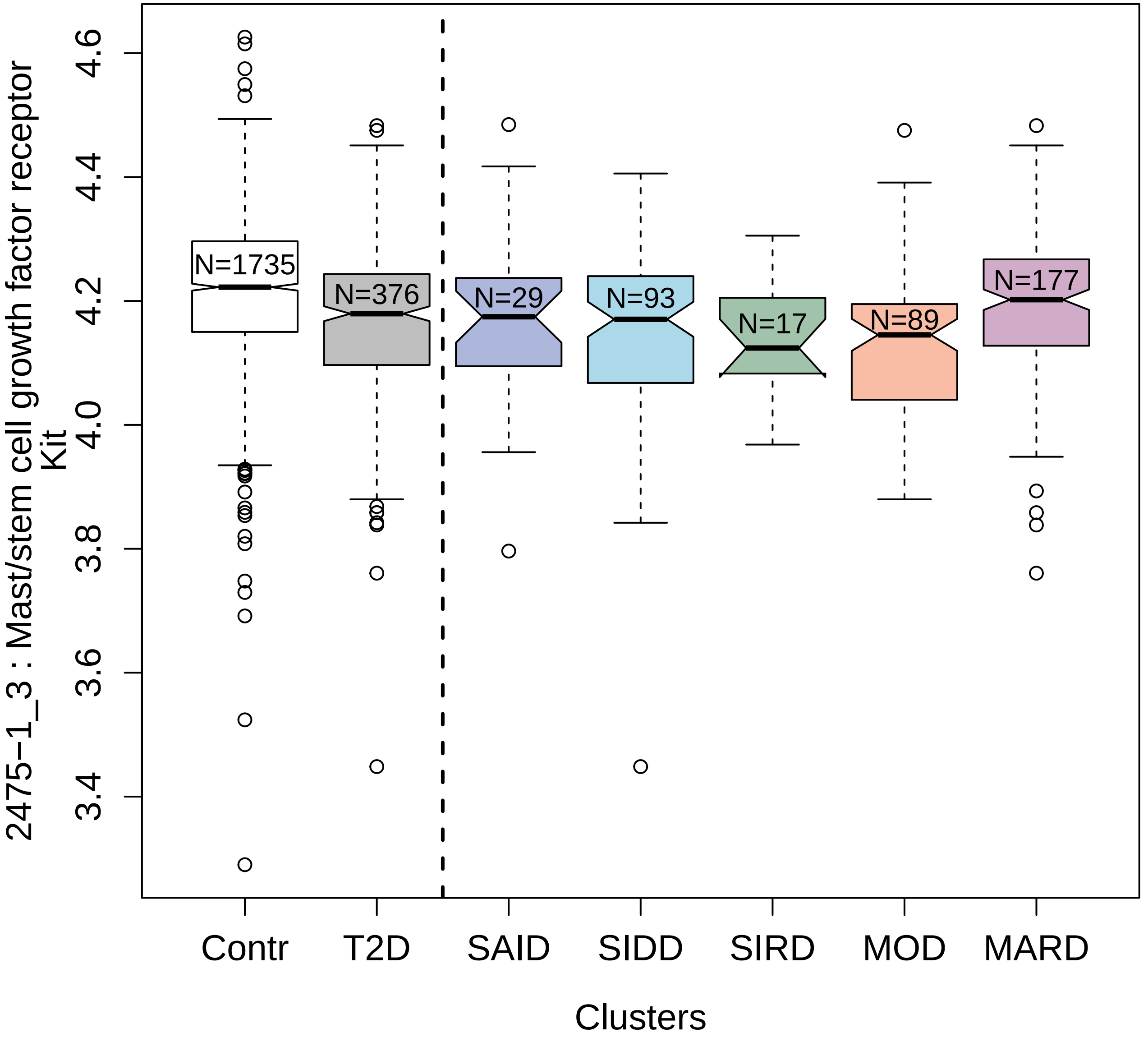

2475−1_3 : Mast/stem cell growth factor receptor
Kit
3.4
3.6
3.8
4.0
4.2
4.4
4.6
N=1735
N=376
N=29
N=93
N=17
N=89
N=177
Contr
T2D
SAID
SIDD
SIRD
MOD
MARD
Clusters

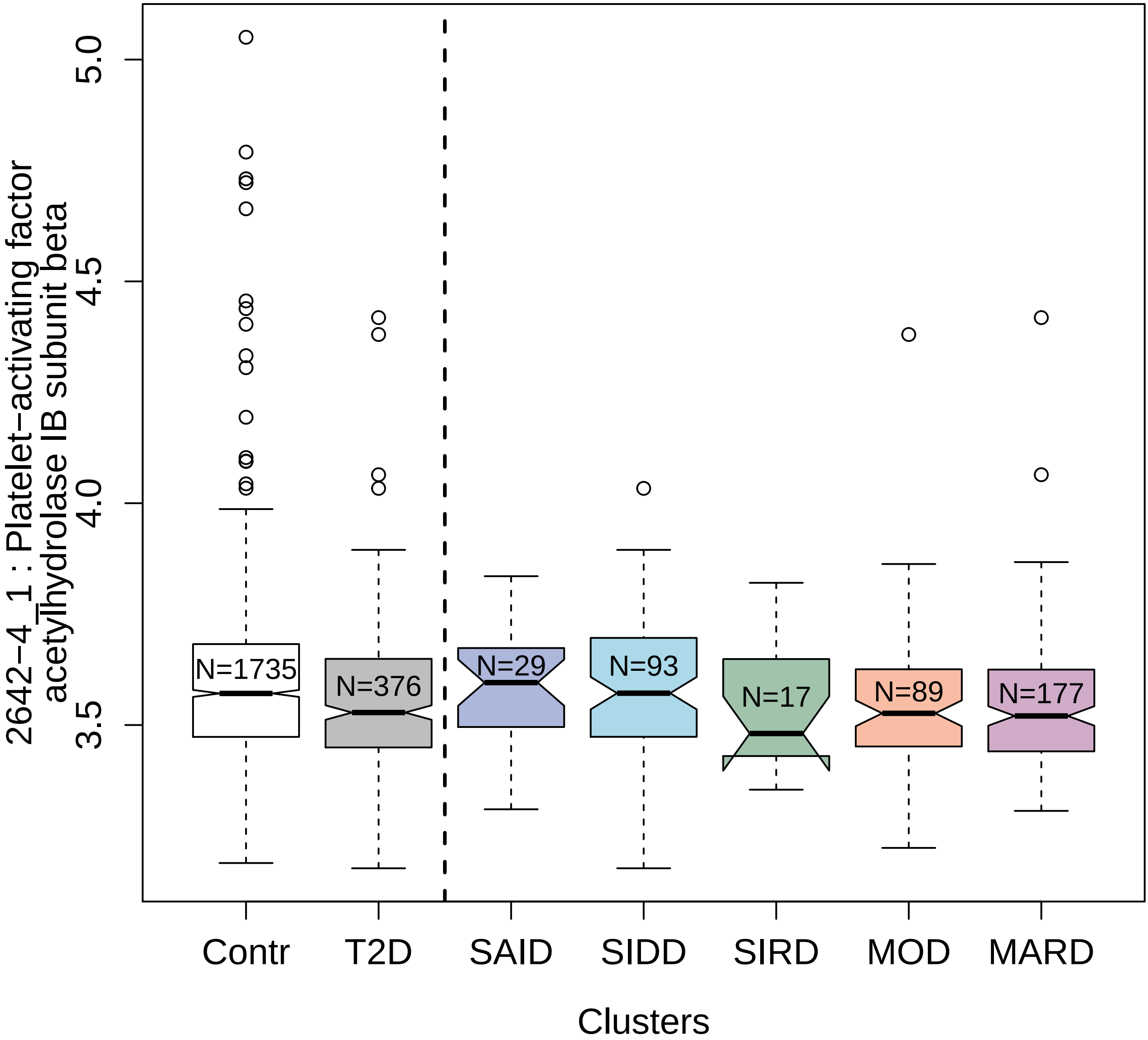

2642−4_1 : Platelet−activating factor acetylhydrolase IB subunit beta
5.0
4.5
4.0
3.5
N=1735
N=376
N=29
N=93
N=17
N=89
N=177
Contr
T2D
SAID
SIDD
SIRD
MOD
MARD
Clusters

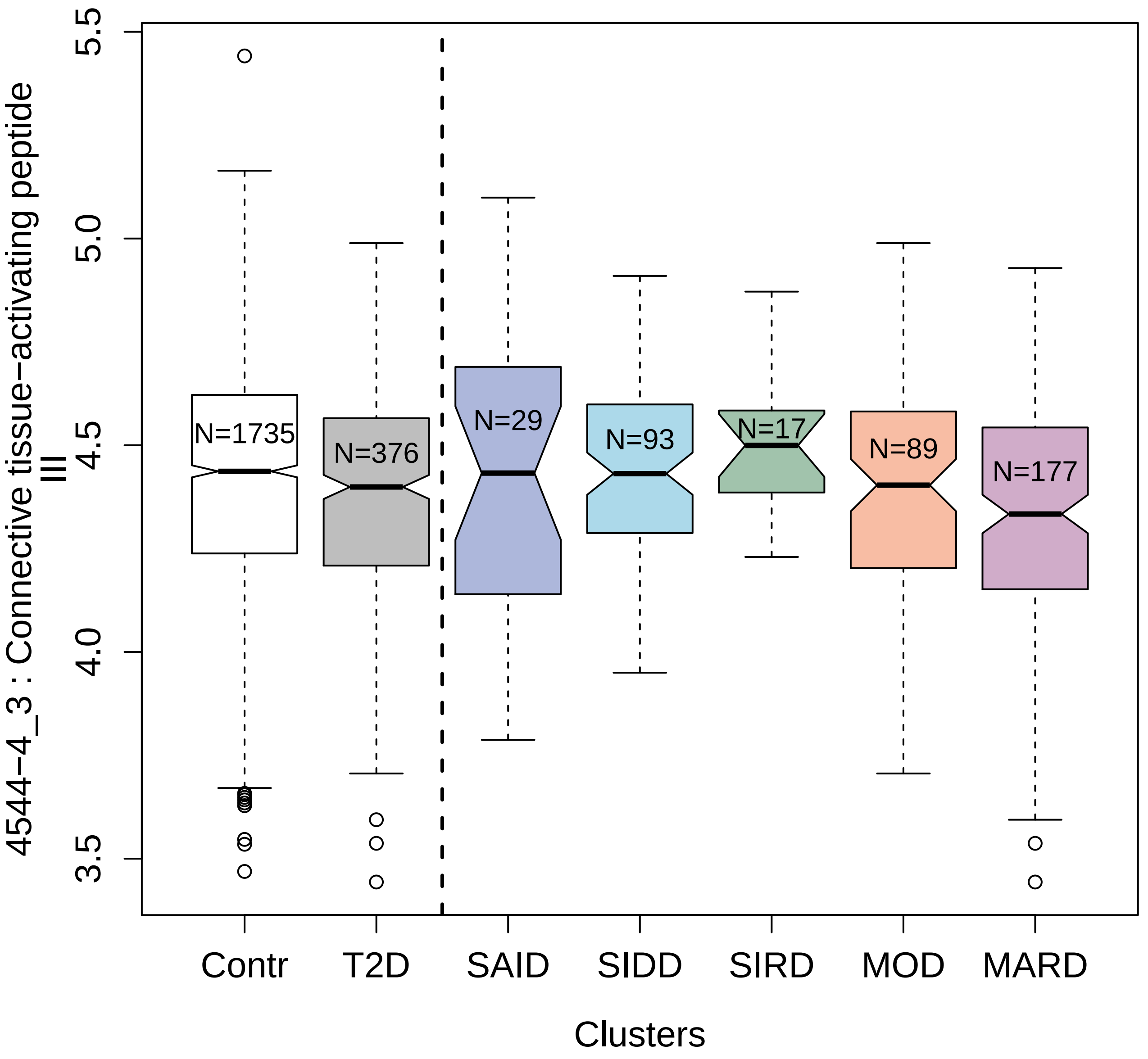

4544−4_3 : Connective tissue−activating peptide III
5.5
5.0
4.5
4.0
3.5
N=1735
N=376
N=29
N=93
N=17
N=89
N=177
Contr
T2D
SAID
SIDD
SIRD
MOD
MARD
Clusters

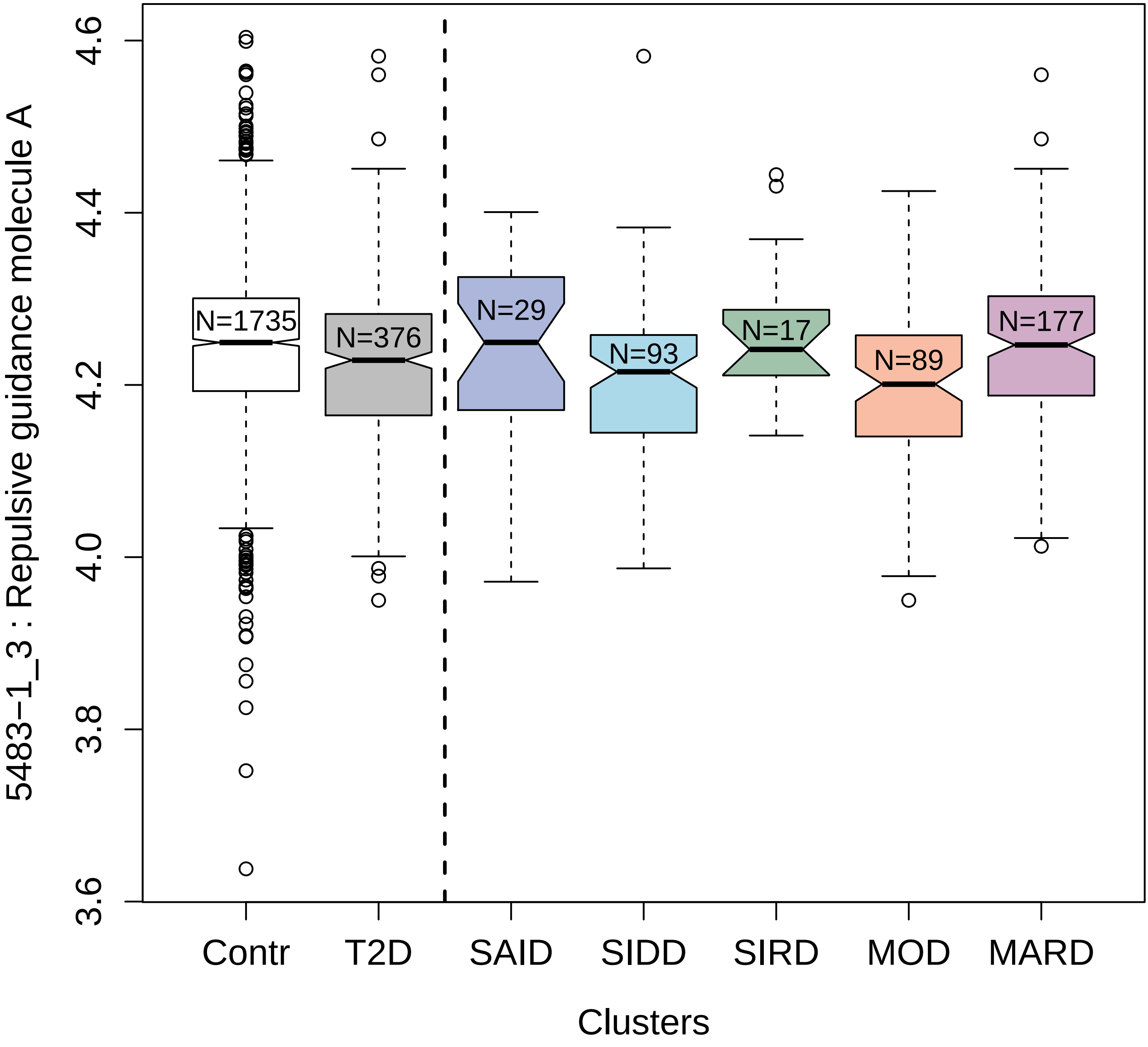

5483−1_3 : Repulsive guidance molecule A
3.6
3.8
4.0
4.2
4.4
4.6
N=1735
N=376
N=29
N=93
N=17
N=89
N=177
Contr
T2D
SAID
SIDD
SIRD
MOD
MARD
Clusters

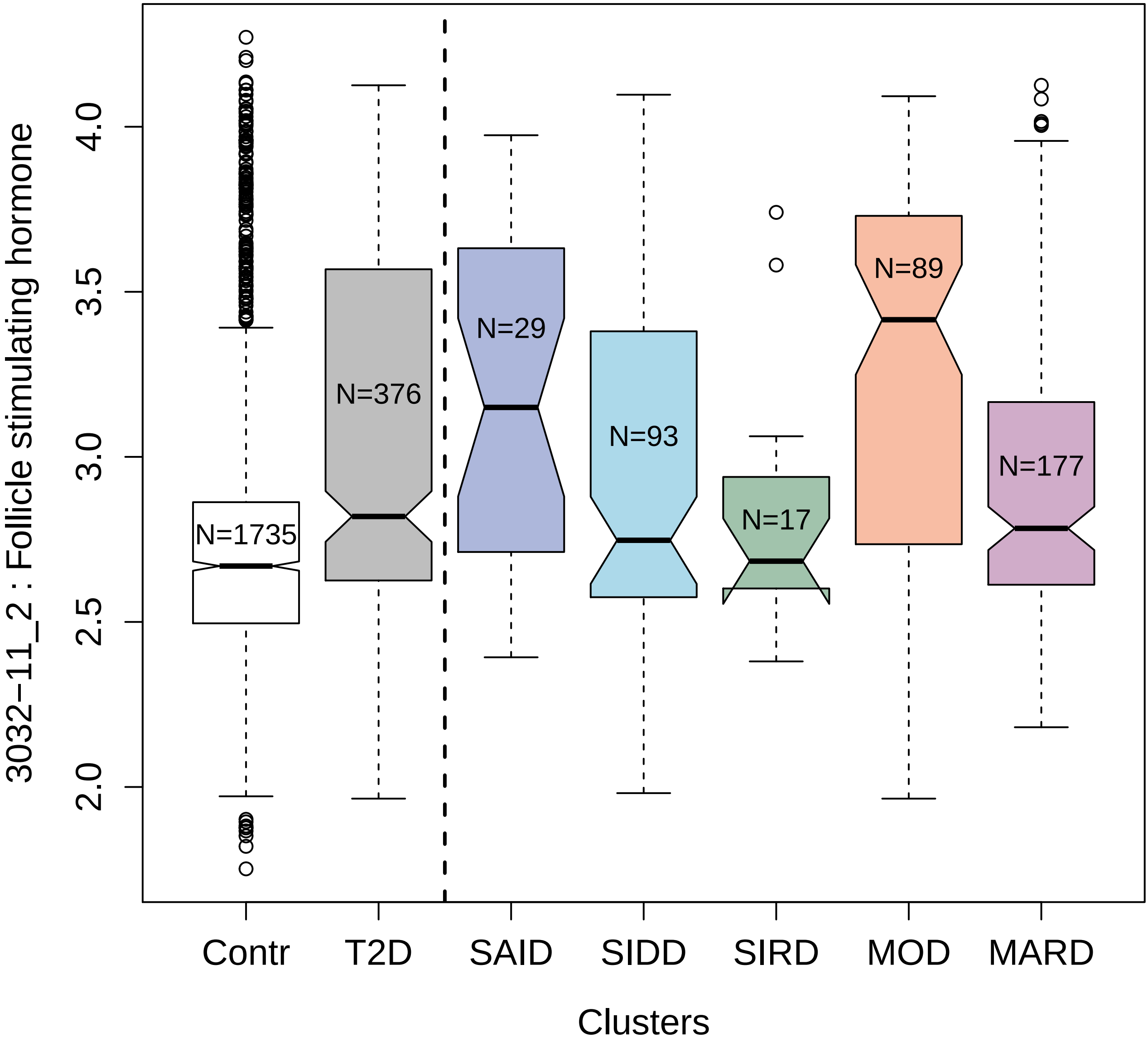

3032−11_2 : Follicle stimulating hormone
2.0
2.5
3.0
3.5
4.0
N=1735
N=376
N=29
N=93
N=17
N=89
N=177
Contr
T2D
SAID
SIDD
SIRD
MOD
MARD
Clusters

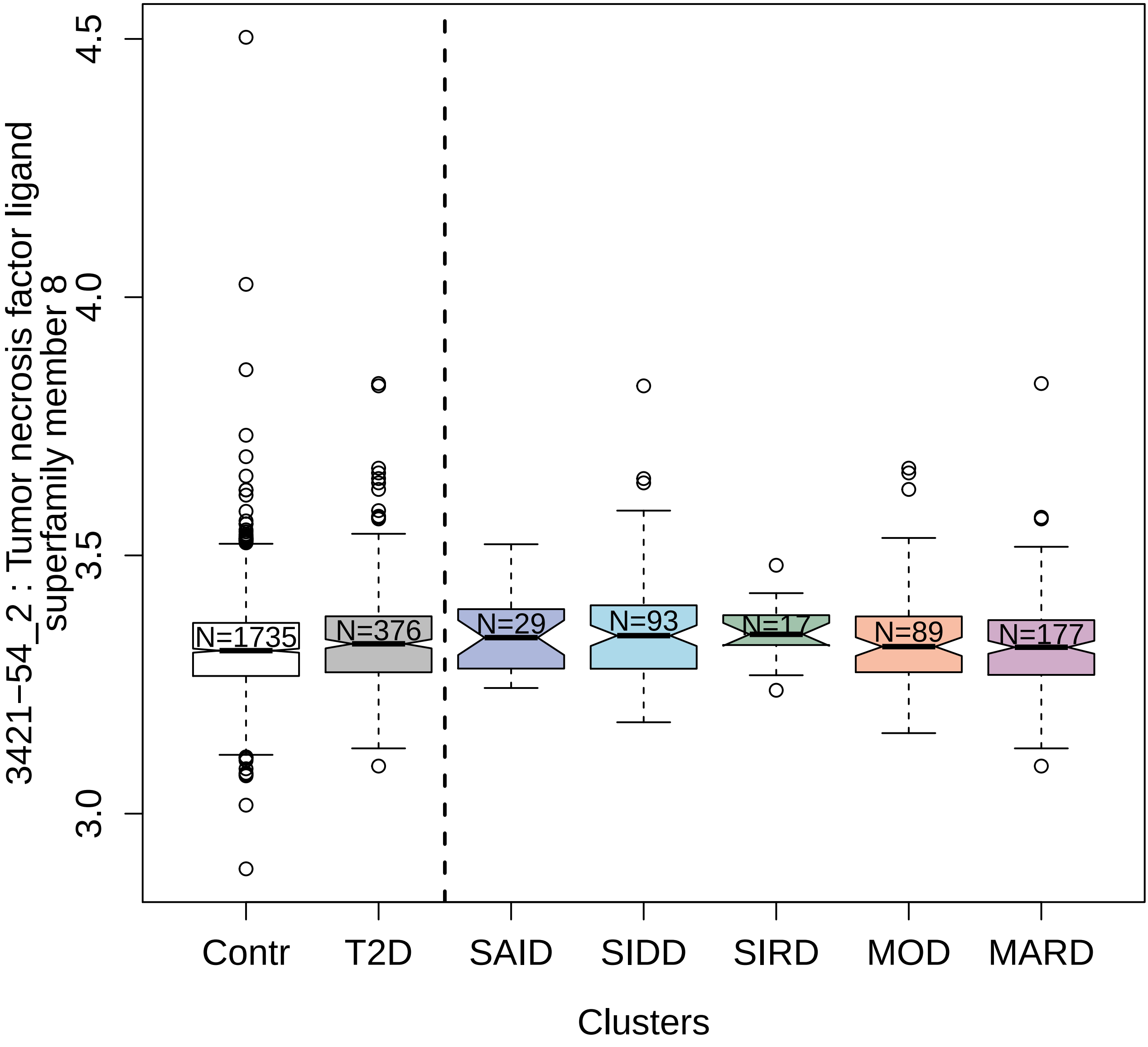

3421−54_2 : Tumor necrosis factor ligand
superfamily member 8
4.5
4.0
3.5
3.0
N=1735
N=376
N=29
N=93
N=17
N=89
N=177
Contr
T2D
SAID
SIDD
SIRD
MOD
MARD
Clusters

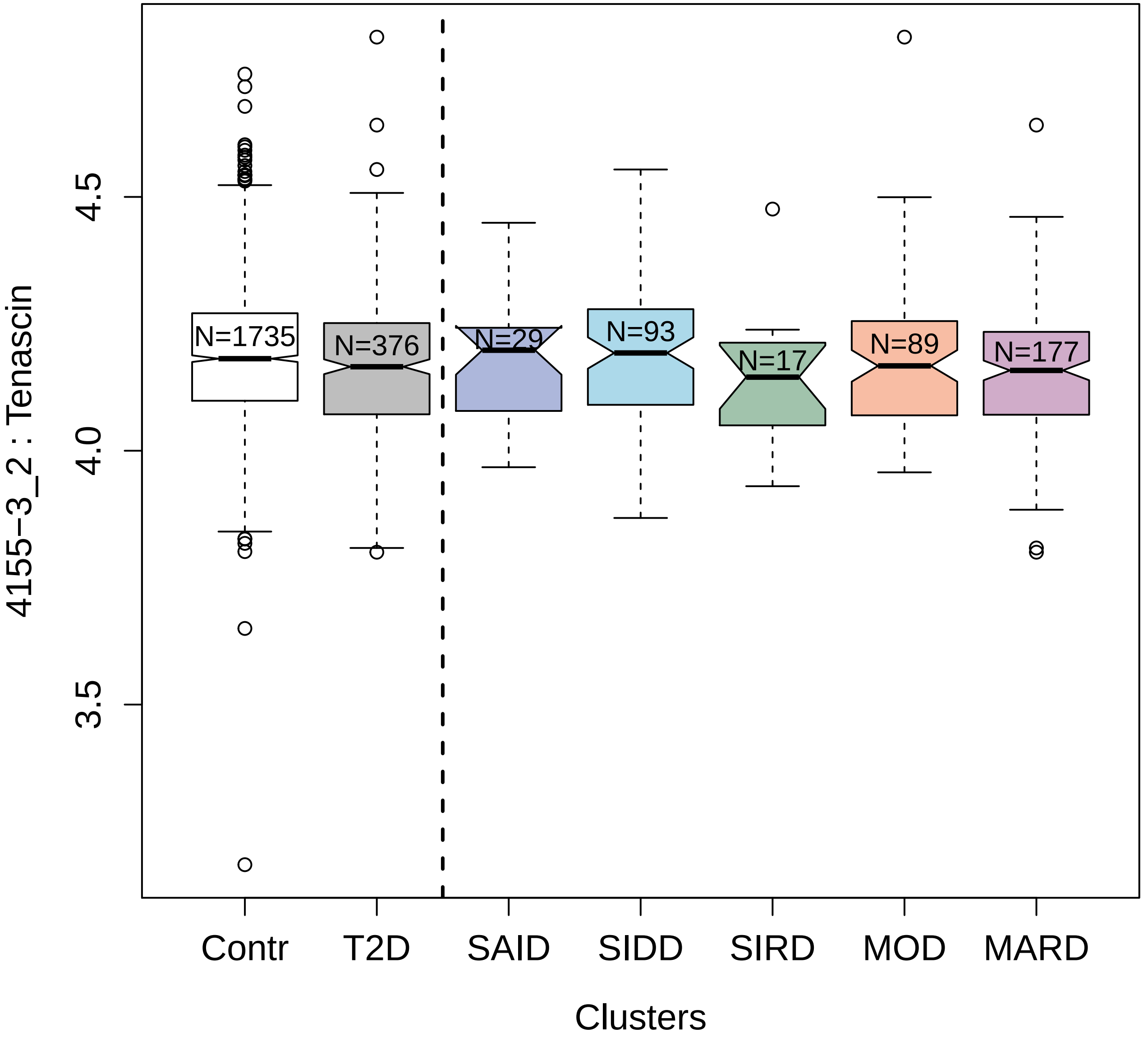

4155−3_2 : Tenascin
3.5
4.0
4.5
N=1735
N=376
N=29
N=93
N=17
N=89
N=177
Contr
T2D
SAID
SIDD
SIRD
MOD
MARD
Clusters

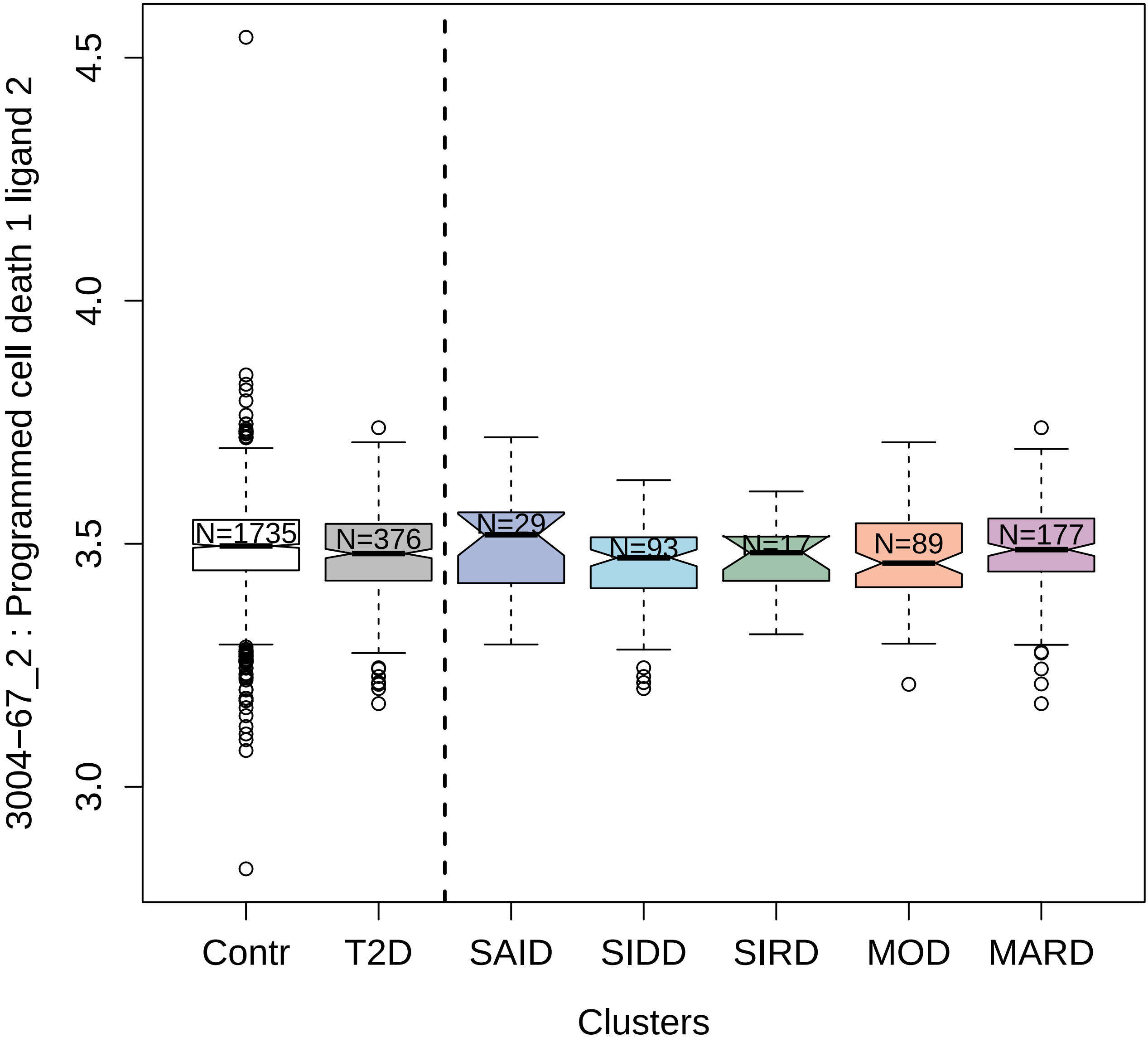
3004−67_2 : Programmed cell death 1 ligand 2
4.5
4.0
3.5
3.0
N=1735
N=376
N=29
N=93
N=17
N=89
N=177
Contr
T2D
SAID
SIDD
SIRD
MOD
MARD
Clusters

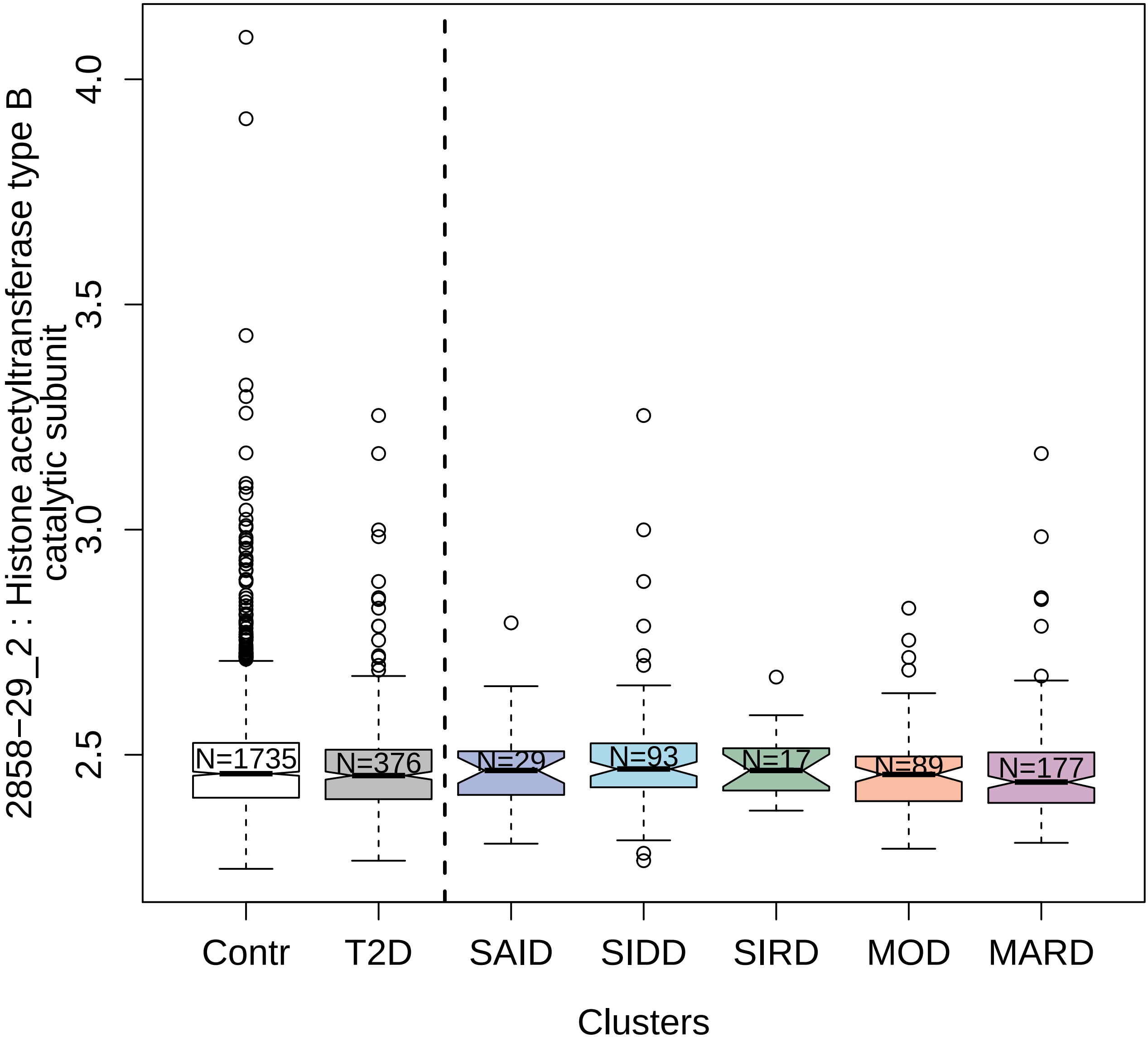
2858−29_2 : Histone acetyltransferase type B catalytic subunit
4.0
3.5
3.0
2.5
N=1735
N=376
N=29
N=93
N=17
N=89
N=177
Contr
T2D
SAID
SIDD
SIRD
MOD
MARD
Clusters

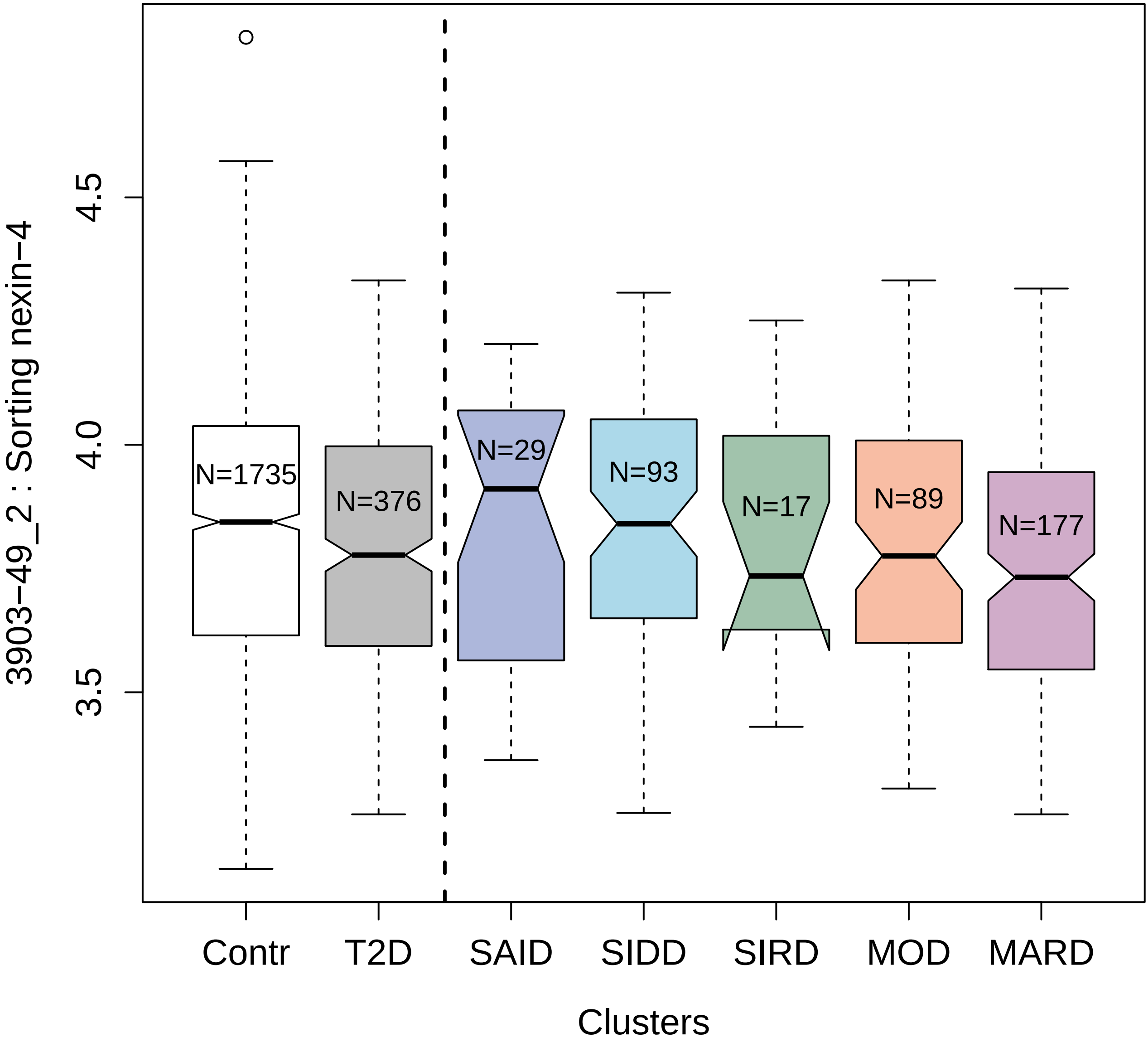

3903−49_2 : Sorting nexin−4
4.5
4.0
3.5
N=1735
N=376
N=29
N=93
N=17
N=89
N=177
Contr
T2D
SAID
SIDD
SIRD
MOD
MARD
Clusters

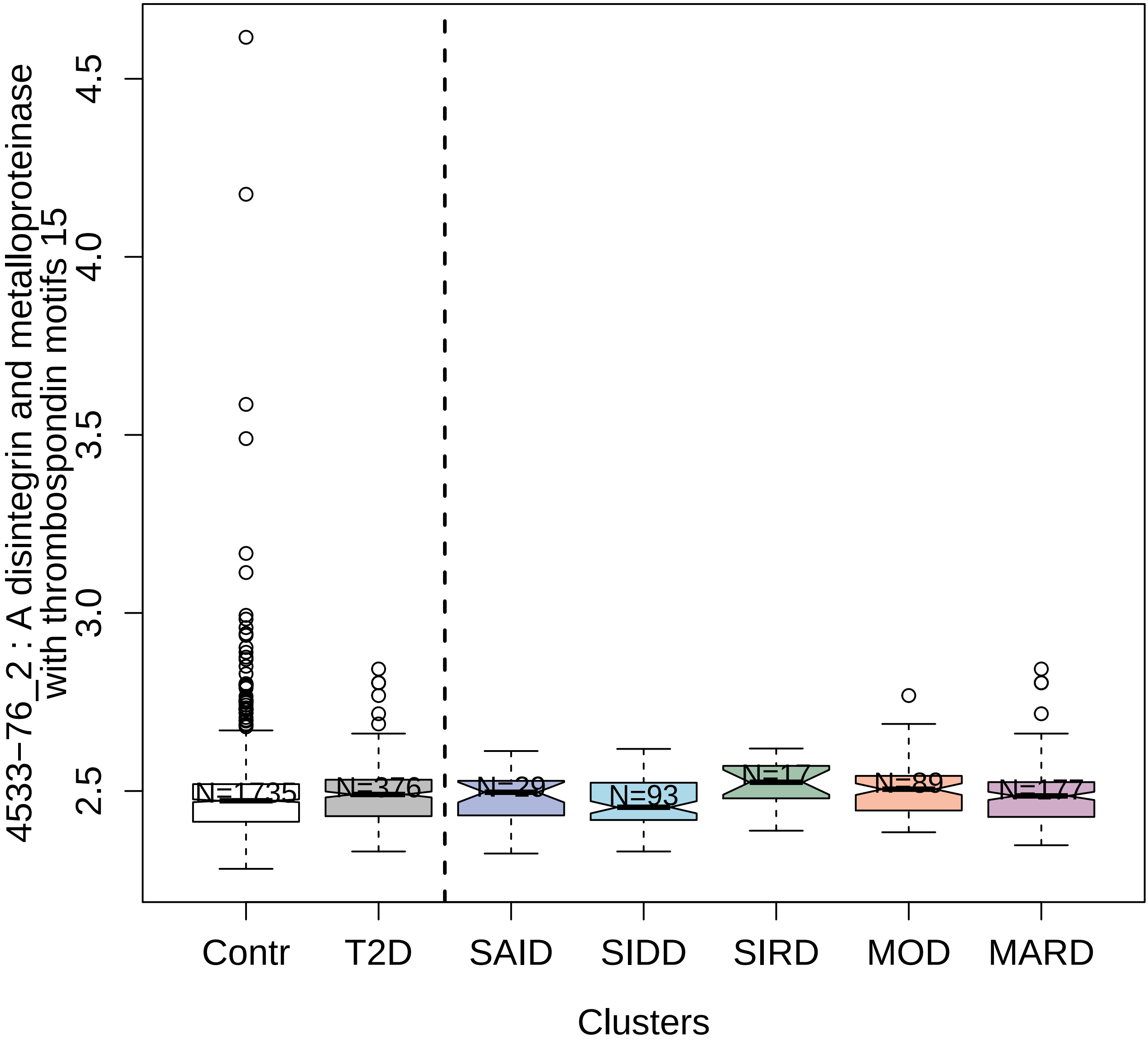

4533−76_2 : A disintegrin and metalloproteinase with thrombospondin motifs 15
4.5
4.0
3.5
3.0
2.5
N=1735
N=376
N=29
N=93
N=17
N=89
N=177
Contr
T2D
SAID
SIDD
SIRD
MOD
MARD
Clusters

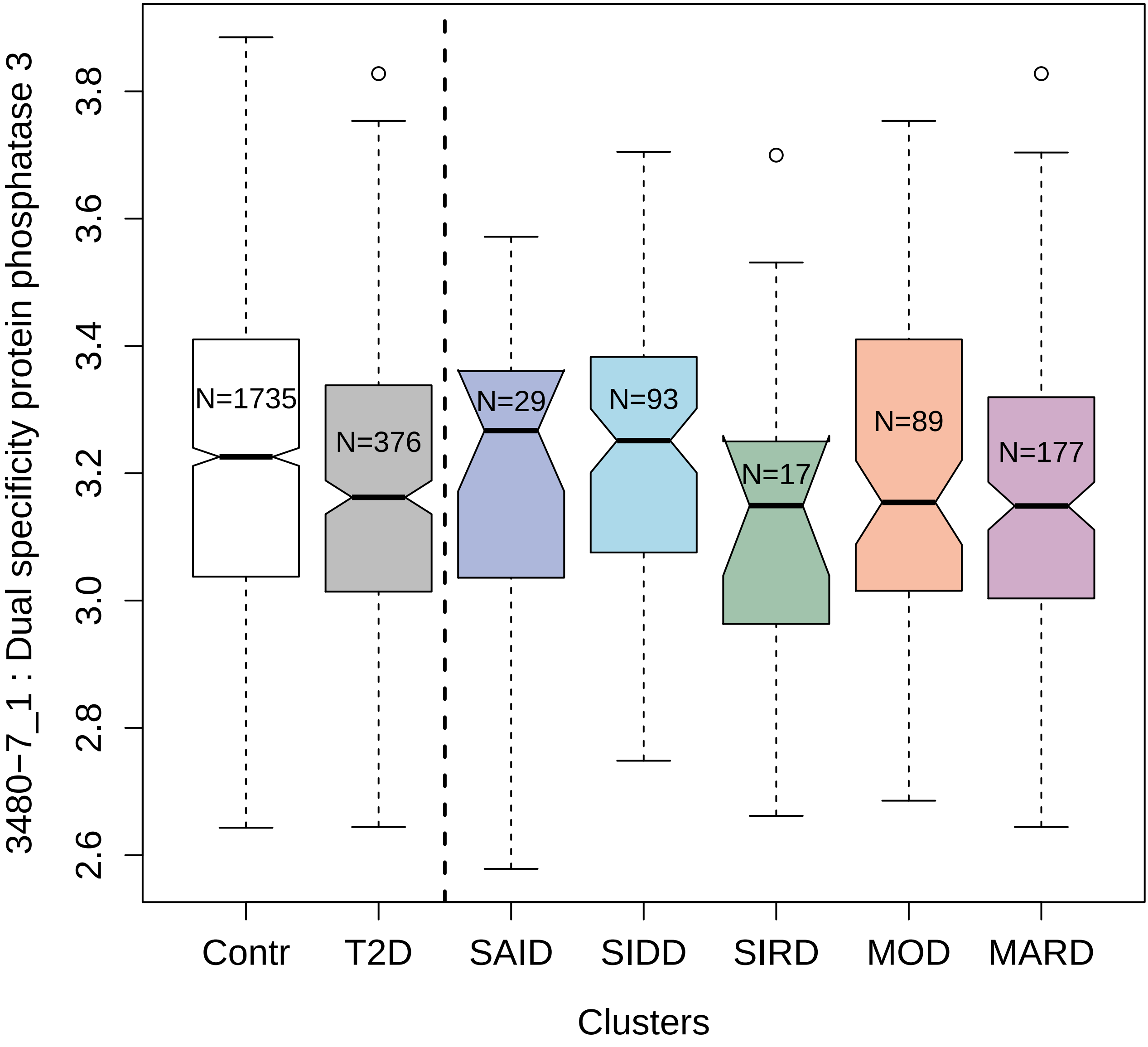
3480−7_1 : Dual specificity protein phosphatase 3
2.6
2.8
3.0
3.2
3.4
3.6
3.8
N=1735
N=376
N=29
N=93
N=17
N=89
N=177
Contr
T2D
SAID
SIDD
SIRD
MOD
MARD
Clusters

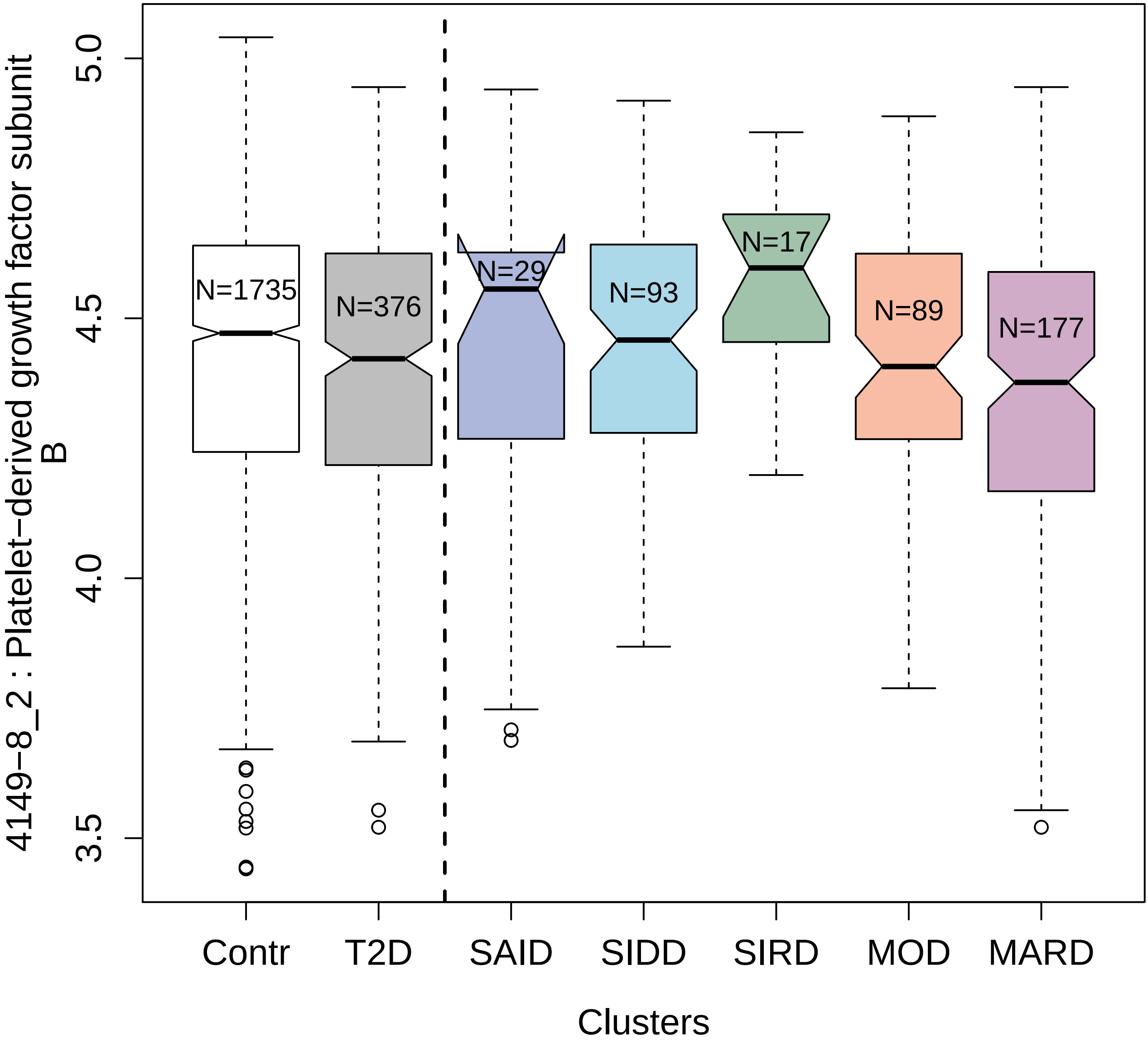
4149−8_2 : Platelet−derived growth factor subunit B
5.0
4.5
4.0
3.5
N=1735
N=376
N=29
N=93
N=17
N=89
N=177
Contr
T2D
SAID
SIDD
SIRD
MOD
MARD
Clusters

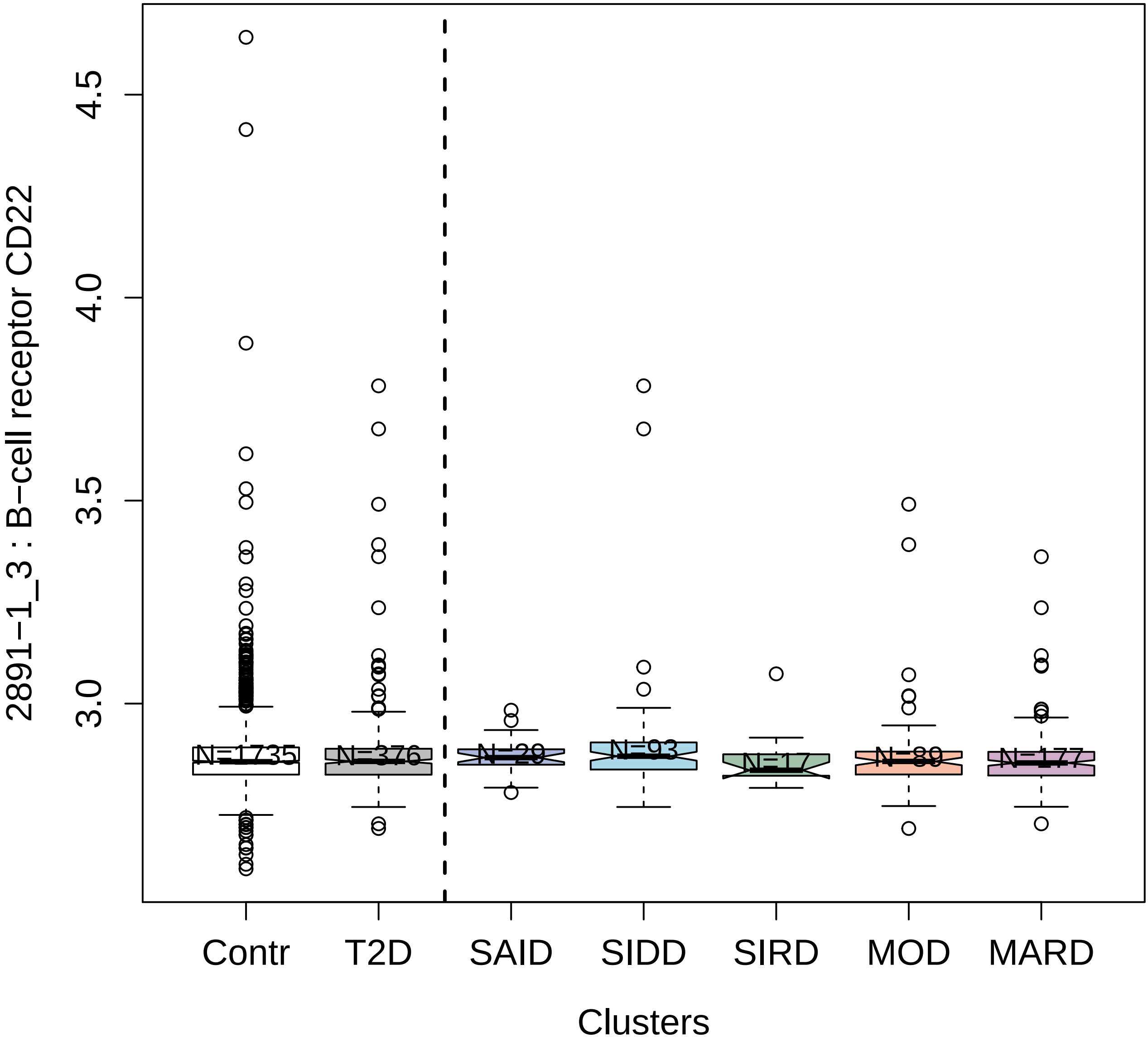

2891−1_3 : B−cell receptor CD22
4.5
4.0
3.5
3.0
N=1735
N=376
N=29
N=93
N=17
N=89
N=177
Contr
T2D
SAID
SIDD
SIRD
MOD
MARD
Clusters

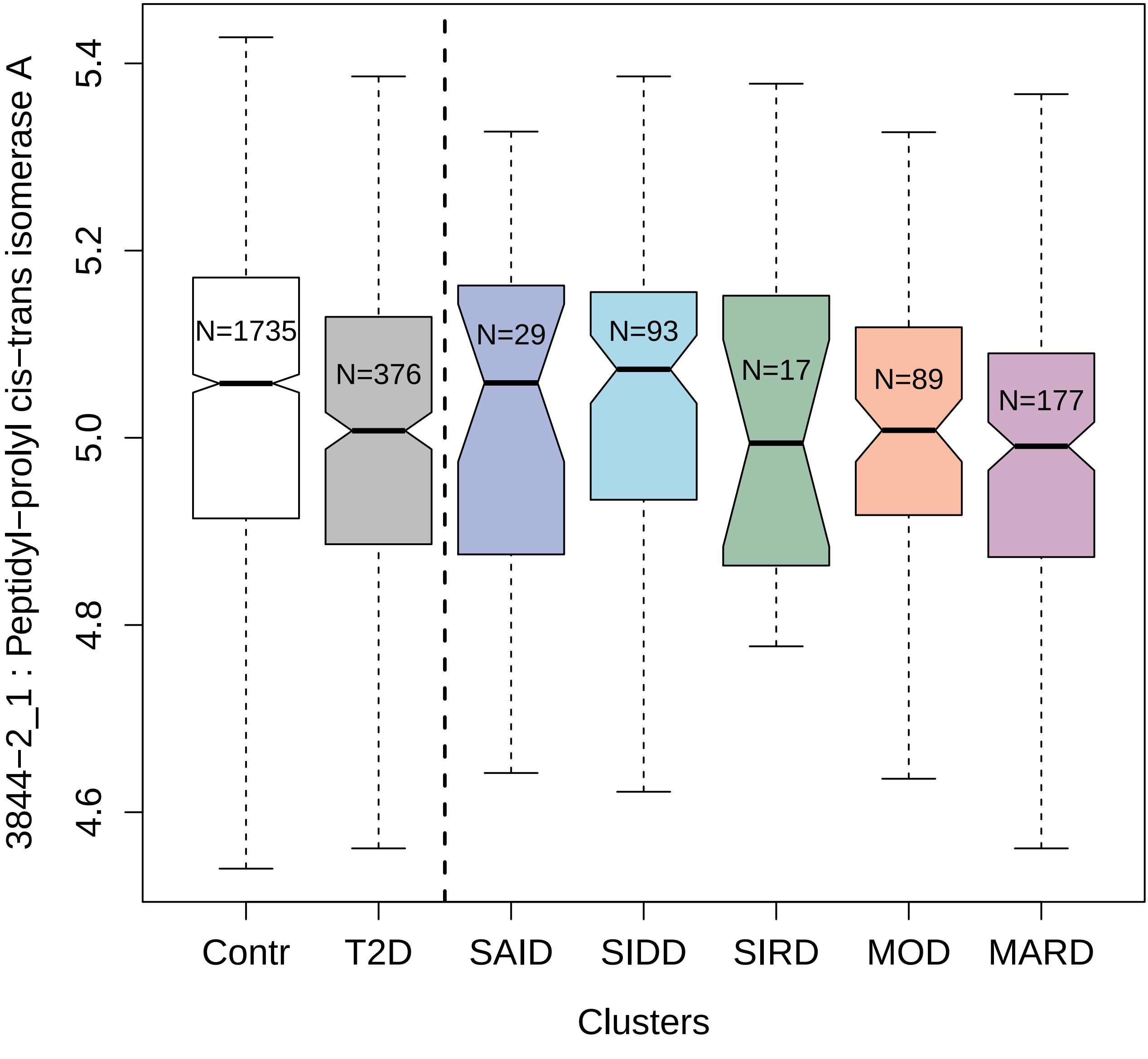

3844−2_1 : Peptidyl−prolyl cis−trans isomerase A
5.4
5.2
5.0
4.8
4.6
N=1735
N=376
N=29
N=93
N=17
N=89
N=177
Contr
T2D
SAID
SIDD
SIRD
MOD
MARD
Clusters

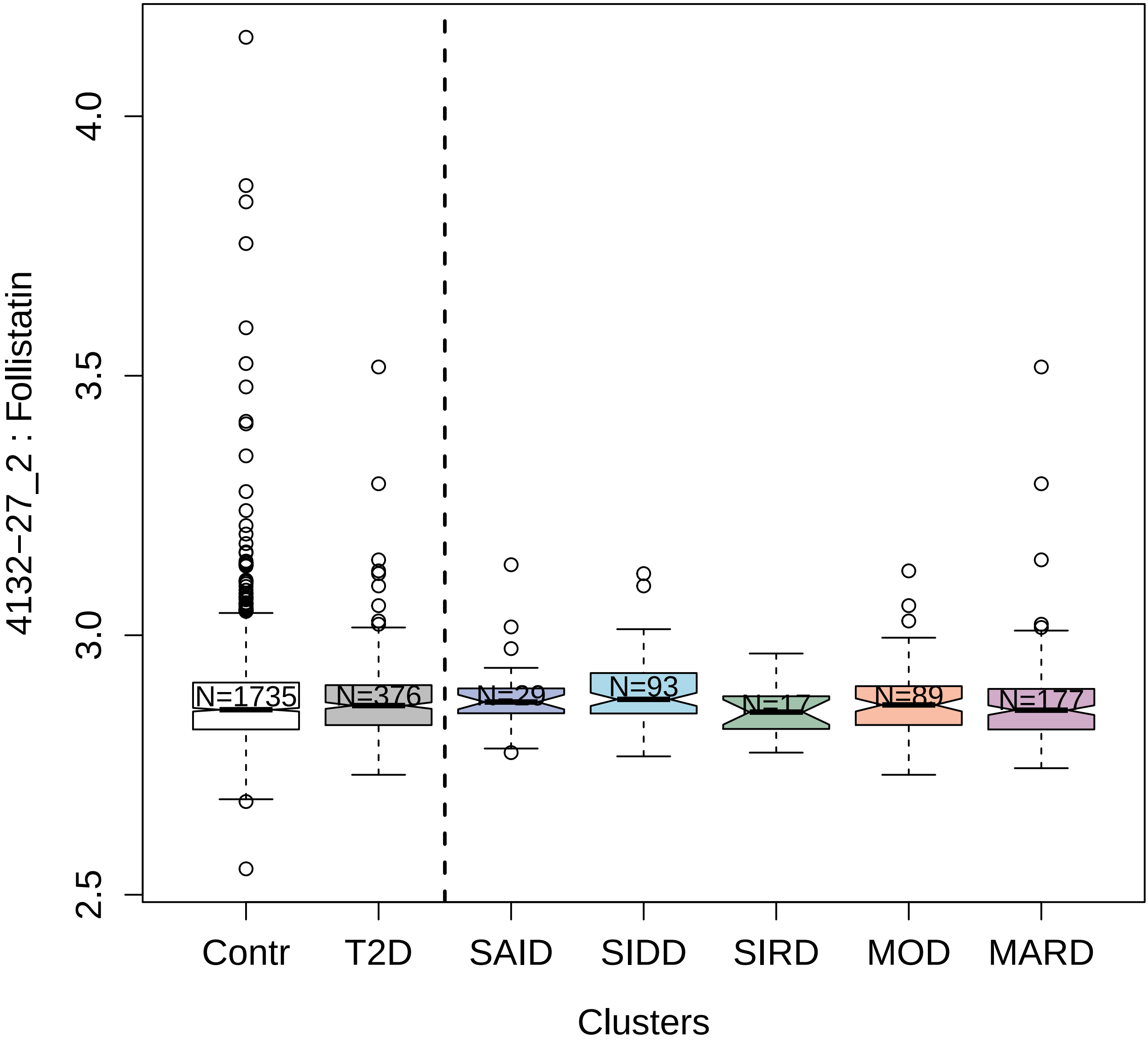
4132−27_2 : Follistatin
4.0
3.5
3.0
2.5
N=1735
N=376
N=29
N=93
N=17
N=89
N=177
Contr
T2D
SAID
SIDD
SIRD
MOD
MARD
Clusters

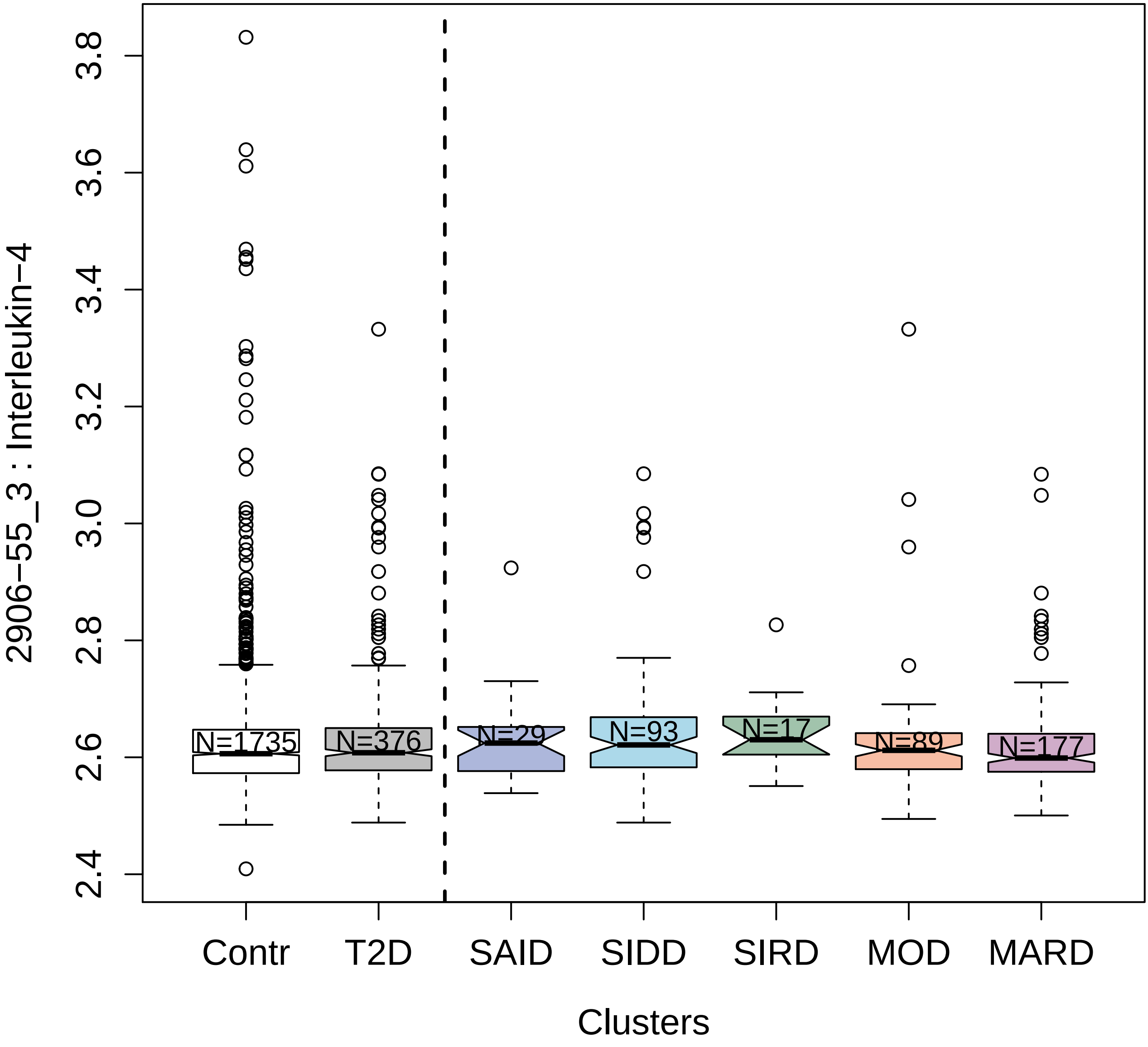

2906−55_3 : Interleukin−4
Clusters
Contr
T2D
SAID
SIDD
SIRD
MOD
MARD
N=1735
N=376
N=29
N=93
N=17
N=89
N=177
2.4
2.6
2.8
3.0
3.2
3.4
3.6
3.8

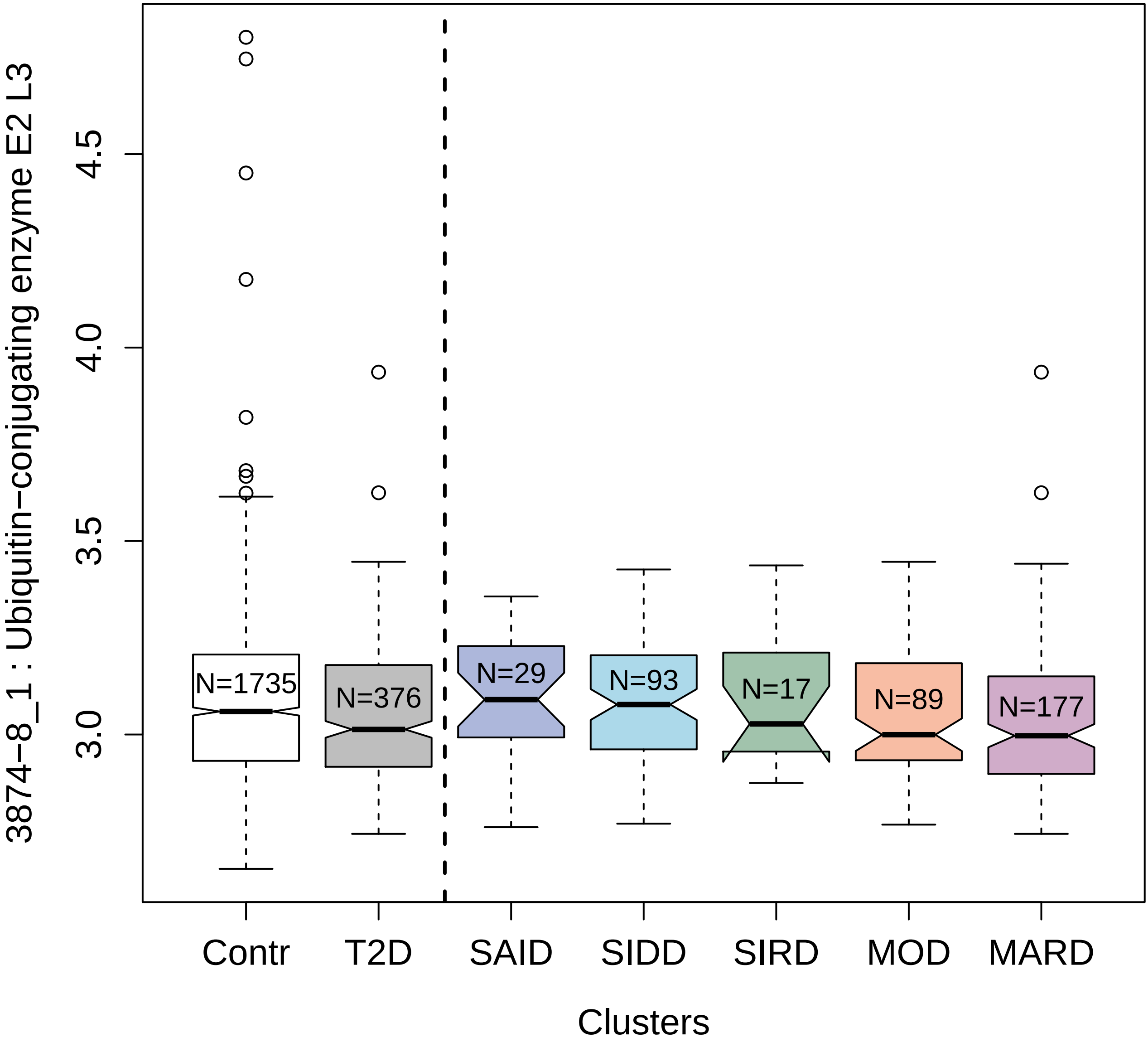
3874−8_1 : Ubiquitin−conjugating enzyme E2 L3
4.5
4.0
3.5
3.0
N=1735
N=376
N=29
N=93
N=17
N=89
N=177
Contr
T2D
SAID
SIDD
SIRD
MOD
MARD
Clusters

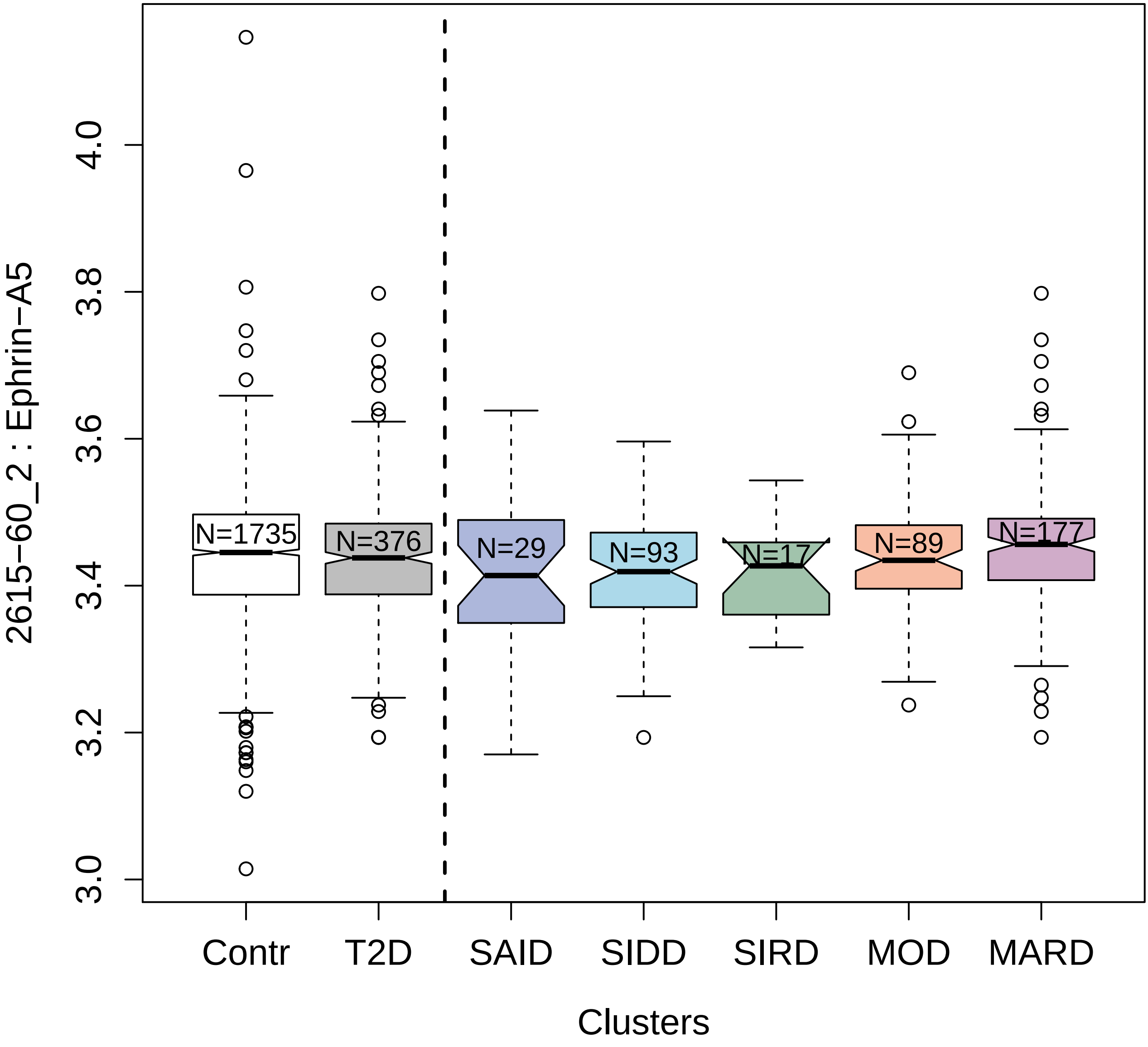

2615−60_2 : Ephrin−A5
4.0
3.8
3.6
3.4
3.2
3.0
N=1735
N=376
N=29
N=93
N=17
N=89
N=177
Contr
T2D
SAID
SIDD
SIRD
MOD
MARD
Clusters

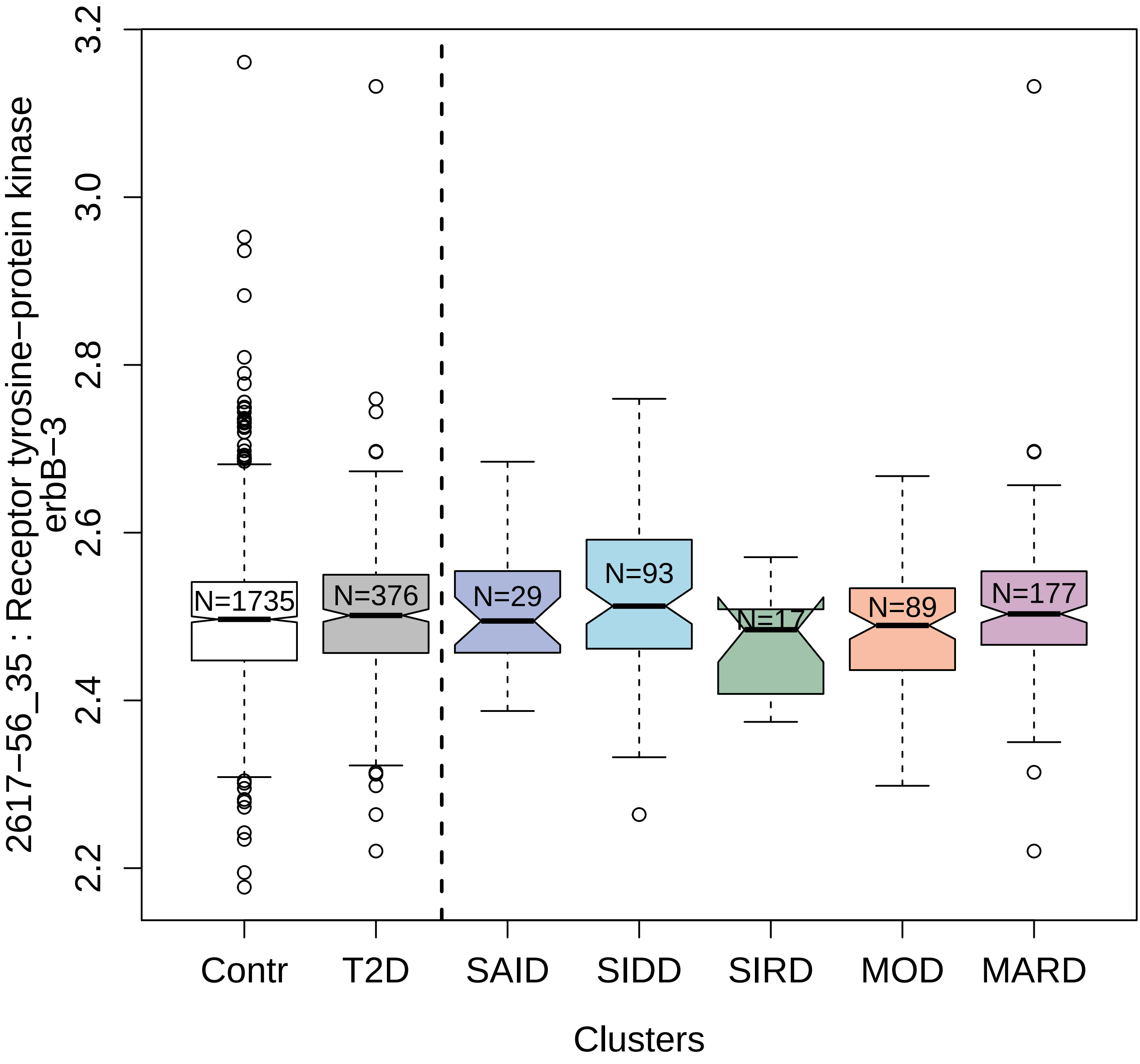

2617−56_35 : Receptor tyrosine−protein kinase erbB−3
3.2
3.0
2.8
2.6
2.4
2.2
N=1735
N=376
N=29
N=93
N=17
N=89
N=177
Contr
T2D
SAID
SIDD
SIRD
MOD
MARD
Clusters

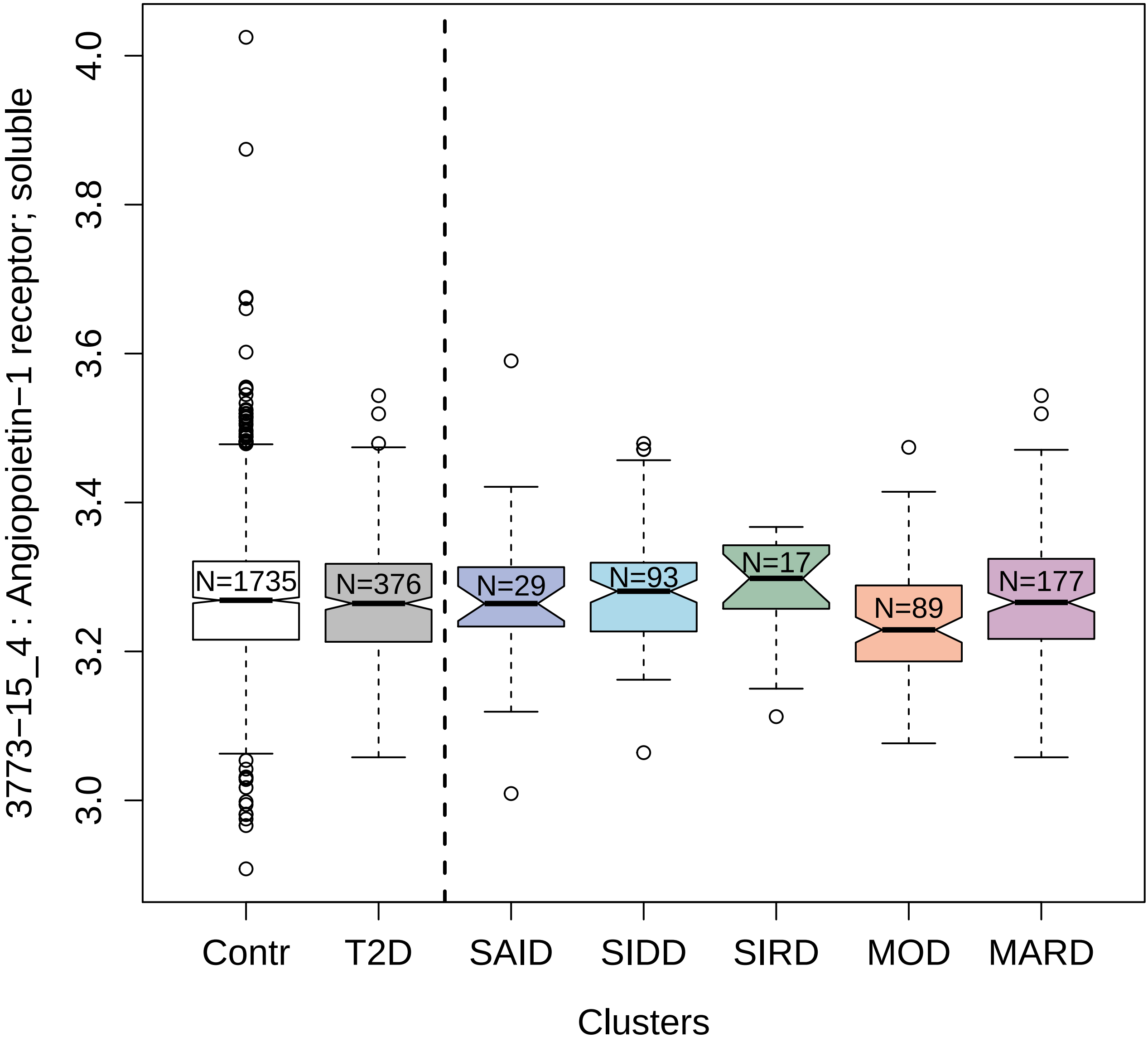
3773−15_4 : Angiopoietin−1 receptor; soluble
4.0
3.8
3.6
3.4
3.2
3.0
N=1735
N=376
N=29
N=93
N=17
N=89
N=177
Contr
T2D
SAID
SIDD
SIRD
MOD
MARD
Clusters

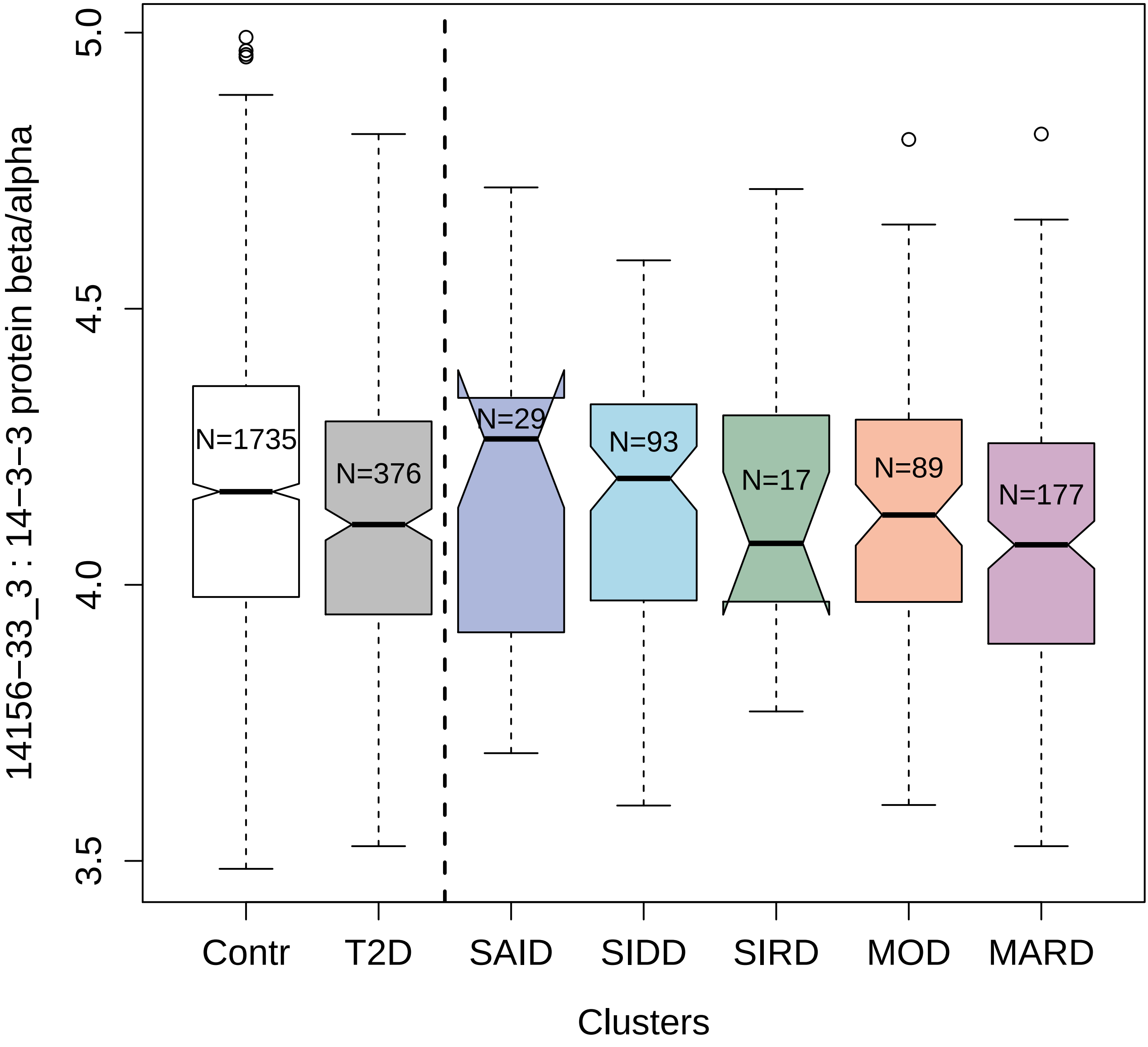

14156−33_3 : 14−3−3 protein beta/alpha
5.0
4.5
4.0
3.5
N=1735
N=376
N=29
N=93
N=17
N=89
N=177
Contr
T2D
SAID
SIDD
SIRD
MOD
MARD
Clusters

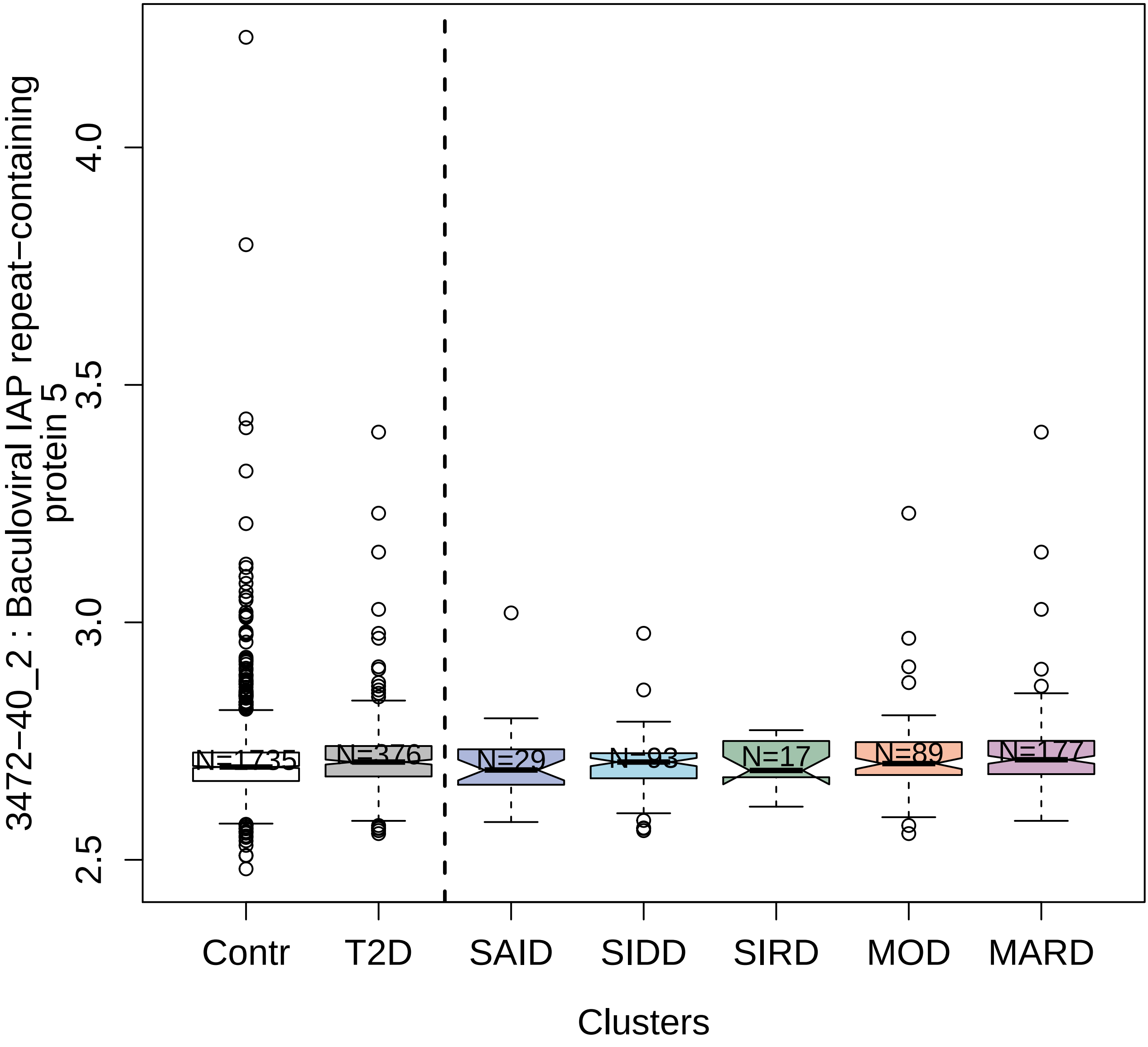
3472−40_2 : Baculoviral IAP repeat−containing protein 5
4.0
3.5
3.0
2.5
N=1735
N=376
N=29
N=93
N=17
N=89
N=177
Contr
T2D
SAID
SIDD
SIRD
MOD
MARD
Clusters

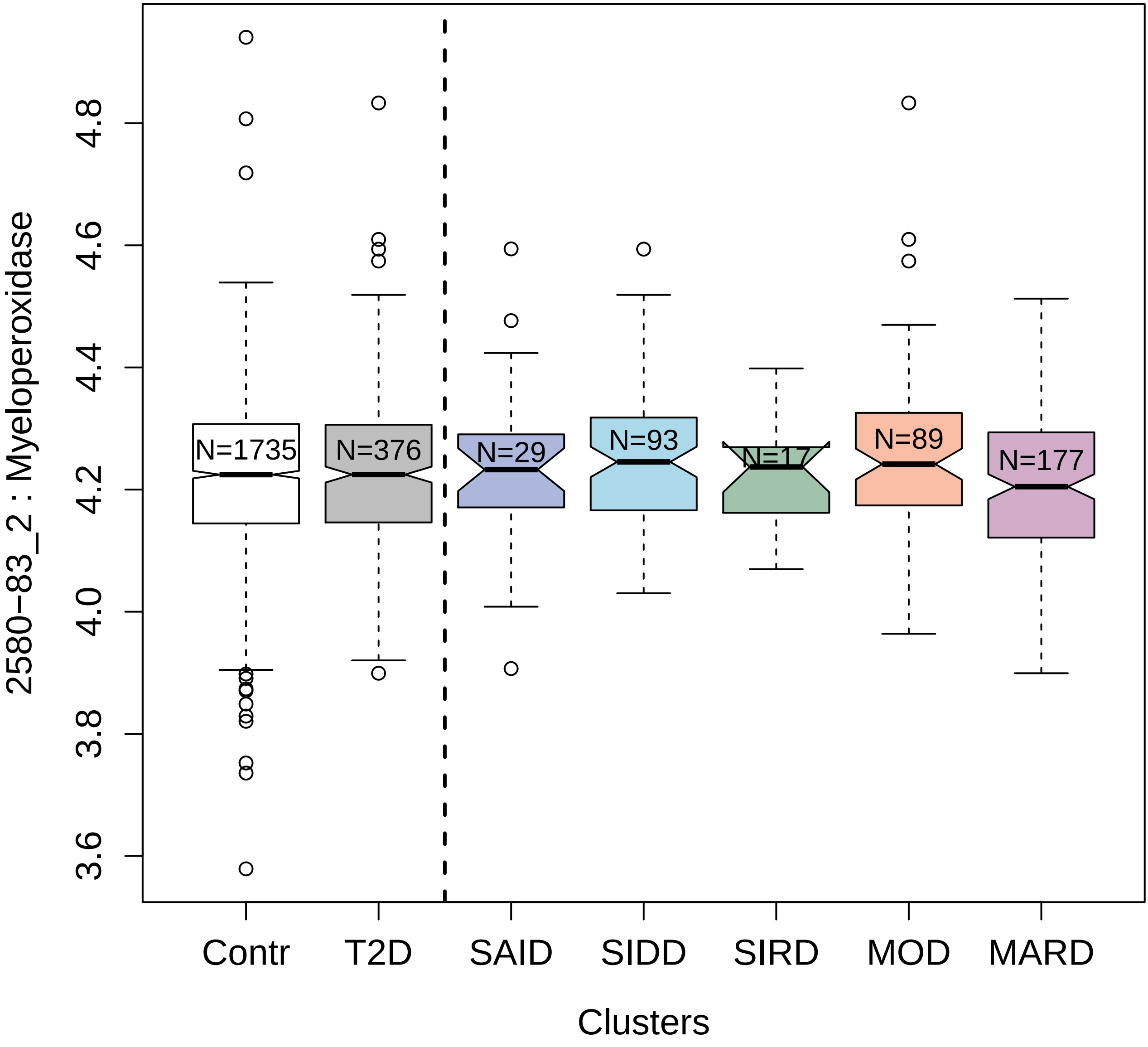

2580−83_2 : Myeloperoxidase
3.6
3.8
4.0
4.2
4.4
4.6
4.8
N=1735
N=376
N=29
N=93
N=17
N=89
N=177
Contr
T2D
SAID
SIDD
SIRD
MOD
MARD
Clusters

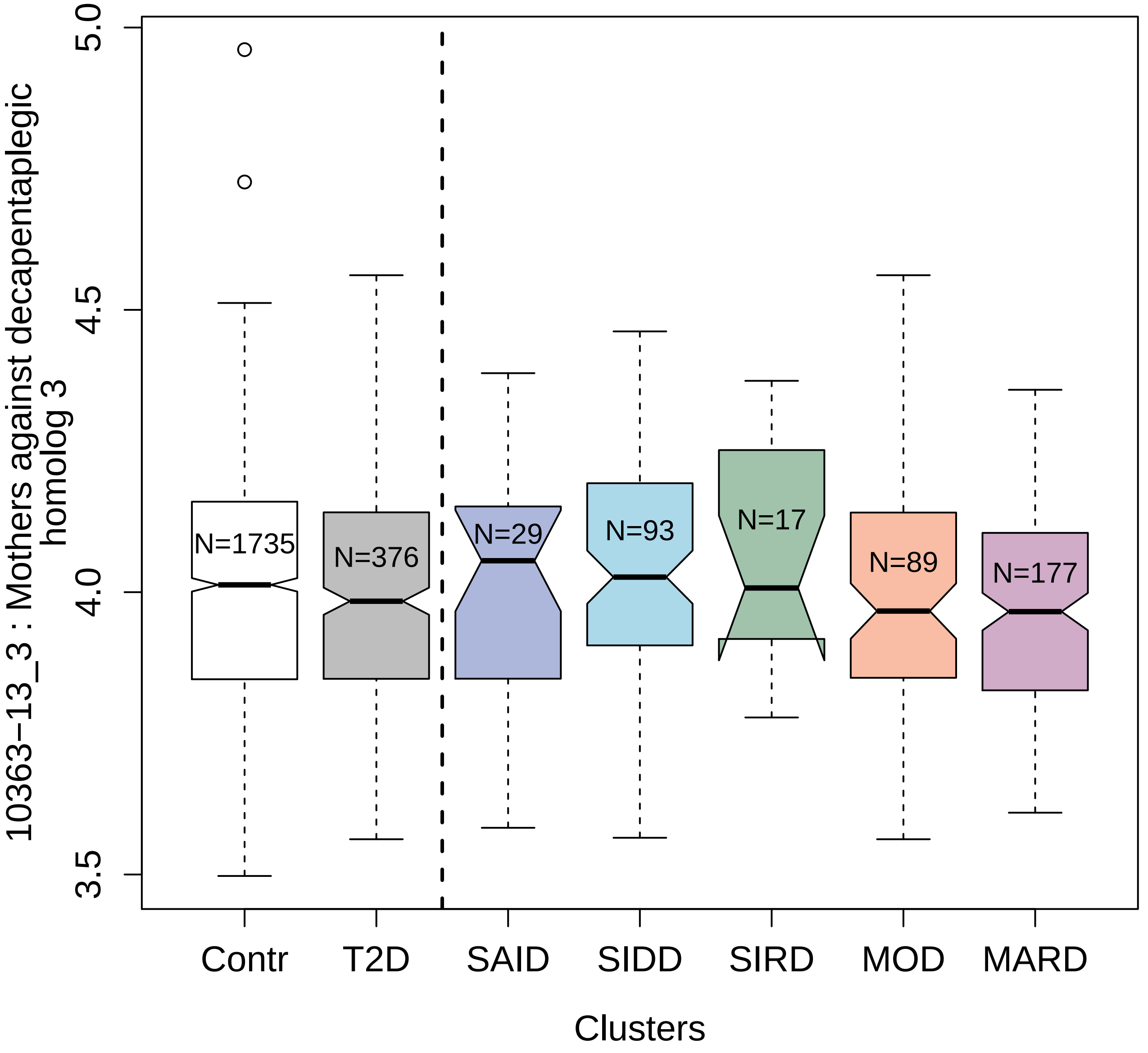
10363−13_3 : Mothers against decapentaplegic homolog 3
5.0
4.5
4.0
3.5
N=1735
N=376
N=29
N=93
N=17
N=89
N=177
Contr
T2D
SAID
SIDD
SIRD
MOD
MARD
Clusters

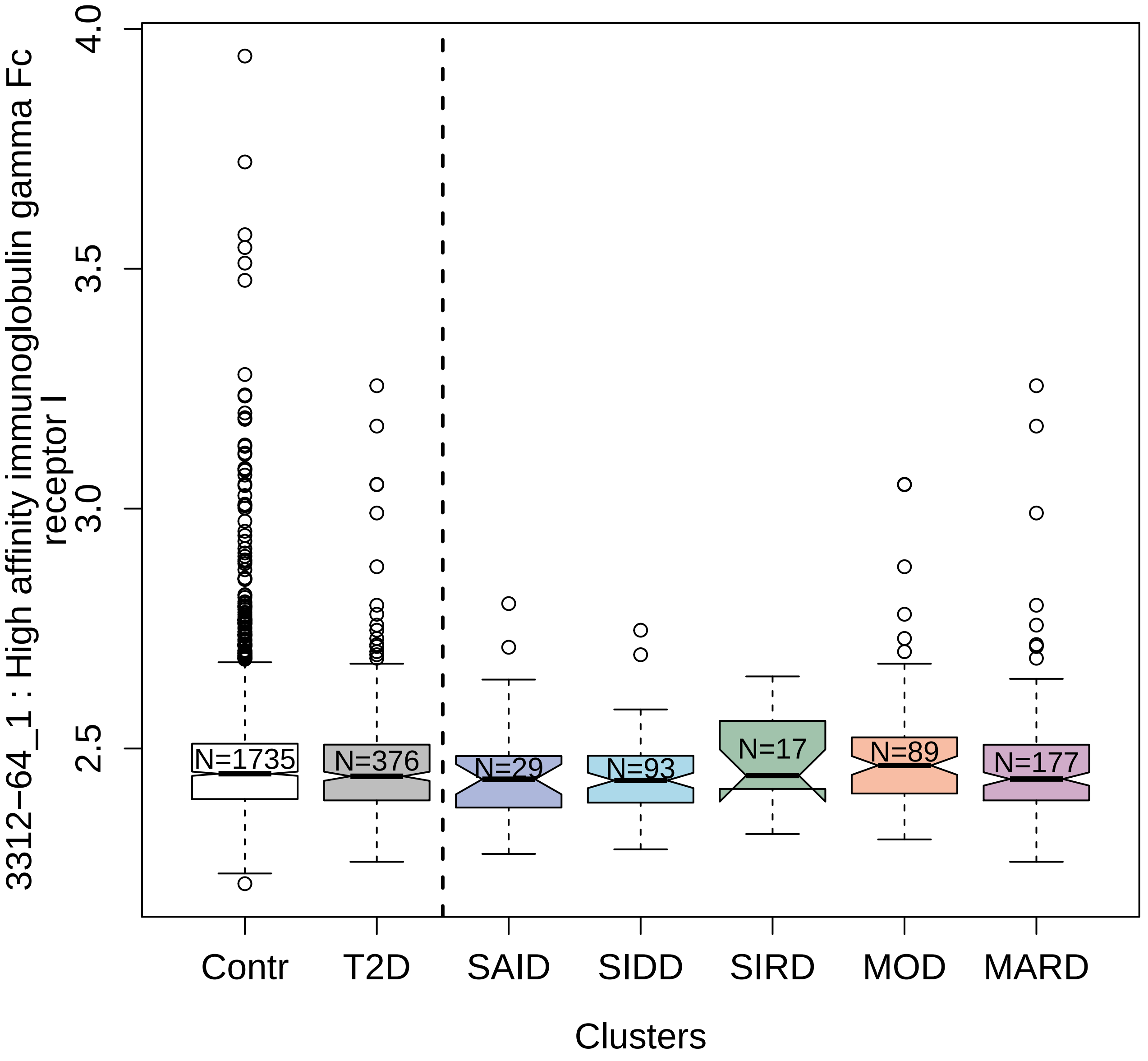

3312−64_1 : High affinity immunoglobulin gamma Fc receptor I
2.5
3.0
3.5
4.0
N=1735
N=376
N=29
N=93
N=17
N=89
N=177
Contr
T2D
SAID
SIDD
SIRD
MOD
MARD
Clusters

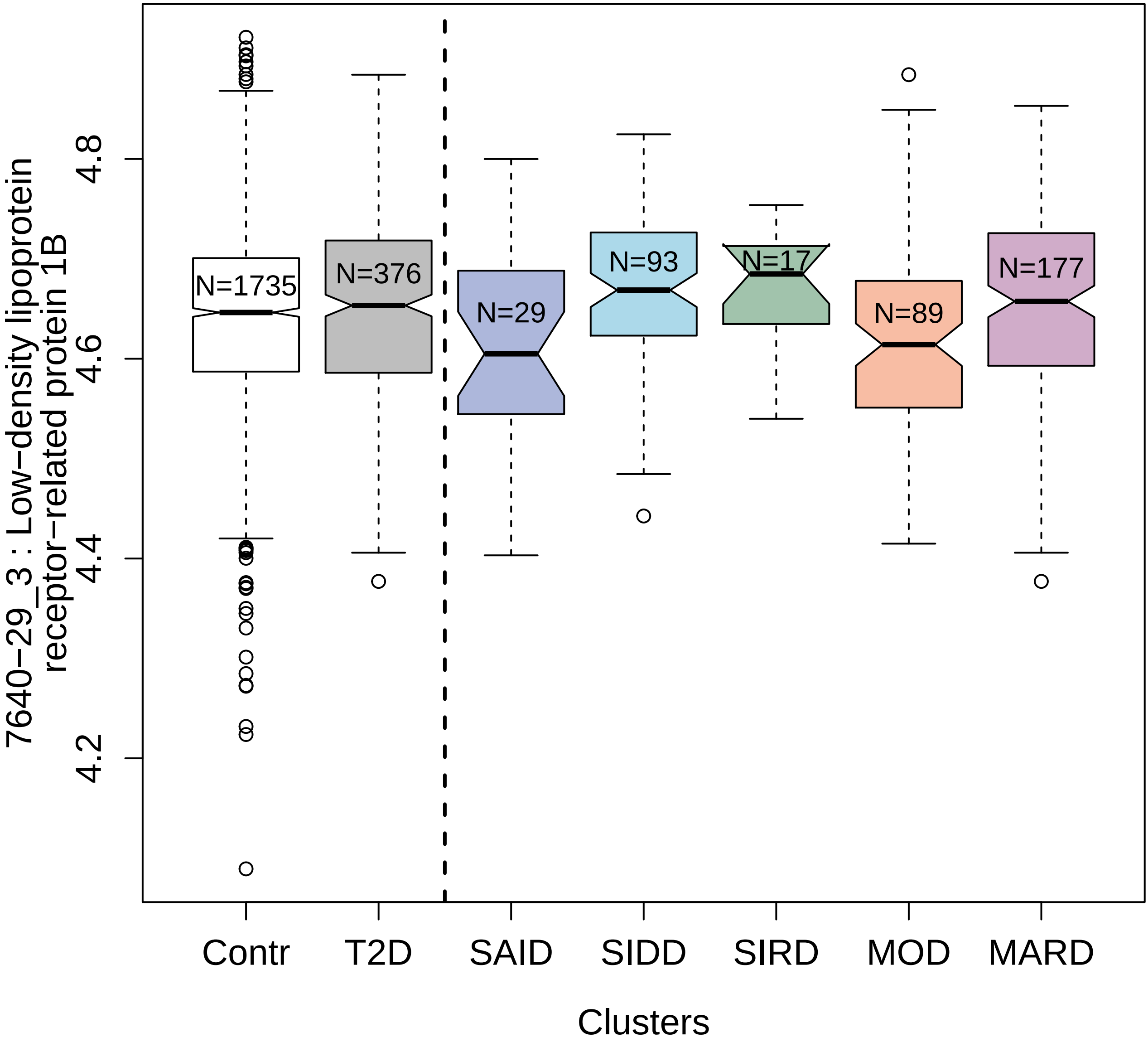
7640−29_3 : Low−density lipoprotein
receptor−related protein 1B
4.2
4.4
4.6
4.8
N=1735
N=376
N=29
N=93
N=17
N=89
N=177
Contr
T2D
SAID
SIDD
SIRD
MOD
MARD
Clusters

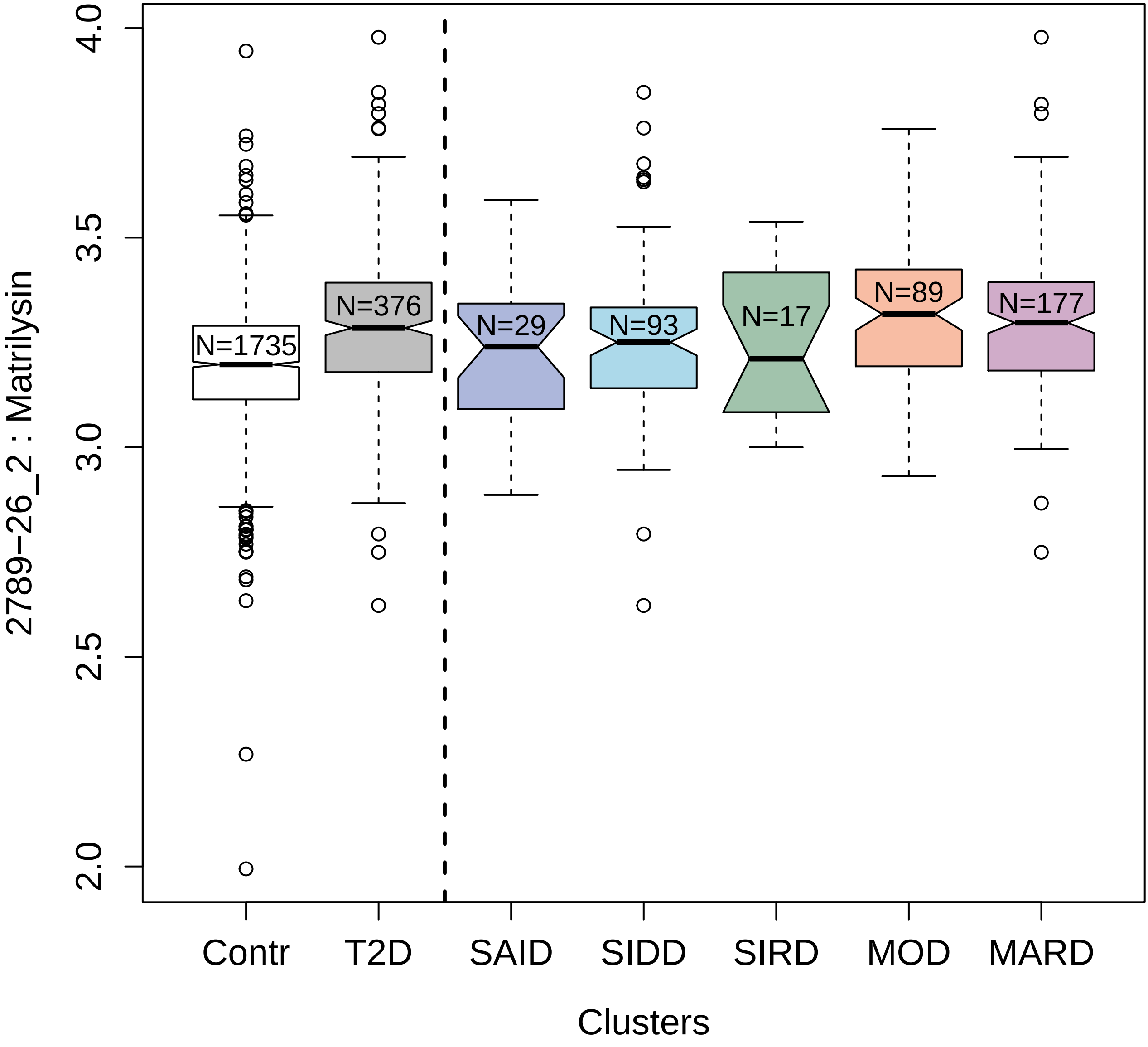

4.0
3.5
3.0
2.5
2.0
2789−26_2 : Matrilysin
N=1735
N=376
N=29
N=93
N=17
N=89
N=177
Contr
T2D
SAID
SIDD
SIRD
MOD
MARD
Clusters

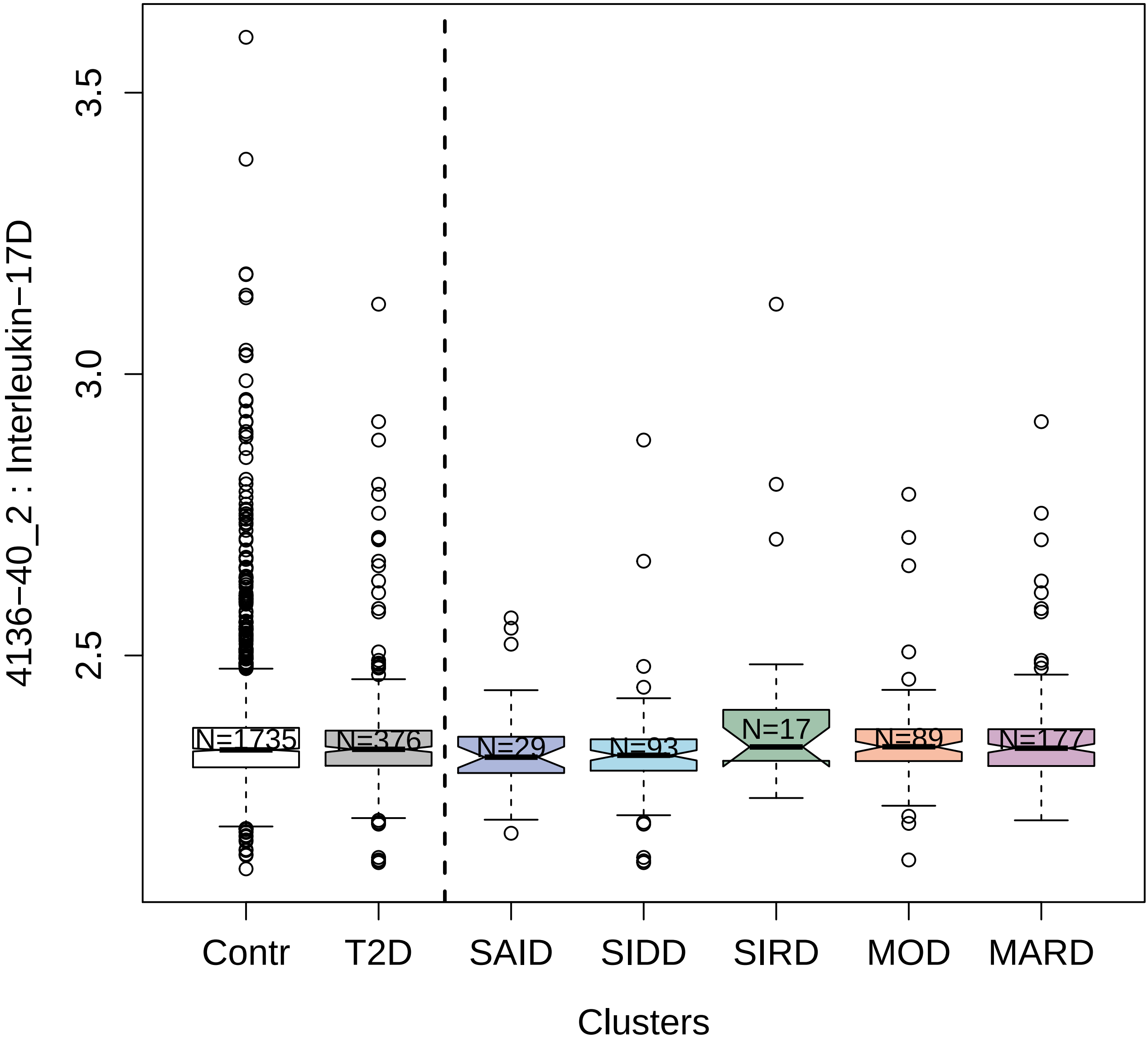

4136−40_2 : Interleukin−17D
3.5
3.0
2.5
N=1735
N=376
N=29
N=93
N=17
N=89
N=177
Contr
T2D
SAID
SIDD
SIRD
MOD
MARD
Clusters

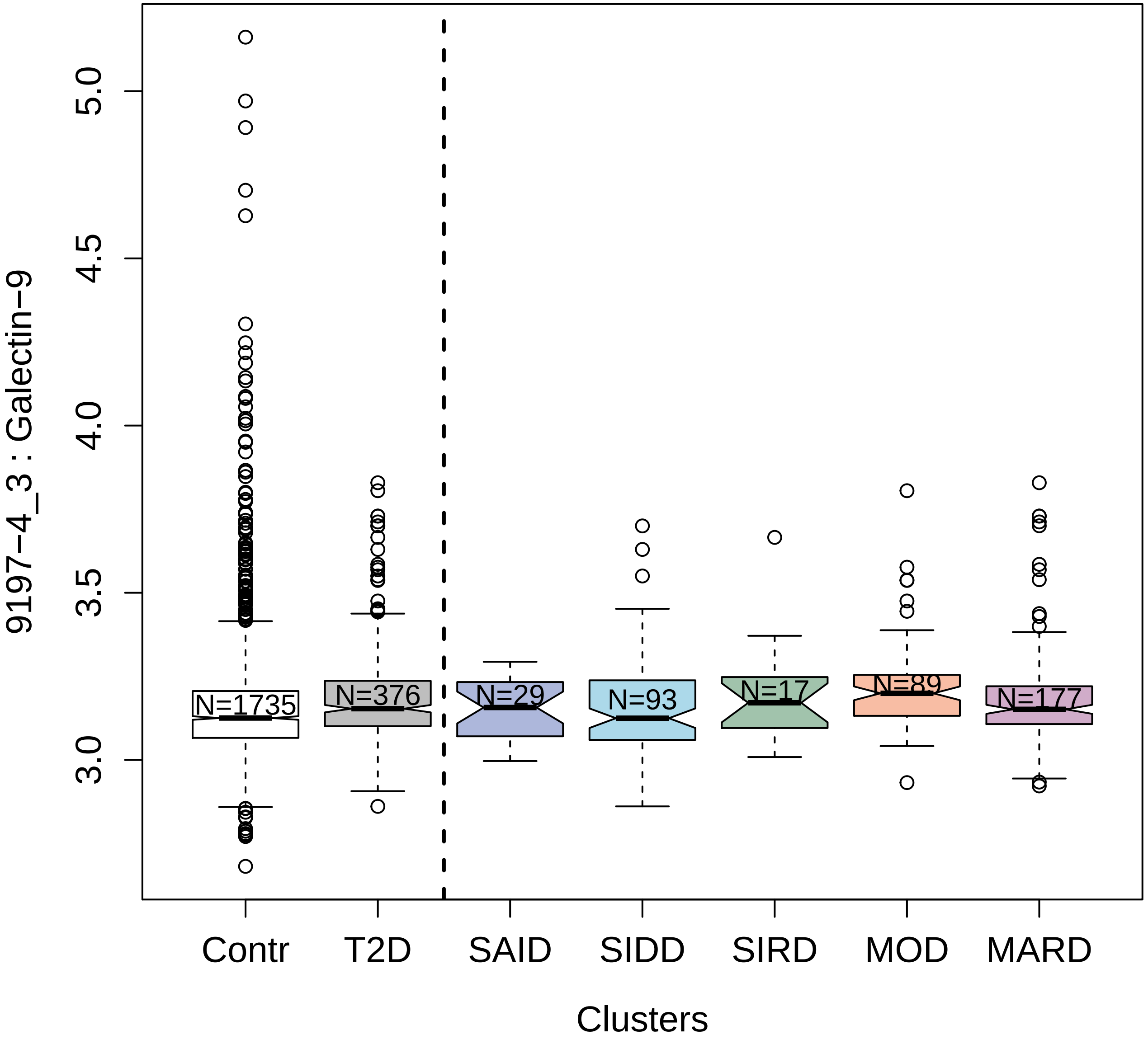

9197−4_3 : Galectin−9
5.0
4.5
4.0
3.5
3.0
N=1735
N=376
N=29
N=93
N=17
N=89
N=177
Contr
T2D
SAID
SIDD
SIRD
MOD
MARD
Clusters

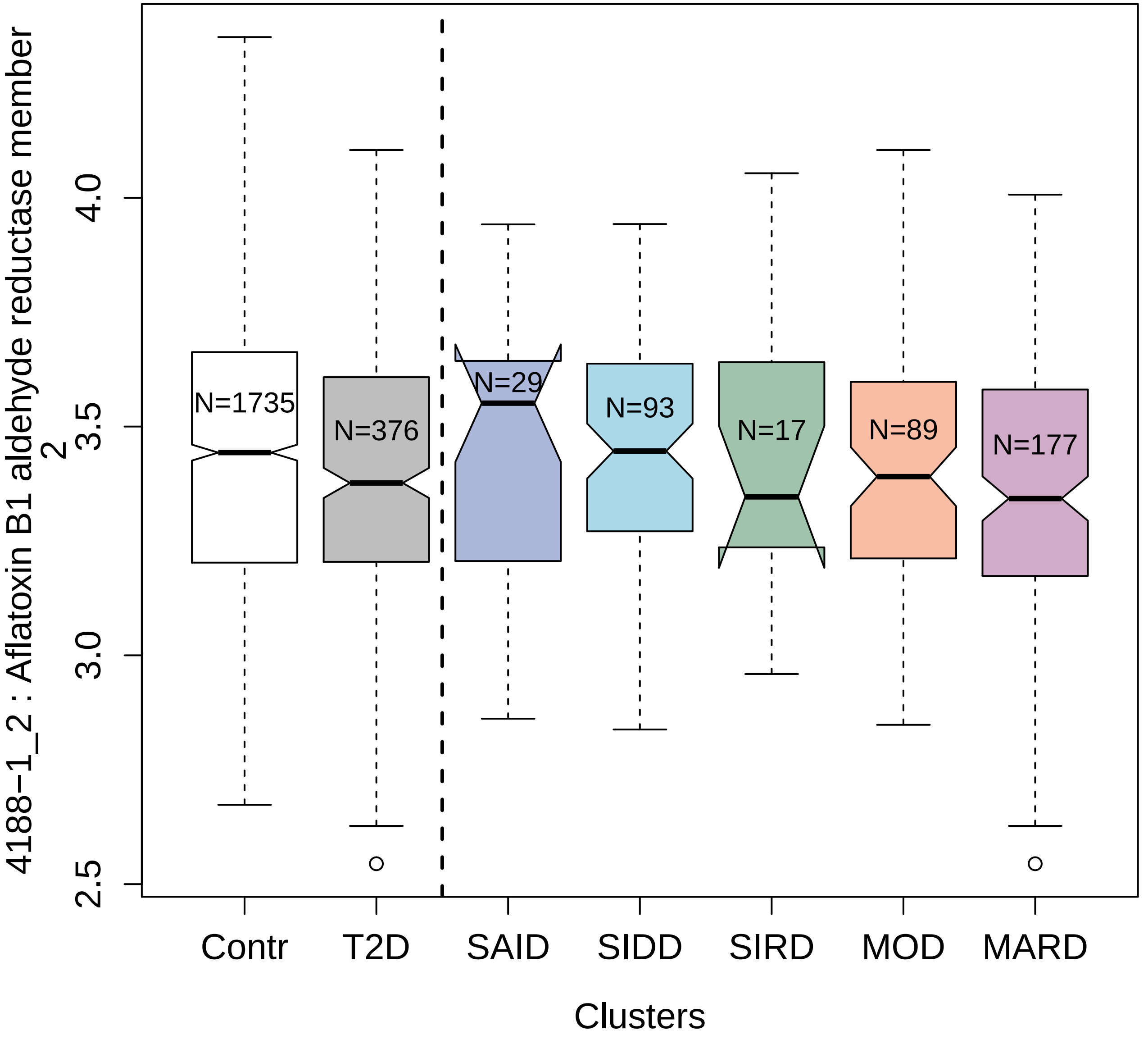

4188−1_2 : Aflatoxin B1 aldehyde reductase member 2
2.5
3.0
3.5
4.0
N=1735
N=376
N=29
N=93
N=17
N=89
N=177
Contr
T2D
SAID
SIDD
SIRD
MOD
MARD
Clusters

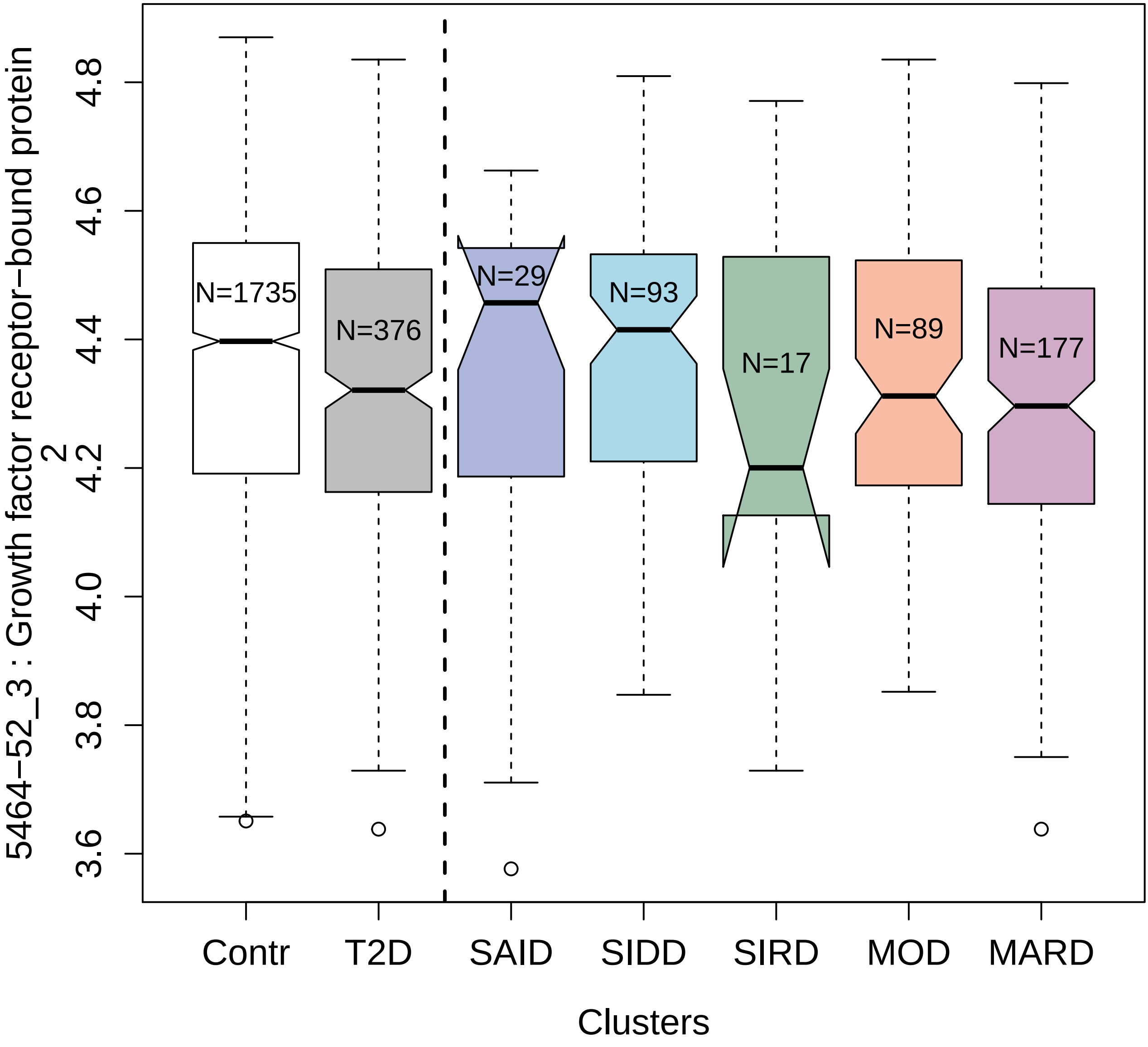
5464−52_3 : Growth factor receptor−bound protein 2
3.6
3.8
4.0
4.2
4.4
4.6
4.8
N=1735
N=376
N=29
N=93
N=17
N=89
N=177
Contr
T2D
SAID
SIDD
SIRD
MOD
MARD
Clusters

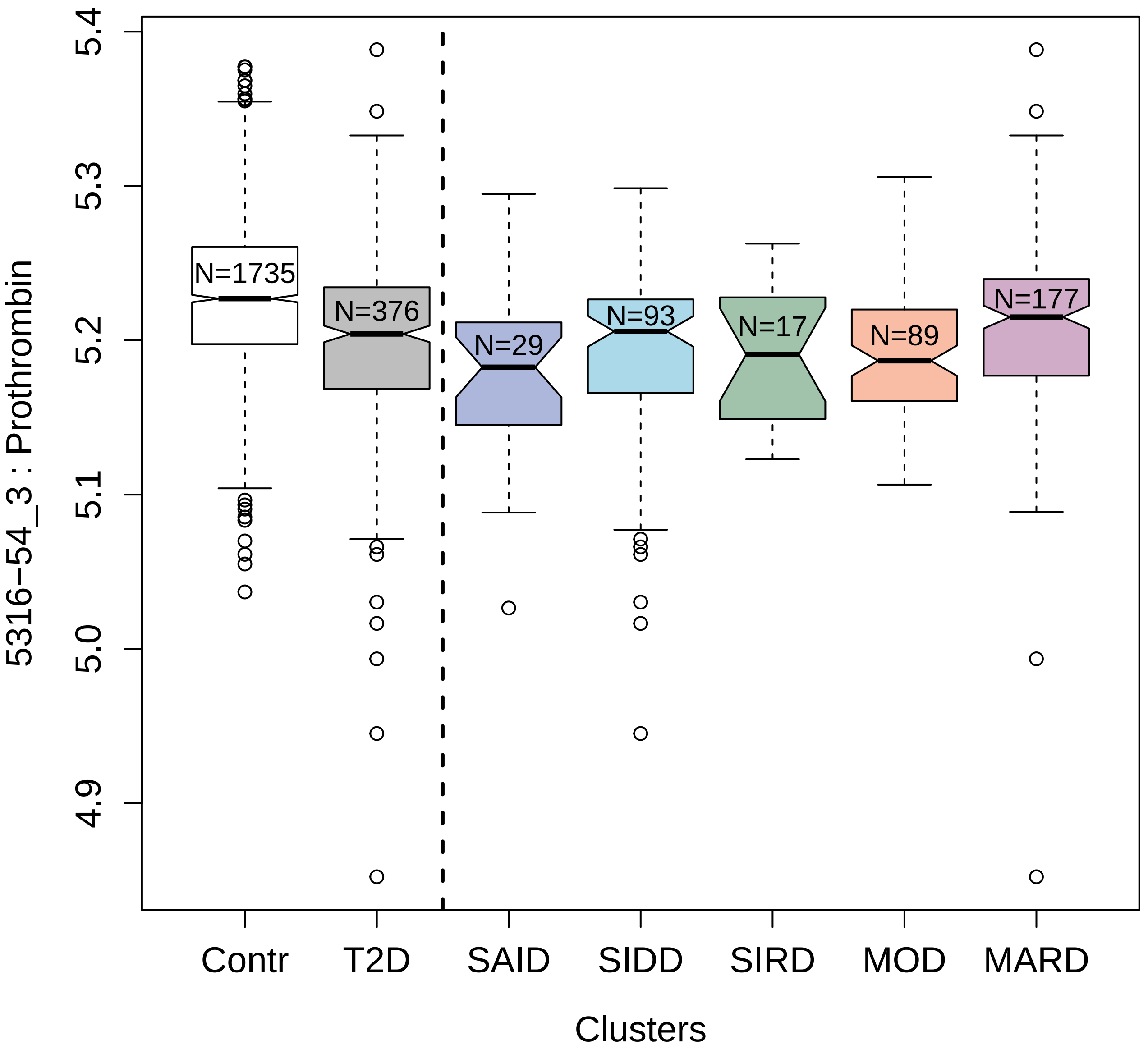
5316−54_3 : Prothrombin
5.4
5.3
5.2
5.1
5.0
4.9
N=1735
N=376
N=29
N=93
N=17
N=89
N=177
Contr
T2D
SAID
SIDD
SIRD
MOD
MARD
Clusters

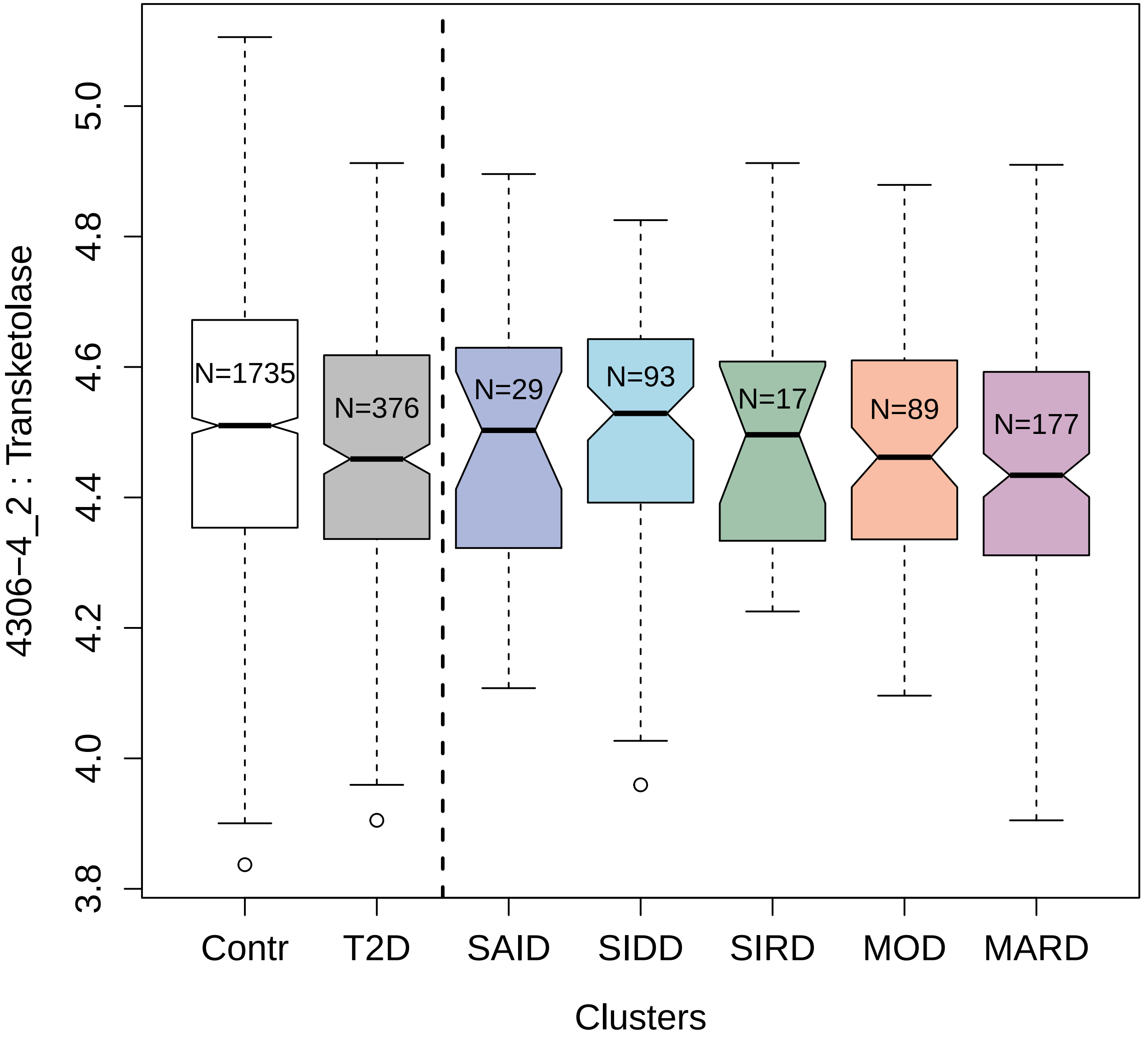

4306−4_2 : Transketolase
3.8
4.0
4.2
4.4
4.6
4.8
5.0
N=1735
N=376
N=29
N=93
N=17
N=89
N=177
Contr
T2D
SAID
SIDD
SIRD
MOD
MARD
Clusters

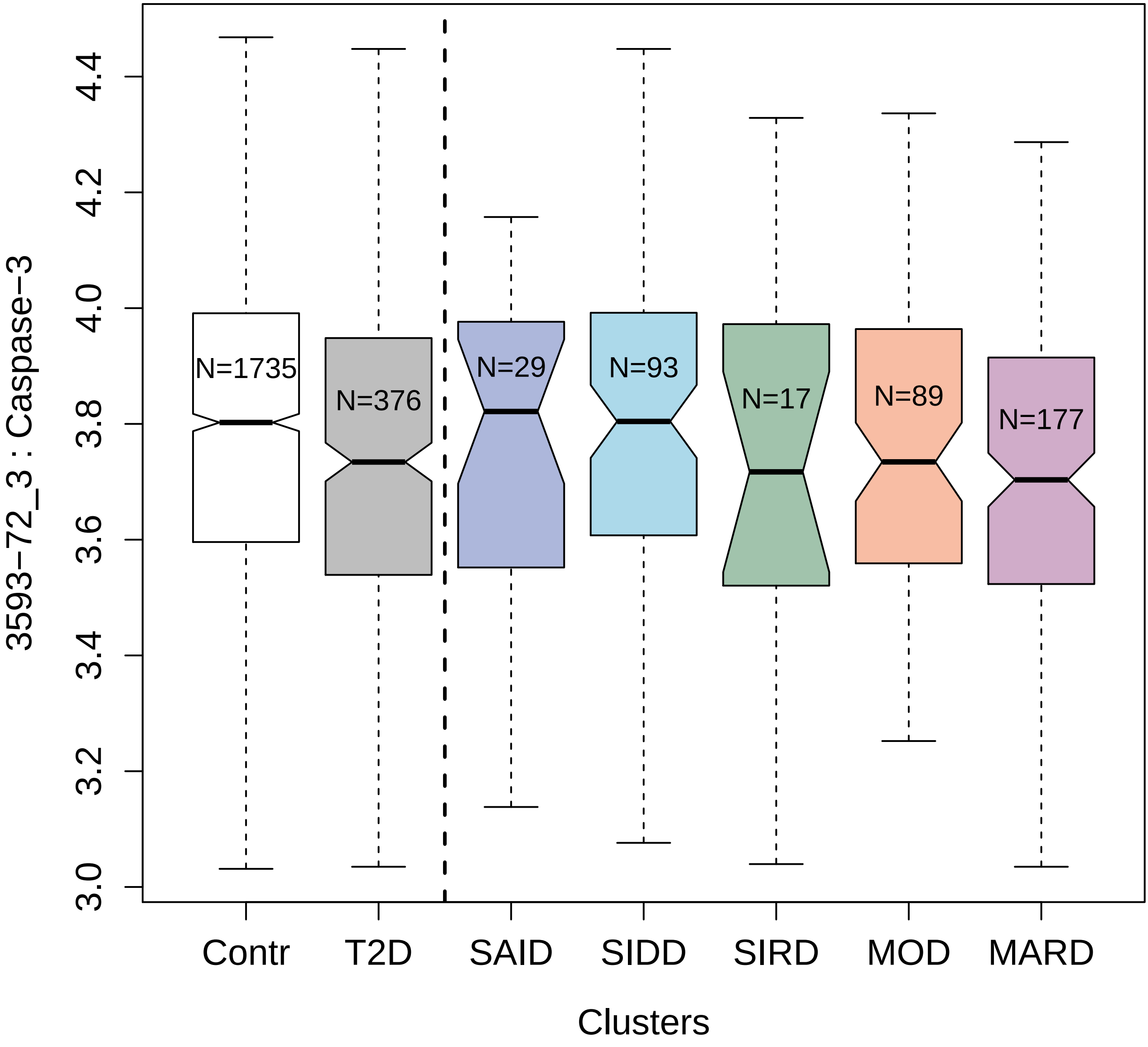

3593−72_3 : Caspase−3
3.0
3.2
3.4
3.6
3.8
4.0
4.2
4.4
N=1735
N=376
N=29
N=93
N=17
N=89
N=177
Contr
T2D
SAID
SIDD
SIRD
MOD
MARD
Clusters

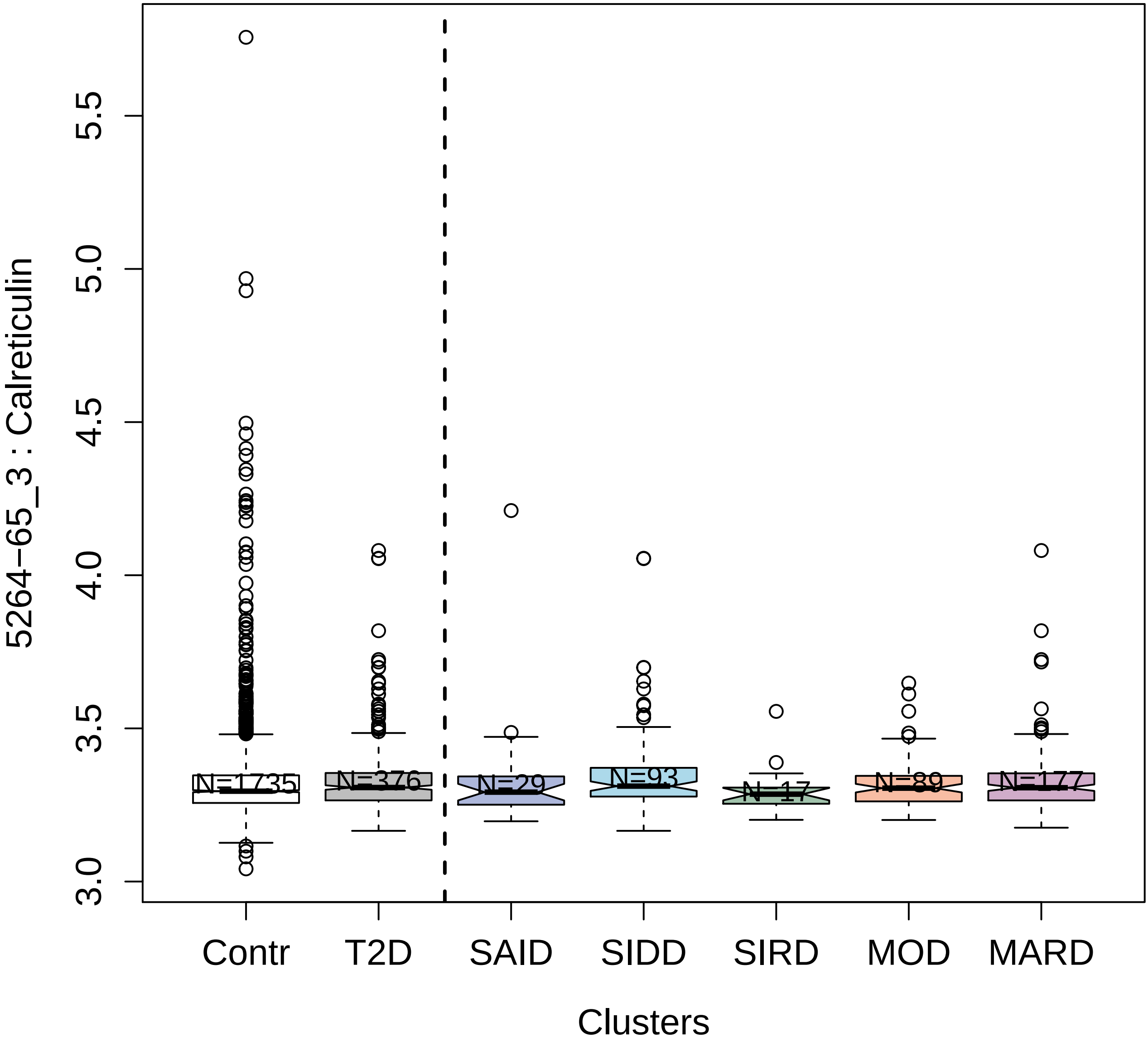

5264−65_3 : Calreticulin
3.0
3.5
4.0
4.5
5.0
5.5
N=1735
N=376
N=29
N=93
N=17
N=89
N=177
Contr
T2D
SAID
SIDD
SIRD
MOD
MARD
Clusters

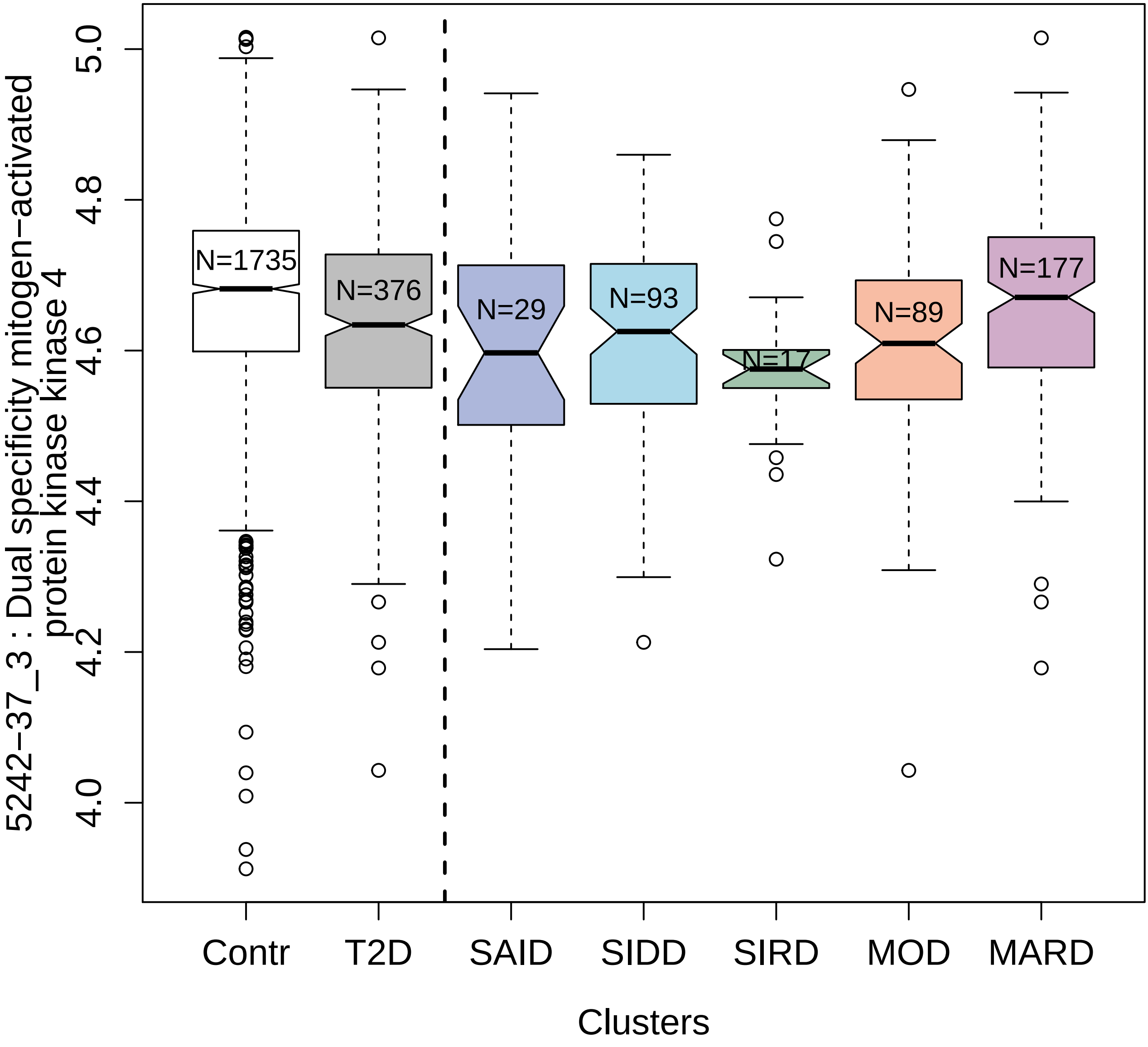
5242−37_3 : Dual specificity mitogen−activated protein kinase kinase 4
5.0
4.8
4.6
4.4
4.2
4.0
N=1735
N=376
N=29
N=93
N=17
N=89
N=177
Contr
T2D
SAID
SIDD
SIRD
MOD
MARD
Clusters

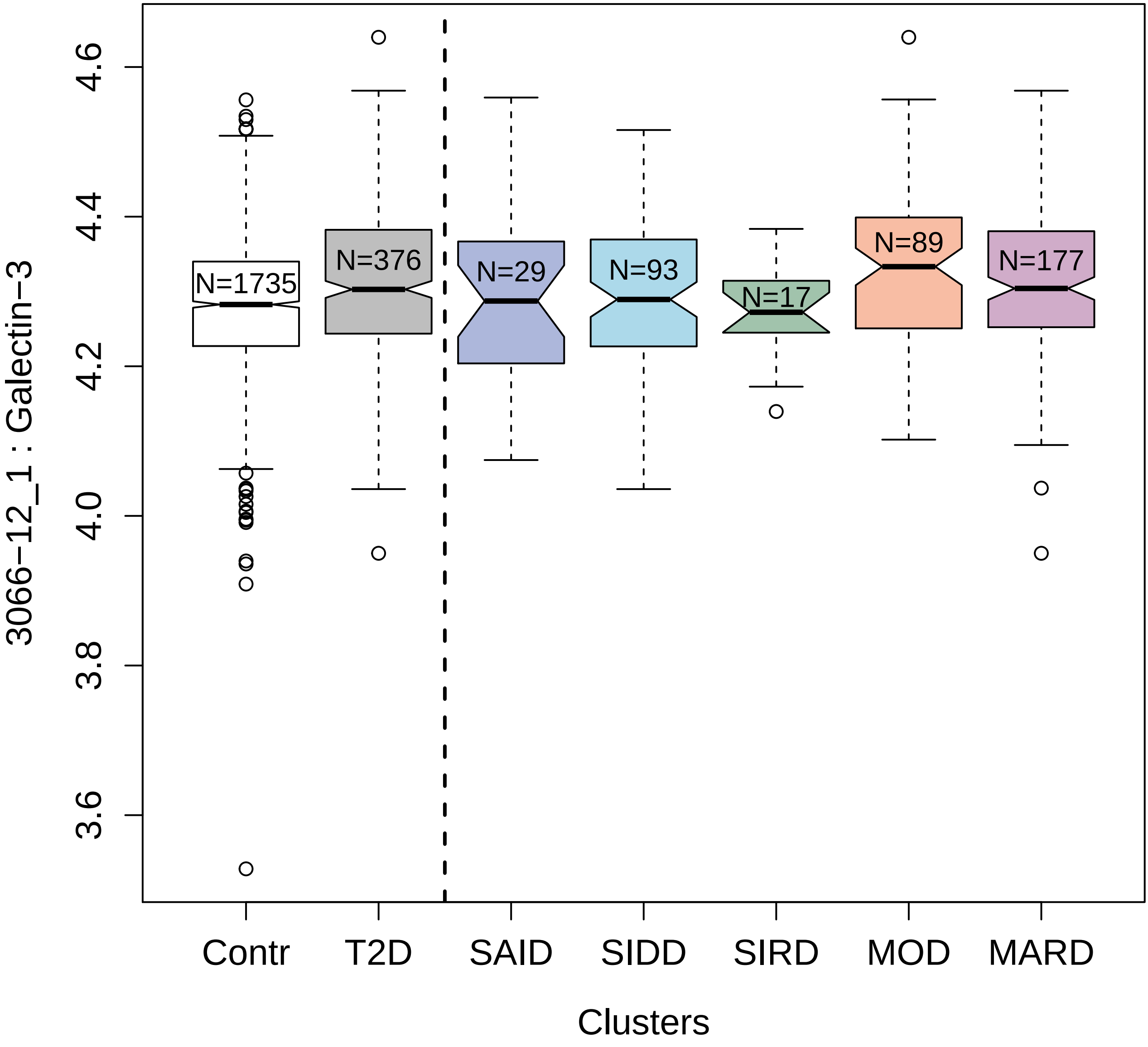
3066−12_1 : Galectin−3
4.6
4.4
4.2
4.0
3.8
3.6
N=1735
N=376
N=29
N=93
N=17
N=89
N=177
Contr
T2D
SAID
SIDD
SIRD
MOD
MARD
Clusters

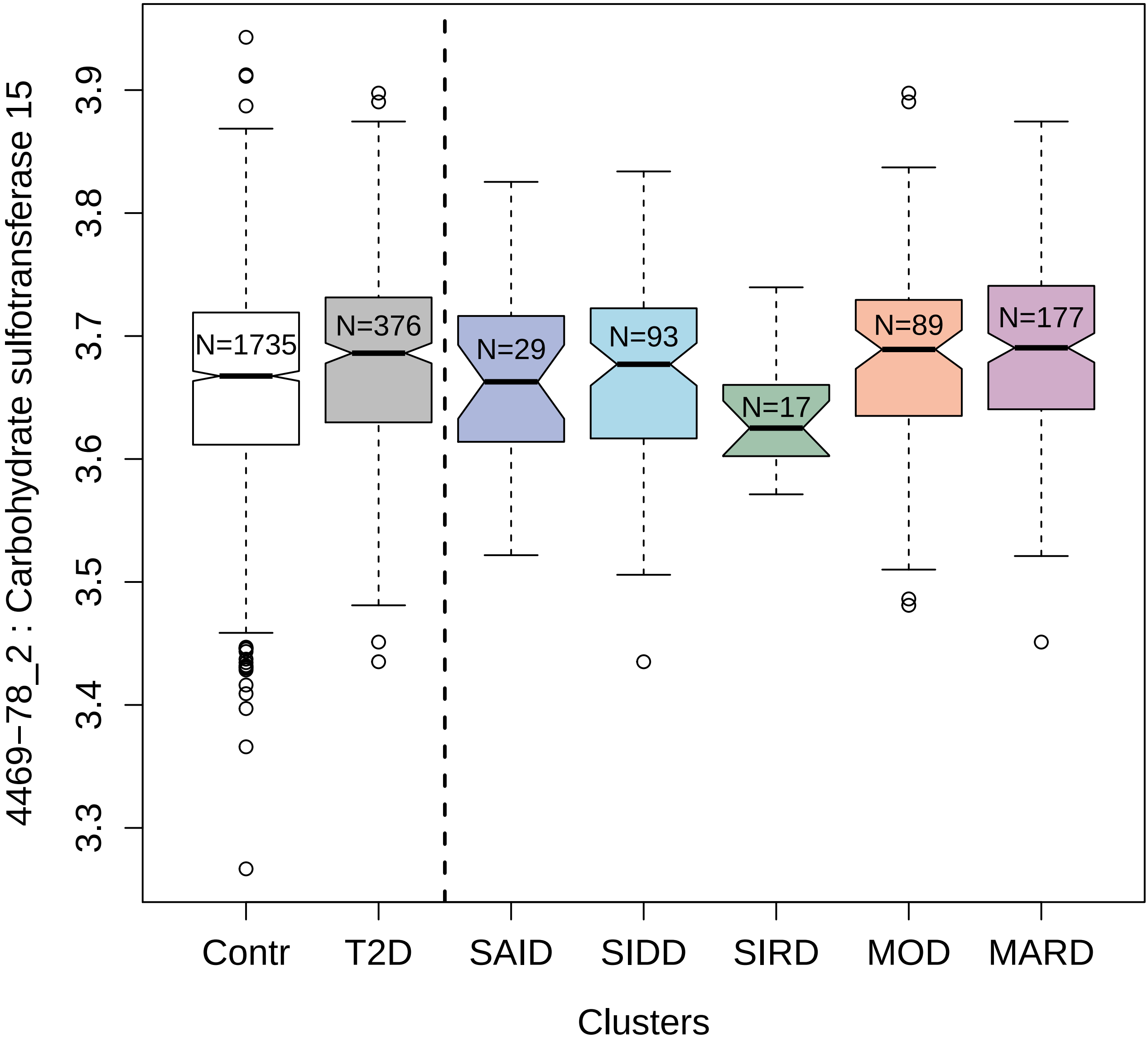
4469−78_2 : Carbohydrate sulfotransferase 15
3.3
3.4
3.5
3.6
3.7
3.8
3.9
N=1735
N=376
N=29
N=93
N=17
N=89
N=177
Contr
T2D
SAID
SIDD
SIRD
MOD
MARD
Clusters

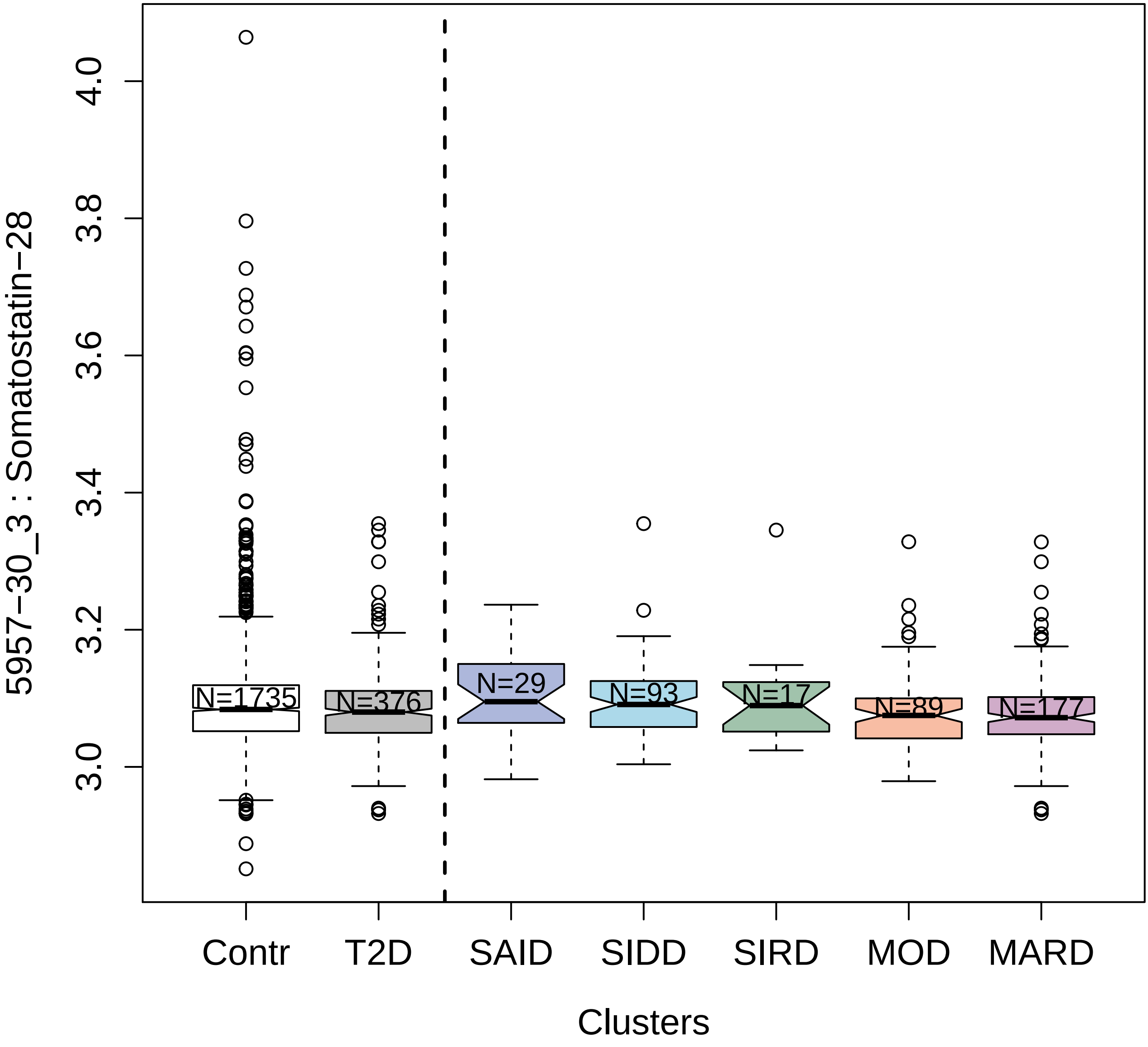

5957−30_3 : Somatostatin−28
4.0
3.8
3.6
3.4
3.2
3.0
N=1735
N=376
N=29
N=93
N=17
N=89
N=177
Contr
T2D
SAID
SIDD
SIRD
MOD
MARD
Clusters

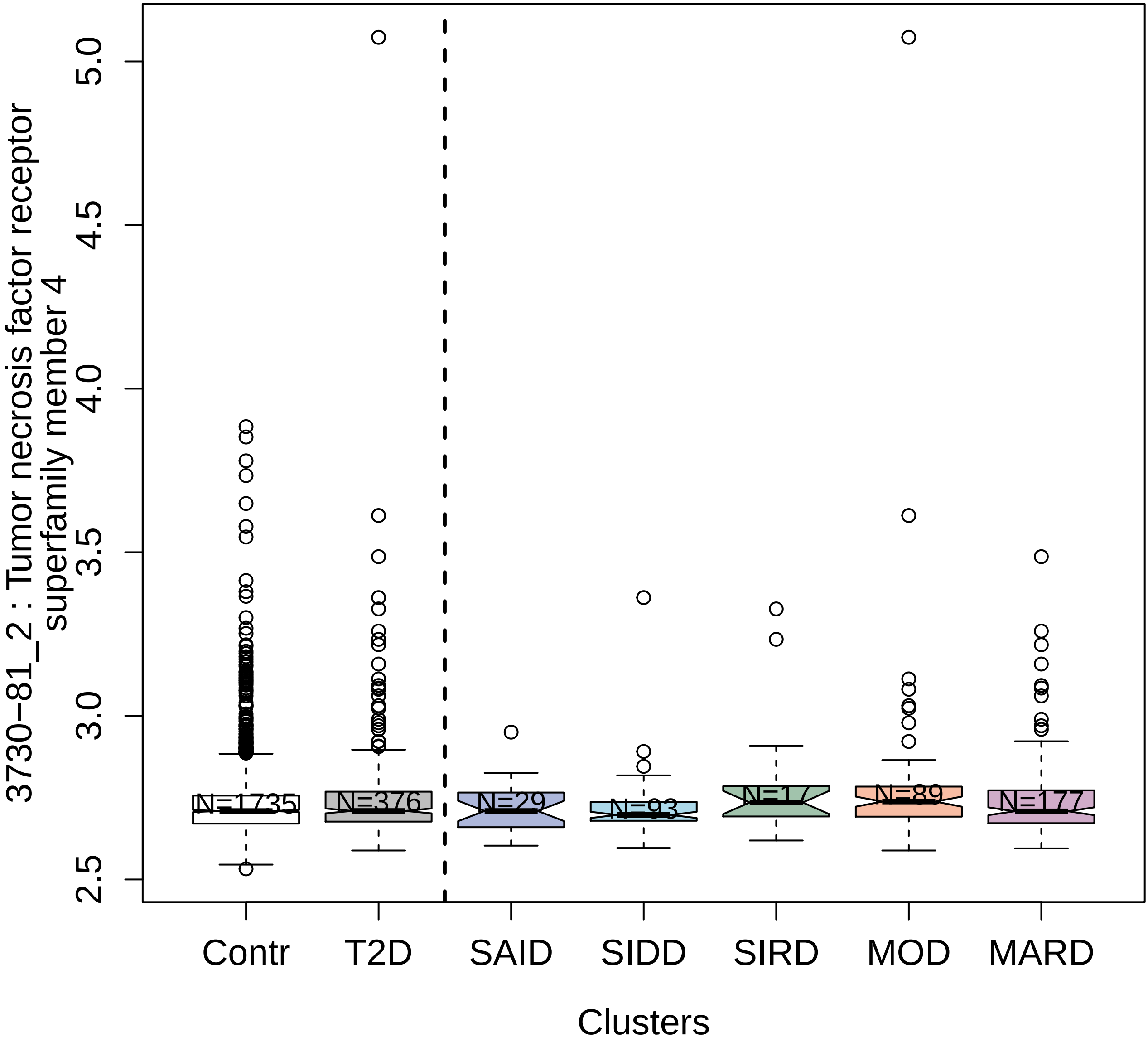
3730−81_2 : Tumor necrosis factor receptor superfamily member 4
2.5
3.0
3.5
4.0
4.5
5.0
N=1735
N=376
N=29
N=93
N=17
N=89
N=177
Contr
T2D
SAID
SIDD
SIRD
MOD
MARD
Clusters

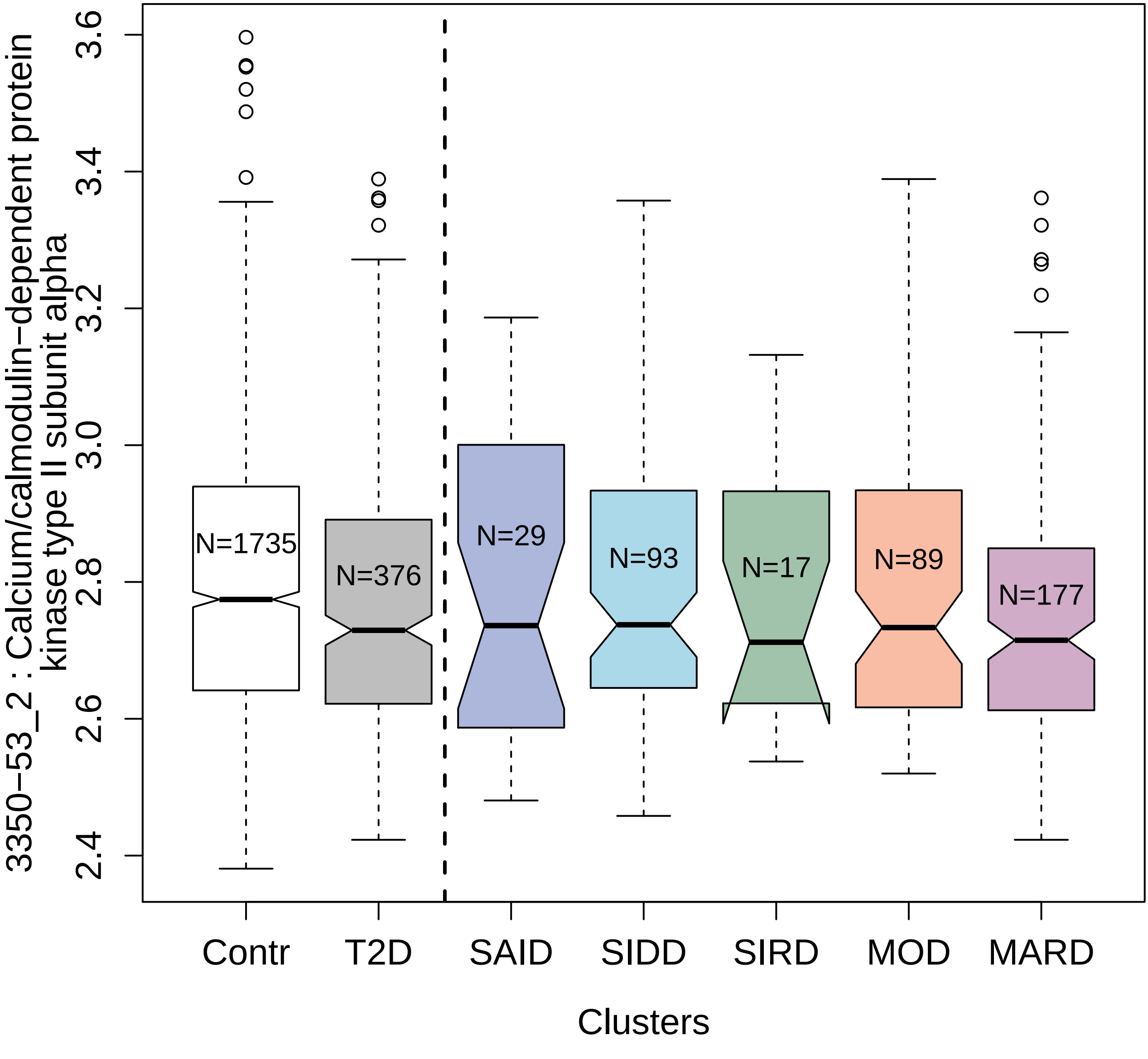
3350−53_2 : Calcium/calmodulin−dependent protein kinase type II subunit alpha
2.4
2.6
2.8
3.0
3.2
3.4
3.6
N=1735
N=376
N=29
N=93
N=17
N=89
N=177
Contr
T2D
SAID
SIDD
SIRD
MOD
MARD
Clusters

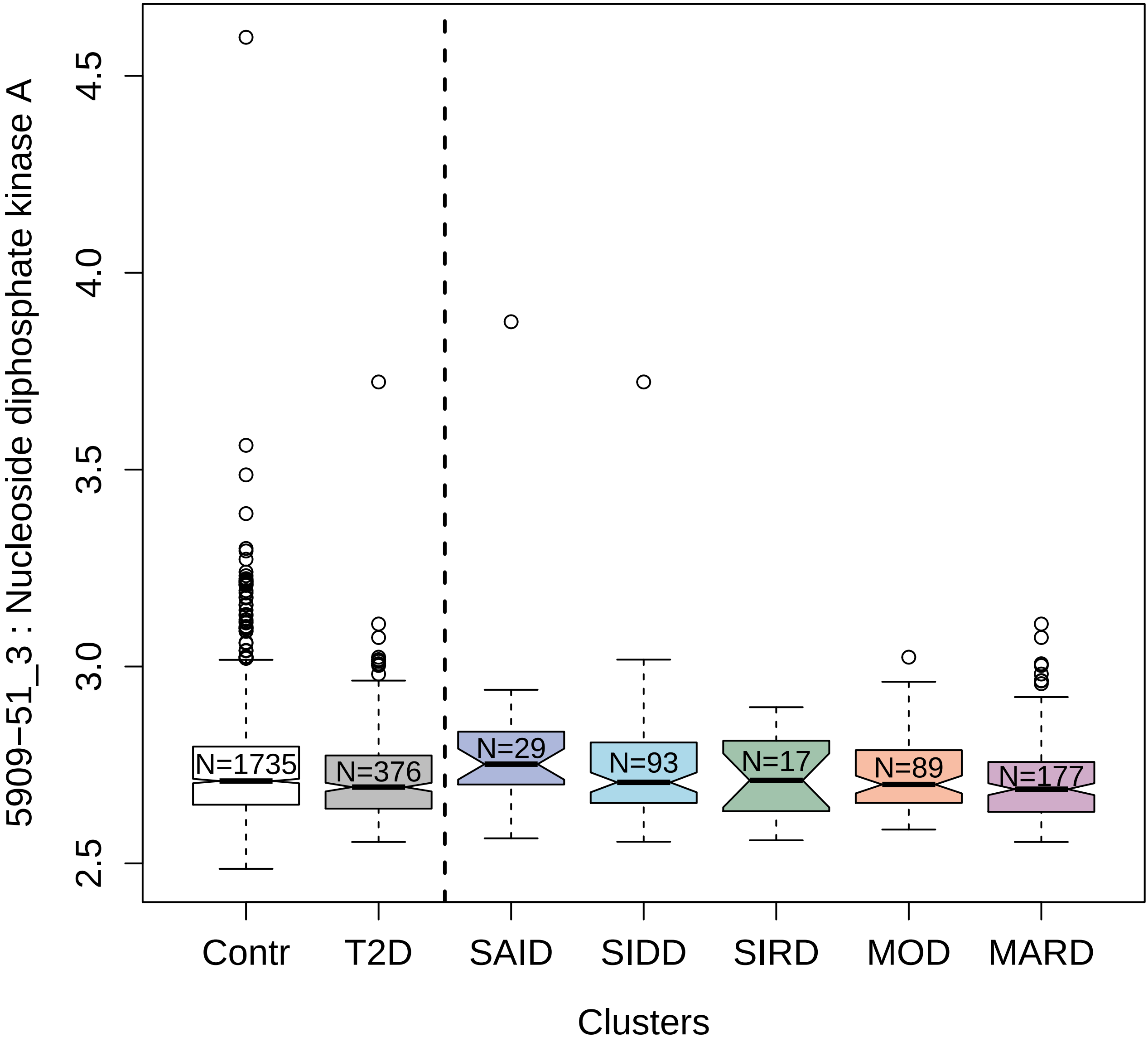
5909−51_3 : Nucleoside diphosphate kinase A
2.5
3.0
3.5
4.0
4.5
N=1735
N=376
N=29
N=93
N=17
N=89
N=177
Contr
T2D
SAID
SIDD
SIRD
MOD
MARD
Clusters

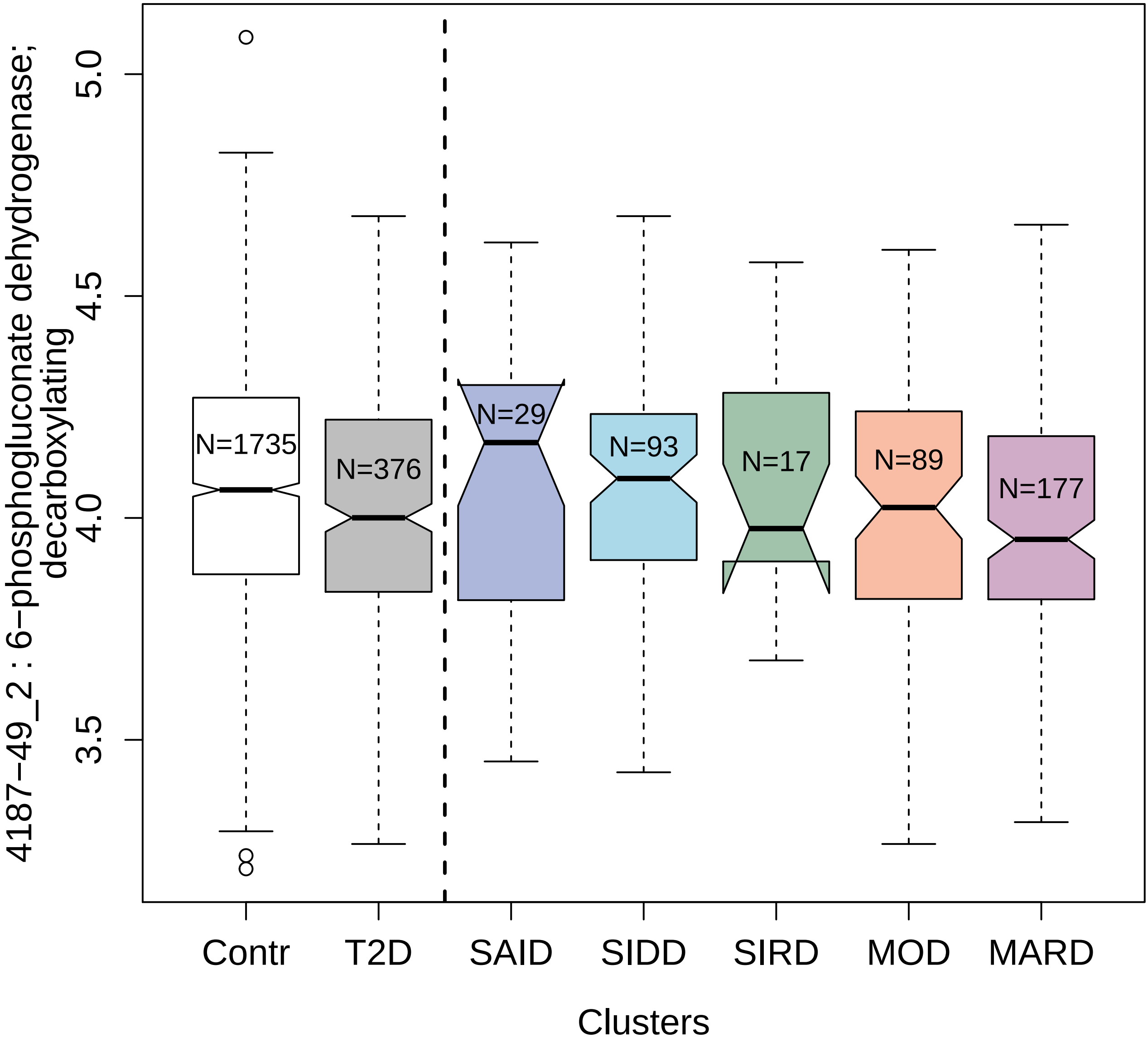

4187−49_2 : 6−phosphogluconate dehydrogenase; decarboxylating
5.0
4.5
4.0
3.5
N=1735
N=376
N=29
N=93
N=17
N=89
N=177
Contr
T2D
SAID
SIDD
SIRD
MOD
MARD
Clusters

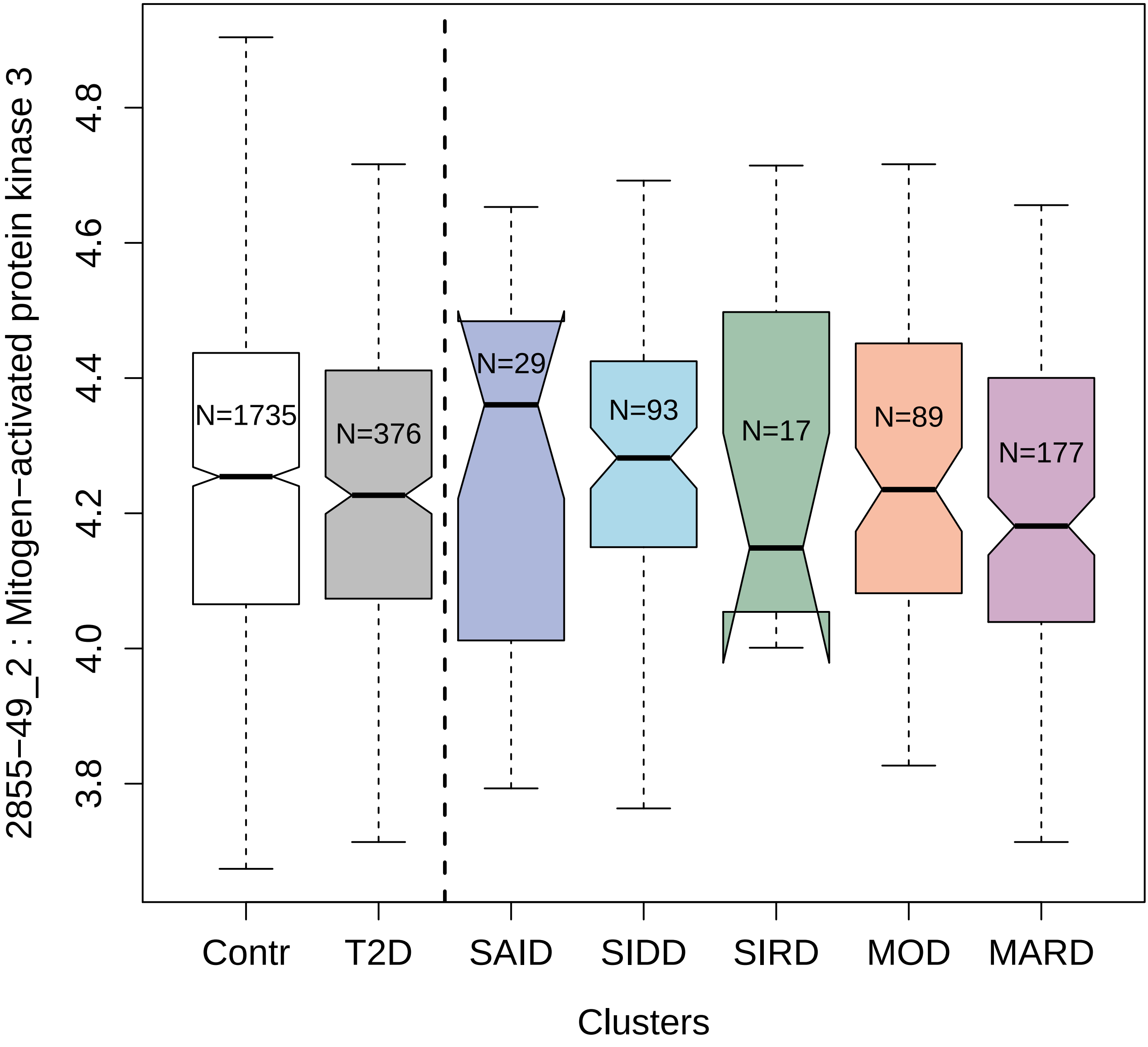
2855−49_2 : Mitogen−activated protein kinase 3
3.8
4.0
4.2
4.4
4.6
4.8
N=1735
N=376
N=29
N=93
N=17
N=89
N=177
Contr
T2D
SAID
SIDD
SIRD
MOD
MARD
Clusters

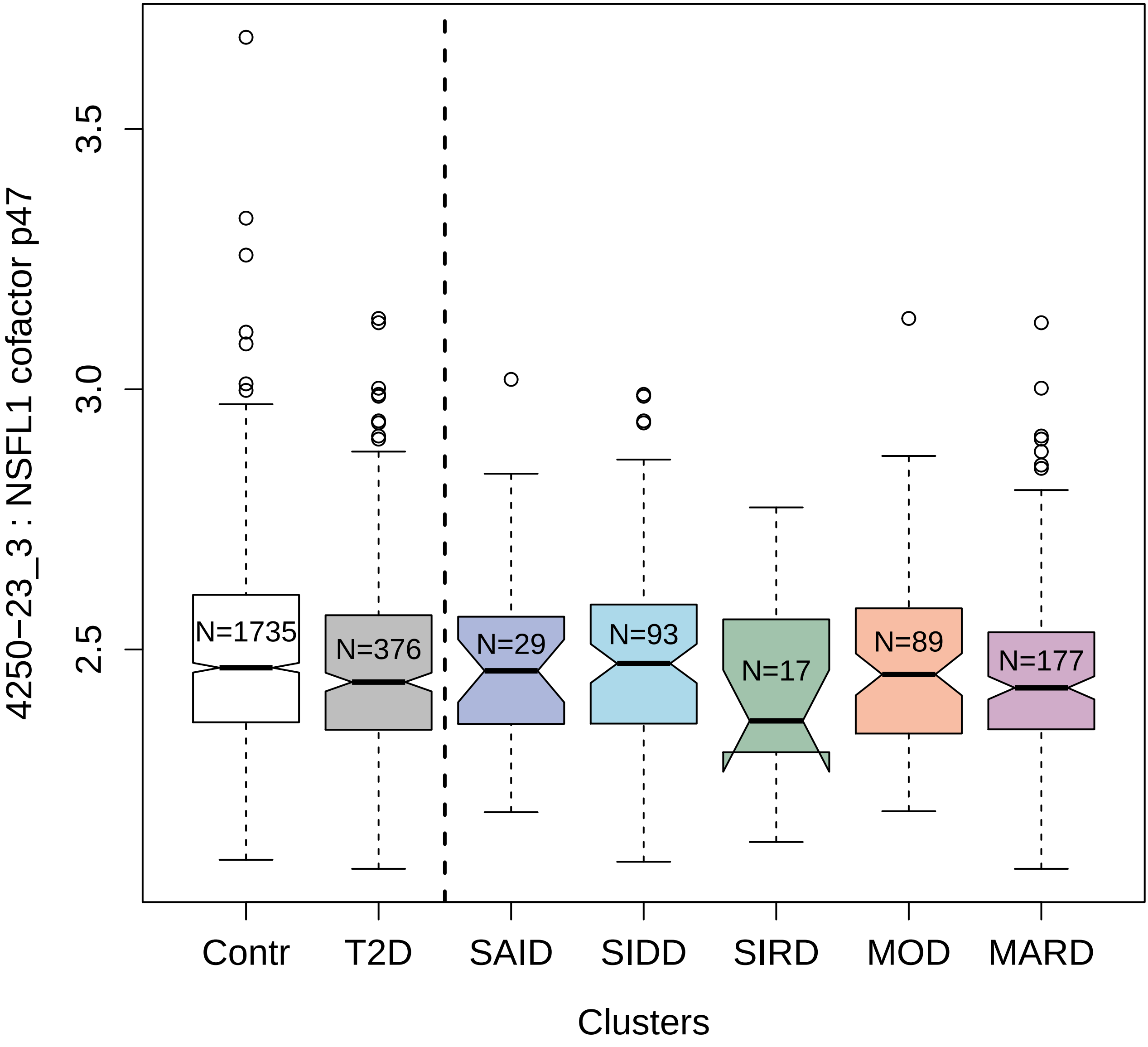

4250−23_3 : NSFL1 cofactor p47
3.5
3.0
2.5
N=1735
N=376
N=29
N=93
N=17
N=89
N=177
Contr
T2D
SAID
SIDD
SIRD
MOD
MARD
Clusters

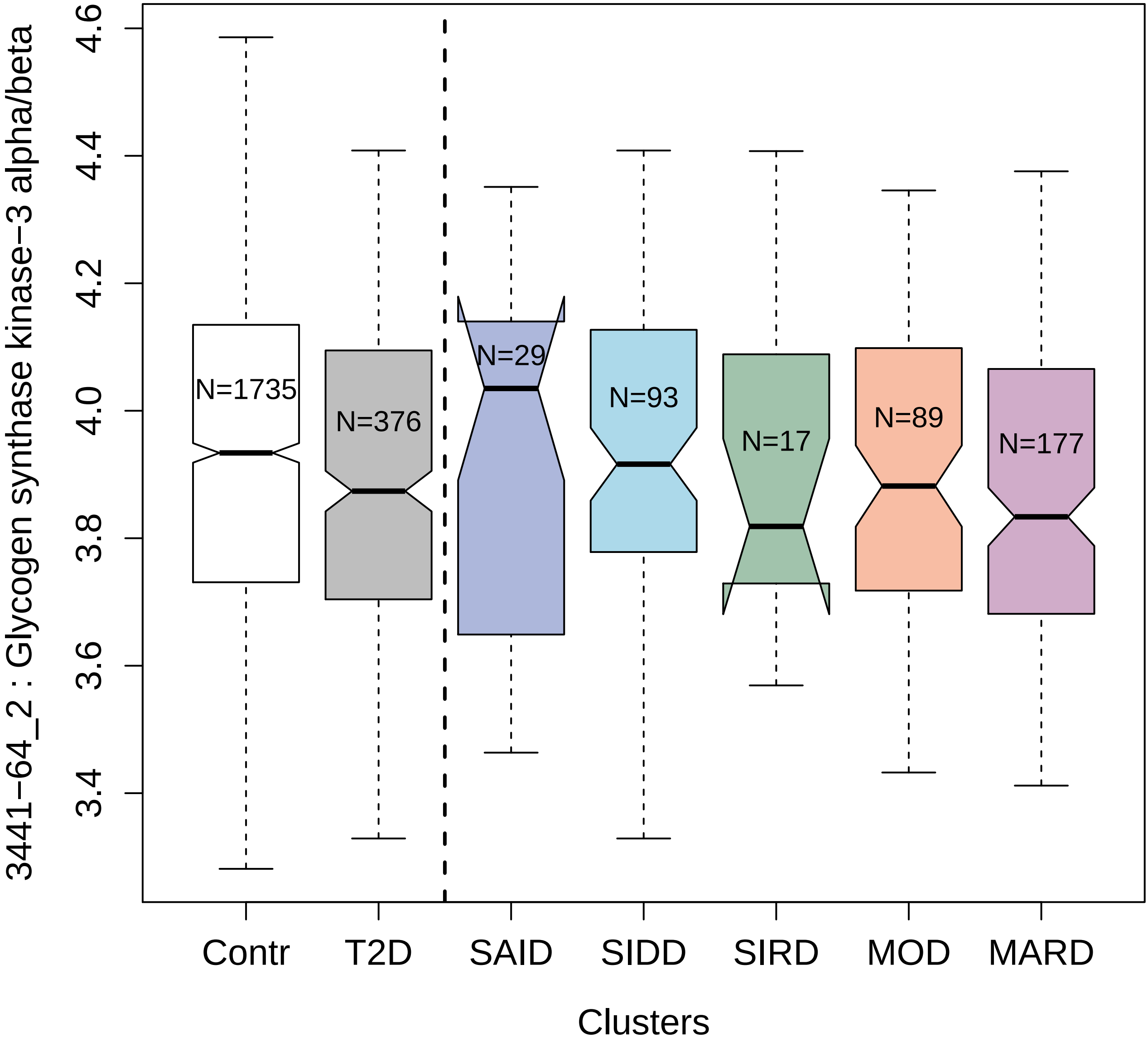

3441−64_2 : Glycogen synthase kinase−3 alpha/beta
4.6
4.4
4.2
4.0
3.8
3.6
3.4
N=1735
N=376
N=29
N=93
N=17
N=89
N=177
Contr
T2D
SAID
SIDD
SIRD
MOD
MARD
Clusters

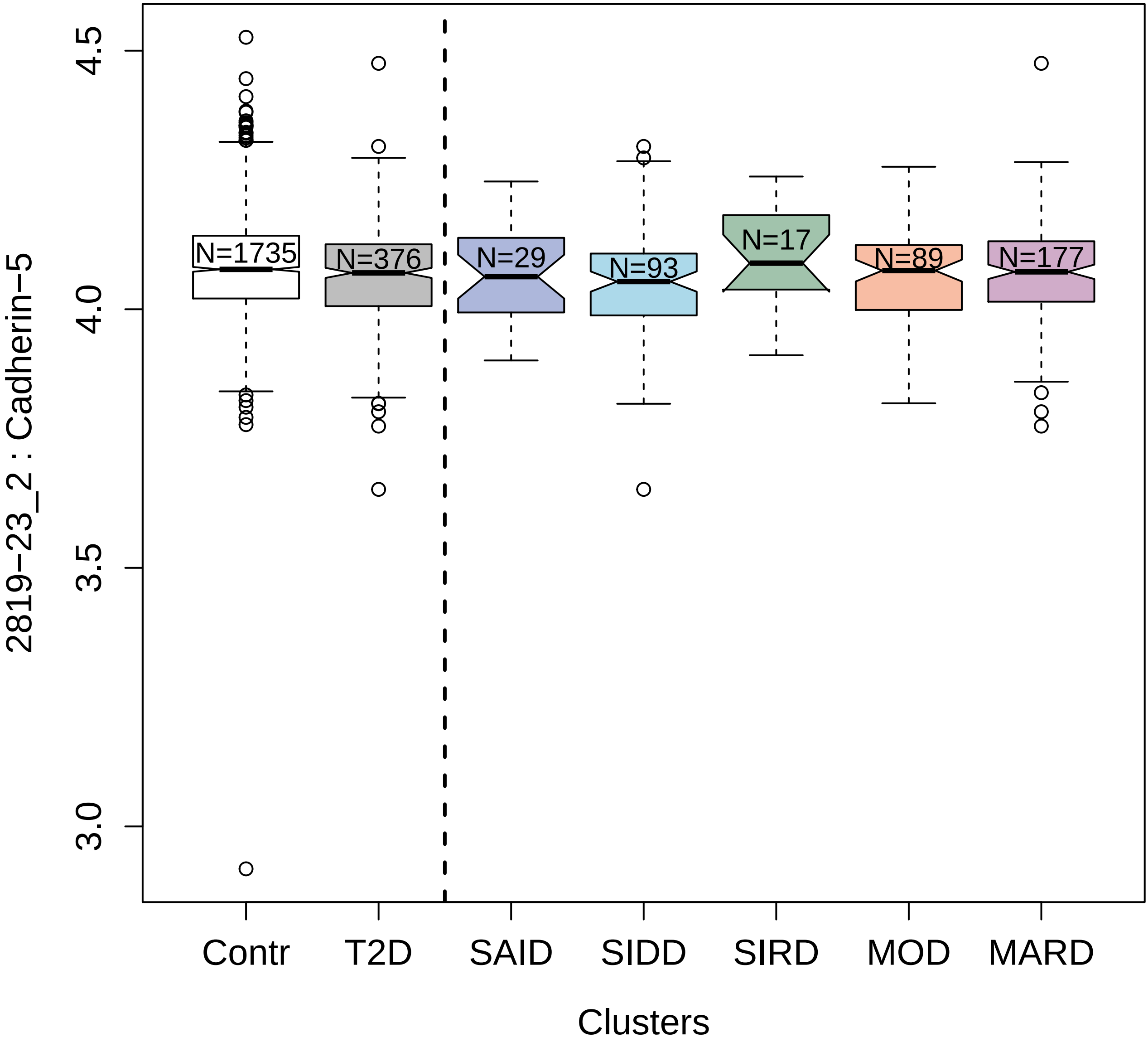
2819−23_2 : Cadherin−5
4.5
4.0
3.5
3.0
N=1735
N=376
N=29
N=93
N=17
N=89
N=177
Contr
T2D
SAID
SIDD
SIRD
MOD
MARD
Clusters

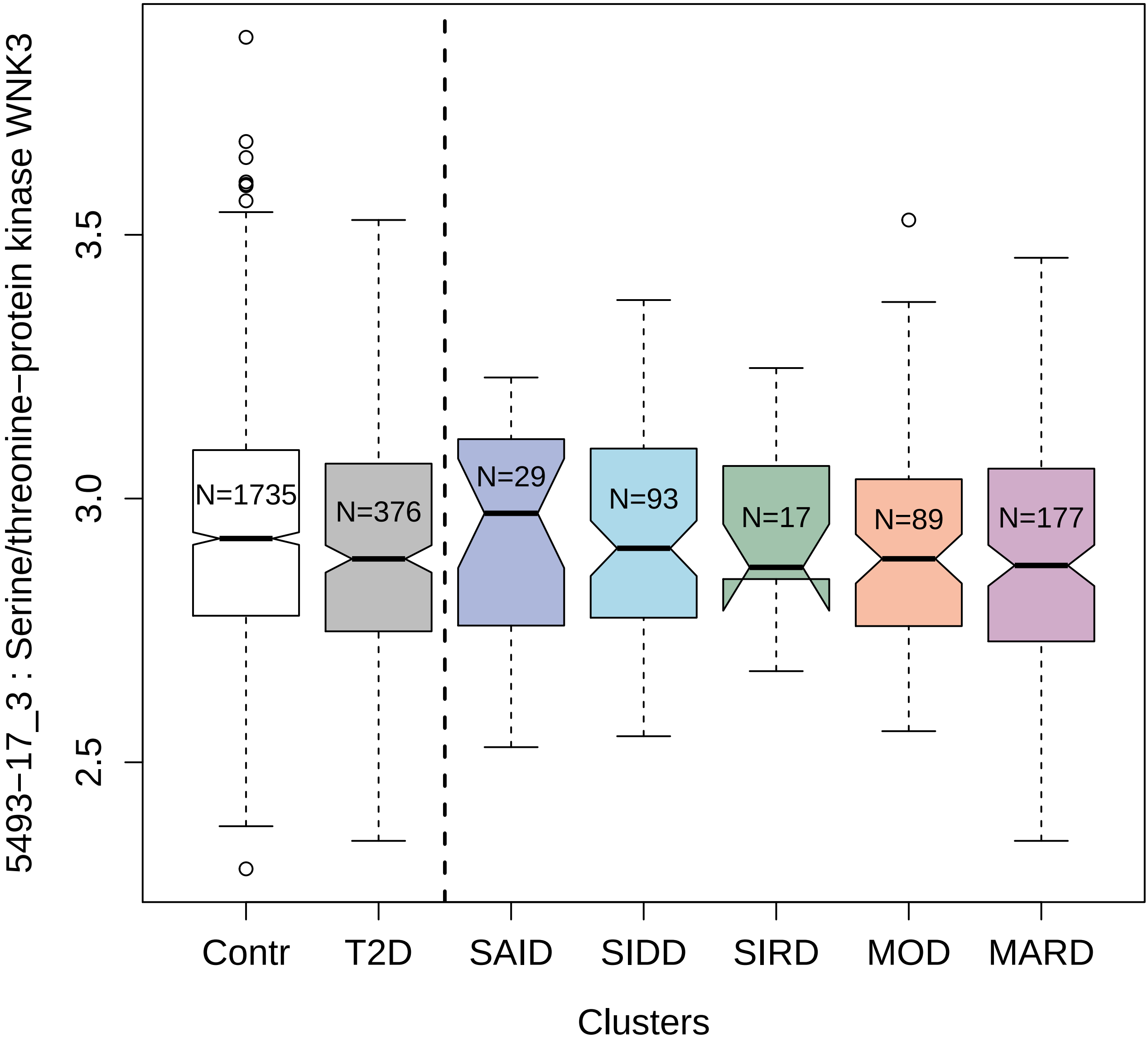
5493−17_3 : Serine/threonine−protein kinase WNK3
3.5
3.0
2.5
N=1735
N=376
N=29
N=93
N=17
N=89
N=177
Contr
T2D
SAID
SIDD
SIRD
MOD
MARD
Clusters

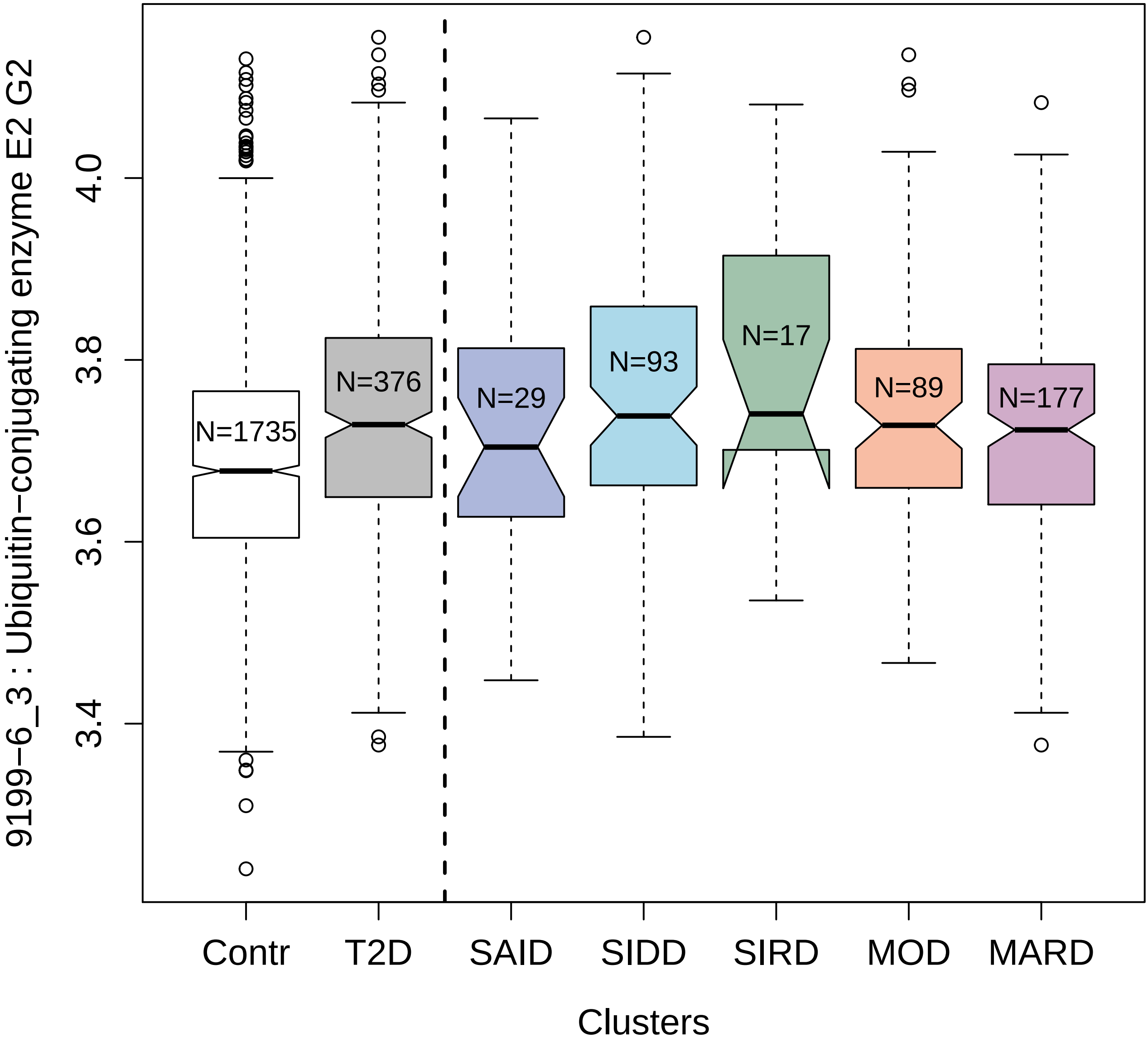
9199−6_3 : Ubiquitin−conjugating enzyme E2 G2
3.4
3.6
3.8
4.0
N=1735
N=376
N=29
N=93
N=17
N=89
N=177
Contr
T2D
SAID
SIDD
SIRD
MOD
MARD
Clusters

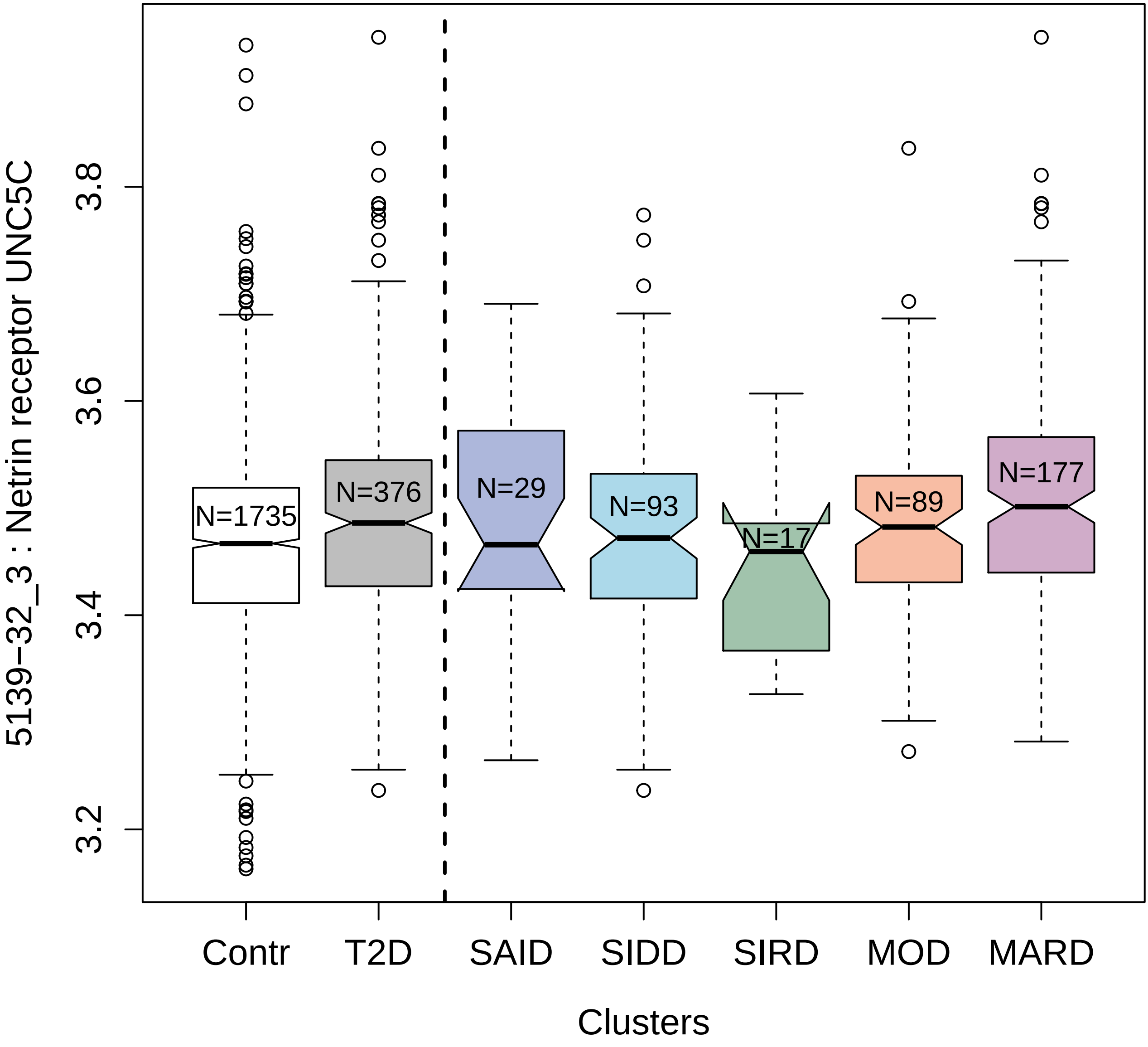
5139−32_3 : Netrin receptor UNC5C
3.2
3.4
3.6
3.8
N=1735
N=376
N=29
N=93
N=17
N=89
N=177
Contr
T2D
SAID
SIDD
SIRD
MOD
MARD
Clusters

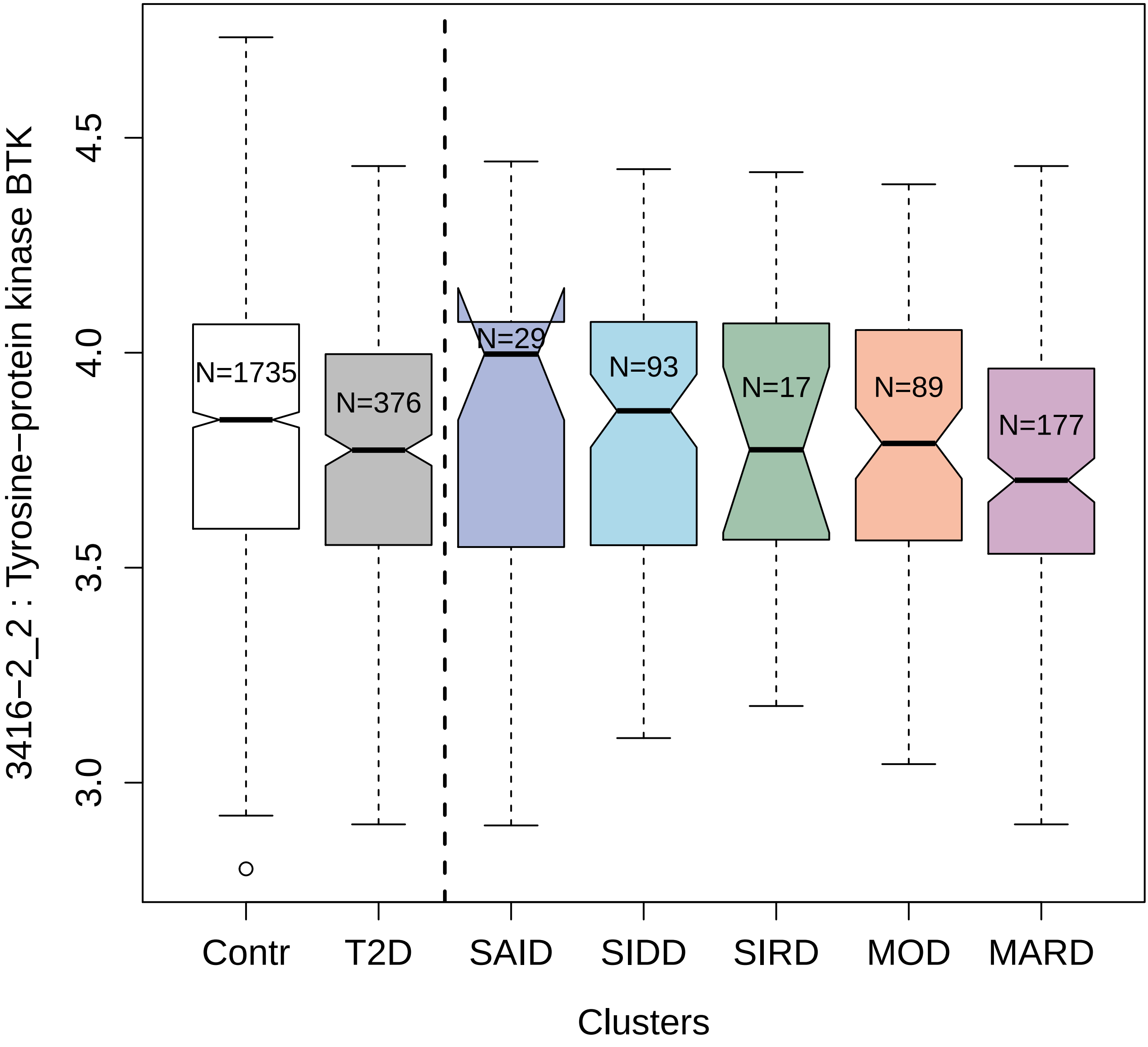
3416−2_2 : Tyrosine−protein kinase BTK
4.5
4.0
3.5
3.0
N=1735
N=376
N=29
N=93
N=17
N=89
N=177
Contr
T2D
SAID
SIDD
SIRD
MOD
MARD
Clusters

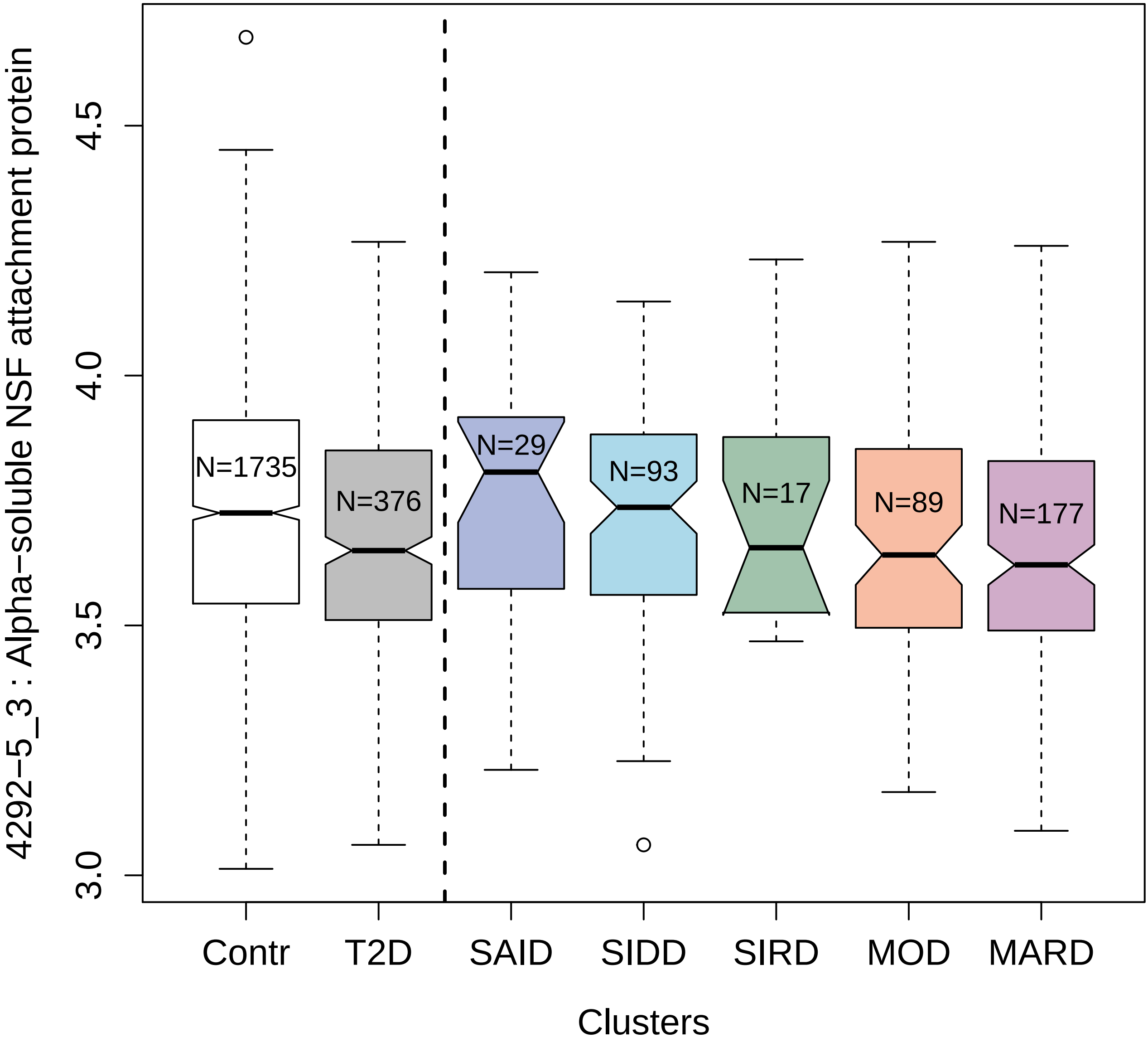
4292−5_3 : Alpha−soluble NSF attachment protein
3.0
3.5
4.0
4.5
N=1735
N=376
N=29
N=93
N=17
N=89
N=177
Contr
T2D
SAID
SIDD
SIRD
MOD
MARD
Clusters

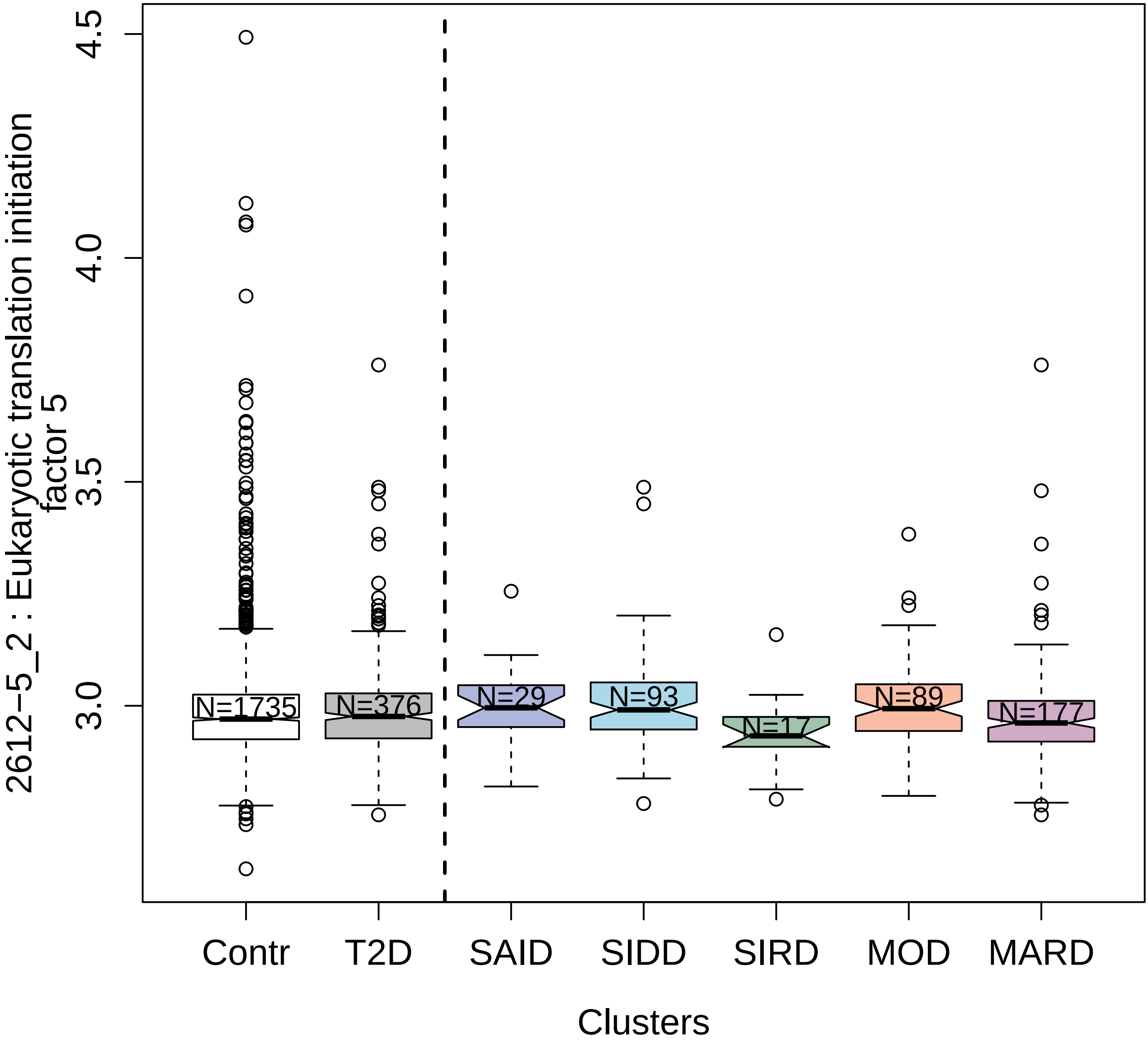
2612−5_2 : Eukaryotic translation initiation factor 5
4.5
4.0
3.5
3.0
N=1735
N=376
N=29
N=93
N=17
N=89
N=177
Contr
T2D
SAID
SIDD
SIRD
MOD
MARD
Clusters

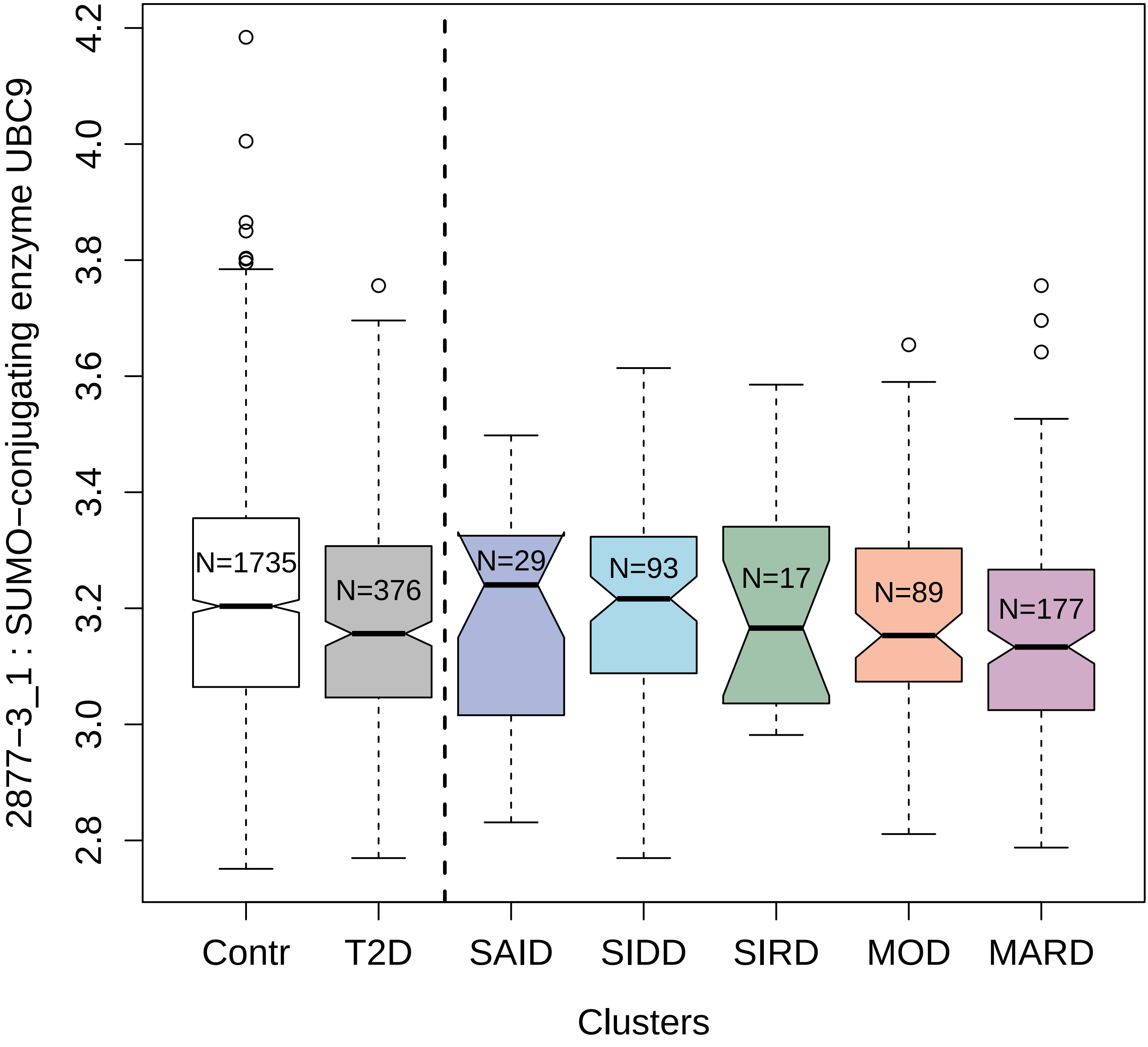

2877−3_1 : SUMO−conjugating enzyme UBC9
4.2
4.0
3.8
3.6
3.4
3.2
3.0
2.8
N=1735
N=376
N=29
N=93
N=17
N=89
N=177
Contr
T2D
SAID
SIDD
SIRD
MOD
MARD
Clusters

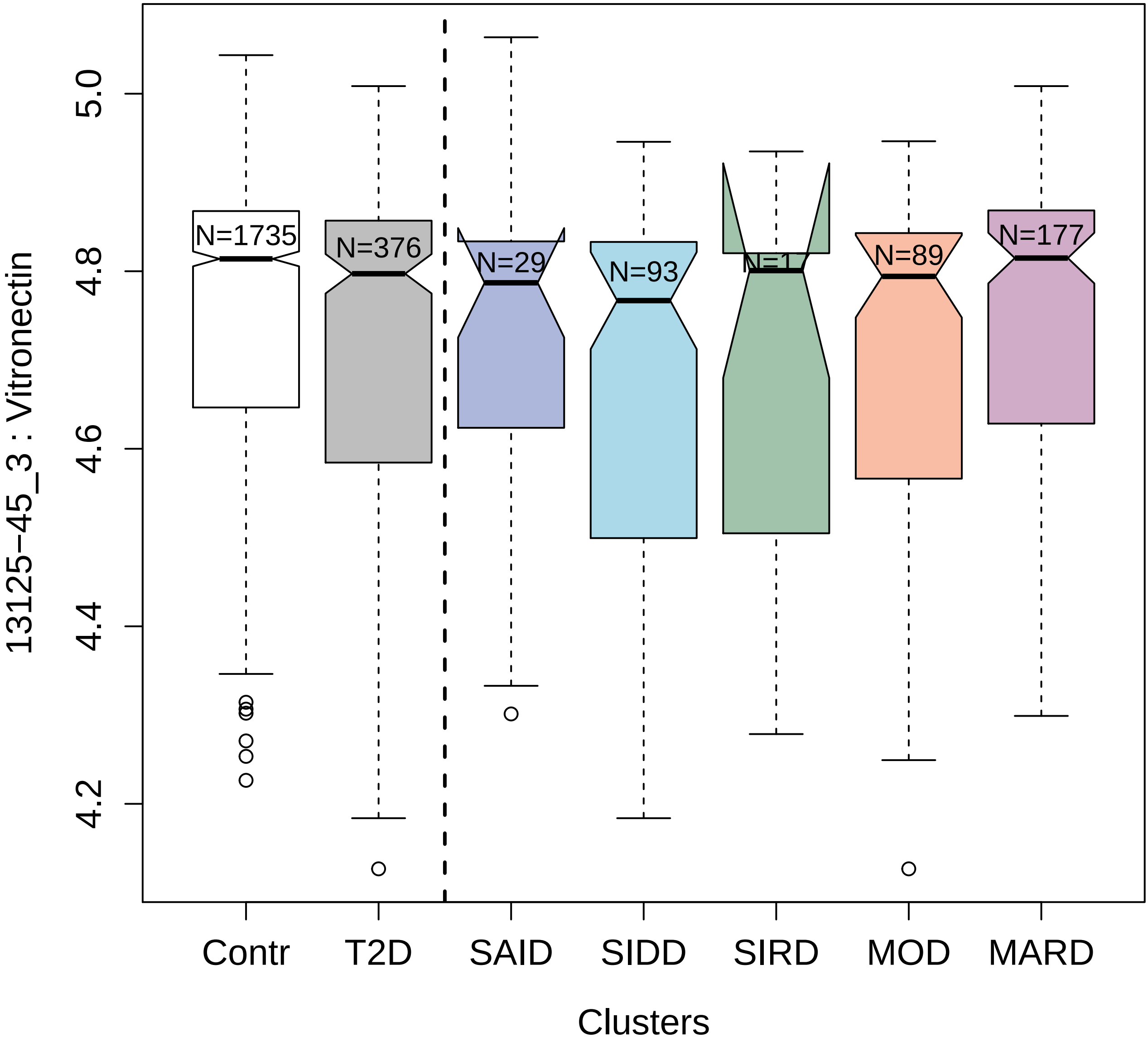

13125−45_3 : Vitronectin
5.0
4.8
4.6
4.4
4.2
N=1735
N=376
N=29
N=93
N=89
N=177
Contr
T2D
SAID
SIDD
SIRD
MOD
MARD
Clusters

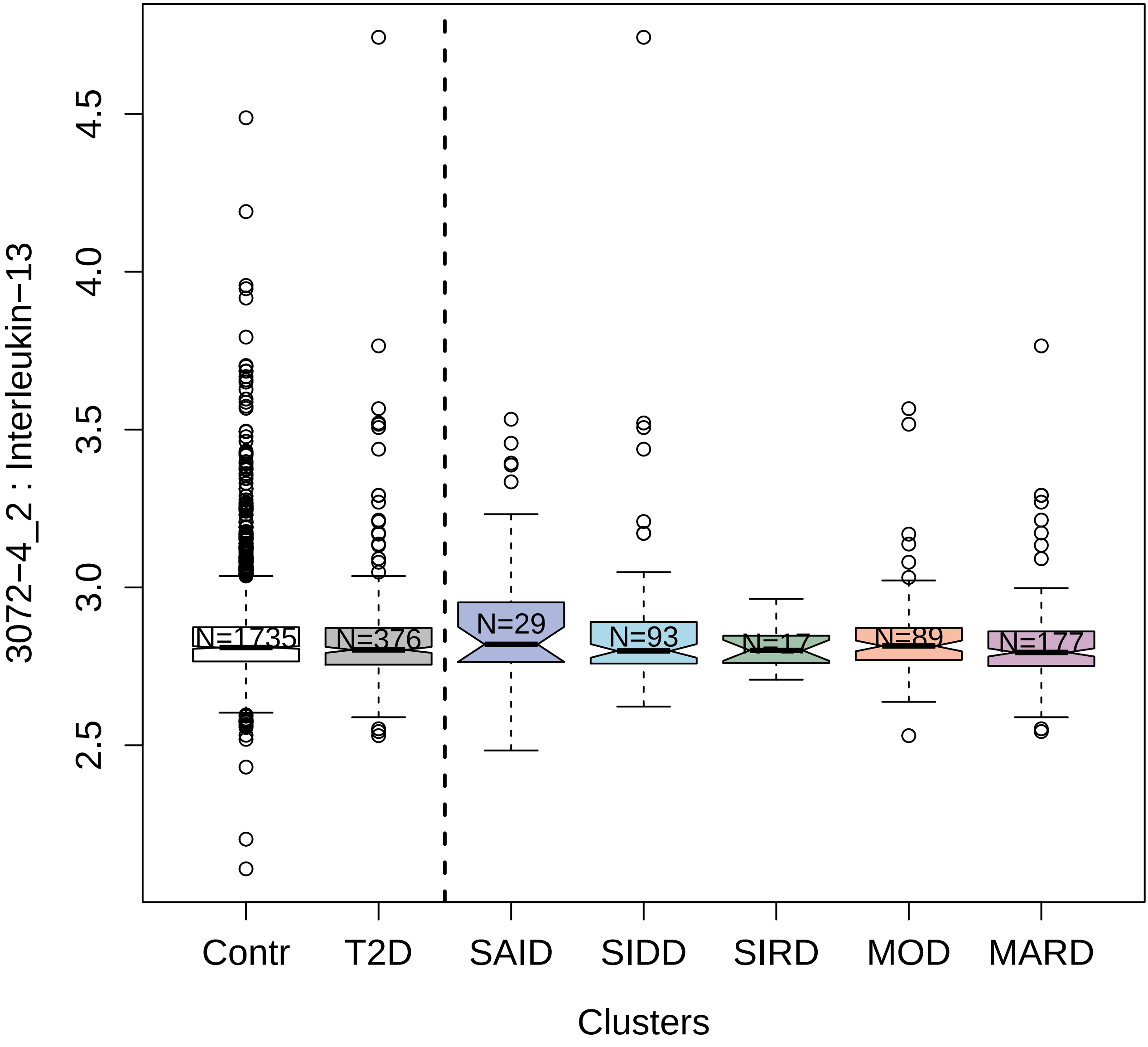
3072−4_2 : Interleukin−13
4.5
4.0
3.5
3.0
2.5
N=1735
N=376
N=29
N=93
N=17
N=89
N=177
Contr
T2D
SAID
SIDD
SIRD
MOD
MARD
Clusters

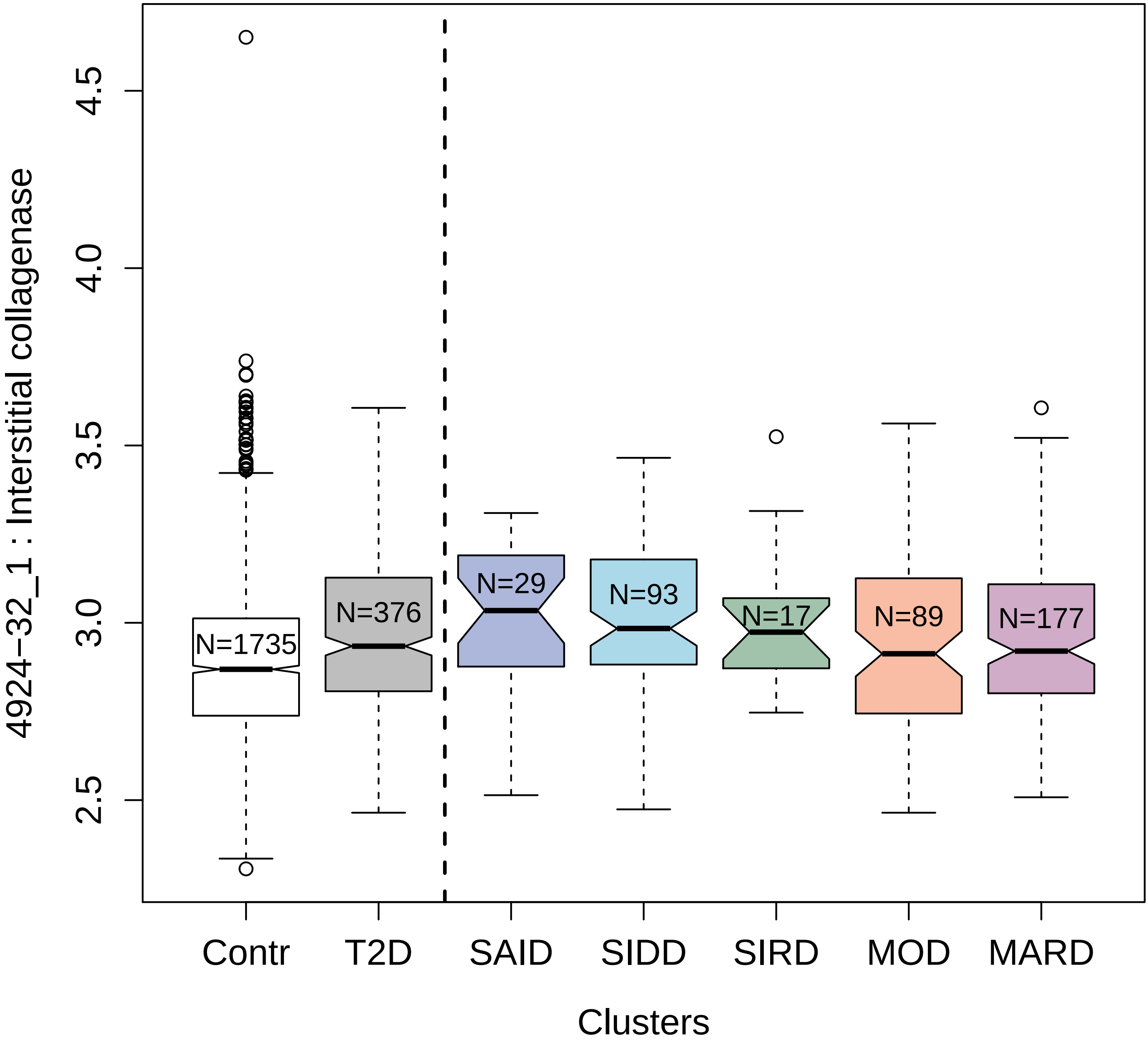

4924−32_1 : Interstitial collagenase
4.5
4.0
3.5
3.0
2.5
N=1735
N=376
N=29
N=93
N=17
N=89
N=177
Contr
T2D
SAID
SIDD
SIRD
MOD
MARD
Clusters

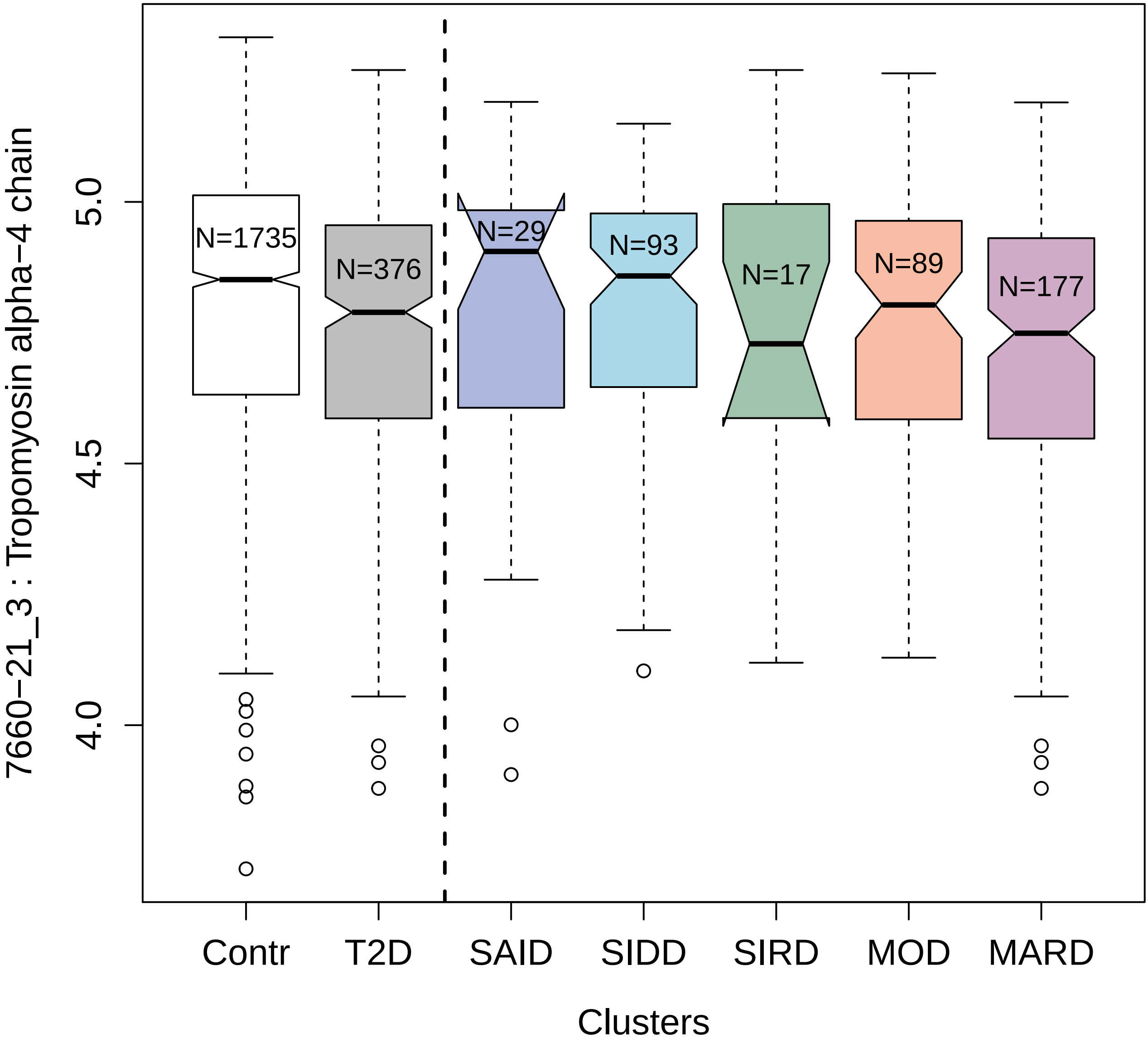
7660−21_3 : Tropomyosin alpha−4 chain
5.0
4.5
4.0
N=1735
N=376
N=29
N=93
N=17
N=89
N=177
Contr
T2D
SAID
SIDD
SIRD
MOD
MARD
Clusters

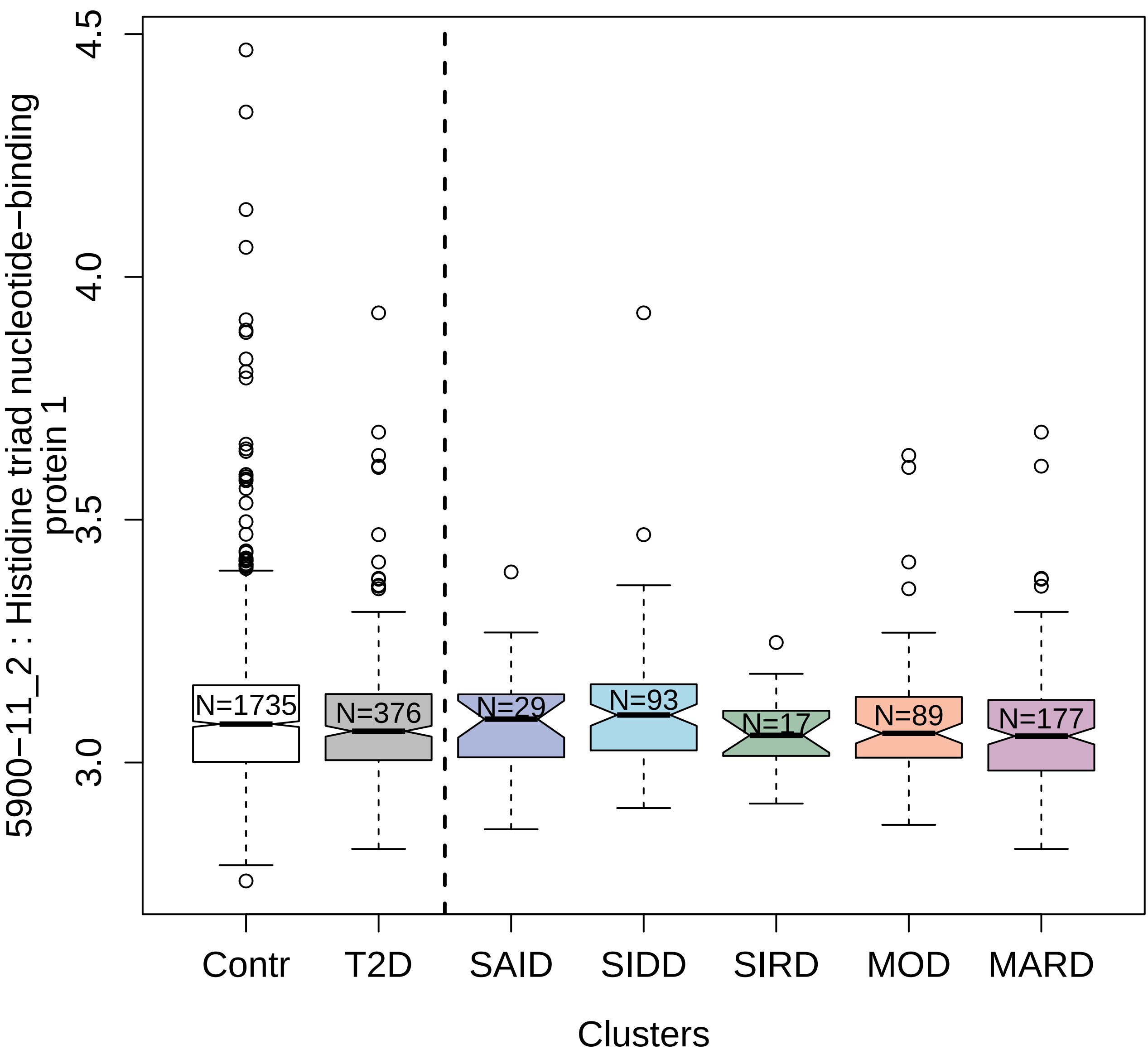
5900−11_2 : Histidine triad nucleotide−binding protein 1
4.5
4.0
3.5
3.0
N=1735
N=376
N=29
N=93
N=17
N=89
N=177
Contr
T2D
SAID
SIDD
SIRD
MOD
MARD
Clusters

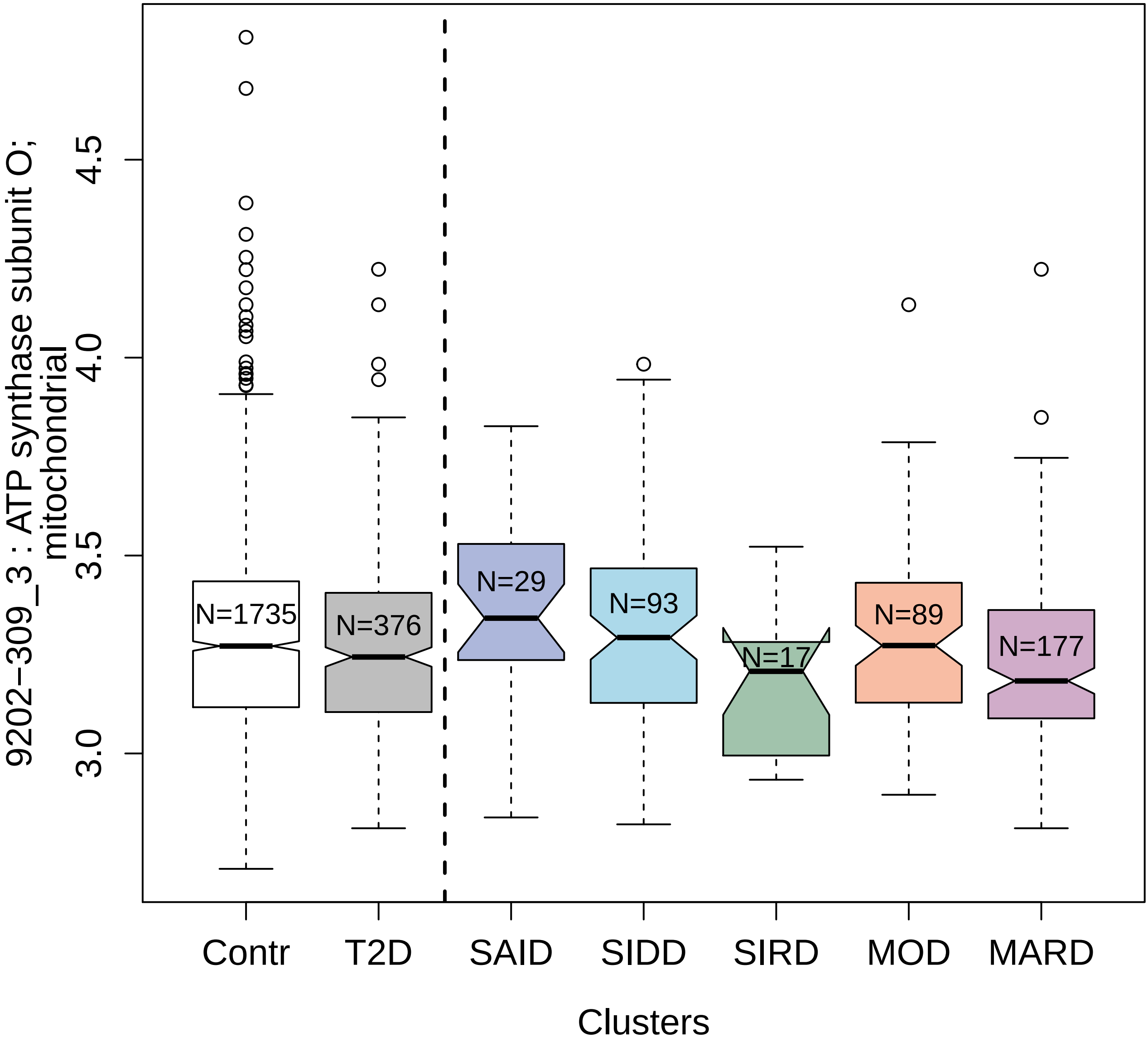

9202−309_3 : ATP synthase subunit O; mitochondrial
4.5
4.0
3.5
3.0
N=1735
N=376
N=29
N=93
N=17
N=89
N=177
Contr
T2D
SAID
SIDD
SIRD
MOD
MARD
Clusters

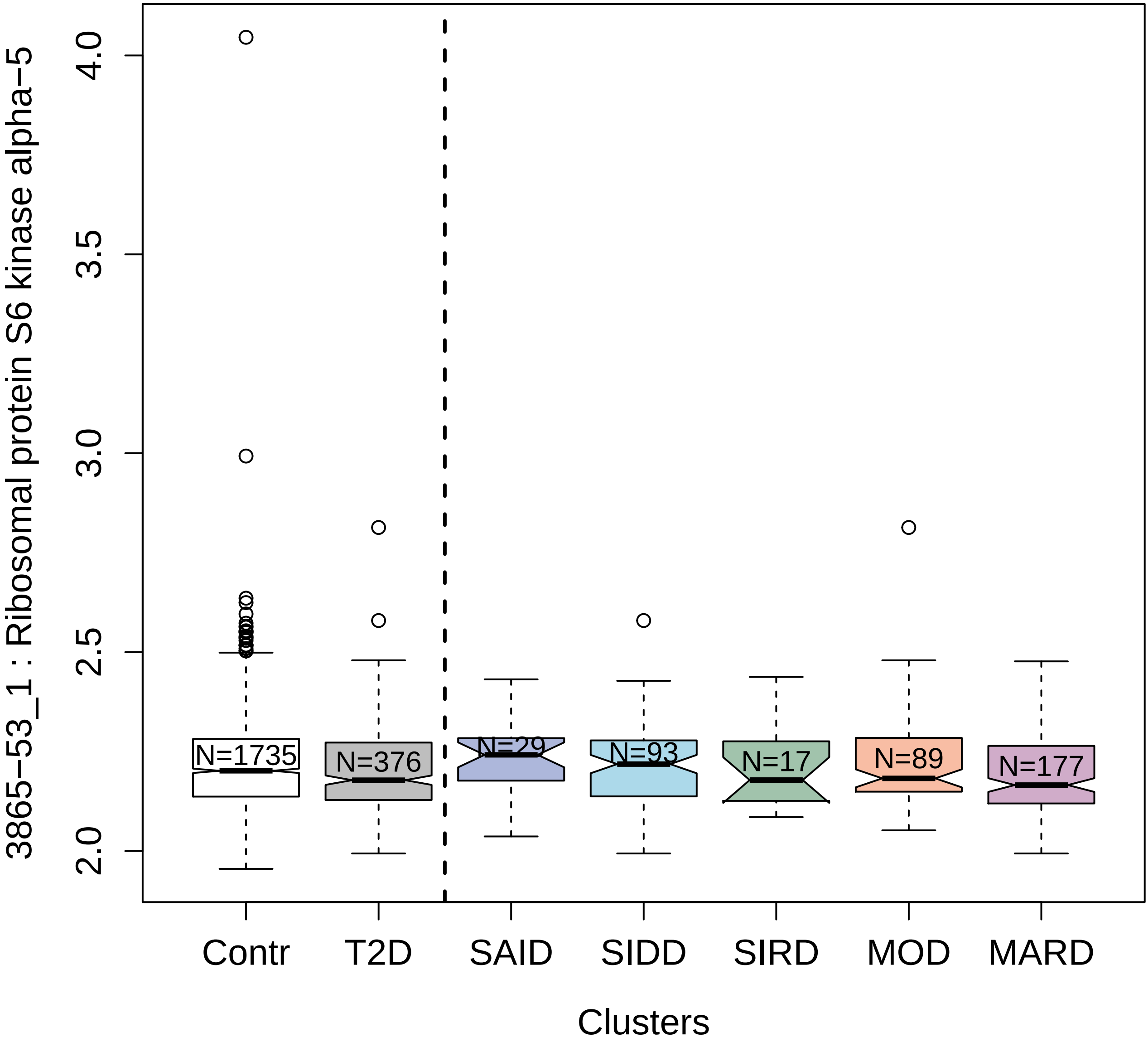

3865−53_1 : Ribosomal protein S6 kinase alpha−5
4.0
3.5
3.0
2.5
2.0
N=1735
N=376
N=29
N=93
N=17
N=89
N=177
Contr
T2D
SAID
SIDD
SIRD
MOD
MARD
Clusters

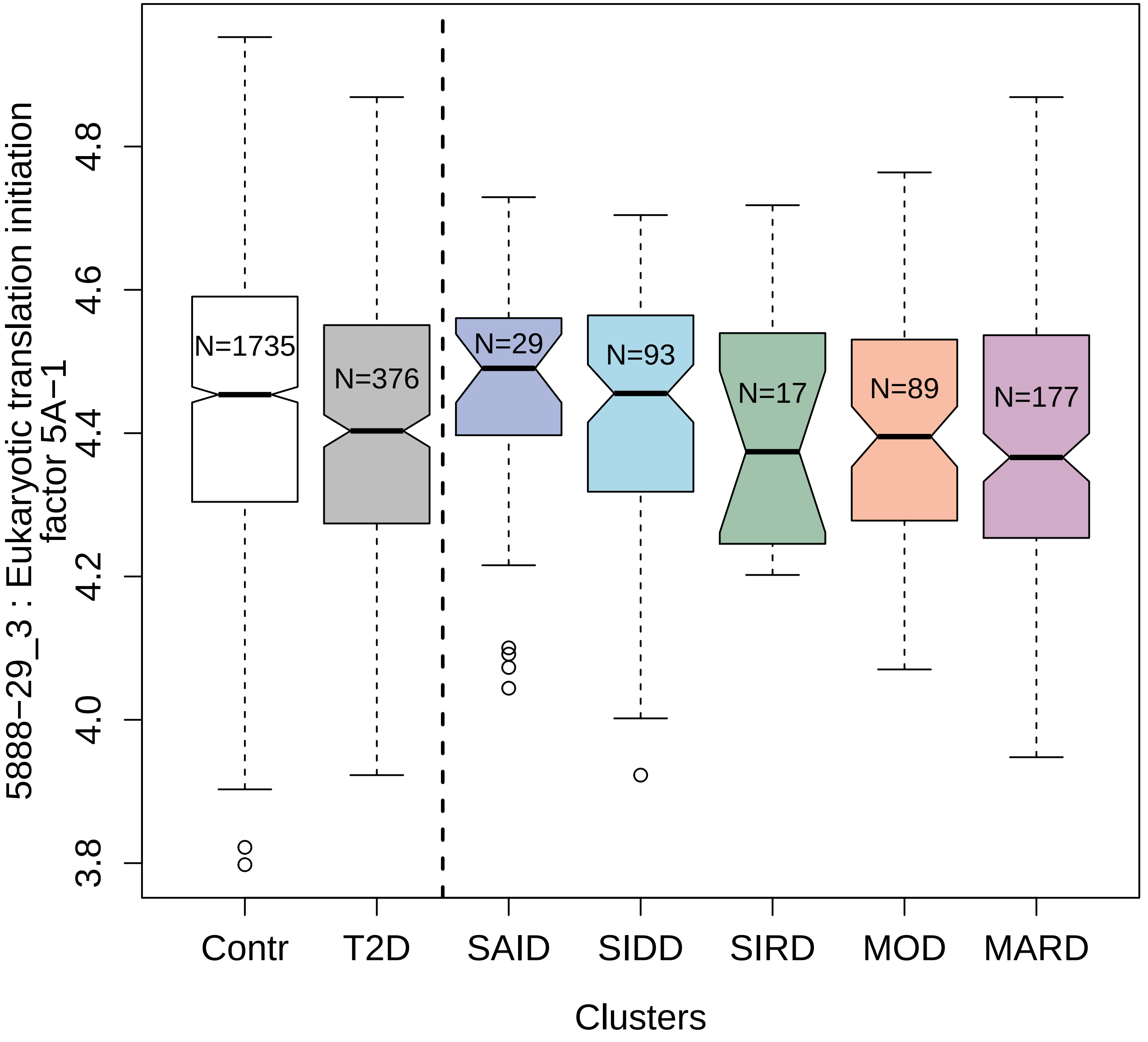
5888−29_3 : Eukaryotic translation initiation factor 5A−1
3.8
4.0
4.2
4.4
4.6
4.8
N=1735
N=376
N=29
N=93
N=17
N=89
N=177
Contr
T2D
SAID
SIDD
SIRD
MOD
MARD
Clusters

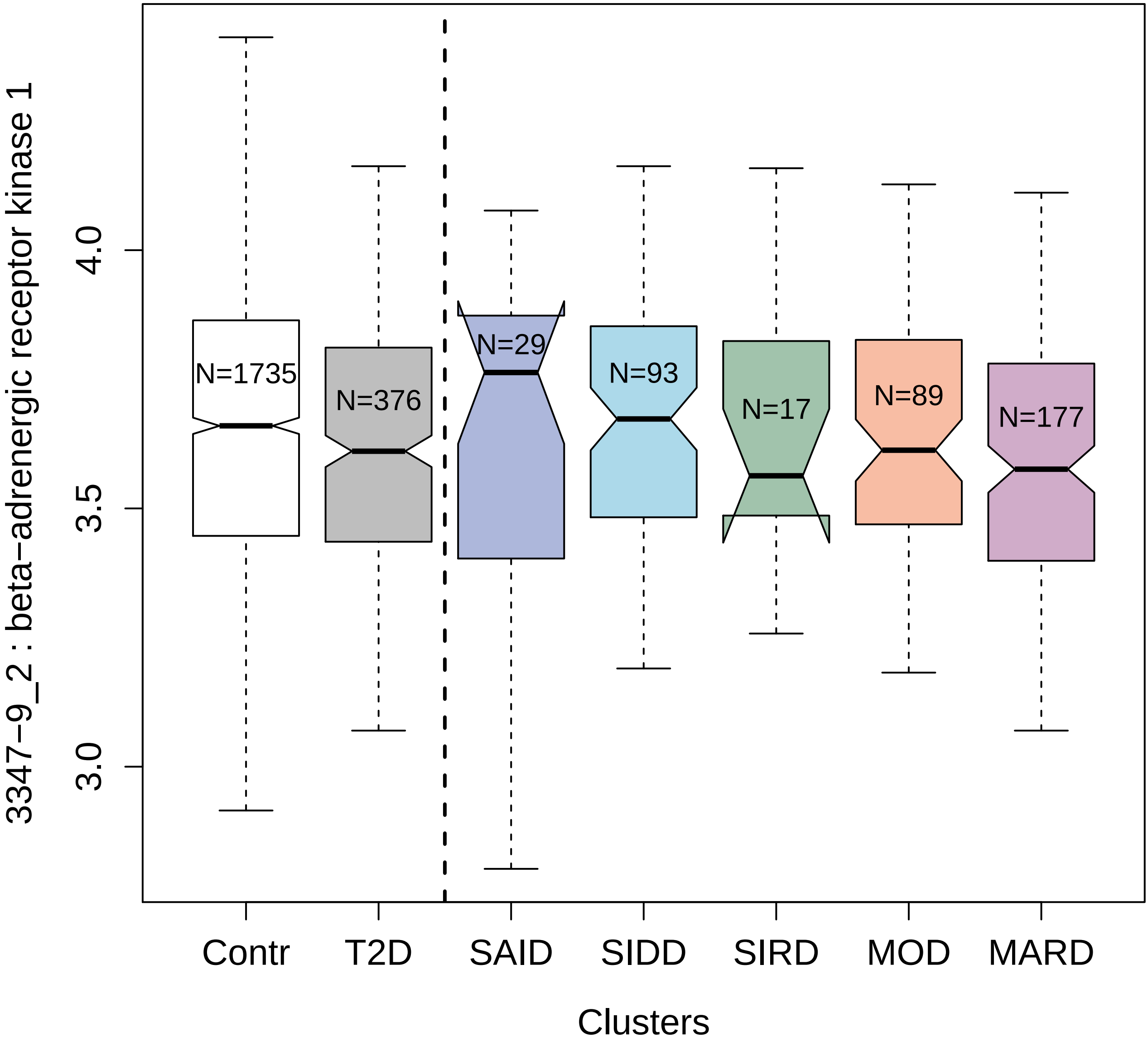

3347−9_2 : beta−adrenergic receptor kinase 1
3.0
3.5
4.0
N=1735
N=376
N=29
N=93
N=17
N=89
N=177
Contr
T2D
SAID
SIDD
SIRD
MOD
MARD
Clusters

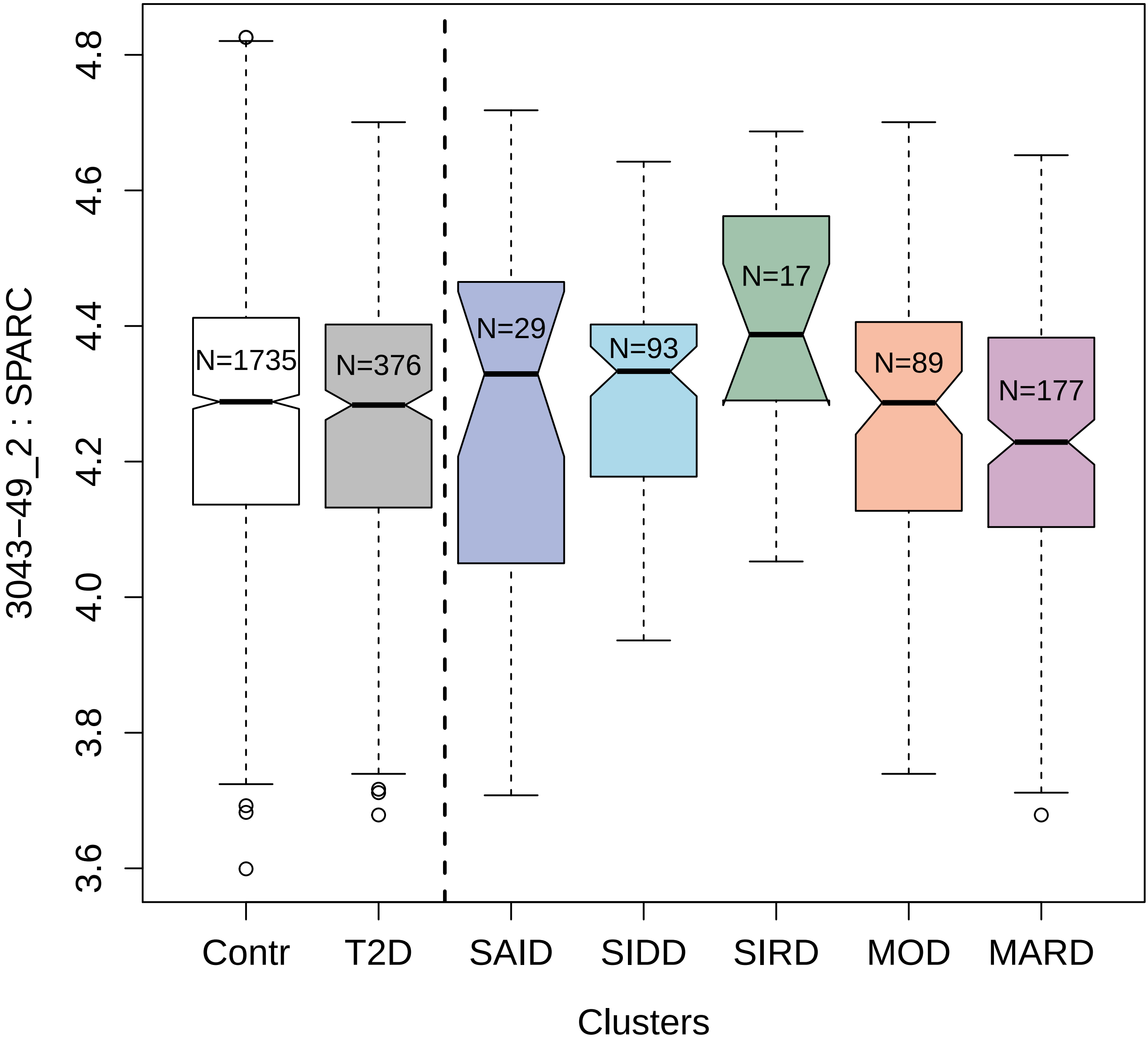
3043−49_2 : SPARC
4.8
4.6
4.4
4.2
4.0
3.8
3.6
N=1735
N=376
N=29
N=93
N=17
N=89
N=177
Contr
T2D
SAID
SIDD
SIRD
MOD
MARD
Clusters

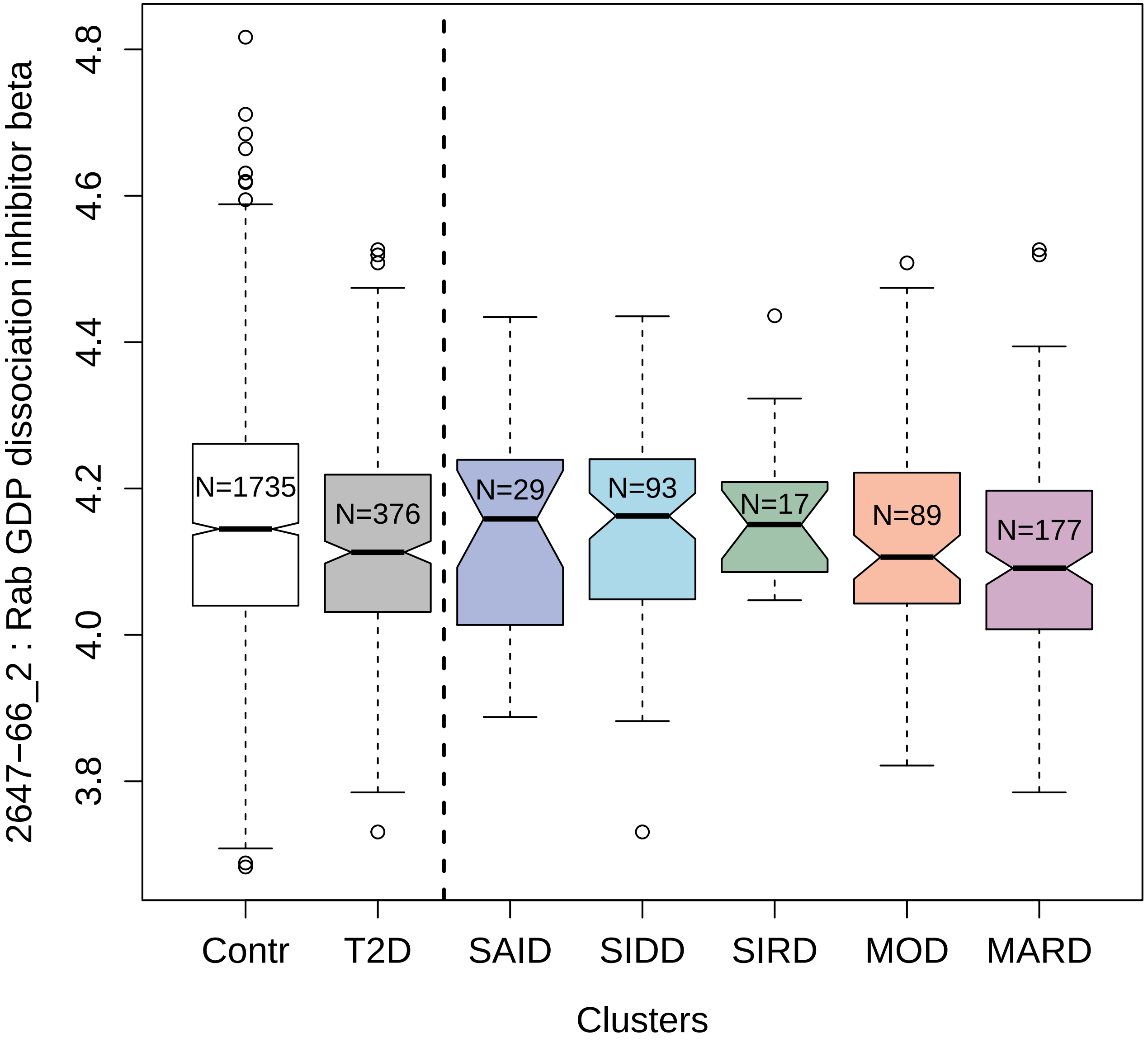
2647−66_2 : Rab GDP dissociation inhibitor beta
4.8
4.6
4.4
4.2
4.0
3.8
N=1735
N=376
N=29
N=93
N=17
N=89
N=177
Contr
T2D
SAID
SIDD
SIRD
MOD
MARD
Clusters

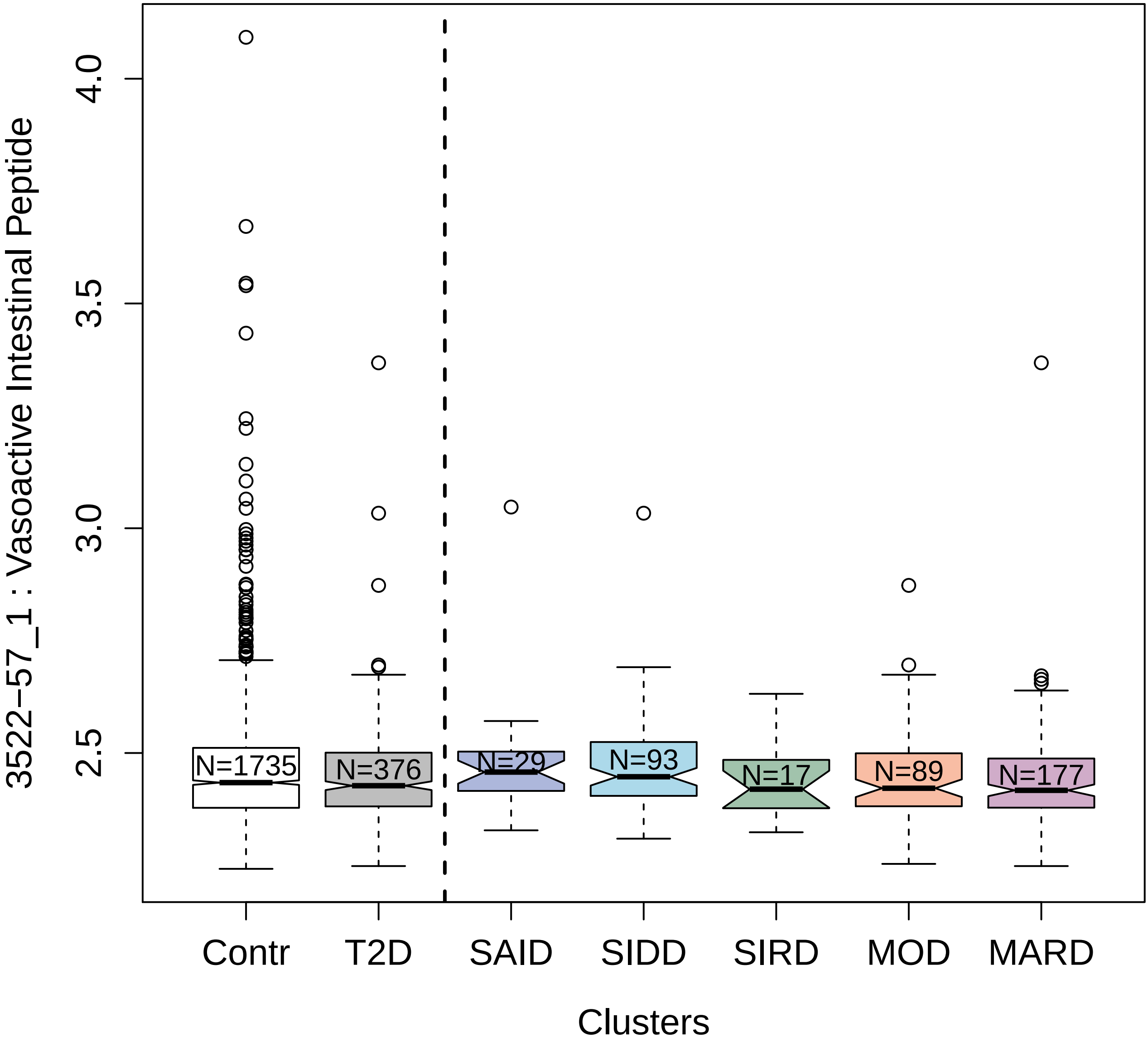
3522−57_1 : Vasoactive Intestinal Peptide
4.0
3.5
3.0
2.5
N=1735
N=376
N=29
N=93
N=17
N=89
N=177
Contr
T2D
SAID
SIDD
SIRD
MOD
MARD
Clusters

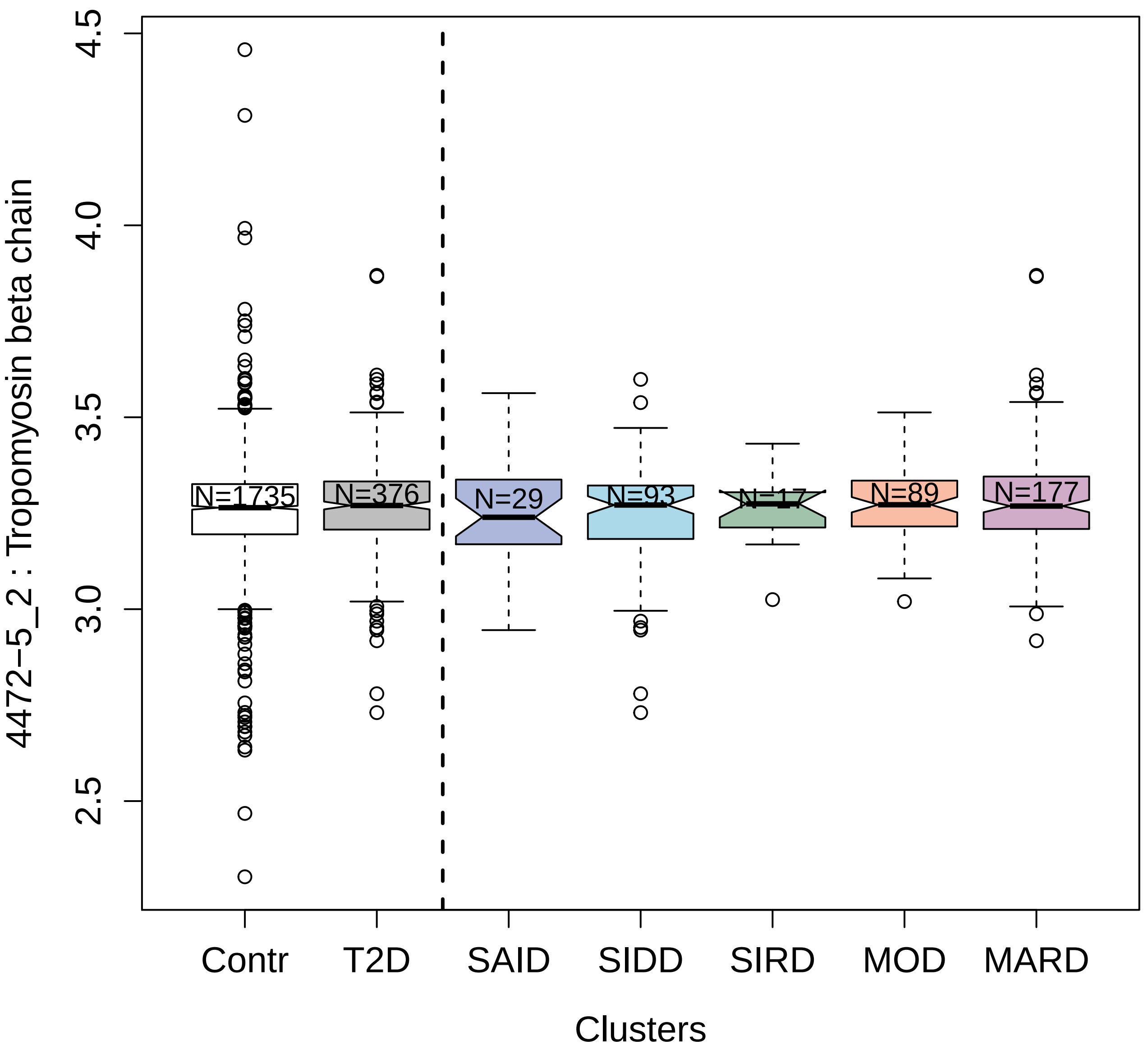
4472−5_2 : Tropomyosin beta chain
4.5
4.0
3.5
3.0
2.5
N=1735
N=376
N=29
N=93
N=17
N=89
N=177
Contr
T2D
SAID
SIDD
SIRD
MOD
MARD
Clusters

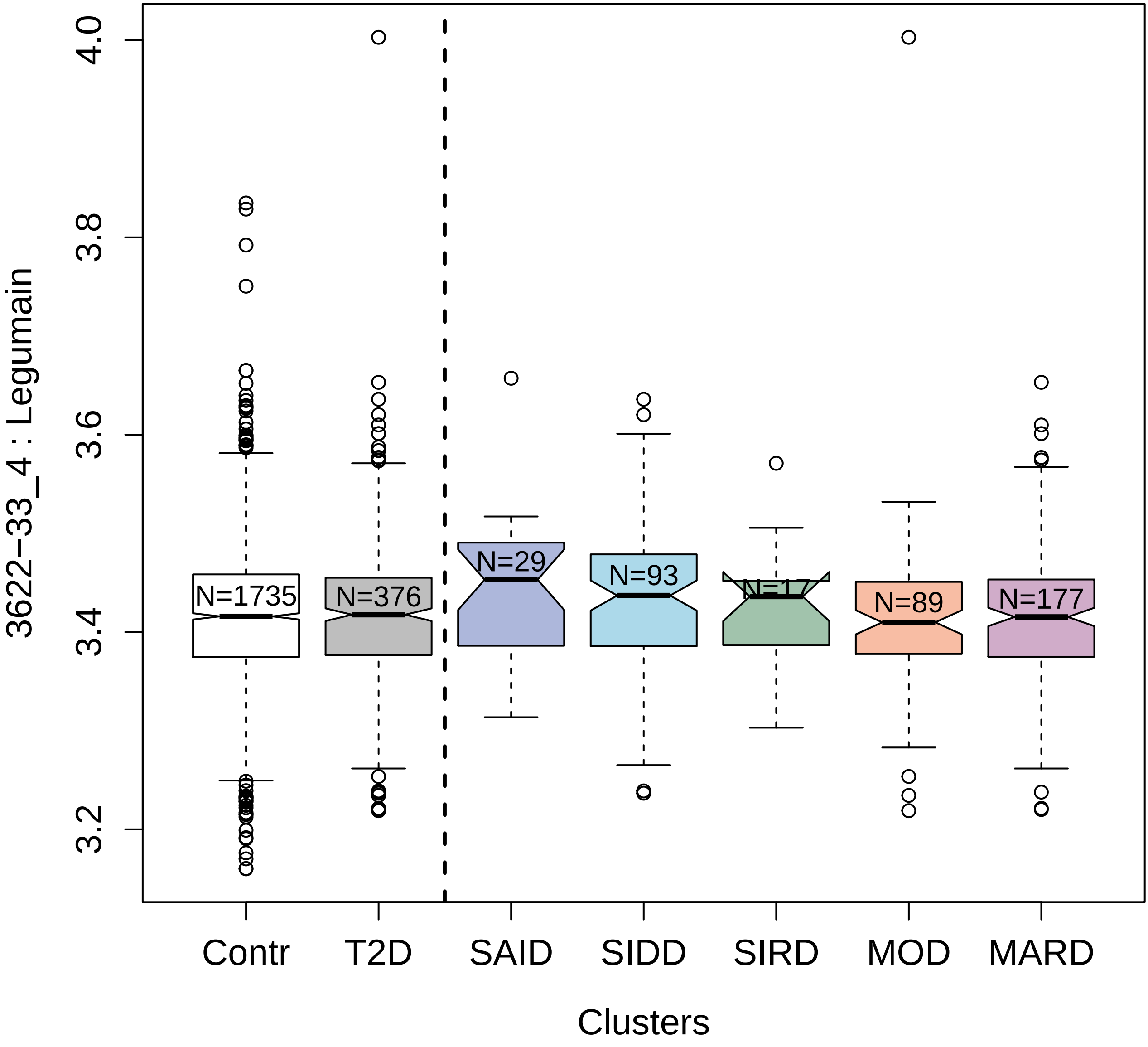
3622−33_4 : Legumain
4.0
3.8
3.6
3.4
3.2
N=1735
N=376
N=29
N=93
N=89
N=177
Contr
T2D
SAID
SIDD
SIRD
MOD
MARD
Clusters

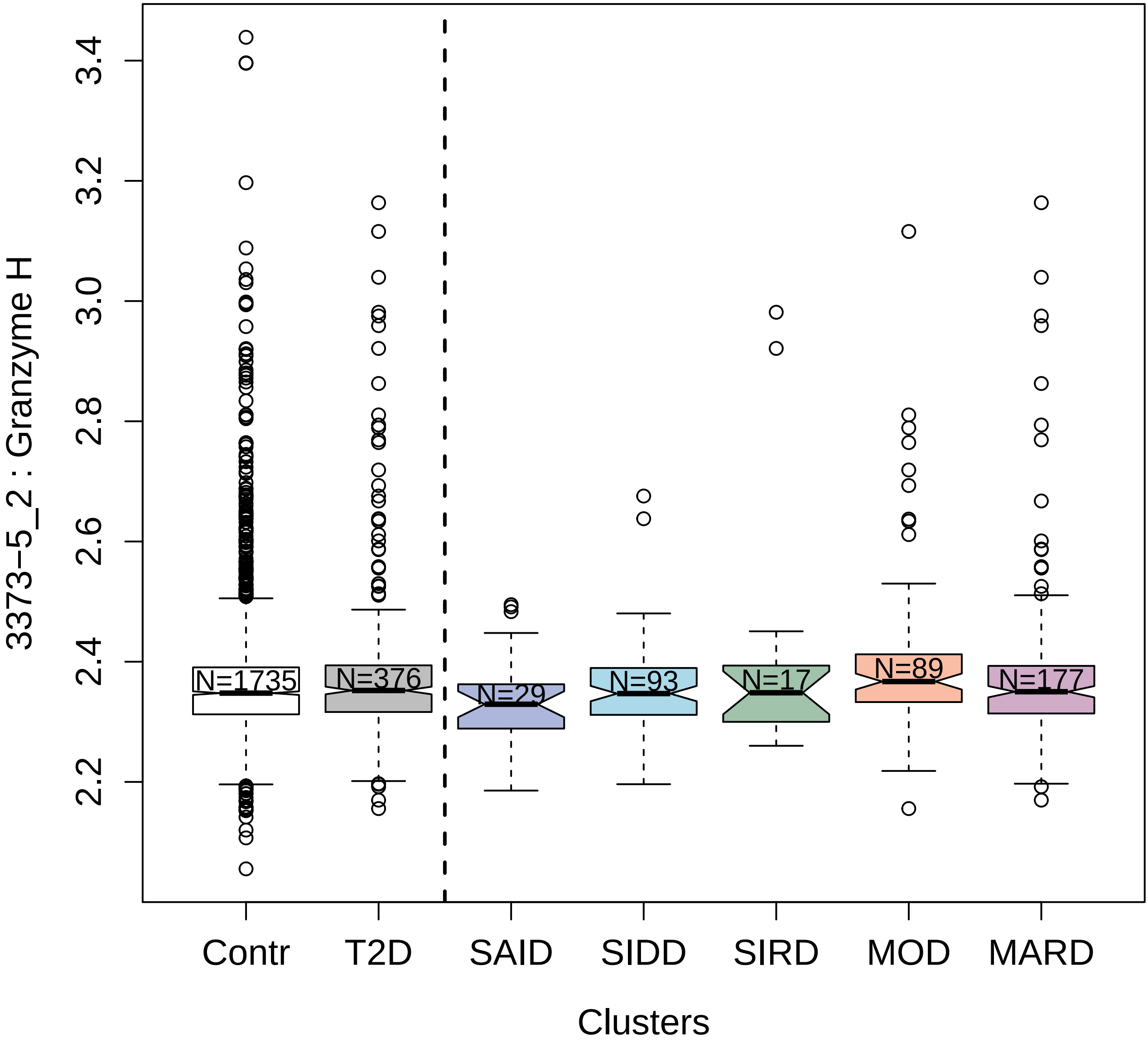
3373−5_2 : Granzyme H
3.4
3.2
3.0
2.8
2.6
2.4
2.2
N=1735
N=376
N=29
N=93
N=17
N=89
N=177
Contr
T2D
SAID
SIDD
SIRD
MOD
MARD
Clusters

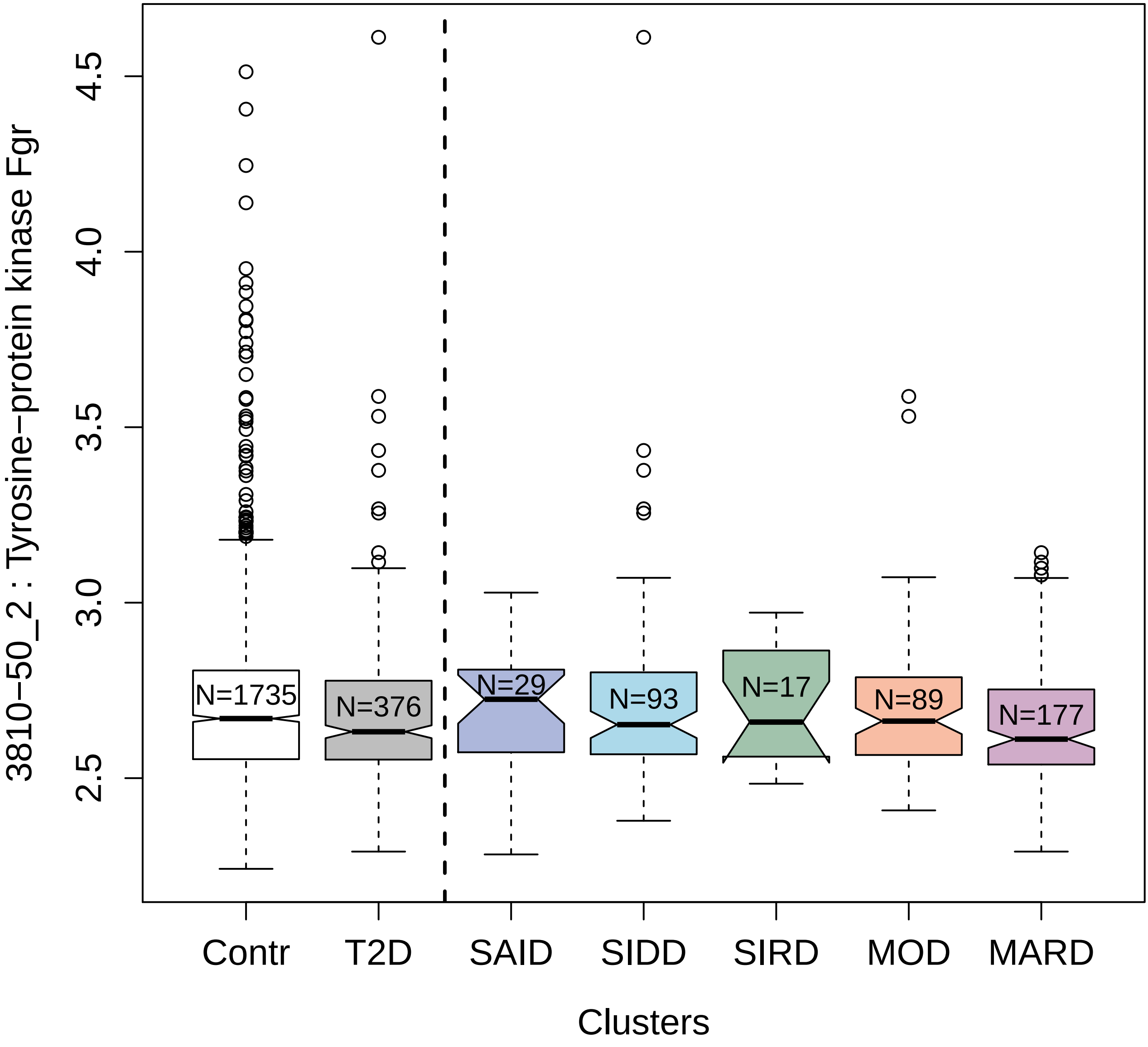
3810−50_2 : Tyrosine−protein kinase Fgr
2.5
3.0
3.5
4.0
4.5
N=1735
N=376
N=29
N=93
N=17
N=89
N=177
Contr
T2D
SAID
SIDD
SIRD
MOD
MARD
Clusters

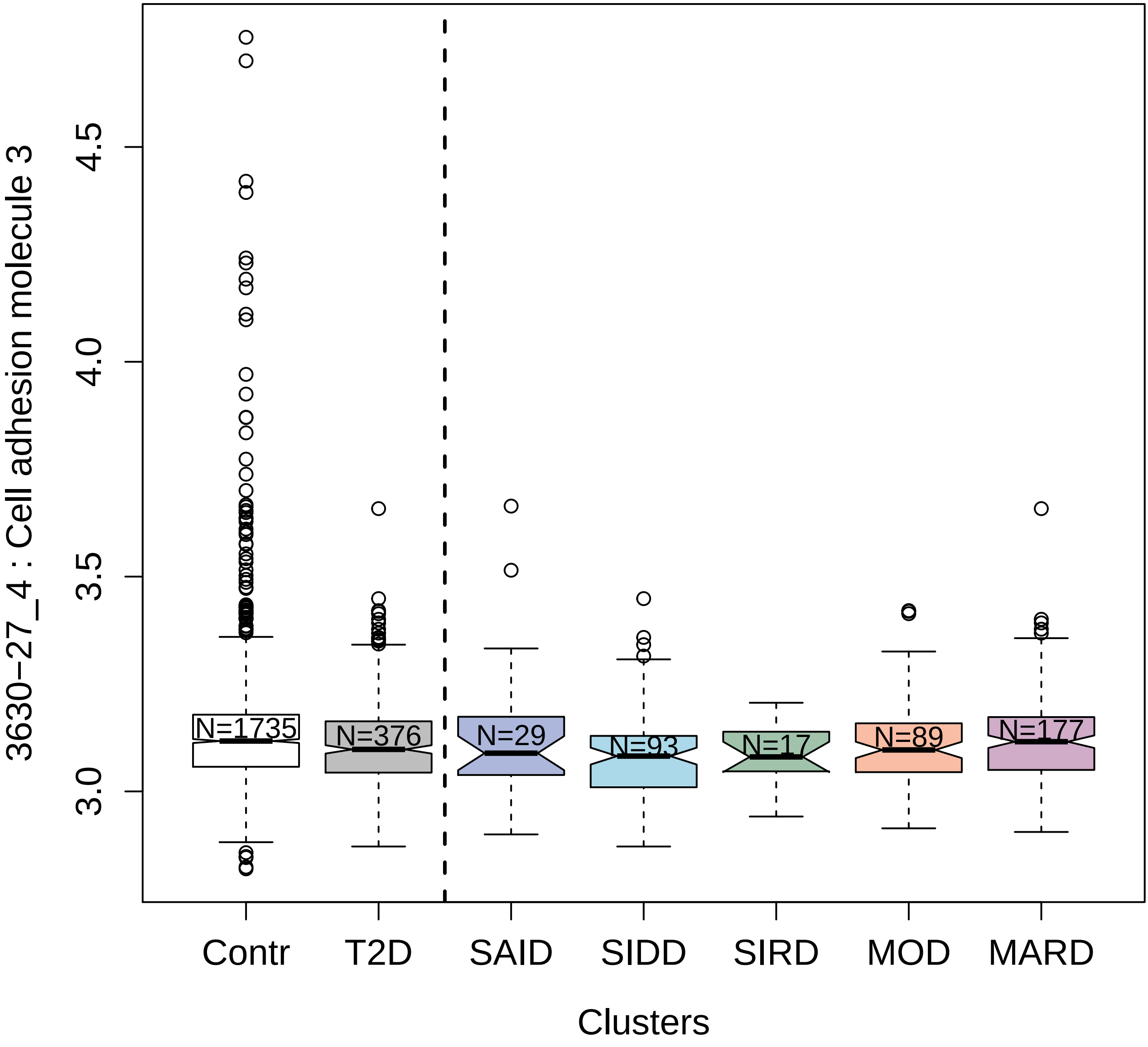

3630−27_4 : Cell adhesion molecule 3
3.0
3.5
4.0
4.5
N=1735
N=376
N=29
N=93
N=17
N=89
N=177
Contr
T2D
SAID
SIDD
SIRD
MOD
MARD
Clusters

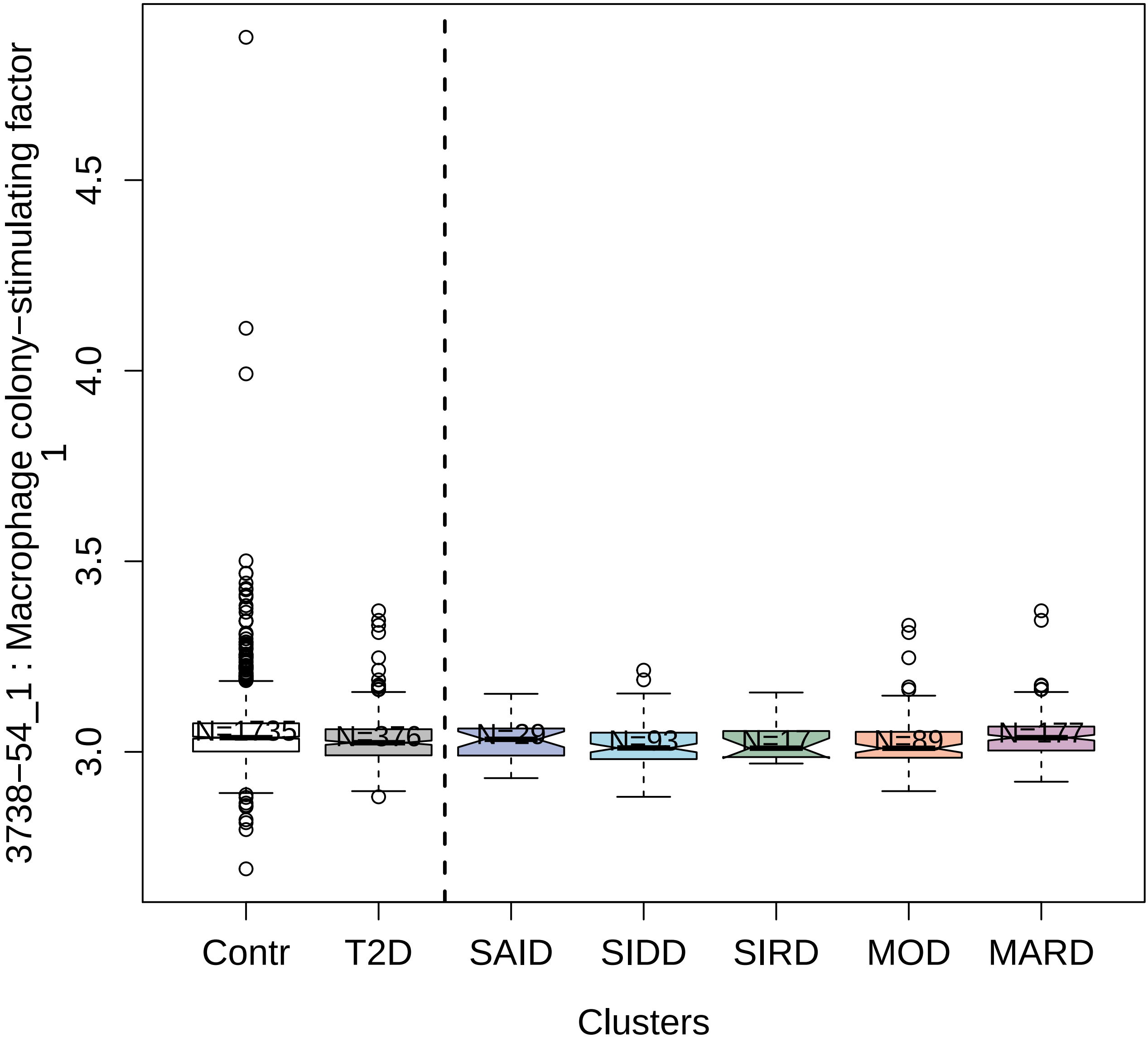
3738−54_1 : Macrophage colony−stimulating factor 1
4.5
4.0
3.5
3.0
N=1735
N=376
N=29
N=93
N=17
N=89
N=177
Contr
T2D
SAID
SIDD
SIRD
MOD
MARD
Clusters

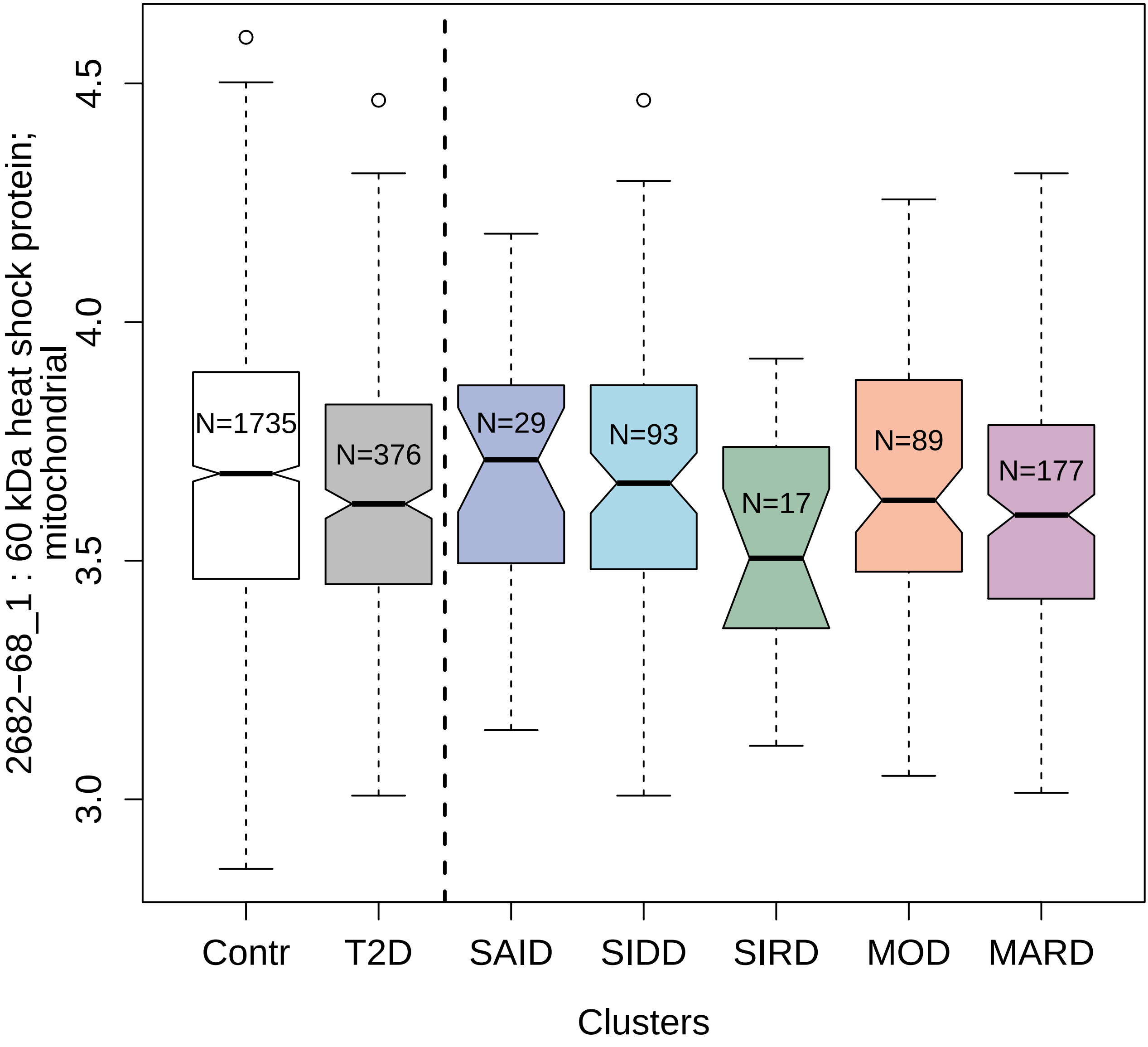

2682−68_1 : 60 kDa heat shock protein;
mitochondrial
4.5
4.0
3.5
3.0
N=1735
N=376
N=29
N=93
N=17
N=89
N=177
Contr
T2D
SAID
SIDD
SIRD
MOD
MARD
Clusters

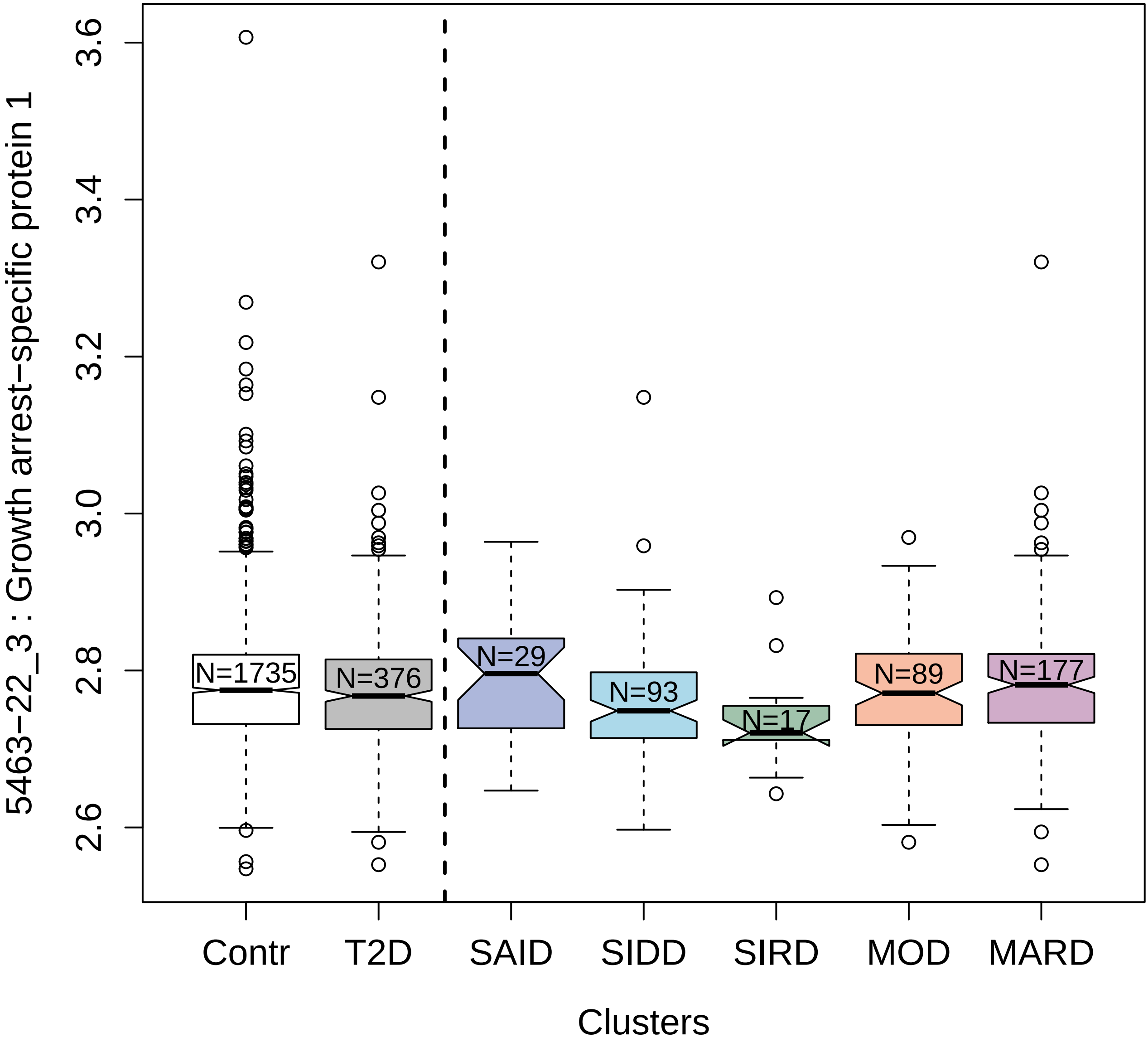

5463−22_3 : Growth arrest−specific protein 1
3.6
3.4
3.2
3.0
2.8
2.6
N=1735
N=376
N=29
N=93
N=17
N=89
N=177
Contr
T2D
SAID
SIDD
SIRD
MOD
MARD
Clusters

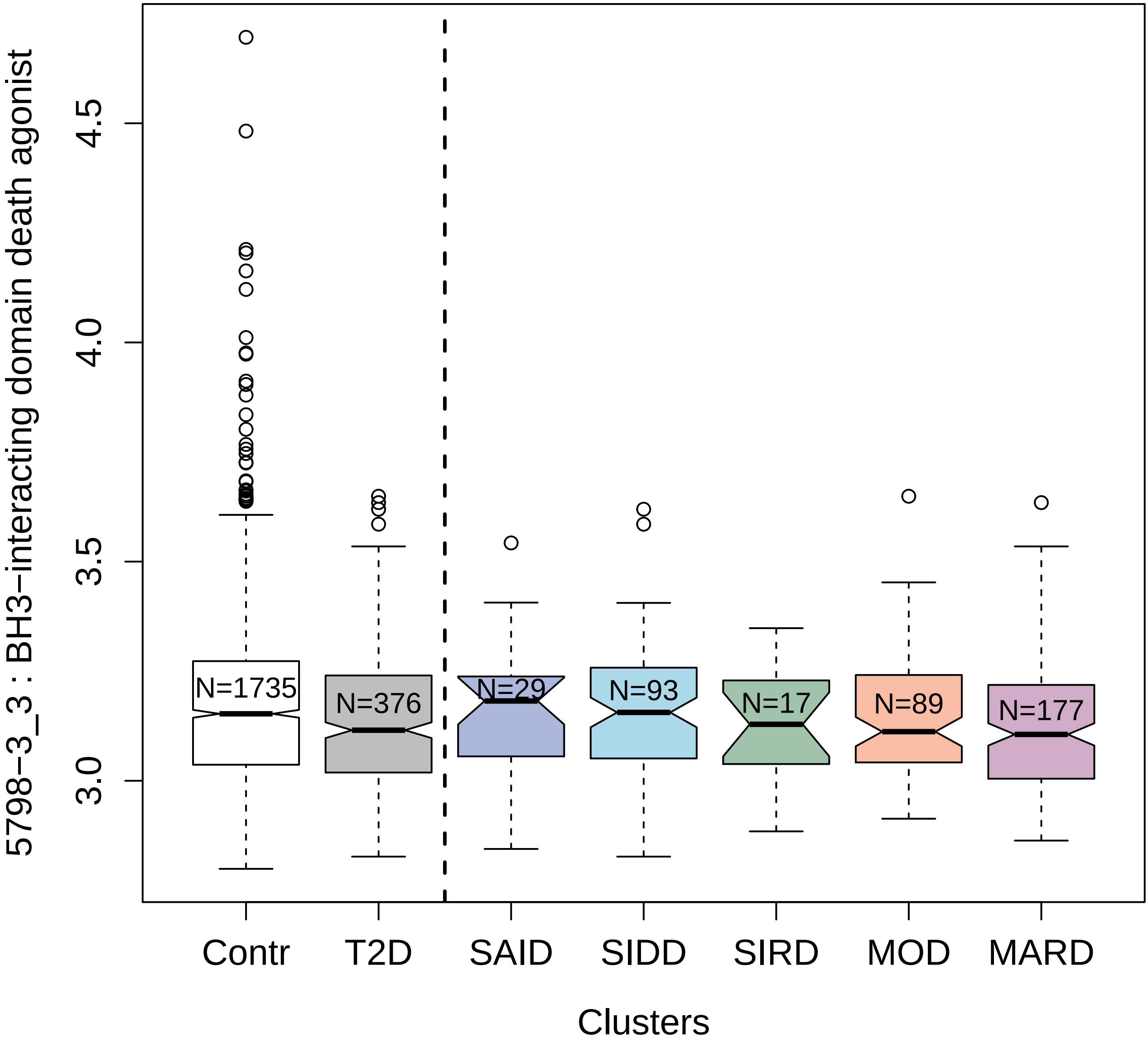
5798−3_3 : BH3−interacting domain death agonist
4.5
4.0
3.5
3.0
N=1735
N=376
N=29
N=93
N=17
N=89
N=177
Contr
T2D
SAID
SIDD
SIRD
MOD
MARD
Clusters

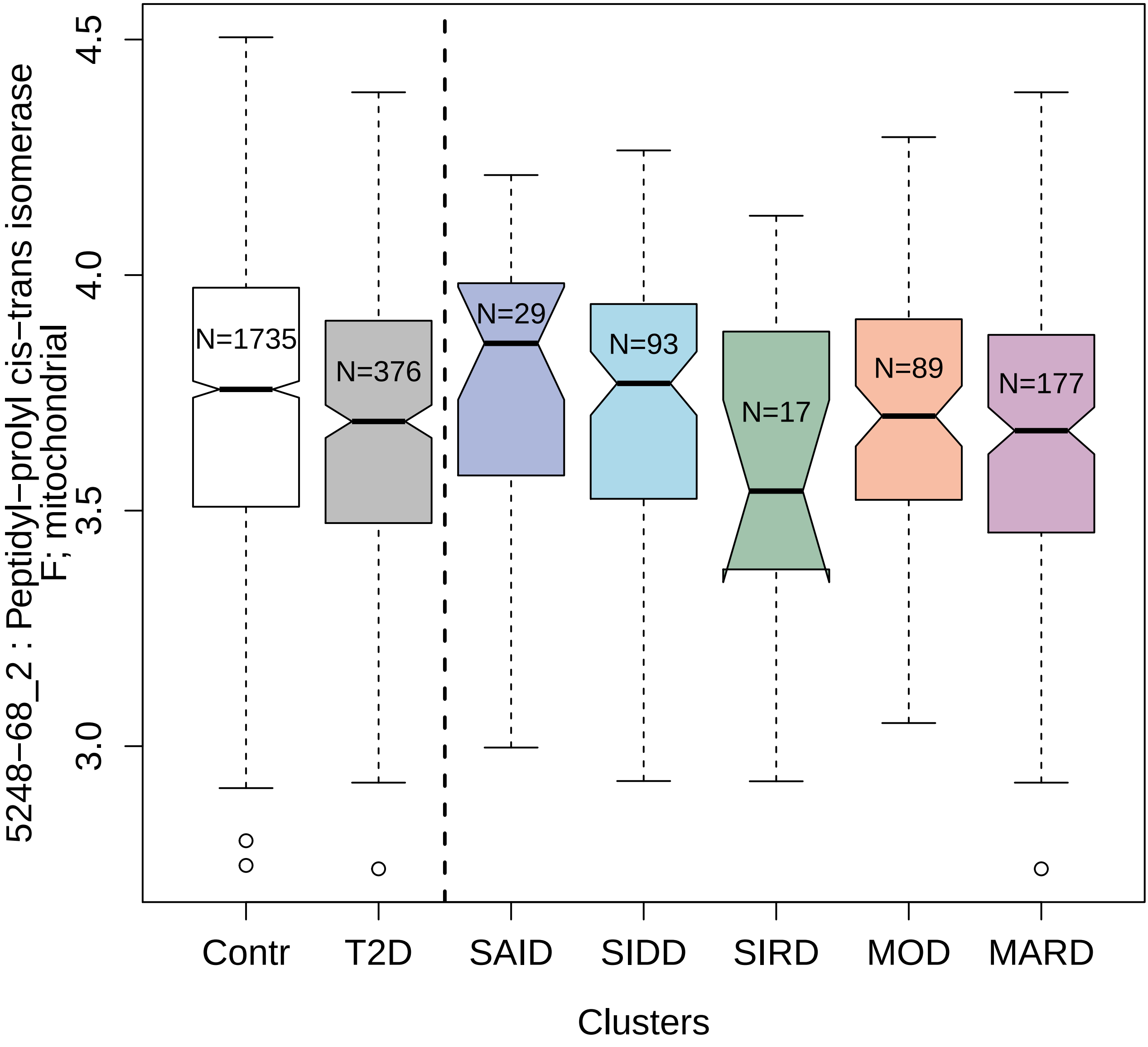

5248−68_2 : Peptidyl−prolyl cis−trans isomerase
F; mitochondrial
4.5
4.0
3.5
3.0
N=1735
N=376
N=29
N=93
N=17
N=89
N=177
Contr
T2D
SAID
SIDD
SIRD
MOD
MARD
Clusters

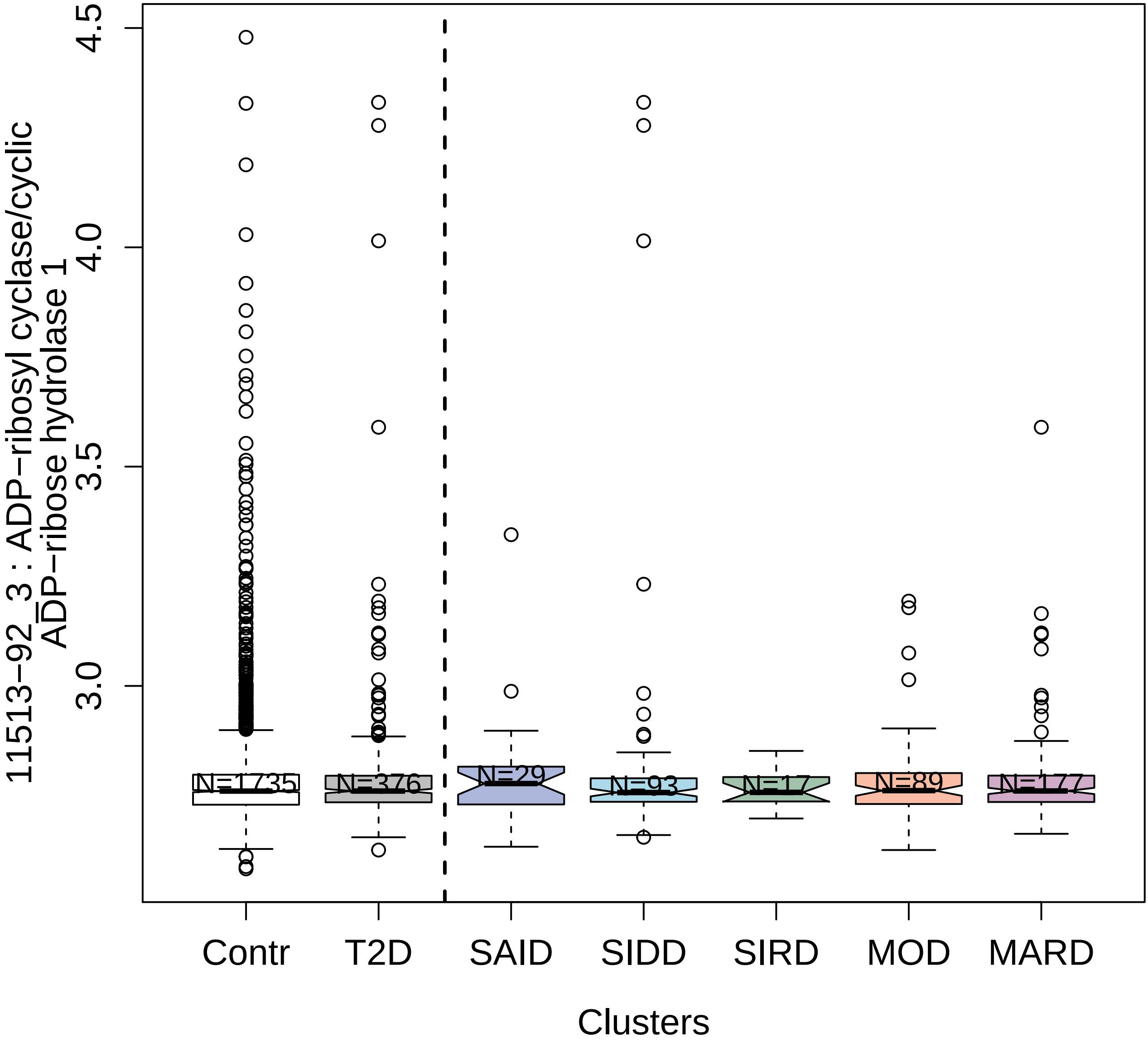

1513−92_3 : ADP−ribosyl cyclase/cyclic ADP−ribose hydrolase 1
4.5
4.0
3.5
3.0
N=1735
N=376
N=29
N=93
N=17
N=89
N=177
Contr
T2D
SAID
SIDD
SIRD
MOD
MARD
Clusters

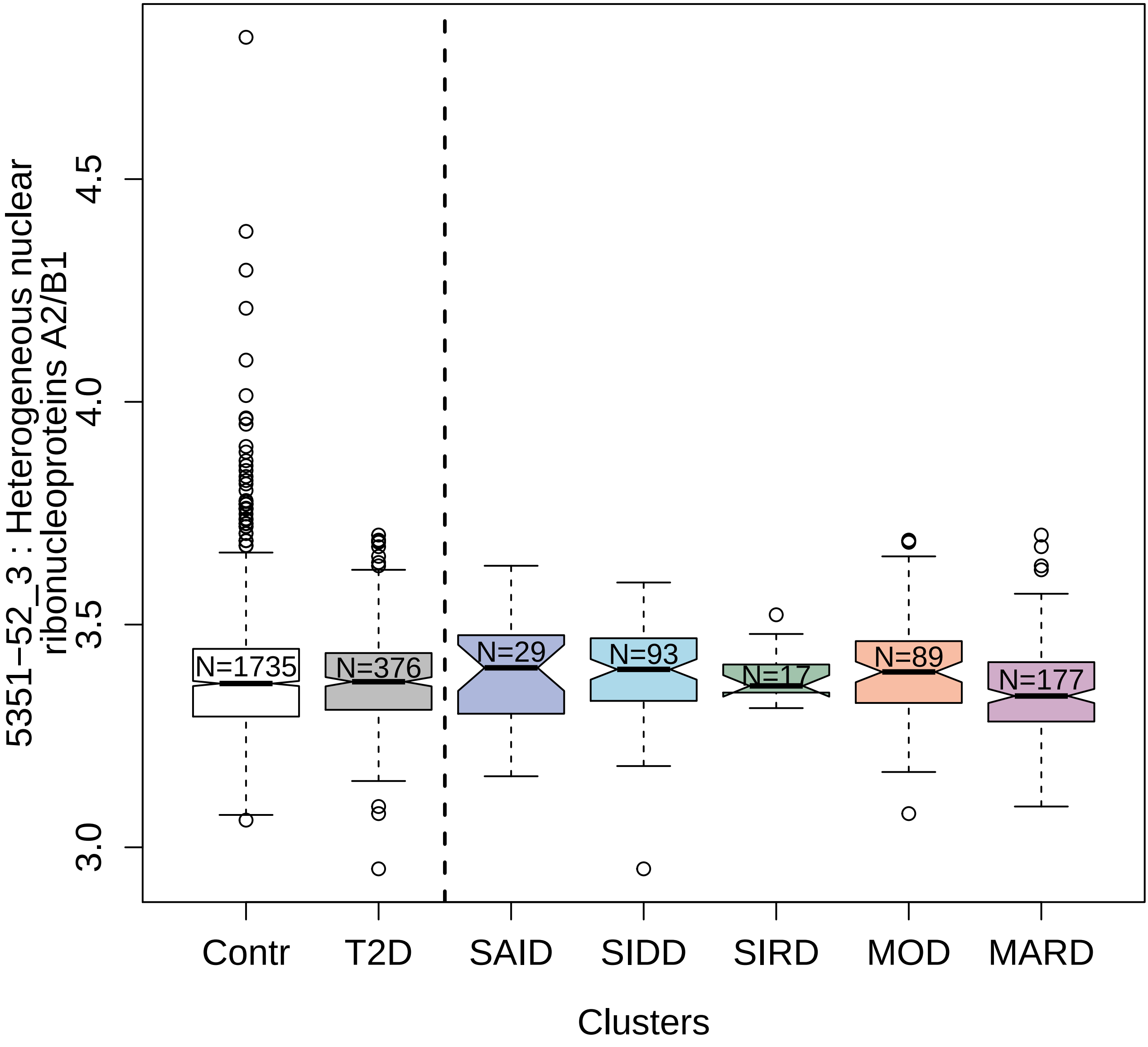

5351−52_3 : Heterogeneous nuclear
ribonucleoproteins A2/B1
3.0
3.5
4.0
4.5
N=1735
N=376
N=29
N=93
N=17
N=89
N=177
Contr
T2D
SAID
SIDD
SIRD
MOD
MARD
Clusters

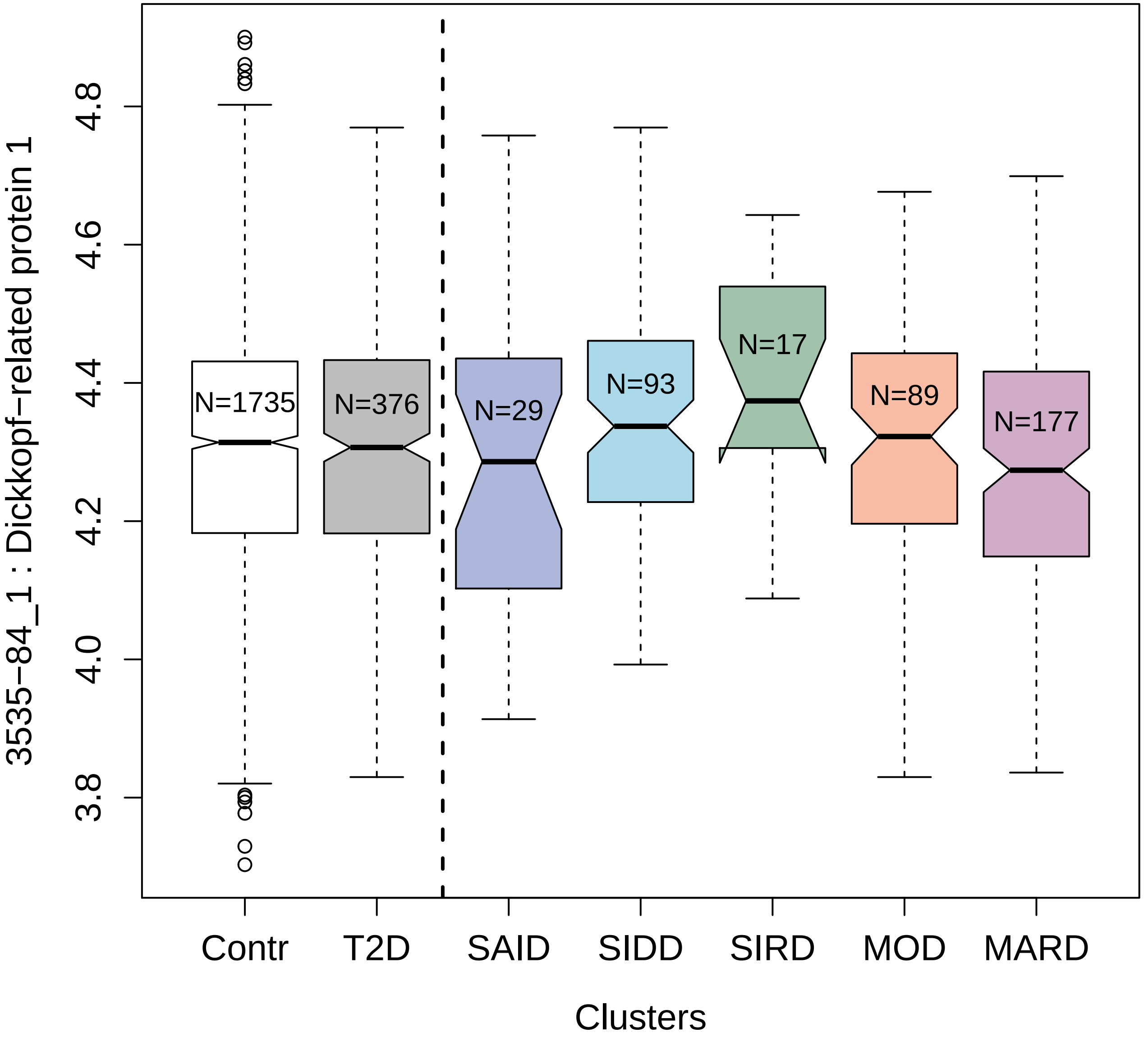
3535−84_1 : Dickkopf−related protein 1
4.8
4.6
4.4
4.2
4.0
3.8
N=1735
N=376
N=29
N=93
N=17
N=89
N=177
Contr
T2D
SAID
SIDD
SIRD
MOD
MARD
Clusters

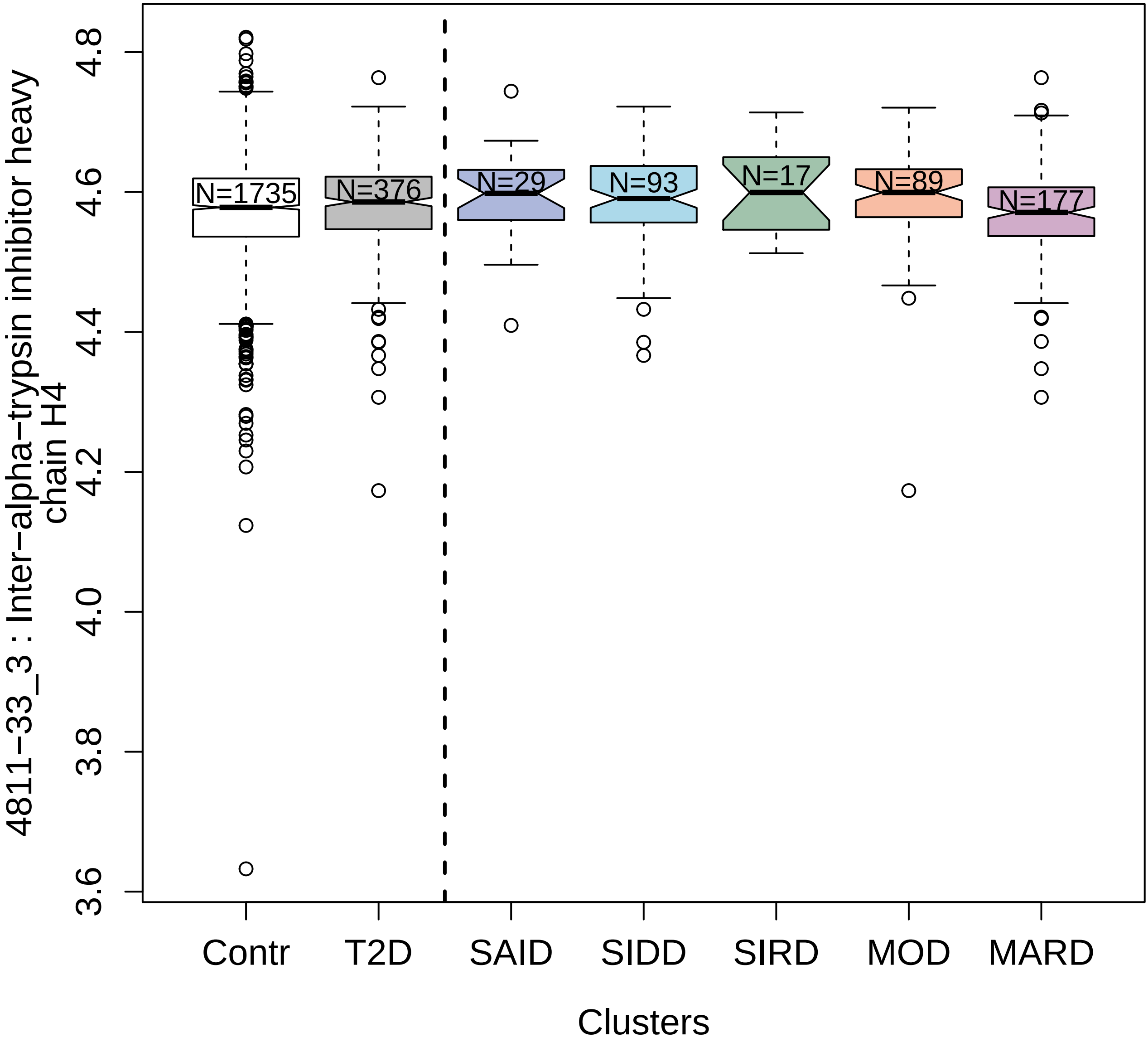

4811−33_3 : Inter−alpha−trypsin inhibitor heavy chain H4
3.6
3.8
4.0
4.2
4.4
4.6
4.8
N=1735
N=376
N=29
N=93
N=17
N=89
N=177
Contr
T2D
SAID
SIDD
SIRD
MOD
MARD
Clusters

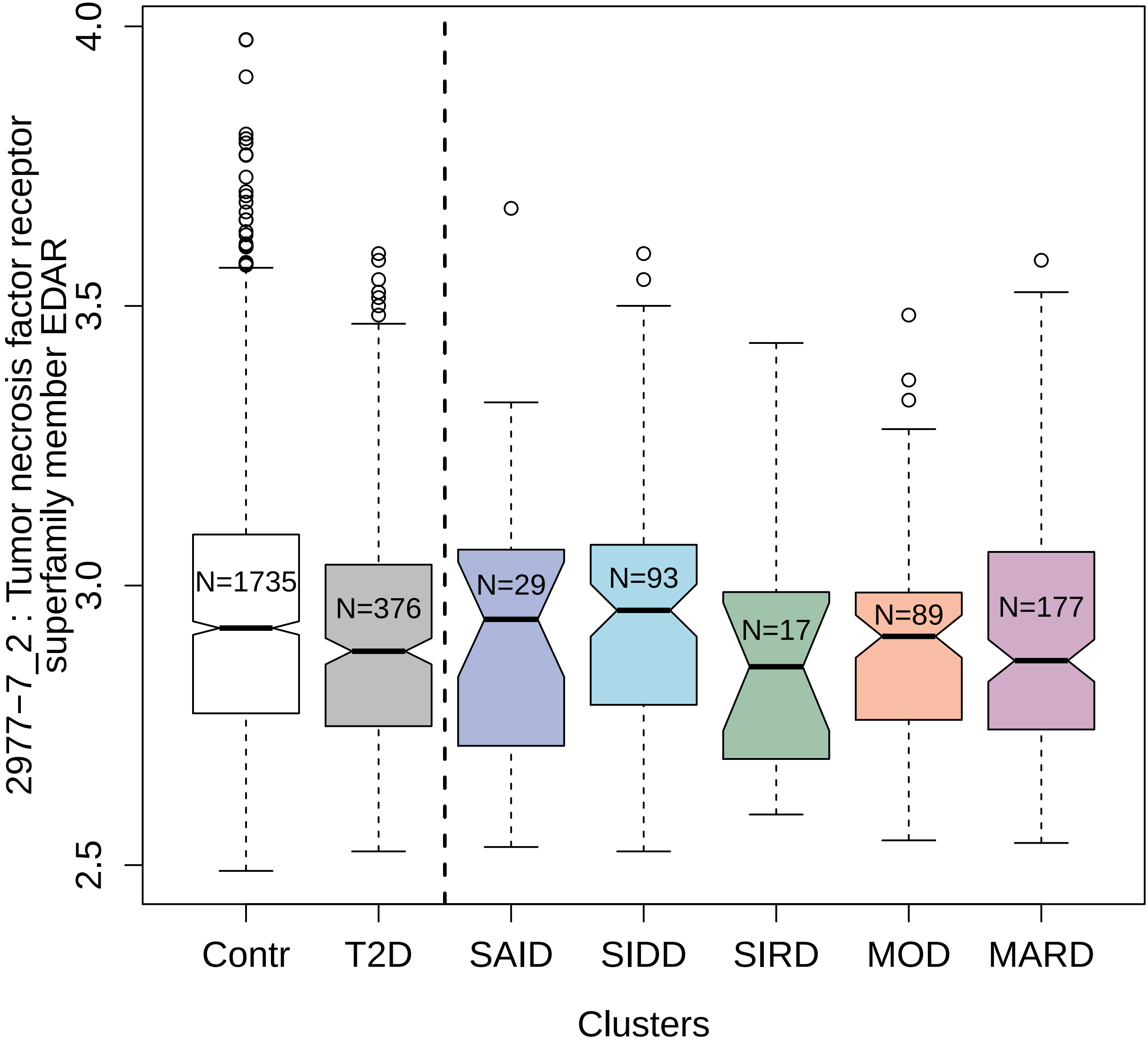
2977−7_2 : Tumor necrosis factor receptor superfamily member EDAR
4.0
3.5
3.0
2.5
N=1735
N=376
N=29
N=93
N=17
N=89
N=177
Contr
T2D
SAID
SIDD
SIRD
MOD
MARD
Clusters

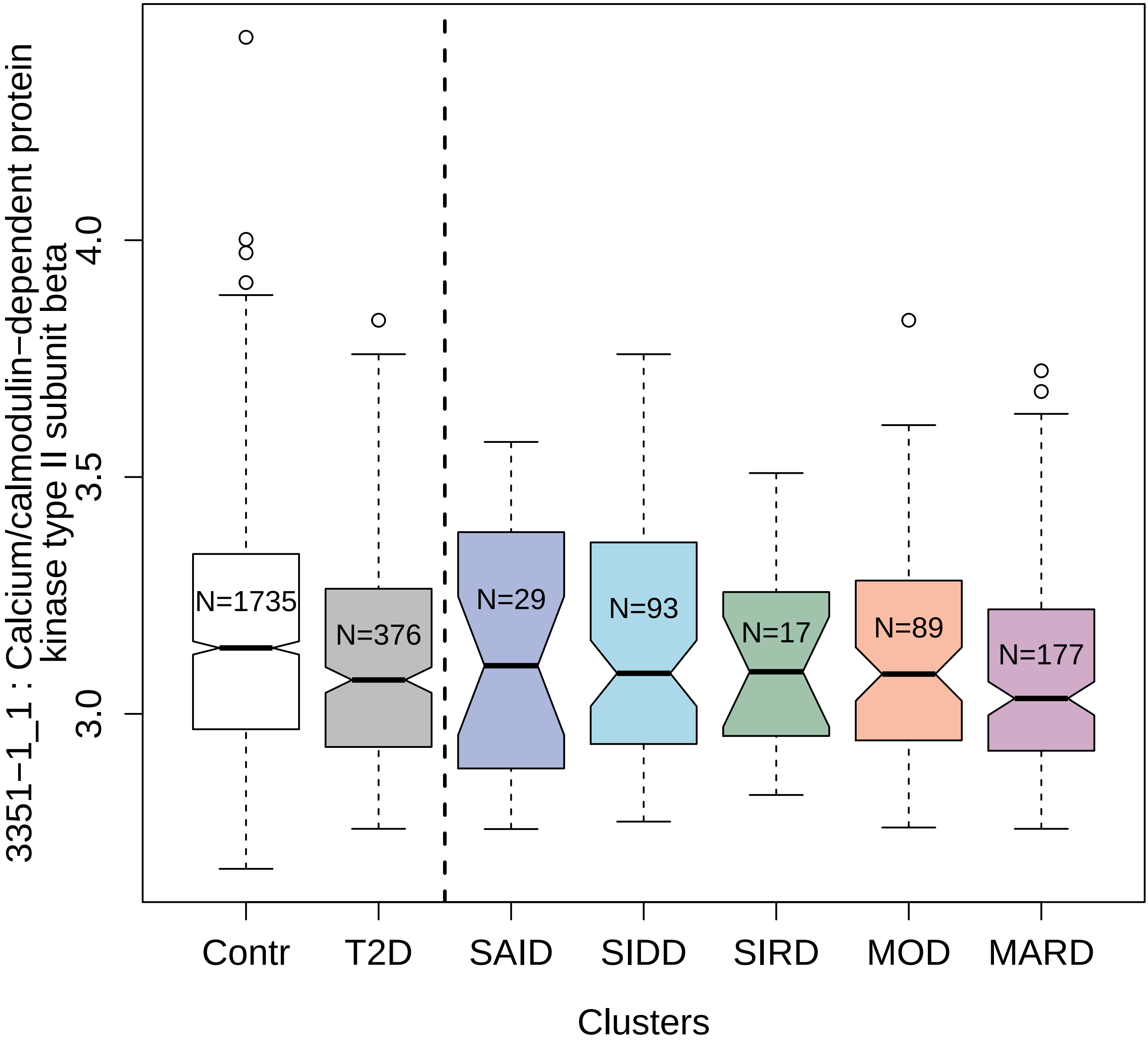
3351−1_1 : Calcium/calmodulin−dependent protein
kinase type II subunit beta
3.0
3.5
4.0
N=1735
N=376
N=29
N=93
N=17
N=89
N=177
Contr
T2D
SAID
SIDD
SIRD
MOD
MARD
Clusters

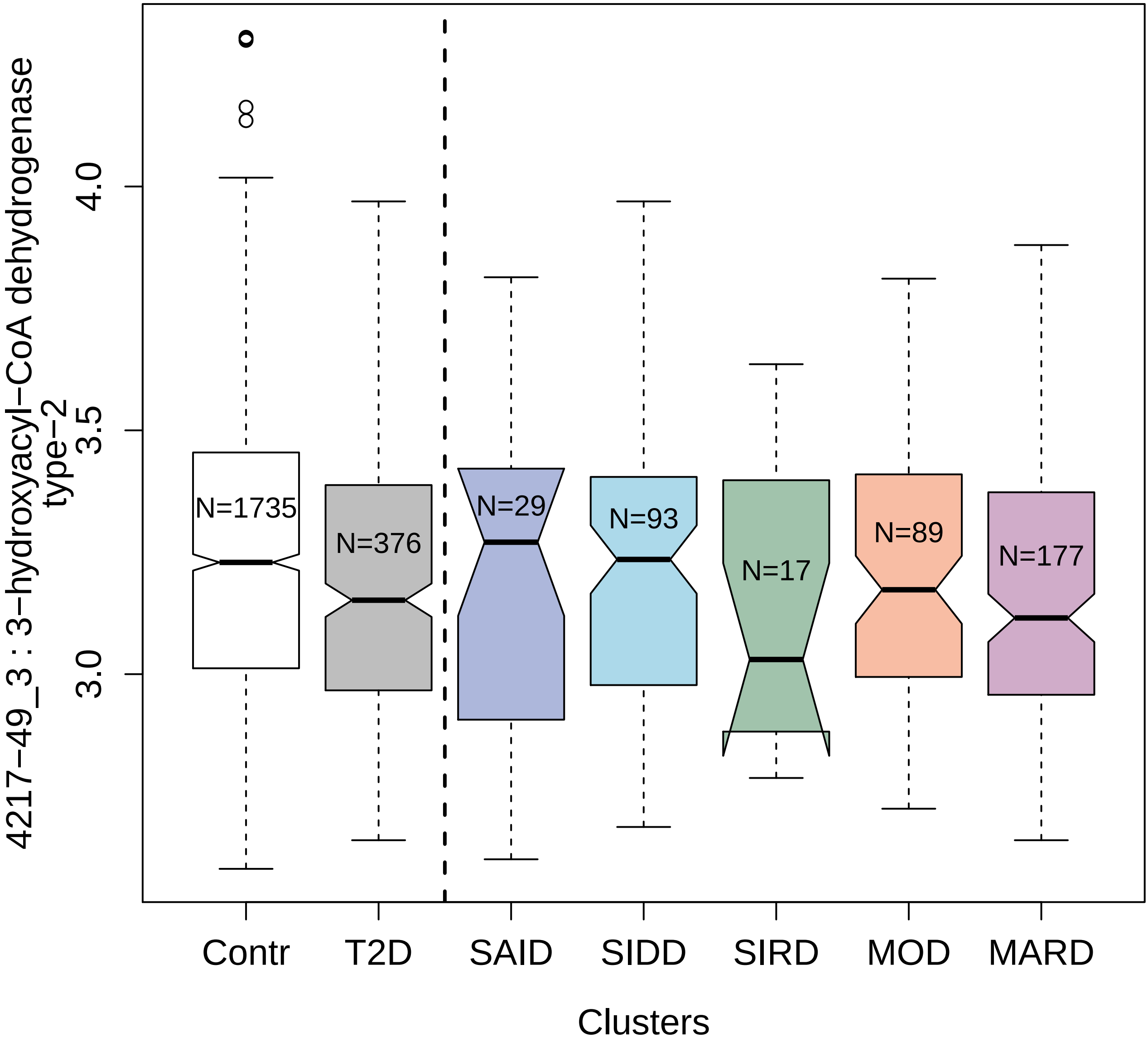
4217−49_3 : 3−hydroxyacyl−CoA dehydrogenase
type−2
4.0
3.5
3.0
N=1735
N=376
N=29
N=93
N=17
N=89
N=177
Contr
T2D
SAID
SIDD
SIRD
MOD
MARD
Clusters

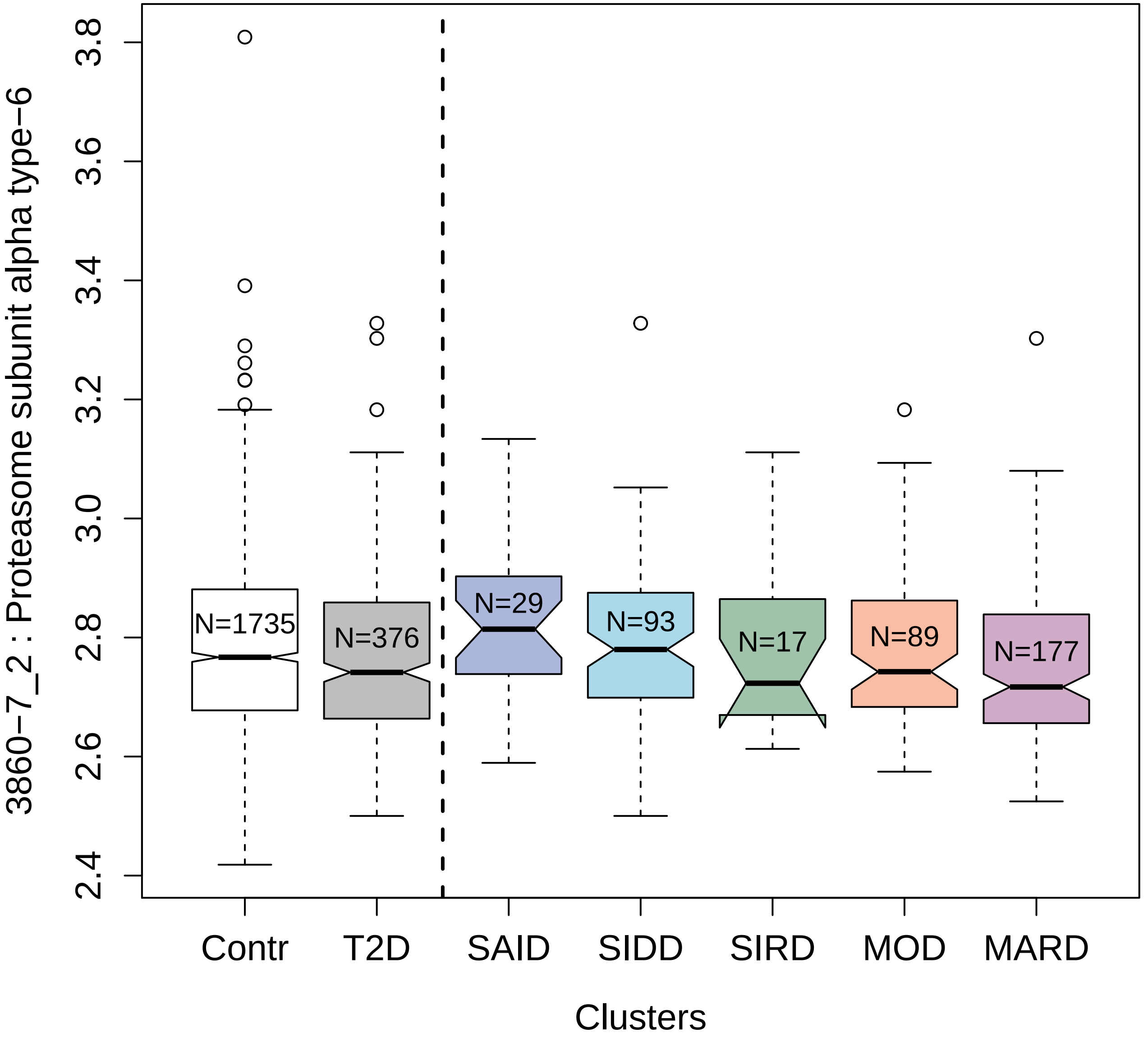
3860−7_2 : Proteasome subunit alpha type−6
2.4
2.6
2.8
3.0
3.2
3.4
3.6
3.8
N=1735
N=376
N=29
N=93
N=17
N=89
N=177
Contr
T2D
SAID
SIDD
SIRD
MOD
MARD
Clusters

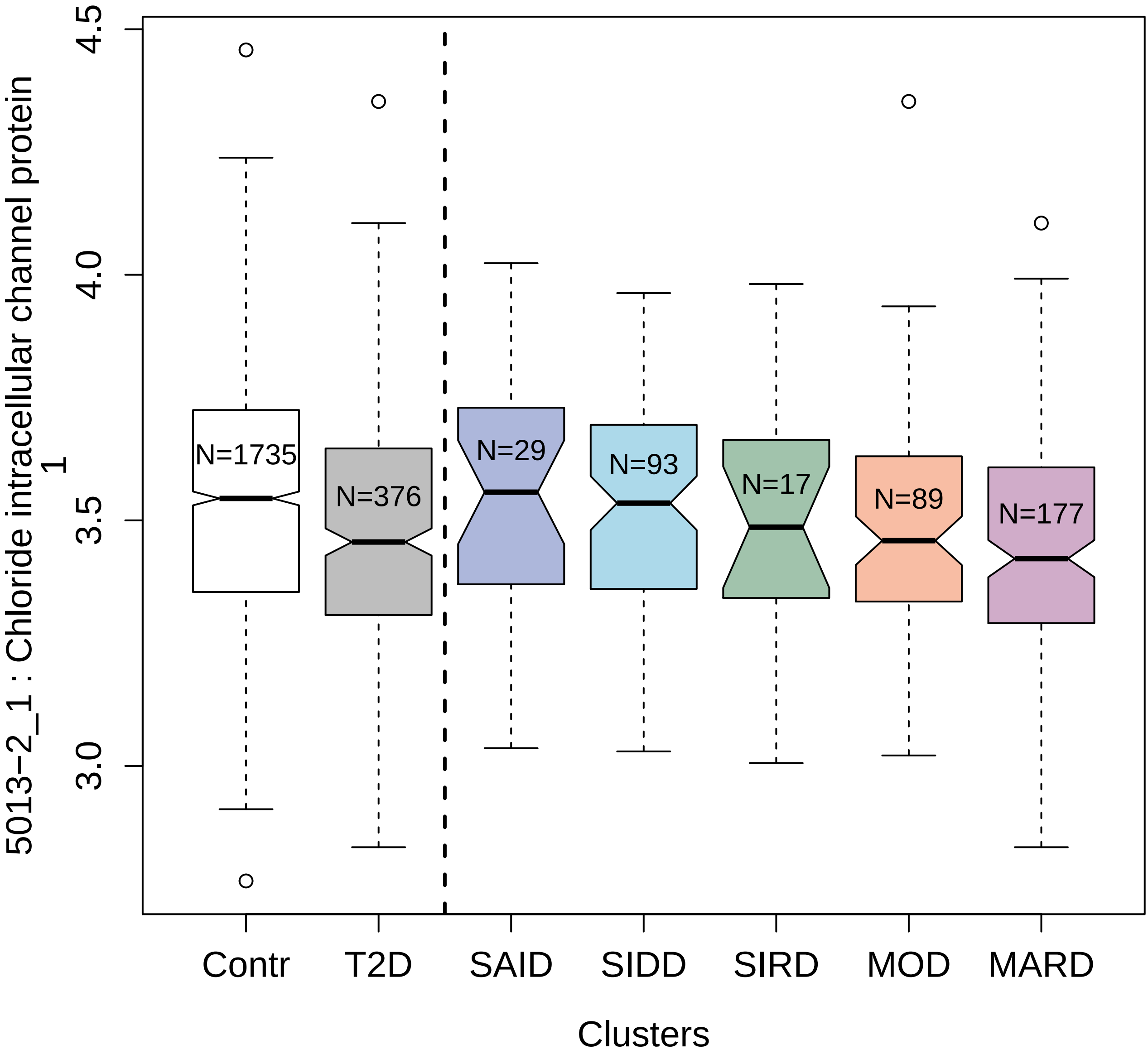

5013−2_1 : Chloride intracellular channel protein 1
4.5
4.0
3.5
3.0
N=1735
N=376
N=29
N=93
N=17
N=89
N=177
Contr
T2D
SAID
SIDD
SIRD
MOD
MARD
Clusters

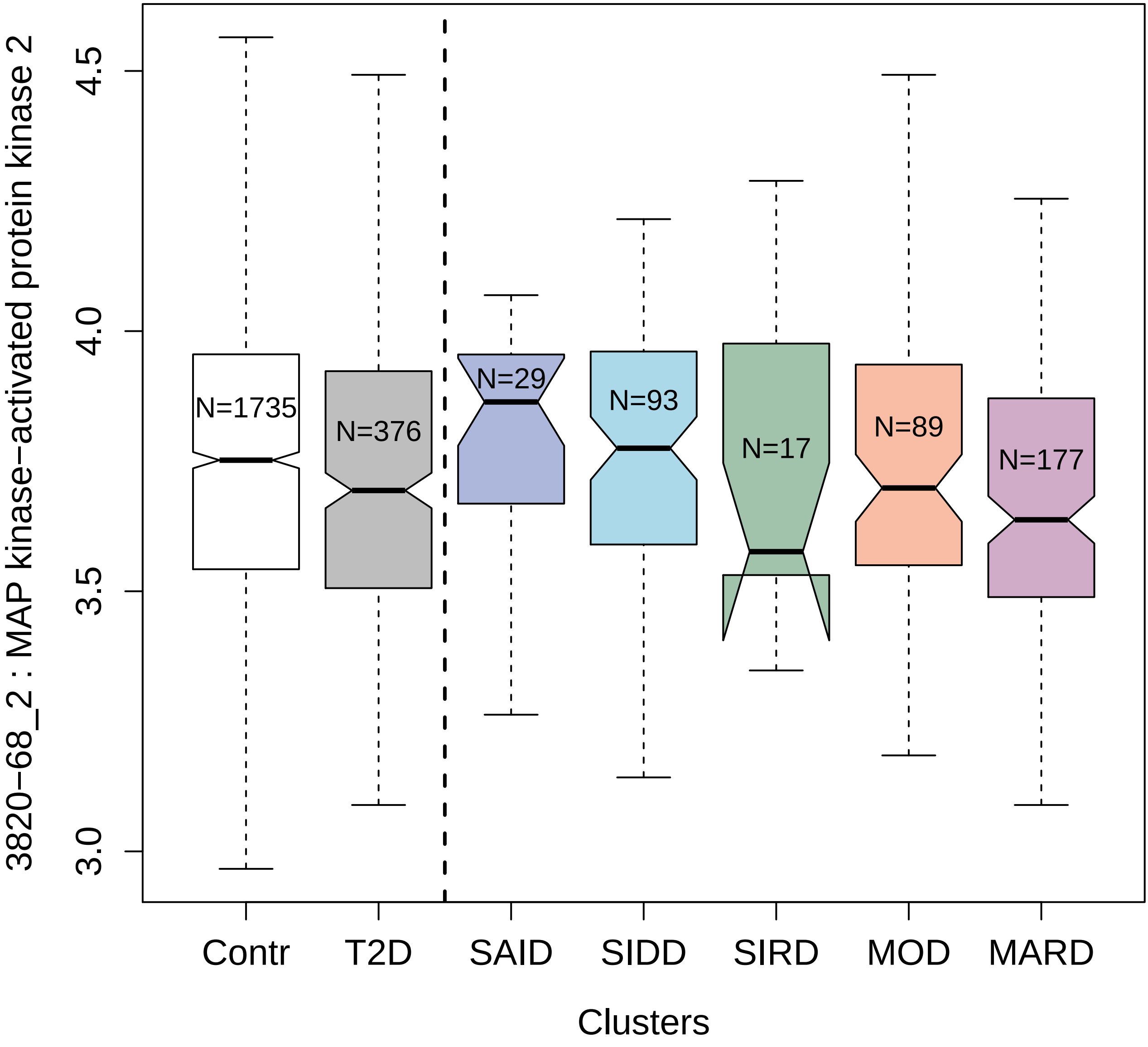

3820−68_2 : MAP kinase−activated protein kinase 2
3.0
3.5
4.0
4.5
N=1735
N=376
N=29
N=93
N=17
N=89
N=177
Contr
T2D
SAID
SIDD
SIRD
MOD
MARD
Clusters

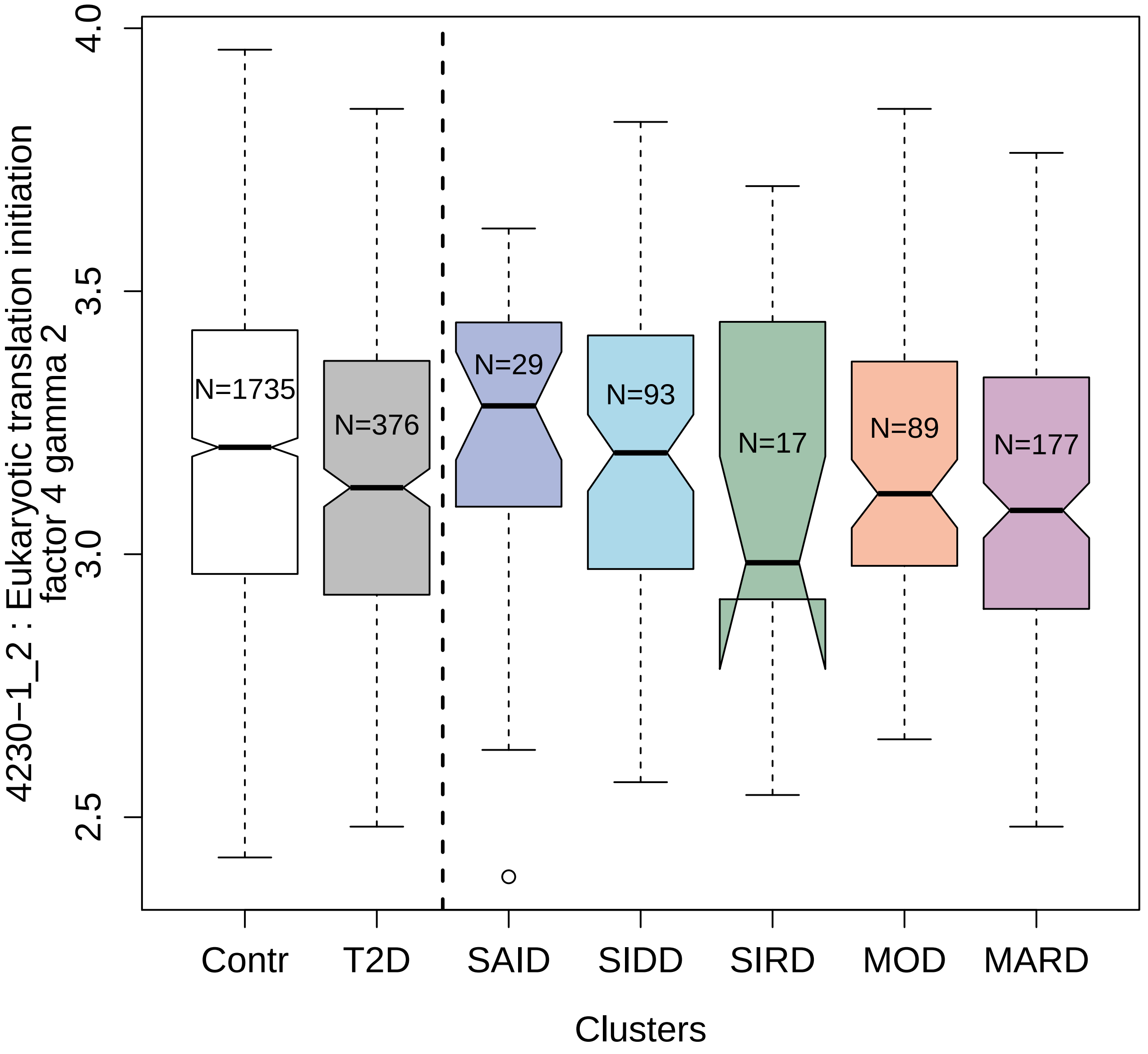

4230−1_2 : Eukaryotic translation initiation
factor 4 gamma 2
4.0
3.5
3.0
2.5
N=1735
N=376
N=29
N=93
N=17
N=89
N=177
Contr
T2D
SAID
SIDD
SIRD
MOD
MARD
Clusters

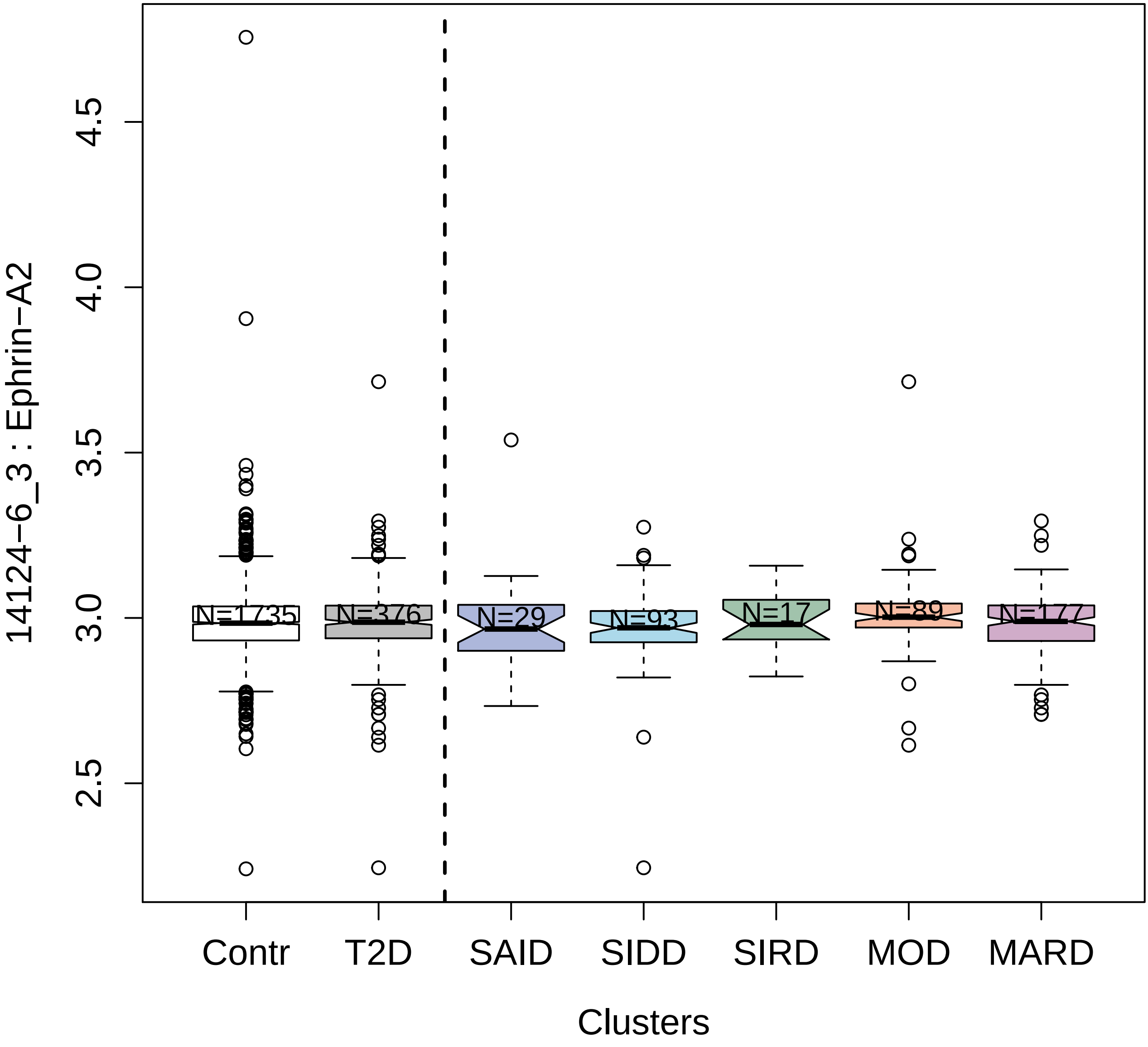
14124−6_3 : Ephrin−A2
4.5
4.0
3.5
3.0
2.5
N=1735
N=376
N=29
N=93
N=17
N=89
N=177
Contr
T2D
SAID
SIDD
SIRD
MOD
MARD
Clusters

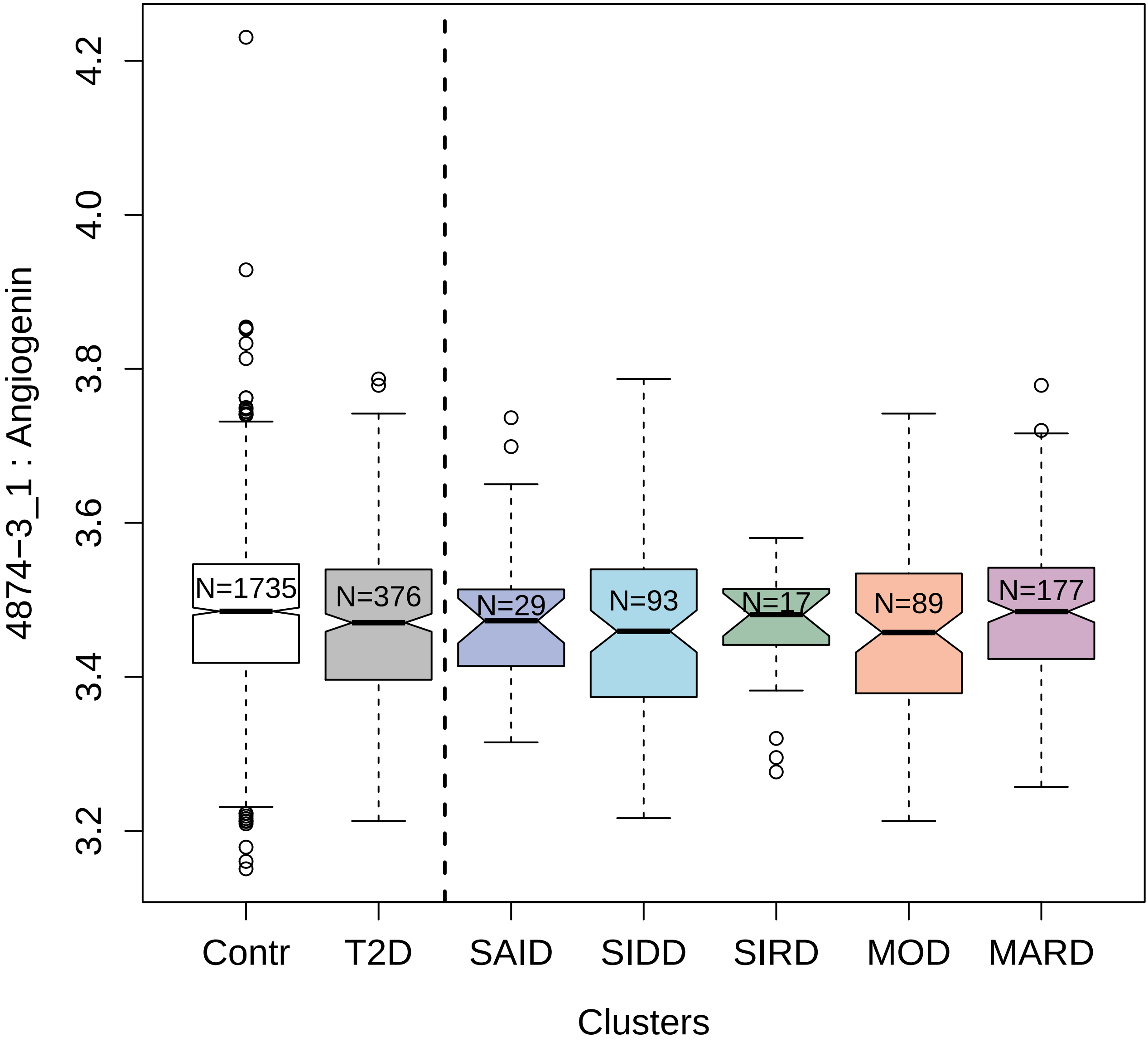
4874−3_1 : Angiogenin
4.2
4.0
3.8
3.6
3.4
3.2
N=1735
N=376
N=29
N=93
N=17
N=89
N=177
Contr
T2D
SAID
SIDD
SIRD
MOD
MARD
Clusters

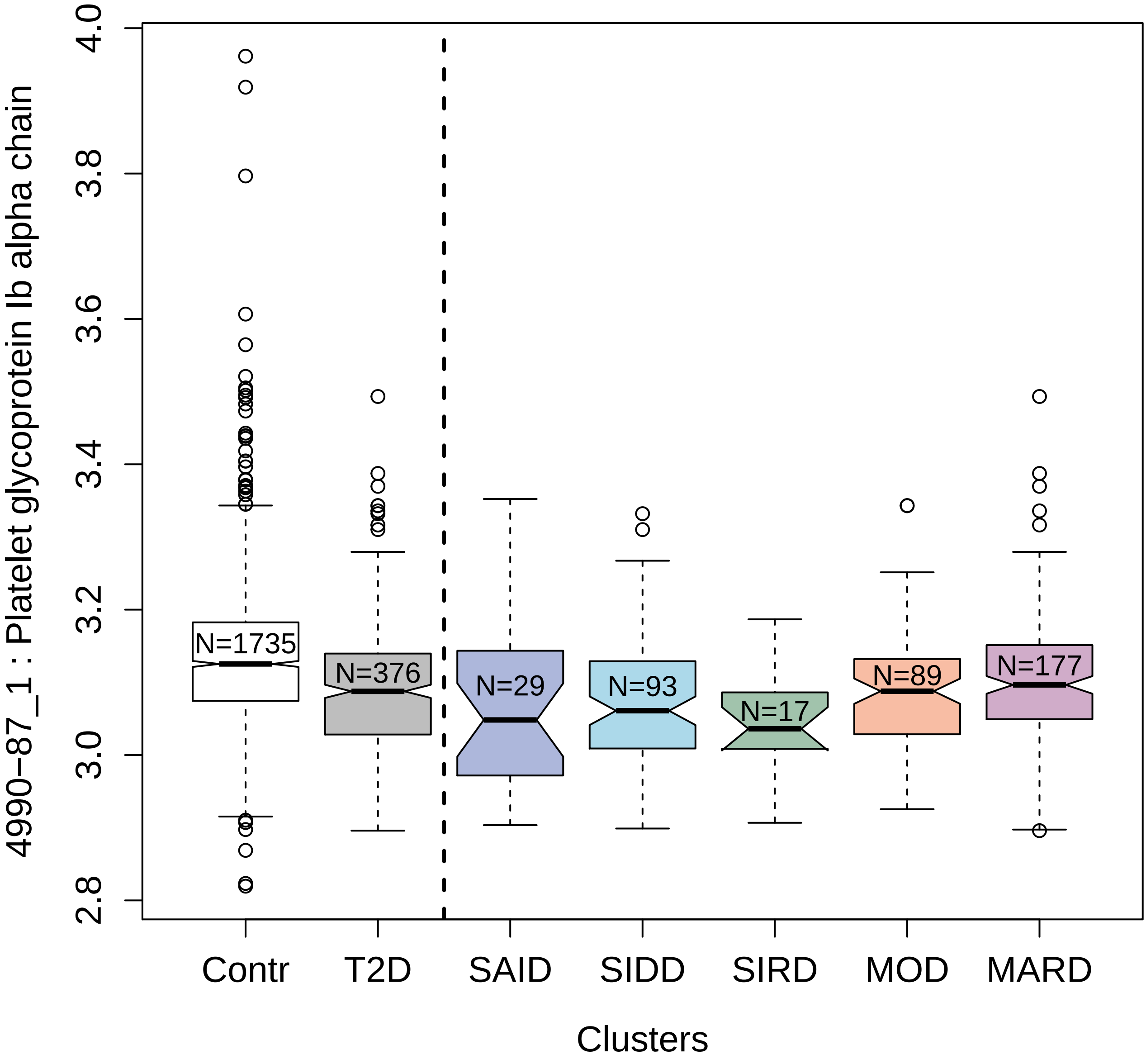
4990−87_1 : Platelet glycoprotein Ib alpha chain
4.0
3.8
3.6
3.4
3.2
3.0
2.8
N=1735
N=376
N=29
N=93
N=17
N=89
N=177
Contr
T2D
SAID
SIDD
SIRD
MOD
MARD
Clusters

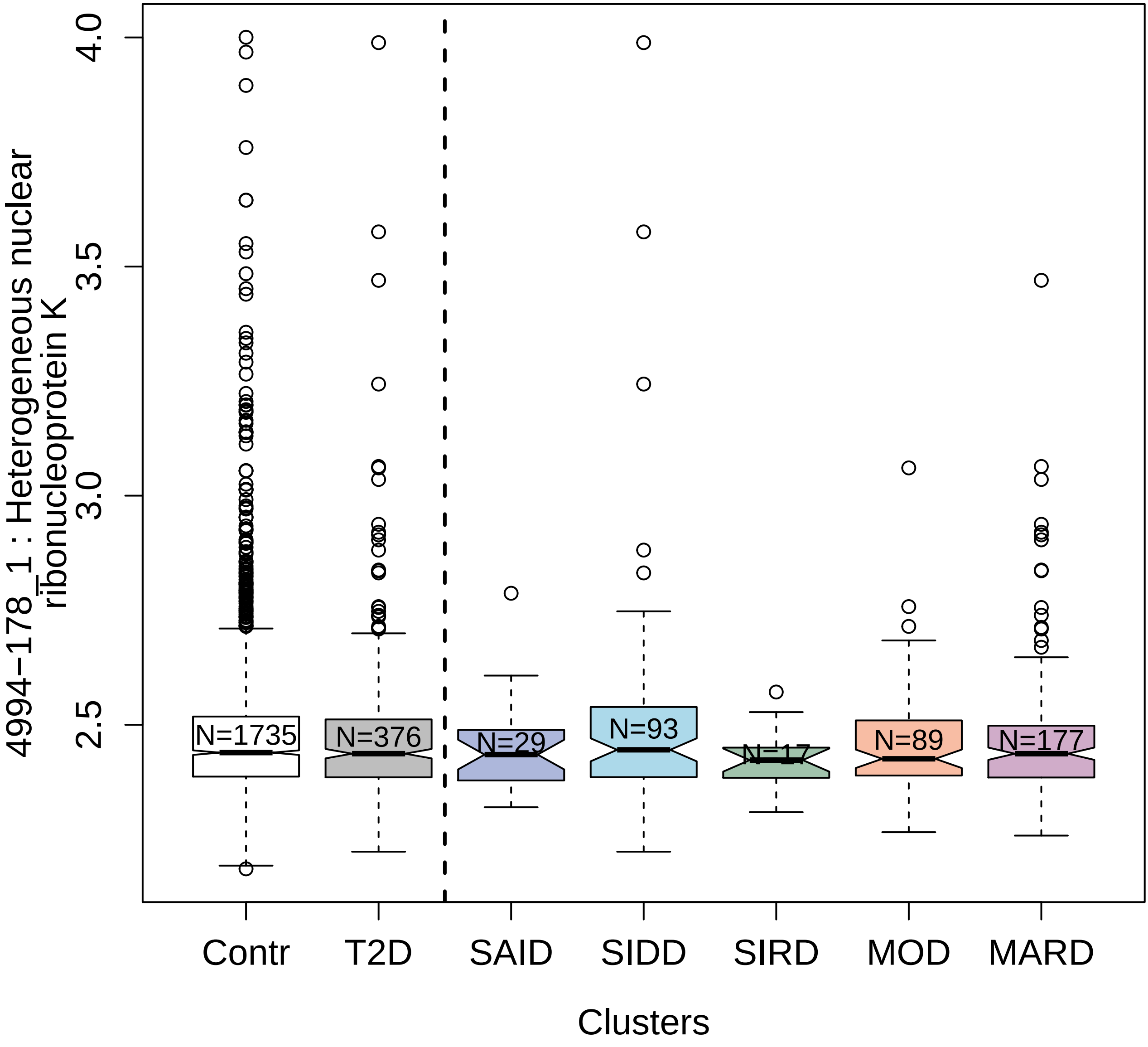

4994−178_1 : Heterogeneous nuclear ribonucleoprotein K
4.0
3.5
3.0
2.5
N=1735
N=376
N=29
N=93
N=17
N=89
N=177
Contr
T2D
SAID
SIDD
SIRD
MOD
MARD
Clusters

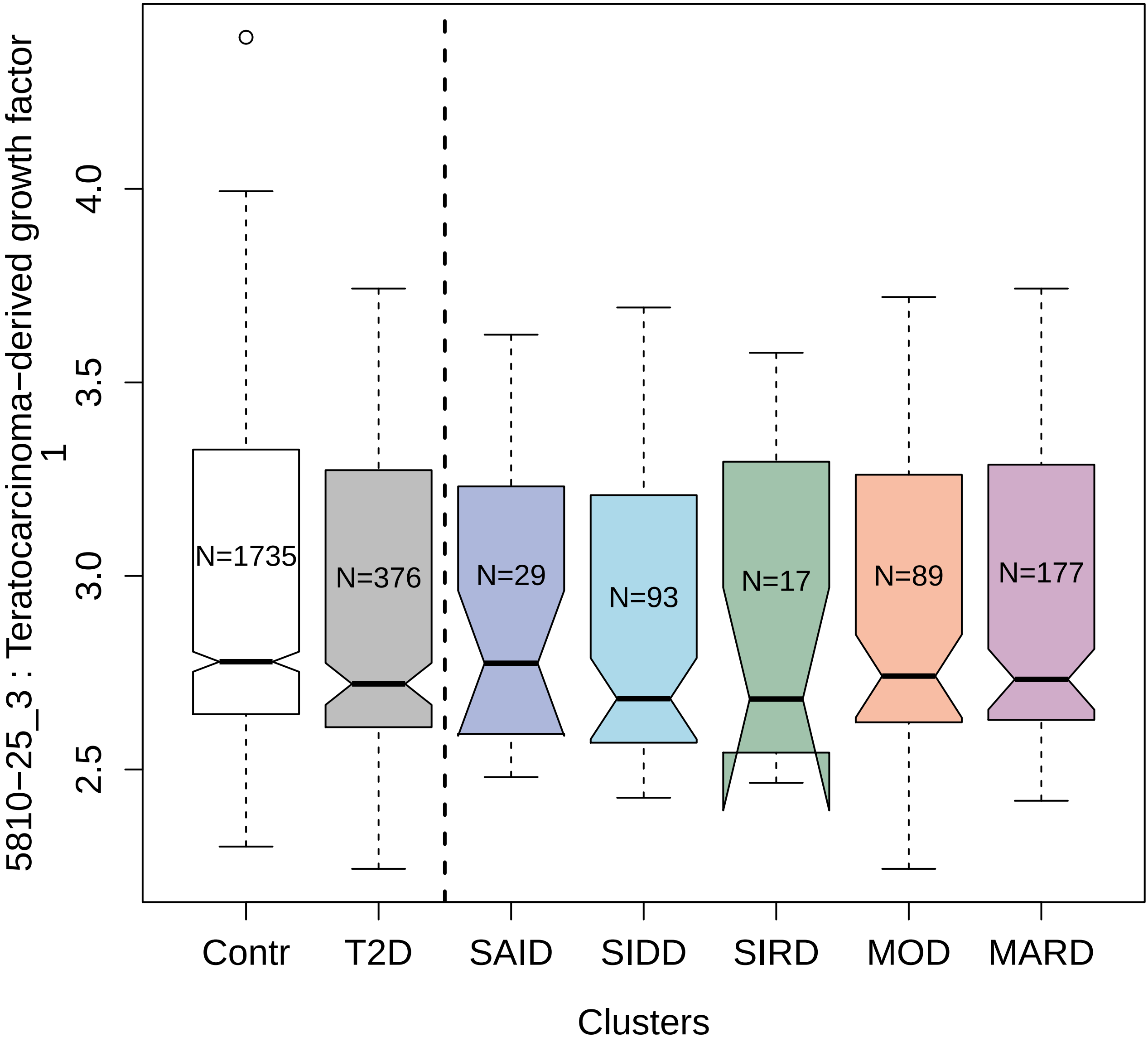
5810−25_3 : Teratocarcinoma−derived growth factor 1
4.0
3.5
3.0
2.5
N=1735
N=376
N=29
N=93
N=17
N=89
N=177
Contr
T2D
SAID
SIDD
SIRD
MOD
MARD
Clusters

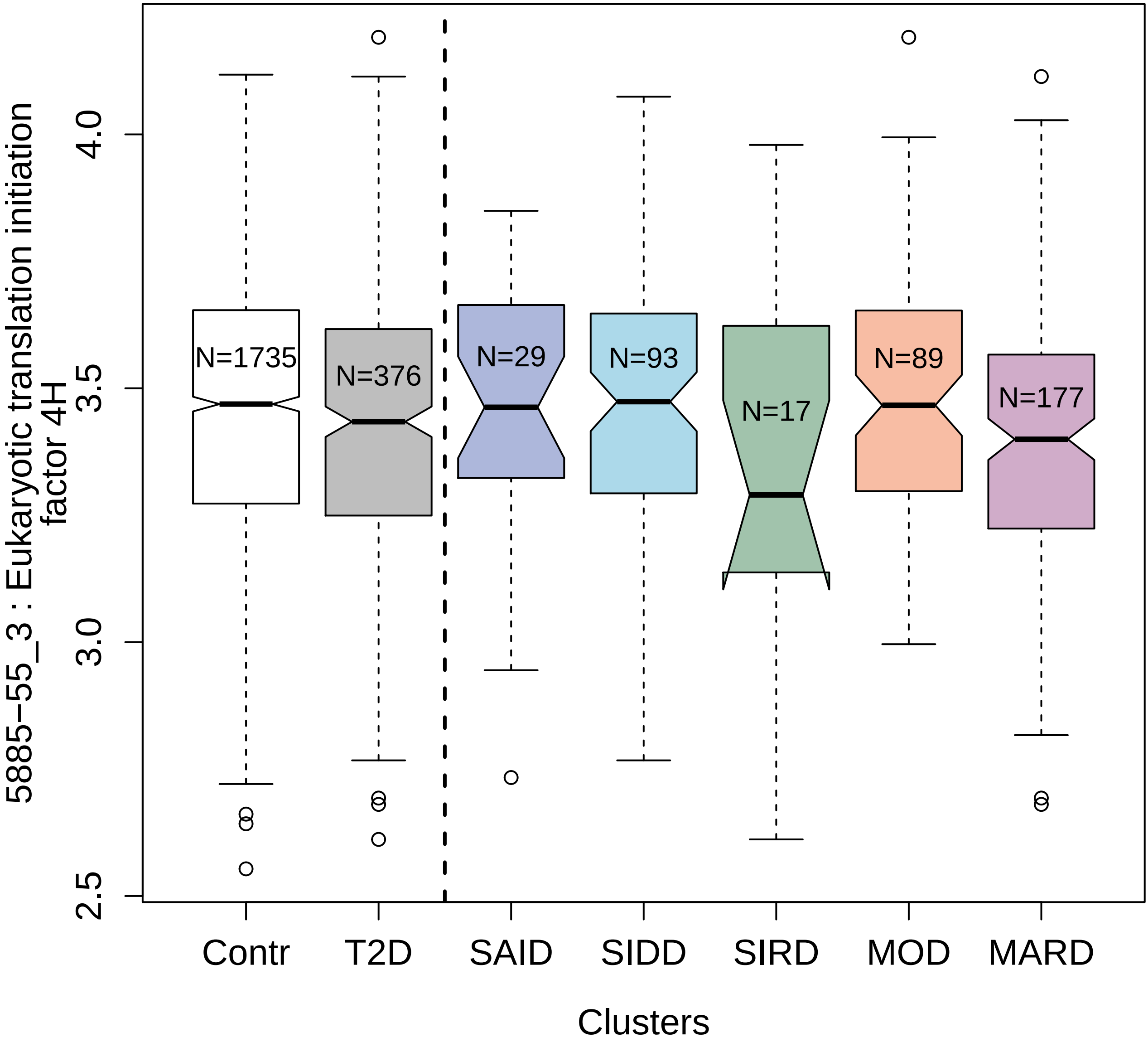

5885−55_3 : Eukaryotic translation initiation
factor 4H
4.0
3.5
3.0
2.5
N=1735
N=376
N=29
N=93
N=17
N=89
N=177
Contr
T2D
SAID
SIDD
SIRD
MOD
MARD
Clusters

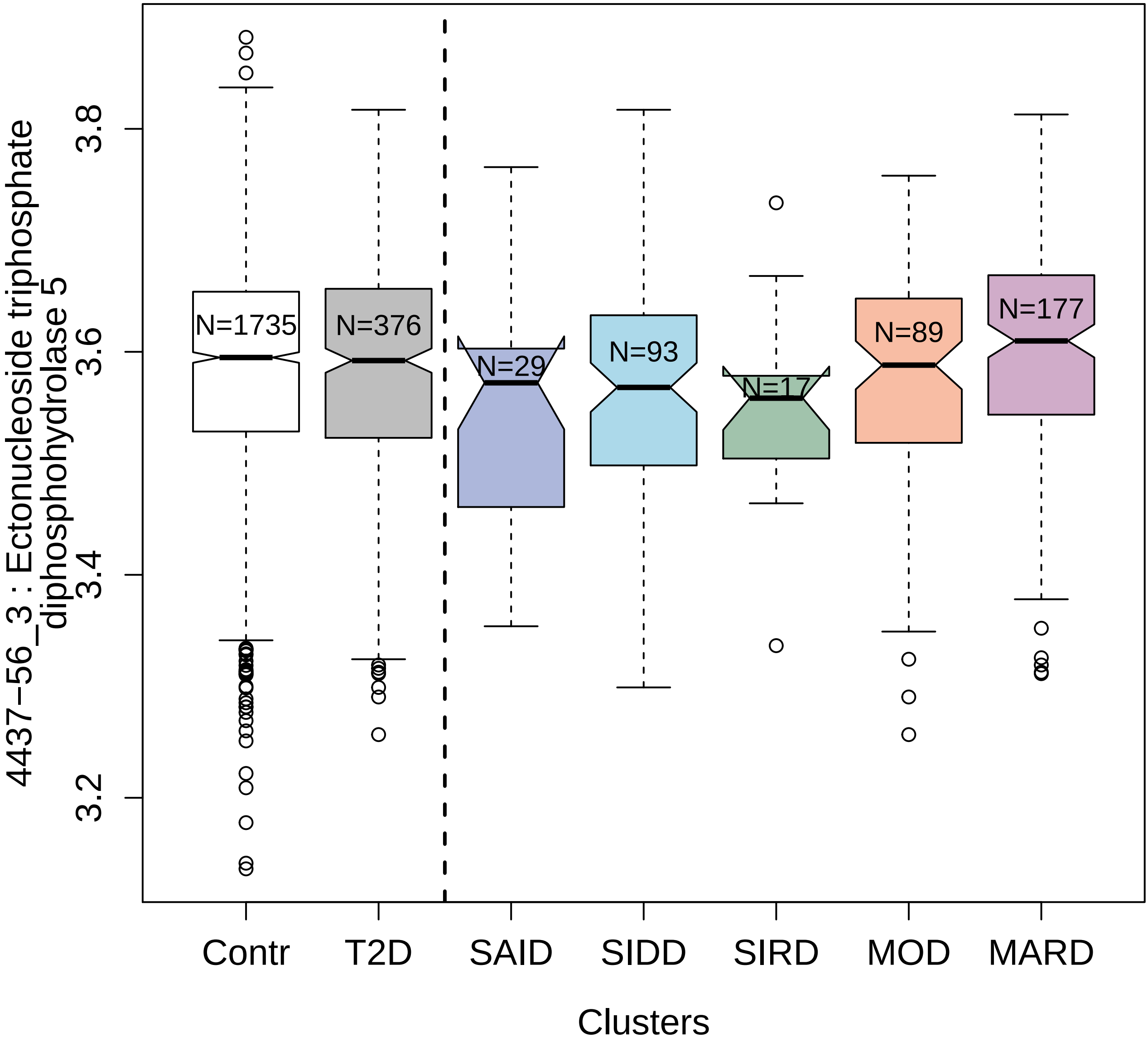
4437−56_3 : Ectonucleoside triphosphate
_diphosphohydrolase 5
3.2
3.4
3.6
3.8
N=1735
N=376
N=29
N=93
N=17
N=89
N=177
Contr
T2D
SAID
SIDD
SIRD
MOD
MARD
Clusters

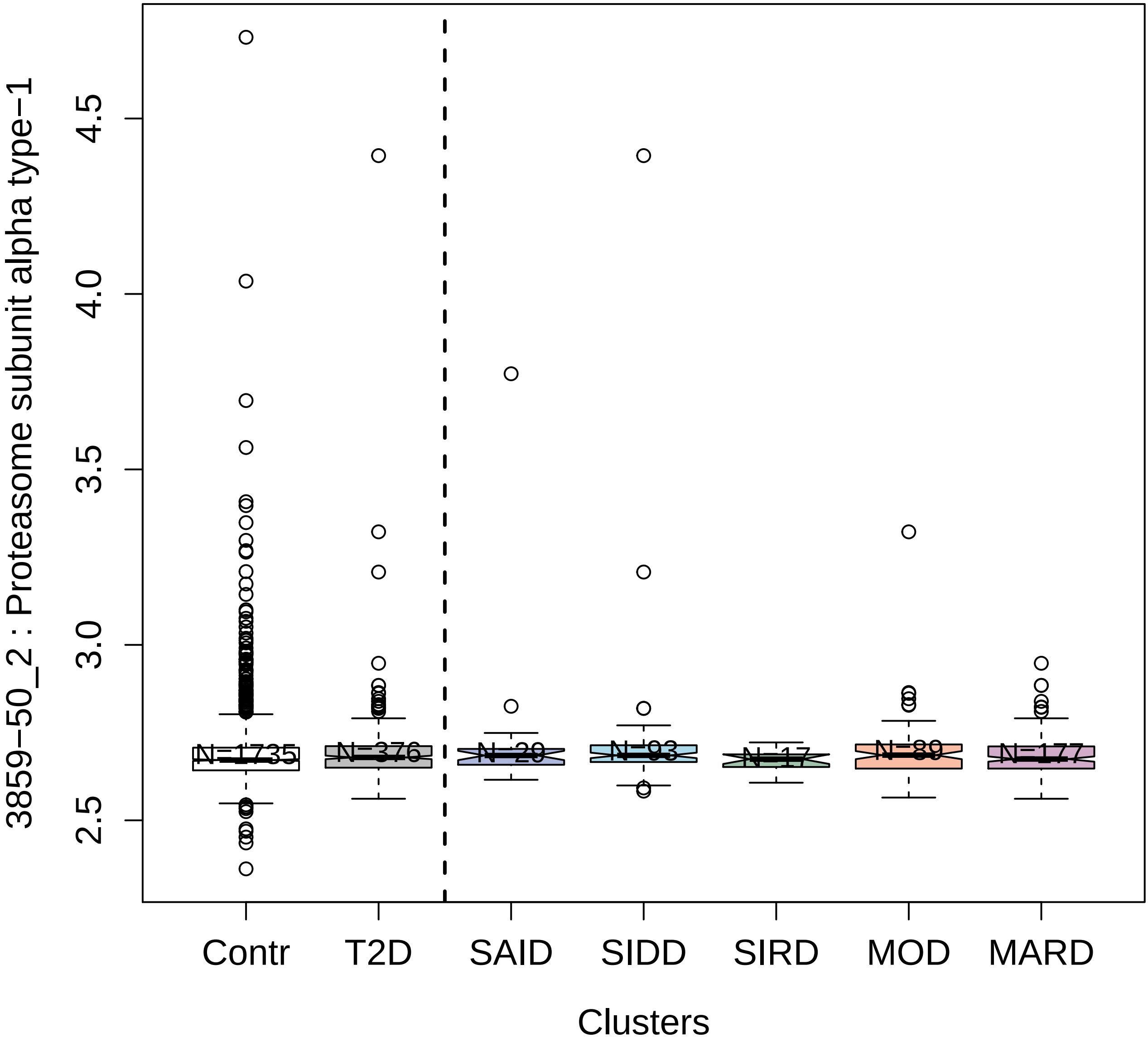
3859−50_2 : Proteasome subunit alpha type−1
2.5
3.0
3.5
4.0
4.5
N=1735
N=376
N=29
N=93
N=17
N=89
N=177
Contr
T2D
SAID
SIDD
SIRD
MOD
MARD
Clusters

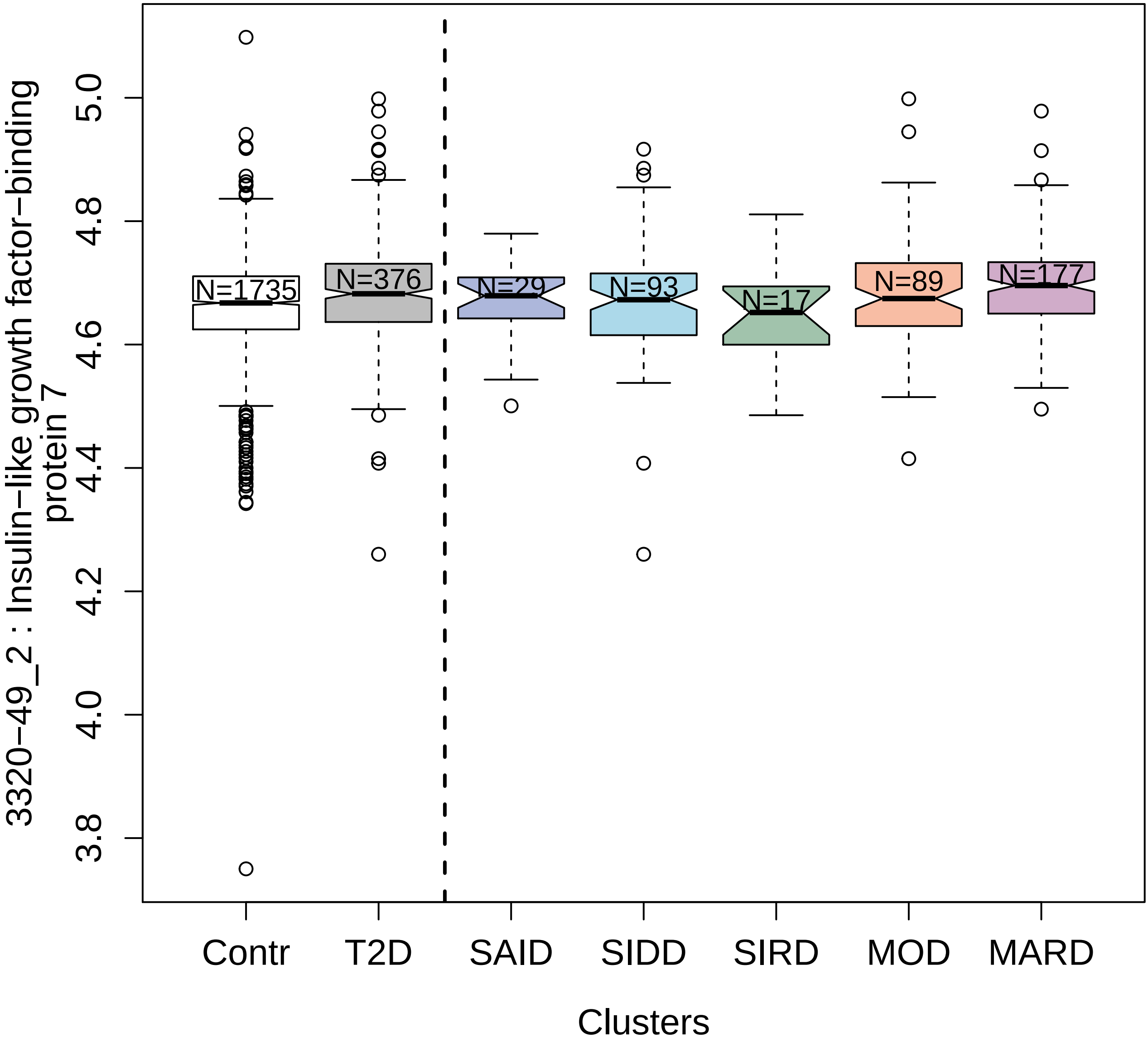
3320−49_2 : Insulin−like growth factor−binding protein 7
5.0
4.8
4.6
4.4
4.2
4.0
3.8
N=1735
N=376
N=29
N=93
N=17
N=89
N=177
Contr
T2D
SAID
SIDD
SIRD
MOD
MARD
Clusters

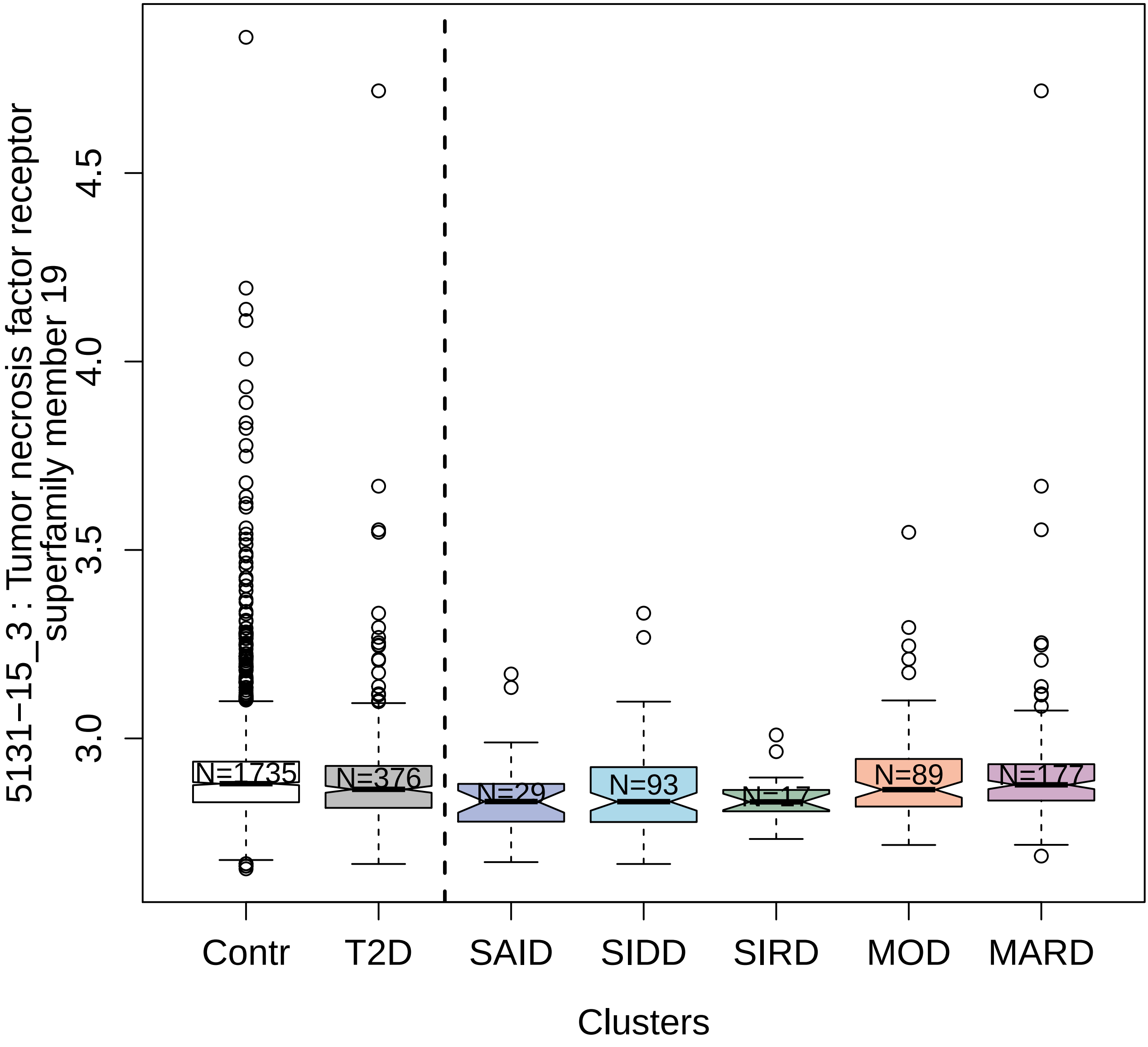
5131−15_3 : Tumor necrosis factor receptor superfamily member 19
3.0
3.5
4.0
4.5
N=1735
N=376
N=29
N=93
N=17
N=89
N=177
Contr
T2D
SAID
SIDD
SIRD
MOD
MARD
Clusters

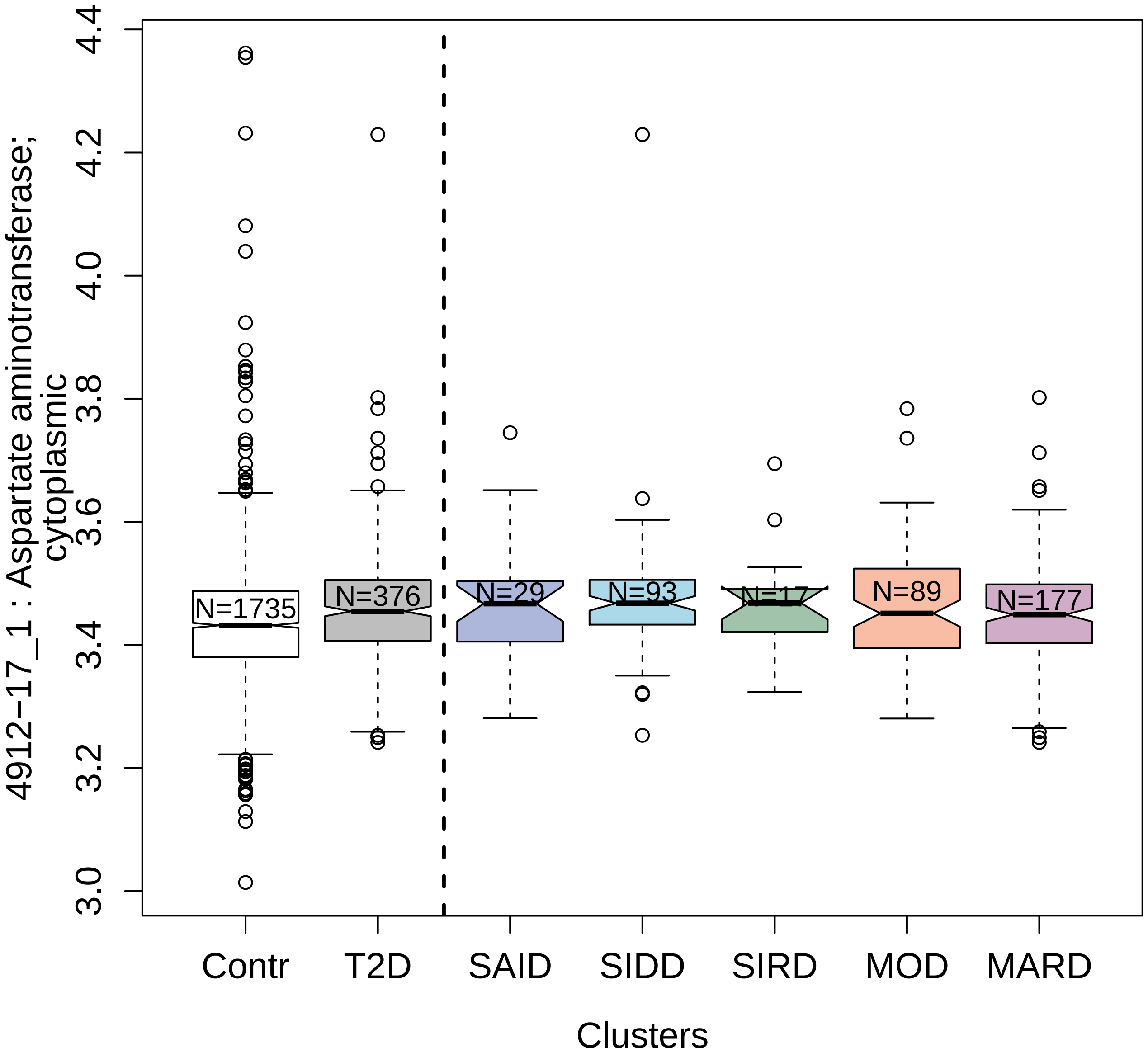

4912−17_1 : Aspartate aminotransferase; cytoplasmic
4.4
4.2
4.0
3.8
3.6
3.4
3.2
3.0
N=1735
N=376
N=29
N=93
N=17
N=89
N=177
Contr
T2D
SAID
SIDD
SIRD
MOD
MARD
Clusters

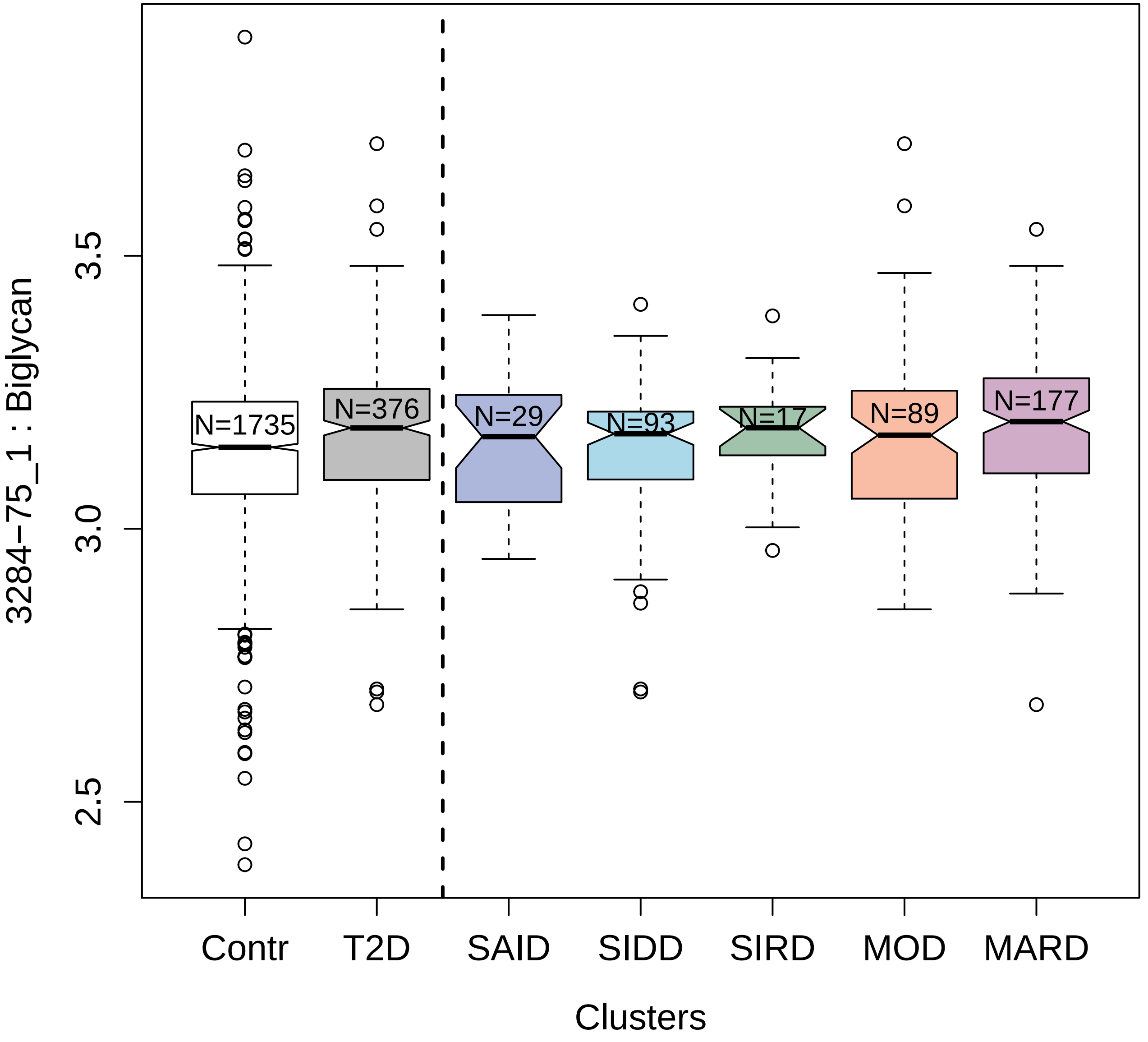
3284−75_1 : Biglycan
3.5
3.0
2.5
N=1735
N=376
N=29
N=93
N=17
N=89
N=177
Contr
T2D
SAID
SIDD
SIRD
MOD
MARD
Clusters

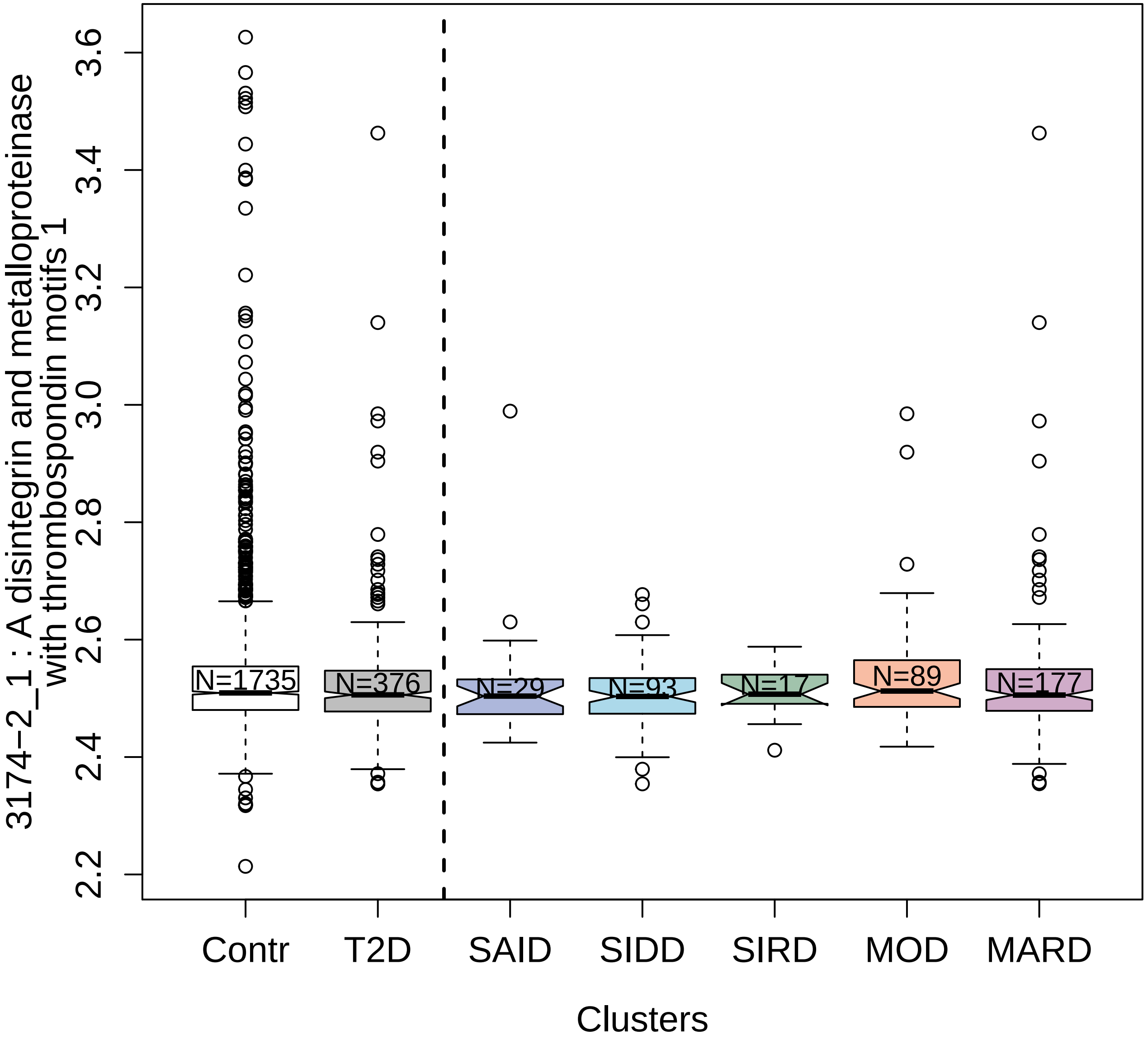
3174−2_1 : A disintegrin and metalloproteinase
with thrombospondin motifs 1
3.6
3.4
3.2
3.0
2.8
2.6
2.4
2.2
N=1735
N=376
N=29
N=93
N=17
N=89
N=177
Contr
T2D
SAID
SIDD
SIRD
MOD
MARD
Clusters

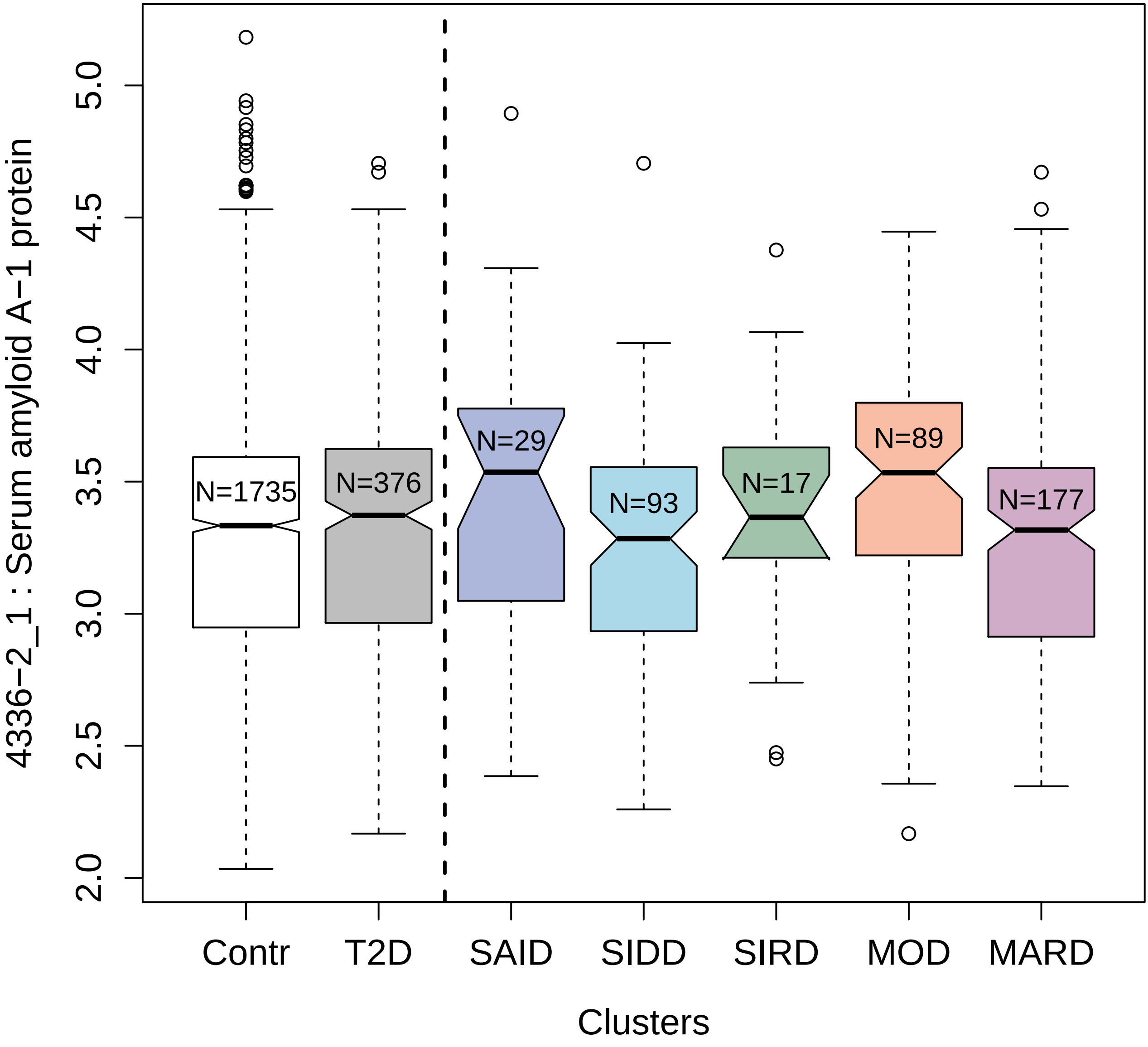
4336−2_1 : Serum amyloid A−1 protein
2.0
2.5
3.0
3.5
4.0
4.5
5.0
N=1735
N=376
N=29
N=93
N=17
N=89
N=177
Contr
T2D
SAID
SIDD
SIRD
MOD
MARD
Clusters

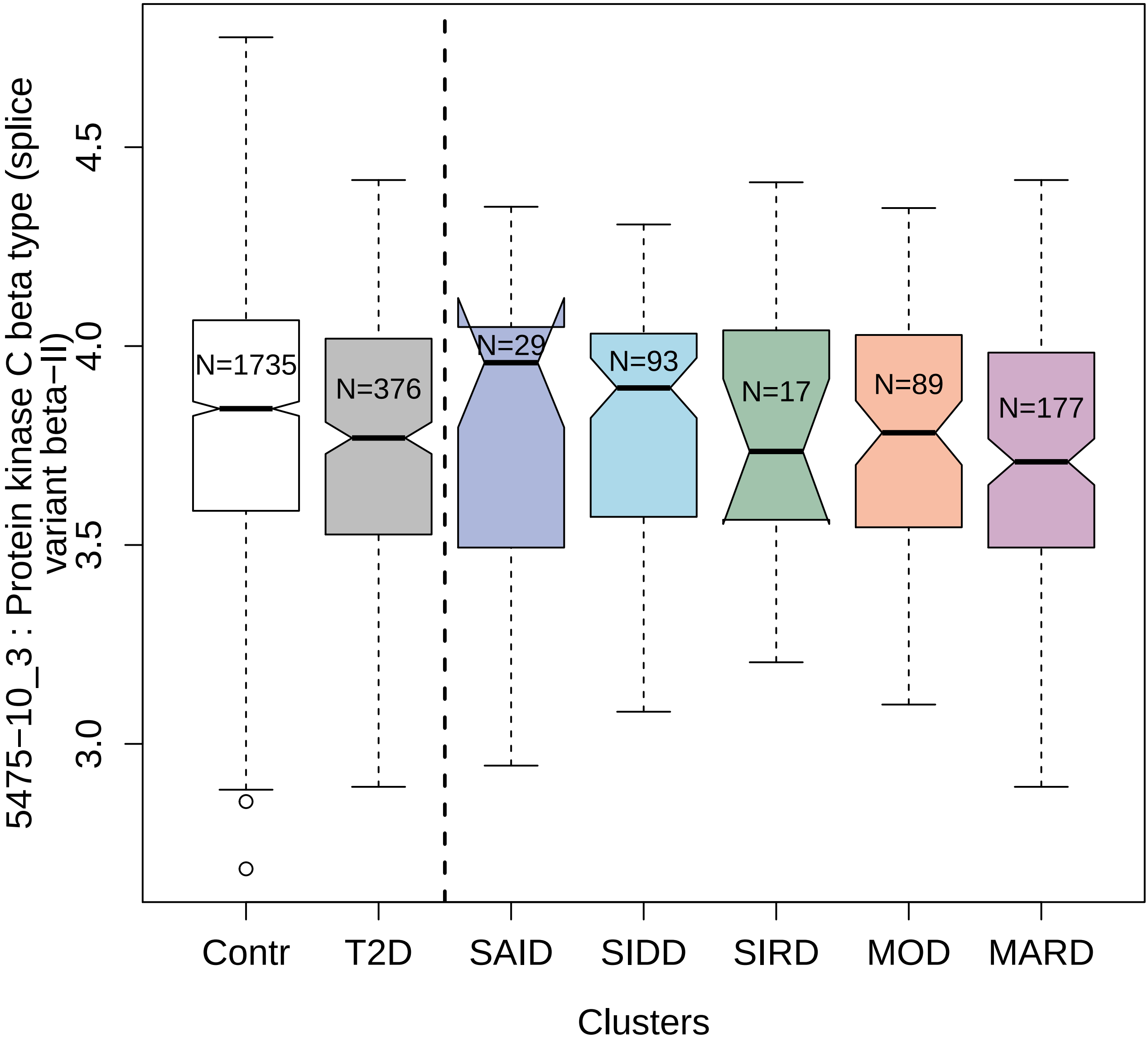

5475−10_3 : Protein kinase C beta type (splice variant beta−II)
3.0
3.5
4.0
4.5
N=1735
N=376
N=29
N=93
N=17
N=89
N=177
Contr
T2D
SAID
SIDD
SIRD
MOD
MARD
Clusters

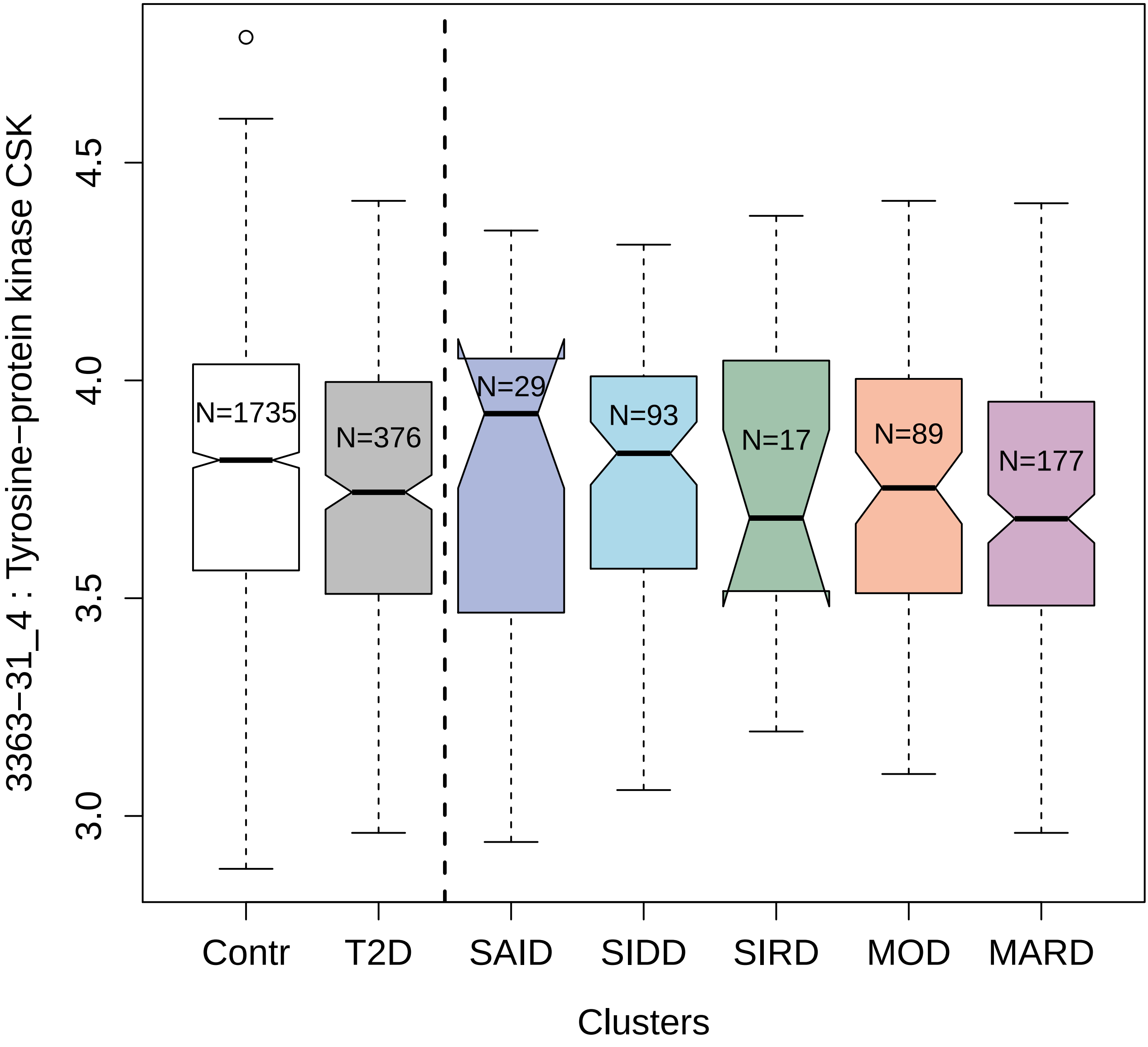

3363−31_4 : Tyrosine−protein kinase CSK
3.0
3.5
4.0
4.5
N=1735
N=376
N=29
N=93
N=17
N=89
N=177
Contr
T2D
SAID
SIDD
SIRD
MOD
MARD
Clusters

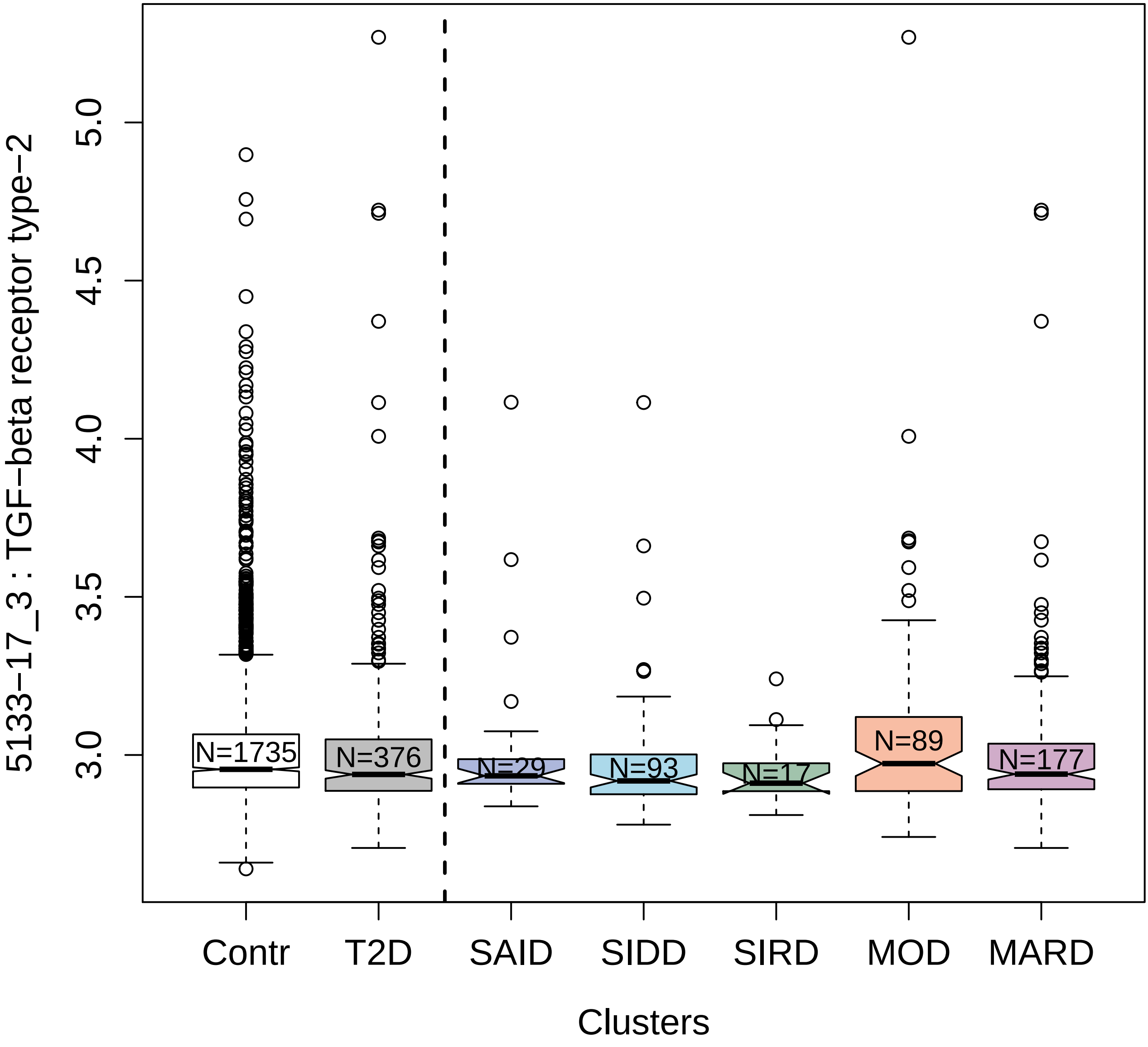

5133−17_3 : TGF−beta receptor type−2
5.0
4.5
4.0
3.5
3.0
N=1735
N=376
N=29
N=93
N=17
N=89
N=177
Contr
T2D
SAID
SIDD
SIRD
MOD
MARD
Clusters

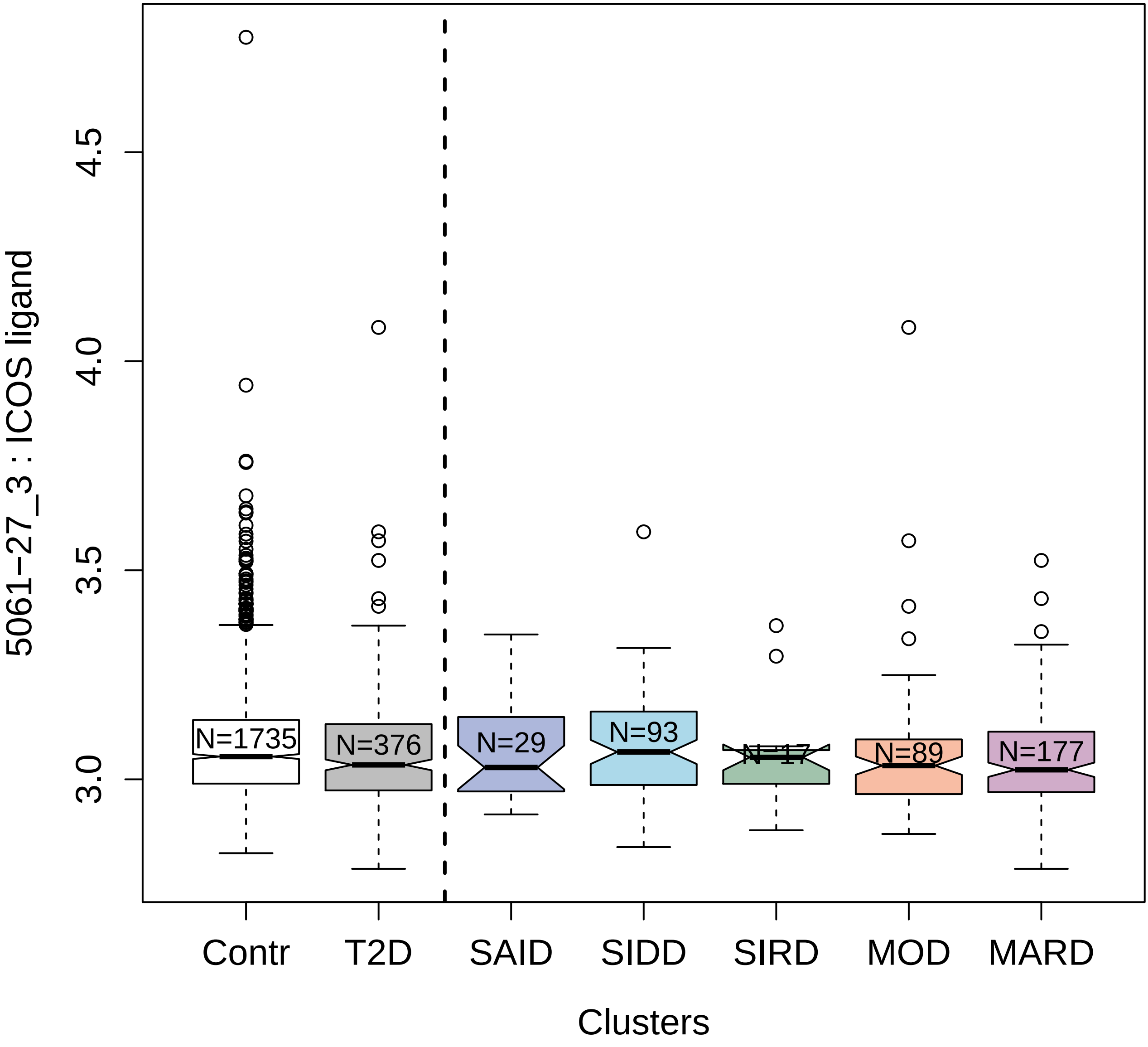
5061−27_3 : ICOS ligand
3.0
3.5
4.0
4.5
N=1735
N=376
N=29
N=93
N=17
N=89
N=177
Contr
T2D
SAID
SIDD
SIRD
MOD
MARD
Clusters

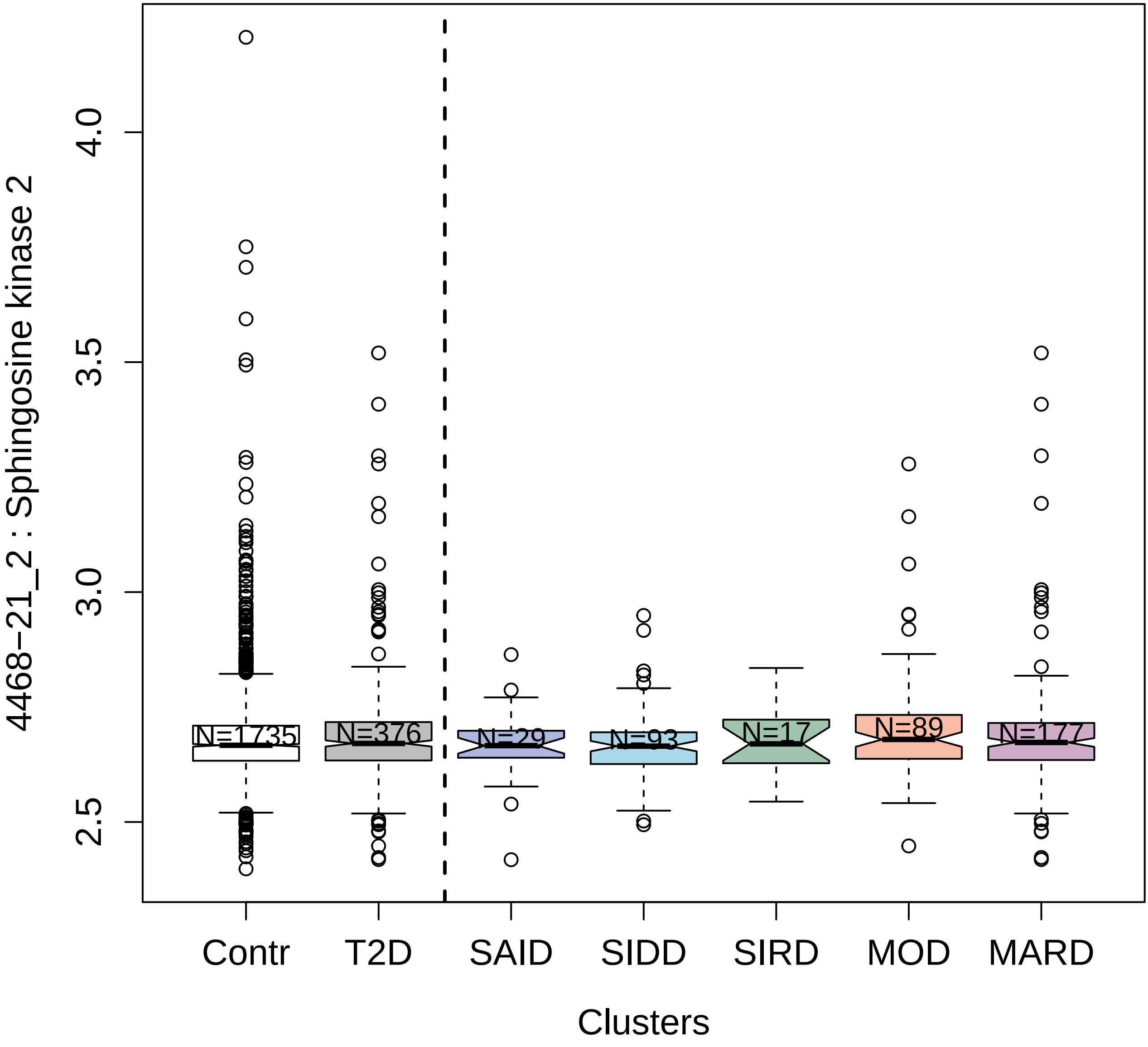
4468−21_2 : Sphingosine kinase 2
4.0
3.5
3.0
2.5
N=1735
N=376
N=29
N=93
N=17
N=89
N=177
Contr
T2D
SAID
SIDD
SIRD
MOD
MARD
Clusters

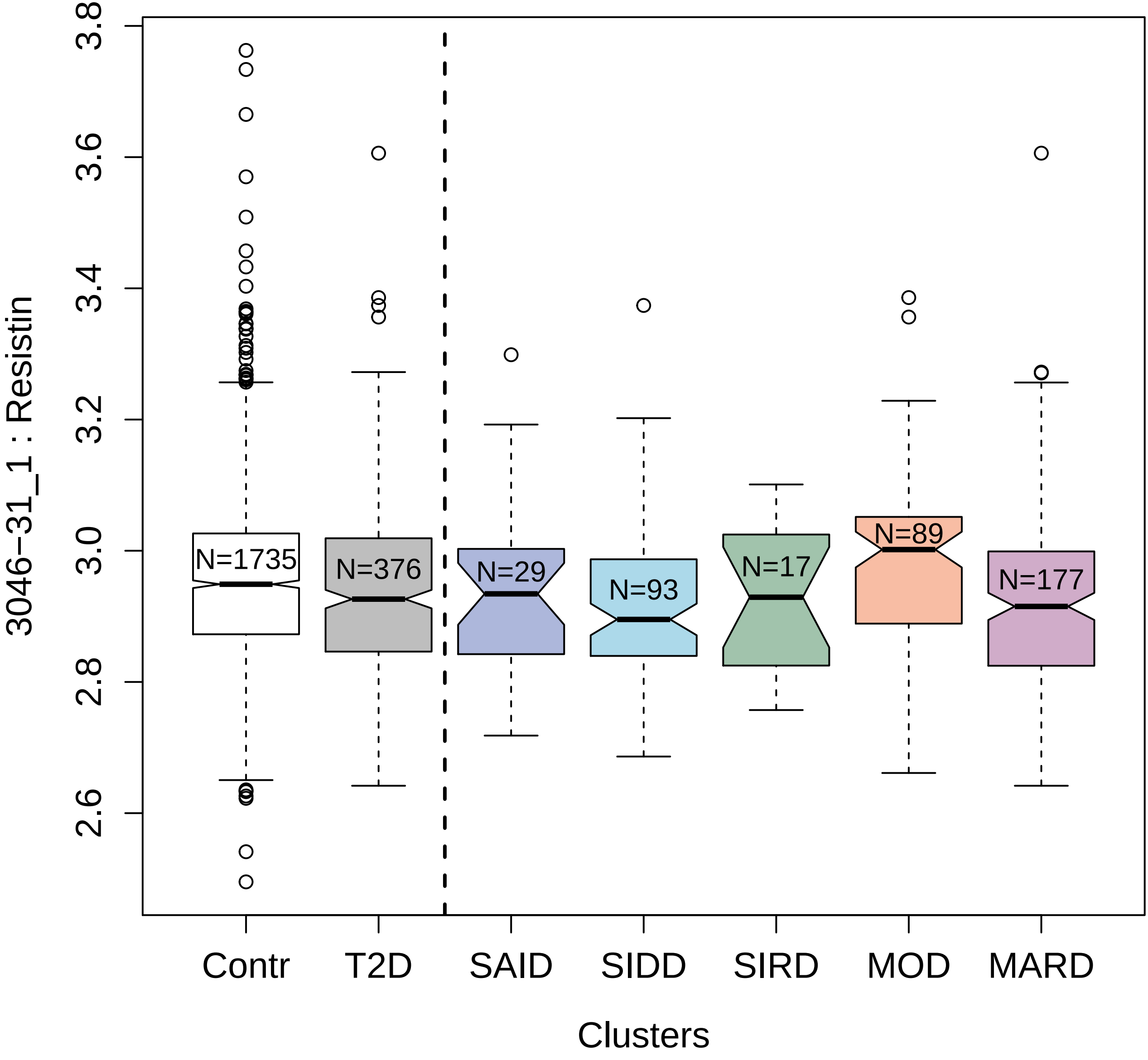

3046−31_1 : Resistin
3.8
3.6
3.4
3.2
3.0
2.8
2.6
N=1735
N=376
N=29
N=93
N=17
N=89
N=177
Contr
T2D
SAID
SIDD
SIRD
MOD
MARD
Clusters

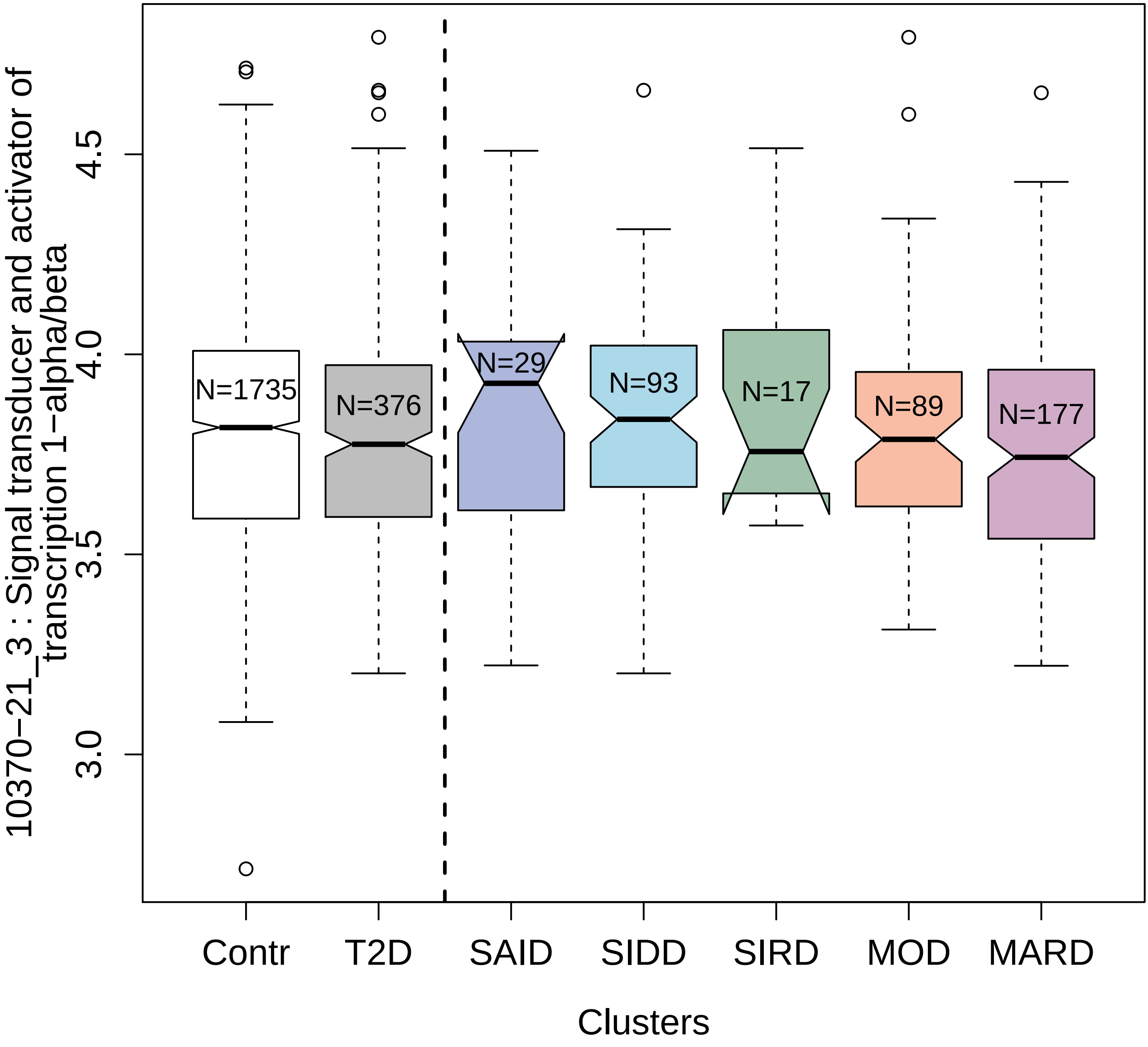

10370−21_3 : Signal transducer and activator of transcription 1−alpha/beta
3.0
3.5
4.0
4.5
N=1735
N=376
N=29
N=93
N=17
N=89
N=177
Contr
T2D
SAID
SIDD
SIRD
MOD
MARD
Clusters

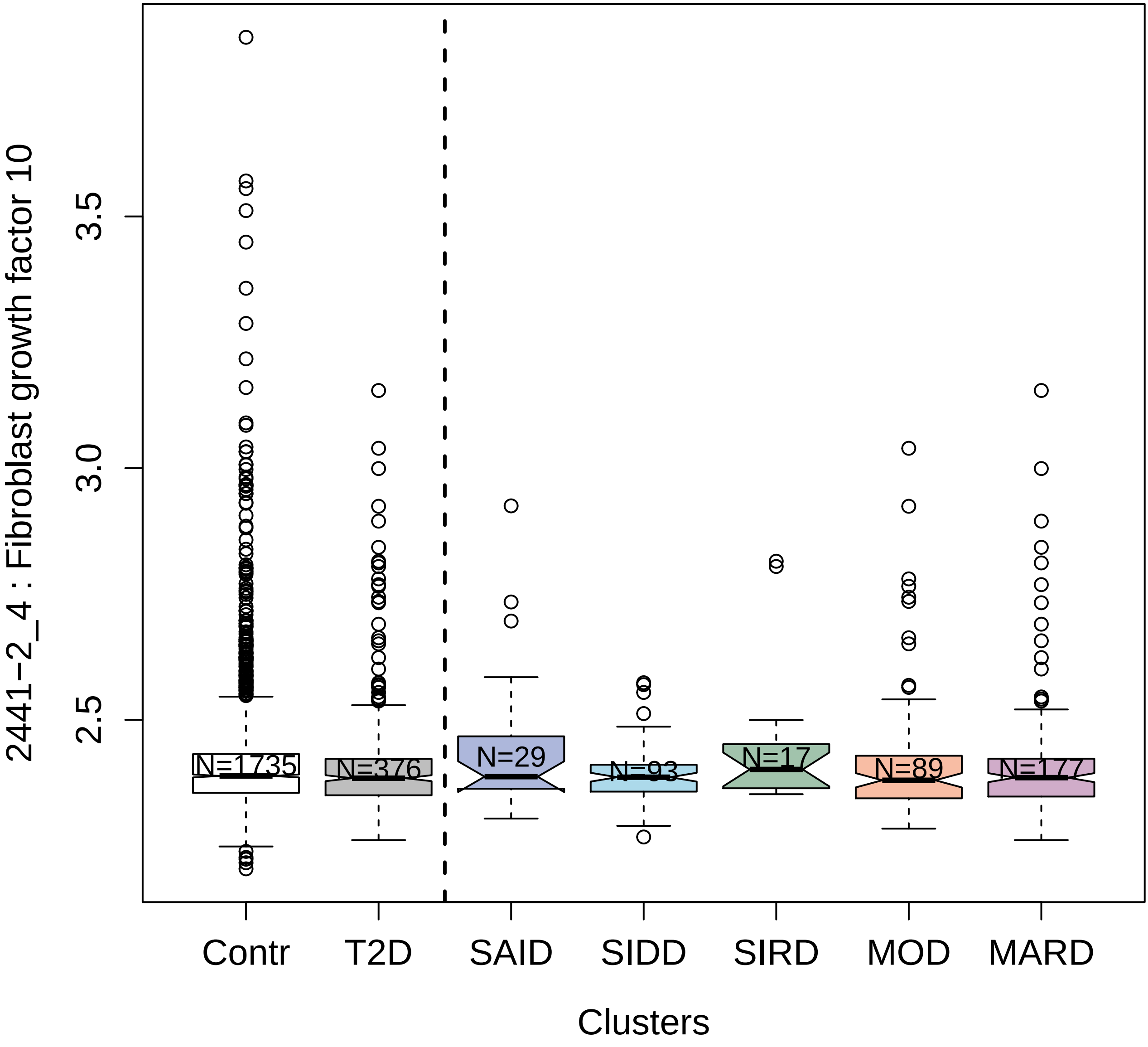

2441−2_4 : Fibroblast growth factor 10
3.5
3.0
2.5
N=1735
N=376
N=29
N=93
N=17
N=89
N=177
Contr
T2D
SAID
SIDD
SIRD
MOD
MARD
Clusters

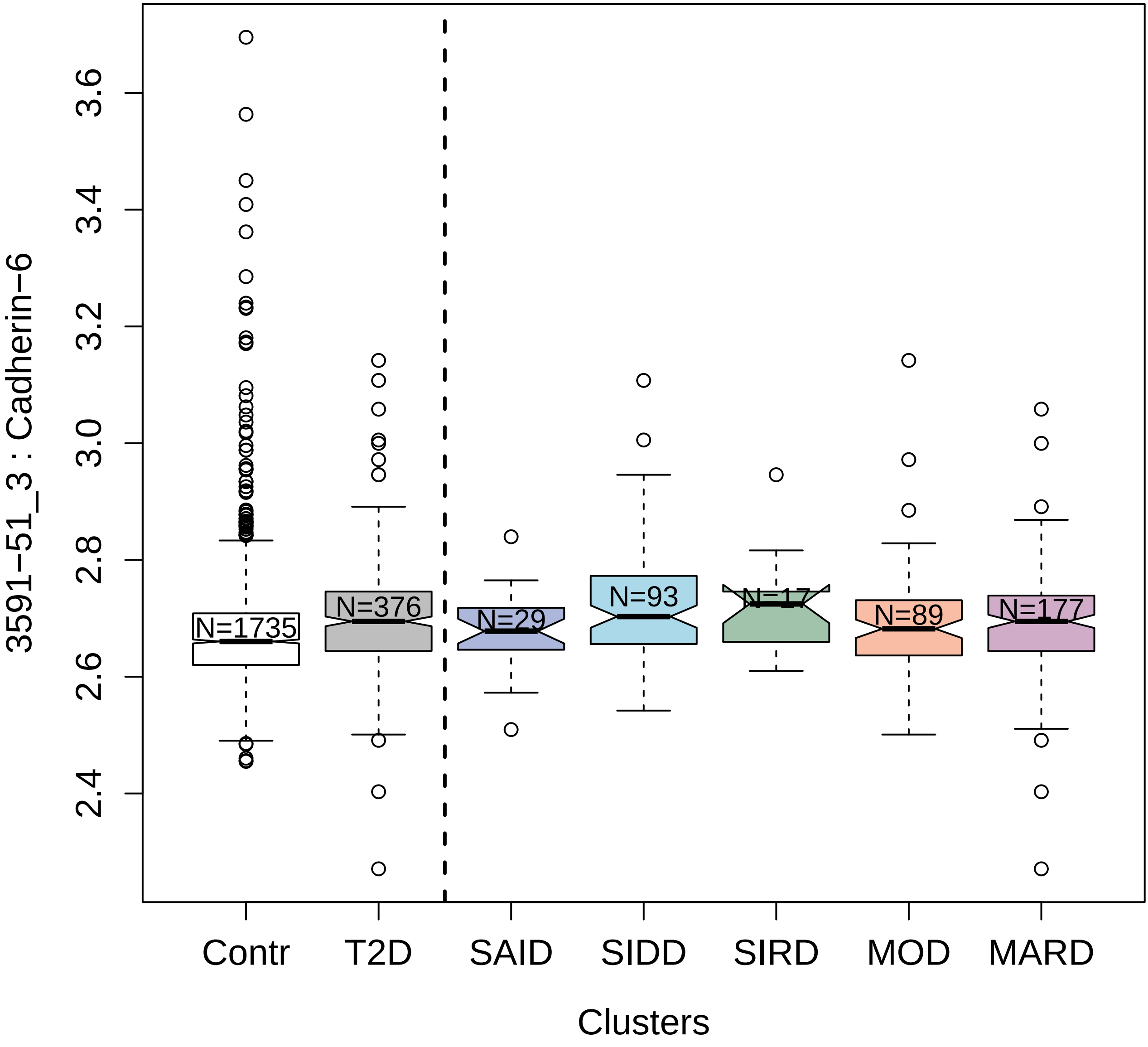
3591−51_3 : Cadherin−6
3.6
3.4
3.2
3.0
2.8
2.6
2.4
N=1735
N=376
N=29
N=93
N=17
N=89
N=177
Contr
T2D
SAID
SIDD
SIRD
MOD
MARD
Clusters

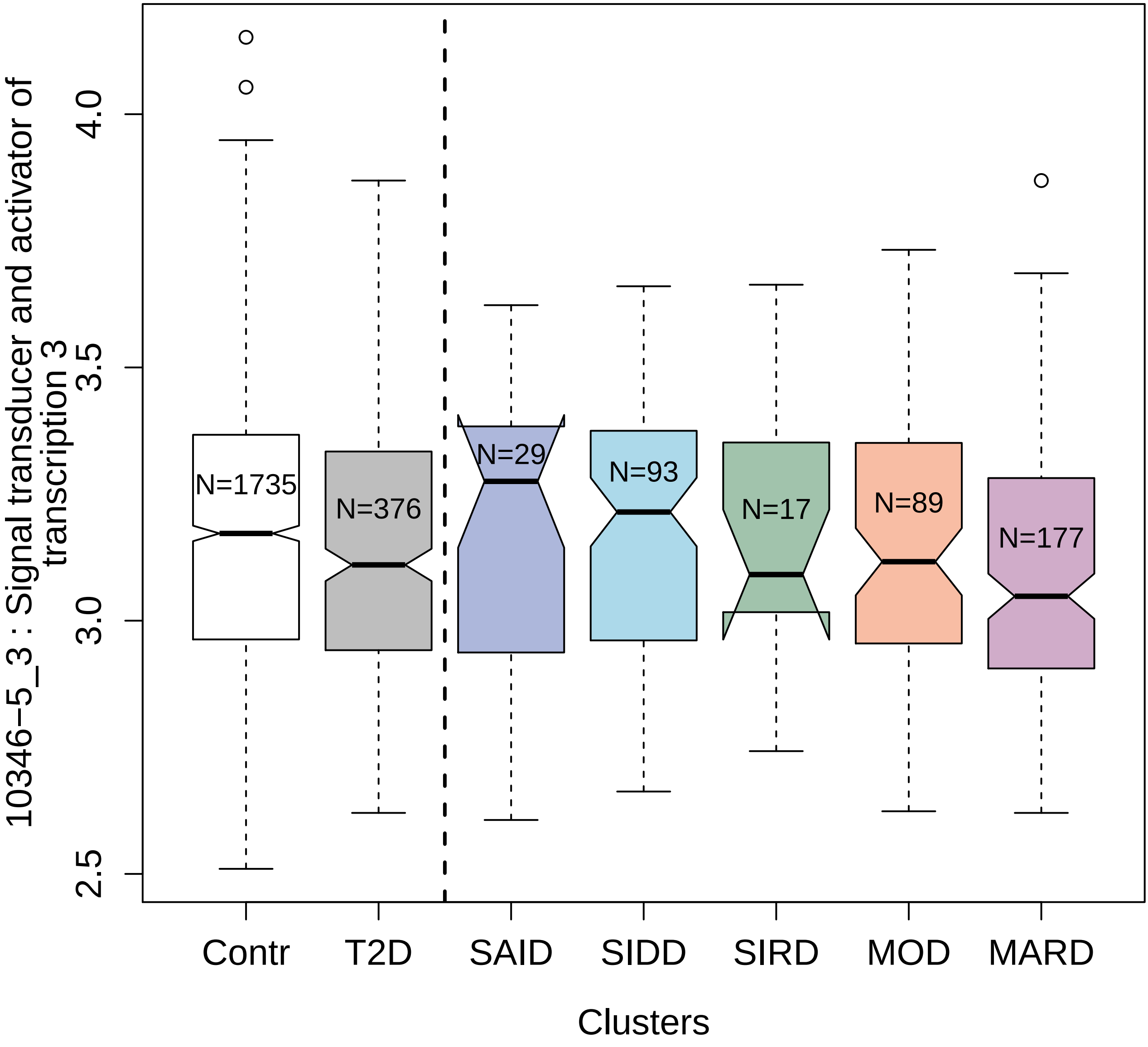
10346−5_3 : Signal transducer and activator of transcription 3
2.5
3.0
3.5
4.0
N=1735
N=376
N=29
N=93
N=17
N=89
N=177
Contr
T2D
SAID
SIDD
SIRD
MOD
MARD
Clusters

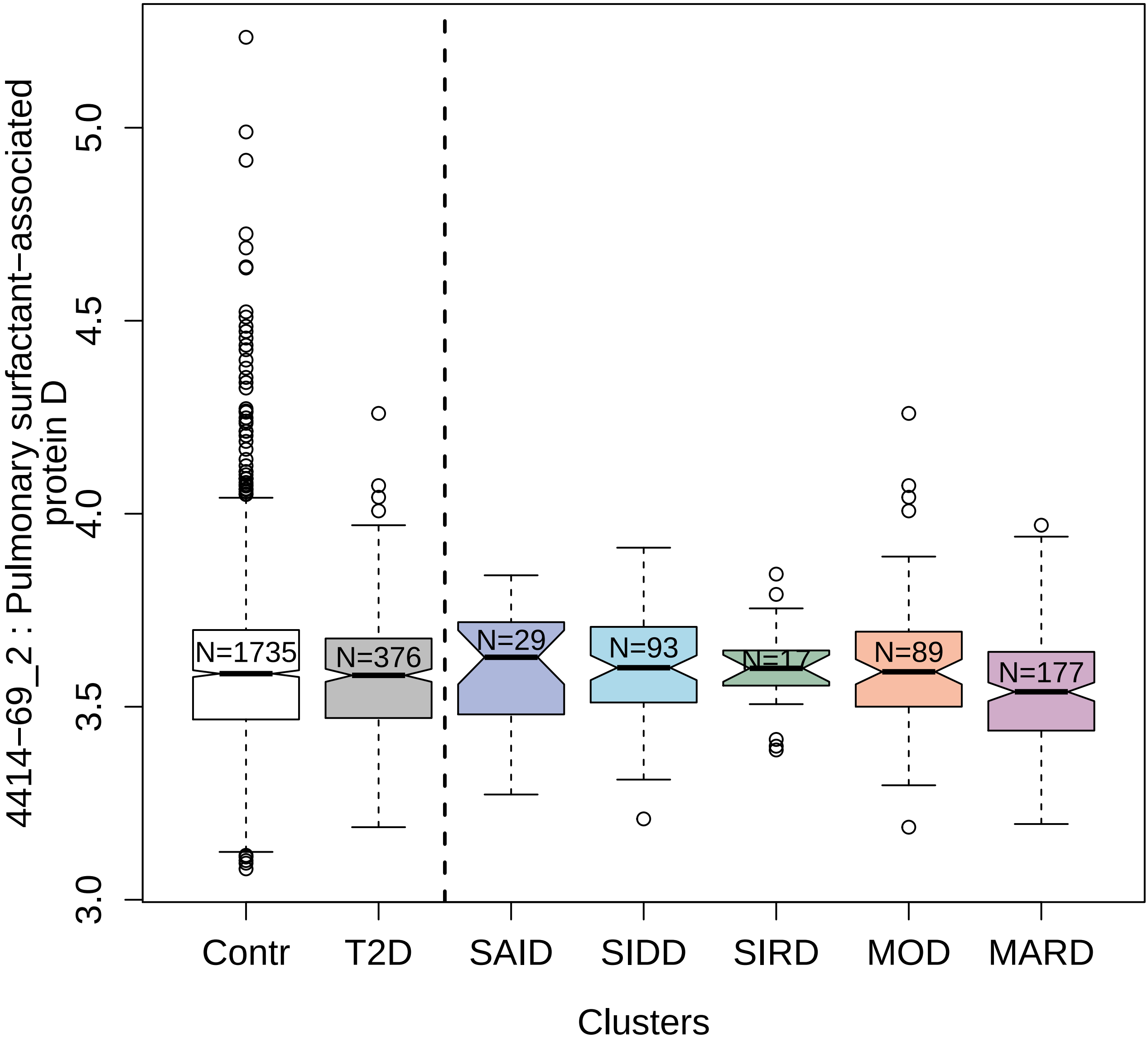

4414−69_2 : Pulmonary surfactant−associated protein D
3.0
3.5
4.0
4.5
5.0
N=1735
N=376
N=29
N=93
N=17
N=89
N=177
Contr
T2D
SAID
SIDD
SIRD
MOD
MARD
Clusters

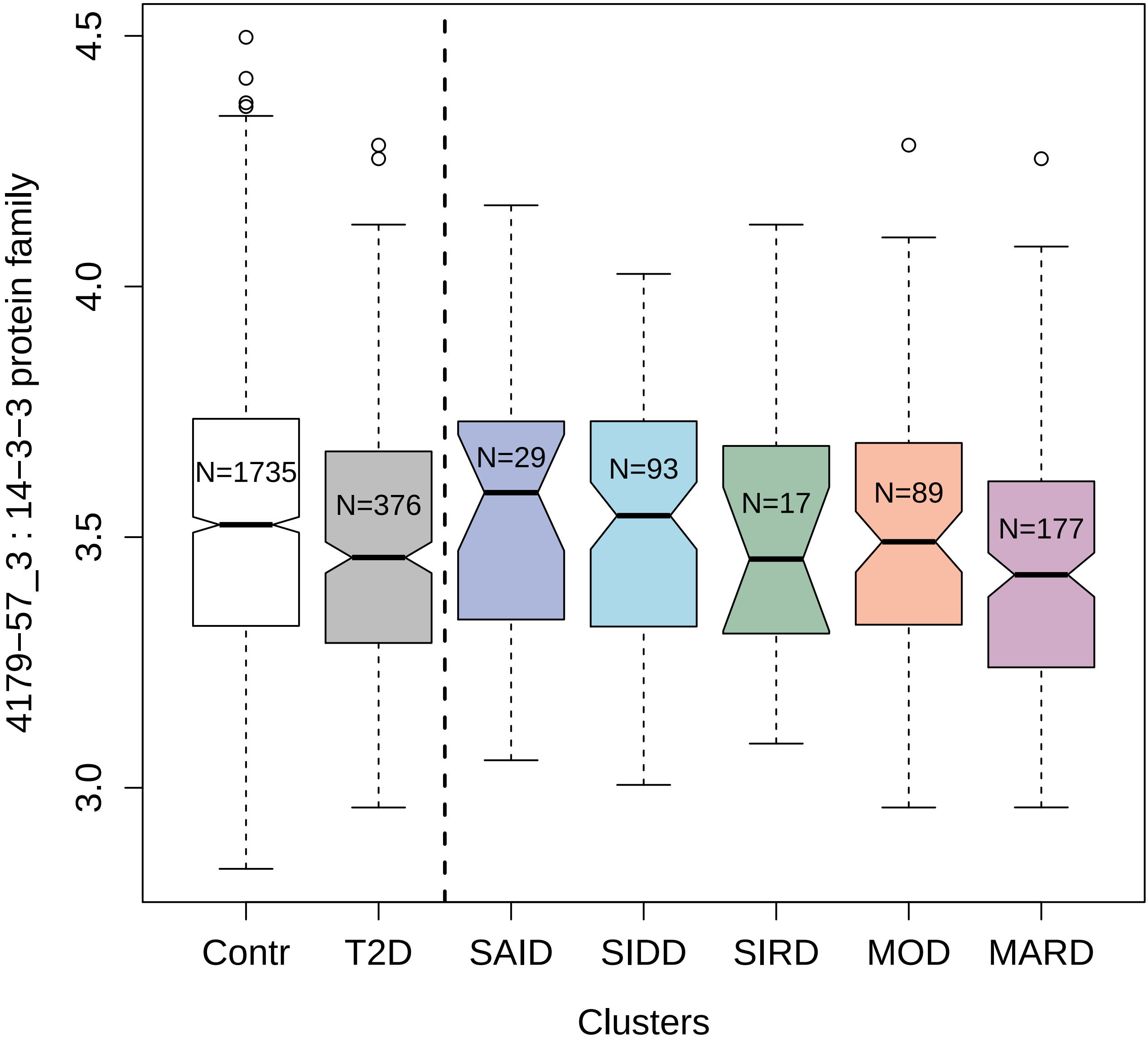

4179−57_3 : 14−3−3 protein family
4.5
4.0
3.5
3.0
N=1735
N=376
N=29
N=93
N=17
N=89
N=177
Contr
T2D
SAID
SIDD
SIRD
MOD
MARD
Clusters

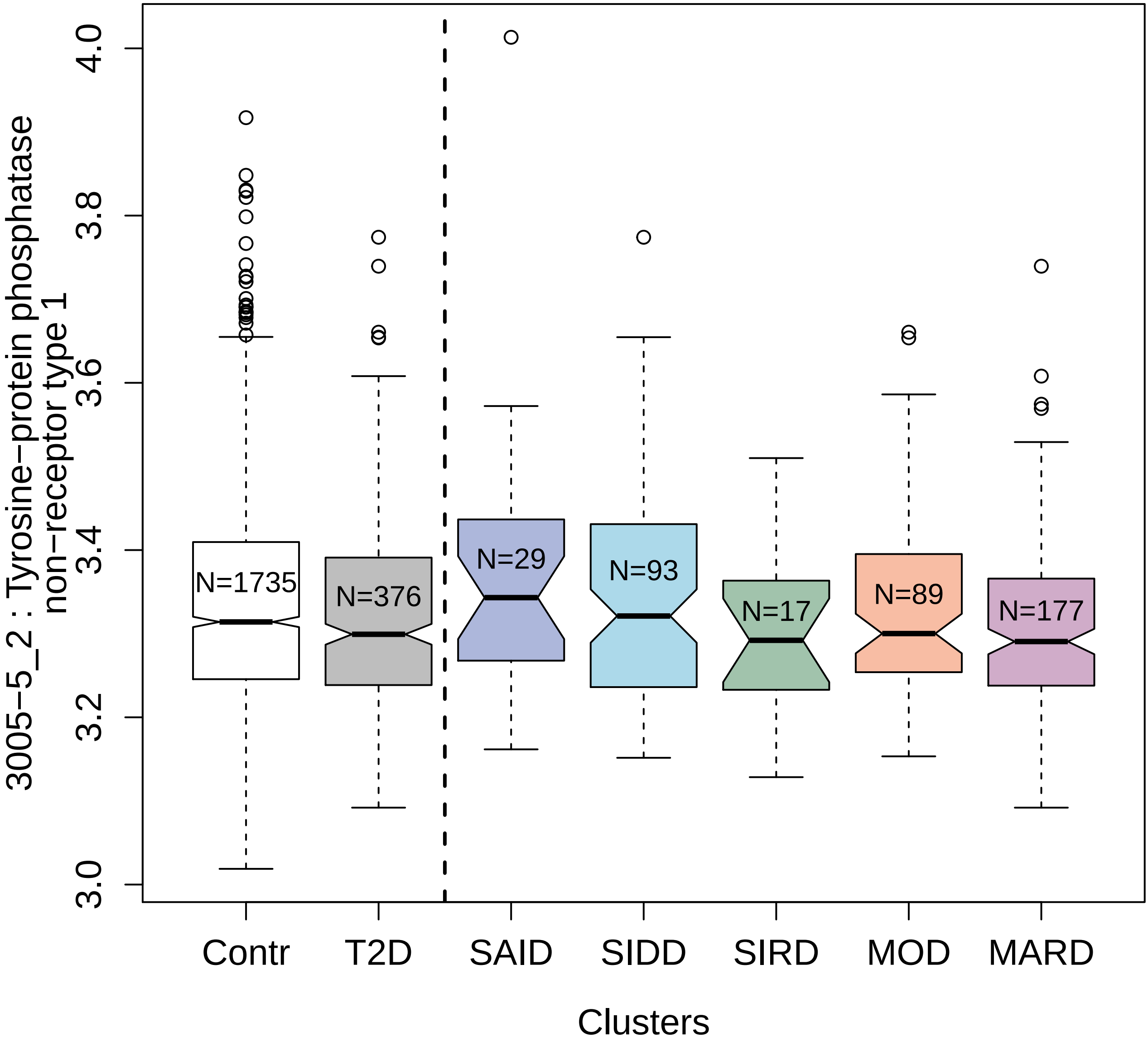
3005−5_2 : Tyrosine−protein phosphatase
non−receptor type 1
4.0
3.8
3.6
3.4
3.2
3.0
N=1735
N=376
N=29
N=93
N=17
N=89
N=177
Contr
T2D
SAID
SIDD
SIRD
MOD
MARD
Clusters

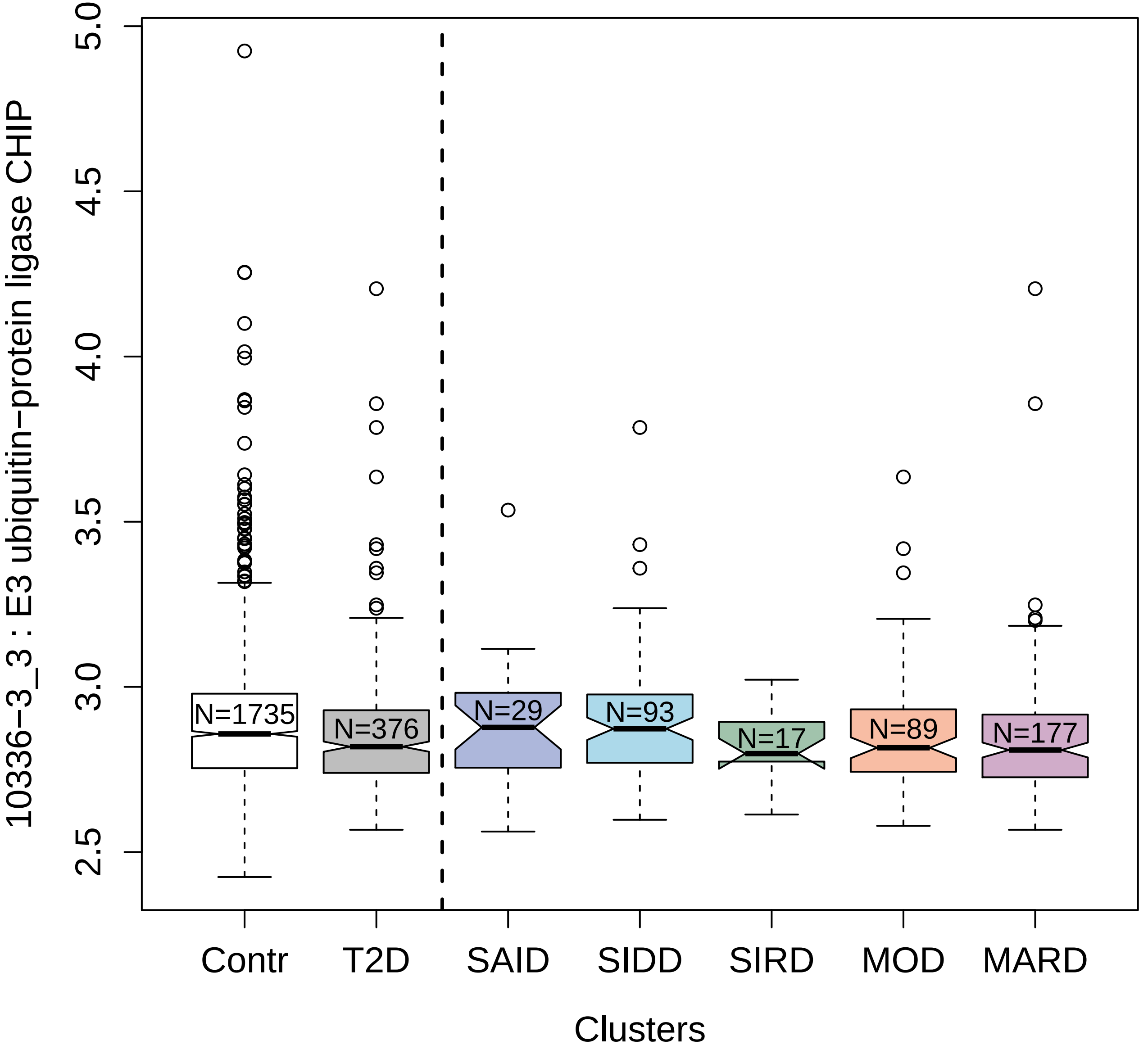

10336−3_3 : E3 ubiquitin−protein ligase CHIP
2.5
3.0
3.5
4.0
4.5
5.0
N=1735
N=376
N=29
N=93
N=17
N=89
N=177
Contr
T2D
SAID
SIDD
SIRD
MOD
MARD
Clusters

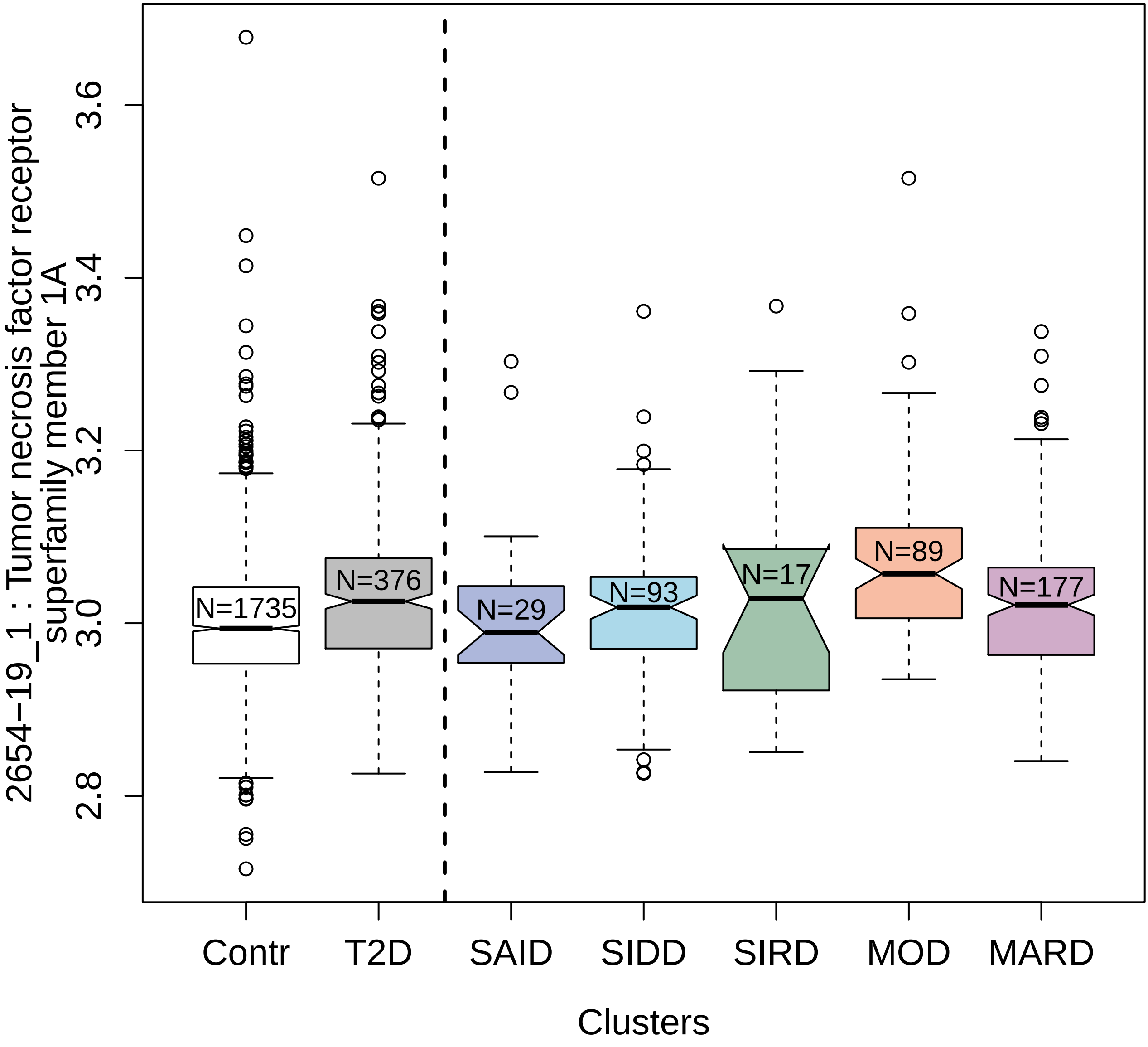

2654−19_1 : Tumor necrosis factor receptor
superfamily member 1A
3.6
3.4
3.2
3.0
2.8
N=1735
N=376
N=29
N=93
N=17
N=89
N=177
Contr
T2D
SAID
SIDD
SIRD
MOD
MARD
Clusters

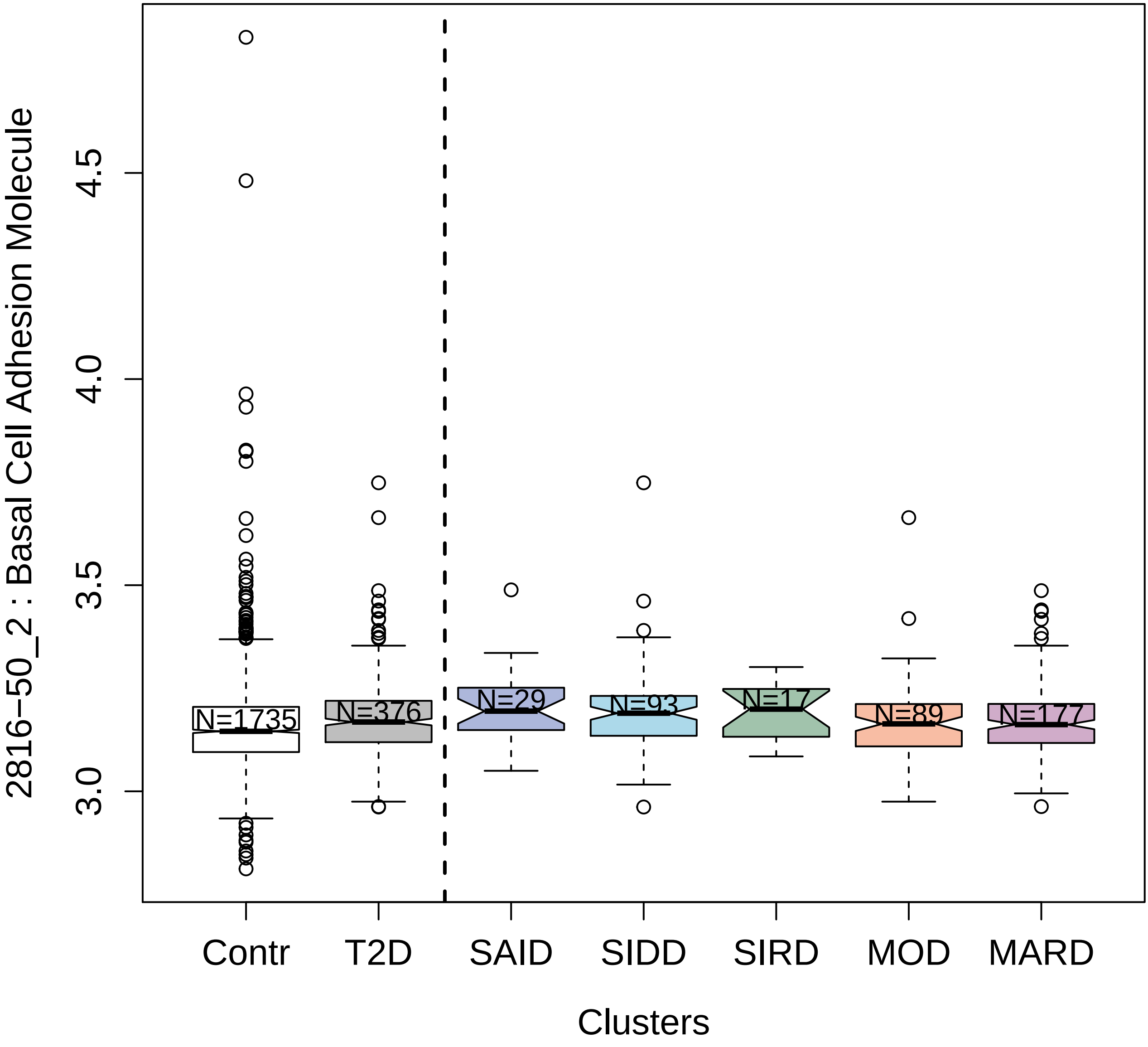
2816−50_2 : Basal Cell Adhesion Molecule
3.0
3.5
4.0
4.5
N=1735
N=376
N=29
N=93
N=17
N=89
N=177
Contr
T2D
SAID
SIDD
SIRD
MOD
MARD
Clusters

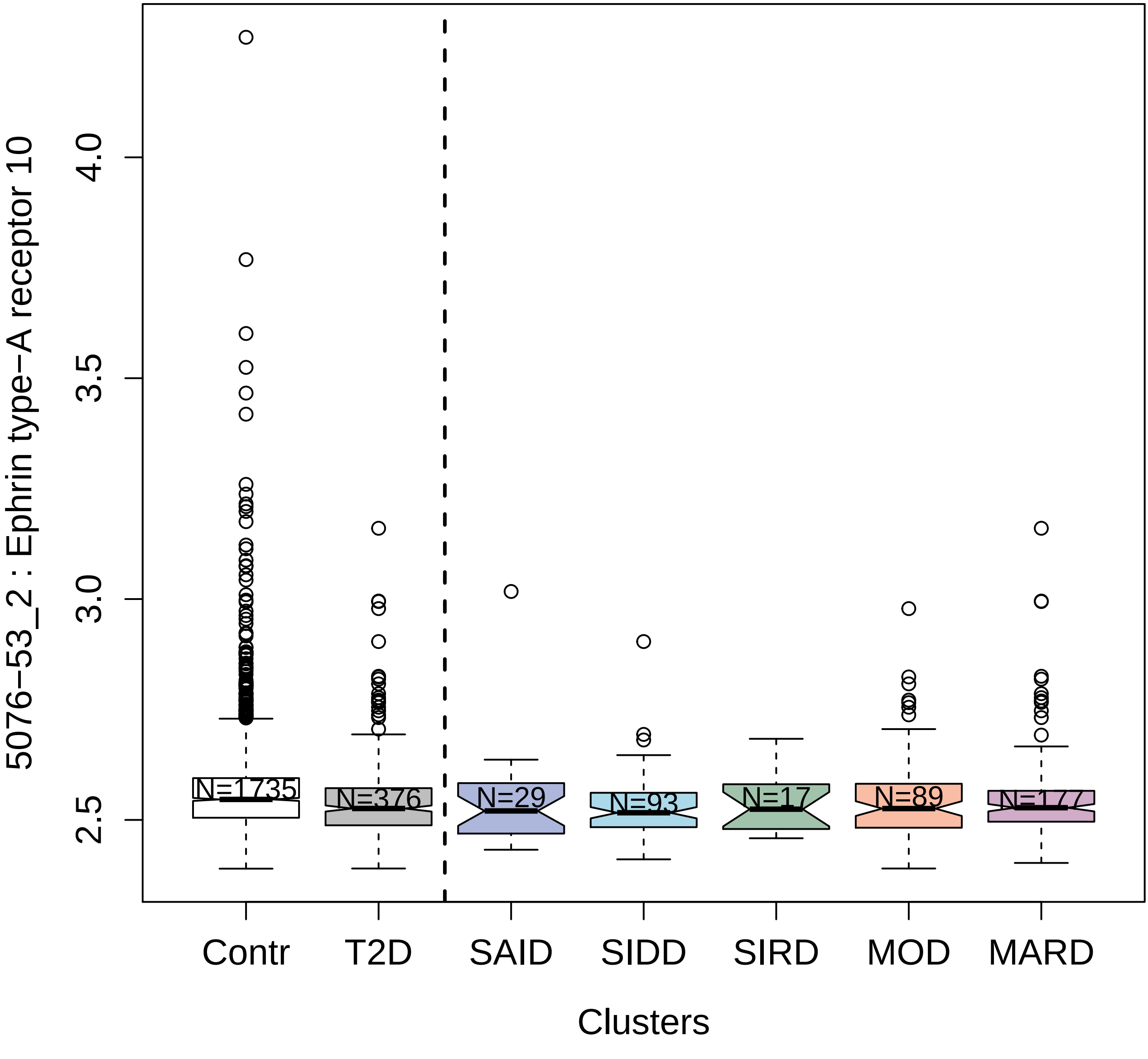

5076−53_2 : Ephrin type−A receptor 10
4.0
3.5
3.0
2.5
N=1735
N=376
N=29
N=93
N=17
N=89
N=177
Contr
T2D
SAID
SIDD
SIRD
MOD
MARD
Clusters

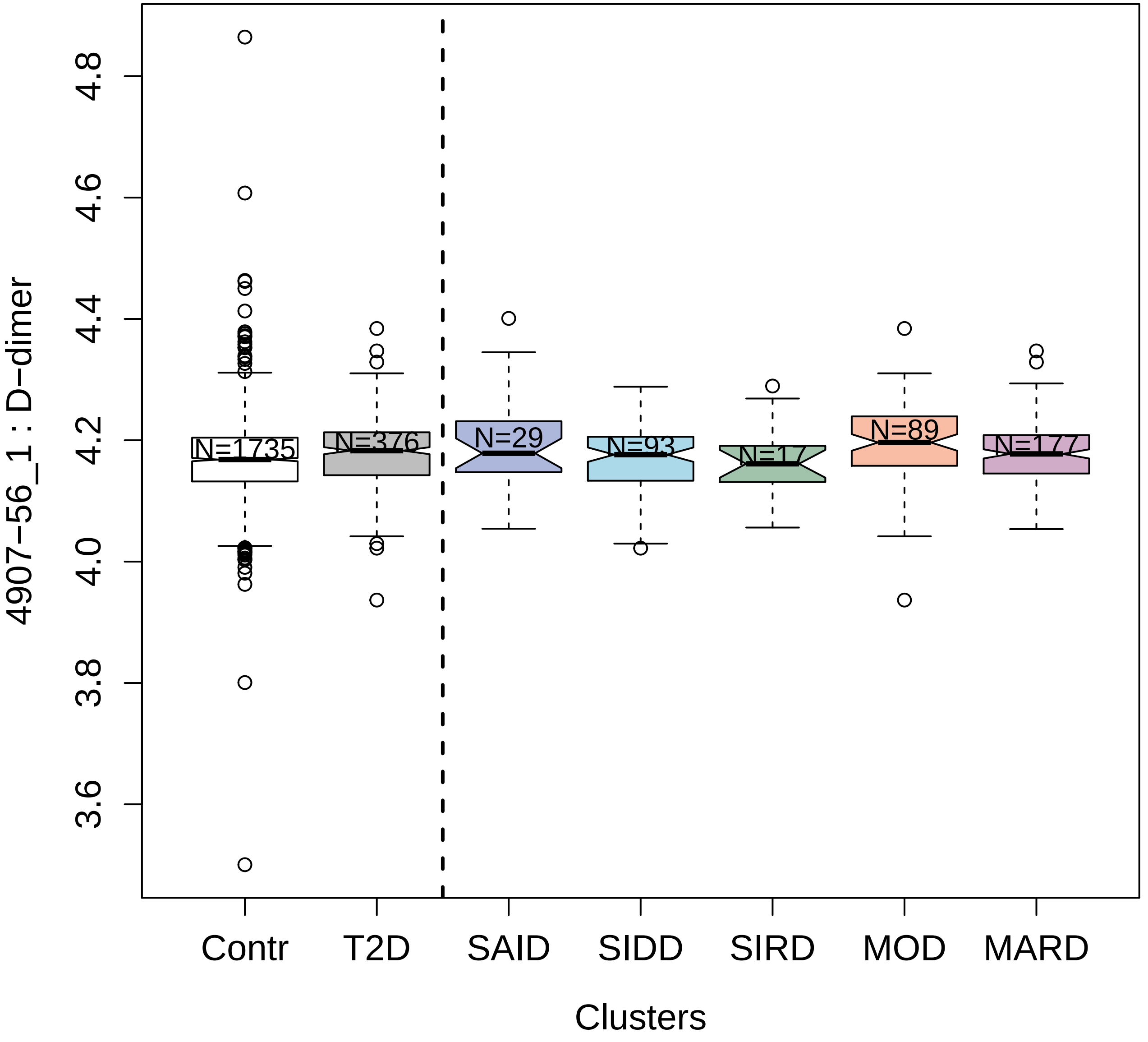
4907−56_1 : D−dimer
4.8
4.6
4.4
4.2
4.0
3.8
3.6
N=1735
N=376
N=29
N=93
N=17
N=89
N=177
Contr
T2D
SAID
SIDD
SIRD
MOD
MARD
Clusters

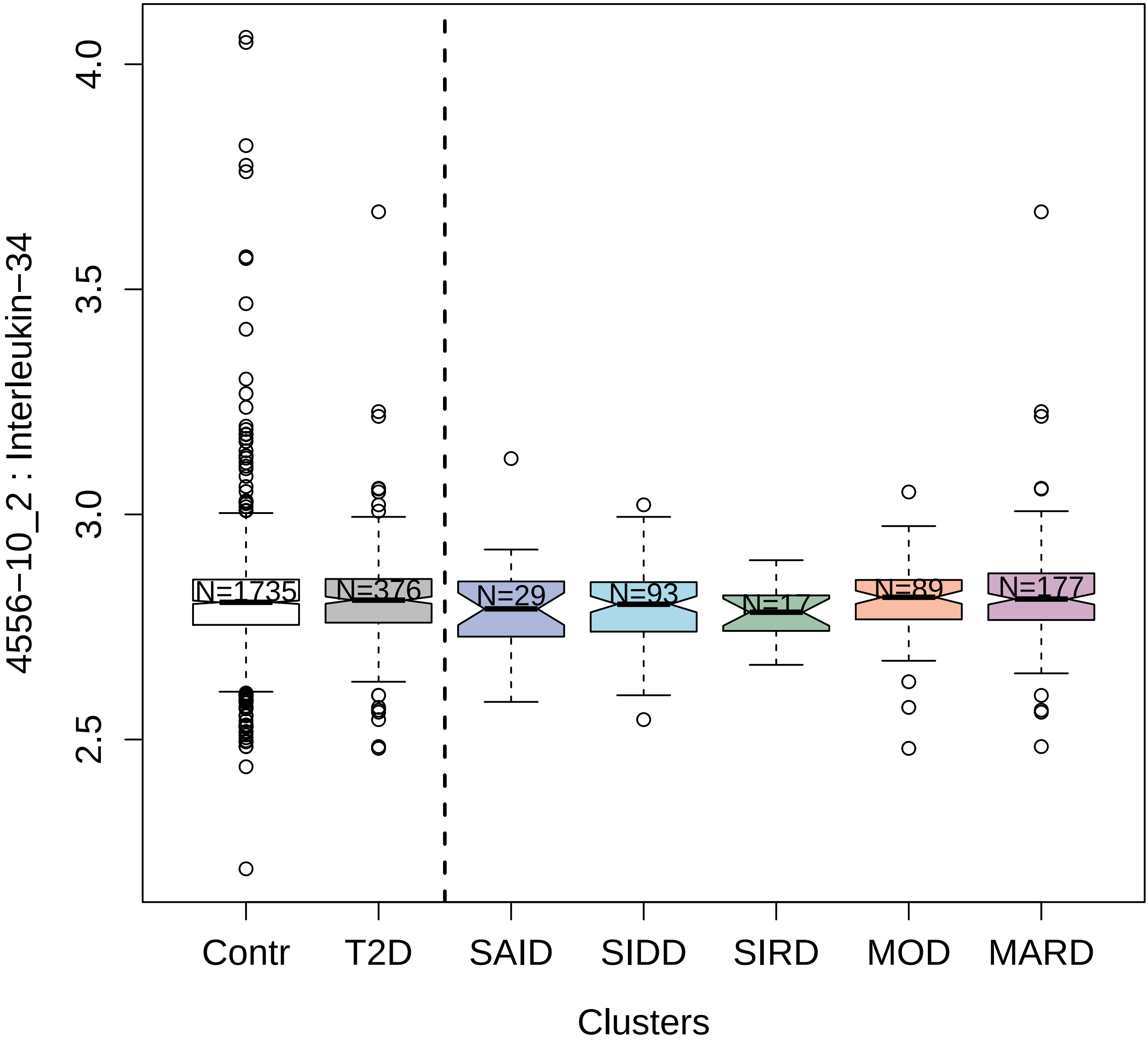
4556−10_2 : Interleukin−34
4.0
3.5
3.0
2.5
N=1735
N=376
N=29
N=93
N=17
N=89
N=177
Contr
T2D
SAID
SIDD
SIRD
MOD
MARD
Clusters

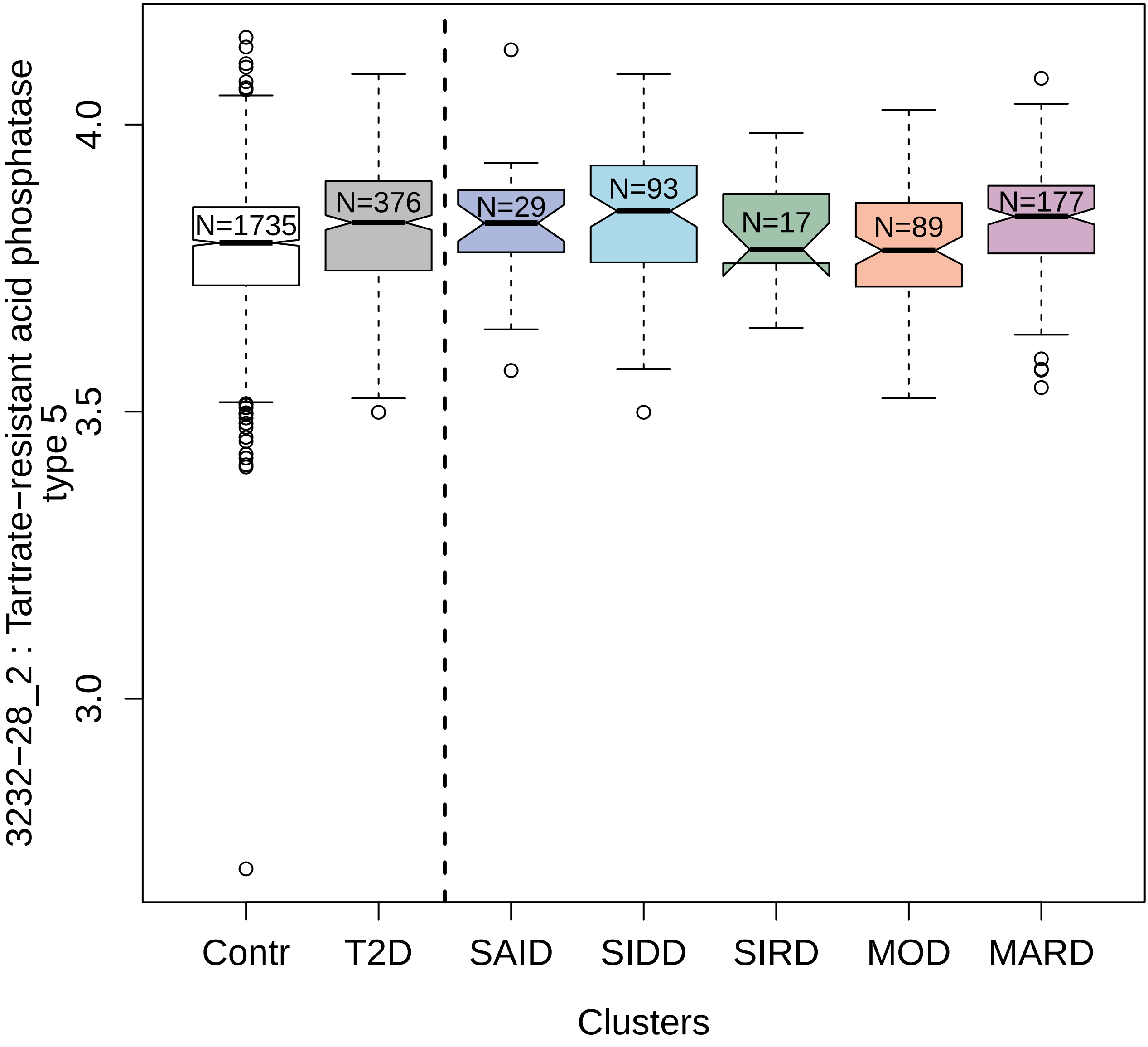

3232−28_2 : Tartrate−resistant acid phosphatase
type 5
4.0
3.5
3.0
N=1735
N=376
N=29
N=93
N=17
N=89
N=177
Contr
T2D
SAID
SIDD
SIRD
MOD
MARD
Clusters

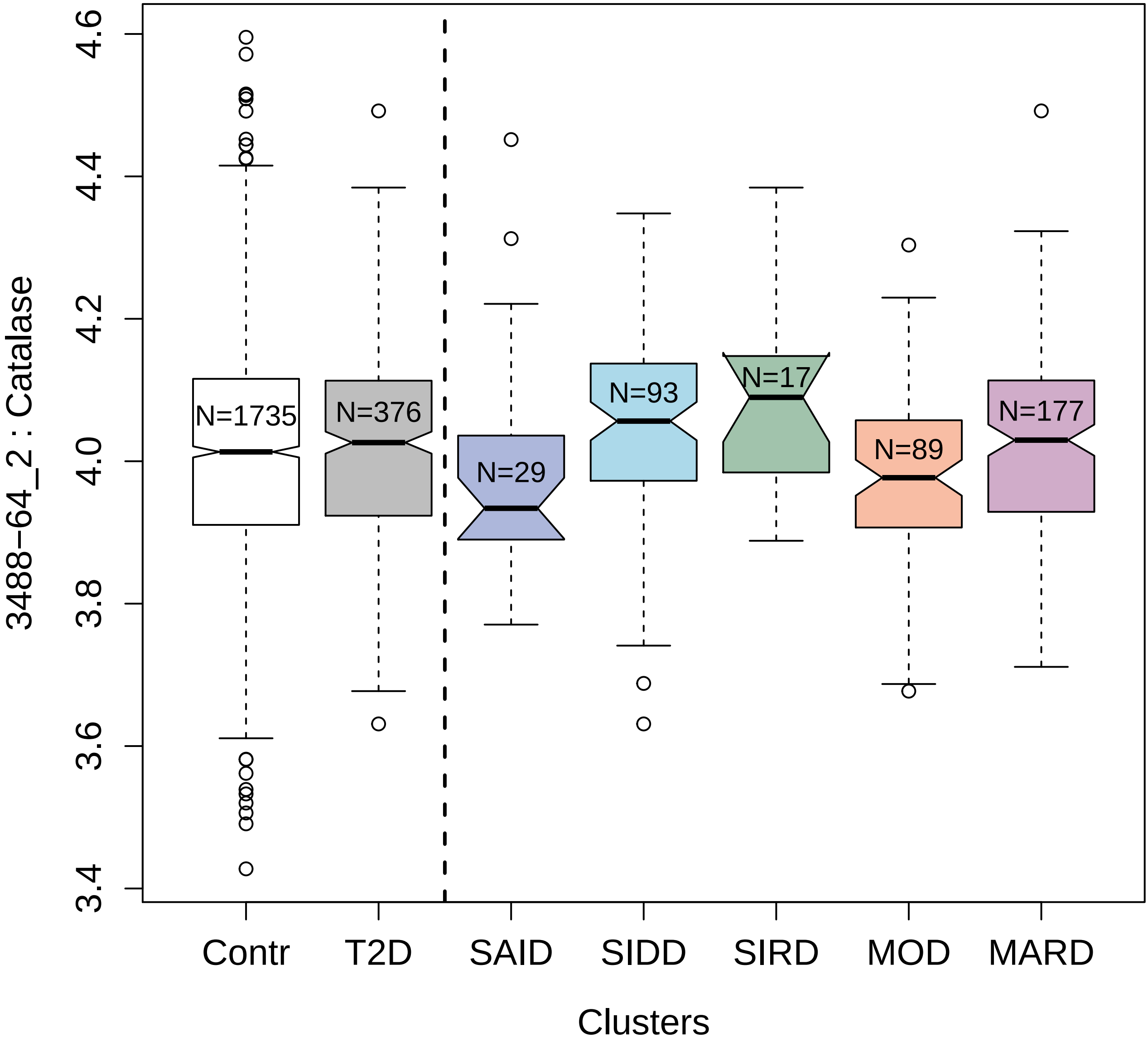

3488−64_2 : Catalase
Clusters
Contr
T2D
SAID
SIDD
SIRD
MOD
MARD
N=1735
N=376
N=29
N=93
N=17
N=89
N=177
3.4
3.6
3.8
4.0
4.2
4.4
4.6

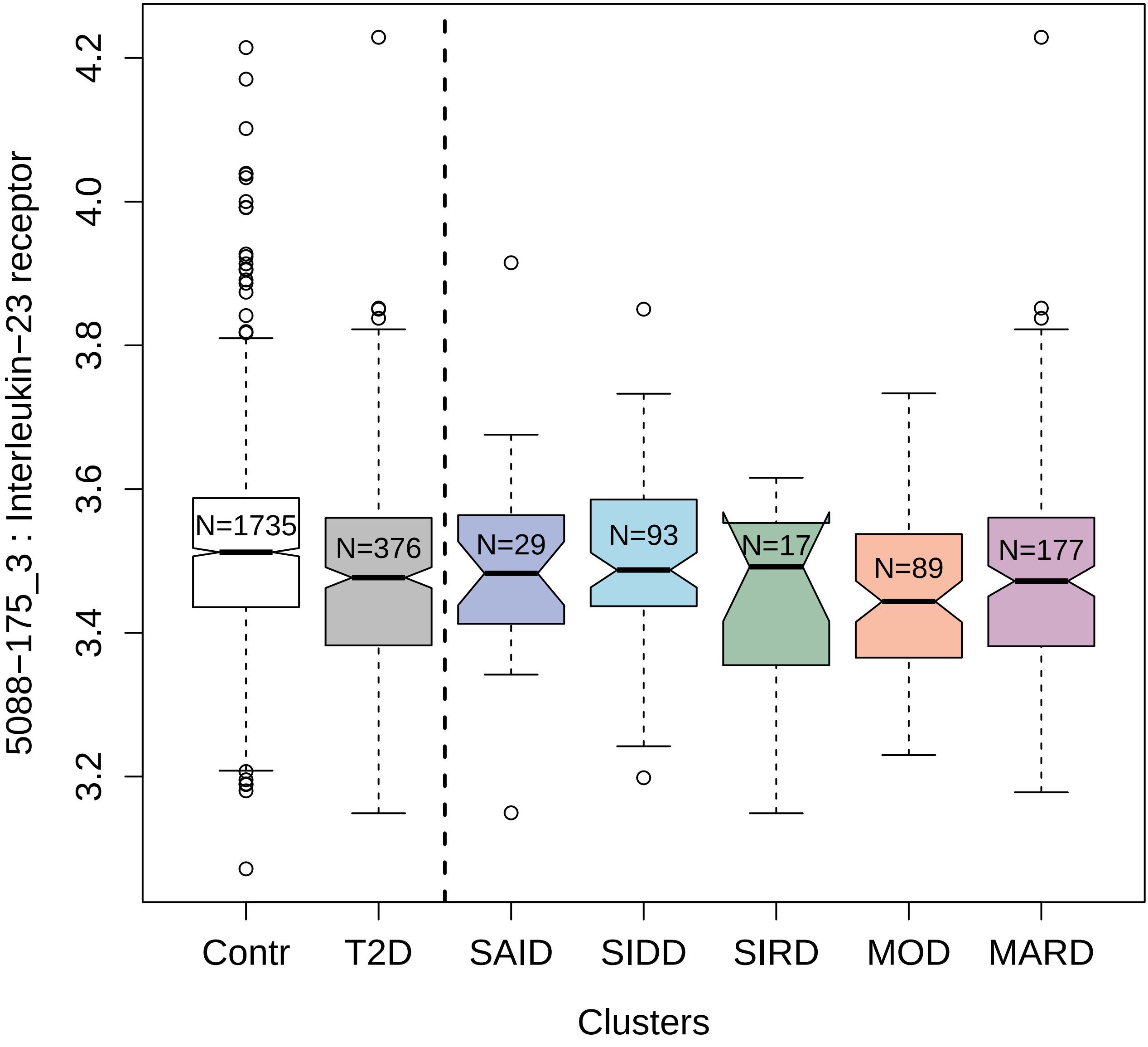
5088−175_3 : Interleukin−23 receptor
4.2
4.0
3.8
3.6
3.4
3.2
N=1735
N=376
N=29
N=93
N=17
N=89
N=177
Contr
T2D
SAID
SIDD
SIRD
MOD
MARD
Clusters

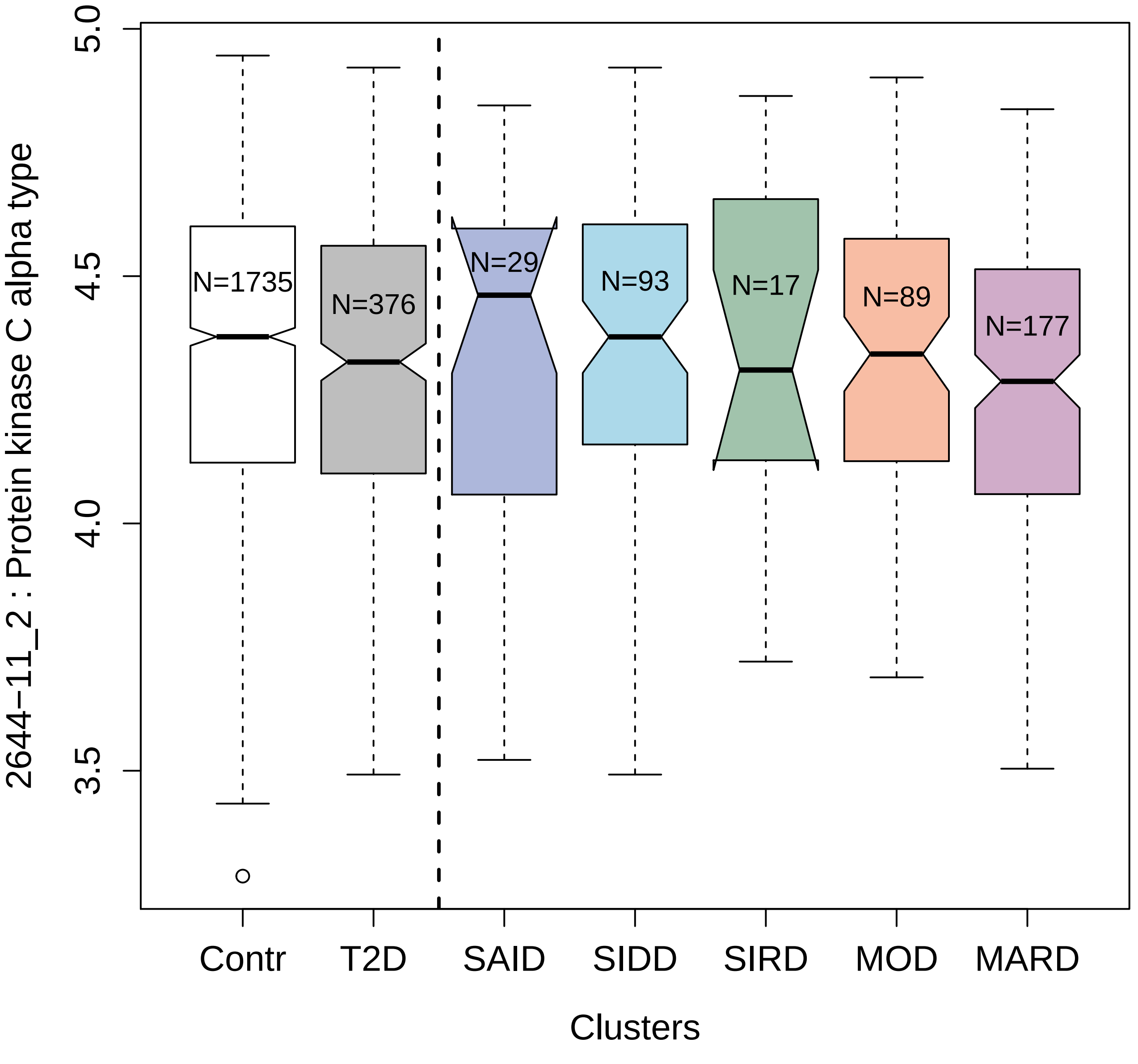

2644−11_2 : Protein kinase C alpha type
5.0
4.5
4.0
3.5
N=1735
N=376
N=29
N=93
N=17
N=89
N=177
Contr
T2D
SAID
SIDD
SIRD
MOD
MARD
Clusters

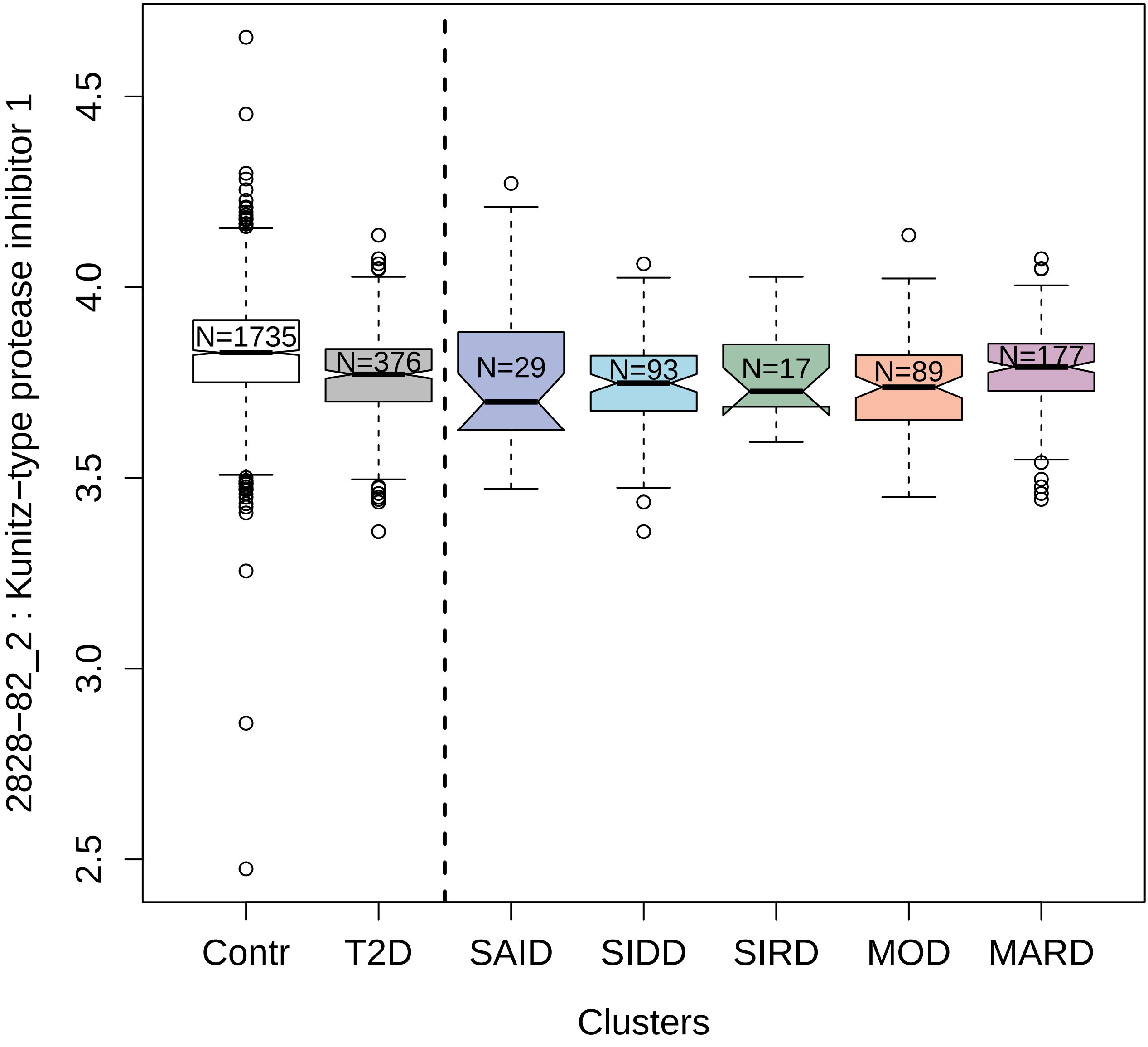
2828−82_2 : Kunitz−type protease inhibitor 1
4.5
4.0
3.5
3.0
2.5
N=1735
N=376
N=29
N=93
N=17
N=89
N=177
Contr
T2D
SAID
SIDD
SIRD
MOD
MARD
Clusters

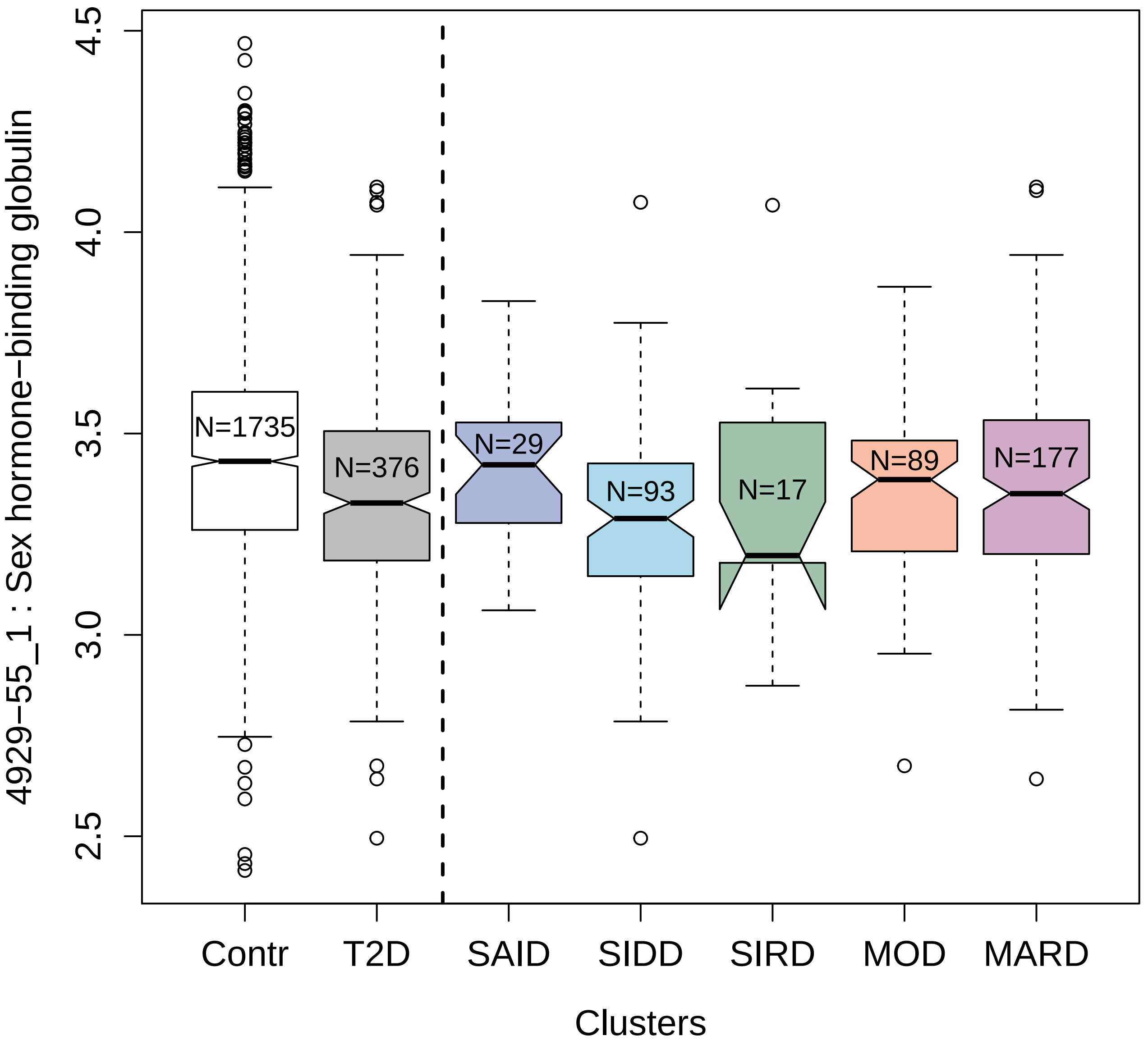
4929−55_1 : Sex hormone−binding globulin
4.5
4.0
3.5
3.0
2.5
N=1735
N=376
N=29
N=93
N=17
N=89
N=177
Contr
T2D
SAID
SIDD
SIRD
MOD
MARD
Clusters

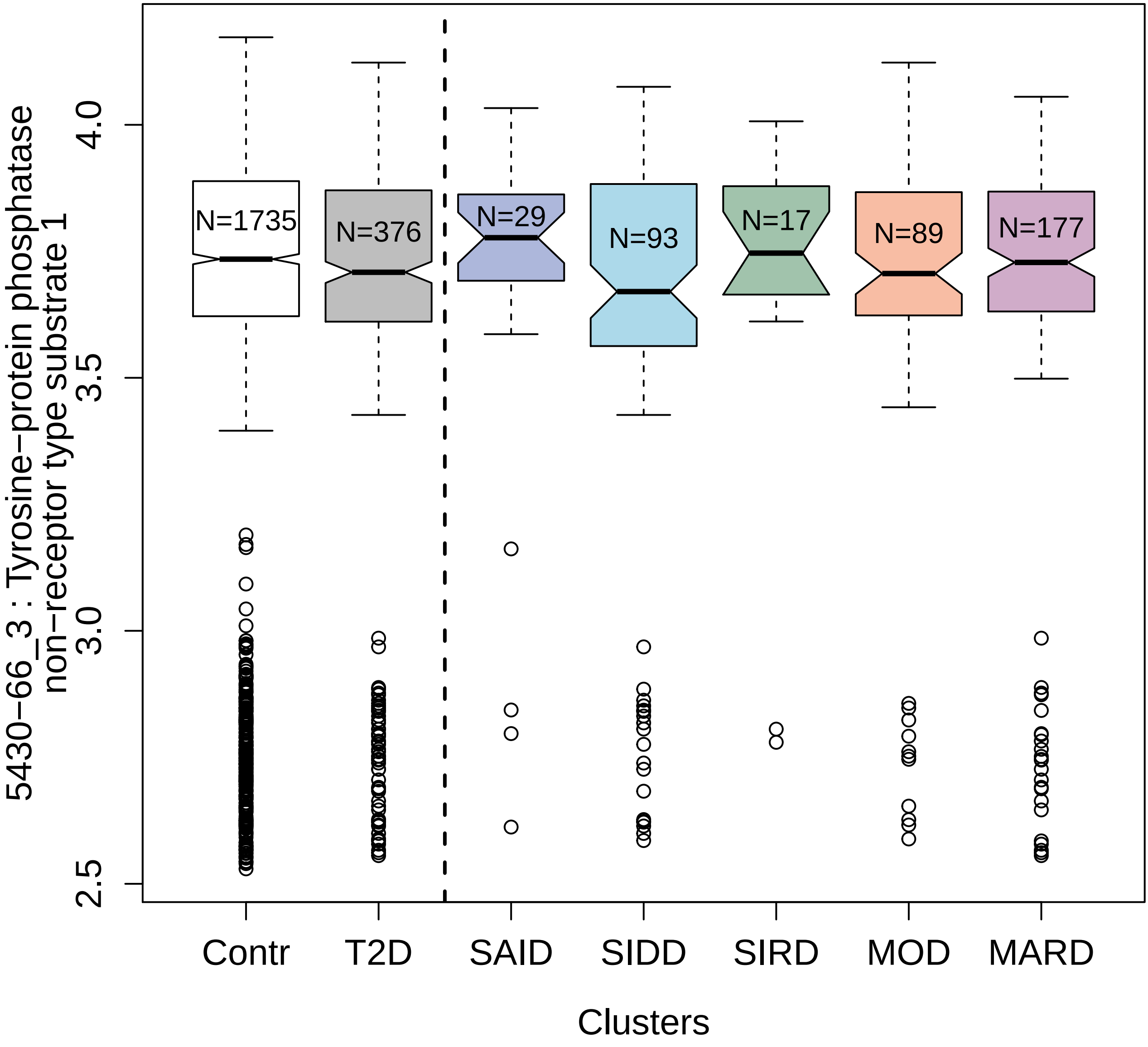
5430−66_3 : Tyrosine−protein phosphatase
non−receptor type substrate 1
2.5
3.0
3.5
4.0
N=1735
N=376
N=29
N=93
N=17
N=89
N=177
Contr
T2D
SAID
SIDD
SIRD
MOD
MARD
Clusters

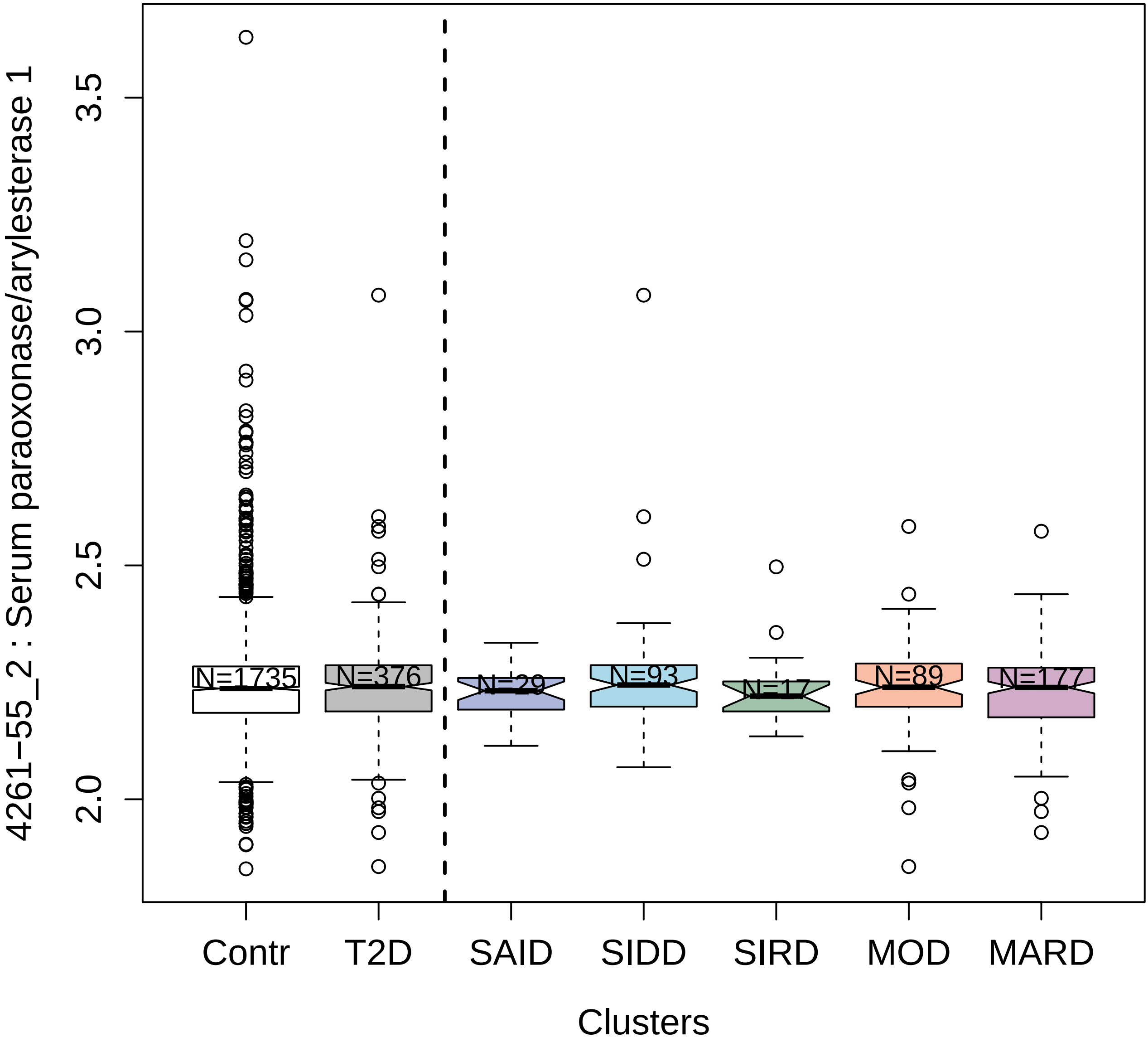

4261−55_2 : Serum paraoxonase/arylesterase 1
2.0
2.5
3.0
3.5
N=1735
N=376
N=29
N=93
N=17
N=89
N=177
Contr
T2D
SAID
SIDD
SIRD
MOD
MARD
Clusters

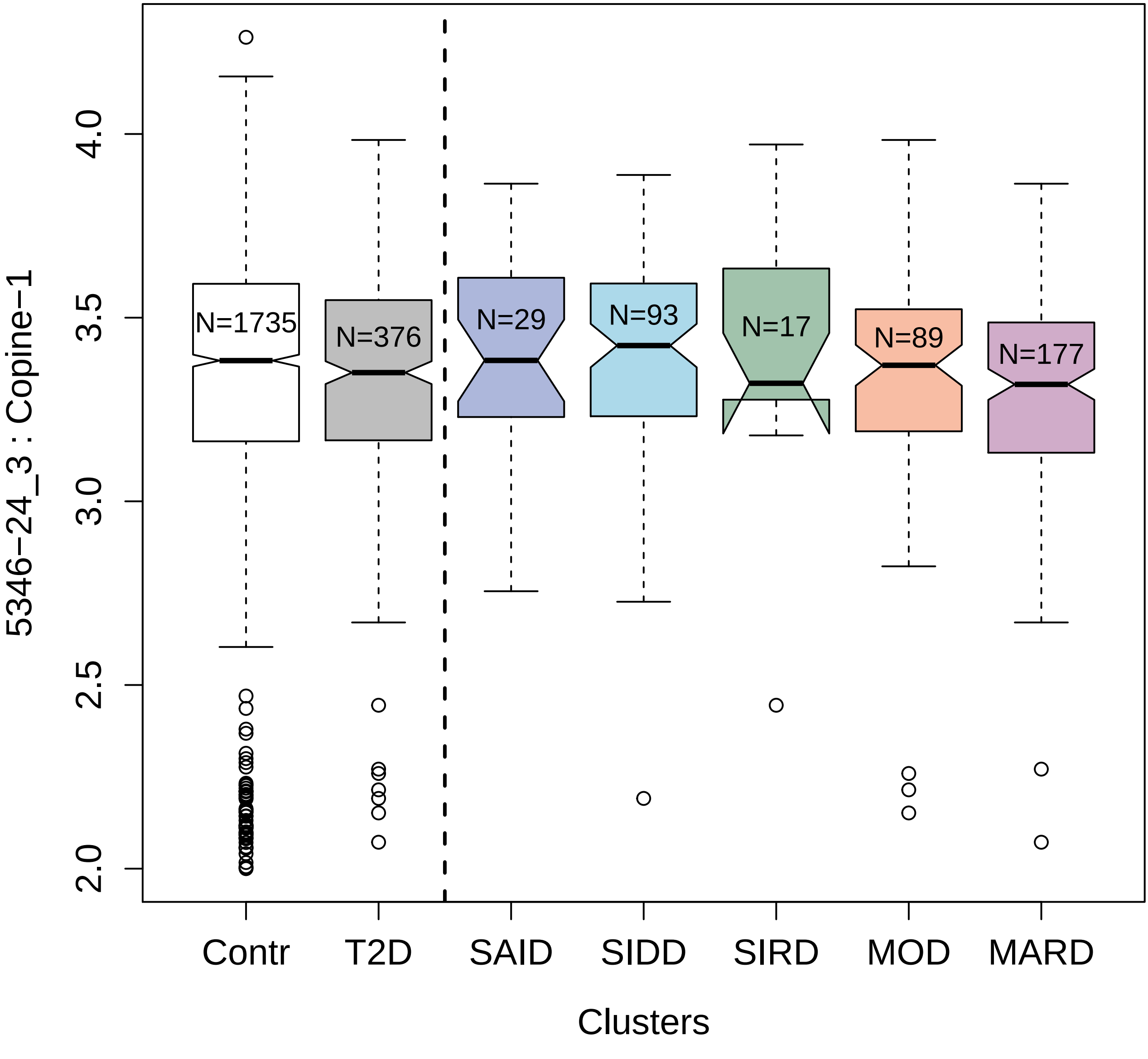

5346−24_3 : Copine−1
4.0
3.5
3.0
2.5
2.0
N=1735
N=376
N=29
N=93
N=17
N=89
N=177
Contr
T2D
SAID
SIDD
SIRD
MOD
MARD
Clusters

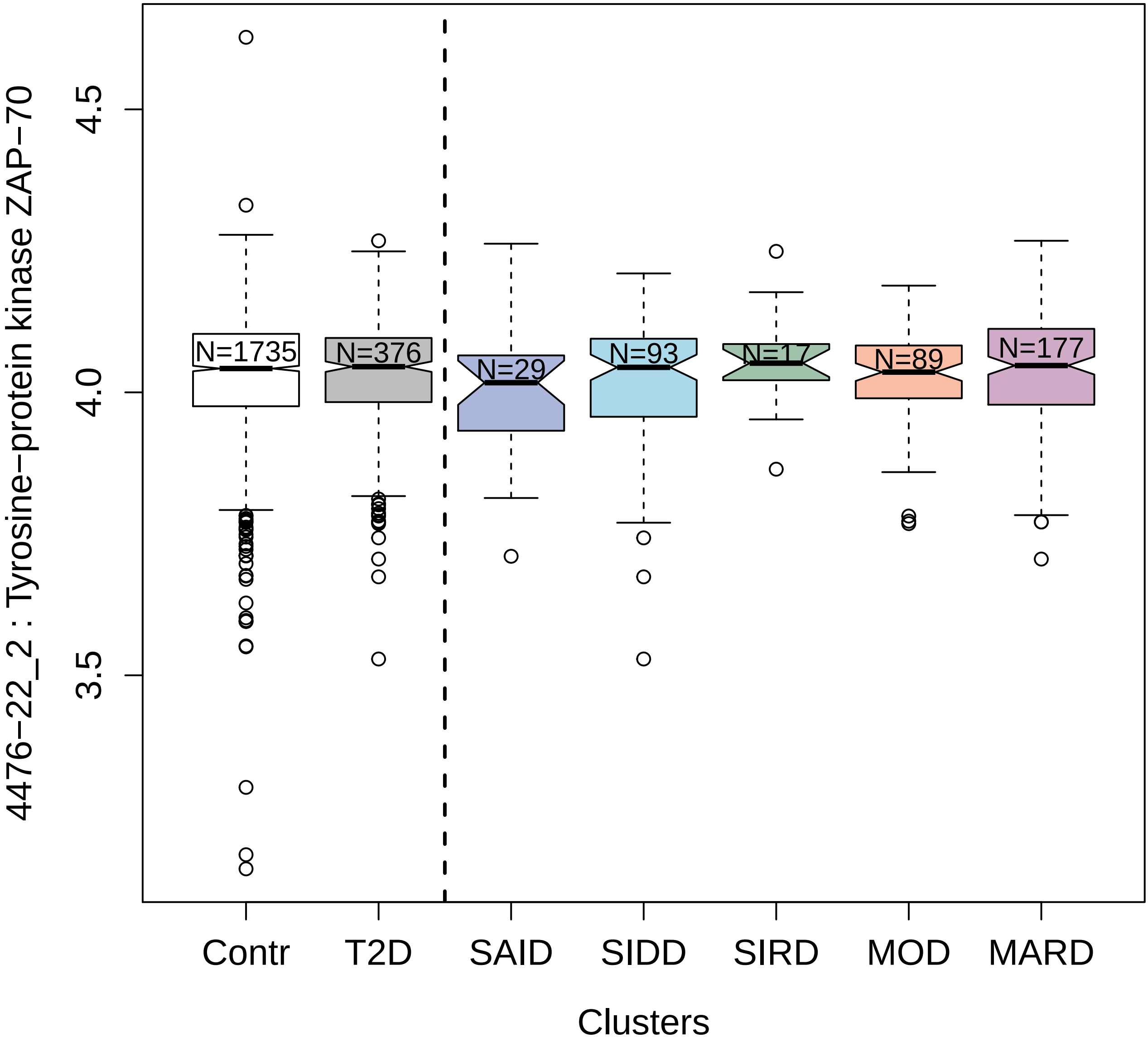
4476−22_2 : Tyrosine−protein kinase ZAP−70
4.5
4.0
3.5
N=1735
N=376
N=29
N=93
N=17
N=89
N=177
Contr
T2D
SAID
SIDD
SIRD
MOD
MARD
Clusters

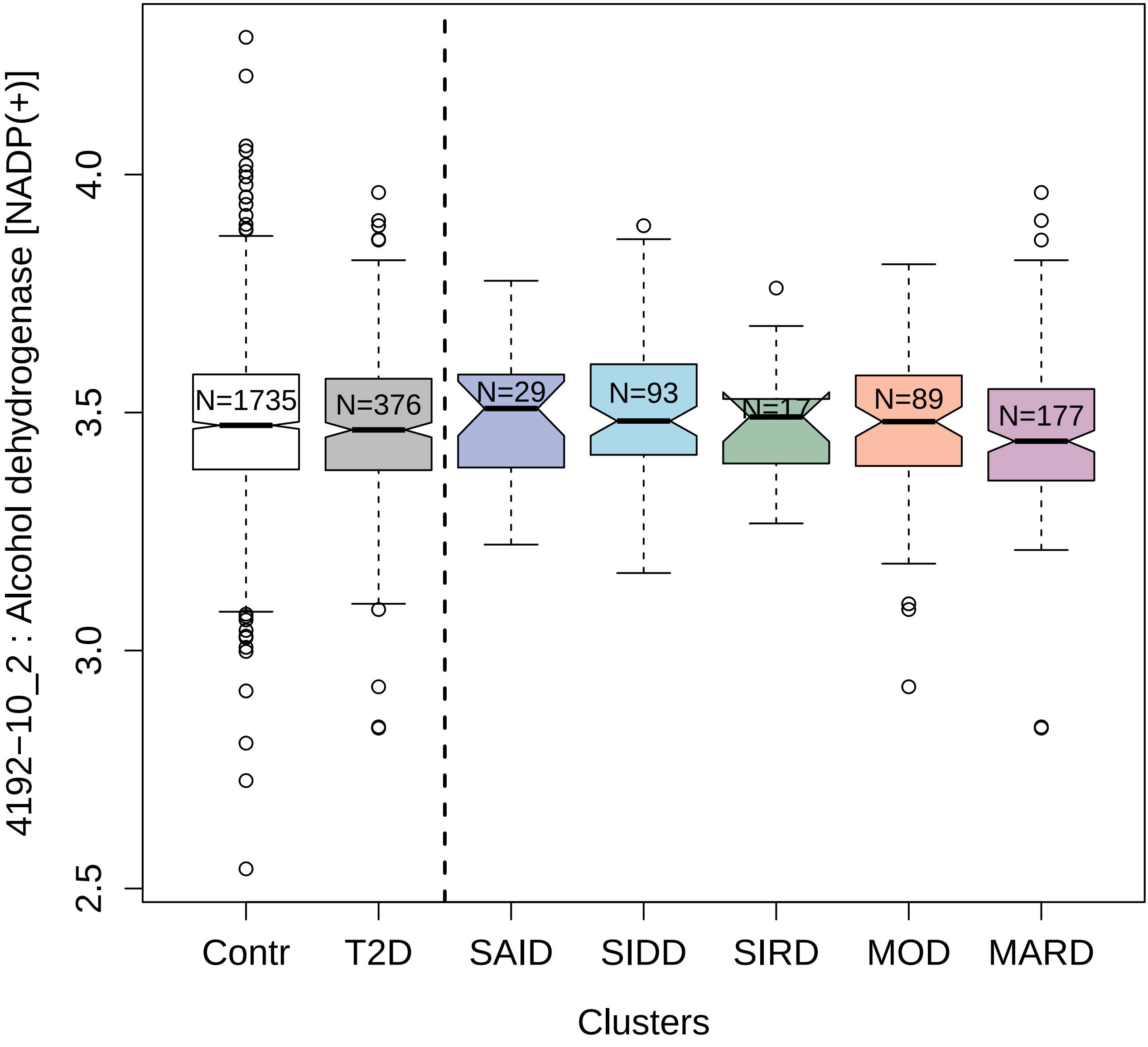

4192−10_2 : Alcohol dehydrogenase [NADP(+)]
2.5
3.0
3.5
4.0
N=1735
N=376
N=29
N=93
N=17
N=89
N=177
Contr
T2D
SAID
SIDD
SIRD
MOD
MARD
Clusters

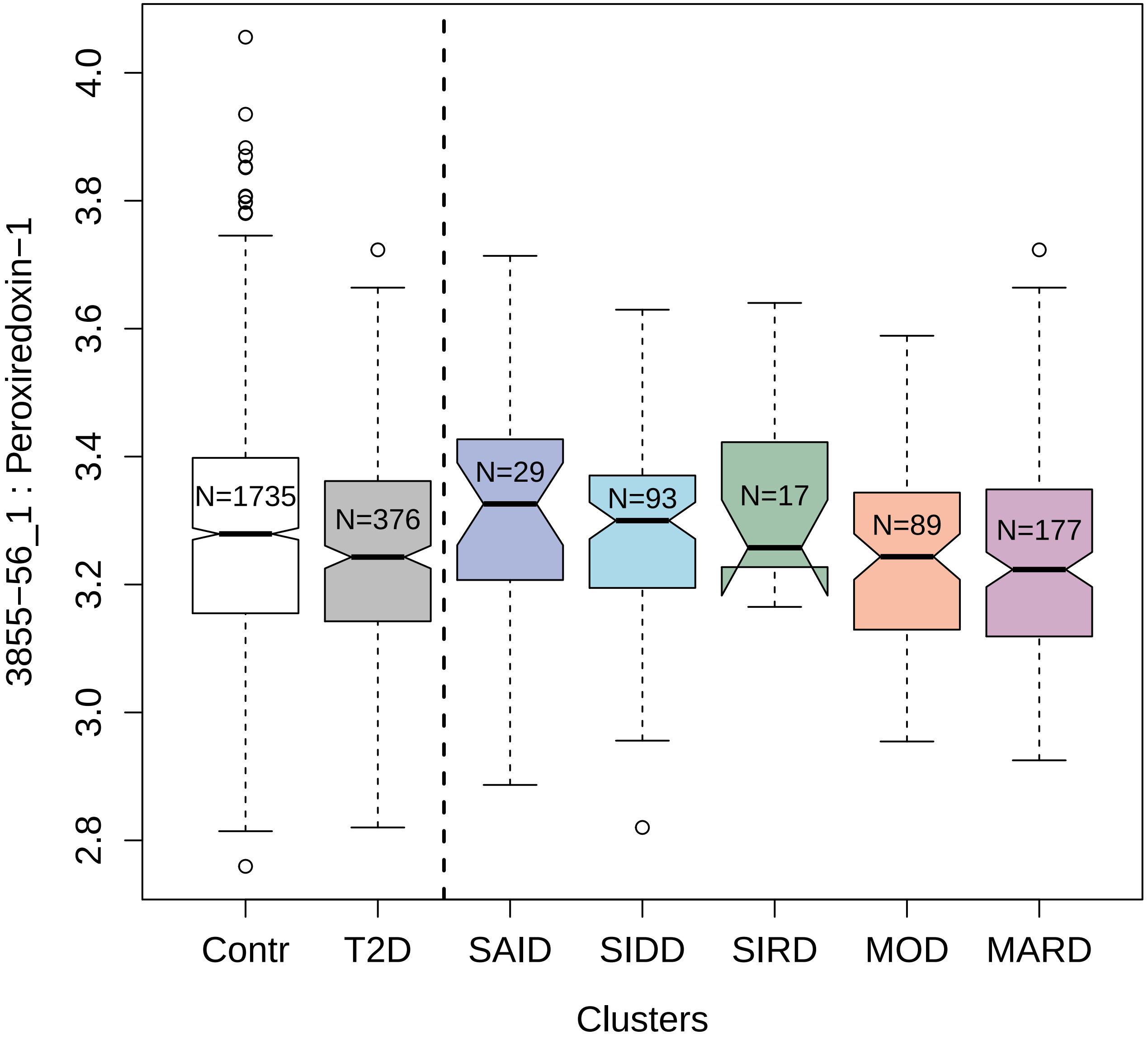

3855−56_1 : Peroxiredoxin−1
4.0
3.8
3.6
3.4
3.2
3.0
2.8
N=1735
N=376
N=29
N=93
N=17
N=89
N=177
Contr
T2D
SAID
SIDD
SIRD
MOD
MARD
Clusters

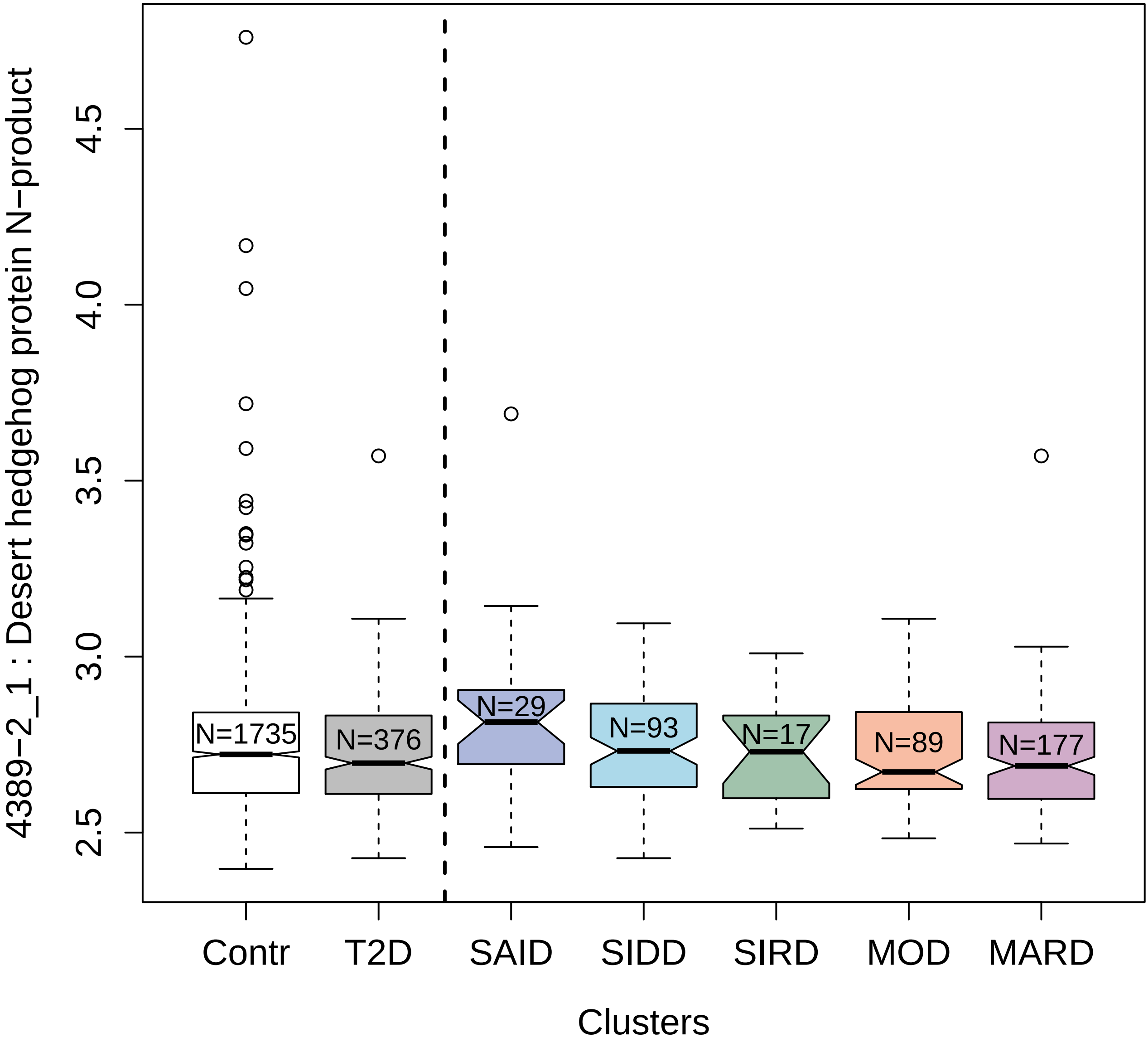
4389−2_1 : Desert hedgehog protein N−product
2.5
3.0
3.5
4.0
4.5
N=1735
N=376
N=29
N=93
N=17
N=89
N=177
Contr
T2D
SAID
SIDD
SIRD
MOD
MARD
Clusters

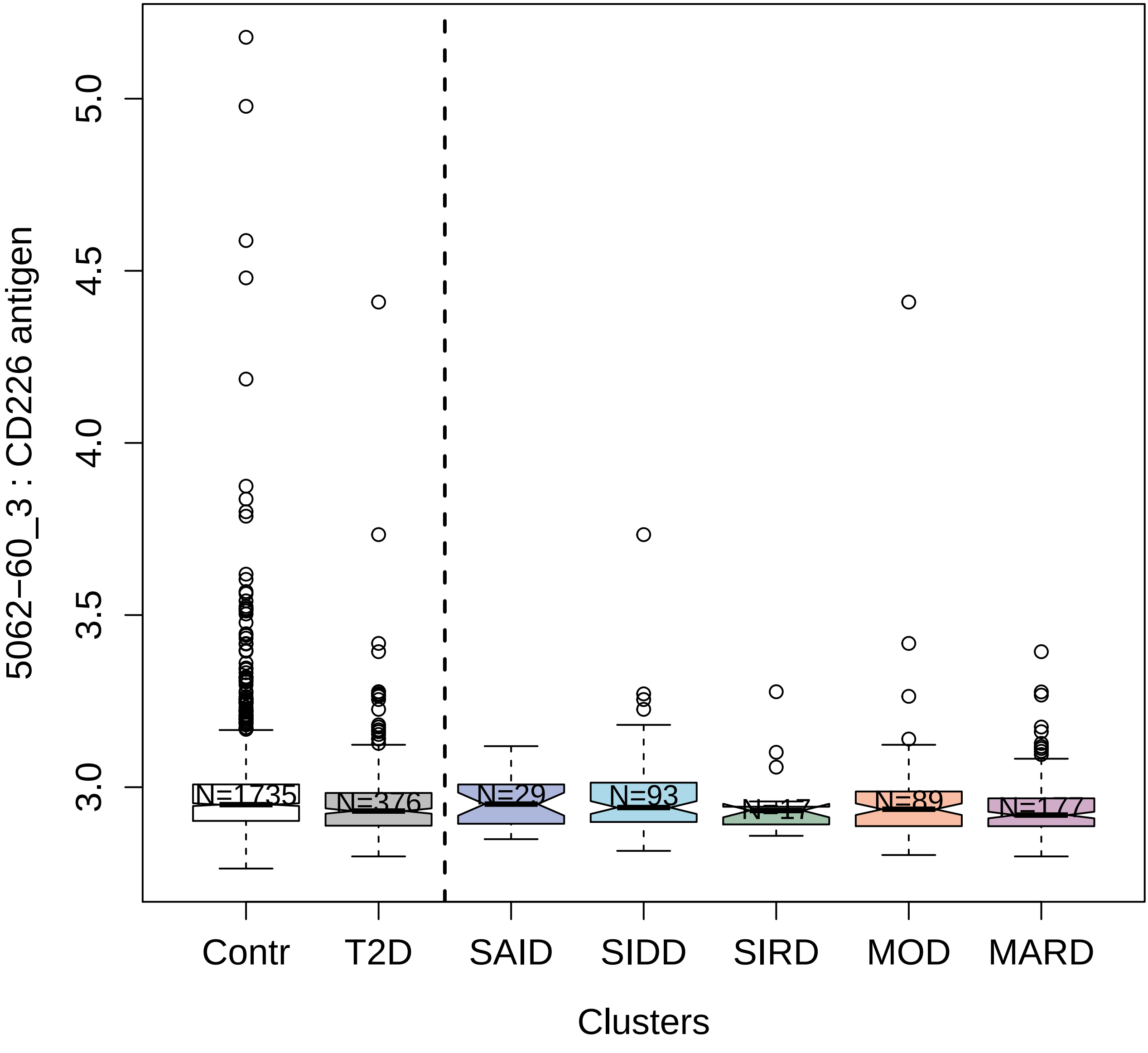

5062−60_3 : CD226 antigen
5.0
4.5
4.0
3.5
3.0
N=1735
N=376
N=29
N=93
N=17
N=89
N=177
Contr
T2D
SAID
SIDD
SIRD
MOD
MARD
Clusters

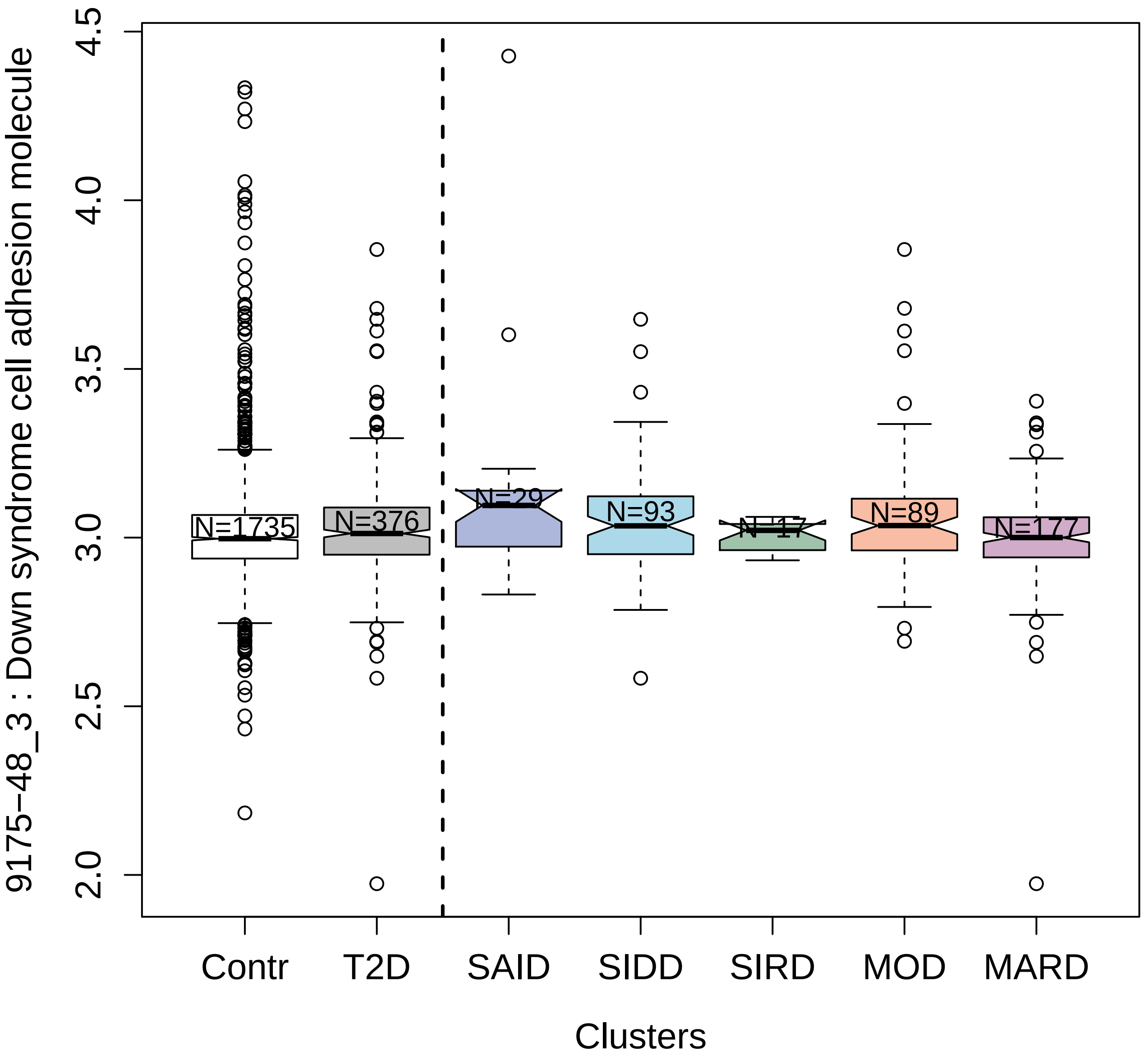
9175−48_3 : Down syndrome cell adhesion molecule
4.5
4.0
3.5
3.0
2.5
2.0
N=1735
N=376
N=29
N=93
N=17
N=89
N=177
Contr
T2D
SAID
SIDD
SIRD
MOD
MARD
Clusters

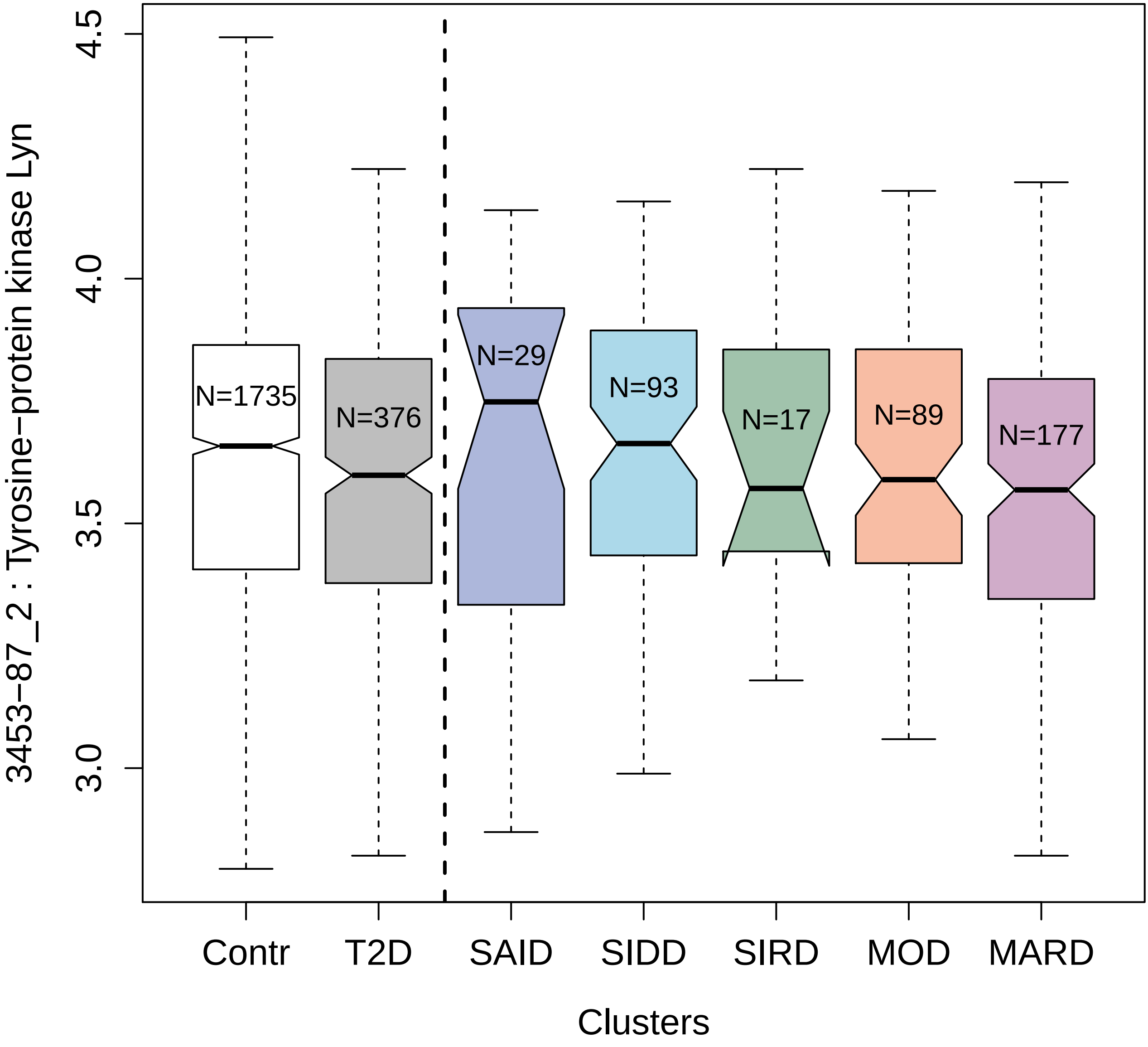

3453−87_2 : Tyrosine−protein kinase Lyn
4.5
4.0
3.5
3.0
N=1735
N=376
N=29
N=93
N=17
N=89
N=177
Contr
T2D
SAID
SIDD
SIRD
MOD
MARD
Clusters

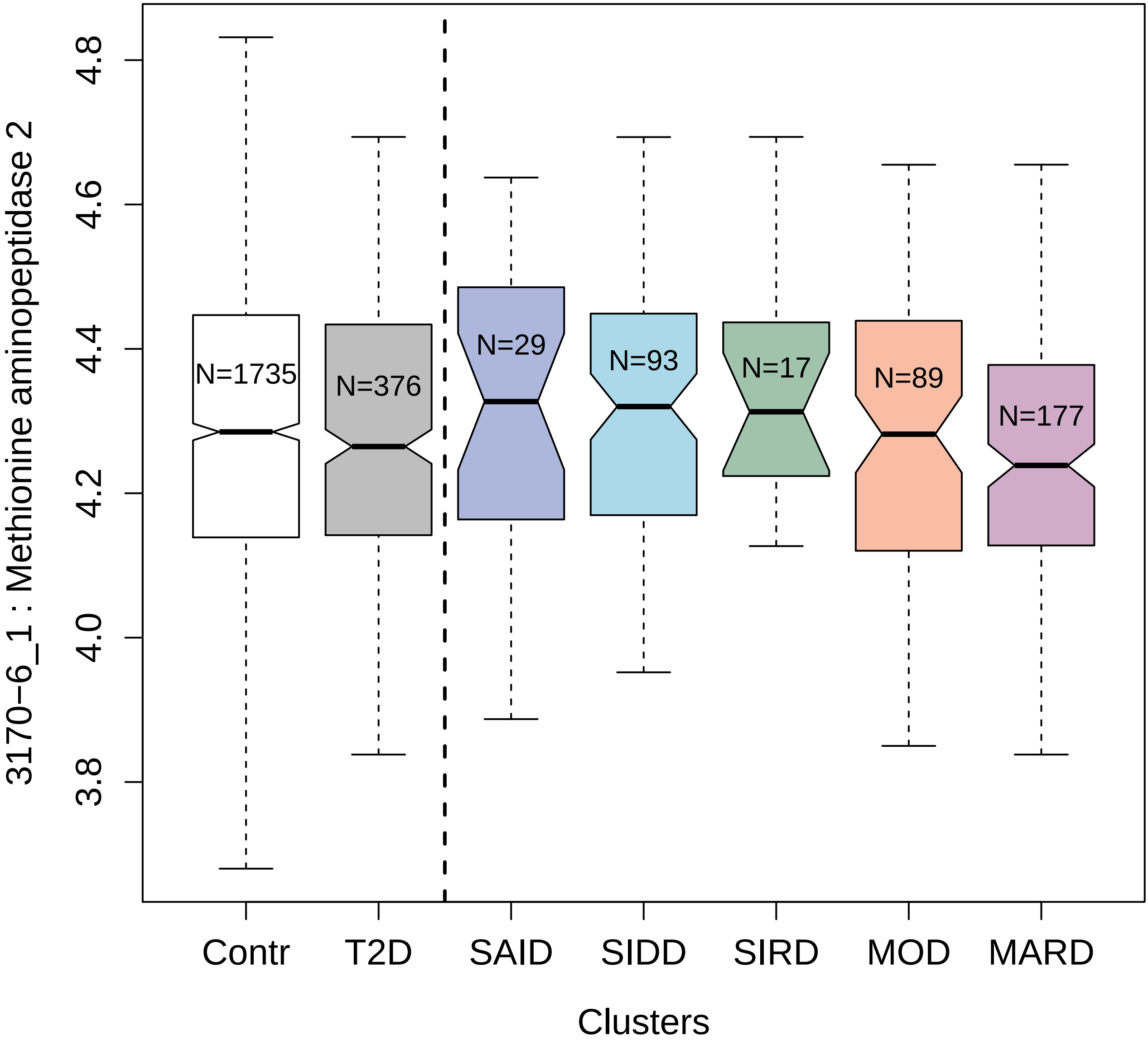

3170−6_1 : Methionine aminopeptidase 2
4.8
4.6
4.4
4.2
4.0
3.8
N=1735
N=376
N=29
N=93
N=17
N=89
N=177
Contr
T2D
SAID
SIDD
SIRD
MOD
MARD
Clusters

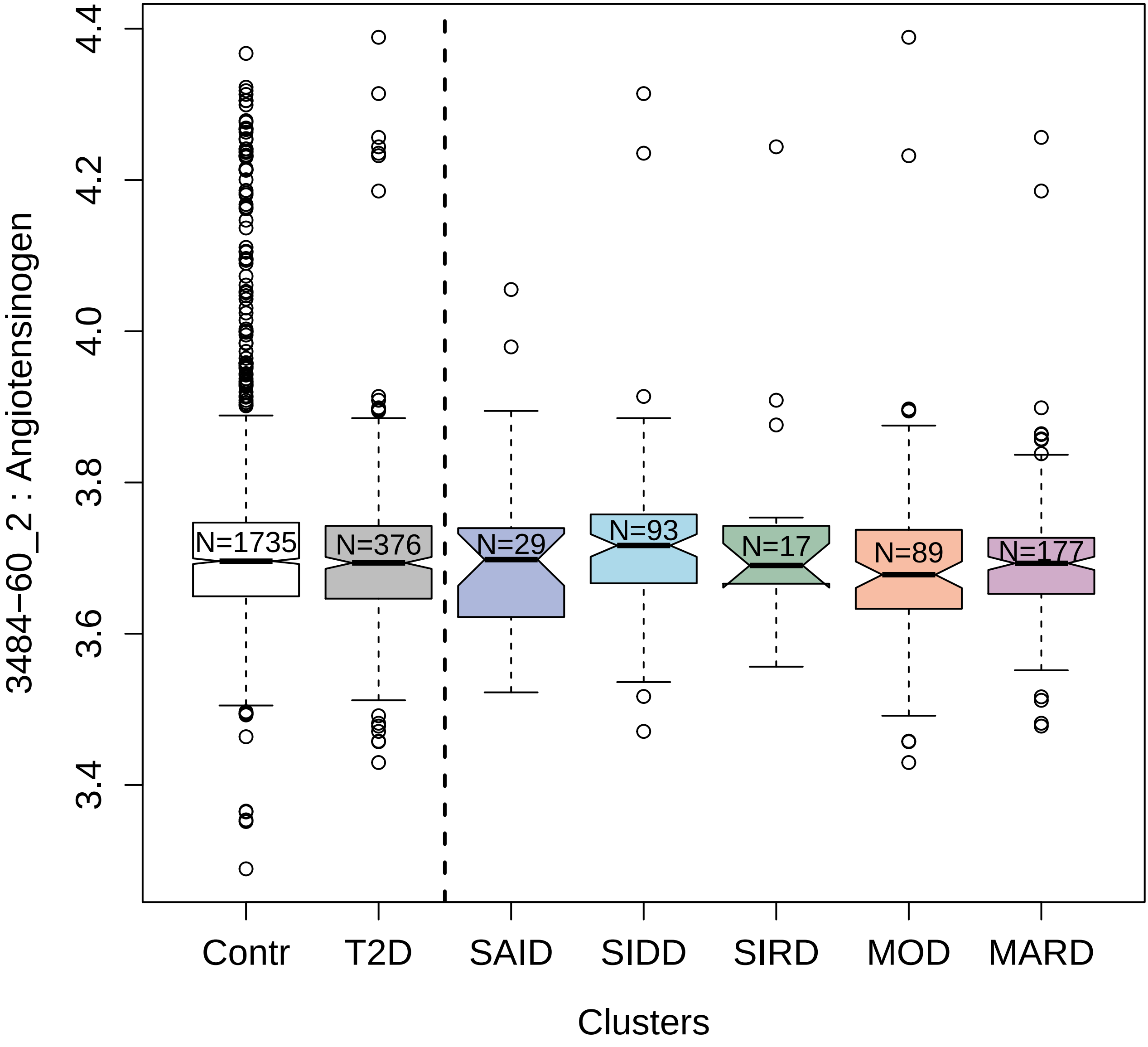

3484−60_2 : Angiotensinogen
4.4
4.2
4.0
3.8
3.6
3.4
N=1735
N=376
N=29
N=93
N=17
N=89
N=177
Contr
T2D
SAID
SIDD
SIRD
MOD
MARD
Clusters

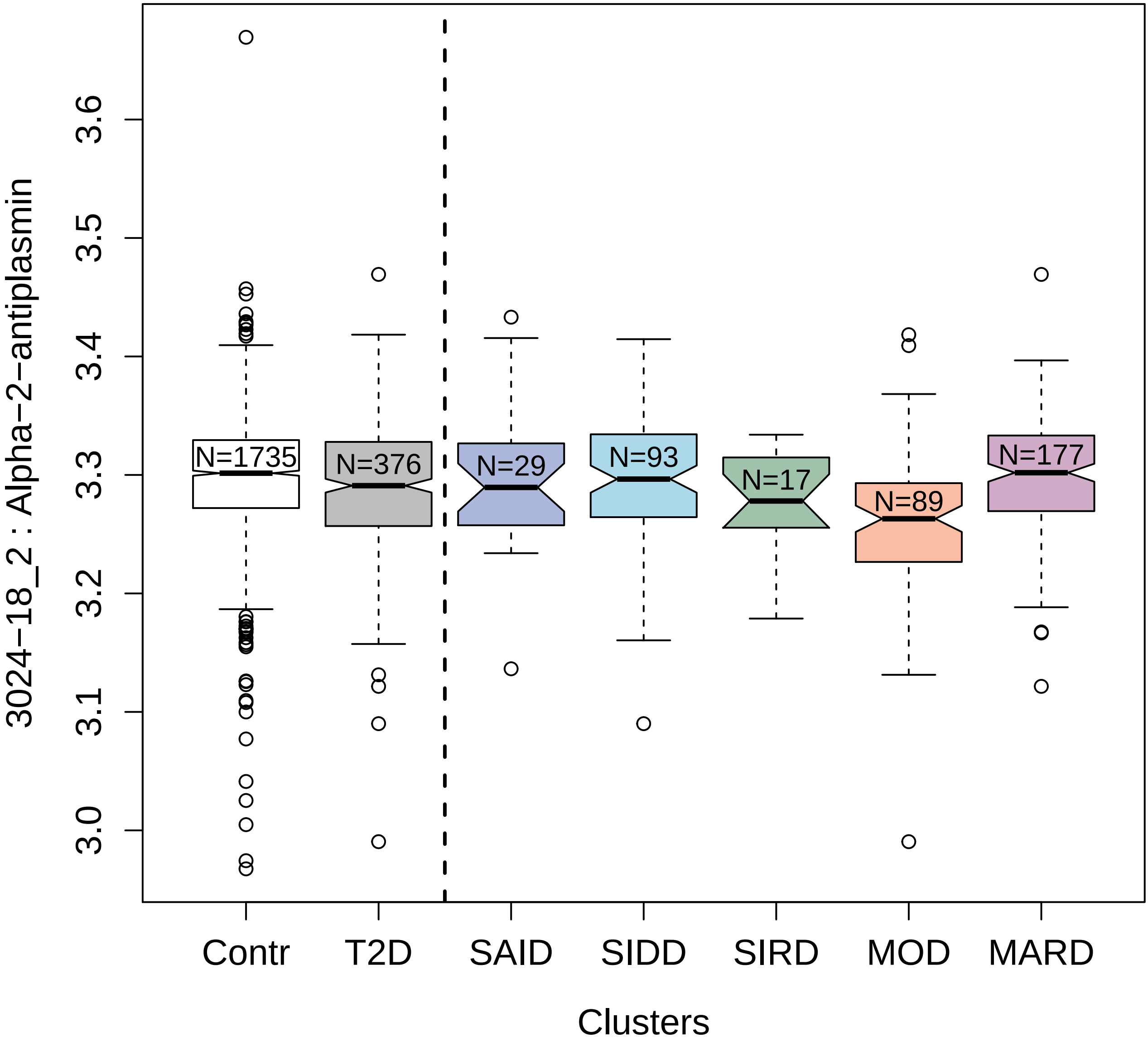
3024−18_2 : Alpha−2−antiplasmin
3.0
3.1
3.2
3.3
3.4
3.5
3.6
N=1735
N=376
N=29
N=93
N=17
N=89
N=177
Contr
T2D
SAID
SIDD
SIRD
MOD
MARD
Clusters

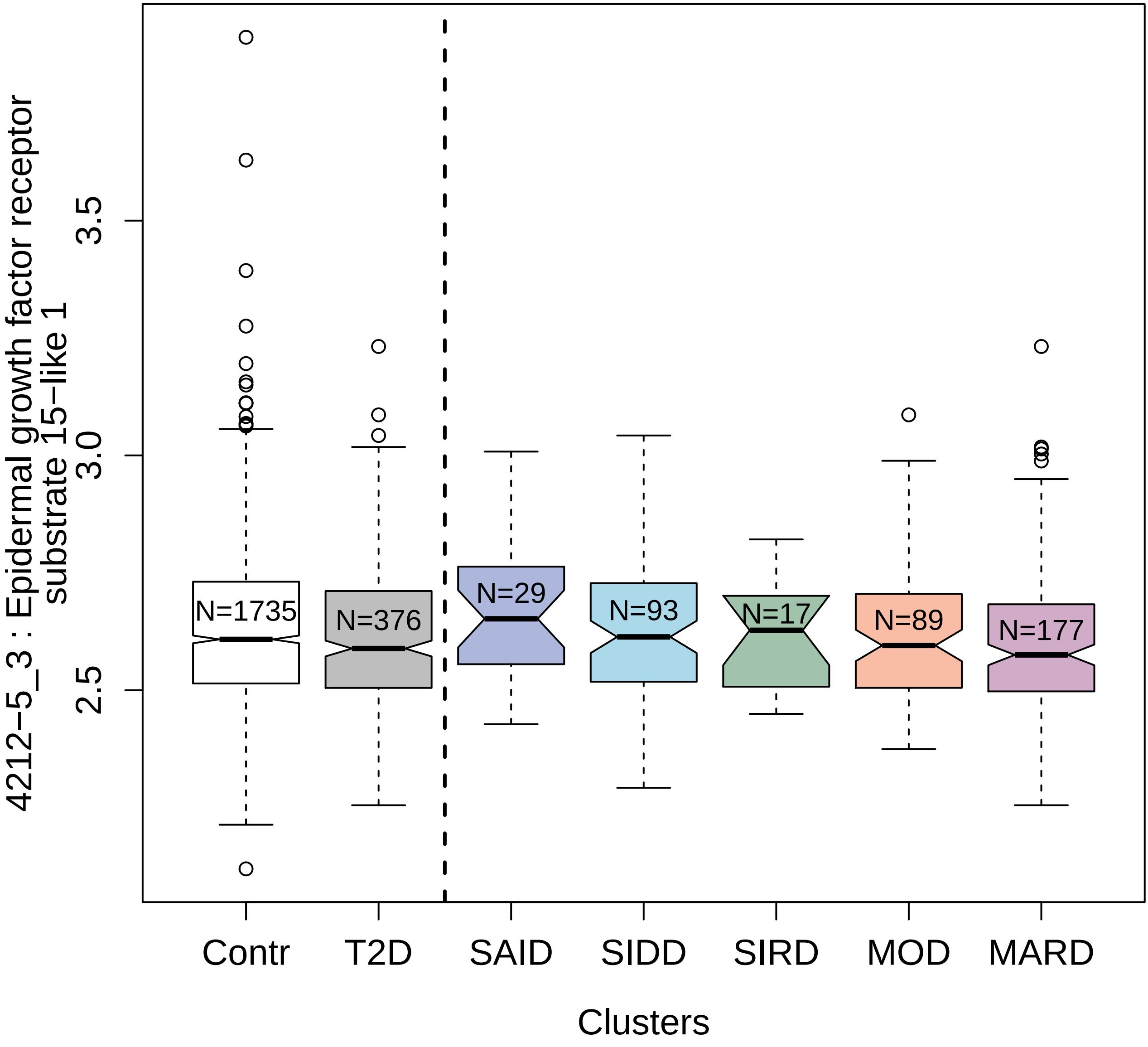
4212−5_3 : Epidermal growth factor receptor
substrate 15−like 1
3.5
3.0
2.5
N=1735
N=376
N=29
N=93
N=17
N=89
N=177
Contr
T2D
SAID
SIDD
SIRD
MOD
MARD
Clusters

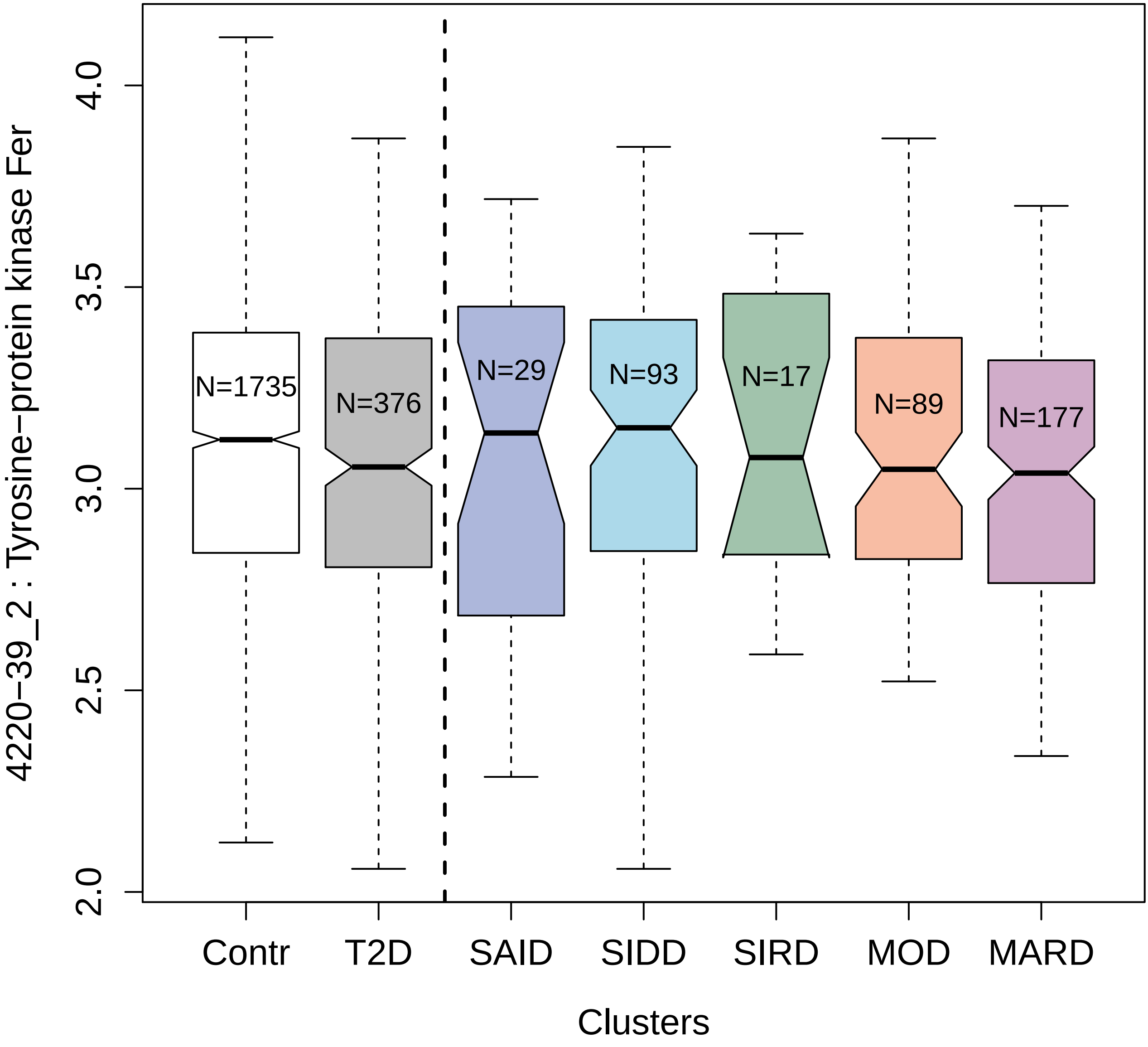
4220−39_2 : Tyrosine−protein kinase Fer
2.0
2.5
3.0
3.5
4.0
N=1735
N=376
N=29
N=93
N=17
N=89
N=177
Contr
T2D
SAID
SIDD
SIRD
MOD
MARD
Clusters

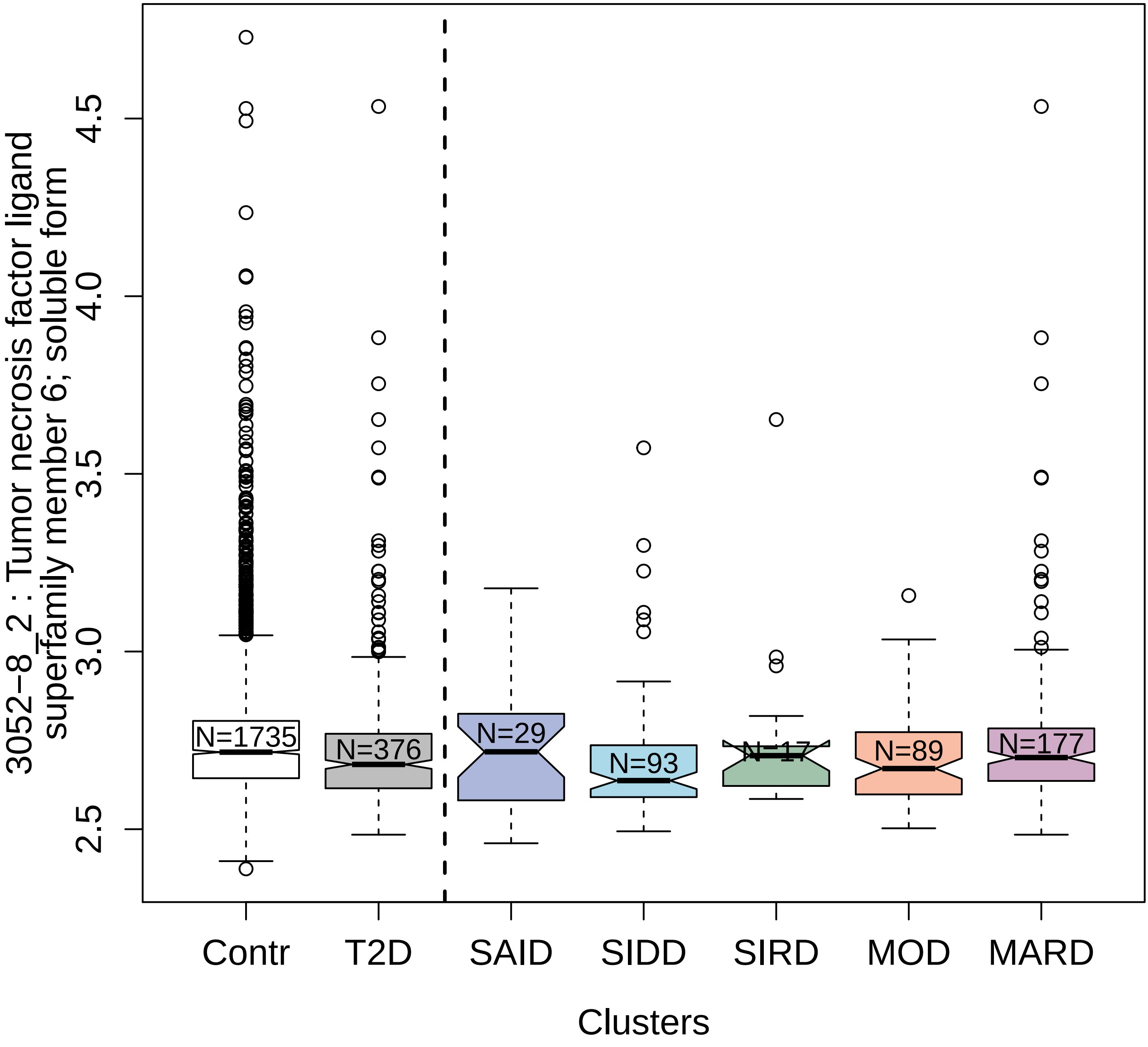
3052−8_2 : Tumor necrosis factor ligand superfamily member 6; soluble form
2.5
3.0
3.5
4.0
4.5
N=1735
N=376
N=29
N=93
N=17
N=89
N=177
Contr
T2D
SAID
SIDD
SIRD
MOD
MARD
Clusters

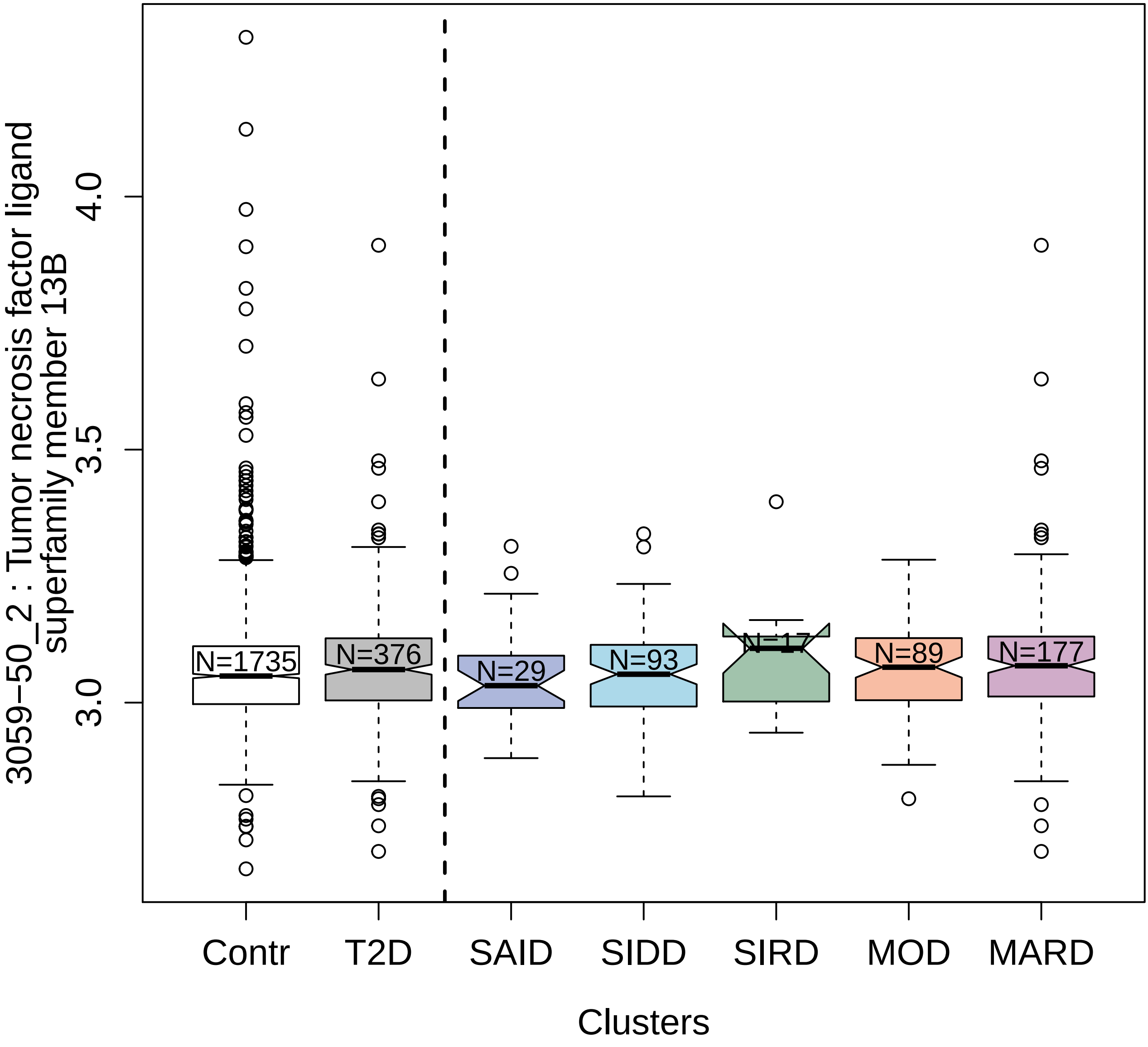

3059−50_2 : Tumor necrosis factor ligand superfamily member 13B
4.0
3.5
3.0
N=1735
N=376
N=29
N=93
N=17
N=89
N=177
Contr
T2D
SAID
SIDD
SIRD
MOD
MARD
Clusters

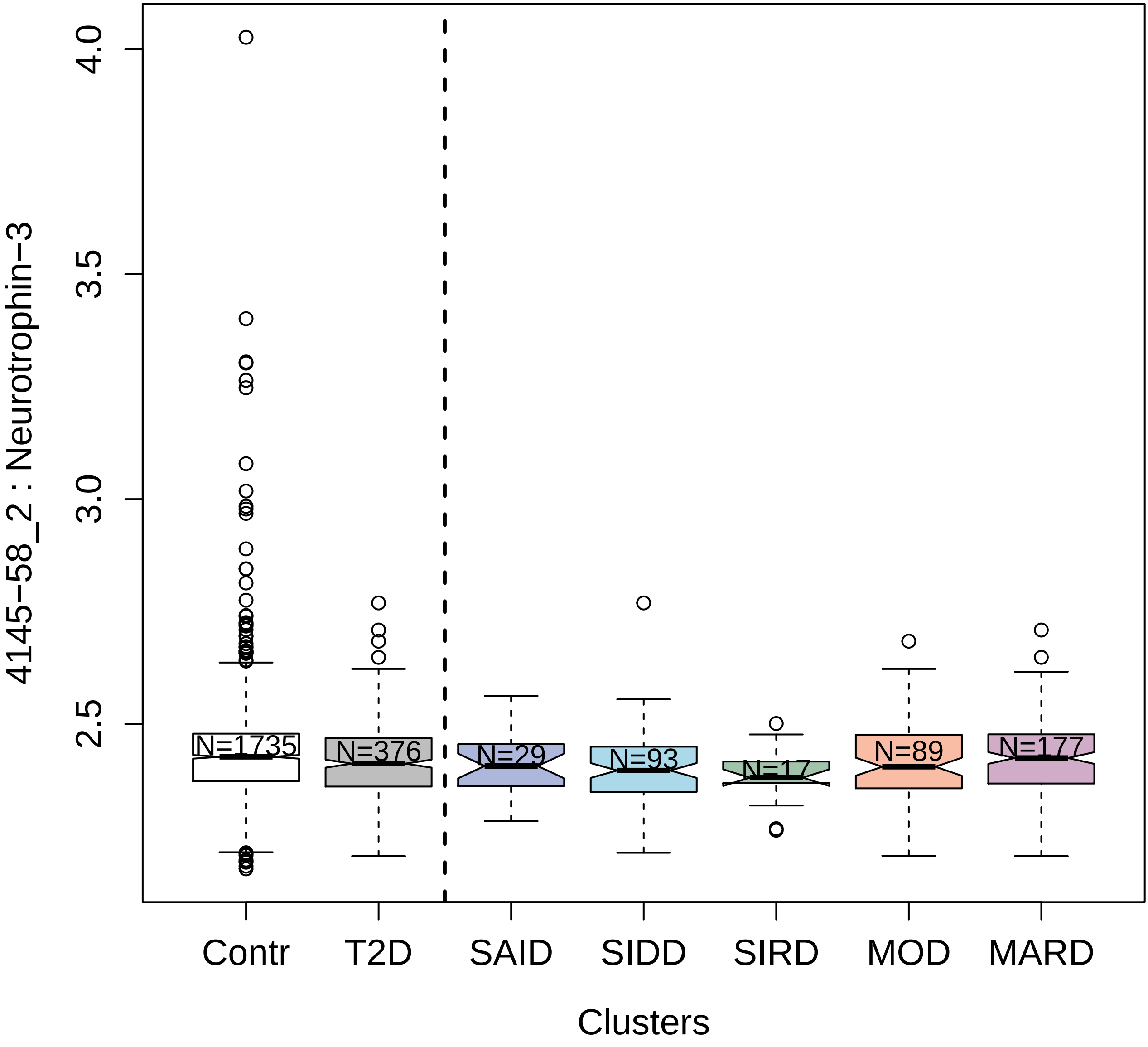

4145−58_2 : Neurotrophin−3
4.0
3.5
3.0
2.5
N=1735
N=376
N=29
N=93
N=17
N=89
N=177
Contr
T2D
SAID
SIDD
SIRD
MOD
MARD
Clusters

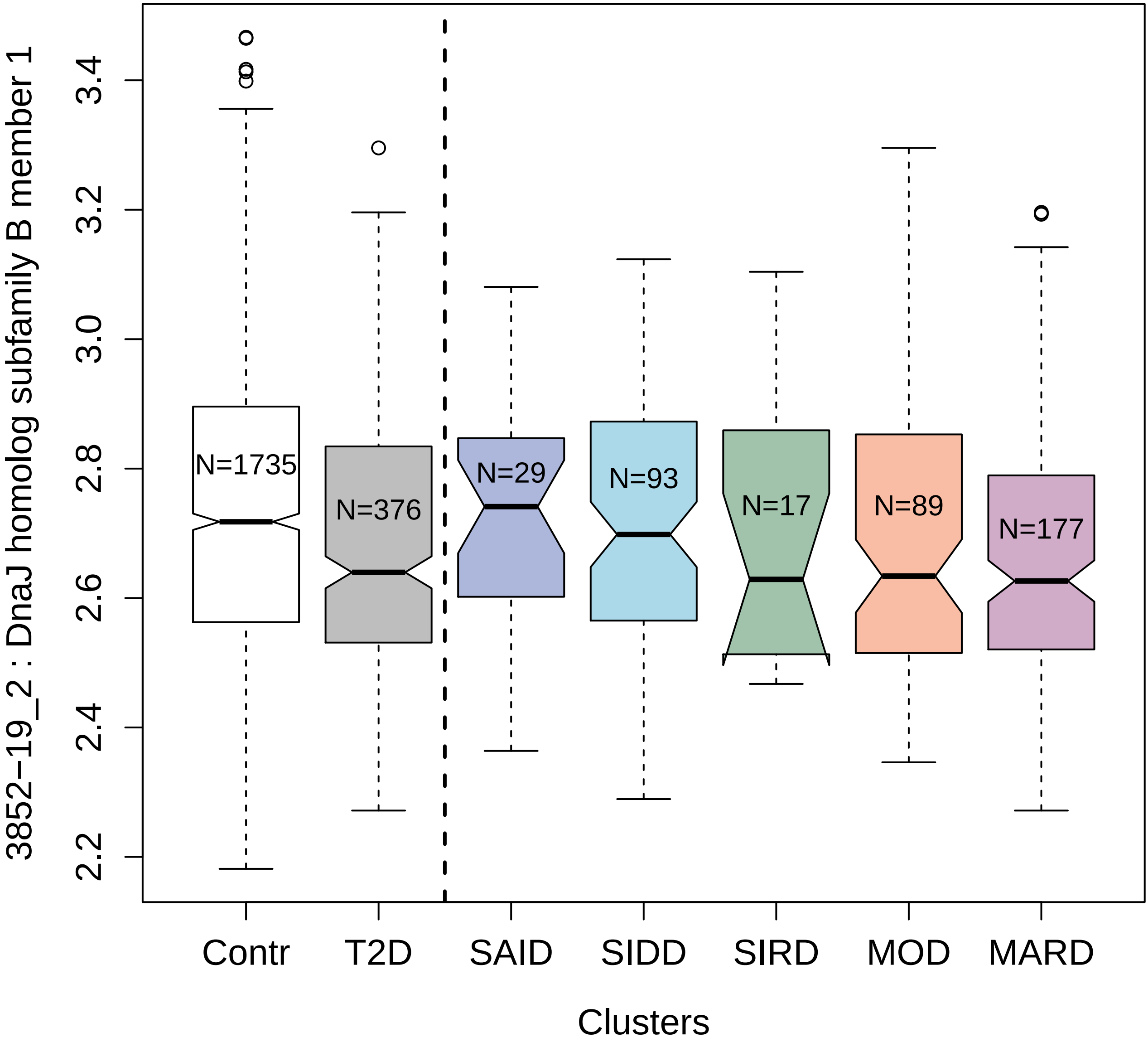
3852−19_2 : DnaJ homolog subfamily B member 1
2.2
2.4
2.6
2.8
3.0
3.2
3.4
N=1735
N=376
N=29
N=93
N=17
N=89
N=177
Contr
T2D
SAID
SIDD
SIRD
MOD
MARD
Clusters

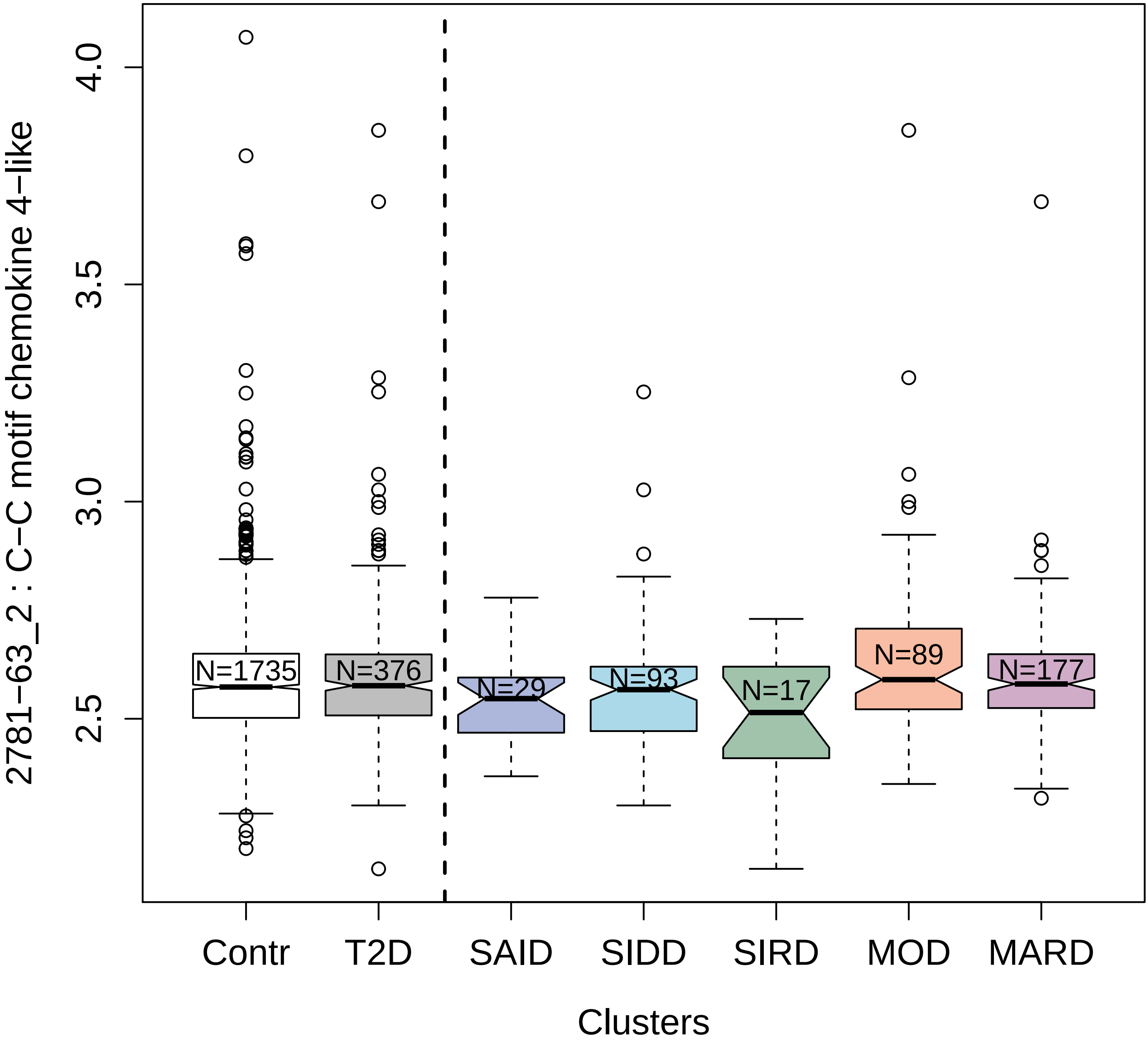

2781−63_2 : C−C motif chemokine 4−like
4.0
3.5
3.0
2.5
N=1735
N=376
N=29
N=93
N=17
N=89
N=177
Contr
T2D
SAID
SIDD
SIRD
MOD
MARD
Clusters

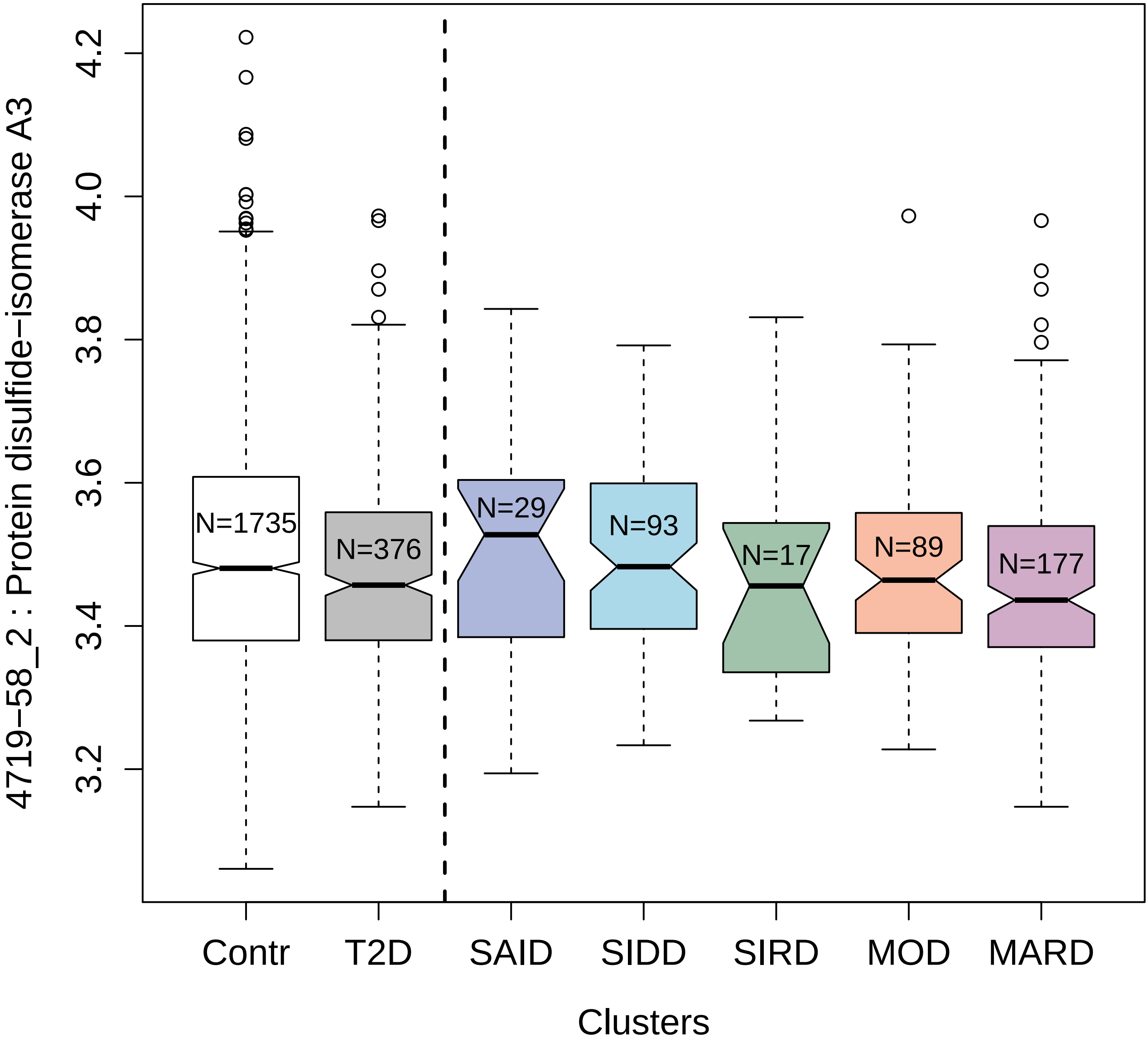

4719−58_2 : Protein disulfide−isomerase A3
3.2
3.4
3.6
3.8
4.0
4.2
N=1735
N=376
N=29
N=93
N=17
N=89
N=177
Contr
T2D
SAID
SIDD
SIRD
MOD
MARD
Clusters

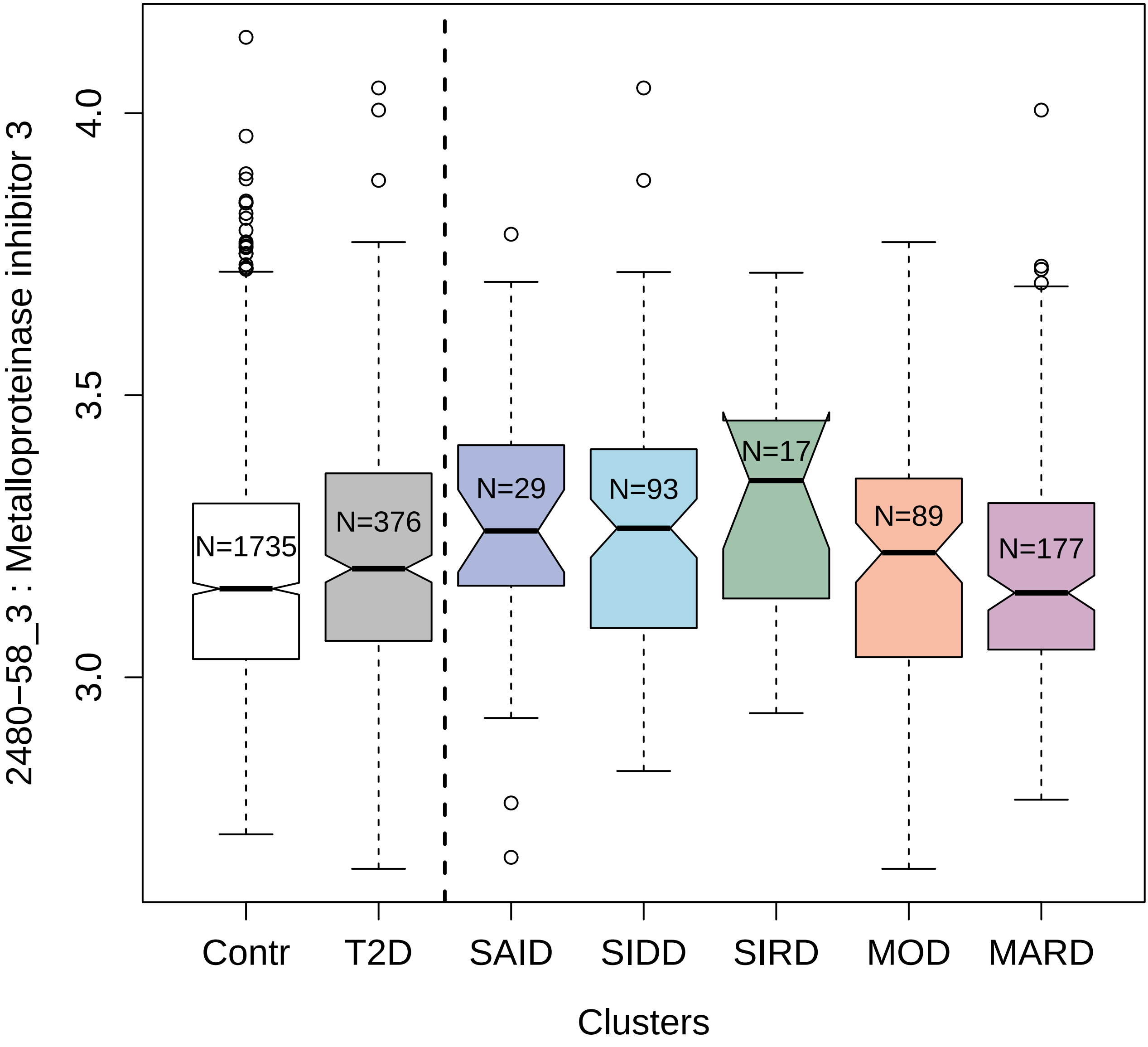
2480−58_3 : Metalloproteinase inhibitor 3
4.0
3.5
3.0
N=1735
N=376
N=29
N=93
N=17
N=89
N=177
Contr
T2D
SAID
SIDD
SIRD
MOD
MARD
Clusters

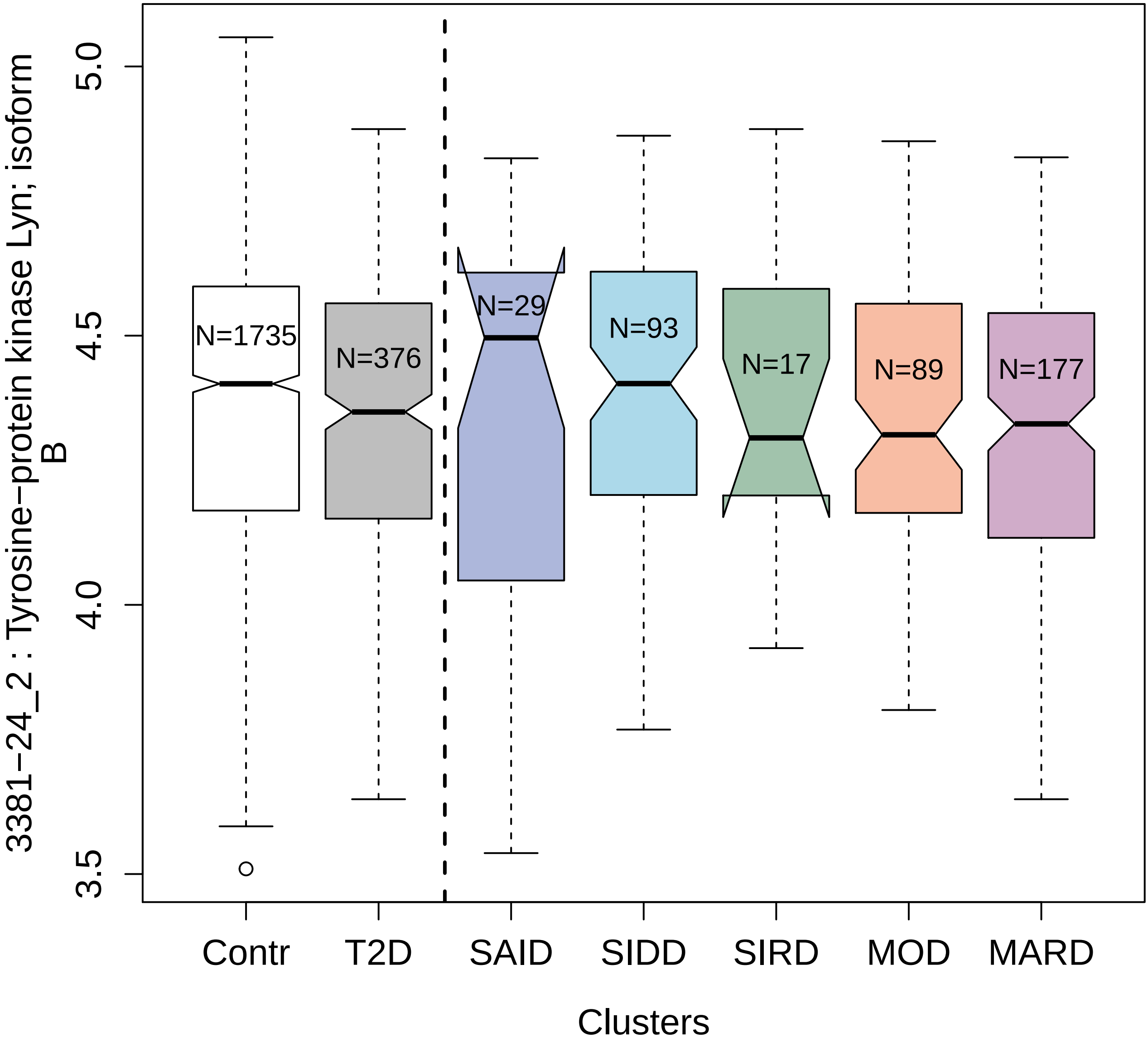
3381−24_2 : Tyrosine−protein kinase Lyn; isoform B
5.0
4.5
4.0
3.5
N=1735
N=376
N=29
N=93
N=17
N=89
N=177
Contr
T2D
SAID
SIDD
SIRD
MOD
MARD
Clusters

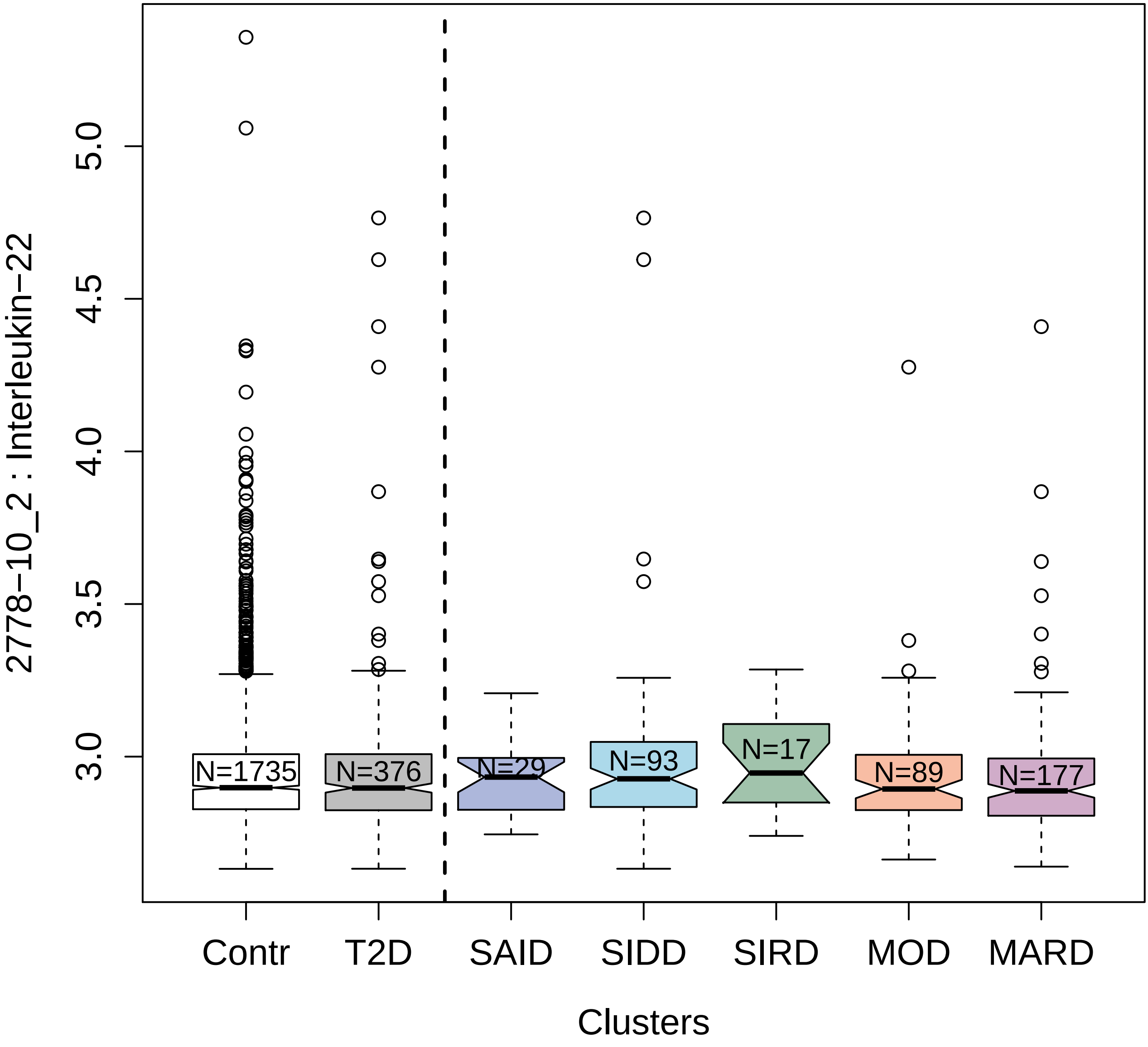

2778−10_2 : Interleukin−22
5.0
4.5
4.0
3.5
3.0
N=1735
N=376
N=29
N=93
N=17
N=89
N=177
Contr
T2D
SAID
SIDD
SIRD
MOD
MARD
Clusters

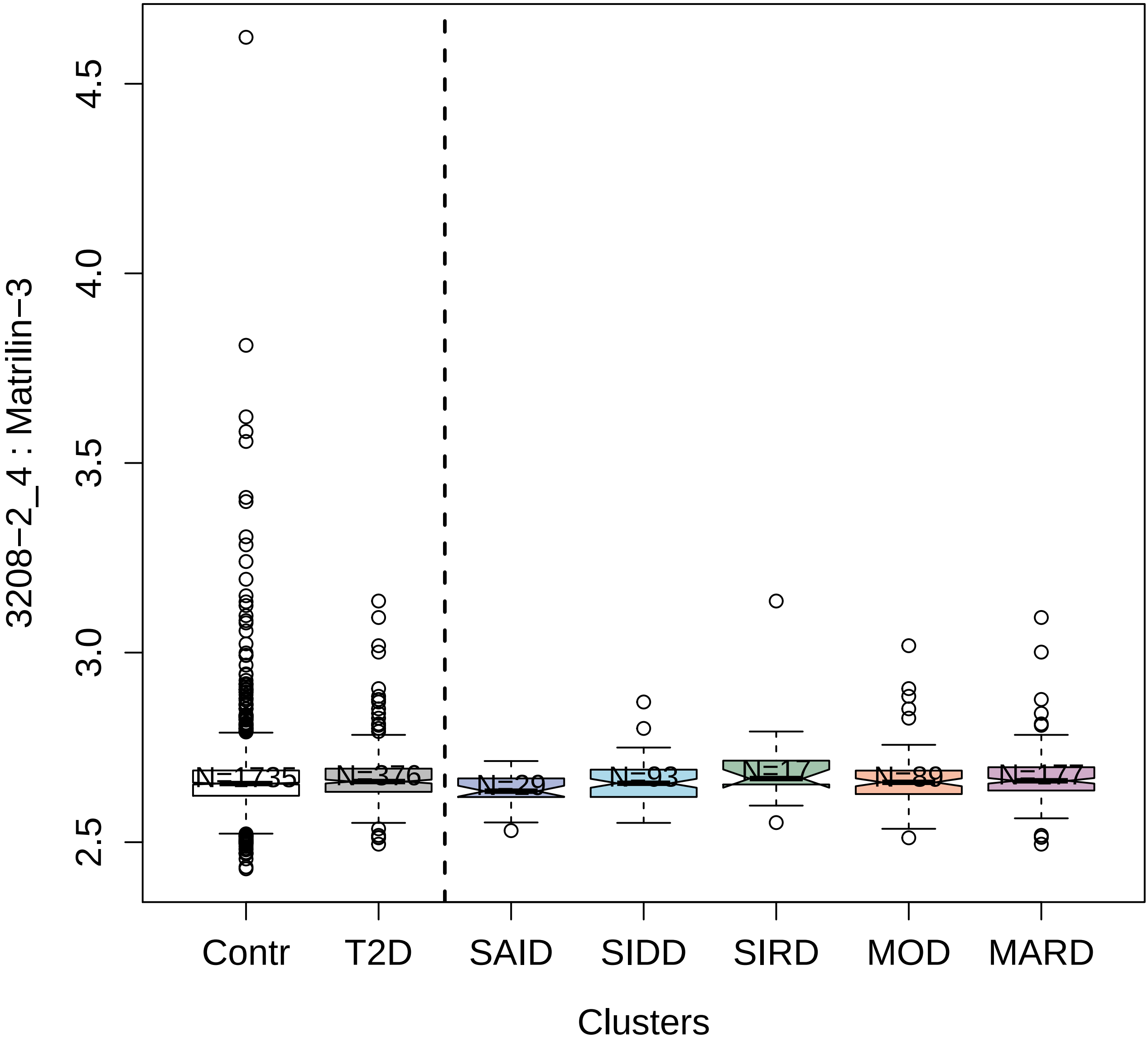

3208−2_4 : Matrilin−3
4.5
4.0
3.5
3.0
2.5
N=1735
N=376
N=29
N=93
N=17
N=89
N=177
Contr
T2D
SAID
SIDD
SIRD
MOD
MARD
Clusters

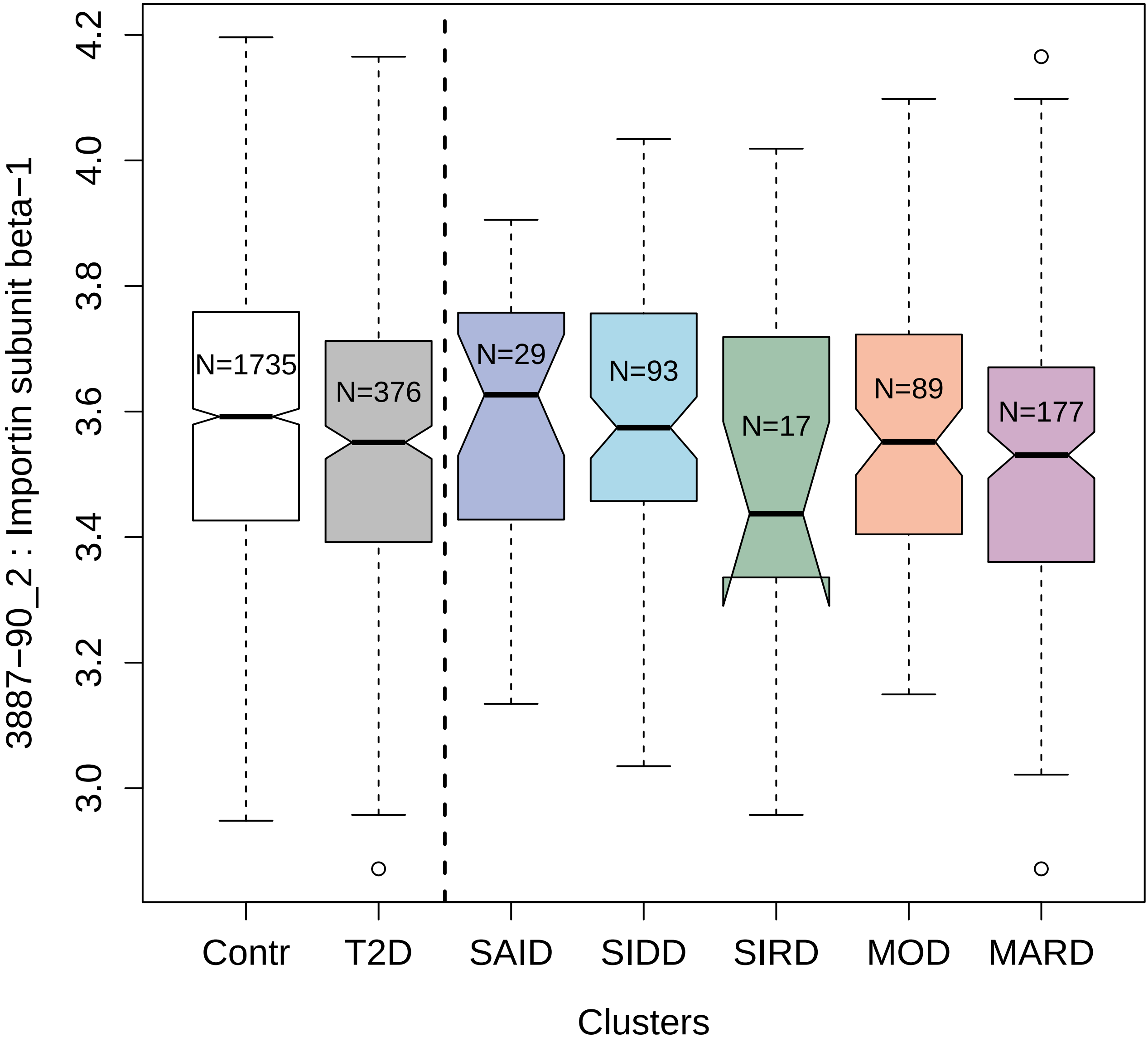
3887−90_2 : Importin subunit beta−1
4.2
4.0
3.8
3.6
3.4
3.2
3.0
N=1735
N=376
N=29
N=93
N=17
N=89
N=177
Contr
T2D
SAID
SIDD
SIRD
MOD
MARD
Clusters

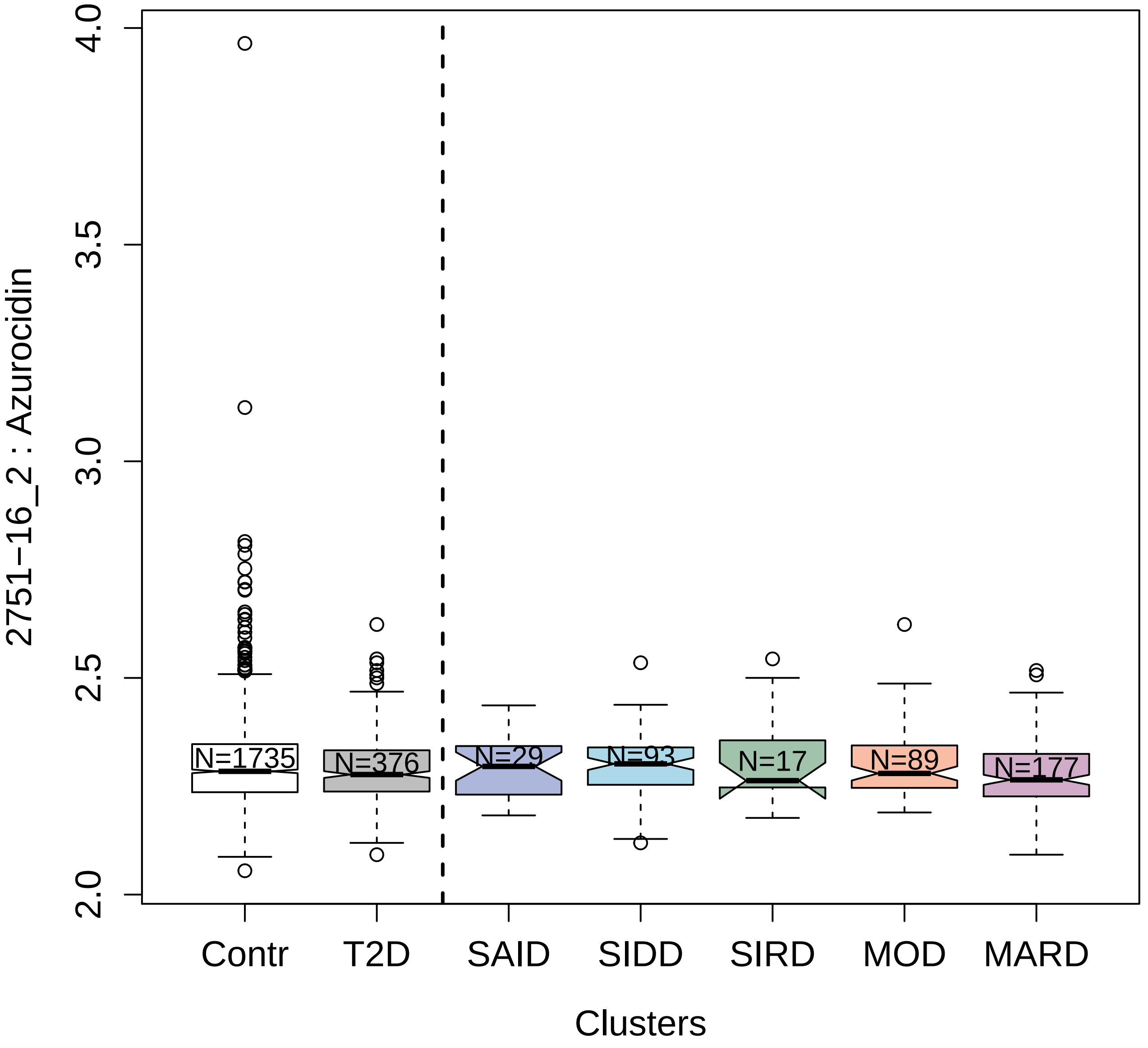

2751−16_2 : Azurocidin
2.0
2.5
3.0
3.5
4.0
N=1735
N=376
N=29
N=93
N=17
N=89
N=177
Contr
T2D
SAID
SIDD
SIRD
MOD
MARD
Clusters

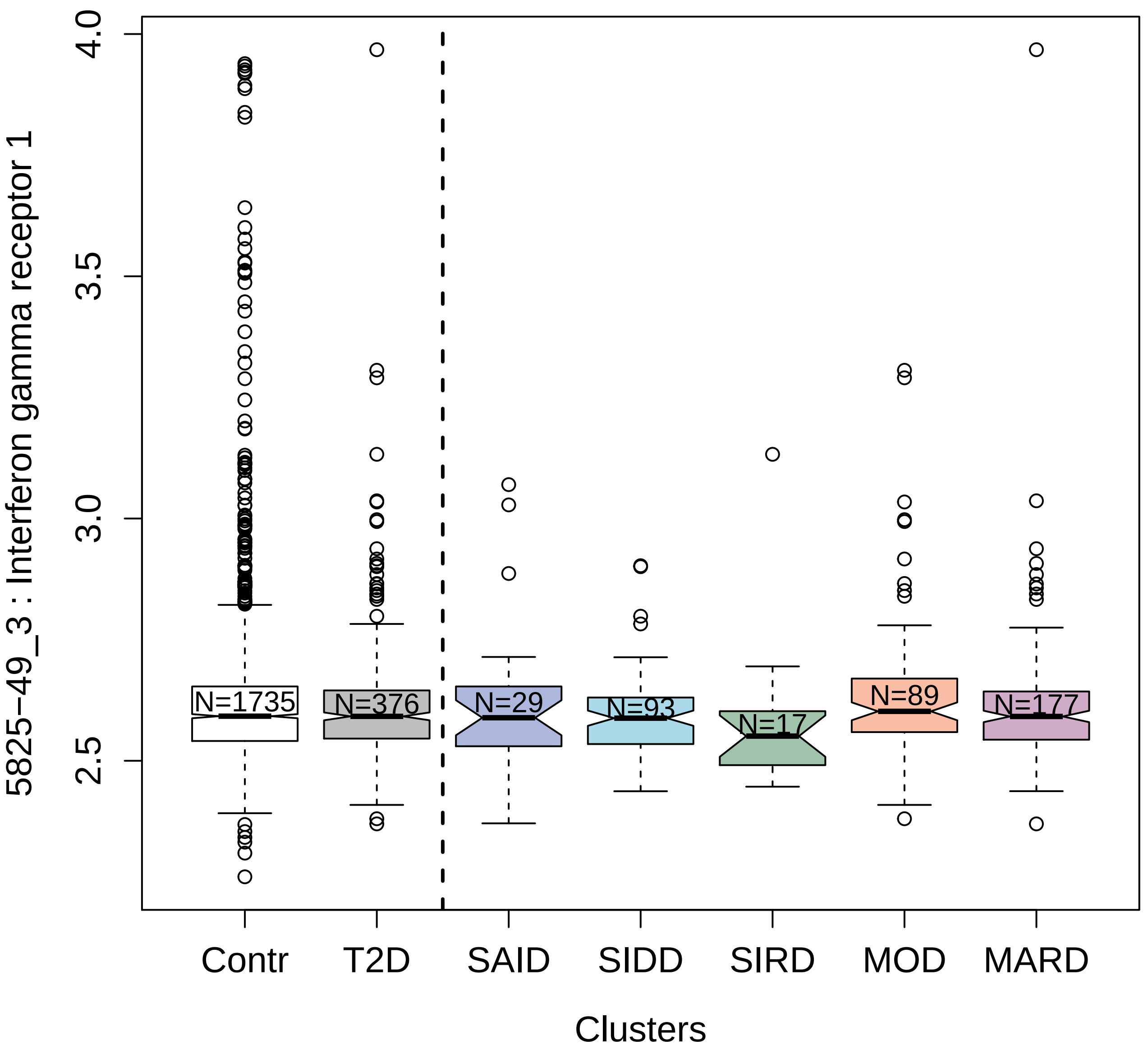
5825−49_3 : Interferon gamma receptor 1
4.0
3.5
3.0
2.5
N=1735
N=376
N=29
N=93
N=17
N=89
N=177
Contr
T2D
SAID
SIDD
SIRD
MOD
MARD
Clusters

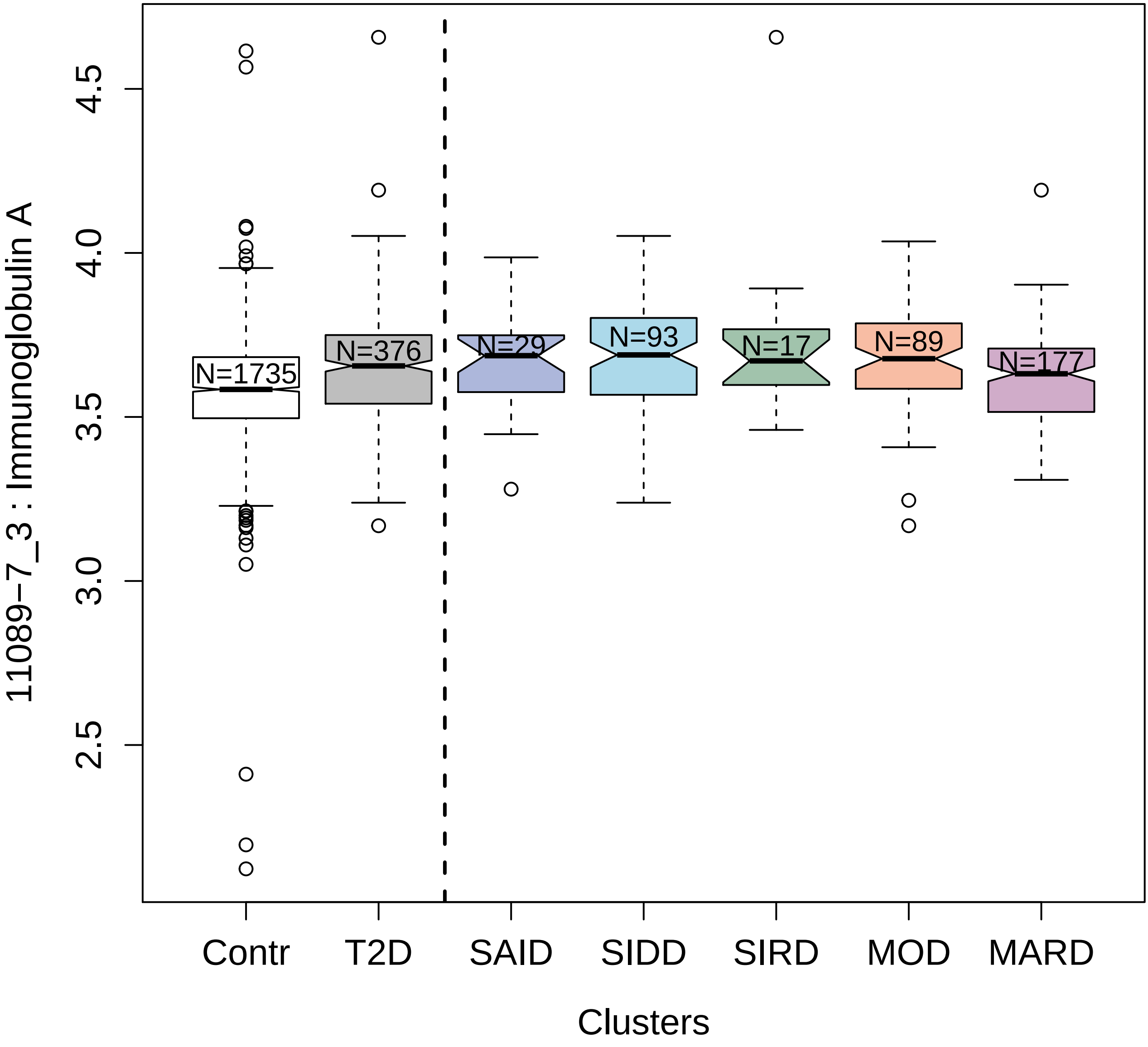
11089−7_3 : Immunoglobulin A
2.5
3.0
3.5
4.0
4.5
N=1735
N=376
N=29
N=93
N=17
N=89
N=177
Contr
T2D
SAID
SIDD
SIRD
MOD
MARD
Clusters

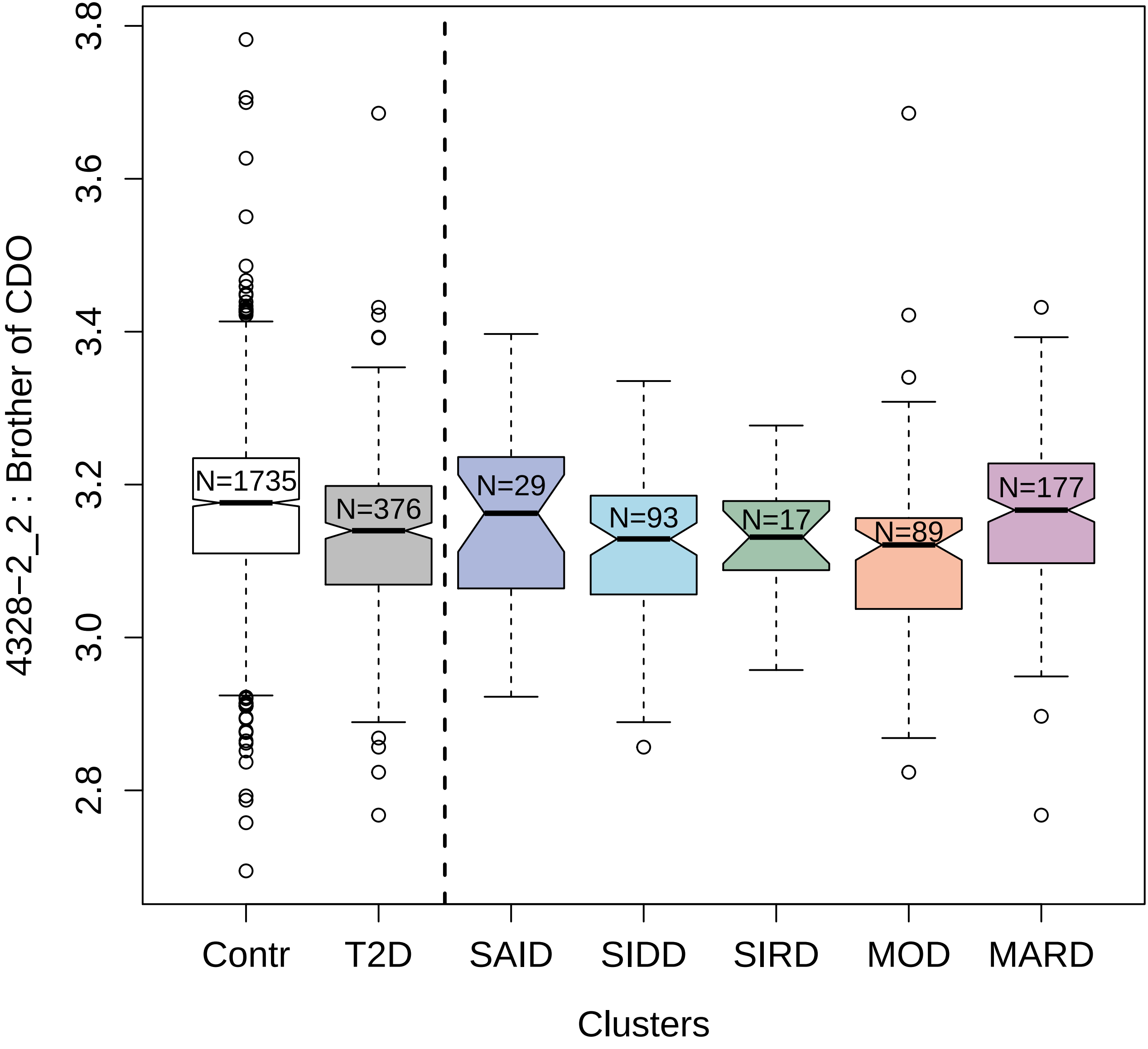

4328−2_2 : Brother of CDO
3.8
3.6
3.4
3.2
3.0
2.8
N=1735
N=376
N=29
N=93
N=17
N=89
N=177
Contr
T2D
SAID
SIDD
SIRD
MOD
MARD
Clusters

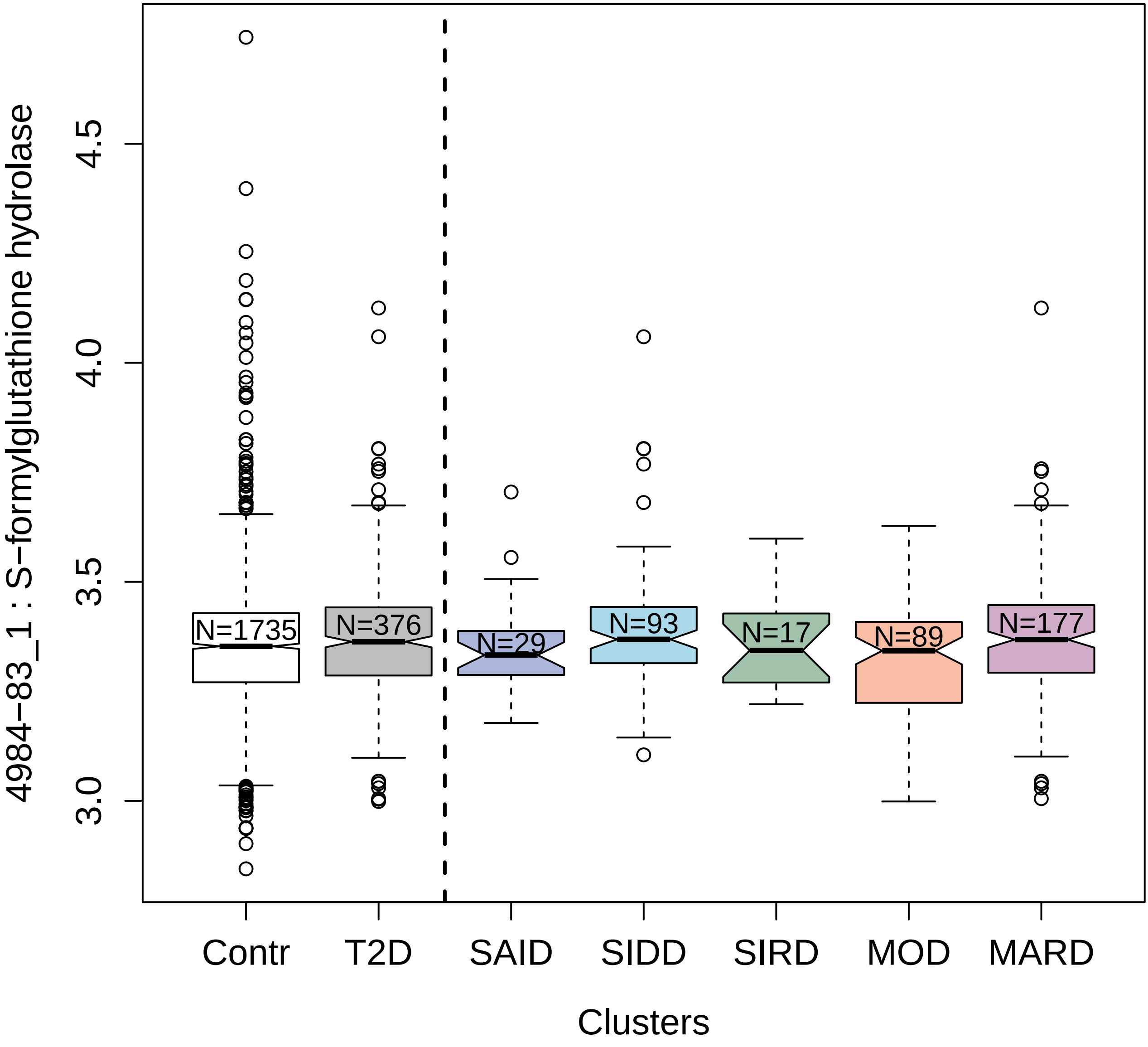

4984−83_1 : S−formylglutathione hydrolase
3.0
3.5
4.0
4.5
N=1735
N=376
N=29
N=93
N=17
N=89
N=177
Contr
T2D
SAID
SIDD
SIRD
MOD
MARD
Clusters

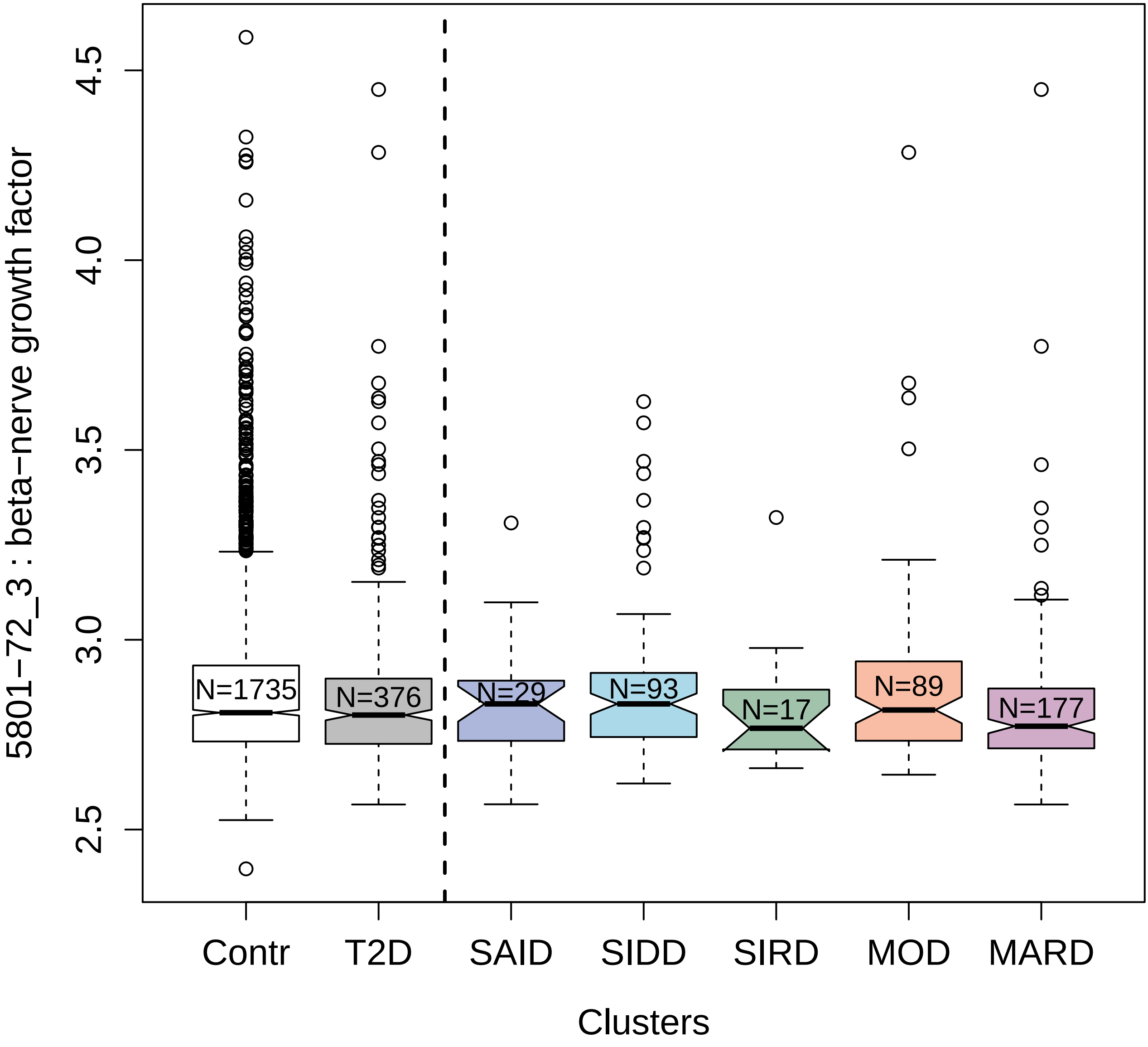
5801−72_3 : beta−nerve growth factor
2.5
3.0
3.5
4.0
4.5
N=1735
N=376
N=29
N=93
N=17
N=89
N=177
Contr
T2D
SAID
SIDD
SIRD
MOD
MARD
Clusters

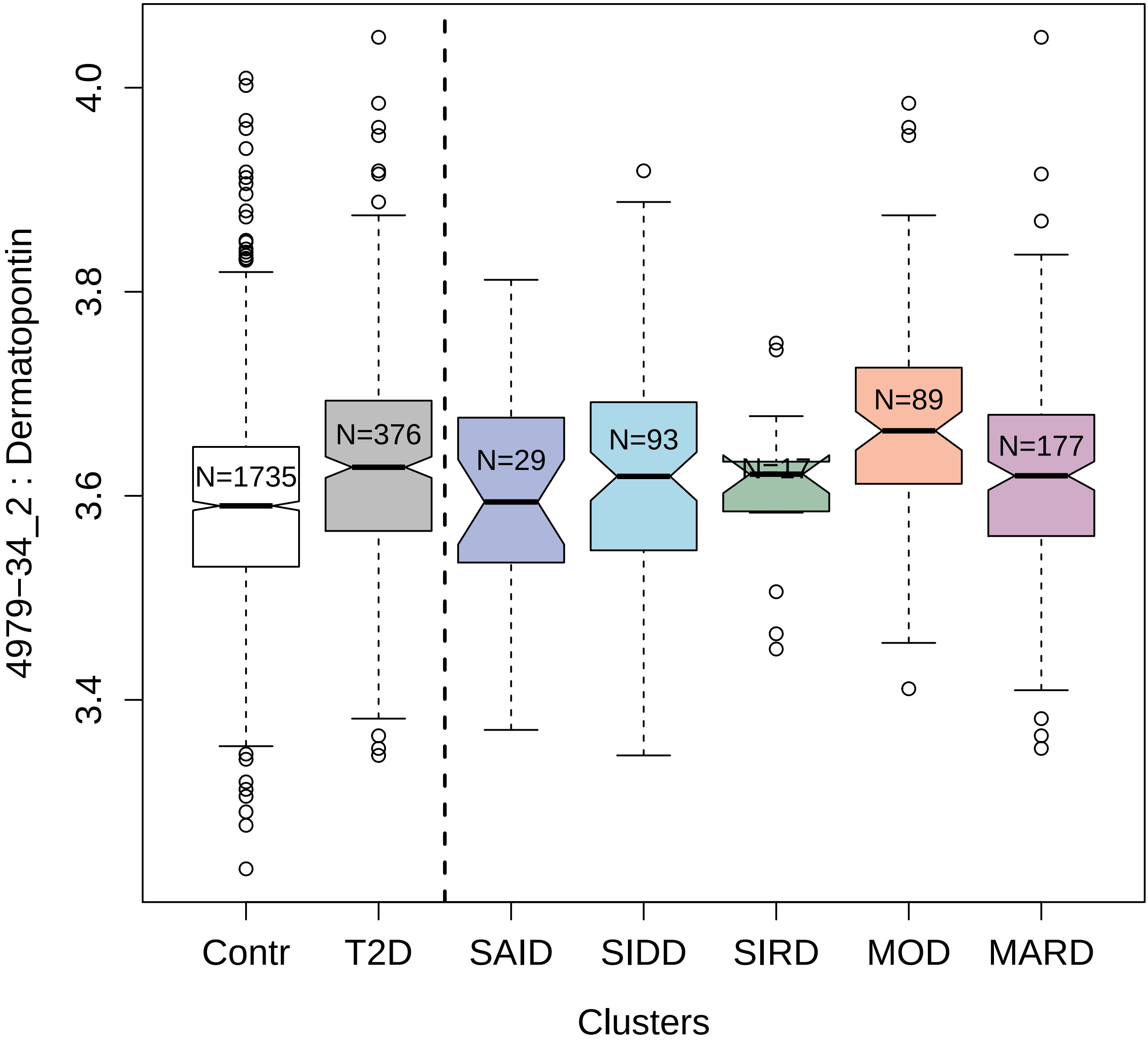

4979−34_2 : Dermatopontin
3.4
3.6
3.8
4.0
N=1735
N=376
N=29
N=93
N=89
N=177
Contr
T2D
SAID
SIDD
SIRD
MOD
MARD
Clusters

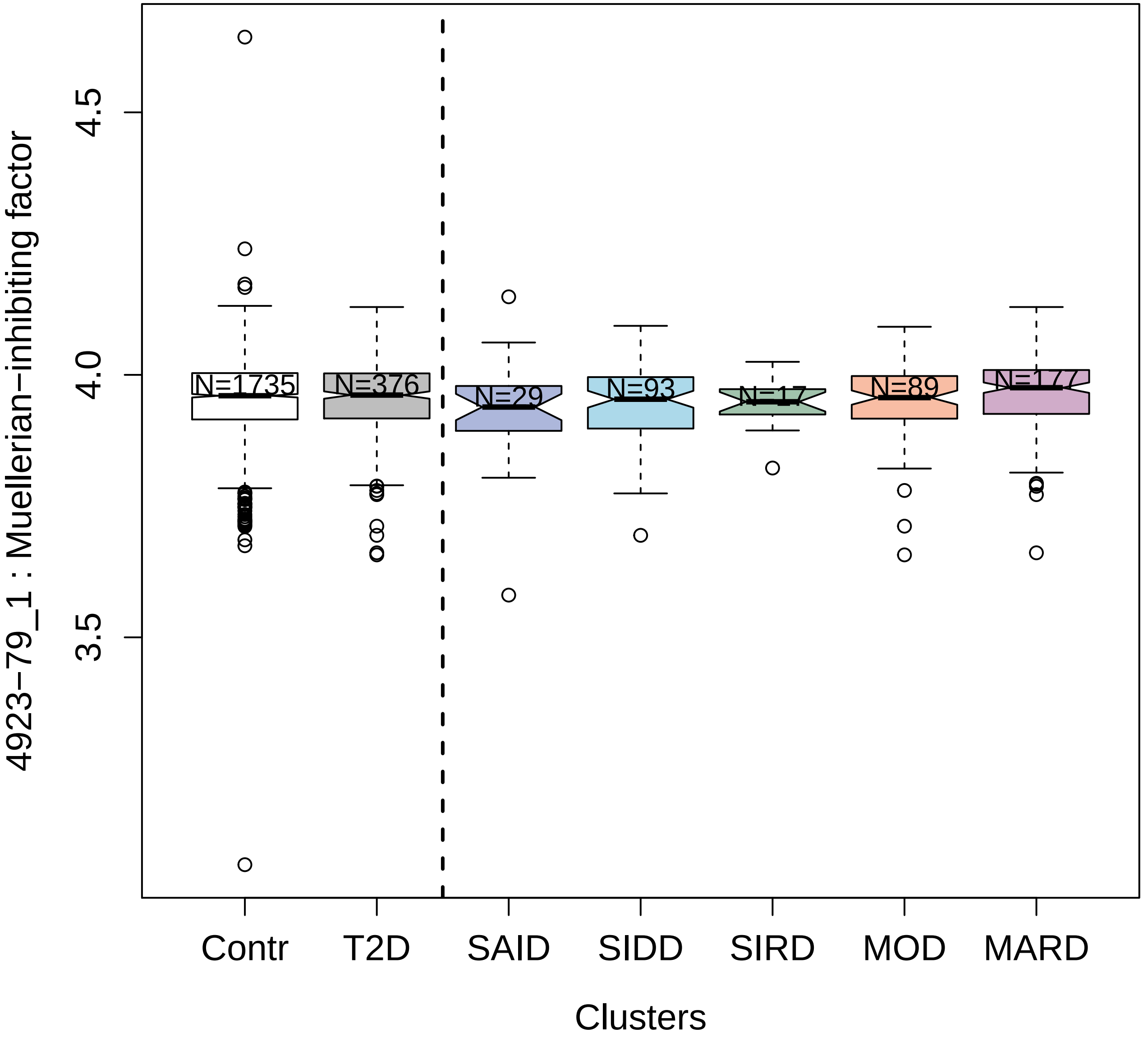

4923−79_1 : Muellerian−inhibiting factor
4.5
4.0
3.5
N=1735
N=376
N=29
N=93
N=17
N=89
N=177
Contr
T2D
SAID
SIDD
SIRD
MOD
MARD
Clusters

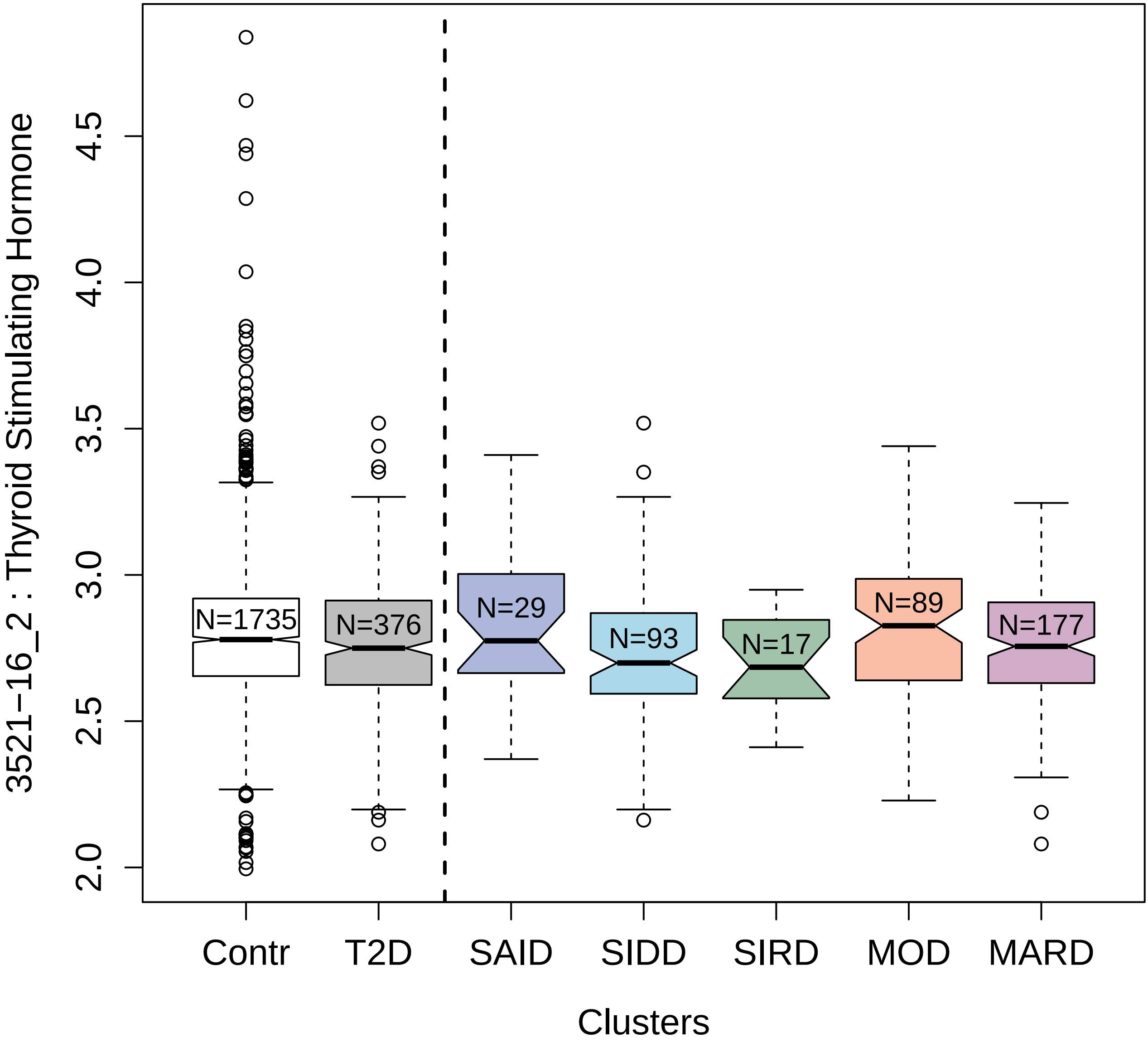
3521−16_2 : Thyroid Stimulating Hormone
2.0
2.5
3.0
3.5
4.0
4.5
N=1735
N=376
N=29
N=93
N=17
N=89
N=177
Contr
T2D
SAID
SIDD
SIRD
MOD
MARD
Clusters

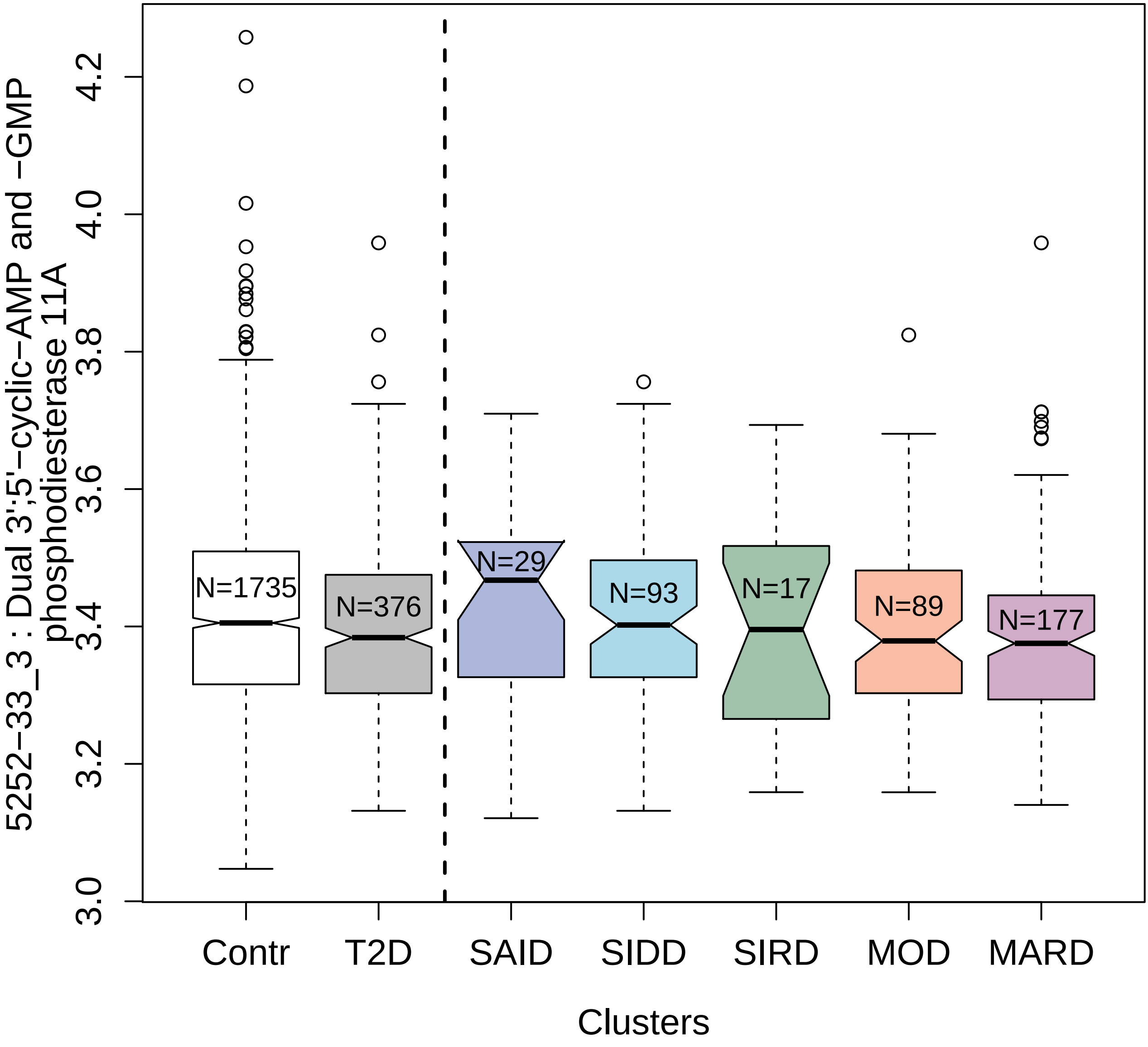

5252−33_3 : Dual 3',5'−cyclic−AMP and −GMP
phosphodiesterase 11A
3.0
3.2
3.4
3.6
3.8
4.0
4.2
N=1735
N=376
N=29
N=93
N=17
N=89
N=177
Contr
T2D
SAID
SIDD
SIRD
MOD
MARD
Clusters

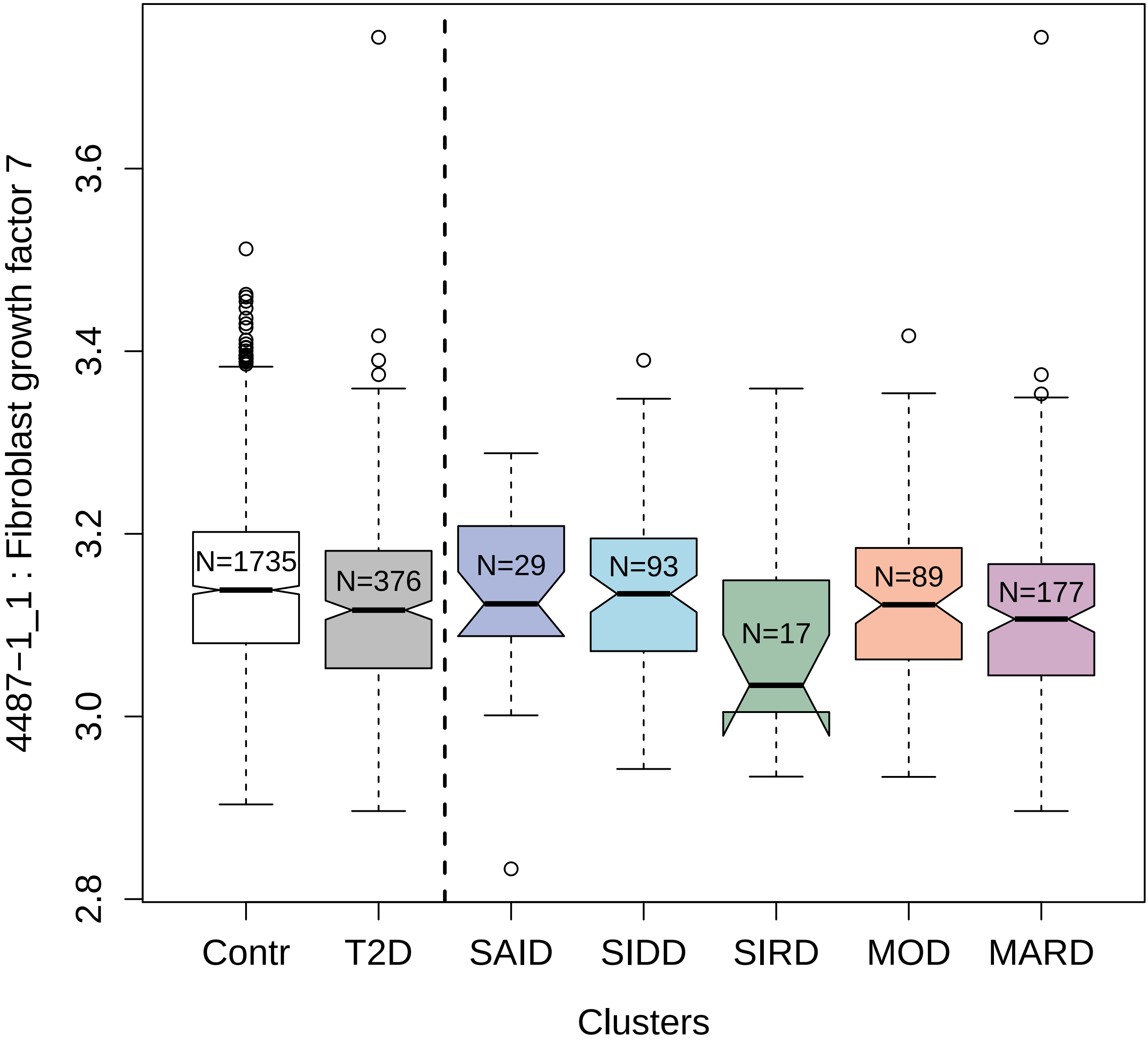
4487−1_1 : Fibroblast growth factor 7
3.6
3.4
3.2
3.0
2.8
N=1735
N=376
N=29
N=93
N=17
N=89
N=177
Contr
T2D
SAID
SIDD
SIRD
MOD
MARD
Clusters

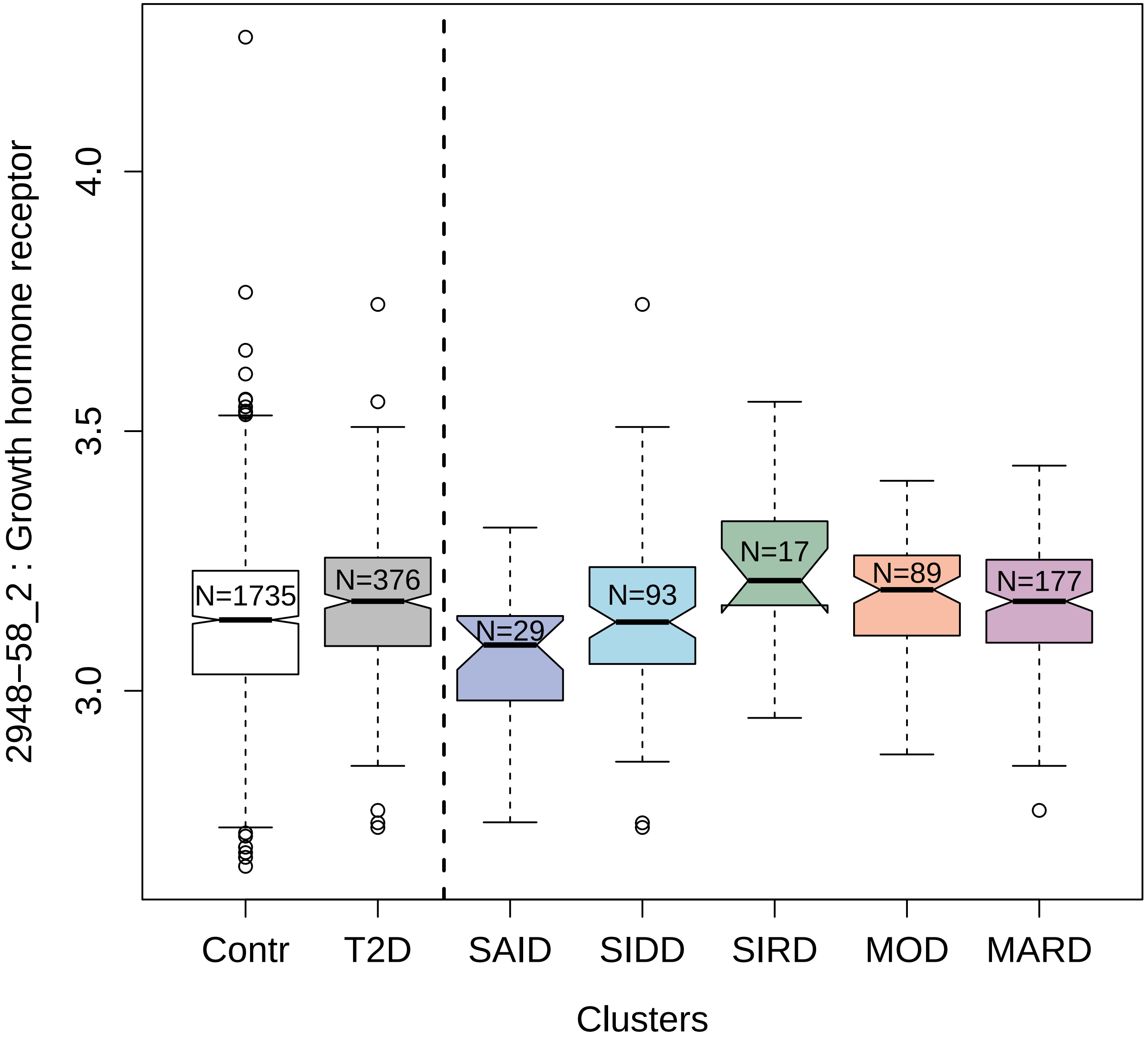
2948−58_2 : Growth hormone receptor
4.0
3.5
3.0
N=1735
N=376
N=29
N=93
N=17
N=89
N=177
Contr
T2D
SAID
SIDD
SIRD
MOD
MARD
Clusters

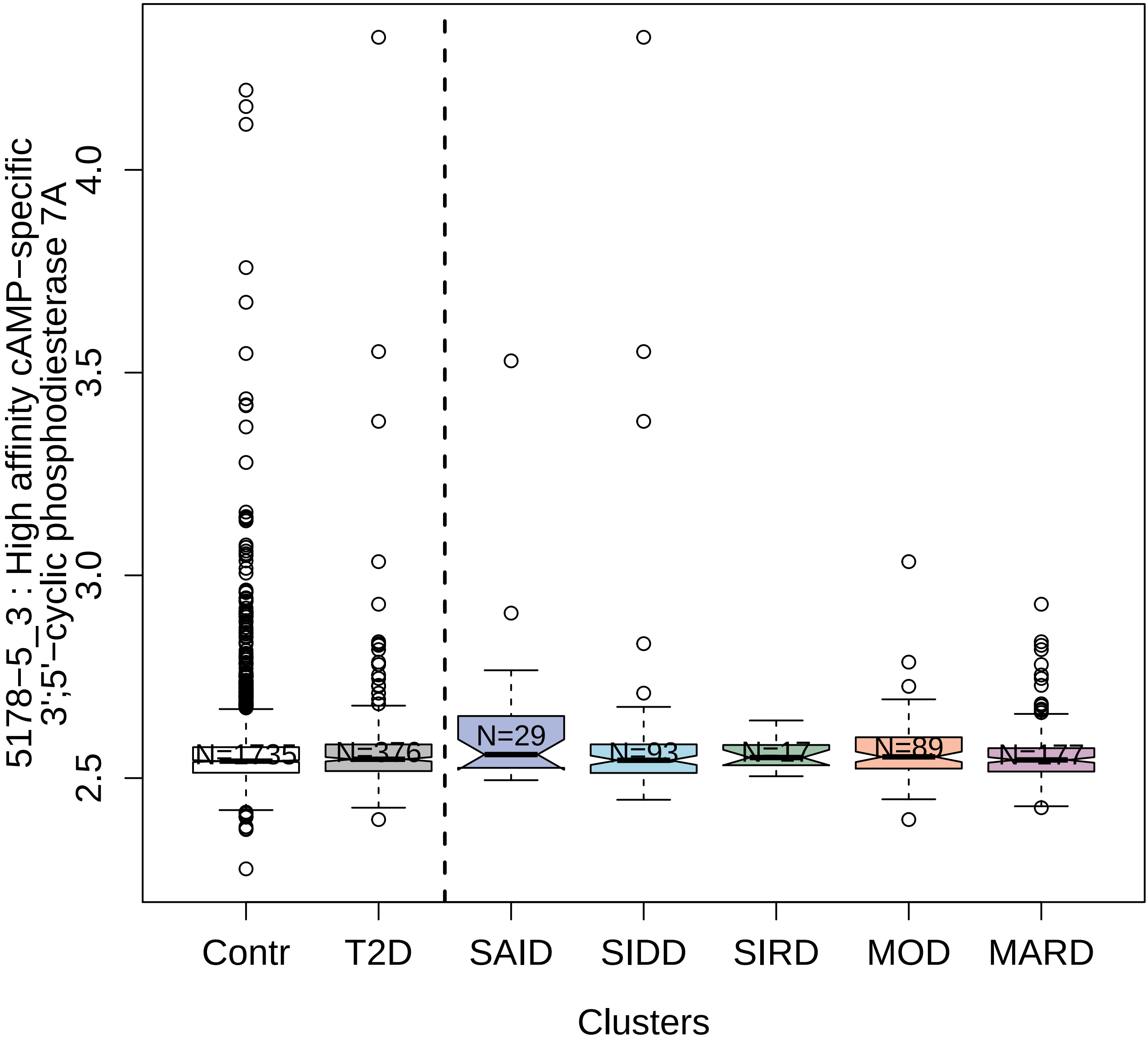

5178−5_3 : High affinity cAMP−specific
3',5'−cyclic phosphodiesterase 7A
2.5
3.0
3.5
4.0
N=1735
N=376
N=29
N=93
N=17
N=89
N=177
Contr
T2D
SAID
SIDD
SIRD
MOD
MARD
Clusters

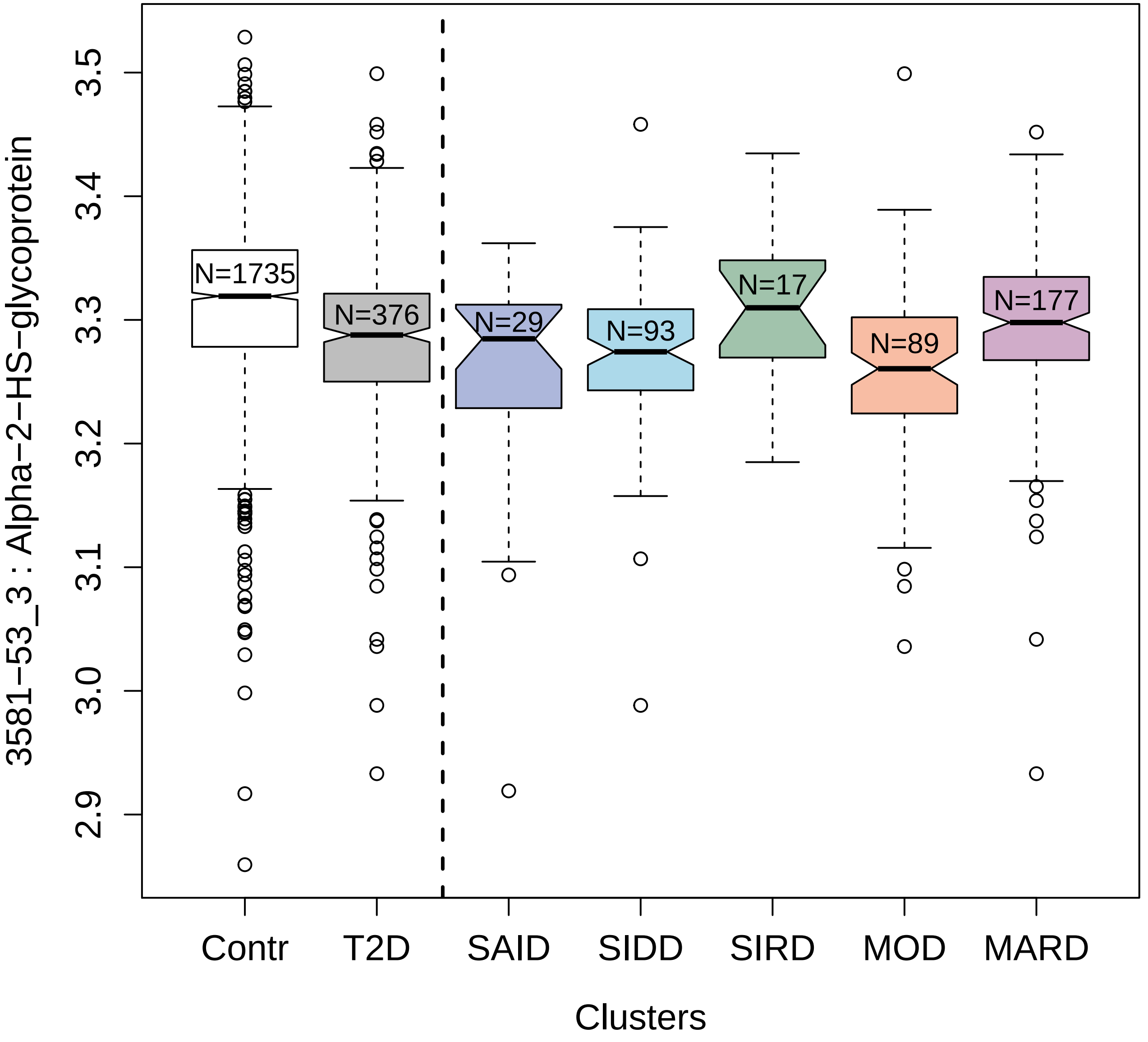

3581−53_3 : Alpha−2−HS−glycoprotein
2.9
3.0
3.1
3.2
3.3
3.4
3.5
N=1735
N=376
N=29
N=93
N=17
N=89
N=177
Contr
T2D
SAID
SIDD
SIRD
MOD
MARD
Clusters

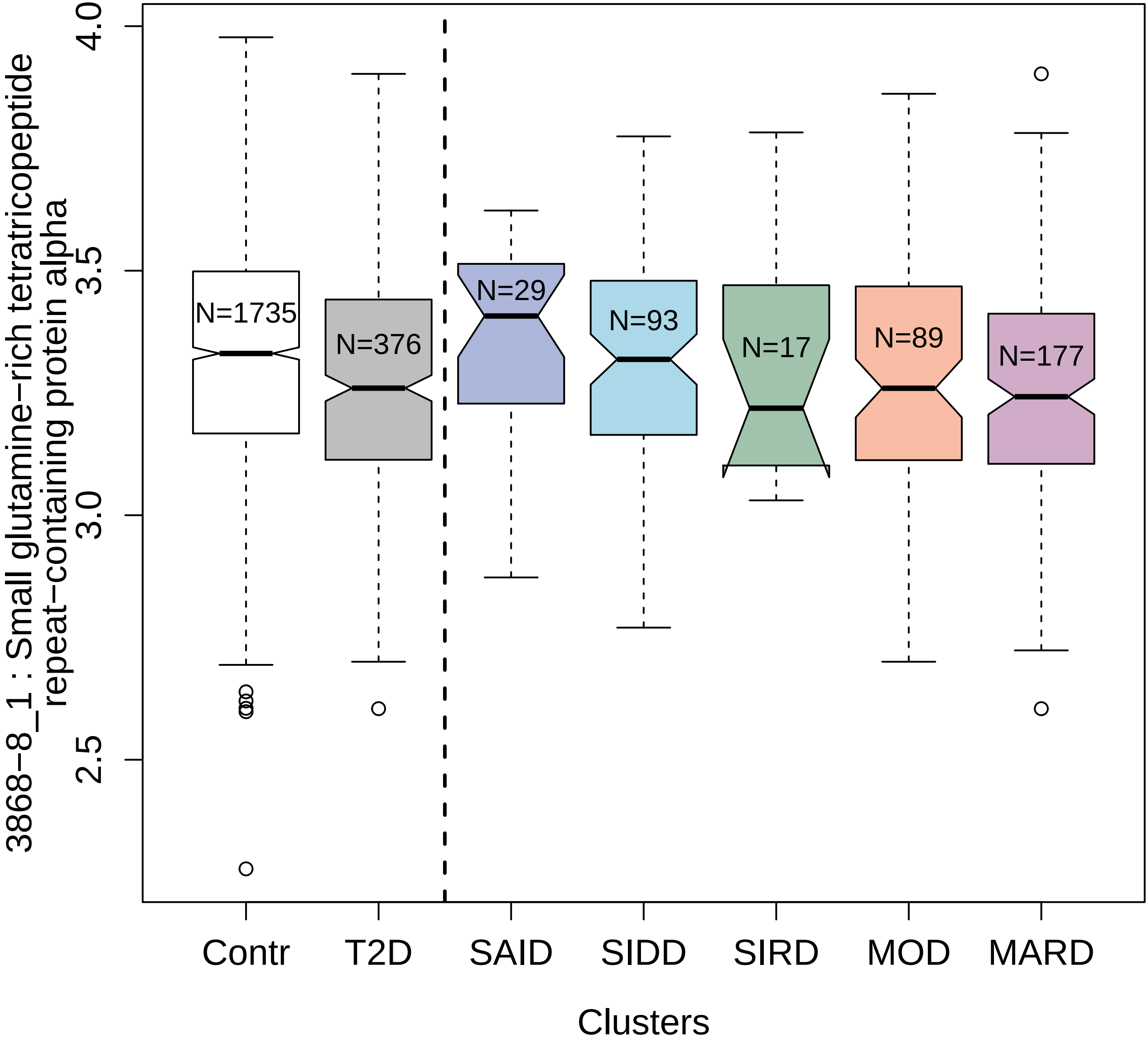

3868−8_1 : Small glutamine−rich tetratricopeptide repeat−containing protein alpha
4.0
3.5
3.0
2.5
N=1735
N=376
N=29
N=93
N=17
N=89
N=177
Contr
T2D
SAID
SIDD
SIRD
MOD
MARD
Clusters

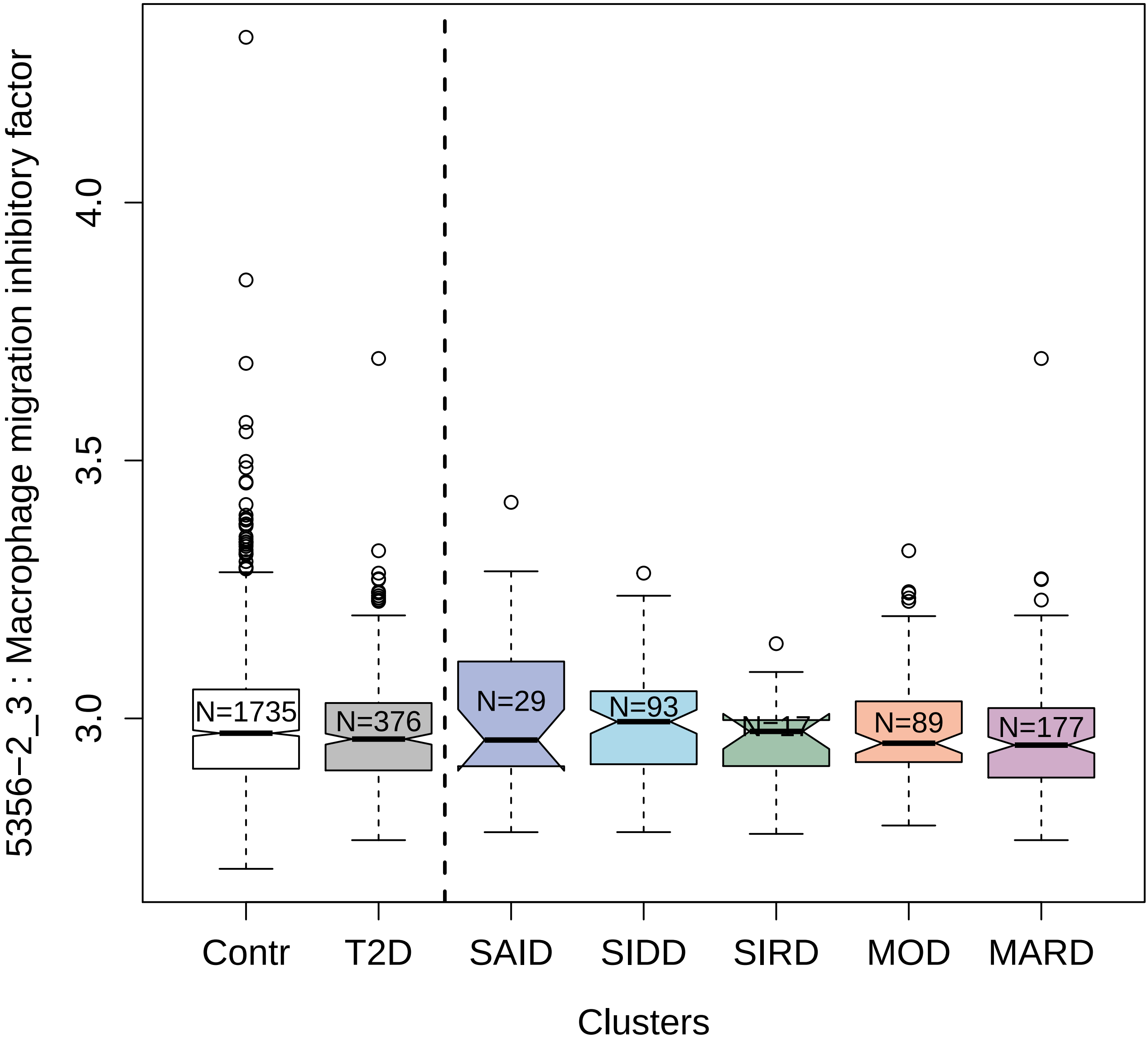

5356−2_3 : Macrophage migration inhibitory factor
4.0
3.5
3.0
N=1735
N=376
N=29
N=93
N=17
N=89
N=177
Contr
T2D
SAID
SIDD
SIRD
MOD
MARD
Clusters

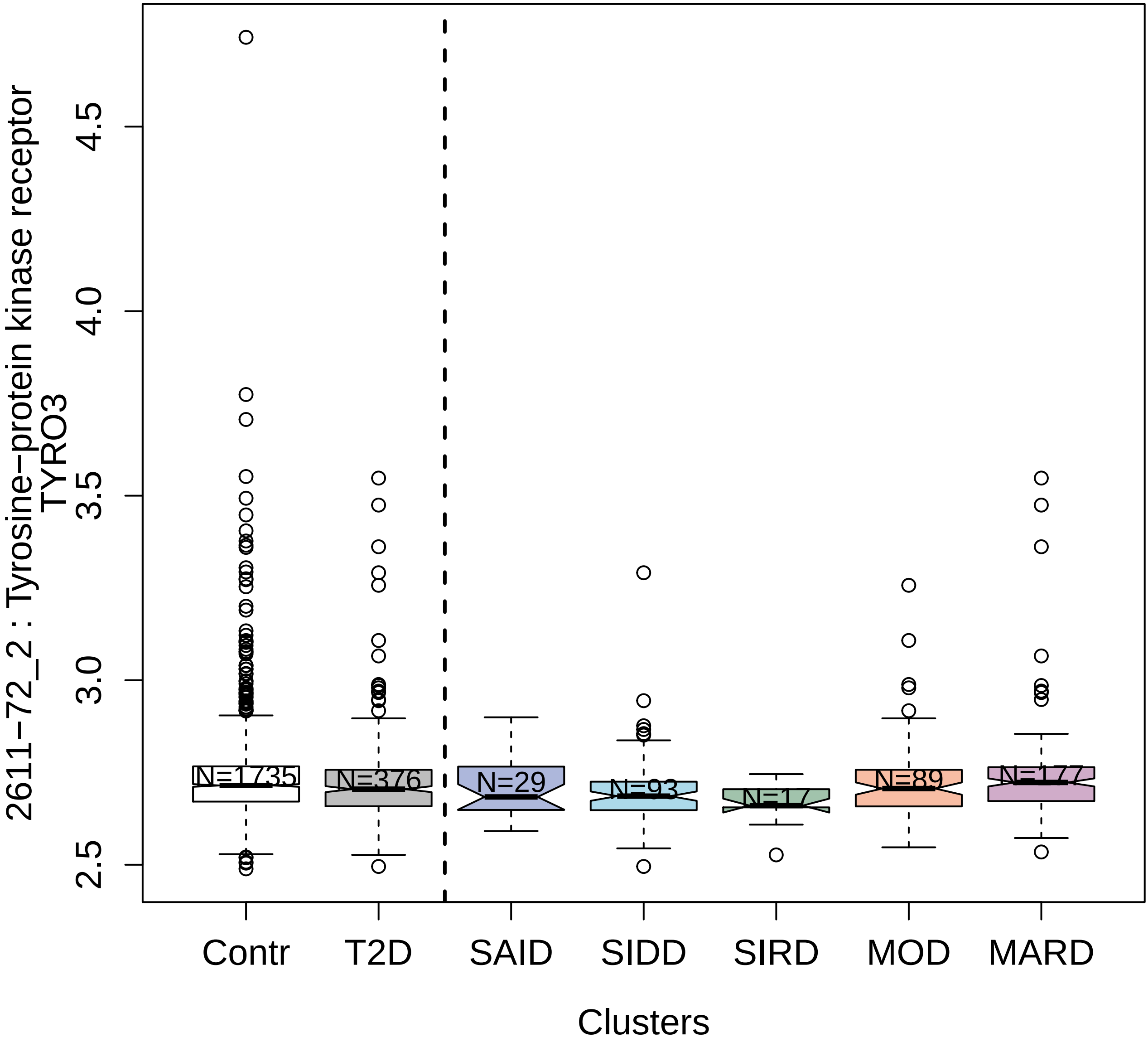

2611−72_2 : Tyrosine−protein kinase receptor
TYRO3
4.5
4.0
3.5
3.0
2.5
N=1735
N=376
N=29
N=93
N=17
N=89
N=177
Contr
T2D
SAID
SIDD
SIRD
MOD
MARD
Clusters

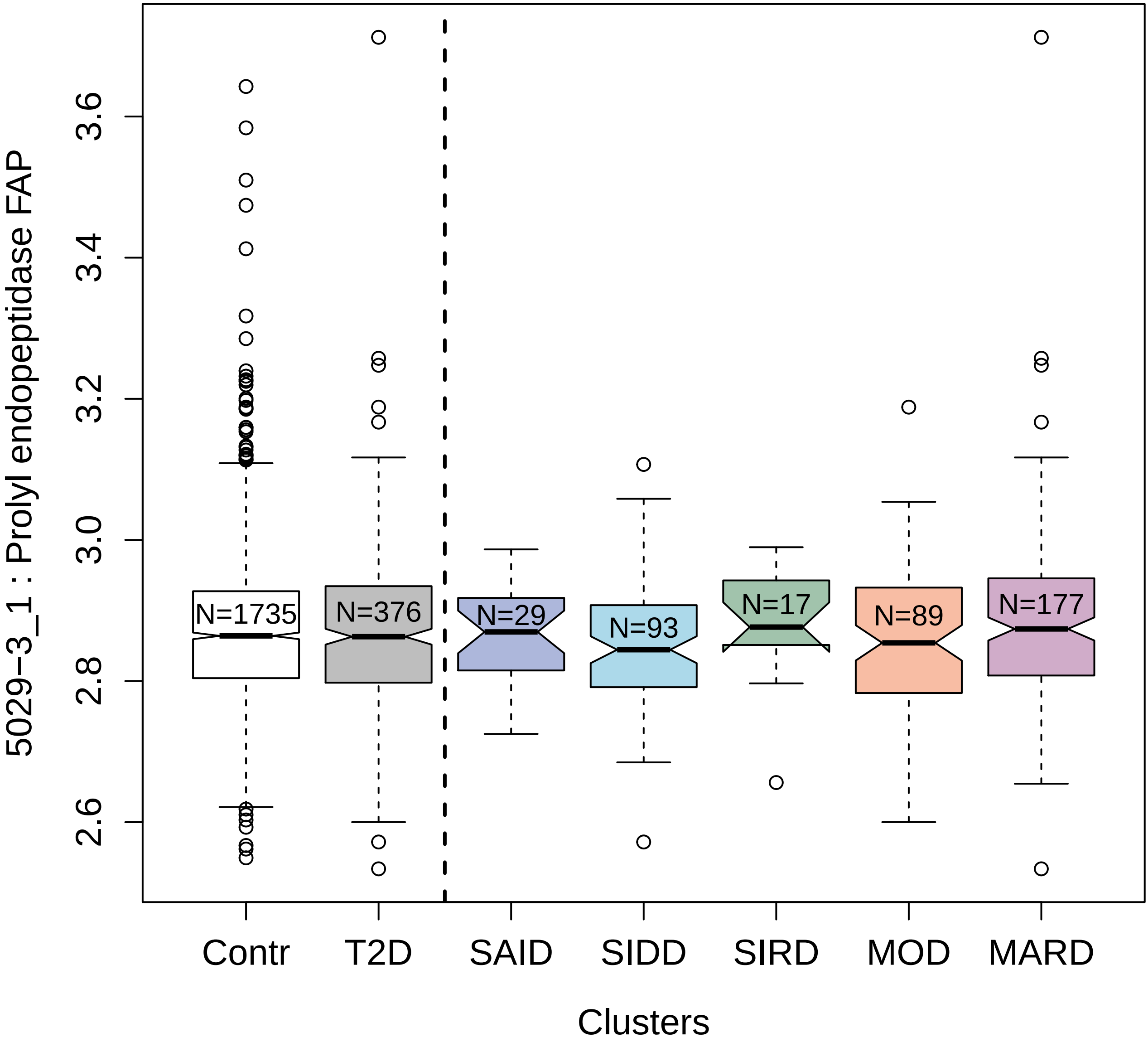

5029−3_1 : Prolyl endopeptidase FAP
2.6
2.8
3.0
3.2
3.4
3.6
N=1735
N=376
N=29
N=93
N=17
N=89
N=177
Contr
T2D
SAID
SIDD
SIRD
MOD
MARD
Clusters

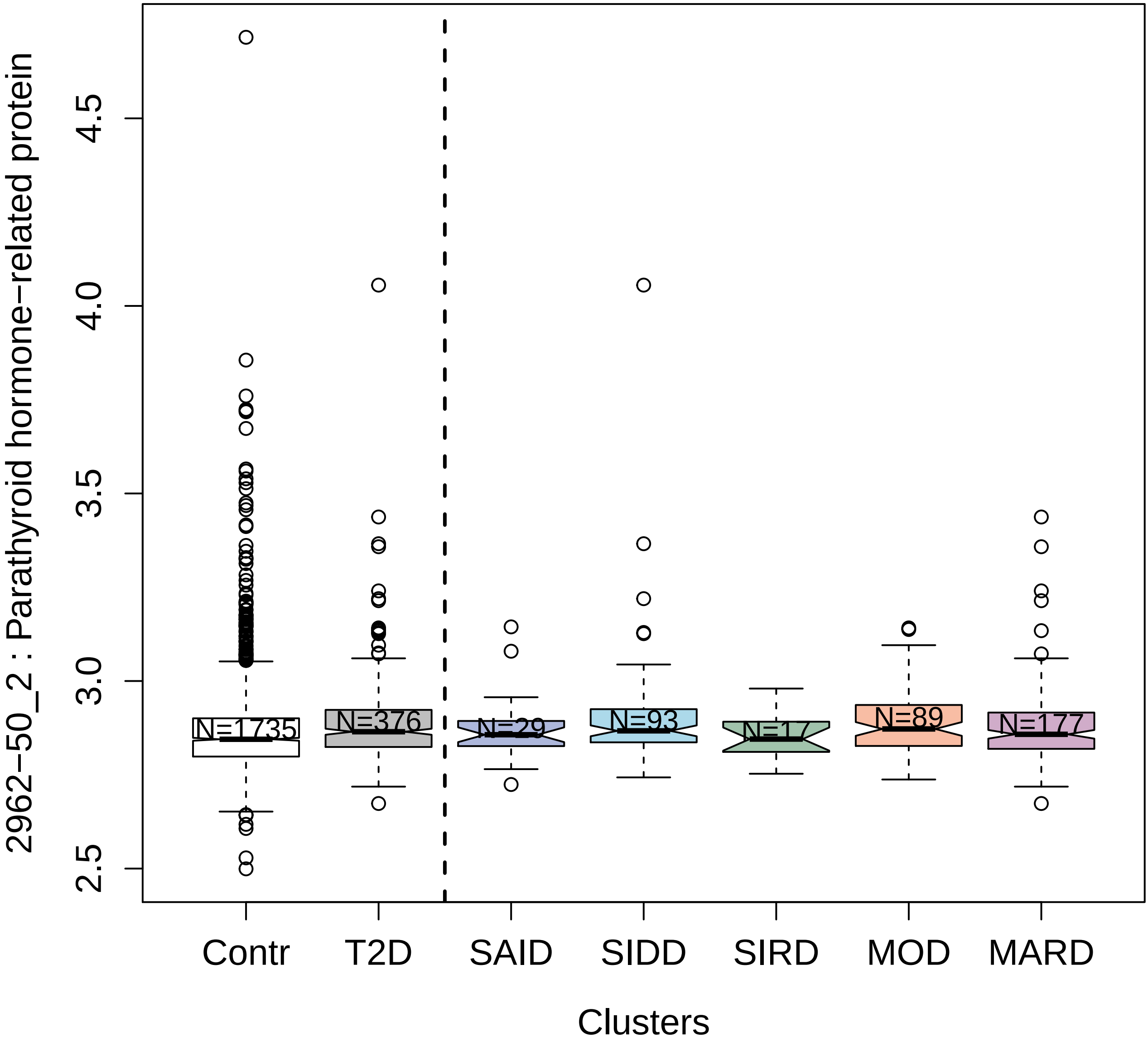
2962−50_2 : Parathyroid hormone−related protein
2.5
3.0
3.5
4.0
4.5
N=1735
N=376
N=29
N=93
N=17
N=89
N=177
Contr
T2D
SAID
SIDD
SIRD
MOD
MARD
Clusters

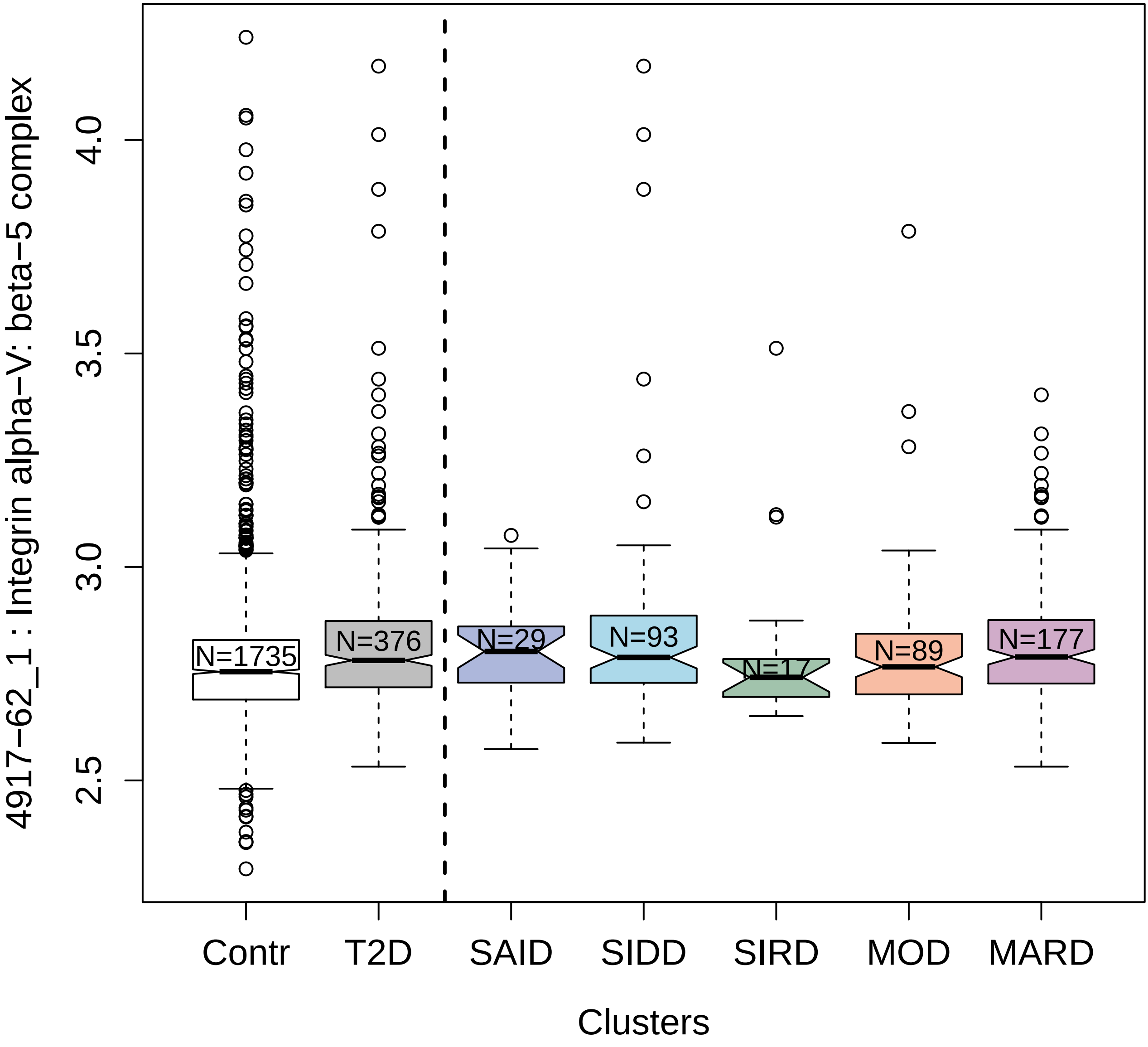

4917−62_1 : Integrin alpha−V: beta−5 complex
2.5
3.0
3.5
4.0
N=1735
N=376
N=29
N=93
N=17
N=89
N=177
Contr
T2D
SAID
SIDD
SIRD
MOD
MARD
Clusters

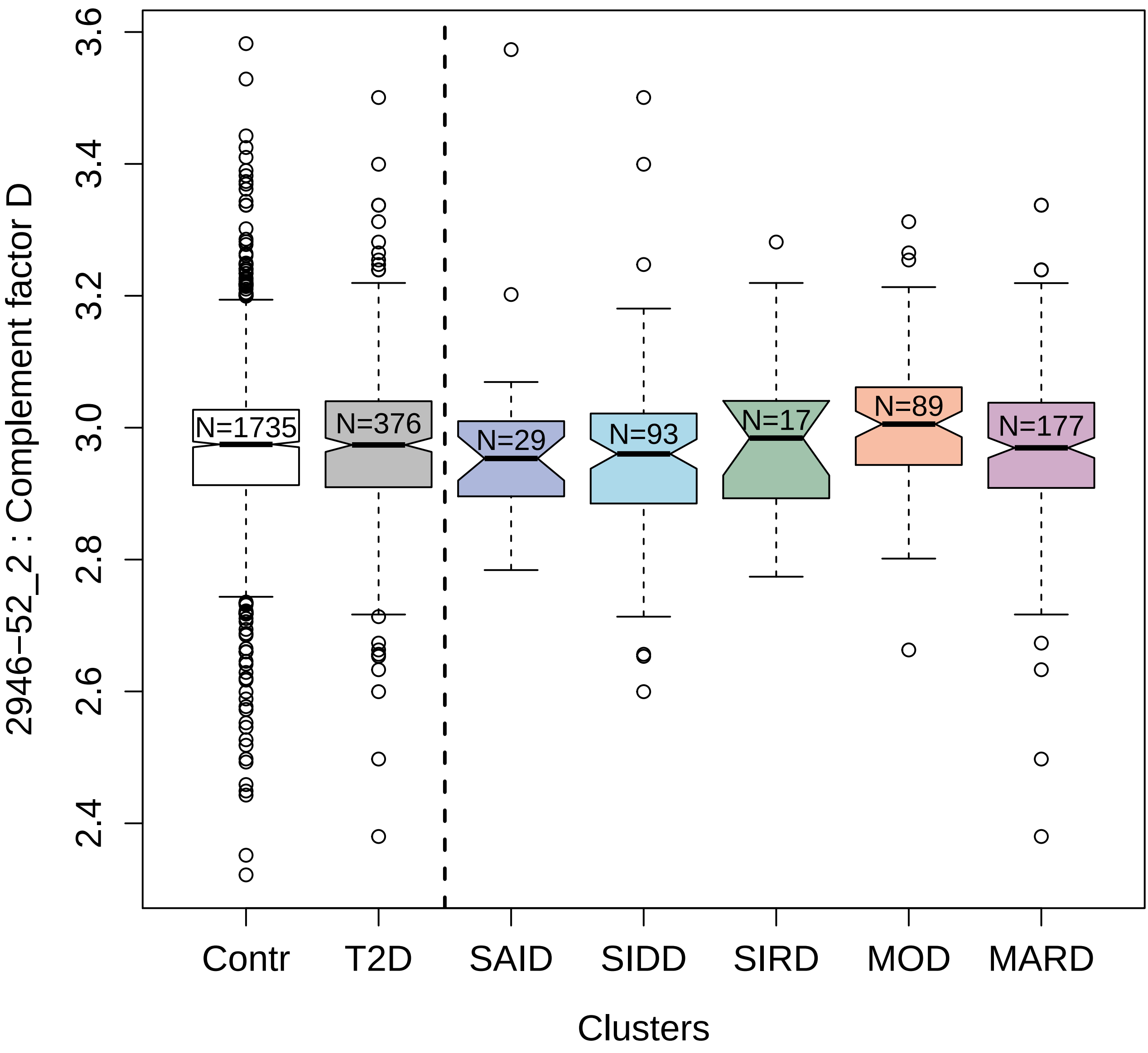

2946−52_2 : Complement factor D
3.6
3.4
3.2
3.0
2.8
2.6
2.4
N=1735
N=376
N=29
N=93
N=17
N=89
N=177
Contr
T2D
SAID
SIDD
SIRD
MOD
MARD
Clusters

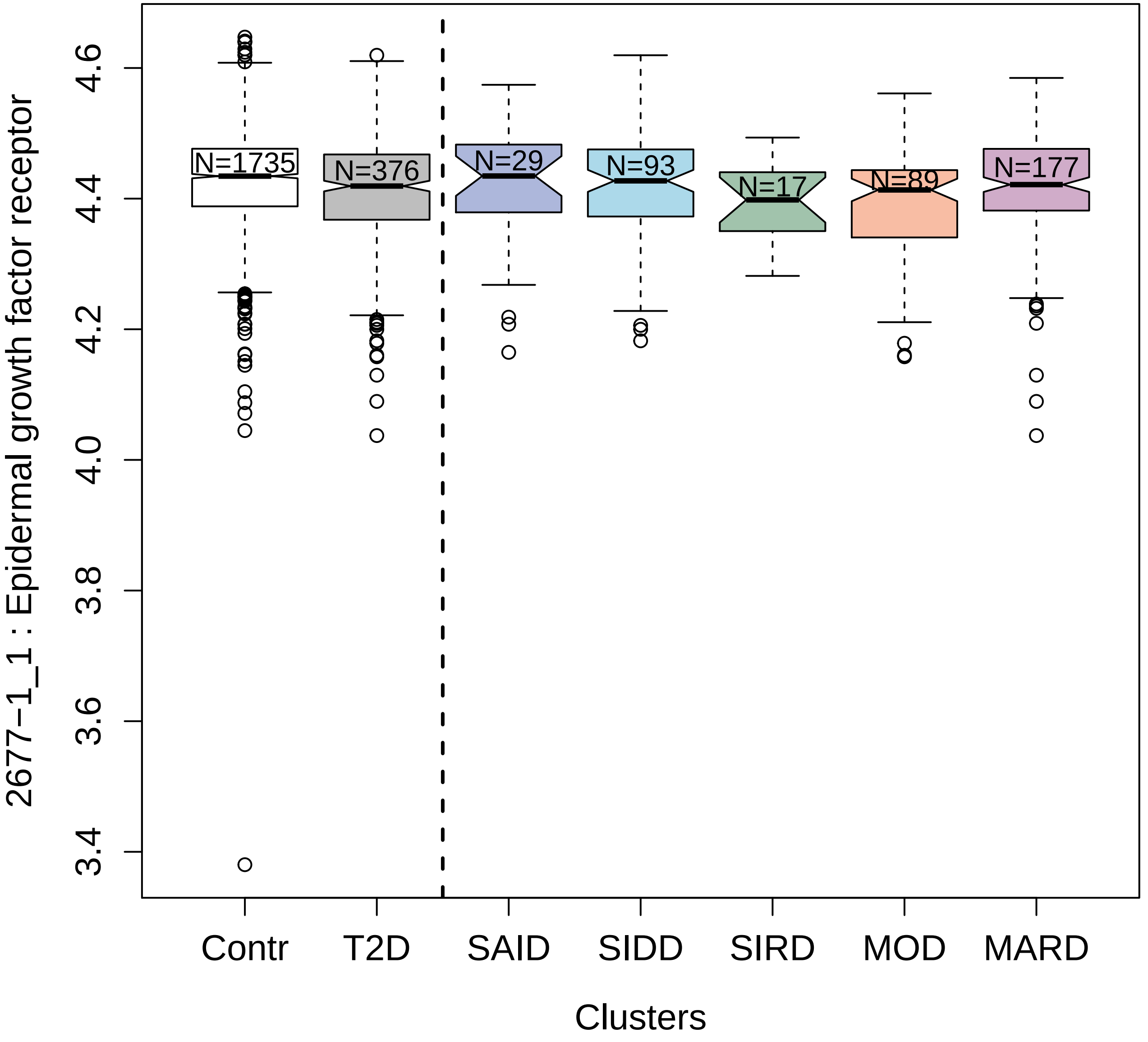

2677−1_1 : Epidermal growth factor receptor
4.6
4.4
4.2
4.0
3.8
3.6
3.4
N=1735
N=376
N=29
N=93
N=17
N=89
N=177
Contr
T2D
SAID
SIDD
SIRD
MOD
MARD
Clusters

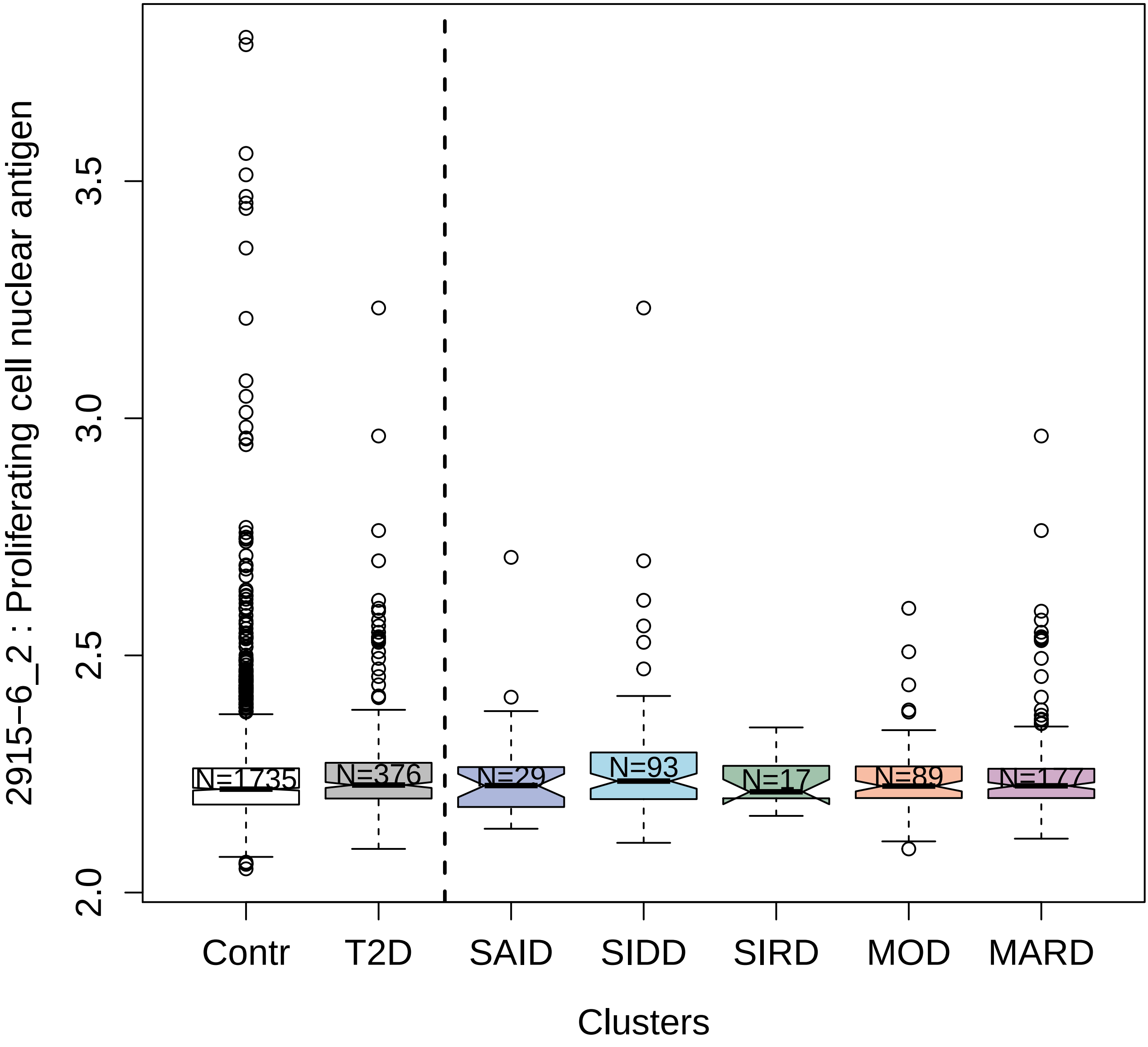

2915−6_2 : Proliferating cell nuclear antigen
2.0
2.5
3.0
3.5
N=1735
N=376
N=29
N=93
N=17
N=89
N=177
Contr
T2D
SAID
SIDD
SIRD
MOD
MARD
Clusters

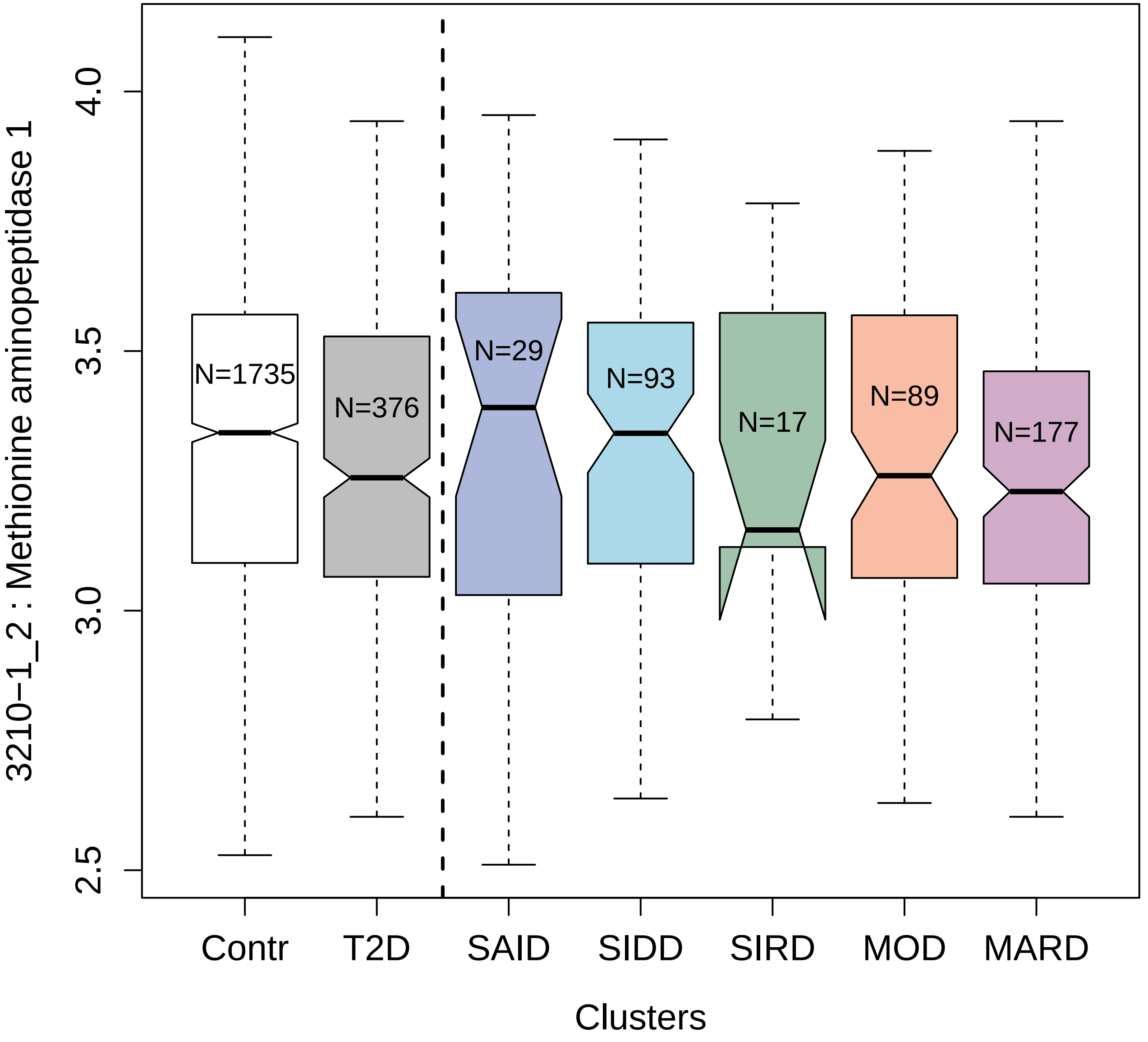

3210−1_2 : Methionine aminopeptidase 1
4.0
3.5
3.0
2.5
N=1735
N=376
N=29
N=93
N=17
N=89
N=177
Contr
T2D
SAID
SIDD
SIRD
MOD
MARD
Clusters

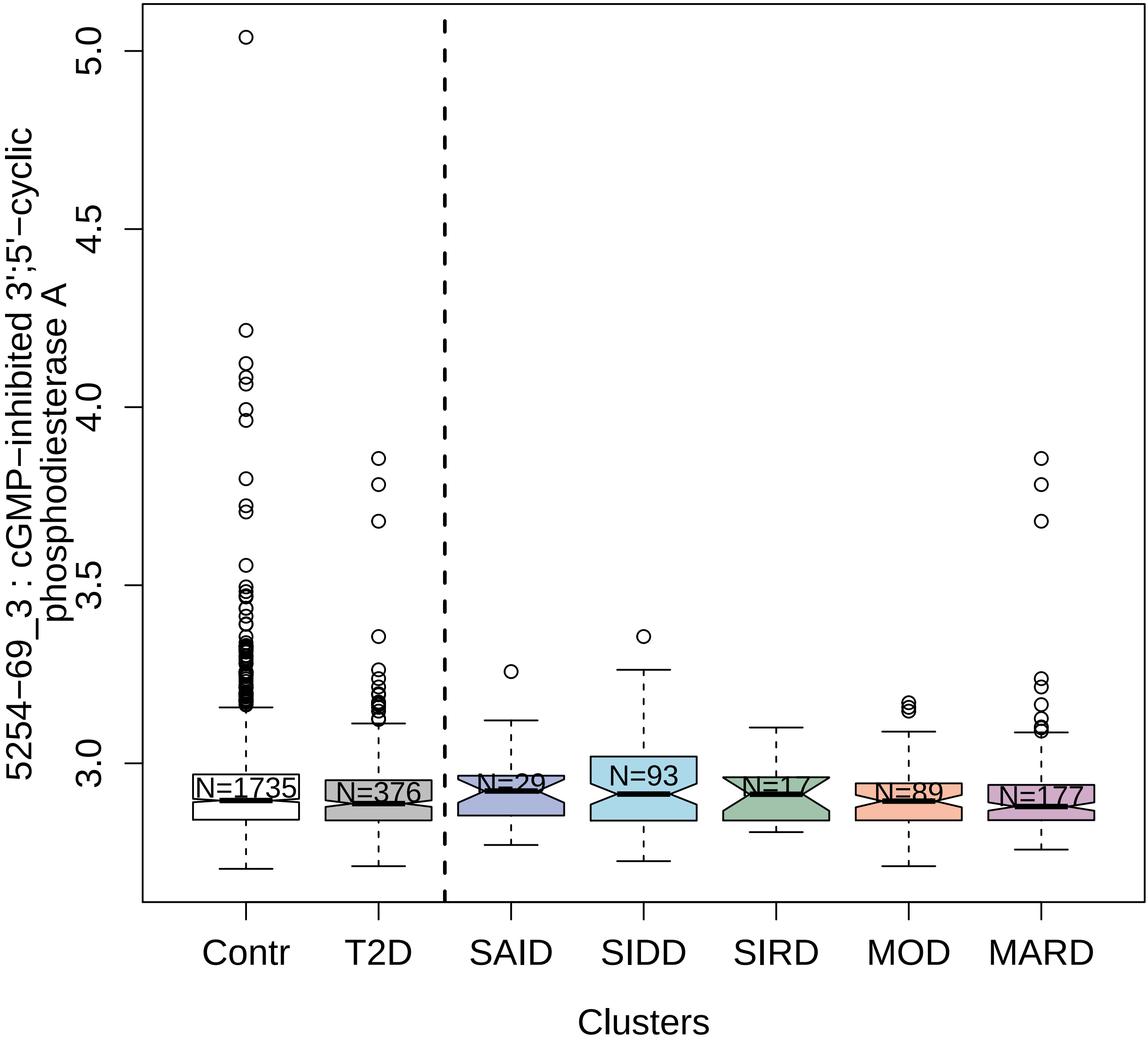
5254−69_3 : cGMP−inhibited 3',5'−cyclic phosphodiesterase A
5.0
4.5
4.0
3.5
3.0
N=1735
N=376
N=29
N=93
N=17
N=89
N=177
Contr
T2D
SAID
SIDD
SIRD
MOD
MARD
Clusters

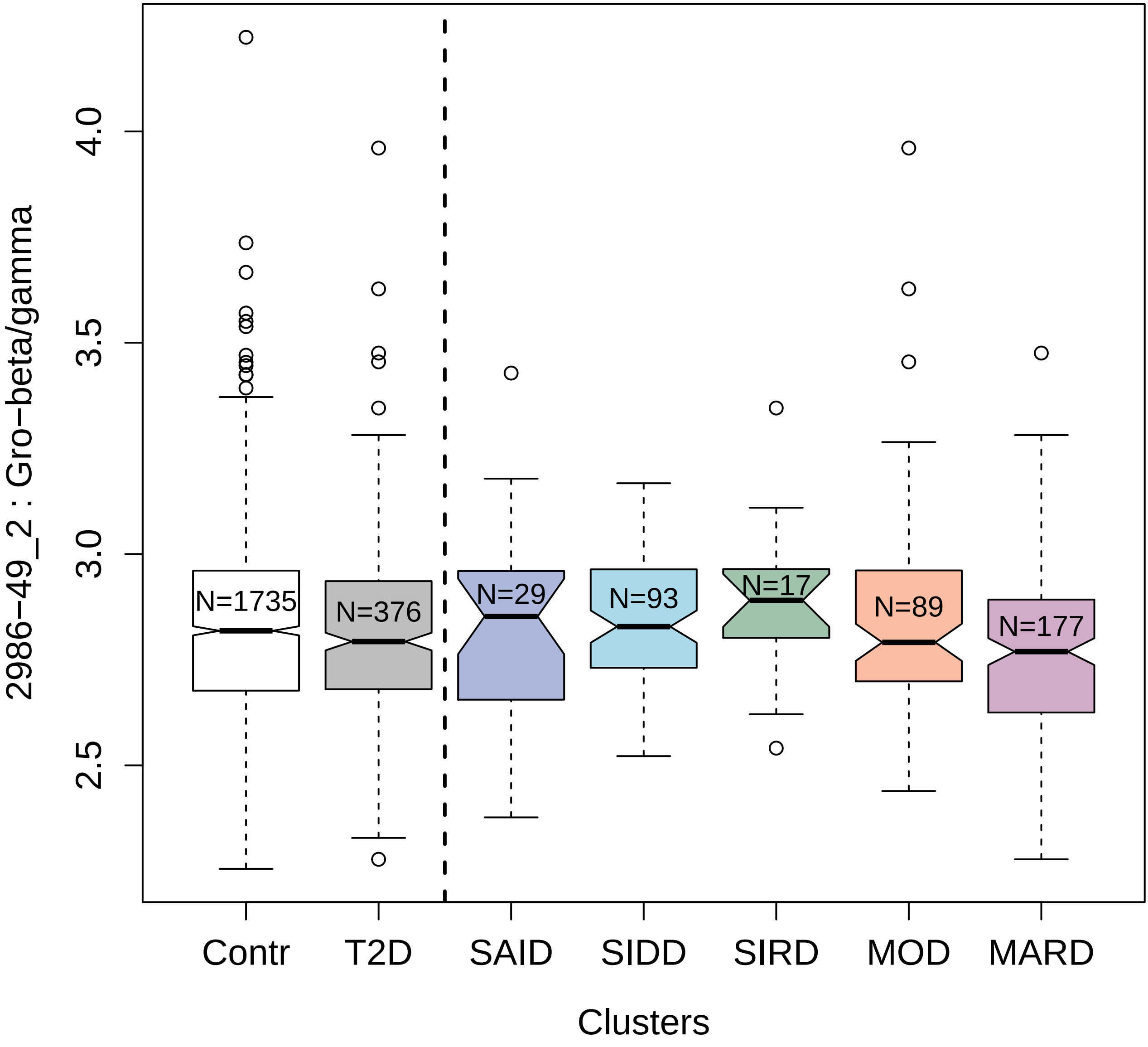
2986−49_2 : Gro−beta/gamma
4.0
3.5
3.0
2.5
N=1735
N=376
N=29
N=93
N=17
N=89
N=177
Contr
T2D
SAID
SIDD
SIRD
MOD
MARD
Clusters

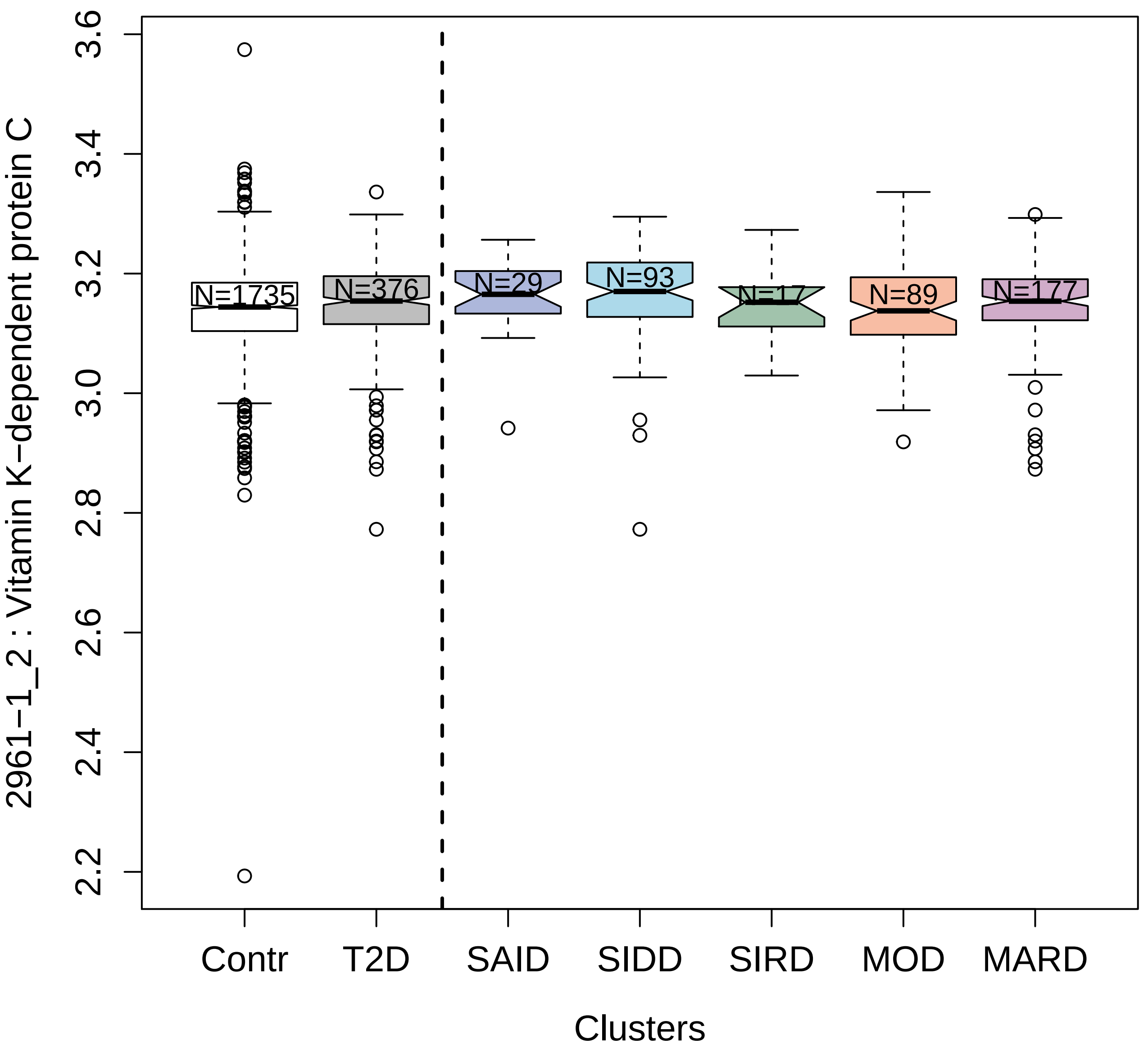

2961−1_2 : Vitamin K−dependent protein C
3.6
3.4
3.2
3.0
2.8
2.6
2.4
2.2
N=1735
N=376
N=29
N=93
N=17
N=89
N=177
Contr
T2D
SAID
SIDD
SIRD
MOD
MARD
Clusters

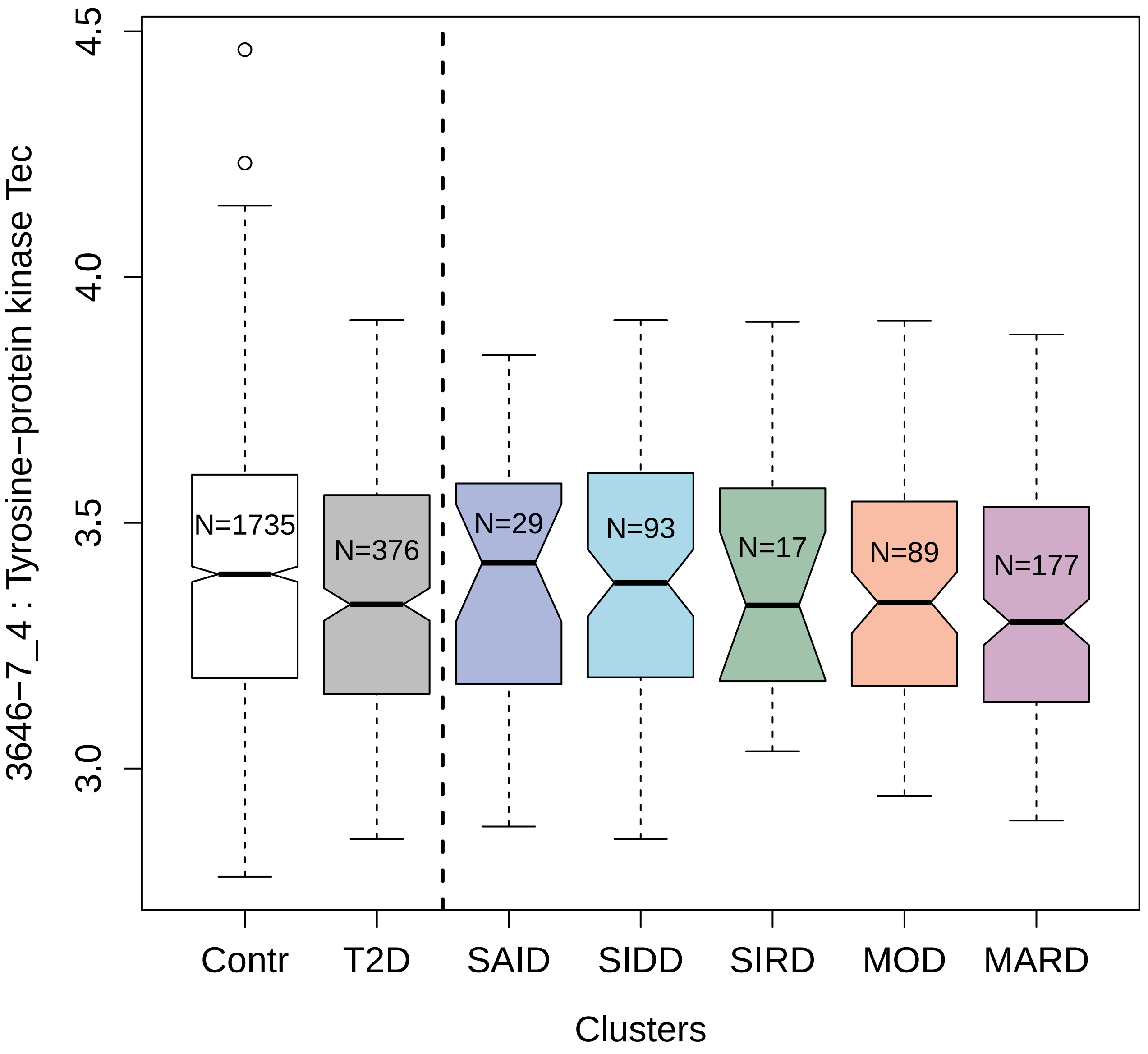
3646−7_4 : Tyrosine−protein kinase Tec
4.5
4.0
3.5
3.0
N=1735
N=376
N=29
N=93
N=17
N=89
N=177
Contr
T2D
SAID
SIDD
SIRD
MOD
MARD
Clusters

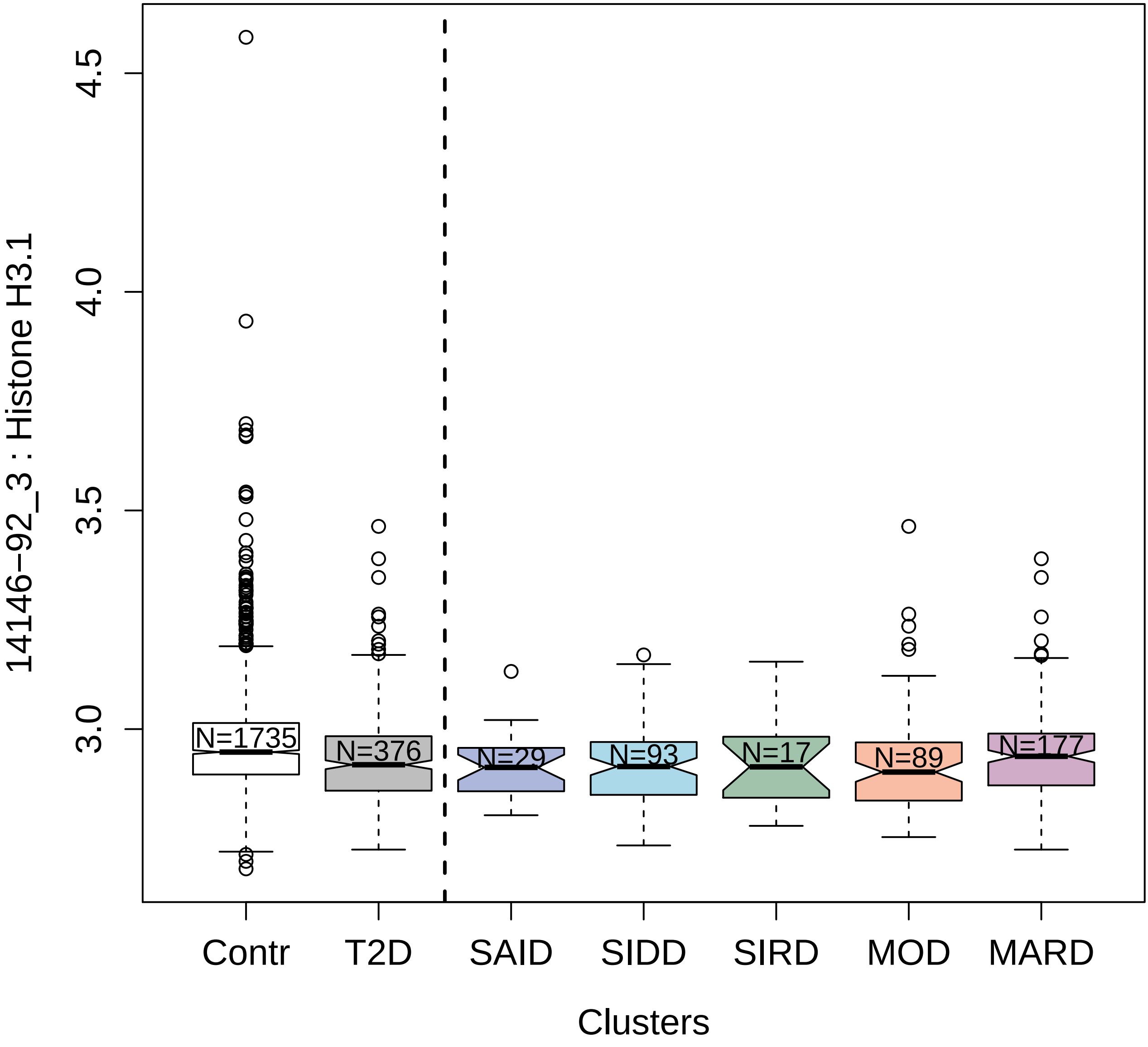
14146−92_3 : Histone H3.1
4.5
4.0
3.5
3.0
N=1735
N=376
N=29
N=93
N=17
N=89
N=177
Contr
T2D
SAID
SIDD
SIRD
MOD
MARD
Clusters

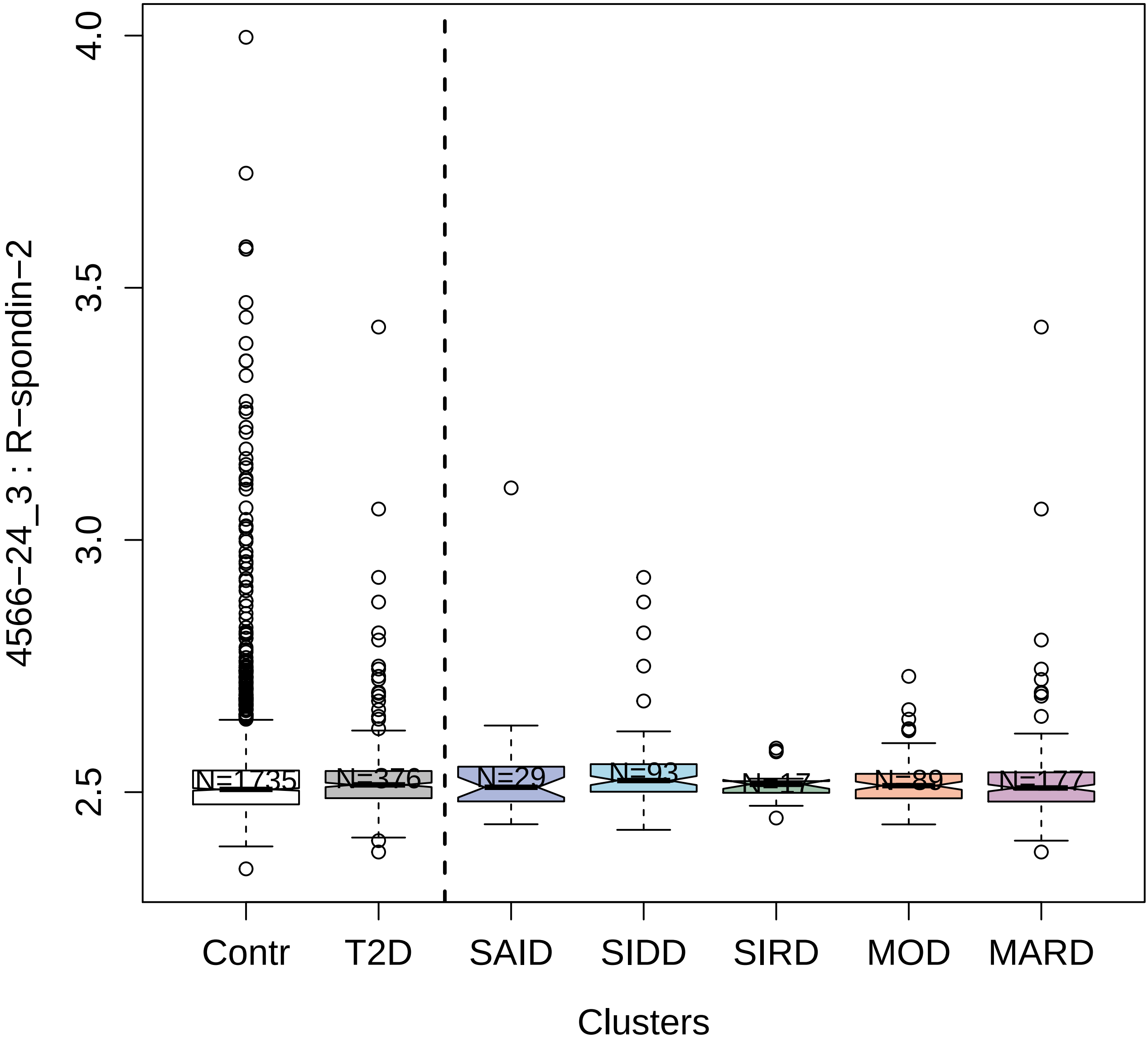

4566−24_3 : R−spondin−2
4.0
3.5
3.0
2.5
N=1735
N=376
N=29
N=93
N=17
N=89
N=177
Contr
T2D
SAID
SIDD
SIRD
MOD
MARD
Clusters

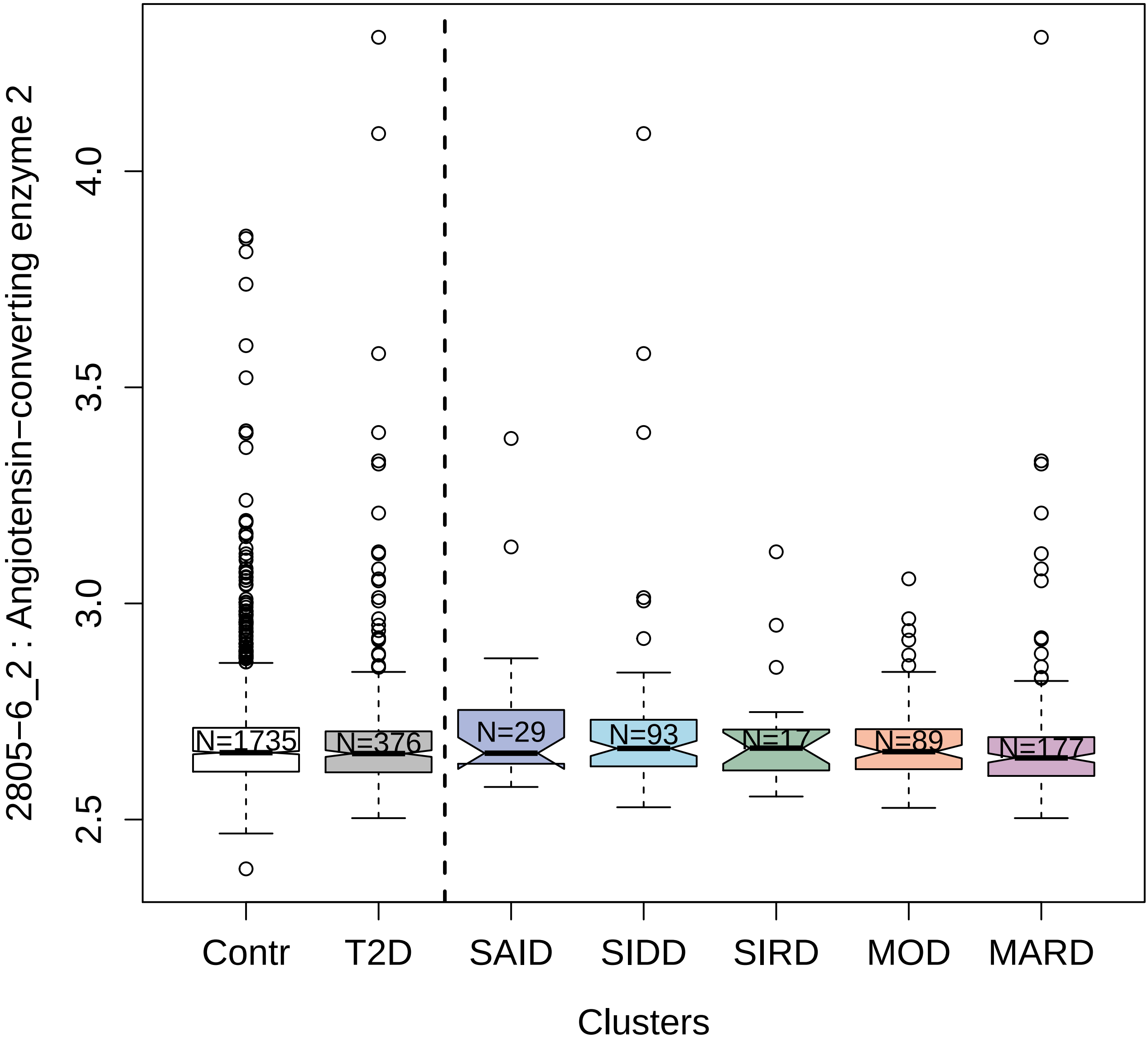

2805−6_2 : Angiotensin−converting enzyme 2
2.5
3.0
3.5
4.0
N=1735
N=376
N=29
N=93
N=17
N=89
N=177
Contr
T2D
SAID
SIDD
SIRD
MOD
MARD
Clusters

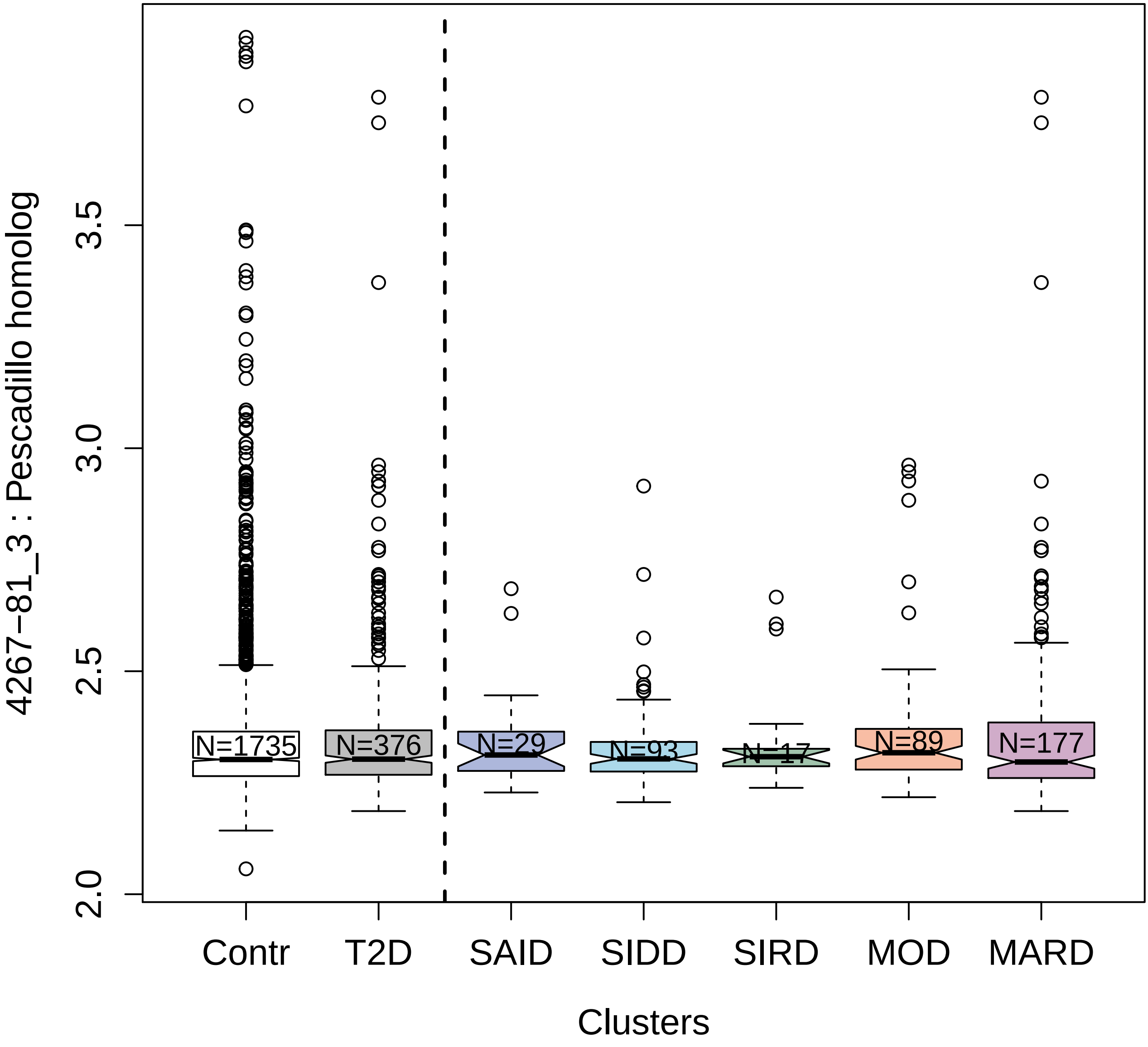

4267−81_3 : Pescadillo homolog
2.0
2.5
3.0
3.5
N=1735
N=376
N=29
N=93
N=17
N=89
N=177
Contr
T2D
SAID
SIDD
SIRD
MOD
MARD
Clusters

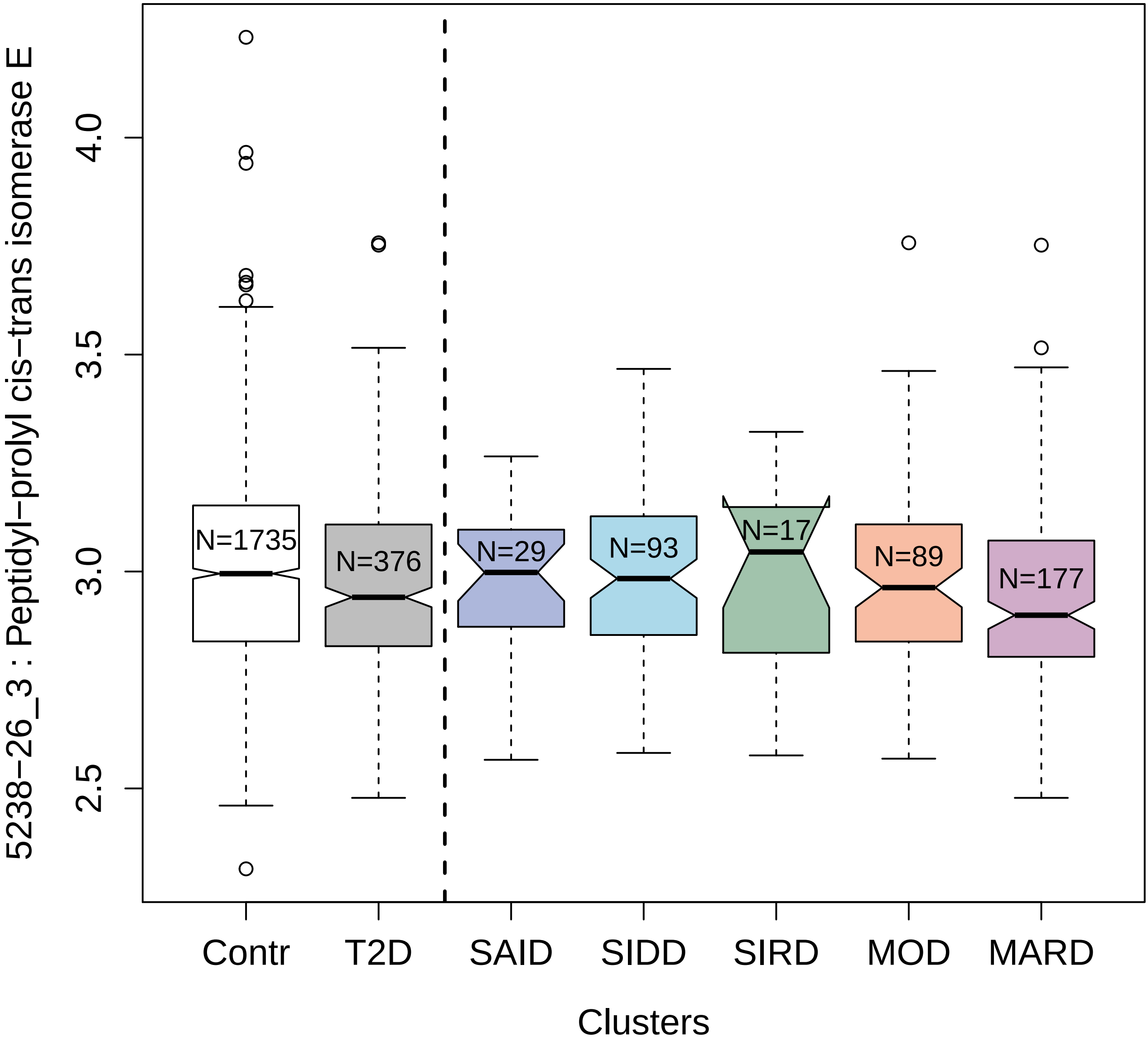
5238−26_3 : Peptidyl−prolyl cis−trans isomerase E
4.0
3.5
3.0
2.5
N=1735
N=376
N=29
N=93
N=17
N=89
N=177
Contr
T2D
SAID
SIDD
SIRD
MOD
MARD
Clusters

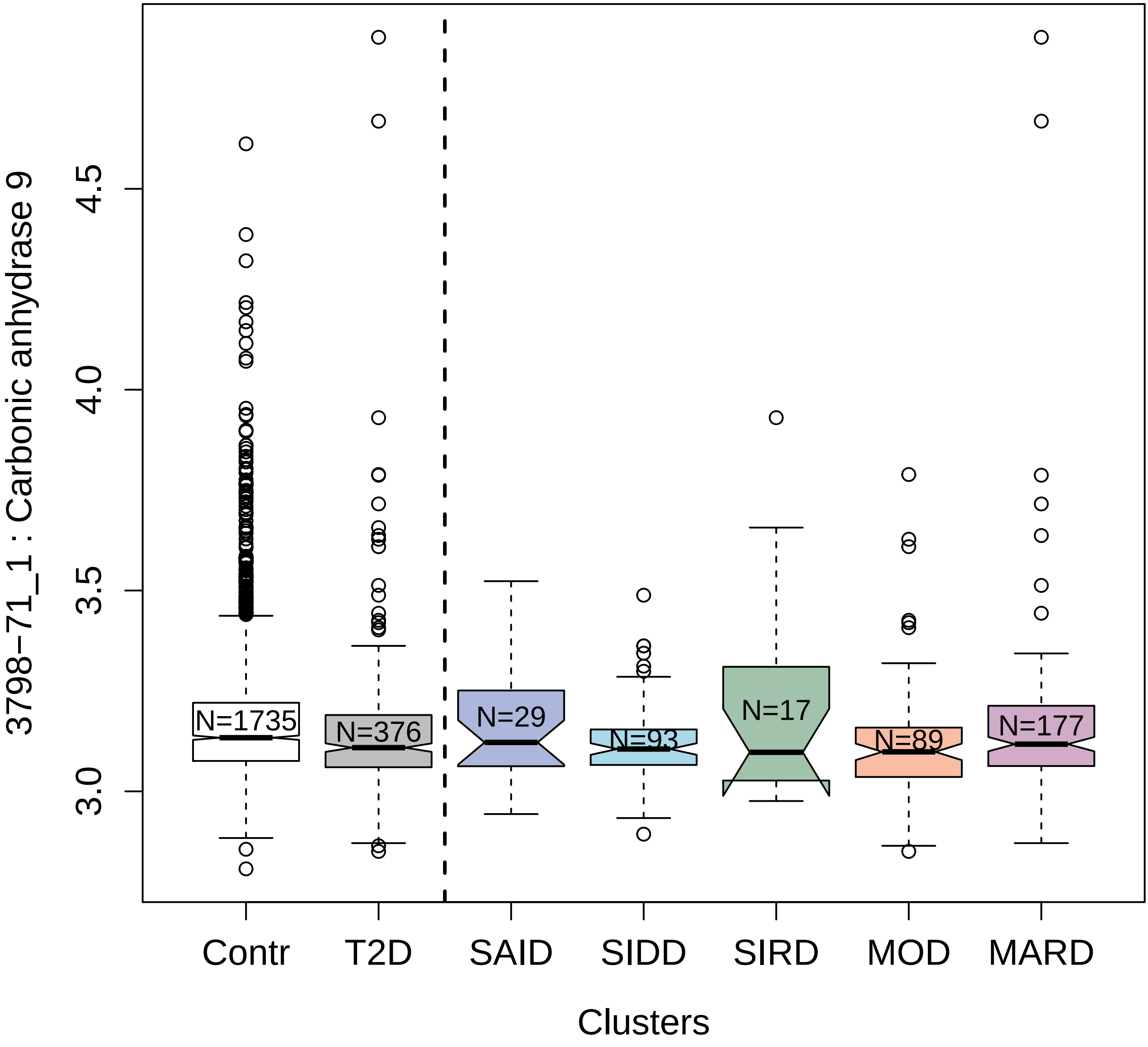
3798−71_1 : Carbonic anhydrase 9
3.0
3.5
4.0
4.5
N=1735
N=376
N=29
N=93
N=17
N=89
N=177
Contr
T2D
SAID
SIDD
SIRD
MOD
MARD
Clusters

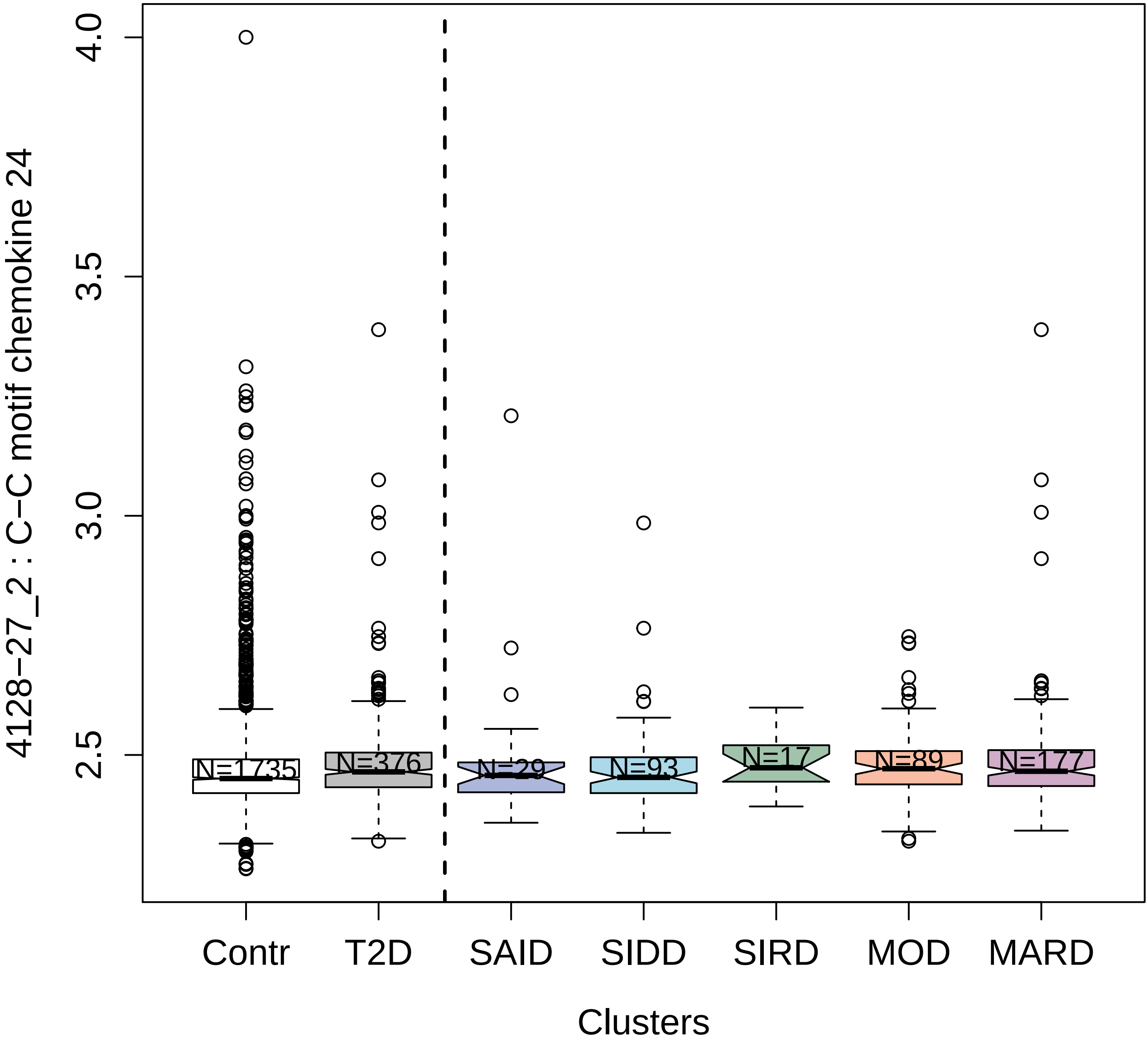

4128−27_2 : C−C motif chemokine 24
4.0
3.5
3.0
2.5
N=1735
N=376
N=29
N=93
N=17
N=89
N=177
Contr
T2D
SAID
SIDD
SIRD
MOD
MARD
Clusters

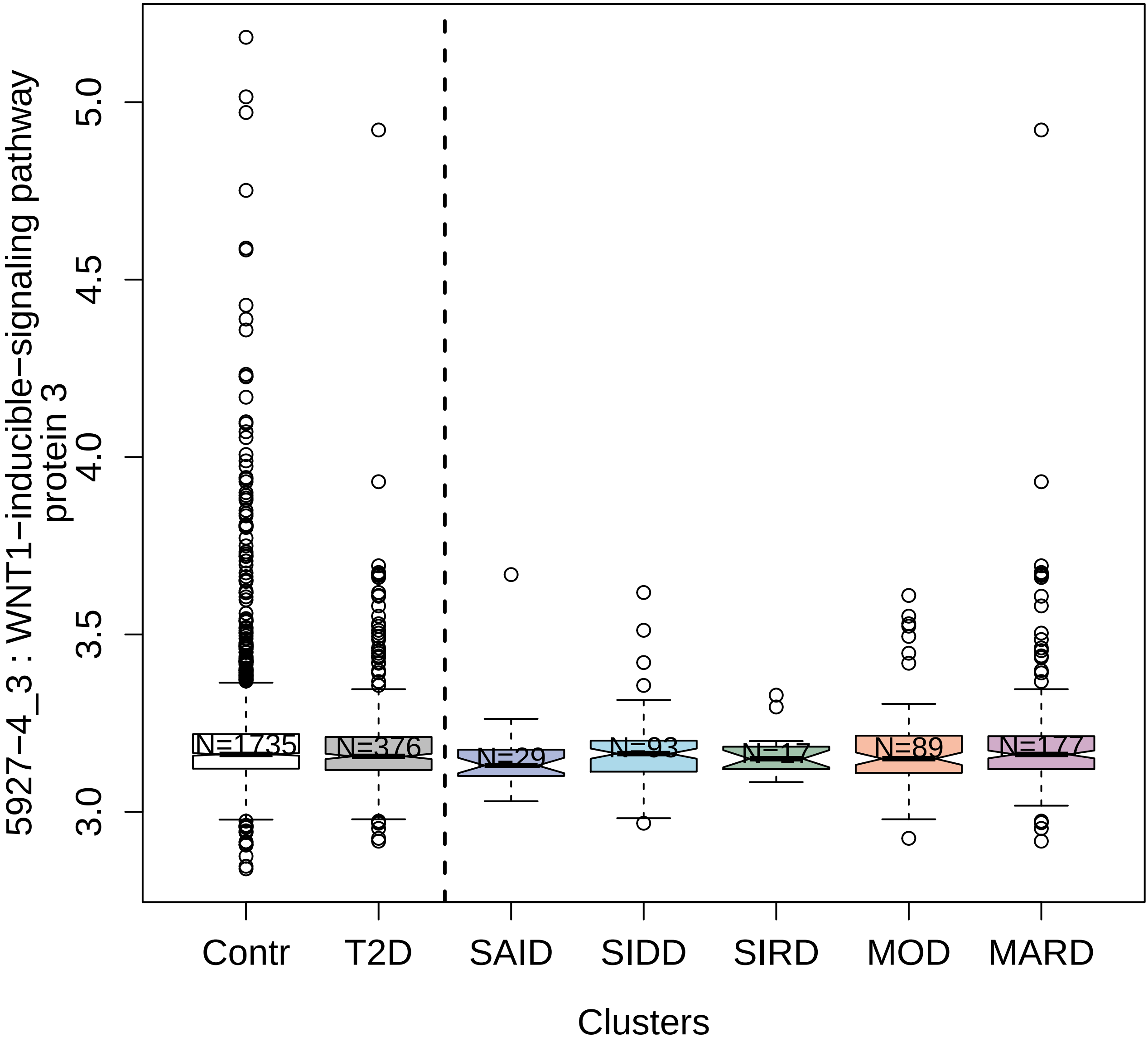
5927−4_3 : WNT1−inducible−signaling pathway
protein 3
5.0
4.5
4.0
3.5
3.0
N=1735
N=376
N=29
N=93
N=17
N=89
N=177
Contr
T2D
SAID
SIDD
SIRD
MOD
MARD
Clusters

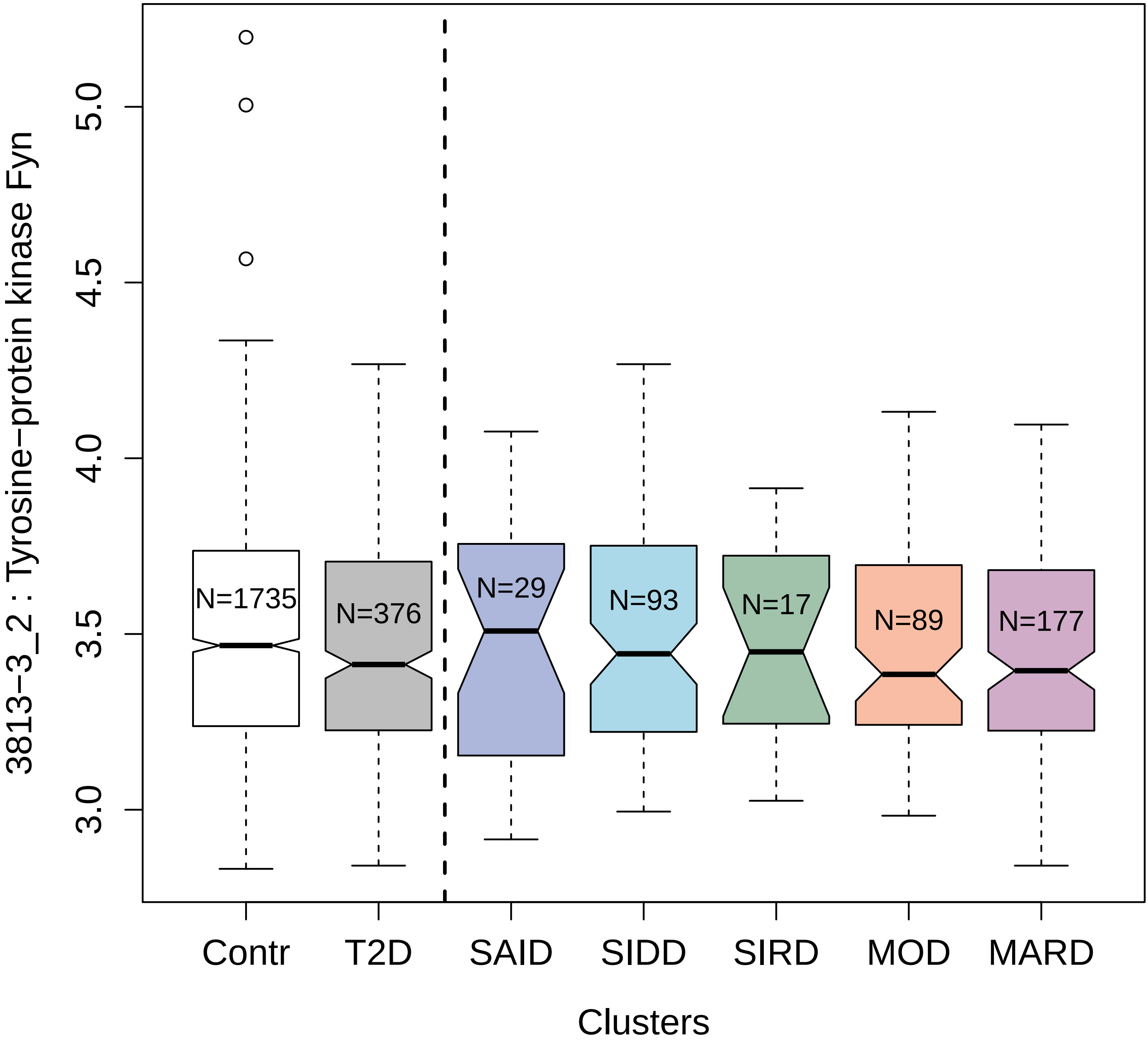

3813−3_2 : Tyrosine−protein kinase Fyn
3.0
3.5
4.0
4.5
5.0
N=1735
N=376
N=29
N=93
N=17
N=89
N=177
Contr
T2D
SAID
SIDD
SIRD
MOD
MARD
Clusters

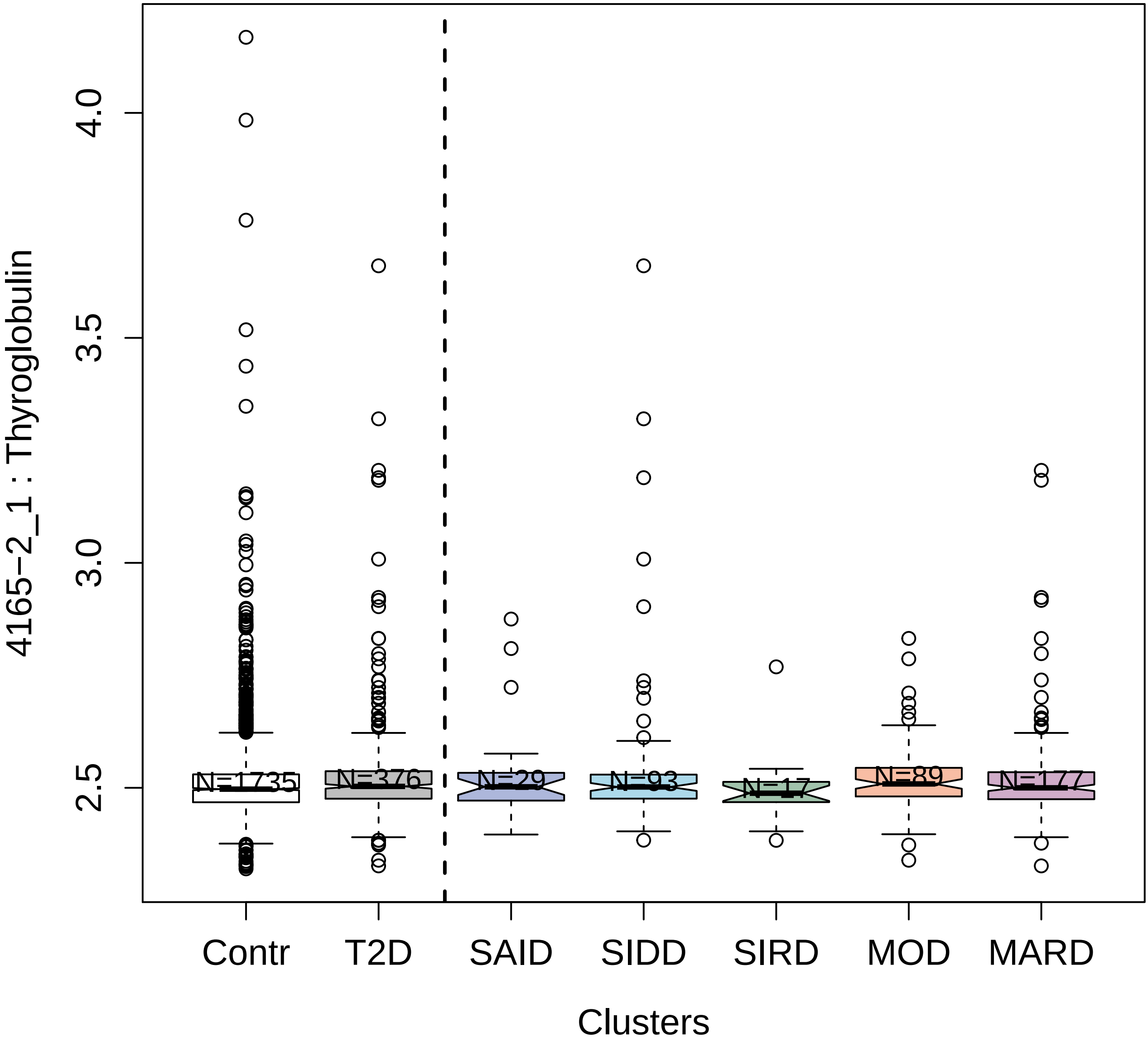
4165−2_1 : Thyroglobulin
4.0
3.5
3.0
2.5
N=1735
N=376
N=29
N=93
N=17
N=89
N=177
Contr
T2D
SAID
SIDD
SIRD
MOD
MARD
Clusters

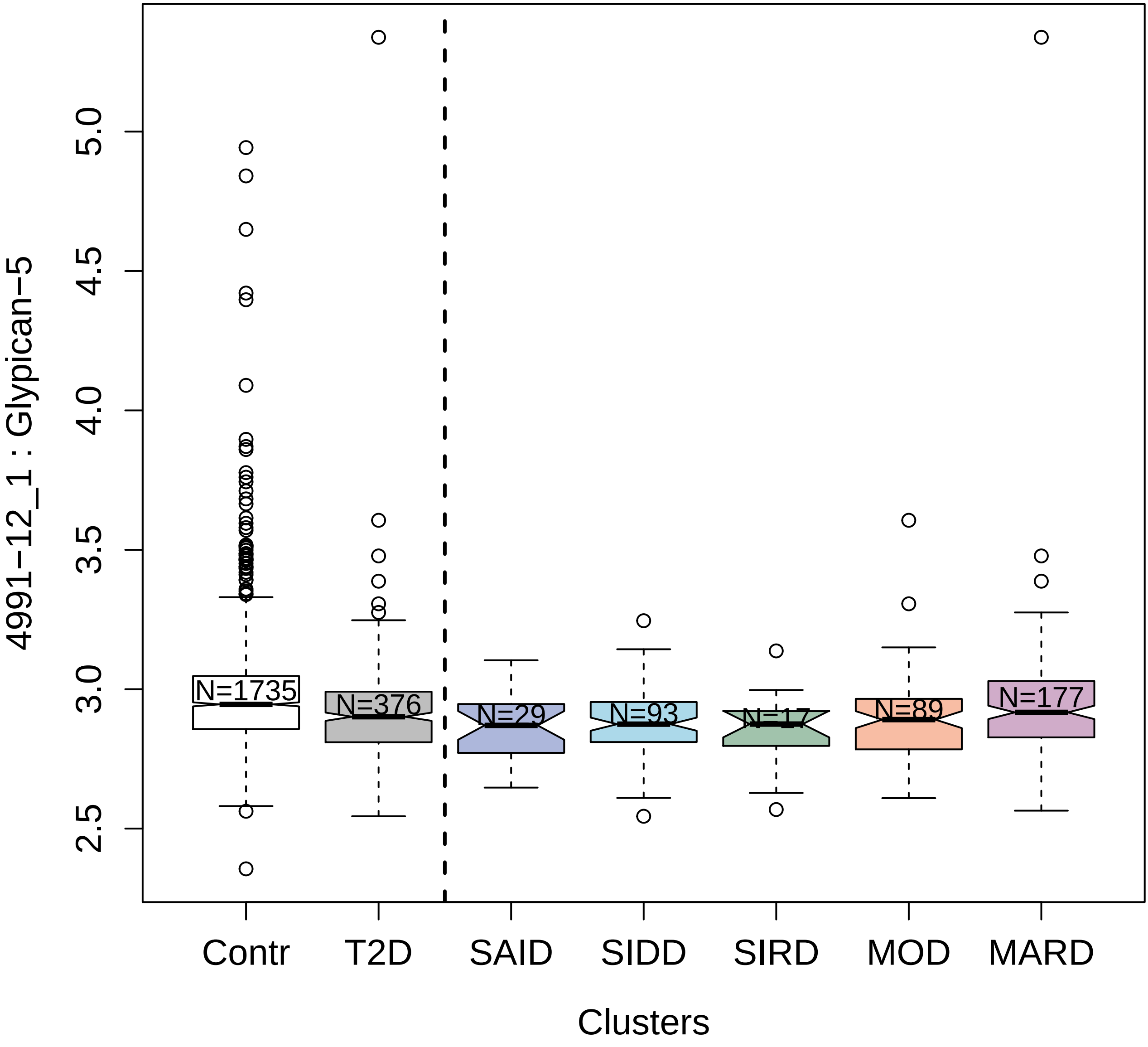
4991−12_1 : Glypican−5
5.0
4.5
4.0
3.5
3.0
2.5
N=1735
N=376
N=29
N=93
N=17
N=89
N=177
Contr
T2D
SAID
SIDD
SIRD
MOD
MARD
Clusters

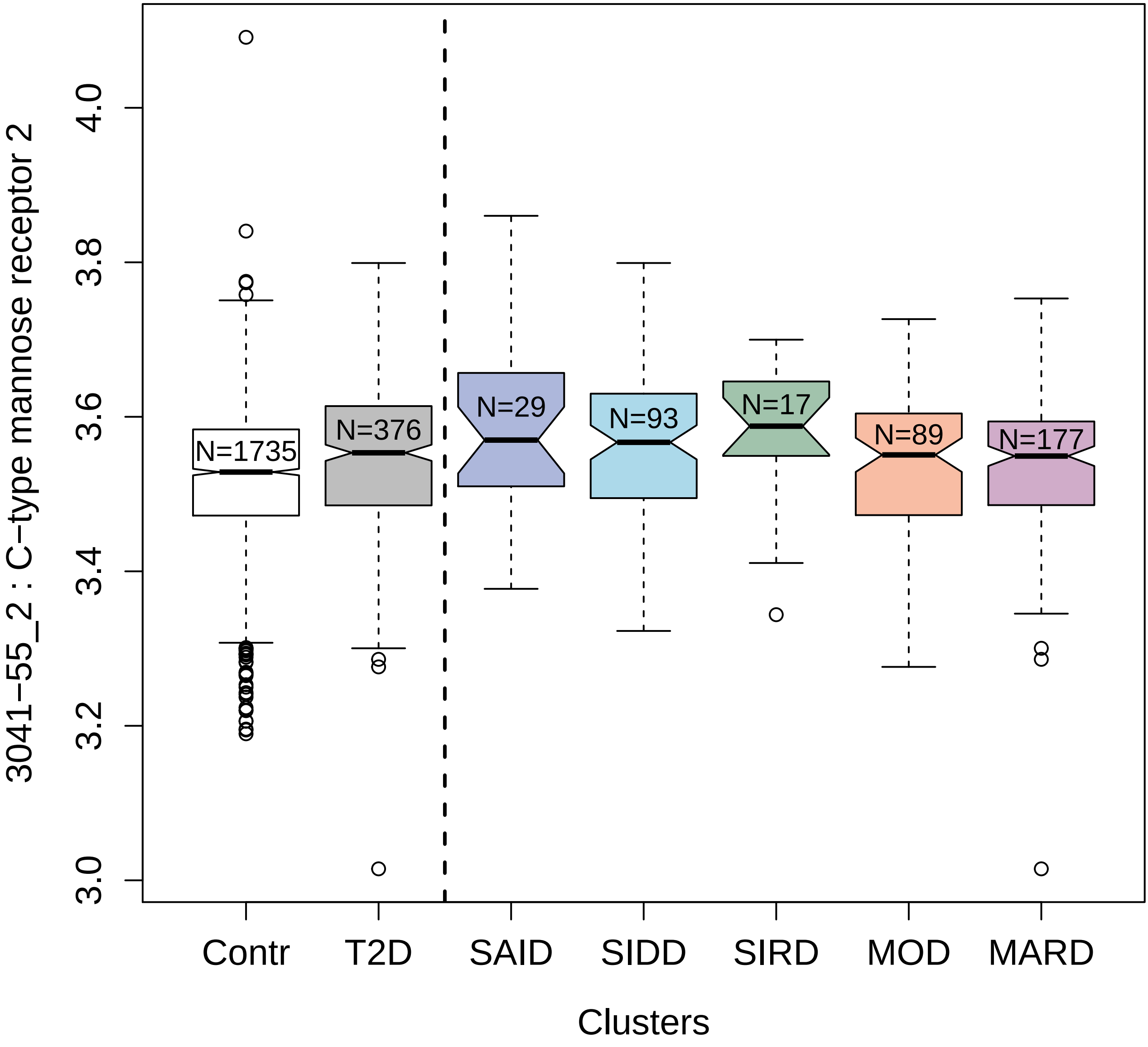

3041−55_2 : C−type mannose receptor 2
4.0
3.8
3.6
3.4
3.2
3.0
N=1735
N=376
N=29
N=93
N=17
N=89
N=177
Contr
T2D
SAID
SIDD
SIRD
MOD
MARD
Clusters

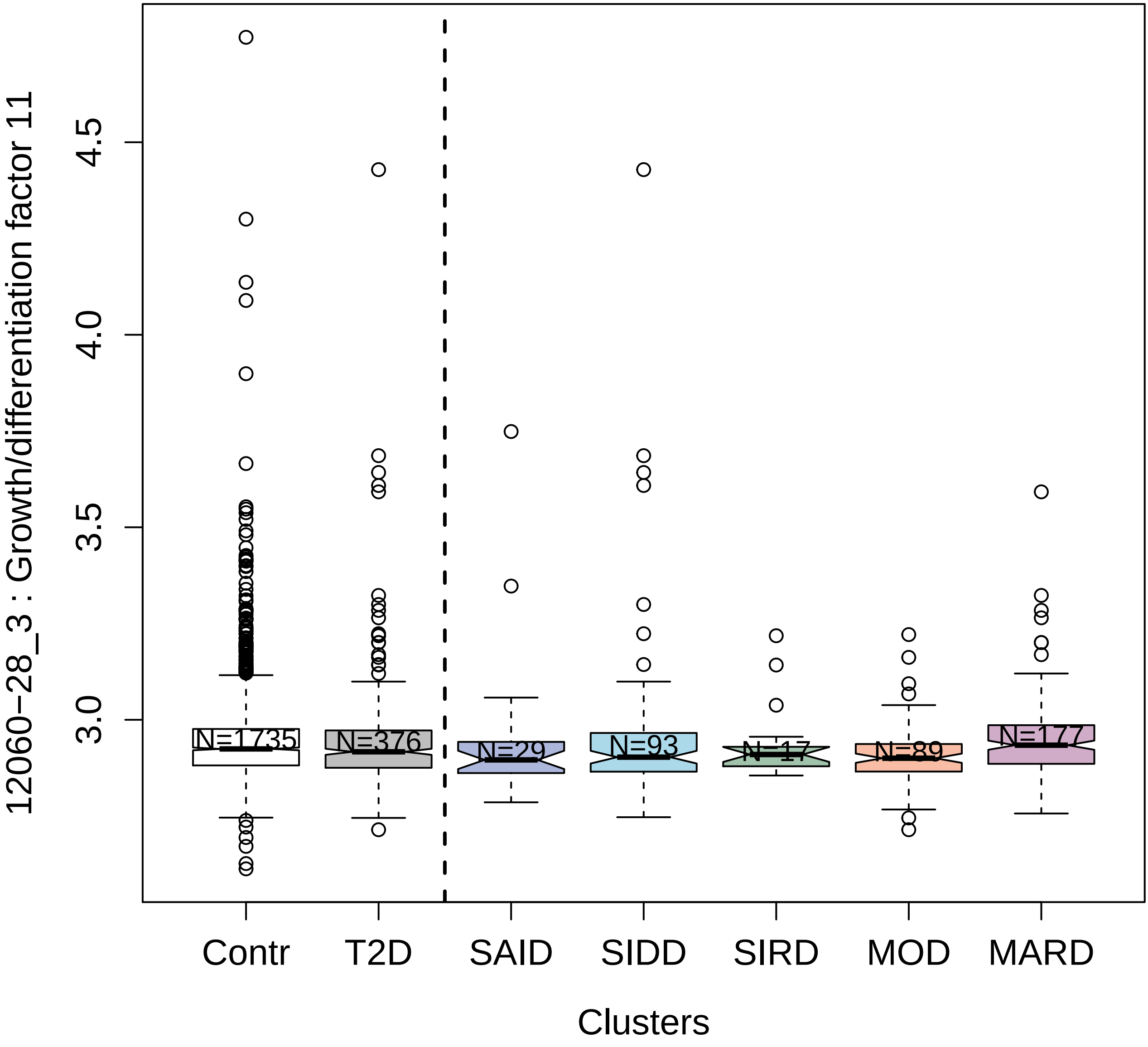
12060−28_3 : Growth/differentiation factor 11
4.5
4.0
3.5
3.0
N=1735
N=376
N=29
N=93
N=17
N=89
N=177
Contr
T2D
SAID
SIDD
SIRD
MOD
MARD
Clusters

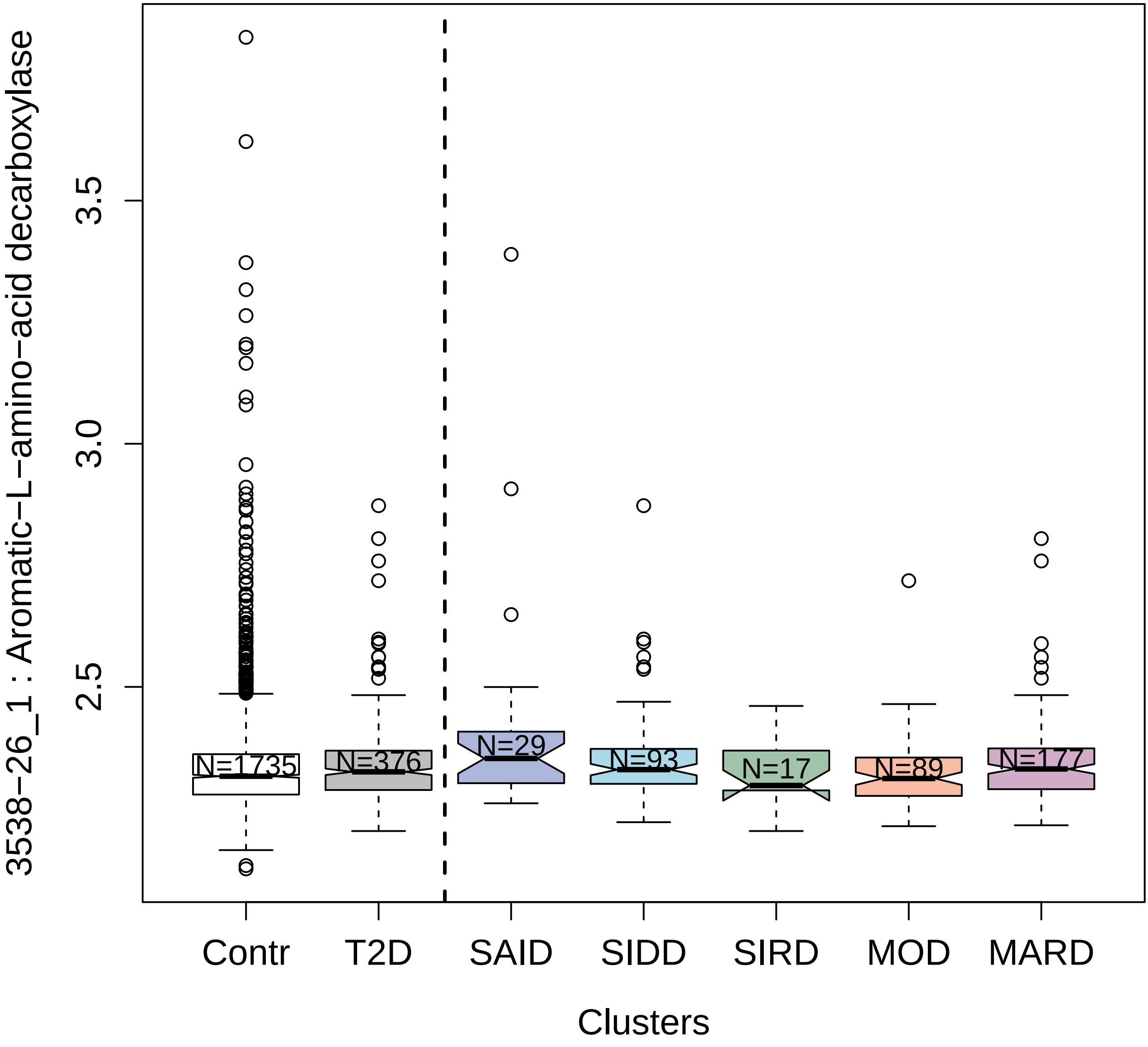
3538−26_1 : Aromatic−L−amino−acid decarboxylase
3.5
3.0
2.5
N=1735
N=376
N=29
N=93
N=17
N=89
N=177
Contr
T2D
SAID
SIDD
SIRD
MOD
MARD
Clusters

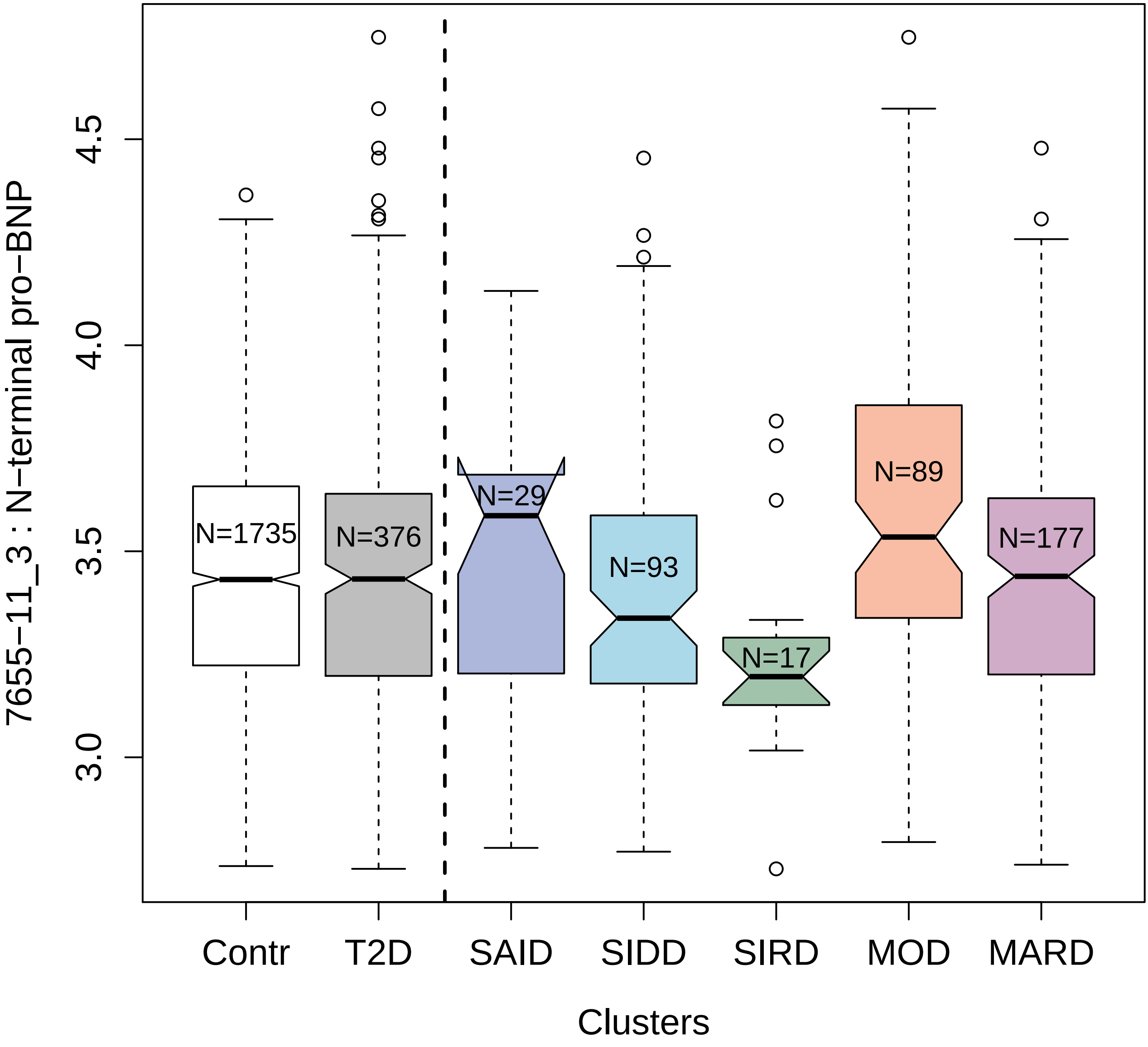

7655−11_3 : N−terminal pro−BNP
3.0
3.5
4.0
4.5
N=1735
N=376
N=29
N=93
N=17
N=89
N=177
Contr
T2D
SAID
SIDD
SIRD
MOD
MARD
Clusters

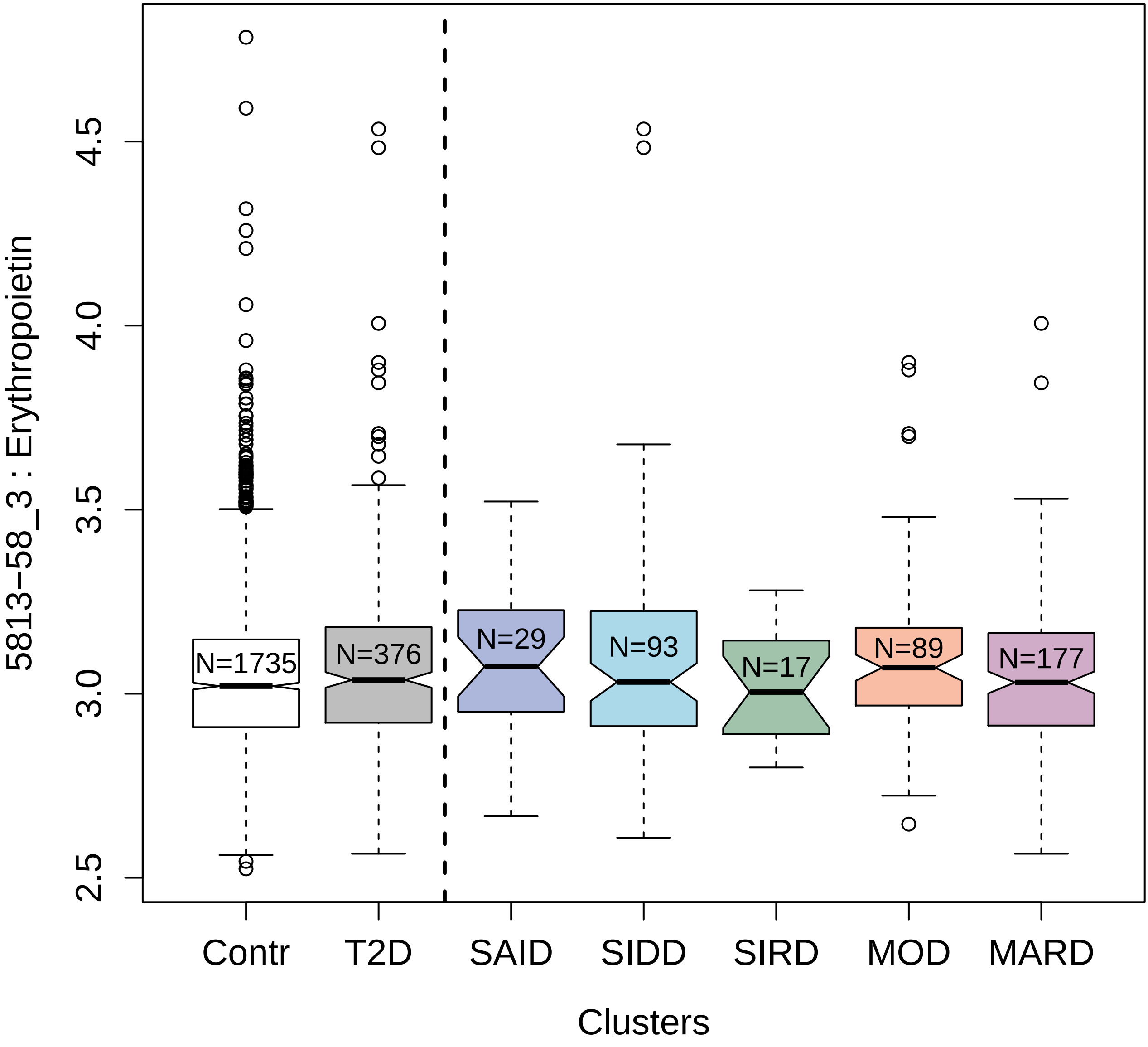

5813−58_3 : Erythropoietin
2.5
3.0
3.5
4.0
4.5
N=1735
N=376
N=29
N=93
N=17
N=89
N=177
Contr
T2D
SAID
SIDD
SIRD
MOD
MARD
Clusters

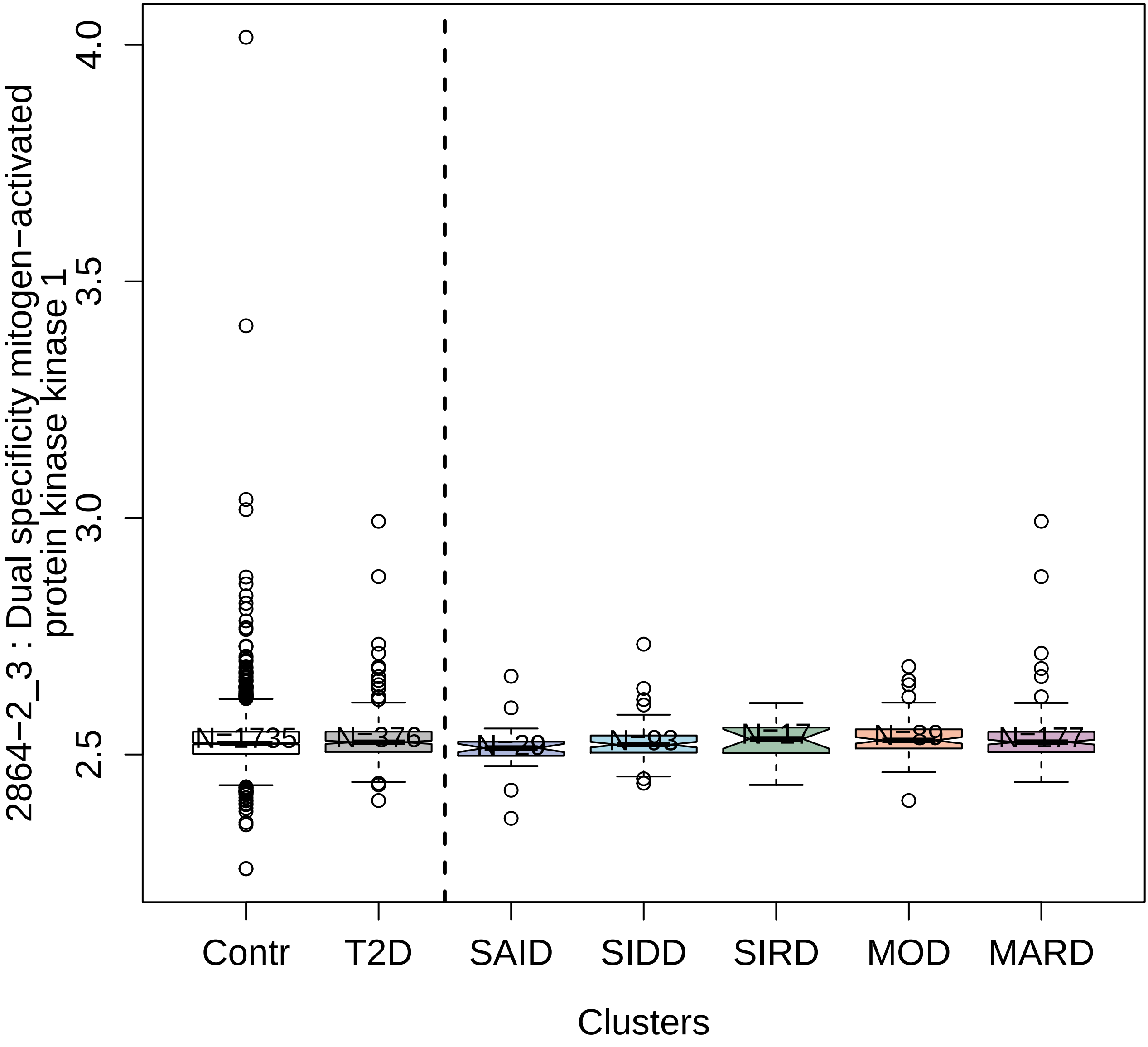
2864−2_3 : Dual specificity mitogen−activated protein kinase kinase 1
4.0
3.5
3.0
2.5
N=1735
N=376
N=29
N=93
N=17
N=89
N=177
Contr
T2D
SAID
SIDD
SIRD
MOD
MARD
Clusters

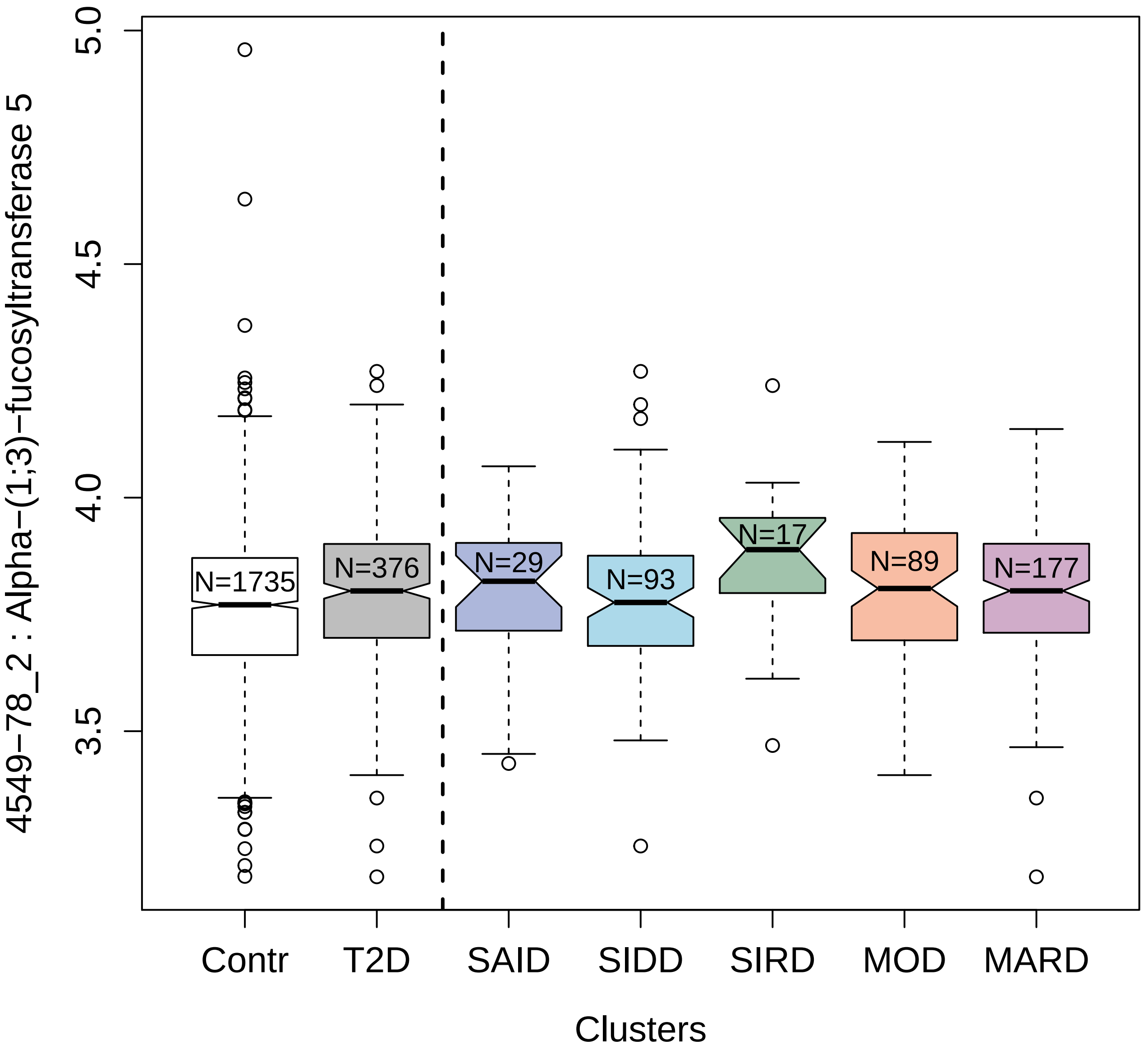

4549−78_2 : Alpha−(1;3)−fucosyltransferase 5
5.0
4.5
4.0
3.5
N=1735
N=376
N=29
N=93
N=17
N=89
N=177
Contr
T2D
SAID
SIDD
SIRD
MOD
MARD
Clusters

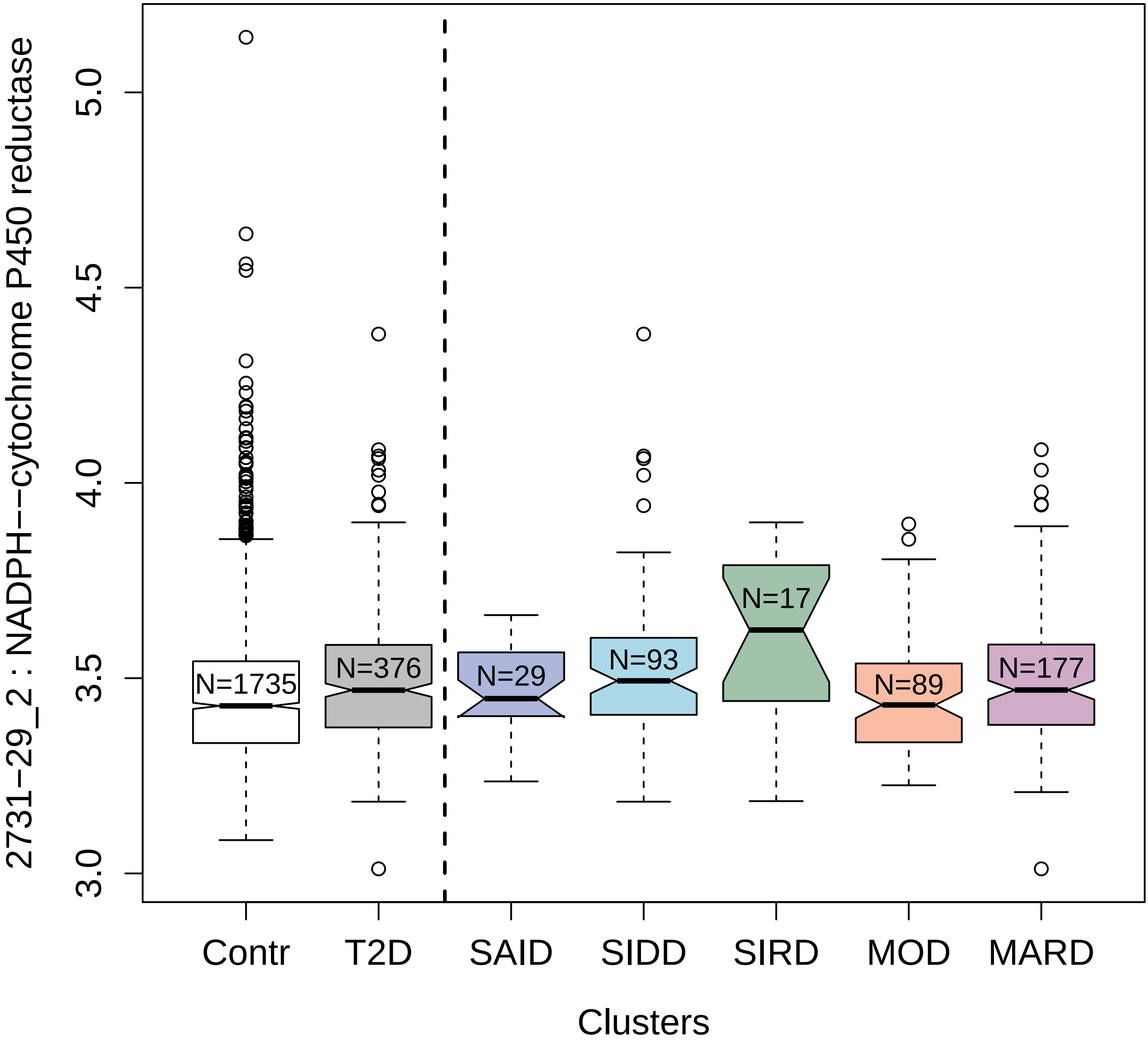
2731−29_2 : NADPH−−cytochrome P450 reductase
3.0
3.5
4.0
4.5
5.0
N=1735
N=376
N=29
N=93
N=17
N=89
N=177
Contr
T2D
SAID
SIDD
SIRD
MOD
MARD
Clusters

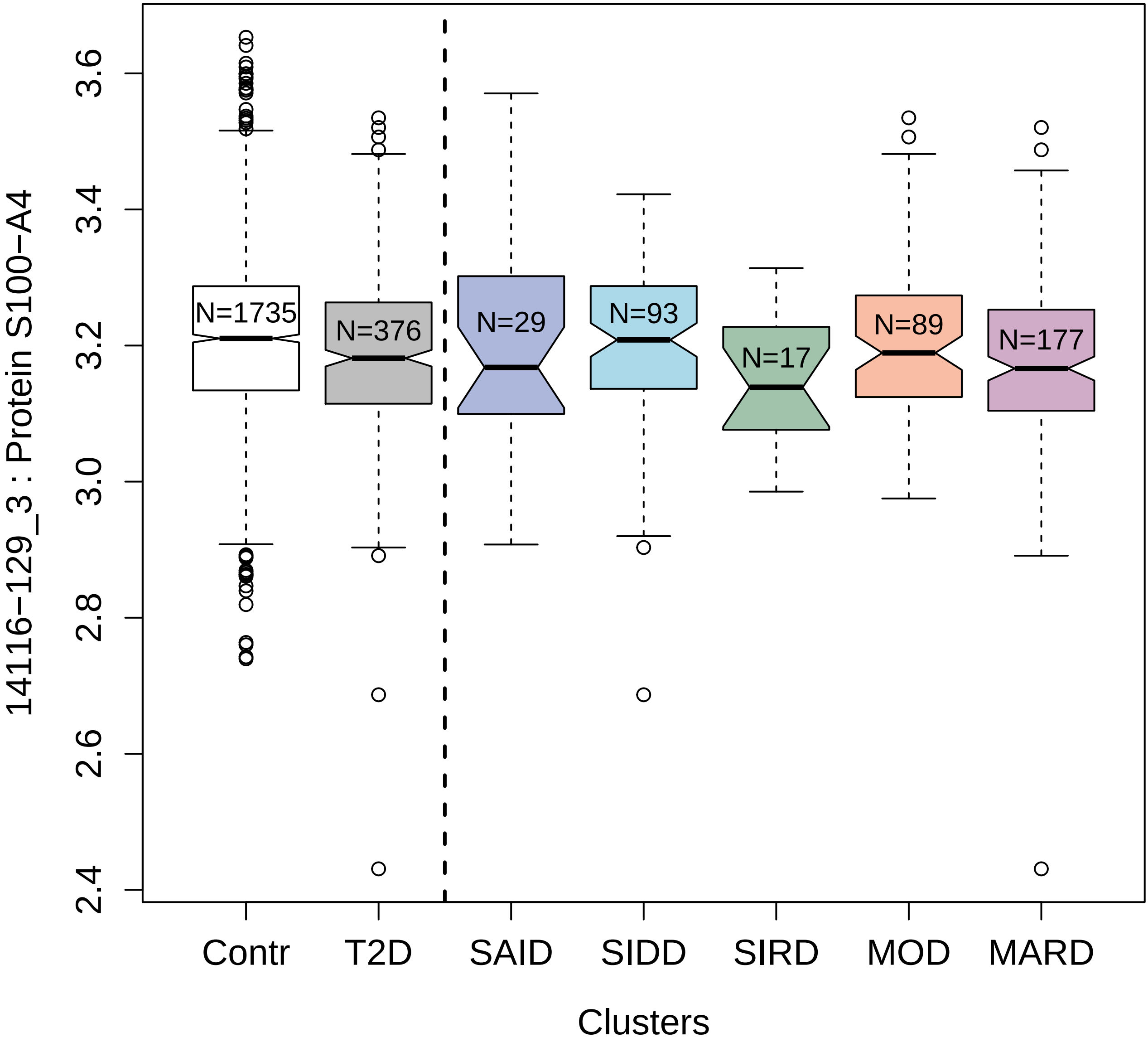

14116−129_3 : Protein S100−A4
3.6
3.4
3.2
3.0
2.8
2.6
2.4
N=1735
N=376
N=29
N=93
N=17
N=89
N=177
Contr
T2D
SAID
SIDD
SIRD
MOD
MARD
Clusters

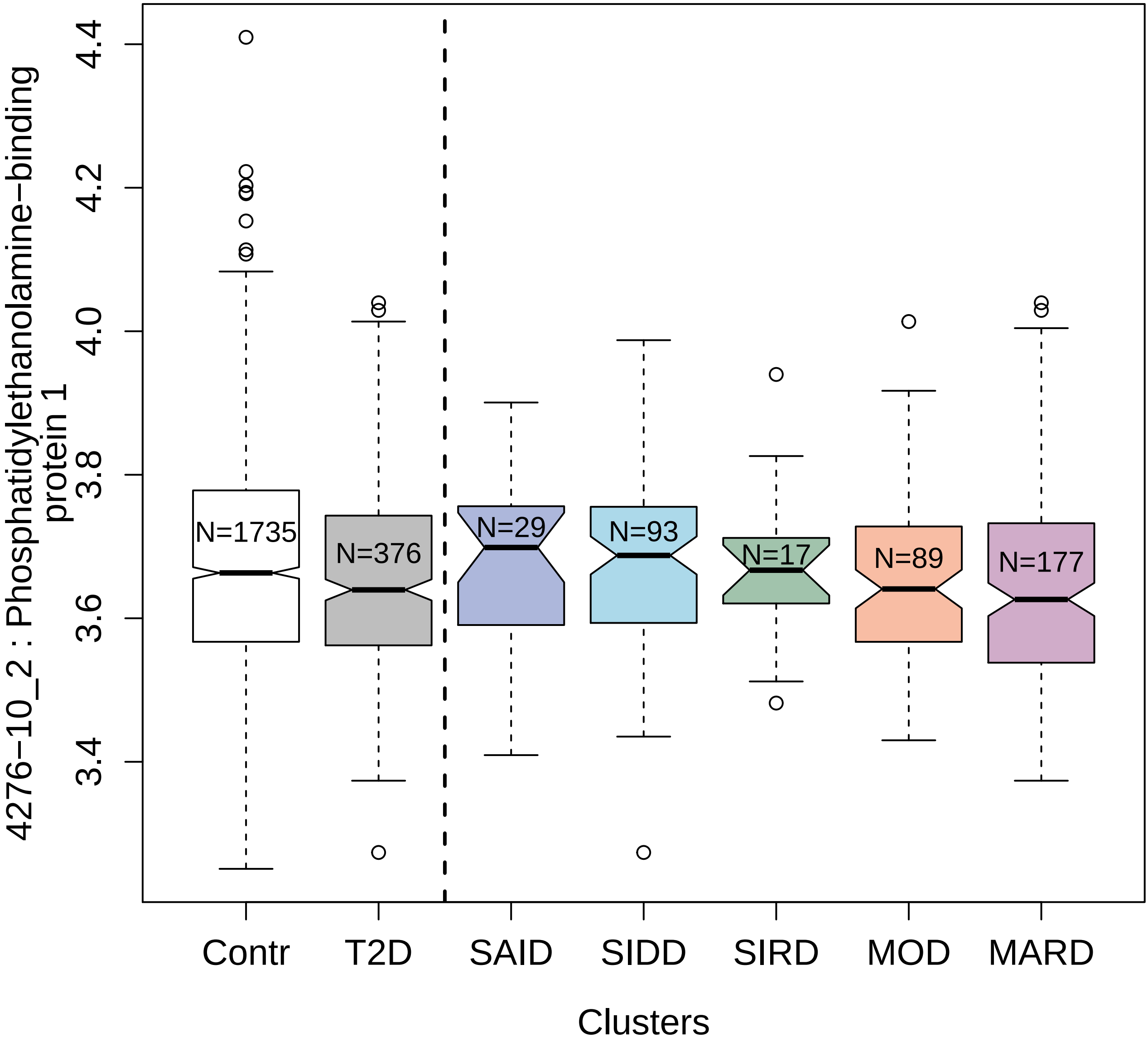
4276−10_2 : Phosphatidylethanolamine−binding protein 1
4.4
4.2
4.0
3.8
3.6
3.4
N=1735
N=376
N=29
N=93
N=17
N=89
N=177
Contr
T2D
SAID
SIDD
SIRD
MOD
MARD
Clusters

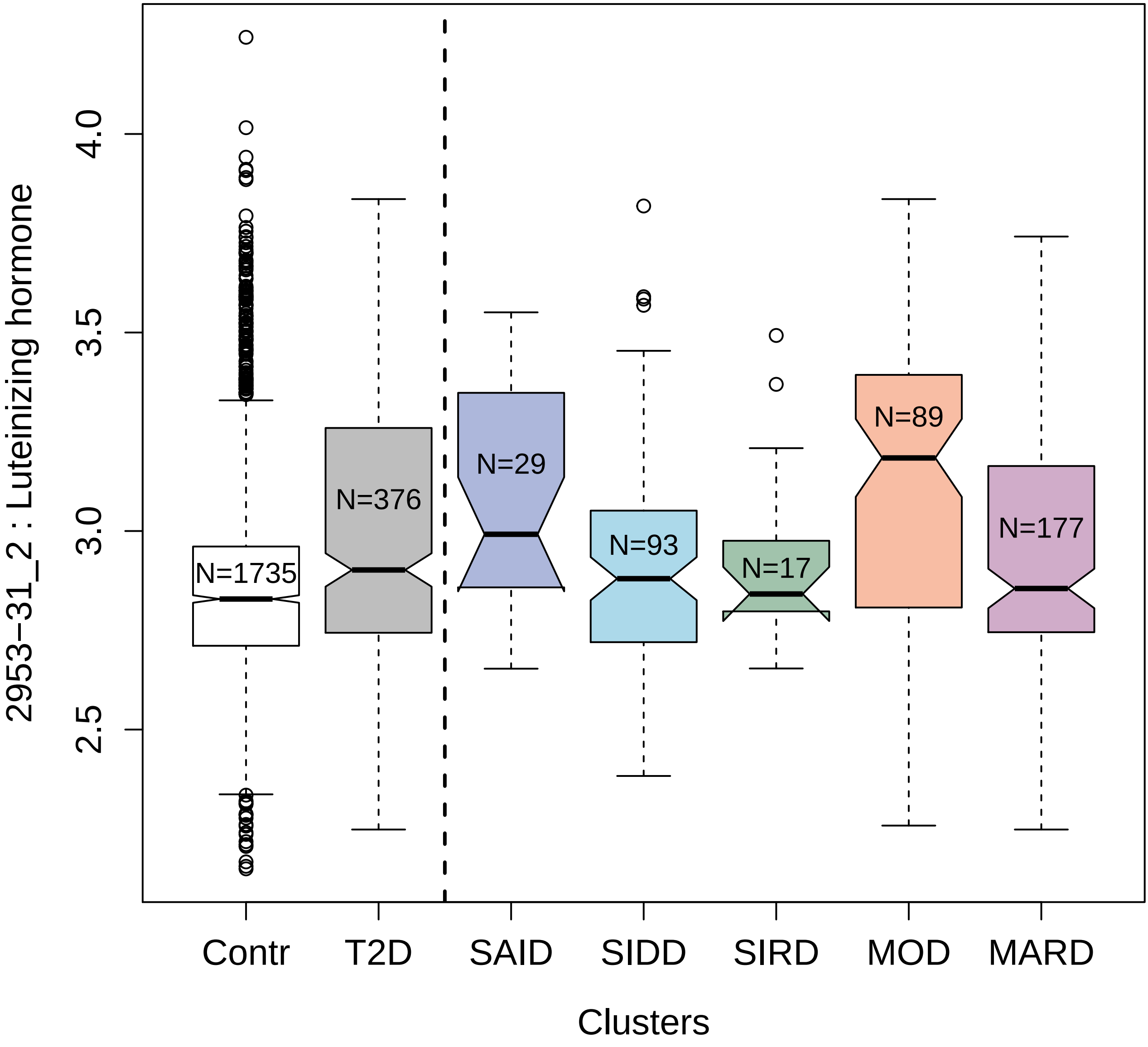

2953−31_2 : Luteinizing hormone
4.0
3.5
3.0
2.5
N=1735
N=376
N=29
N=93
N=17
N=89
N=177
Contr
T2D
SAID
SIDD
SIRD
MOD
MARD
Clusters

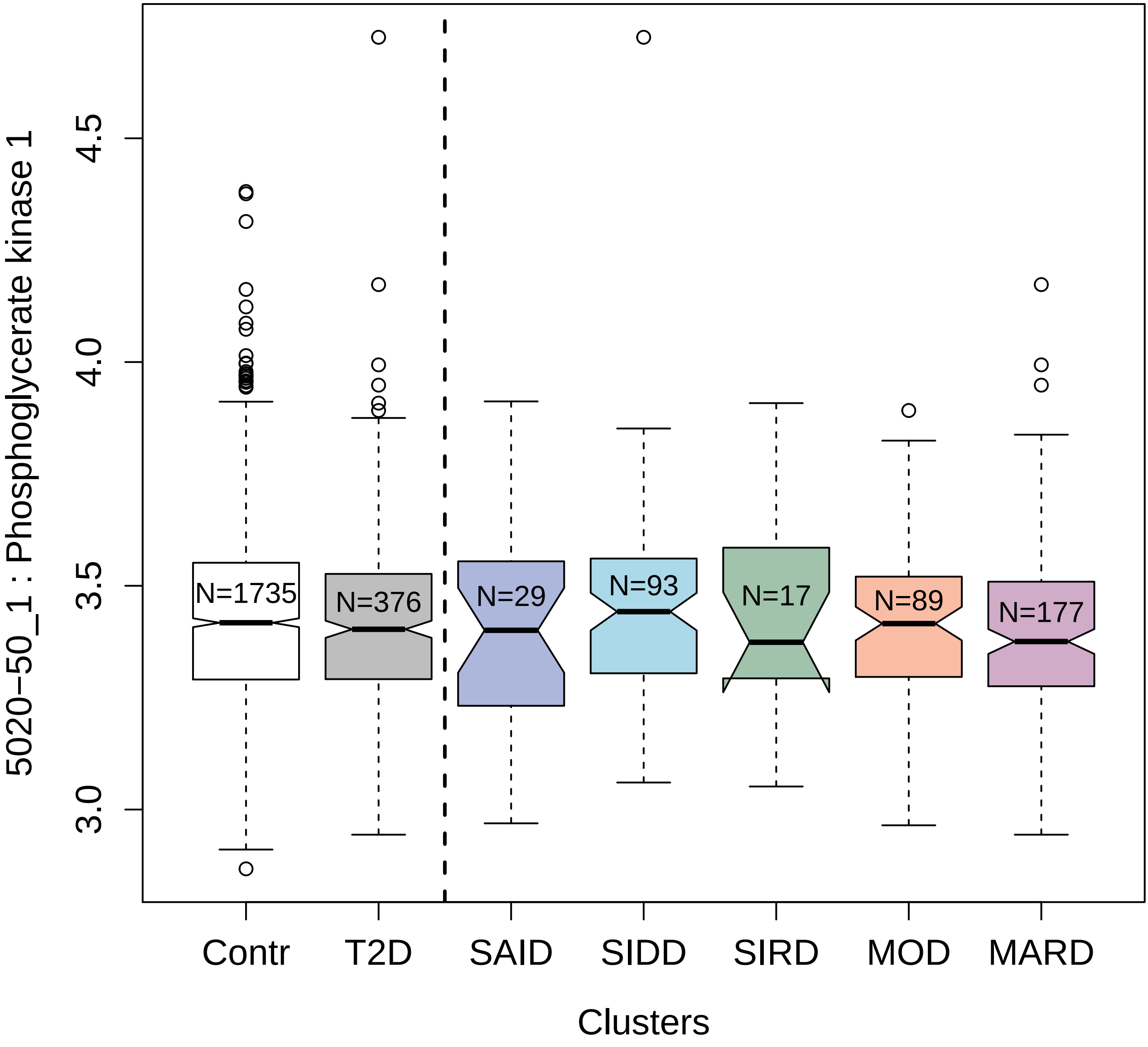

5020−50_1 : Phosphoglycerate kinase 1
3.0
3.5
4.0
4.5
N=1735
N=376
N=29
N=93
N=17
N=89
N=177
Contr
T2D
SAID
SIDD
SIRD
MOD
MARD
Clusters

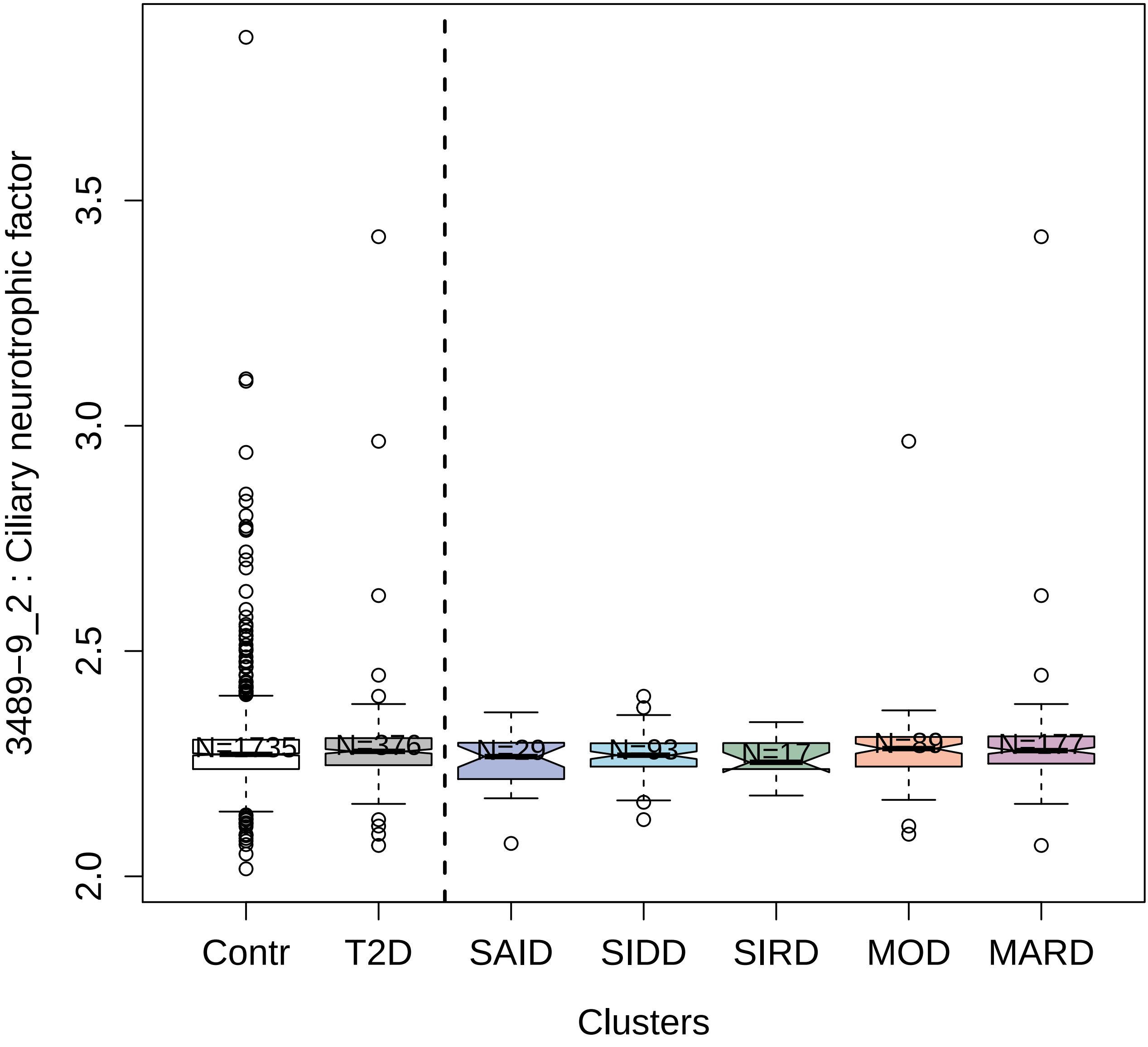
3489−9_2 : Ciliary neurotrophic factor
2.0
2.5
3.0
3.5
N=1735
N=376
N=29
N=93
N=17
N=89
N=177
Contr
T2D
SAID
SIDD
SIRD
MOD
MARD
Clusters

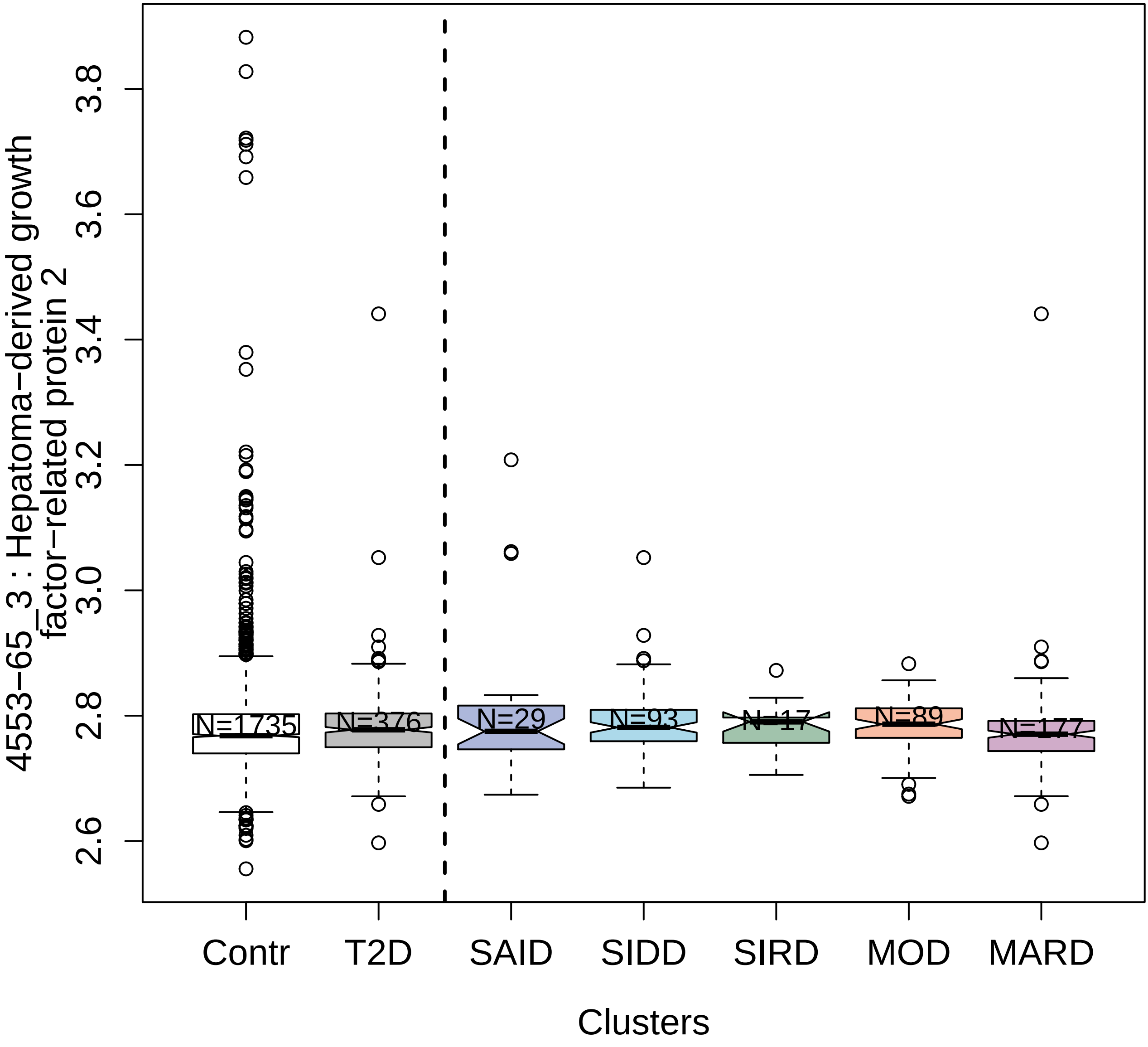
4553−65_3 : Hepatoma−derived growth
factor−related protein 2
3.8
3.6
3.4
3.2
3.0
2.8
2.6
N=1735
N=376
N=29
N=93
N=17
N=89
N=177
Contr
T2D
SAID
SIDD
SIRD
MOD
MARD
Clusters

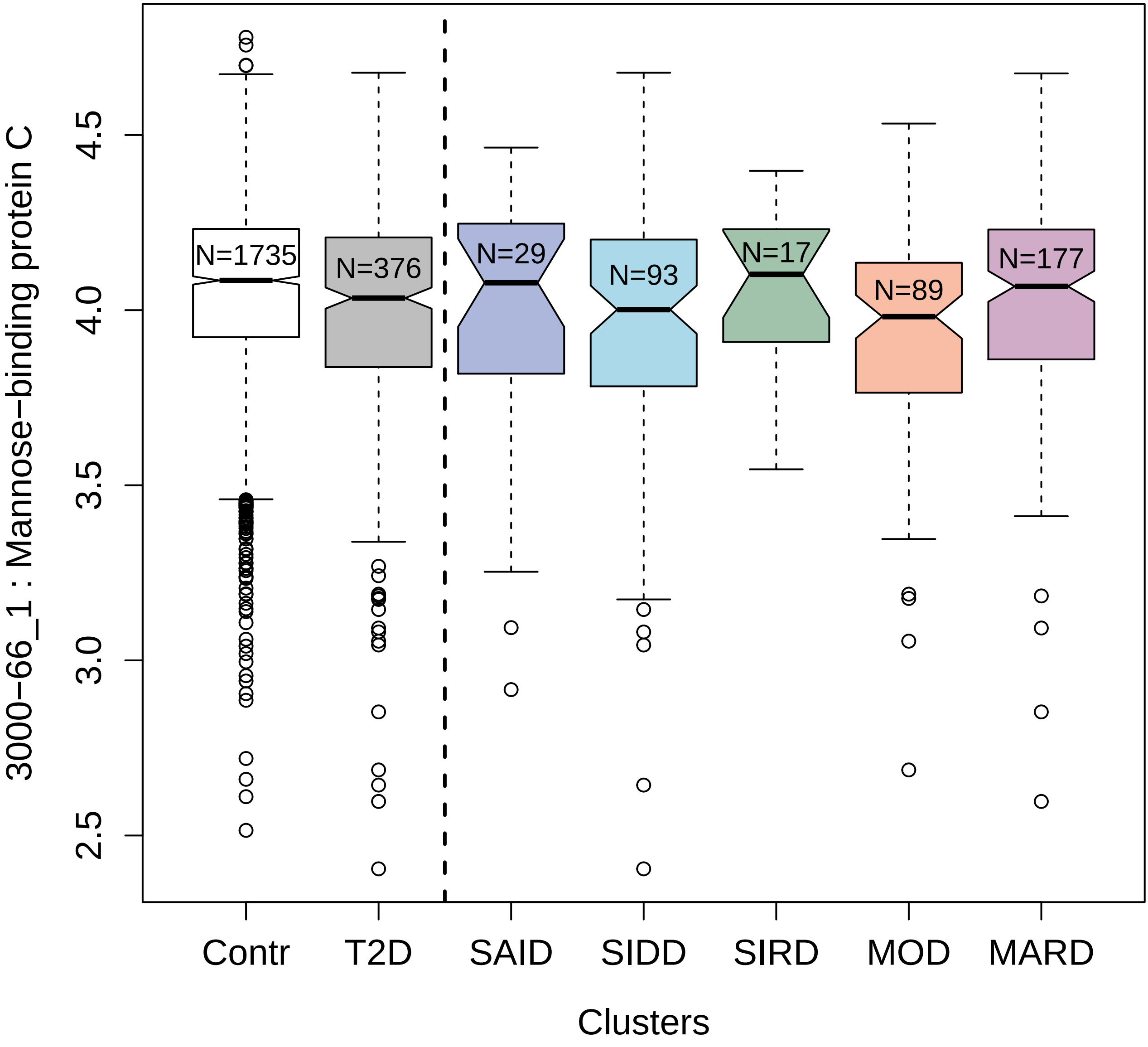
3000−66_1 : Mannose−binding protein C
4.5
4.0
3.5
3.0
2.5
N=1735
N=376
N=29
N=93
N=17
N=89
N=177
Contr
T2D
SAID
SIDD
SIRD
MOD
MARD
Clusters

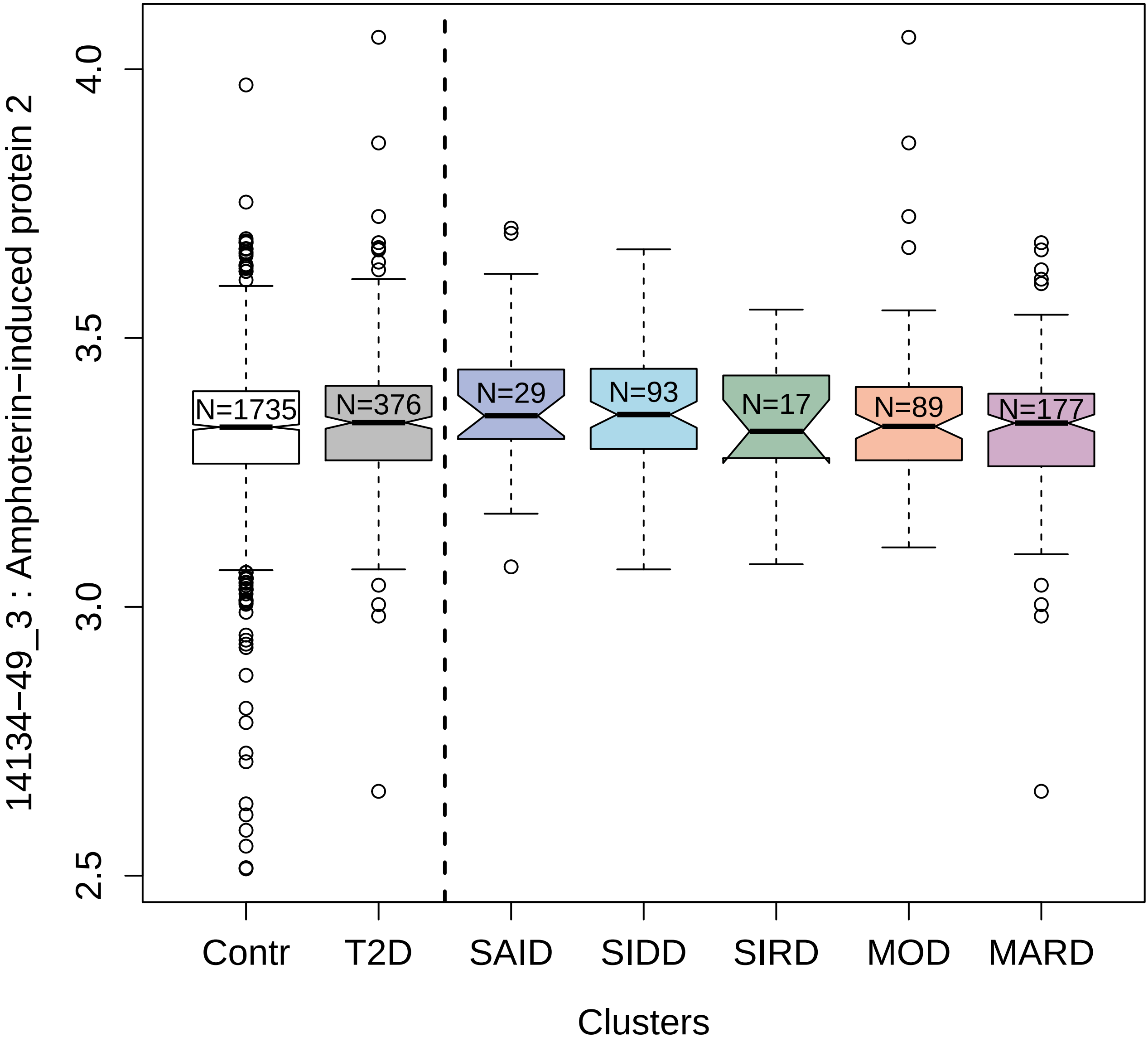
14134−49_3 : Amphoterin−induced protein 2
4.0
3.5
3.0
2.5
N=1735
N=376
N=29
N=93
N=17
N=89
N=177
Contr
T2D
SAID
SIDD
SIRD
MOD
MARD
Clusters

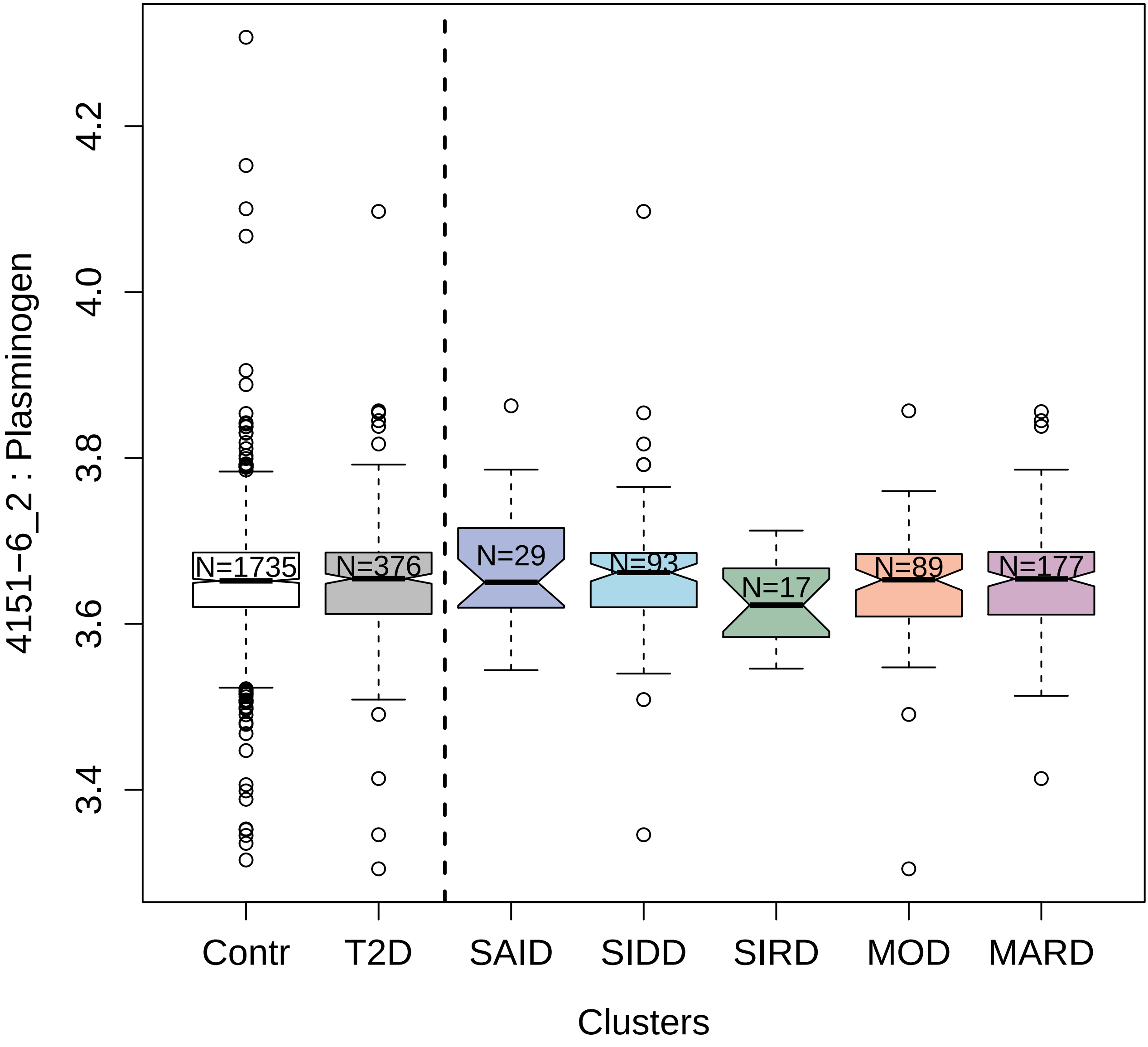

4151−6_2 : Plasminogen
3.4
3.6
3.8
4.0
4.2
N=1735
N=376
N=29
N=93
N=17
N=89
N=177
Contr
T2D
SAID
SIDD
SIRD
MOD
MARD
Clusters

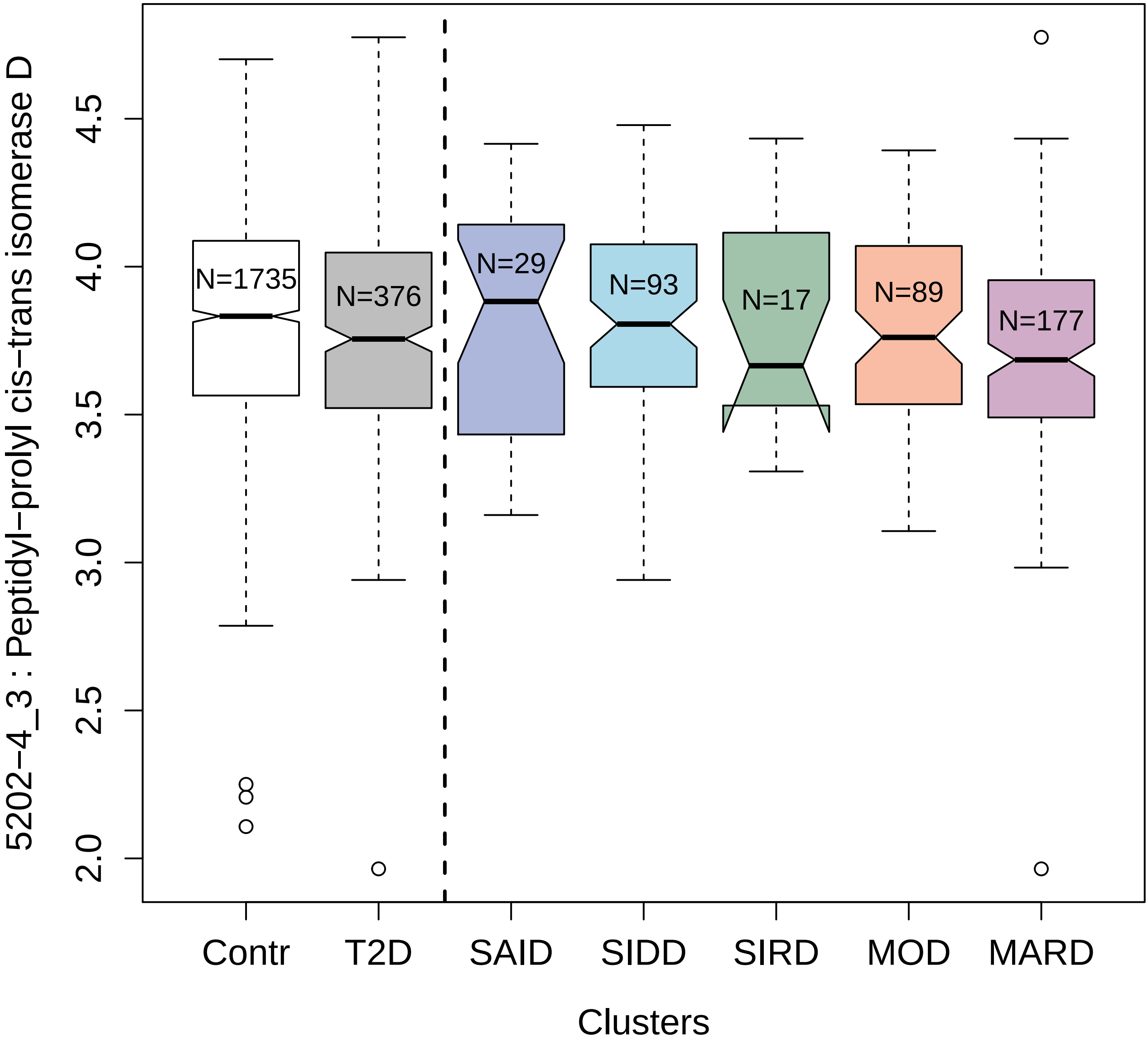
5202−4_3 : Peptidyl−prolyl cis−trans isomerase D
Clusters
Contr
T2D
SAID
SIDD
SIRD
MOD
MARD
N=1735
N=376
N=29
N=93
N=17
N=89
N=177
2.0
2.5
3.0
3.5
4.0
4.5

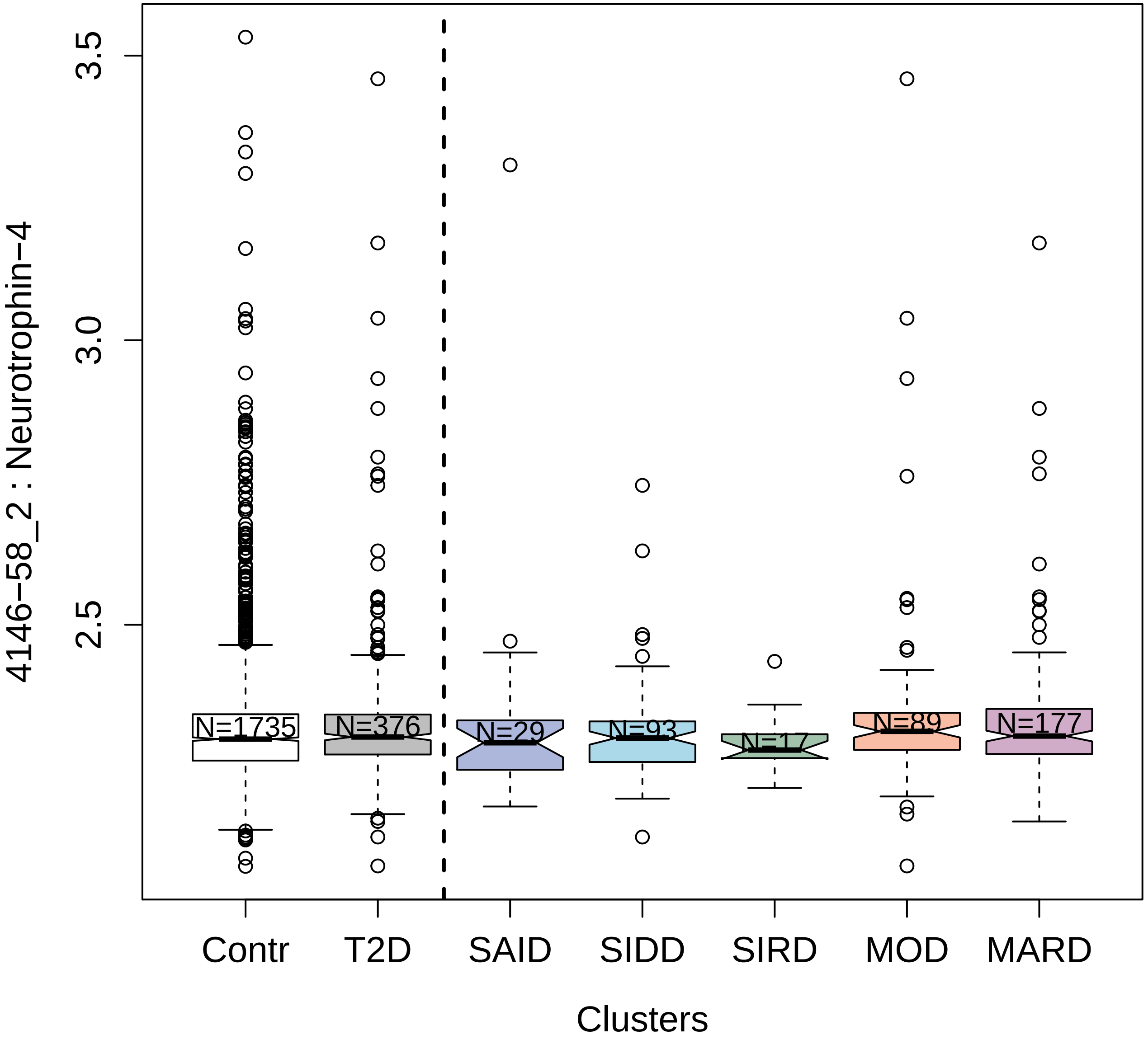
4146−58_2 : Neurotrophin−4
3.5
3.0
2.5
N=1735
N=376
N=29
N=93
N=17
N=89
N=177
Contr
T2D
SAID
SIDD
SIRD
MOD
MARD
Clusters

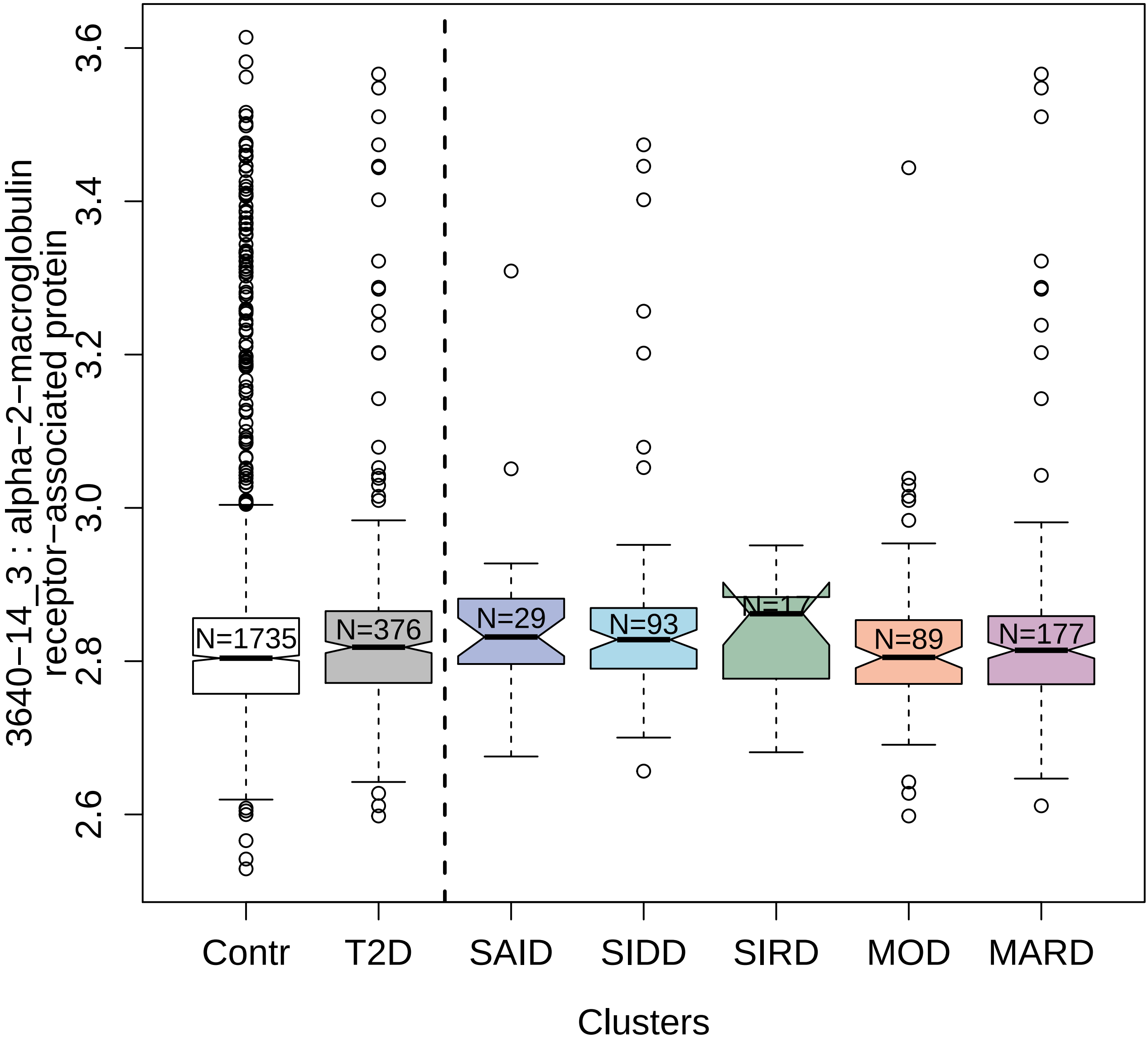
3640−14_3 : alpha−2−macroglobulin
receptor−associated protein
2.6
2.8
3.0
3.2
3.4
3.6
N=1735
N=376
N=29
N=93
N=89
N=177
Contr
T2D
SAID
SIDD
SIRD
MOD
MARD
Clusters

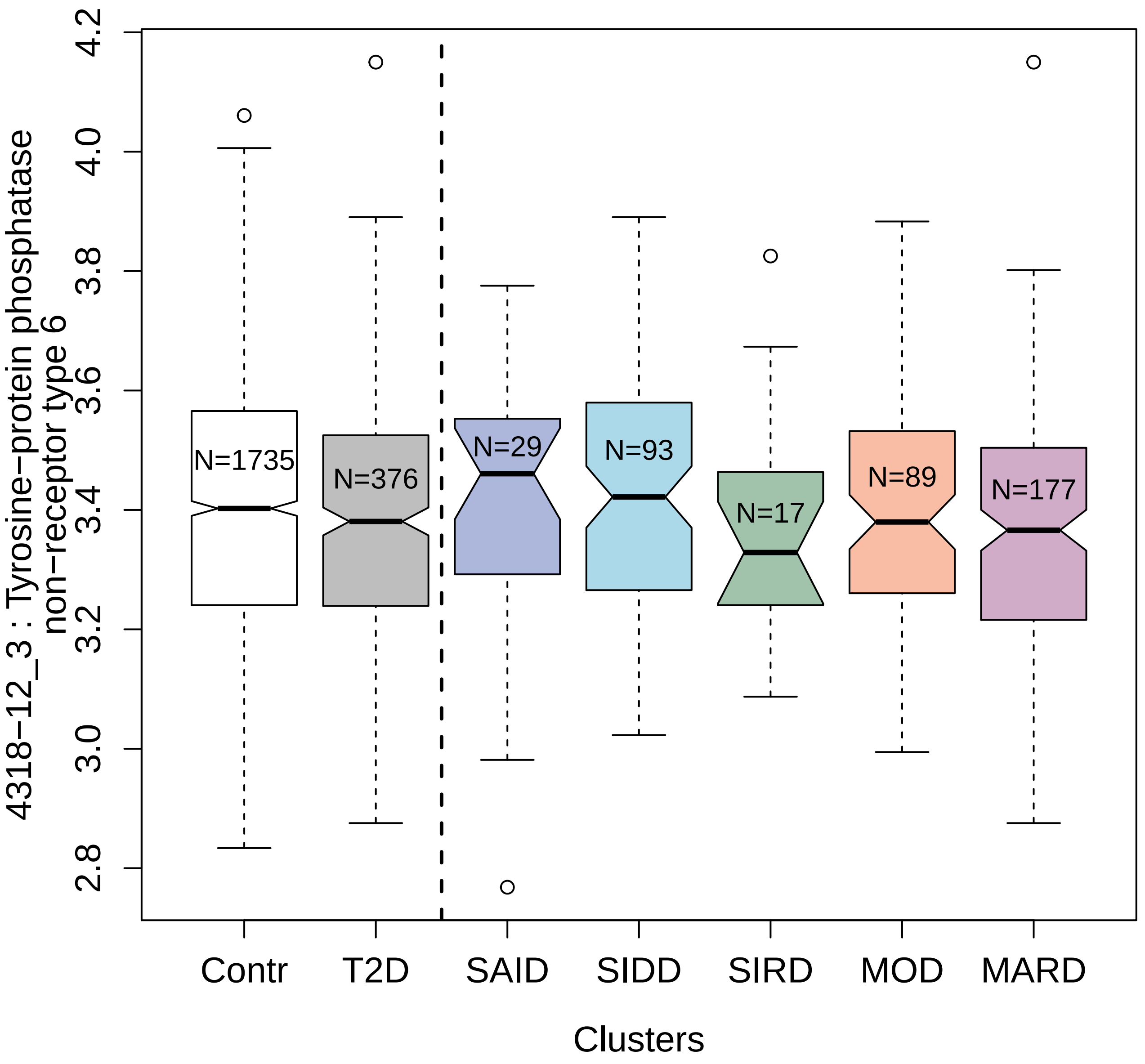
4318−12_3 : Tyrosine−protein phosphatase
non−receptor type 6
4.2
4.0
3.8
3.6
3.4
3.2
3.0
2.8
N=1735
N=376
N=29
N=93
N=17
N=89
N=177
Contr
T2D
SAID
SIDD
SIRD
MOD
MARD
Clusters

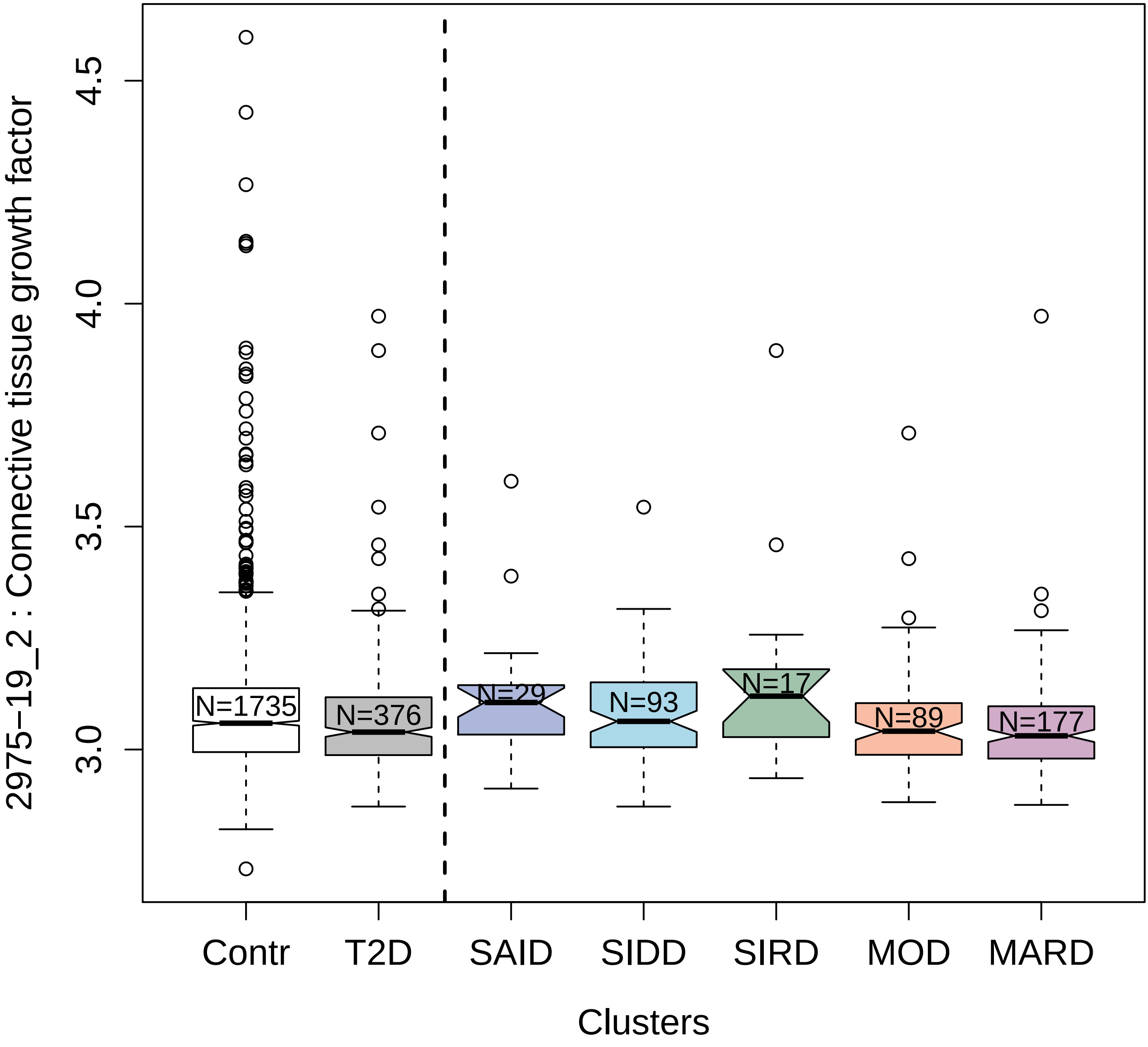

2975−19_2 : Connective tissue growth factor
3.0
3.5
4.0
4.5
N=1735
N=376
N=29
N=93
N=17
N=89
N=177
Contr
T2D
SAID
SIDD
SIRD
MOD
MARD
Clusters

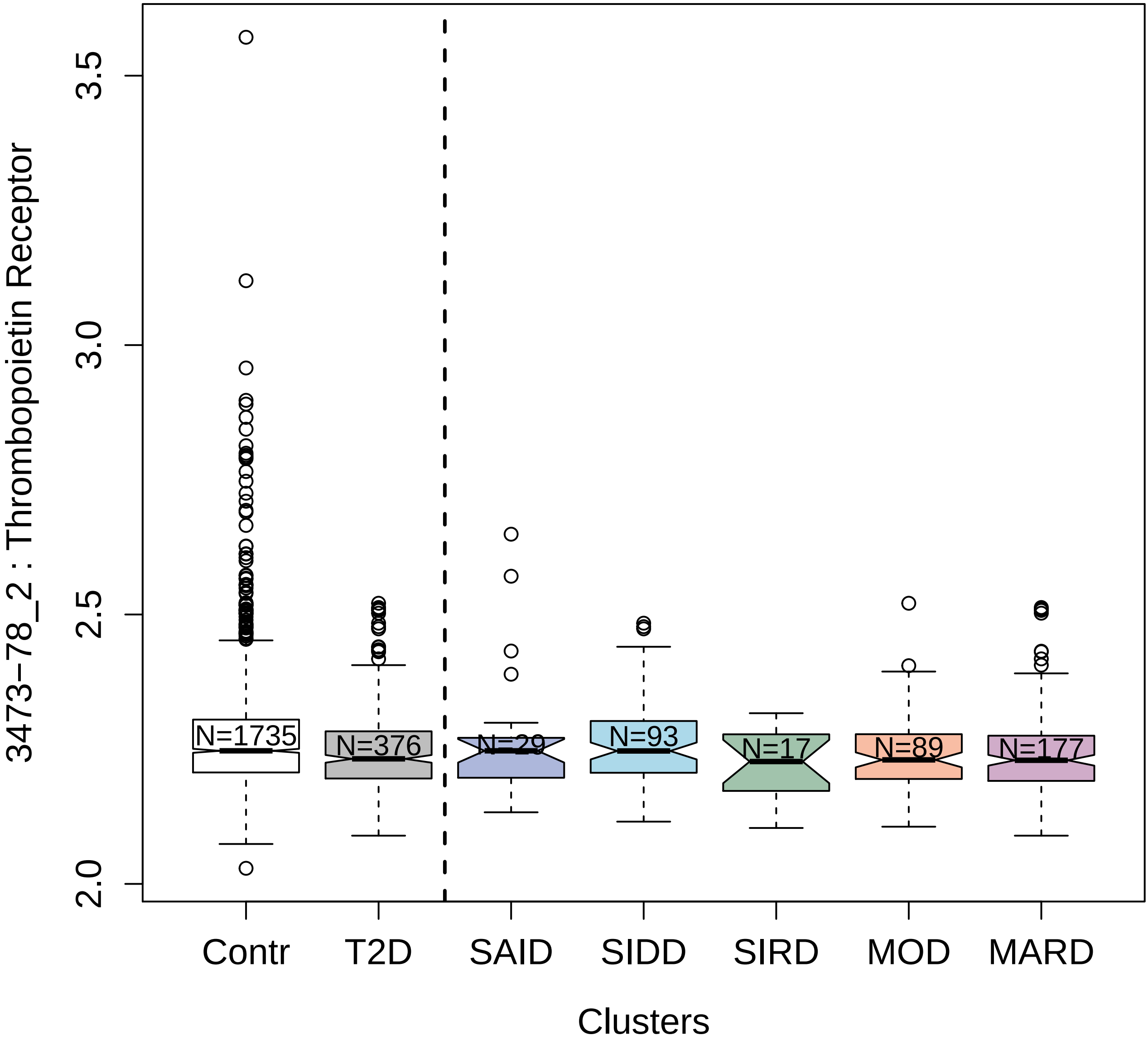

3473−78_2 : Thrombopoietin Receptor
3.5
3.0
2.5
2.0
N=1735
N=376
N=29
N=93
N=17
N=89
N=177
Contr
T2D
SAID
SIDD
SIRD
MOD
MARD
Clusters

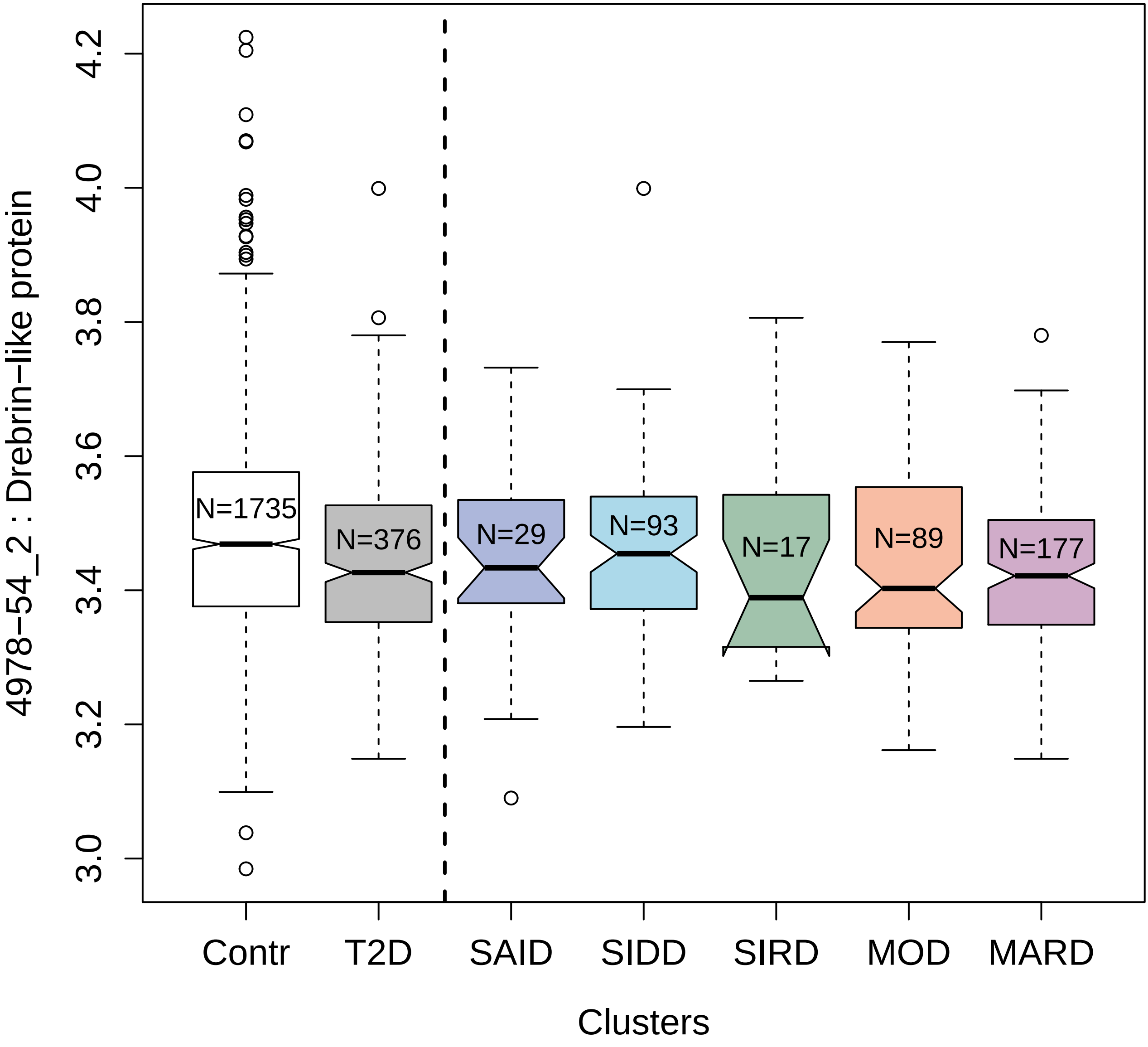

4978−54_2 : Drebrin−like protein
4.2
4.0
3.8
3.6
3.4
3.2
3.0
N=1735
N=376
N=29
N=93
N=17
N=89
N=177
Contr
T2D
SAID
SIDD
SIRD
MOD
MARD
Clusters

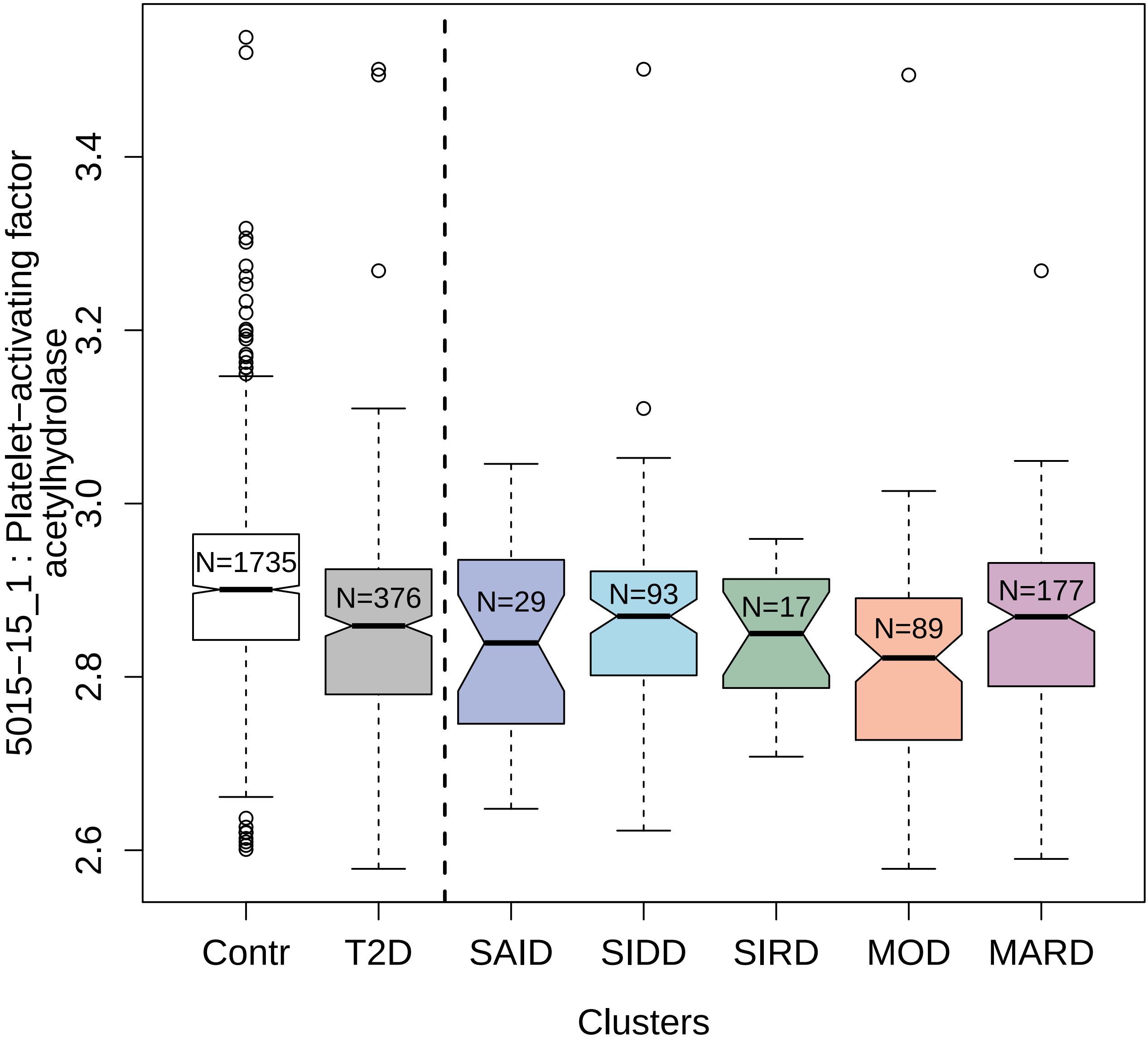

5015−15_1 : Platelet−activating factor acetylhydrolase
2.6
2.8
3.0
3.2
3.4
N=1735
N=376
N=29
N=93
N=17
N=89
N=177
Contr
T2D
SAID
SIDD
SIRD
MOD
MARD
Clusters

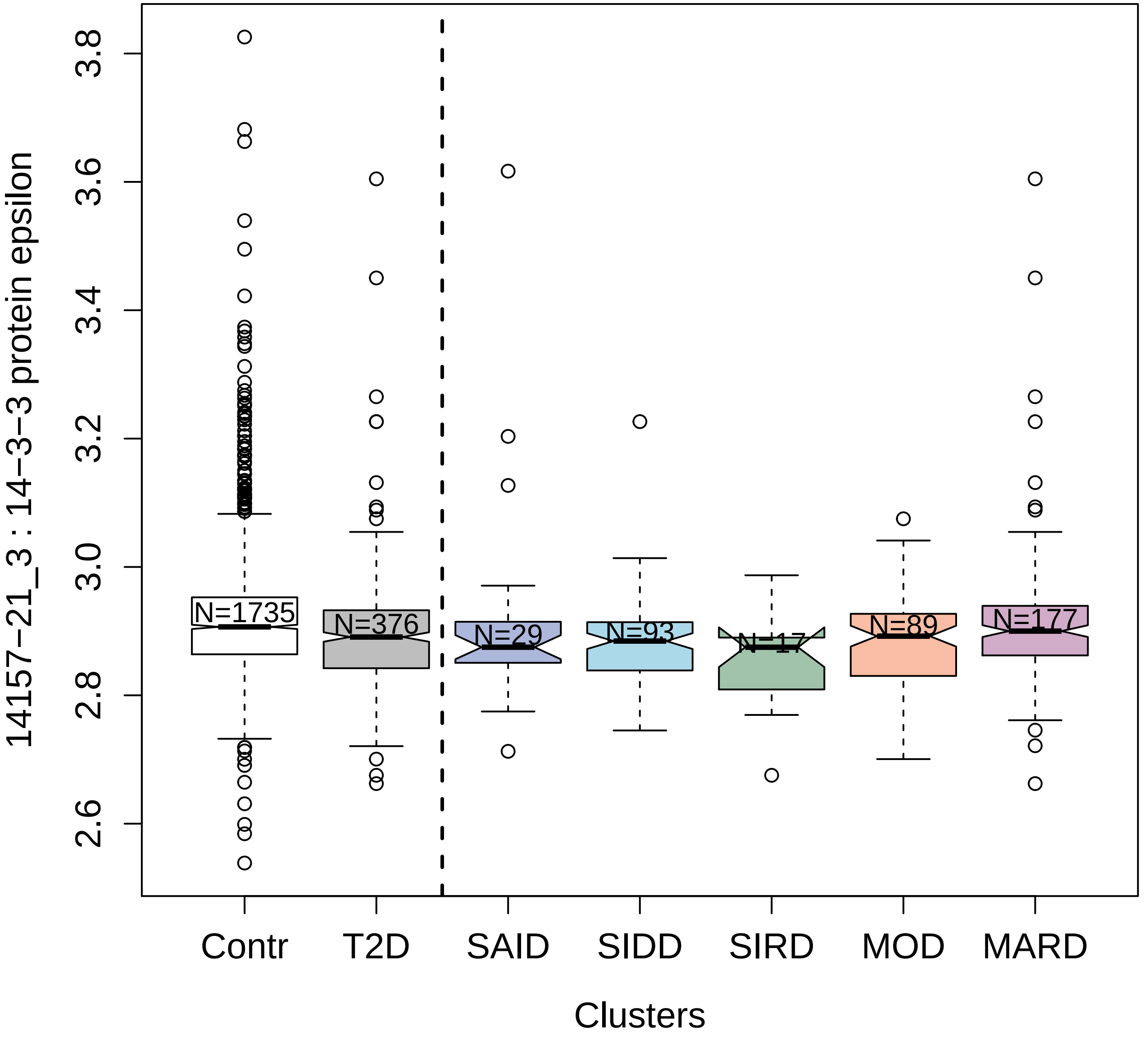

14157−21_3 : 14−3−3 protein epsilon
2.6
2.8
3.0
3.2
3.4
3.6
3.8
N=1735
N=376
N=29
N=93
N=17
N=89
N=177
Contr
T2D
SAID
SIDD
SIRD
MOD
MARD
Clusters

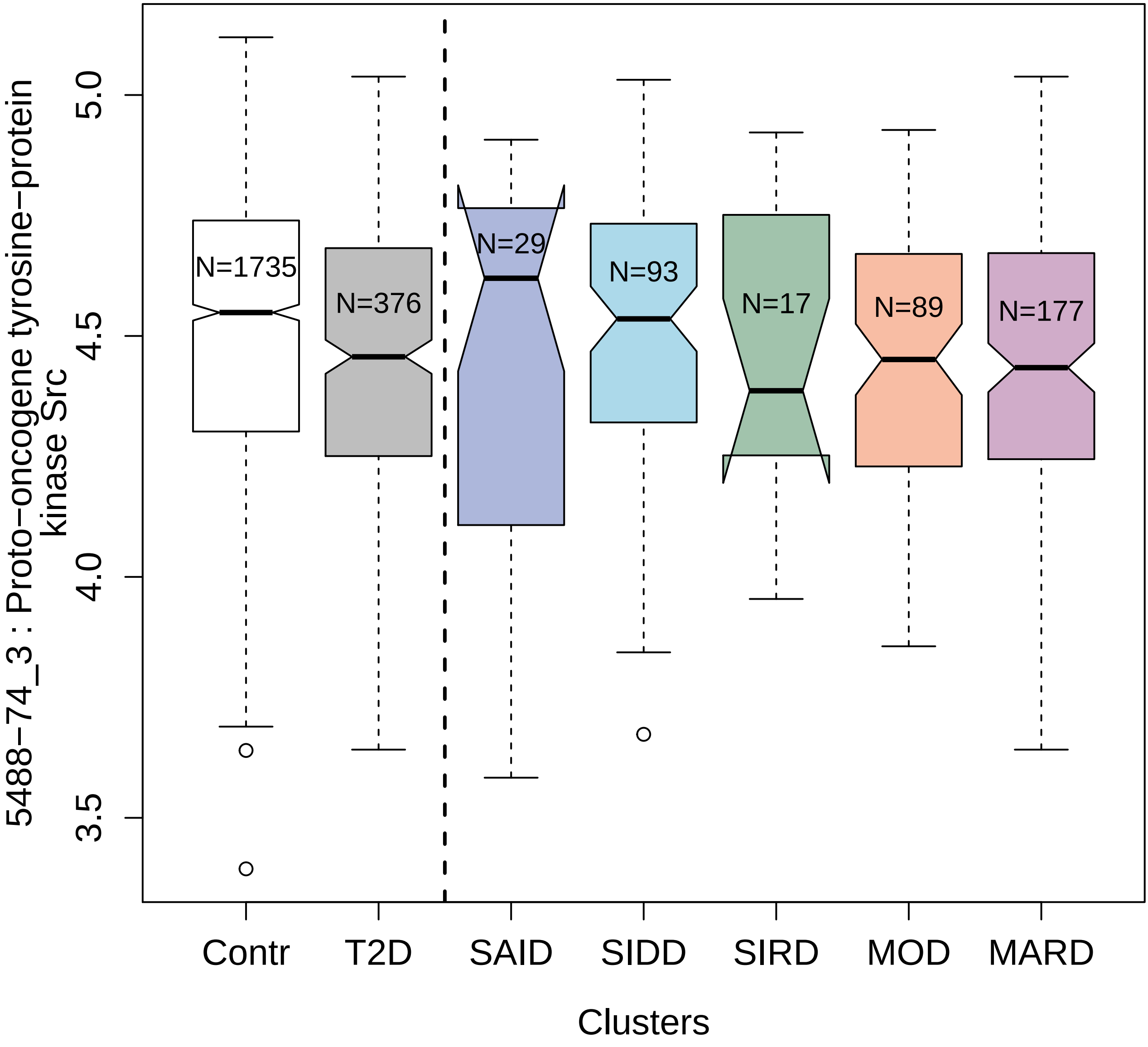

5488−74_3 : Proto−oncogene tyrosine−protein kinase Src
3.5
4.0
4.5
5.0
N=1735
N=376
N=29
N=93
N=17
N=89
N=177
Contr
T2D
SAID
SIDD
SIRD
MOD
MARD
Clusters

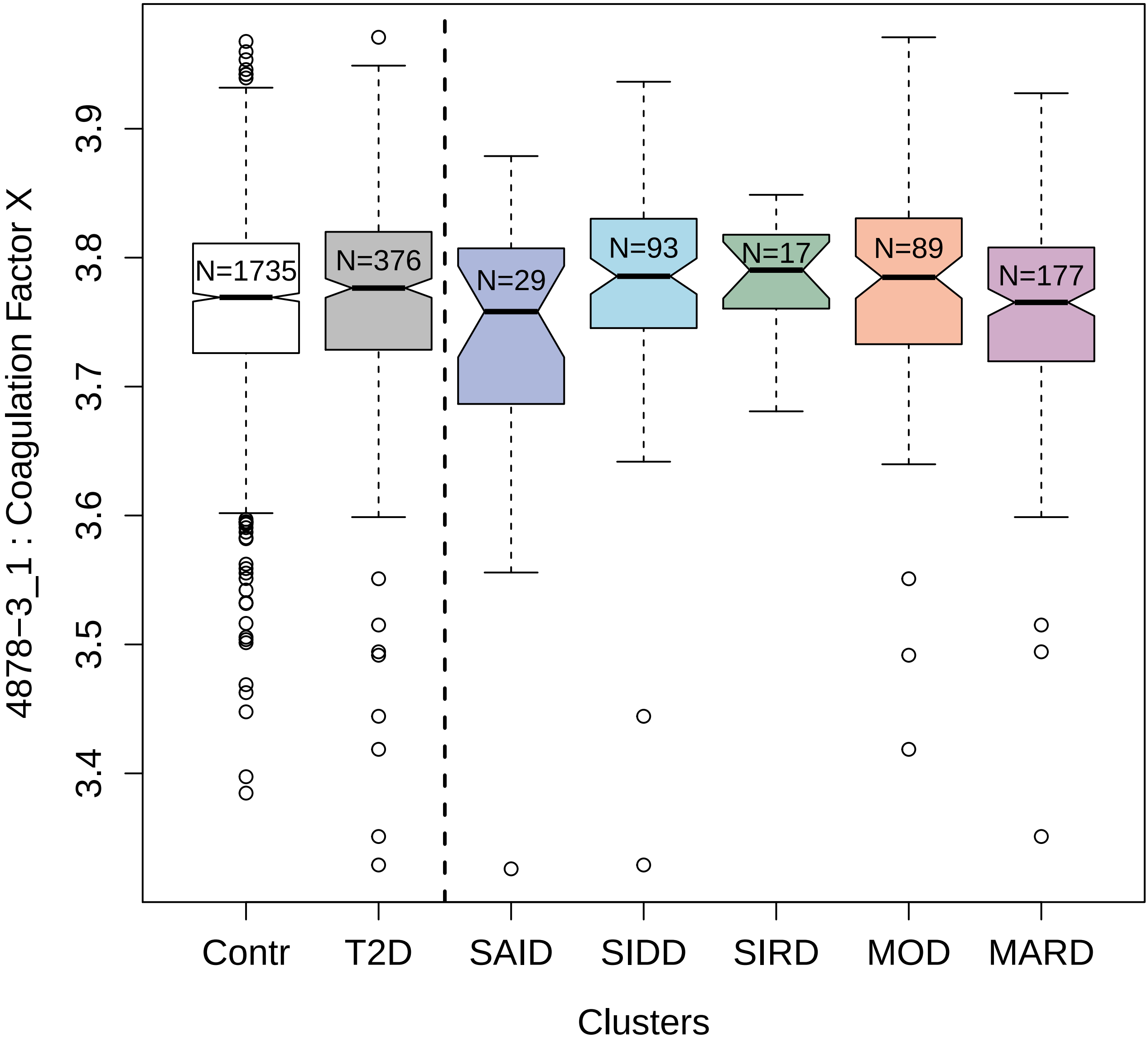
4878−3_1 : Coagulation Factor X
3.4
3.5
3.6
3.7
3.8
3.9
N=1735
N=376
N=29
N=93
N=17
N=89
N=177
Contr
T2D
SAID
SIDD
SIRD
MOD
MARD
Clusters

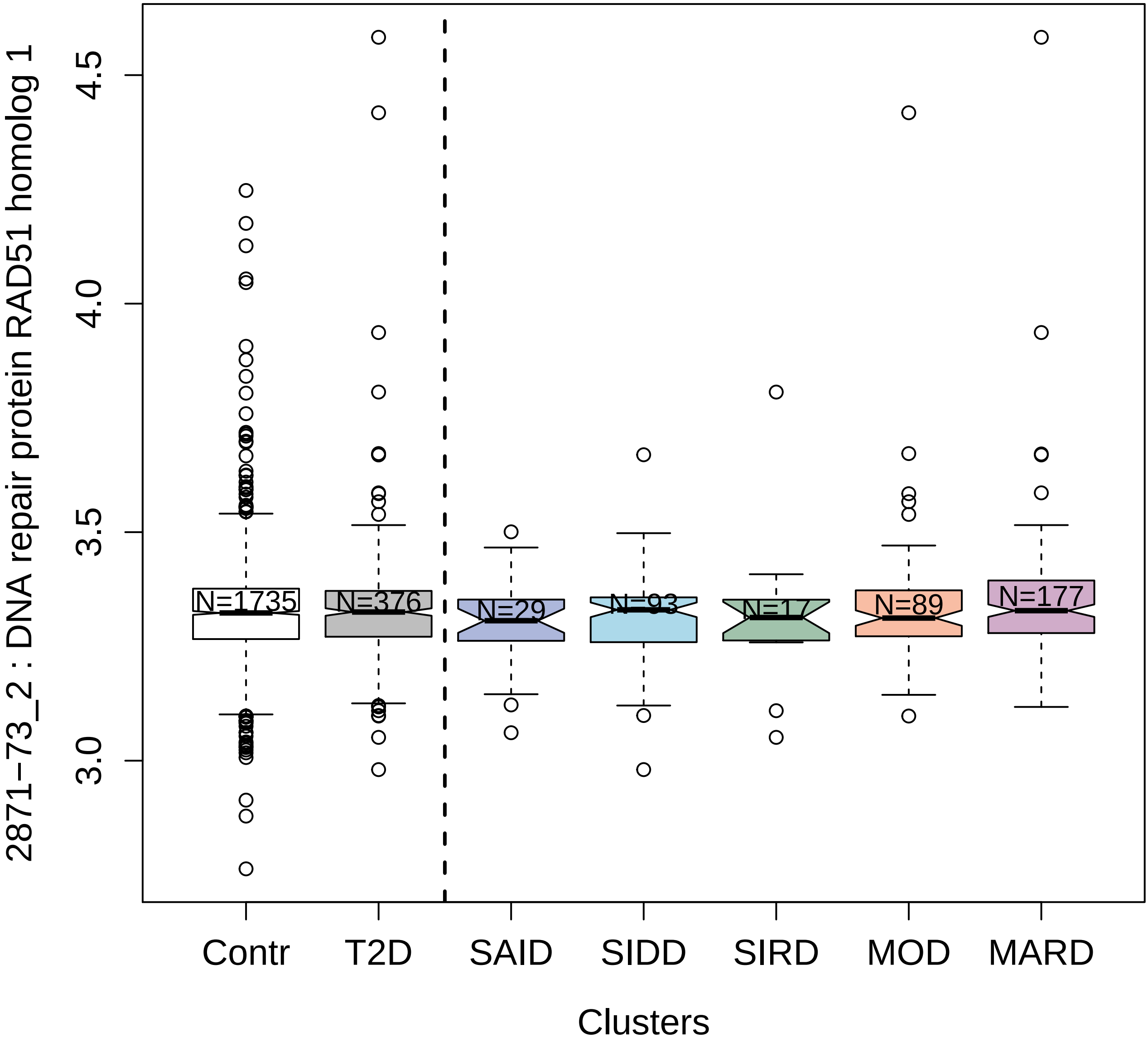
2871−73_2 : DNA repair protein RAD51 homolog 1
3.0
3.5
4.0
4.5
N=1735
N=376
N=29
N=93
N=17
N=89
N=177
Contr
T2D
SAID
SIDD
SIRD
MOD
MARD
Clusters

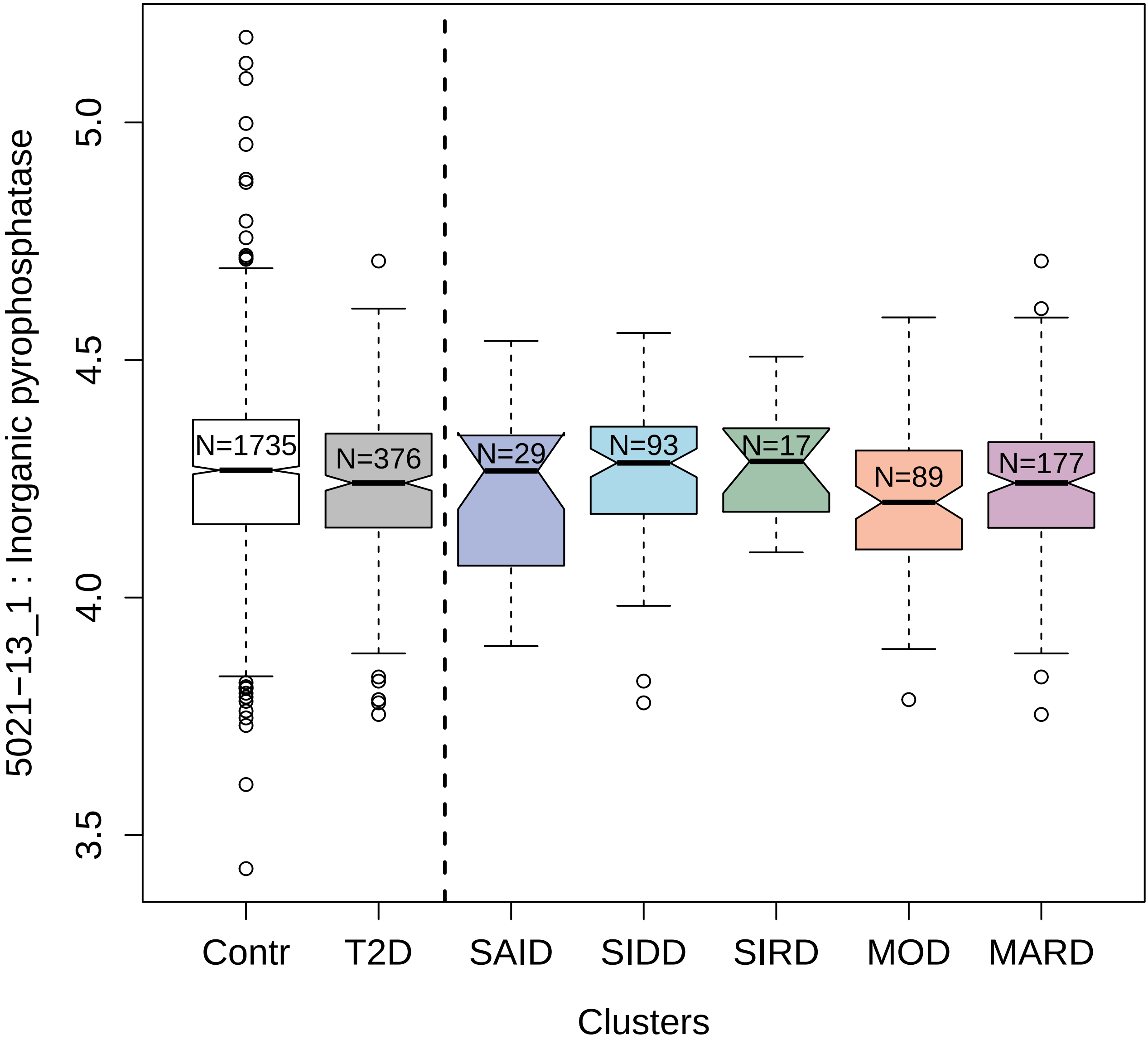

5021−13_1 : Inorganic pyrophosphatase
3.5
4.0
4.5
5.0
N=1735
N=376
N=29
N=93
N=17
N=89
N=177
Contr
T2D
SAID
SIDD
SIRD
MOD
MARD
Clusters

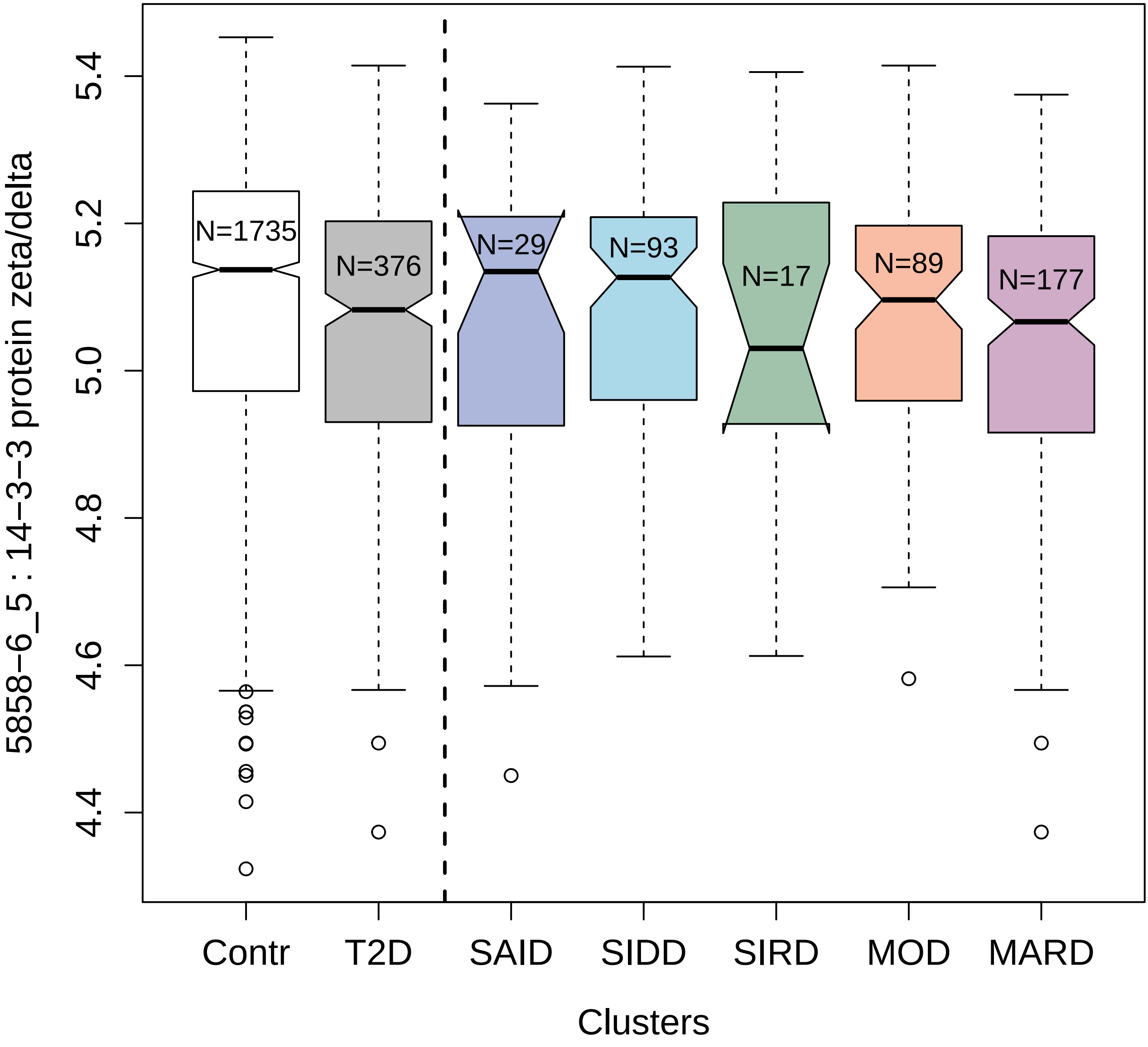

5858−6_5 : 14−3−3 protein zeta/delta
5.4
5.2
5.0
4.8
4.6
4.4
N=1735
N=376
N=29
N=93
N=17
N=89
N=177
Contr
T2D
SAID
SIDD
SIRD
MOD
MARD
Clusters

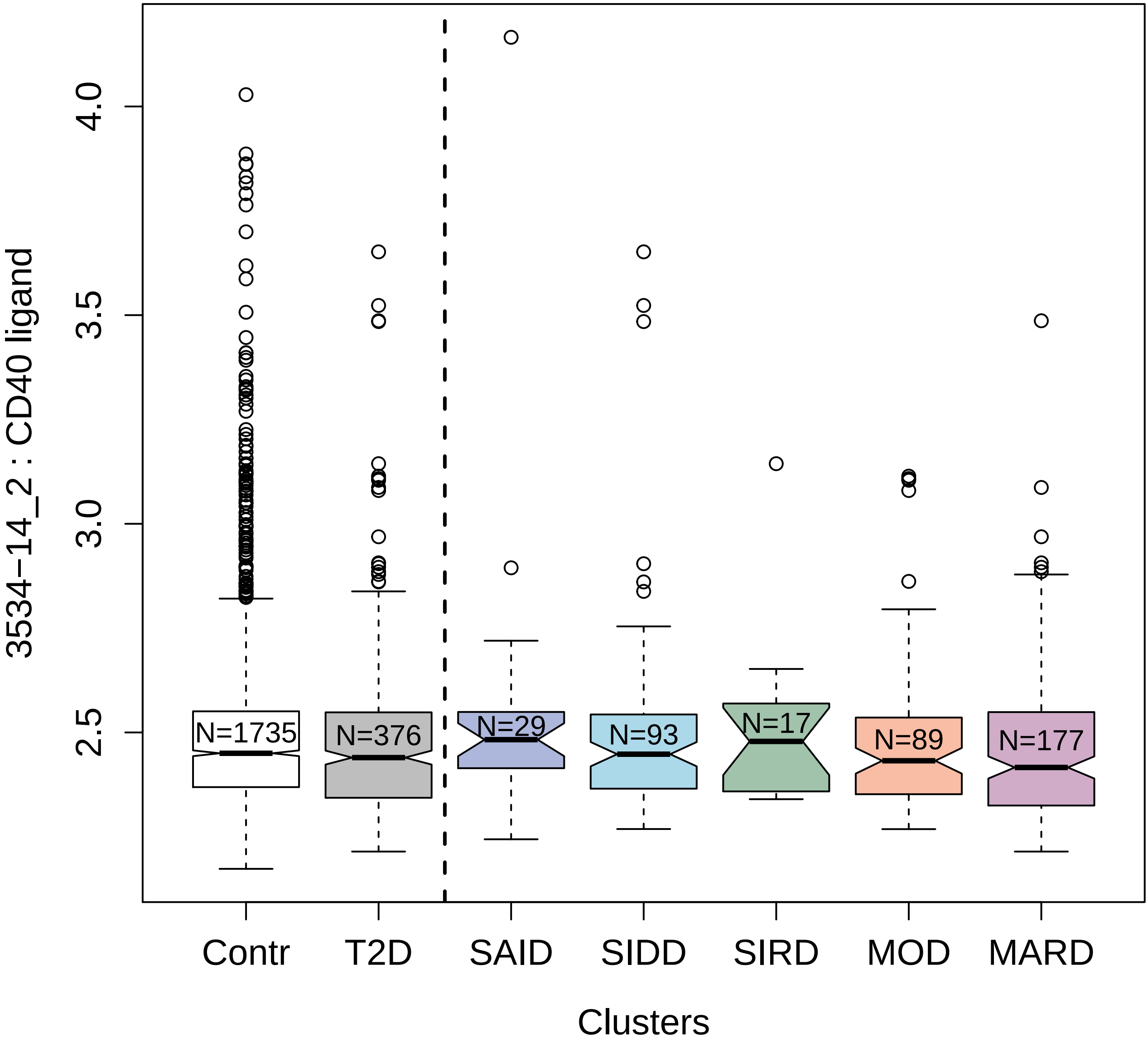
3534−14_2 : CD40 ligand
4.0
3.5
3.0
2.5
N=1735
N=376
N=29
N=93
N=17
N=89
N=177
Contr
T2D
SAID
SIDD
SIRD
MOD
MARD
Clusters

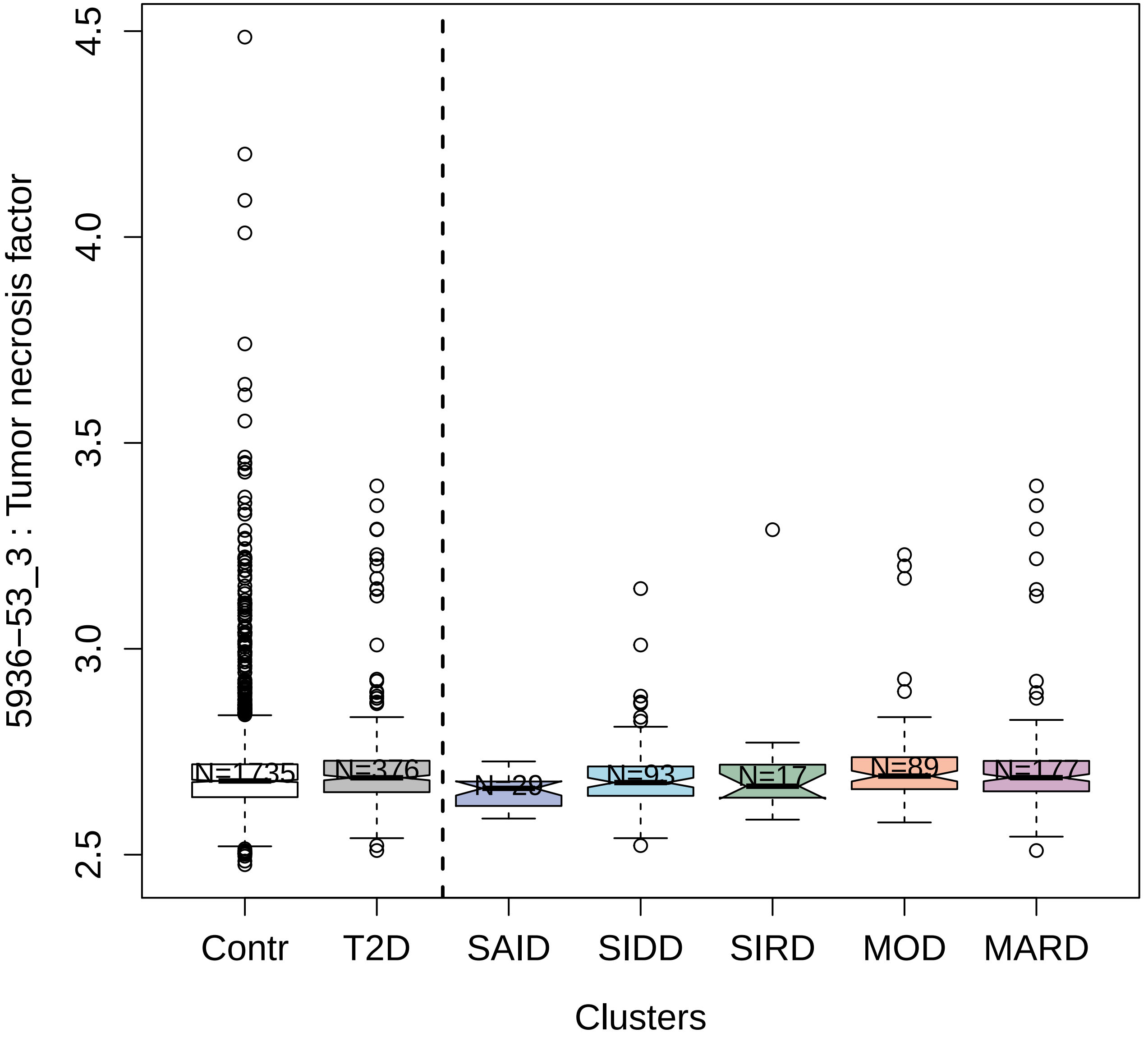
5936−53_3 : Tumor necrosis factor
4.5
4.0
3.5
3.0
2.5
N=1735
N=376
N=20
N=93
N=17
N=89
N=177
Contr
T2D
SAID
SIDD
SIRD
MOD
MARD
Clusters

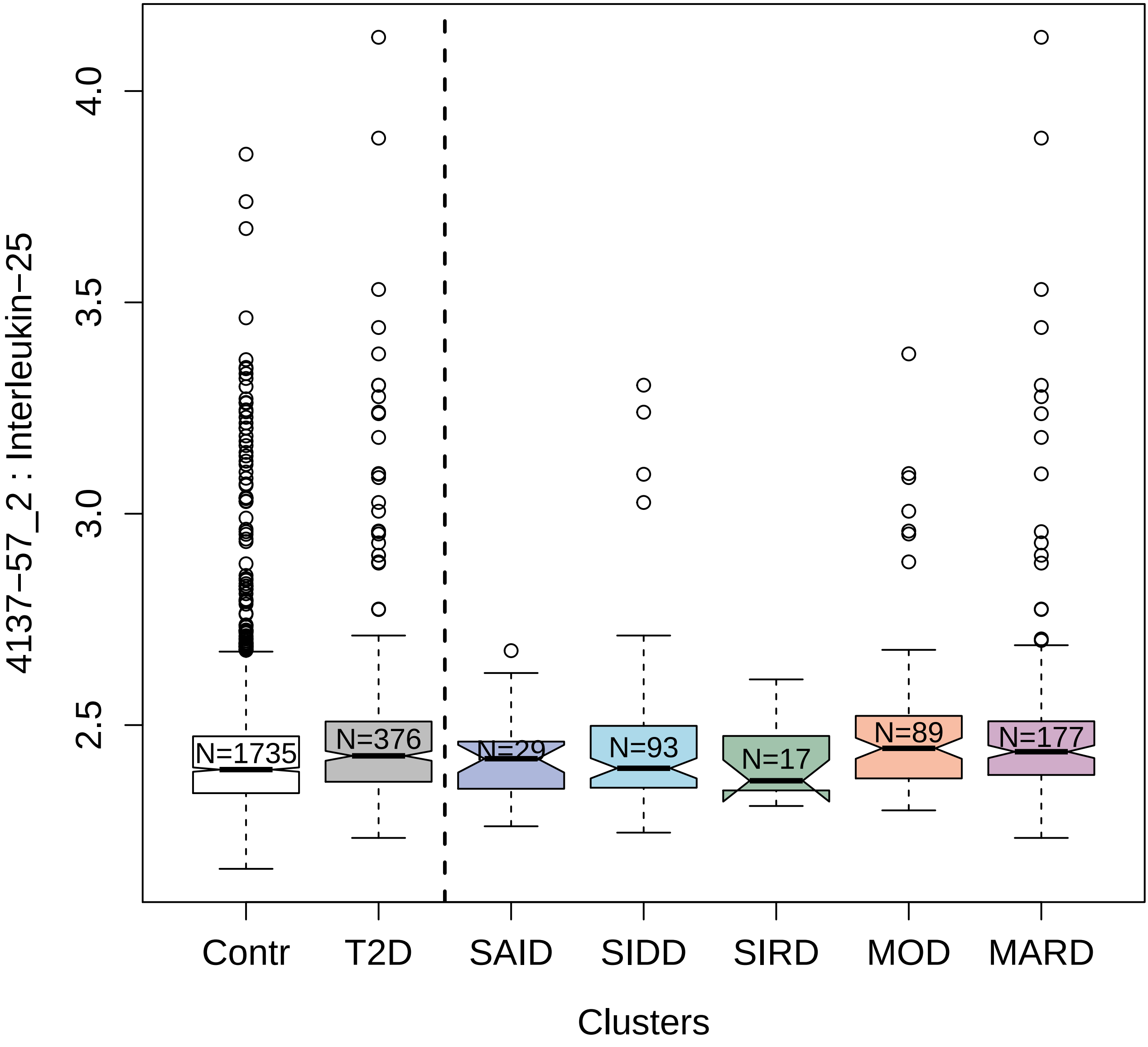
4137−57_2 : Interleukin−25
4.0
3.5
3.0
2.5
N=1735
N=376
N=29
N=93
N=17
N=89
N=177
Contr
T2D
SAID
SIDD
SIRD
MOD
MARD
Clusters

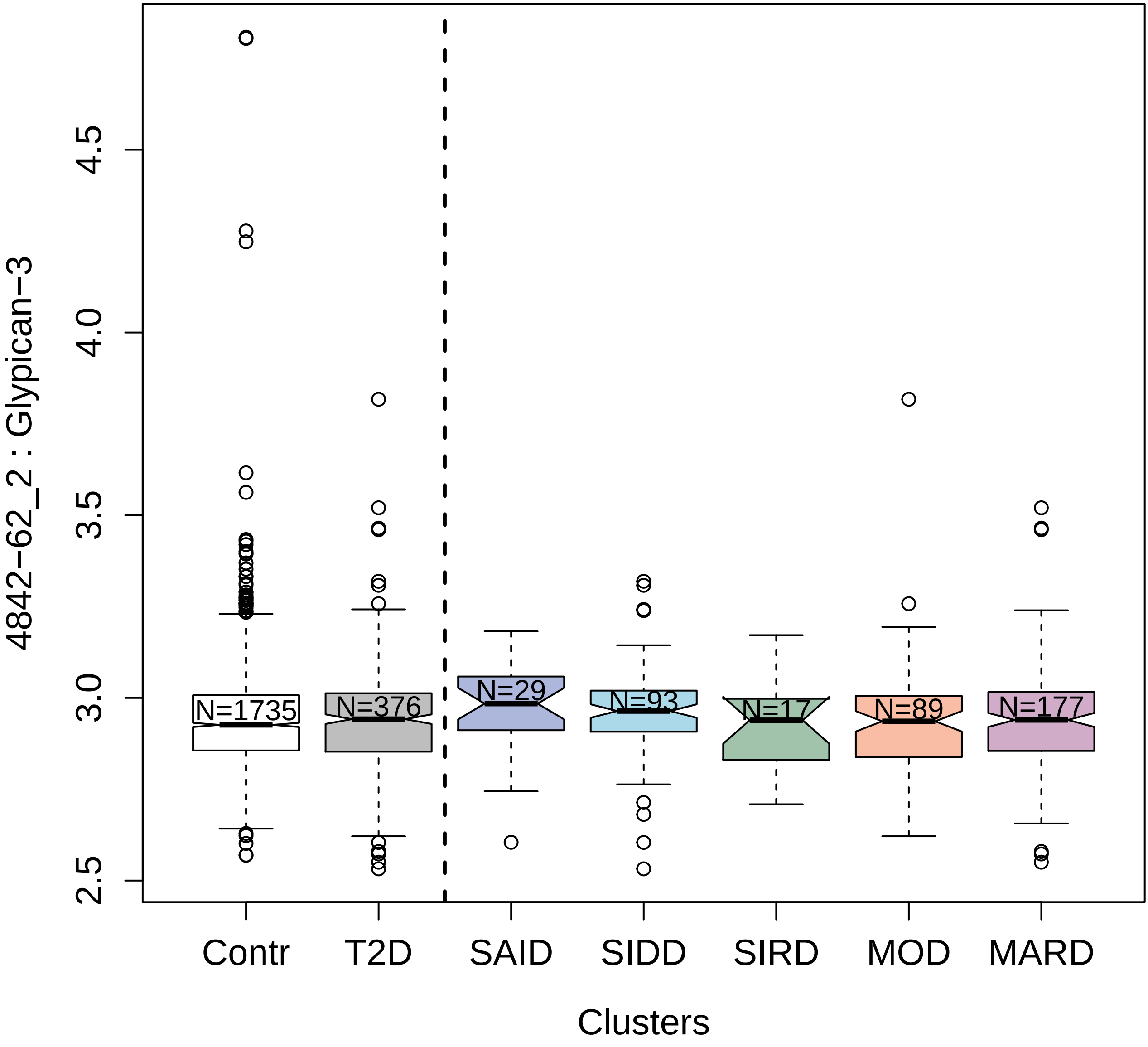

4842−62_2 : Glypican−3
4.5
4.0
3.5
3.0
2.5
N=1735
N=376
N=29
N=93
N=17
N=89
N=177
Contr
T2D
SAID
SIDD
SIRD
MOD
MARD
Clusters

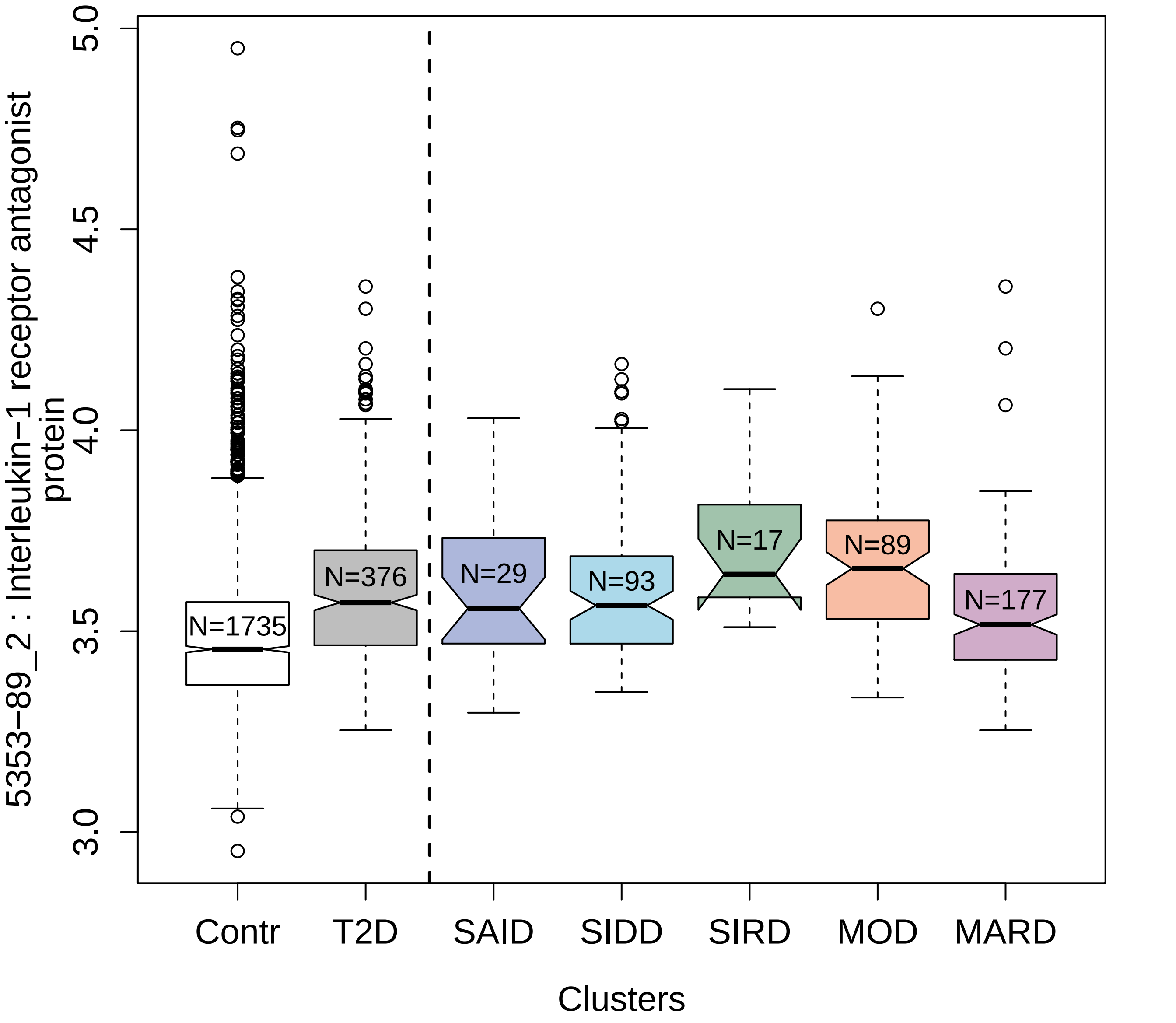
5353−89_2 : Interleukin−1 receptor antagonist protein
5.0
4.5
4.0
3.5
3.0
N=1735
N=376
N=29
N=93
N=17
N=89
N=177
Contr
T2D
SAID
SIDD
SIRD
MOD
MARD
Clusters

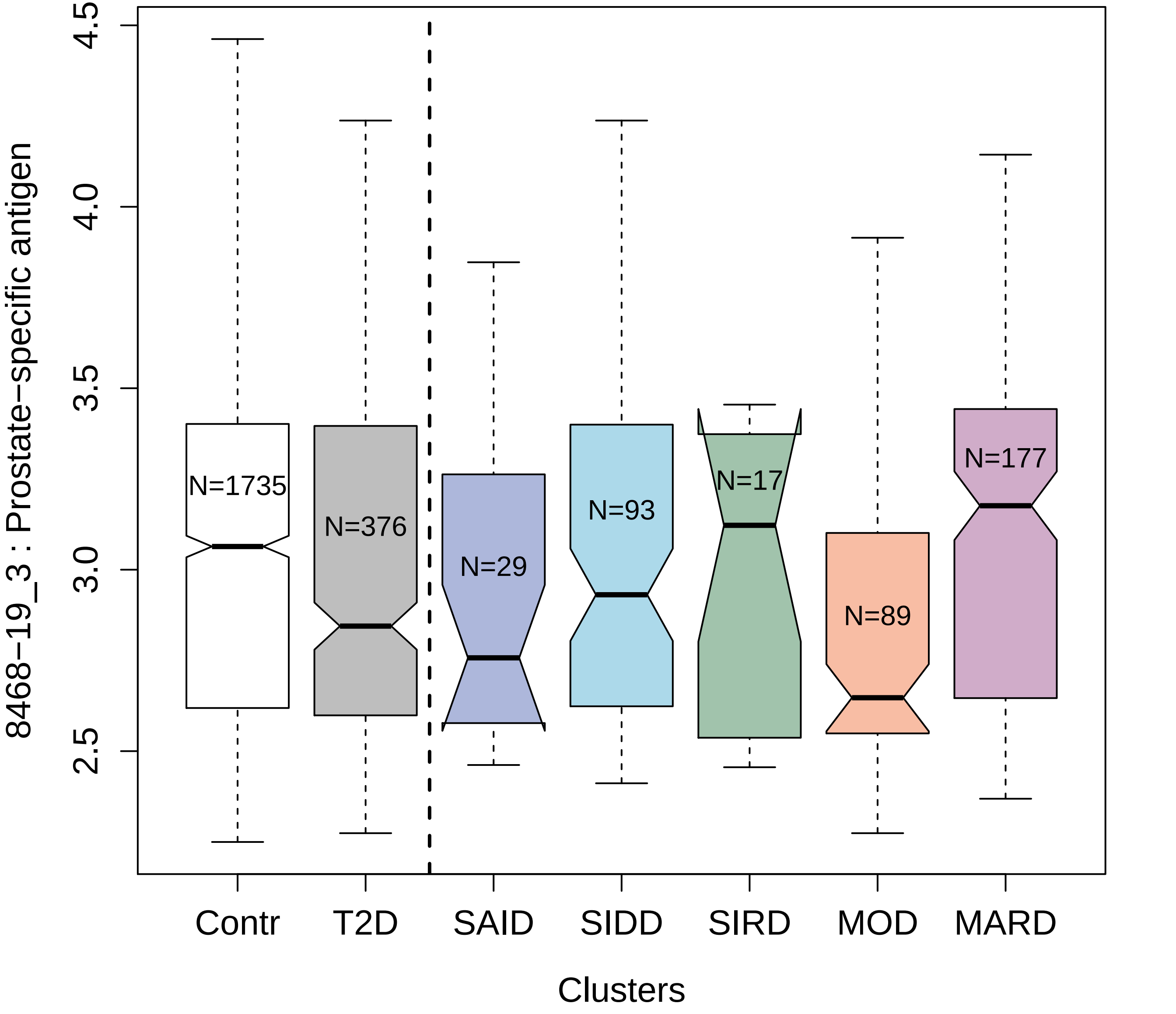

8468−19_3 : Prostate−specific antigen
4.5
4.0
3.5
3.0
2.5
N=1735
N=376
N=29
N=93
N=17
N=89
N=177
Contr
T2D
SAID
SIDD
SIRD
MOD
MARD
Clusters

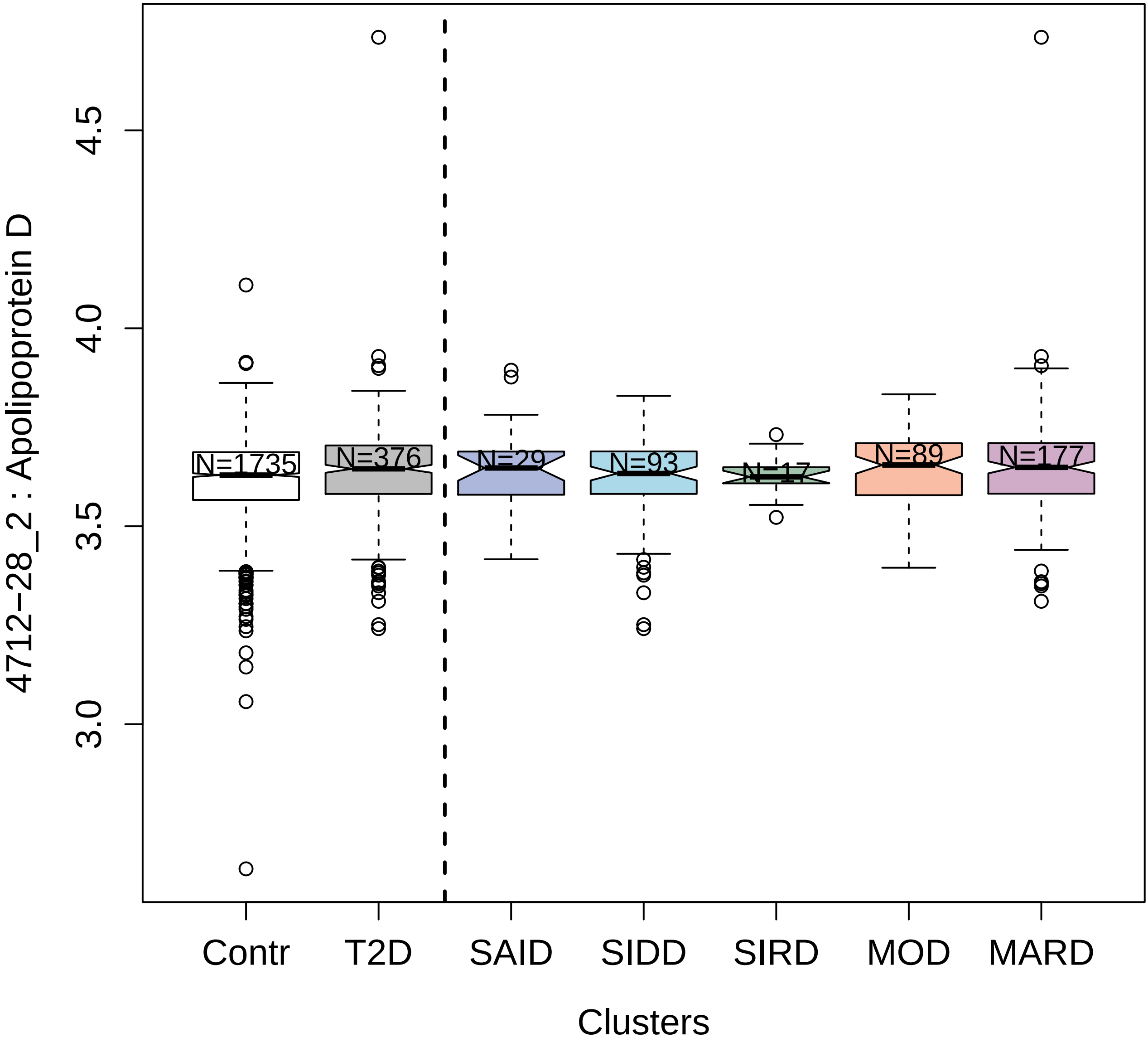

4712−28_2 : Apolipoprotein D
4.5
4.0
3.5
3.0
N=1735
N=376
N=29
N=93
N=17
N=89
N=177
Contr
T2D
SAID
SIDD
SIRD
MOD
MARD
Clusters

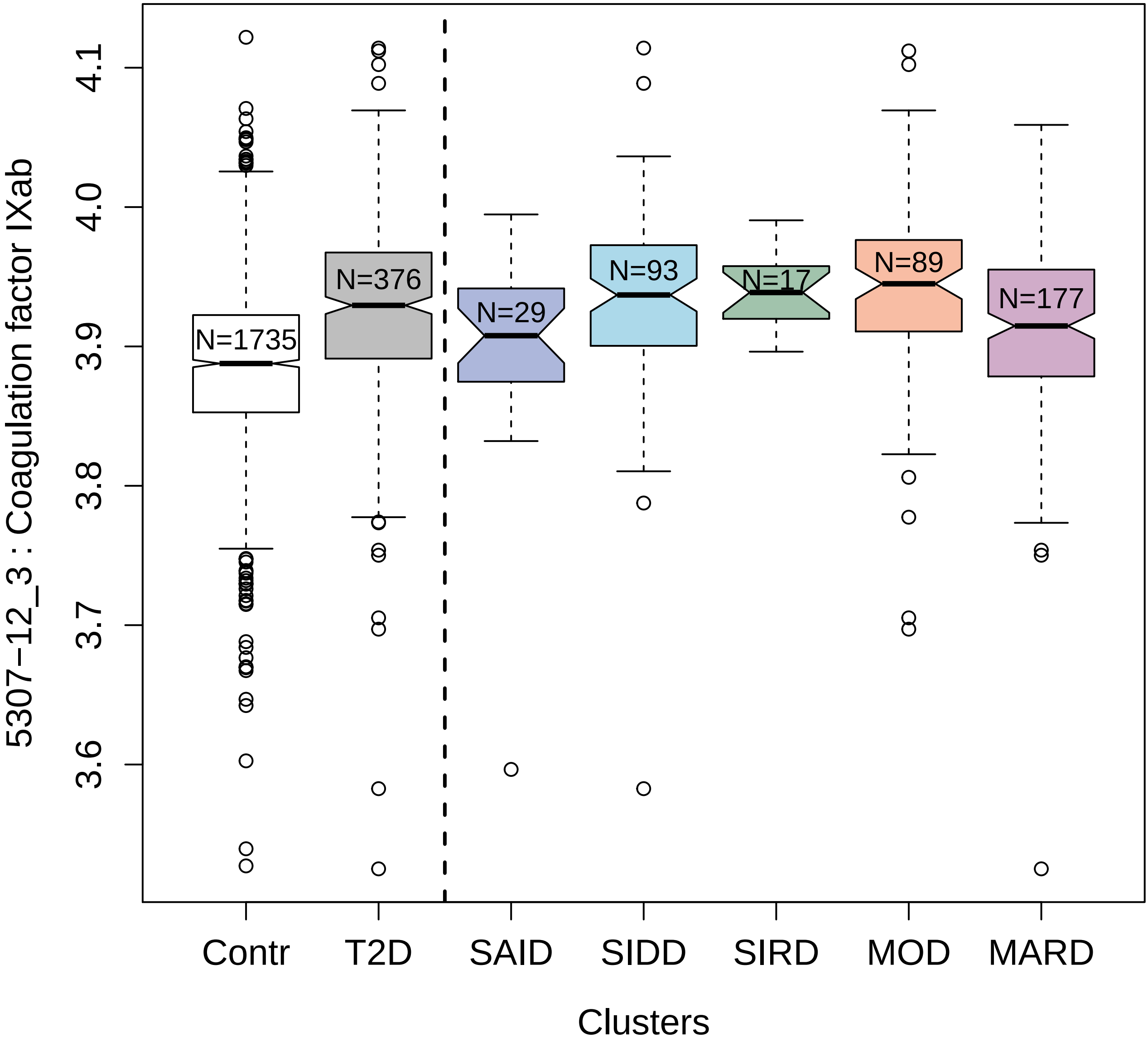

5307−12_3 : Coagulation factor IXab
4.1
4.0
3.9
3.8
3.7
3.6
N=1735
N=376
N=29
N=93
N=17
N=89
N=177
Contr
T2D
SAID
SIDD
SIRD
MOD
MARD
Clusters

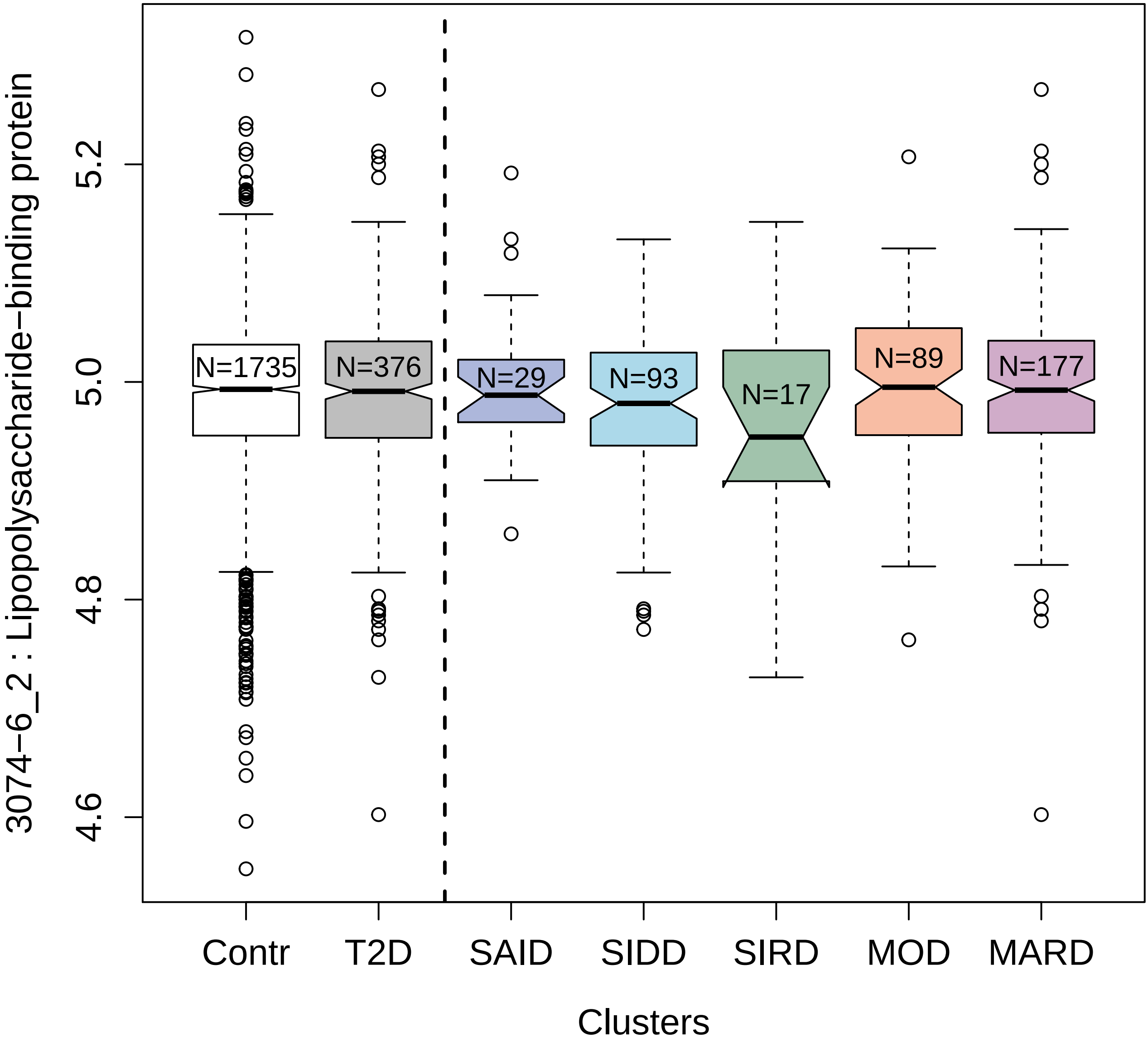

3074−6_2 : Lipopolysaccharide−binding protein
5.2
5.0
4.8
4.6
N=1735
N=376
N=29
N=93
N=17
N=89
N=177
Contr
T2D
SAID
SIDD
SIRD
MOD
MARD
Clusters

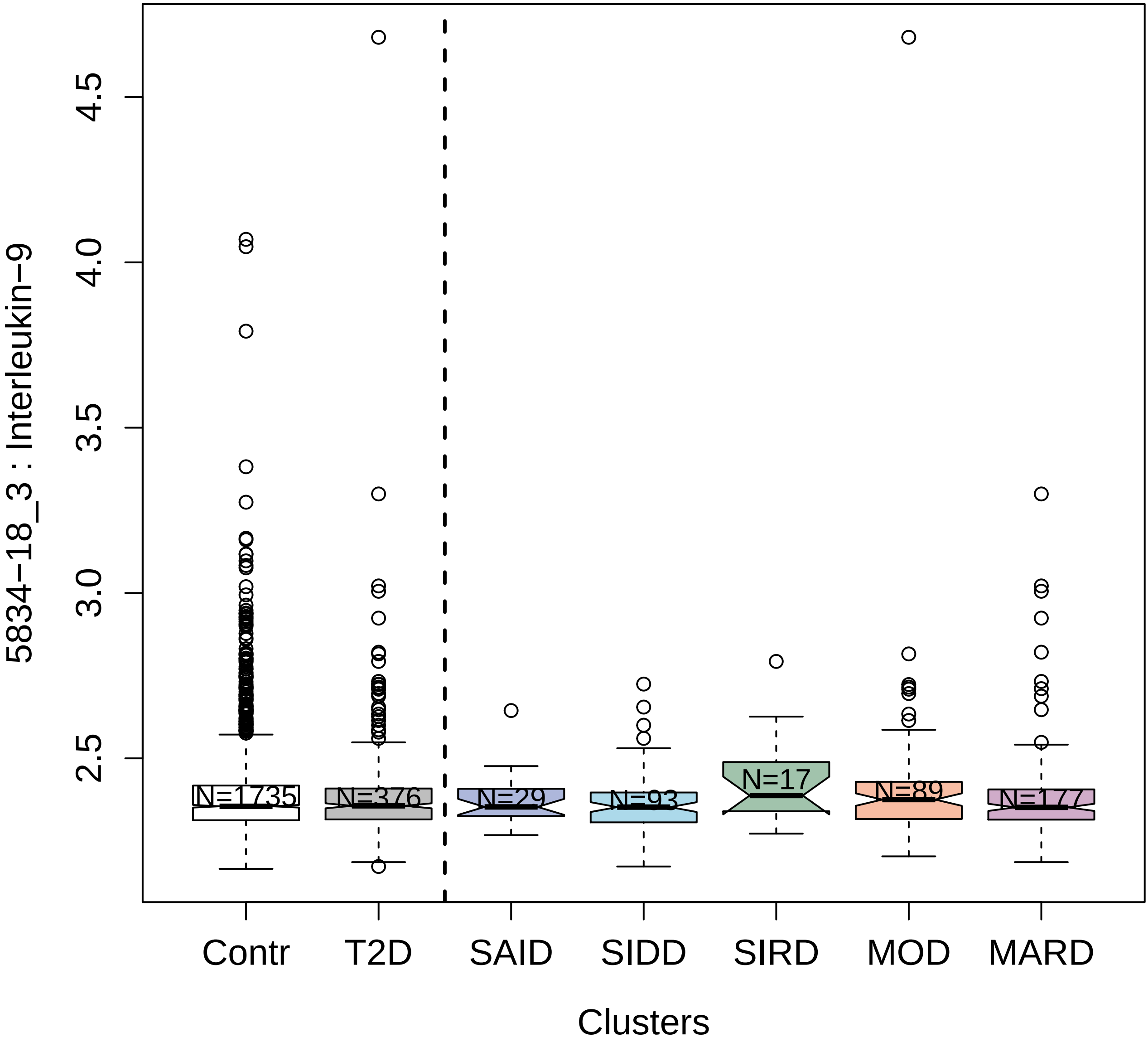
5834−18_3 : Interleukin−9
4.5
4.0
3.5
3.0
2.5
N=1735
N=376
N=29
N=93
N=17
N=89
N=177
Contr
T2D
SAID
SIDD
SIRD
MOD
MARD
Clusters

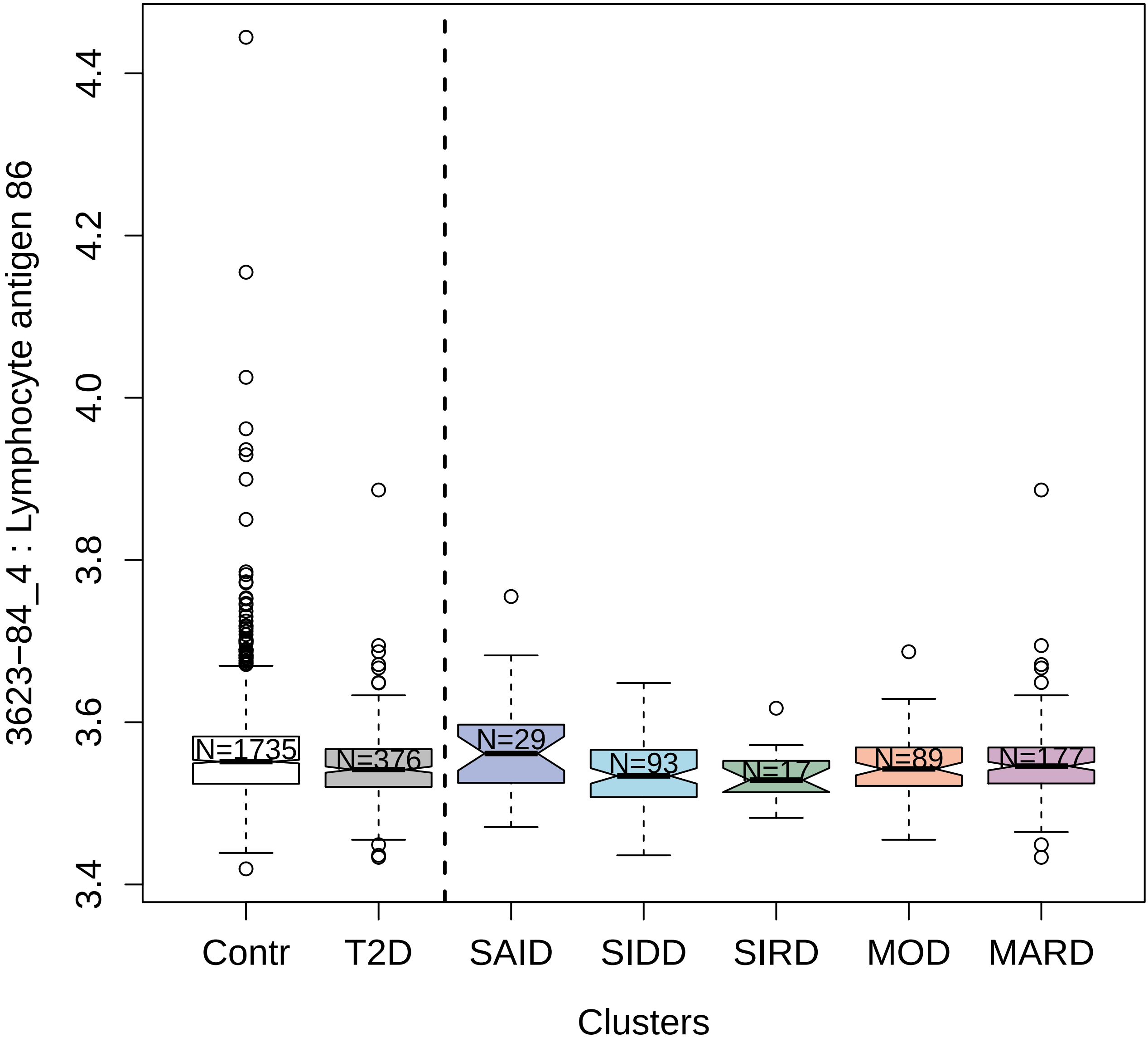
3623−84_4 : Lymphocyte antigen 86
4.4
4.2
4.0
3.8
3.6
3.4
N=1735
N=376
N=29
N=93
N=17
N=89
N=177
Contr
T2D
SAID
SIDD
SIRD
MOD
MARD
Clusters

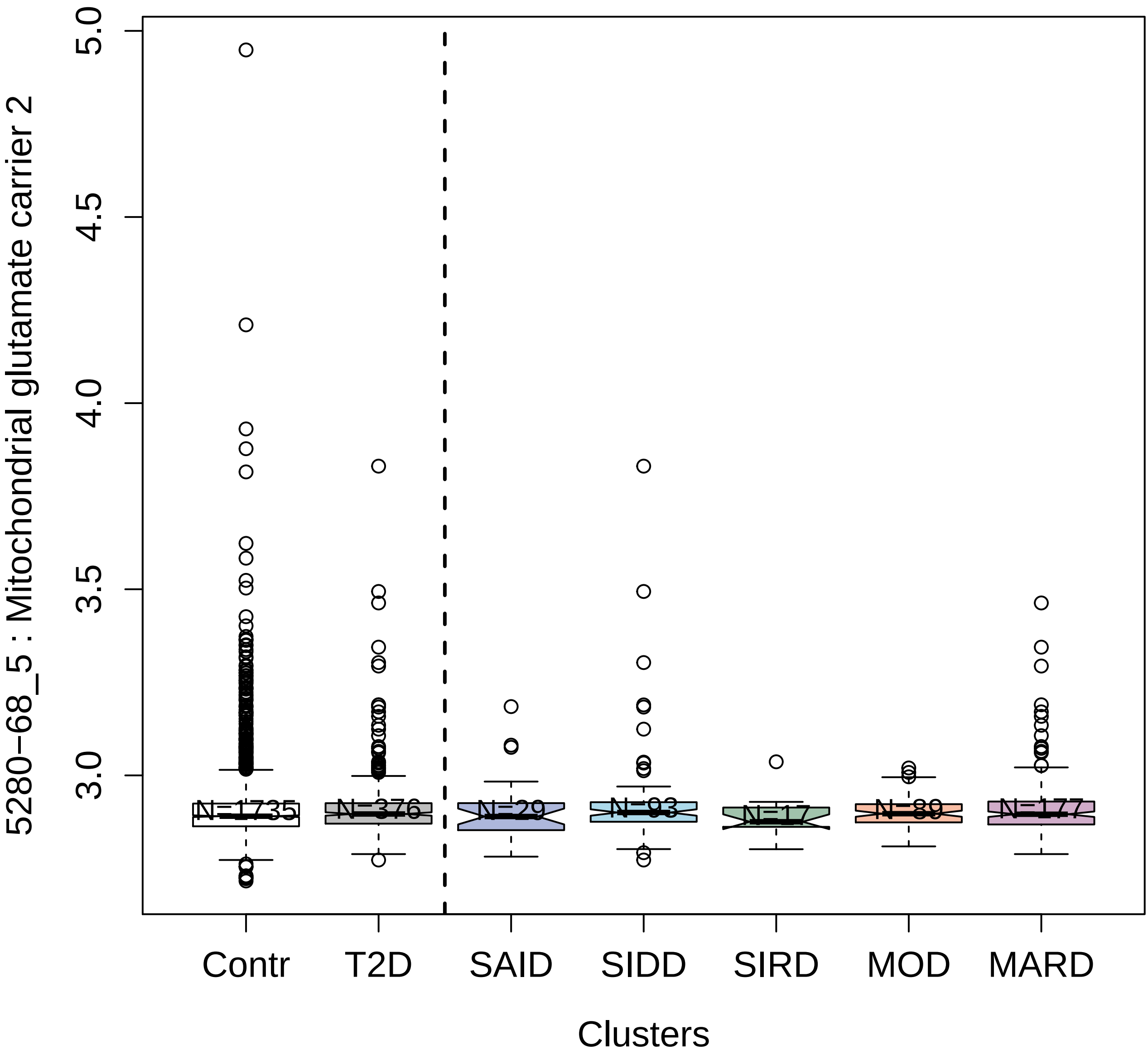

5280−68_5 : Mitochondrial glutamate carrier 2
5.0
4.5
4.0
3.5
3.0
N=1735
N=376
N=29
N=93
N=17
N=89
N=177
Contr
T2D
SAID
SIDD
SIRD
MOD
MARD
Clusters

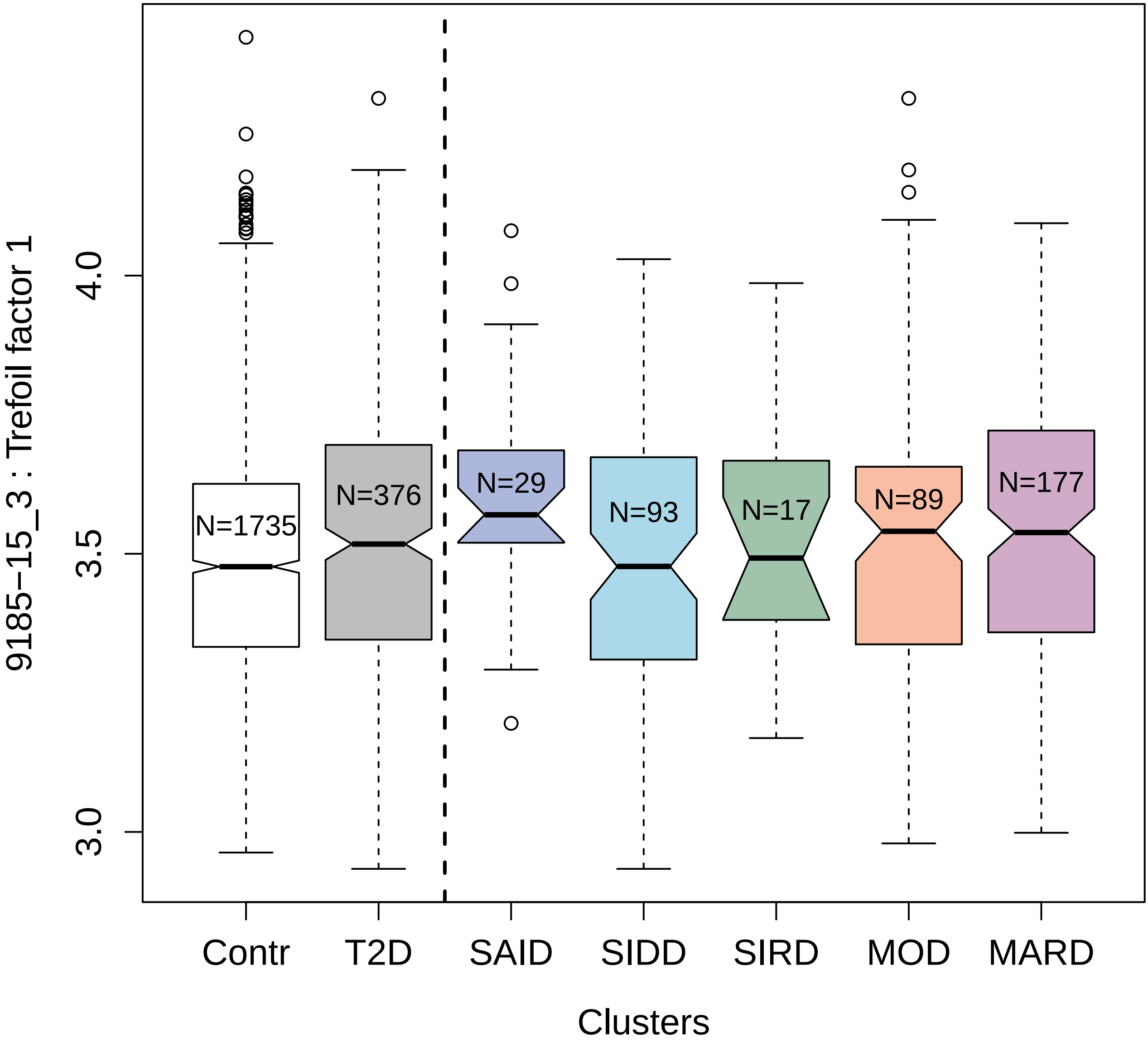

9185−15_3 : Trefoil factor 1
3.0
3.5
4.0
N=1735
N=376
N=29
N=93
N=17
N=89
N=177
Contr
T2D
SAID
SIDD
SIRD
MOD
MARD
Clusters

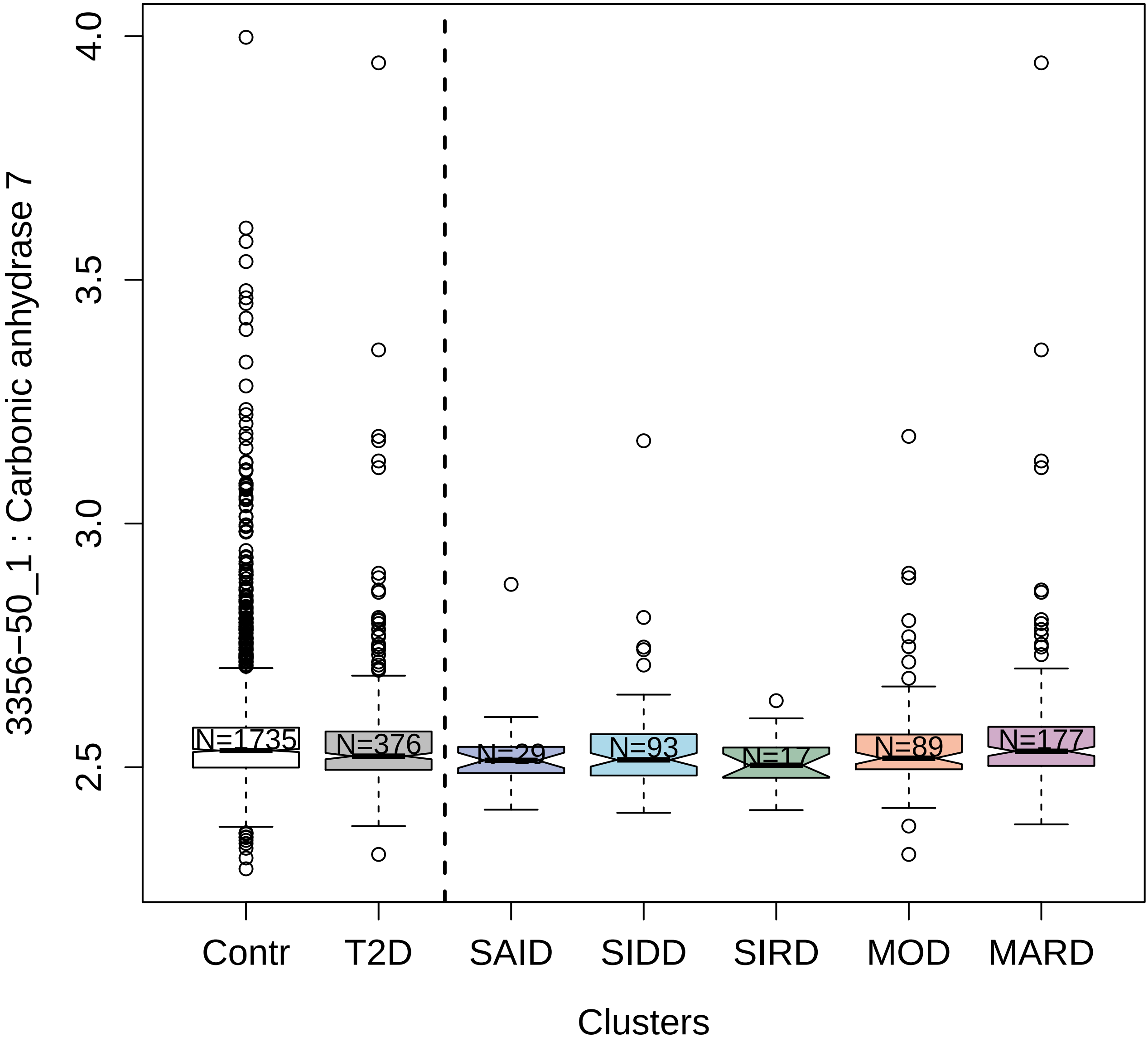
3356−50_1 : Carbonic anhydrase 7
4.0
3.5
3.0
2.5
N=1735
N=376
N=29
N=93
N=17
N=89
N=177
Contr
T2D
SAID
SIDD
SIRD
MOD
MARD
Clusters

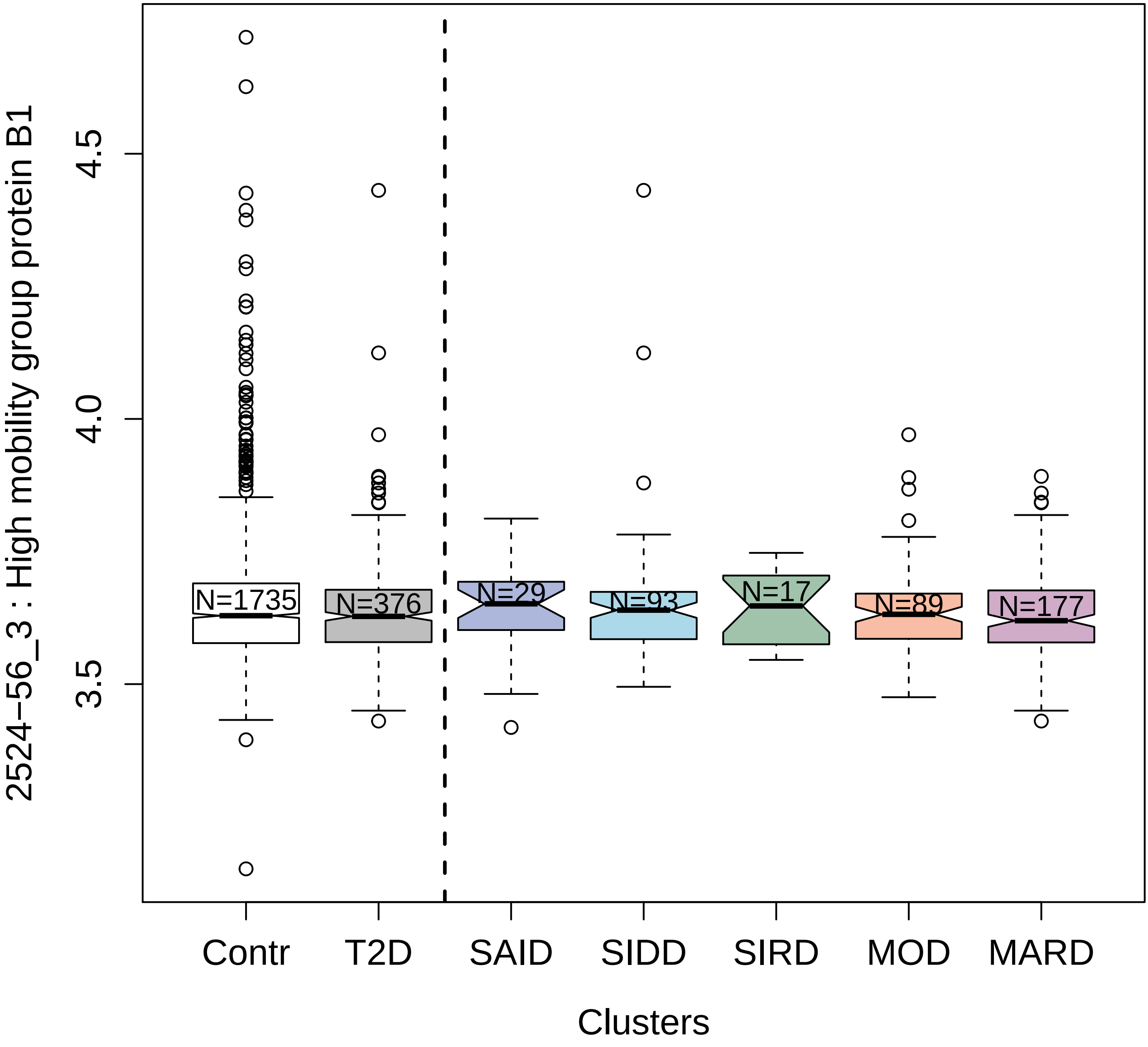

2524−56_3 : High mobility group protein B1
4.5
4.0
3.5
N=1735
N=376
N=29
N=93
N=17
N=89
N=177
Contr
T2D
SAID
SIDD
SIRD
MOD
MARD
Clusters

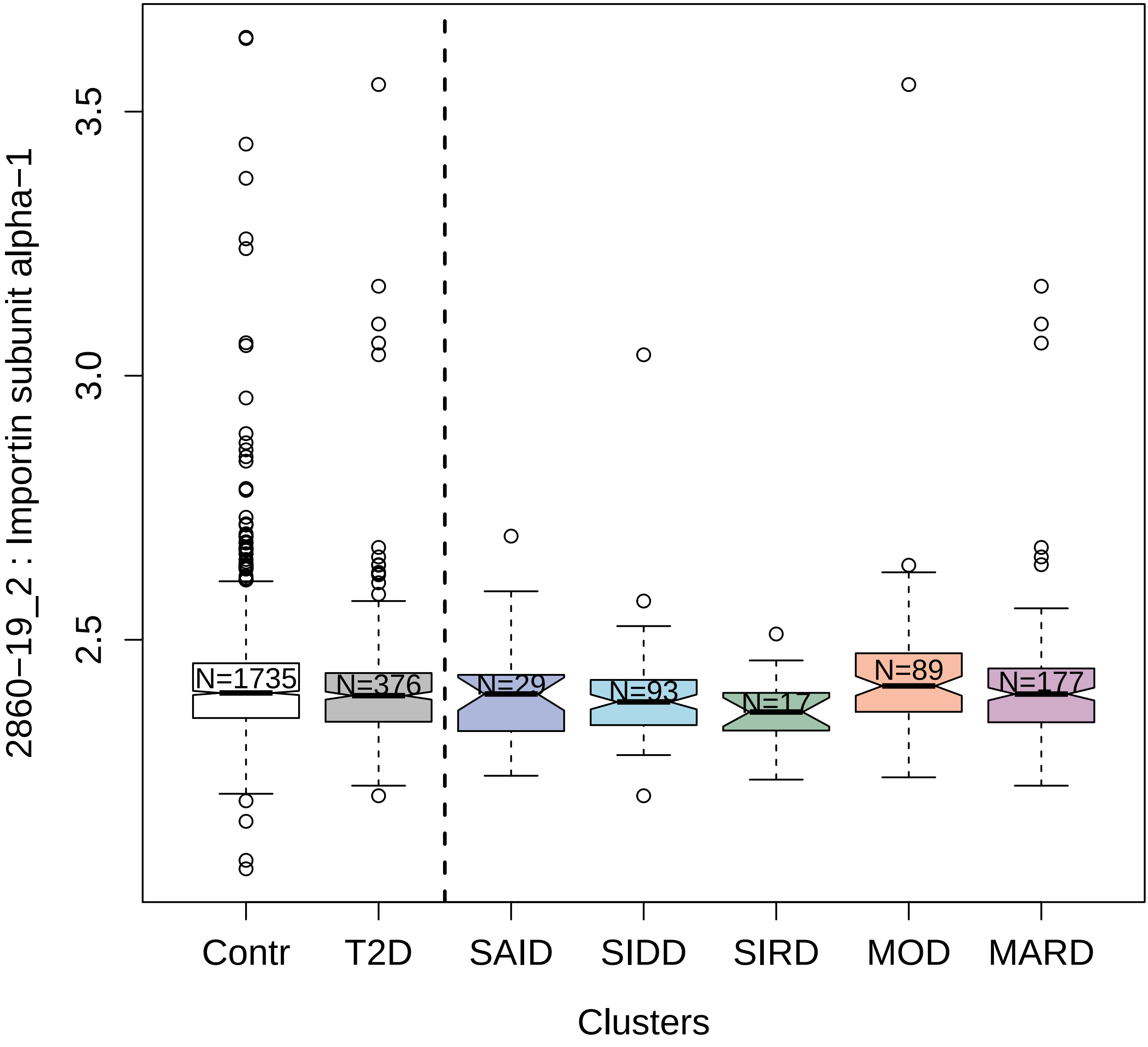

2860−19_2 : Importin subunit alpha−1
3.5
3.0
2.5
N=1735
N=376
N=29
N=93
N=17
N=89
N=177
Contr
T2D
SAID
SIDD
SIRD
MOD
MARD
Clusters

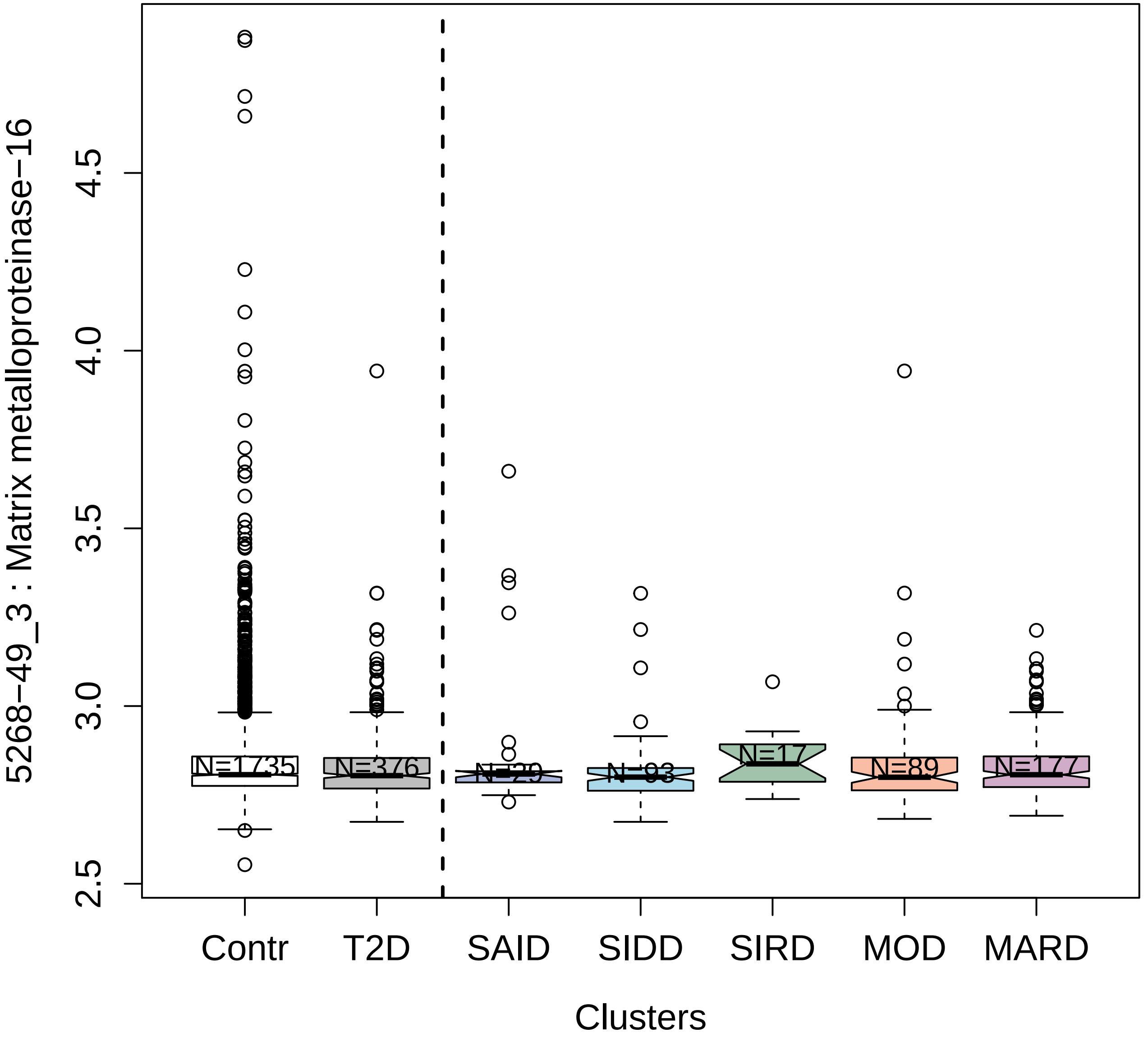

5268−49_3 : Matrix metalloproteinase−16
2.5
3.0
3.5
4.0
4.5
N=1735
N=376
N=29
N=93
N=17
N=89
N=177
Contr
T2D
SAID
SIDD
SIRD
MOD
MARD
Clusters

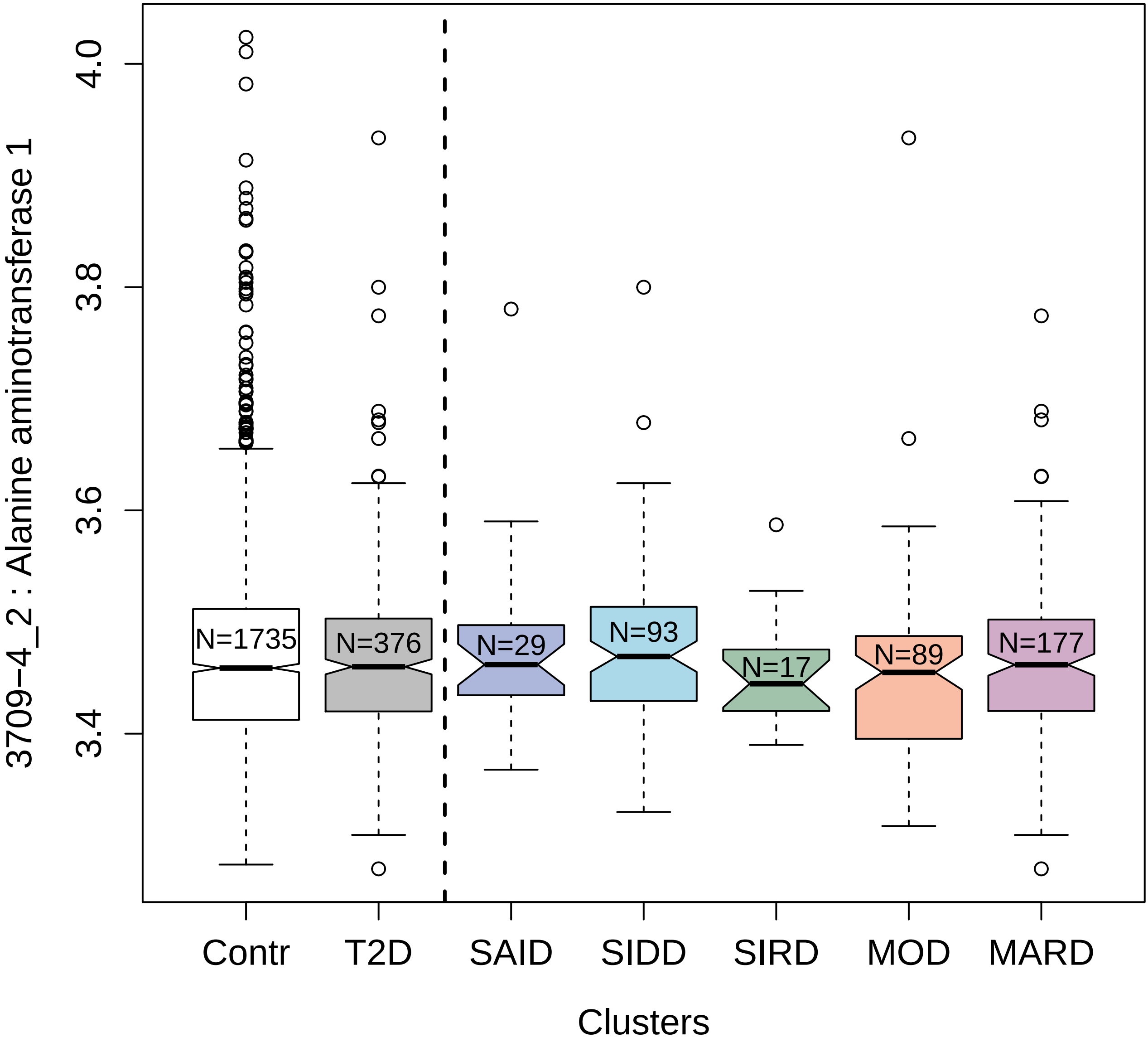

3709−4_2 : Alanine aminotransferase 1
4.0
3.8
3.6
3.4
N=1735
N=376
N=29
N=93
N=17
N=89
N=177
Contr
T2D
SAID
SIDD
SIRD
MOD
MARD
Clusters

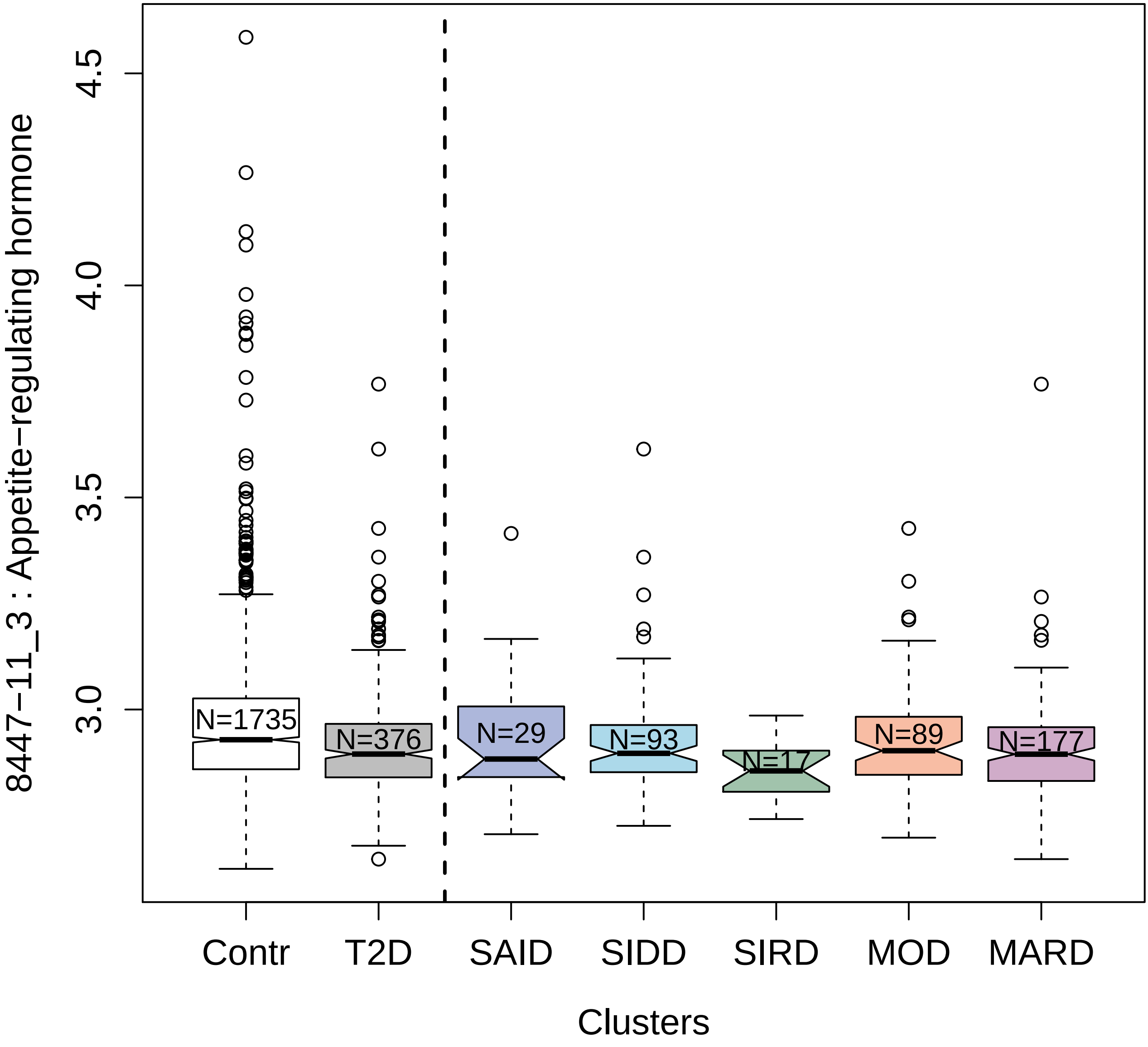

8447−11_3 : Appetite−regulating hormone
4.5
4.0
3.5
3.0
N=1735
N=376
N=29
N=93
N=17
N=89
N=177
Contr
T2D
SAID
SIDD
SIRD
MOD
MARD
Clusters

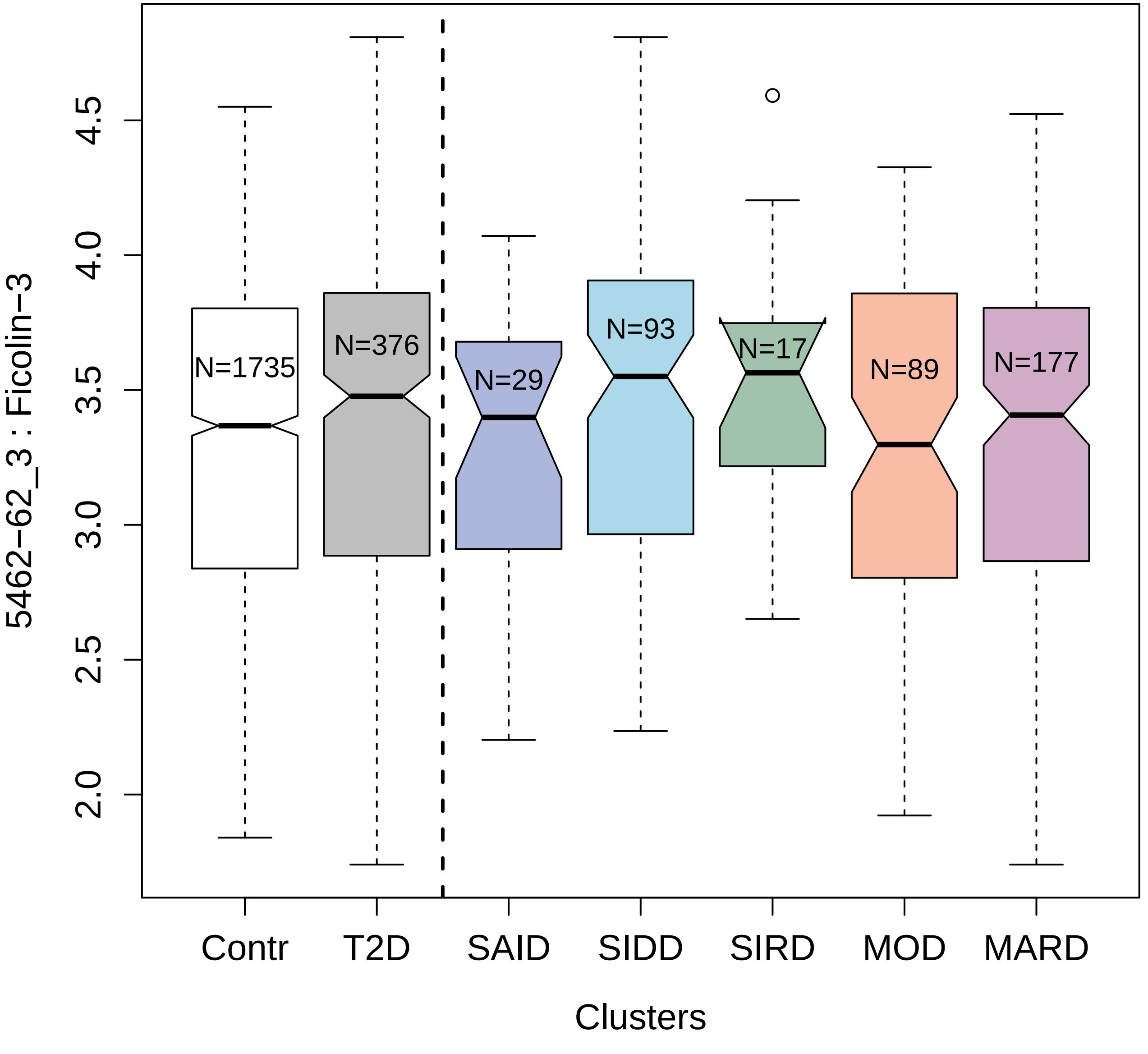

5462−62_3 : Ficolin−3
2.0
2.5
3.0
3.5
4.0
4.5
N=1735
N=376
N=29
N=93
N=17
N=89
N=177
Contr
T2D
SAID
SIDD
SIRD
MOD
MARD
Clusters

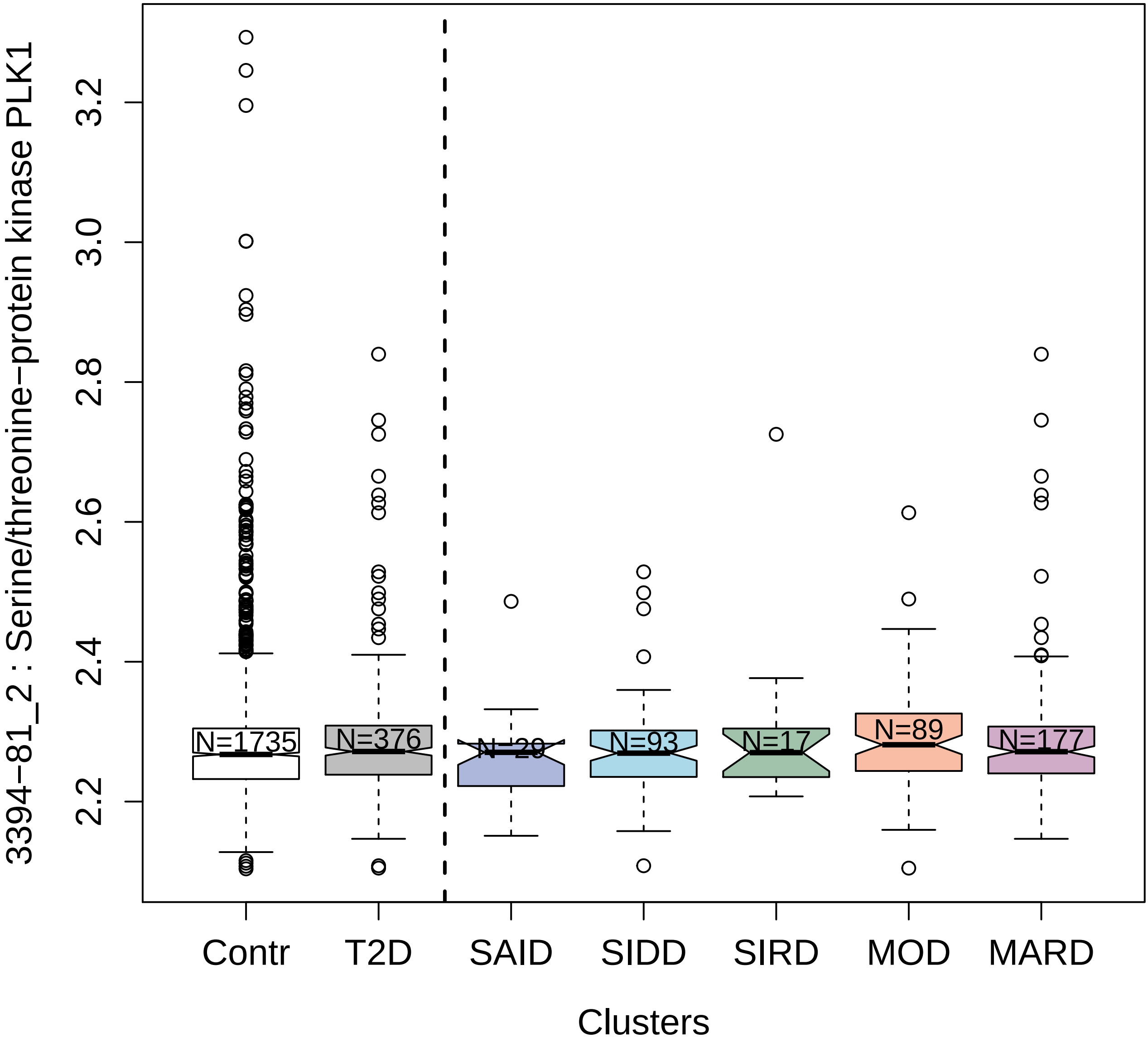
3394−81_2 : Serine/threonine−protein kinase PLK1
2.2
2.4
2.6
2.8
3.0
3.2
N=1735
N=376
N=29
N=93
N=17
N=89
N=177
Contr
T2D
SAID
SIDD
SIRD
MOD
MARD
Clusters

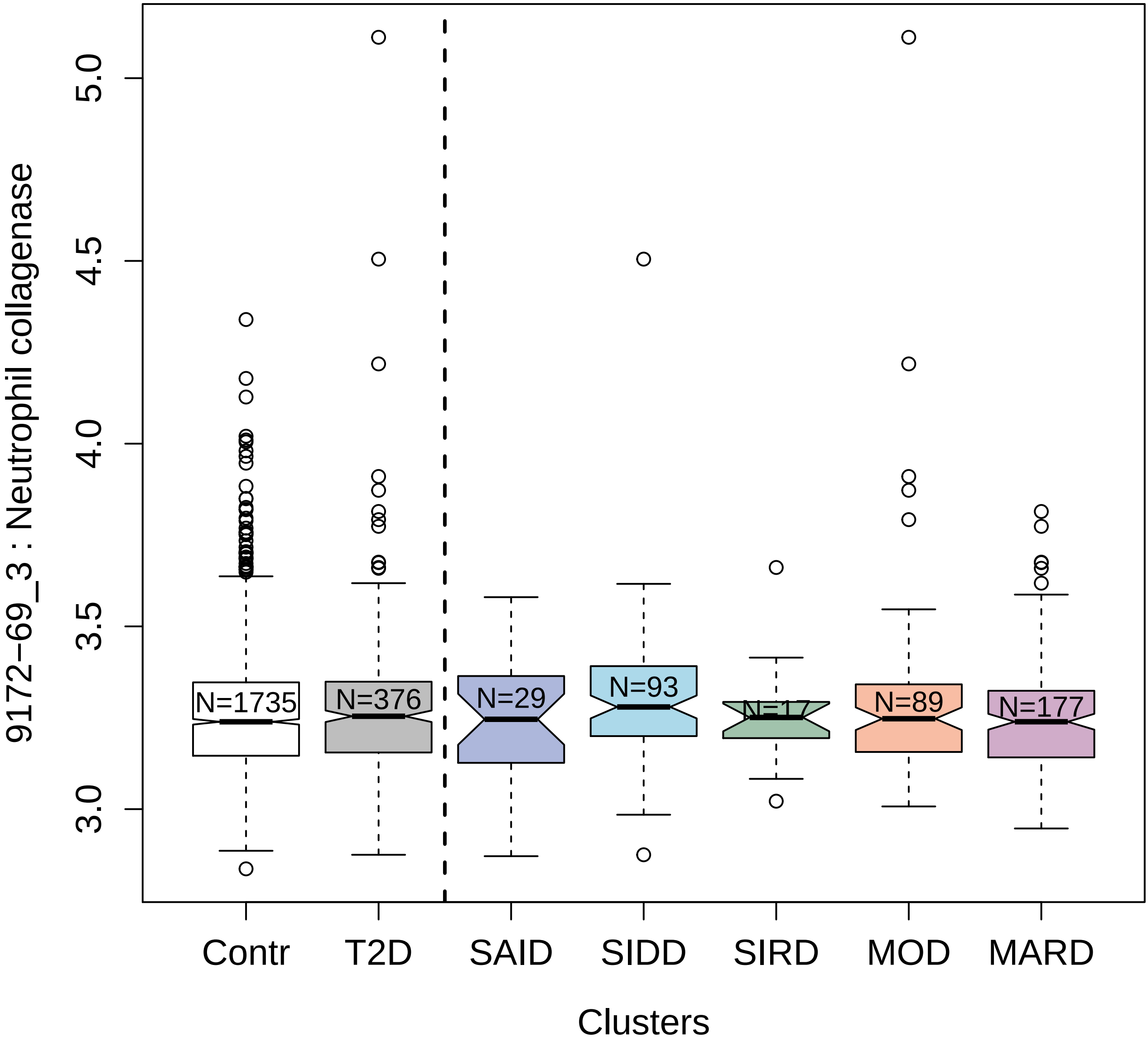
9172−69_3 : Neutrophil collagenase
5.0
4.5
4.0
3.5
3.0
N=1735
N=376
N=29
N=93
N=17
N=89
N=177
Contr
T2D
SAID
SIDD
SIRD
MOD
MARD
Clusters

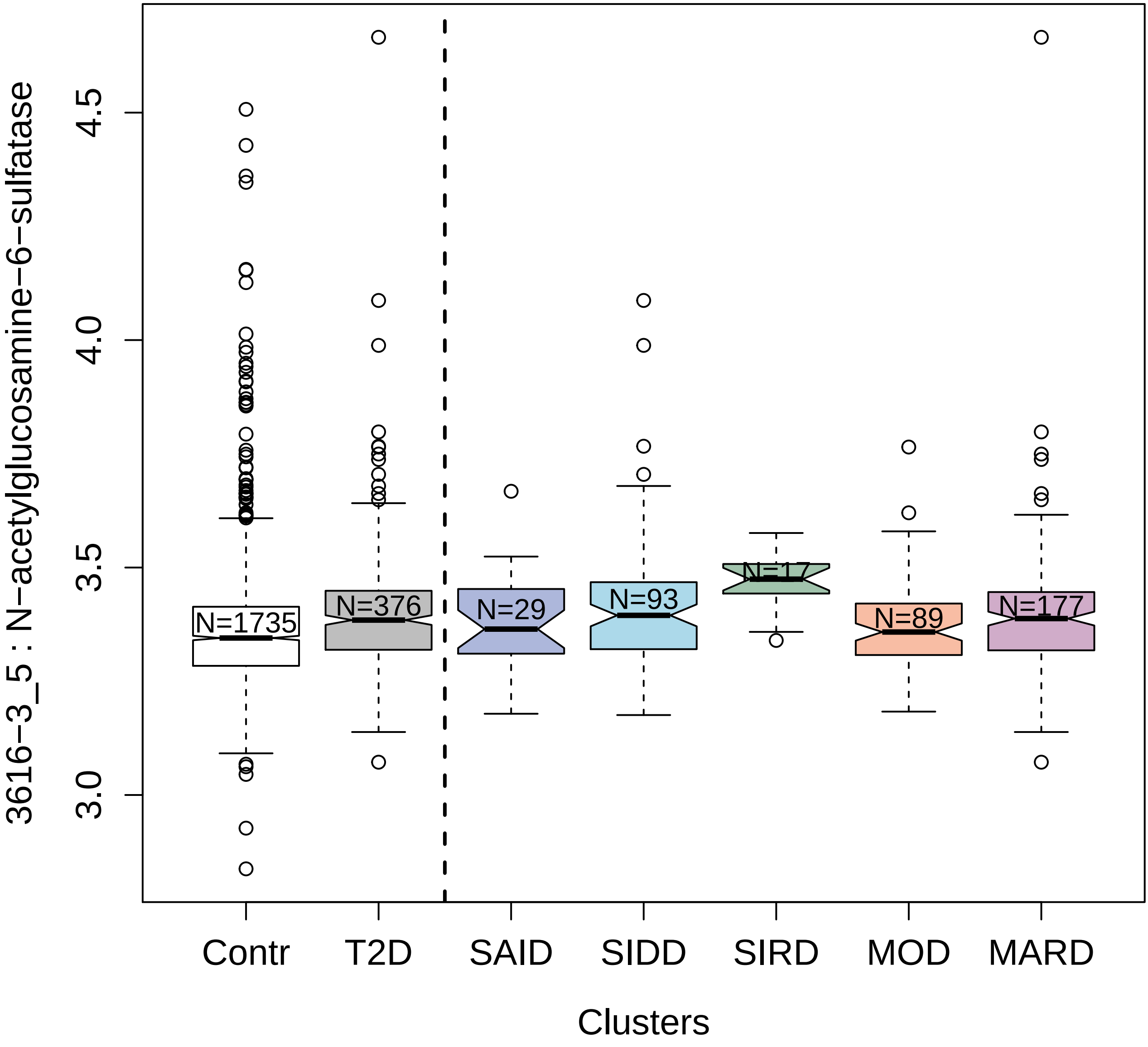

3616−3_5 : N−acetylglucosamine−6−sulfatase
3.0
3.5
4.0
4.5
N=1735
N=376
N=29
N=93
N=17
N=89
N=177
Contr
T2D
SAID
SIDD
SIRD
MOD
MARD
Clusters

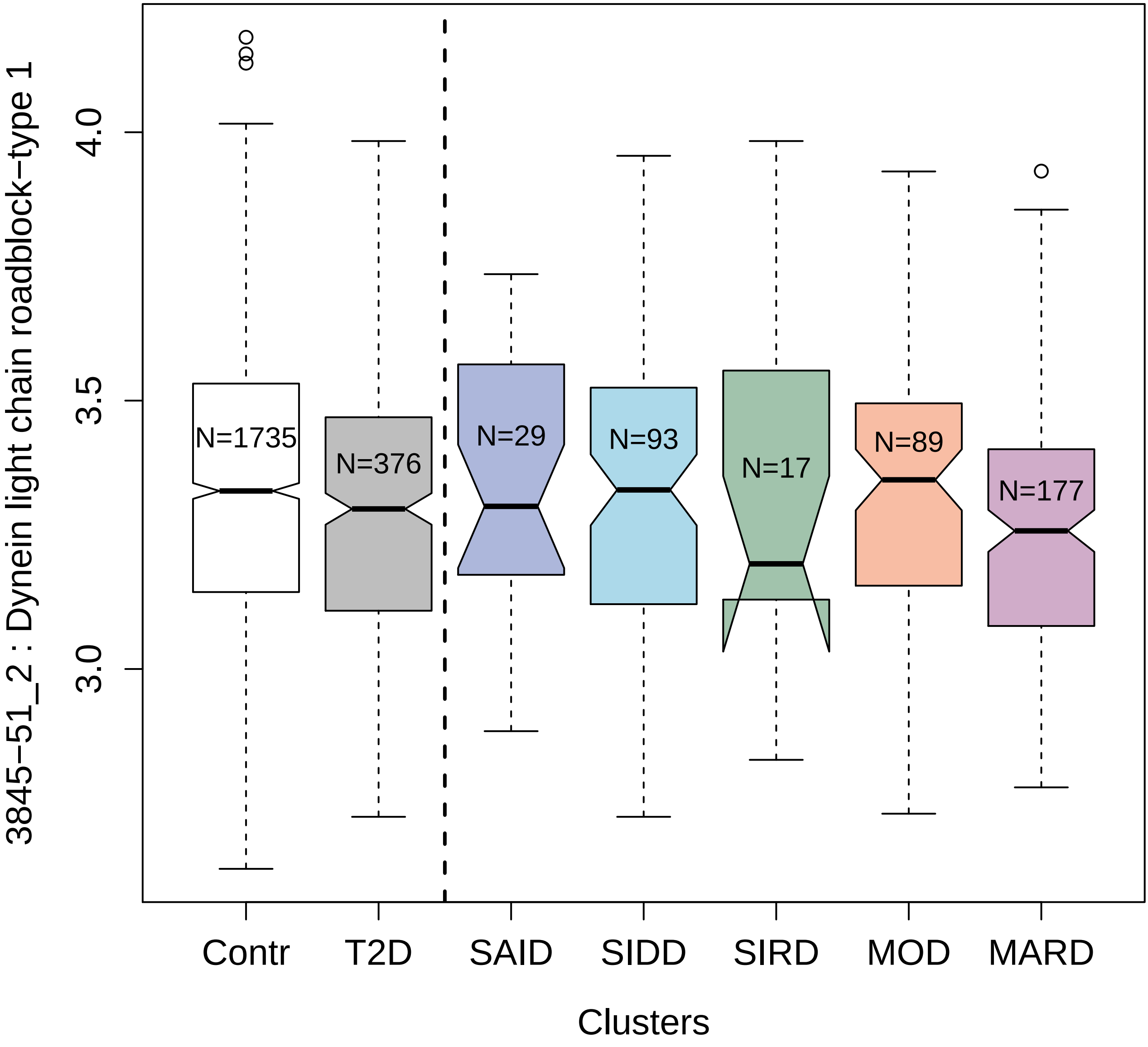
3845−51_2 : Dynein light chain roadblock−type 1
4.0
3.5
3.0
N=1735
N=376
N=29
N=93
N=17
N=89
N=177
Contr
T2D
SAID
SIDD
SIRD
MOD
MARD
Clusters

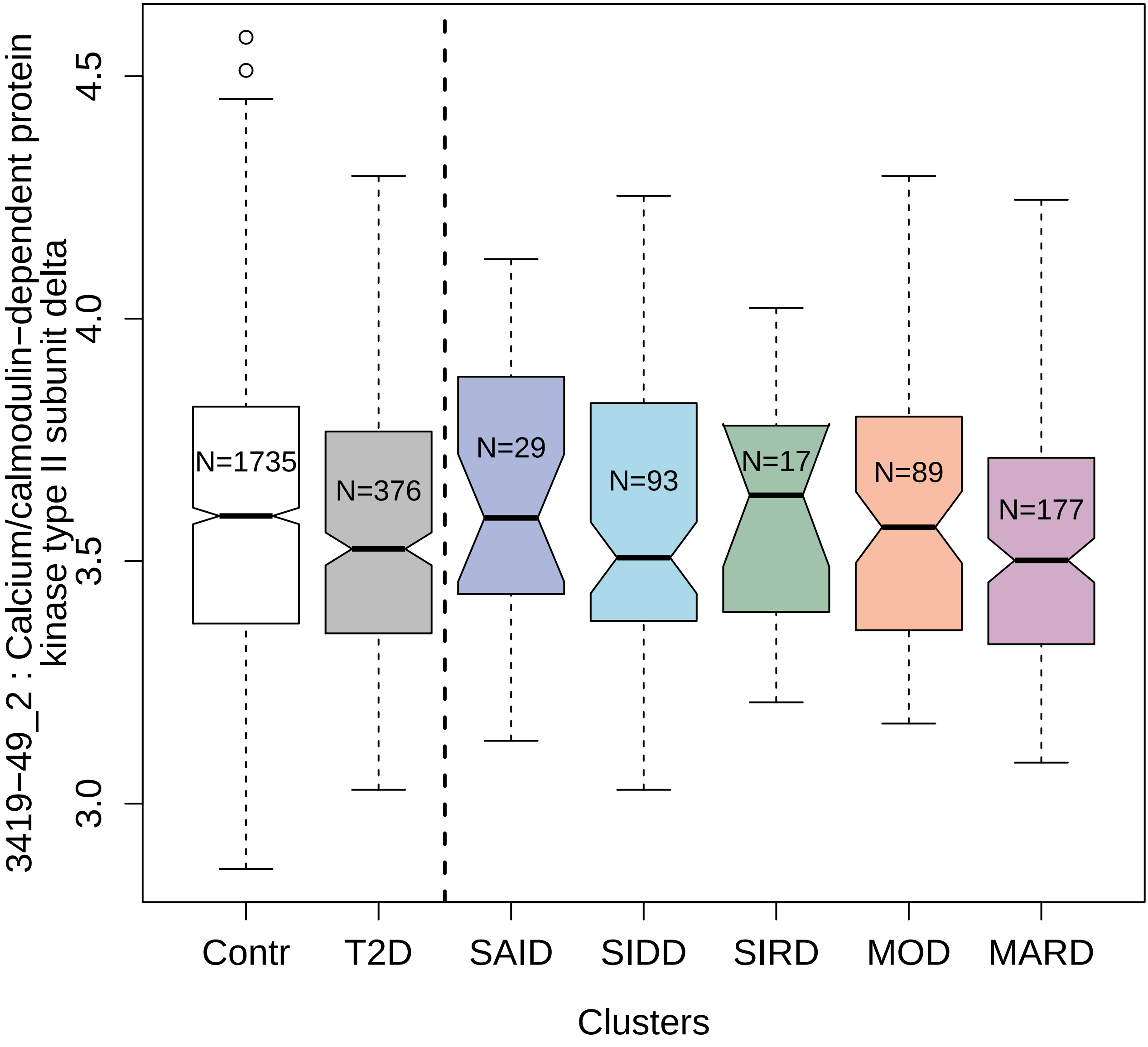
3419−49_2 : Calcium/calmodulin−dependent protein
kinase type II subunit delta
4.5
4.0
3.5
3.0
N=1735
N=376
N=29
N=93
N=17
N=89
N=177
Contr
T2D
SAID
SIDD
SIRD
MOD
MARD
Clusters

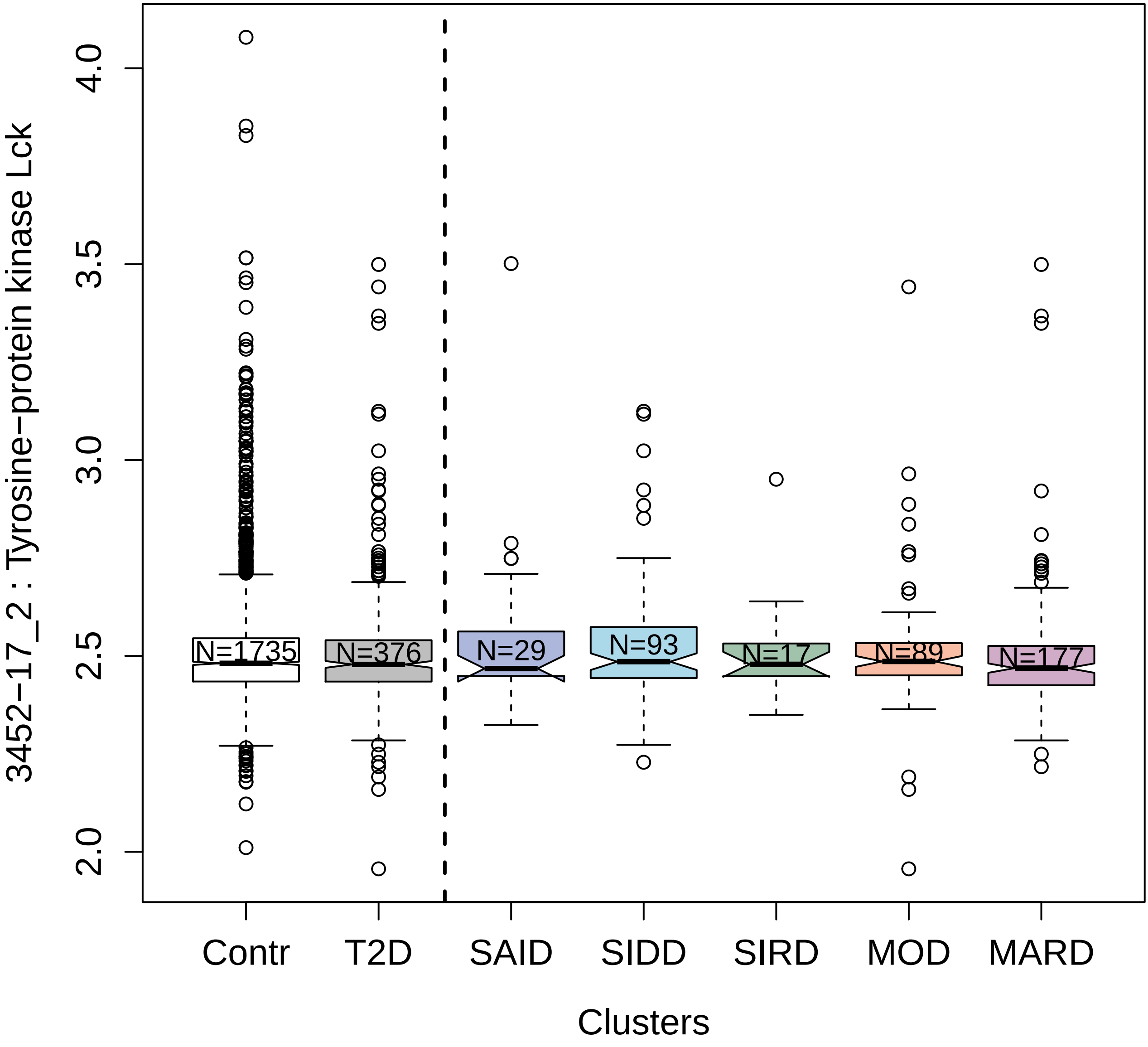
3452−17_2 : Tyrosine−protein kinase Lck
4.0
3.5
3.0
2.5
2.0
N=1735
N=376
N=29
N=93
N=17
N=89
N=177
Contr
T2D
SAID
SIDD
SIRD
MOD
MARD
Clusters

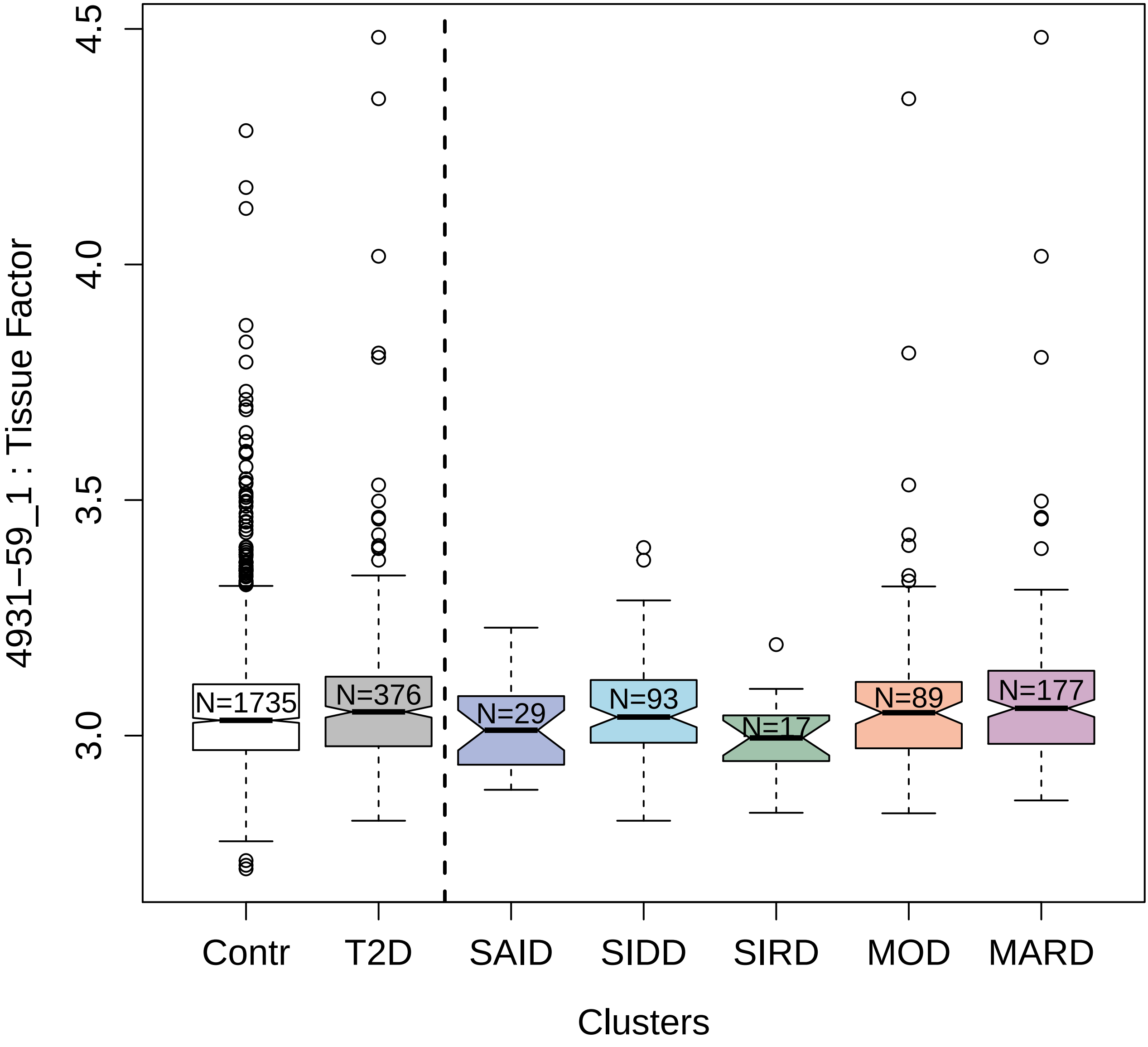

4931−59_1 : Tissue Factor
4.5
4.0
3.5
3.0
N=1735
N=376
N=29
N=93
N=17
N=89
N=177
Contr
T2D
SAID
SIDD
SIRD
MOD
MARD
Clusters

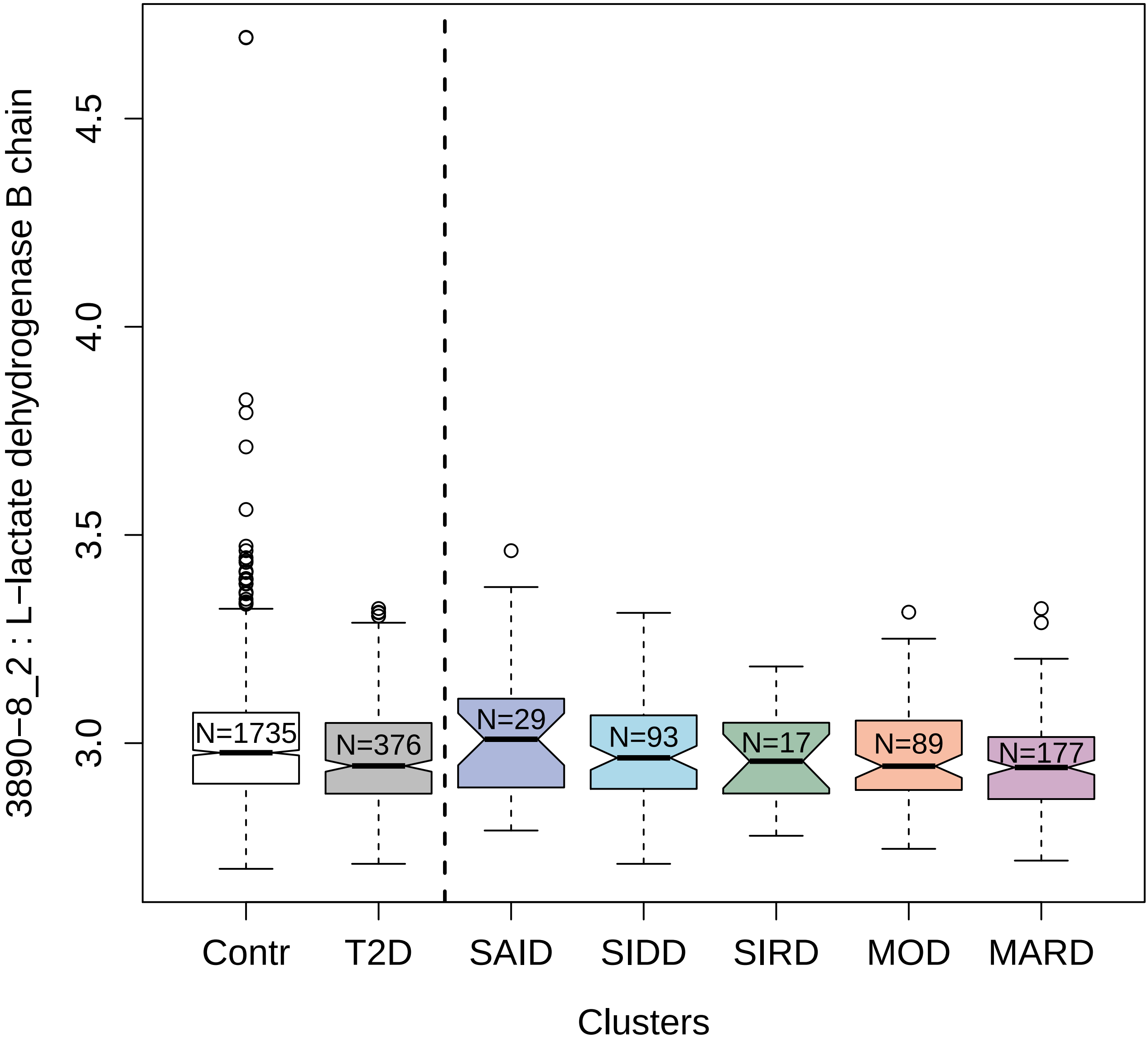
3890−8_2 : L−lactate dehydrogenase B chain
4.5
4.0
3.5
3.0
N=1735
N=376
N=29
N=93
N=17
N=89
N=177
Contr
T2D
SAID
SIDD
SIRD
MOD
MARD
Clusters

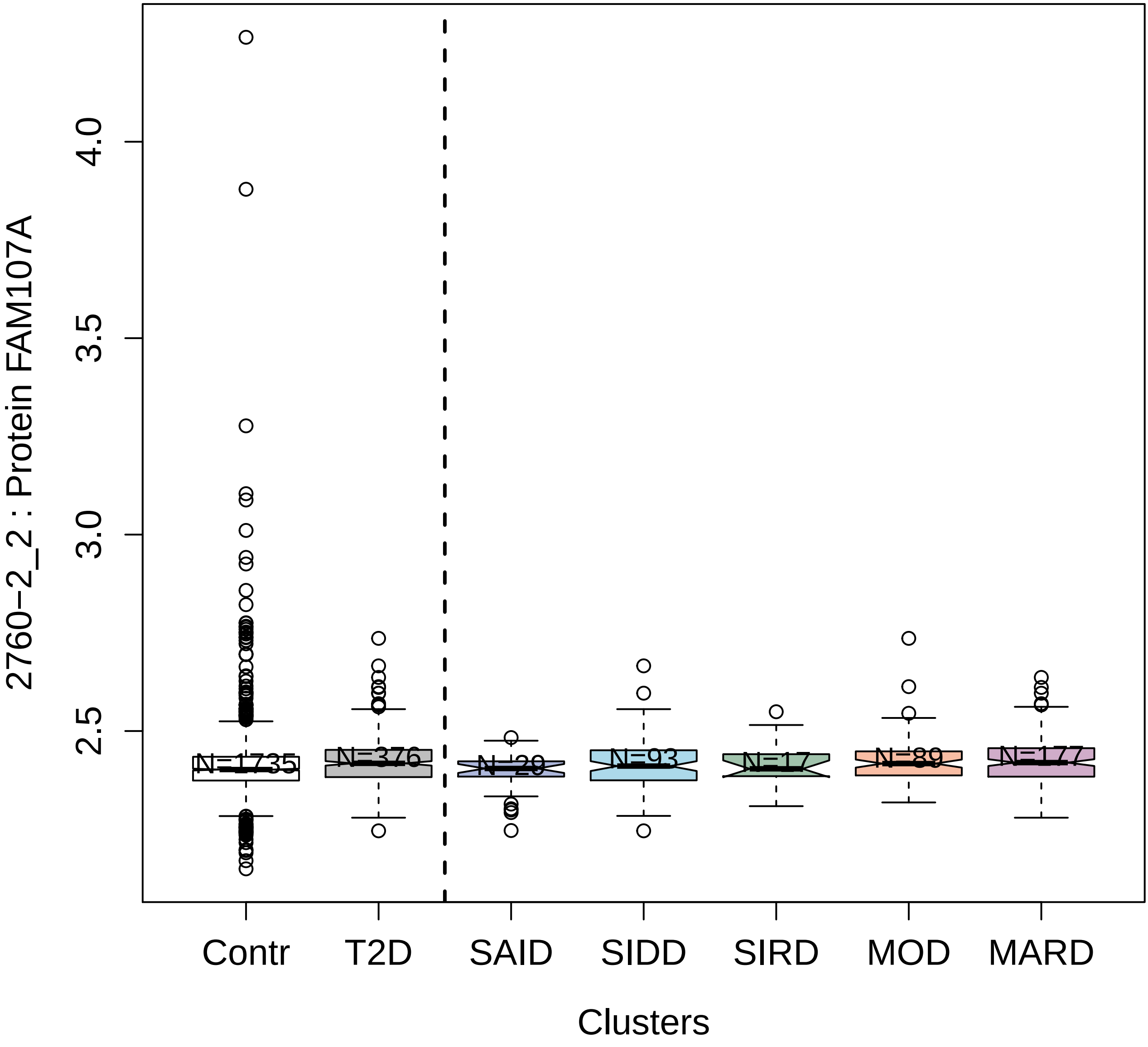

2760−2_2 : Protein FAM107A
4.0
3.5
3.0
2.5
N=1735
N=376
N=20
N=93
N=17
N=89
N=177
Contr
T2D
SAID
SIDD
SIRD
MOD
MARD
Clusters

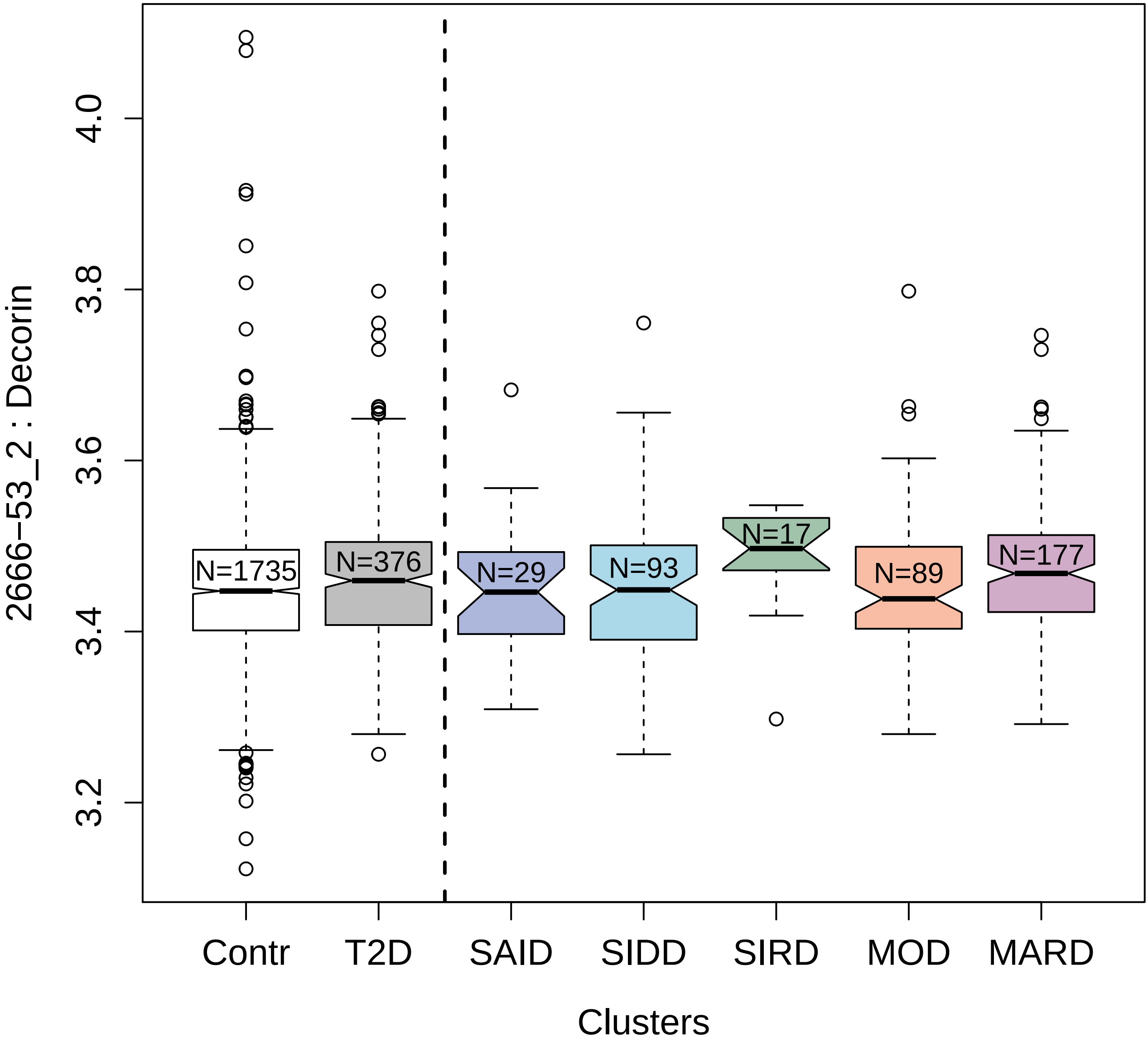

2666−53_2 : Decorin
4.0
3.8
3.6
3.4
3.2
N=1735
N=376
N=29
N=93
N=17
N=89
N=177
Contr
T2D
SAID
SIDD
SIRD
MOD
MARD
Clusters

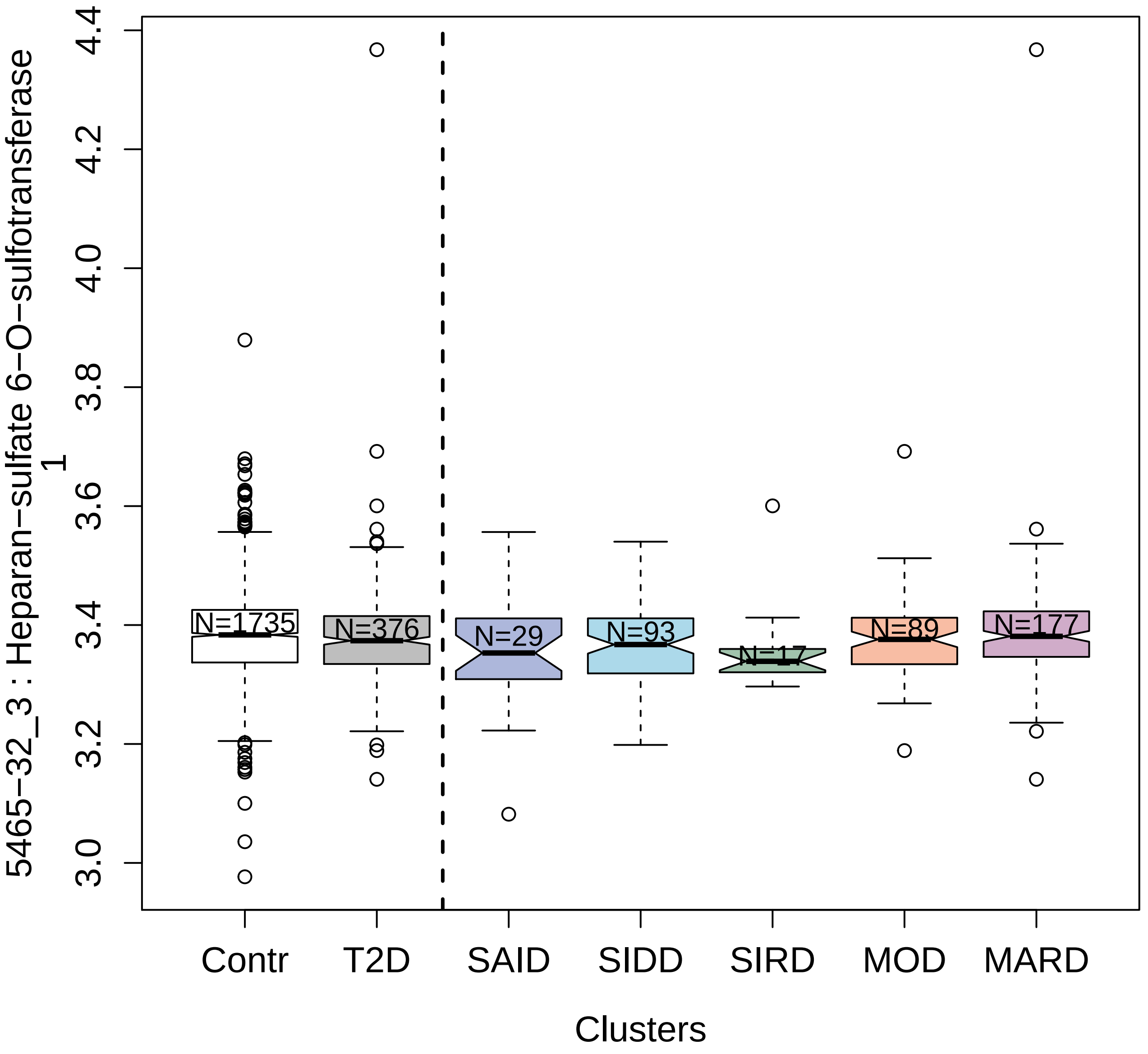

5465−32_3 : Heparan−sulfate 6−O−sulfotransferase 1
3.0
3.2
3.4
3.6
3.8
4.0
4.2
4.4
N=1735
N=376
N=29
N=93
N=17
N=89
N=177
Contr
T2D
SAID
SIDD
SIRD
MOD
MARD
Clusters

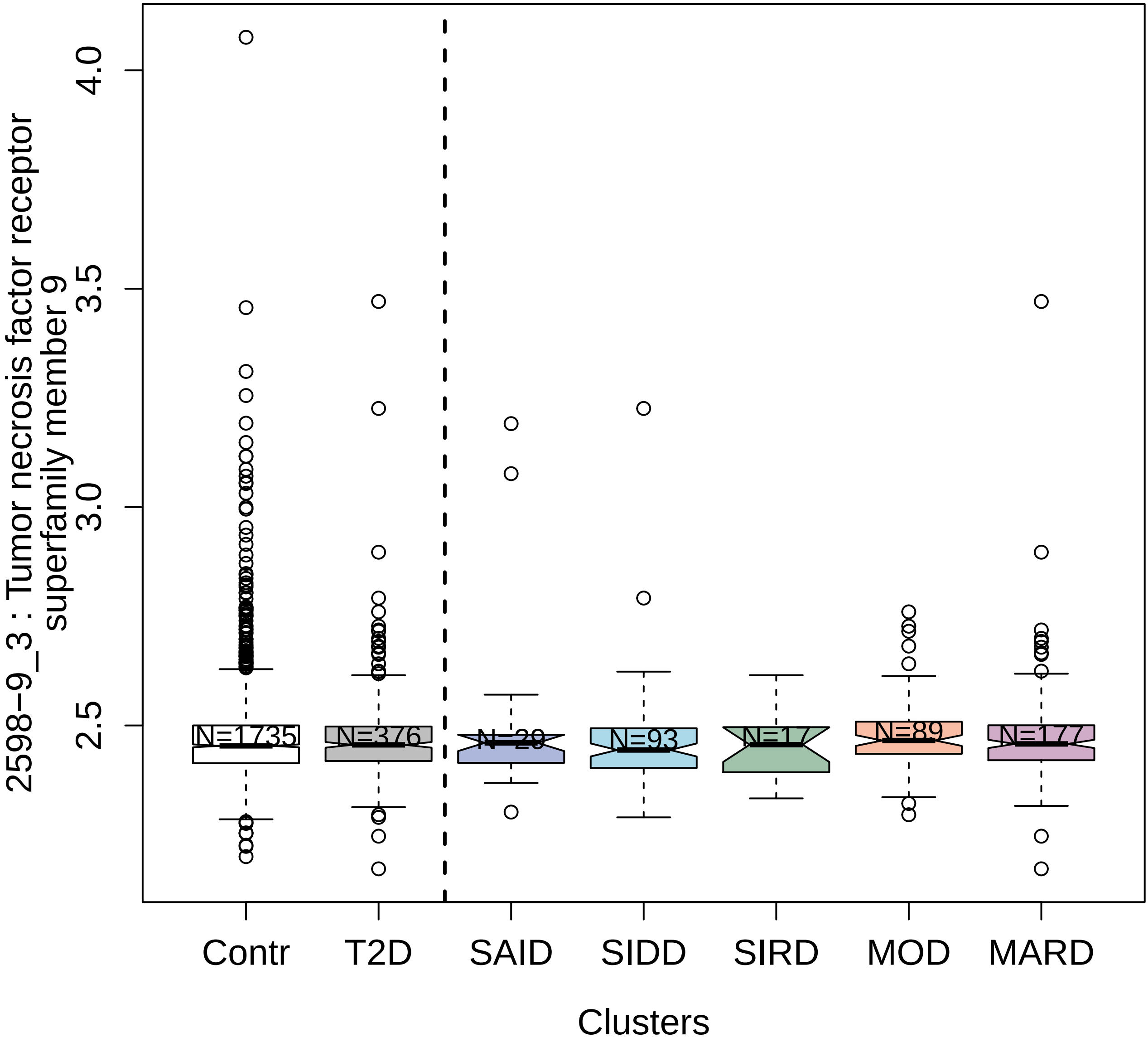
2598−9_3 : Tumor necrosis factor receptor
superfamily member 9
4.0
3.5
3.0
2.5
N=1735
N=376
N=29
N=93
N=17
N=89
N=177
Contr
T2D
SAID
SIDD
SIRD
MOD
MARD
Clusters

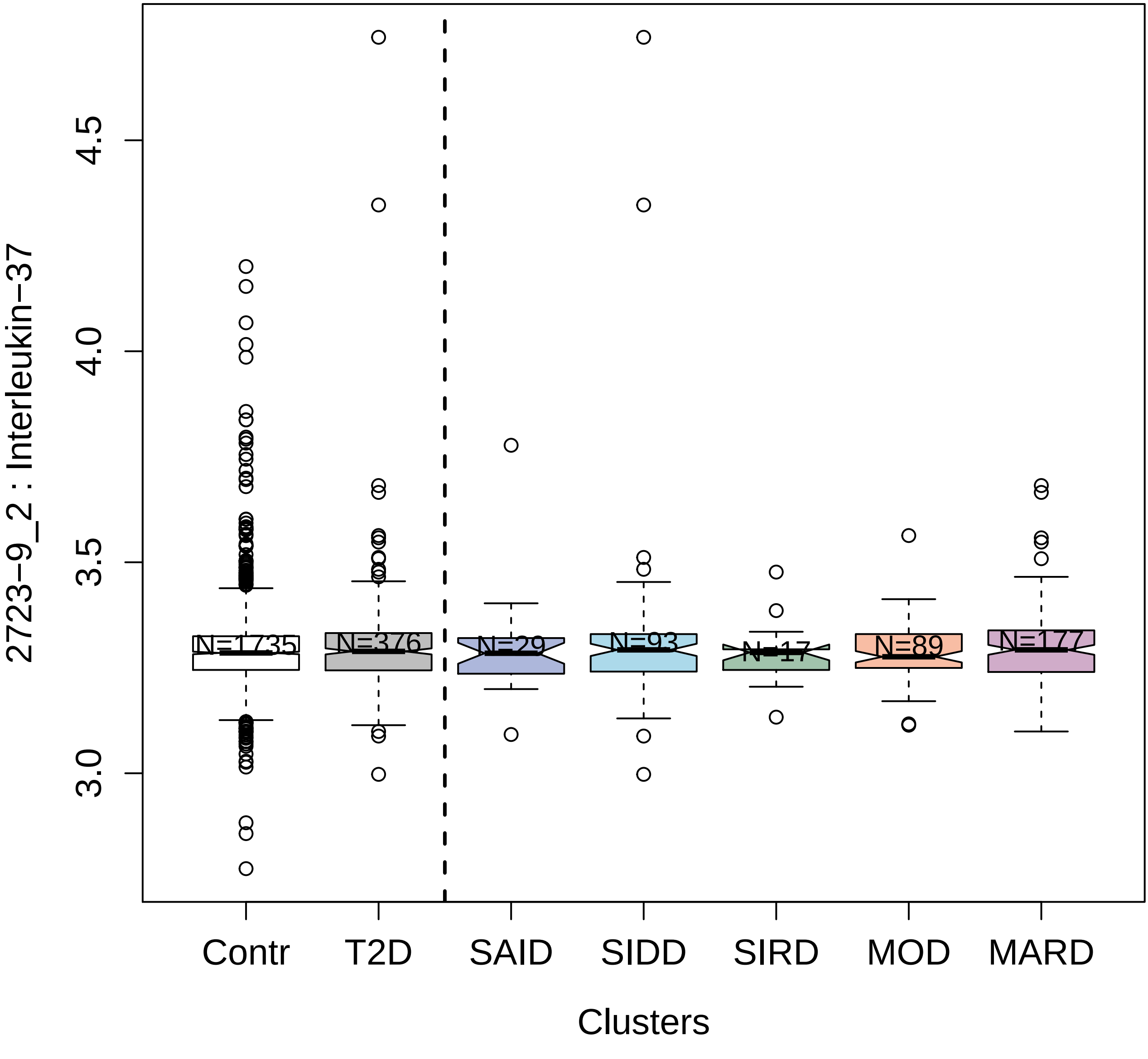
2723−9_2 : Interleukin−37
3.0
3.5
4.0
4.5
N=1735
N=376
N=29
N=93
N=17
N=89
N=177
Contr
T2D
SAID
SIDD
SIRD
MOD
MARD
Clusters

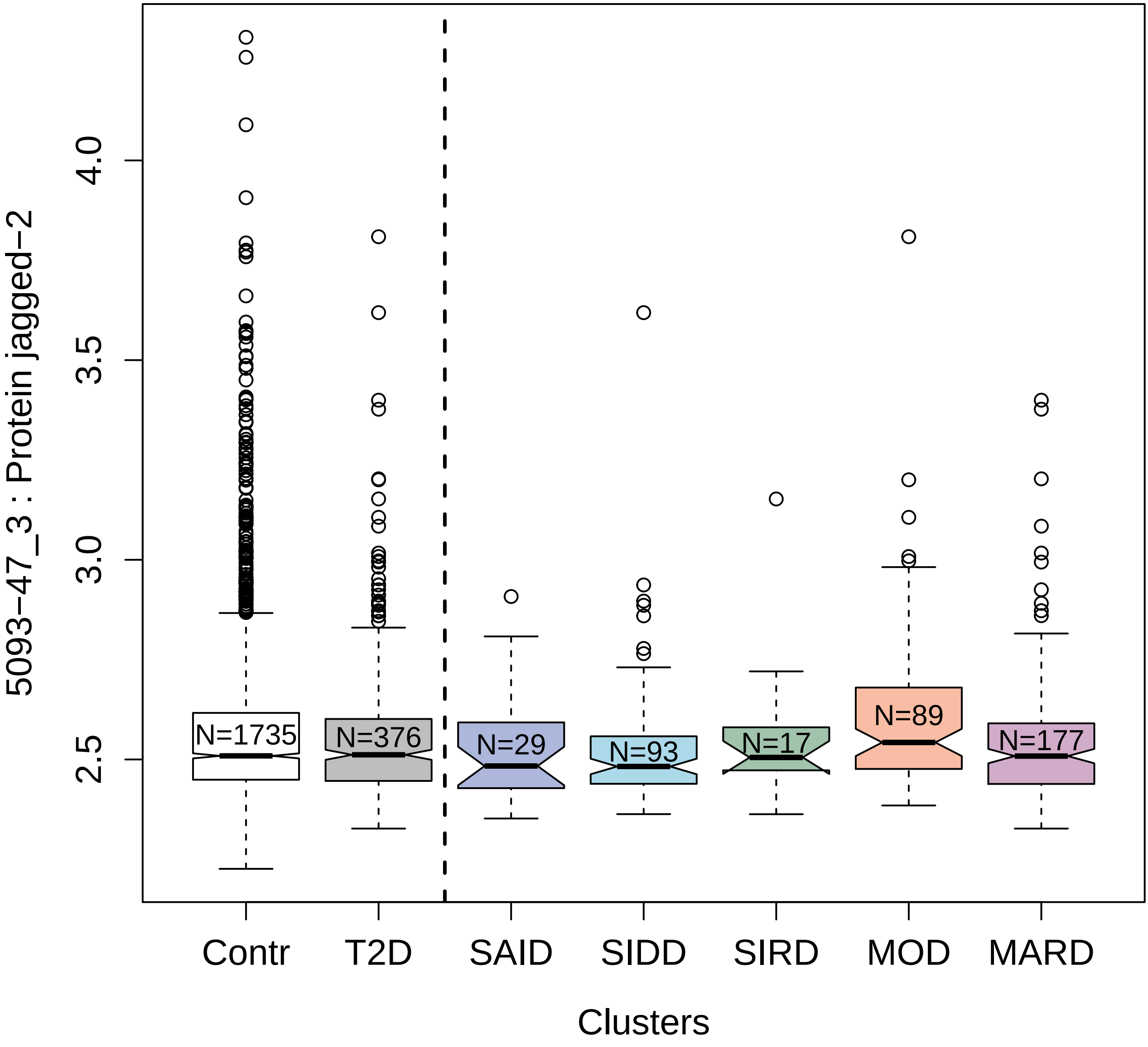

5093−47_3 : Protein jagged−2
2.5
3.0
3.5
4.0
N=1735
N=376
N=29
N=93
N=17
N=89
N=177
Contr
T2D
SAID
SIDD
SIRD
MOD
MARD
Clusters

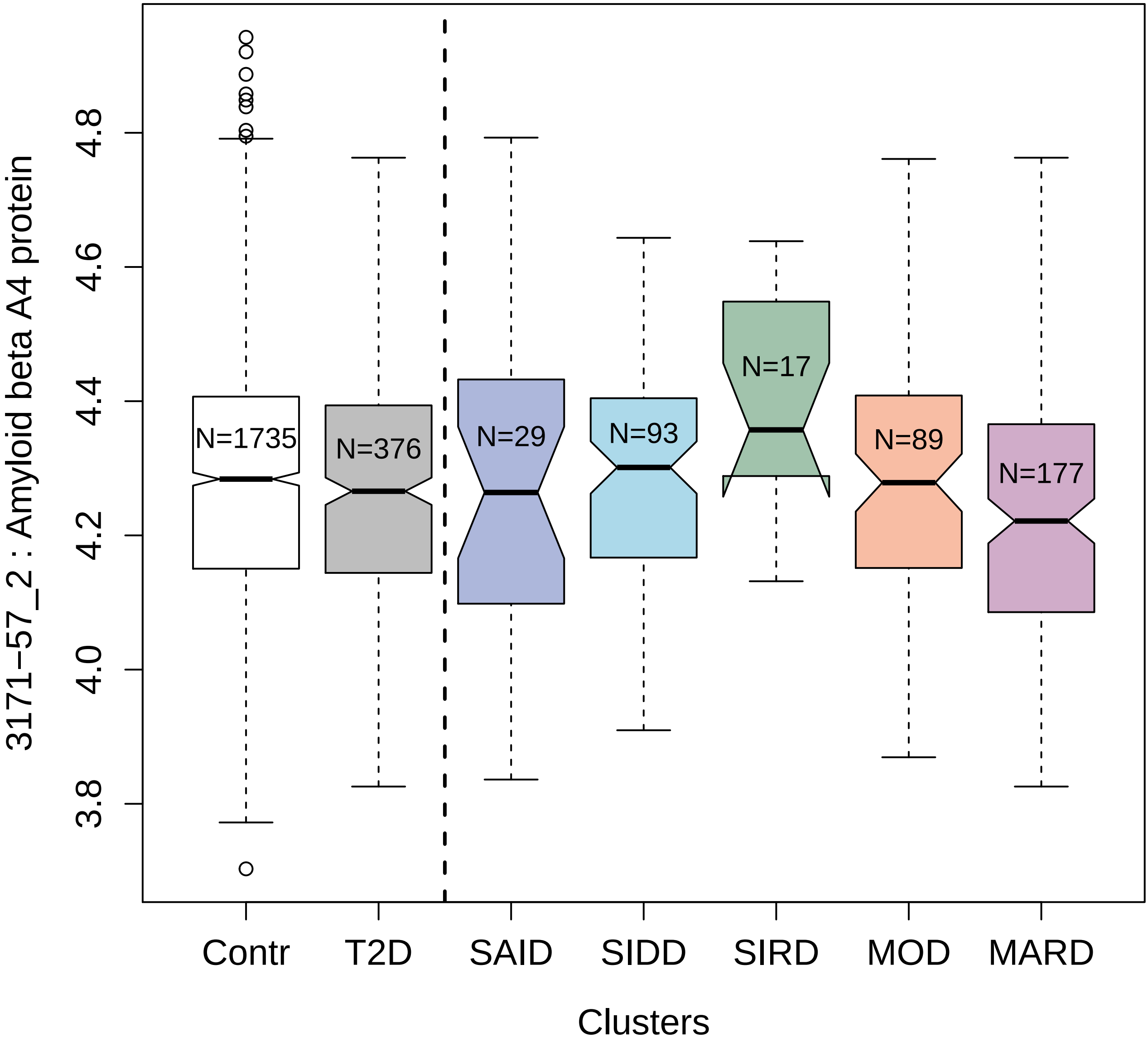

3171−57_2 : Amyloid beta A4 protein
3.8
4.0
4.2
4.4
4.6
4.8
N=1735
N=376
N=29
N=93
N=17
N=89
N=177
Contr
T2D
SAID
SIDD
SIRD
MOD
MARD
Clusters

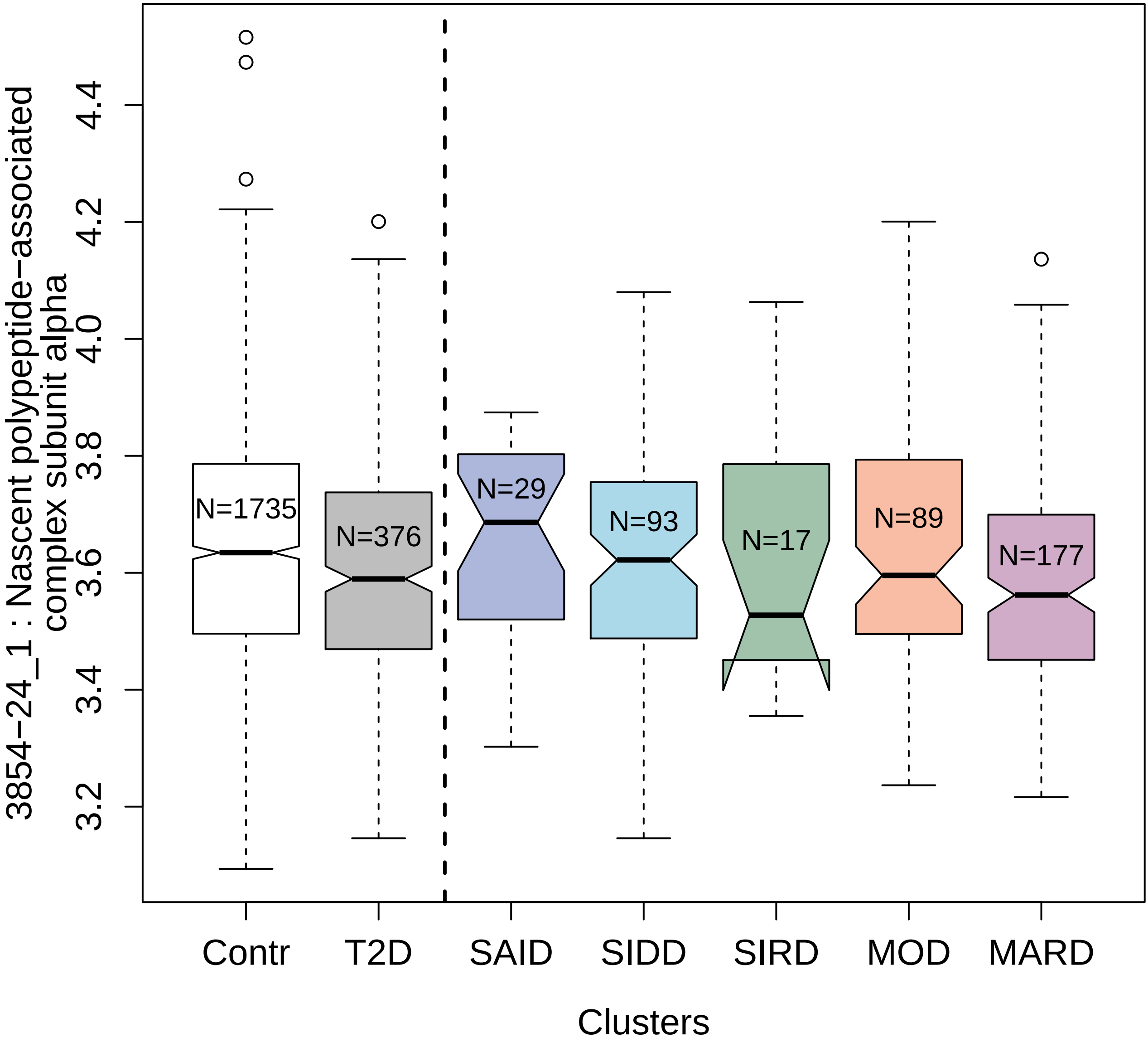
3854−24_1 : Nascent polypeptide−associated complex subunit alpha
3.2
3.4
3.6
3.8
4.0
4.2
4.4
N=1735
N=376
N=29
N=93
N=17
N=89
N=177
Contr
T2D
SAID
SIDD
SIRD
MOD
MARD
Clusters

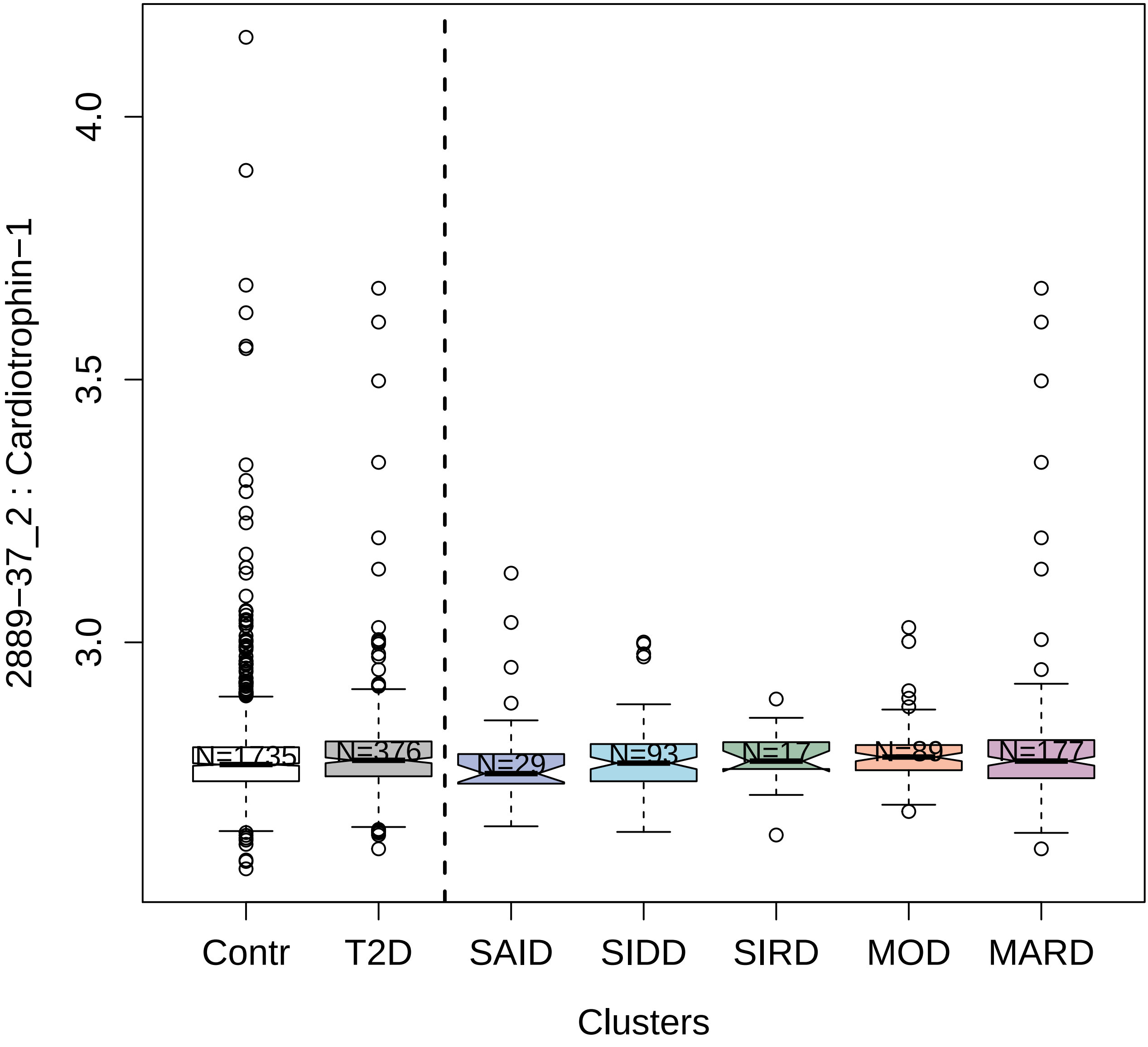
2889−37_2 : Cardiotrophin−1
4.0
3.5
3.0
N=1735
N=376
N=29
N=93
N=17
N=89
N=177
Contr
T2D
SAID
SIDD
SIRD
MOD
MARD
Clusters

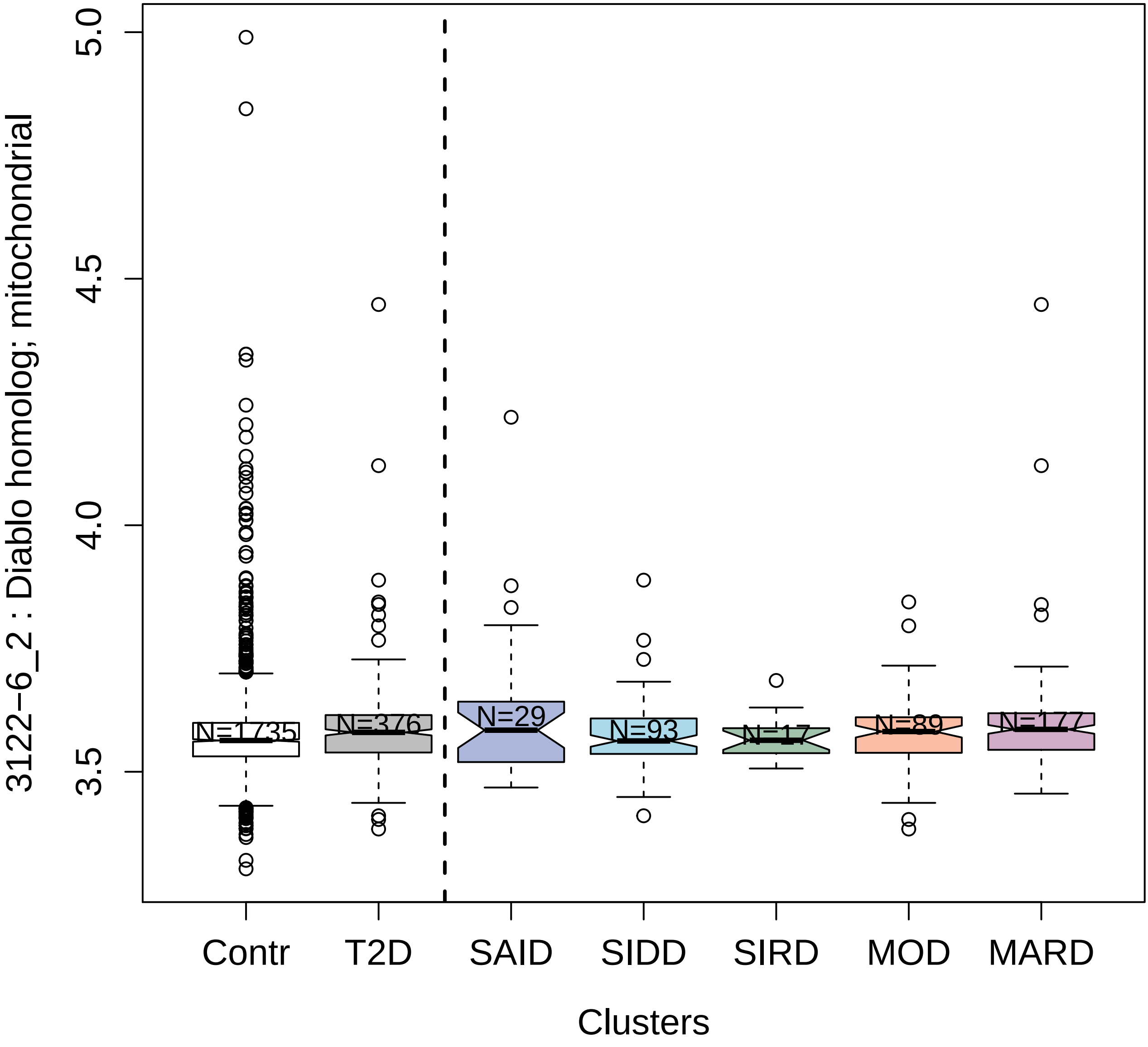

3122−6_2 : Diablo homolog; mitochondrial
5.0
4.5
4.0
3.5
N=1735
N=376
N=29
N=93
N=17
N=89
N=177
Contr
T2D
SAID
SIDD
SIRD
MOD
MARD
Clusters

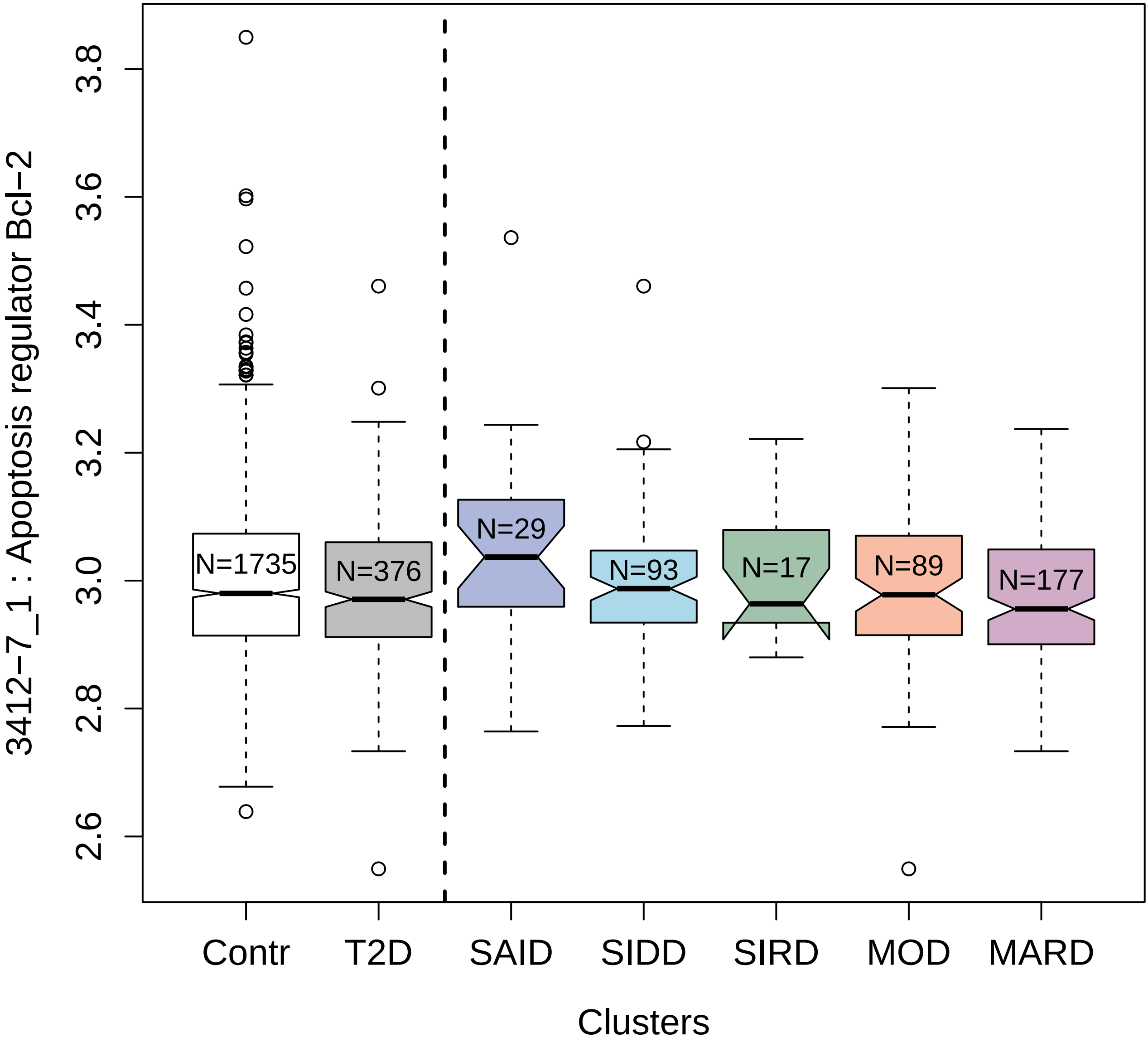

3412−7_1 : Apoptosis regulator Bcl−2
3.8
3.6
3.4
3.2
3.0
2.8
2.6
N=1735
N=376
N=29
N=93
N=17
N=89
N=177
Contr
T2D
SAID
SIDD
SIRD
MOD
MARD
Clusters

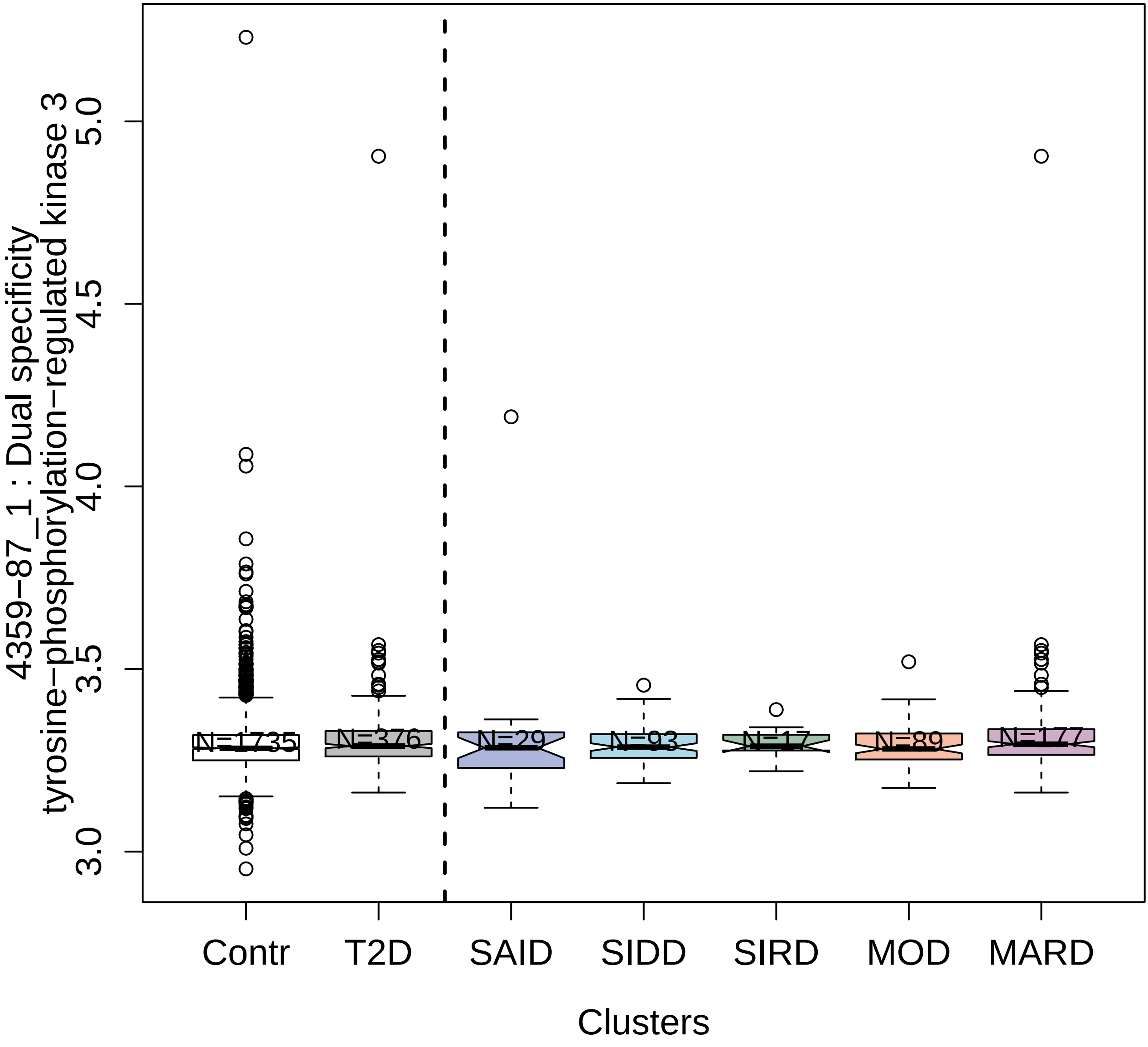
4359−87_1 : Dual specificity
tyrosine−phosphorylation−regulated kinase 3
3.0
3.5
4.0
4.5
5.0
N=1735
N=376
N=29
N=93
N=17
N=89
N=177
Contr
T2D
SAID
SIDD
SIRD
MOD
MARD
Clusters

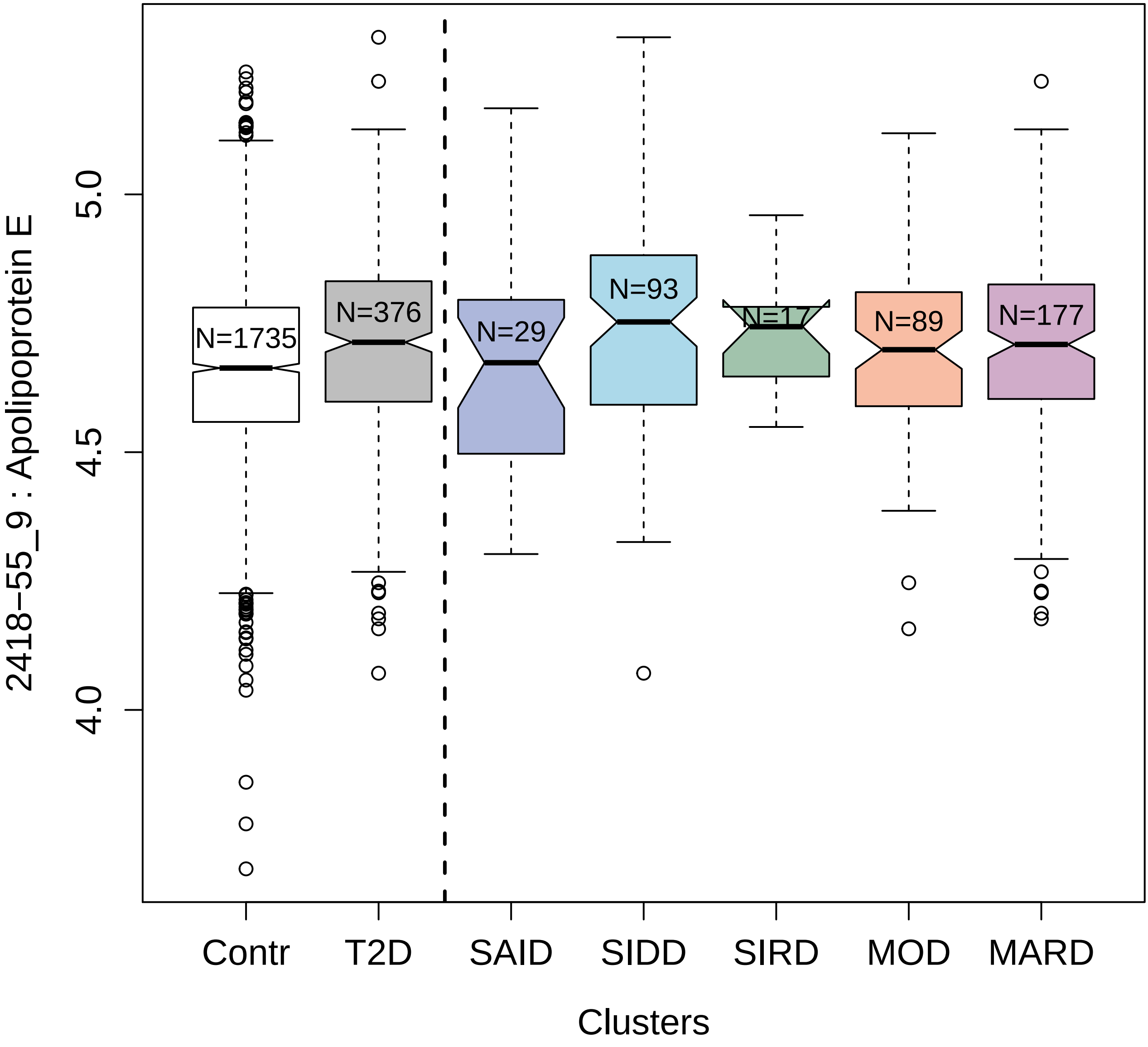
2418−55_9 : Apolipoprotein E
5.0
4.5
4.0
N=1735
N=376
N=29
N=93
N=17
N=89
N=177
Contr
T2D
SAID
SIDD
SIRD
MOD
MARD
Clusters

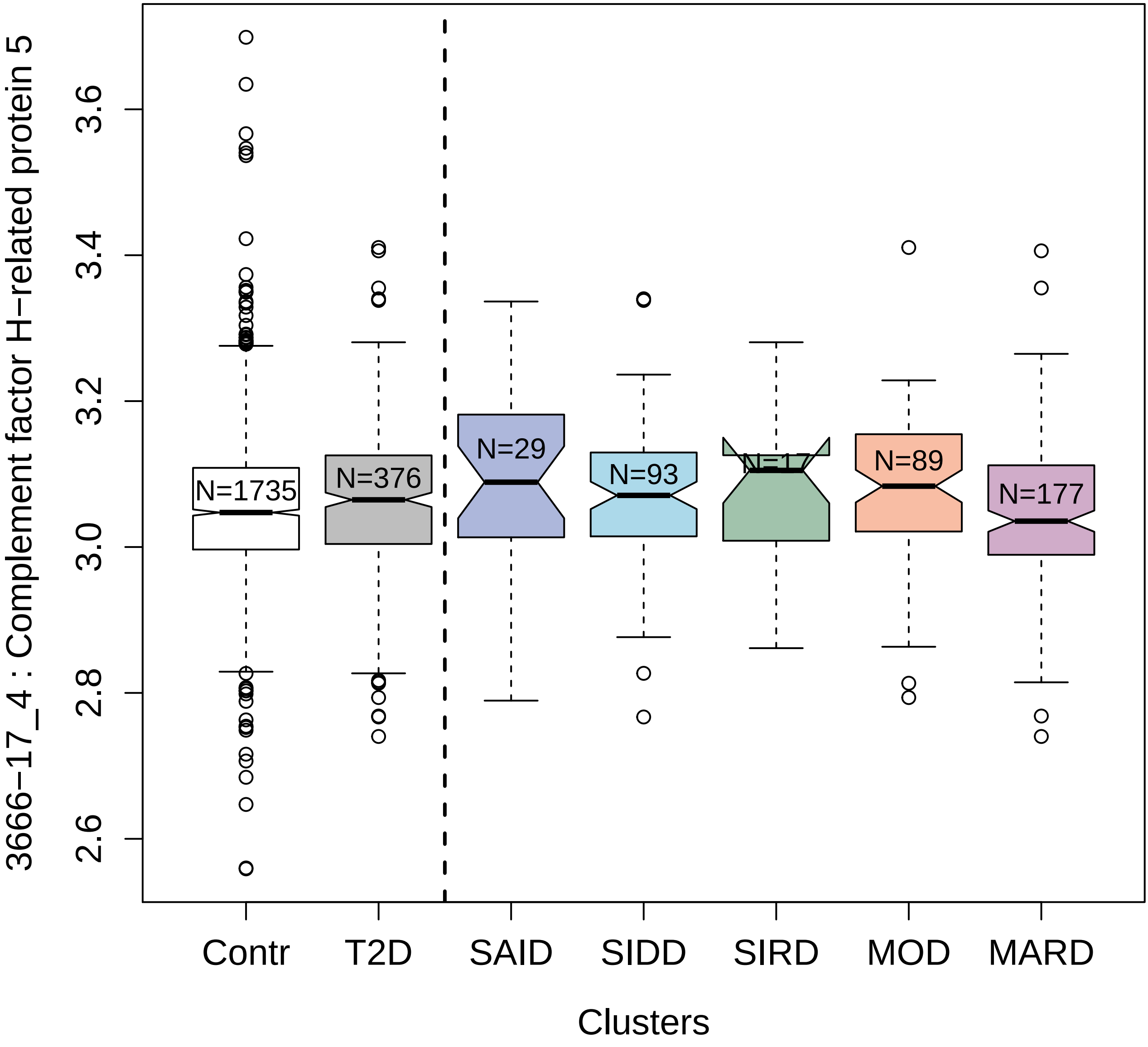

3666−17_4 : Complement factor H−related protein 5
2.6
2.8
3.0
3.2
3.4
3.6
N=1735
N=376
N=29
N=93
N=89
N=177
Contr
T2D
SAID
SIDD
SIRD
MOD
MARD
Clusters

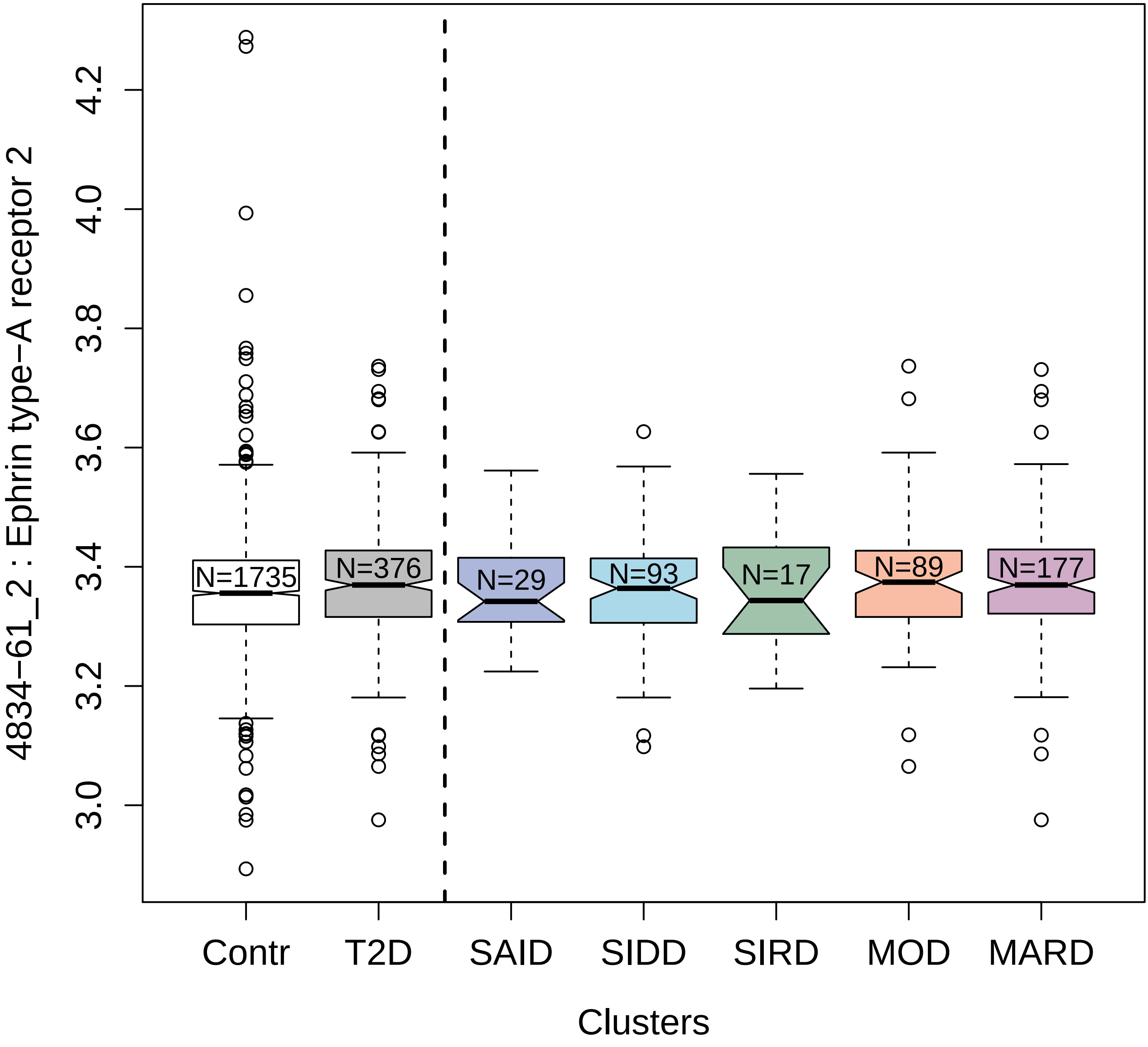

4834−61_2 : Ephrin type−A receptor 2
4.2
4.0
3.8
3.6
3.4
3.2
3.0
N=1735
N=376
N=29
N=93
N=17
N=89
N=177
Contr
T2D
SAID
SIDD
SIRD
MOD
MARD
Clusters

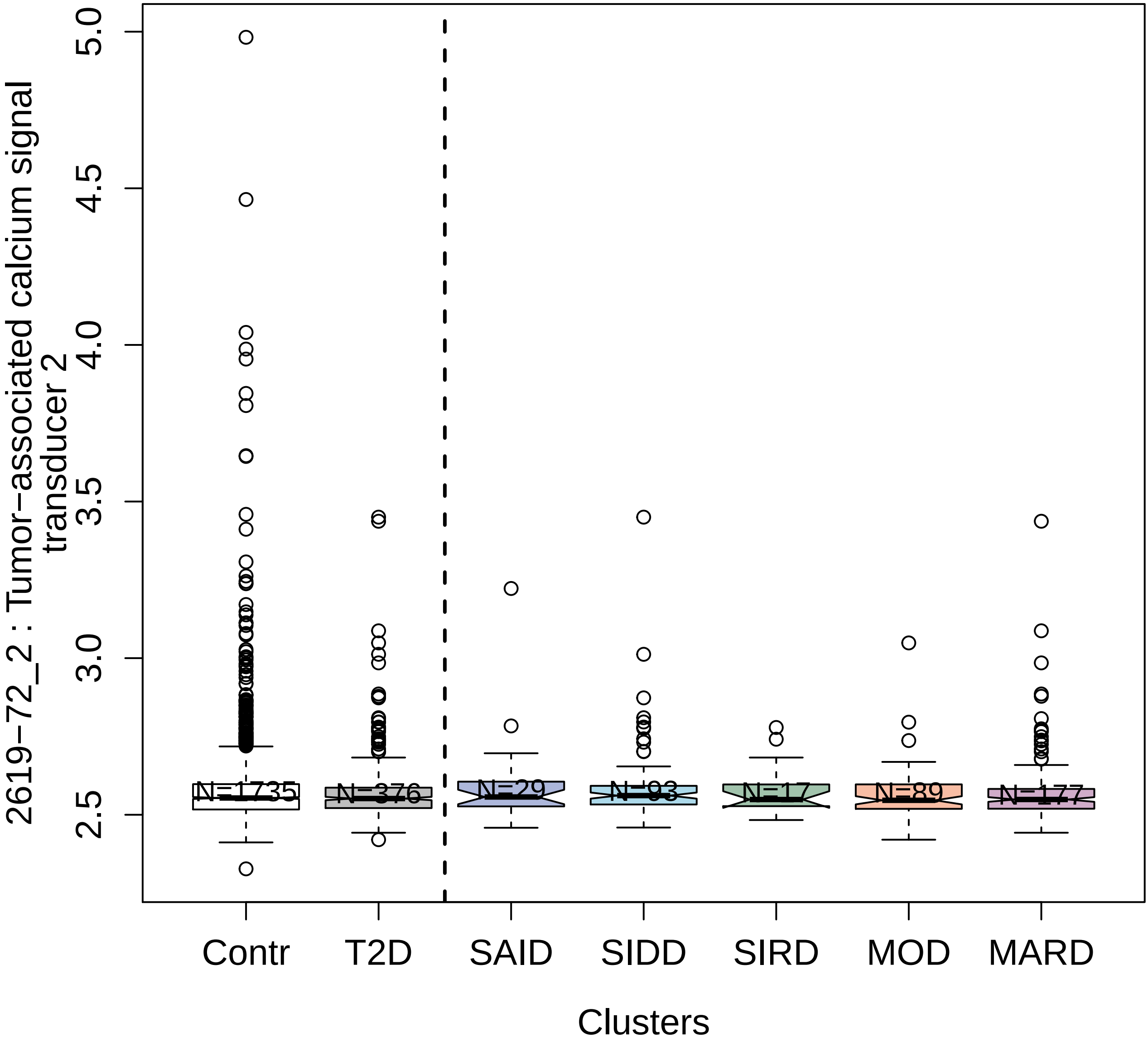
2619−72_2 : Tumor−associated calcium signal transducer 2
Clusters
Contr
T2D
SAID
SIDD
SIRD
MOD
MARD
N=1735
N=376
N=29
N=93
N=17
N=89
N=177
2.5
3.0
3.5
4.0
4.5
5.0

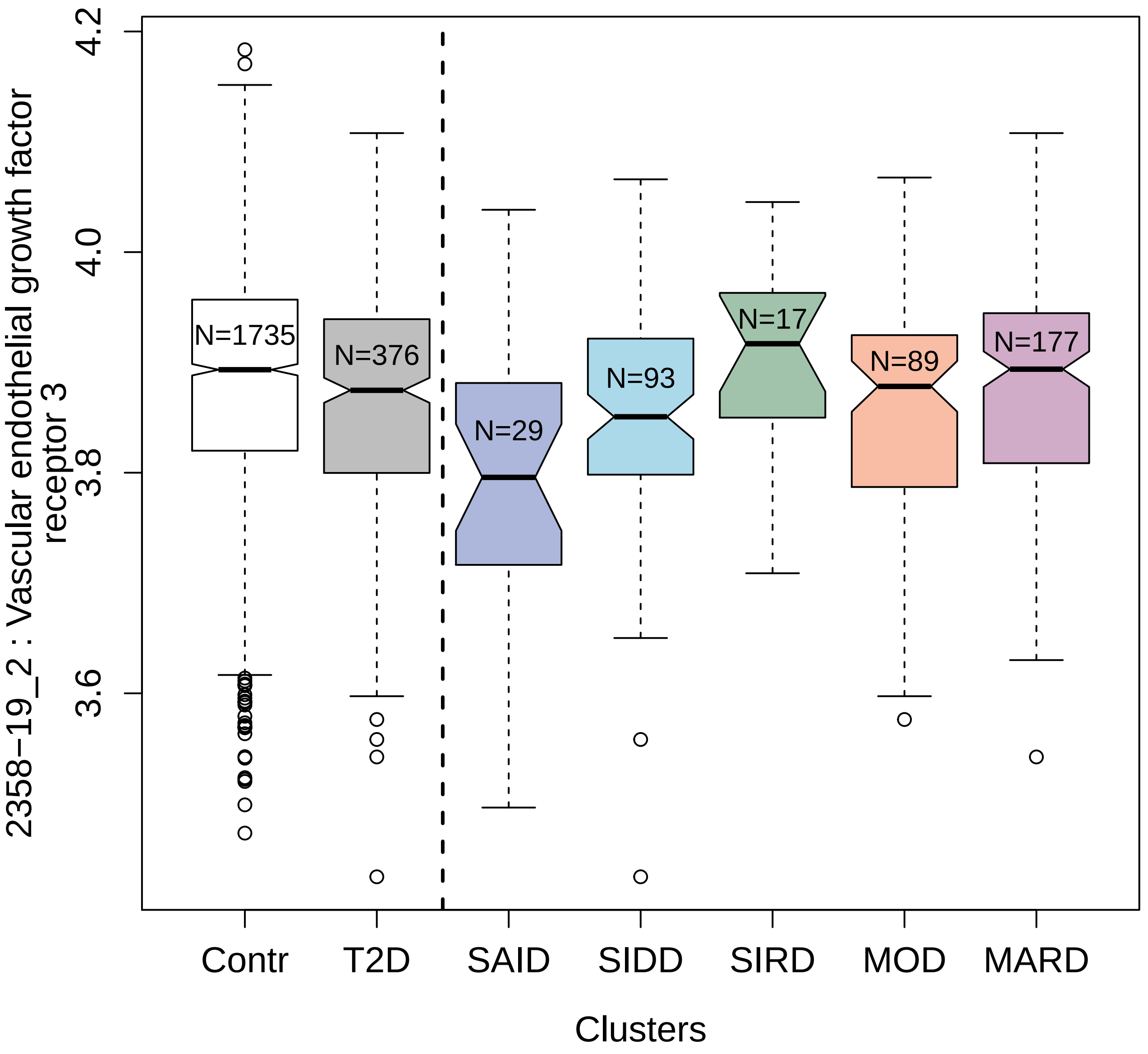

2358−19_2 : Vascular endothelial growth factor receptor 3
4.2
4.0
3.8
3.6
N=1735
N=376
N=29
N=93
N=17
N=89
N=177
Contr
T2D
SAID
SIDD
SIRD
MOD
MARD
Clusters

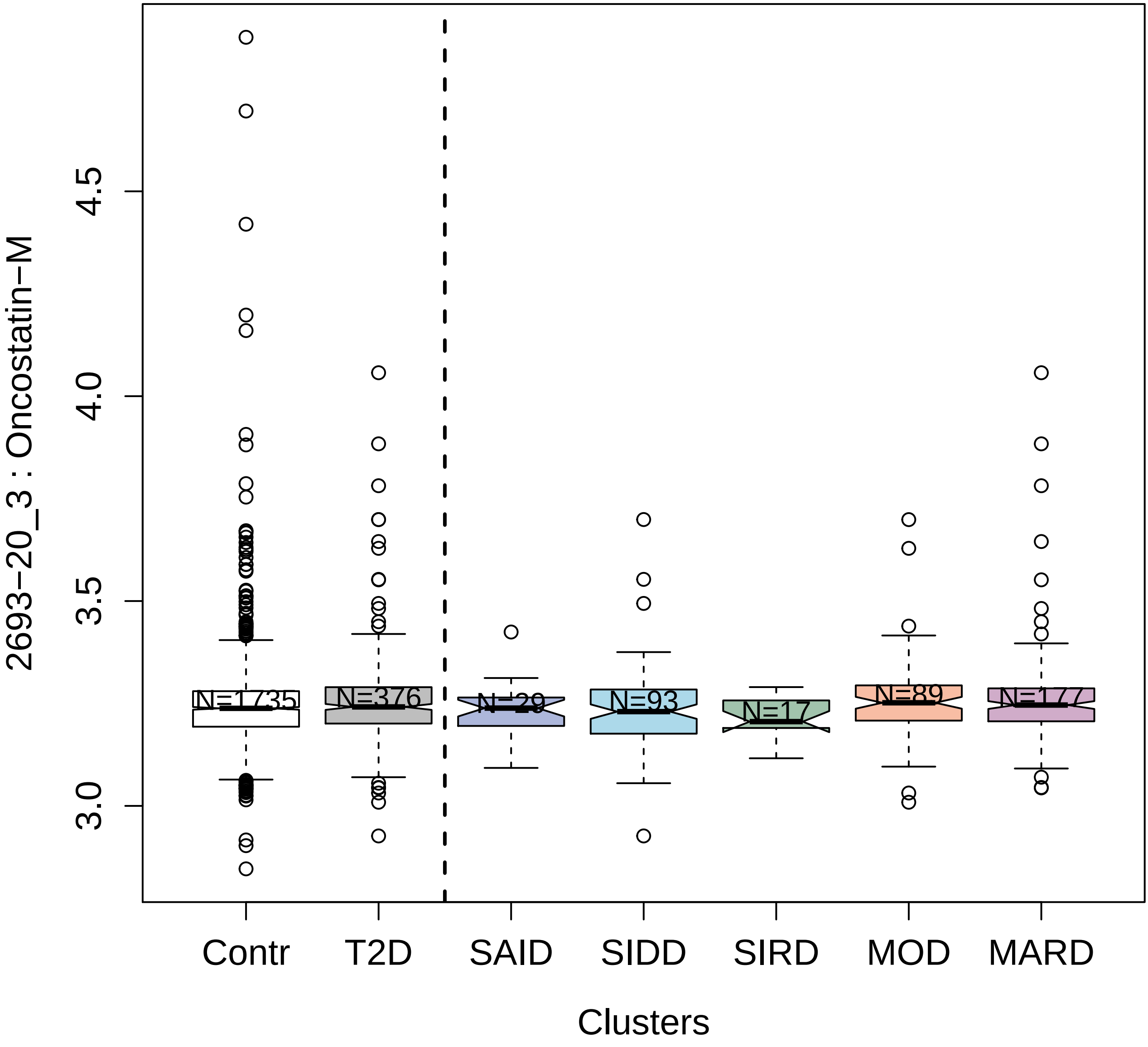
2693−20_3 : Oncostatin−M
4.5
4.0
3.5
3.0
N=1735
N=376
N=29
N=93
N=17
N=89
N=177
Contr
T2D
SAID
SIDD
SIRD
MOD
MARD
Clusters

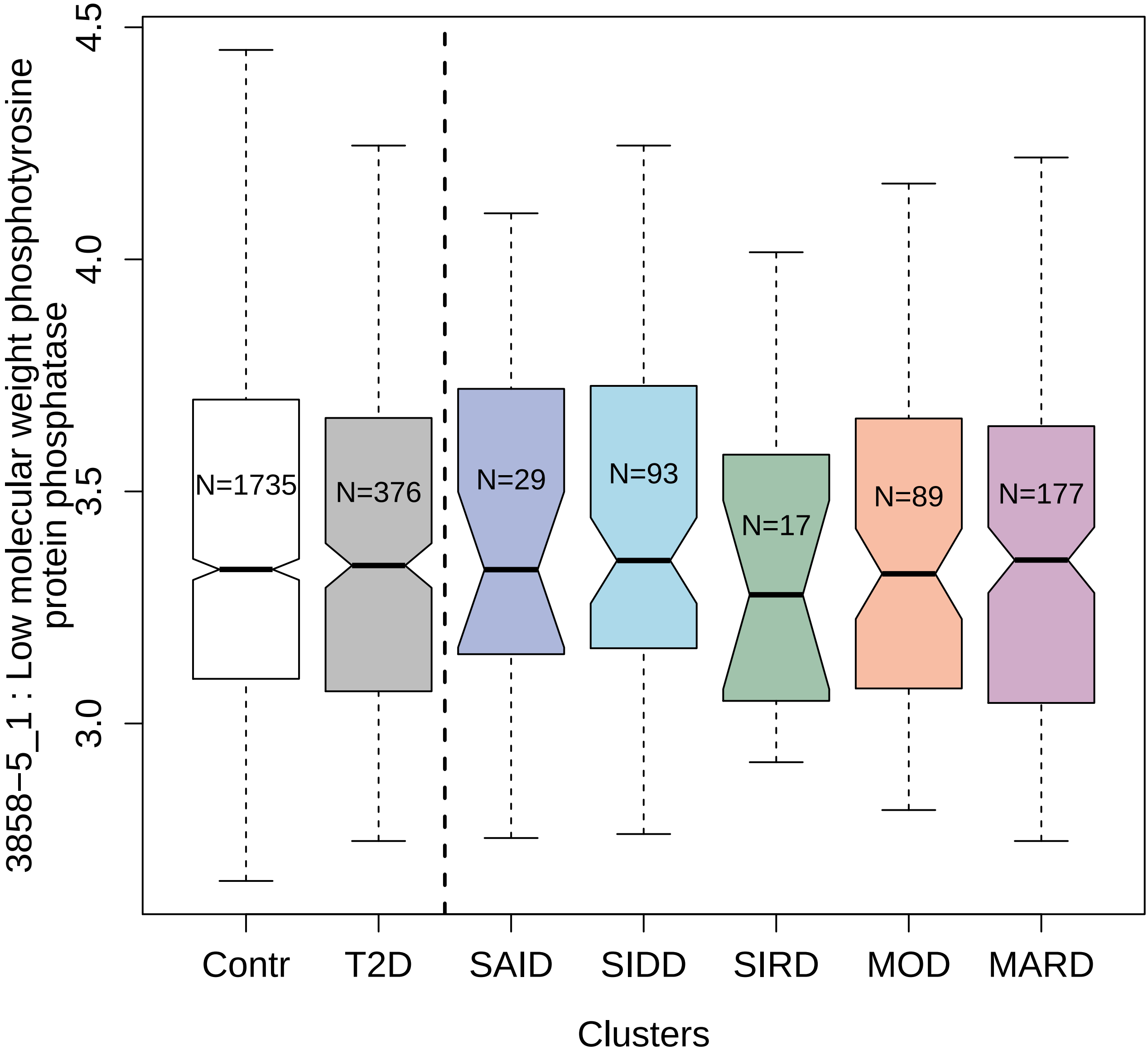
3858–5_1 : Low molecular weight phosphotyrosine protein phosphatase
4.5
4.0
3.5
3.0
N=1735
N=376
N=29
N=93
N=17
N=89
N=177
Contr
T2D
SAID
SIDD
SIRD
MOD
MARD
Clusters

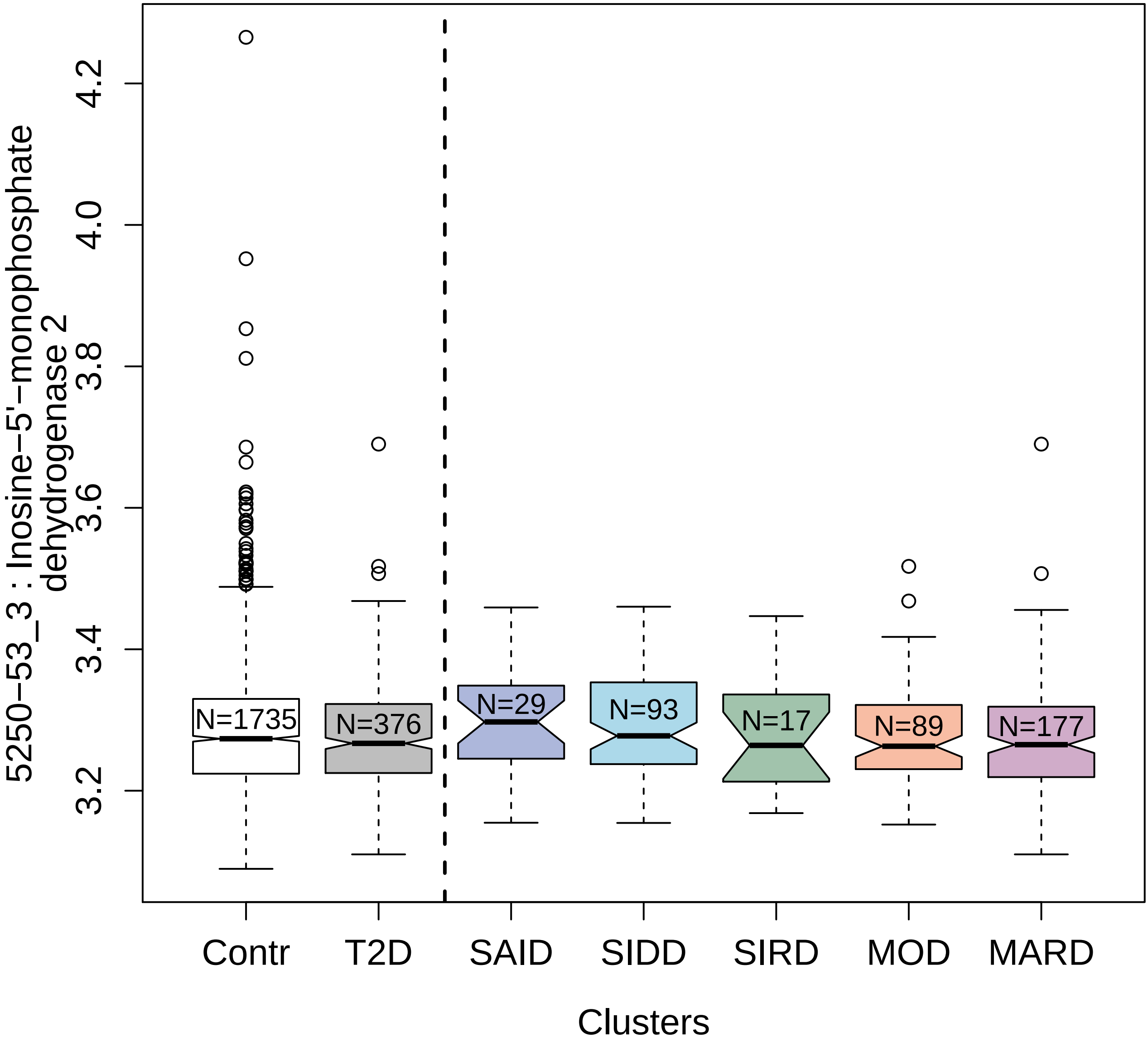

5250−53_3 : Inosine−5'−monophosphate dehydrogenase 2
3.2
3.4
3.6
3.8
4.0
4.2
N=1735
N=376
N=29
N=93
N=17
N=89
N=177
Contr
T2D
SAID
SIDD
SIRD
MOD
MARD
Clusters

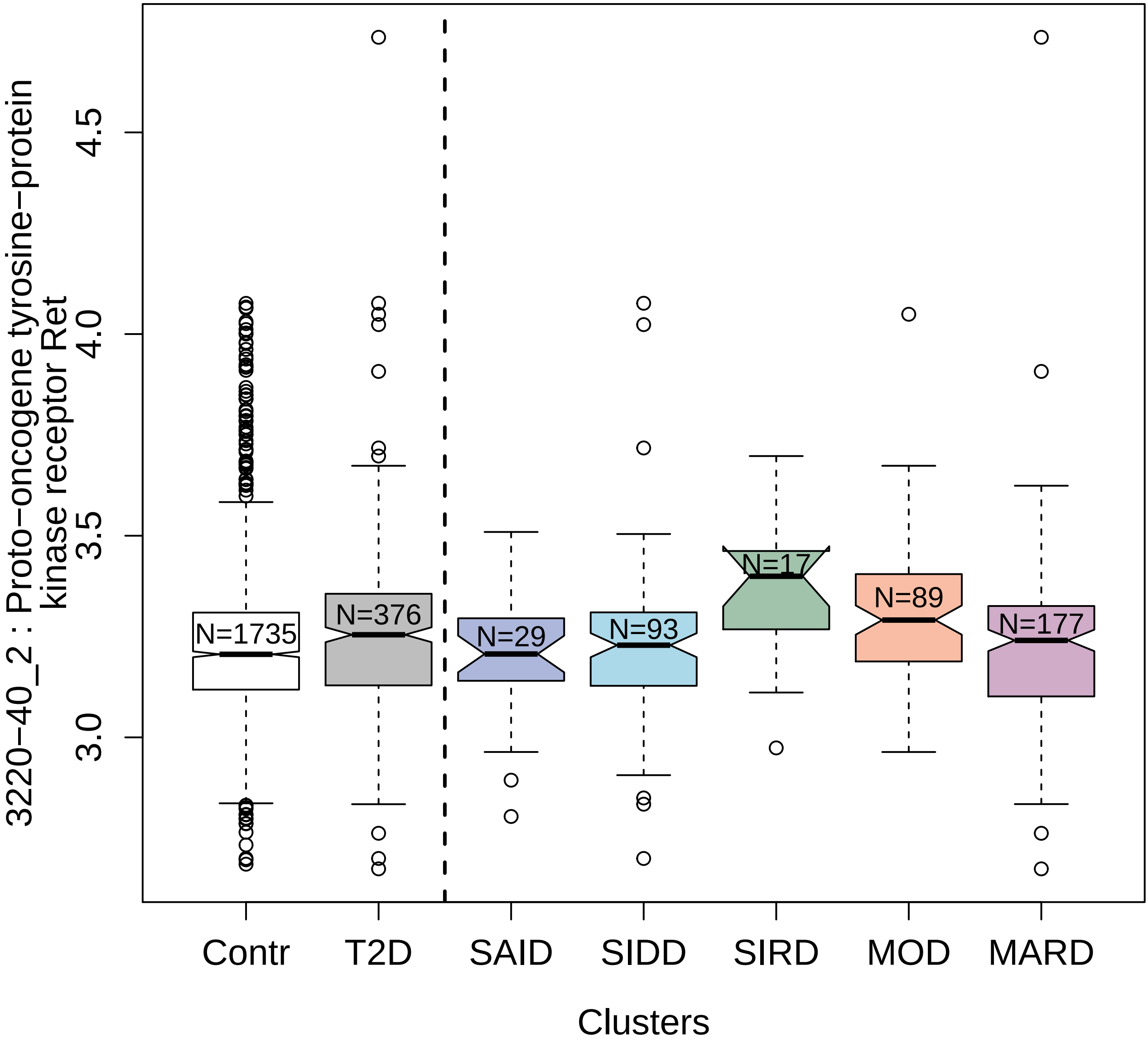

3220−40_2 : Proto−oncogene tyrosine−protein
kinase receptor Ret
4.5
4.0
3.5
3.0
N=1735
N=376
N=29
N=93
N=17
N=89
N=177
Contr
T2D
SAID
SIDD
SIRD
MOD
MARD
Clusters

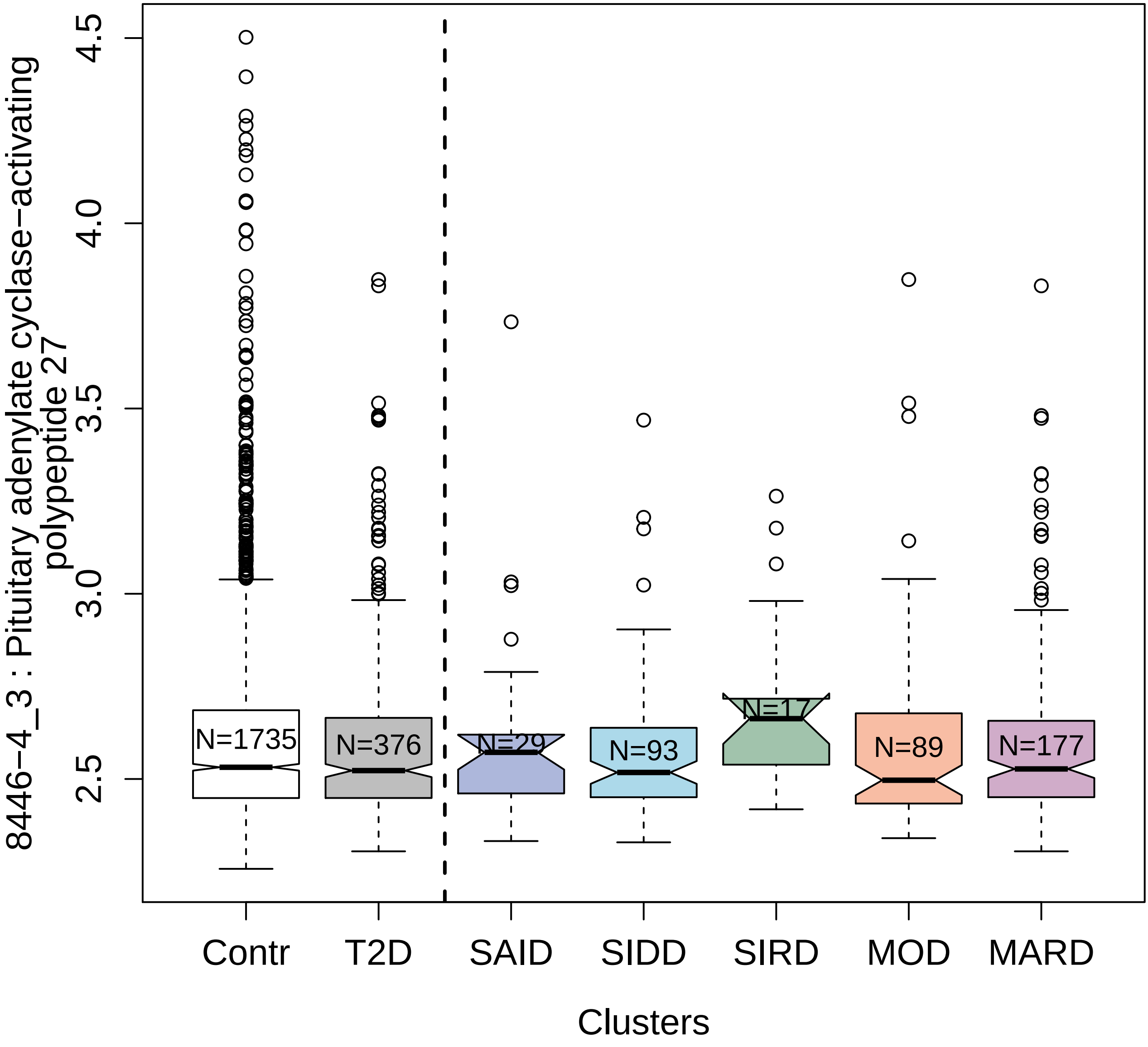
8446−4_3 : Pituitary adenylate cyclase−activating polypeptide 27
4.5
4.0
3.5
3.0
2.5
N=1735
N=376
N=29
N=93
N=17
N=89
N=177
Contr
T2D
SAID
SIDD
SIRD
MOD
MARD
Clusters

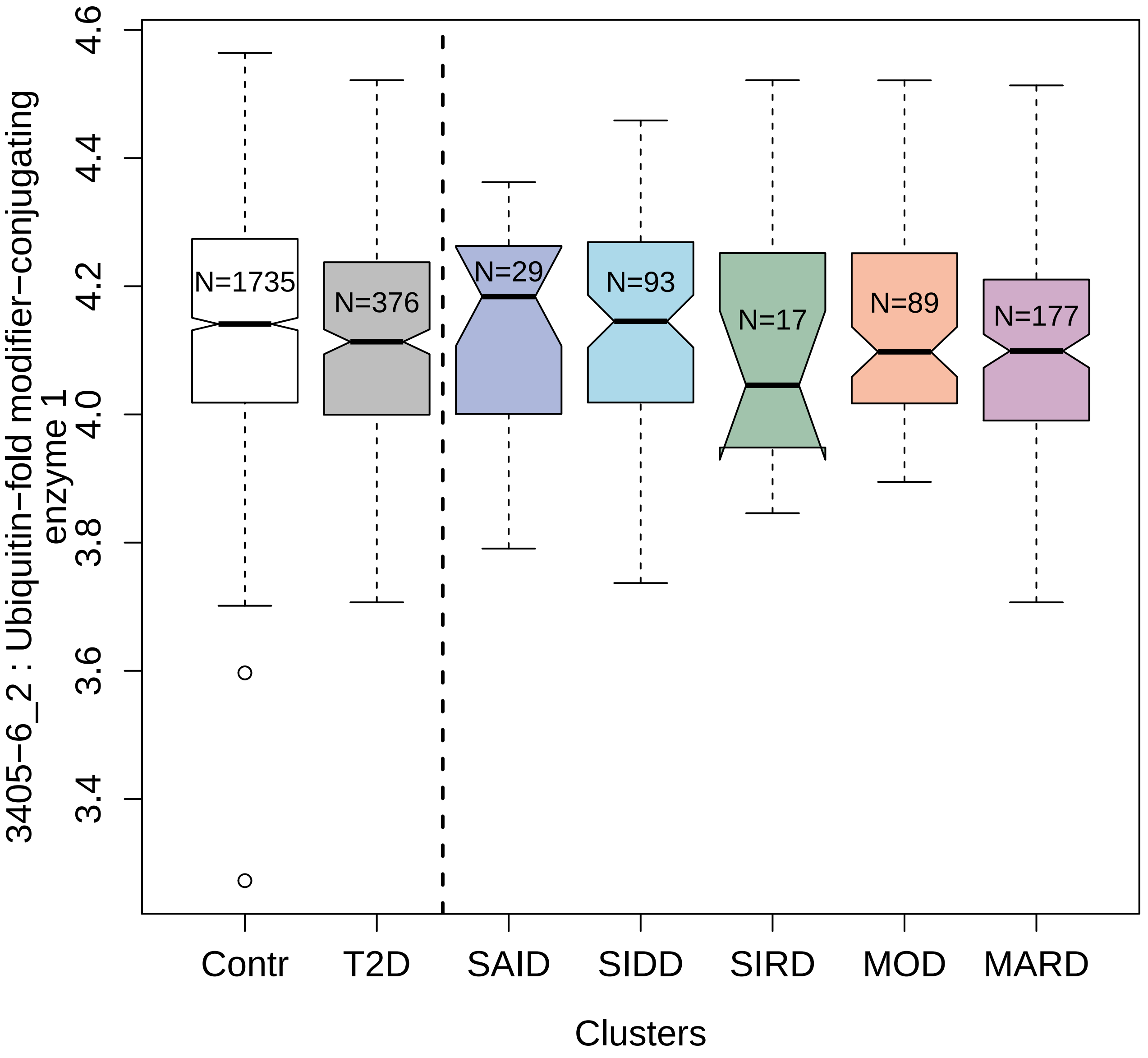
3405−6_2 : Ubiquitin−fold modifier−conjugating enzyme 1
Clusters
Contr
T2D
SAID
SIDD
SIRD
MOD
MARD
N=1735
N=376
N=29
N=93
N=17
N=89
N=177
3.4
3.6
3.8
4.0
4.2
4.4
4.6

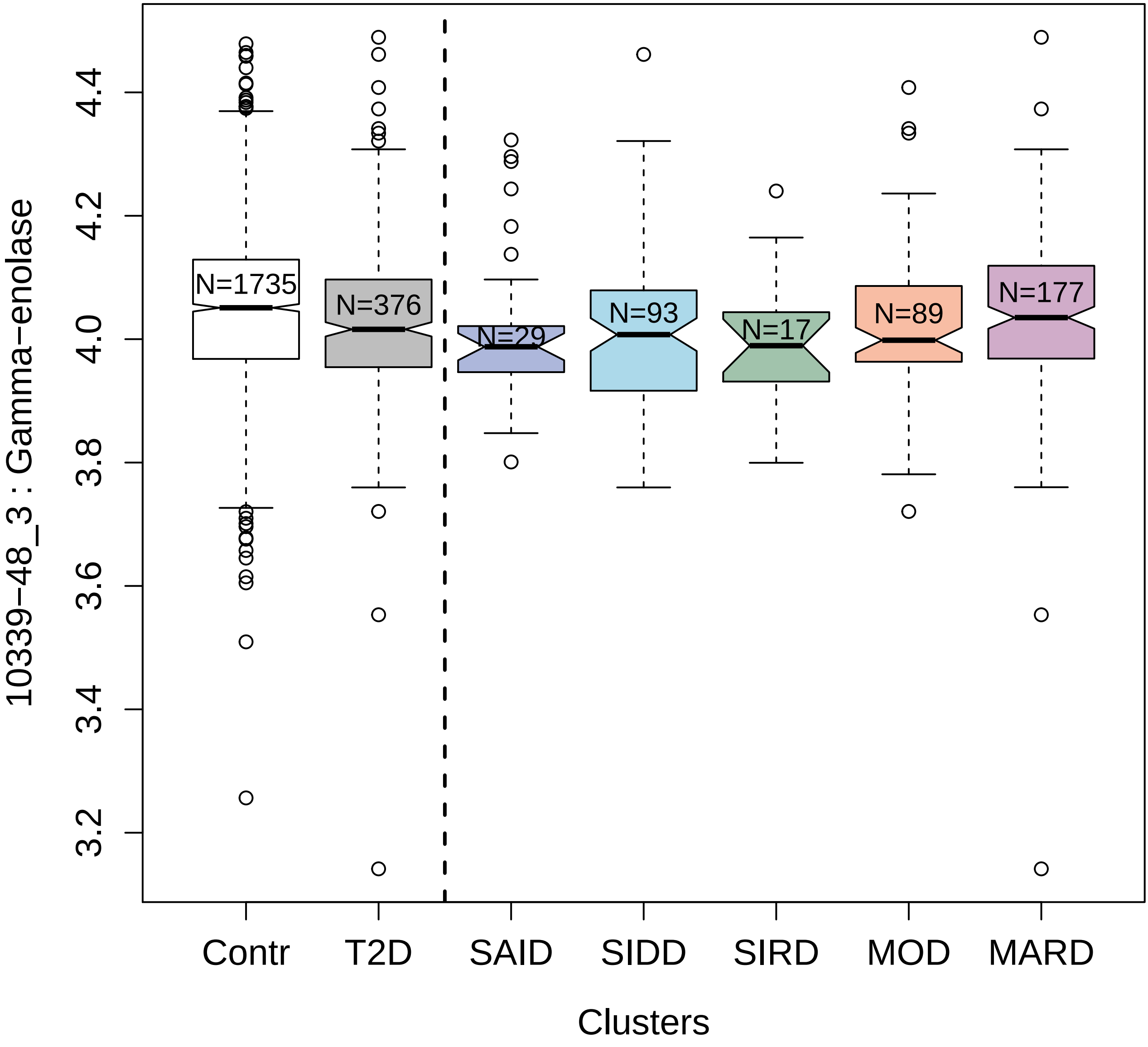

10339−48_3 : Gamma−enolase
Clusters
Contr
T2D
SAID
SIDD
SIRD
MOD
MARD
N=1735
N=376
N=29
N=93
N=17
N=89
N=177
3.2
3.4
3.6
3.8
4.0
4.2
4.4

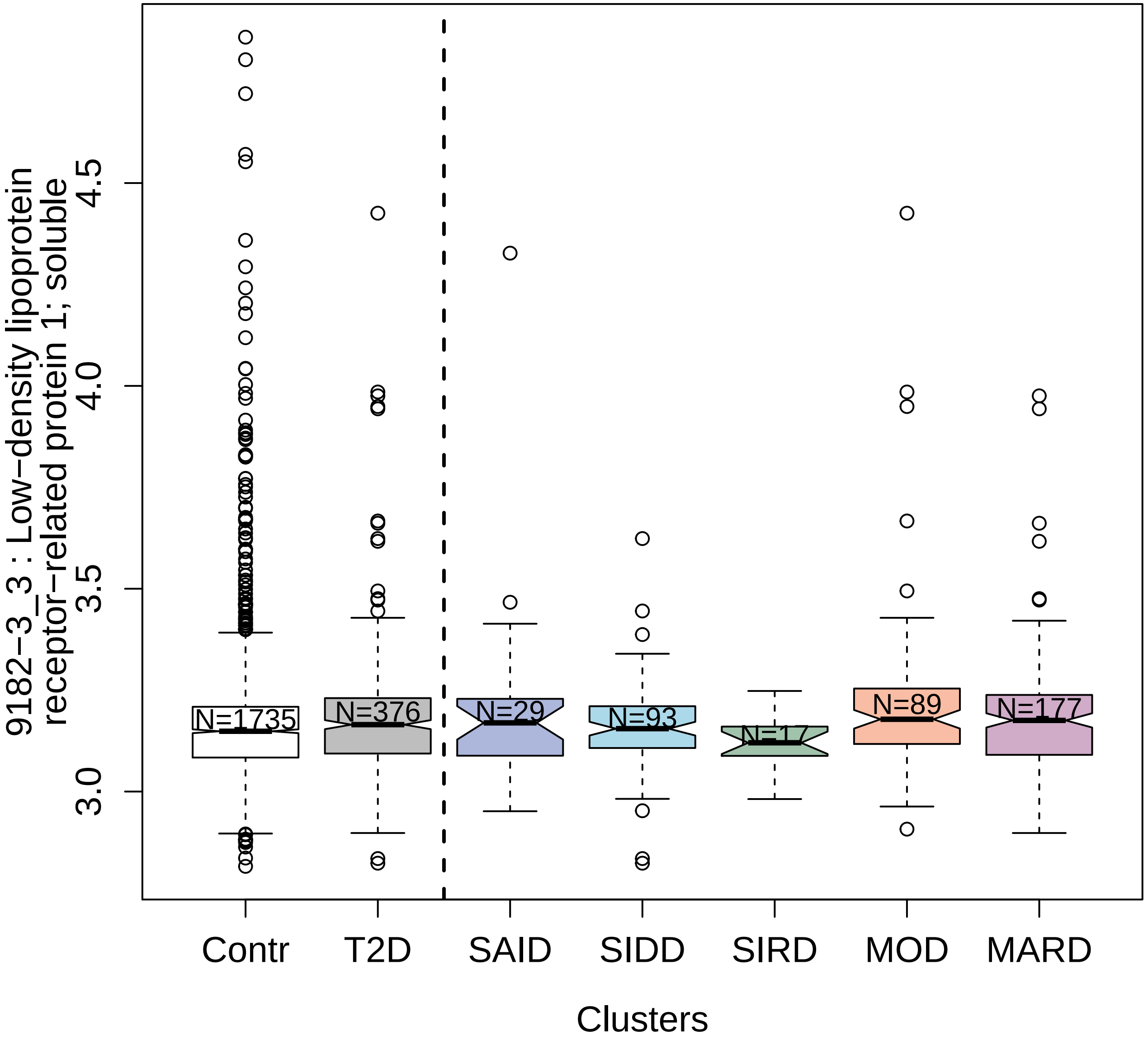

9182−3_3 : Low−density lipoprotein receptor−related protein 1; soluble
3.0
3.5
4.0
4.5
N=1735
N=376
N=29
N=93
N=17
N=89
N=177
Contr
T2D
SAID
SIDD
SIRD
MOD
MARD
Clusters

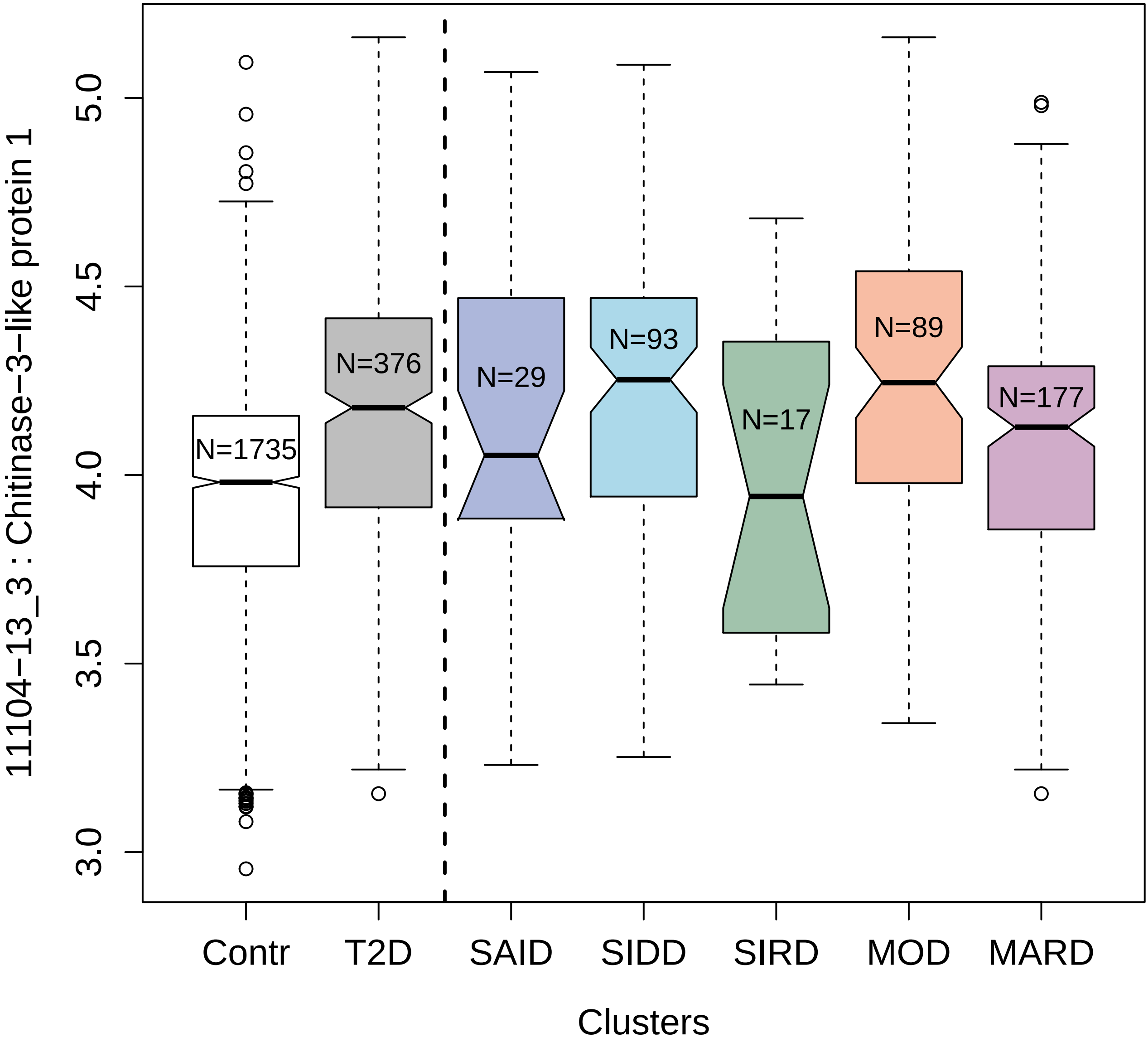
11104−13_3 : Chitinase−3−like protein 1
5.0
4.5
4.0
3.5
3.0
N=1735
N=376
N=29
N=93
N=17
N=89
N=177
Contr
T2D
SAID
SIDD
SIRD
MOD
MARD
Clusters

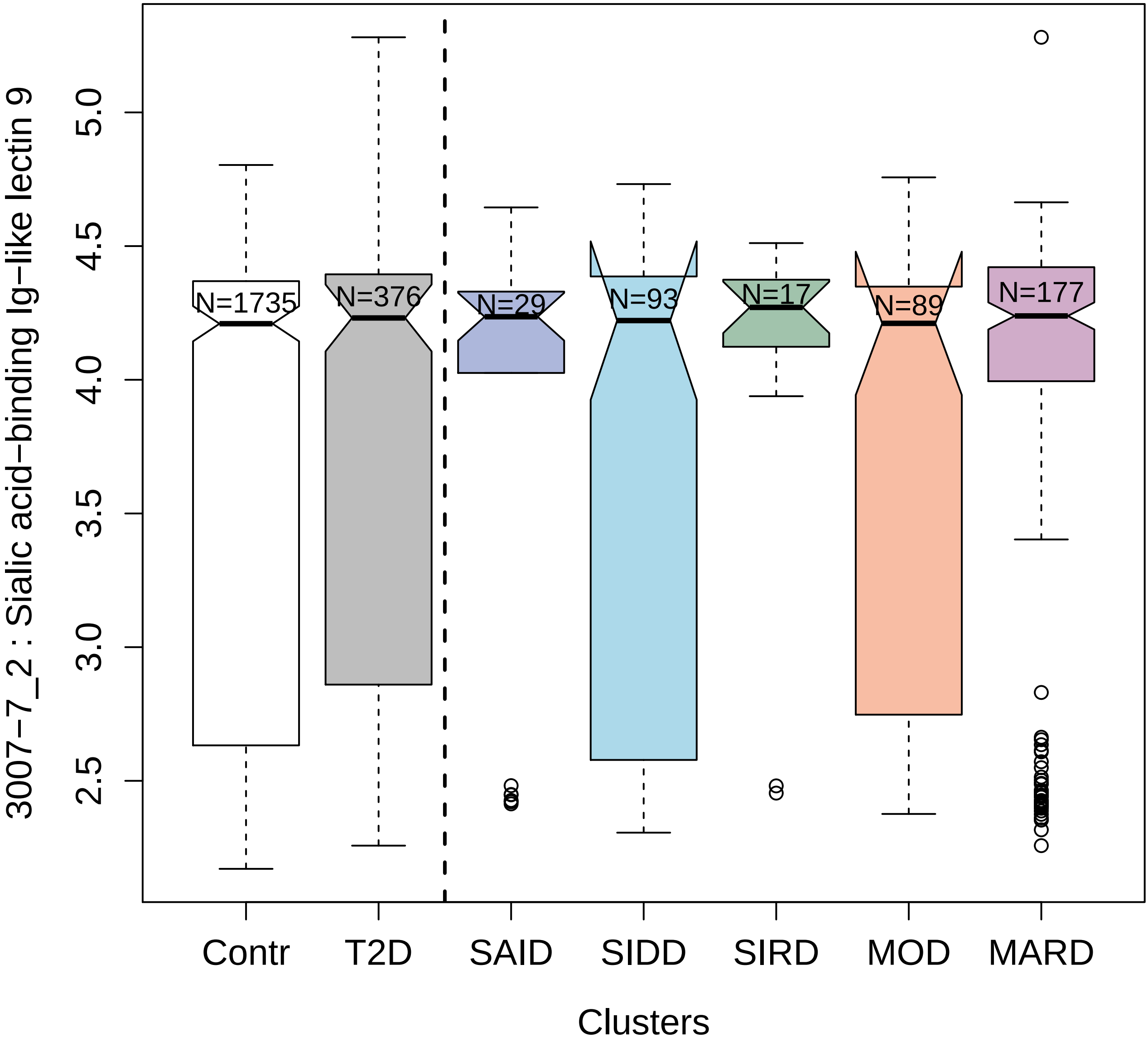

3007−7_2 : Sialic acid−binding Ig−like lectin 9
2.5
3.0
3.5
4.0
4.5
5.0
N=1735
N=376
N=29
N=93
N=17
N=89
N=177
Contr
T2D
SAID
SIDD
SIRD
MOD
MARD
Clusters

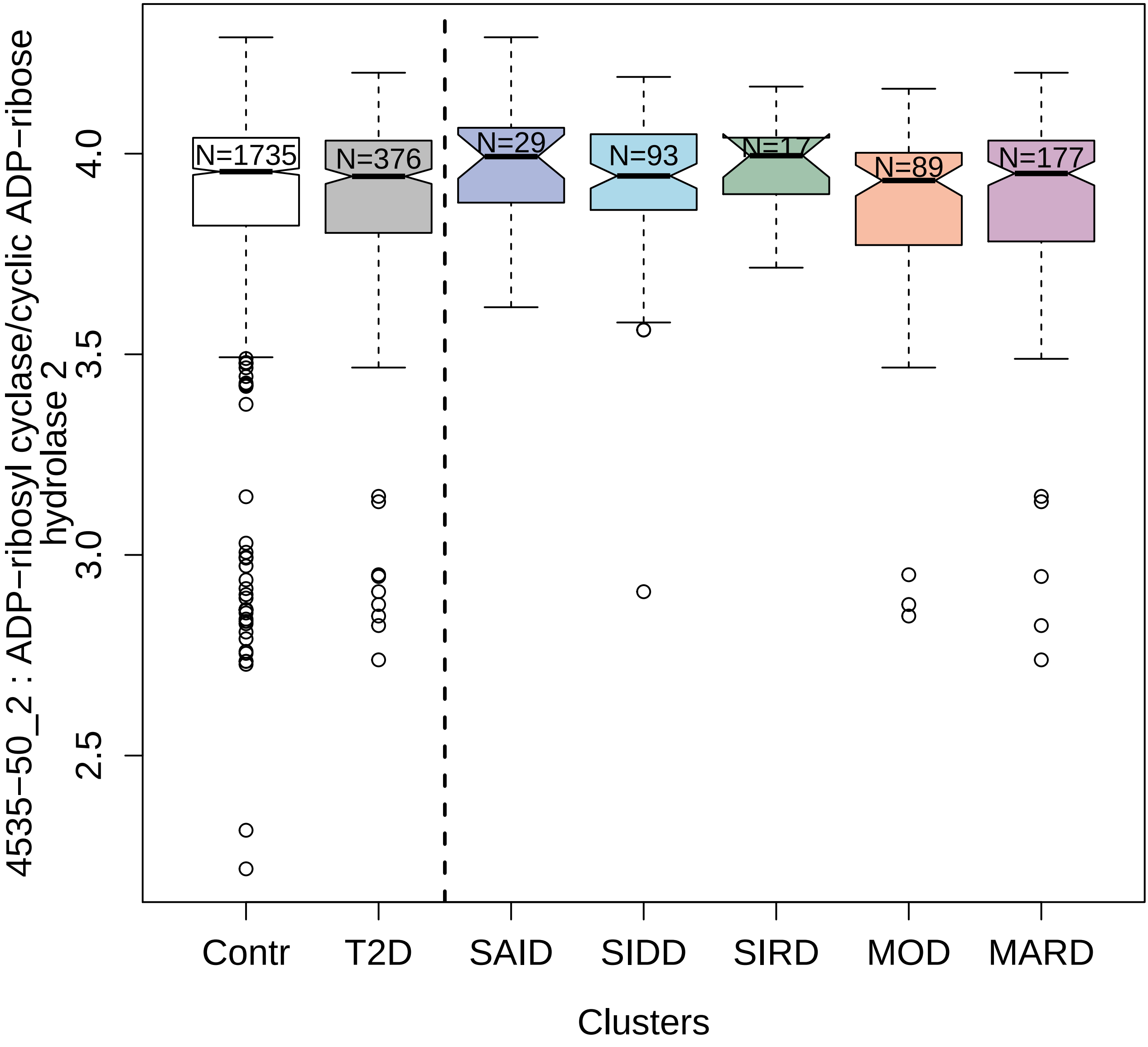
4535−50_2 : ADP−ribosyl cyclase/cyclic ADP−ribose hydrolase 2
4.0
3.5
3.0
2.5
N=1735
N=376
N=29
N=93
N=17
N=89
N=177
Contr
T2D
SAID
SIDD
SIRD
MOD
MARD
Clusters

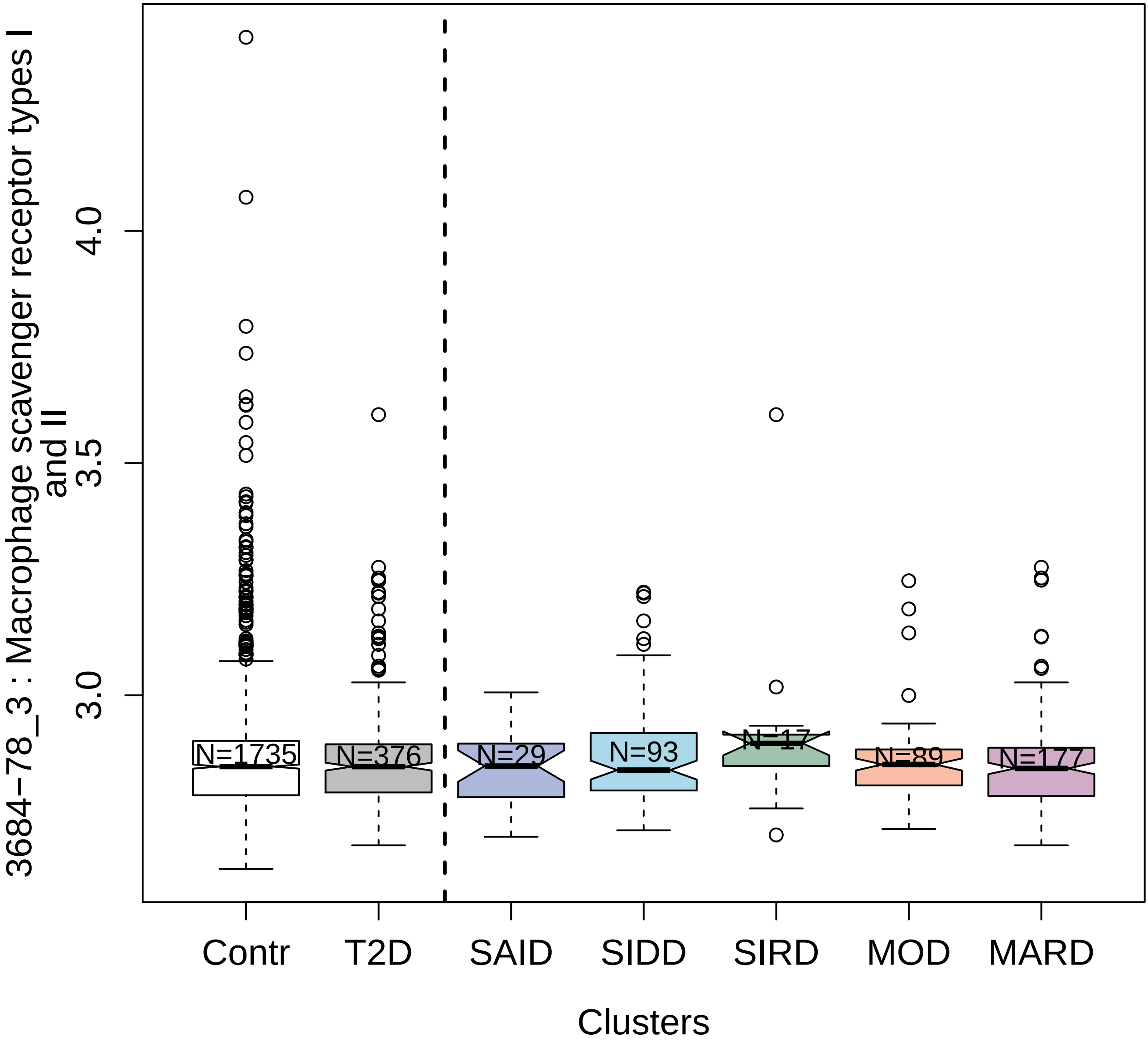

3684−78_3 : Macrophage scavenger receptor types I and II
4.0
3.5
3.0
N=1735
N=376
N=29
N=93
N=17
N=89
N=177
Contr
T2D
SAID
SIDD
SIRD
MOD
MARD
Clusters

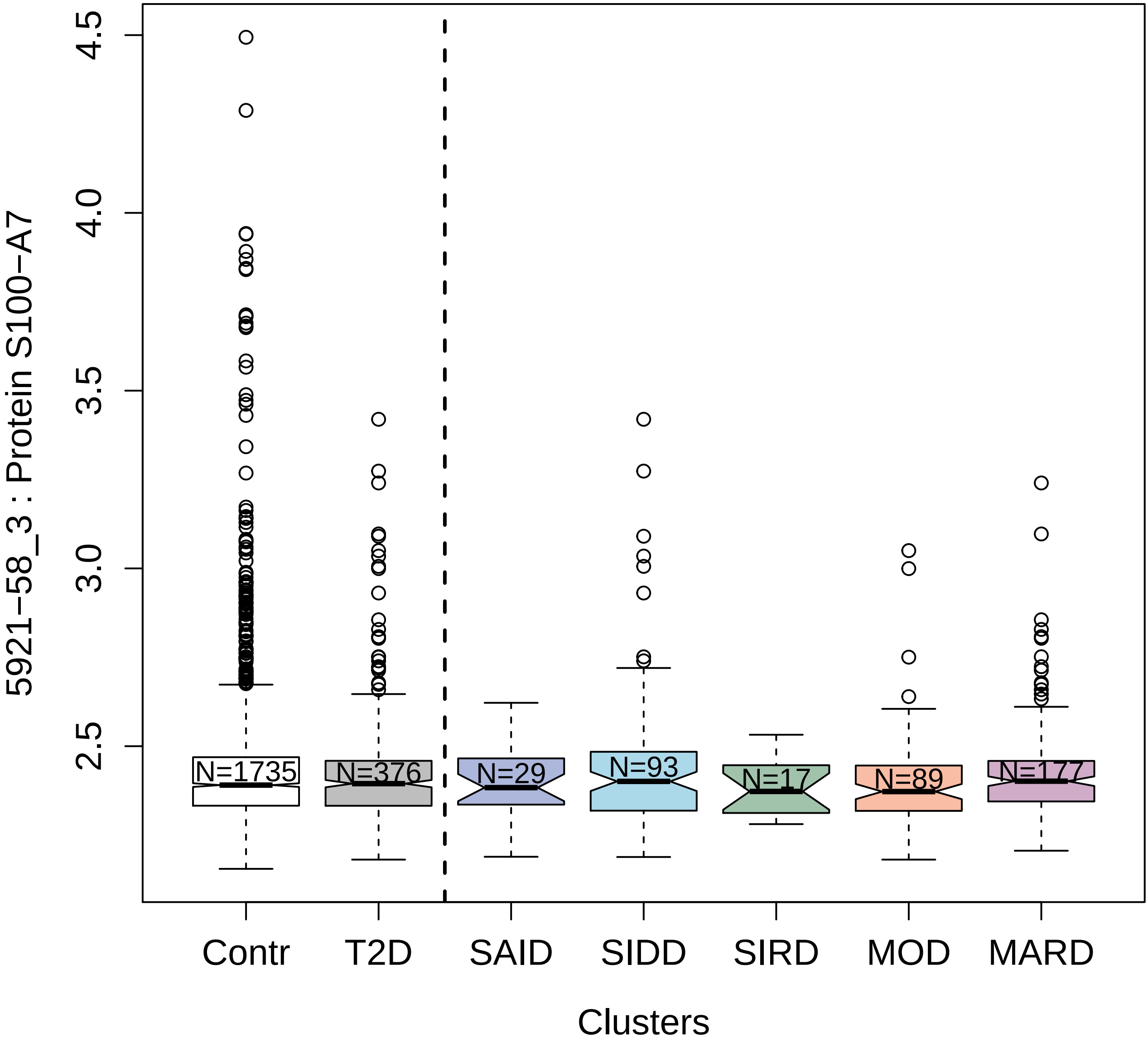

5921−58_3 : Protein S100−A7
4.5
4.0
3.5
3.0
2.5
N=1735
N=376
N=29
N=93
N=17
N=89
N=177
Contr
T2D
SAID
SIDD
SIRD
MOD
MARD
Clusters

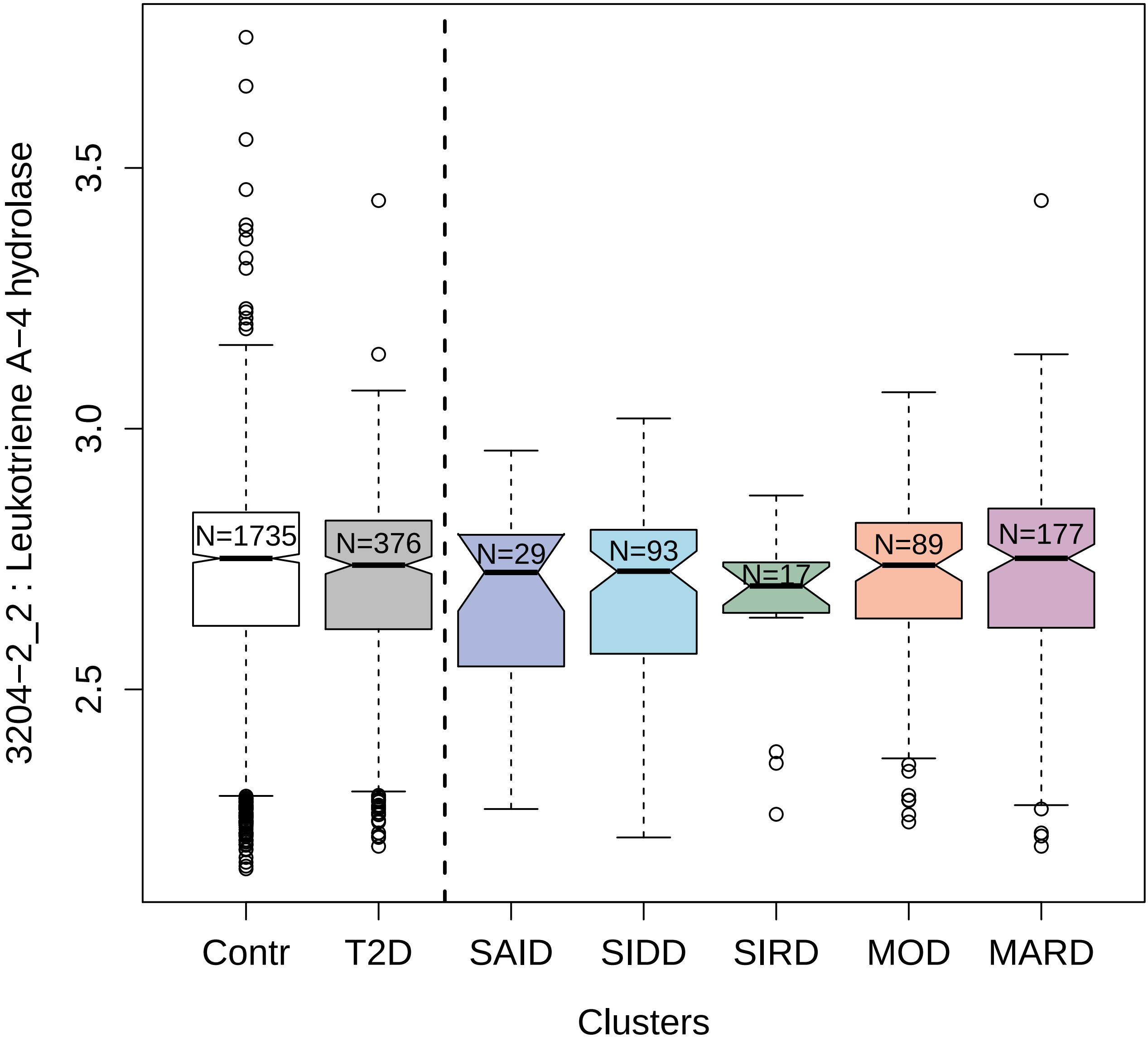
3204−2_2 : Leukotriene A−4 hydrolase
3.5
3.0
2.5
N=1735
N=376
N=29
N=93
N=17
N=89
N=177
Contr
T2D
SAID
SIDD
SIRD
MOD
MARD
Clusters

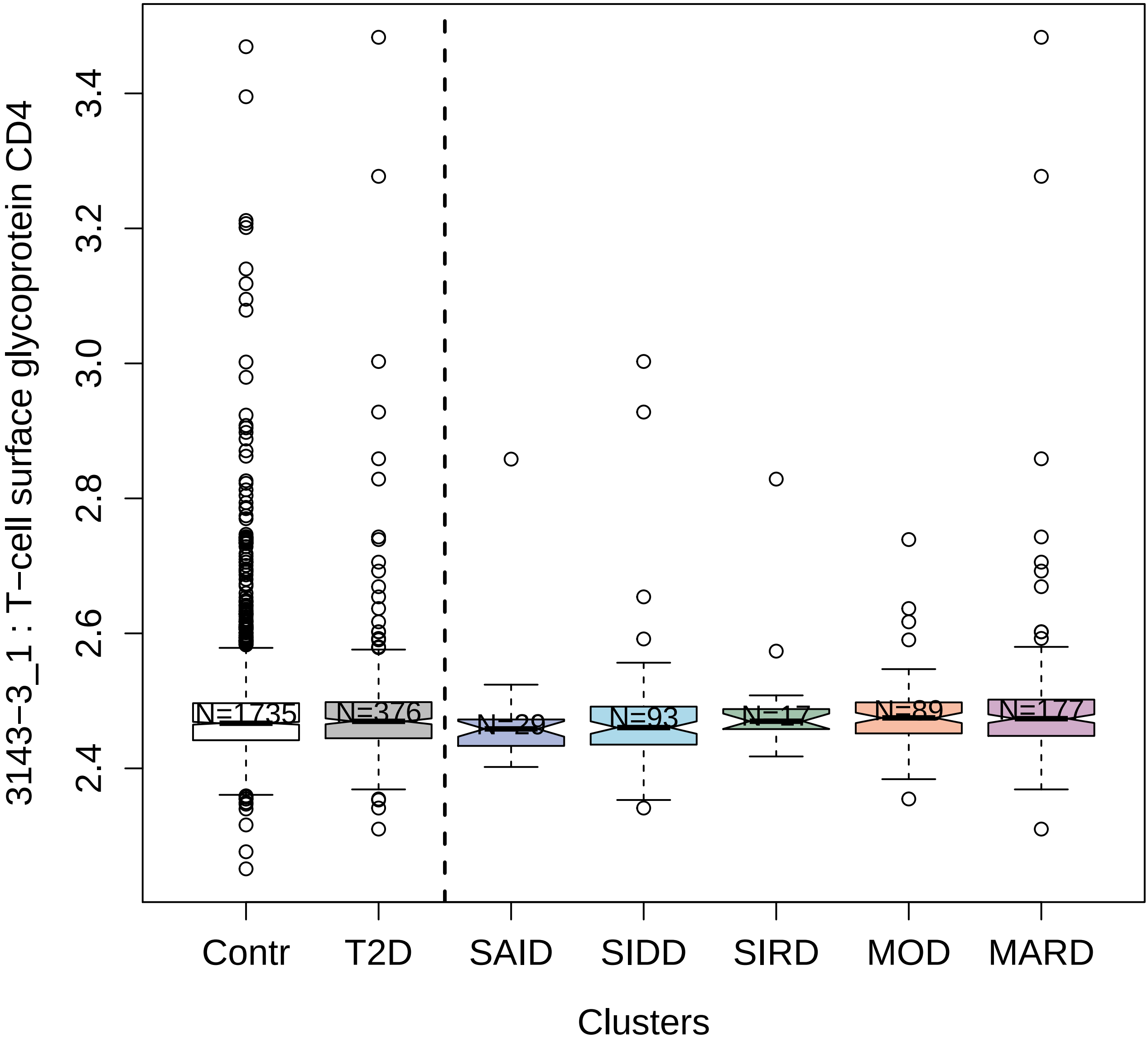
3143−3_1 : T−cell surface glycoprotein CD4
3.4
3.2
3.0
2.8
2.6
2.4
N=1735
N=376
N=29
N=93
N=17
N=89
N=177
Contr
T2D
SAID
SIDD
SIRD
MOD
MARD
Clusters

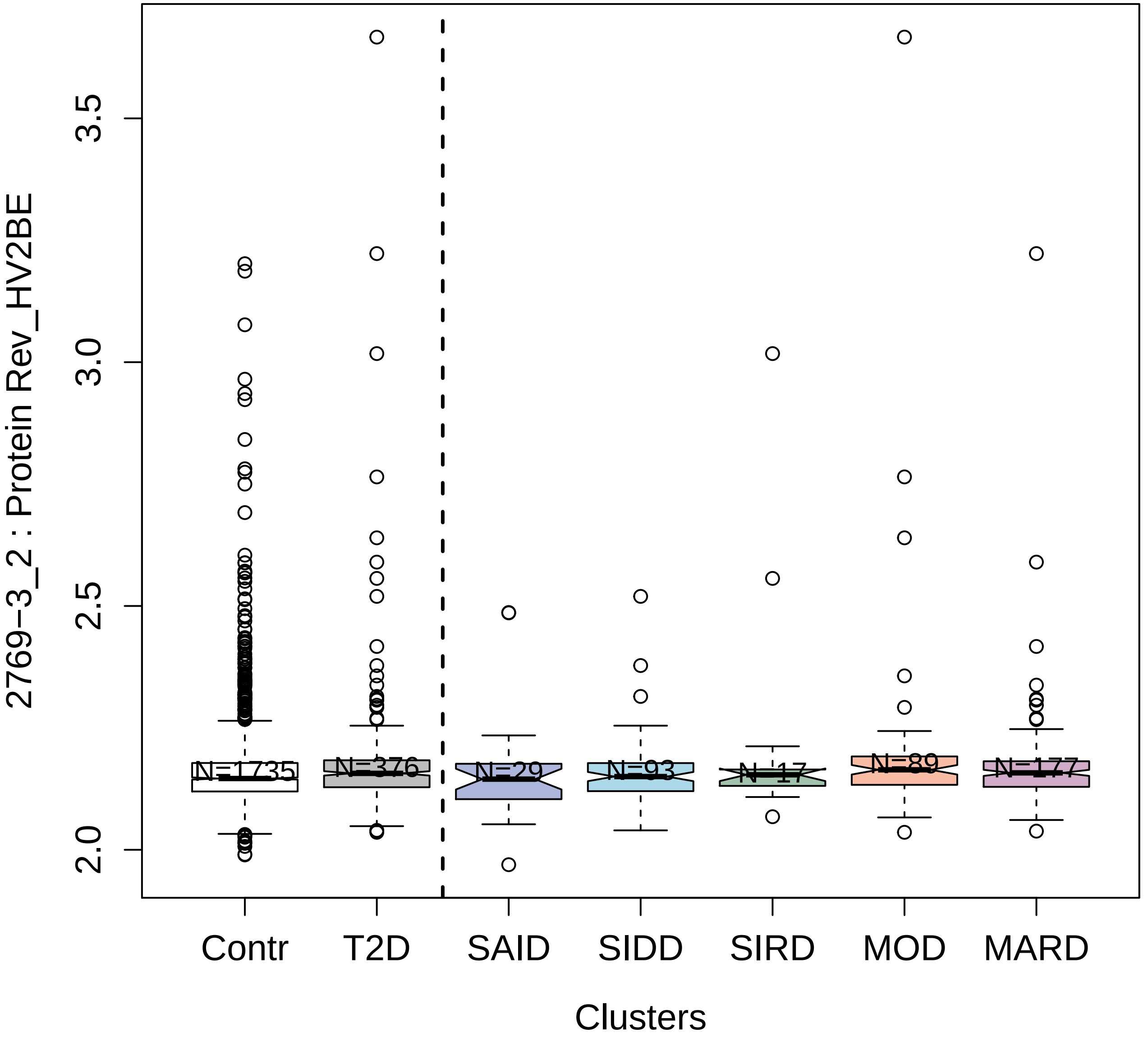

2769−3_2 : Protein Rev_HV2BE
3.5
3.0
2.5
2.0
N=1735
N=376
N=29
N=93
N=17
N=89
N=177
Contr
T2D
SAID
SIDD
SIRD
MOD
MARD
Clusters

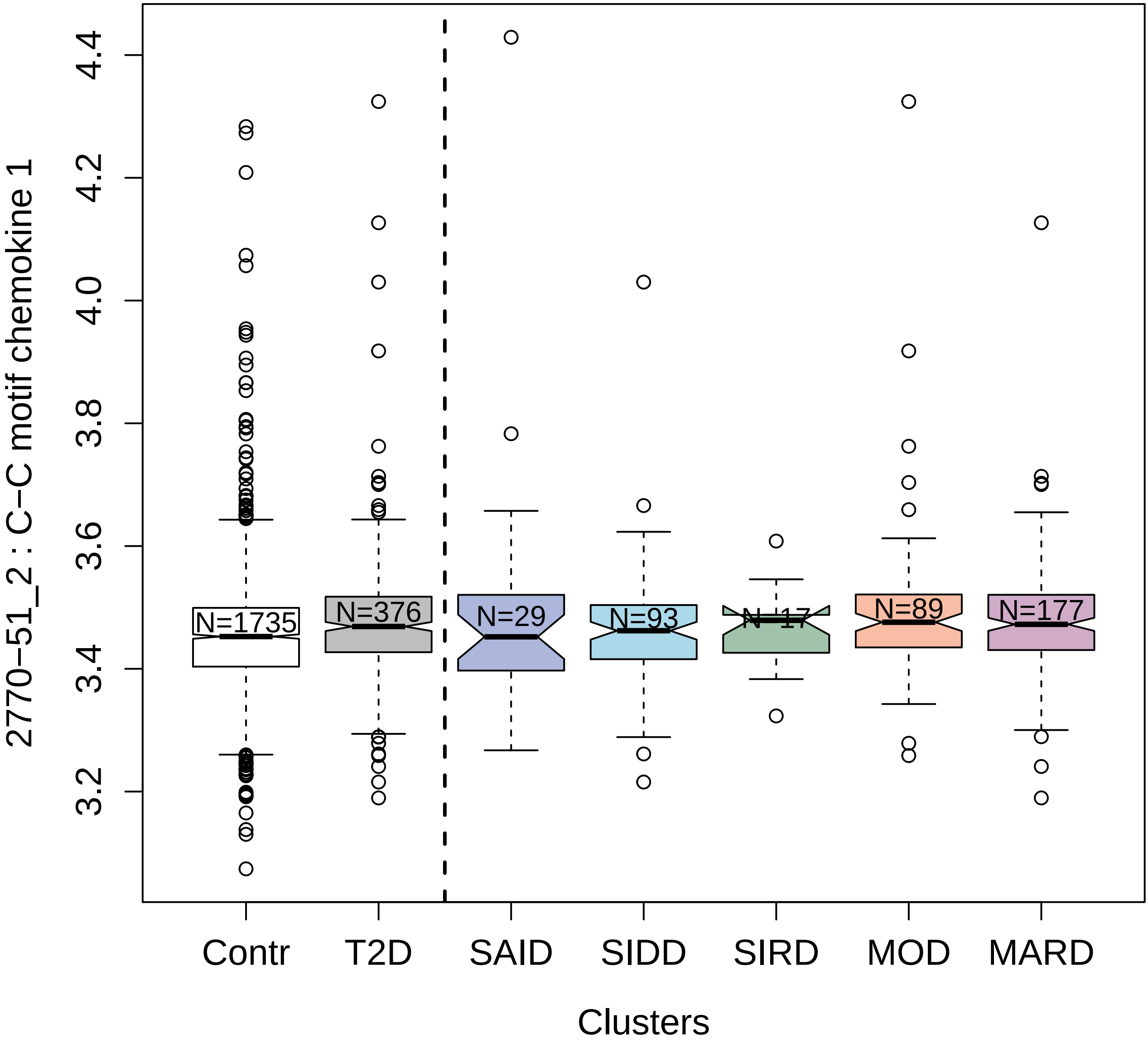

2770−51_2 : C−C motif chemokine 1
3.2
3.4
3.6
3.8
4.0
4.2
4.4
N=1735
N=376
N=29
N=93
N=17
N=89
N=177
Contr
T2D
SAID
SIDD
SIRD
MOD
MARD
Clusters

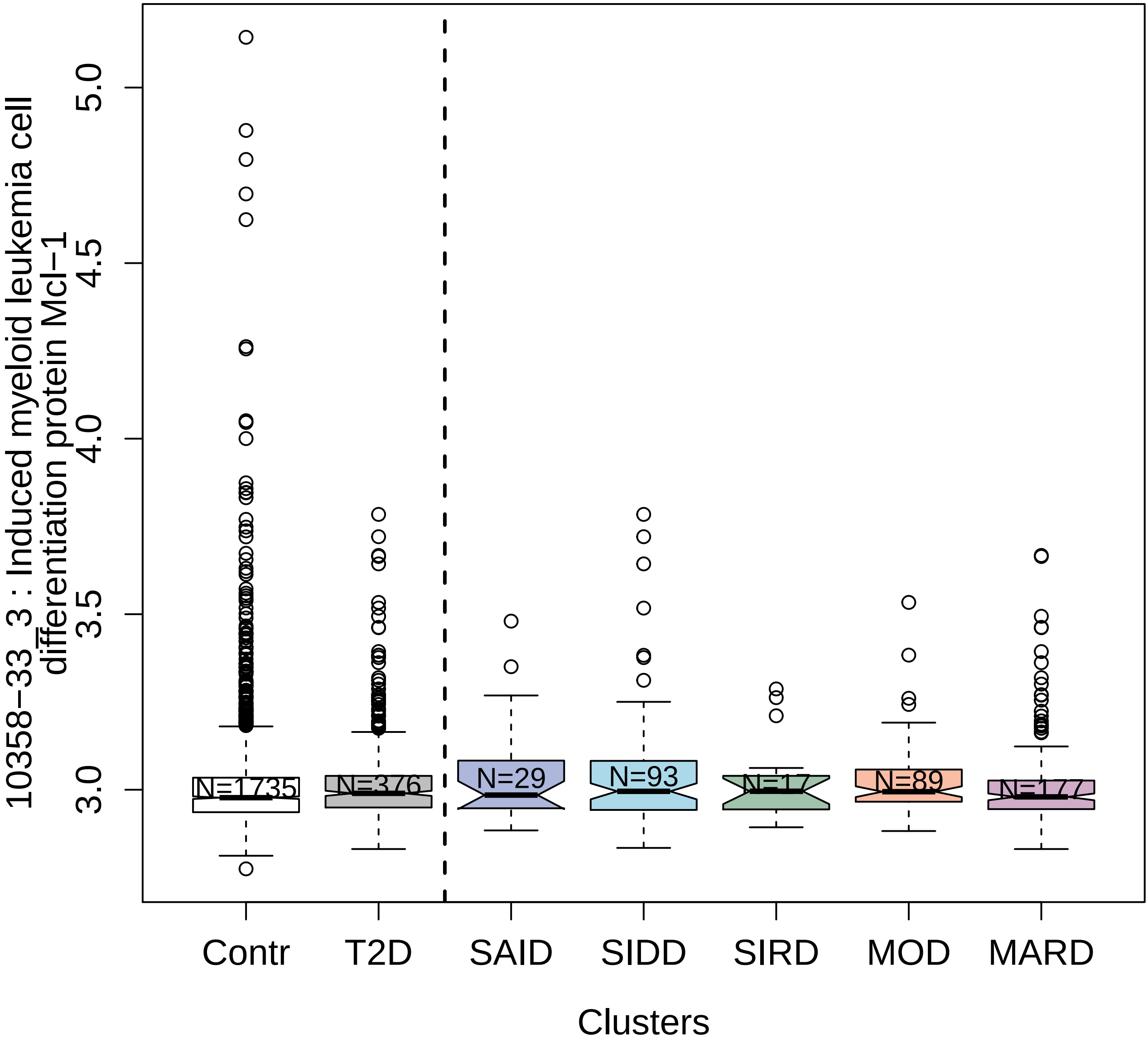
10358−33_3 : Induced myeloid leukemia cell
differentiation protein Mcl−1
5.0
4.5
4.0
3.5
3.0
N=1735
N=376
N=29
N=93
N=17
N=89
N=177
Contr
T2D
SAID
SIDD
SIRD
MOD
MARD
Clusters

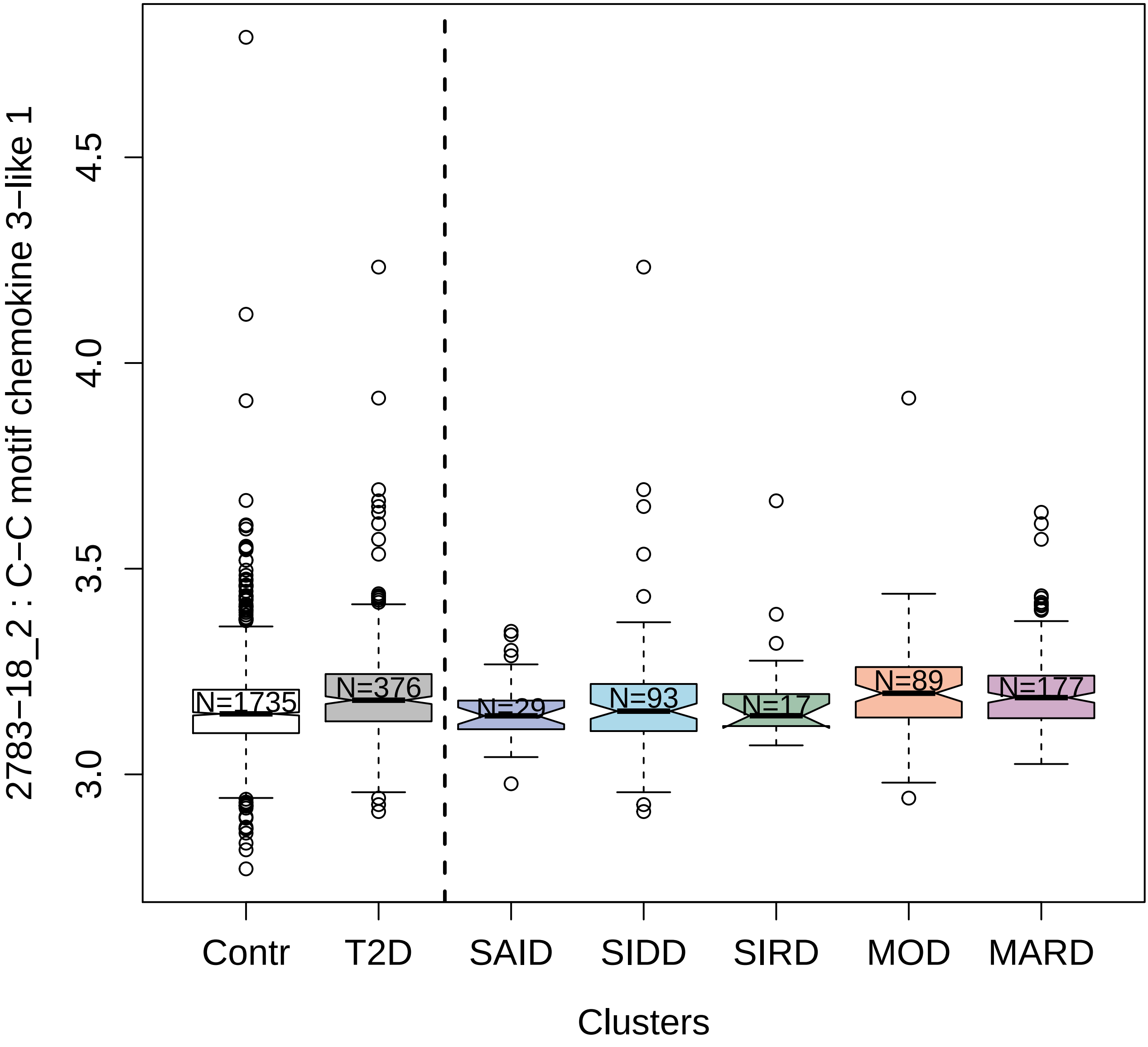

2783−18_2 : C−C motif chemokine 3−like 1
4.5
4.0
3.5
3.0
N=1735
N=376
N=29
N=93
N=17
N=89
N=177
Contr
T2D
SAID
SIDD
SIRD
MOD
MARD
Clusters

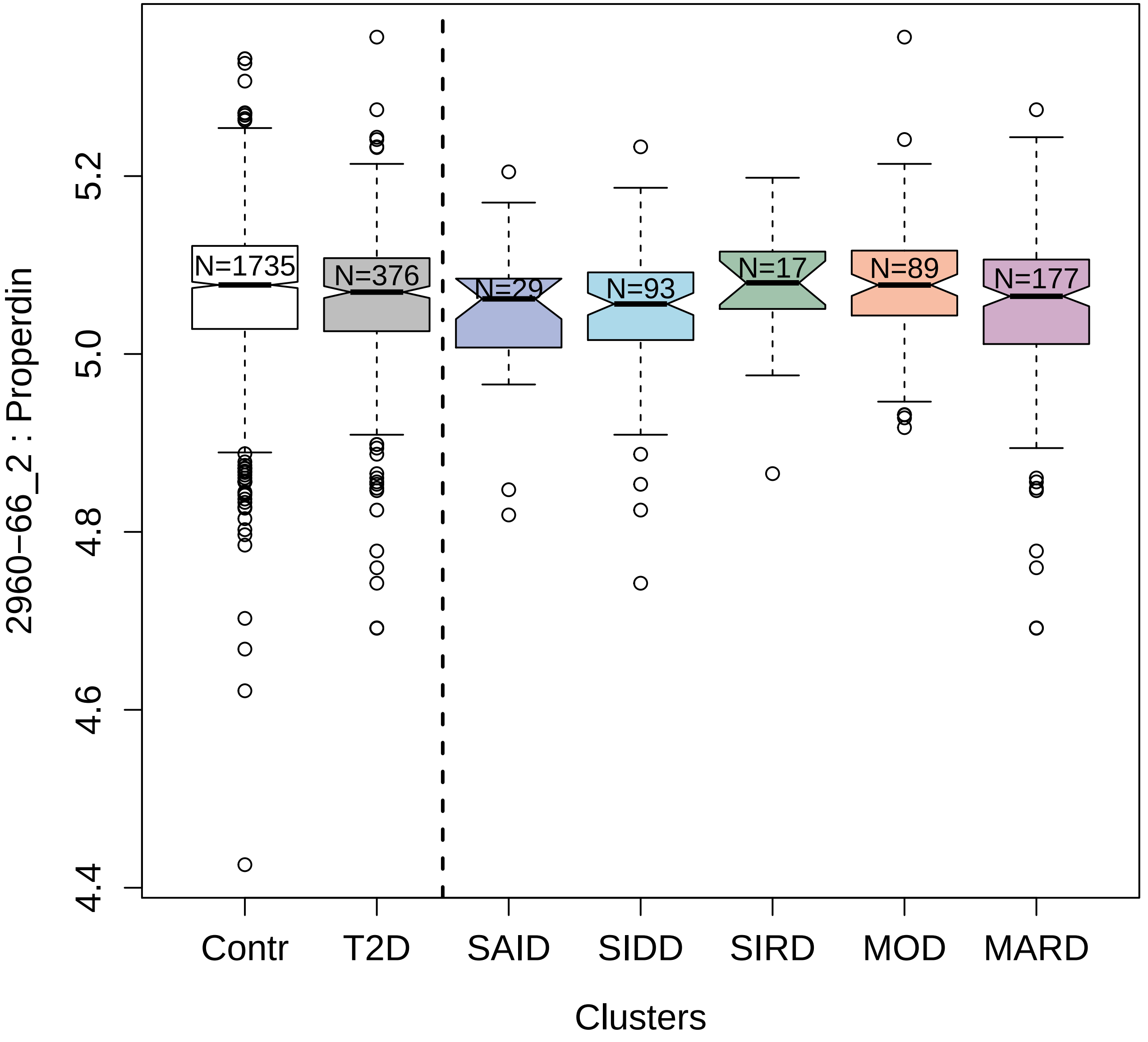

2960−66_2 : Properdin
5.2
5.0
4.8
4.6
4.4
N=1735
N=376
N=29
N=93
N=17
N=89
N=177
Contr
T2D
SAID
SIDD
SIRD
MOD
MARD
Clusters

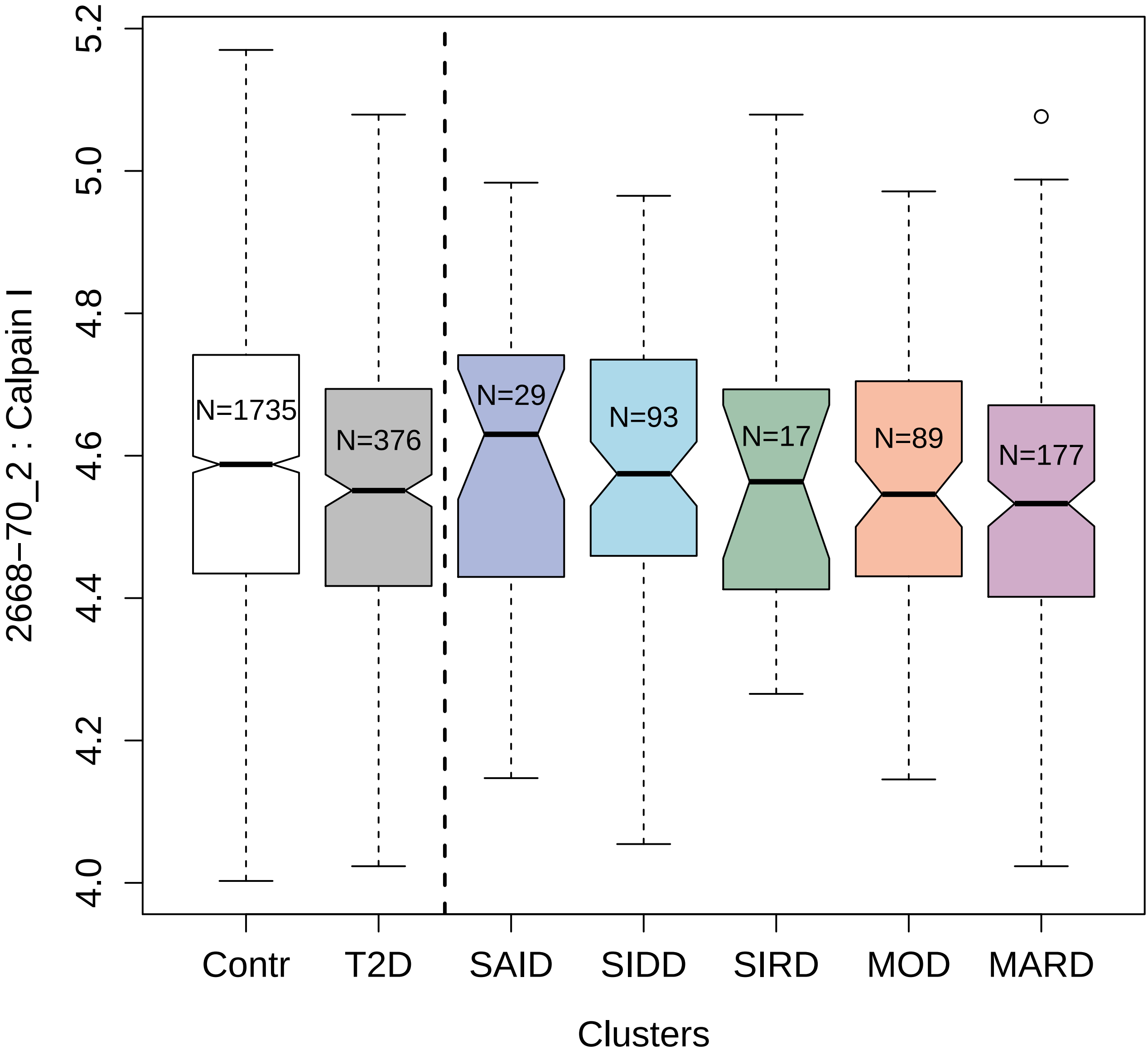

2668−70_2 : Calpain I
5.2
5.0
4.8
4.6
4.4
4.2
4.0
N=1735
N=376
N=29
N=93
N=17
N=89
N=177
Contr
T2D
SAID
SIDD
SIRD
MOD
MARD
Clusters

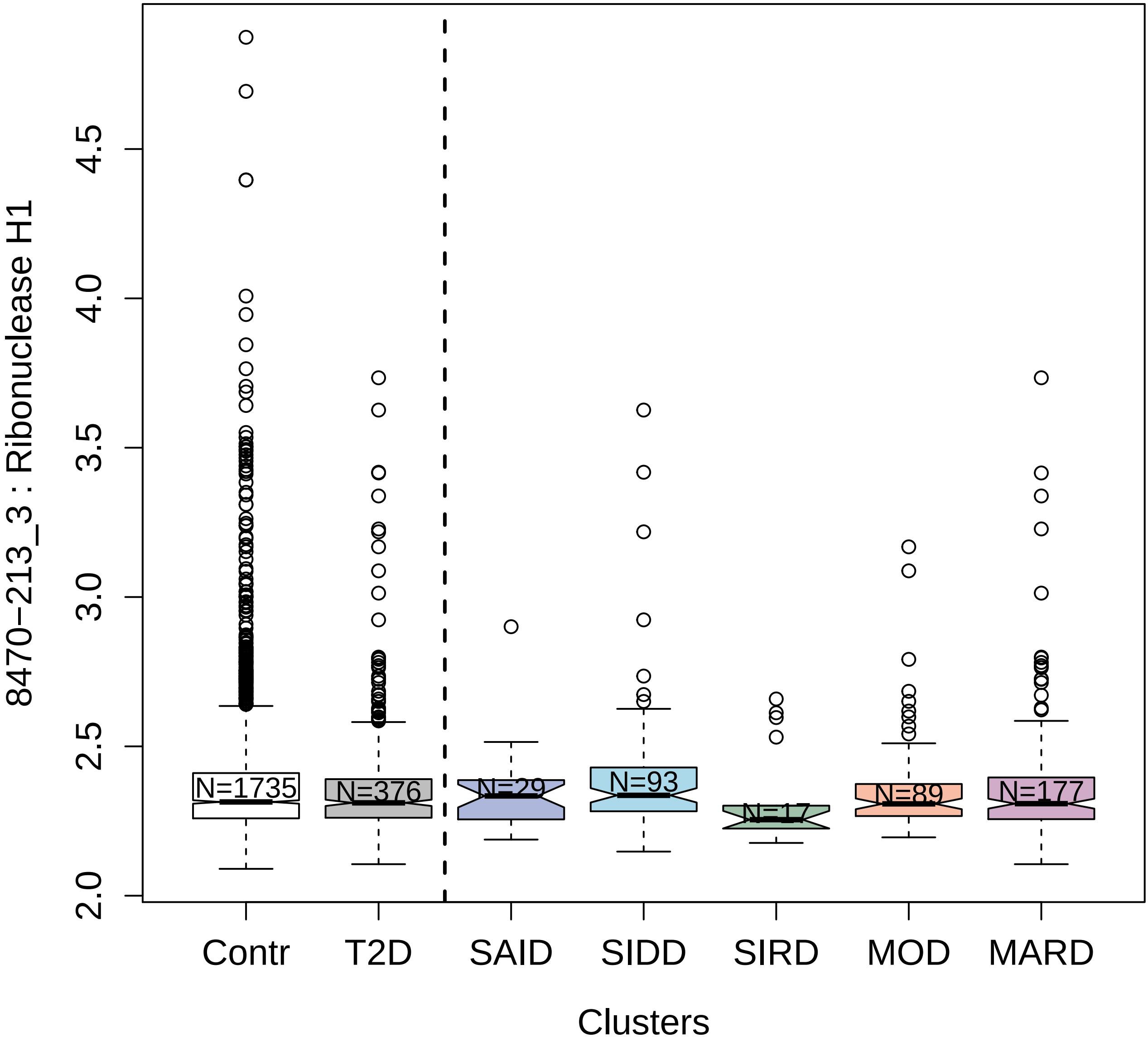
8470−213_3 : Ribonuclease H1
2.0
2.5
3.0
3.5
4.0
4.5
N=1735
N=376
N=29
N=93
N=17
N=89
N=177
Contr
T2D
SAID
SIDD
SIRD
MOD
MARD
Clusters

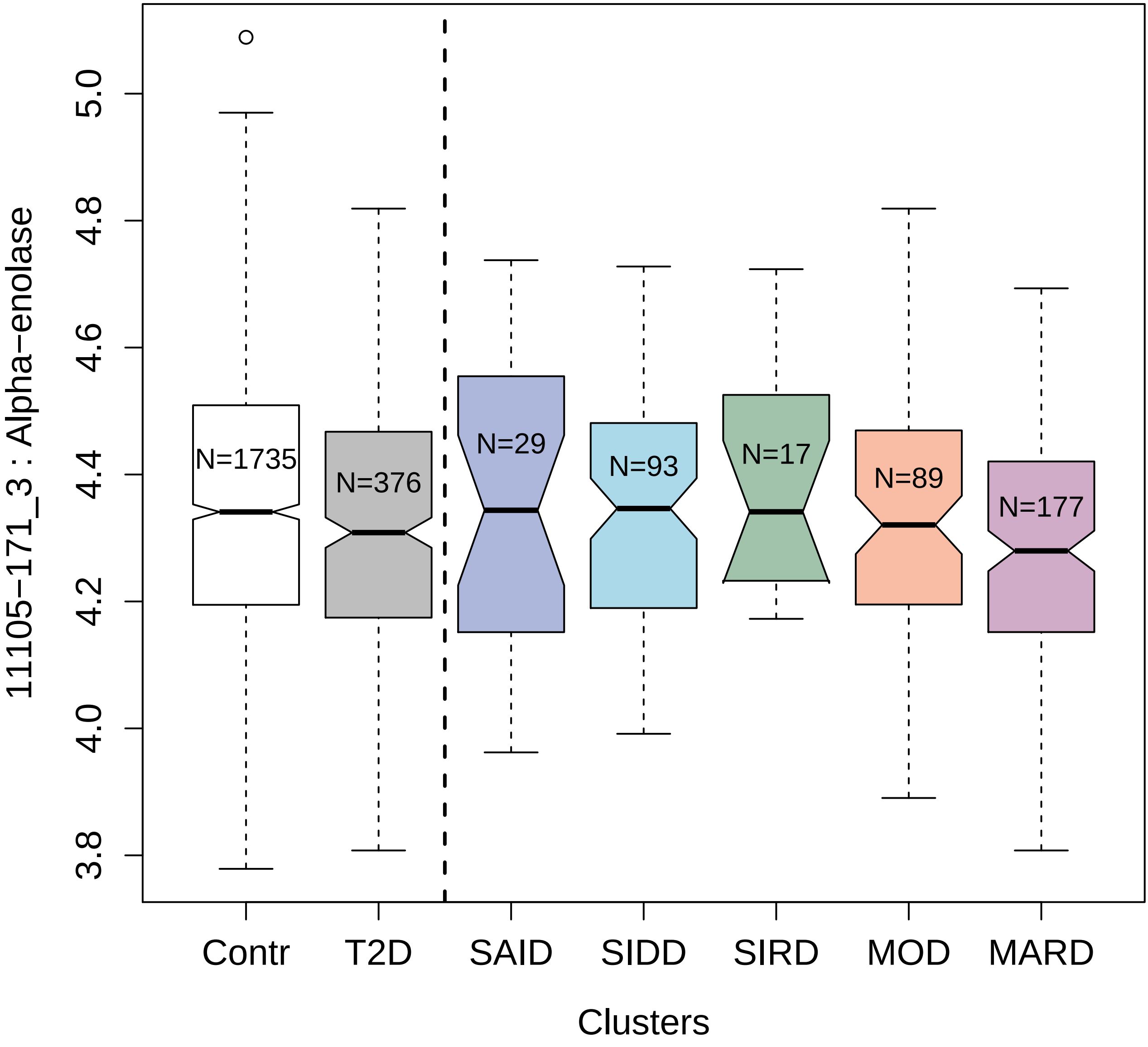

11105−171_3 : Alpha−enolase
3.8
4.0
4.2
4.4
4.6
4.8
5.0
N=1735
N=376
N=29
N=93
N=17
N=89
N=177
Contr
T2D
SAID
SIDD
SIRD
MOD
MARD
Clusters

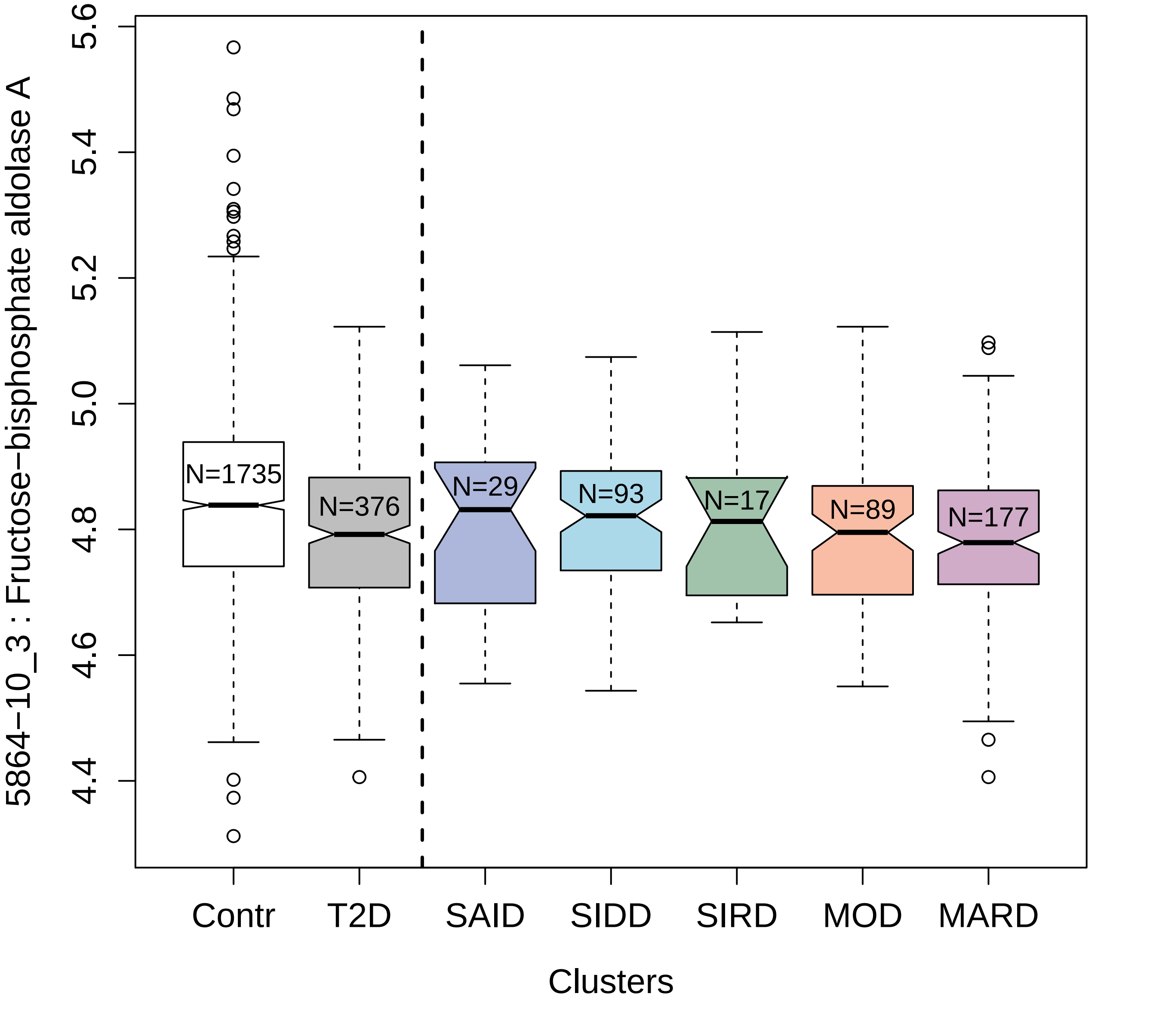
5864−10_3 : Fructose−bisphosphate aldolase A
5.6
5.4
5.2
5.0
4.8
4.6
4.4
N=1735
N=376
N=29
N=93
N=17
N=89
N=177
Contr
T2D
SAID
SIDD
SIRD
MOD
MARD
Clusters

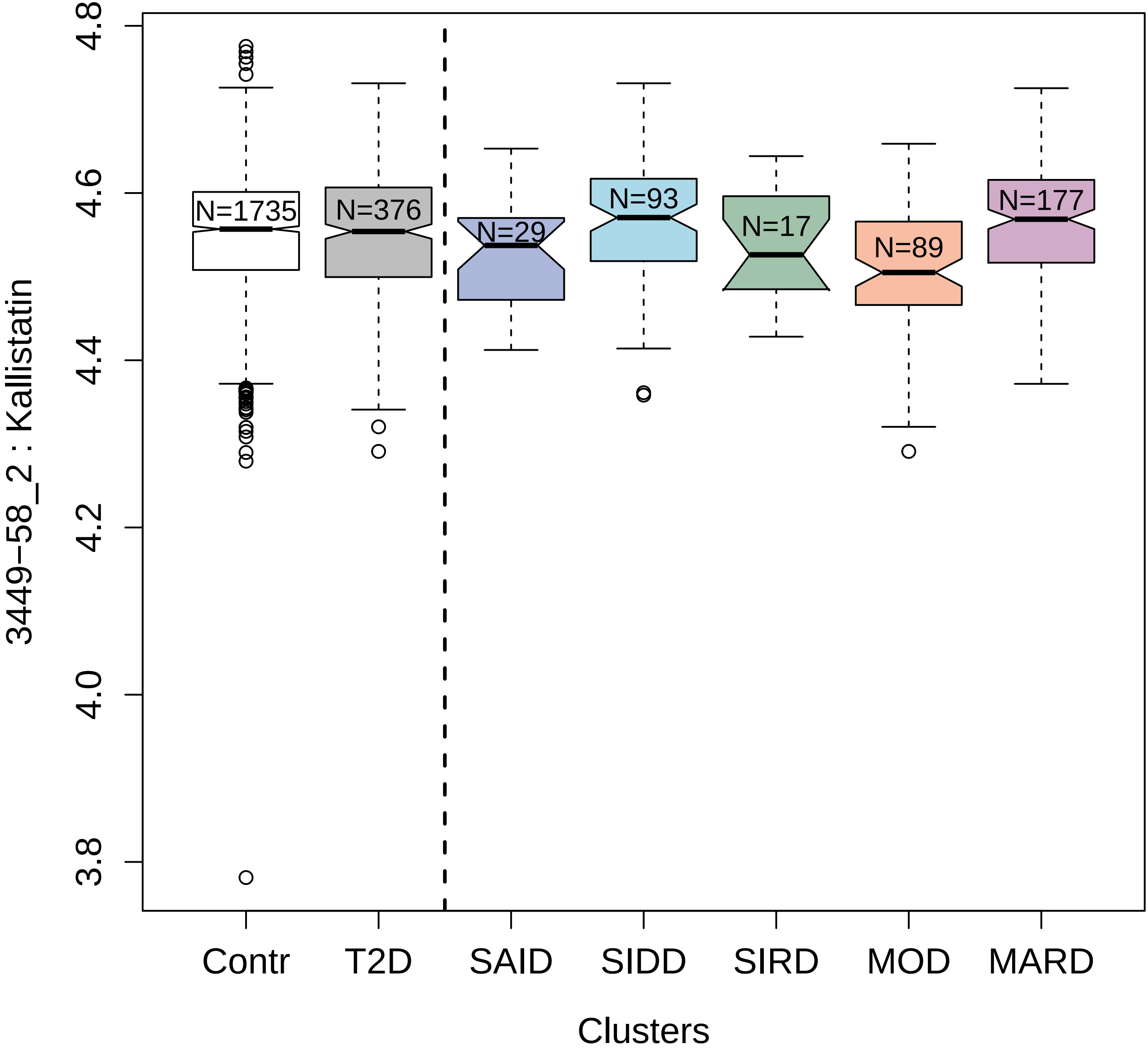
3449−58_2 : Kallistatin
4.8
4.6
4.4
4.2
4.0
3.8
N=1735
N=376
N=29
N=93
N=17
N=89
N=177
Contr
T2D
SAID
SIDD
SIRD
MOD
MARD
Clusters

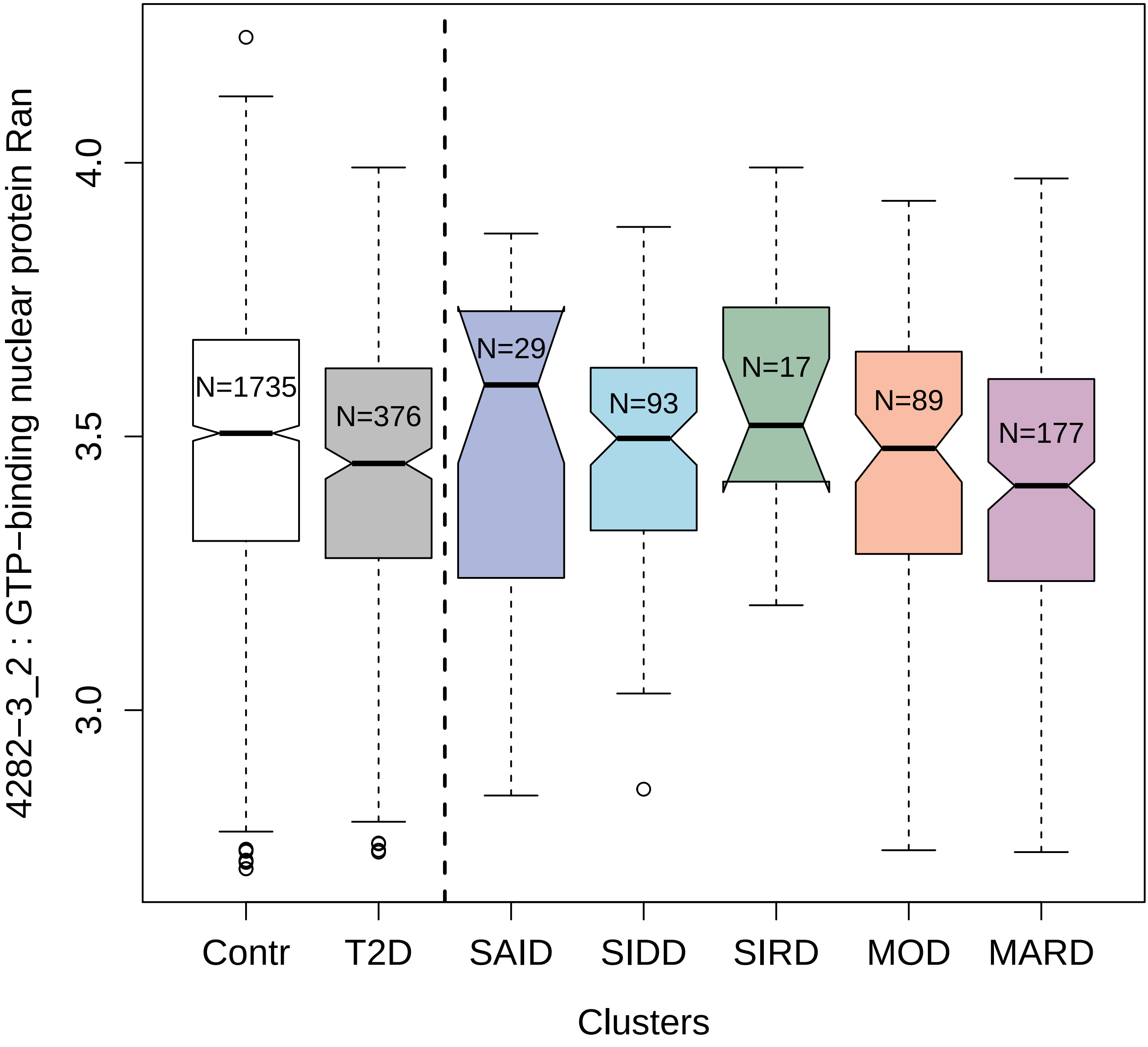
4282−3_2 : GTP−binding nuclear protein Ran
4.0
3.5
3.0
N=1735
N=376
N=29
N=93
N=17
N=89
N=177
Contr
T2D
SAID
SIDD
SIRD
MOD
MARD
Clusters

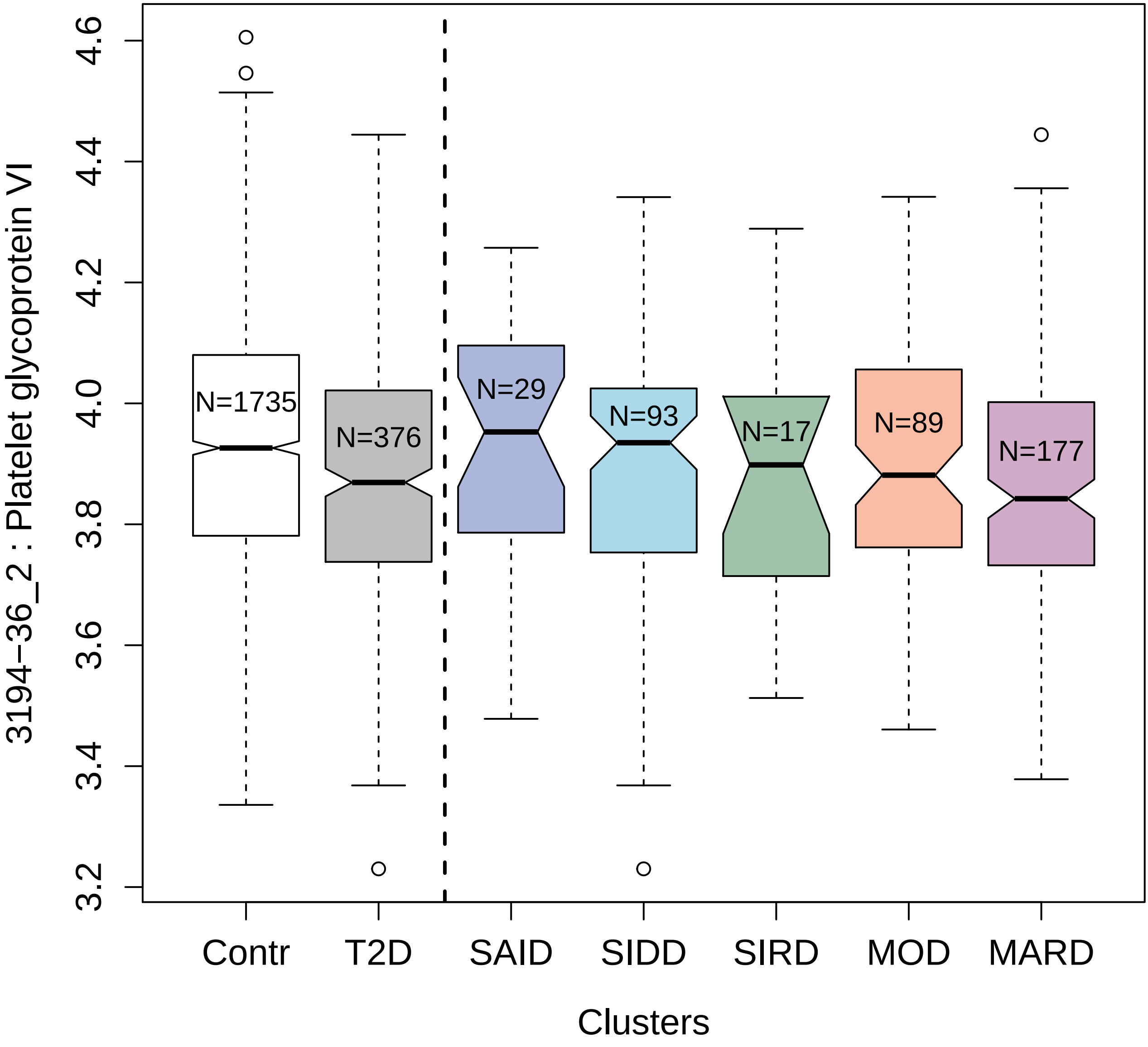

3194−36_2 : Platelet glycoprotein VI
4.6
4.4
4.2
4.0
3.8
3.6
3.4
3.2
N=1735
N=376
N=29
N=93
N=17
N=89
N=177
Contr
T2D
SAID
SIDD
SIRD
MOD
MARD
Clusters

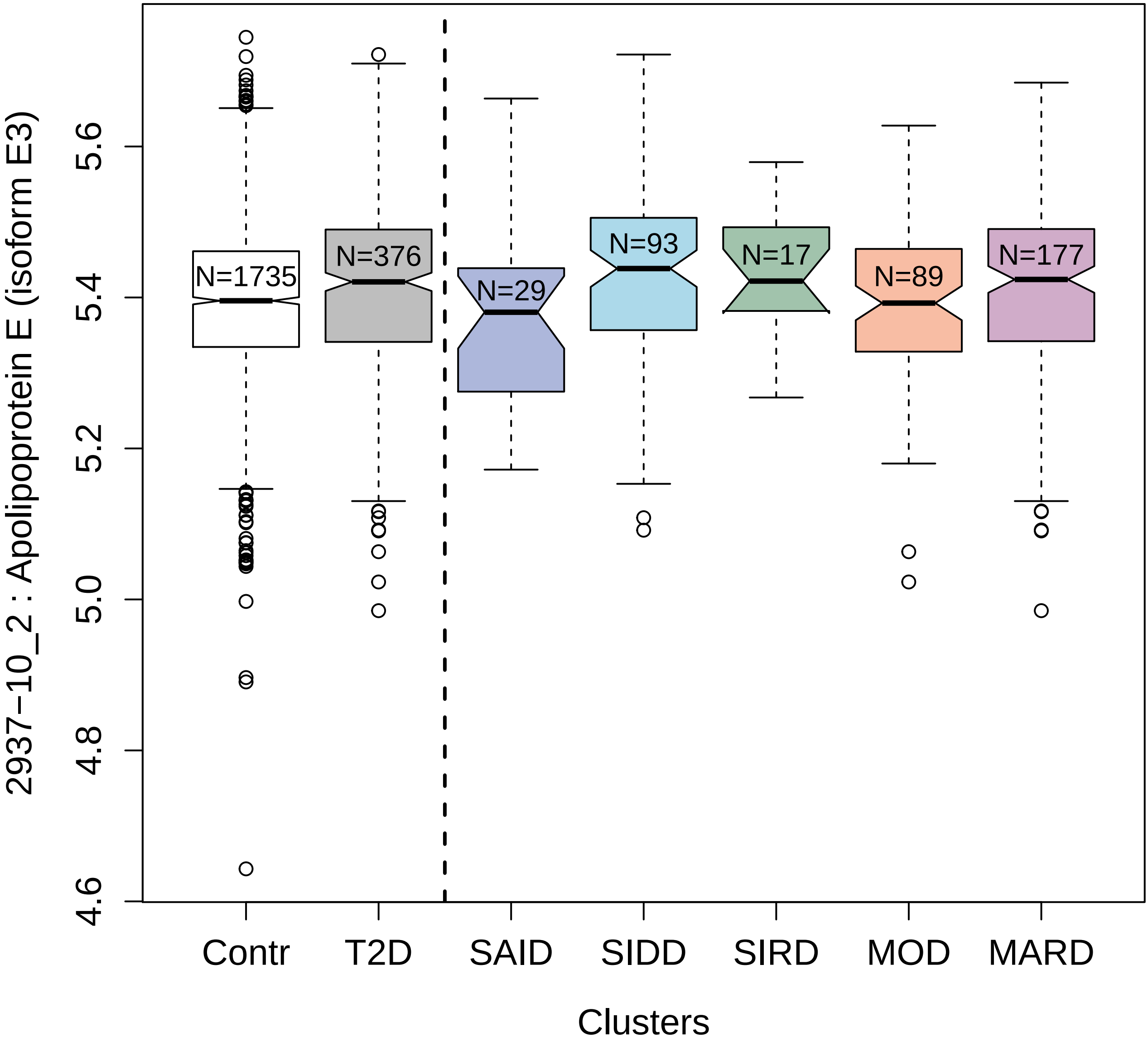

2937−10_2 : Apolipoprotein E (isoform E3)
4.6
4.8
5.0
5.2
5.4
5.6
N=1735
N=376
N=29
N=93
N=17
N=89
N=177
Contr
T2D
SAID
SIDD
SIRD
MOD
MARD
Clusters

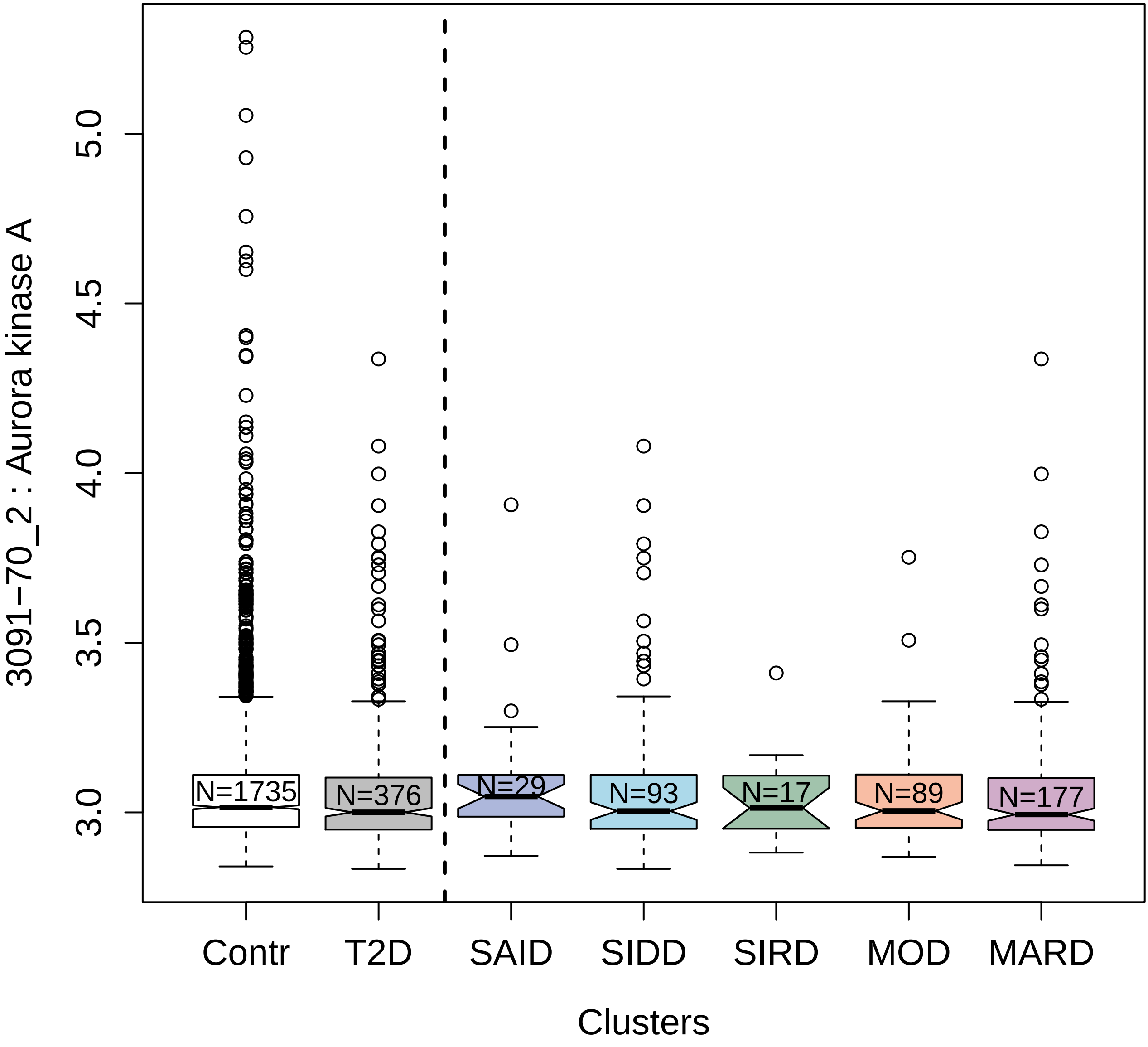

3091−70_2 : Aurora kinase A
5.0
4.5
4.0
3.5
3.0
N=1735
N=376
N=29
N=93
N=17
N=89
N=177
Contr
T2D
SAID
SIDD
SIRD
MOD
MARD
Clusters

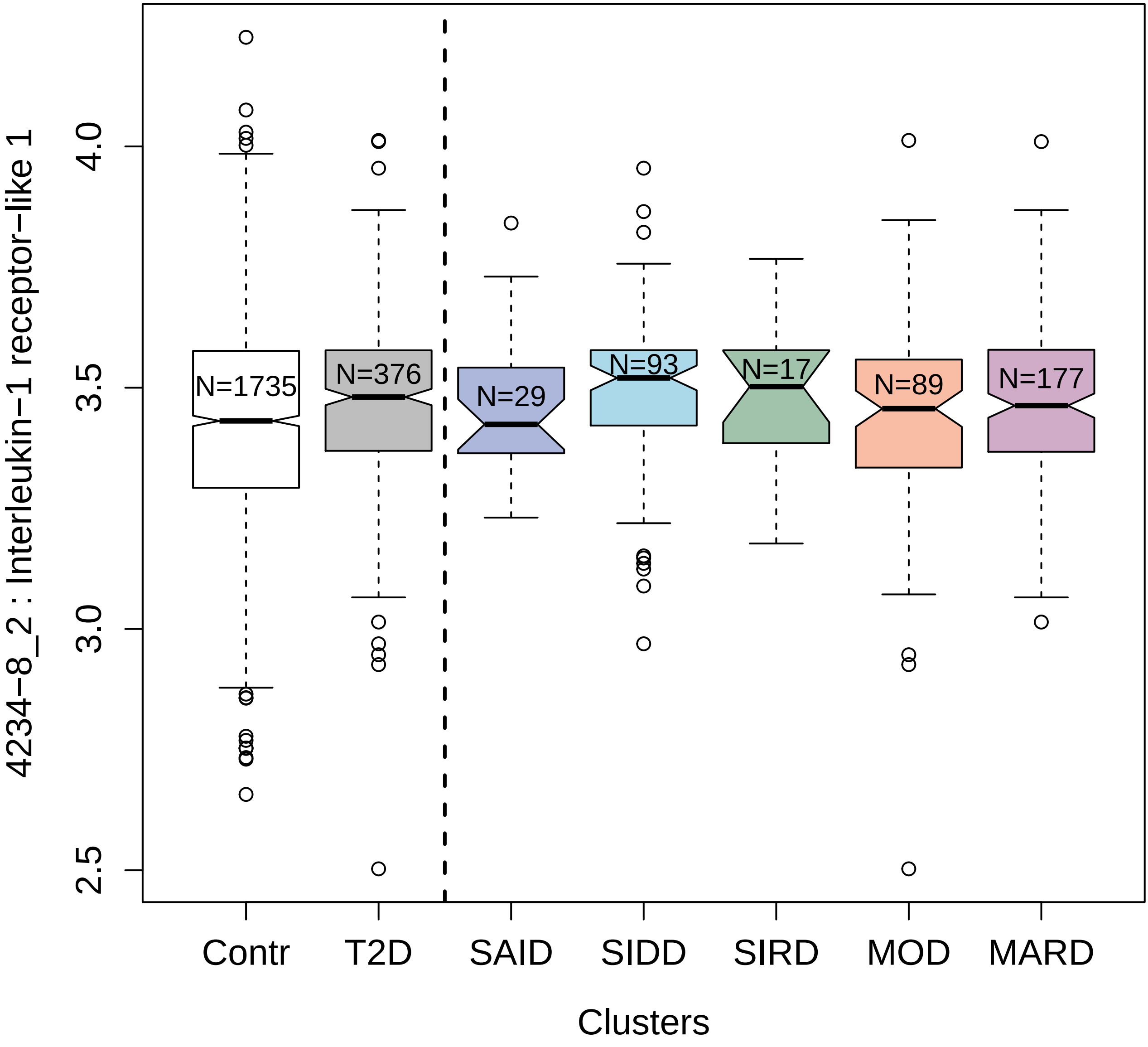

4234−8_2 : Interleukin−1 receptor−like 1
2.5
3.0
3.5
4.0
N=1735
N=376
N=29
N=93
N=17
N=89
N=177
Contr
T2D
SAID
SIDD
SIRD
MOD
MARD
Clusters

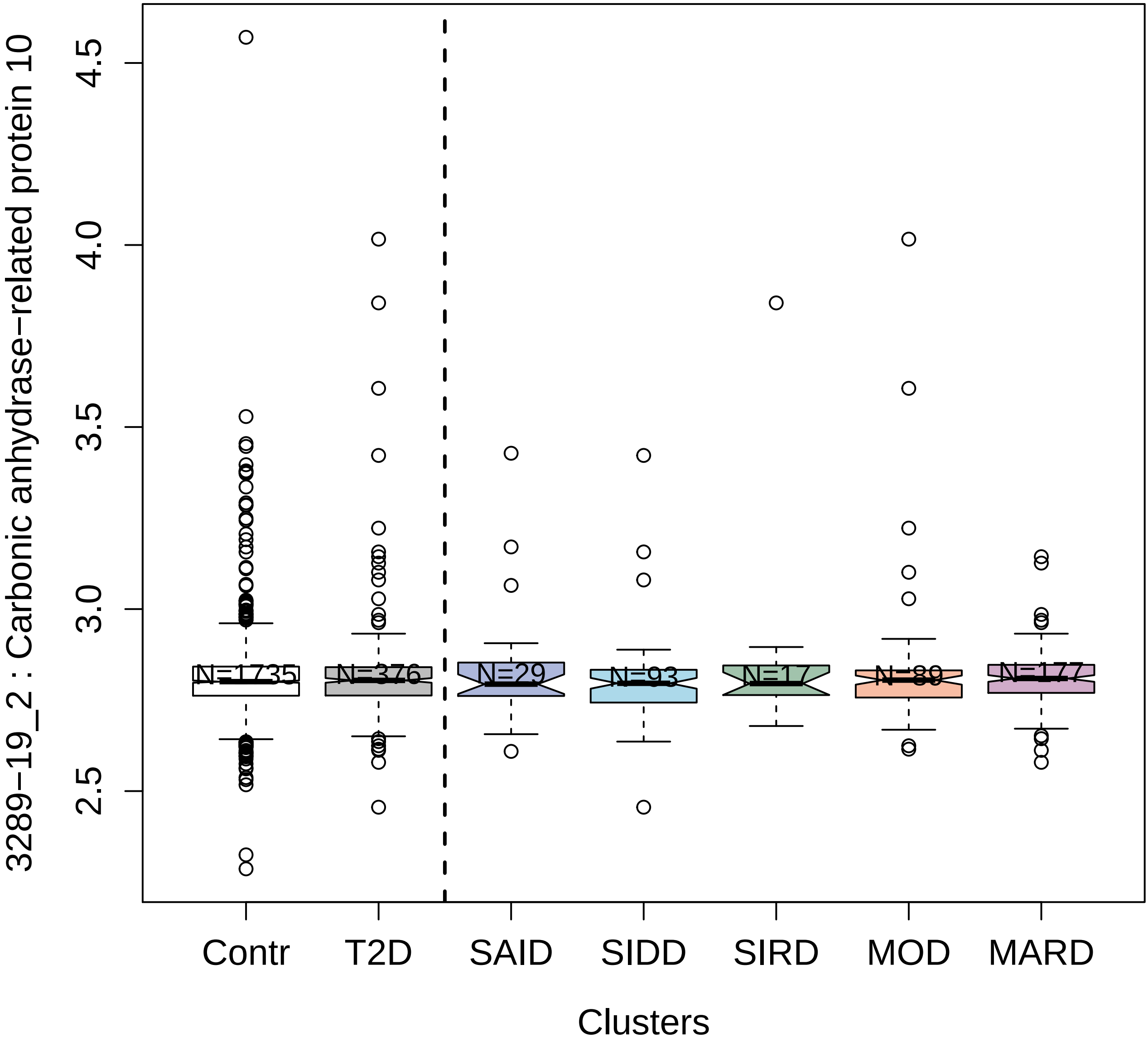
3289−19_2 : Carbonic anhydrase−related protein 10
Clusters
Contr
T2D
SAID
SIDD
SIRD
MOD
MARD
N=1735
N=376
N=29
N=93
N=17
N=89
N=177
2.5
3.0
3.5
4.0
4.5

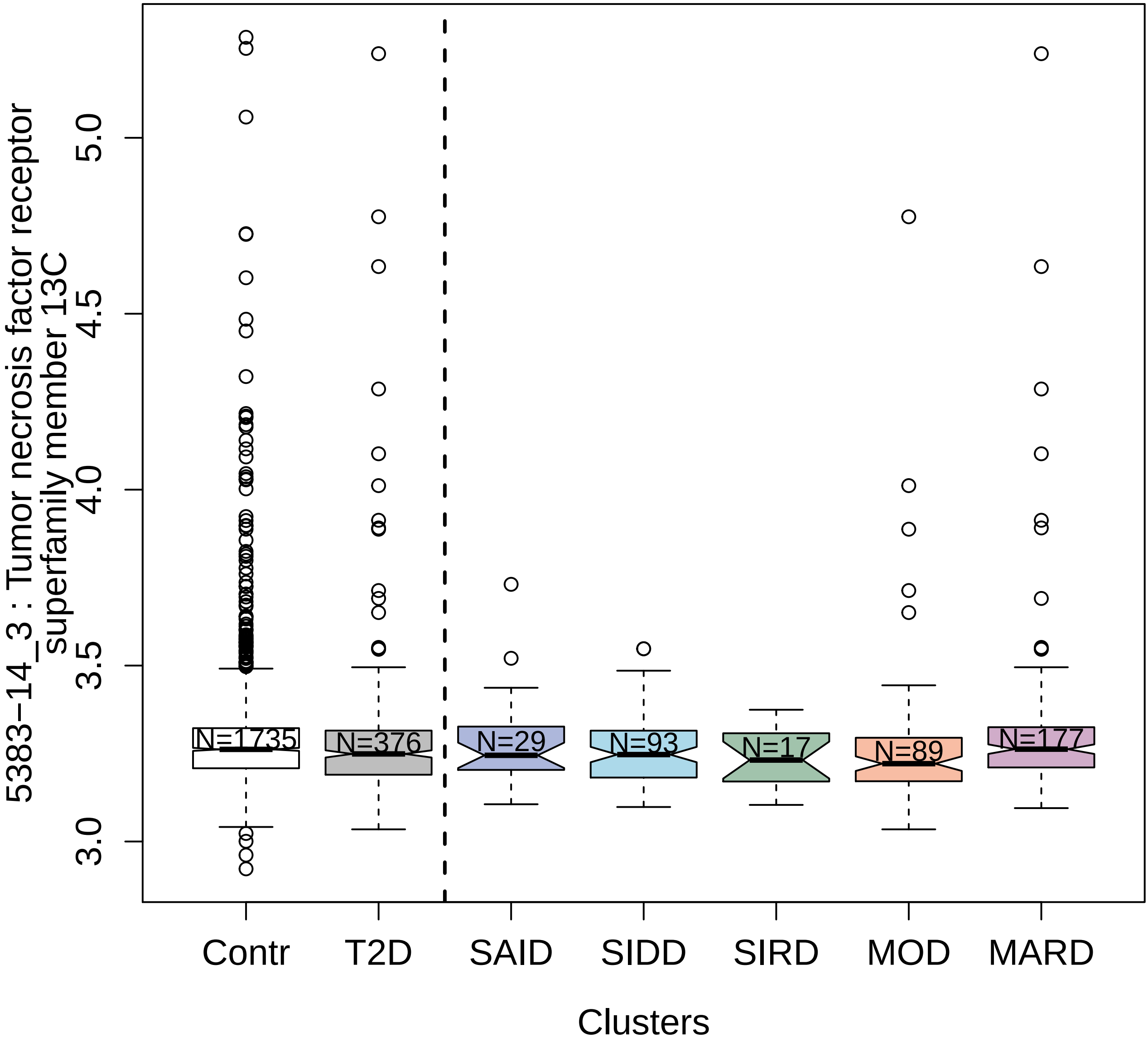

5383−14_3 : Tumor necrosis factor receptor superfamily member 13C
3.0
3.5
4.0
4.5
5.0
N=1735
N=376
N=29
N=93
N=17
N=89
N=177
Contr
T2D
SAID
SIDD
SIRD
MOD
MARD
Clusters

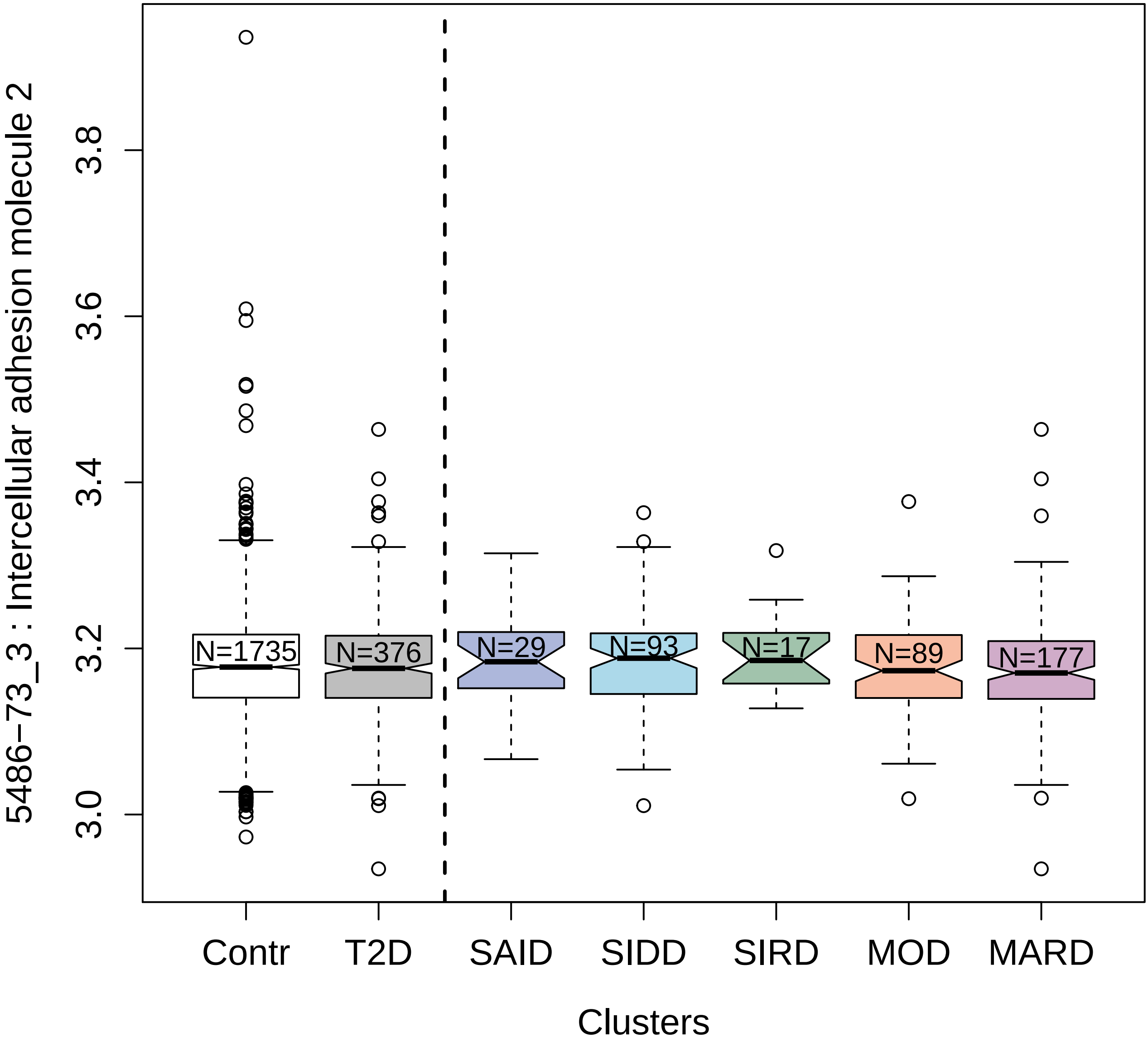

5486−73_3 : Intercellular adhesion molecule 2
3.0
3.2
3.4
3.6
3.8
N=1735
N=376
N=29
N=93
N=17
N=89
N=177
Contr
T2D
SAID
SIDD
SIRD
MOD
MARD
Clusters

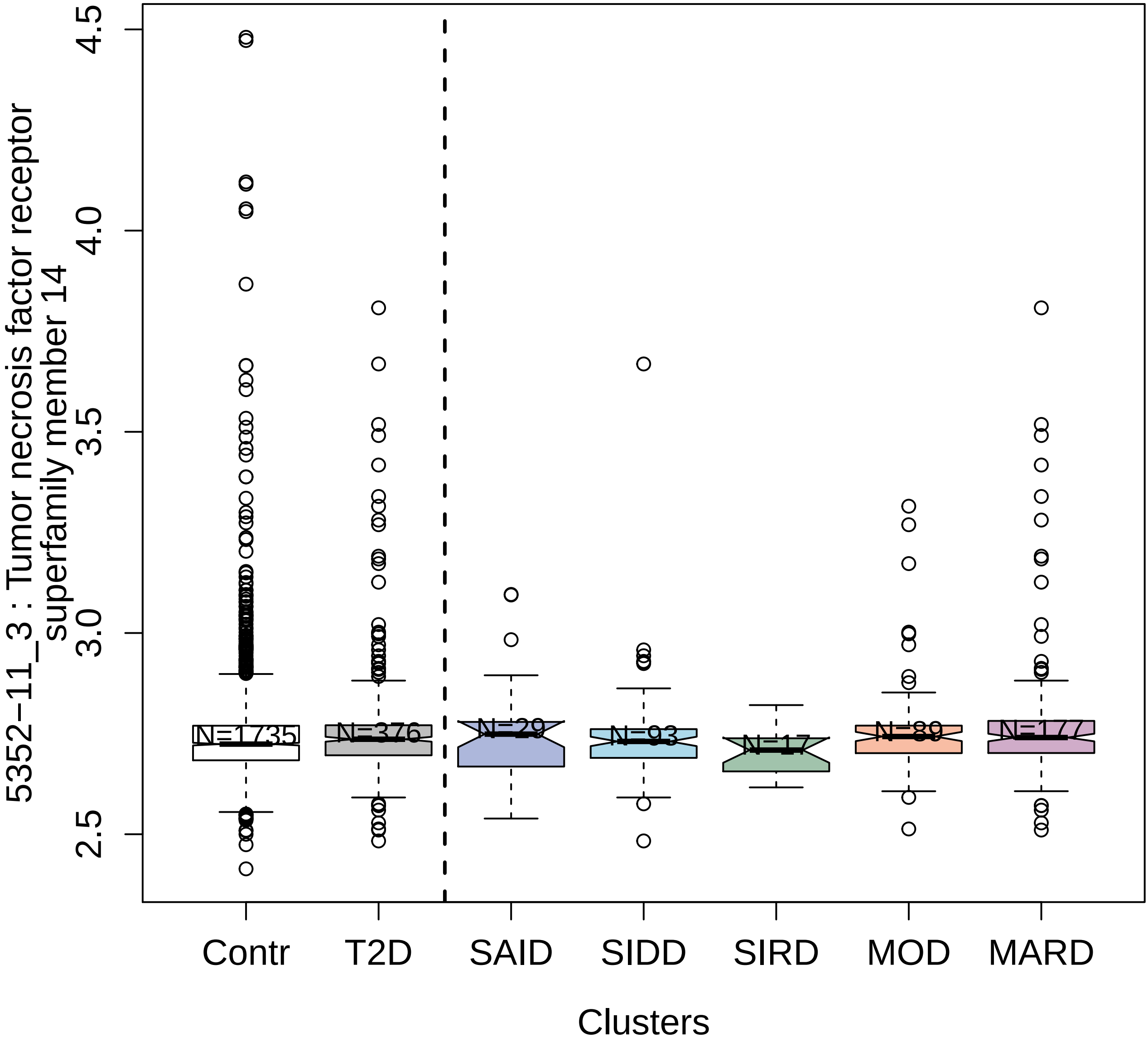

5352−11_3 : Tumor necrosis factor receptor superfamily member 14
4.5
4.0
3.5
3.0
2.5
N=1735
N=376
N=29
N=93
N=17
N=89
N=177
Contr
T2D
SAID
SIDD
SIRD
MOD
MARD
Clusters

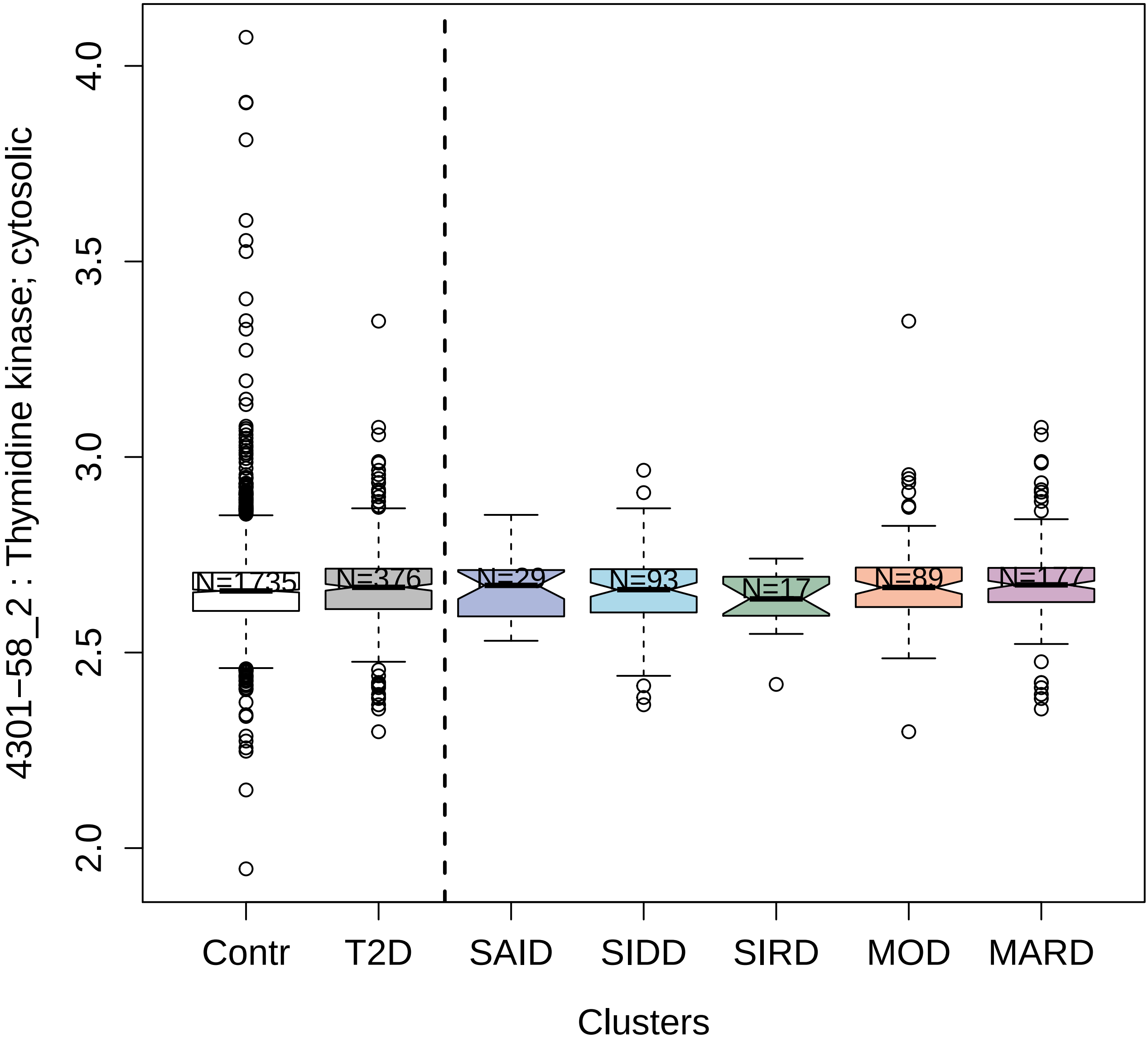

4301−58_2 : Thymidine kinase; cytosolic
2.0
2.5
3.0
3.5
4.0
N=1735
N=376
N=29
N=93
N=17
N=89
N=177
Contr
T2D
SAID
SIDD
SIRD
MOD
MARD
Clusters

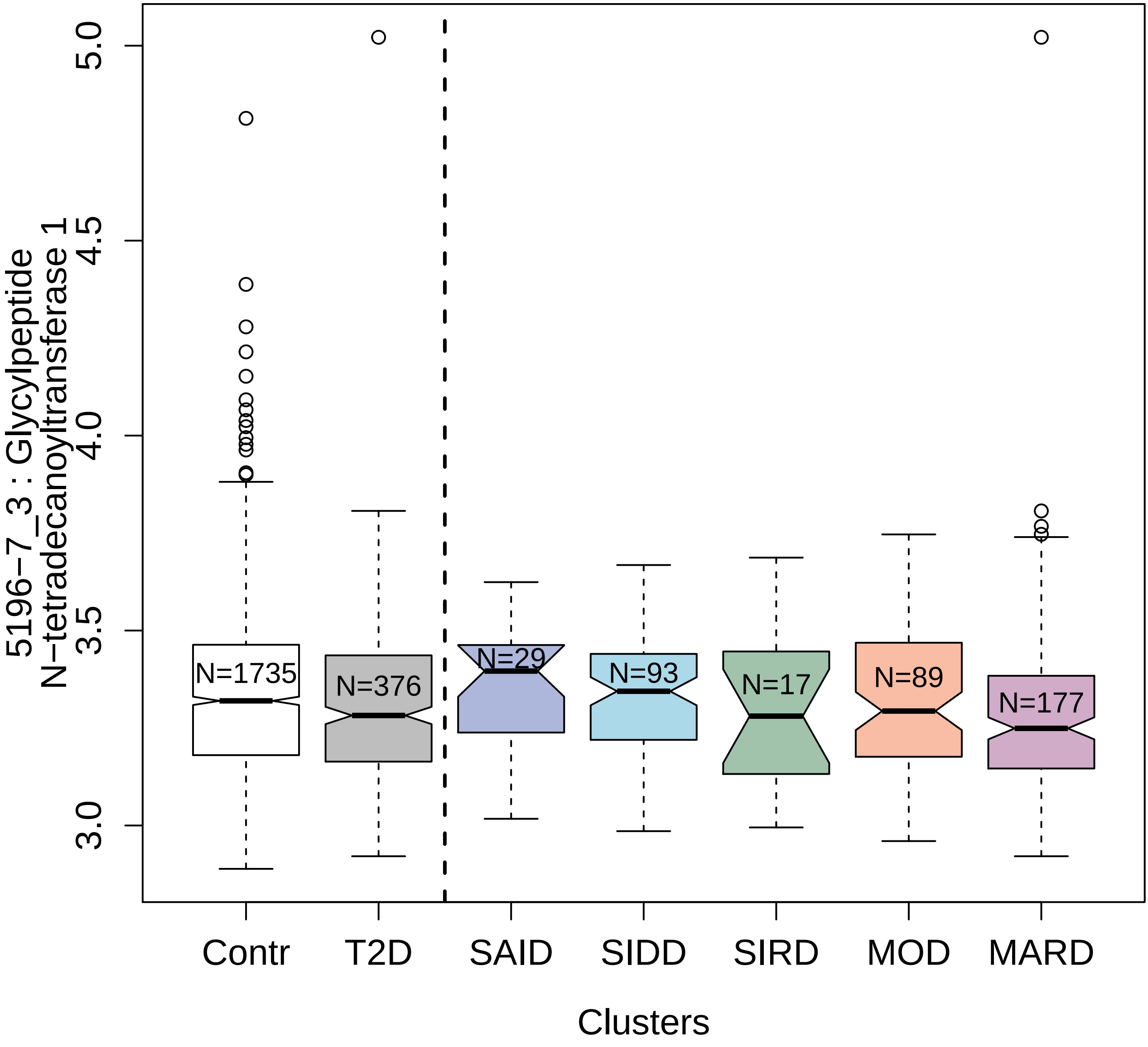

5196−7_3 : Glycylpeptide
N−tetradecanoyltransferase 1
5.0
4.5
4.0
3.5
3.0
N=1735
N=376
N=29
N=93
N=17
N=89
N=177
Contr
T2D
SAID
SIDD
SIRD
MOD
MARD
Clusters

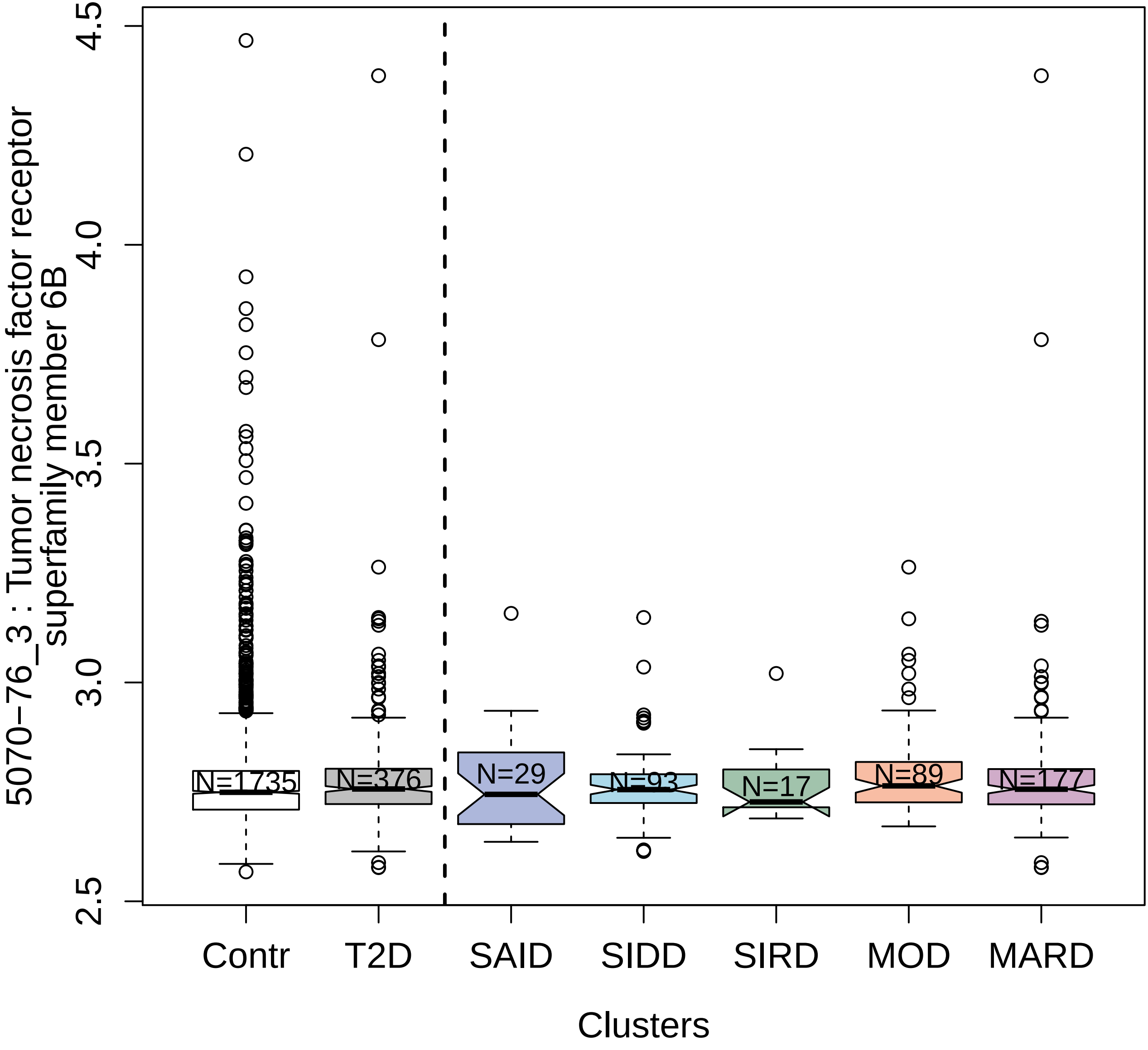

5070−76_3 : Tumor necrosis factor receptor superfamily member 6B
2.5
3.0
3.5
4.0
4.5
N=1735
N=376
N=29
N=93
N=17
N=89
N=177
Contr
T2D
SAID
SIDD
SIRD
MOD
MARD
Clusters

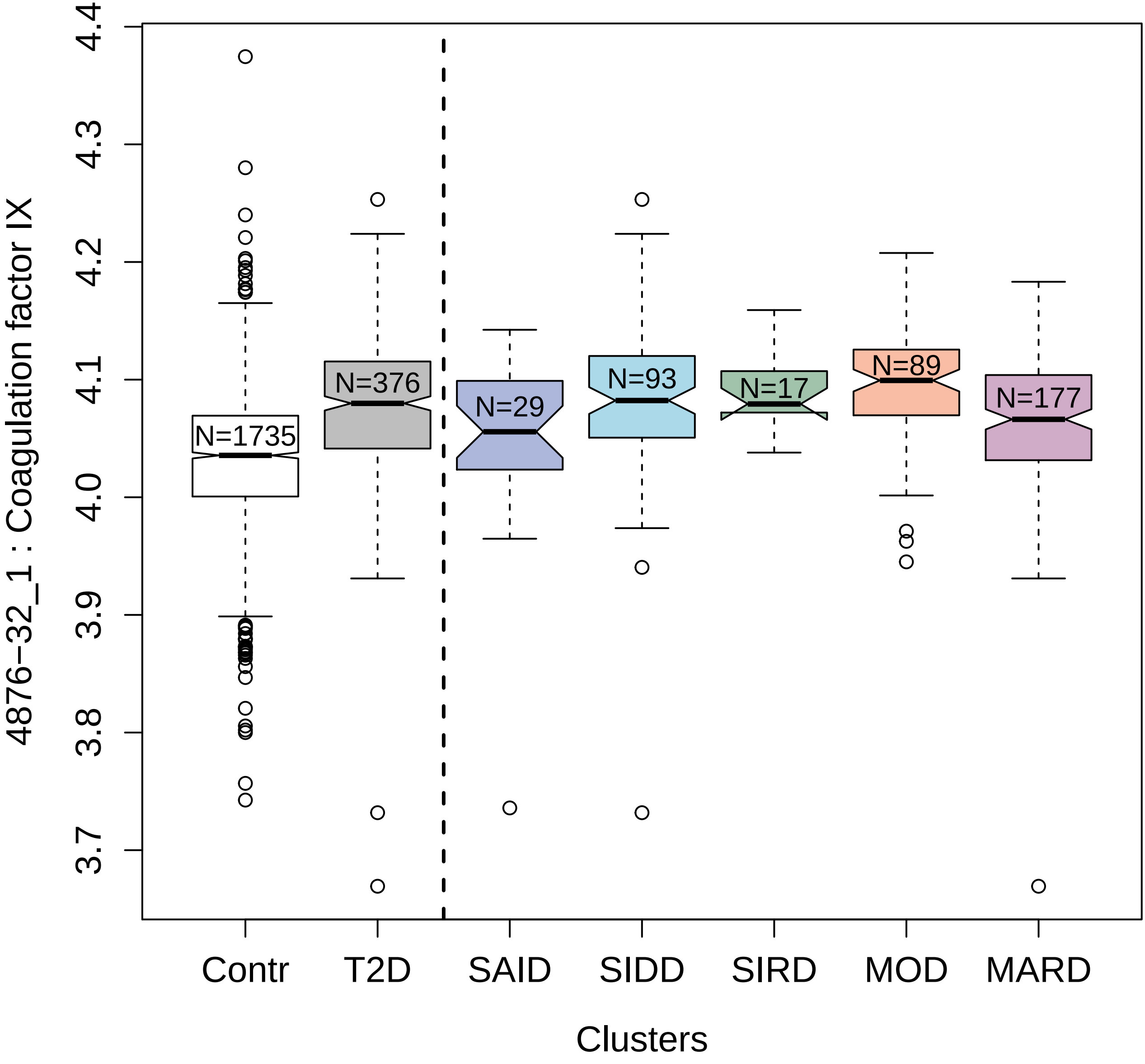
4876−32_1 : Coagulation factor IX
Clusters
Contr
T2D
SAID
SIDD
SIRD
MOD
MARD
N=1735
N=376
N=29
N=93
N=17
N=89
N=177
3.7
3.8
3.9
4.0
4.1
4.2
4.3
4.4

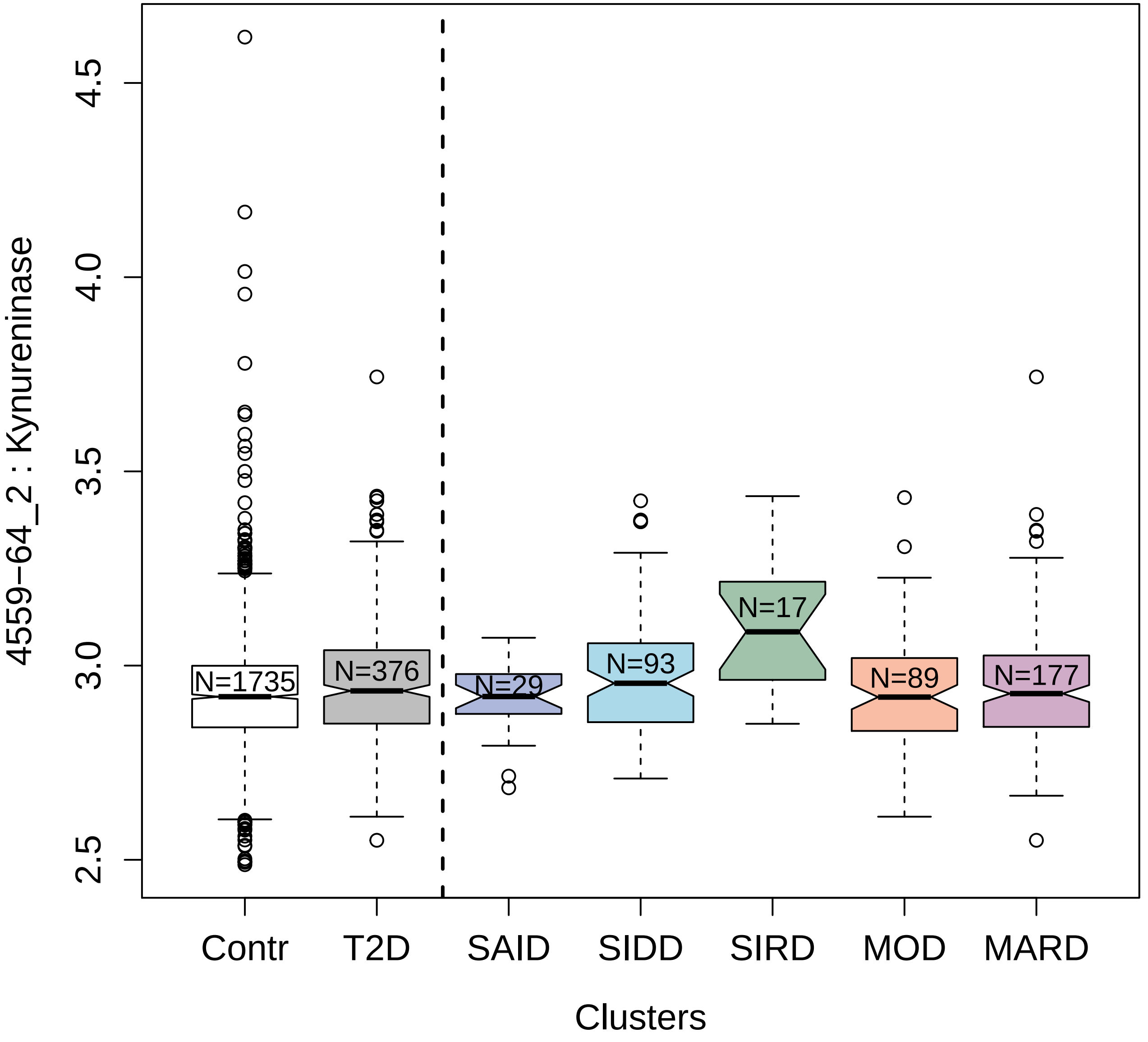

4559−64_2 : Kynureninase
2.5
3.0
3.5
4.0
4.5
N=1735
N=376
N=29
N=93
N=17
N=89
N=177
Contr
T2D
SAID
SIDD
SIRD
MOD
MARD
Clusters

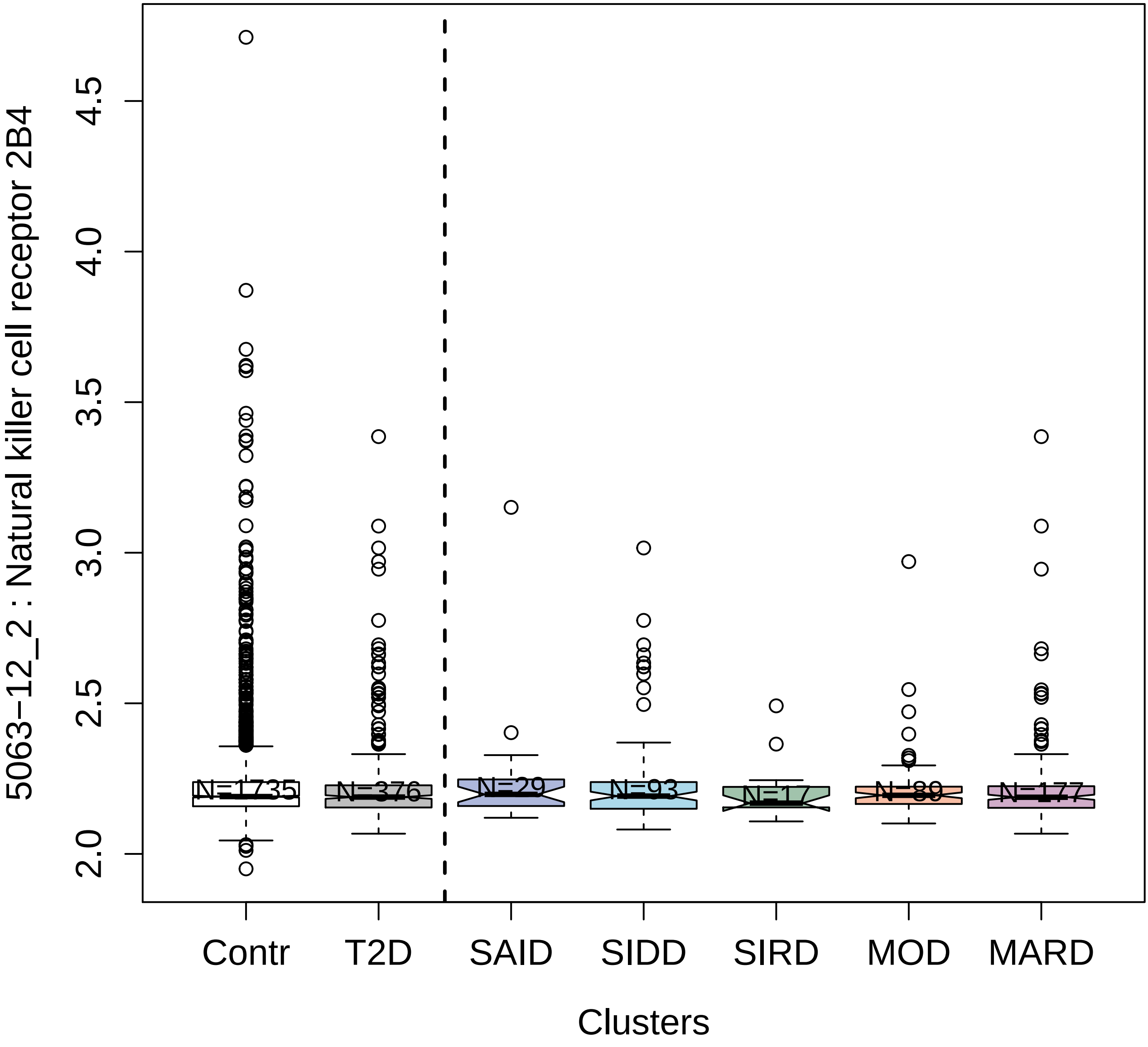
5063−12_2 : Natural killer cell receptor 2B4
Clusters
Contr
T2D
SAID
SIDD
SIRD
MOD
MARD
N=1735
N=376
N=29
N=93
N=17
N=89
N=177
2.0
2.5
3.0
3.5
4.0
4.5

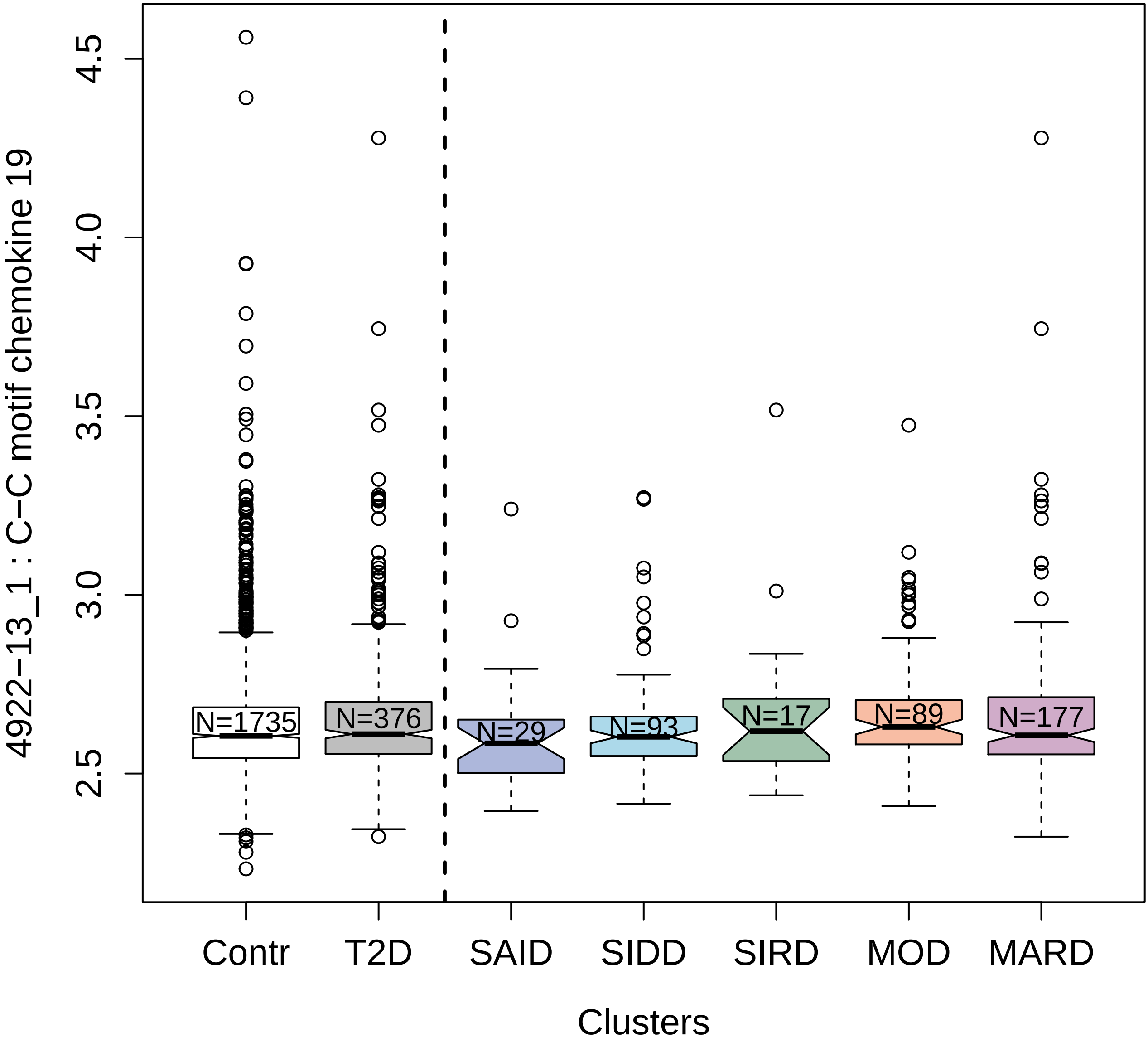

4922−13_1 : C−C motif chemokine 19
4.5
4.0
3.5
3.0
2.5
N=1735
N=376
N=29
N=93
N=17
N=89
N=177
Contr
T2D
SAID
SIDD
SIRD
MOD
MARD
Clusters

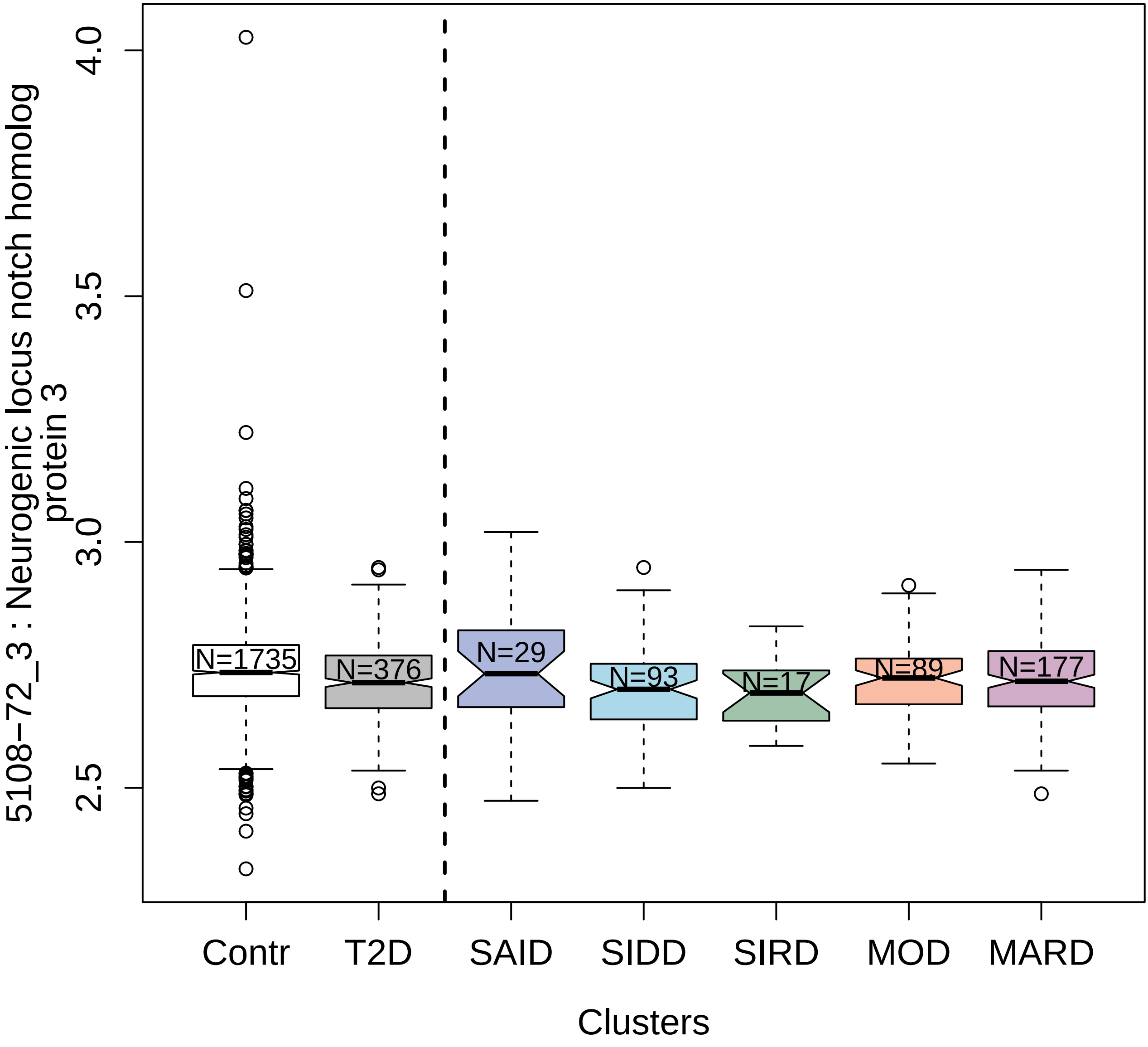
5108−72_3 : Neurogenic locus notch homolog protein 3
4.0
3.5
3.0
2.5
N=1735
N=376
N=29
N=93
N=17
N=89
N=177
Contr
T2D
SAID
SIDD
SIRD
MOD
MARD
Clusters

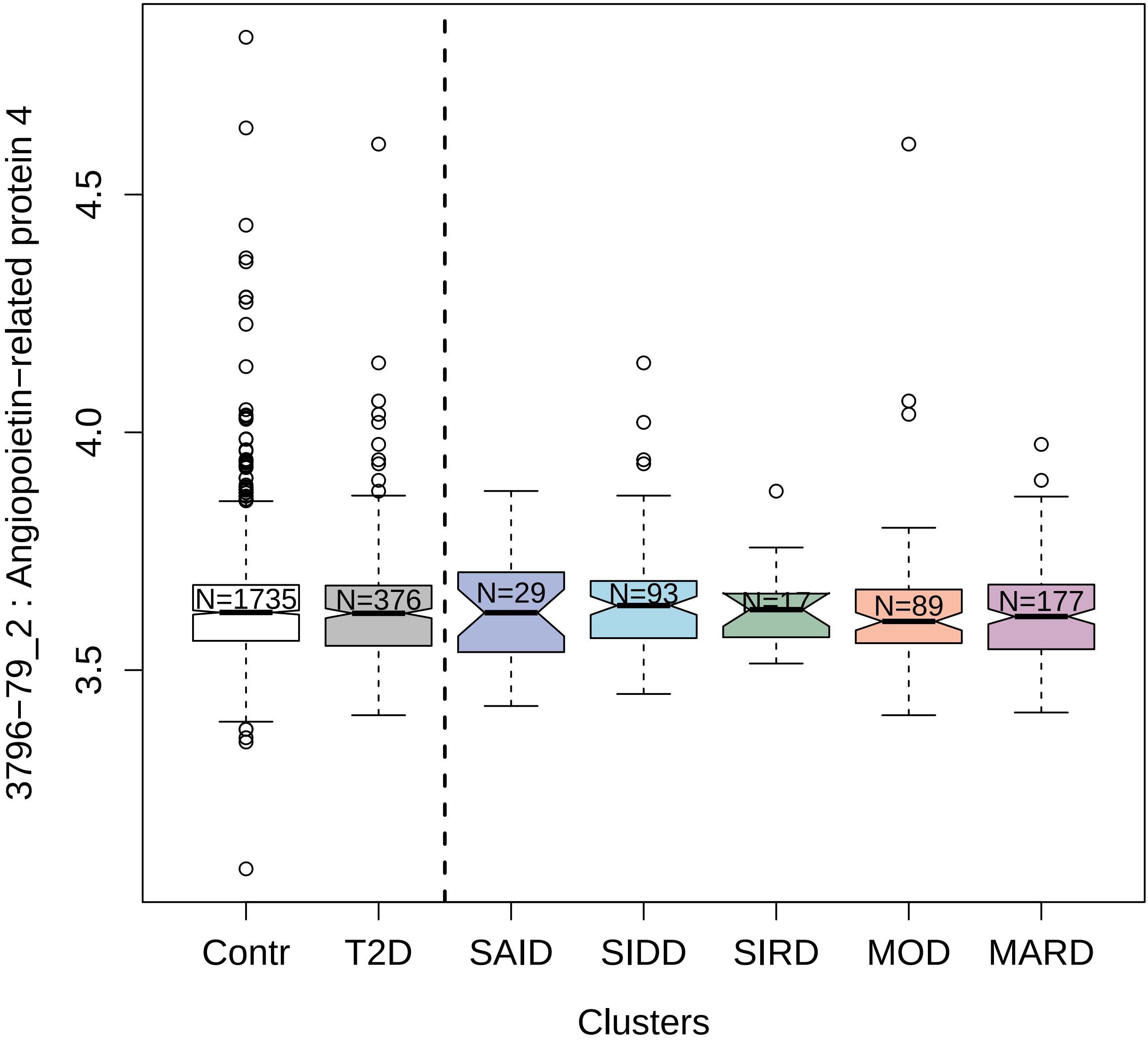

3796−79_2 : Angiopoietin−related protein 4
4.5
4.0
3.5
N=1735
N=376
N=29
N=93
N=17
N=89
N=177
Contr
T2D
SAID
SIDD
SIRD
MOD
MARD
Clusters

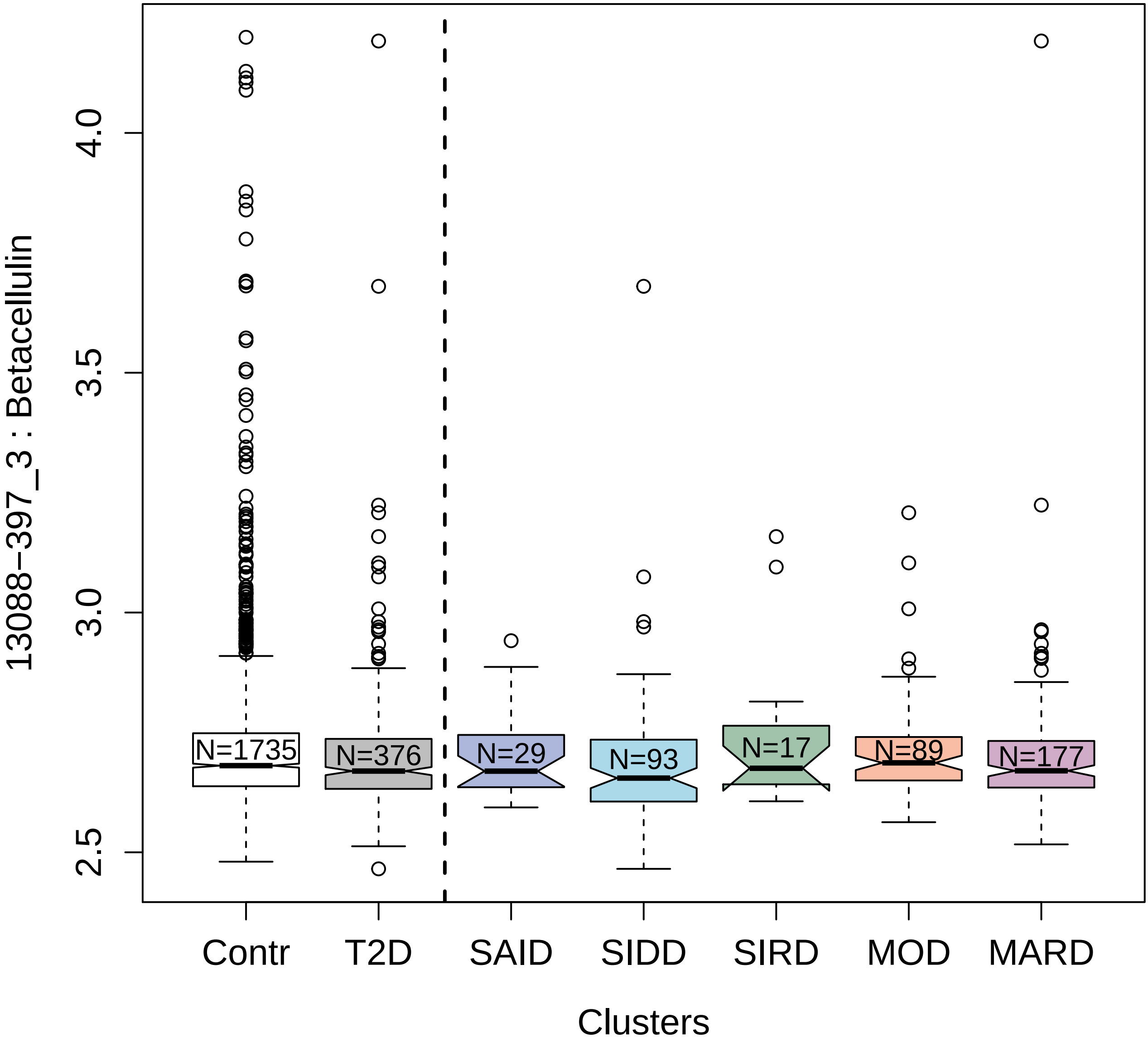

13088−397_3 : Betacellulin
2.5
3.0
3.5
4.0
N=1735
N=376
N=29
N=93
N=17
N=89
N=177
Contr
T2D
SAID
SIDD
SIRD
MOD
MARD
Clusters

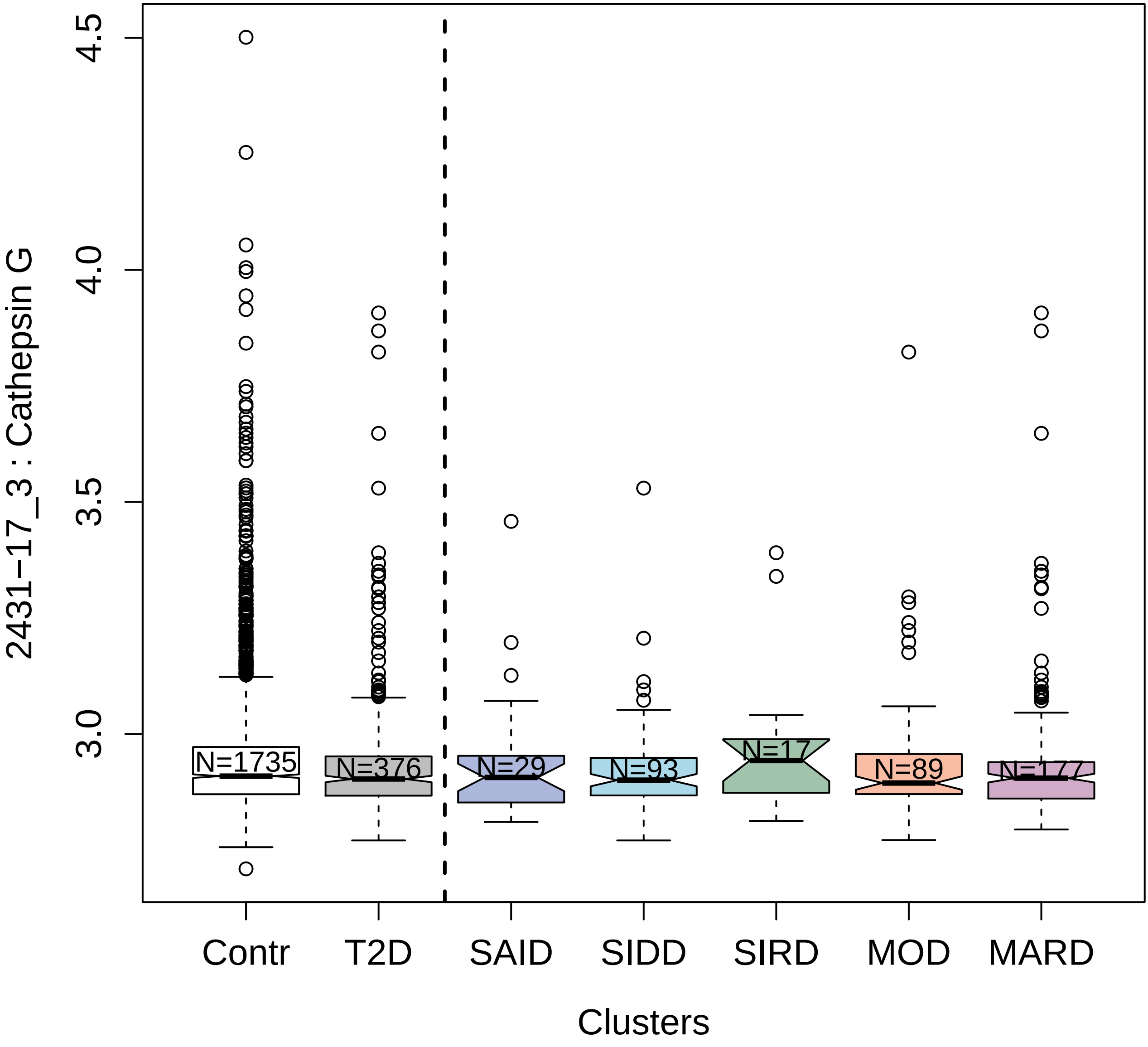
2431−17_3 : Cathepsin G
4.5
4.0
3.5
3.0
N=1735
N=376
N=29
N=93
N=17
N=89
N=177
Contr
T2D
SAID
SIDD
SIRD
MOD
MARD
Clusters

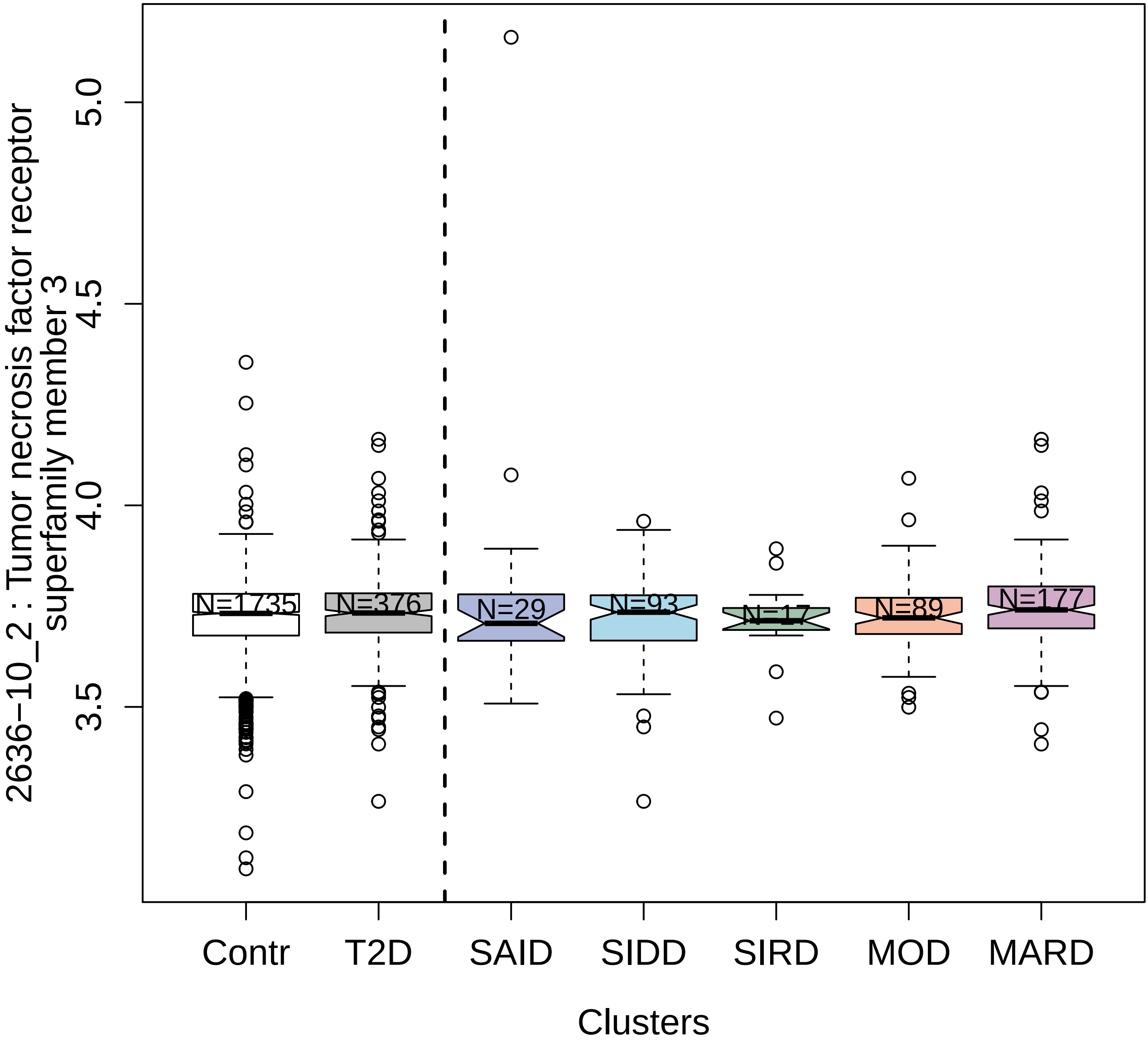
2636−10_2 : Tumor necrosis factor receptor superfamily member 3
5.0
4.5
4.0
3.5
N=1735
N=376
N=29
N=93
N=17
N=89
N=177
Contr
T2D
SAID
SIDD
SIRD
MOD
MARD
Clusters

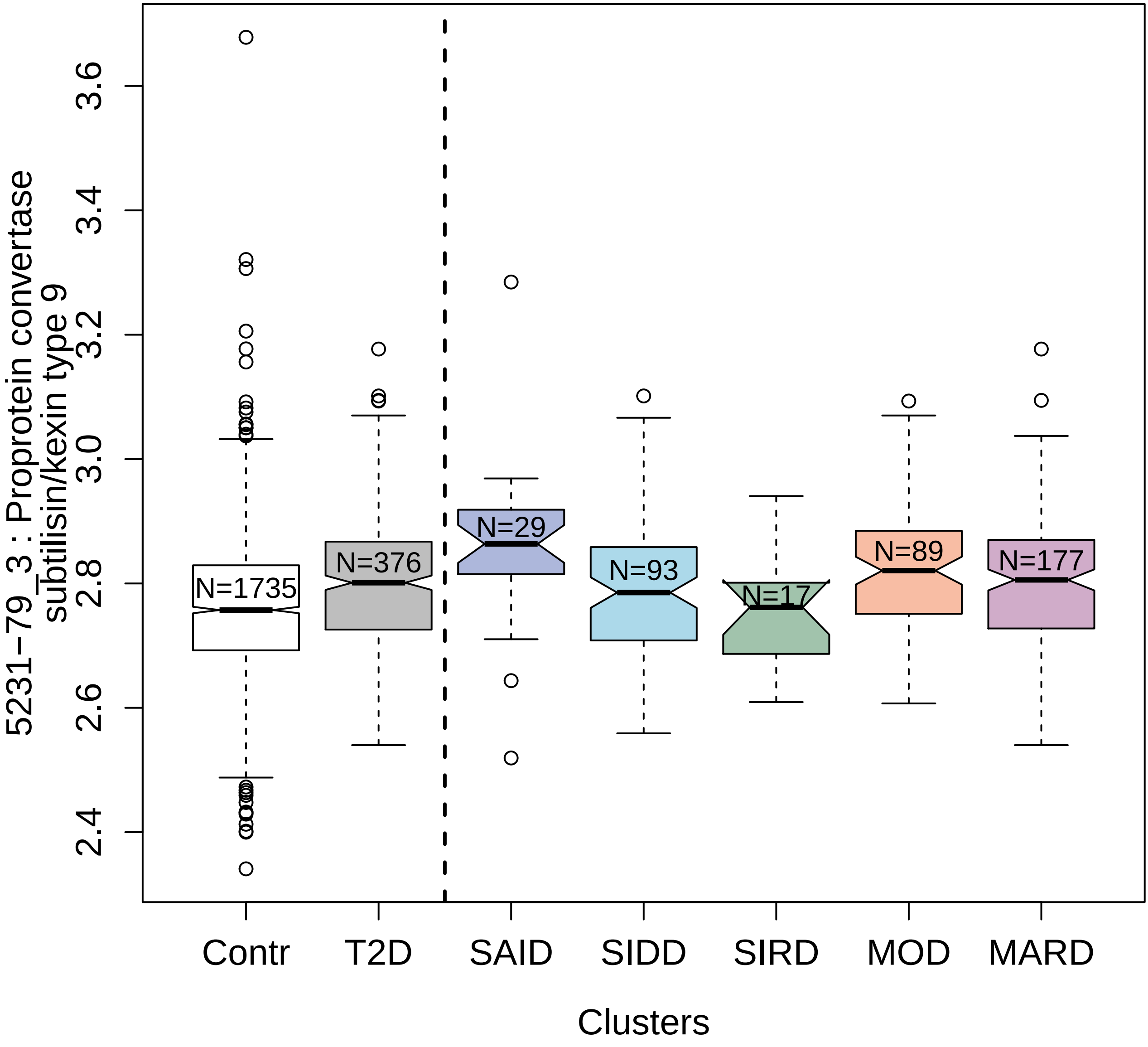

5231−79_3 : Proprotein convertase subtilisin/kexin type 9
3.6
3.4
3.2
3.0
2.8
2.6
2.4
N=1735
N=376
N=29
N=93
N=17
N=89
N=177
Contr
T2D
SAID
SIDD
SIRD
MOD
MARD
Clusters

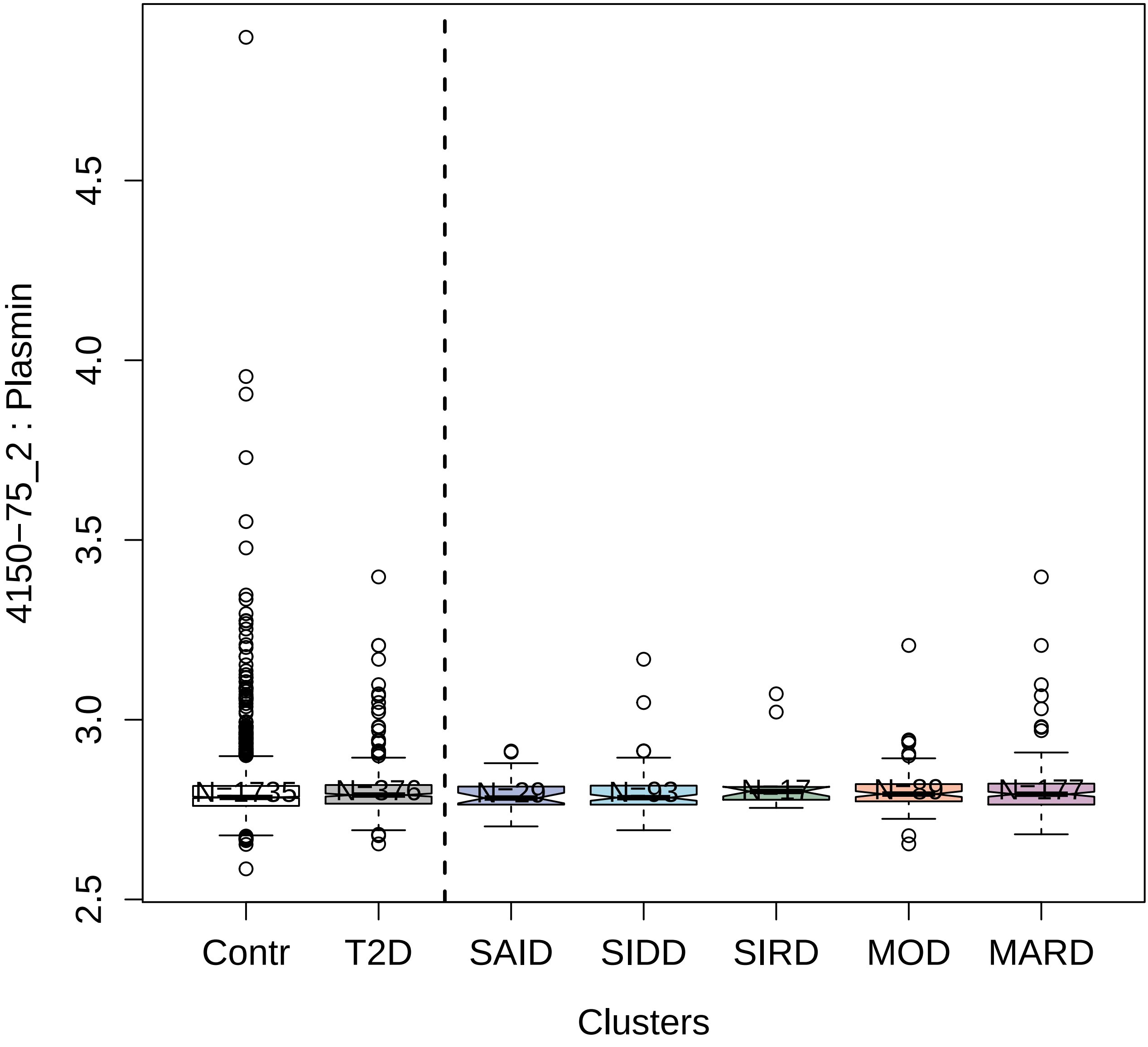

4150−75_2 : Plasmin
2.5
3.0
3.5
4.0
4.5
N=1735
N=376
N=29
N=93
N=17
N=89
N=177
Contr
T2D
SAID
SIDD
SIRD
MOD
MARD
Clusters

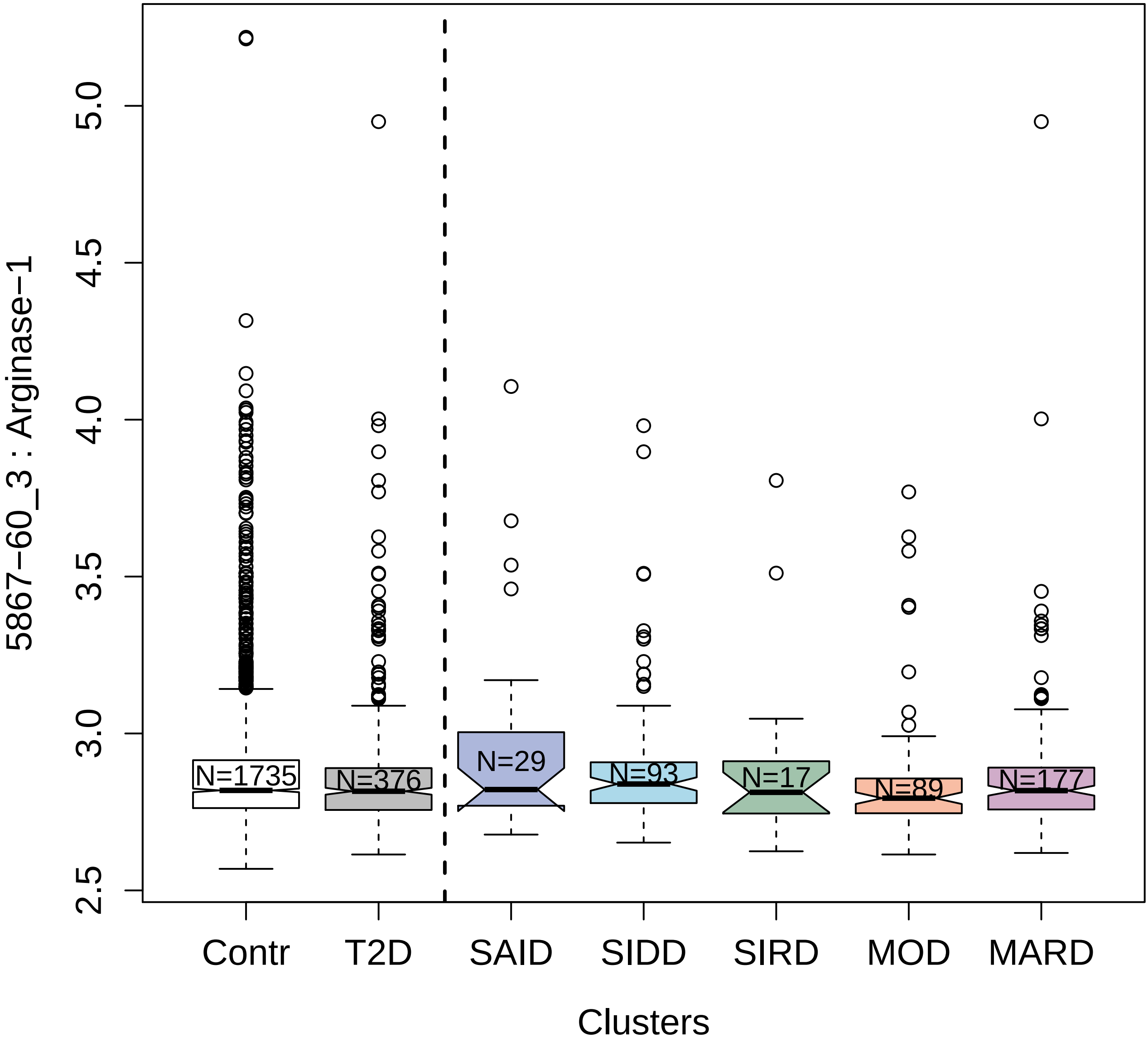
5867−60_3 : Arginase−1
5.0
4.5
4.0
3.5
3.0
2.5
N=1735
N=376
N=29
N=93
N=17
N=89
N=177
Contr
T2D
SAID
SIDD
SIRD
MOD
MARD
Clusters

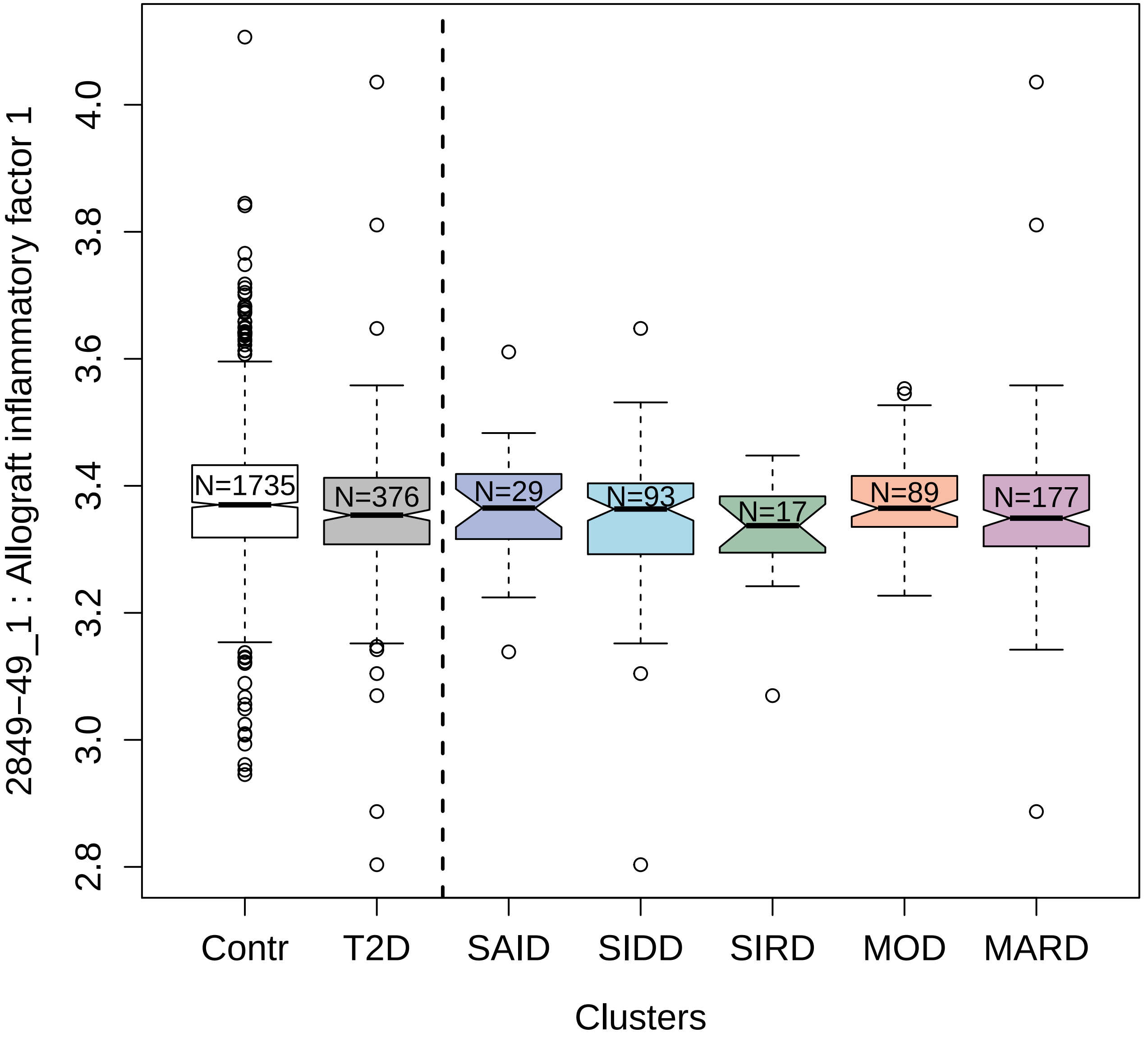
2849−49_1 : Allograft inflammatory factor 1
4.0
3.8
3.6
3.4
3.2
3.0
2.8
N=1735
N=376
N=29
N=93
N=17
N=89
N=177
Contr
T2D
SAID
SIDD
SIRD
MOD
MARD
Clusters

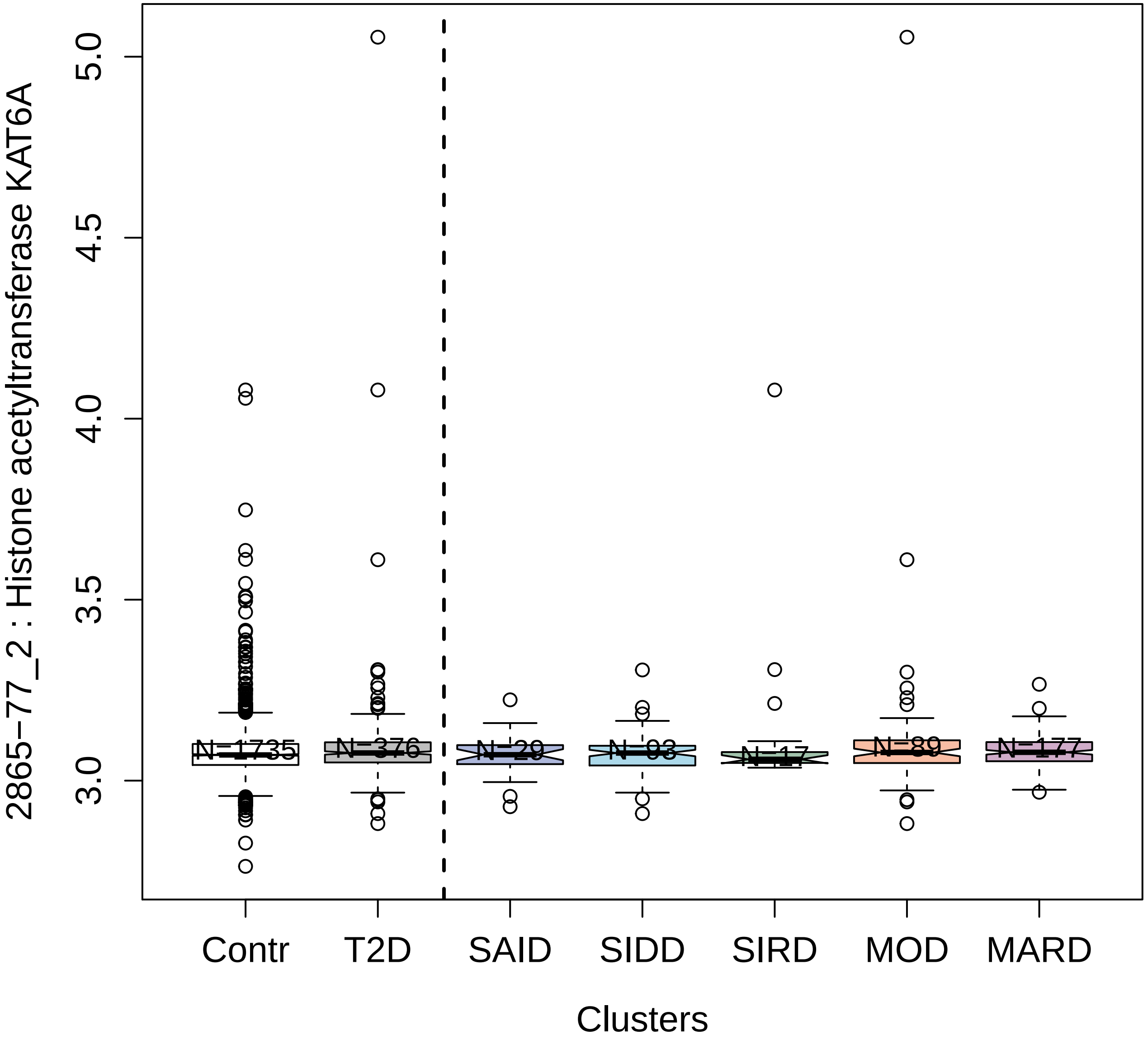

2865−77_2 : Histone acetyltransferase KAT6A
5.0
4.5
4.0
3.5
3.0
N=1735
N=376
N=29
N=93
N=17
N=89
N=177
Contr
T2D
SAID
SIDD
SIRD
MOD
MARD
Clusters

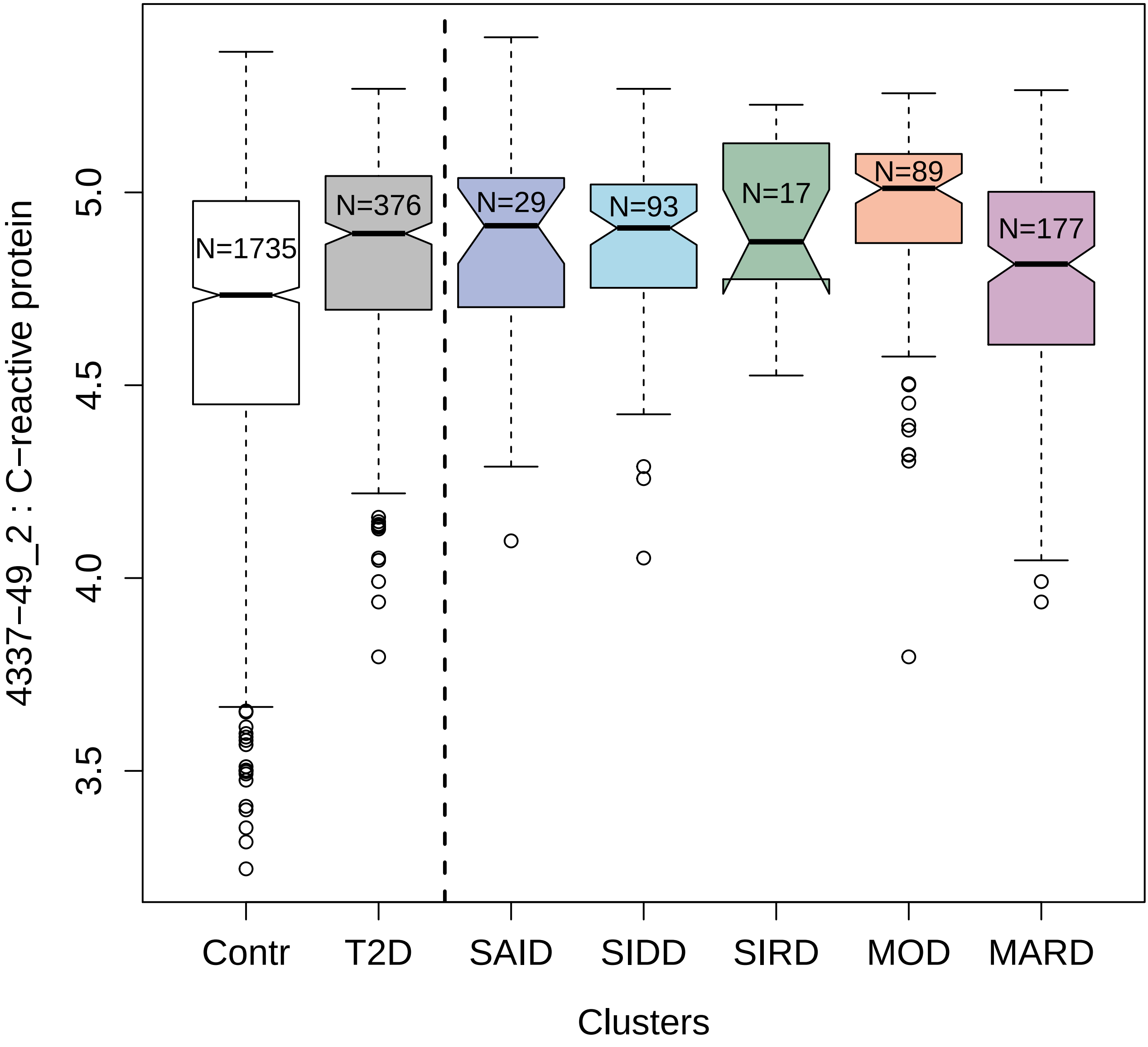
4337−49_2 : C−reactive protein
5.0
4.5
4.0
3.5
N=1735
N=376
N=29
N=93
N=17
N=89
N=177
Contr
T2D
SAID
SIDD
SIRD
MOD
MARD
Clusters

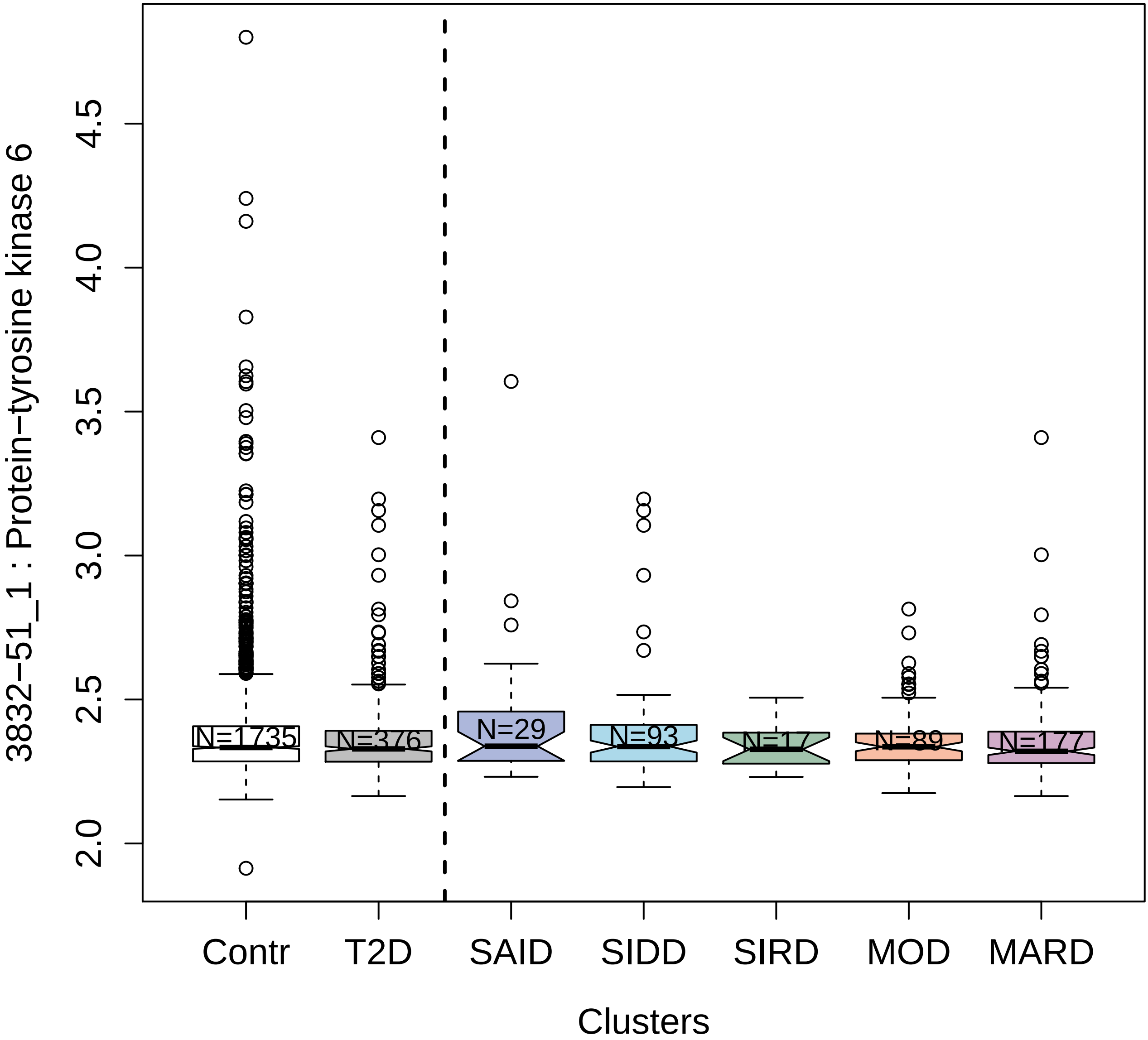
3832−51_1 : Protein−tyrosine kinase 6
2.0
2.5
3.0
3.5
4.0
4.5
N=1735
N=376
N=29
N=93
N=17
N=89
N=177
Contr
T2D
SAID
SIDD
SIRD
MOD
MARD
Clusters

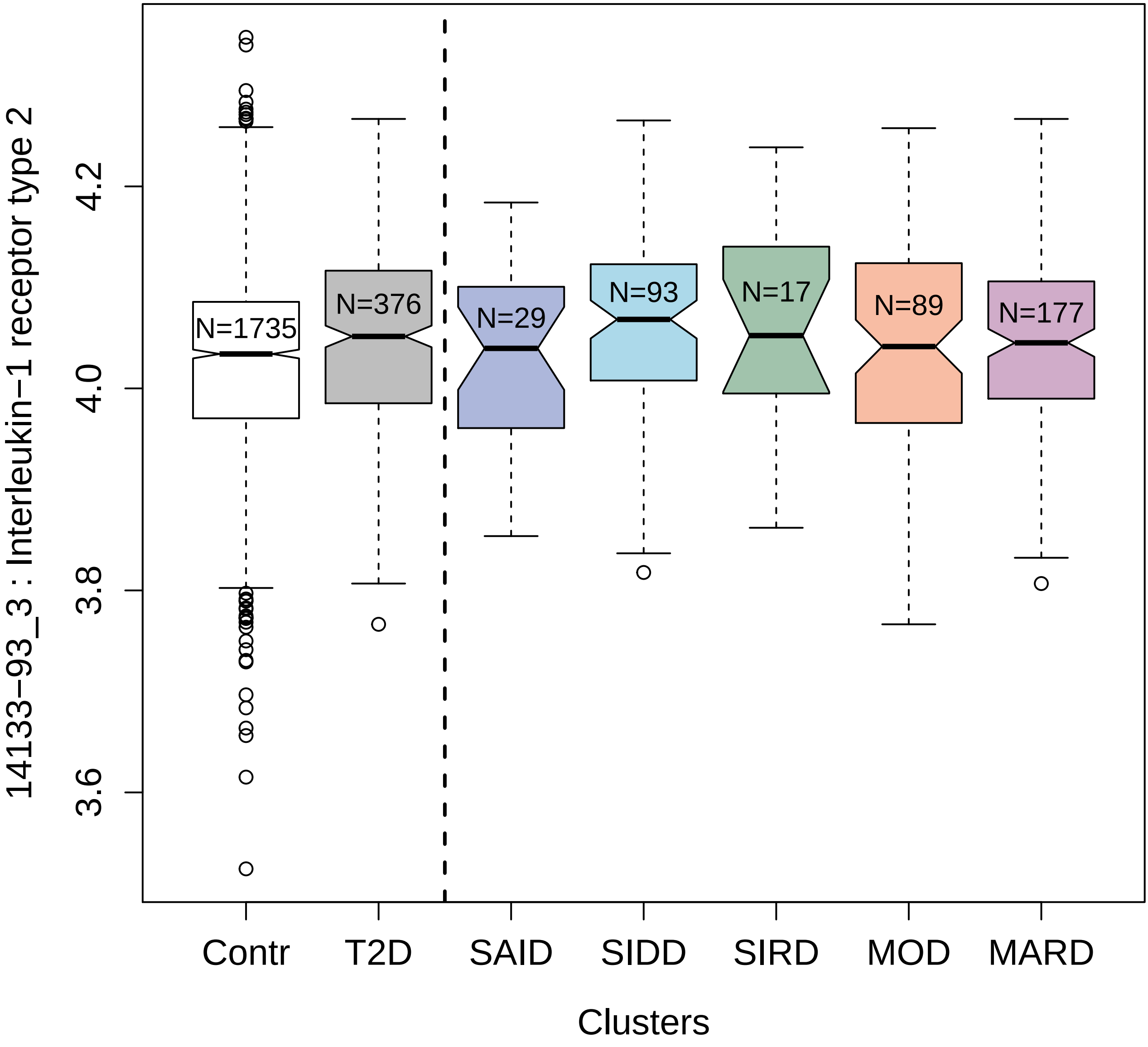
14133−93_3 : Interleukin−1 receptor type 2
4.2
4.0
3.8
3.6
N=1735
N=376
N=29
N=93
N=17
N=89
N=177
Contr
T2D
SAID
SIDD
SIRD
MOD
MARD
Clusters

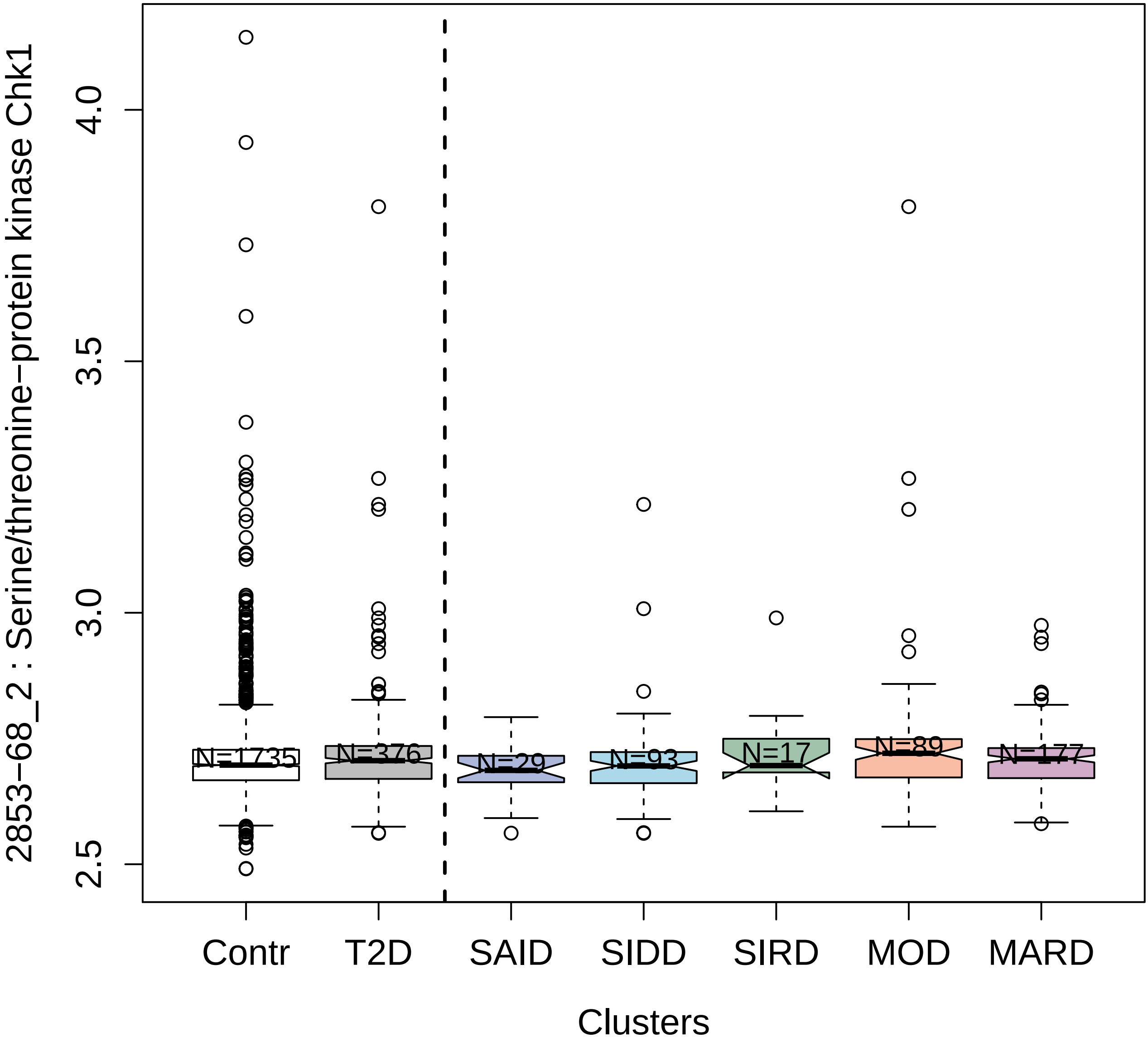
2853−68_2 : Serine/threonine−protein kinase Chk1
2.5
3.0
3.5
4.0
N=1735
N=376
N=29
N=93
N=17
N=89
N=177
Contr
T2D
SAID
SIDD
SIRD
MOD
MARD
Clusters

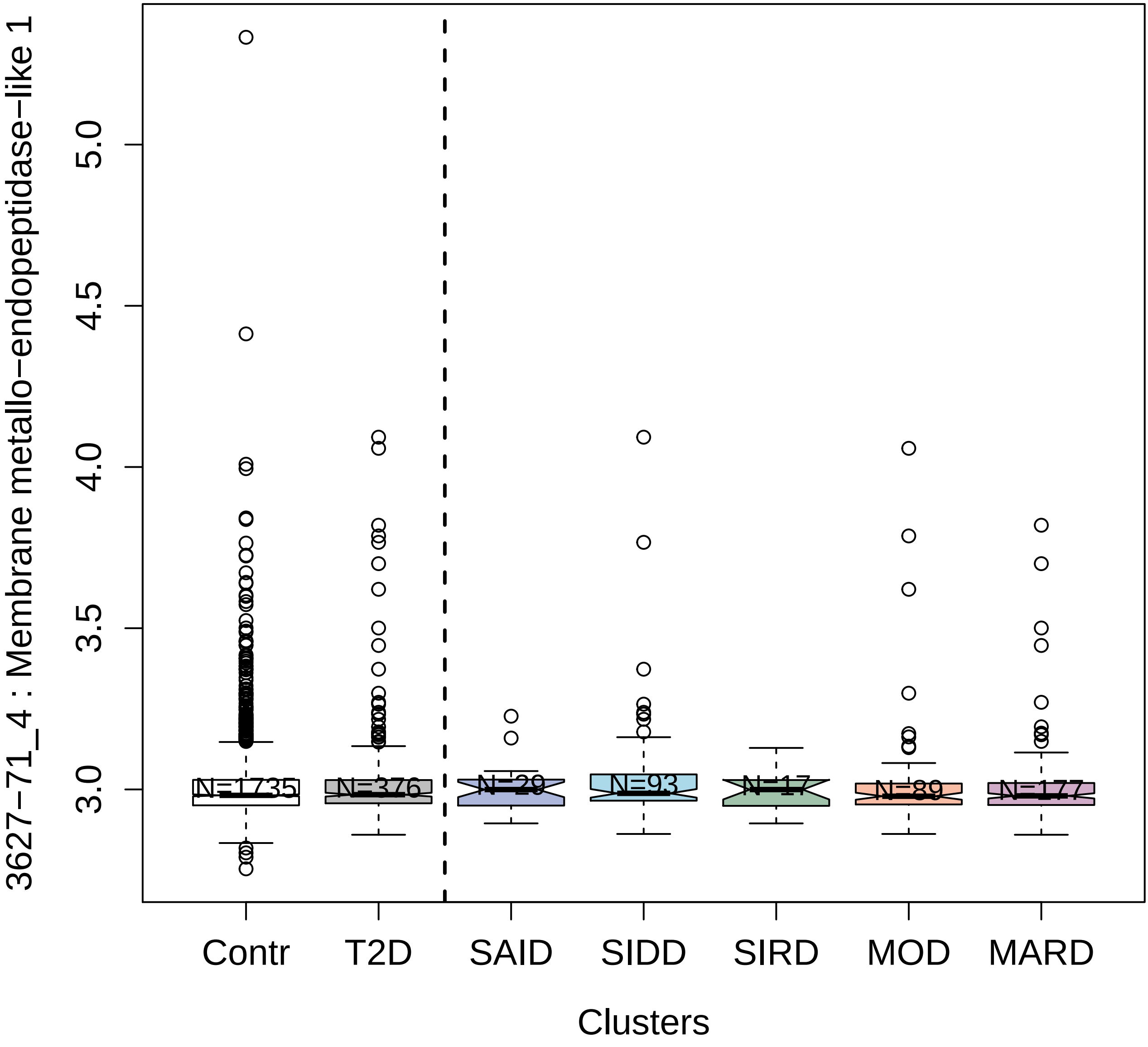

3627−71_4 : Membrane metallo−endopeptidase−like 1
3.0
3.5
4.0
4.5
5.0
N=1735
N=376
N=29
N=93
N=17
N=89
N=177
Contr
T2D
SAID
SIDD
SIRD
MOD
MARD
Clusters

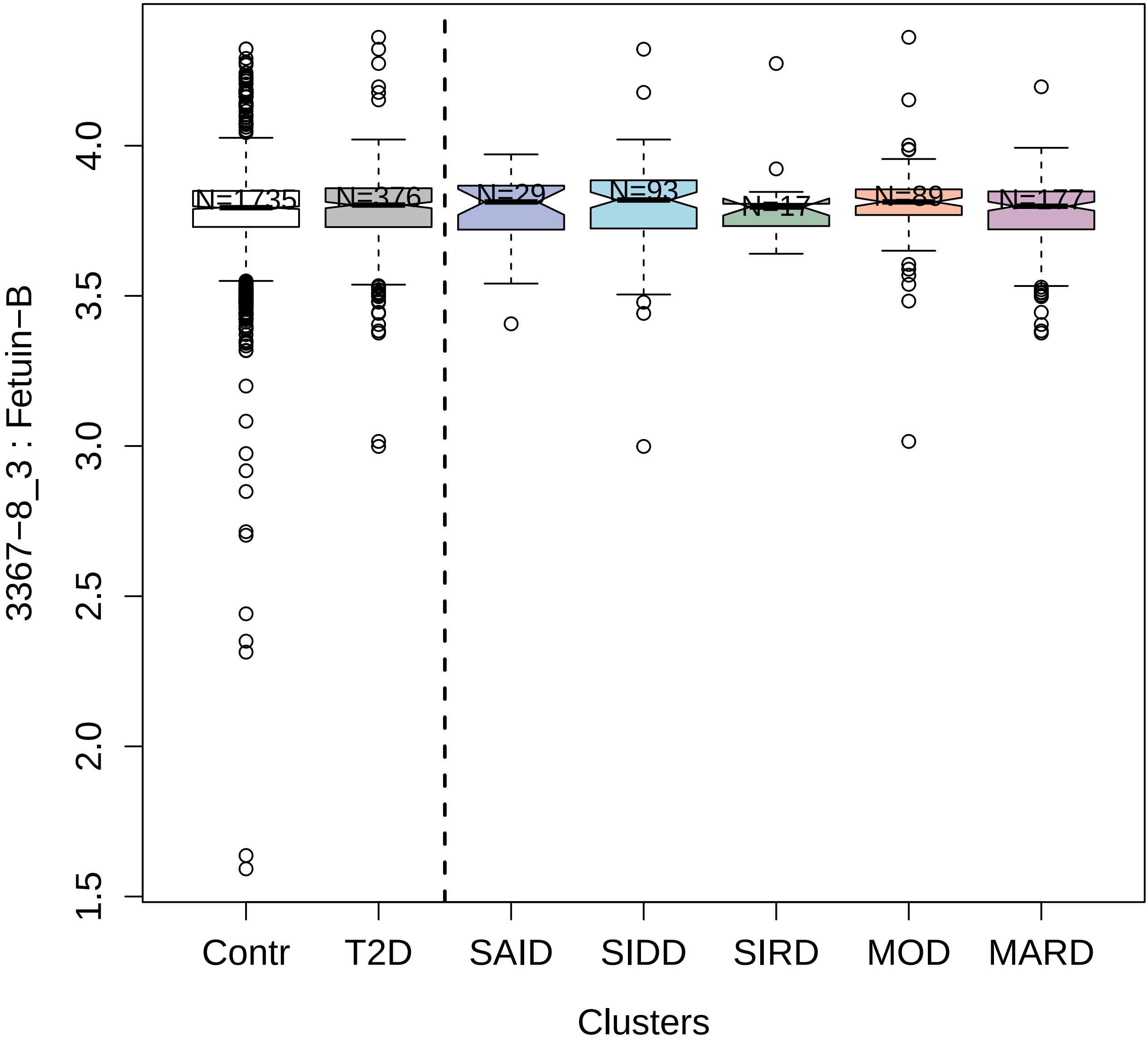

3367−8_3 : Fetuin−B
1.5
2.0
2.5
3.0
3.5
4.0
N=1735
N=376
N=29
N=93
N=17
N=89
N=177
Contr
T2D
SAID
SIDD
SIRD
MOD
MARD
Clusters

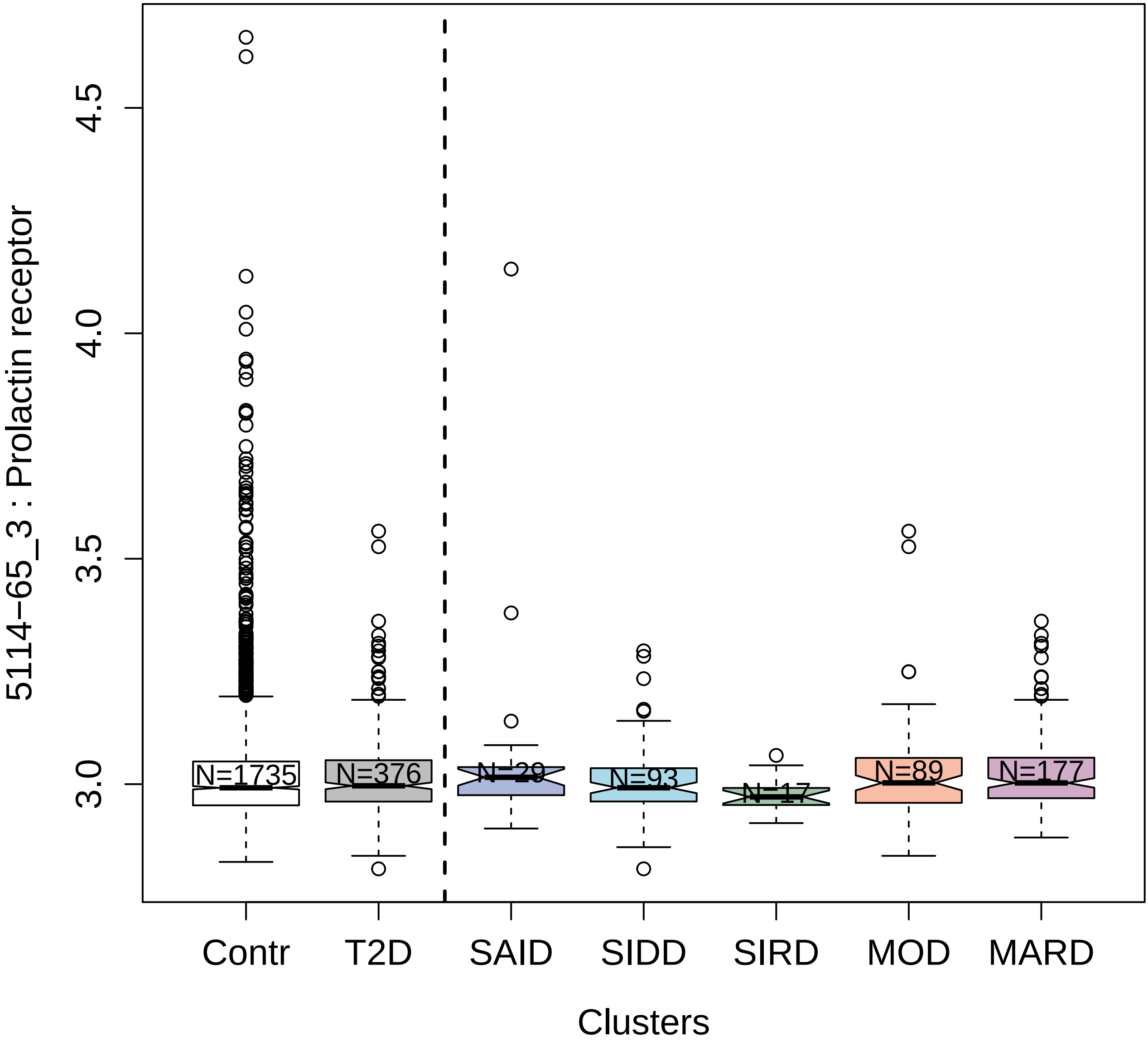
5114−65_3 : Prolactin receptor
4.5
4.0
3.5
3.0
N=1735
N=376
N=29
N=93
N=17
N=89
N=177
Contr
T2D
SAID
SIDD
SIRD
MOD
MARD
Clusters

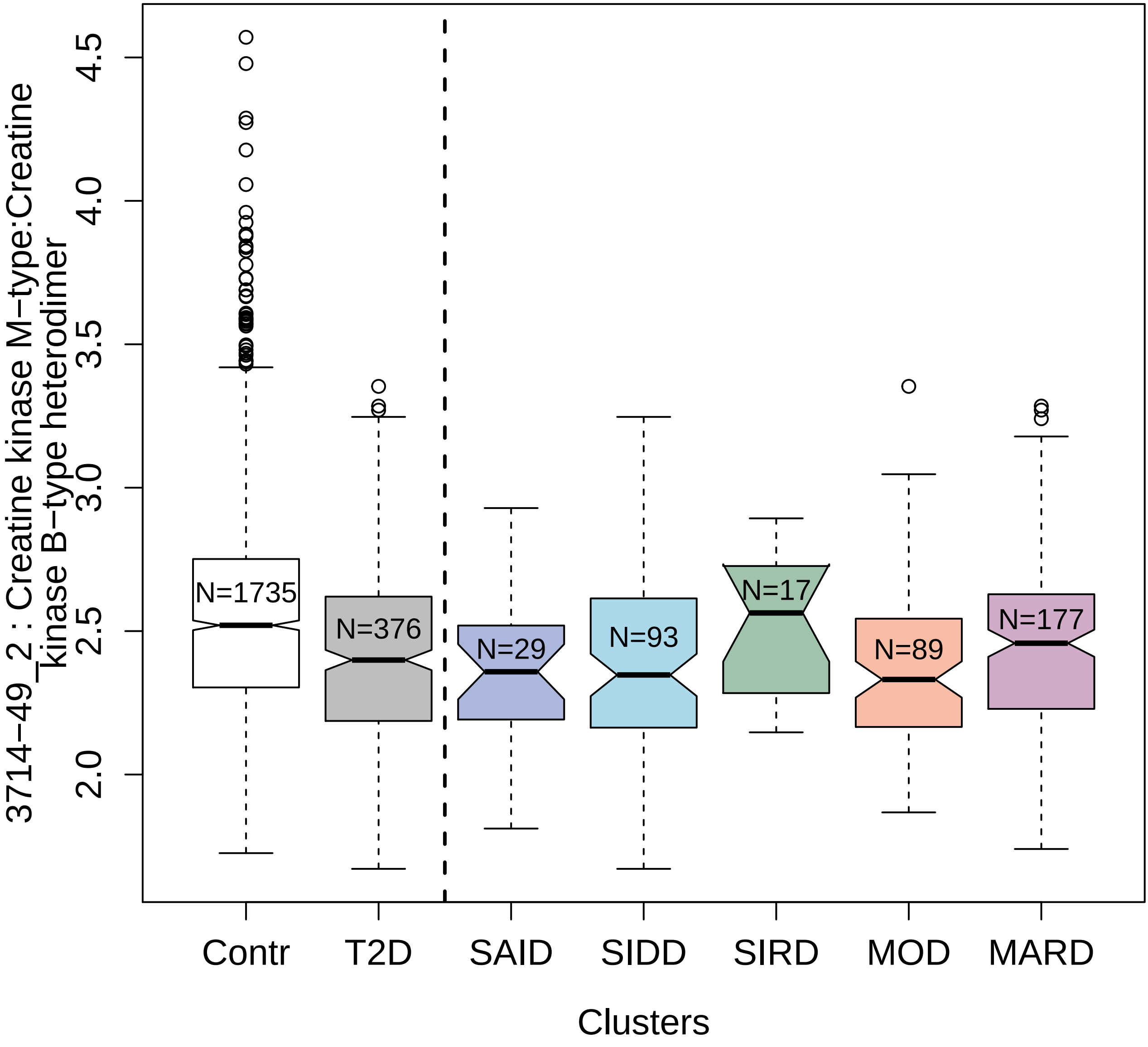

3714−49_2 : Creatine kinase M−type:Creatine kinase B−type heterodimer
4.5
4.0
3.5
3.0
2.5
2.0
N=1735
N=376
N=29
N=93
N=17
N=89
N=177
Contr
T2D
SAID
SIDD
SIRD
MOD
MARD
Clusters

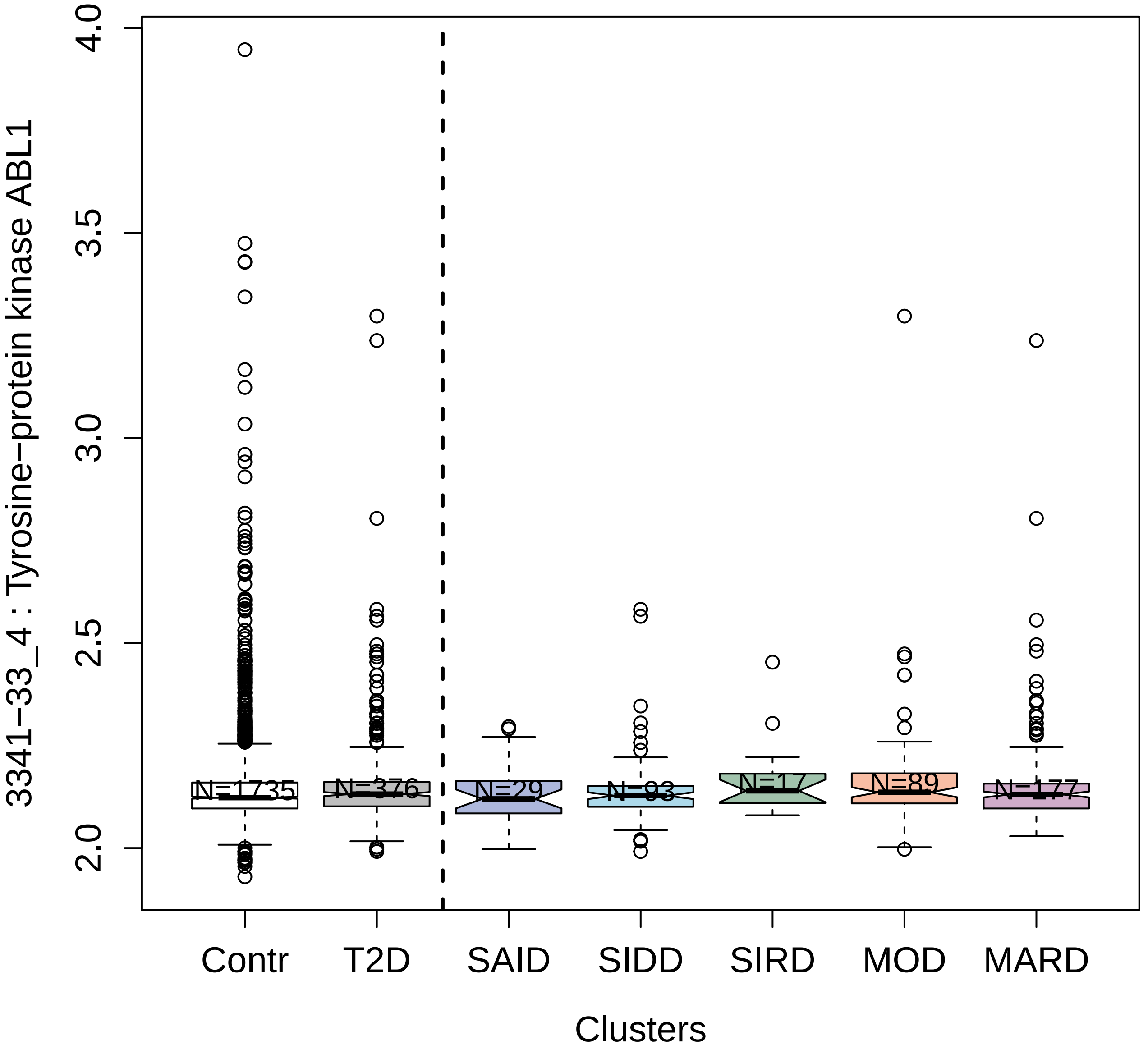

3341−33_4 : Tyrosine−protein kinase ABL1
4.0
3.5
3.0
2.5
2.0
N=1735
N=376
N=29
N=93
N=17
N=89
N=177
Contr
T2D
SAID
SIDD
SIRD
MOD
MARD
Clusters

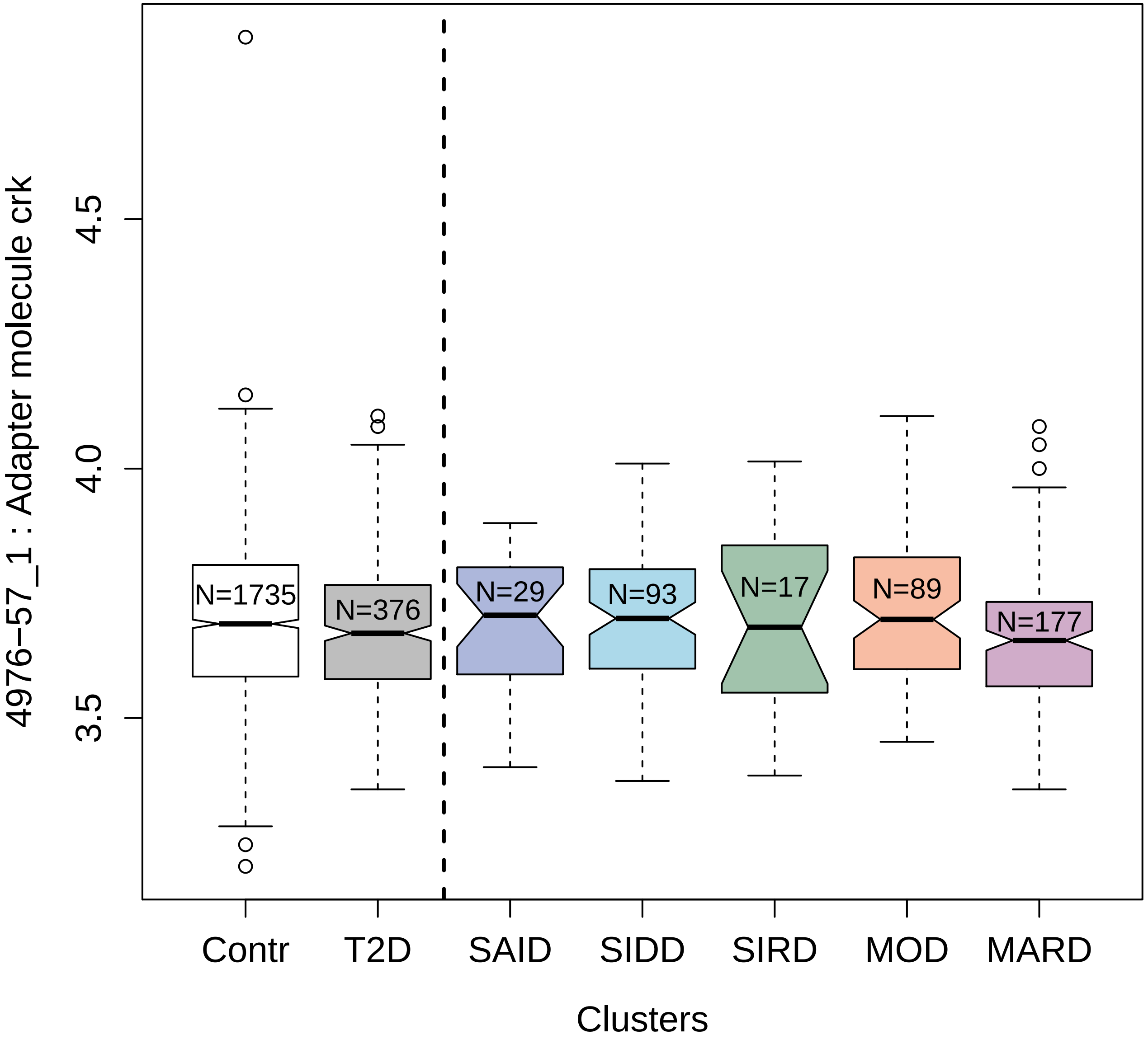

4976−57_1 : Adapter molecule crk
3.5
4.0
4.5
N=1735
N=376
N=29
N=93
N=17
N=89
N=177
Contr
T2D
SAID
SIDD
SIRD
MOD
MARD
Clusters

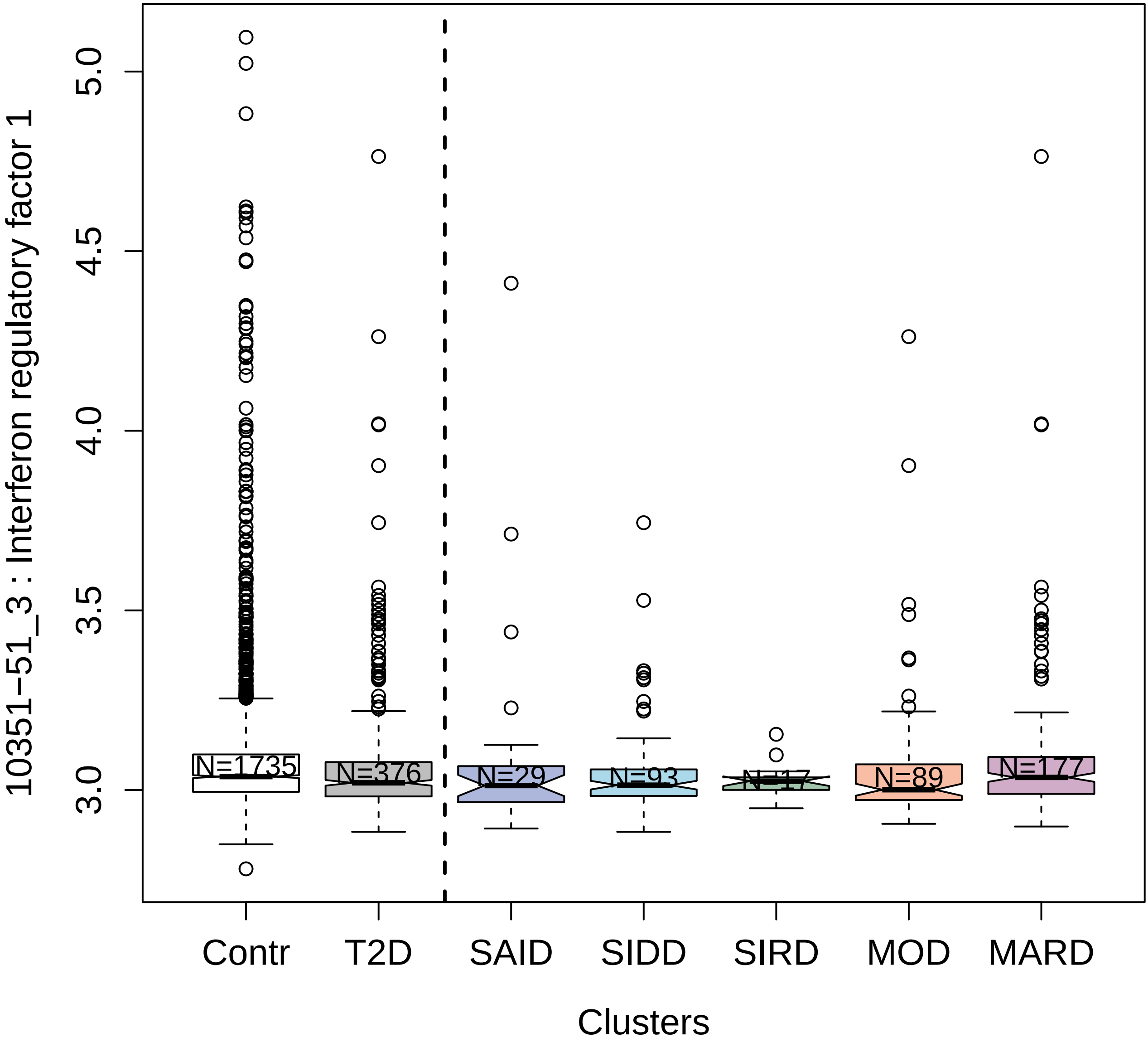

10351−51_3 : Interferon regulatory factor 1
5.0
4.5
4.0
3.5
3.0
N=1735
N=376
N=29
N=93
N=17
N=89
N=177
Contr
T2D
SAID
SIDD
SIRD
MOD
MARD
Clusters

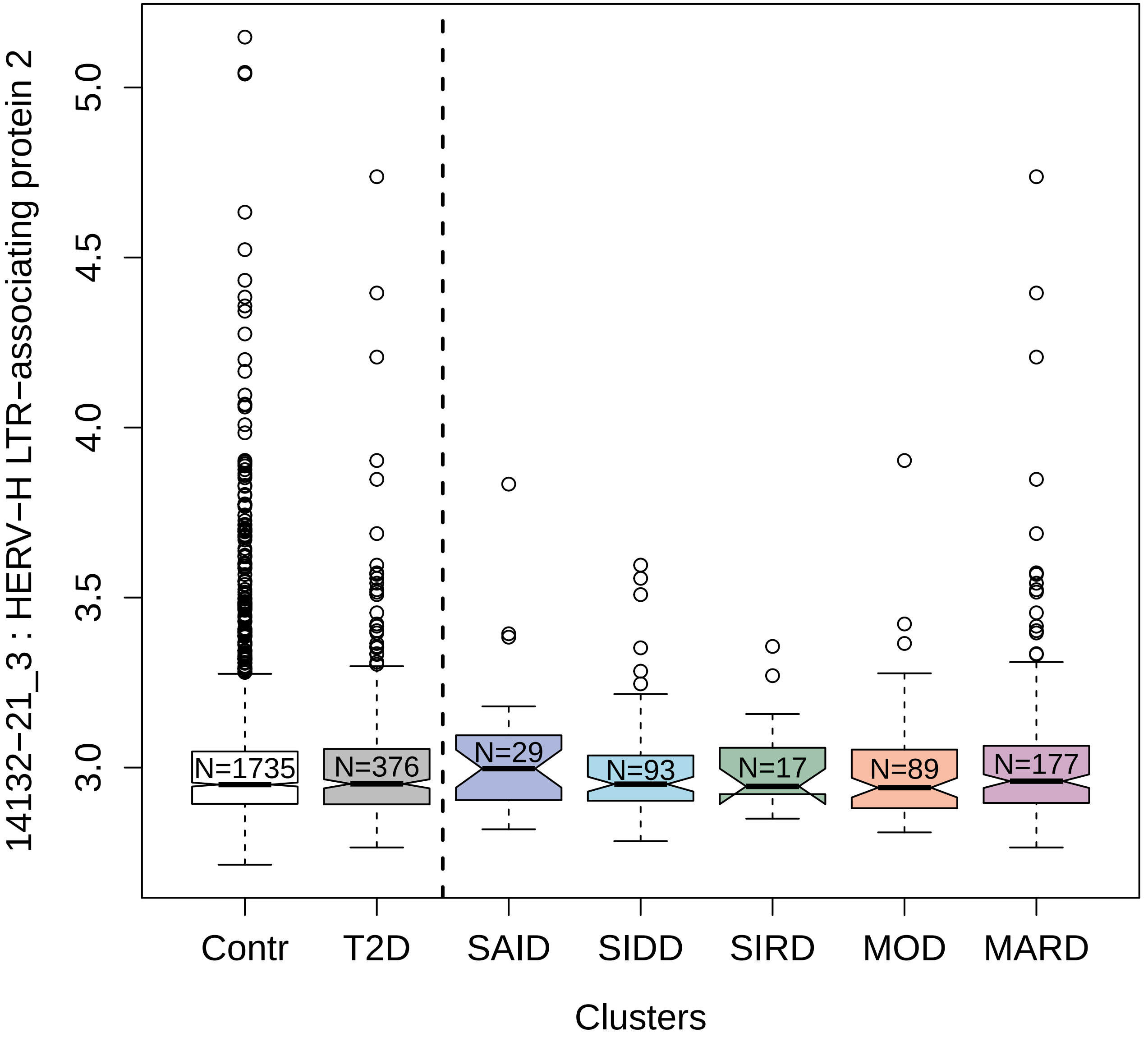

14132−21_3 : HERV−H LTR−associating protein 2
5.0
4.5
4.0
3.5
3.0
N=1735
N=376
N=29
N=93
N=17
N=89
N=177
Contr
T2D
SAID
SIDD
SIRD
MOD
MARD
Clusters

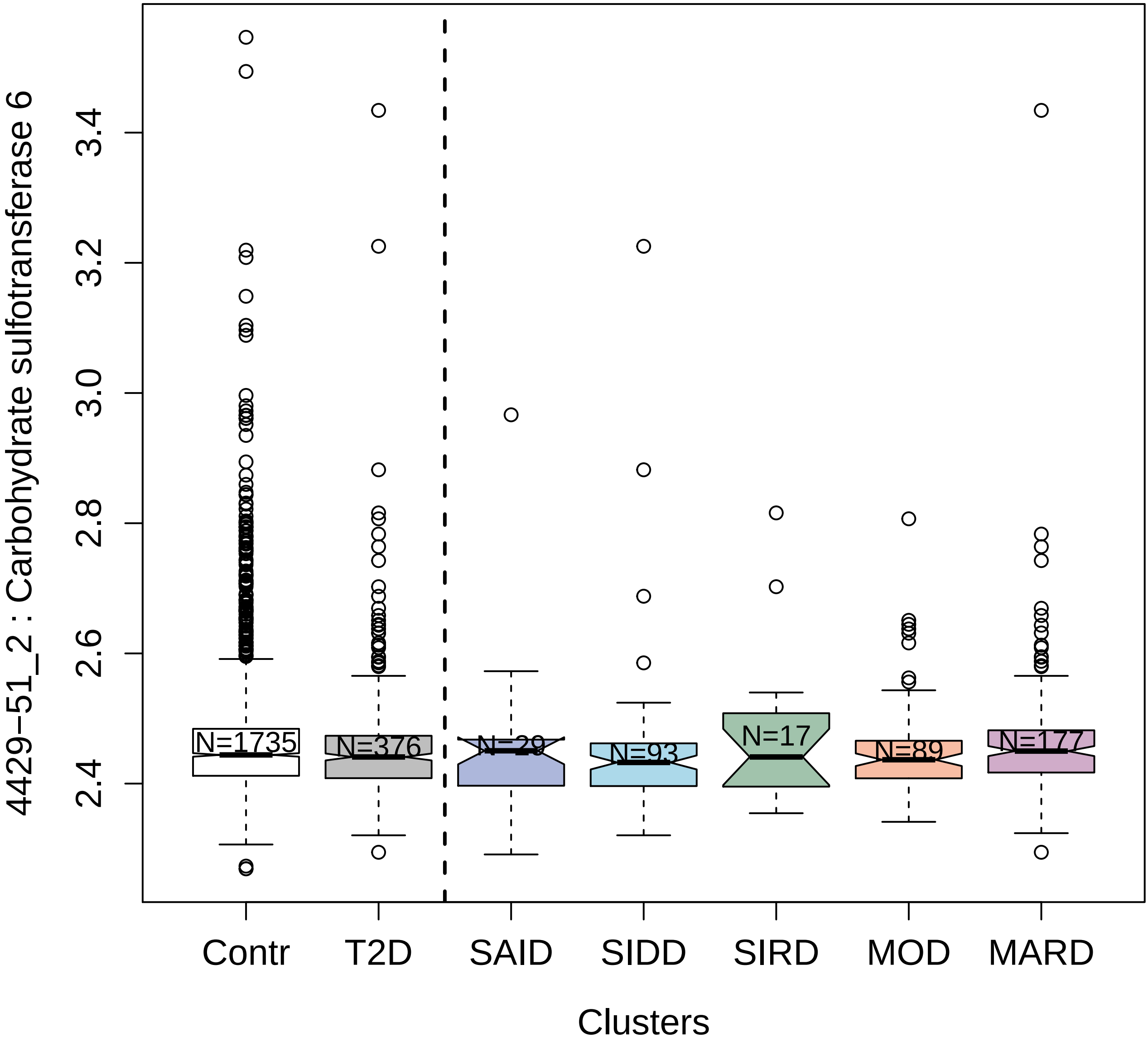

4429−51_2 : Carbohydrate sulfotransferase 6
2.4
2.6
2.8
3.0
3.2
3.4
N=1735
N=376
N=29
N=93
N=17
N=89
N=177
Contr
T2D
SAID
SIDD
SIRD
MOD
MARD
Clusters

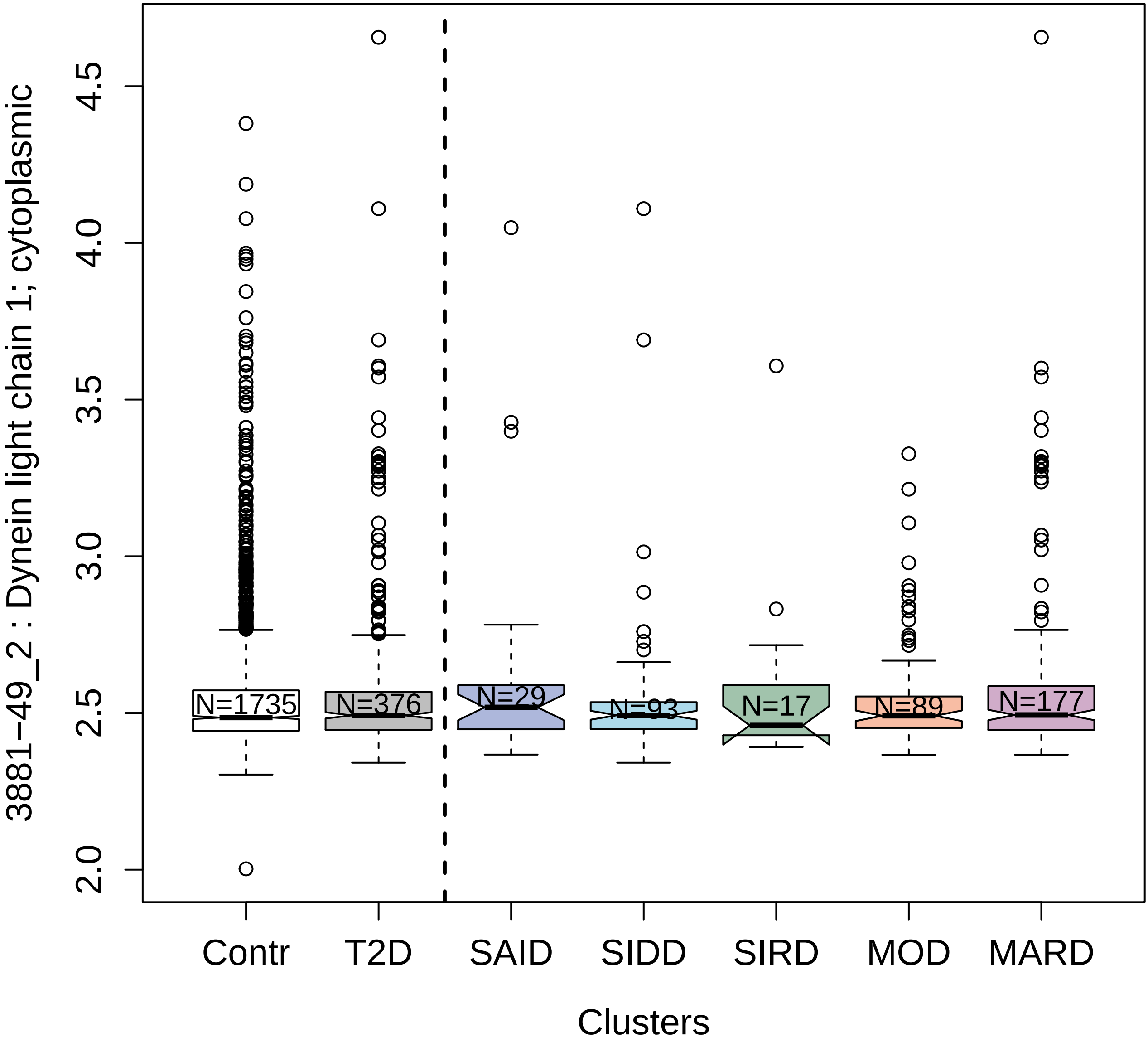
3881−49_2 : Dynein light chain 1; cytoplasmic
2.0
2.5
3.0
3.5
4.0
4.5
N=1735
N=376
N=29
N=93
N=17
N=89
N=177
Contr
T2D
SAID
SIDD
SIRD
MOD
MARD
Clusters

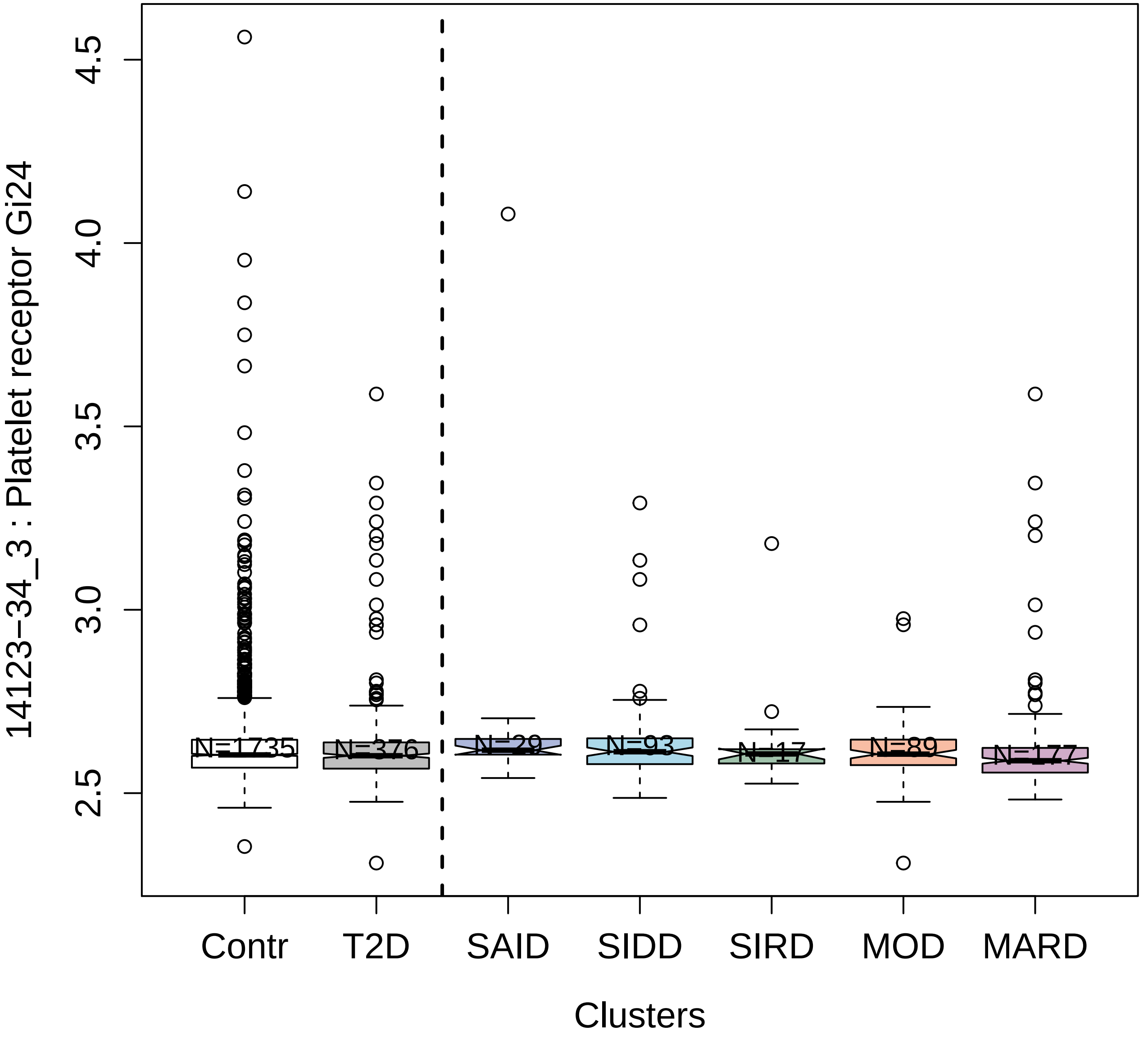

14123−34_3 : Platelet receptor Gi24
4.5
4.0
3.5
3.0
2.5
N=1735
N=376
N=29
N=93
N=17
N=89
N=177
Contr
T2D
SAID
SIDD
SIRD
MOD
MARD
Clusters

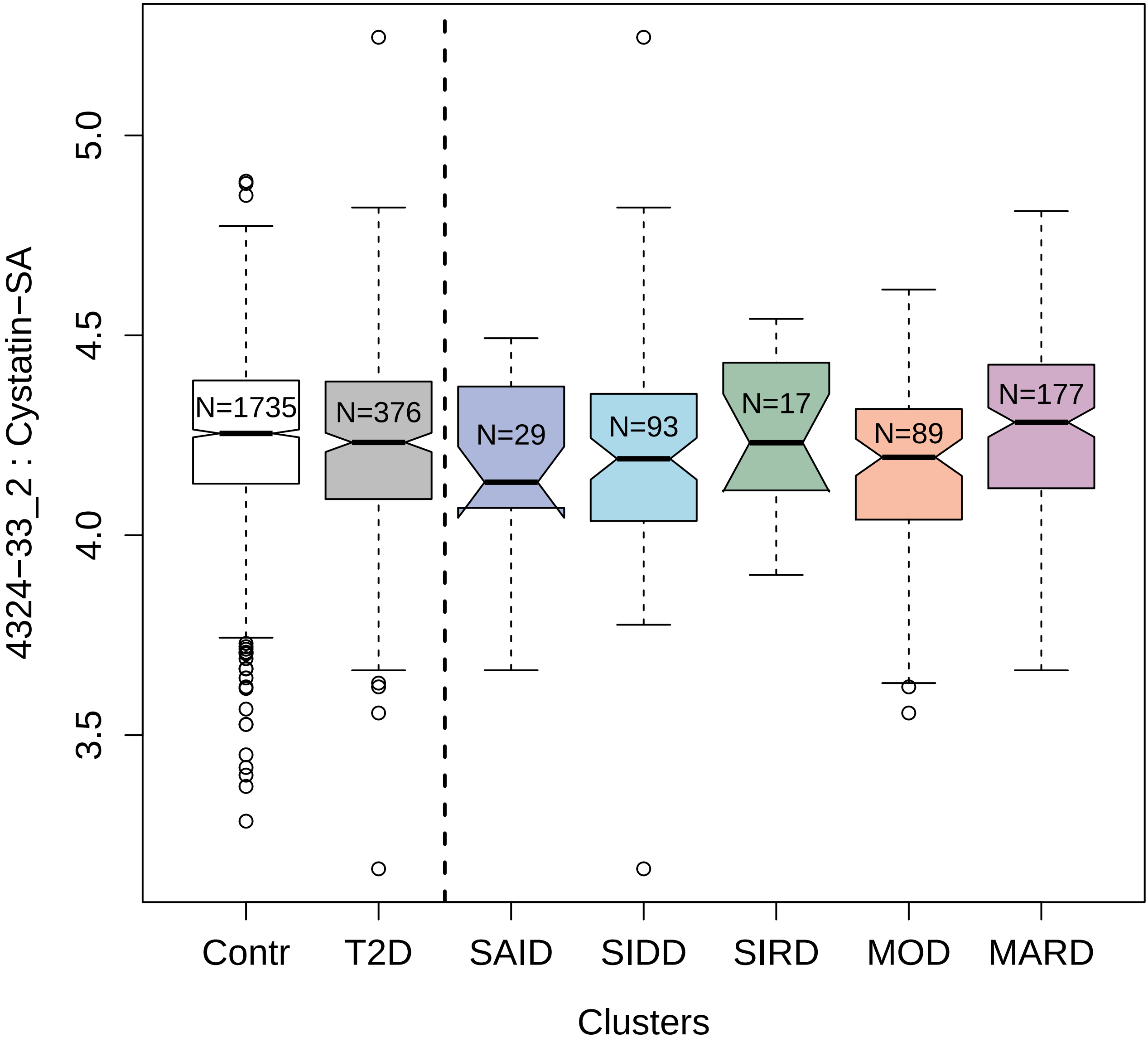

4324−33_2 : Cystatin−SA
5.0
4.5
4.0
3.5
N=1735
N=376
N=29
N=93
N=17
N=89
N=177
Contr
T2D
SAID
SIDD
SIRD
MOD
MARD
Clusters

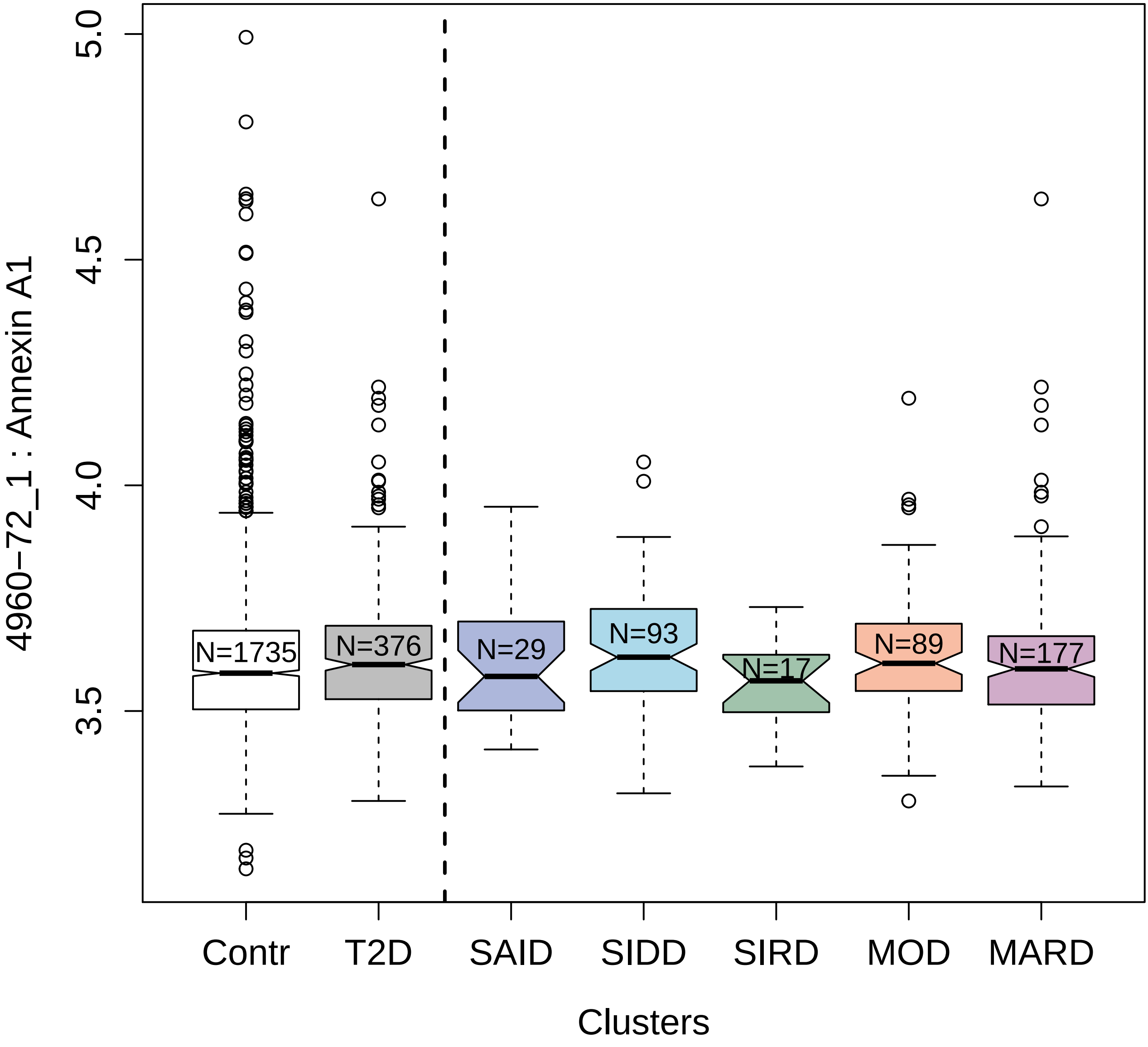
4960−72_1 : Annexin A1
5.0
4.5
4.0
3.5
N=1735
N=376
N=29
N=93
N=17
N=89
N=177
Contr
T2D
SAID
SIDD
SIRD
MOD
MARD
Clusters

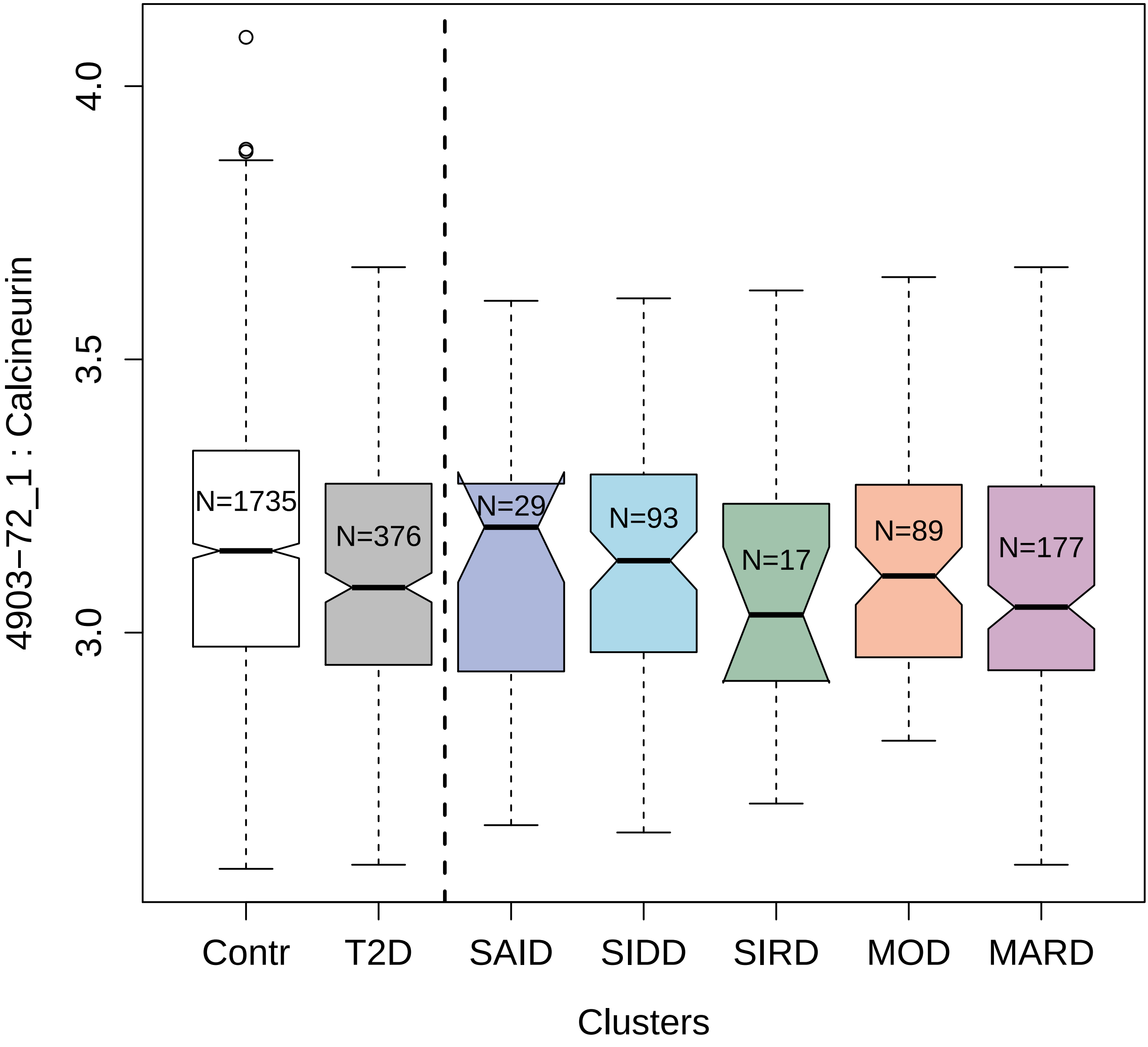
4903−72_1 : Calcineurin
4.0
3.5
3.0
N=1735
N=376
N=29
N=93
N=17
N=89
N=177
Contr
T2D
SAID
SIDD
SIRD
MOD
MARD
Clusters

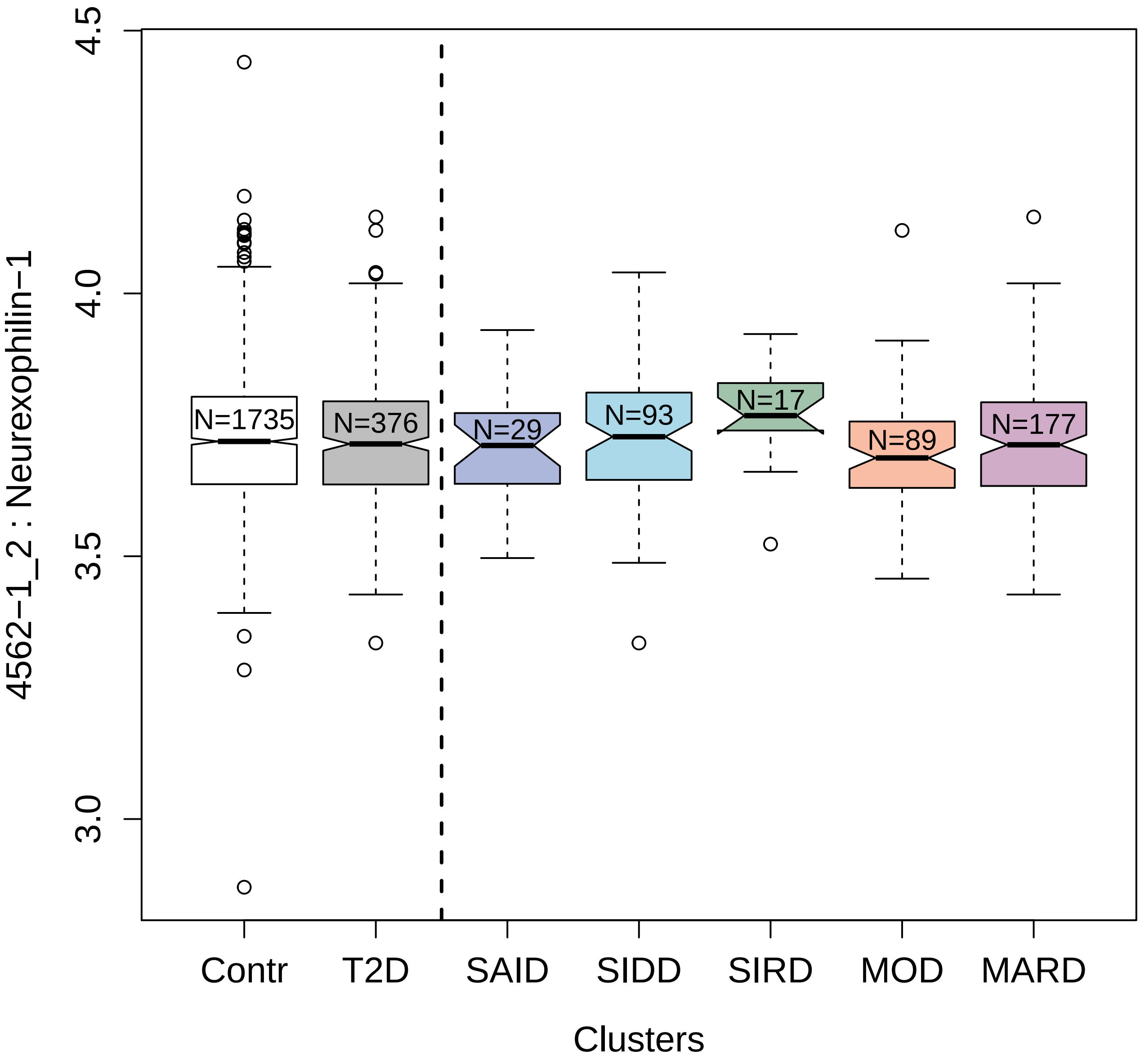

4562−1_2 : Neurexophilin−1
4.5
4.0
3.5
3.0
N=1735
N=376
N=29
N=93
N=17
N=89
N=177
Contr
T2D
SAID
SIDD
SIRD
MOD
MARD
Clusters

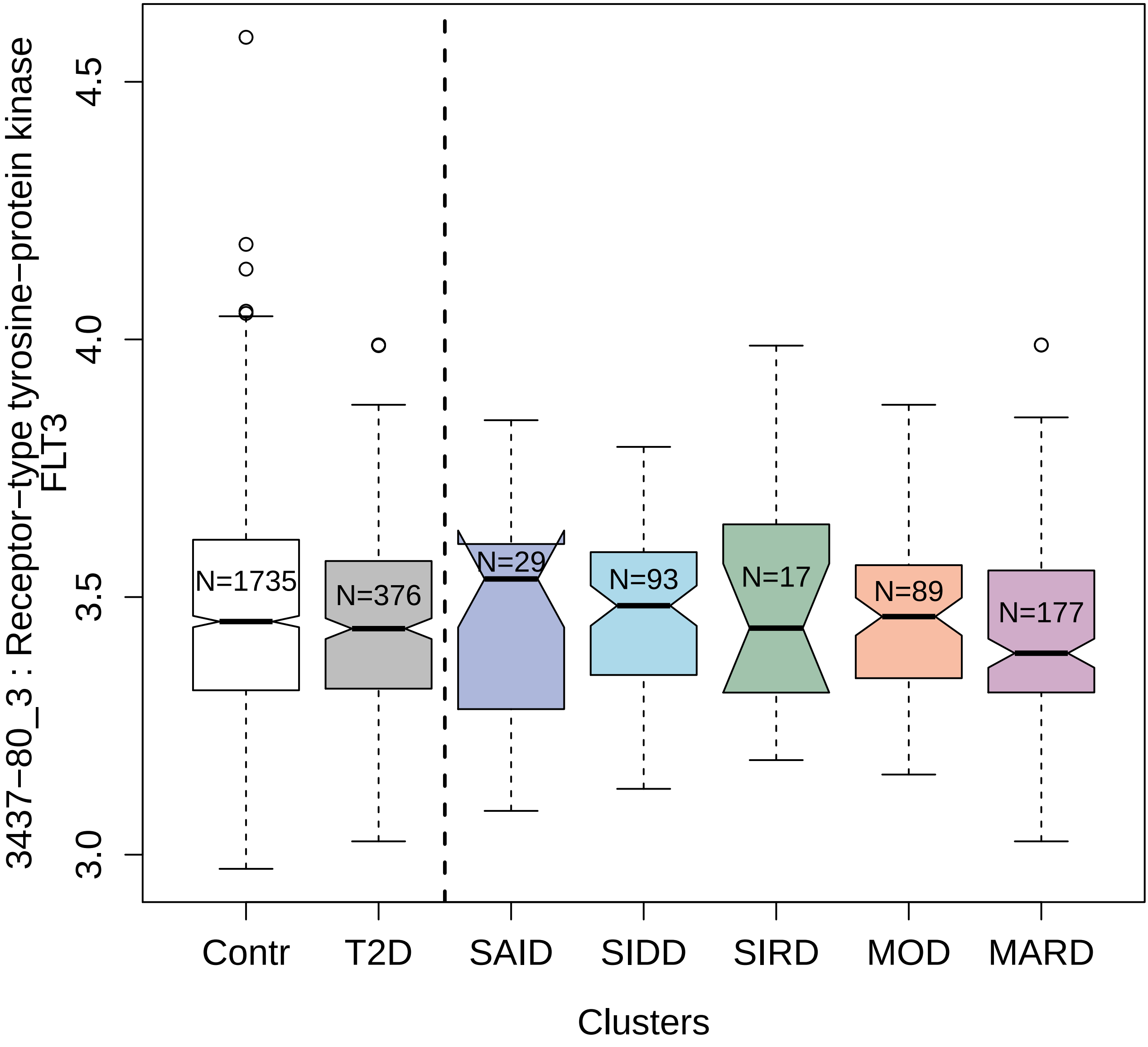

3437−80_3 : Receptor−type tyrosine−protein kinase
FLT3
4.5
4.0
3.5
3.0
N=1735
N=376
N=29
N=93
N=17
N=89
N=177
Contr
T2D
SAID
SIDD
SIRD
MOD
MARD
Clusters

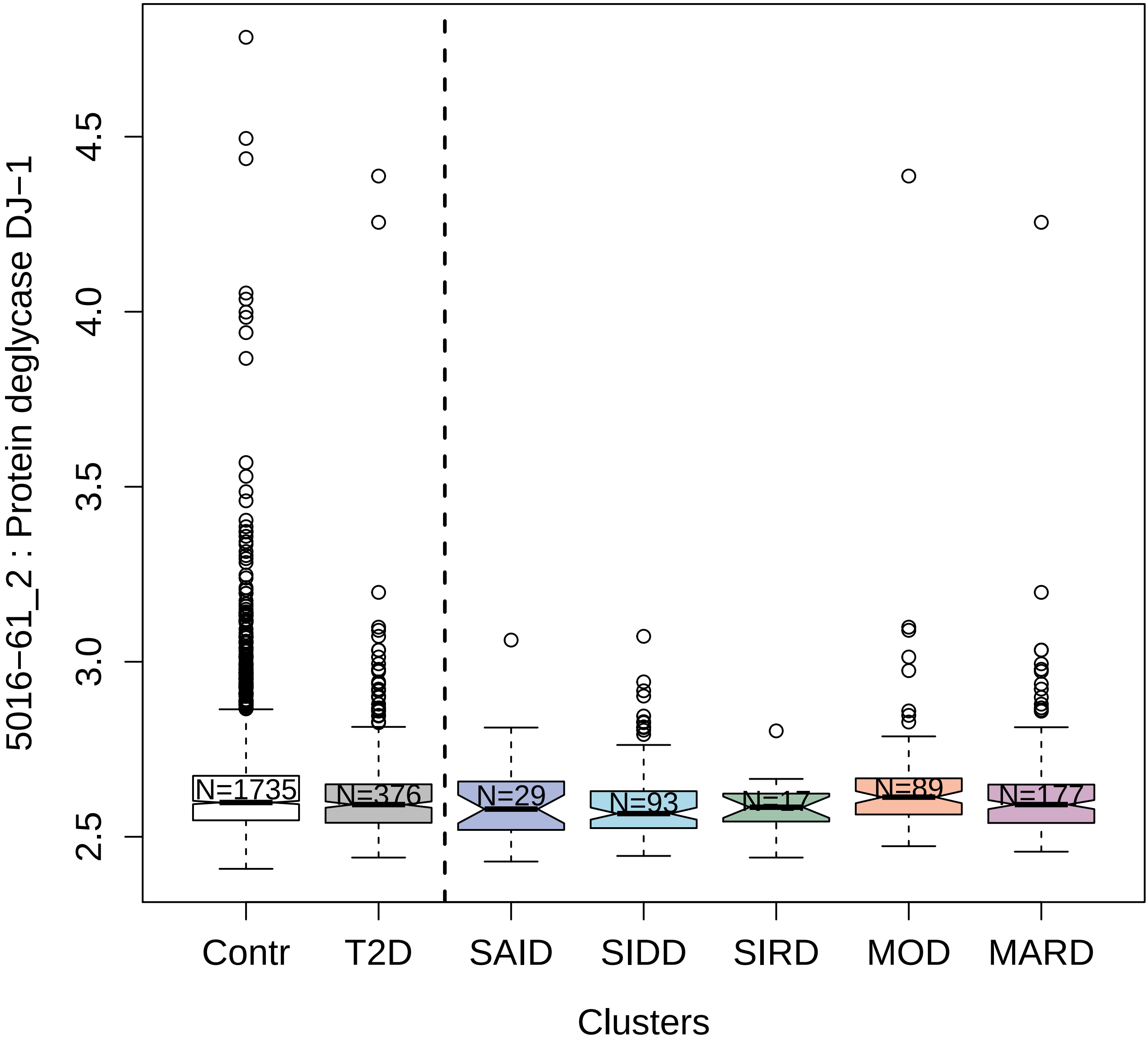

5016−61_2 : Protein deglycase DJ−1
4.5
4.0
3.5
3.0
2.5
N=1735
N=376
N=29
N=93
N=17
N=89
N=177
Contr
T2D
SAID
SIDD
SIRD
MOD
MARD
Clusters

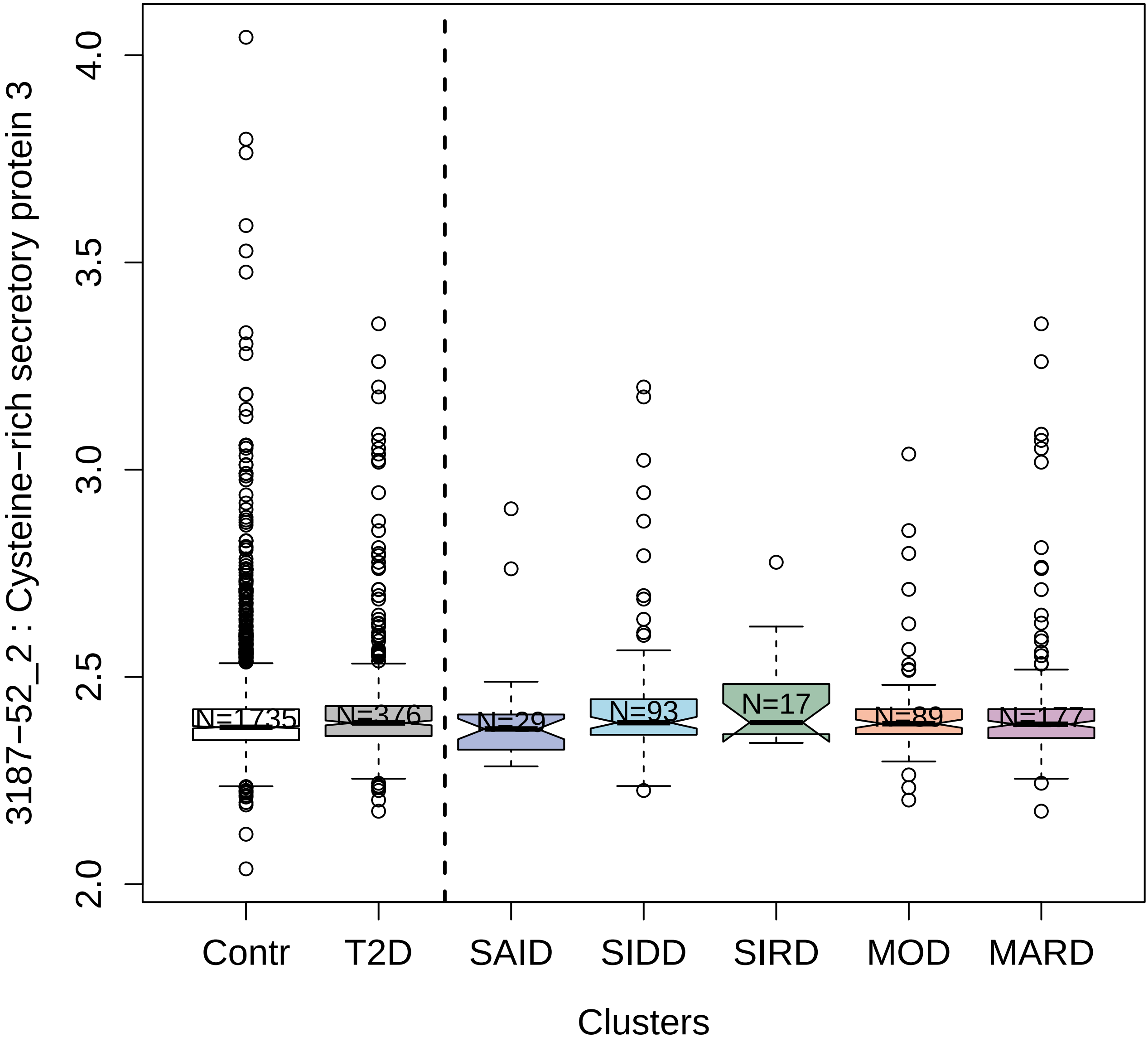
3187−52_2 : Cysteine−rich secretory protein 3
2.0
2.5
3.0
3.5
4.0
N=1735
N=376
N=29
N=93
N=17
N=89
N=177
Contr
T2D
SAID
SIDD
SIRD
MOD
MARD
Clusters

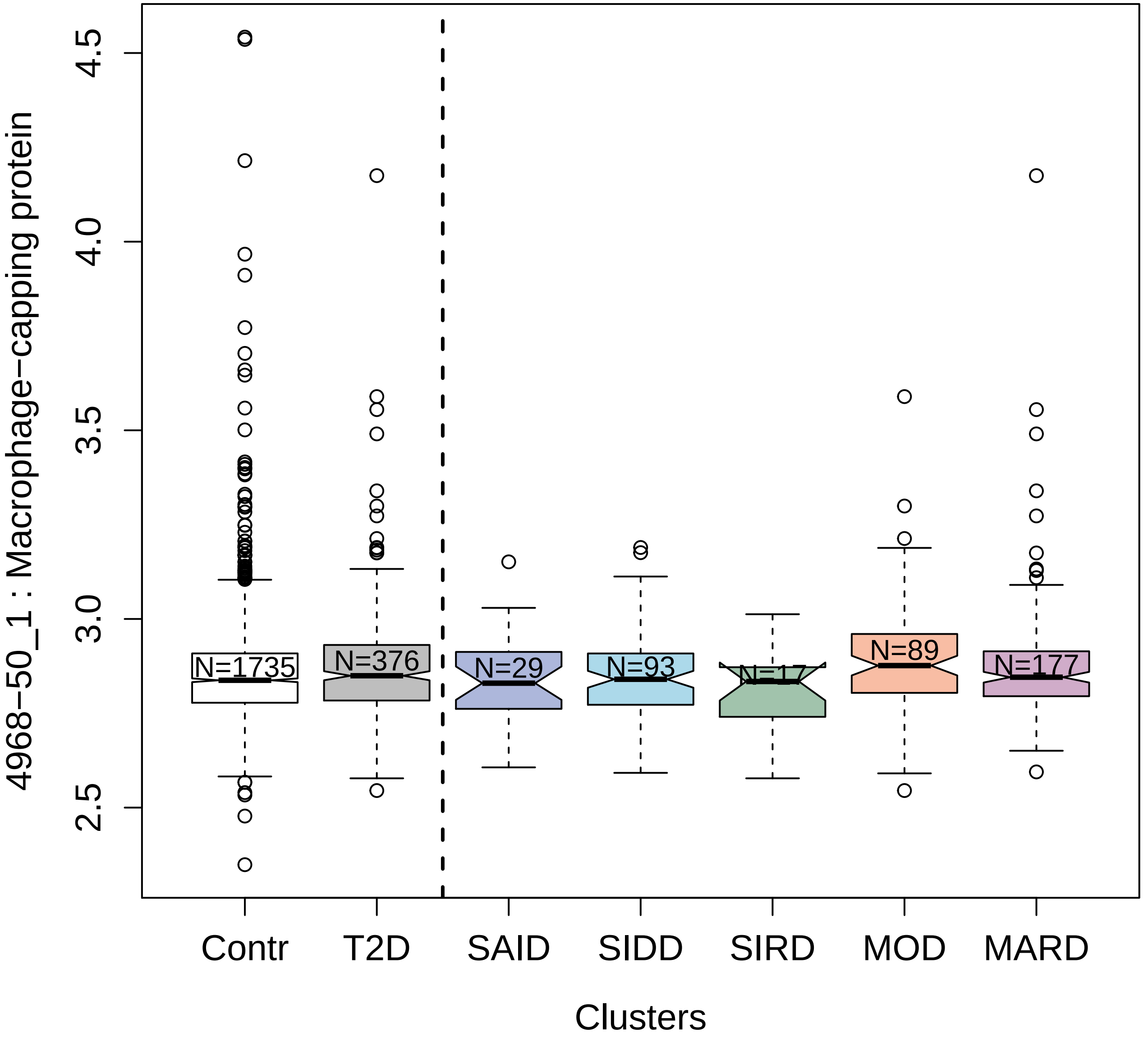

4968−50_1 : Macrophage−capping protein
2.5
3.0
3.5
4.0
4.5
N=1735
N=376
N=29
N=93
N=17
N=89
N=177
Contr
T2D
SAID
SIDD
SIRD
MOD
MARD
Clusters

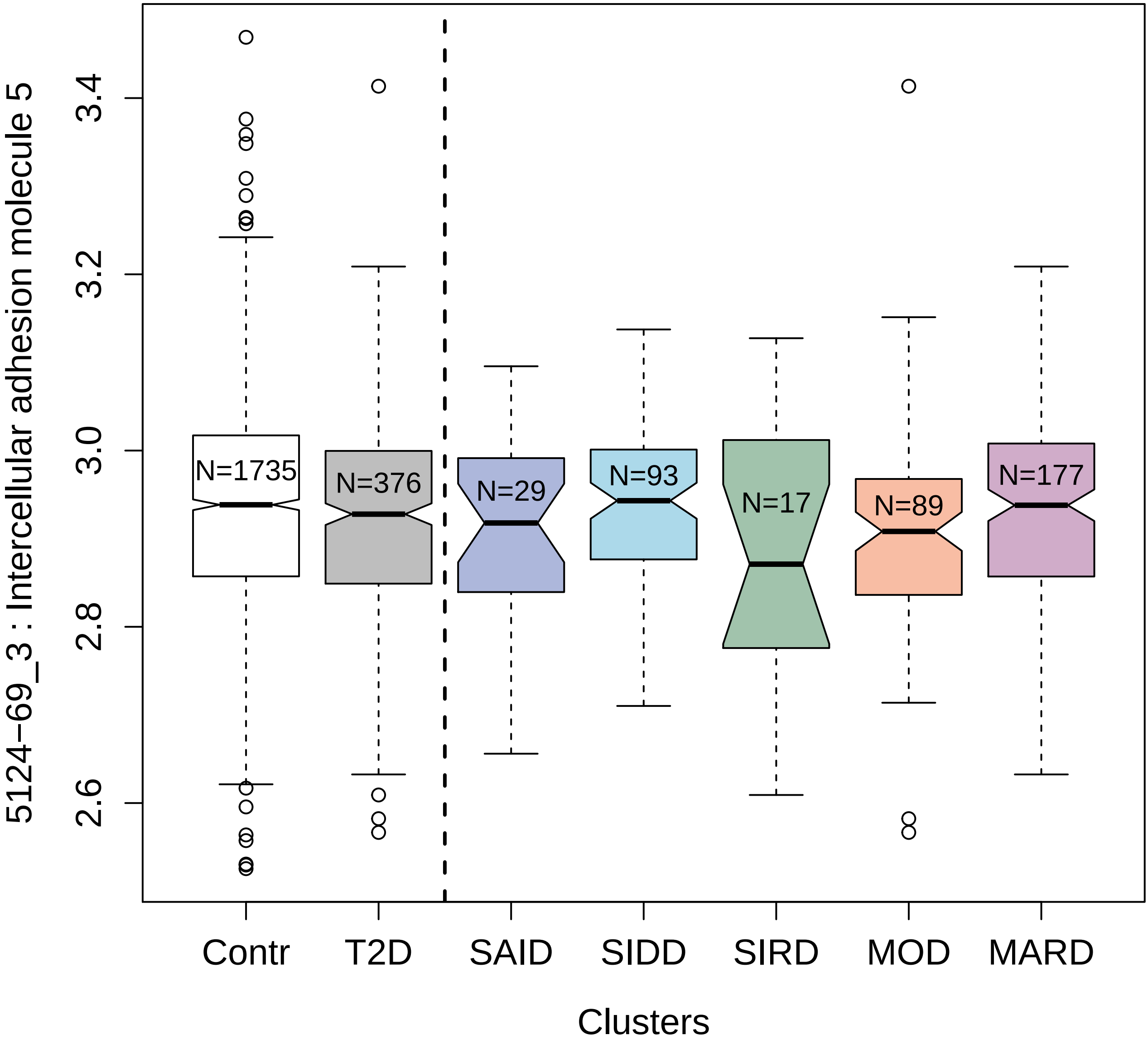

5124−69_3 : Intercellular adhesion molecule 5
3.4
3.2
3.0
2.8
2.6
N=1735
N=376
N=29
N=93
N=17
N=89
N=177
Contr
T2D
SAID
SIDD
SIRD
MOD
MARD
Clusters

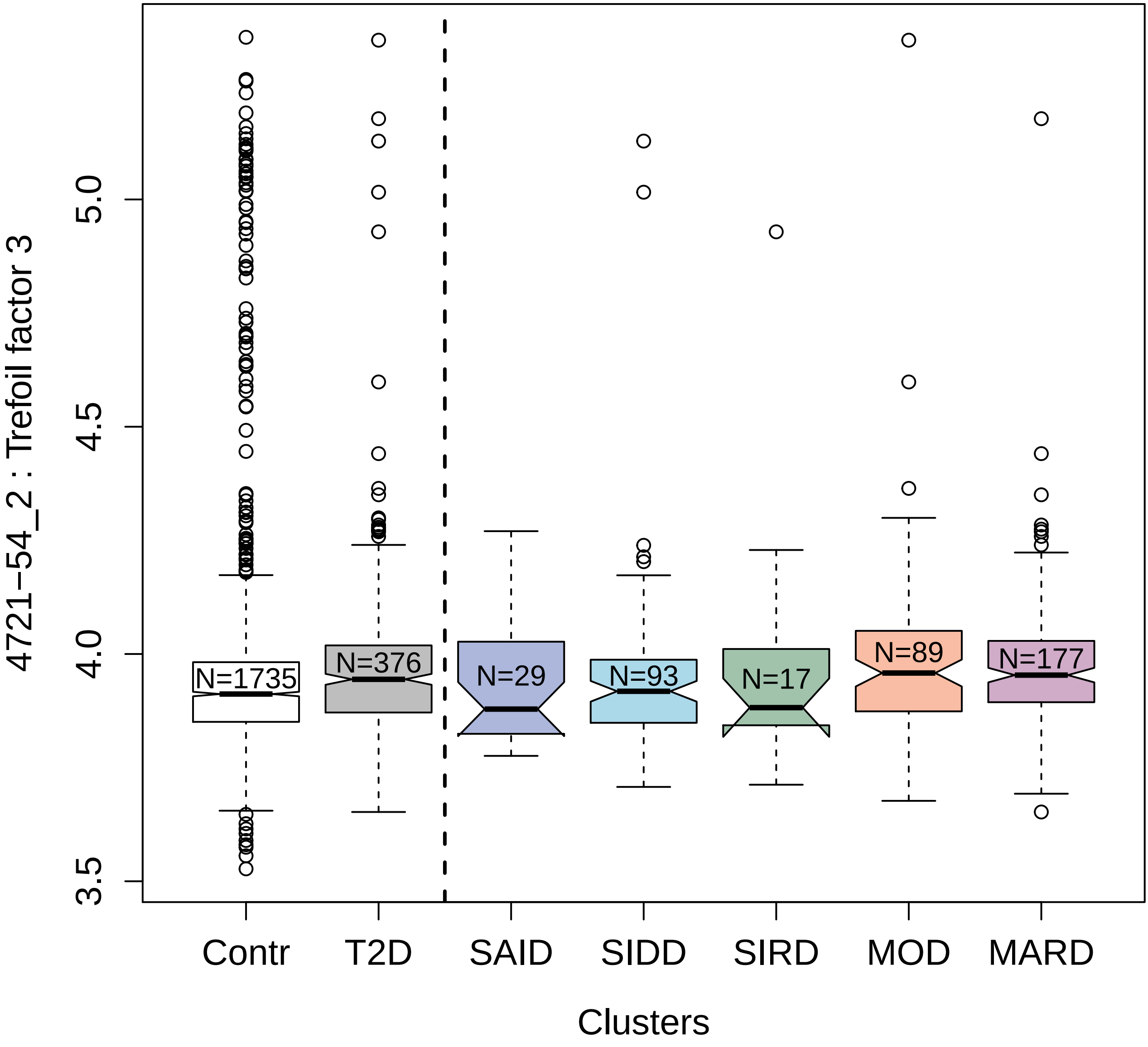
4721−54_2 : Trefoil factor 3
3.5
4.0
4.5
5.0
N=1735
N=376
N=29
N=93
N=17
N=89
N=177
Contr
T2D
SAID
SIDD
SIRD
MOD
MARD
Clusters

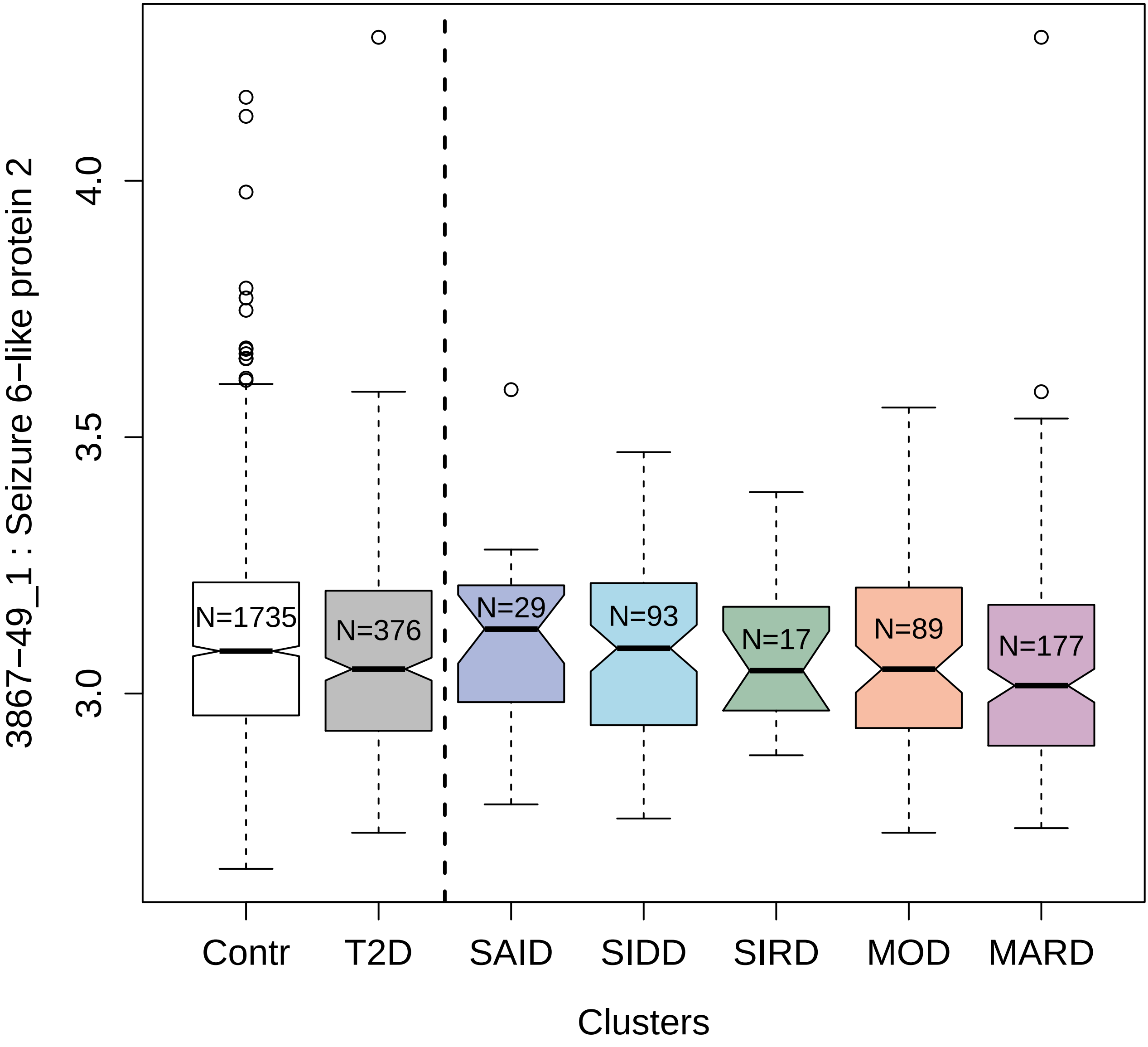

3867−49_1 : Seizure 6−like protein 2
4.0
3.5
3.0
N=1735
N=376
N=29
N=93
N=17
N=89
N=177
Contr
T2D
SAID
SIDD
SIRD
MOD
MARD
Clusters

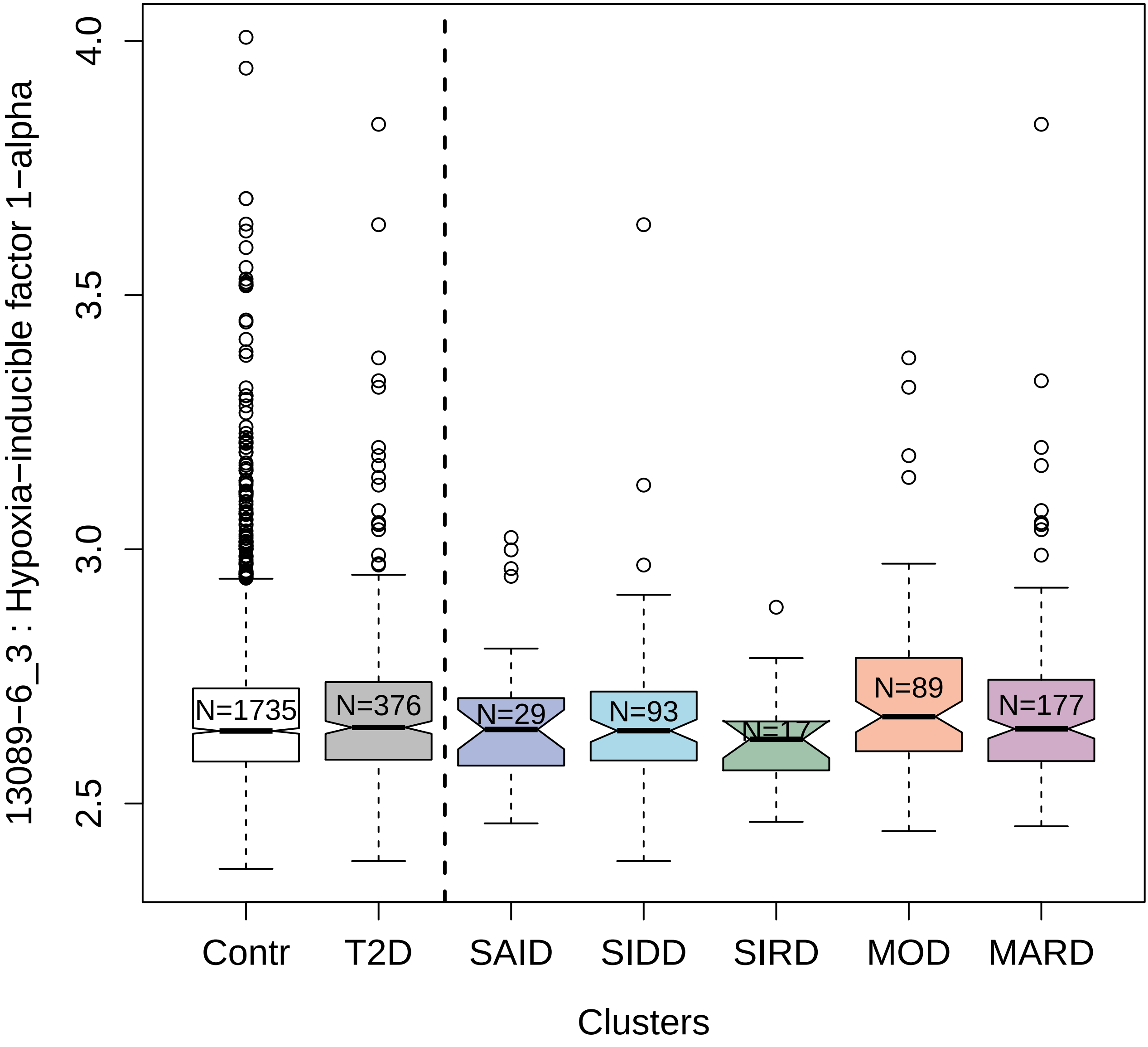

13089−6_3 : Hypoxia−inducible factor 1−alpha
4.0
3.5
3.0
2.5
N=1735
N=376
N=29
N=93
N=89
N=177
Contr
T2D
SAID
SIDD
SIRD
MOD
MARD
Clusters

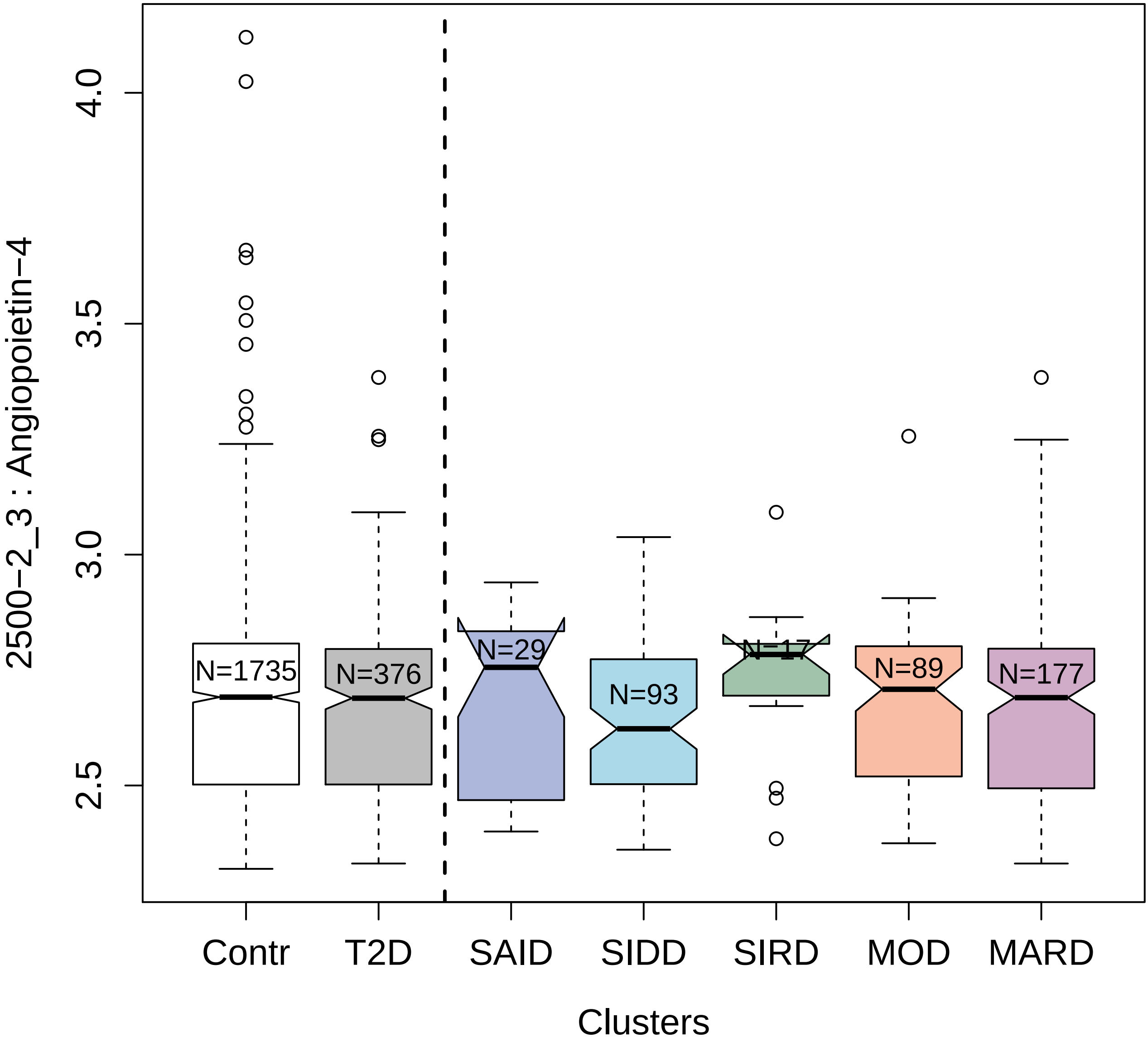
2500−2_3 : Angiopoietin−4
4.0
3.5
3.0
2.5
N=1735
N=376
N=29
N=93
N=17
N=89
N=177
Contr
T2D
SAID
SIDD
SIRD
MOD
MARD
Clusters

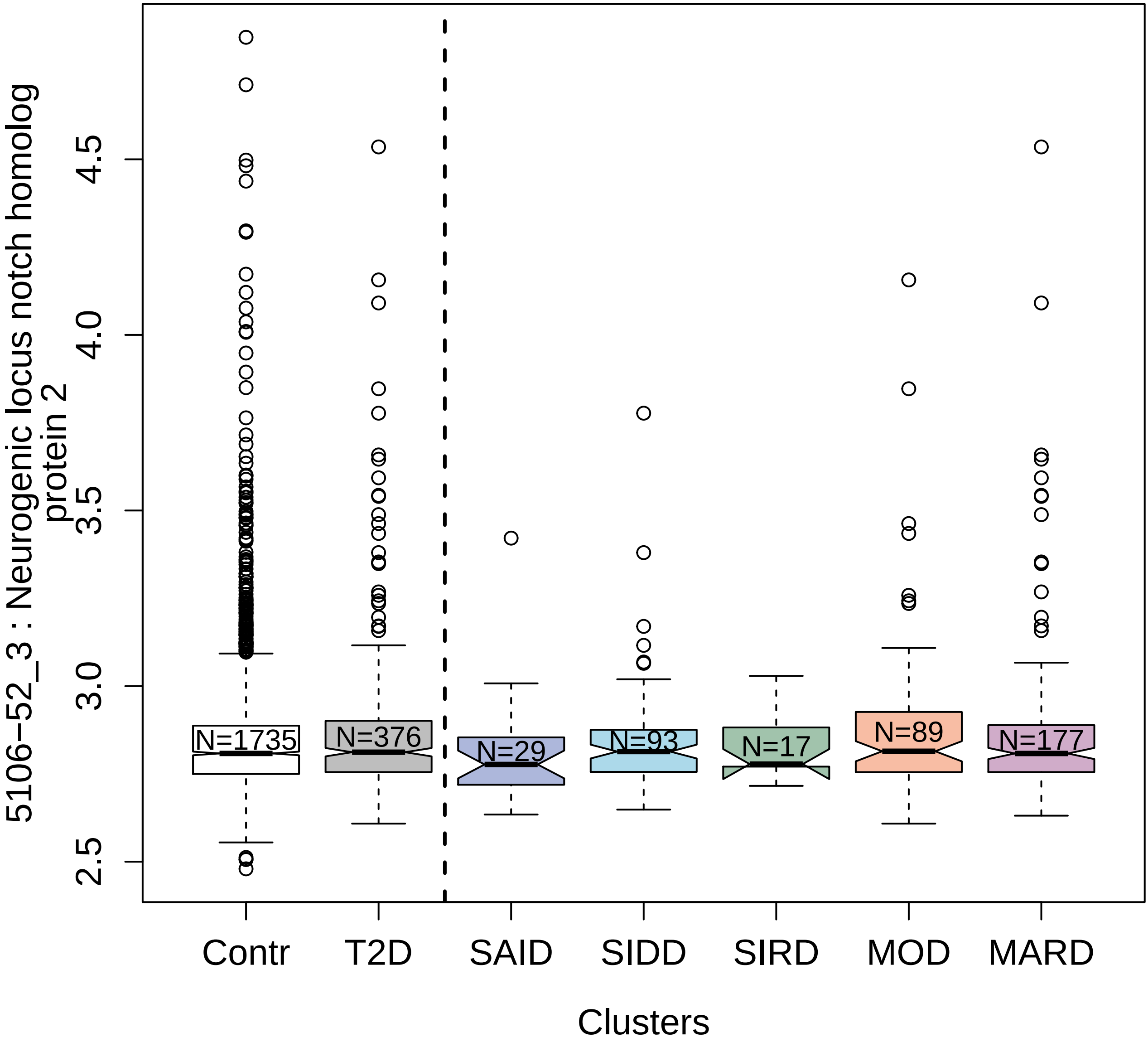
5106−52_3 : Neurogenic locus notch homolog protein 2
2.5
3.0
3.5
4.0
4.5
N=1735
N=376
N=29
N=93
N=17
N=89
N=177
Contr
T2D
SAID
SIDD
SIRD
MOD
MARD
Clusters

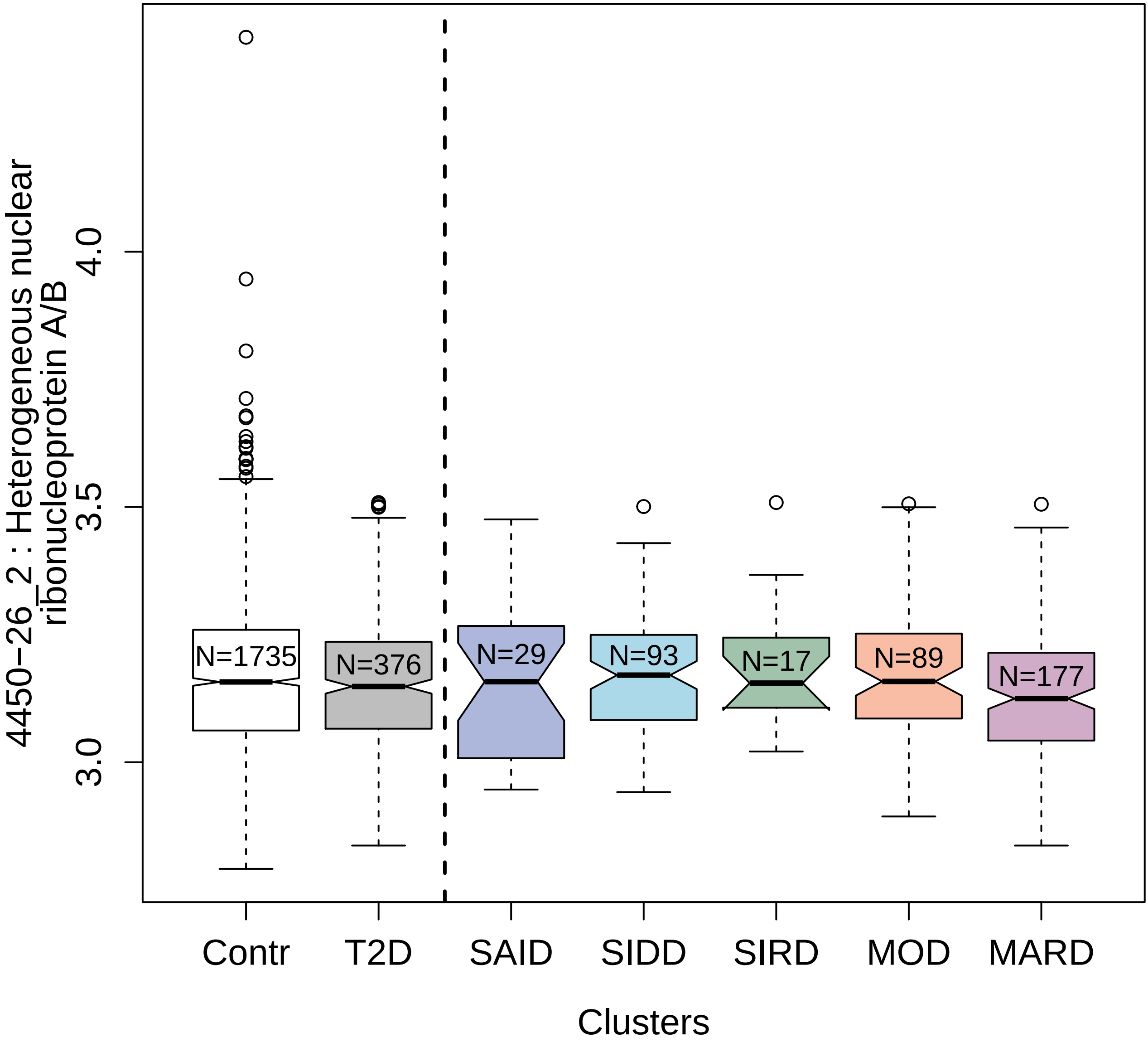
4450−26_2 : Heterogeneous nuclear ribonucleoprotein A/B
3.0
3.5
4.0
N=1735
N=376
N=29
N=93
N=17
N=89
N=177
Contr
T2D
SAID
SIDD
SIRD
MOD
MARD
Clusters

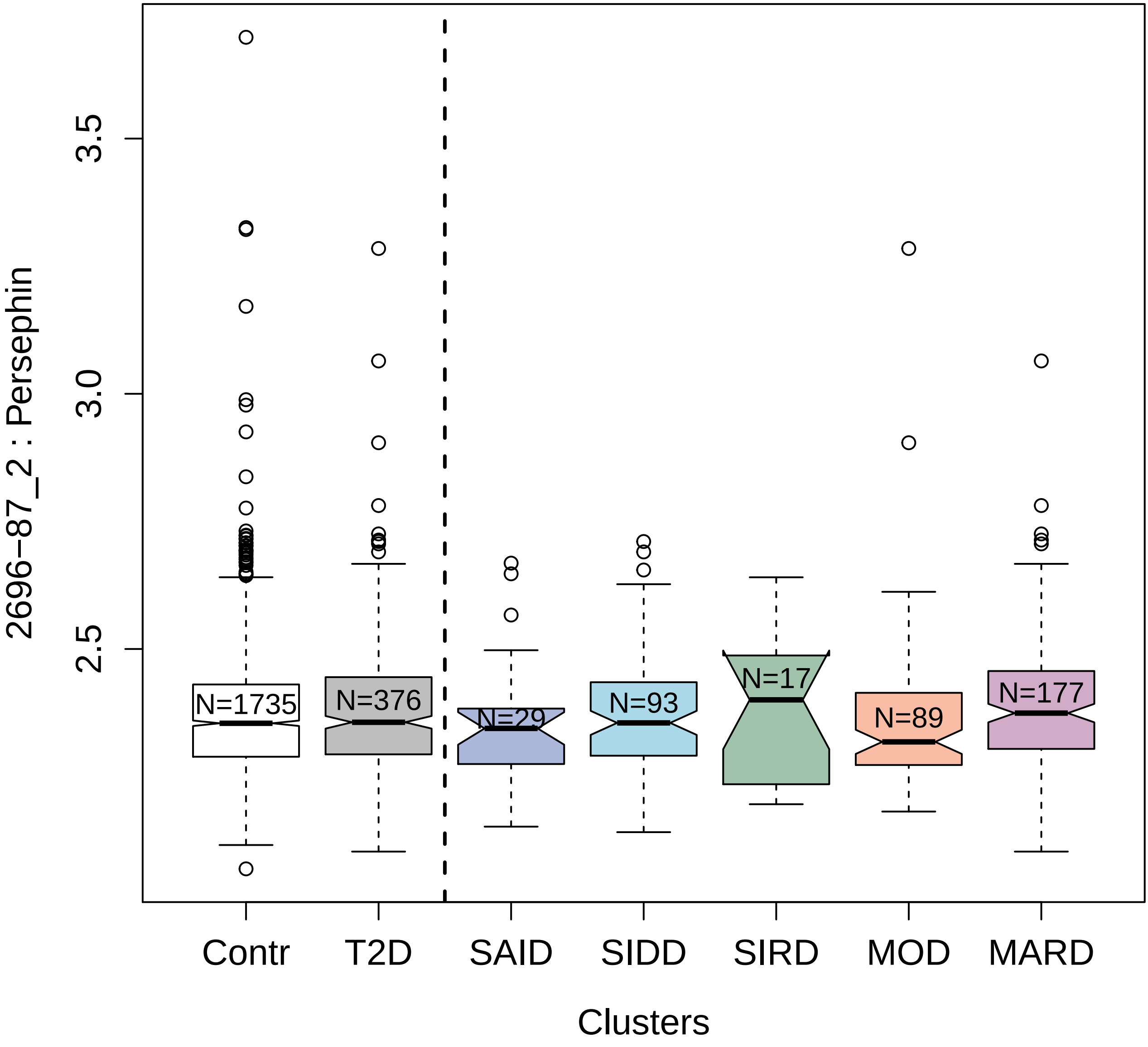

2696−87_2 : Persephin
3.5
3.0
2.5
N=1735
N=376
N=29
N=93
N=17
N=89
N=177
Contr
T2D
SAID
SIDD
SIRD
MOD
MARD
Clusters

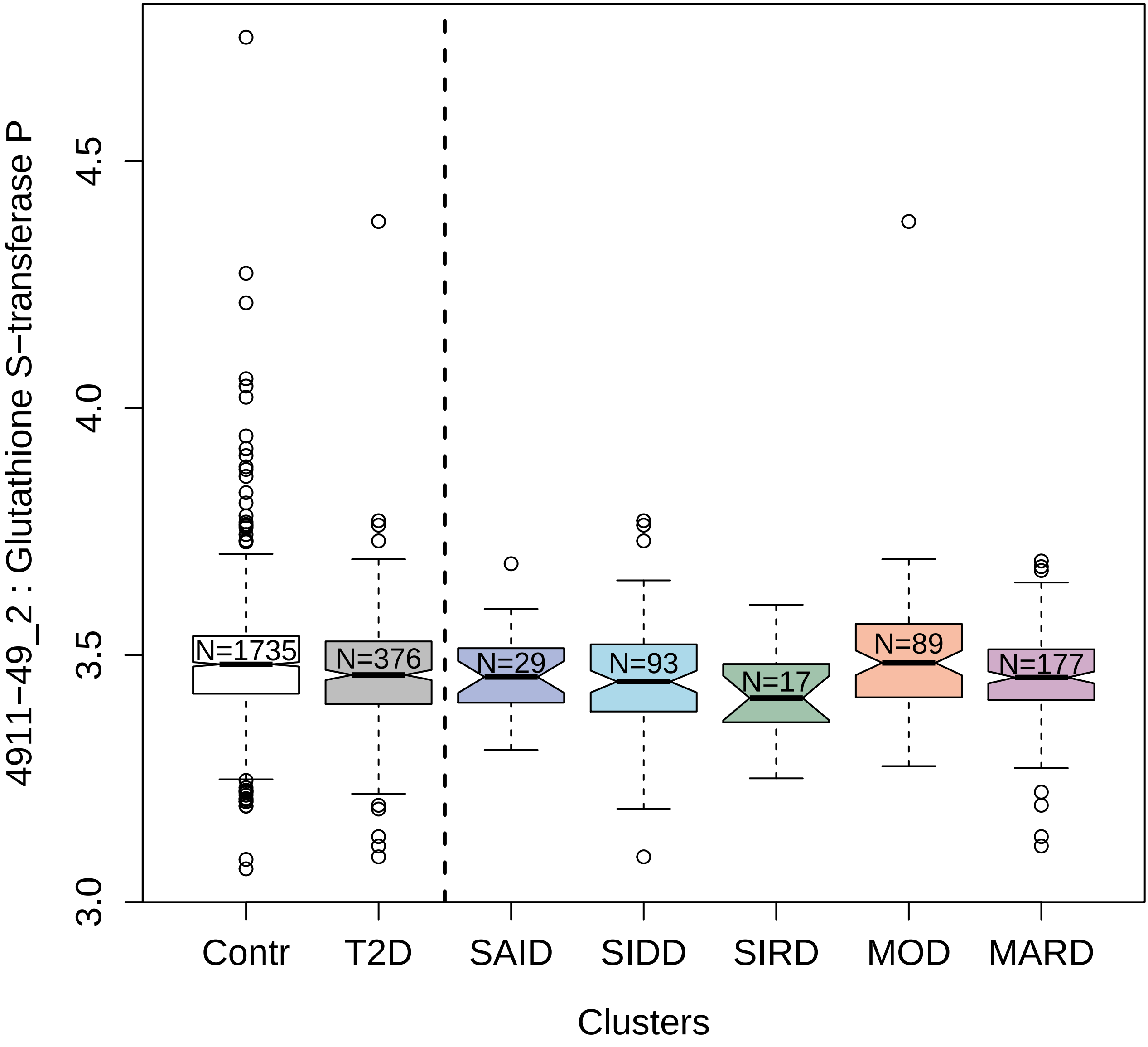
4911−49_2 : Glutathione S−transferase P
3.0
3.5
4.0
4.5
N=1735
N=376
N=29
N=93
N=17
N=89
N=177
Contr
T2D
SAID
SIDD
SIRD
MOD
MARD
Clusters

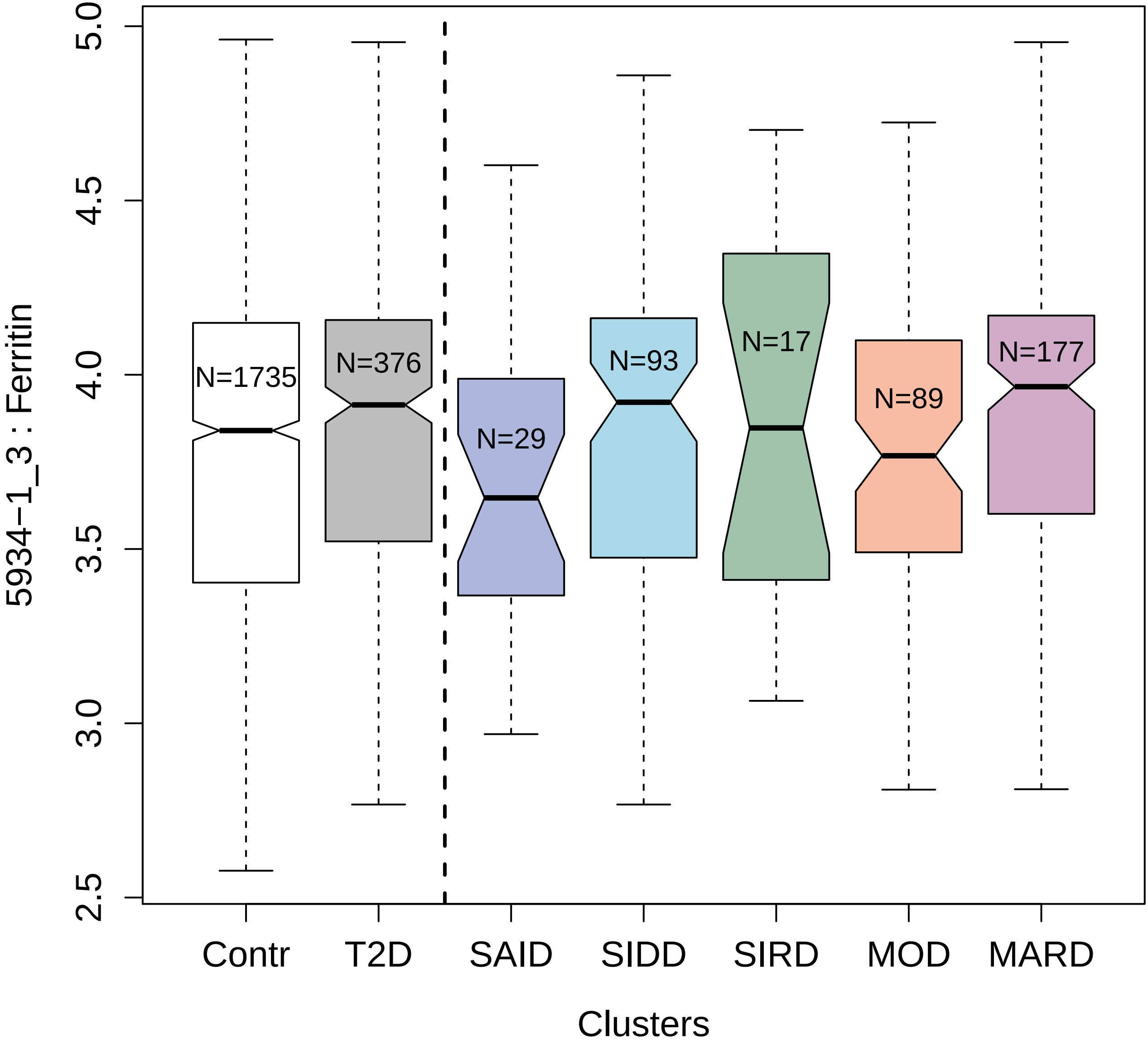

5934−1_3 : Ferritin
5.0
4.5
4.0
3.5
3.0
2.5
N=1735
N=376
N=29
N=93
N=17
N=89
N=177
Contr
T2D
SAID
SIDD
SIRD
MOD
MARD
Clusters

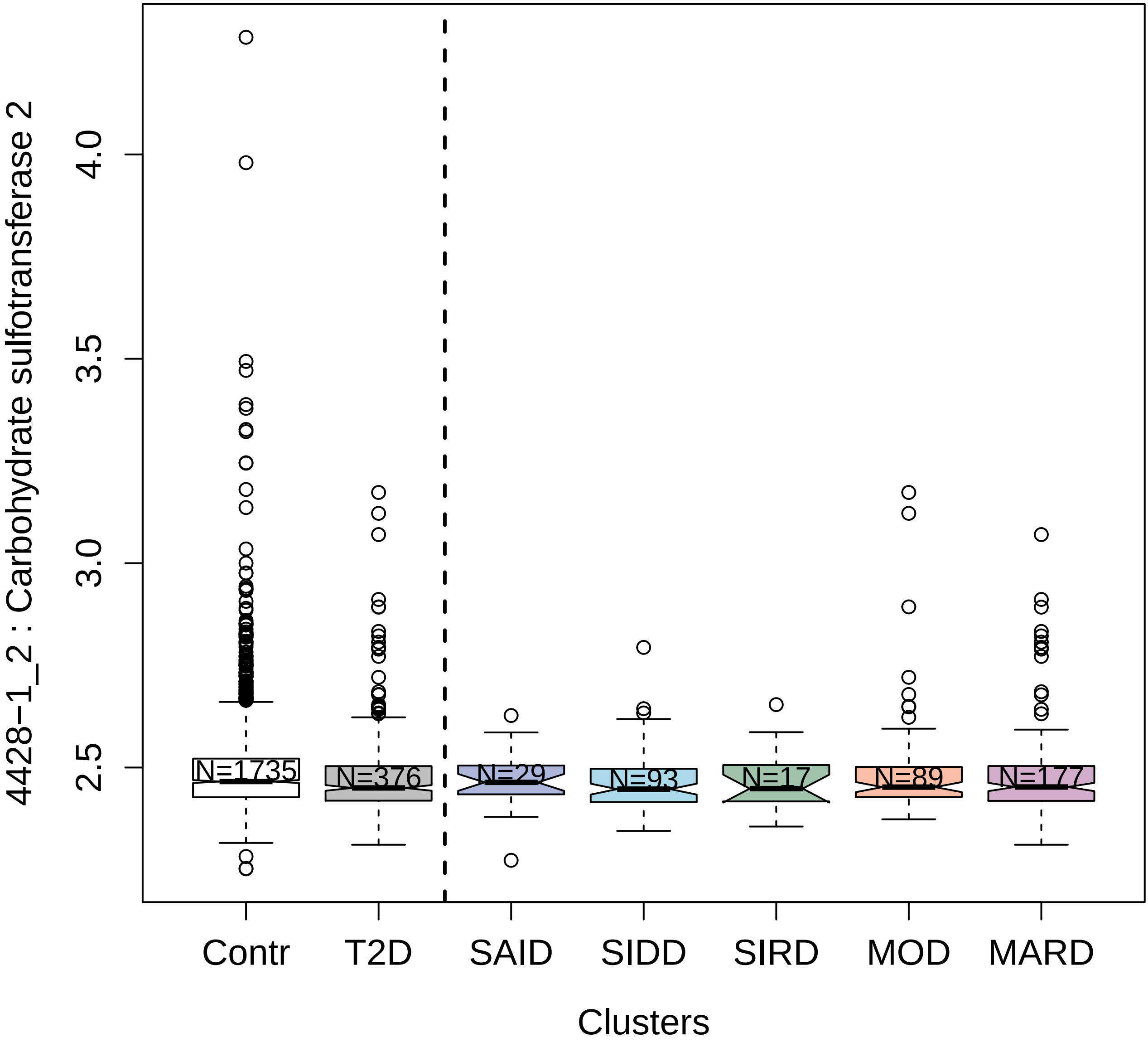

4428−1_2 : Carbohydrate sulfotransferase 2
4.0
3.5
3.0
2.5
N=1735
N=376
N=29
N=93
N=17
N=89
N=177
Contr
T2D
SAID
SIDD
SIRD
MOD
MARD
Clusters

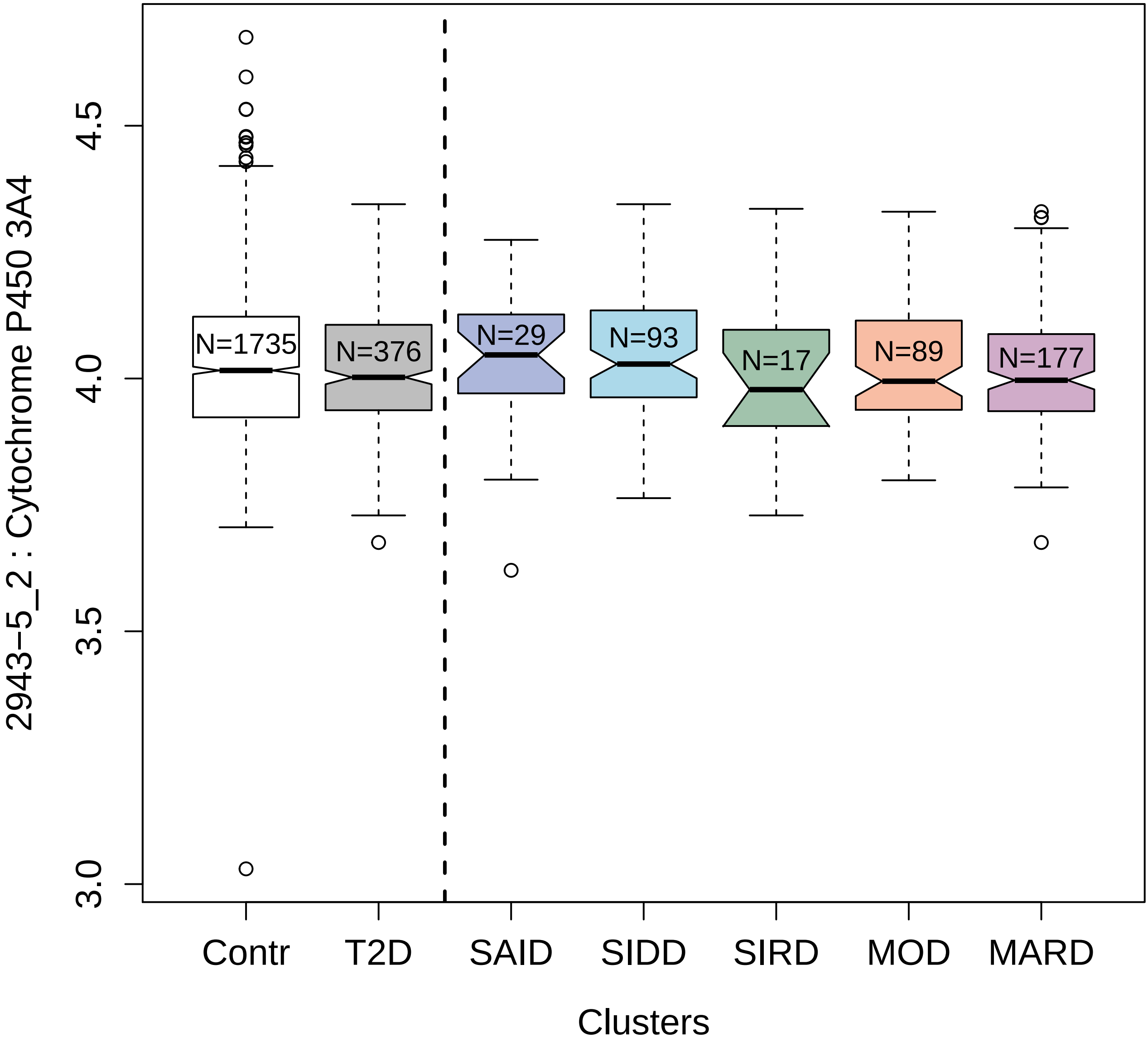
N=1735
N=376
N=29
N=93
N=17
N=89
N=177
4.5
4.0
3.5
3.0
2943−5_2 : Cytochrome P450 3A4
Contr
T2D
SAID
SIDD
SIRD
MOD
MARD
Clusters

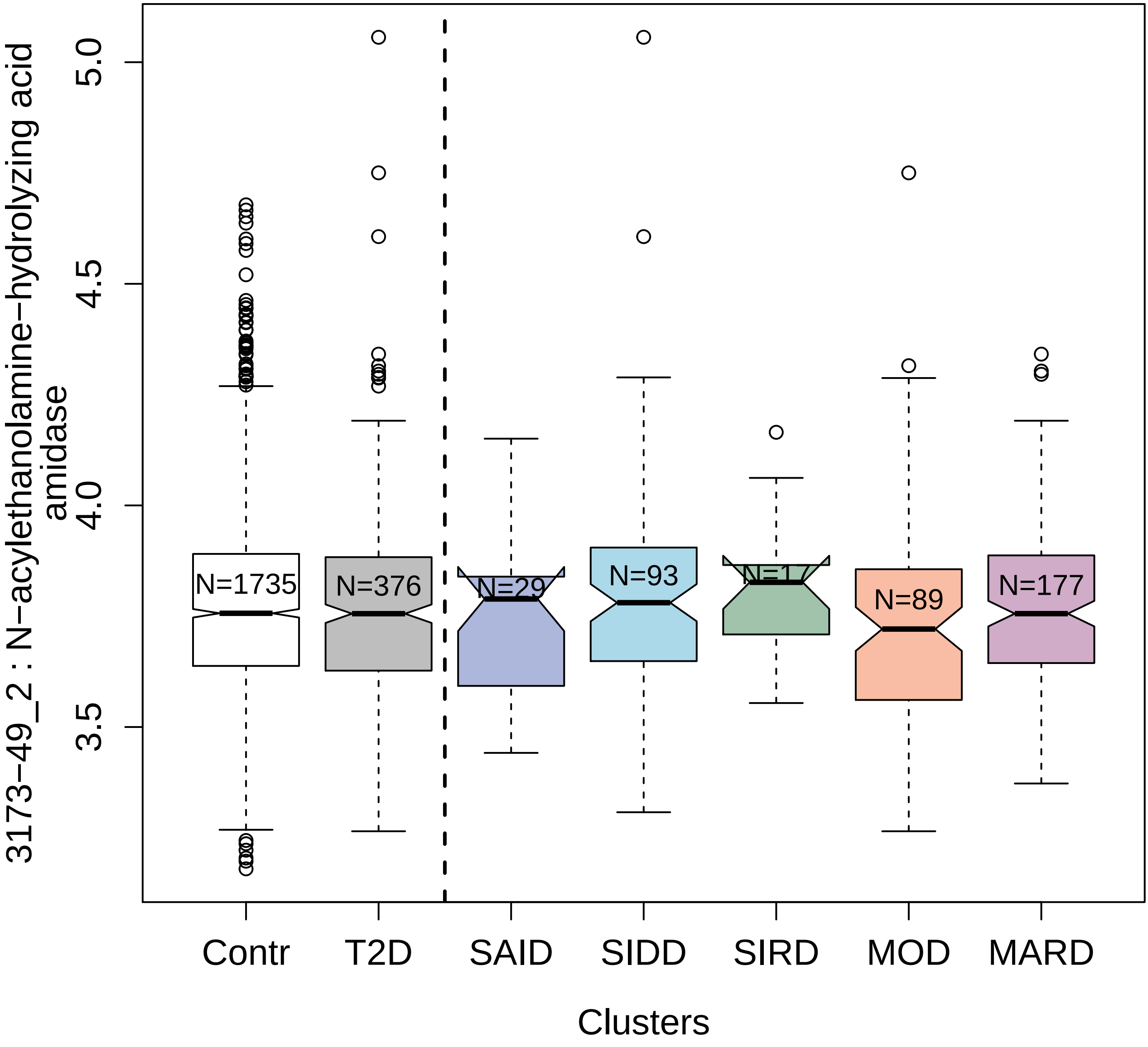

3173−49_2 : N−acylethanolamine−hydrolyzing acid amidase
5.0
4.5
4.0
3.5
N=1735
N=376
N=29
N=93
N=17
N=89
N=177
Contr
T2D
SAID
SIDD
SIRD
MOD
MARD
Clusters

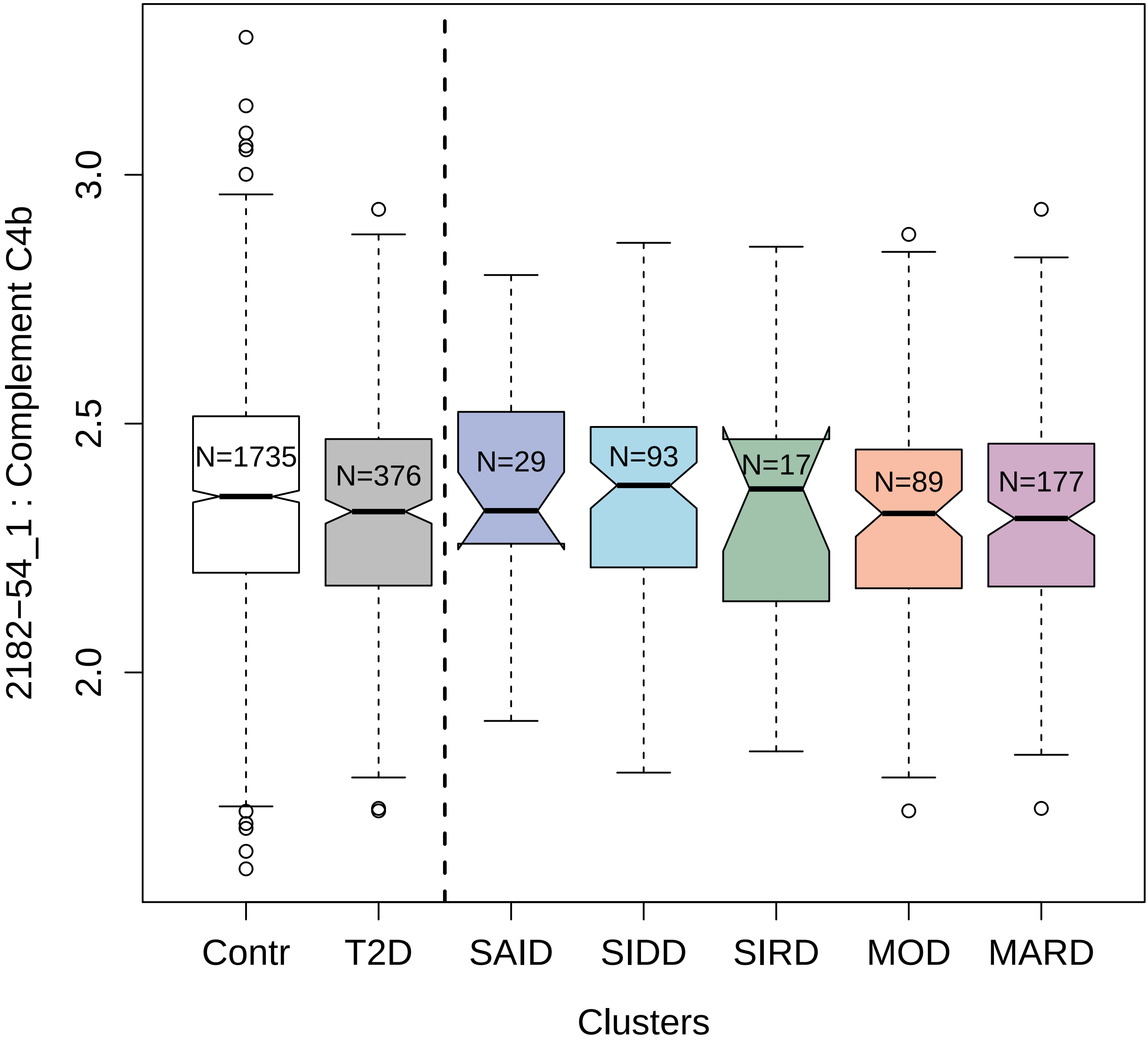
2182−54_1 : Complement C4b
3.0
2.5
2.0
N=1735
N=376
N=29
N=93
N=17
N=89
N=177
Contr
T2D
SAID
SIDD
SIRD
MOD
MARD
Clusters

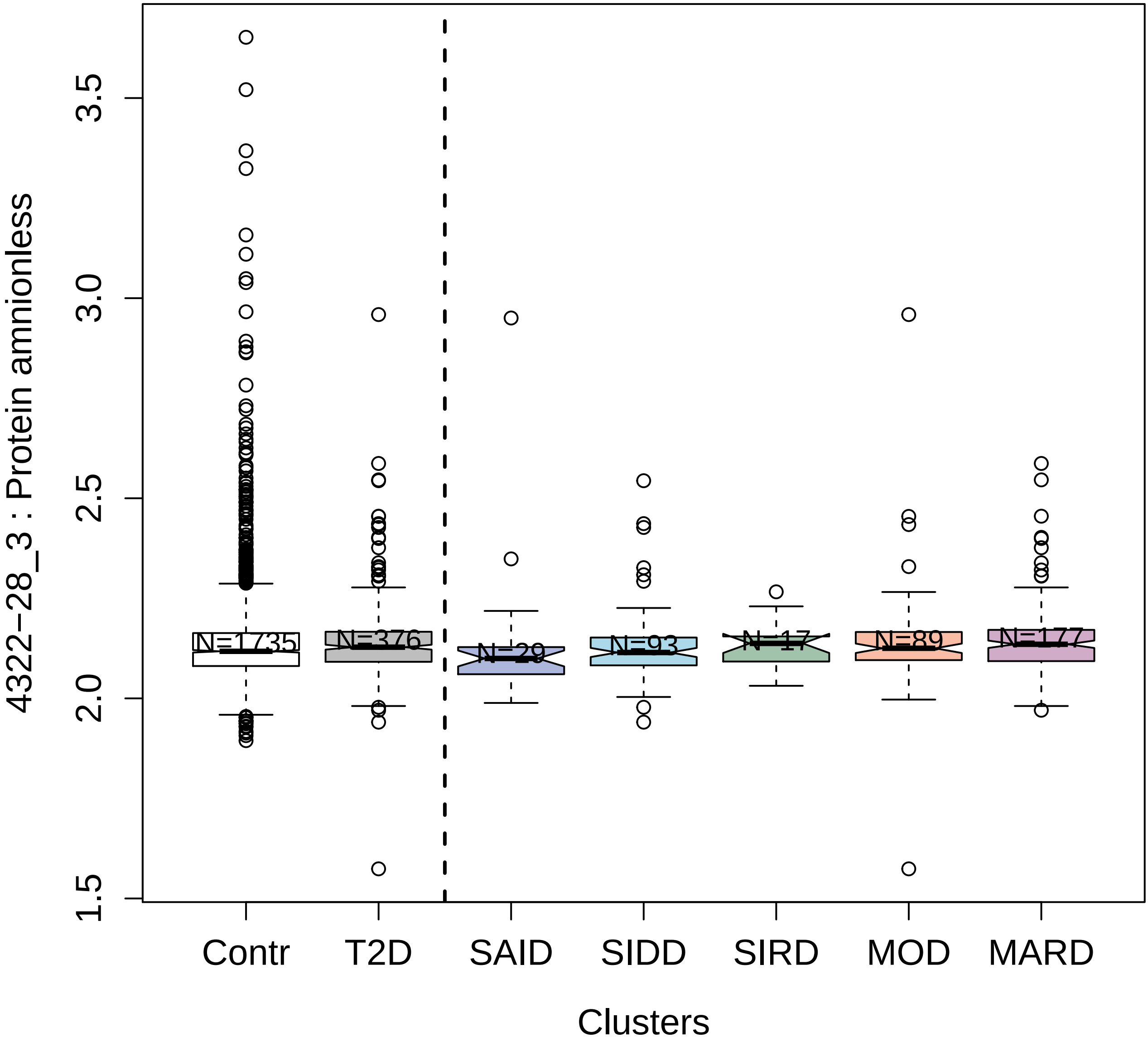
4322−28_3 : Protein amnionless
1.5
2.0
2.5
3.0
3.5
N=1735
N=376
N=29
N=93
N=17
N=89
N=177
Contr
T2D
SAID
SIDD
SIRD
MOD
MARD
Clusters

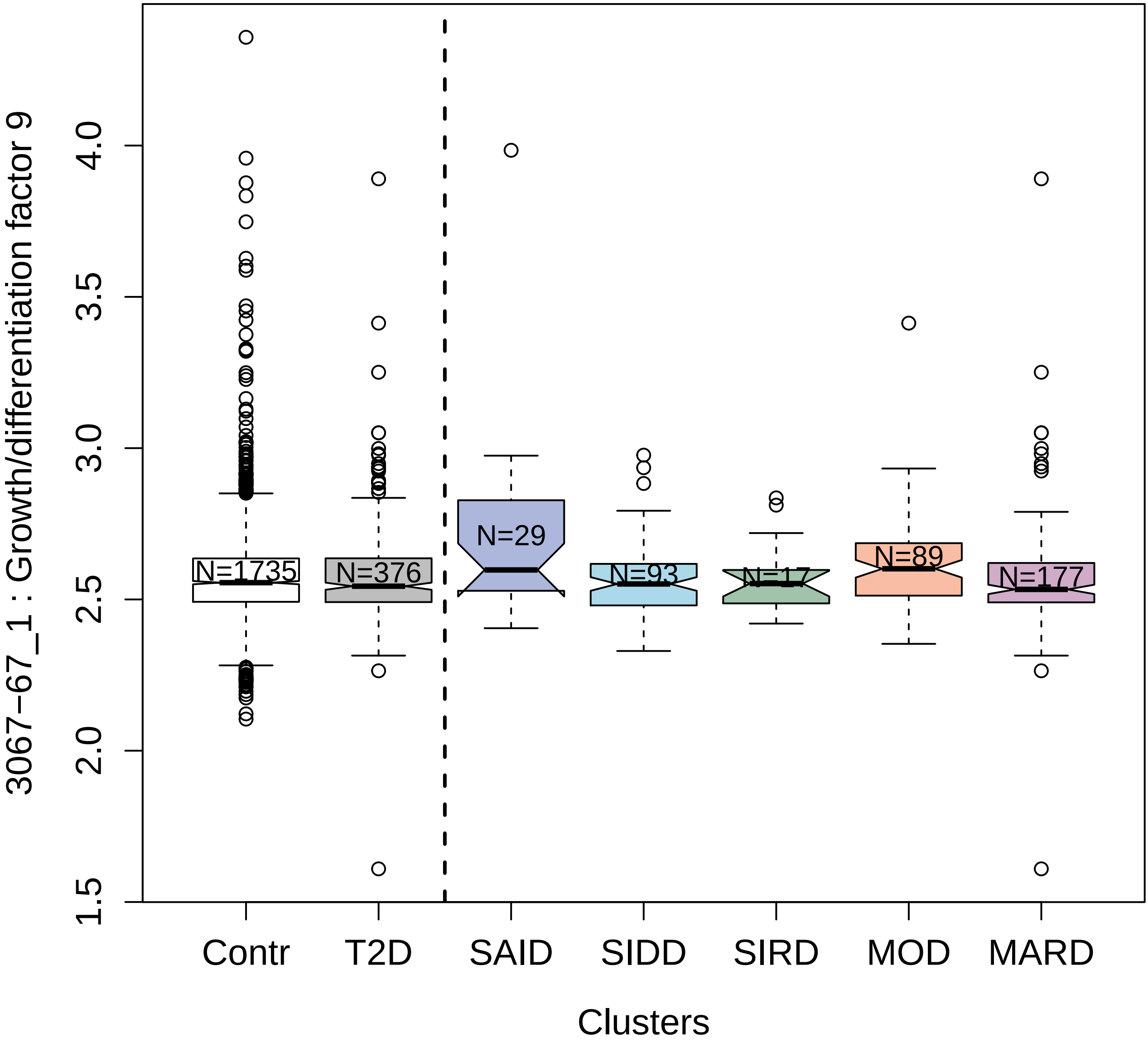
3067−67_1 : Growth/differentiation factor 9
1.5
2.0
2.5
3.0
3.5
4.0
N=1735
N=376
N=29
N=93
N=17
N=89
N=177
Contr
T2D
SAID
SIDD
SIRD
MOD
MARD
Clusters

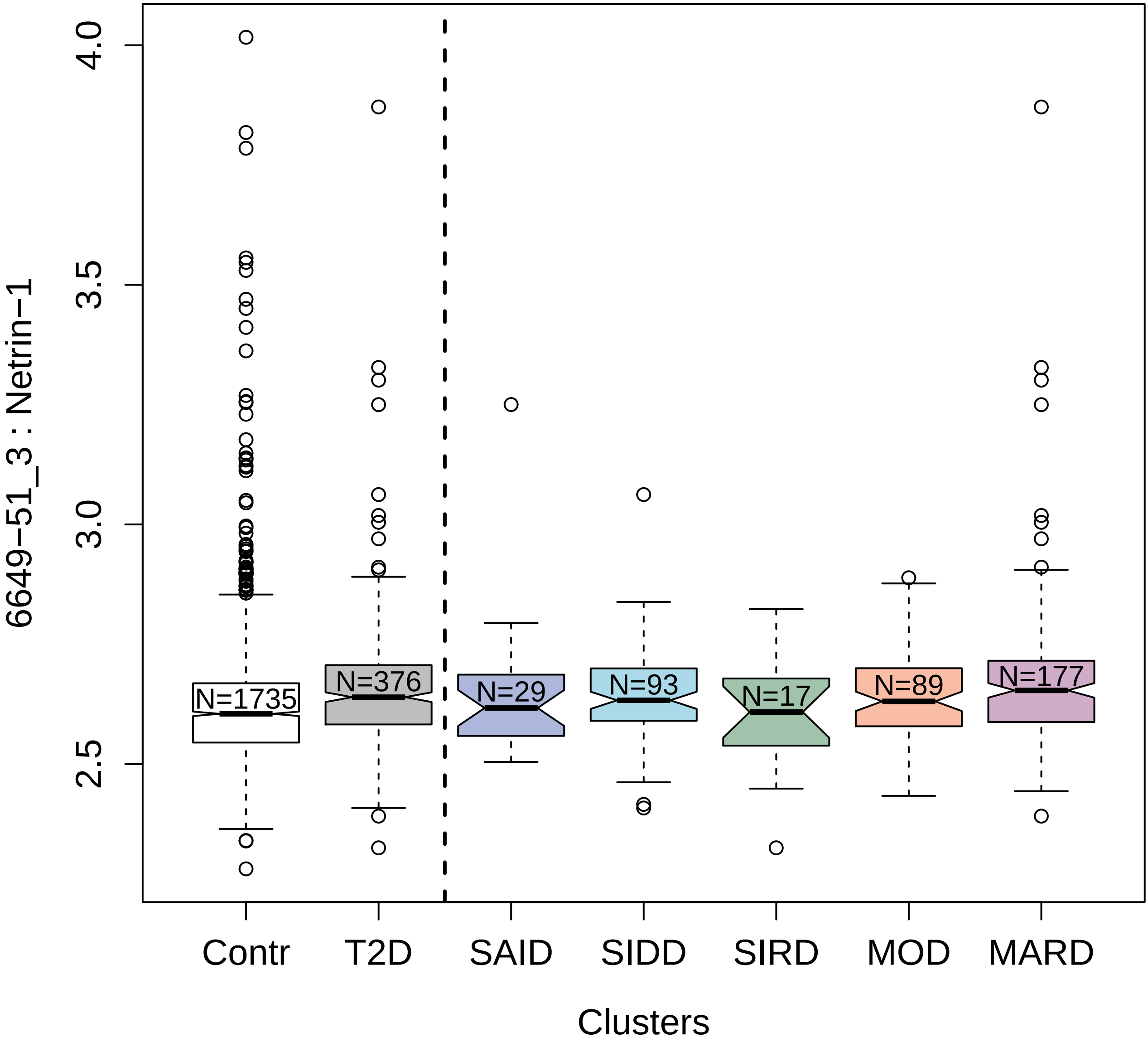
6649−51_3 : Netrin−1
4.0
3.5
3.0
2.5
N=1735
N=376
N=29
N=93
N=17
N=89
N=177
Contr
T2D
SAID
SIDD
SIRD
MOD
MARD
Clusters

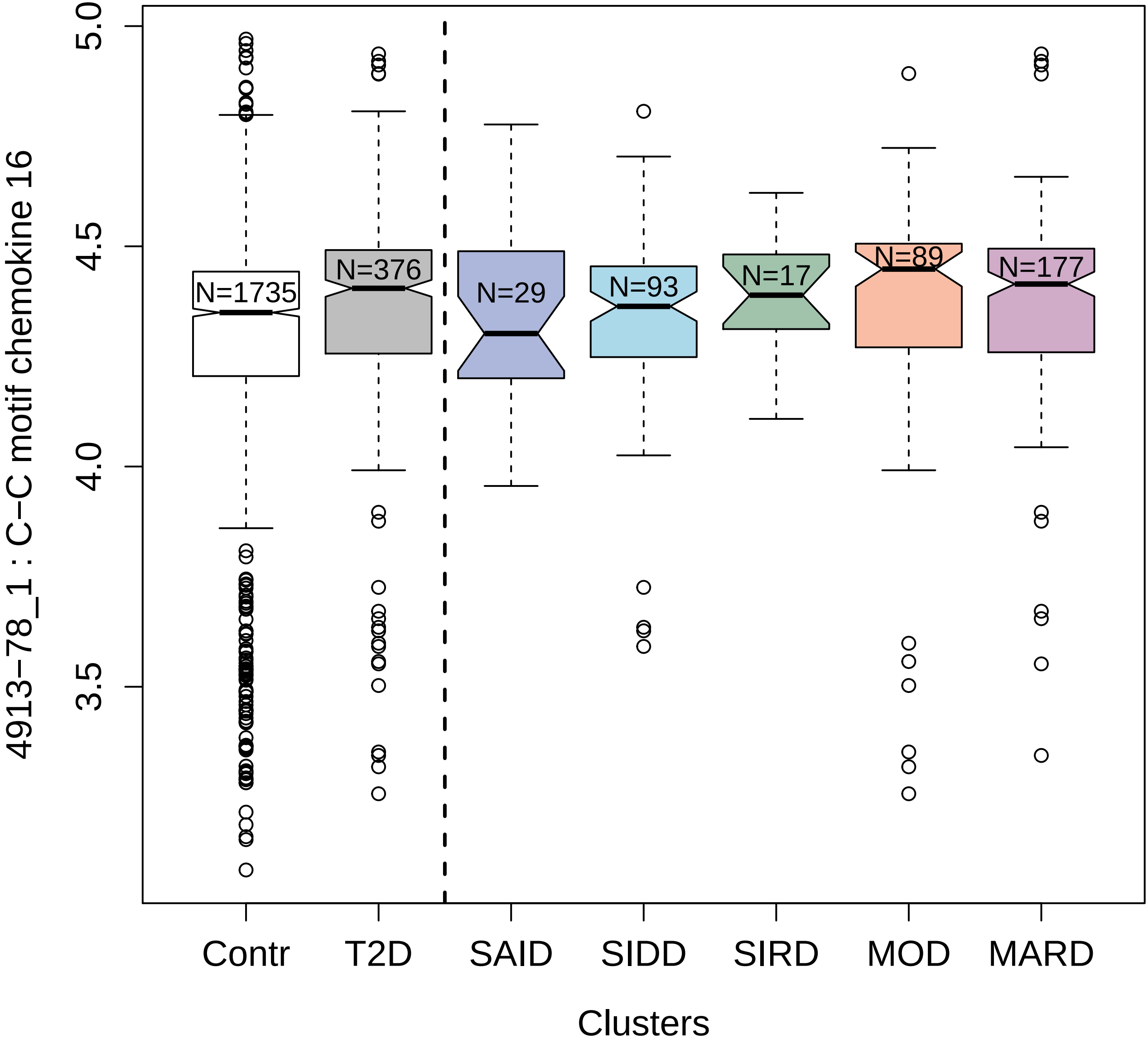
4913−78_1 : C−C motif chemokine 16
5.0
4.5
4.0
3.5
N=1735
N=376
N=29
N=93
N=17
N=89
N=177
Contr
T2D
SAID
SIDD
SIRD
MOD
MARD
Clusters

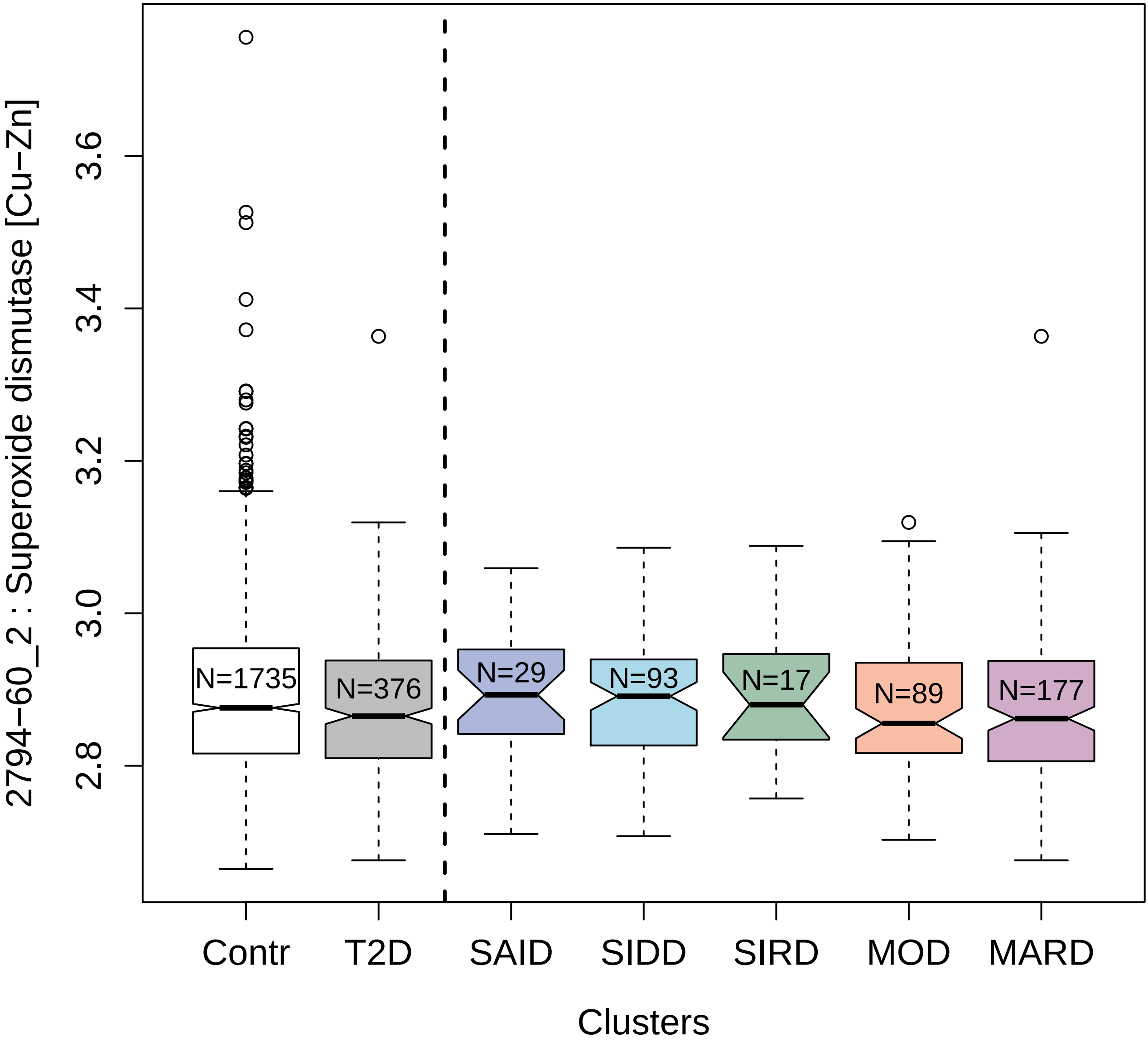

2794−60_2 : Superoxide dismutase [Cu−Zn]
3.6
3.4
3.2
3.0
2.8
N=1735
N=376
N=29
N=93
N=17
N=89
N=177
Contr
T2D
SAID
SIDD
SIRD
MOD
MARD
Clusters

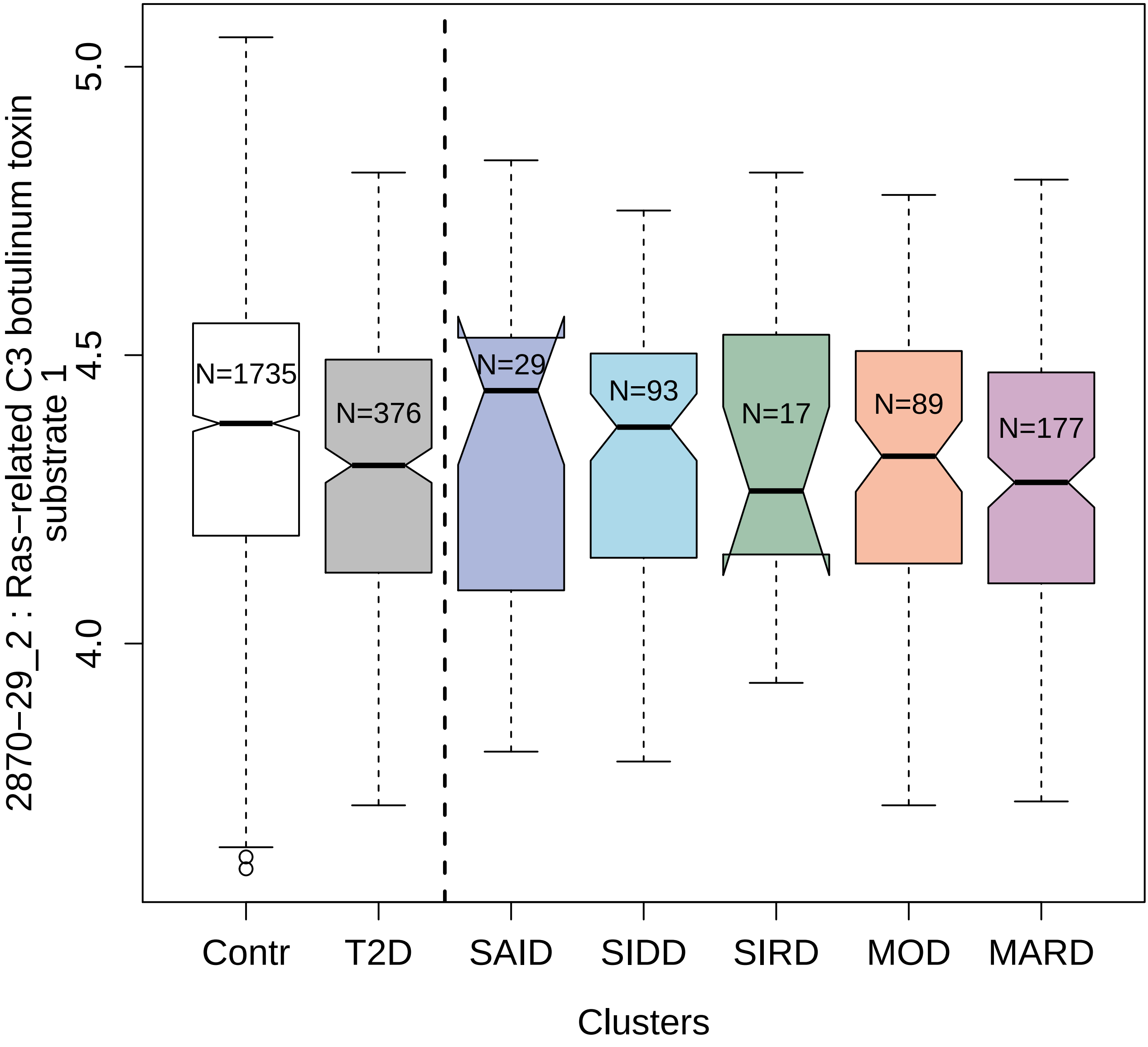
2870−29_2 : Ras−related C3 botulinum toxin substrate 1
5.0
4.5
4.0
N=1735
N=376
N=29
N=93
N=17
N=89
N=177
Contr
T2D
SAID
SIDD
SIRD
MOD
MARD
Clusters

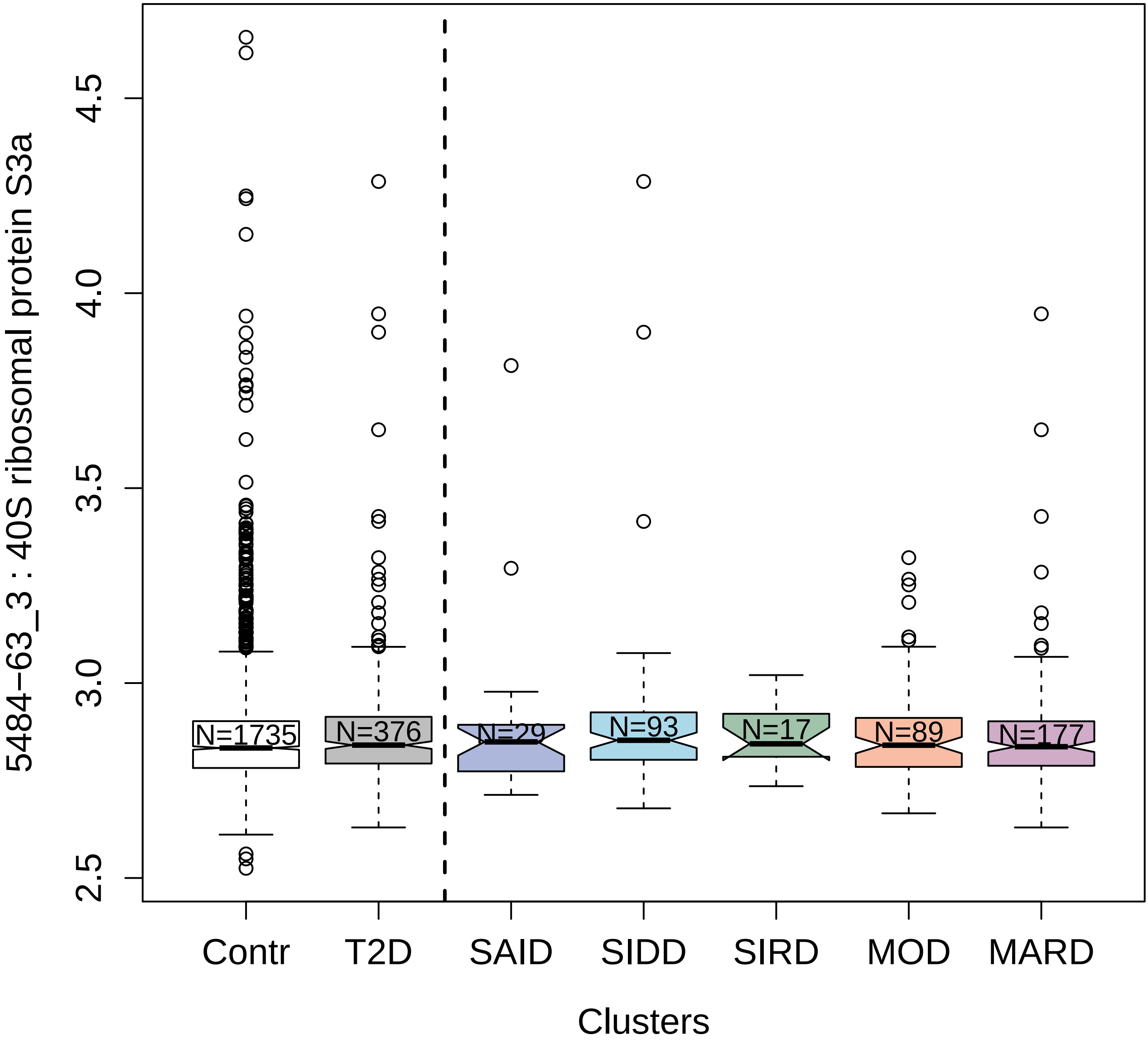

5484−63_3 : 40S ribosomal protein S3a
2.5
3.0
3.5
4.0
4.5
N=1735
N=376
N=29
N=93
N=17
N=89
N=177
Contr
T2D
SAID
SIDD
SIRD
MOD
MARD
Clusters

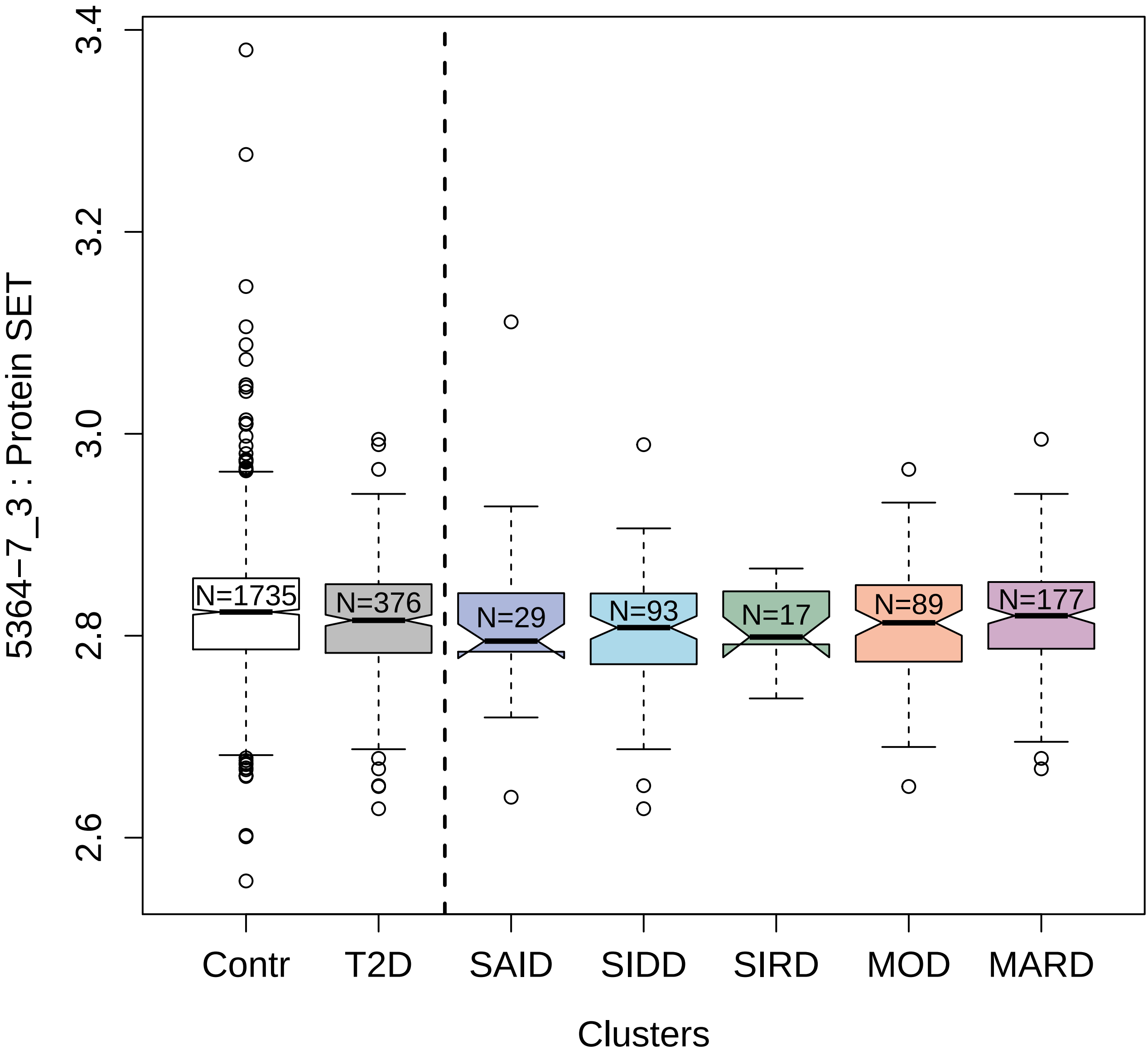
5364−7_3 : Protein SET
3.4
3.2
3.0
2.8
2.6
N=1735
N=376
N=29
N=93
N=17
N=89
N=177
Contr
T2D
SAID
SIDD
SIRD
MOD
MARD
Clusters

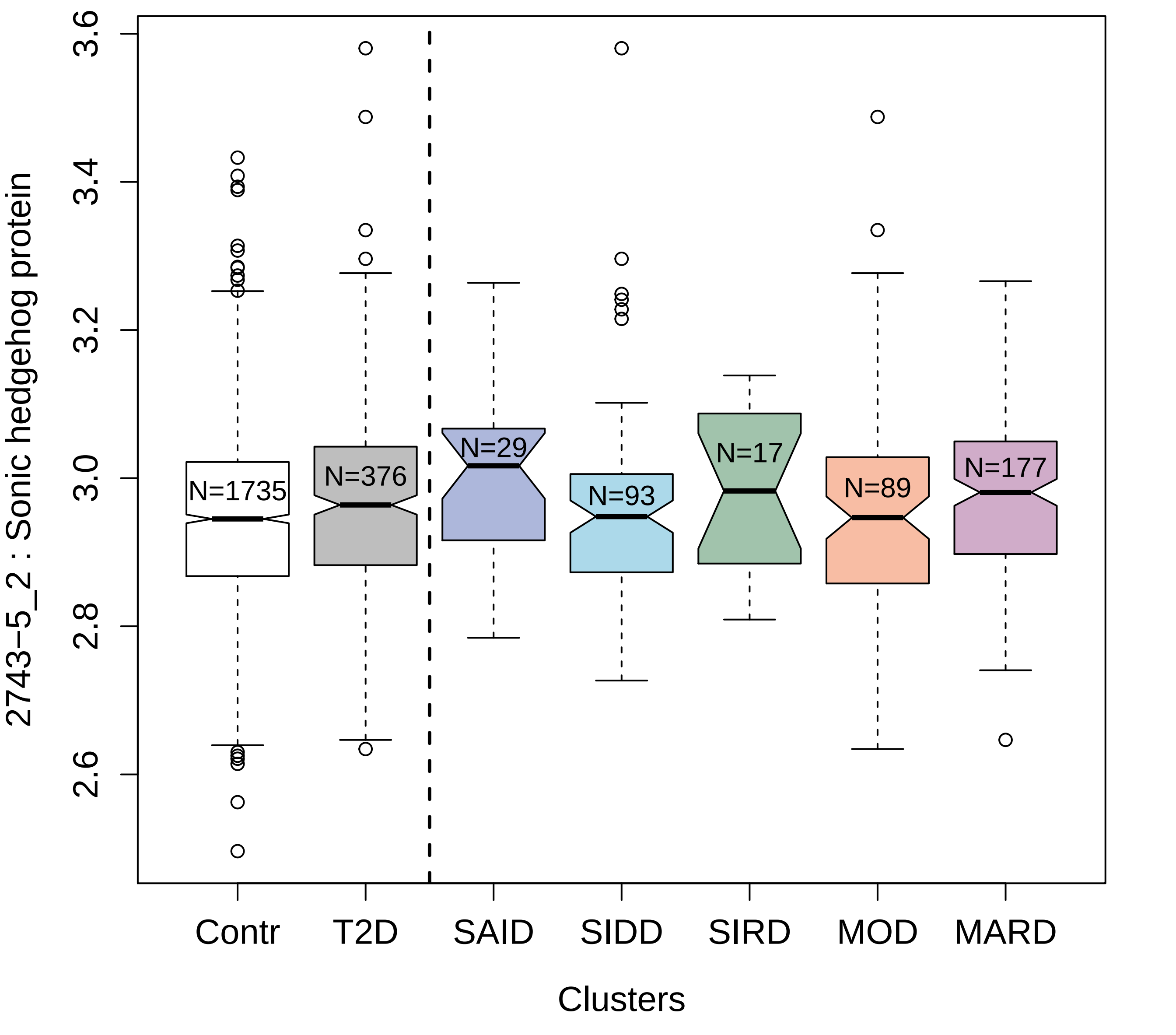
2743−5_2 : Sonic hedgehog protein
3.6
3.4
3.2
3.0
2.8
2.6
N=1735
N=376
N=29
N=93
N=17
N=89
N=177
Contr
T2D
SAID
SIDD
SIRD
MOD
MARD
Clusters

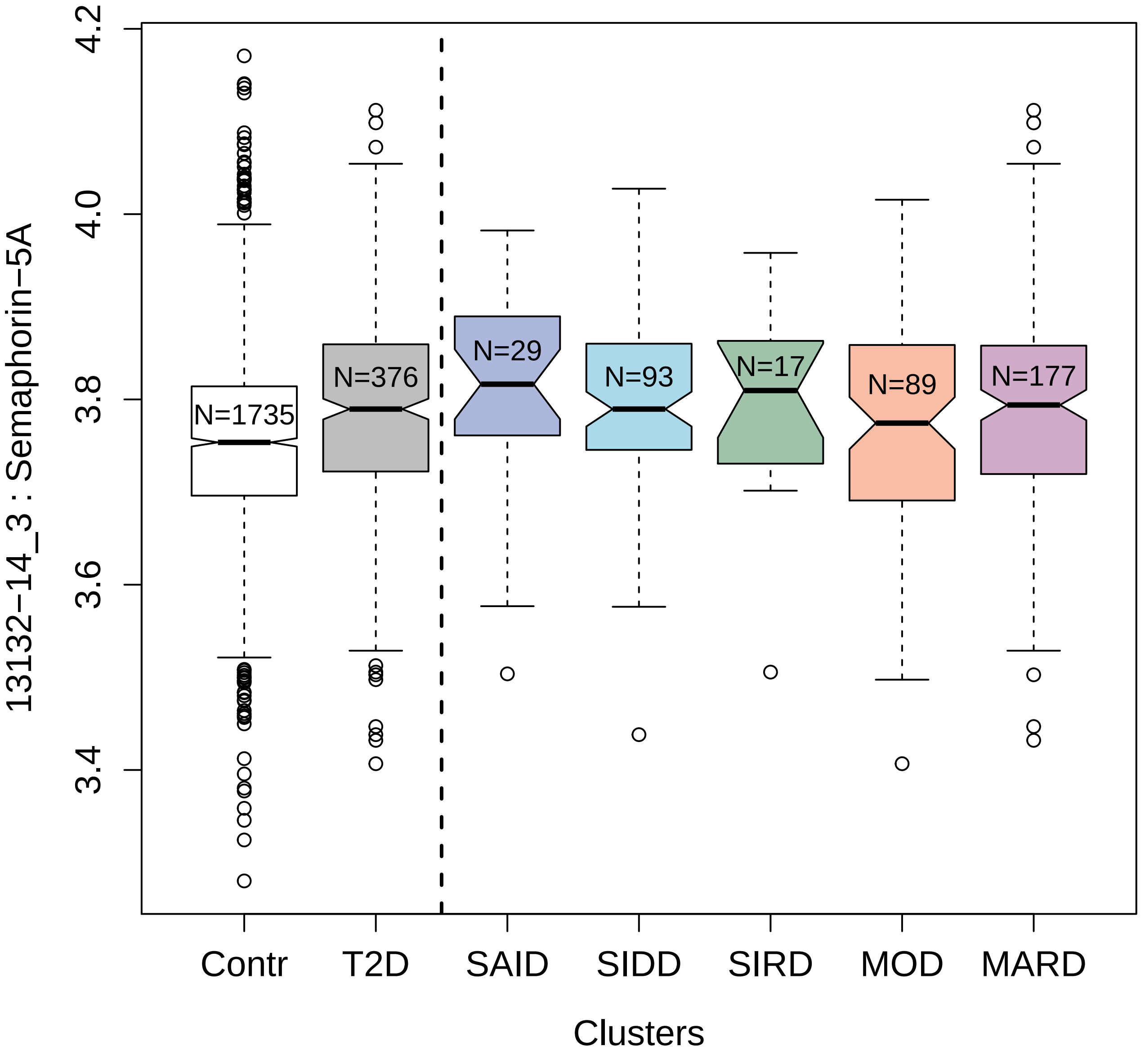

13132−14_3 : Semaphorin−5A
4.2
4.0
3.8
3.6
3.4
N=1735
N=376
N=29
N=93
N=17
N=89
N=177
Contr
T2D
SAID
SIDD
SIRD
MOD
MARD
Clusters

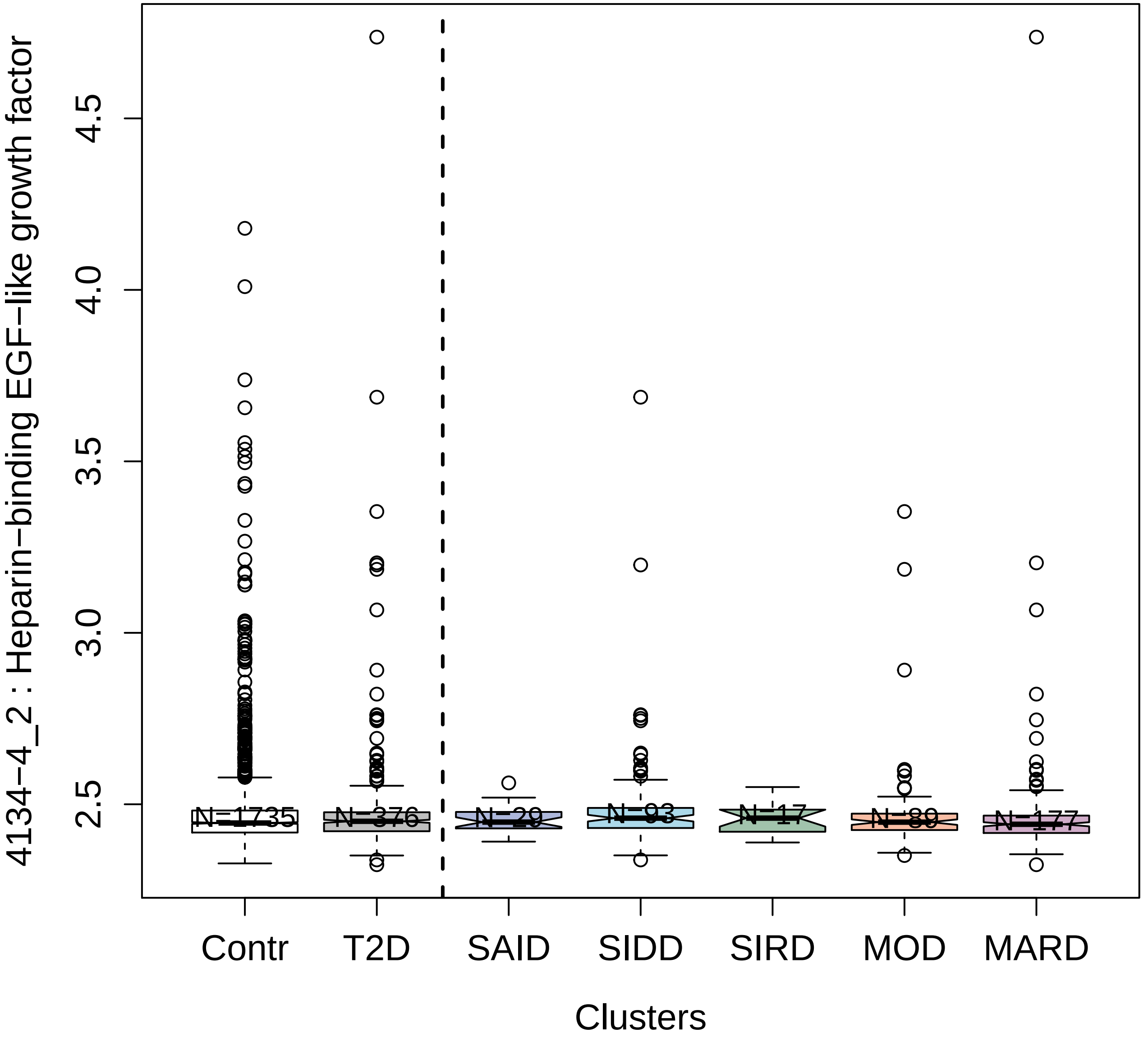

4134−4_2 : Heparin−binding EGF−like growth factor
4.5
4.0
3.5
3.0
2.5
N=1735
N=376
N=29
N=93
N=17
N=89
N=177
Contr
T2D
SAID
SIDD
SIRD
MOD
MARD
Clusters

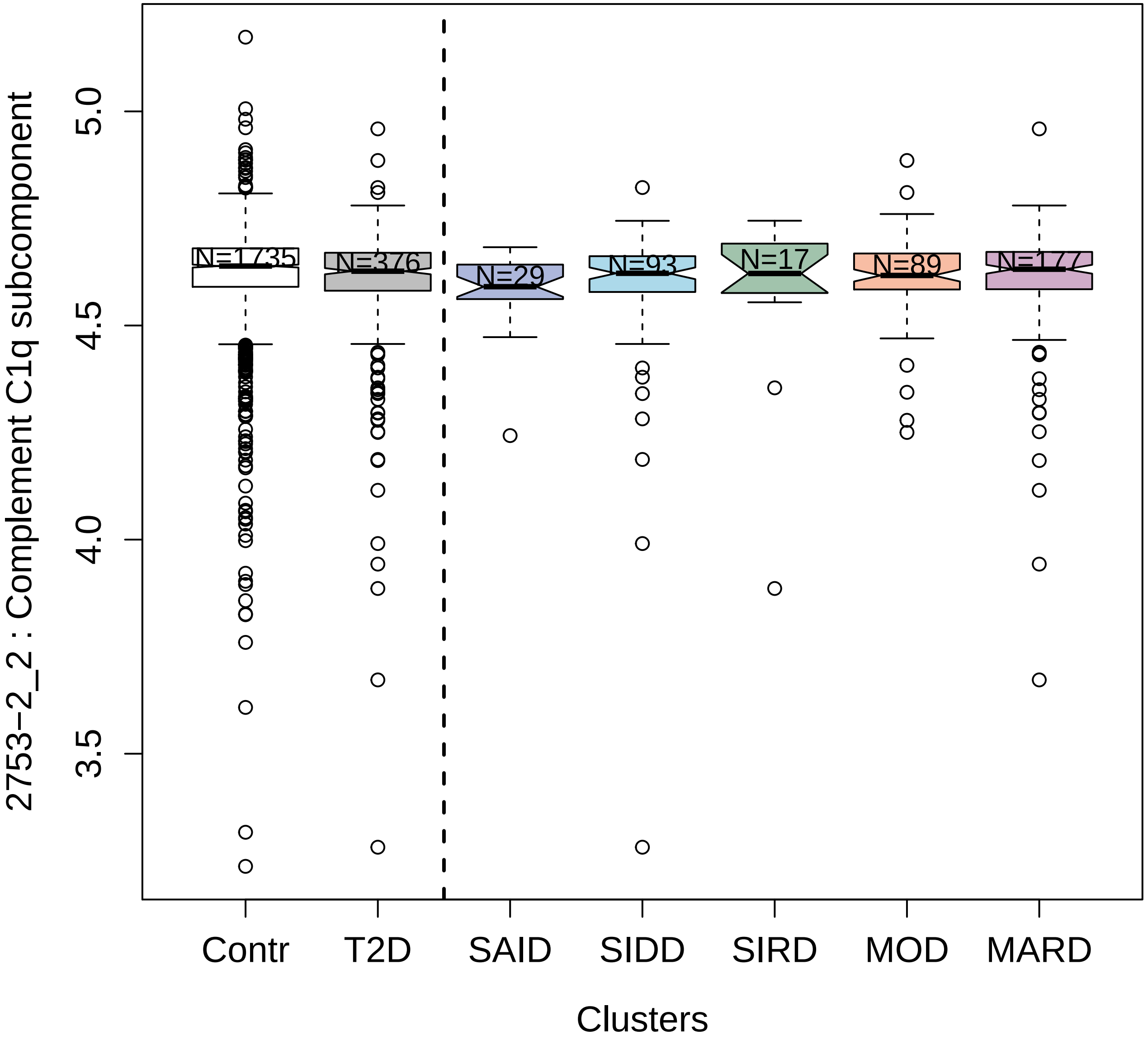
2753−2_2 : Complement C1q subcomponent
5.0
4.5
4.0
3.5
N=1735
N=376
N=29
N=93
N=17
N=89
N=177
Contr
T2D
SAID
SIDD
SIRD
MOD
MARD
Clusters

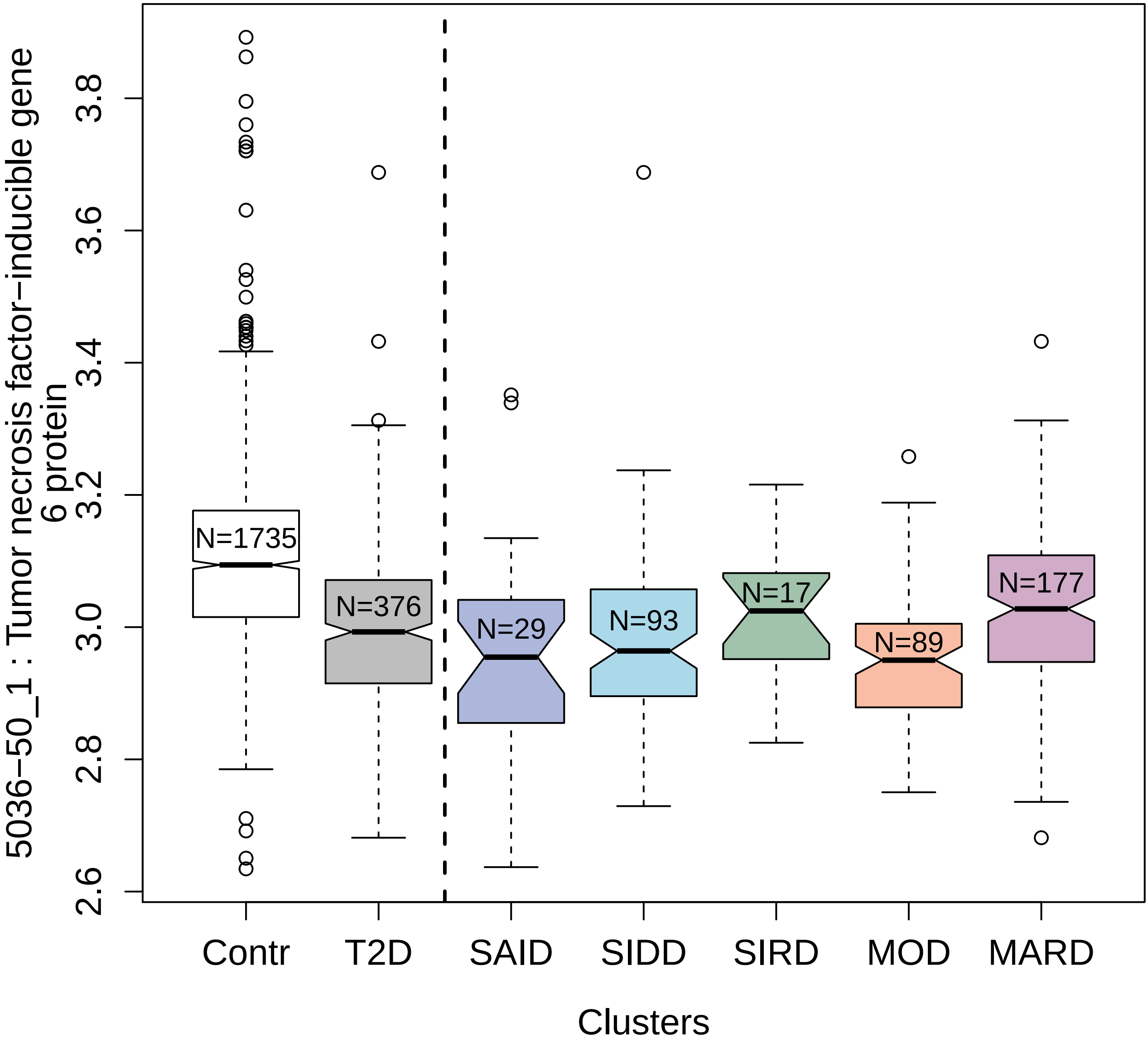
5036−50_1 : Tumor necrosis factor−inducible gene 6 protein
3.8
3.6
3.4
3.2
3.0
2.8
2.6
N=1735
N=376
N=29
N=93
N=17
N=89
N=177
Contr
T2D
SAID
SIDD
SIRD
MOD
MARD
Clusters

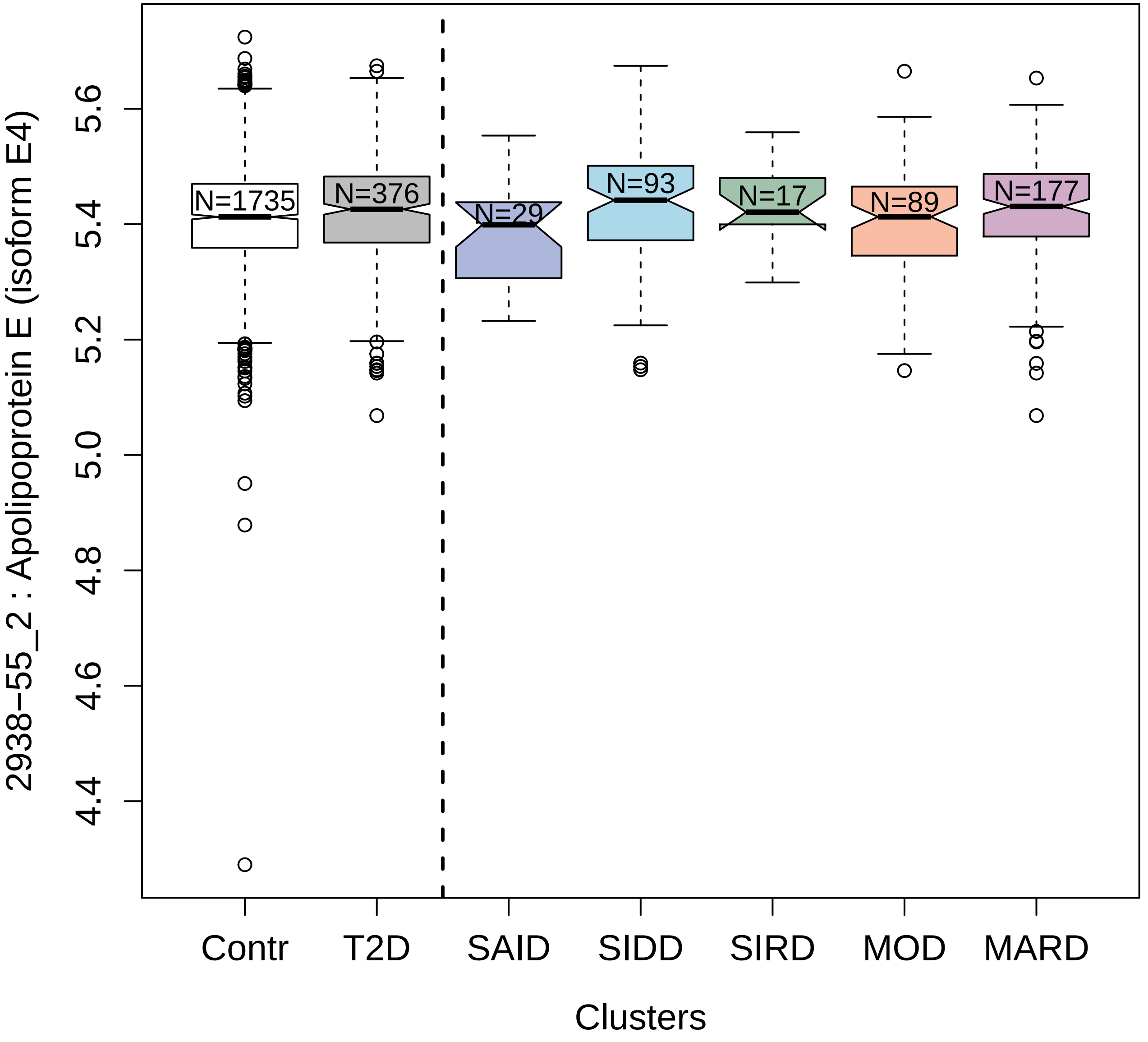

N=1735
N=376
N=29
N=93
N=17
N=89
N=177
5.6
5.4
5.2
5.0
4.8
4.6
4.4
2938−55_2 : Apolipoprotein E (isoform E4)
Contr
T2D
SAID
SIDD
SIRD
MOD
MARD
Clusters

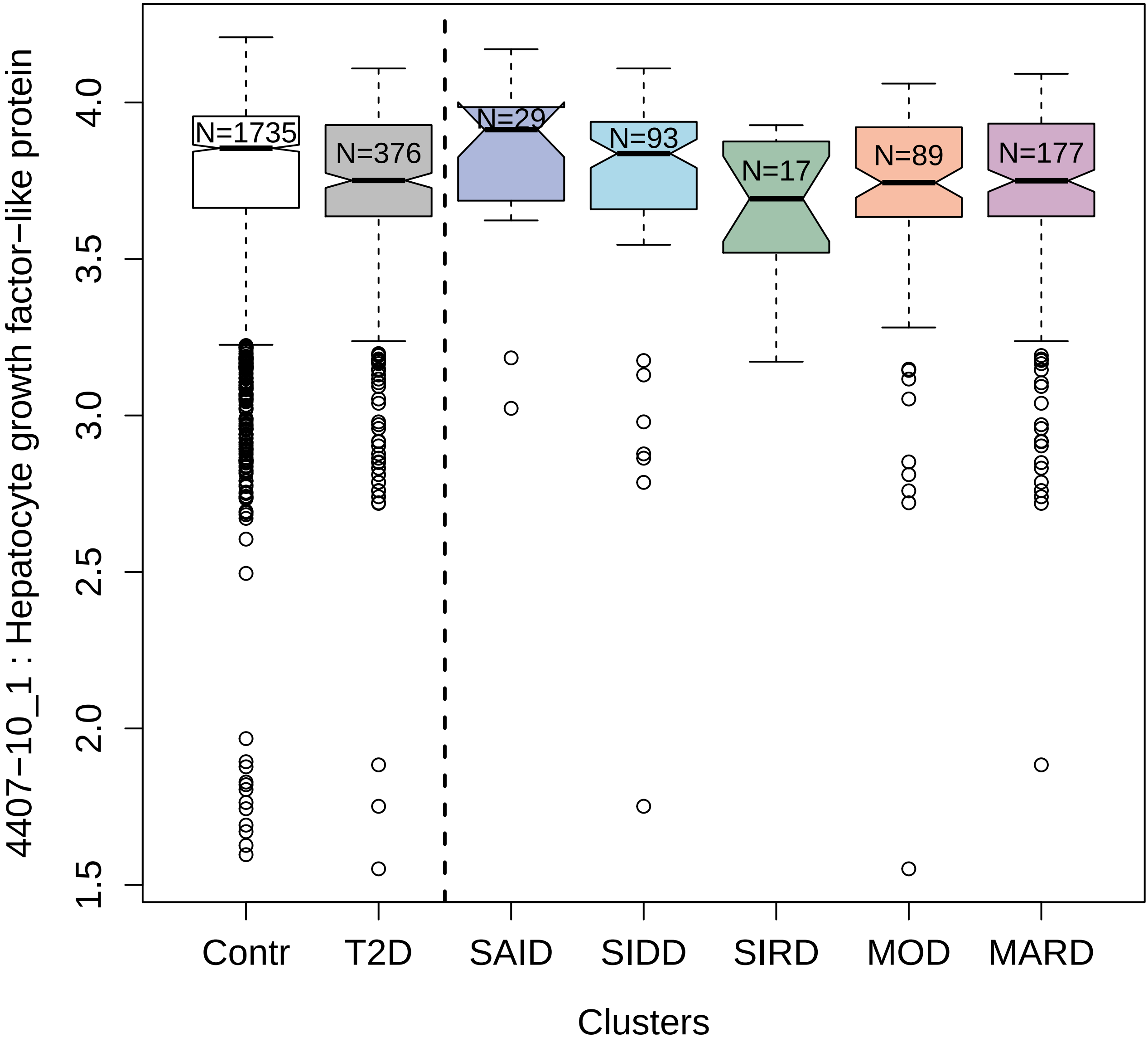
4407−10_1 : Hepatocyte growth factor−like protein
1.5
2.0
2.5
3.0
3.5
4.0
N=1735
N=376
N=29
N=93
N=17
N=89
N=177
Contr
T2D
SAID
SIDD
SIRD
MOD
MARD
Clusters

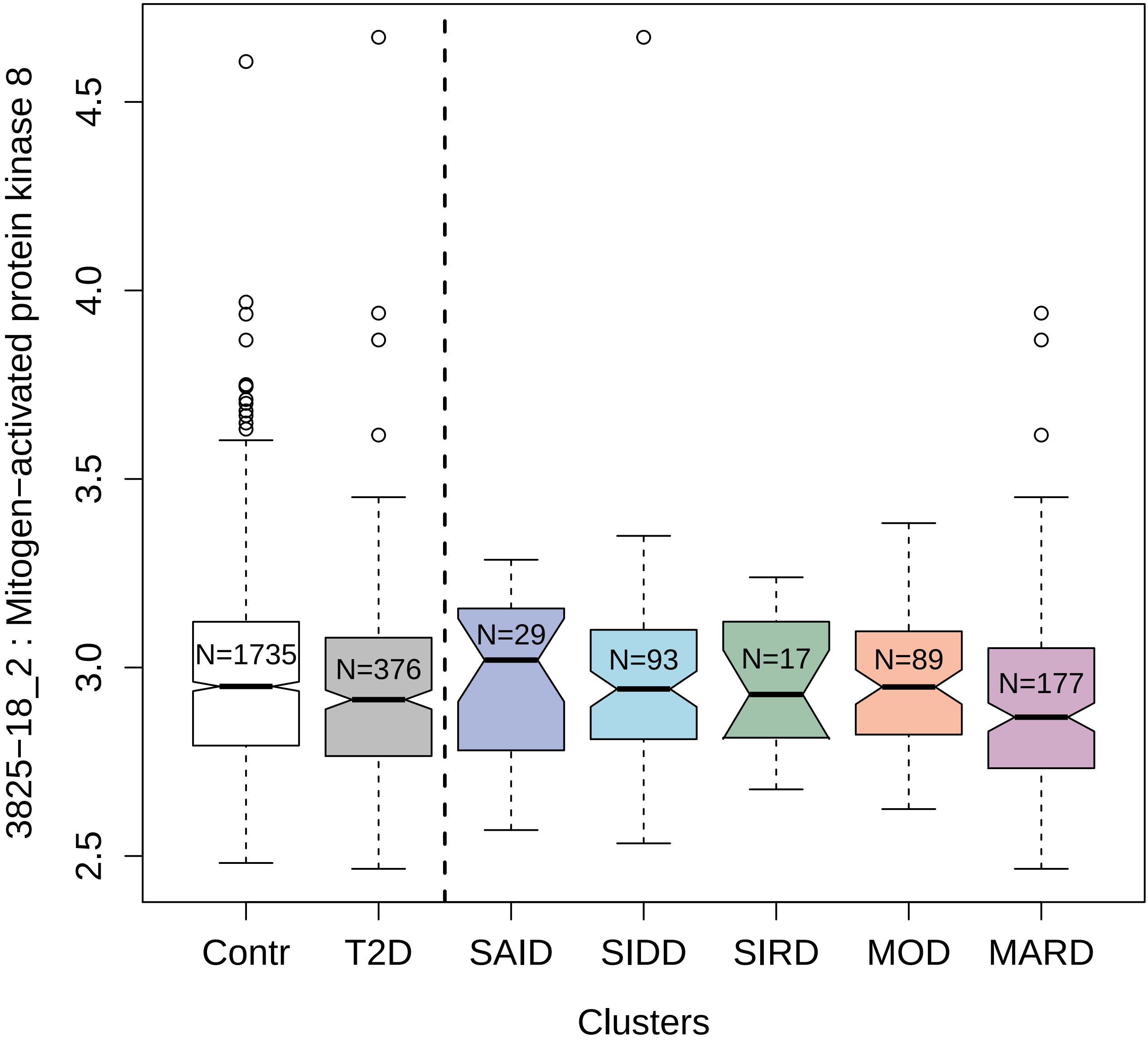

3825−18_2 : Mitogen−activated protein kinase 8
2.5
3.0
3.5
4.0
4.5
N=1735
N=376
N=29
N=93
N=17
N=89
N=177
Contr
T2D
SAID
SIDD
SIRD
MOD
MARD
Clusters

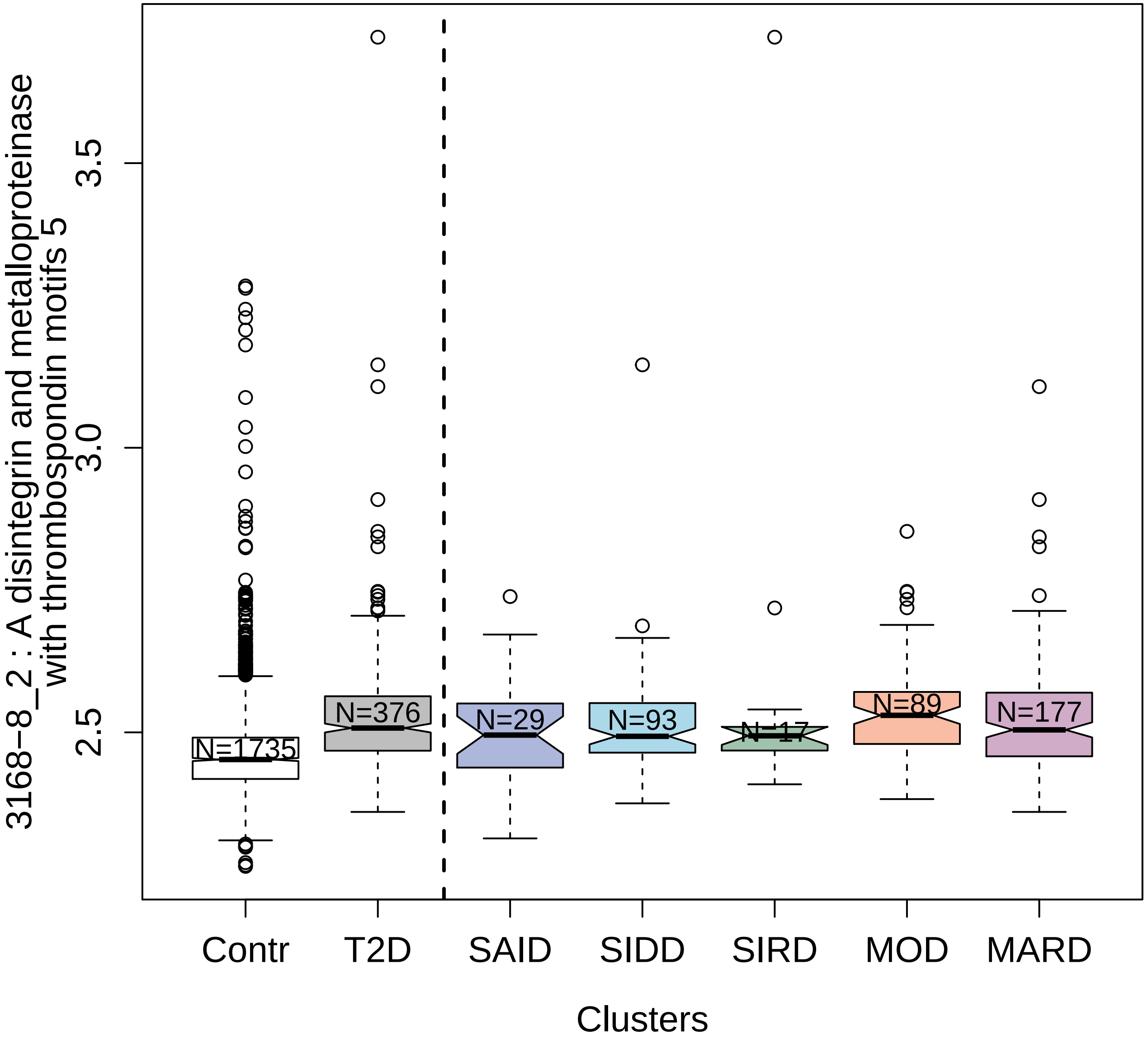
3168−8_2 : A disintegrin and metalloproteinase
with thrombospondin motifs 5
3.5
3.0
2.5
N=1735
N=376
N=29
N=93
N=17
N=89
N=177
Contr
T2D
SAID
SIDD
SIRD
MOD
MARD
Clusters

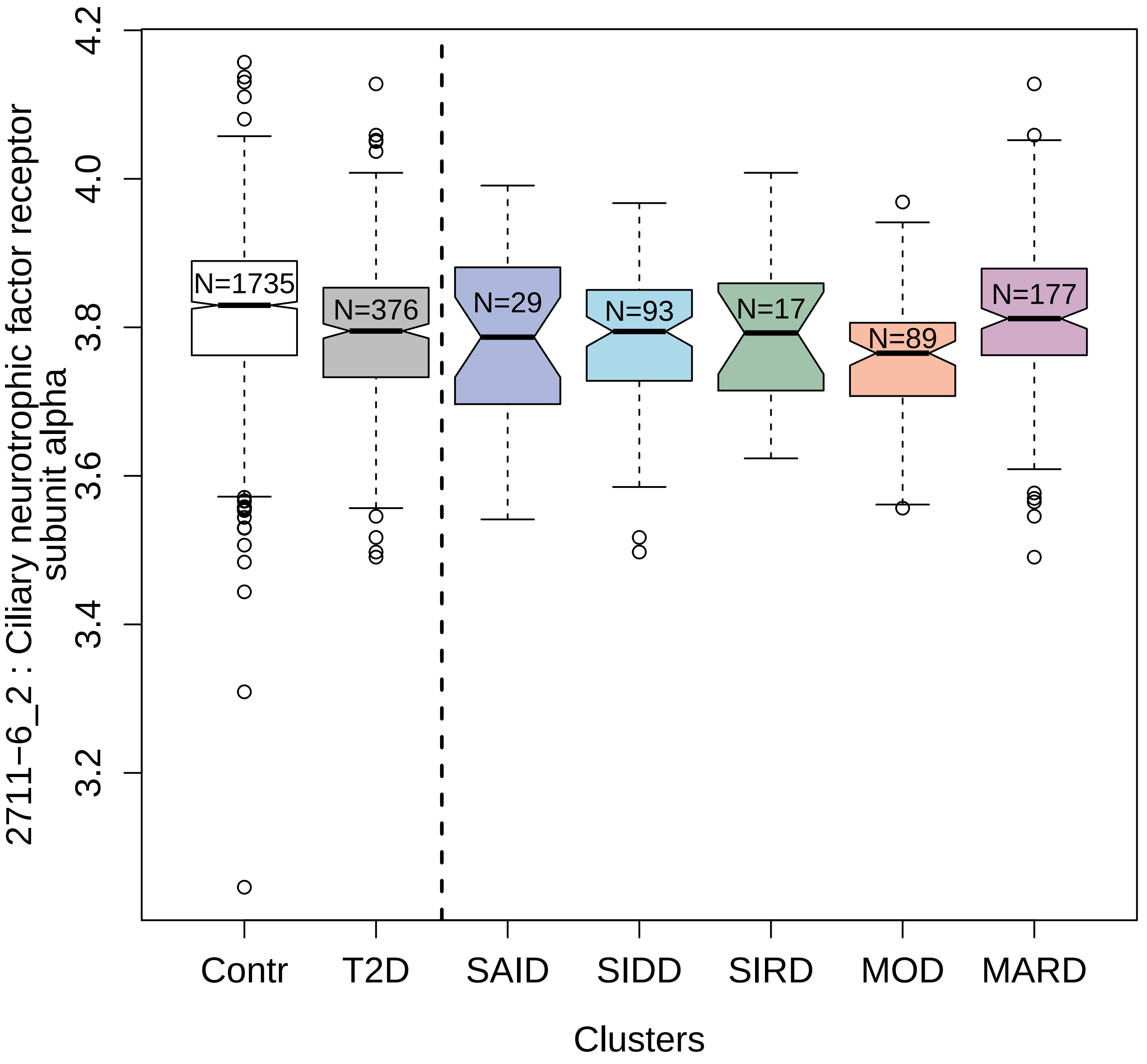
2711−6_2 : Ciliary neurotrophic factor receptor subunit alpha
4.2
4.0
3.8
3.6
3.4
3.2
N=1735
N=376
N=29
N=93
N=17
N=89
N=177
Contr
T2D
SAID
SIDD
SIRD
MOD
MARD
Clusters

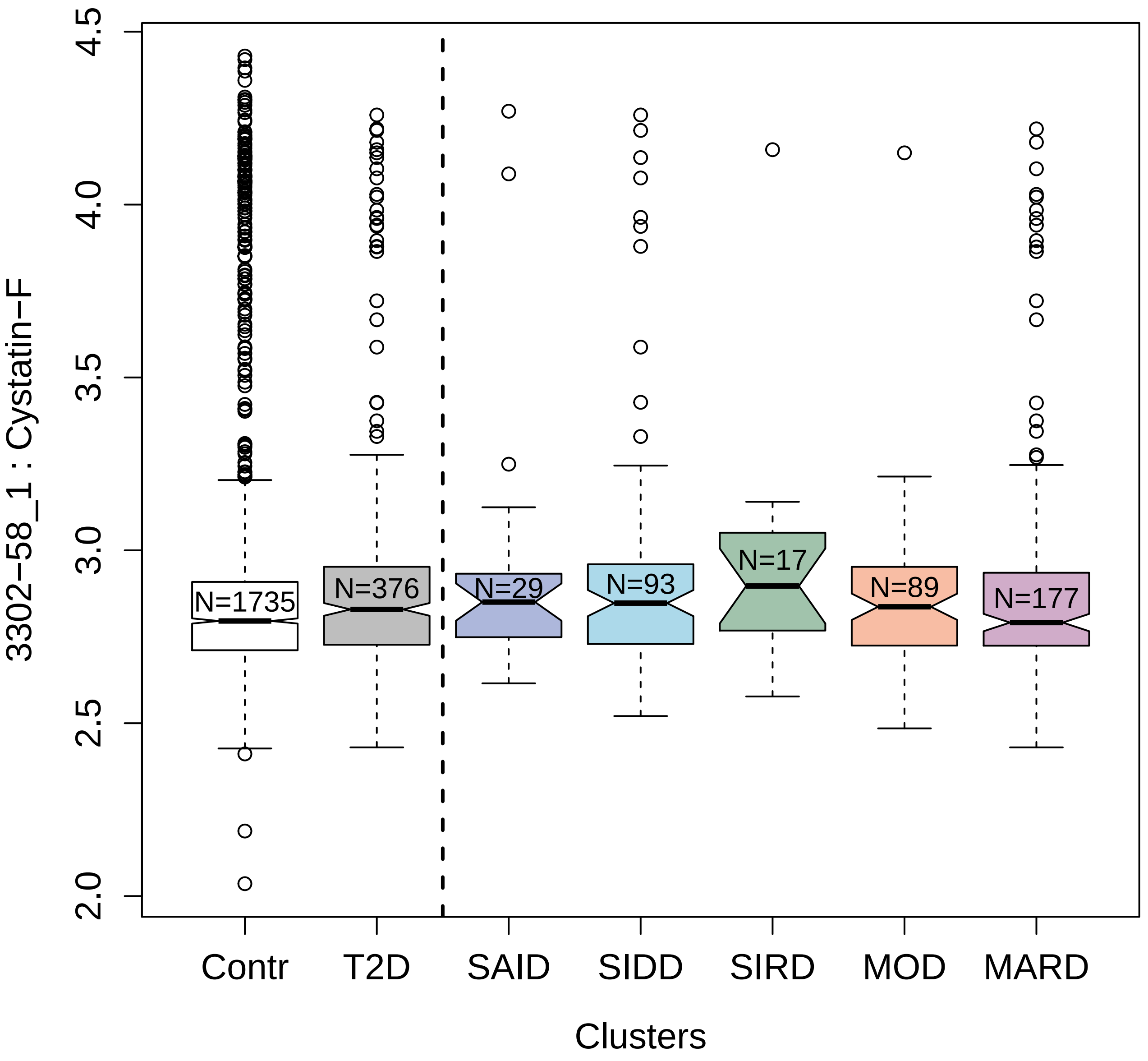

3302−58_1 : Cystatin−F
4.5
4.0
3.5
3.0
2.5
2.0
N=1735
N=376
N=29
N=93
N=17
N=89
N=177
Contr
T2D
SAID
SIDD
SIRD
MOD
MARD
Clusters

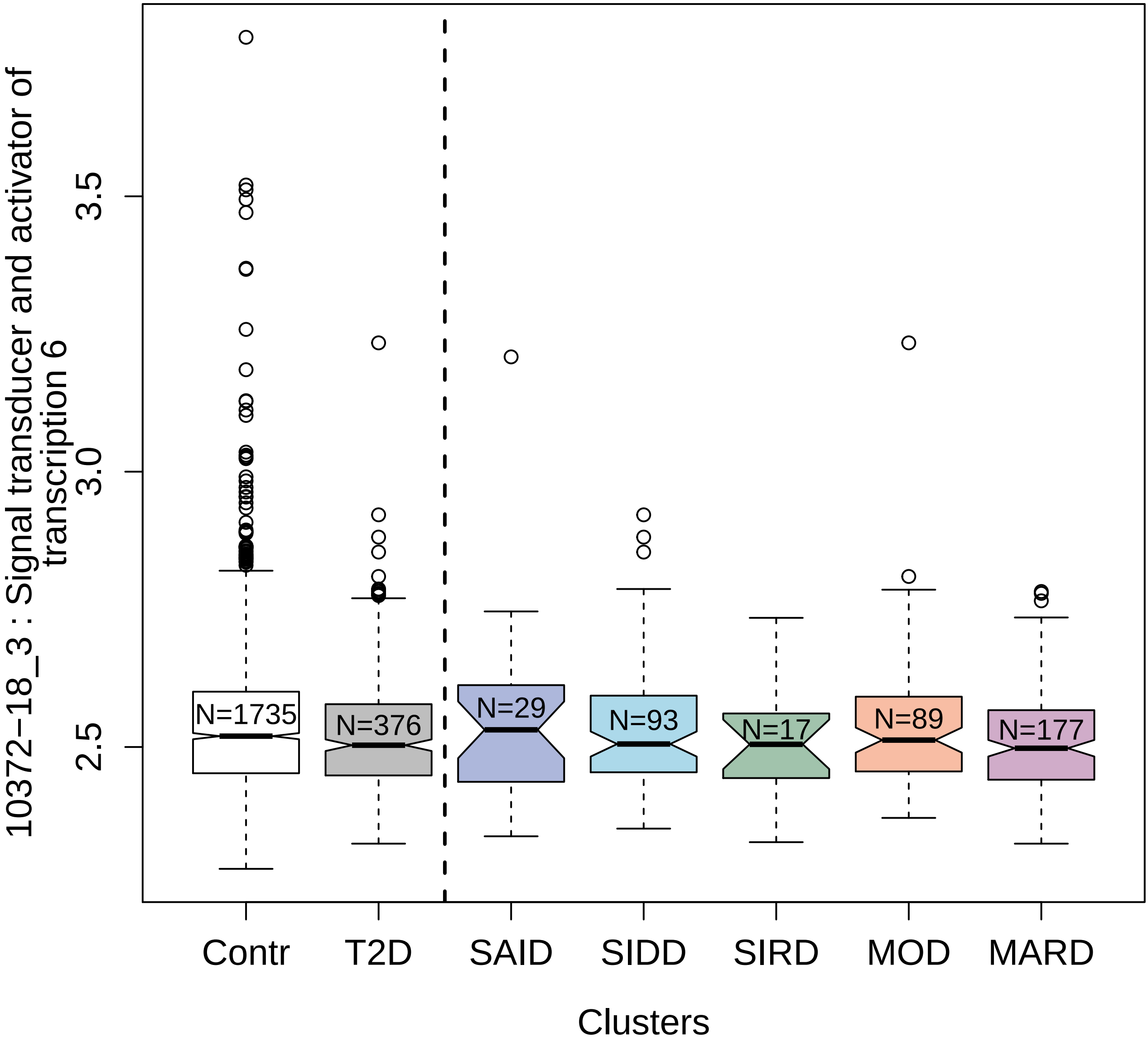
10372−18_3 : Signal transducer and activator of transcription 6
3.5
3.0
2.5
N=1735
N=376
N=29
N=93
N=17
N=89
N=177
Contr
T2D
SAID
SIDD
SIRD
MOD
MARD
Clusters

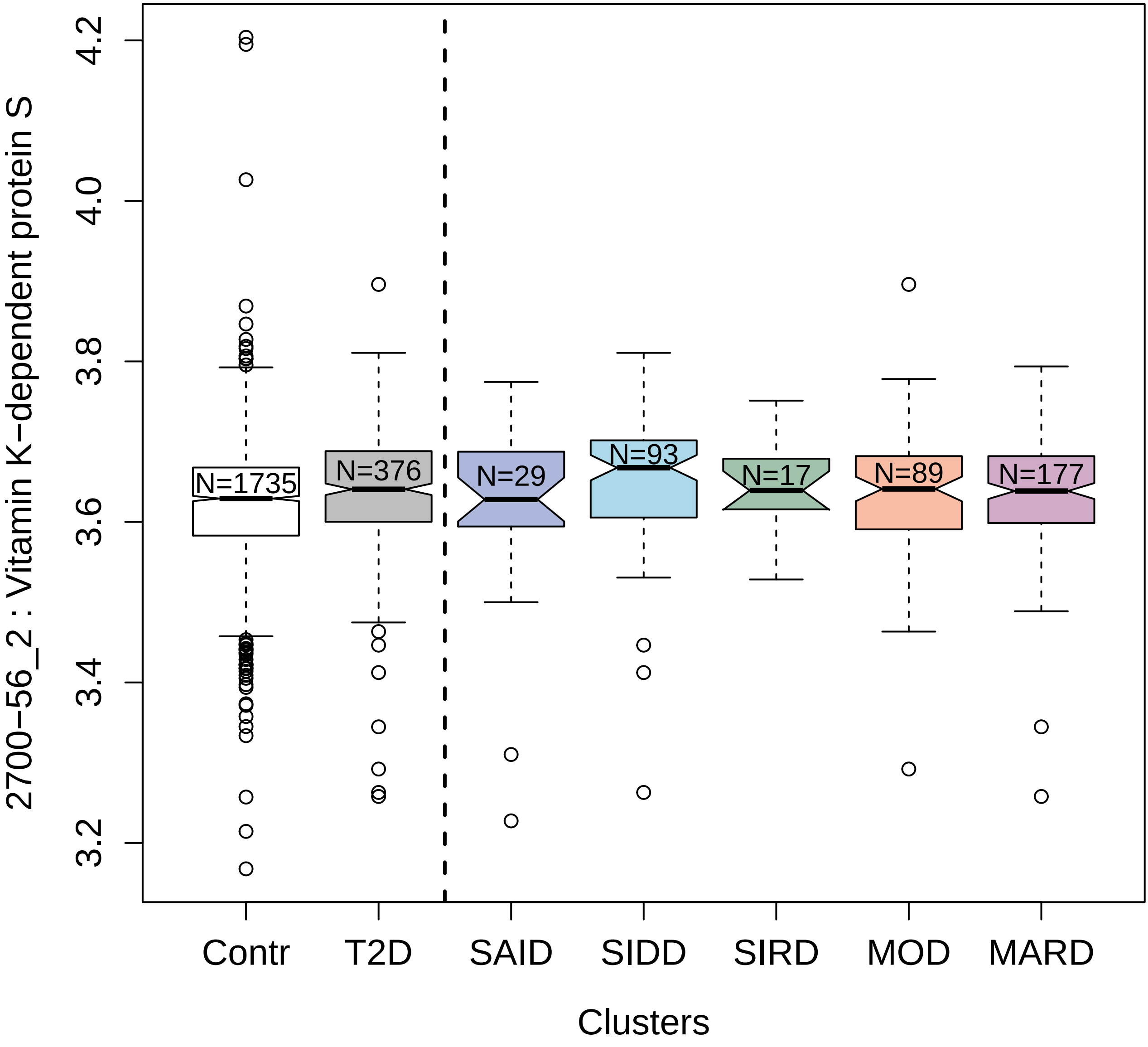

2700−56_2 : Vitamin K−dependent protein S
4.2
4.0
3.8
3.6
3.4
3.2
N=1735
N=376
N=29
N=93
N=17
N=89
N=177
Contr
T2D
SAID
SIDD
SIRD
MOD
MARD
Clusters

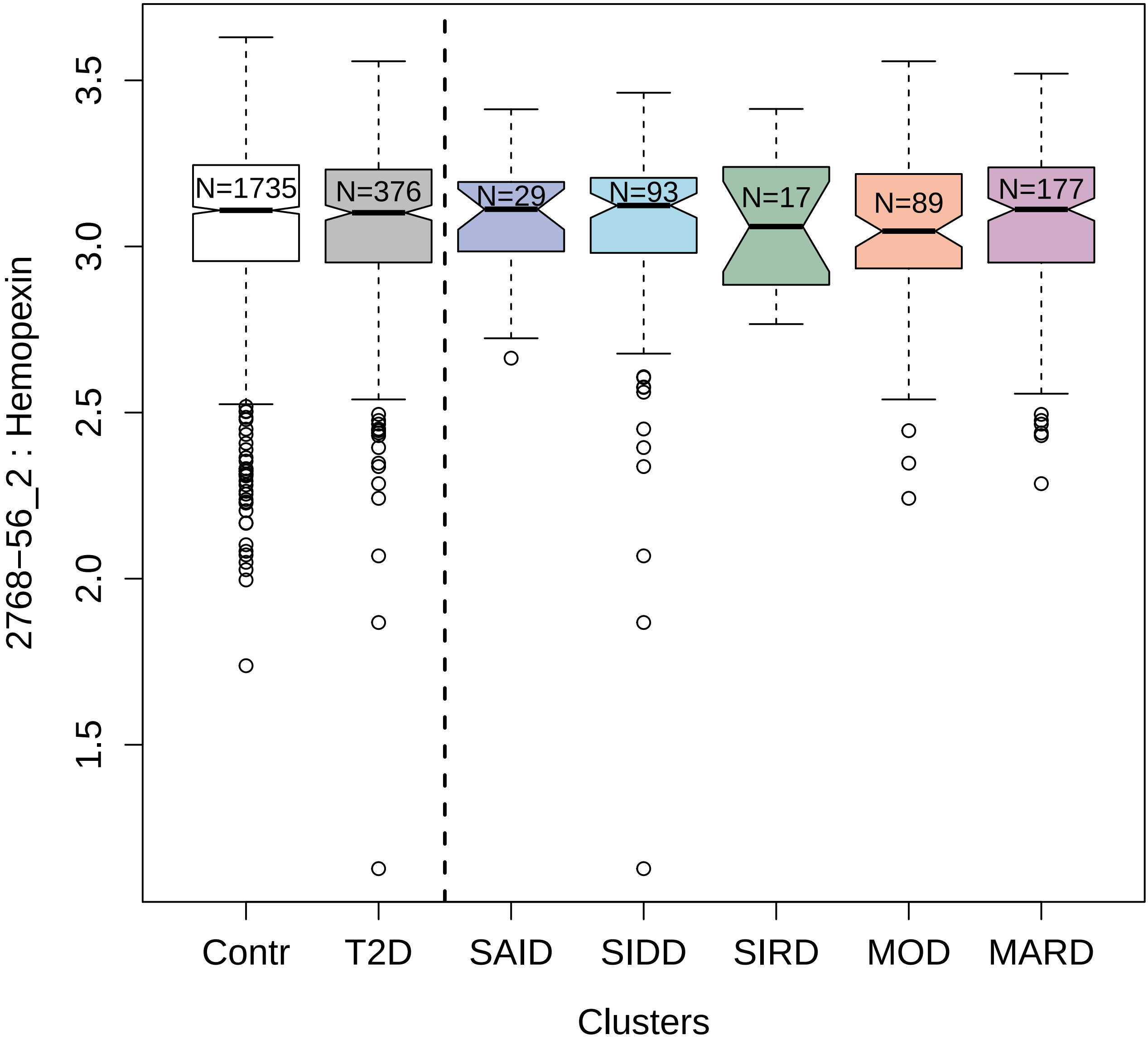

2768−56_2 : Hemopexin
3.5
3.0
2.5
2.0
1.5
N=1735
N=376
N=29
N=93
N=17
N=89
N=177
Contr
T2D
SAID
SIDD
SIRD
MOD
MARD
Clusters

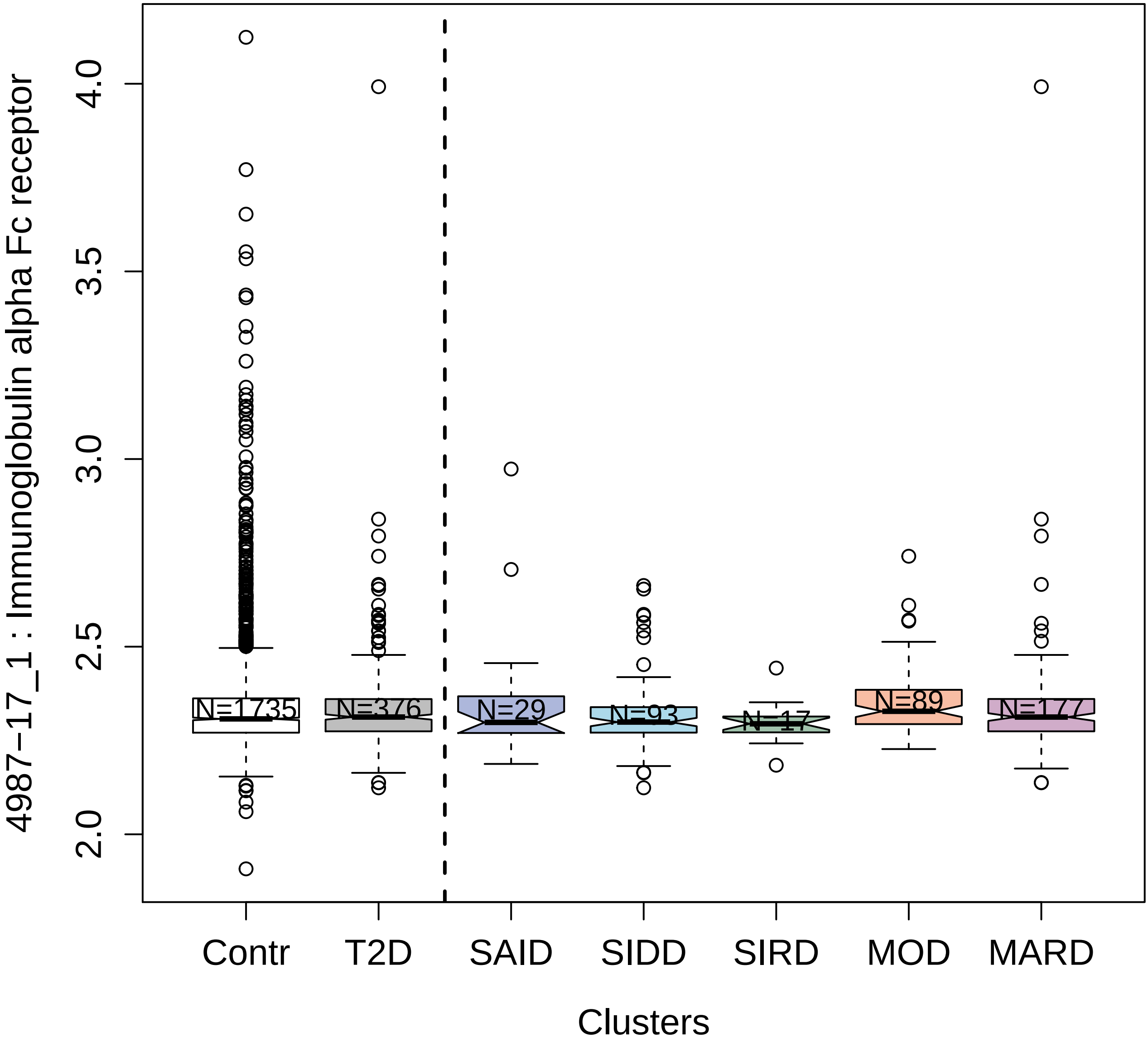

4987−17_1 : Immunoglobulin alpha Fc receptor
4.0
3.5
3.0
2.5
2.0
N=1735
N=376
N=29
N=93
N=17
N=89
N=177
Contr
T2D
SAID
SIDD
SIRD
MOD
MARD
Clusters

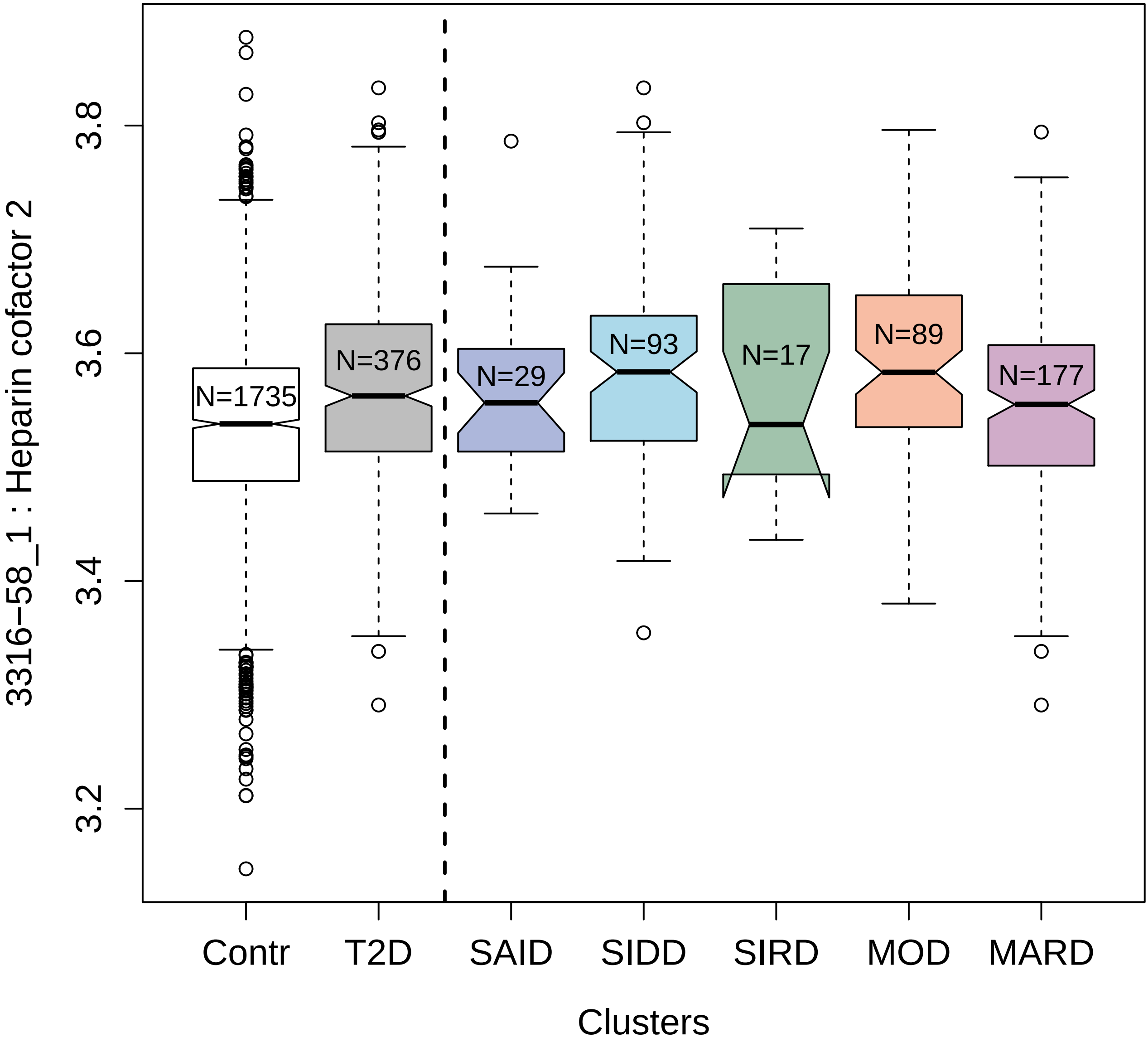

3316−58_1 : Heparin cofactor 2
3.2
3.4
3.6
3.8
N=1735
N=376
N=29
N=93
N=17
N=89
N=177
Contr
T2D
SAID
SIDD
SIRD
MOD
MARD
Clusters

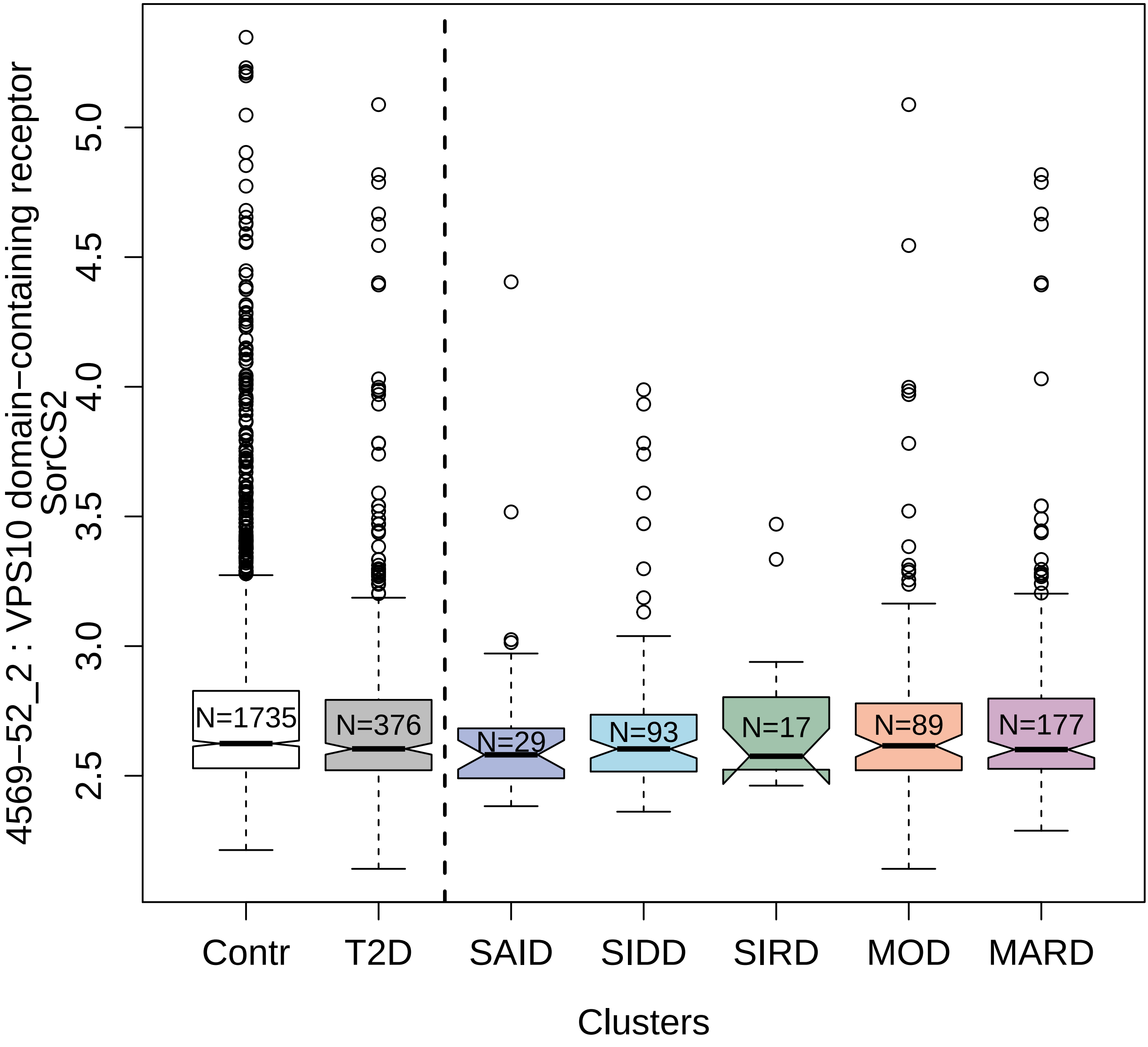
4569−52_2 : VPS10 domain−containing receptor
SorCS2
5.0
4.5
4.0
3.5
3.0
2.5
N=1735
N=376
N=29
N=93
N=17
N=89
N=177
Contr
T2D
SAID
SIDD
SIRD
MOD
MARD
Clusters

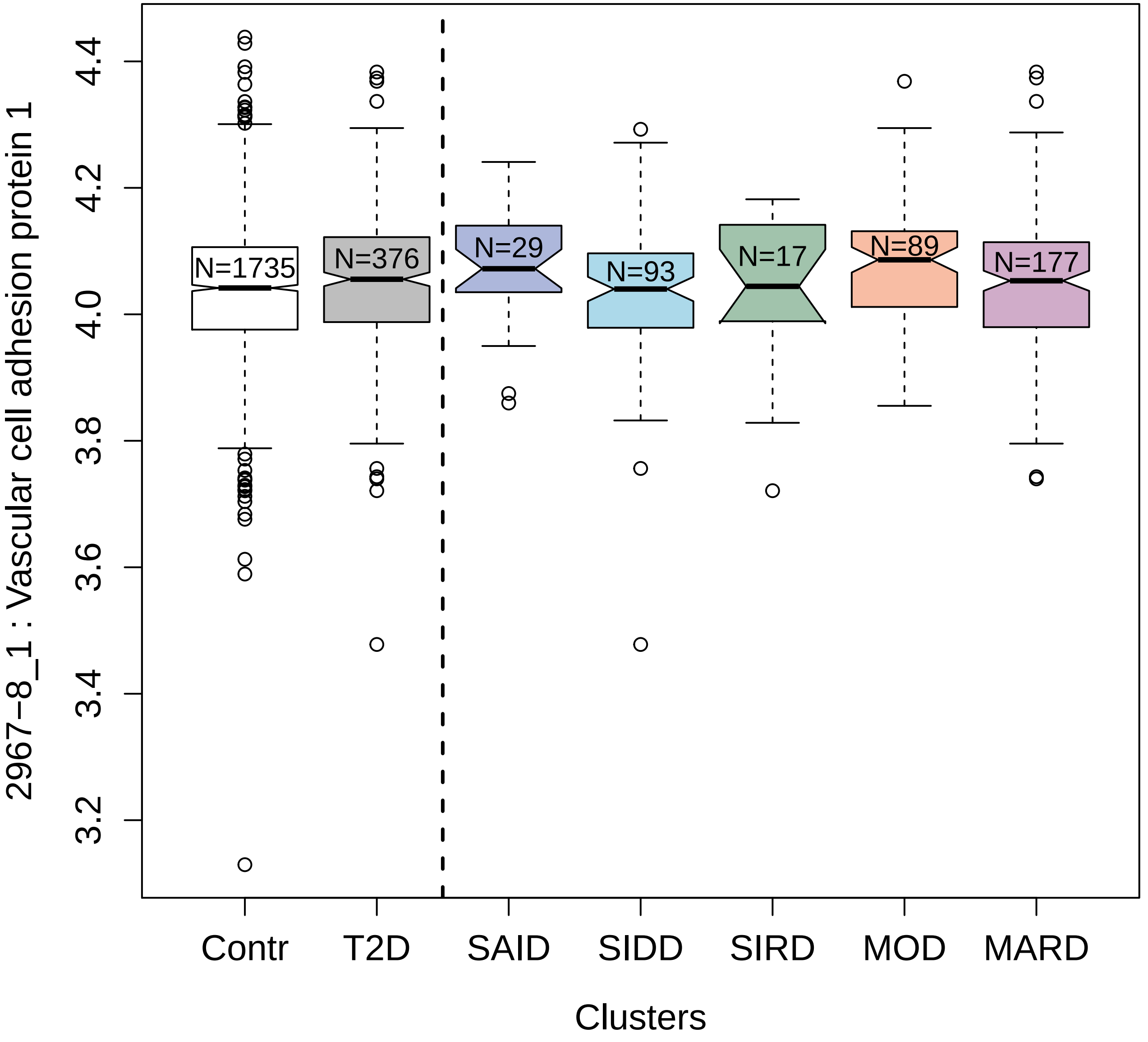

2967−8_1 : Vascular cell adhesion protein 1
3.2
3.4
3.6
3.8
4.0
4.2
4.4
N=1735
N=376
N=29
N=93
N=17
N=89
N=177
Contr
T2D
SAID
SIDD
SIRD
MOD
MARD
Clusters

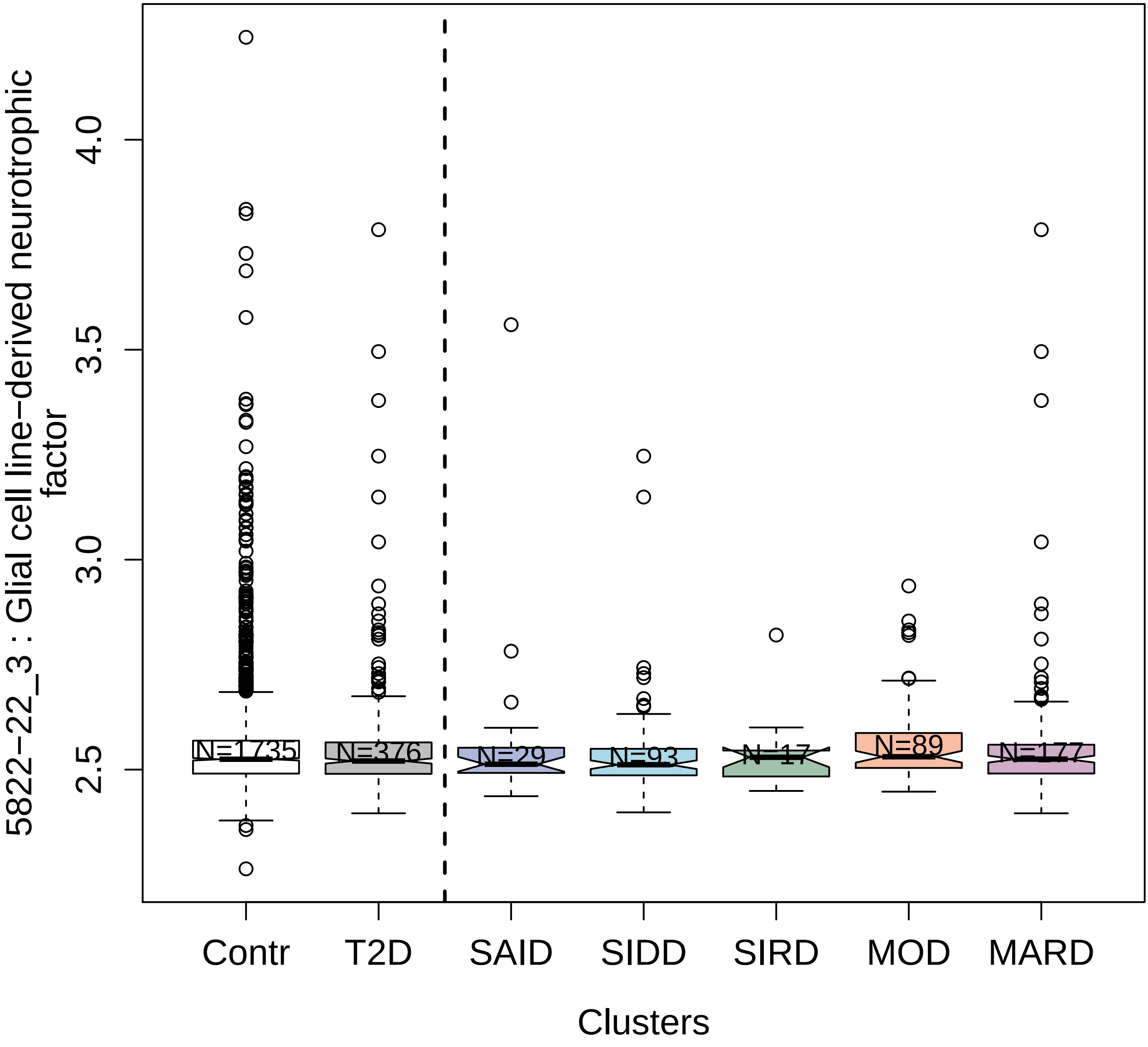

5822−22_3 : Glial cell line−derived neurotrophic factor
4.0
3.5
3.0
2.5
N=1735
N=376
N=29
N=93
N=17
N=89
N=177
Contr
T2D
SAID
SIDD
SIRD
MOD
MARD
Clusters

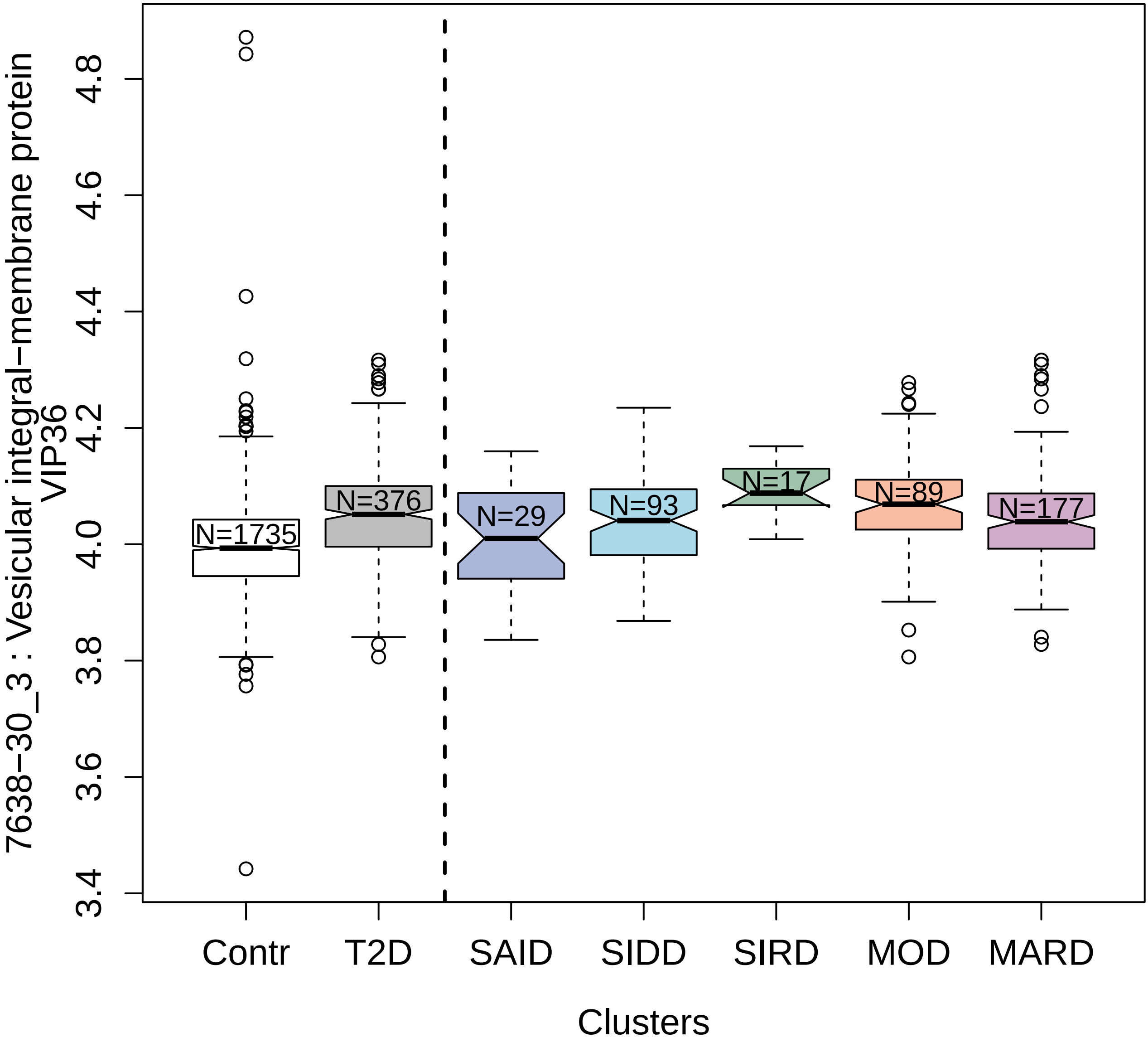

7638−30_3 : Vesicular integral−membrane protein
VIP36
4.8
4.6
4.4
4.2
4.0
3.8
3.6
3.4
N=1735
N=376
N=29
N=93
N=17
N=89
N=177
Contr
T2D
SAID
SIDD
SIRD
MOD
MARD
Clusters

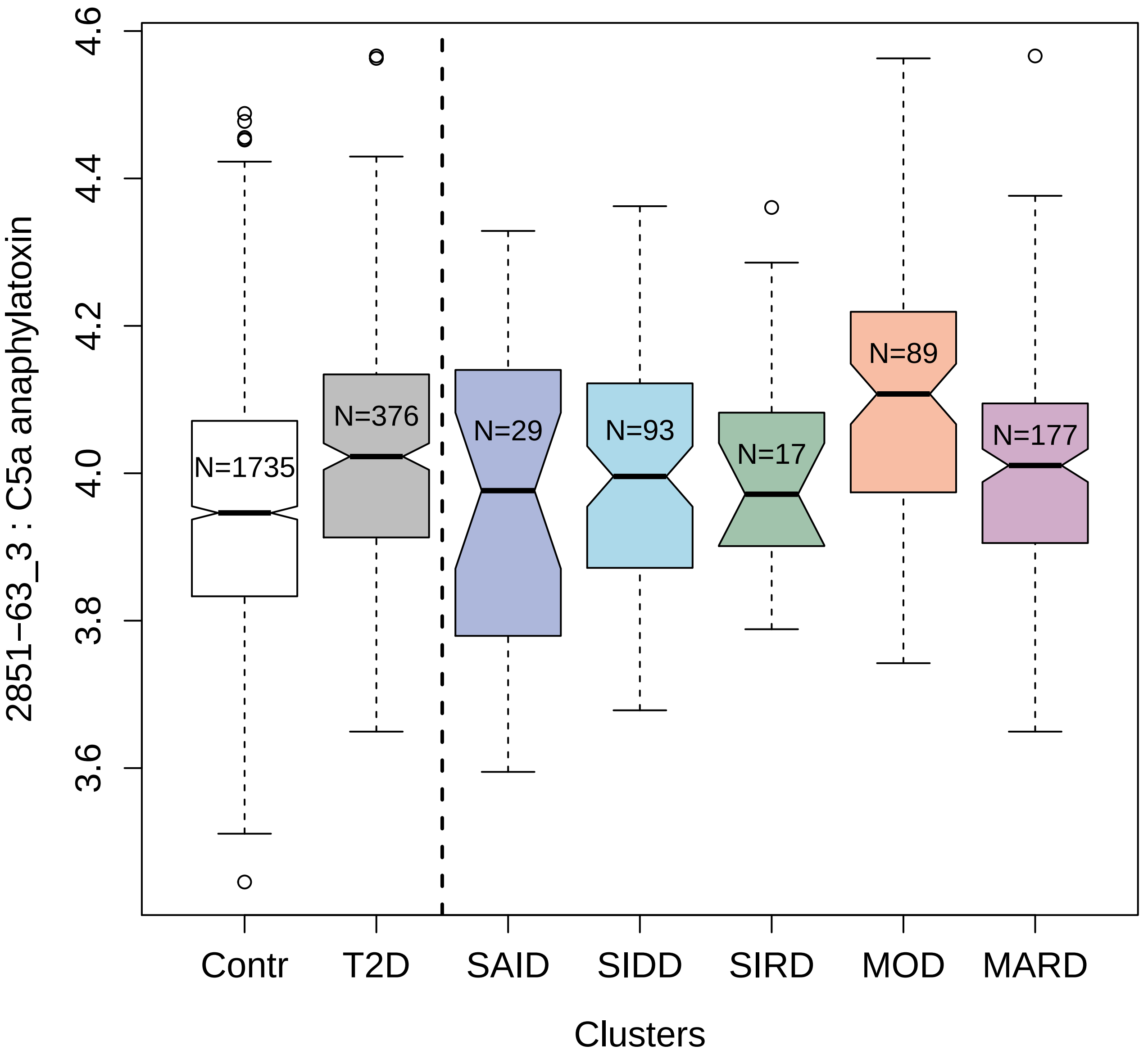

2851−63_3 : C5a anaphylatoxin
4.6
4.4
4.2
4.0
3.8
3.6
N=1735
N=376
N=29
N=93
N=17
N=89
N=177
Contr
T2D
SAID
SIDD
SIRD
MOD
MARD
Clusters

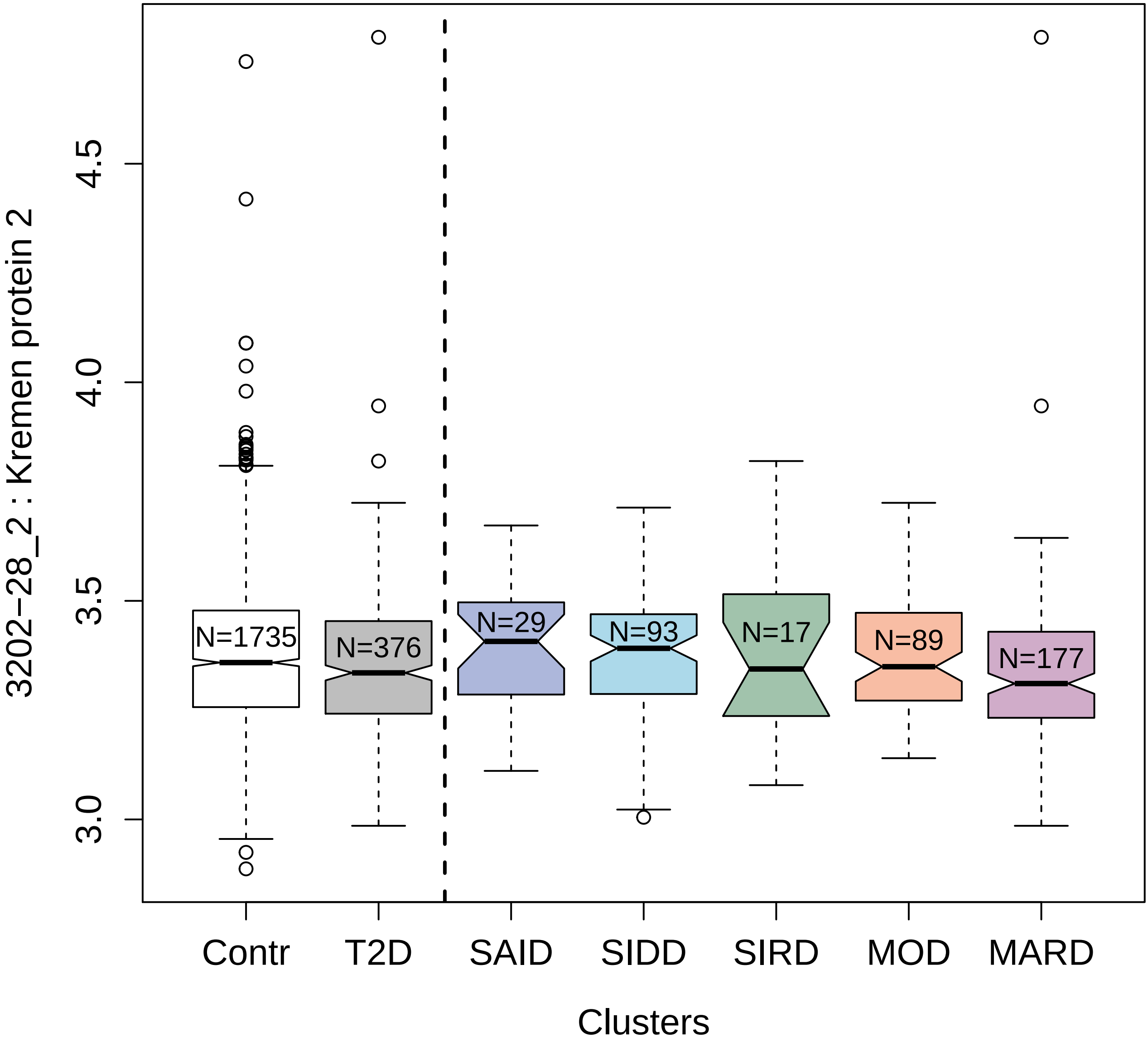
3202−28_2 : Kremen protein 2
3.0
3.5
4.0
4.5
N=1735
N=376
N=29
N=93
N=17
N=89
N=177
Contr
T2D
SAID
SIDD
SIRD
MOD
MARD
Clusters

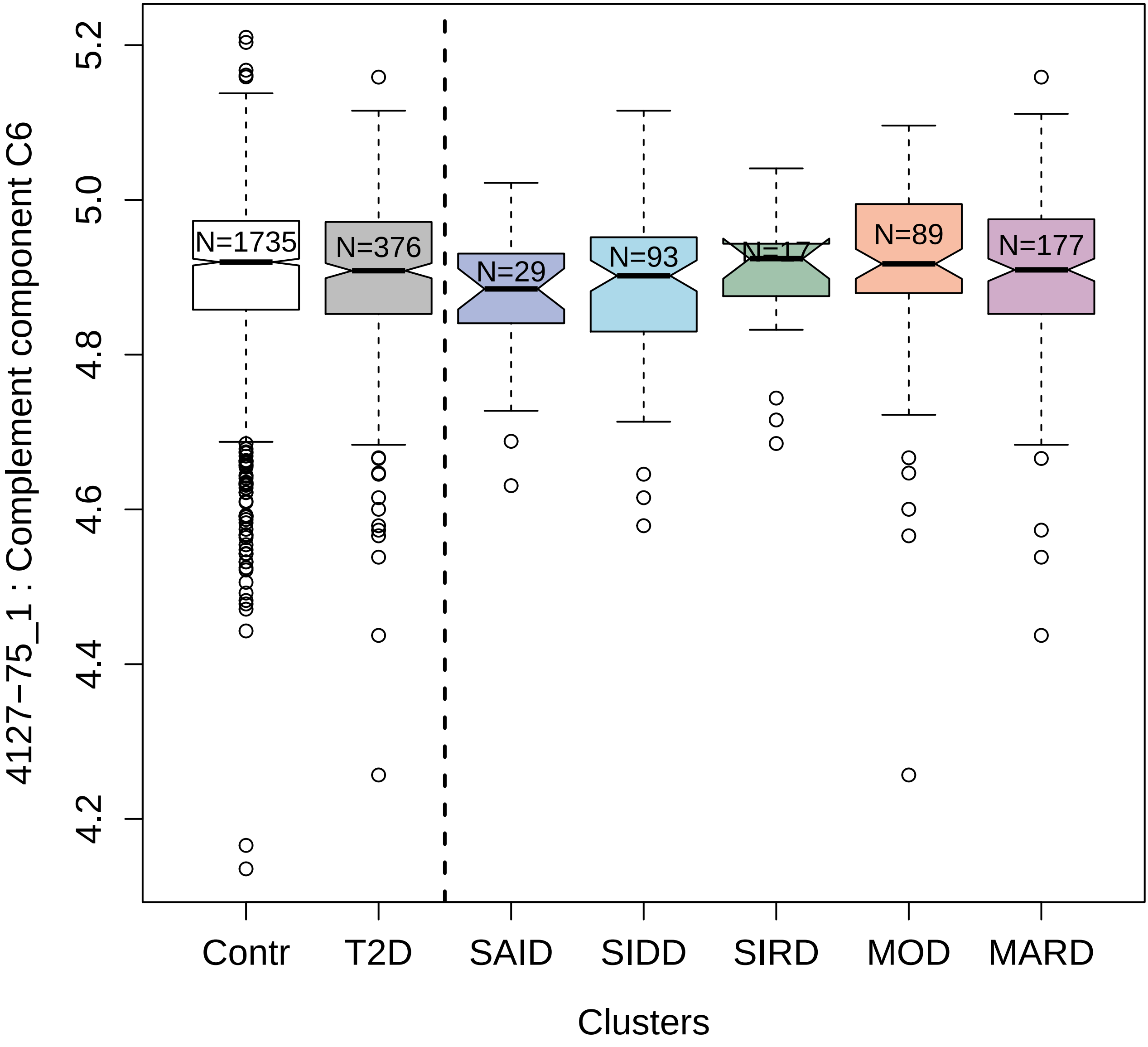

4127−75_1 : Complement component C6
5.2
5.0
4.8
4.6
4.4
4.2
N=1735
N=376
N=29
N=93
N=17
N=89
N=177
Contr
T2D
SAID
SIDD
SIRD
MOD
MARD
Clusters

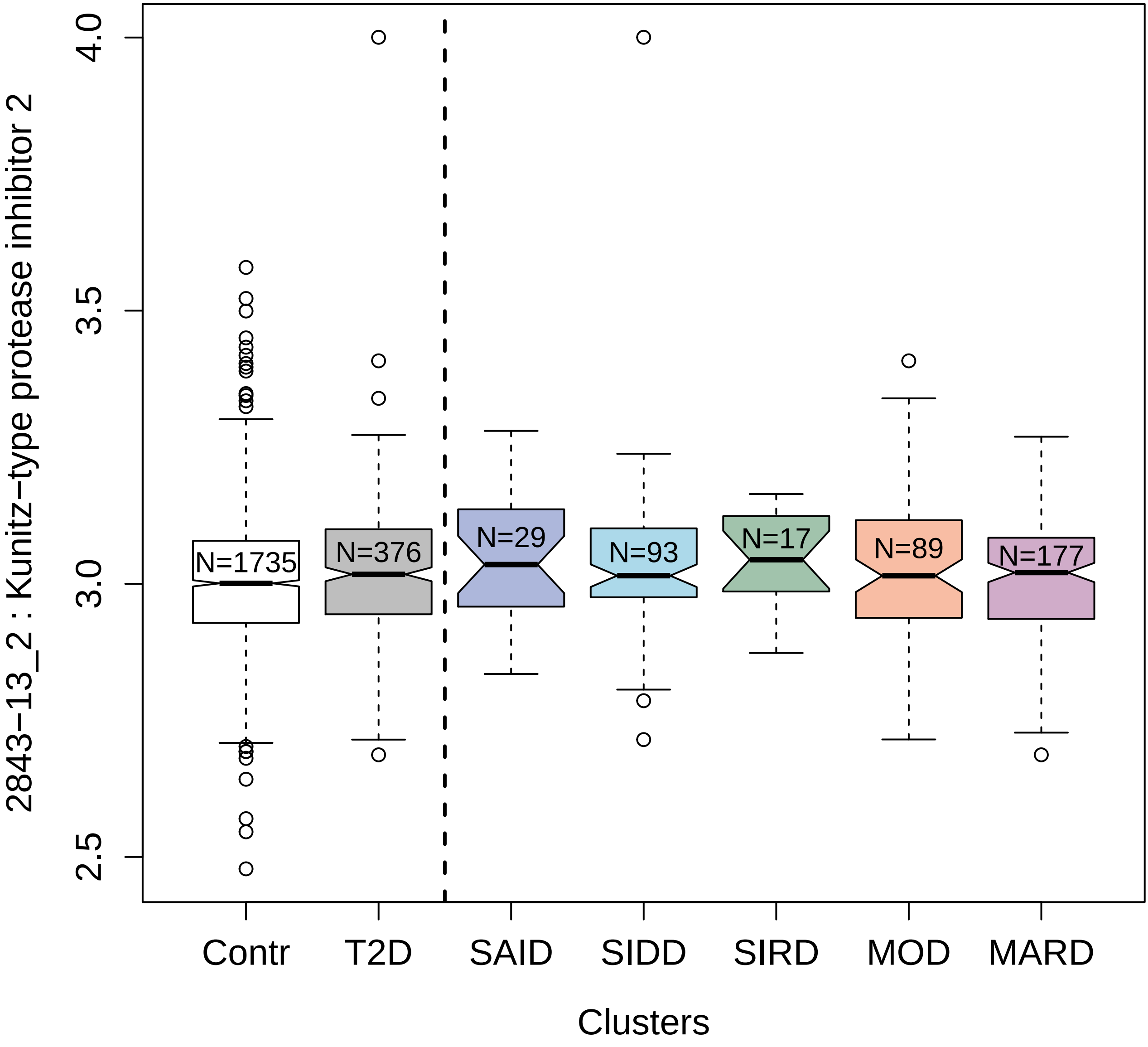
2843−13_2 : Kunitz−type protease inhibitor 2
2.5
3.0
3.5
4.0
N=1735
N=376
N=29
N=93
N=17
N=89
N=177
Contr
T2D
SAID
SIDD
SIRD
MOD
MARD
Clusters

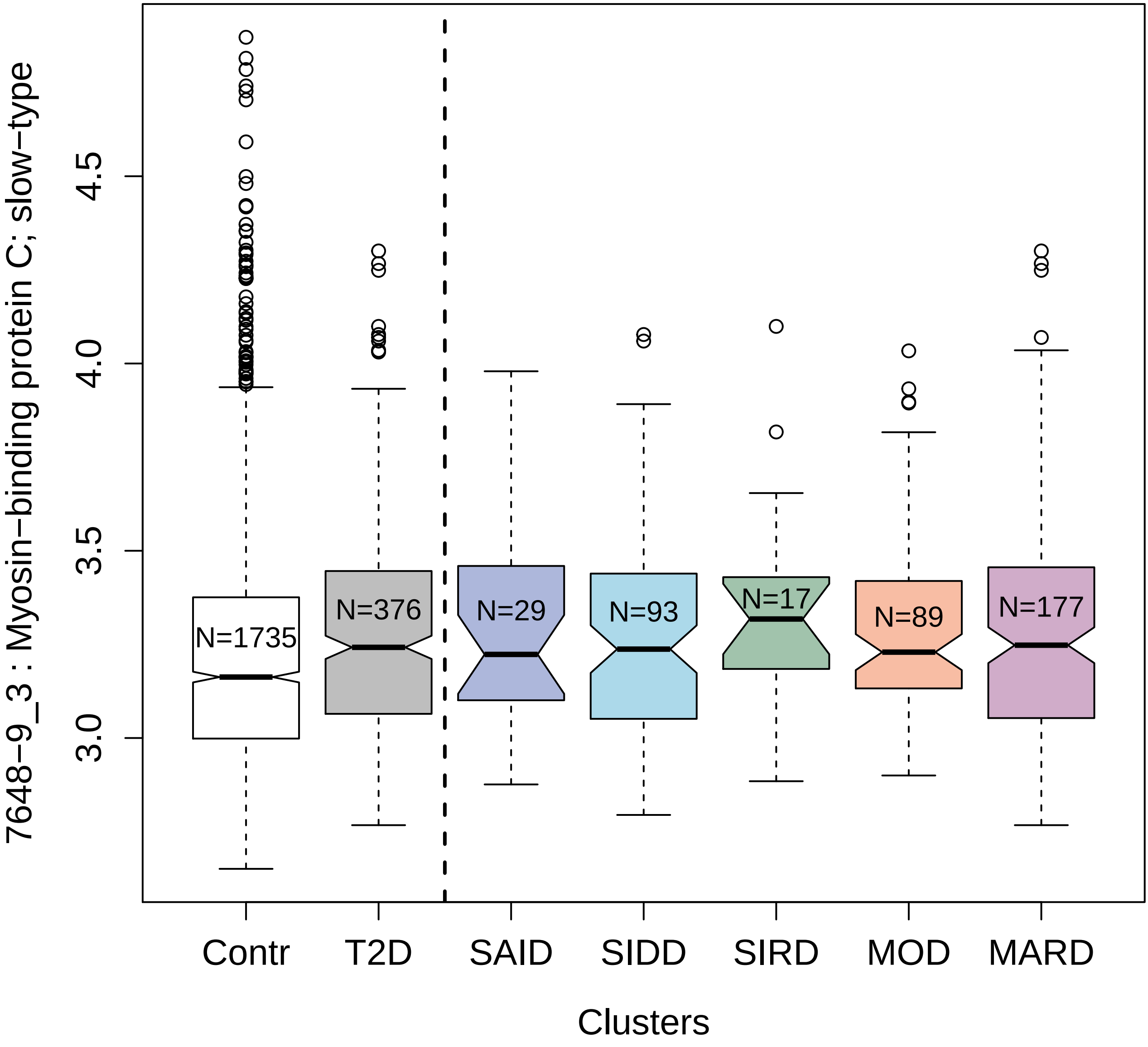

7648−9_3 : Myosin−binding protein C; slow−type
3.0
3.5
4.0
4.5
N=1735
N=376
N=29
N=93
N=17
N=89
N=177
Contr
T2D
SAID
SIDD
SIRD
MOD
MARD
Clusters

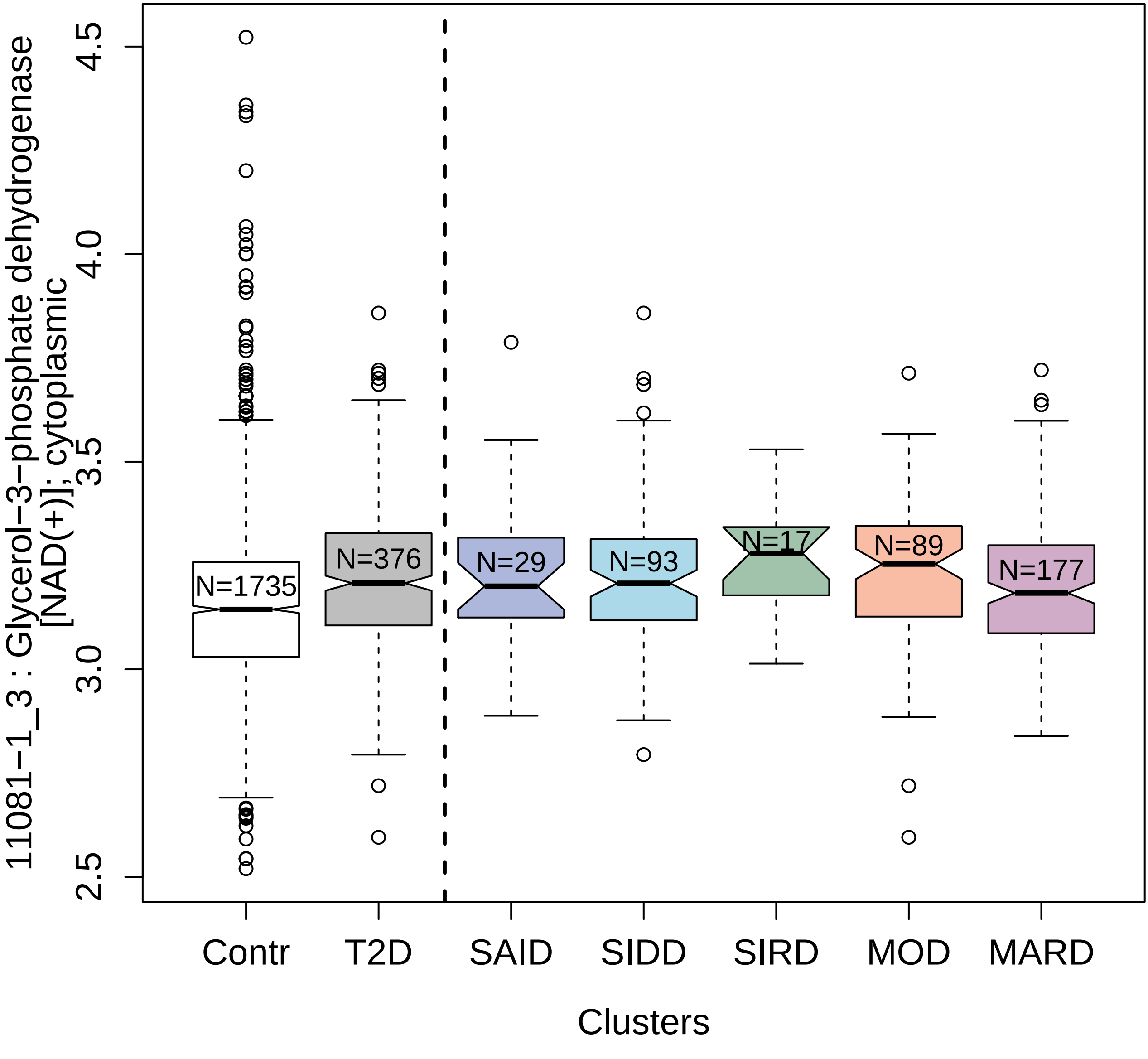

11081−1_3 : Glycerol−3−phosphate dehydrogenase [NAD(+)], cytoplasmic
2.5
3.0
3.5
4.0
4.5
N=1735
N=376
N=29
N=93
N=17
N=89
N=177
Contr
T2D
SAID
SIDD
SIRD
MOD
MARD
Clusters

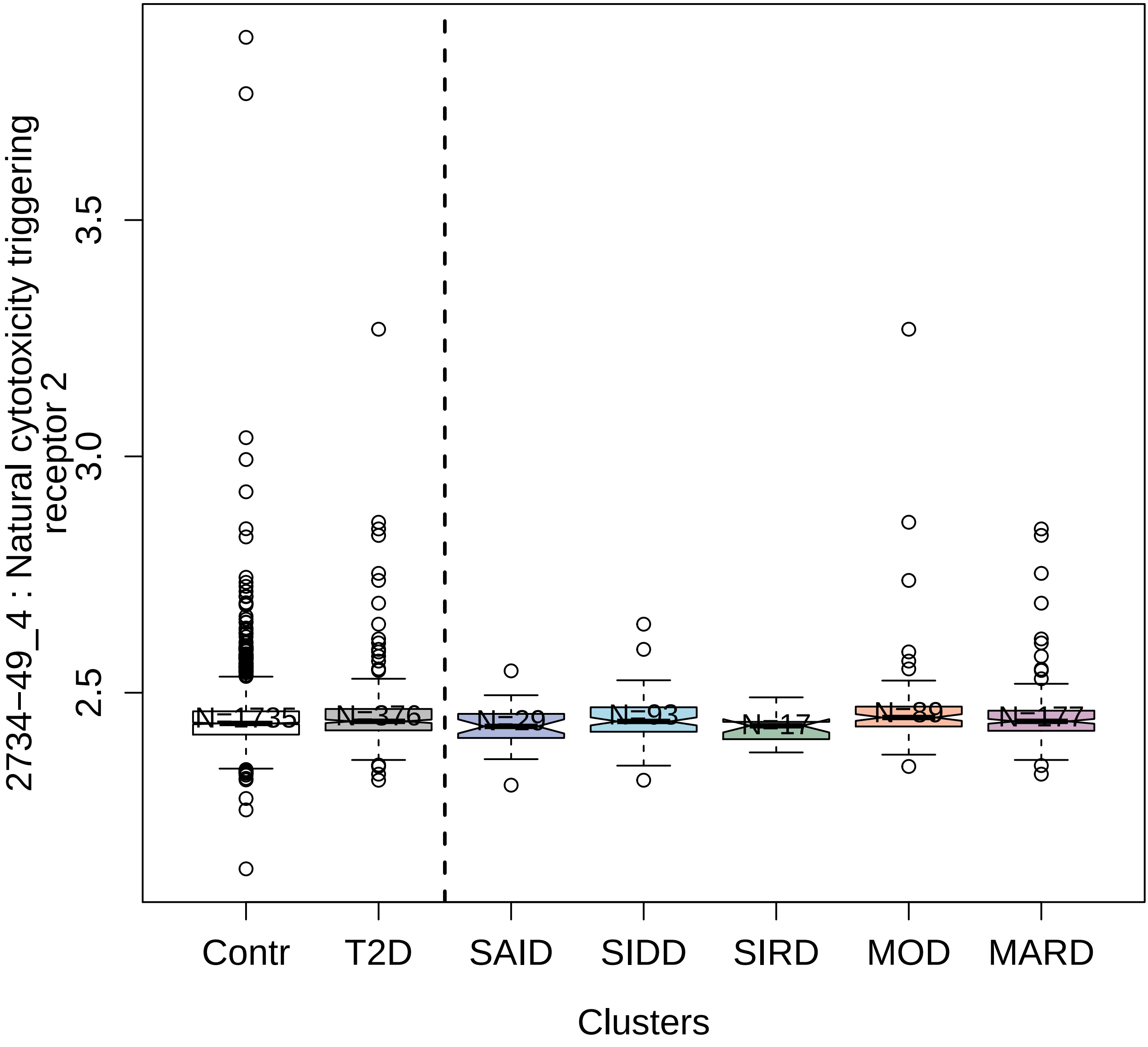
2734−49_4 : Natural cytotoxicity triggering receptor 2
3.5
3.0
2.5
N=1735
N=376
N=29
N=93
N=17
N=89
N=177
Contr
T2D
SAID
SIDD
SIRD
MOD
MARD
Clusters

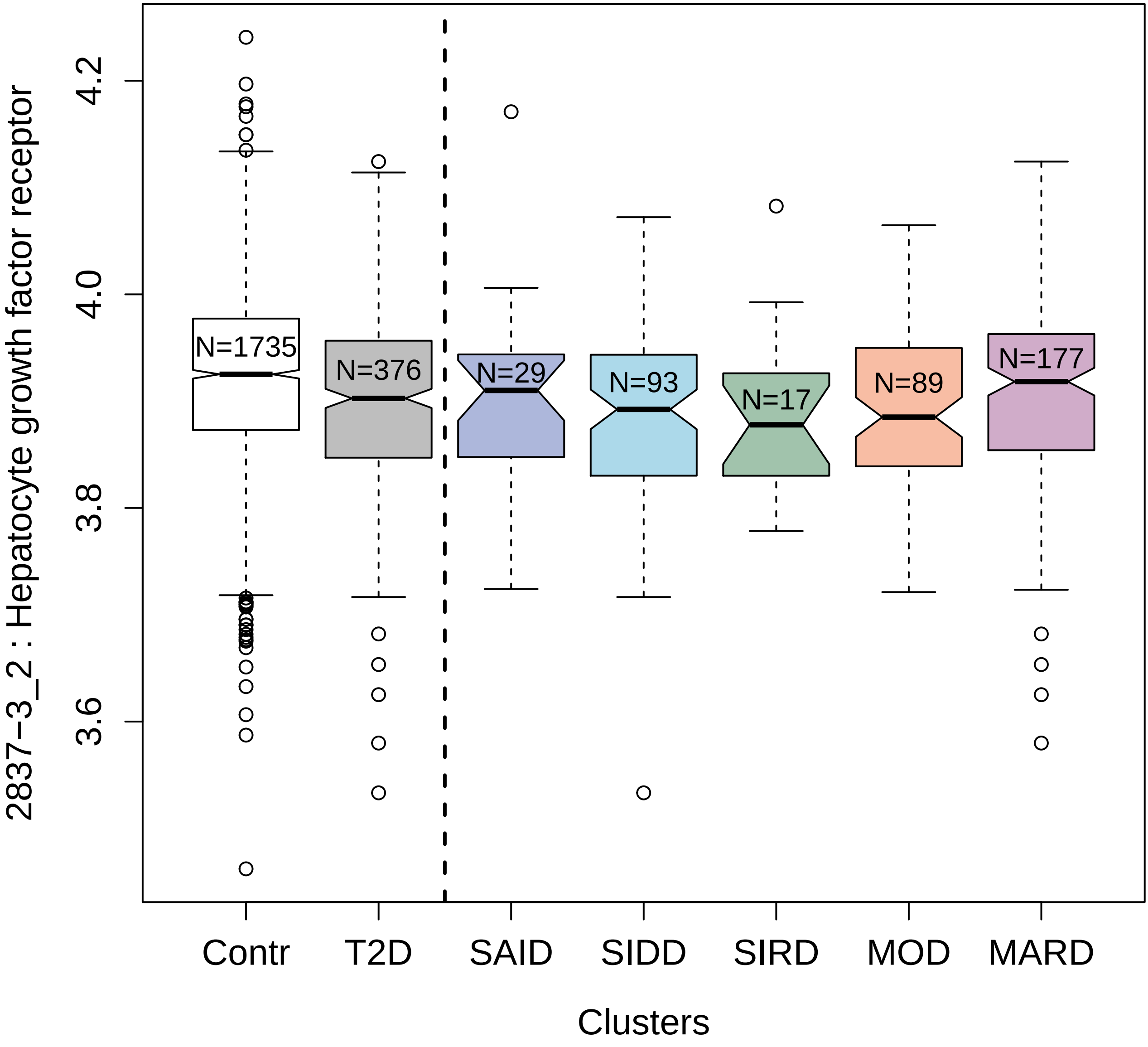

2837−3_2 : Hepatocyte growth factor receptor
4.2
4.0
3.8
3.6
N=1735
N=376
N=29
N=93
N=17
N=89
N=177
Contr
T2D
SAID
SIDD
SIRD
MOD
MARD
Clusters

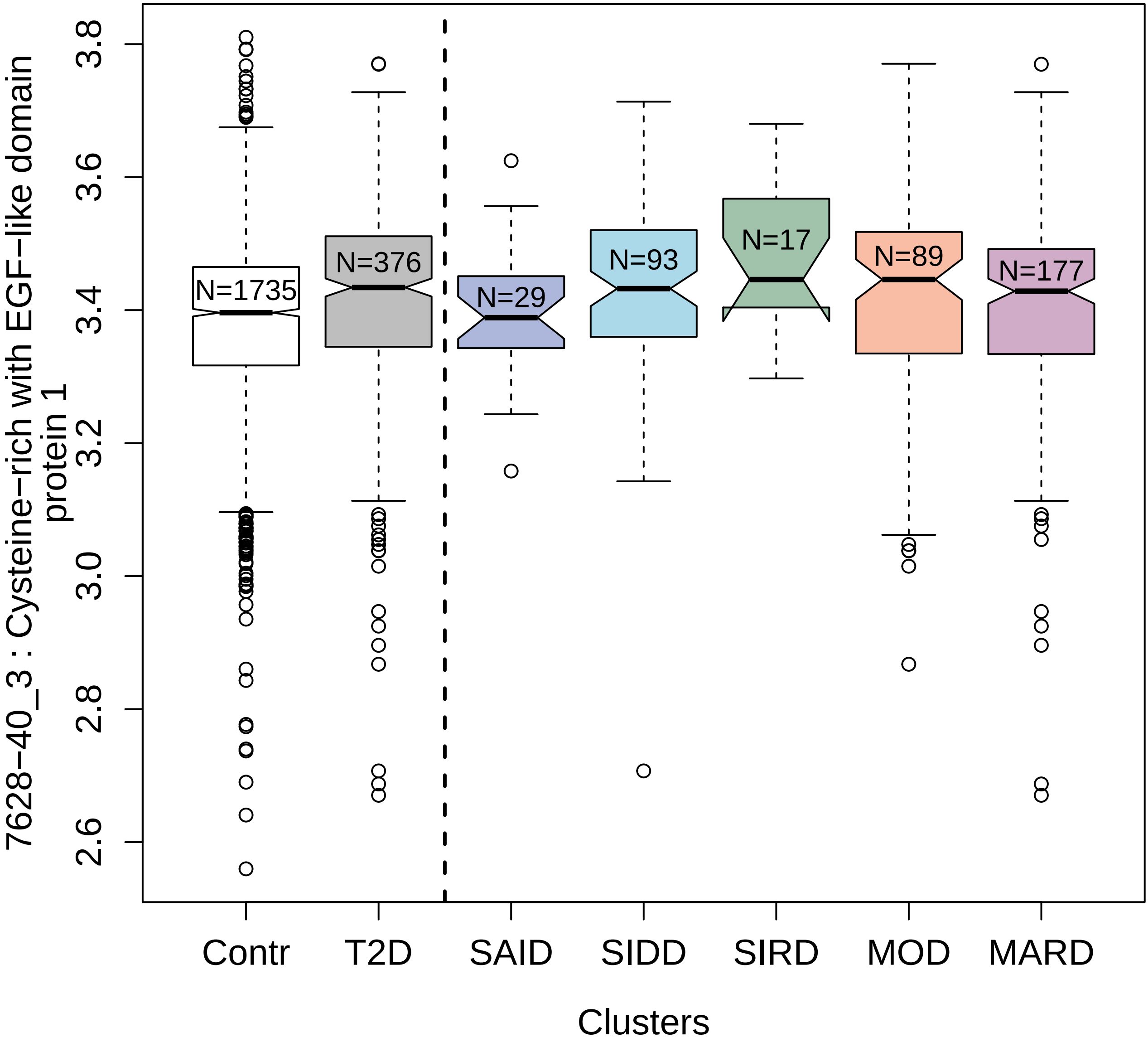

7628−40_3 : Cysteine−rich with EGF−like domain protein 1
3.8
3.6
3.4
3.2
3.0
2.8
2.6
N=1735
N=376
N=29
N=93
N=17
N=89
N=177
Contr
T2D
SAID
SIDD
SIRD
MOD
MARD
Clusters

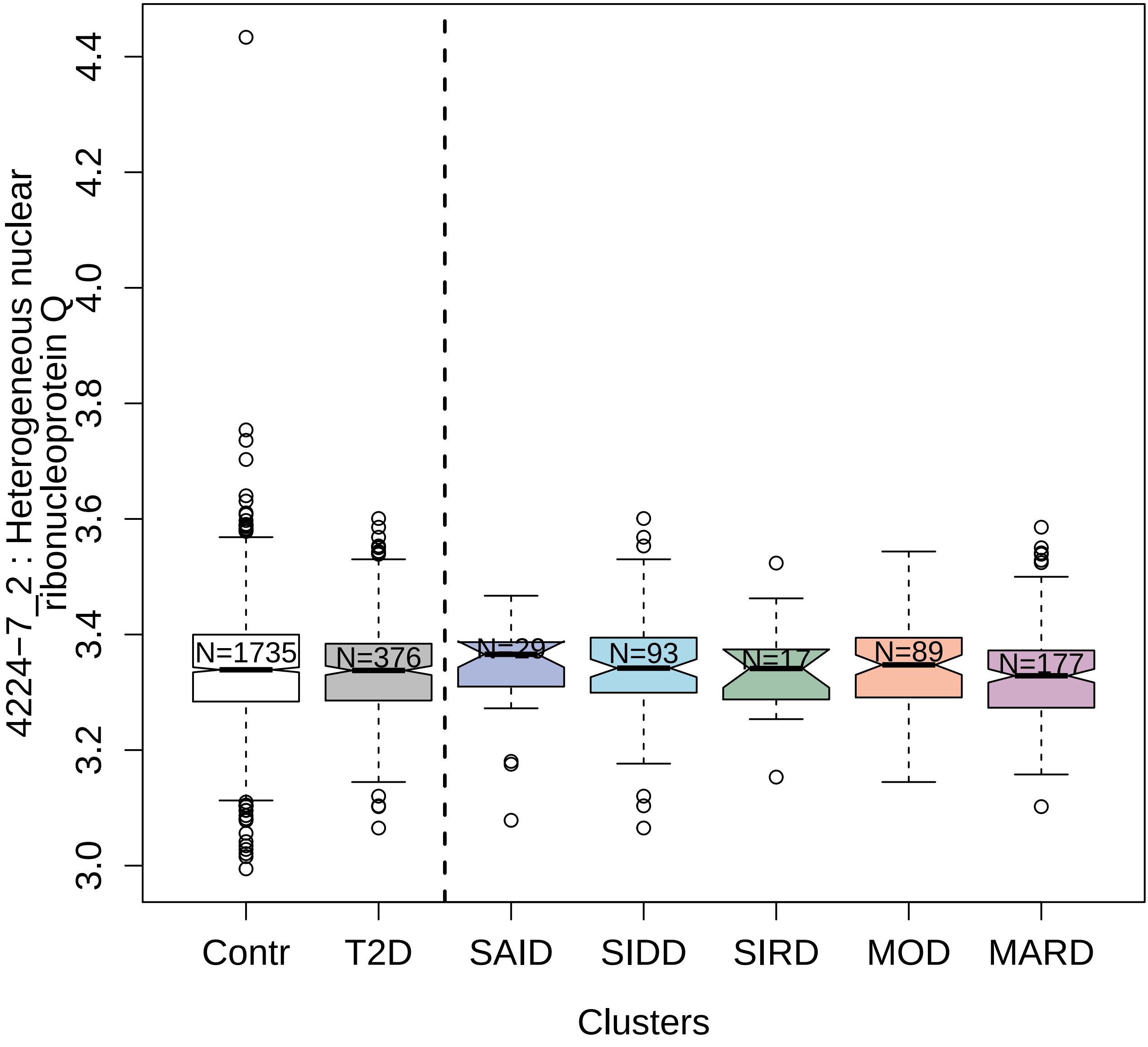

4224−7_2 : Heterogeneous nuclear ribonucleoprotein Q
4.4
4.2
4.0
3.8
3.6
3.4
3.2
3.0
N=1735
N=376
N=29
N=93
N=17
N=89
N=177
Contr
T2D
SAID
SIDD
SIRD
MOD
MARD
Clusters

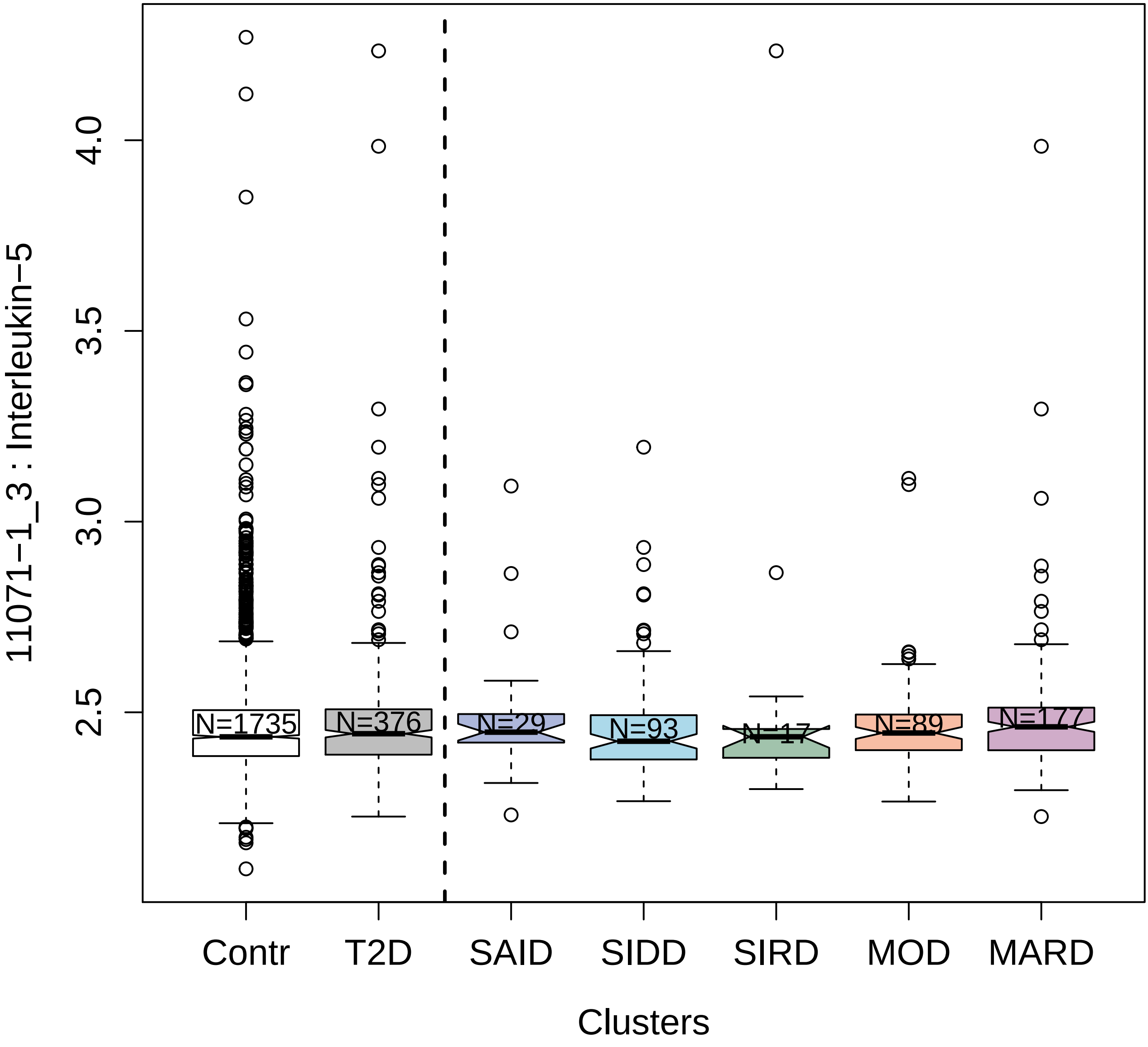
11071−1_3 : Interleukin−5
4.0
3.5
3.0
2.5
N=1735
N=376
N=29
N=93
N=17
N=89
N=177
Contr
T2D
SAID
SIDD
SIRD
MOD
MARD
Clusters

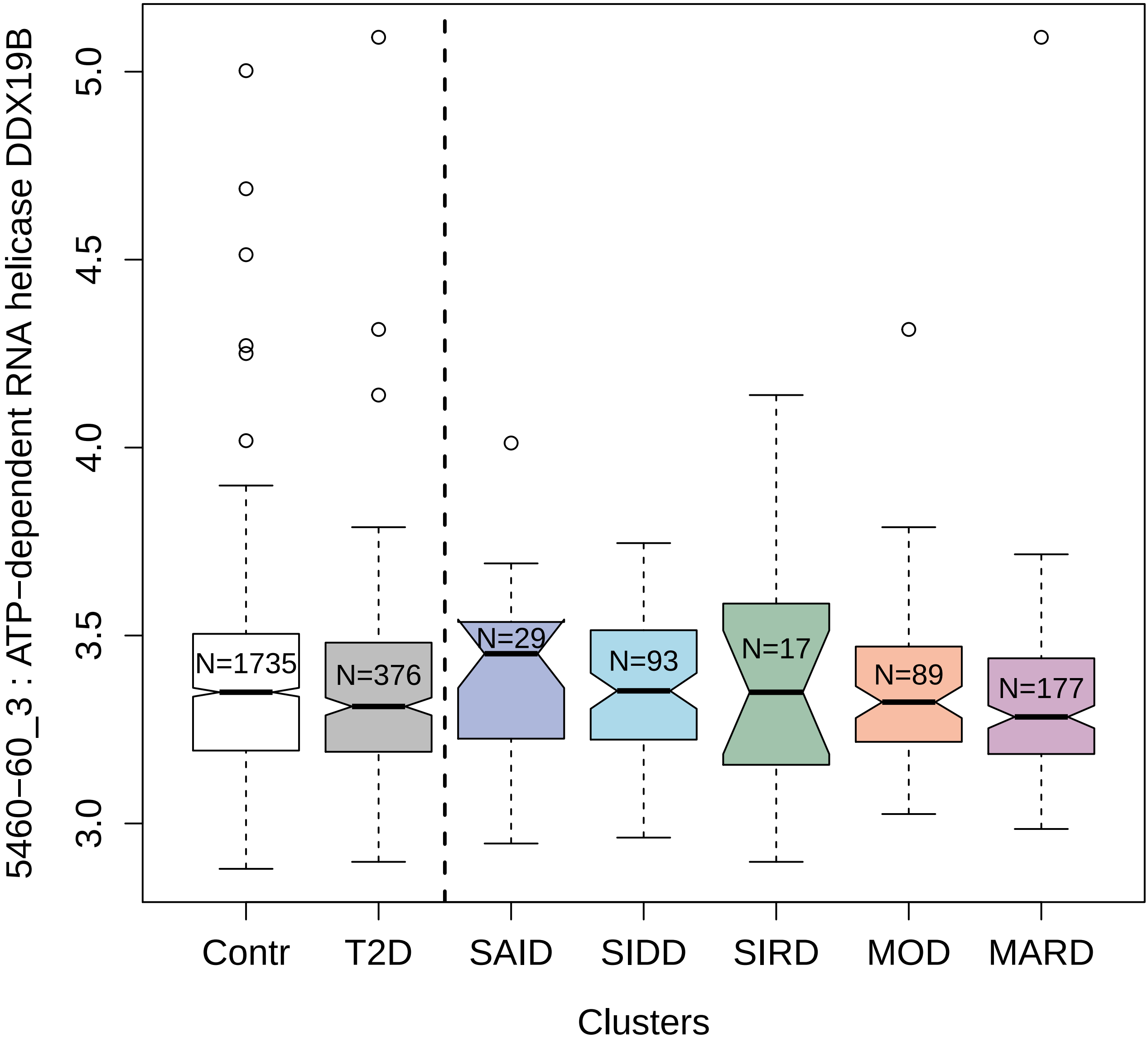
5460−60_3 : ATP−dependent RNA helicase DDX19B
3.0
3.5
4.0
4.5
5.0
N=1735
N=376
N=29
N=93
N=17
N=89
N=177
Contr
T2D
SAID
SIDD
SIRD
MOD
MARD
Clusters

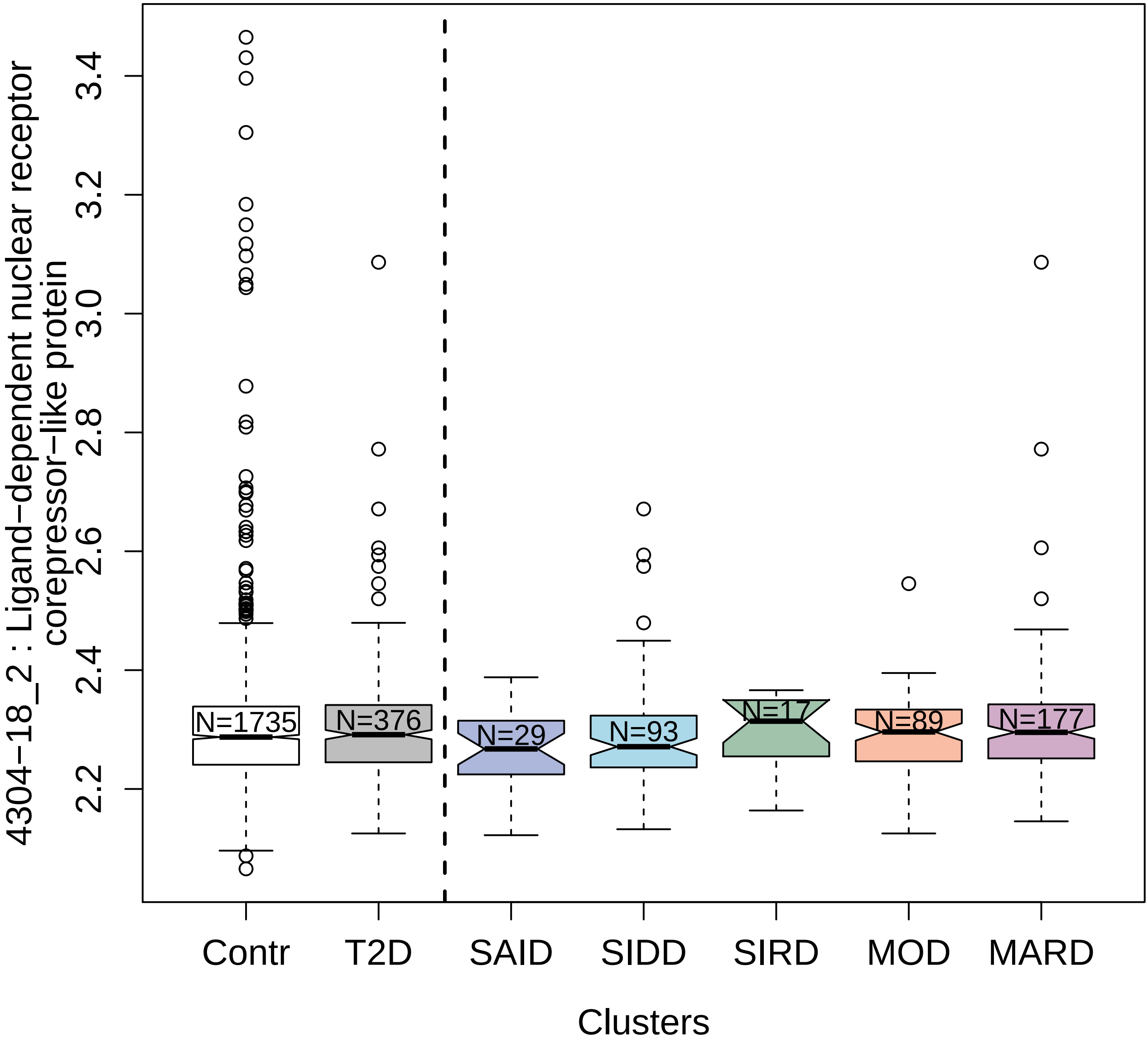

4304−18_2 : Ligand−dependent nuclear receptor corepressor−like protein
2.2
2.4
2.6
2.8
3.0
3.2
3.4
N=1735
N=376
N=29
N=93
N=17
N=89
N=177
Contr
T2D
SAID
SIDD
SIRD
MOD
MARD
Clusters

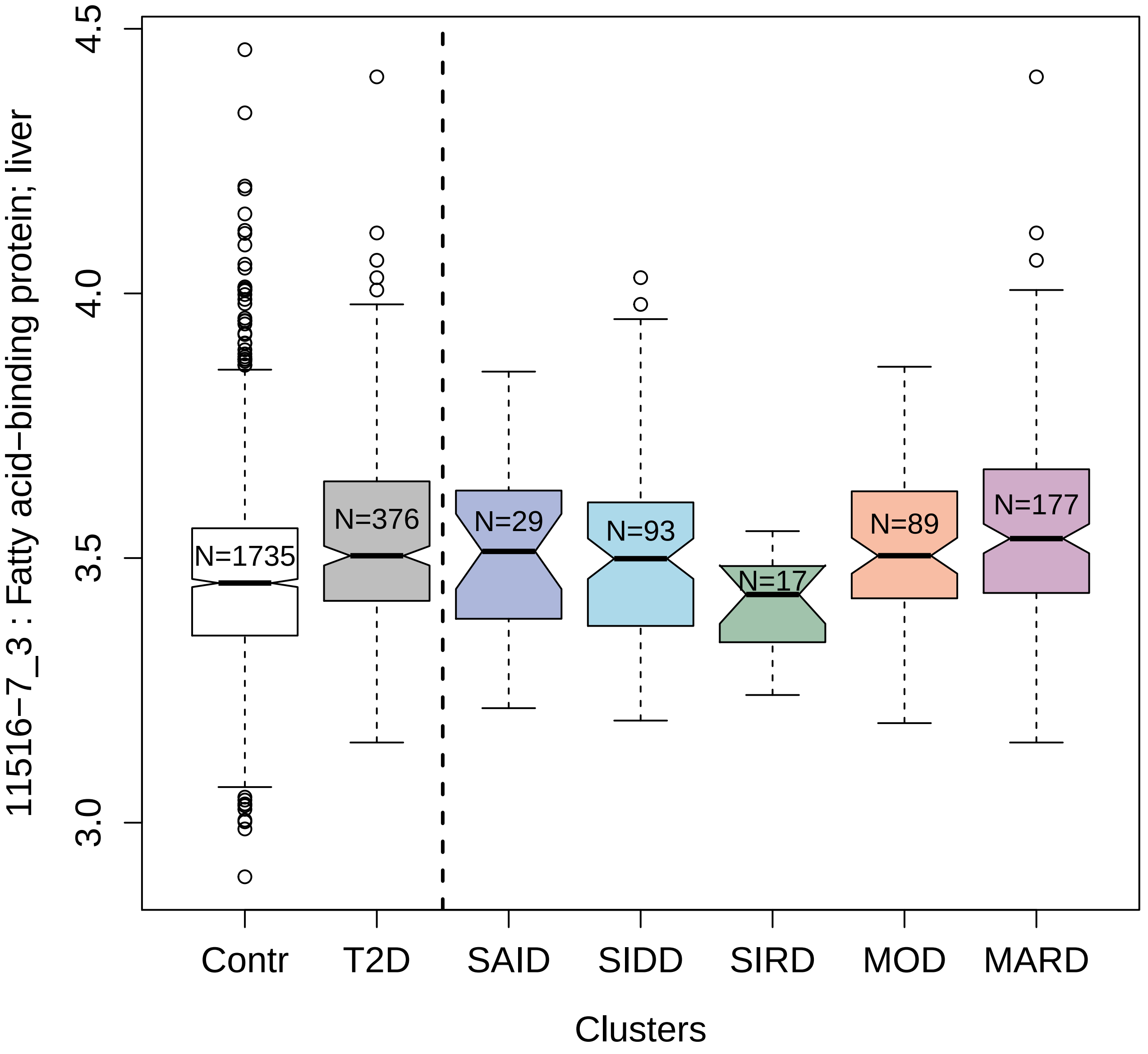
11516−7_3 : Fatty acid−binding protein; liver
4.5
4.0
3.5
3.0
N=1735
N=376
N=29
N=93
N=17
N=89
N=177
Contr
T2D
SAID
SIDD
SIRD
MOD
MARD
Clusters

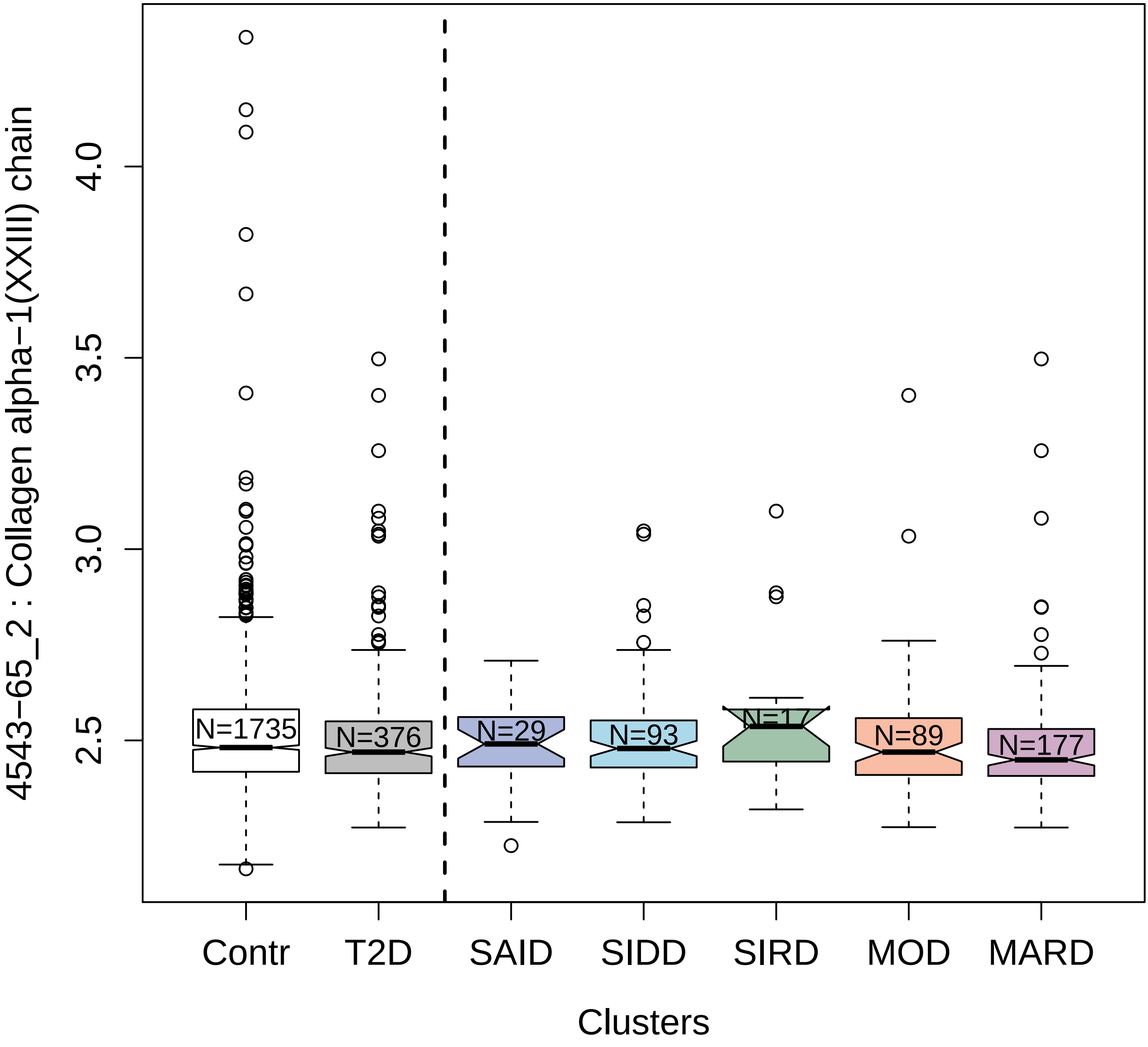
4543−65_2 : Collagen alpha−1(XXIII) chain
4.0
3.5
3.0
2.5
N=1735
N=376
N=29
N=93
N=17
N=89
N=177
Contr
T2D
SAID
SIDD
SIRD
MOD
MARD
Clusters

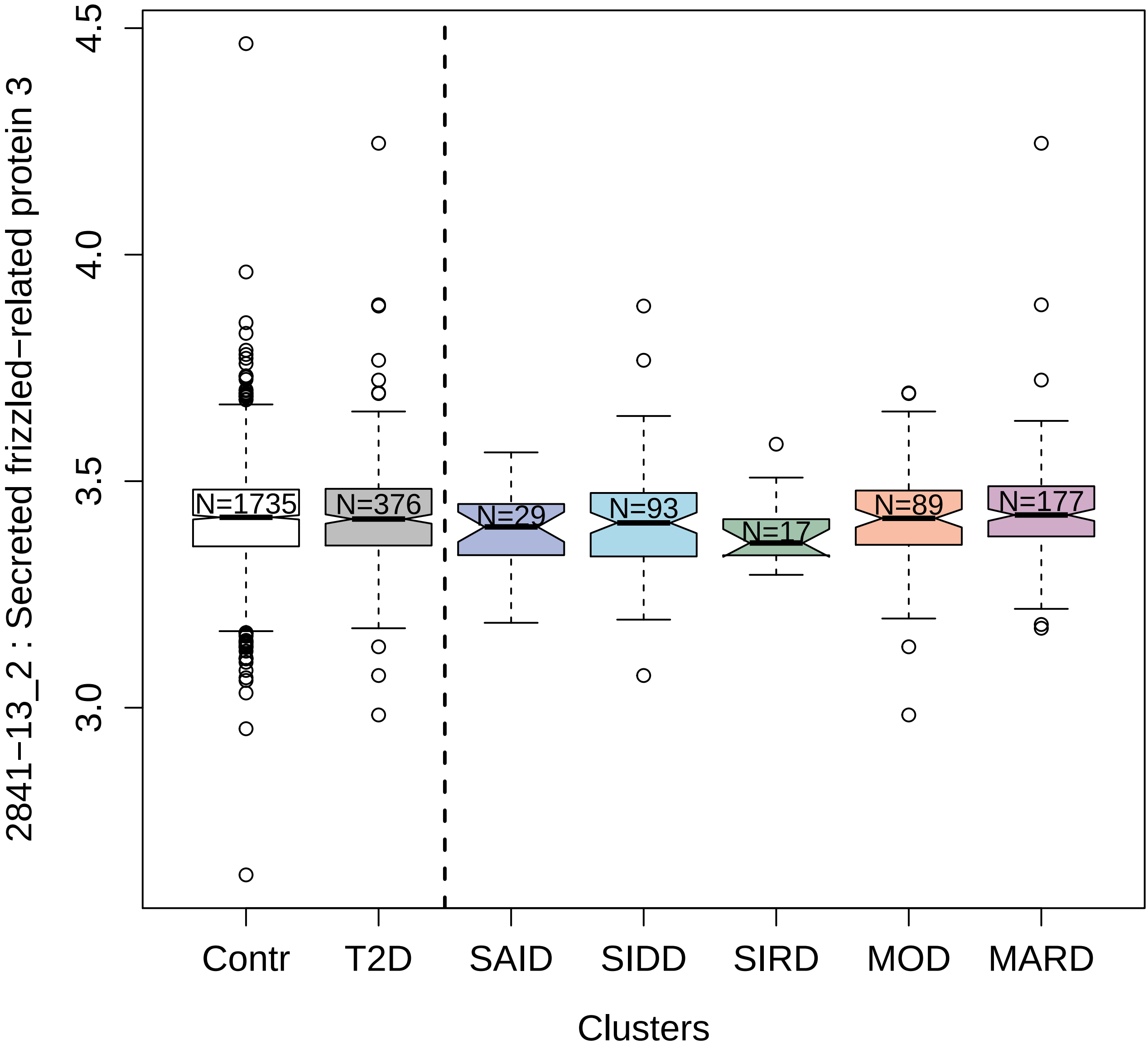
2841−13_2 : Secreted frizzled−related protein 3
4.5
4.0
3.5
3.0
N=1735
N=376
N=29
N=93
N=17
N=89
N=177
Contr
T2D
SAID
SIDD
SIRD
MOD
MARD
Clusters

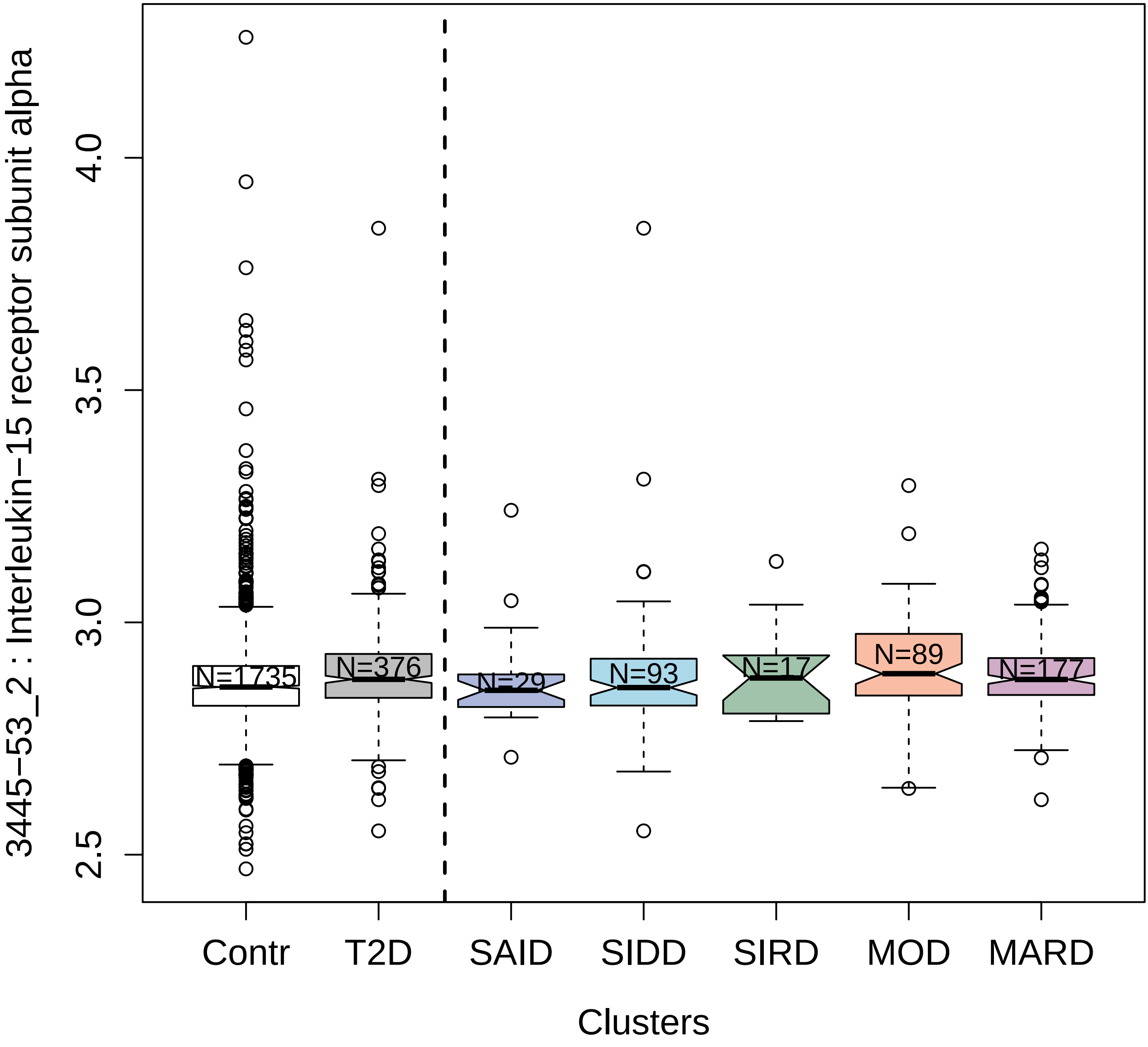
3445−53_2 : Interleukin−15 receptor subunit alpha
2.5
3.0
3.5
4.0
N=1735
N=376
N=29
N=93
N=17
N=89
N=177
Contr
T2D
SAID
SIDD
SIRD
MOD
MARD
Clusters

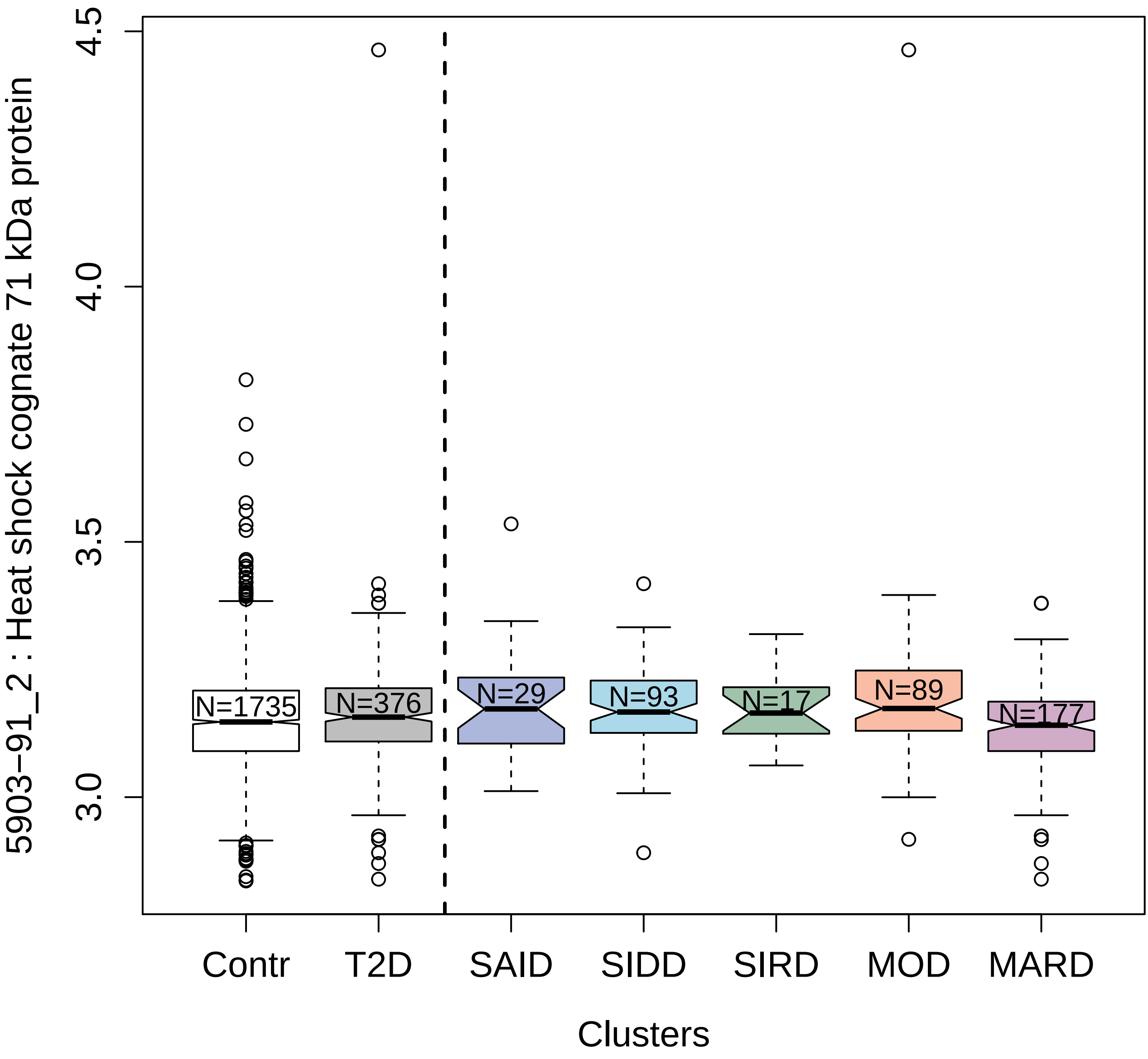
5903−91_2 : Heat shock cognate 71 kDa protein
3.0
3.5
4.0
4.5
N=1735
N=376
N=29
N=93
N=17
N=89
N=177
Contr
T2D
SAID
SIDD
SIRD
MOD
MARD
Clusters

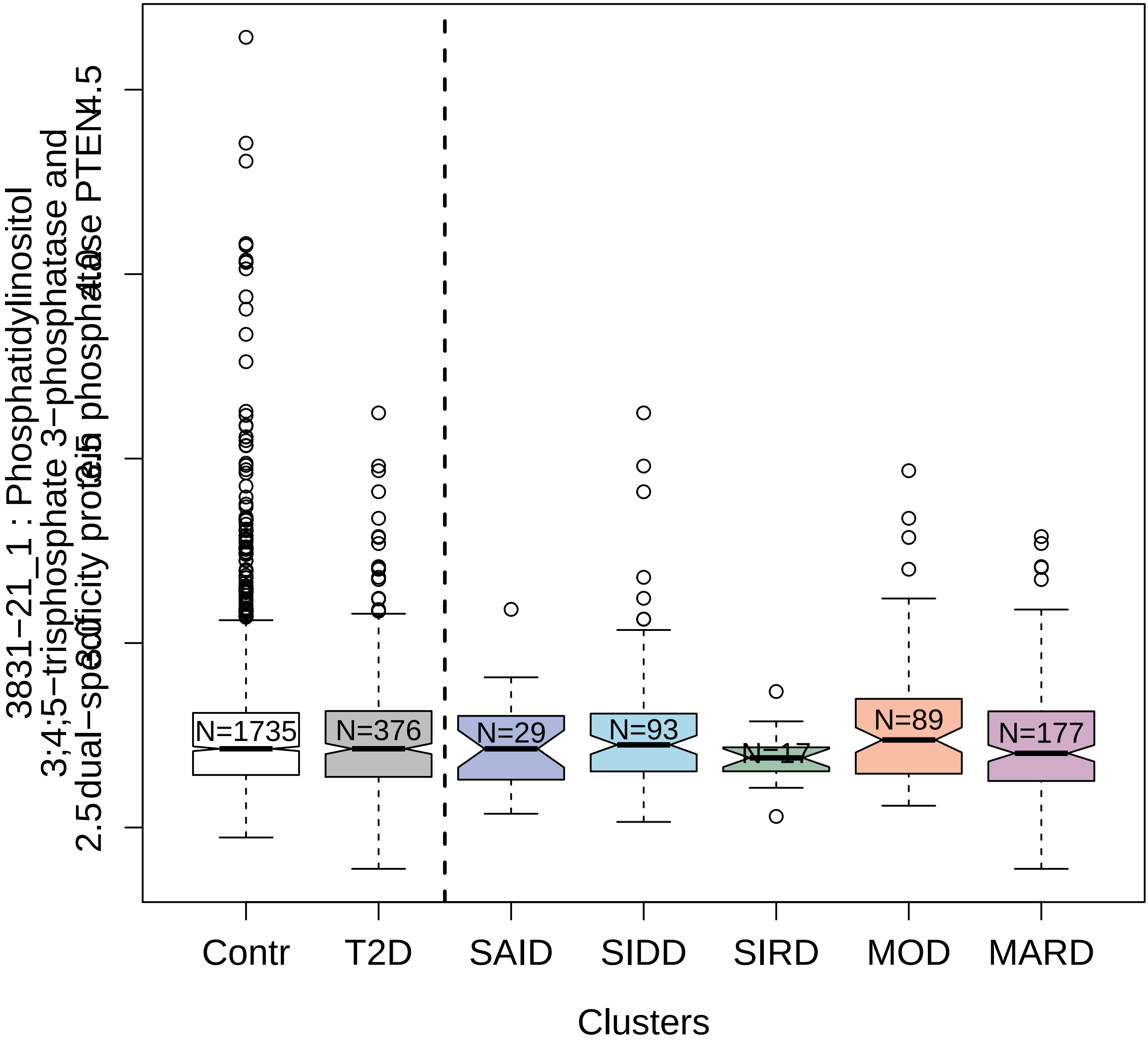

3831−21_1 : Phosphatidylinositol 3,4,5−trisphosphate 3−phosphatase and dual−specificity protein phosphatase PTEN
2.5
3.0
3.5
4.0
4.5
N=1735
N=376
N=29
N=93
N=17
N=89
N=177
Contr
T2D
SAID
SIDD
SIRD
MOD
MARD
Clusters

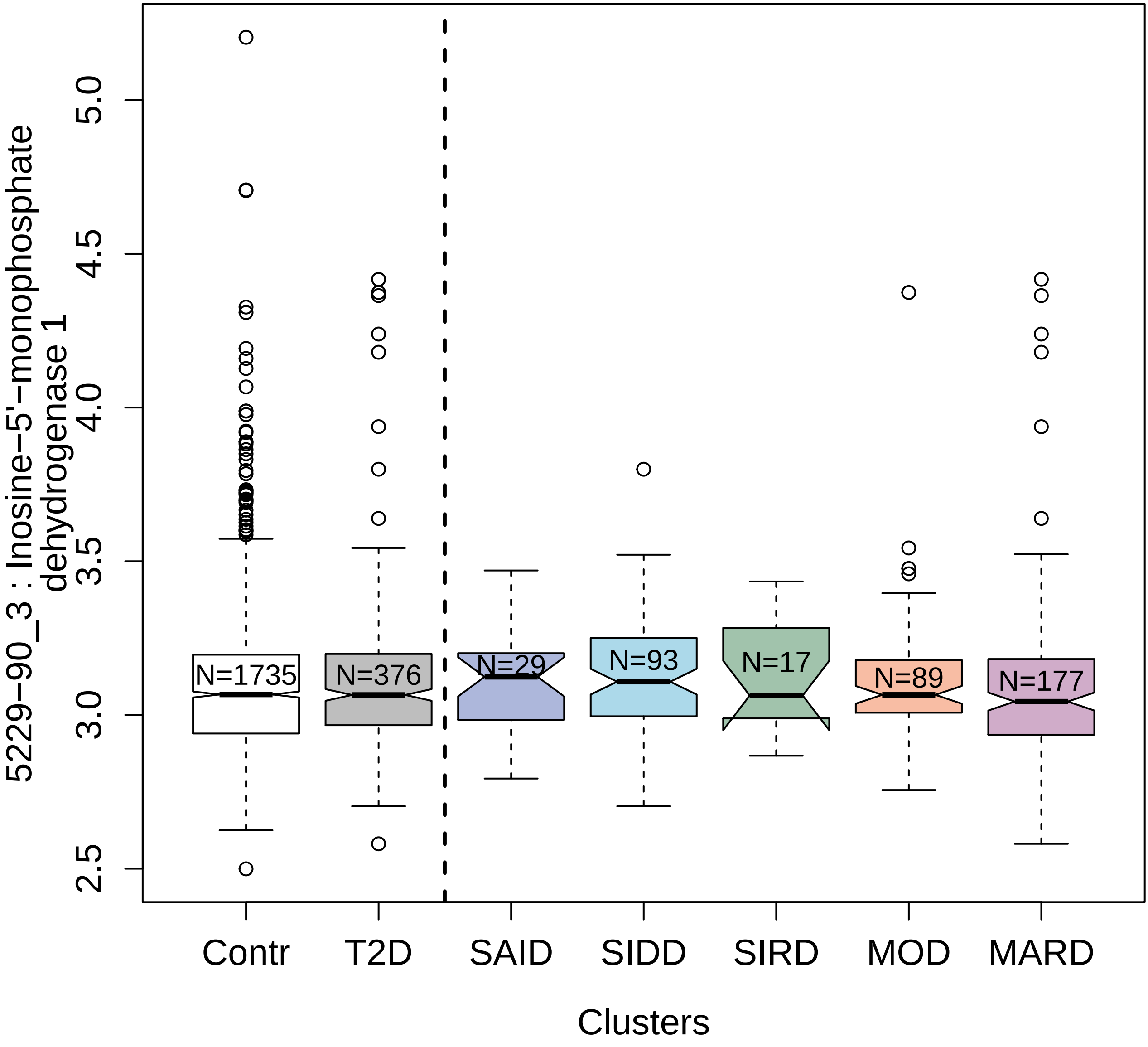

5229−90_3 : Inosine−5'−monophosphate dehydrogenase 1
2.5
3.0
3.5
4.0
4.5
5.0
N=1735
N=376
N=29
N=93
N=17
N=89
N=177
Contr
T2D
SAID
SIDD
SIRD
MOD
MARD
Clusters

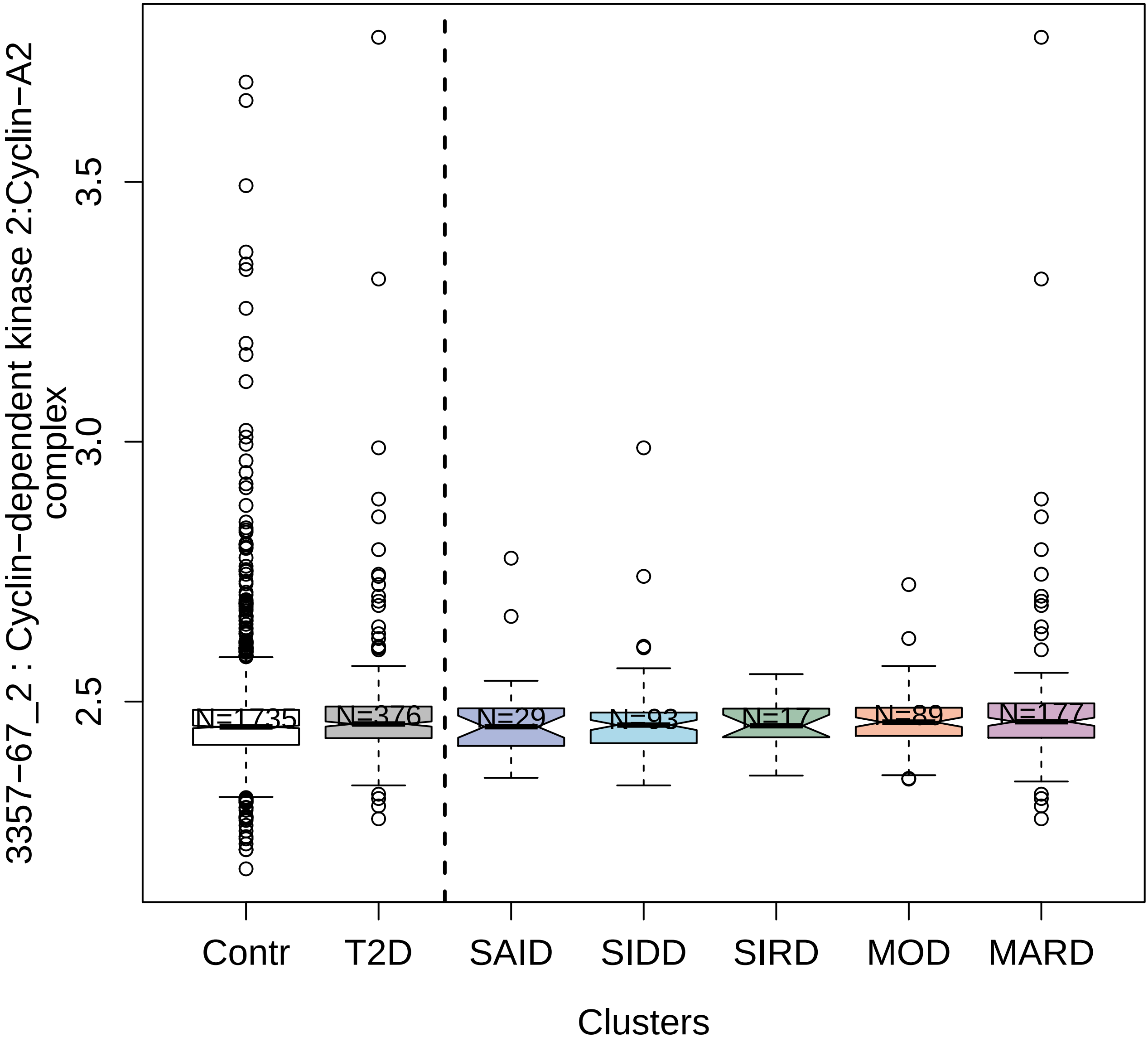
3357−67_2 : Cyclin−dependent kinase 2:Cyclin−A2 complex
3.5
3.0
2.5
N=1735
N=376
N=29
N=93
N=17
N=89
N=177
Contr
T2D
SAID
SIDD
SIRD
MOD
MARD
Clusters

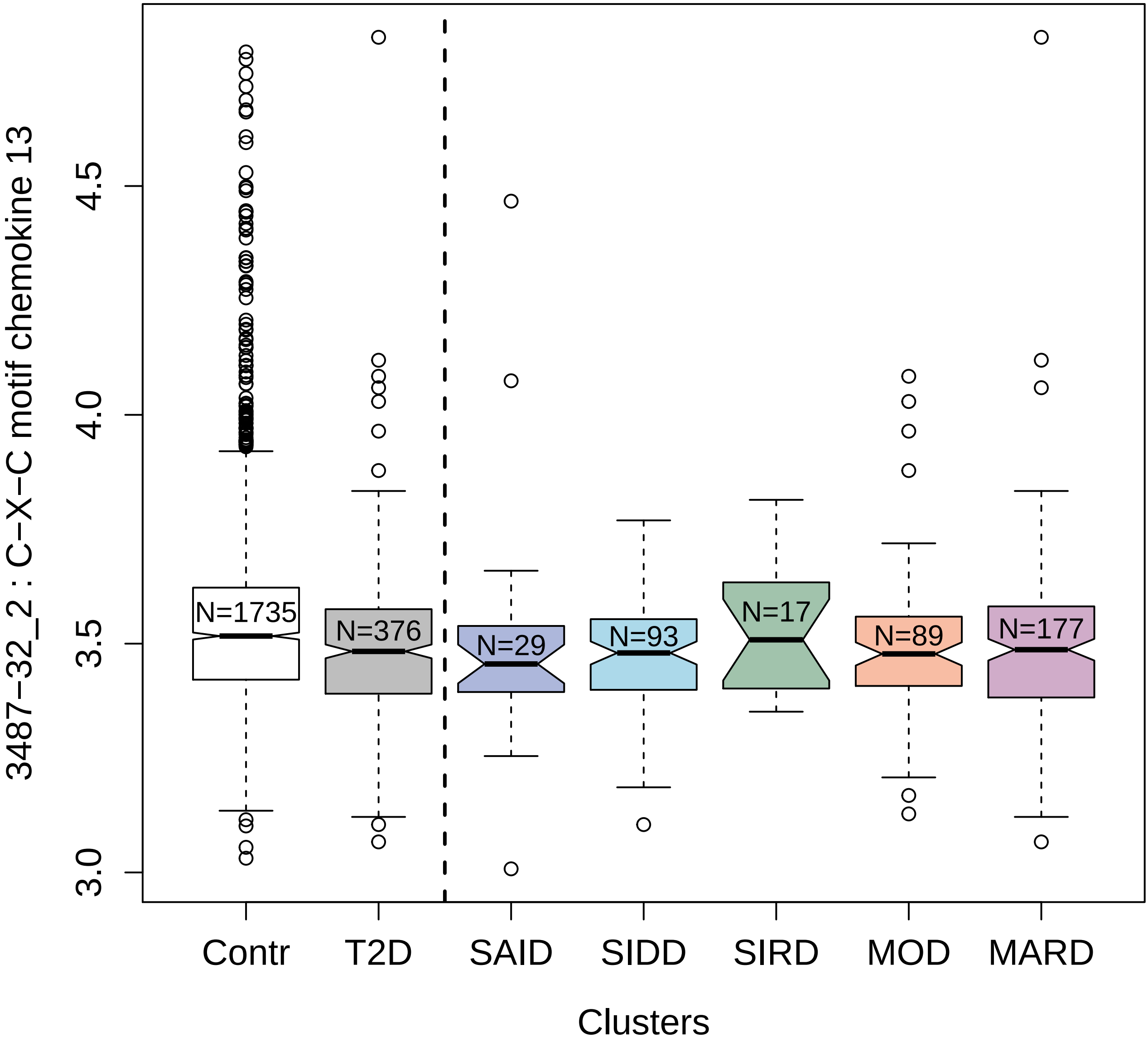
3487−32_2 : C−X−C motif chemokine 13
3.0
3.5
4.0
4.5
N=1735
N=376
N=29
N=93
N=17
N=89
N=177
Contr
T2D
SAID
SIDD
SIRD
MOD
MARD
Clusters

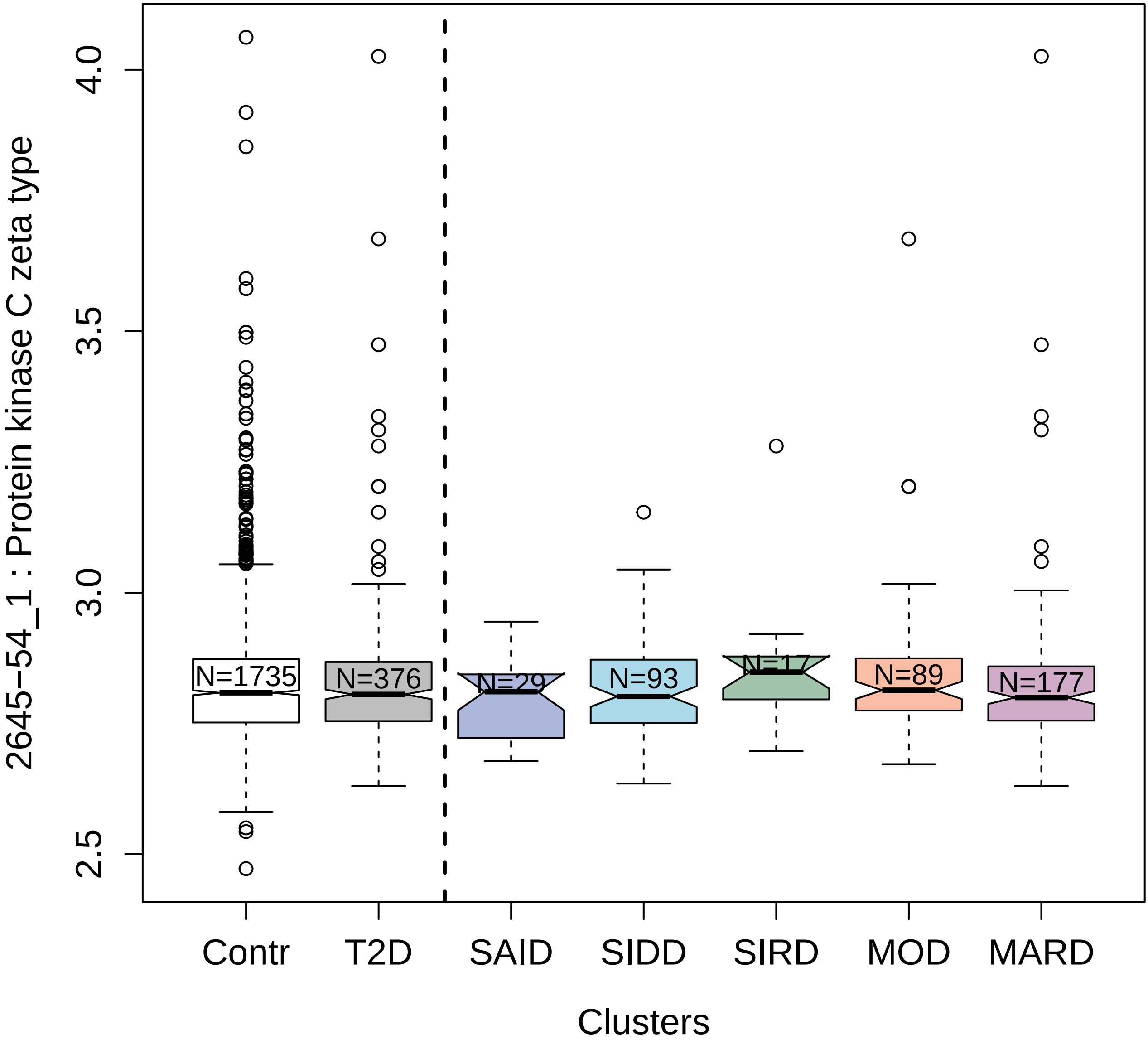
2645−54_1 : Protein kinase C zeta type
4.0
3.5
3.0
2.5
N=1735
N=376
N=29
N=93
N=17
N=89
N=177
Contr
T2D
SAID
SIDD
SIRD
MOD
MARD
Clusters

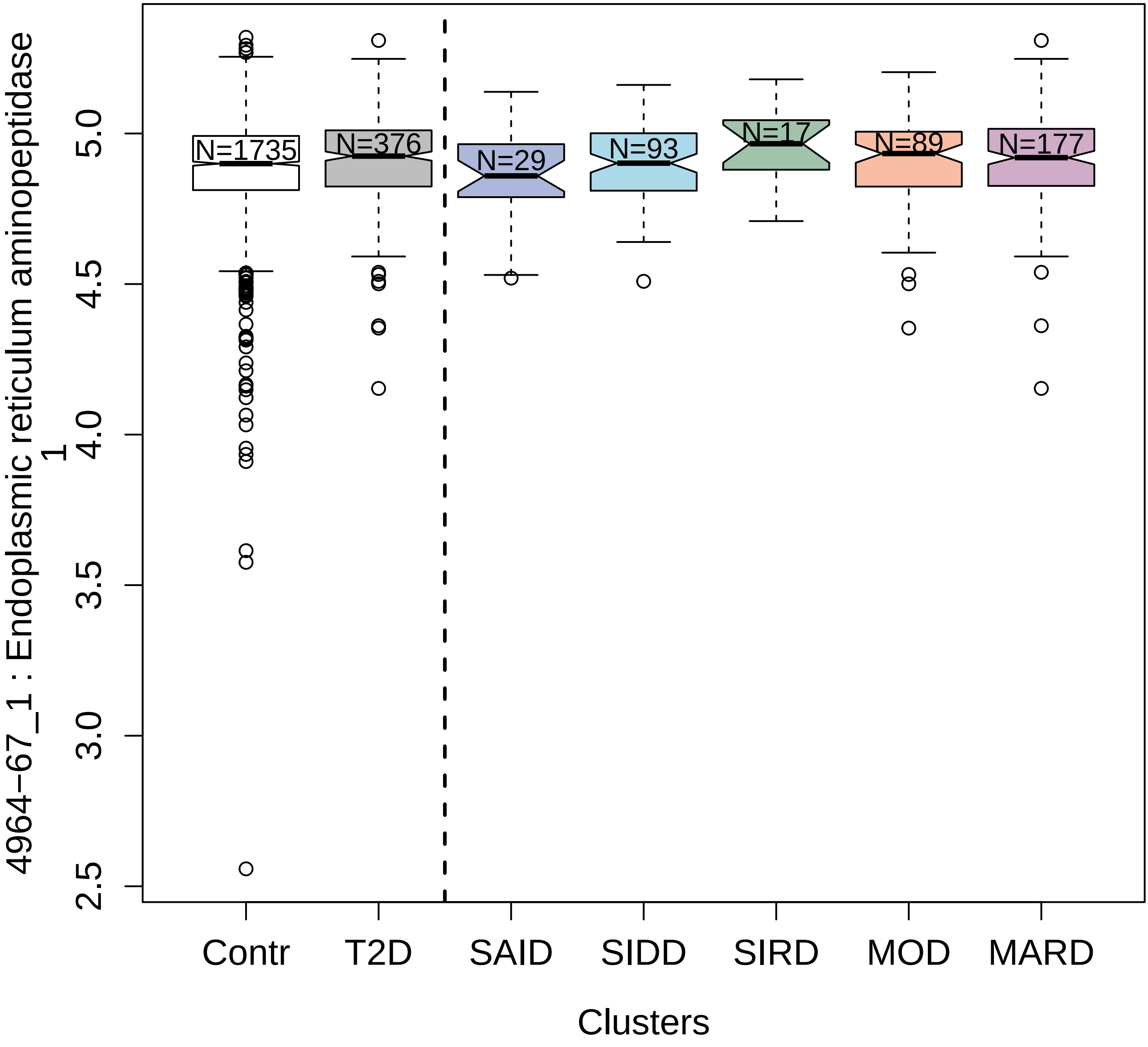

4964−67_1 : Endoplasmic reticulum aminopeptidase 1
2.5
3.0
3.5
4.0
4.5
5.0
N=1735
N=376
N=29
N=93
N=17
N=89
N=177
Contr
T2D
SAID
SIDD
SIRD
MOD
MARD
Clusters

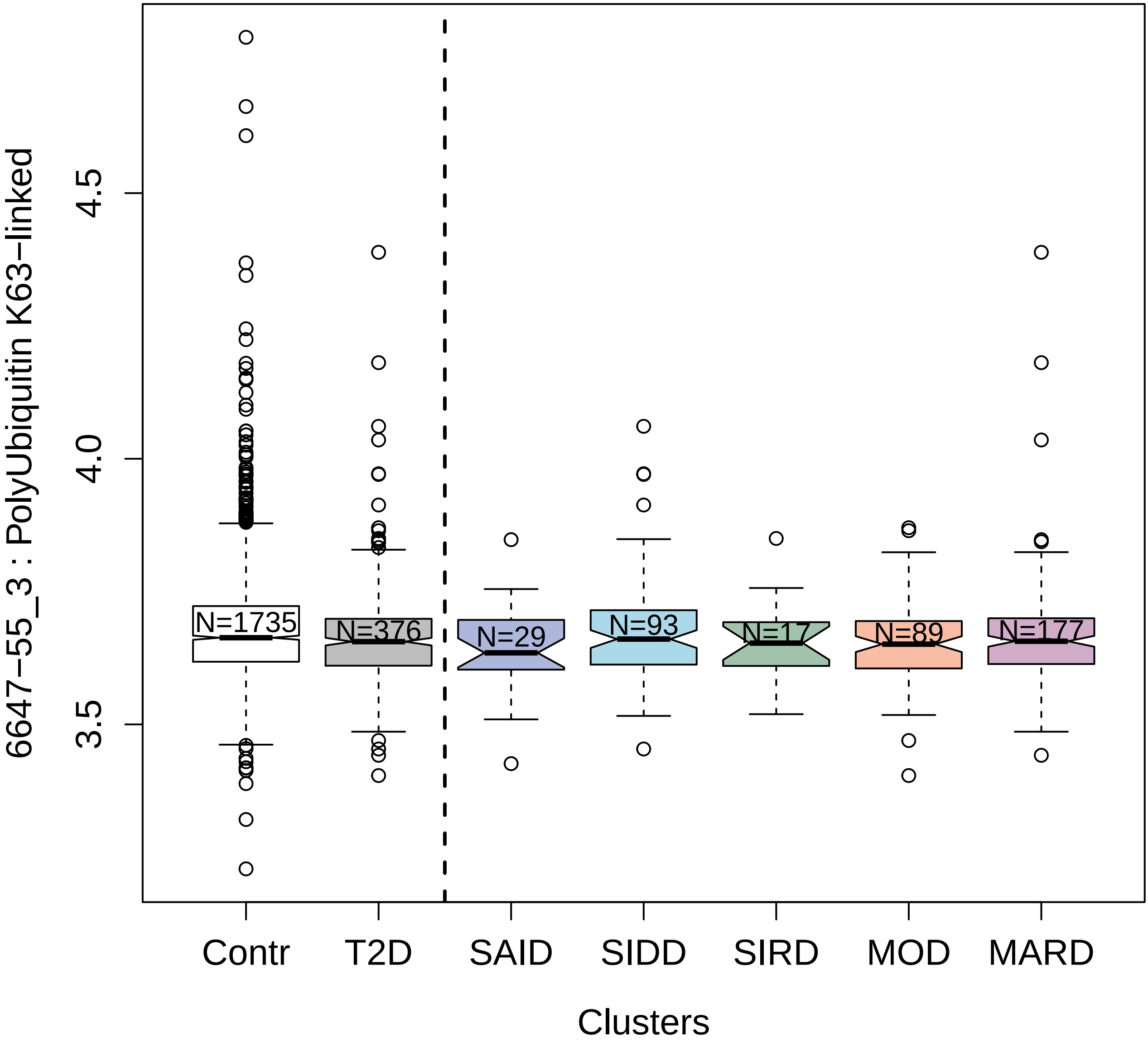
6647−55_3 : PolyUbiquitin K63−linked
4.5
4.0
3.5
N=1735
N=376
N=29
N=93
N=17
N=89
N=177
Contr
T2D
SAID
SIDD
SIRD
MOD
MARD
Clusters

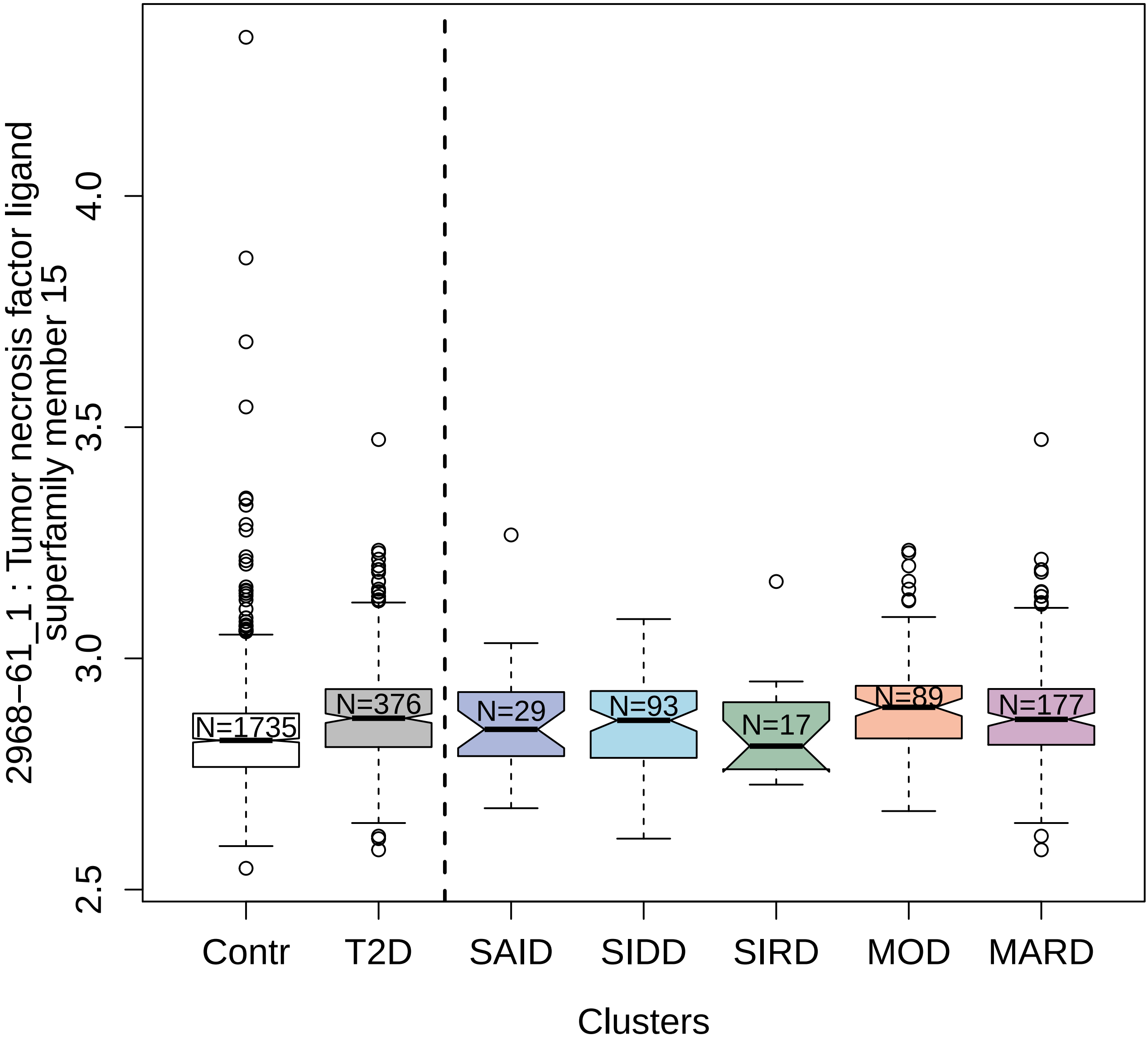

2968−61_1 : Tumor necrosis factor ligand superfamily member 15
2.5
3.0
3.5
4.0
N=1735
N=376
N=29
N=93
N=17
N=89
N=177
Contr
T2D
SAID
SIDD
SIRD
MOD
MARD
Clusters

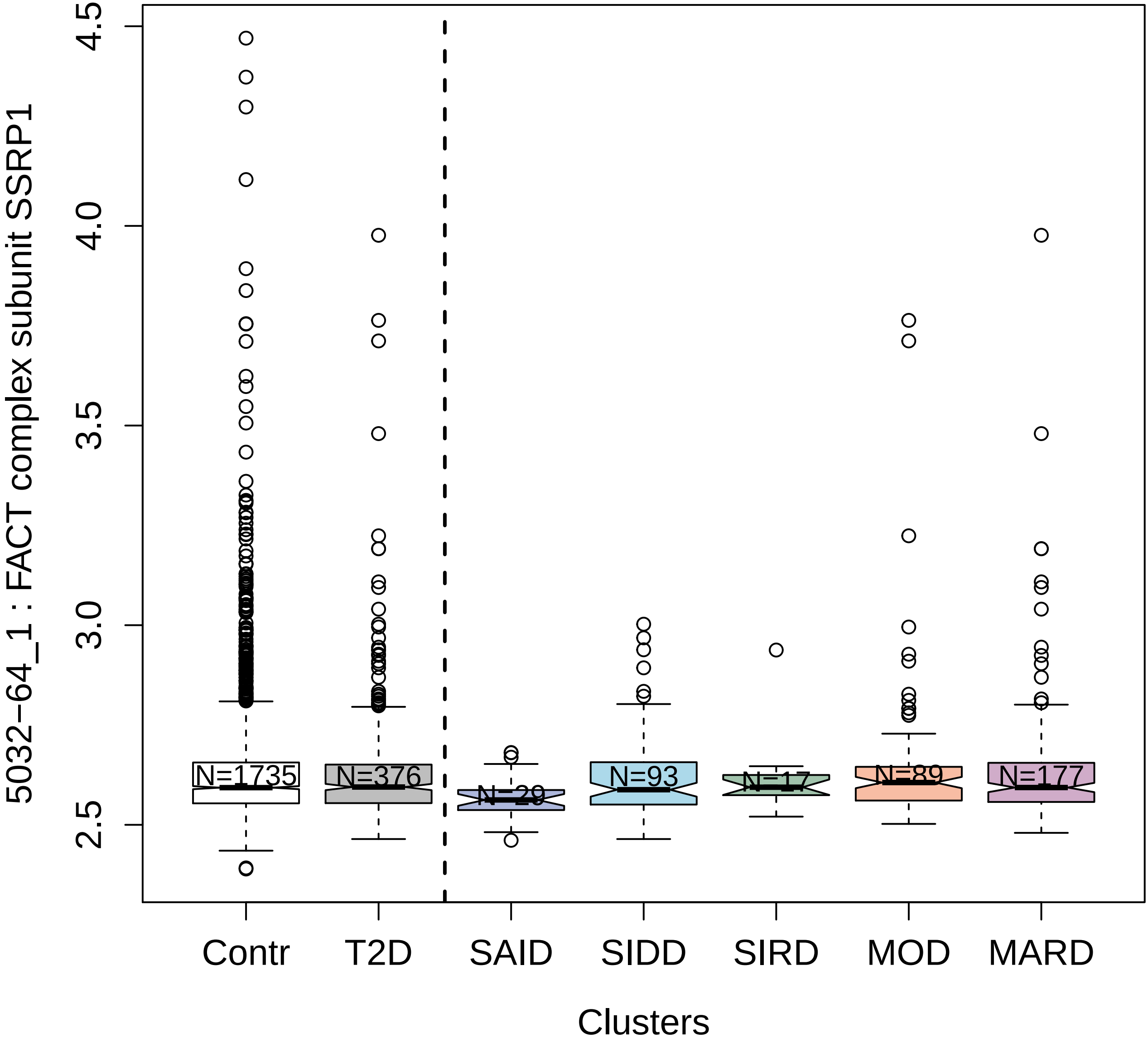
5032−64_1 : FACT complex subunit SSRP1
4.5
4.0
3.5
3.0
2.5
N=1735
N=376
N=29
N=93
N=17
N=89
N=177
Contr
T2D
SAID
SIDD
SIRD
MOD
MARD
Clusters

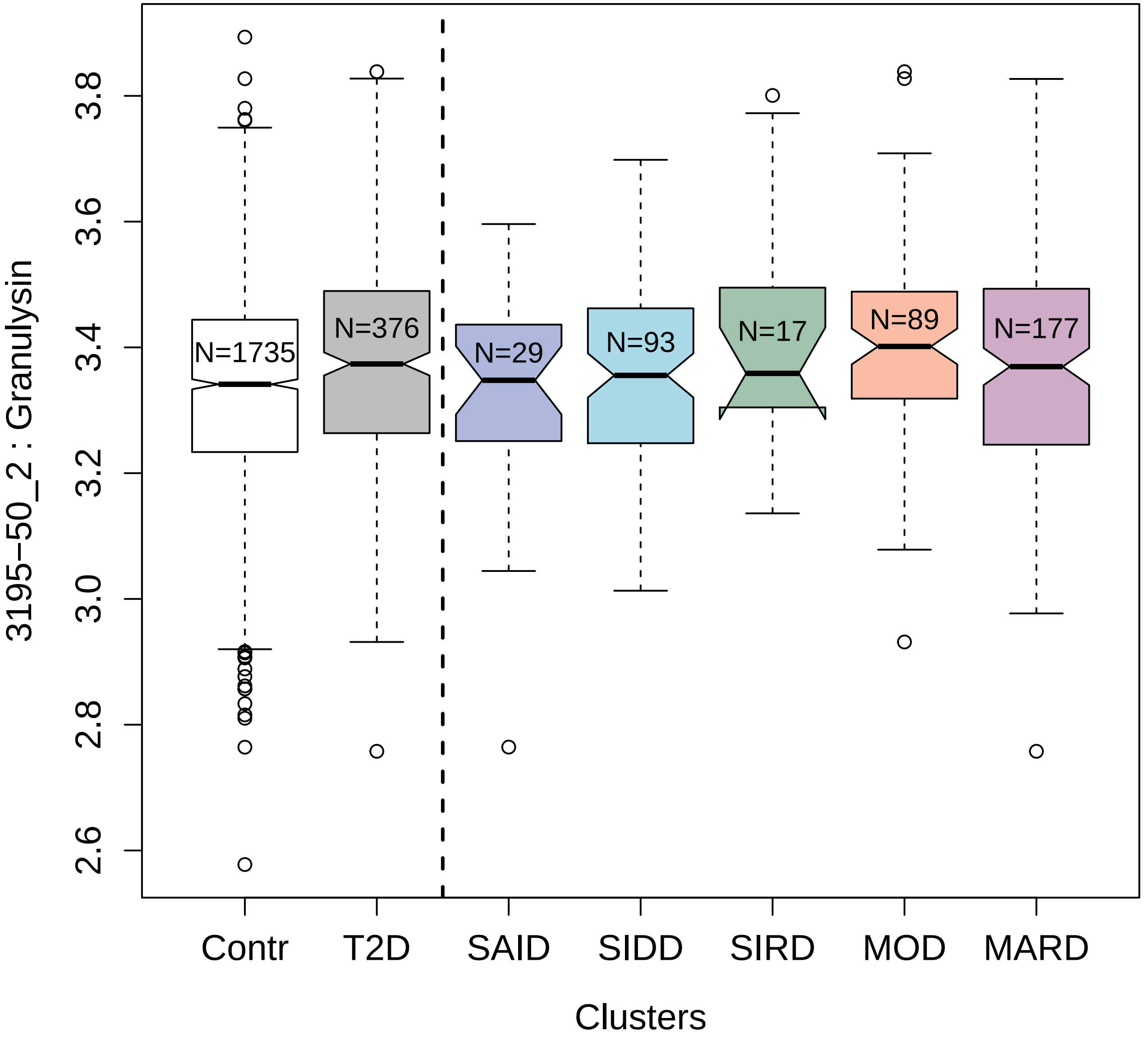

3195−50_2 : Granulysin
2.6
2.8
3.0
3.2
3.4
3.6
3.8
N=1735
N=376
N=29
N=93
N=17
N=89
N=177
Contr
T2D
SAID
SIDD
SIRD
MOD
MARD
Clusters

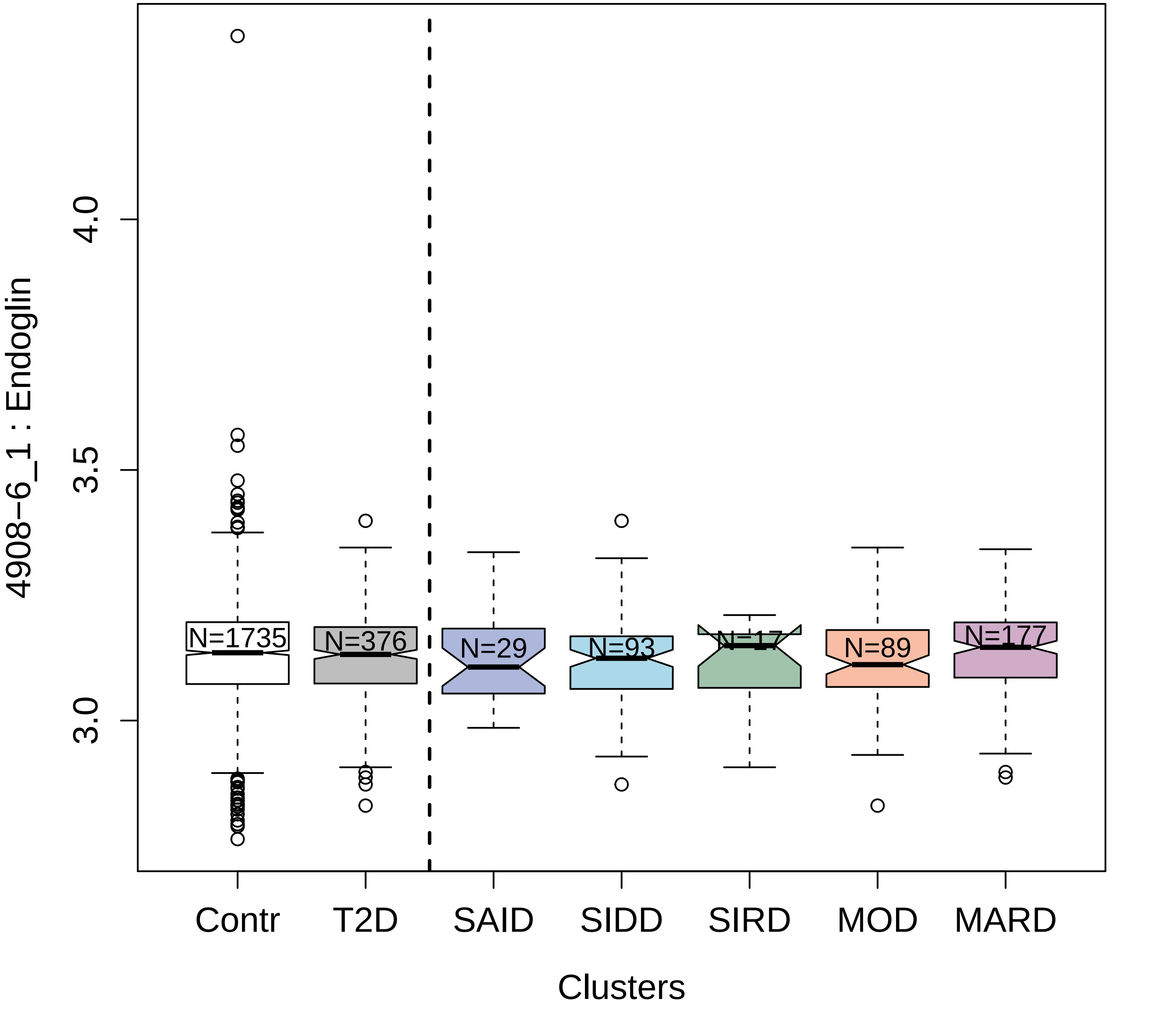

4908−6_1 : Endoglin
4.0
3.5
3.0
N=1735
N=376
N=29
N=93
N=17
N=89
N=177
Contr
T2D
SAID
SIDD
SIRD
MOD
MARD
Clusters

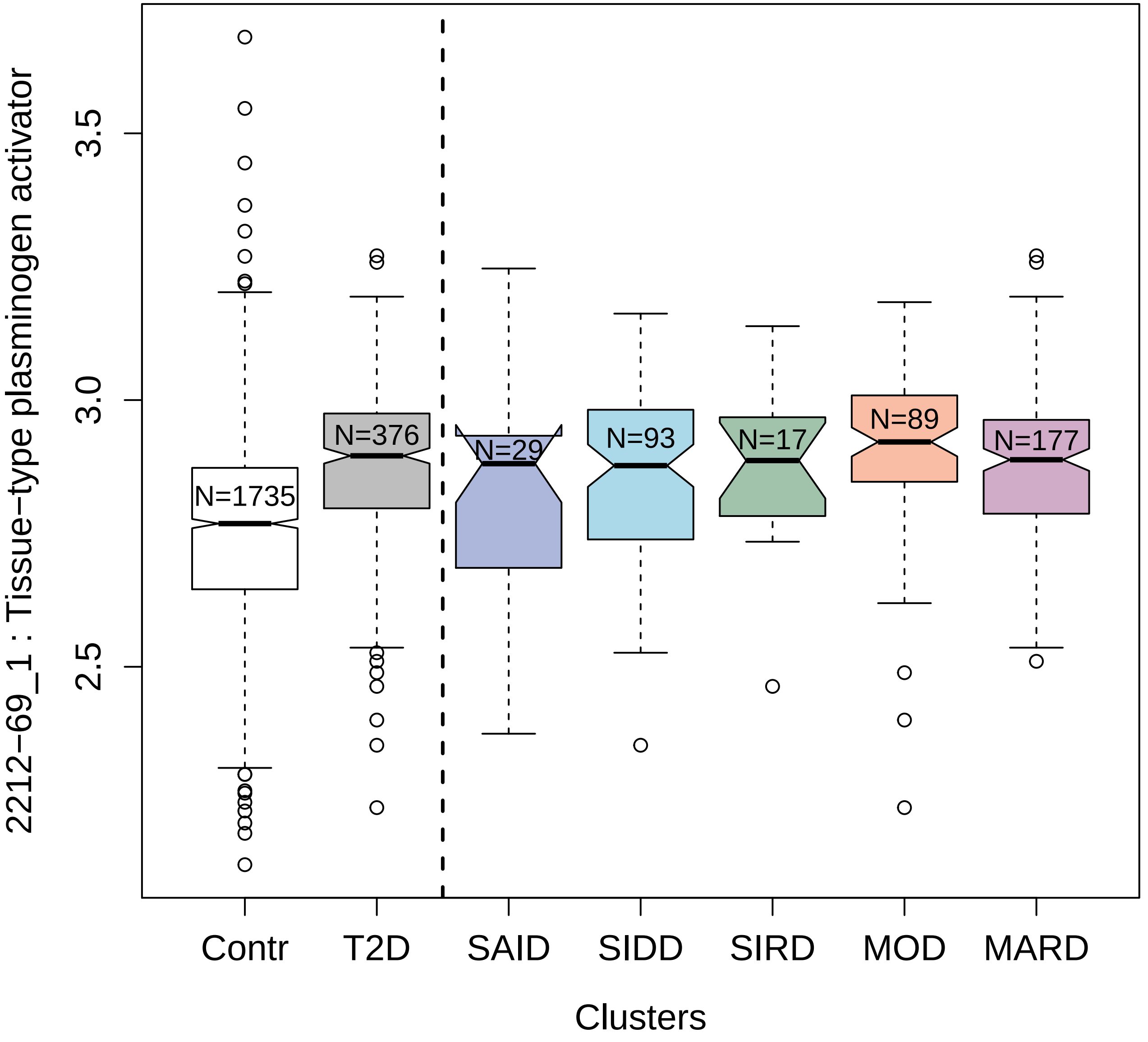
2212−69_1 : Tissue−type plasminogen activator
3.5
3.0
2.5
N=1735
N=376
N=29
N=93
N=17
N=89
N=177
Contr
T2D
SAID
SIDD
SIRD
MOD
MARD
Clusters

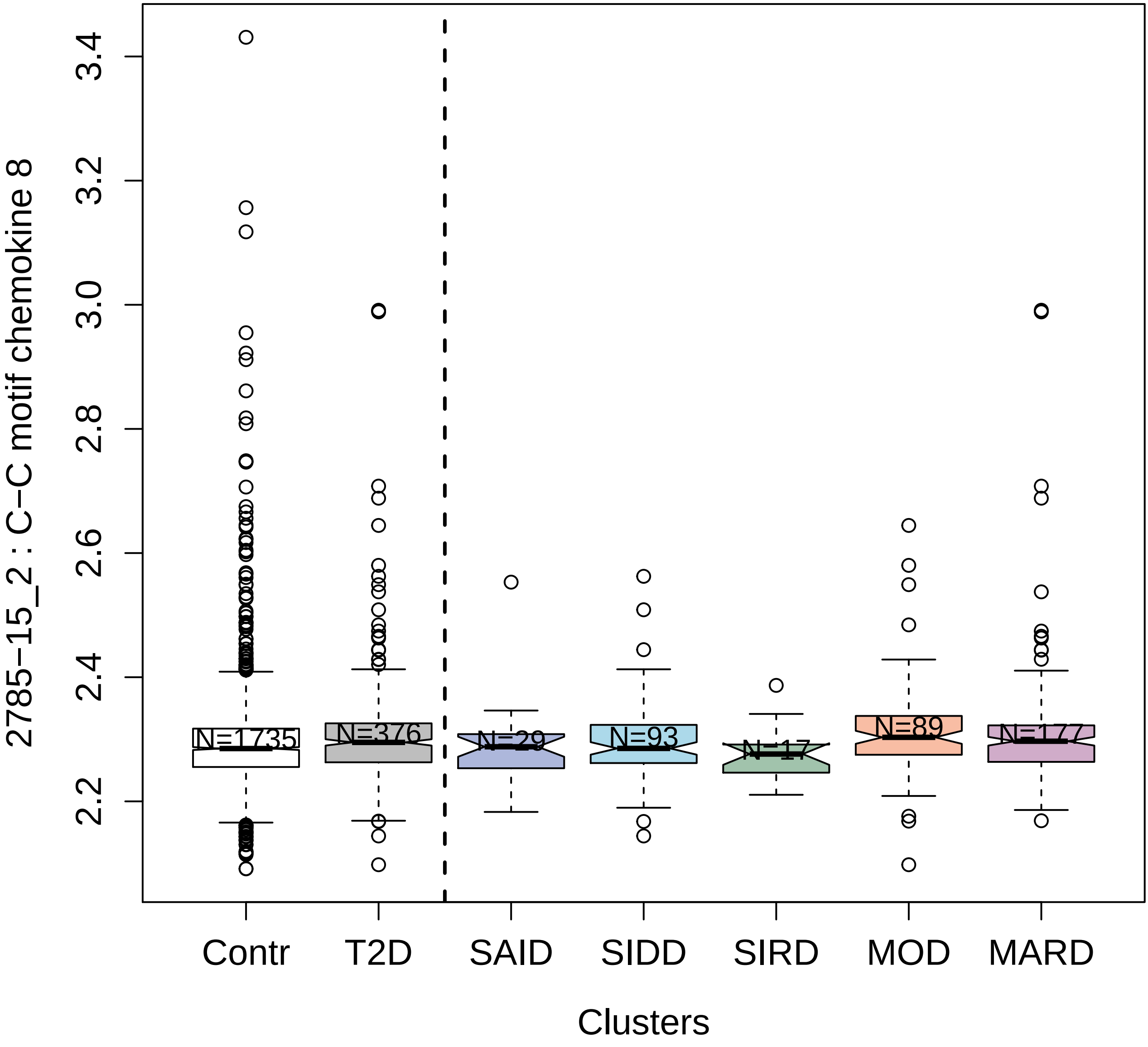

2785−15_2 : C−C motif chemokine 8
3.4
3.2
3.0
2.8
2.6
2.4
2.2
N=1735
N=376
N=29
N=93
N=17
N=89
N=177
Contr
T2D
SAID
SIDD
SIRD
MOD
MARD
Clusters

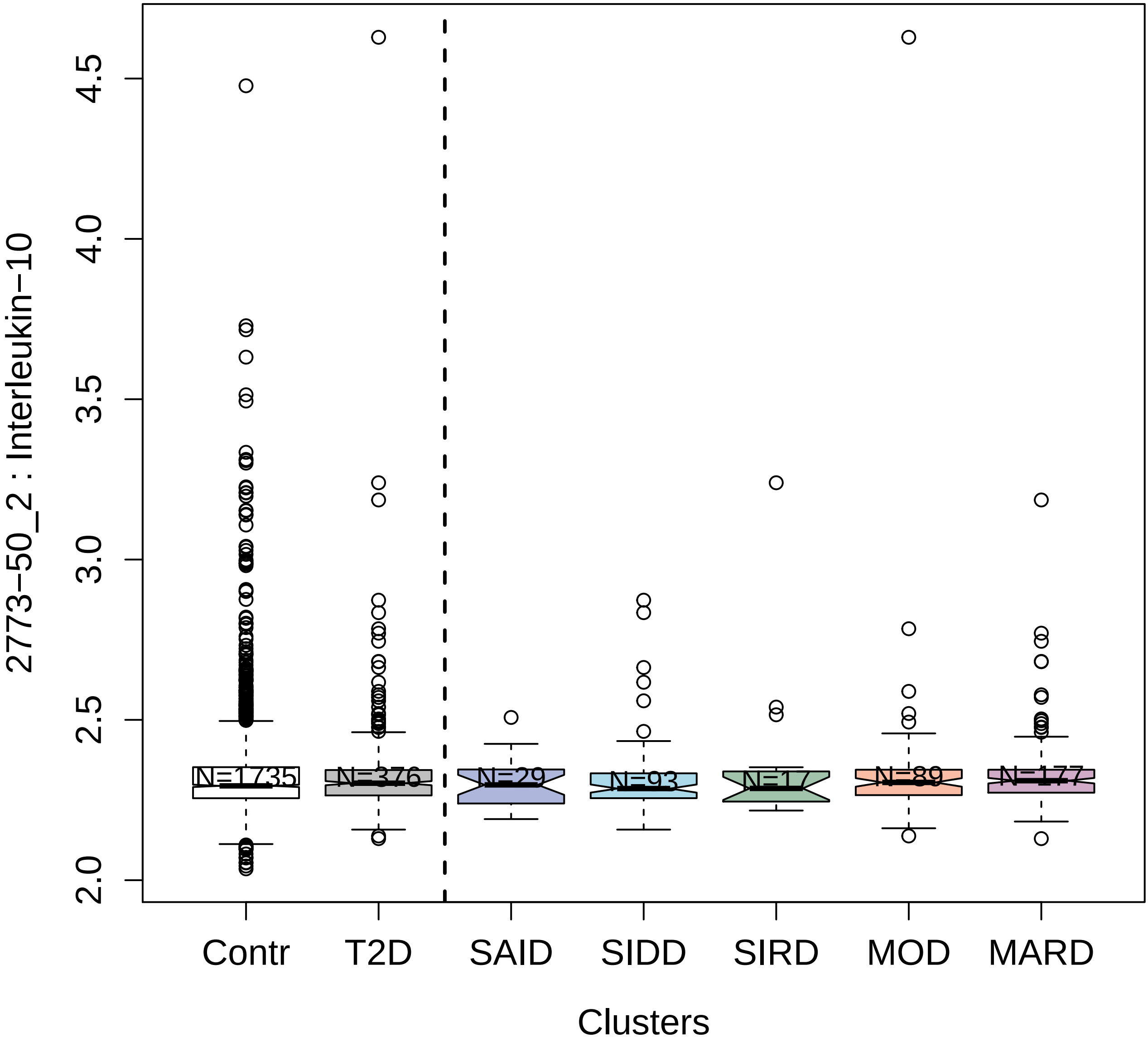

2773−50_2 : Interleukin−10
4.5
4.0
3.5
3.0
2.5
2.0
N=1735
N=376
N=29
N=93
N=17
N=89
N=177
Contr
T2D
SAID
SIDD
SIRD
MOD
MARD
Clusters

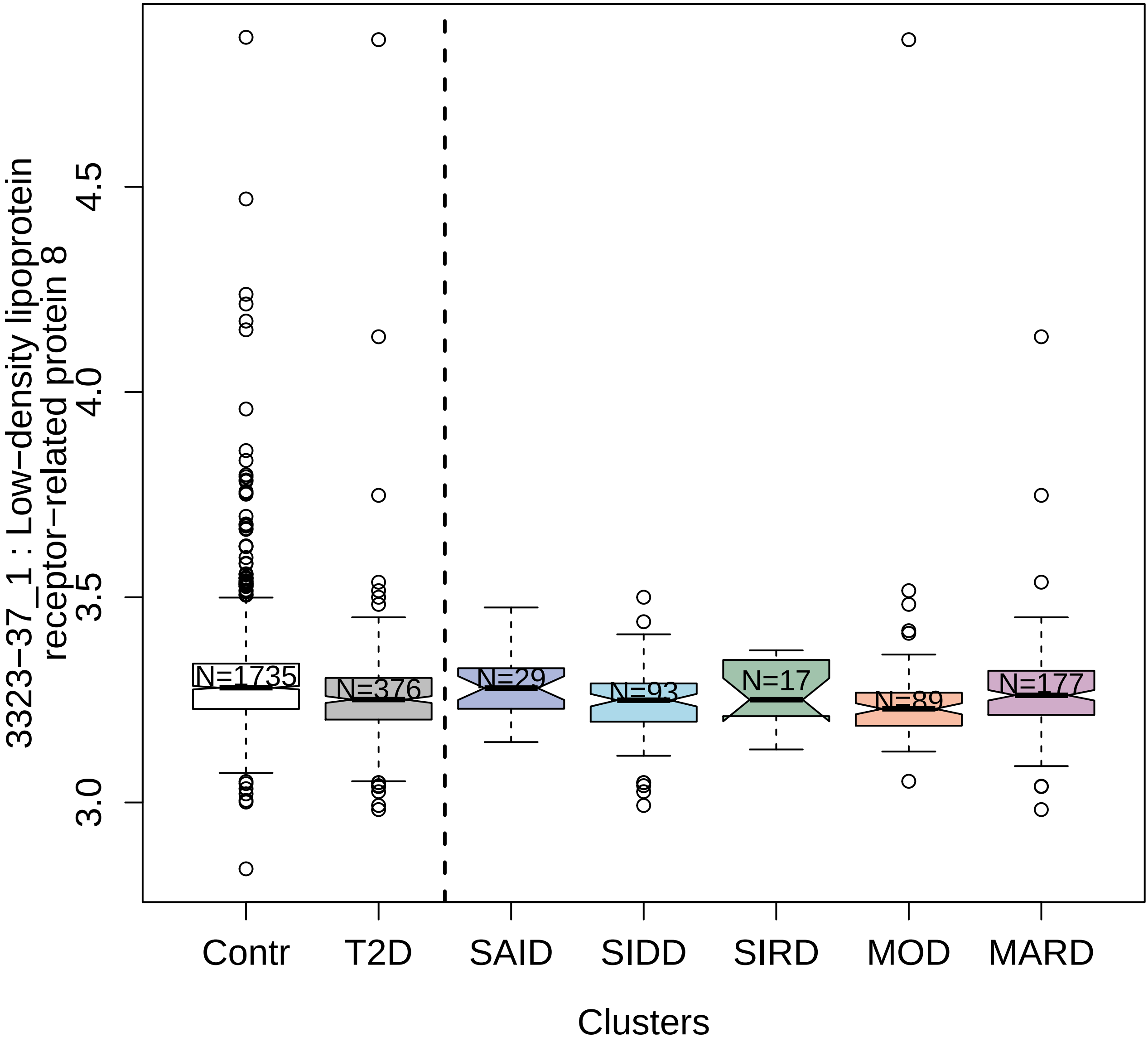

3323−37_1 : Low−density lipoprotein
receptor−related protein 8
3.0
3.5
4.0
4.5
N=1735
N=376
N=29
N=93
N=17
N=89
N=177
Contr
T2D
SAID
SIDD
SIRD
MOD
MARD
Clusters

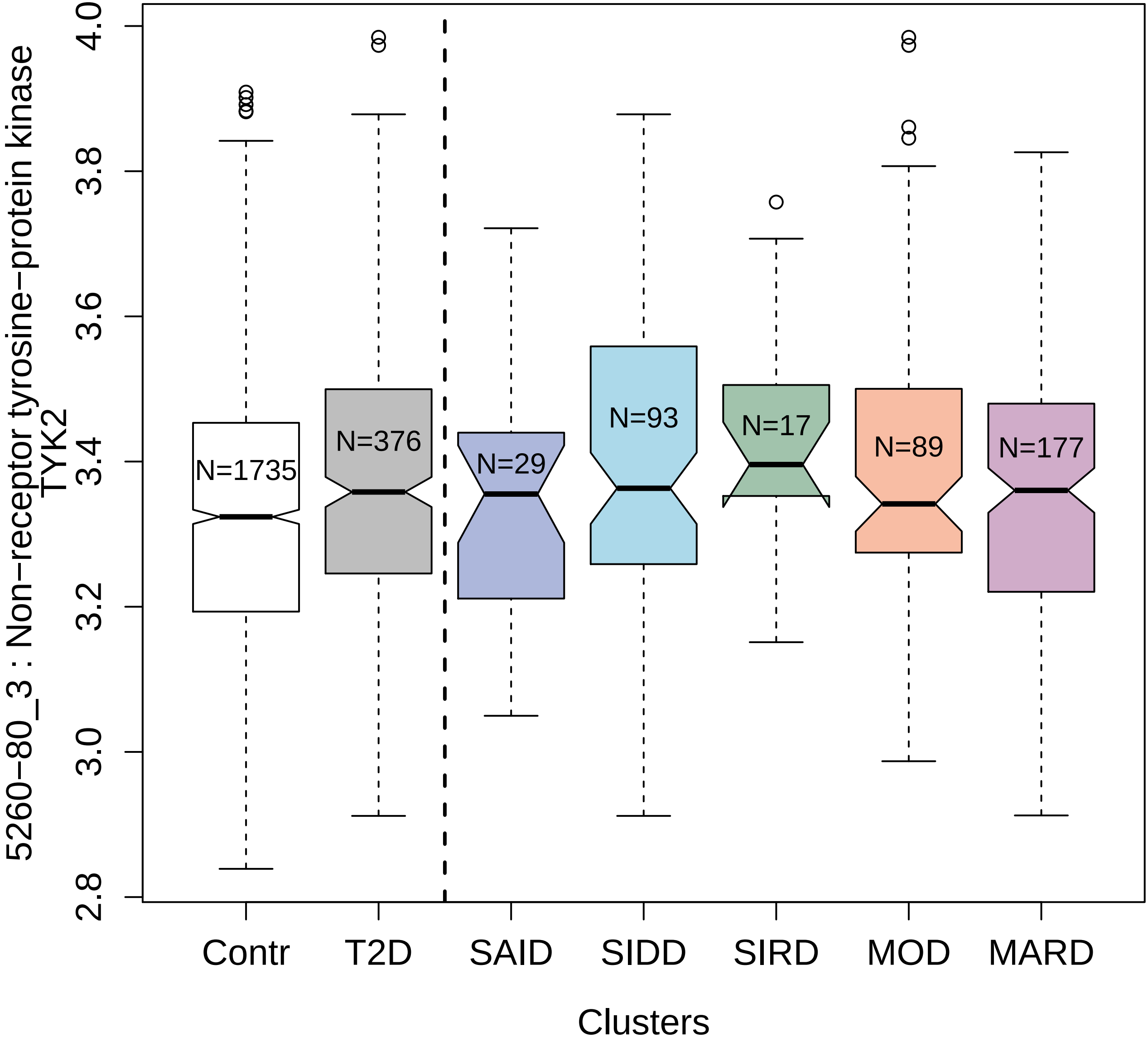
5260−80_3 : Non−receptor tyrosine−protein kinase
TYK2
4.0
3.8
3.6
3.4
3.2
3.0
2.8
N=1735
N=376
N=29
N=93
N=17
N=89
N=177
Contr
T2D
SAID
SIDD
SIRD
MOD
MARD
Clusters

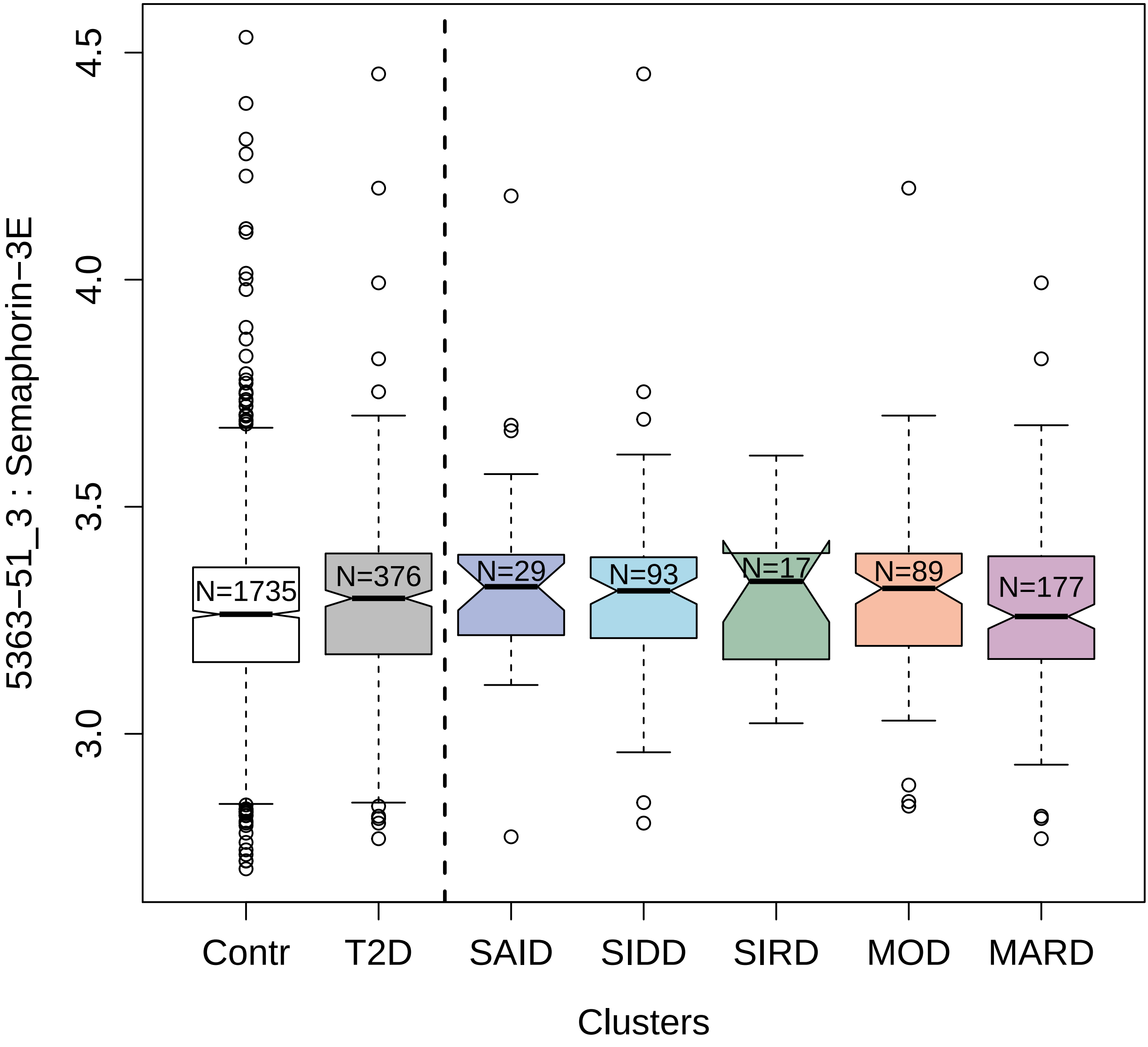
5363−51_3 : Semaphorin−3E
4.5
4.0
3.5
3.0
N=1735
N=376
N=29
N=93
N=17
N=89
N=177
Contr
T2D
SAID
SIDD
SIRD
MOD
MARD
Clusters

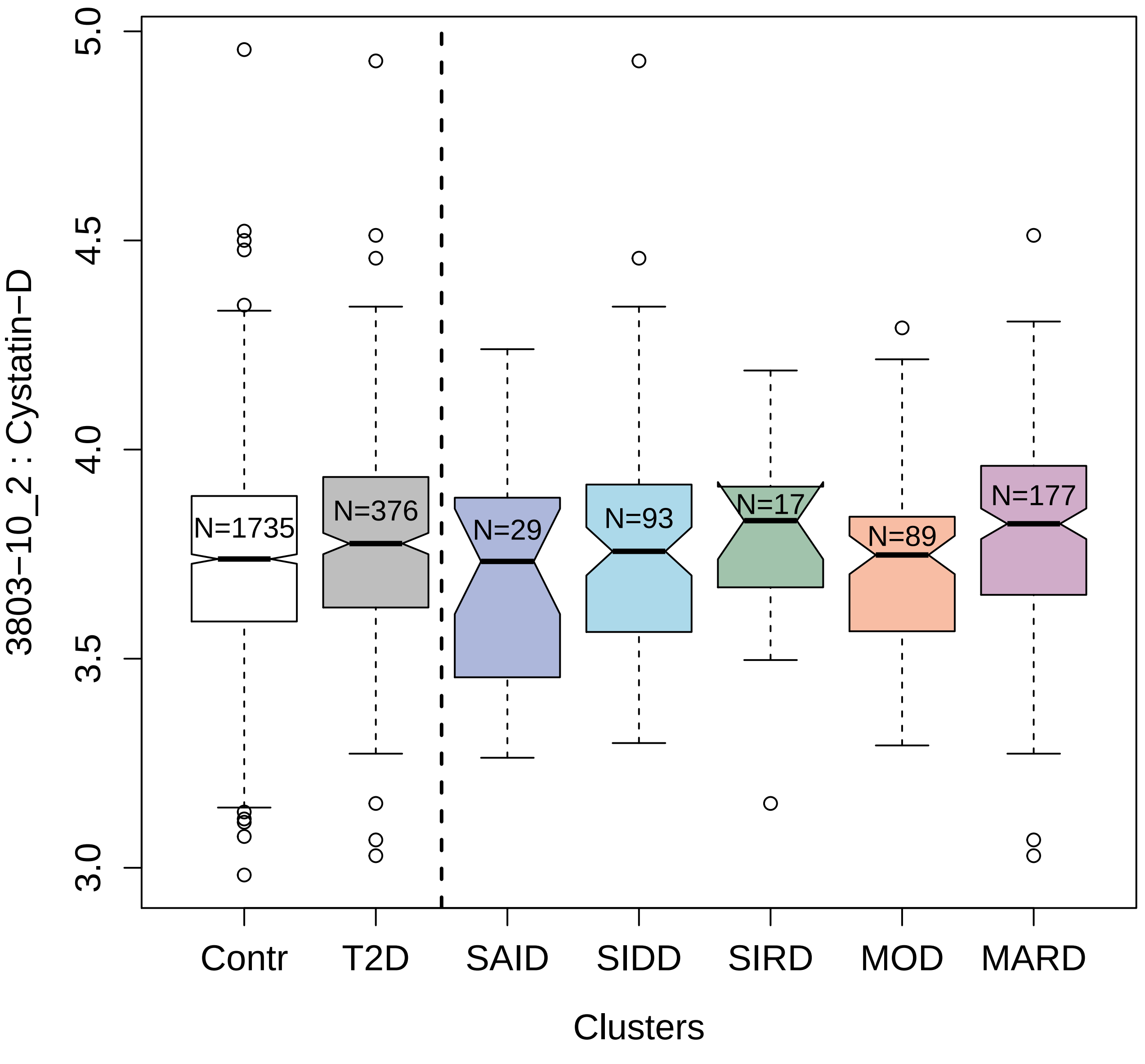
3803−10_2 : Cystatin−D
5.0
4.5
4.0
3.5
3.0
N=1735
N=376
N=29
N=93
N=17
N=89
N=177
Contr
T2D
SAID
SIDD
SIRD
MOD
MARD
Clusters

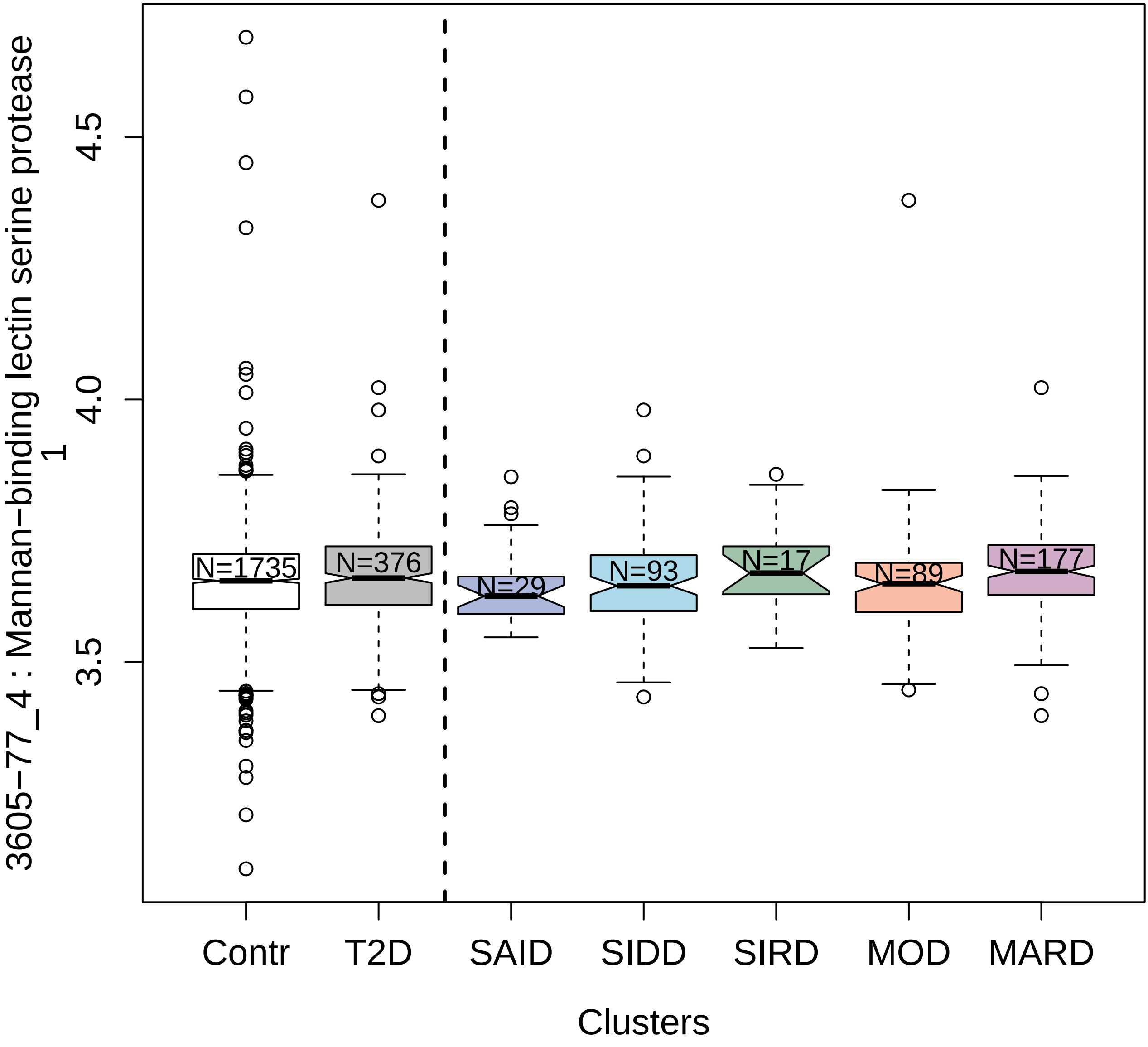
3605−77_4 : Mannan−binding lectin serine protease 1
4.5
4.0
3.5
N=1735
N=376
N=29
N=93
N=17
N=89
N=177
Contr
T2D
SAID
SIDD
SIRD
MOD
MARD
Clusters

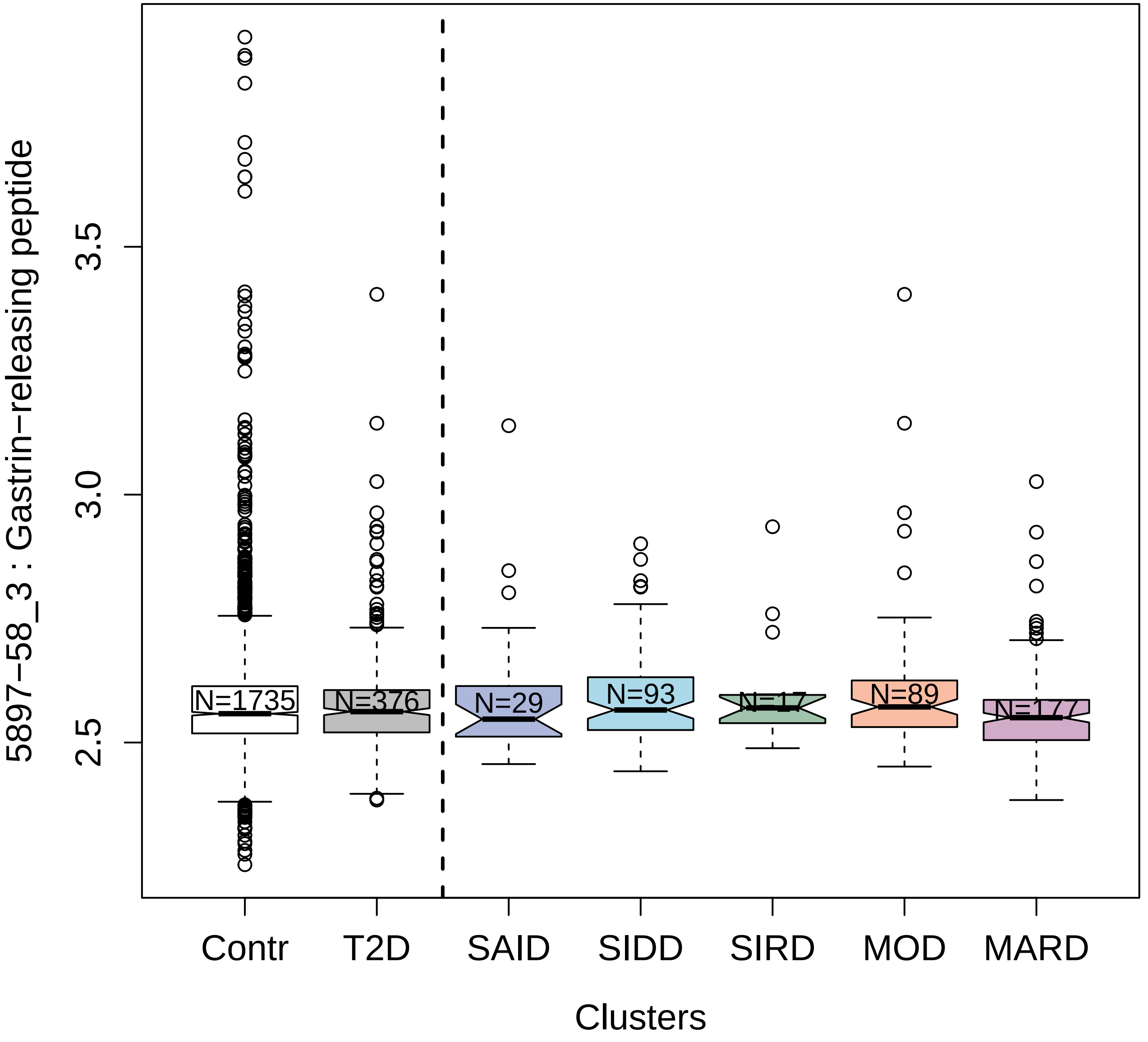
5897−58_3 : Gastrin−releasing peptide
2.5
3.0
3.5
N=1735
N=376
N=29
N=93
N=17
N=89
N=177
Contr
T2D
SAID
SIDD
SIRD
MOD
MARD
Clusters

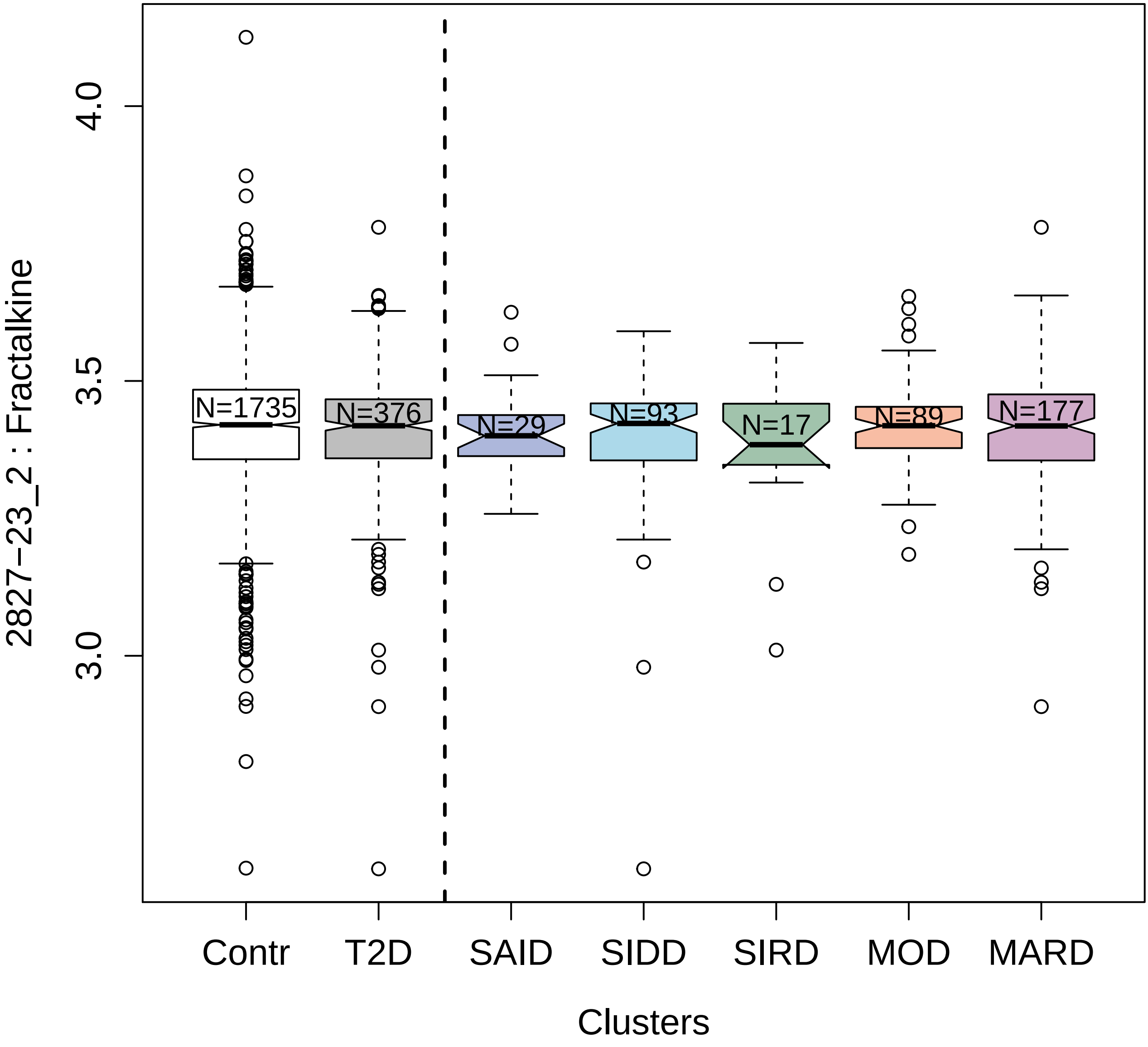

2827−23_2 : Fractalkine
4.0
3.5
3.0
N=1735
N=376
N=29
N=93
N=17
N=89
N=177
Contr
T2D
SAID
SIDD
SIRD
MOD
MARD
Clusters

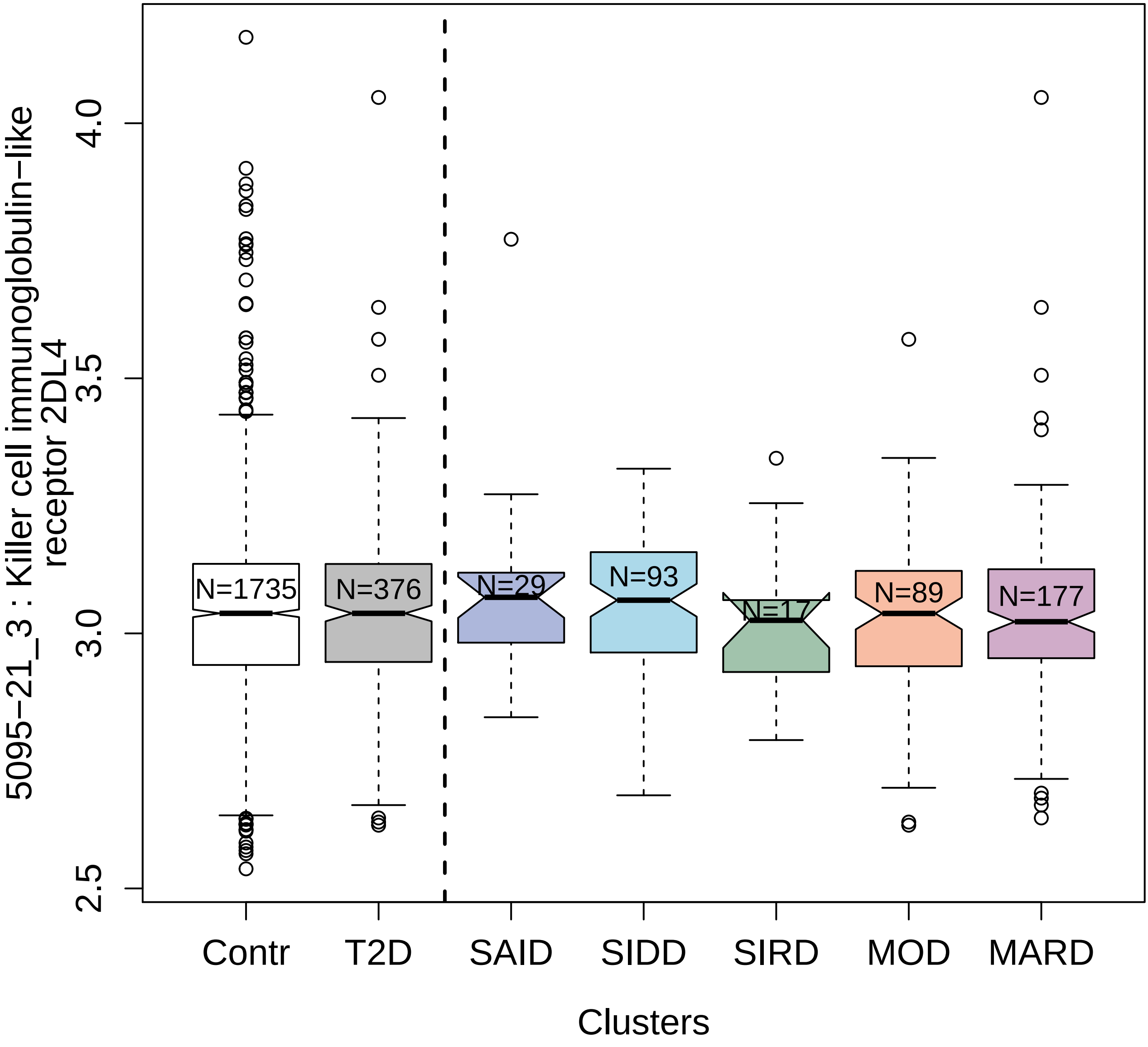
5095−21_3 : Killer cell immunoglobulin−like receptor 2DL4
4.0
3.5
3.0
2.5
N=1735
N=376
N=29
N=93
N=17
N=89
N=177
Contr
T2D
SAID
SIDD
SIRD
MOD
MARD
Clusters

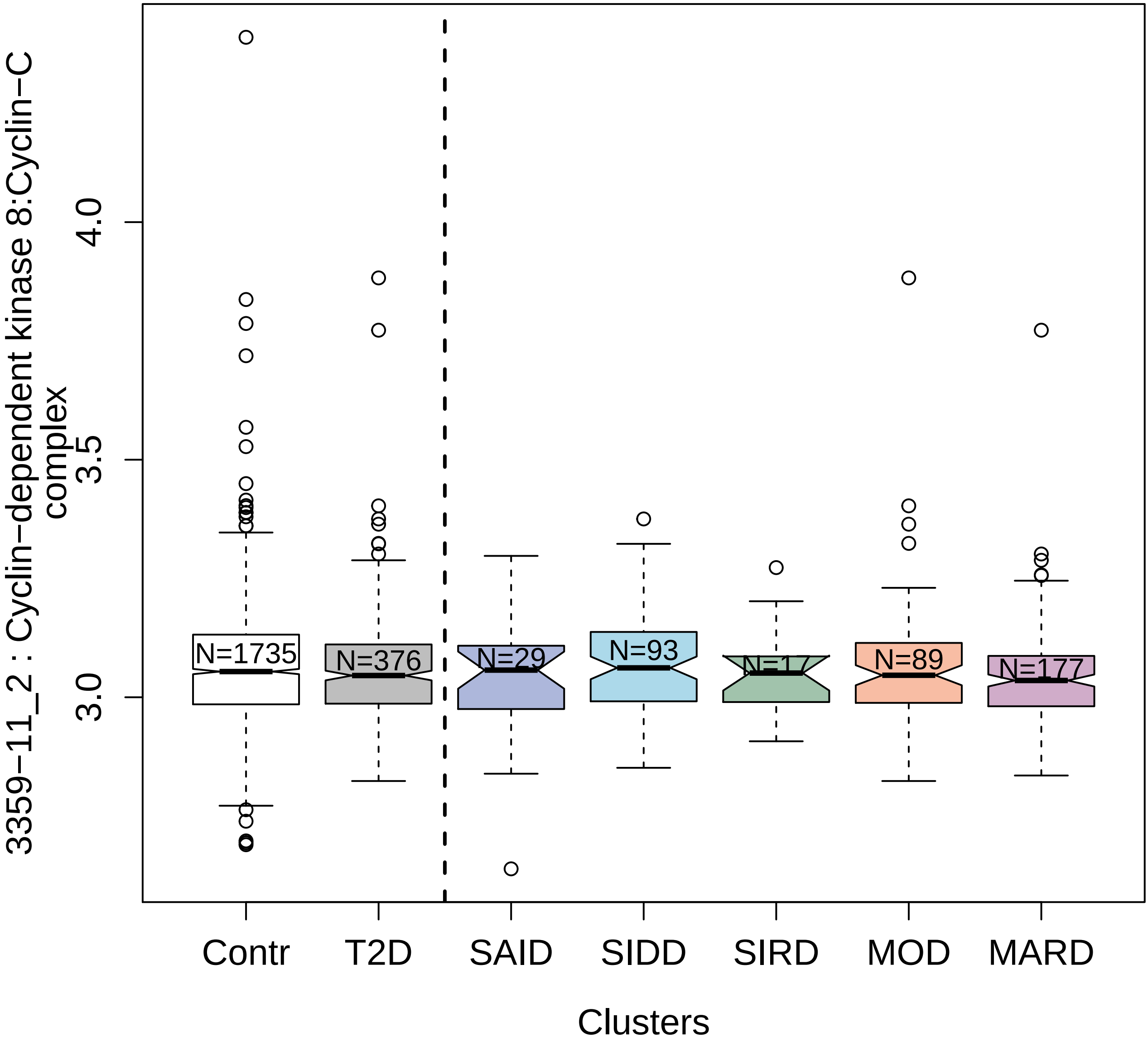

3359−11_2 : Cyclin−dependent kinase 8:Cyclin−C complex
3.0
3.5
4.0
N=1735
N=376
N=29
N=93
N=89
N=177
Contr
T2D
SAID
SIDD
SIRD
MOD
MARD
Clusters

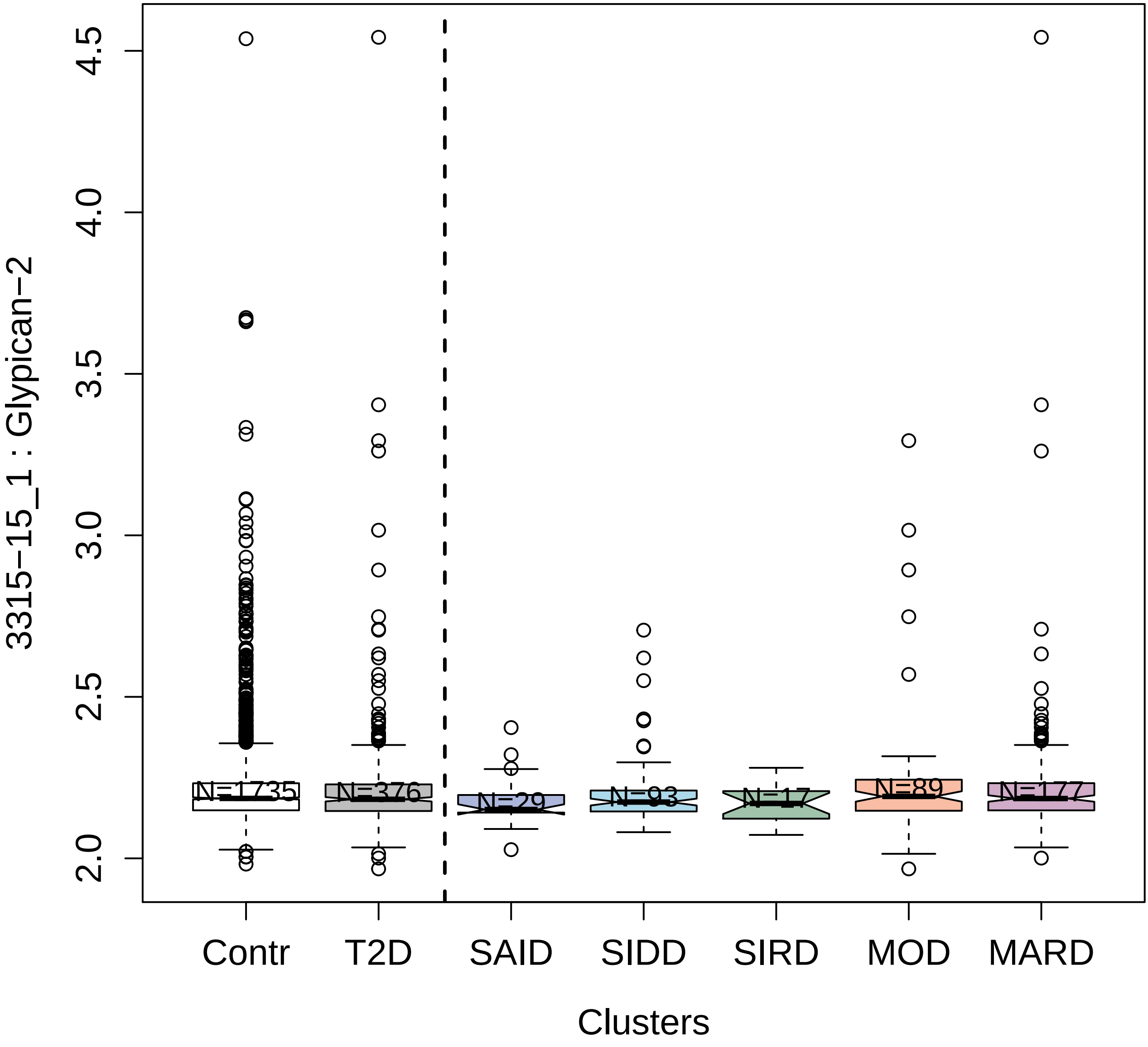

3315−15_1 : Glypican−2
2.0
2.5
3.0
3.5
4.0
4.5
N=1735
N=376
N=29
N=93
N=17
N=89
N=177
Contr
T2D
SAID
SIDD
SIRD
MOD
MARD
Clusters

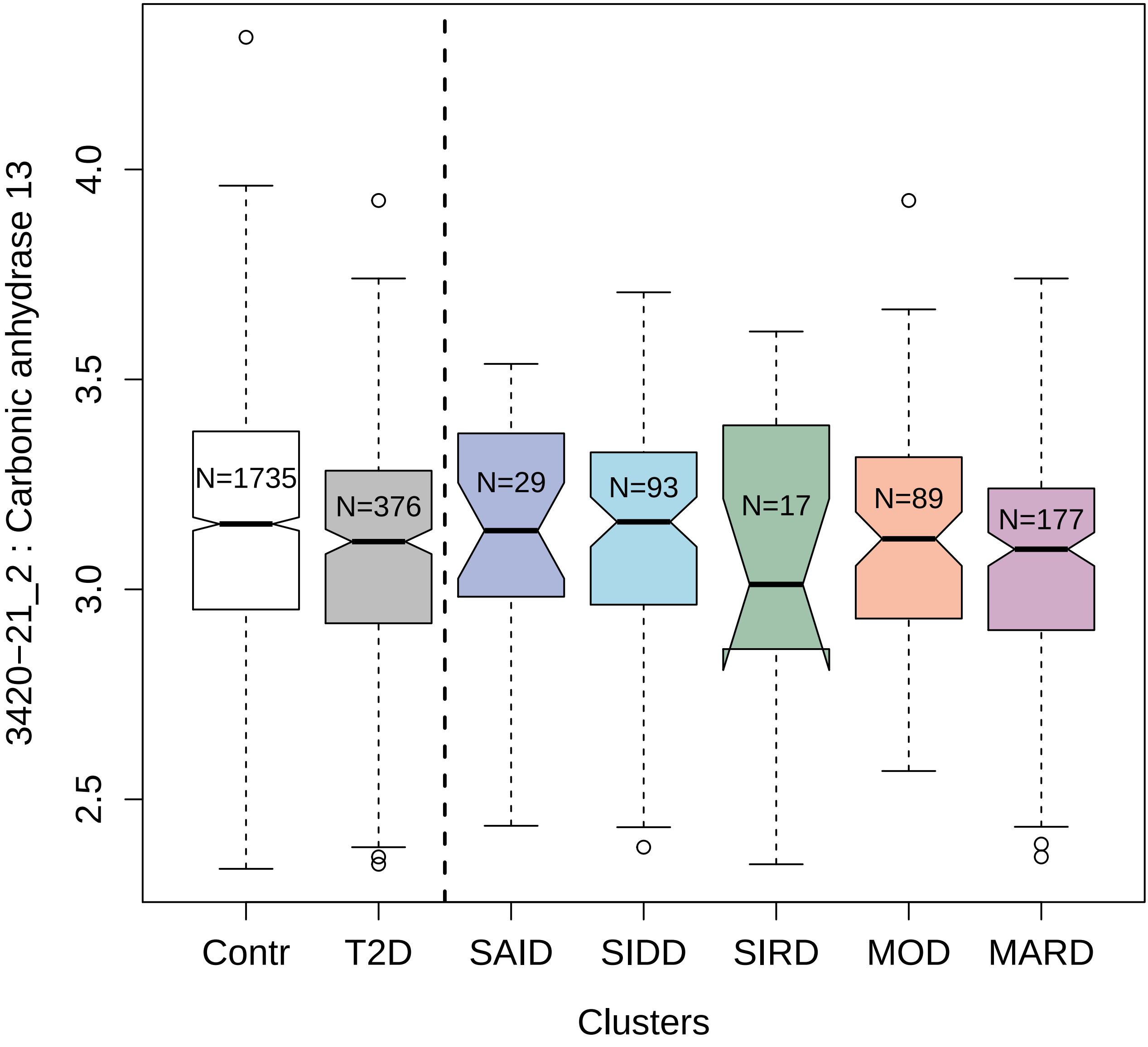
3420−21_2 : Carbonic anhydrase 13
2.5
3.0
3.5
4.0
N=1735
N=376
N=29
N=93
N=17
N=89
N=177
Contr
T2D
SAID
SIDD
SIRD
MOD
MARD
Clusters

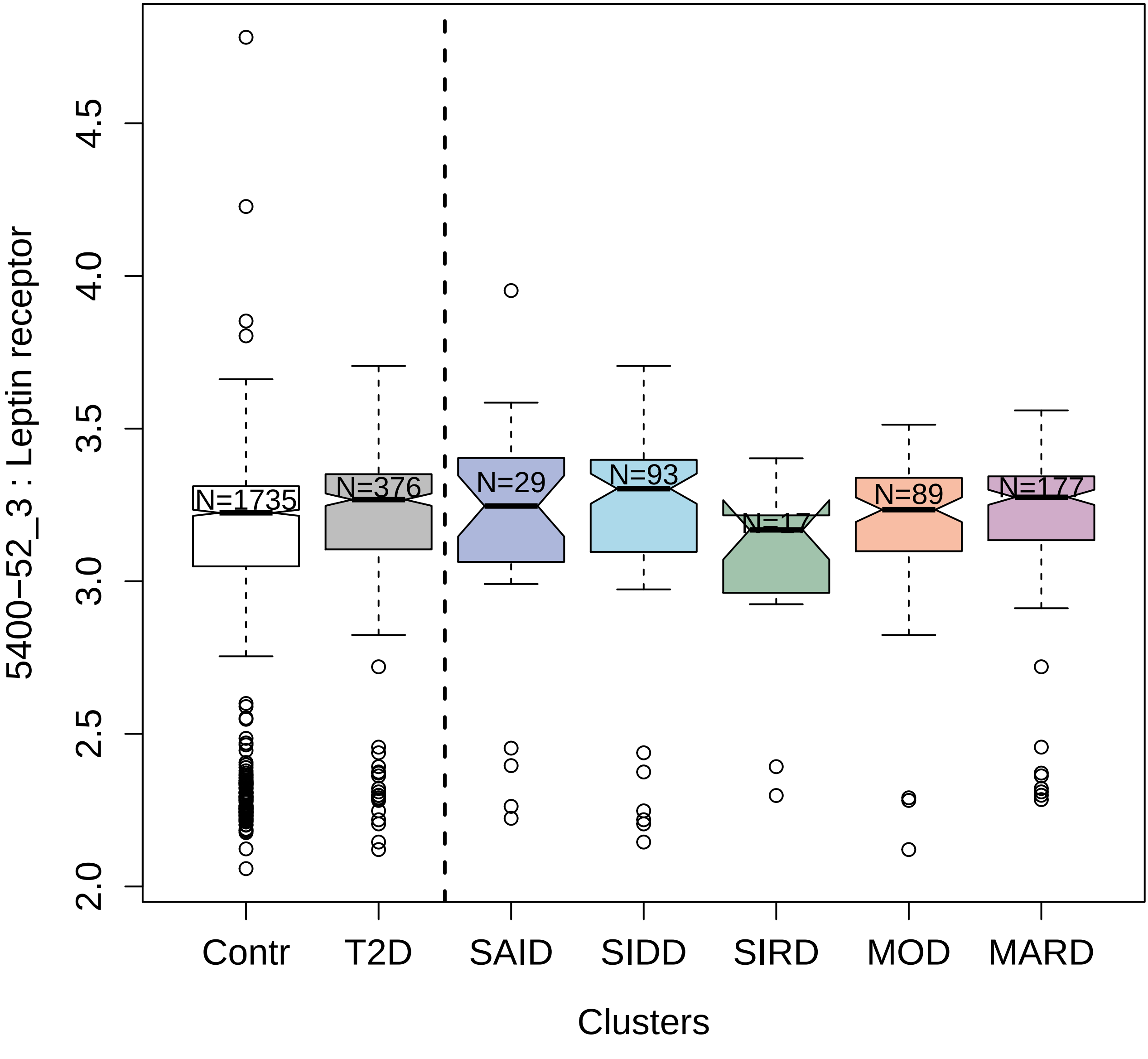

5400−52_3 : Leptin receptor
2.0
2.5
3.0
3.5
4.0
4.5
N=1735
N=376
N=29
N=93
N=89
N=177
Contr
T2D
SAID
SIDD
SIRD
MOD
MARD
Clusters

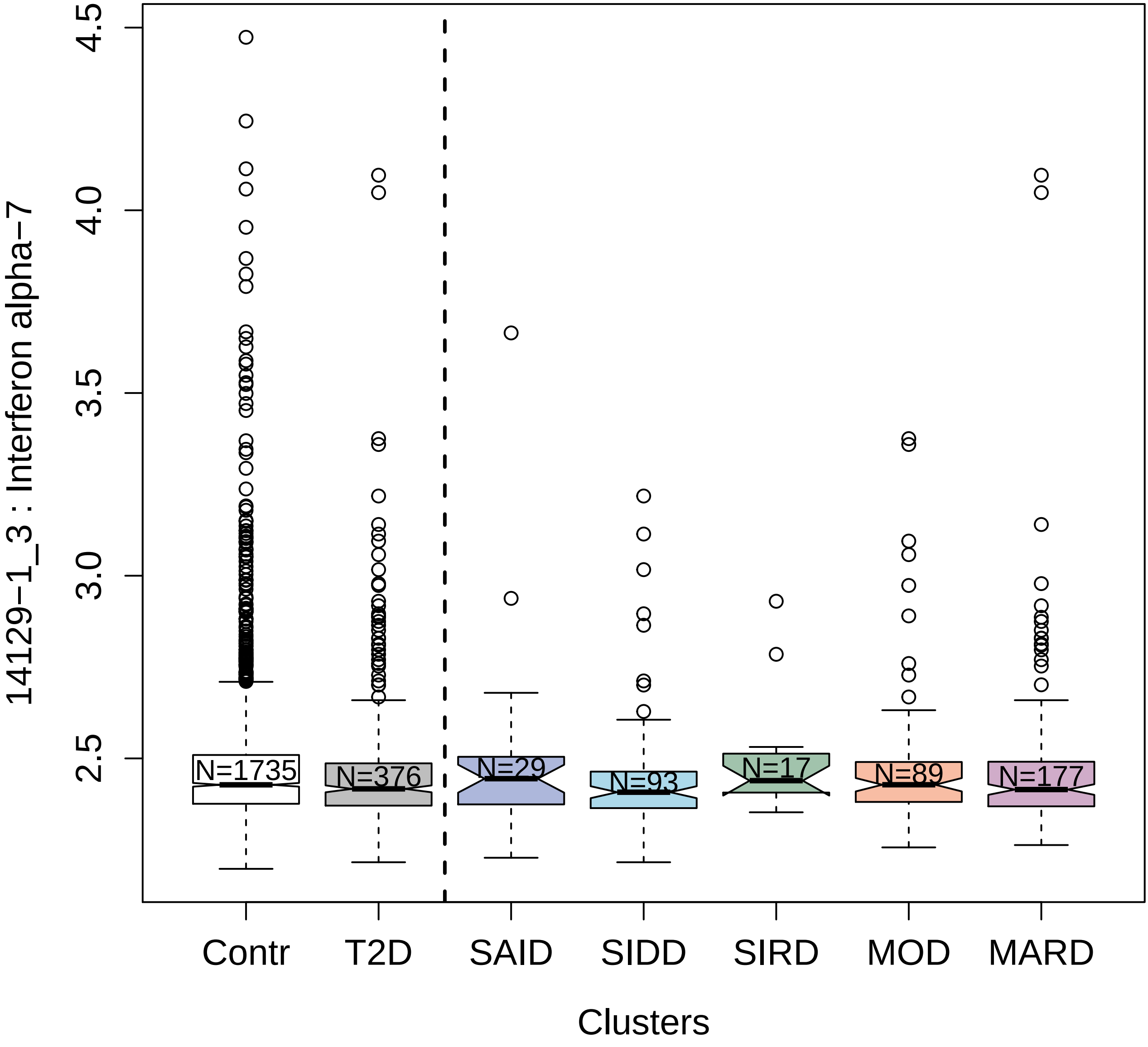
14129−1_3 : Interferon alpha−7
4.5
4.0
3.5
3.0
2.5
N=1735
N=376
N=29
N=93
N=17
N=89
N=177
Contr
T2D
SAID
SIDD
SIRD
MOD
MARD
Clusters

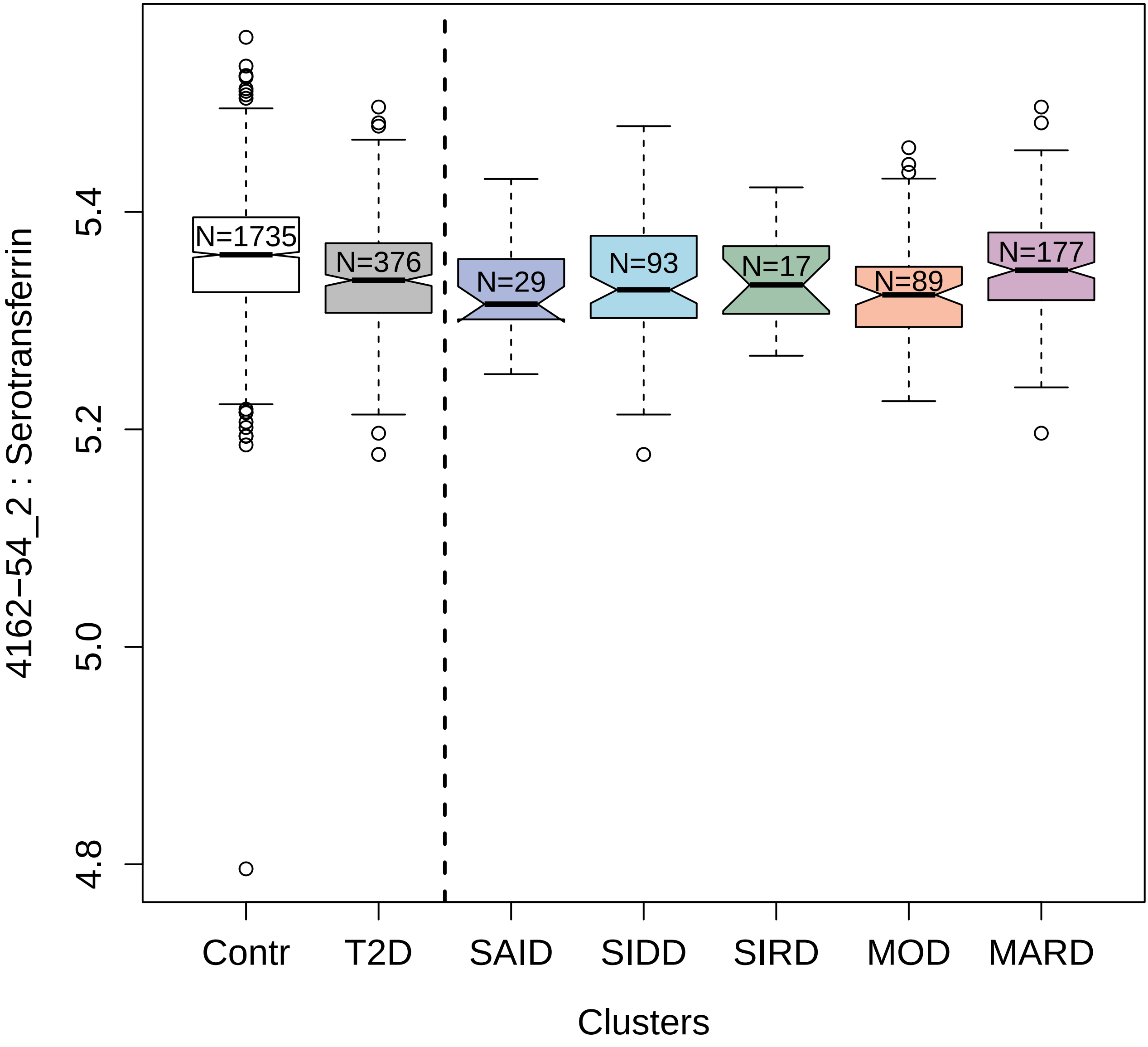

4162−54_2 : Serotransferrin
4.8
5.0
5.2
5.4
N=1735
N=376
N=29
N=93
N=17
N=89
N=177
Contr
T2D
SAID
SIDD
SIRD
MOD
MARD
Clusters

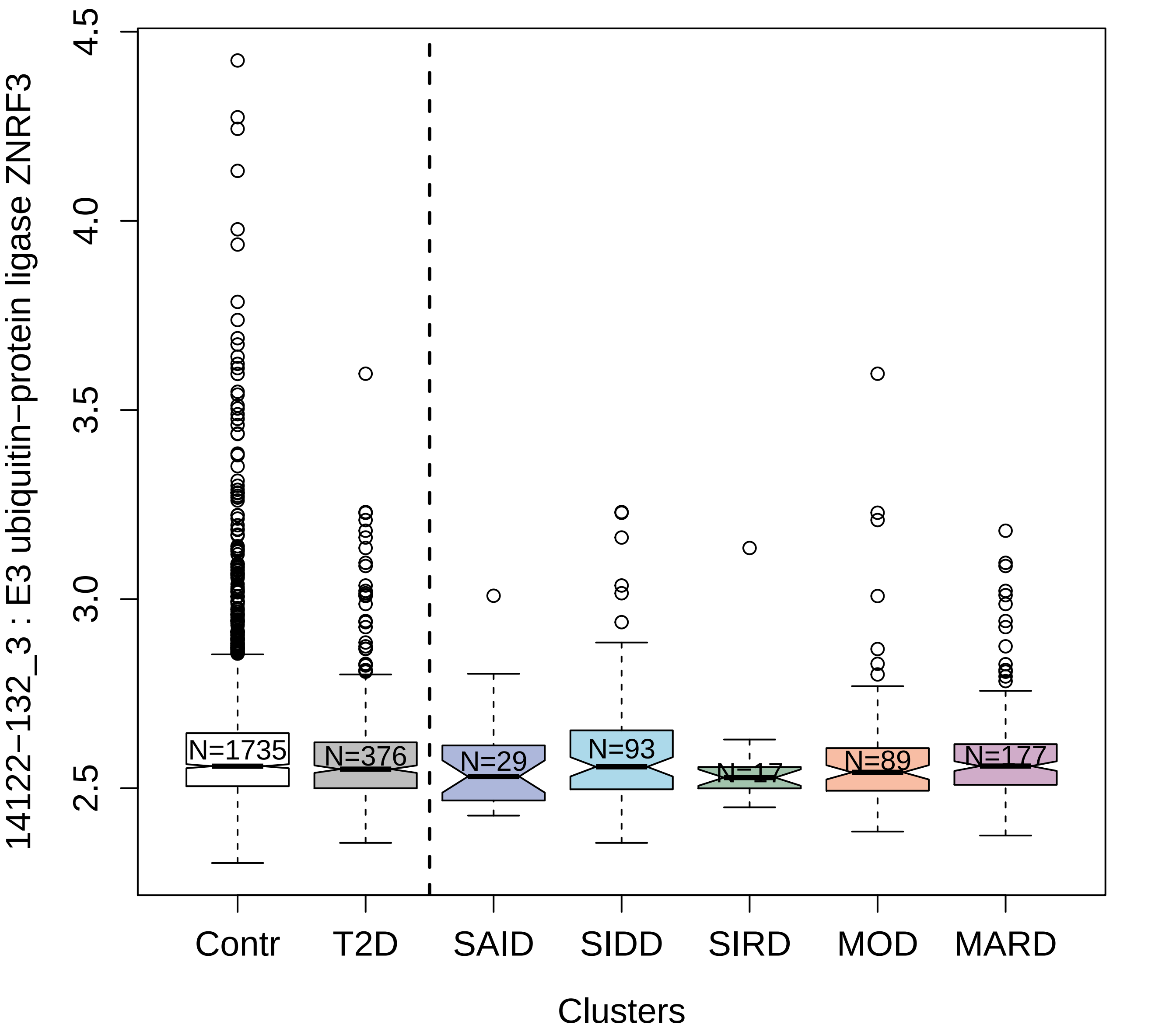

14122−132_3 : E3 ubiquitin−protein ligase ZNRF3
4.5
4.0
3.5
3.0
2.5
N=1735
N=376
N=29
N=93
N=17
N=89
N=177
Contr
T2D
SAID
SIDD
SIRD
MOD
MARD
Clusters

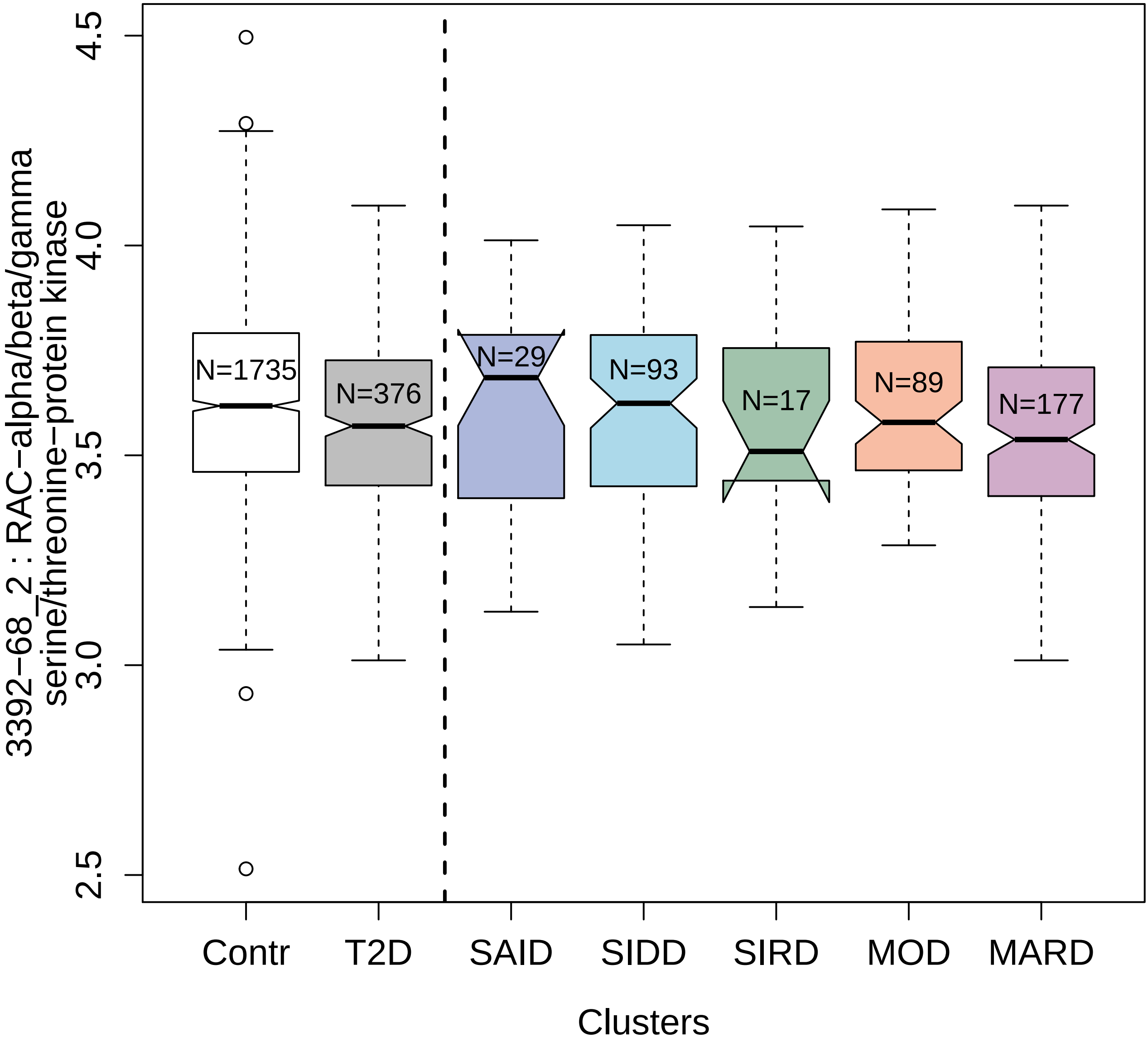
3392−68_2 : RAC−alpha/beta/gamma
serine/threonine−protein kinase
4.5
4.0
3.5
3.0
2.5
N=1735
N=376
N=29
N=93
N=17
N=89
N=177
Contr
T2D
SAID
SIDD
SIRD
MOD
MARD
Clusters

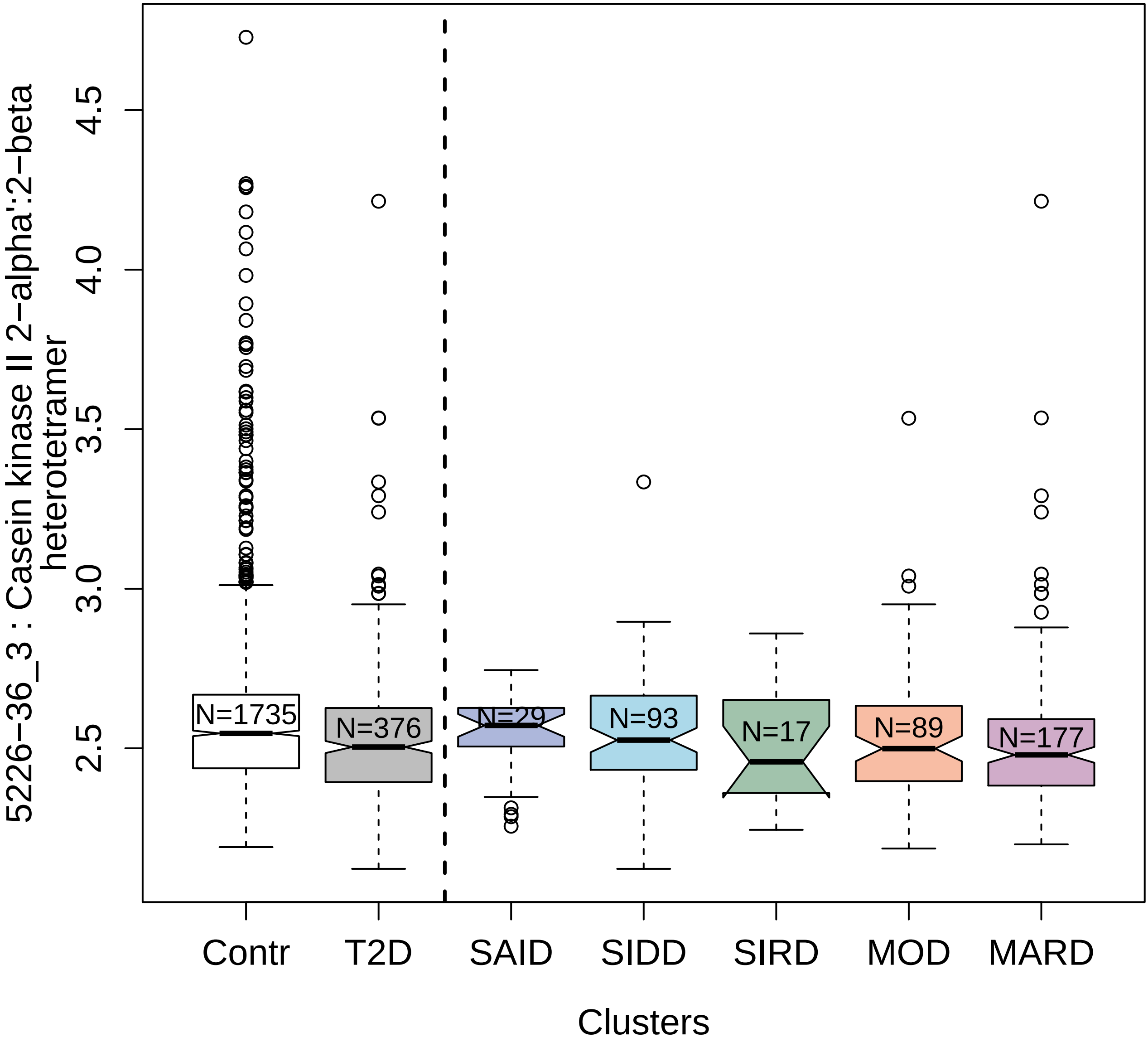

5226−36_3 : Casein kinase II 2−alpha':2−beta heterotetramer
4.5
4.0
3.5
3.0
2.5
N=1735
N=376
N=29
N=93
N=17
N=89
N=177
Contr
T2D
SAID
SIDD
SIRD
MOD
MARD
Clusters

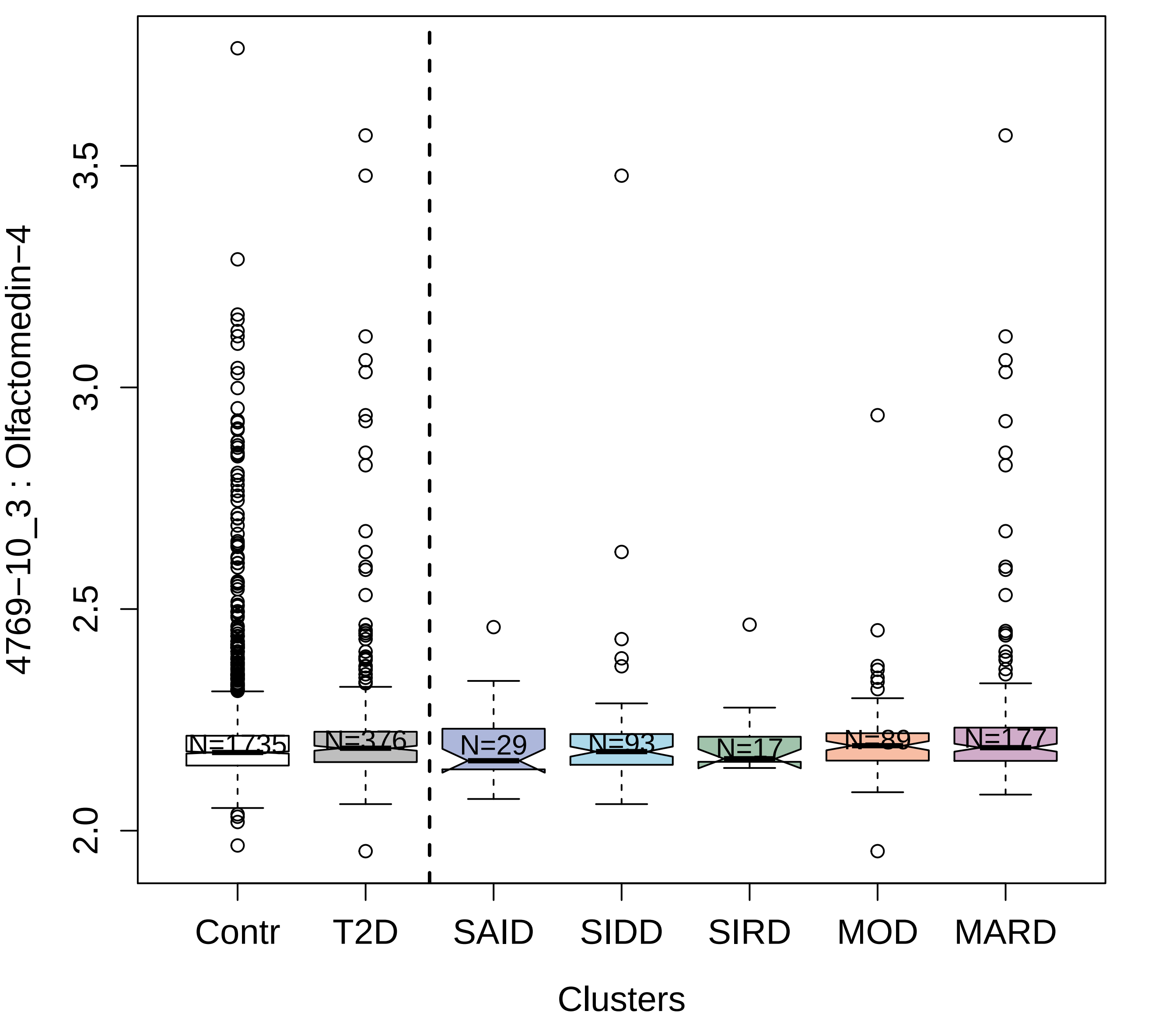

4769−10_3 : Olfactomedin−4
2.0
2.5
3.0
3.5
N=1735
N=376
N=29
N=93
N=17
N=89
N=177
Contr
T2D
SAID
SIDD
SIRD
MOD
MARD
Clusters

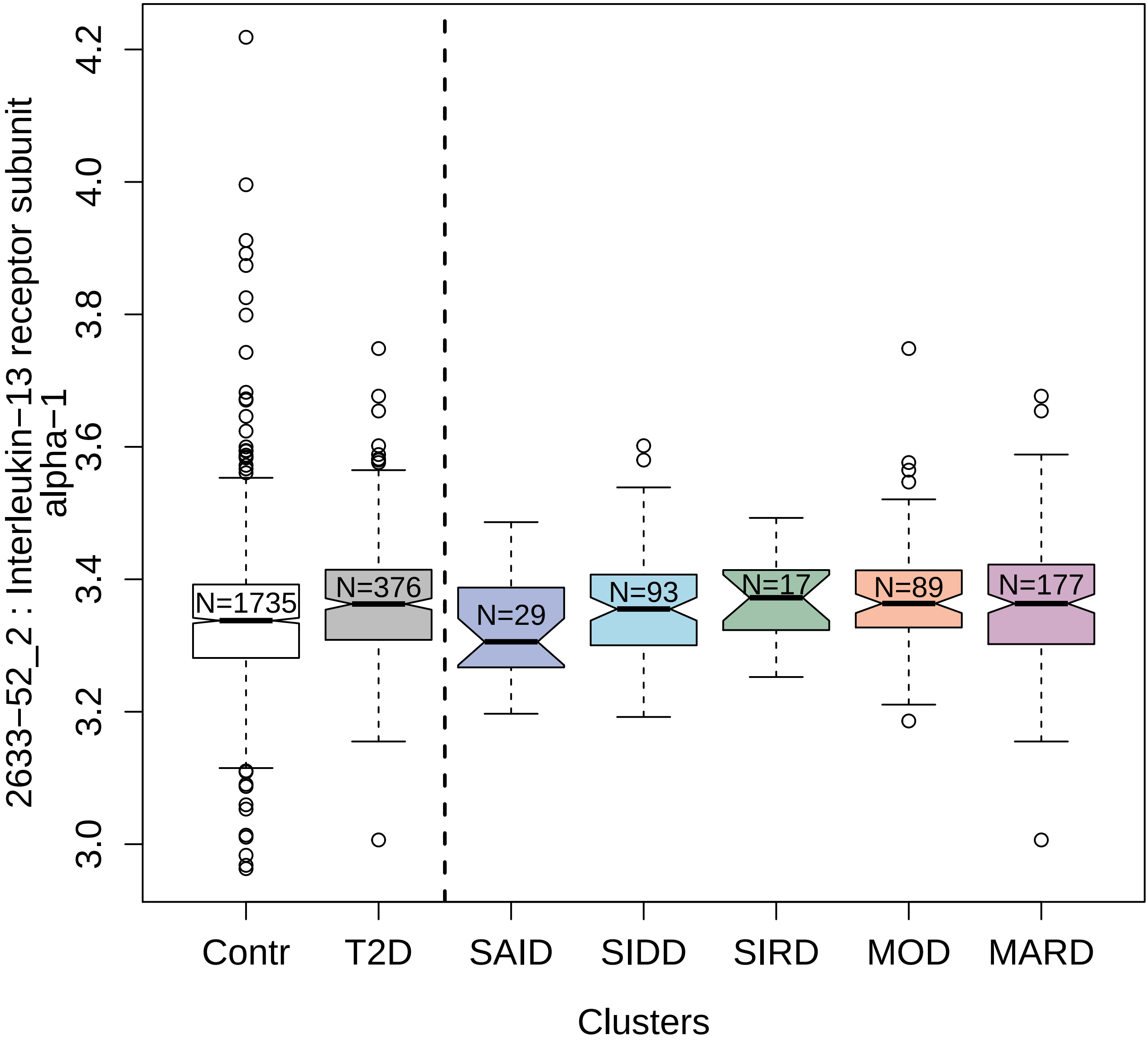

2633−52_2 : Interleukin−13 receptor subunit alpha−1
4.2
4.0
3.8
3.6
3.4
3.2
3.0
N=1735
N=376
N=29
N=93
N=17
N=89
N=177
Contr
T2D
SAID
SIDD
SIRD
MOD
MARD
Clusters

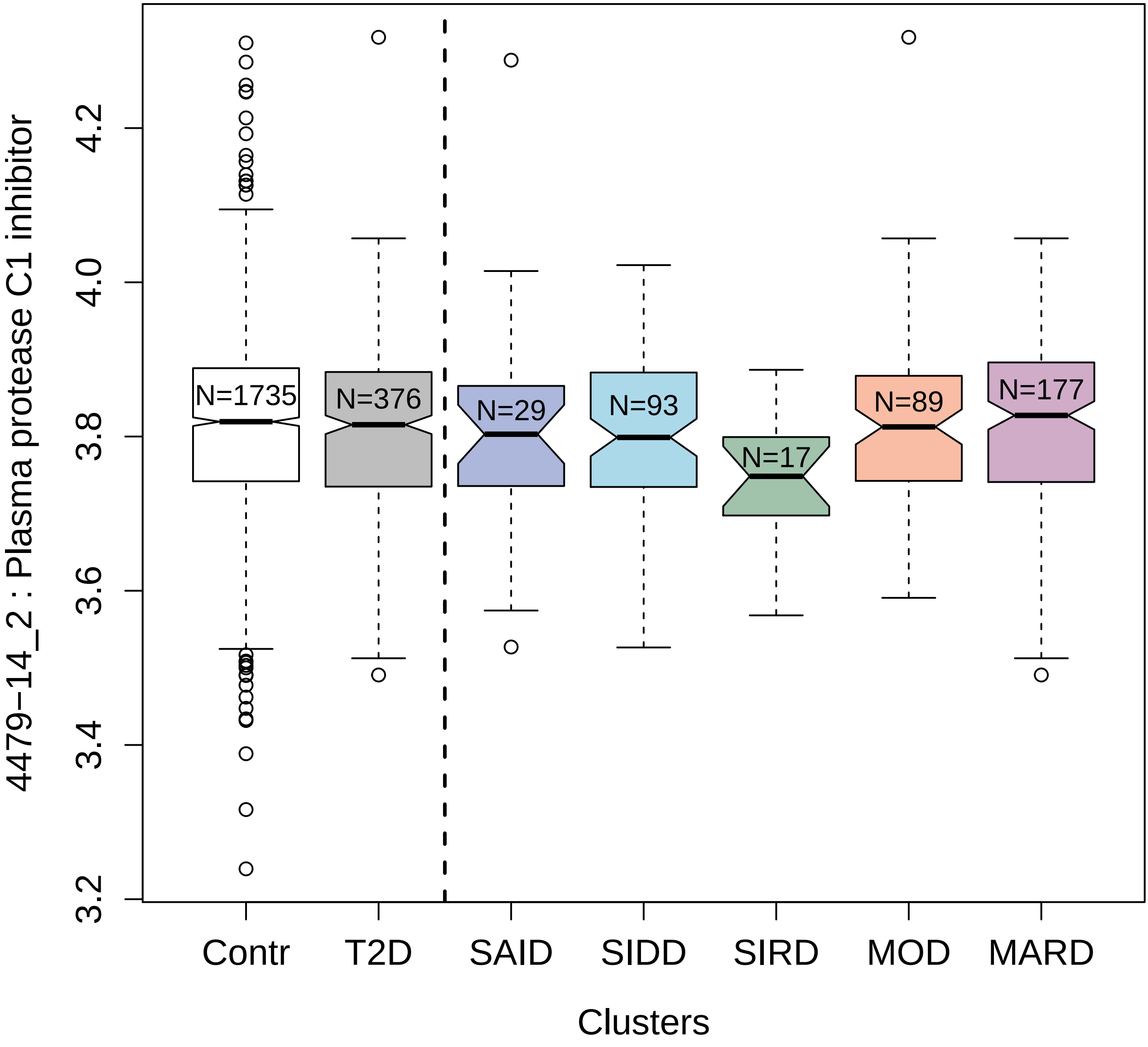
4479−14_2 : Plasma protease C1 inhibitor
3.2
3.4
3.6
3.8
4.0
4.2
N=1735
N=376
N=29
N=93
N=17
N=89
N=177
Contr
T2D
SAID
SIDD
SIRD
MOD
MARD
Clusters

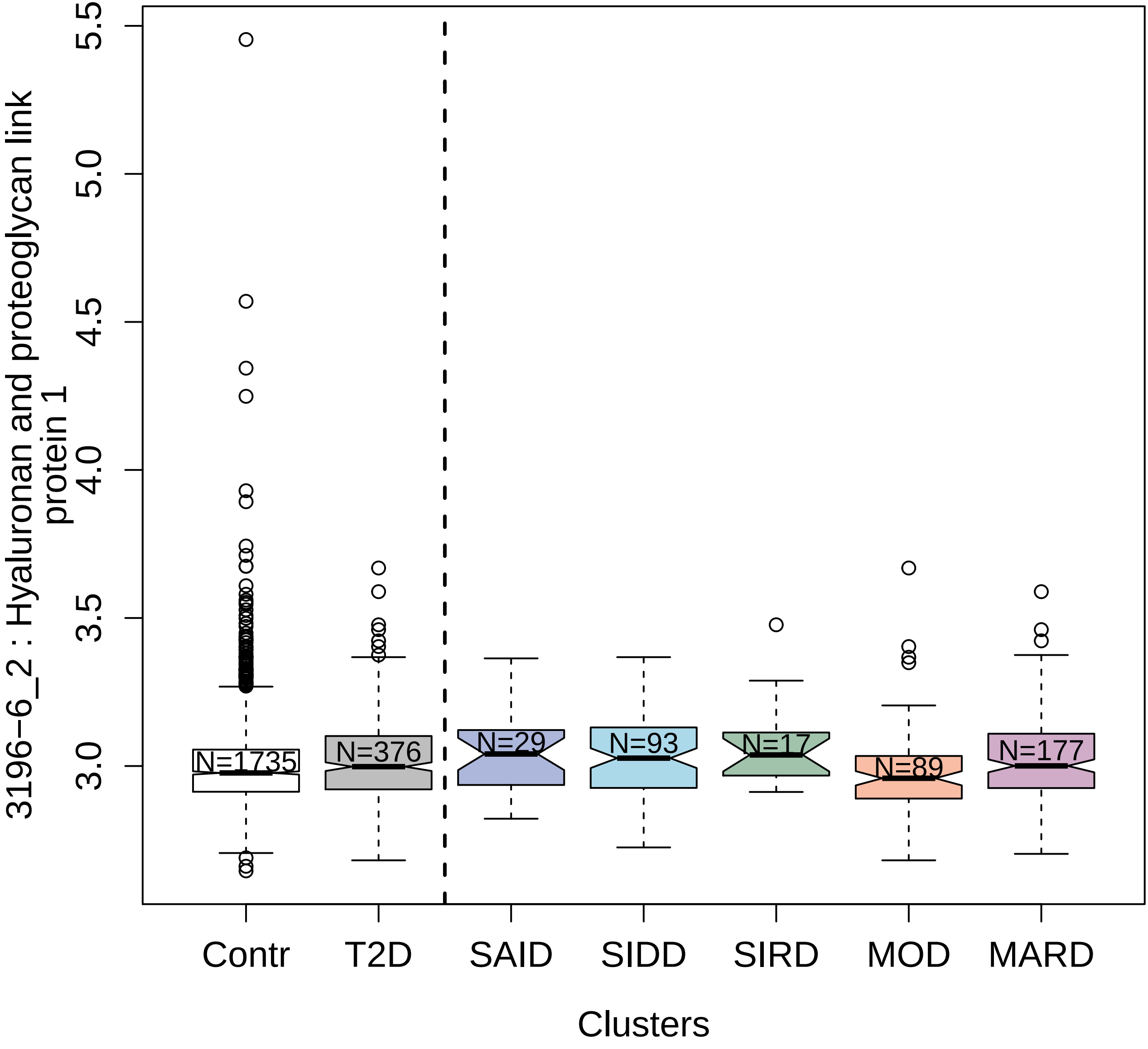

3196−6_2 : Hyaluronan and proteoglycan link protein 1
5.5
5.0
4.5
4.0
3.5
3.0
N=1735
N=376
N=29
N=93
N=17
N=89
N=177
Contr
T2D
SAID
SIDD
SIRD
MOD
MARD
Clusters

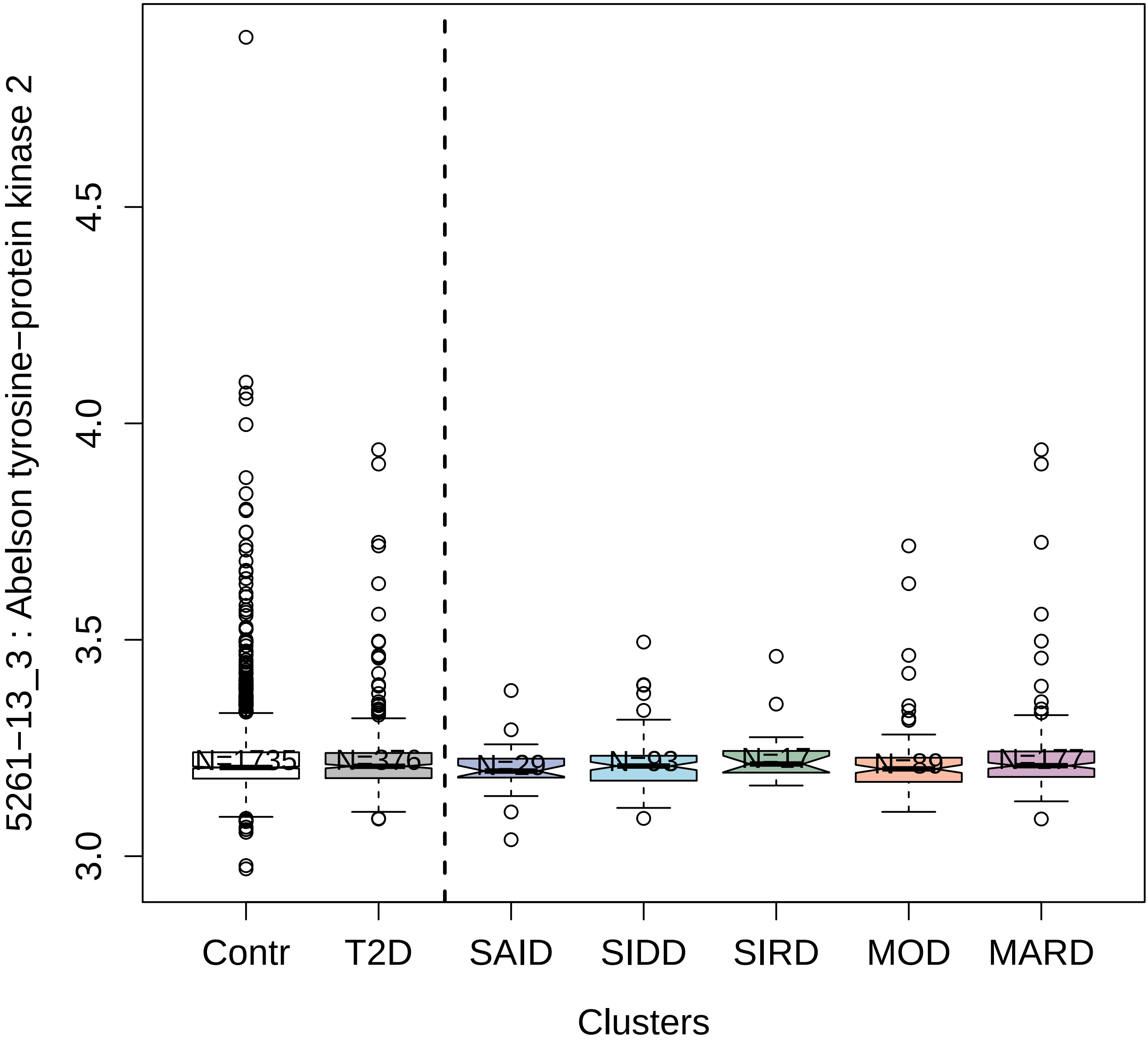
5261−13_3 : Abelson tyrosine−protein kinase 2
3.0
3.5
4.0
4.5
N=1735
N=376
N=29
N=93
N=17
N=89
N=177
Contr
T2D
SAID
SIDD
SIRD
MOD
MARD
Clusters

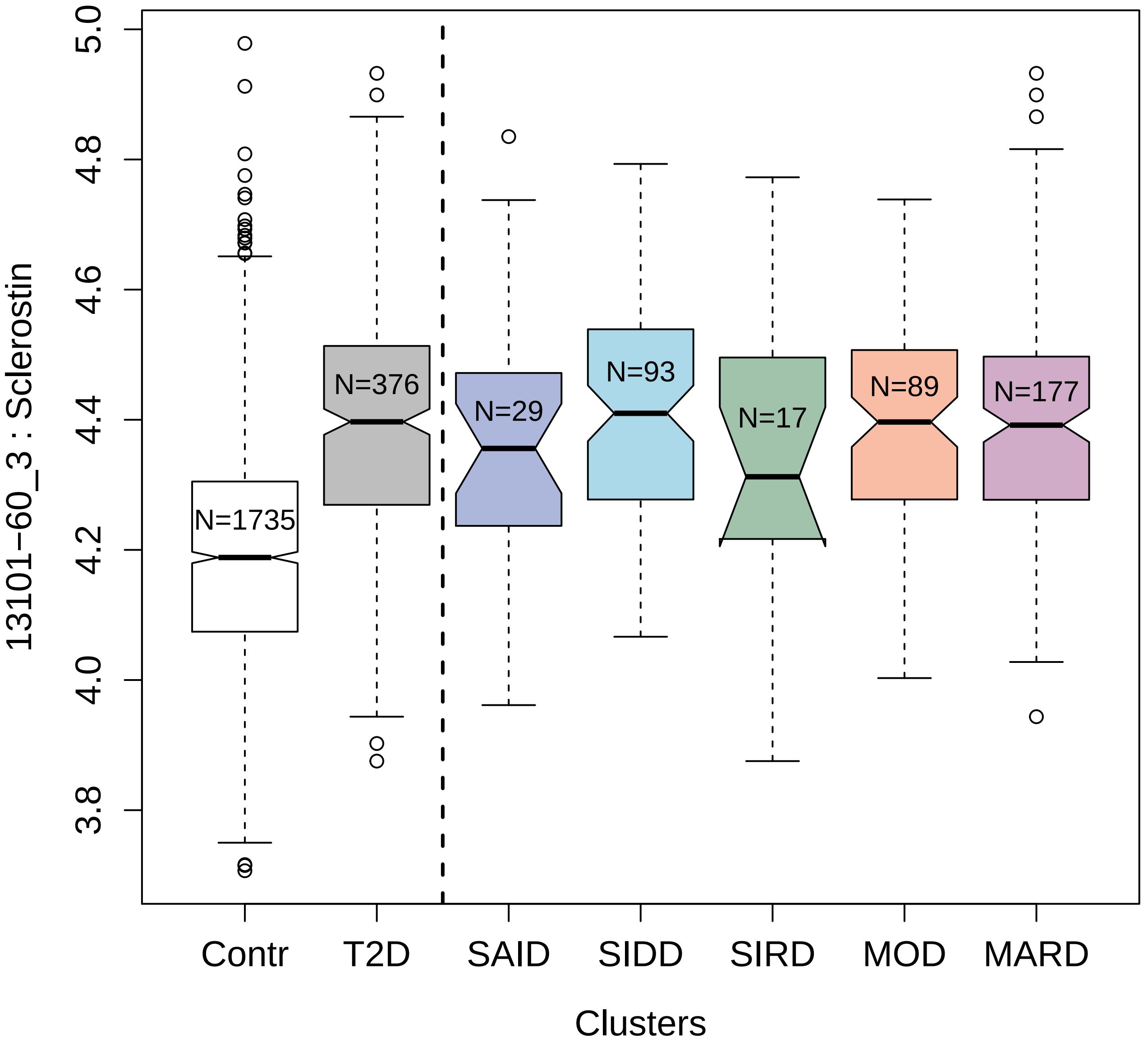

13101−60_3 : Sclerostin
5.0
4.8
4.6
4.4
4.2
4.0
3.8
N=1735
N=376
N=29
N=93
N=17
N=89
N=177
Contr
T2D
SAID
SIDD
SIRD
MOD
MARD
Clusters

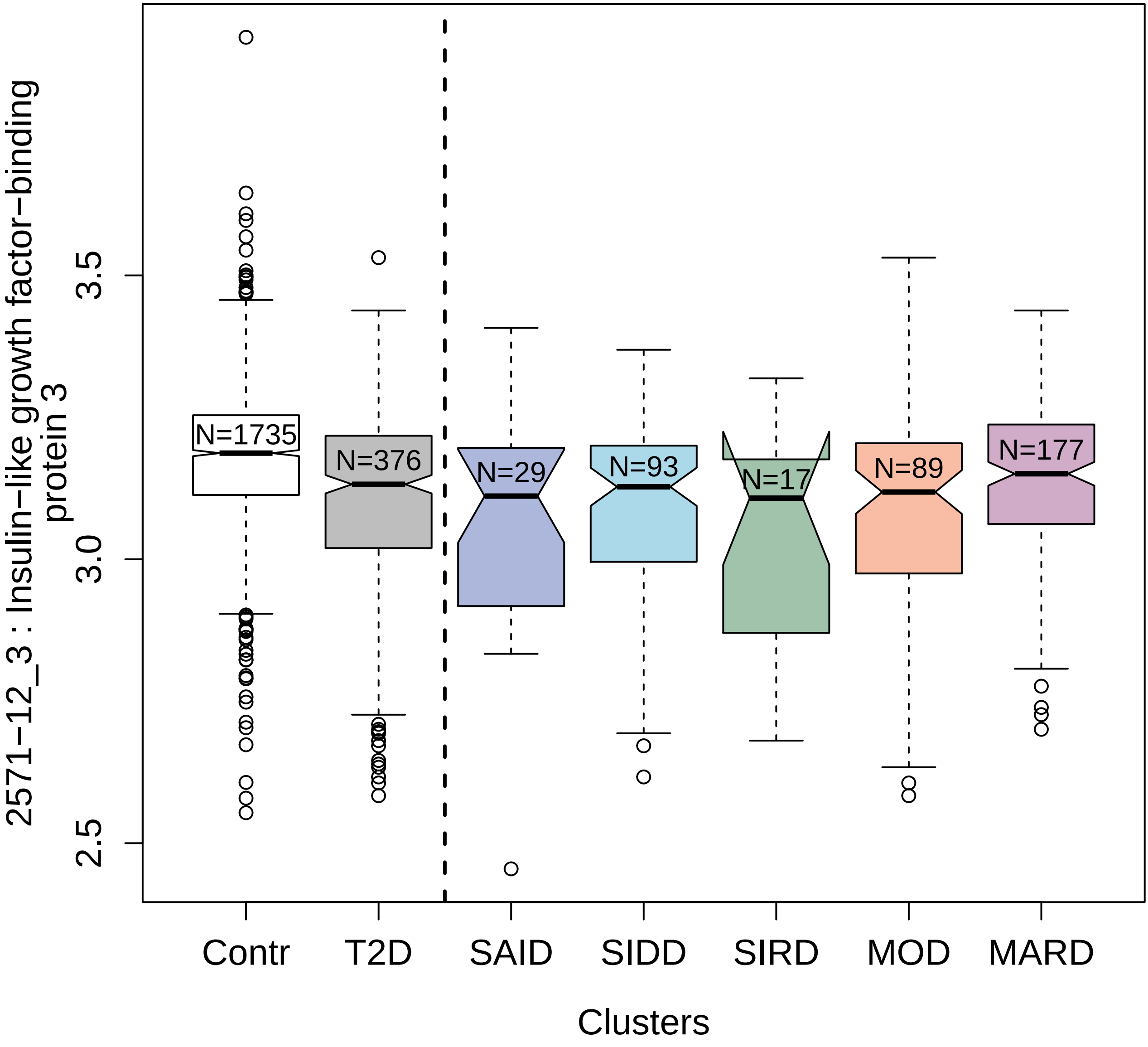
2571−12_3 : Insulin−like growth factor−binding protein 3
2.5
3.0
3.5
N=1735
N=376
N=29
N=93
N=17
N=89
N=177
Contr
T2D
SAID
SIDD
SIRD
MOD
MARD
Clusters

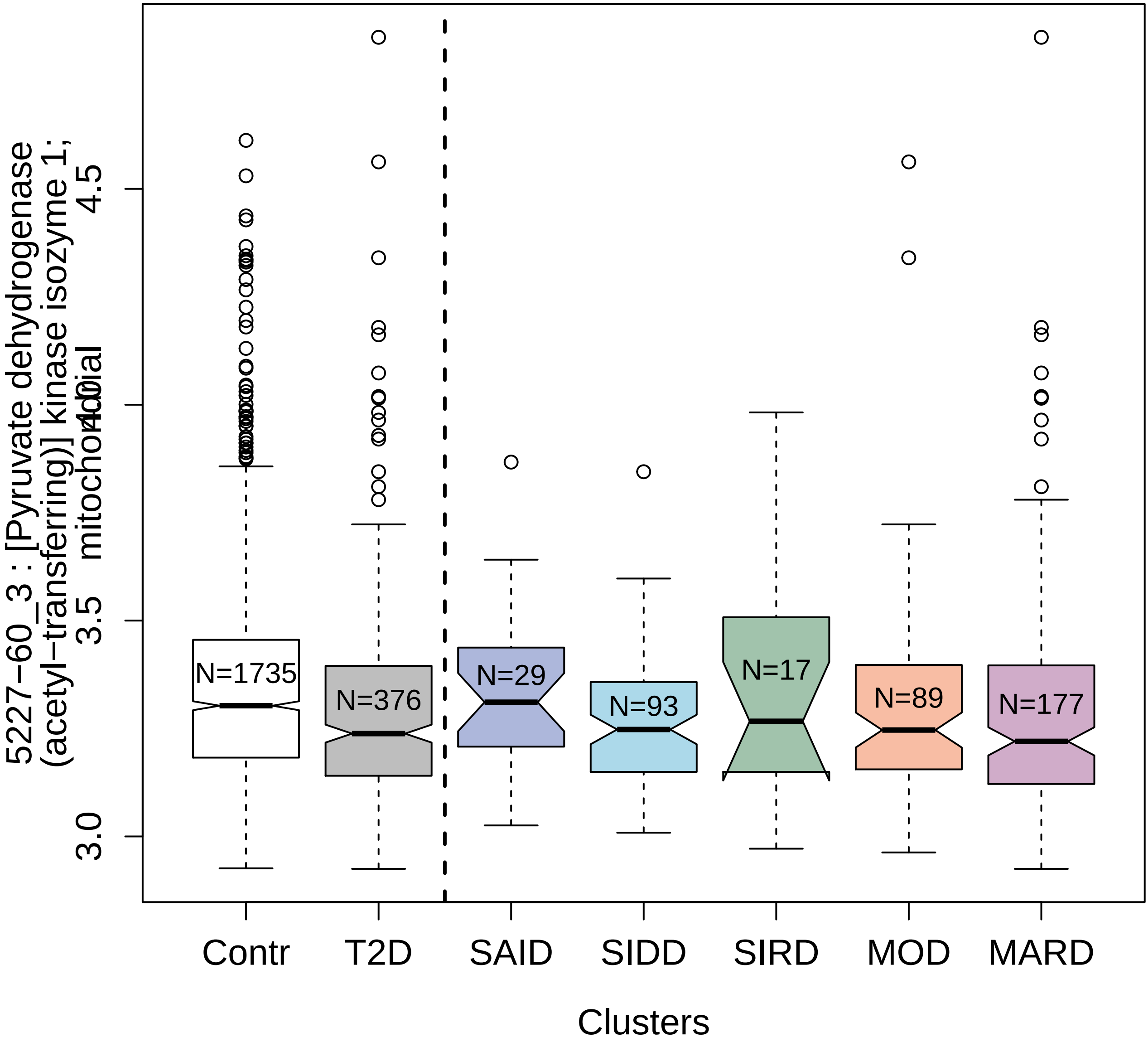

5227−60_3 : [Pyruvate dehydrogenase (acetyl−transferring)] kinase isozyme 1, mitochondrial
3.0
3.5
4.0
4.5
N=1735
N=376
N=29
N=93
N=17
N=89
N=177
Contr
T2D
SAID
SIDD
SIRD
MOD
MARD
Clusters

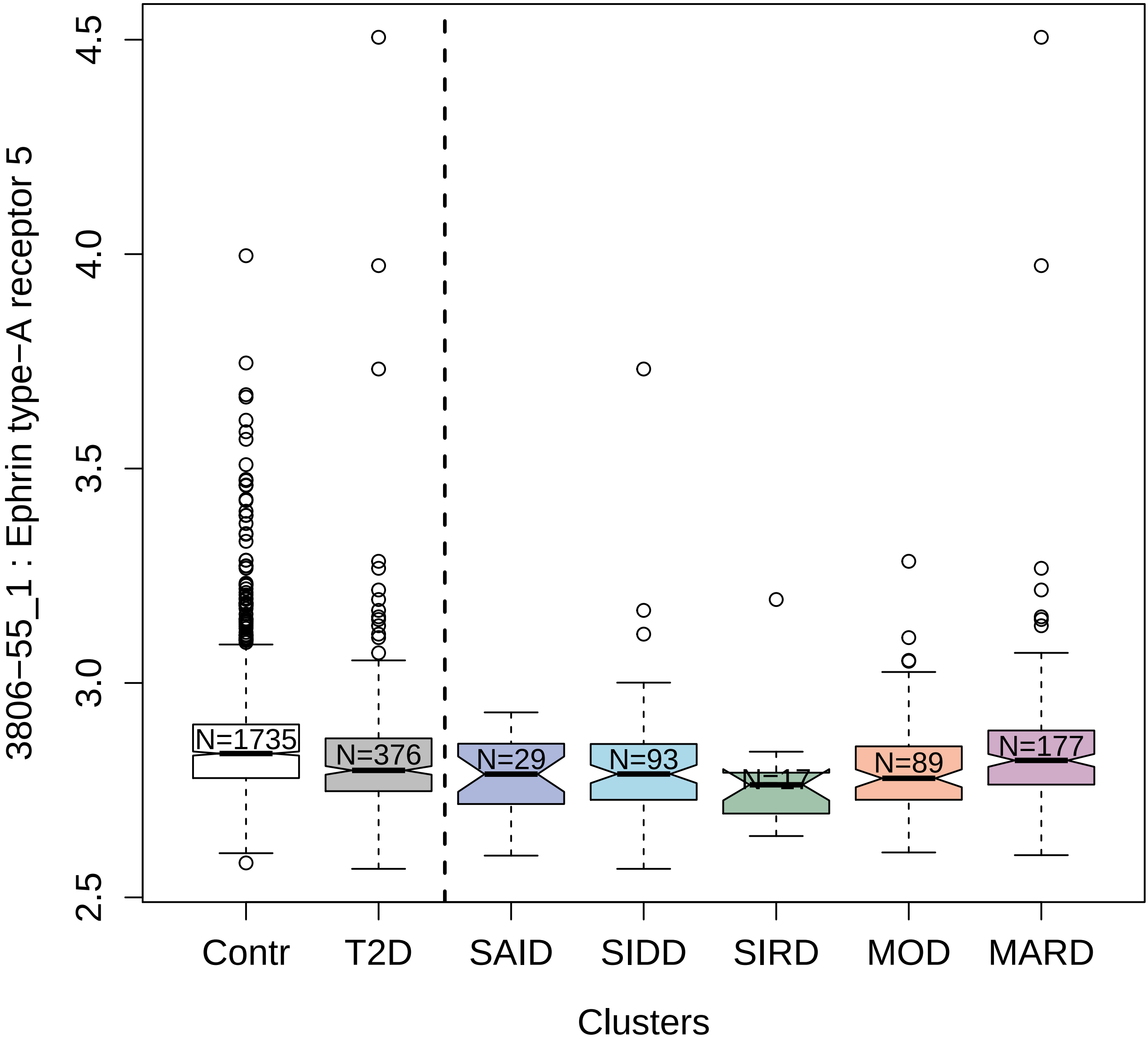
3806−55_1 : Ephrin type−A receptor 5
2.5
3.0
3.5
4.0
4.5
N=1735
N=376
N=29
N=93
N=17
N=89
N=177
Contr
T2D
SAID
SIDD
SIRD
MOD
MARD
Clusters

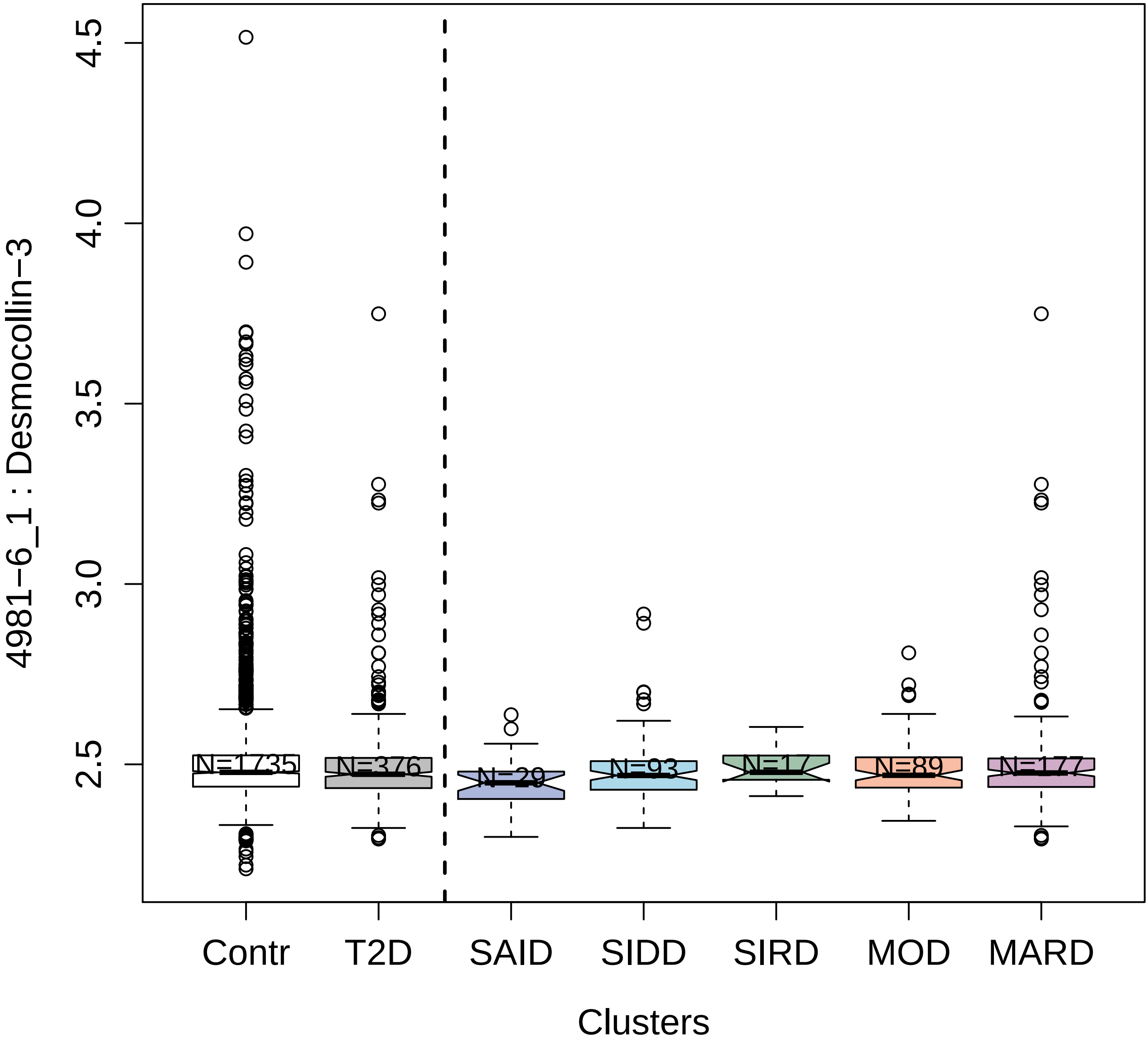

4981−6_1 : Desmocollin−3
4.5
4.0
3.5
3.0
2.5
N=1735
N=376
N=29
N=93
N=17
N=89
N=177
Contr
T2D
SAID
SIDD
SIRD
MOD
MARD
Clusters

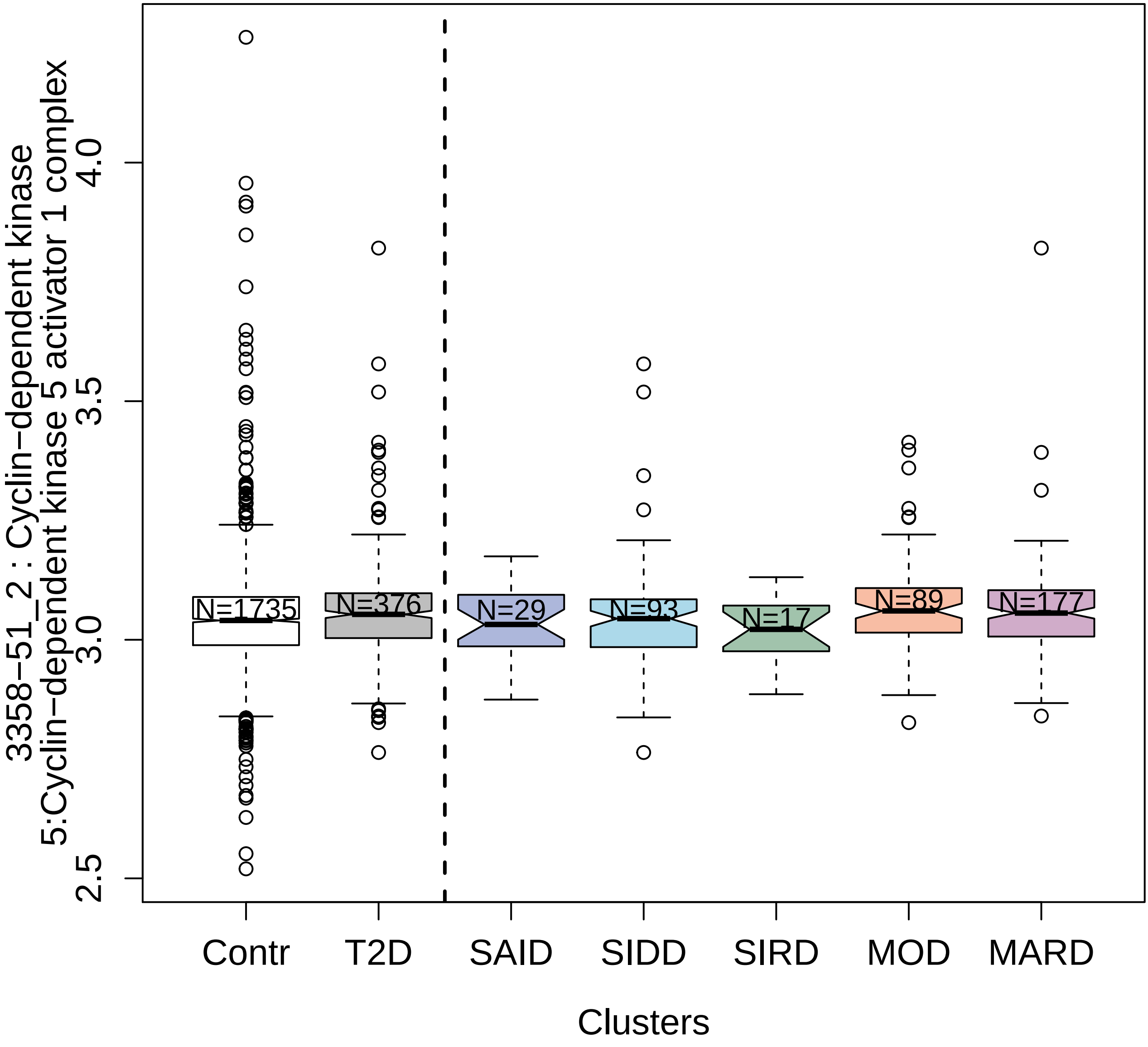

3358−51_2 : Cyclin−dependent kinase
5:Cyclin−dependent kinase 5 activator 1 complex
2.5
3.0
3.5
4.0
N=1735
N=376
N=29
N=93
N=17
N=89
N=177
Contr
T2D
SAID
SIDD
SIRD
MOD
MARD
Clusters

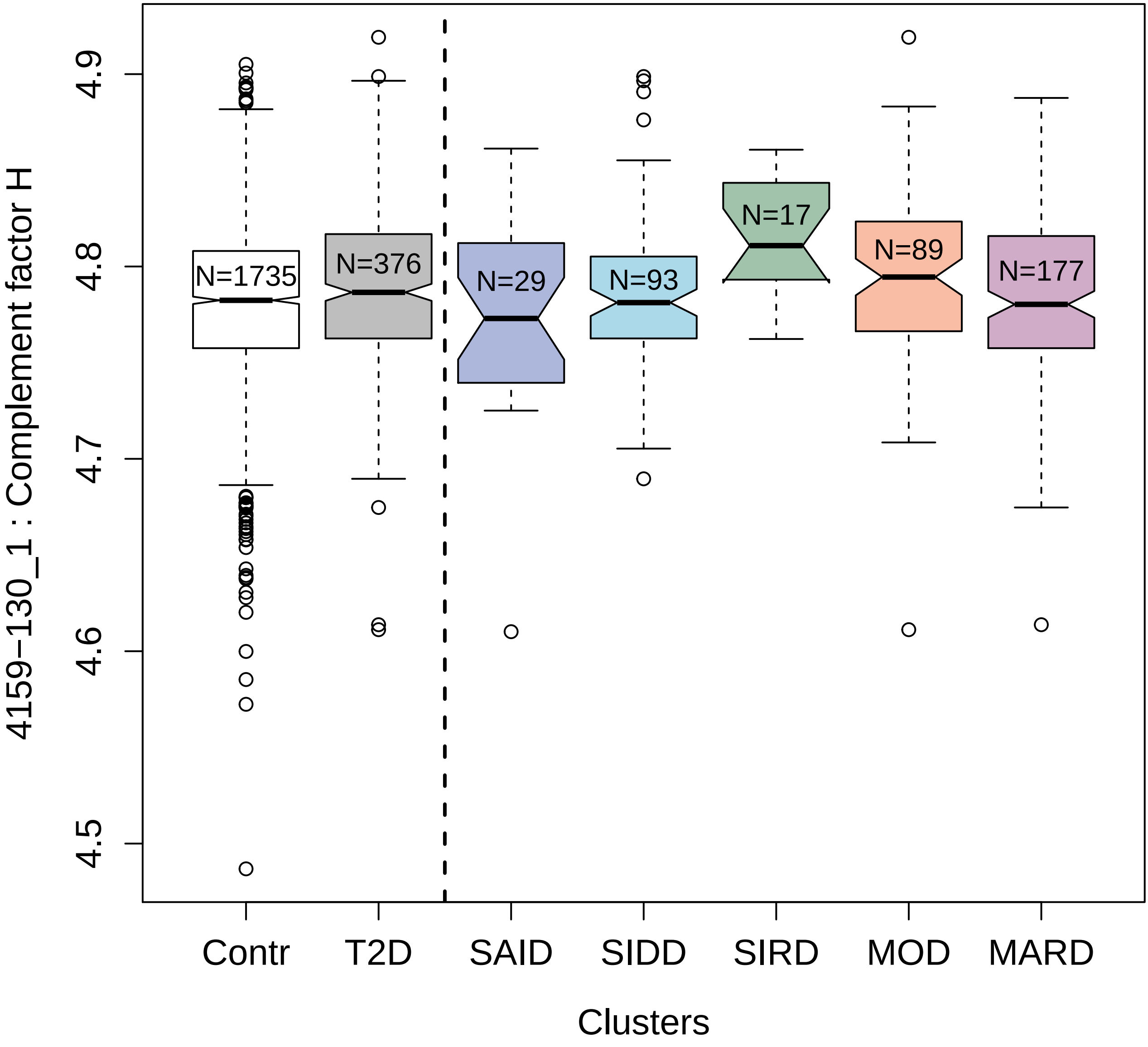

4159−130_1 : Complement factor H
4.9
4.8
4.7
4.6
4.5
N=1735
N=376
N=29
N=93
N=17
N=89
N=177
Contr
T2D
SAID
SIDD
SIRD
MOD
MARD
Clusters

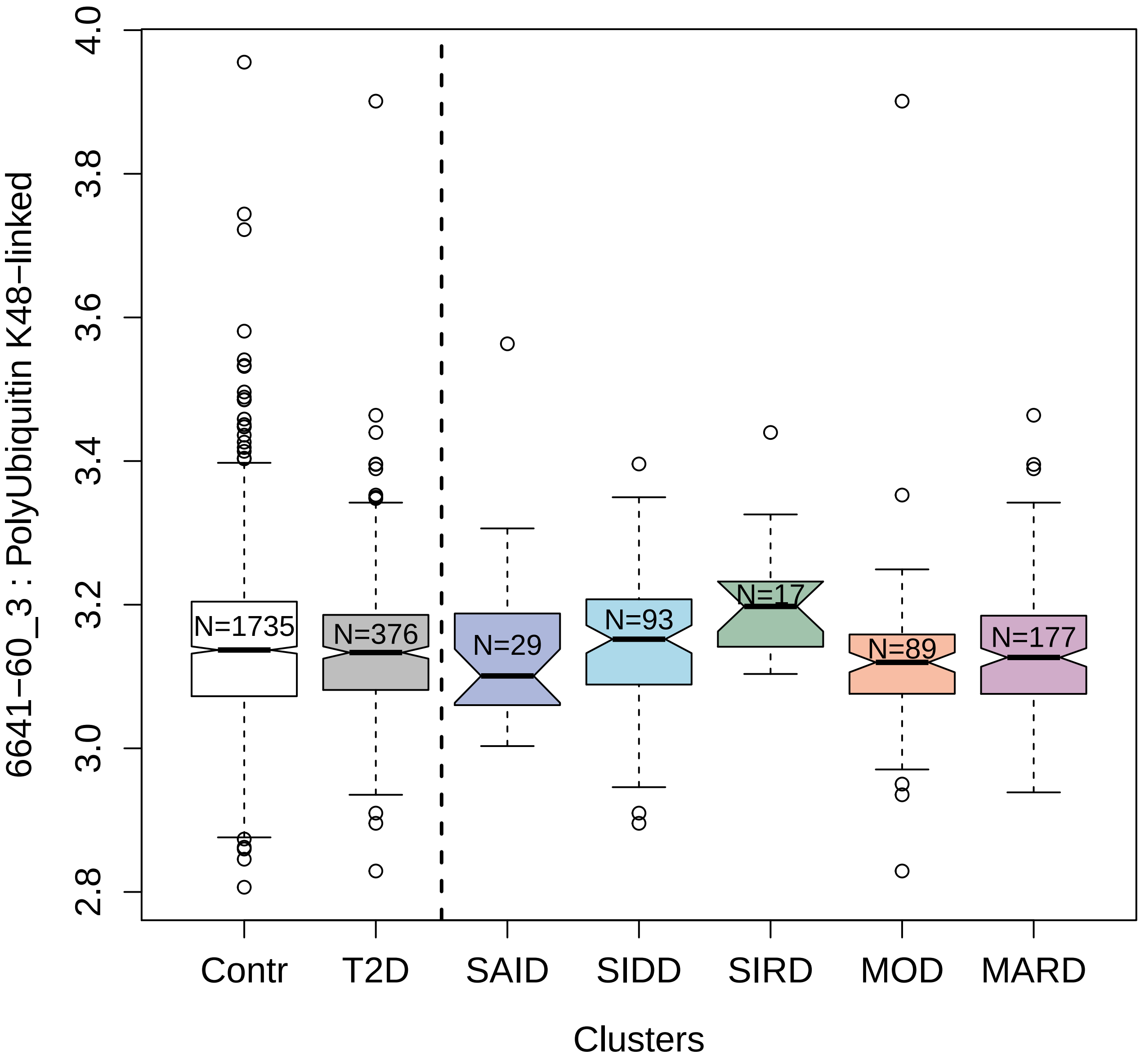
6641−60_3 : PolyUbiquitin K48−linked
4.0
3.8
3.6
3.4
3.2
3.0
2.8
N=1735
N=376
N=29
N=93
N=17
N=89
N=177
Contr
T2D
SAID
SIDD
SIRD
MOD
MARD
Clusters

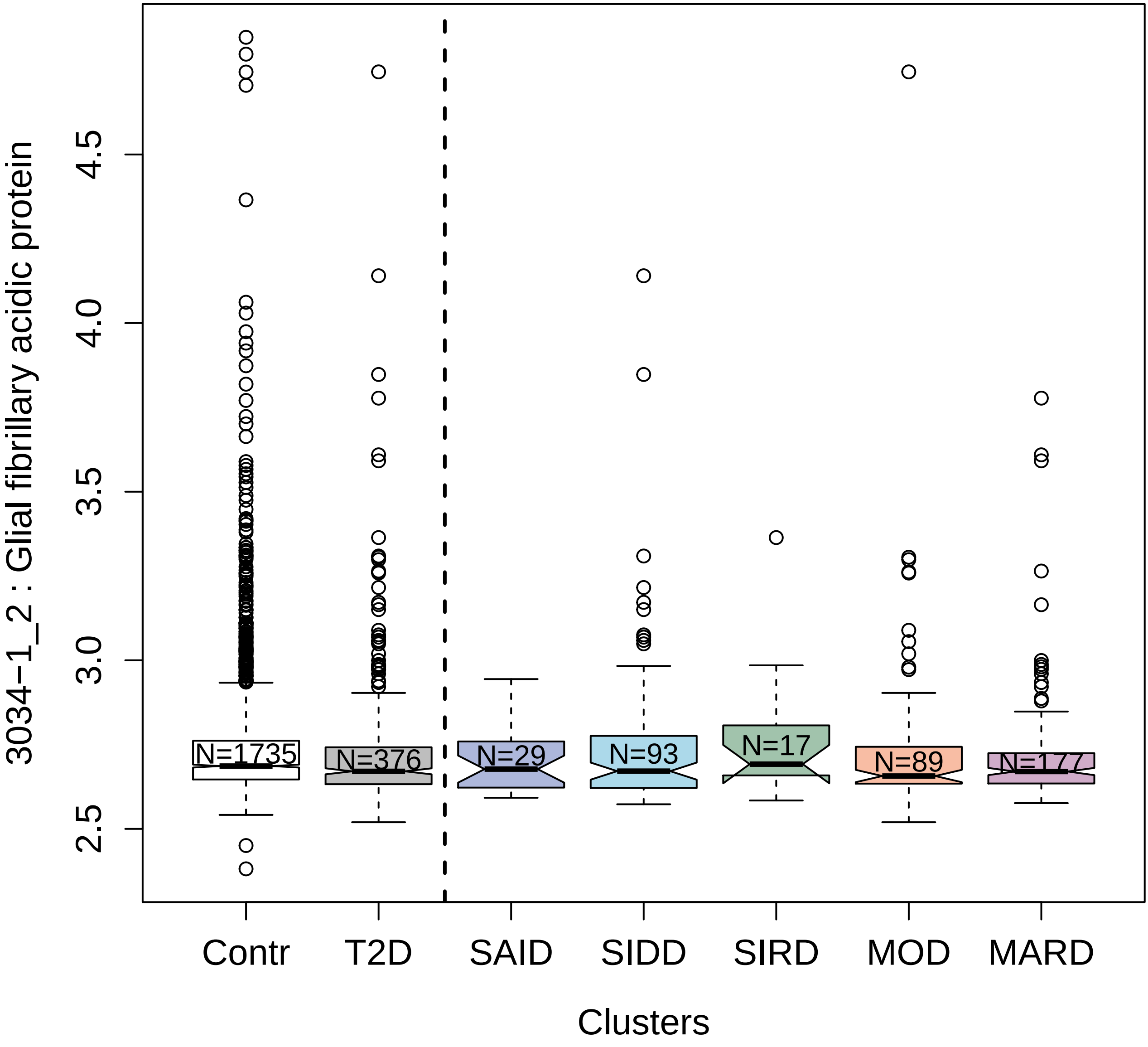
3034−1_2 : Glial fibrillary acidic protein
2.5
3.0
3.5
4.0
4.5
N=1735
N=376
N=29
N=93
N=17
N=89
N=177
Contr
T2D
SAID
SIDD
SIRD
MOD
MARD
Clusters

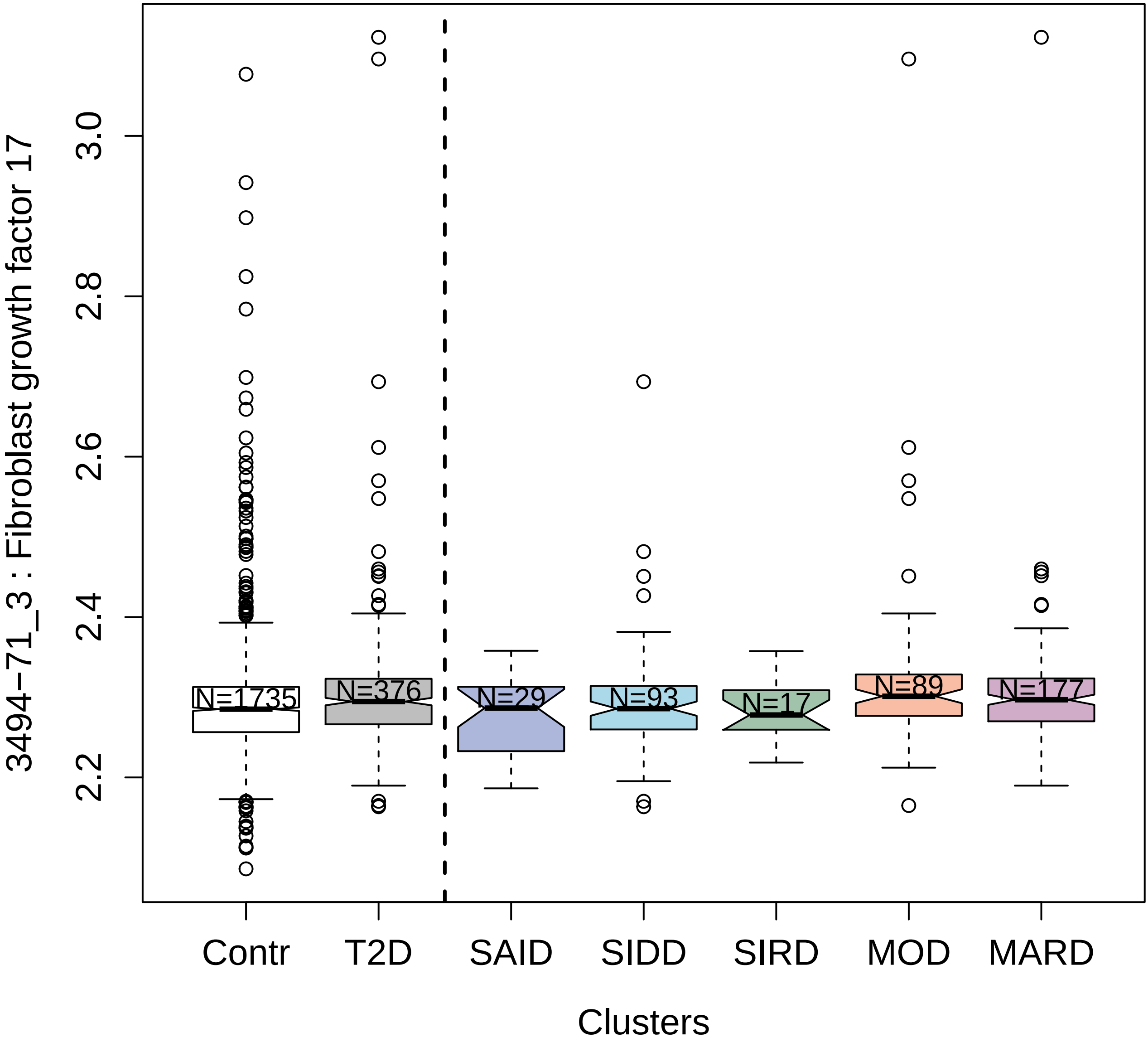
3494−71_3 : Fibroblast growth factor 17
2.2
2.4
2.6
2.8
3.0
N=1735
N=376
N=29
N=93
N=17
N=89
N=177
Contr
T2D
SAID
SIDD
SIRD
MOD
MARD
Clusters

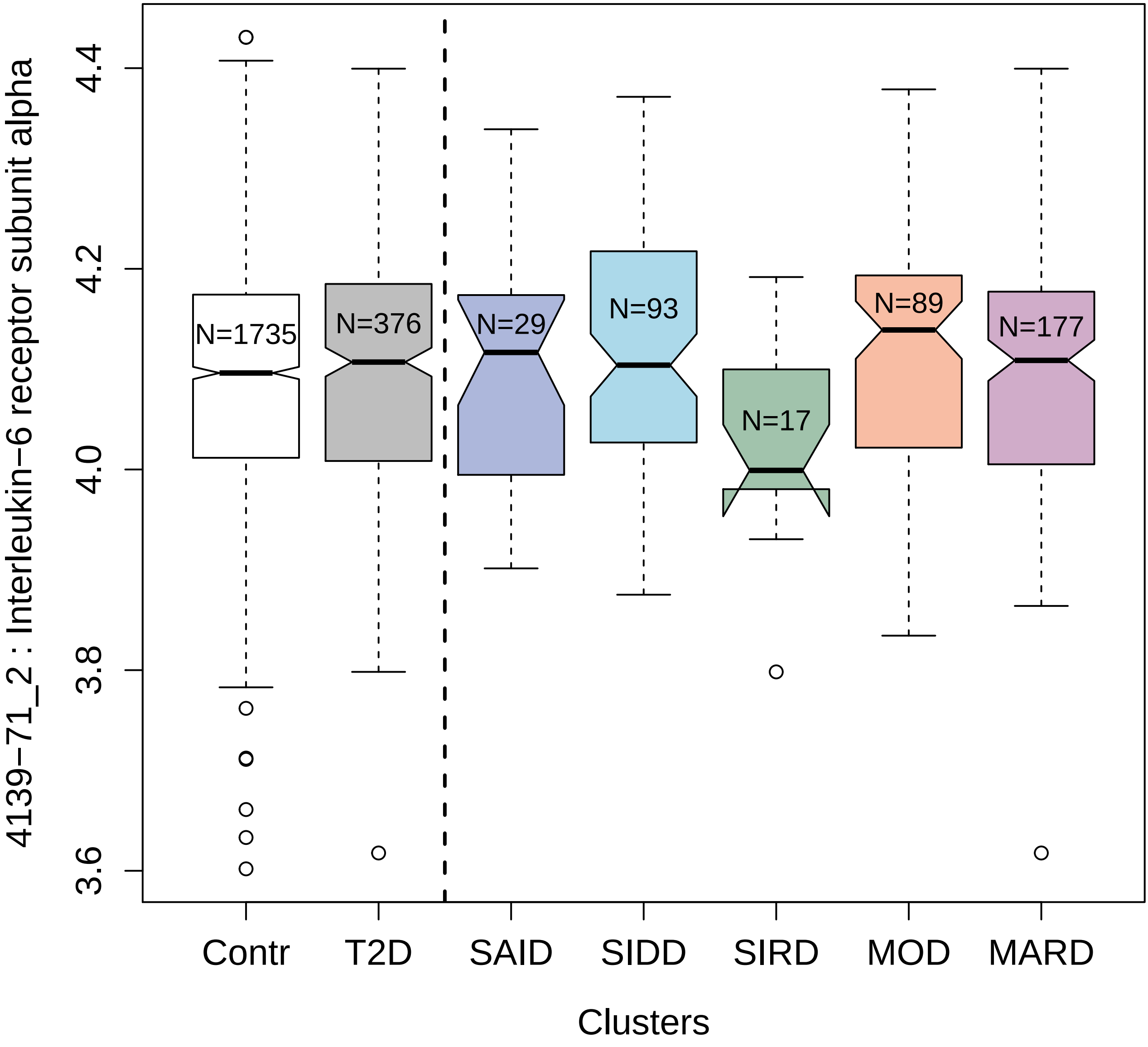

4139−71_2 : Interleukin−6 receptor subunit alpha
3.6
3.8
4.0
4.2
4.4
N=1735
N=376
N=29
N=93
N=17
N=89
N=177
Contr
T2D
SAID
SIDD
SIRD
MOD
MARD
Clusters

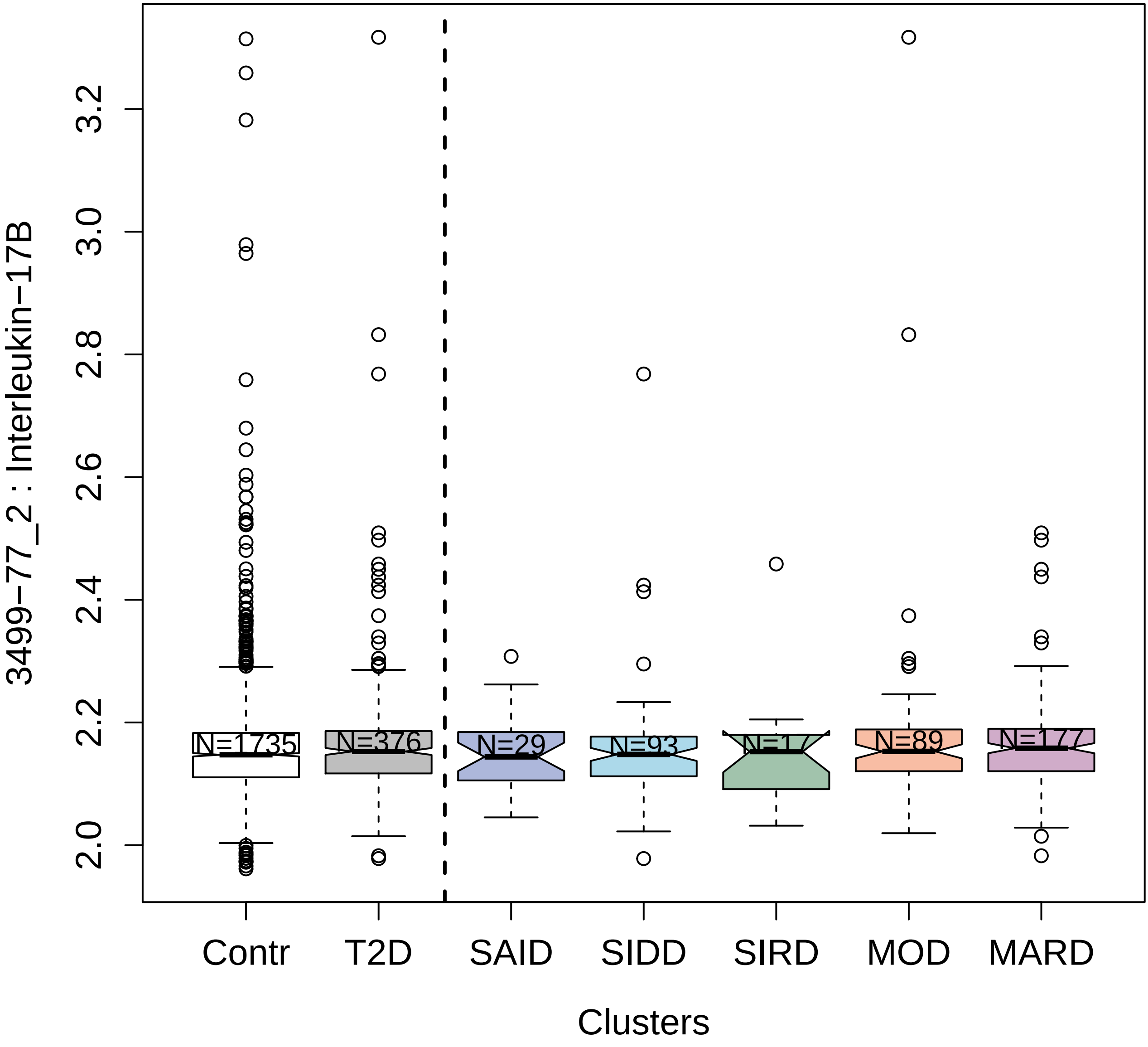
3499−77_2 : Interleukin−17B
3.2
3.0
2.8
2.6
2.4
2.2
2.0
N=1735
N=376
N=29
N=93
N=17
N=89
N=177
Contr
T2D
SAID
SIDD
SIRD
MOD
MARD
Clusters

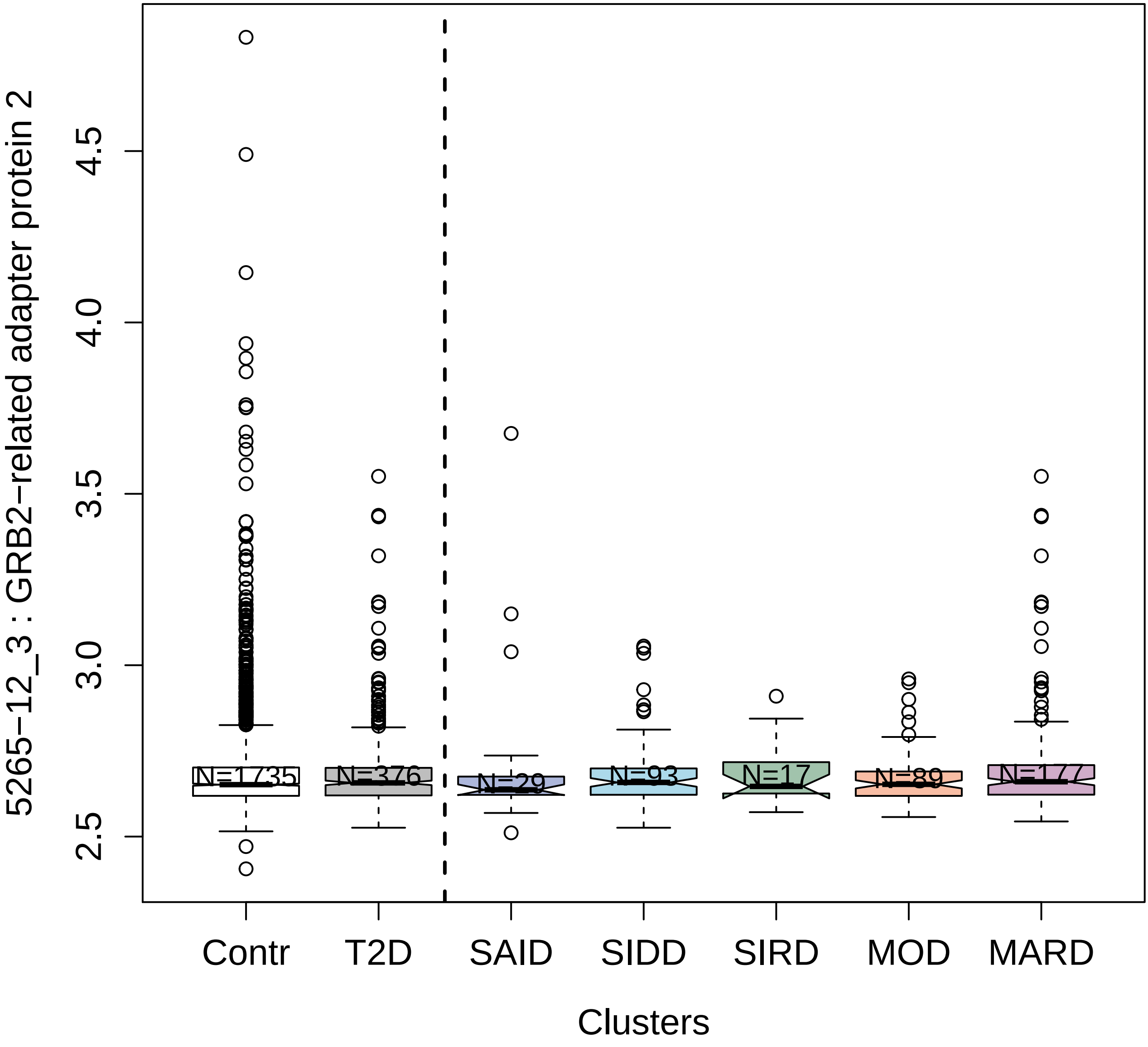
5265−12_3 : GRB2−related adapter protein 2
2.5
3.0
3.5
4.0
4.5
N=1735
N=376
N=29
N=93
N=17
N=89
N=177
Contr
T2D
SAID
SIDD
SIRD
MOD
MARD
Clusters

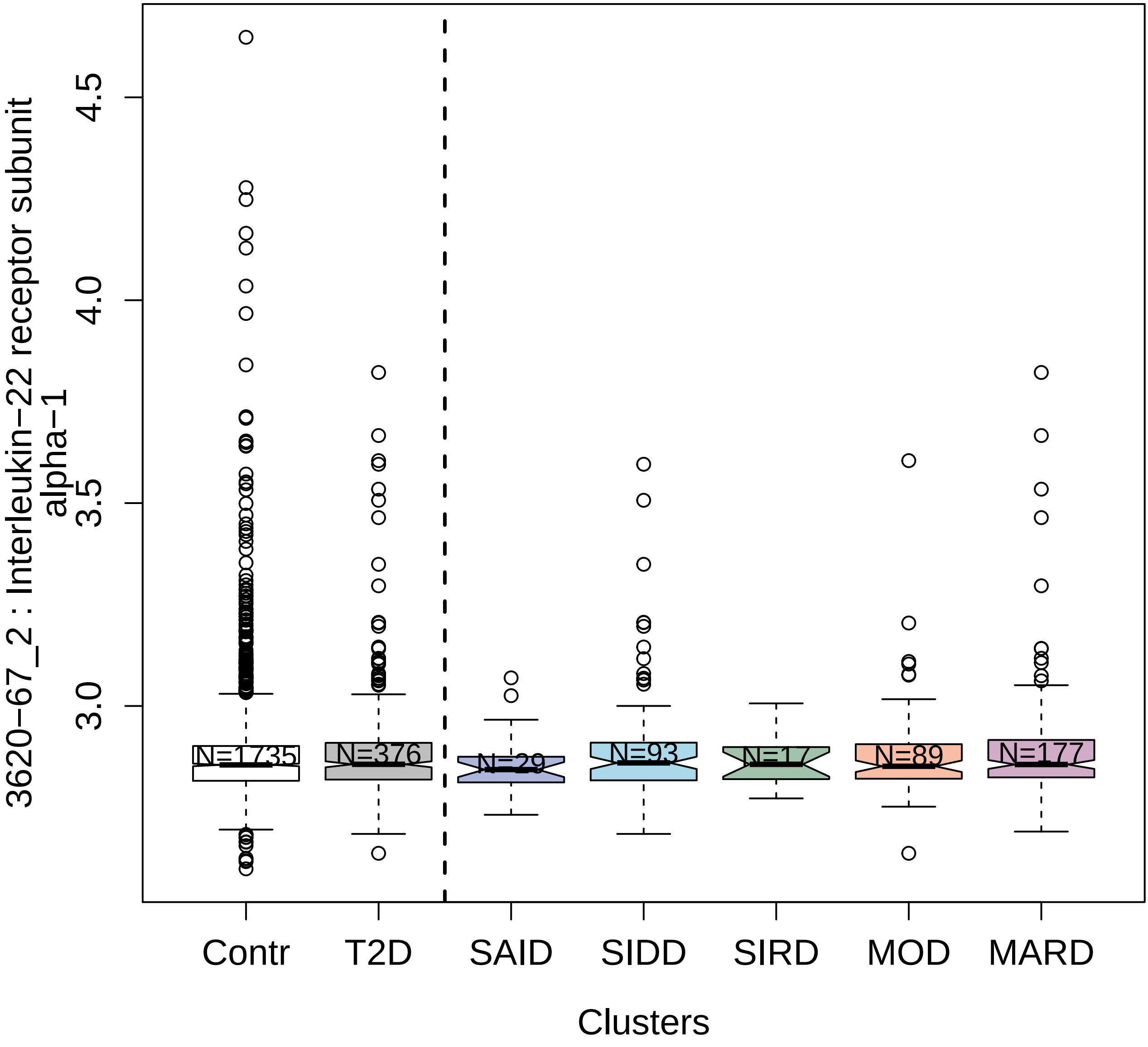

3620−67_2 : Interleukin−22 receptor subunit alpha−1
4.5
4.0
3.5
3.0
N=1735
N=376
N=29
N=93
N=17
N=89
N=177
Contr
T2D
SAID
SIDD
SIRD
MOD
MARD
Clusters

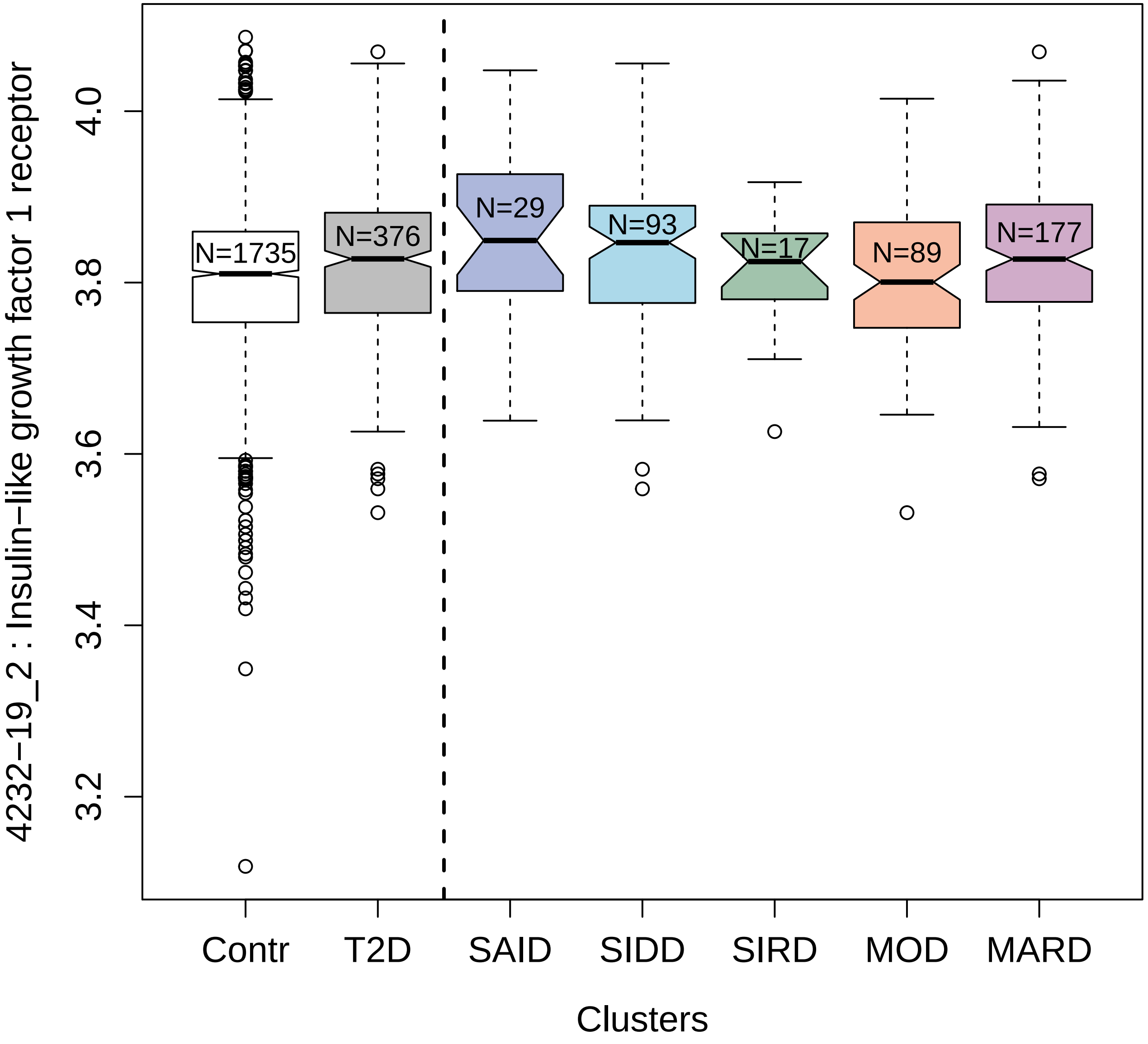
4232−19_2 : Insulin−like growth factor 1 receptor
3.2
3.4
3.6
3.8
4.0
N=1735
N=376
N=29
N=93
N=17
N=89
N=177
Contr
T2D
SAID
SIDD
SIRD
MOD
MARD
Clusters

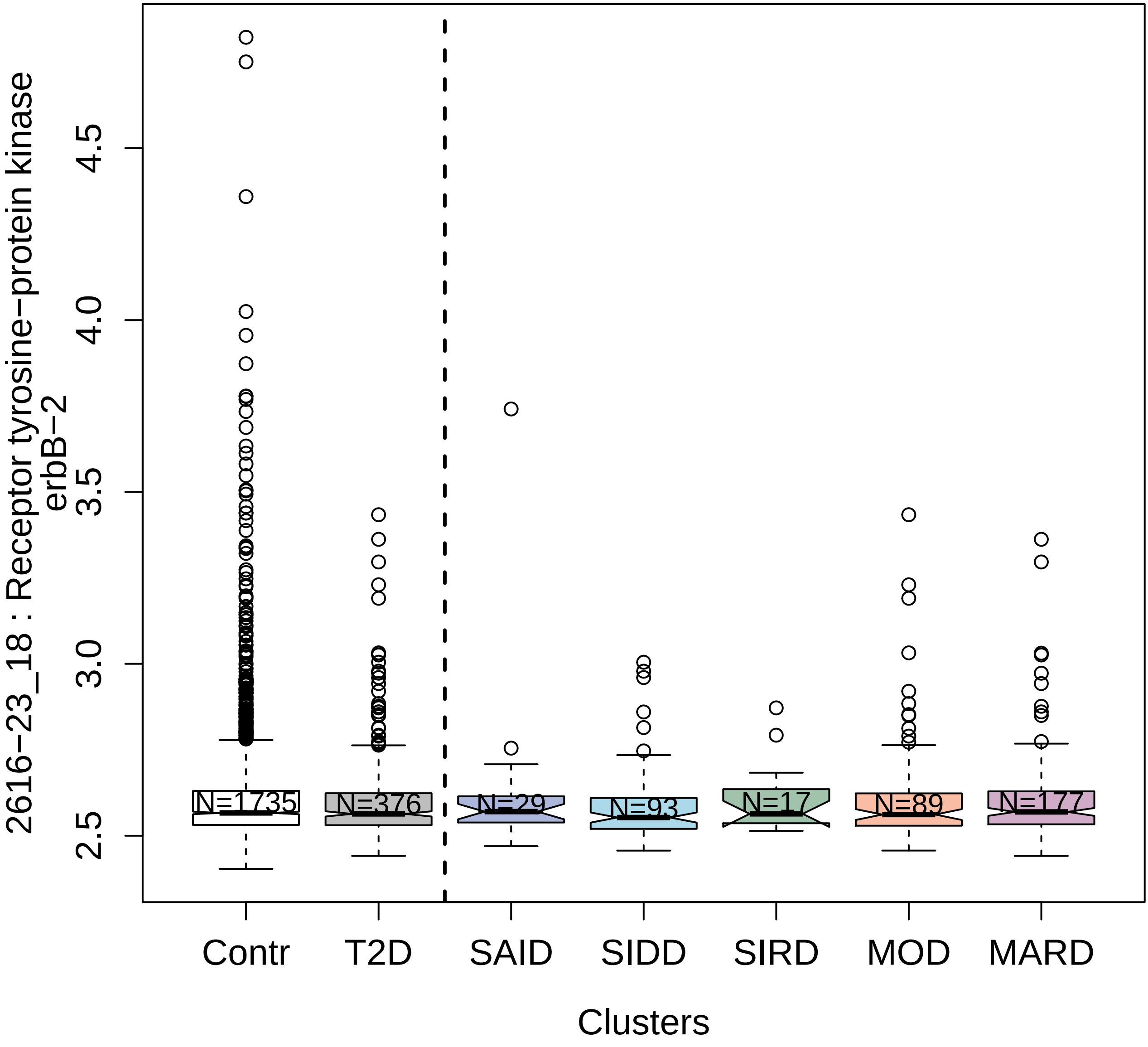

2616−23_18 : Receptor tyrosine−protein kinase
erbB−2
4.5
4.0
3.5
3.0
2.5
N=1735
N=376
N=29
N=93
N=17
N=89
N=177
Contr
T2D
SAID
SIDD
SIRD
MOD
MARD
Clusters

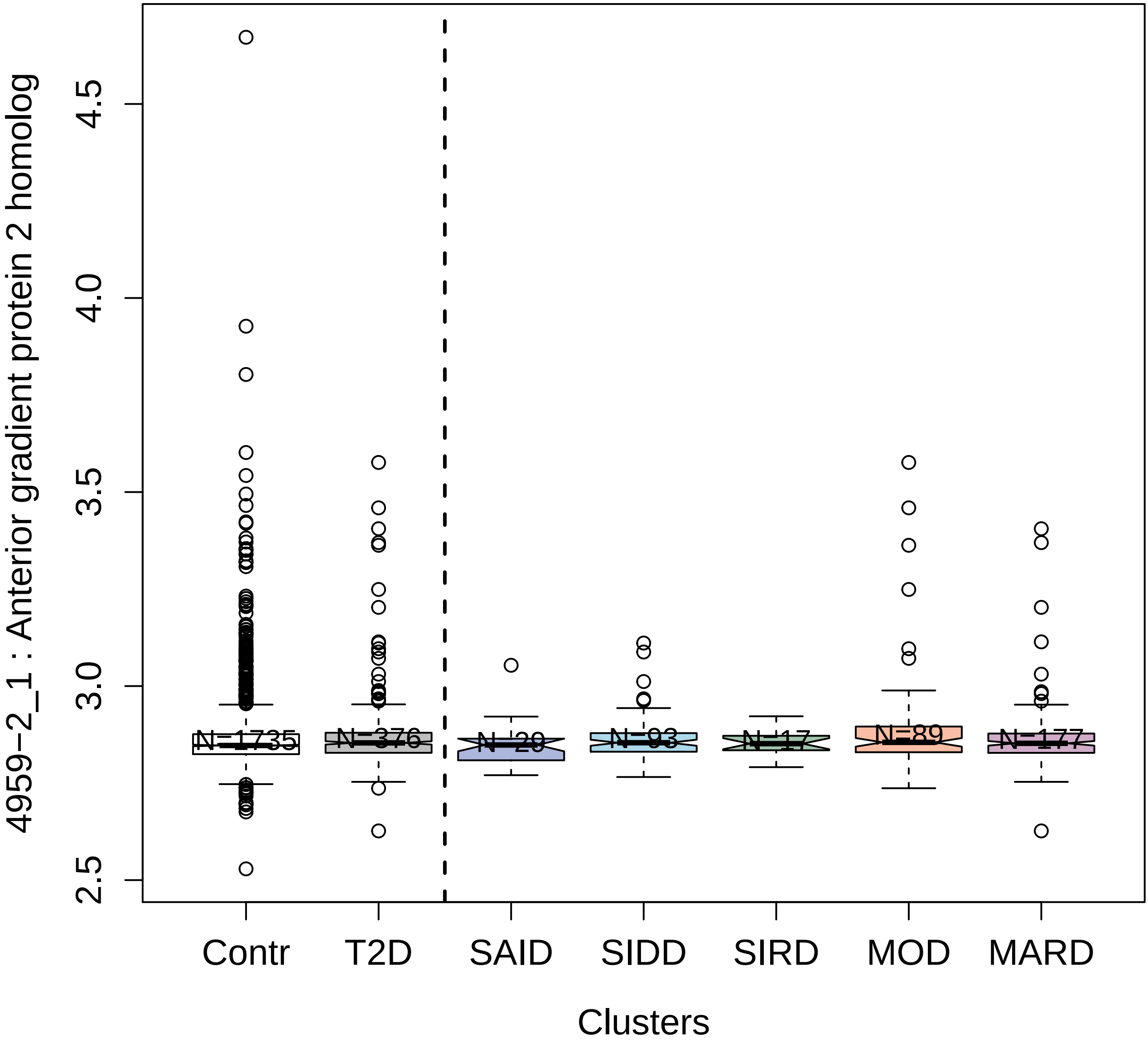

4959−2_1 : Anterior gradient protein 2 homolog
2.5
3.0
3.5
4.0
4.5
N=1735
N=376
N=29
N=93
N=17
N=89
N=177
Contr
T2D
SAID
SIDD
SIRD
MOD
MARD
Clusters

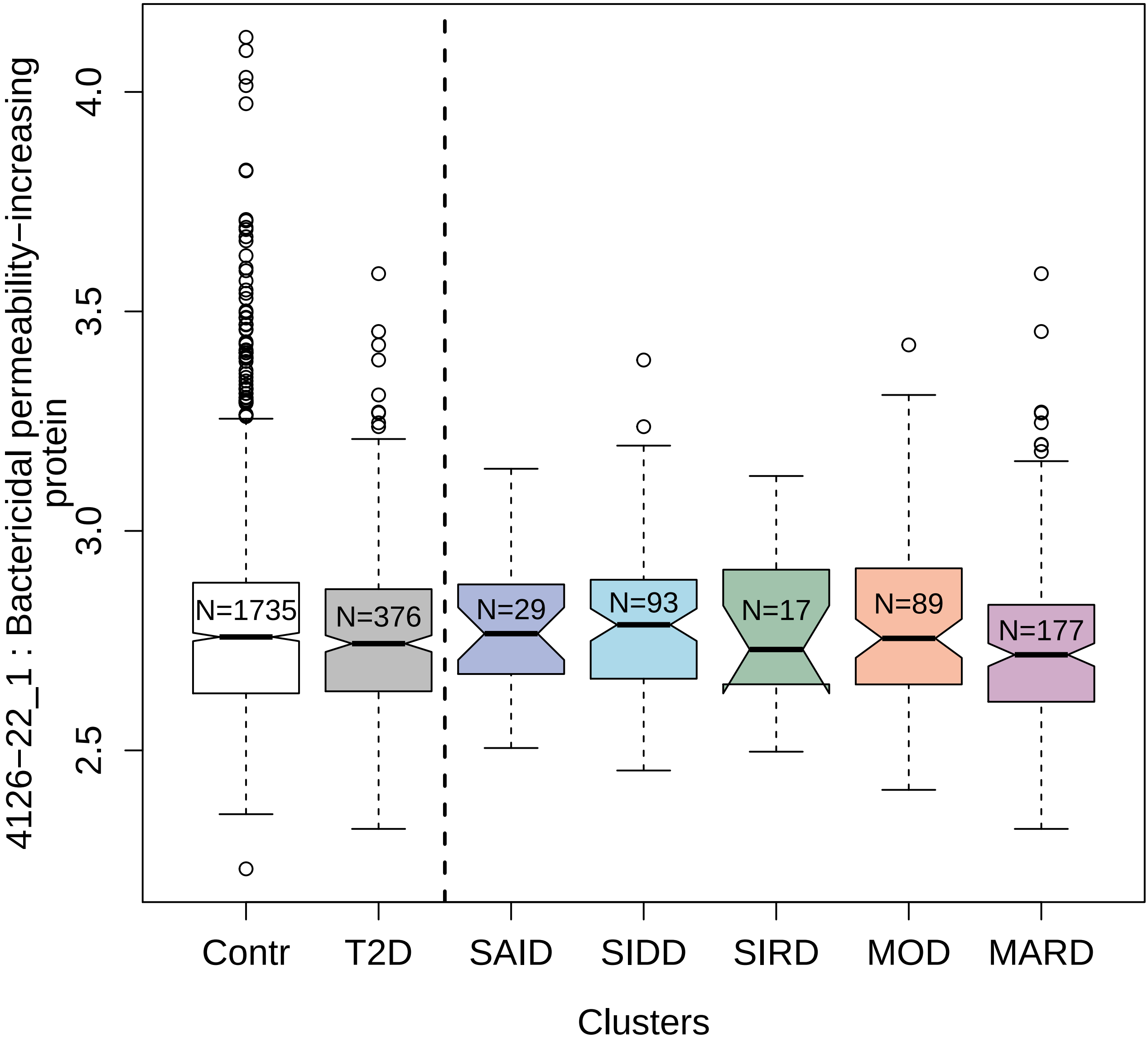
4126−22_1 : Bactericidal permeability−increasing protein
4.0
3.5
3.0
2.5
N=1735
N=376
N=29
N=93
N=17
N=89
N=177
Contr
T2D
SAID
SIDD
SIRD
MOD
MARD
Clusters

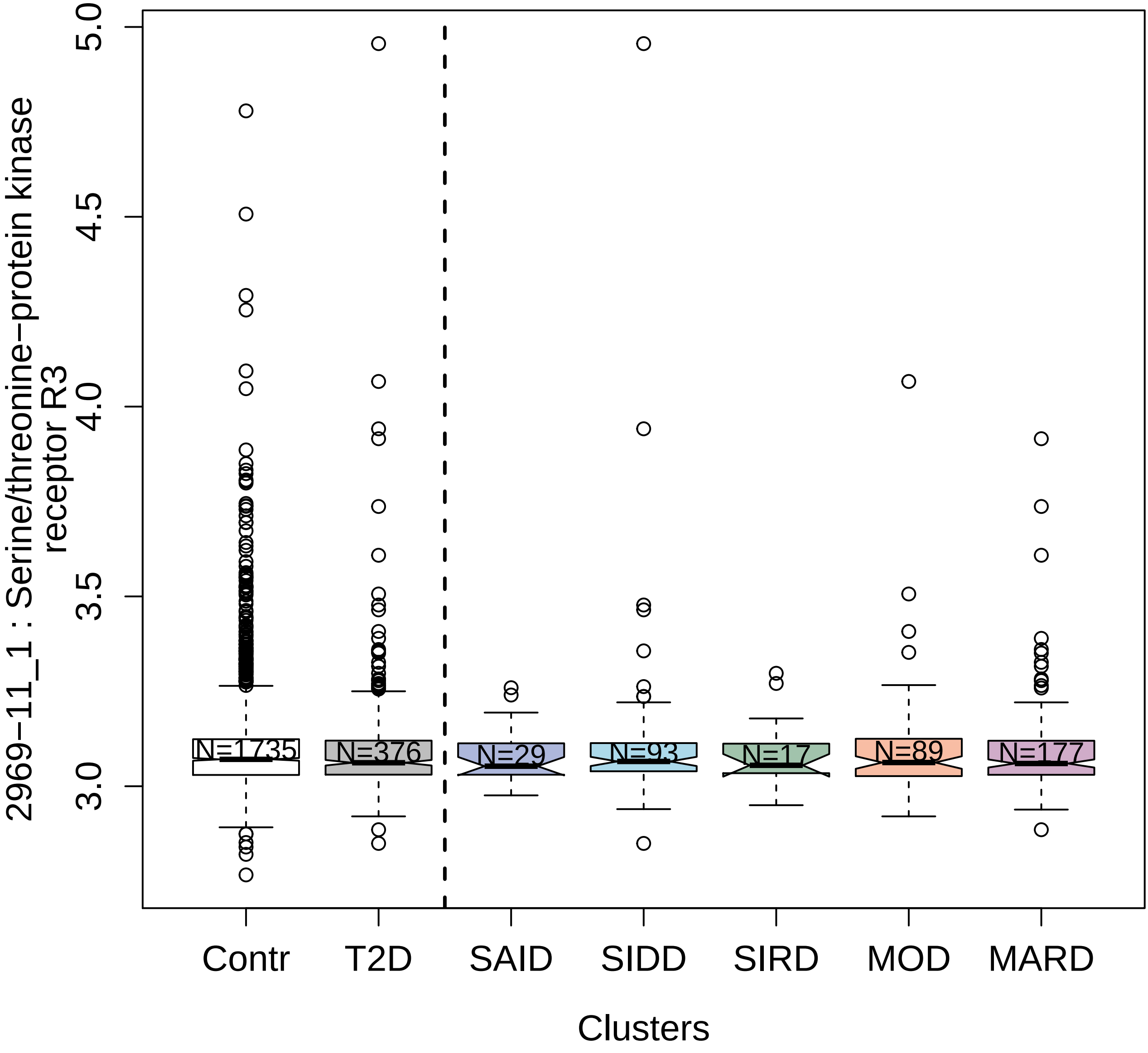

2969−11_1 : Serine/threonine−protein kinase
receptor R3
5.0
4.5
4.0
3.5
3.0
N=1735
N=376
N=29
N=93
N=17
N=89
N=177
Contr
T2D
SAID
SIDD
SIRD
MOD
MARD
Clusters

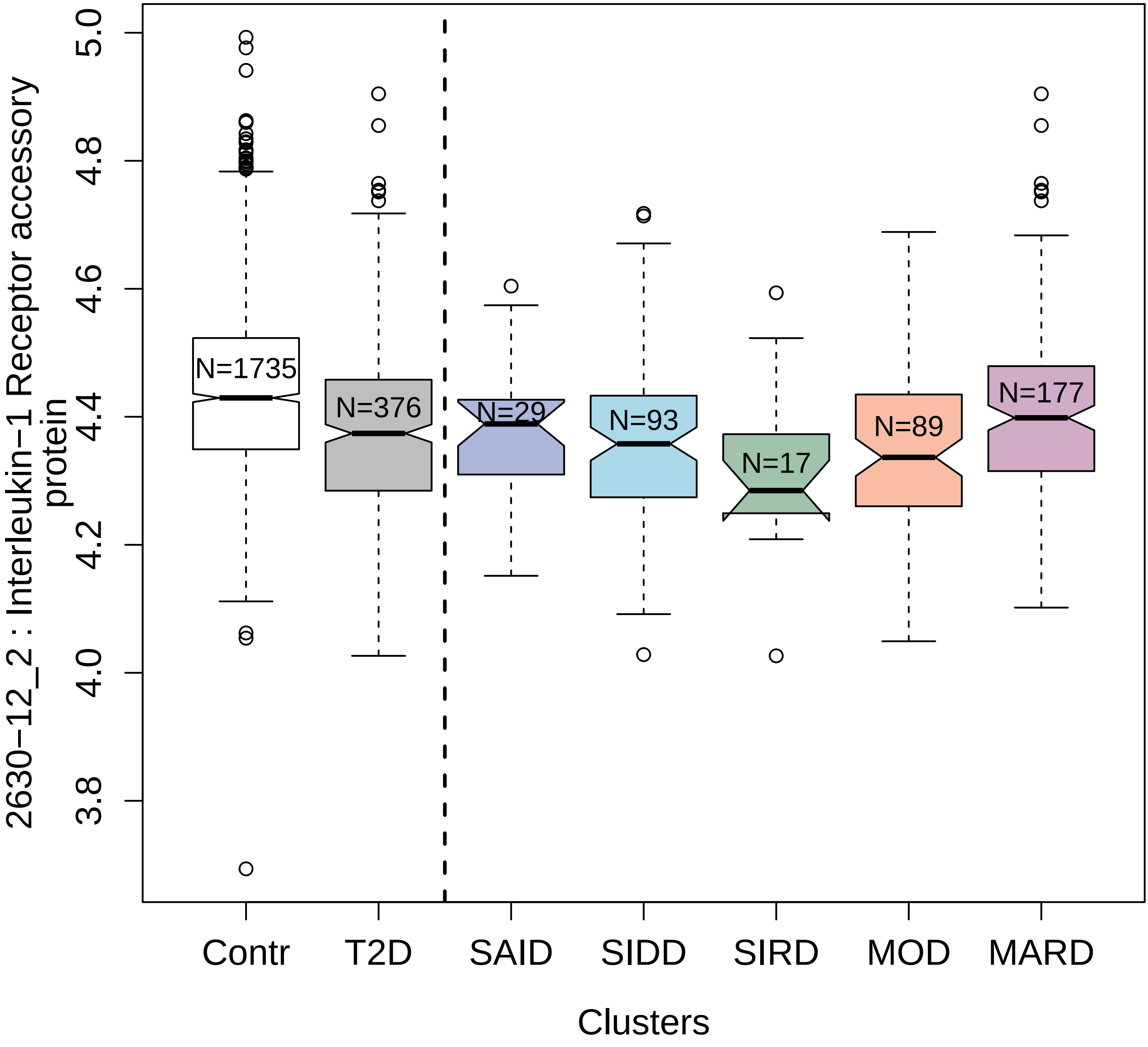
2630−12_2 : Interleukin−1 Receptor accessory protein
5.0
4.8
4.6
4.4
4.2
4.0
3.8
N=1735
N=376
N=29
N=93
N=17
N=89
N=177
Contr
T2D
SAID
SIDD
SIRD
MOD
MARD
Clusters

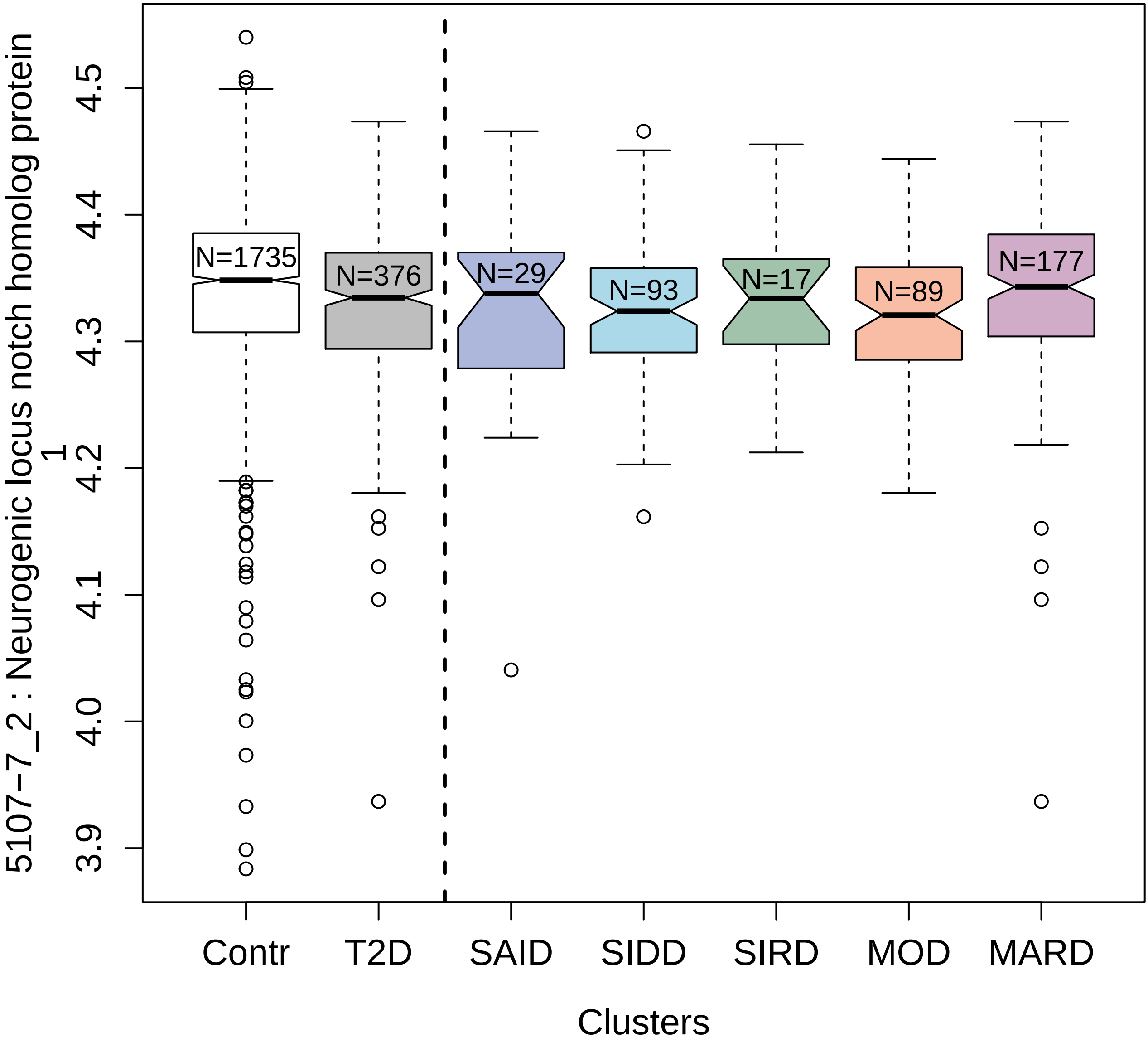
5107−7_2 : Neurogenic locus notch homolog protein 1
3.9
4.0
4.1
4.2
4.3
4.4
4.5
N=1735
N=376
N=29
N=93
N=17
N=89
N=177
Contr
T2D
SAID
SIDD
SIRD
MOD
MARD
Clusters

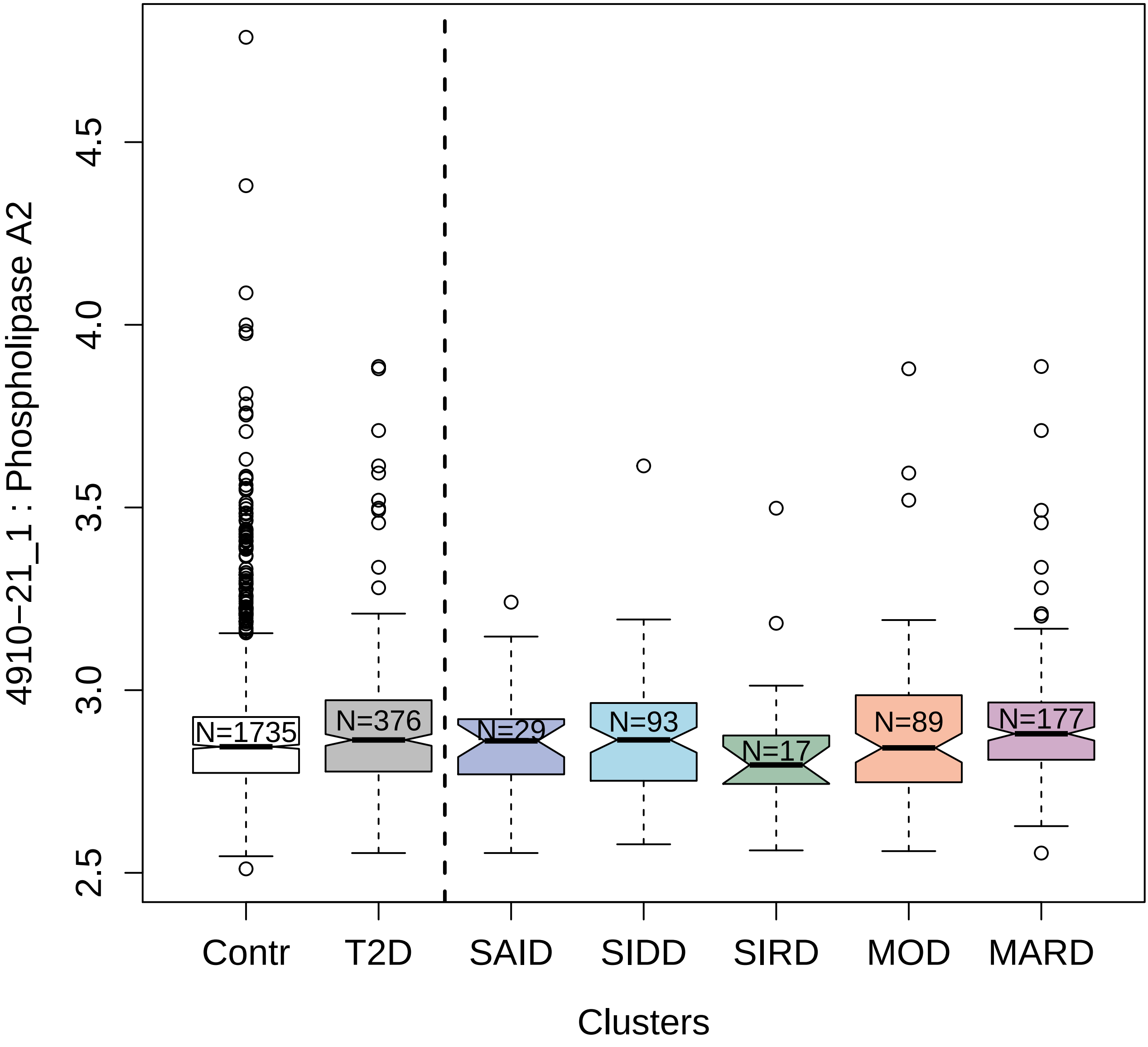
4910−21_1 : Phosphoplipase A2
2.5
3.0
3.5
4.0
4.5
N=1735
N=376
N=29
N=93
N=17
N=89
N=177
Contr
T2D
SAID
SIDD
SIRD
MOD
MARD
Clusters

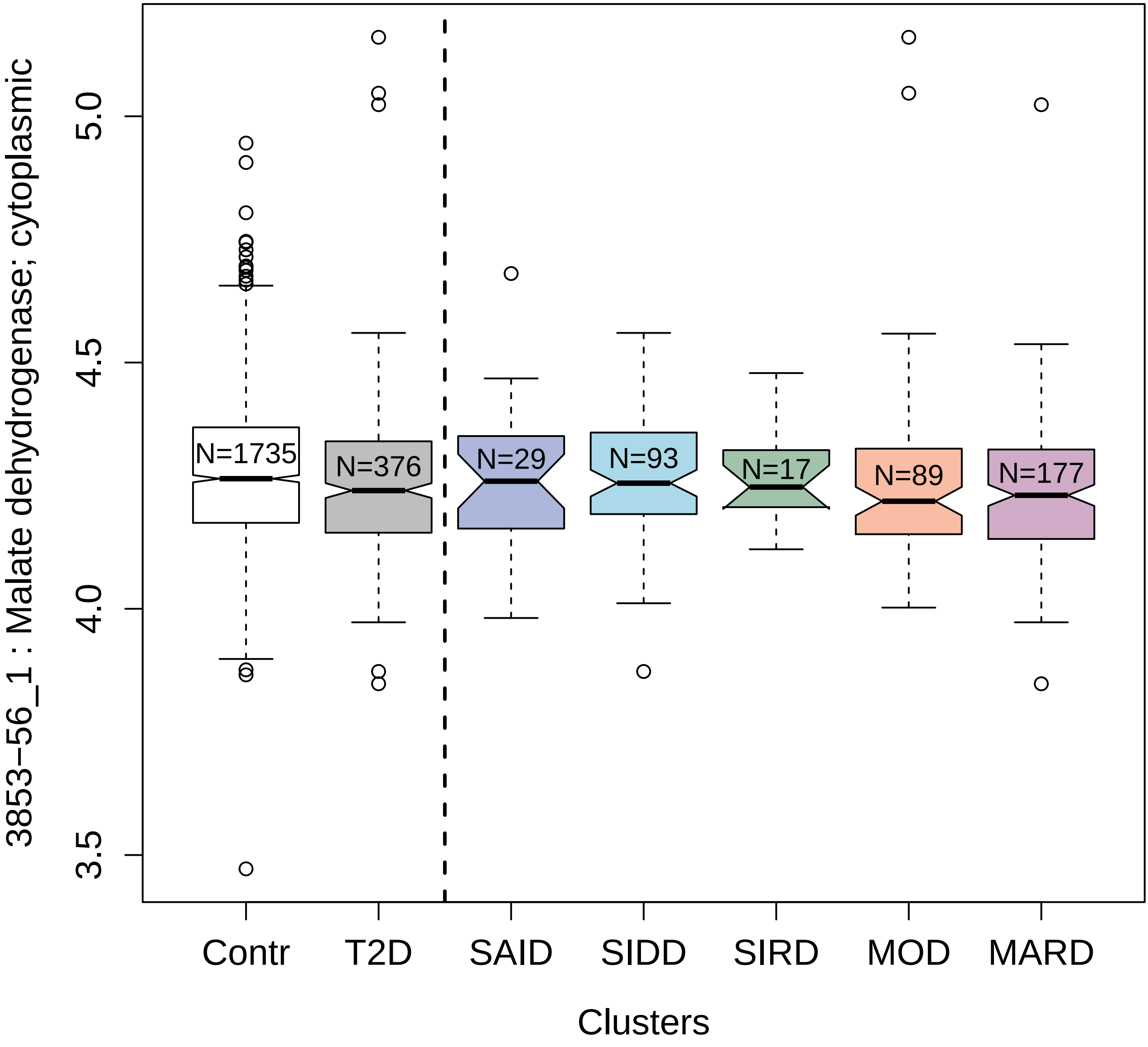
3853−56_1 : Malate dehydrogenase; cytoplasmic
5.0
4.5
4.0
3.5
N=1735
N=376
N=29
N=93
N=17
N=89
N=177
Contr
T2D
SAID
SIDD
SIRD
MOD
MARD
Clusters

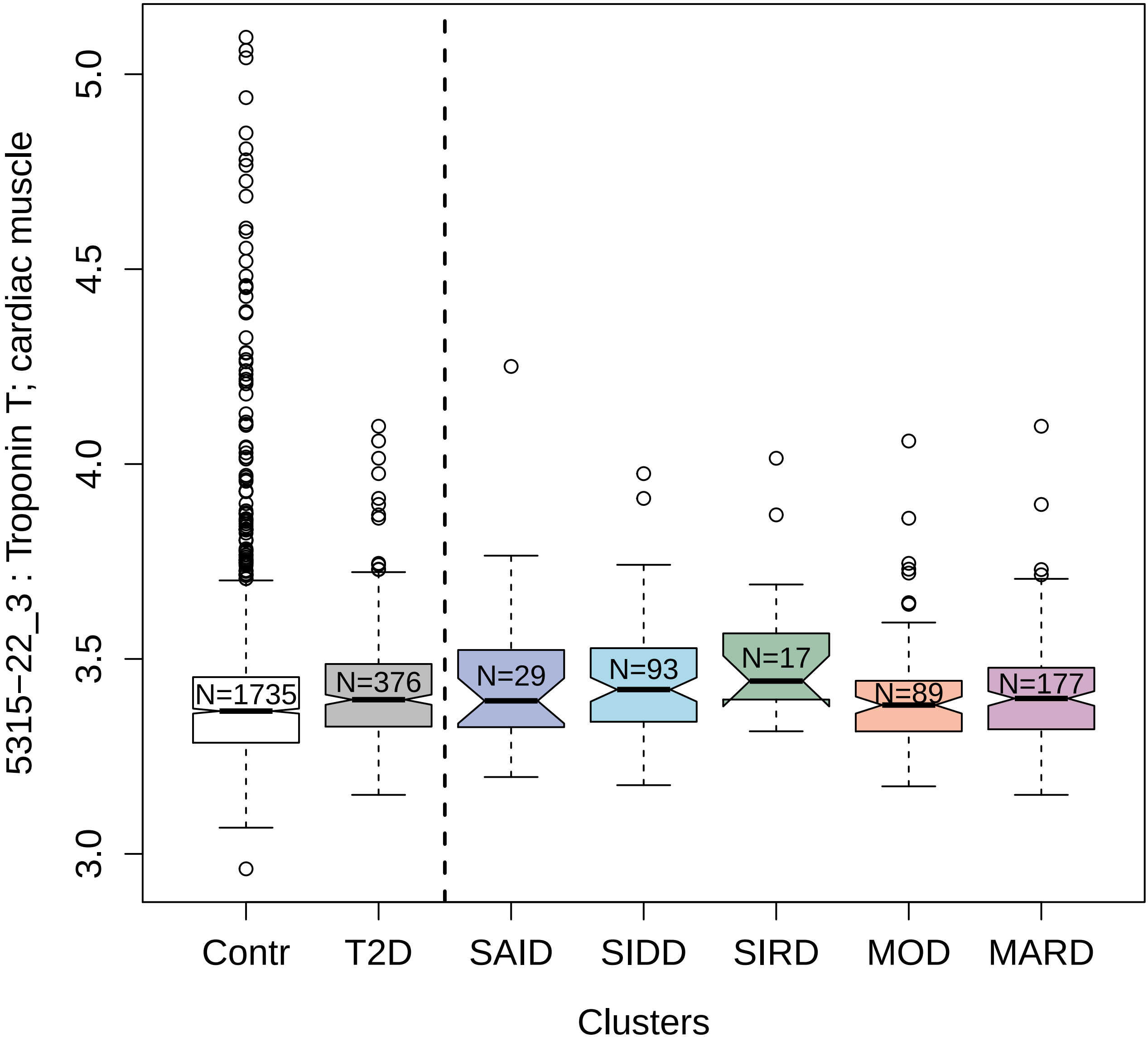

5315−22_3 : Troponin T; cardiac muscle
3.0
3.5
4.0
4.5
5.0
N=1735
N=376
N=29
N=93
N=17
N=89
N=177
Contr
T2D
SAID
SIDD
SIRD
MOD
MARD
Clusters

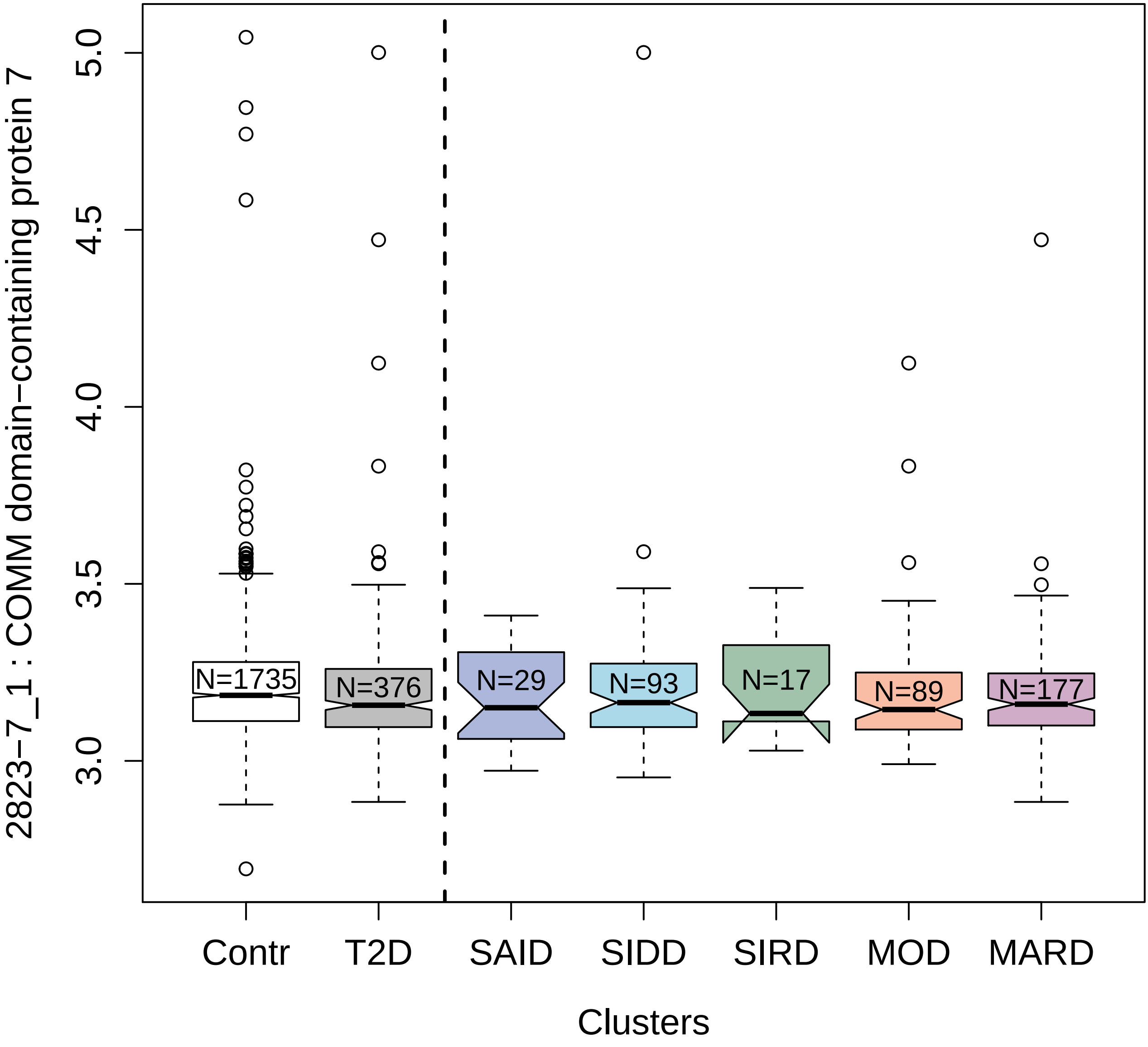

2823−7_1 : COMM domain−containing protein 7
5.0
4.5
4.0
3.5
3.0
N=1735
N=376
N=29
N=93
N=17
N=89
N=177
Contr
T2D
SAID
SIDD
SIRD
MOD
MARD
Clusters

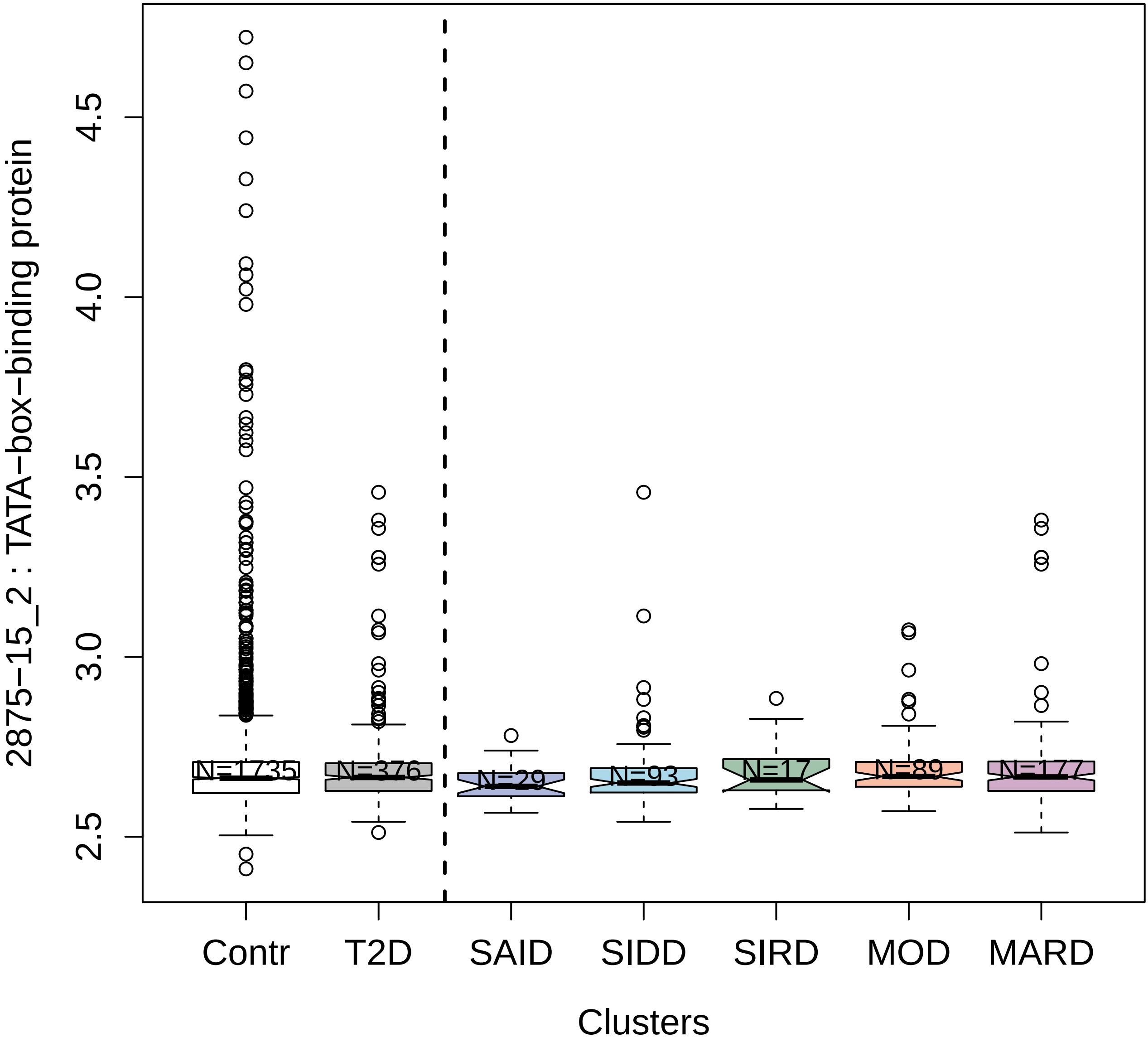
2875−15_2 : TATA−box−binding protein
4.5
4.0
3.5
3.0
2.5
N=1735
N=376
N=29
N=93
N=17
N=89
N=177
Contr
T2D
SAID
SIDD
SIRD
MOD
MARD
Clusters

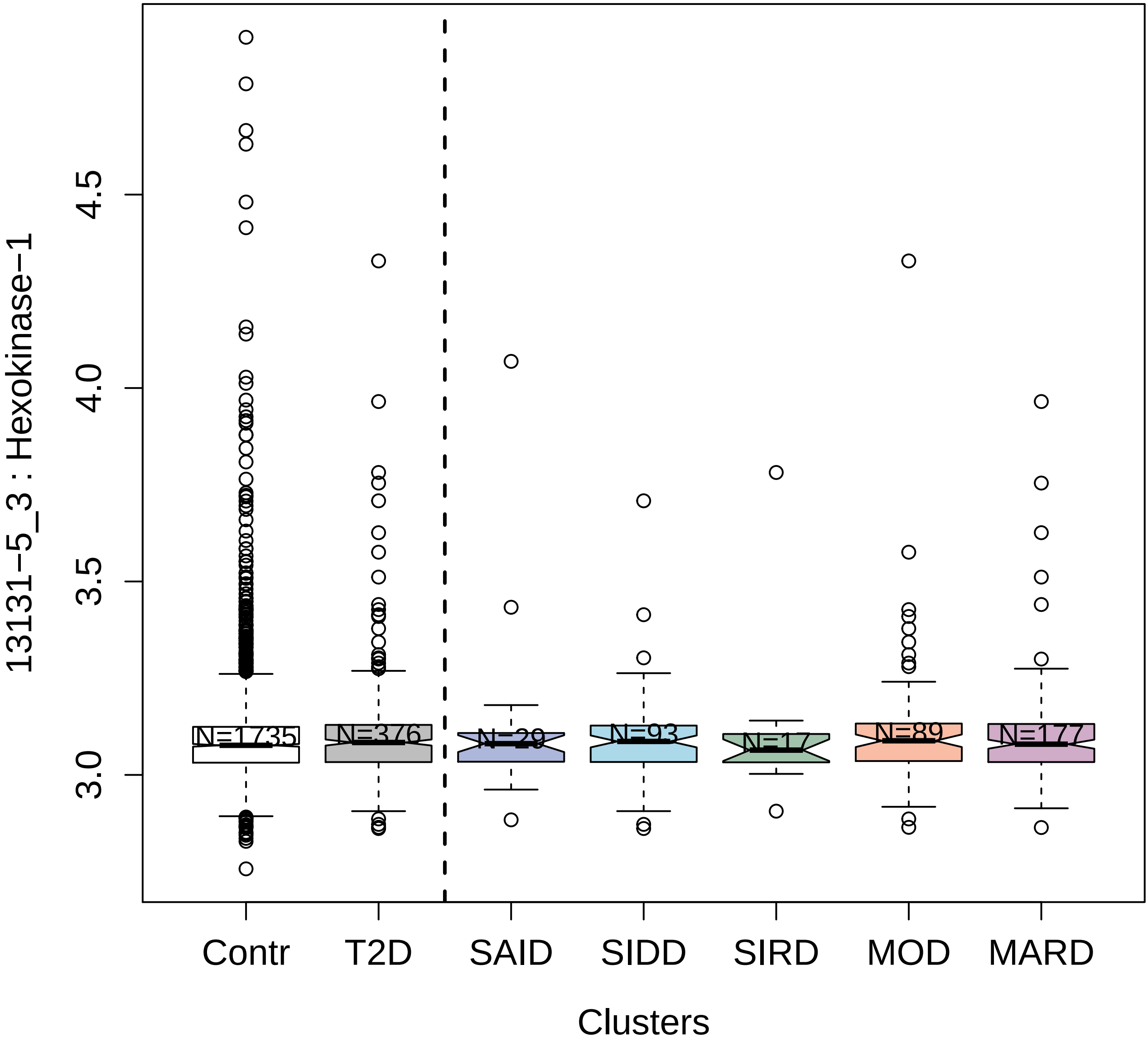

13131−5_3 : Hexokinase−1
3.0
3.5
4.0
4.5
N=1735
N=376
N=29
N=93
N=17
N=89
N=177
Contr
T2D
SAID
SIDD
SIRD
MOD
MARD
Clusters

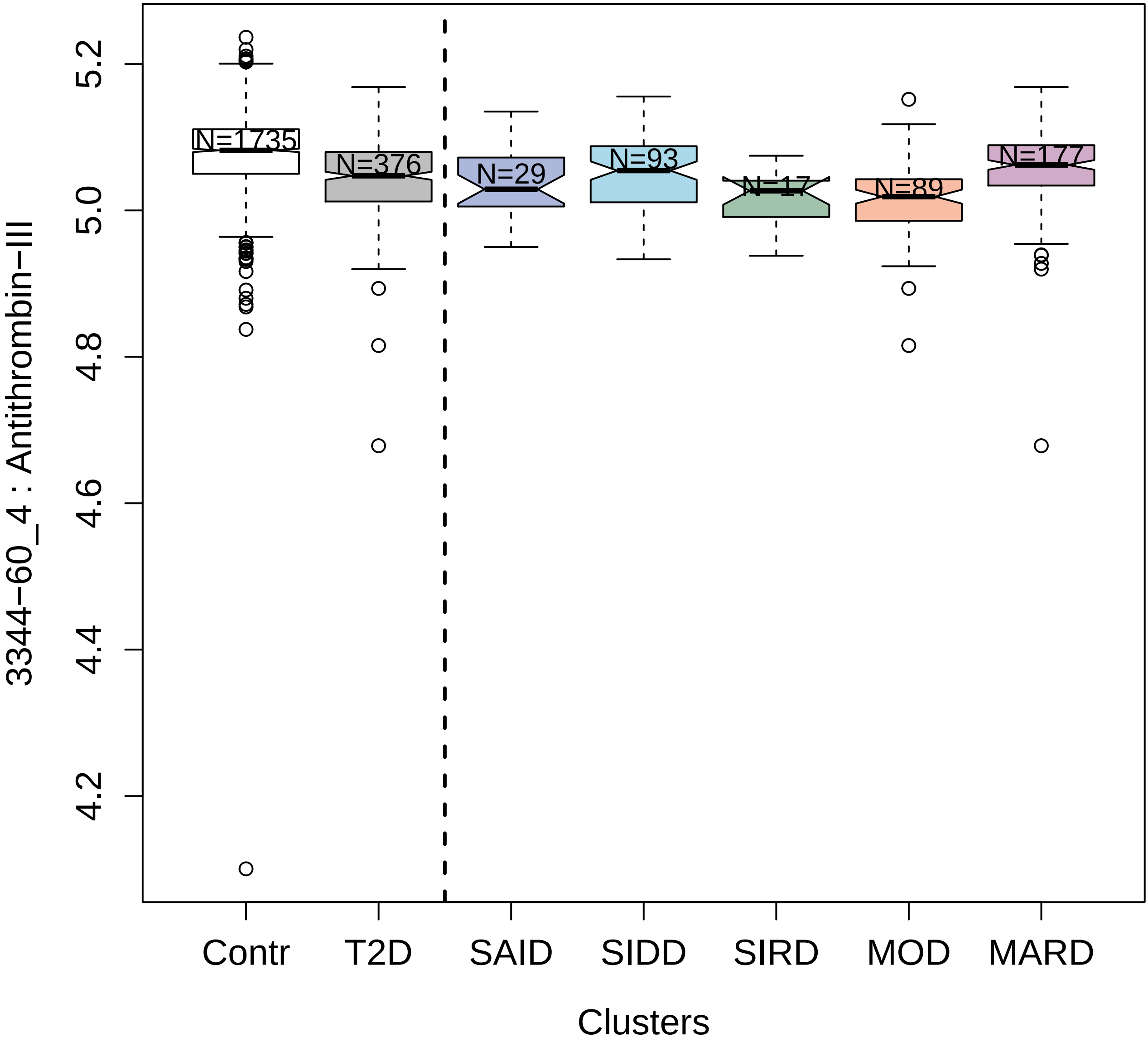

3344−60_4 : Antithrombin−III
5.2
5.0
4.8
4.6
4.4
4.2
N=1735
N=376
N=29
N=93
N=17
N=89
N=177
Contr
T2D
SAID
SIDD
SIRD
MOD
MARD
Clusters

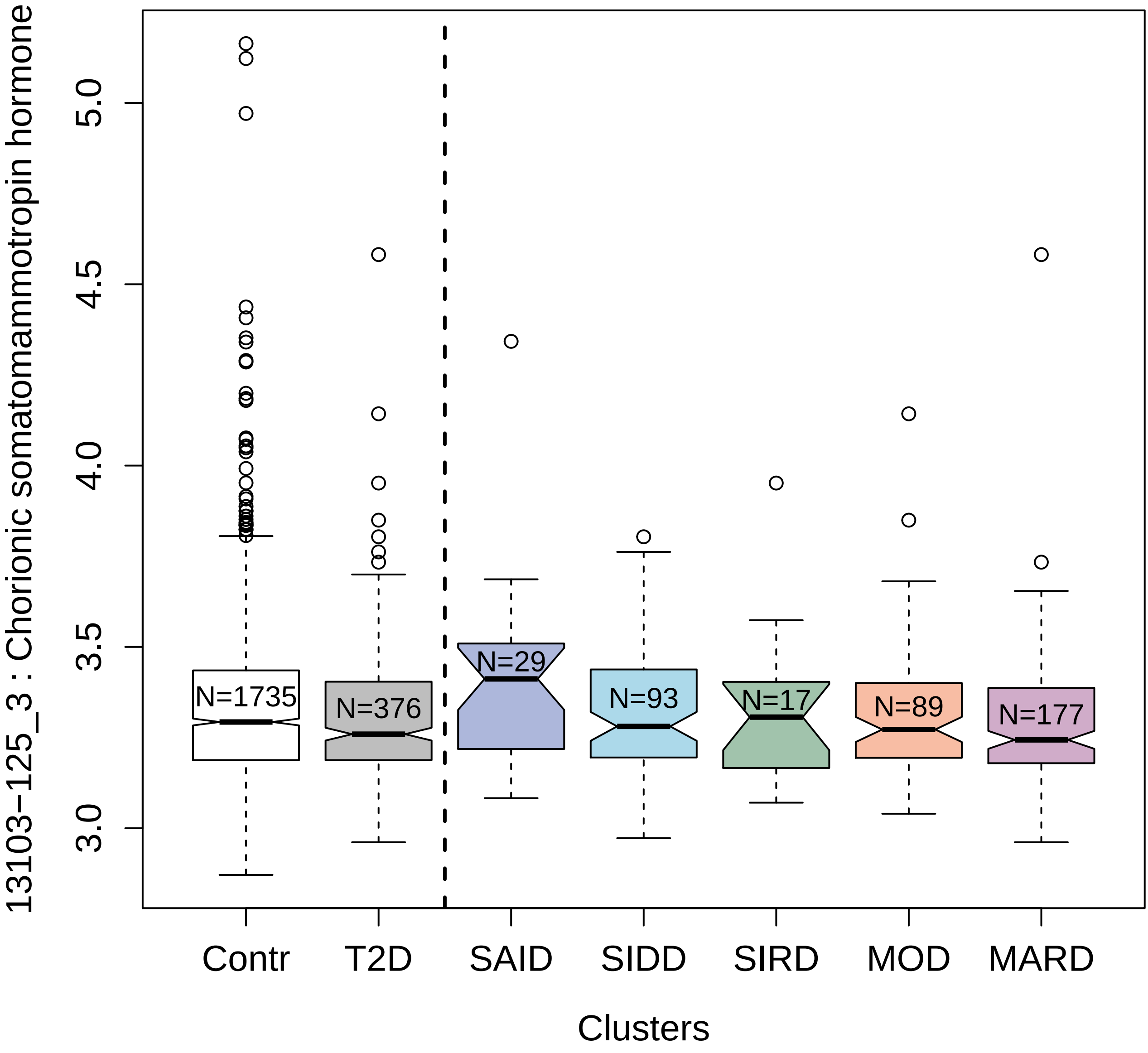
13103−125_3 : Chorionic somatomammotropin hormone
5.0
4.5
4.0
3.5
3.0
N=1735
N=376
N=29
N=93
N=17
N=89
N=177
Contr
T2D
SAID
SIDD
SIRD
MOD
MARD
Clusters

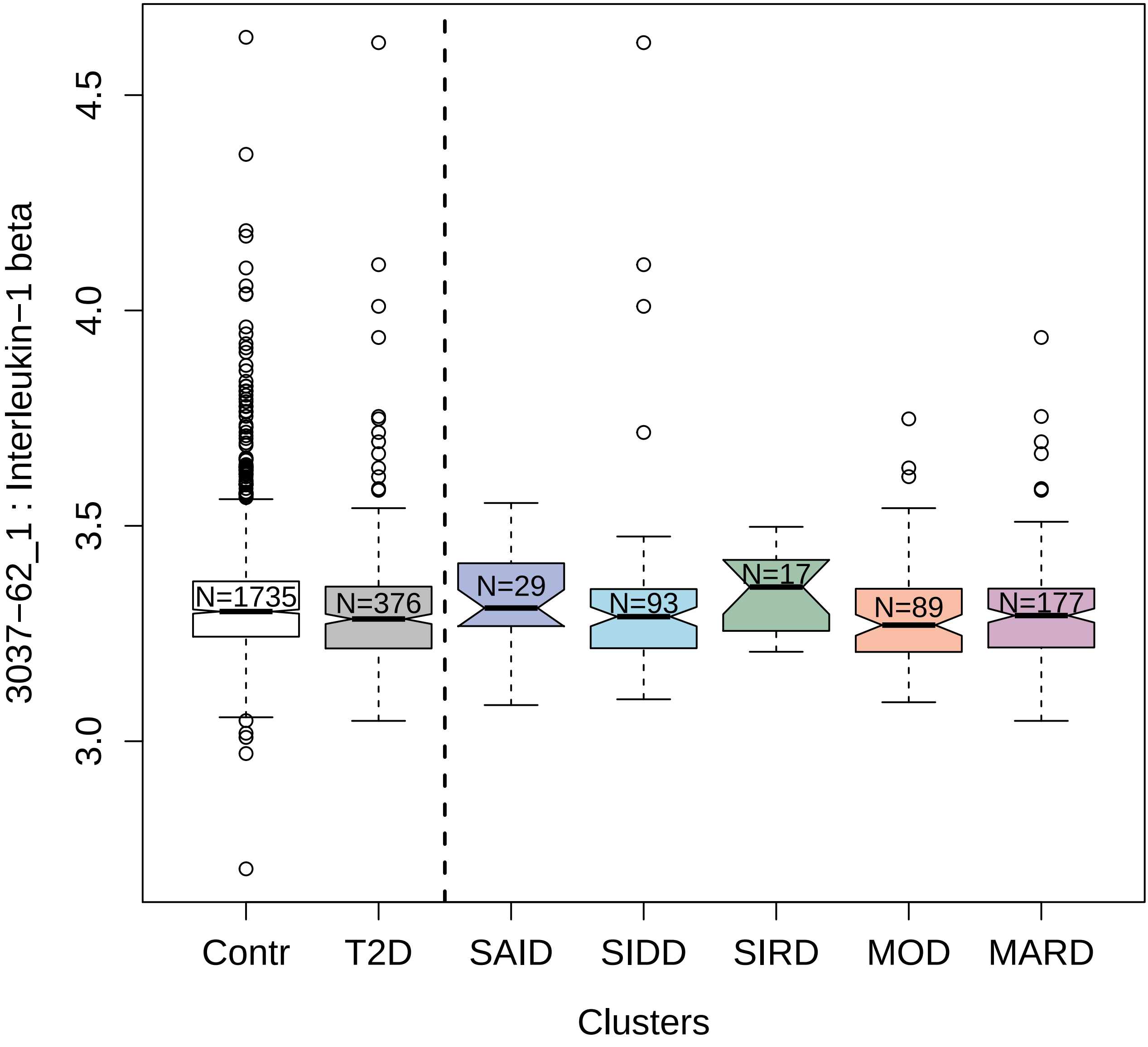
3037−62_1 : Interleukin−1 beta
4.5
4.0
3.5
3.0
N=1735
N=376
N=29
N=93
N=17
N=89
N=177
Contr
T2D
SAID
SIDD
SIRD
MOD
MARD
Clusters

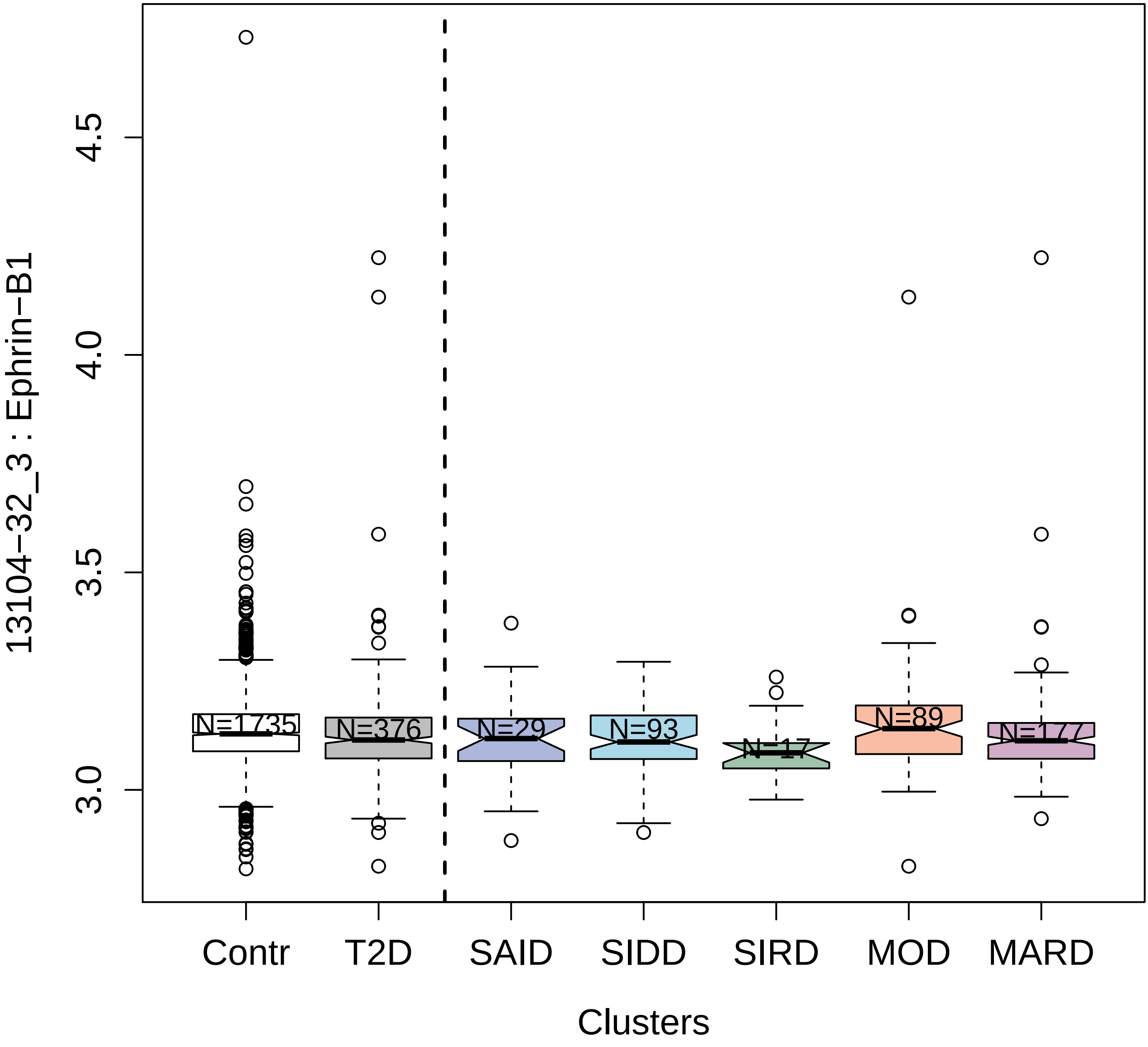
13104−32_3 : Ephrin−B1
3.0
3.5
4.0
4.5
N=1735
N=376
N=29
N=93
N=17
N=89
N=177
Contr
T2D
SAID
SIDD
SIRD
MOD
MARD
Clusters

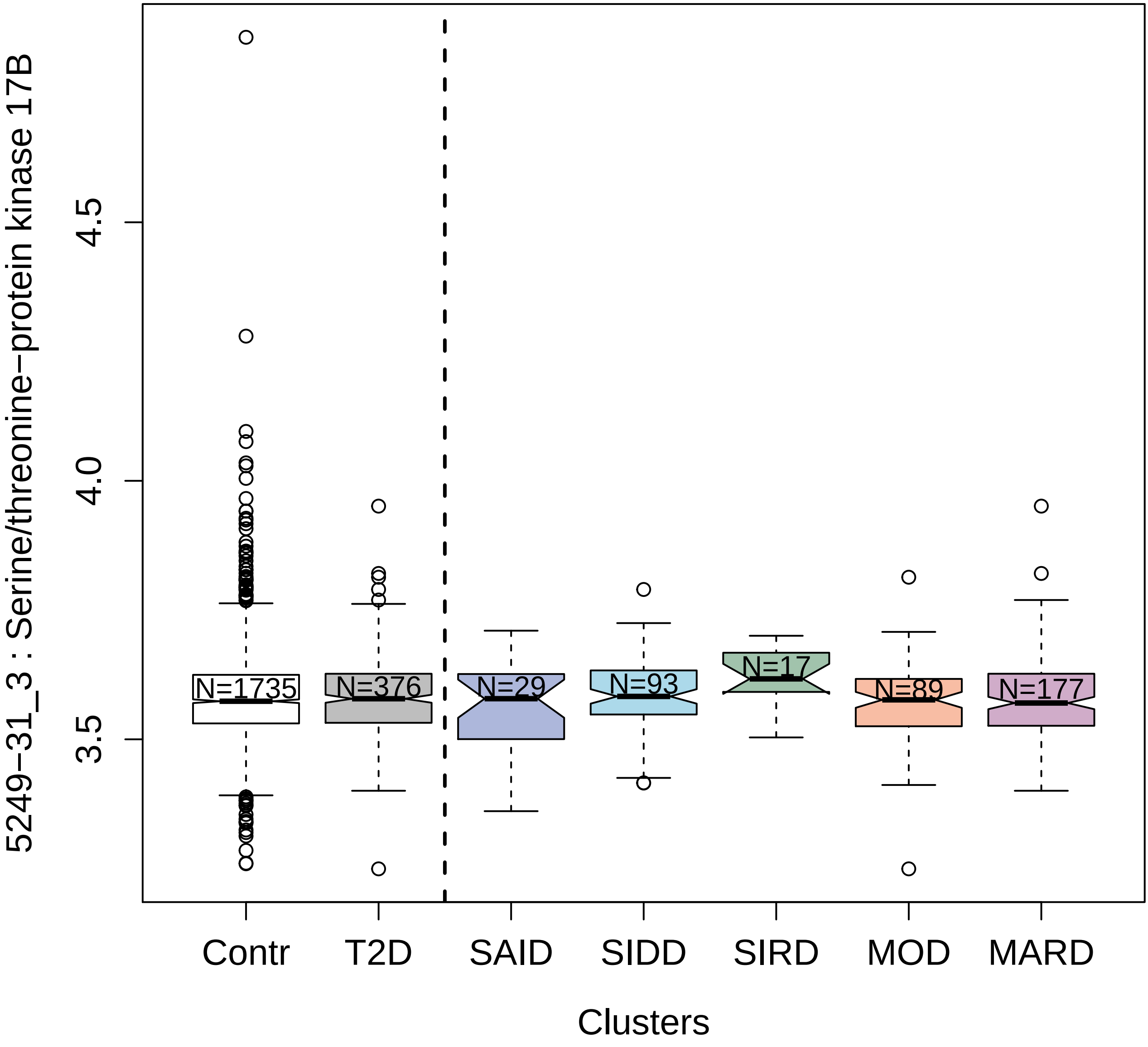

5249−31_3 : Serine/threonine−protein kinase 17B
4.5
4.0
3.5
N=1735
N=376
N=29
N=93
N=17
N=89
N=177
Contr
T2D
SAID
SIDD
SIRD
MOD
MARD
Clusters

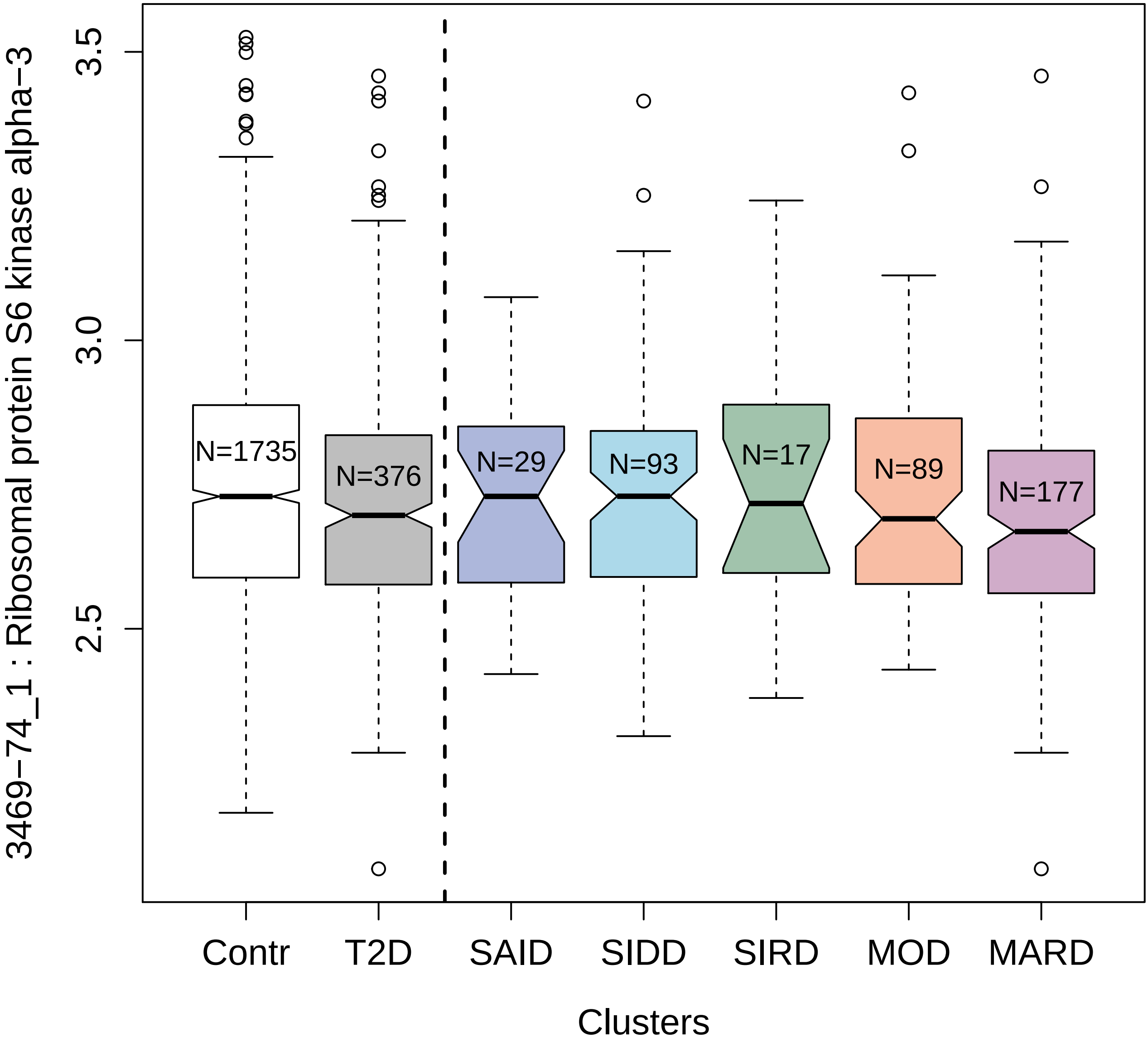
3469−74_1 : Ribosomal protein S6 kinase alpha−3
3.5
3.0
2.5
N=1735
N=376
N=29
N=93
N=17
N=89
N=177
Contr
T2D
SAID
SIDD
SIRD
MOD
MARD
Clusters

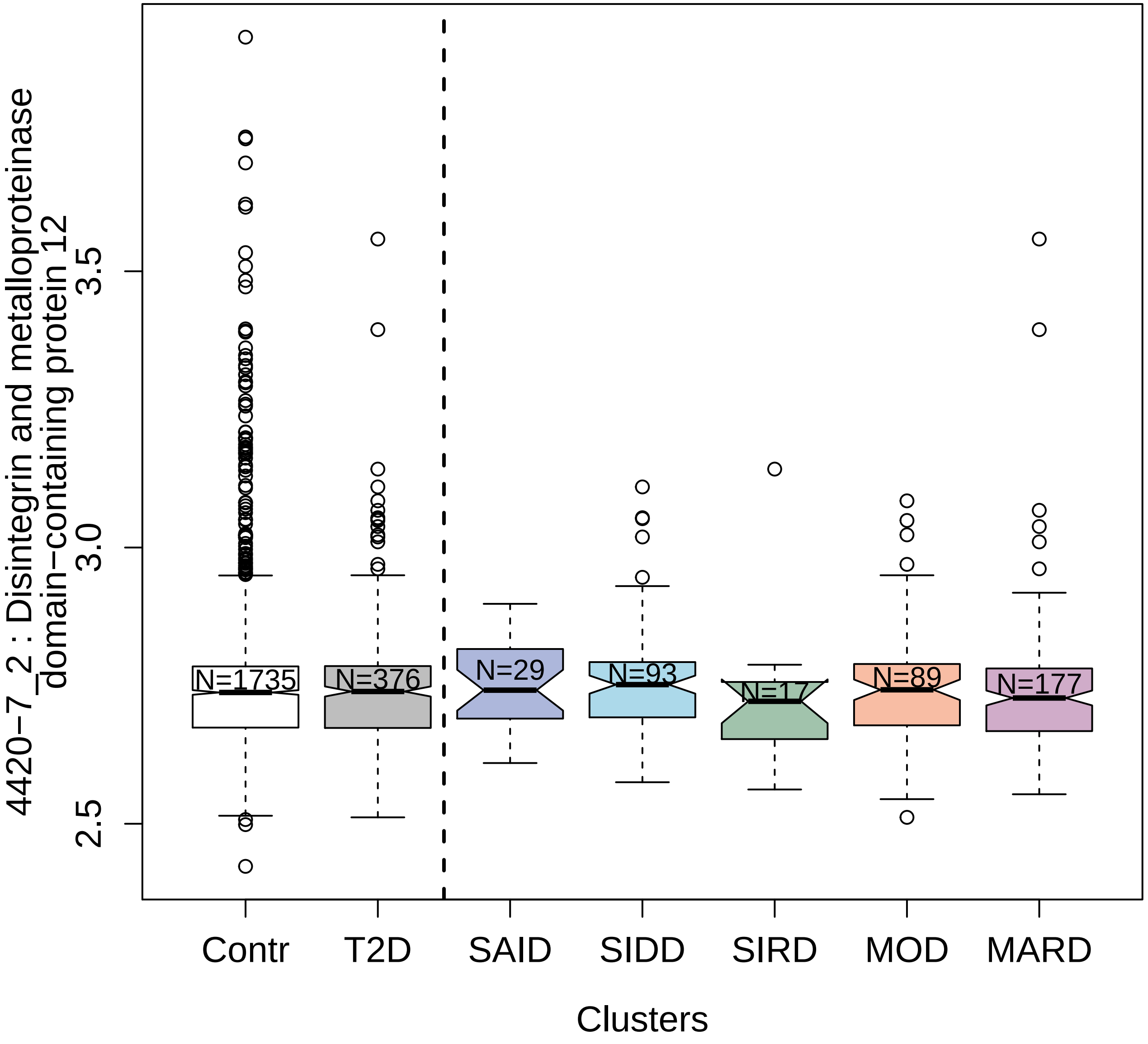
4420−7_2 : Disintegrin and metalloproteinase
domain−containing protein 12
3.5
3.0
2.5
N=1735
N=376
N=29
N=93
N=17
N=89
N=177
Contr
T2D
SAID
SIDD
SIRD
MOD
MARD
Clusters

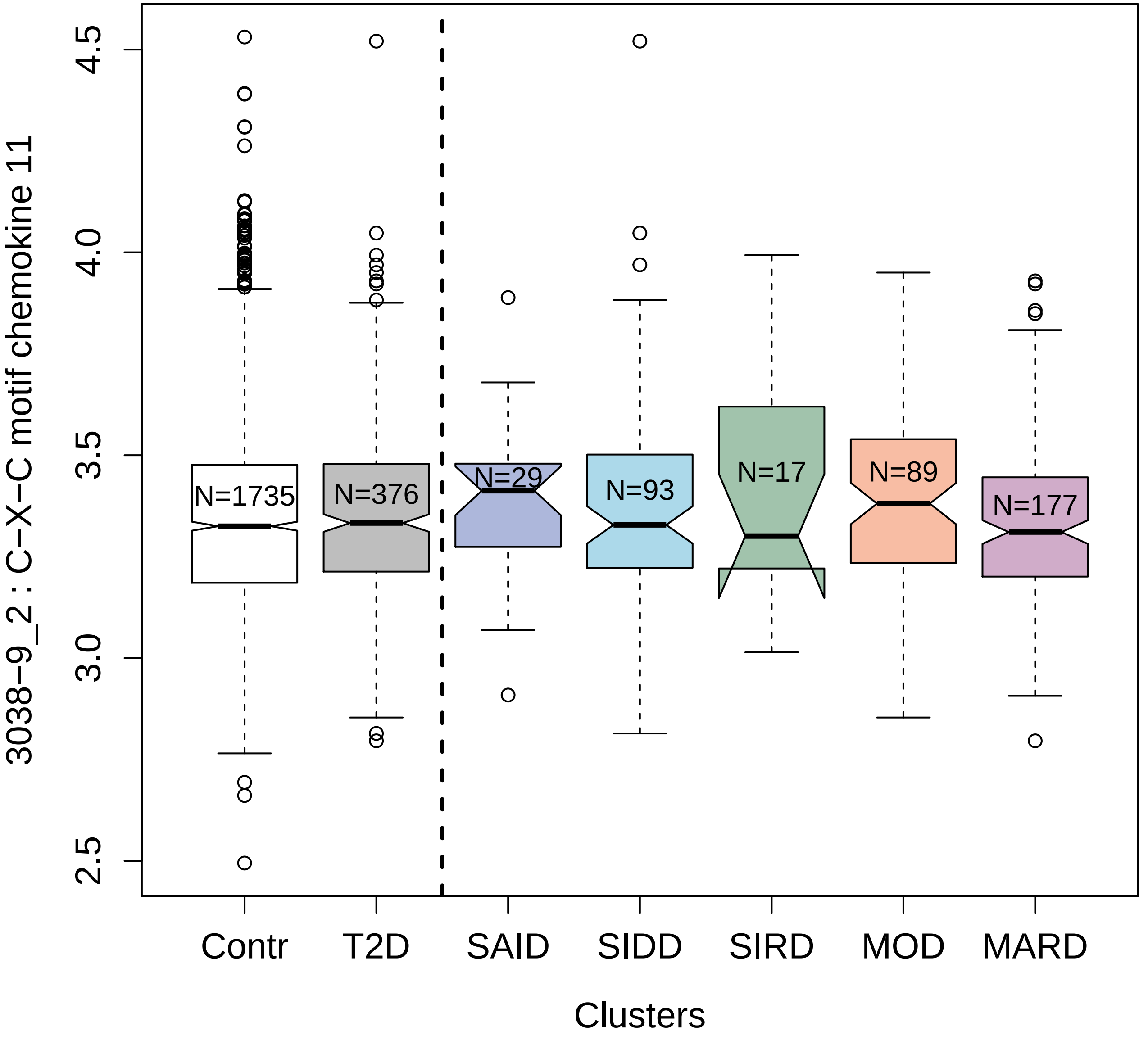
3038−9_2 : C−X−C motif chemokine 11
Clusters
Contr
T2D
SAID
SIDD
SIRD
MOD
MARD
N=1735
N=376
N=29
N=93
N=17
N=89
N=177
2.5
3.0
3.5
4.0
4.5

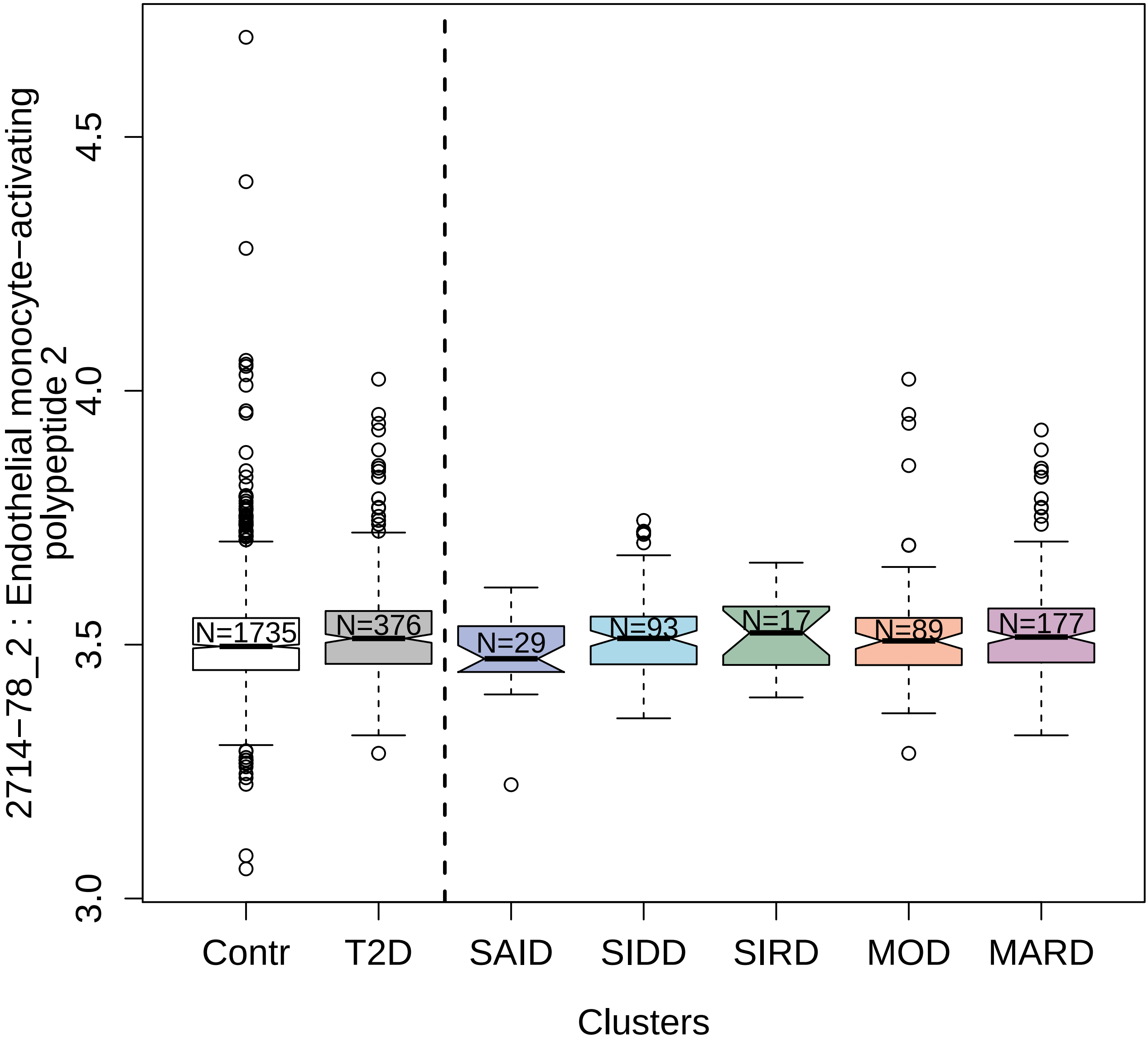

2714−78_2 : Endothelial monocyte−activating polypeptide 2
3.0
3.5
4.0
4.5
N=1735
N=376
N=29
N=93
N=17
N=89
N=177
Contr
T2D
SAID
SIDD
SIRD
MOD
MARD
Clusters

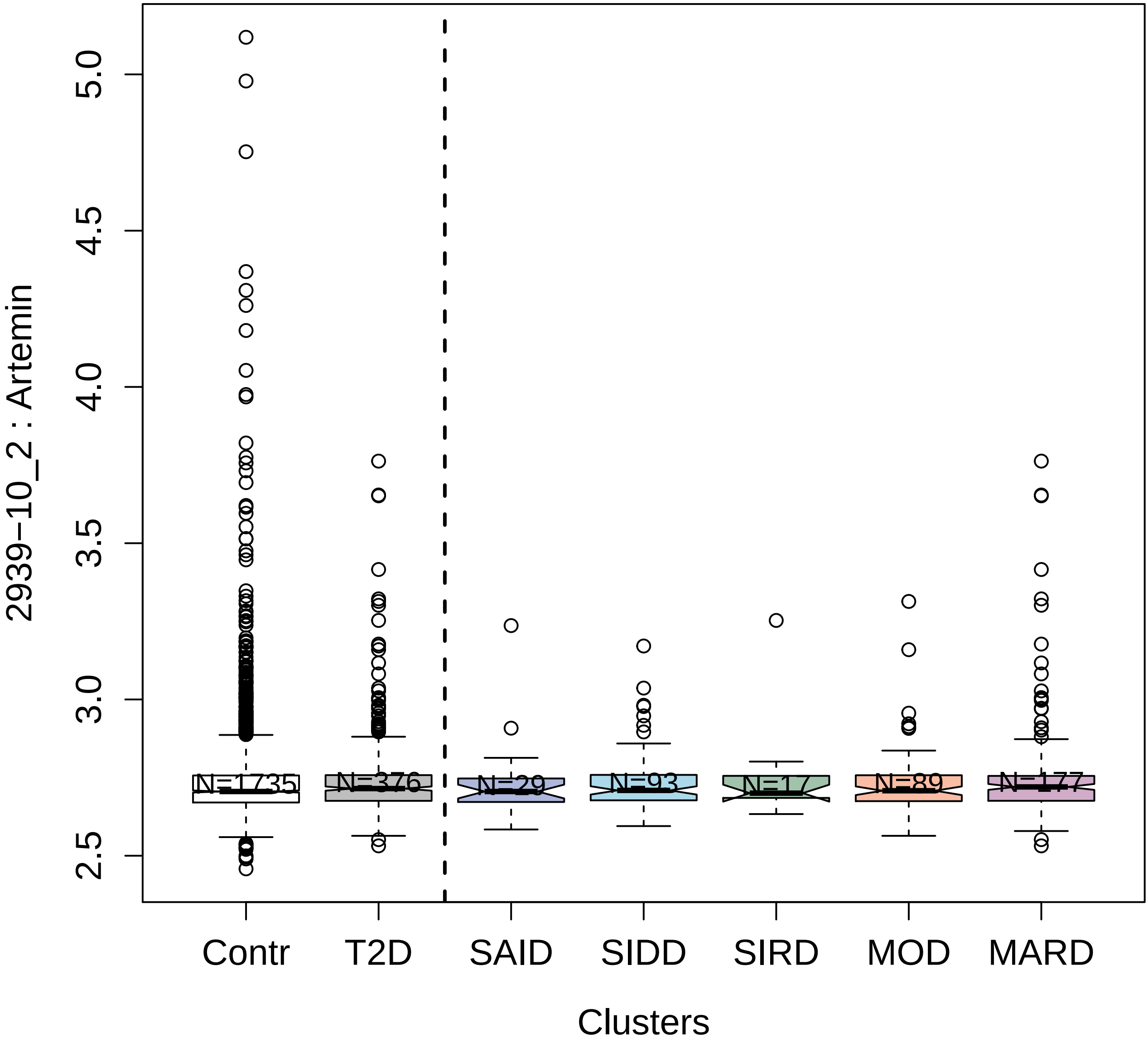

2939−10_2 : Artemin
5.0
4.5
4.0
3.5
3.0
2.5
N=1735
N=376
N=29
N=93
N=17
N=89
N=177
Contr
T2D
SAID
SIDD
SIRD
MOD
MARD
Clusters

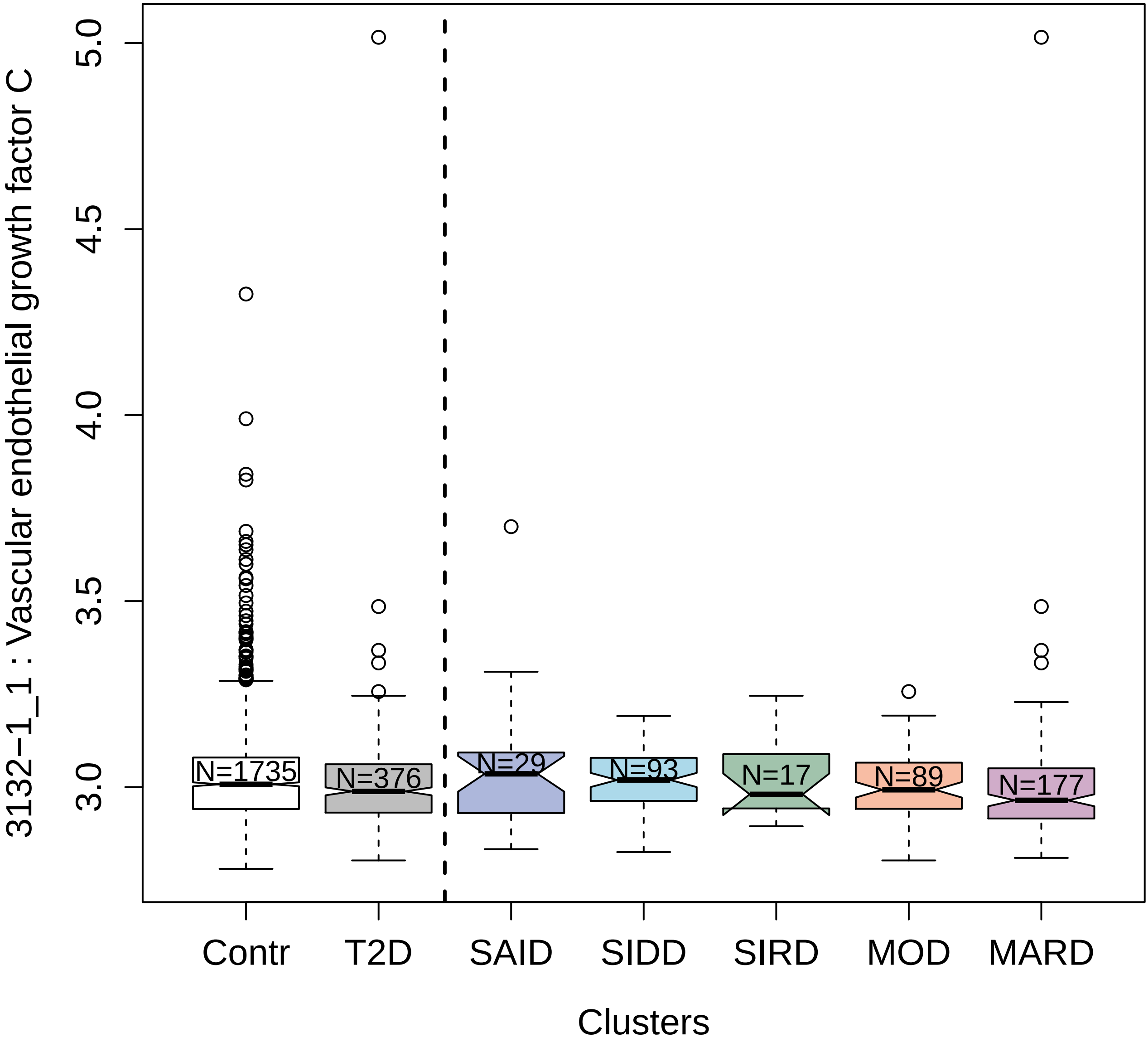

3132−1_1 : Vascular endothelial growth factor C
5.0
4.5
4.0
3.5
3.0
N=1735
N=376
N=29
N=93
N=17
N=89
N=177
Contr
T2D
SAID
SIDD
SIRD
MOD
MARD
Clusters

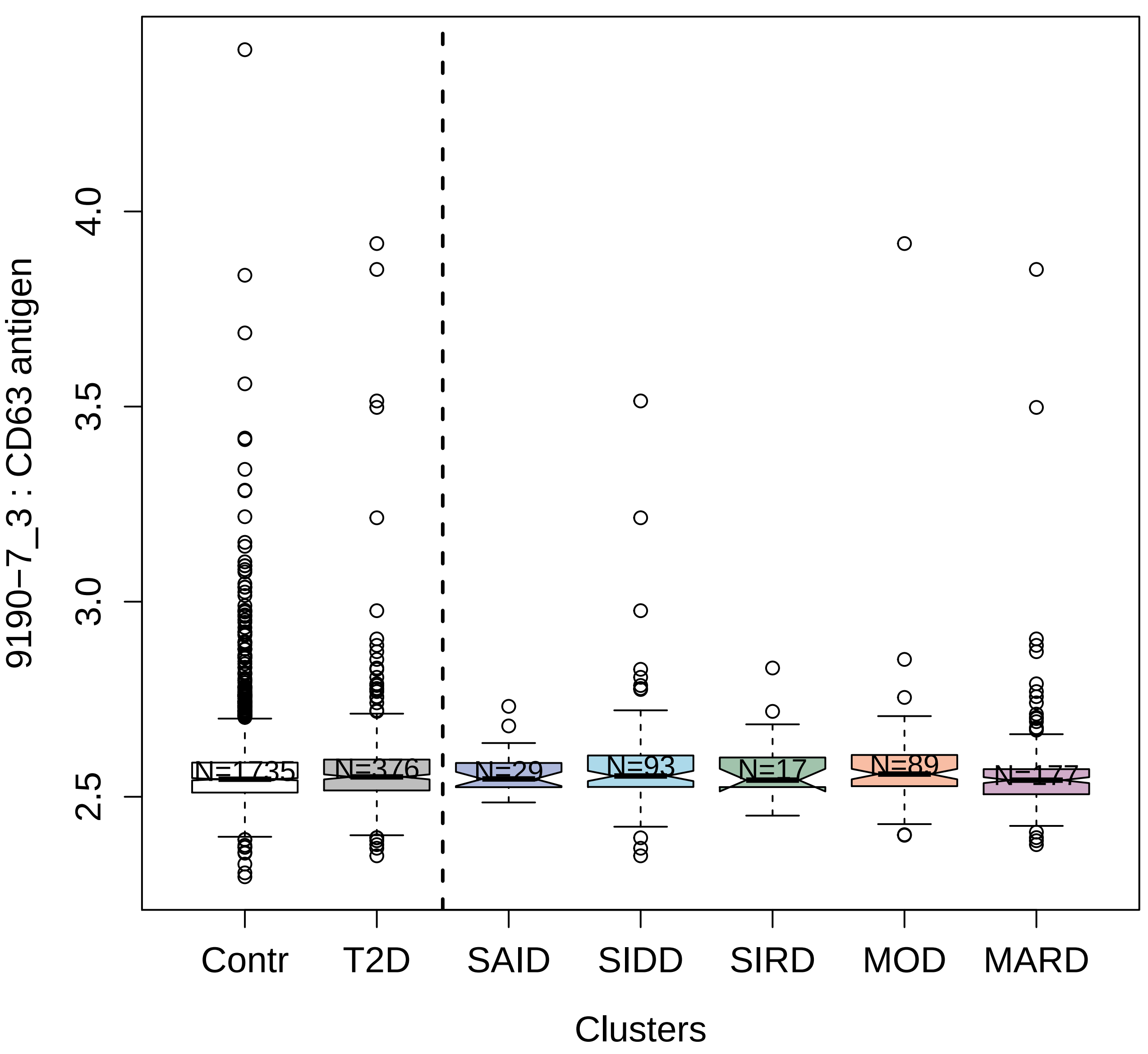

9190−7_3 : CD63 antigen
4.0
3.5
3.0
2.5
N=1735
N=376
N=29
N=93
N=17
N=89
N=177
Contr
T2D
SAID
SIDD
SIRD
MOD
MARD
Clusters

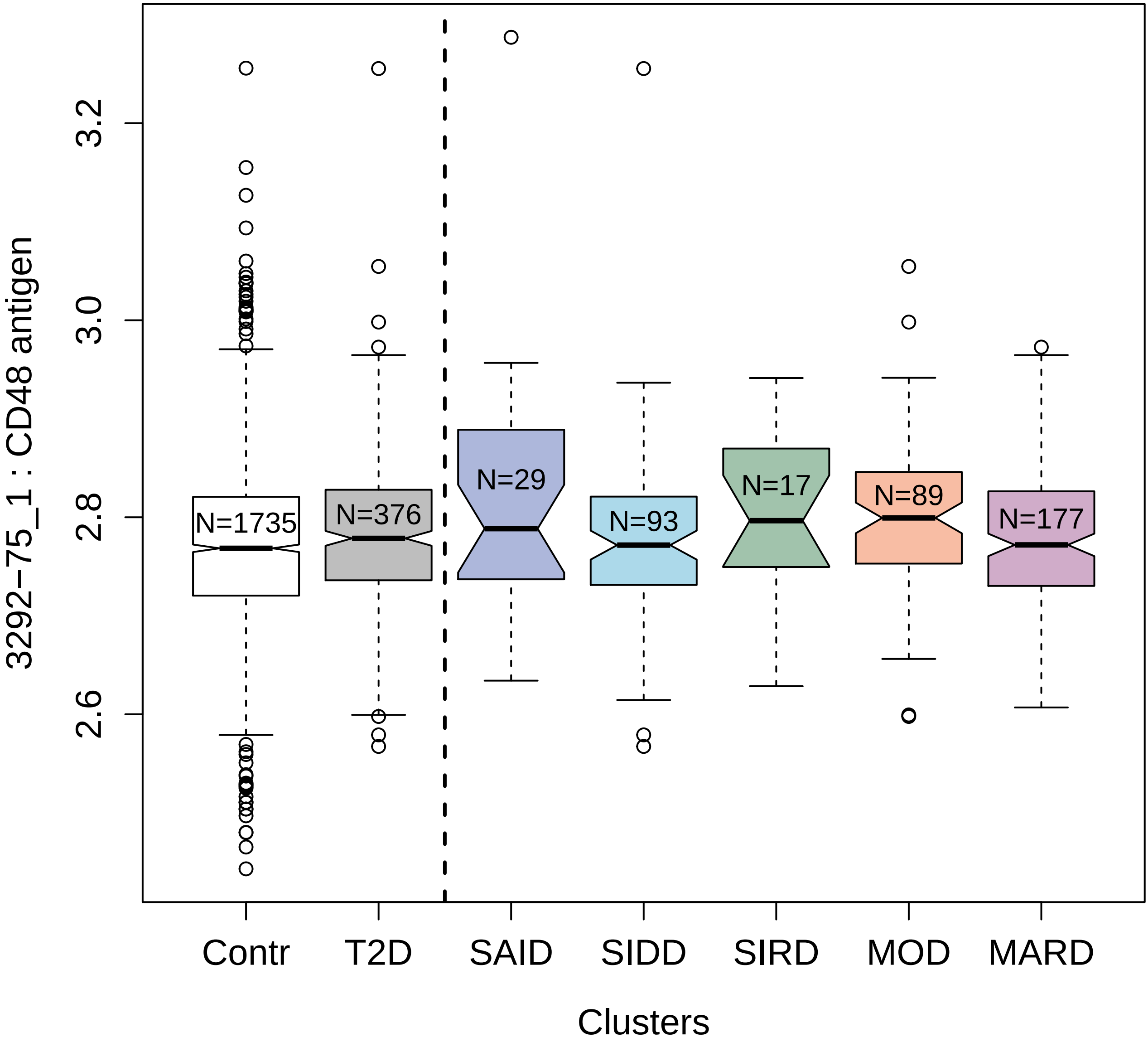
3292−75_1 : CD48 antigen
2.6
2.8
3.0
3.2
N=1735
N=376
N=29
N=93
N=17
N=89
N=177
Contr
T2D
SAID
SIDD
SIRD
MOD
MARD
Clusters

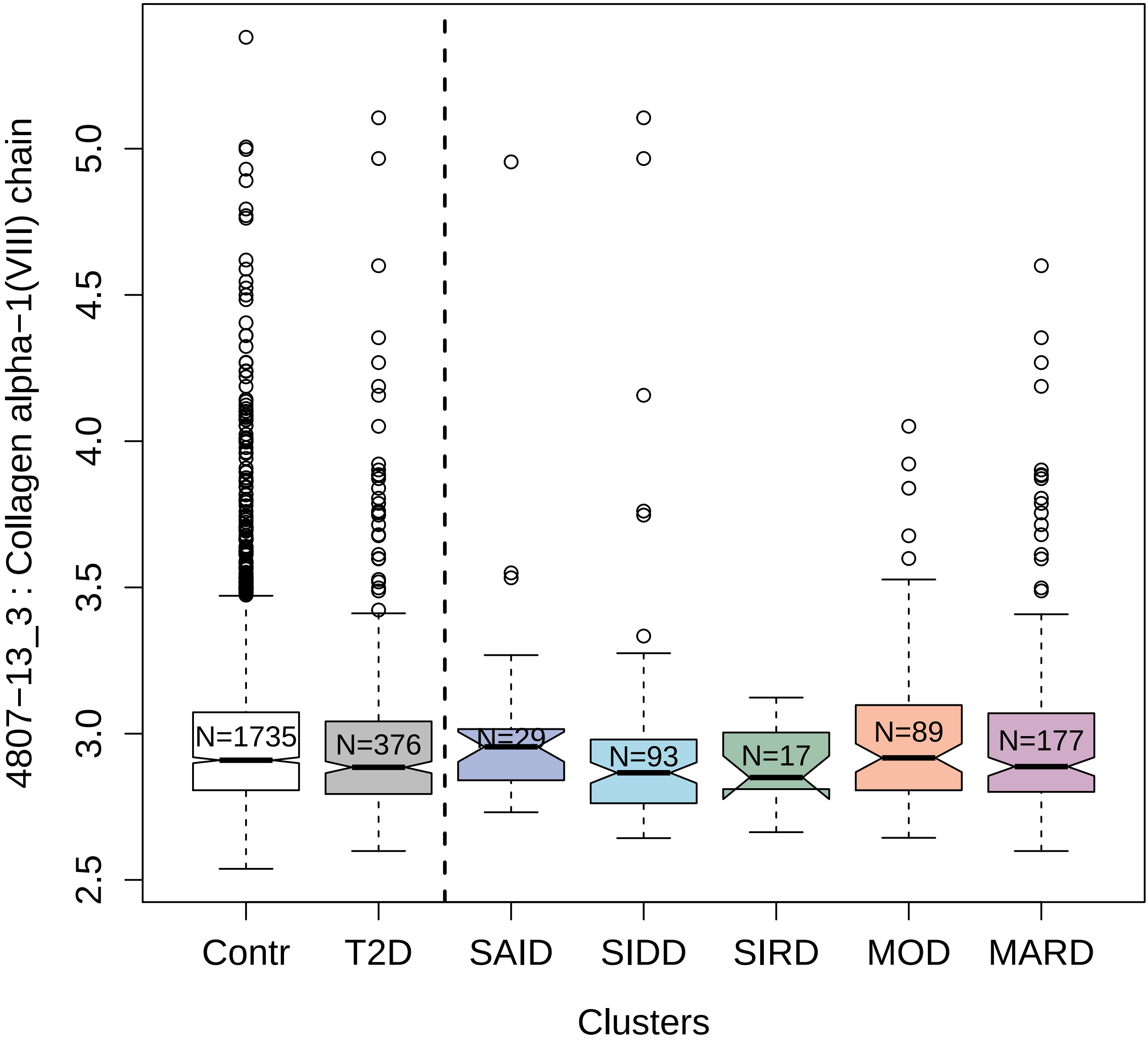

4807−13_3 : Collagen alpha−1(VIII) chain
2.5
3.0
3.5
4.0
4.5
5.0
N=1735
N=376
N=29
N=93
N=17
N=89
N=177
Contr
T2D
SAID
SIDD
SIRD
MOD
MARD
Clusters

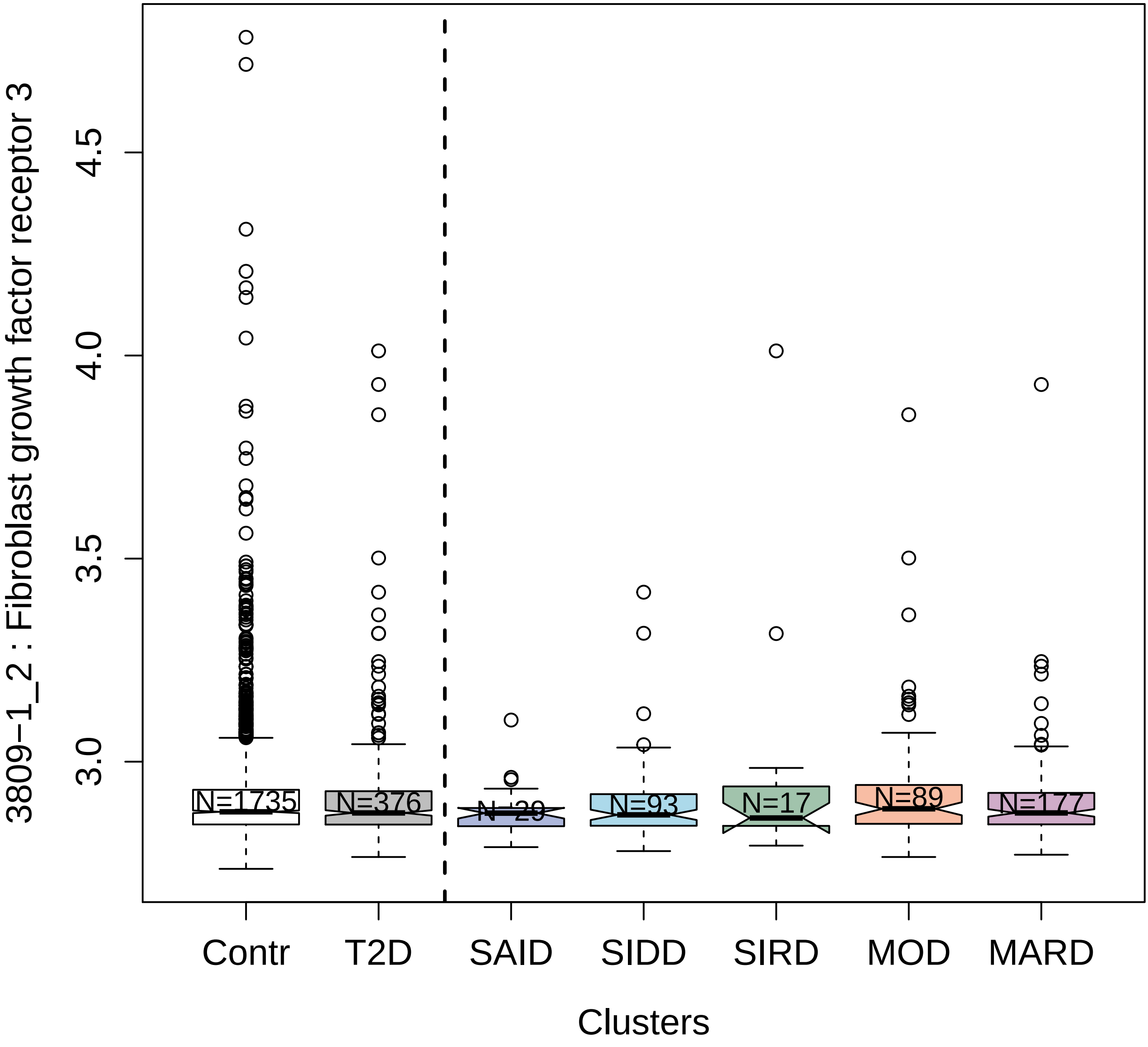
3809−1_2 : Fibroblast growth factor receptor 3
4.5
4.0
3.5
3.0
N=1735
N=376
N=29
N=93
N=17
N=89
N=177
Contr
T2D
SAID
SIDD
SIRD
MOD
MARD
Clusters

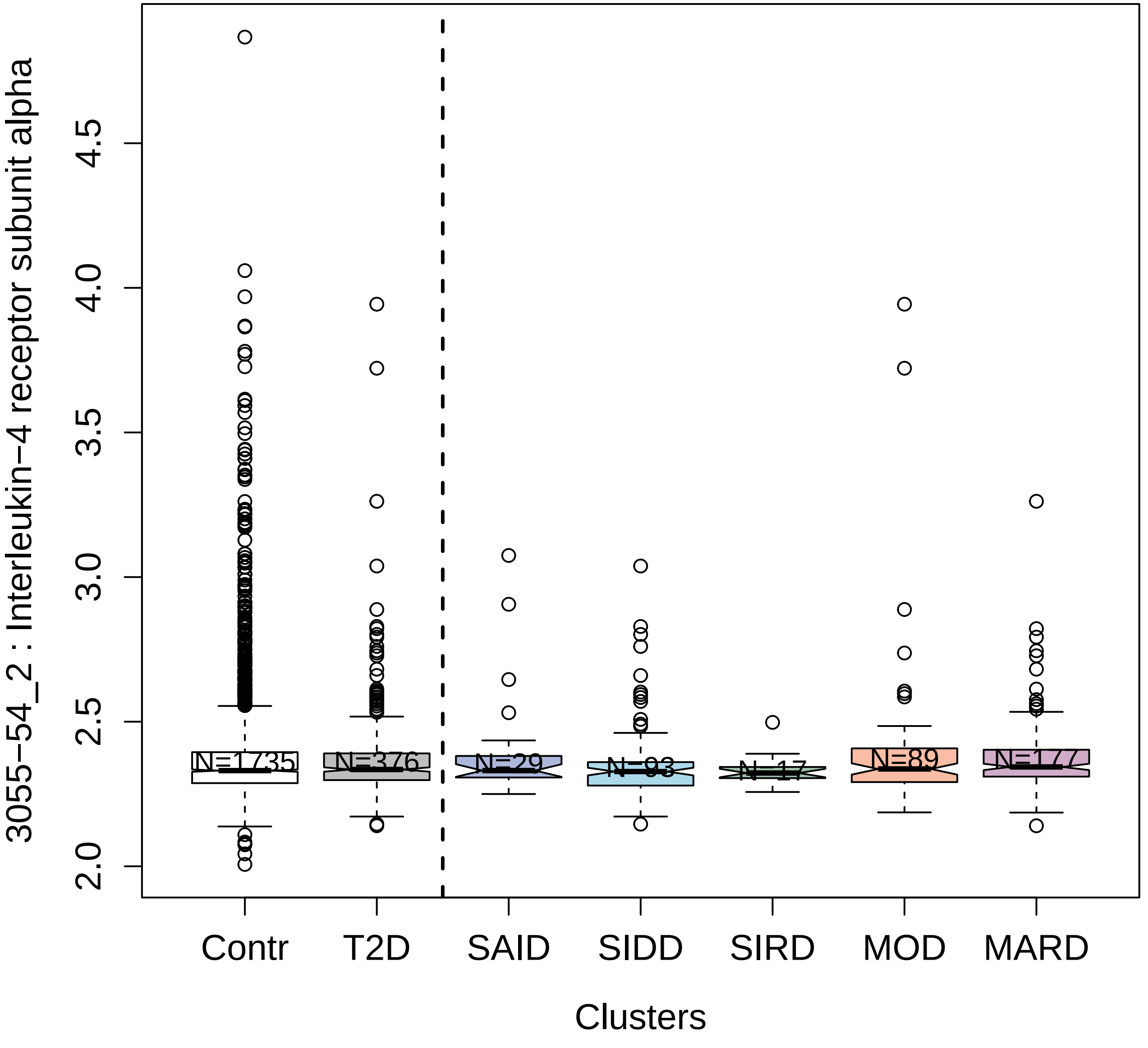

3055−54_2 : Interleukin−4 receptor subunit alpha
2.0
2.5
3.0
3.5
4.0
4.5
N=1735
N=376
N=29
N=89
N=177
Contr
T2D
SAID
SIDD
SIRD
MOD
MARD
Clusters

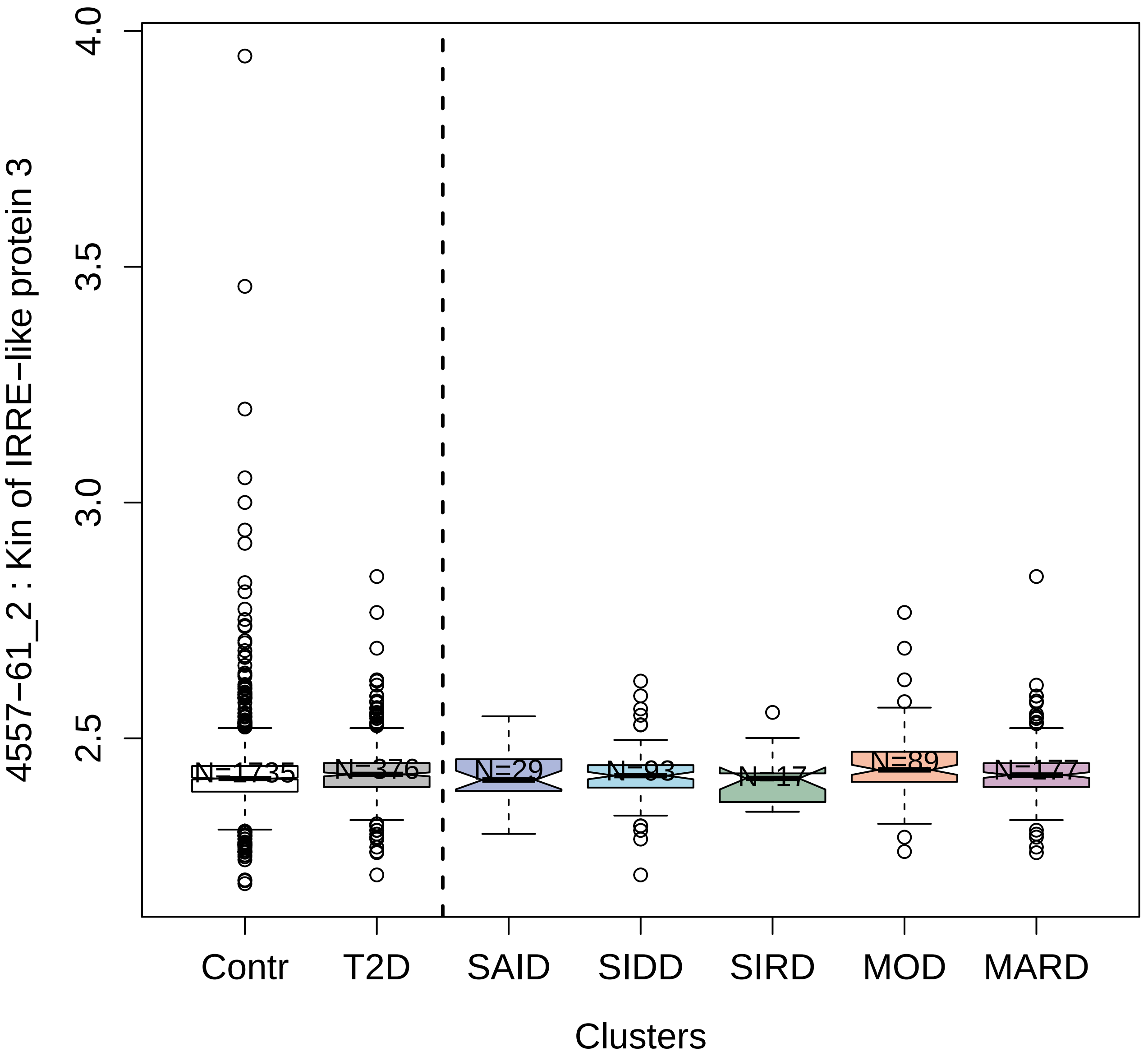
4557−61_2 : Kin of IRRE−like protein 3
4.0
3.5
3.0
2.5
N=1735
N=376
N=29
N=93
N=17
N=89
N=177
Contr
T2D
SAID
SIDD
SIRD
MOD
MARD
Clusters

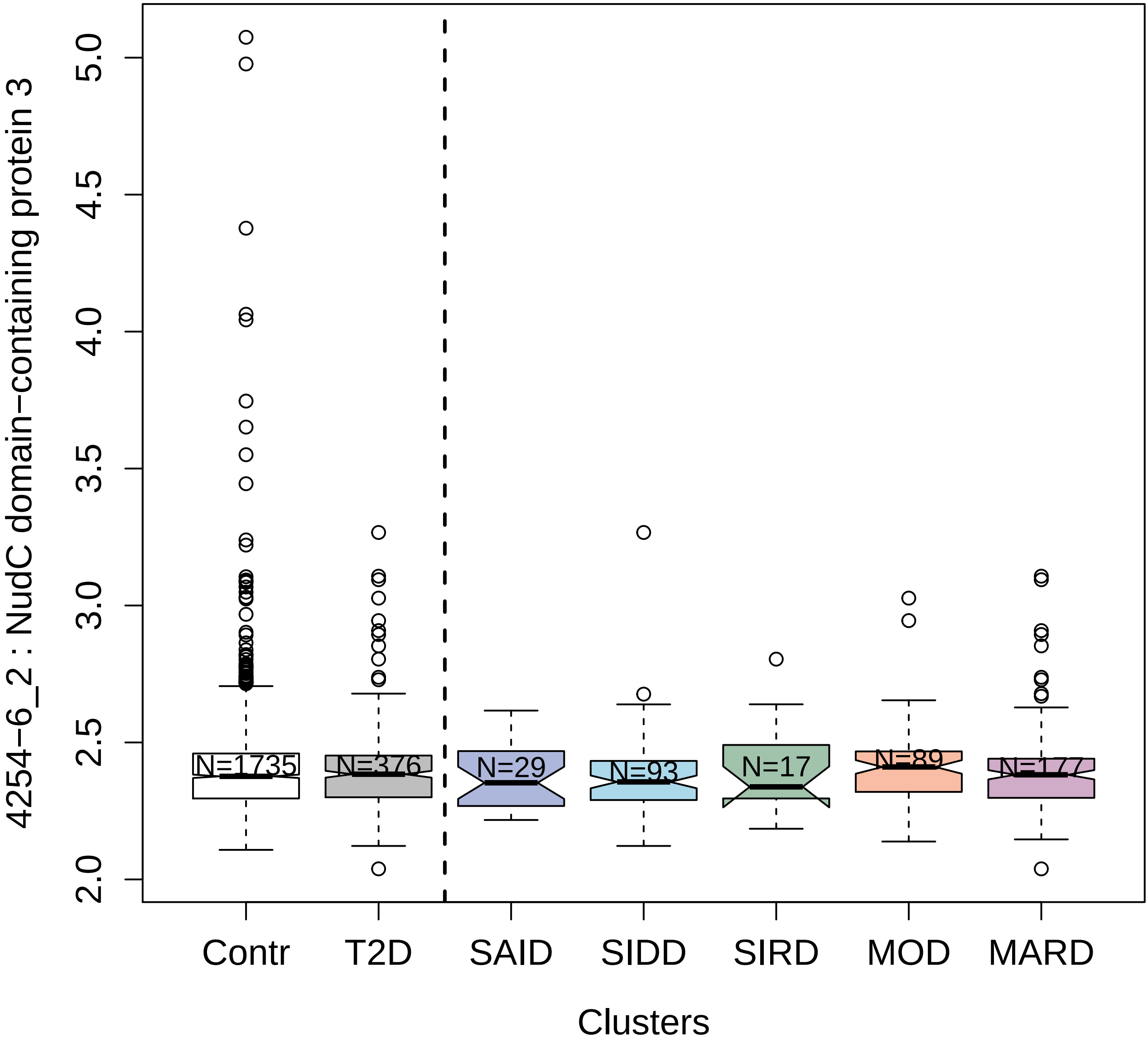

4254−6_2 : NudC domain−containing protein 3
5.0
4.5
4.0
3.5
3.0
2.5
2.0
N=1735
N=376
N=29
N=93
N=17
N=89
N=177
Contr
T2D
SAID
SIDD
SIRD
MOD
MARD
Clusters

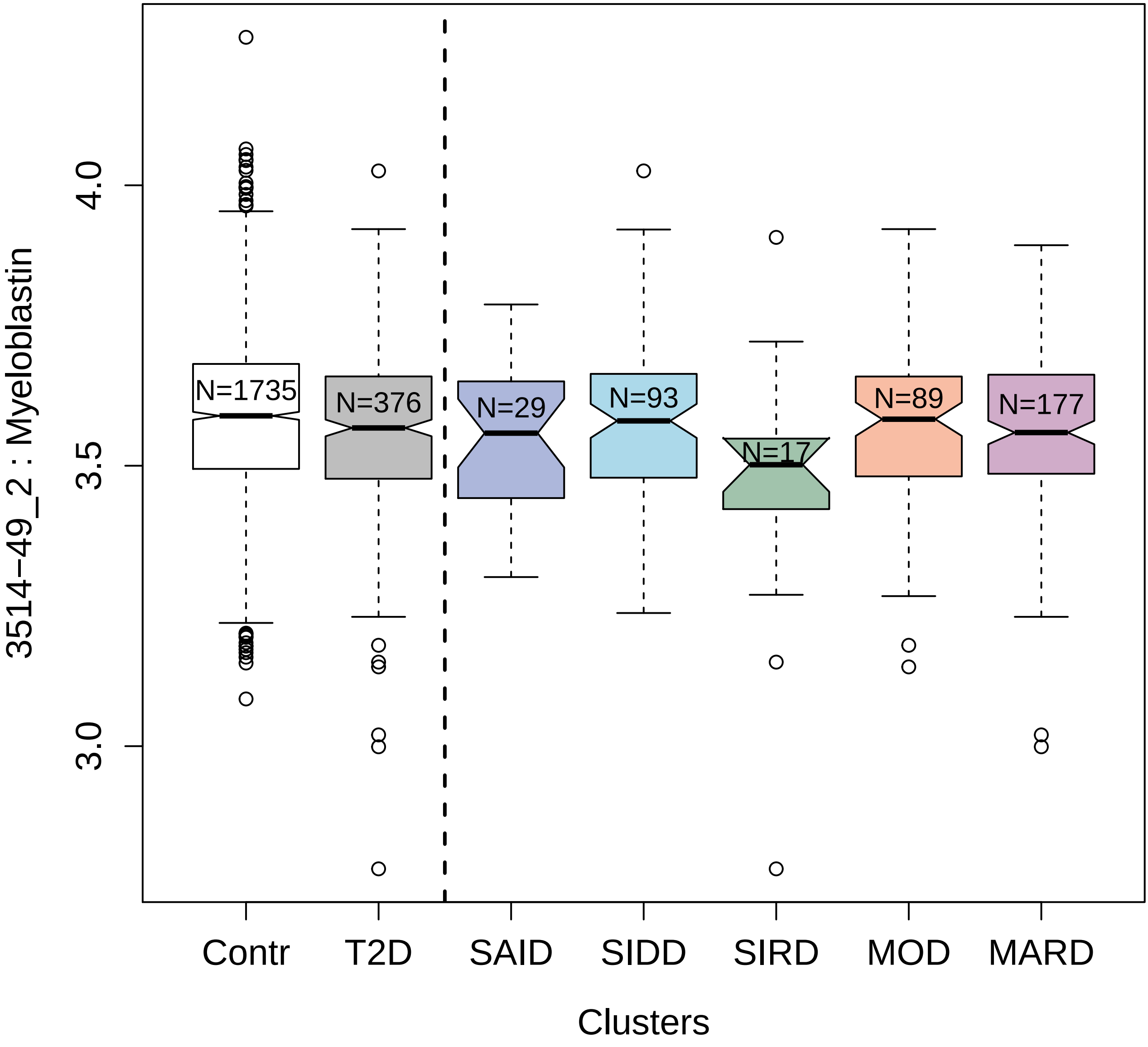

3514−49_2 : Myeloblastin
4.0
3.5
3.0
N=1735
N=376
N=29
N=93
N=17
N=89
N=177
Contr
T2D
SAID
SIDD
SIRD
MOD
MARD
Clusters

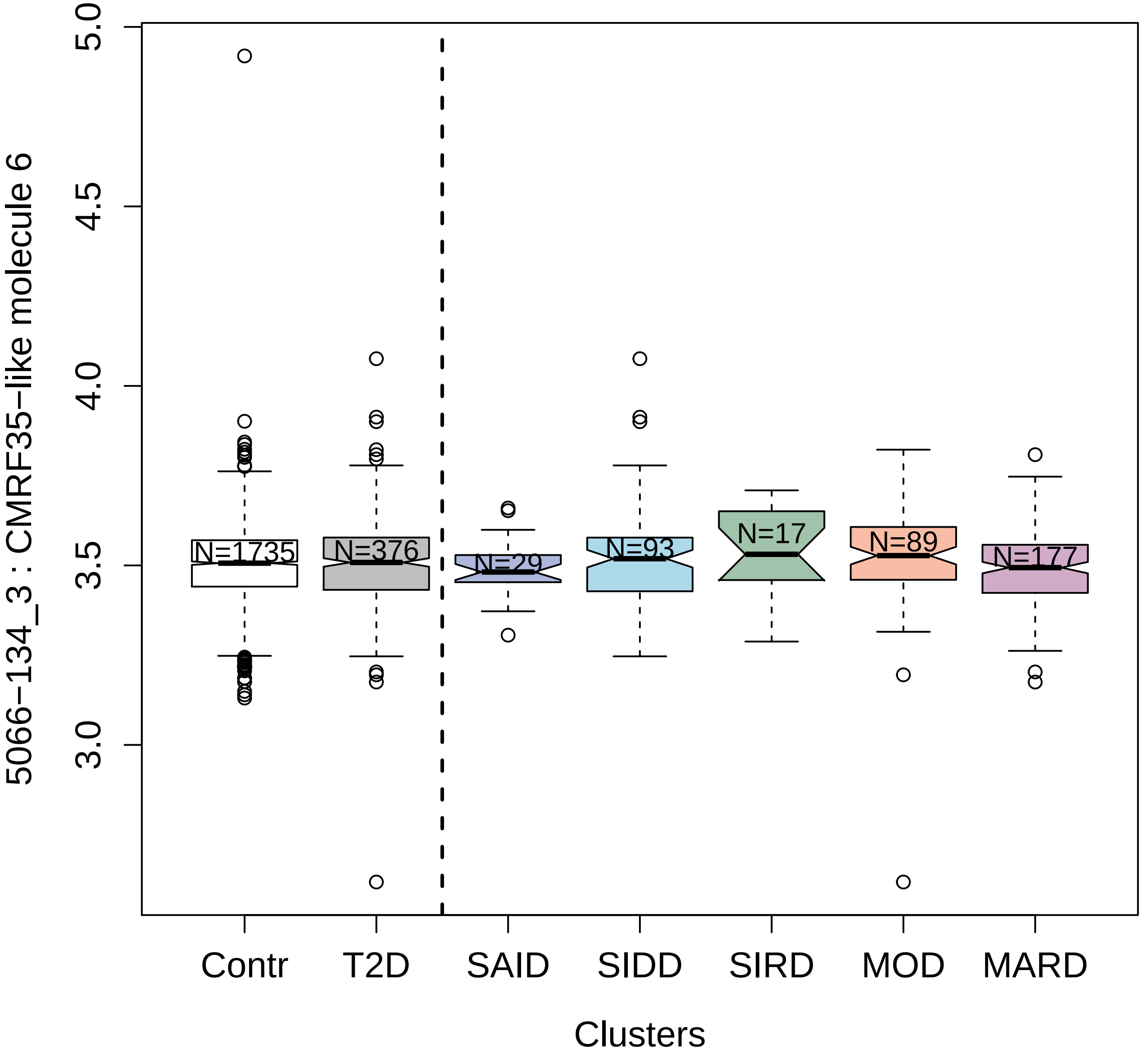

5066−134_3 : CMRF35−like molecule 6
5.0
4.5
4.0
3.5
3.0
N=1735
N=376
N=29
N=93
N=17
N=89
N=177
Contr
T2D
SAID
SIDD
SIRD
MOD
MARD
Clusters

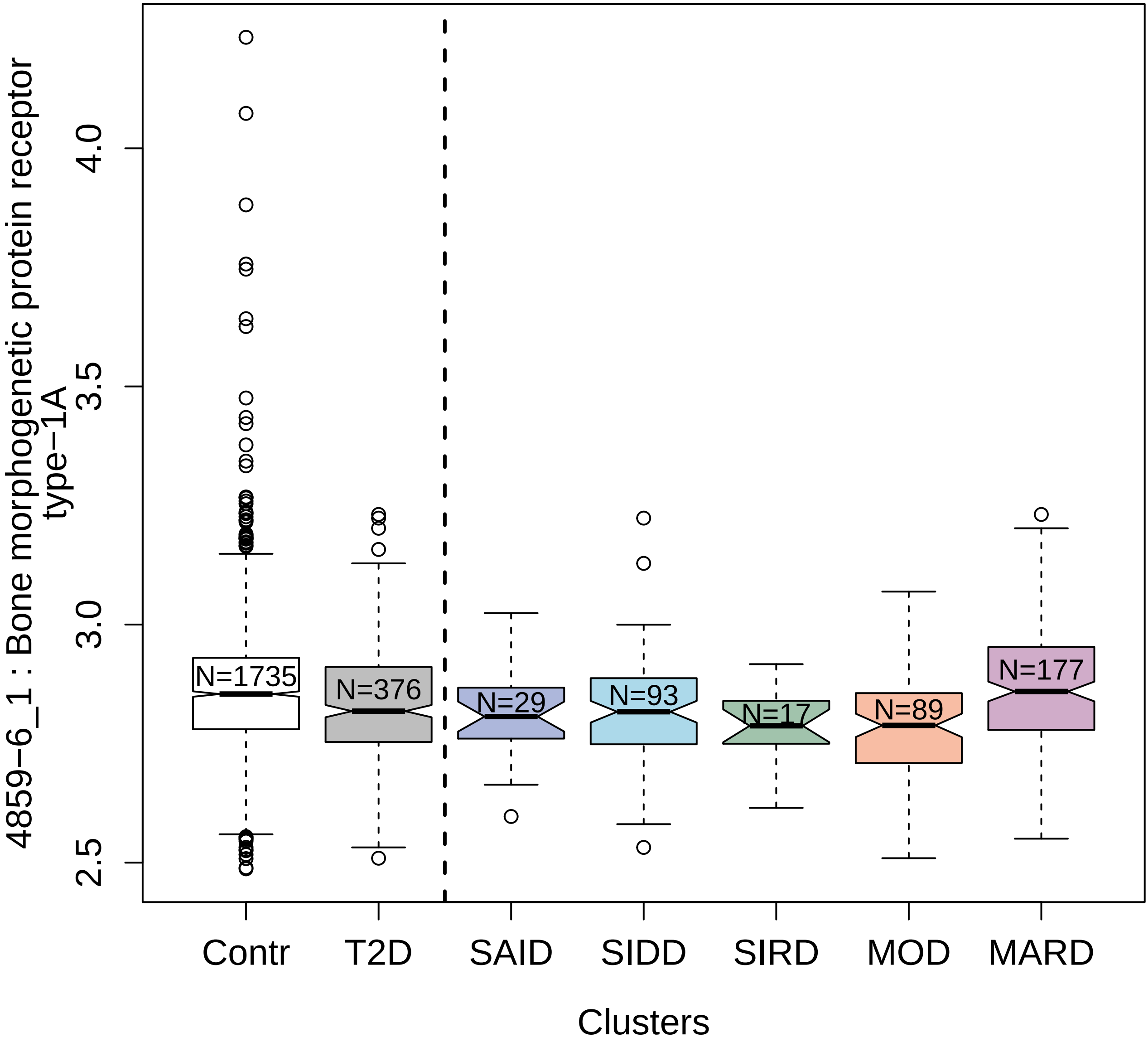

4859−6_1 : Bone morphogenetic protein receptor type−1A
4.0
3.5
3.0
2.5
N=1735
N=376
N=29
N=93
N=17
N=89
N=177
Contr
T2D
SAID
SIDD
SIRD
MOD
MARD
Clusters

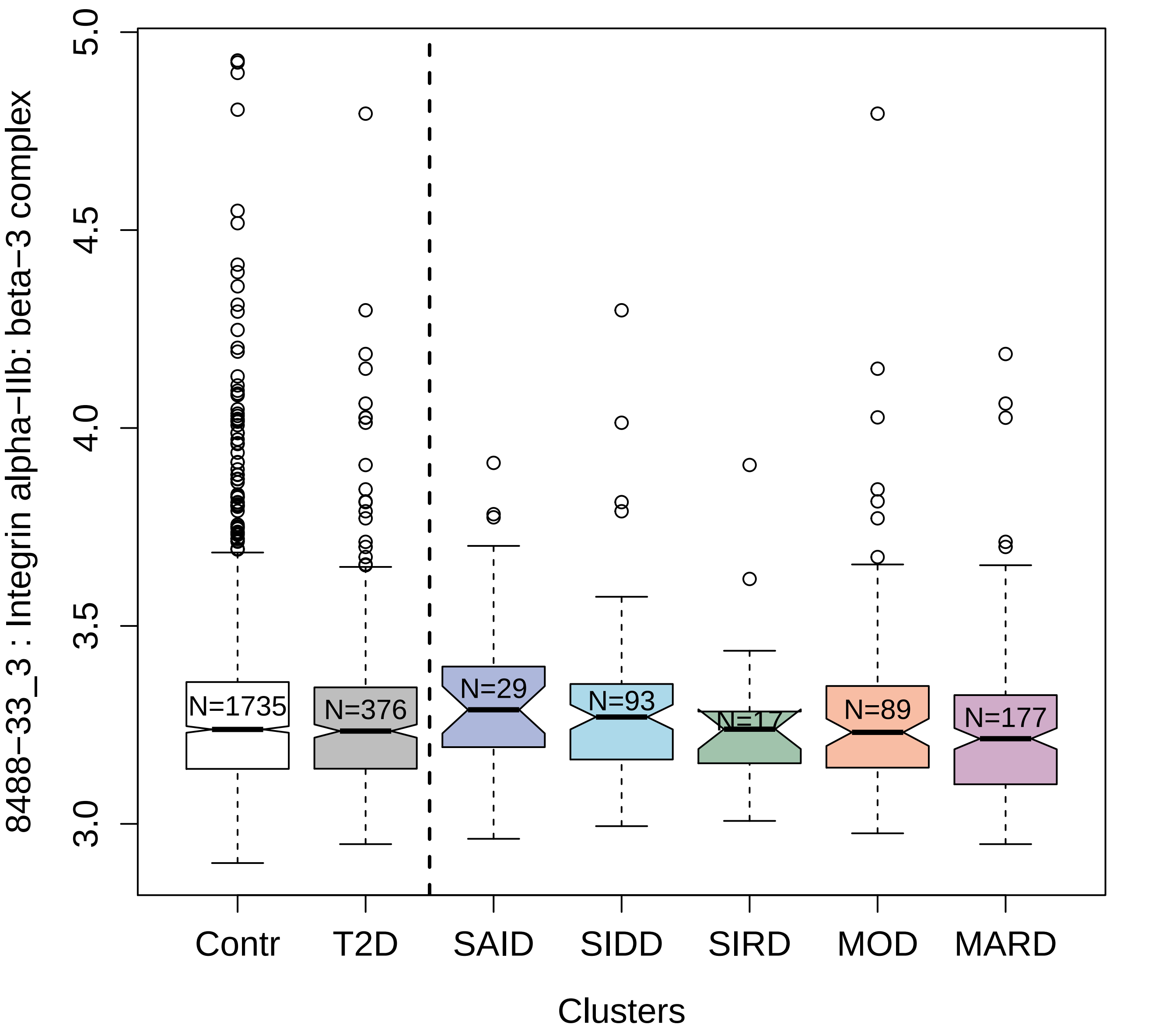
8488−33_3 : Integrin alpha−IIb: beta−3 complex
5.0
4.5
4.0
3.5
3.0
N=1735
N=376
N=29
N=93
N=17
N=89
N=177
Contr
T2D
SAID
SIDD
SIRD
MOD
MARD
Clusters

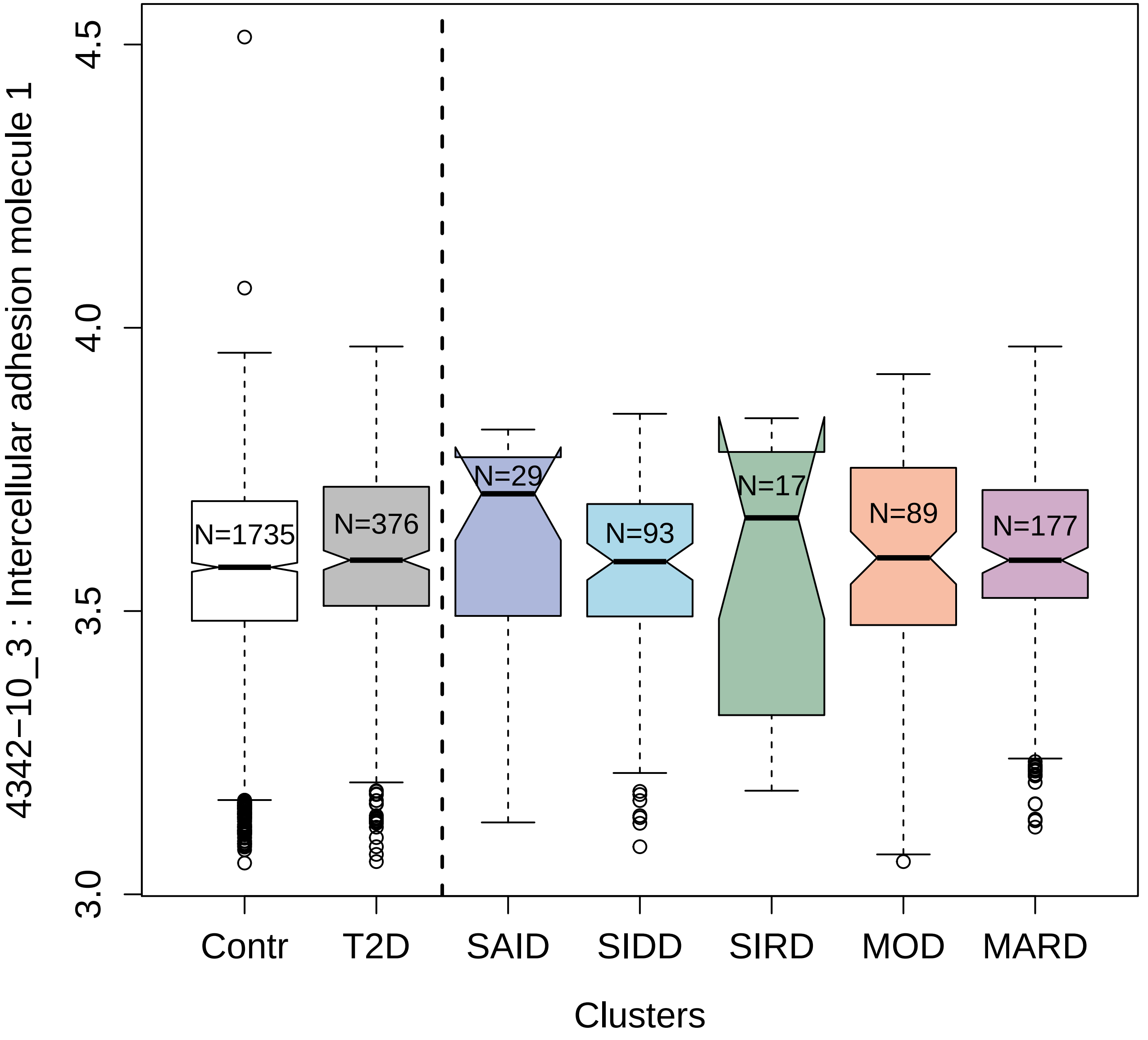

4342−10_3 : Intercellular adhesion molecule 1
3.0
3.5
4.0
4.5
N=1735
N=376
N=29
N=93
N=17
N=89
N=177
Contr
T2D
SAID
SIDD
SIRD
MOD
MARD
Clusters

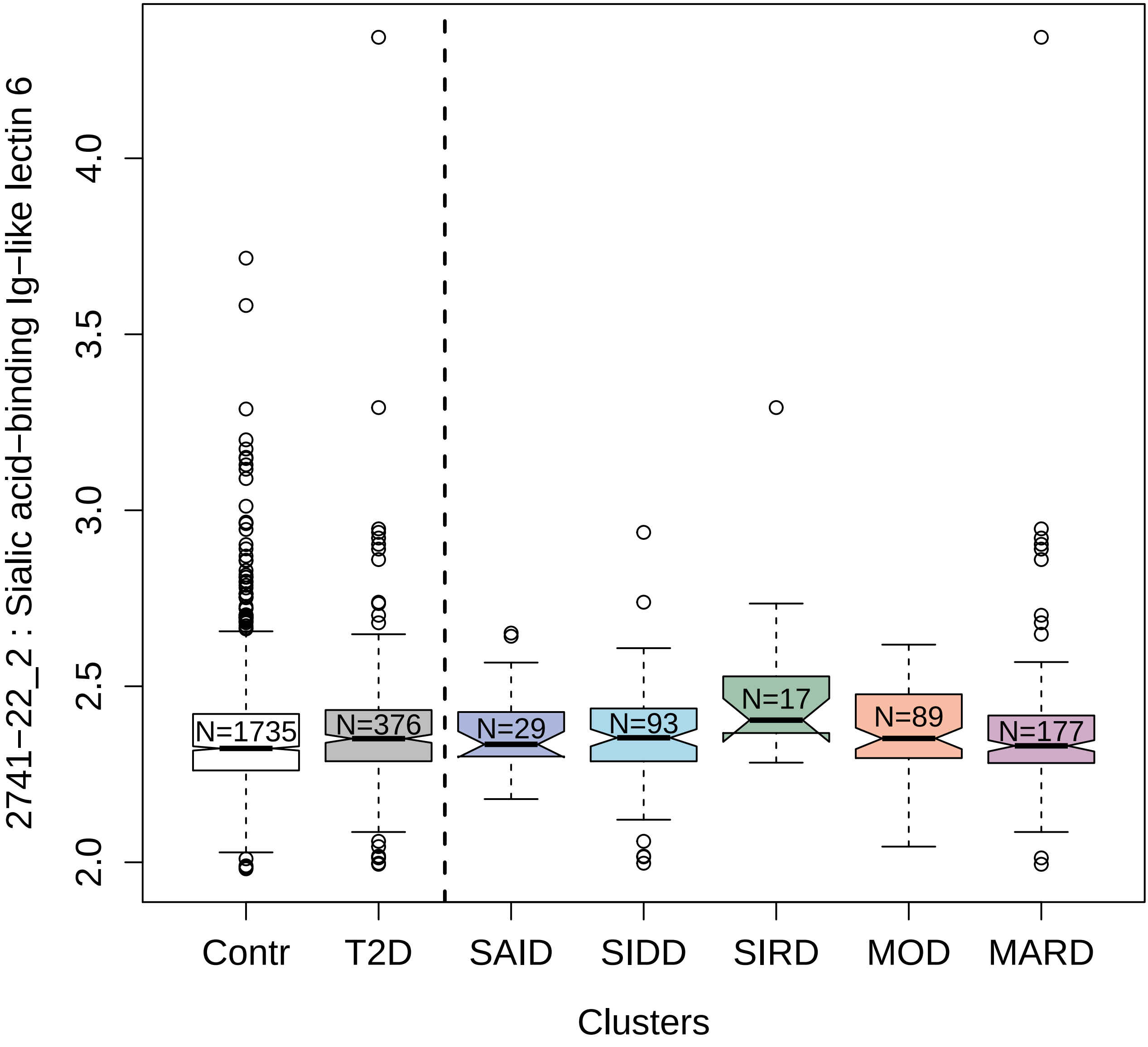
2741−22_2 : Sialic acid−binding Ig−like lectin 6
4.0
3.5
3.0
2.5
2.0
N=1735
N=376
N=29
N=93
N=17
N=89
N=177
Contr
T2D
SAID
SIDD
SIRD
MOD
MARD
Clusters

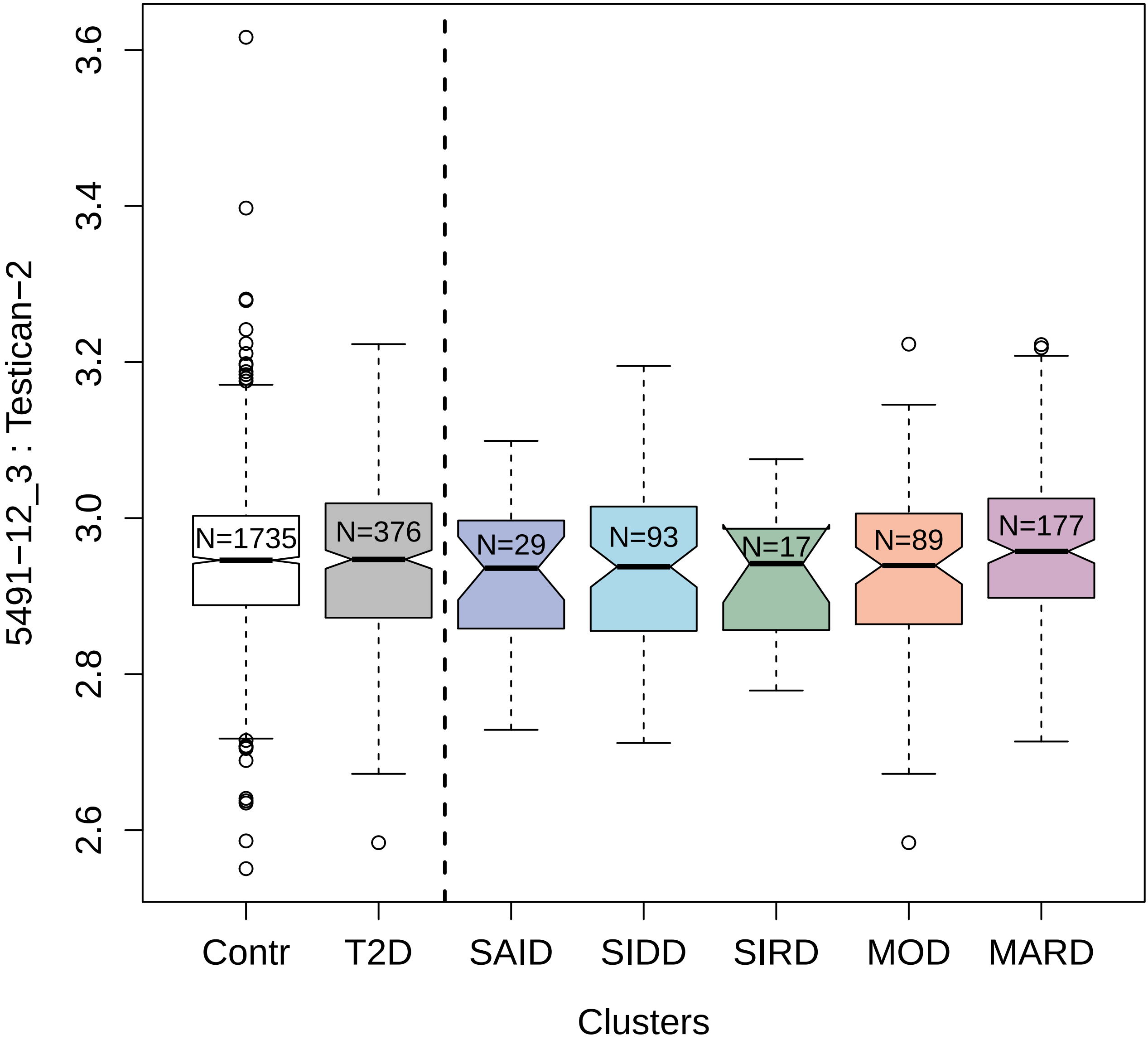

5491−12_3 : Testican−2
3.6
3.4
3.2
3.0
2.8
2.6
N=1735
N=376
N=29
N=93
N=17
N=89
N=177
Contr
T2D
SAID
SIDD
SIRD
MOD
MARD
Clusters

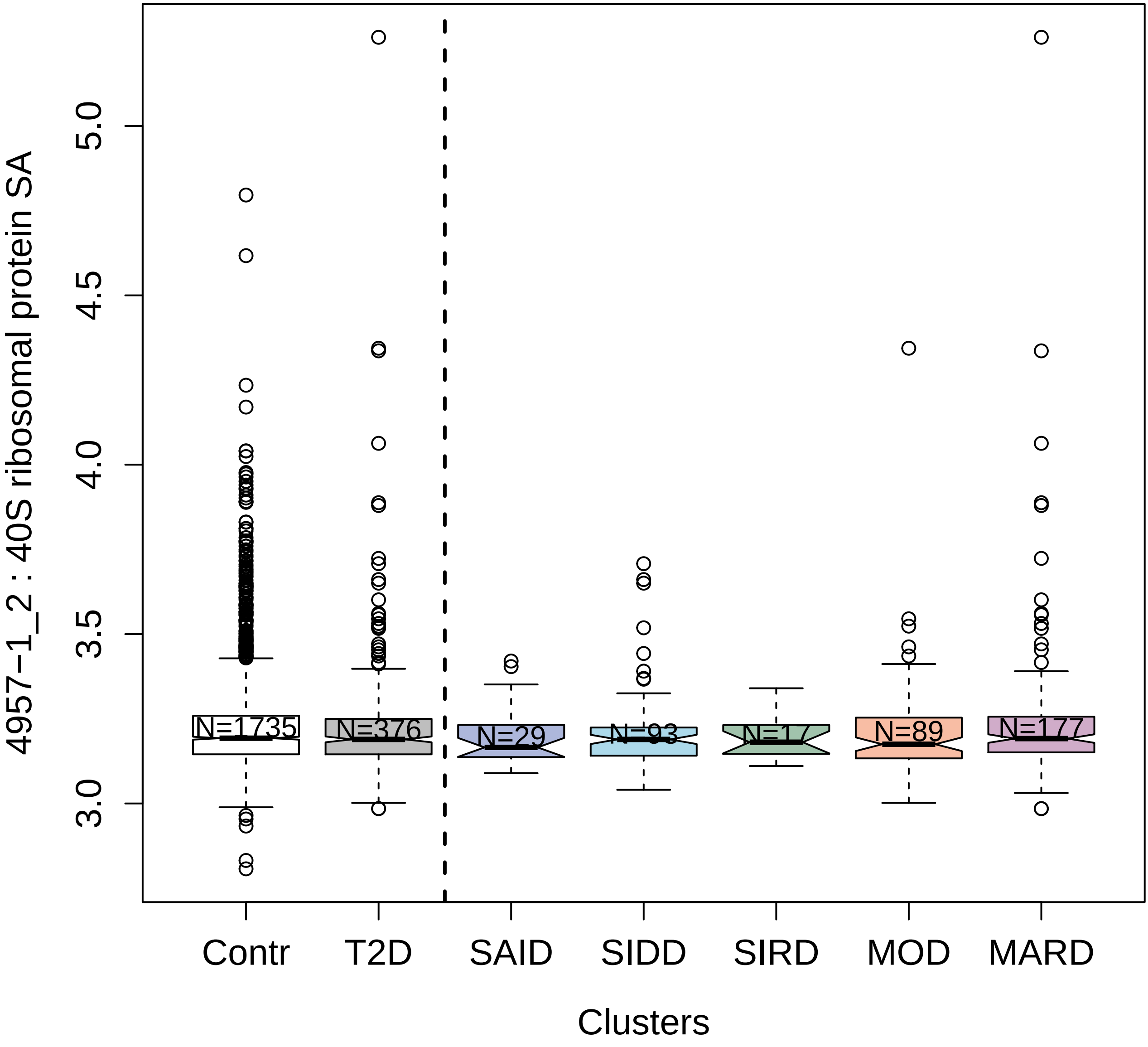

4957−1_2 : 40S ribosomal protein SA
3.0
3.5
4.0
4.5
5.0
N=1735
N=376
N=29
N=93
N=17
N=89
N=177
Contr
T2D
SAID
SIDD
SIRD
MOD
MARD
Clusters

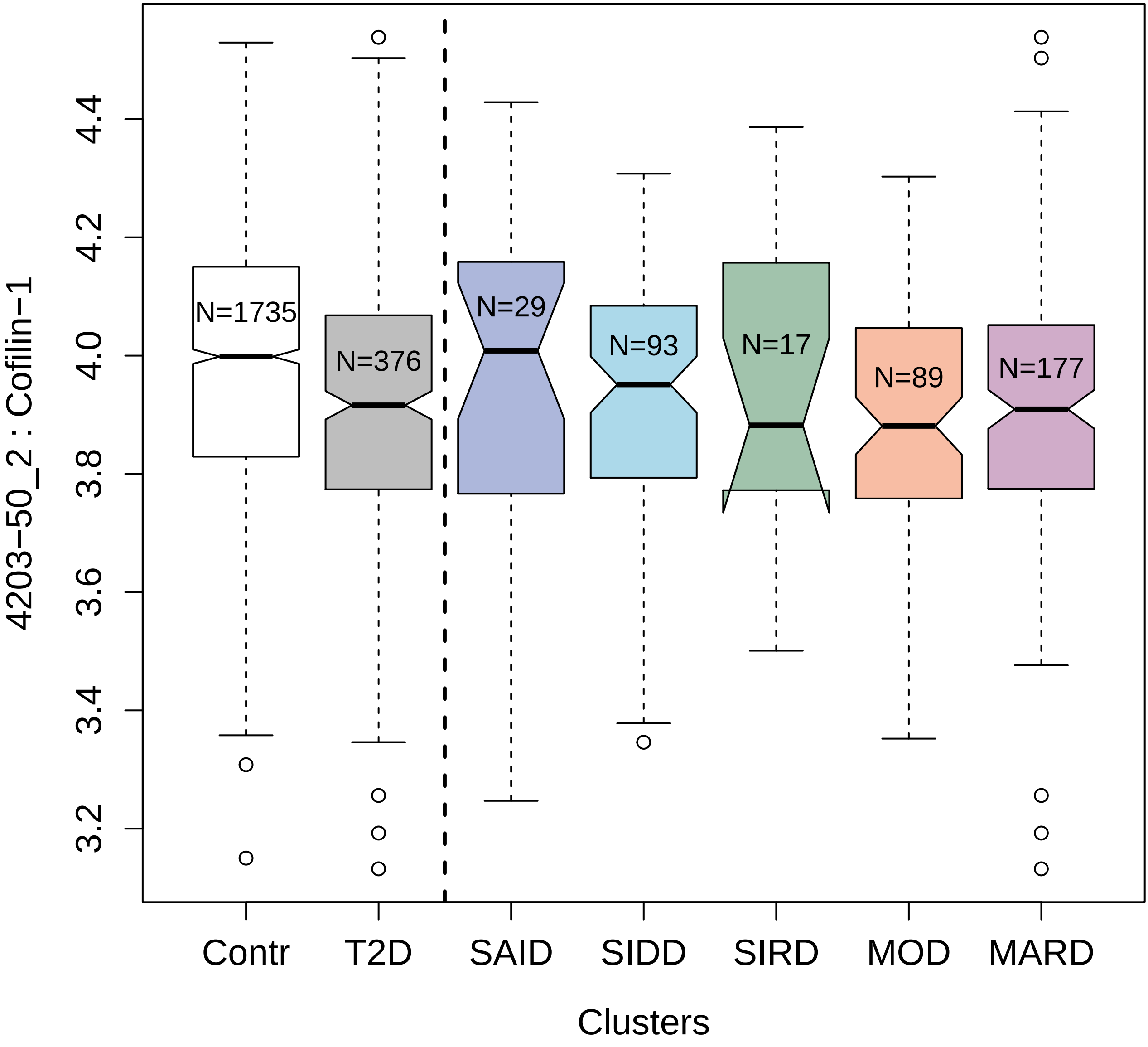
4203−50_2 : Cofilin−1
3.2
3.4
3.6
3.8
4.0
4.2
4.4
N=1735
N=376
N=29
N=93
N=17
N=89
N=177
Contr
T2D
SAID
SIDD
SIRD
MOD
MARD
Clusters

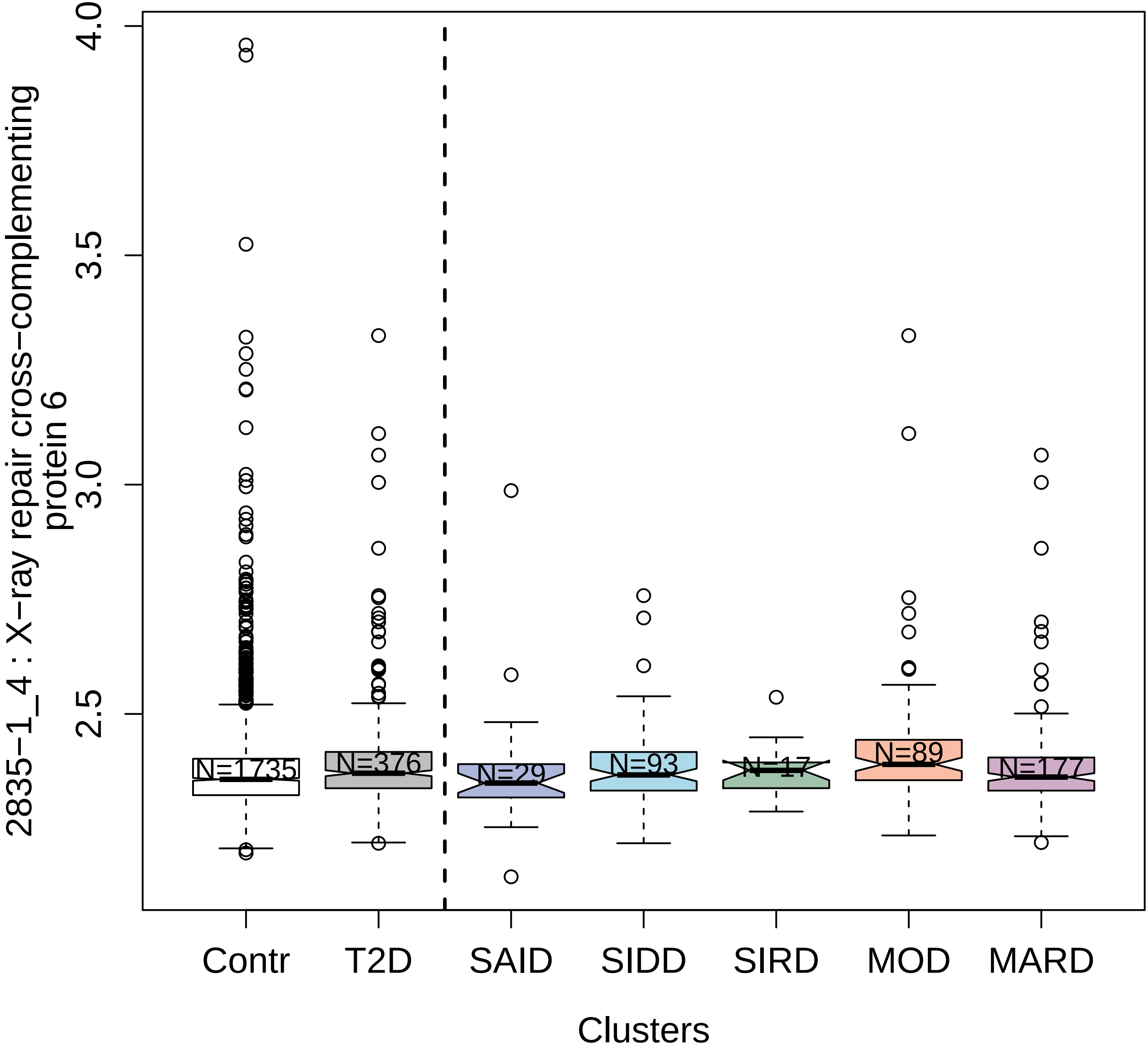

2835−1_4 : X−ray repair cross−complementing
protein 6
4.0
3.5
3.0
2.5
N=1735
N=376
N=29
N=93
N=17
N=89
N=177
Contr
T2D
SAID
SIDD
SIRD
MOD
MARD
Clusters

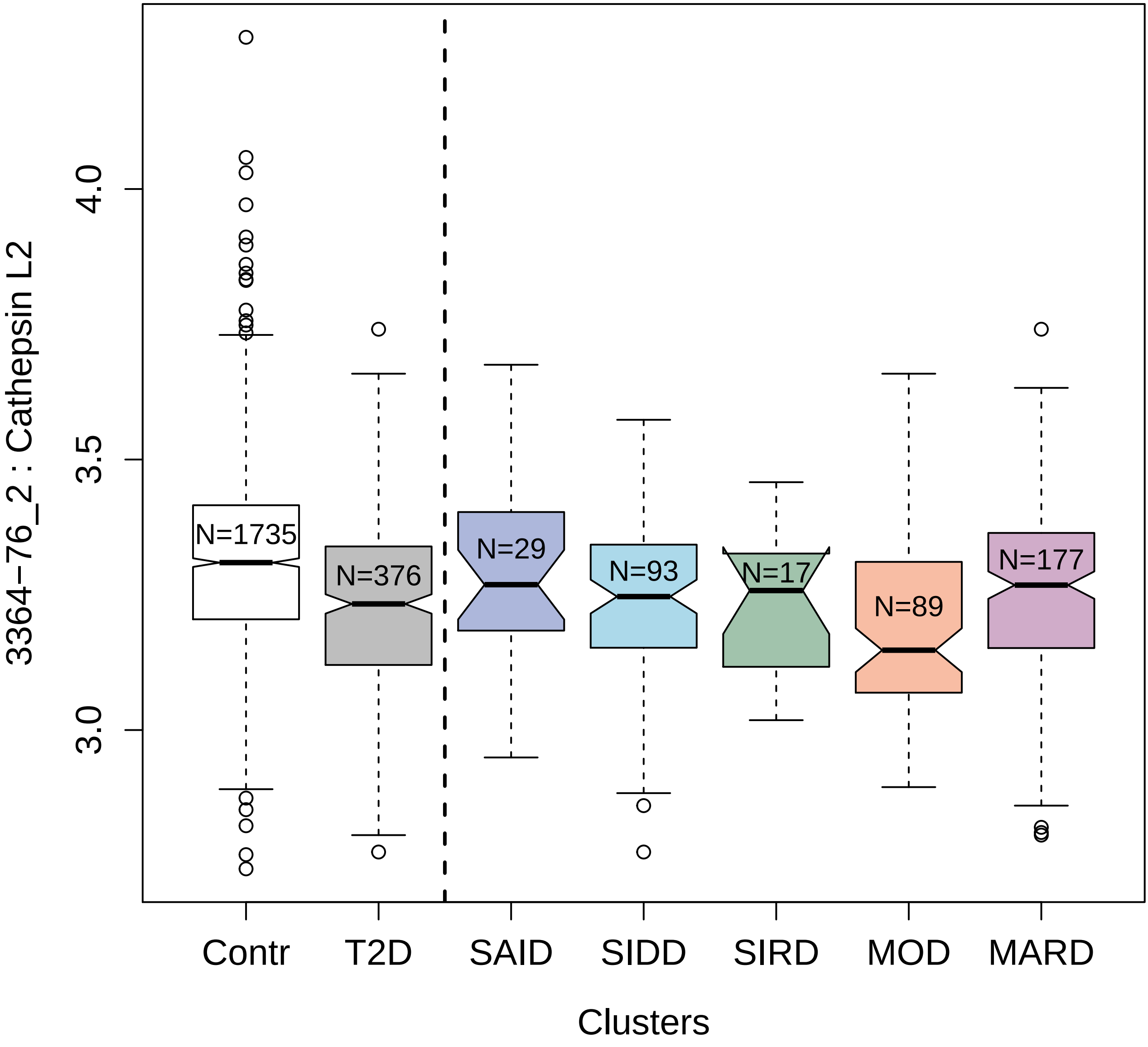

3364−76_2 : Cathepsin L2
4.0
3.5
3.0
N=1735
N=376
N=29
N=93
N=17
N=89
N=177
Contr
T2D
SAID
SIDD
SIRD
MOD
MARD
Clusters

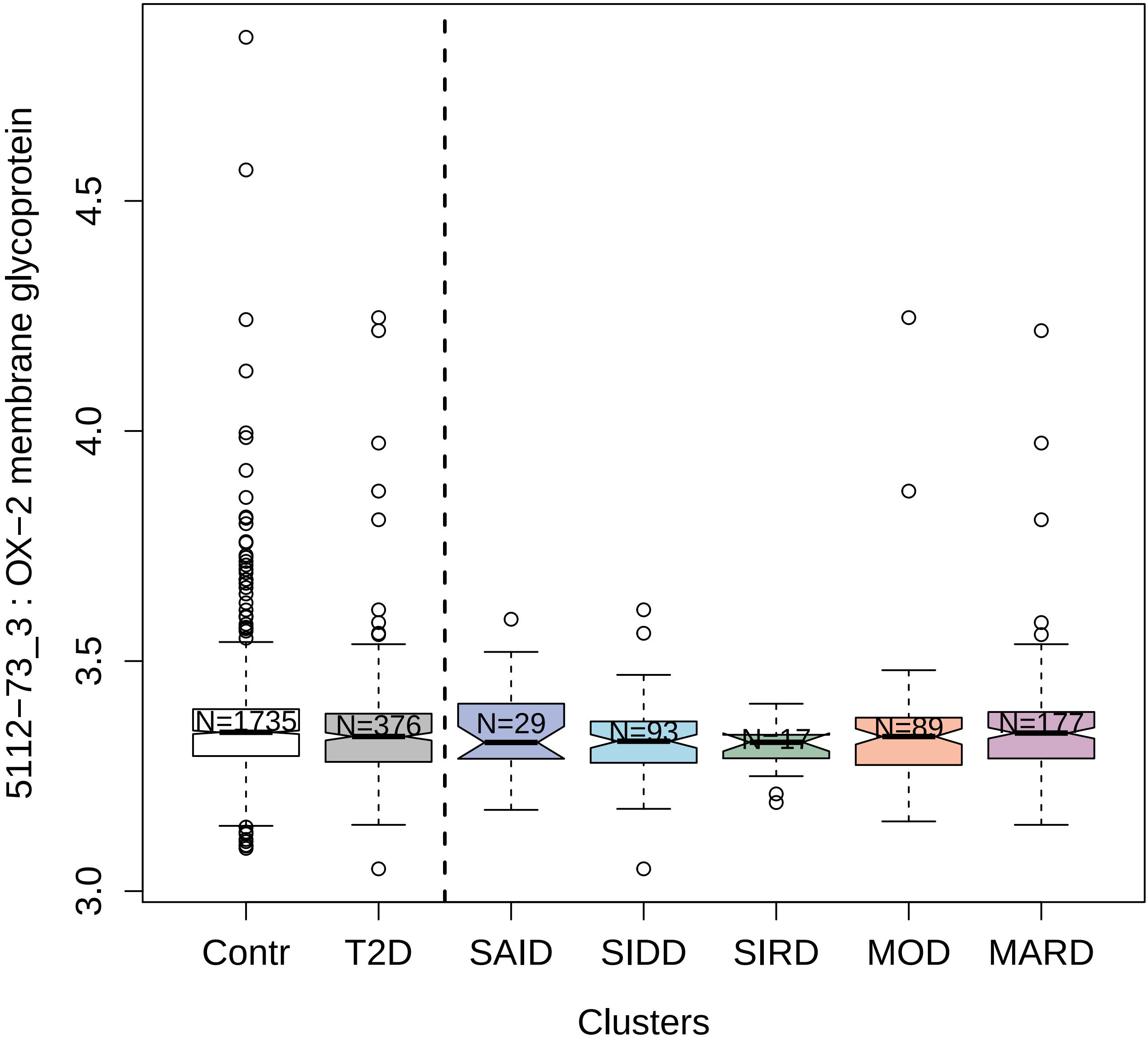
5112−73_3 : OX−2 membrane glycoprotein
4.5
4.0
3.5
3.0
N=1735
N=376
N=29
N=93
N=17
N=89
N=177
Contr
T2D
SAID
SIDD
SIRD
MOD
MARD
Clusters

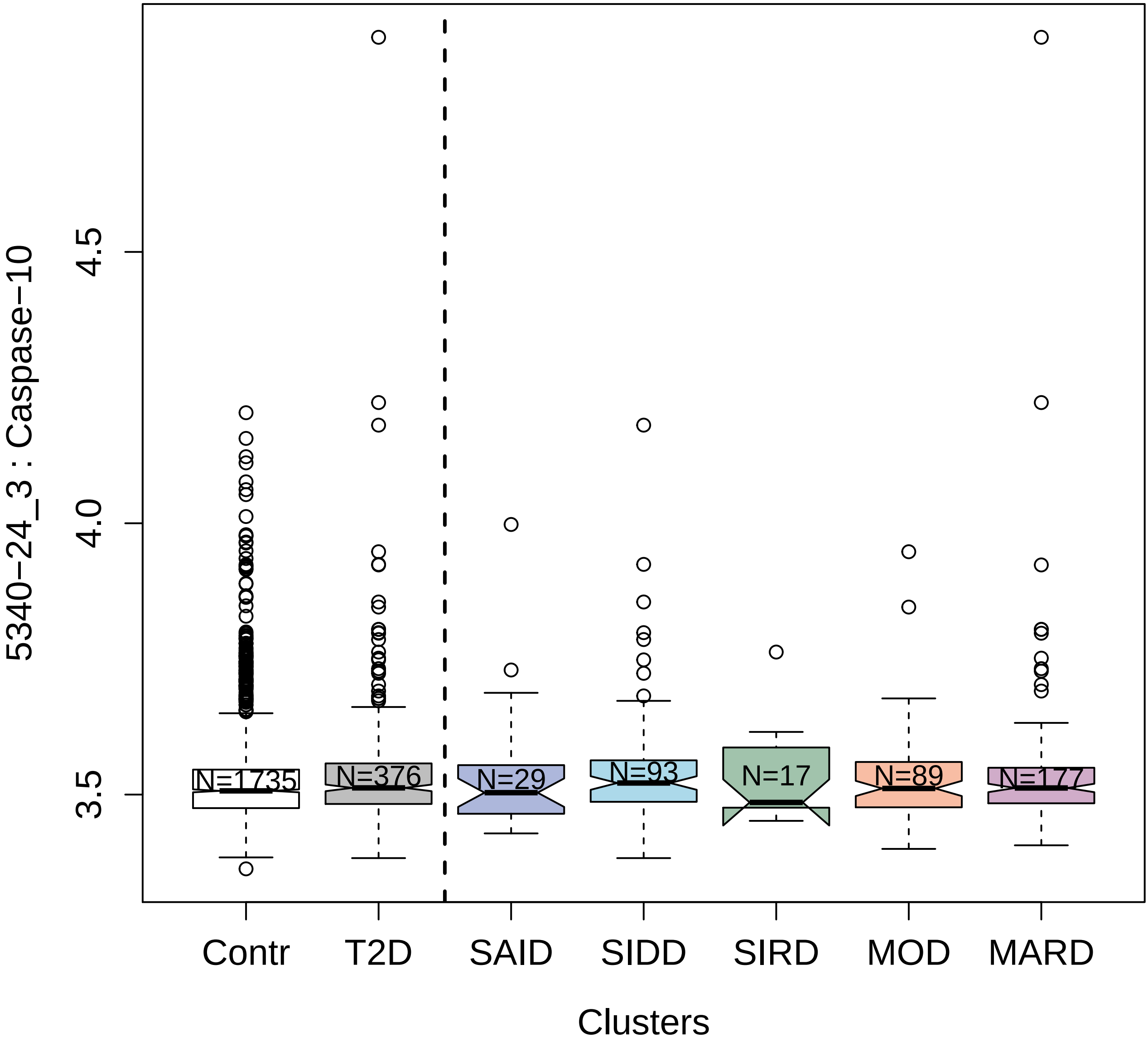

5340−24_3 : Caspase−10
3.5
4.0
4.5
N=1735
N=376
N=29
N=93
N=17
N=89
N=177
Contr
T2D
SAID
SIDD
SIRD
MOD
MARD
Clusters

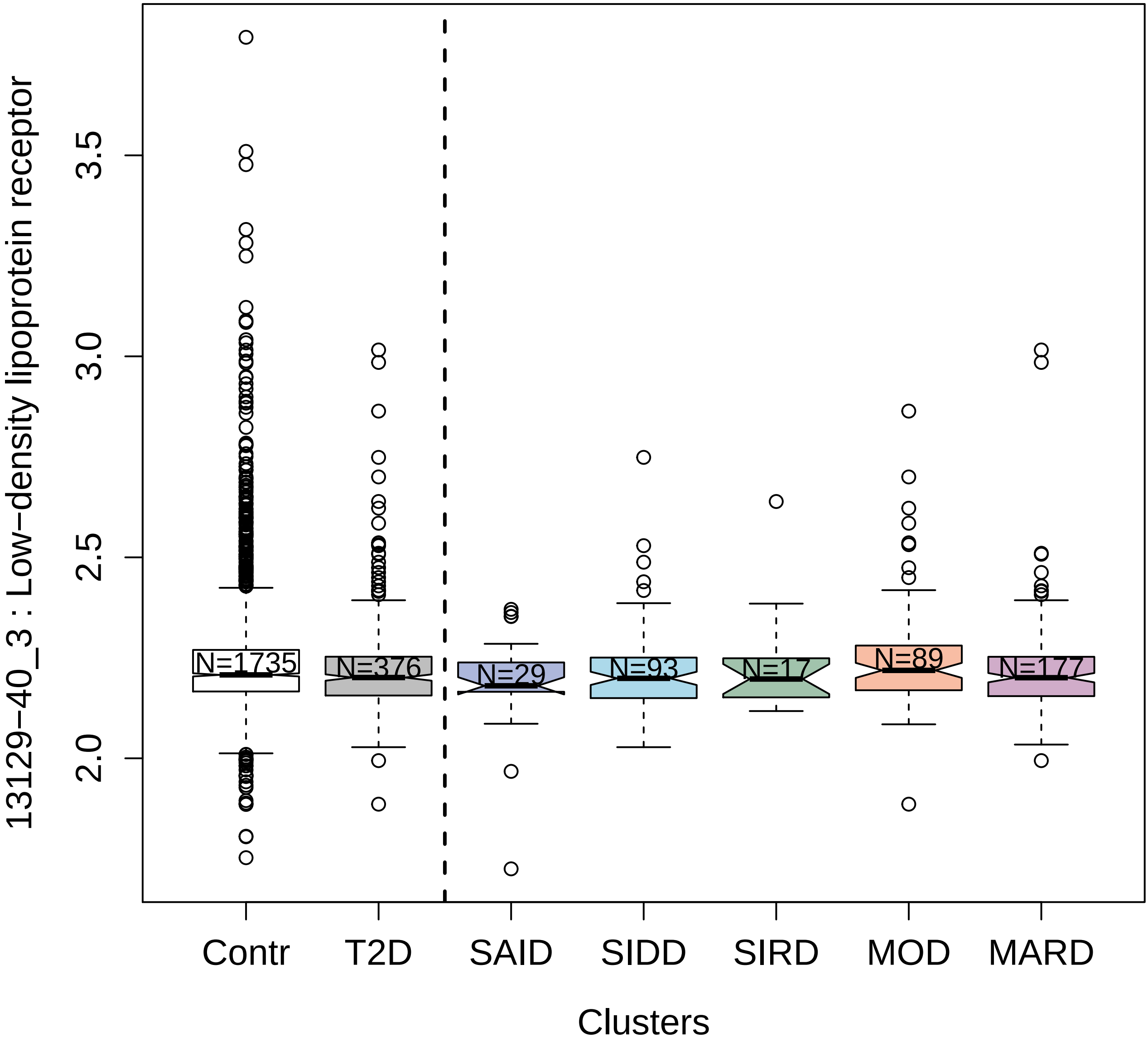
13129−40_3 : Low−density lipoprotein receptor
3.5
3.0
2.5
2.0
N=1735
N=376
N=29
N=93
N=17
N=89
N=177
Contr
T2D
SAID
SIDD
SIRD
MOD
MARD
Clusters

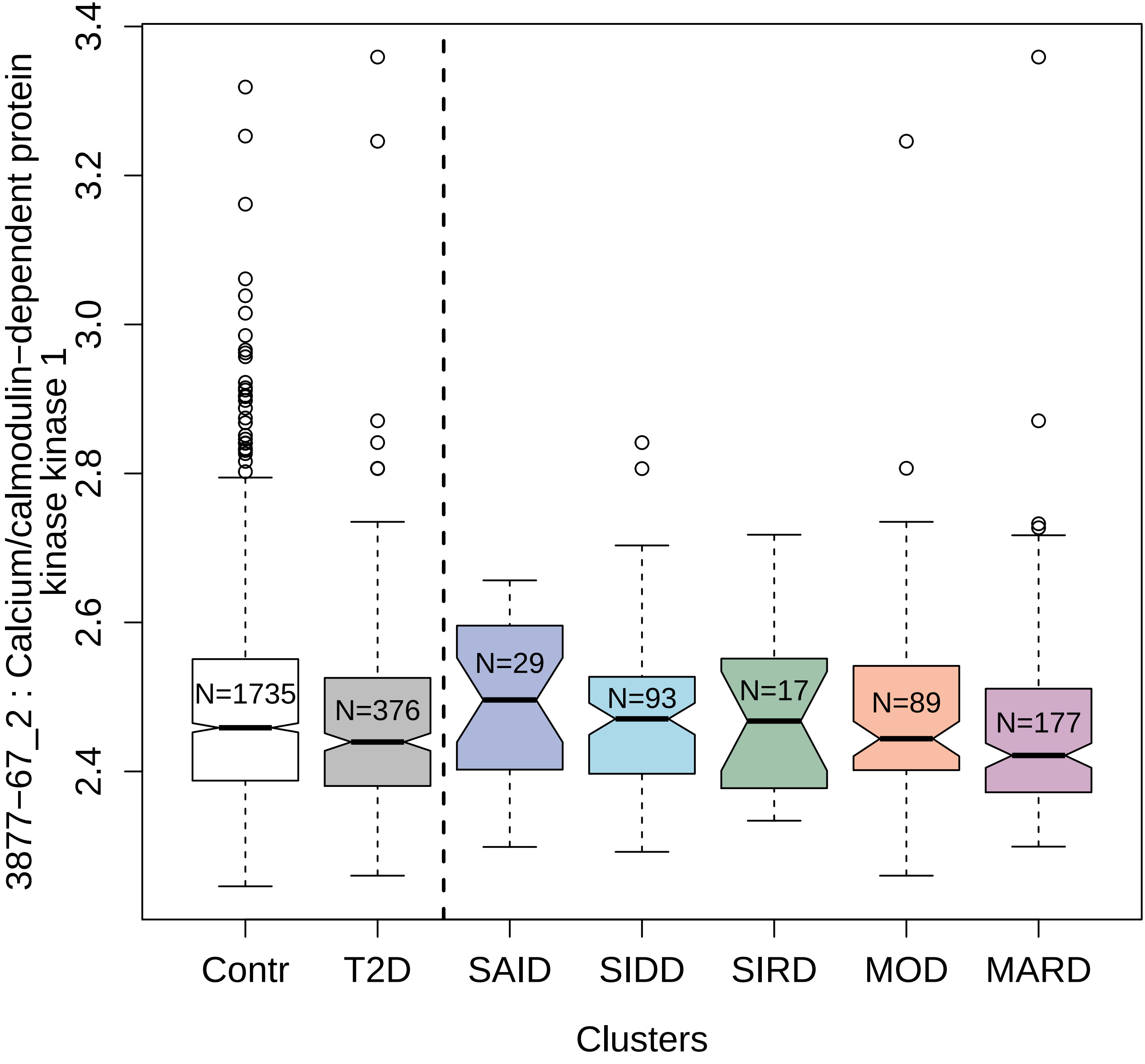

3877−67_2 : Calcium/calmodulin−dependent protein
kinase kinase 1
3.4
3.2
3.0
2.8
2.6
2.4
N=1735
N=376
N=29
N=93
N=17
N=89
N=177
Contr
T2D
SAID
SIDD
SIRD
MOD
MARD
Clusters

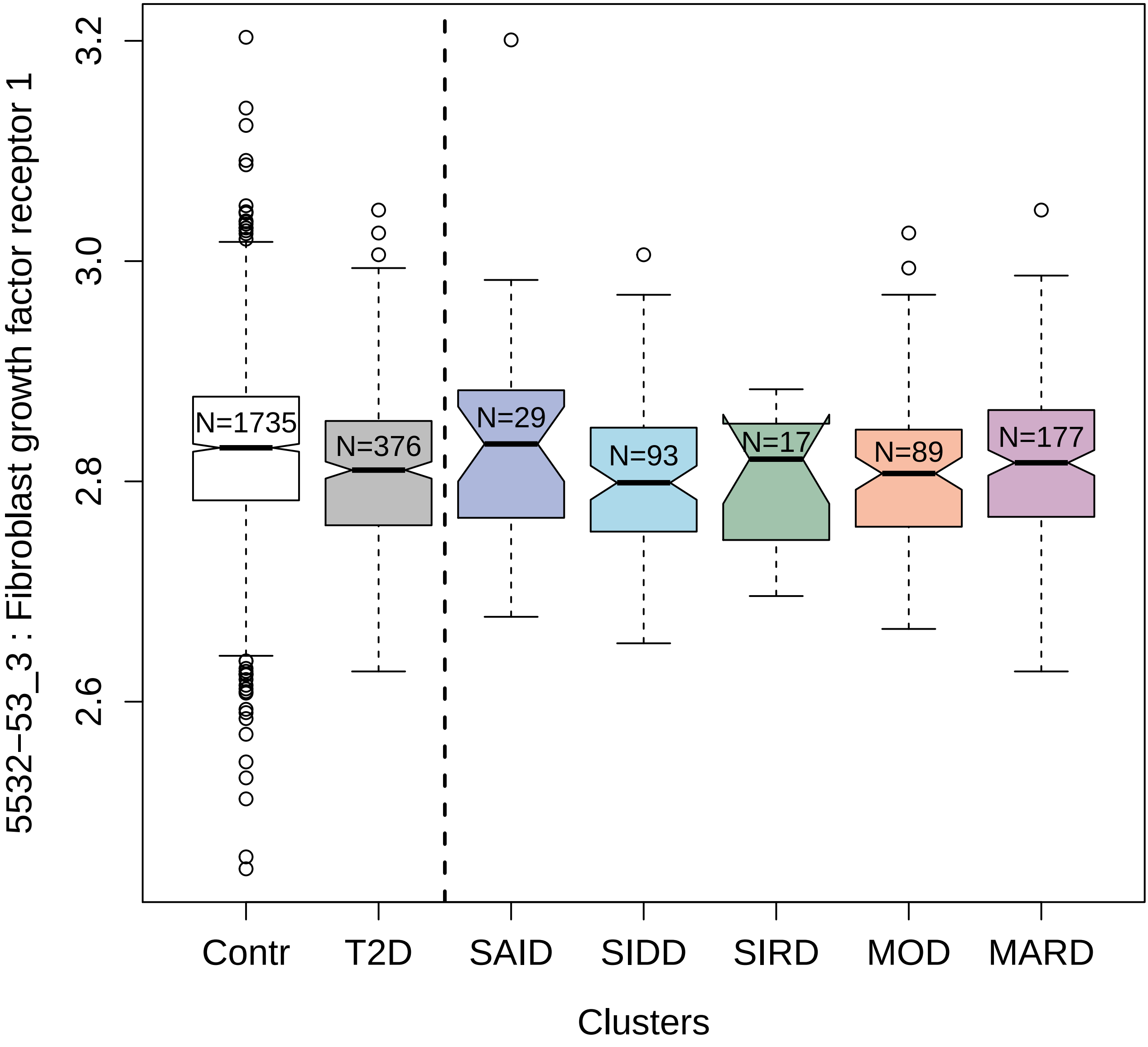
5532−53_3 : Fibroblast growth factor receptor 1
3.2
3.0
2.8
2.6
N=1735
N=376
N=29
N=93
N=17
N=89
N=177
Contr
T2D
SAID
SIDD
SIRD
MOD
MARD
Clusters

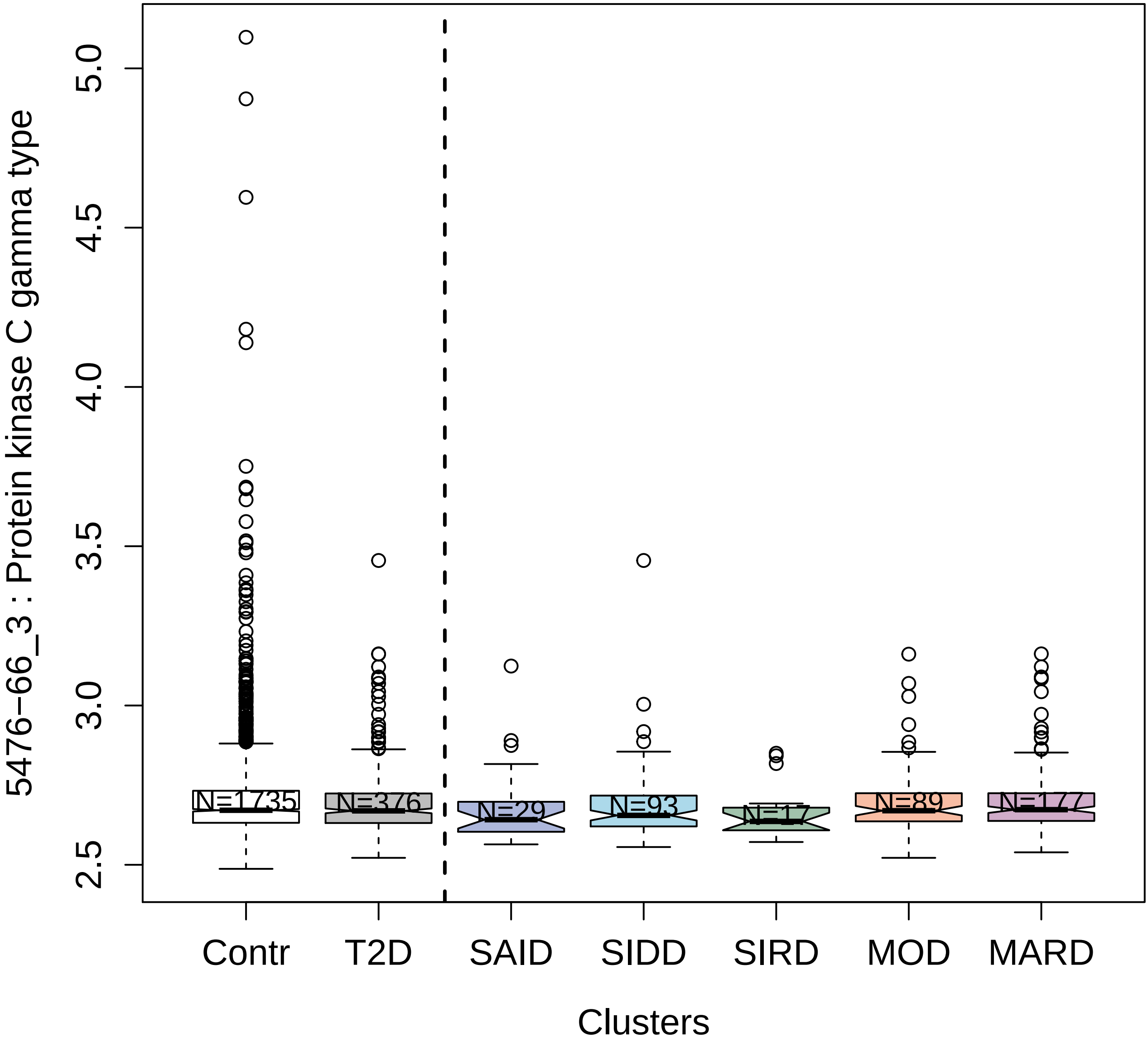
5476−66_3 : Protein kinase C gamma type
5.0
4.5
4.0
3.5
3.0
2.5
N=1735
N=376
N=29
N=93
N=17
N=89
N=177
Contr
T2D
SAID
SIDD
SIRD
MOD
MARD
Clusters

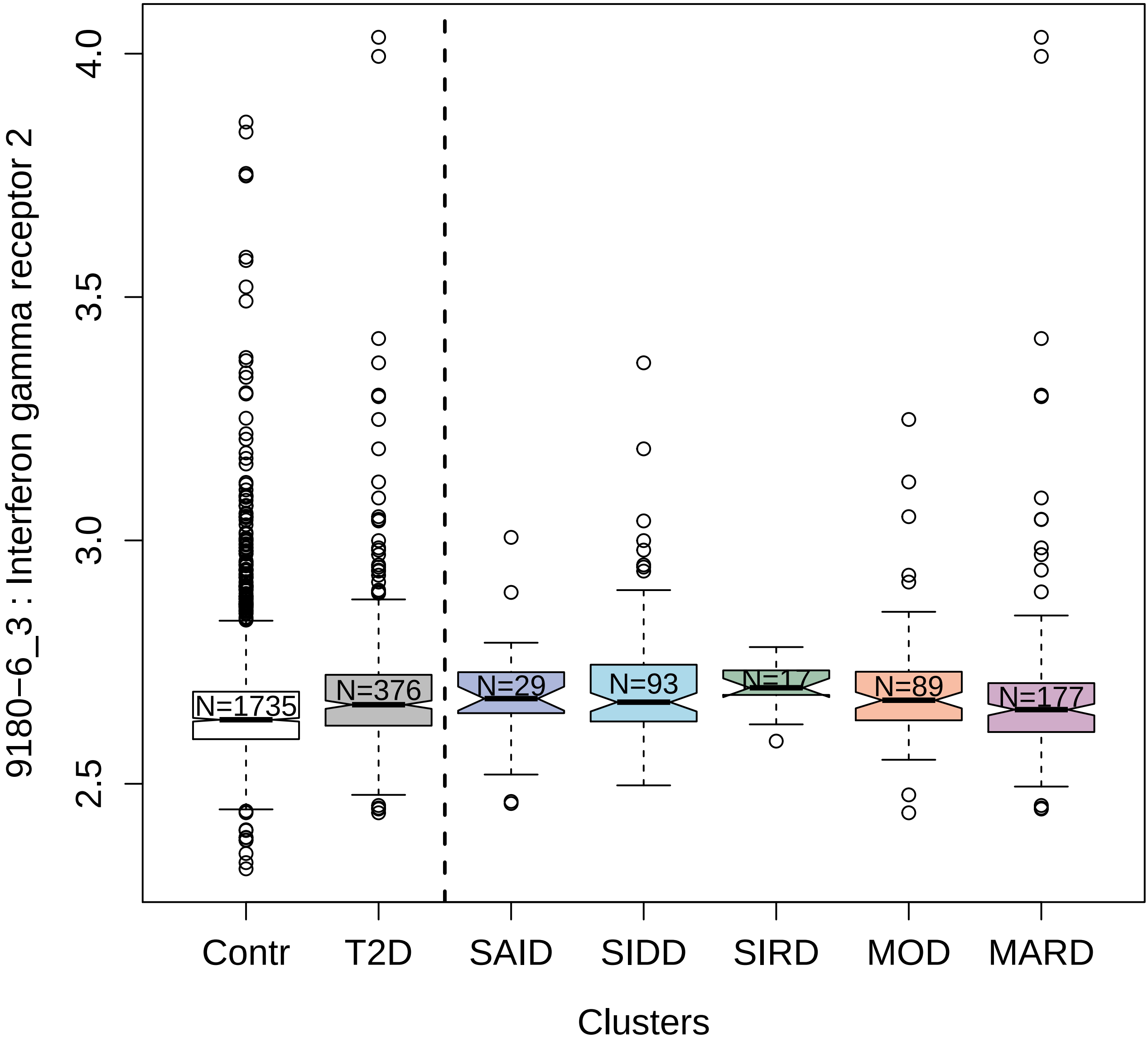
9180−6_3 : Interferon gamma receptor 2
4.0
3.5
3.0
2.5
N=1735
N=376
N=29
N=93
N=17
N=89
N=177
Contr
T2D
SAID
SIDD
SIRD
MOD
MARD
Clusters

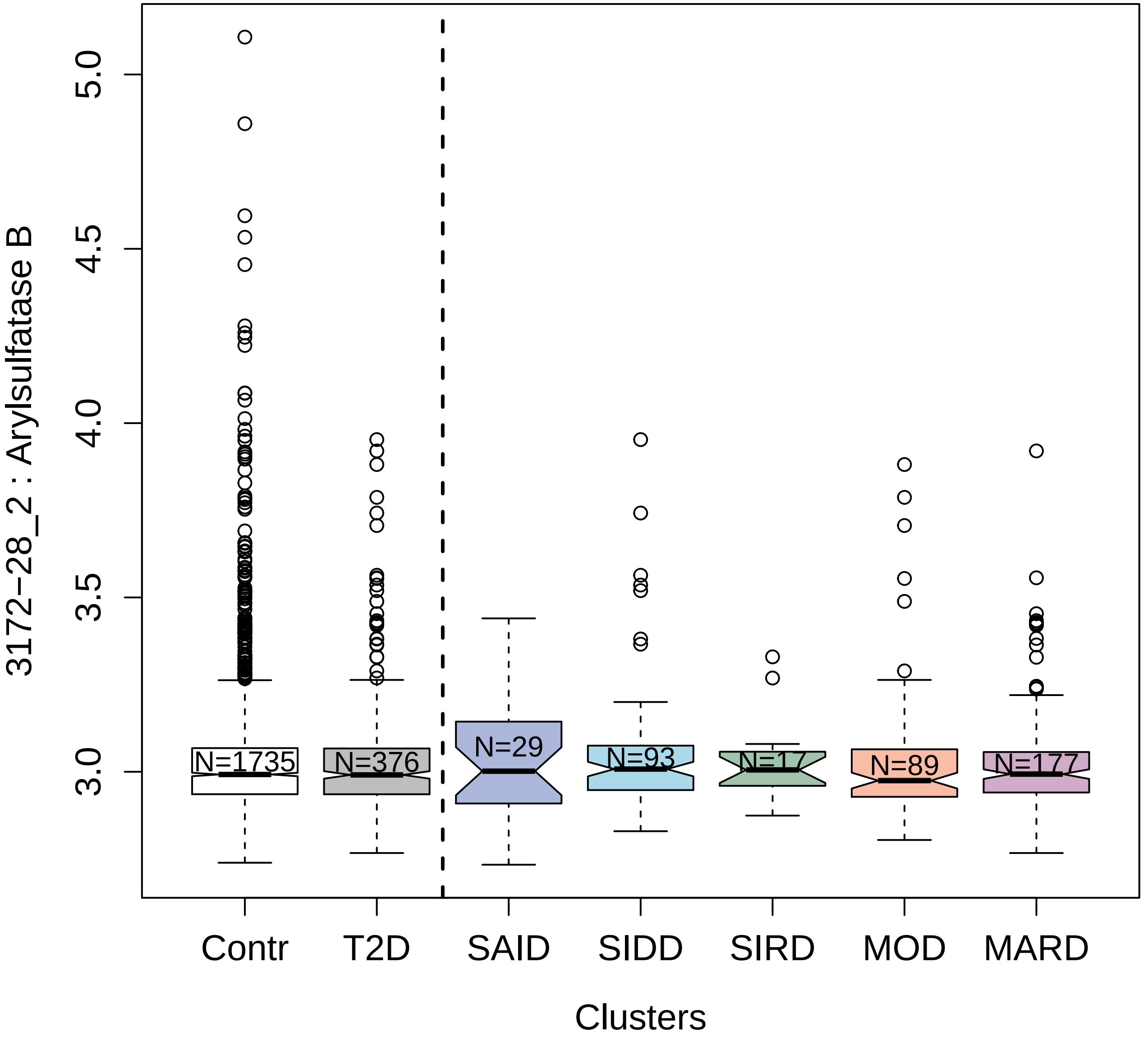

3172−28_2 : Arylsulfatase B
5.0
4.5
4.0
3.5
3.0
N=1735
N=376
N=29
N=93
N=17
N=89
N=177
Contr
T2D
SAID
SIDD
SIRD
MOD
MARD
Clusters

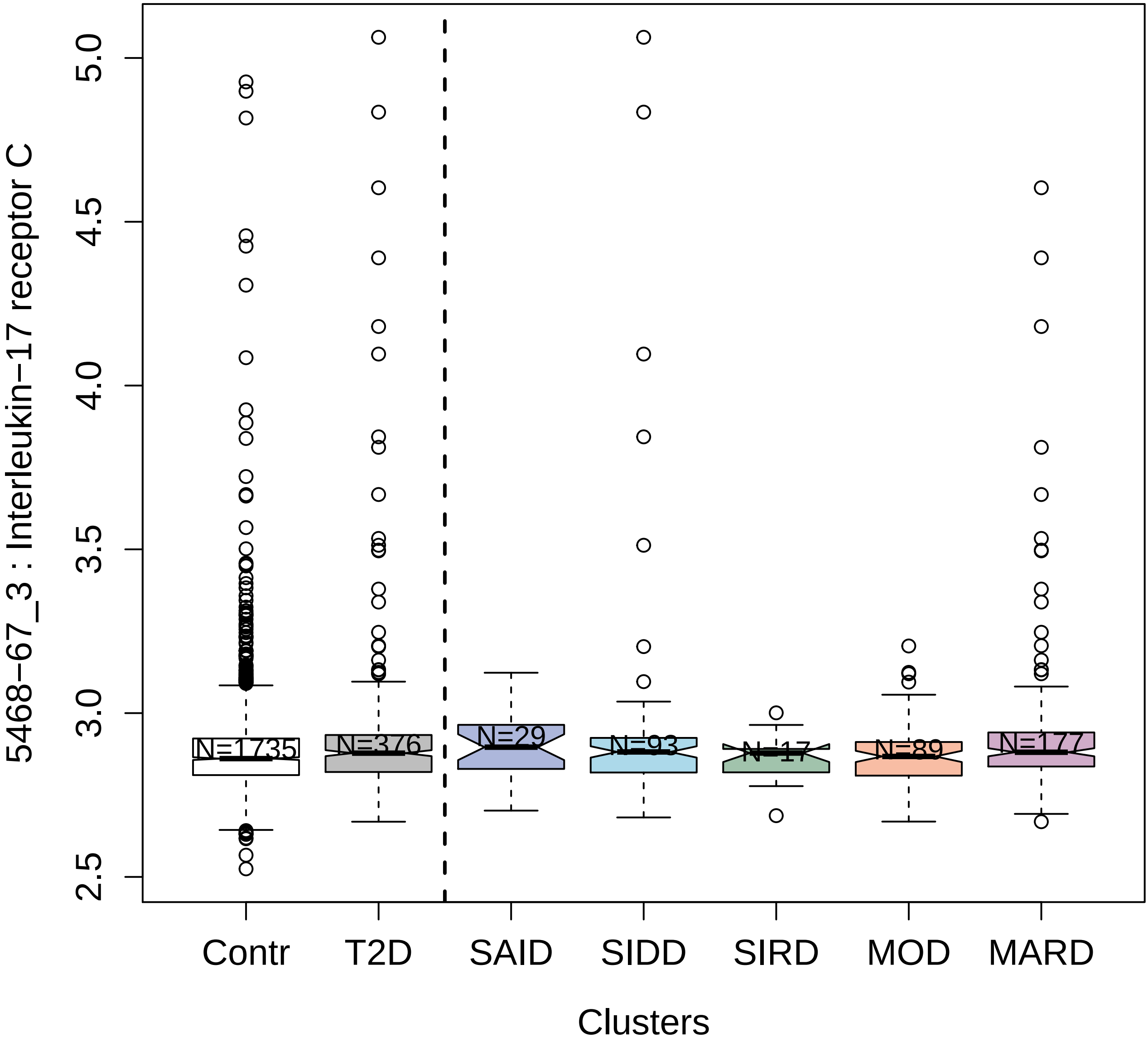
5468−67_3 : Interleukin−17 receptor C
2.5
3.0
3.5
4.0
4.5
5.0
N=1735
N=376
N=29
N=93
N=17
N=89
N=177
Contr
T2D
SAID
SIDD
SIRD
MOD
MARD
Clusters

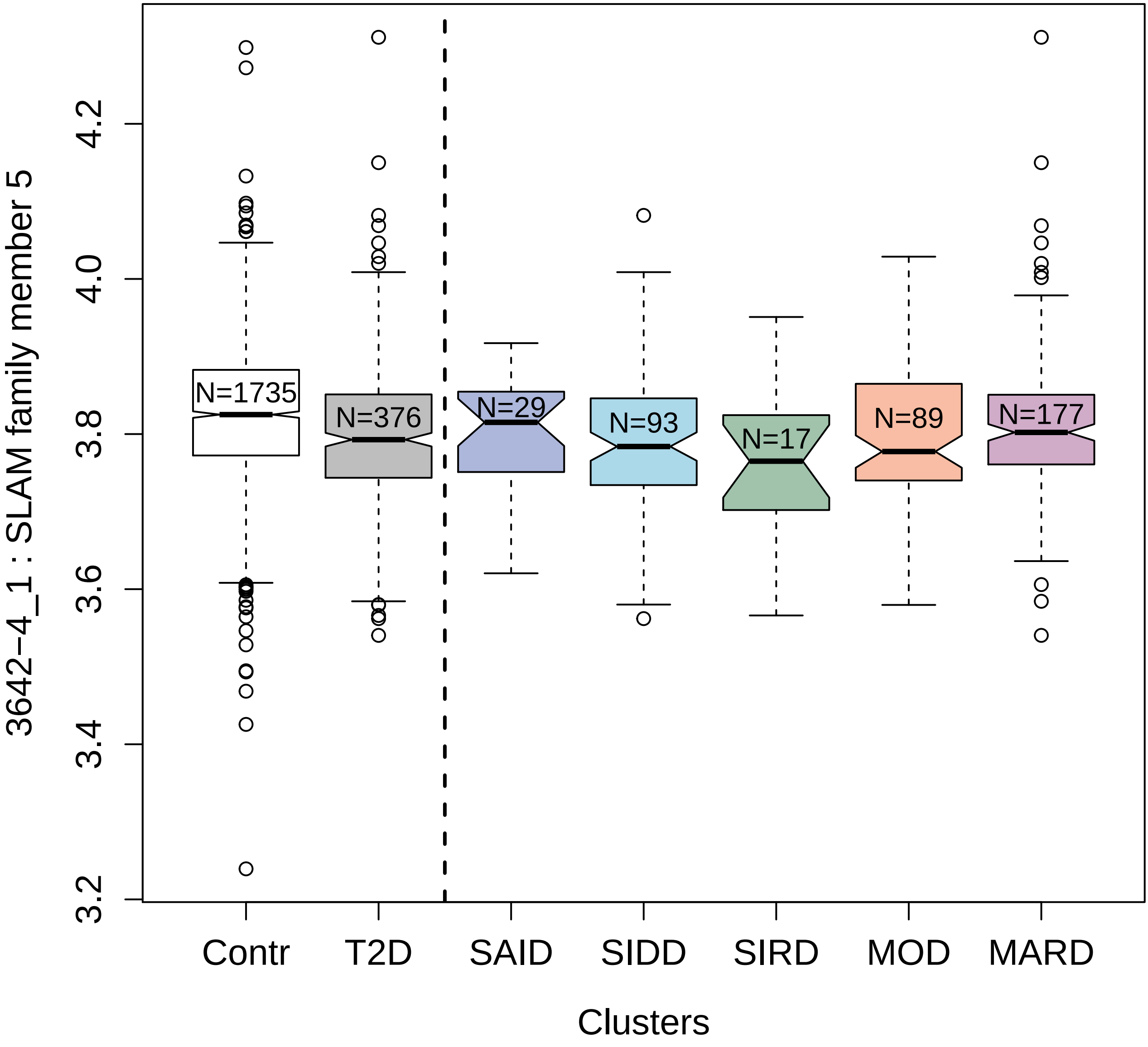

3642−4_1 : SLAM family member 5
4.2
4.0
3.8
3.6
3.4
3.2
N=1735
N=376
N=29
N=93
N=17
N=89
N=177
Contr
T2D
SAID
SIDD
SIRD
MOD
MARD
Clusters

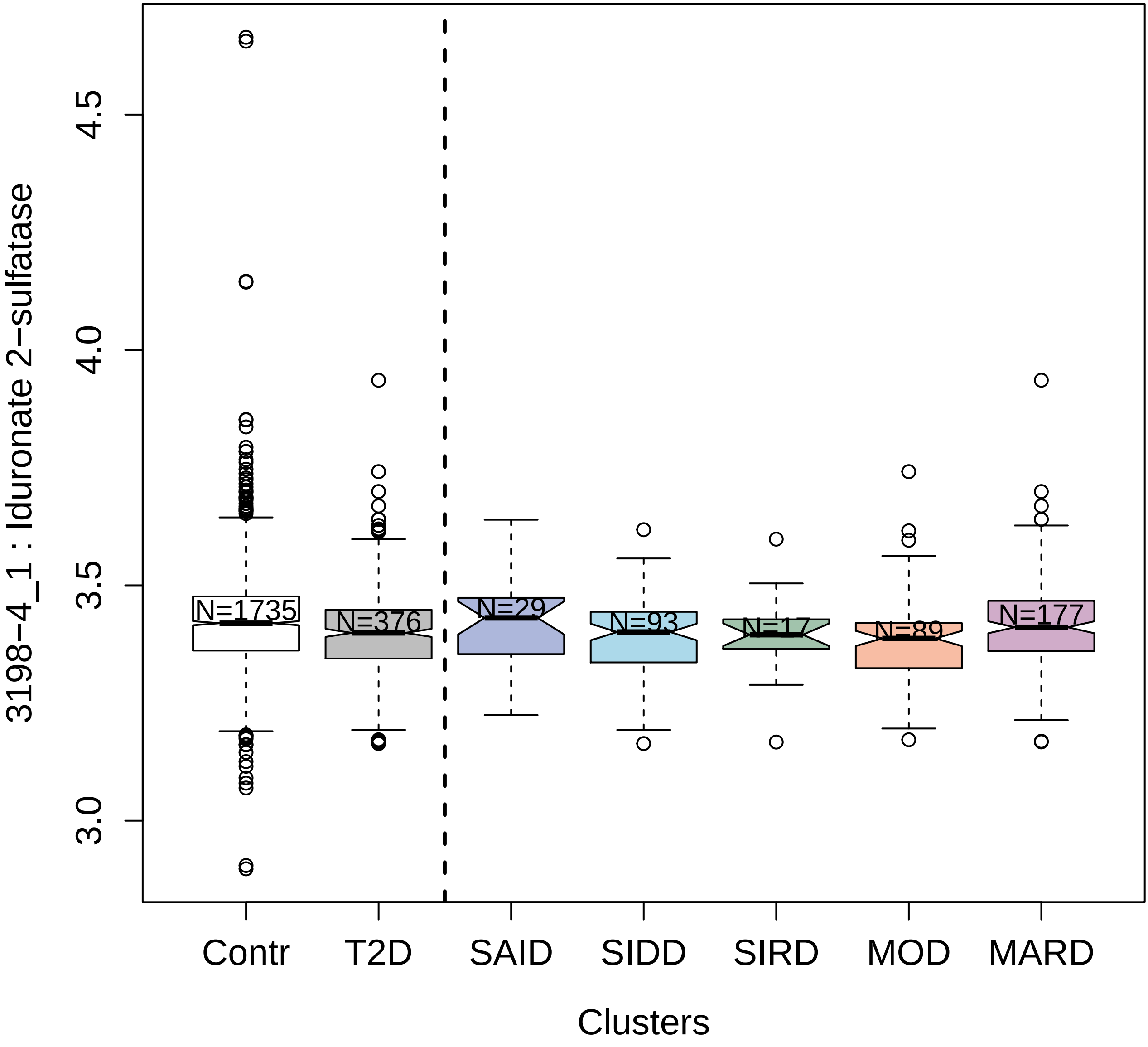

3198−4_1 : Iduronate 2−sulfatase
4.5
4.0
3.5
3.0
N=1735
N=376
N=29
N=93
N=17
N=89
N=177
Contr
T2D
SAID
SIDD
SIRD
MOD
MARD
Clusters

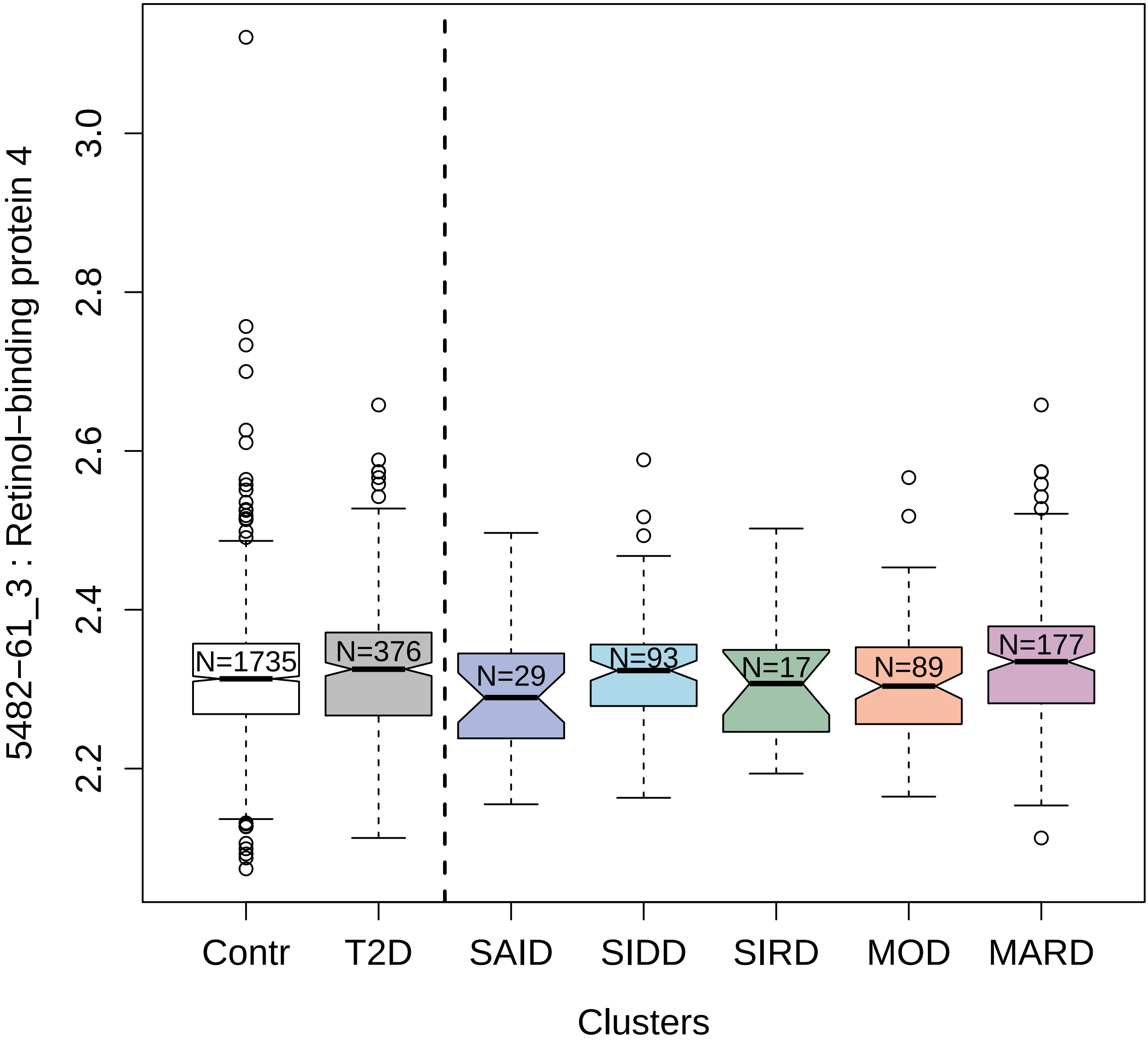
5482−61_3 : Retinol−binding protein 4
3.0
2.8
2.6
2.4
2.2
N=1735
N=376
N=29
N=93
N=17
N=89
N=177
Contr
T2D
SAID
SIDD
SIRD
MOD
MARD
Clusters

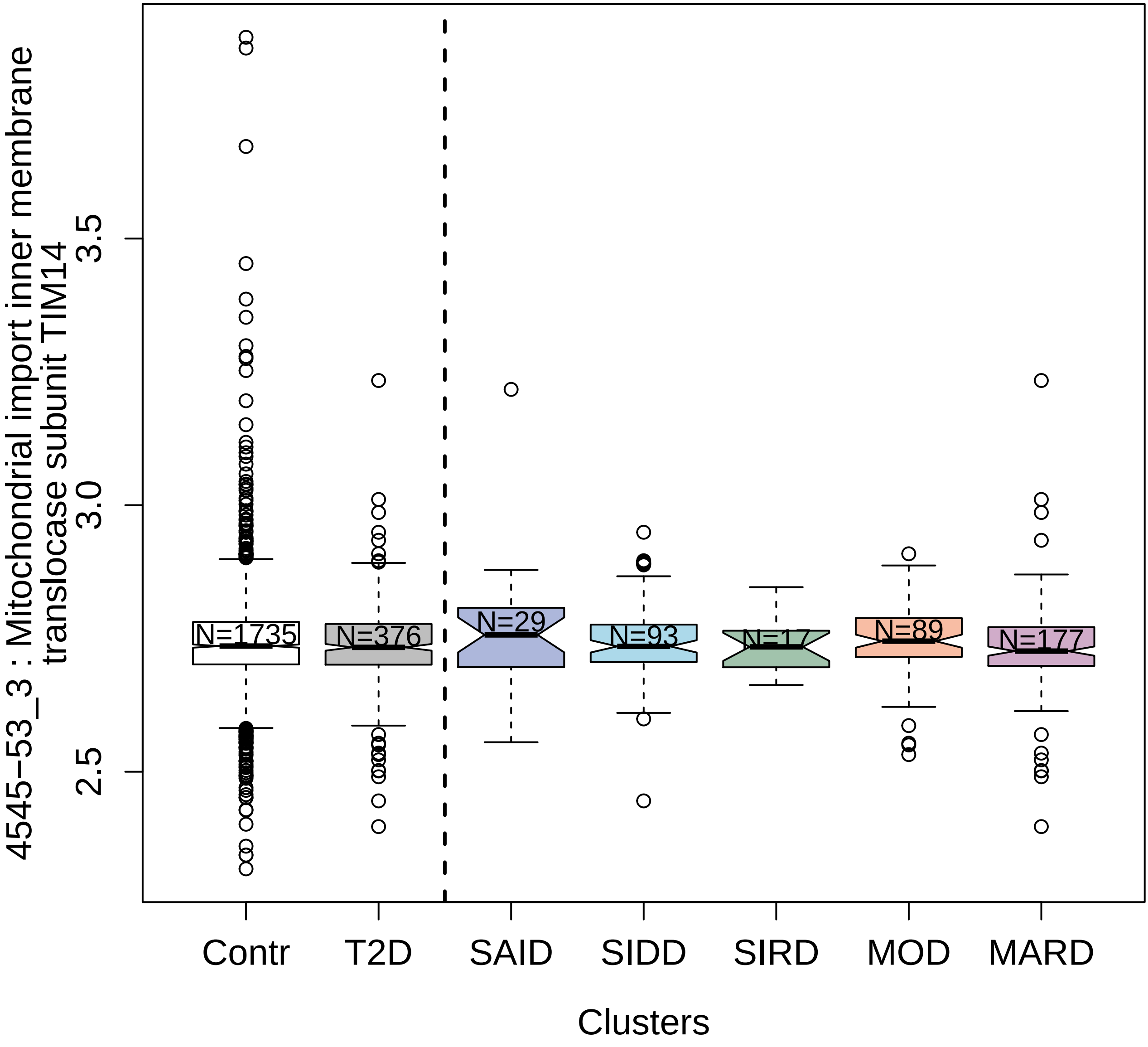

4545−53_3 : Mitochondrial import inner membrane translocase subunit TIM14
3.5
3.0
2.5
N=1735
N=376
N=29
N=93
N=17
N=89
N=177
Contr
T2D
SAID
SIDD
SIRD
MOD
MARD
Clusters

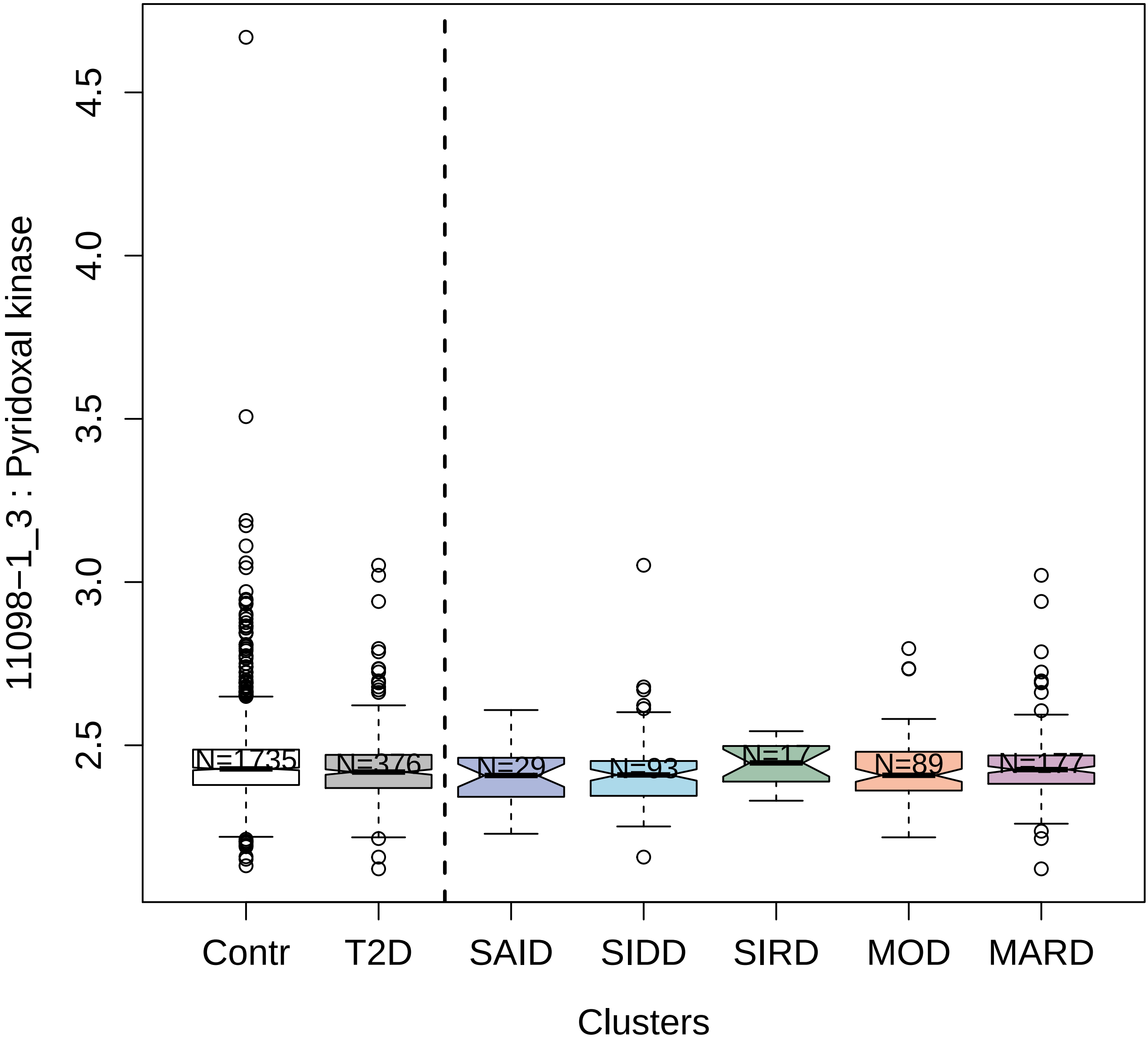

11098−1_3 : Pyridoxal kinase
4.5
4.0
3.5
3.0
2.5
N=1735
N=376
N=29
N=93
N=17
N=89
N=177
Contr
T2D
SAID
SIDD
SIRD
MOD
MARD
Clusters

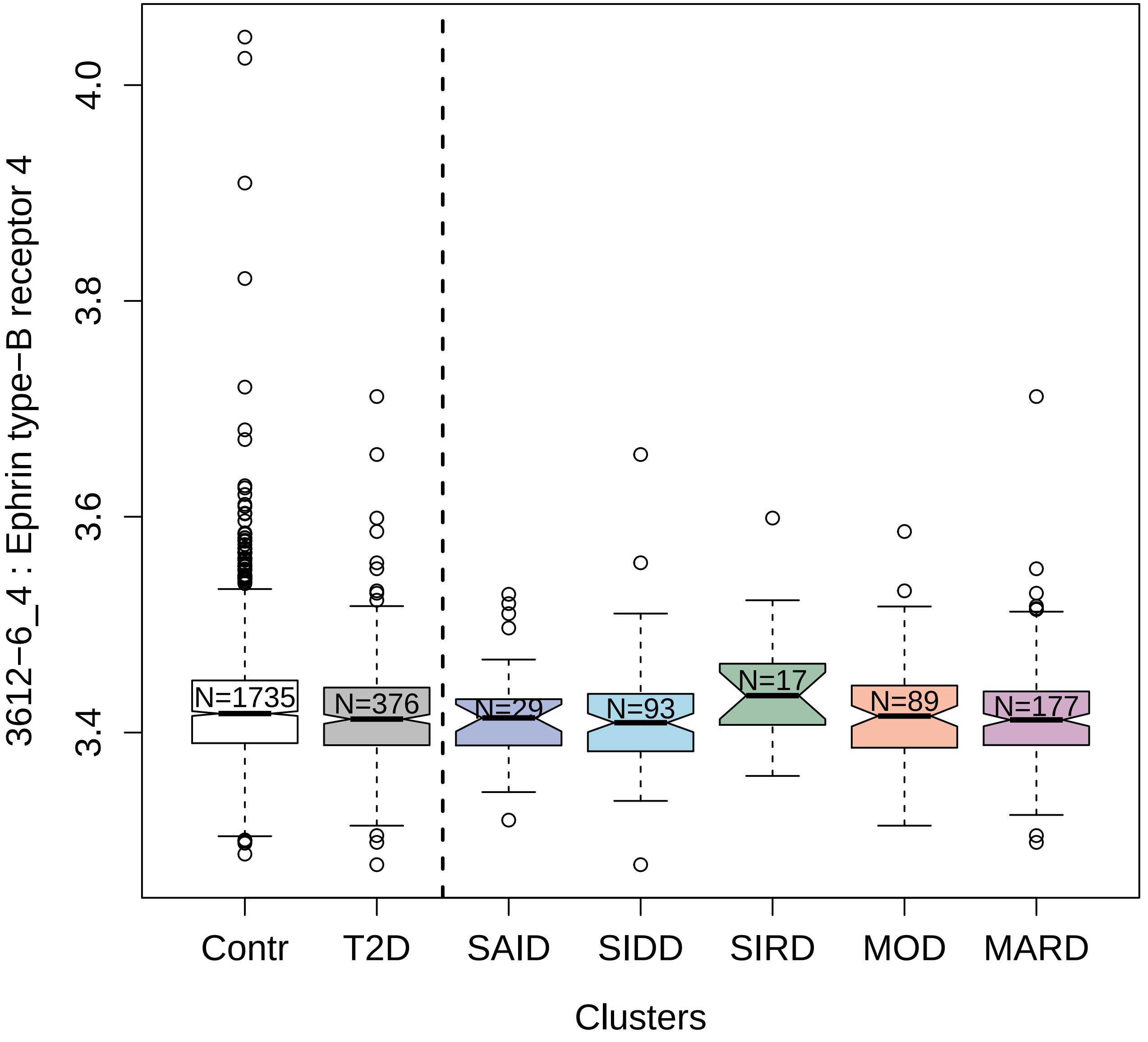

3612−6_4 : Ephrin type−B receptor 4
4.0
3.8
3.6
3.4
N=1735
N=376
N=29
N=93
N=17
N=89
N=177
Contr
T2D
SAID
SIDD
SIRD
MOD
MARD
Clusters

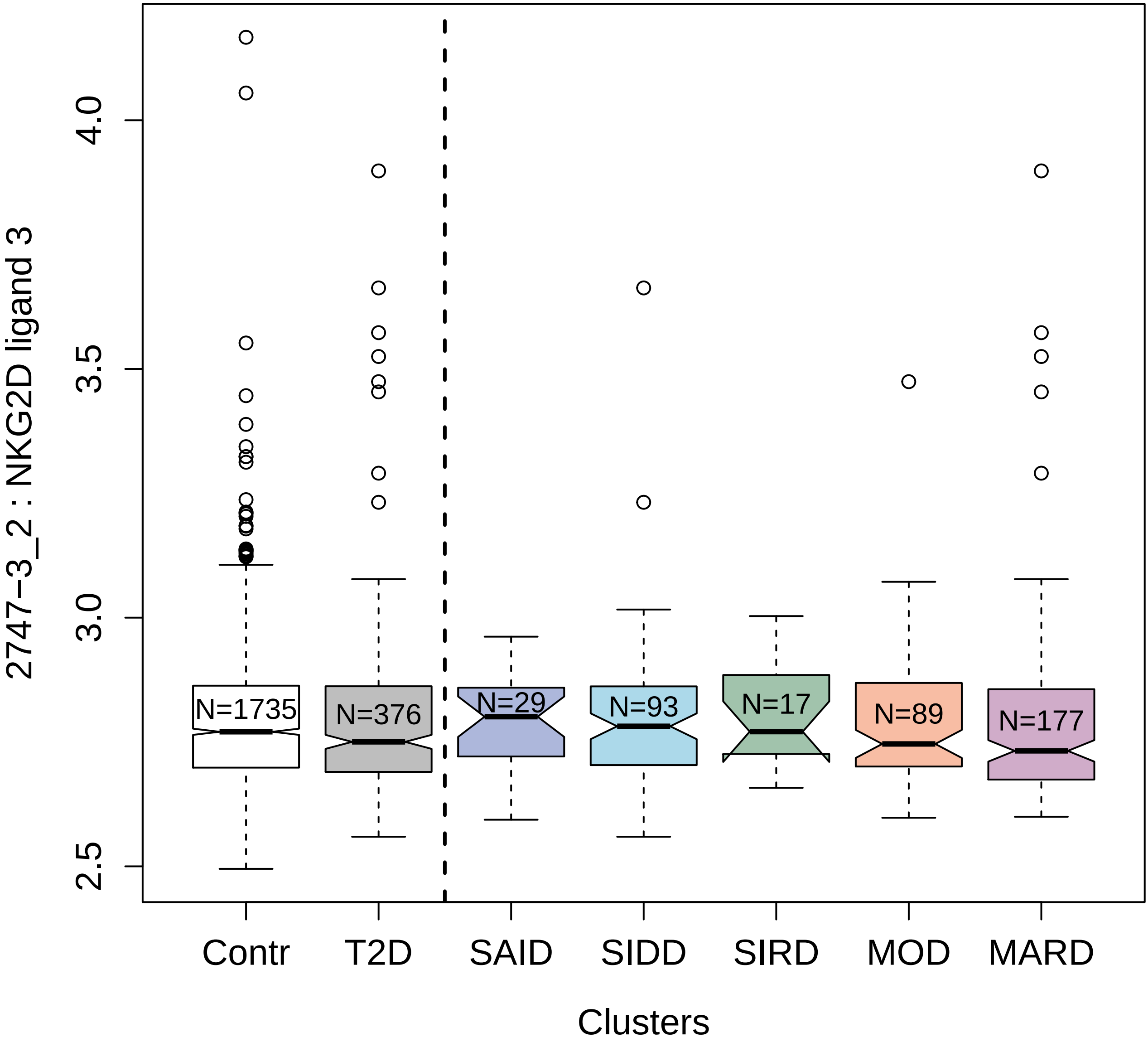
2747−3_2 : NKG2D ligand 3
2.5
3.0
3.5
4.0
N=1735
N=376
N=29
N=93
N=17
N=89
N=177
Contr
T2D
SAID
SIDD
SIRD
MOD
MARD
Clusters

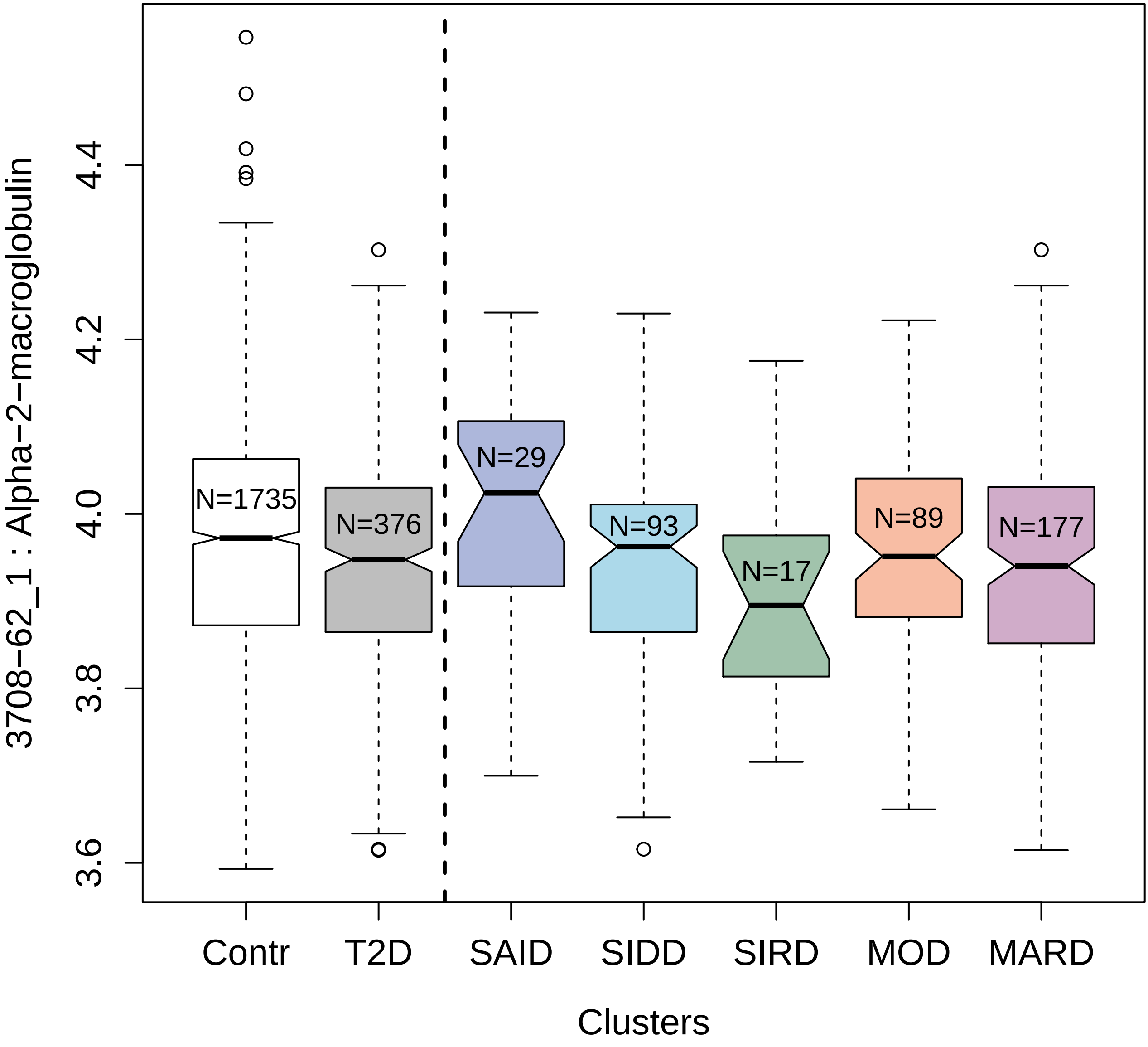

3708−62_1 : Alpha−2−macroglobulin
3.6
3.8
4.0
4.2
4.4
N=1735
N=376
N=29
N=93
N=17
N=89
N=177
Contr
T2D
SAID
SIDD
SIRD
MOD
MARD
Clusters

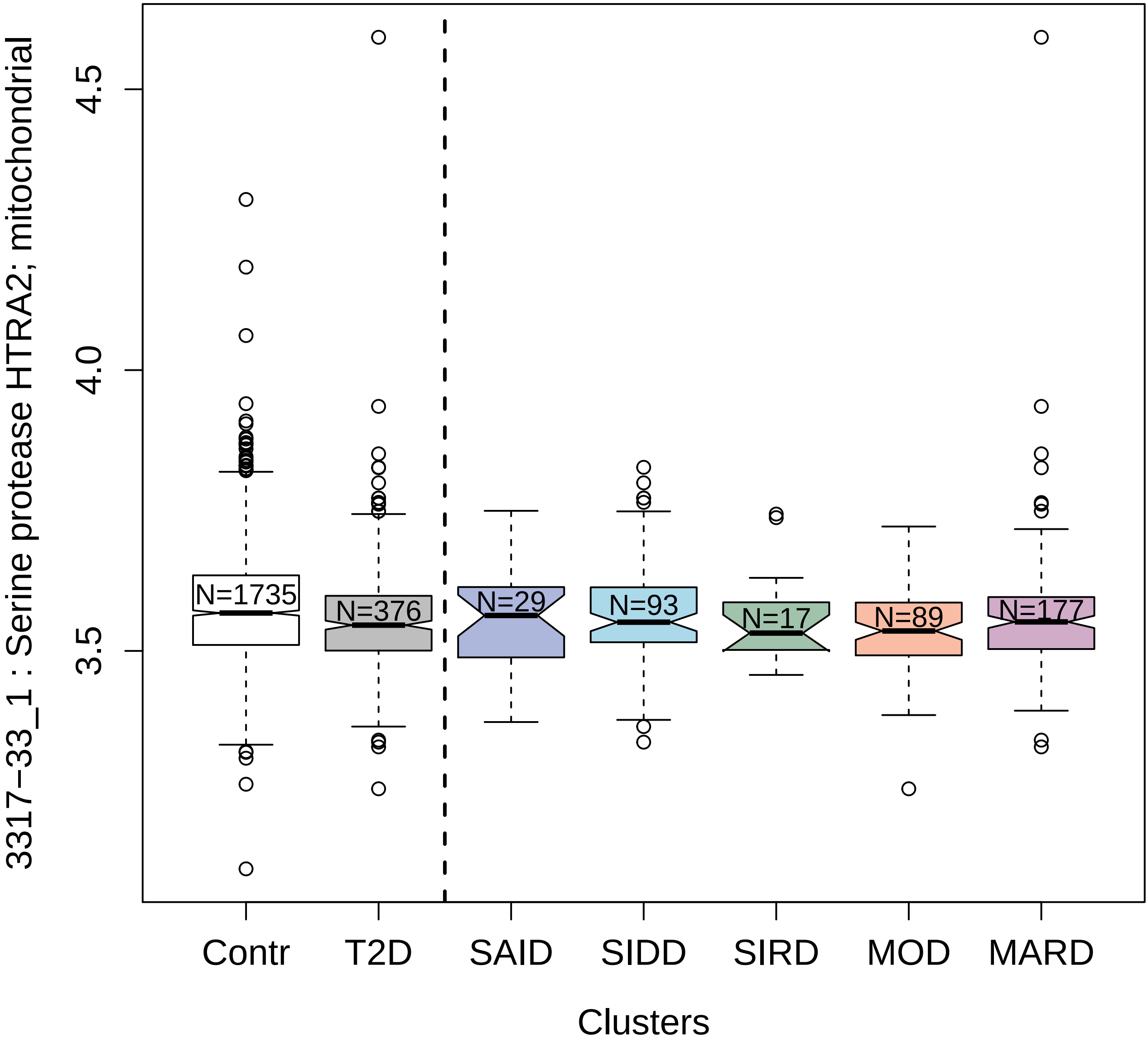
3317−33_1 : Serine protease HTRA2; mitochondrial
4.5
4.0
3.5
N=1735
N=376
N=29
N=93
N=17
N=89
N=177
Contr
T2D
SAID
SIDD
SIRD
MOD
MARD
Clusters

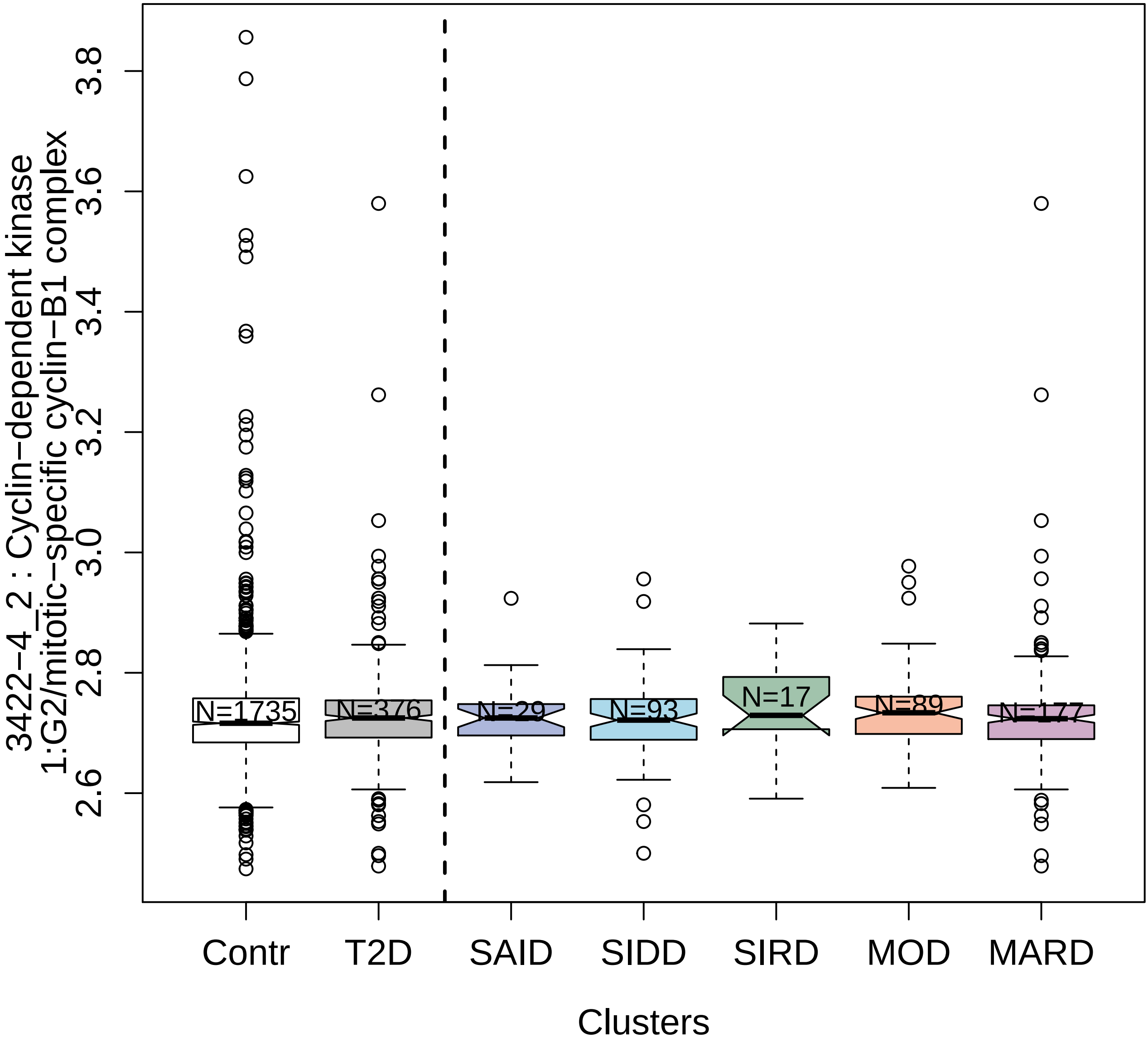

3422−4_2 : Cyclin−dependent kinase
1:G2/mitotic−specific cyclin−B1 complex
3.8
3.6
3.4
3.2
3.0
2.8
2.6
N=1735
N=376
N=29
N=93
N=17
N=89
N=177
Contr
T2D
SAID
SIDD
SIRD
MOD
MARD
Clusters

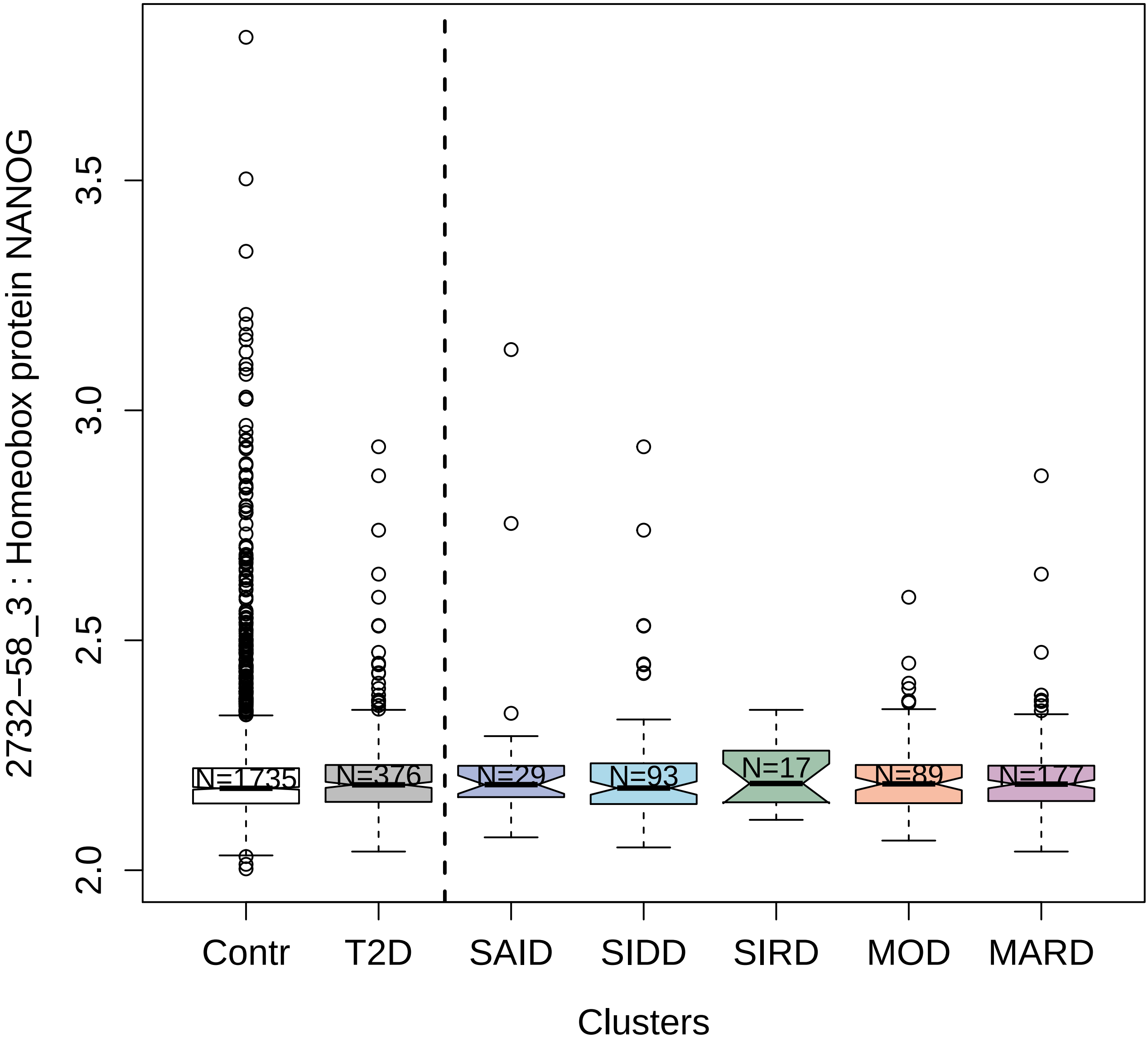

2732−58_3 : Homeobox protein NANOG
3.5
3.0
2.5
2.0
N=1735
N=376
N=29
N=93
N=17
N=89
N=177
Contr
T2D
SAID
SIDD
SIRD
MOD
MARD
Clusters

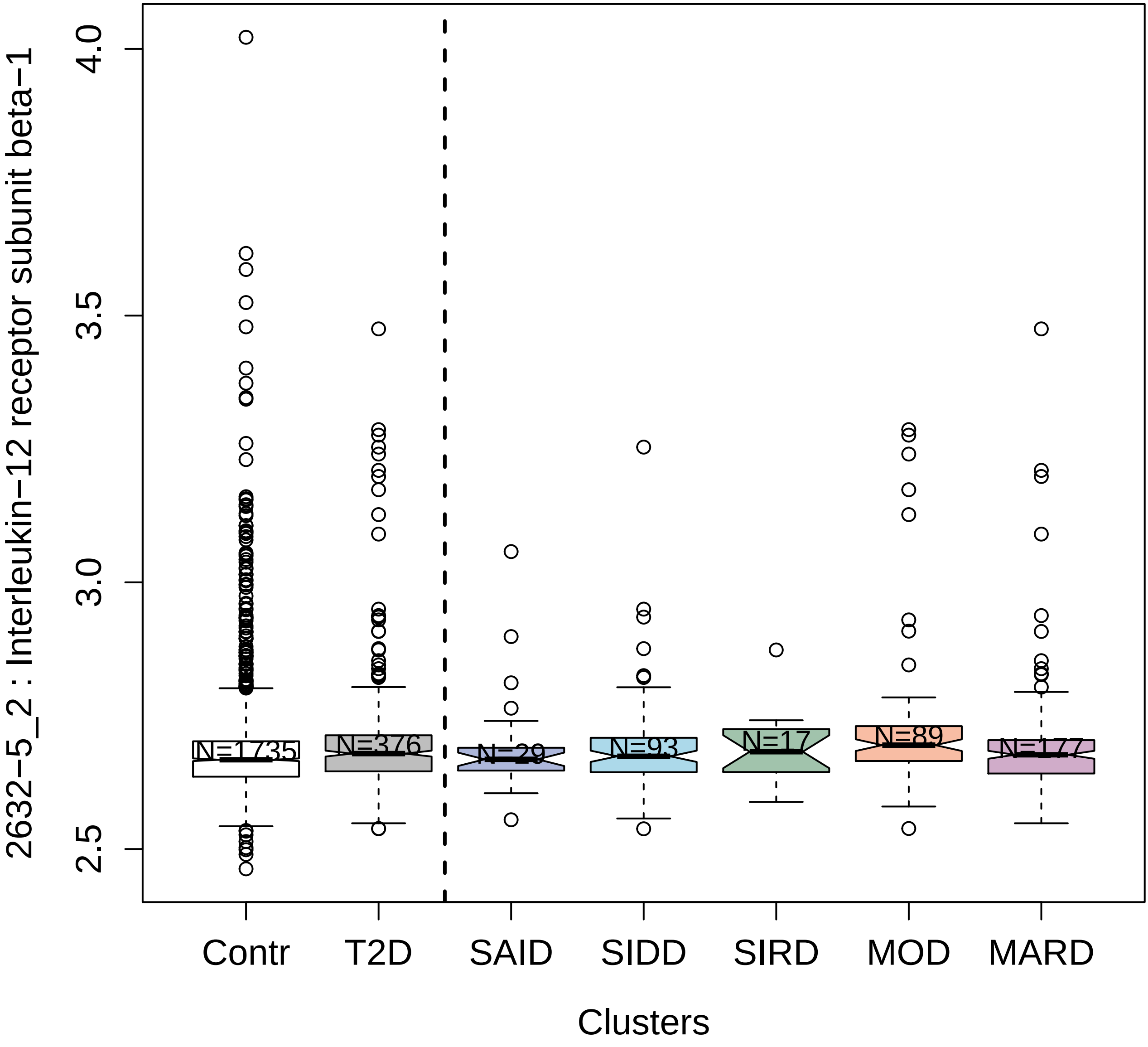

2632−5_2 : Interleukin−12 receptor subunit beta−1
4.0
3.5
3.0
2.5
N=1735
N=376
N=29
N=93
N=17
N=89
N=177
Contr
T2D
SAID
SIDD
SIRD
MOD
MARD
Clusters

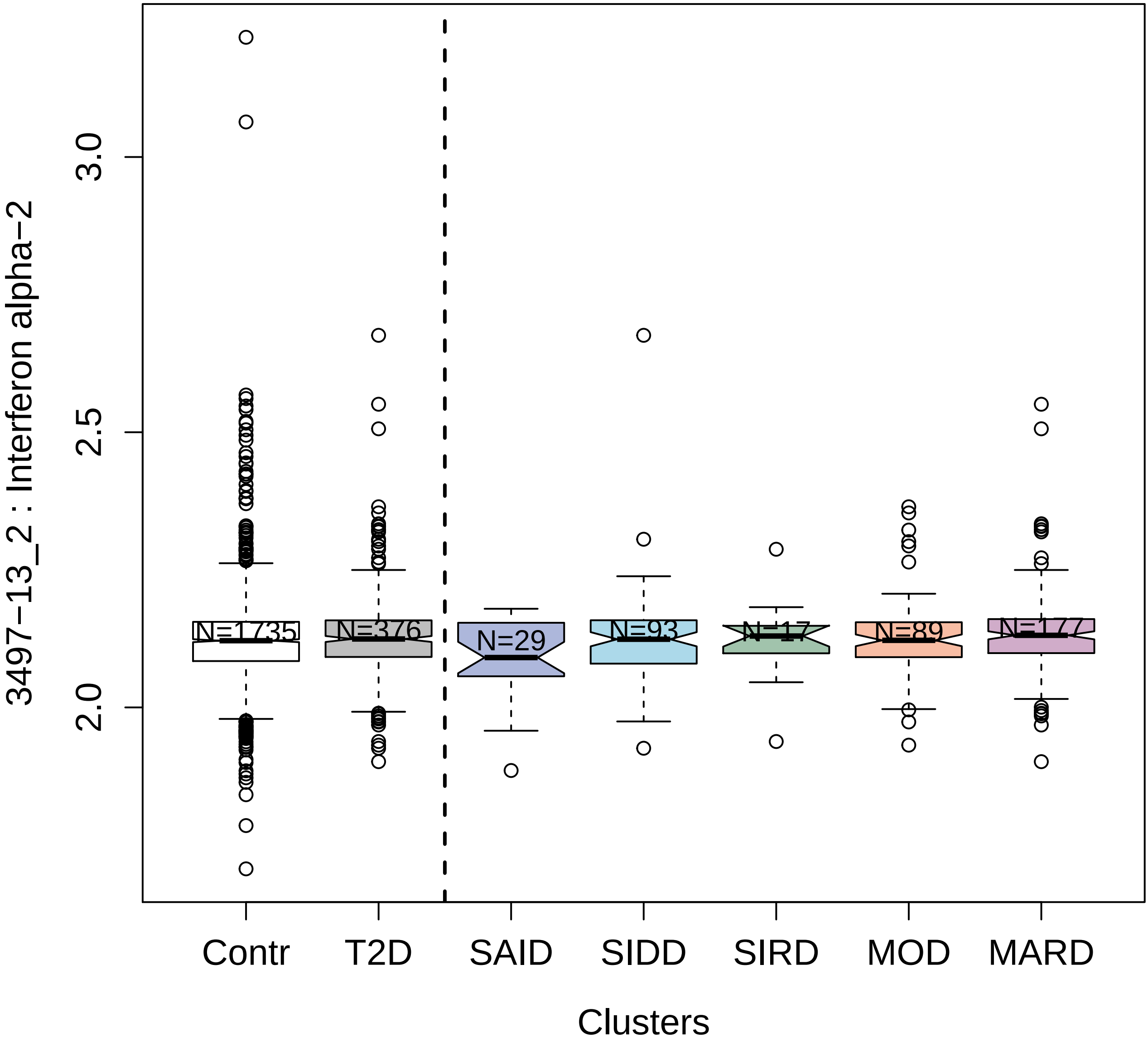
3497−13_2 : Interferon alpha−2
3.0
2.5
2.0
N=1735
N=376
N=29
N=93
N=17
N=89
N=177
Contr
T2D
SAID
SIDD
SIRD
MOD
MARD
Clusters

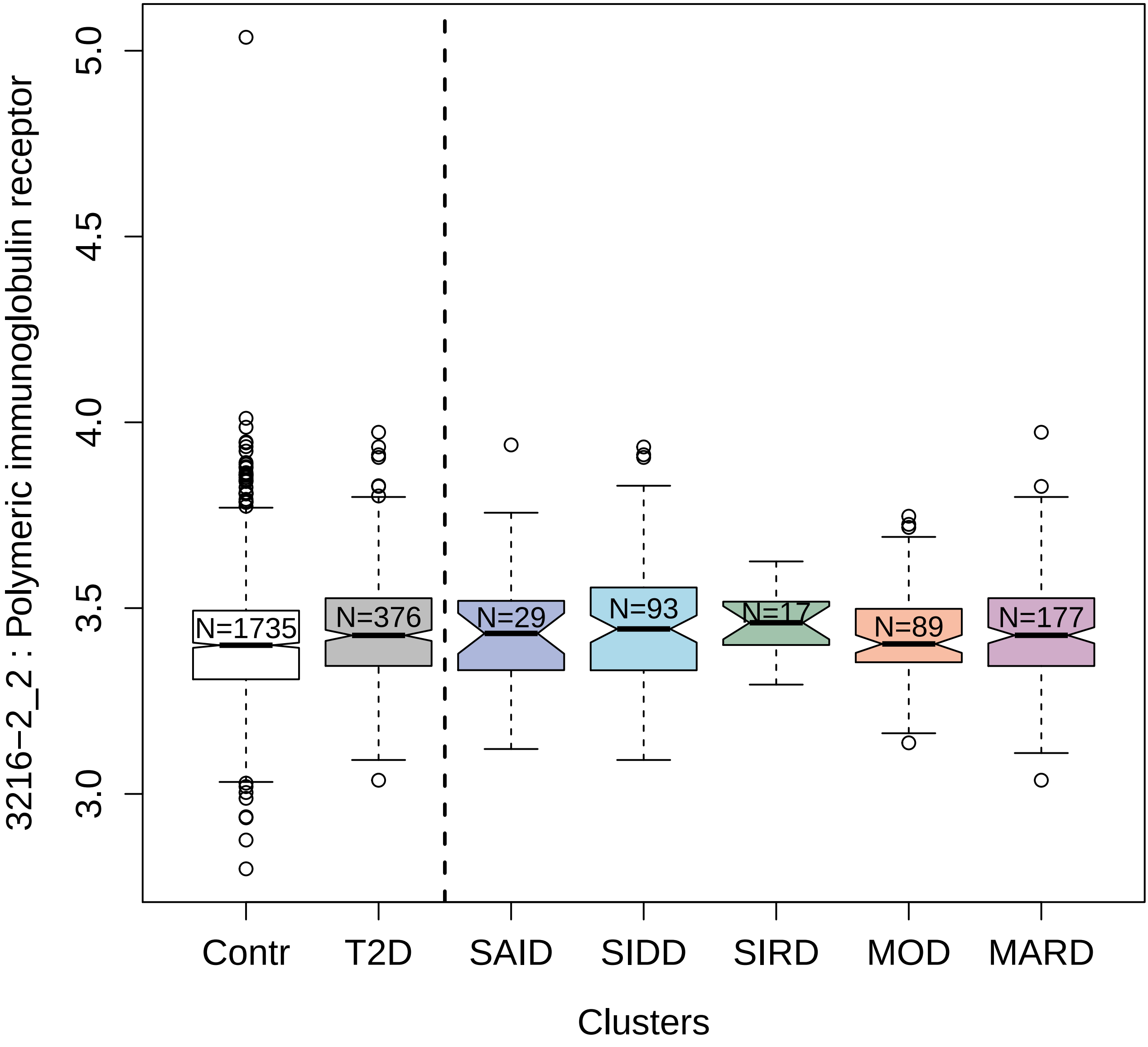
3216−2_2 : Polymeric immunoglobulin receptor
5.0
4.5
4.0
3.5
3.0
N=1735
N=376
N=29
N=93
N=17
N=89
N=177
Contr
T2D
SAID
SIDD
SIRD
MOD
MARD
Clusters

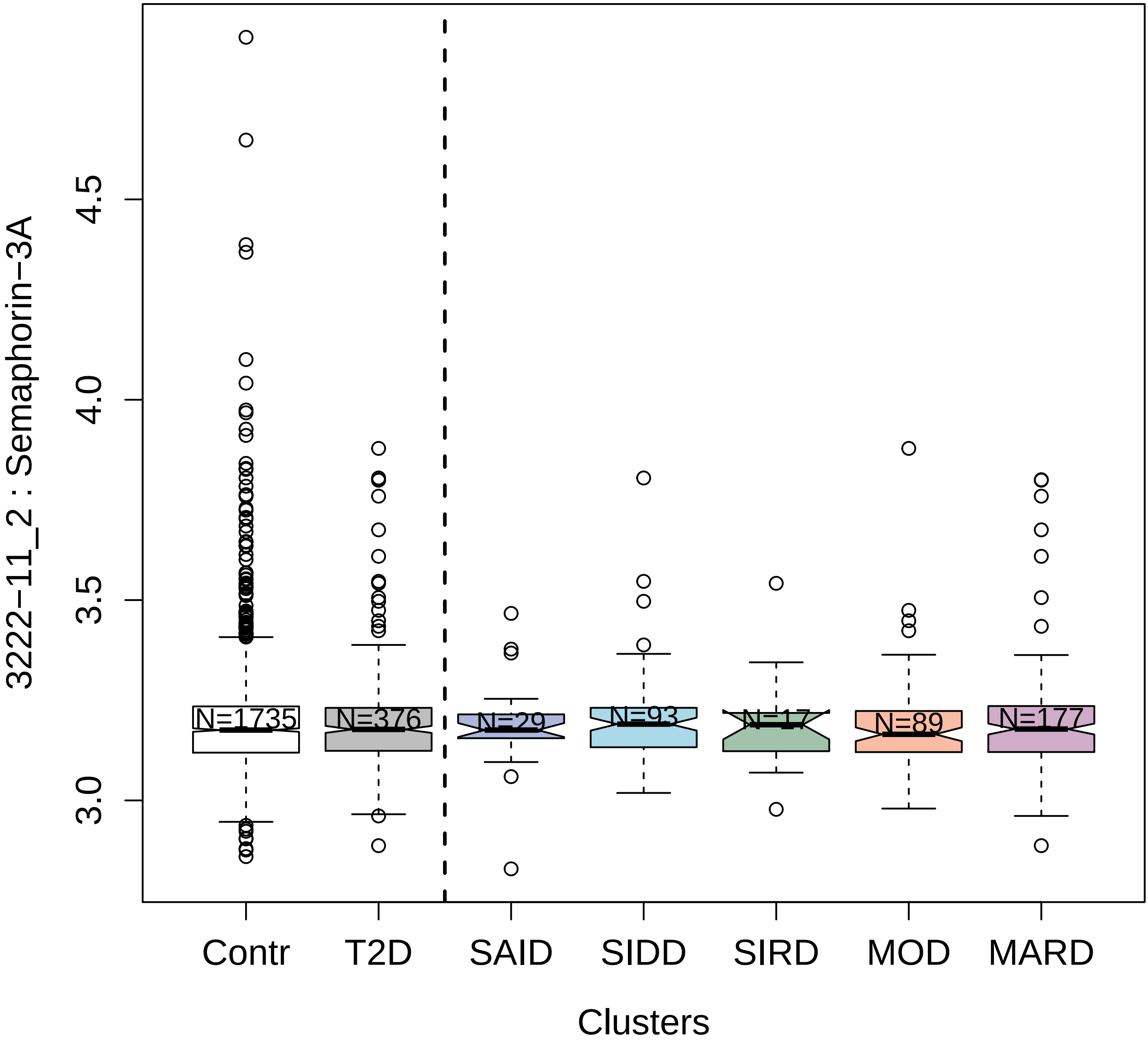

3222−11_2 : Semaphorin−3A
4.5
4.0
3.5
3.0
N=1735
N=376
N=29
N=93
N=17
N=89
N=177
Contr
T2D
SAID
SIDD
SIRD
MOD
MARD
Clusters

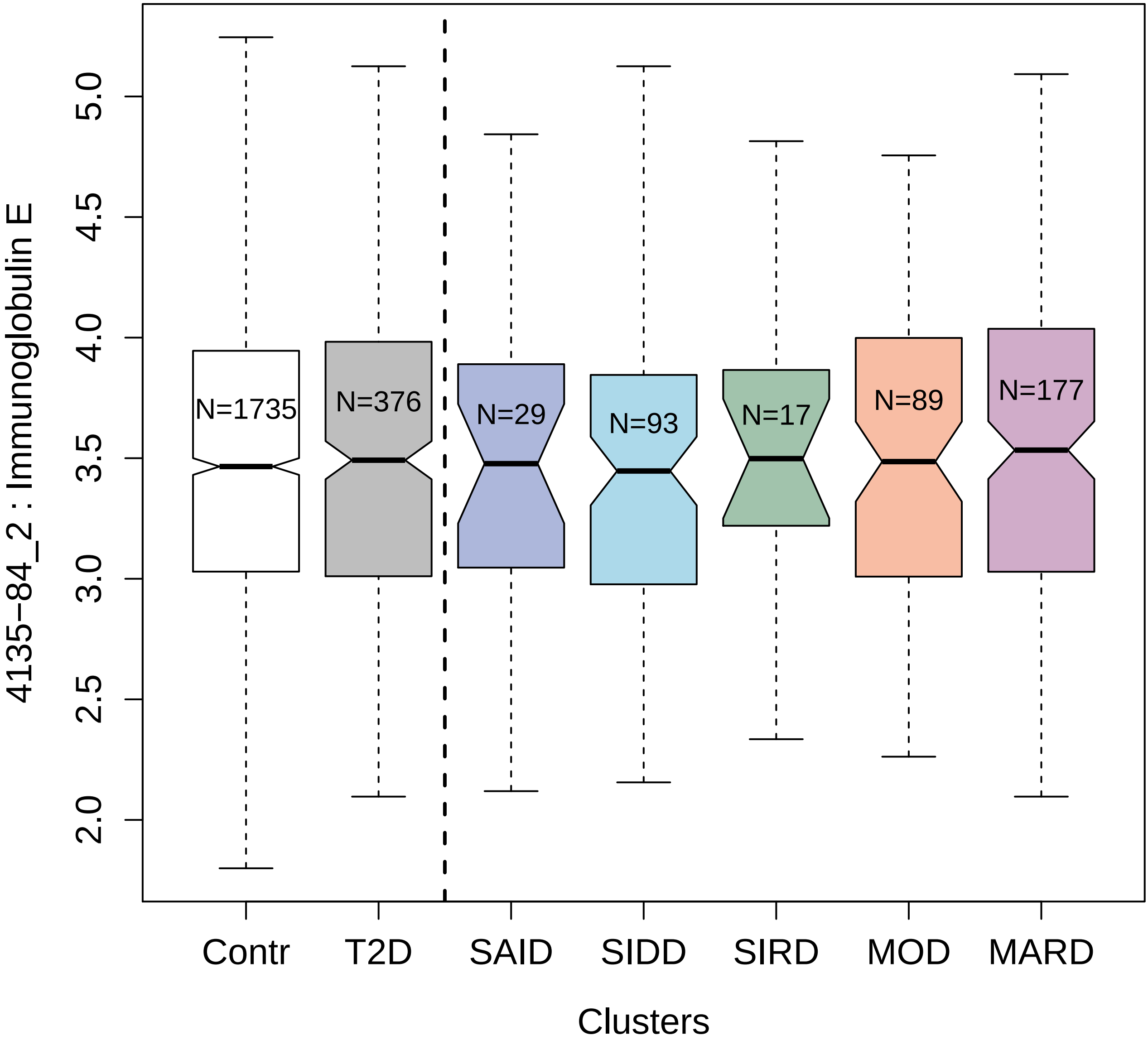

4135−84_2 : Immunoglobulin E
2.0
2.5
3.0
3.5
4.0
4.5
5.0
N=1735
N=376
N=29
N=93
N=17
N=89
N=177
Contr
T2D
SAID
SIDD
SIRD
MOD
MARD
Clusters

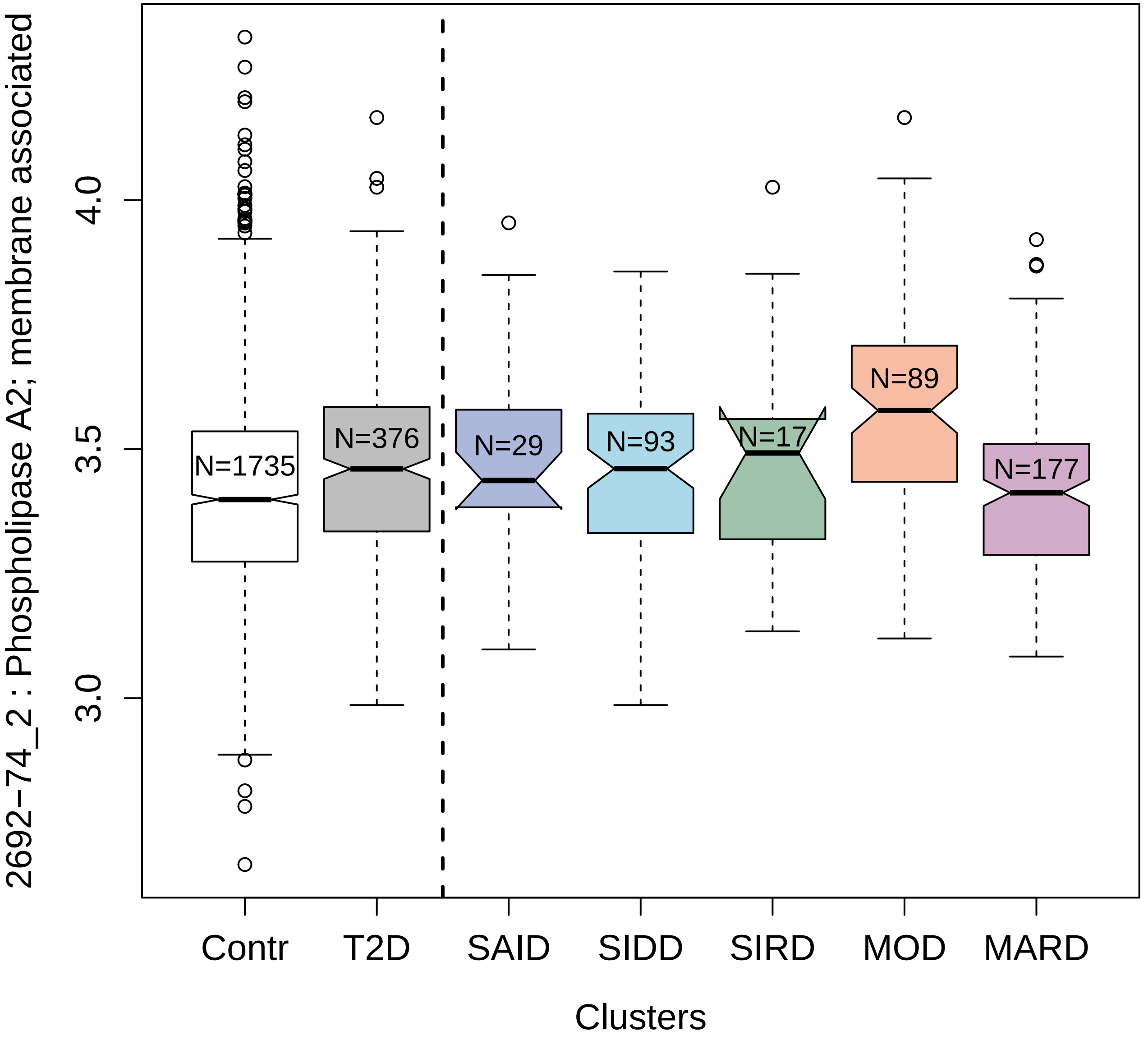
2692−74_2 : Phospholipase A2; membrane associated
4.0
3.5
3.0
N=1735
N=376
N=29
N=93
N=17
N=89
N=177
Contr
T2D
SAID
SIDD
SIRD
MOD
MARD
Clusters

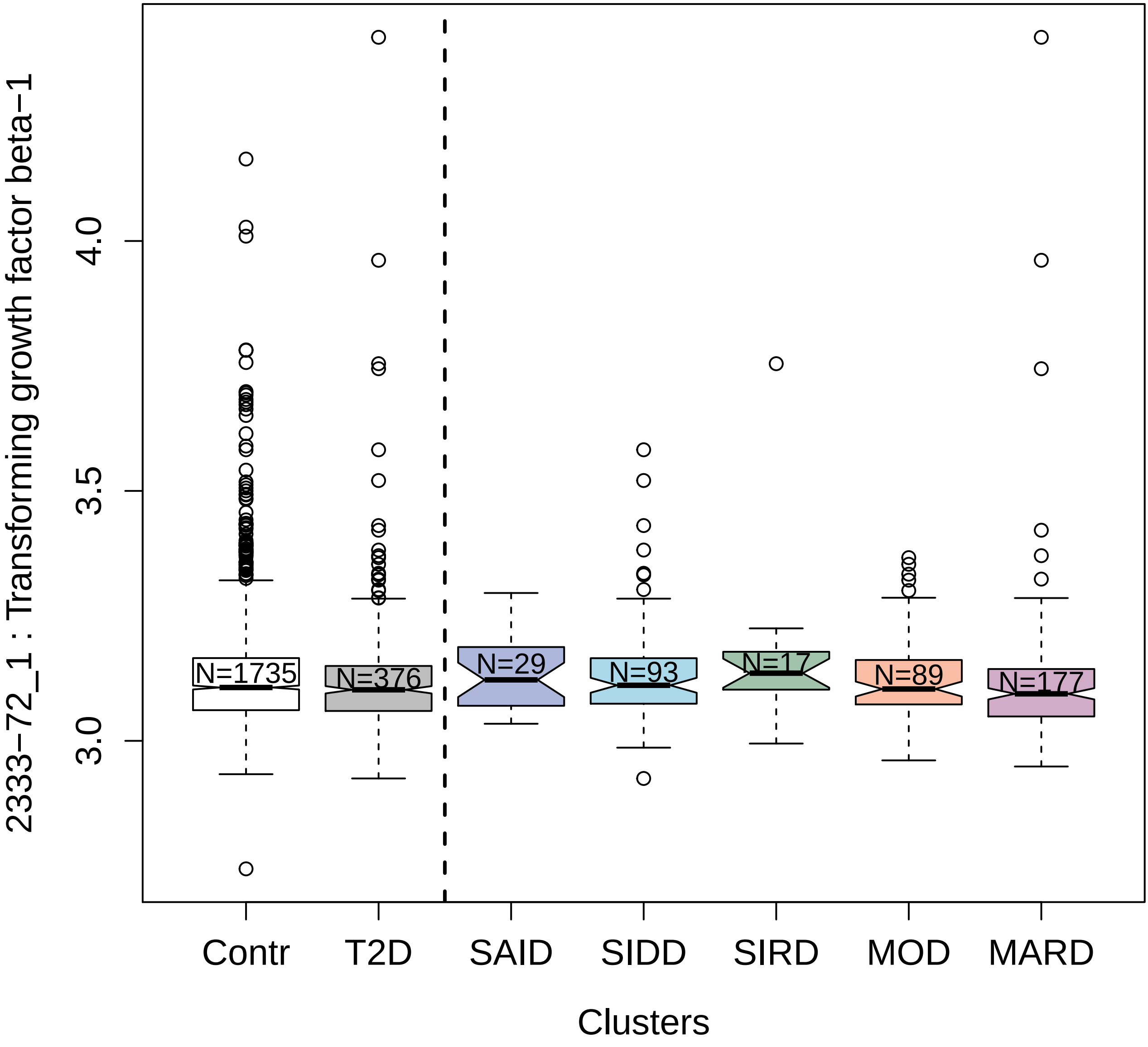
2333−72_1 : Transforming growth factor beta−1
4.0
3.5
3.0
N=1735
N=376
N=29
N=93
N=17
N=89
N=177
Contr
T2D
SAID
SIDD
SIRD
MOD
MARD
Clusters

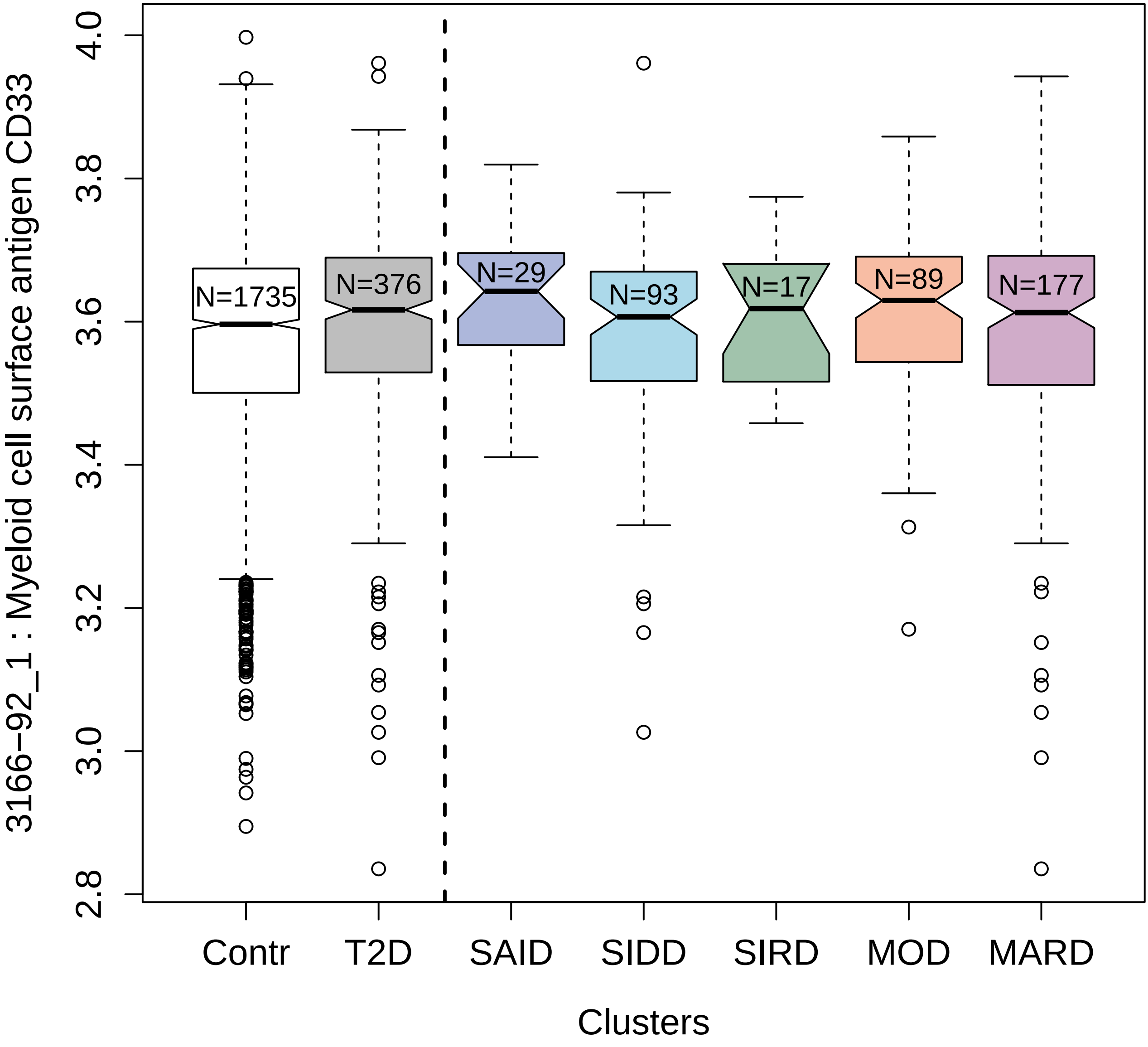

3166−92_1 : Myeloid cell surface antigen CD33
4.0
3.8
3.6
3.4
3.2
3.0
2.8
N=1735
N=376
N=29
N=93
N=17
N=89
N=177
Contr
T2D
SAID
SIDD
SIRD
MOD
MARD
Clusters

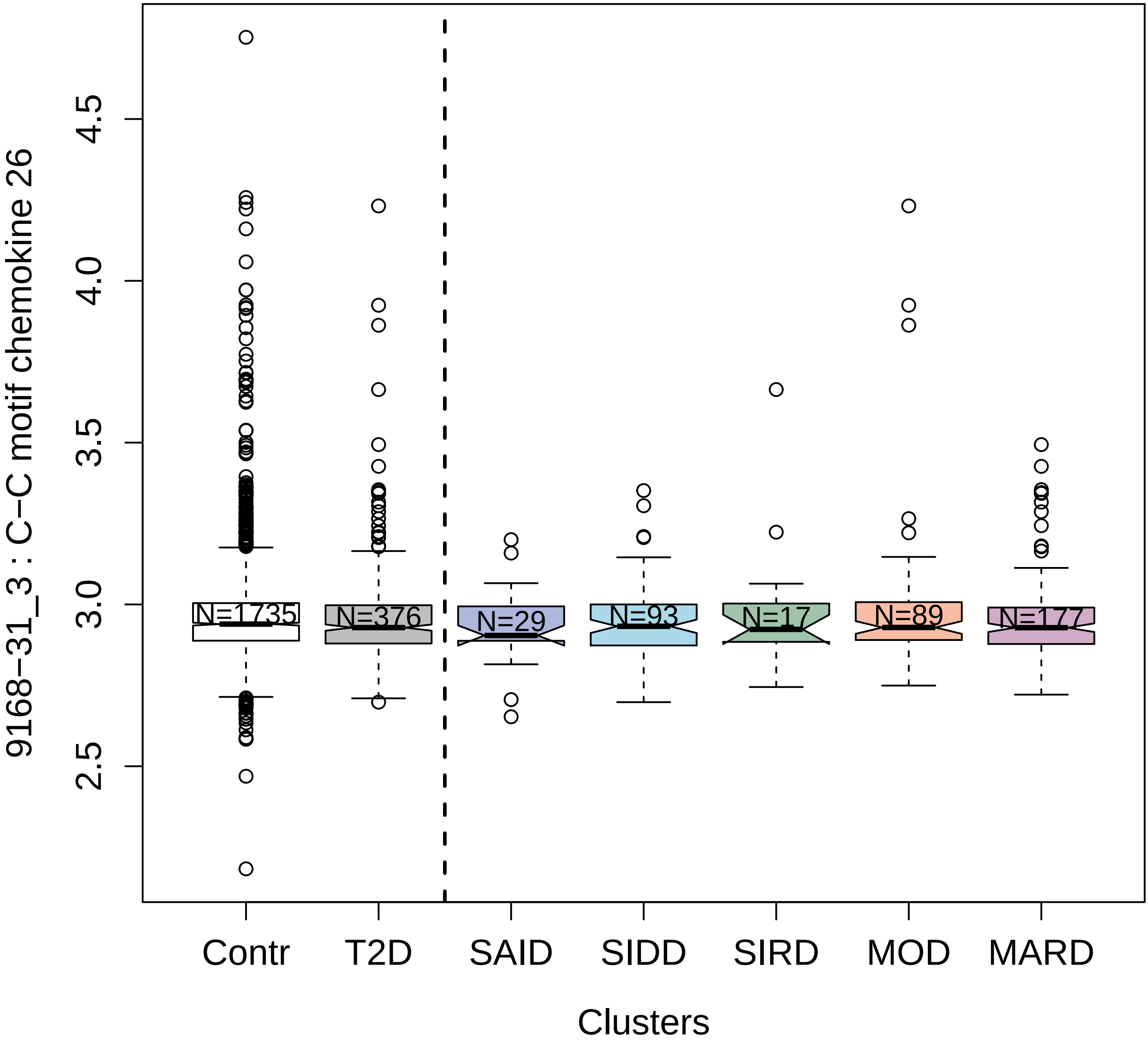

9168−31_3 : C−C motif chemokine 26
2.5
3.0
3.5
4.0
4.5
N=1735
N=376
N=29
N=93
N=17
N=89
N=177
Contr
T2D
SAID
SIDD
SIRD
MOD
MARD
Clusters

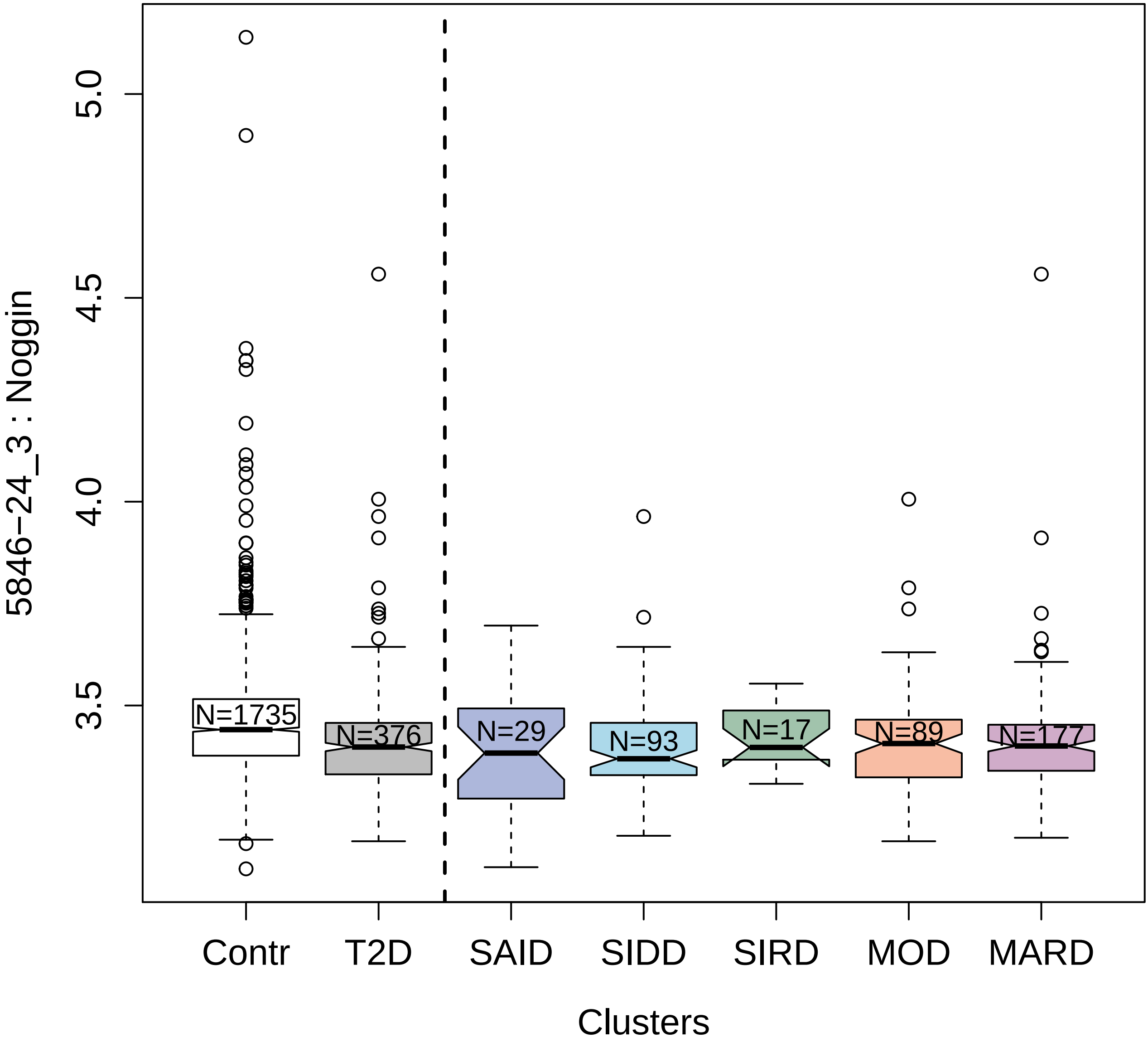
5846−24_3 : Noggin
5.0
4.5
4.0
3.5
N=1735
N=376
N=29
N=93
N=17
N=89
N=177
Contr
T2D
SAID
SIDD
SIRD
MOD
MARD
Clusters

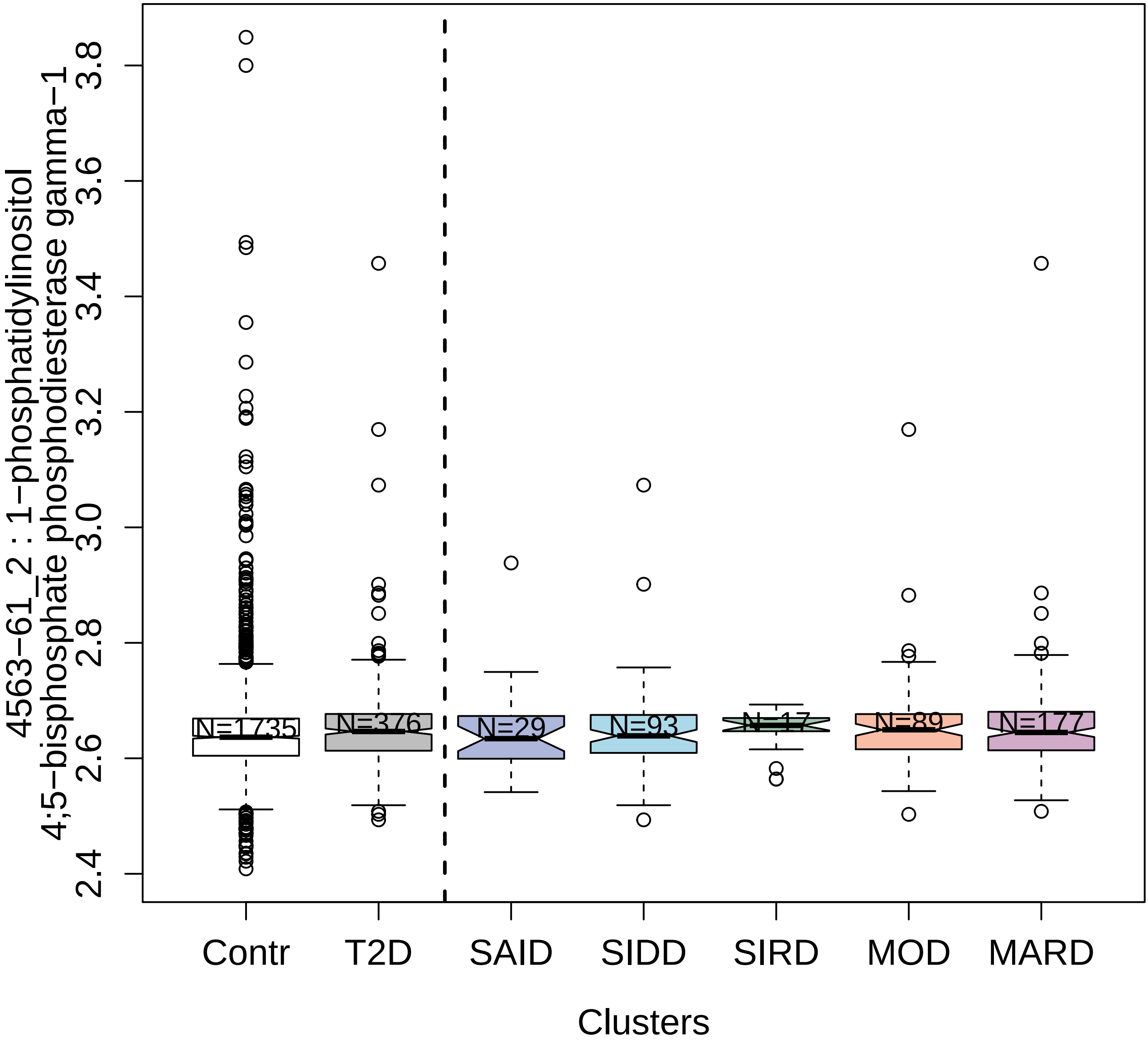
4563−61_2 : 1−phosphatidylinositol
4;5−bisphosphate phosphodiesterase gamma−1
3.8
3.6
3.4
3.2
3.0
2.8
2.6
2.4
N=1735
N=376
N=29
N=93
N=17
N=89
N=177
Contr
T2D
SAID
SIDD
SIRD
MOD
MARD
Clusters

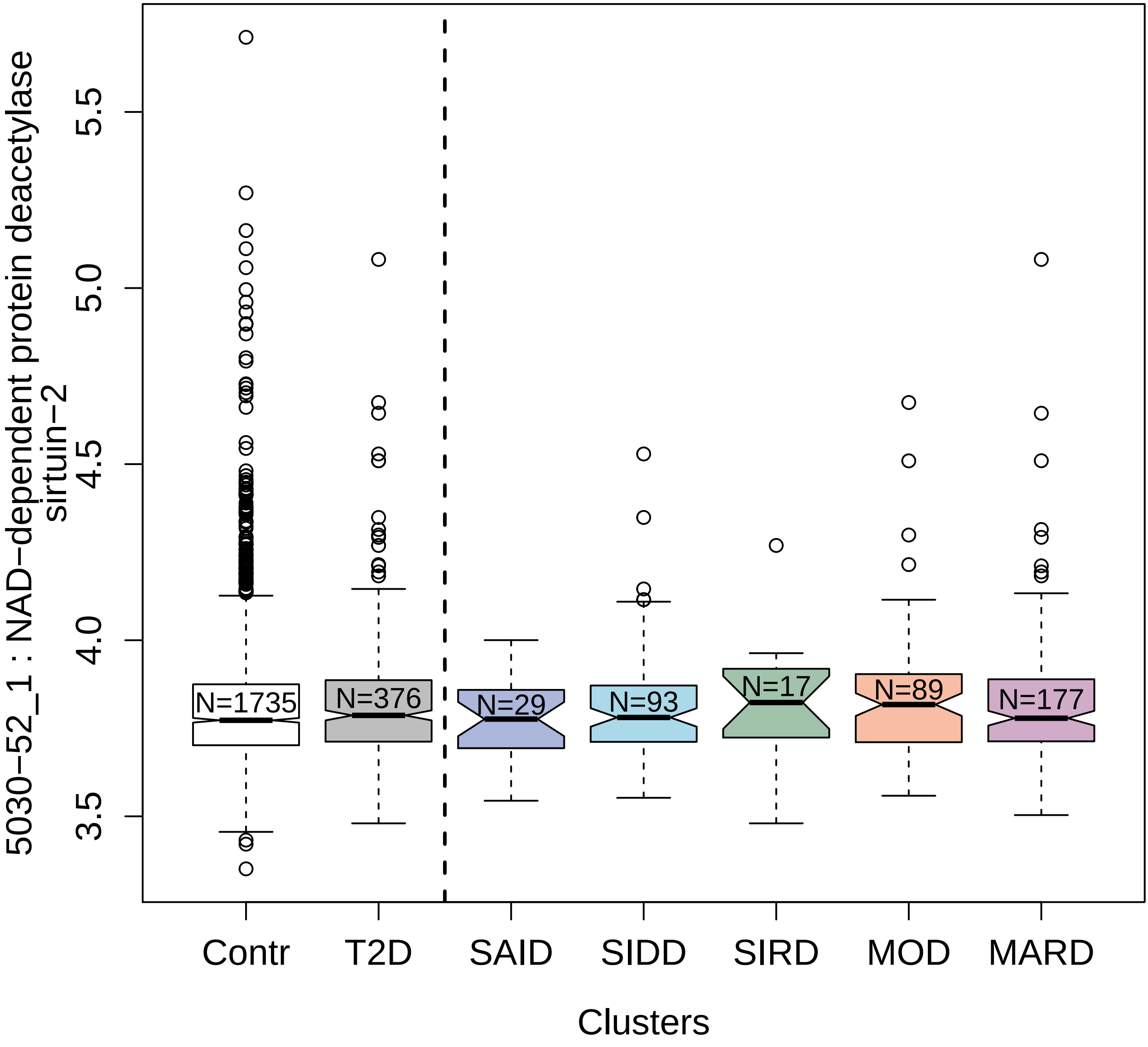
5030−52_1 : NAD−dependent protein deacetylase
sirtuin−2
5.5
5.0
4.5
4.0
3.5
N=1735
N=376
N=29
N=93
N=17
N=89
N=177
Contr
T2D
SAID
SIDD
SIRD
MOD
MARD
Clusters

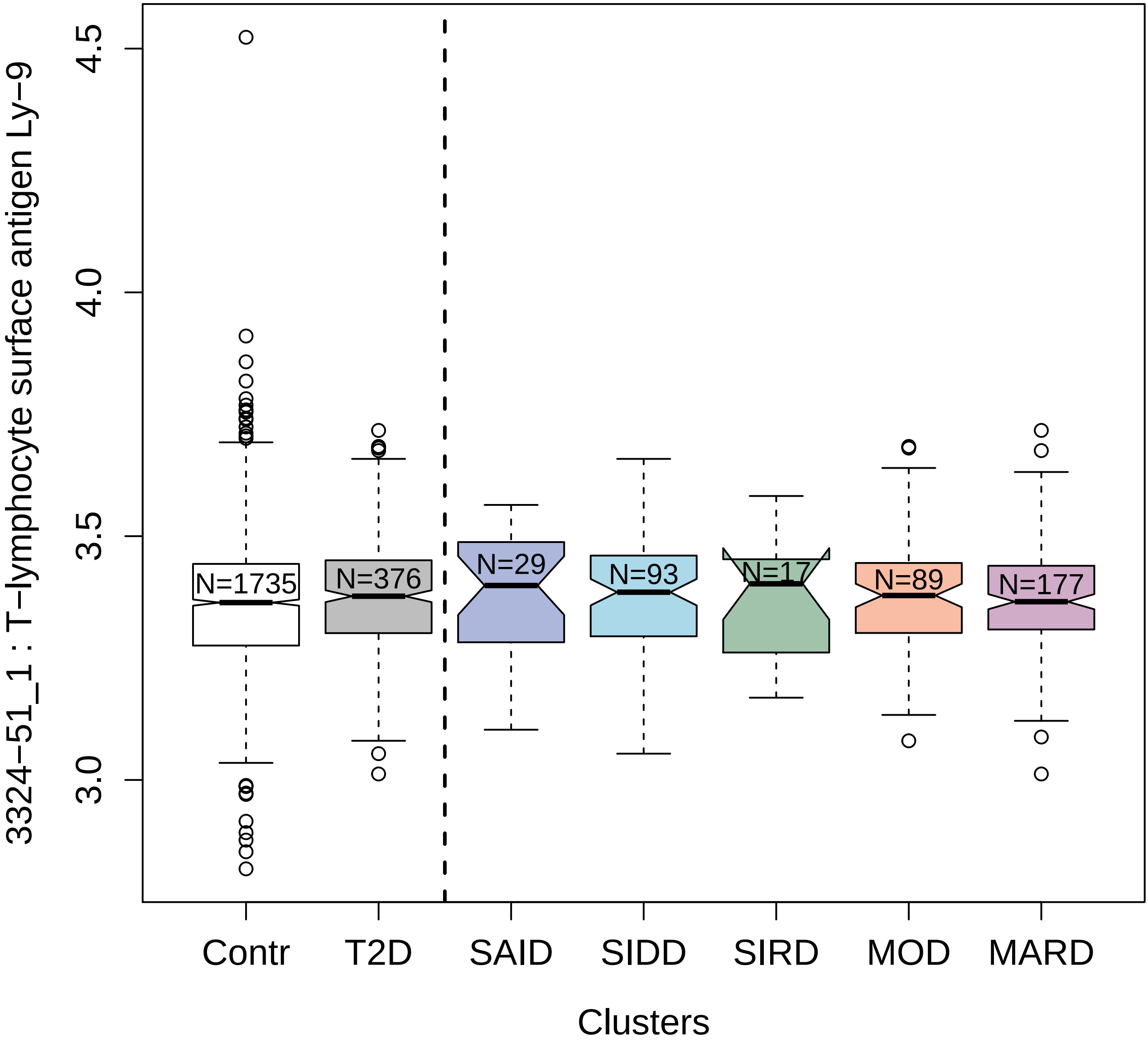
3324−51_1 : T−lymphocyte surface antigen Ly−9
4.5
4.0
3.5
3.0
N=1735
N=376
N=29
N=93
N=17
N=89
N=177
Contr
T2D
SAID
SIDD
SIRD
MOD
MARD
Clusters

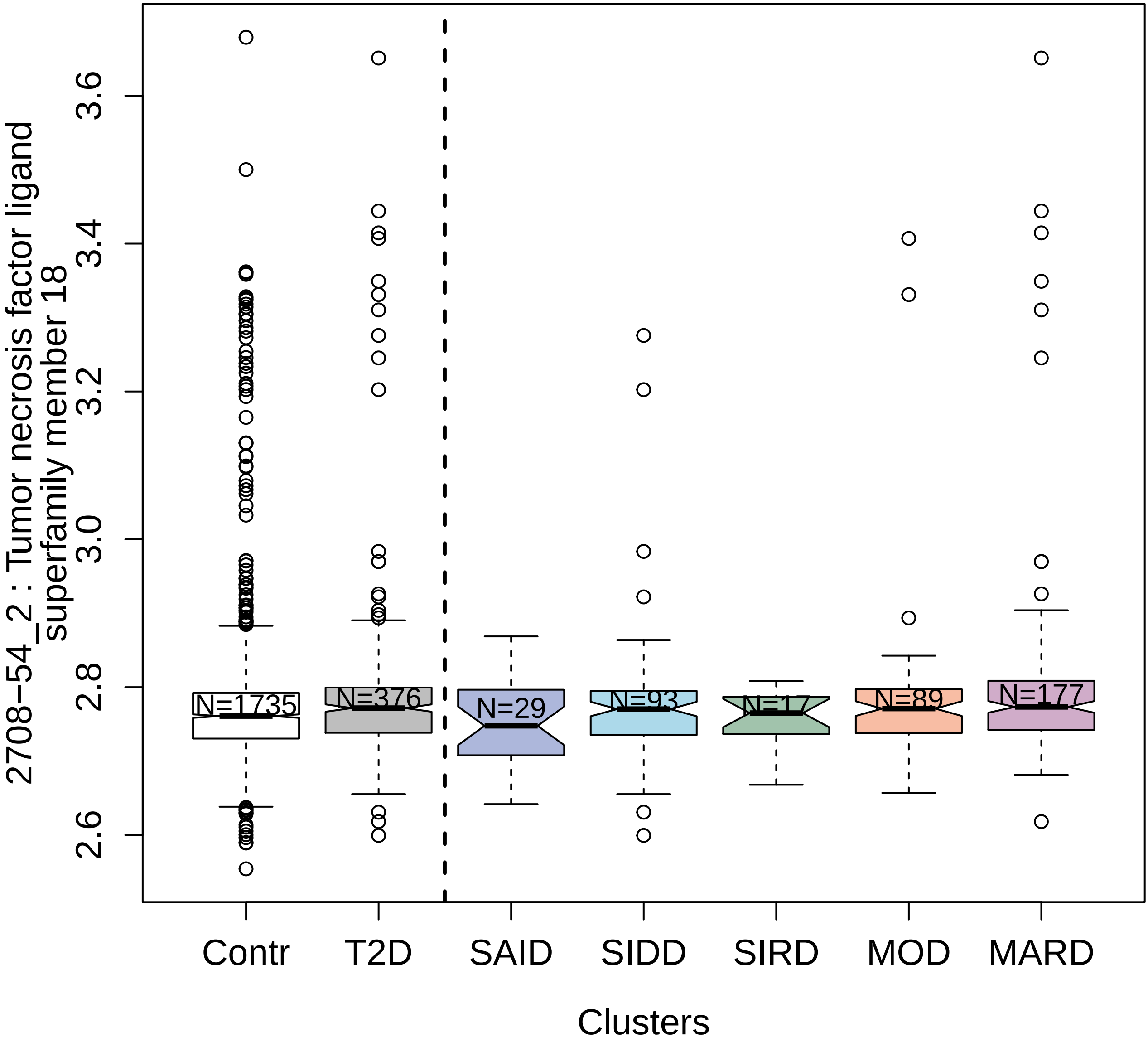

2708−54_2 : Tumor necrosis factor ligand superfamily member 18
2.6
2.8
3.0
3.2
3.4
3.6
N=1735
N=376
N=29
N=93
N=17
N=89
N=177
Contr
T2D
SAID
SIDD
SIRD
MOD
MARD
Clusters

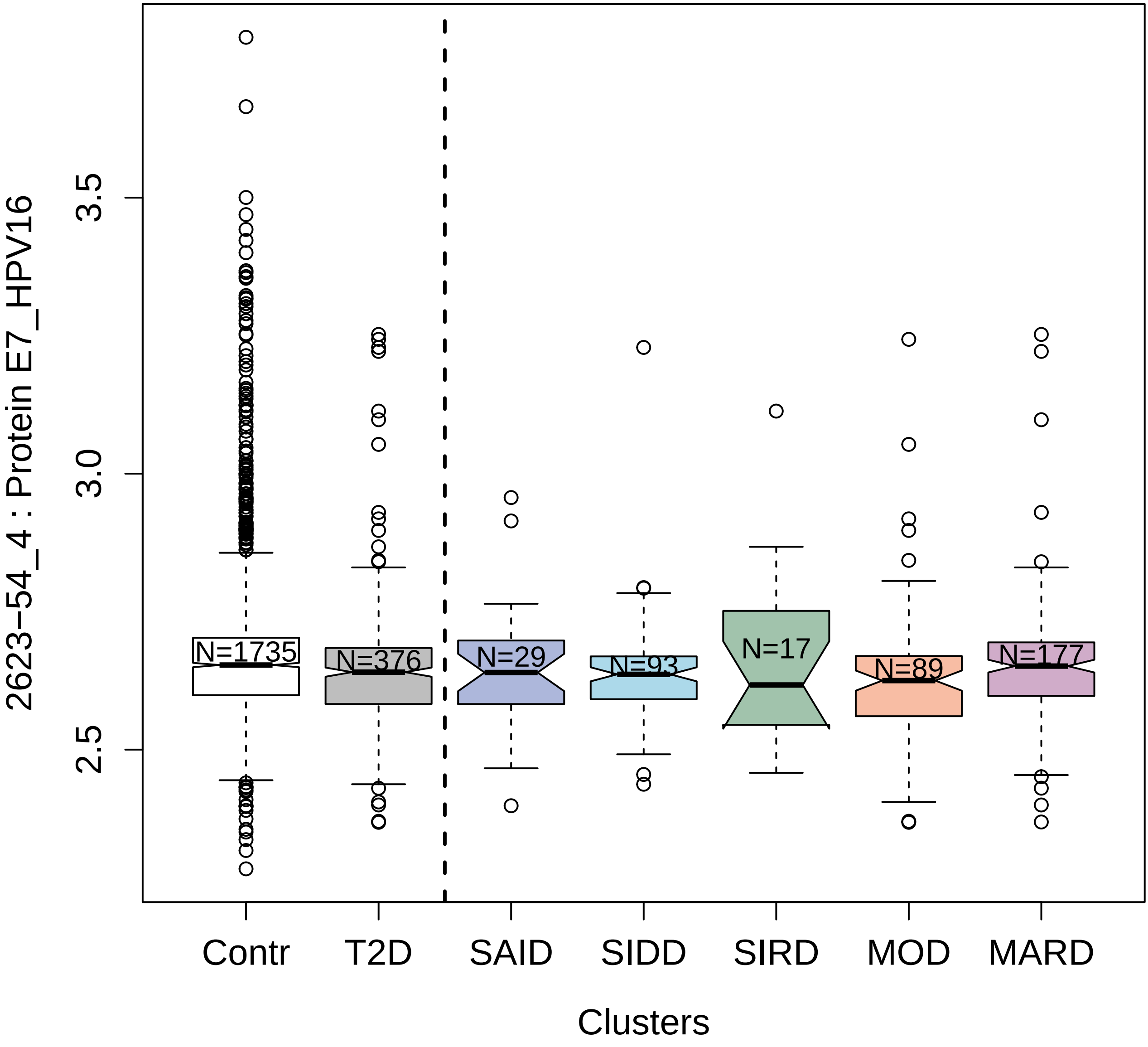

2623−54_4 : Protein E7_HPV16
3.5
3.0
2.5
N=1735
N=376
N=29
N=93
N=17
N=89
N=177
Contr
T2D
SAID
SIDD
SIRD
MOD
MARD
Clusters

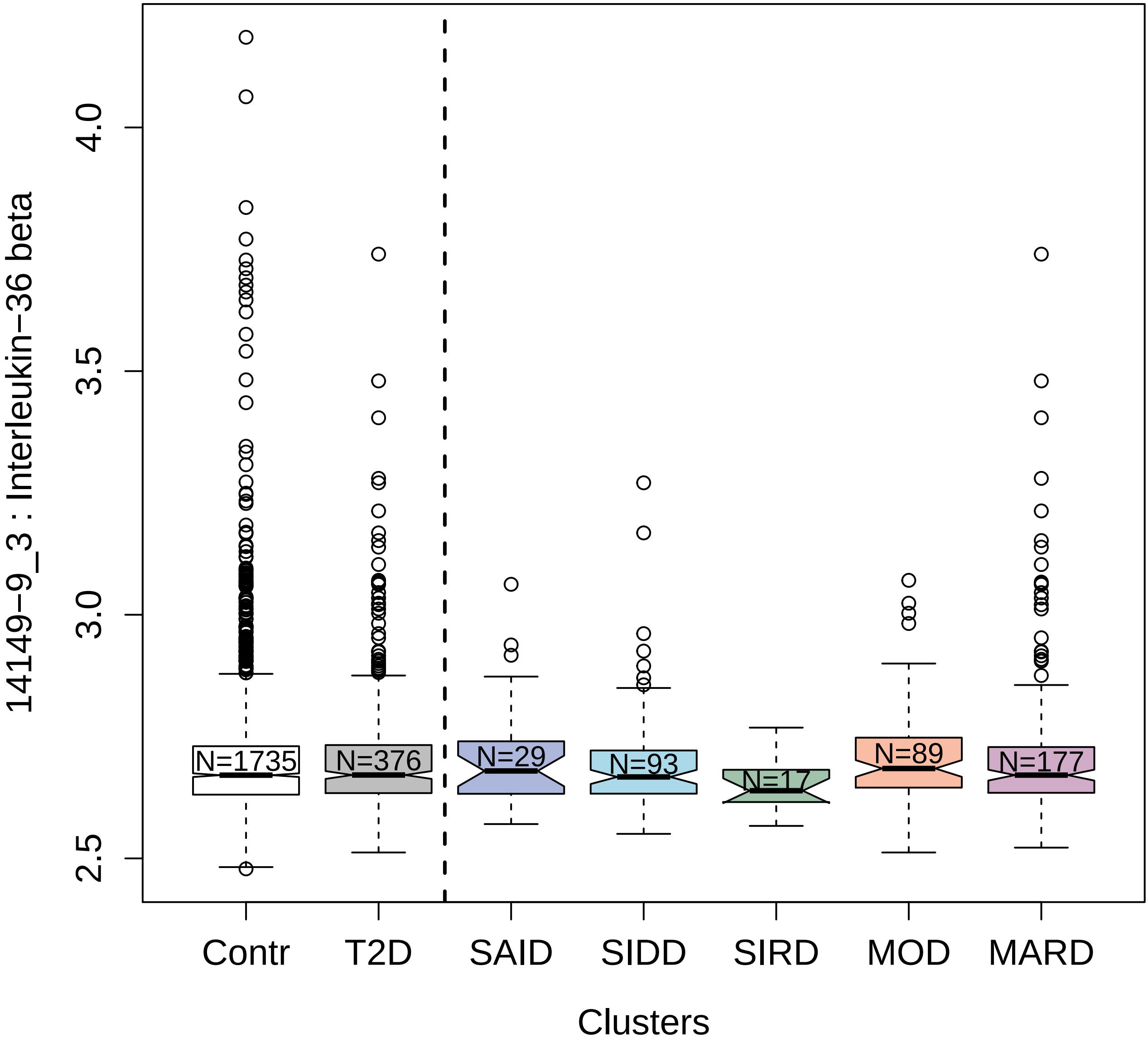
14149−9_3 : Interleukin−36 beta
4.0
3.5
3.0
2.5
N=1735
N=376
N=29
N=93
N=17
N=89
N=177
Contr
T2D
SAID
SIDD
SIRD
MOD
MARD
Clusters

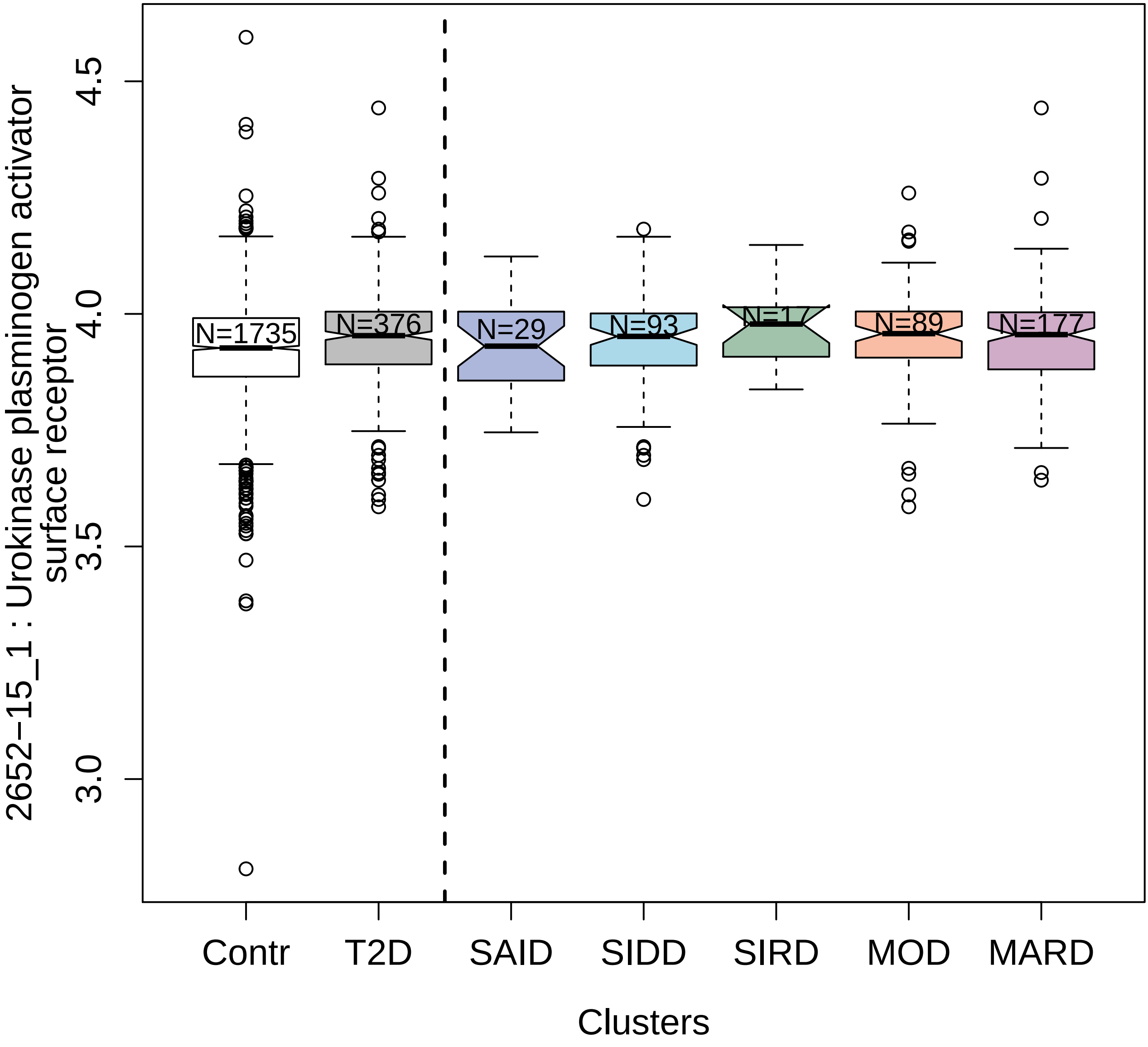

2652−15_1 : Urokinase plasminogen activator surface receptor
4.5
4.0
3.5
3.0
N=1735
N=376
N=29
N=93
N=17
N=89
N=177
Contr
T2D
SAID
SIDD
SIRD
MOD
MARD
Clusters

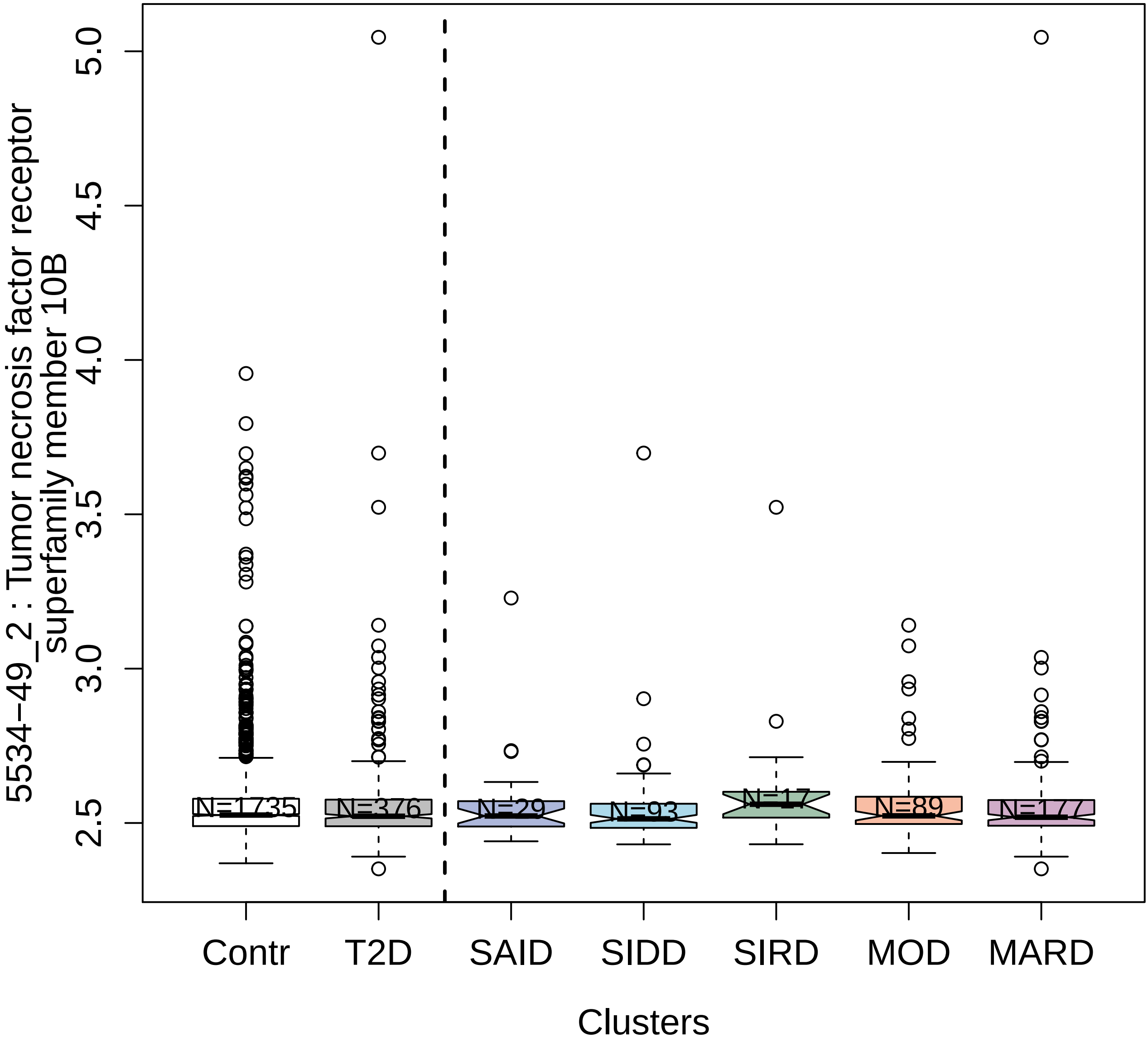
5534−49_2 : Tumor necrosis factor receptor
superfamily member 10B
5.0
4.5
4.0
3.5
3.0
2.5
N=1735
N=376
N=29
N=93
N=17
N=89
N=177
Contr
T2D
SAID
SIDD
SIRD
MOD
MARD
Clusters

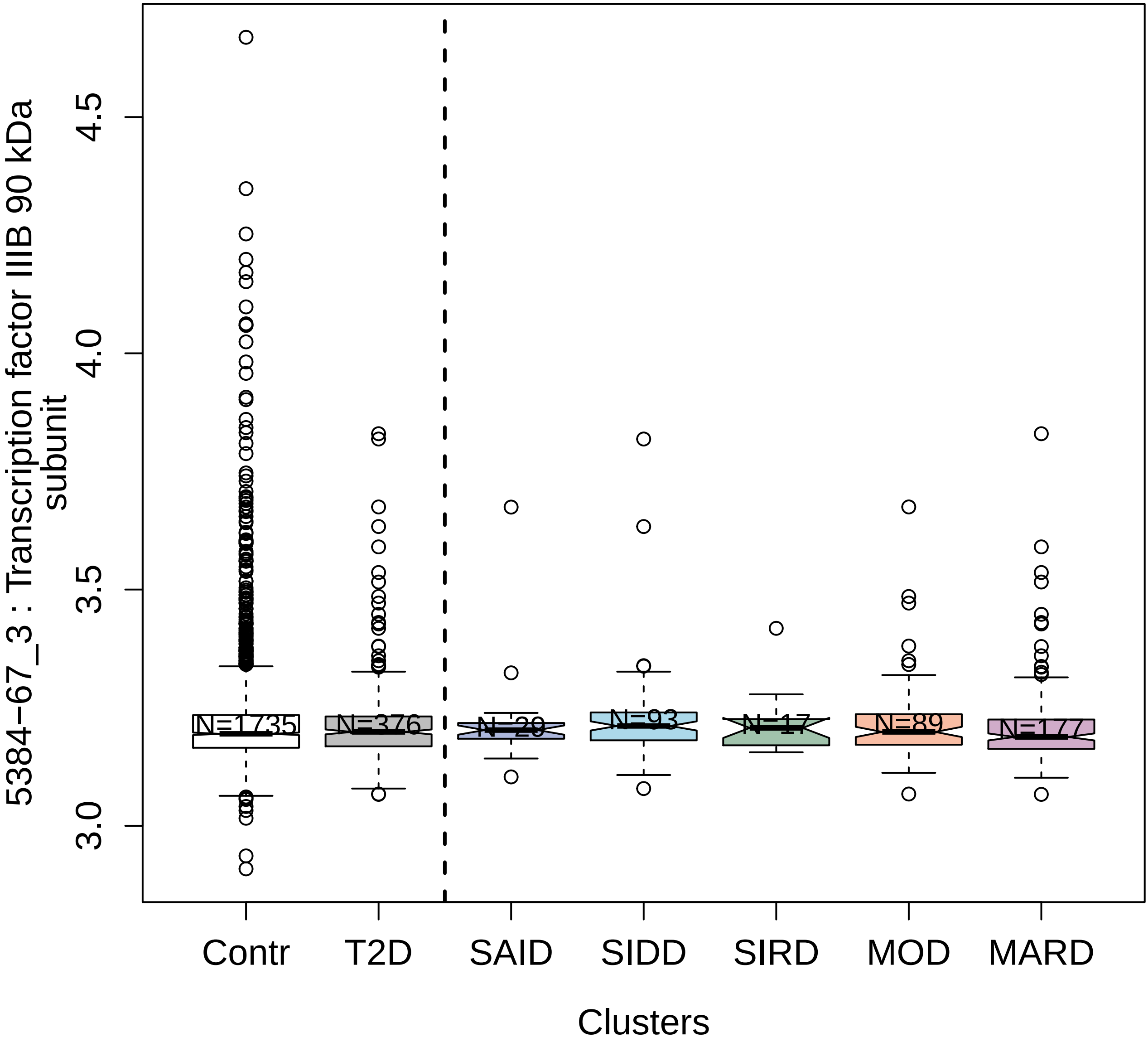
5384−67_3 : Transcription factor IIIB 90 kDa subunit
3.0
3.5
4.0
4.5
N=1735
N=376
N=20
N=93
N=17
N=89
N=177
Contr
T2D
SAID
SIDD
SIRD
MOD
MARD
Clusters

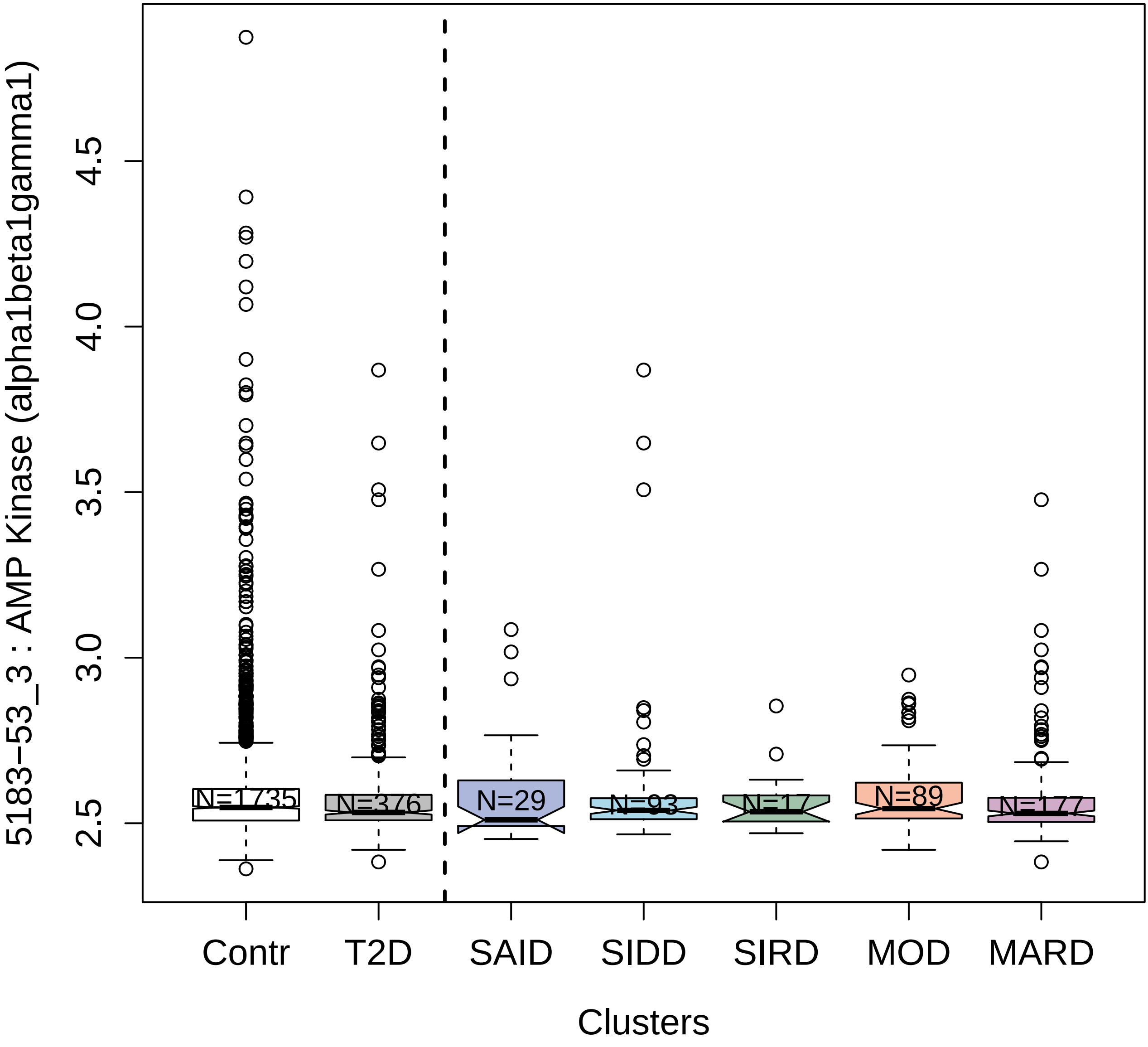
5183−53_3 : AMP Kinase (alpha1beta1gamma1)
2.5
3.0
3.5
4.0
4.5
N=1735
N=376
N=29
N=93
N=17
N=89
N=177
Contr
T2D
SAID
SIDD
SIRD
MOD
MARD
Clusters

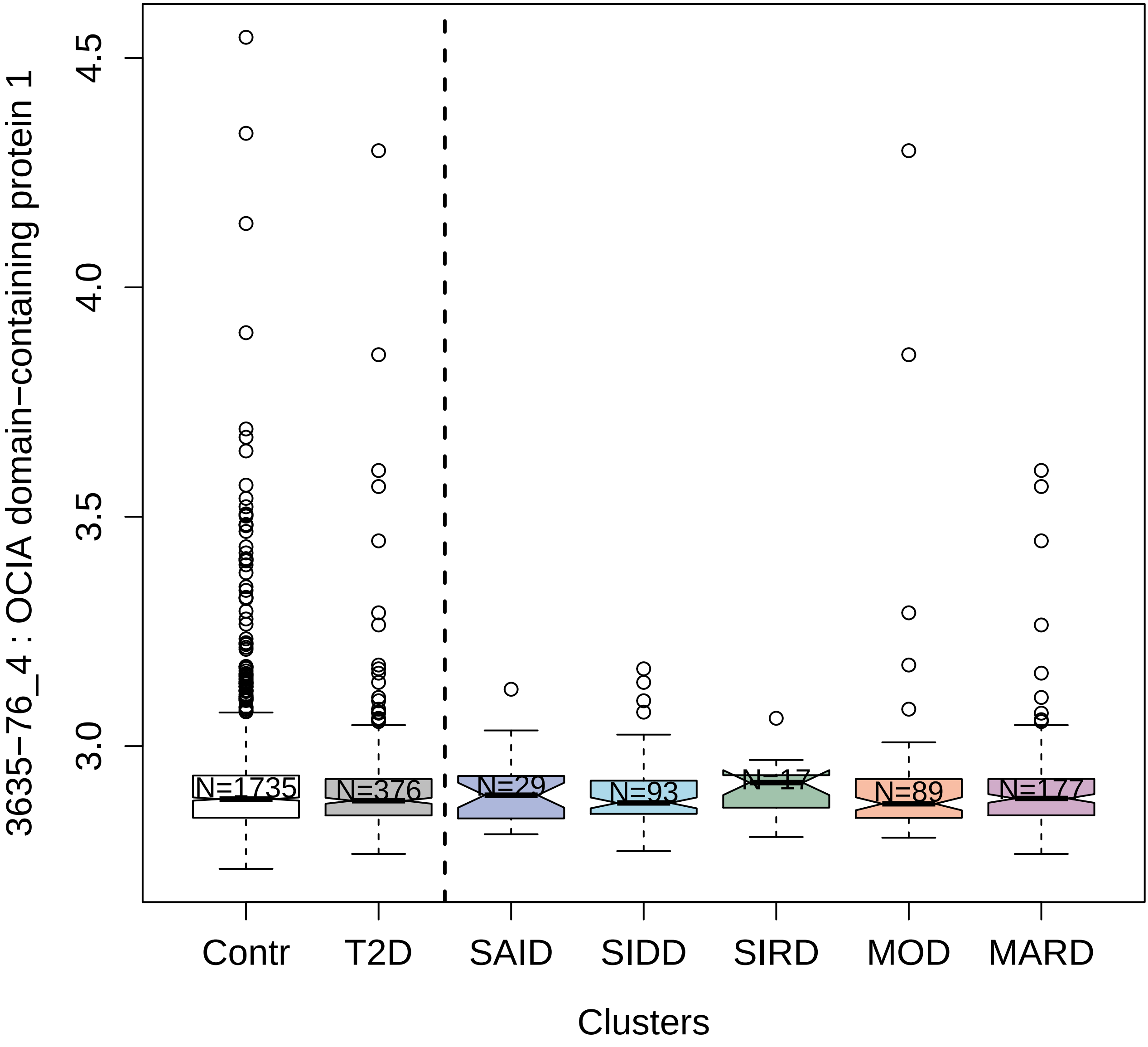

3635−76_4 : OCIA domain−containing protein 1
4.5
4.0
3.5
3.0
N=1735
N=376
N=29
N=93
N=17
N=89
N=177
Contr
T2D
SAID
SIDD
SIRD
MOD
MARD
Clusters

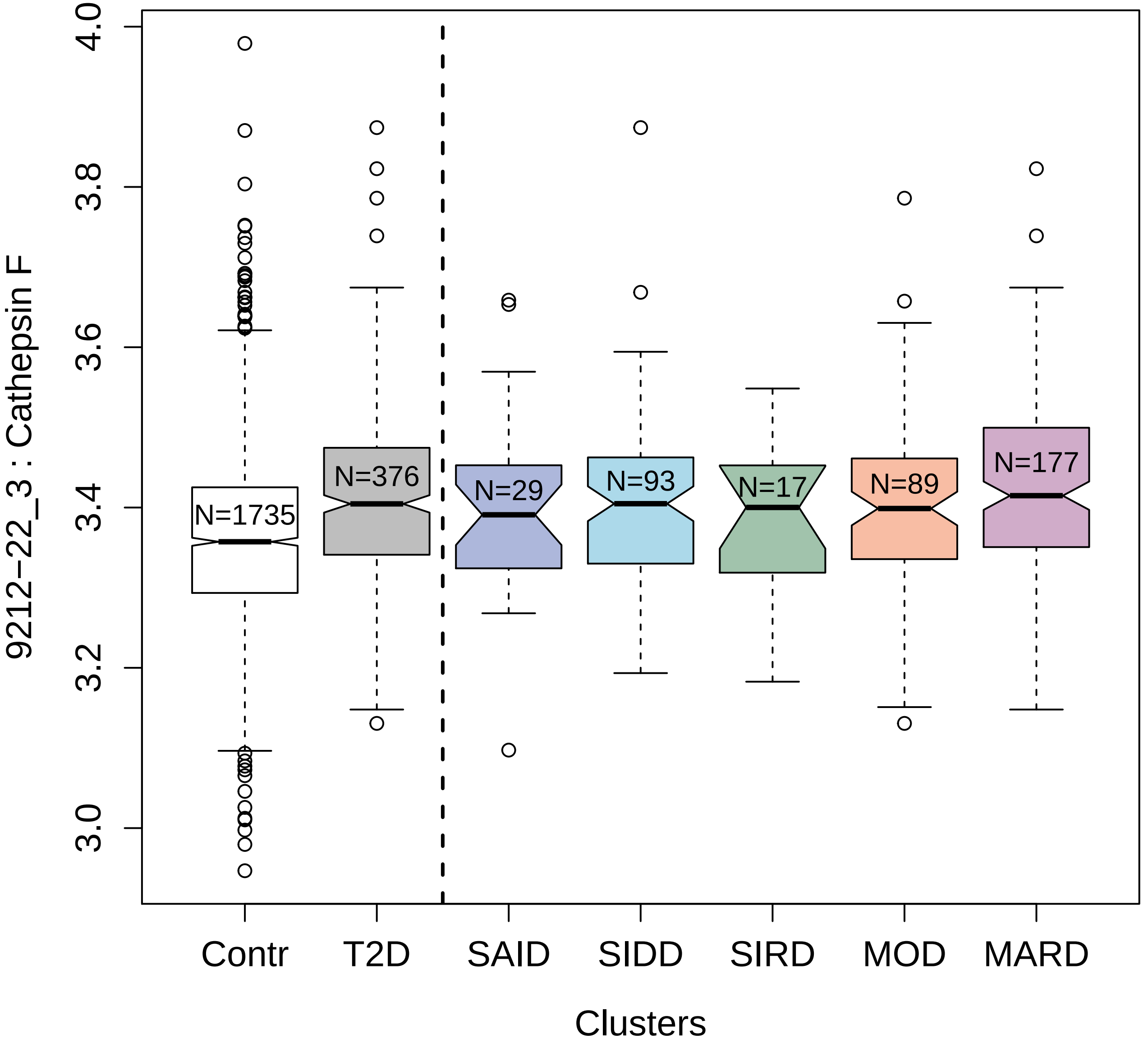

9212−22_3 : Cathepsin F
4.0
3.8
3.6
3.4
3.2
3.0
N=1735
N=376
N=29
N=93
N=17
N=89
N=177
Contr
T2D
SAID
SIDD
SIRD
MOD
MARD
Clusters

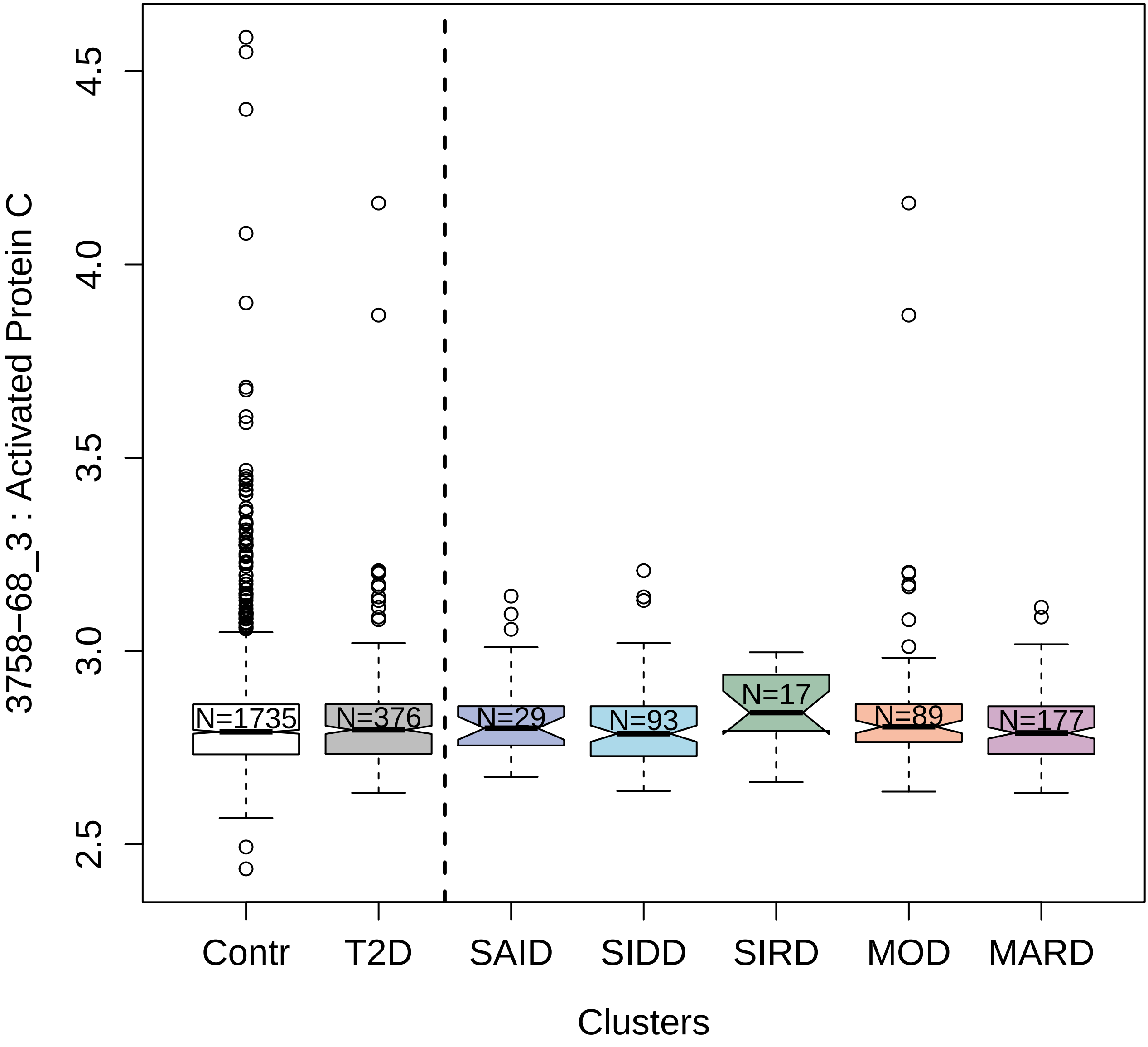
3758−68_3 : Activated Protein C
4.5
4.0
3.5
3.0
2.5
N=1735
N=376
N=29
N=93
N=17
N=89
N=177
Contr
T2D
SAID
SIDD
SIRD
MOD
MARD
Clusters

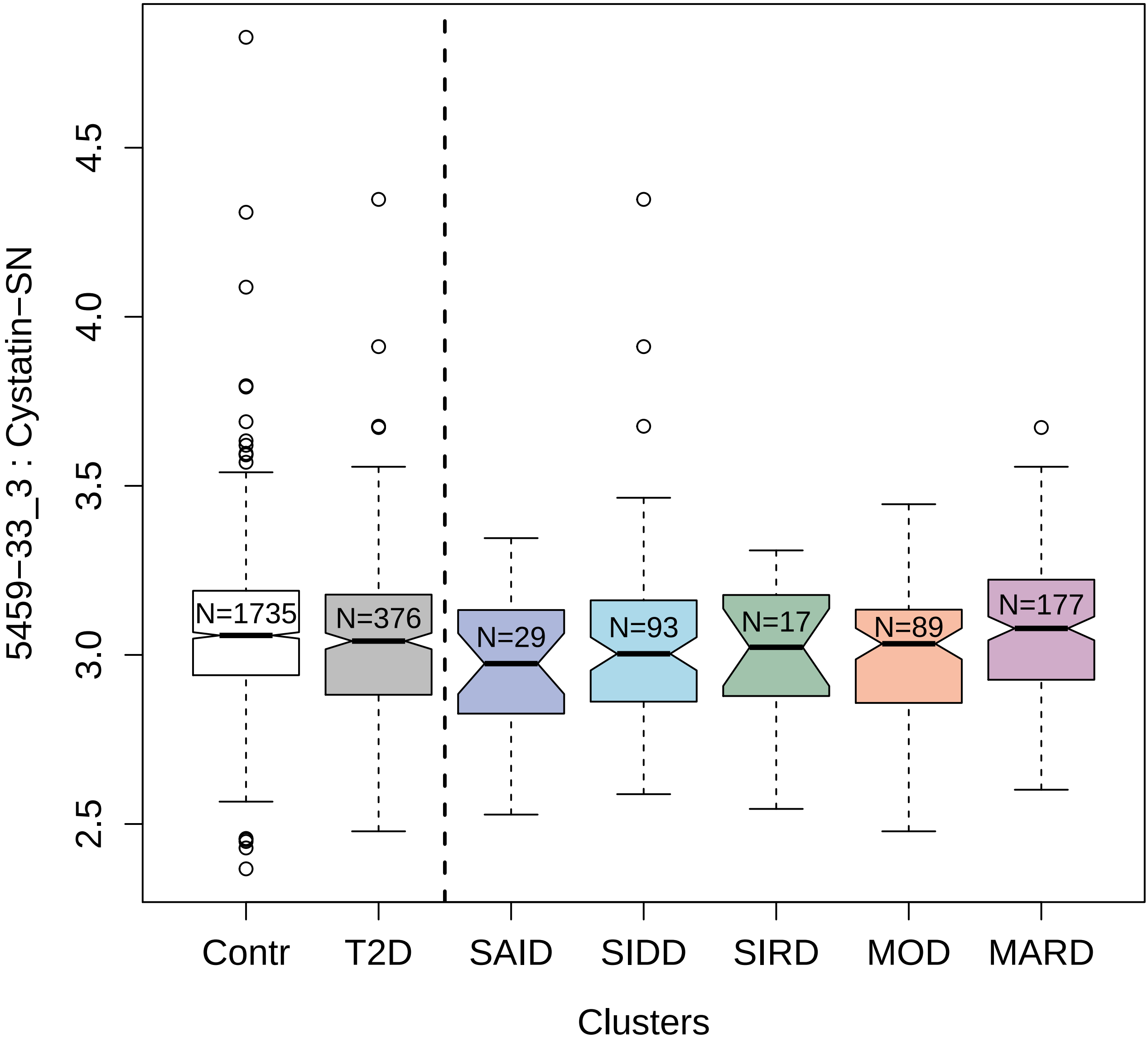

5459−33_3 : Cystatin−SN
4.5
4.0
3.5
3.0
2.5
N=1735
N=376
N=29
N=93
N=17
N=89
N=177
Contr
T2D
SAID
SIDD
SIRD
MOD
MARD
Clusters

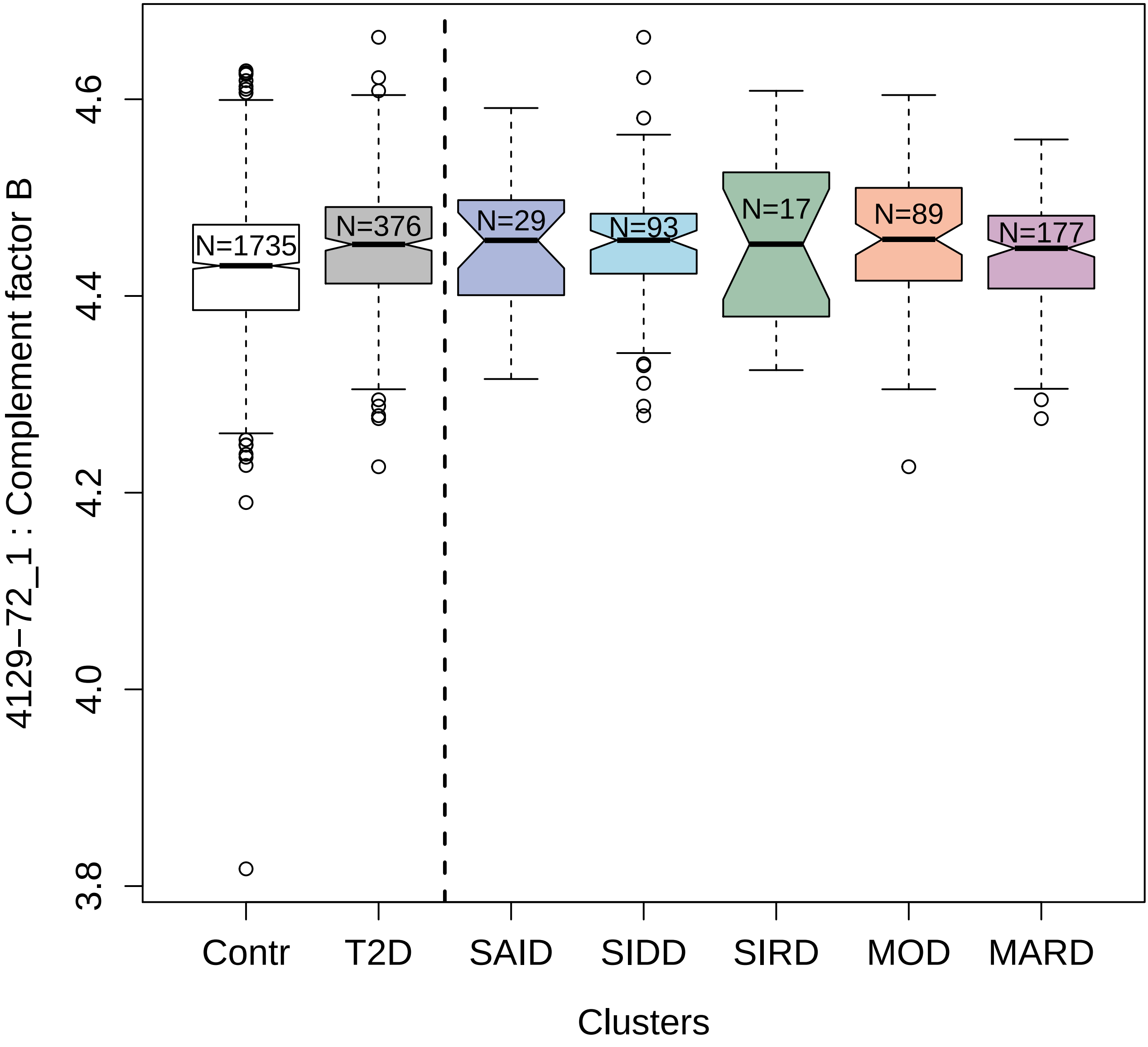
4129−72_1 : Complement factor B
4.6
4.4
4.2
4.0
3.8
N=1735
N=376
N=29
N=93
N=17
N=89
N=177
Contr
T2D
SAID
SIDD
SIRD
MOD
MARD
Clusters

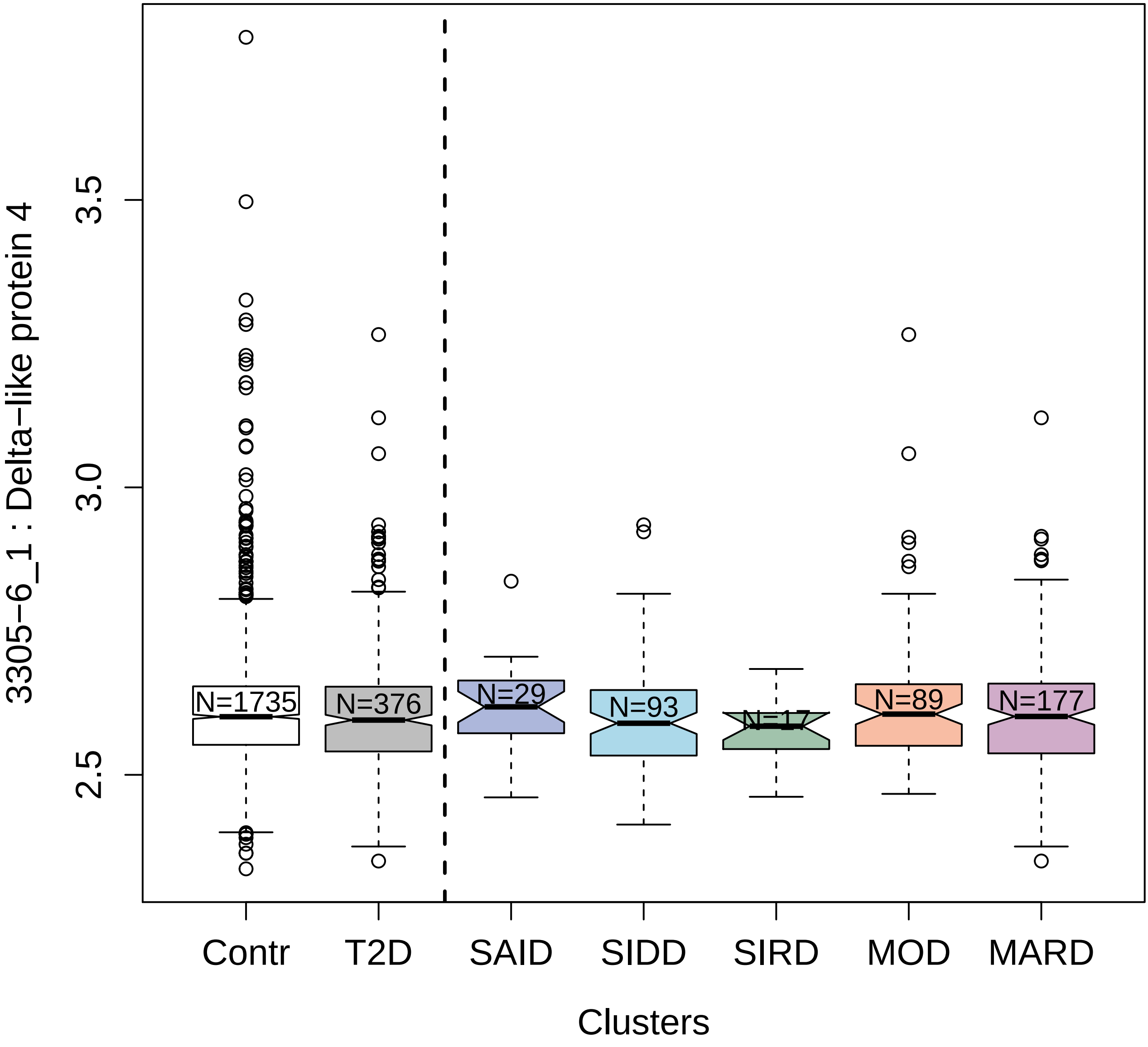

3305−6_1 : Delta−like protein 4
3.5
3.0
2.5
N=1735
N=376
N=29
N=93
N=17
N=89
N=177
Contr
T2D
SAID
SIDD
SIRD
MOD
MARD
Clusters

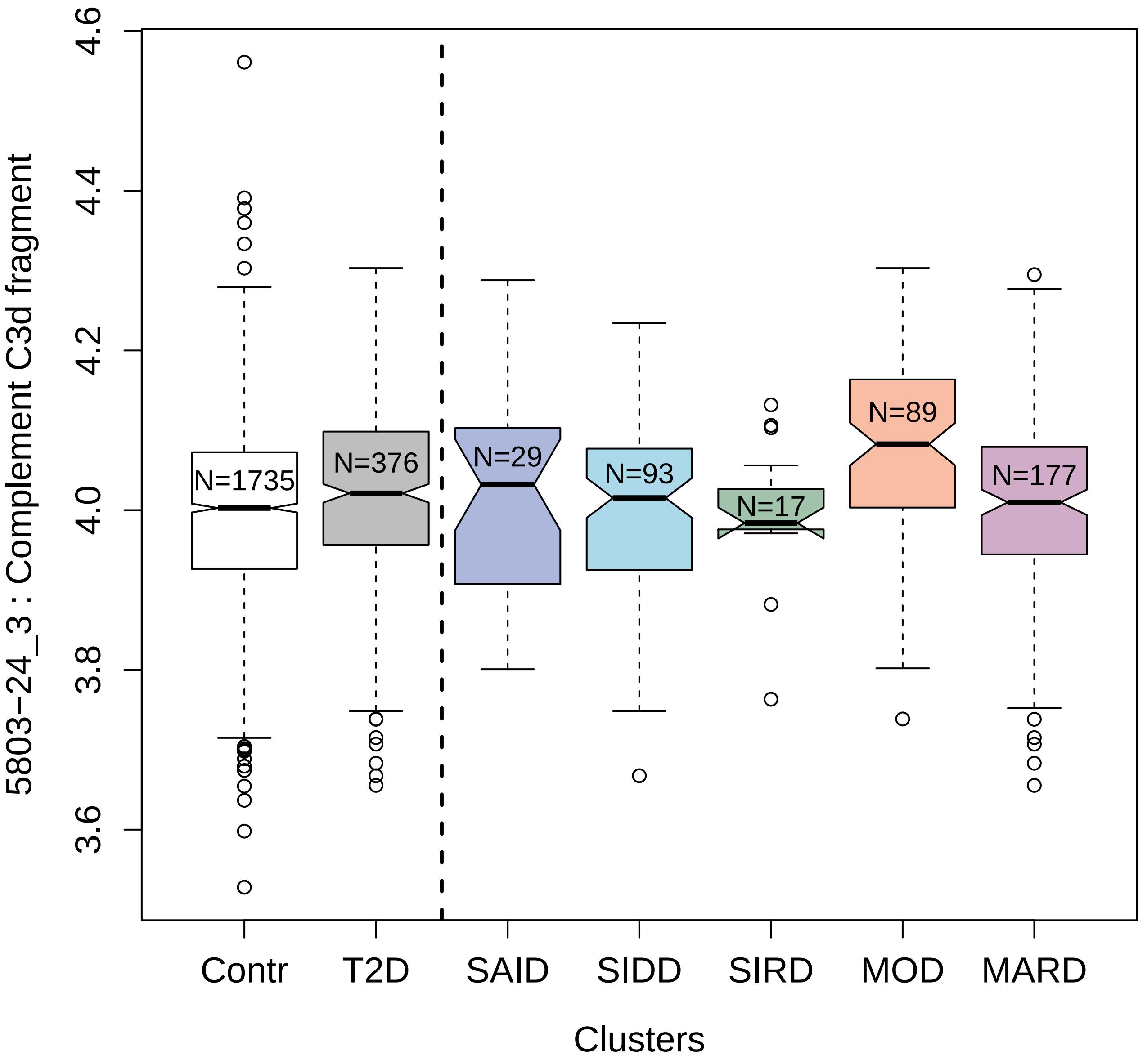
5803−24_3 : Complement C3d fragment
4.6
4.4
4.2
4.0
3.8
3.6
N=1735
N=376
N=29
N=93
N=17
N=89
N=177
Contr
T2D
SAID
SIDD
SIRD
MOD
MARD
Clusters

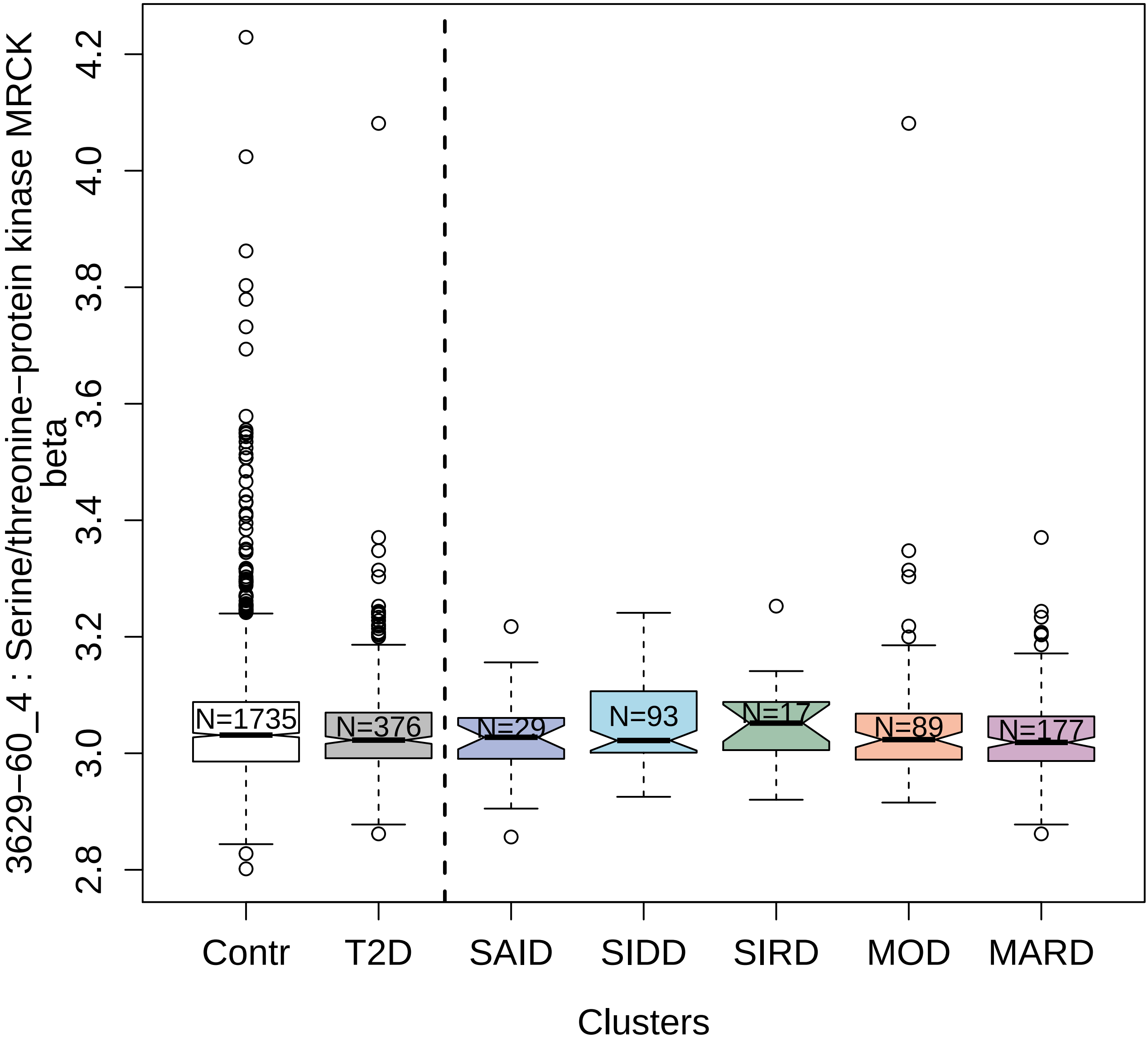
3629−60_4 : Serine/threonine−protein kinase MRCK beta
4.2
4.0
3.8
3.6
3.4
3.2
3.0
2.8
N=1735
N=376
N=29
N=93
N=17
N=89
N=177
Contr
T2D
SAID
SIDD
SIRD
MOD
MARD
Clusters

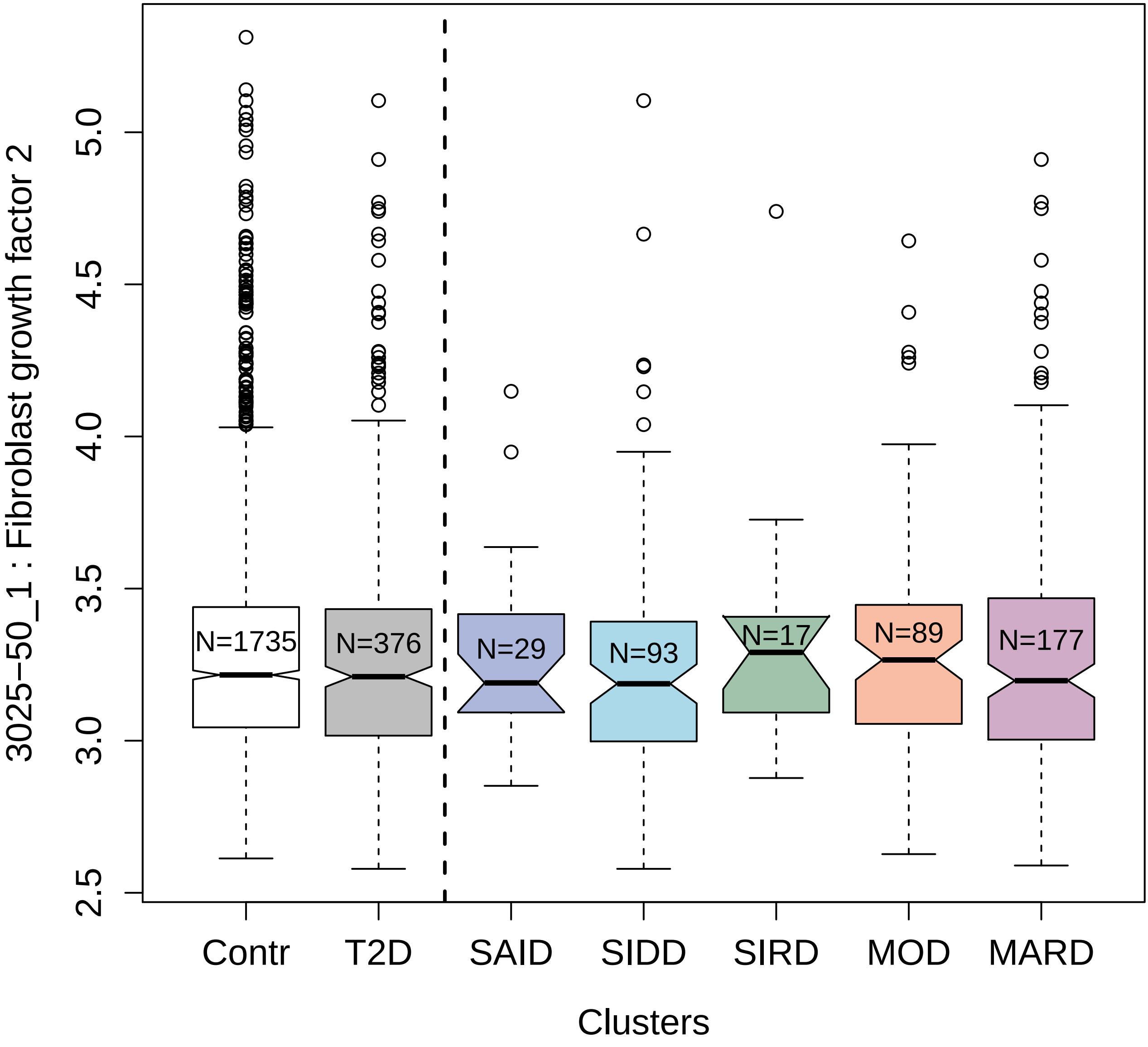
3025−50_1 : Fibroblast growth factor 2
2.5
3.0
3.5
4.0
4.5
5.0
N=1735
N=376
N=29
N=93
N=17
N=89
N=177
Contr
T2D
SAID
SIDD
SIRD
MOD
MARD
Clusters

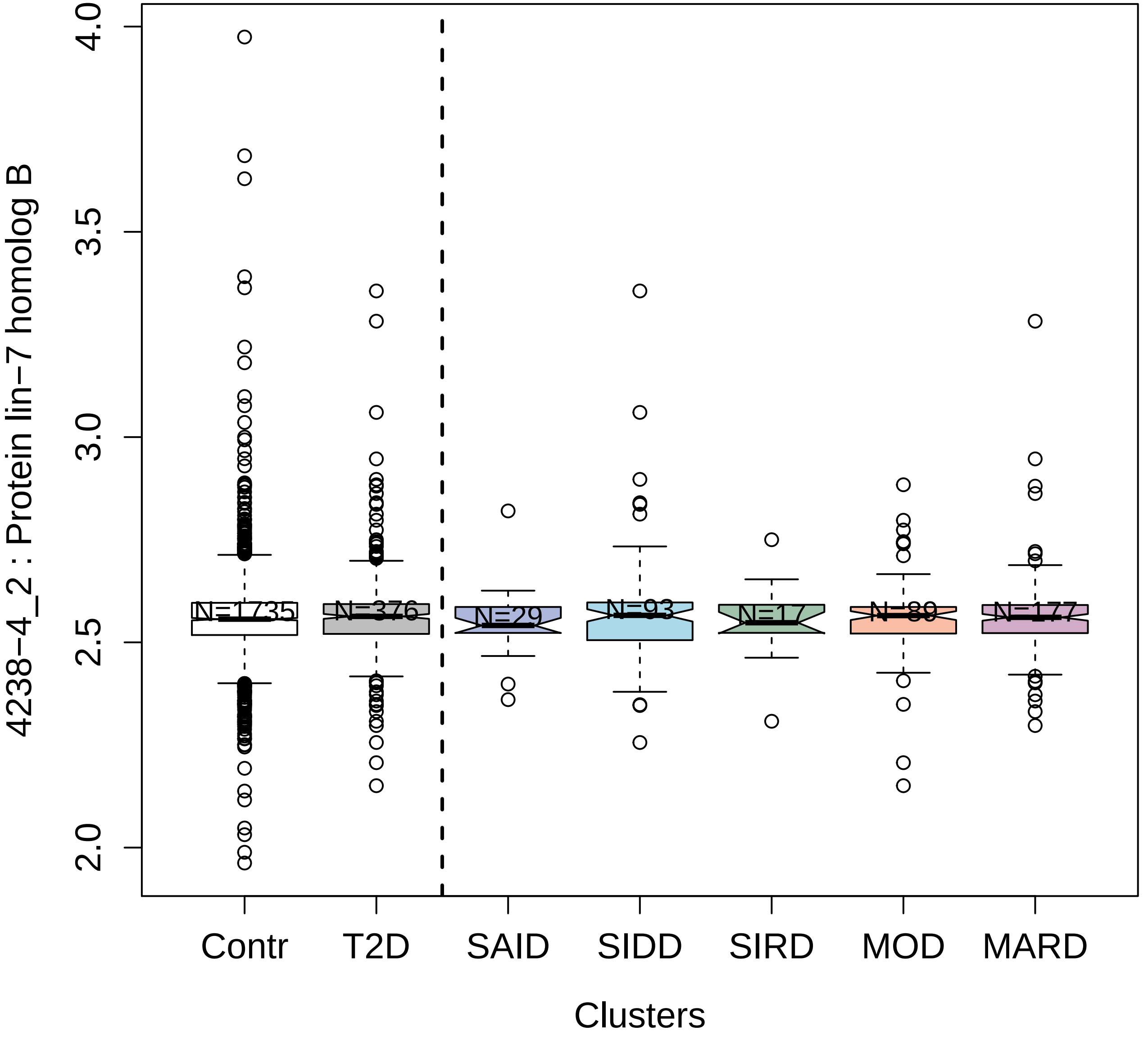

4238−4_2 : Protein lin−7 homolog B
4.0
3.5
3.0
2.5
2.0
N=1735
N=376
N=29
N=93
N=17
N=89
N=177
Contr
T2D
SAID
SIDD
SIRD
MOD
MARD
Clusters

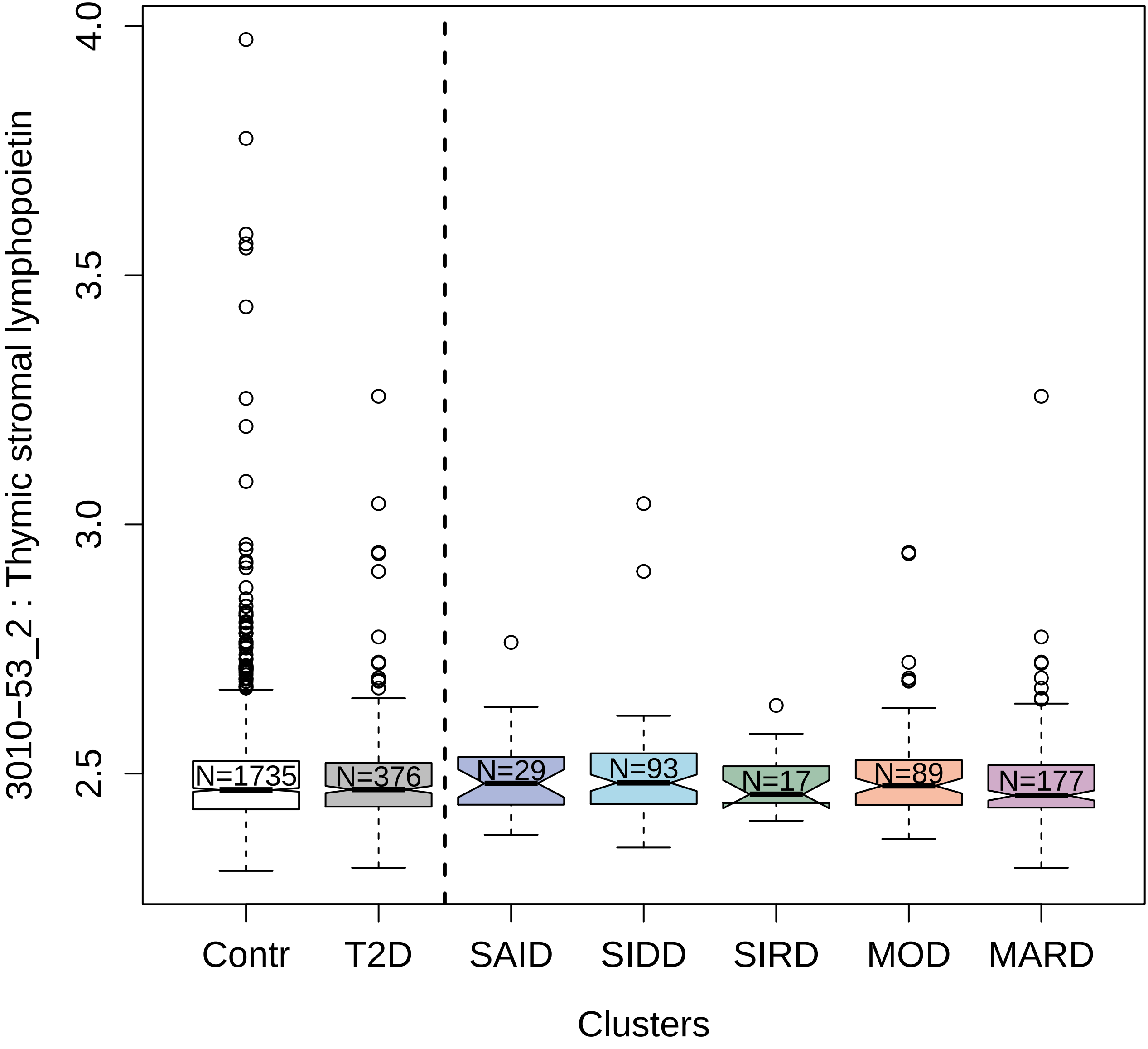
3010−53_2 : Thymic stromal lymphopoietin
4.0
3.5
3.0
2.5
N=1735
N=376
N=29
N=93
N=17
N=89
N=177
Contr
T2D
SAID
SIDD
SIRD
MOD
MARD
Clusters

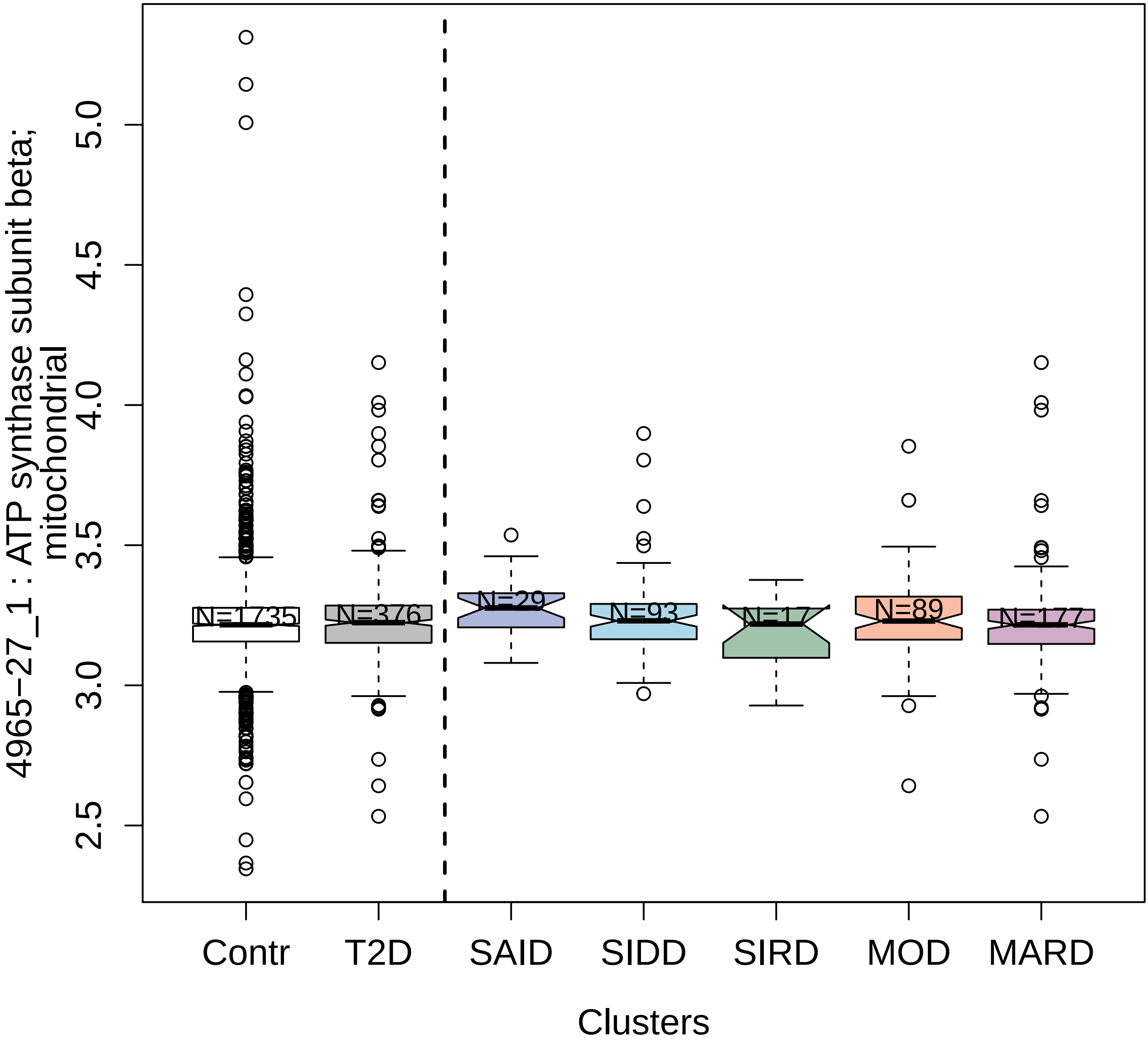

4965−27_1 : ATP synthase subunit beta; mitochondrial
2.5
3.0
3.5
4.0
4.5
5.0
N=1735
N=376
N=29
N=93
N=17
N=89
N=177
Contr
T2D
SAID
SIDD
SIRD
MOD
MARD
Clusters

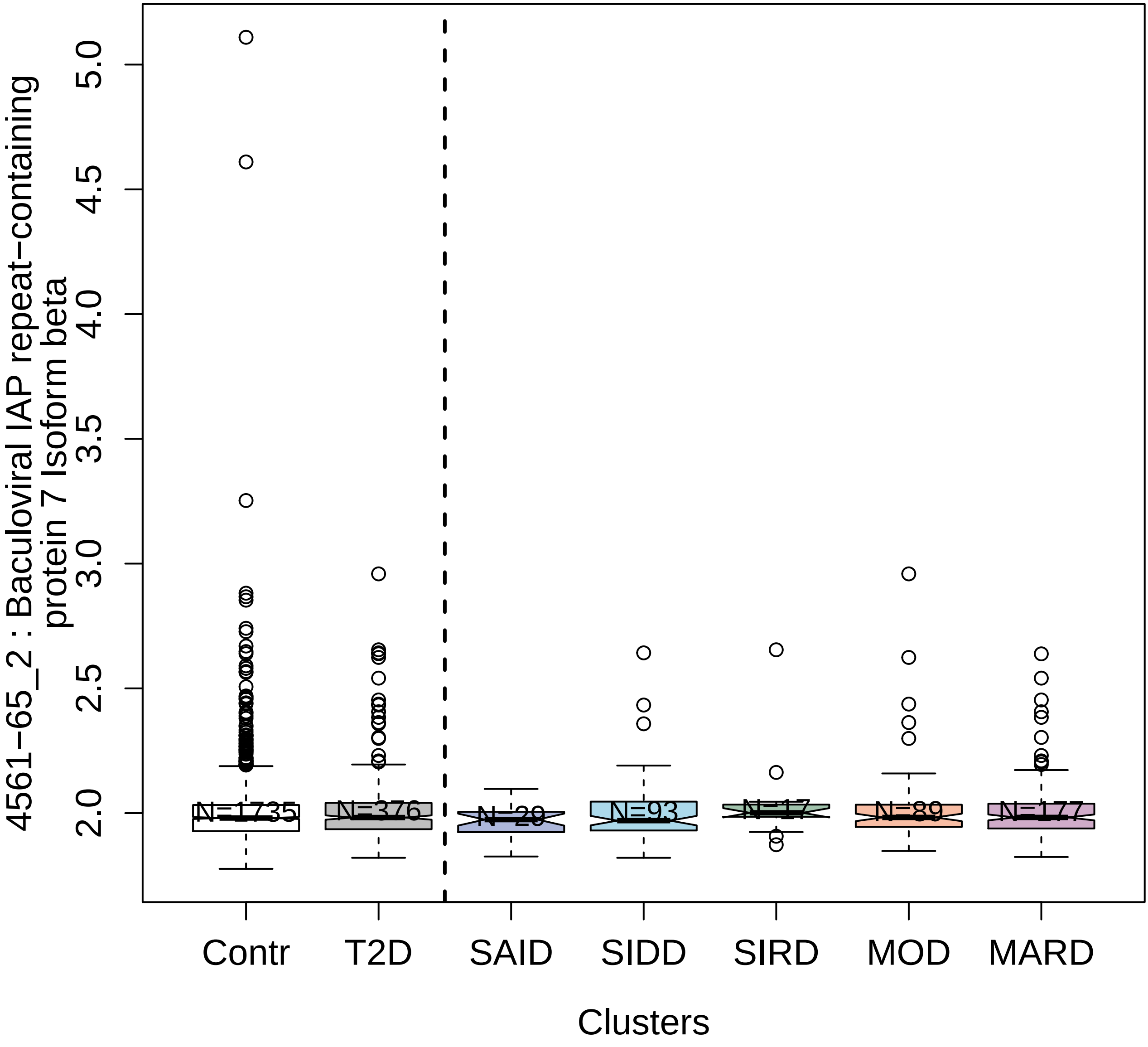
4561−65_2 : Baculoviral IAP repeat−containing protein 7 Isoform beta
5.0
4.5
4.0
3.5
3.0
2.5
2.0
N=1735
N=376
N=29
N=93
N=17
N=89
N=177
Contr
T2D
SAID
SIDD
SIRD
MOD
MARD
Clusters

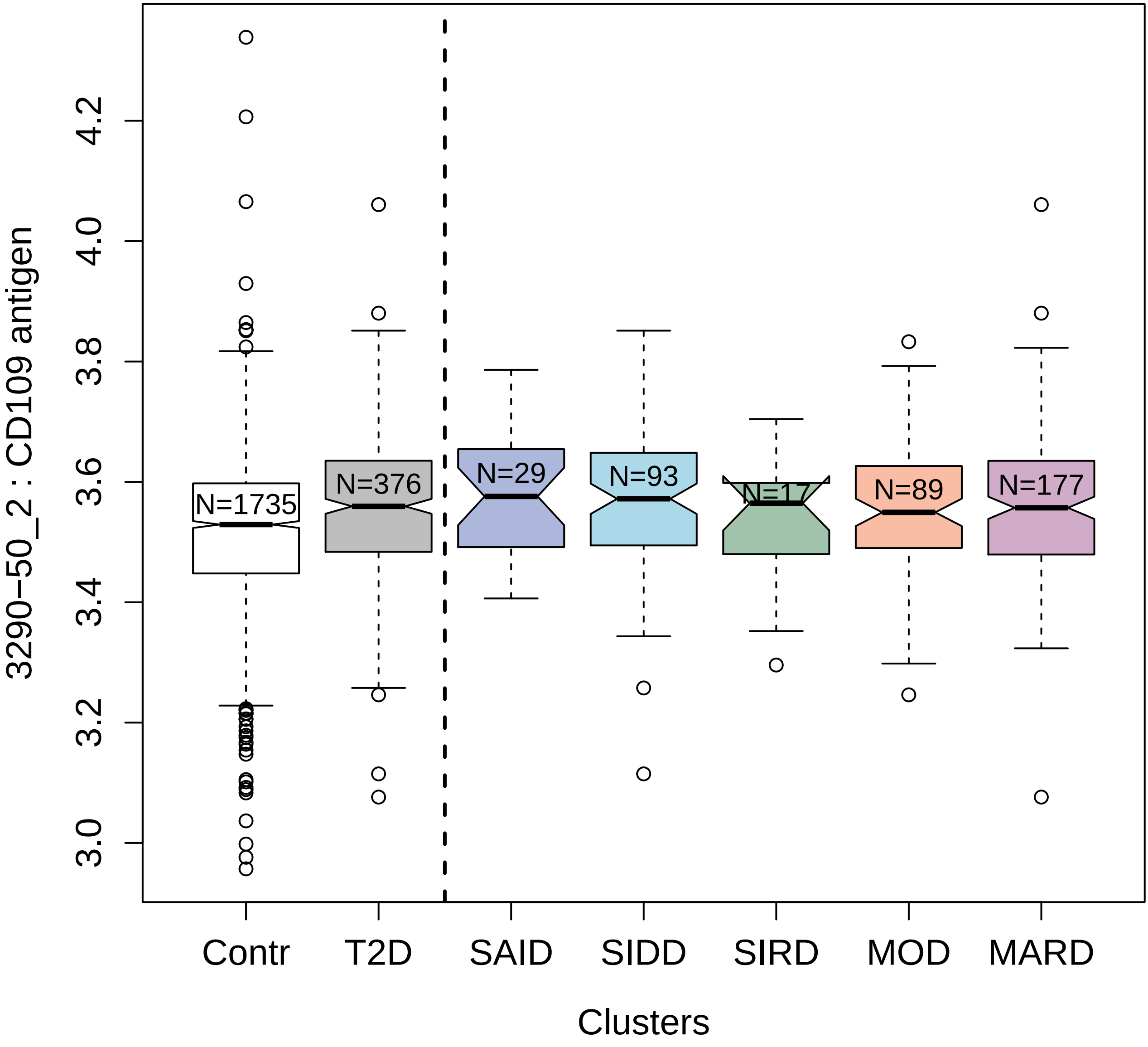
3290−50_2 : CD109 antigen
4.2
4.0
3.8
3.6
3.4
3.2
3.0
N=1735
N=376
N=29
N=93
N=17
N=89
N=177
Contr
T2D
SAID
SIDD
SIRD
MOD
MARD
Clusters

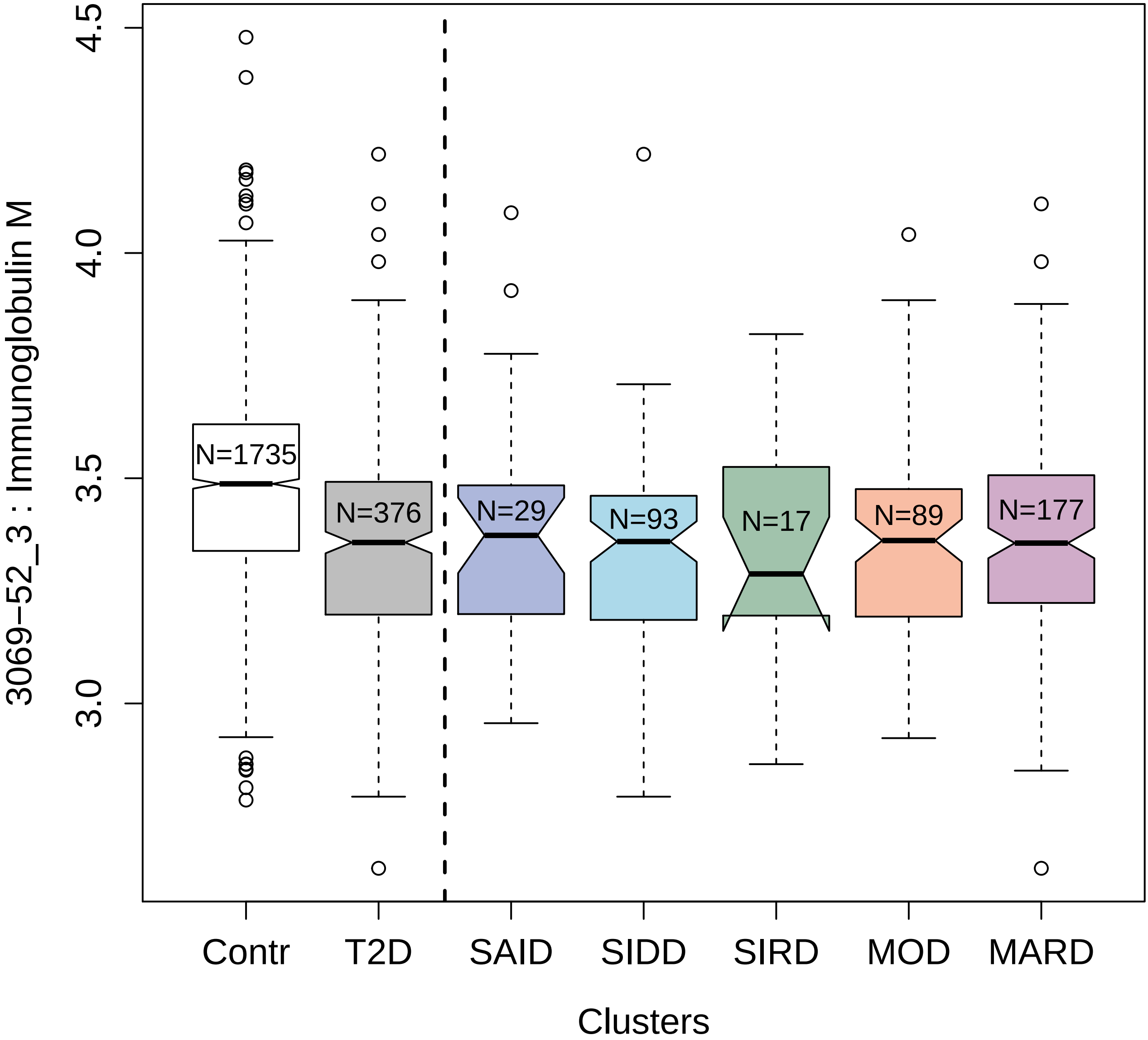
3069−52_3 : Immunoglobulin M
4.5
4.0
3.5
3.0
N=1735
N=376
N=29
N=93
N=17
N=89
N=177
Contr
T2D
SAID
SIDD
SIRD
MOD
MARD
Clusters

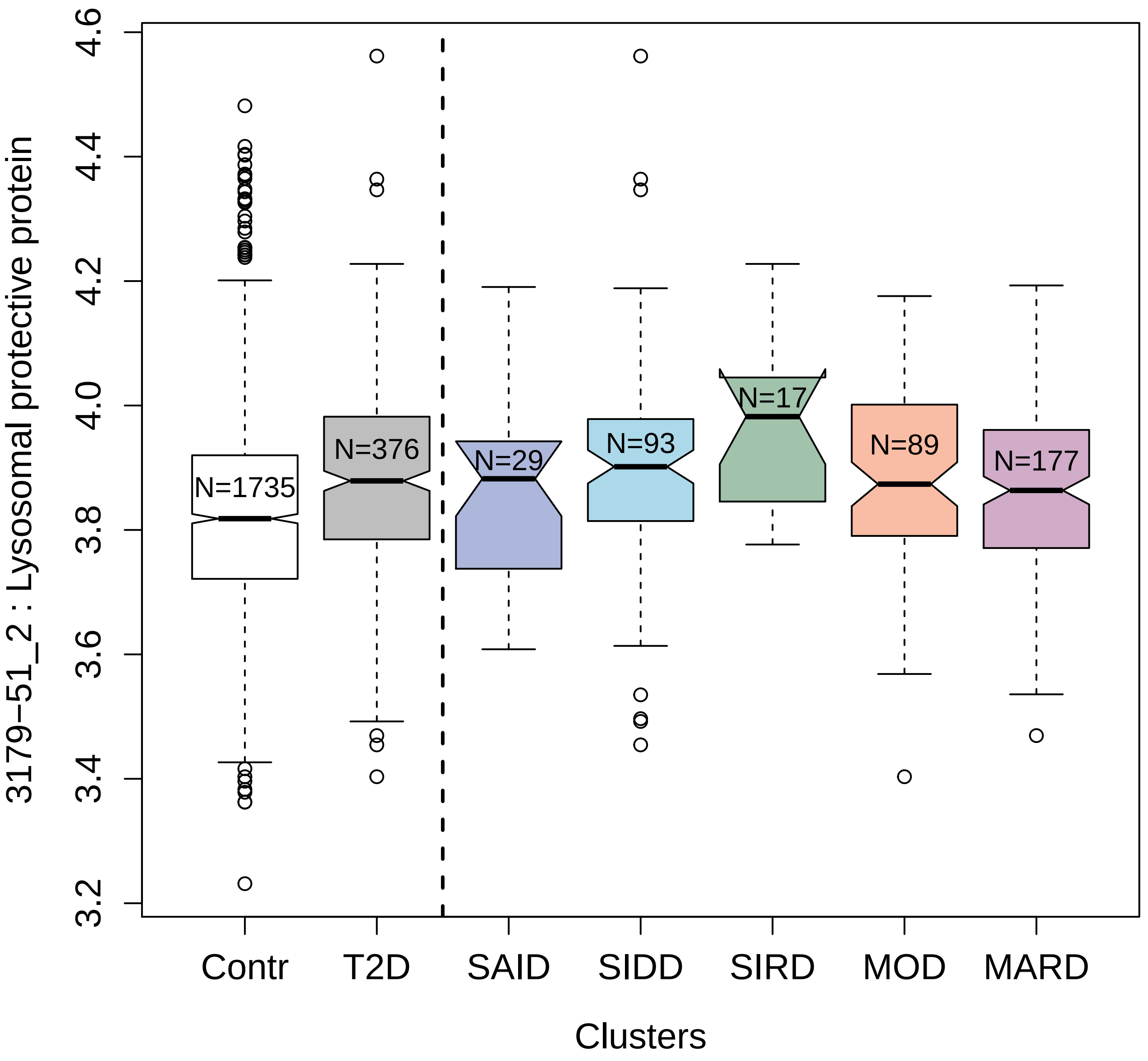

3179–51_2 : Lysosomal protective protein
3.2
3.4
3.6
3.8
4.0
4.2
4.4
4.6
N=1735
N=376
N=29
N=93
N=17
N=89
N=177
Contr
T2D
SAID
SIDD
SIRD
MOD
MARD
Clusters

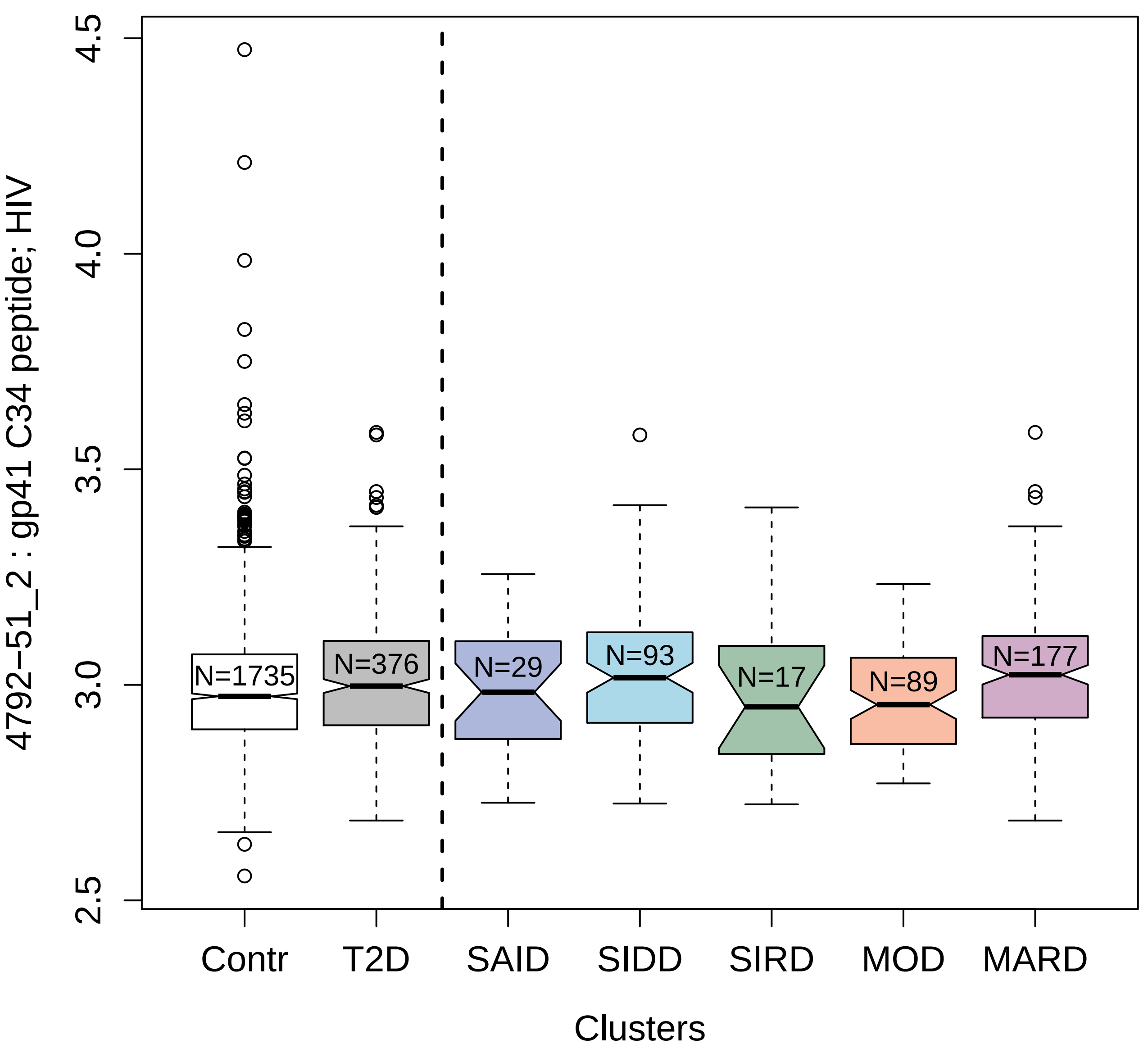
4792−51_2 : gp41 C34 peptide; HIV
4.5
4.0
3.5
3.0
2.5
N=1735
N=376
N=29
N=93
N=17
N=89
N=177
Contr
T2D
SAID
SIDD
SIRD
MOD
MARD
Clusters

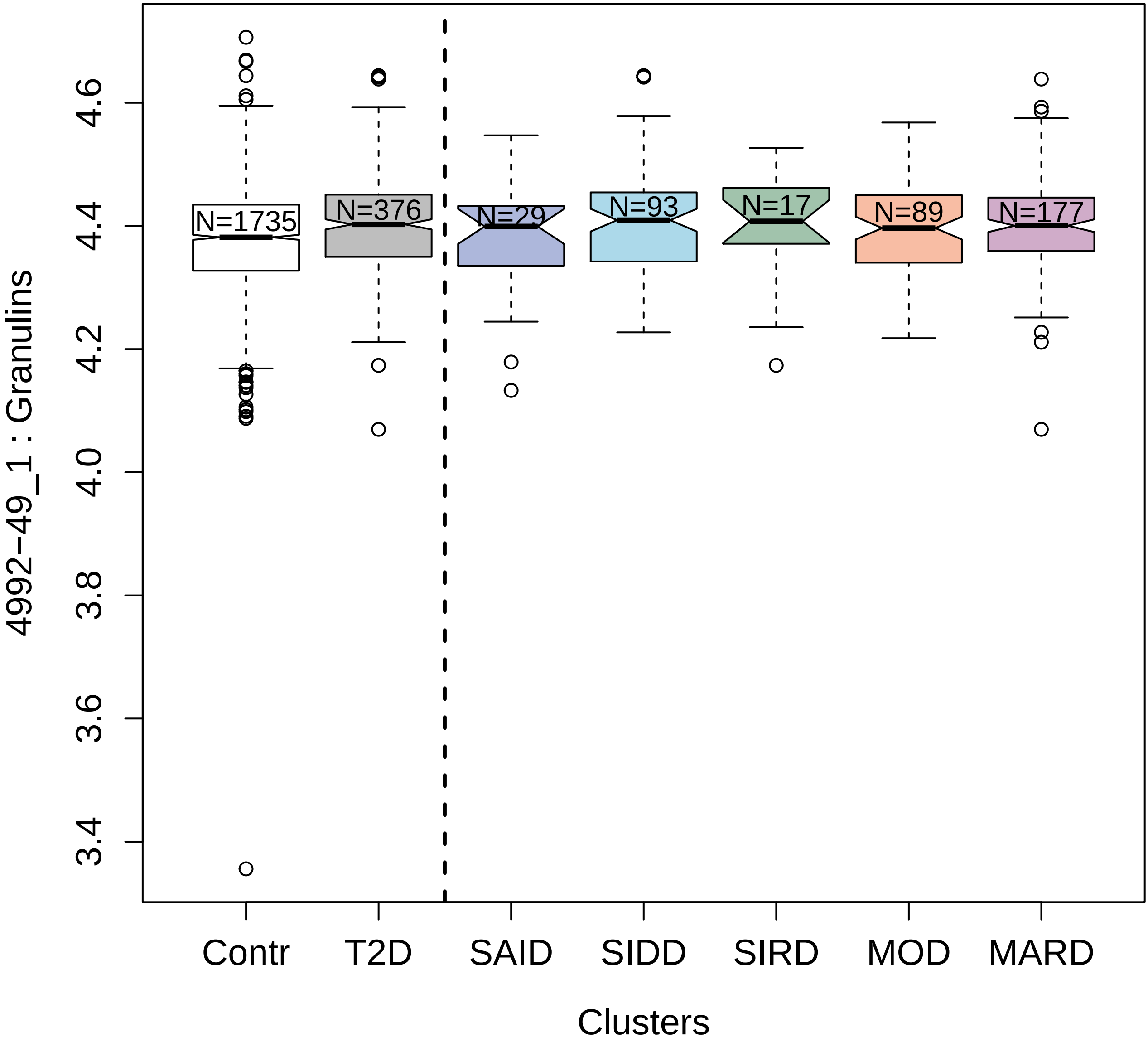

4992−49_1 : Granulins
Clusters
Contr
T2D
SAID
SIDD
SIRD
MOD
MARD
N=1735
N=376
N=29
N=93
N=17
N=89
N=177
3.4
3.6
3.8
4.0
4.2
4.4
4.6

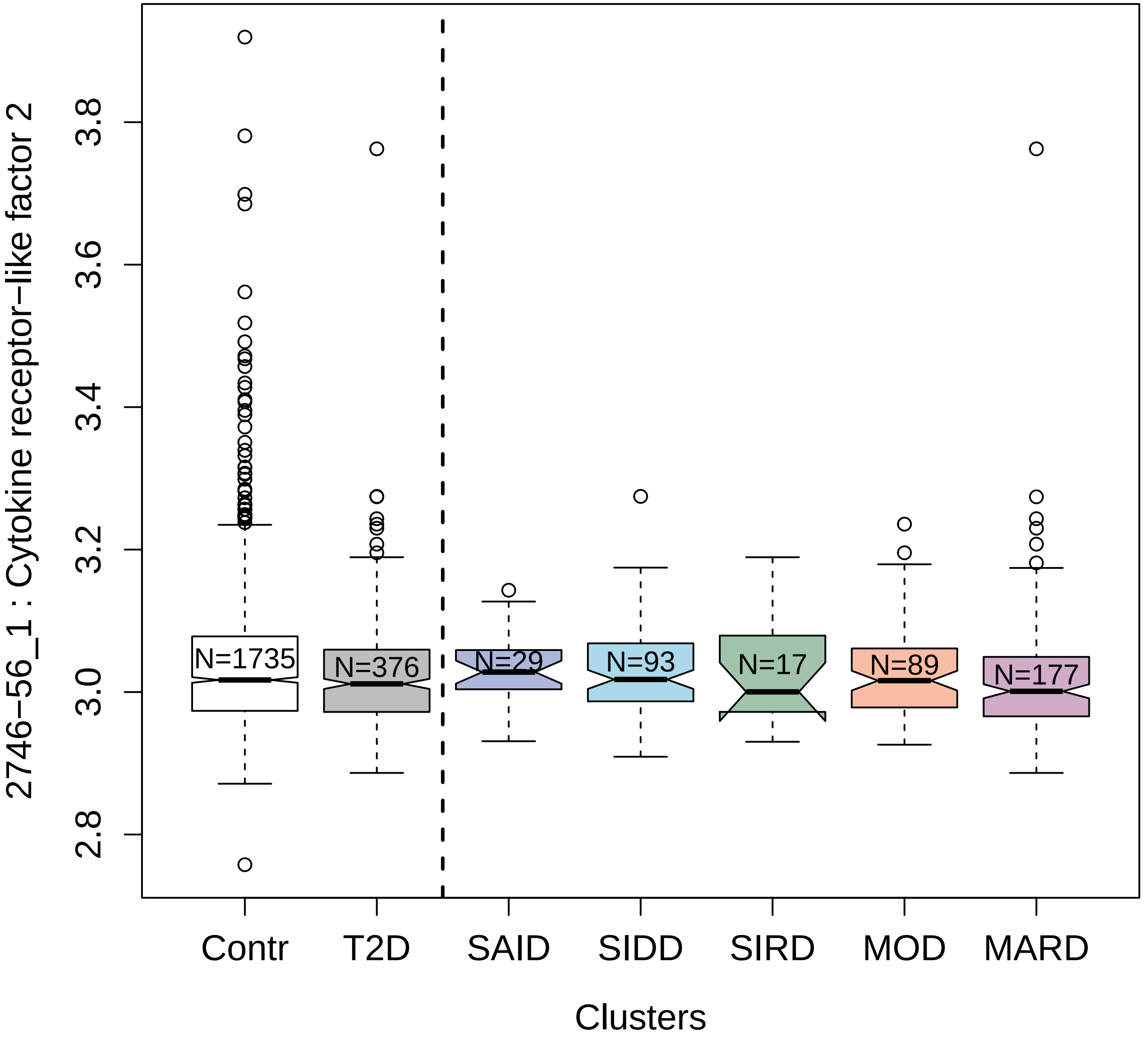

2746−56_1 : Cytokine receptor−like factor 2
2.8
3.0
3.2
3.4
3.6
3.8
N=1735
N=376
N=29
N=93
N=17
N=89
N=177
Contr
T2D
SAID
SIDD
SIRD
MOD
MARD
Clusters

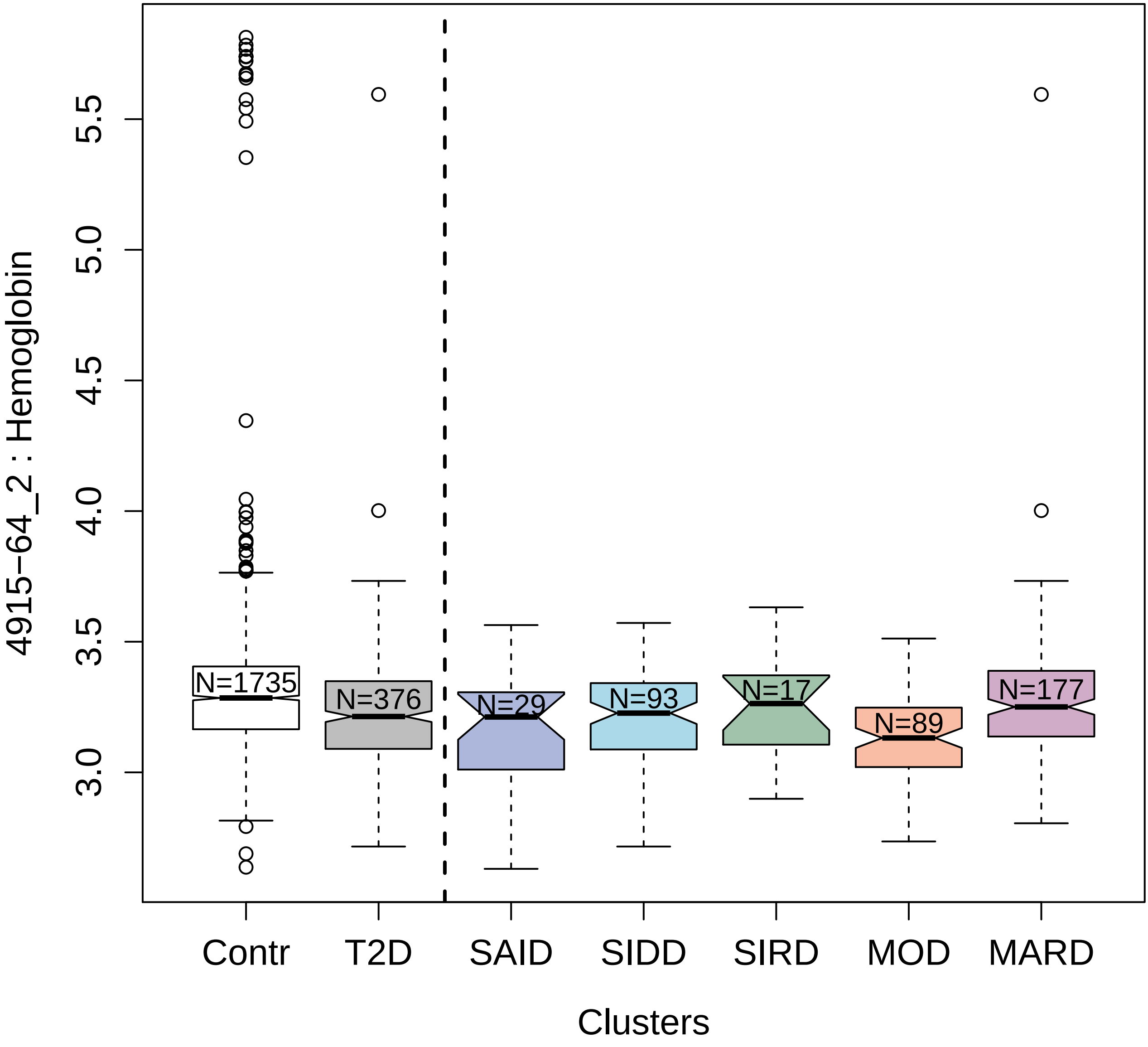
4915−64_2 : Hemoglobin
5.5
5.0
4.5
4.0
3.5
3.0
N=1735
N=376
N=29
N=93
N=17
N=89
N=177
Contr
T2D
SAID
SIDD
SIRD
MOD
MARD
Clusters

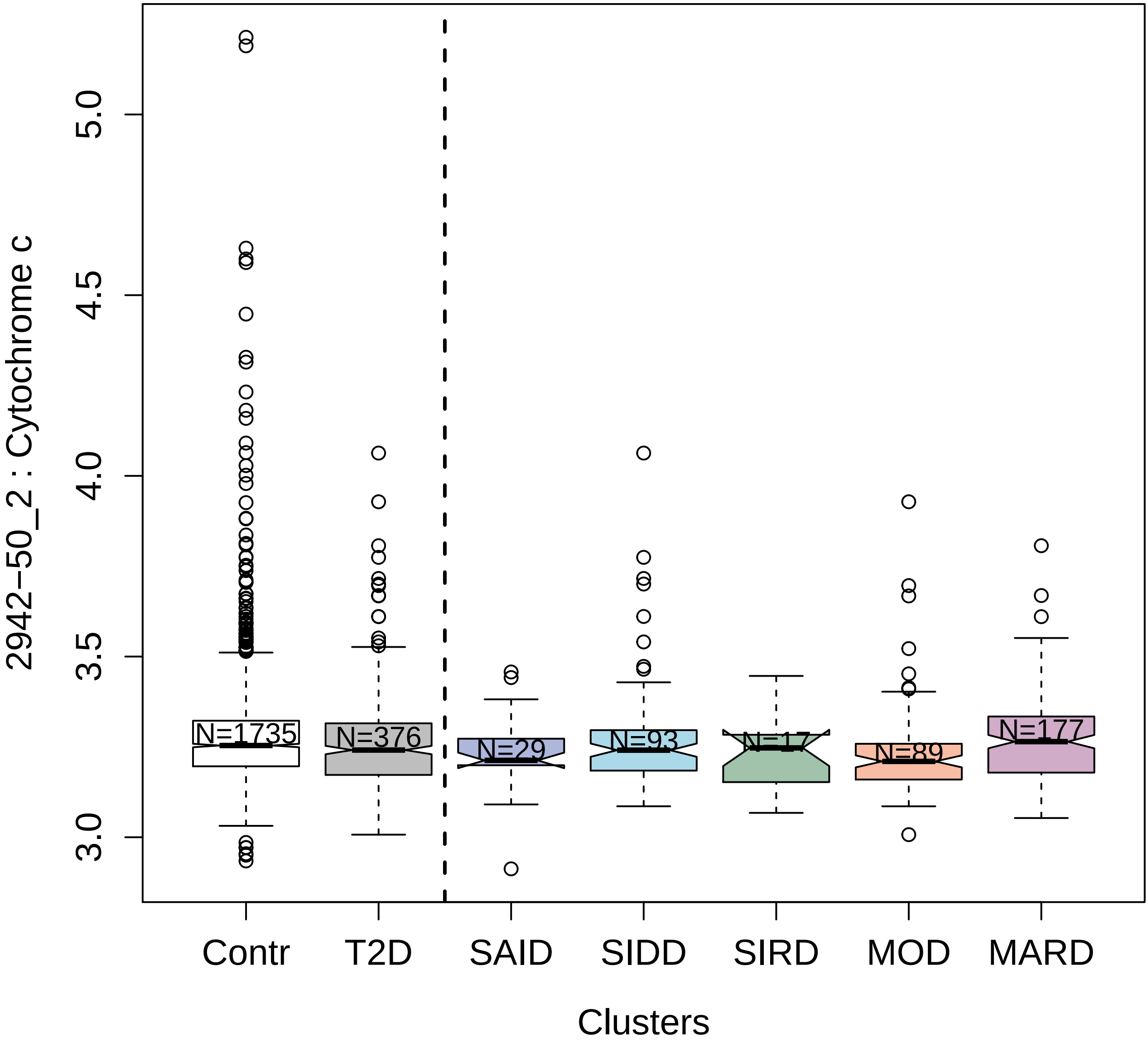

2942−50_2 : Cytochrome c
5.0
4.5
4.0
3.5
3.0
N=1735
N=376
N=29
N=93
N=17
N=89
N=177
Contr
T2D
SAID
SIDD
SIRD
MOD
MARD
Clusters

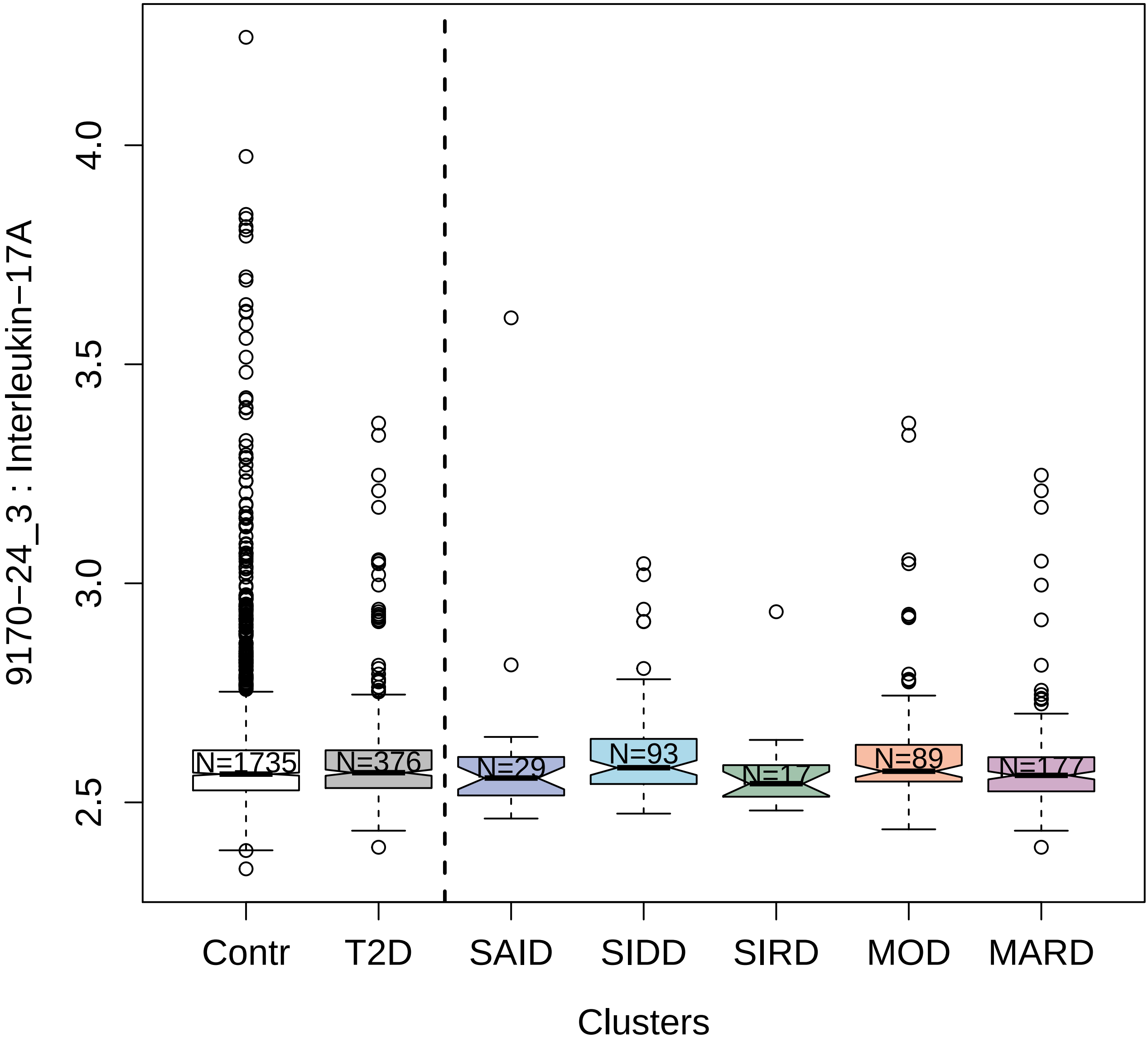

9170−24_3 : Interleukin−17A
4.0
3.5
3.0
2.5
N=1735
N=376
N=29
N=93
N=17
N=89
N=177
Contr
T2D
SAID
SIDD
SIRD
MOD
MARD
Clusters

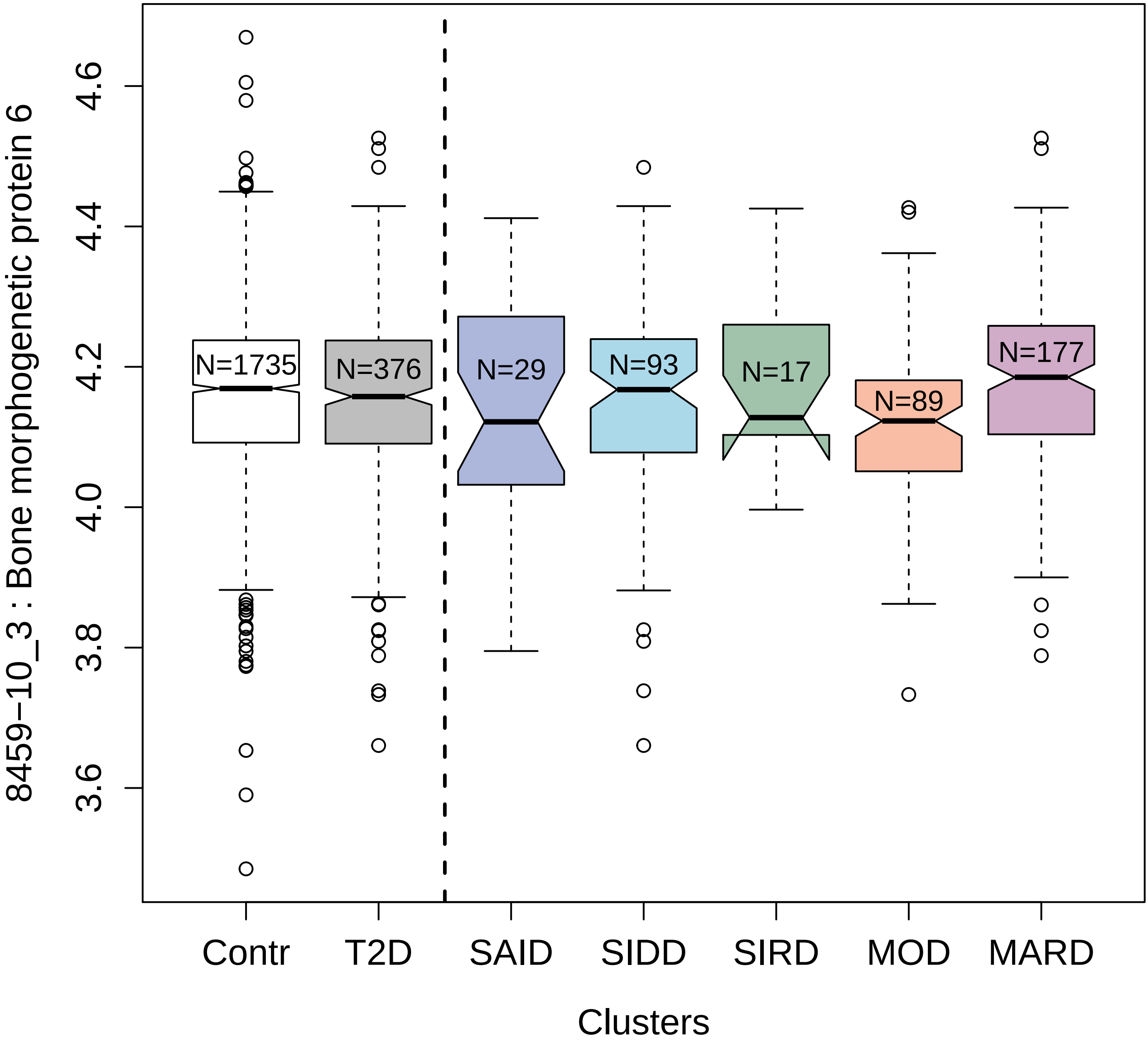

8459−10_3 : Bone morphogenetic protein 6
3.6
3.8
4.0
4.2
4.4
4.6
N=1735
N=376
N=29
N=93
N=17
N=89
N=177
Contr
T2D
SAID
SIDD
SIRD
MOD
MARD
Clusters

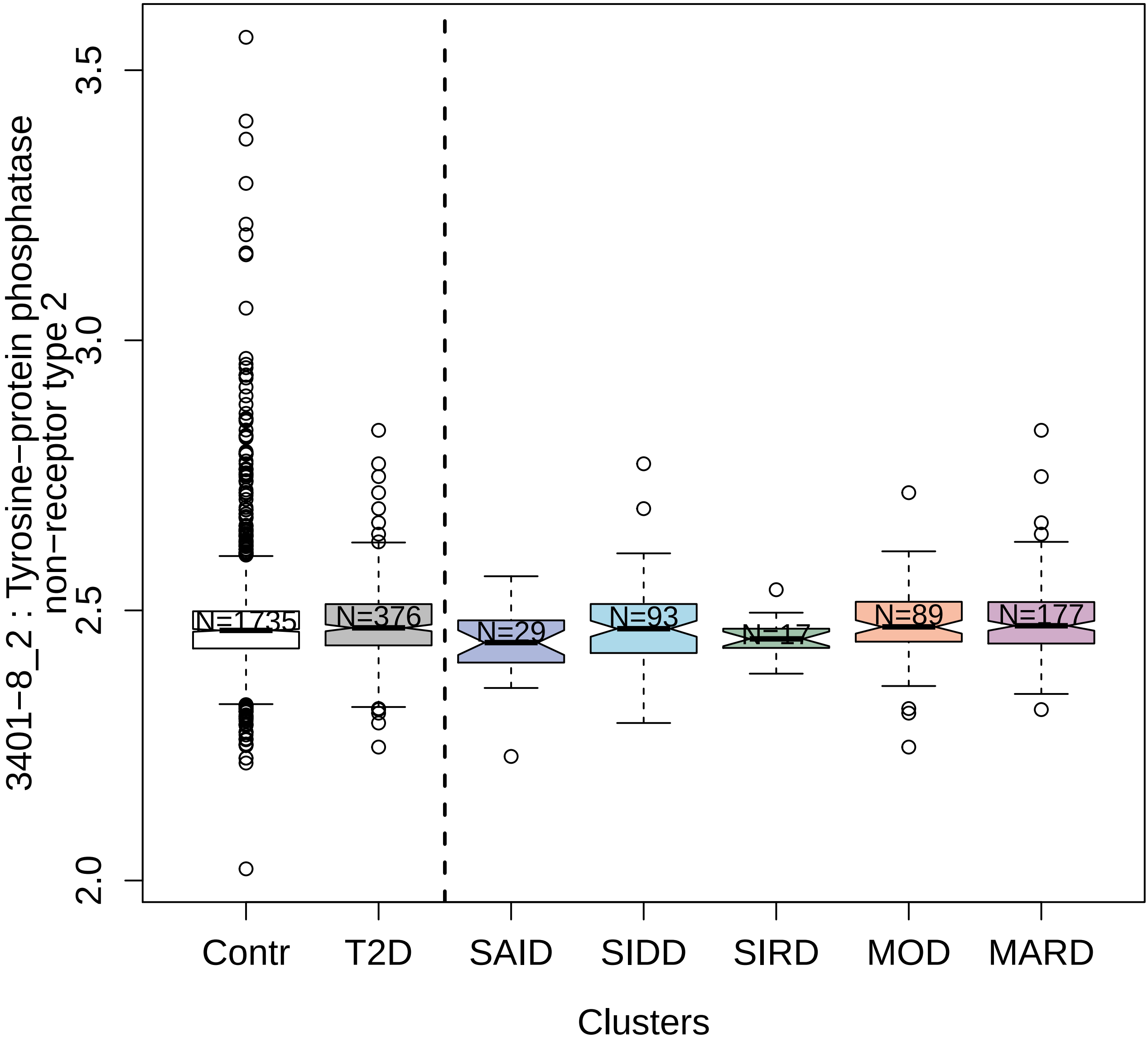

3401−8_2 : Tyrosine−protein phosphatase
non−receptor type 2
2.0
2.5
3.0
3.5
N=1735
N=376
N=29
N=93
N=17
N=89
N=177
Contr
T2D
SAID
SIDD
SIRD
MOD
MARD
Clusters

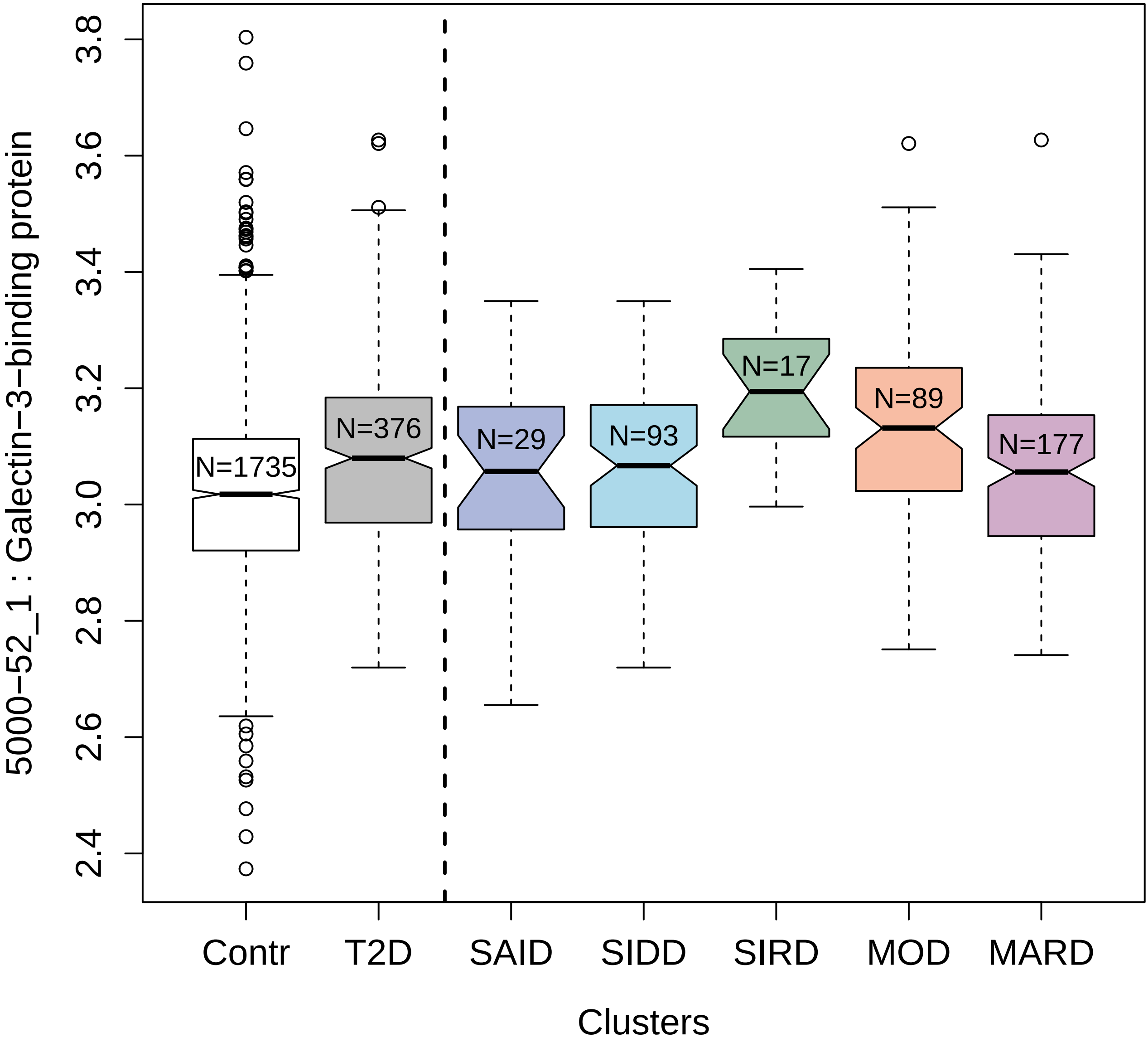

5000−52_1 : Galectin−3−binding protein
3.8
3.6
3.4
3.2
3.0
2.8
2.6
2.4
N=1735
N=376
N=29
N=93
N=17
N=89
N=177
Contr
T2D
SAID
SIDD
SIRD
MOD
MARD
Clusters

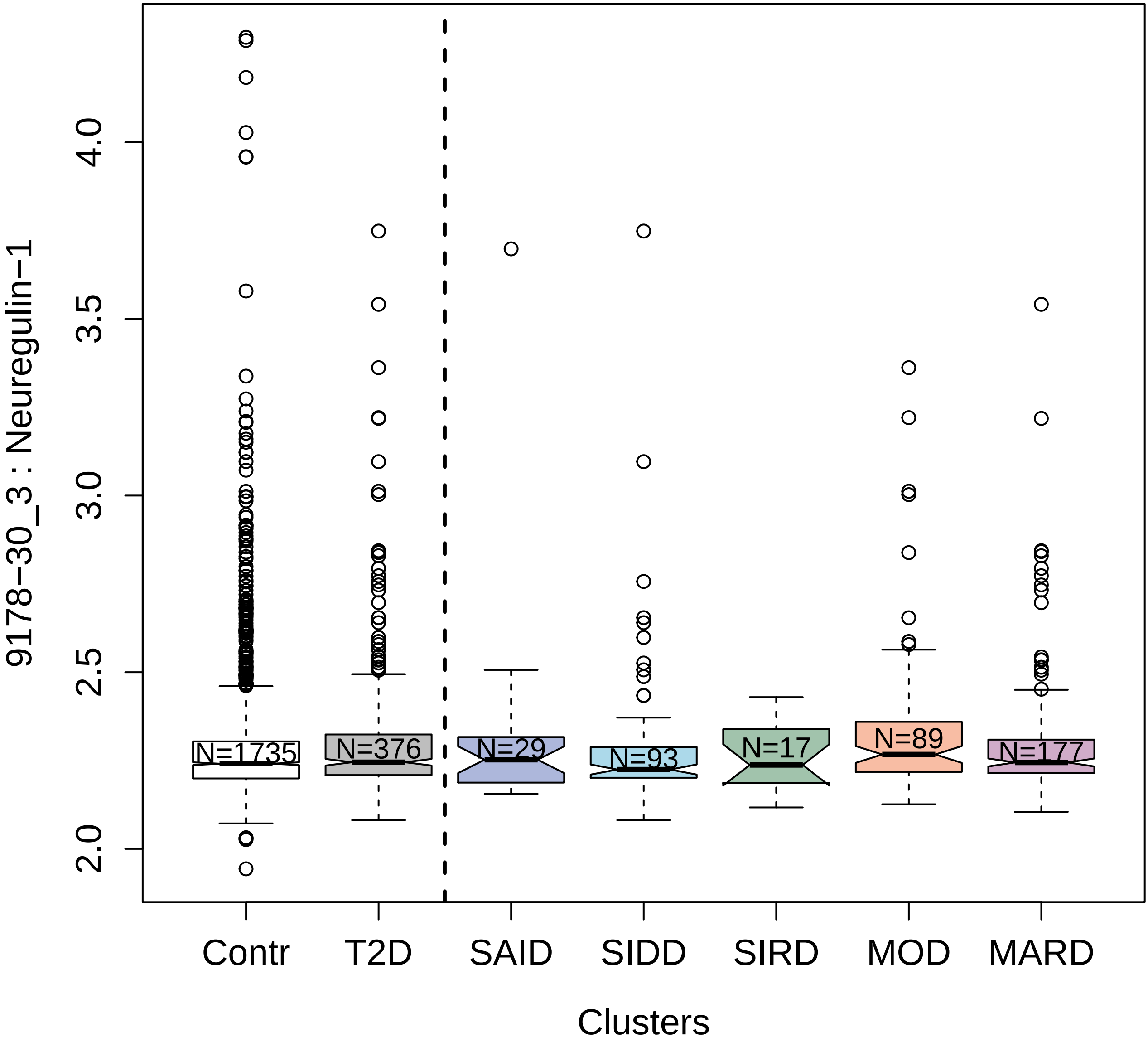
9178−30_3 : Neuregulin−1
2.0
2.5
3.0
3.5
4.0
N=1735
N=376
N=29
N=93
N=17
N=89
N=177
Contr
T2D
SAID
SIDD
SIRD
MOD
MARD
Clusters

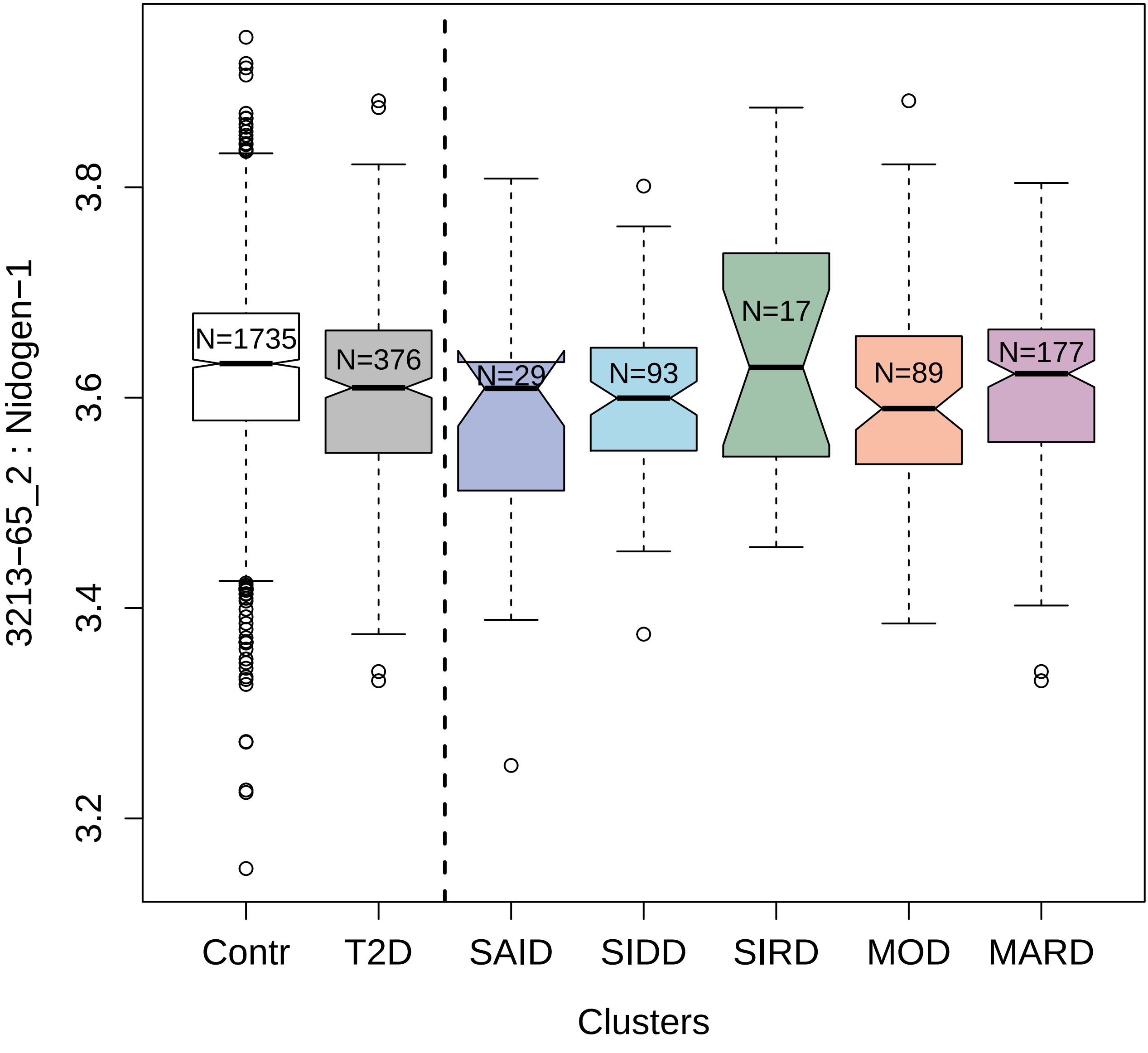

3213−65_2 : Nidogen−1
3.2
3.4
3.6
3.8
N=1735
N=376
N=29
N=93
N=17
N=89
N=177
Contr
T2D
SAID
SIDD
SIRD
MOD
MARD
Clusters

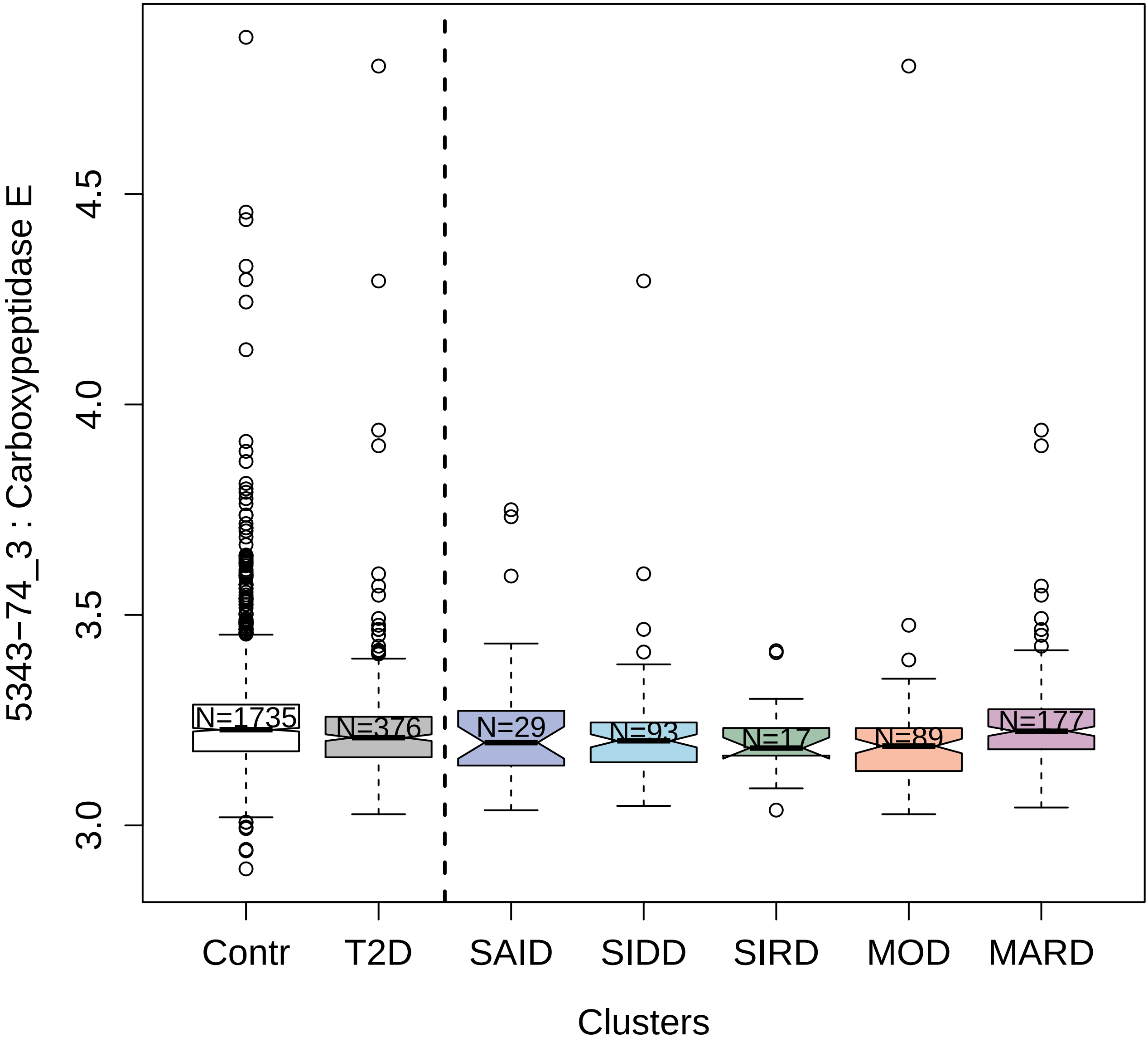
5343−74_3 : Carboxypeptidase E
4.5
4.0
3.5
3.0
N=1735
N=376
N=29
N=93
N=17
N=89
N=177
Contr
T2D
SAID
SIDD
SIRD
MOD
MARD
Clusters

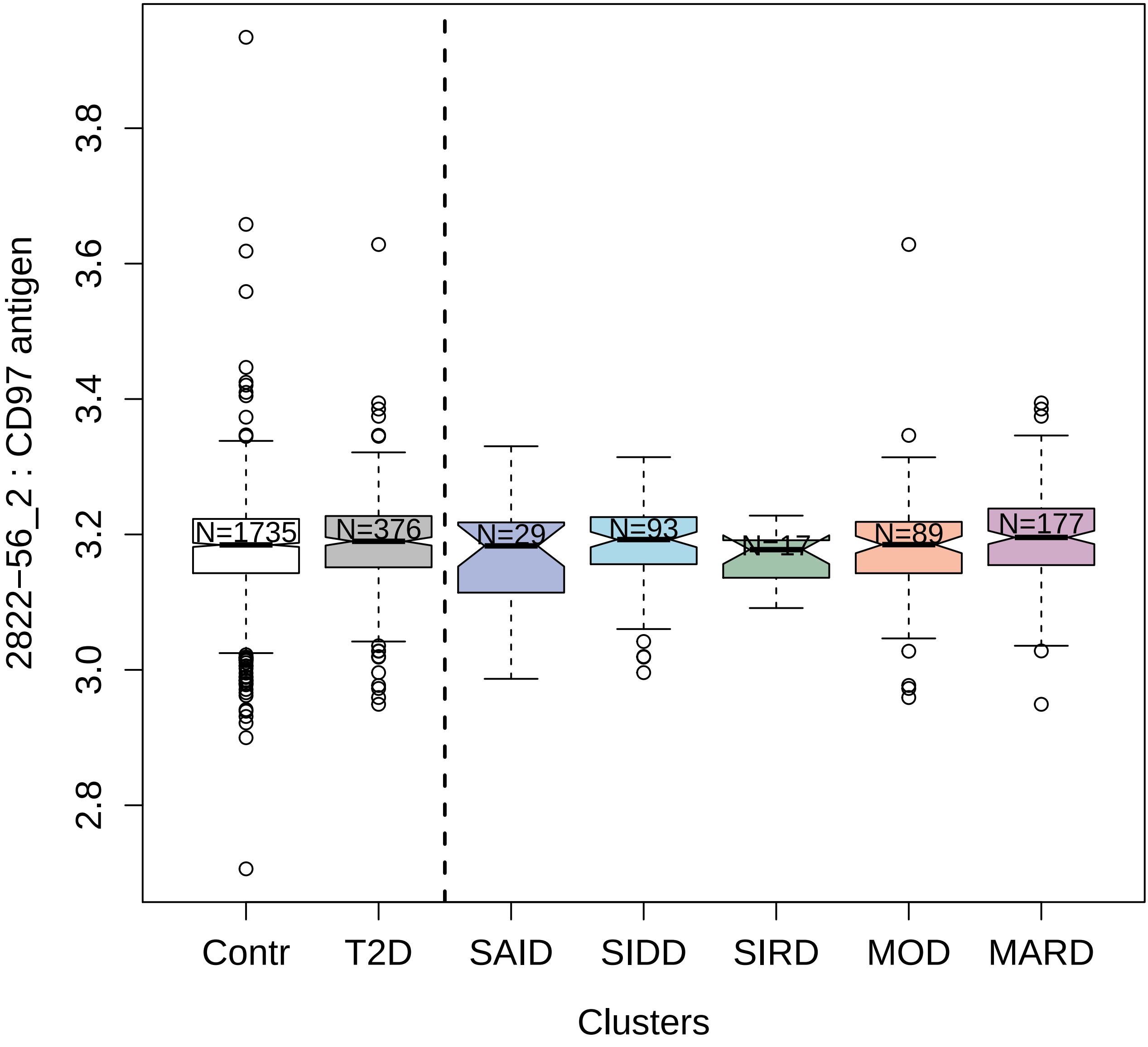

2822−56_2 : CD97 antigen
2.8
3.0
3.2
3.4
3.6
3.8
N=1735
N=376
N=29
N=93
N=17
N=89
N=177
Contr
T2D
SAID
SIDD
SIRD
MOD
MARD
Clusters

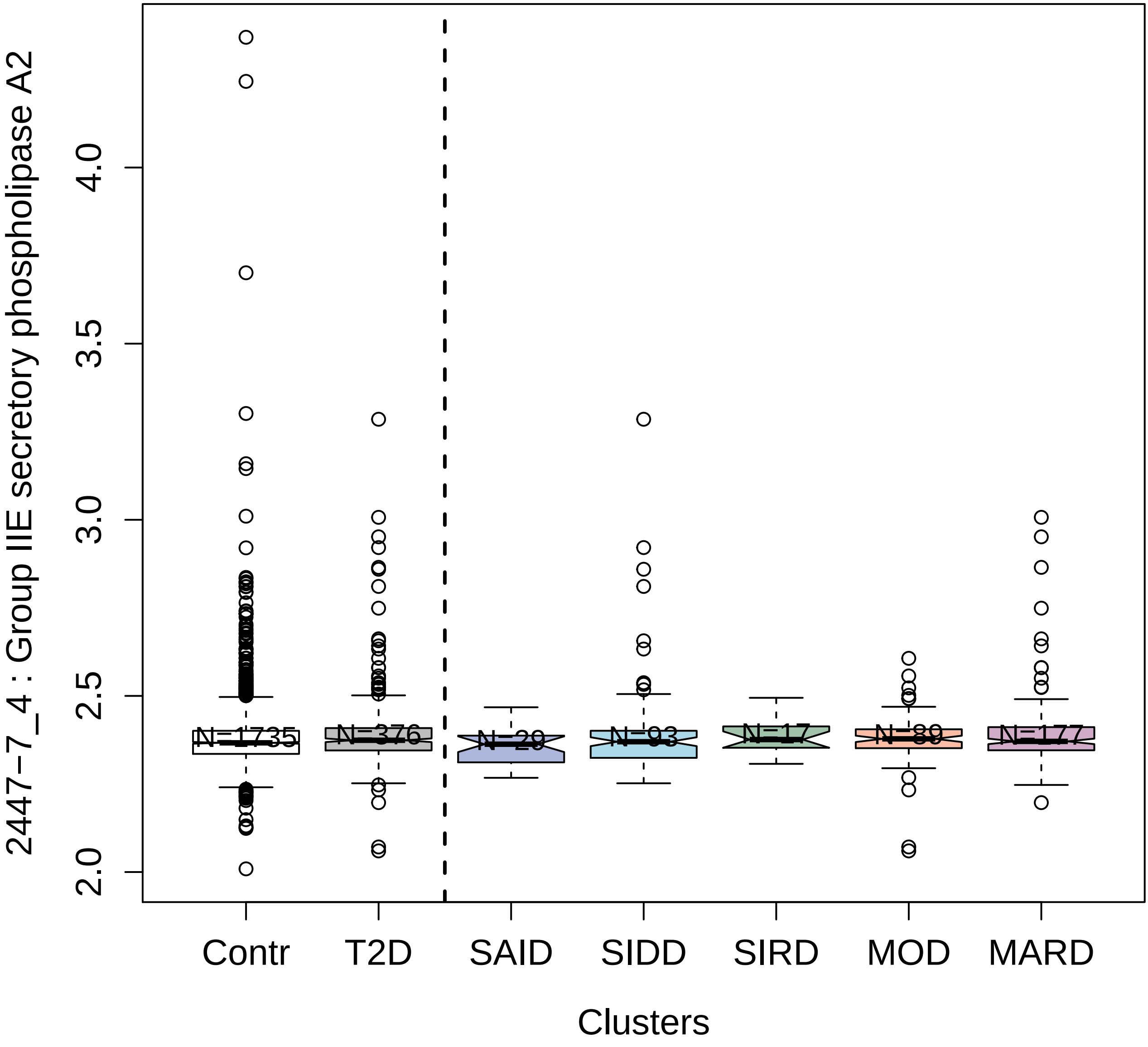
2447−7_4 : Group IIE secretory phospholipase A2
2.0
2.5
3.0
3.5
4.0
N=1735
N=376
N=29
N=93
N=17
N=89
N=177
Contr
T2D
SAID
SIDD
SIRD
MOD
MARD
Clusters

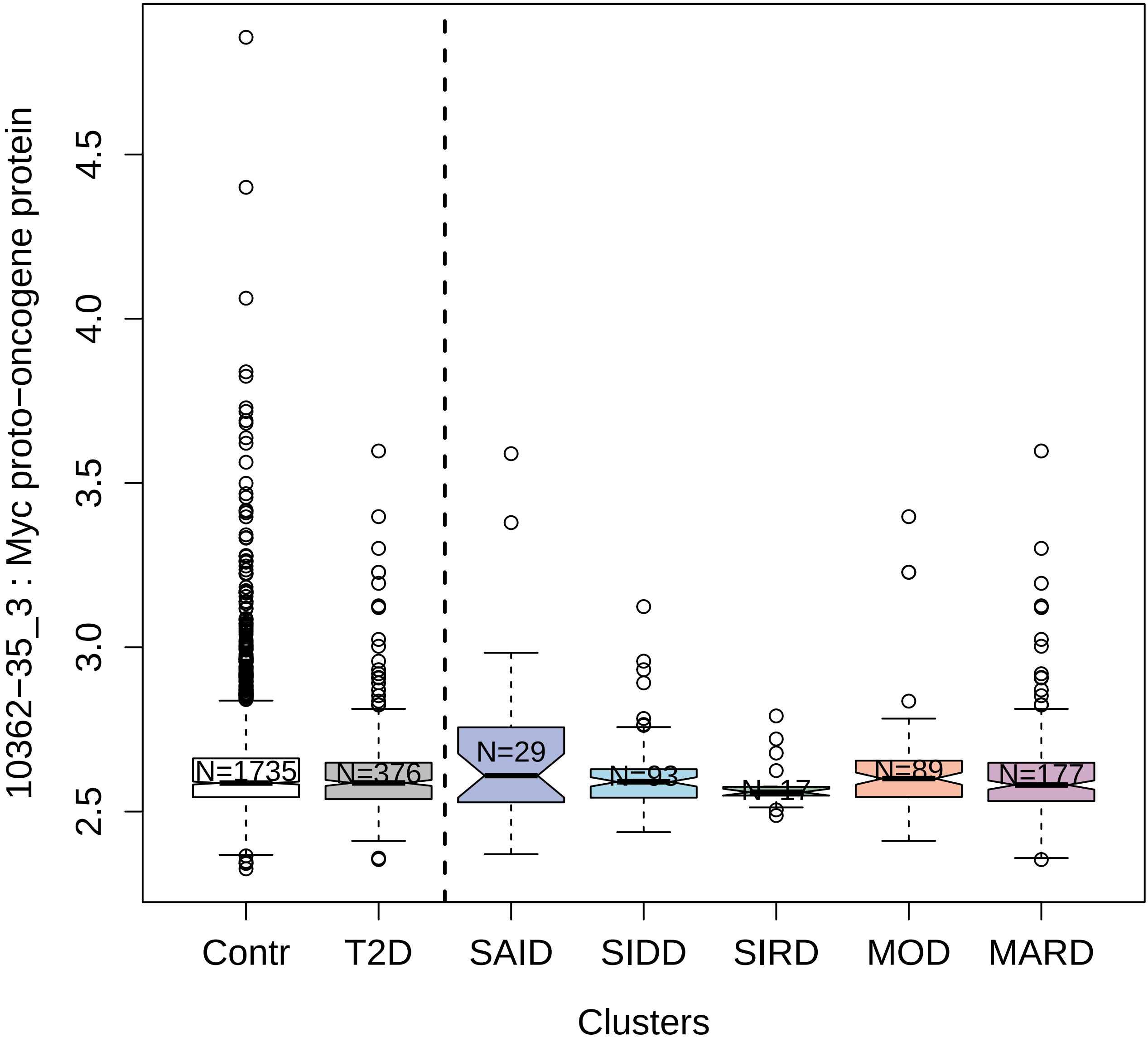
10362−35_3 : Myc proto−oncogene protein
4.5
4.0
3.5
3.0
2.5
N=1735
N=376
N=29
N=93
N=17
N=89
N=177
Contr
T2D
SAID
SIDD
SIRD
MOD
MARD
Clusters

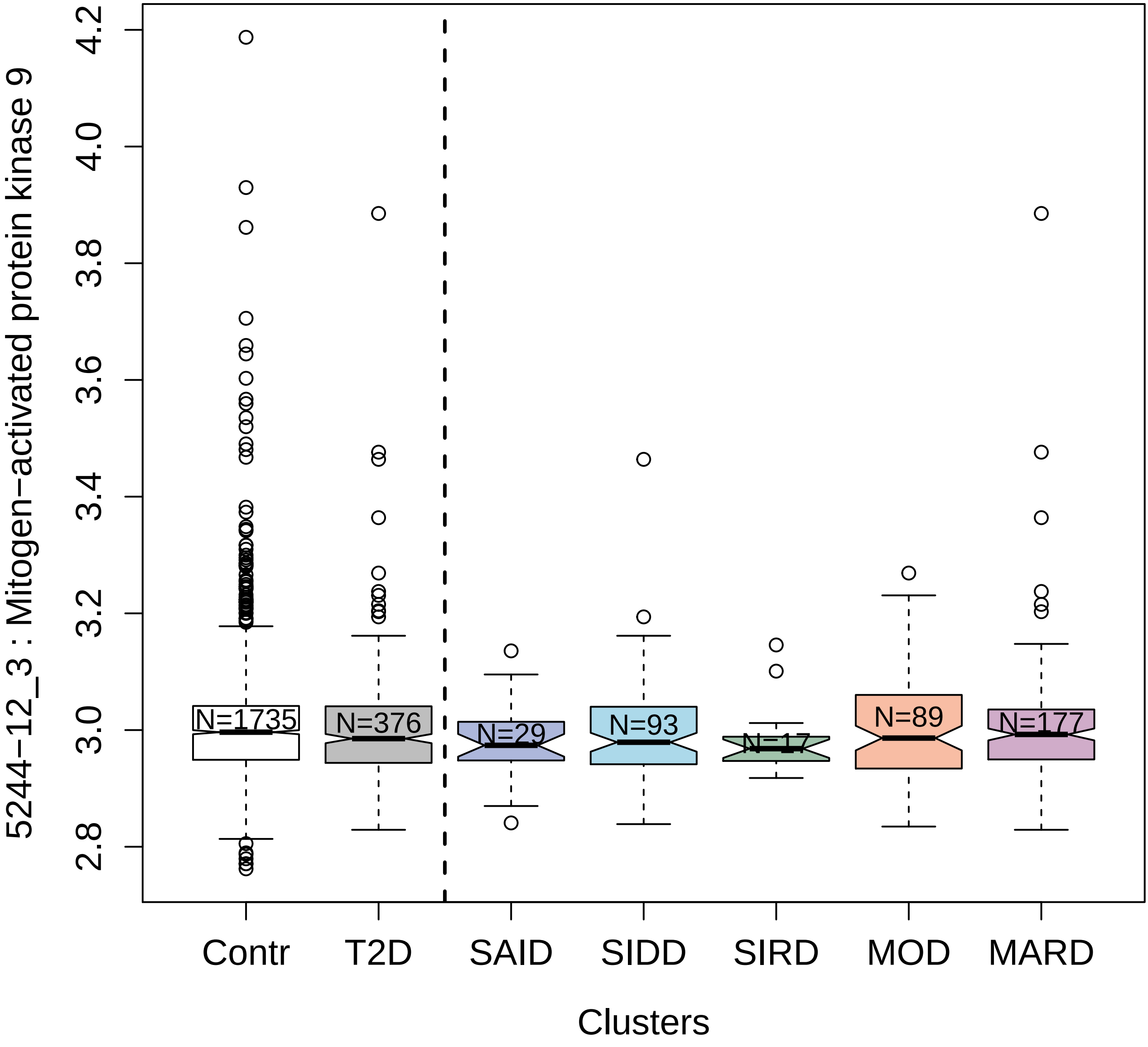

5244−12_3 : Mitogen−activated protein kinase 9
2.8
3.0
3.2
3.4
3.6
3.8
4.0
4.2
N=1735
N=376
N=29
N=93
N=17
N=89
N=177
Contr
T2D
SAID
SIDD
SIRD
MOD
MARD
Clusters

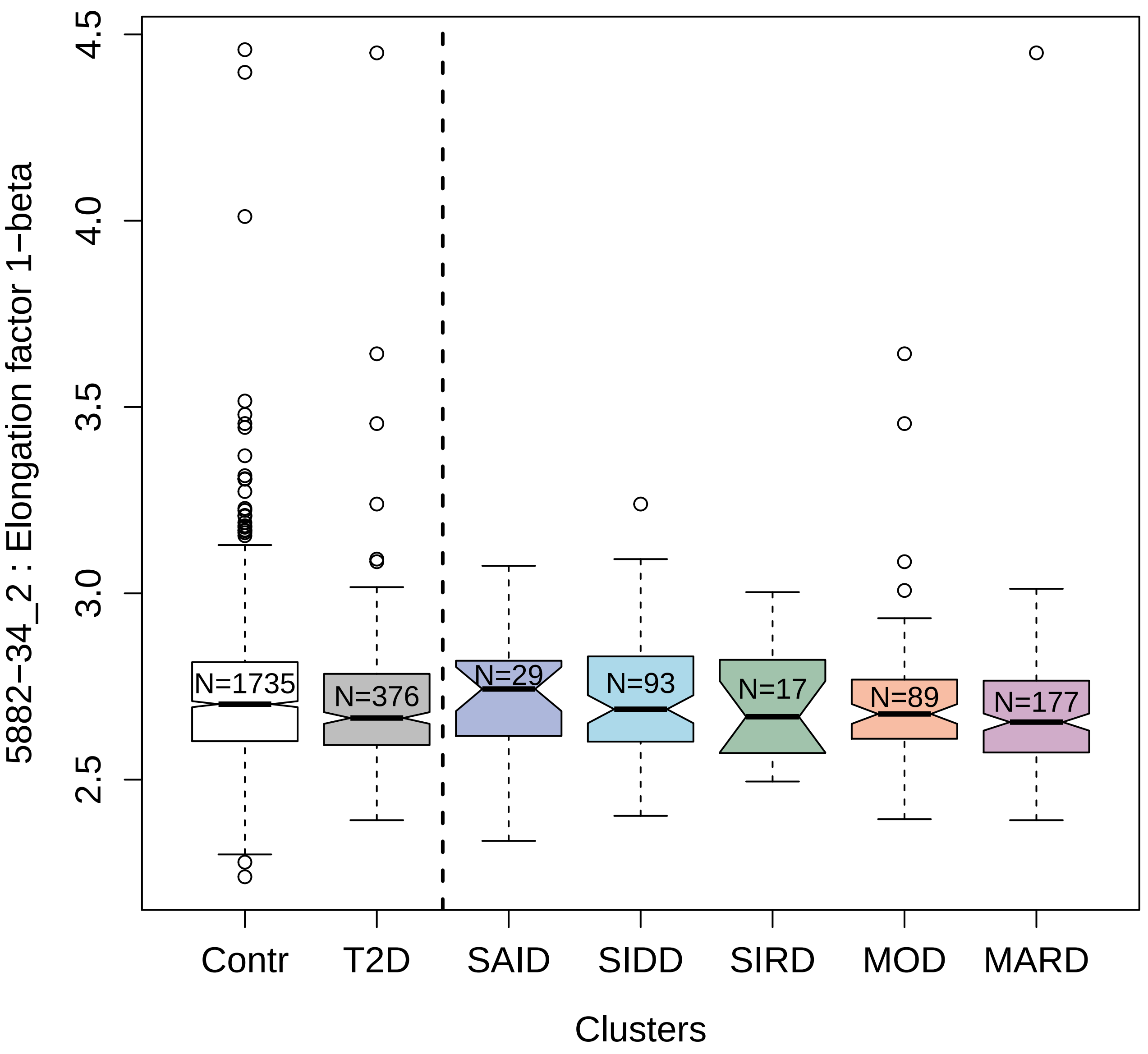
5882−34_2 : Elongation factor 1−beta
4.5
4.0
3.5
3.0
2.5
N=1735
N=376
N=29
N=93
N=17
N=89
N=177
Contr
T2D
SAID
SIDD
SIRD
MOD
MARD
Clusters

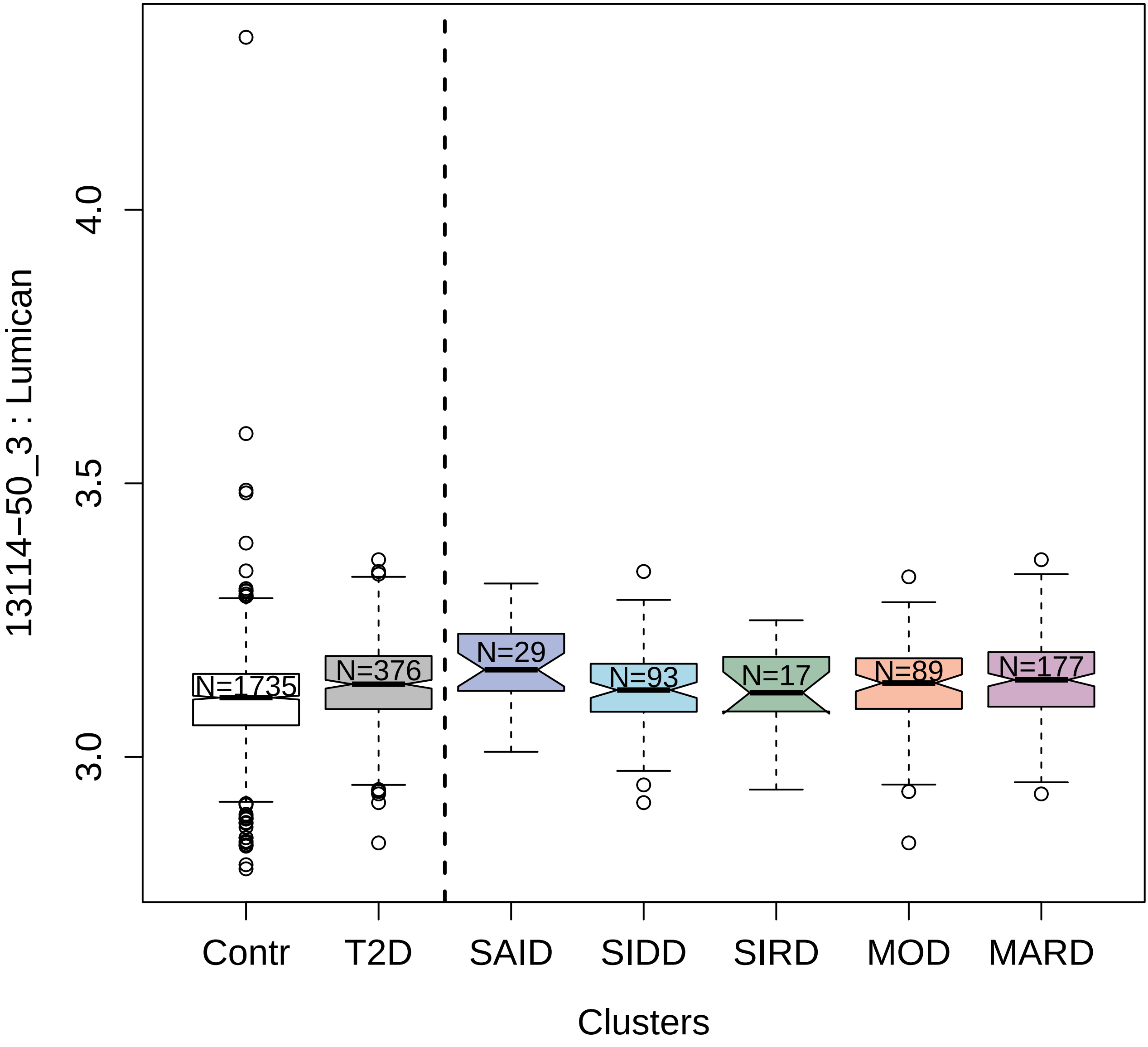

13114−50_3 : Lumican
4.0
3.5
3.0
N=1735
N=376
N=29
N=93
N=17
N=89
N=177
Contr
T2D
SAID
SIDD
SIRD
MOD
MARD
Clusters

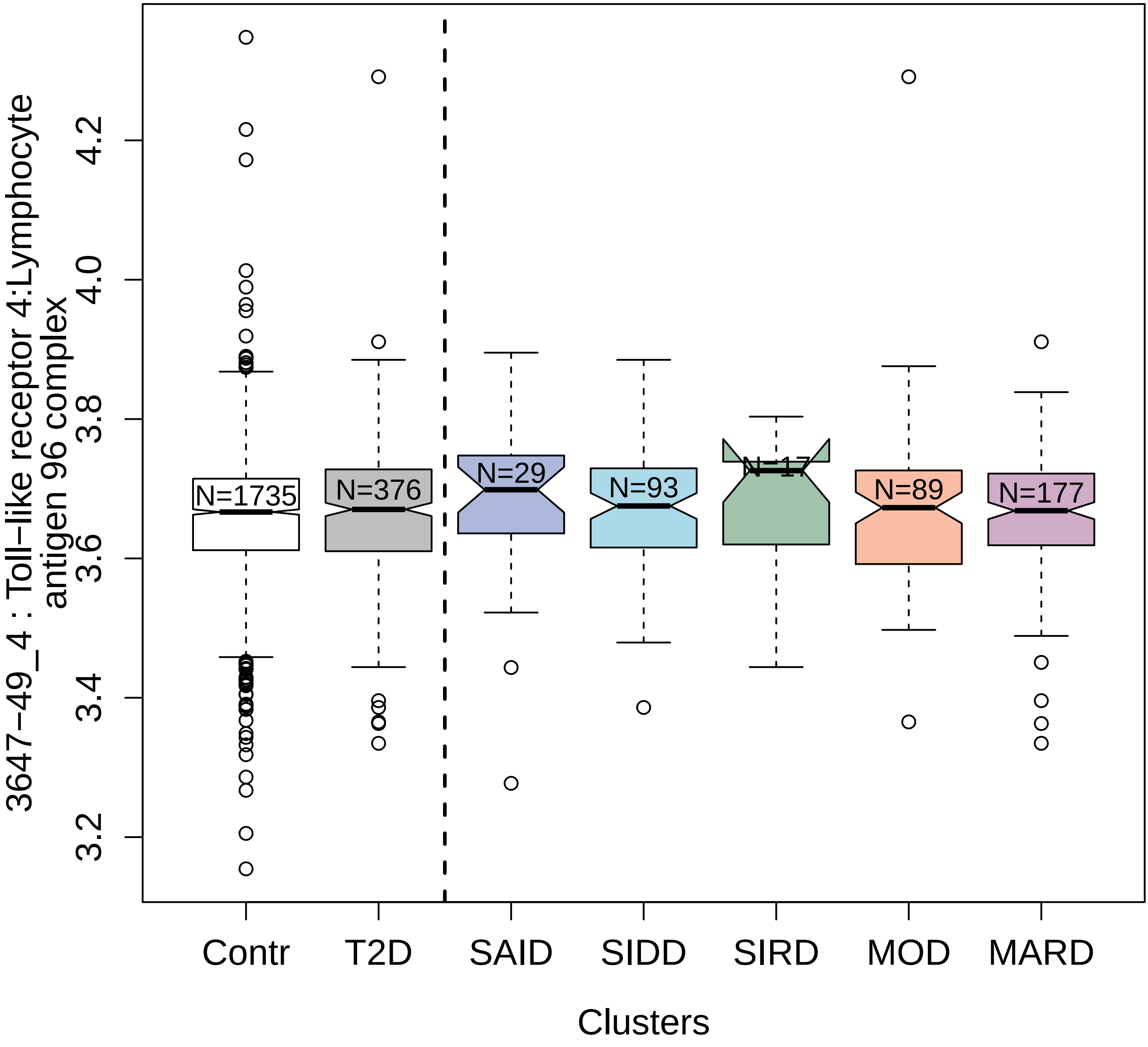

3647−49_4 : Toll−like receptor 4:Lymphocyte antigen 96 complex
3.2
3.4
3.6
3.8
4.0
4.2
N=1735
N=376
N=29
N=93
N=17
N=89
N=177
Contr
T2D
SAID
SIDD
SIRD
MOD
MARD
Clusters

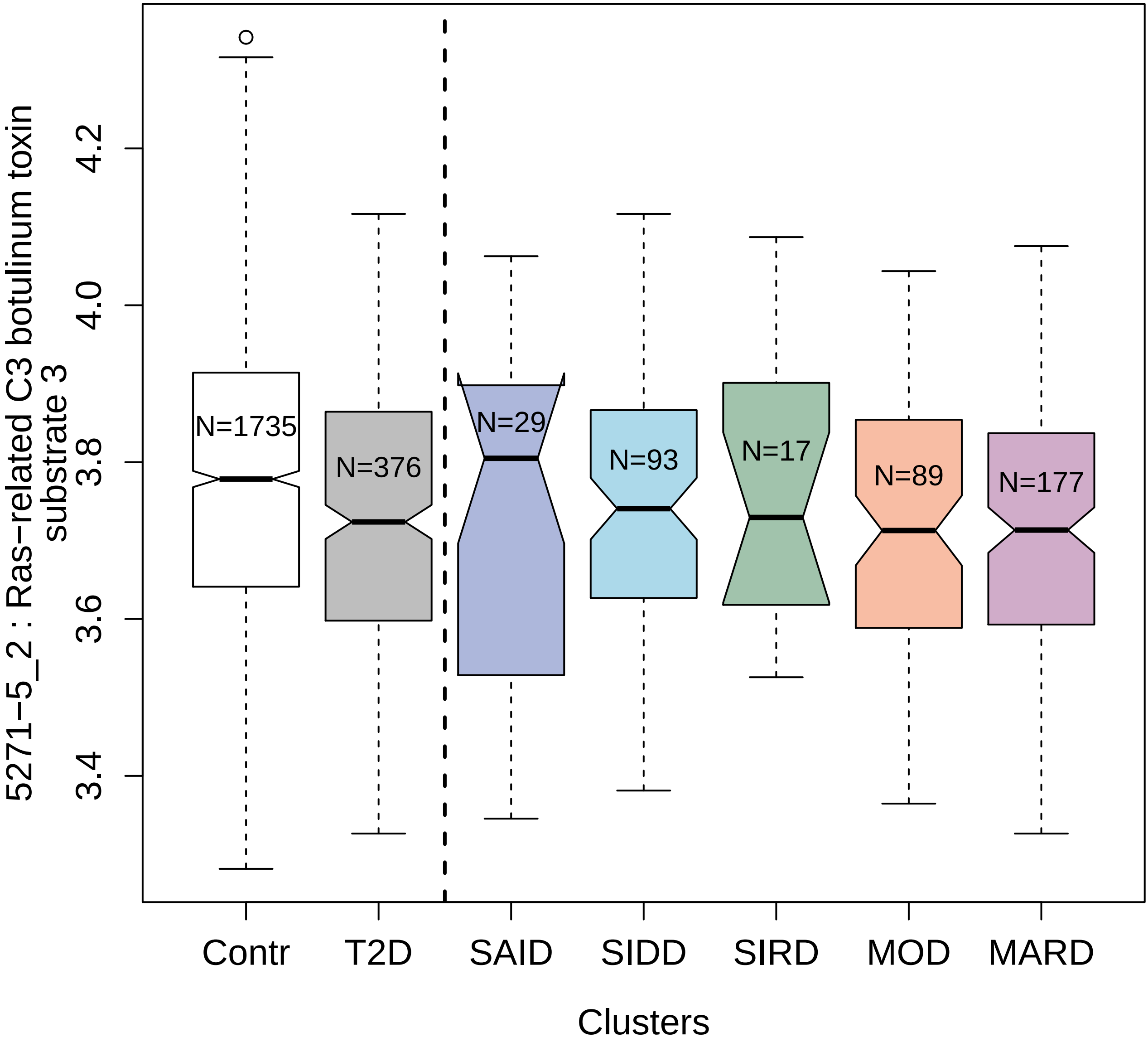
5271−5_2 : Ras−related C3 botulinum toxin substrate 3
3.4
3.6
3.8
4.0
4.2
N=1735
N=376
N=29
N=93
N=17
N=89
N=177
Contr
T2D
SAID
SIDD
SIRD
MOD
MARD
Clusters

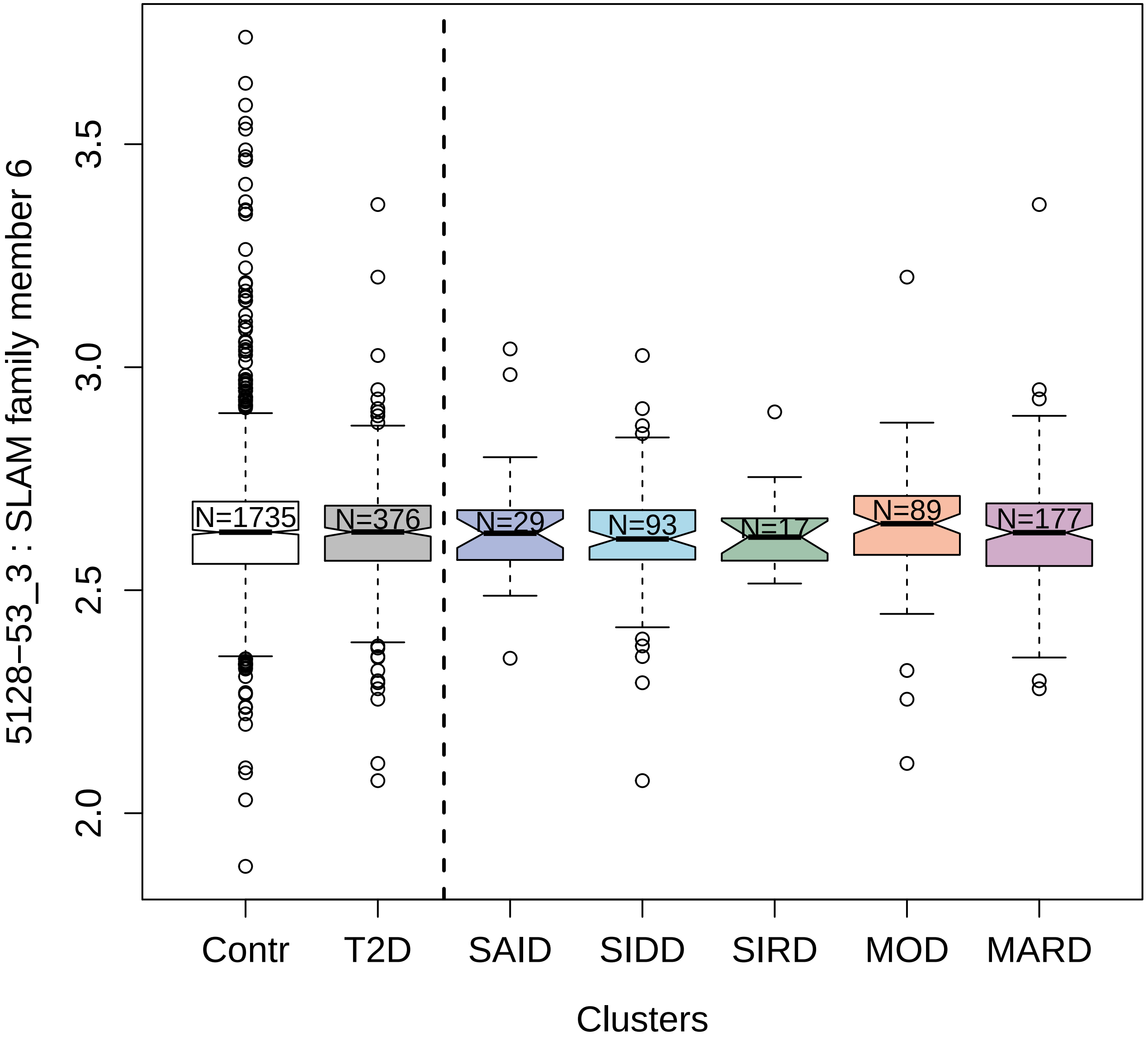

5128−53_3 : SLAM family member 6
2.0
2.5
3.0
3.5
N=1735
N=376
N=29
N=93
N=17
N=89
N=177
Contr
T2D
SAID
SIDD
SIRD
MOD
MARD
Clusters

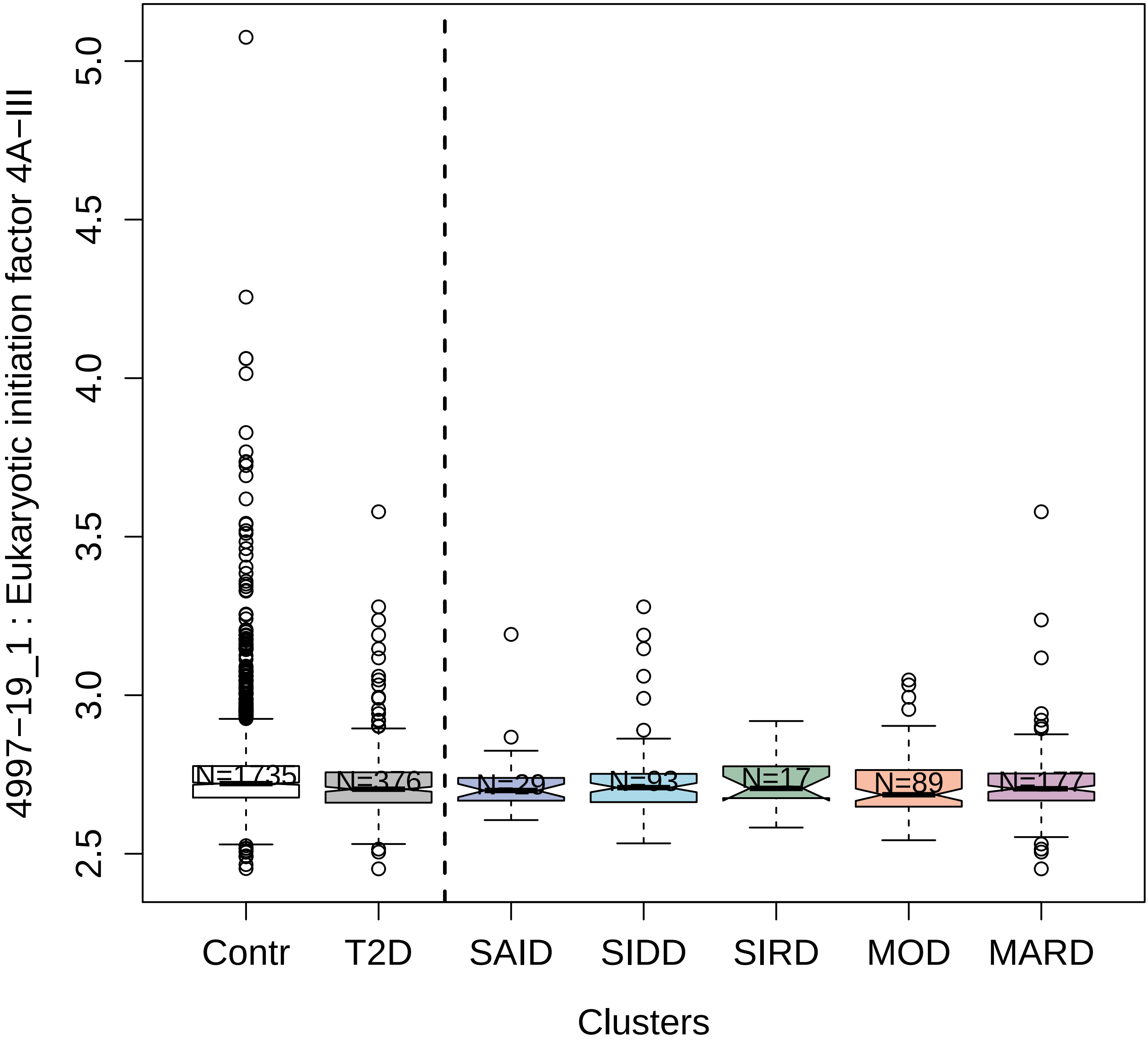
4997−19_1 : Eukaryotic initiation factor 4A−III
5.0
4.5
4.0
3.5
3.0
2.5
N=1735
N=376
N=29
N=93
N=17
N=89
N=177
Contr
T2D
SAID
SIDD
SIRD
MOD
MARD
Clusters

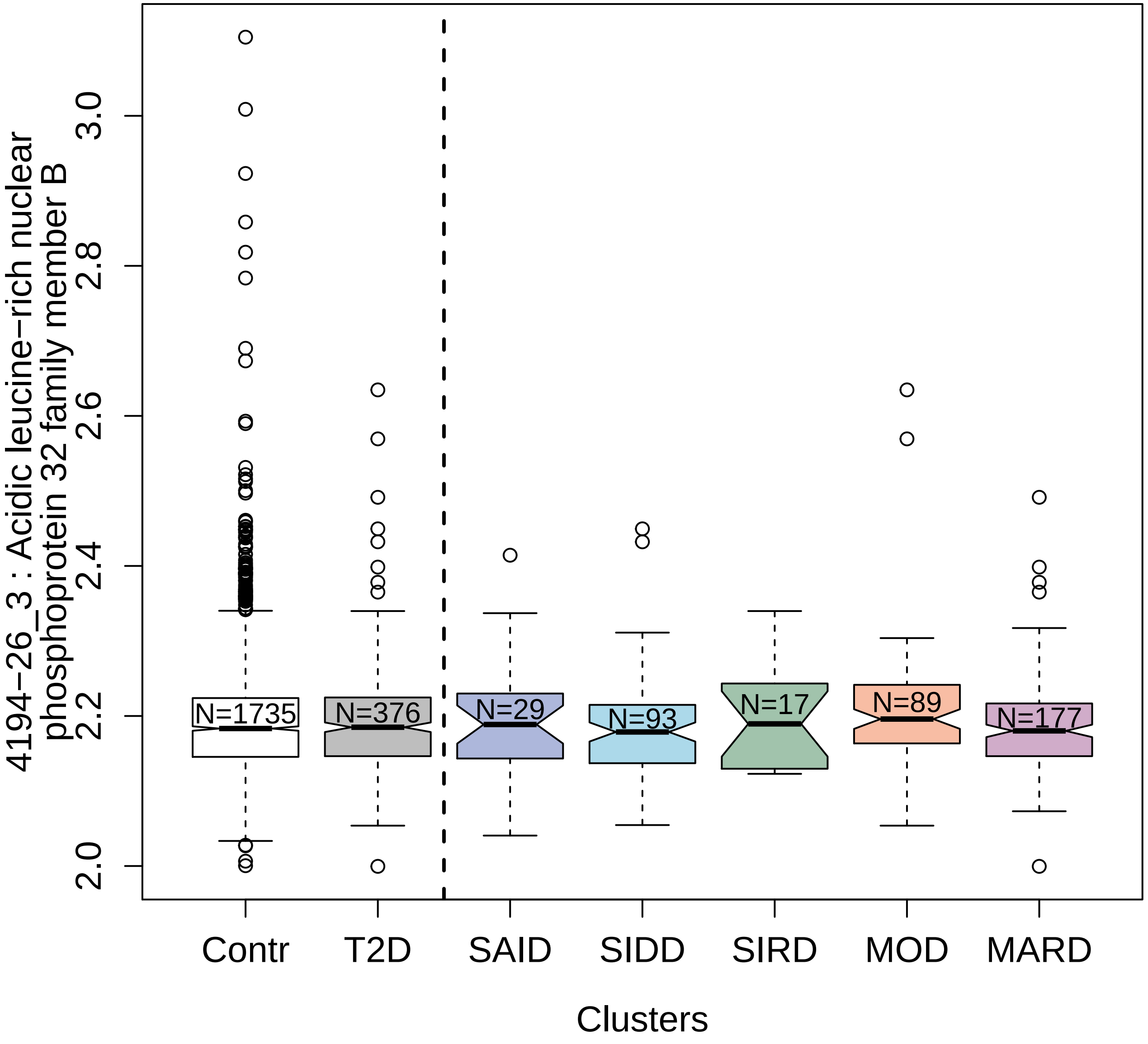
4194−26_3 : Acidic leucine−rich nuclear
phosphoprotein 32 family member B
2.0
2.2
2.4
2.6
2.8
3.0
N=1735
N=376
N=29
N=93
N=17
N=89
N=177
Contr
T2D
SAID
SIDD
SIRD
MOD
MARD
Clusters

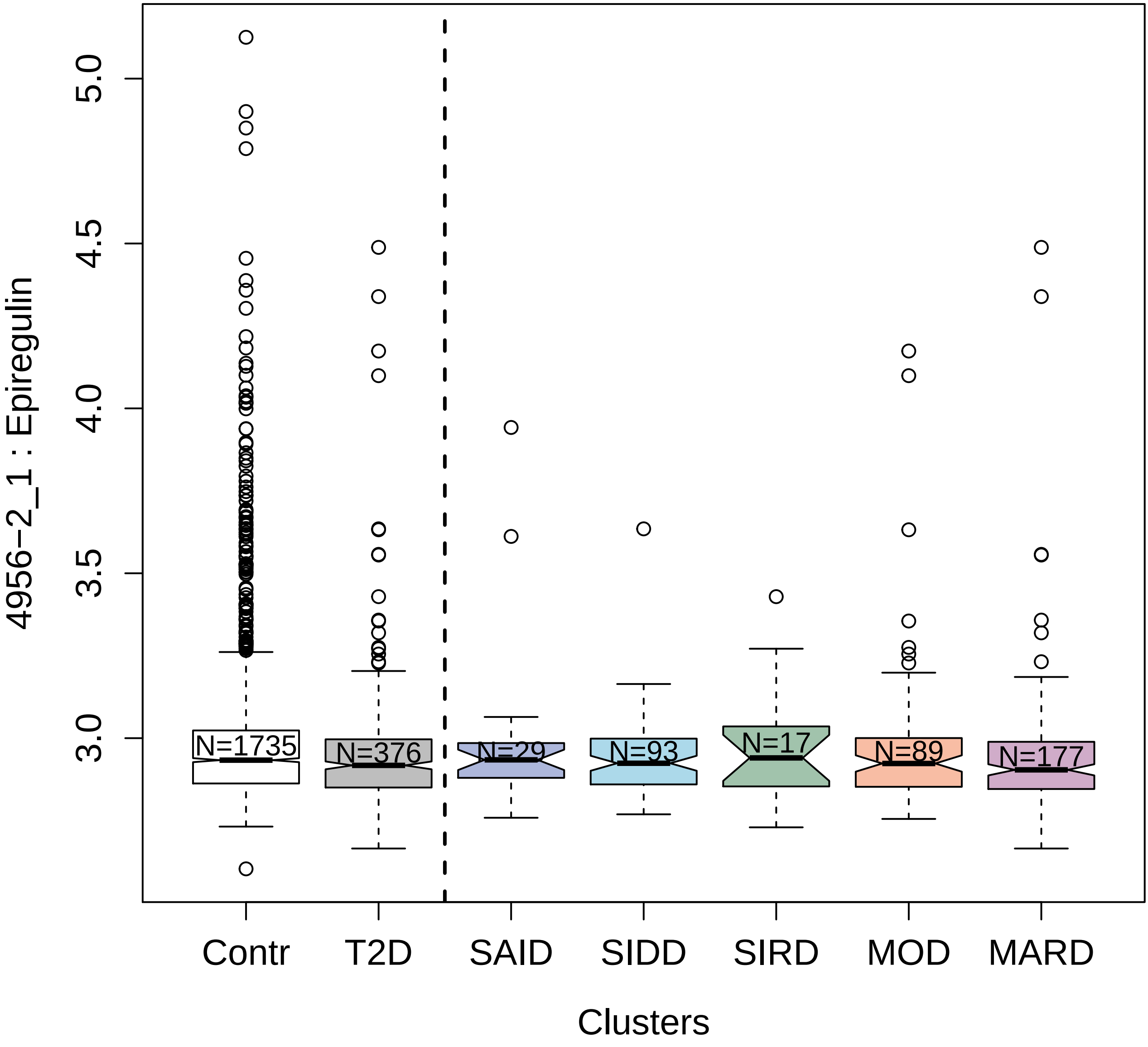
4956−2_1 : Epiregulin
5.0
4.5
4.0
3.5
3.0
N=1735
N=376
N=29
N=93
N=17
N=89
N=177
Contr
T2D
SAID
SIDD
SIRD
MOD
MARD
Clusters

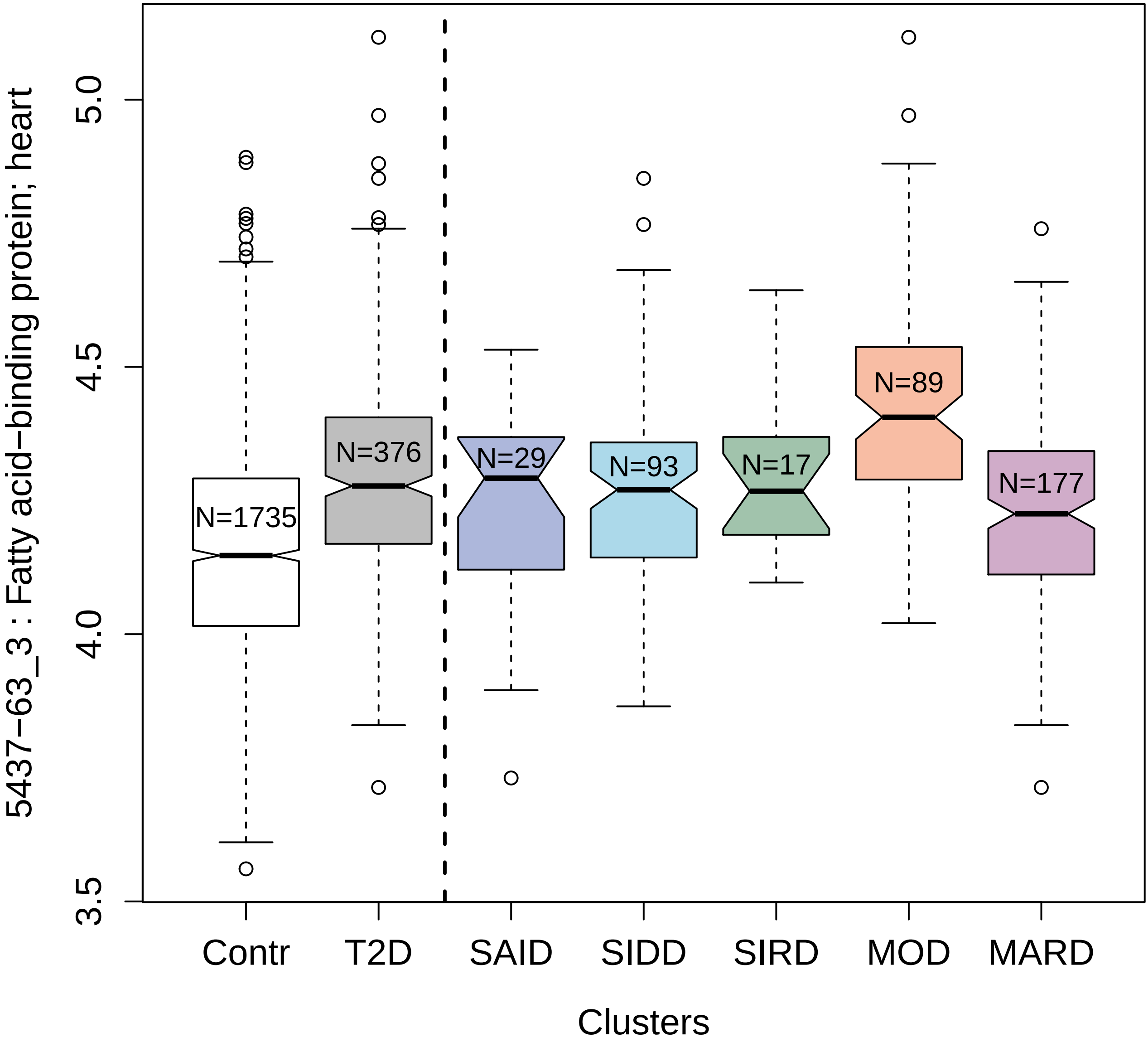

5437−63_3 : Fatty acid−binding protein; heart
3.5
4.0
4.5
5.0
N=1735
N=376
N=29
N=93
N=17
N=89
N=177
Contr
T2D
SAID
SIDD
SIRD
MOD
MARD
Clusters

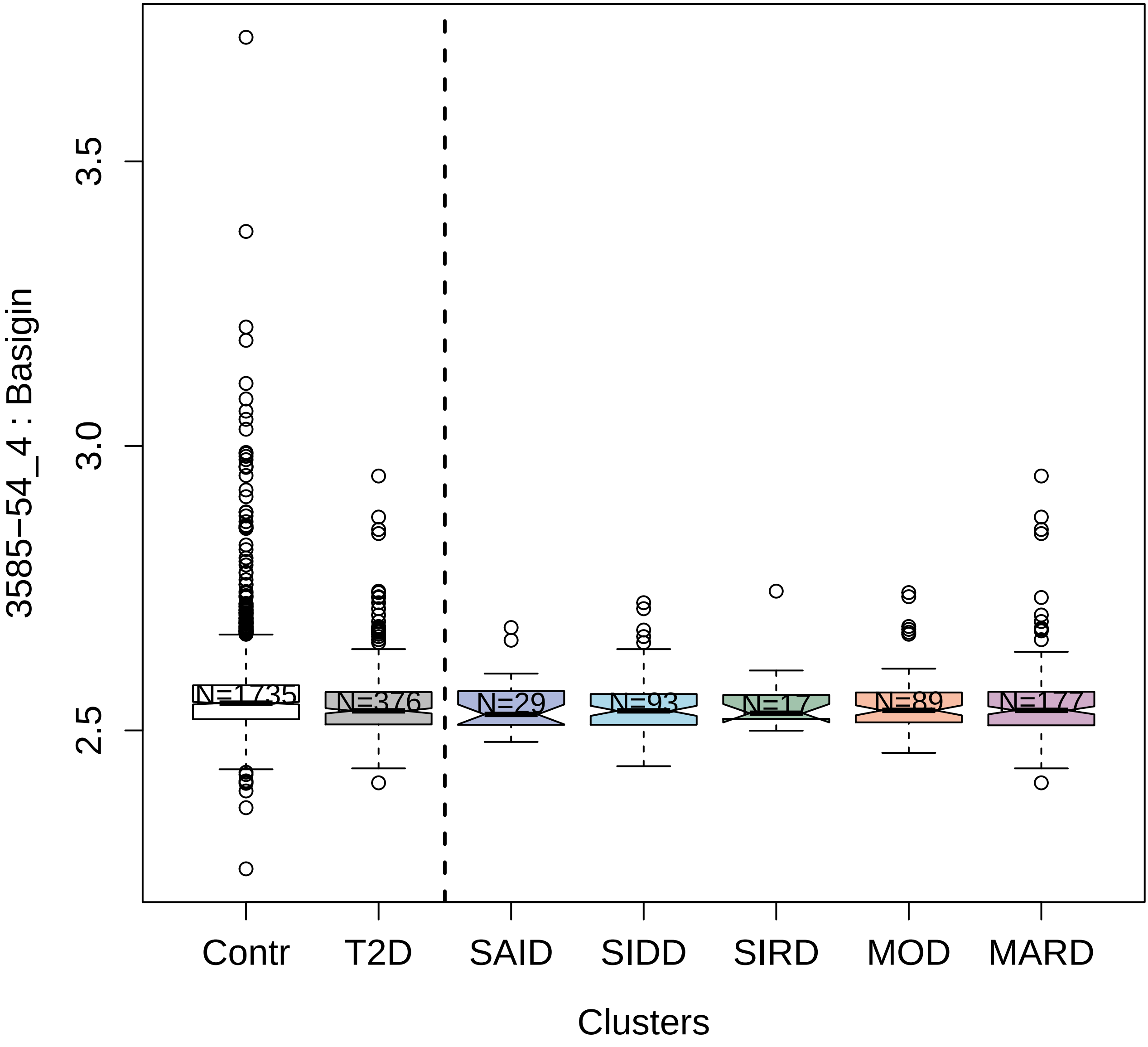

3585−54_4 : Basigin
3.5
3.0
2.5
N=1735
N=376
N=29
N=93
N=17
N=89
N=177
Contr
T2D
SAID
SIDD
SIRD
MOD
MARD
Clusters

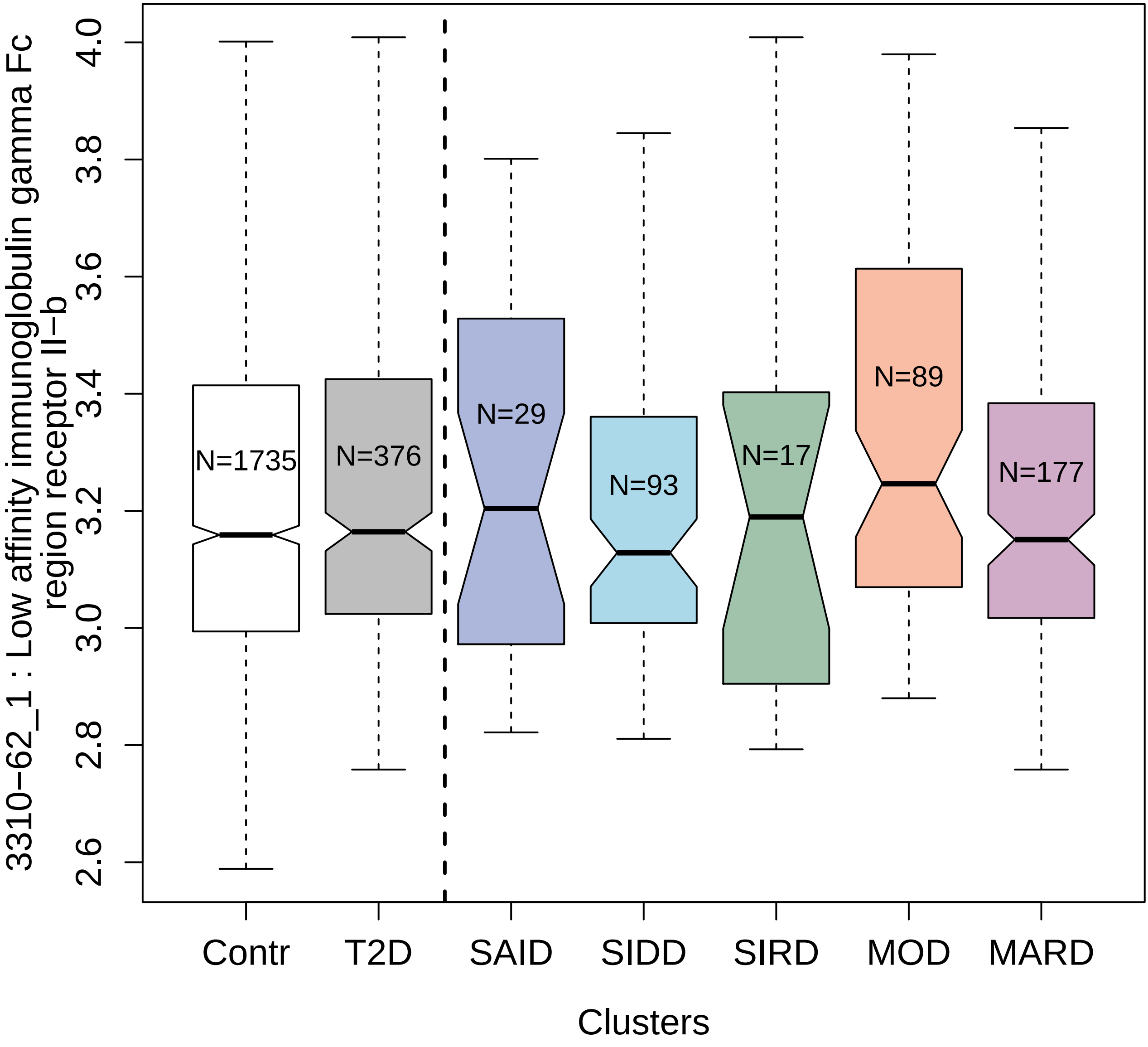
3310−62_1 : Low affinity immunoglobulin gamma Fc region receptor II−b
2.6
2.8
3.0
3.2
3.4
3.6
3.8
4.0
N=1735
N=376
N=29
N=93
N=17
N=89
N=177
Contr
T2D
SAID
SIDD
SIRD
MOD
MARD
Clusters

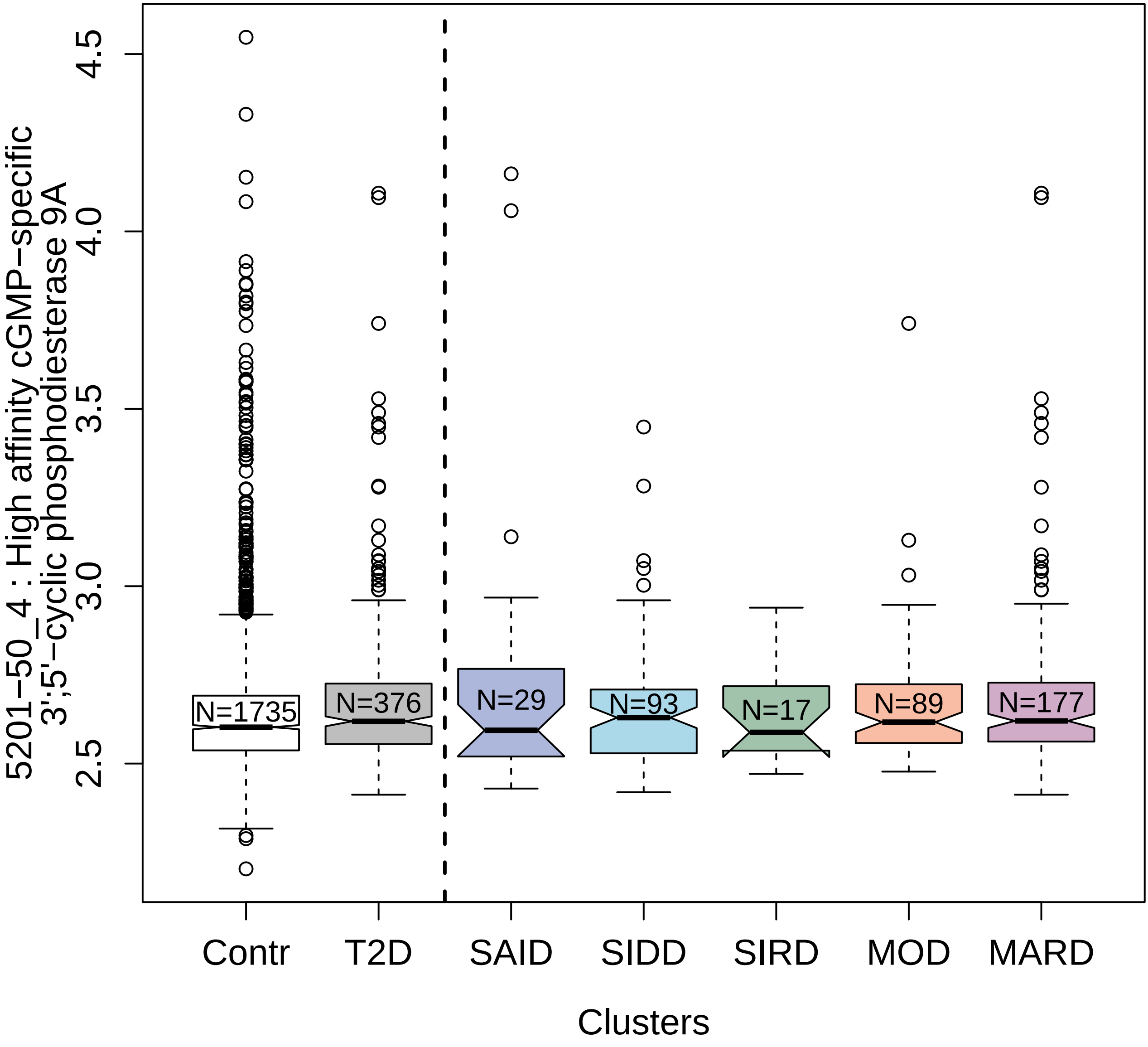

5201−50_4 : High affinity cGMP−specific
3',5'−cyclic phosphodiesterase 9A
2.5
3.0
3.5
4.0
4.5
N=1735
N=376
N=29
N=93
N=17
N=89
N=177
Contr
T2D
SAID
SIDD
SIRD
MOD
MARD
Clusters

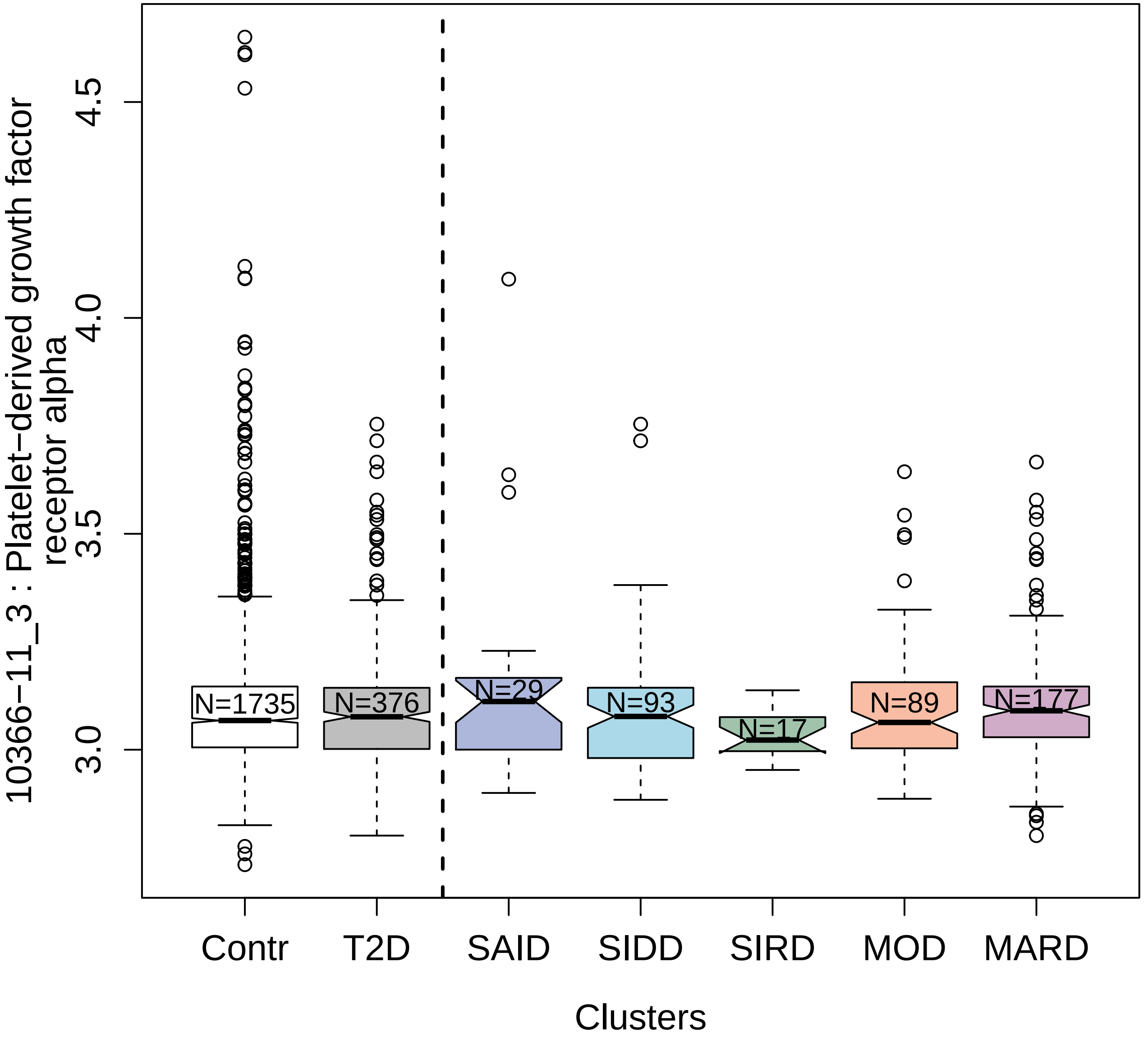
10366−11_3 : Platelet−derived growth factor receptor alpha
3.0
3.5
4.0
4.5
N=1735
N=376
N=29
N=93
N=17
N=89
N=177
Contr
T2D
SAID
SIDD
SIRD
MOD
MARD
Clusters

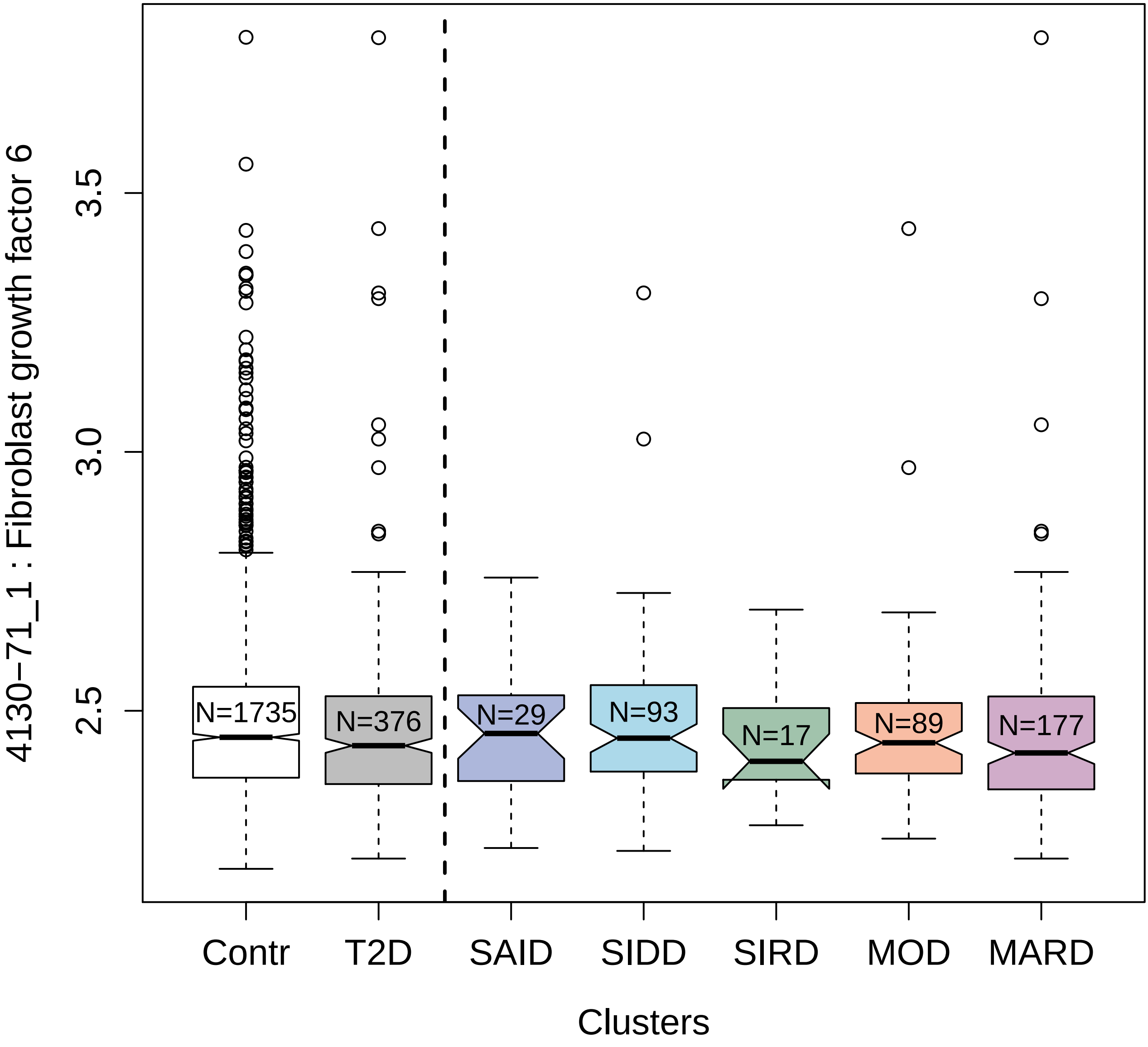

4130−71_1 : Fibroblast growth factor 6
3.5
3.0
2.5
N=1735
N=376
N=29
N=93
N=17
N=89
N=177
Contr
T2D
SAID
SIDD
SIRD
MOD
MARD
Clusters

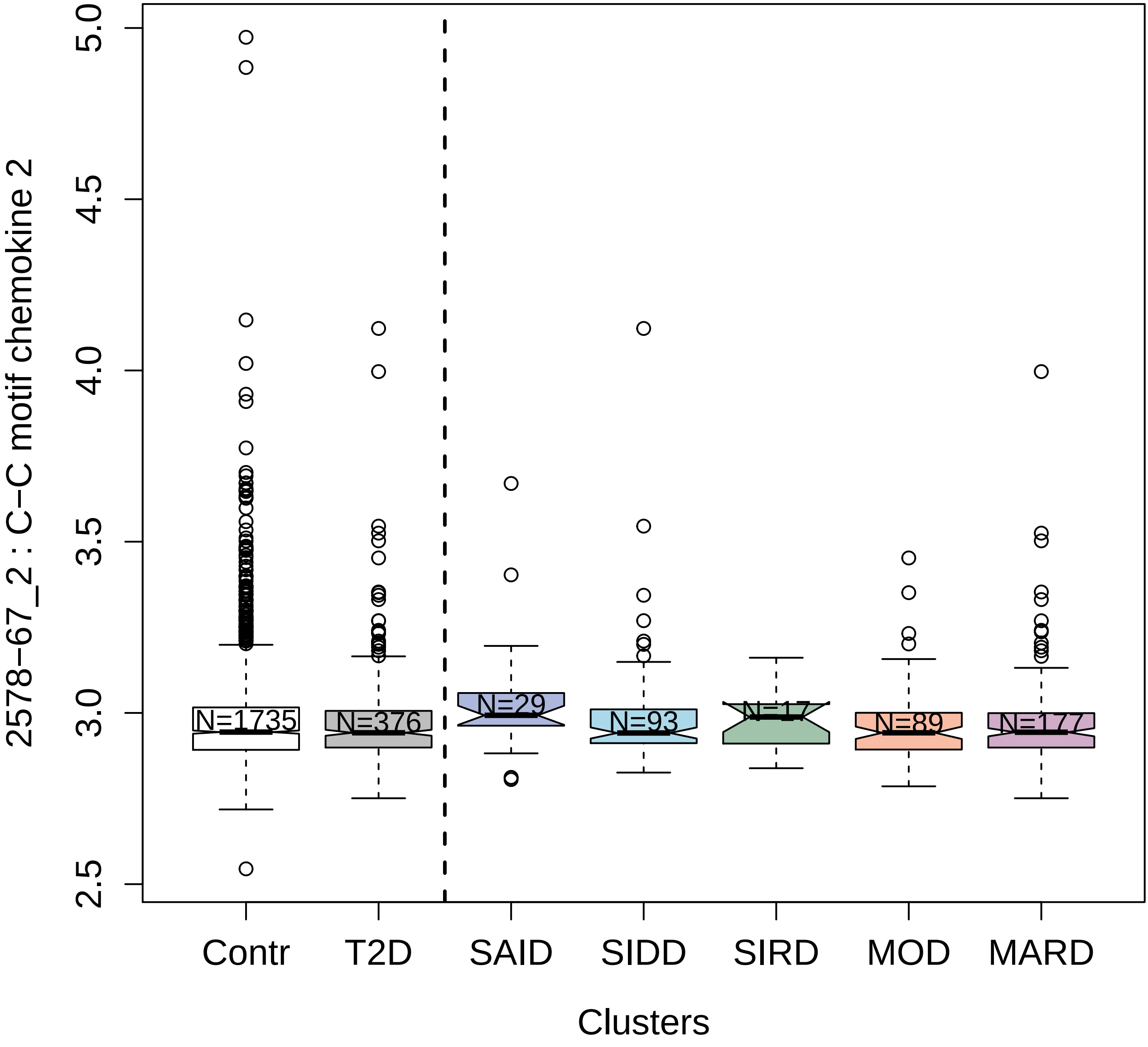
2578−67_2 : C−C motif chemokine 2
5.0
4.5
4.0
3.5
3.0
2.5
N=1735
N=376
N=29
N=93
N=17
N=89
N=177
Contr
T2D
SAID
SIDD
SIRD
MOD
MARD
Clusters

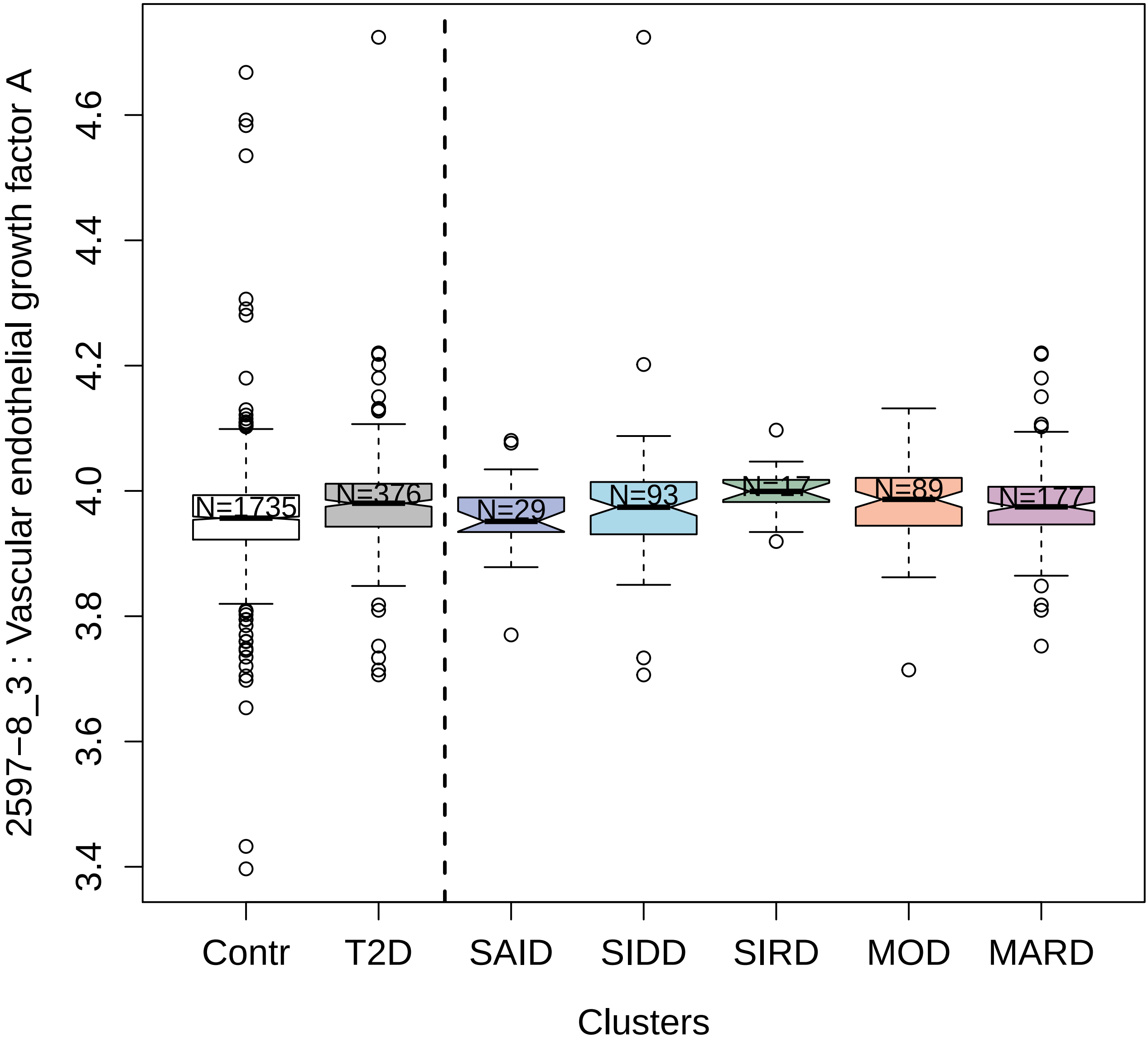
2597−8_3 : Vascular endothelial growth factor A
3.4
3.6
3.8
4.0
4.2
4.4
4.6
N=1735
N=376
N=29
N=93
N=17
N=89
N=177
Contr
T2D
SAID
SIDD
SIRD
MOD
MARD
Clusters

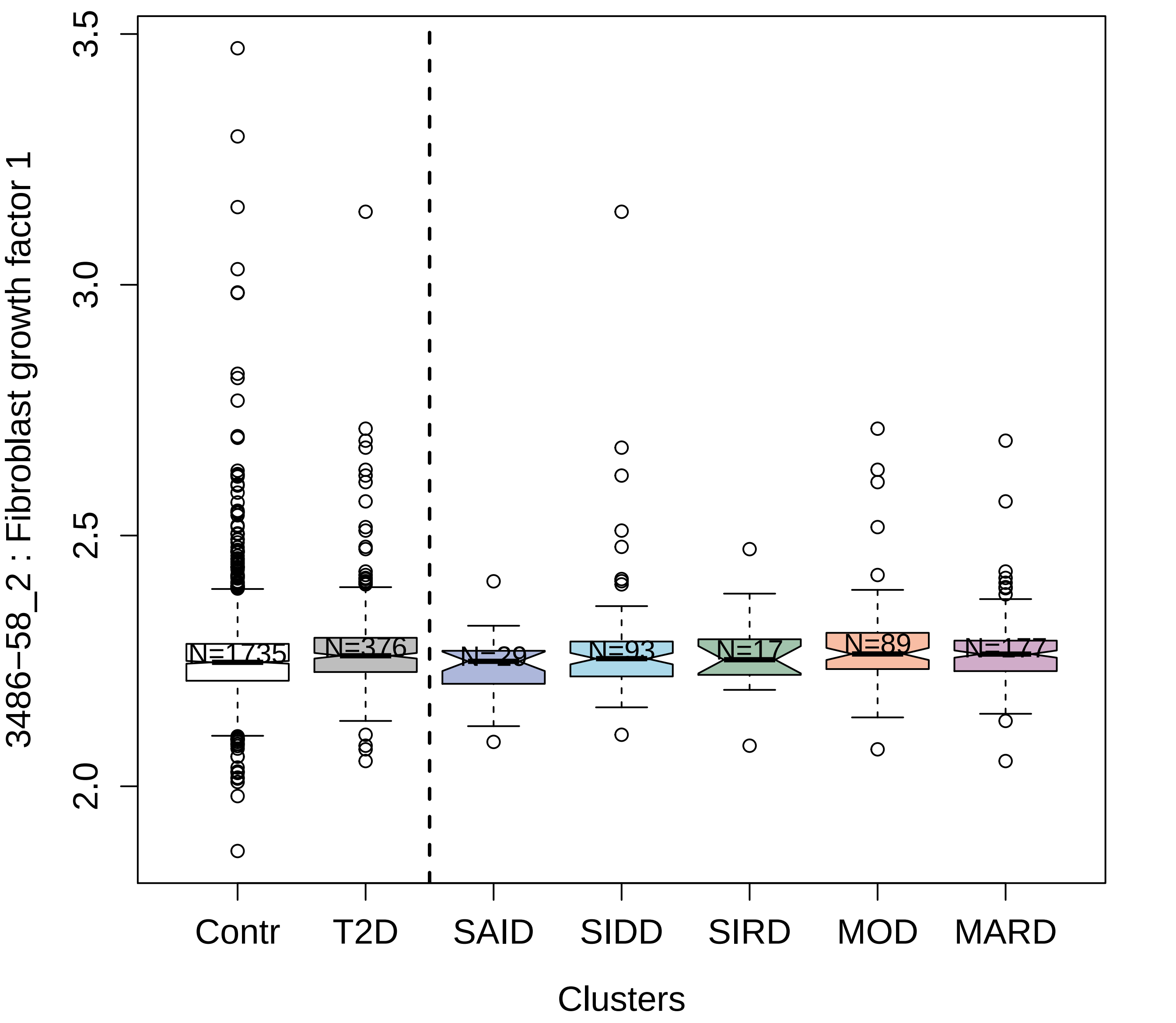
3486−58_2 : Fibroblast growth factor 1
3.5
3.0
2.5
2.0
N=1735
N=376
N=29
N=93
N=17
N=89
N=177
Contr
T2D
SAID
SIDD
SIRD
MOD
MARD
Clusters

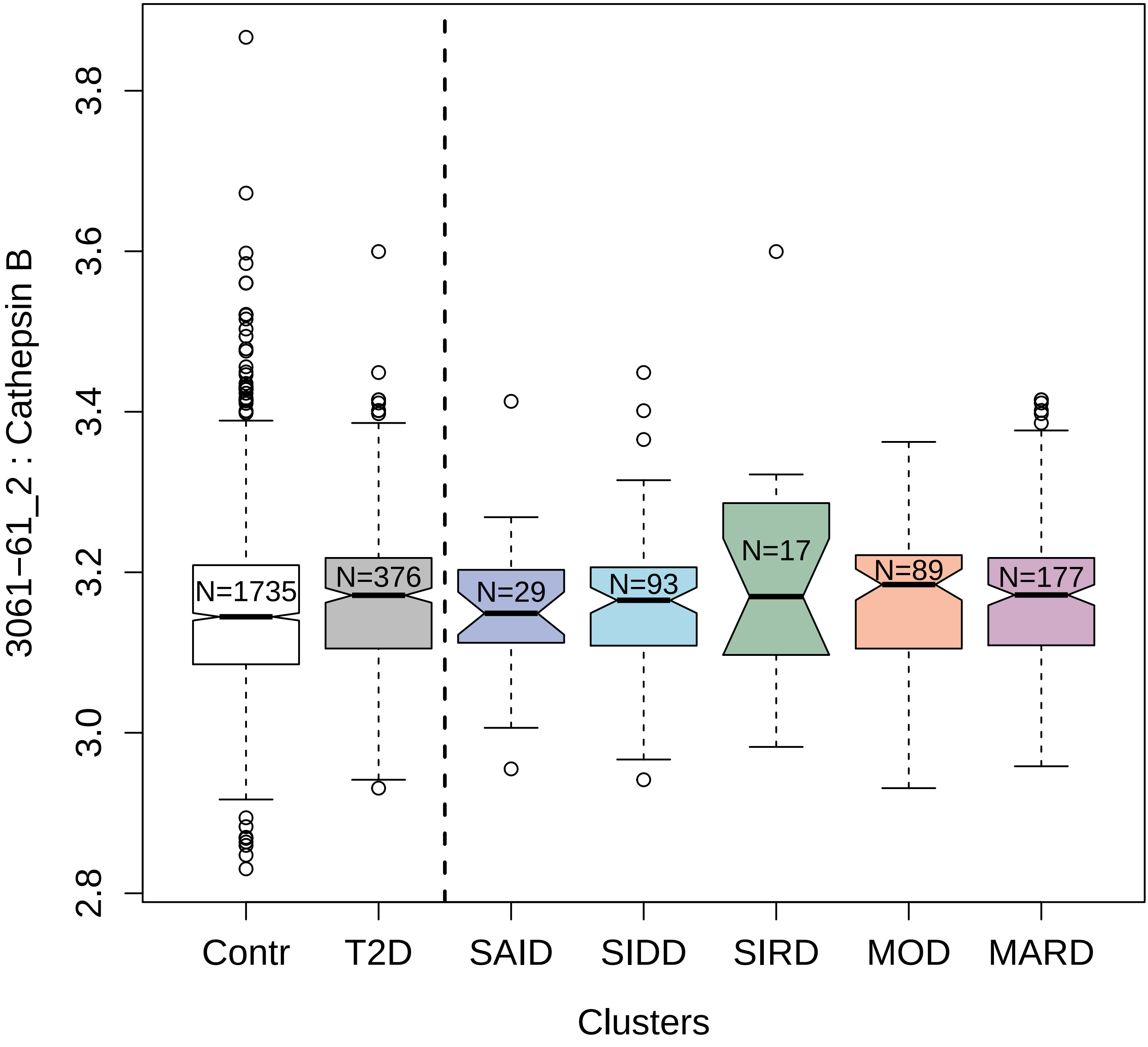
3061−61_2 : Cathepsin B
2.8
3.0
3.2
3.4
3.6
3.8
N=1735
N=376
N=29
N=93
N=17
N=89
N=177
Contr
T2D
SAID
SIDD
SIRD
MOD
MARD
Clusters

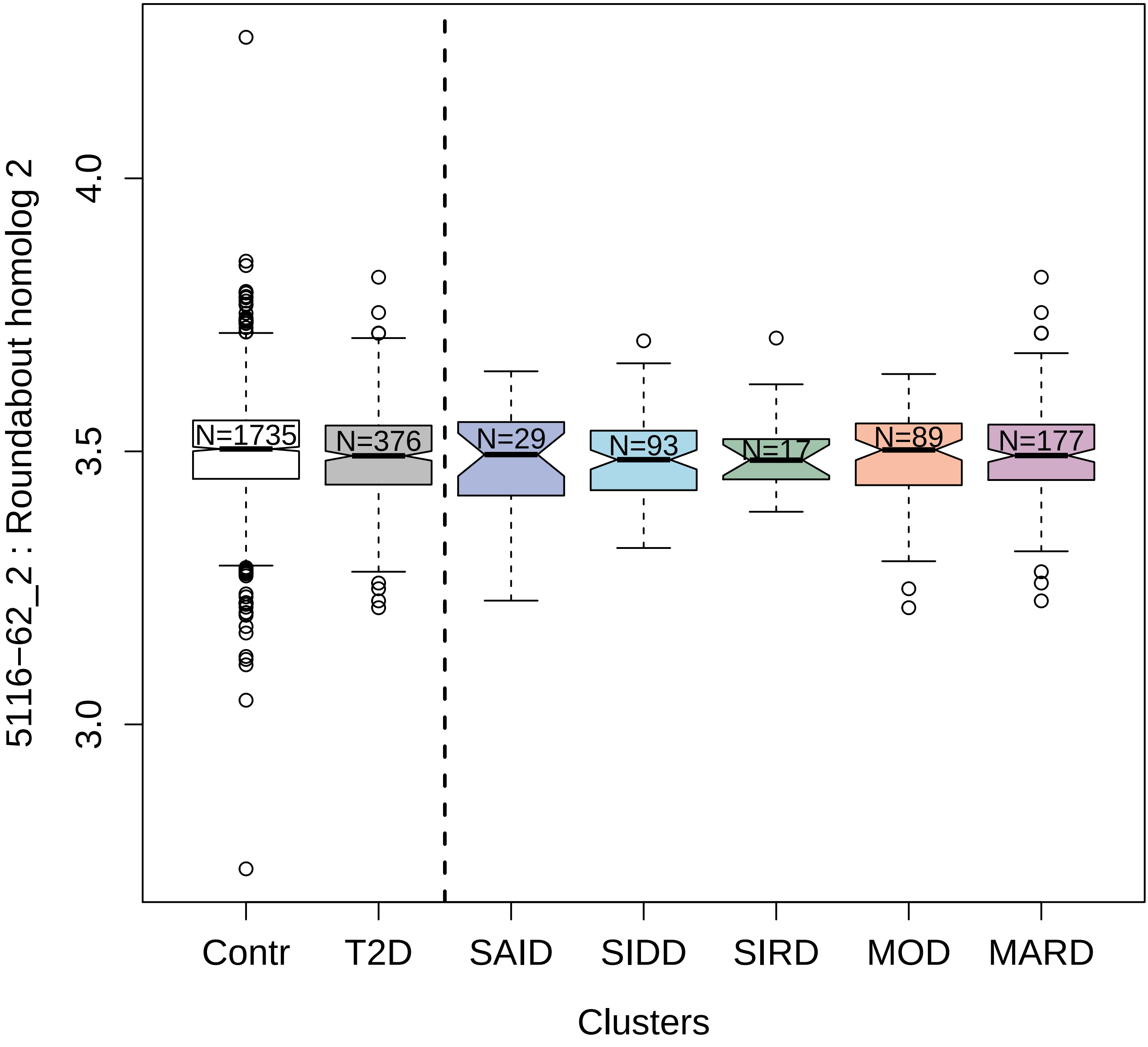

5116−62_2 : Roundabout homolog 2
4.0
3.5
3.0
N=1735
N=376
N=29
N=93
N=17
N=89
N=177
Contr
T2D
SAID
SIDD
SIRD
MOD
MARD
Clusters

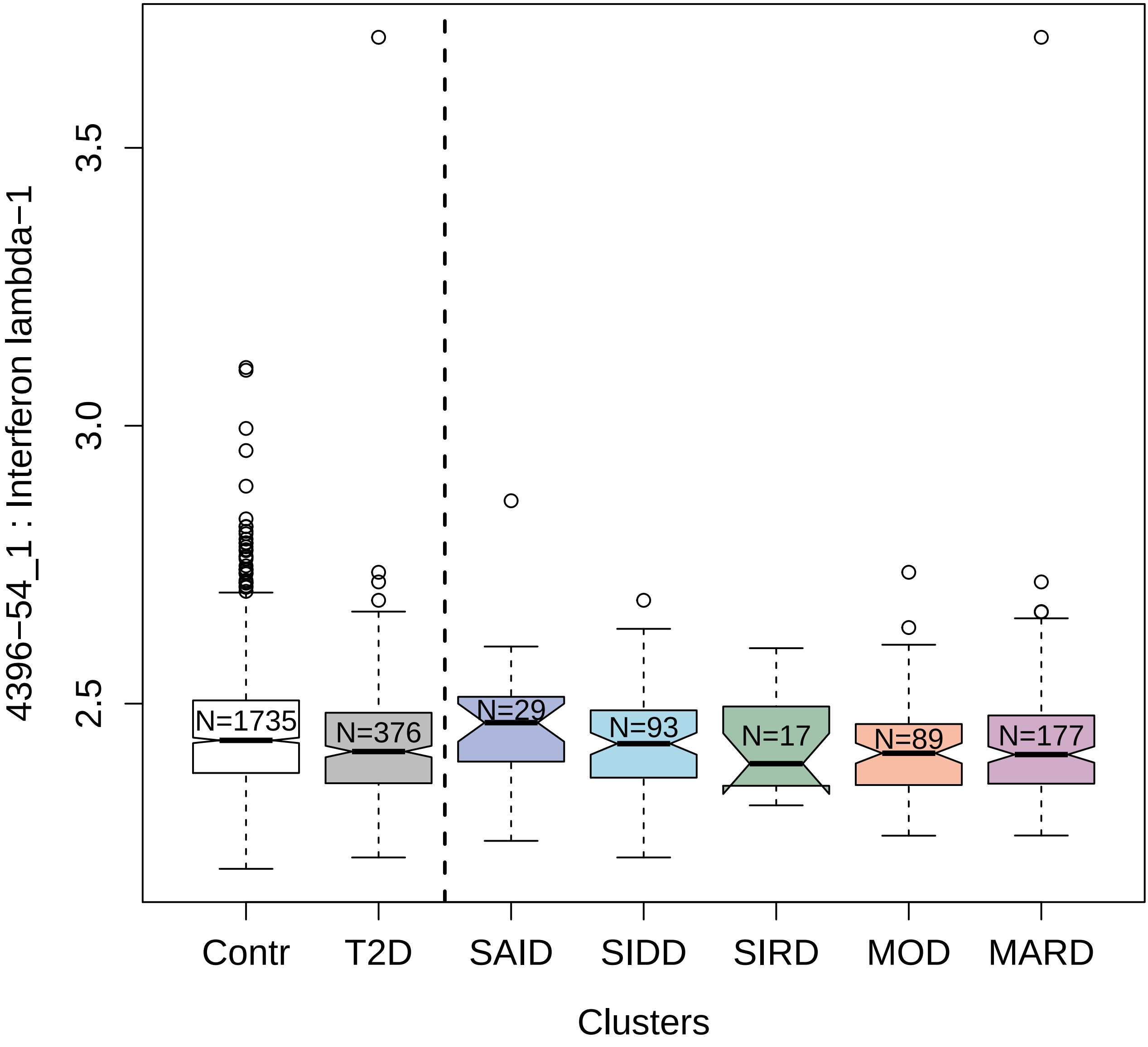
4396−54_1 : Interferon lambda−1
3.5
3.0
2.5
N=1735
N=376
N=29
N=93
N=17
N=89
N=177
Contr
T2D
SAID
SIDD
SIRD
MOD
MARD
Clusters

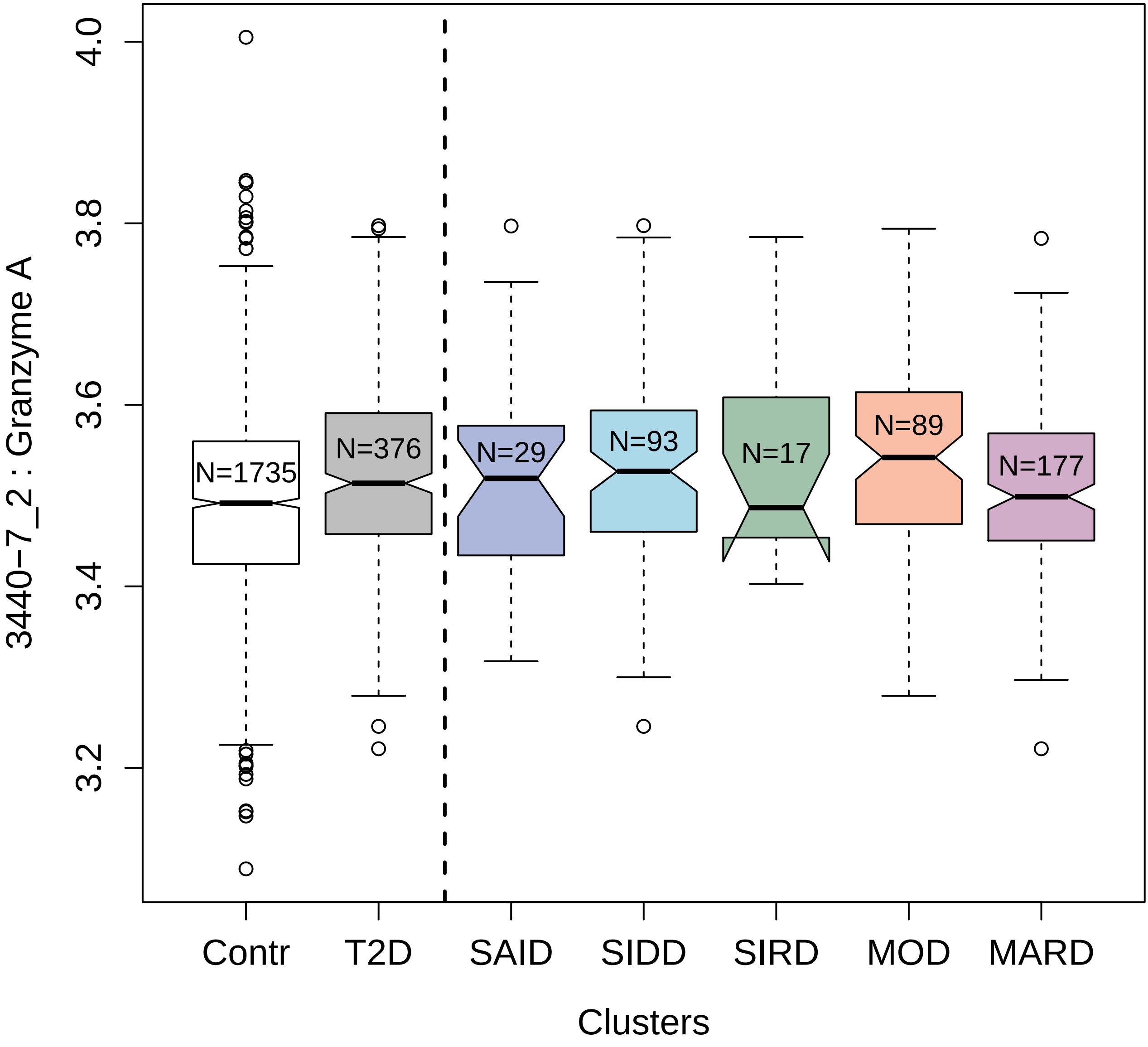

3440−7_2 : Granzyme A
4.0
3.8
3.6
3.4
3.2
N=1735
N=376
N=29
N=93
N=17
N=89
N=177
Contr
T2D
SAID
SIDD
SIRD
MOD
MARD
Clusters

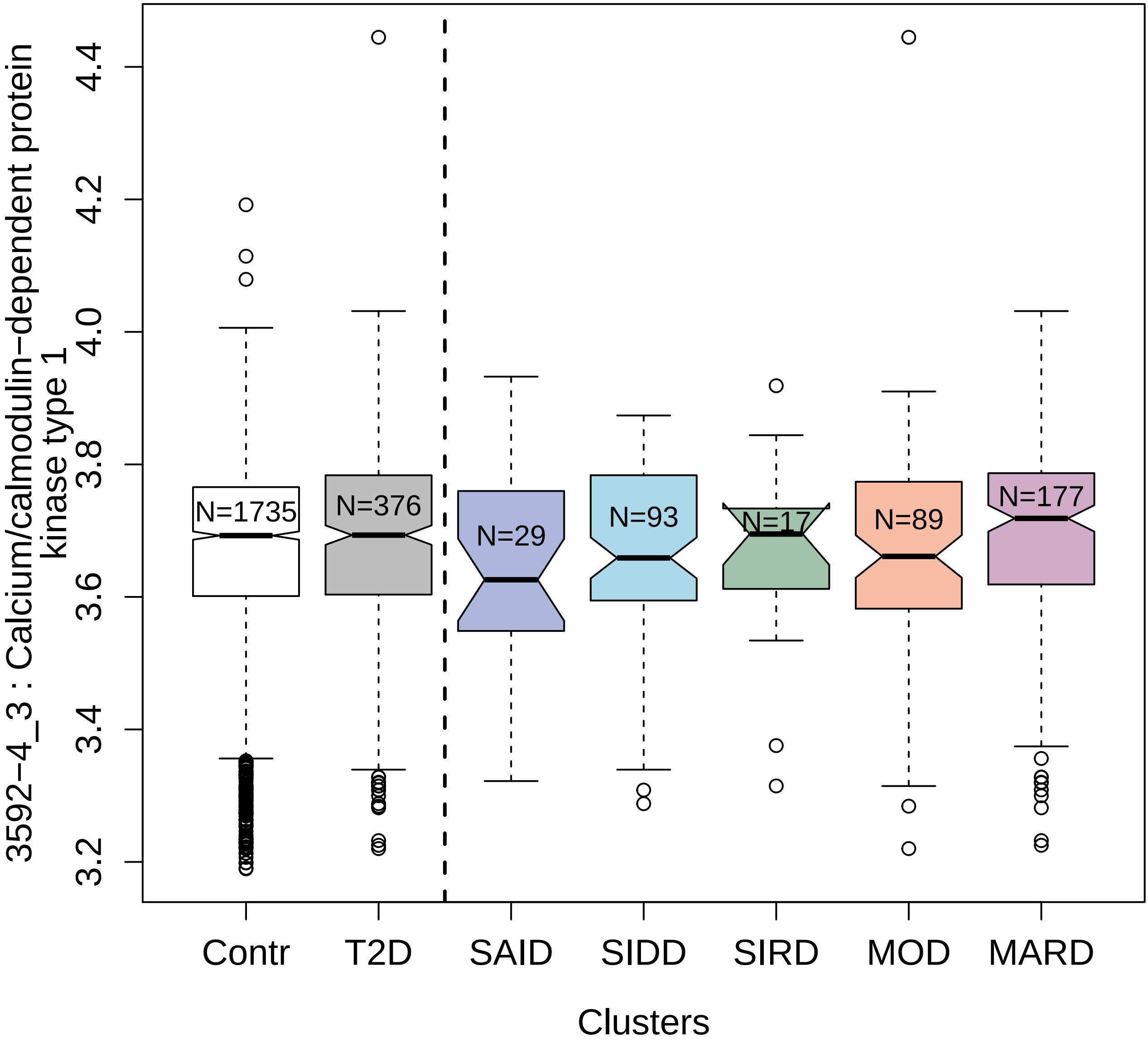
3592−4_3 : Calcium/calmodulin−dependent protein kinase type 1
3.2
3.4
3.6
3.8
4.0
4.2
4.4
N=1735
N=376
N=29
N=93
N=17
N=89
N=177
Contr
T2D
SAID
SIDD
SIRD
MOD
MARD
Clusters

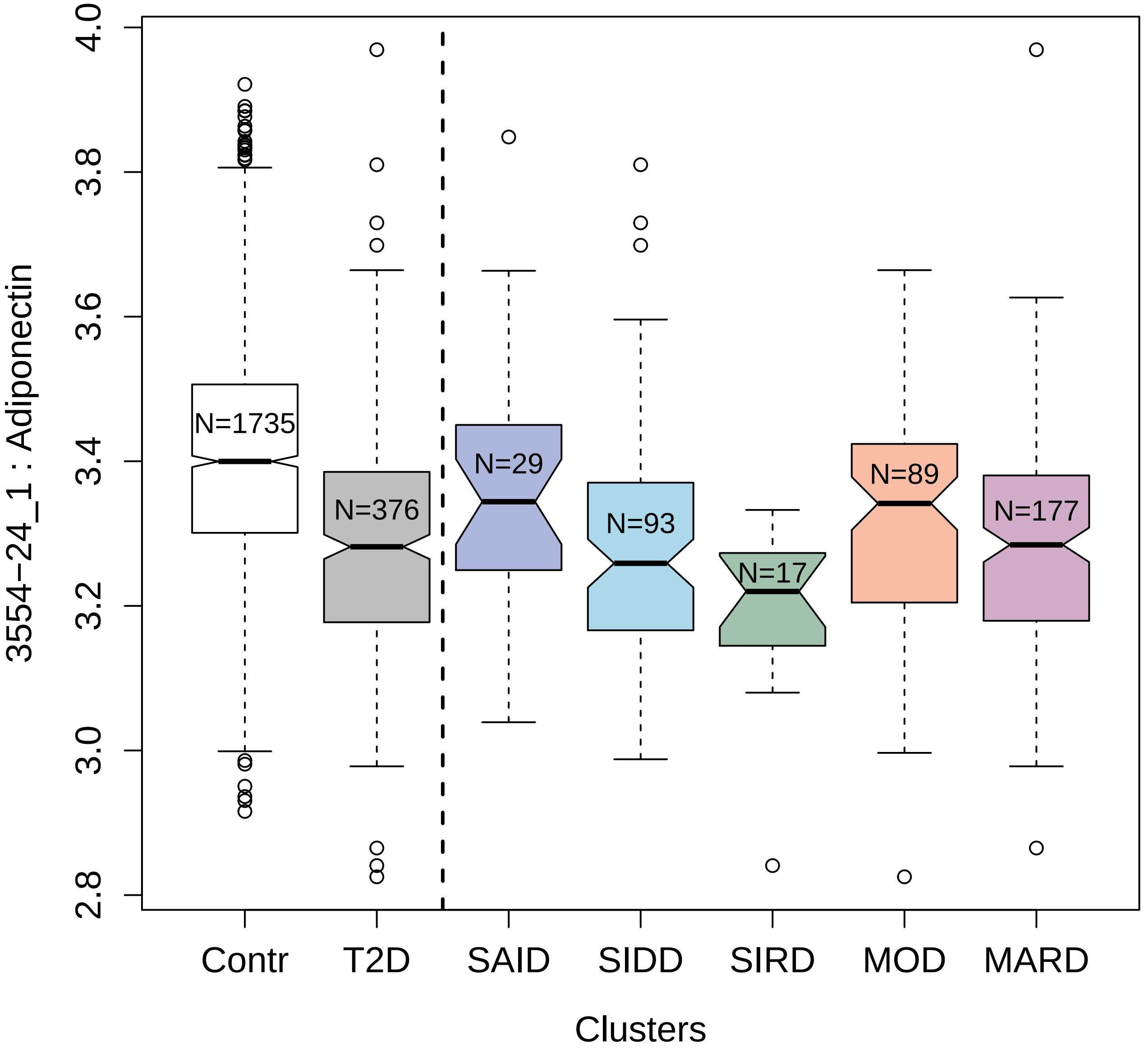
3554−24_1 : Adiponectin
4.0
3.8
3.6
3.4
3.2
3.0
2.8
N=1735
N=376
N=29
N=93
N=17
N=89
N=177
Contr
T2D
SAID
SIDD
SIRD
MOD
MARD
Clusters

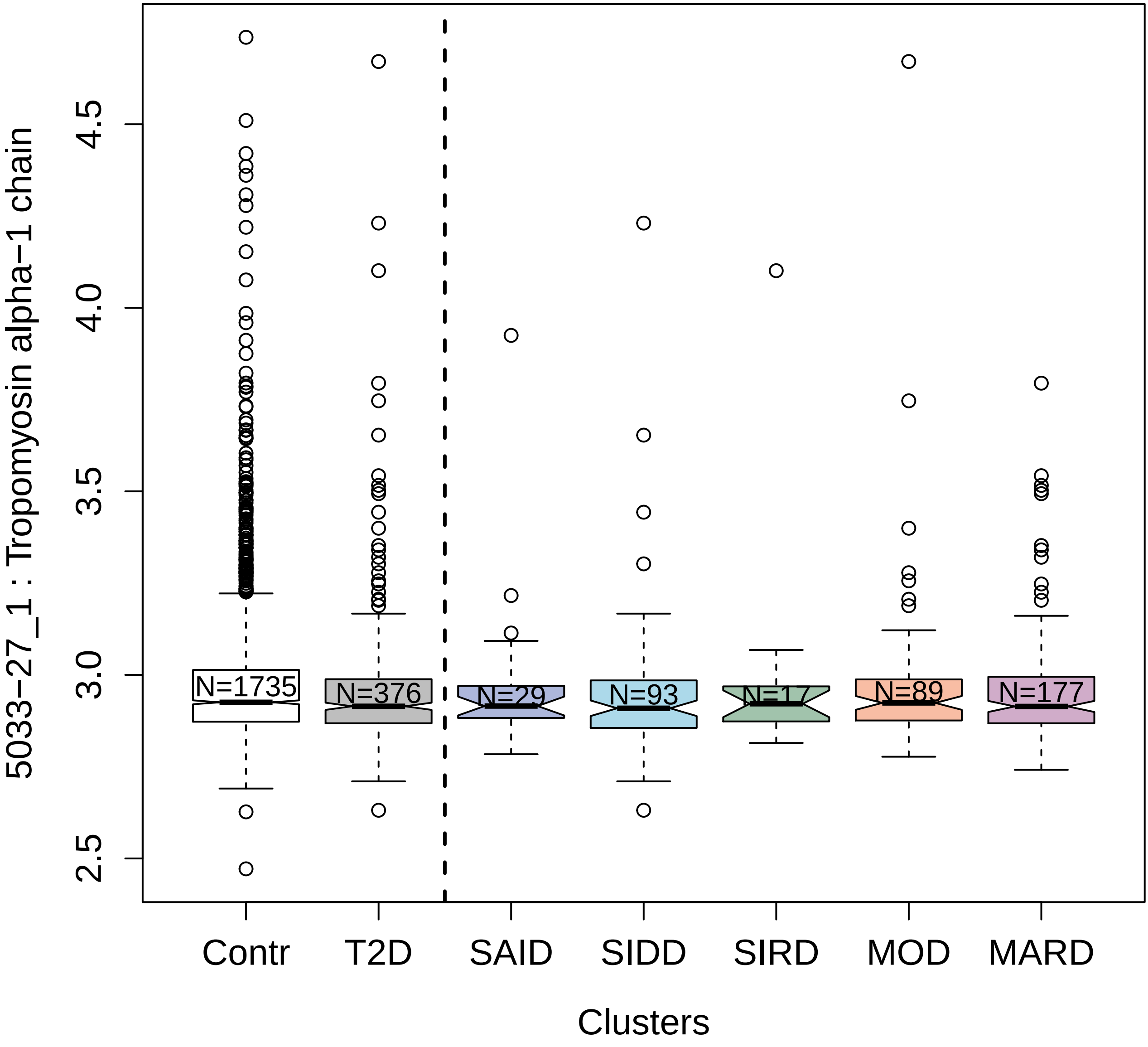
5033−27_1 : Tropomyosin alpha−1 chain
2.5
3.0
3.5
4.0
4.5
N=1735
N=376
N=29
N=93
N=17
N=89
N=177
Contr
T2D
SAID
SIDD
SIRD
MOD
MARD
Clusters

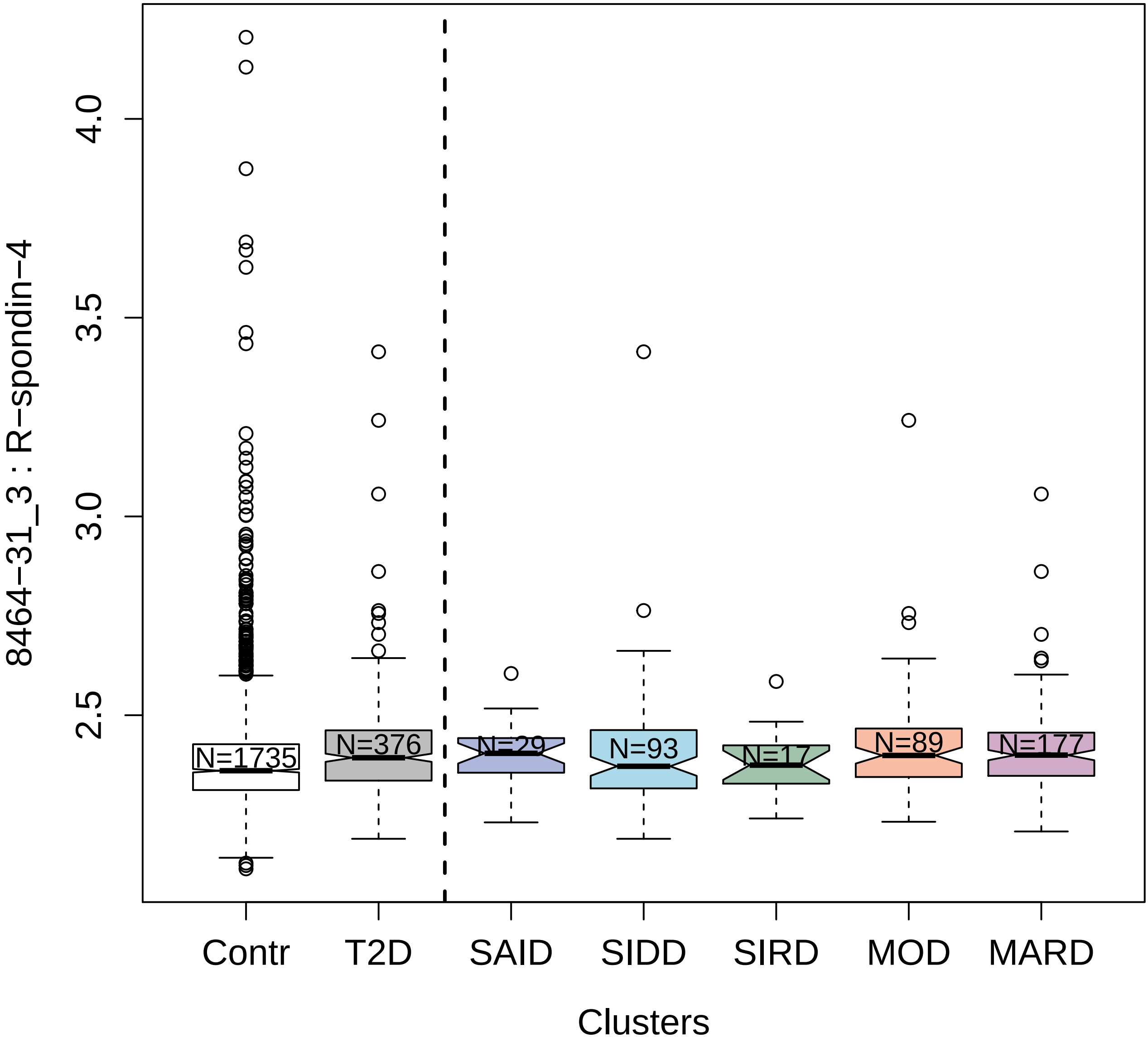
8464−31_3 : R−spondin−4
4.0
3.5
3.0
2.5
N=1735
N=376
N=29
N=93
N=17
N=89
N=177
Contr
T2D
SAID
SIDD
SIRD
MOD
MARD
Clusters

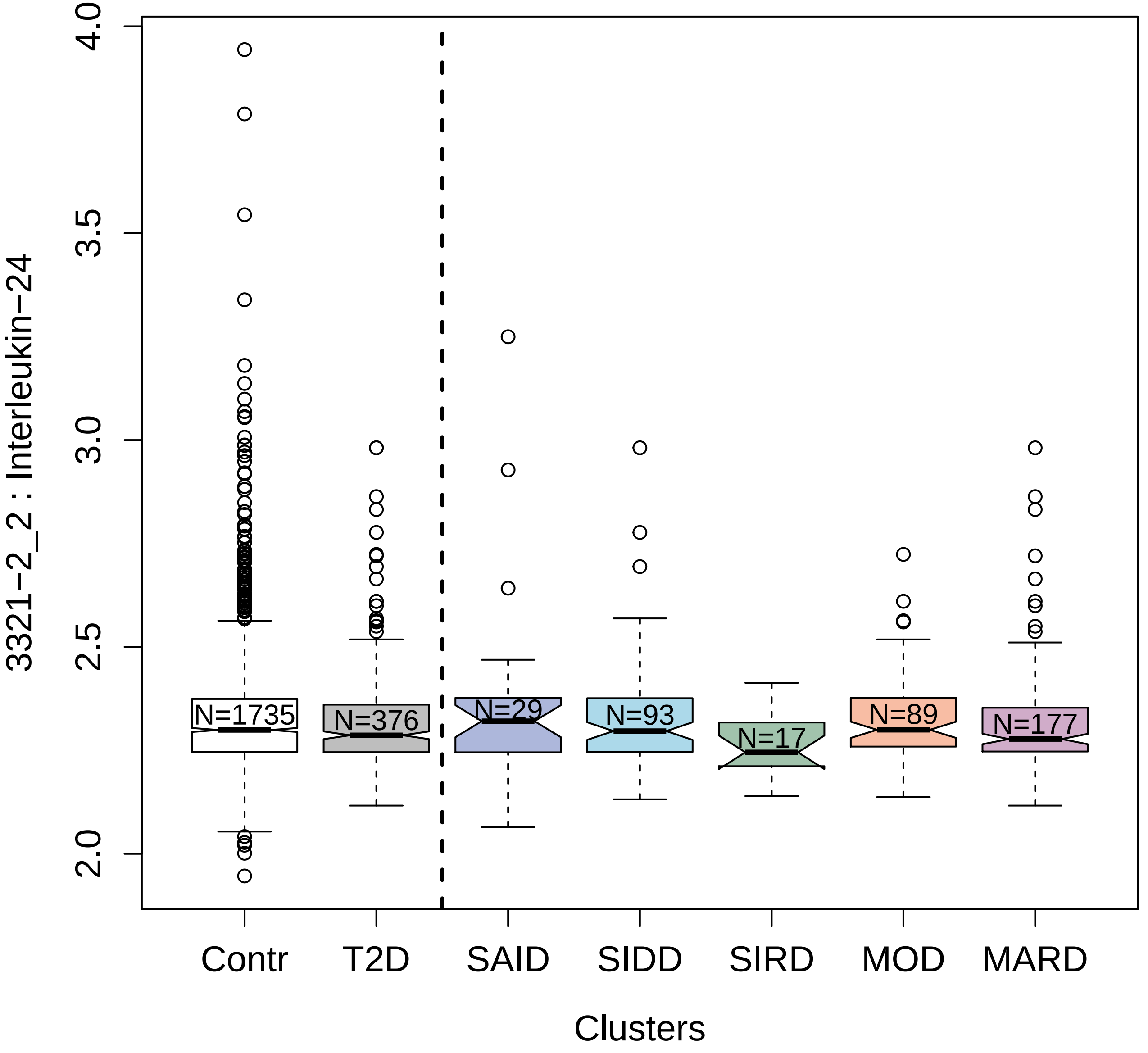
3321−2_2 : Interleukin−24
4.0
3.5
3.0
2.5
2.0
N=1735
N=376
N=29
N=93
N=17
N=89
N=177
Contr
T2D
SAID
SIDD
SIRD
MOD
MARD
Clusters

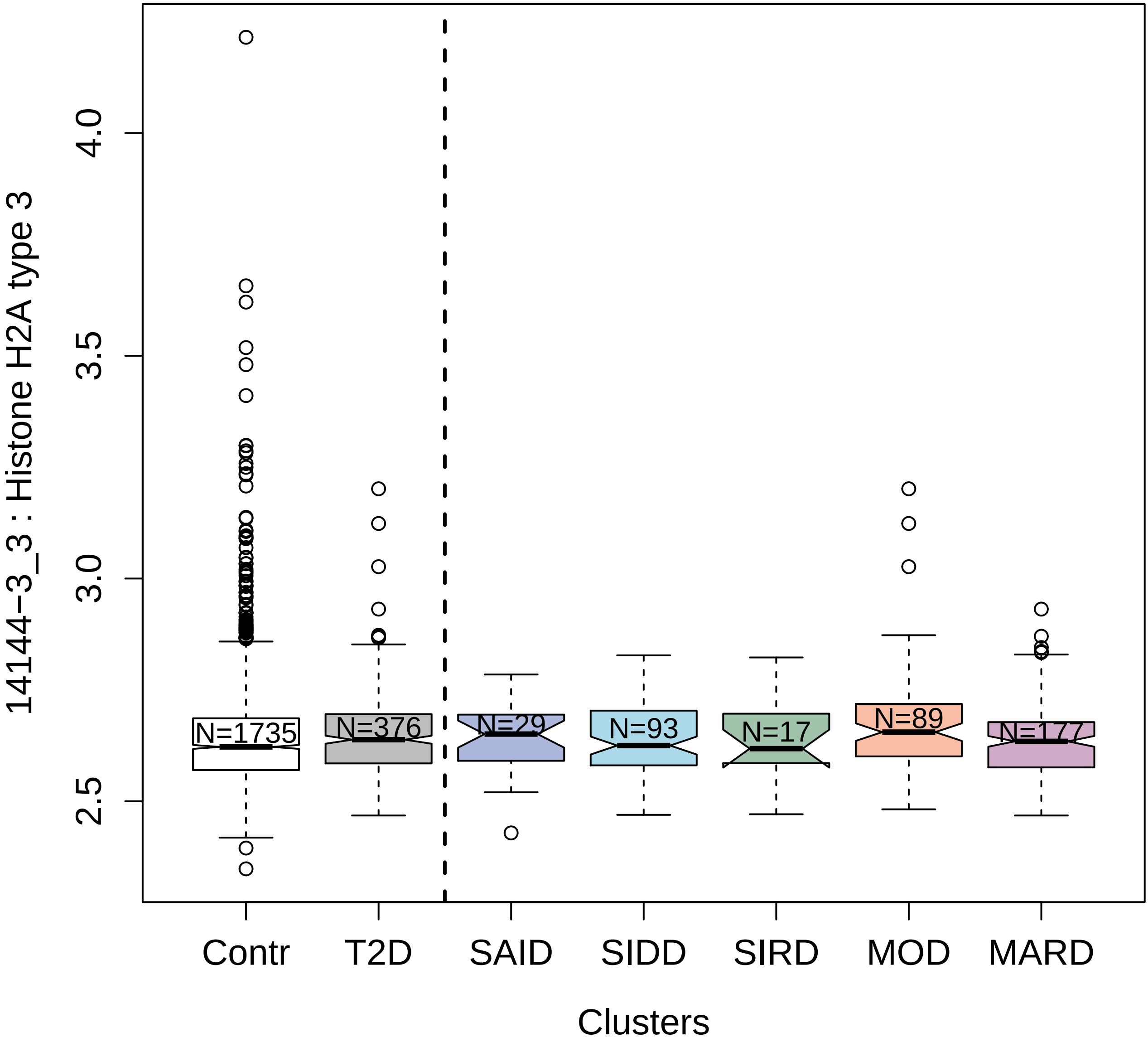

14144−3_3 : Histone H2A type 3
4.0
3.5
3.0
2.5
N=1735
N=376
N=29
N=93
N=17
N=89
N=177
Contr
T2D
SAID
SIDD
SIRD
MOD
MARD
Clusters

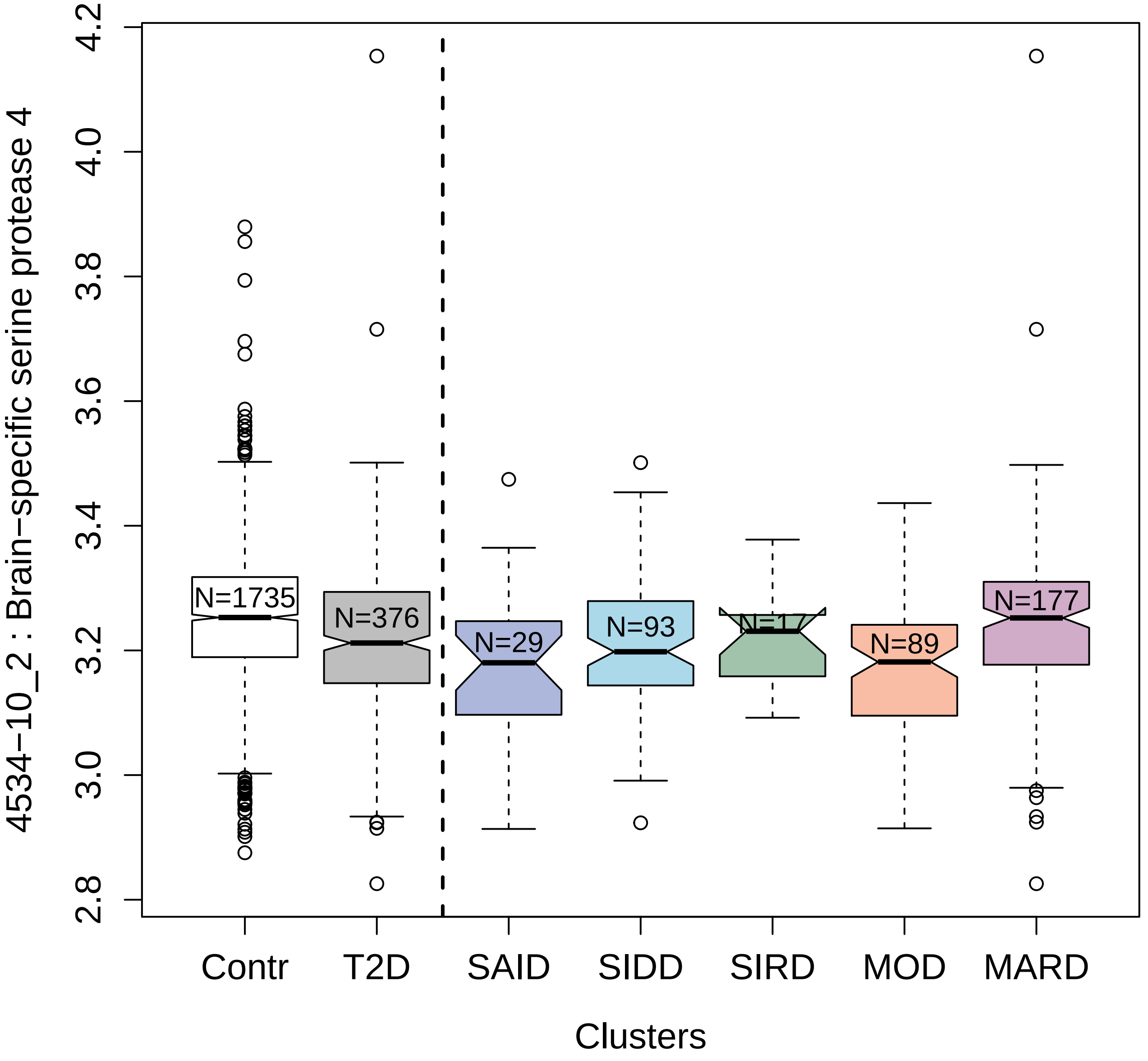
4534−10_2 : Brain−specific serine protease 4
2.8
3.0
3.2
3.4
3.6
3.8
4.0
4.2
N=1735
N=376
N=29
N=93
N=17
N=89
N=177
Contr
T2D
SAID
SIDD
SIRD
MOD
MARD
Clusters

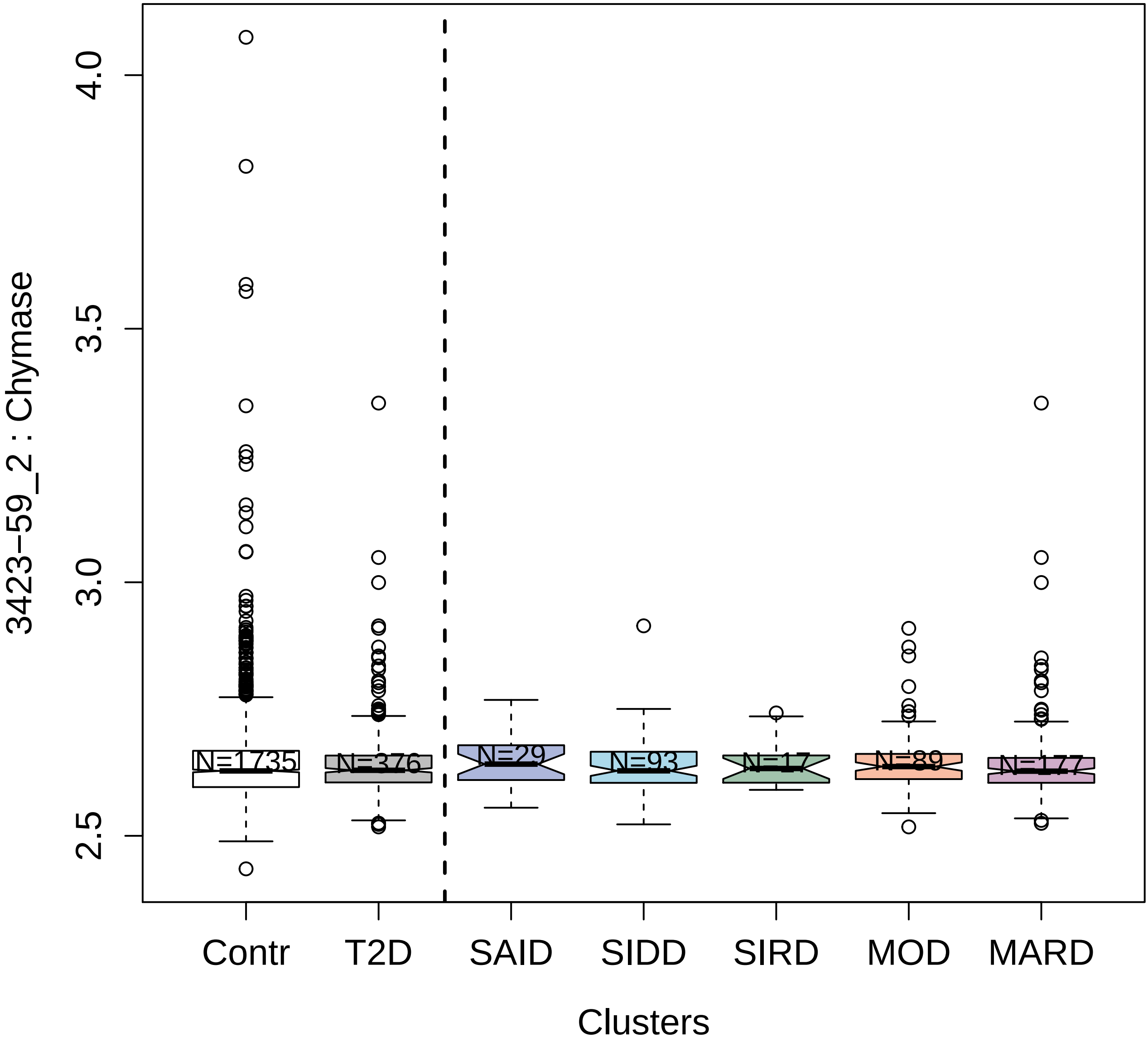

3423−59_2 : Chymase
4.0
3.5
3.0
2.5
N=1735
N=376
N=29
N=93
N=17
N=89
N=177
Contr
T2D
SAID
SIDD
SIRD
MOD
MARD
Clusters

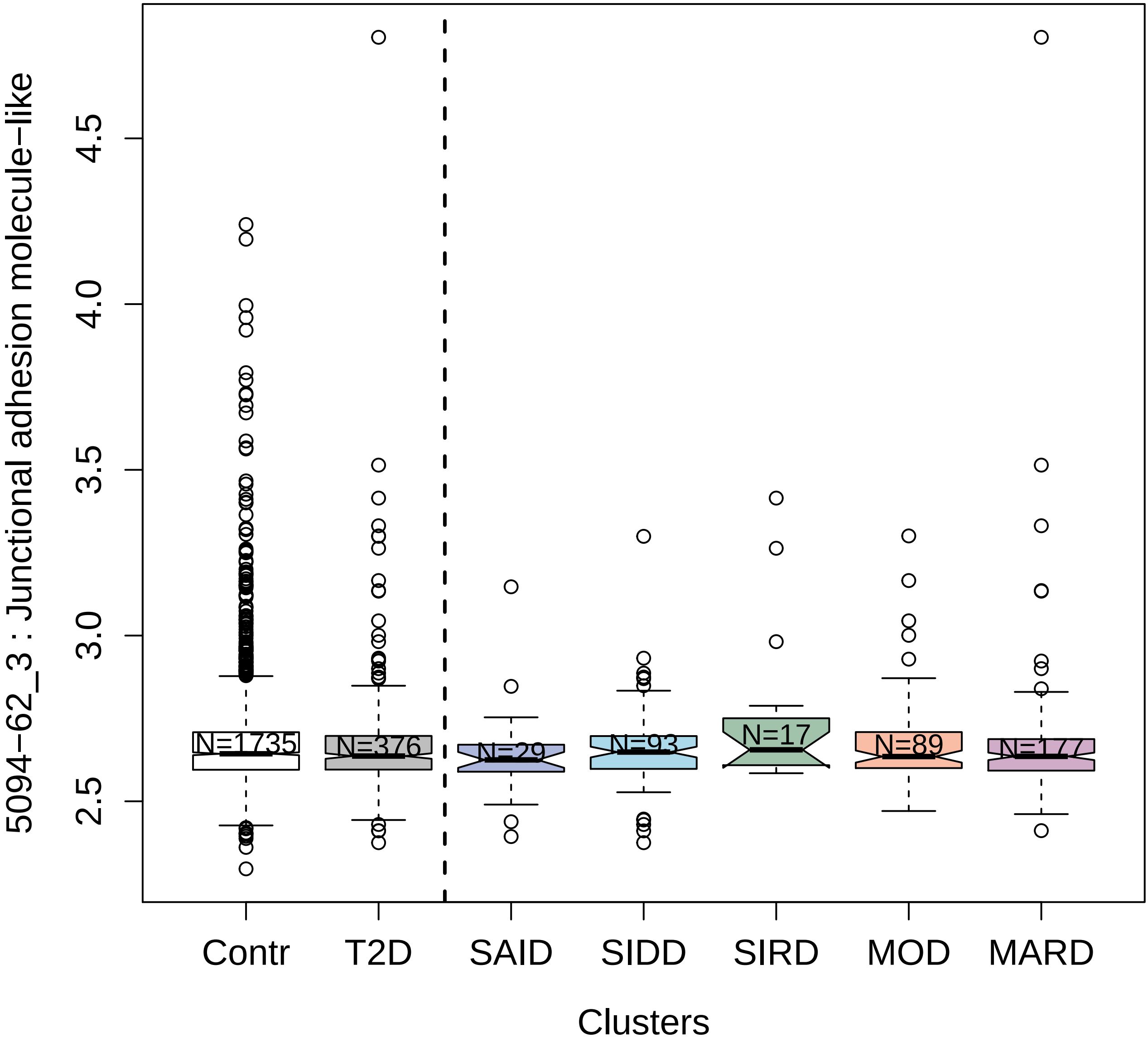

5094−62_3 : Junctional adhesion molecule−like
2.5
3.0
3.5
4.0
4.5
N=1735
N=376
N=29
N=93
N=17
N=89
N=177
Contr
T2D
SAID
SIDD
SIRD
MOD
MARD
Clusters

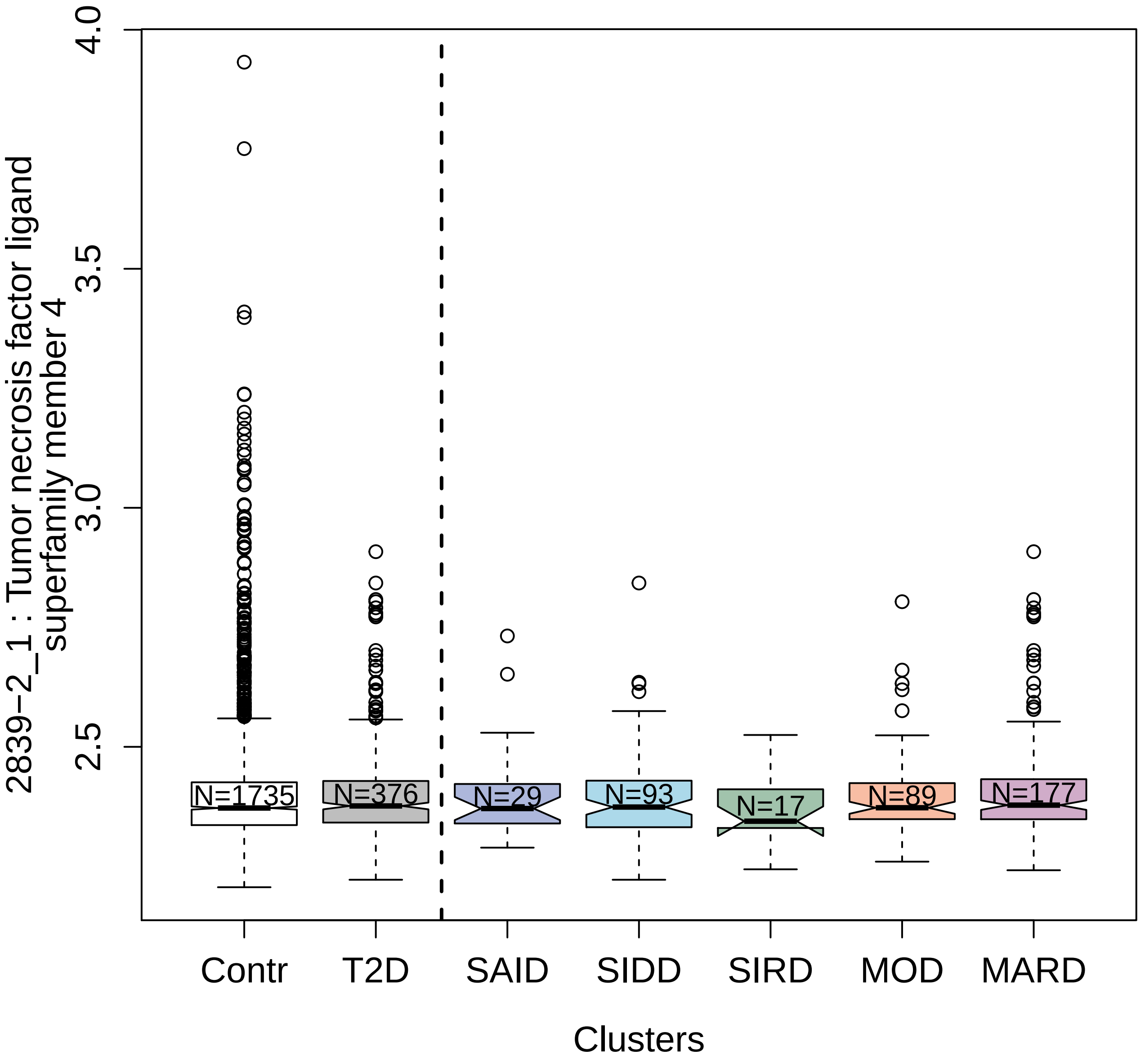
2839−2_1 : Tumor necrosis factor ligand
superfamily member 4
4.0
3.5
3.0
2.5
N=1735
N=376
N=29
N=93
N=17
N=89
N=177
Contr
T2D
SAID
SIDD
SIRD
MOD
MARD
Clusters

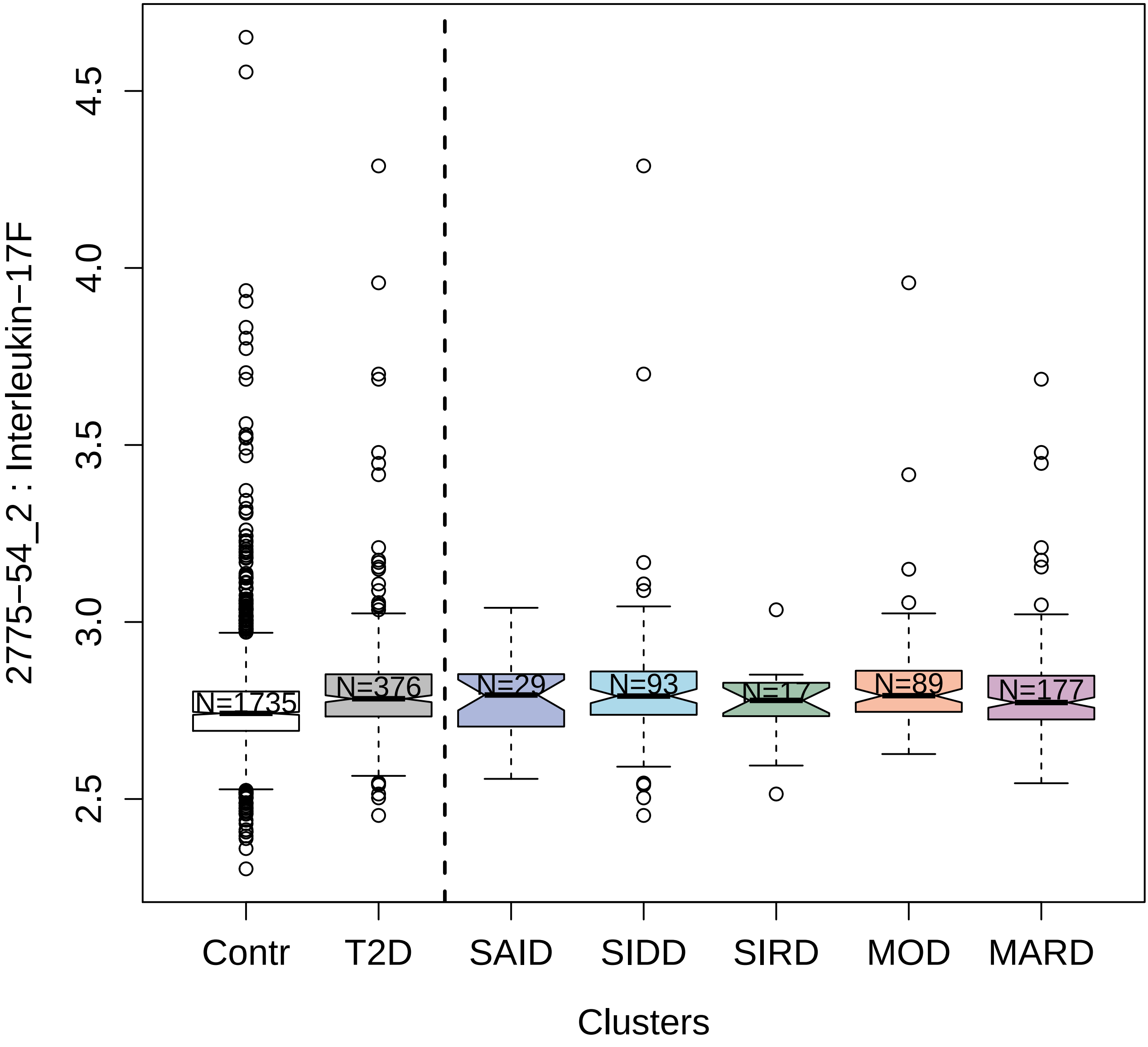
2775−54_2 : Interleukin−17F
4.5
4.0
3.5
3.0
2.5
N=1735
N=376
N=29
N=93
N=17
N=89
N=177
Contr
T2D
SAID
SIDD
SIRD
MOD
MARD
Clusters

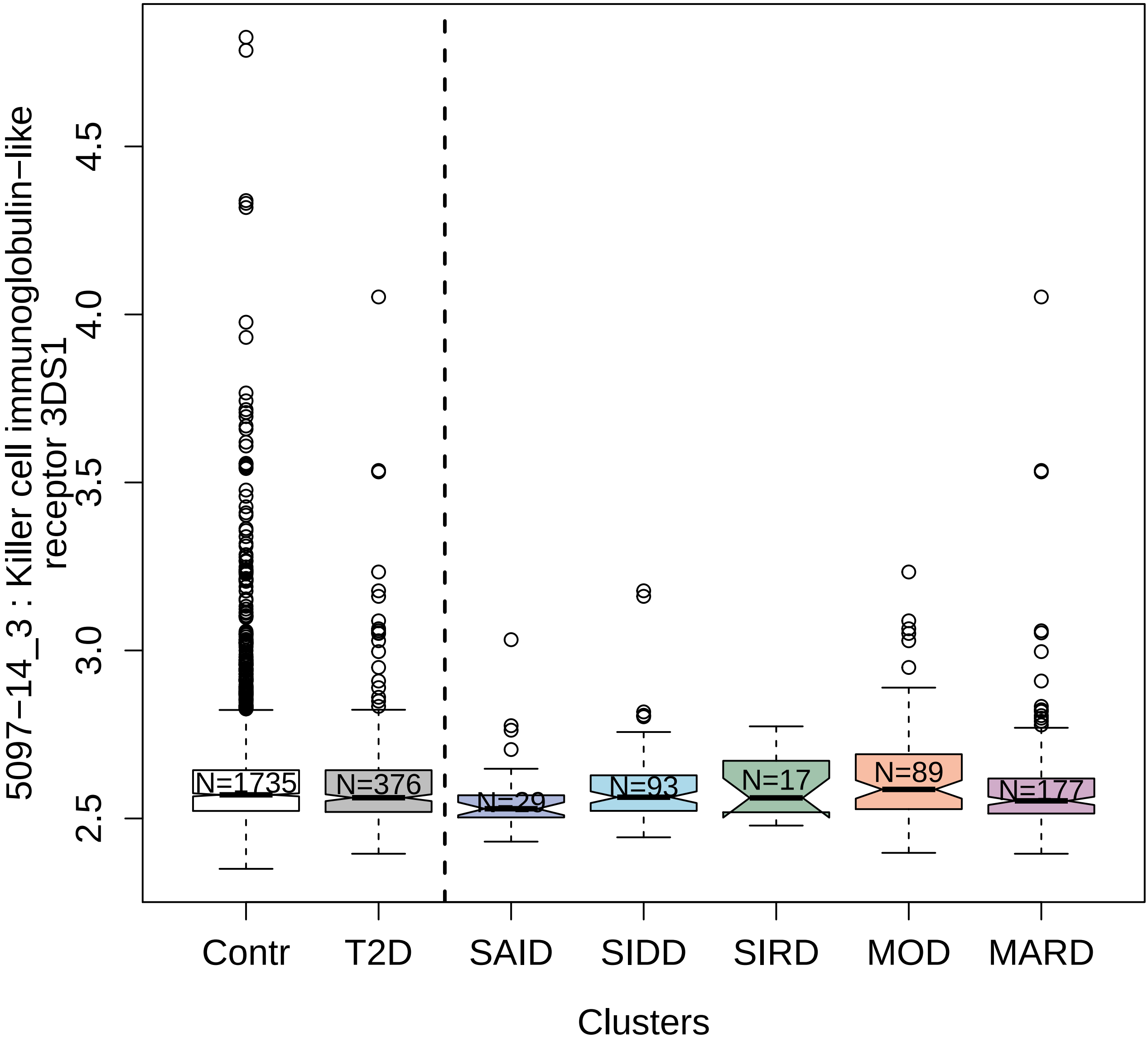
5097−14_3 : Killer cell immunoglobulin−like
receptor 3DS1
4.5
4.0
3.5
3.0
2.5
N=1735
N=376
N=29
N=93
N=17
N=89
N=177
Contr
T2D
SAID
SIDD
SIRD
MOD
MARD
Clusters

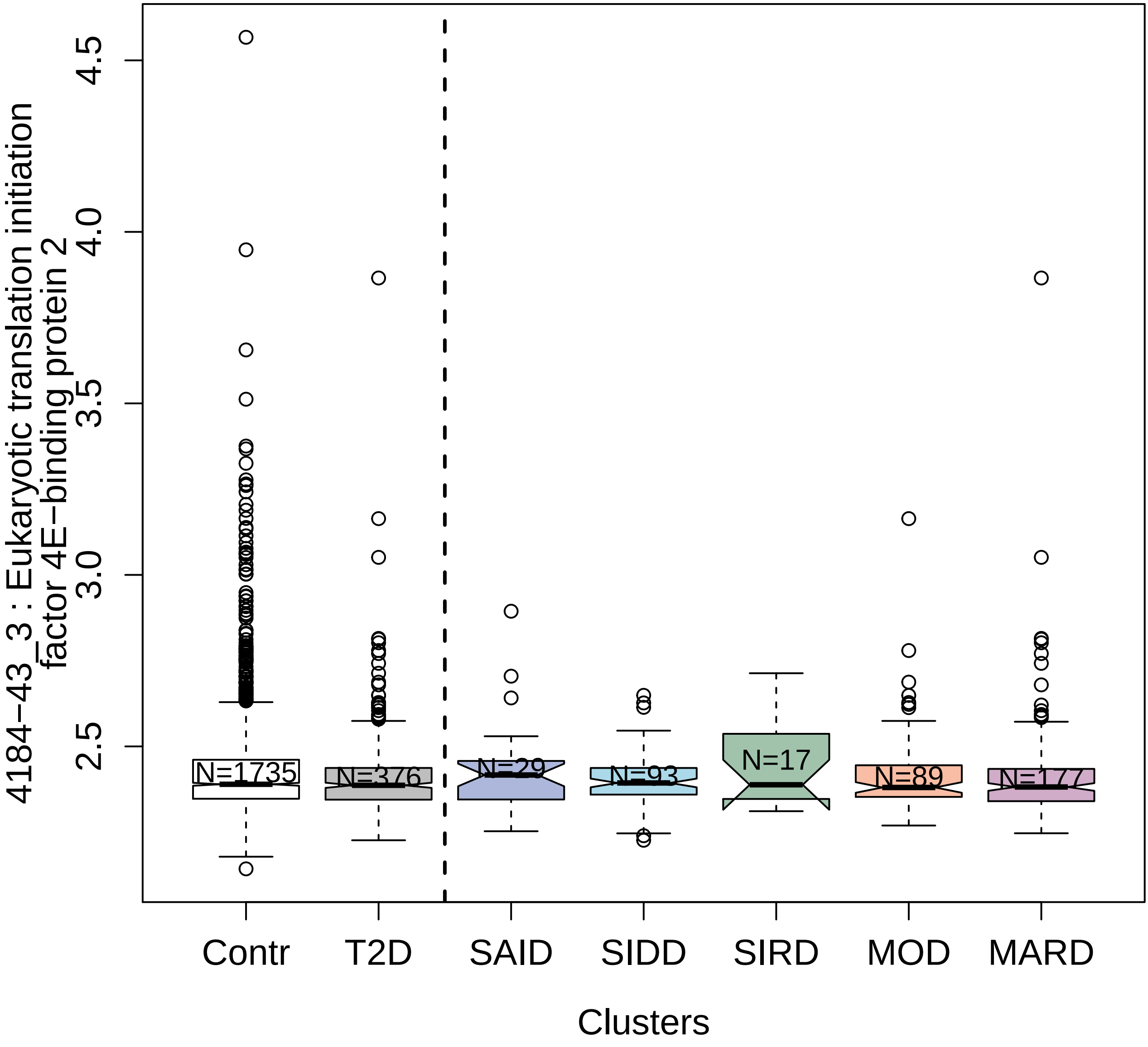
4184−43_3 : Eukaryotic translation initiation
factor 4E−binding protein 2
4.5
4.0
3.5
3.0
2.5
N=1735
N=376
N=29
N=93
N=17
N=89
N=177
Contr
T2D
SAID
SIDD
SIRD
MOD
MARD
Clusters

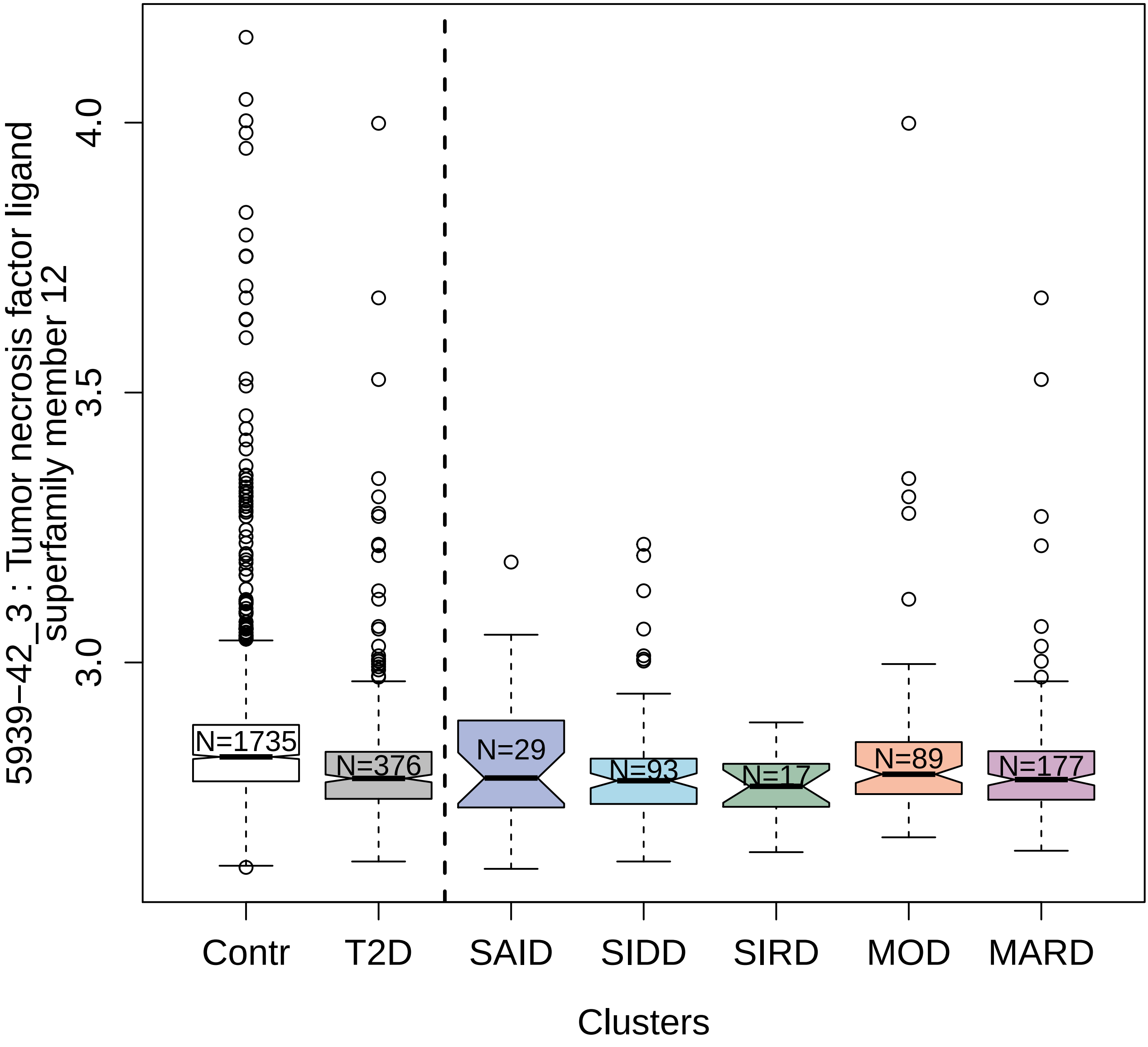

5939−42_3 : Tumor necrosis factor ligand superfamily member 12
4.0
3.5
3.0
N=1735
N=376
N=29
N=93
N=17
N=89
N=177
Contr
T2D
SAID
SIDD
SIRD
MOD
MARD
Clusters

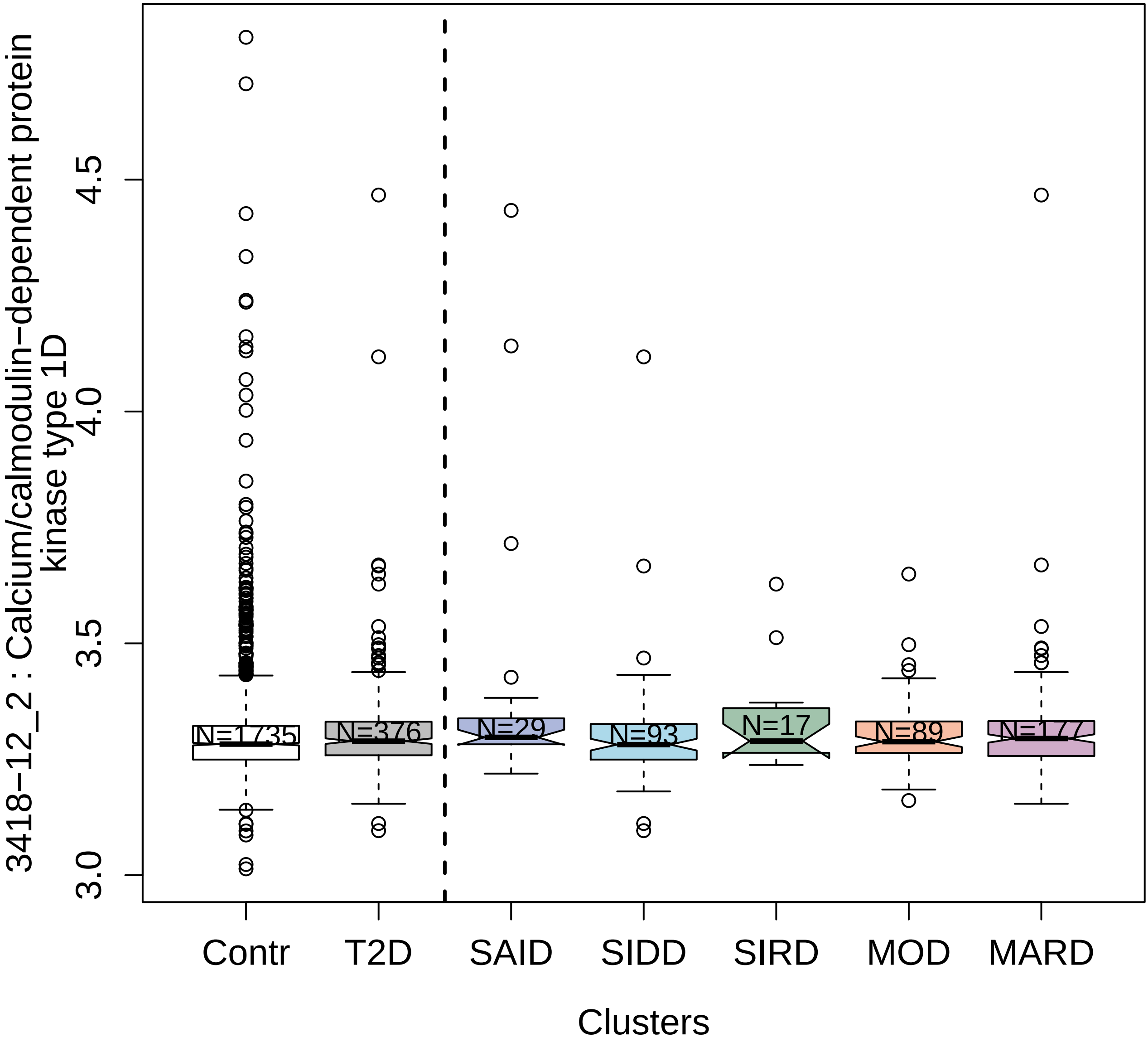
3418−12_2 : Calcium/calmodulin−dependent protein kinase type 1D
4.5
4.0
3.5
3.0
N=1735
N=376
N=29
N=93
N=17
N=89
N=177
Contr
T2D
SAID
SIDD
SIRD
MOD
MARD
Clusters

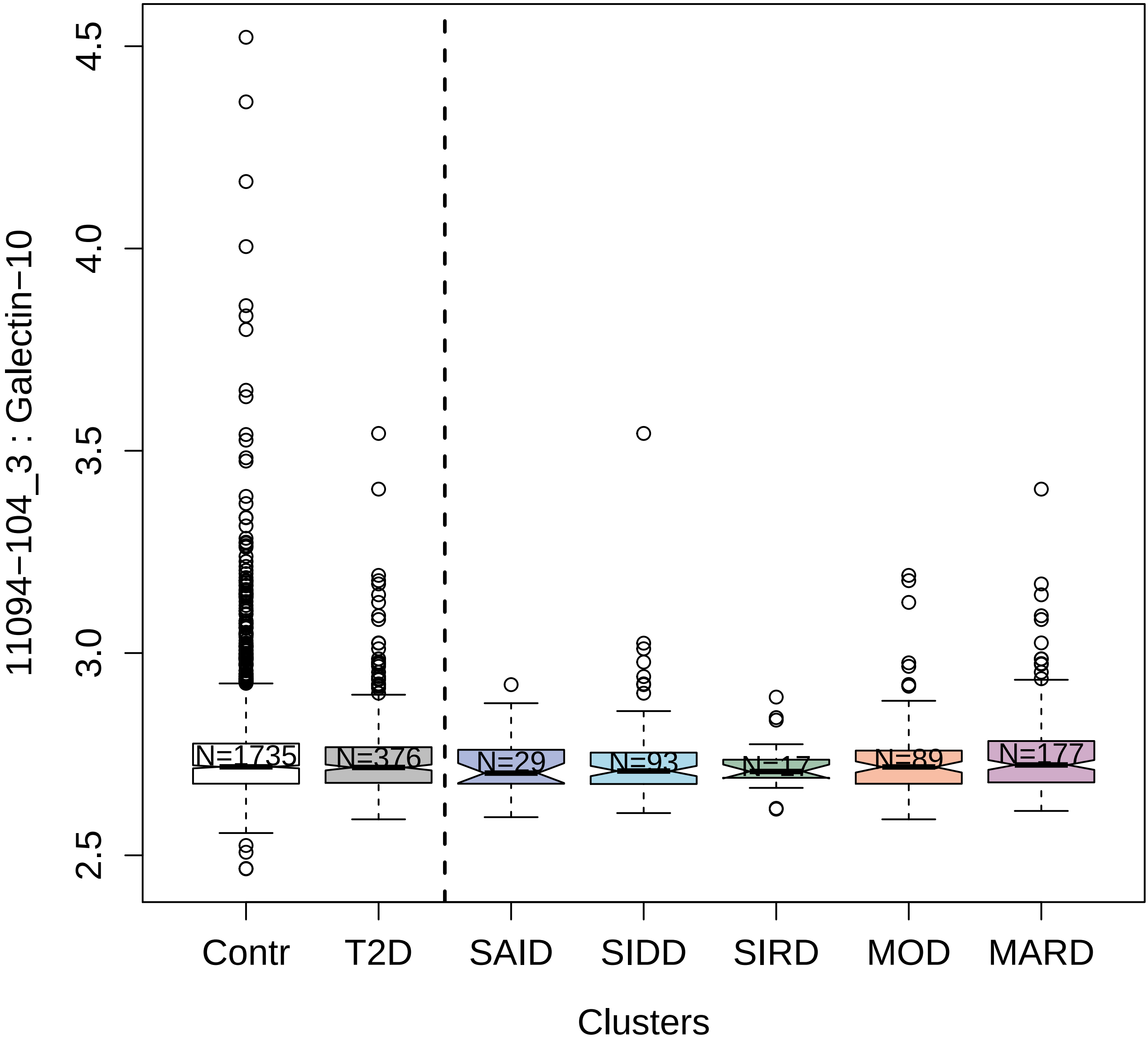
11094−104_3 : Galectin−10
4.5
4.0
3.5
3.0
2.5
N=1735
N=376
N=29
N=93
N=17
N=89
N=177
Contr
T2D
SAID
SIDD
SIRD
MOD
MARD
Clusters

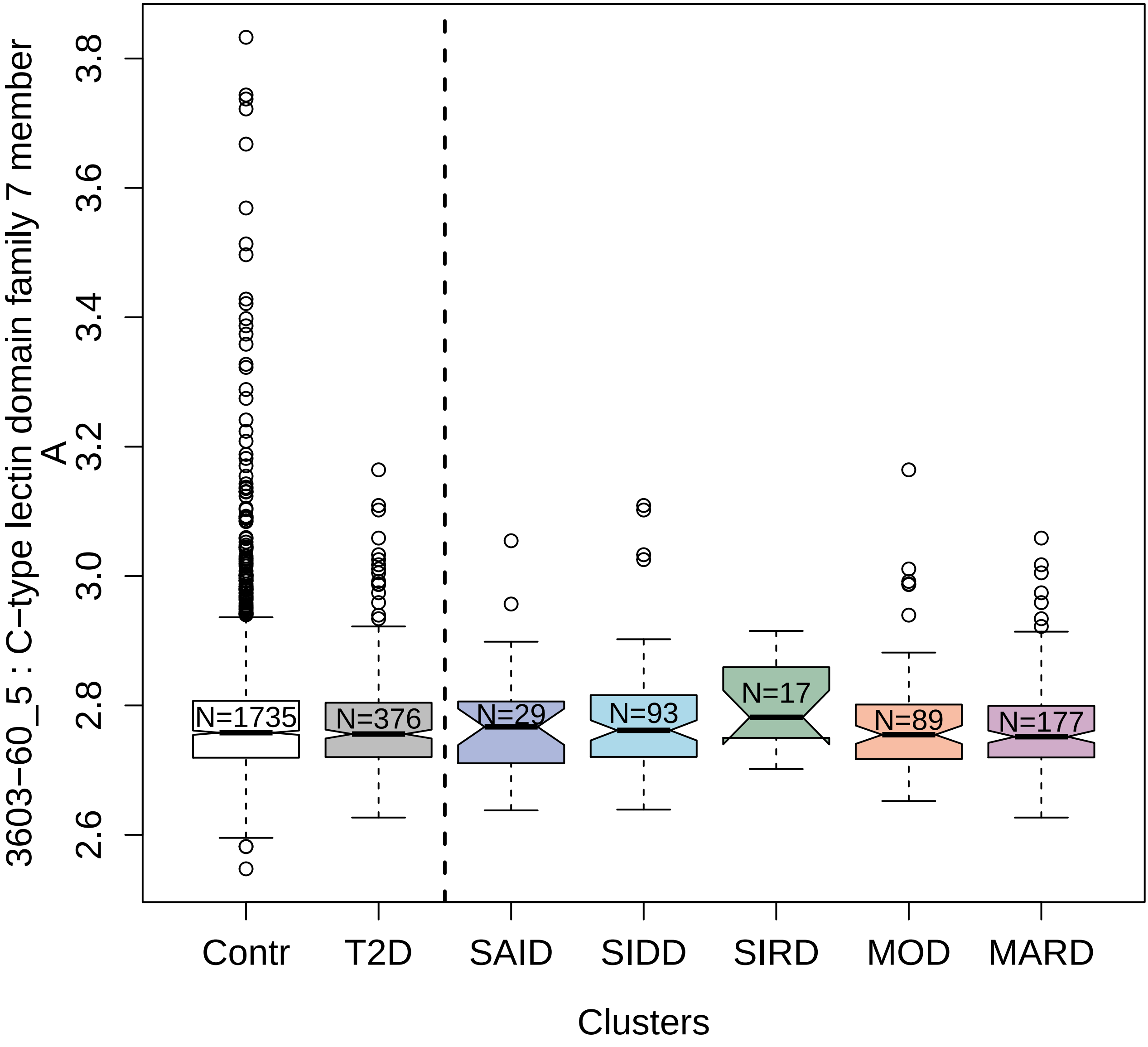

3603−60_5 : C−type lectin domain family 7 member A
3.8
3.6
3.4
3.2
3.0
2.8
2.6
N=1735
N=376
N=29
N=93
N=17
N=89
N=177
Contr
T2D
SAID
SIDD
SIRD
MOD
MARD
Clusters

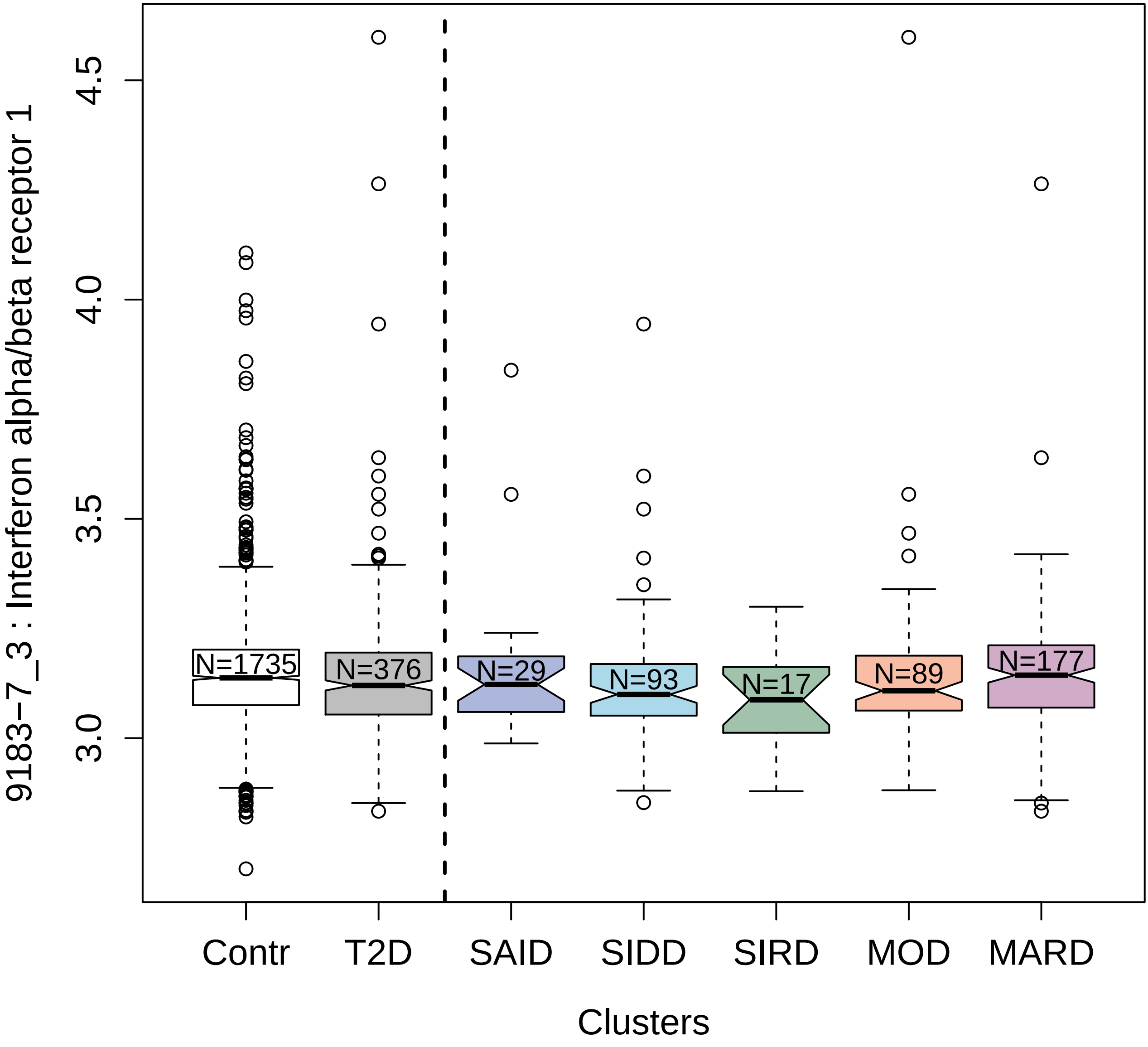
9183−7_3 : Interferon alpha/beta receptor 1
4.5
4.0
3.5
3.0
N=1735
N=376
N=29
N=93
N=17
N=89
N=177
Contr
T2D
SAID
SIDD
SIRD
MOD
MARD
Clusters

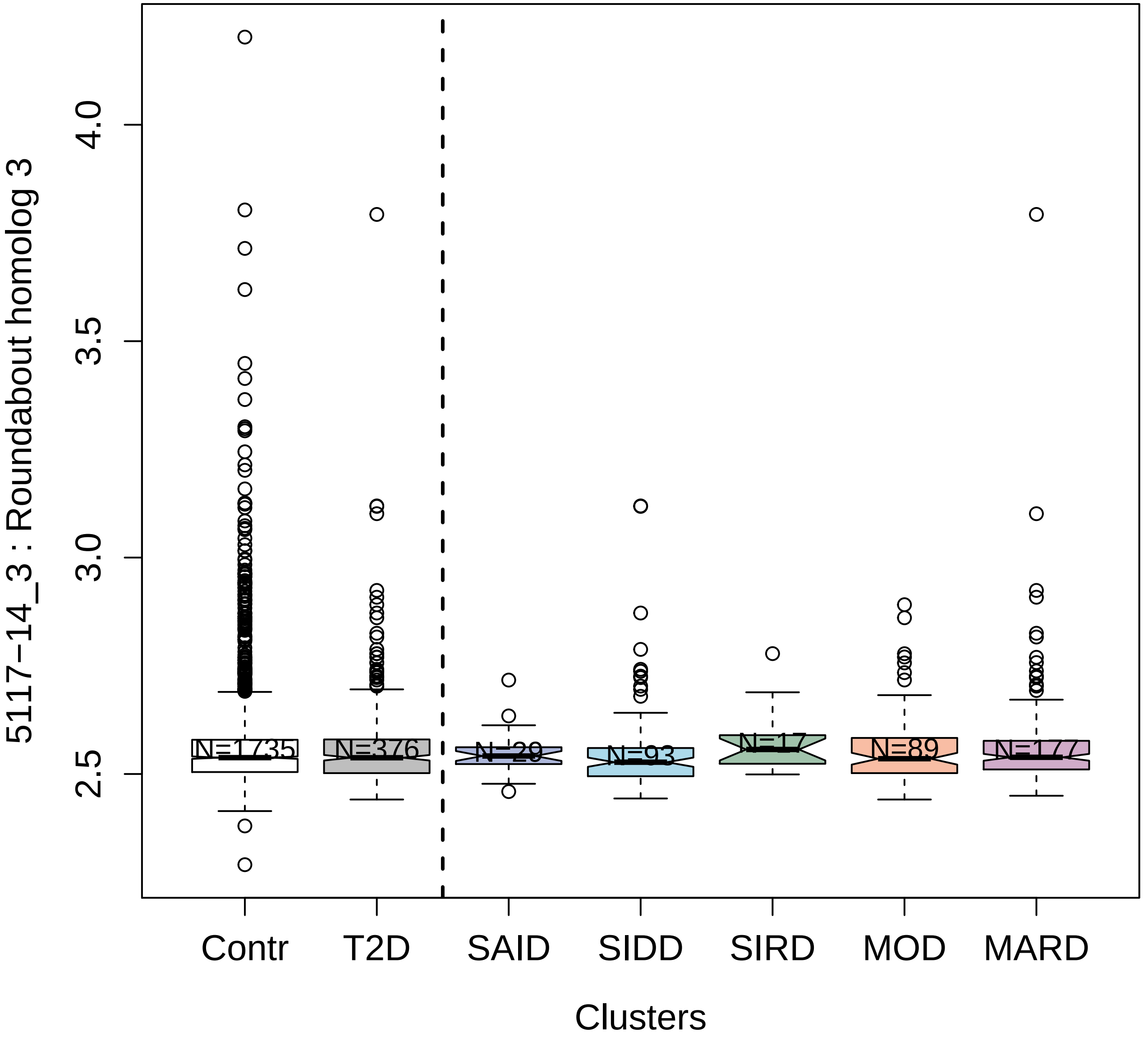

5117−14_3 : Roundabout homolog 3
2.5
3.0
3.5
4.0
N=1735
N=376
N=29
N=93
N=17
N=89
N=177
Contr
T2D
SAID
SIDD
SIRD
MOD
MARD
Clusters

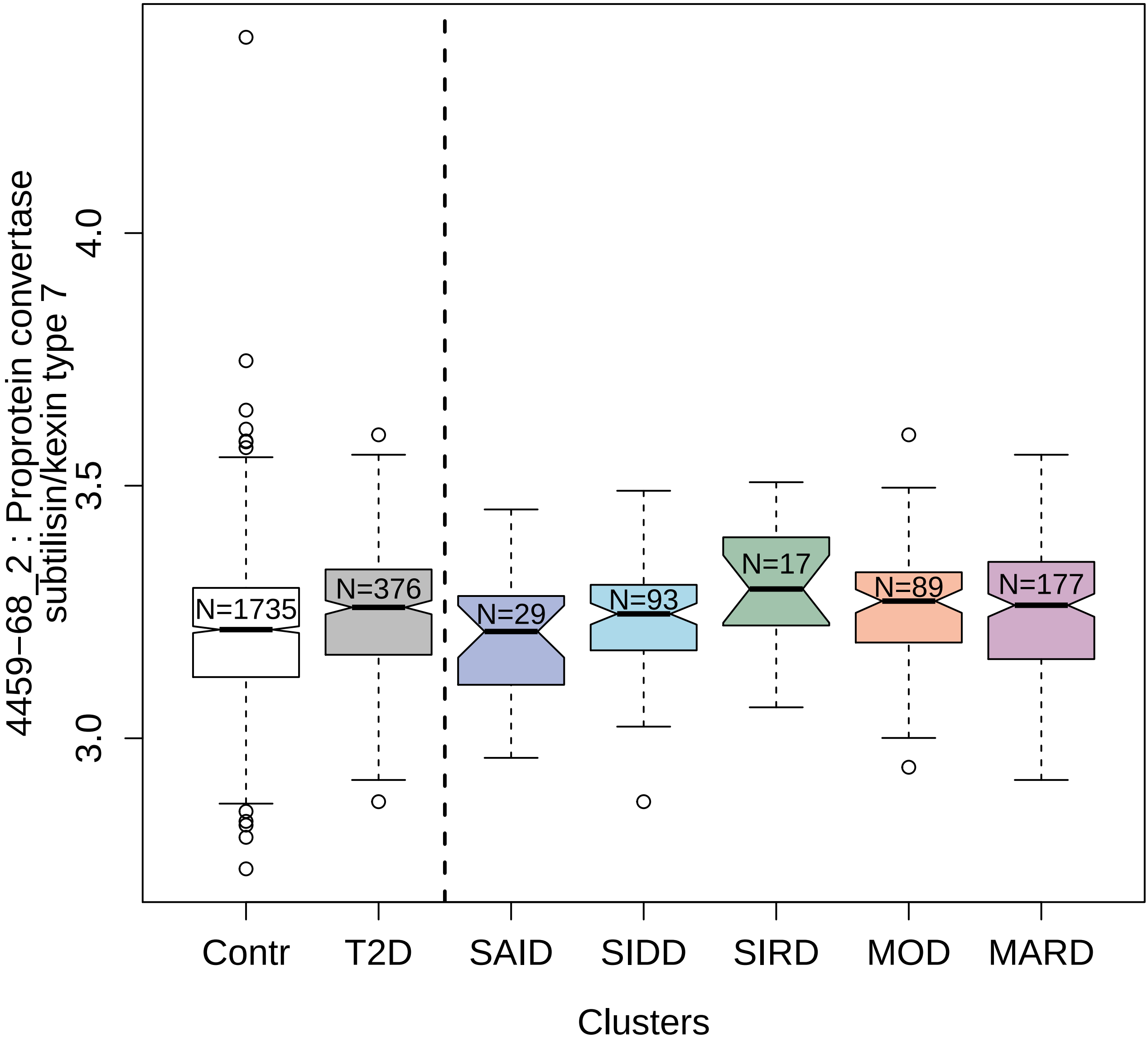
4459−68_2 : Proprotein convertase
subtilisin/kexin type 7
3.0
3.5
4.0
N=1735
N=376
N=29
N=93
N=17
N=89
N=177
Contr
T2D
SAID
SIDD
SIRD
MOD
MARD
Clusters

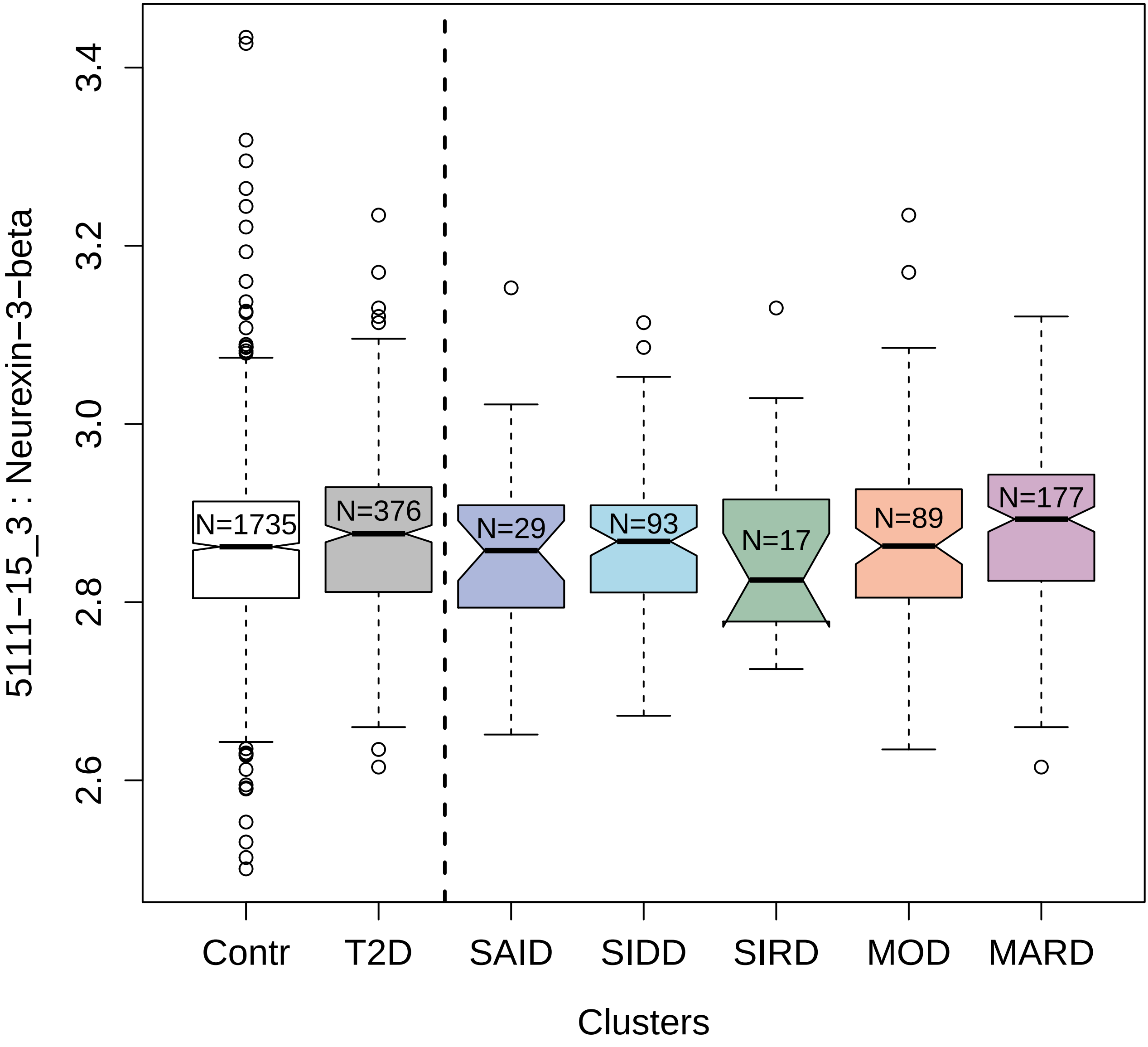
5111−15_3 : Neurexin−3−beta
3.4
3.2
3.0
2.8
2.6
N=1735
N=376
N=29
N=93
N=17
N=89
N=177
Contr
T2D
SAID
SIDD
SIRD
MOD
MARD
Clusters

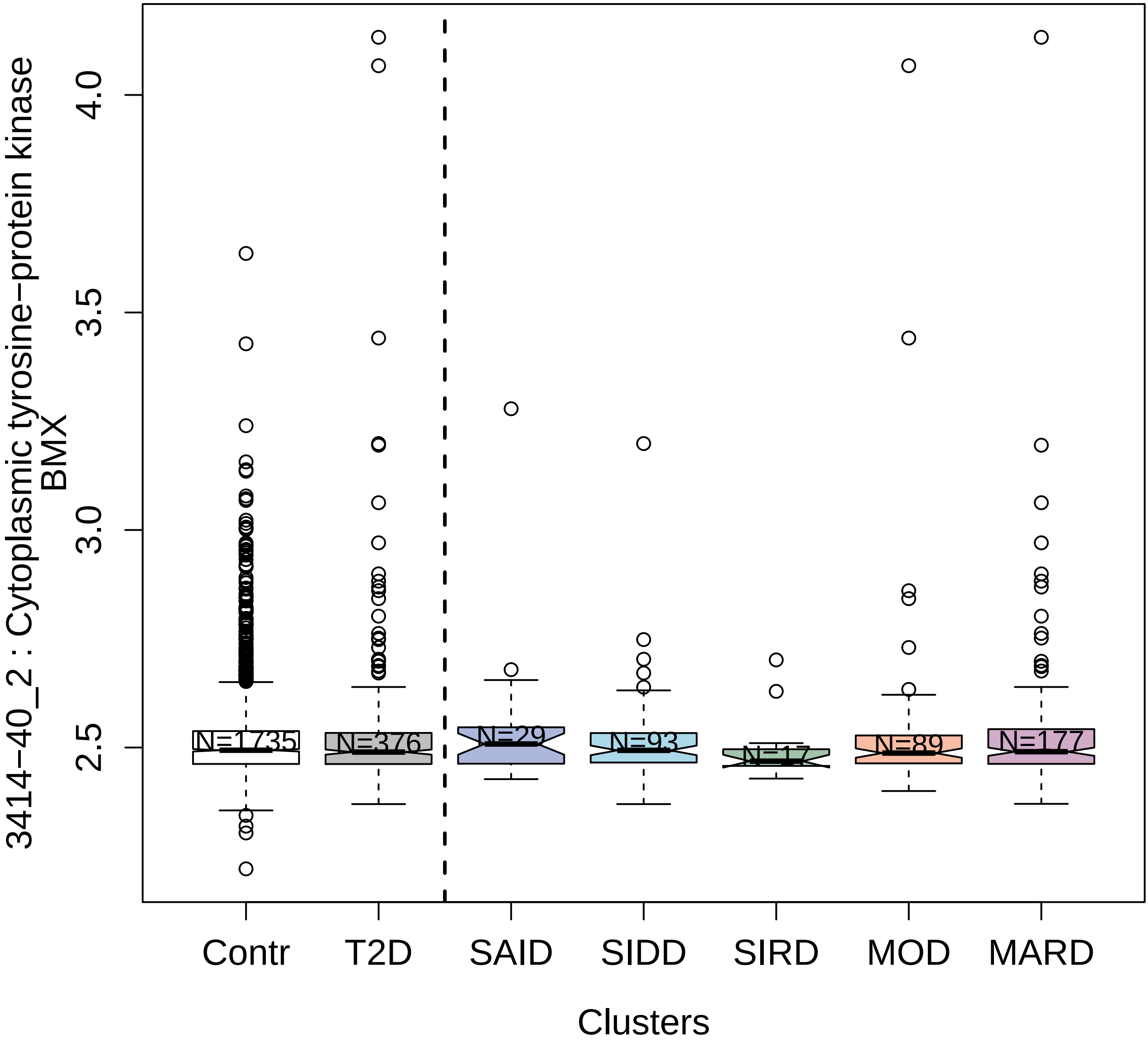
3414−40_2 : Cytoplasmic tyrosine−protein kinase
BMX
2.5
3.0
3.5
4.0
N=1735
N=376
N=29
N=93
N=17
N=89
N=177
Contr
T2D
SAID
SIDD
SIRD
MOD
MARD
Clusters

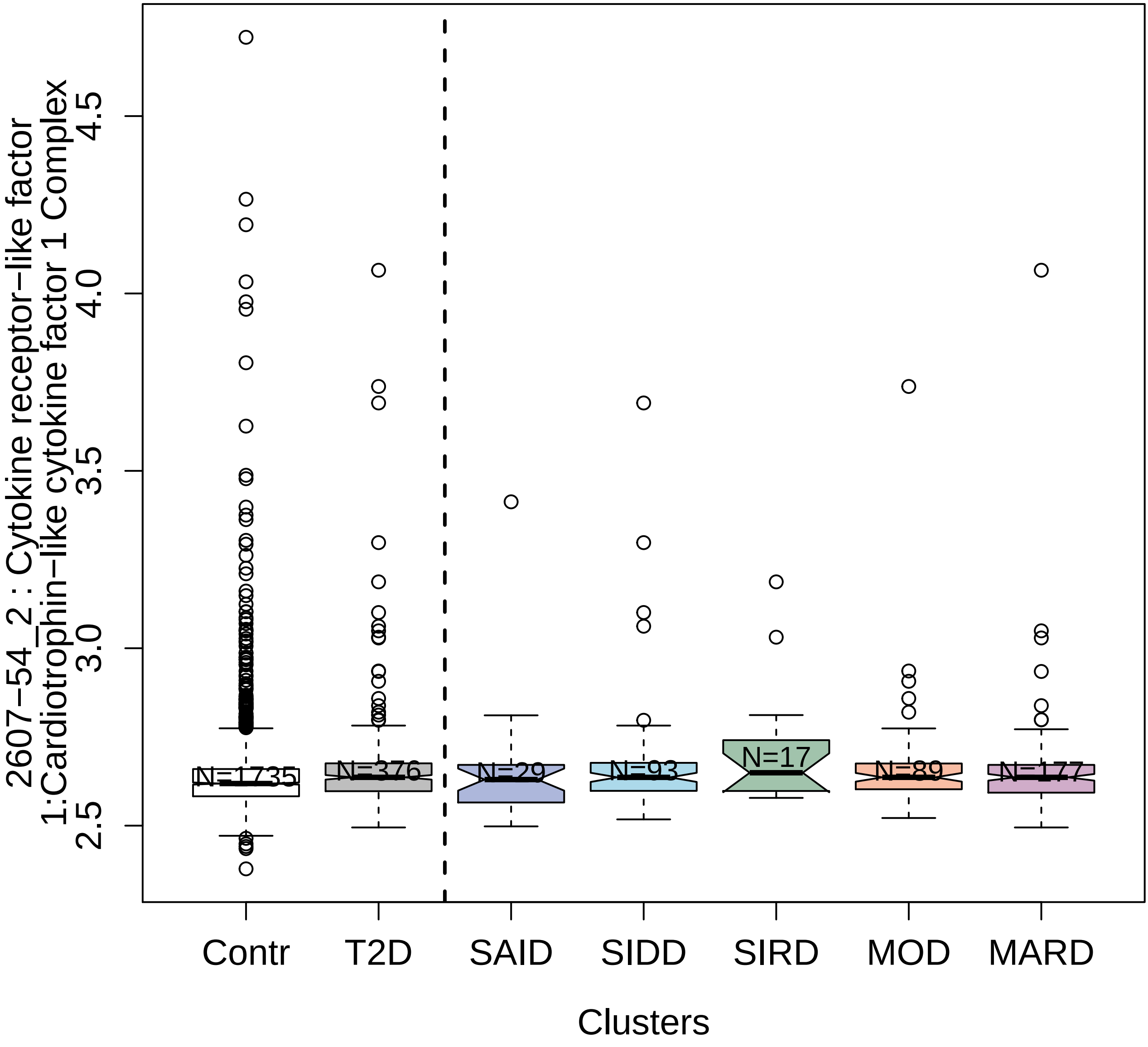
2607−54_2 : Cytokine receptor−like factor
1:Cardiotrophin−like cytokine factor 1 Complex
2.5
3.0
3.5
4.0
4.5
N=1735
N=376
N=29
N=93
N=17
N=89
N=177
Contr
T2D
SAID
SIDD
SIRD
MOD
MARD
Clusters

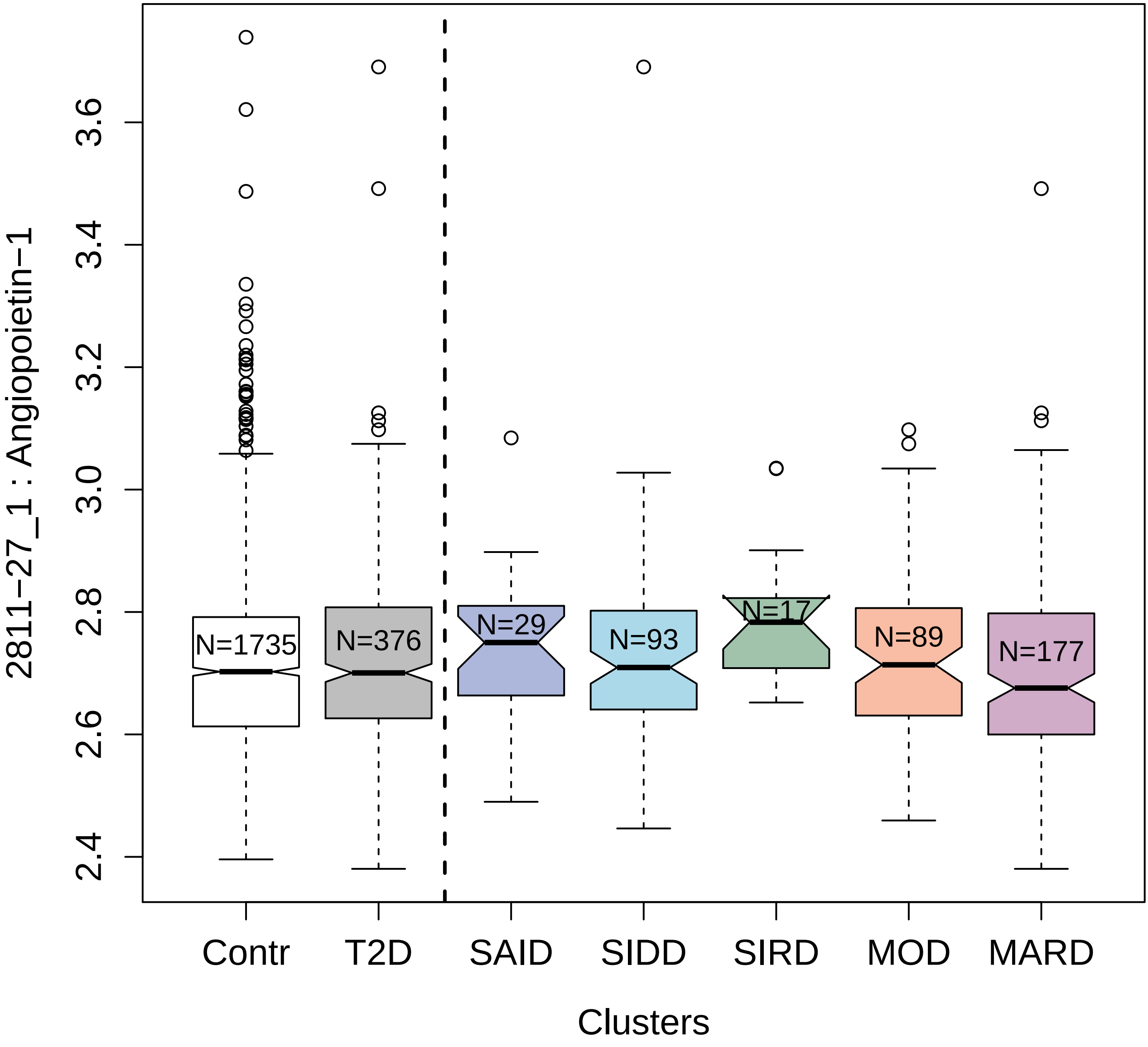

2811−27_1 : Angiopoietin−1
3.6
3.4
3.2
3.0
2.8
2.6
2.4
N=1735
N=376
N=29
N=93
N=17
N=89
N=177
Contr
T2D
SAID
SIDD
SIRD
MOD
MARD
Clusters

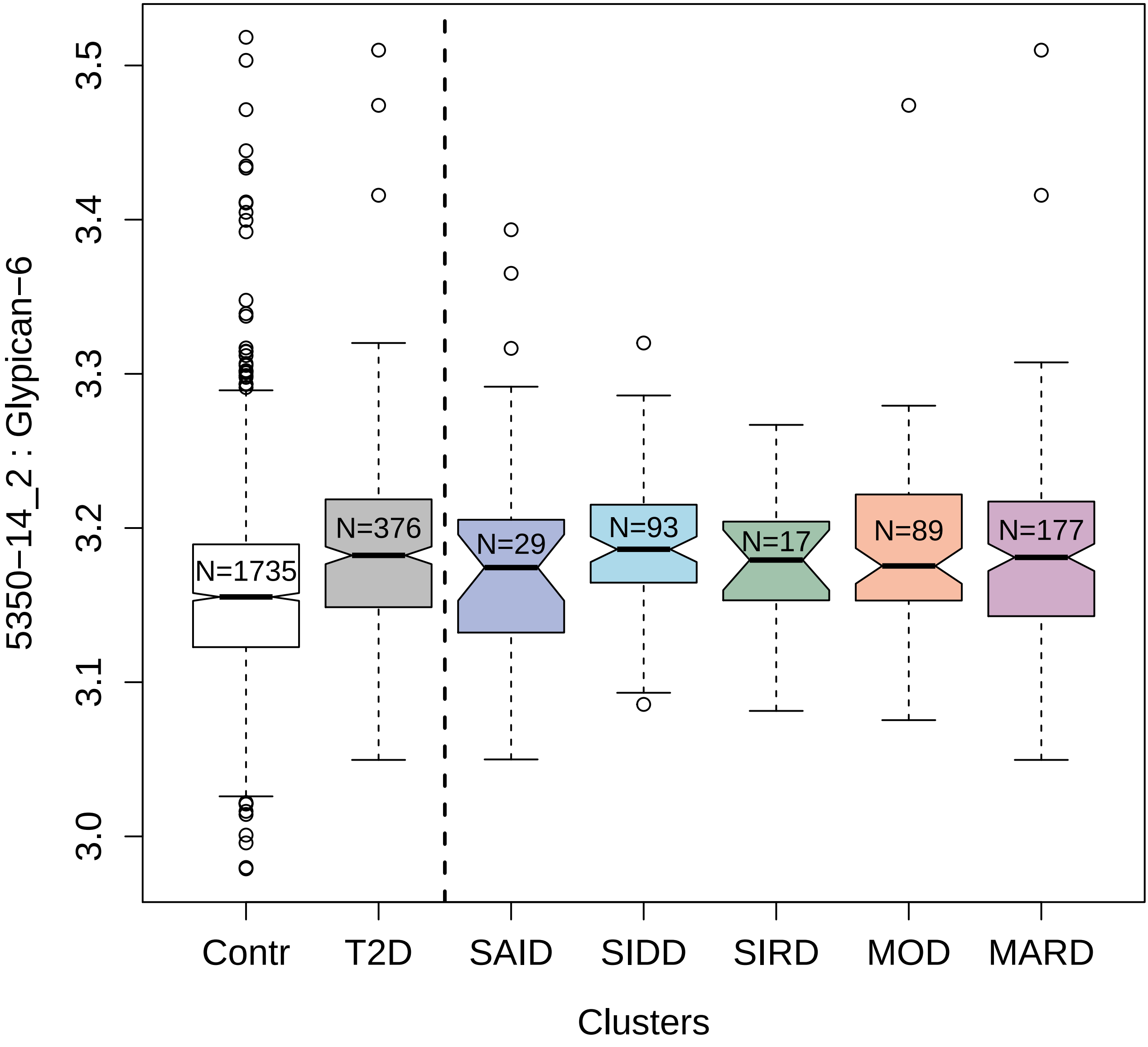

5350−14_2 : Glypican−6
3.0
3.1
3.2
3.3
3.4
3.5
N=1735
N=376
N=29
N=93
N=17
N=89
N=177
Contr
T2D
SAID
SIDD
SIRD
MOD
MARD
Clusters

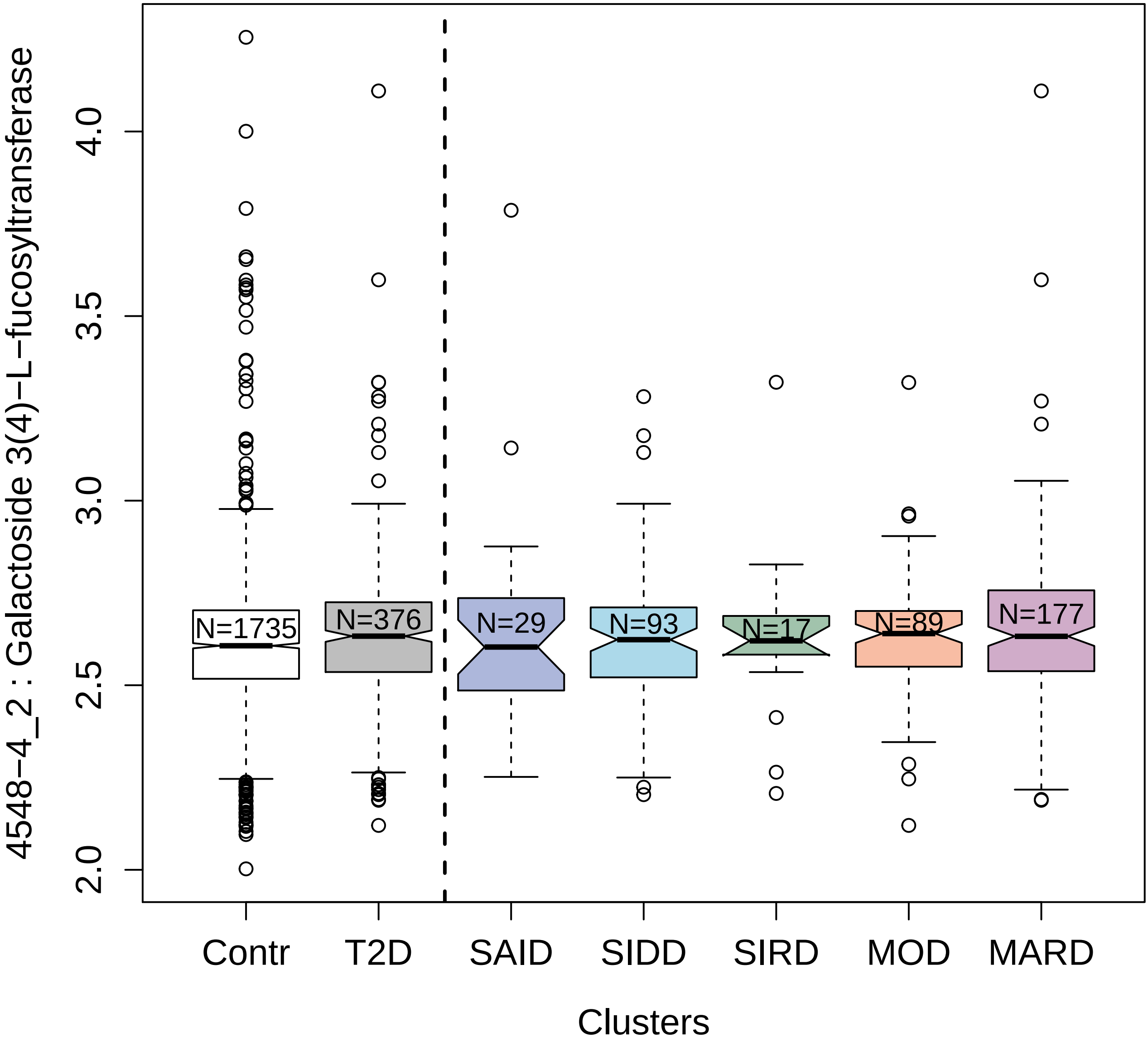

4548−4_2 : Galactoside 3(4)−L−fucosyltransferase
2.0
2.5
3.0
3.5
4.0
N=1735
N=376
N=29
N=93
N=17
N=89
N=177
Contr
T2D
SAID
SIDD
SIRD
MOD
MARD
Clusters

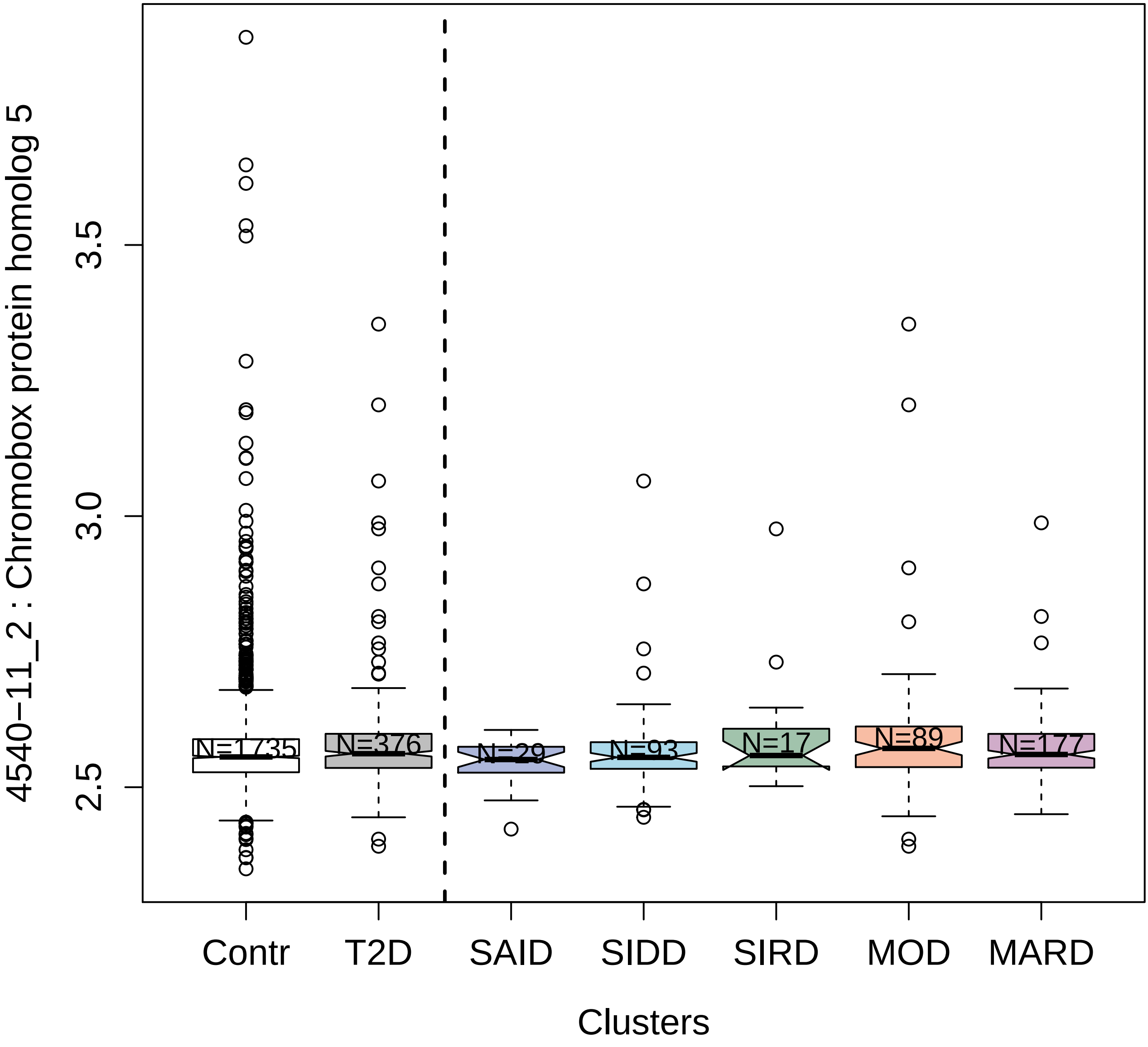

4540−11_2 : Chromobox protein homolog 5
3.5
3.0
2.5
N=1735
N=376
N=29
N=93
N=17
N=89
N=177
Contr
T2D
SAID
SIDD
SIRD
MOD
MARD
Clusters

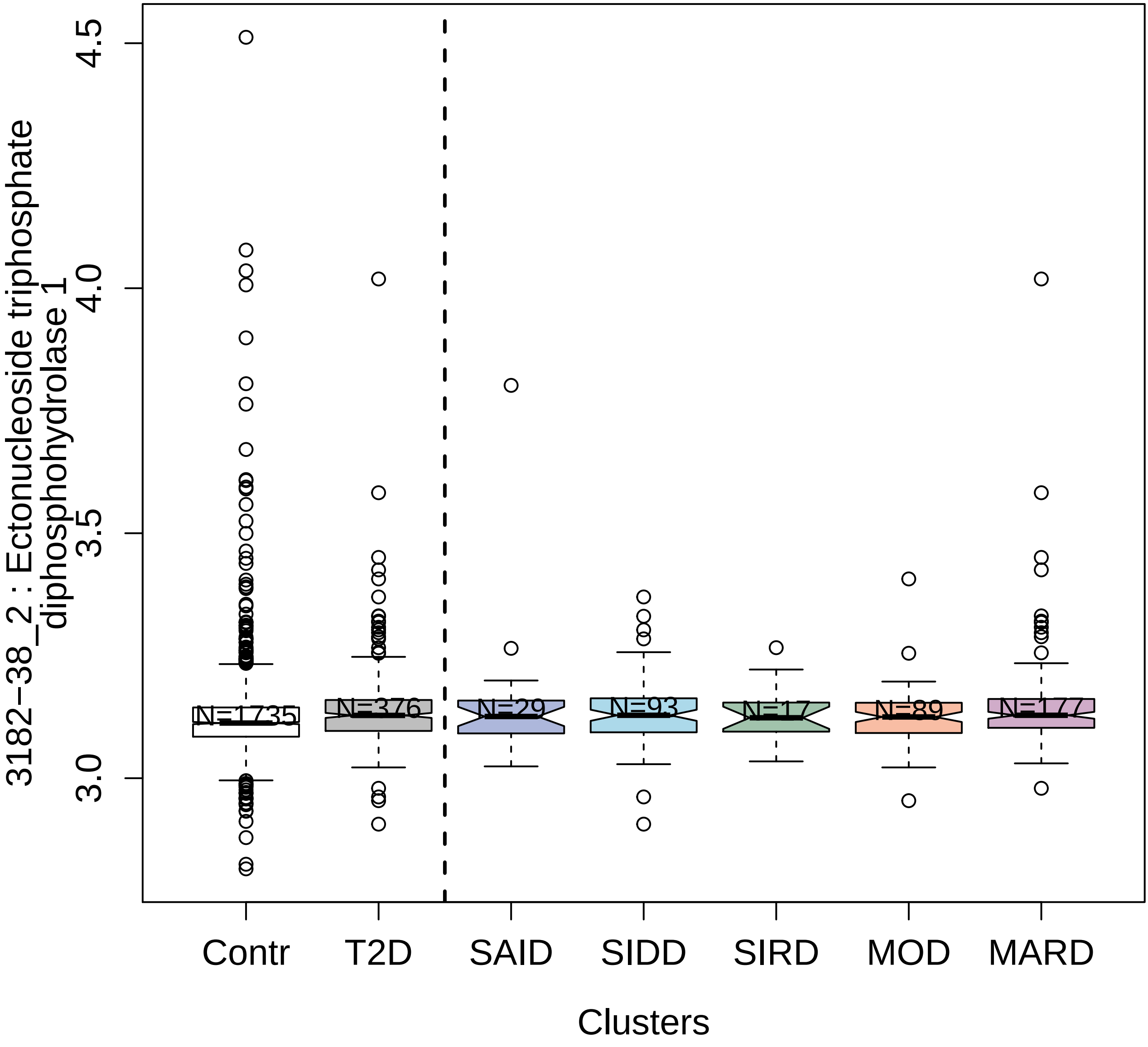

3182−38_2 : Ectonucleoside triphosphate
_diphosphohydrolase 1
4.5
4.0
3.5
3.0
N=1735
N=376
N=29
N=93
N=17
N=89
N=177
Contr
T2D
SAID
SIDD
SIRD
MOD
MARD
Clusters

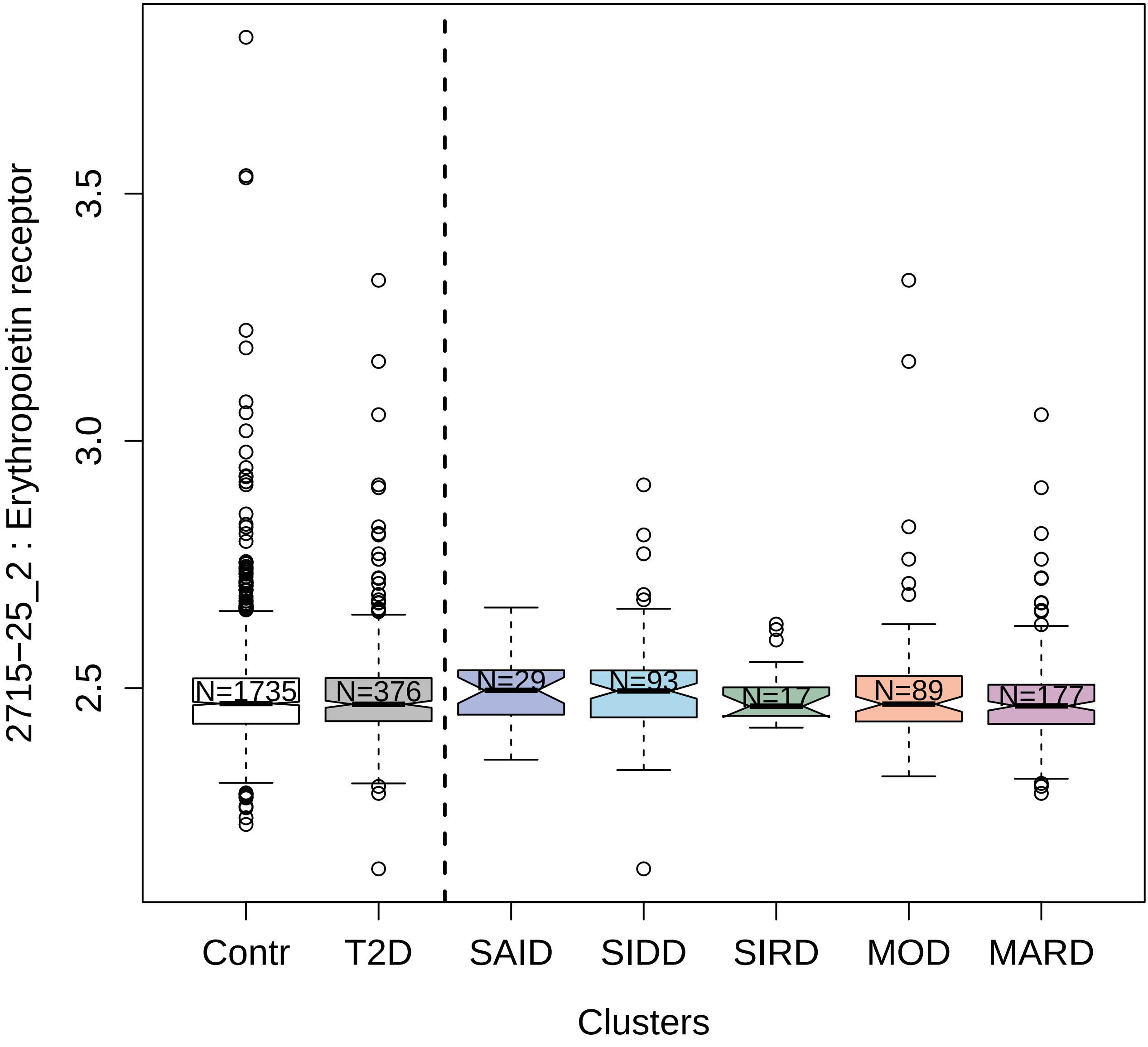
2715−25_2 : Erythropoietin receptor
3.5
3.0
2.5
N=1735
N=376
N=29
N=93
N=17
N=89
N=177
Contr
T2D
SAID
SIDD
SIRD
MOD
MARD
Clusters

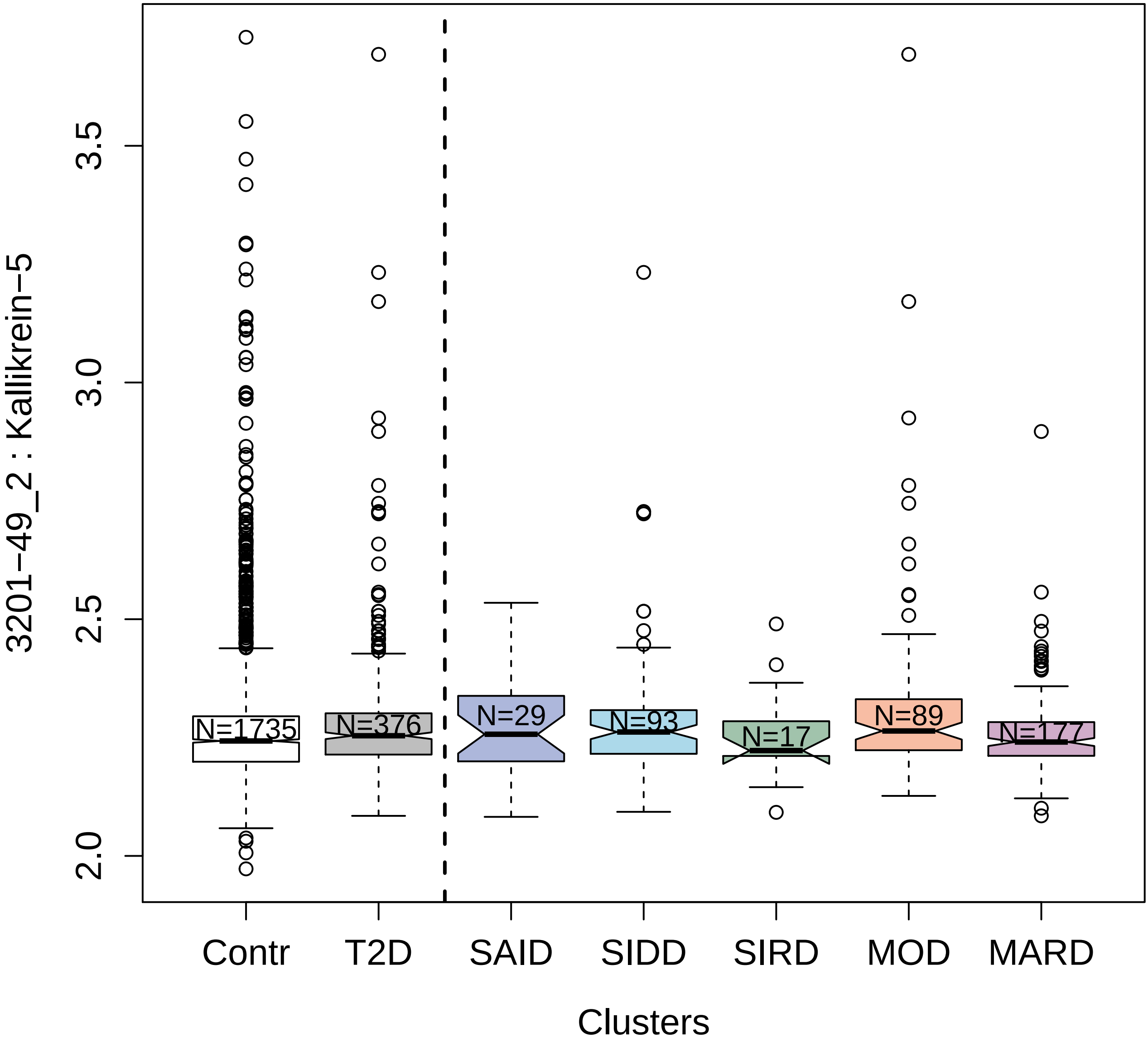
3201−49_2 : Kallikrein−5
2.0
2.5
3.0
3.5
N=1735
N=376
N=29
N=93
N=17
N=89
N=177
Contr
T2D
SAID
SIDD
SIRD
MOD
MARD
Clusters

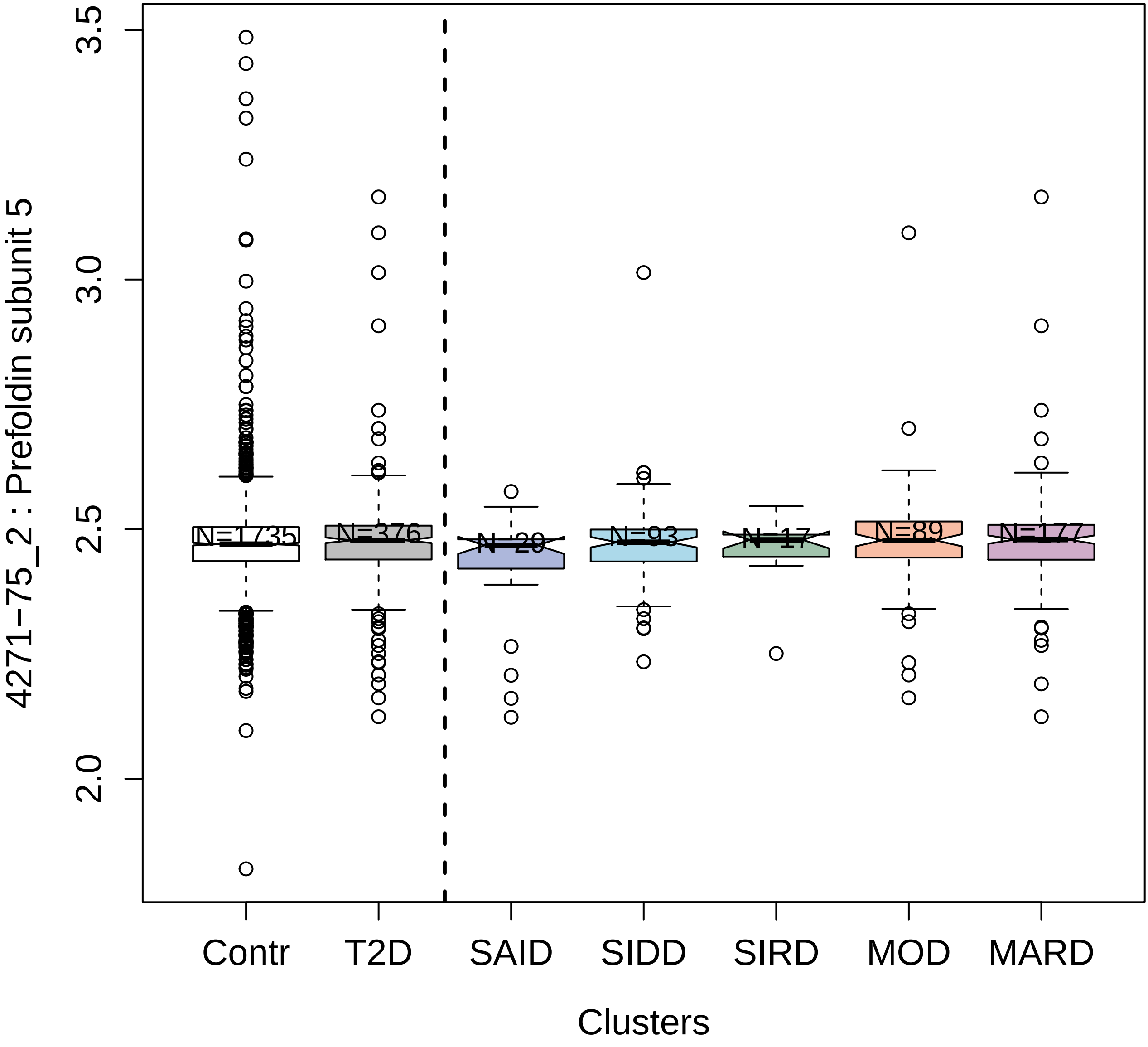
4271−75_2 : Prefoldin subunit 5
3.5
3.0
2.5
2.0
N=1735
N=376
N=29
N=93
N=17
N=89
N=177
Contr
T2D
SAID
SIDD
SIRD
MOD
MARD
Clusters

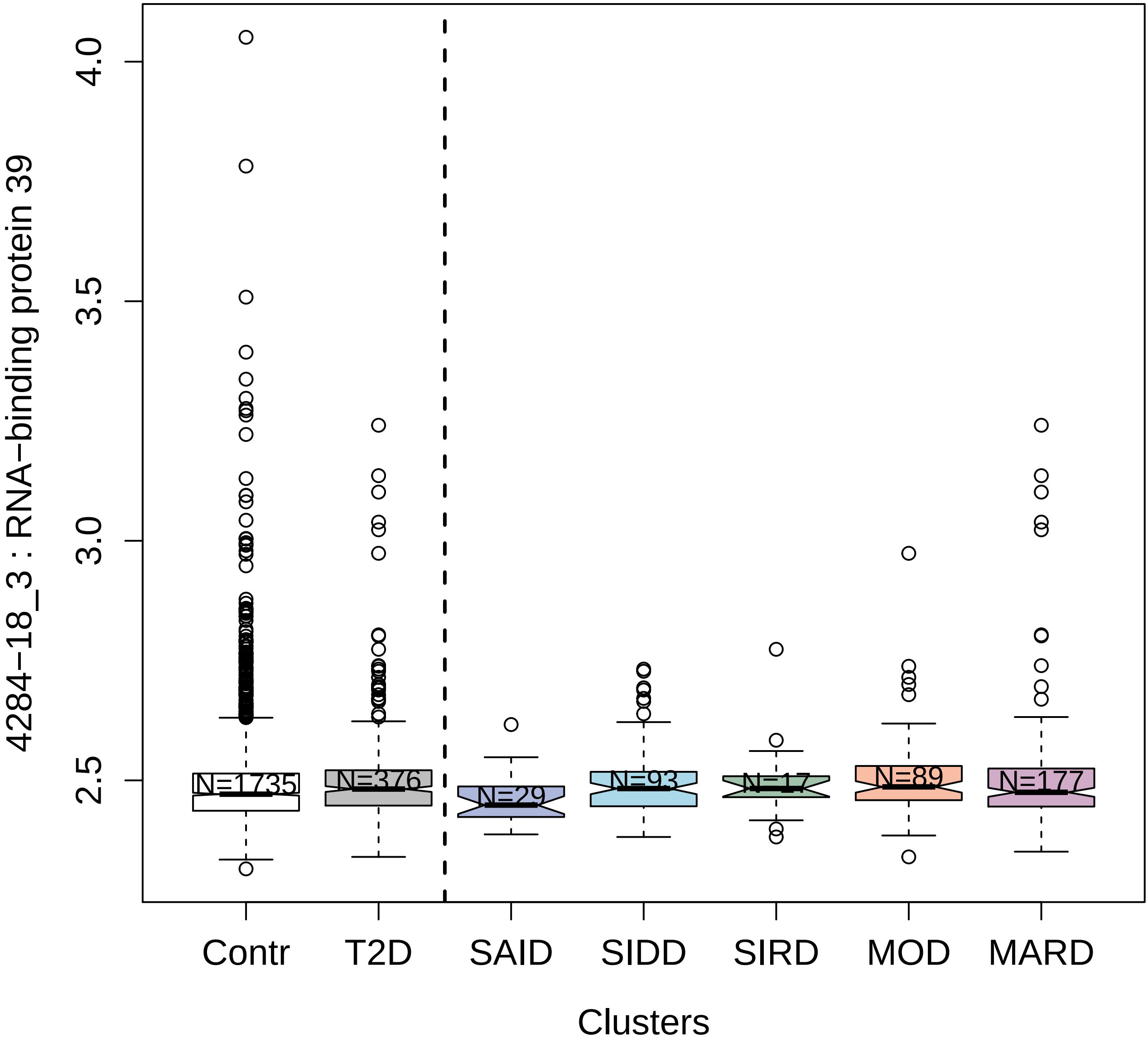
4284−18_3 : RNA−binding protein 39
4.0
3.5
3.0
2.5
N=1735
N=376
N=29
N=93
N=17
N=89
N=177
Contr
T2D
SAID
SIDD
SIRD
MOD
MARD
Clusters

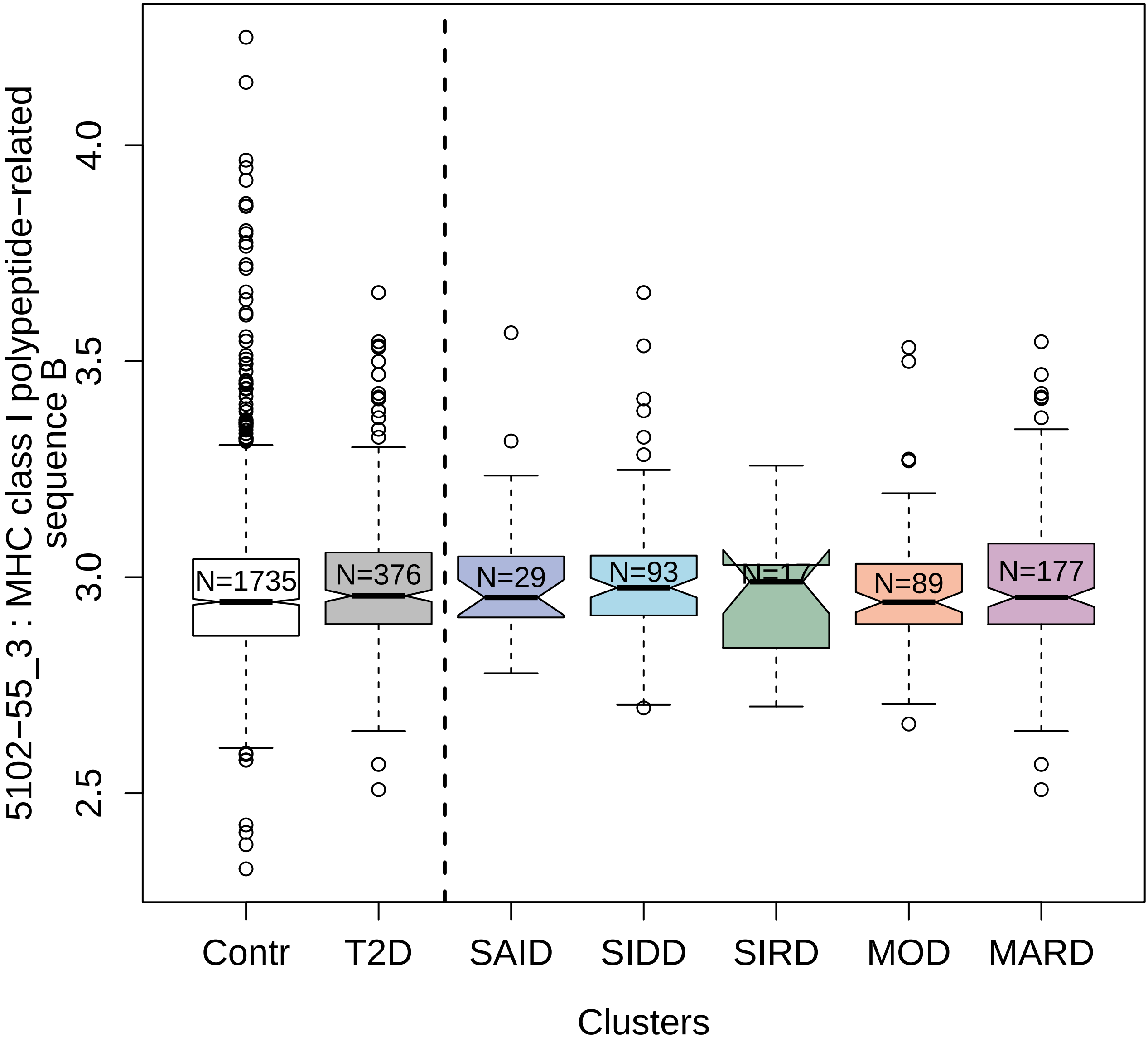

5102−55_3 : MHC class I polypeptide−related sequence B
4.0
3.5
3.0
2.5
N=1735
N=376
N=29
N=93
N=89
N=177
Contr
T2D
SAID
SIDD
SIRD
MOD
MARD
Clusters

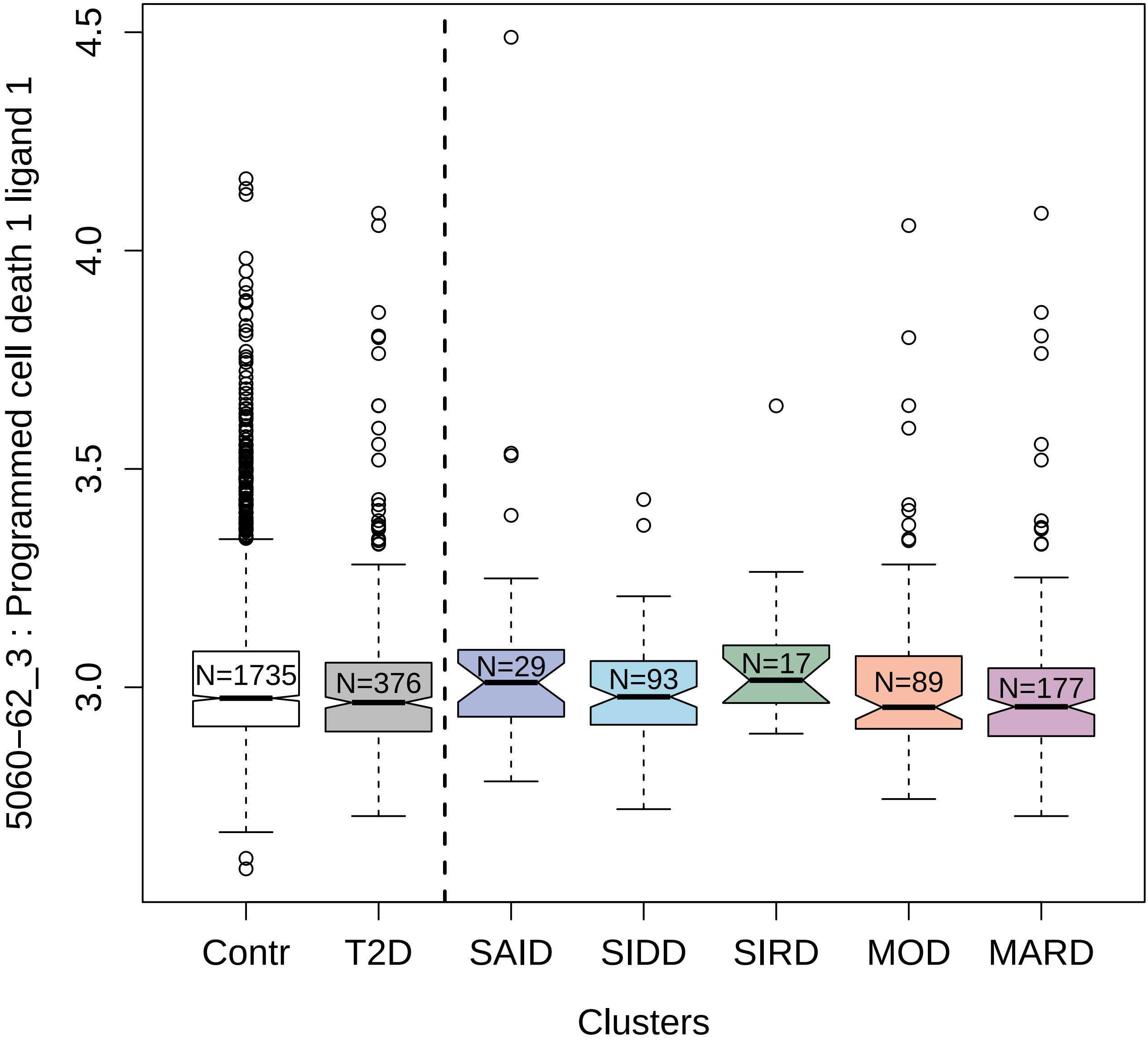

5060−62_3 : Programmed cell death 1 ligand 1
4.5
4.0
3.5
3.0
N=1735
N=376
N=29
N=93
N=17
N=89
N=177
Contr
T2D
SAID
SIDD
SIRD
MOD
MARD
Clusters

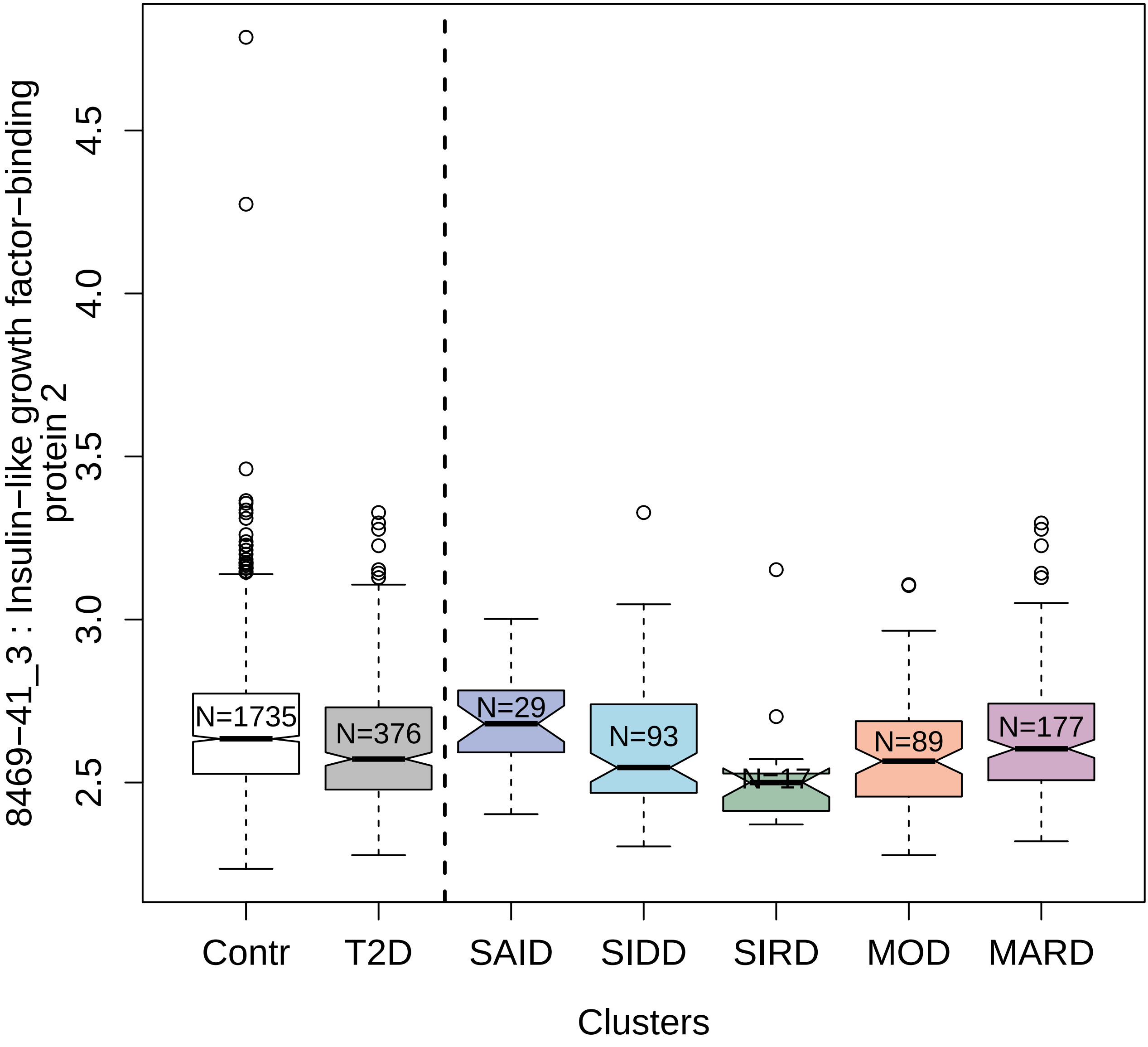

8469−41_3 : Insulin−like growth factor−binding protein 2
4.5
4.0
3.5
3.0
2.5
N=1735
N=376
N=29
N=93
N=17
N=89
N=177
Contr
T2D
SAID
SIDD
SIRD
MOD
MARD
Clusters

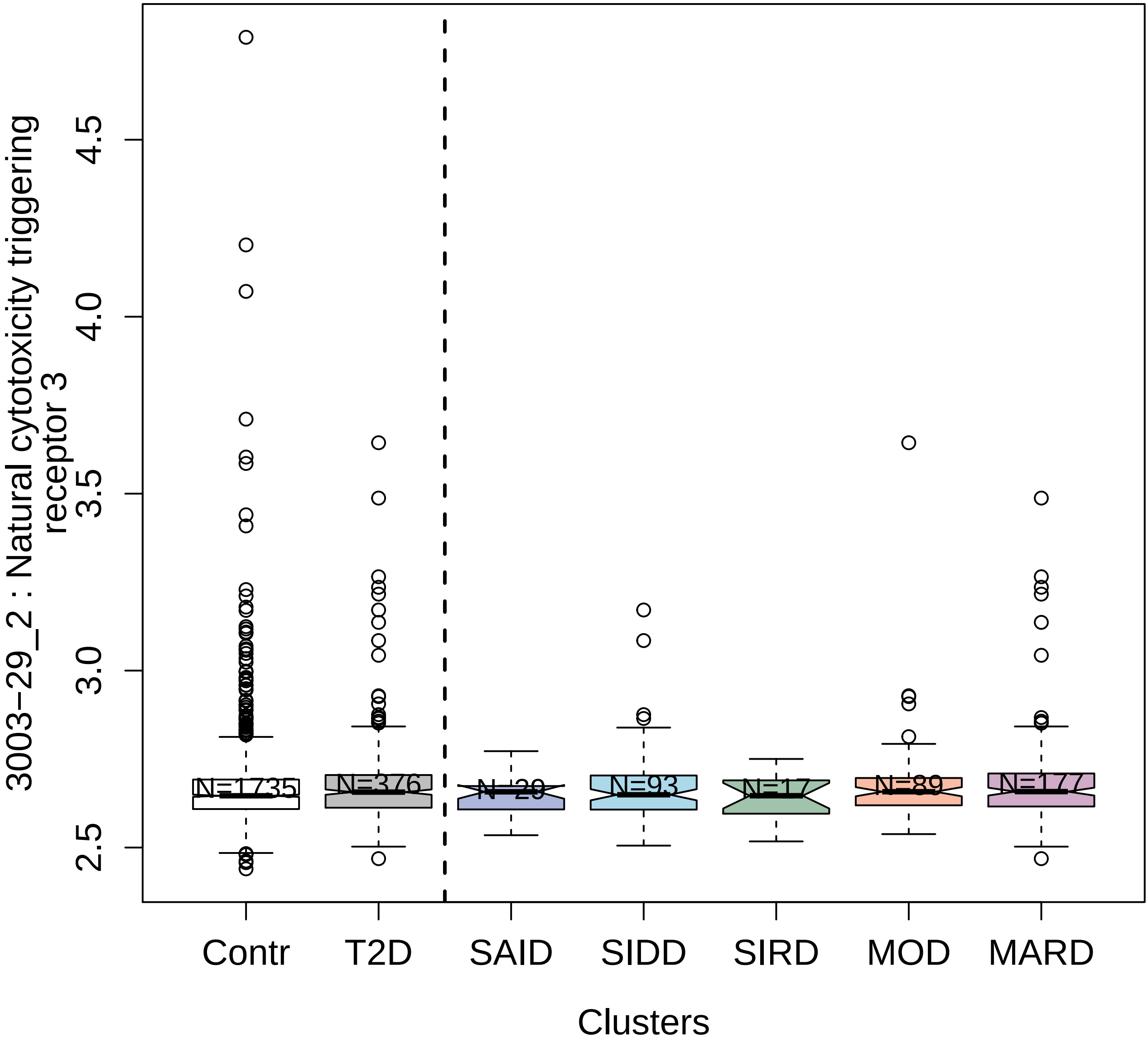
3003−29_2 : Natural cytotoxicity triggering receptor 3
2.5
3.0
3.5
4.0
4.5
N=1735
N=376
N=29
N=93
N=17
N=89
N=177
Contr
T2D
SAID
SIDD
SIRD
MOD
MARD
Clusters

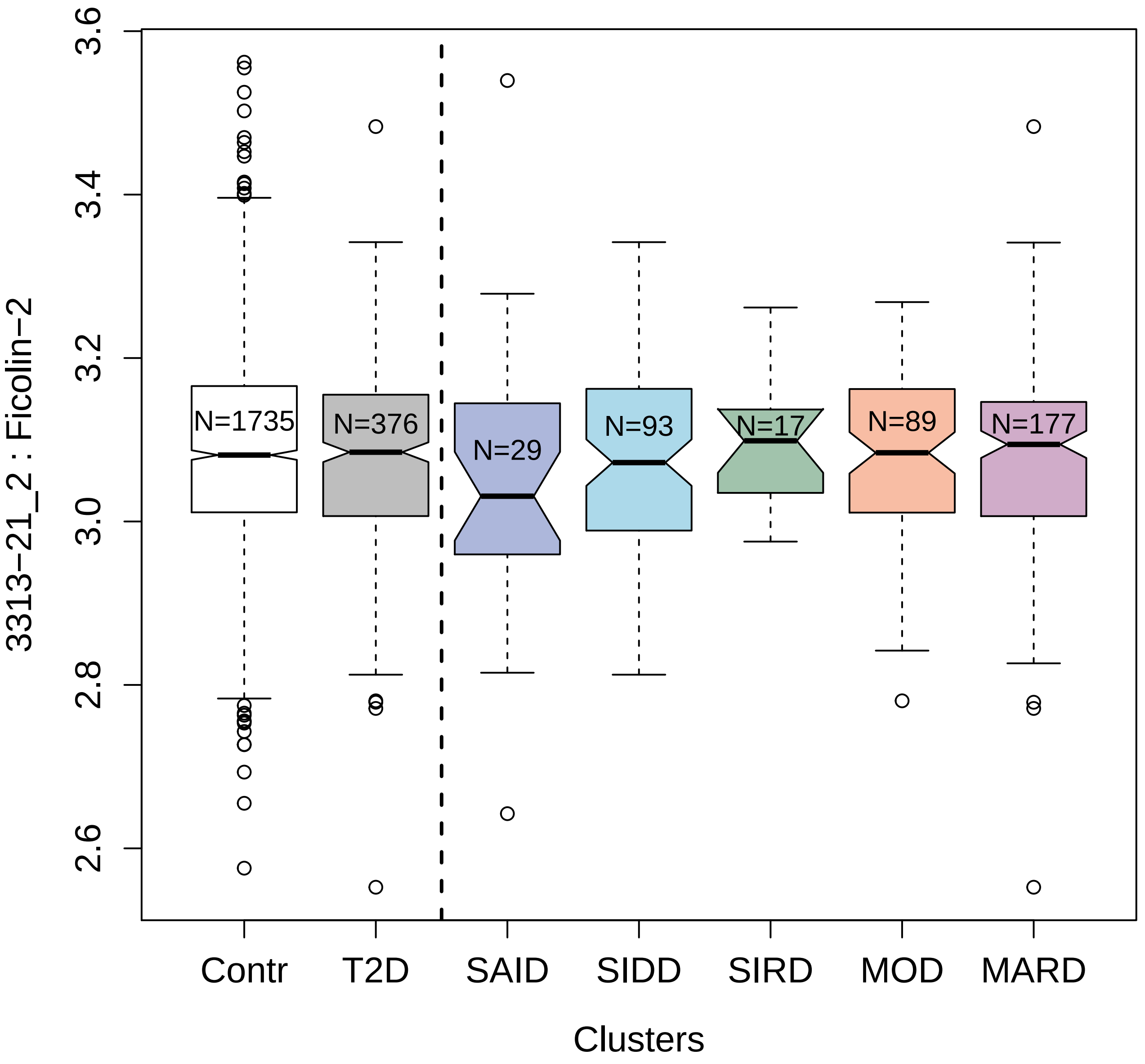
3313−21_2 : Ficolin−2
3.6
3.4
3.2
3.0
2.8
2.6
N=1735
N=376
N=29
N=93
N=17
N=89
N=177
Contr
T2D
SAID
SIDD
SIRD
MOD
MARD
Clusters

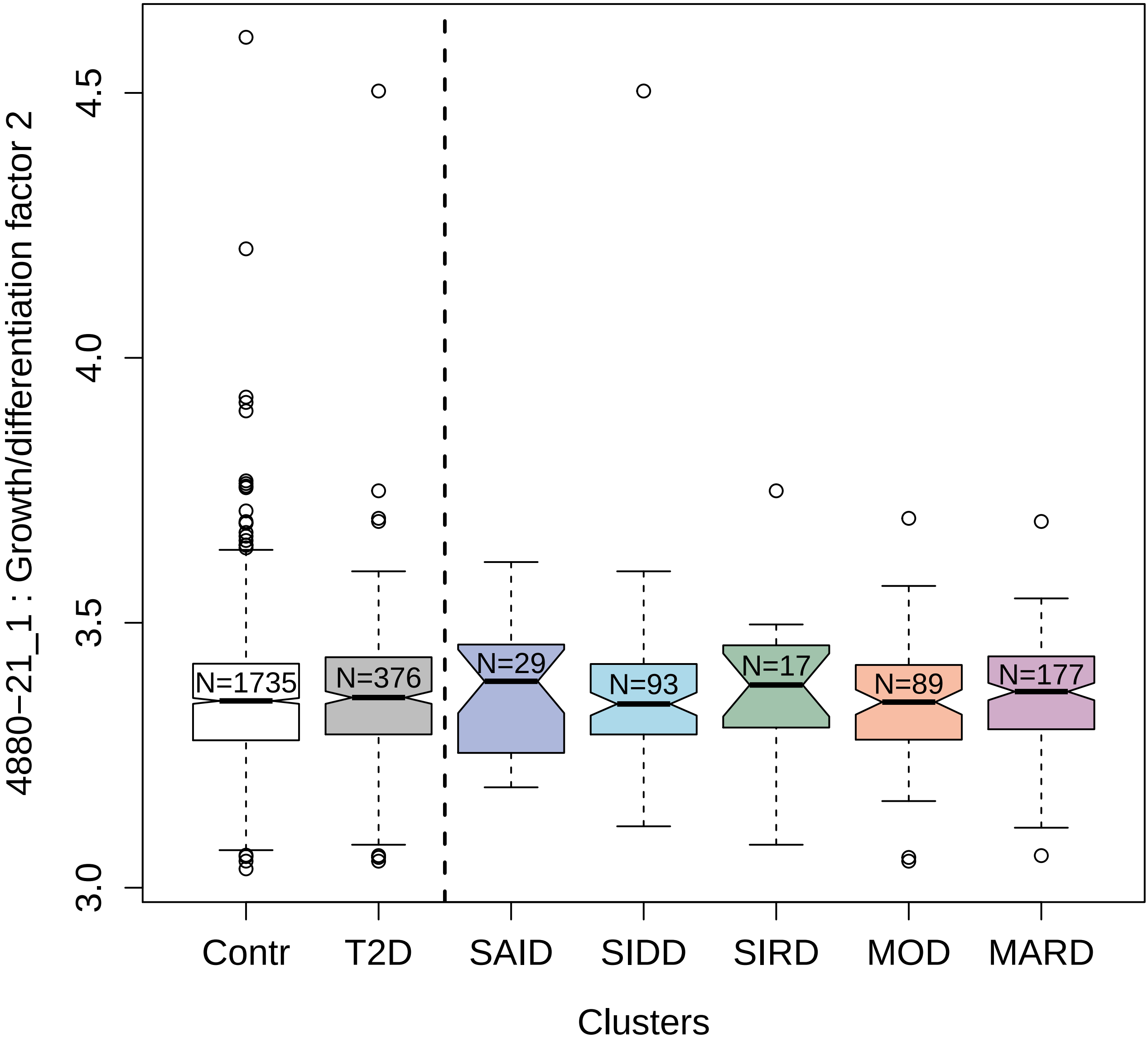
4880−21_1 : Growth/differentiation factor 2
3.0
3.5
4.0
4.5
N=1735
N=376
N=29
N=93
N=17
N=89
N=177
Contr
T2D
SAID
SIDD
SIRD
MOD
MARD
Clusters

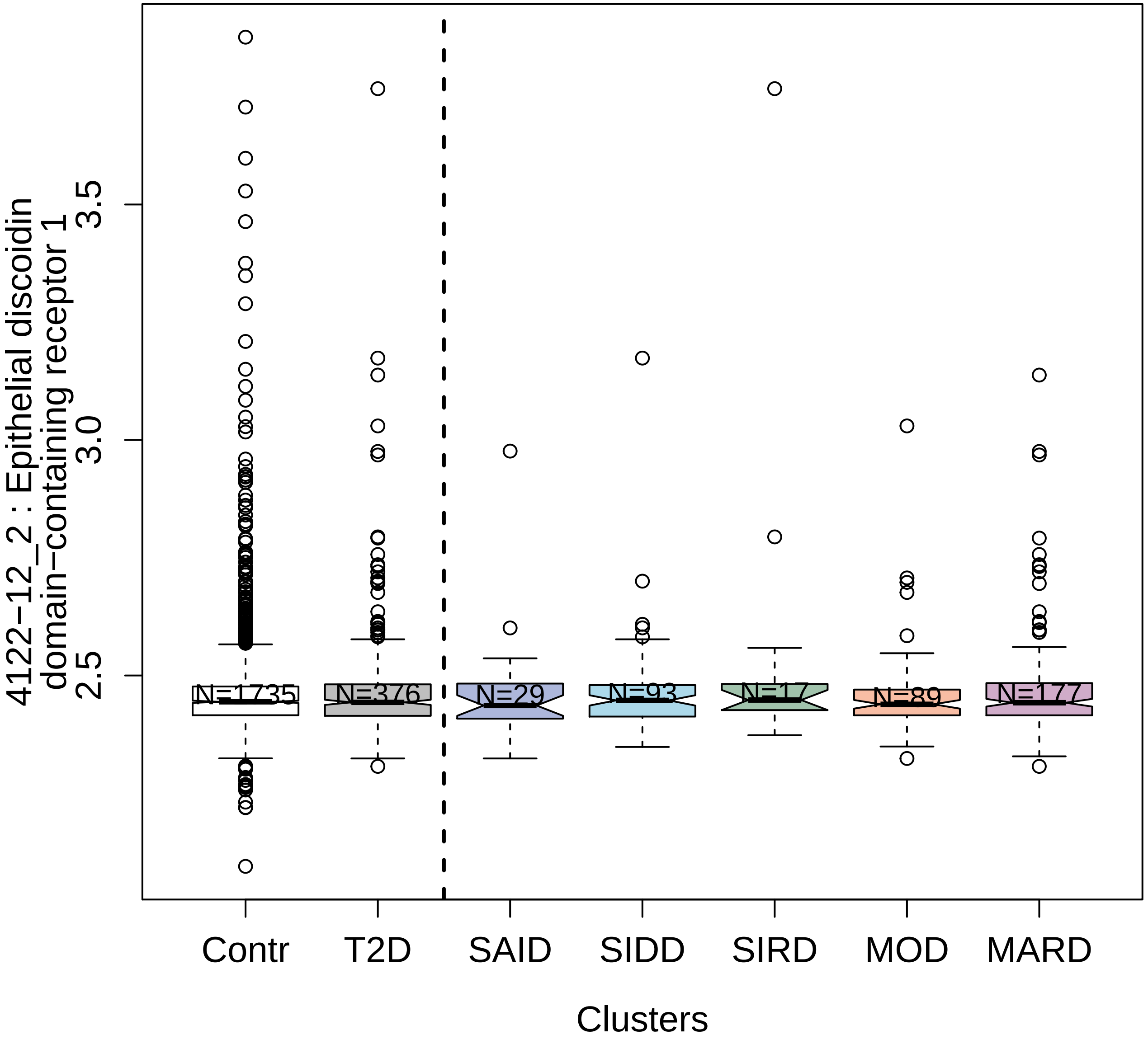
4122−12_2 : Epithelial discoidin
domain−containing receptor 1
2.5
3.0
3.5
N=1735
N=376
N=29
N=93
N=17
N=89
N=177
Contr
T2D
SAID
SIDD
SIRD
MOD
MARD
Clusters

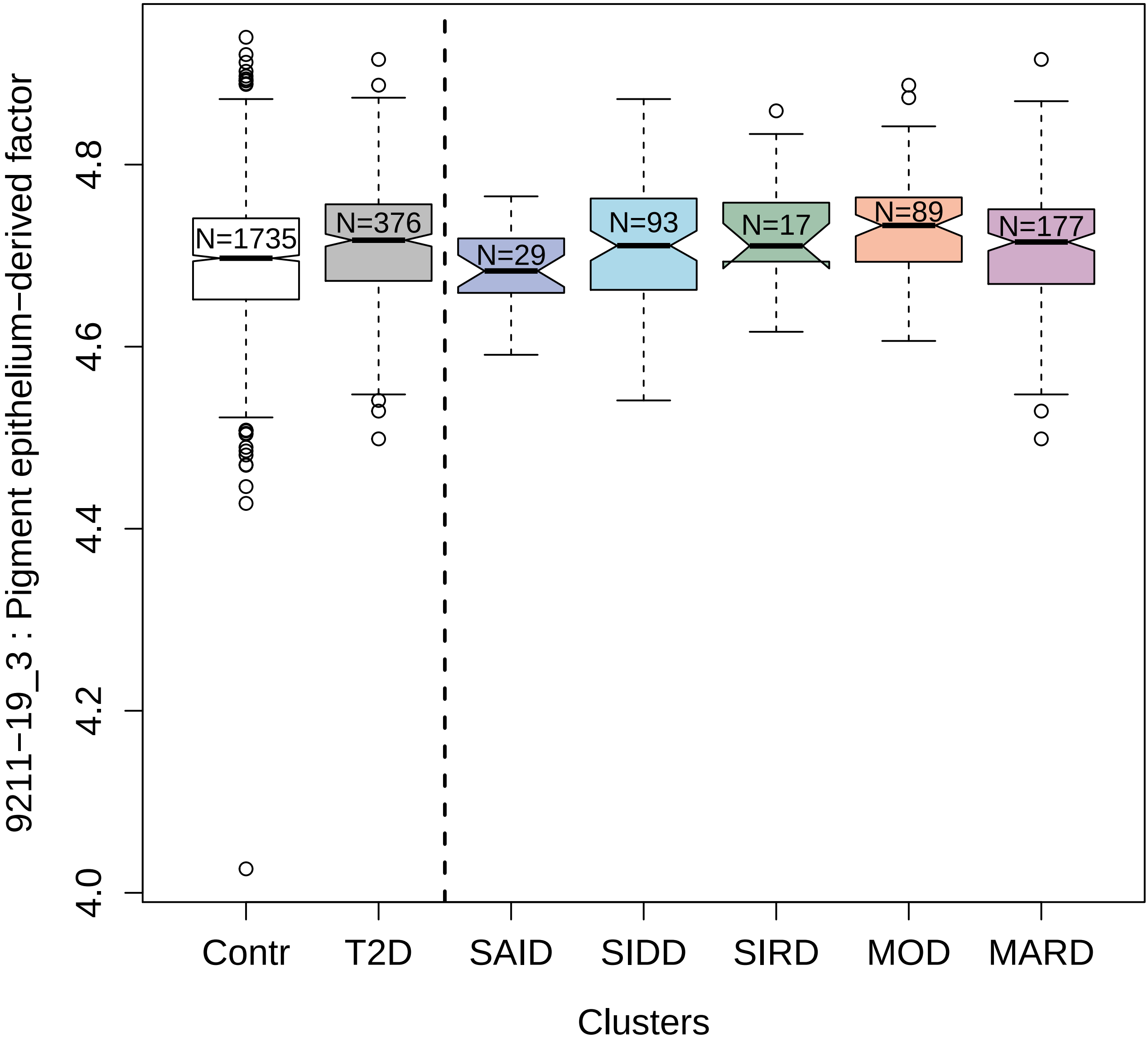

N=1735
N=376
N=29
N=93
N=17
N=89
N=177
4.0
4.2
4.4
4.6
4.8
9211−19_3 : Pigment epithelium−derived factor
Contr
T2D
SAID
SIDD
SIRD
MOD
MARD
Clusters

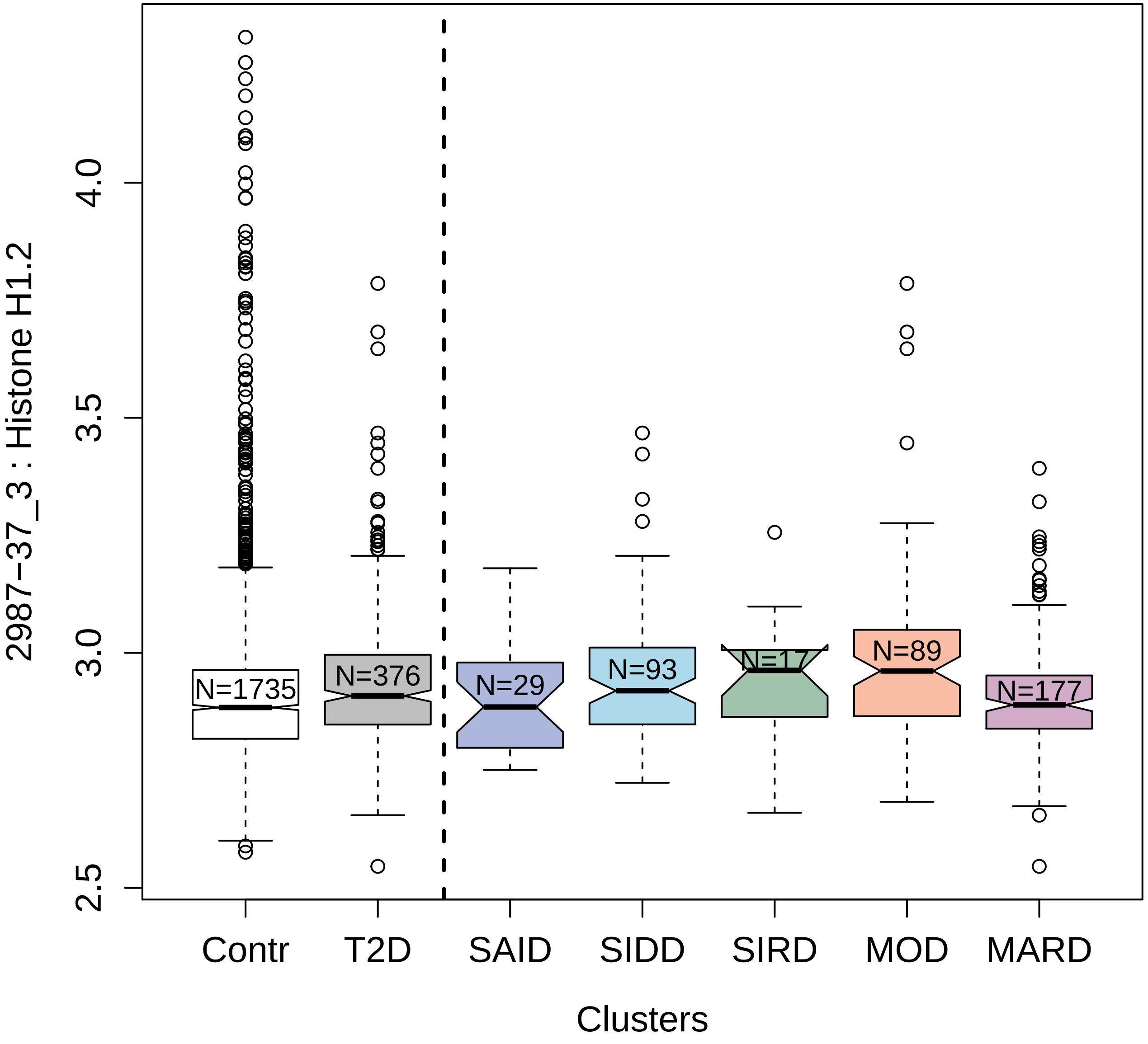
2987−37_3 : Histone H1.2
4.0
3.5
3.0
2.5
N=1735
N=376
N=29
N=93
N=17
N=89
N=177
Contr
T2D
SAID
SIDD
SIRD
MOD
MARD
Clusters

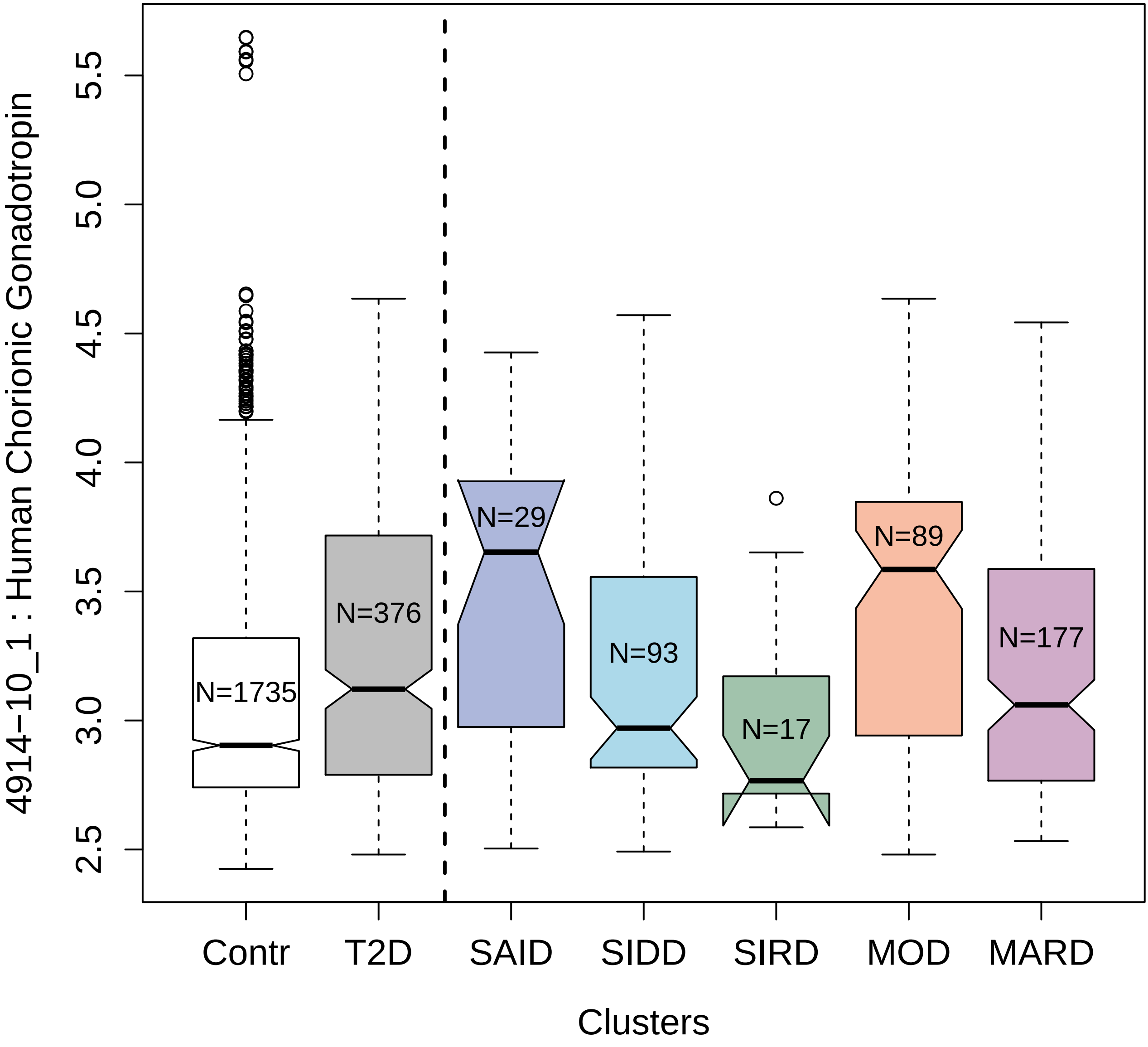

4914−10_1 : Human Chorionic Gonadotropin
2.5
3.0
3.5
4.0
4.5
5.0
5.5
N=1735
N=376
N=29
N=93
N=17
N=89
N=177
Contr
T2D
SAID
SIDD
SIRD
MOD
MARD
Clusters

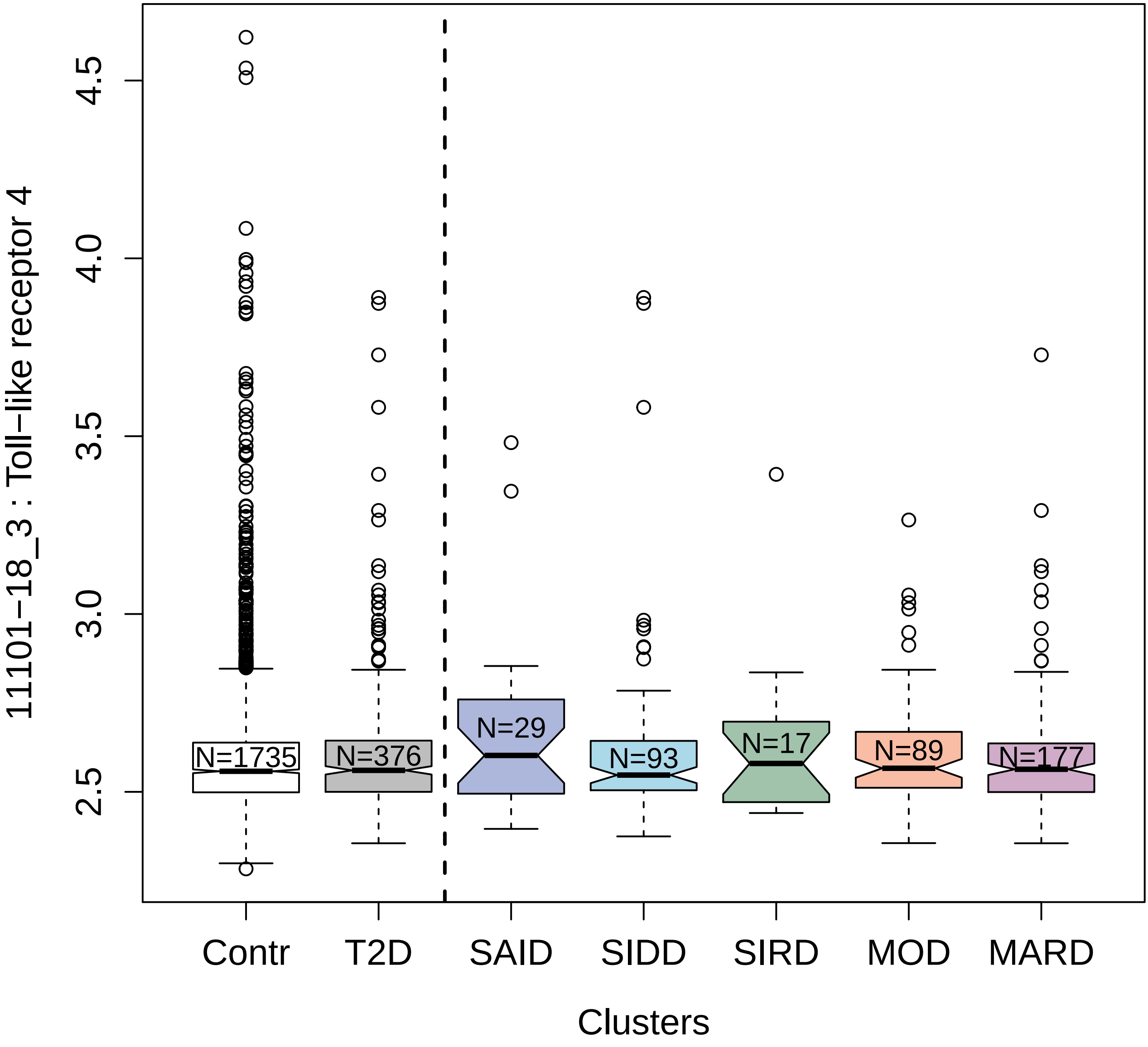

11101−18_3 : Toll−like receptor 4
4.5
4.0
3.5
3.0
2.5
N=1735
N=376
N=29
N=93
N=17
N=89
N=177
Contr
T2D
SAID
SIDD
SIRD
MOD
MARD
Clusters

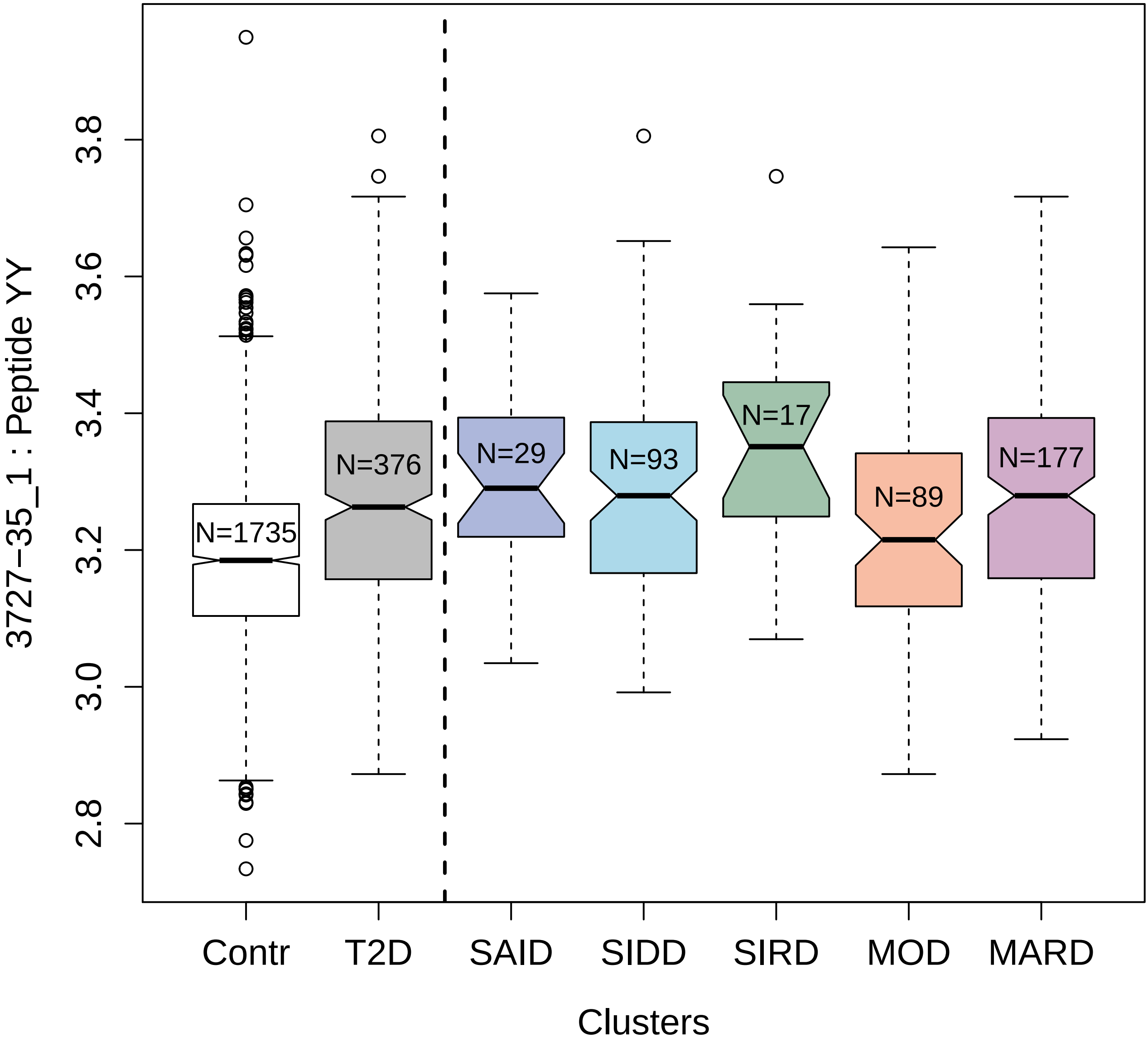
3727−35_1 : Peptide YY
3.8
3.6
3.4
3.2
3.0
2.8
N=1735
N=376
N=29
N=93
N=17
N=89
N=177
Contr
T2D
SAID
SIDD
SIRD
MOD
MARD
Clusters

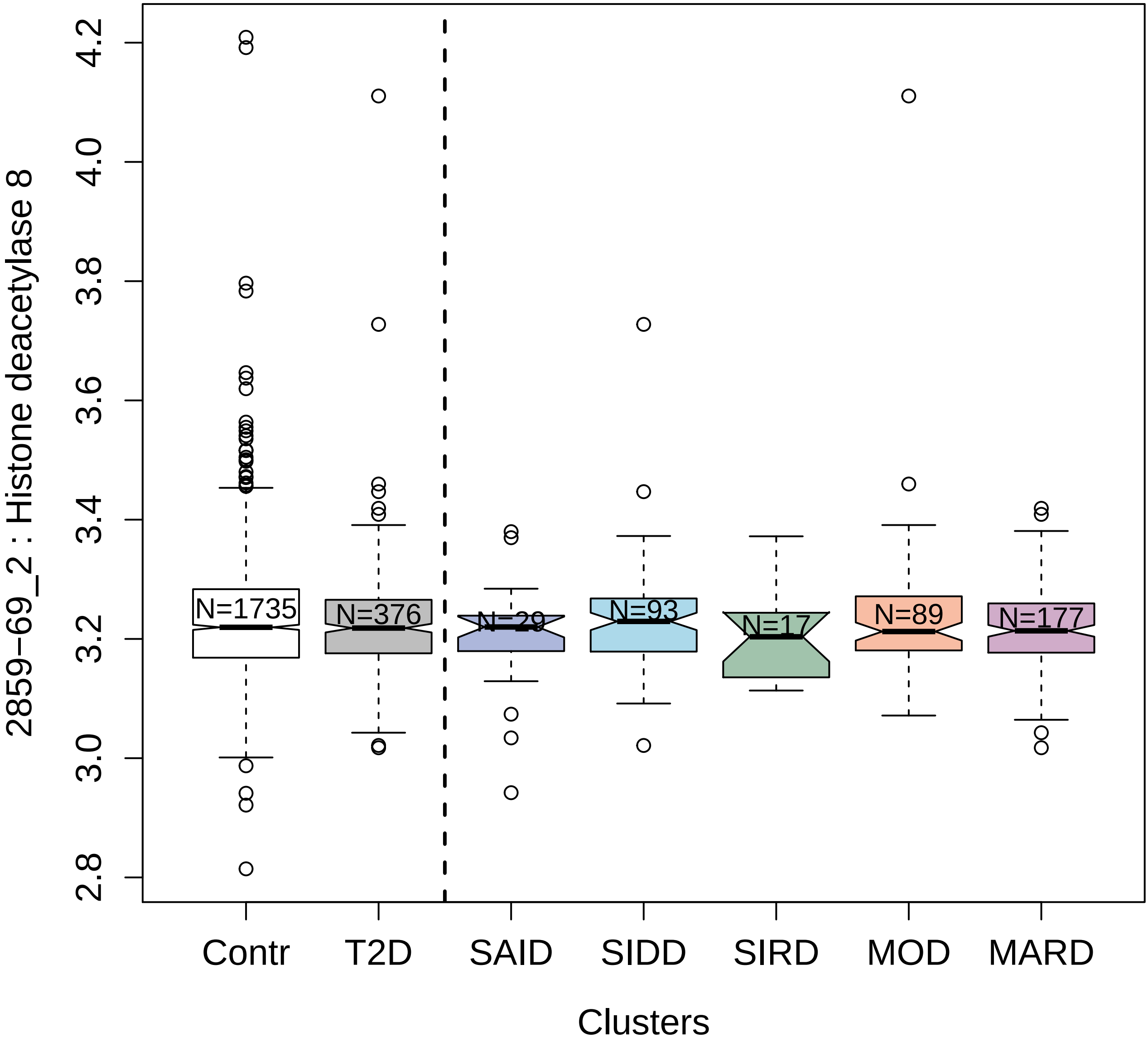

2859−69_2 : Histone deacetylase 8
2.8
3.0
3.2
3.4
3.6
3.8
4.0
4.2
N=1735
N=376
N=29
N=93
N=17
N=89
N=177
Contr
T2D
SAID
SIDD
SIRD
MOD
MARD
Clusters

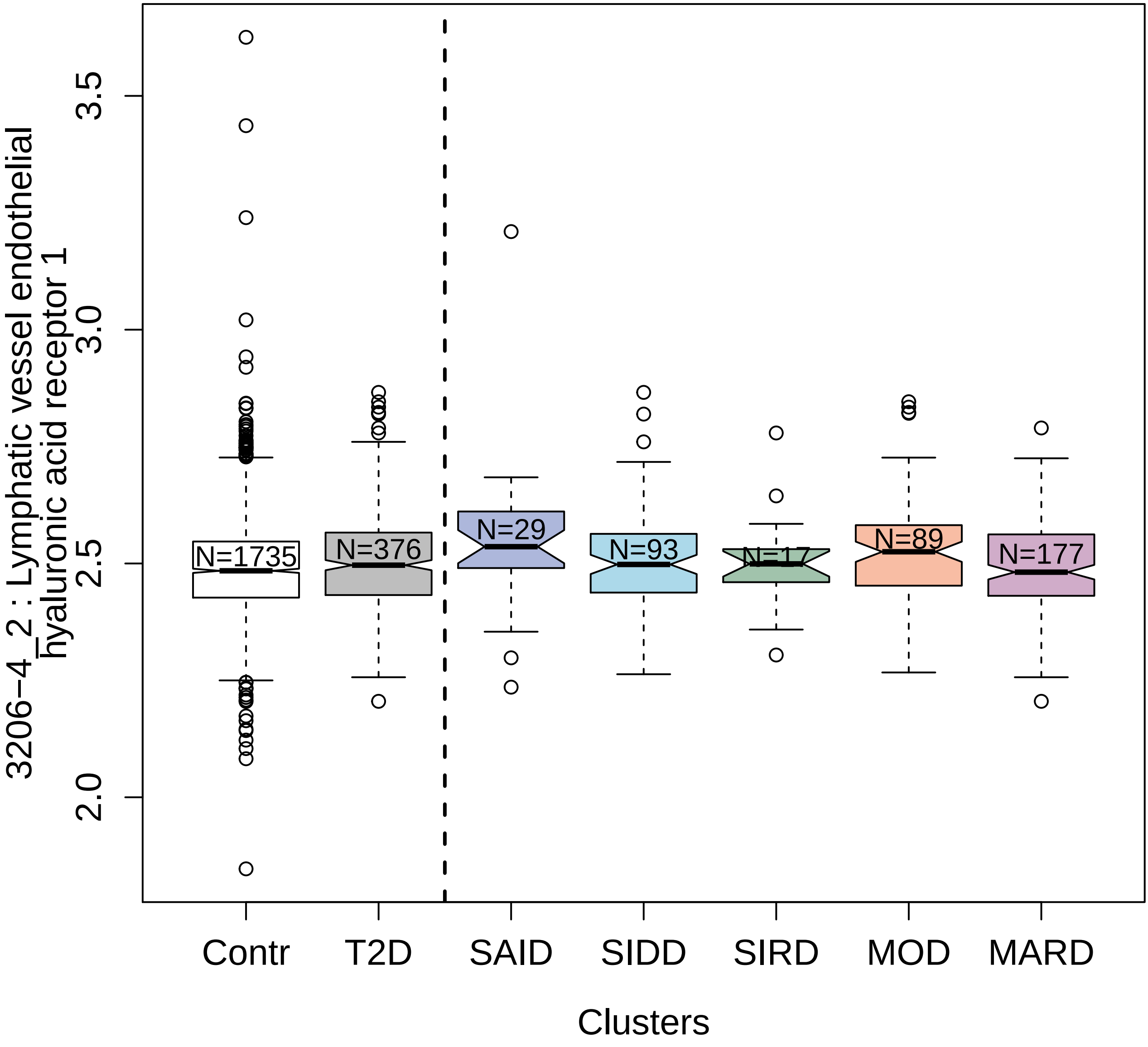

3206−4_2 : Lymphatic vessel endothelial hyaluronic acid receptor 1
3.5
3.0
2.5
2.0
N=1735
N=376
N=29
N=93
N=17
N=89
N=177
Contr
T2D
SAID
SIDD
SIRD
MOD
MARD
Clusters

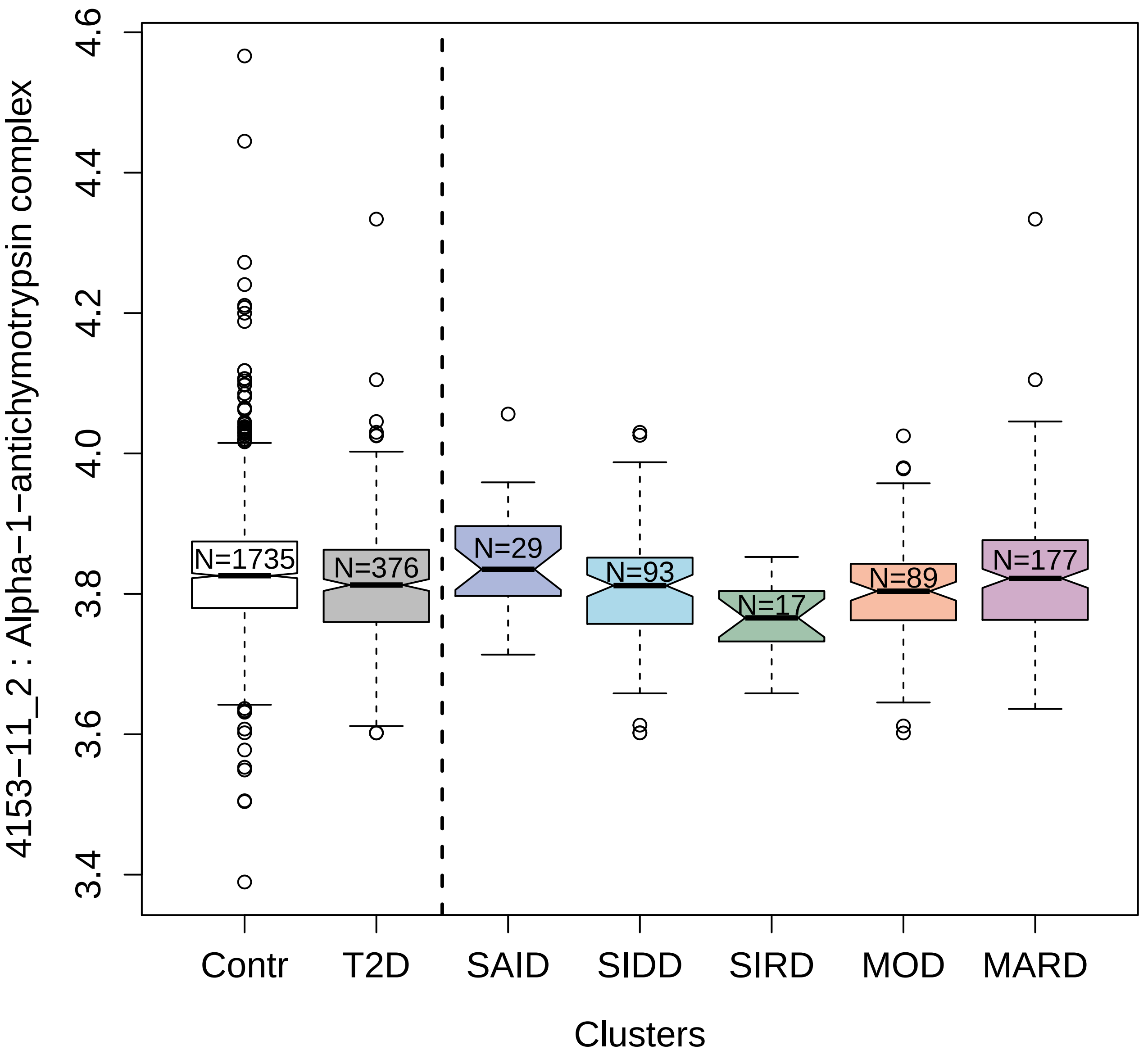

4153−11_2 : Alpha−1−antichymotrypsin complex
4.6
4.4
4.2
4.0
3.8
3.6
3.4
N=1735
N=376
N=29
N=93
N=17
N=89
N=177
Contr
T2D
SAID
SIDD
SIRD
MOD
MARD
Clusters

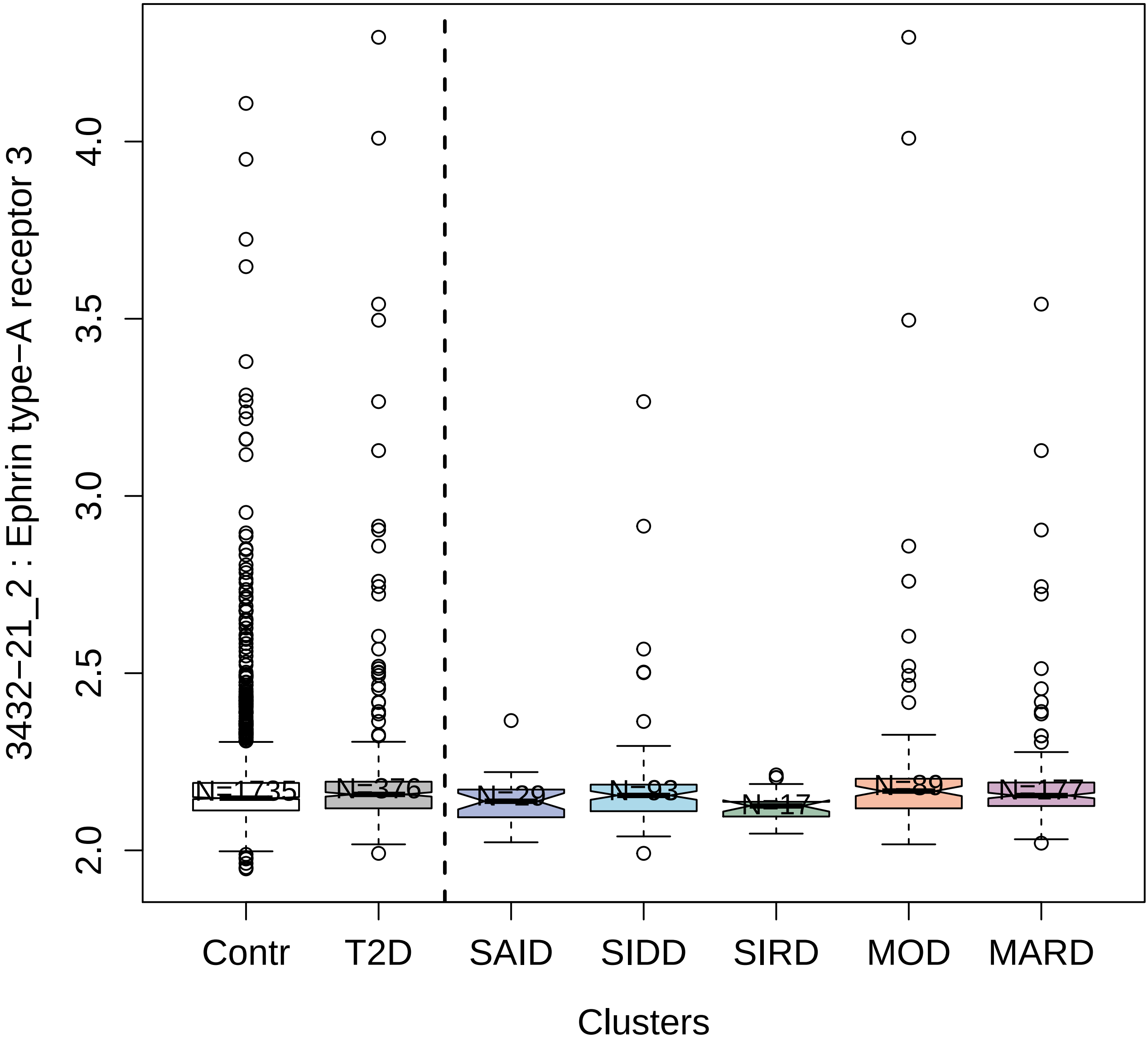
3432−21_2 : Ephrin type−A receptor 3
2.0
2.5
3.0
3.5
4.0
N=1735
N=376
N=29
N=93
N=17
N=89
N=177
Contr
T2D
SAID
SIDD
SIRD
MOD
MARD
Clusters

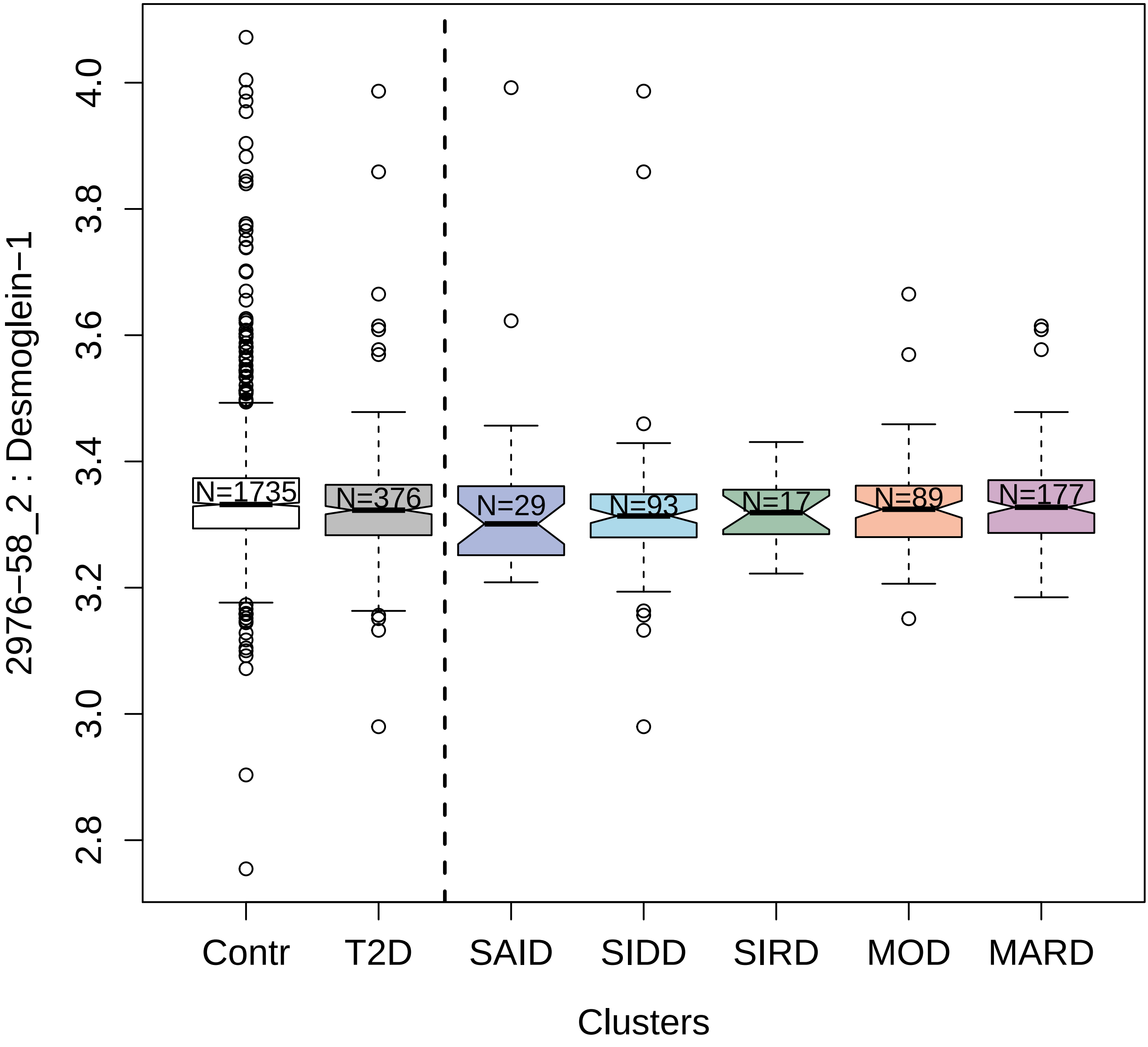

2976−58_2 : Desmoglein−1
4.0
3.8
3.6
3.4
3.2
3.0
2.8
N=1735
N=376
N=29
N=93
N=17
N=89
N=177
Contr
T2D
SAID
SIDD
SIRD
MOD
MARD
Clusters

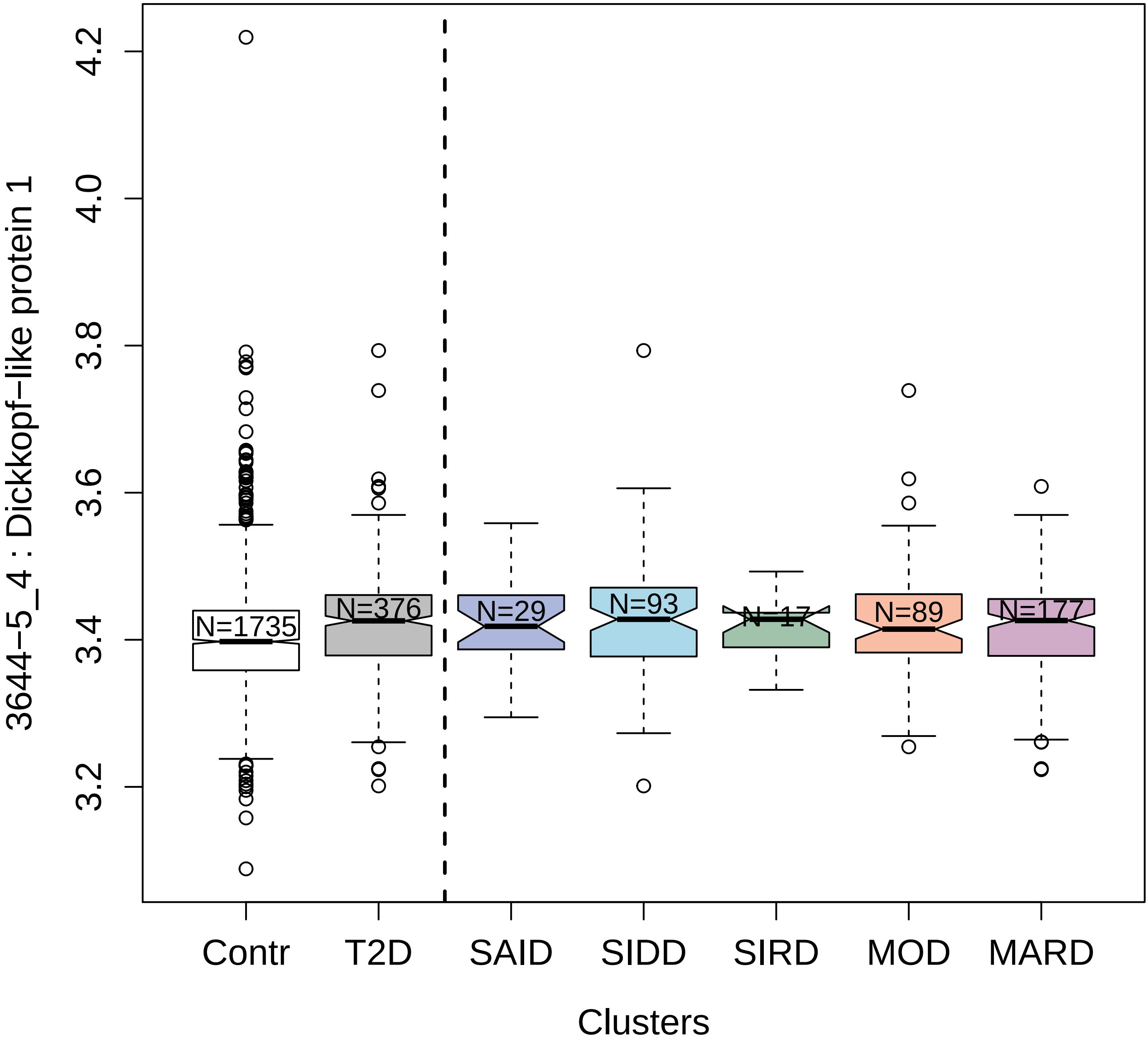

3644−5_4 : Dickkopf−like protein 1
4.2
4.0
3.8
3.6
3.4
3.2
N=1735
N=376
N=29
N=93
N=17
N=89
N=177
Contr
T2D
SAID
SIDD
SIRD
MOD
MARD
Clusters

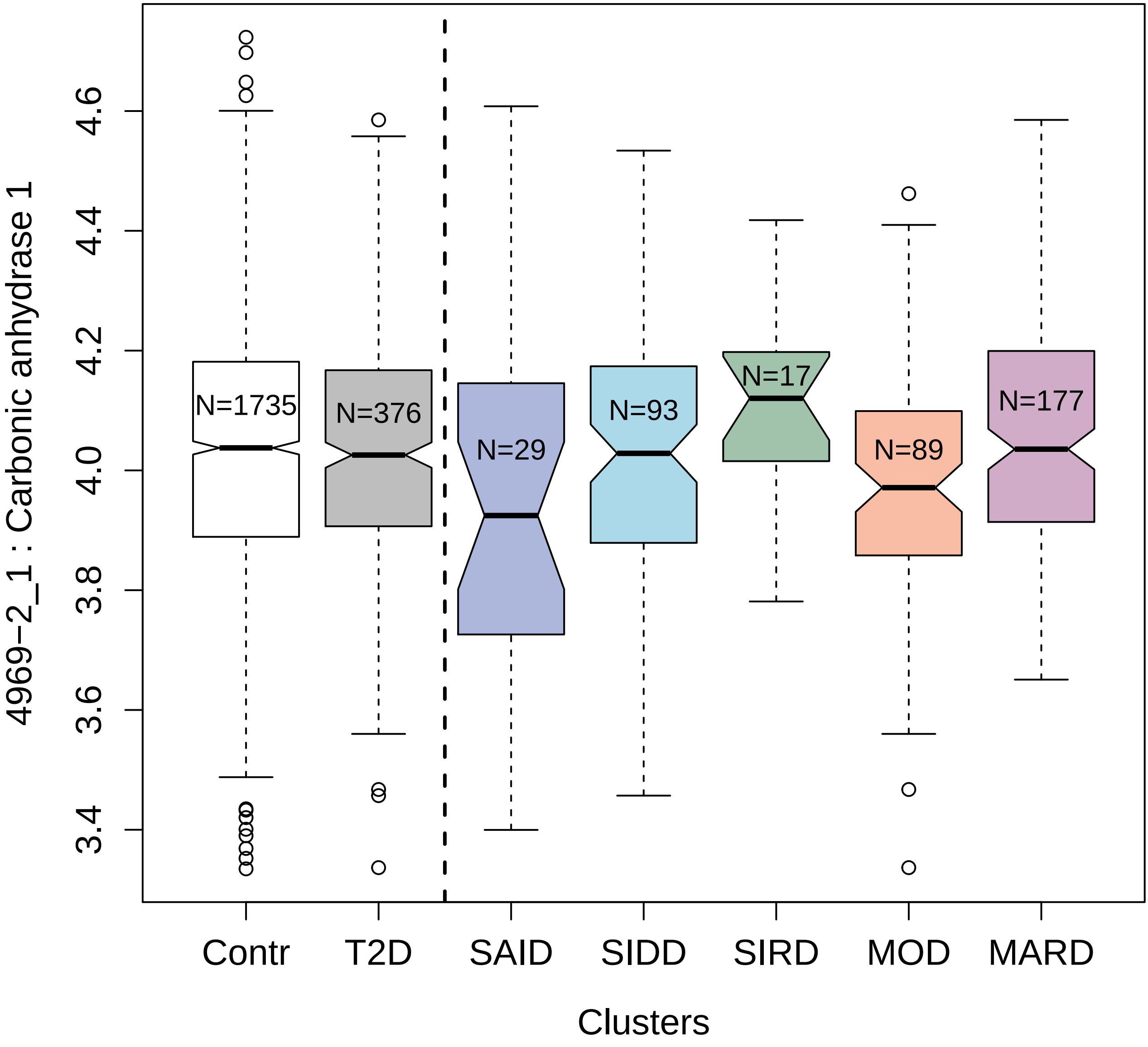
4969−2_1 : Carbonic anhydrase 1
3.4
3.6
3.8
4.0
4.2
4.4
4.6
N=1735
N=376
N=29
N=93
N=17
N=89
N=177
Contr
T2D
SAID
SIDD
SIRD
MOD
MARD
Clusters

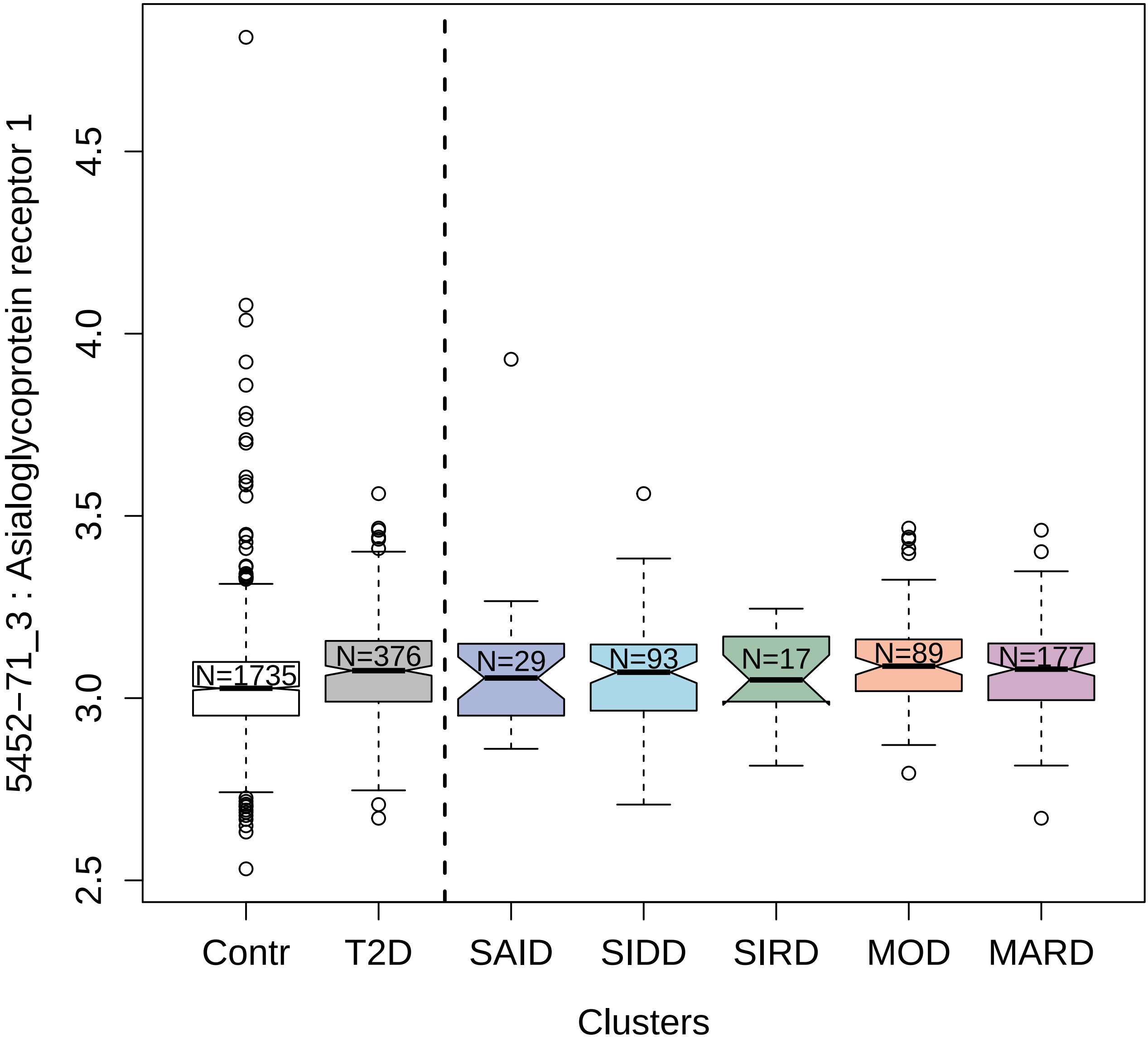

5452−71_3 : Asialoglycoprotein receptor 1
2.5
3.0
3.5
4.0
4.5
N=1735
N=376
N=29
N=93
N=17
N=89
N=177
Contr
T2D
SAID
SIDD
SIRD
MOD
MARD
Clusters

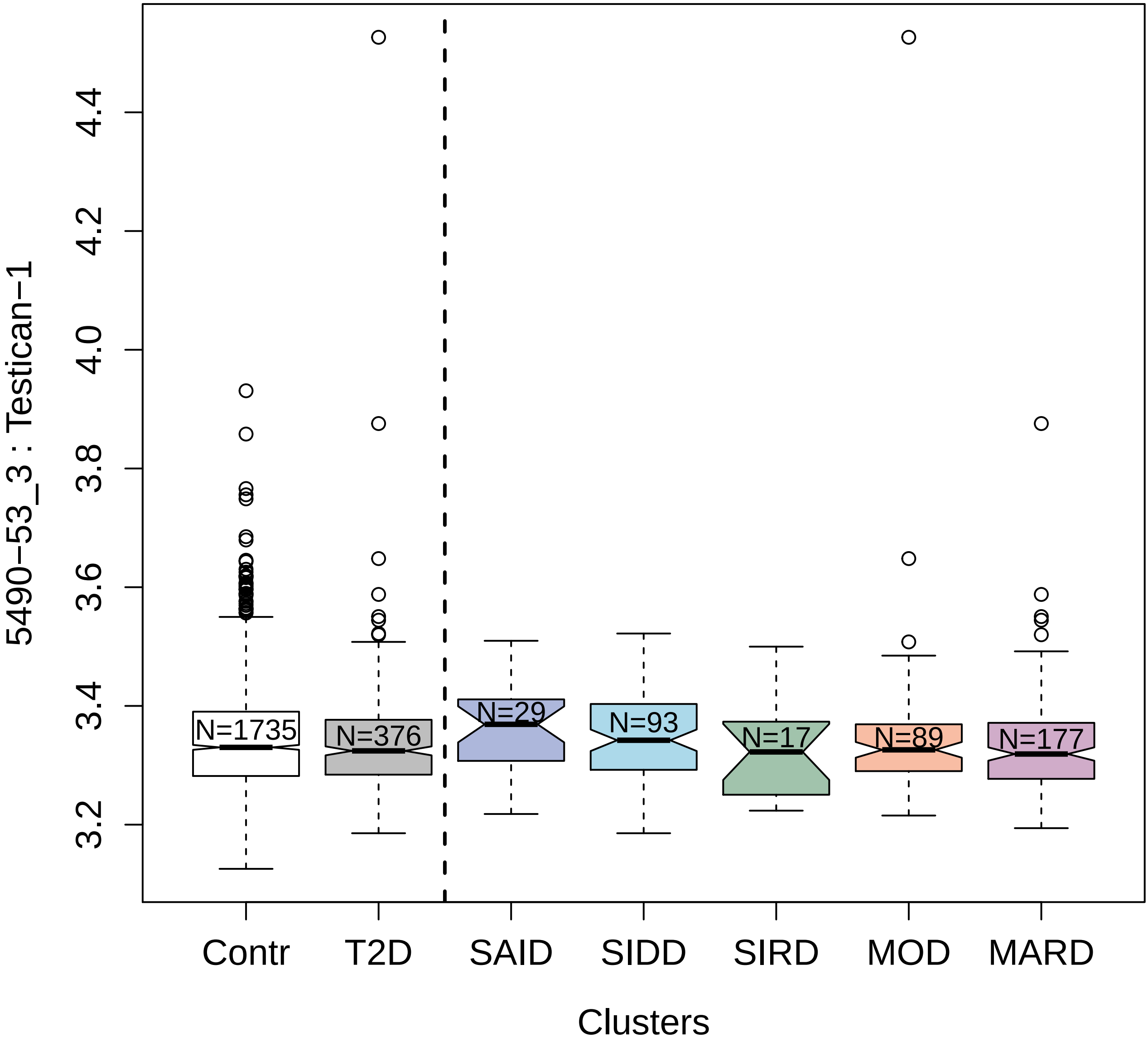

5490−53_3 : Testican−1
4.4
4.2
4.0
3.8
3.6
3.4
3.2
N=1735
N=376
N=29
N=93
N=17
N=89
N=177
Contr
T2D
SAID
SIDD
SIRD
MOD
MARD
Clusters

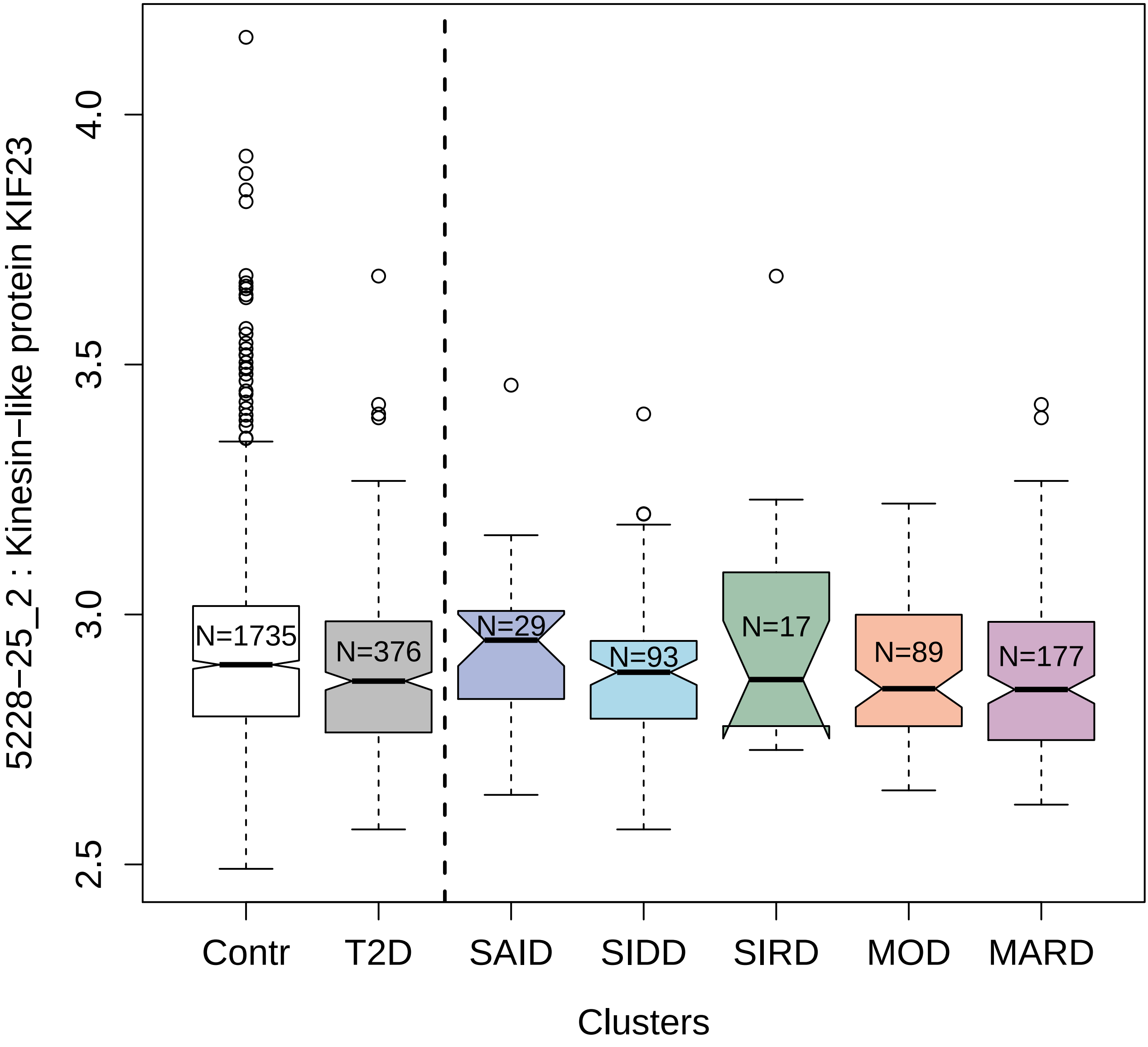
5228−25_2 : Kinesin−like protein KIF23
2.5
3.0
3.5
4.0
N=1735
N=376
N=29
N=93
N=17
N=89
N=177
Contr
T2D
SAID
SIDD
SIRD
MOD
MARD
Clusters

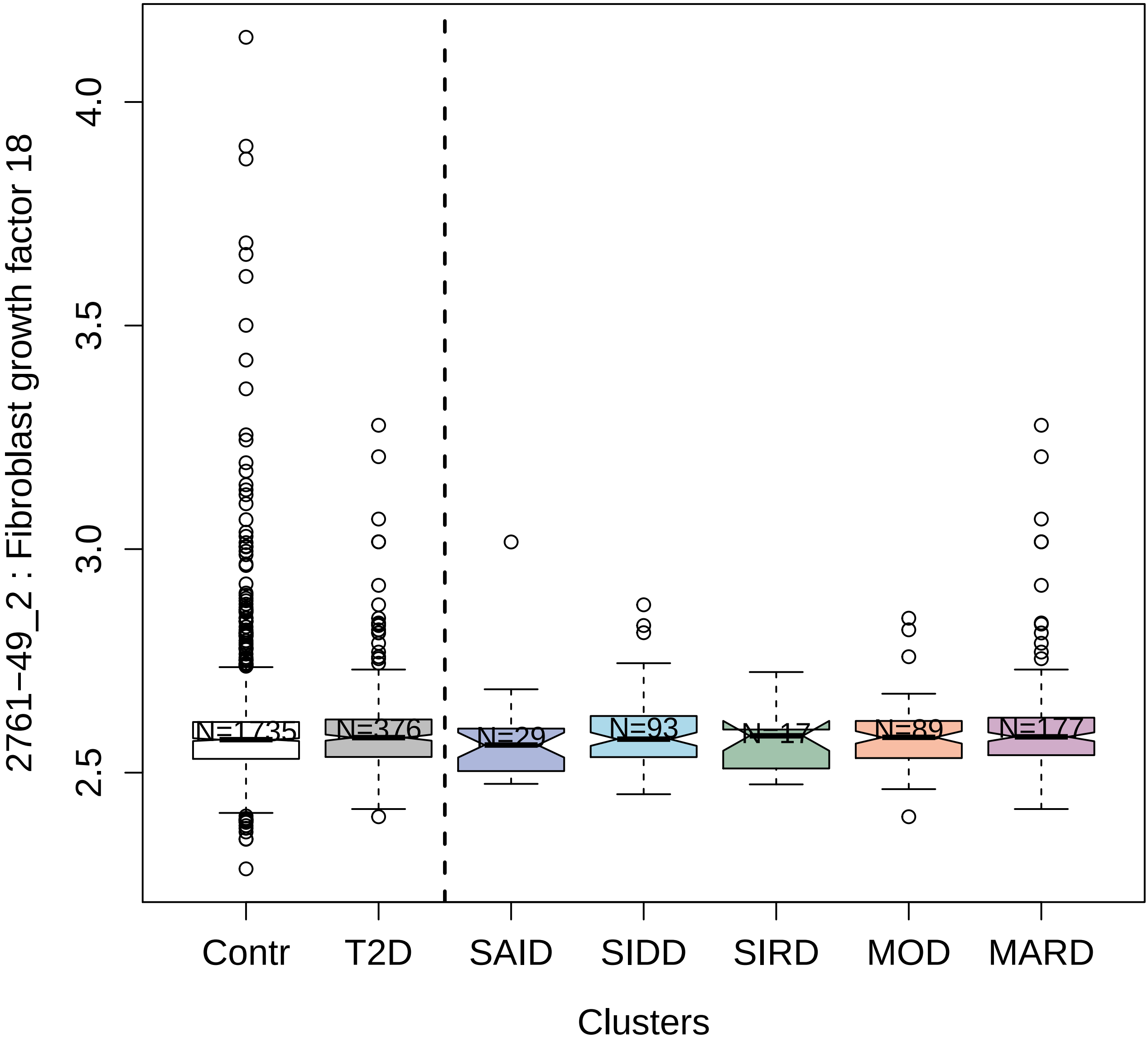
2761−49_2 : Fibroblast growth factor 18
4.0
3.5
3.0
2.5
N=1735
N=376
N=29
N=93
N=17
N=89
N=177
Contr
T2D
SAID
SIDD
SIRD
MOD
MARD
Clusters

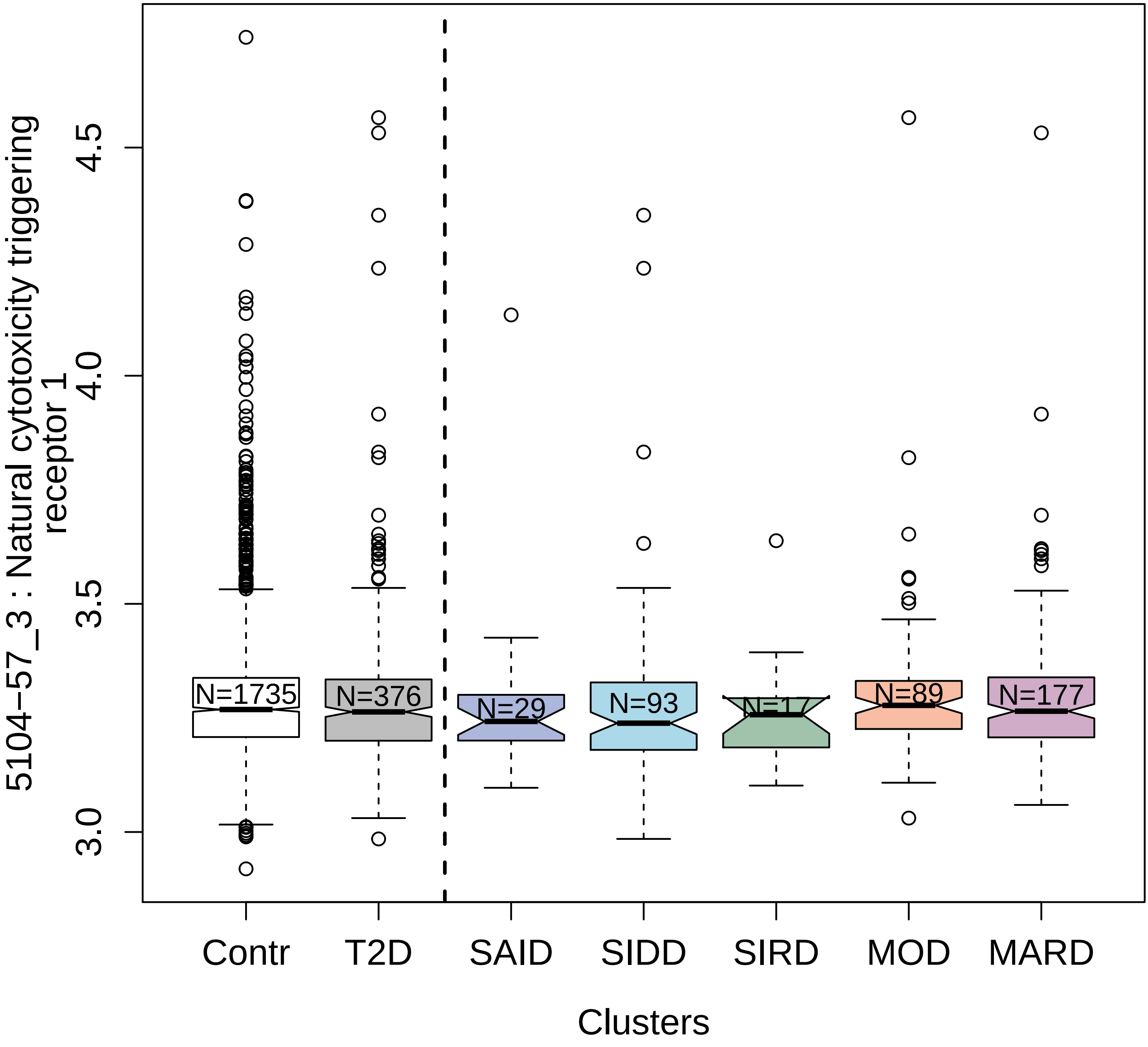
5104−57_3 : Natural cytotoxicity triggering receptor 1
3.0
3.5
4.0
4.5
N=1735
N=376
N=29
N=93
N=17
N=89
N=177
Contr
T2D
SAID
SIDD
SIRD
MOD
MARD
Clusters

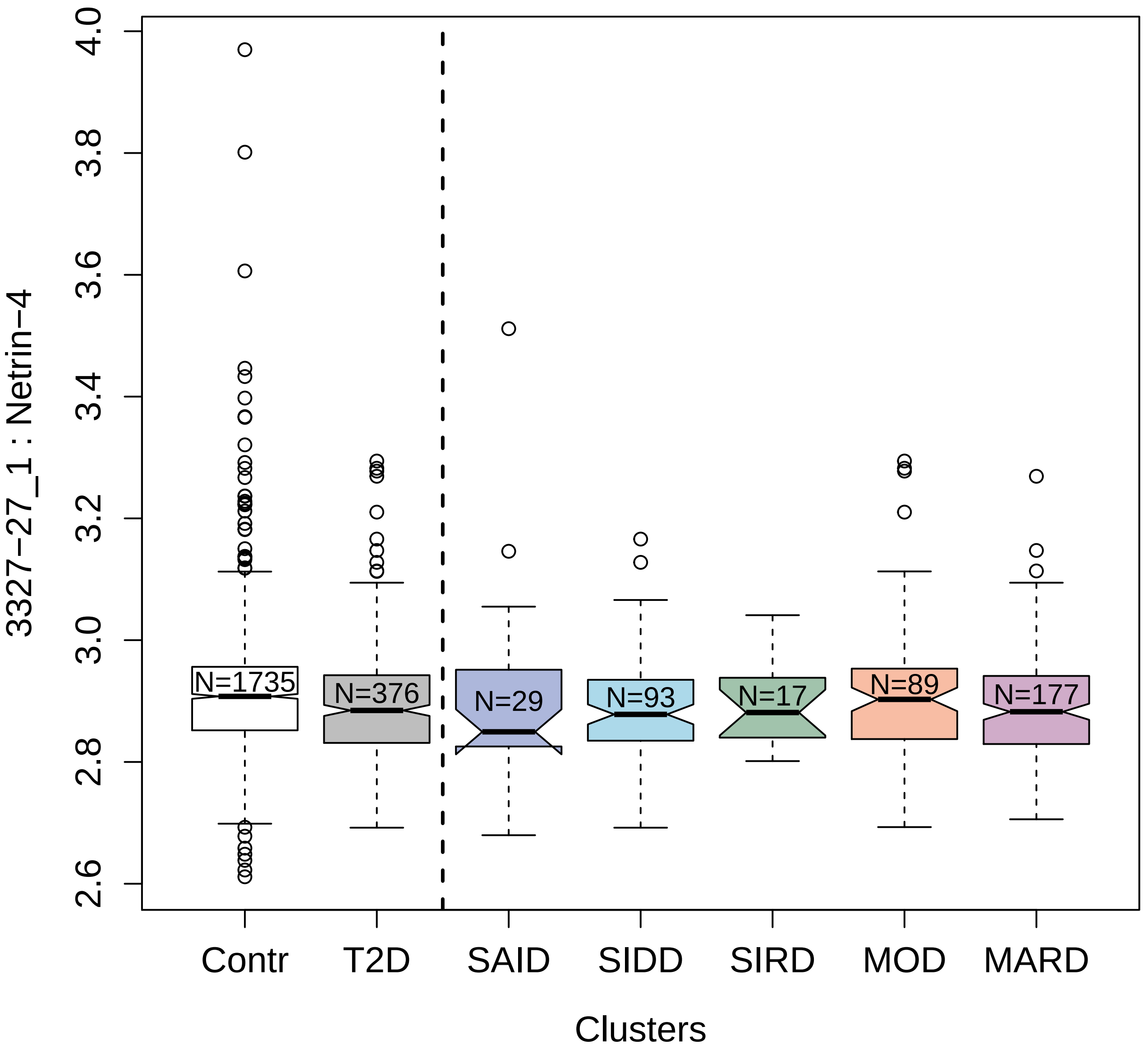
3327−27_1 : Netrin−4
4.0
3.8
3.6
3.4
3.2
3.0
2.8
2.6
N=1735
N=376
N=29
N=93
N=17
N=89
N=177
Contr
T2D
SAID
SIDD
SIRD
MOD
MARD
Clusters

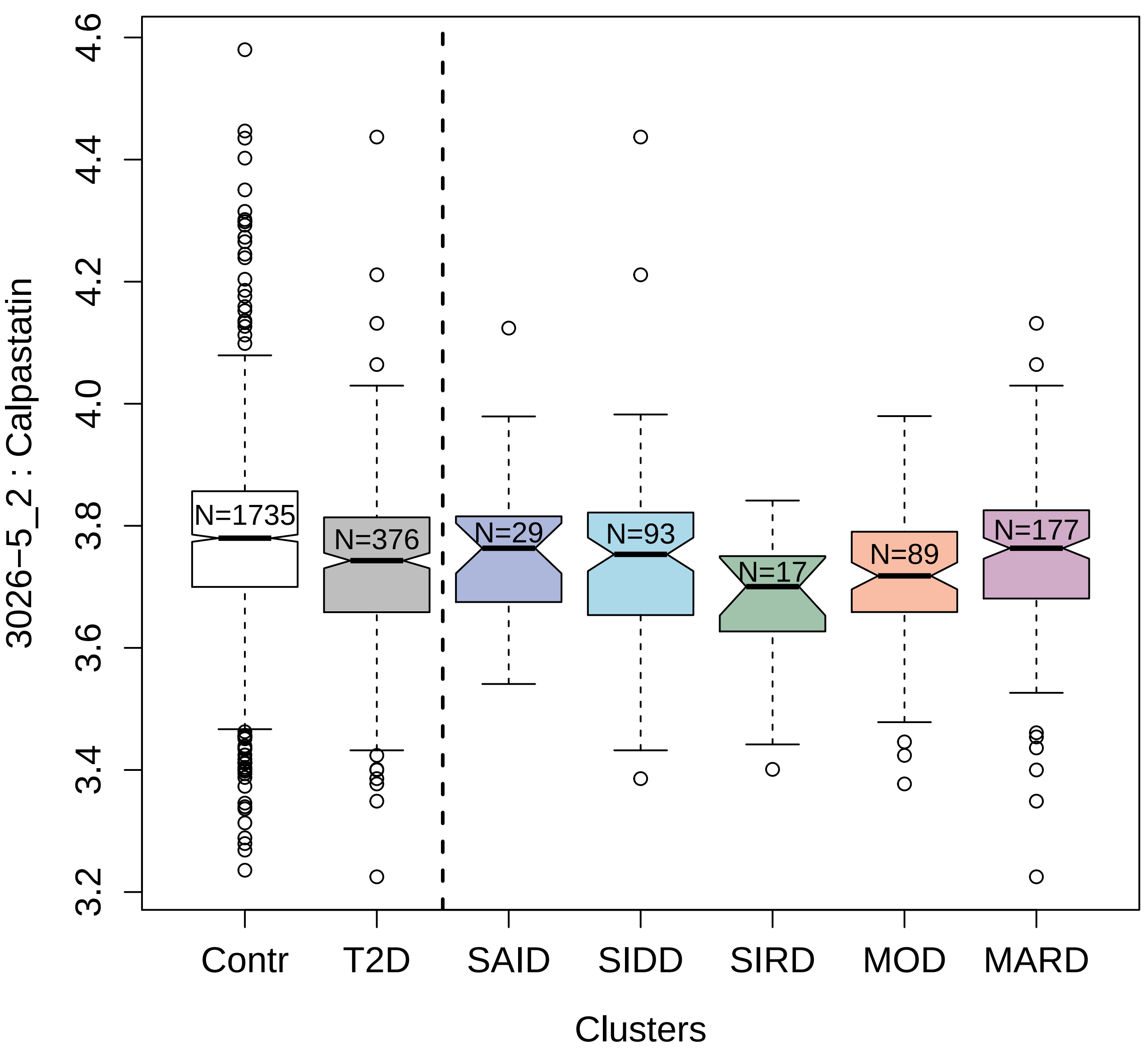
3026−5_2 : Calpastatin
4.6
4.4
4.2
4.0
3.8
3.6
3.4
3.2
N=1735
N=376
N=29
N=93
N=17
N=89
N=177
Contr
T2D
SAID
SIDD
SIRD
MOD
MARD
Clusters

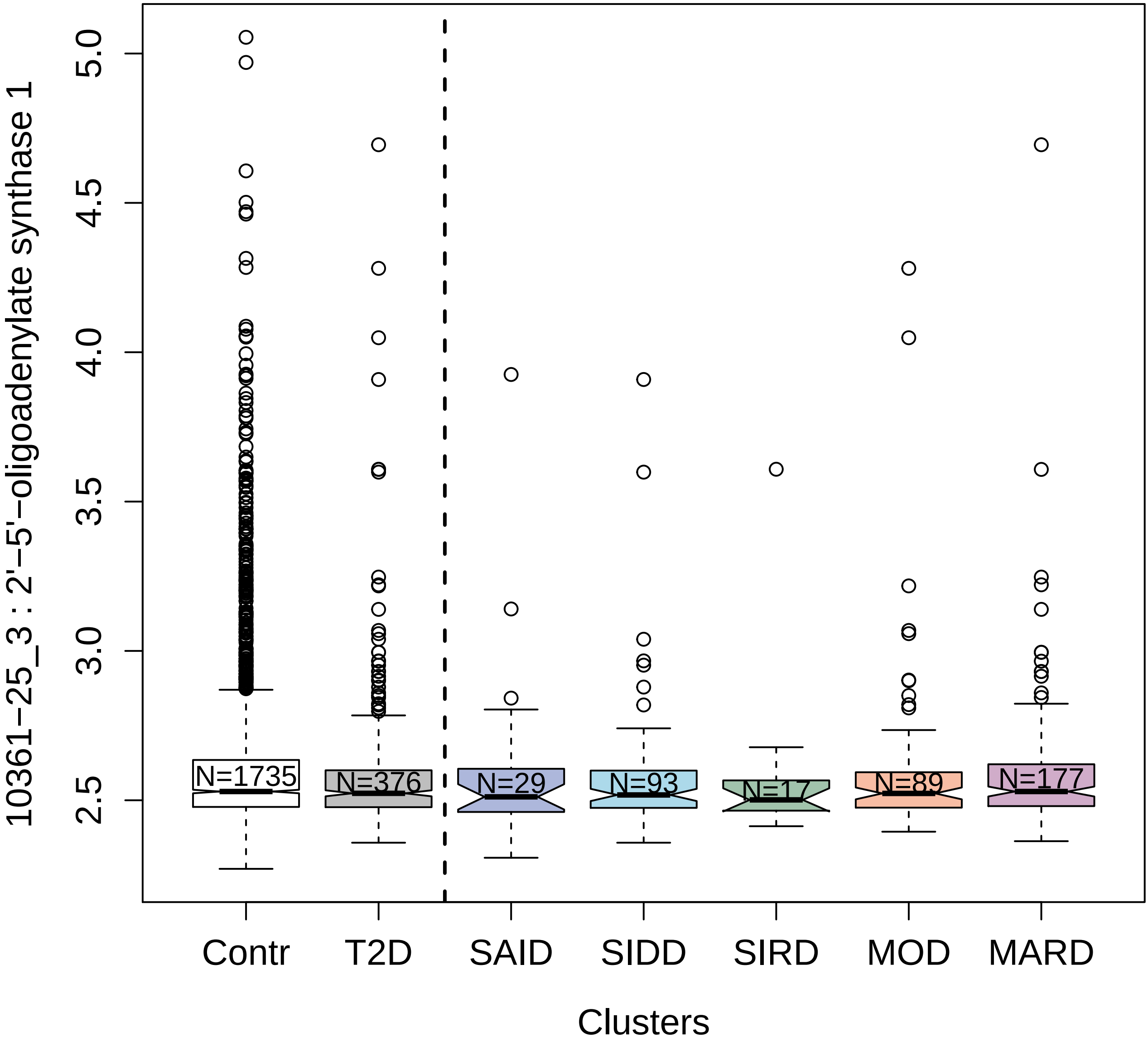

10361−25_3 : 2'−5'−oligoadenylate synthase 1
5.0
4.5
4.0
3.5
3.0
2.5
N=1735
N=376
N=29
N=93
N=17
N=89
N=177
Contr
T2D
SAID
SIDD
SIRD
MOD
MARD
Clusters

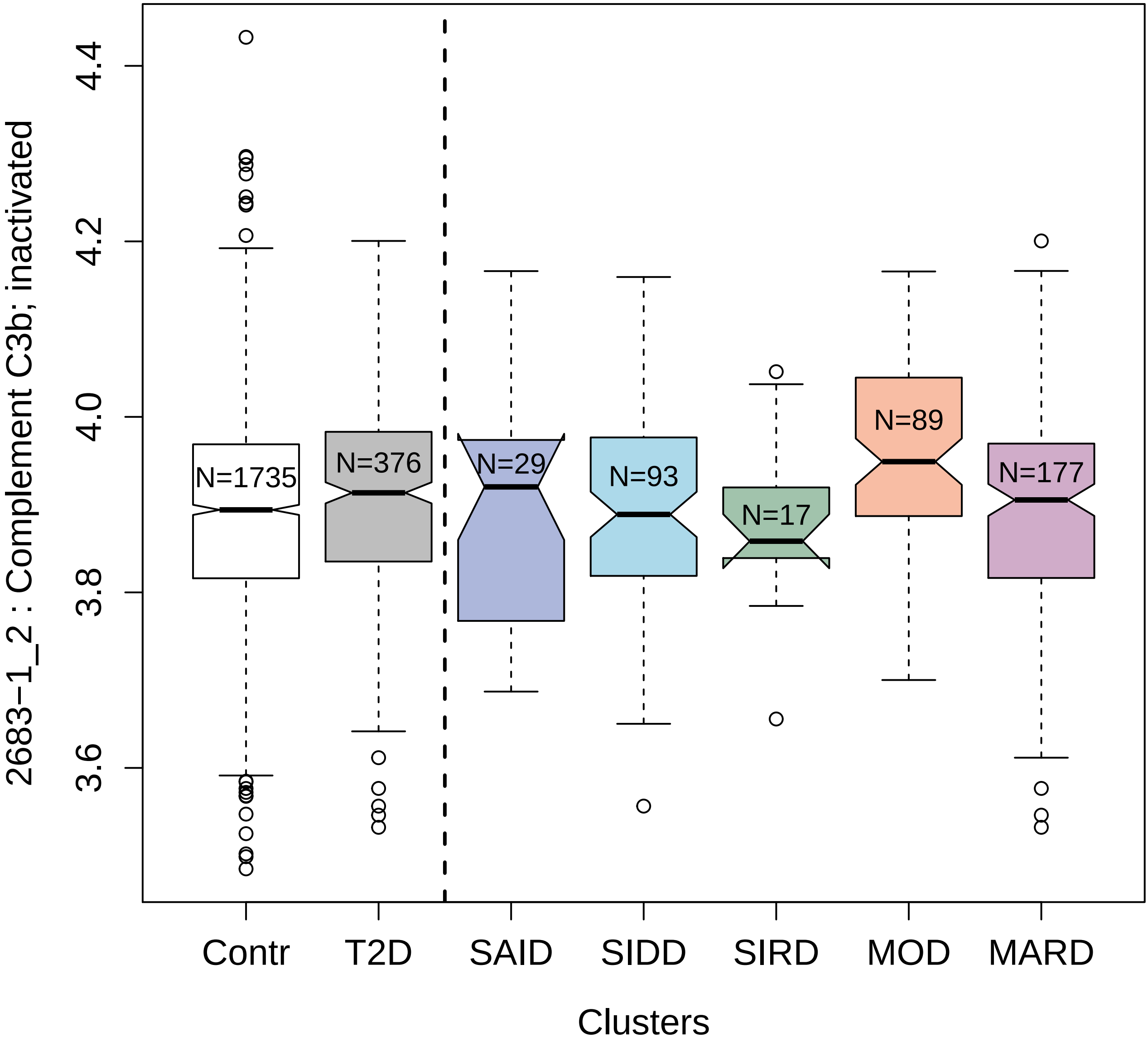

2683−1_2 : Complement C3b; inactivated
4.4
4.2
4.0
3.8
3.6
N=1735
N=376
N=29
N=93
N=17
N=89
N=177
Contr
T2D
SAID
SIDD
SIRD
MOD
MARD
Clusters

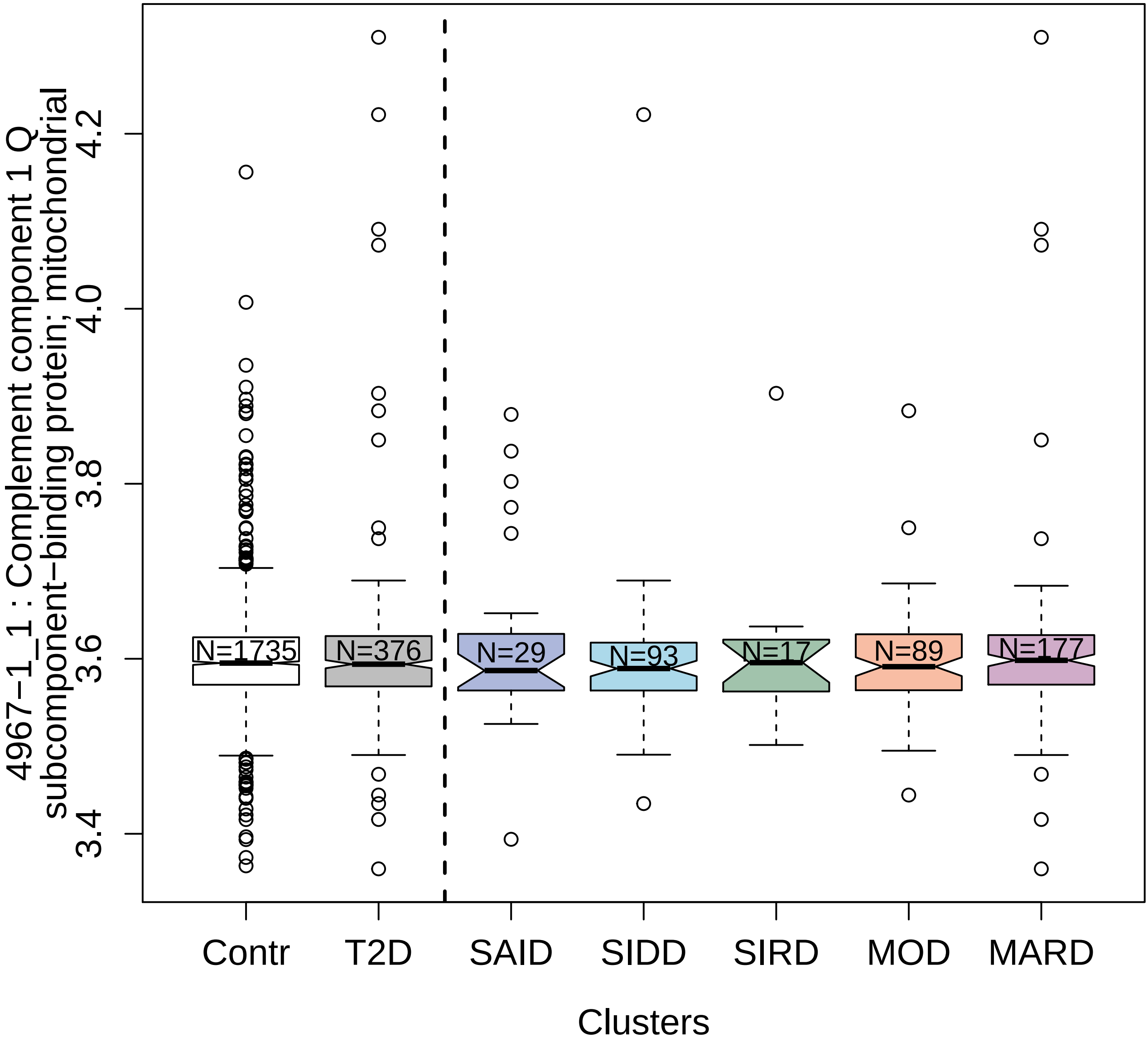
4967−1_1 : Complement component 1 Q subcomponent−binding protein; mitochondrial
3.4
3.6
3.8
4.0
4.2
N=1735
N=376
N=29
N=93
N=17
N=89
N=177
Contr
T2D
SAID
SIDD
SIRD
MOD
MARD
Clusters

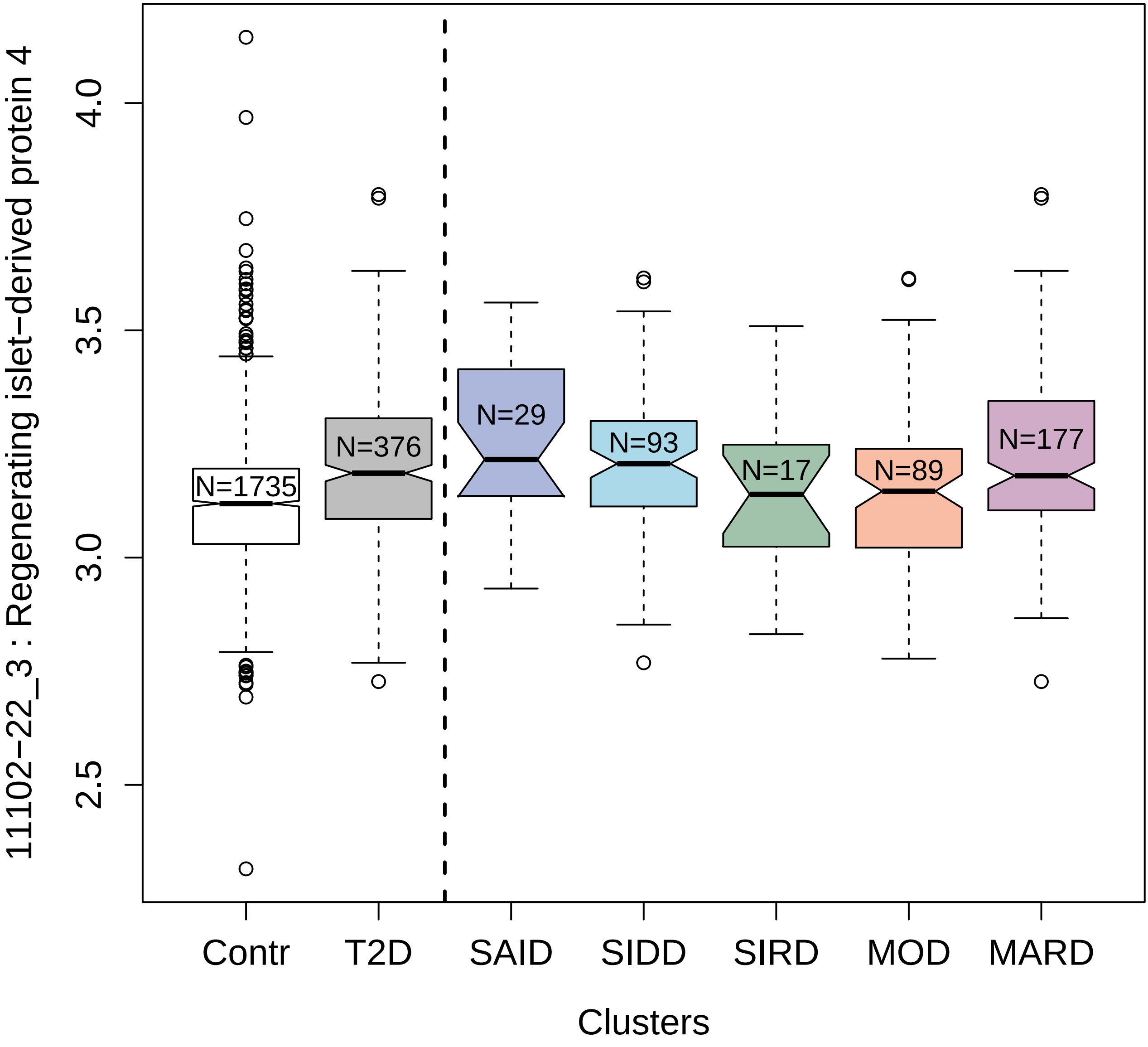

11102−22_3 : Regenerating islet−derived protein 4
2.5
3.0
3.5
4.0
N=1735
N=376
N=29
N=93
N=17
N=89
N=177
Contr
T2D
SAID
SIDD
SIRD
MOD
MARD
Clusters

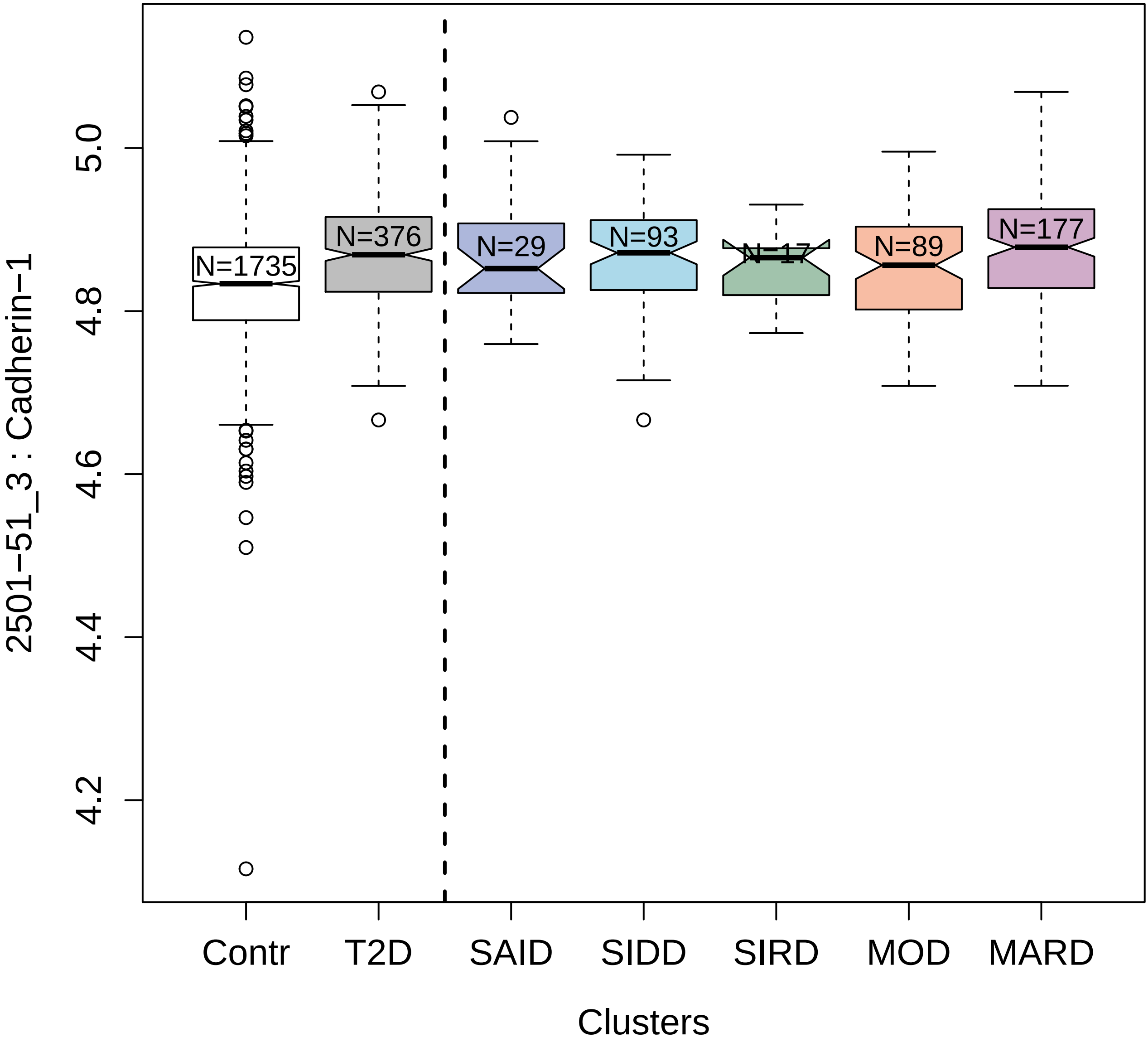
2501−51_3 : Cadherin−1
4.2
4.4
4.6
4.8
5.0
N=1735
N=376
N=29
N=93
N=89
N=177
Contr
T2D
SAID
SIDD
SIRD
MOD
MARD
Clusters

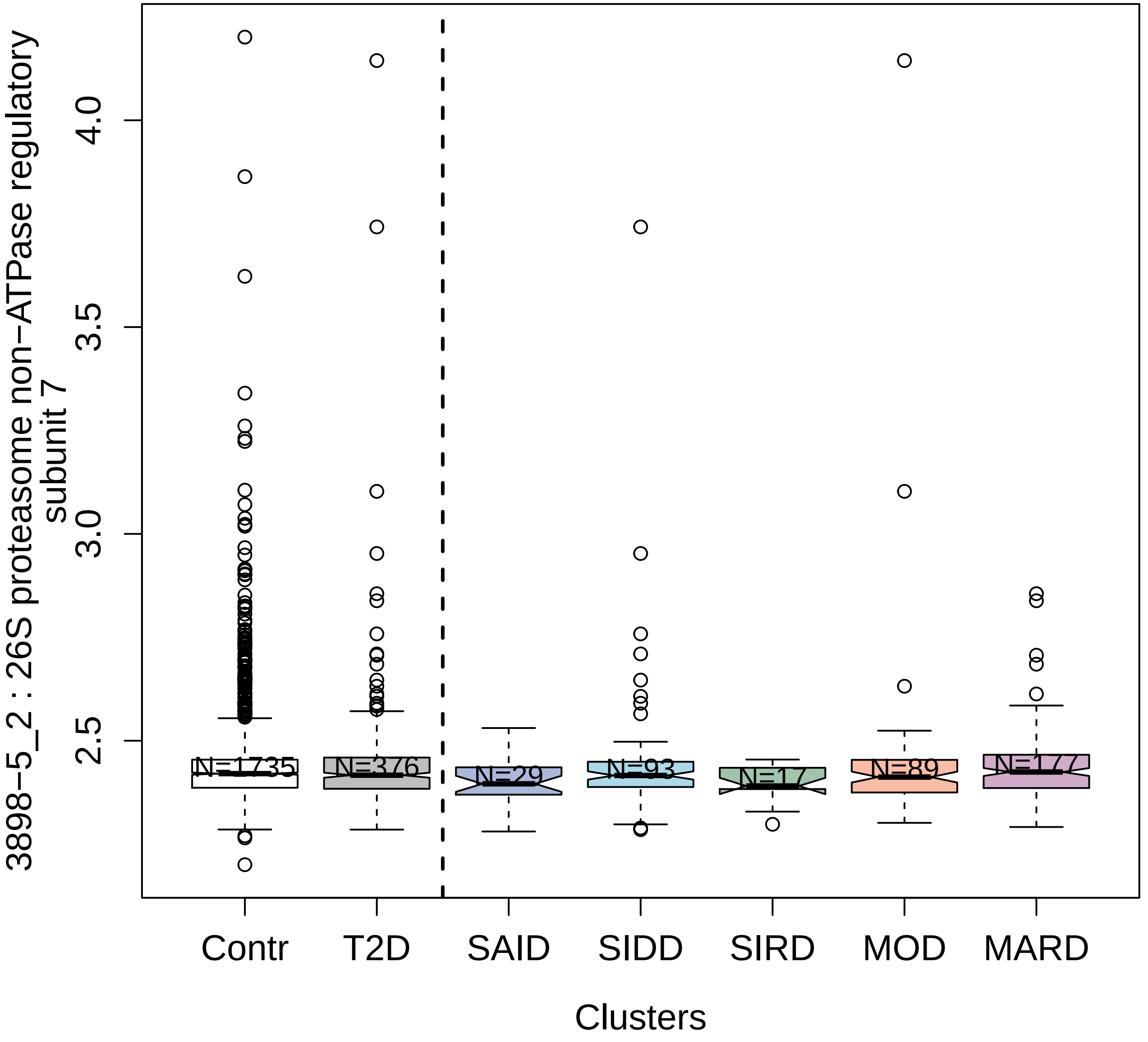
3898−5_2 : 26S proteasome non−ATPase regulatory subunit 7
4.0
3.5
3.0
2.5
N=1735
N=376
N=29
N=93
N=17
N=89
N=177
Contr
T2D
SAID
SIDD
SIRD
MOD
MARD
Clusters

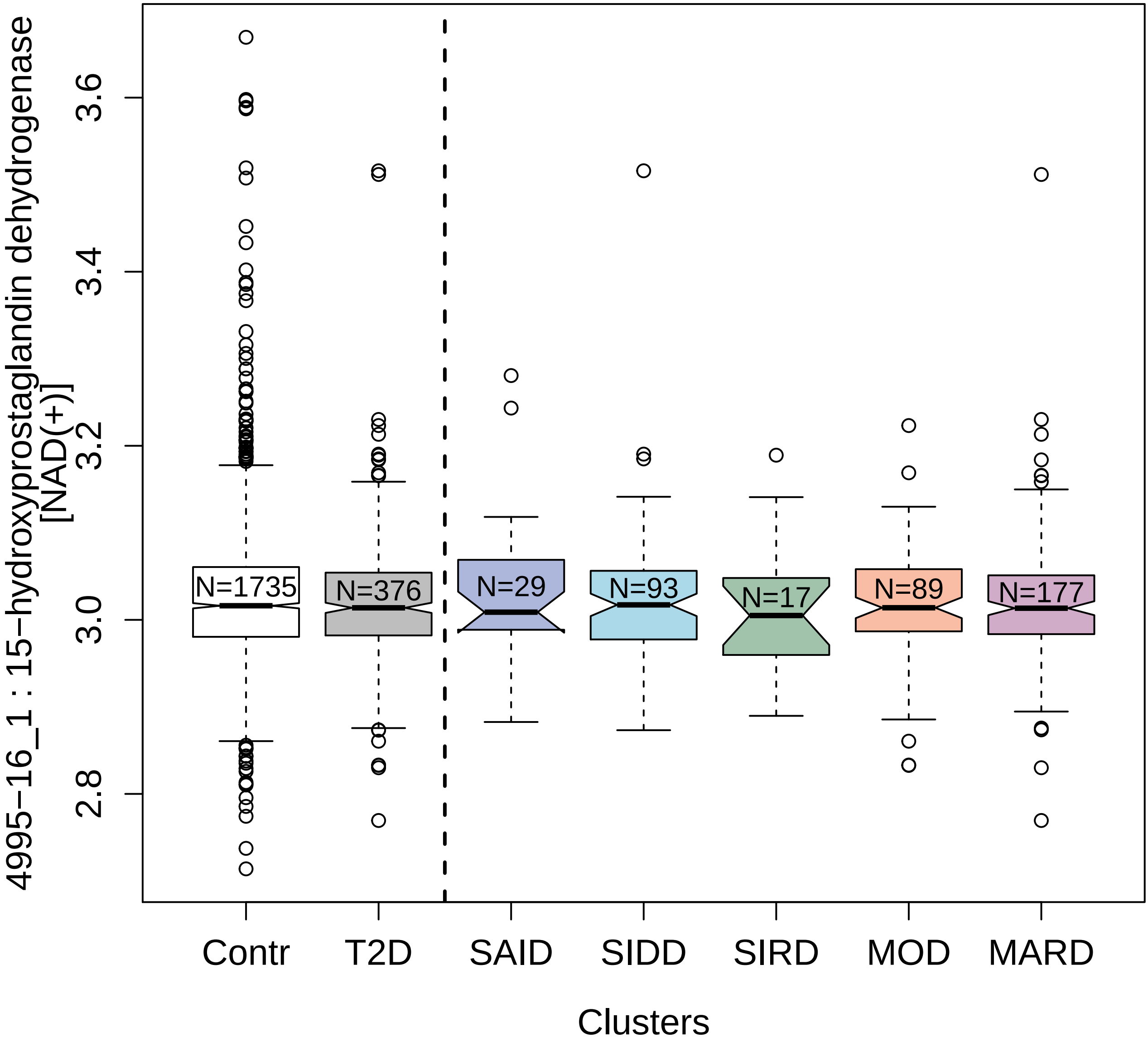

4995−16_1 : 15−hydroxyprostaglandin dehydrogenase [NAD(+)]
3.6
3.4
3.2
3.0
2.8
N=1735
N=376
N=29
N=93
N=17
N=89
N=177
Contr
T2D
SAID
SIDD
SIRD
MOD
MARD
Clusters

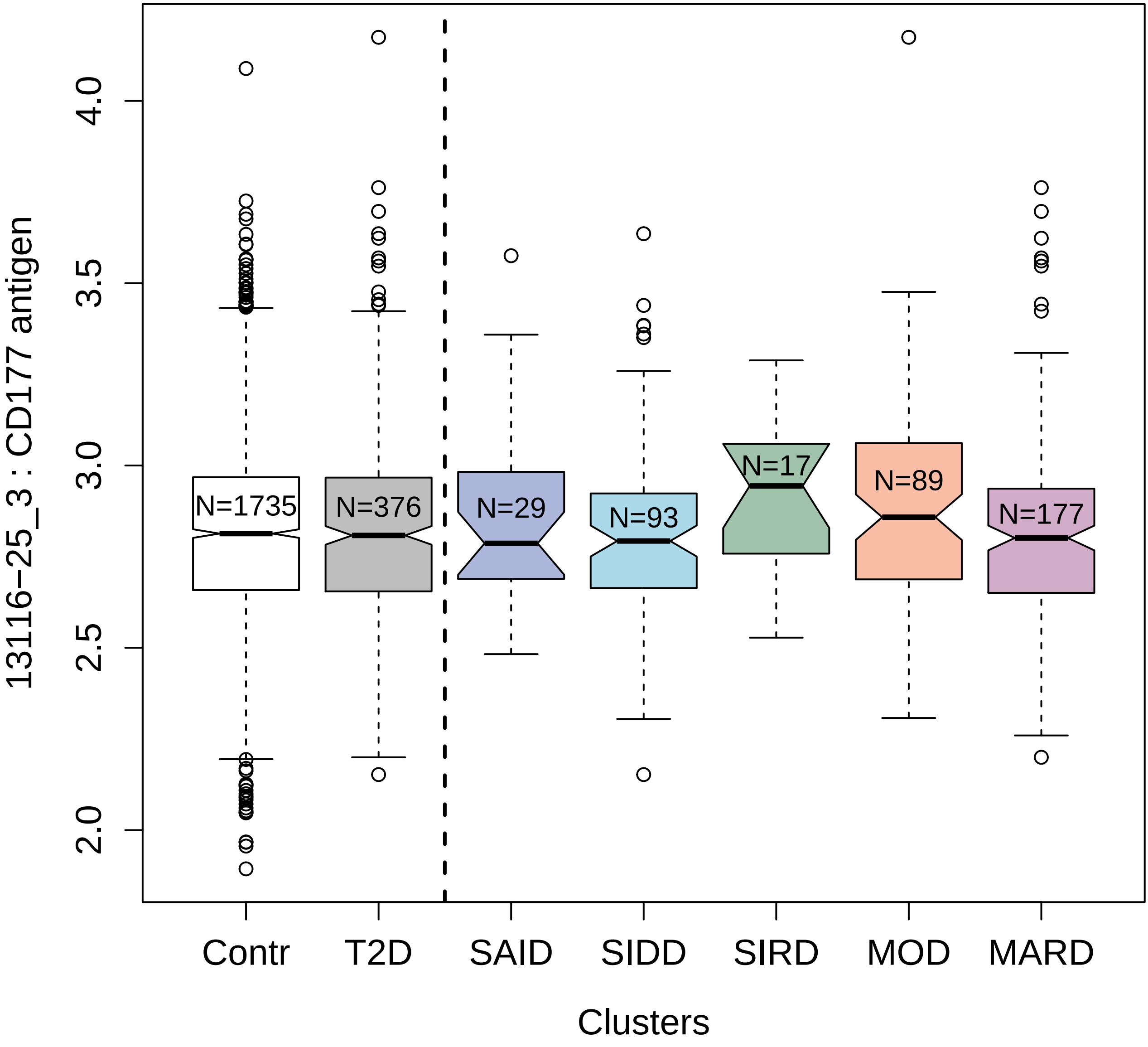

13116−25_3 : CD177 antigen
4.0
3.5
3.0
2.5
2.0
N=1735
N=376
N=29
N=93
N=17
N=89
N=177
Contr
T2D
SAID
SIDD
SIRD
MOD
MARD
Clusters

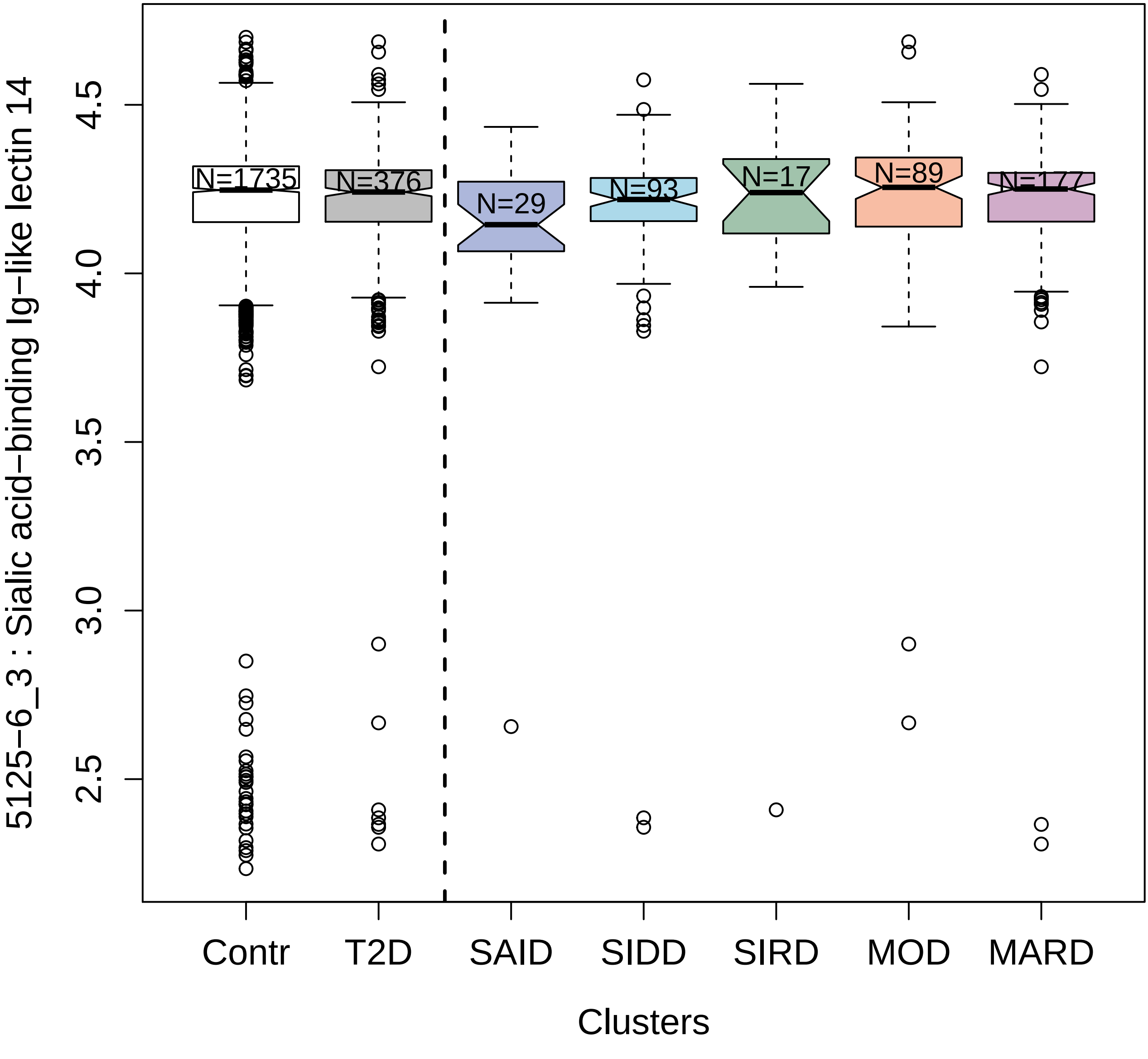

5125−6_3 : Sialic acid−binding Ig−like lectin 14
4.5
4.0
3.5
3.0
2.5
N=1735
N=376
N=29
N=93
N=17
N=89
N=177
Contr
T2D
SAID
SIDD
SIRD
MOD
MARD
Clusters

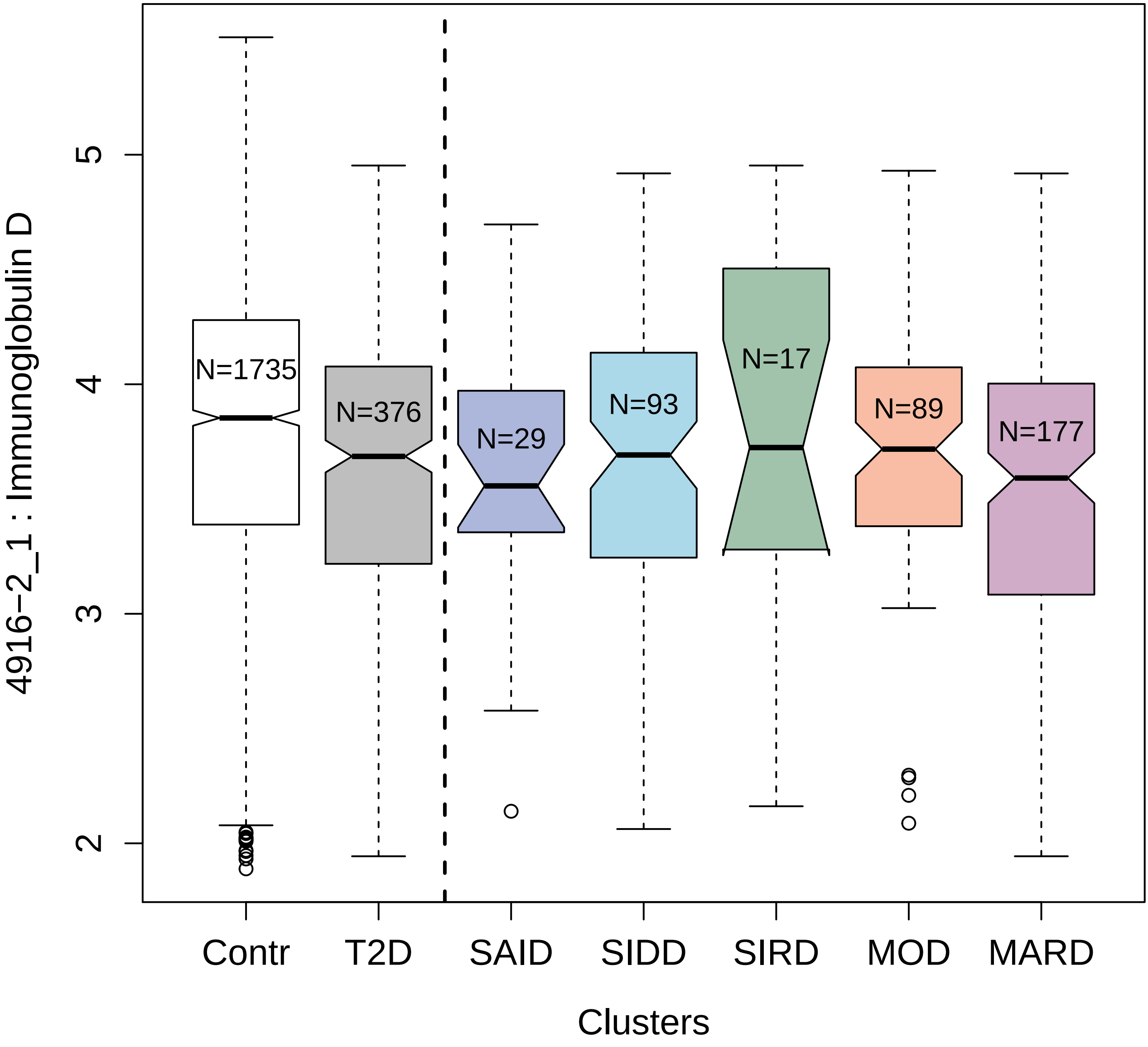

4916−2_1 : Immunoglobulin D
5
4
3
2
N=1735
N=376
N=29
N=93
N=17
N=89
N=177
Contr
T2D
SAID
SIDD
SIRD
MOD
MARD
Clusters

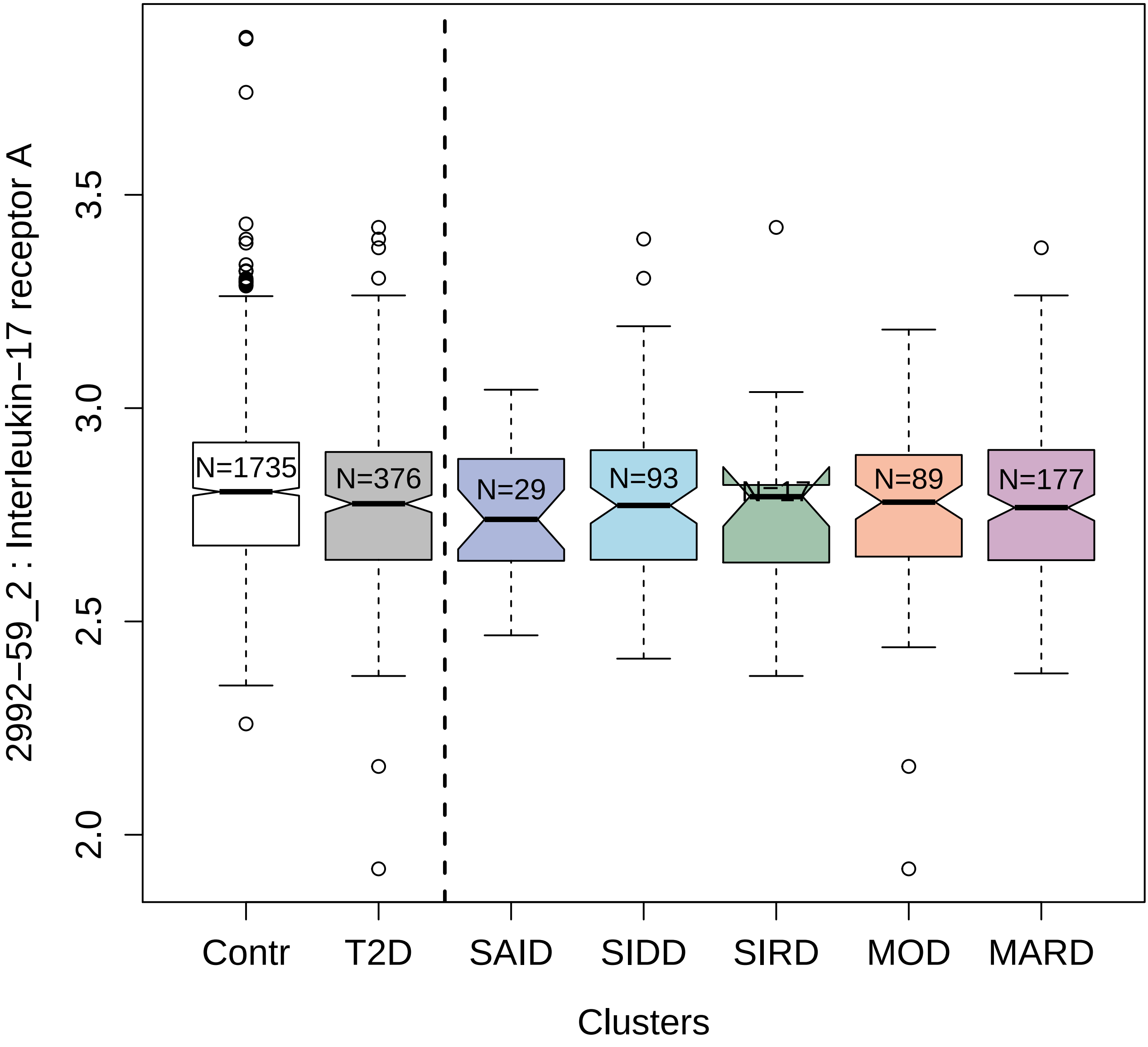

2992−59_2 : Interleukin−17 receptor A
3.5
3.0
2.5
2.0
N=1735
N=376
N=29
N=93
N=17
N=89
N=177
Contr
T2D
SAID
SIDD
SIRD
MOD
MARD
Clusters

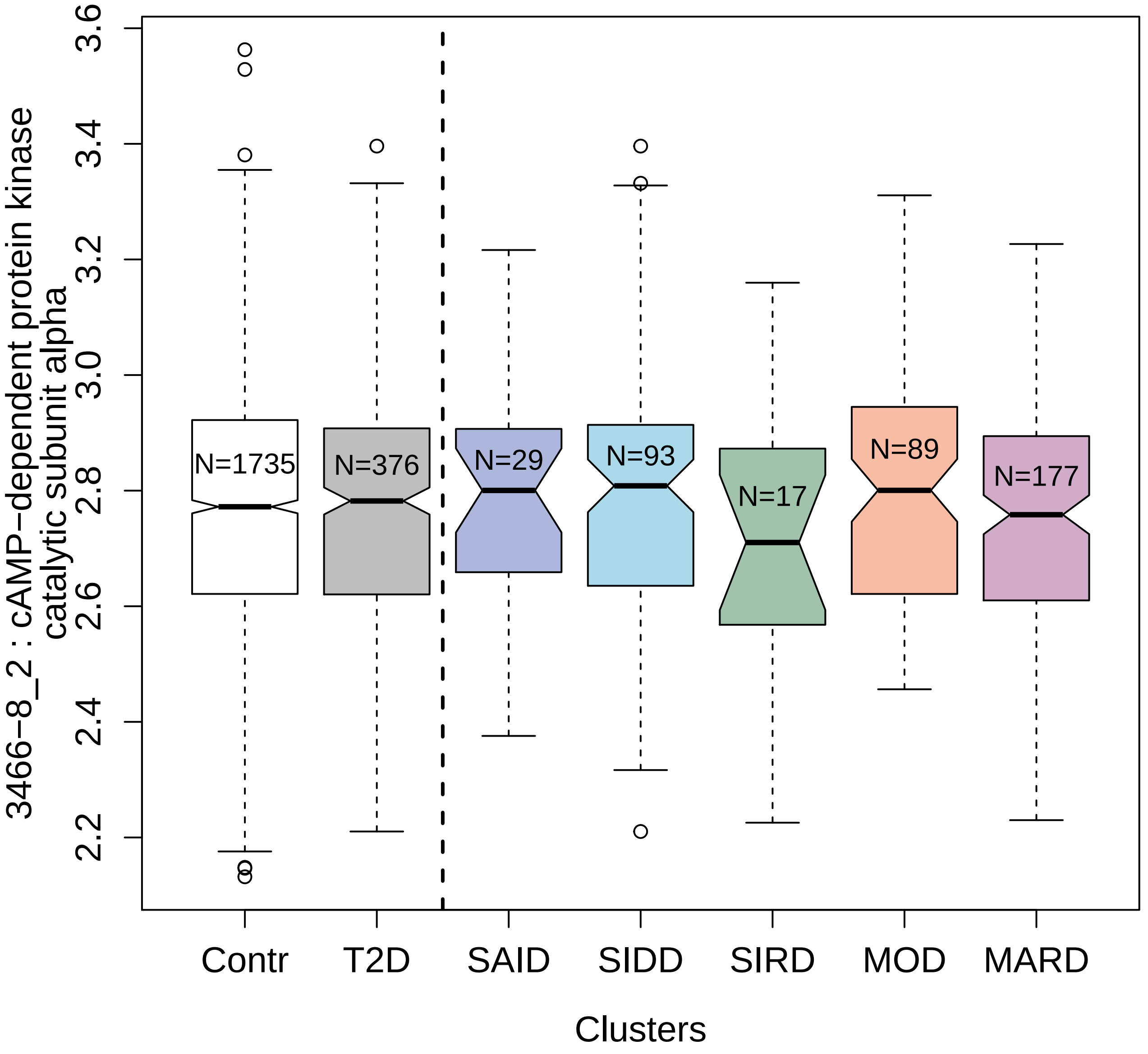

3466−8_2 : cAMP−dependent protein kinase
catalytic subunit alpha
3.6
3.4
3.2
3.0
2.8
2.6
2.4
2.2
N=1735
N=376
N=29
N=93
N=17
N=89
N=177
Contr
T2D
SAID
SIDD
SIRD
MOD
MARD
Clusters

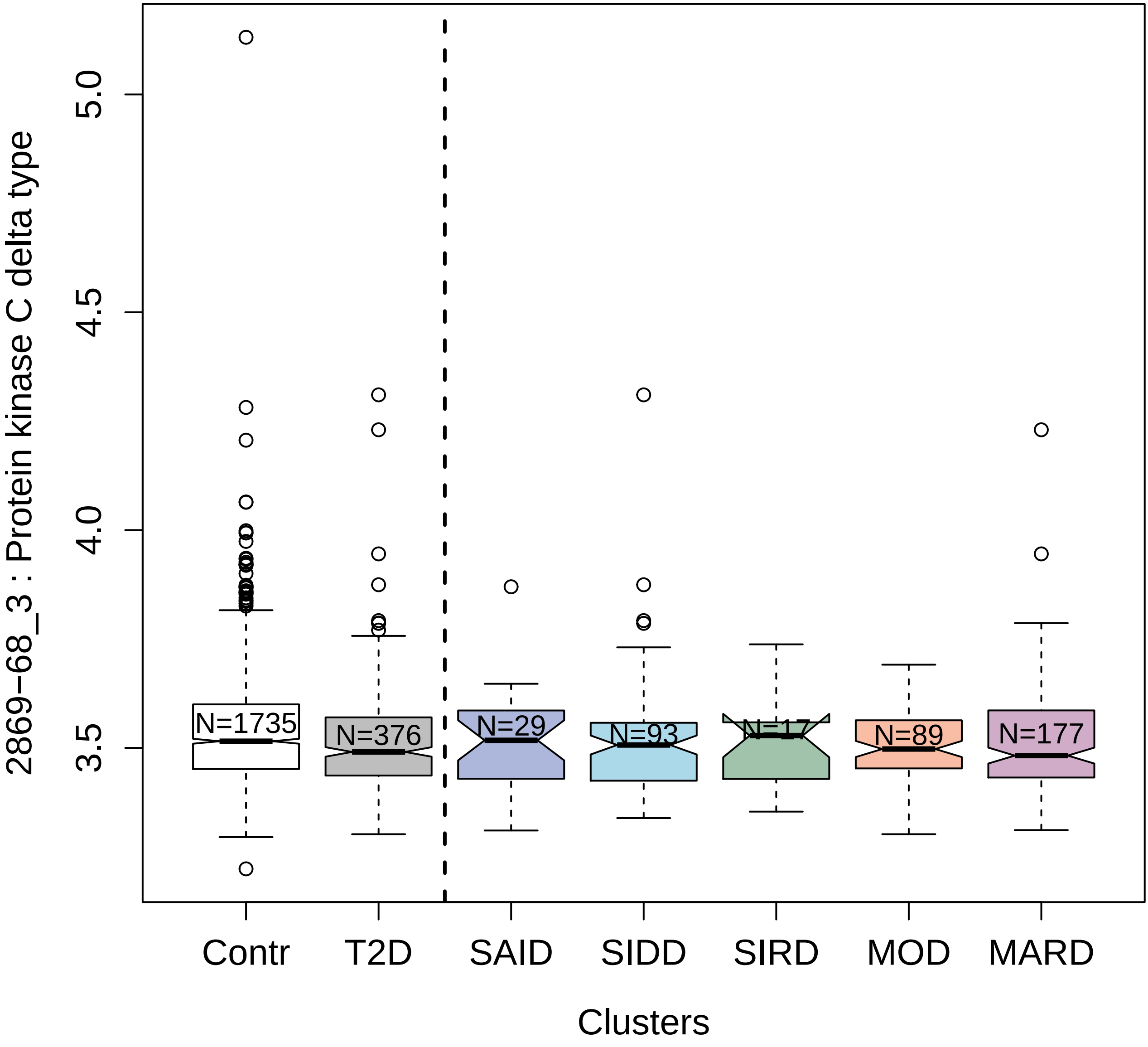

2869−68_3 : Protein kinase C delta type
5.0
4.5
4.0
3.5
N=1735
N=376
N=29
N=93
N=17
N=89
N=177
Contr
T2D
SAID
SIDD
SIRD
MOD
MARD
Clusters

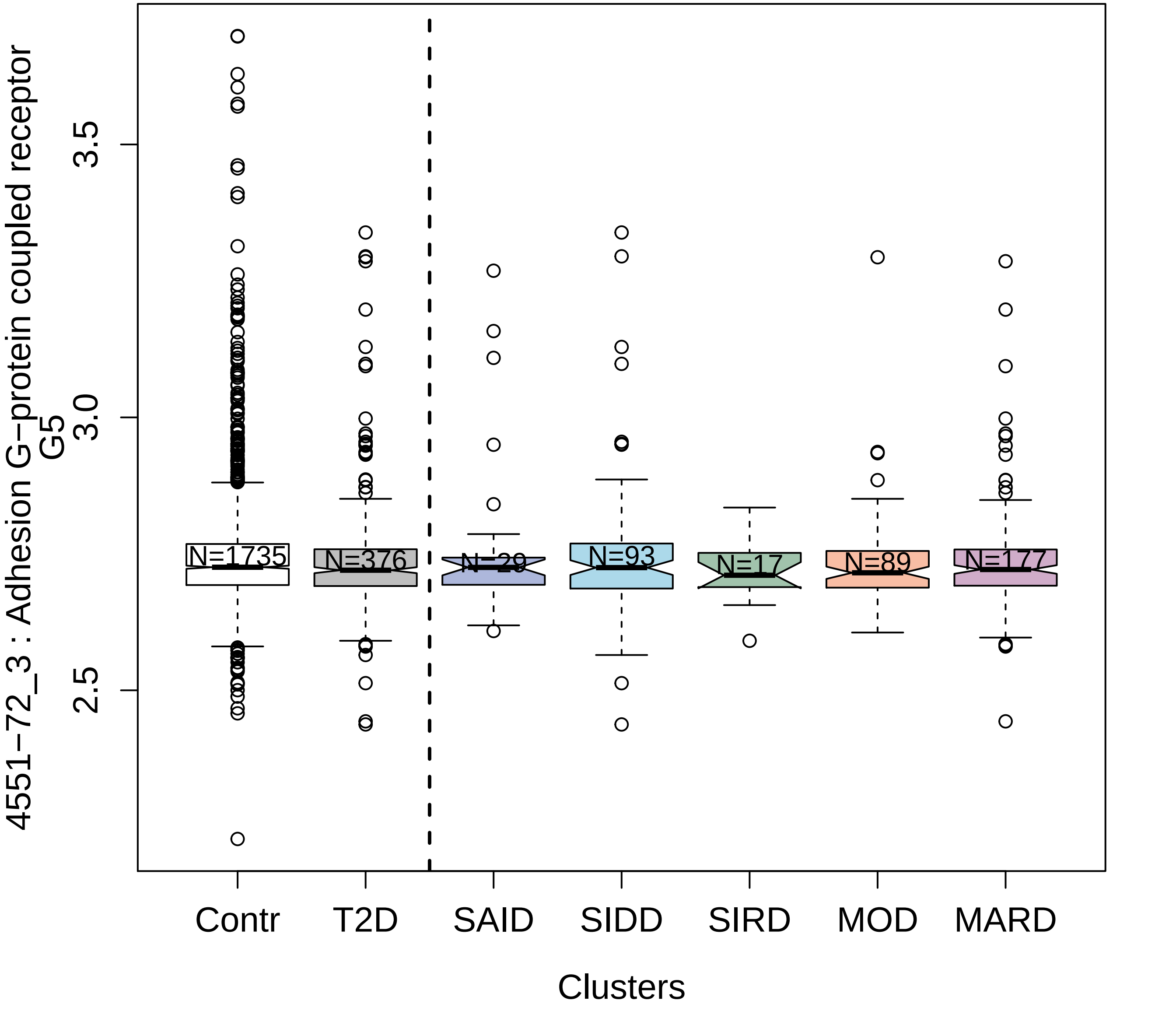
4551−72_3 : Adhesion G−protein coupled receptor G5
3.5
3.0
2.5
N=1735
N=376
N=29
N=93
N=17
N=89
N=177
Contr
T2D
SAID
SIDD
SIRD
MOD
MARD
Clusters

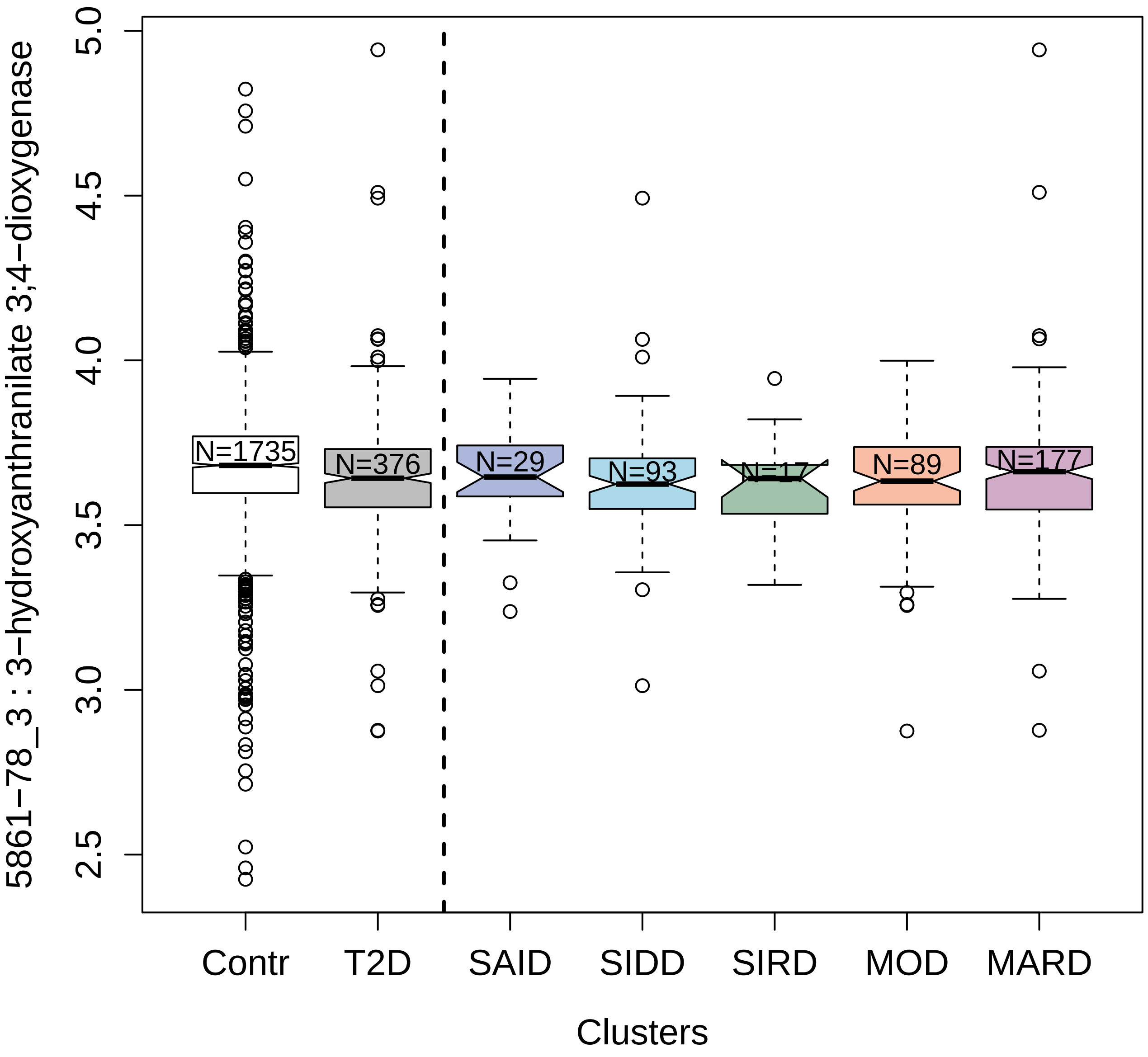

5861−78_3 : 3−hydroxyanthranilate 3;4−dioxygenase
5.0
4.5
4.0
3.5
3.0
2.5
N=1735
N=376
N=29
N=93
N=17
N=89
N=177
Contr
T2D
SAID
SIDD
SIRD
MOD
MARD
Clusters

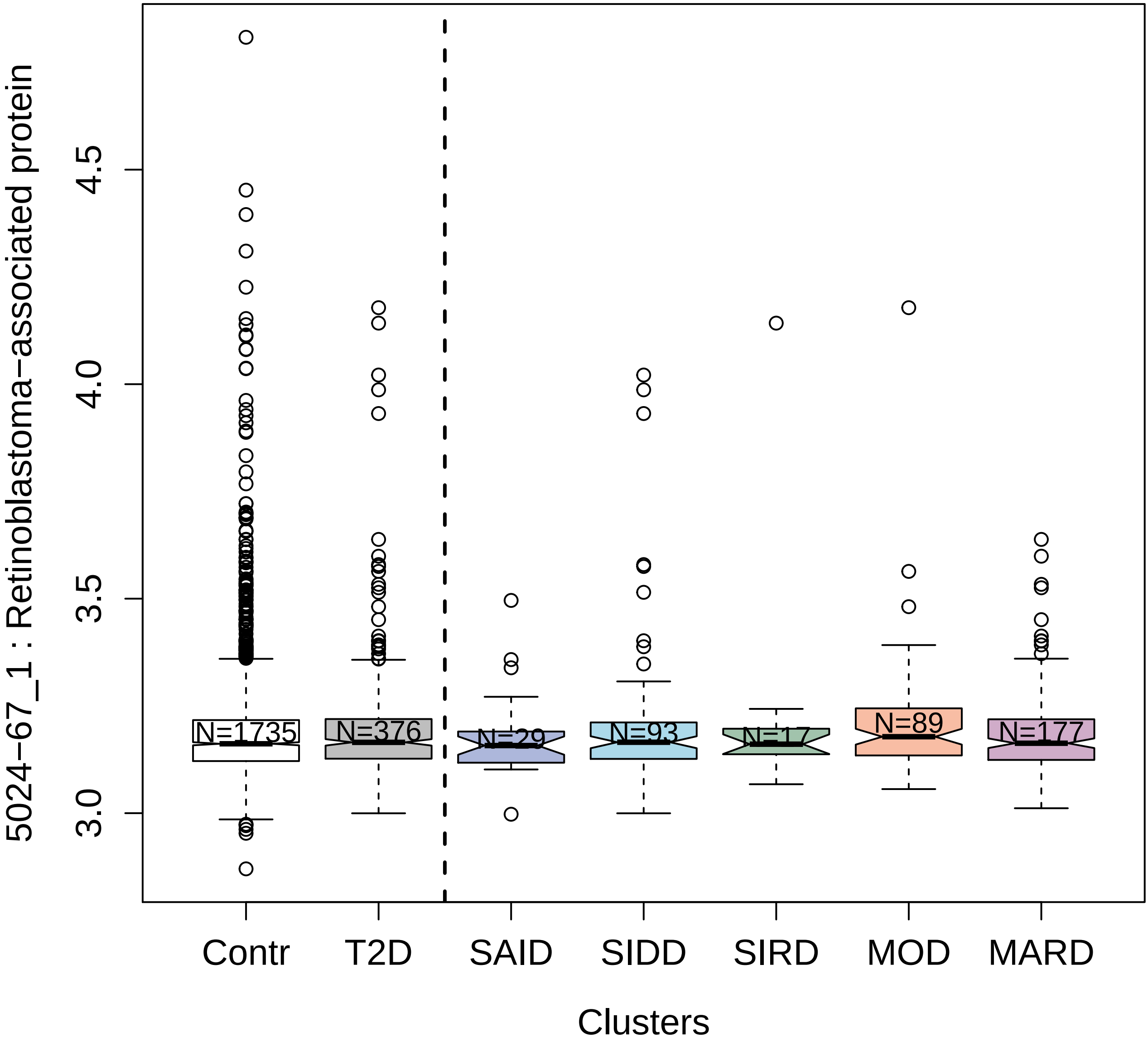
5024−67_1 : Retinoblastoma−associated protein
3.0
3.5
4.0
4.5
N=1735
N=376
N=29
N=93
N=17
N=89
N=177
Contr
T2D
SAID
SIDD
SIRD
MOD
MARD
Clusters

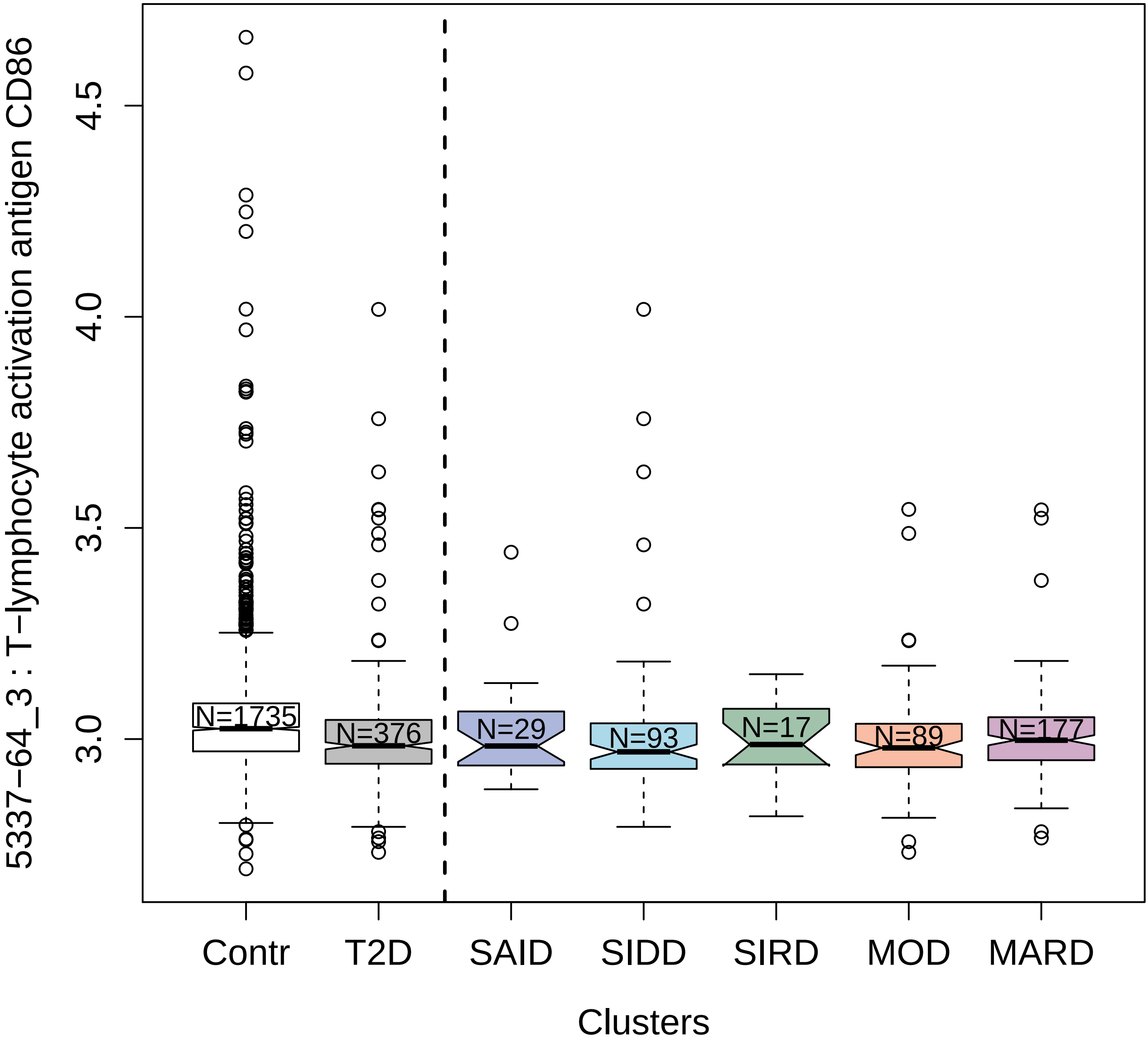

5337−64_3 : T−lymphocyte activation antigen CD86
4.5
4.0
3.5
3.0
N=1735
N=376
N=29
N=93
N=17
N=89
N=177
Contr
T2D
SAID
SIDD
SIRD
MOD
MARD
Clusters

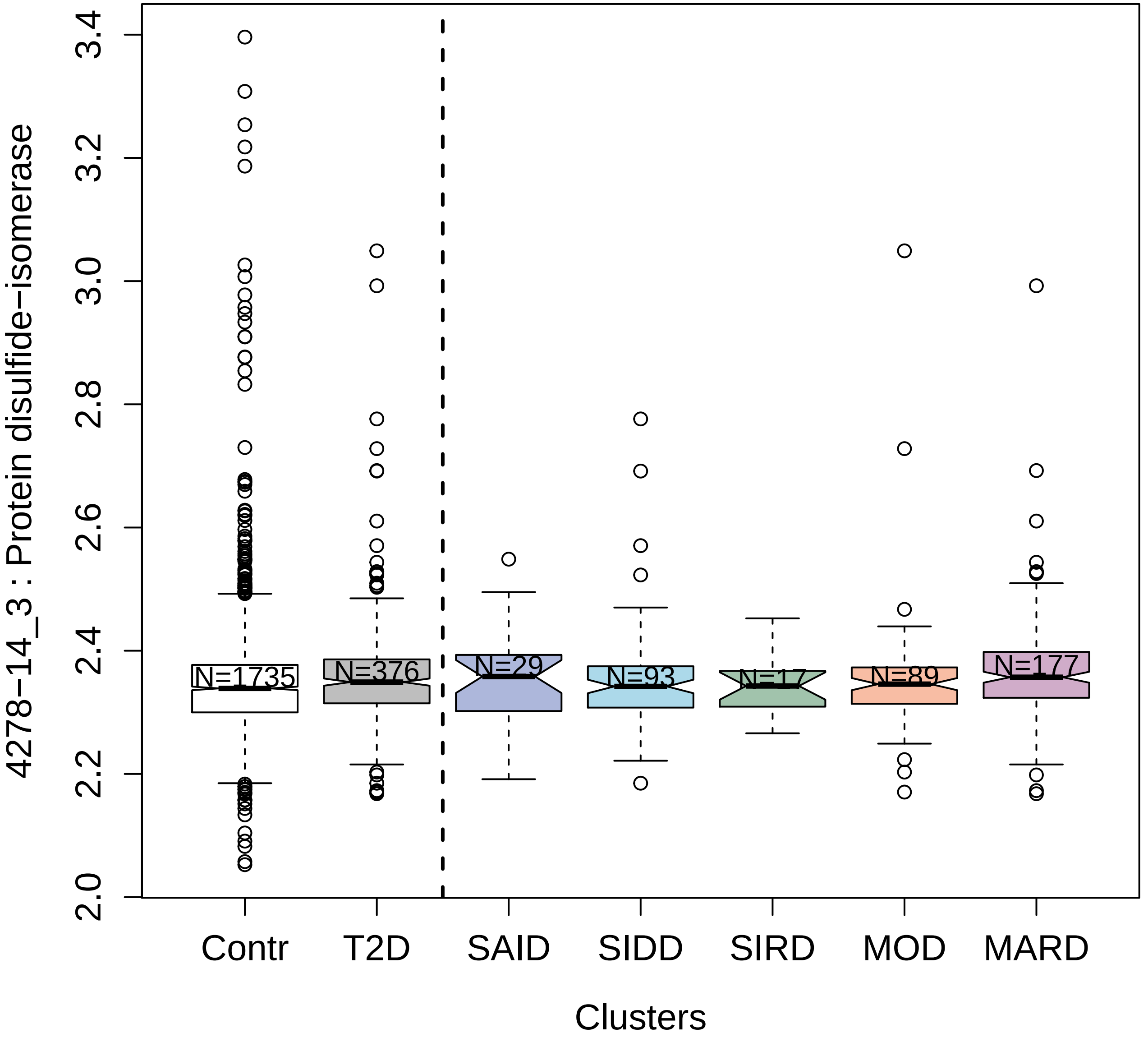

4278−14_3 : Protein disulfide−isomerase
2.0
2.2
2.4
2.6
2.8
3.0
3.2
3.4
N=1735
N=376
N=29
N=93
N=17
N=89
N=177
Contr
T2D
SAID
SIDD
SIRD
MOD
MARD
Clusters

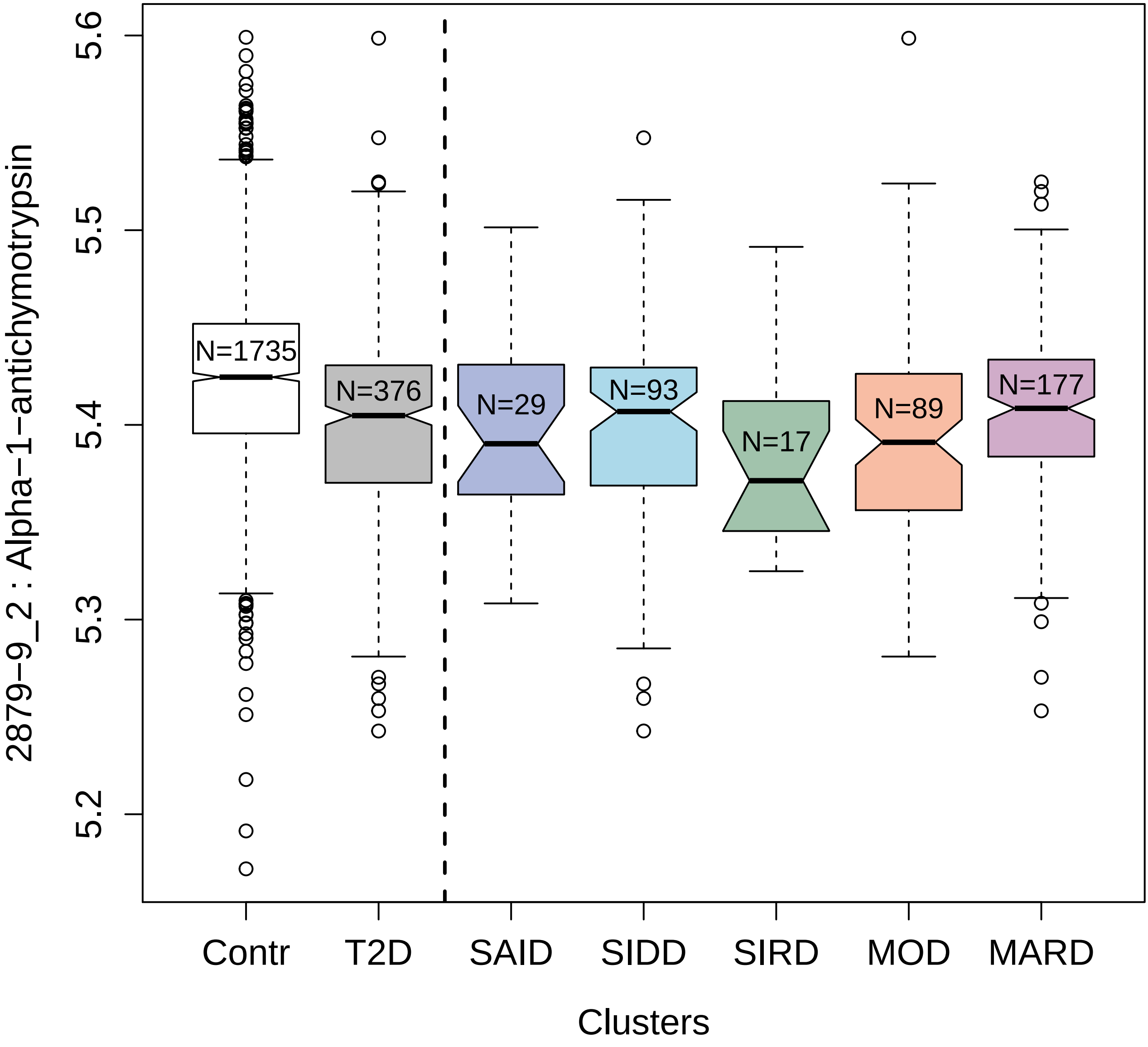
2879−9_2 : Alpha−1−antichymotrypsin
5.2
5.3
5.4
5.5
5.6
N=1735
N=376
N=29
N=93
N=17
N=89
N=177
Contr
T2D
SAID
SIDD
SIRD
MOD
MARD
Clusters

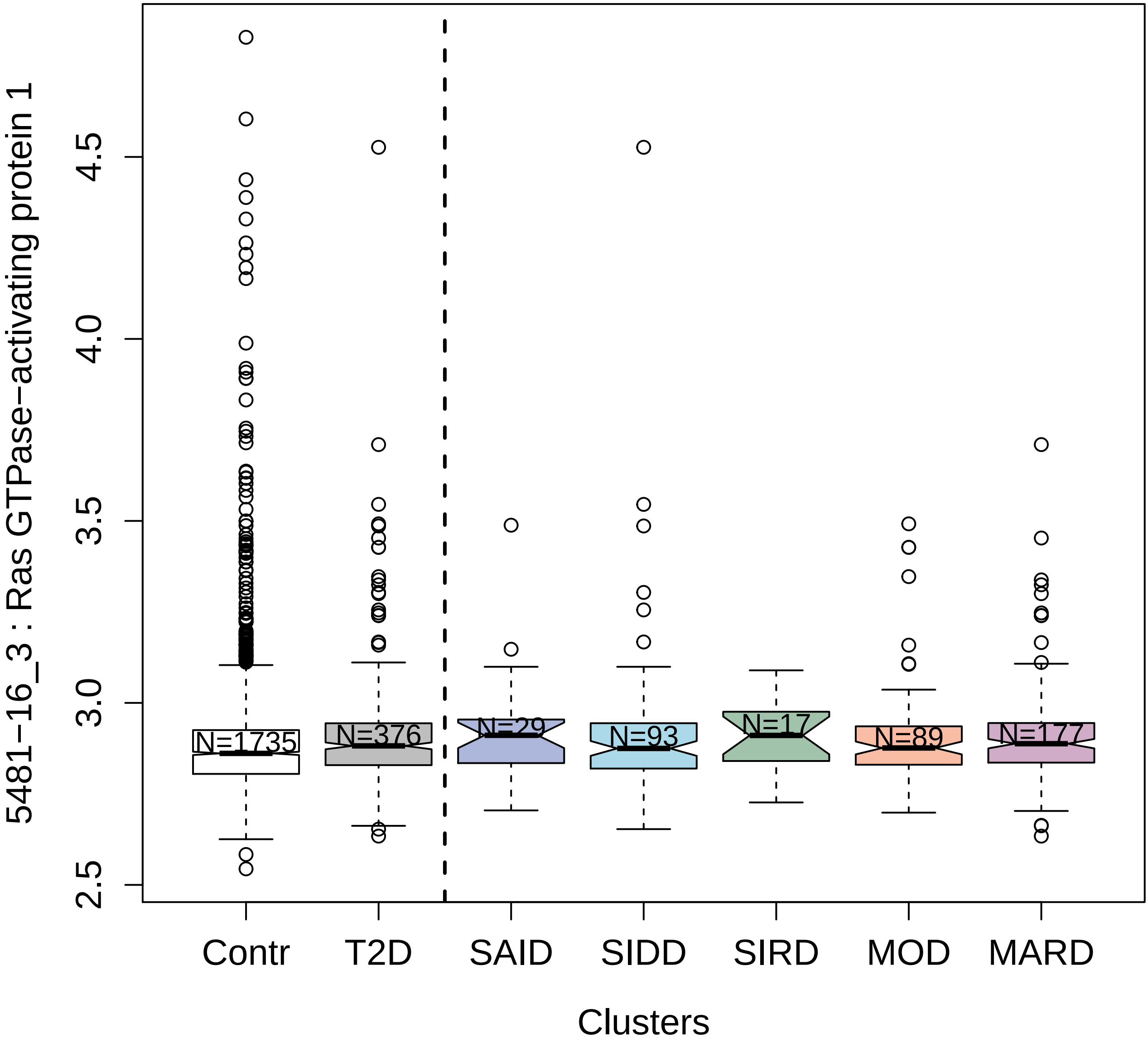

5481−16_3 : Ras GTPase−activating protein 1
2.5
3.0
3.5
4.0
4.5
N=1735
N=376
N=29
N=93
N=17
N=89
N=177
Contr
T2D
SAID
SIDD
SIRD
MOD
MARD
Clusters

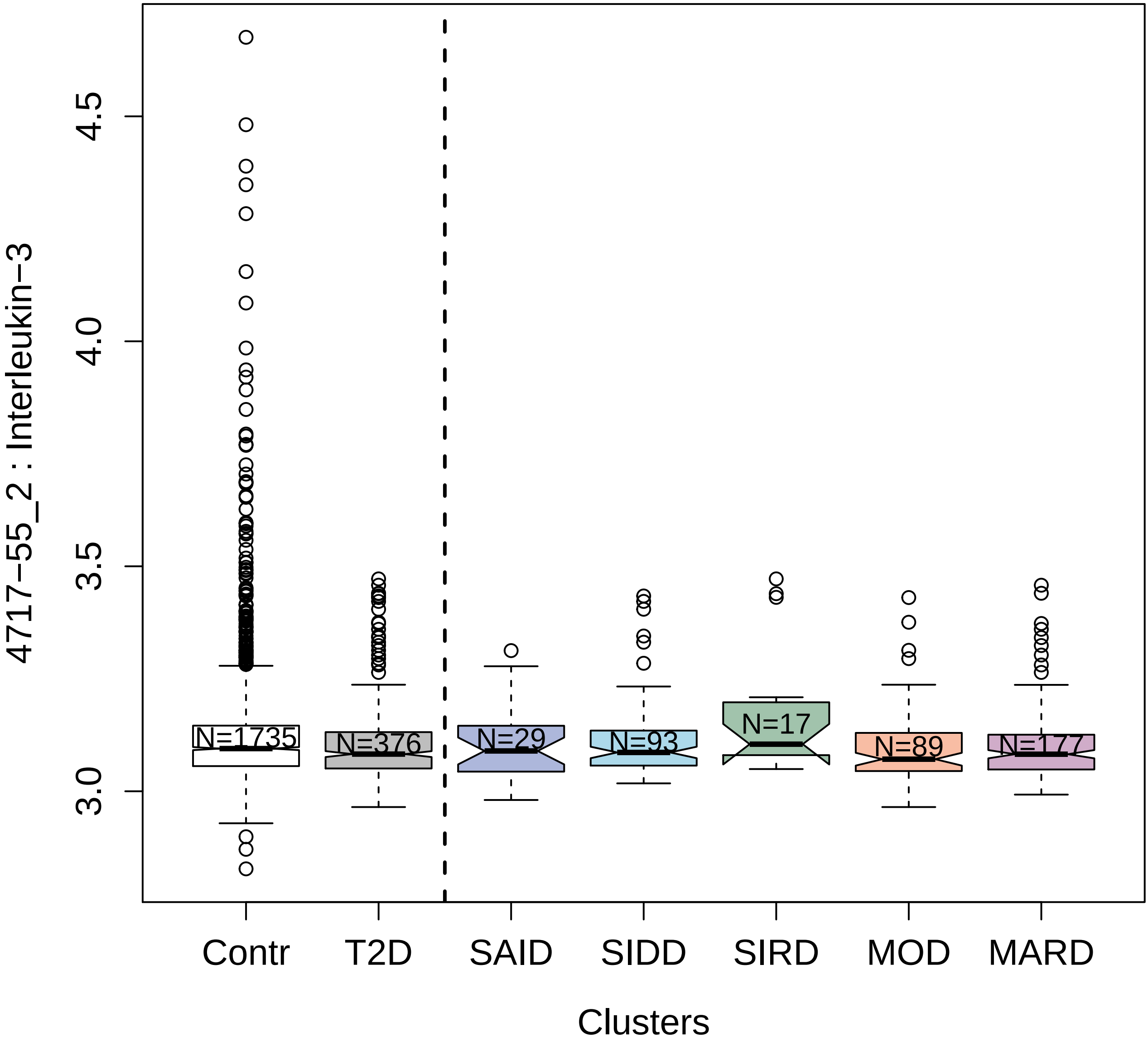

4717−55_2 : Interleukin−3
3.0
3.5
4.0
4.5
N=1735
N=376
N=29
N=93
N=17
N=89
N=177
Contr
T2D
SAID
SIDD
SIRD
MOD
MARD
Clusters

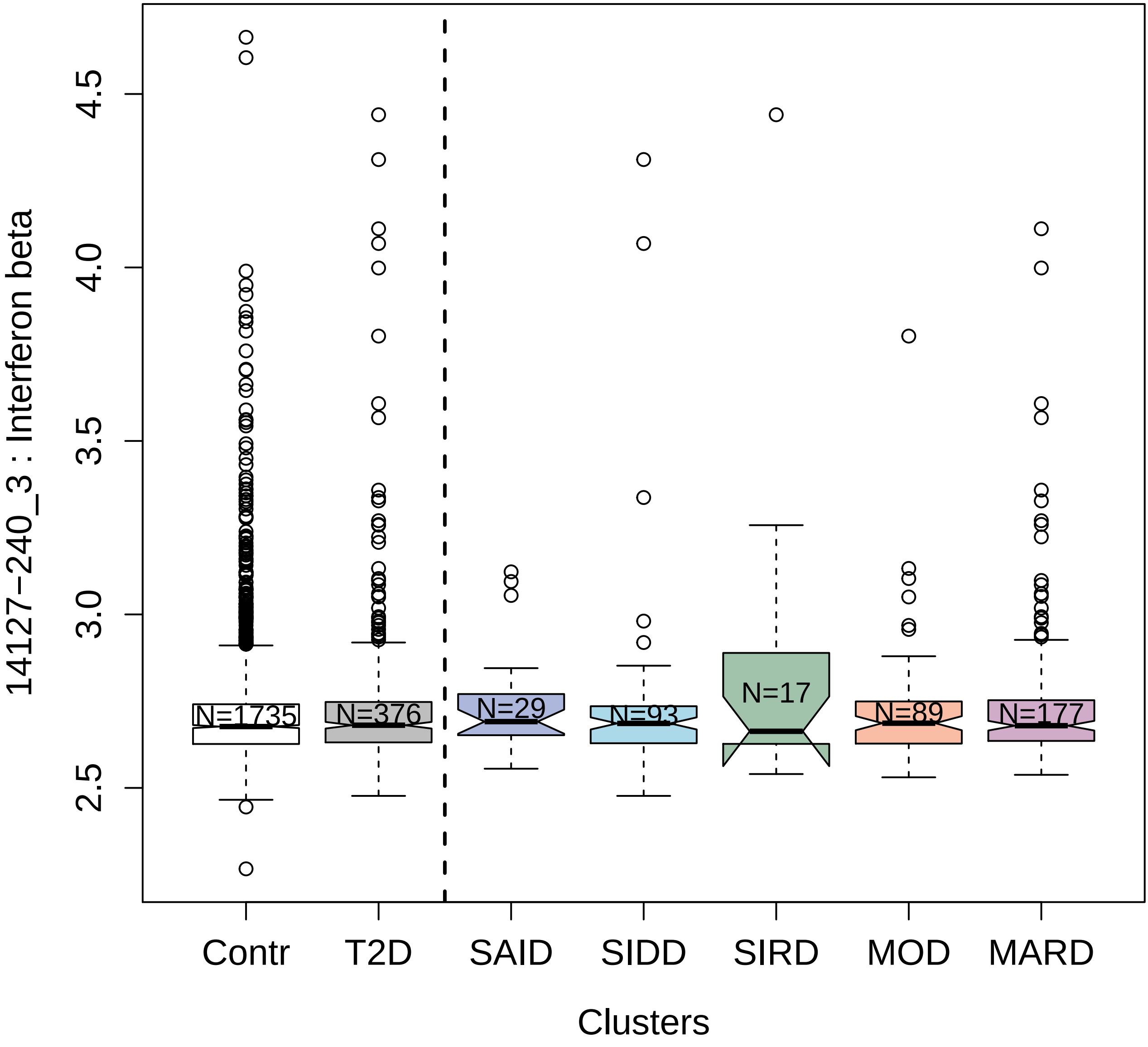

14127−240_3 : Interferon beta
4.5
4.0
3.5
3.0
2.5
N=1735
N=376
N=29
N=93
N=17
N=89
N=177
Contr
T2D
SAID
SIDD
SIRD
MOD
MARD
Clusters

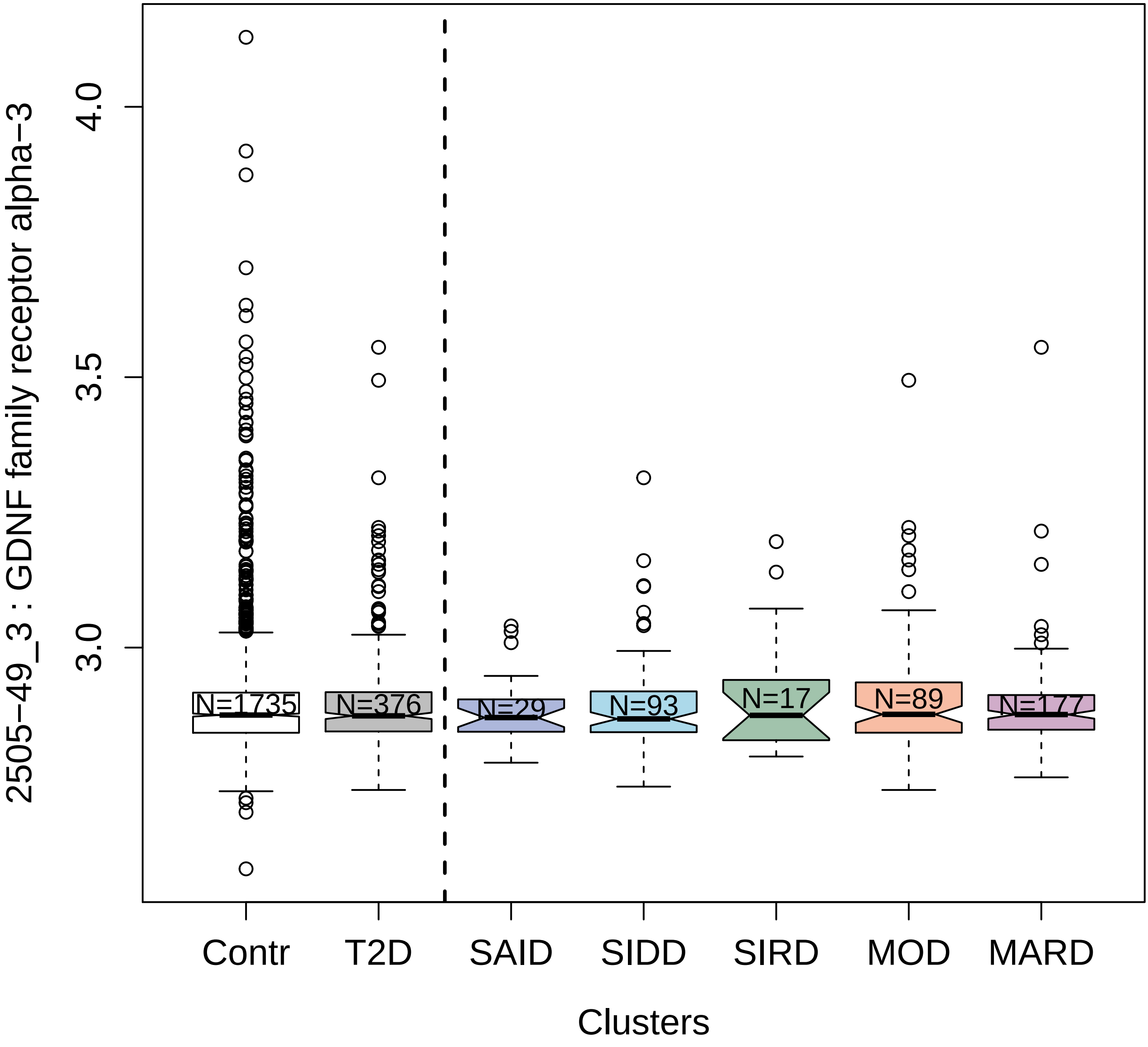

2505−49_3 : GDNF family receptor alpha−3
4.0
3.5
3.0
N=1735
N=376
N=29
N=93
N=17
N=89
N=177
Contr
T2D
SAID
SIDD
SIRD
MOD
MARD
Clusters

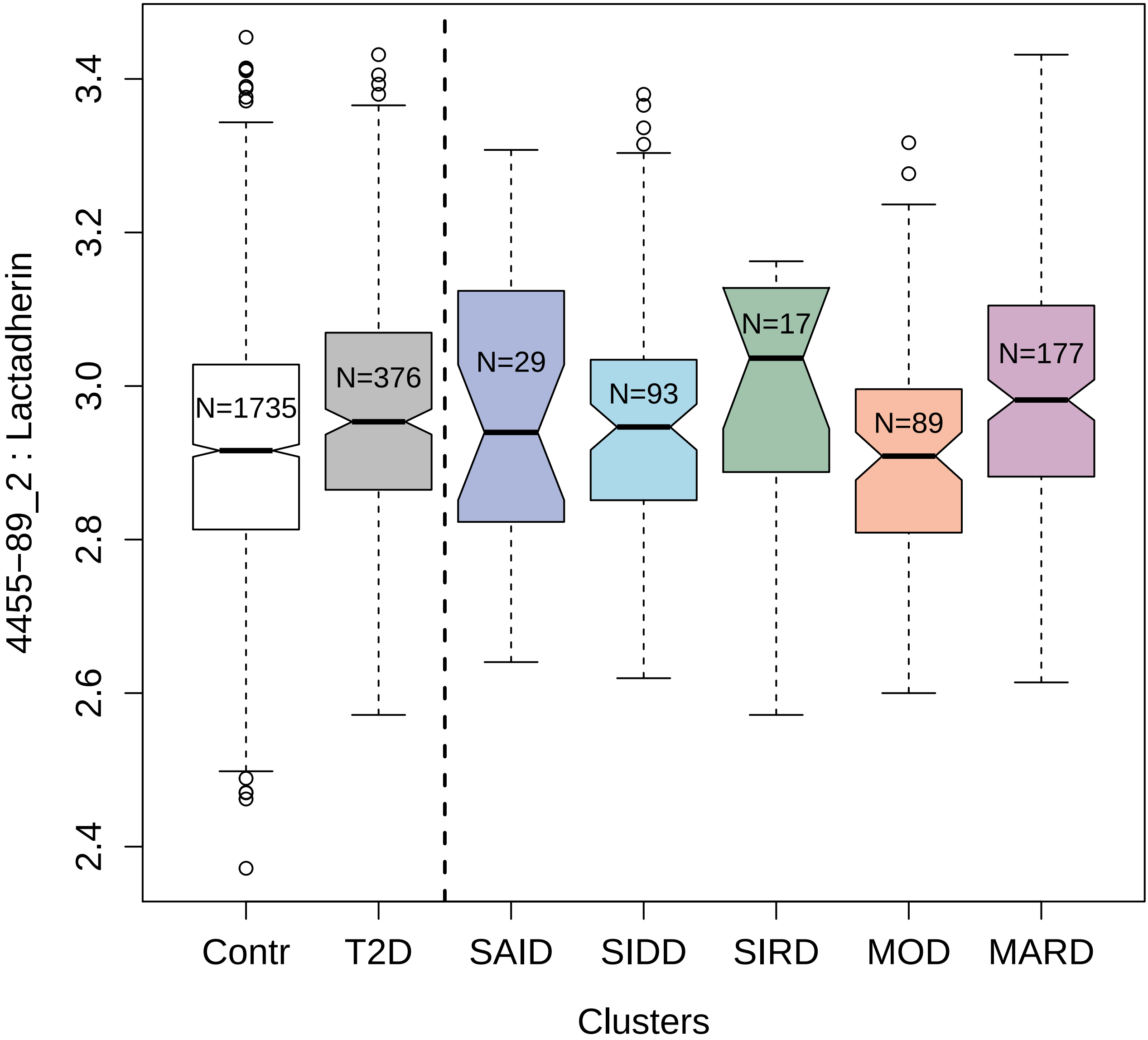

4455−89_2 : Lactadherin
3.4
3.2
3.0
2.8
2.6
2.4
N=1735
N=376
N=29
N=93
N=17
N=89
N=177
Contr
T2D
SAID
SIDD
SIRD
MOD
MARD
Clusters

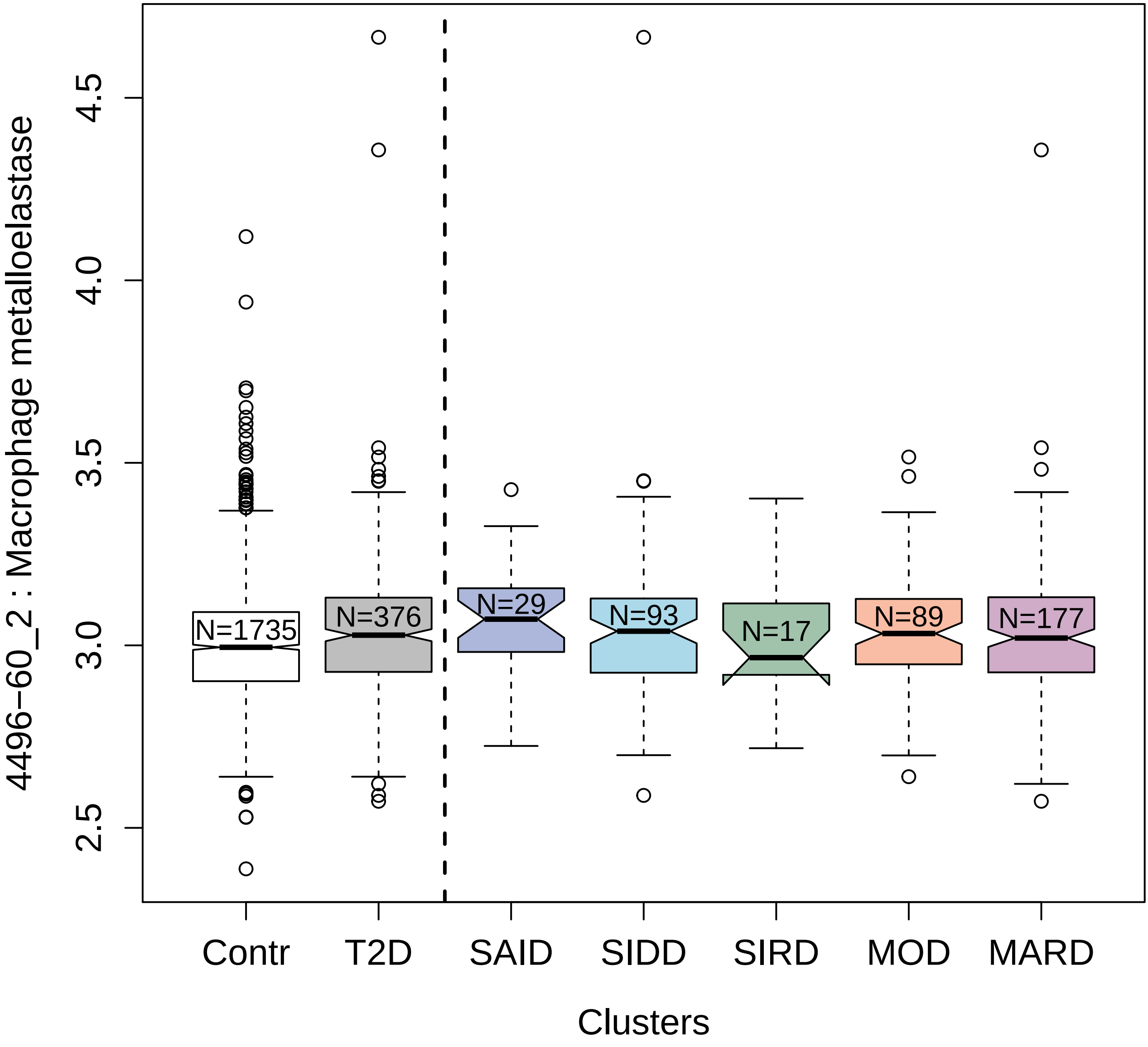
4496−60_2 : Macrophage metalloelastase
2.5
3.0
3.5
4.0
4.5
N=1735
N=376
N=29
N=93
N=17
N=89
N=177
Contr
T2D
SAID
SIDD
SIRD
MOD
MARD
Clusters

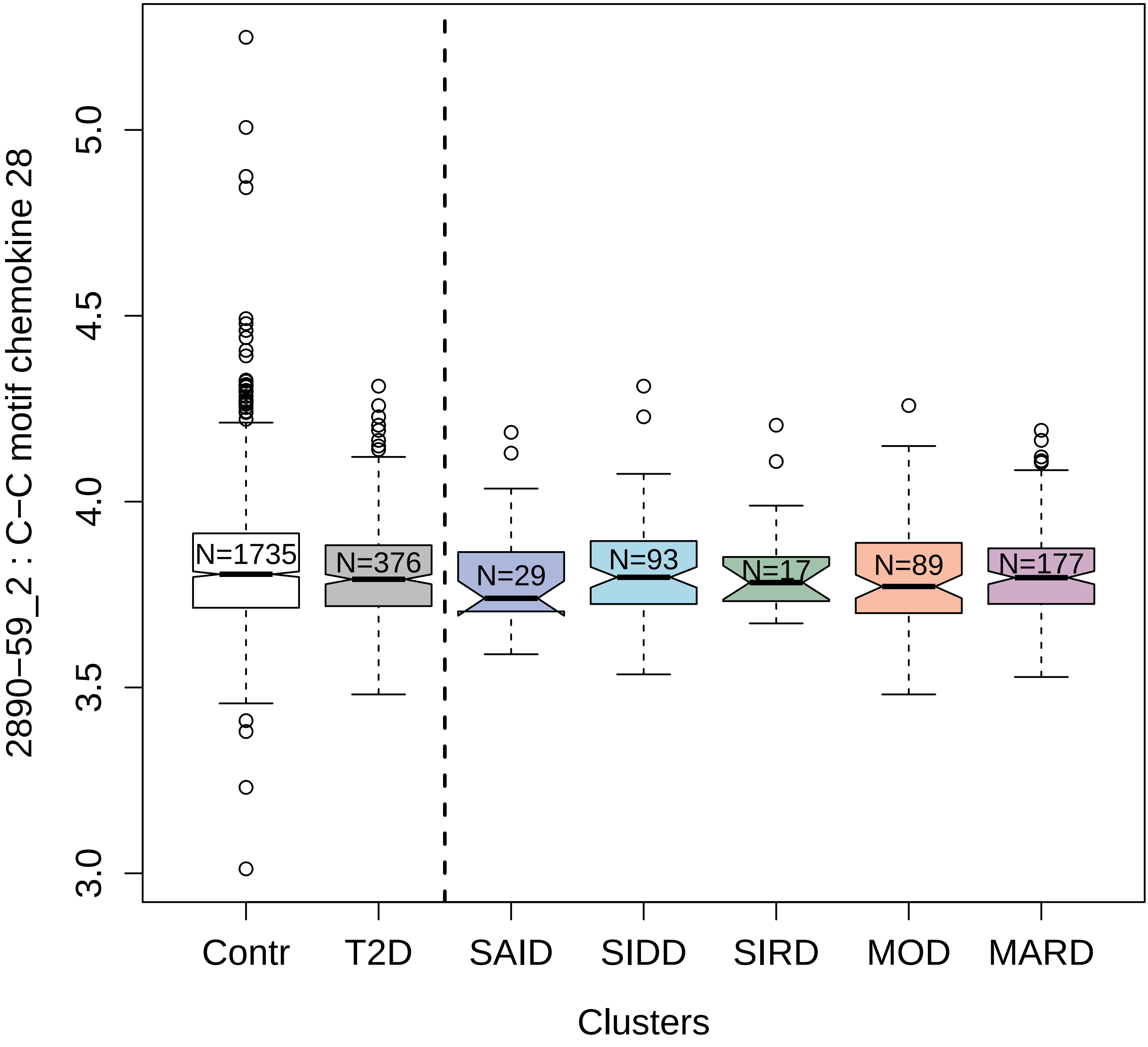

2890−59_2 : C−C motif chemokine 28
5.0
4.5
4.0
3.5
3.0
N=1735
N=376
N=29
N=93
N=17
N=89
N=177
Contr
T2D
SAID
SIDD
SIRD
MOD
MARD
Clusters

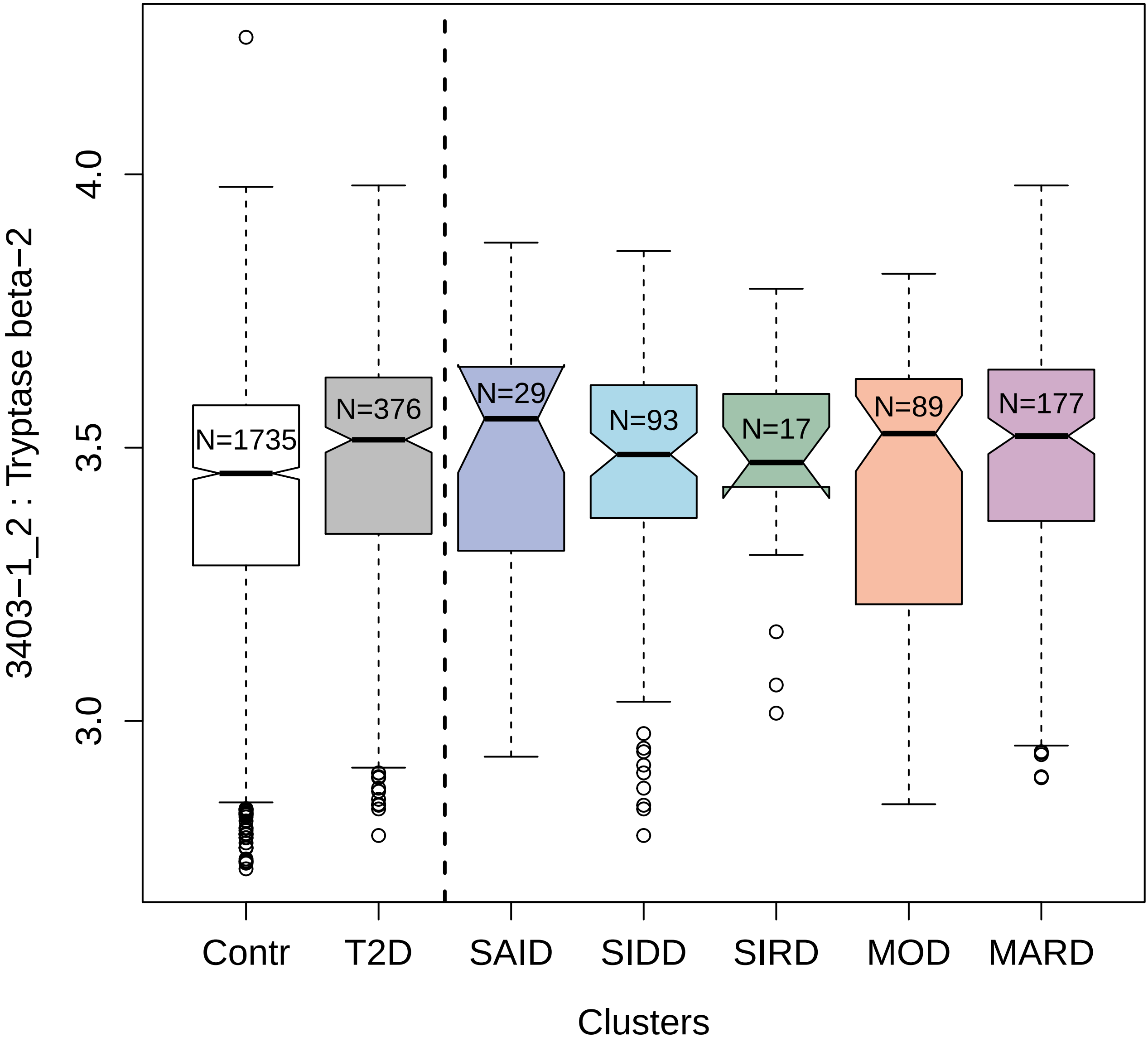
3403−1_2 : Tryptase beta−2
4.0
3.5
3.0
N=1735
N=376
N=29
N=93
N=17
N=89
N=177
Contr
T2D
SAID
SIDD
SIRD
MOD
MARD
Clusters

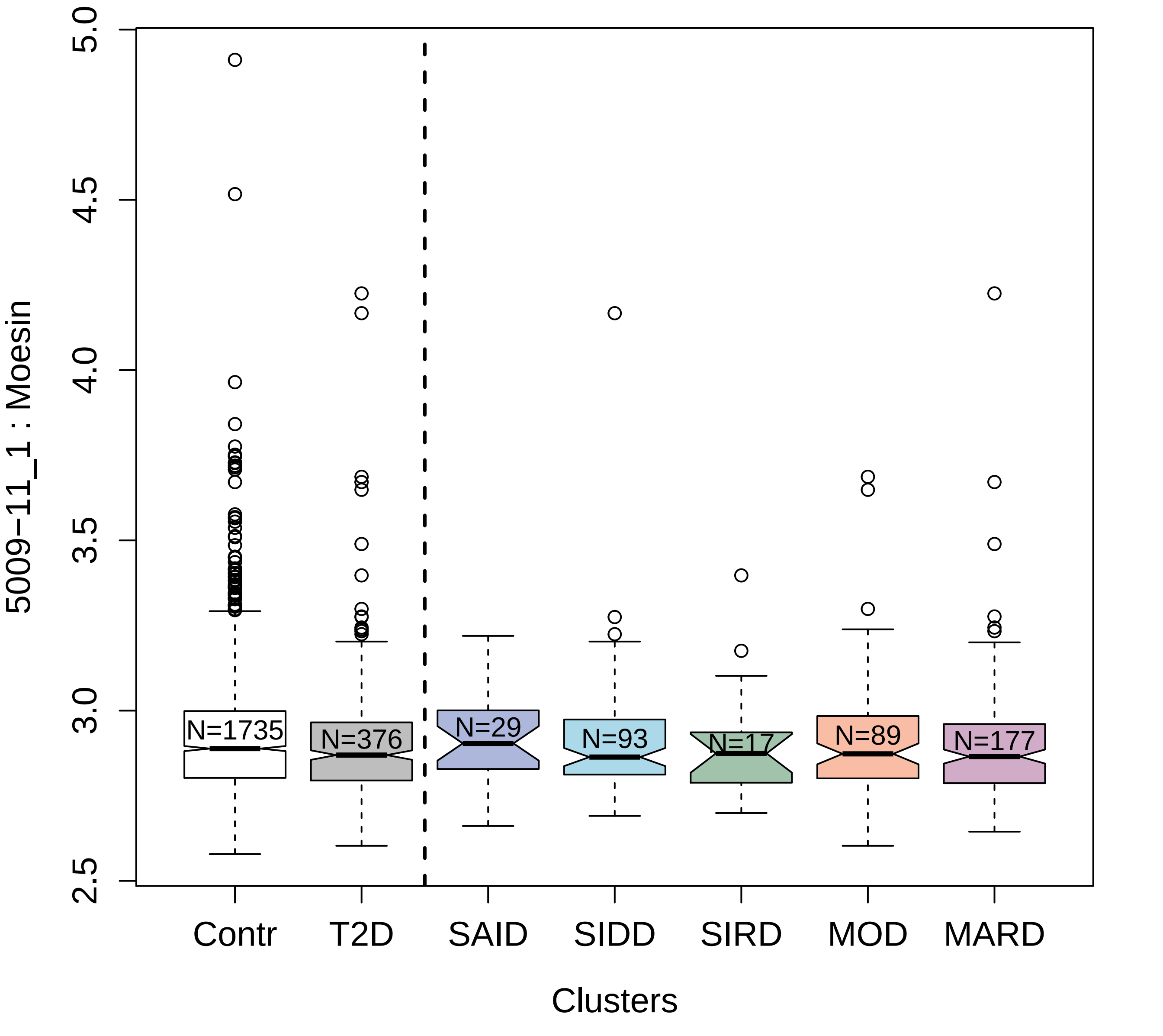
5009−11_1 : Moesin
5.0
4.5
4.0
3.5
3.0
2.5
N=1735
N=376
N=29
N=93
N=17
N=89
N=177
Contr
T2D
SAID
SIDD
SIRD
MOD
MARD
Clusters

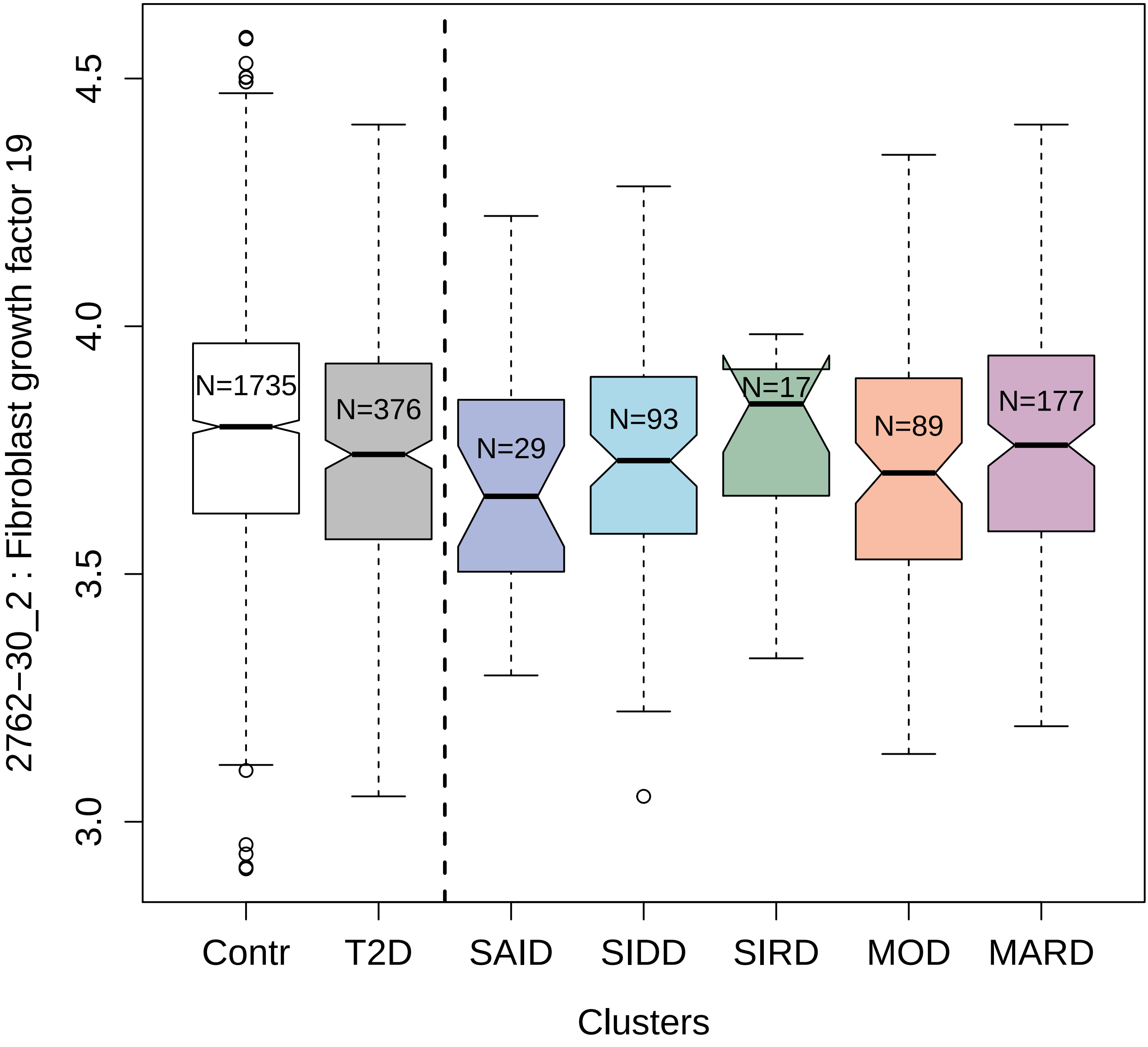

2762−30_2 : Fibroblast growth factor 19
3.0
3.5
4.0
4.5
N=1735
N=376
N=29
N=93
N=17
N=89
N=177
Contr
T2D
SAID
SIDD
SIRD
MOD
MARD
Clusters

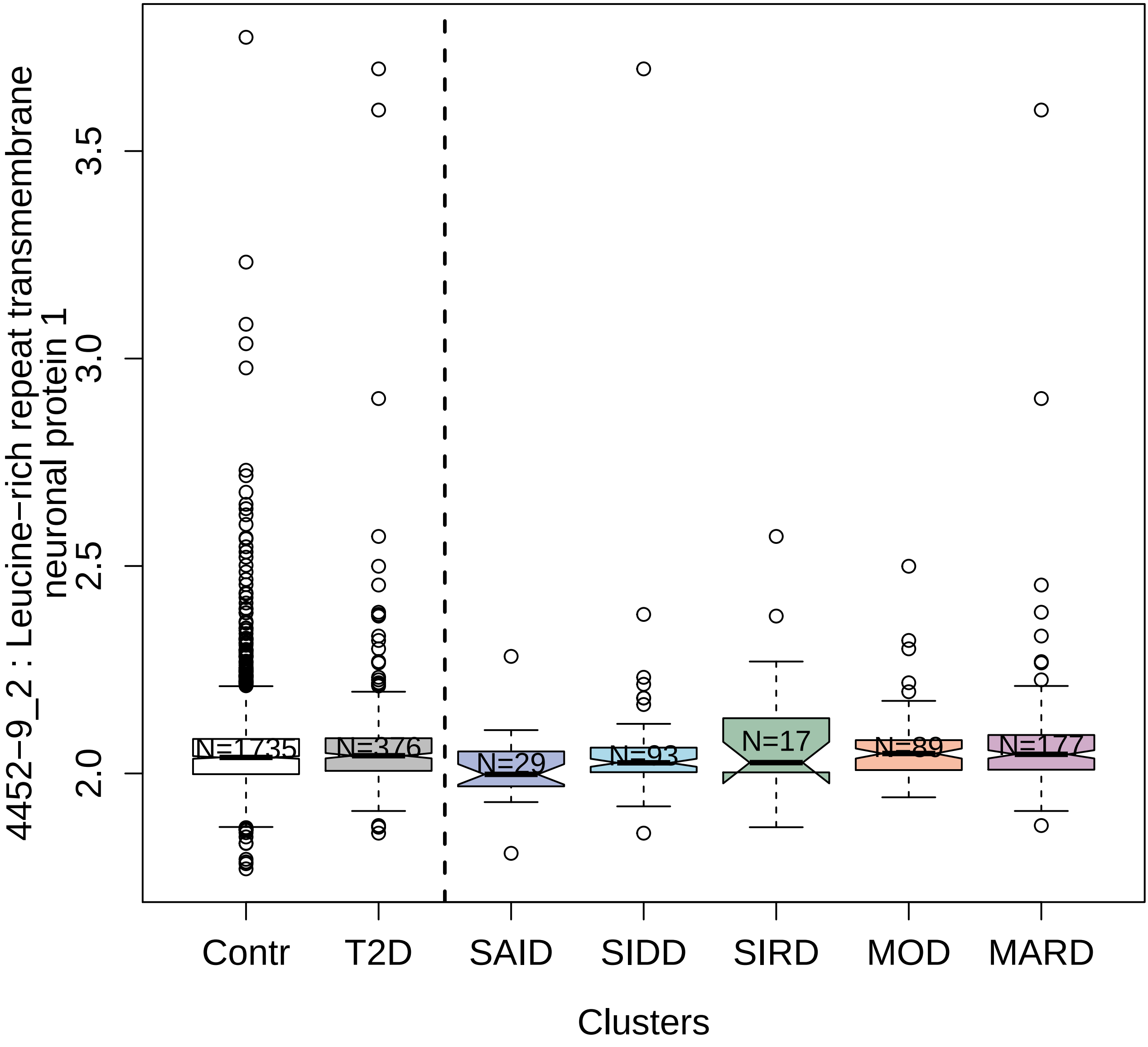

4452−9_2 : Leucine−rich repeat transmembrane
neuronal protein 1
2.0
2.5
3.0
3.5
N=1735
N=376
N=29
N=93
N=17
N=89
N=177
Contr
T2D
SAID
SIDD
SIRD
MOD
MARD
Clusters

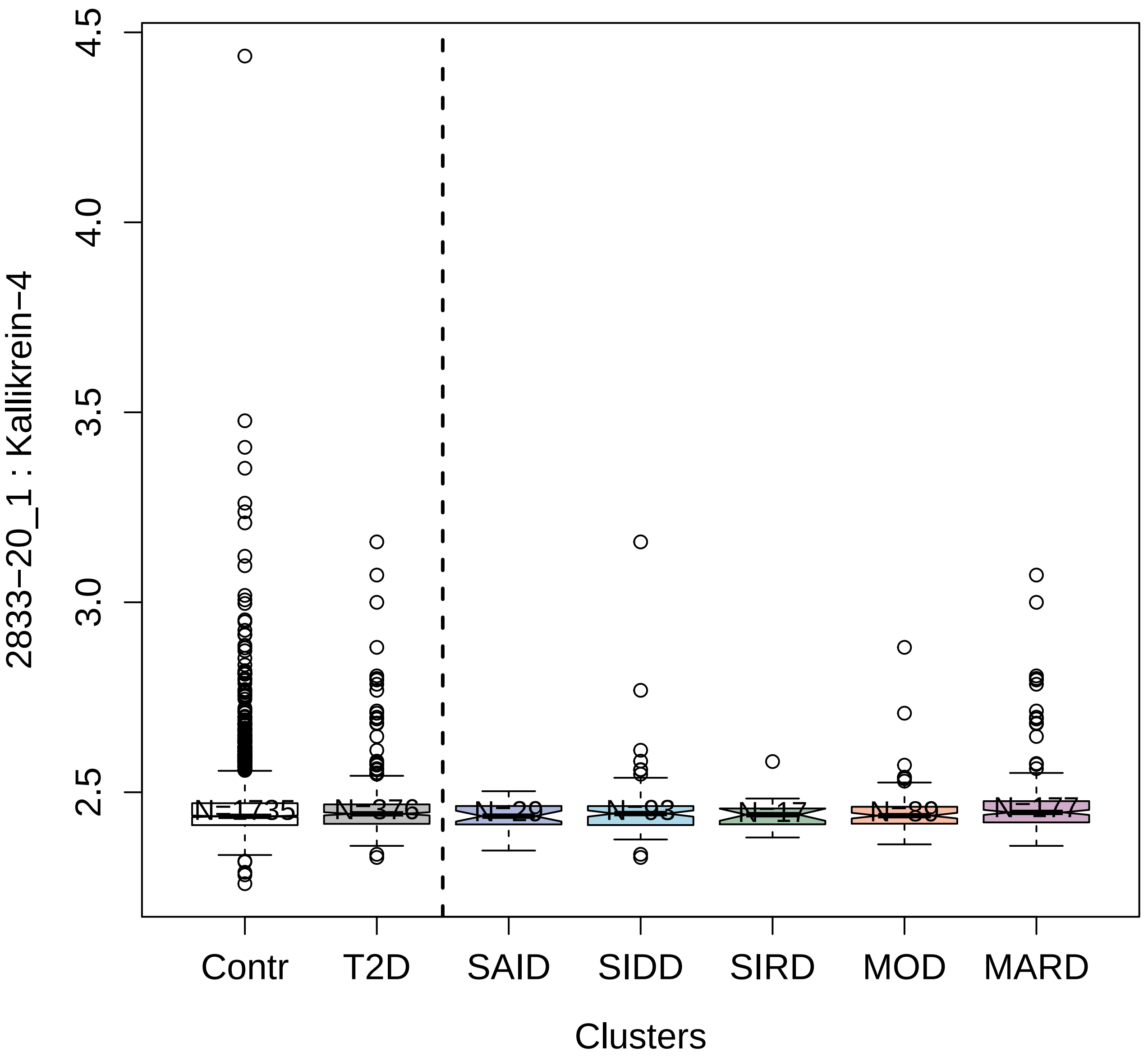

2833−20_1 : Kallikrein−4
4.5
4.0
3.5
3.0
2.5
N=1735
N=376
N=29
N=93
N=17
N=89
N=177
Contr
T2D
SAID
SIDD
SIRD
MOD
MARD
Clusters

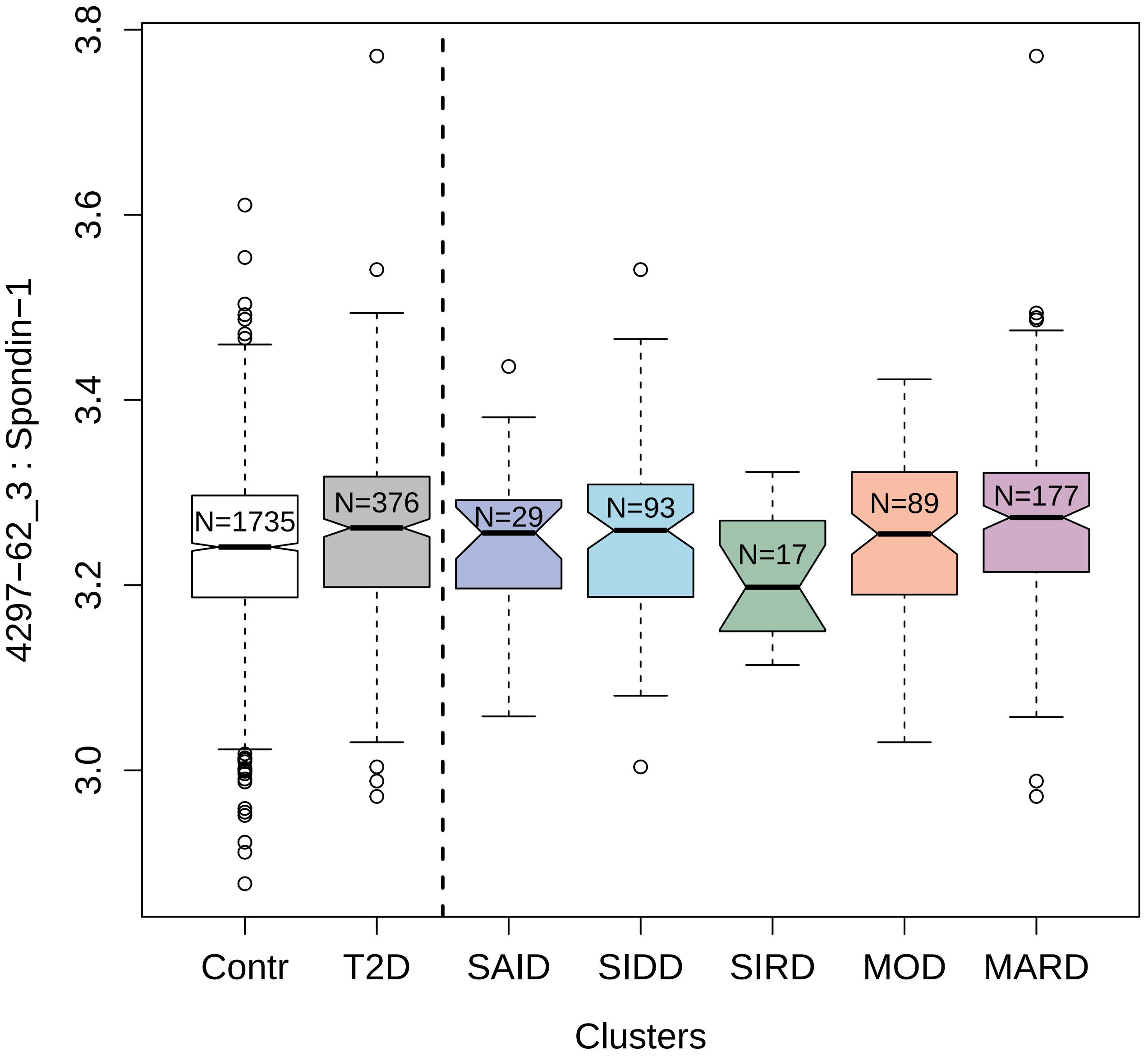

4297−62_3 : Spondin−1
3.8
3.6
3.4
3.2
3.0
N=1735
N=376
N=29
N=93
N=17
N=89
N=177
Contr
T2D
SAID
SIDD
SIRD
MOD
MARD
Clusters

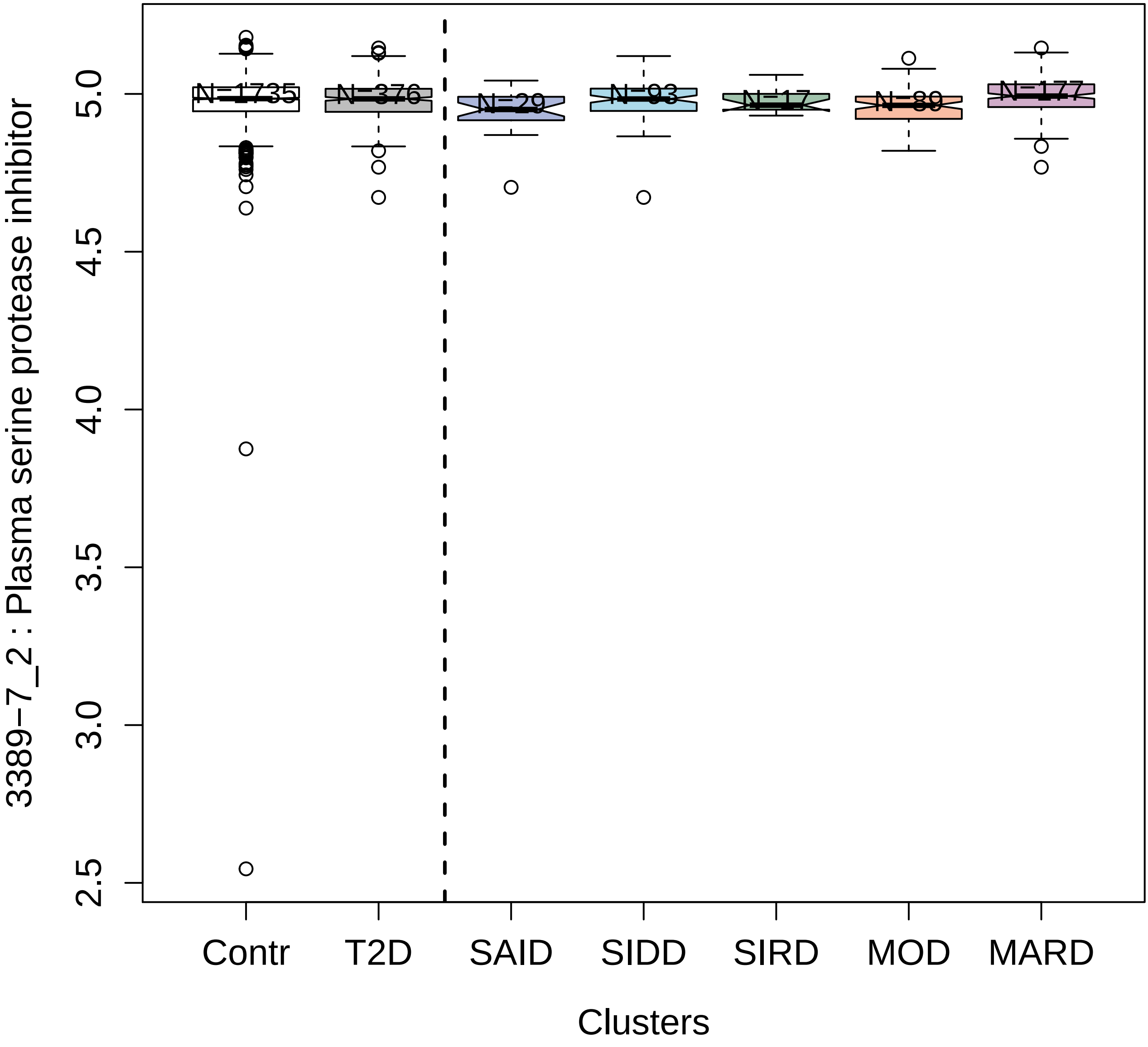

3389−7_2 : Plasma serine protease inhibitor
5.0
4.5
4.0
3.5
3.0
2.5
N=1735
N=376
N=29
N=93
N=17
N=89
N=177
Contr
T2D
SAID
SIDD
SIRD
MOD
MARD
Clusters

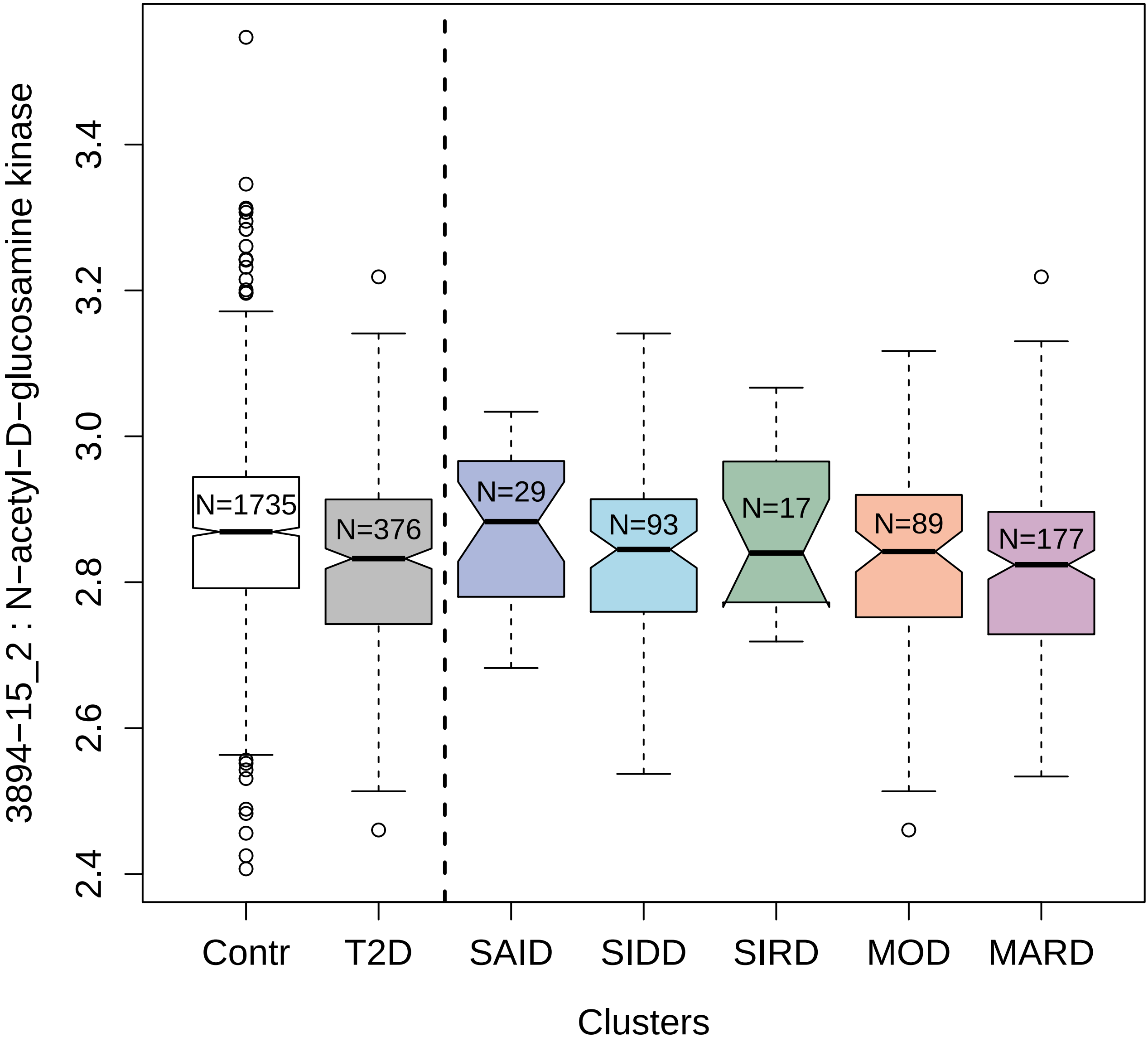

3894−15_2 : N−acetyl−D−glucosamine kinase
2.4
2.6
2.8
3.0
3.2
3.4
N=1735
N=376
N=29
N=93
N=17
N=89
N=177
Contr
T2D
SAID
SIDD
SIRD
MOD
MARD
Clusters

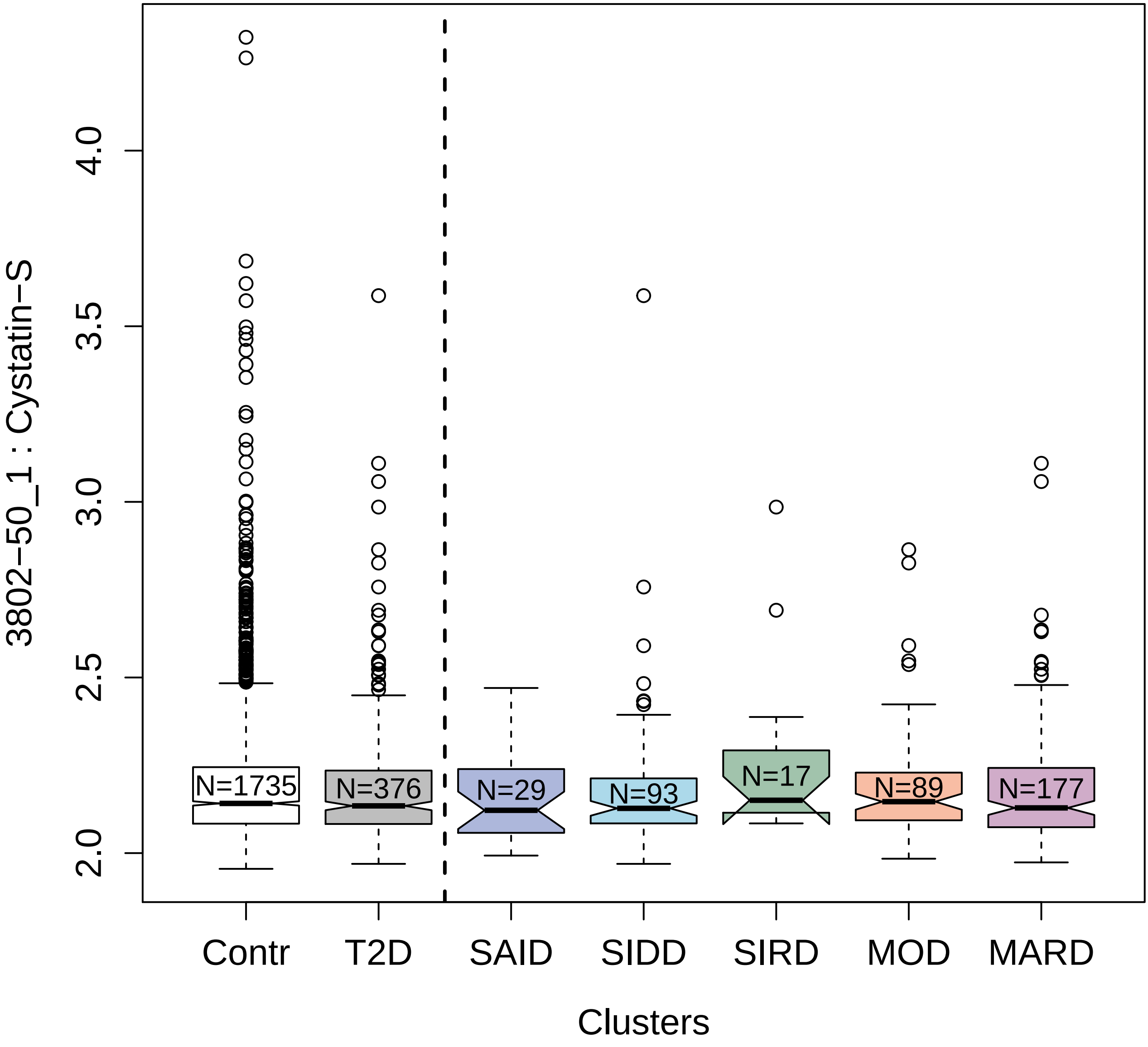
3802−50_1 : Cystatin−S
2.0
2.5
3.0
3.5
4.0
N=1735
N=376
N=29
N=93
N=17
N=89
N=177
Contr
T2D
SAID
SIDD
SIRD
MOD
MARD
Clusters

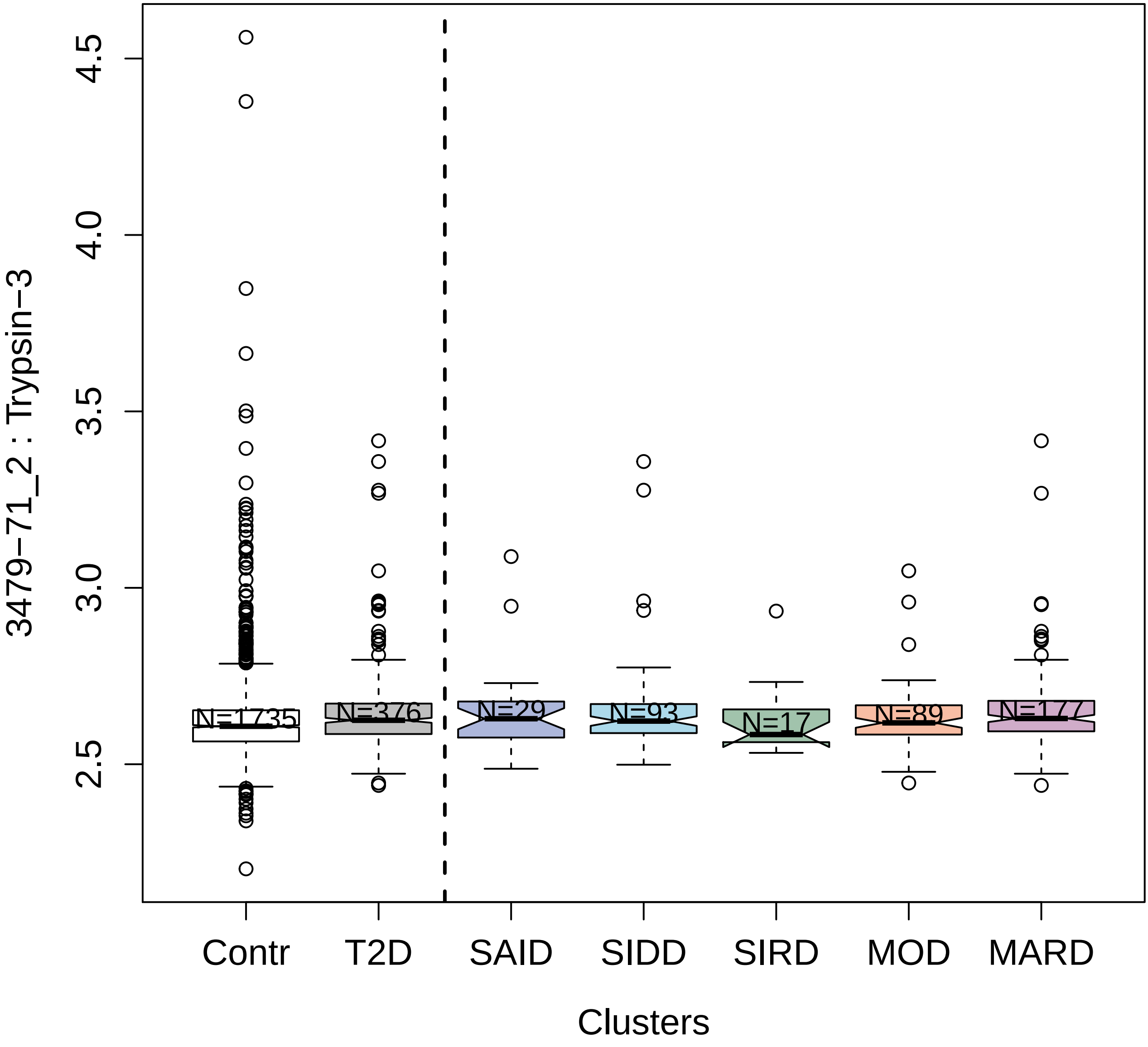

3479−71_2 : Trypsin−3
4.5
4.0
3.5
3.0
2.5
N=1735
N=376
N=29
N=93
N=17
N=89
N=177
Contr
T2D
SAID
SIDD
SIRD
MOD
MARD
Clusters

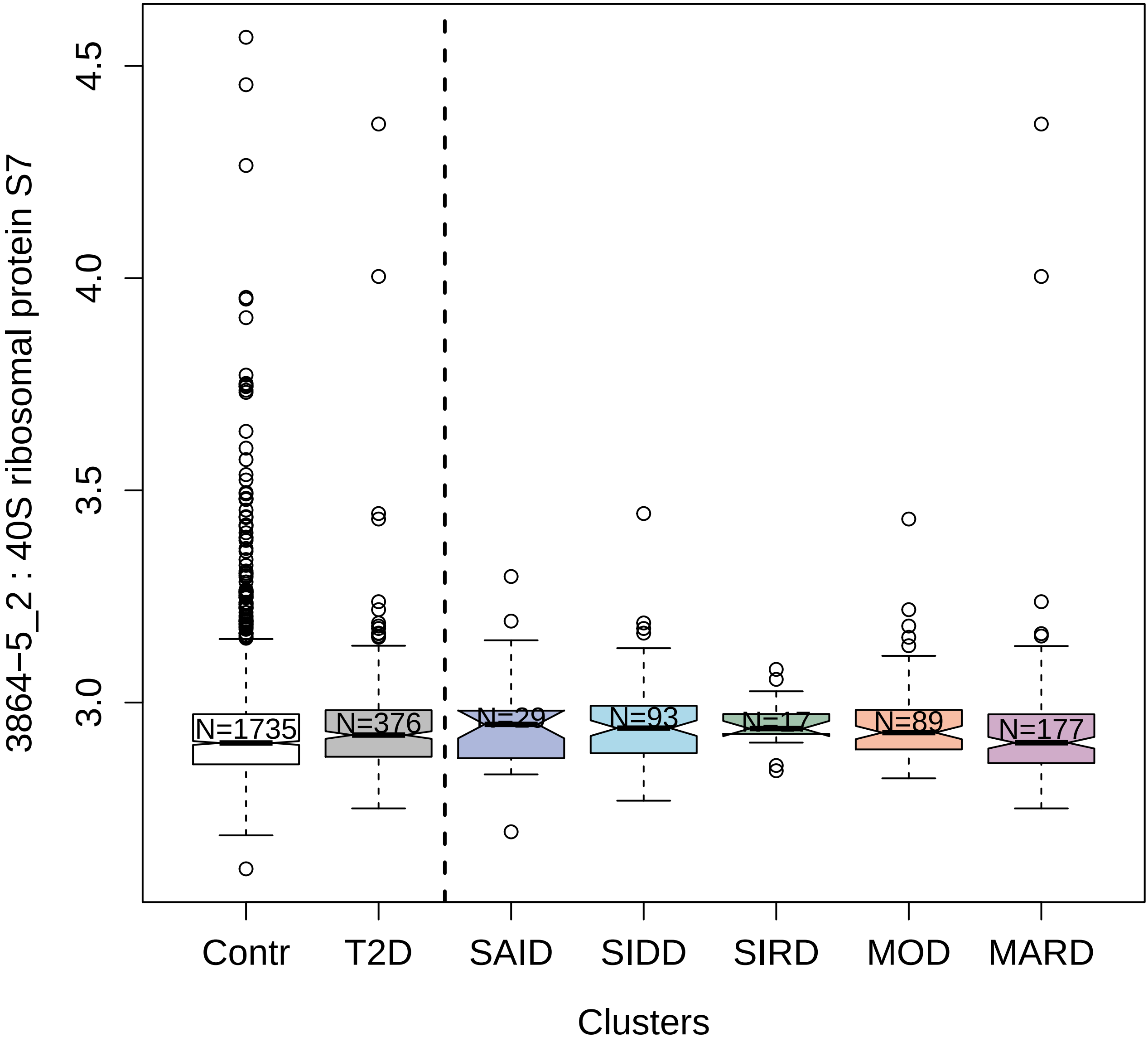
3864−5_2 : 40S ribosomal protein S7
4.5
4.0
3.5
3.0
N=1735
N=376
N=29
N=93
N=17
N=89
N=177
Contr
T2D
SAID
SIDD
SIRD
MOD
MARD
Clusters

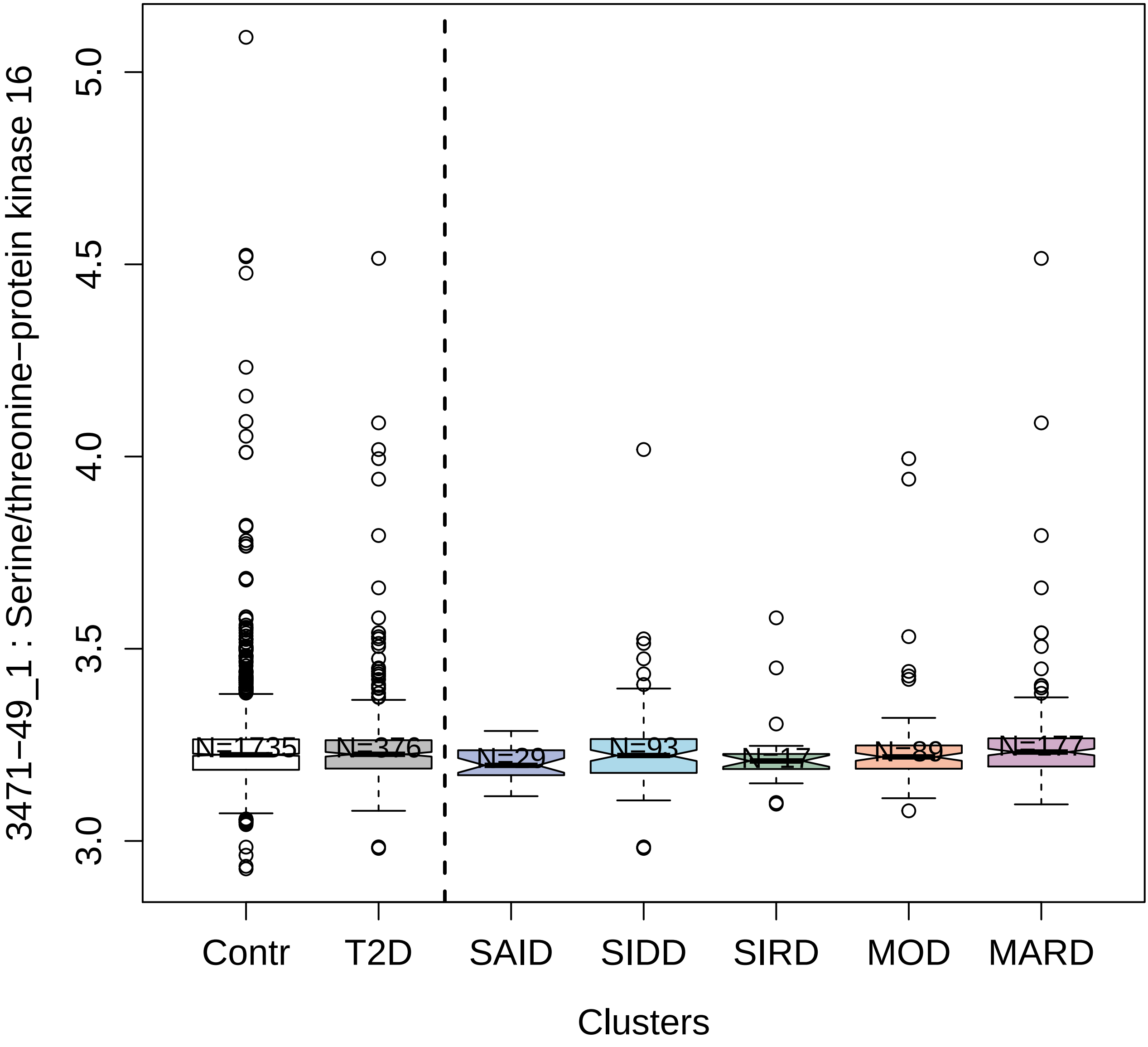
3471−49_1 : Serine/threonine−protein kinase 16
5.0
4.5
4.0
3.5
3.0
N=1735
N=376
N=29
N=93
N=17
N=89
N=177
Contr
T2D
SAID
SIDD
SIRD
MOD
MARD
Clusters

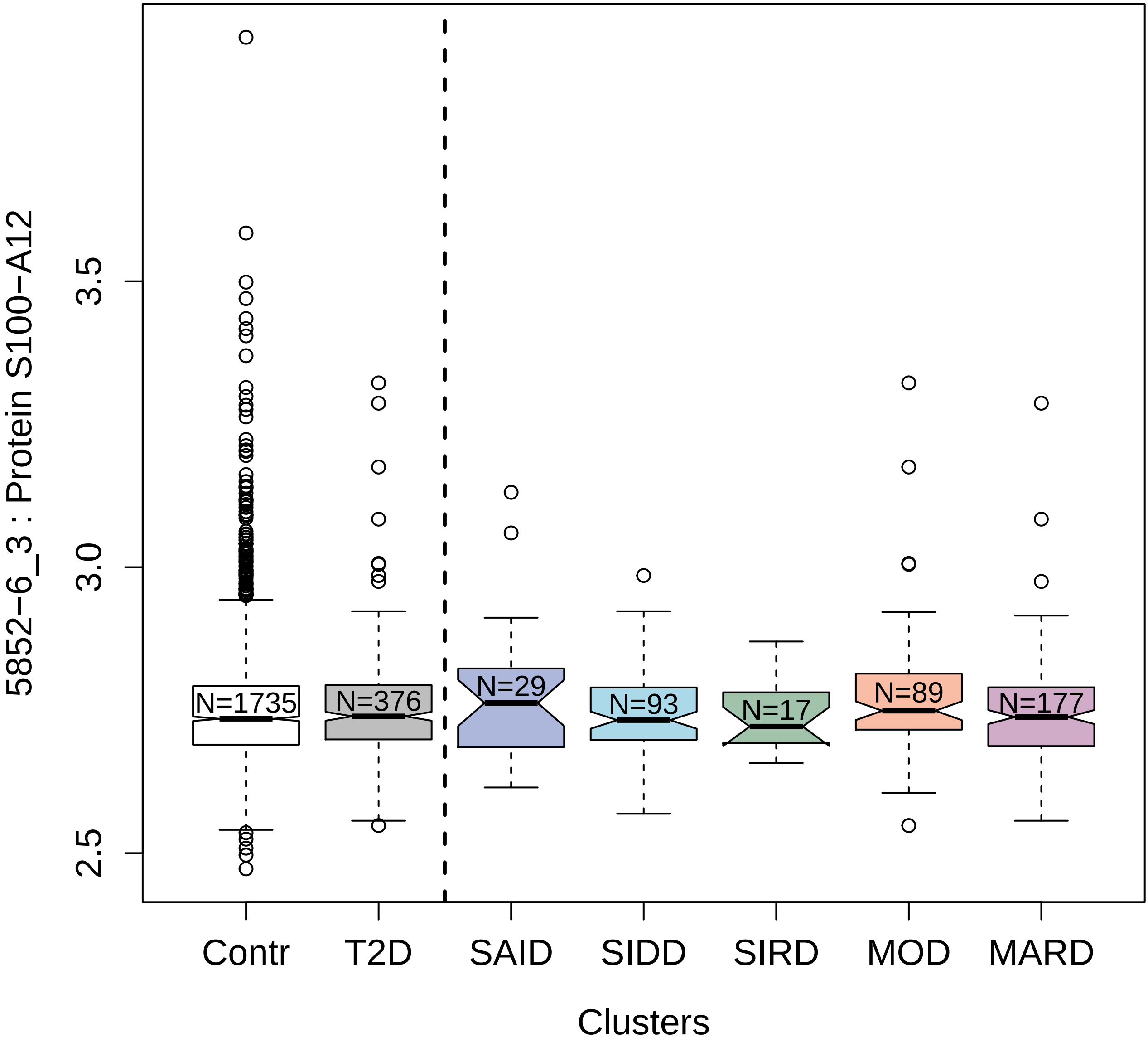
5852−6_3 : Protein S100−A12
3.5
3.0
2.5
N=1735
N=376
N=29
N=93
N=17
N=89
N=177
Contr
T2D
SAID
SIDD
SIRD
MOD
MARD
Clusters

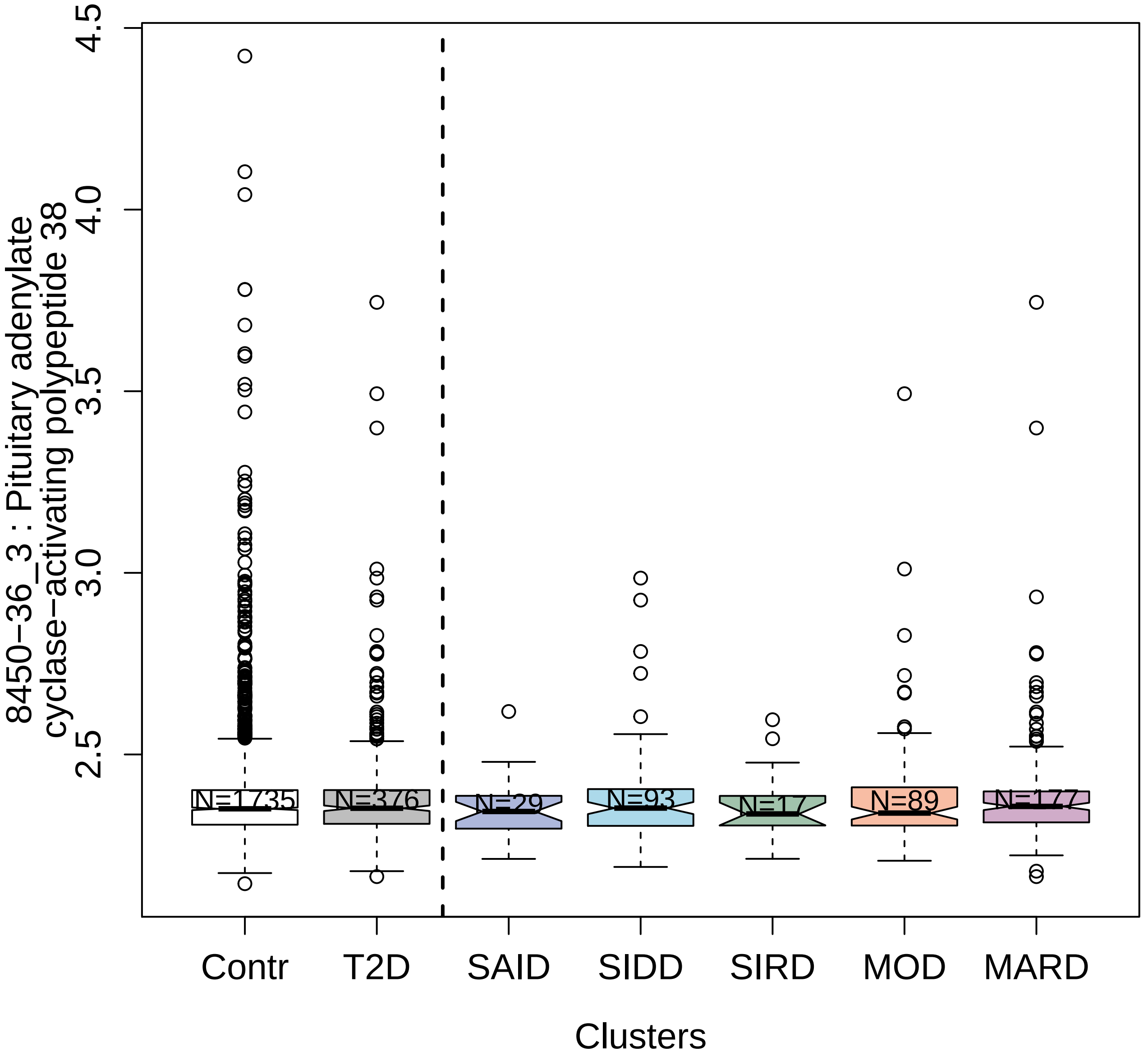

8450−36_3 : Pituitary adenylate
cyclase−activating polypeptide 38
4.5
4.0
3.5
3.0
2.5
N=1735
N=376
N=29
N=93
N=17
N=89
N=177
Contr
T2D
SAID
SIDD
SIRD
MOD
MARD
Clusters

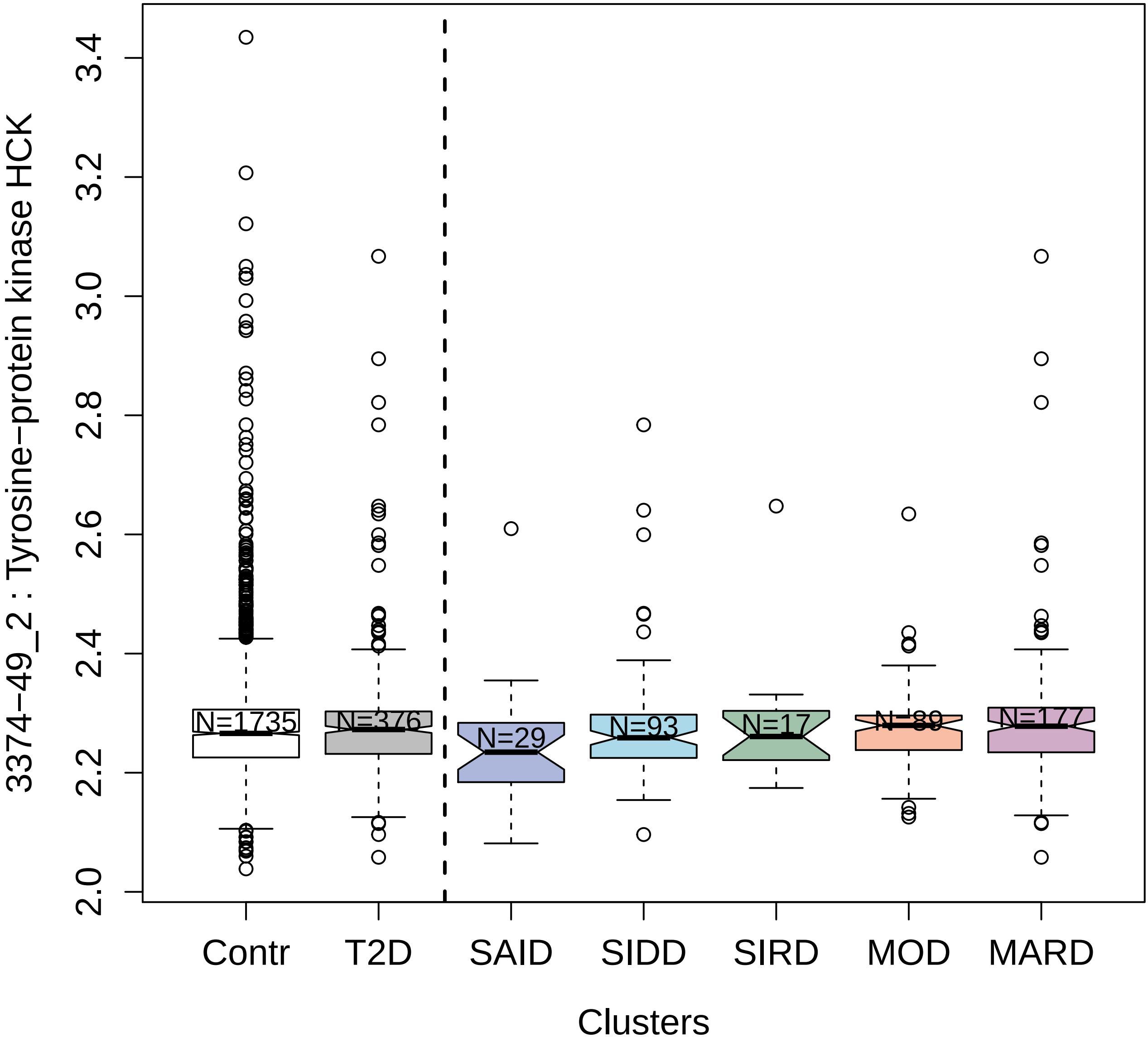

3374−49_2 : Tyrosine−protein kinase HCK
2.0
2.2
2.4
2.6
2.8
3.0
3.2
3.4
N=1735
N=376
N=29
N=93
N=17
N=89
N=177
Contr
T2D
SAID
SIDD
SIRD
MOD
MARD
Clusters

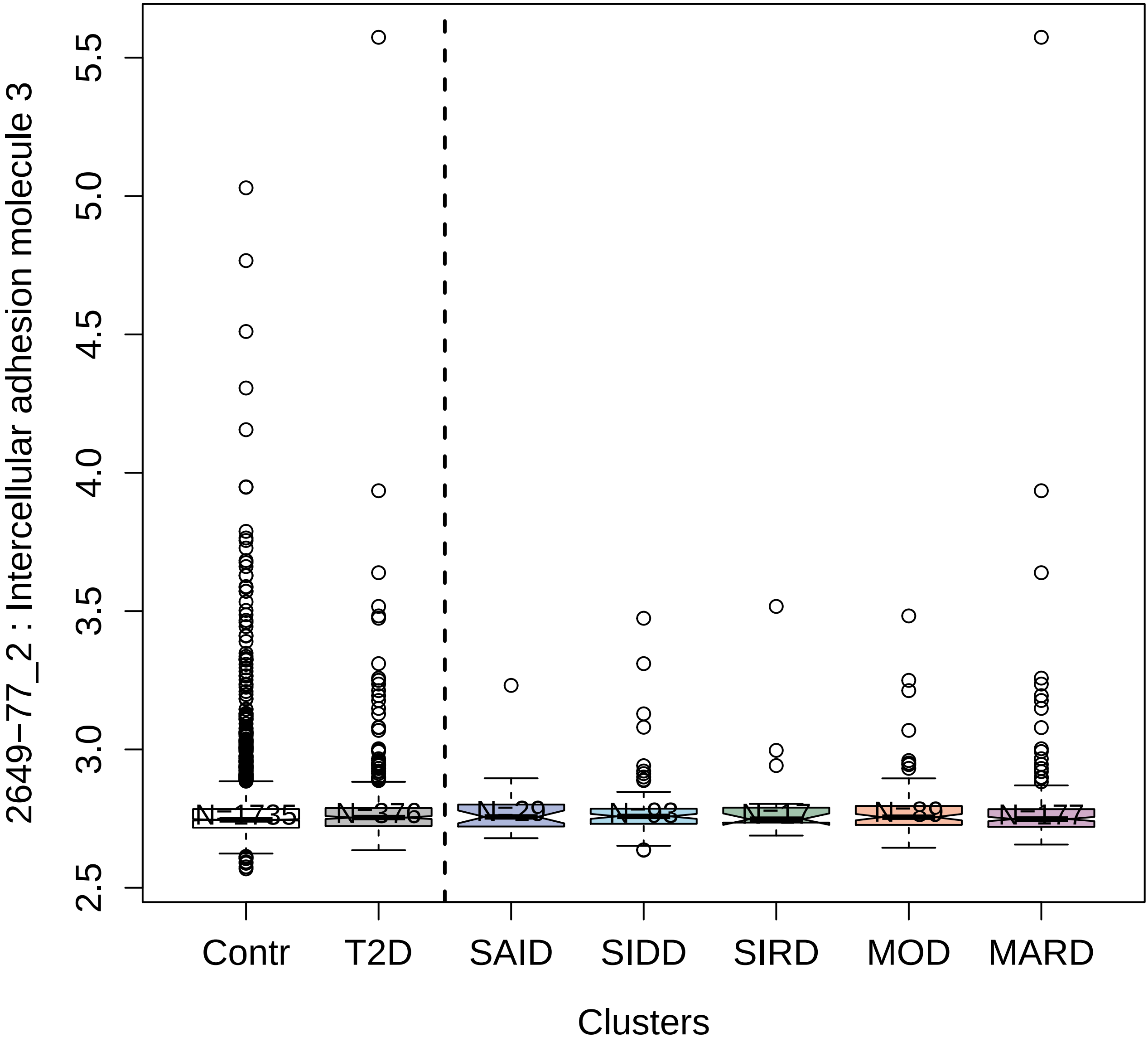
2649−77_2 : Intercellular adhesion molecule 3
Clusters
Contr
T2D
SAID
SIDD
SIRD
MOD
MARD
2.5
3.0
3.5
4.0
4.5
5.0
5.5
N=1735
N=376
N=29
N=89
N=177

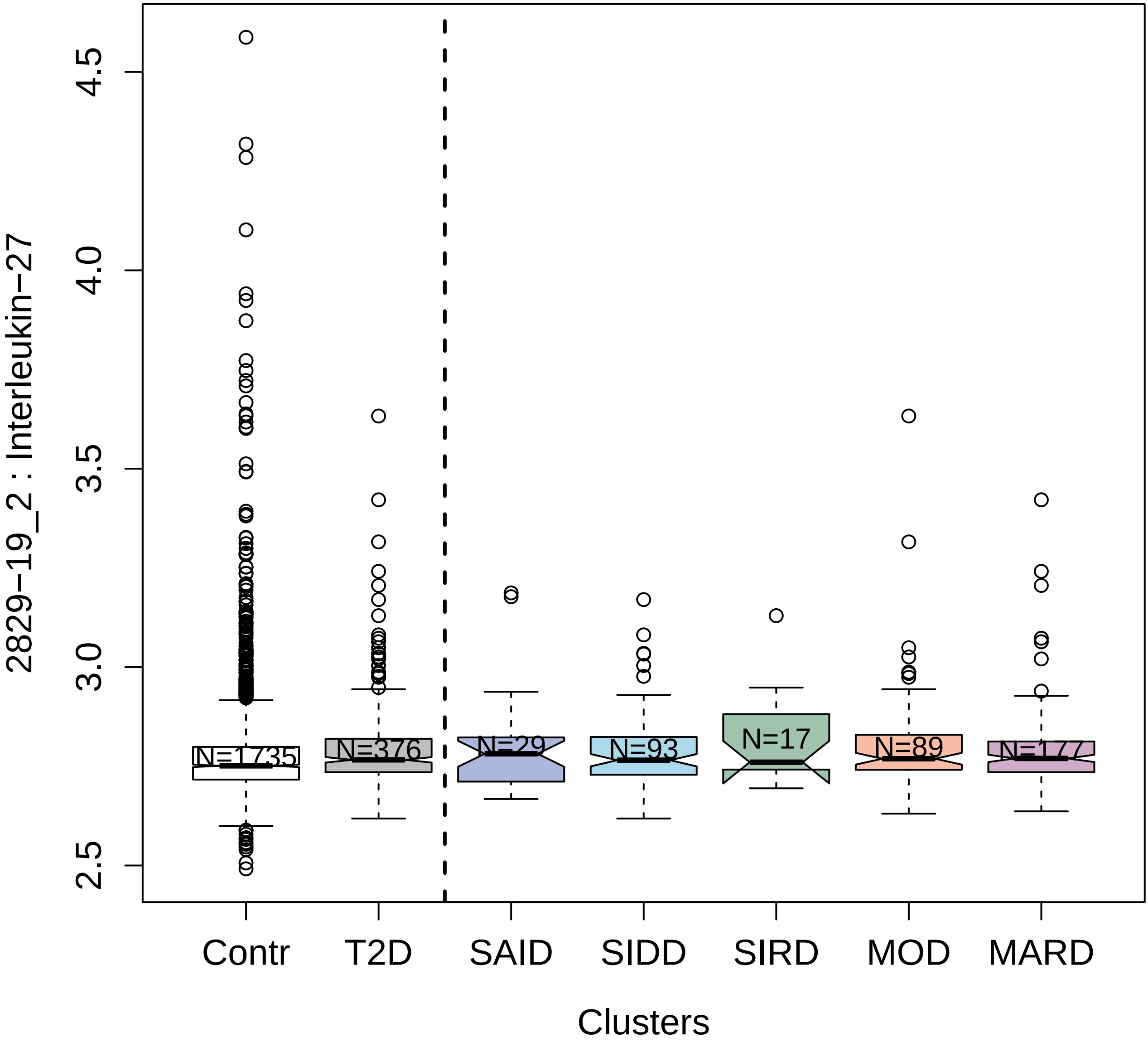
2829−19_2 : Interleukin−27
4.5
4.0
3.5
3.0
2.5
N=1735
N=376
N=29
N=93
N=17
N=89
N=177
Contr
T2D
SAID
SIDD
SIRD
MOD
MARD
Clusters

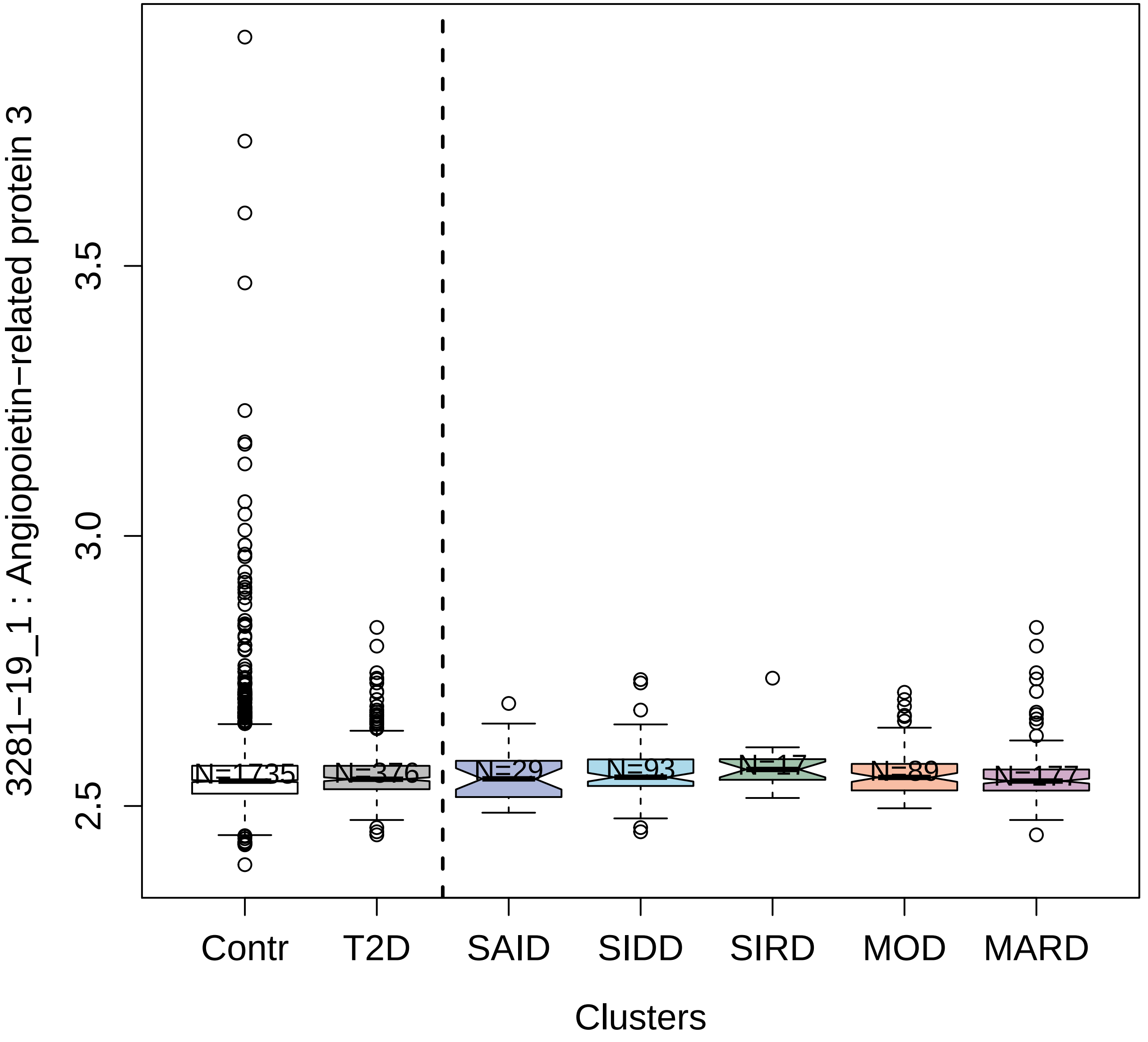

3281−19_1 : Angiopoietin−related protein 3
3.5
3.0
2.5
N=1735
N=376
N=29
N=93
N=17
N=89
N=177
Contr
T2D
SAID
SIDD
SIRD
MOD
MARD
Clusters

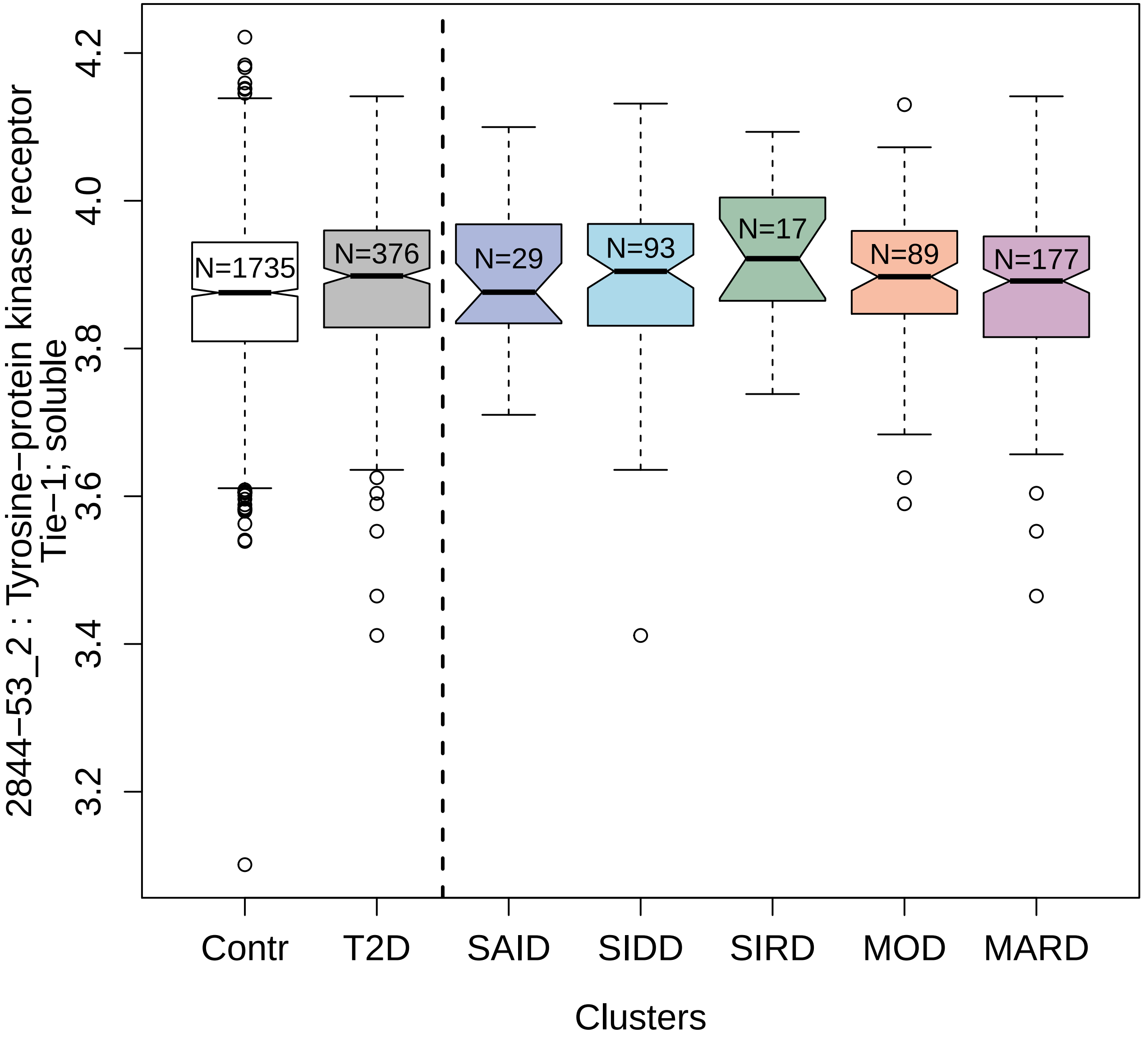

2844−53_2 : Tyrosine−protein kinase receptor
Tie−1; soluble
4.2
4.0
3.8
3.6
3.4
3.2
N=1735
N=376
N=29
N=93
N=17
N=89
N=177
Contr
T2D
SAID
SIDD
SIRD
MOD
MARD
Clusters

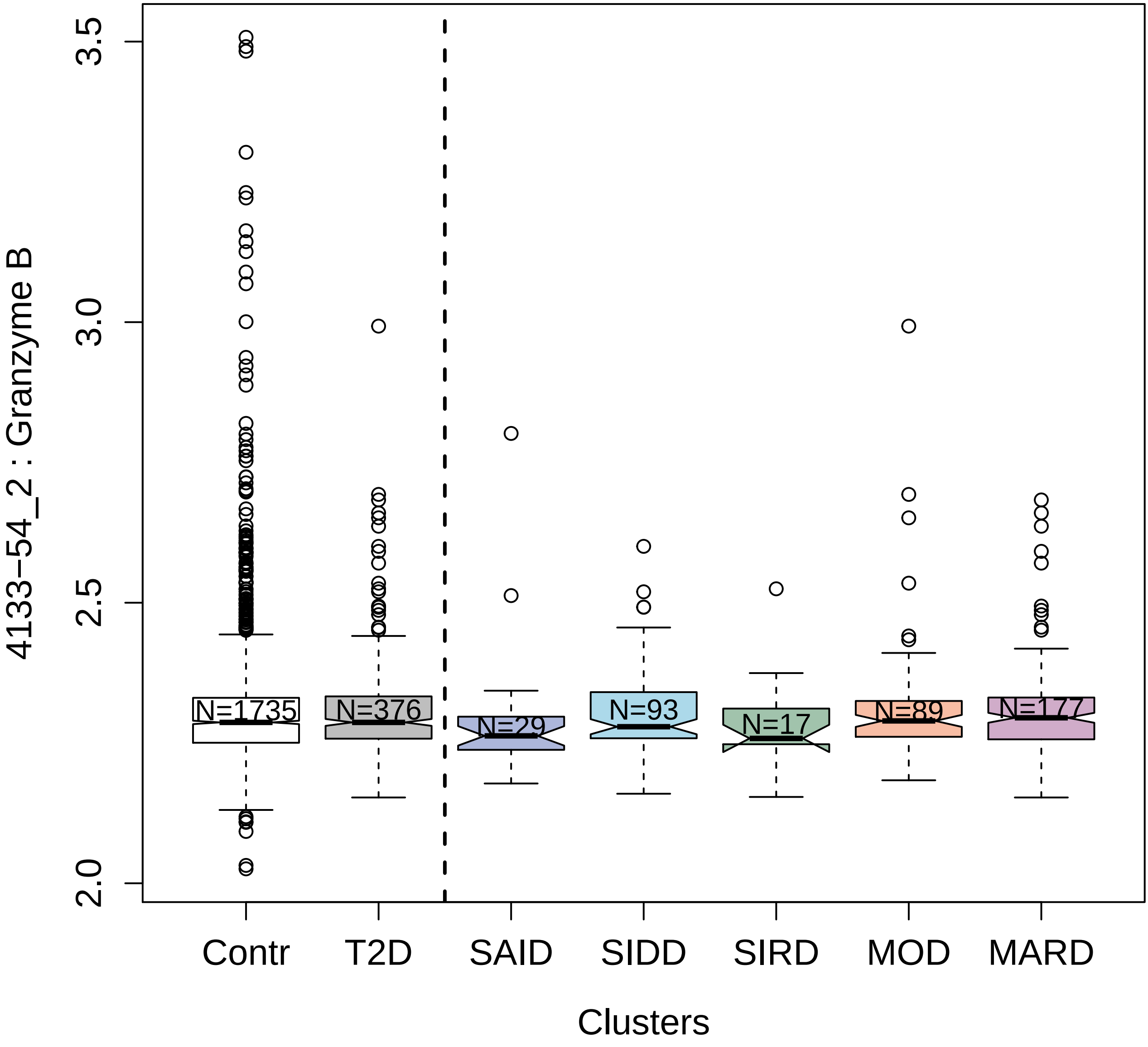

4133−54_2 : Granzyme B
3.5
3.0
2.5
2.0
N=1735
N=376
N=29
N=93
N=17
N=89
N=177
Contr
T2D
SAID
SIDD
SIRD
MOD
MARD
Clusters

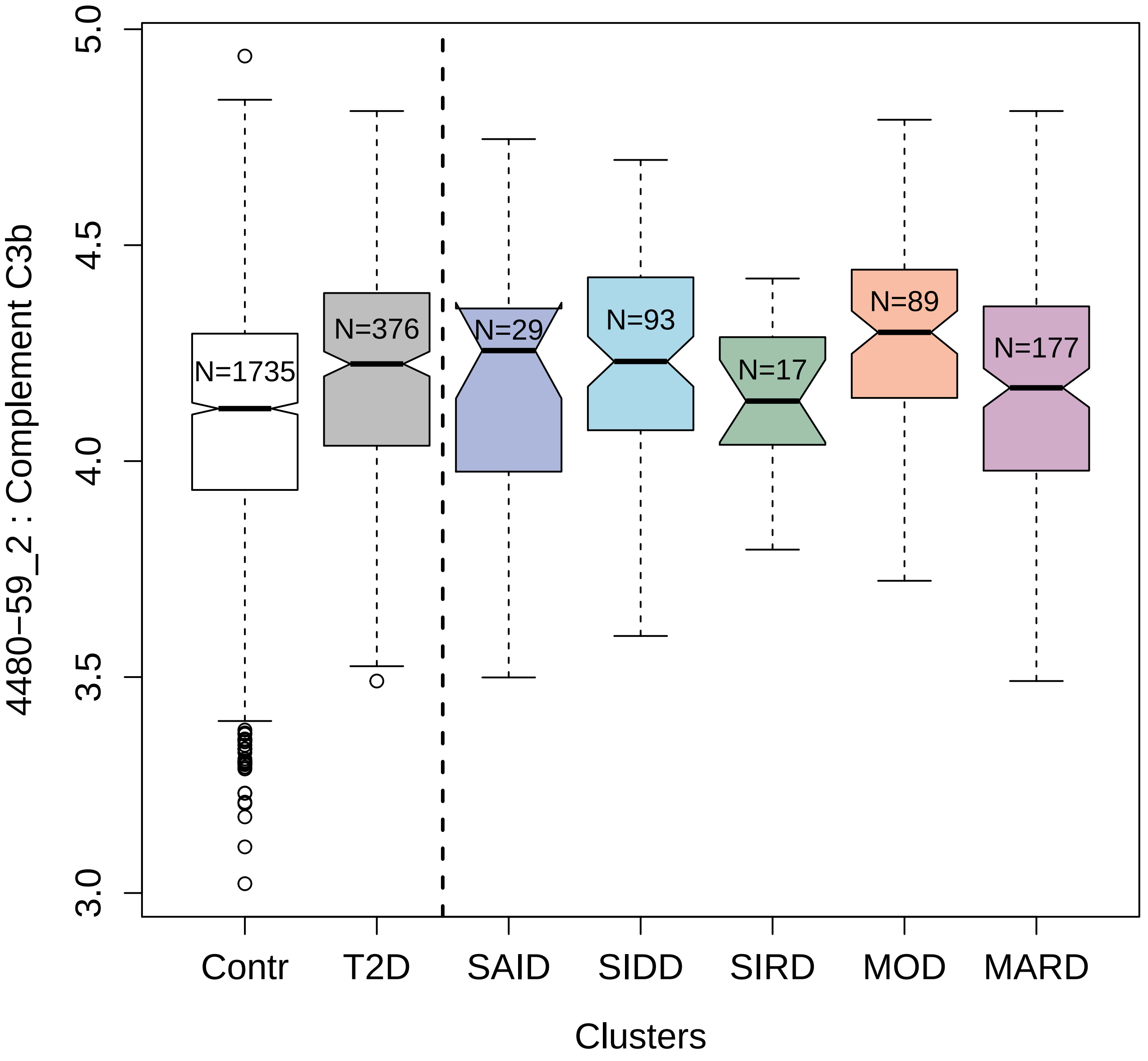
4480−59_2 : Complement C3b
5.0
4.5
4.0
3.5
3.0
N=1735
N=376
N=29
N=93
N=17
N=89
N=177
Contr
T2D
SAID
SIDD
SIRD
MOD
MARD
Clusters

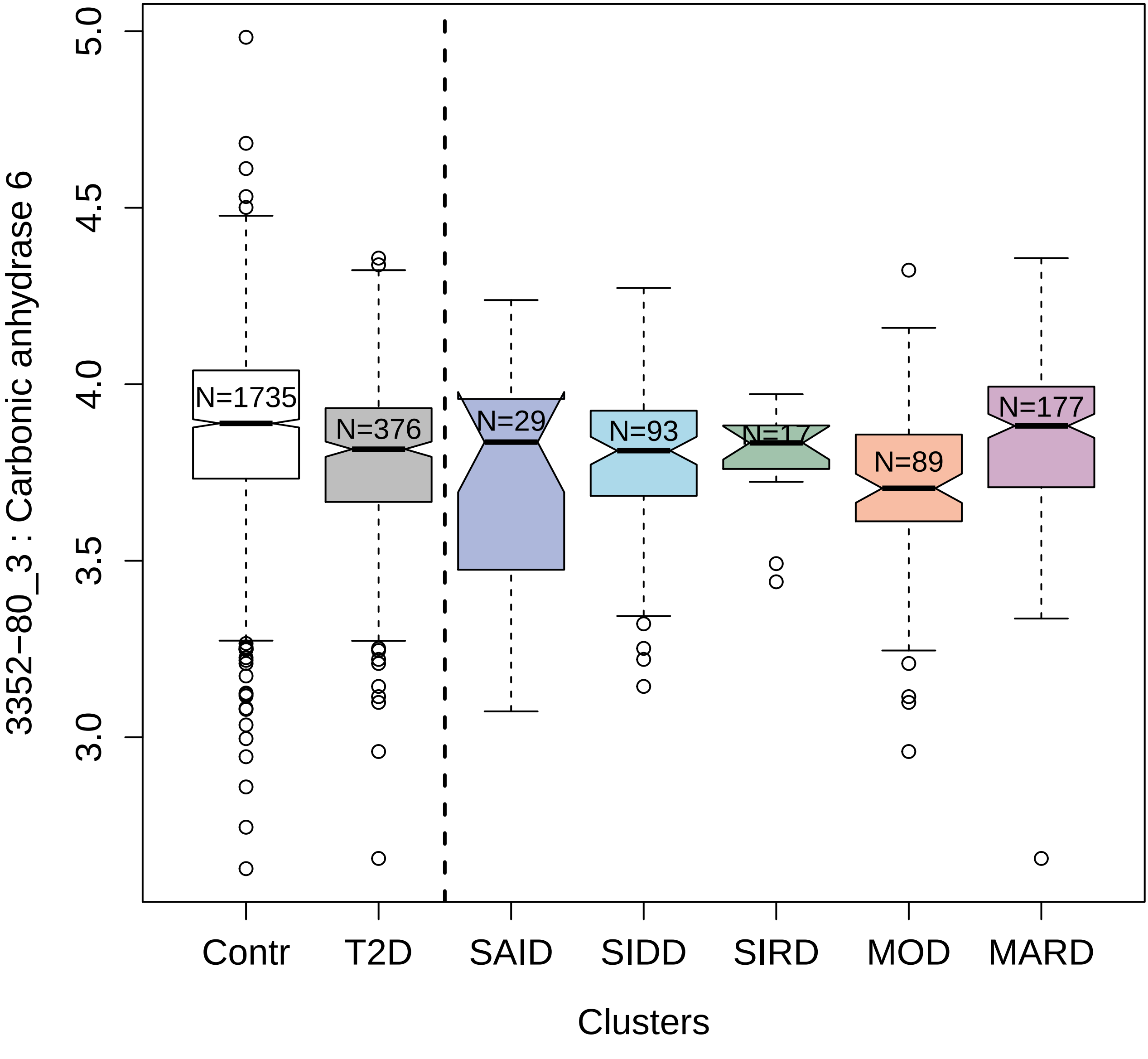
3352−80_3 : Carbonic anhydrase 6
5.0
4.5
4.0
3.5
3.0
N=1735
N=376
N=29
N=93
N=17
N=89
N=177
Contr
T2D
SAID
SIDD
SIRD
MOD
MARD
Clusters

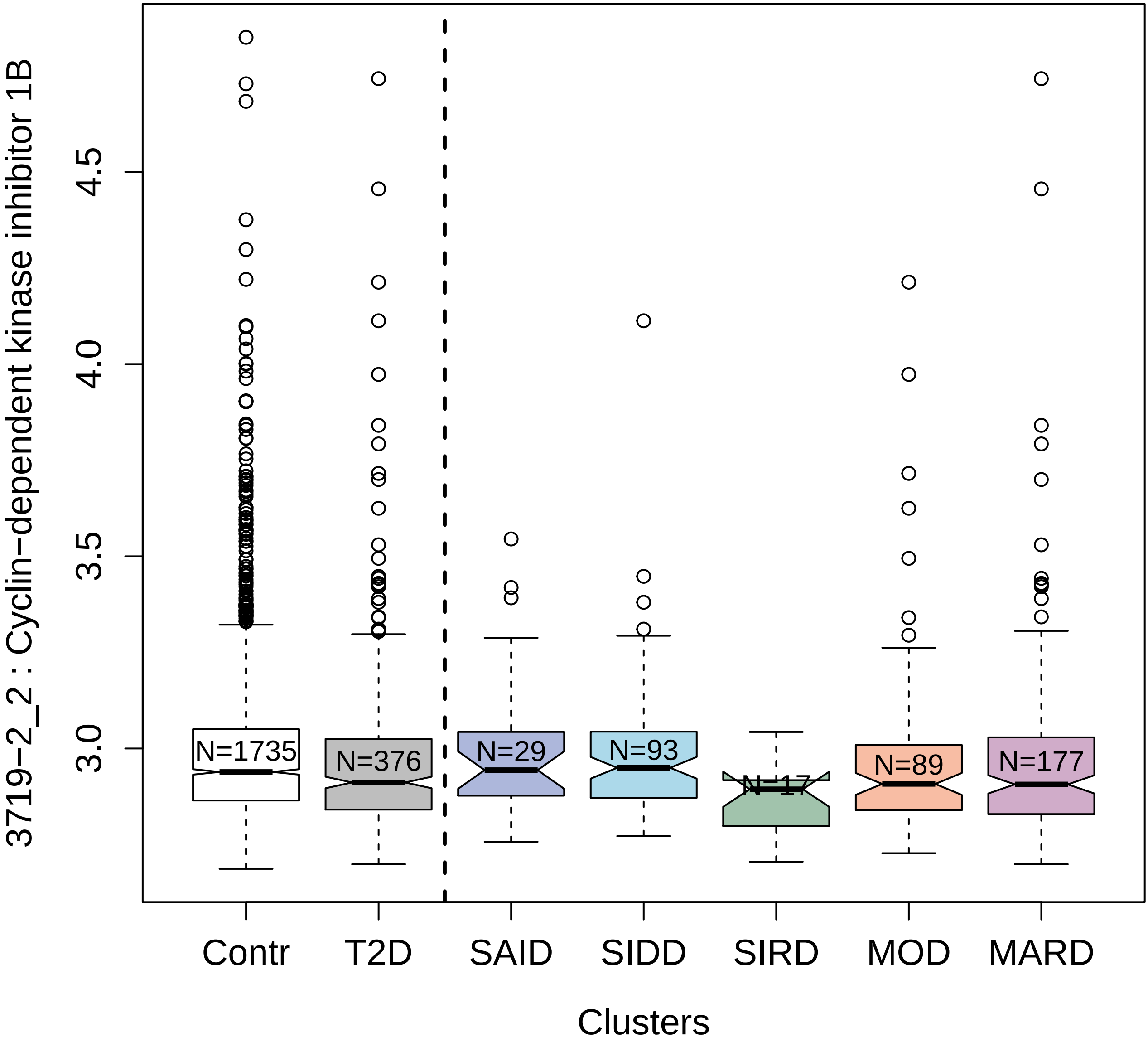

3719−2_2 : Cyclin−dependent kinase inhibitor 1B
3.0
3.5
4.0
4.5
N=1735
N=376
N=29
N=93
N=17
N=89
N=177
Contr
T2D
SAID
SIDD
SIRD
MOD
MARD
Clusters

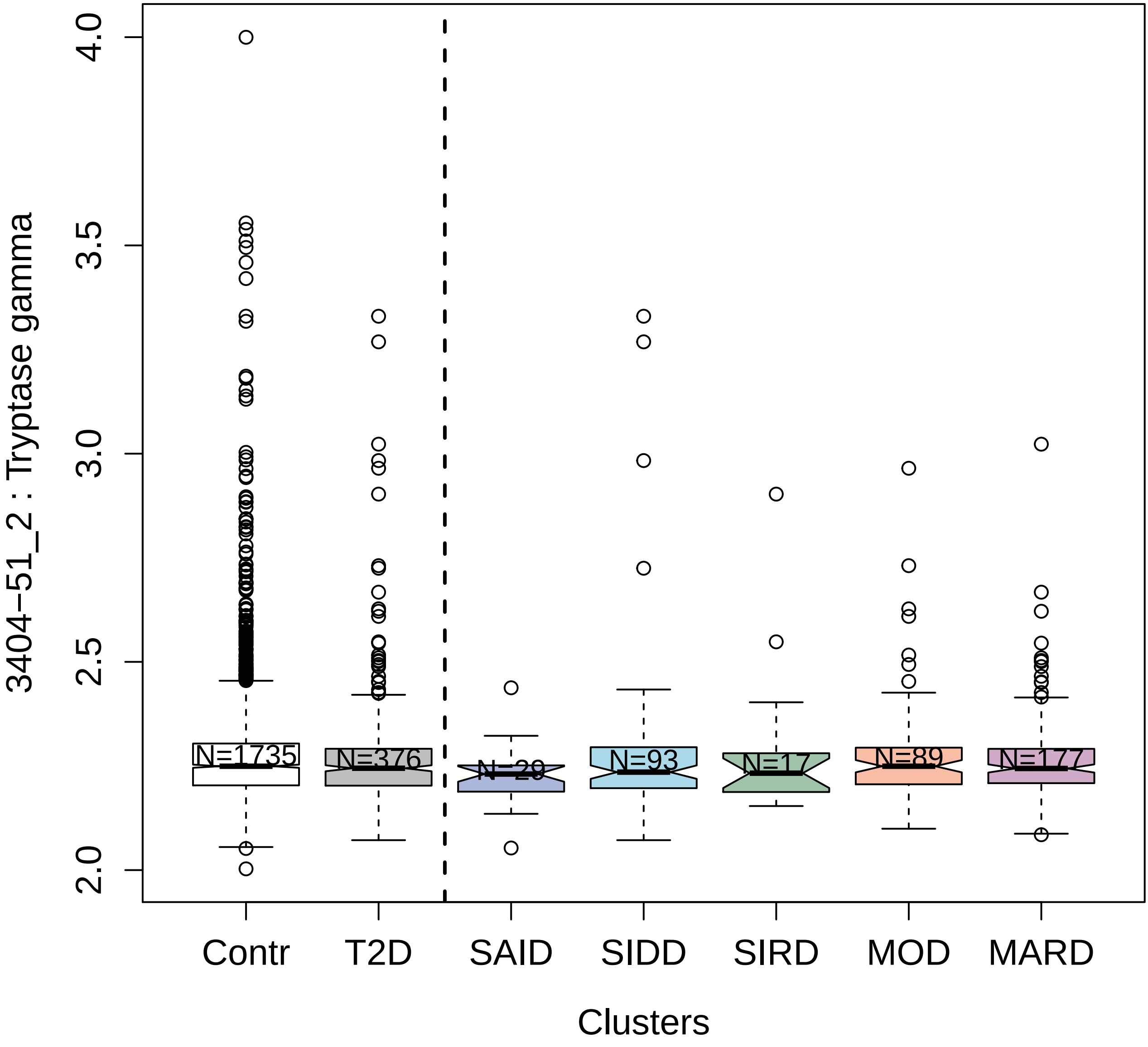

3404−51_2 : Tryptase gamma
4.0
3.5
3.0
2.5
2.0
N=1735
N=376
N=29
N=93
N=17
N=89
N=177
Contr
T2D
SAID
SIDD
SIRD
MOD
MARD
Clusters

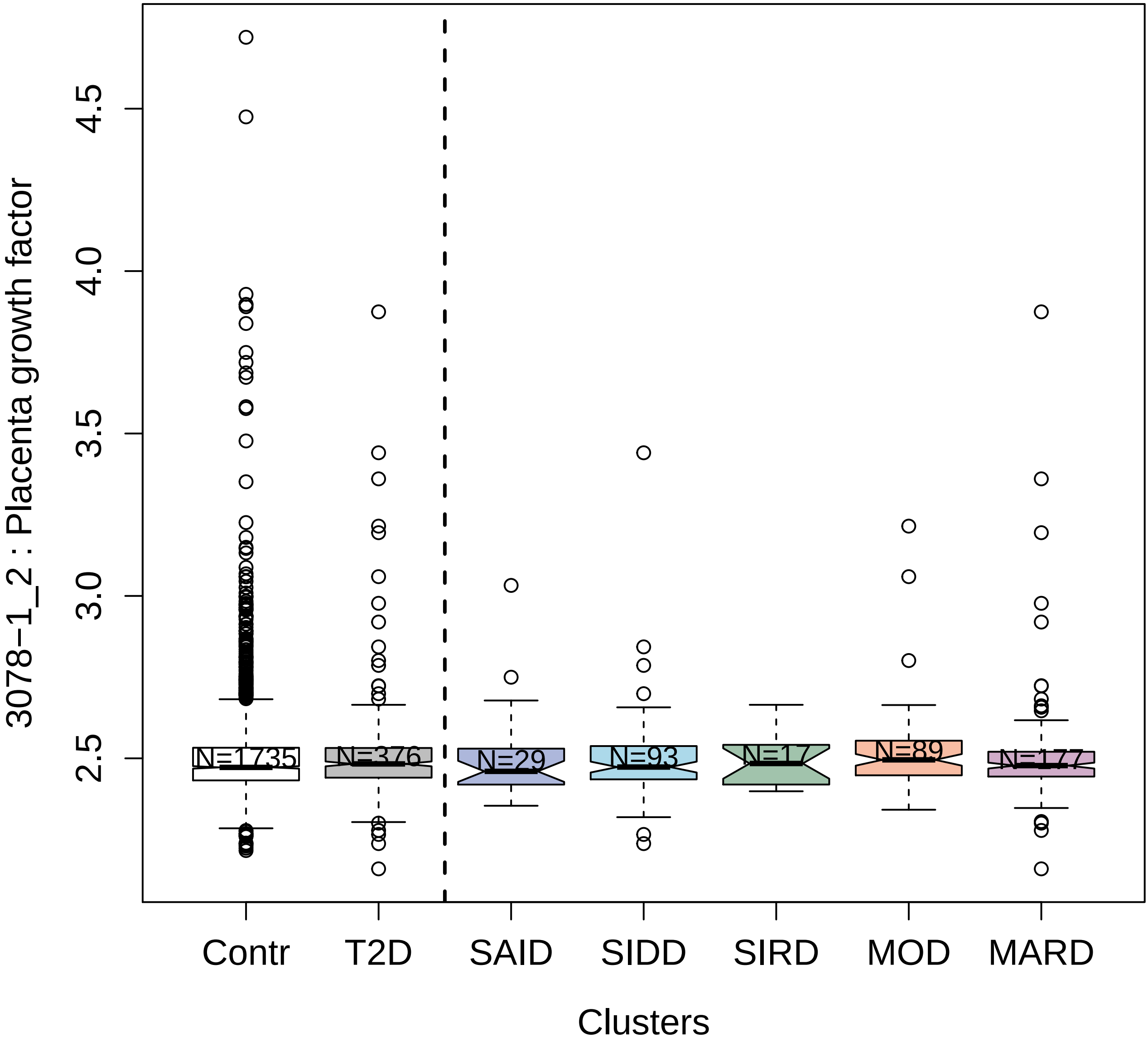
3078−1_2 : Placenta growth factor
4.5
4.0
3.5
3.0
2.5
N=1735
N=376
N=29
N=93
N=17
N=89
N=177
Contr
T2D
SAID
SIDD
SIRD
MOD
MARD
Clusters

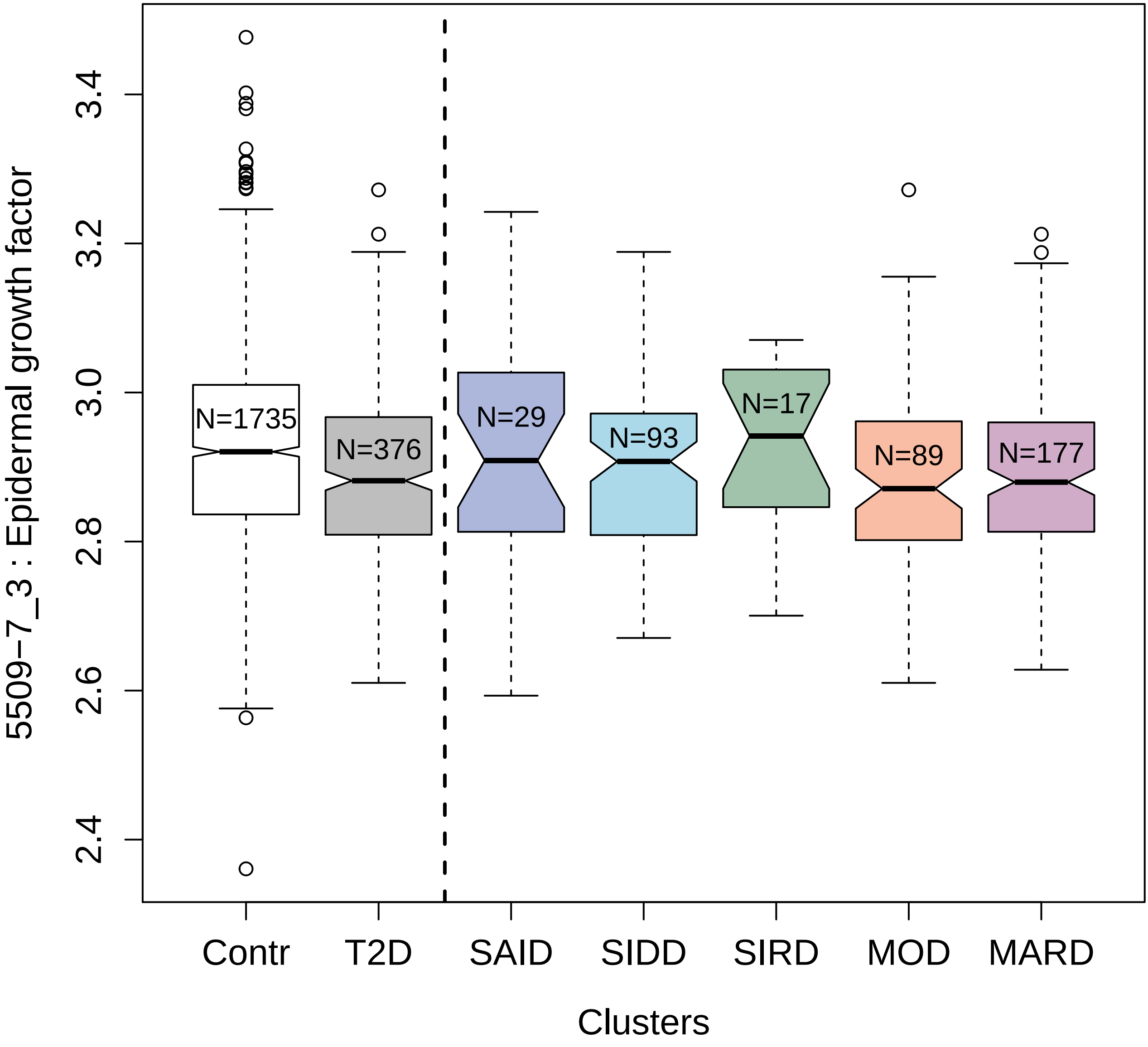

5509−7_3 : Epidermal growth factor
2.4
2.6
2.8
3.0
3.2
3.4
N=1735
N=376
N=29
N=93
N=17
N=89
N=177
Contr
T2D
SAID
SIDD
SIRD
MOD
MARD
Clusters

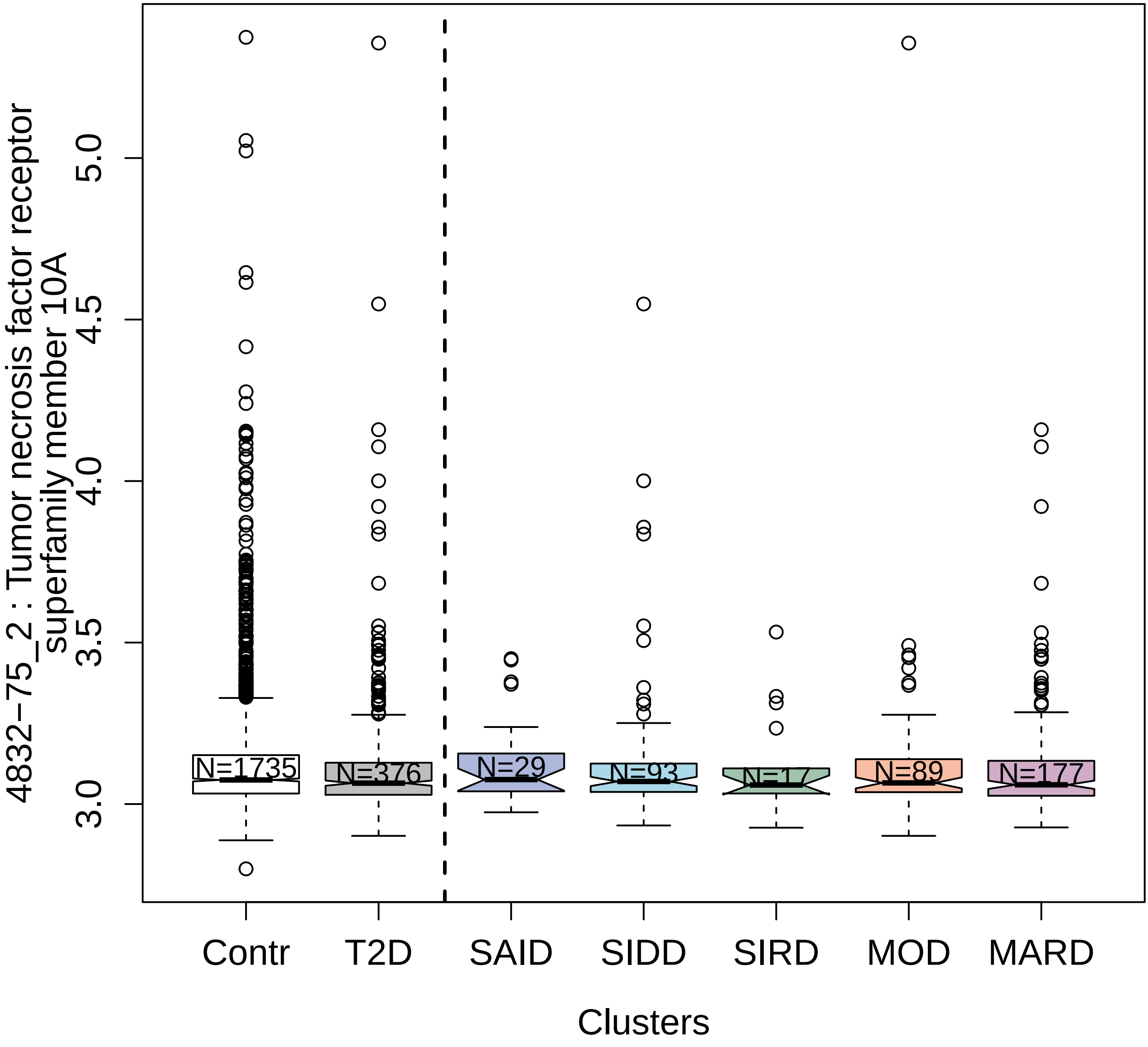

4832−75_2 : Tumor necrosis factor receptor superfamily member 10A
3.0
3.5
4.0
4.5
5.0
N=1735
N=376
N=29
N=93
N=17
N=89
N=177
Contr
T2D
SAID
SIDD
SIRD
MOD
MARD
Clusters

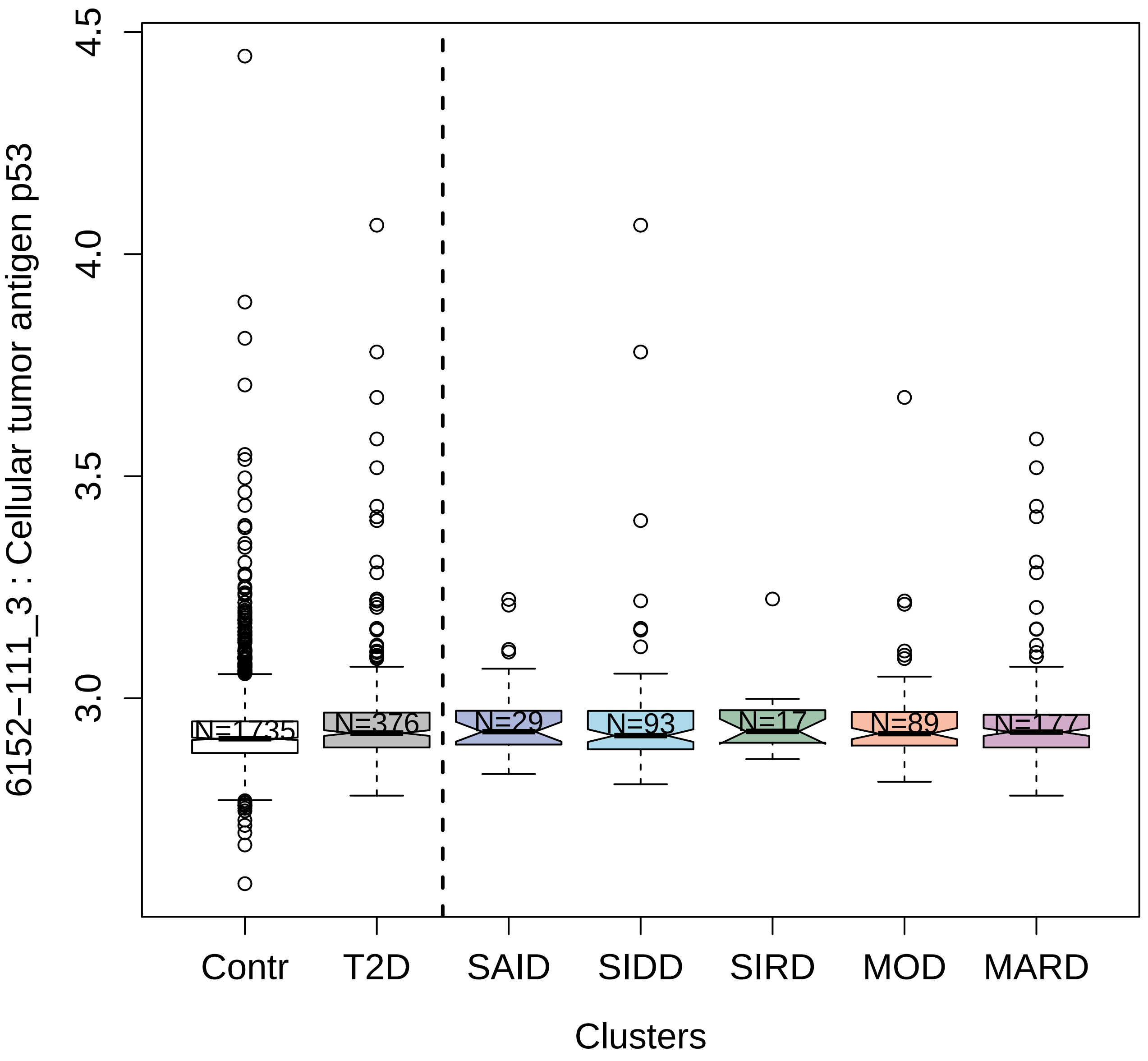
6152−111_3 : Cellular tumor antigen p53
4.5
4.0
3.5
3.0
N=1735
N=376
N=29
N=93
N=17
N=89
N=177
Contr
T2D
SAID
SIDD
SIRD
MOD
MARD
Clusters

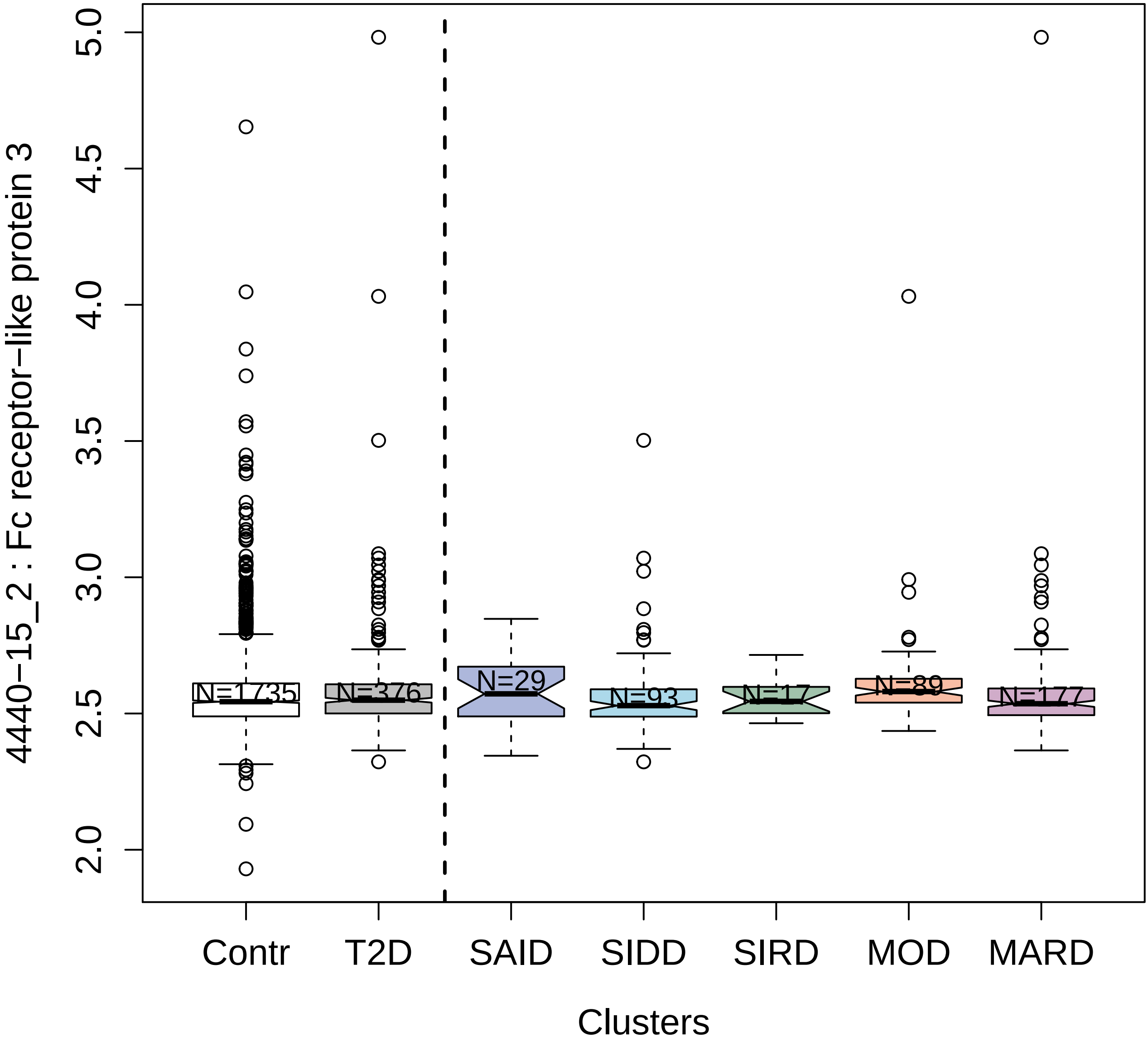
4440−15_2 : Fc receptor−like protein 3
5.0
4.5
4.0
3.5
3.0
2.5
2.0
N=1735
N=376
N=29
N=93
N=17
N=89
N=177
Contr
T2D
SAID
SIDD
SIRD
MOD
MARD
Clusters

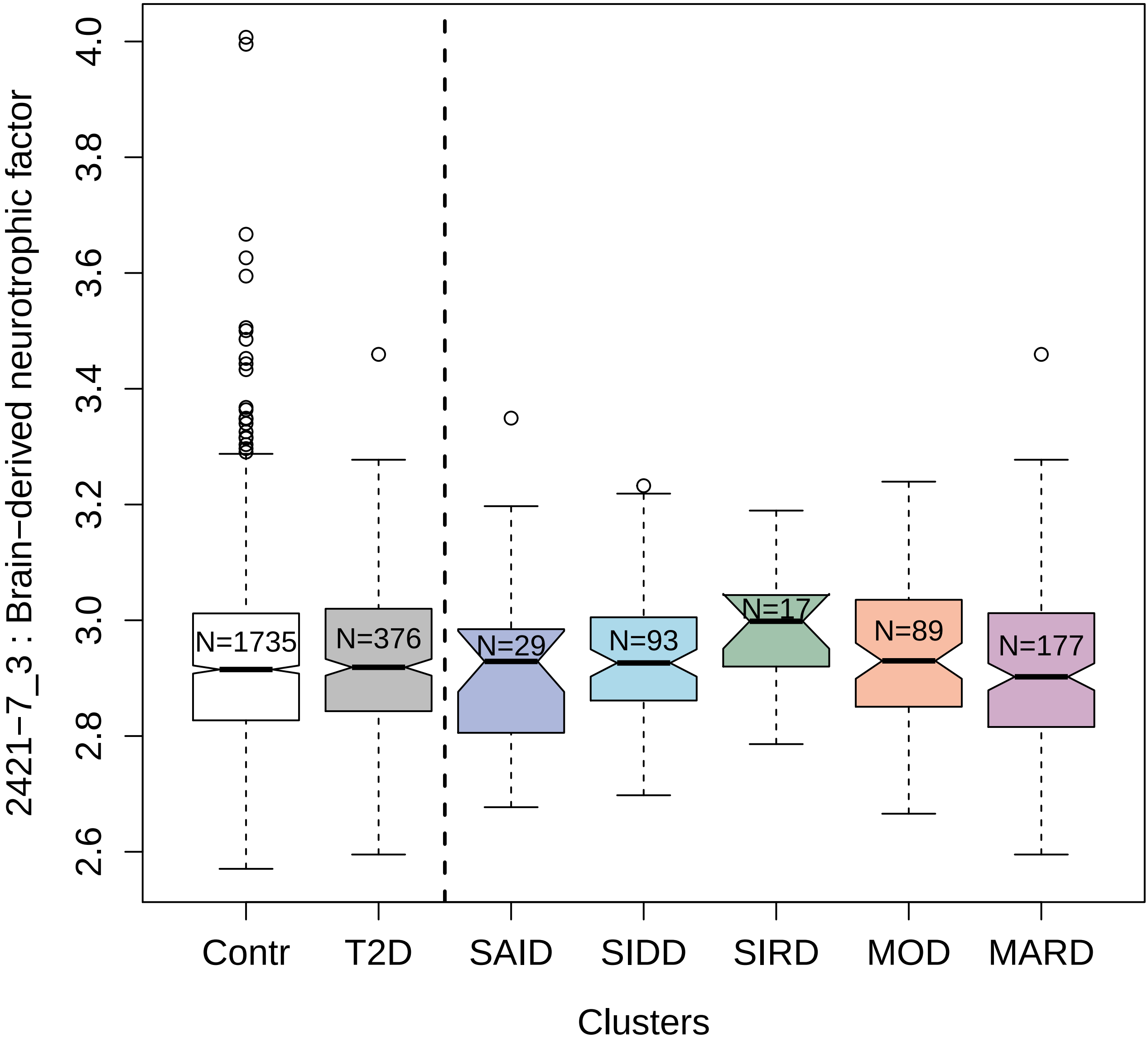
2421−7_3 : Brain−derived neurotrophic factor
4.0
3.8
3.6
3.4
3.2
3.0
2.8
2.6
N=1735
N=376
N=29
N=93
N=17
N=89
N=177
Contr
T2D
SAID
SIDD
SIRD
MOD
MARD
Clusters

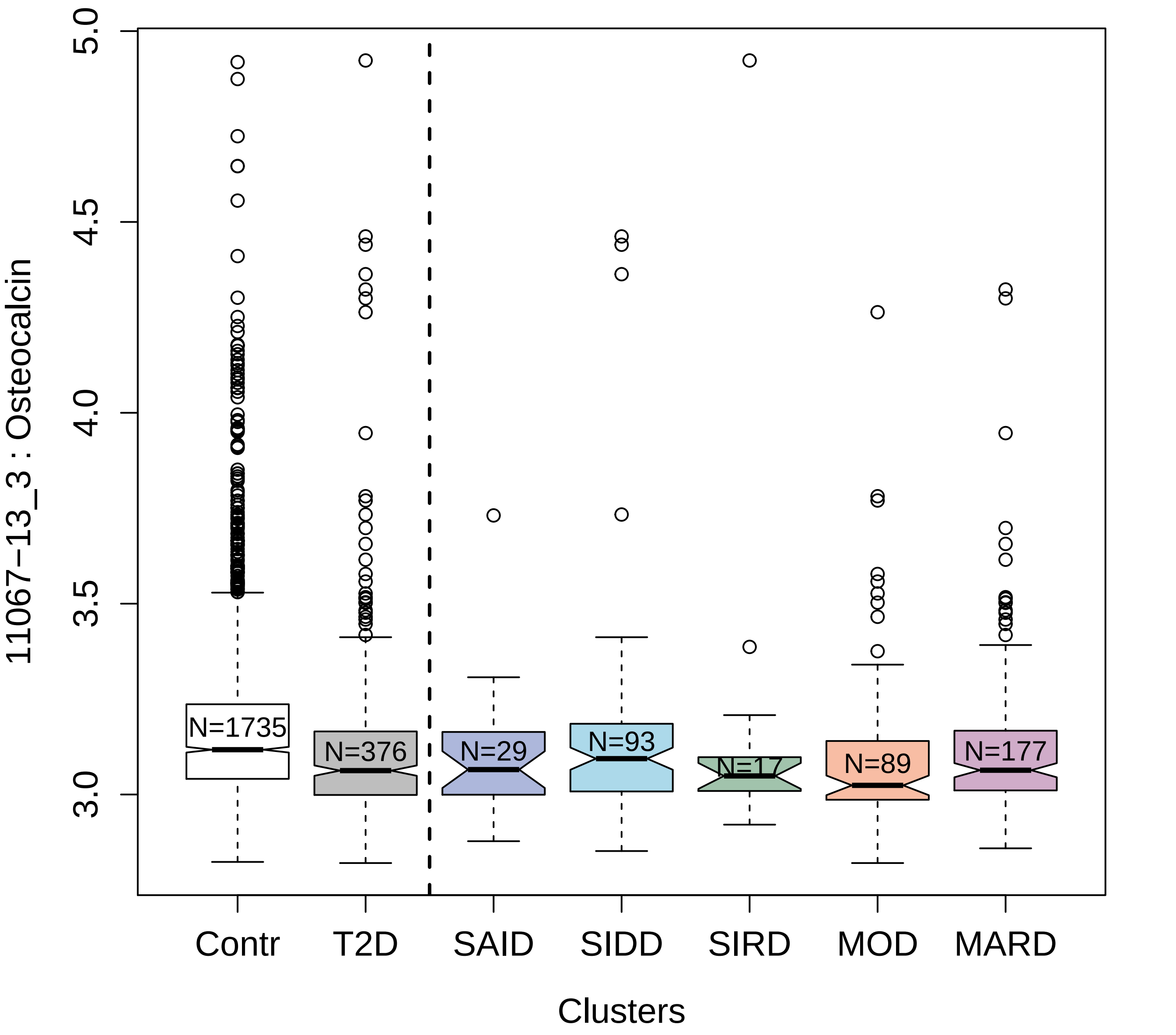

11067−13_3 : Osteocalcin
5.0
4.5
4.0
3.5
3.0
N=1735
N=376
N=29
N=93
N=17
N=89
N=177
Contr
T2D
SAID
SIDD
SIRD
MOD
MARD
Clusters

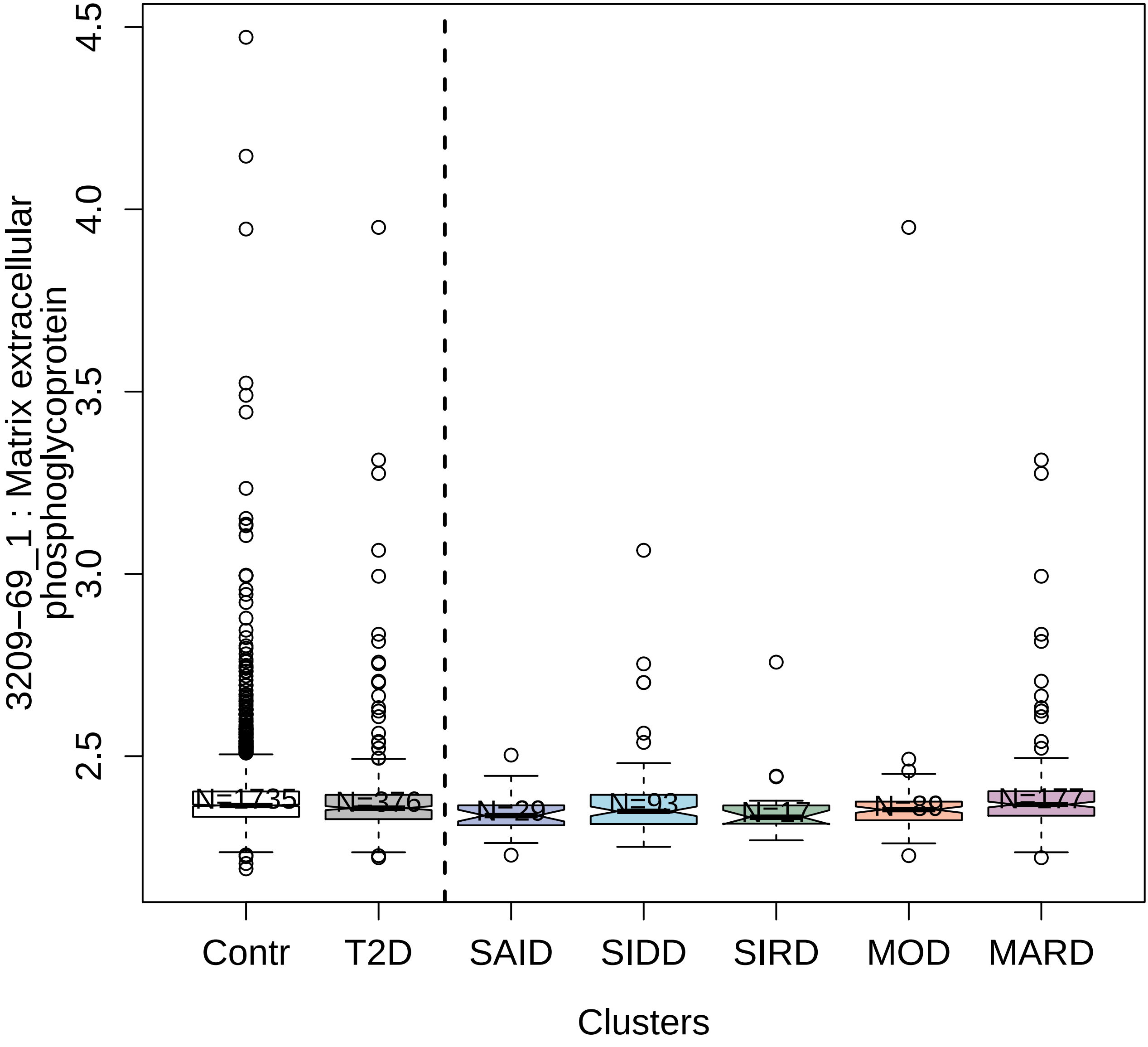

3209−69_1 : Matrix extracellular phosphoglycoprotein
4.5
4.0
3.5
3.0
2.5
N=1735
N=376
N=29
N=93
N=17
N=89
N=177
Contr
T2D
SAID
SIDD
SIRD
MOD
MARD
Clusters

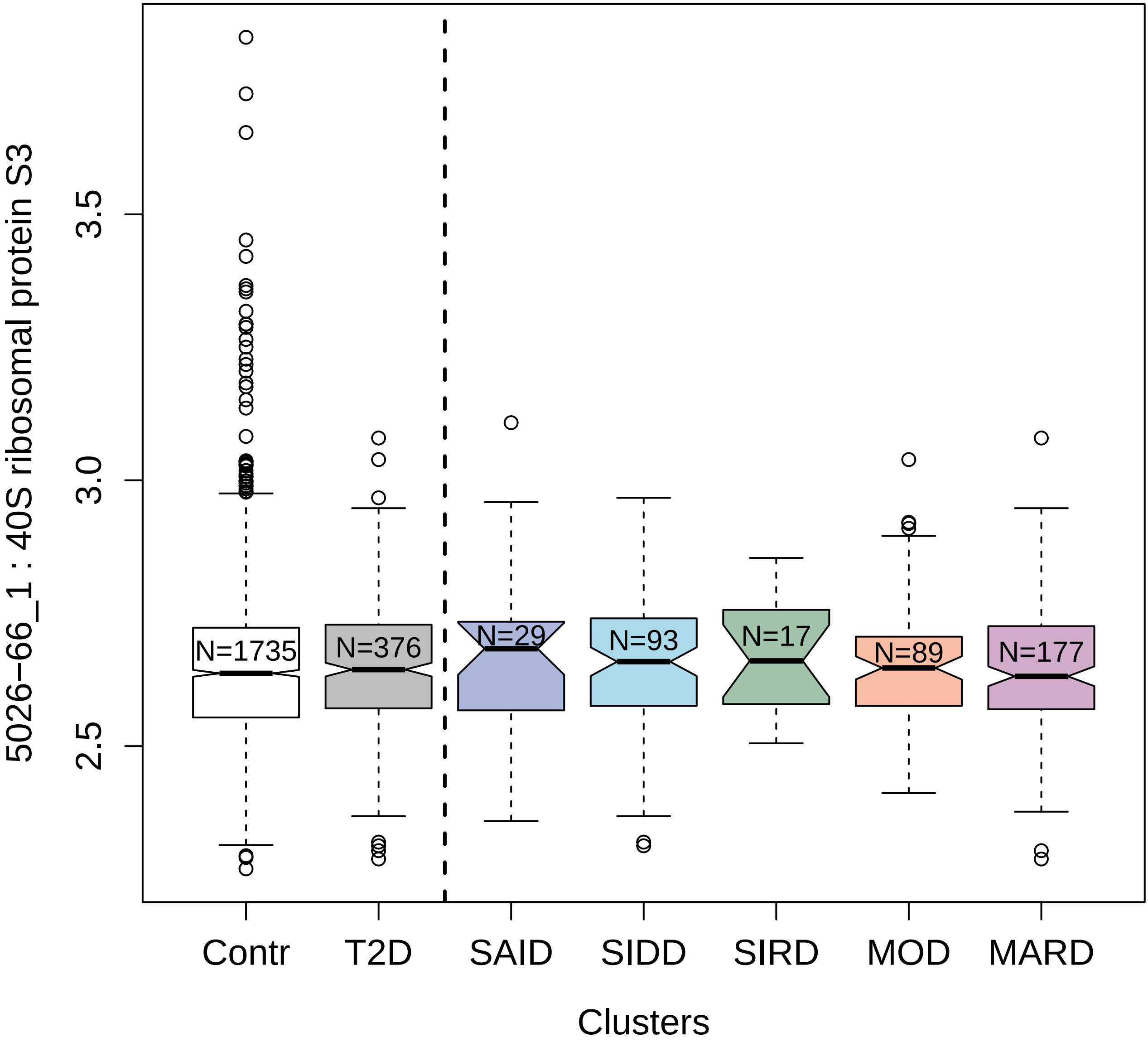
5026−66_1 : 40S ribosomal protein S3
3.5
3.0
2.5
N=1735
N=376
N=29
N=93
N=17
N=89
N=177
Contr
T2D
SAID
SIDD
SIRD
MOD
MARD
Clusters

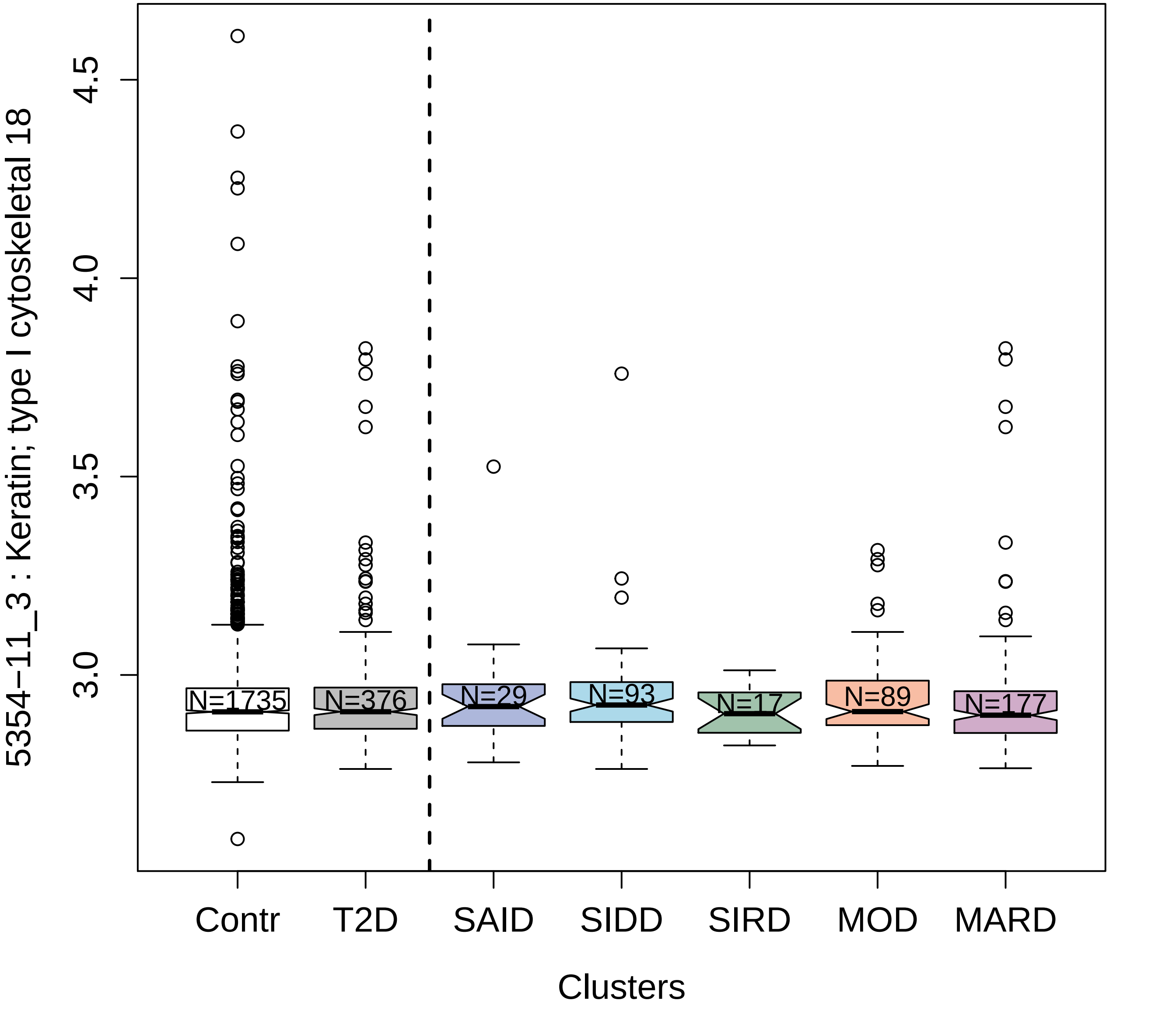

5354−11_3 : Keratin; type I cytoskeletal 18
4.5
4.0
3.5
3.0
N=1735
N=376
N=29
N=93
N=17
N=89
N=177
Contr
T2D
SAID
SIDD
SIRD
MOD
MARD
Clusters

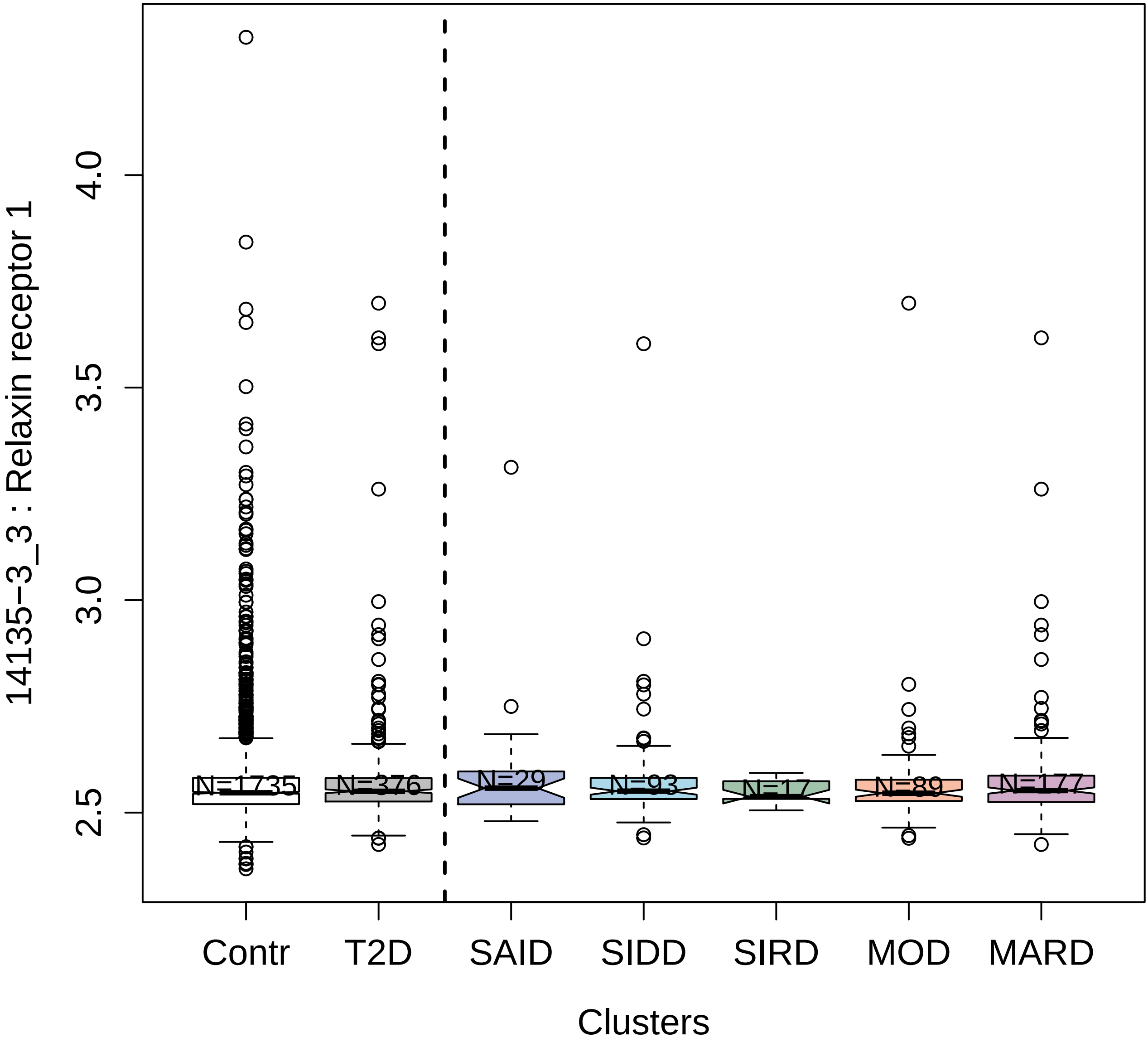

14135−3_3 : Relaxin receptor 1
2.5
3.0
3.5
4.0
N=1735
N=376
N=29
N=93
N=17
N=89
N=177
Contr
T2D
SAID
SIDD
SIRD
MOD
MARD
Clusters

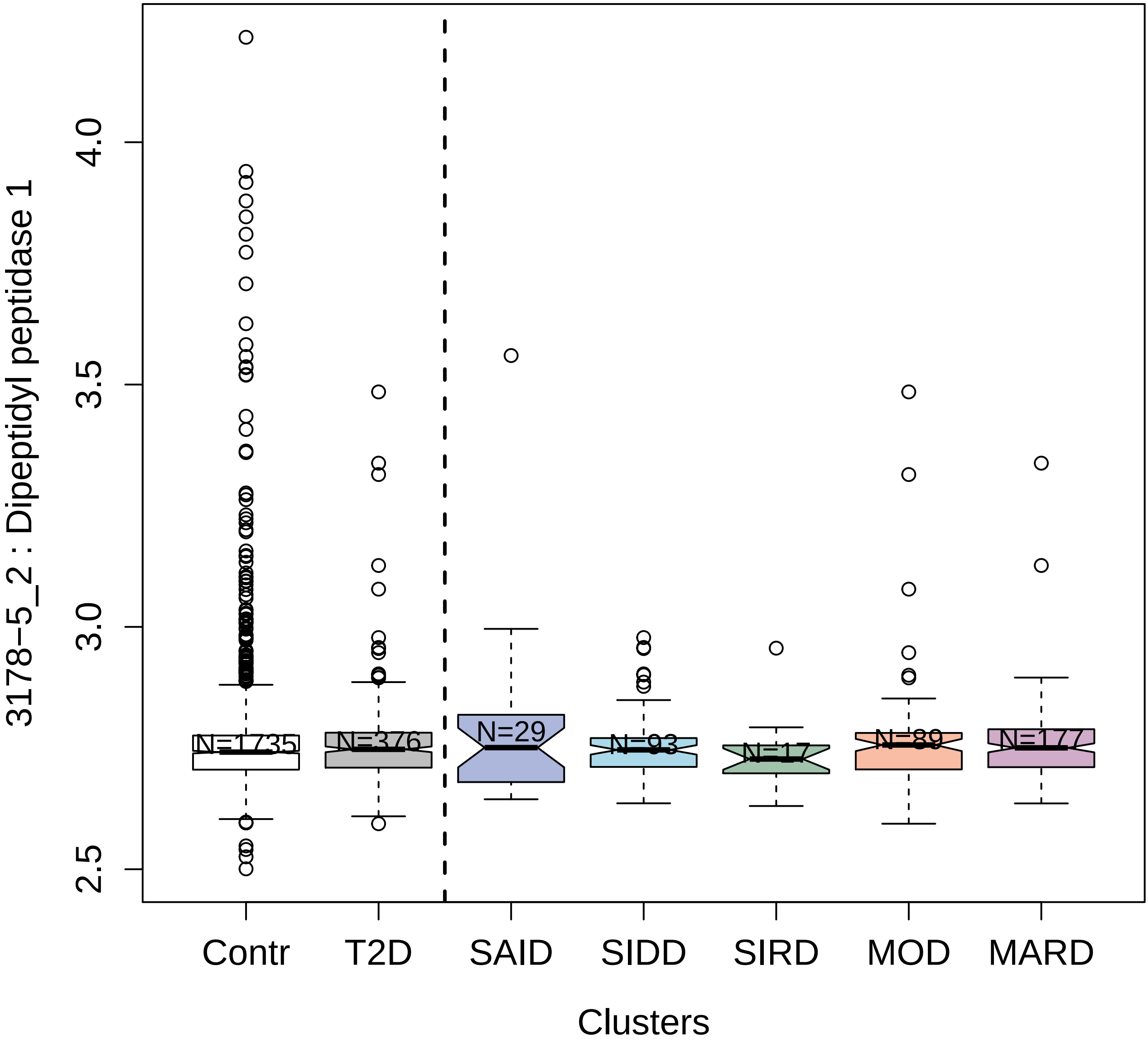

3178−5_2 : Dipeptidyl peptidase 1
4.0
3.5
3.0
2.5
N=1735
N=376
N=29
N=93
N=17
N=89
N=177
Contr
T2D
SAID
SIDD
SIRD
MOD
MARD
Clusters

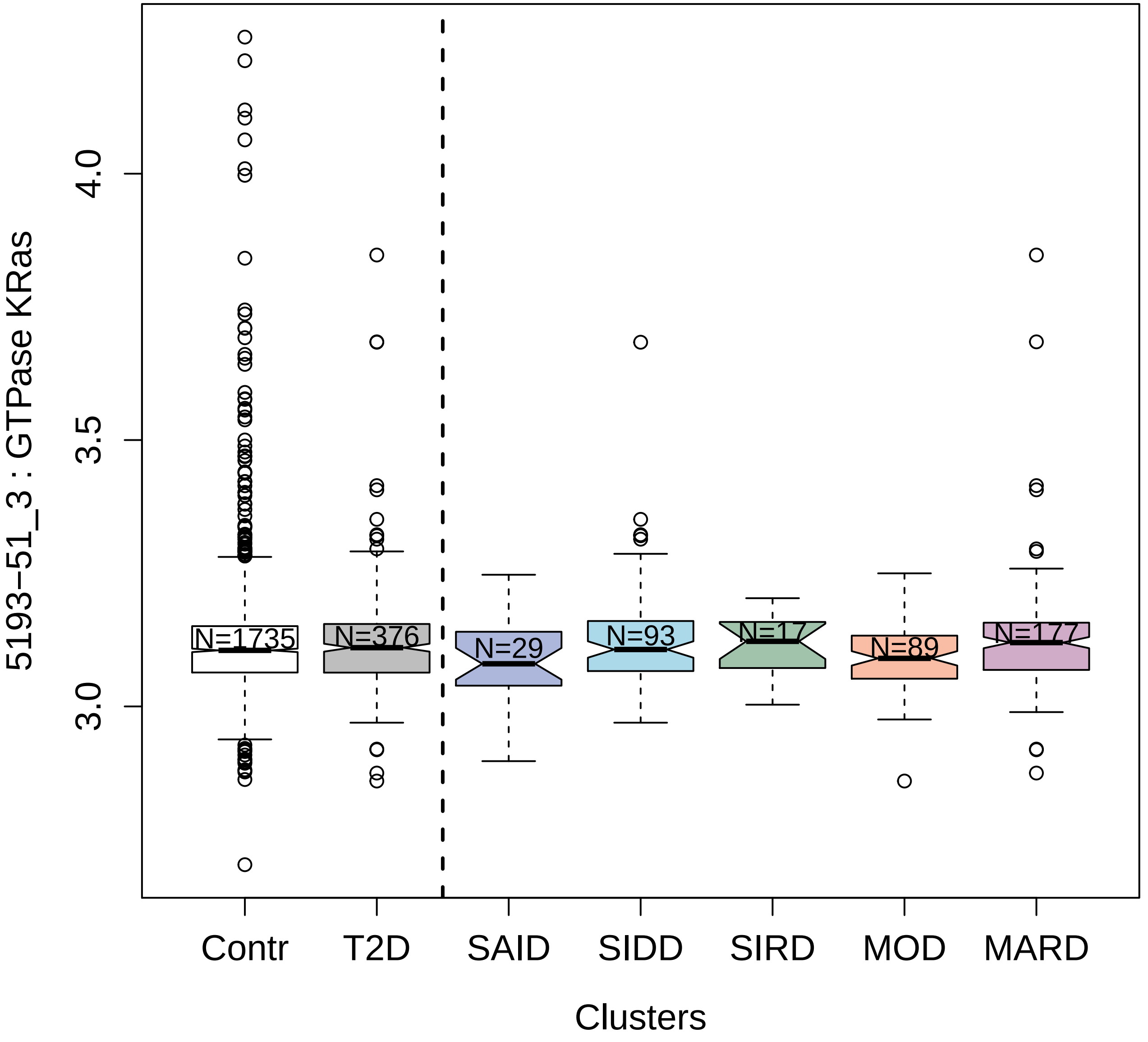

5193−51_3 : GTPase KRas
4.0
3.5
3.0
N=1735
N=376
N=29
N=93
N=17
N=89
N=177
Contr
T2D
SAID
SIDD
SIRD
MOD
MARD
Clusters

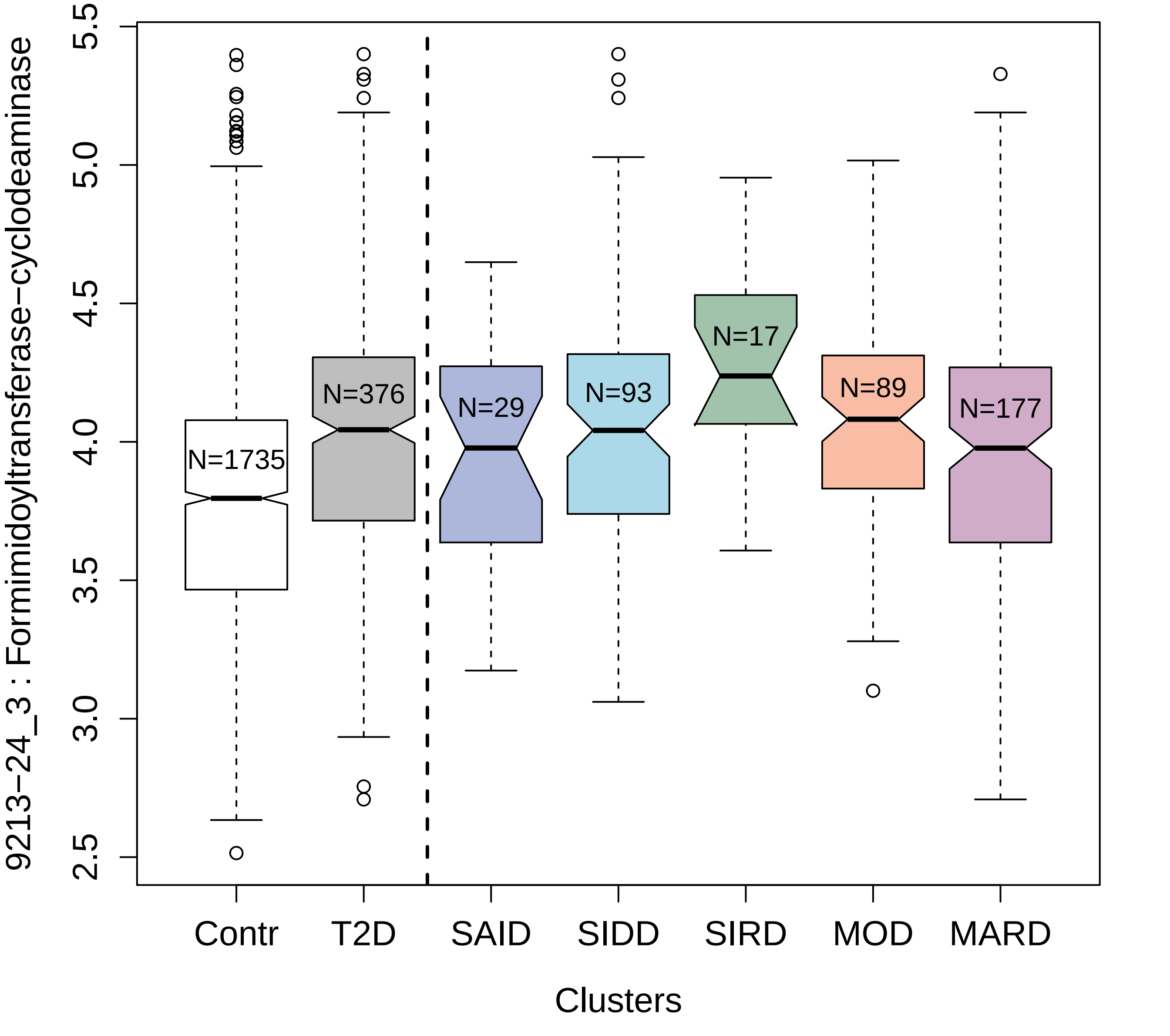

9213−24_3 : Formimidoyltransferase−cyclodeaminase
5.5
5.0
4.5
4.0
3.5
3.0
2.5
N=1735
N=376
N=29
N=93
N=17
N=89
N=177
Contr
T2D
SAID
SIDD
SIRD
MOD
MARD
Clusters

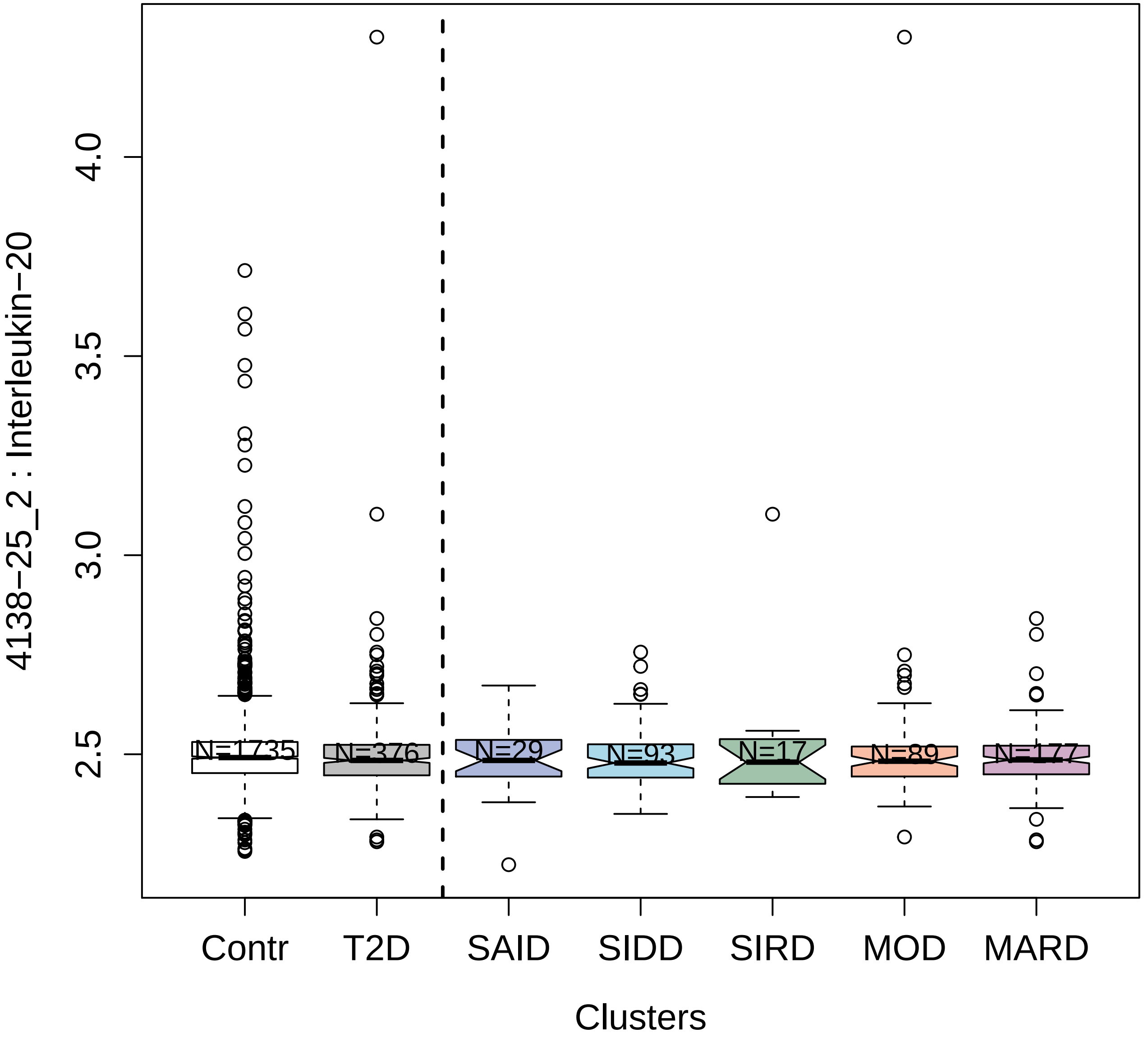

4138−25_2 : Interleukin−20
4.0
3.5
3.0
2.5
N=1735
N=376
N=29
N=93
N=17
N=89
N=177
Contr
T2D
SAID
SIDD
SIRD
MOD
MARD
Clusters

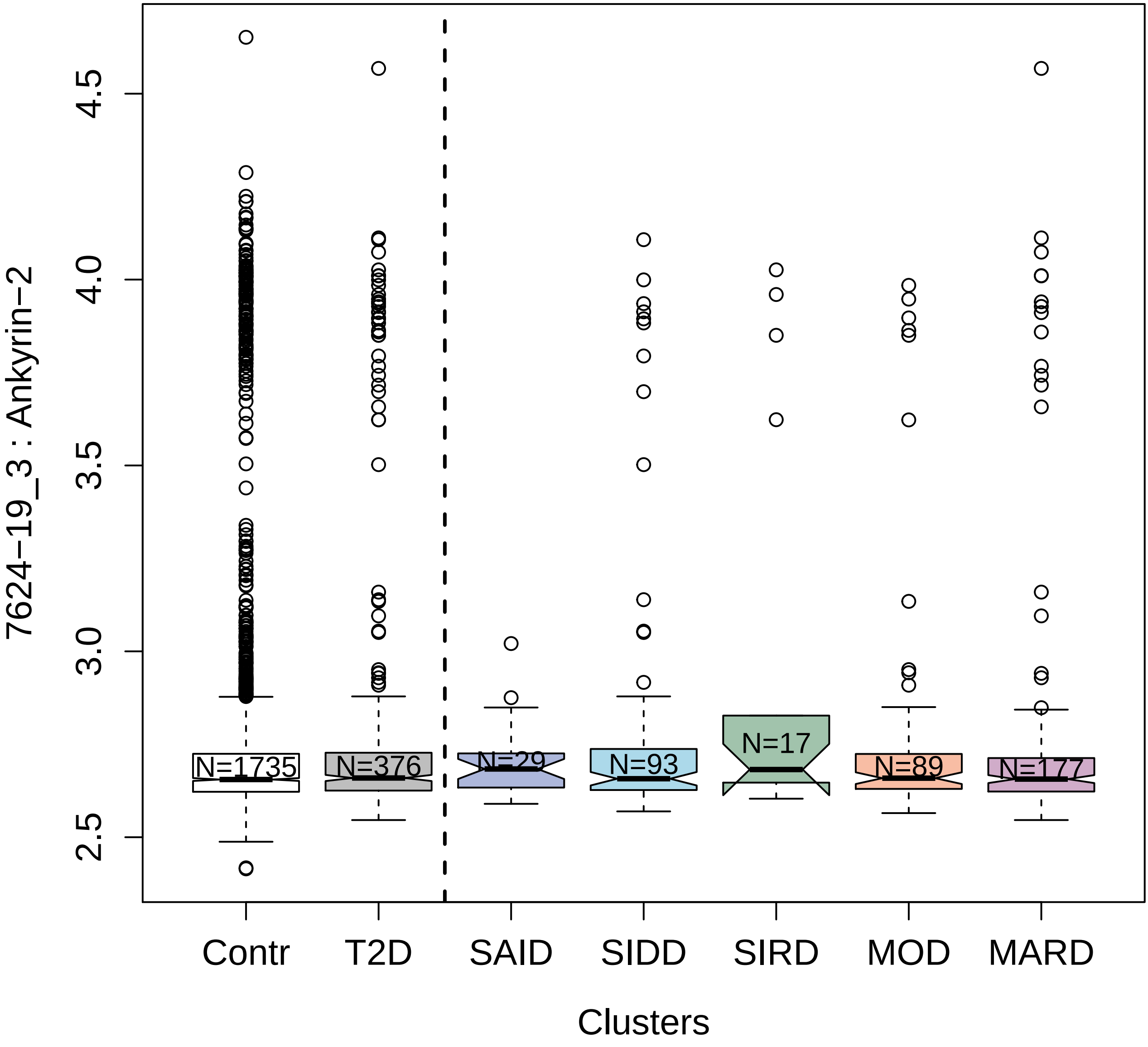
7624−19_3 : Ankyrin−2
2.5
3.0
3.5
4.0
4.5
N=1735
N=376
N=29
N=93
N=17
N=89
N=177
Contr
T2D
SAID
SIDD
SIRD
MOD
MARD
Clusters

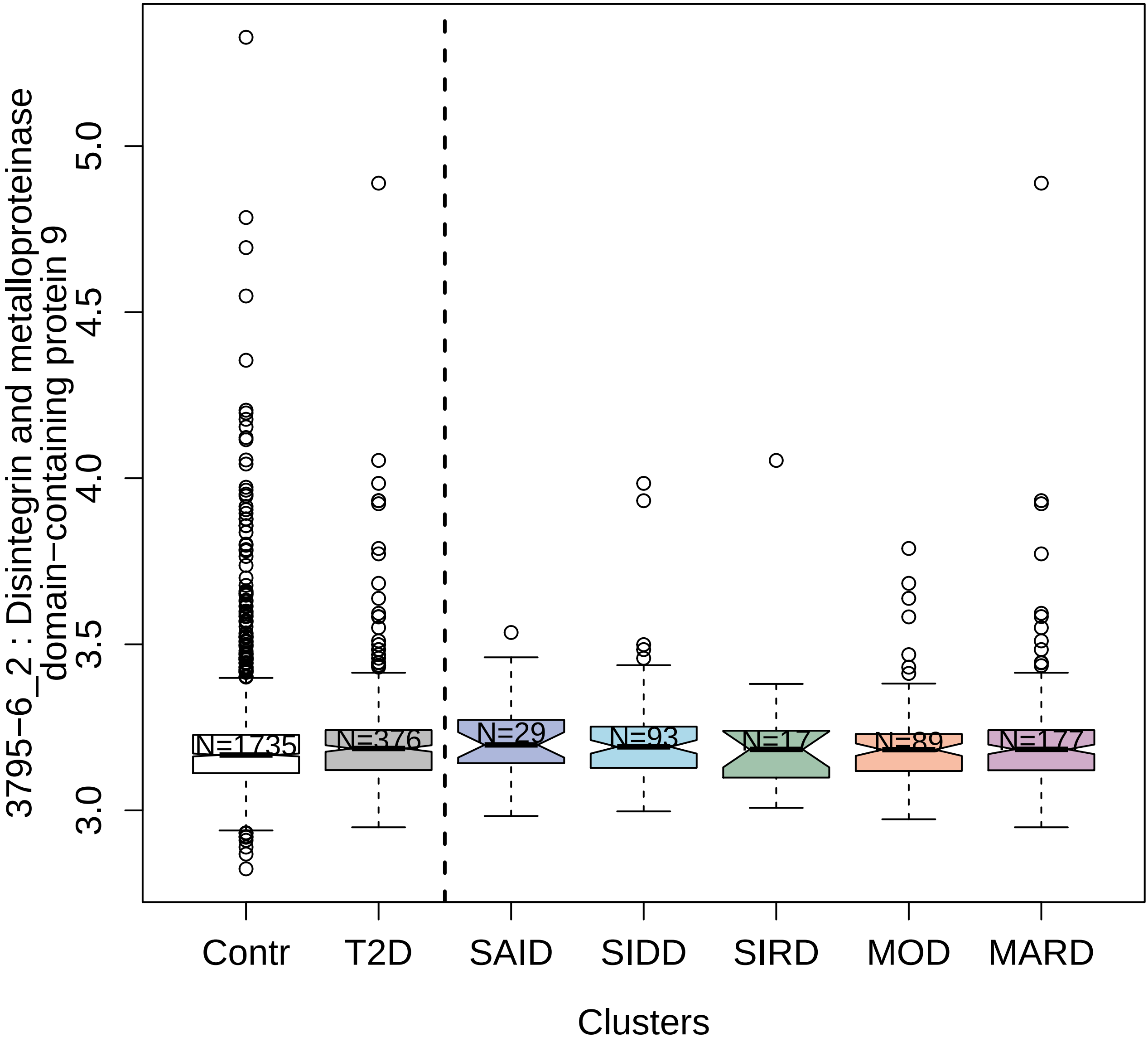
3795−6_2 : Disintegrin and metalloproteinase domain−containing protein 9
5.0
4.5
4.0
3.5
3.0
N=1735
N=376
N=29
N=93
N=17
N=89
N=177
Contr
T2D
SAID
SIDD
SIRD
MOD
MARD
Clusters

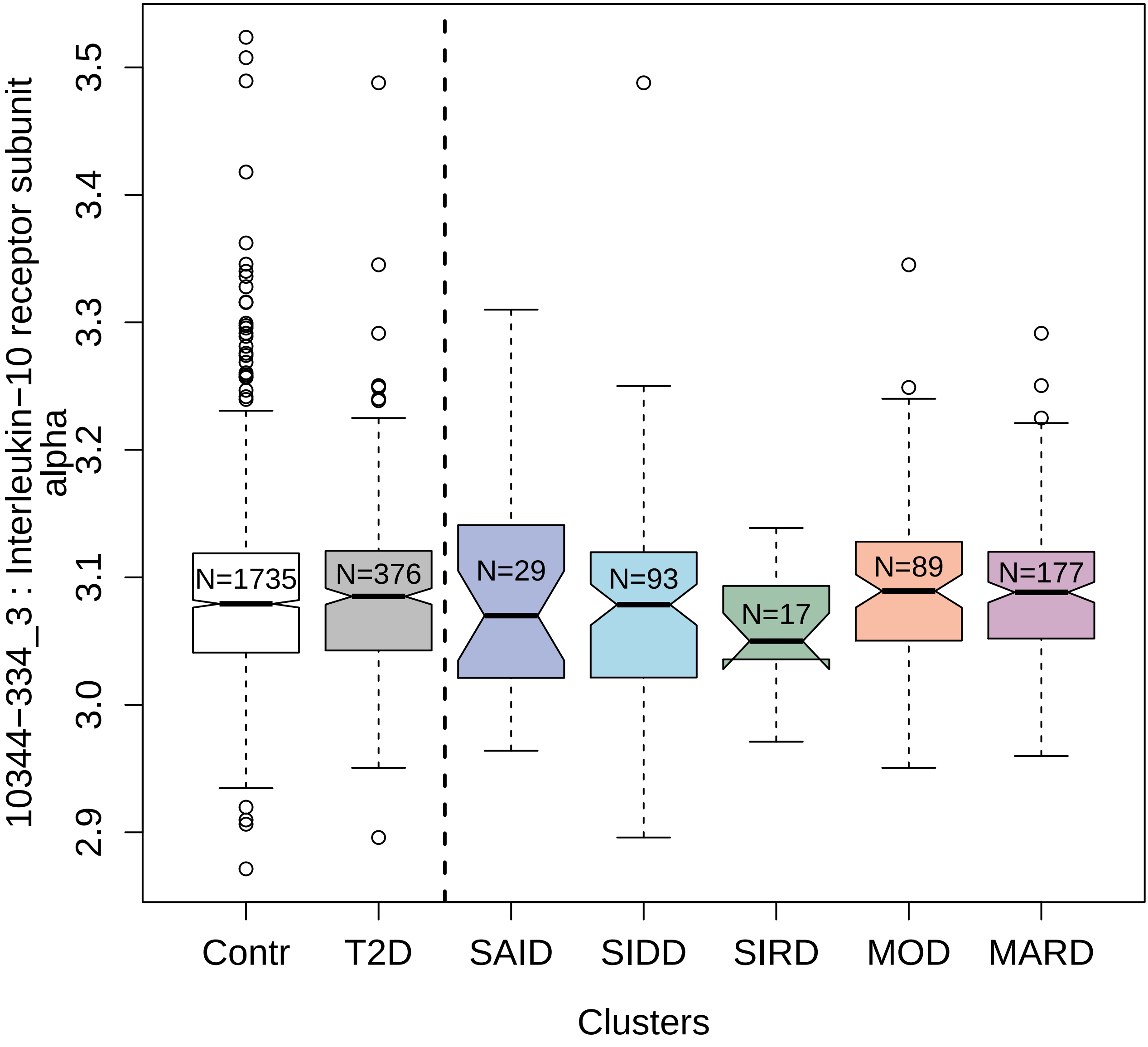

10344−334_3 : Interleukin−10 receptor subunit alpha
2.9
3.0
3.1
3.2
3.3
3.4
3.5
N=1735
N=376
N=29
N=93
N=17
N=89
N=177
Contr
T2D
SAID
SIDD
SIRD
MOD
MARD
Clusters

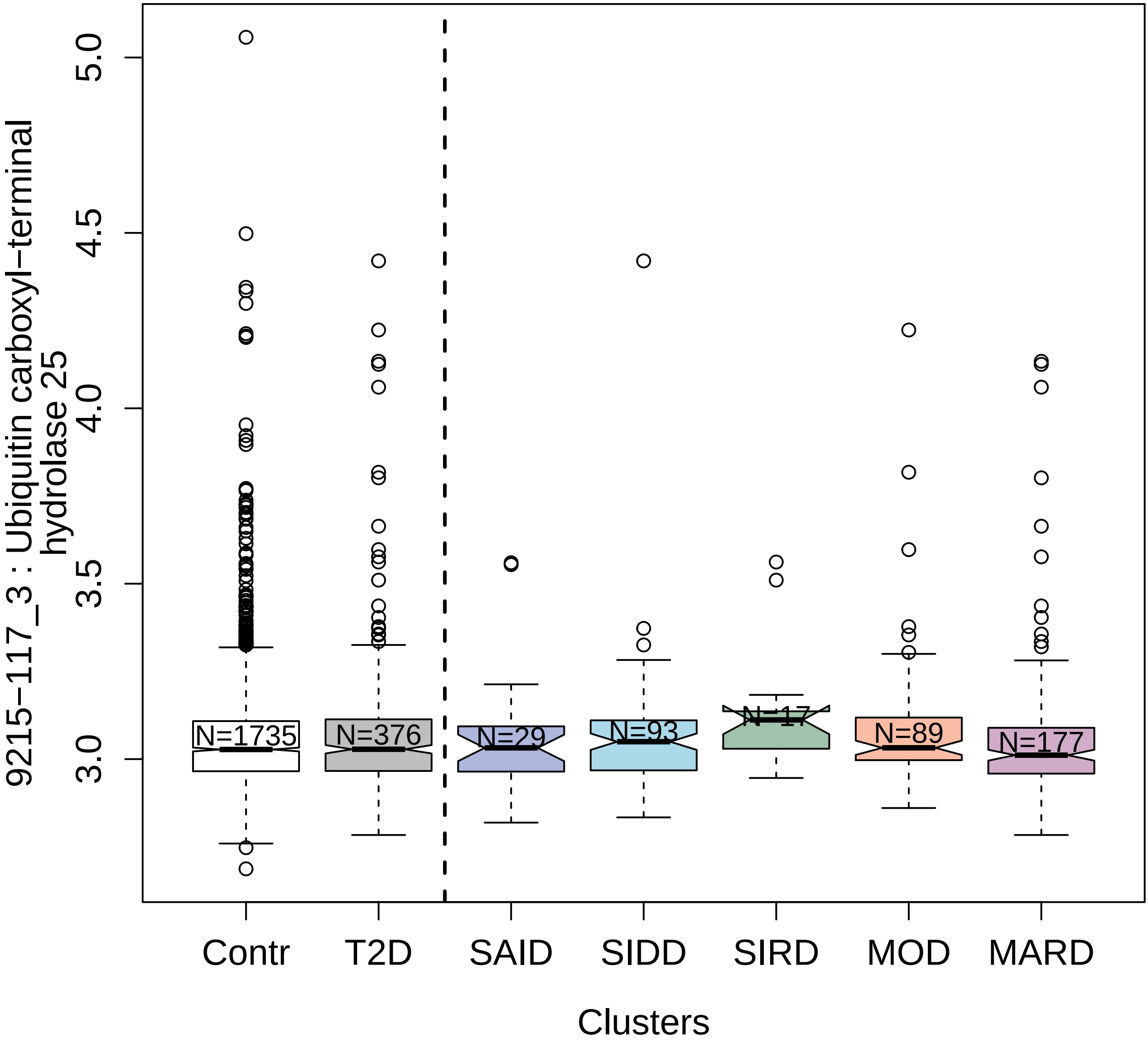

9215−117_3 : Ubiquitin carboxyl−terminal hydrolase 25
5.0
4.5
4.0
3.5
3.0
N=1735
N=376
N=29
N=93
N=17
N=89
N=177
Contr
T2D
SAID
SIDD
SIRD
MOD
MARD
Clusters

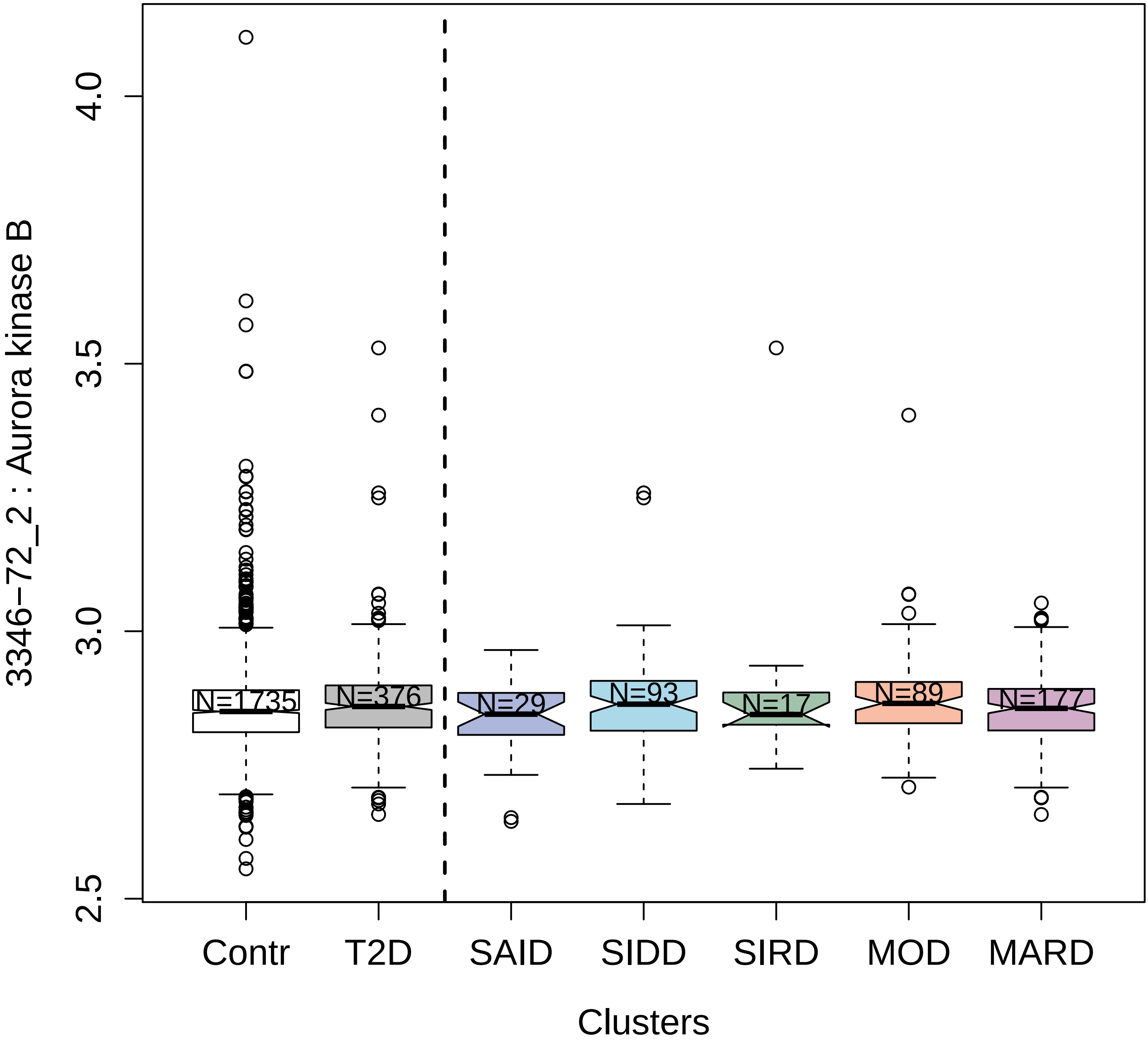

3346−72_2 : Aurora kinase B
4.0
3.5
3.0
2.5
N=1735
N=376
N=29
N=93
N=17
N=89
N=177
Contr
T2D
SAID
SIDD
SIRD
MOD
MARD
Clusters

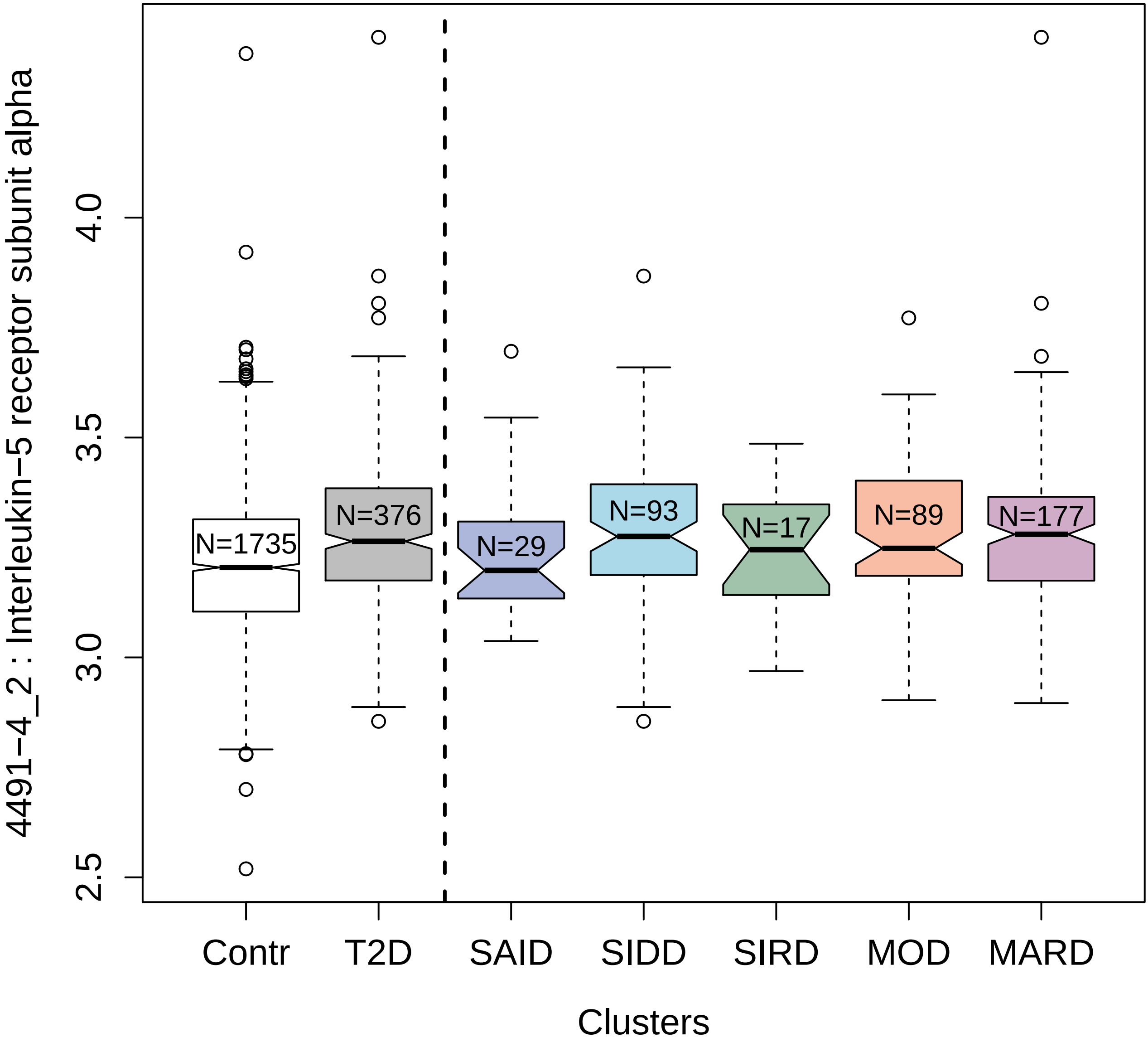

4491−4_2 : Interleukin−5 receptor subunit alpha
Clusters
Contr
T2D
SAID
SIDD
SIRD
MOD
MARD
N=1735
N=376
N=29
N=93
N=17
N=89
N=177
2.5
3.0
3.5
4.0

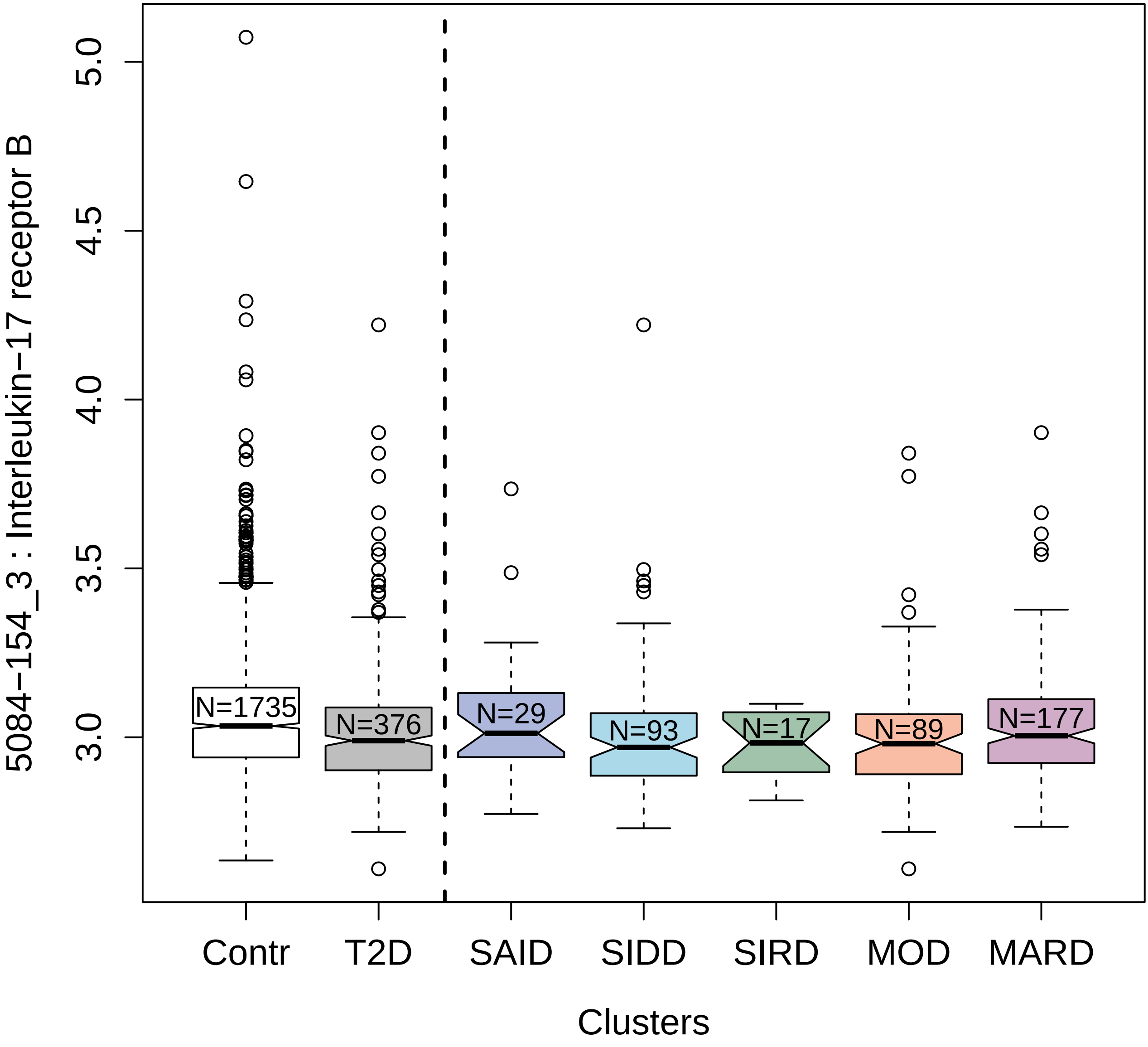
5084−154_3 : Interleukin−17 receptor B
5.0
4.5
4.0
3.5
3.0
N=1735
N=376
N=29
N=93
N=17
N=89
N=177
Contr
T2D
SAID
SIDD
SIRD
MOD
MARD
Clusters

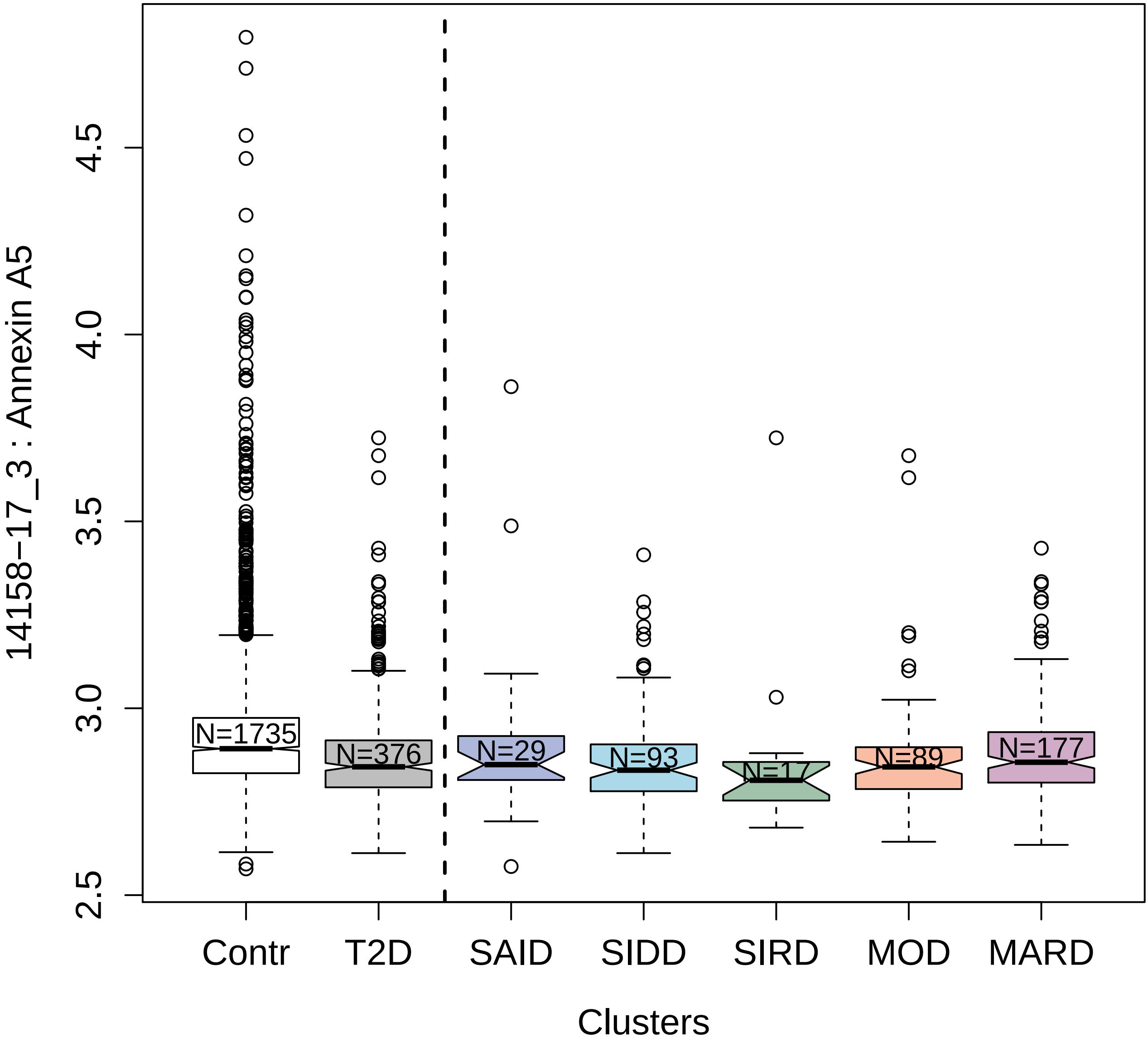

14158−17_3 : Annexin A5
4.5
4.0
3.5
3.0
2.5
N=1735
N=376
N=29
N=93
N=17
N=89
N=177
Contr
T2D
SAID
SIDD
SIRD
MOD
MARD
Clusters

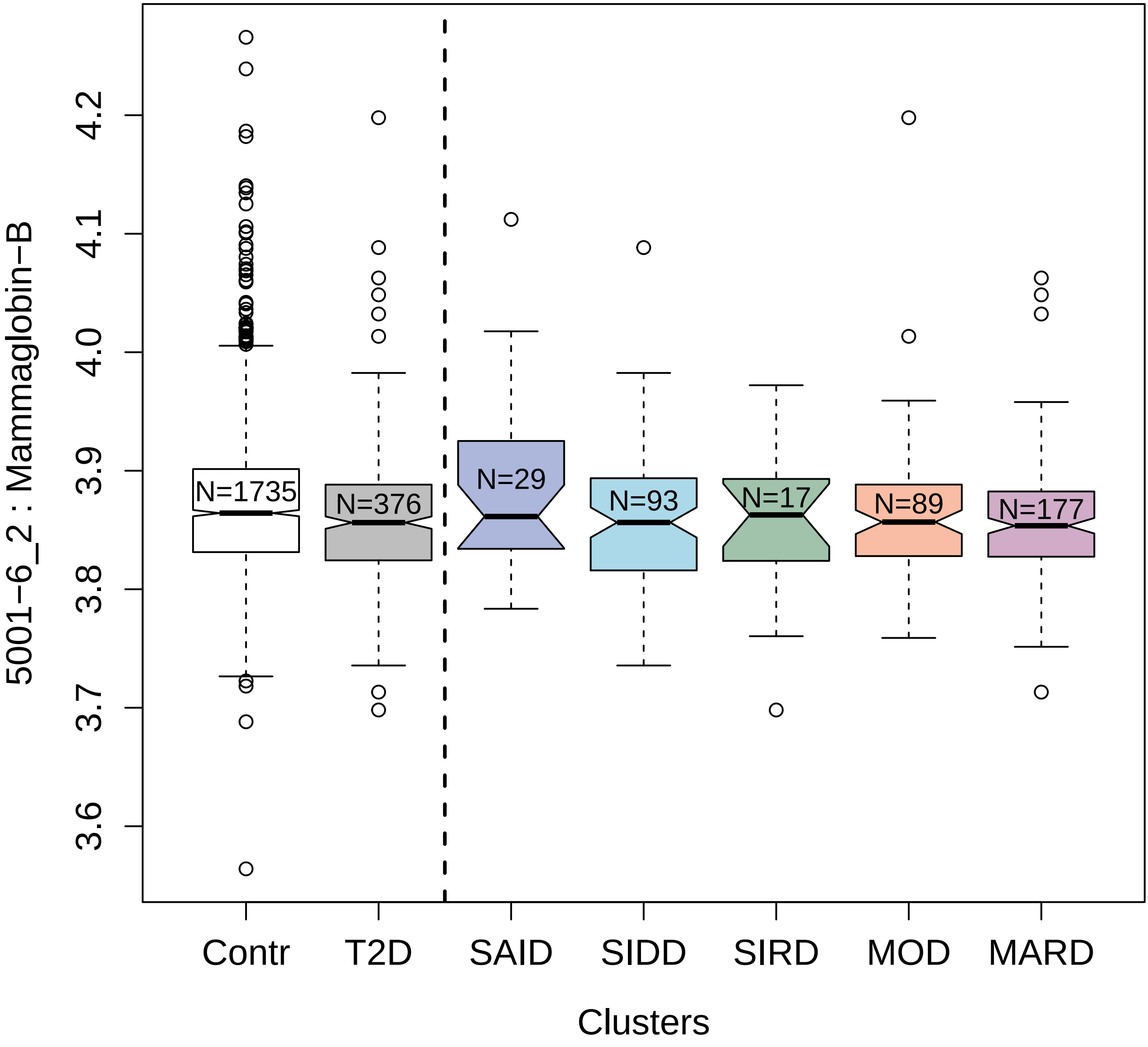

5001−6_2 : Mammaglobin−B
3.6
3.7
3.8
3.9
4.0
4.1
4.2
N=1735
N=376
N=29
N=93
N=17
N=89
N=177
Contr
T2D
SAID
SIDD
SIRD
MOD
MARD
Clusters

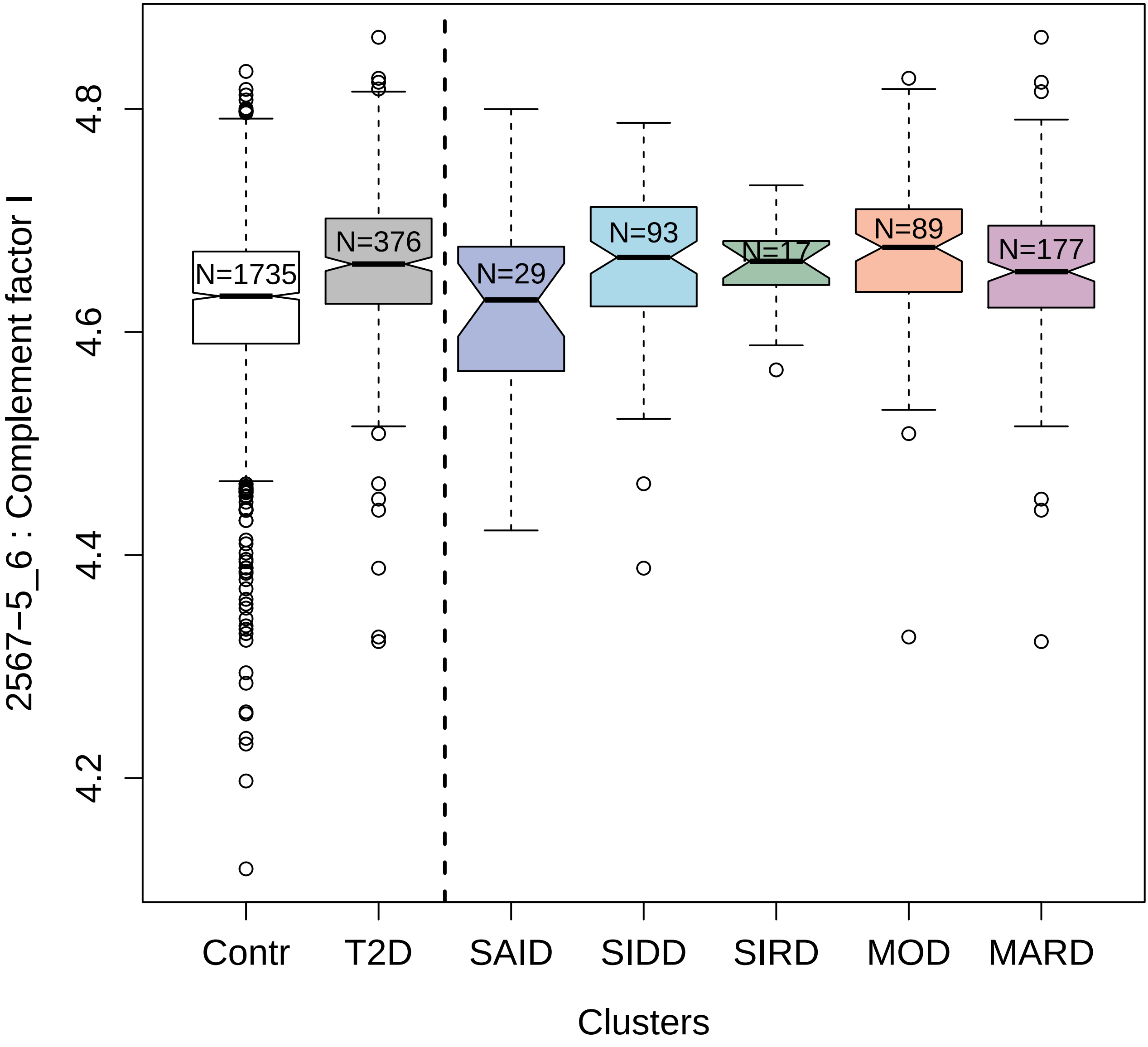
2567−5_6 : Complement factor I
4.2
4.4
4.6
4.8
N=1735
N=376
N=29
N=93
N=17
N=89
N=177
Contr
T2D
SAID
SIDD
SIRD
MOD
MARD
Clusters

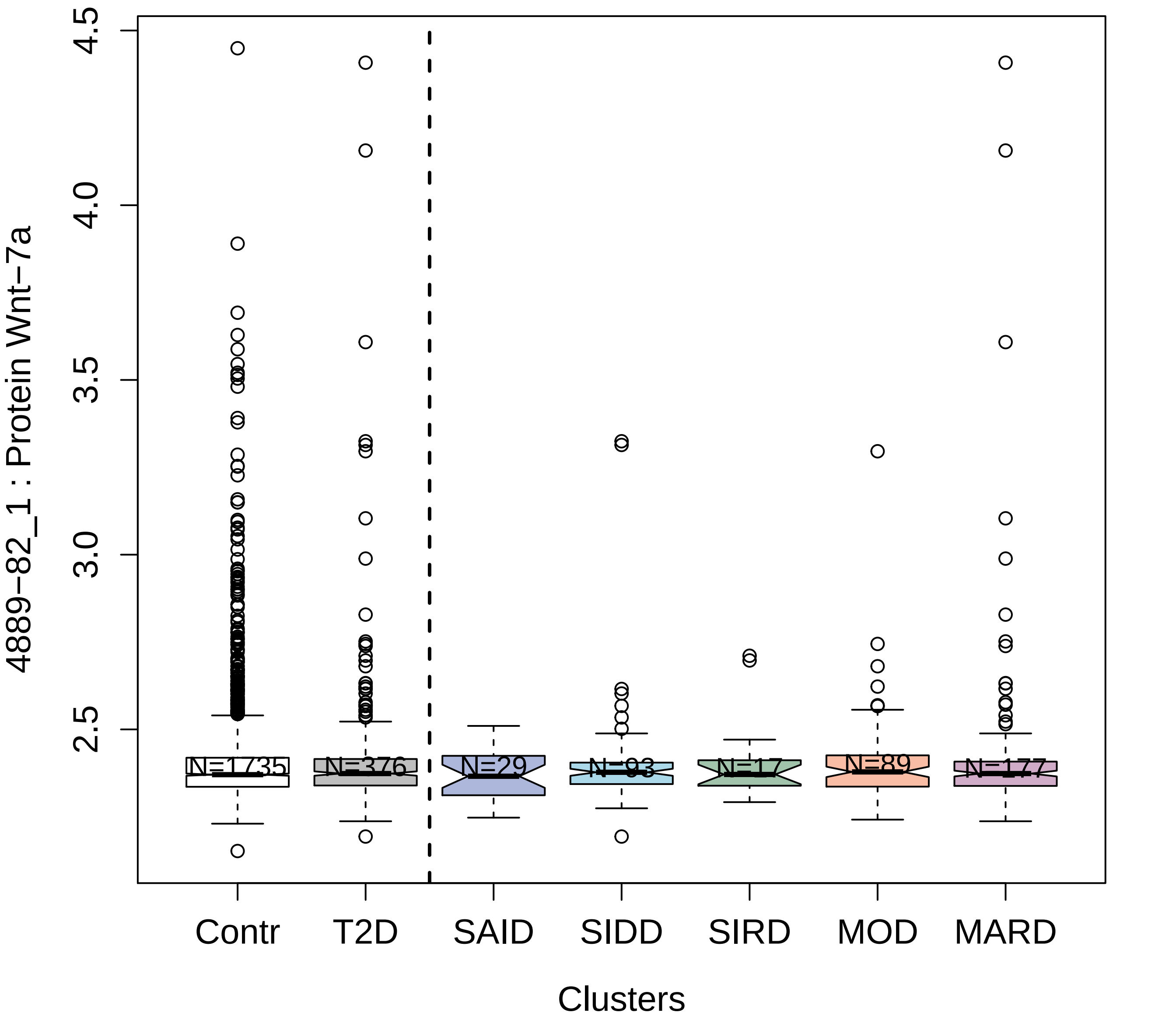
4889−82_1 : Protein Wnt−7a
4.5
4.0
3.5
3.0
2.5
N=1735
N=376
N=29
N=93
N=17
N=89
N=177
Contr
T2D
SAID
SIDD
SIRD
MOD
MARD
Clusters

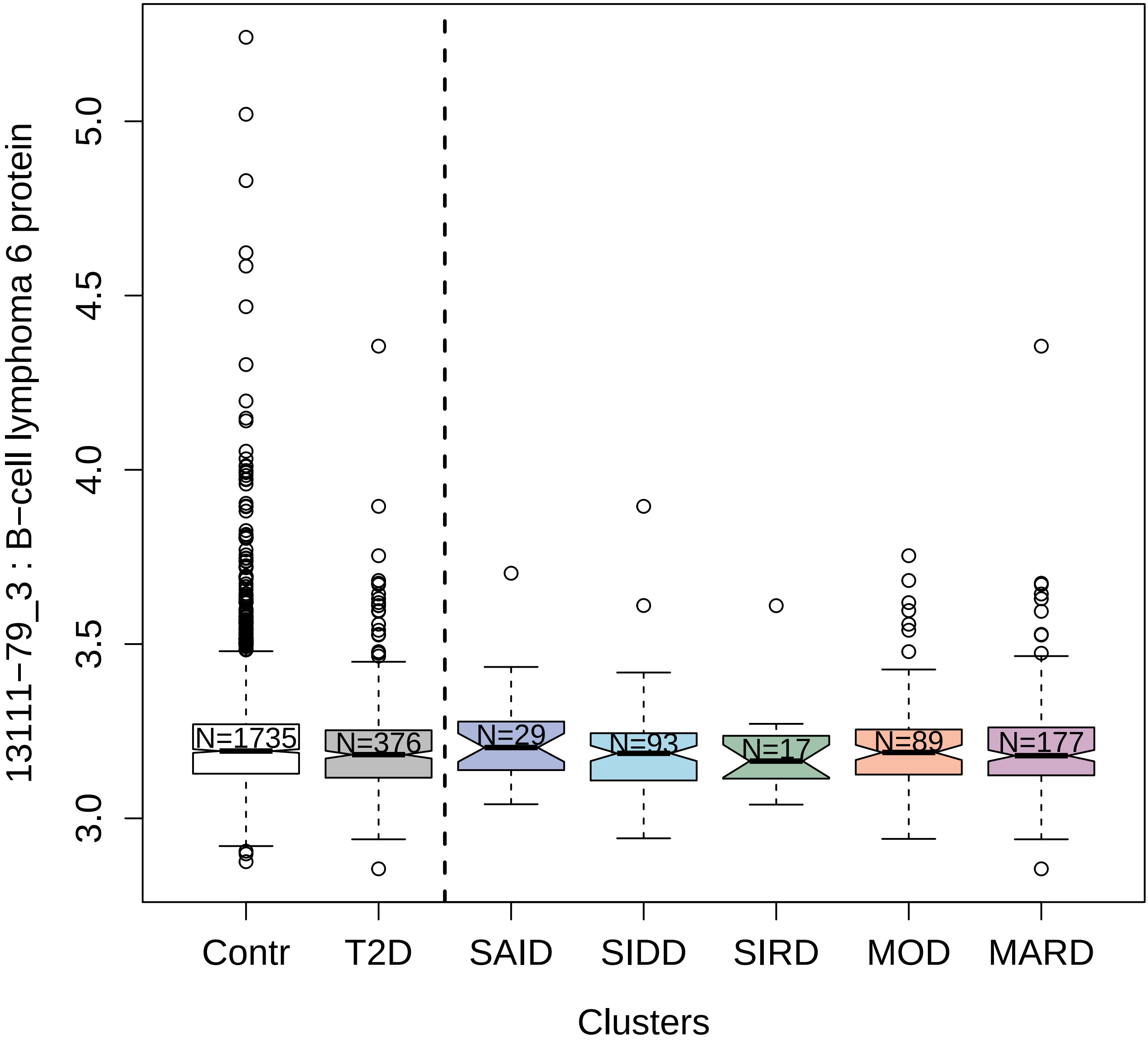

13111−79_3 : B−cell lymphoma 6 protein
3.0
3.5
4.0
4.5
5.0
N=1735
N=376
N=29
N=93
N=17
N=89
N=177
Contr
T2D
SAID
SIDD
SIRD
MOD
MARD
Clusters

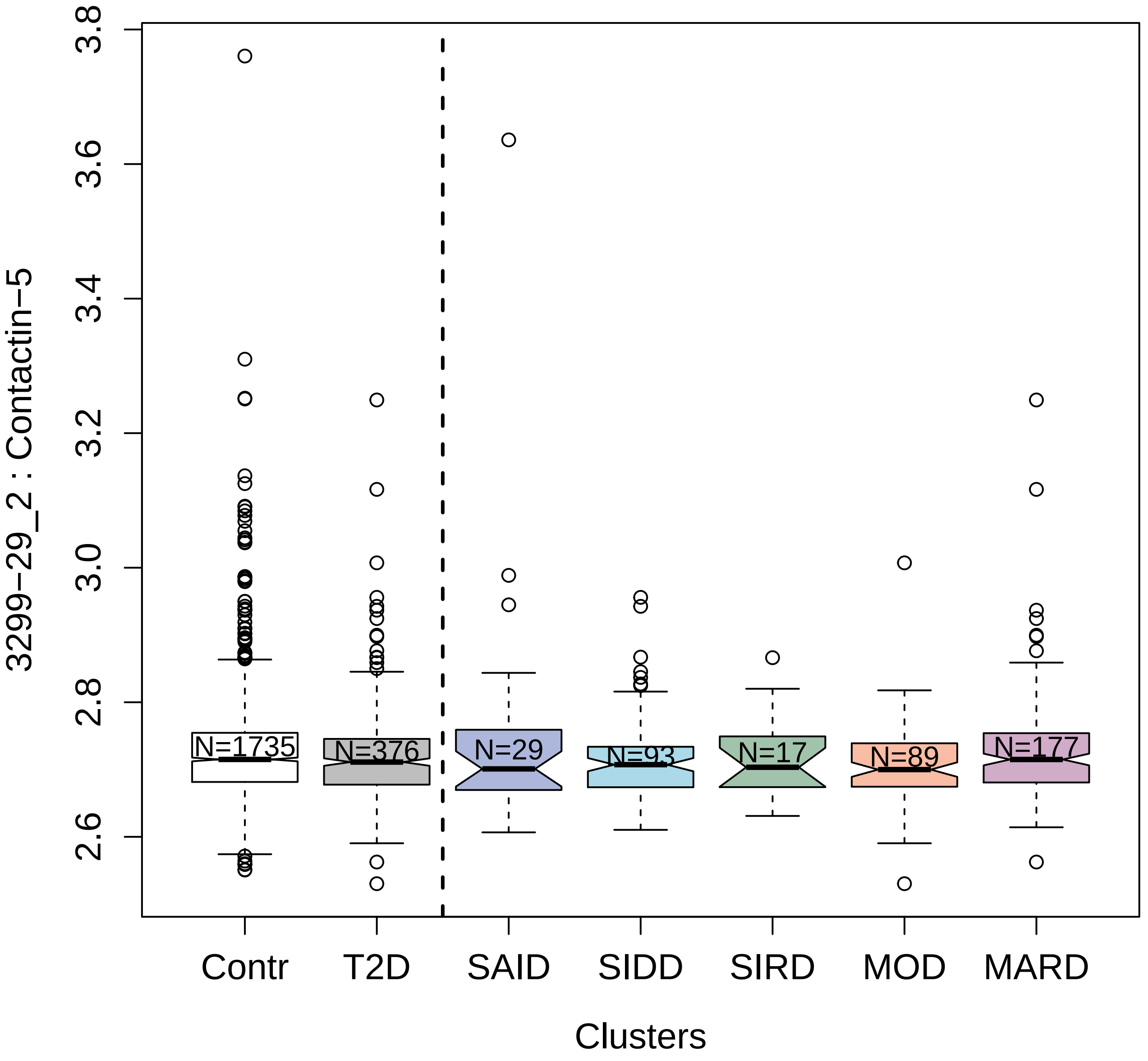
3299−29_2 : Contactin−5
3.8
3.6
3.4
3.2
3.0
2.8
2.6
N=1735
N=376
N=29
N=93
N=17
N=89
N=177
Contr
T2D
SAID
SIDD
SIRD
MOD
MARD
Clusters

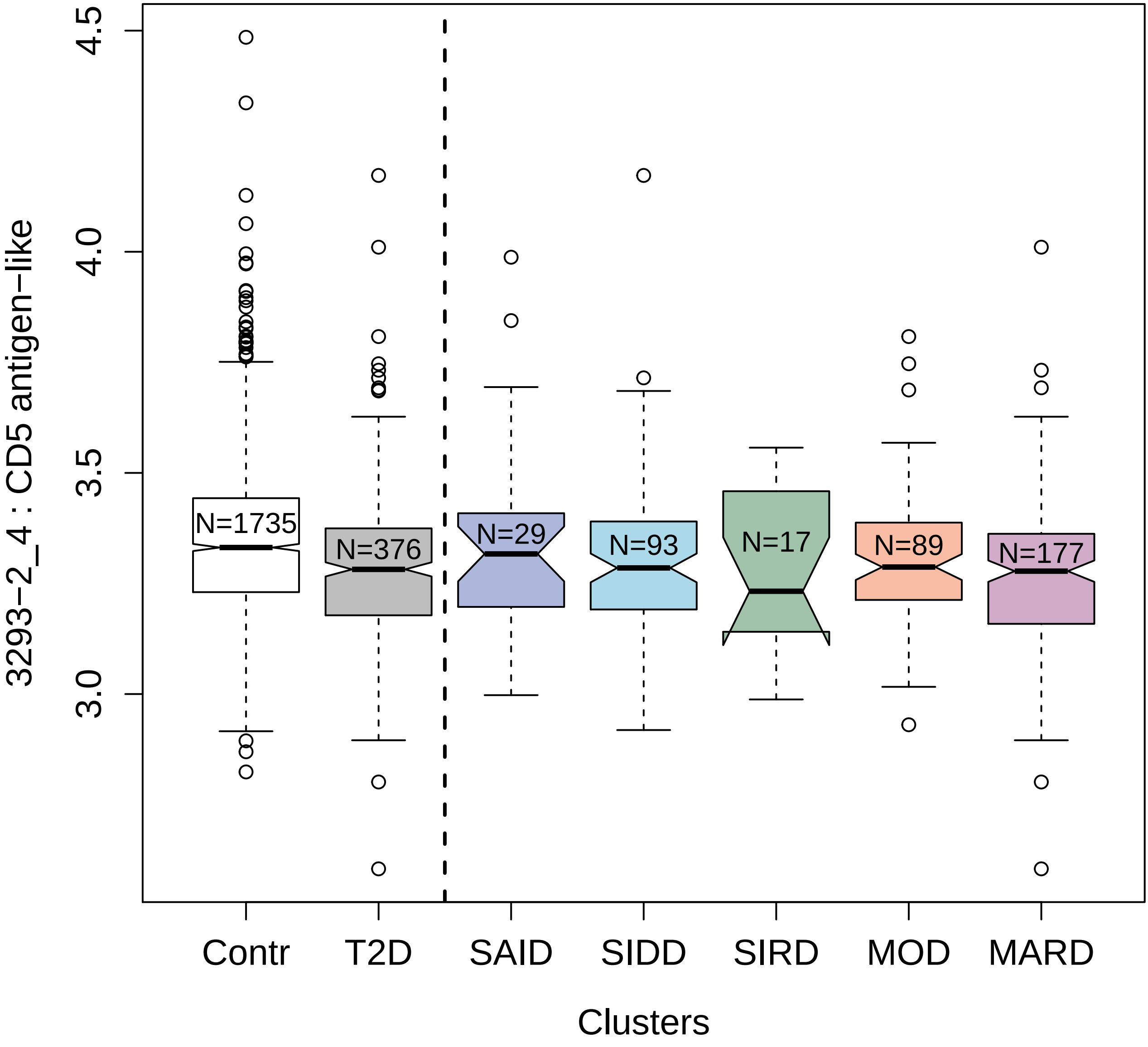
3293−2_4 : CD5 antigen−like
4.5
4.0
3.5
3.0
N=1735
N=376
N=29
N=93
N=17
N=89
N=177
Contr
T2D
SAID
SIDD
SIRD
MOD
MARD
Clusters

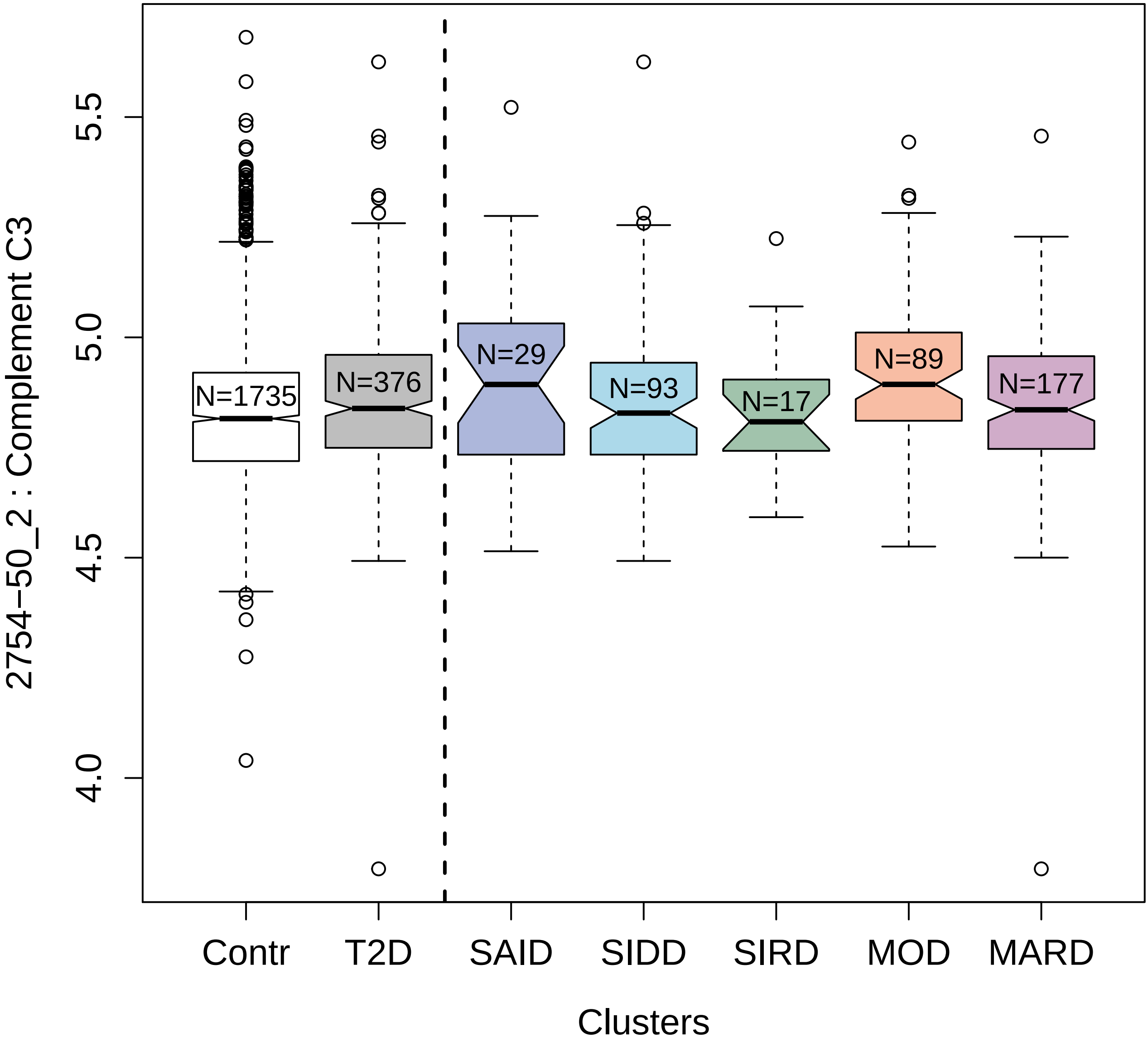

2754−50_2 : Complement C3
5.5
5.0
4.5
4.0
N=1735
N=376
N=29
N=93
N=17
N=89
N=177
Contr
T2D
SAID
SIDD
SIRD
MOD
MARD
Clusters

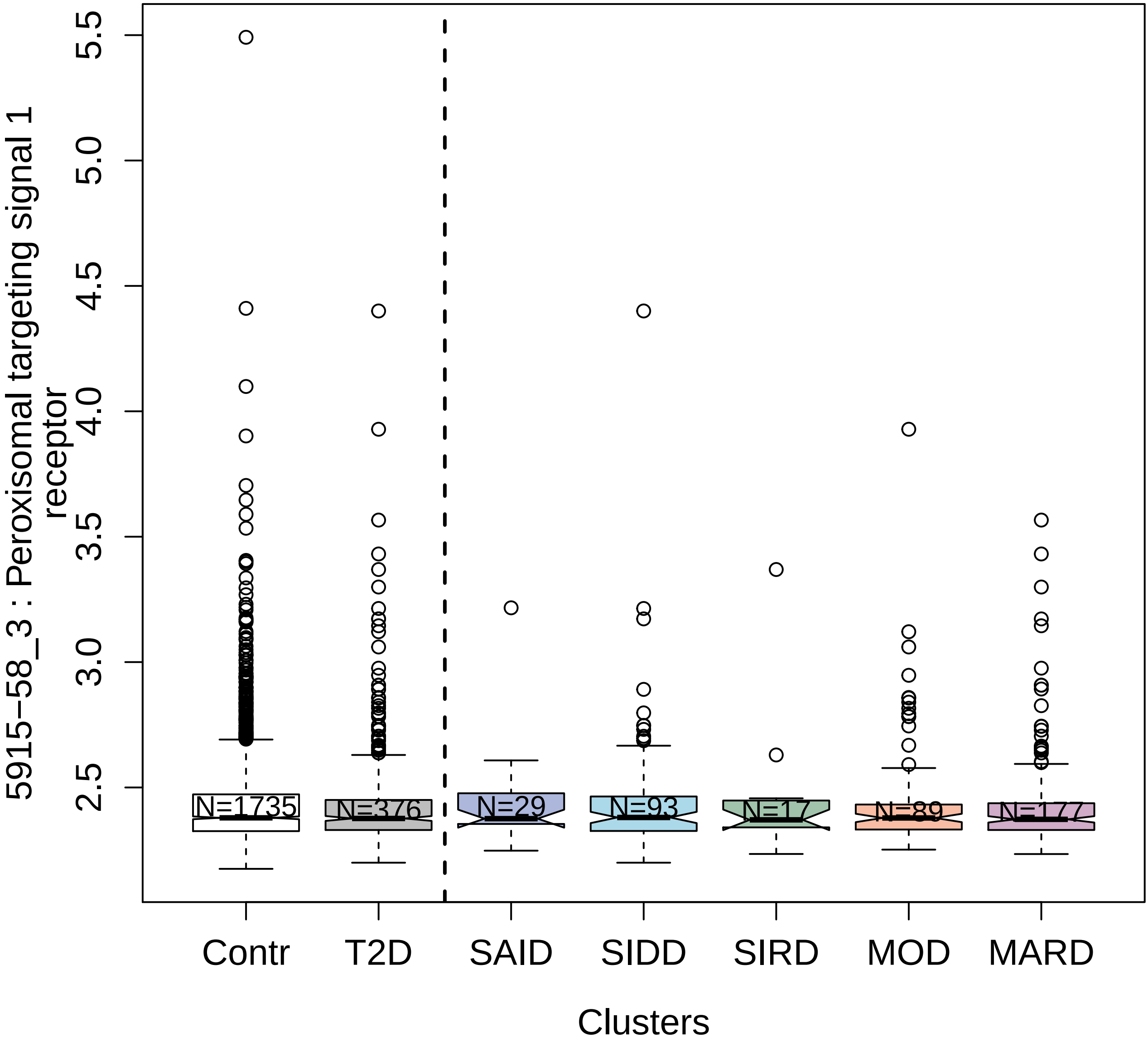
5915−58_3 : Peroxisomal targeting signal 1 receptor
5.5
5.0
4.5
4.0
3.5
3.0
2.5
N=1735
N=376
N=29
N=93
N=17
N=89
N=177
Contr
T2D
SAID
SIDD
SIRD
MOD
MARD
Clusters

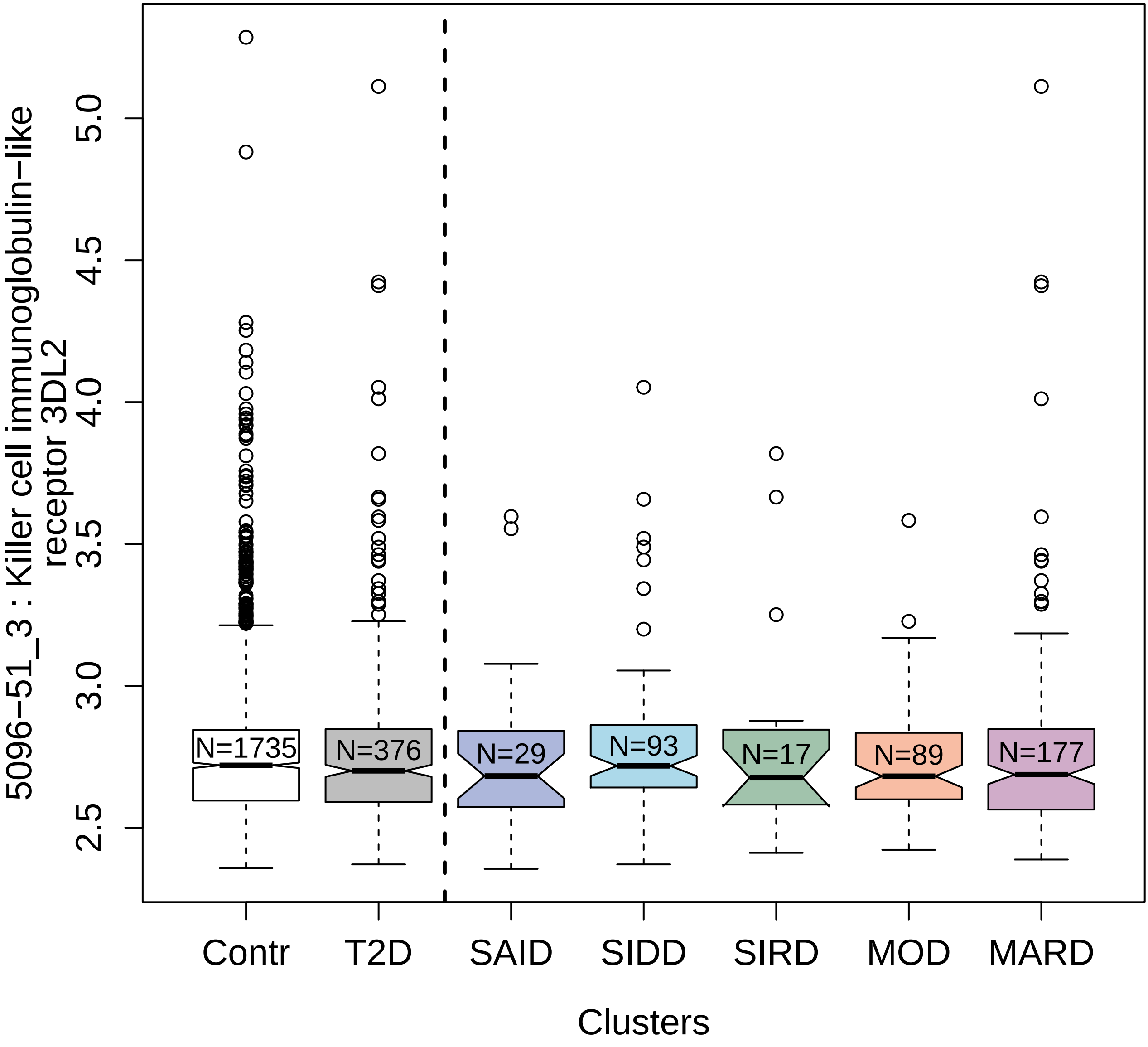

5096−51_3 : Killer cell immunoglobulin−like receptor 3DL2
5.0
4.5
4.0
3.5
3.0
2.5
N=1735
N=376
N=29
N=93
N=17
N=89
N=177
Contr
T2D
SAID
SIDD
SIRD
MOD
MARD
Clusters

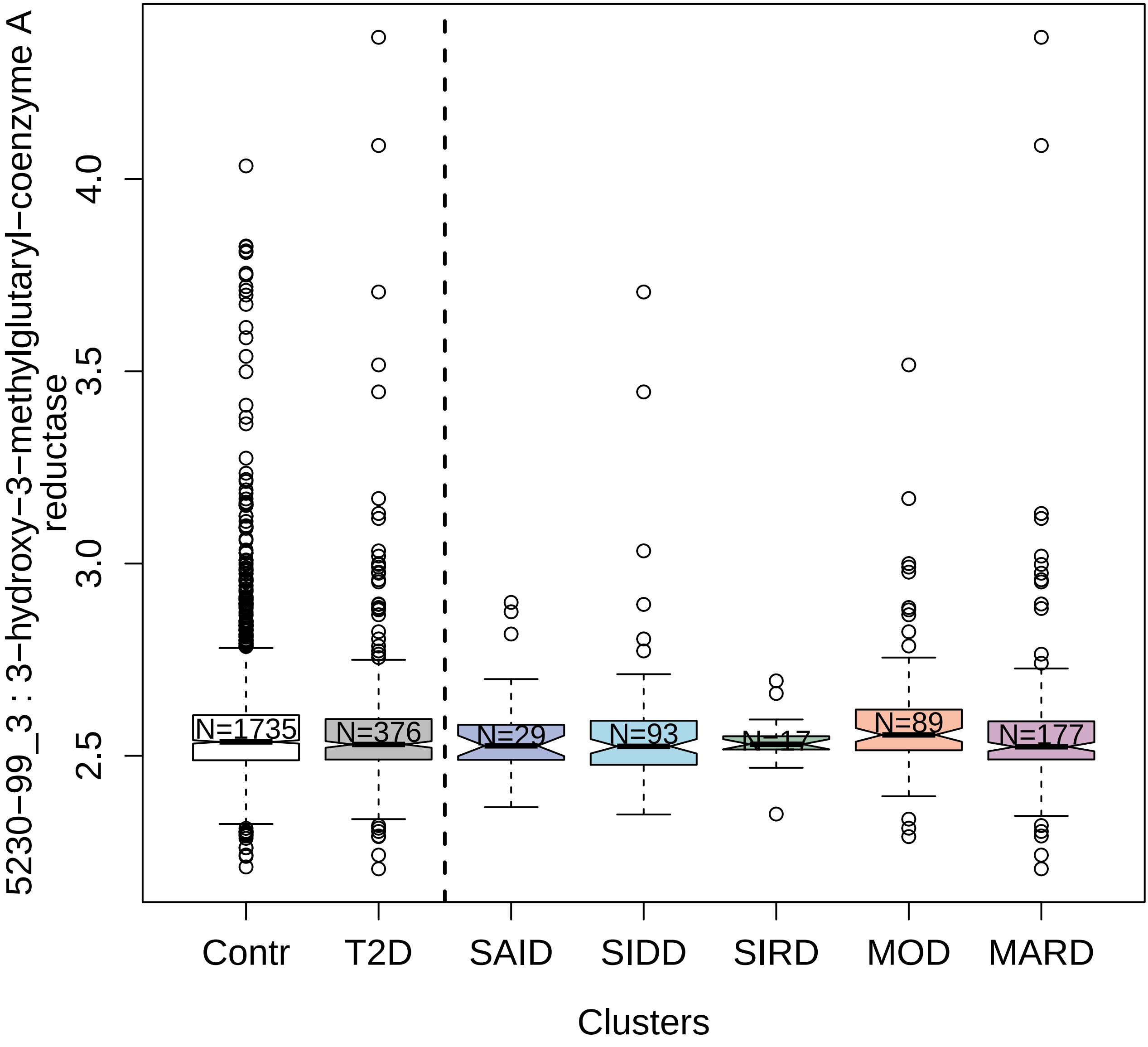
5230−99_3 : 3−hydroxy−3−methylglutaryl−coenzyme A reductase
2.5
3.0
3.5
4.0
N=1735
N=376
N=29
N=93
N=17
N=89
N=177
Contr
T2D
SAID
SIDD
SIRD
MOD
MARD
Clusters

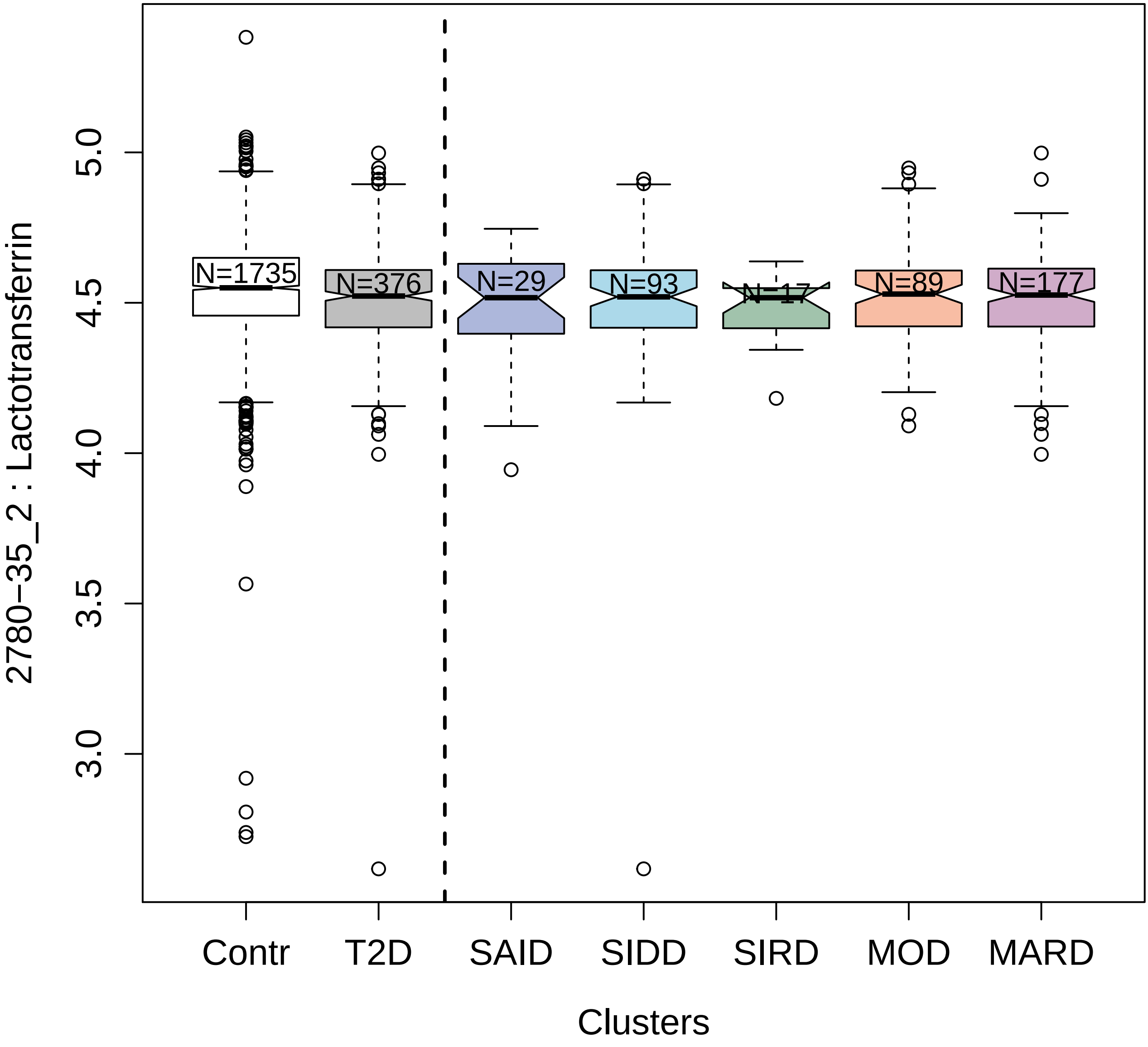

2780−35_2 : Lactotransferrin
5.0
4.5
4.0
3.5
3.0
N=1735
N=376
N=29
N=93
N=17
N=89
N=177
Contr
T2D
SAID
SIDD
SIRD
MOD
MARD
Clusters

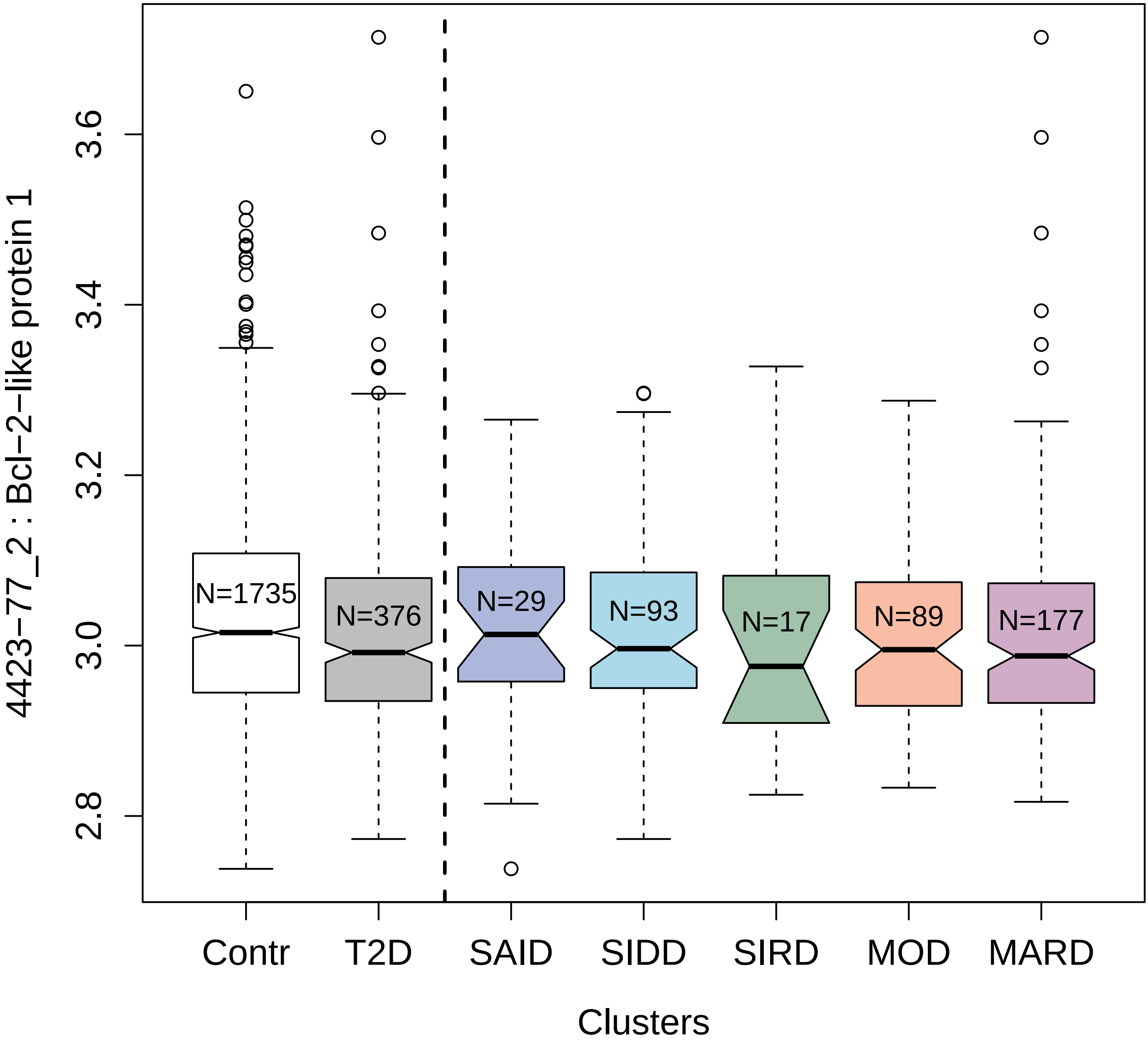

4423−77_2 : Bcl−2−like protein 1
2.8
3.0
3.2
3.4
3.6
N=1735
N=376
N=29
N=93
N=17
N=89
N=177
Contr
T2D
SAID
SIDD
SIRD
MOD
MARD
Clusters

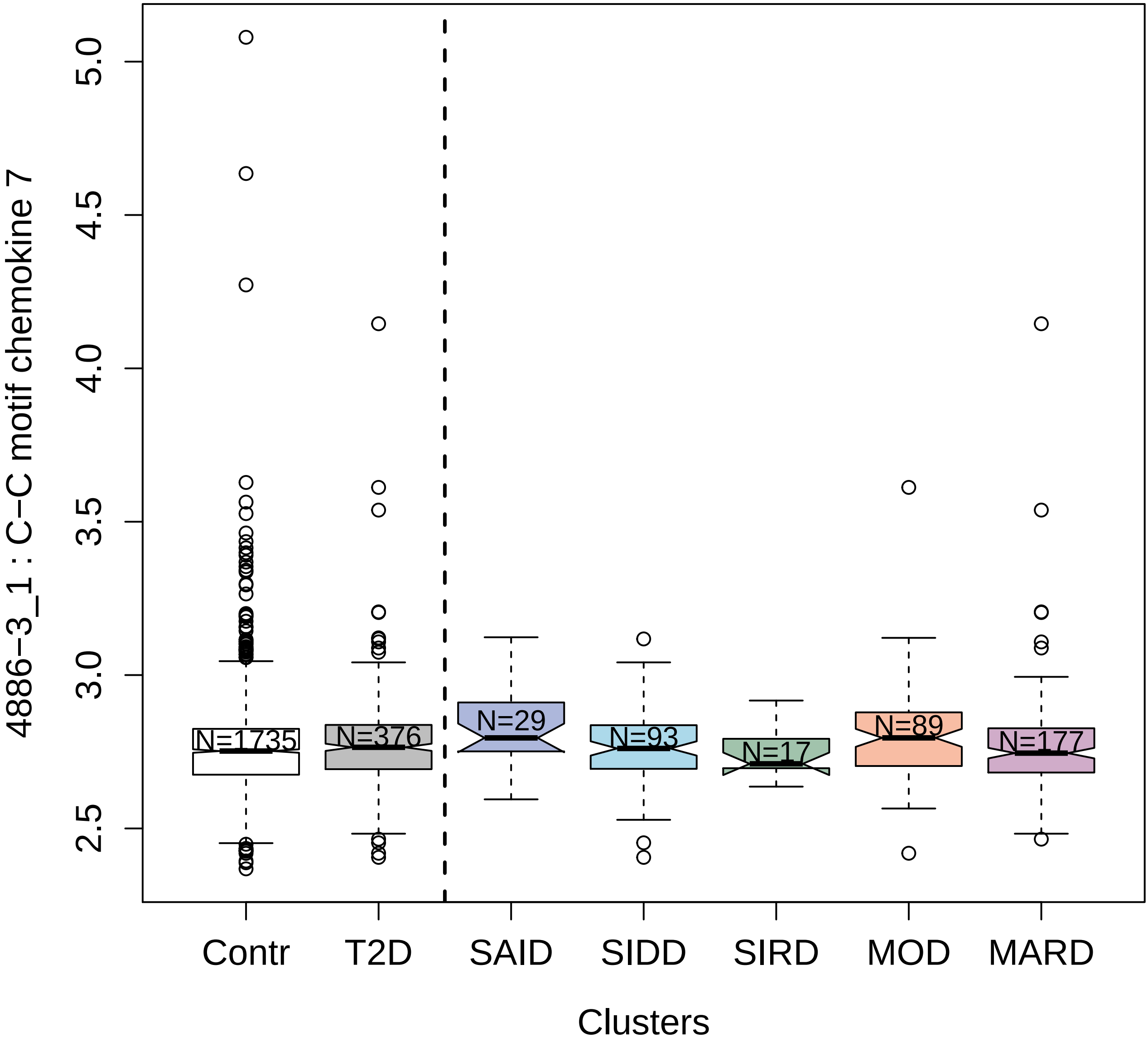

4886−3_1 : C−C motif chemokine 7
5.0
4.5
4.0
3.5
3.0
2.5
N=1735
N=376
N=29
N=93
N=17
N=89
N=177
Contr
T2D
SAID
SIDD
SIRD
MOD
MARD
Clusters

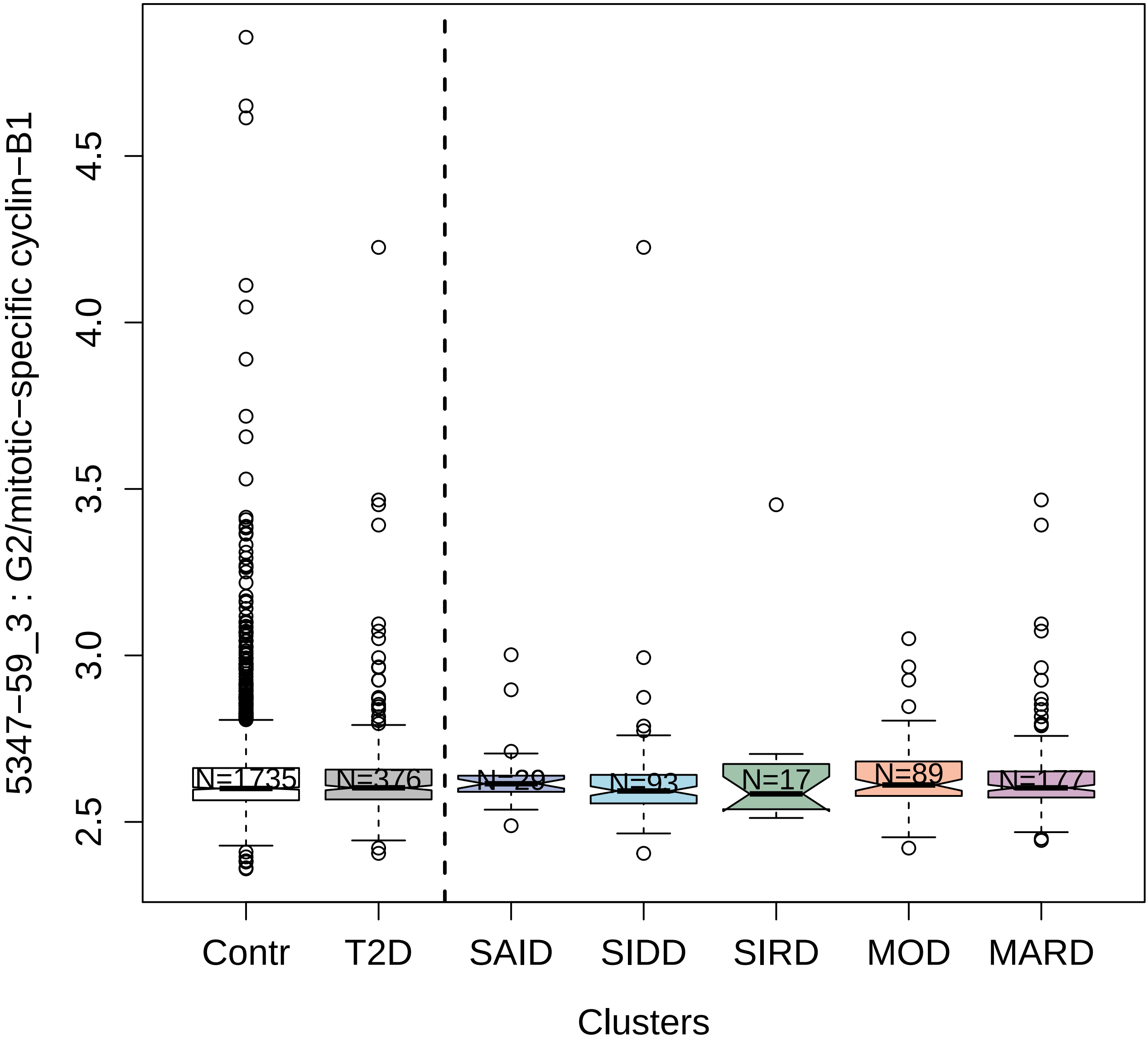

5347−59_3 : G2/mitotic−specific cyclin−B1
2.5
3.0
3.5
4.0
4.5
N=1735
N=376
N=29
N=93
N=17
N=89
N=177
Contr
T2D
SAID
SIDD
SIRD
MOD
MARD
Clusters

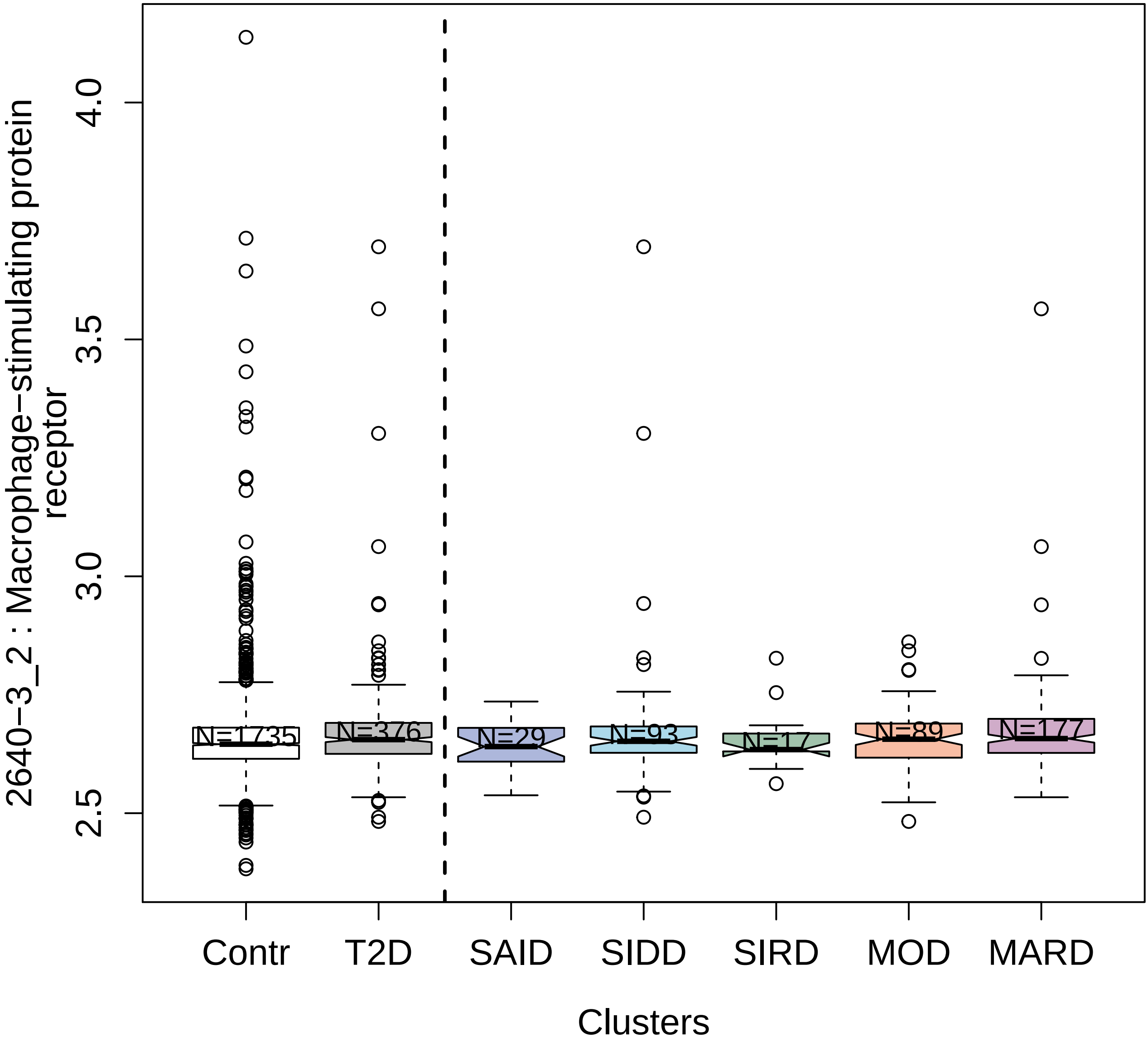
2640−3_2 : Macrophage−stimulating protein receptor
4.0
3.5
3.0
2.5
N=1735
N=376
N=29
N=93
N=17
N=89
N=177
Contr
T2D
SAID
SIDD
SIRD
MOD
MARD
Clusters

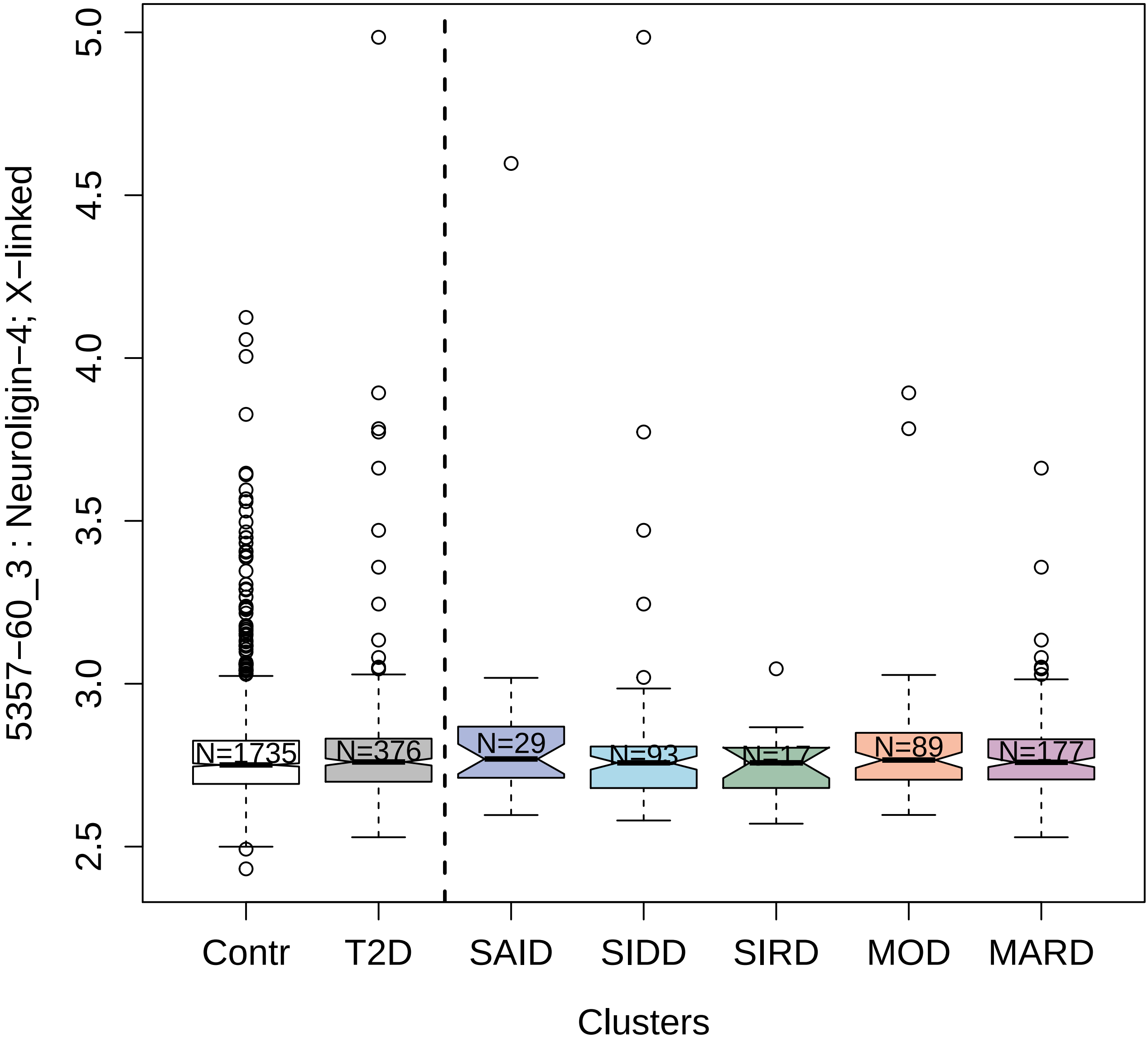

5357−60_3 : Neuroligin−4; X−linked
5.0
4.5
4.0
3.5
3.0
2.5
N=1735
N=376
N=29
N=93
N=17
N=89
N=177
Contr
T2D
SAID
SIDD
SIRD
MOD
MARD
Clusters

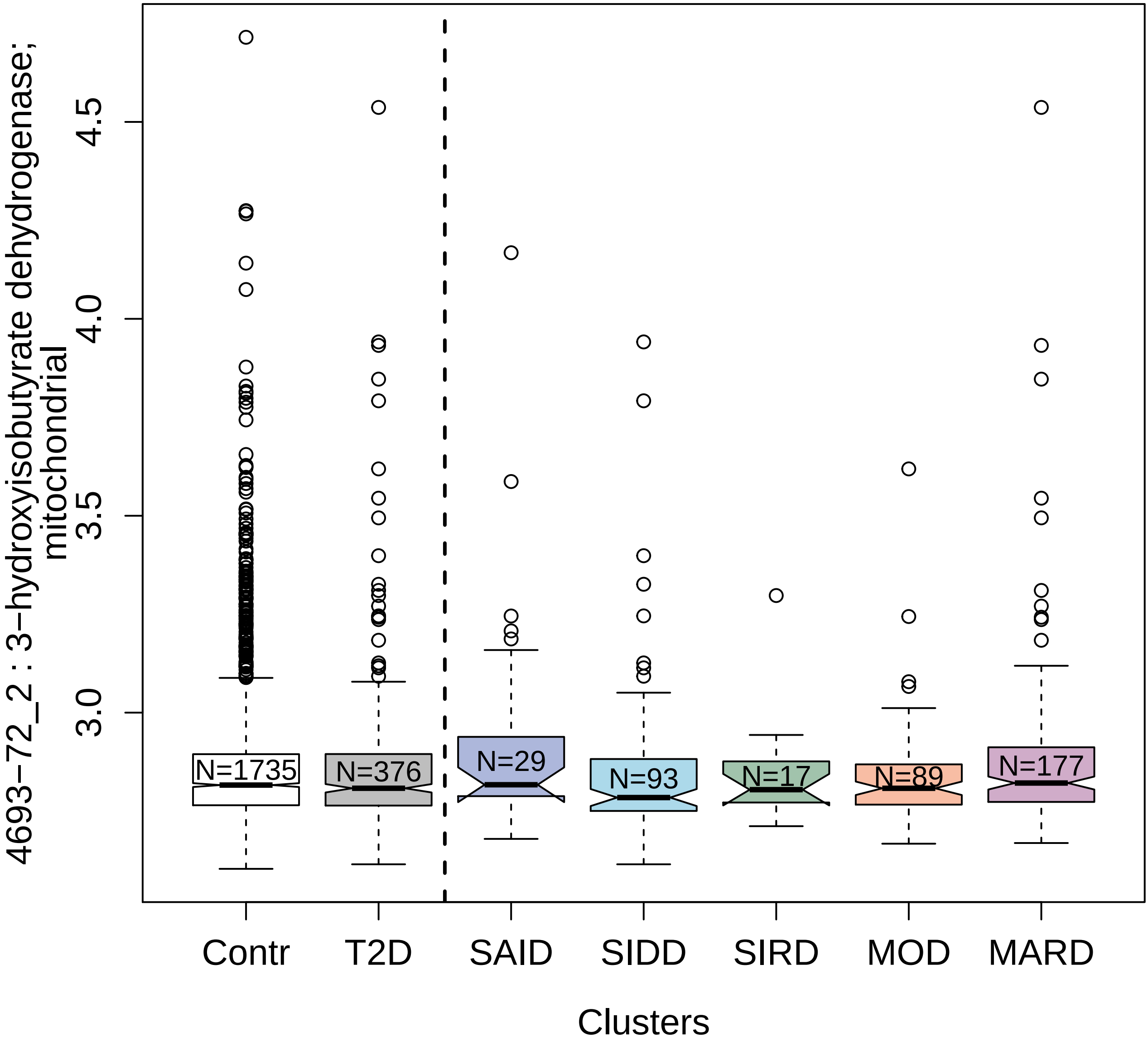
4693−72_2 : 3−hydroxyisobutyrate dehydrogenase; mitochondrial
3.0
3.5
4.0
4.5
N=1735
N=376
N=29
N=93
N=17
N=89
N=177
Contr
T2D
SAID
SIDD
SIRD
MOD
MARD
Clusters

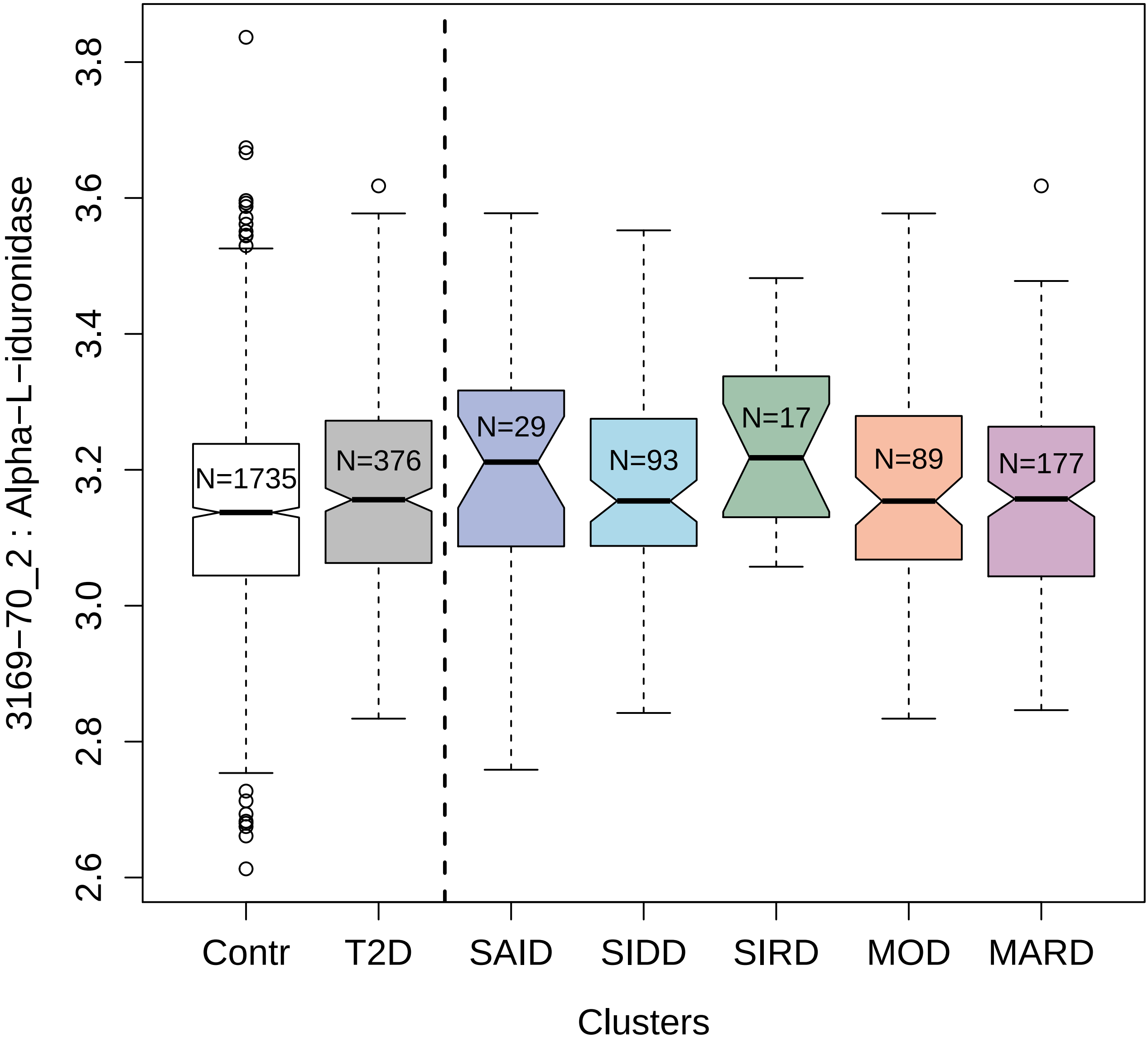
3169−70_2 : Alpha−L−iduronidase
3.8
3.6
3.4
3.2
3.0
2.8
2.6
N=1735
N=376
N=29
N=93
N=17
N=89
N=177
Contr
T2D
SAID
SIDD
SIRD
MOD
MARD
Clusters

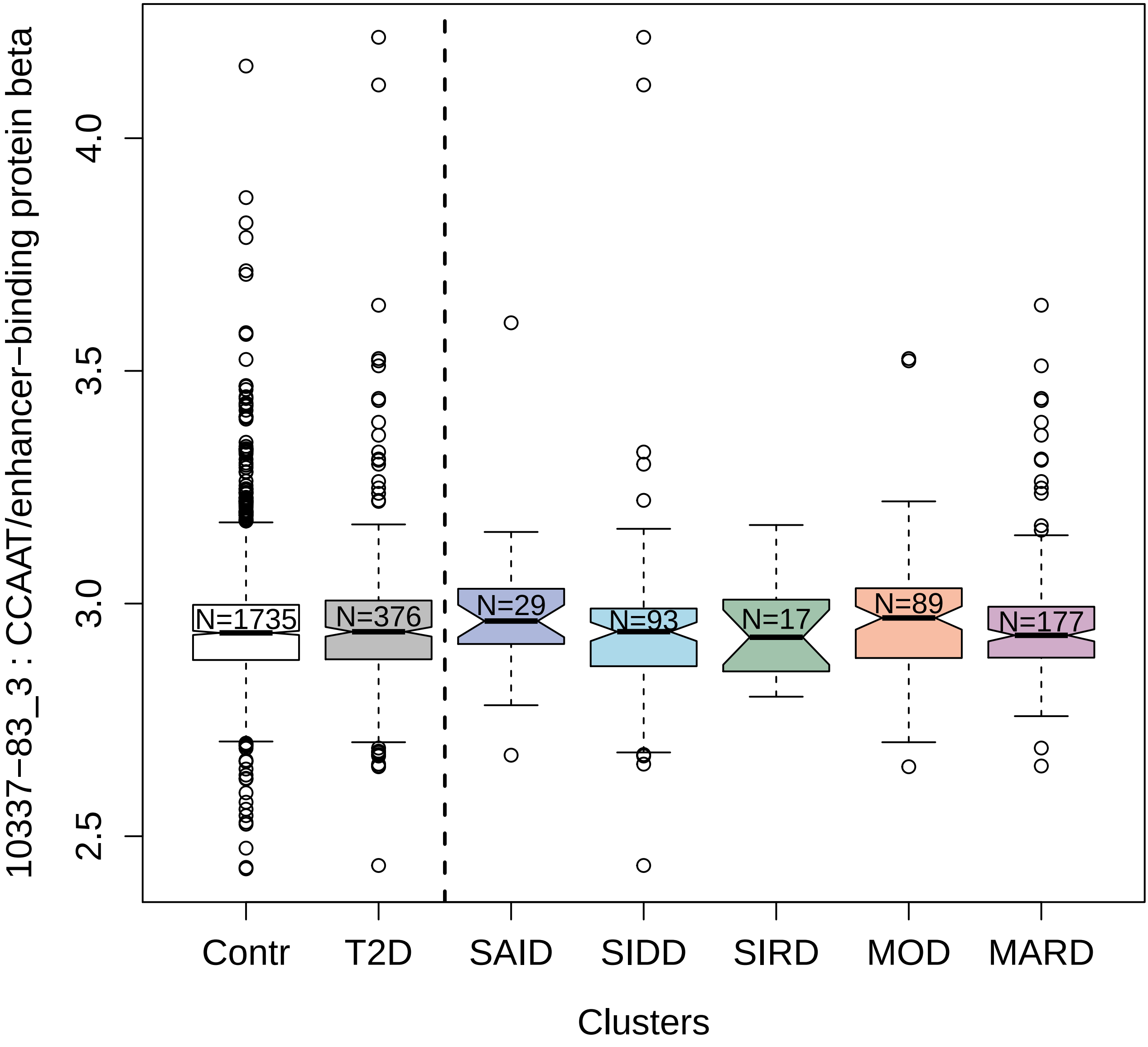
10337−83_3 : CCAAT/enhancer−binding protein beta
4.0
3.5
3.0
2.5
N=1735
N=376
N=29
N=93
N=17
N=89
N=177
Contr
T2D
SAID
SIDD
SIRD
MOD
MARD
Clusters

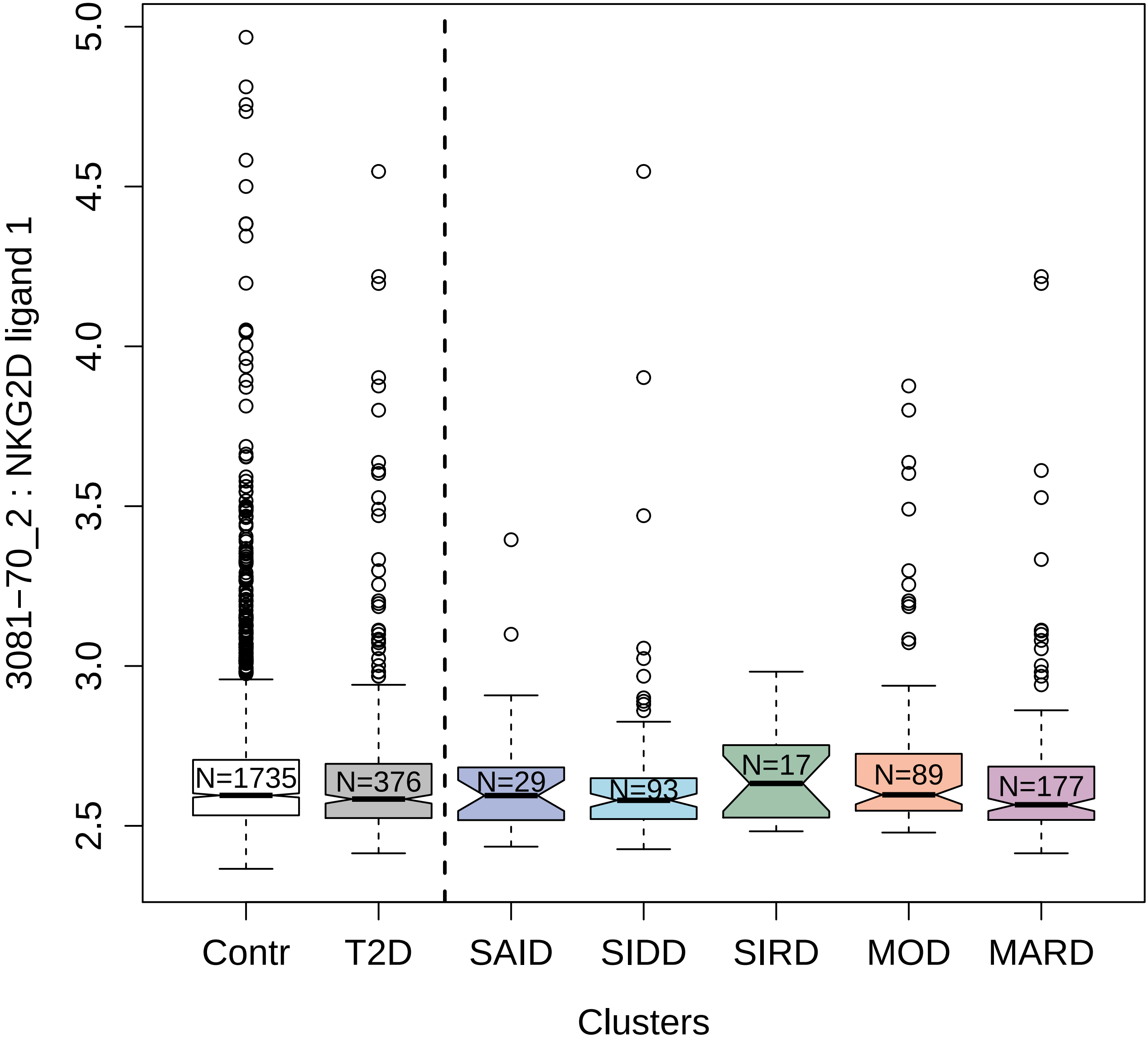

3081−70_2 : NKG2D ligand 1
5.0
4.5
4.0
3.5
3.0
2.5
N=1735
N=376
N=29
N=93
N=17
N=89
N=177
Contr
T2D
SAID
SIDD
SIRD
MOD
MARD
Clusters

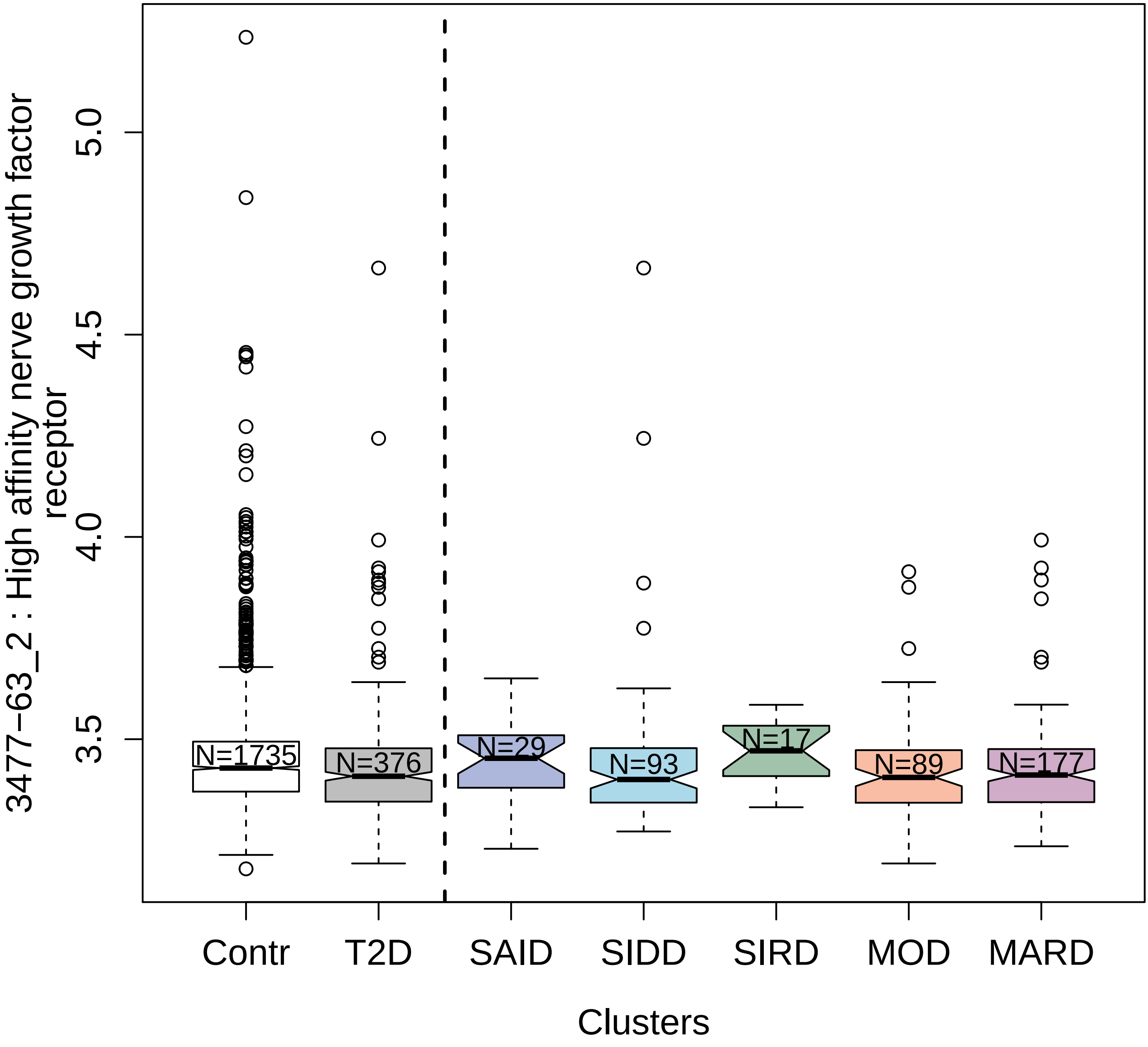
3477−63_2 : High affinity nerve growth factor receptor
5.0
4.5
4.0
3.5
N=1735
N=376
N=29
N=93
N=17
N=89
N=177
Contr
T2D
SAID
SIDD
SIRD
MOD
MARD
Clusters

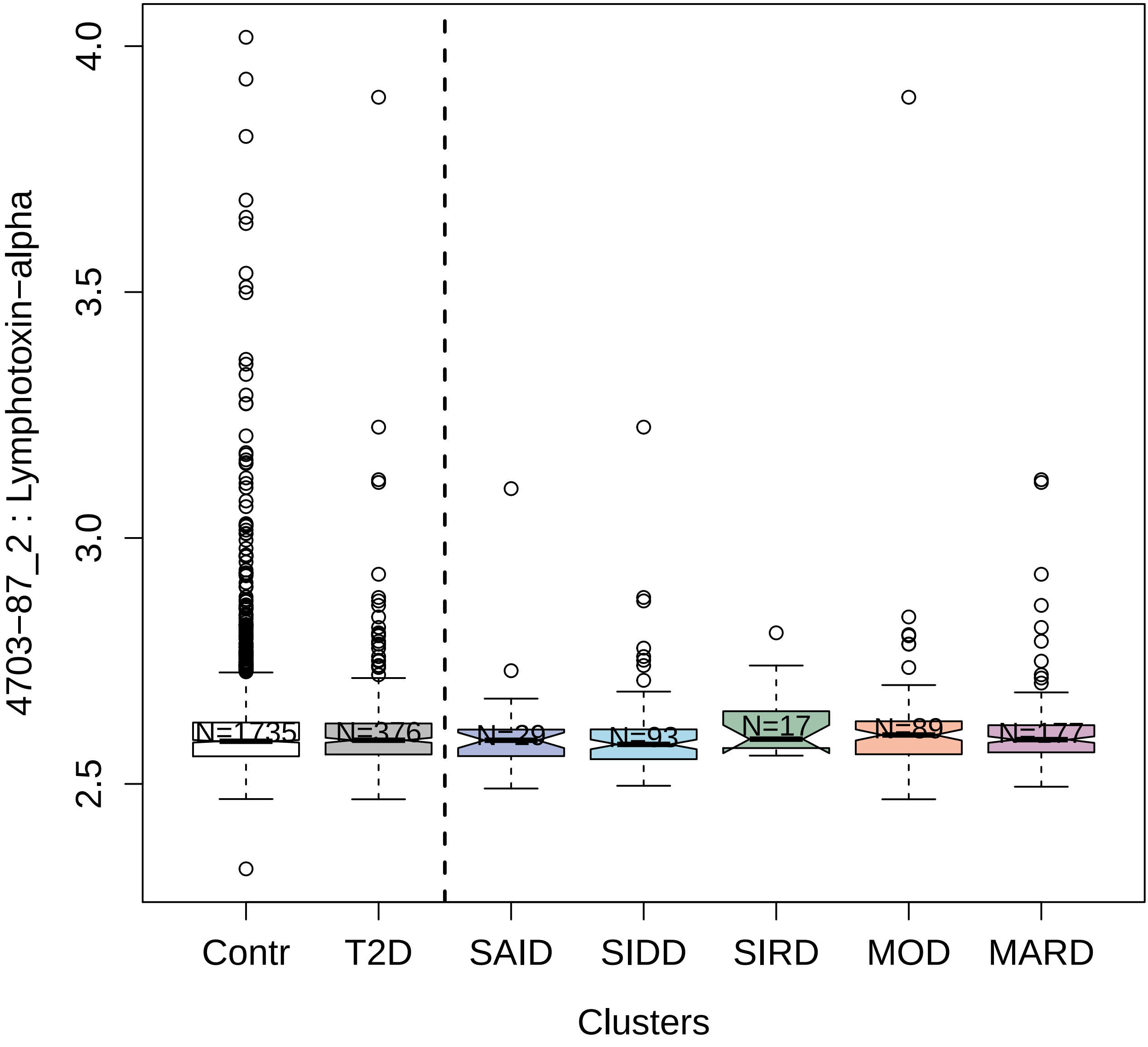

4703−87_2 : Lymphotoxin−alpha
4.0
3.5
3.0
2.5
N=1735
N=376
N=29
N=93
N=17
N=89
N=177
Contr
T2D
SAID
SIDD
SIRD
MOD
MARD
Clusters

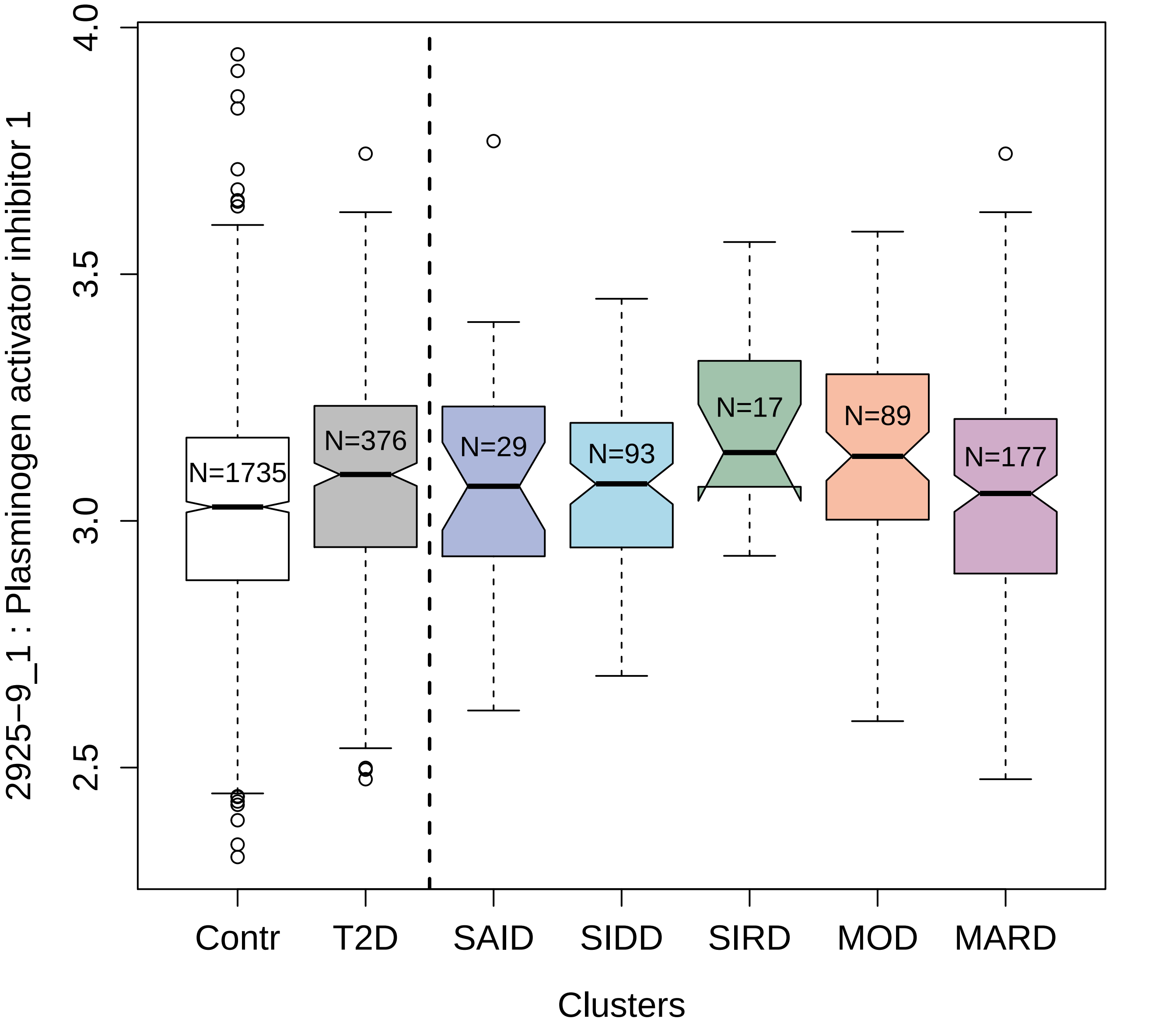

2925−9_1 : Plasminogen activator inhibitor 1
4.0
3.5
3.0
2.5
N=1735
N=376
N=29
N=93
N=17
N=89
N=177
Contr
T2D
SAID
SIDD
SIRD
MOD
MARD
Clusters

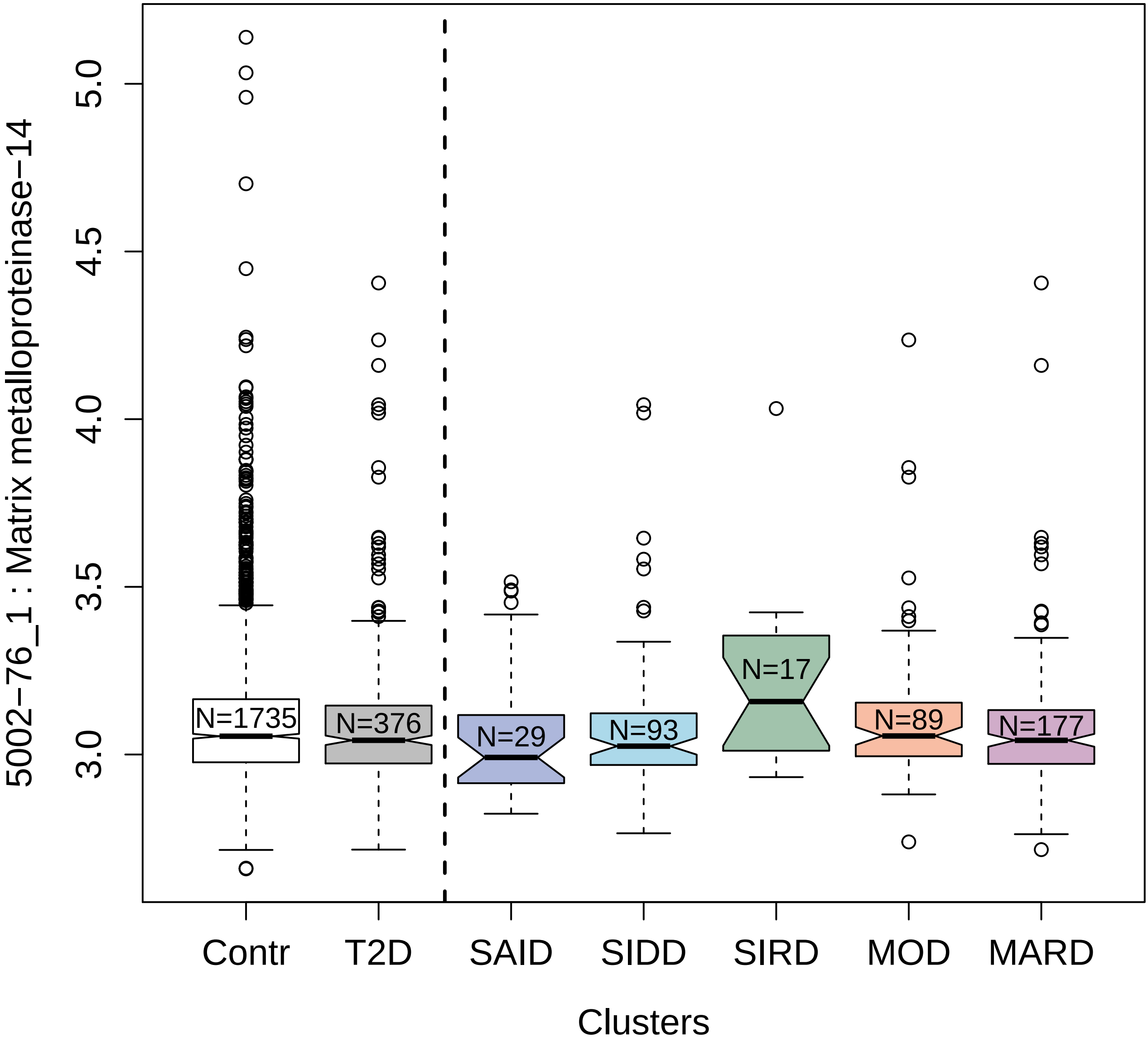
5002−76_1 : Matrix metalloproteinase−14
5.0
4.5
4.0
3.5
3.0
N=1735
N=376
N=29
N=93
N=17
N=89
N=177
Contr
T2D
SAID
SIDD
SIRD
MOD
MARD
Clusters

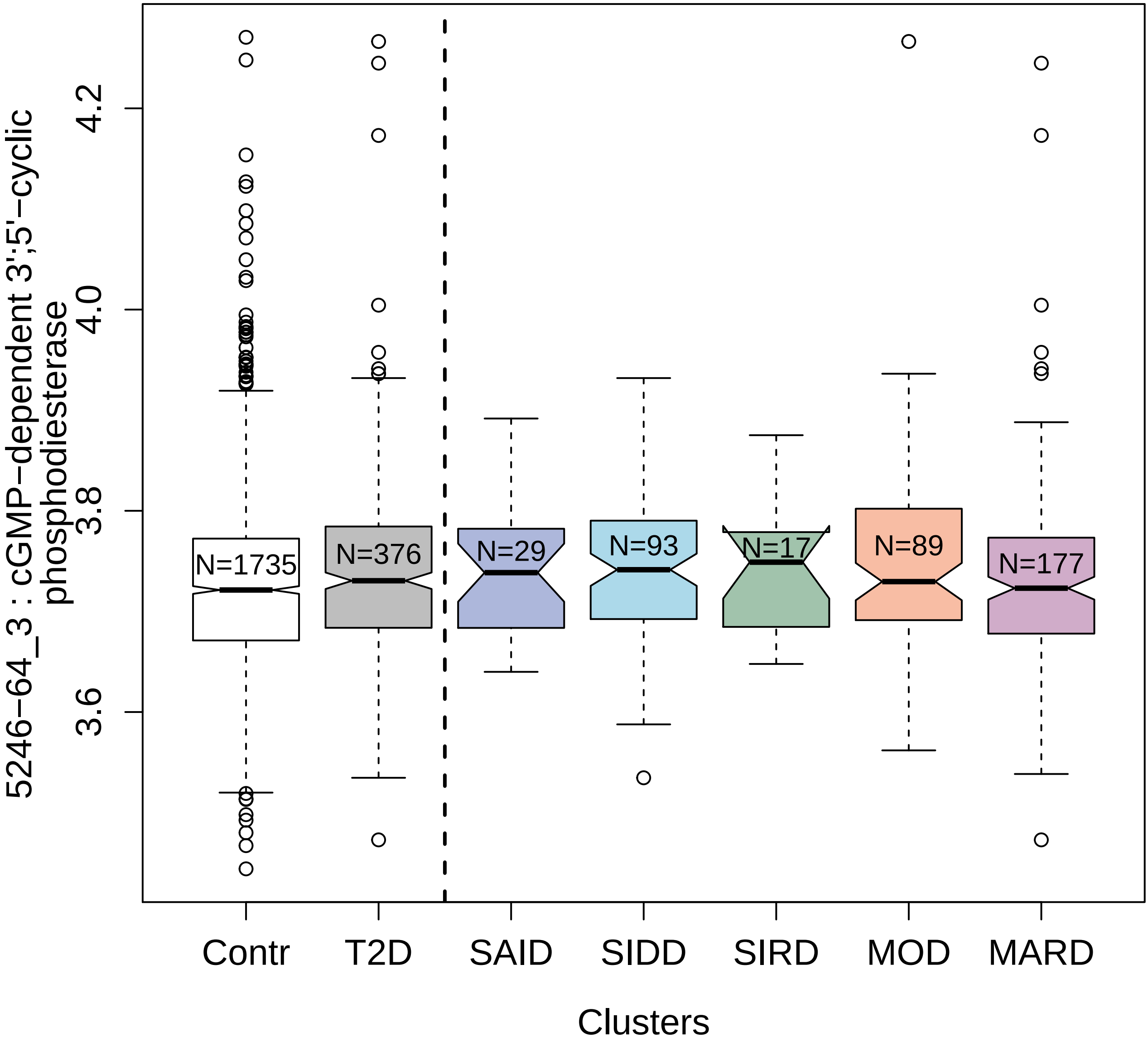
5246−64_3 : cGMP−dependent 3';5'−cyclic phosphodiesterase
4.2
4.0
3.8
3.6
N=1735
N=376
N=29
N=93
N=17
N=89
N=177
Contr
T2D
SAID
SIDD
SIRD
MOD
MARD
Clusters

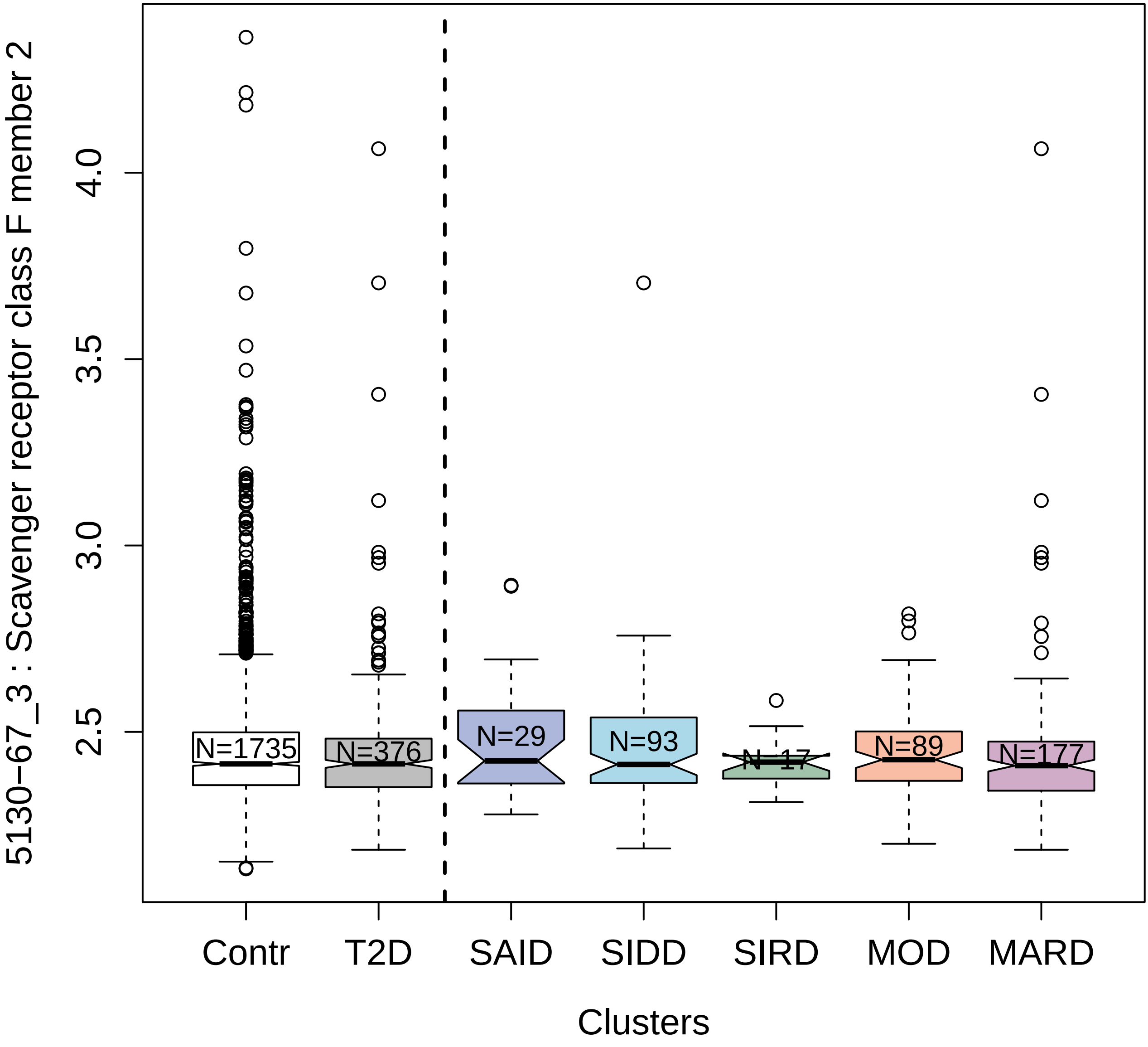

5130−67_3 : Scavenger receptor class F member 2
2.5
3.0
3.5
4.0
N=1735
N=376
N=29
N=93
N=17
N=89
N=177
Contr
T2D
SAID
SIDD
SIRD
MOD
MARD
Clusters

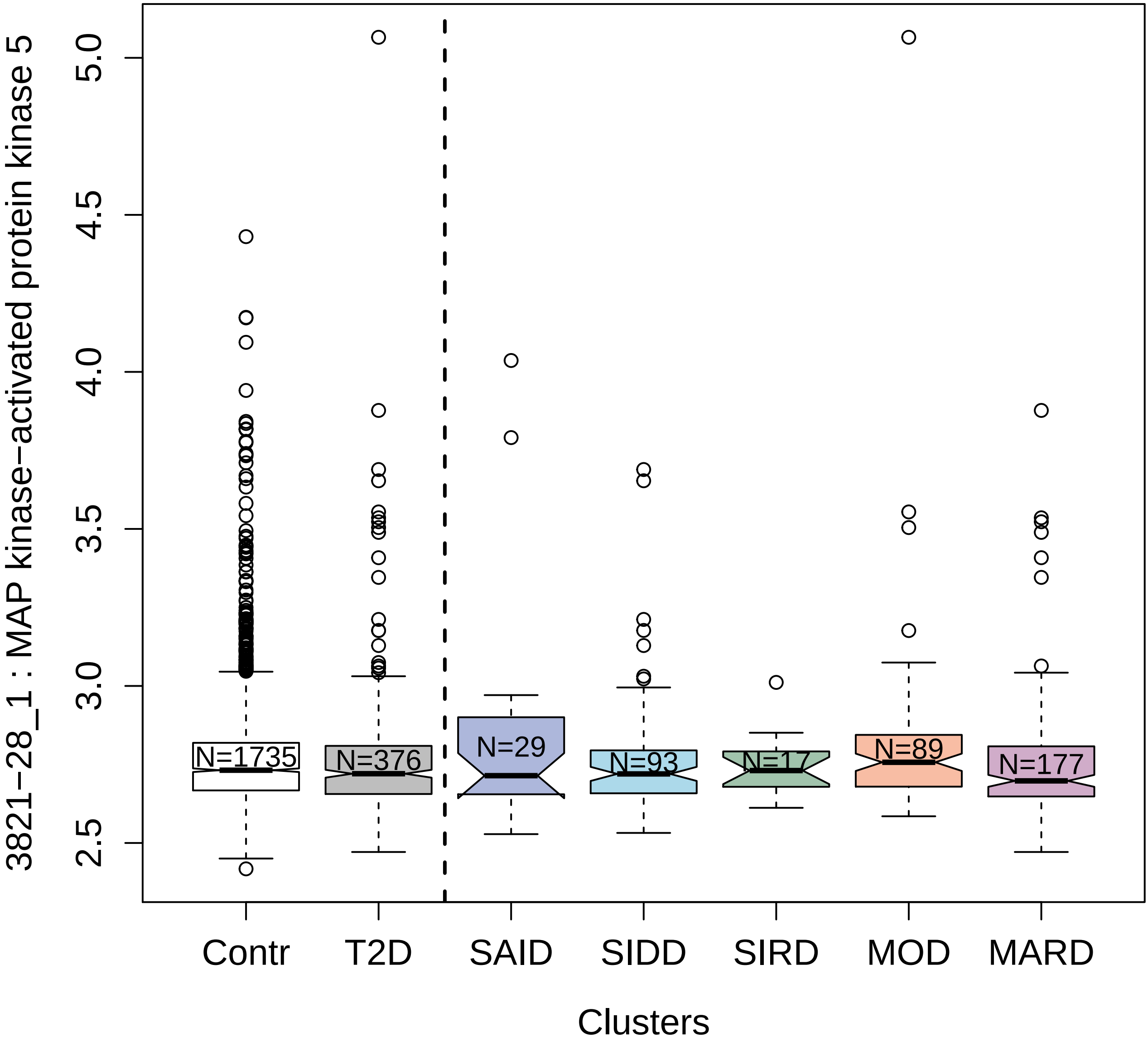

3821−28_1 : MAP kinase−activated protein kinase 5
5.0
4.5
4.0
3.5
3.0
2.5
N=1735
N=376
N=29
N=93
N=17
N=89
N=177
Contr
T2D
SAID
SIDD
SIRD
MOD
MARD
Clusters

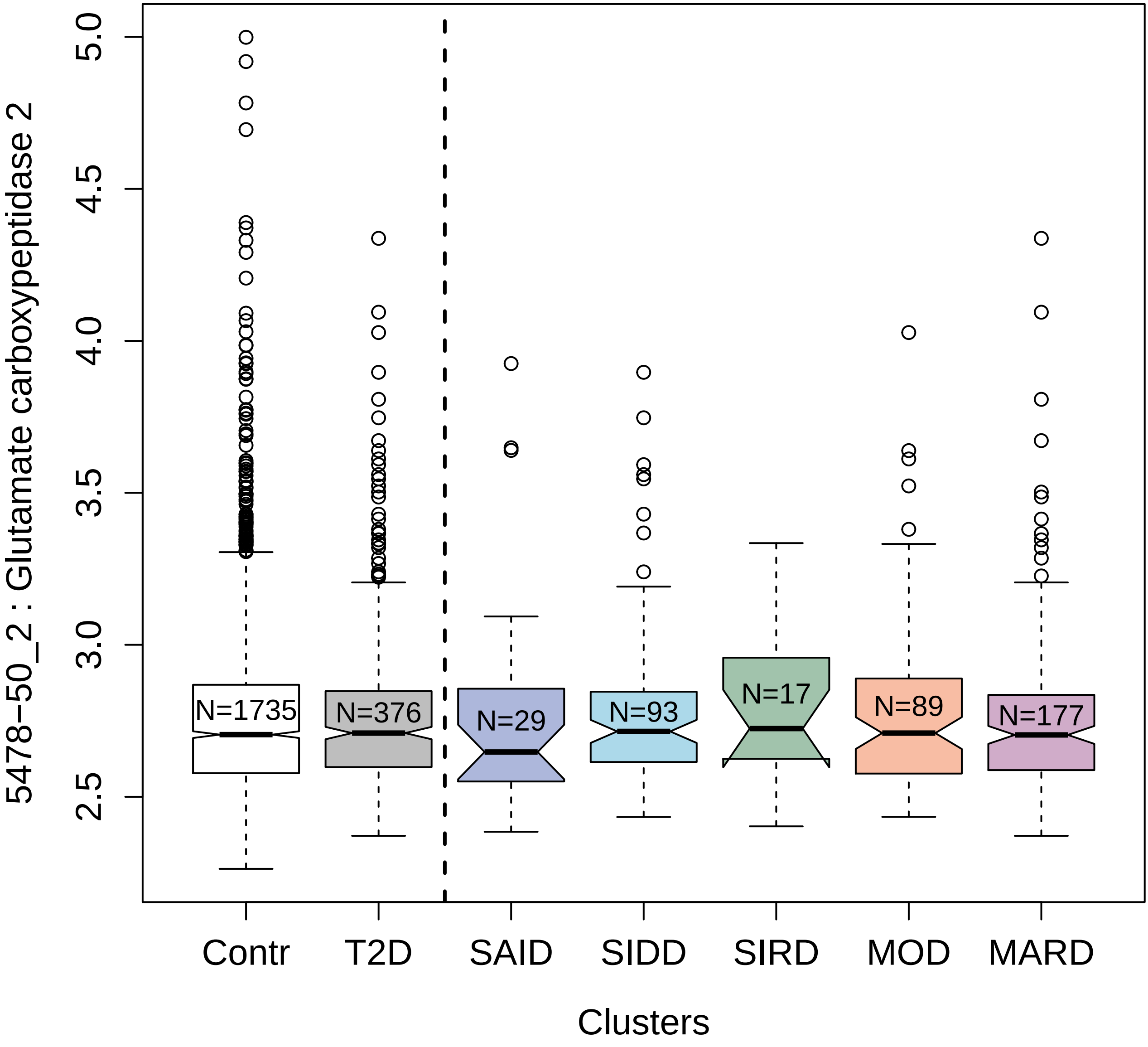

5478−50_2 : Glutamate carboxypeptidase 2
5.0
4.5
4.0
3.5
3.0
2.5
N=1735
N=376
N=29
N=93
N=17
N=89
N=177
Contr
T2D
SAID
SIDD
SIRD
MOD
MARD
Clusters

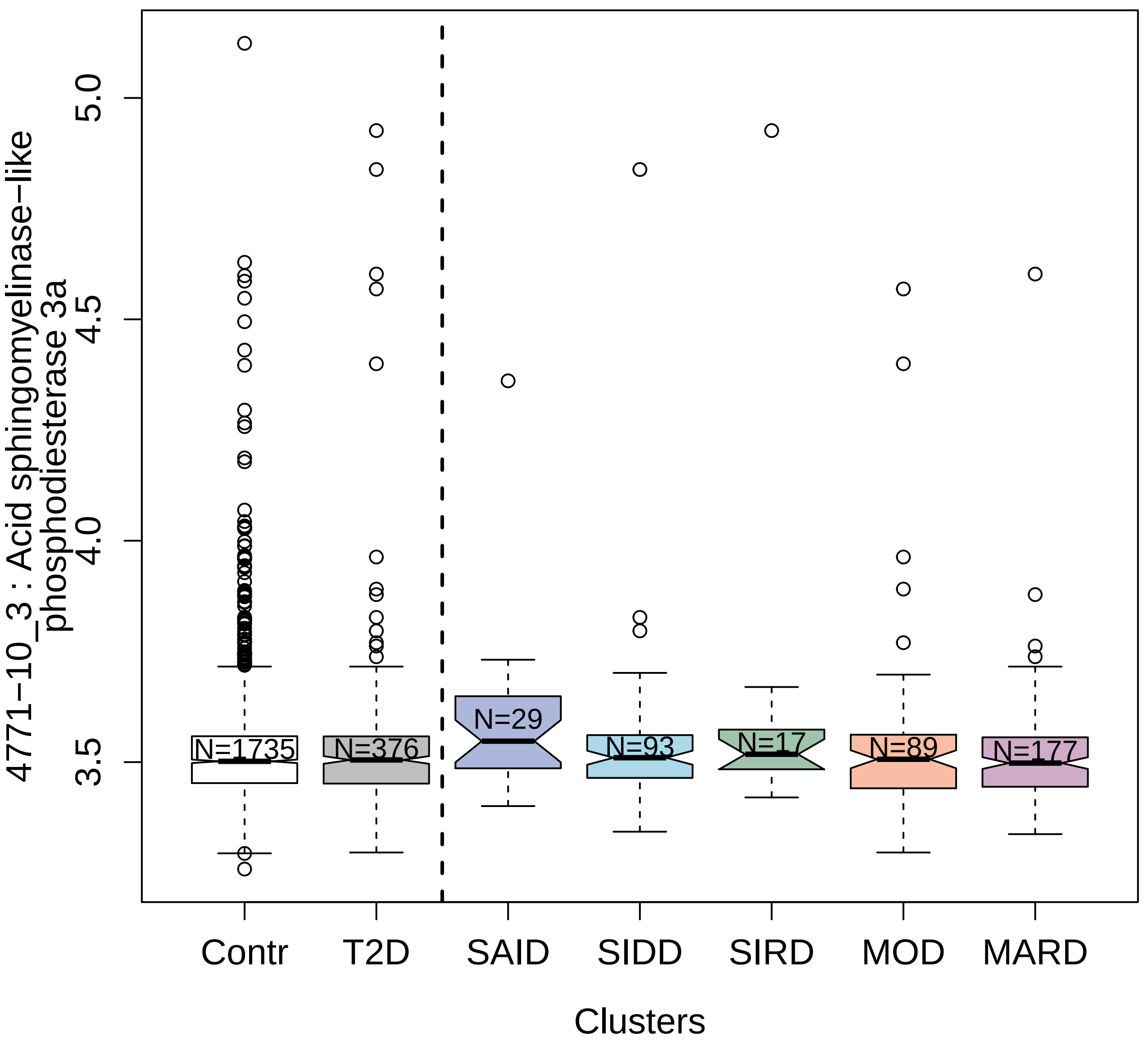

4771−10_3 : Acid sphingomyelinase−like
phosphodiesterase 3a
5.0
4.5
4.0
3.5
N=1735
N=376
N=29
N=93
N=17
N=89
N=177
Contr
T2D
SAID
SIDD
SIRD
MOD
MARD
Clusters

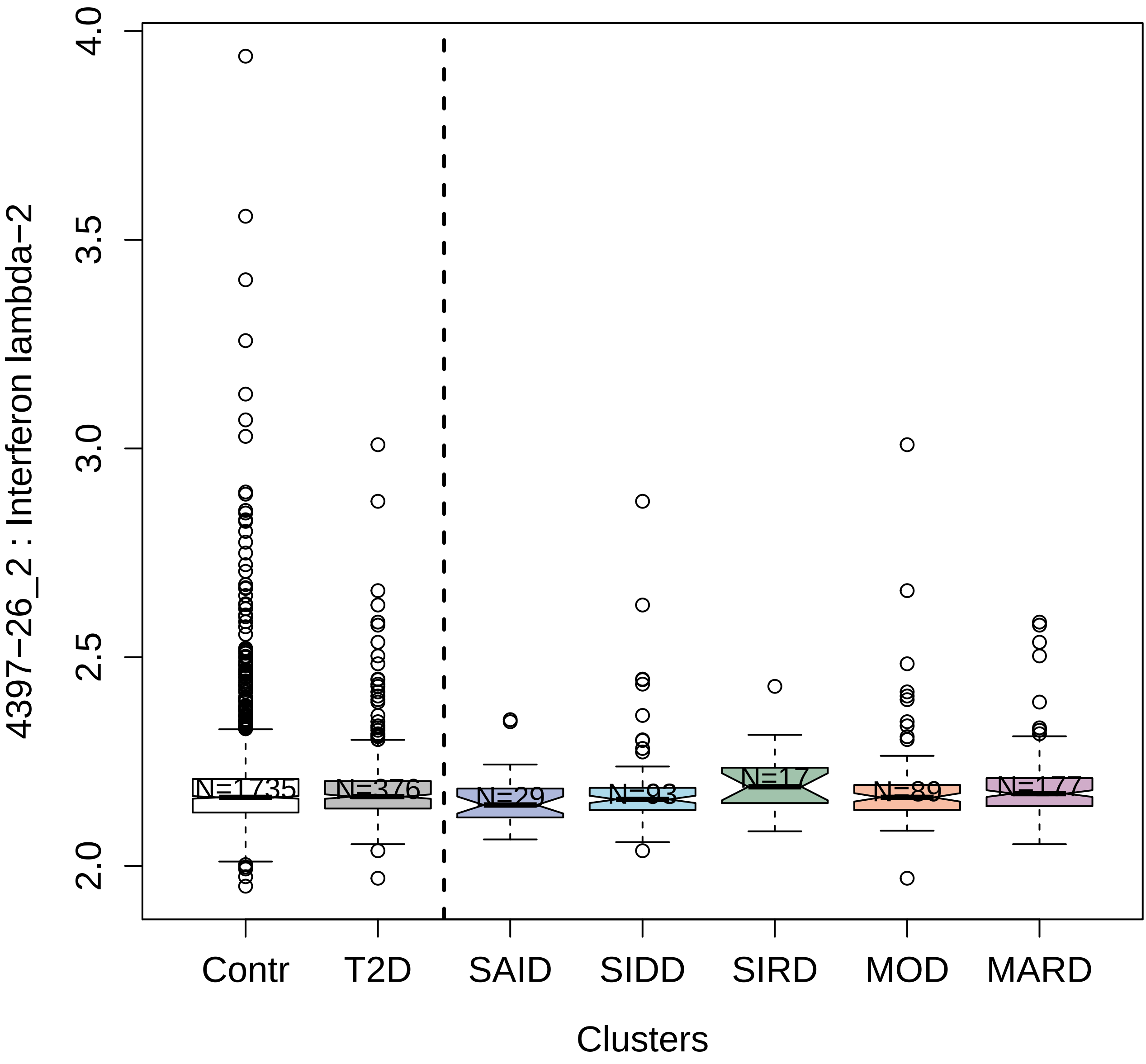

4397−26_2 : Interferon lambda−2
4.0
3.5
3.0
2.5
2.0
N=1735
N=376
N=29
N=93
N=17
N=89
N=177
Contr
T2D
SAID
SIDD
SIRD
MOD
MARD
Clusters

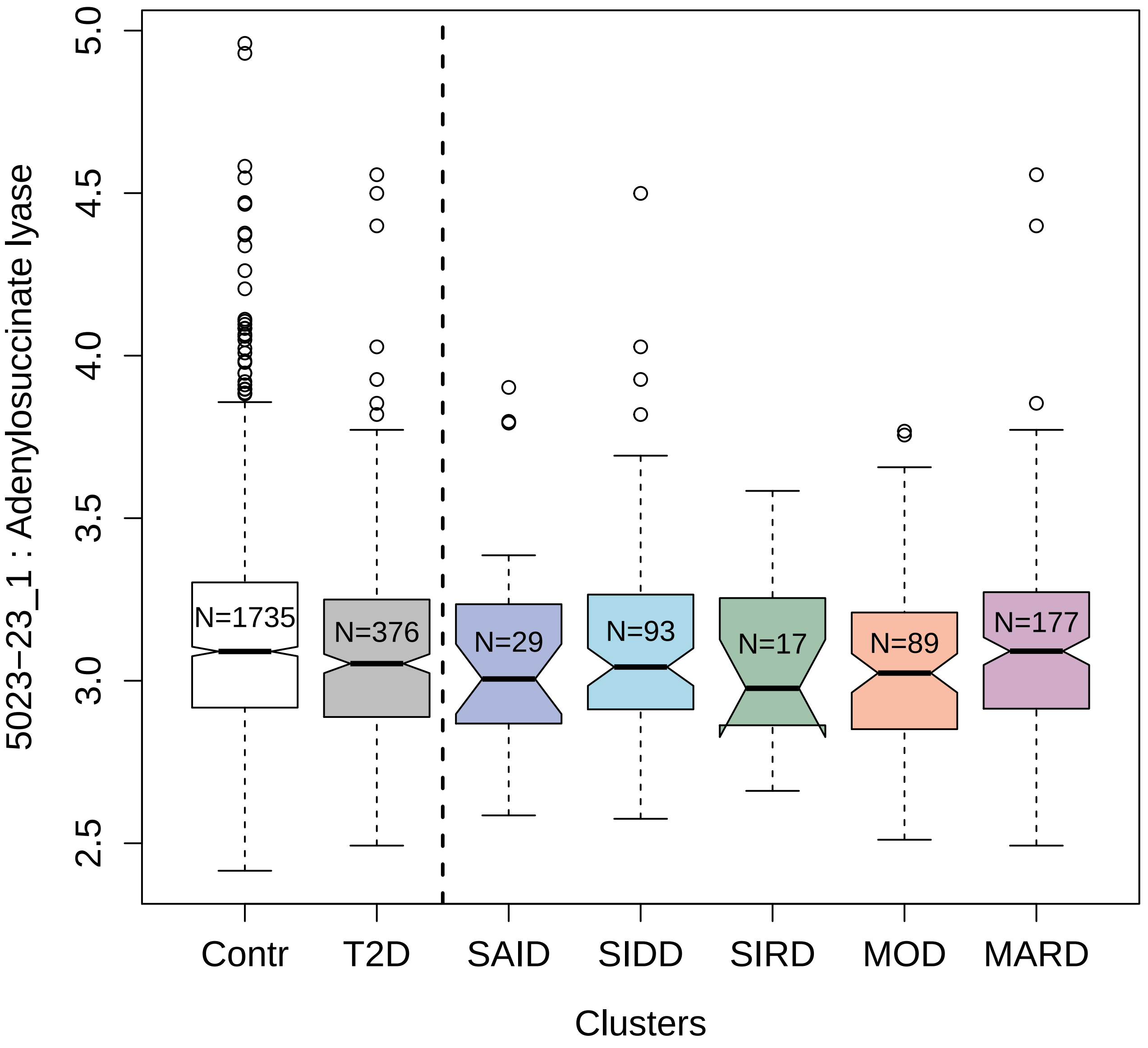
5023−23_1 : Adenylosuccinate lyase
5.0
4.5
4.0
3.5
3.0
2.5
N=1735
N=376
N=29
N=93
N=17
N=89
N=177
Contr
T2D
SAID
SIDD
SIRD
MOD
MARD
Clusters

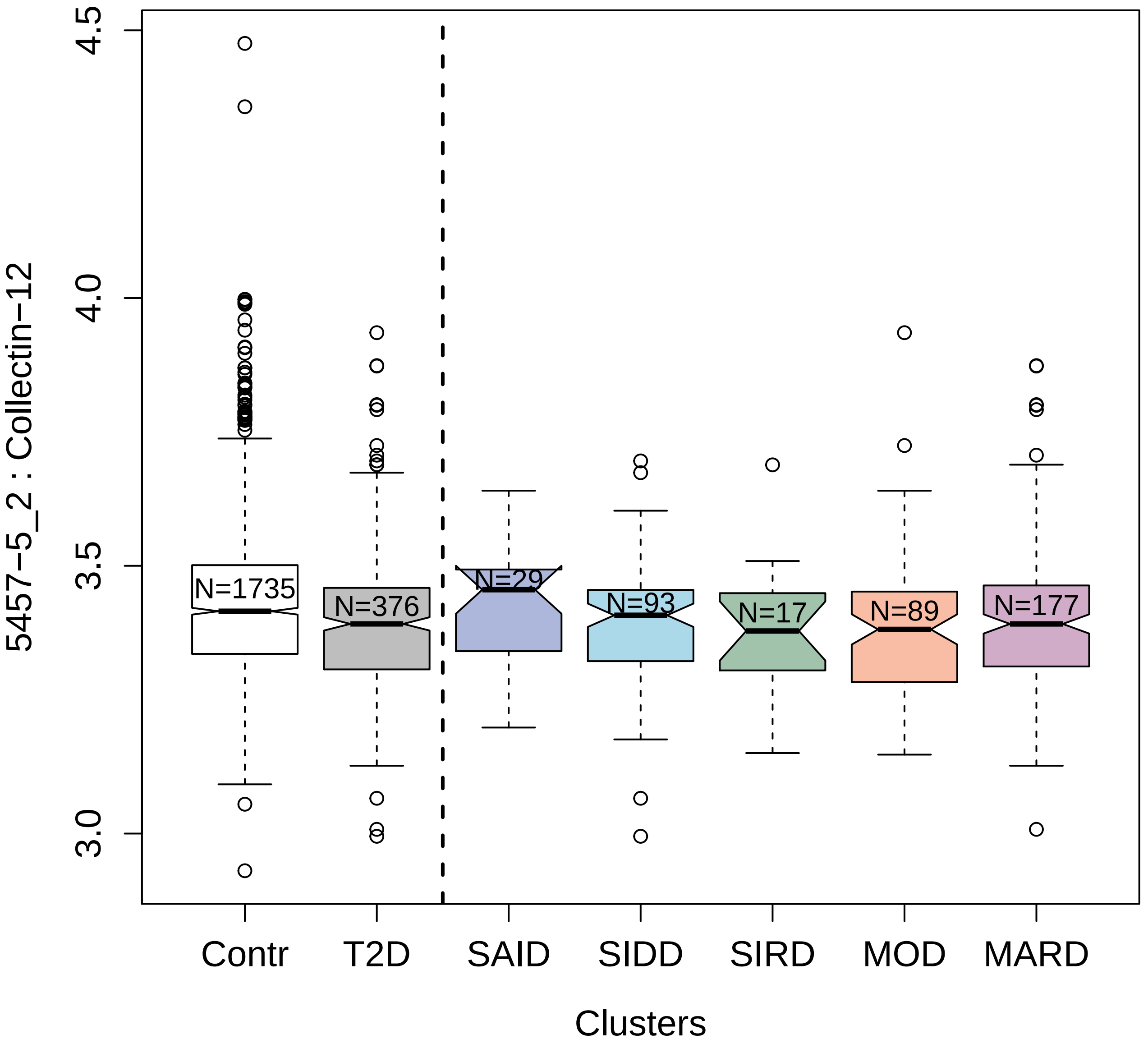

5457−5_2 : Collectin−12
4.5
4.0
3.5
3.0
N=1735
N=376
N=29
N=93
N=17
N=89
N=177
Contr
T2D
SAID
SIDD
SIRD
MOD
MARD
Clusters

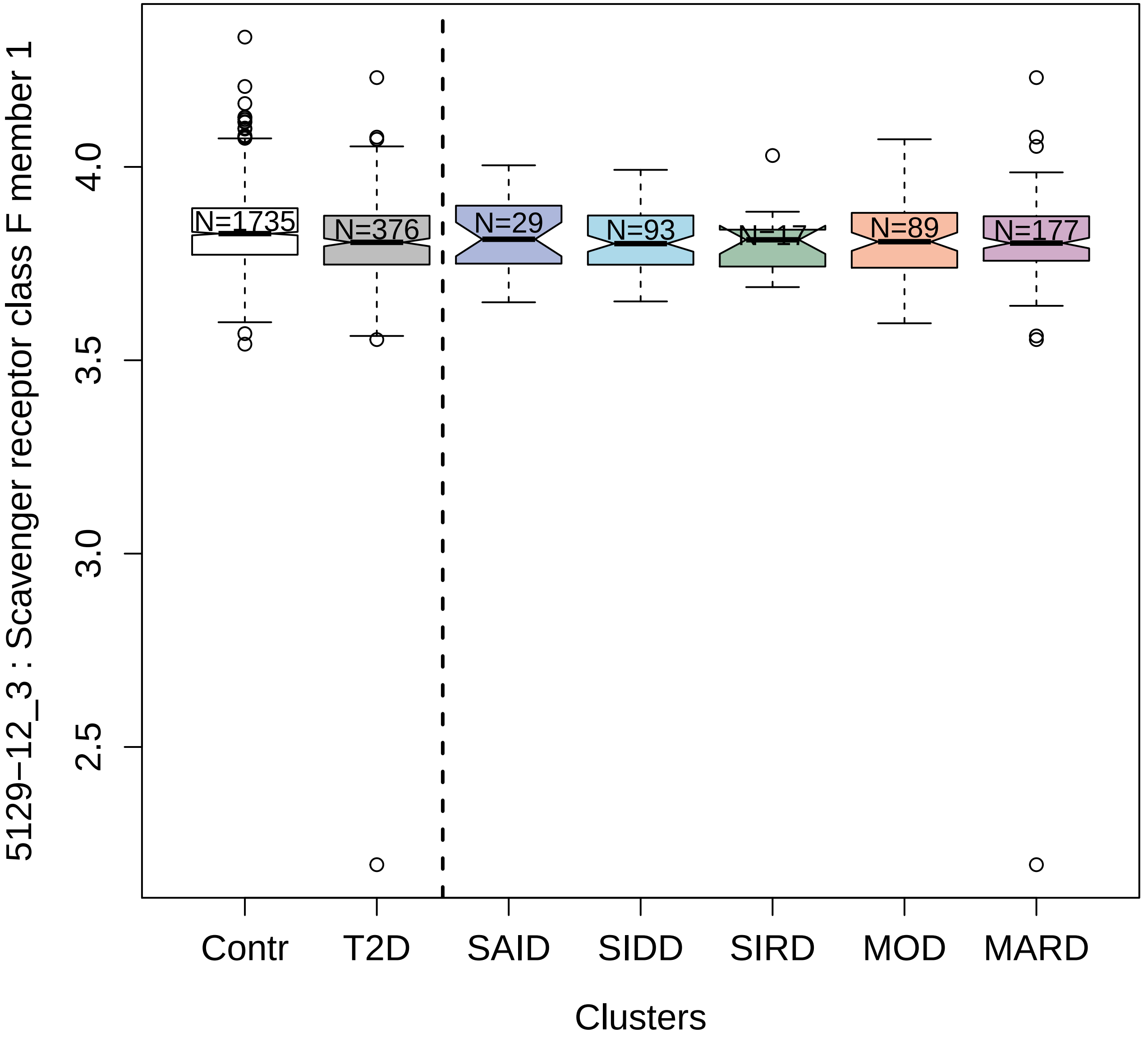

5129−12_3 : Scavenger receptor class F member 1
4.0
3.5
3.0
2.5
N=1735
N=376
N=29
N=93
N=17
N=89
N=177
Contr
T2D
SAID
SIDD
SIRD
MOD
MARD
Clusters

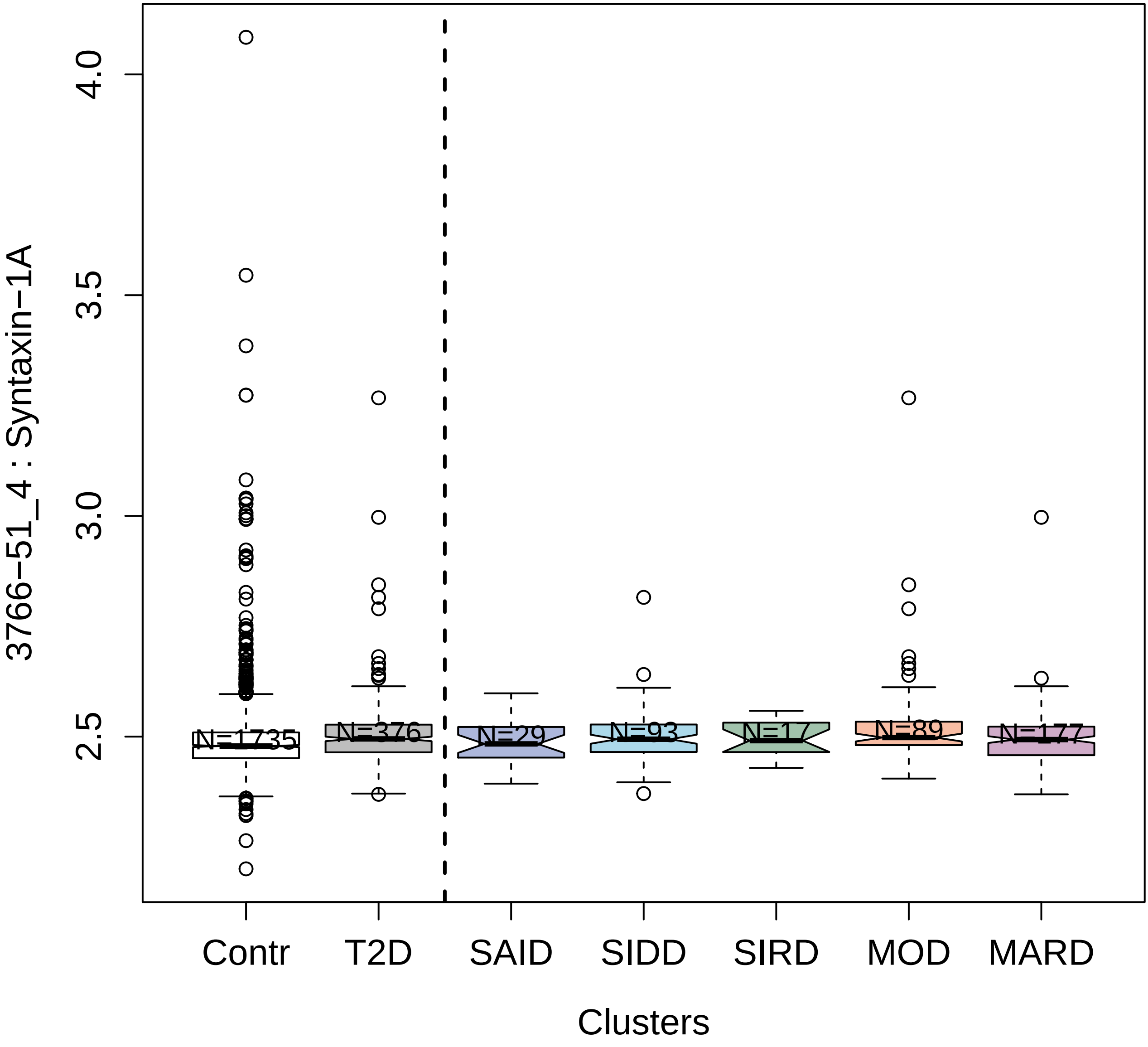
3766−51_4 : Syntaxin−1A
4.0
3.5
3.0
2.5
N=1735
N=376
N=29
N=93
N=17
N=89
N=177
Contr
T2D
SAID
SIDD
SIRD
MOD
MARD
Clusters

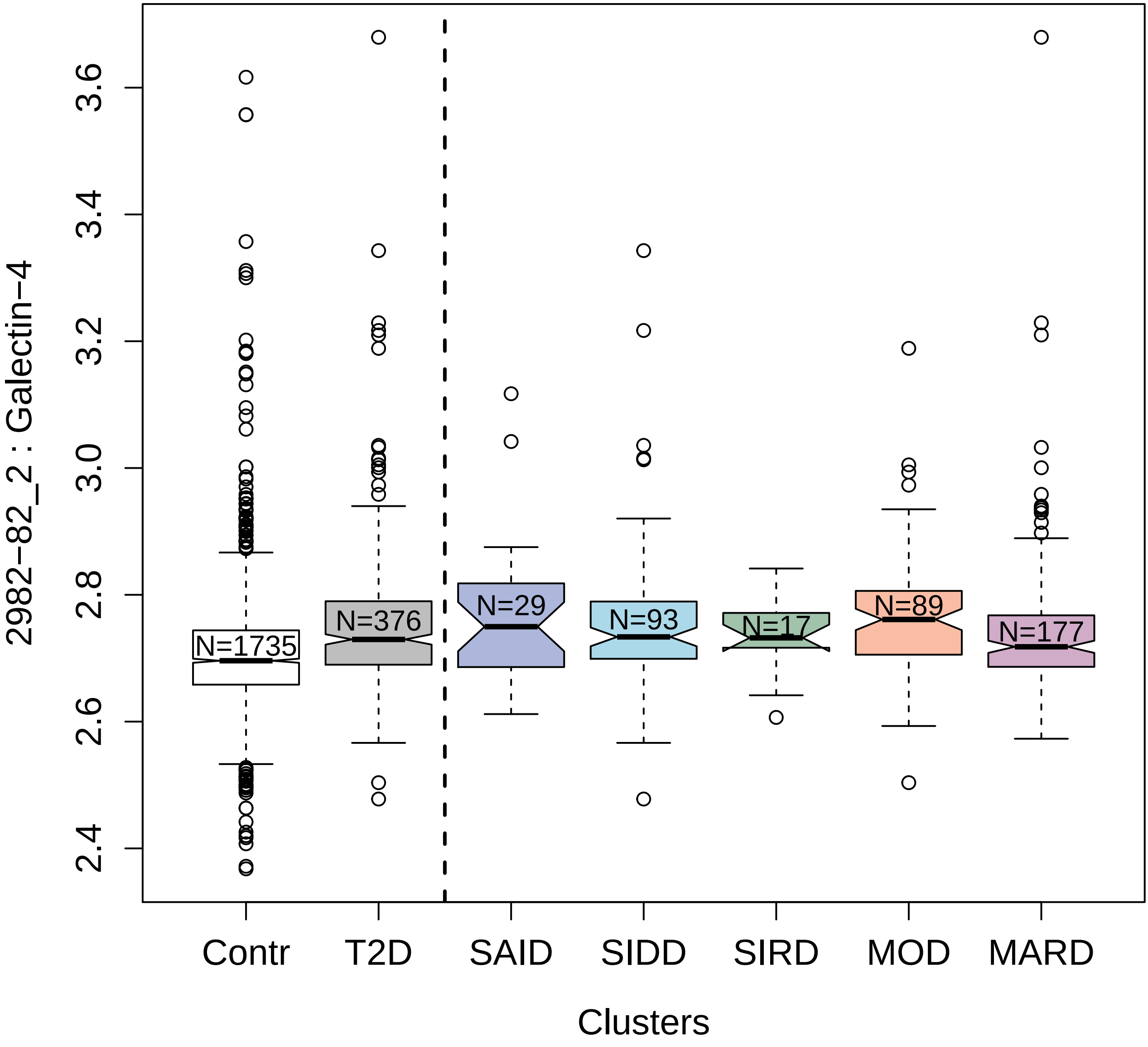
2982−82_2 : Galectin−4
3.6
3.4
3.2
3.0
2.8
2.6
2.4
N=1735
N=376
N=29
N=93
N=17
N=89
N=177
Contr
T2D
SAID
SIDD
SIRD
MOD
MARD
Clusters

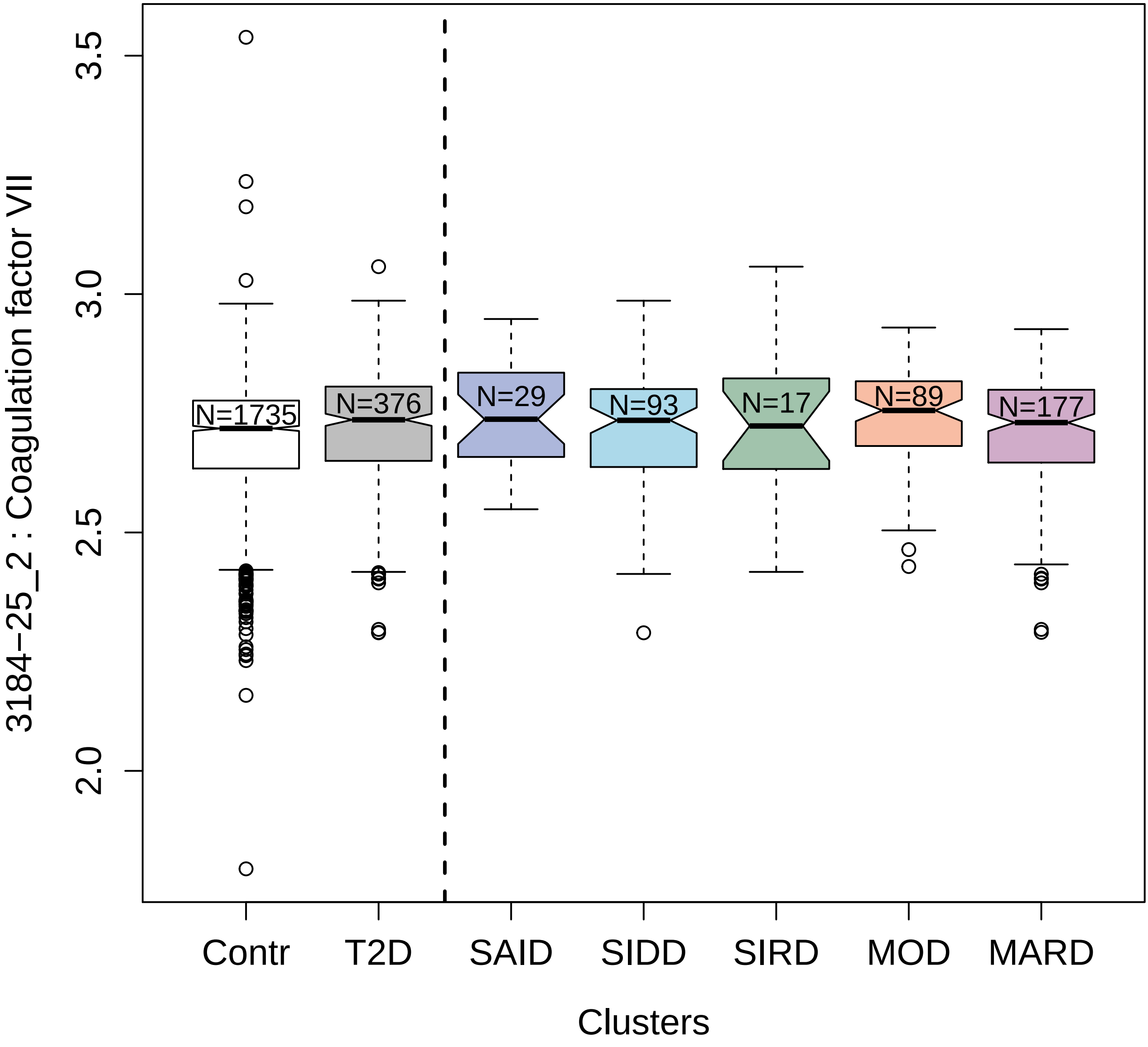
3184−25_2 : Coagulation factor VII
3.5
3.0
2.5
2.0
N=1735
N=376
N=29
N=93
N=17
N=89
N=177
Contr
T2D
SAID
SIDD
SIRD
MOD
MARD
Clusters

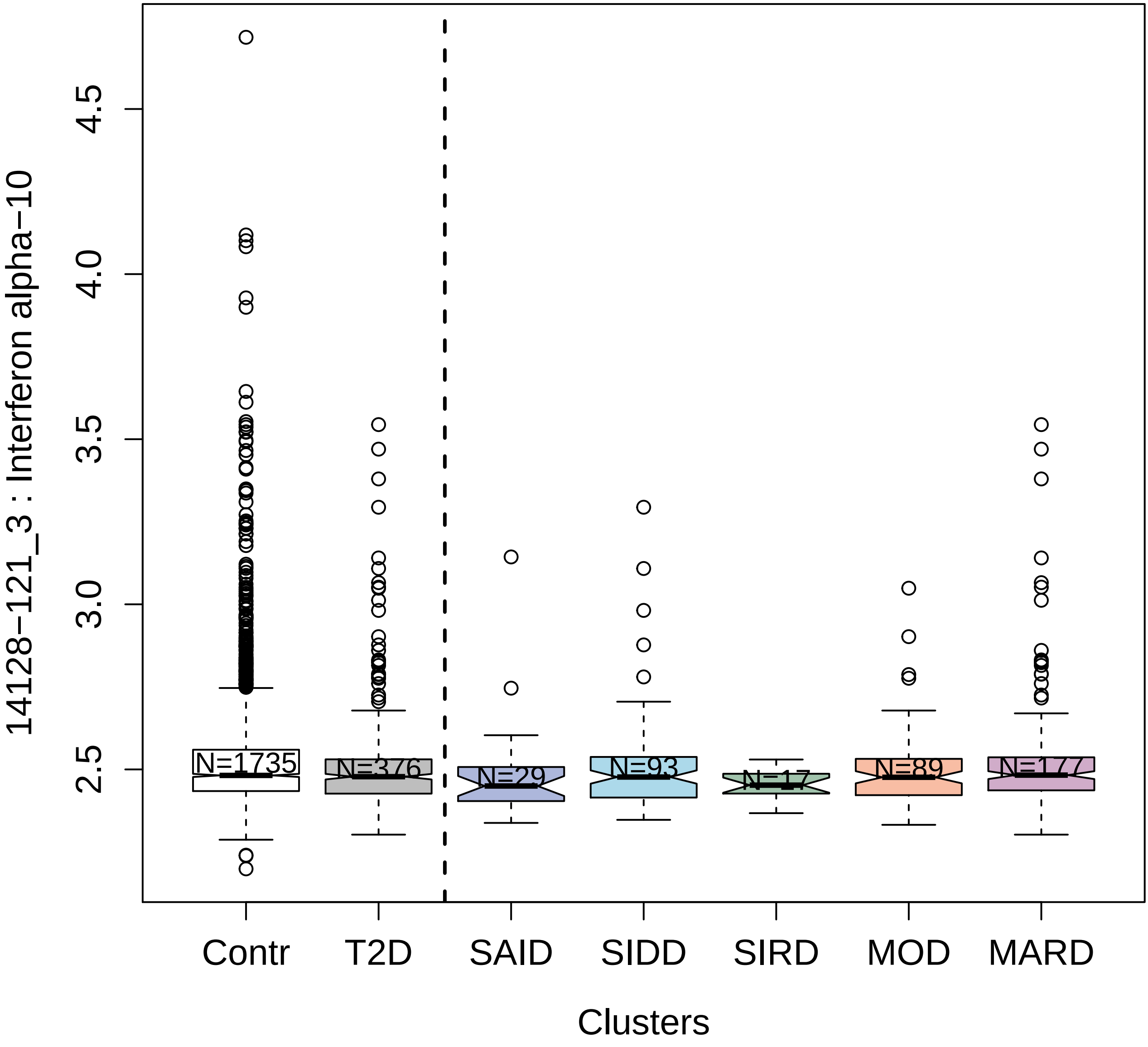
14128−121_3 : Interferon alpha−10
4.5
4.0
3.5
3.0
2.5
N=1735
N=376
N=29
N=93
N=17
N=89
N=177
Contr
T2D
SAID
SIDD
SIRD
MOD
MARD
Clusters

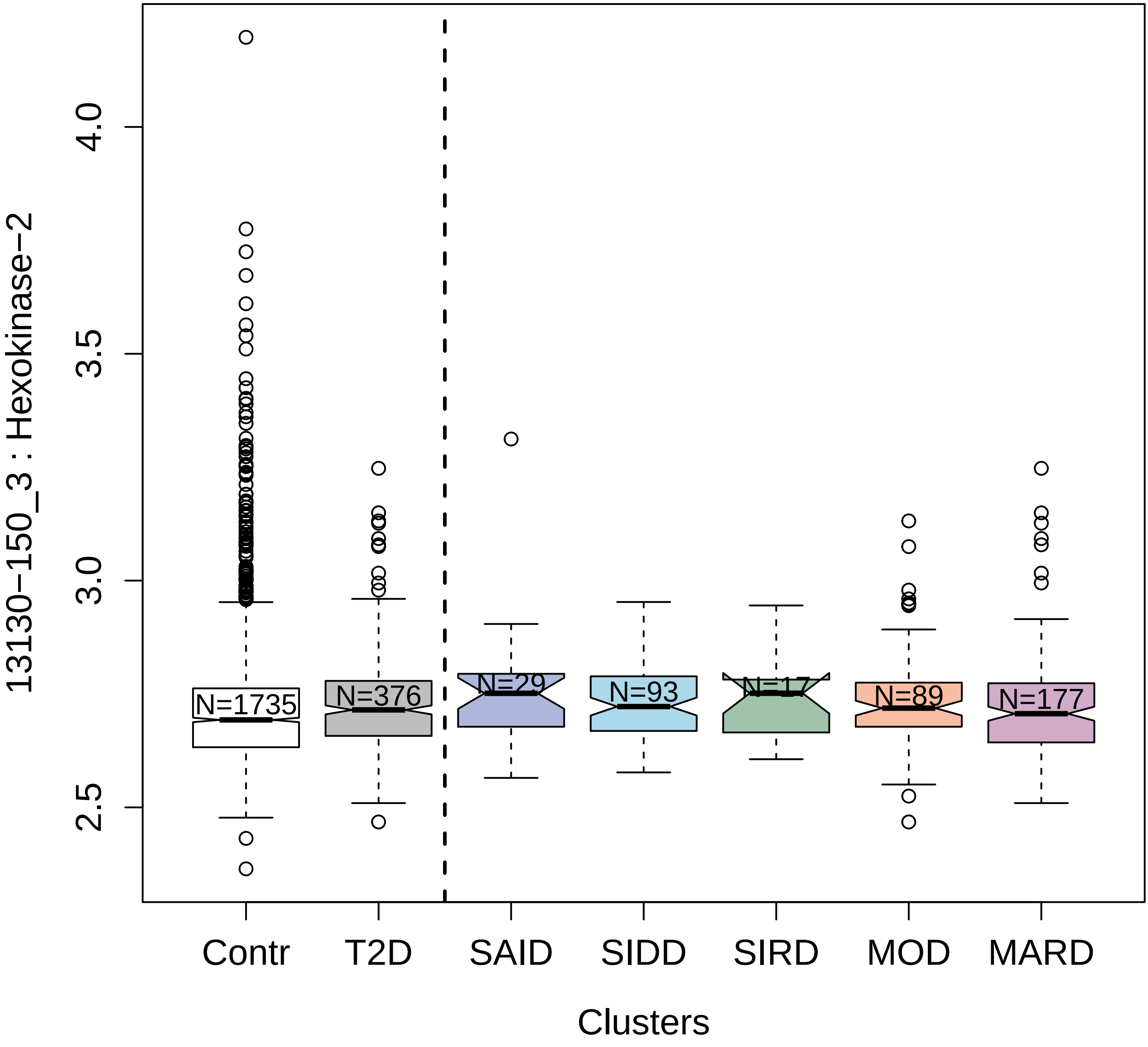

13130−150_3 : Hexokinase−2
4.0
3.5
3.0
2.5
N=1735
N=376
N=29
N=93
N=17
N=89
N=177
Contr
T2D
SAID
SIDD
SIRD
MOD
MARD
Clusters

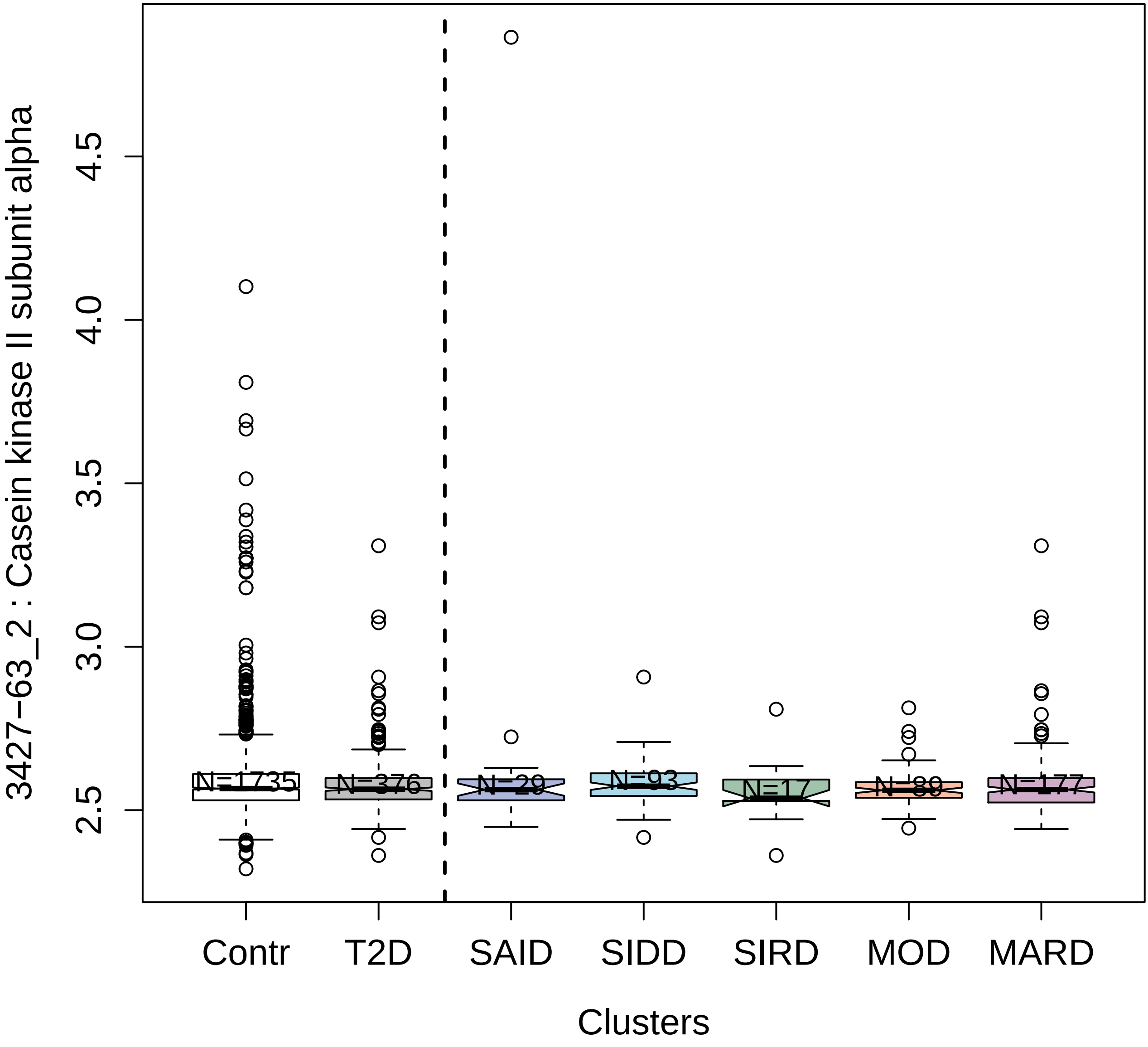

3427−63_2 : Casein kinase II subunit alpha
4.5
4.0
3.5
3.0
2.5
N=1735
N=376
N=29
N=93
N=17
N=89
N=177
Contr
T2D
SAID
SIDD
SIRD
MOD
MARD
Clusters

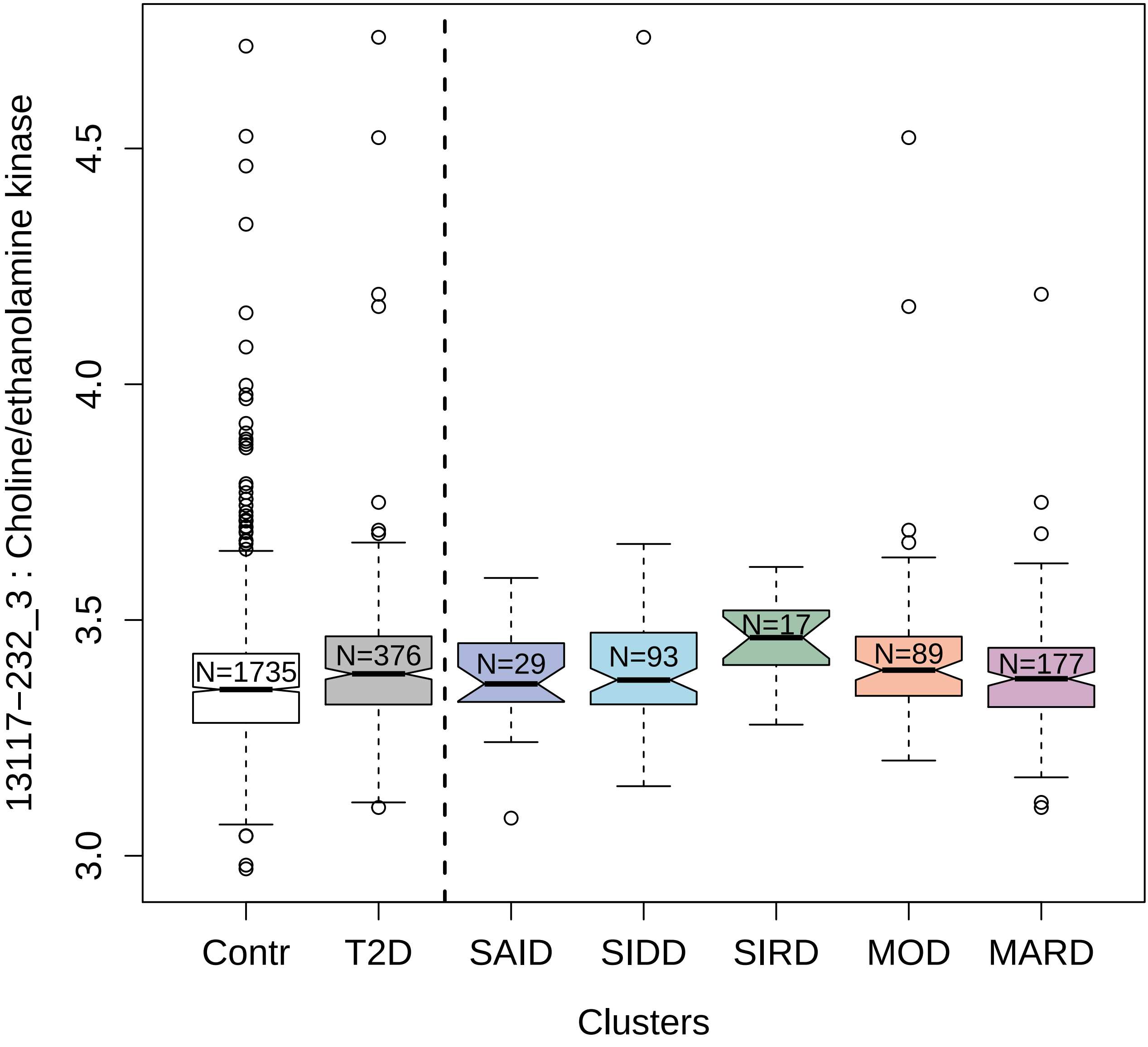

13117−232_3 : Choline/ethanolamine kinase
3.0
3.5
4.0
4.5
N=1735
N=376
N=29
N=93
N=17
N=89
N=177
Contr
T2D
SAID
SIDD
SIRD
MOD
MARD
Clusters

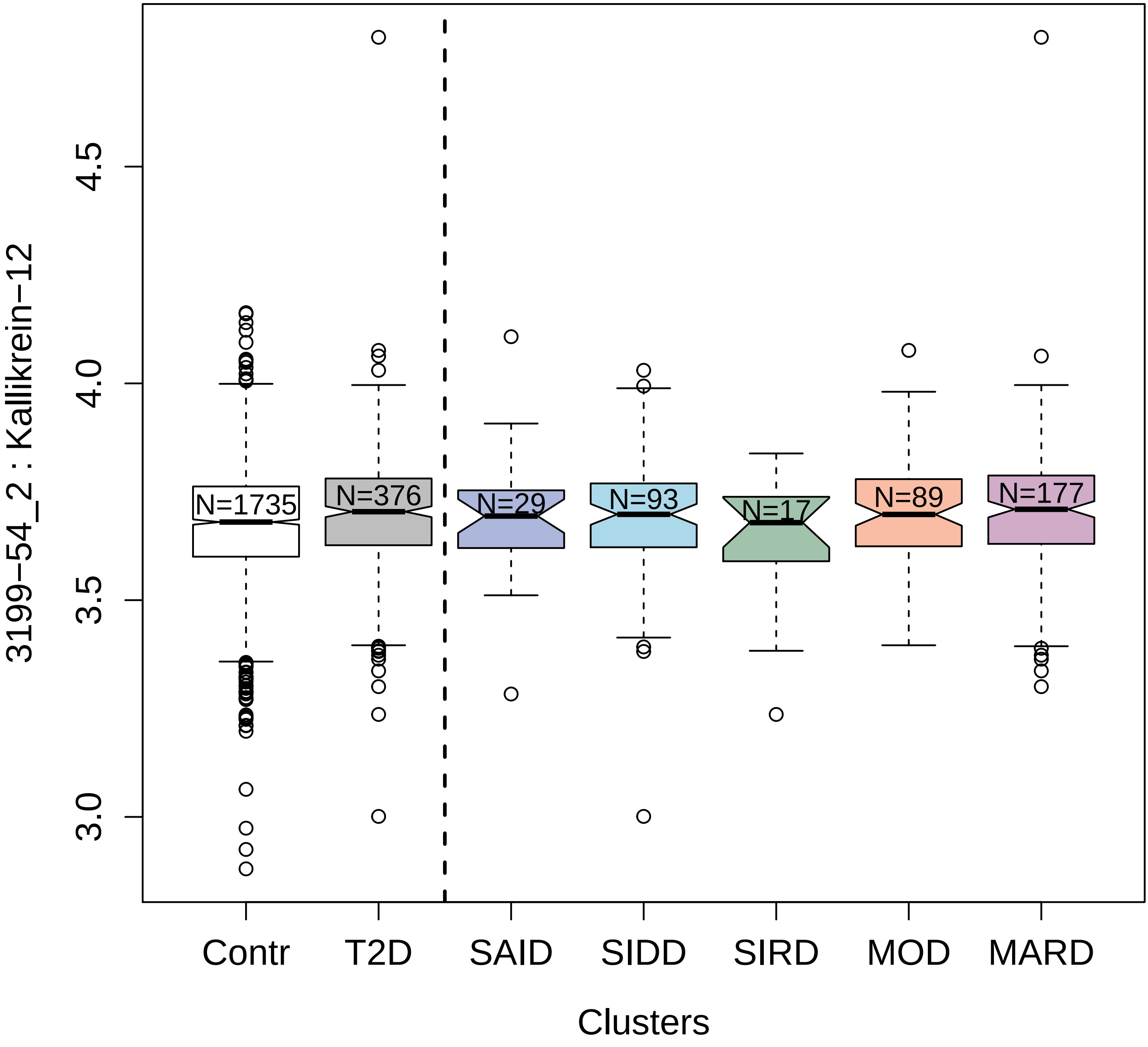
3199−54_2 : Kallikrein−12
4.5
4.0
3.5
3.0
N=1735
N=376
N=29
N=93
N=17
N=89
N=177
Contr
T2D
SAID
SIDD
SIRD
MOD
MARD
Clusters

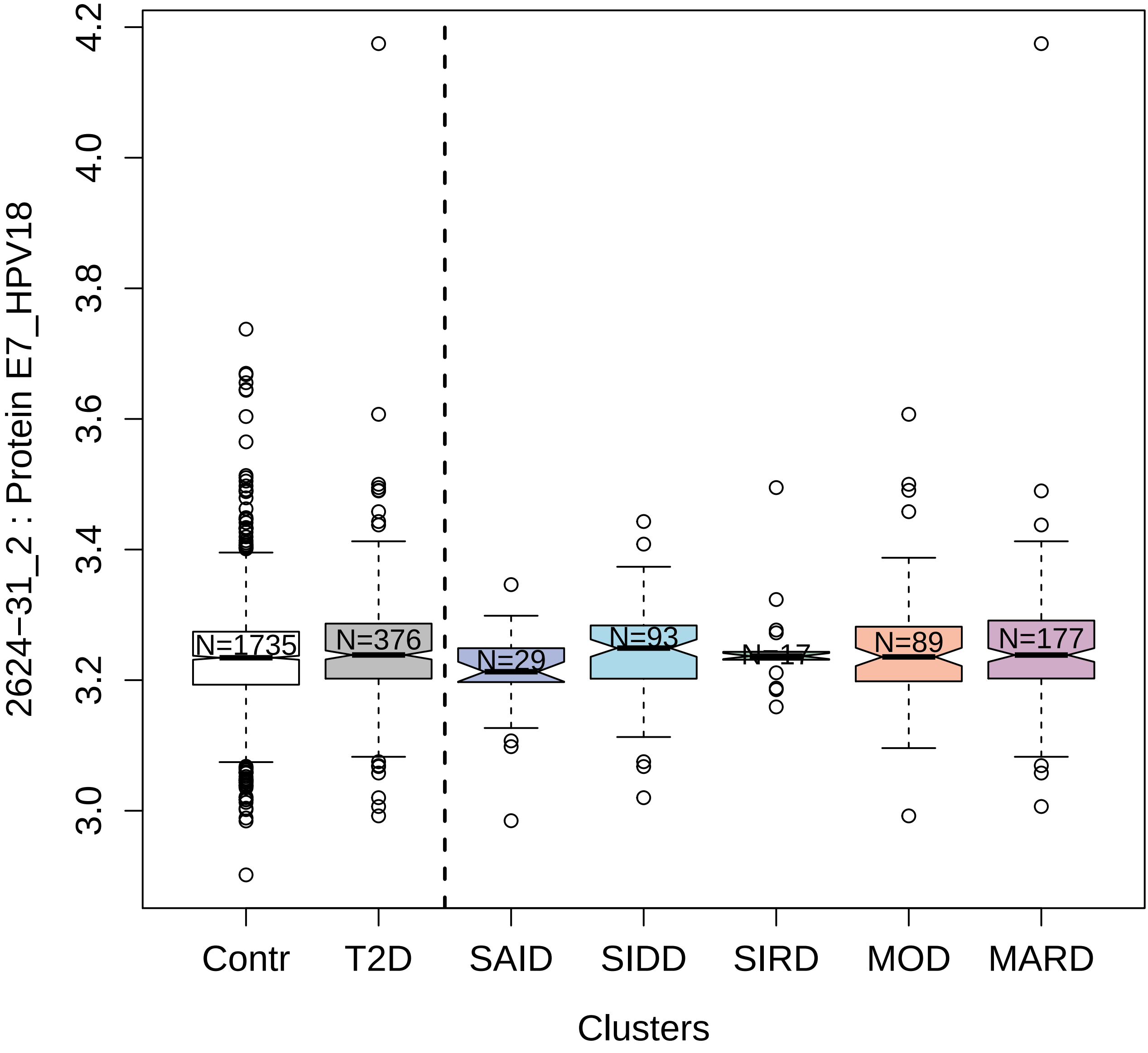

2624−31_2 : Protein E7_HPV18
4.2
4.0
3.8
3.6
3.4
3.2
3.0
N=1735
N=376
N=29
N=93
N=17
N=89
N=177
Contr
T2D
SAID
SIDD
SIRD
MOD
MARD
Clusters

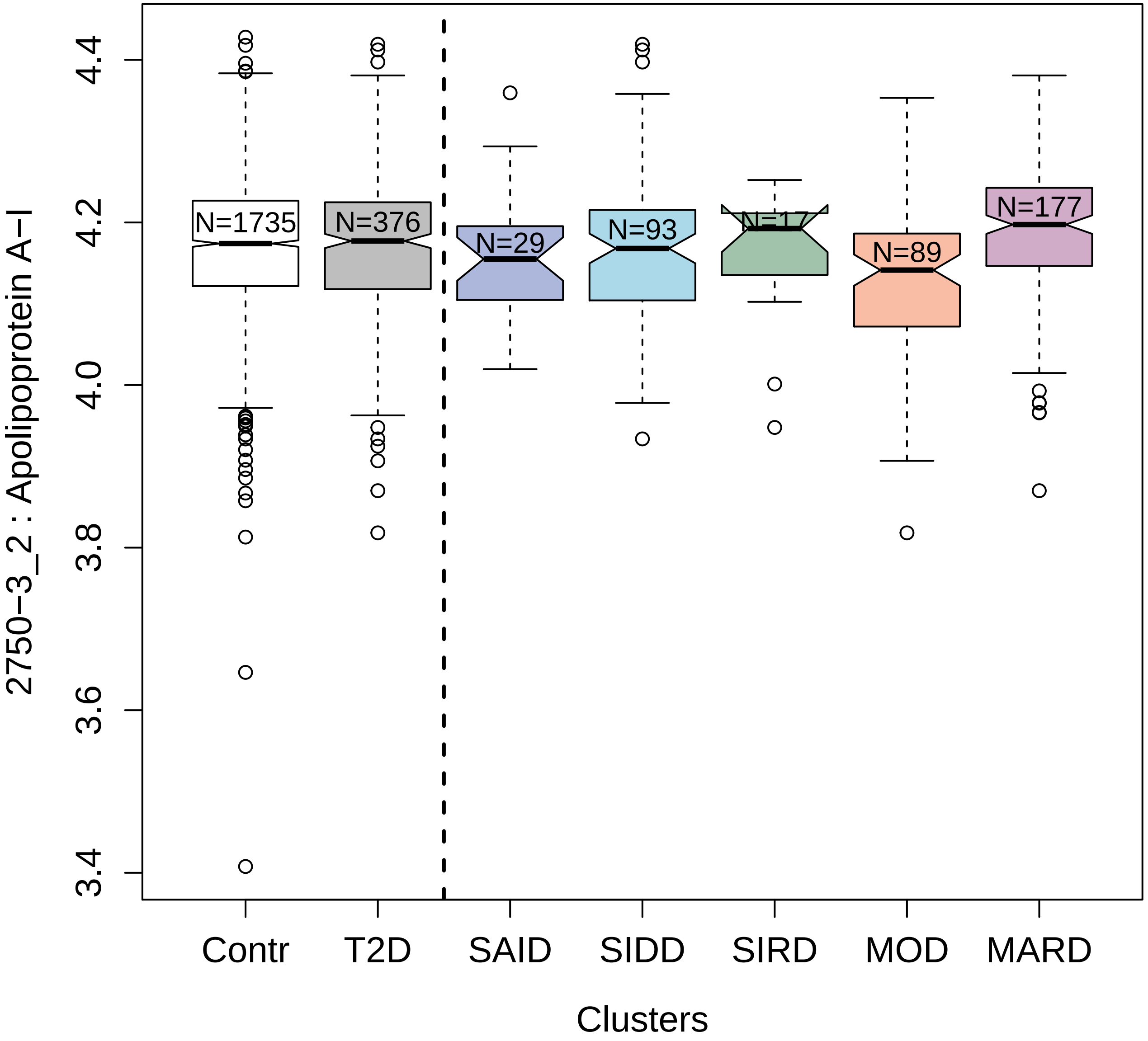

2750−3_2 : Apolipoprotein A−I
3.4
3.6
3.8
4.0
4.2
4.4
N=1735
N=376
N=29
N=93
N=17
N=89
N=177
Contr
T2D
SAID
SIDD
SIRD
MOD
MARD
Clusters

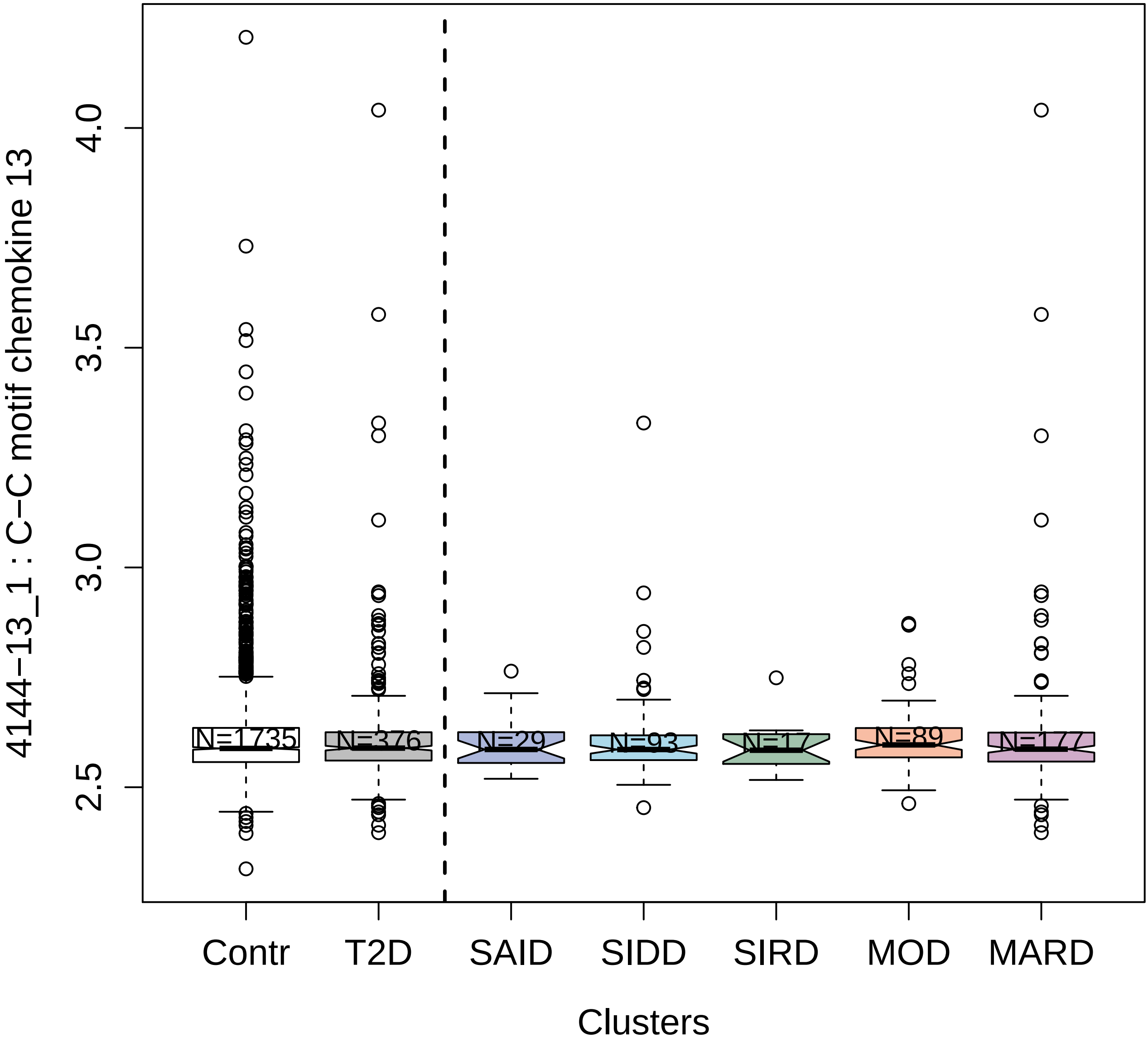
4144−13_1 : C−C motif chemokine 13
4.0
3.5
3.0
2.5
N=1735
N=376
N=29
N=93
N=17
N=89
N=177
Contr
T2D
SAID
SIDD
SIRD
MOD
MARD
Clusters

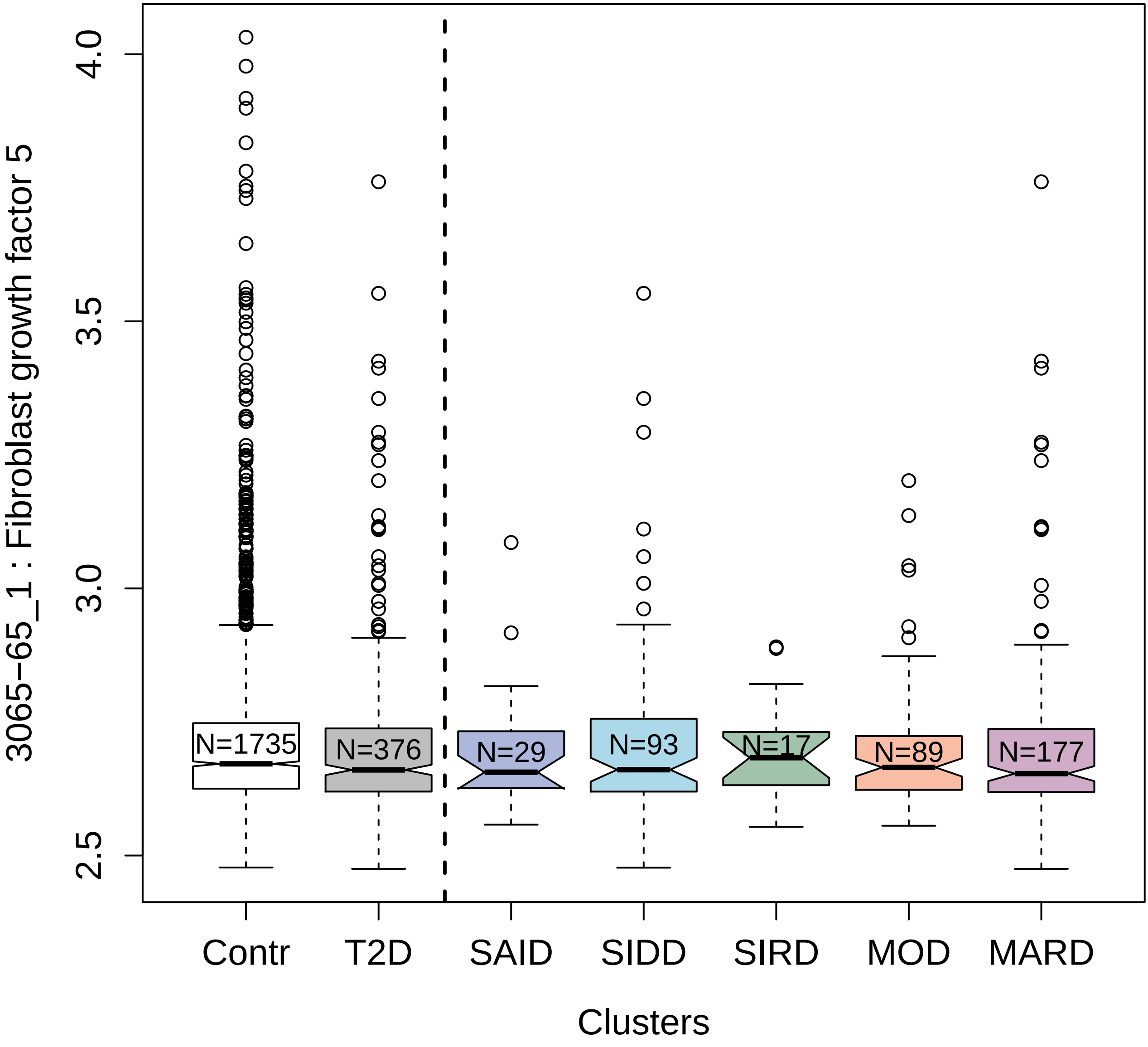
3065−65_1 : Fibroblast growth factor 5
4.0
3.5
3.0
2.5
N=1735
N=376
N=29
N=93
N=17
N=89
N=177
Contr
T2D
SAID
SIDD
SIRD
MOD
MARD
Clusters

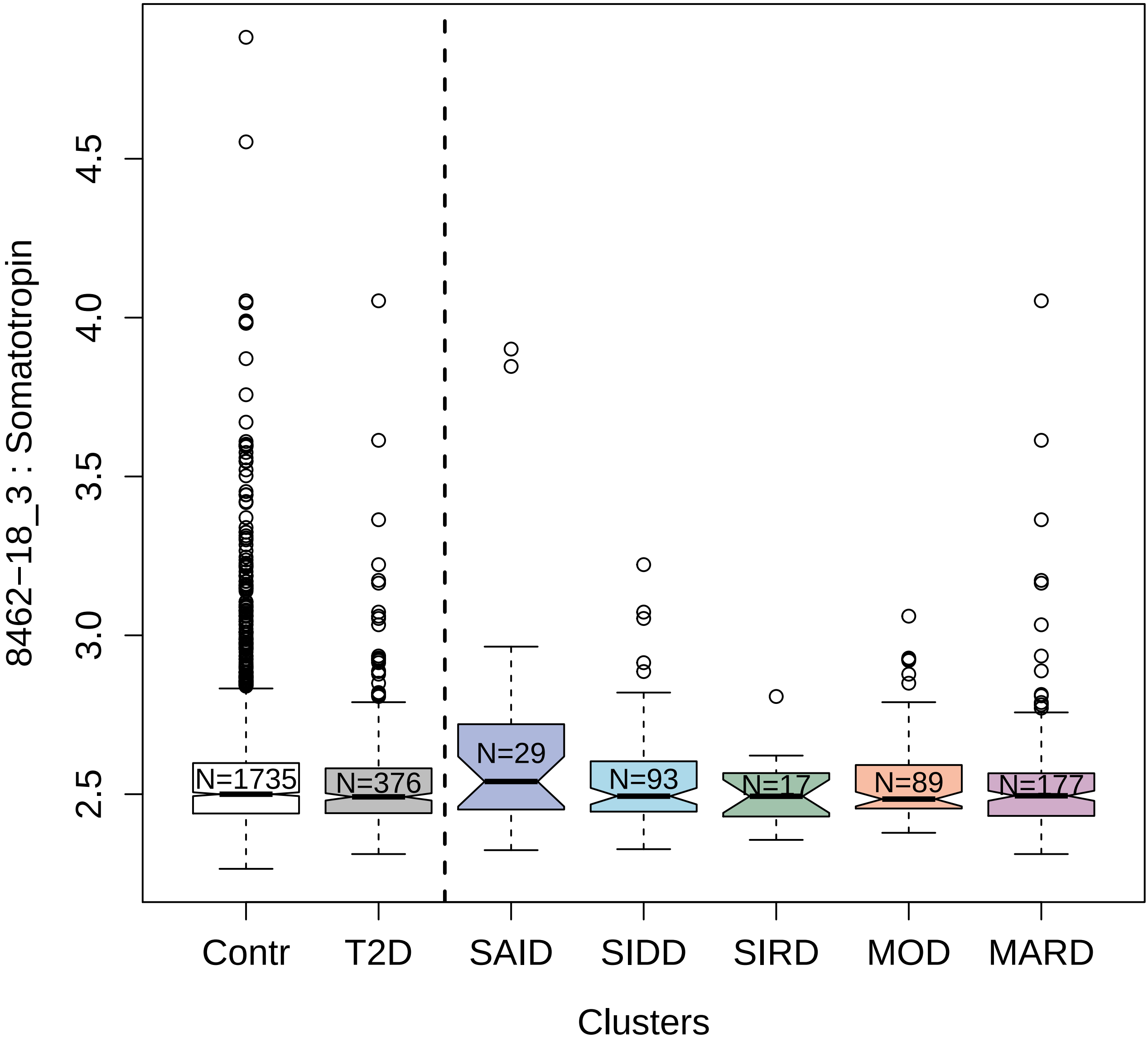

8462−18_3 : Somatotropin
4.5
4.0
3.5
3.0
2.5
N=1735
N=376
N=29
N=93
N=17
N=89
N=177
Contr
T2D
SAID
SIDD
SIRD
MOD
MARD
Clusters

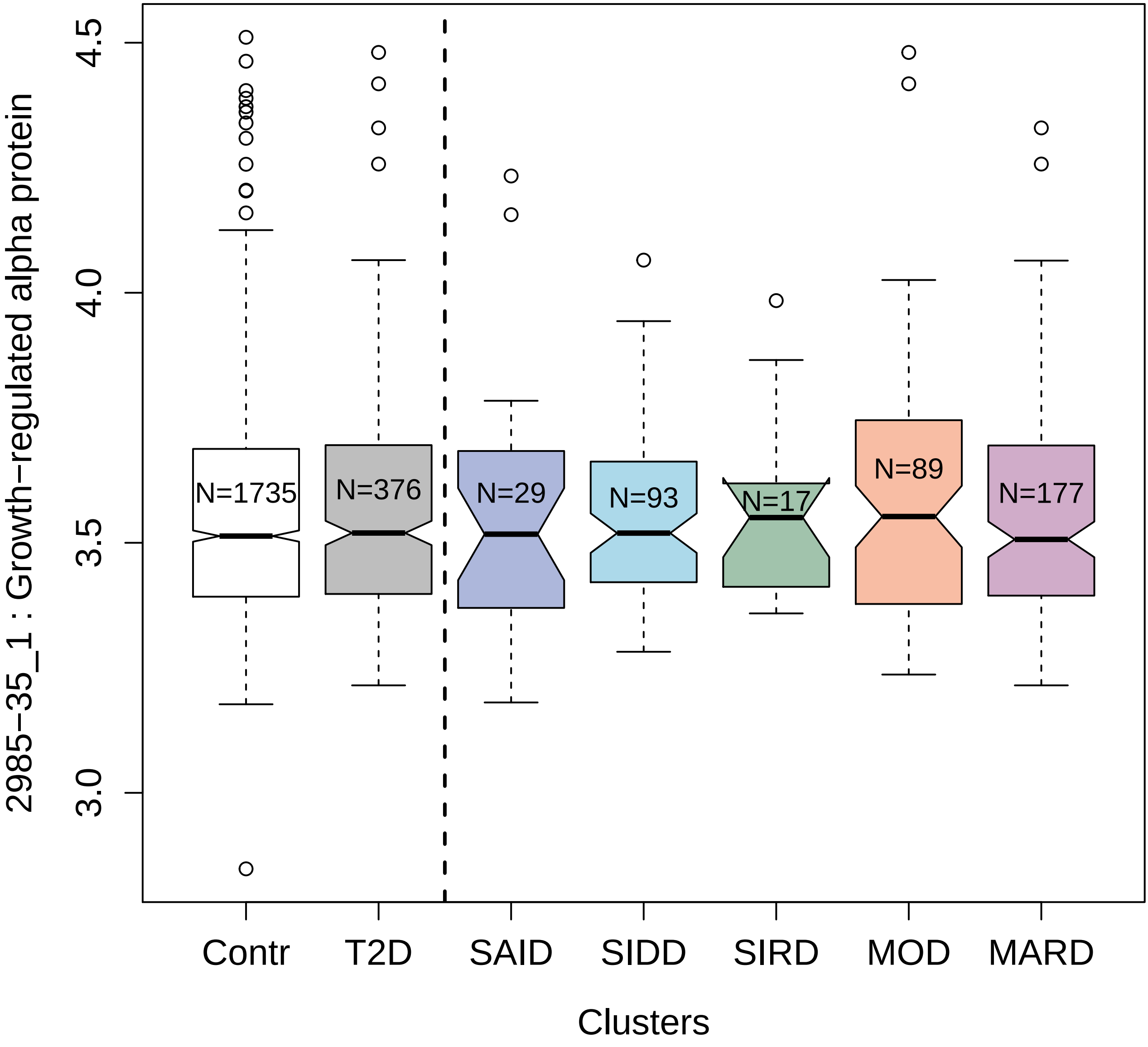
2985−35_1 : Growth−regulated alpha protein
4.5
4.0
3.5
3.0
N=1735
N=376
N=29
N=93
N=17
N=89
N=177
Contr
T2D
SAID
SIDD
SIRD
MOD
MARD
Clusters

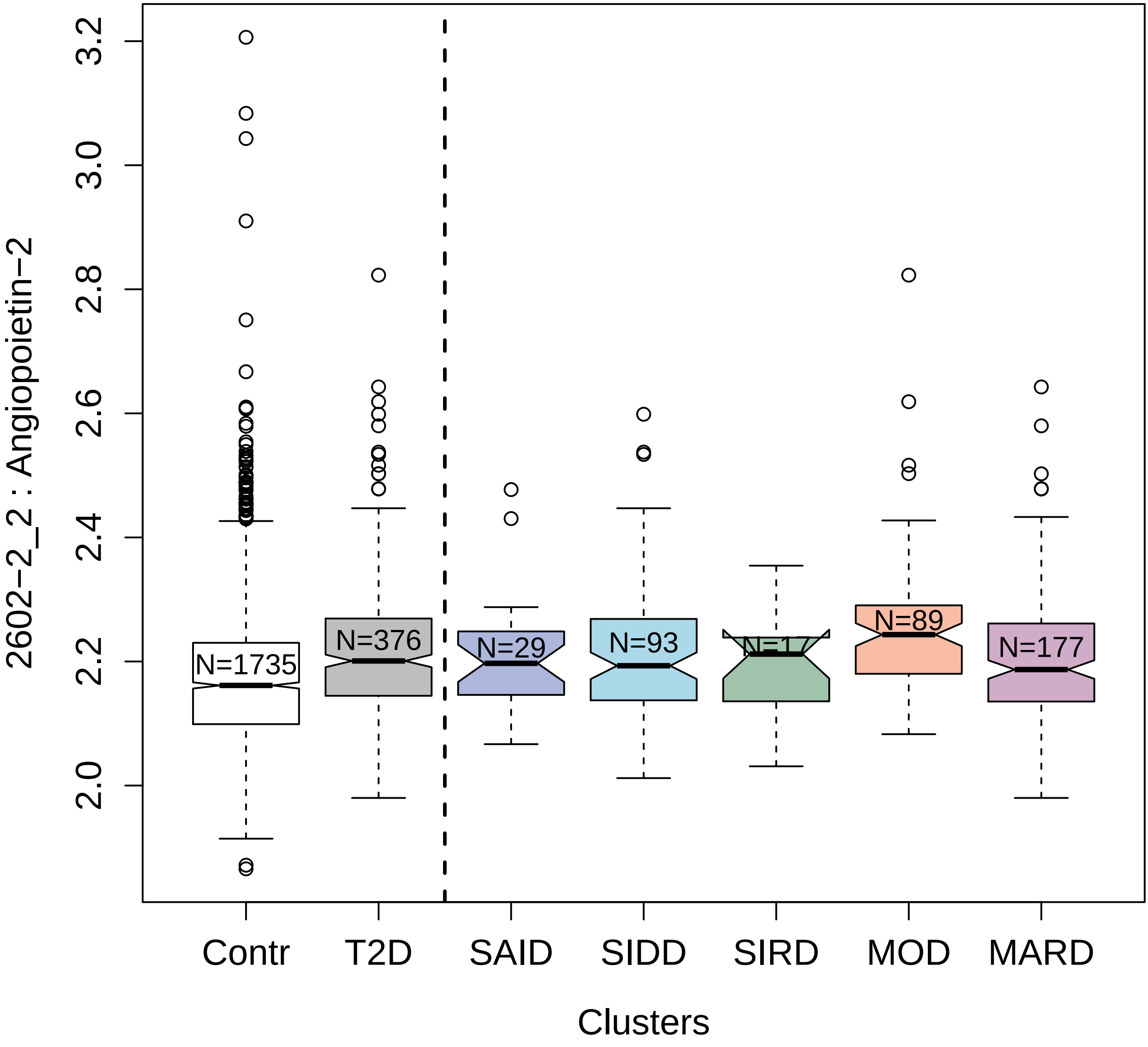

2602−2_2 : Angiopoietin−2
3.2
3.0
2.8
2.6
2.4
2.2
2.0
N=1735
N=376
N=29
N=93
N=89
N=177
Contr
T2D
SAID
SIDD
SIRD
MOD
MARD
Clusters

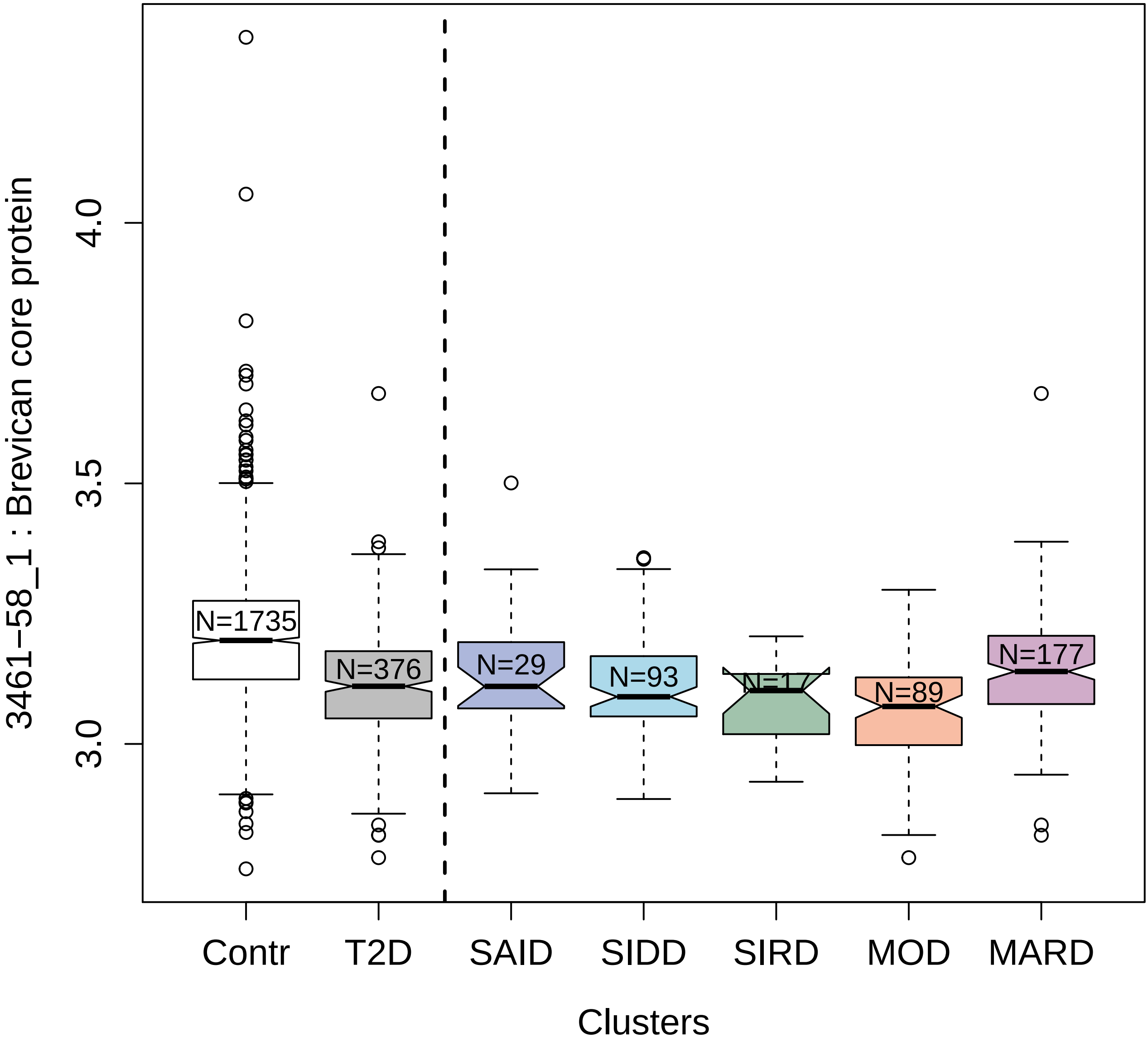

3461−58_1 : Brevican core protein
4.0
3.5
3.0
N=1735
N=376
N=29
N=93
N=89
N=177
Contr
T2D
SAID
SIDD
SIRD
MOD
MARD
Clusters

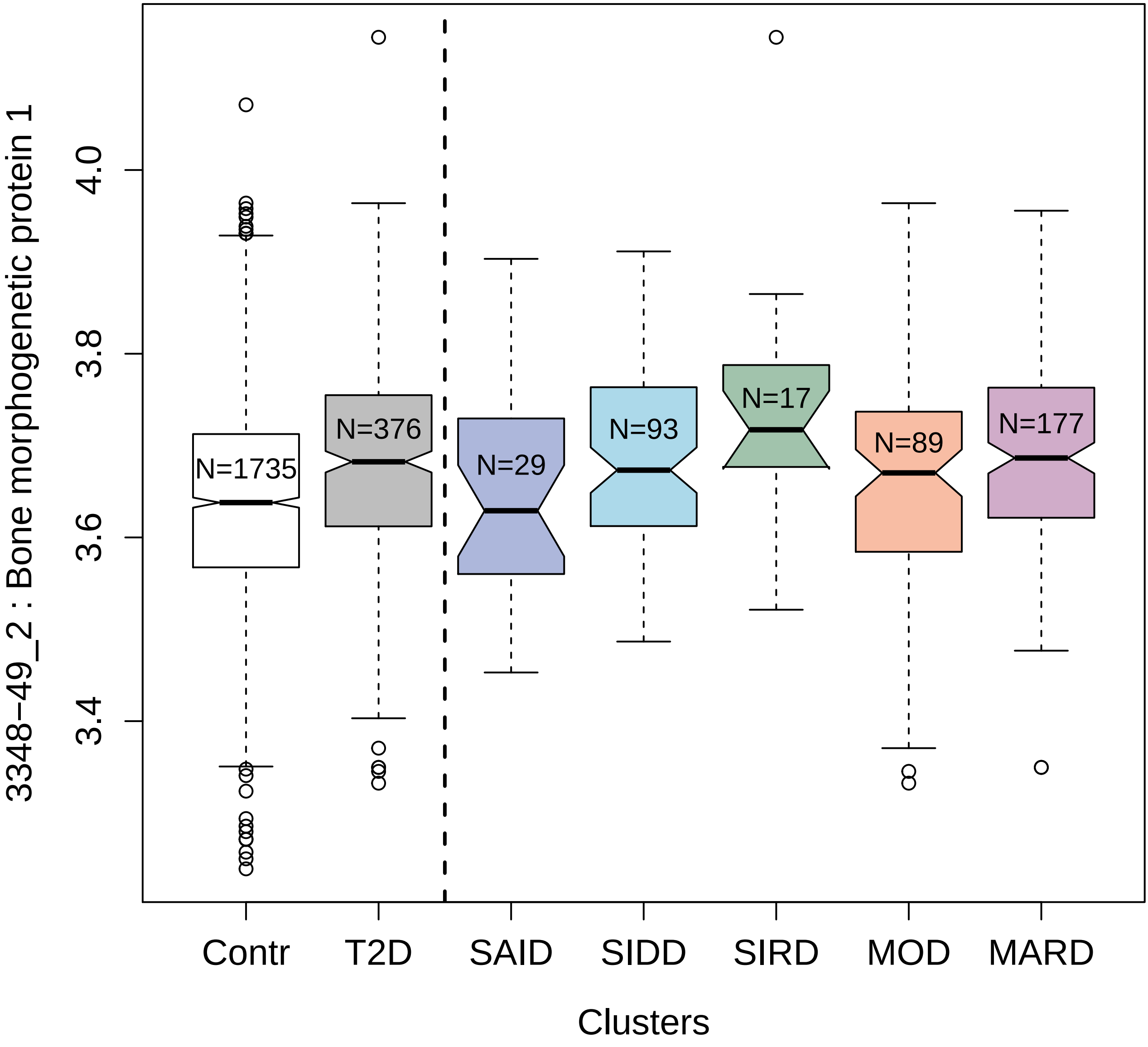

3348−49_2 : Bone morphogenetic protein 1
4.0
3.8
3.6
3.4
N=1735
N=376
N=29
N=93
N=17
N=89
N=177
Contr
T2D
SAID
SIDD
SIRD
MOD
MARD
Clusters

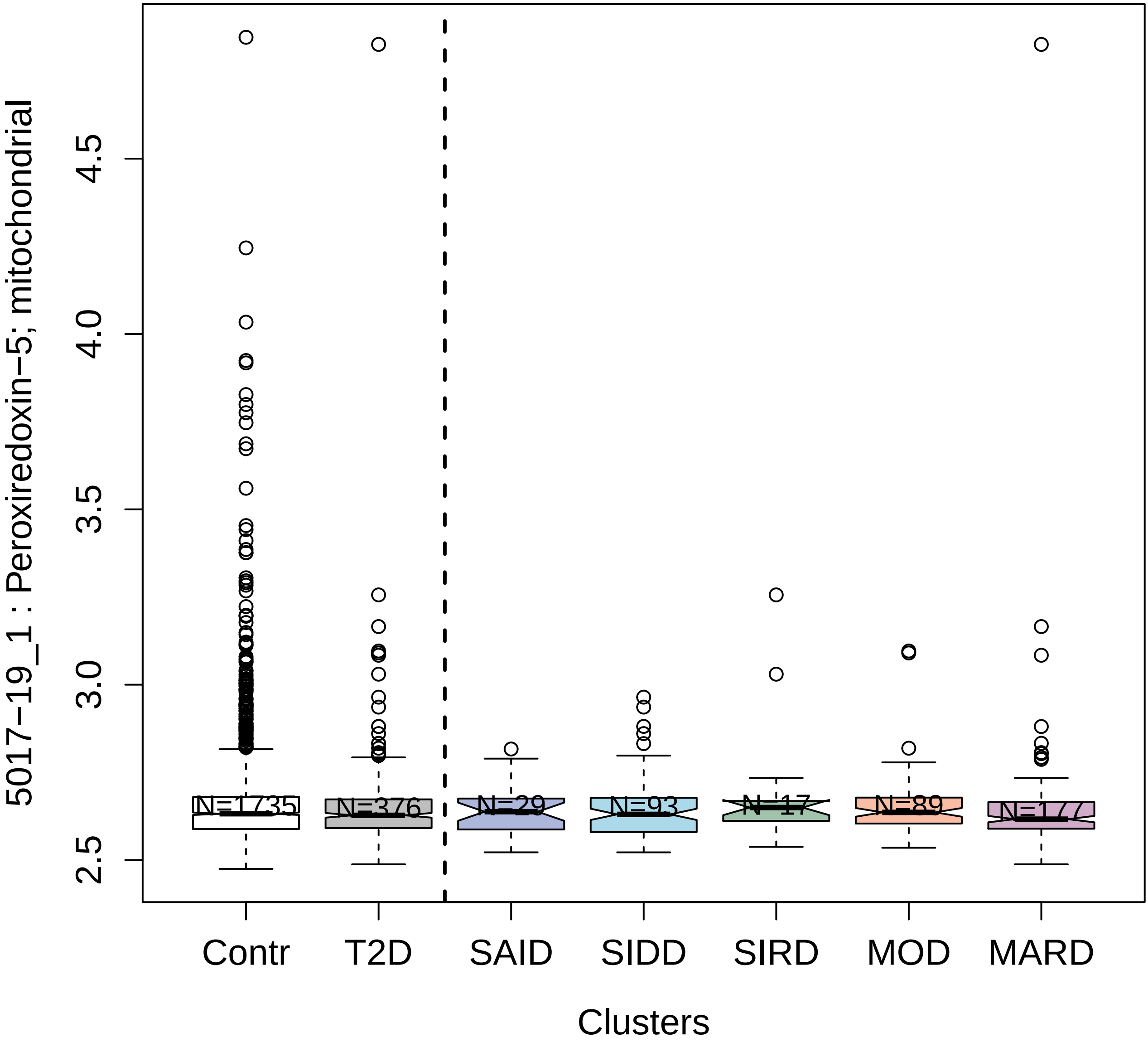

5017−19_1 : Peroxiredoxin−5; mitochondrial
2.5
3.0
3.5
4.0
4.5
N=1735
N=376
N=29
N=93
N=17
N=89
N=177
Contr
T2D
SAID
SIDD
SIRD
MOD
MARD
Clusters

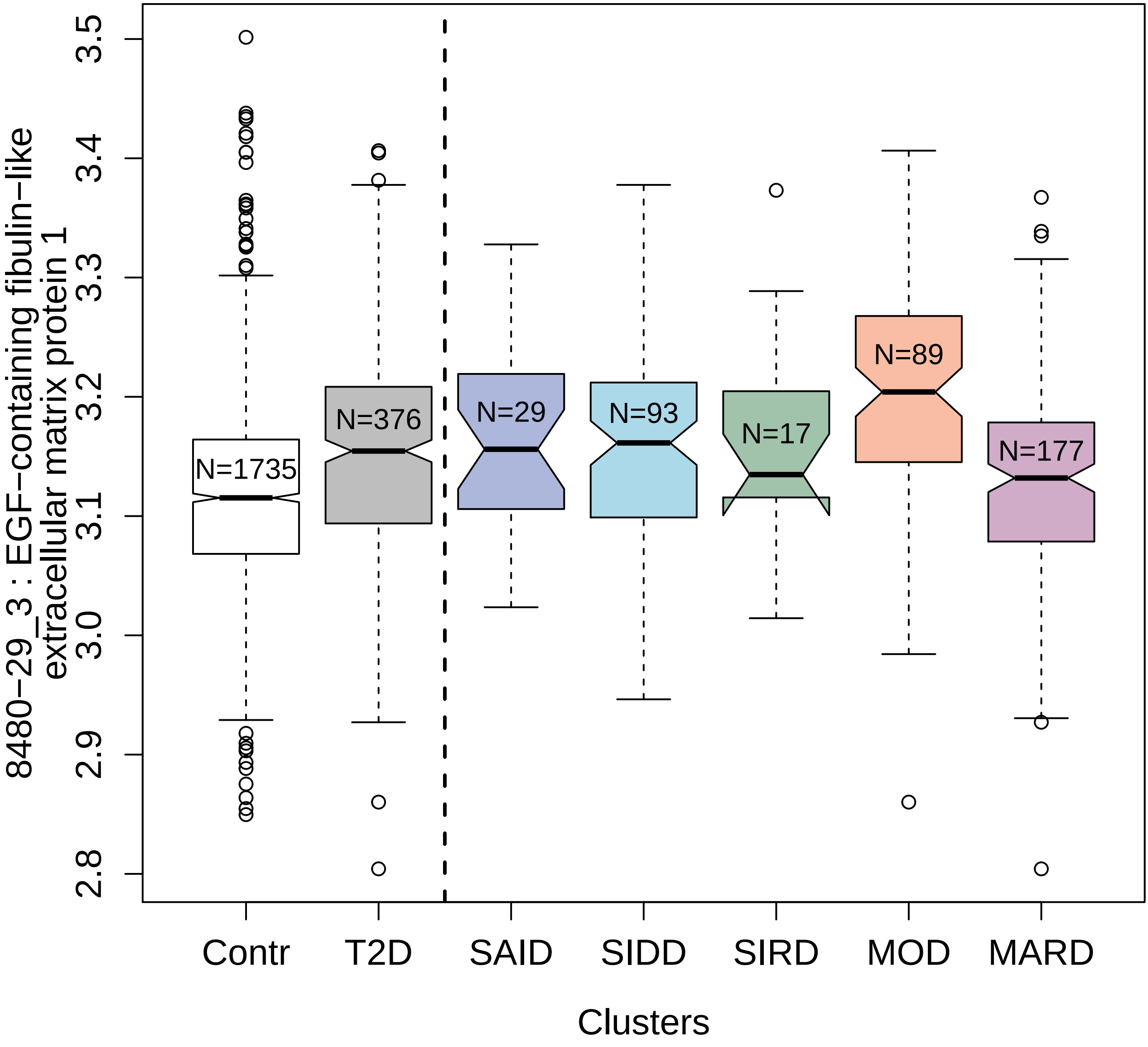
8480−29_3 : EGF−containing fibulin−like extracellular matrix protein 1
2.8
2.9
3.0
3.1
3.2
3.3
3.4
3.5
N=1735
N=376
N=29
N=93
N=17
N=89
N=177
Contr
T2D
SAID
SIDD
SIRD
MOD
MARD
Clusters

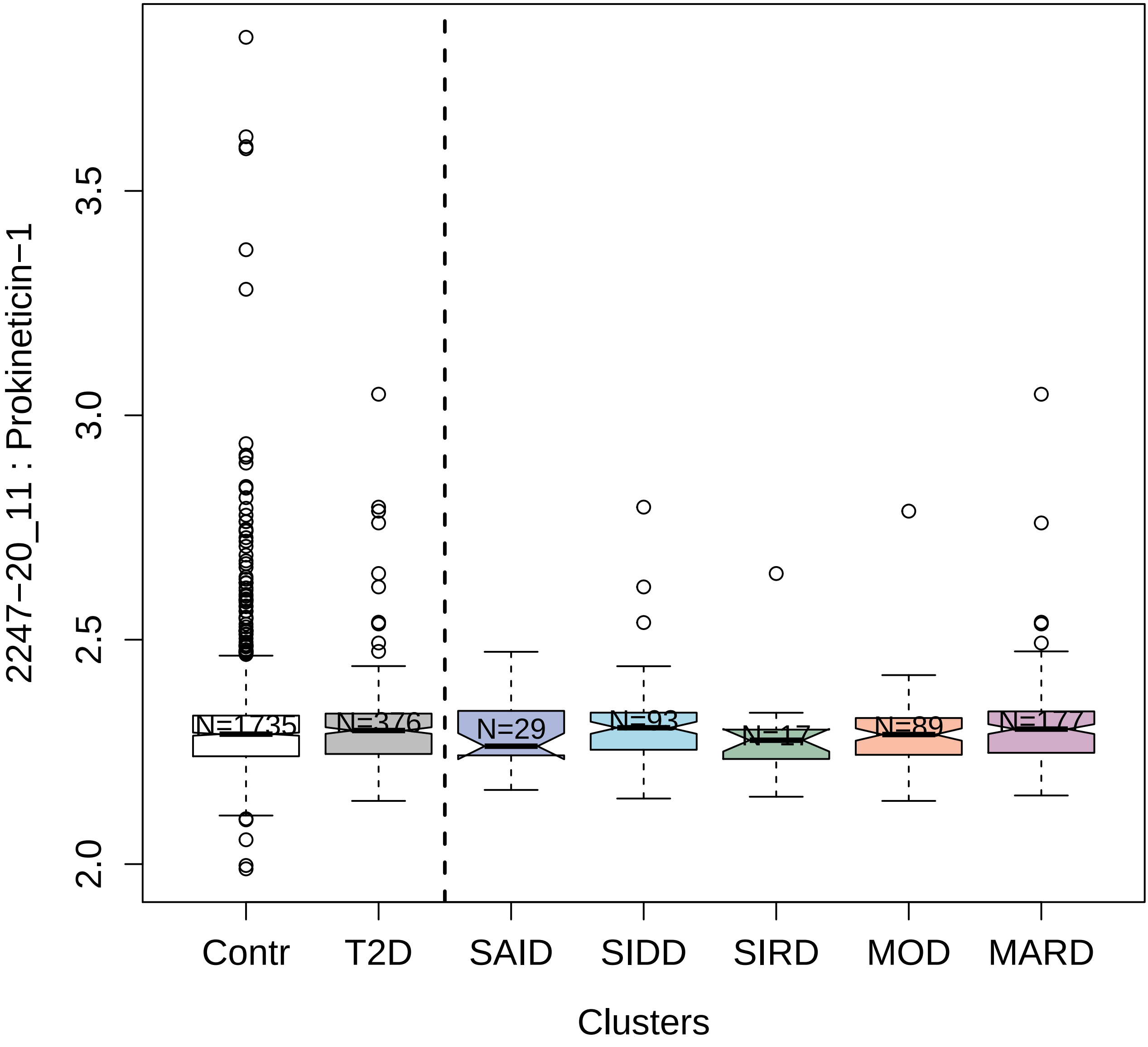

2247−20_11 : Prokineticin−1
2.0
2.5
3.0
3.5
N=1735
N=376
N=29
N=93
N=17
N=89
N=177
Contr
T2D
SAID
SIDD
SIRD
MOD
MARD
Clusters

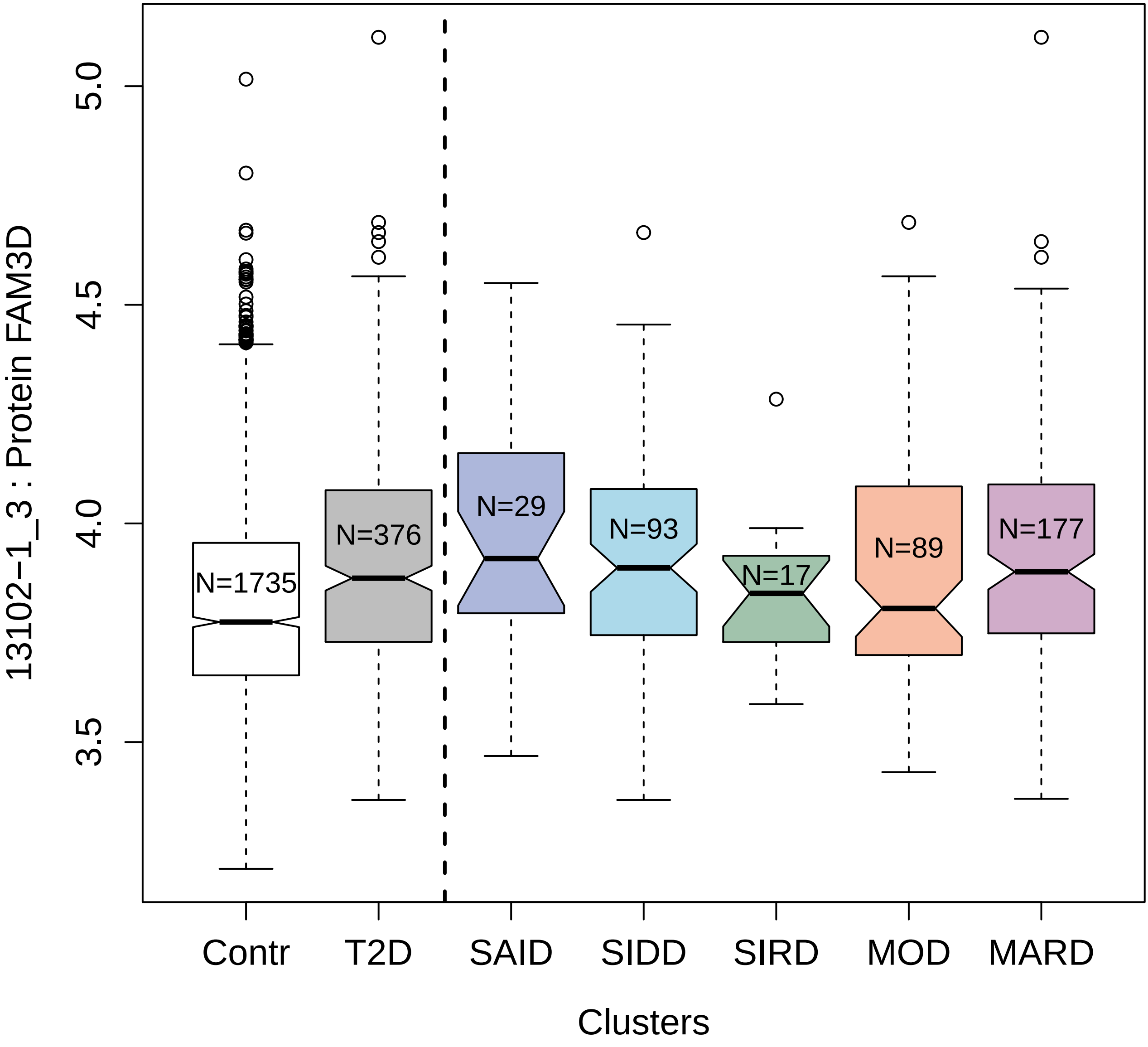

13102−1_3 : Protein FAM3D
5.0
4.5
4.0
3.5
N=1735
N=376
N=29
N=93
N=17
N=89
N=177
Contr
T2D
SAID
SIDD
SIRD
MOD
MARD
Clusters

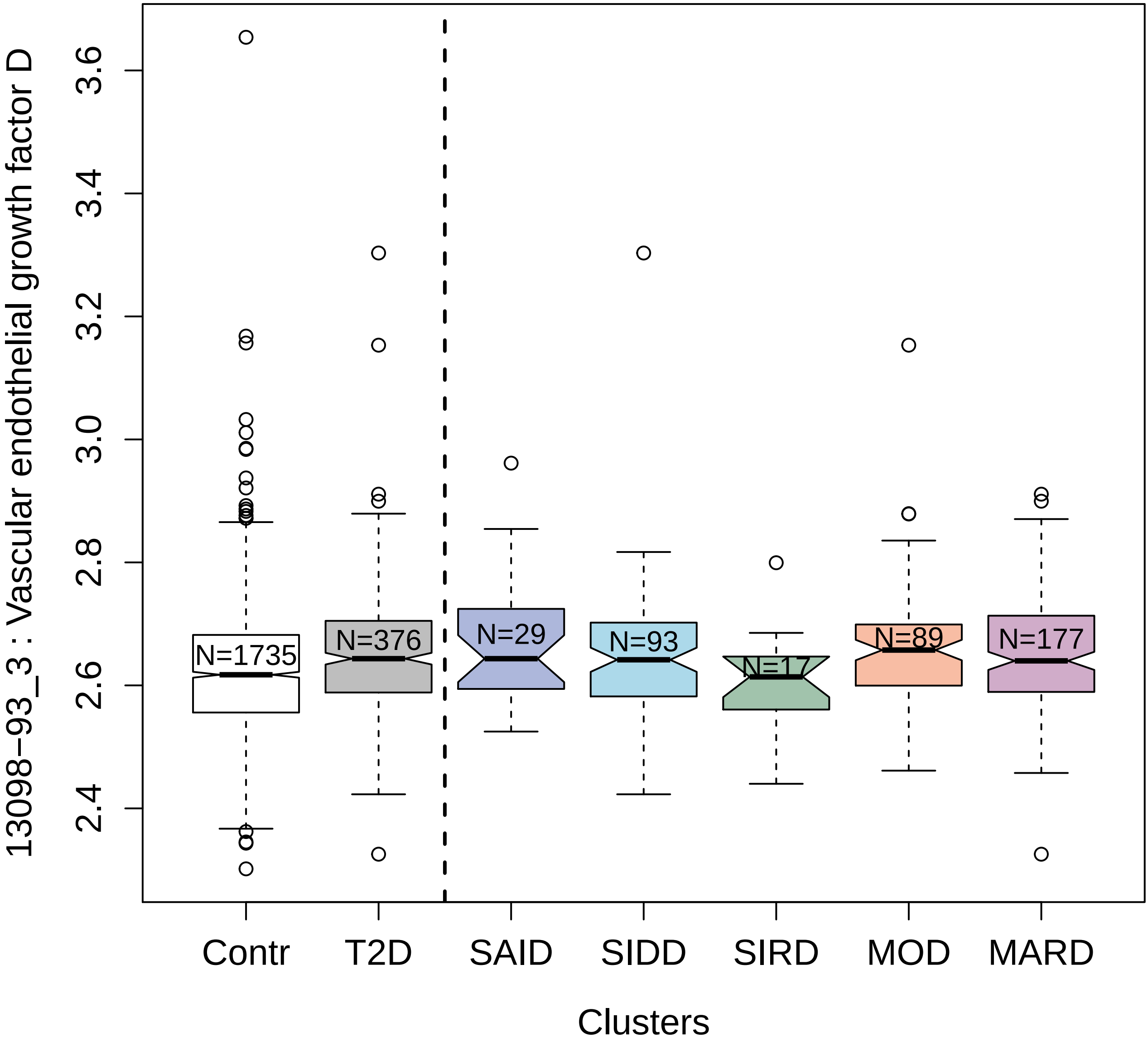

13098−93_3 : Vascular endothelial growth factor D
3.6
3.4
3.2
3.0
2.8
2.6
2.4
N=1735
N=376
N=29
N=93
N=17
N=89
N=177
Contr
T2D
SAID
SIDD
SIRD
MOD
MARD
Clusters

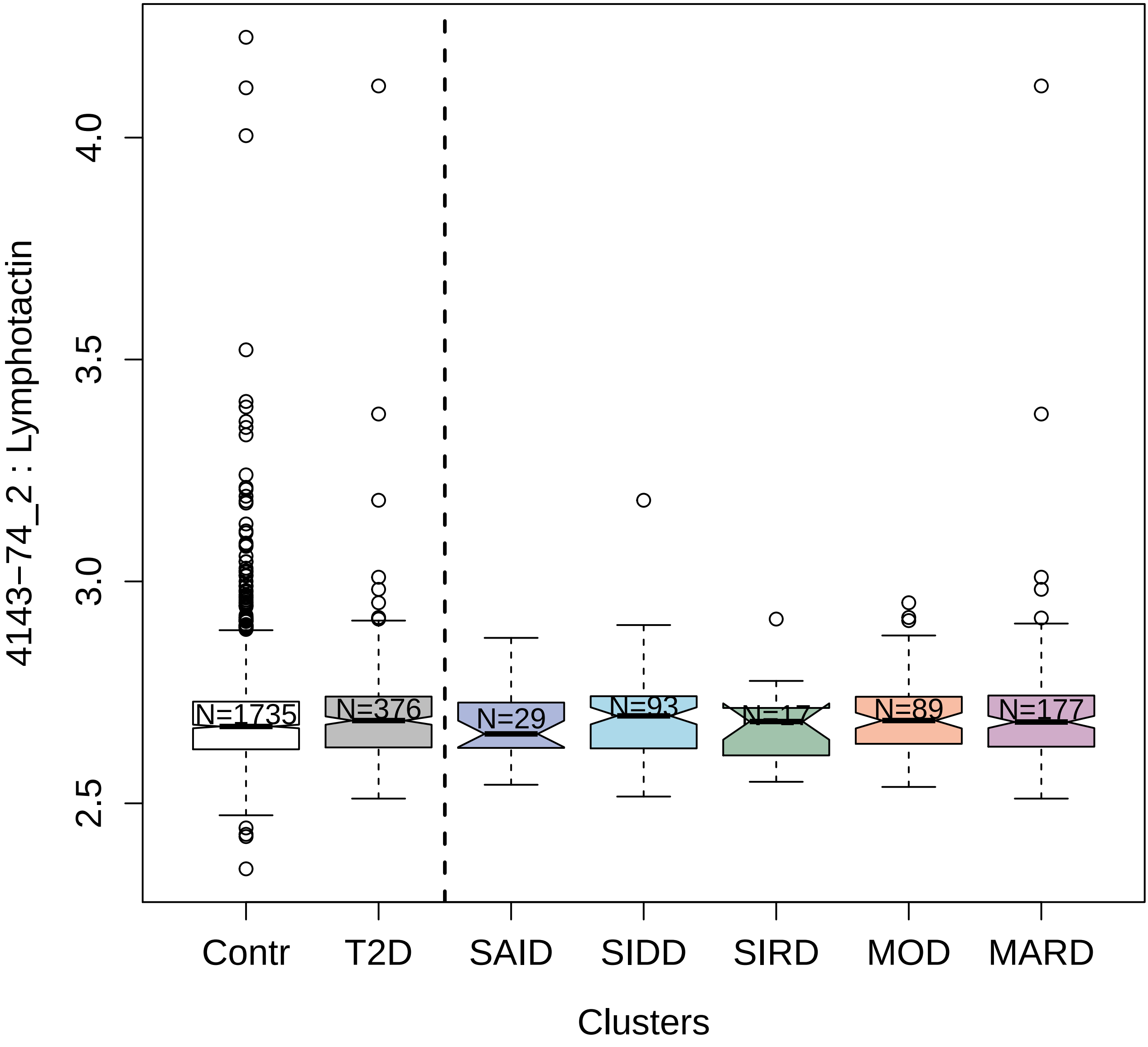

4143−74_2 : Lymphotactin
4.0
3.5
3.0
2.5
N=1735
N=376
N=29
N=93
N=17
N=89
N=177
Contr
T2D
SAID
SIDD
SIRD
MOD
MARD
Clusters

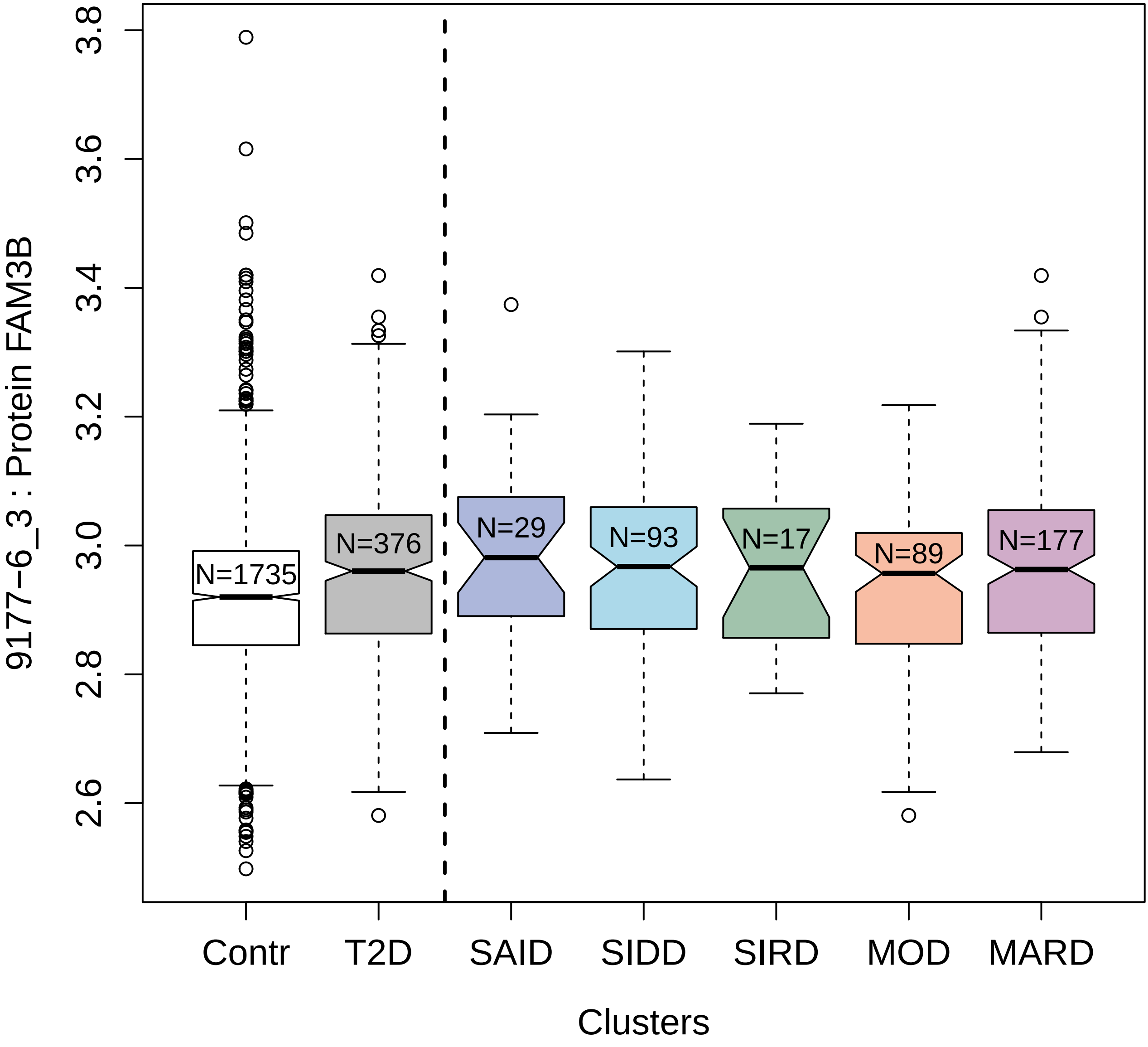

9177−6_3 : Protein FAM3B
3.8
3.6
3.4
3.2
3.0
2.8
2.6
N=1735
N=376
N=29
N=93
N=17
N=89
N=177
Contr
T2D
SAID
SIDD
SIRD
MOD
MARD
Clusters

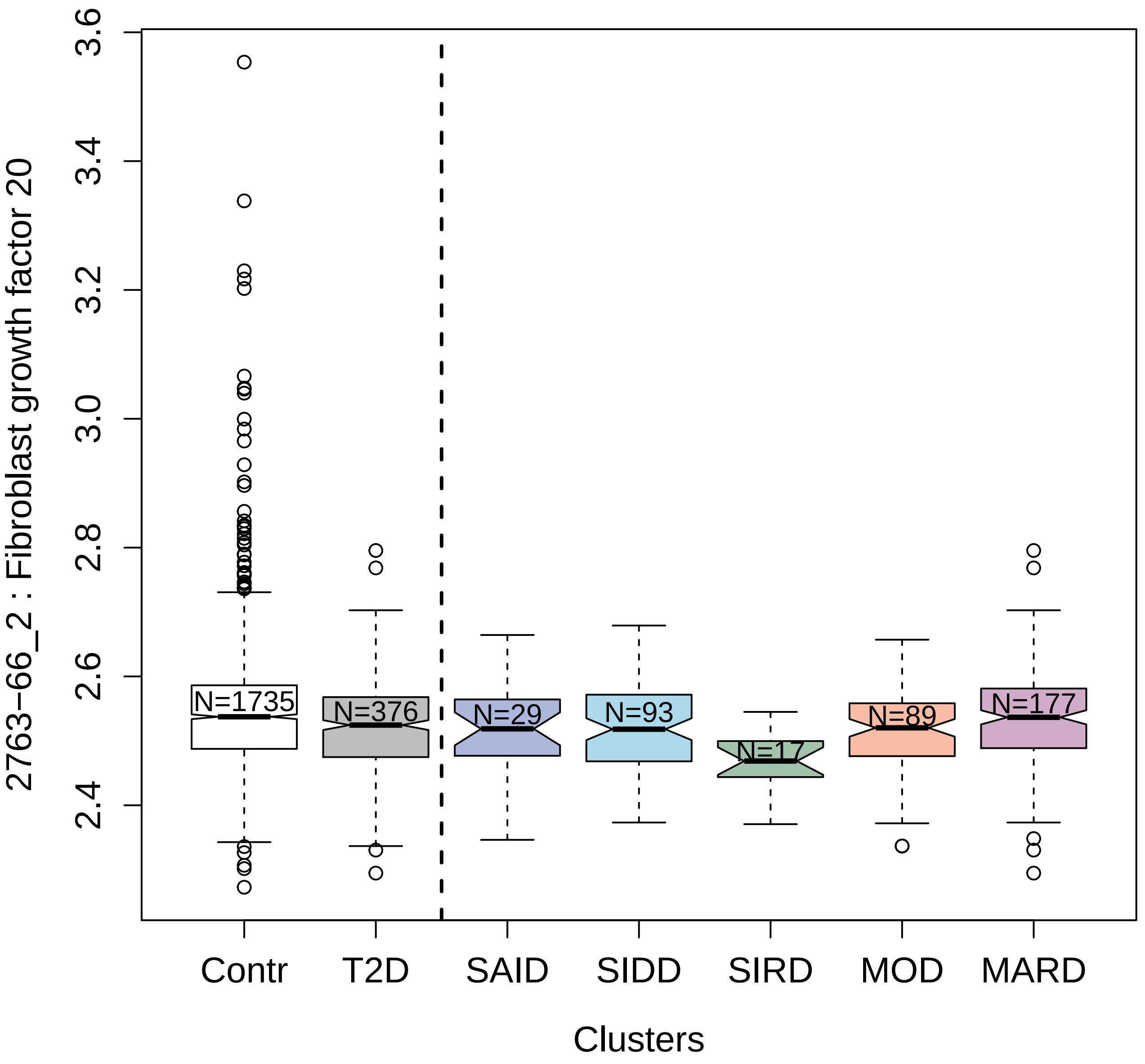

2763−66_2 : Fibroblast growth factor 20
3.6
3.4
3.2
3.0
2.8
2.6
2.4
N=1735
N=376
N=29
N=93
N=17
N=89
N=177
Contr
T2D
SAID
SIDD
SIRD
MOD
MARD
Clusters

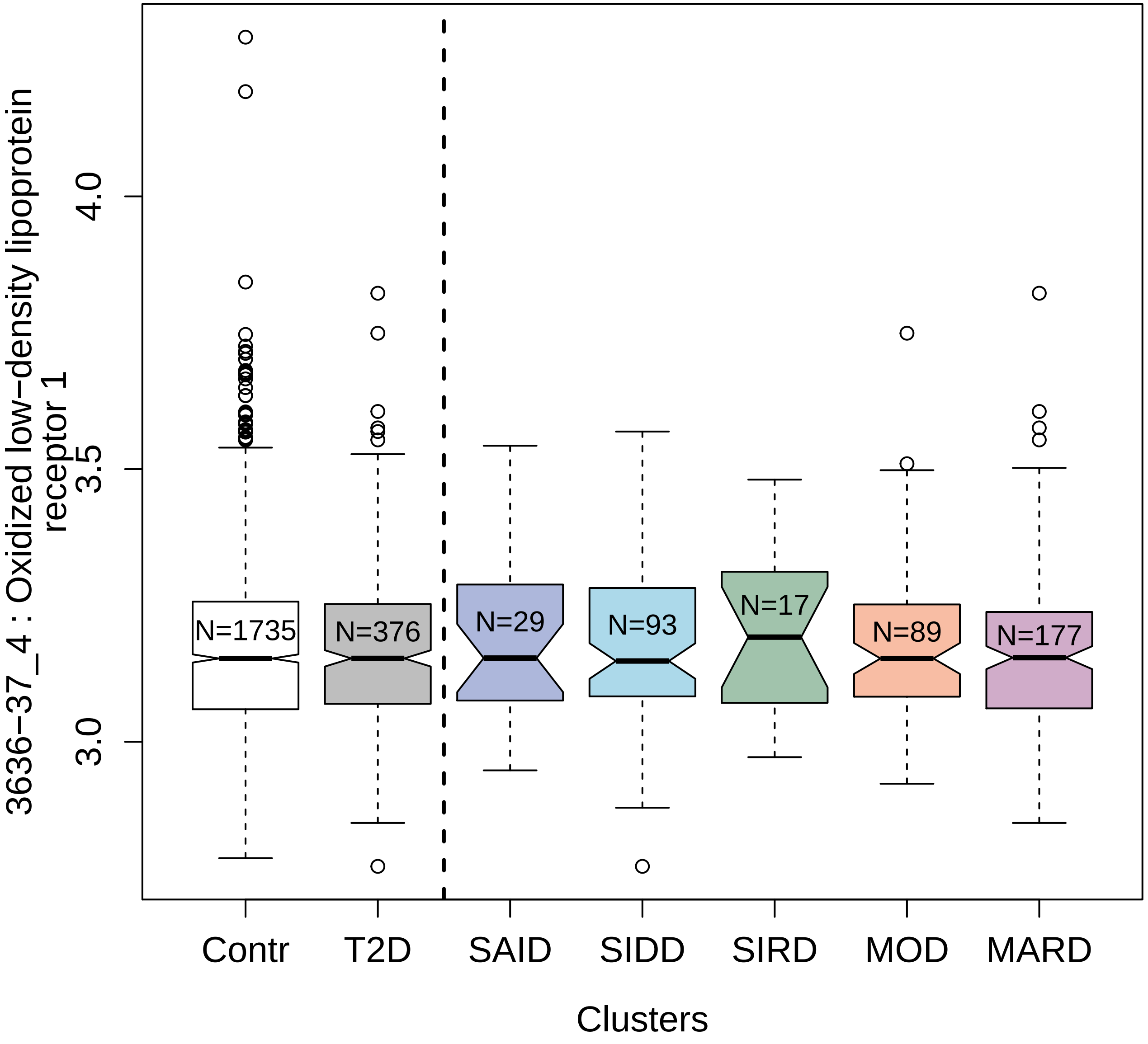

3636−37_4 : Oxidized low−density lipoprotein receptor 1
3.0
3.5
4.0
N=1735
N=376
N=29
N=93
N=17
N=89
N=177
Contr
T2D
SAID
SIDD
SIRD
MOD
MARD
Clusters

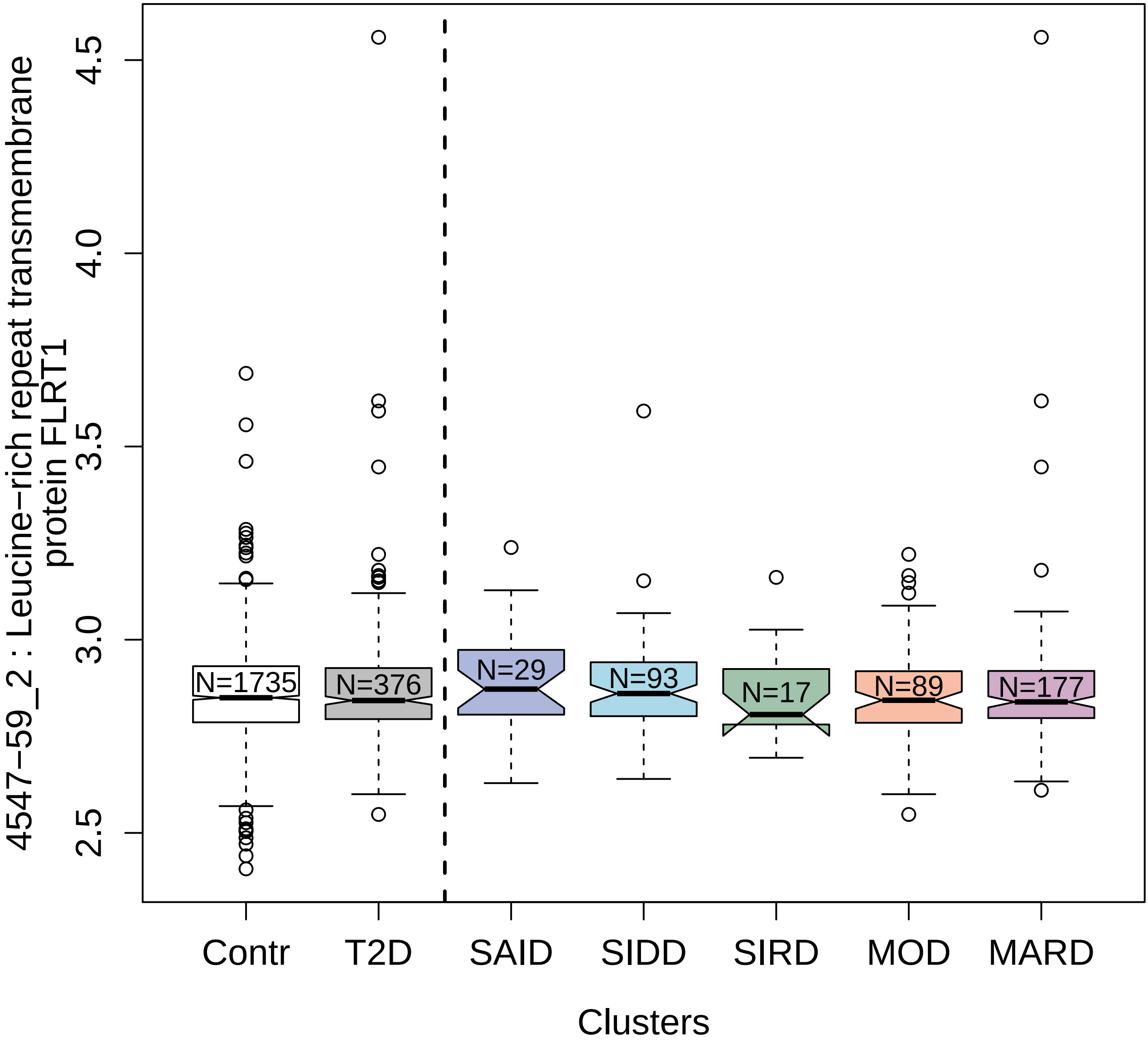
4547−59_2 : Leucine−rich repeat transmembrane protein FLRT1
2.5
3.0
3.5
4.0
4.5
N=1735
N=376
N=29
N=93
N=17
N=89
N=177
Contr
T2D
SAID
SIDD
SIRD
MOD
MARD
Clusters

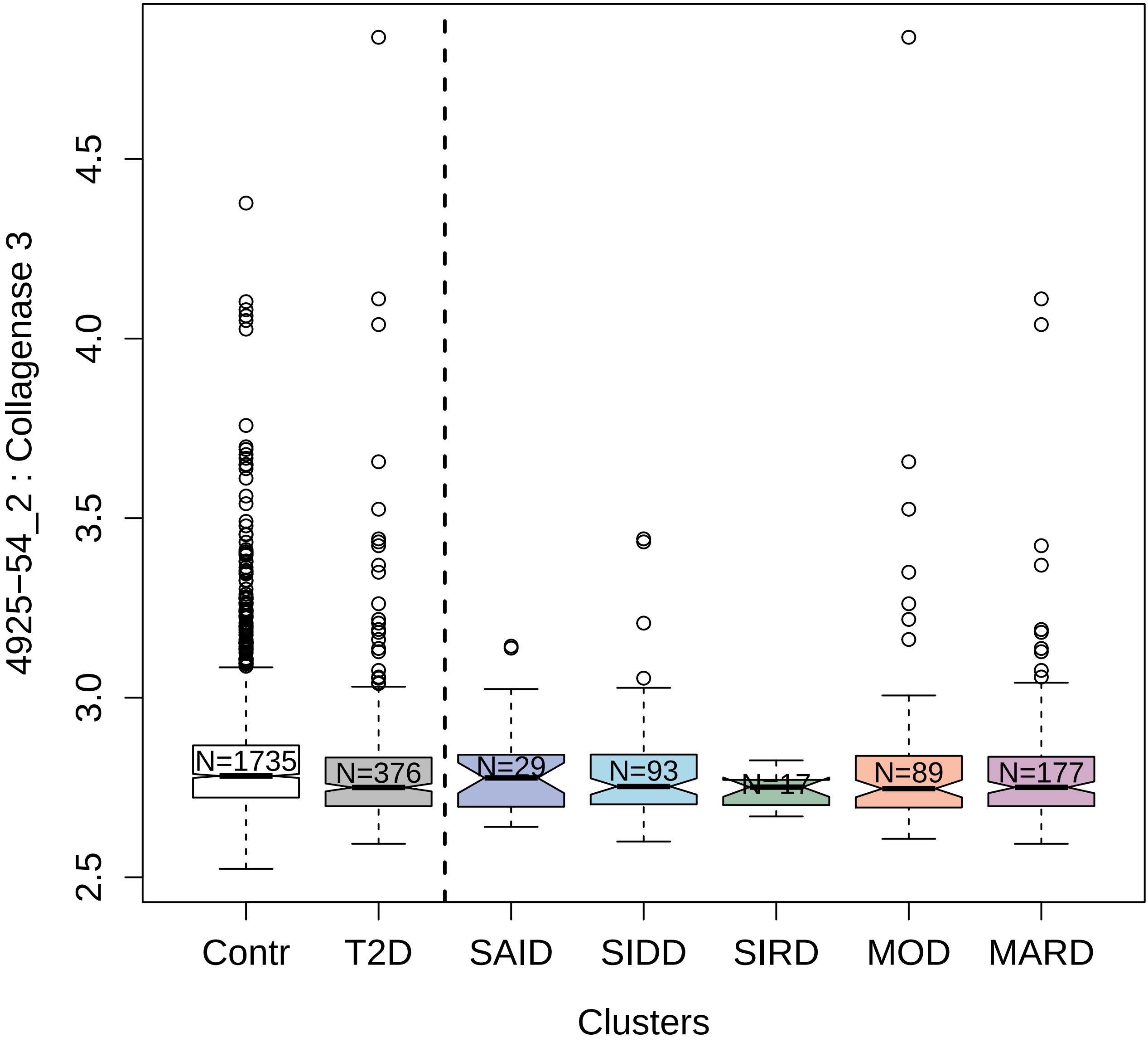
4925–54_2 : Collagenase 3
2.5
3.0
3.5
4.0
4.5
N=1735
N=376
N=29
N=93
N=17
N=89
N=177
Contr
T2D
SAID
SIDD
SIRD
MOD
MARD
Clusters

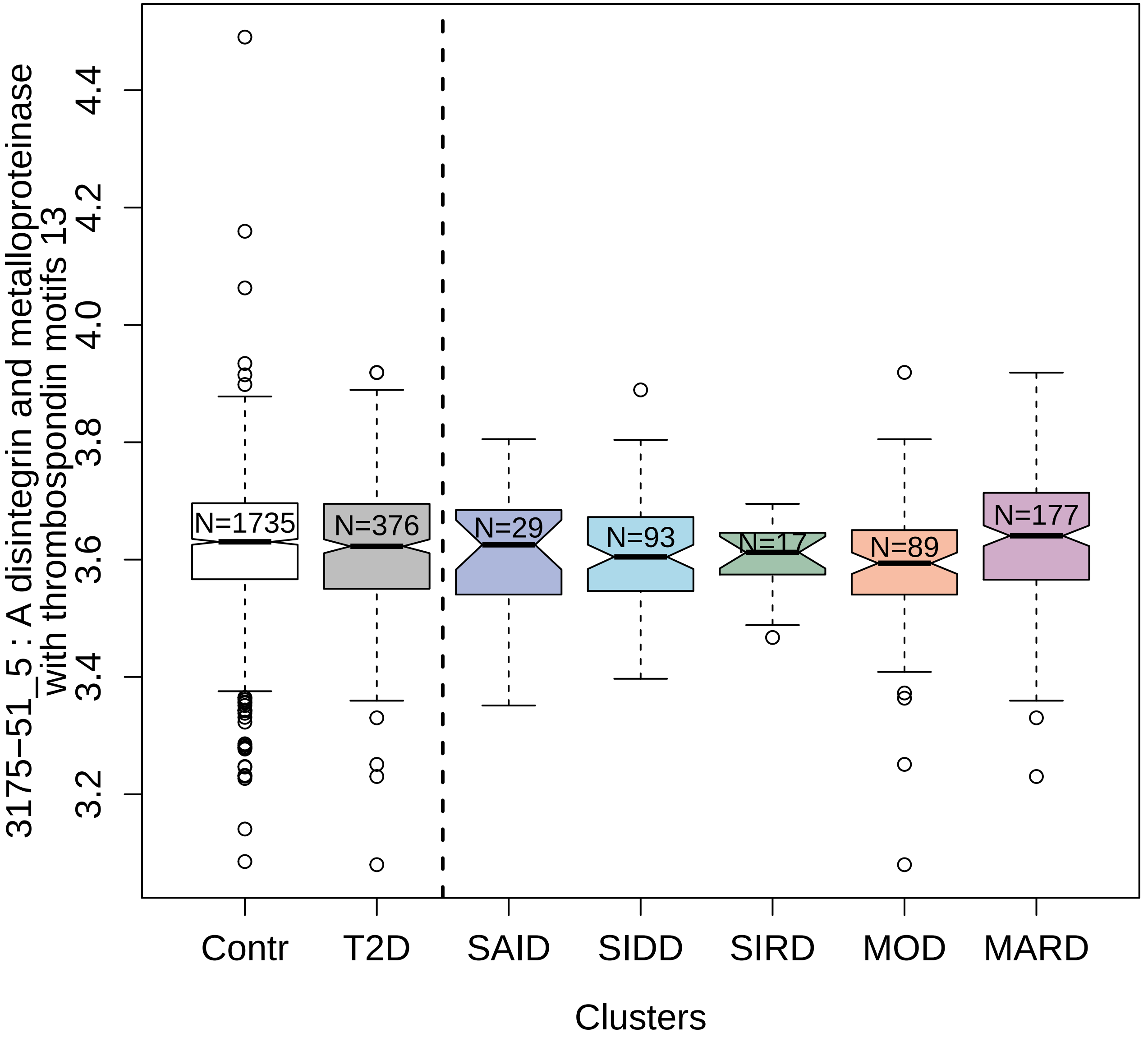
3175−51_5 : A disintegrin and metalloproteinase
with thrombospondin motifs 13
4.4
4.2
4.0
3.8
3.6
3.4
3.2
N=1735
N=376
N=29
N=93
N=17
N=89
N=177
Contr
T2D
SAID
SIDD
SIRD
MOD
MARD
Clusters

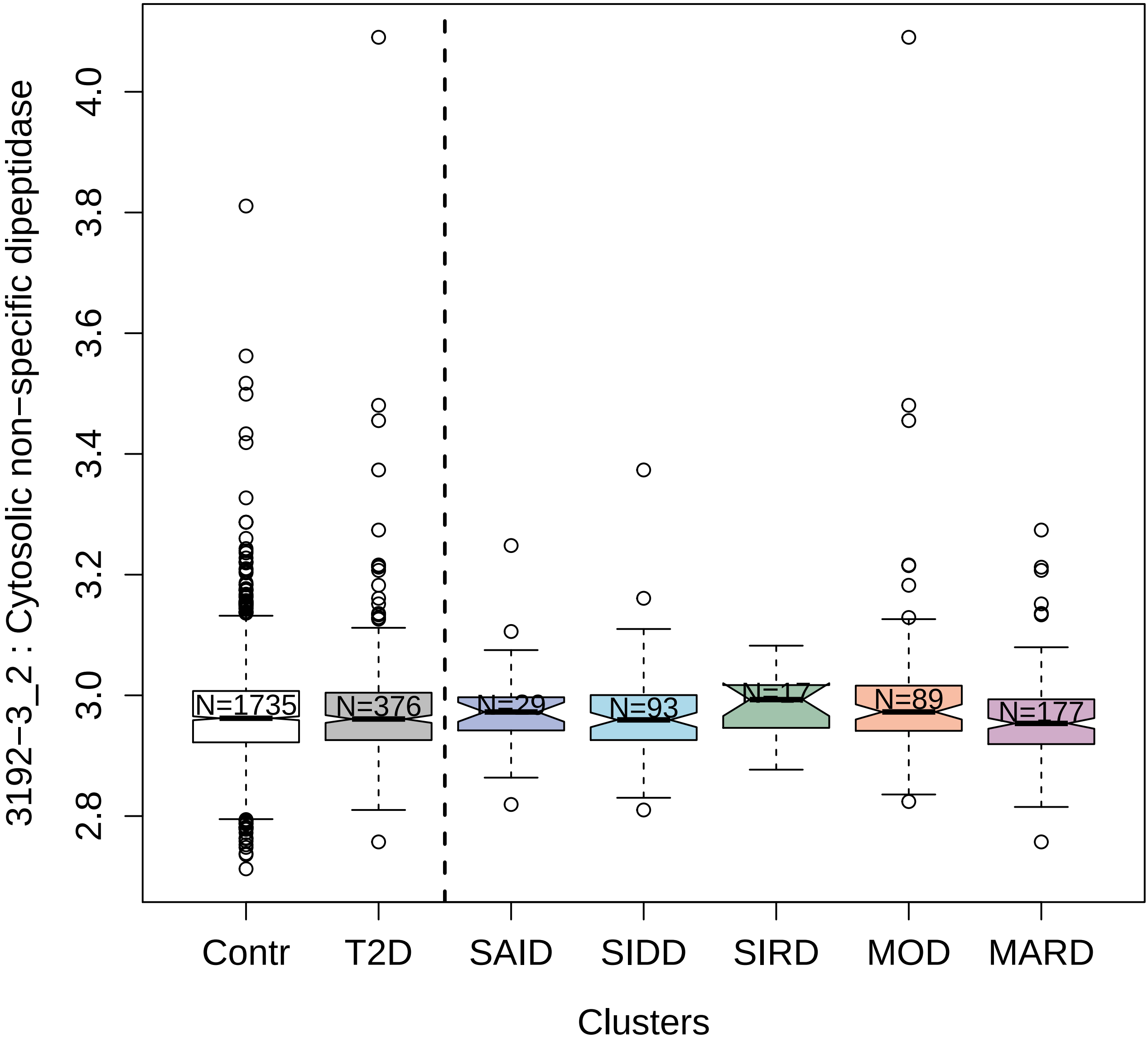
3192−3_2 : Cytosolic non−specific dipeptidase
2.8
3.0
3.2
3.4
3.6
3.8
4.0
N=1735
N=376
N=29
N=93
N=17
N=89
N=177
Contr
T2D
SAID
SIDD
SIRD
MOD
MARD
Clusters

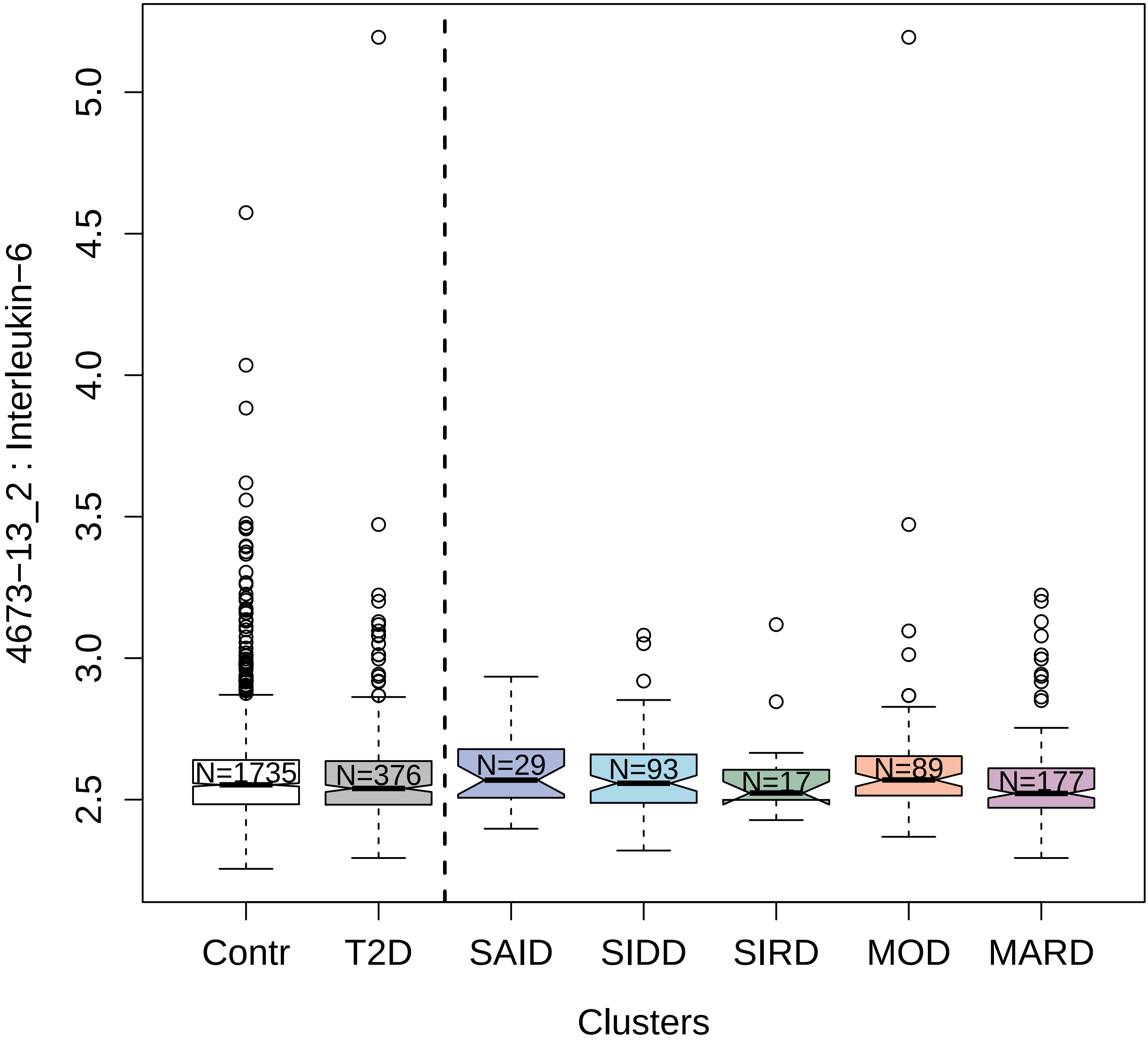

4673−13_2 : Interleukin−6
5.0
4.5
4.0
3.5
3.0
2.5
N=1735
N=376
N=29
N=93
N=17
N=89
N=177
Contr
T2D
SAID
SIDD
SIRD
MOD
MARD
Clusters

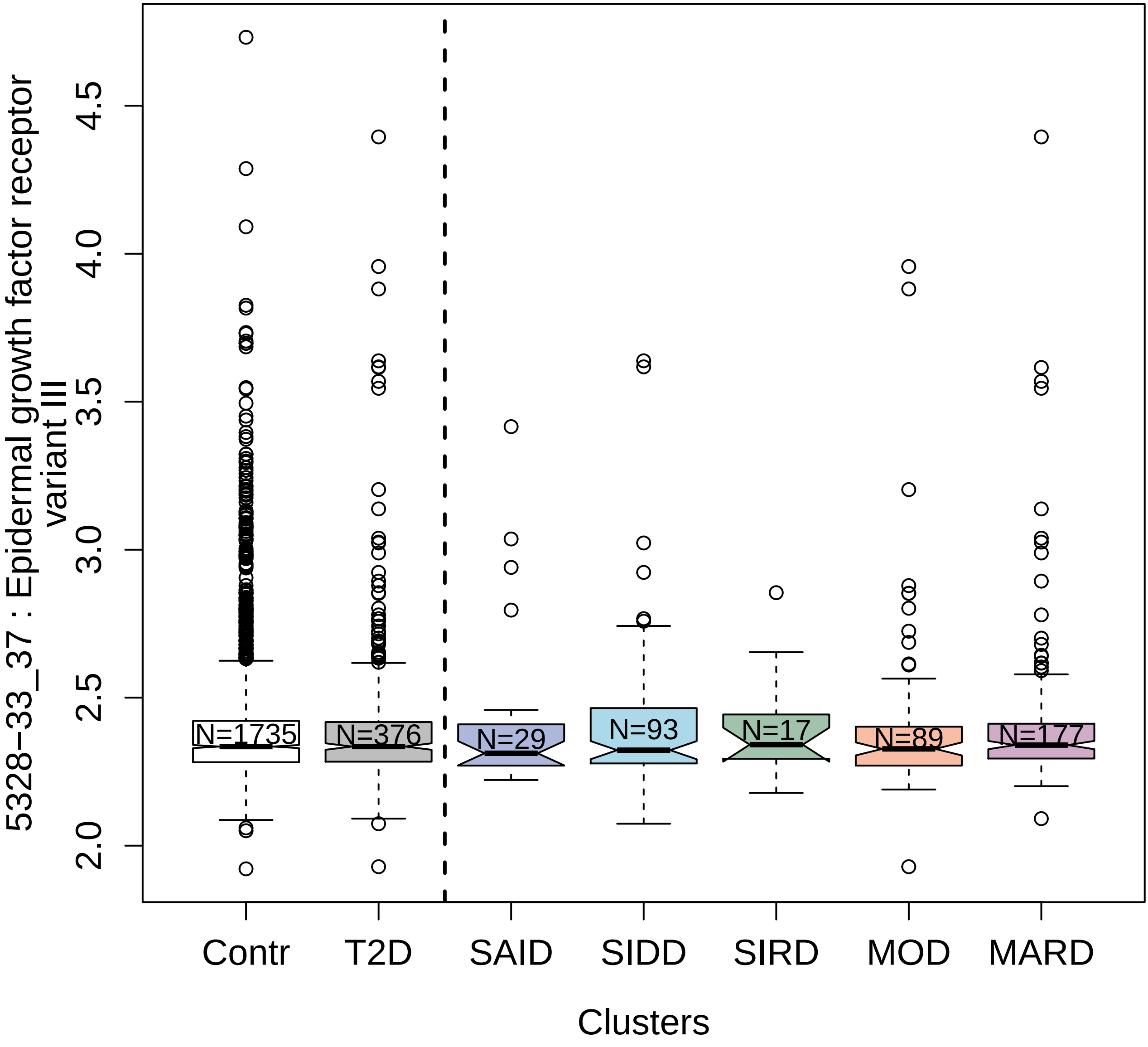
5328−33_37 : Epidermal growth factor receptor variant III
4.5
4.0
3.5
3.0
2.5
2.0
N=1735
N=376
N=29
N=93
N=17
N=89
N=177
Contr
T2D
SAID
SIDD
SIRD
MOD
MARD
Clusters

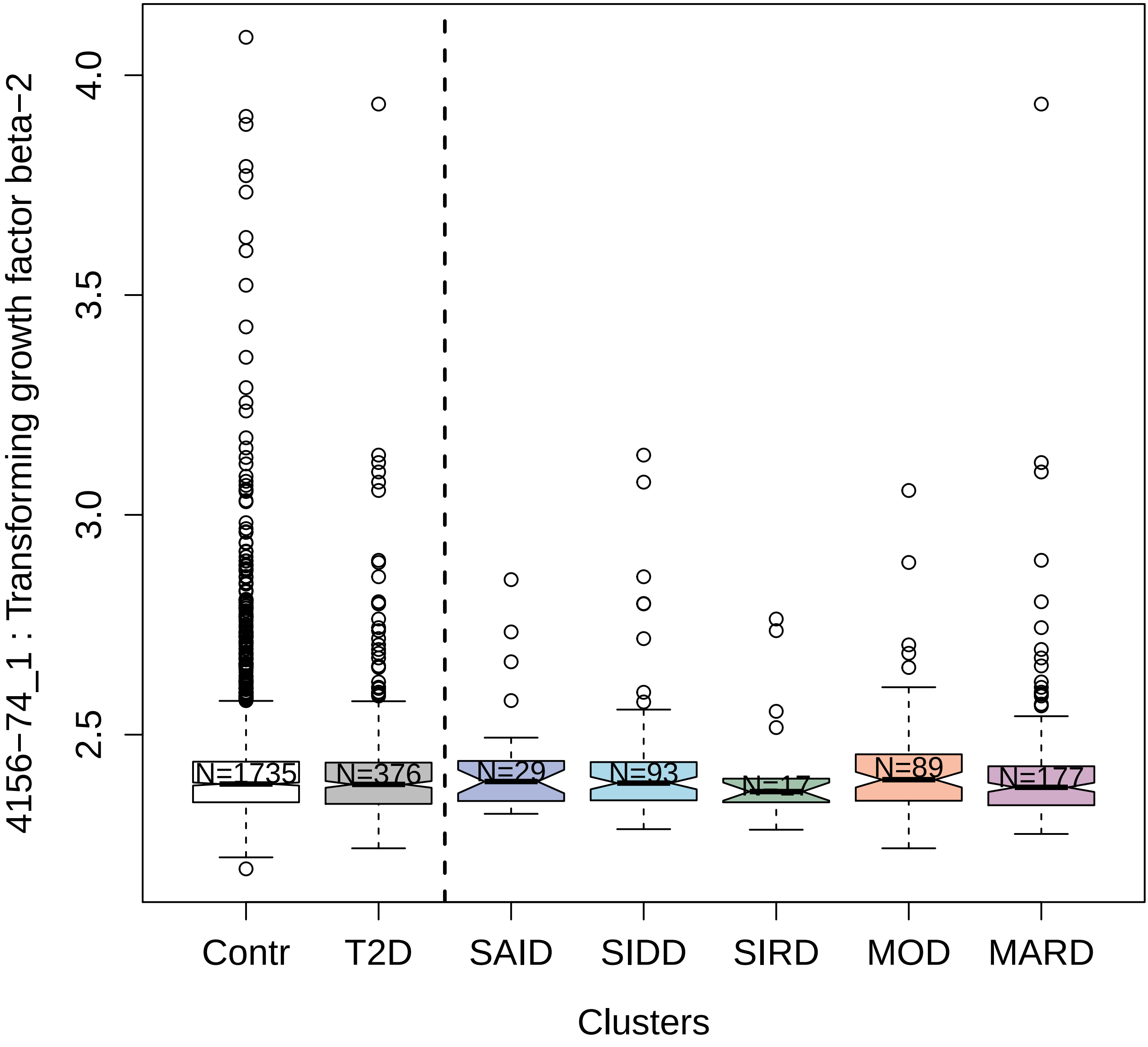

4156−74_1 : Transforming growth factor beta−2
4.0
3.5
3.0
2.5
N=1735
N=376
N=29
N=93
N=17
N=89
N=177
Contr
T2D
SAID
SIDD
SIRD
MOD
MARD
Clusters

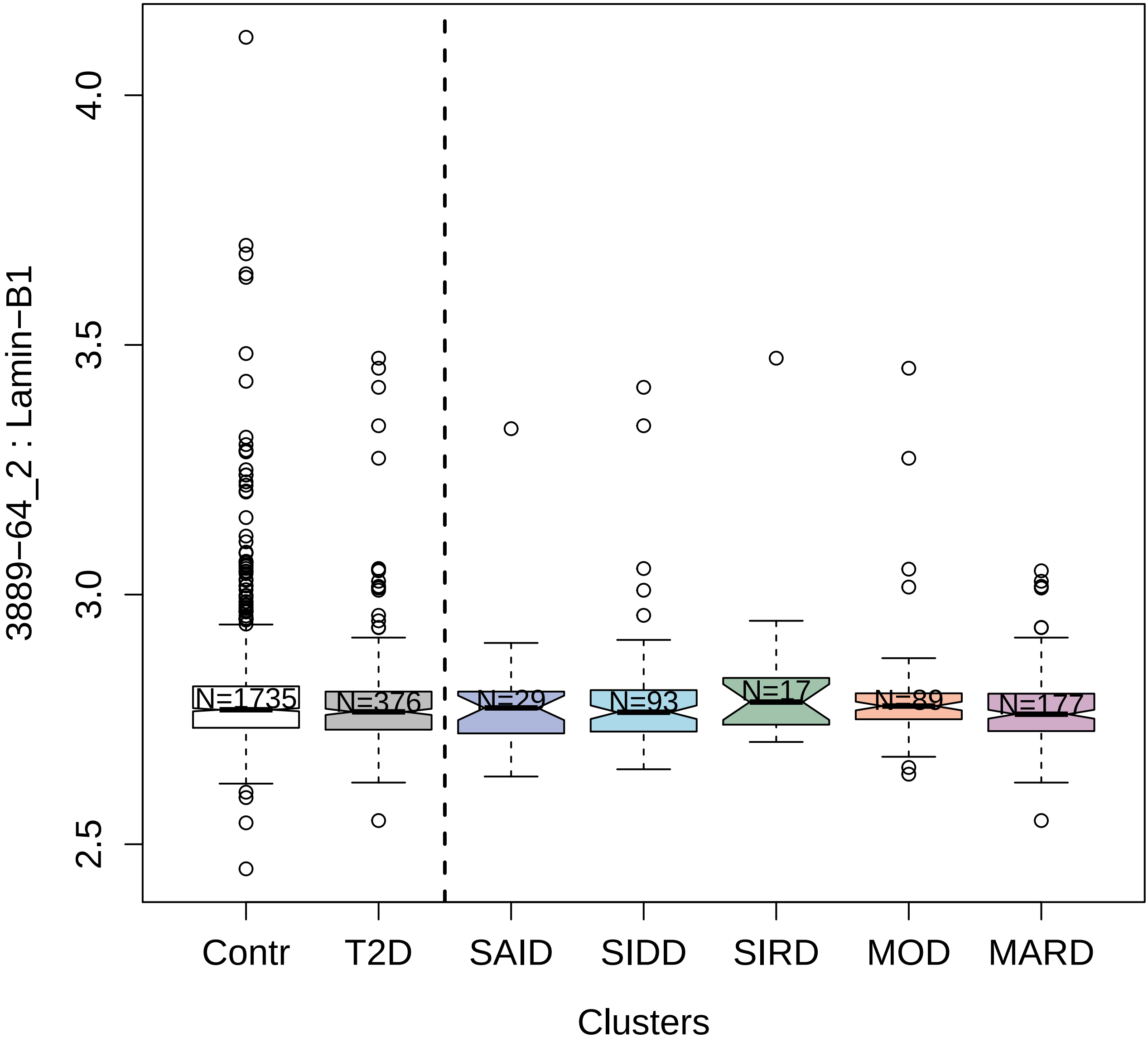

3889−64_2 : Lamin−B1
4.0
3.5
3.0
2.5
N=1735
N=376
N=29
N=93
N=17
N=89
N=177
Contr
T2D
SAID
SIDD
SIRD
MOD
MARD
Clusters

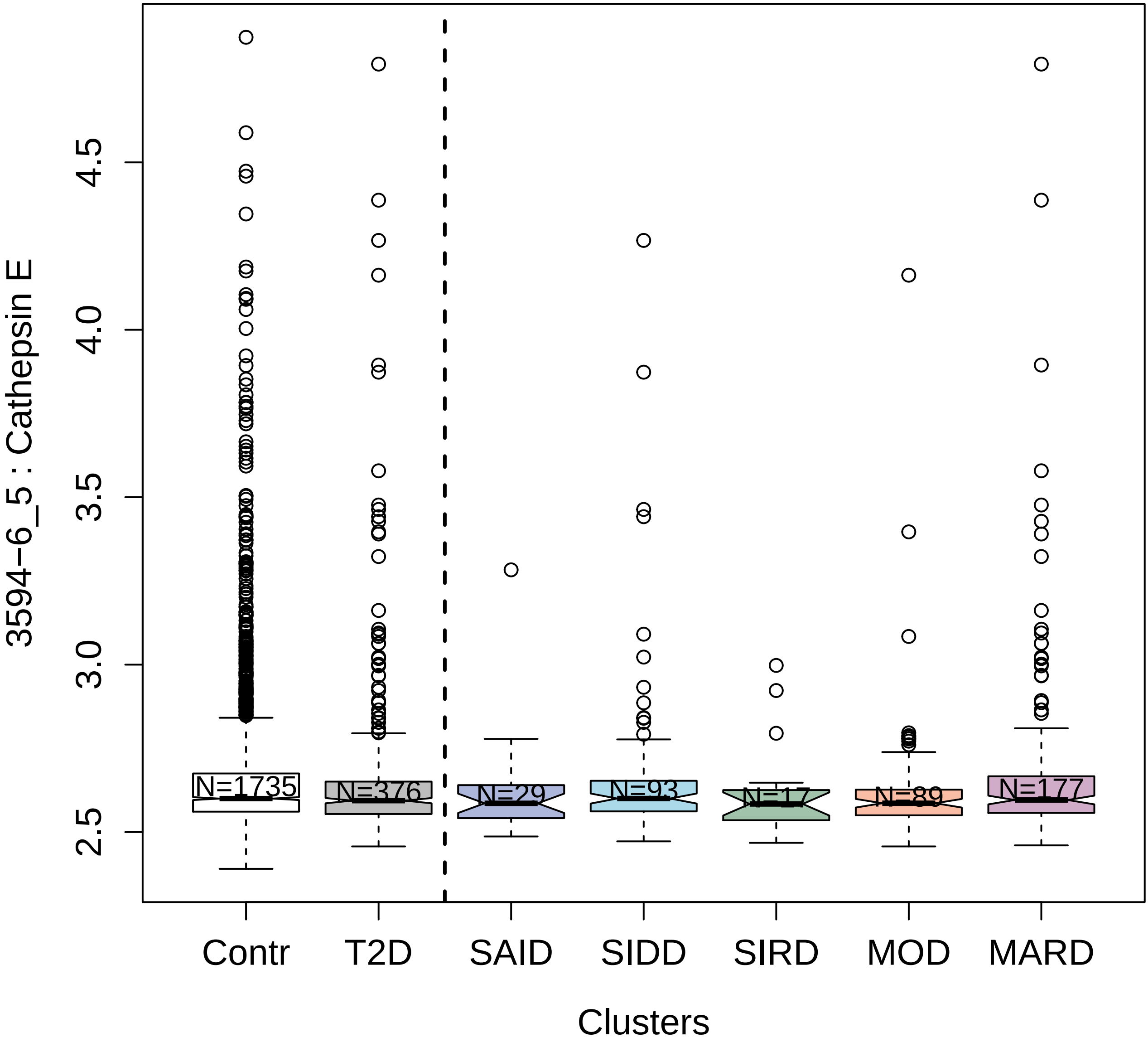
3594−6_5 : Cathepsin E
2.5
3.0
3.5
4.0
4.5
N=1735
N=376
N=29
N=93
N=17
N=89
N=177
Contr
T2D
SAID
SIDD
SIRD
MOD
MARD
Clusters

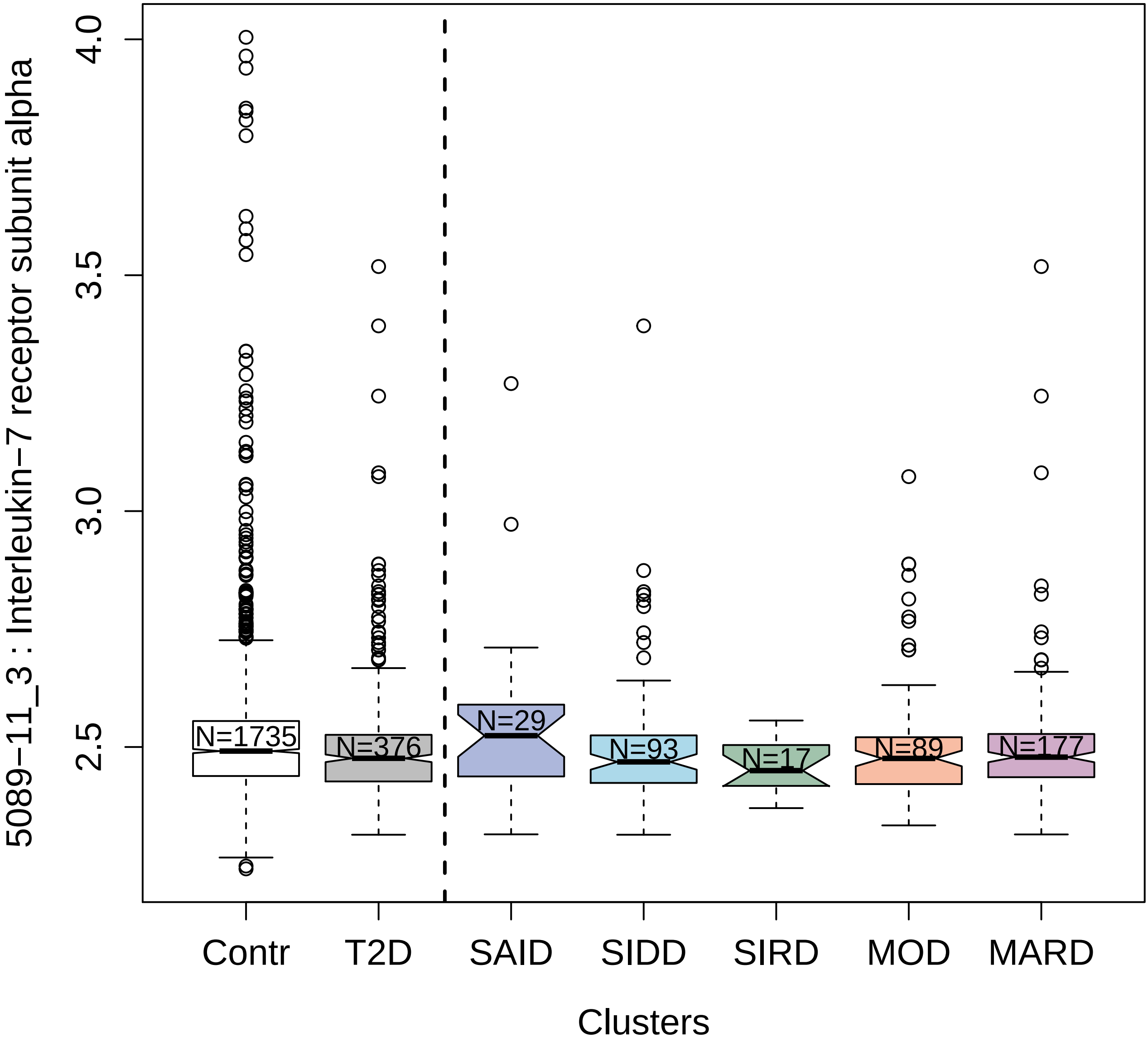

5089−11_3 : Interleukin−7 receptor subunit alpha
4.0
3.5
3.0
2.5
N=1735
N=376
N=29
N=93
N=17
N=89
N=177
Contr
T2D
SAID
SIDD
SIRD
MOD
MARD
Clusters

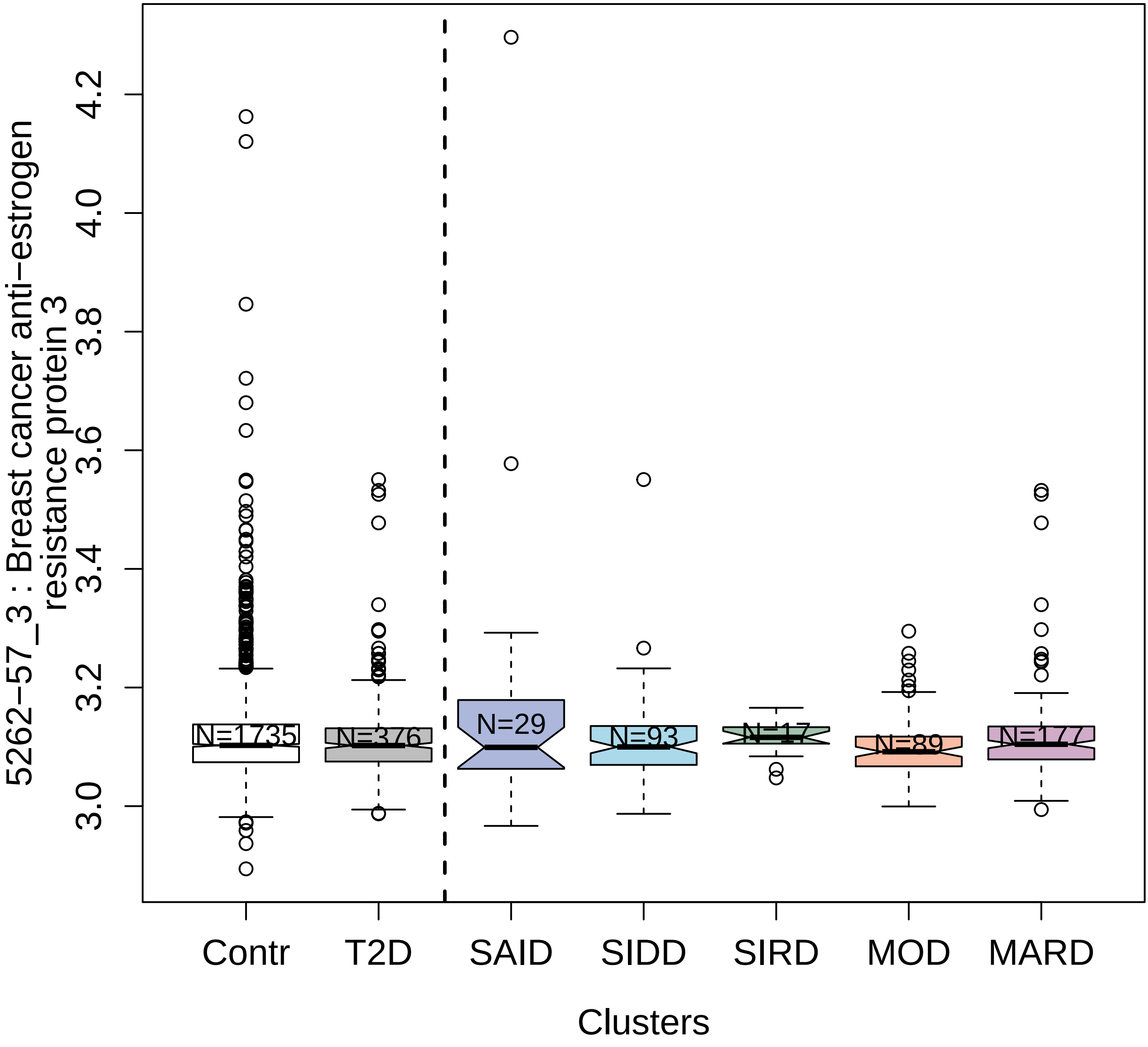
5262−57_3 : Breast cancer anti−estrogen resistance protein 3
3.0
3.2
3.4
3.6
3.8
4.0
4.2
N=1735
N=376
N=29
N=93
N=17
N=89
N=177
Contr
T2D
SAID
SIDD
SIRD
MOD
MARD
Clusters

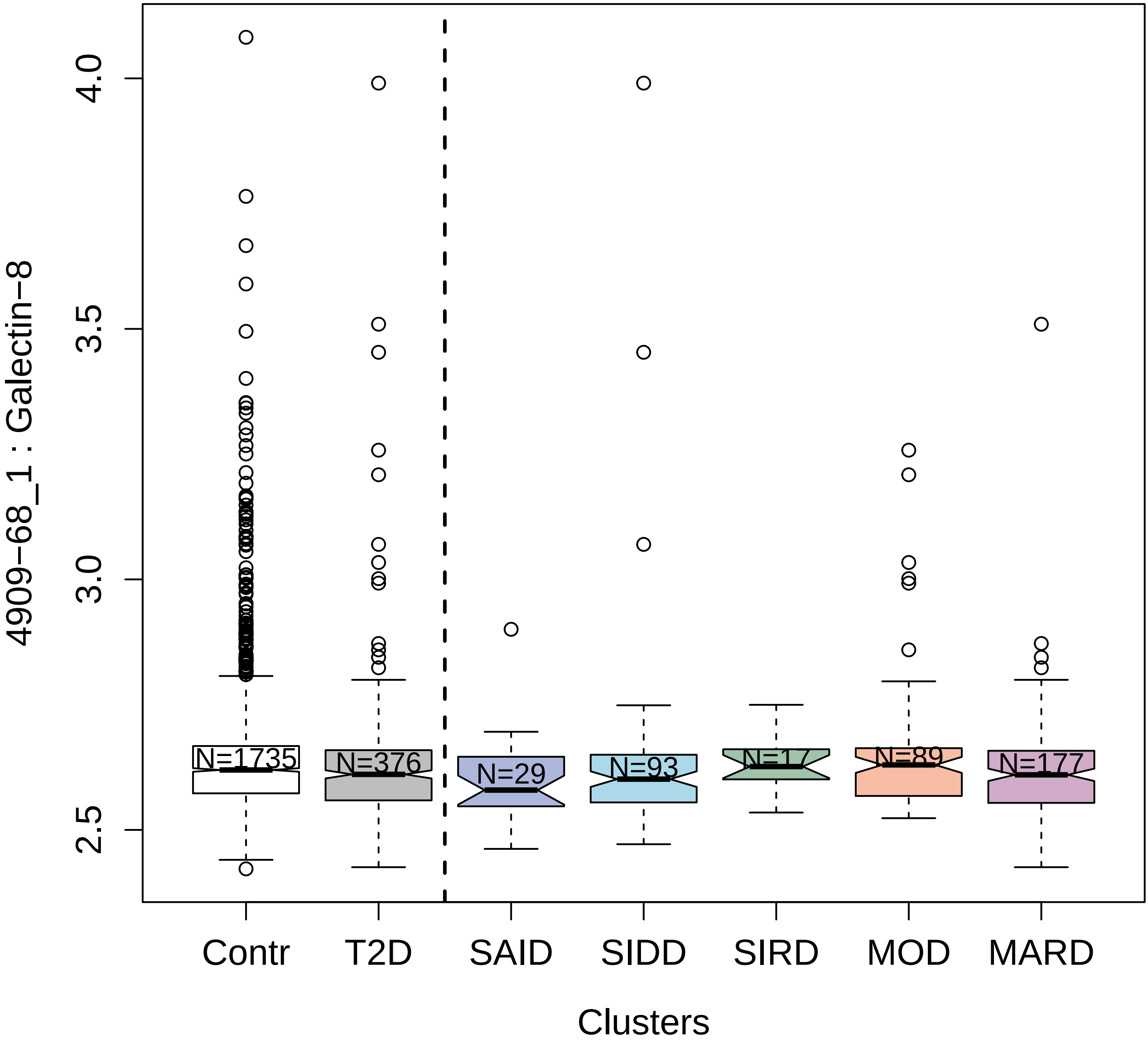
4909−68_1 : Galectin−8
4.0
3.5
3.0
2.5
N=1735
N=376
N=29
N=93
N=17
N=89
N=177
Contr
T2D
SAID
SIDD
SIRD
MOD
MARD
Clusters

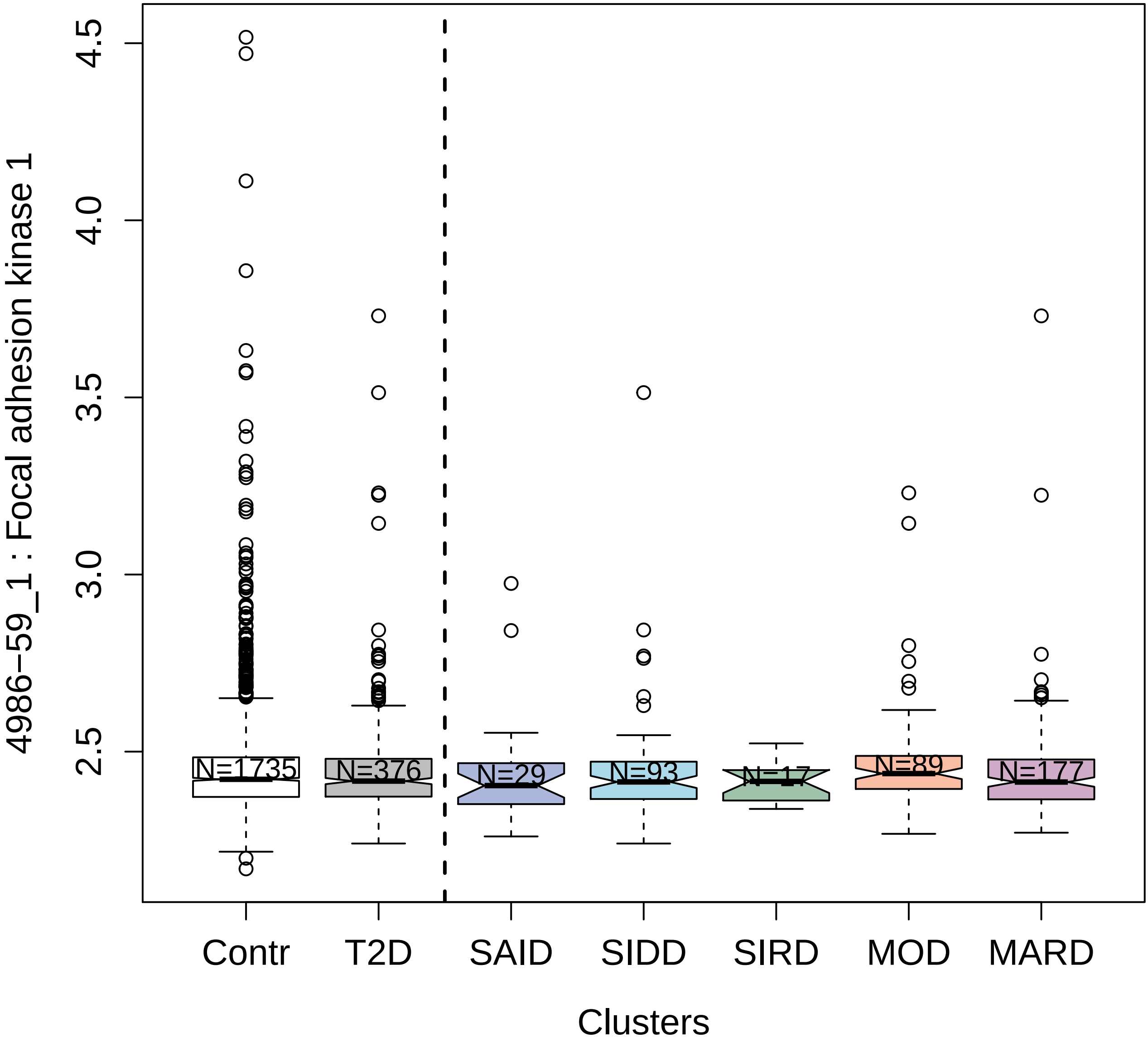

4986−59_1 : Focal adhesion kinase 1
4.5
4.0
3.5
3.0
2.5
N=1735
N=376
N=29
N=93
N=17
N=89
N=177
Contr
T2D
SAID
SIDD
SIRD
MOD
MARD
Clusters

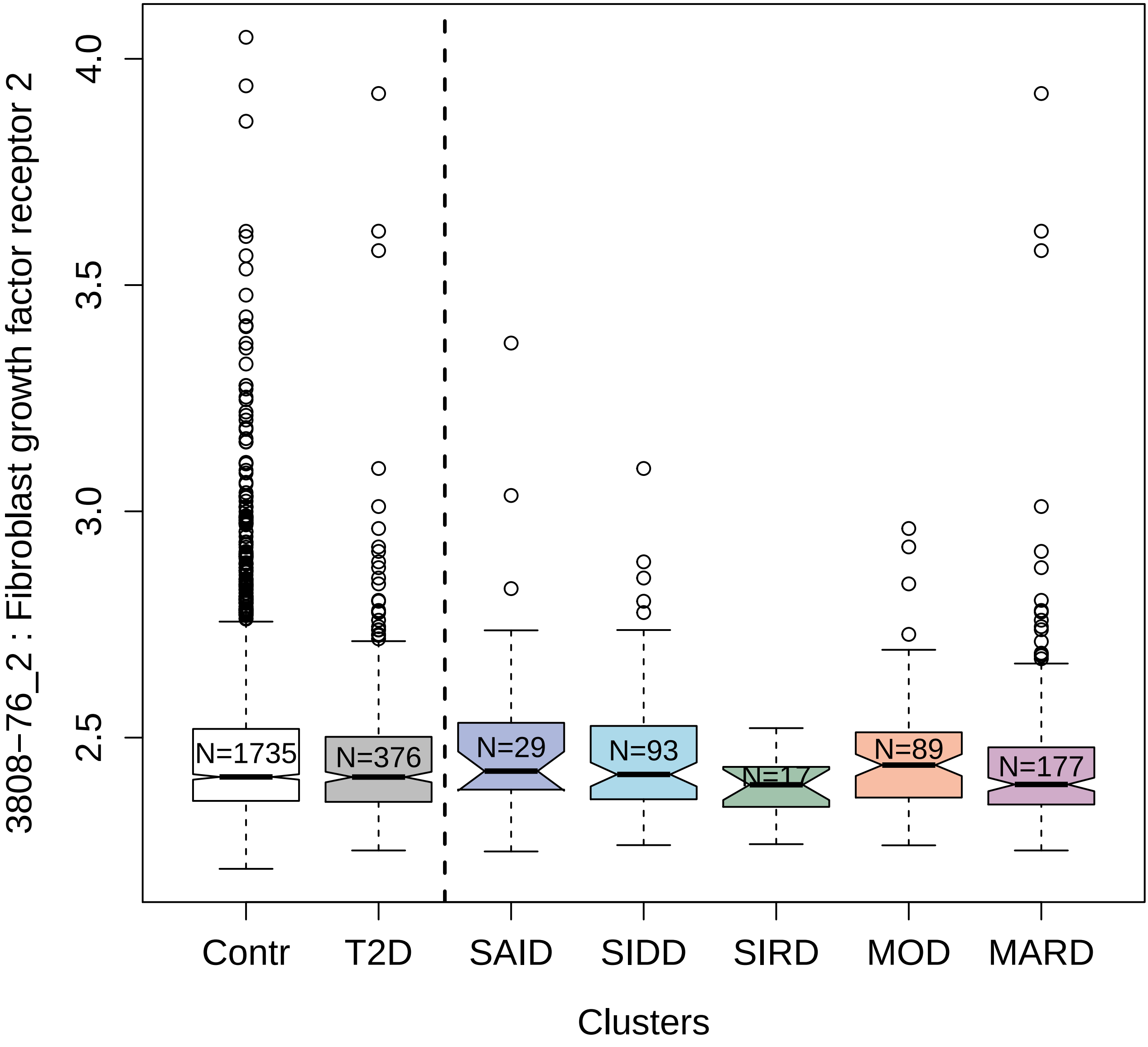

3808−76_2 : Fibroblast growth factor receptor 2
4.0
3.5
3.0
2.5
N=1735
N=376
N=29
N=93
N=89
N=177
Contr
T2D
SAID
SIDD
SIRD
MOD
MARD
Clusters

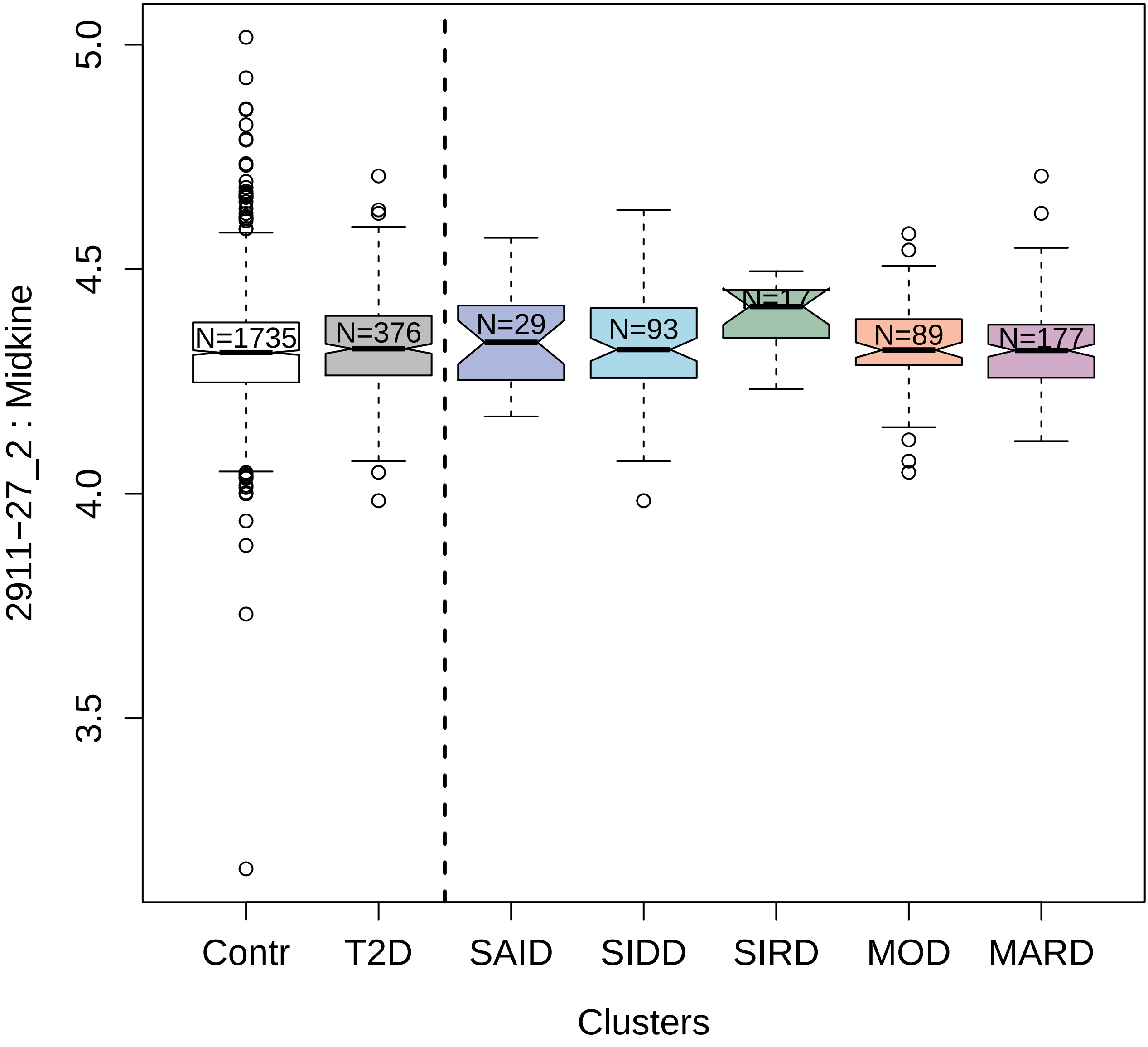

2911−27_2 : Midkine
5.0
4.5
4.0
3.5
N=1735
N=376
N=29
N=93
N=17
N=89
N=177
Contr
T2D
SAID
SIDD
SIRD
MOD
MARD
Clusters

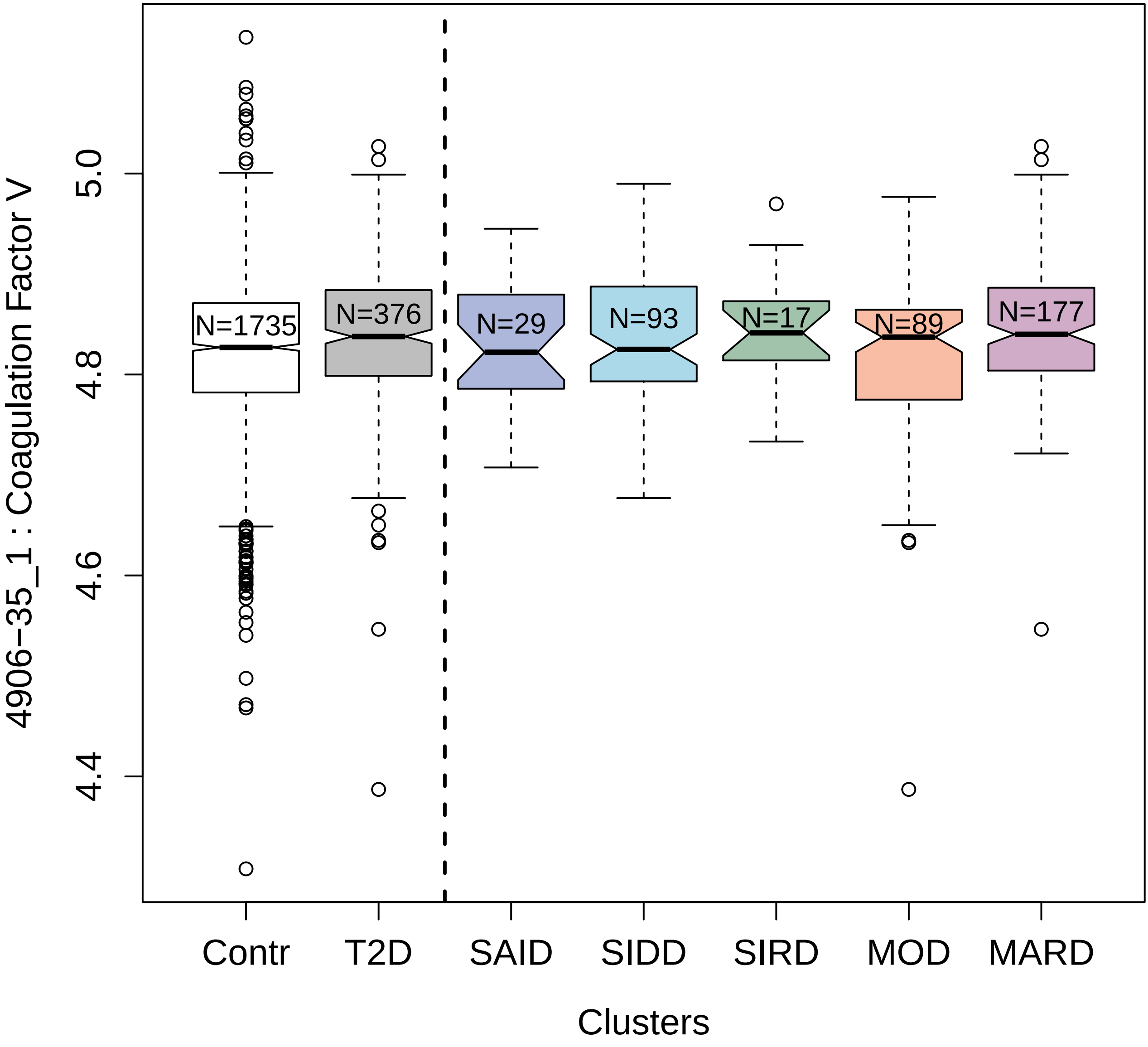

4906−35_1 : Coagulation Factor V
5.0
4.8
4.6
4.4
N=1735
N=376
N=29
N=93
N=17
N=89
N=177
Contr
T2D
SAID
SIDD
SIRD
MOD
MARD
Clusters

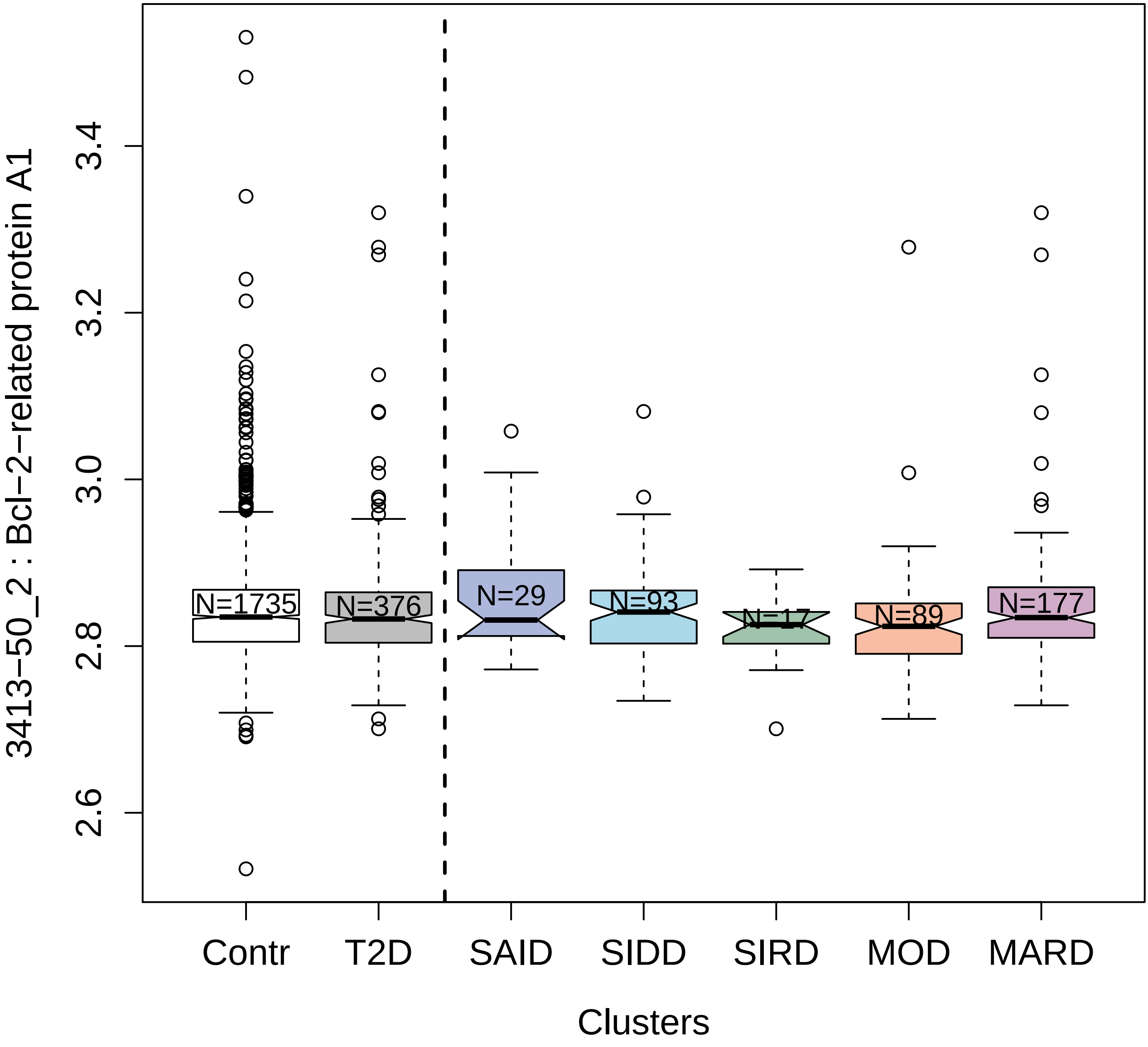

3413−50_2 : Bcl−2−related protein A1
3.4
3.2
3.0
2.8
2.6
N=1735
N=376
N=29
N=93
N=17
N=89
N=177
Contr
T2D
SAID
SIDD
SIRD
MOD
MARD
Clusters

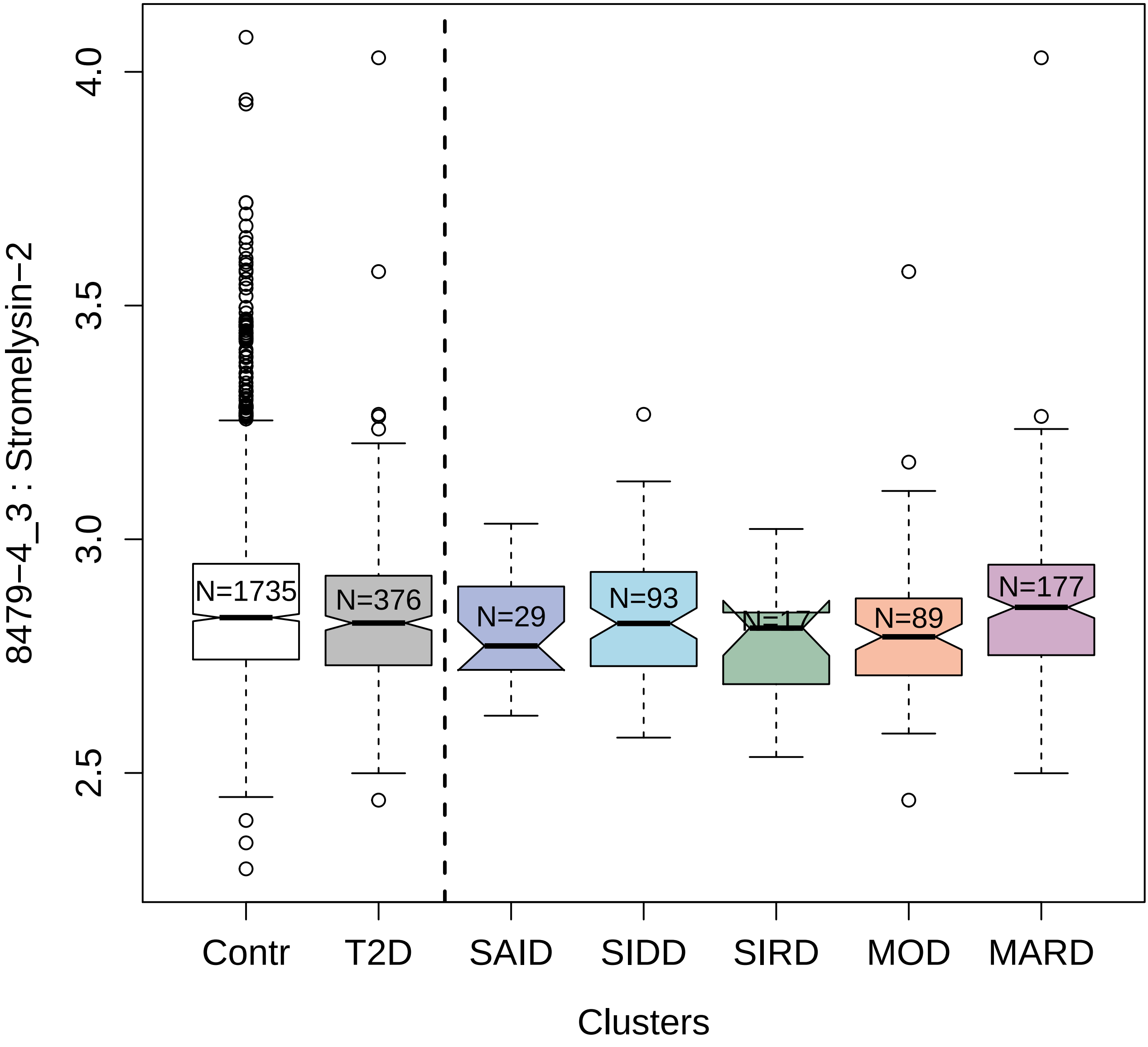

8479−4_3 : Stromelysin−2
4.0
3.5
3.0
2.5
N=1735
N=376
N=29
N=93
N=89
N=177
Contr
T2D
SAID
SIDD
SIRD
MOD
MARD
Clusters

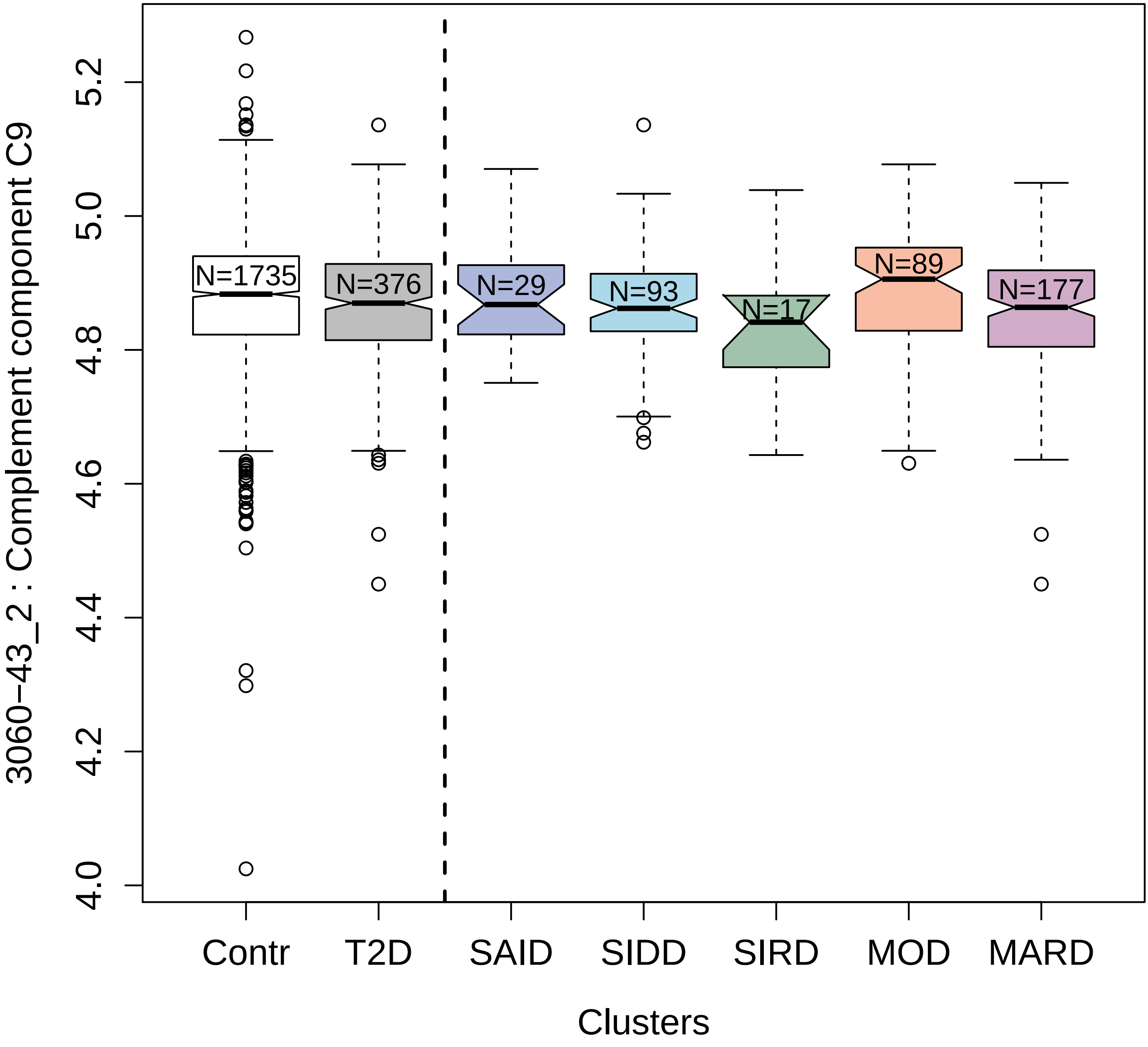
3060−43_2 : Complement component C9
5.2
5.0
4.8
4.6
4.4
4.2
4.0
N=1735
N=376
N=29
N=93
N=17
N=89
N=177
Contr
T2D
SAID
SIDD
SIRD
MOD
MARD
Clusters

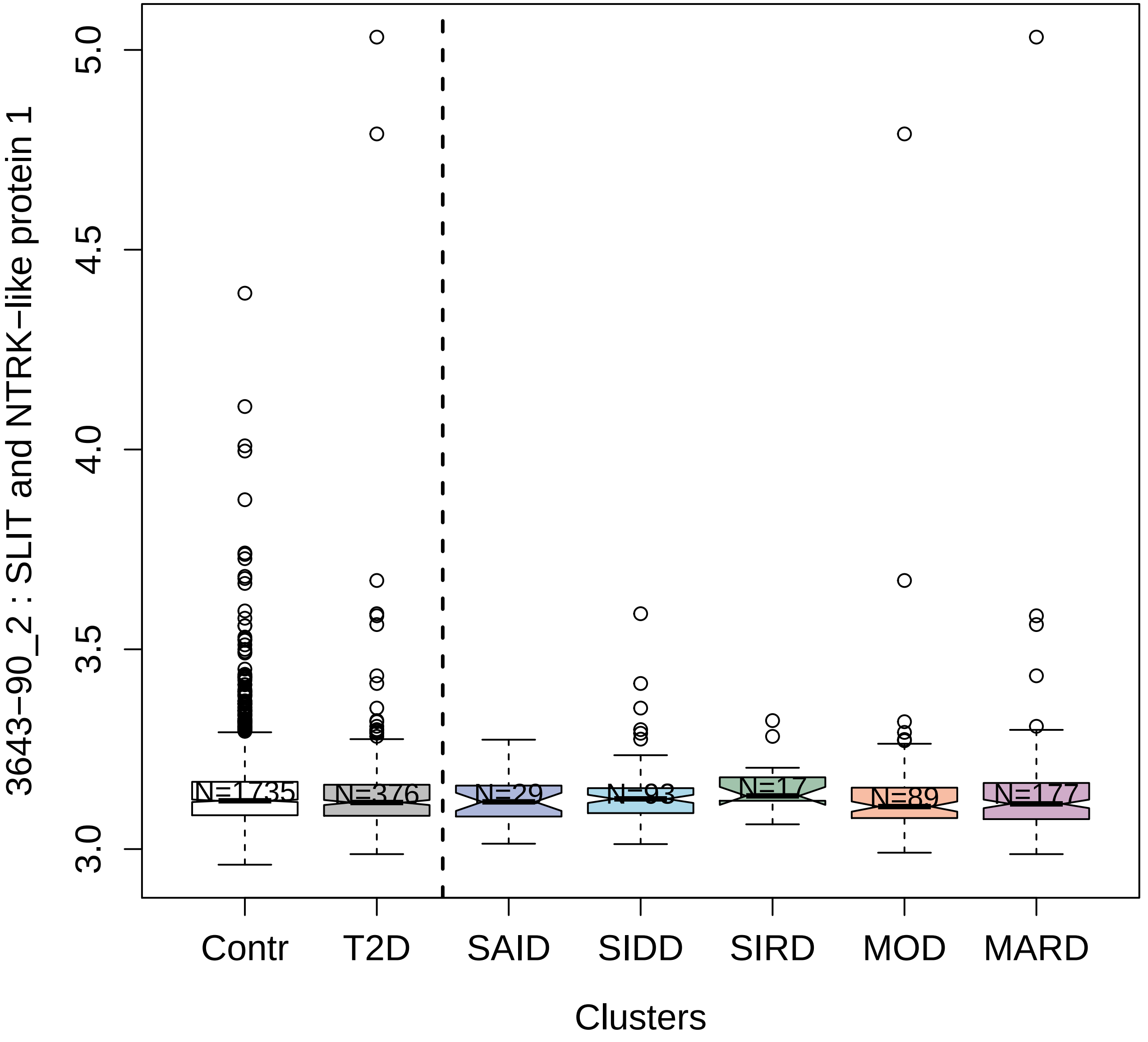

3643−90_2 : SLIT and NTRK−like protein 1
5.0
4.5
4.0
3.5
3.0
N=1735
N=376
N=29
N=93
N=17
N=89
N=177
Contr
T2D
SAID
SIDD
SIRD
MOD
MARD
Clusters

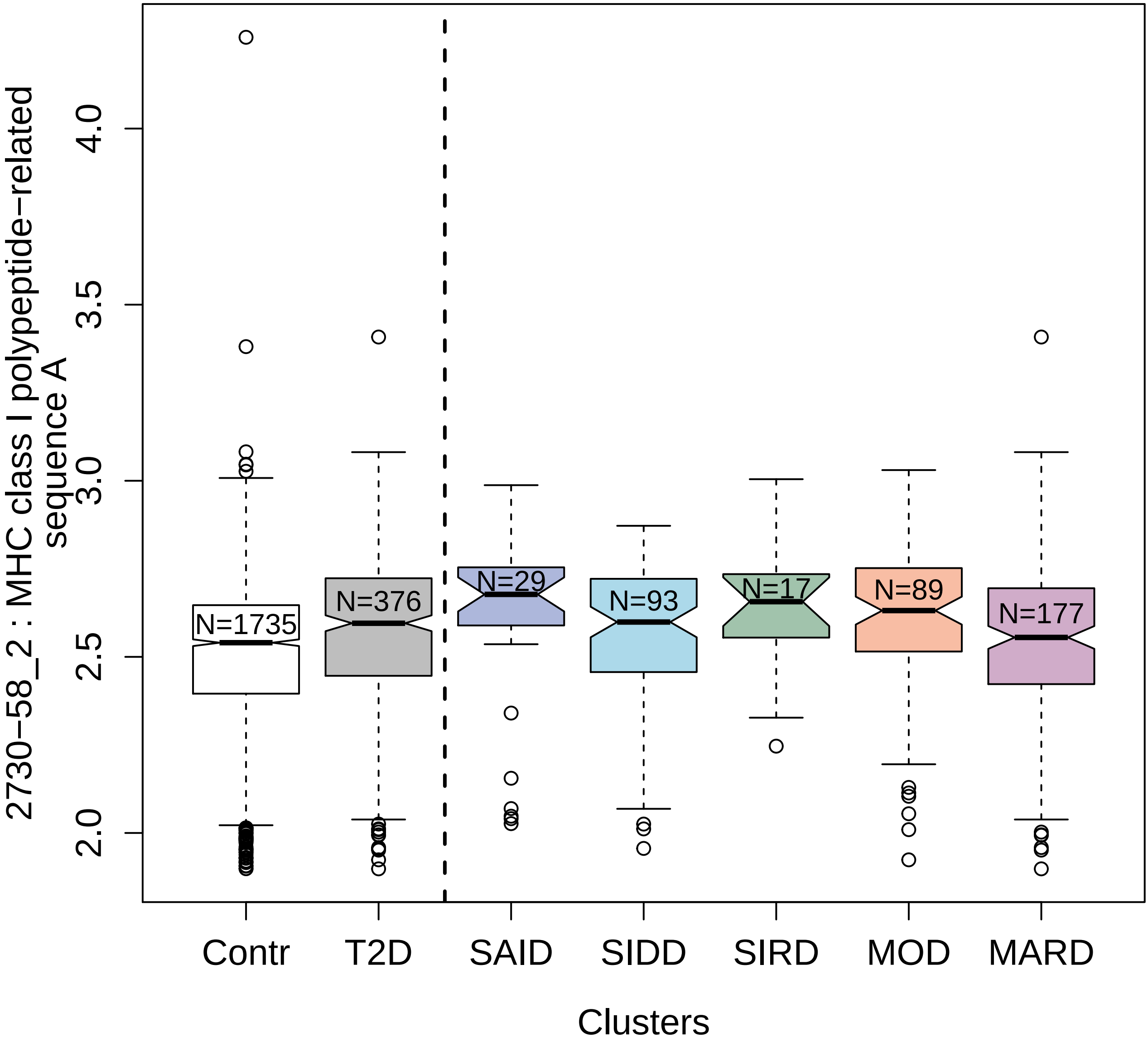

2730−58_2 : MHC class I polypeptide−related sequence A
Clusters
Contr
T2D
SAID
SIDD
SIRD
MOD
MARD
N=1735
N=376
N=29
N=93
N=17
N=89
N=177
2.0
2.5
3.0
3.5
4.0

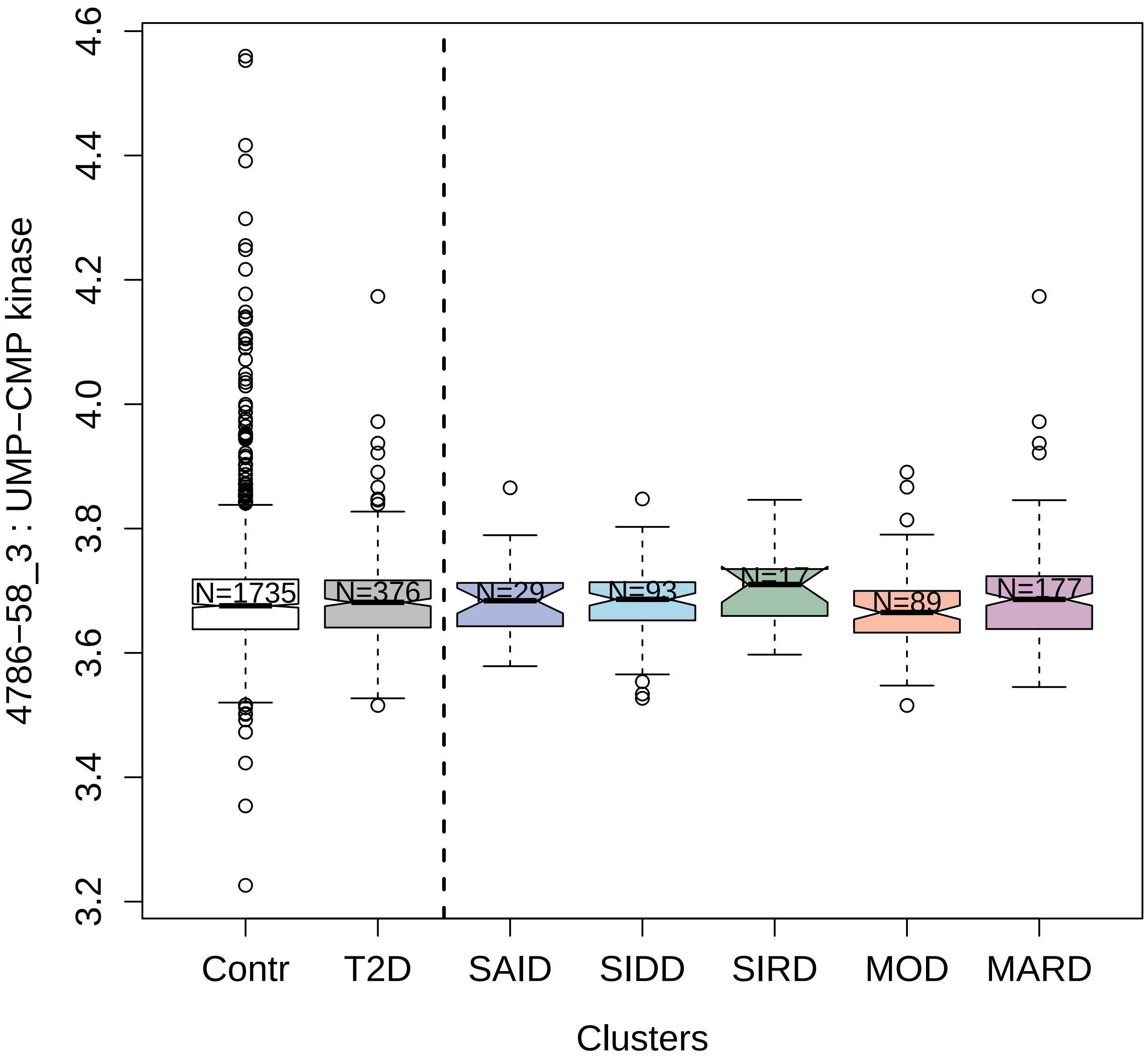
4786−58_3 : UMP−CMP kinase
3.2
3.4
3.6
3.8
4.0
4.2
4.4
4.6
N=1735
N=376
N=29
N=93
N=17
N=89
N=177
Contr
T2D
SAID
SIDD
SIRD
MOD
MARD
Clusters

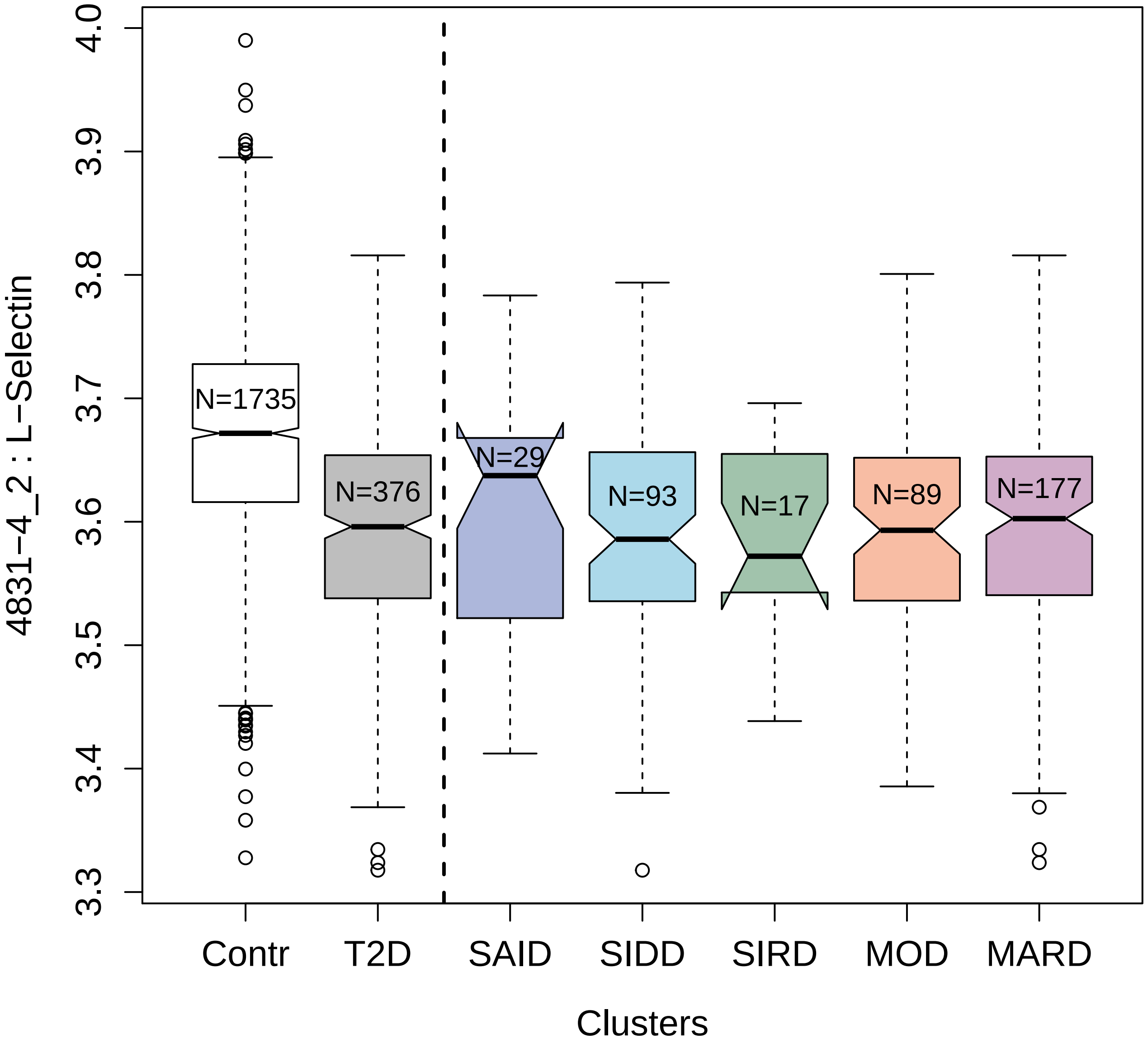

4831−4_2 : L−Selectin
4.0
3.9
3.8
3.7
3.6
3.5
3.4
3.3
N=1735
N=376
N=29
N=93
N=17
N=89
N=177
Contr
T2D
SAID
SIDD
SIRD
MOD
MARD
Clusters

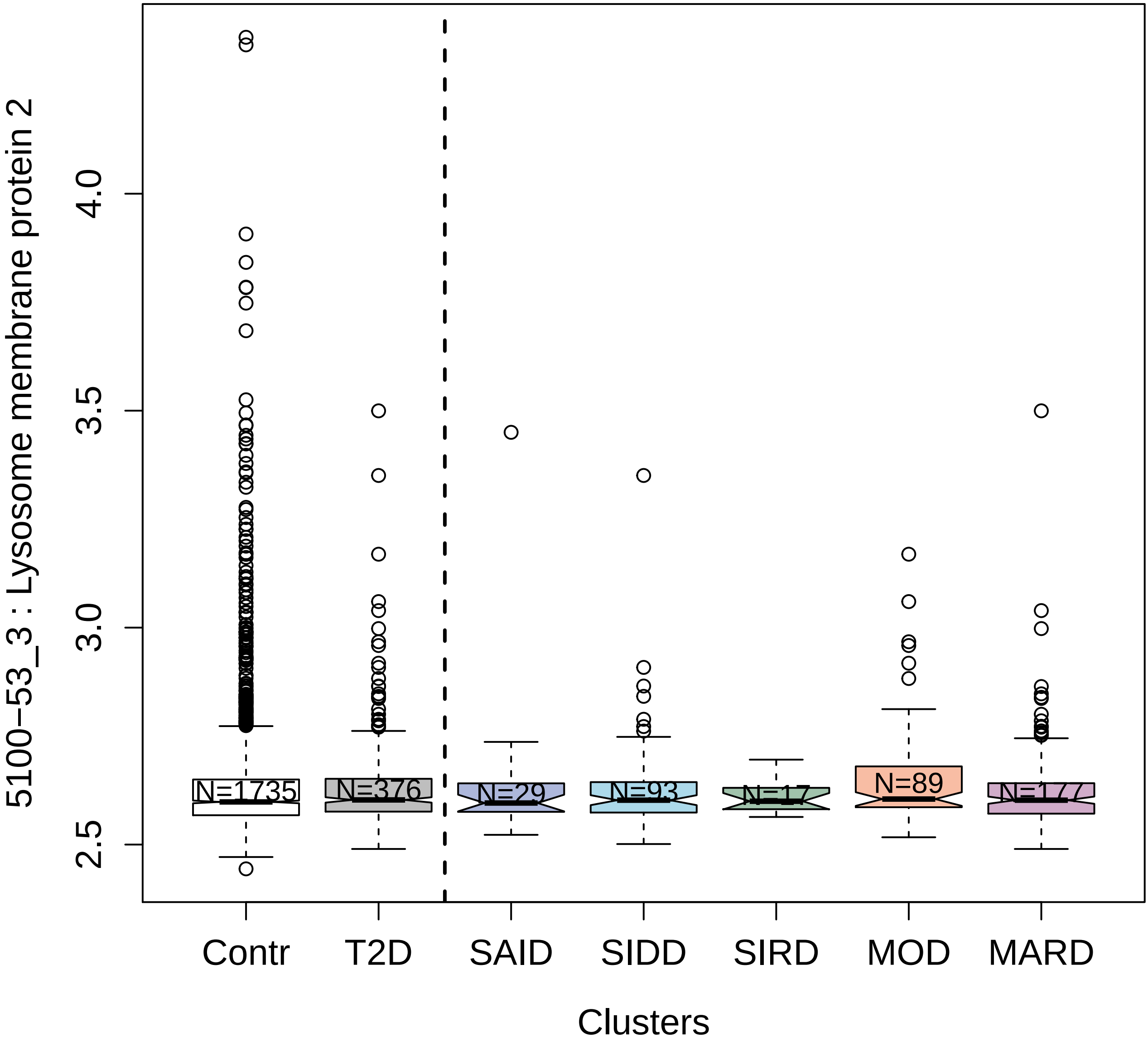

5100−53_3 : Lysosome membrane protein 2
4.0
3.5
3.0
2.5
N=1735
N=376
N=29
N=93
N=17
N=89
N=177
Contr
T2D
SAID
SIDD
SIRD
MOD
MARD
Clusters

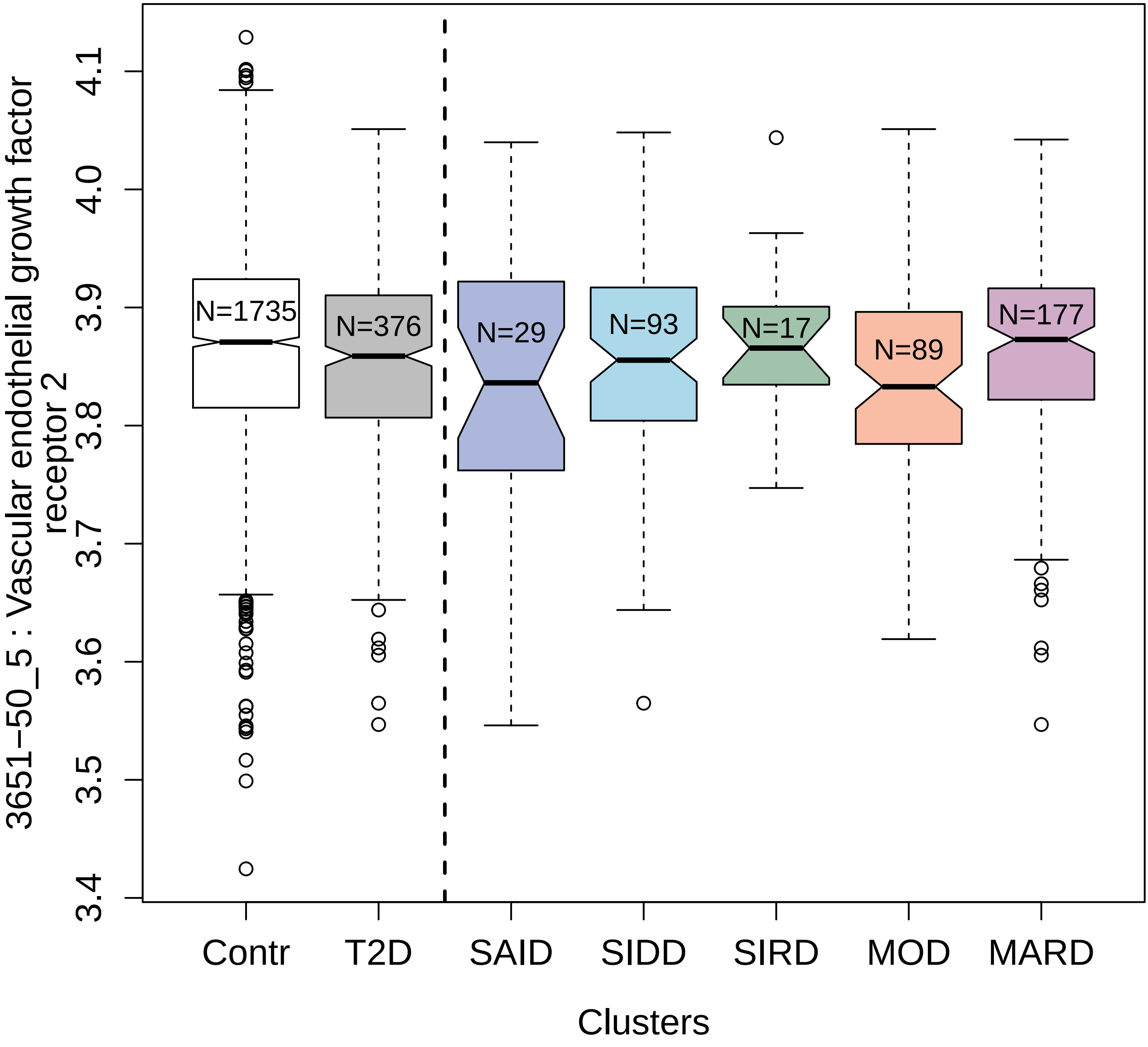
3651−50_5 : Vascular endothelial growth factor receptor 2
4.1
4.0
3.9
3.8
3.7
3.6
3.5
3.4
N=1735
N=376
N=29
N=93
N=17
N=89
N=177
Contr
T2D
SAID
SIDD
SIRD
MOD
MARD
Clusters

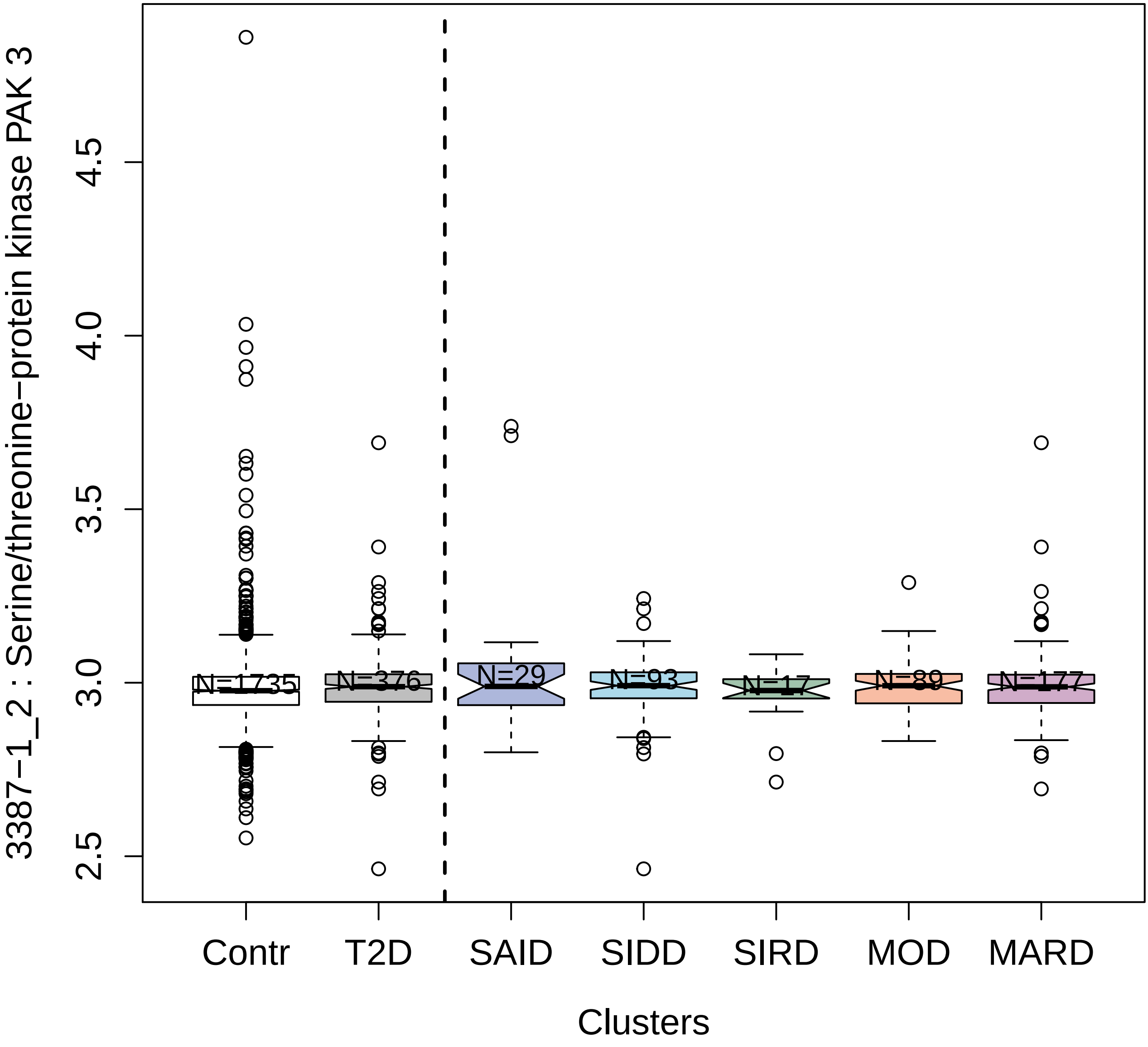

3387−1_2 : Serine/threonine−protein kinase PAK 3
2.5
3.0
3.5
4.0
4.5
N=1735
N=376
N=29
N=93
N=17
N=89
N=177
Contr
T2D
SAID
SIDD
SIRD
MOD
MARD
Clusters

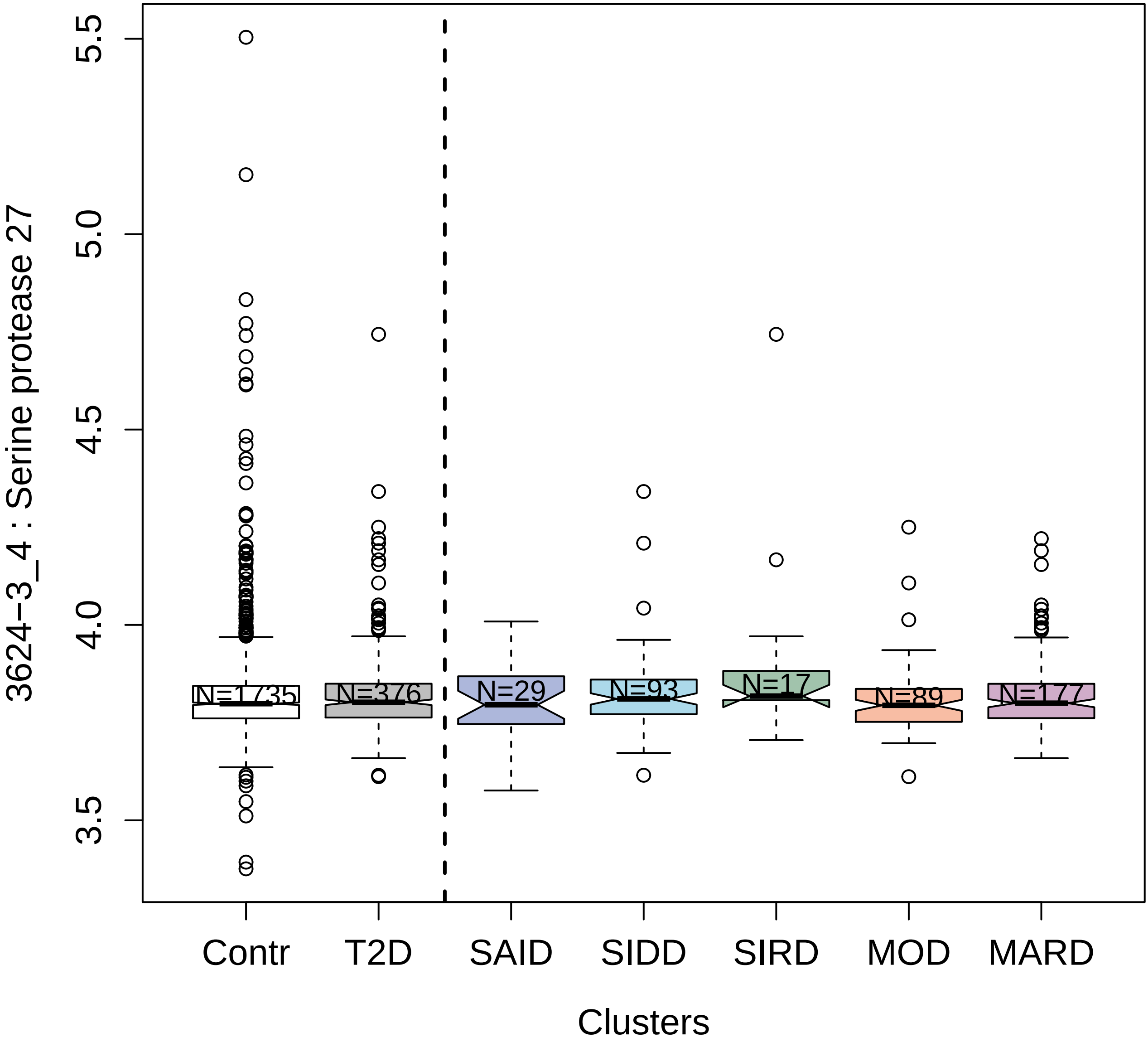

3624−3_4 : Serine protease 27
5.5
5.0
4.5
4.0
3.5
N=1735
N=376
N=29
N=93
N=17
N=89
N=177
Contr
T2D
SAID
SIDD
SIRD
MOD
MARD
Clusters

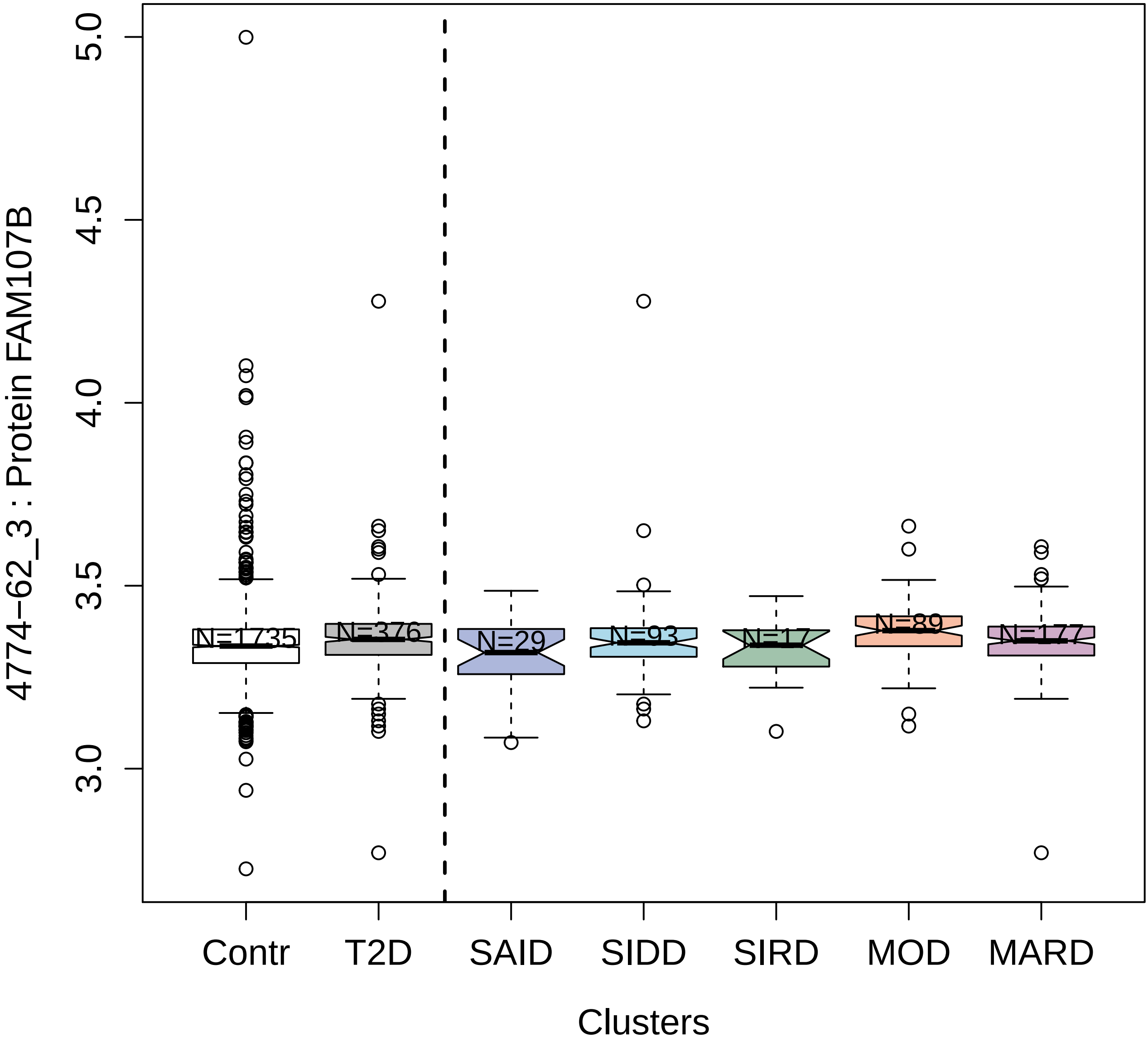

4774−62_3 : Protein FAM107B
5.0
4.5
4.0
3.5
3.0
N=1735
N=376
N=29
N=93
N=17
N=89
N=177
Contr
T2D
SAID
SIDD
SIRD
MOD
MARD
Clusters

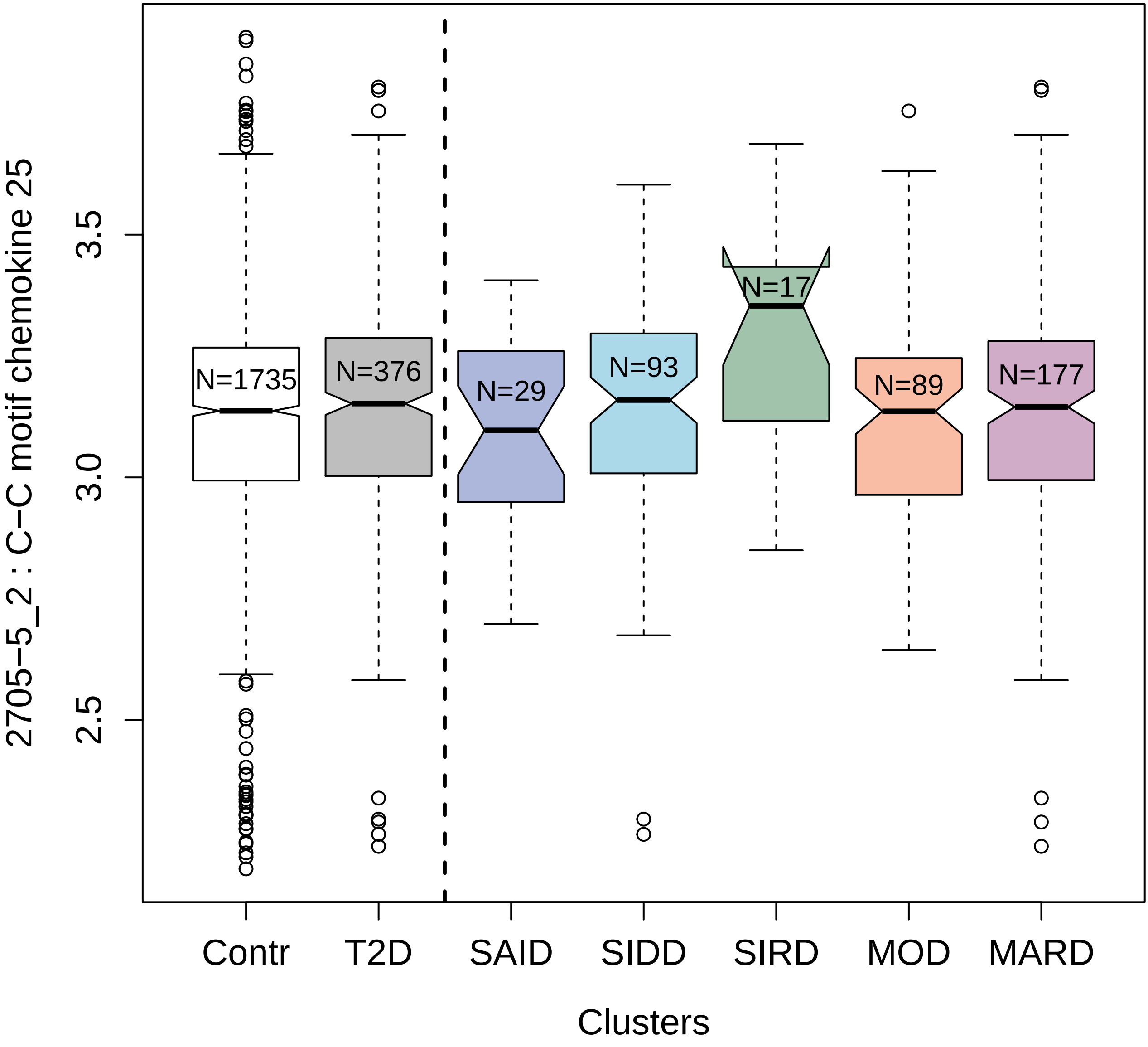

2705−5_2 : C−C motif chemokine 25
3.5
3.0
2.5
N=1735
N=376
N=29
N=93
N=17
N=89
N=177
Contr
T2D
SAID
SIDD
SIRD
MOD
MARD
Clusters

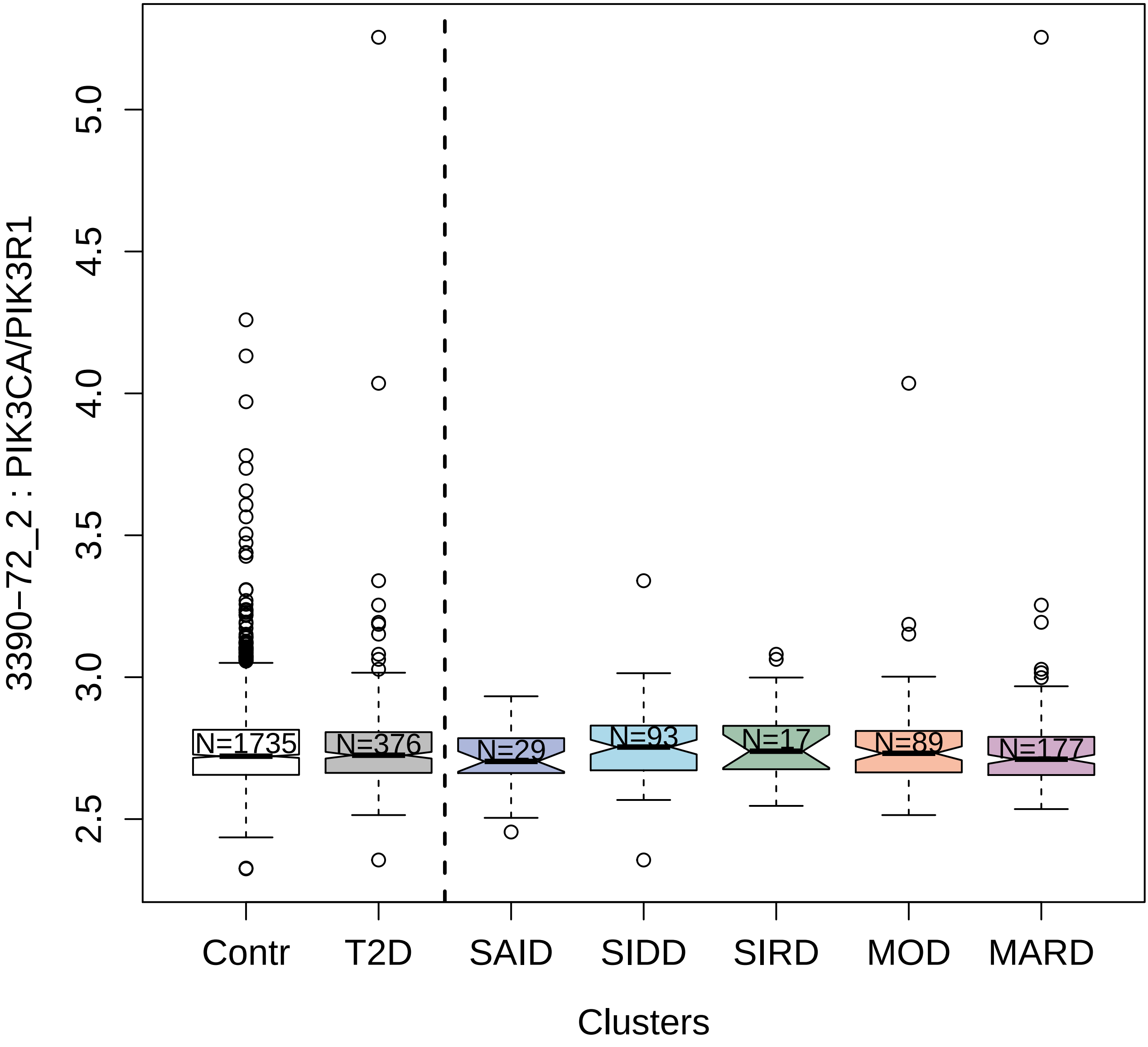

3390−72_2 : PIK3CA/PIK3R1
5.0
4.5
4.0
3.5
3.0
2.5
N=1735
N=376
N=29
N=93
N=17
N=89
N=177
Contr
T2D
SAID
SIDD
SIRD
MOD
MARD
Clusters

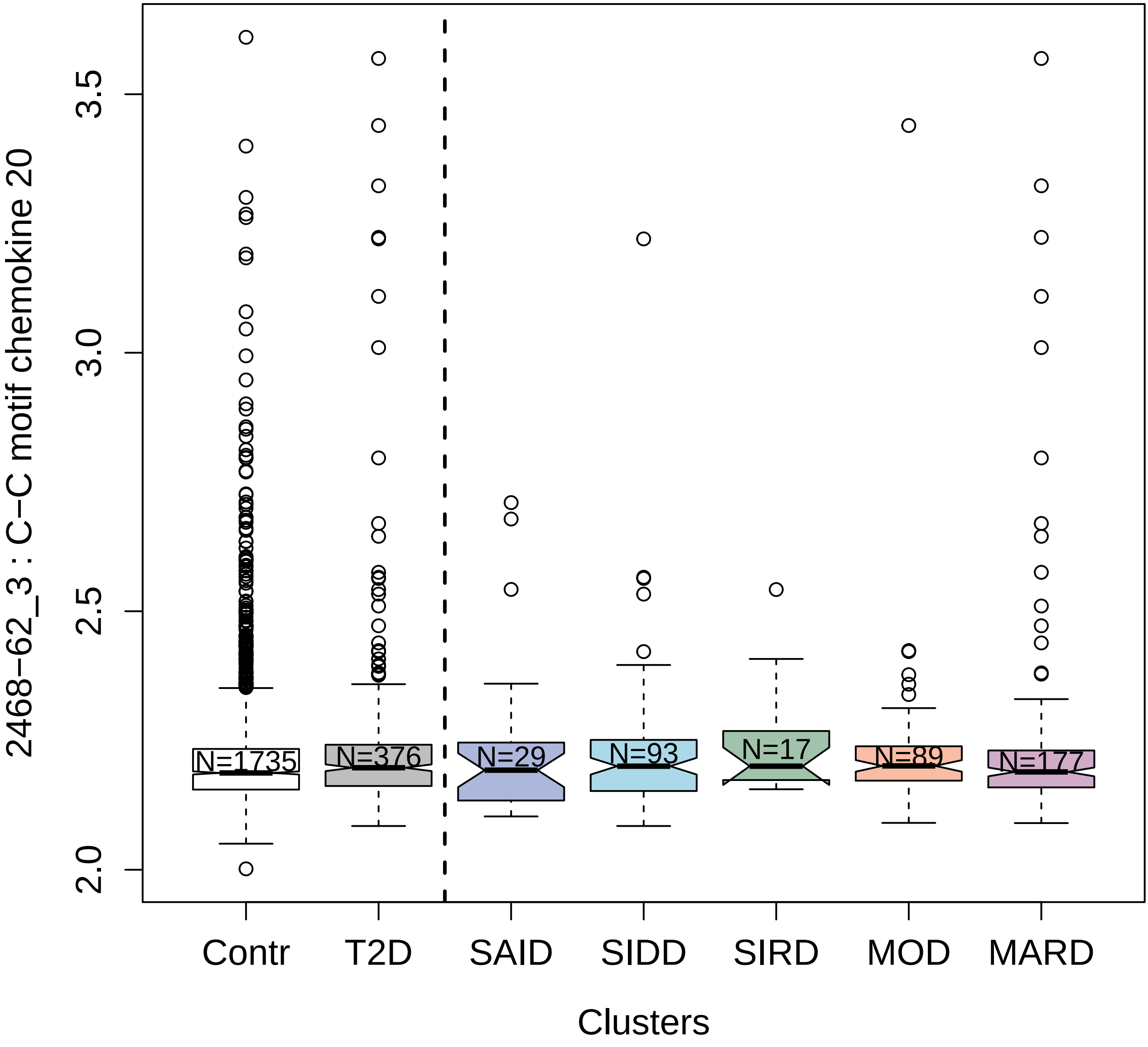
2468−62_3 : C−C motif chemokine 20
2.0
2.5
3.0
3.5
N=1735
N=376
N=29
N=93
N=17
N=89
N=177
Contr
T2D
SAID
SIDD
SIRD
MOD
MARD
Clusters

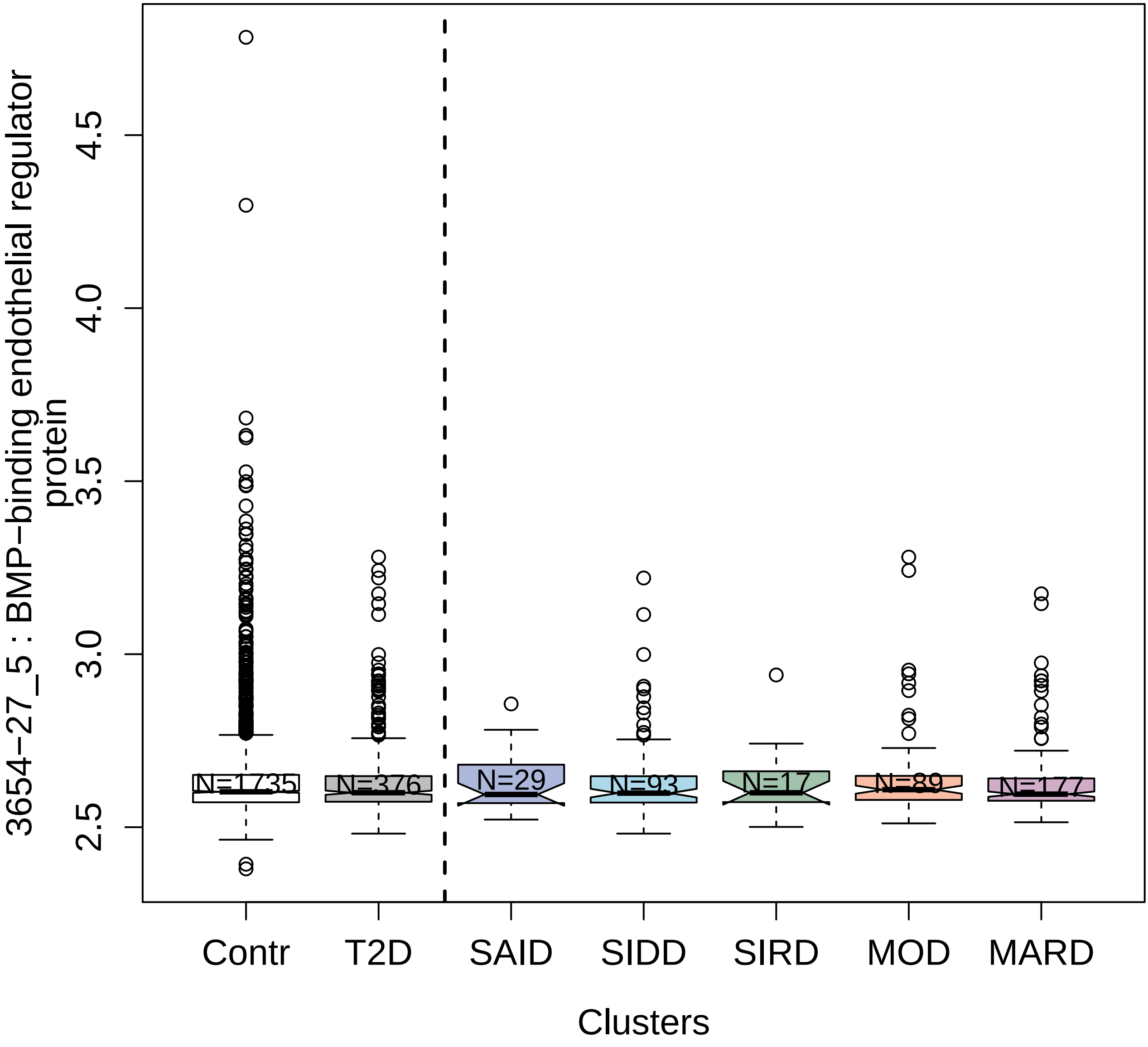

3654−27_5 : BMP−binding endothelial regulator protein
2.5
3.0
3.5
4.0
4.5
N=1735
N=376
N=29
N=93
N=17
N=89
N=177
Contr
T2D
SAID
SIDD
SIRD
MOD
MARD
Clusters

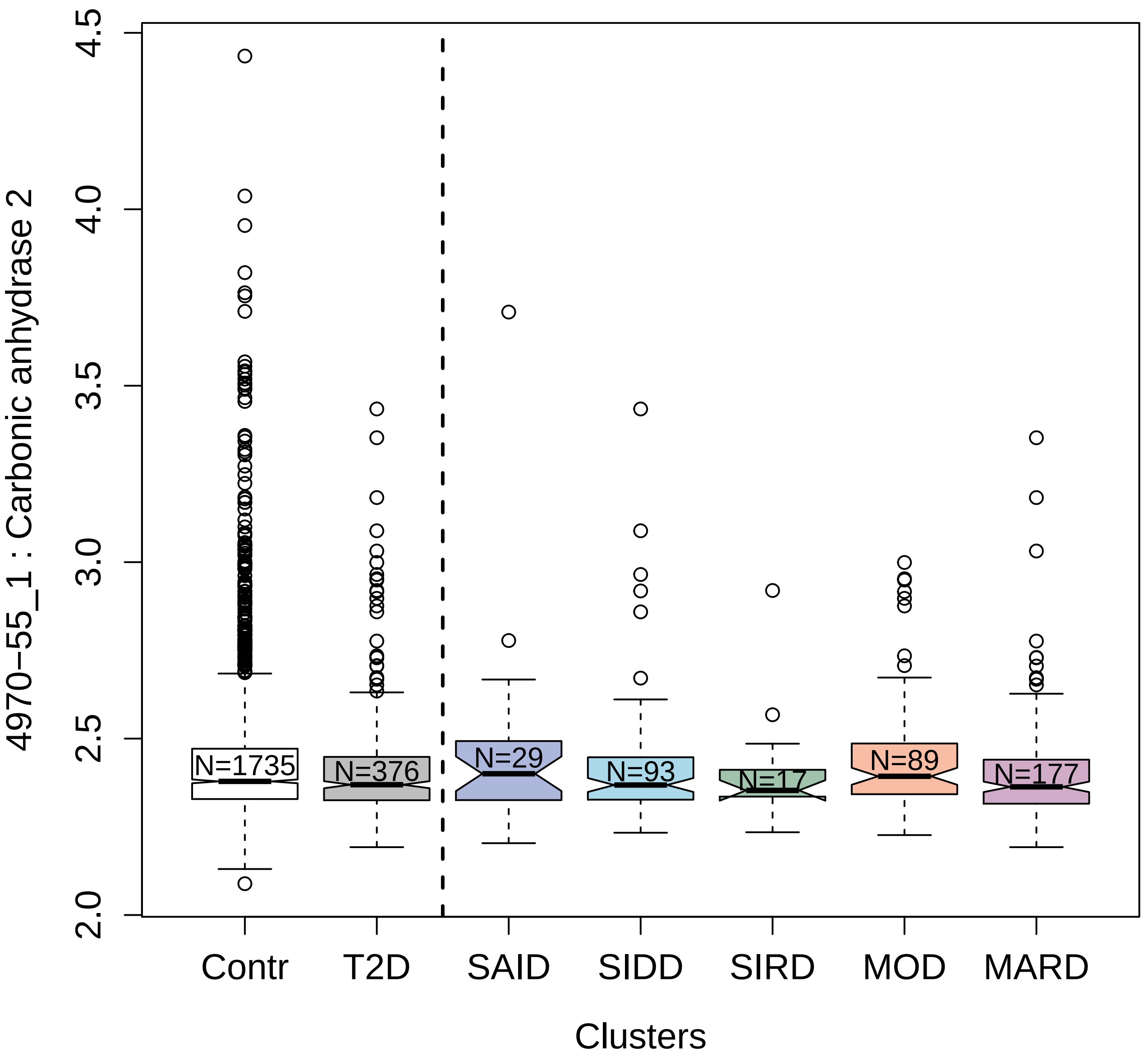
4970−55_1 : Carbonic anhydrase 2
2.0
2.5
3.0
3.5
4.0
4.5
N=1735
N=376
N=29
N=93
N=17
N=89
N=177
Contr
T2D
SAID
SIDD
SIRD
MOD
MARD
Clusters

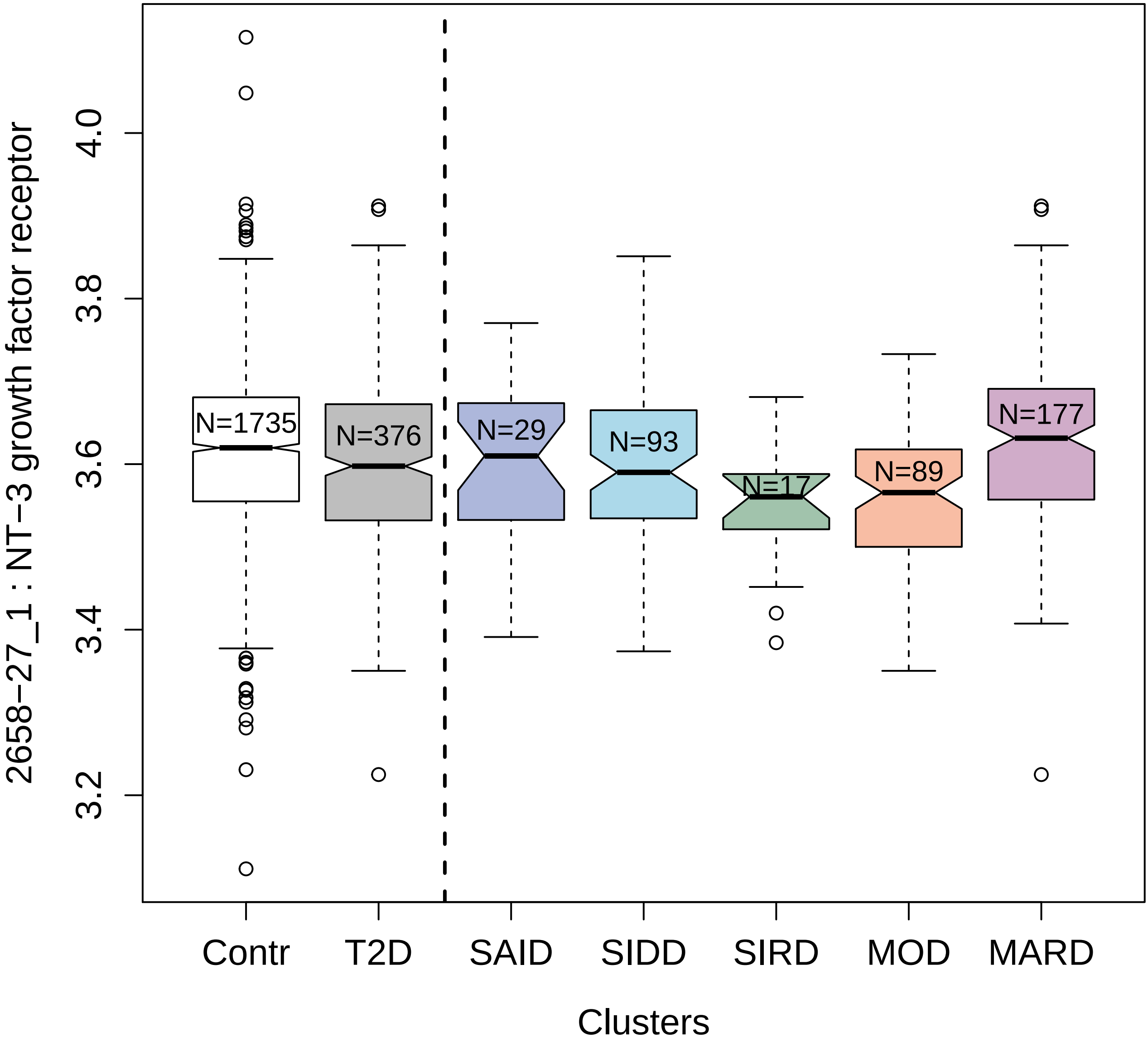
2658−27_1 : NT−3 growth factor receptor
3.2
3.4
3.6
3.8
4.0
N=1735
N=376
N=29
N=93
N=17
N=89
N=177
Contr
T2D
SAID
SIDD
SIRD
MOD
MARD
Clusters

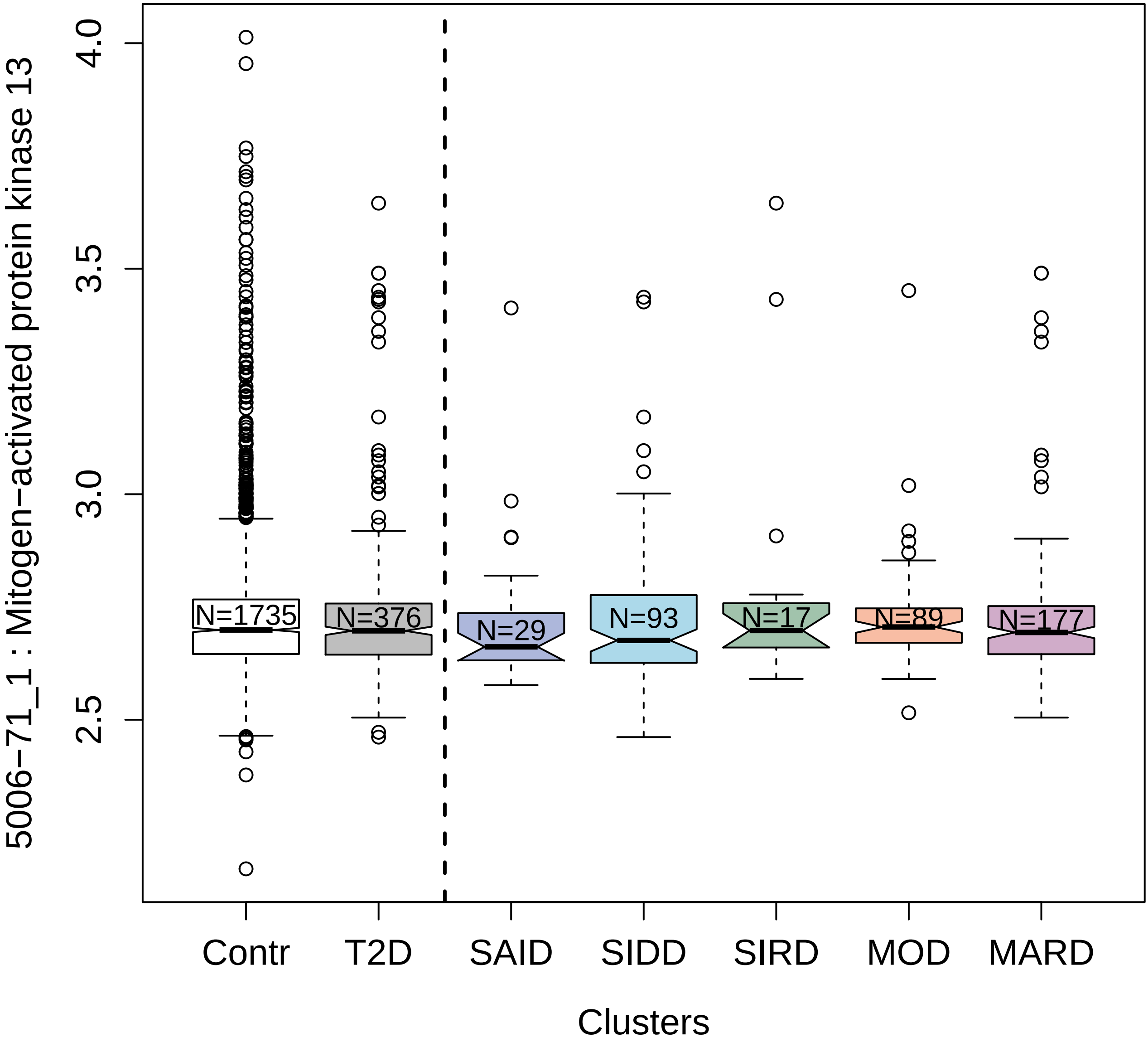

5006−71_1 : Mitogen−activated protein kinase 13
4.0
3.5
3.0
2.5
N=1735
N=376
N=29
N=93
N=17
N=89
N=177
Contr
T2D
SAID
SIDD
SIRD
MOD
MARD
Clusters

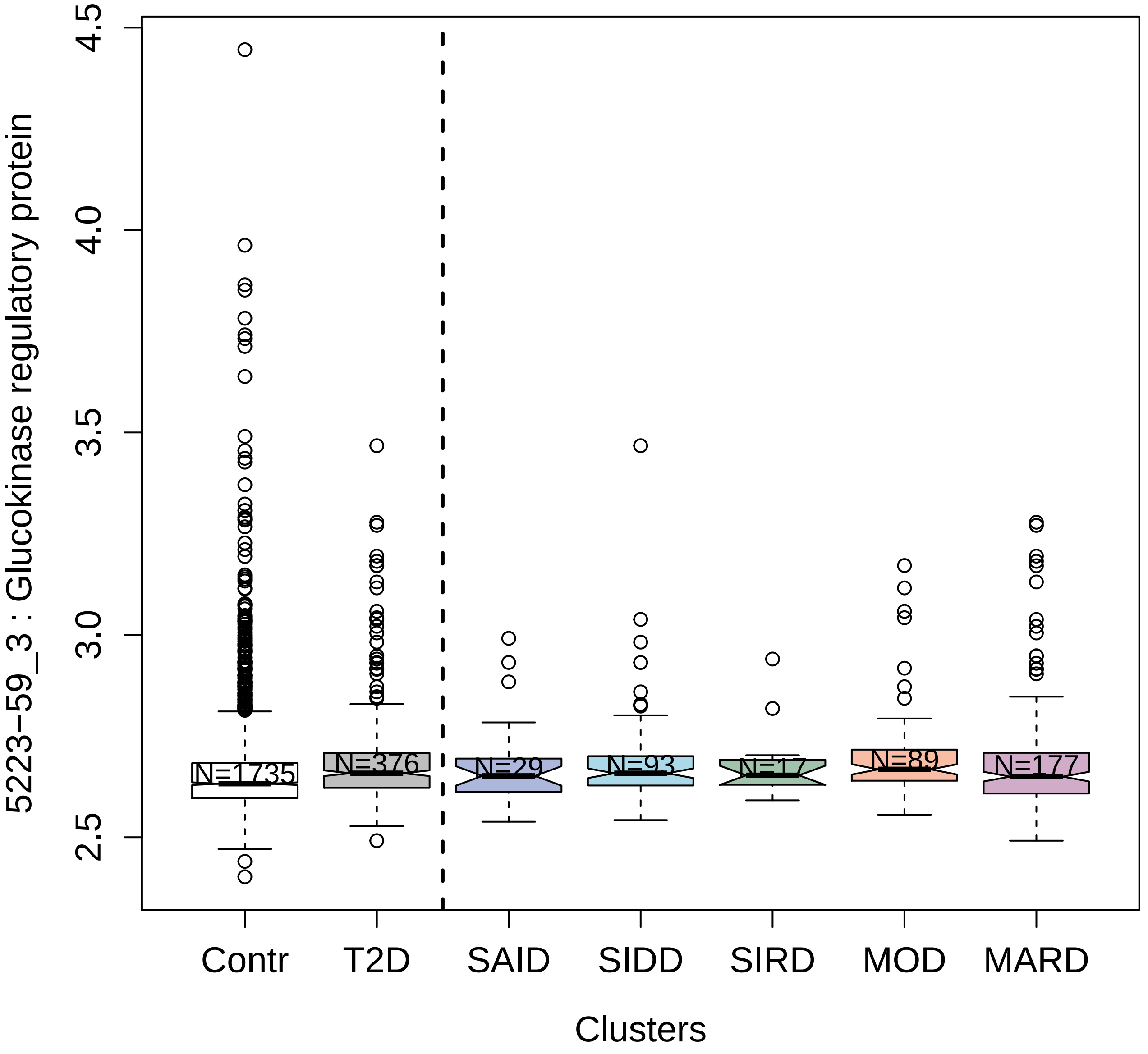

5223−59_3 : Glucokinase regulatory protein
4.5
4.0
3.5
3.0
2.5
N=1735
N=376
N=29
N=93
N=17
N=89
N=177
Contr
T2D
SAID
SIDD
SIRD
MOD
MARD
Clusters

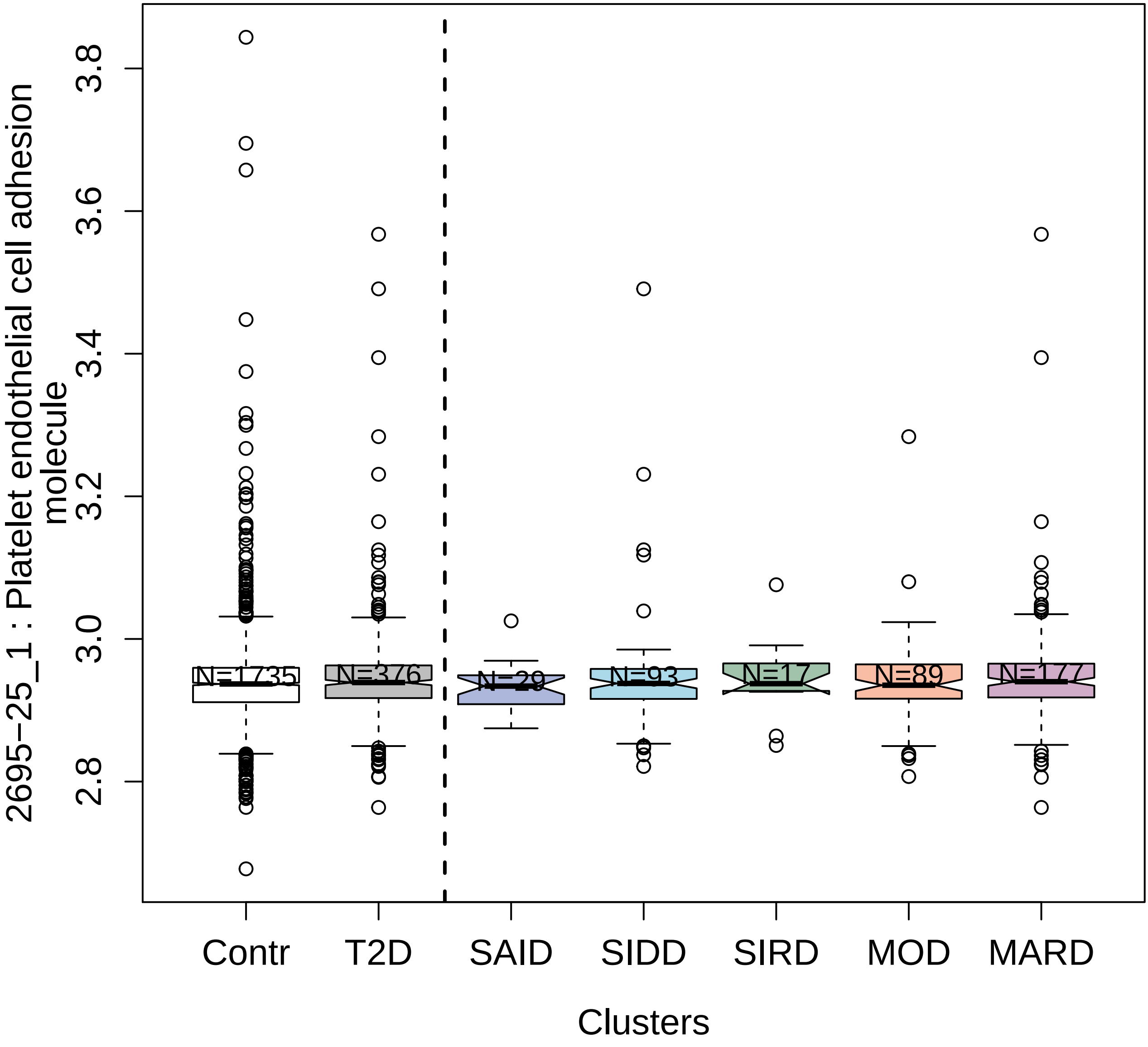

2695−25_1 : Platelet endothelial cell adhesion molecule
3.8
3.6
3.4
3.2
3.0
2.8
N=1735
N=376
N=29
N=93
N=17
N=89
N=177
Contr
T2D
SAID
SIDD
SIRD
MOD
MARD
Clusters

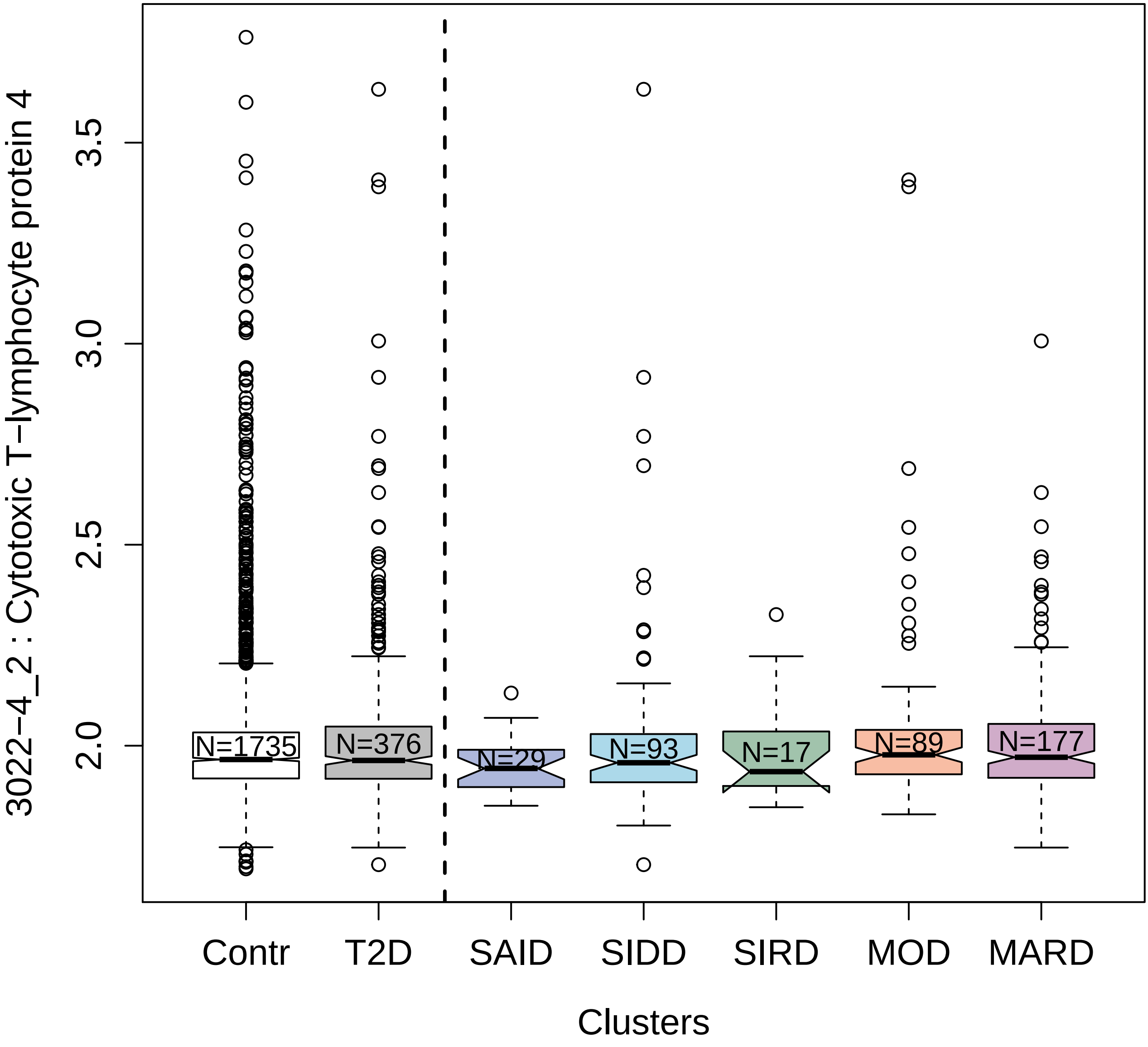
3022−4_2 : Cytotoxic T−lymphocyte protein 4
3.5
3.0
2.5
2.0
N=1735
N=376
N=29
N=93
N=17
N=89
N=177
Contr
T2D
SAID
SIDD
SIRD
MOD
MARD
Clusters

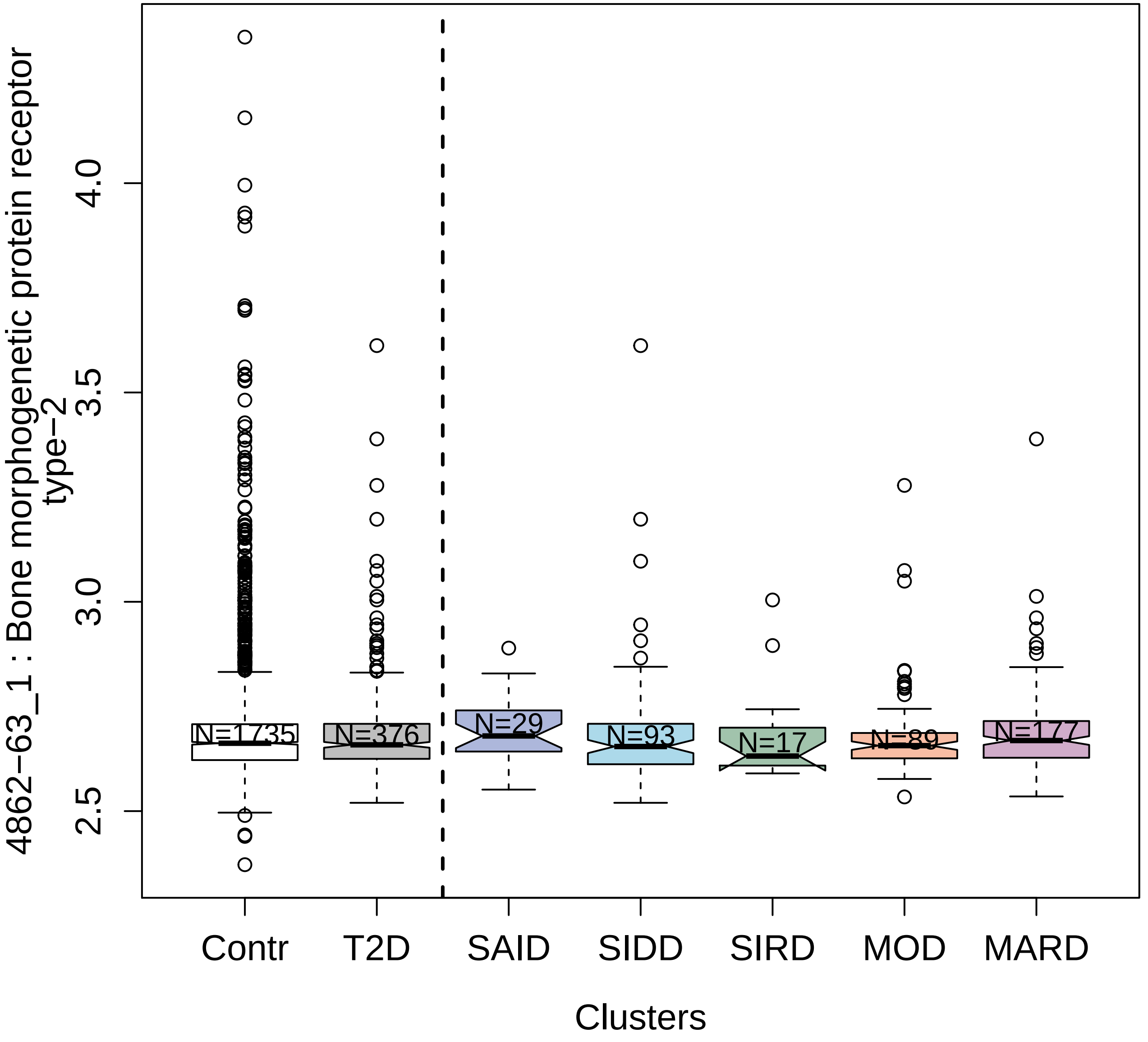

4862−63_1 : Bone morphogenetic protein receptor type−2
2.5
3.0
3.5
4.0
N=1735
N=376
N=29
N=93
N=17
N=89
N=177
Contr
T2D
SAID
SIDD
SIRD
MOD
MARD
Clusters

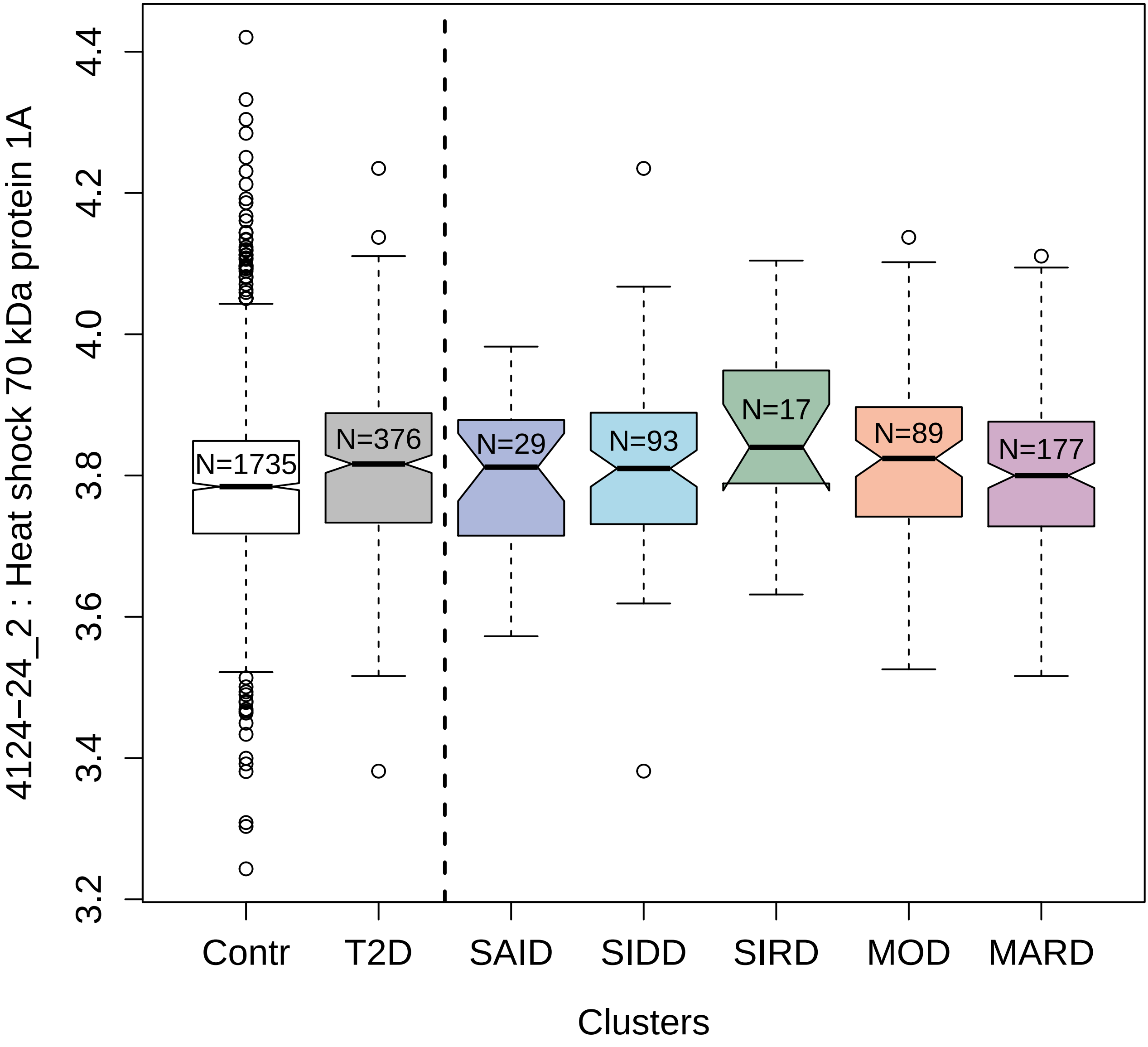

4124−24_2 : Heat shock 70 kDa protein 1A
4.4
4.2
4.0
3.8
3.6
3.4
3.2
N=1735
N=376
N=29
N=93
N=17
N=89
N=177
Contr
T2D
SAID
SIDD
SIRD
MOD
MARD
Clusters

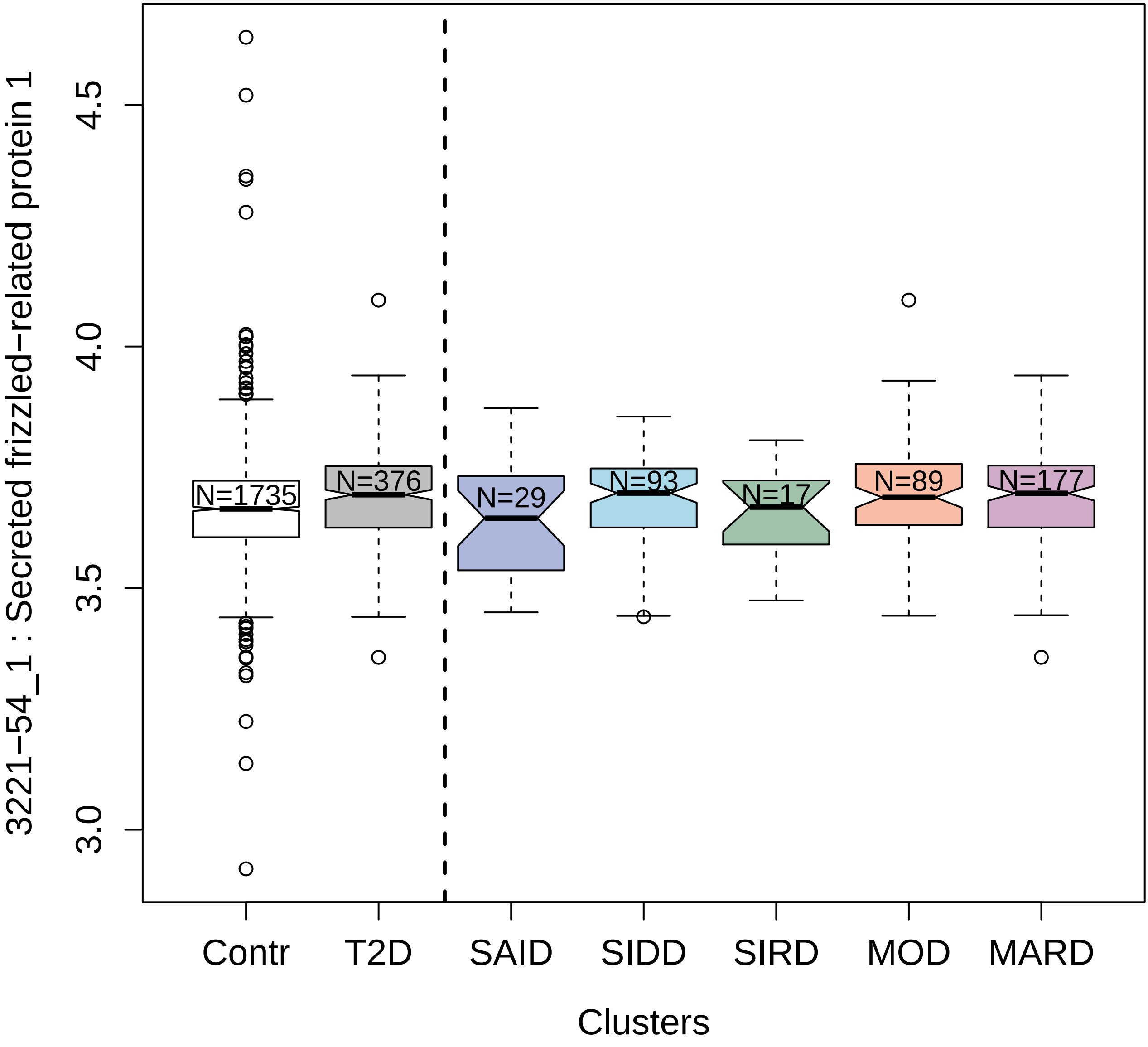
3221−54_1 : Secreted frizzled−related protein 1
3.0
3.5
4.0
4.5
N=1735
N=376
N=29
N=93
N=17
N=89
N=177
Contr
T2D
SAID
SIDD
SIRD
MOD
MARD
Clusters

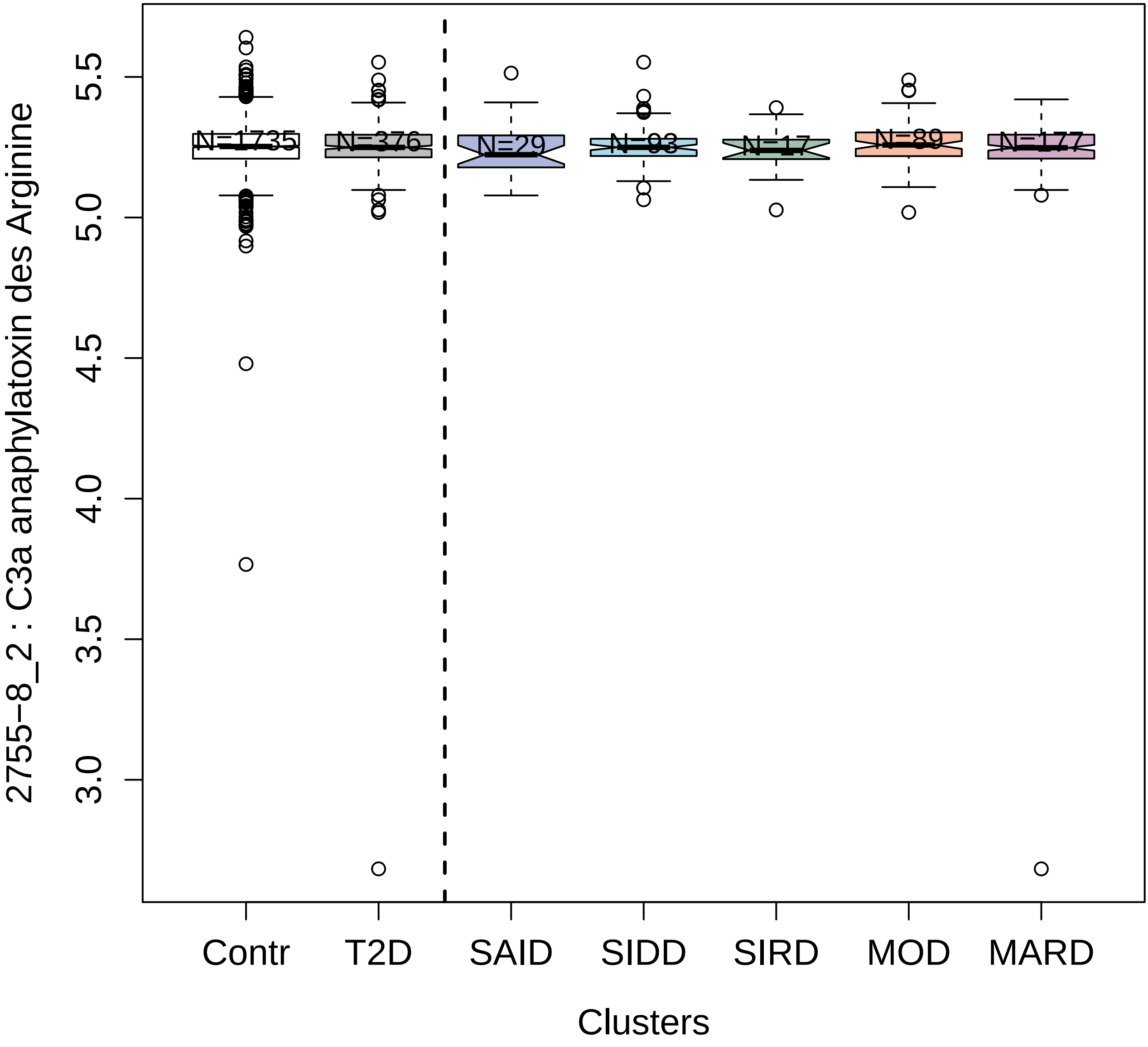

2755−8_2 : C3a anaphylatoxin des Arginine
5.5
5.0
4.5
4.0
3.5
3.0
N=1735
N=376
N=29
N=93
N=17
N=89
N=177
Contr
T2D
SAID
SIDD
SIRD
MOD
MARD
Clusters

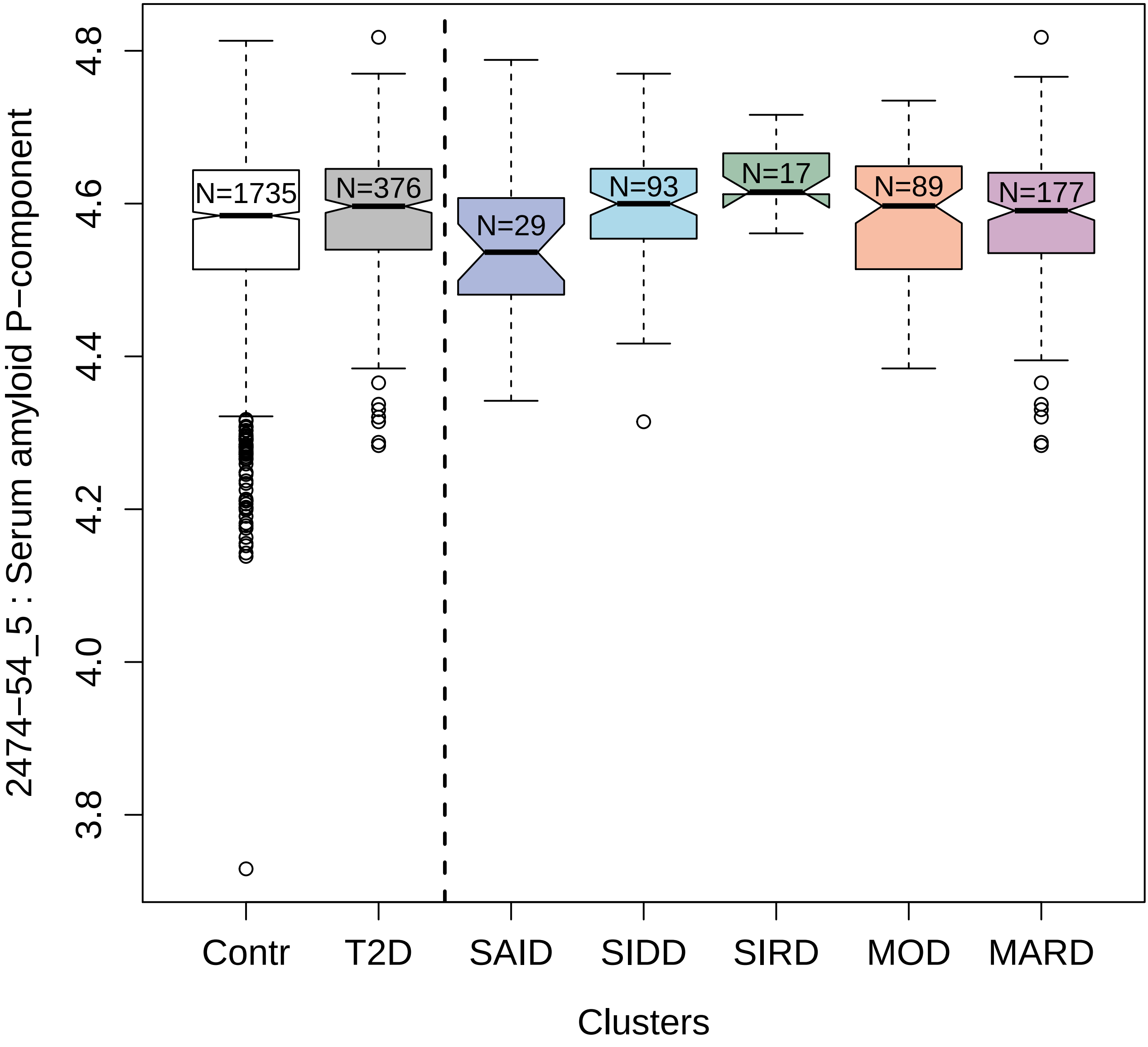

2474−54_5 : Serum amyloid P−component
4.8
4.6
4.4
4.2
4.0
3.8
N=1735
N=376
N=29
N=93
N=17
N=89
N=177
Contr
T2D
SAID
SIDD
SIRD
MOD
MARD
Clusters

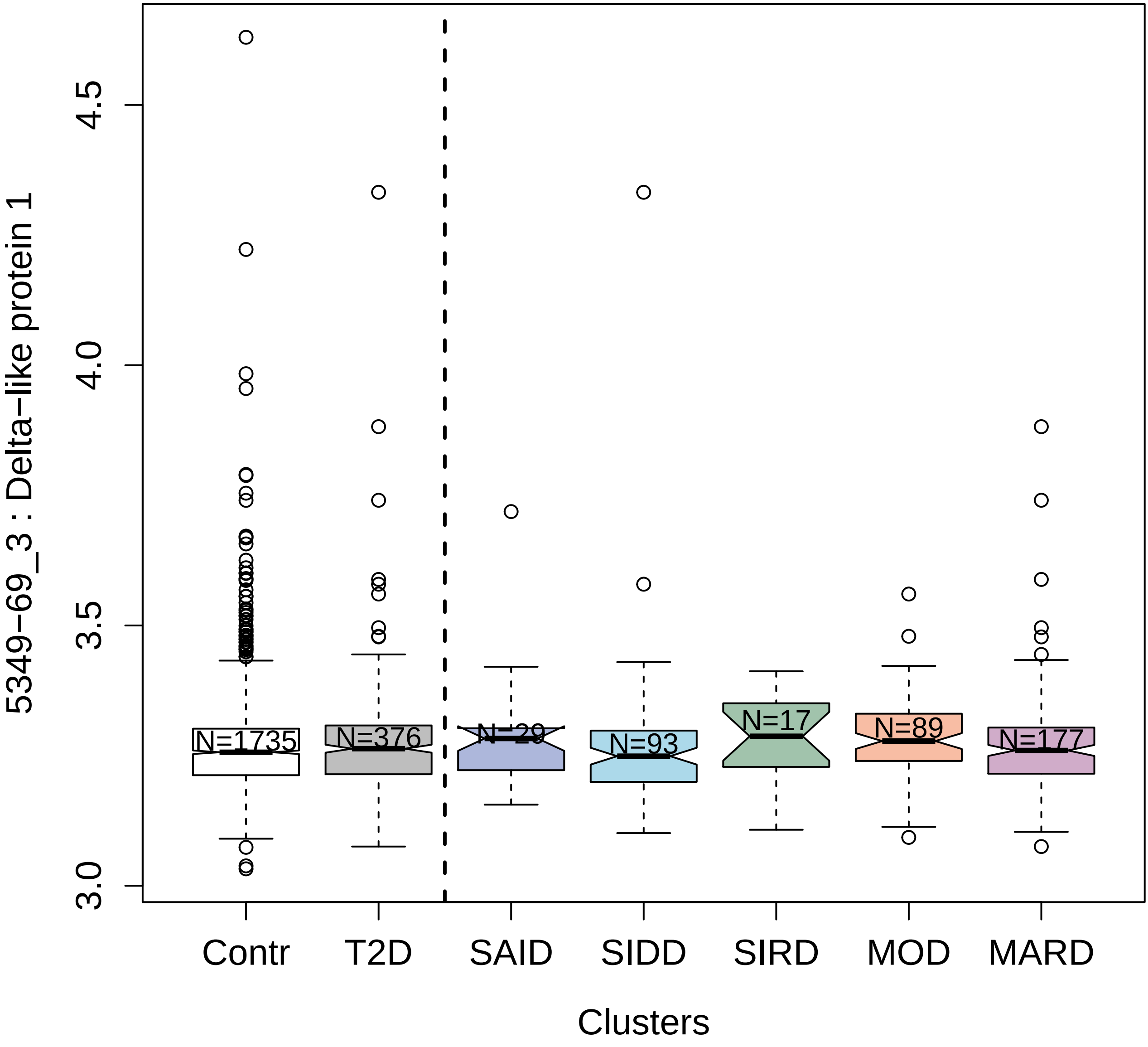

5349−69_3 : Delta−like protein 1
3.0
3.5
4.0
4.5
N=1735
N=376
N=29
N=93
N=17
N=89
N=177
Contr
T2D
SAID
SIDD
SIRD
MOD
MARD
Clusters

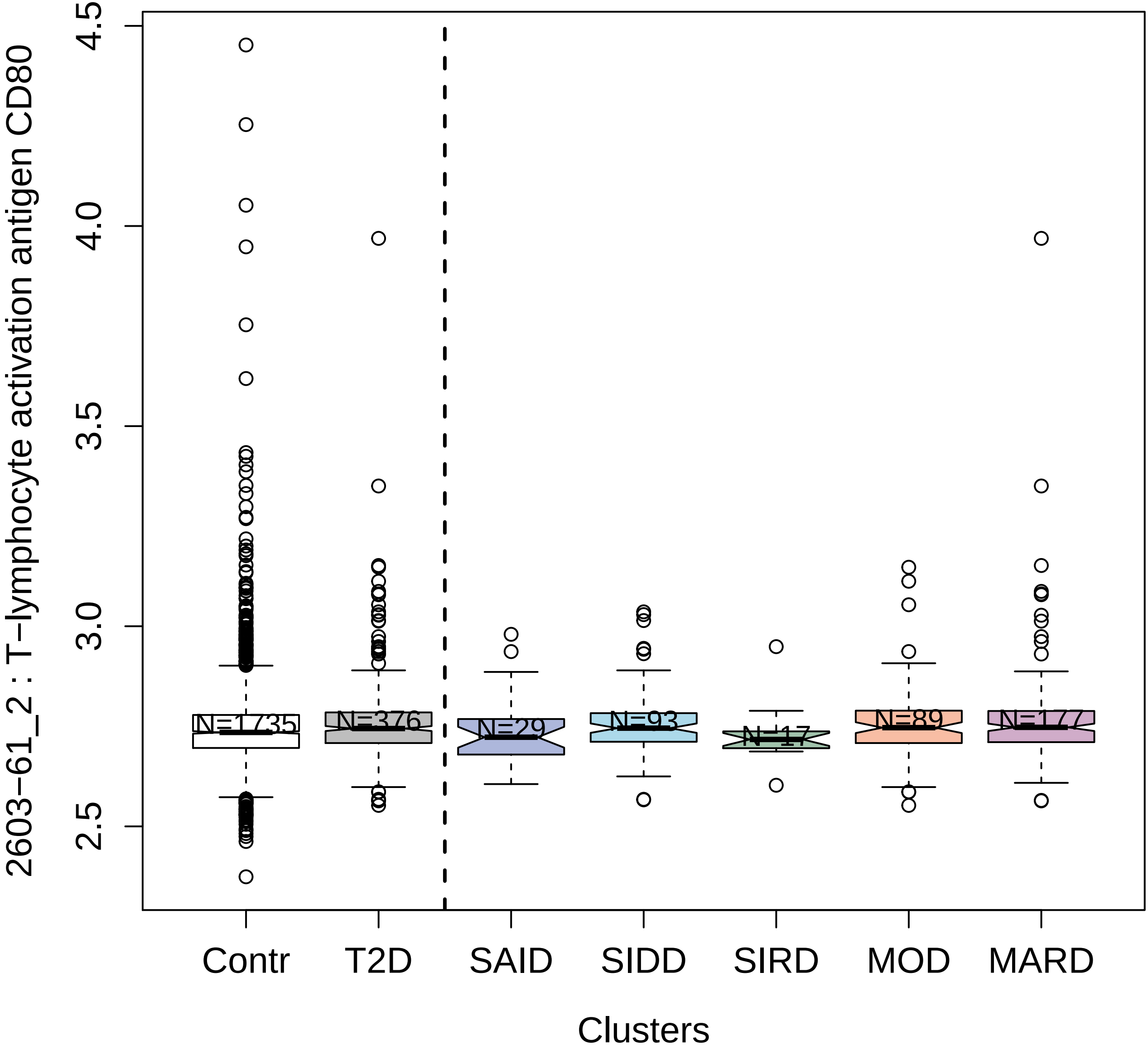

2603−61_2 : T−lymphocyte activation antigen CD80
4.5
4.0
3.5
3.0
2.5
N=1735
N=376
N=29
N=93
N=17
N=89
N=177
Contr
T2D
SAID
SIDD
SIRD
MOD
MARD
Clusters

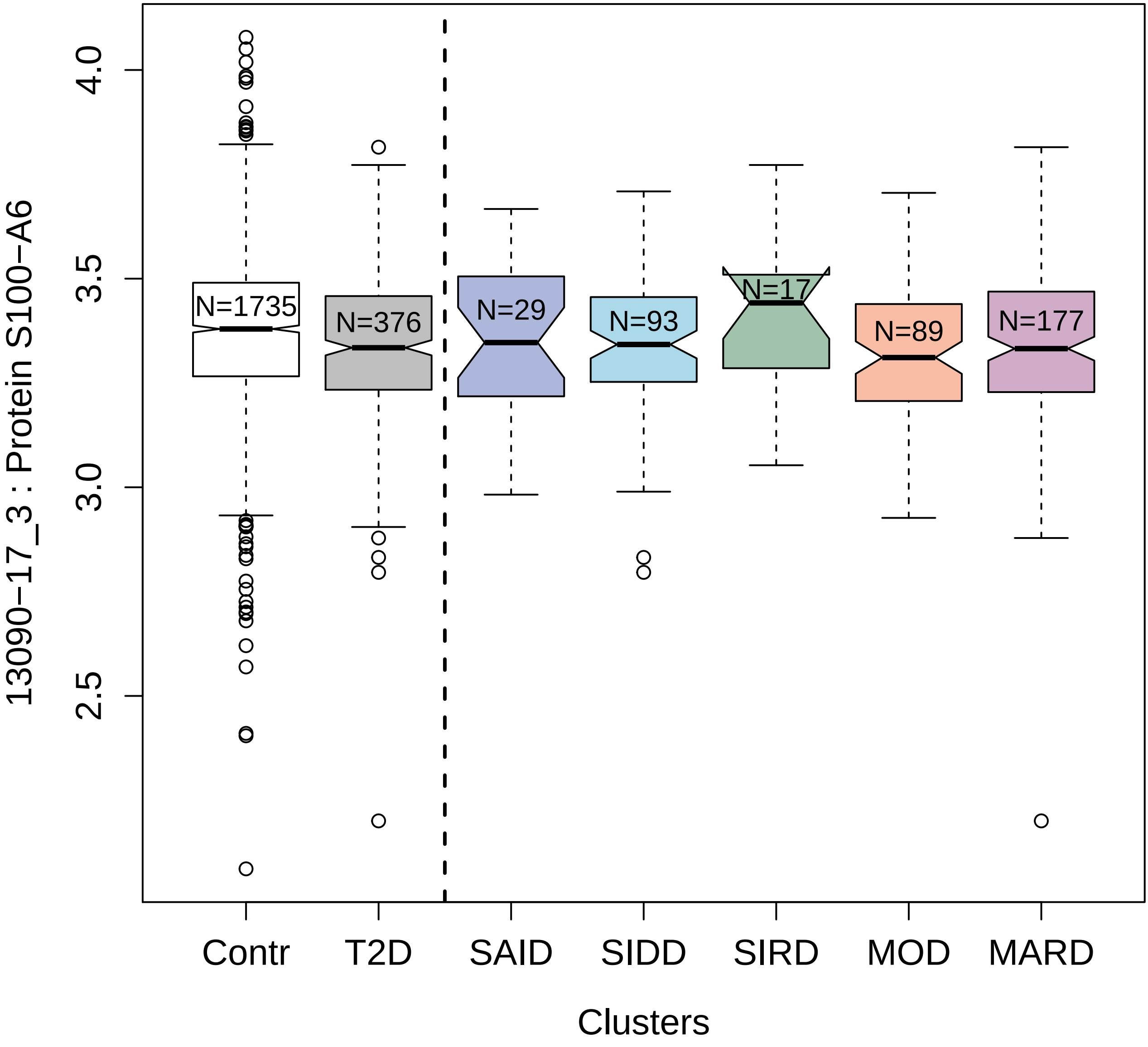
13090−17_3 : Protein S100−A6
4.0
3.5
3.0
2.5
N=1735
N=376
N=29
N=93
N=17
N=89
N=177
Contr
T2D
SAID
SIDD
SIRD
MOD
MARD
Clusters

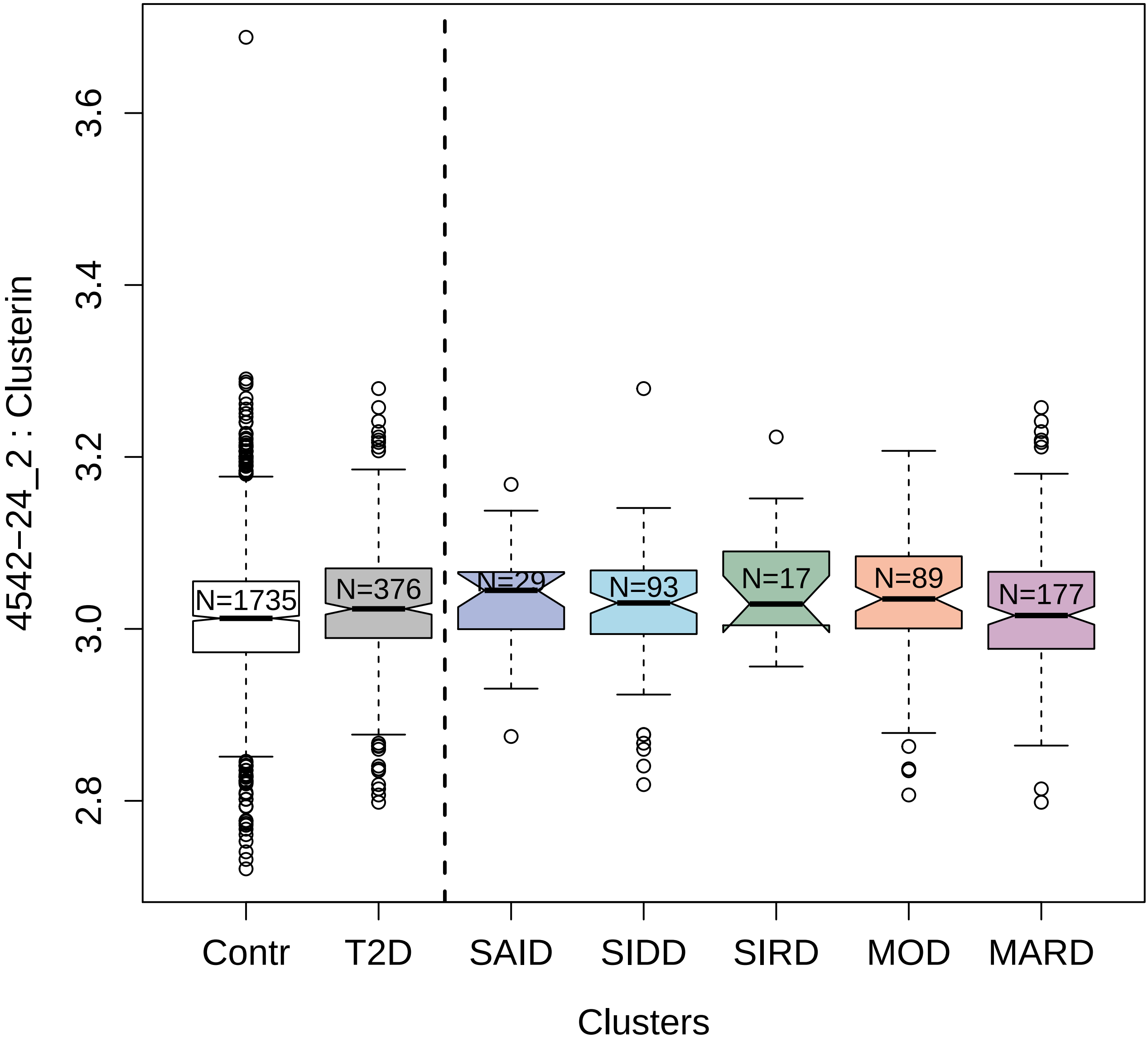

4542−24_2 : Clusterin
3.6
3.4
3.2
3.0
2.8
N=1735
N=376
N=29
N=93
N=17
N=89
N=177
Contr
T2D
SAID
SIDD
SIRD
MOD
MARD
Clusters

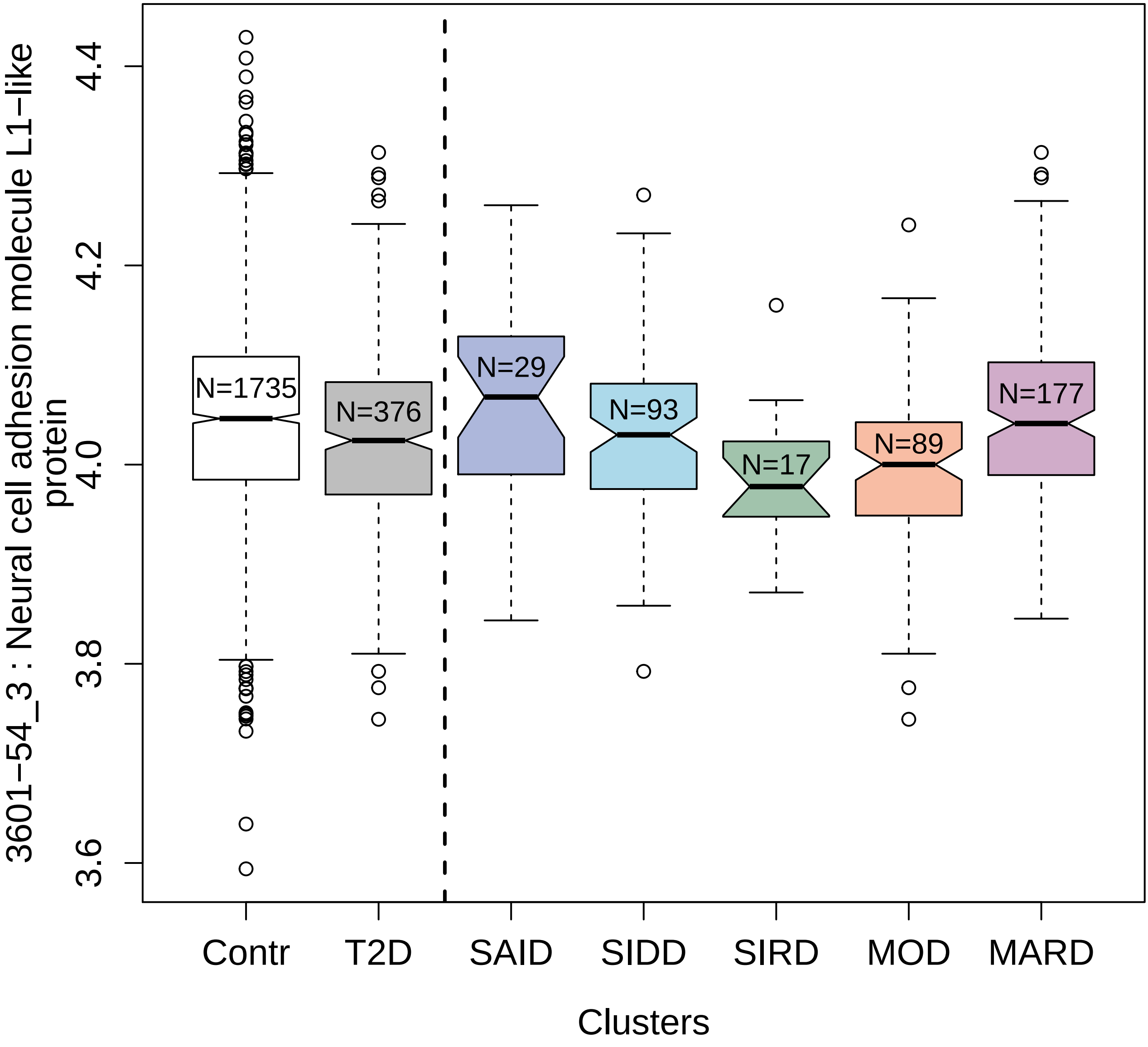

3601−54_3 : Neural cell adhesion molecule L1−like protein
4.4
4.2
4.0
3.8
3.6
N=1735
N=376
N=29
N=93
N=17
N=89
N=177
Contr
T2D
SAID
SIDD
SIRD
MOD
MARD
Clusters

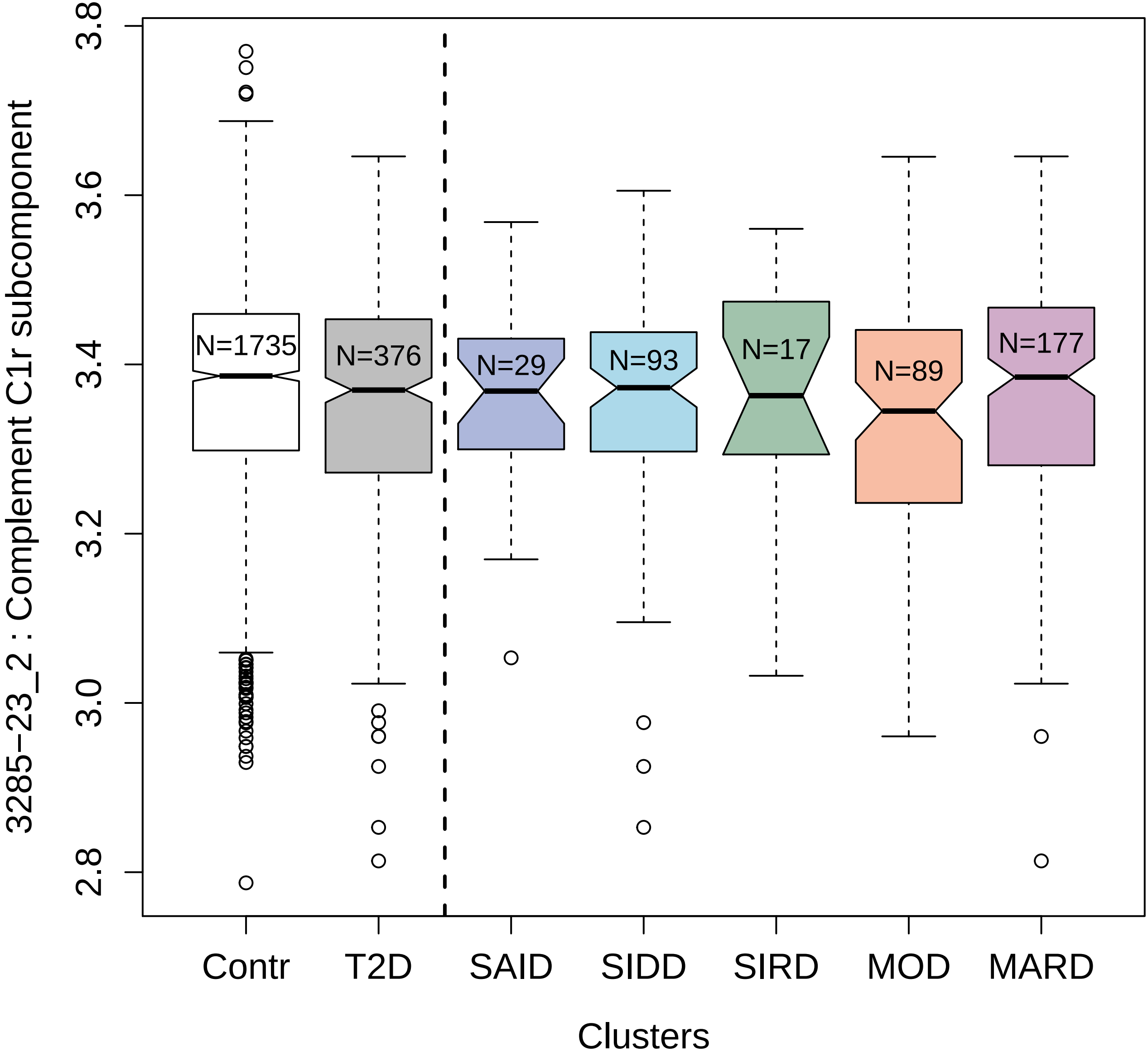

3285−23_2 : Complement C1r subcomponent
3.8
3.6
3.4
3.2
3.0
2.8
N=1735
N=376
N=29
N=93
N=17
N=89
N=177
Contr
T2D
SAID
SIDD
SIRD
MOD
MARD
Clusters

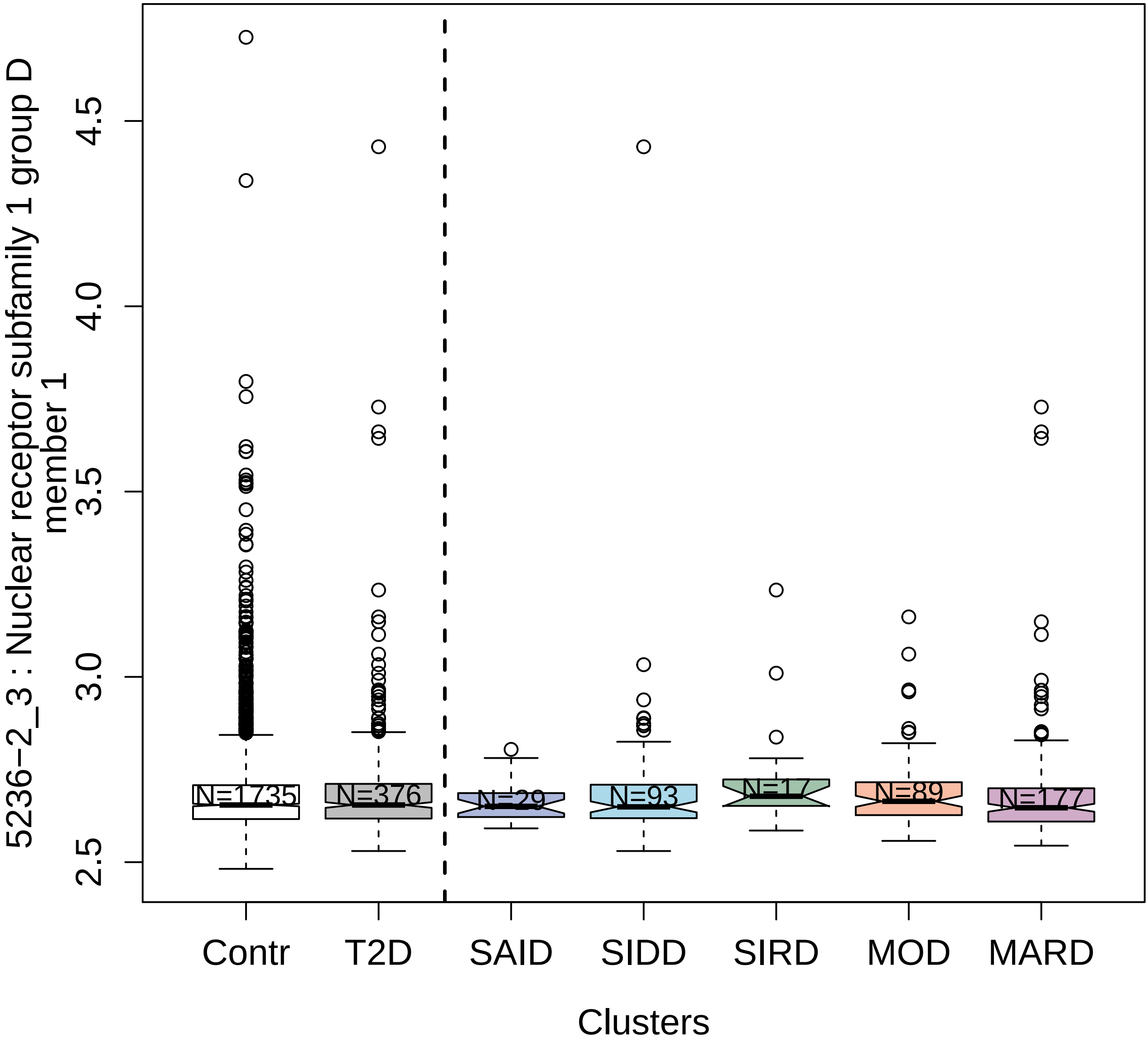

5236−2_3 : Nuclear receptor subfamily 1 group D member 1
2.5
3.0
3.5
4.0
4.5
N=1735
N=376
N=29
N=93
N=17
N=89
N=177
Contr
T2D
SAID
SIDD
SIRD
MOD
MARD
Clusters

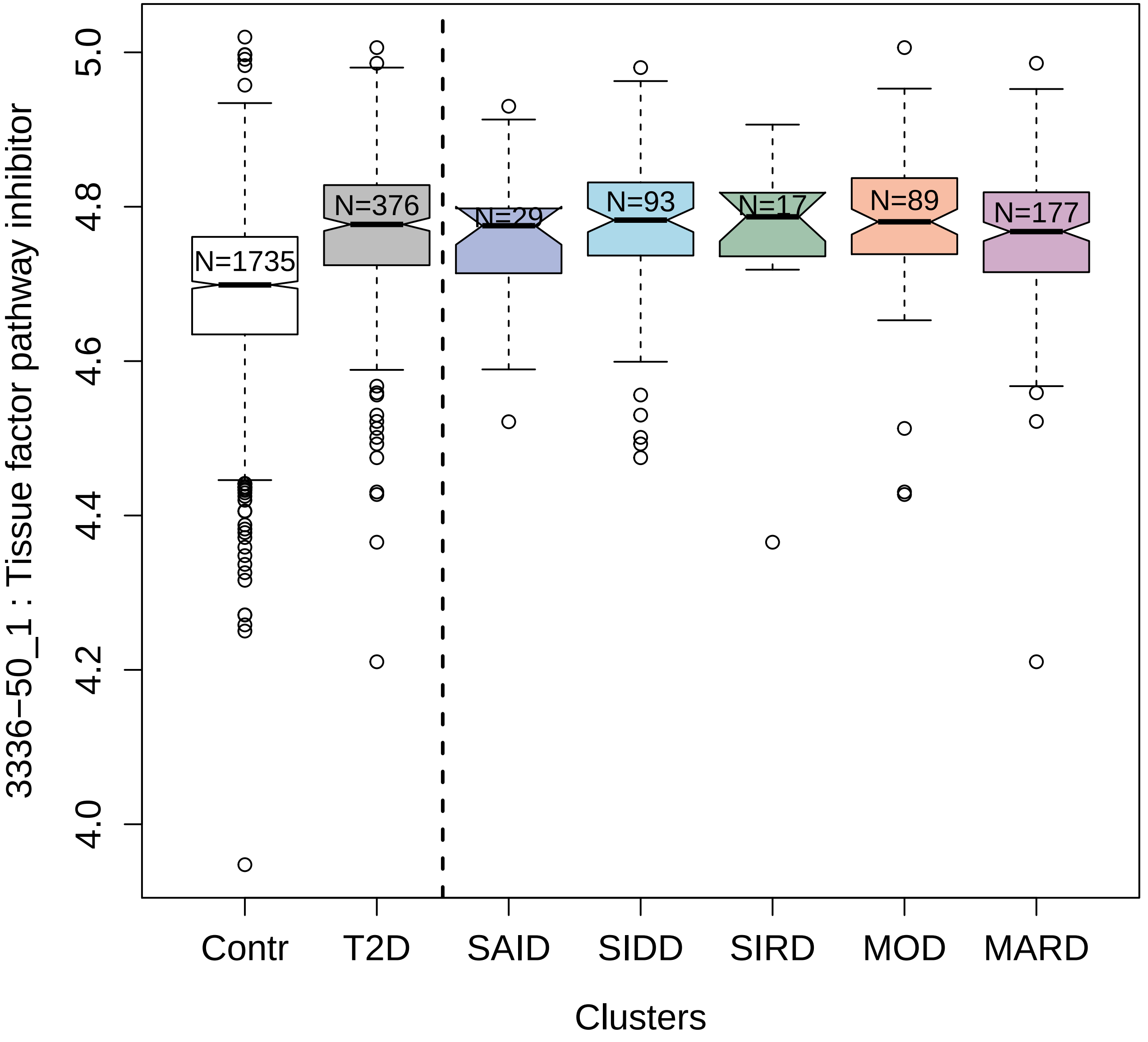

3336−50_1 : Tissue factor pathway inhibitor
5.0
4.8
4.6
4.4
4.2
4.0
N=1735
N=376
N=29
N=93
N=17
N=89
N=177
Contr
T2D
SAID
SIDD
SIRD
MOD
MARD
Clusters

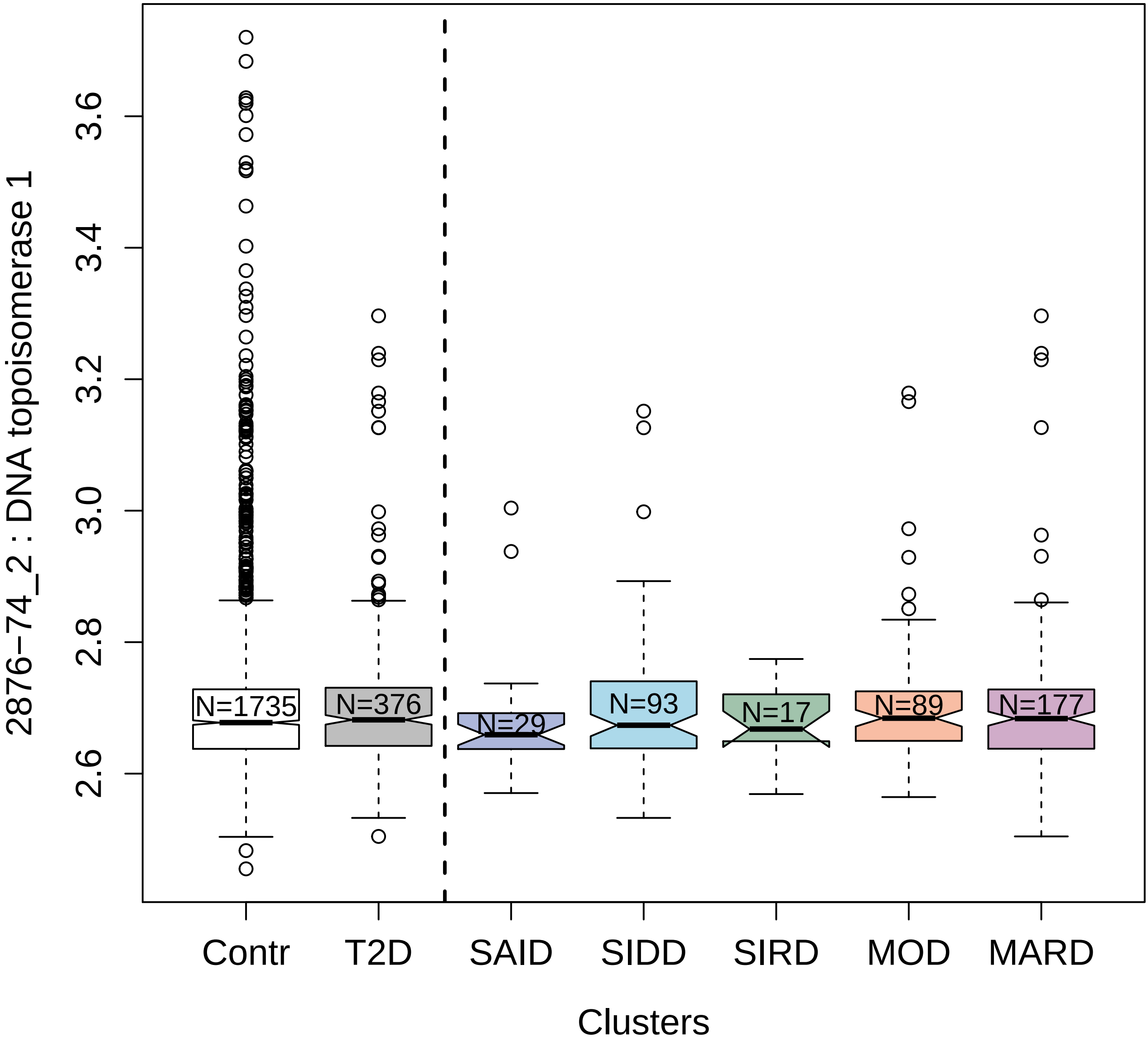

2876−74_2 : DNA topoisomerase 1
2.6
2.8
3.0
3.2
3.4
3.6
N=1735
N=376
N=29
N=93
N=17
N=89
N=177
Contr
T2D
SAID
SIDD
SIRD
MOD
MARD
Clusters

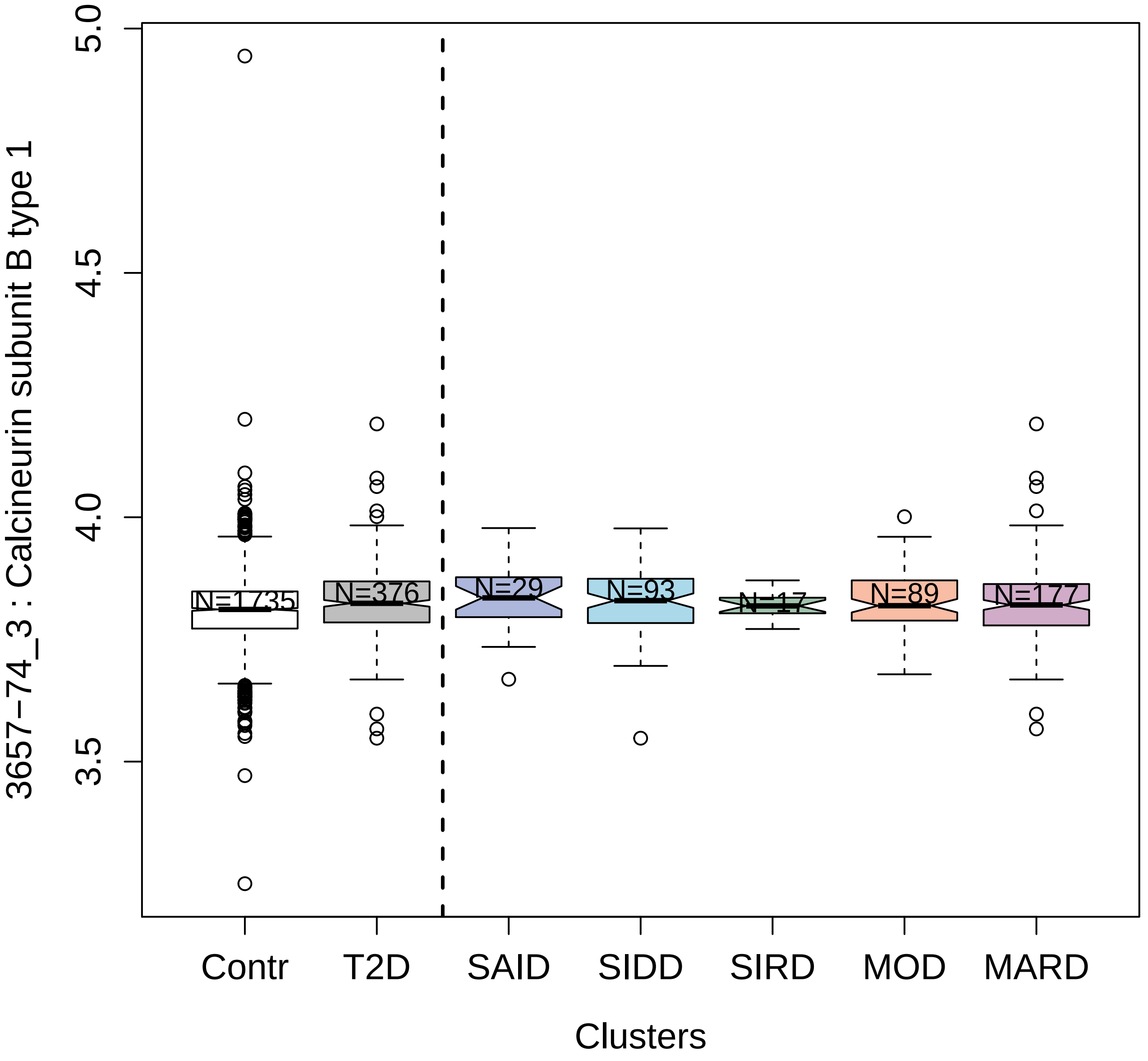

3657−74_3 : Calcineurin subunit B type 1
5.0
4.5
4.0
3.5
N=1735
N=376
N=29
N=93
N=17
N=89
N=177
Contr
T2D
SAID
SIDD
SIRD
MOD
MARD
Clusters

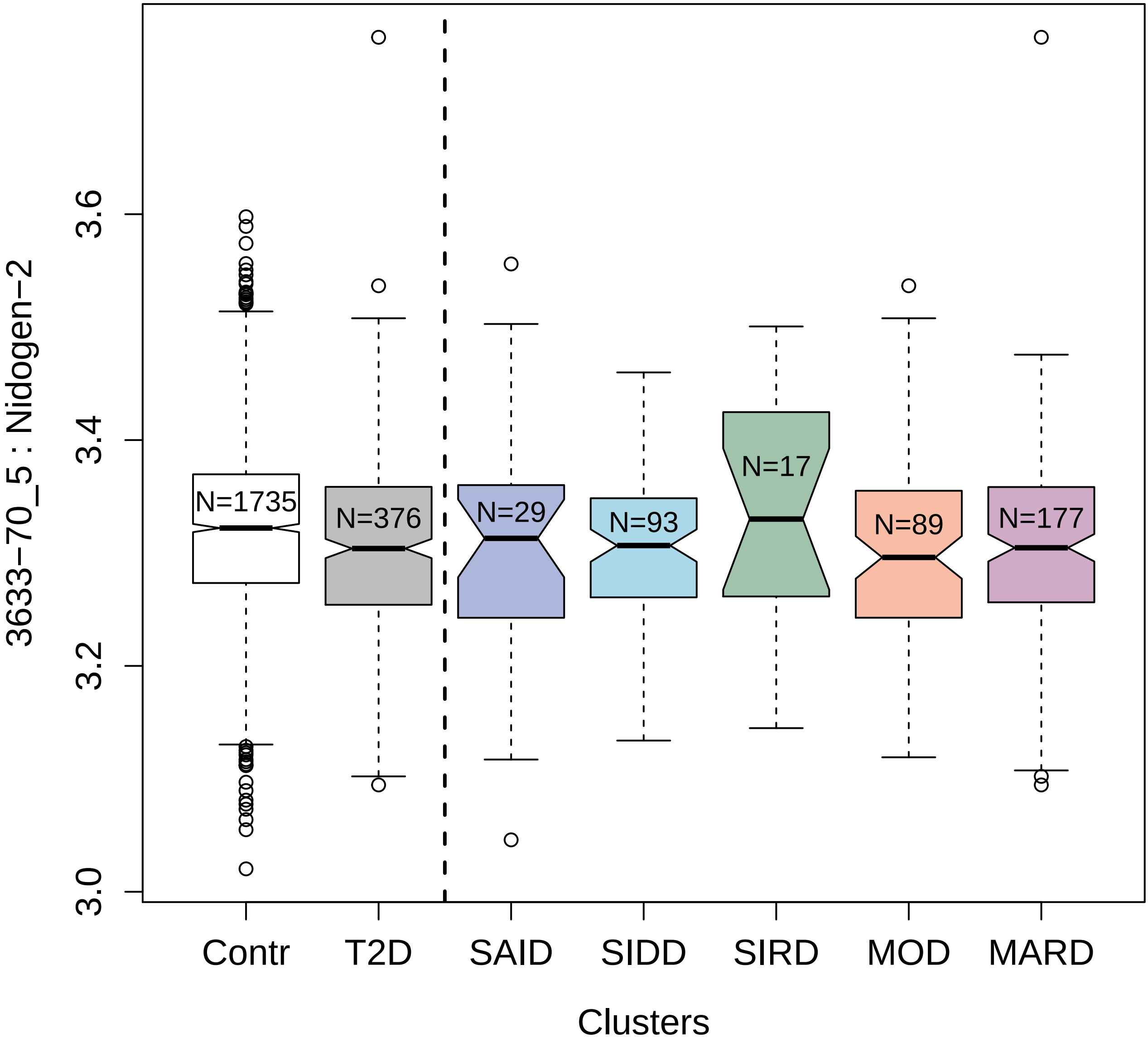
3633−70_5 : Nidogen−2
3.0
3.2
3.4
3.6
N=1735
N=376
N=29
N=93
N=17
N=89
N=177
Contr
T2D
SAID
SIDD
SIRD
MOD
MARD
Clusters

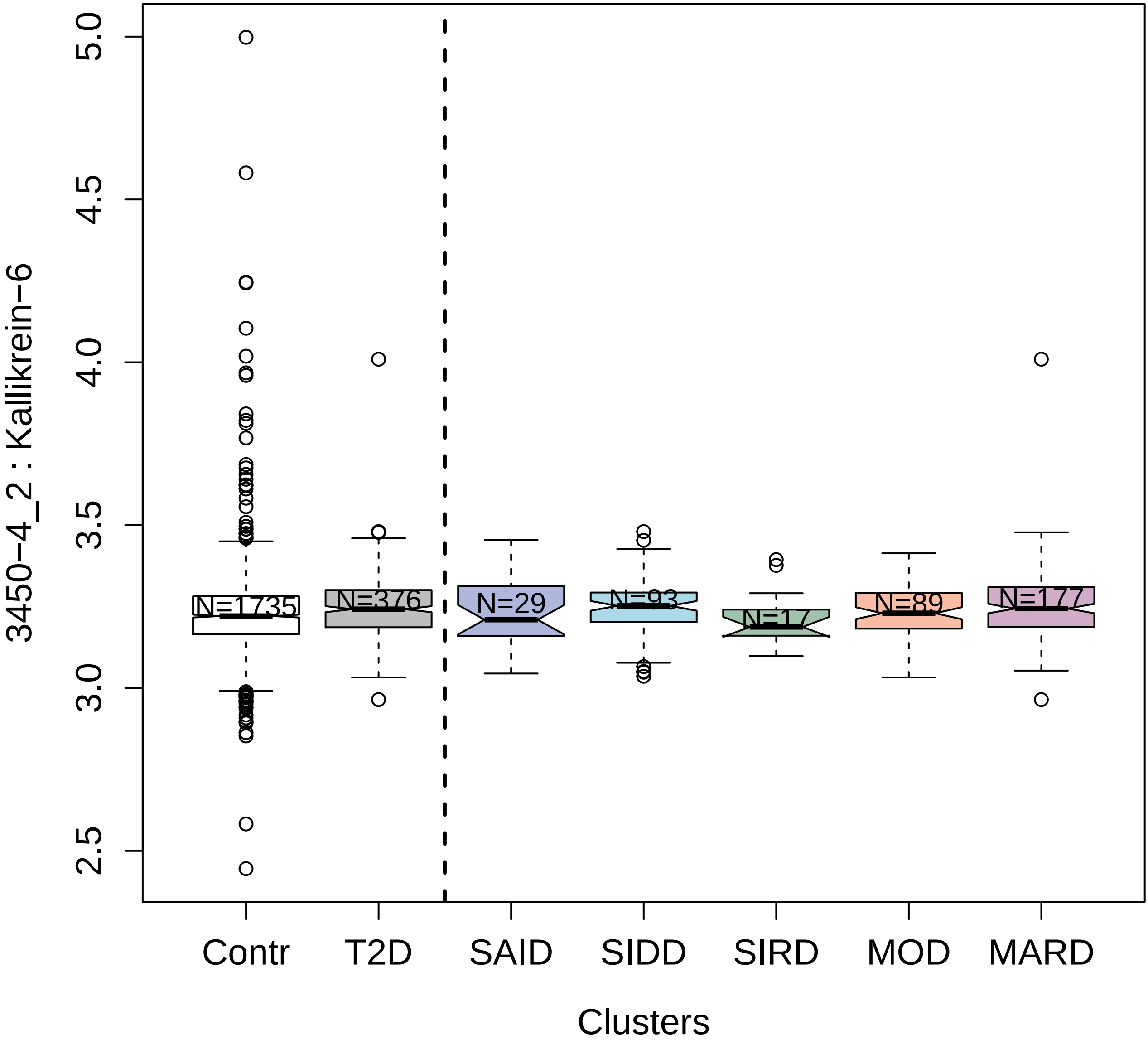
3450−4_2 : Kallikrein−6
5.0
4.5
4.0
3.5
3.0
2.5
N=1735
N=376
N=29
N=93
N=17
N=89
N=177
Contr
T2D
SAID
SIDD
SIRD
MOD
MARD
Clusters

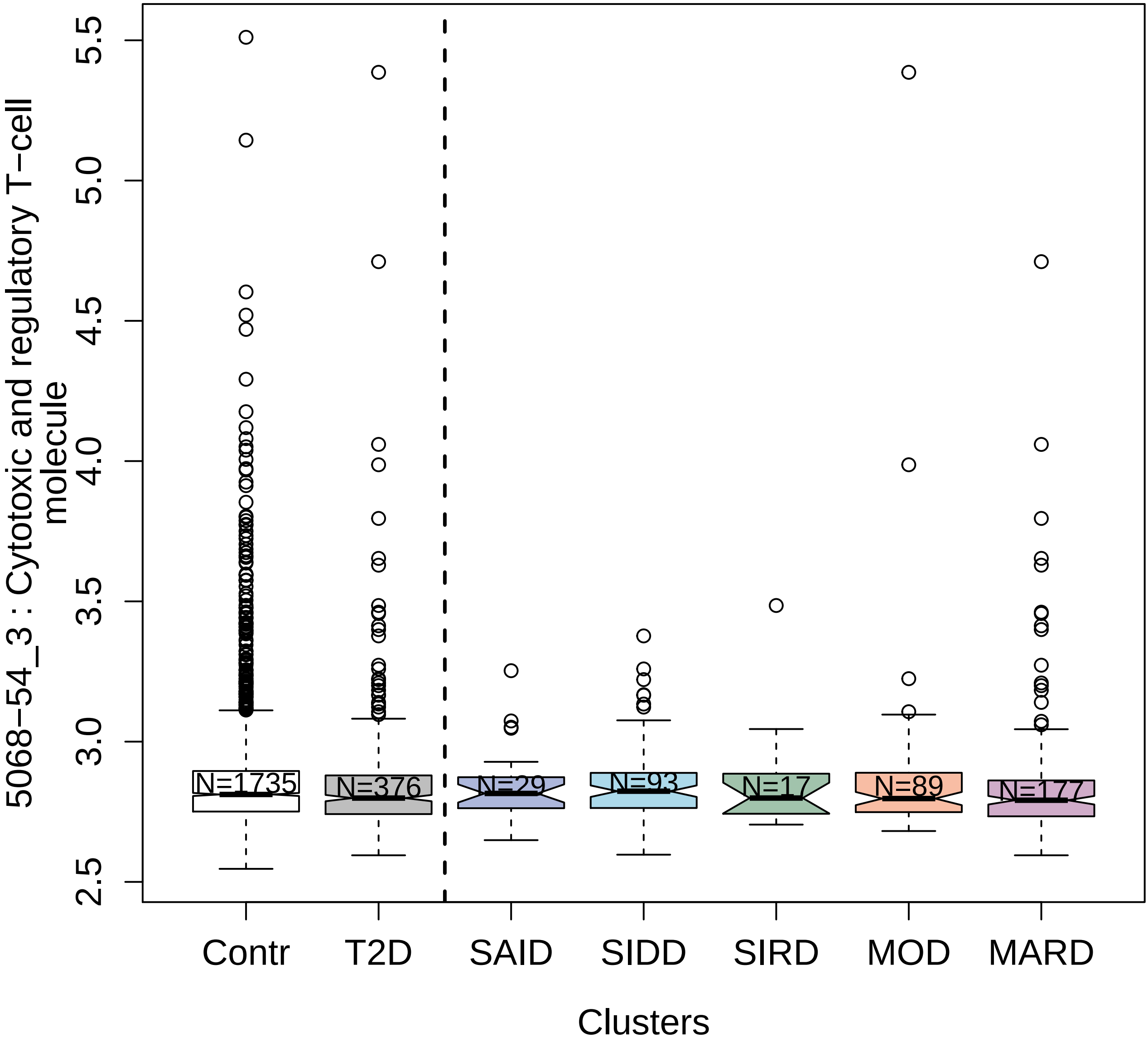
5068−54_3 : Cytotoxic and regulatory T−cell molecule
5.5
5.0
4.5
4.0
3.5
3.0
2.5
N=1735
N=376
N=29
N=93
N=17
N=89
N=177
Contr
T2D
SAID
SIDD
SIRD
MOD
MARD
Clusters

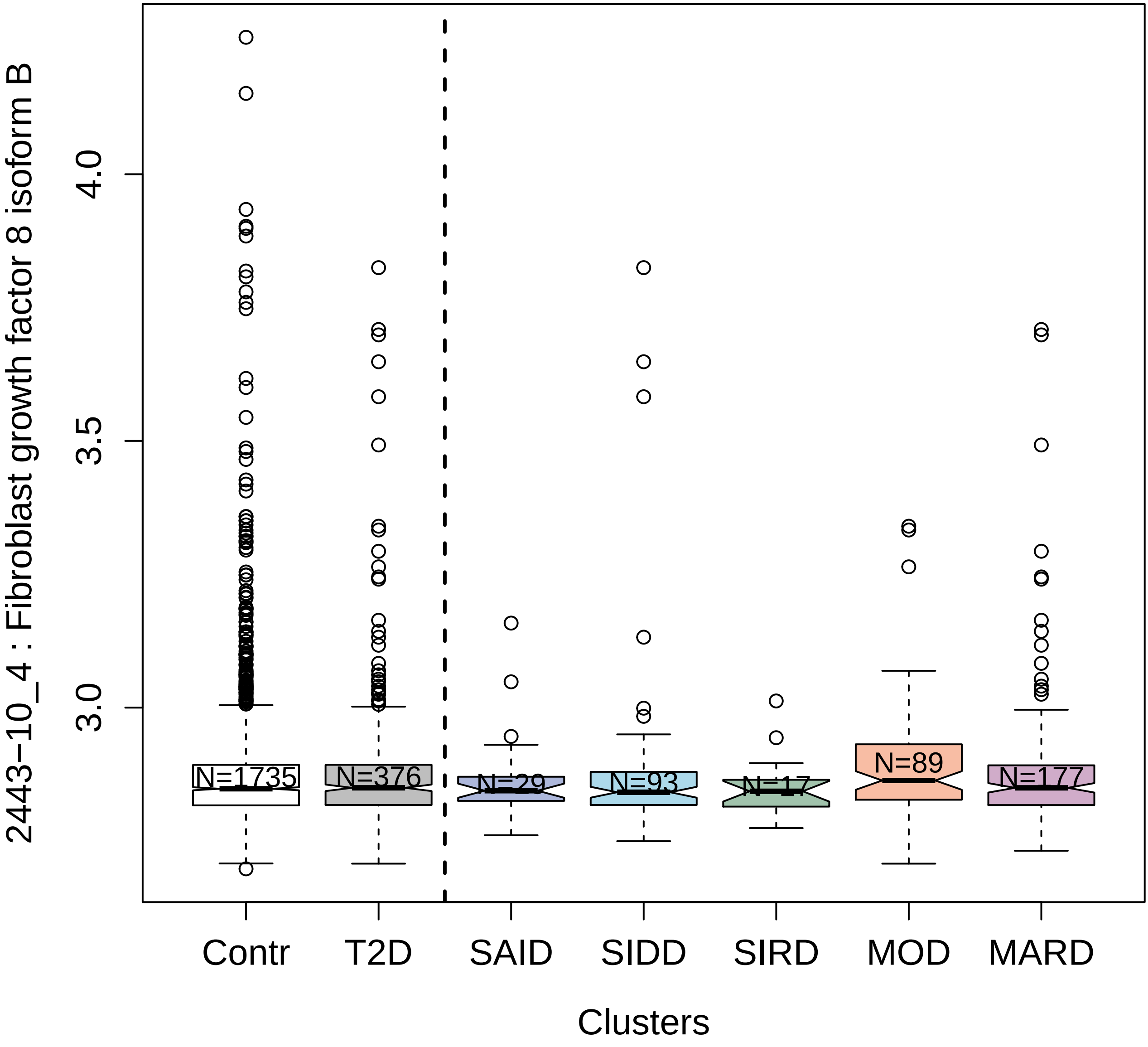

2443−10_4 : Fibroblast growth factor 8 isoform B
4.0
3.5
3.0
N=1735
N=376
N=29
N=93
N=17
N=89
N=177
Contr
T2D
SAID
SIDD
SIRD
MOD
MARD
Clusters

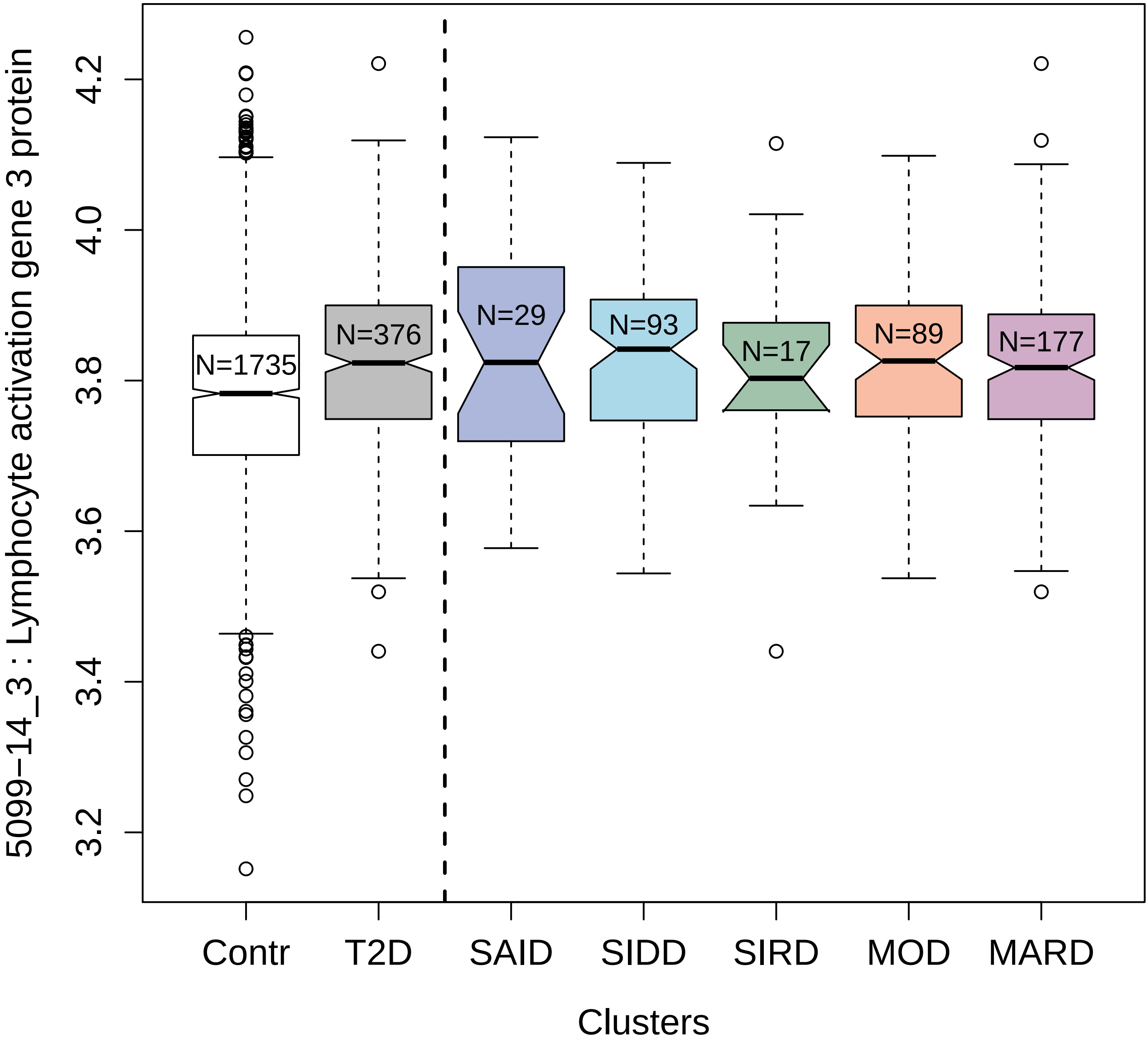
5099−14_3 : Lymphocyte activation gene 3 protein
3.2
3.4
3.6
3.8
4.0
4.2
N=1735
N=376
N=29
N=93
N=17
N=89
N=177
Contr
T2D
SAID
SIDD
SIRD
MOD
MARD
Clusters

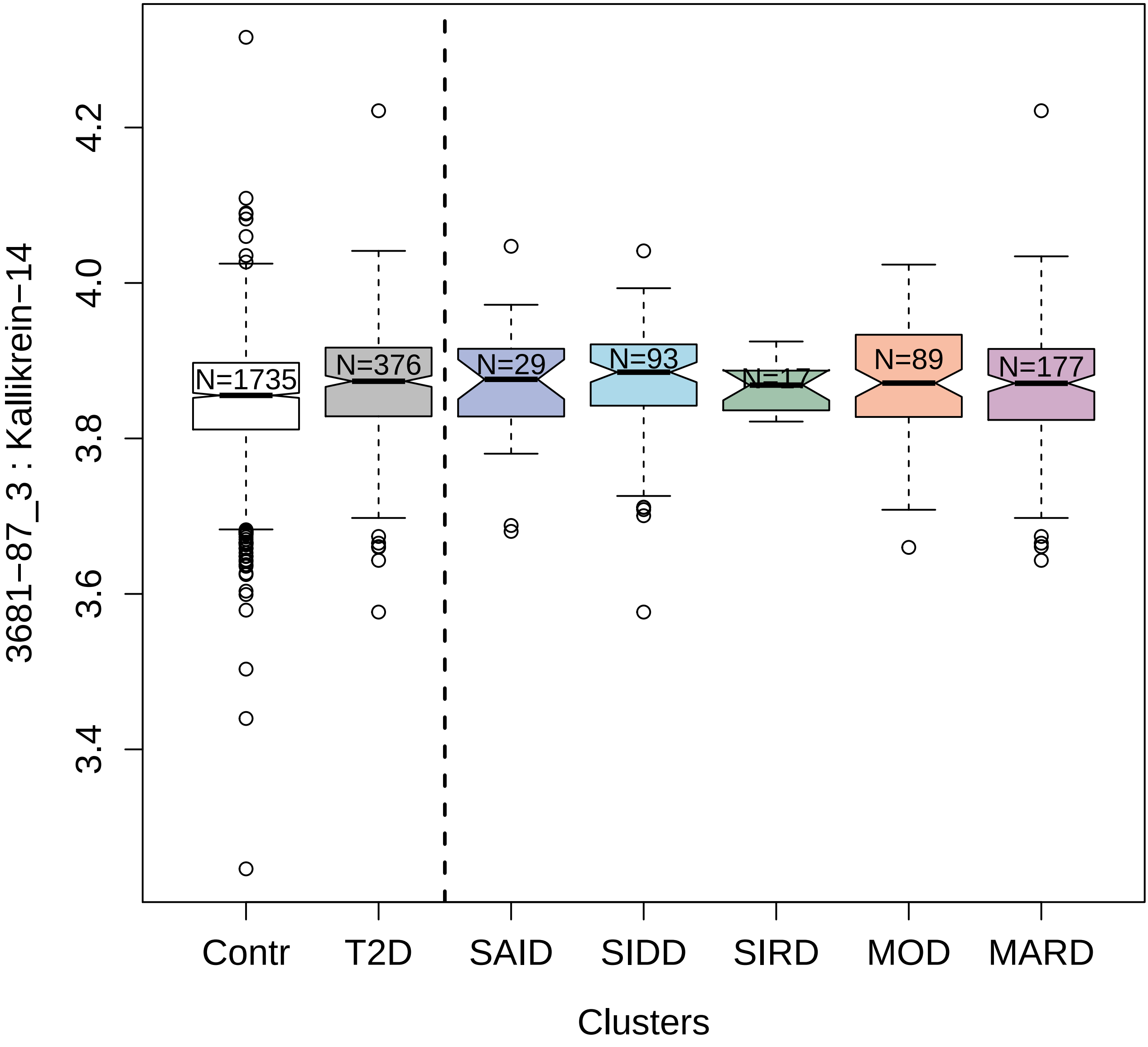

3681−87_3 : Kallikrein−14
4.2
4.0
3.8
3.6
3.4
N=1735
N=376
N=29
N=93
N=17
N=89
N=177
Contr
T2D
SAID
SIDD
SIRD
MOD
MARD
Clusters

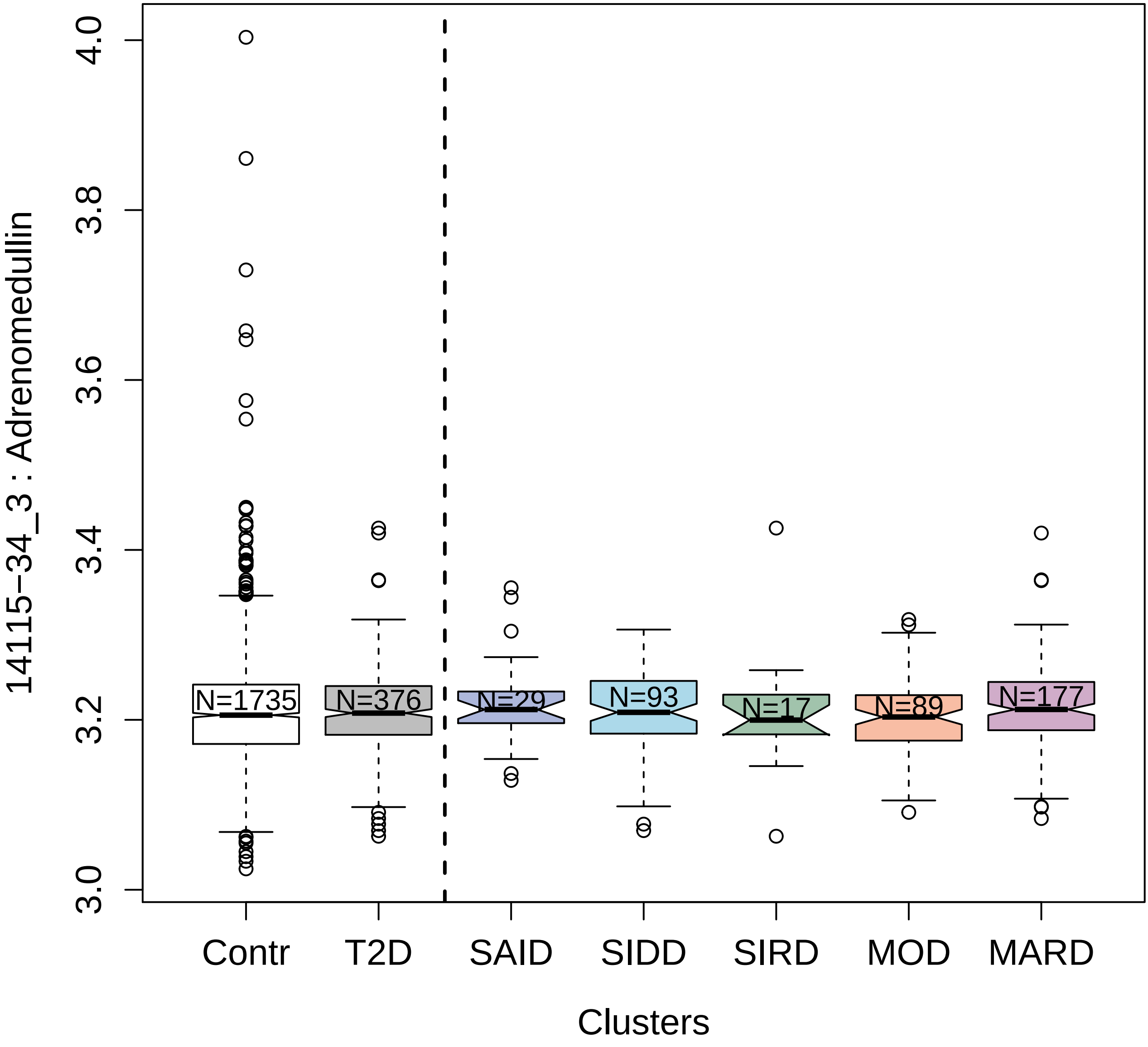

14115−34_3 : Adrenomedullin
4.0
3.8
3.6
3.4
3.2
3.0
N=1735
N=376
N=29
N=93
N=17
N=89
N=177
Contr
T2D
SAID
SIDD
SIRD
MOD
MARD
Clusters

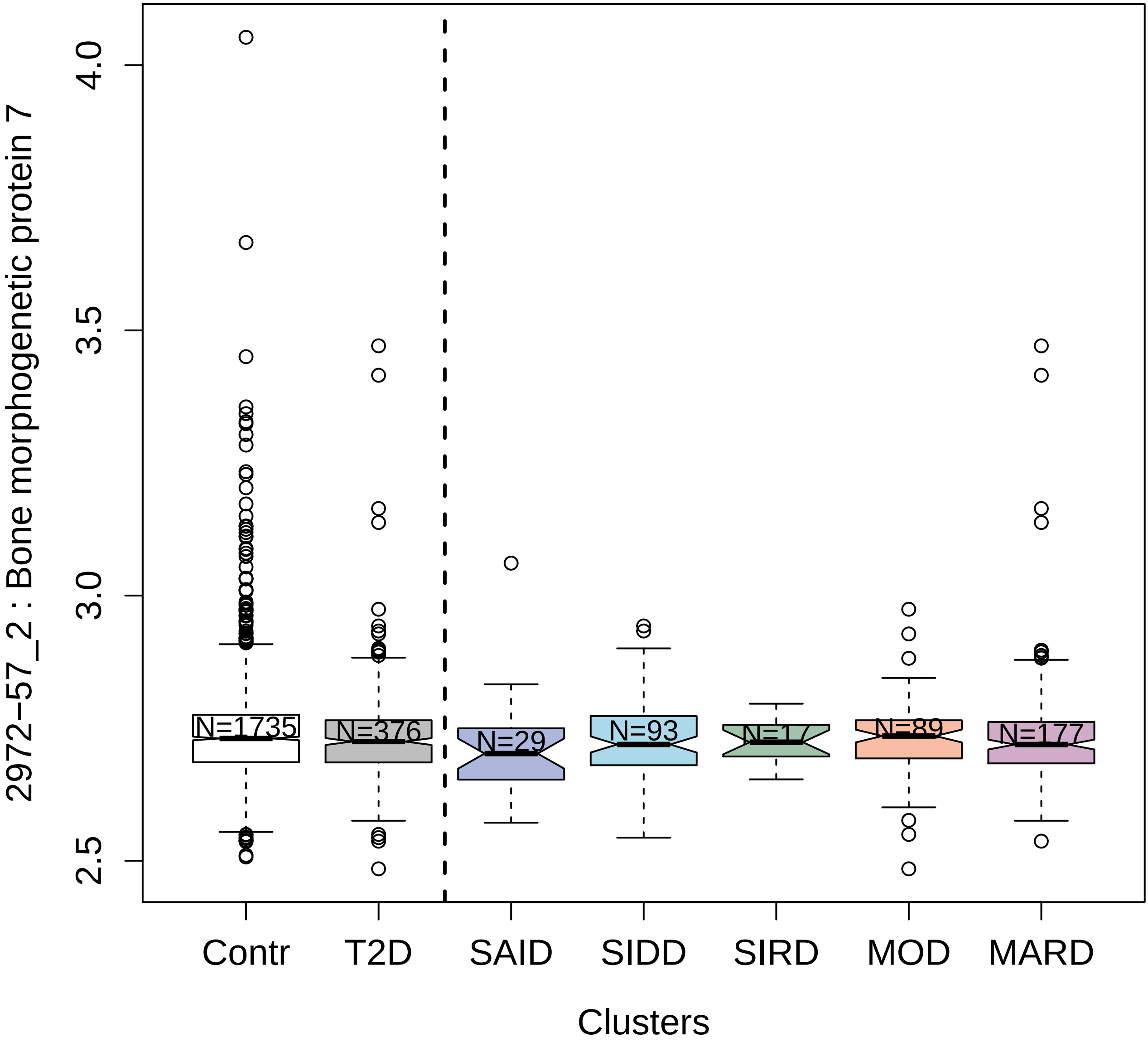
2972−57_2 : Bone morphogenetic protein 7
4.0
3.5
3.0
2.5
N=1735
N=376
N=29
N=93
N=17
N=89
N=177
Contr
T2D
SAID
SIDD
SIRD
MOD
MARD
Clusters

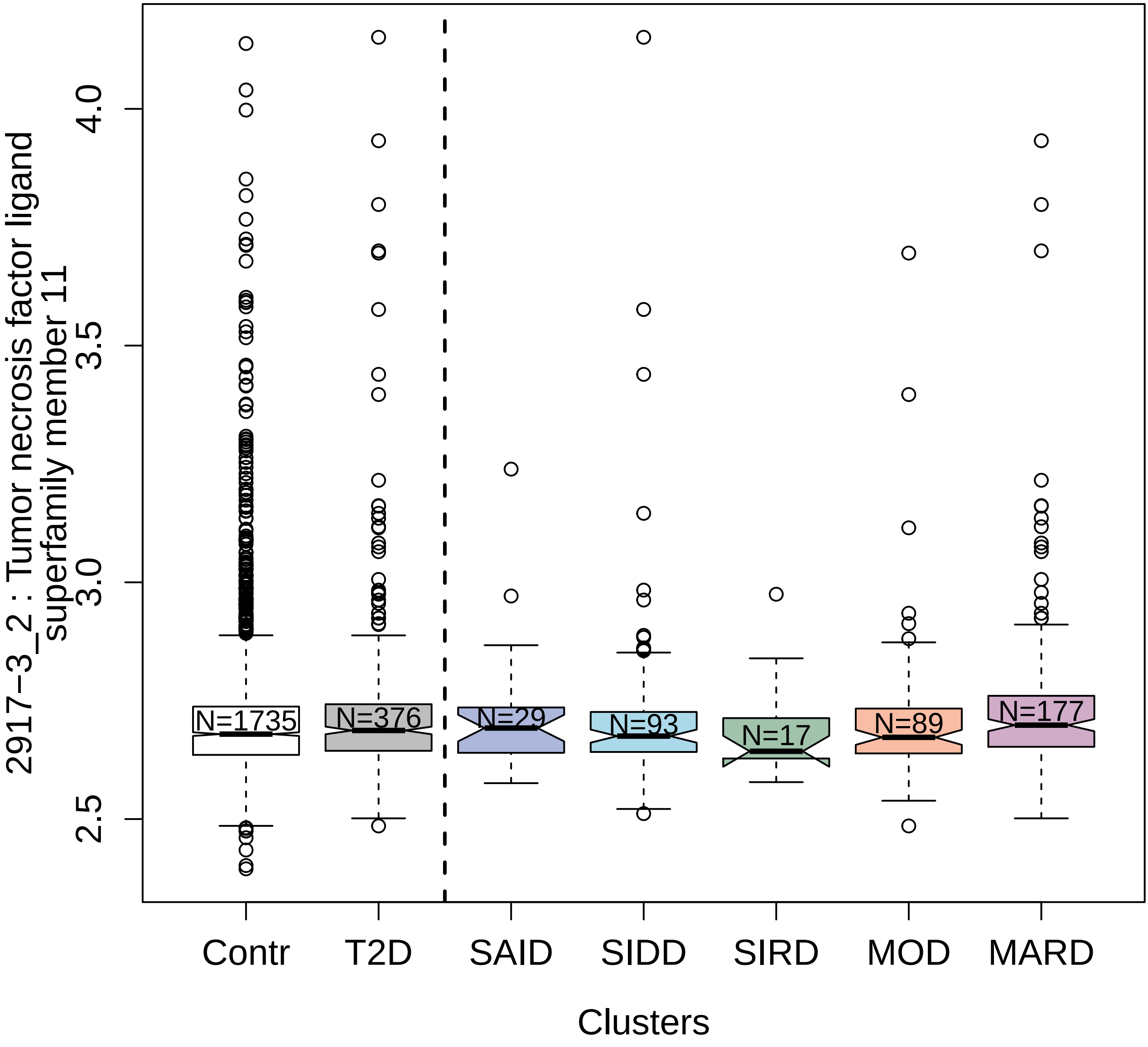

2917−3_2 : Tumor necrosis factor ligand superfamily member 11
2.5
3.0
3.5
4.0
N=1735
N=376
N=29
N=93
N=17
N=89
N=177
Contr
T2D
SAID
SIDD
SIRD
MOD
MARD
Clusters

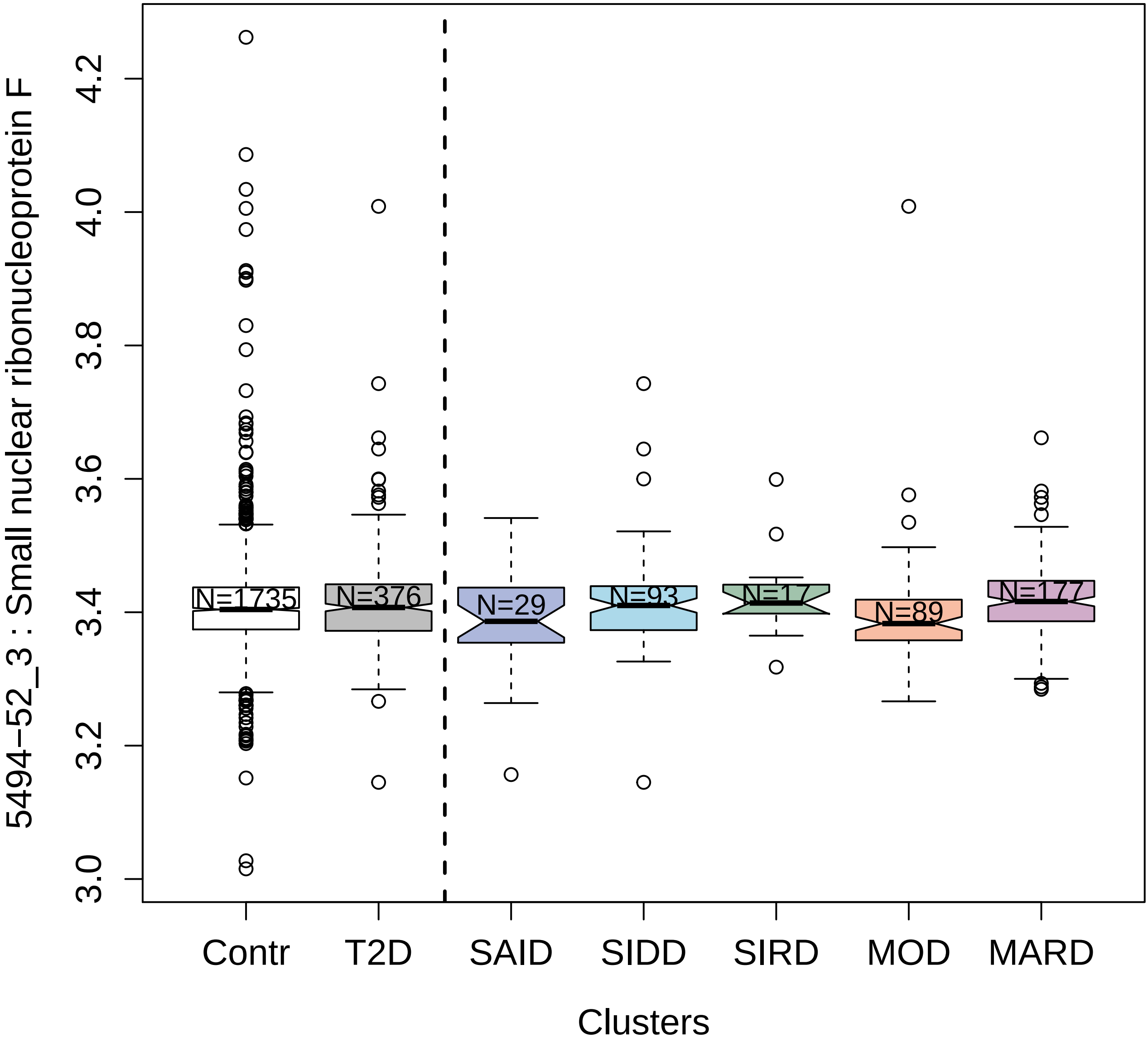
5494−52_3 : Small nuclear ribonucleoprotein F
3.0
3.2
3.4
3.6
3.8
4.0
4.2
N=1735
N=376
N=29
N=93
N=17
N=89
N=177
Contr
T2D
SAID
SIDD
SIRD
MOD
MARD
Clusters

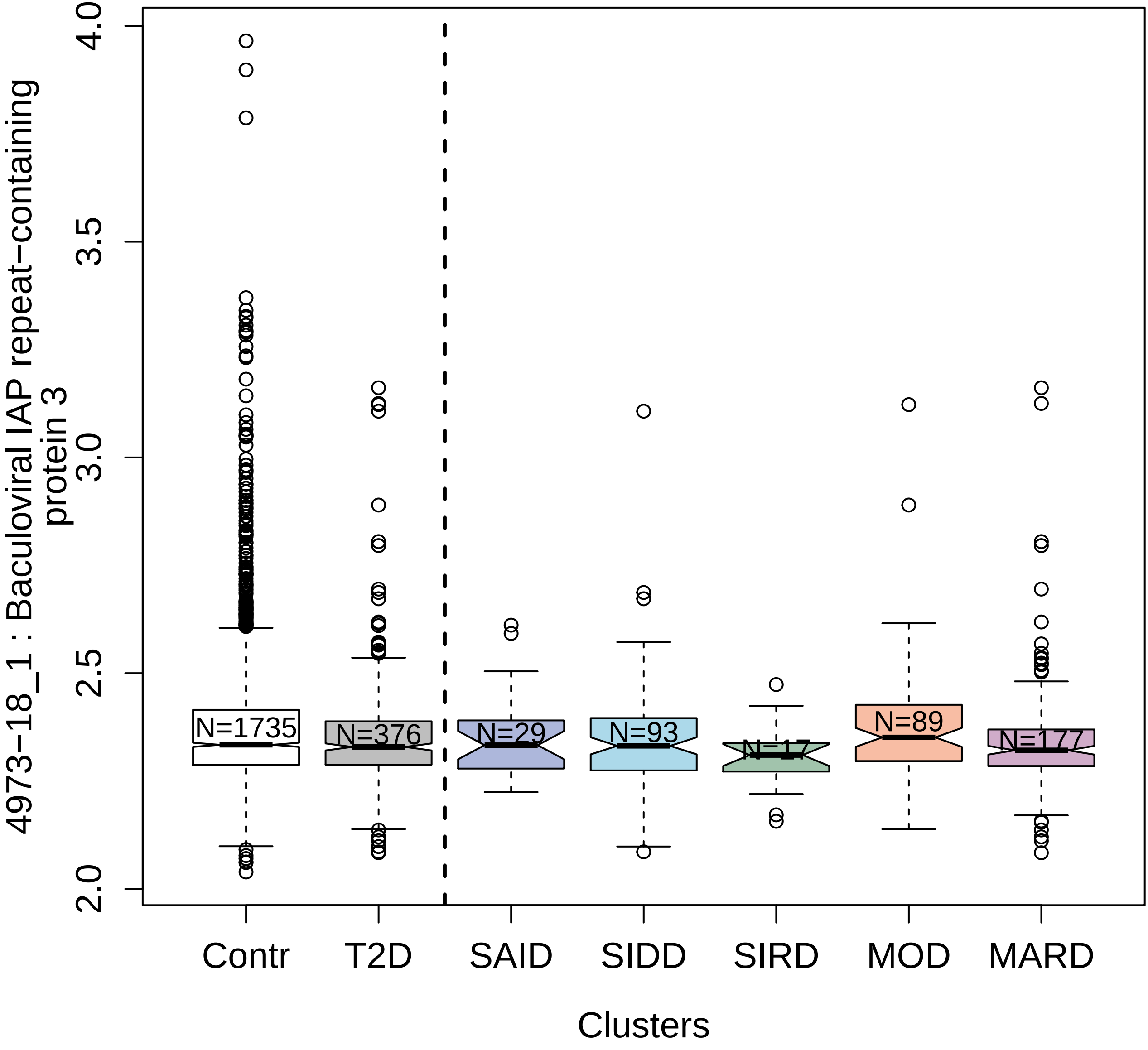

4973−18_1 : Baculoviral IAP repeat−containing protein 3
4.0
3.5
3.0
2.5
2.0
N=1735
N=376
N=29
N=93
N=17
N=89
N=177
Contr
T2D
SAID
SIDD
SIRD
MOD
MARD
Clusters

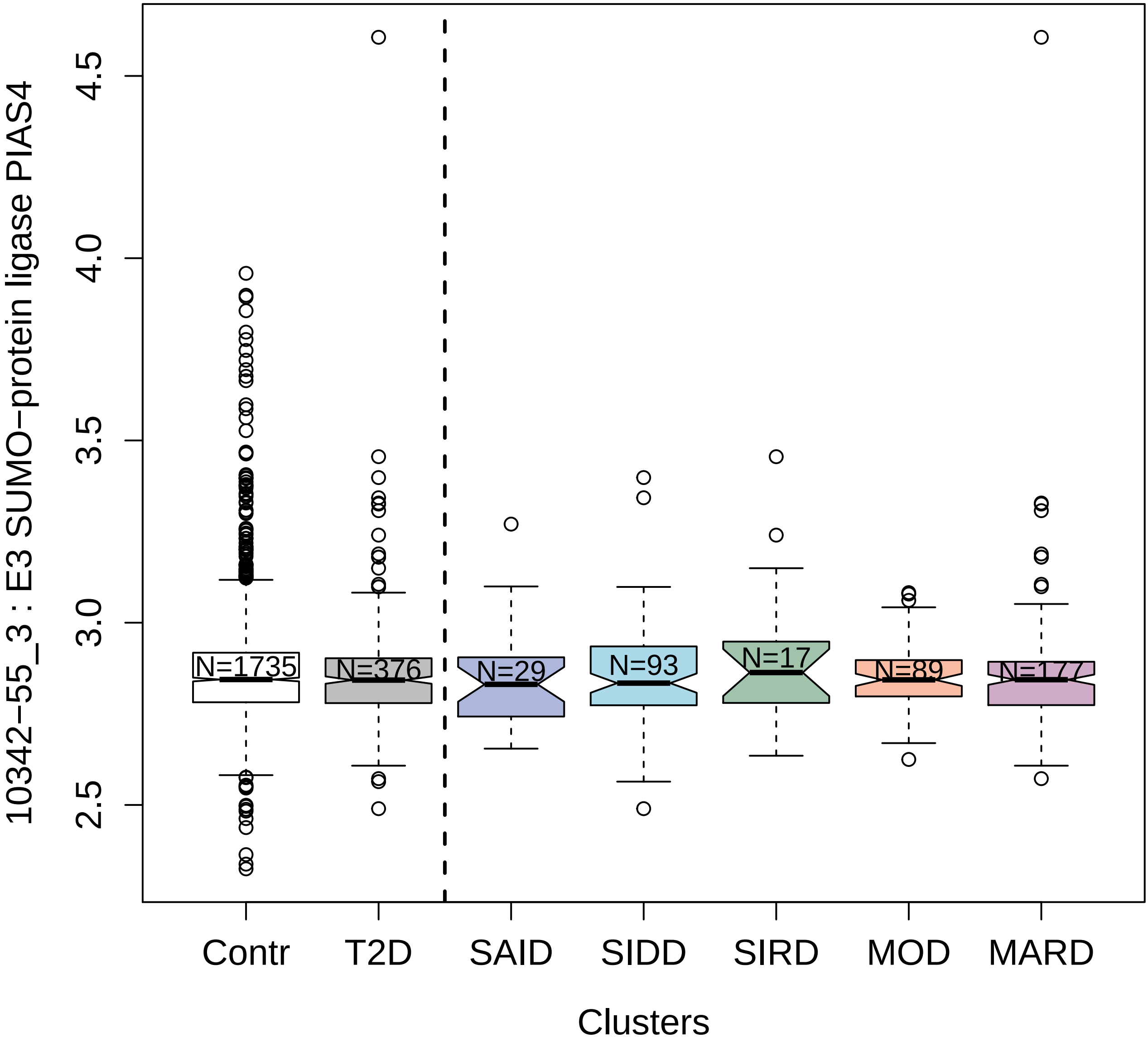

10342−55_3 : E3 SUMO−protein ligase PIAS4
2.5
3.0
3.5
4.0
4.5
N=1735
N=376
N=29
N=93
N=17
N=89
N=177
Contr
T2D
SAID
SIDD
SIRD
MOD
MARD
Clusters

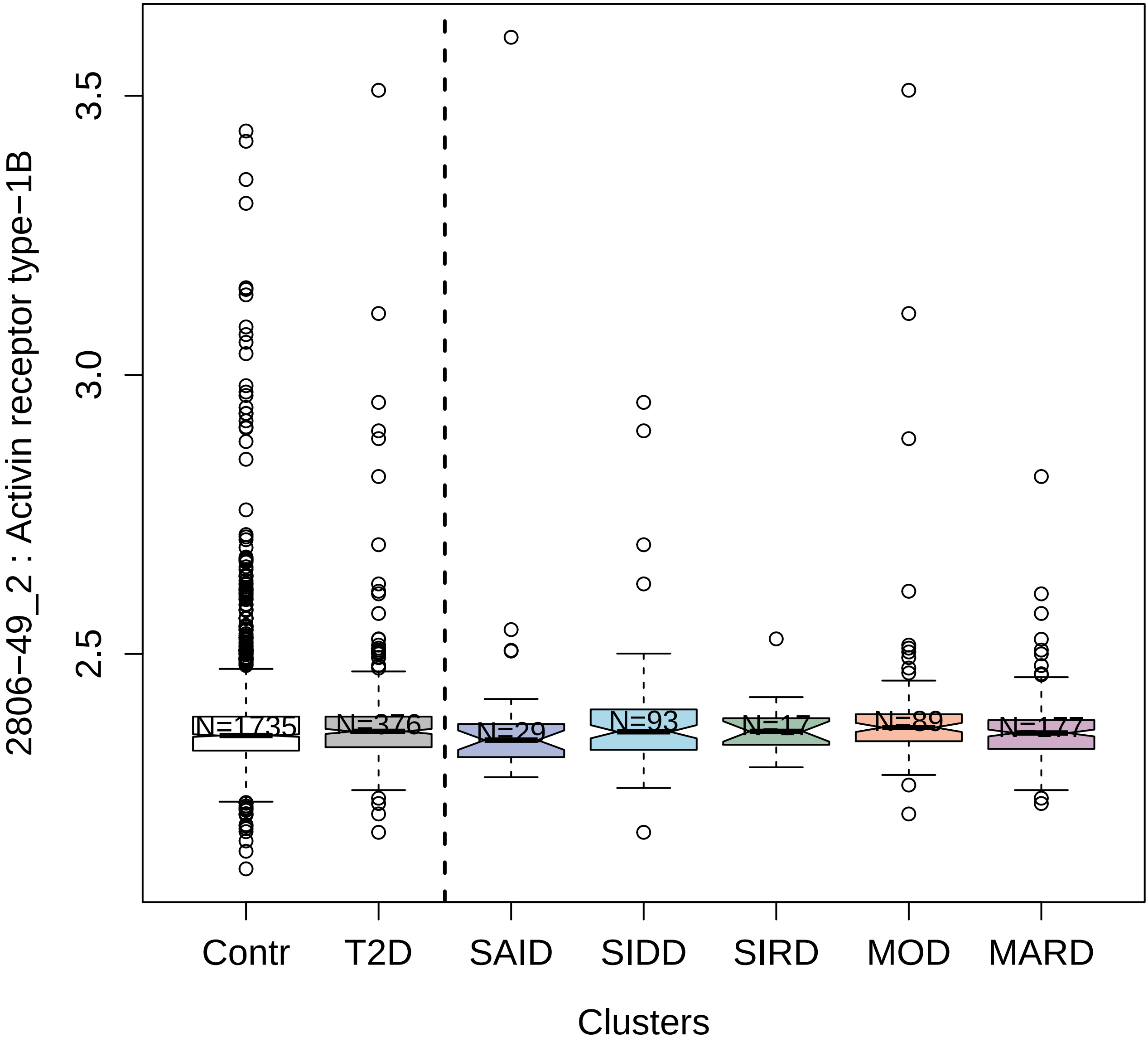

2806−49_2 : Activin receptor type−1B
3.5
3.0
2.5
N=1735
N=376
N=29
N=93
N=17
N=89
N=177
Contr
T2D
SAID
SIDD
SIRD
MOD
MARD
Clusters

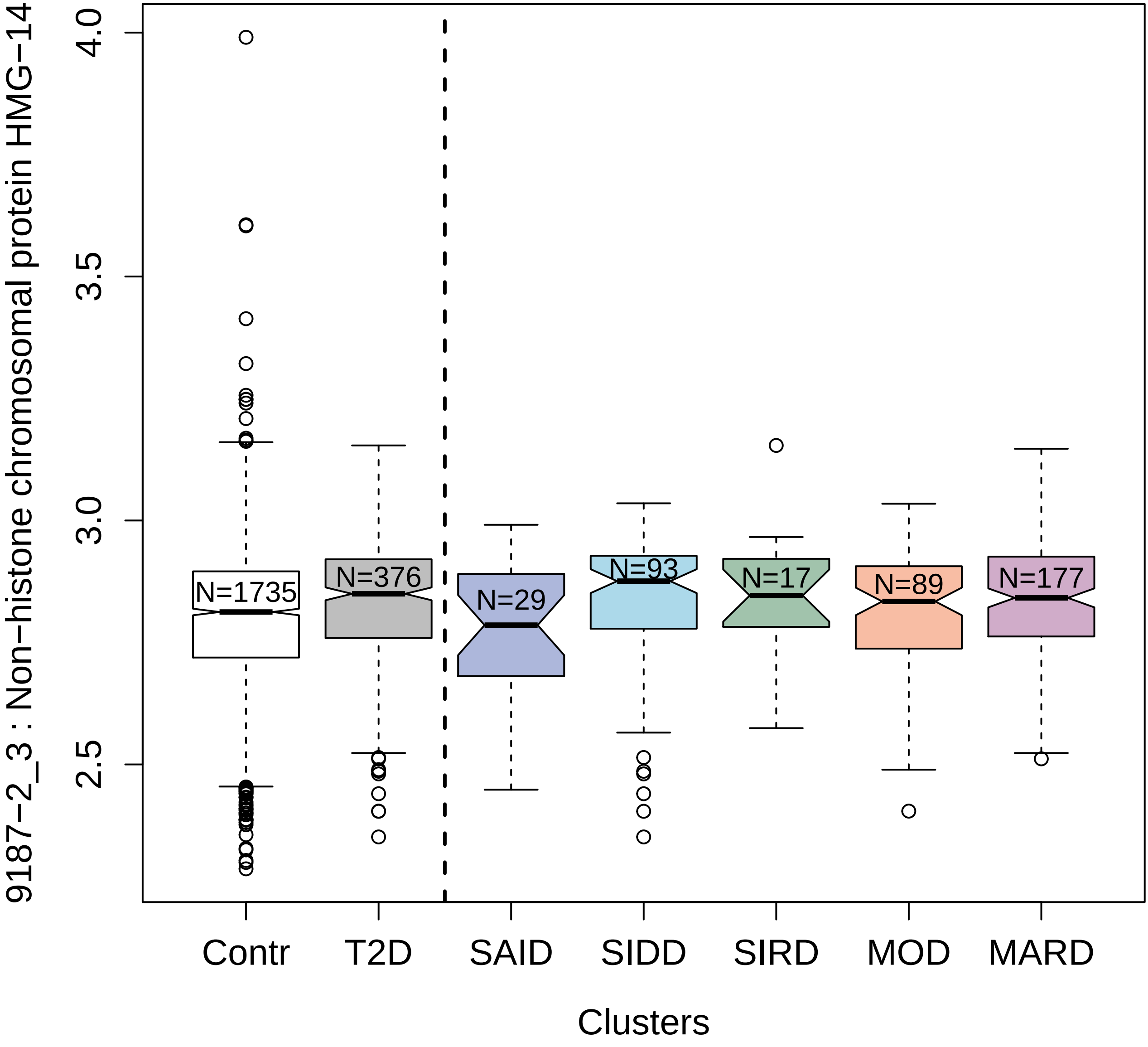

9187−2_3 : Non−histone chromosomal protein HMG−14
4.0
3.5
3.0
2.5
N=1735
N=376
N=29
N=93
N=17
N=89
N=177
Contr
T2D
SAID
SIDD
SIRD
MOD
MARD
Clusters

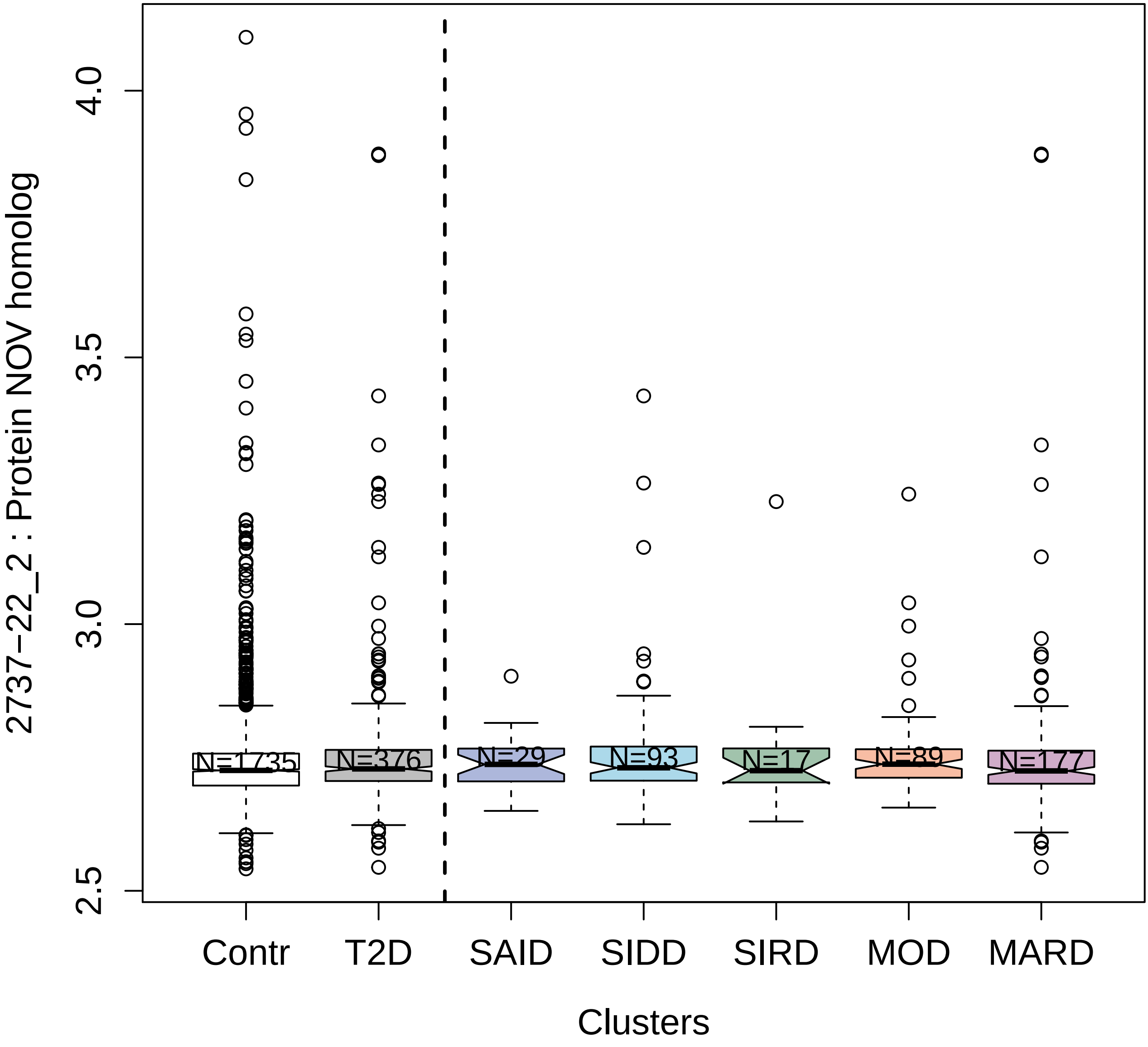

2737−22_2 : Protein NOV homolog
4.0
3.5
3.0
2.5
N=1735
N=376
N=29
N=93
N=17
N=89
N=177
Contr
T2D
SAID
SIDD
SIRD
MOD
MARD
Clusters

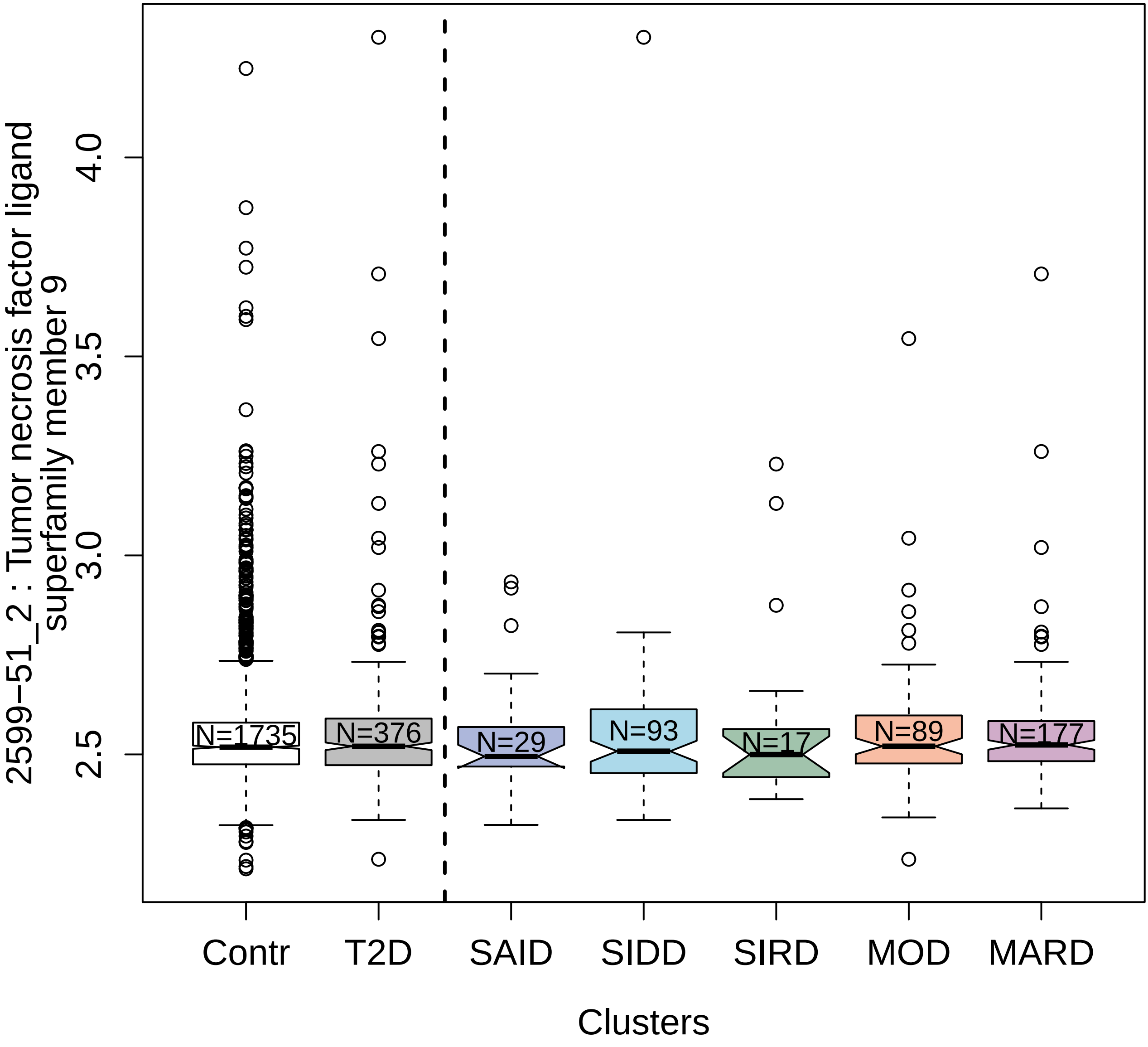

2599−51_2 : Tumor necrosis factor ligand superfamily member 9
2.5
3.0
3.5
4.0
N=1735
N=376
N=29
N=93
N=17
N=89
N=177
Contr
T2D
SAID
SIDD
SIRD
MOD
MARD
Clusters

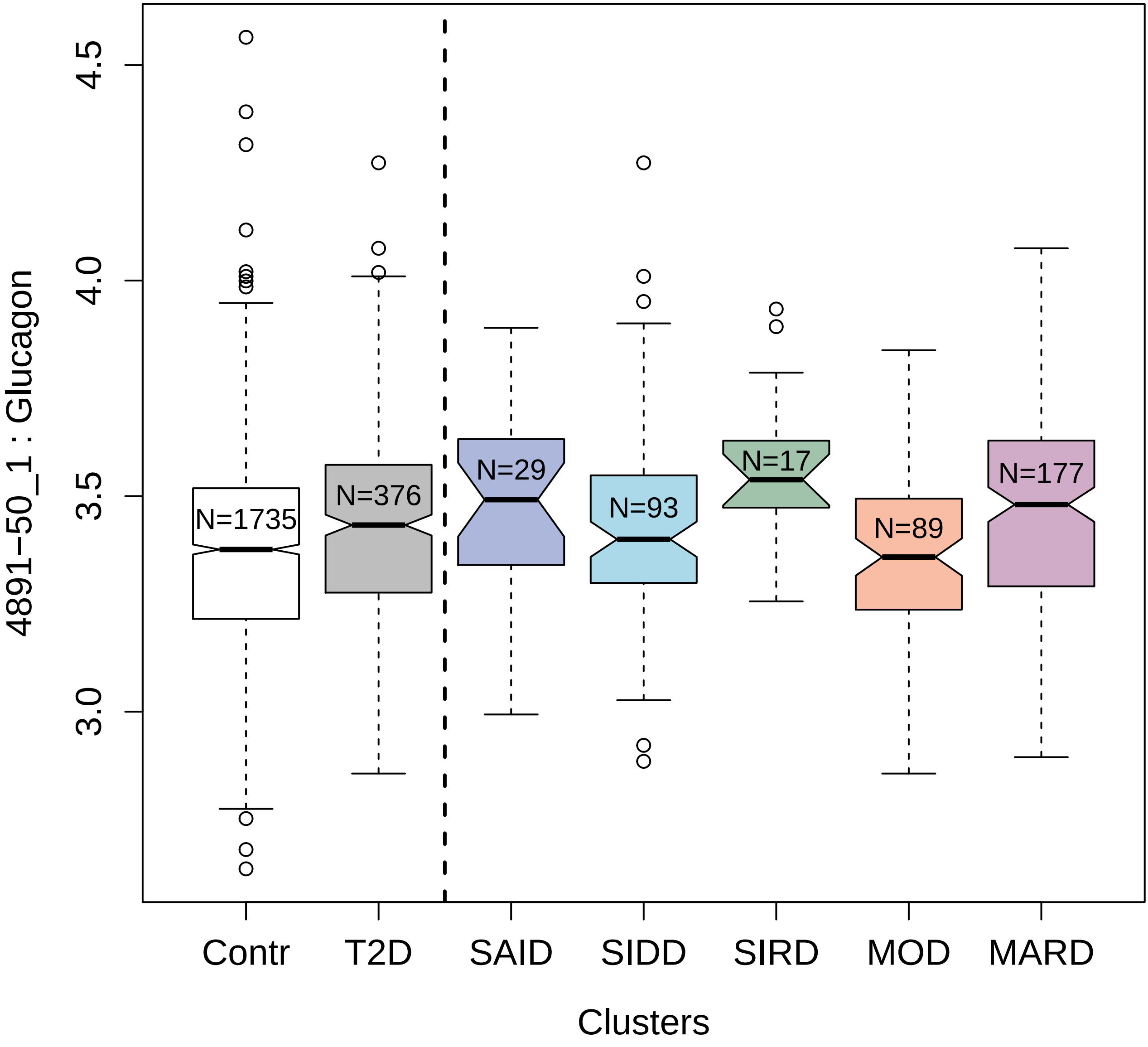
4891−50_1 : Glucagon
4.5
4.0
3.5
3.0
N=1735
N=376
N=29
N=93
N=17
N=89
N=177
Contr
T2D
SAID
SIDD
SIRD
MOD
MARD
Clusters

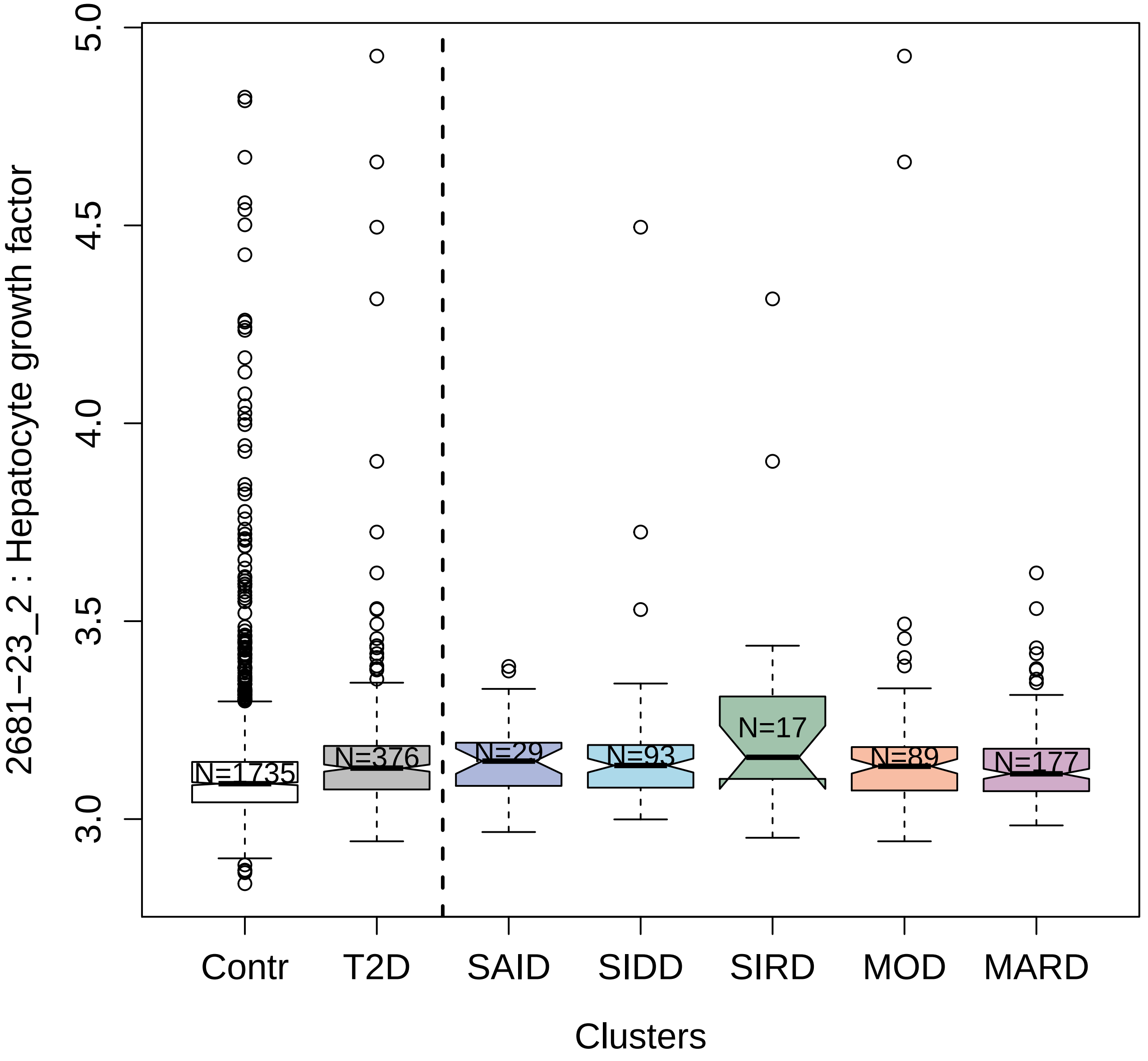
2681−23_2 : Hepatocyte growth factor
5.0
4.5
4.0
3.5
3.0
N=1735
N=376
N=29
N=93
N=17
N=89
N=177
Contr
T2D
SAID
SIDD
SIRD
MOD
MARD
Clusters

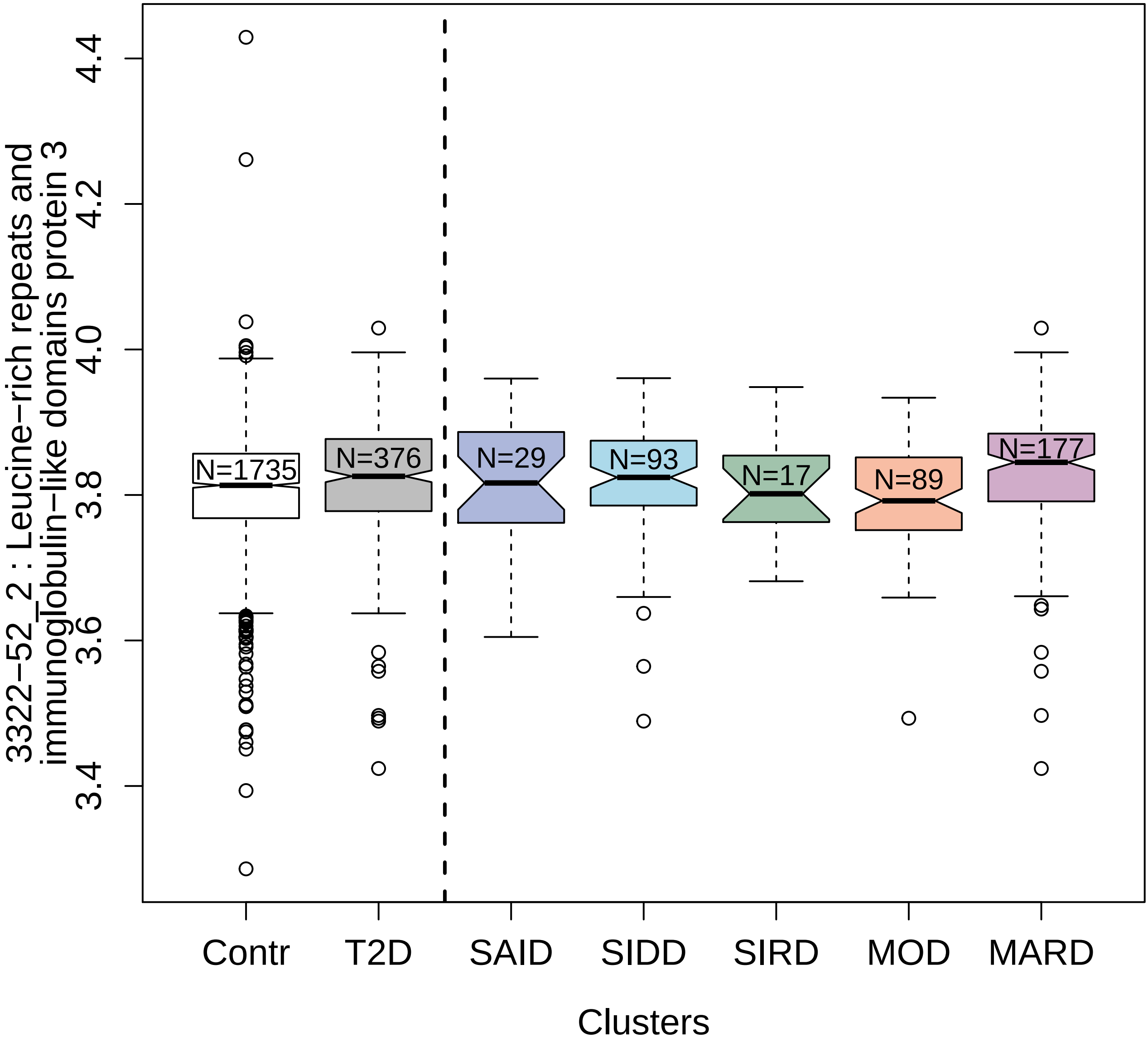

3322−52_2 : Leucine−rich repeats and immunoglobulin−like domains protein 3
3.4
3.6
3.8
4.0
4.2
4.4
N=1735
N=376
N=29
N=93
N=17
N=89
N=177
Contr
T2D
SAID
SIDD
SIRD
MOD
MARD
Clusters

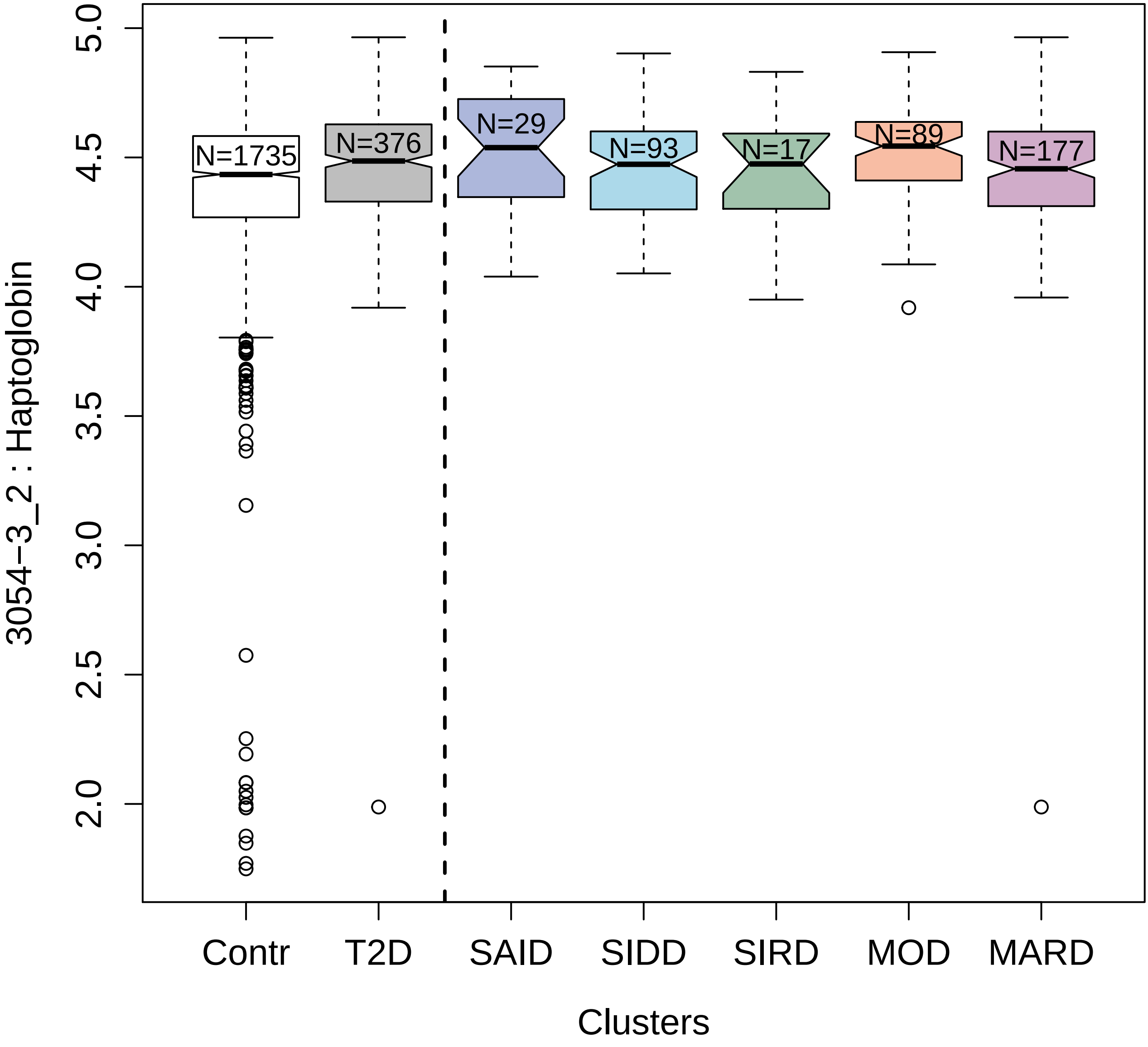
3054−3_2 : Haptoglobin
5.0
4.5
4.0
3.5
3.0
2.5
2.0
N=1735
N=376
N=29
N=93
N=17
N=89
N=177
Contr
T2D
SAID
SIDD
SIRD
MOD
MARD
Clusters

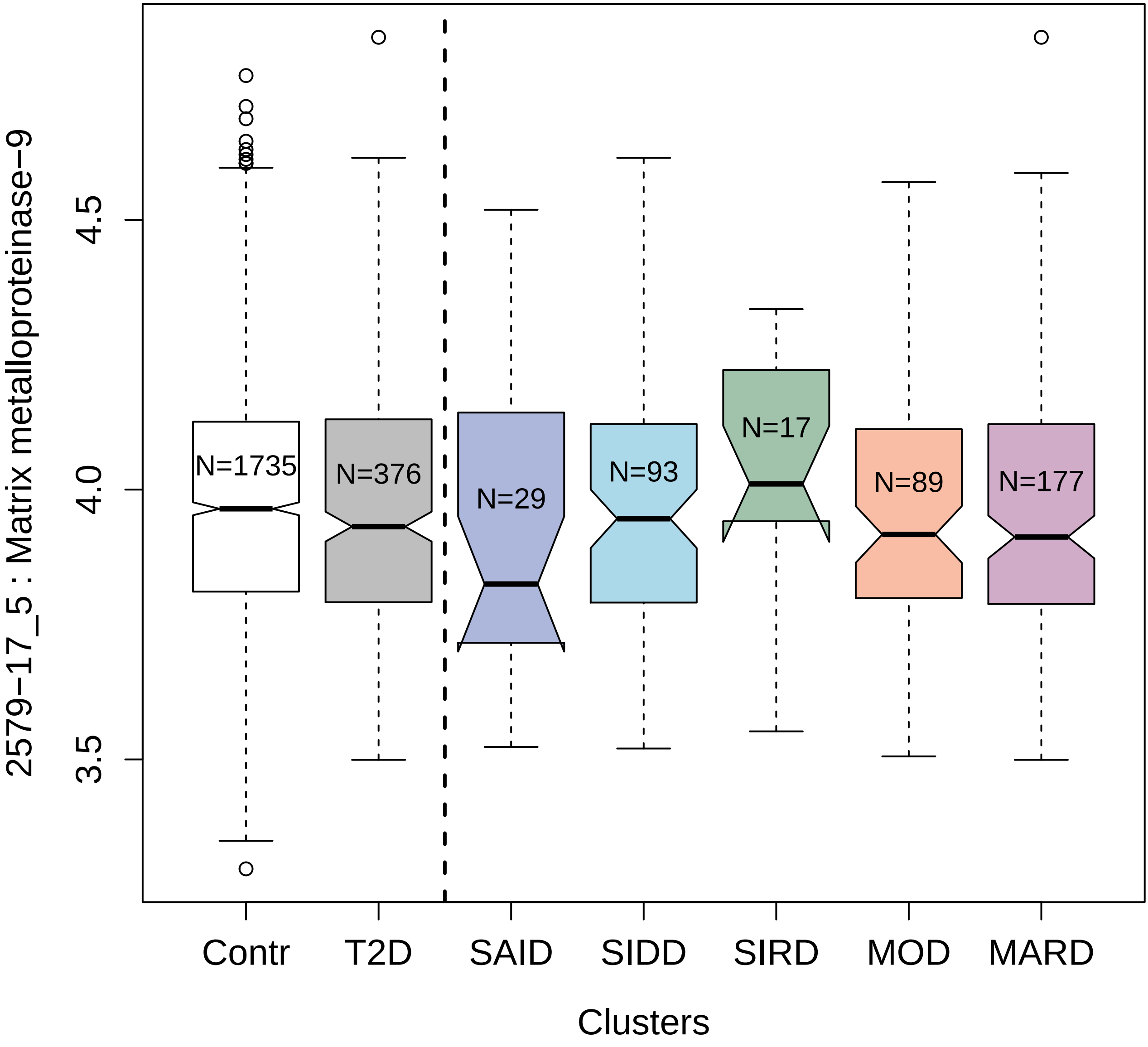
2579−17_5 : Matrix metalloproteinase−9
4.5
4.0
3.5
N=1735
N=376
N=29
N=93
N=17
N=89
N=177
Contr
T2D
SAID
SIDD
SIRD
MOD
MARD
Clusters

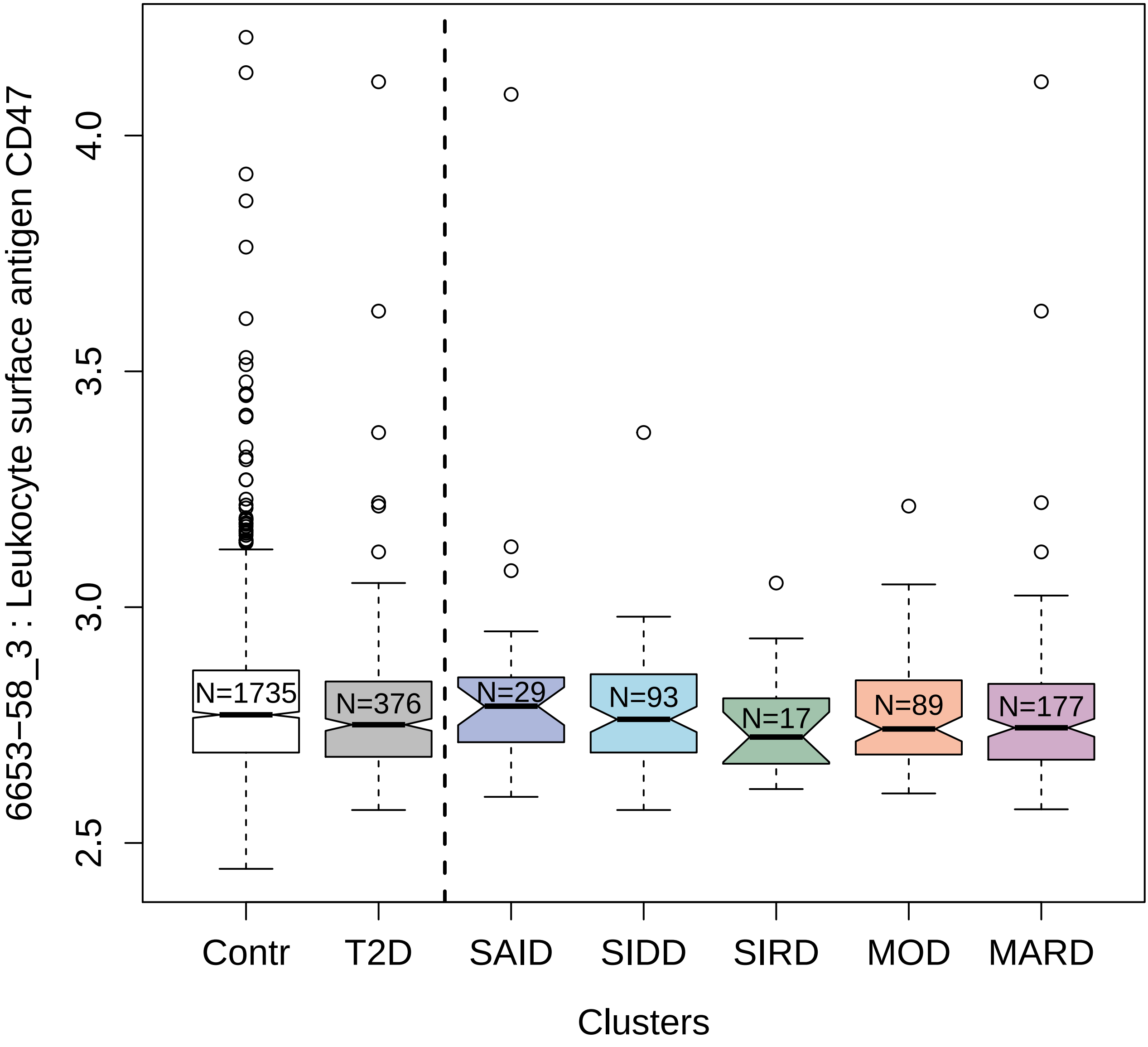

6653−58_3 : Leukocyte surface antigen CD47
4.0
3.5
3.0
2.5
N=1735
N=376
N=29
N=93
N=17
N=89
N=177
Contr
T2D
SAID
SIDD
SIRD
MOD
MARD
Clusters

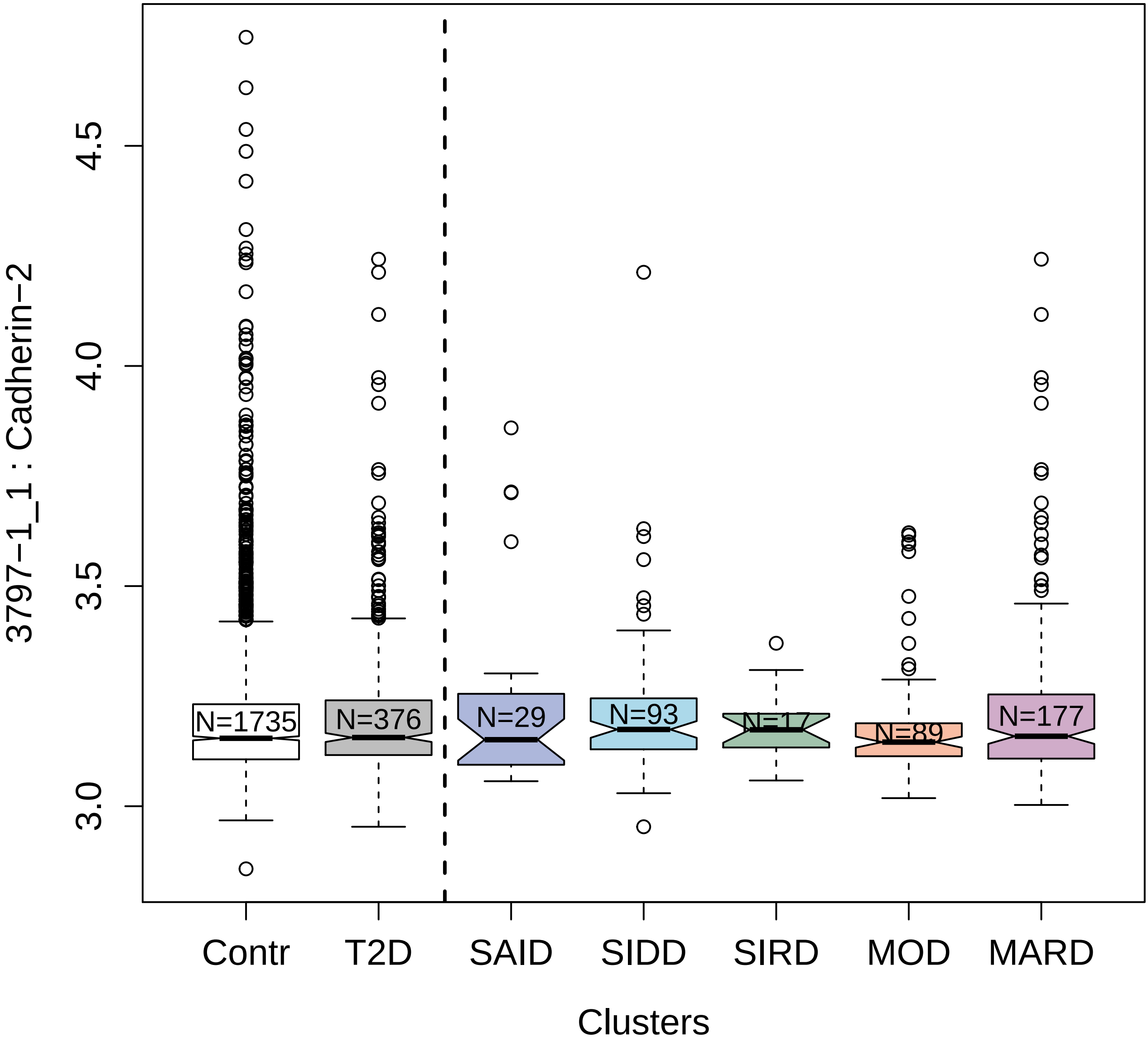

3797−1_1 : Cadherin−2
4.5
4.0
3.5
3.0
N=1735
N=376
N=29
N=93
N=17
N=89
N=177
Contr
T2D
SAID
SIDD
SIRD
MOD
MARD
Clusters

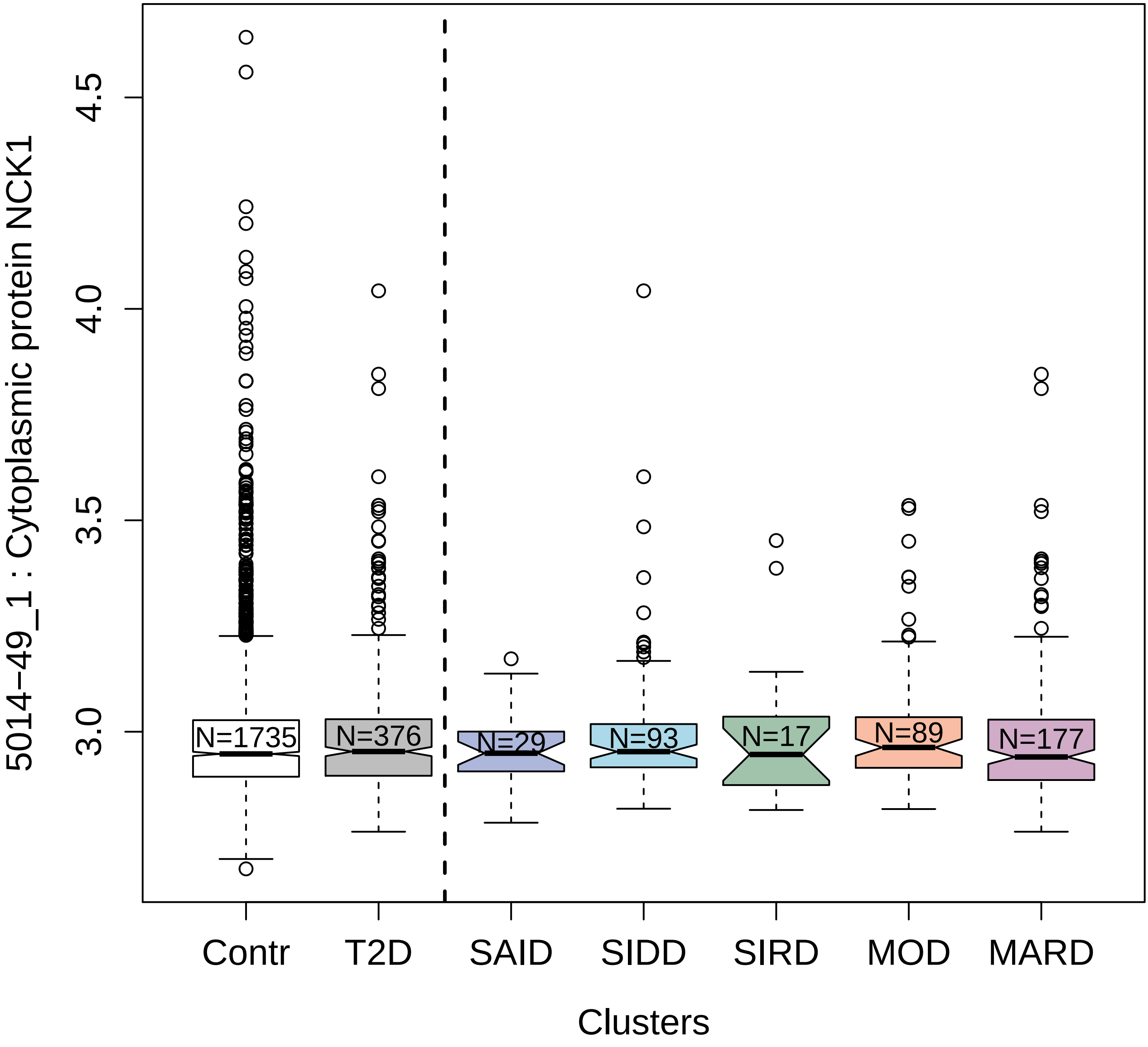

5014−49_1 : Cytoplasmic protein NCK1
4.5
4.0
3.5
3.0
N=1735
N=376
N=29
N=93
N=17
N=89
N=177
Contr
T2D
SAID
SIDD
SIRD
MOD
MARD
Clusters

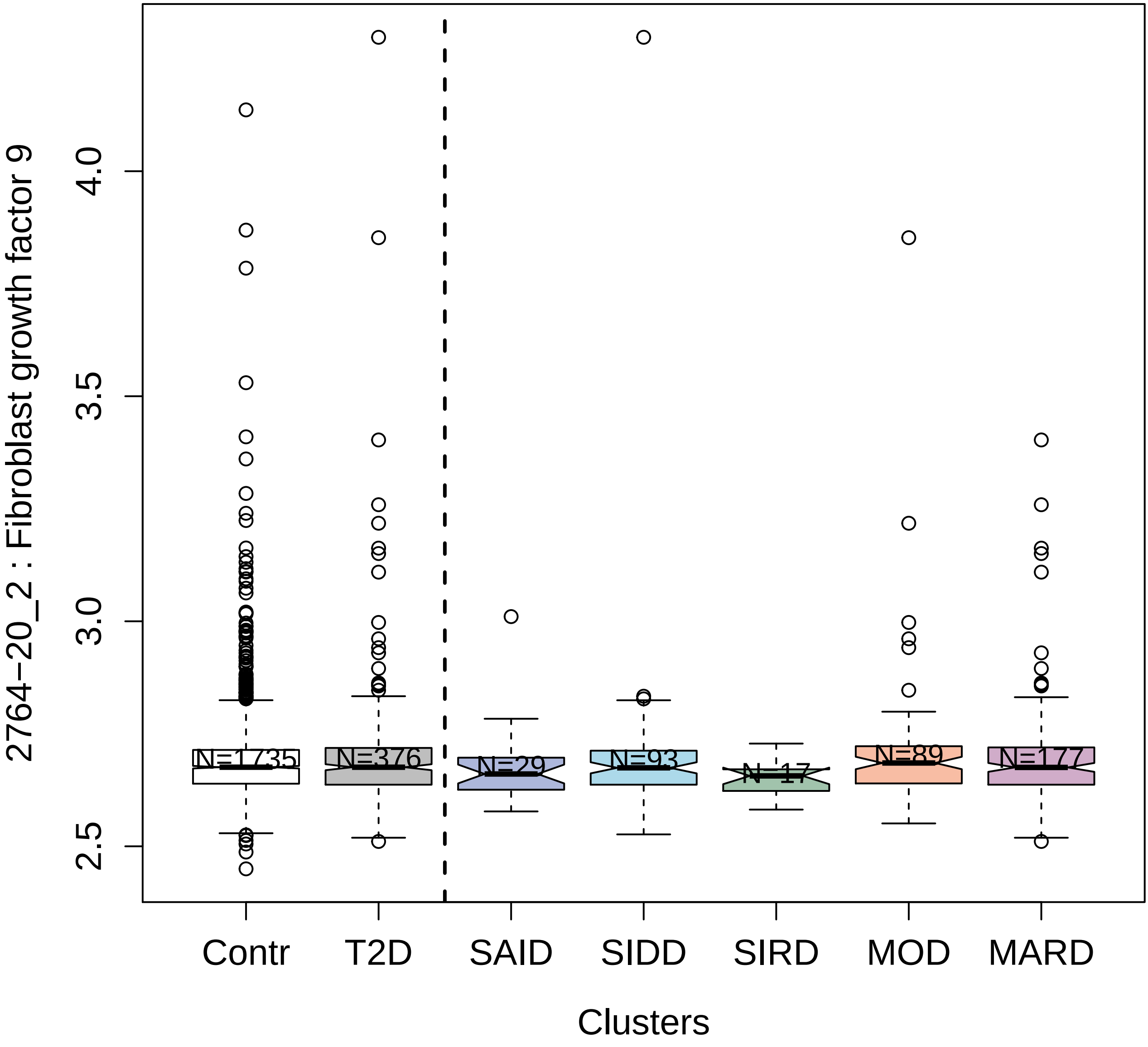

2764−20_2 : Fibroblast growth factor 9
2.5
3.0
3.5
4.0
N=1735
N=376
N=29
N=93
N=17
N=89
N=177
Contr
T2D
SAID
SIDD
SIRD
MOD
MARD
Clusters

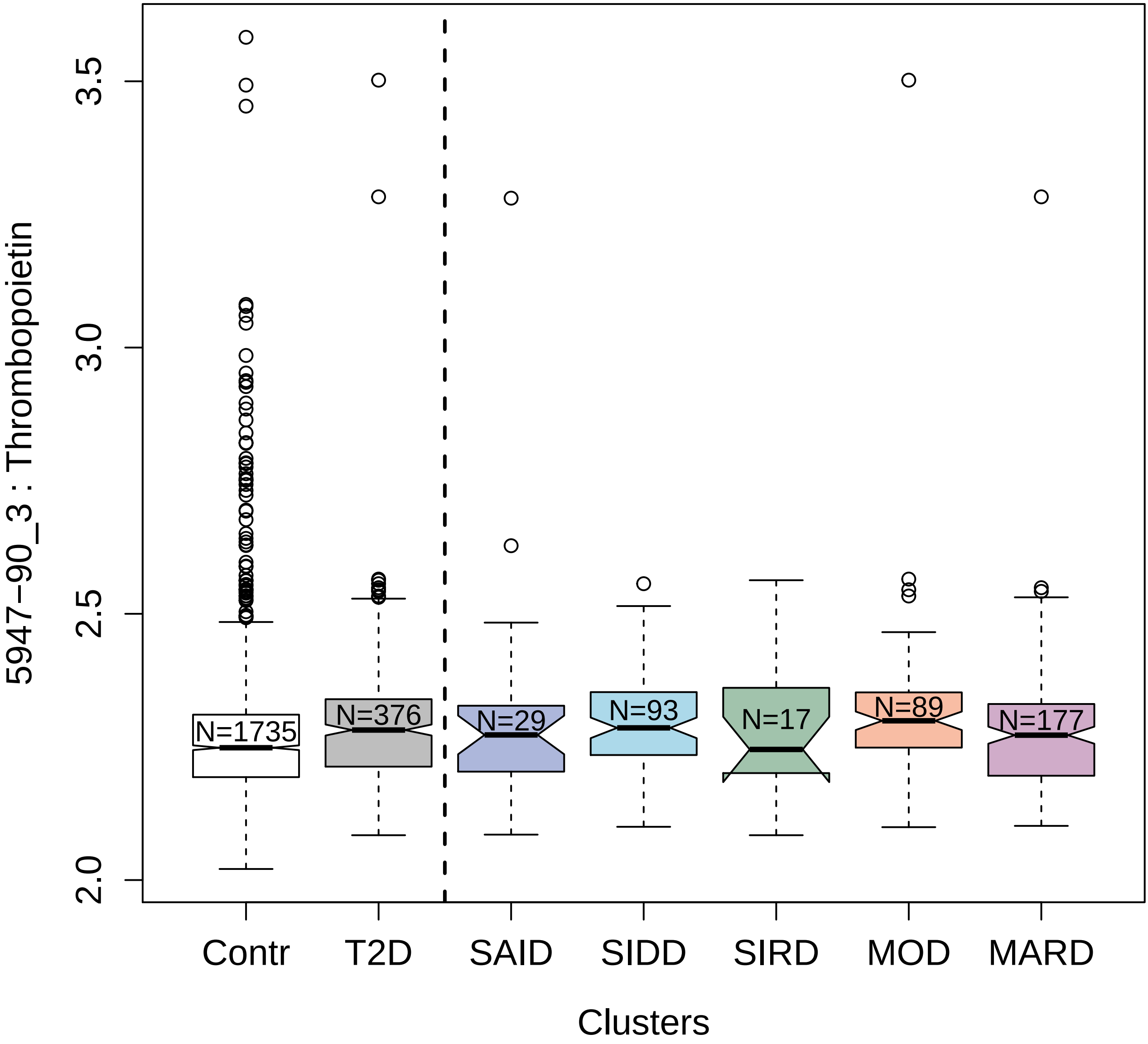

5947−90_3 : Thrombopoietin
2.0
2.5
3.0
3.5
N=1735
N=376
N=29
N=93
N=17
N=89
N=177
Contr
T2D
SAID
SIDD
SIRD
MOD
MARD
Clusters

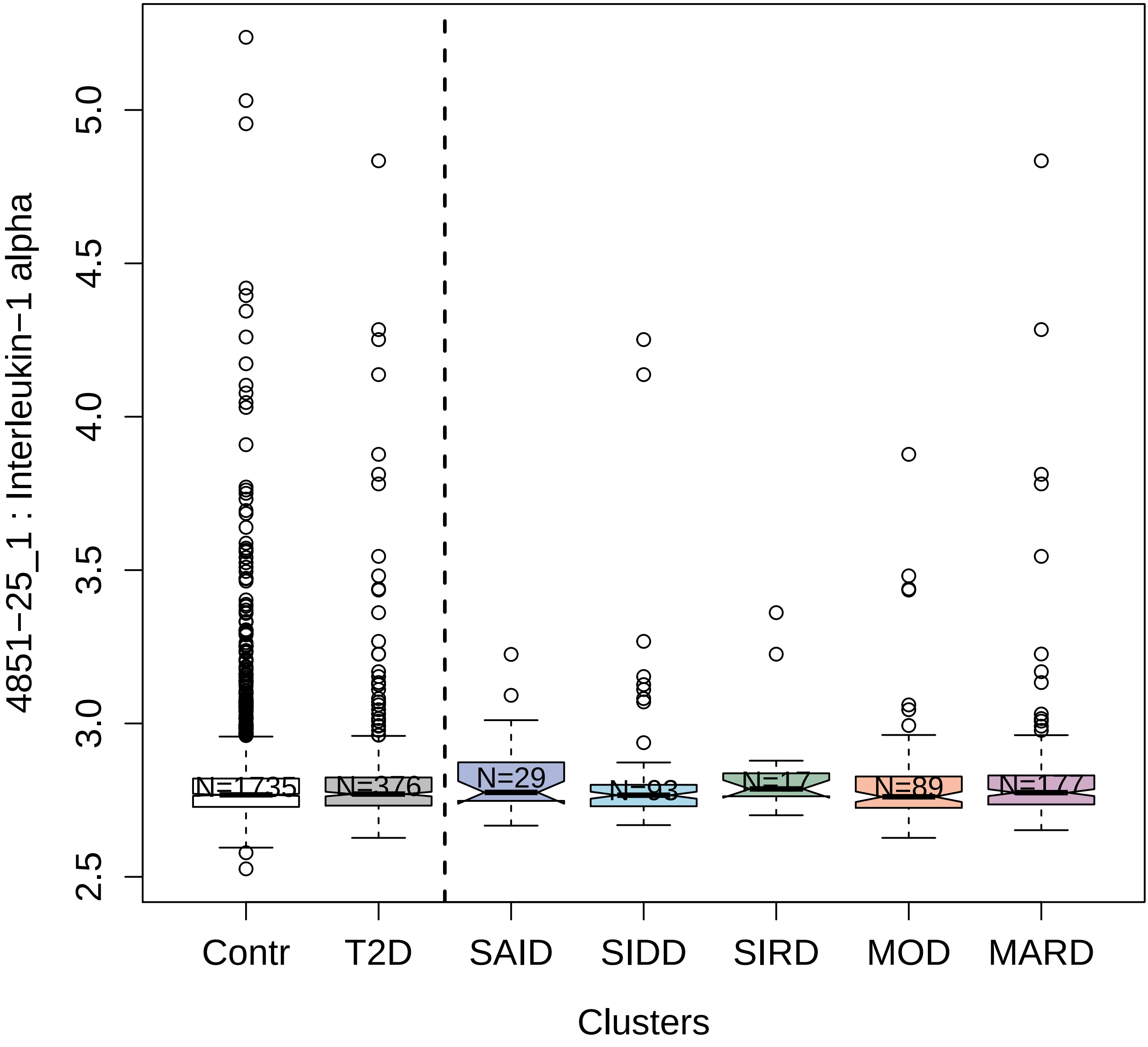
4851−25_1 : Interleukin−1 alpha
2.5
3.0
3.5
4.0
4.5
5.0
Contr
T2D
SAID
SIDD
SIRD
MOD
MARD
Clusters
N=1735
N=376
N=29
N=93
N=17
N=89
N=177

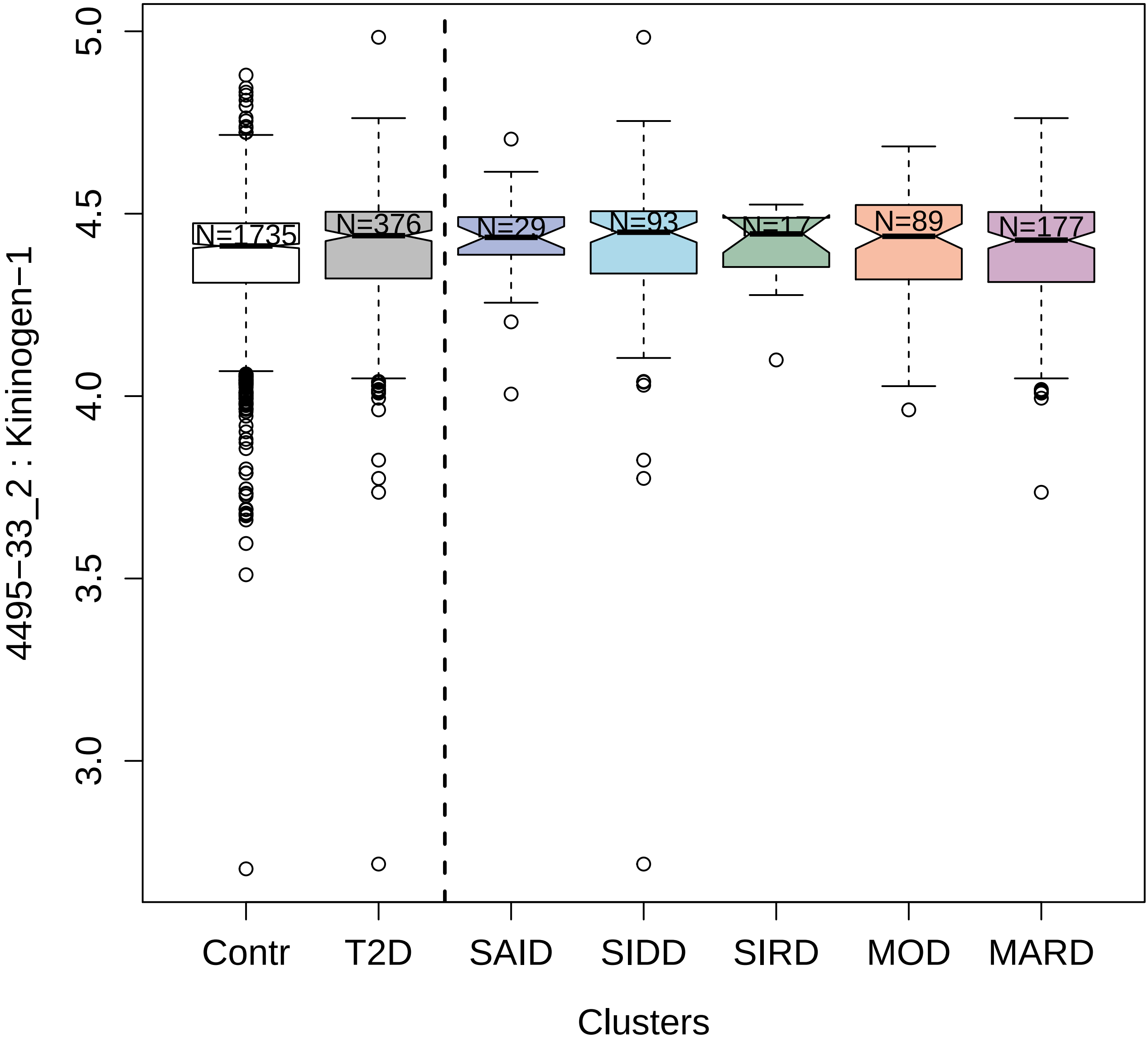

4495−33_2 : Kininogen−1
5.0
4.5
4.0
3.5
3.0
N=1735
N=376
N=29
N=93
N=17
N=89
N=177
Contr
T2D
SAID
SIDD
SIRD
MOD
MARD
Clusters

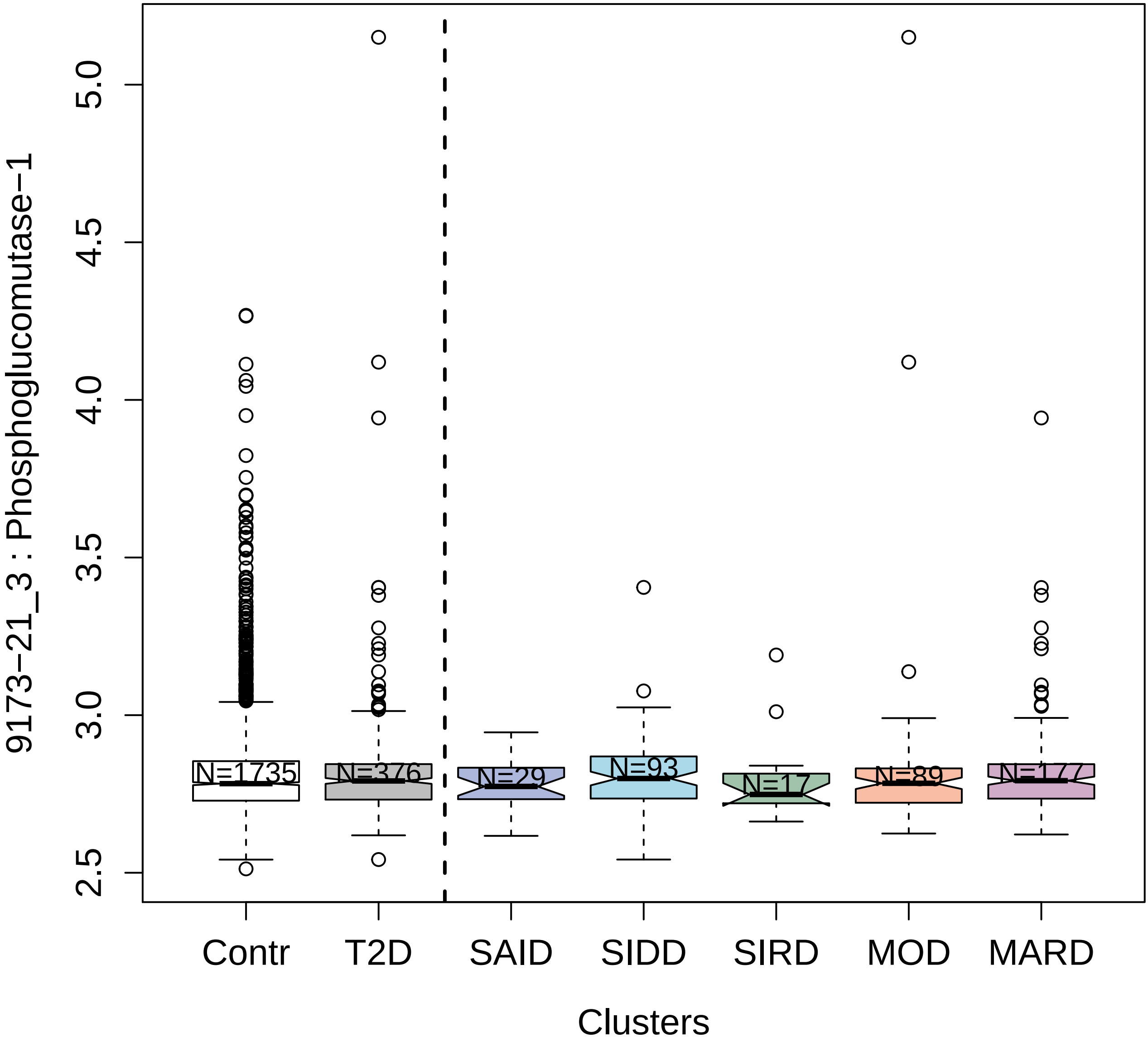

9173−21_3 : Phosphoglucomutase−1
5.0
4.5
4.0
3.5
3.0
2.5
N=1735
N=376
N=29
N=93
N=17
N=89
N=177
Contr
T2D
SAID
SIDD
SIRD
MOD
MARD
Clusters

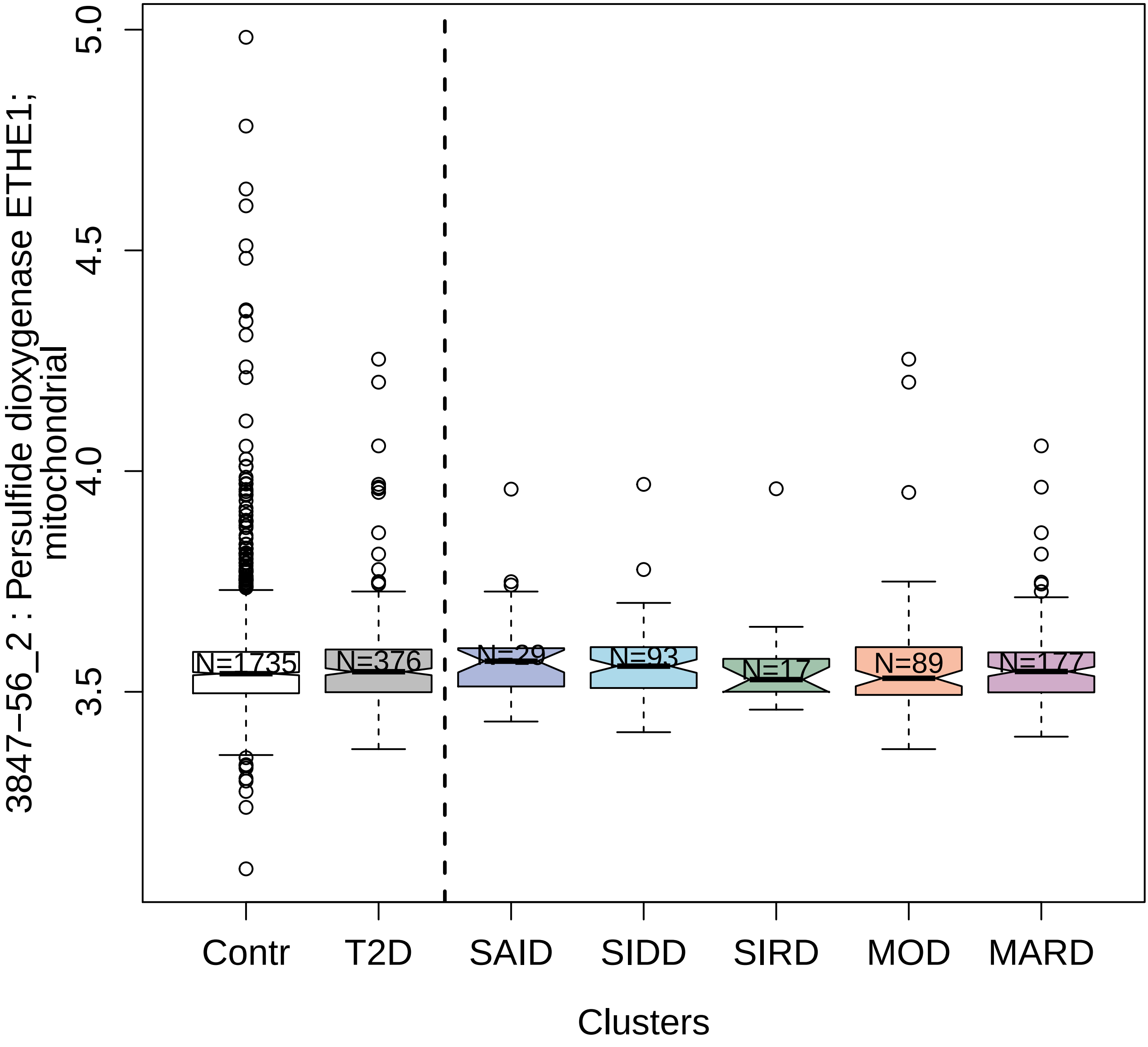
3847−56_2 : Persulfide dioxygenase ETHE1; mitochondrial
5.0
4.5
4.0
3.5
N=1735
N=376
N=29
N=93
N=17
N=89
N=177
Contr
T2D
SAID
SIDD
SIRD
MOD
MARD
Clusters

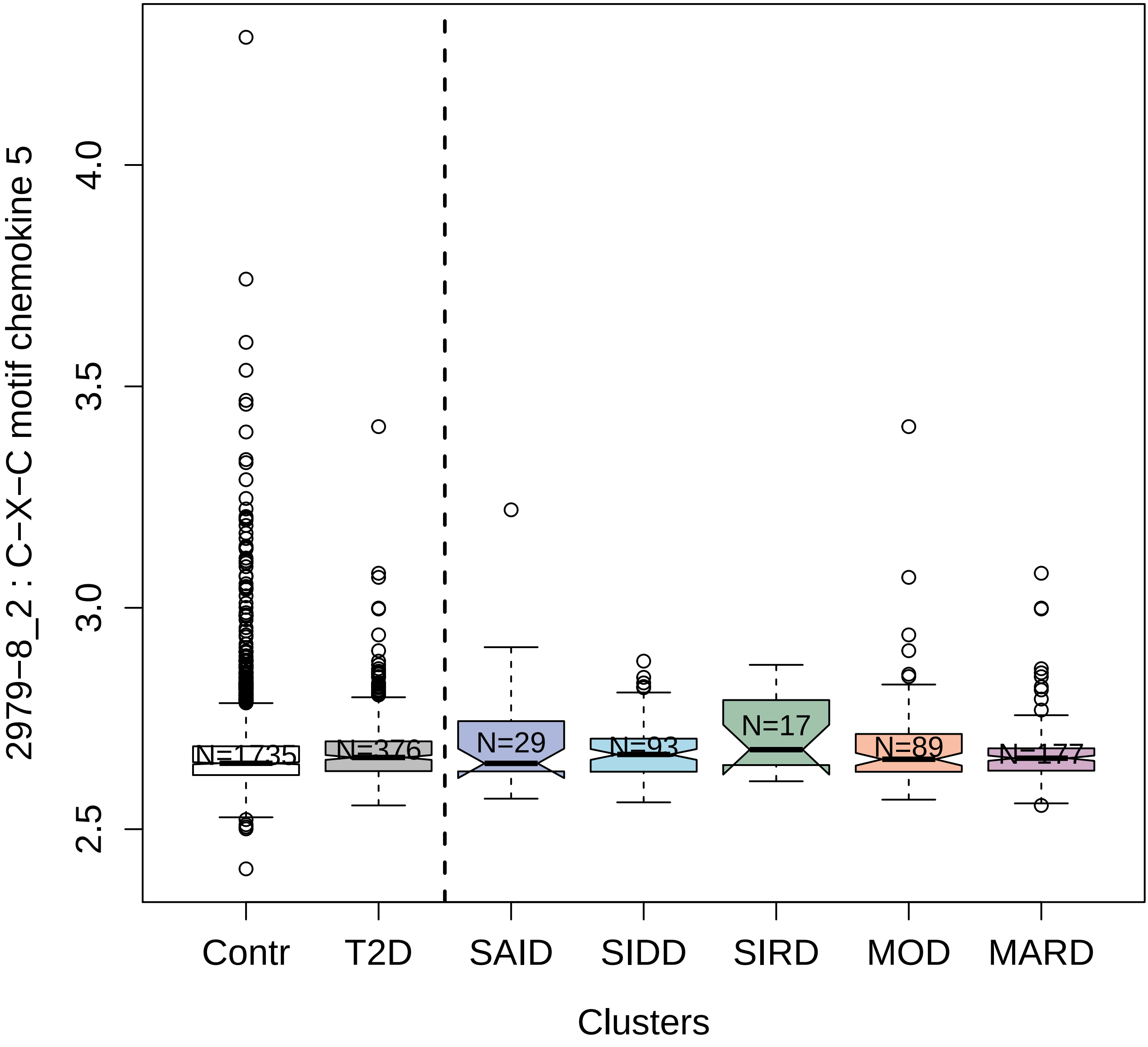

2979−8_2 : C−X−C motif chemokine 5
4.0
3.5
3.0
2.5
N=1735
N=376
N=29
N=93
N=17
N=89
N=177
Contr
T2D
SAID
SIDD
SIRD
MOD
MARD
Clusters

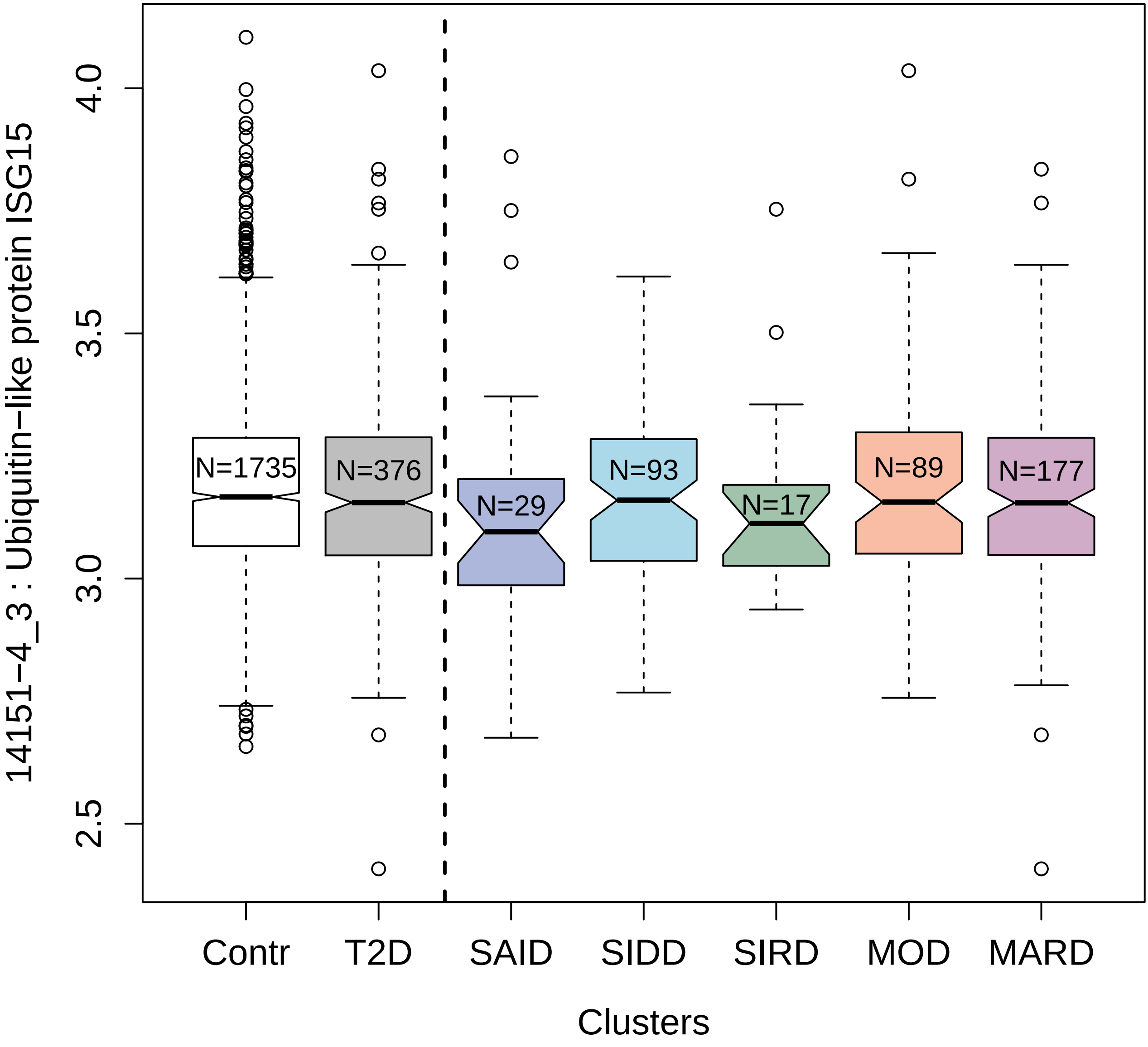

14151−4_3 : Ubiquitin−like protein ISG15
4.0
3.5
3.0
2.5
N=1735
N=376
N=29
N=93
N=17
N=89
N=177
Contr
T2D
SAID
SIDD
SIRD
MOD
MARD
Clusters

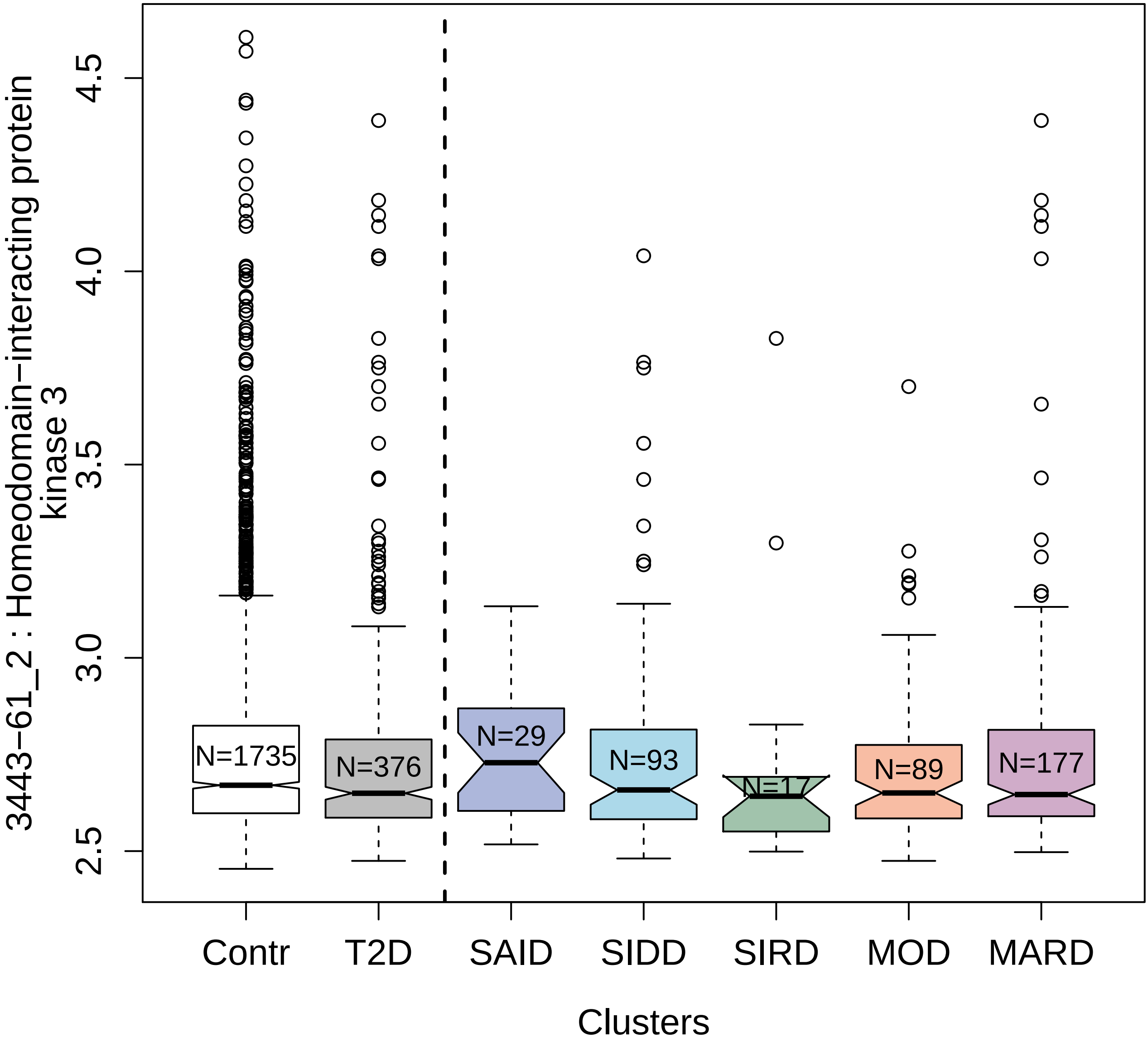

3443−61_2 : Homeodomain−interacting protein kinase 3
2.5
3.0
3.5
4.0
4.5
N=1735
N=376
N=29
N=93
N=89
N=177
Contr
T2D
SAID
SIDD
SIRD
MOD
MARD
Clusters

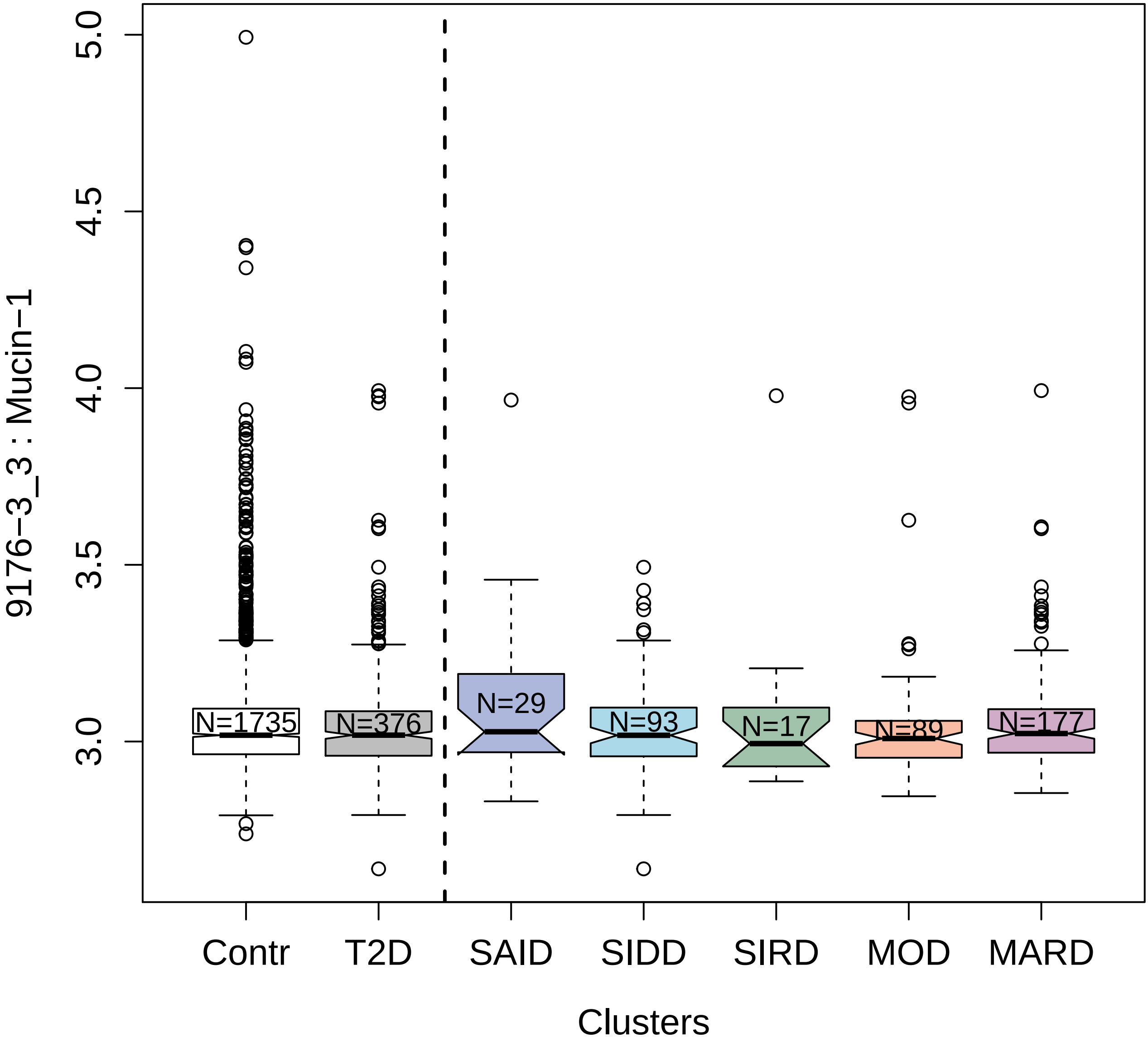

9176−3_3 : Mucin−1
5.0
4.5
4.0
3.5
3.0
N=1735
N=376
N=29
N=93
N=17
N=89
N=177
Contr
T2D
SAID
SIDD
SIRD
MOD
MARD
Clusters

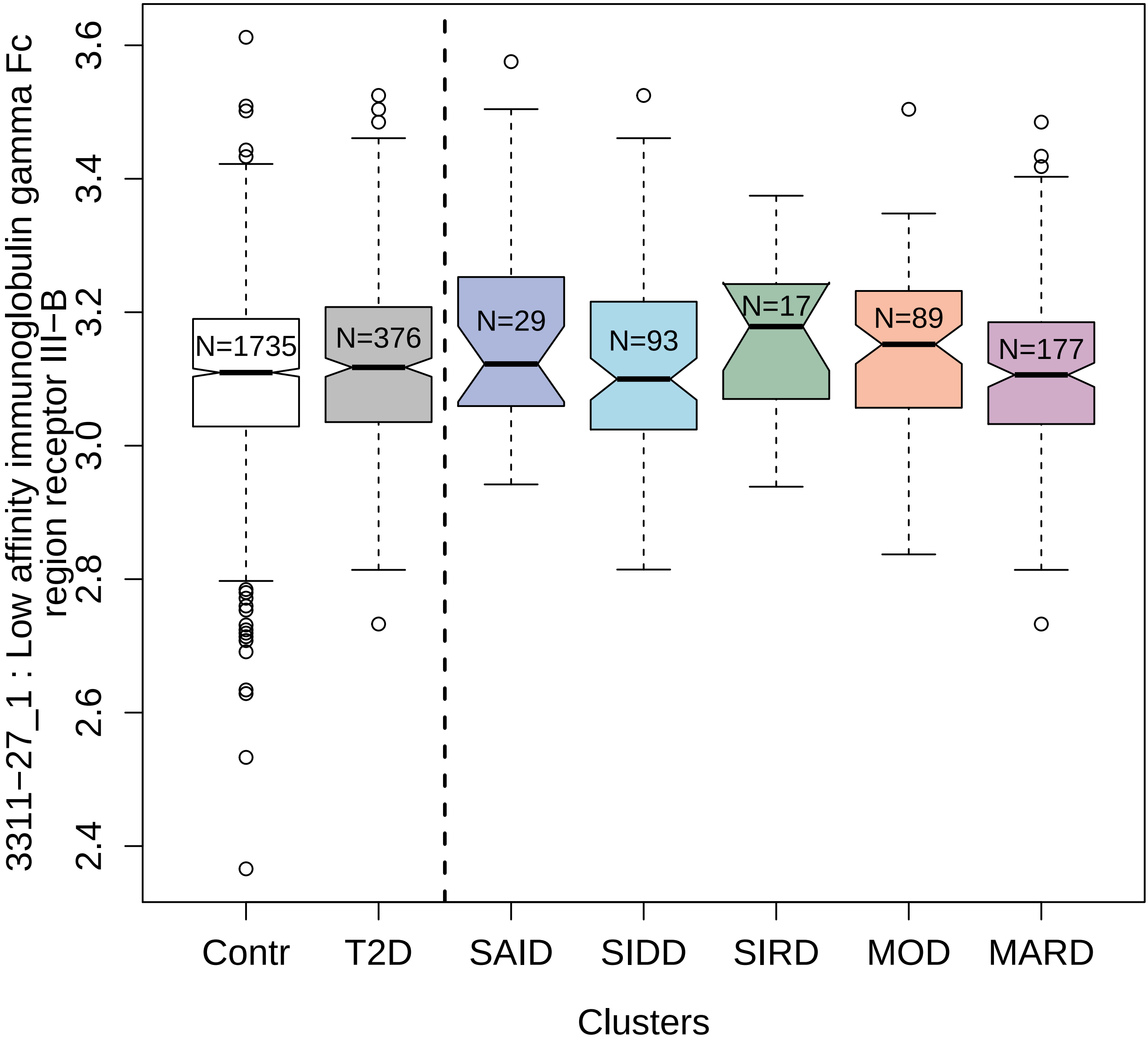

3311−27_1 : Low affinity immunoglobulin gamma Fc region receptor III−B
2.4
2.6
2.8
3.0
3.2
3.4
3.6
N=1735
N=376
N=29
N=93
N=17
N=89
N=177
Contr
T2D
SAID
SIDD
SIRD
MOD
MARD
Clusters

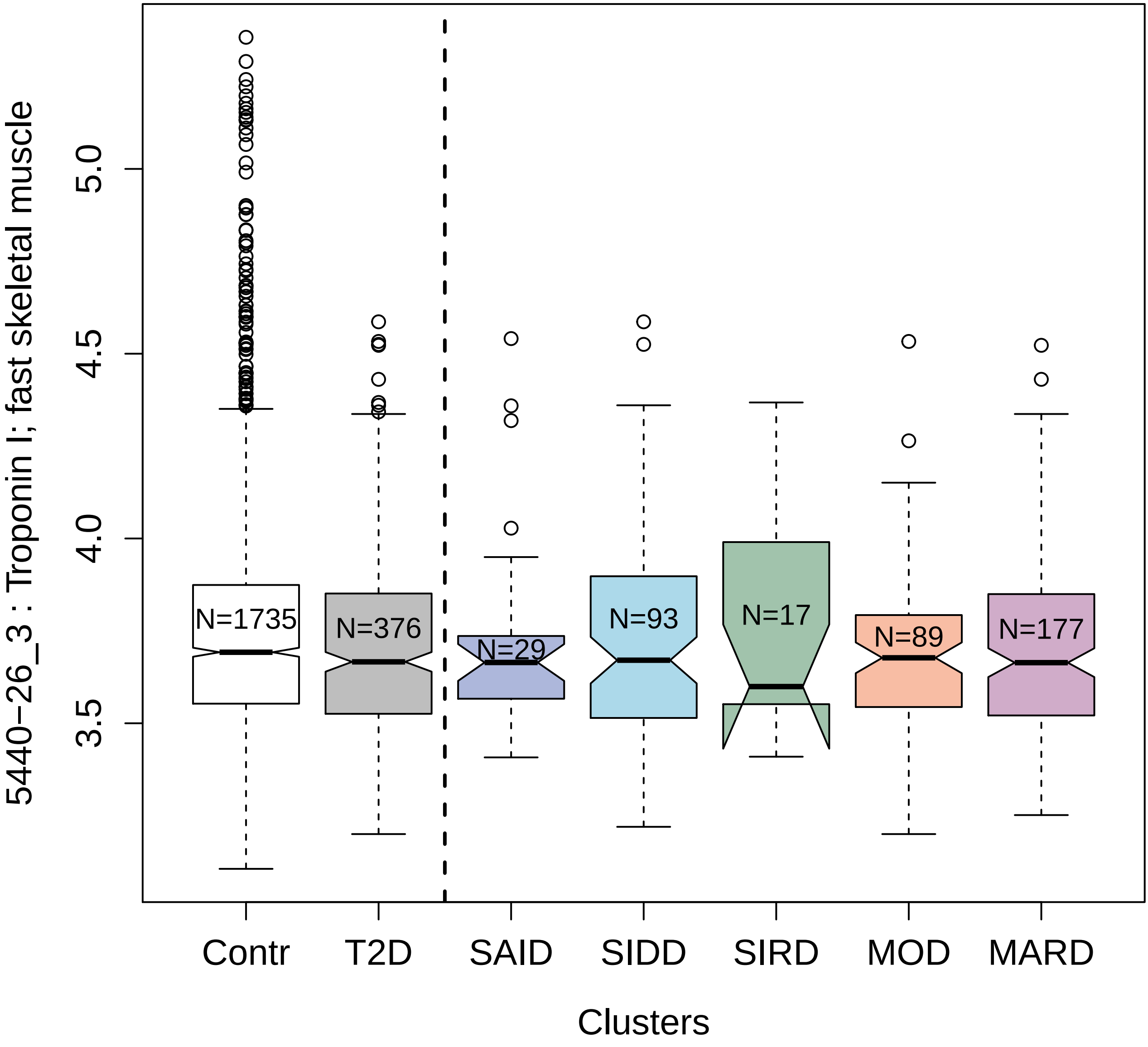

5440−26_3 : Troponin I; fast skeletal muscle
3.5
4.0
4.5
5.0
N=1735
N=376
N=29
N=93
N=17
N=89
N=177
Contr
T2D
SAID
SIDD
SIRD
MOD
MARD
Clusters

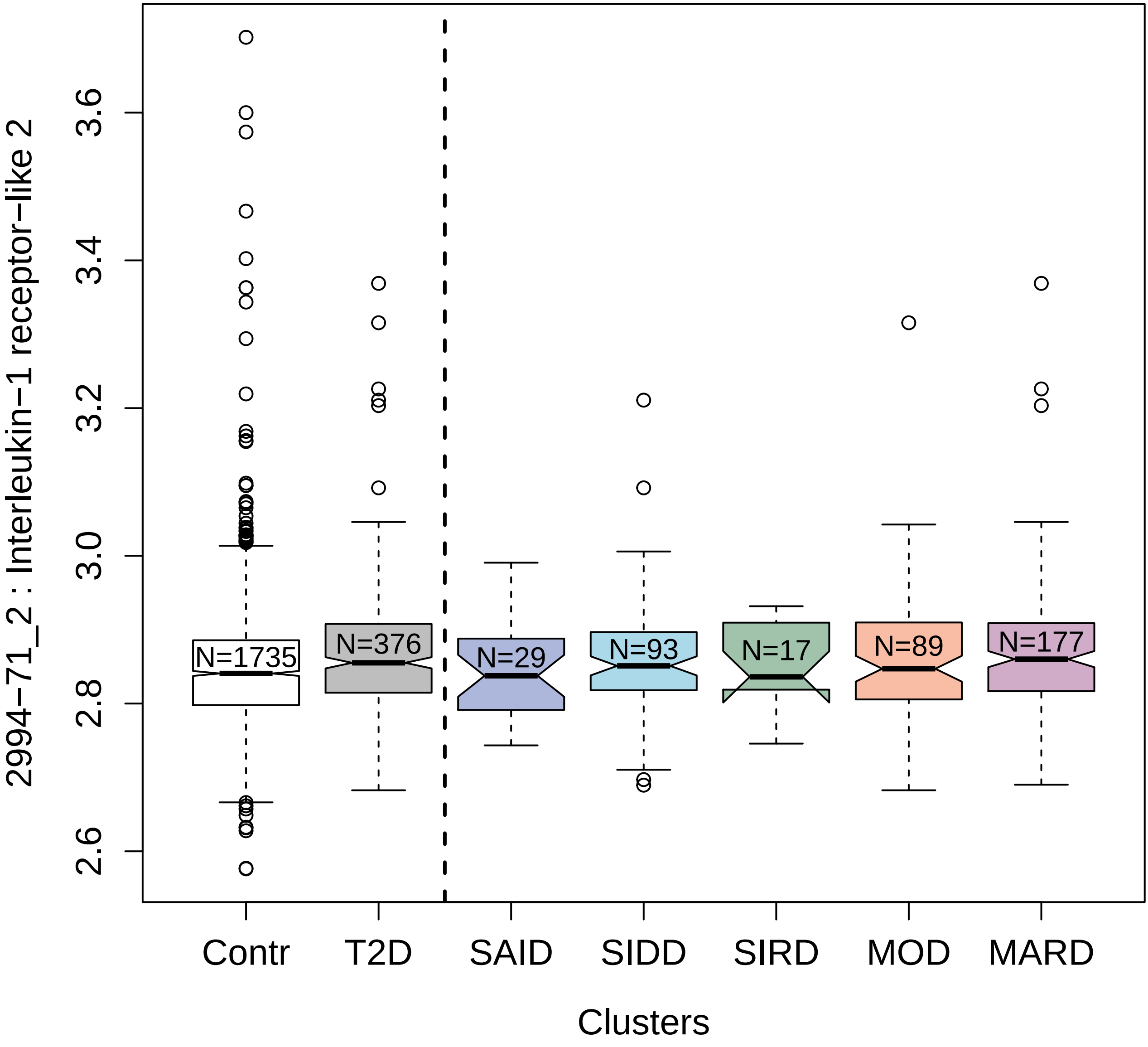

2994−71_2 : Interleukin−1 receptor−like 2
2.6
2.8
3.0
3.2
3.4
3.6
N=1735
N=376
N=29
N=93
N=17
N=89
N=177
Contr
T2D
SAID
SIDD
SIRD
MOD
MARD
Clusters

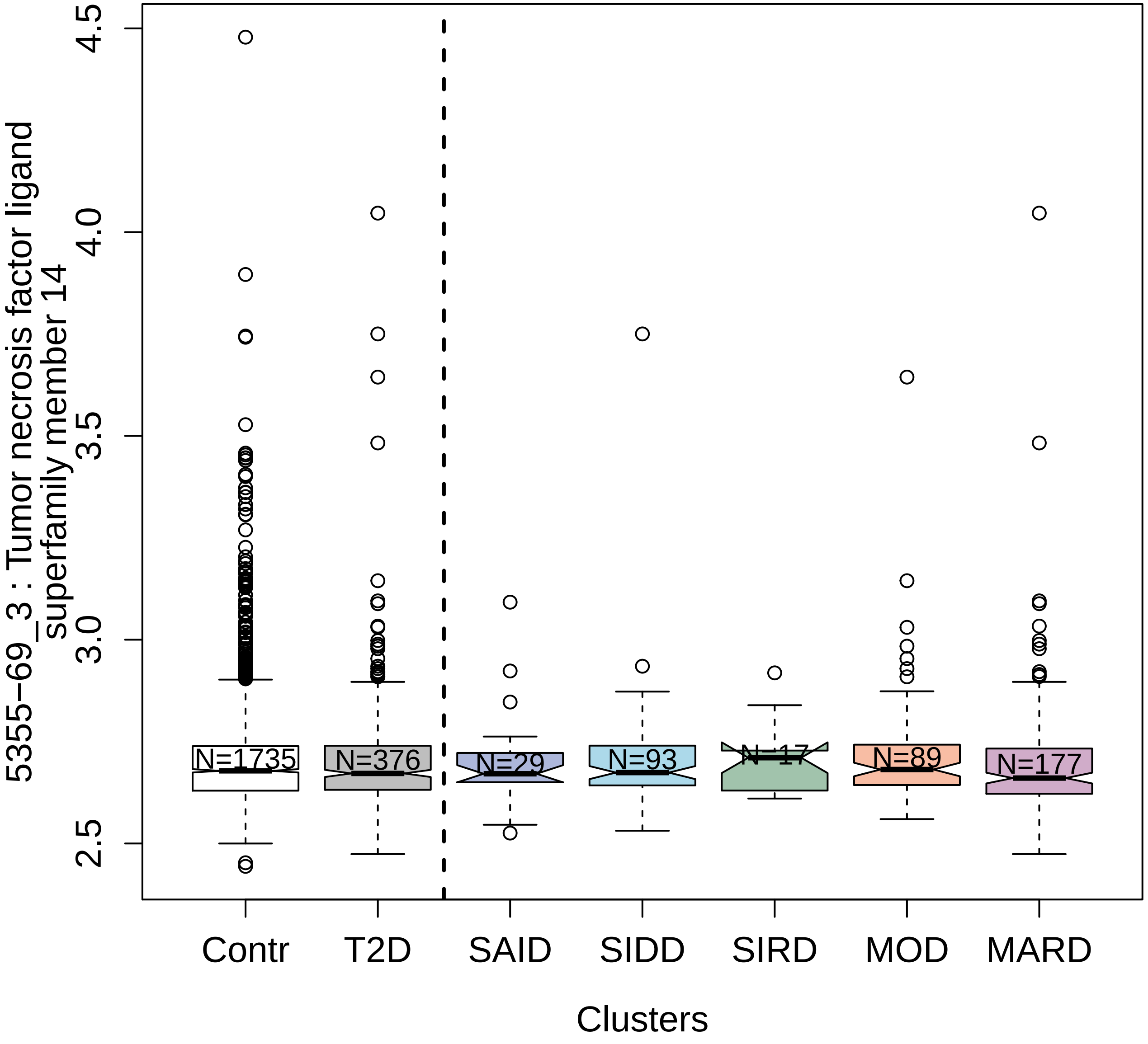
5355−69_3 : Tumor necrosis factor ligand superfamily member 14
Clusters
Contr
T2D
SAID
SIDD
SIRD
MOD
MARD
N=1735
N=376
N=29
N=93
N=17
N=89
N=177
2.5
3.0
3.5
4.0
4.5

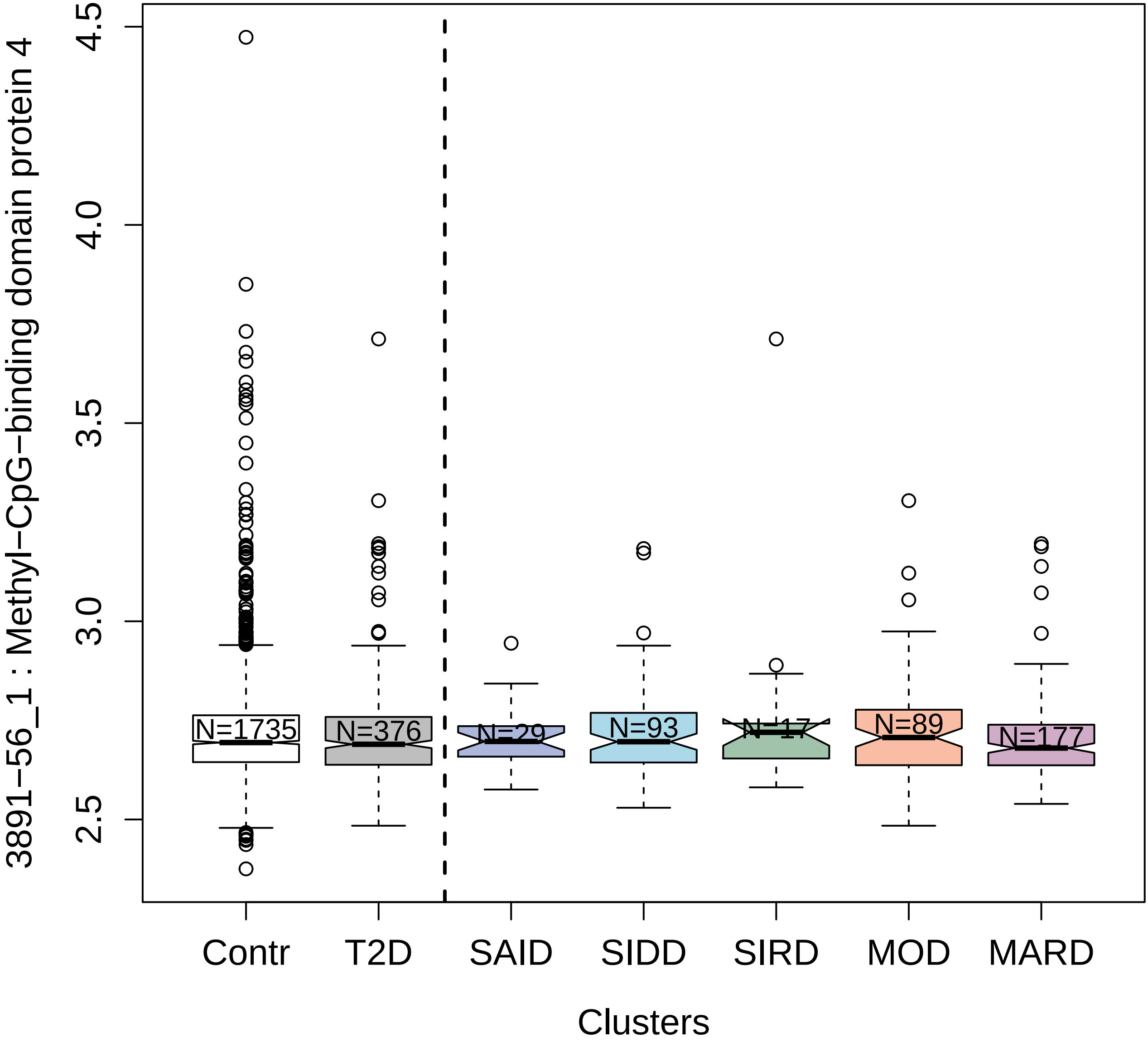

3891−56_1 : Methyl−CpG−binding domain protein 4
4.5
4.0
3.5
3.0
2.5
N=1735
N=376
N=29
N=93
N=17
N=89
N=177
Contr
T2D
SAID
SIDD
SIRD
MOD
MARD
Clusters

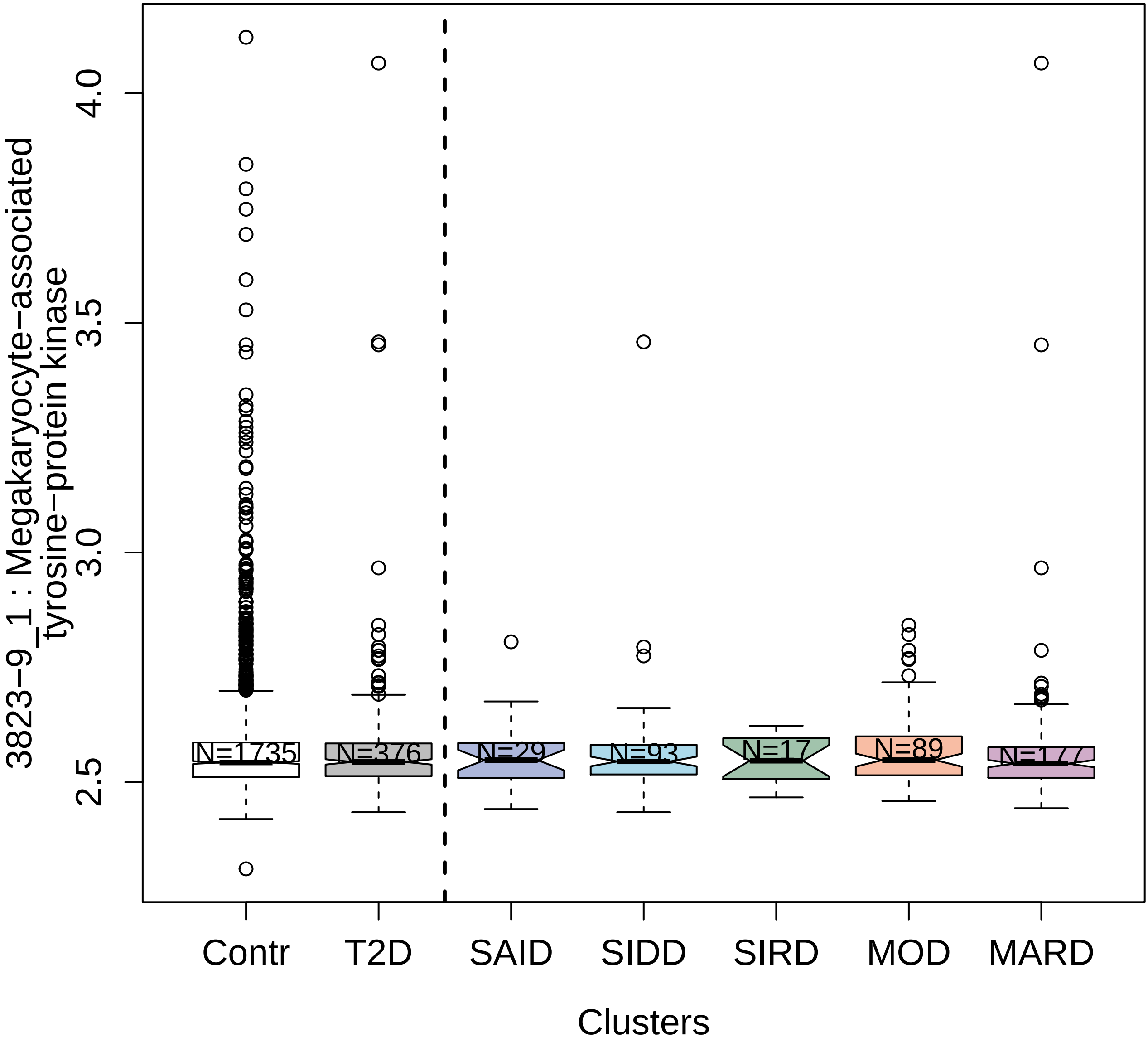

3823−9_1 : Megakaryocyte−associated
tyrosine−protein kinase
2.5
3.0
3.5
4.0
N=1735
N=376
N=29
N=93
N=17
N=89
N=177
Contr
T2D
SAID
SIDD
SIRD
MOD
MARD
Clusters

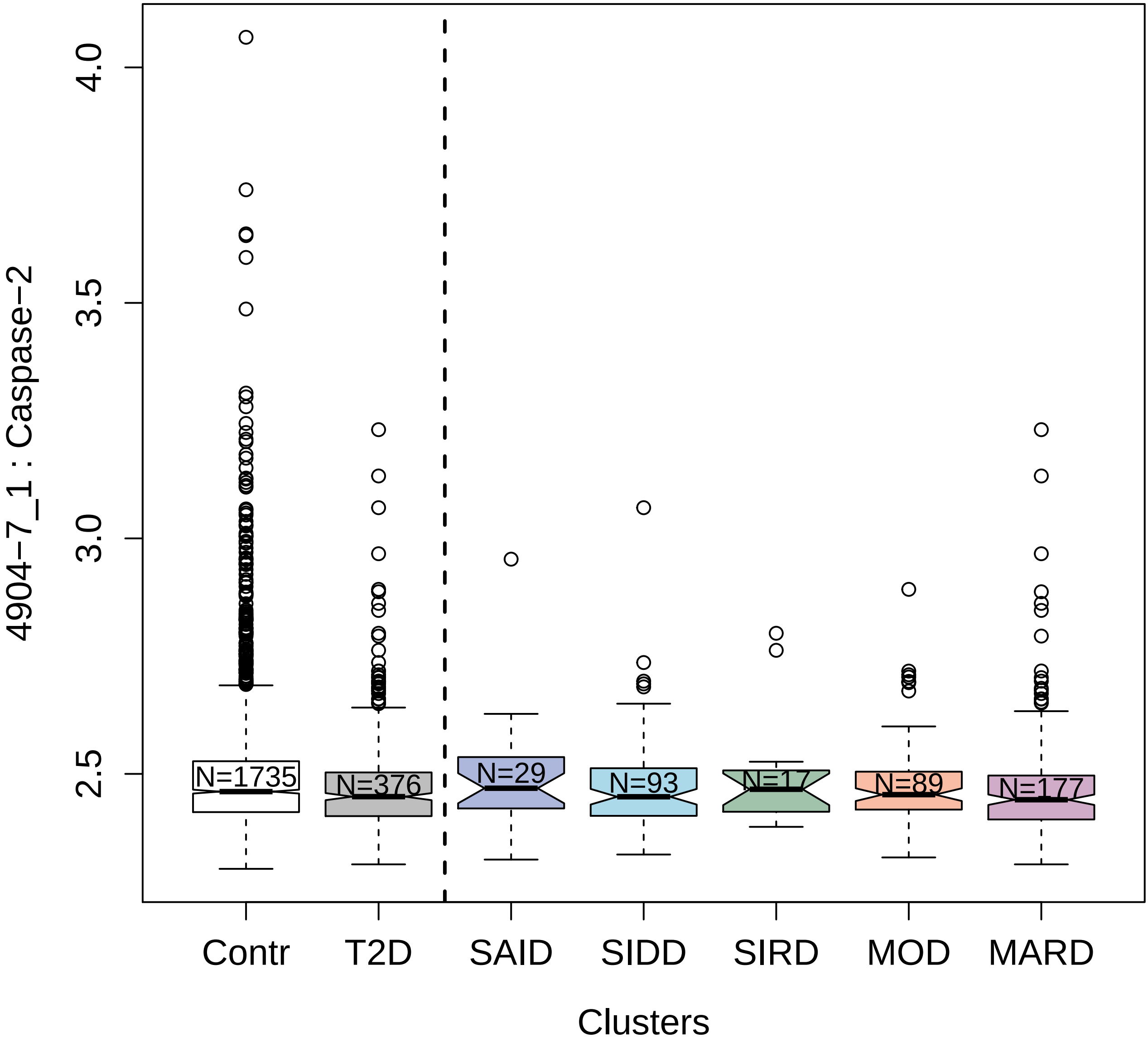

4904−7_1 : Caspase−2
4.0
3.5
3.0
2.5
N=1735
N=376
N=29
N=93
N=17
N=89
N=177
Contr
T2D
SAID
SIDD
SIRD
MOD
MARD
Clusters

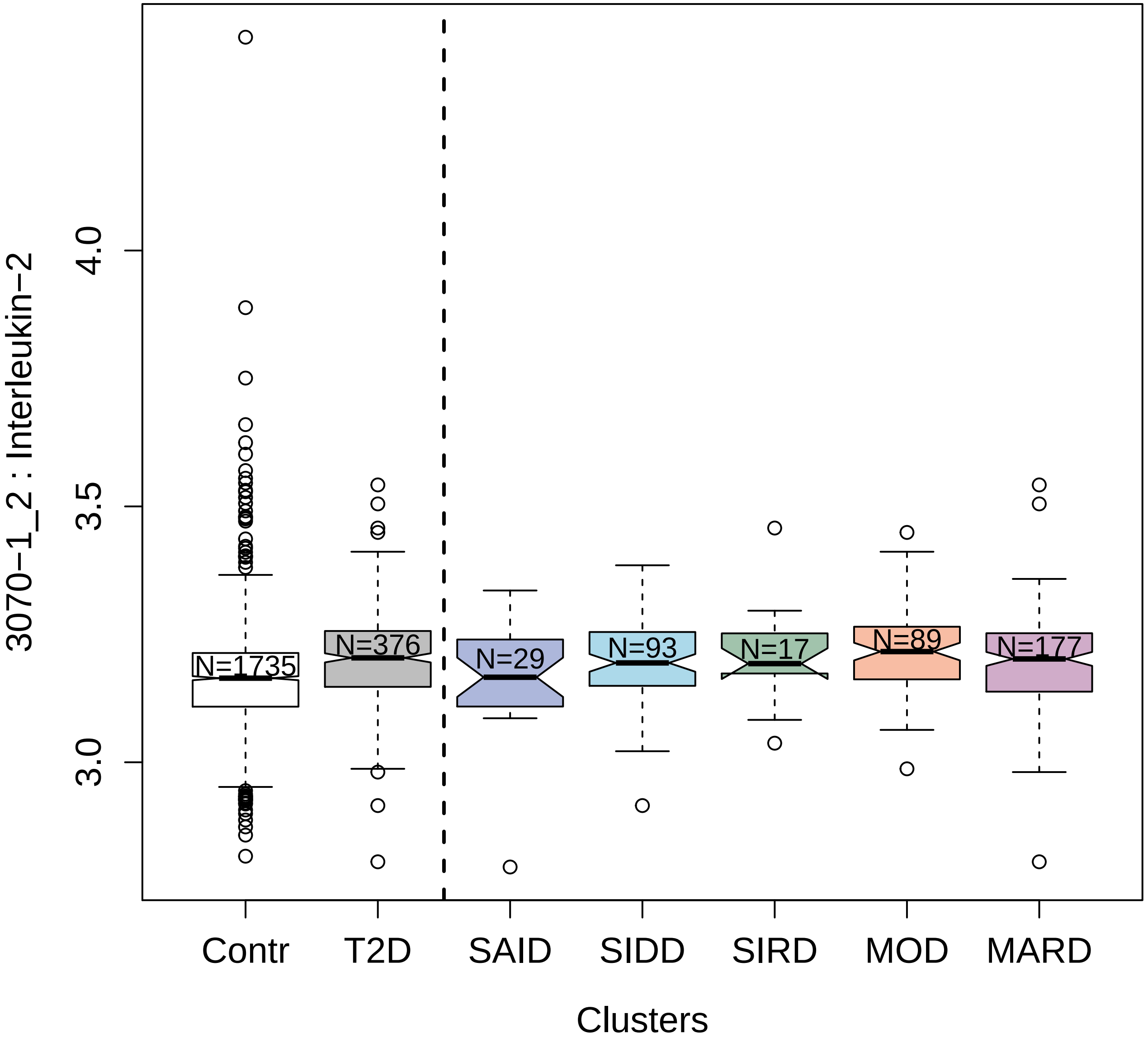

3070−1_2 : Interleukin−2
4.0
3.5
3.0
N=1735
N=376
N=29
N=93
N=17
N=89
N=177
Contr
T2D
SAID
SIDD
SIRD
MOD
MARD
Clusters

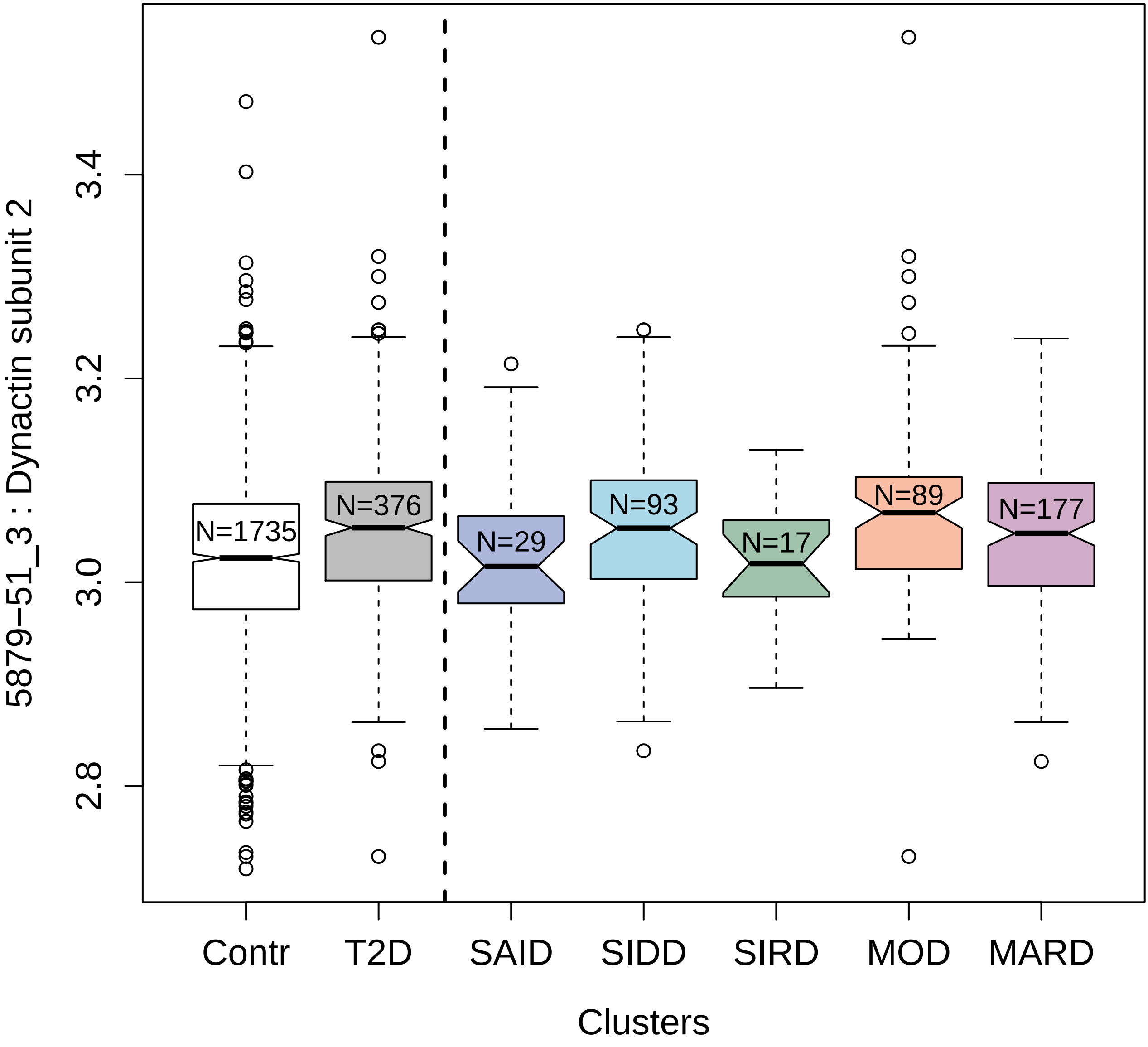
5879−51_3 : Dynactin subunit 2
3.4
3.2
3.0
2.8
N=1735
N=376
N=29
N=93
N=17
N=89
N=177
Contr
T2D
SAID
SIDD
SIRD
MOD
MARD
Clusters

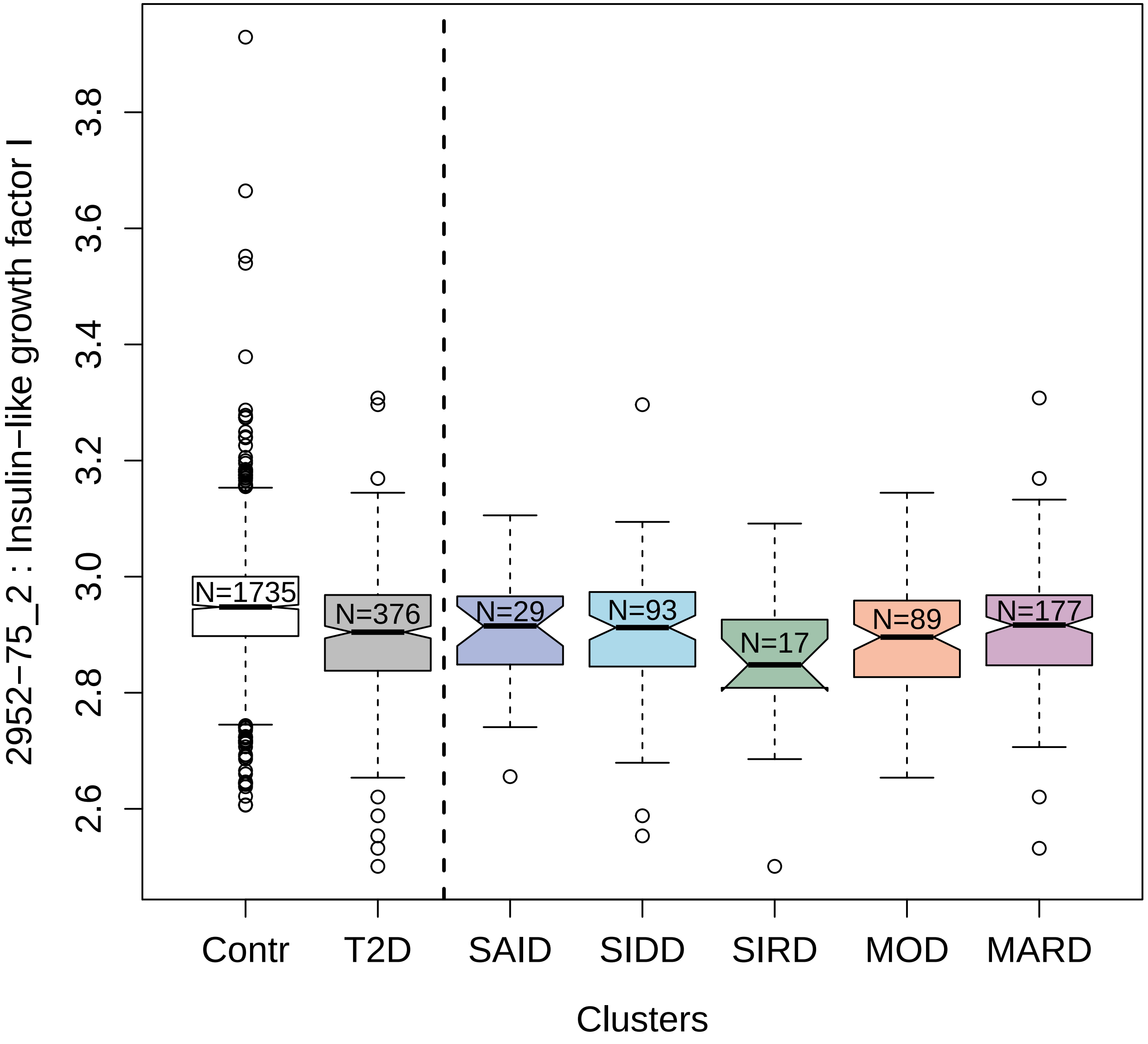

2952−75_2 : Insulin−like growth factor I
3.8
3.6
3.4
3.2
3.0
2.8
2.6
N=1735
N=376
N=29
N=93
N=17
N=89
N=177
Contr
T2D
SAID
SIDD
SIRD
MOD
MARD
Clusters

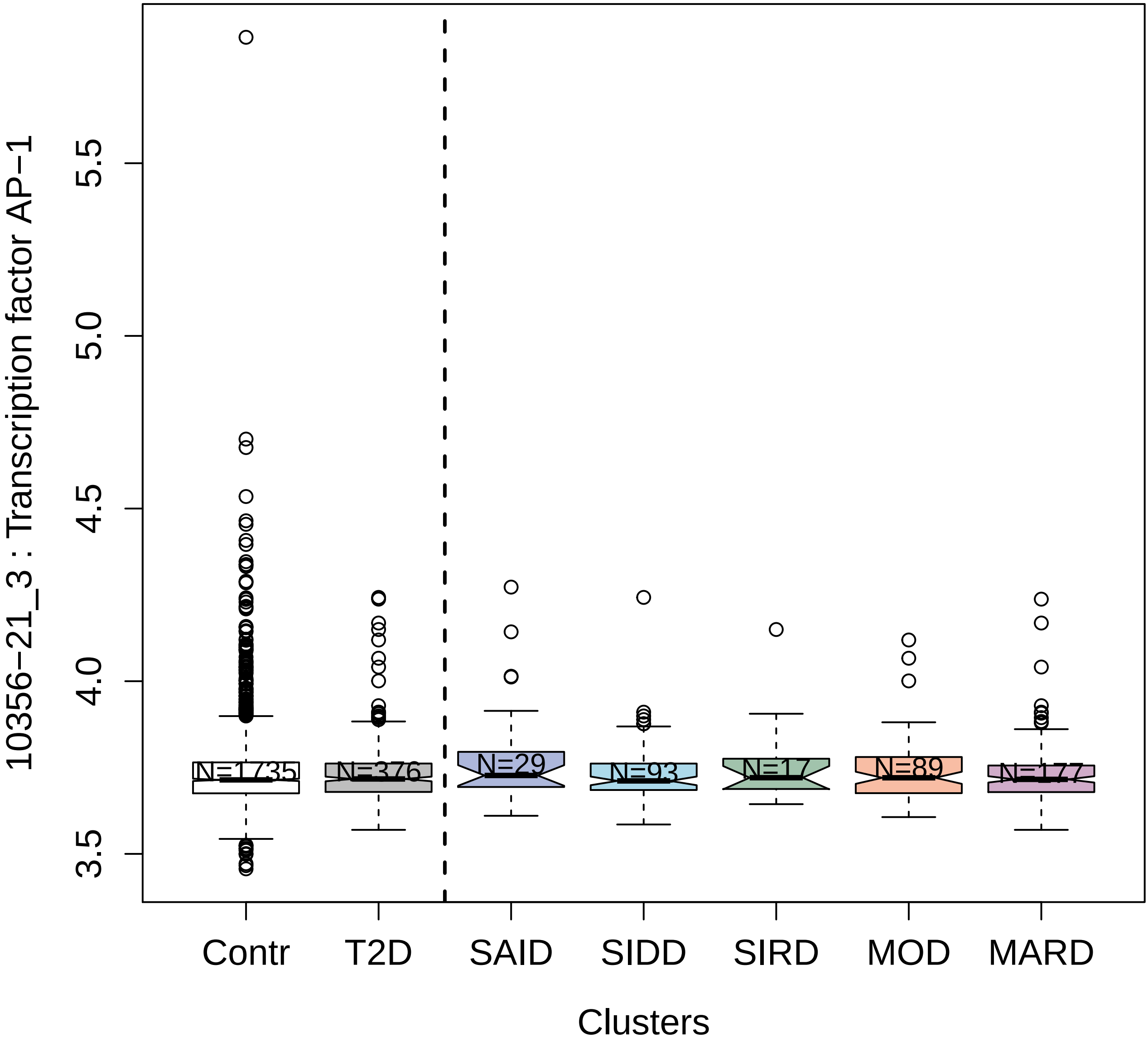
10356−21_3 : Transcription factor AP−1
3.5
4.0
4.5
5.0
5.5
N=1735
N=376
N=29
N=93
N=17
N=89
N=177
Contr
T2D
SAID
SIDD
SIRD
MOD
MARD
Clusters

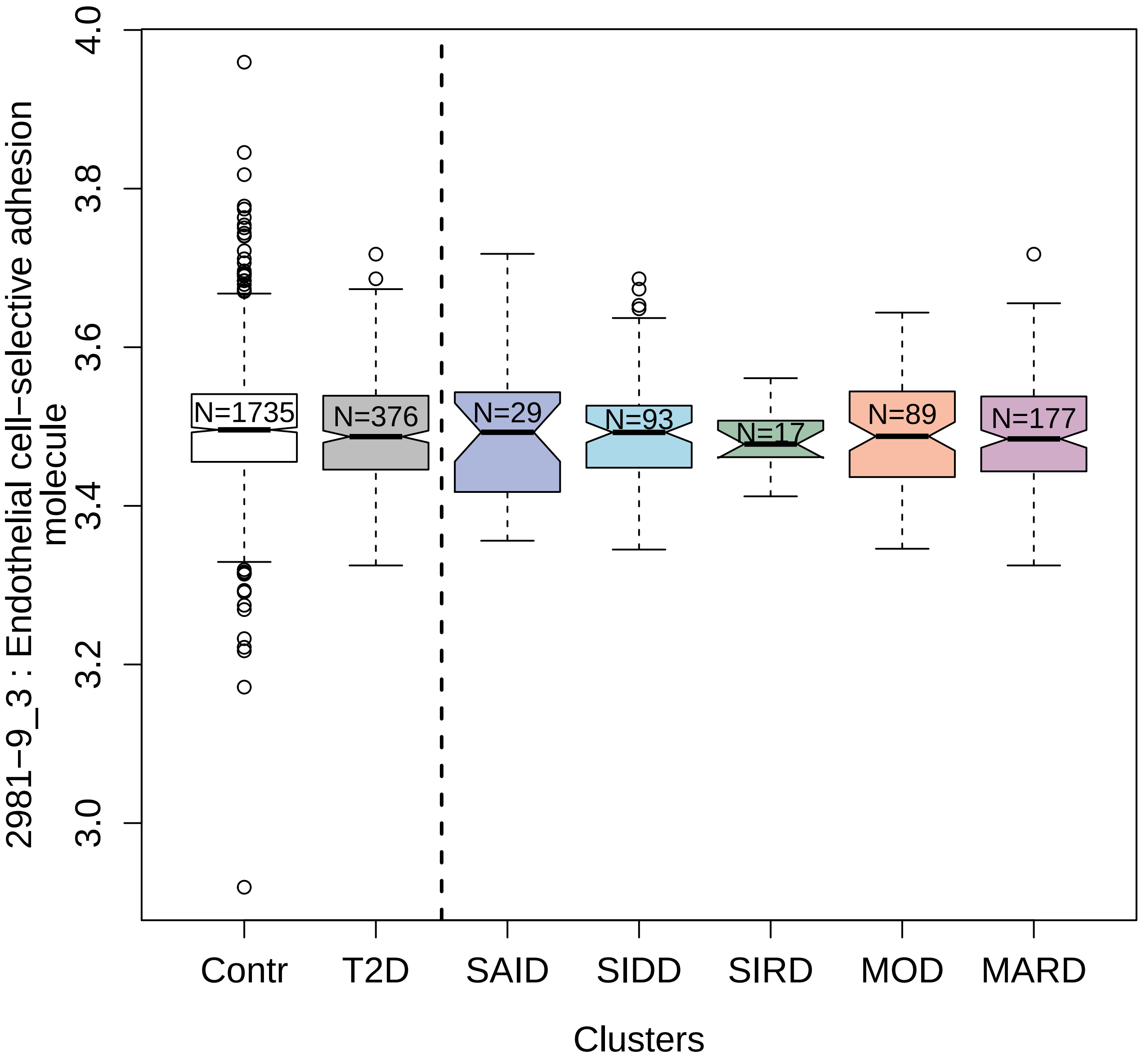

2981−9_3 : Endothelial cell−selective adhesion molecule
4.0
3.8
3.6
3.4
3.2
3.0
N=1735
N=376
N=29
N=93
N=17
N=89
N=177
Contr
T2D
SAID
SIDD
SIRD
MOD
MARD
Clusters

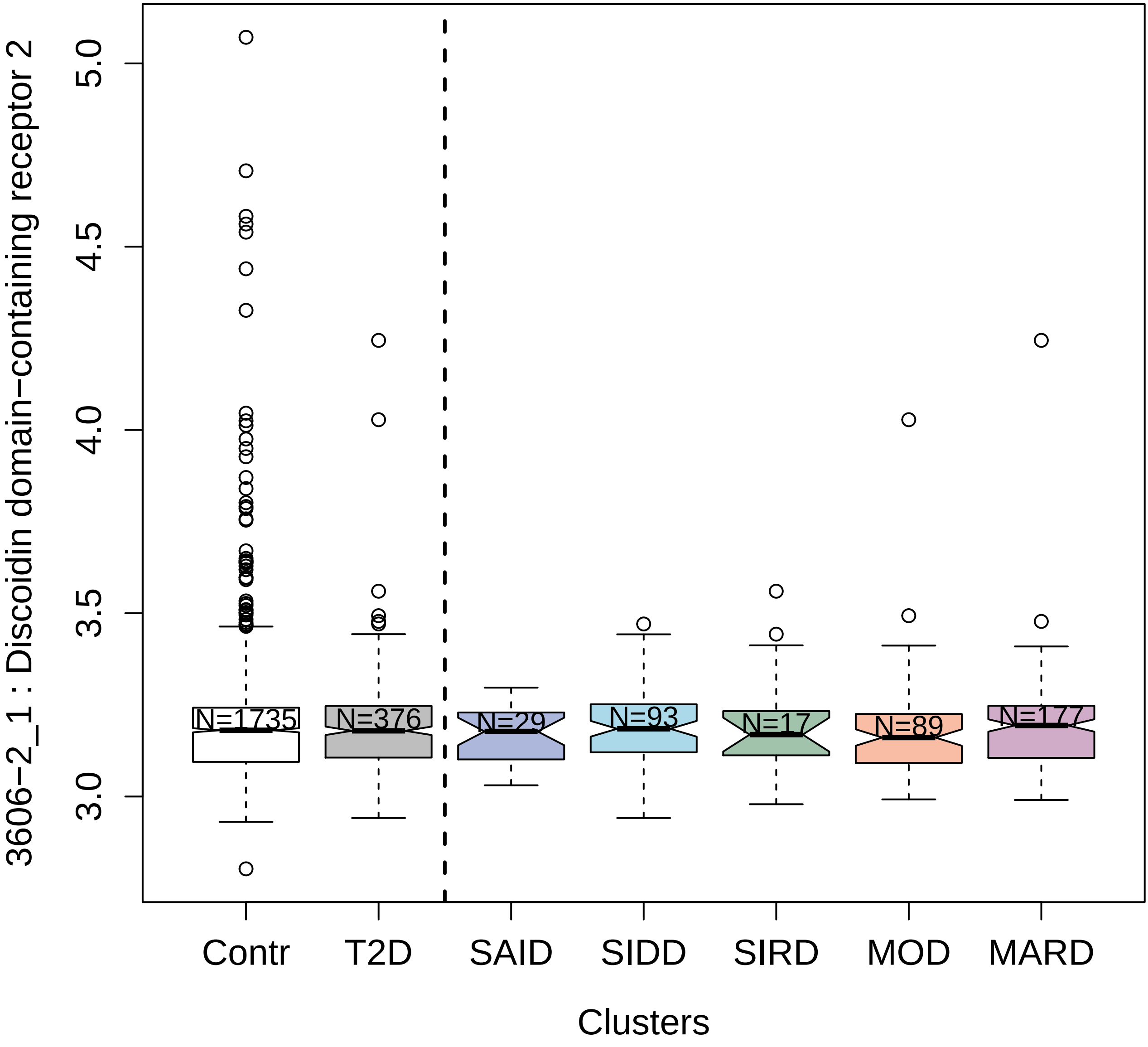
3606−2_1 : Discoidin domain−containing receptor 2
5.0
4.5
4.0
3.5
3.0
N=1735
N=376
N=29
N=93
N=17
N=89
N=177
Contr
T2D
SAID
SIDD
SIRD
MOD
MARD
Clusters

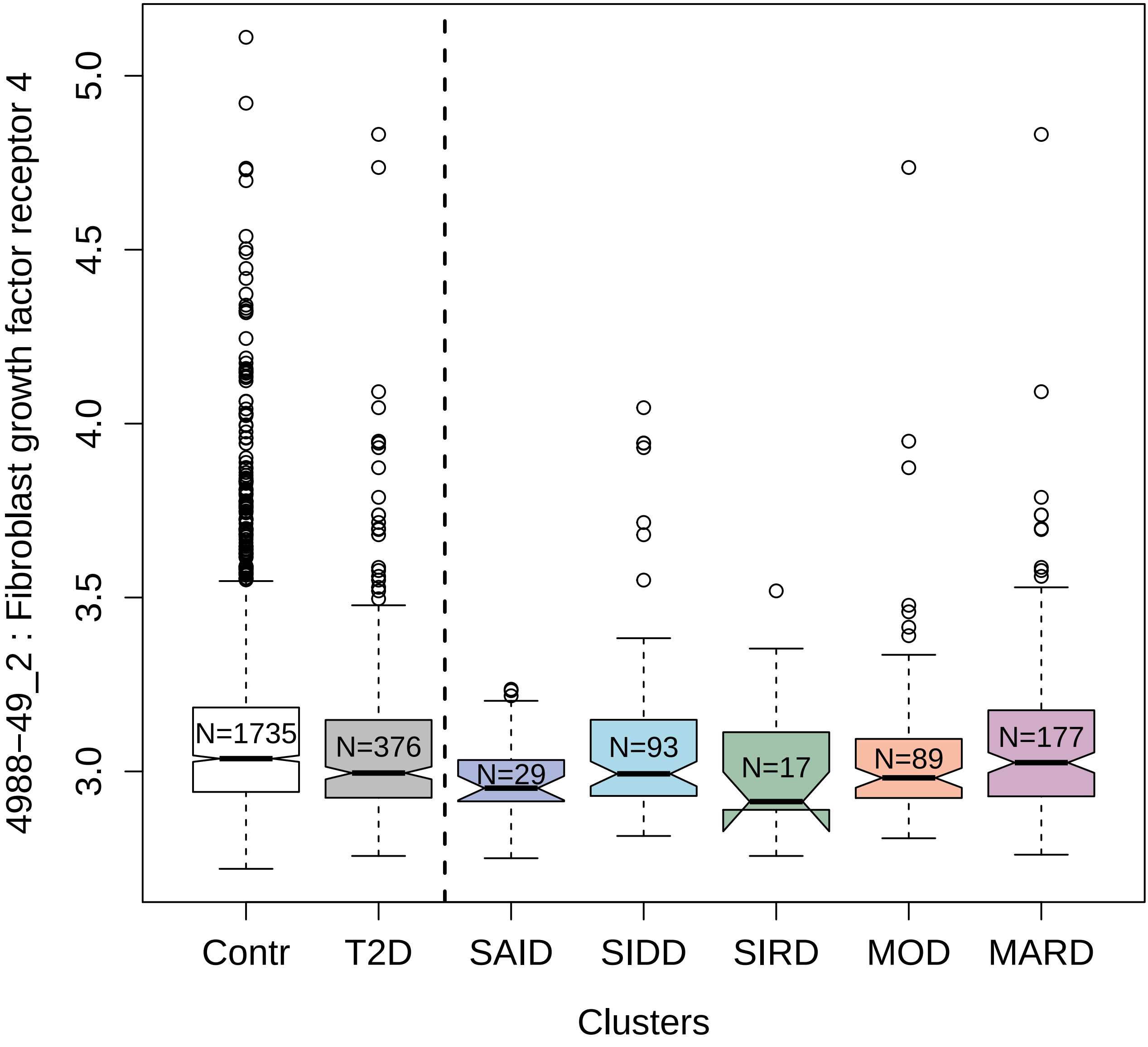

4988−49_2 : Fibroblast growth factor receptor 4
3.0
3.5
4.0
4.5
5.0
N=1735
N=376
N=29
N=93
N=17
N=89
N=177
Contr
T2D
SAID
SIDD
SIRD
MOD
MARD
Clusters

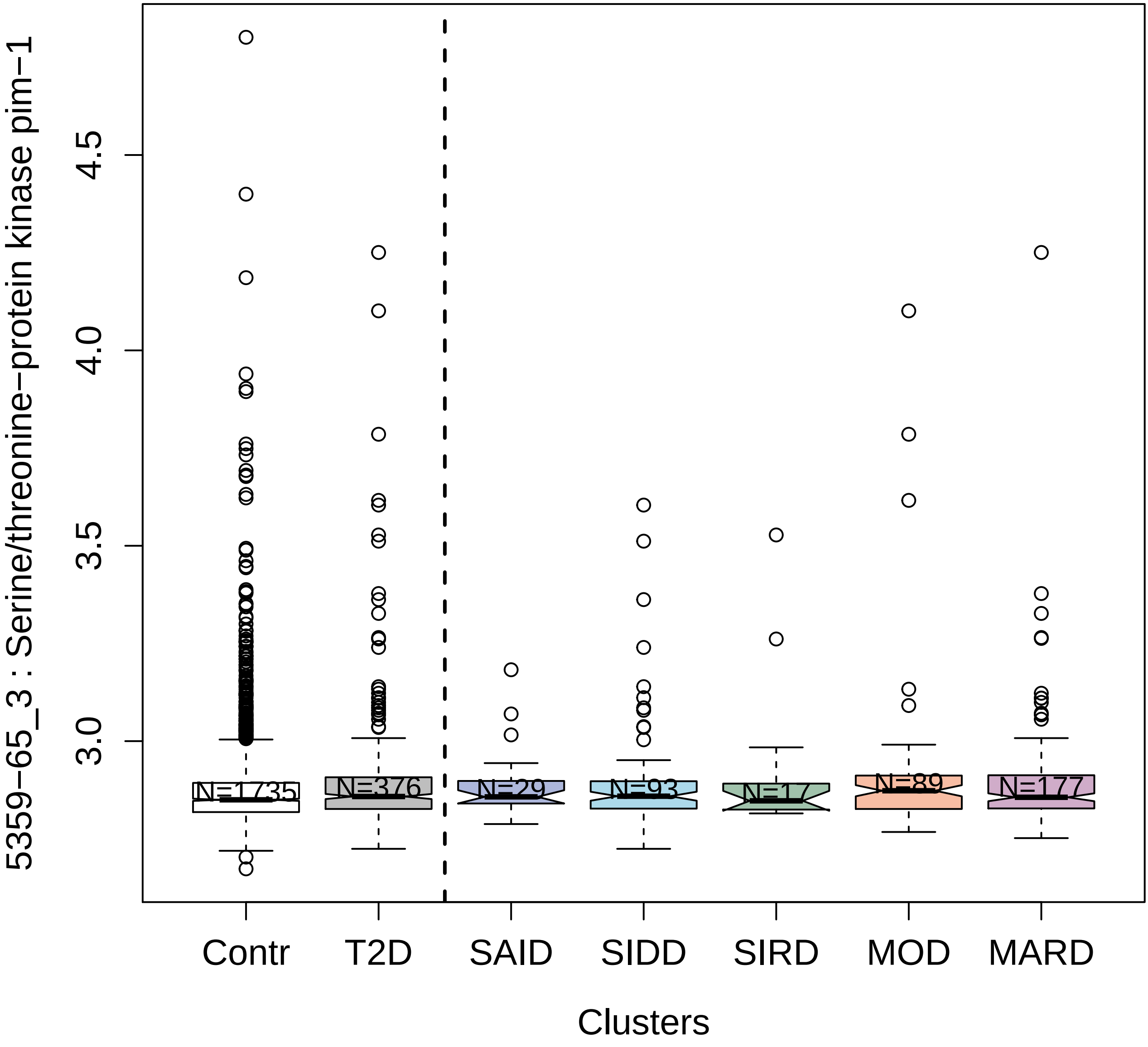
5359−65_3 : Serine/threonine−protein kinase pim−1
3.0
3.5
4.0
4.5
N=1735
N=376
N=29
N=93
N=17
N=89
N=177
Contr
T2D
SAID
SIDD
SIRD
MOD
MARD
Clusters

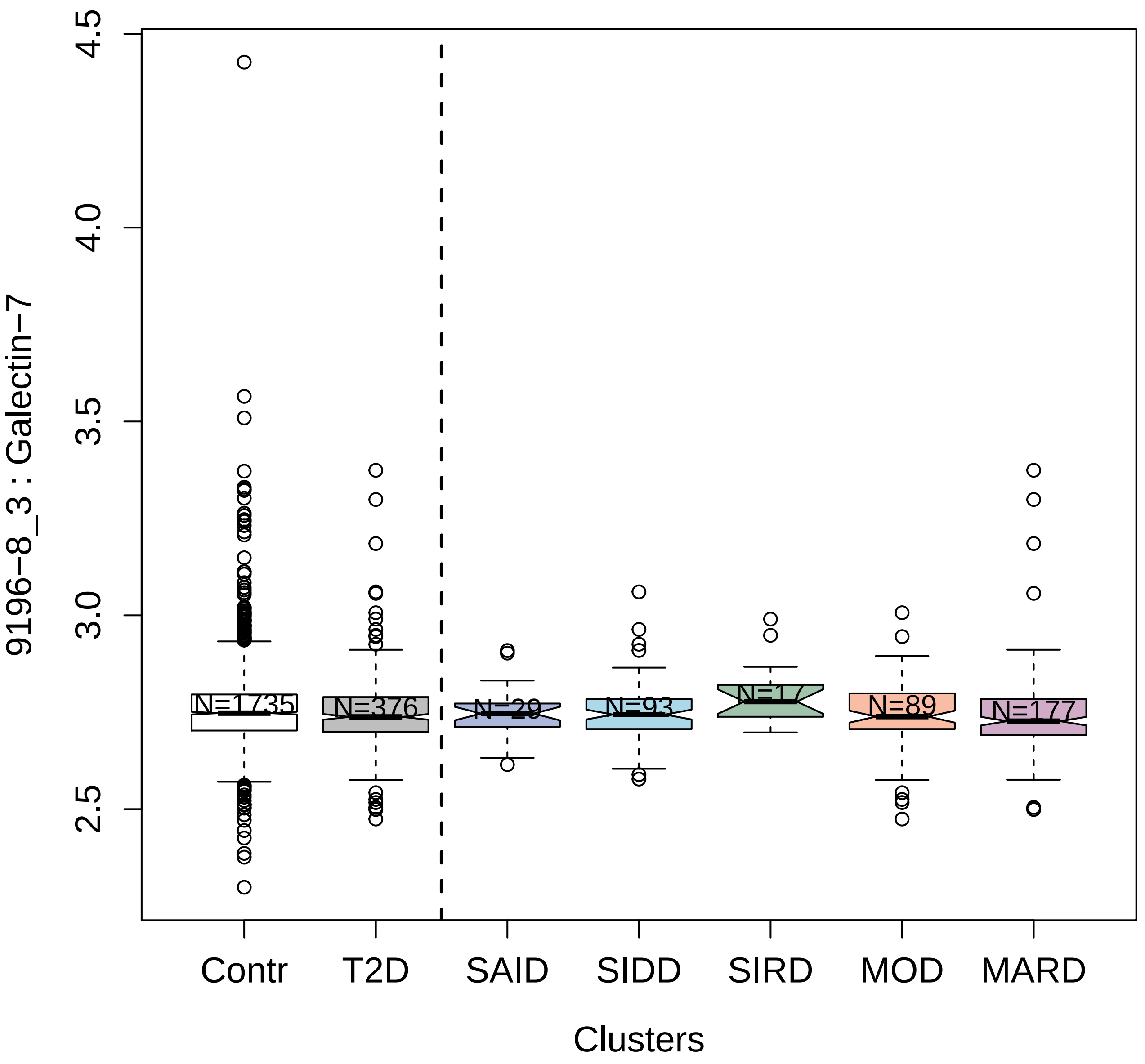

9196−8_3 : Galectin−7
4.5
4.0
3.5
3.0
2.5
N=1735
N=376
N=29
N=93
N=17
N=89
N=177
Contr
T2D
SAID
SIDD
SIRD
MOD
MARD
Clusters

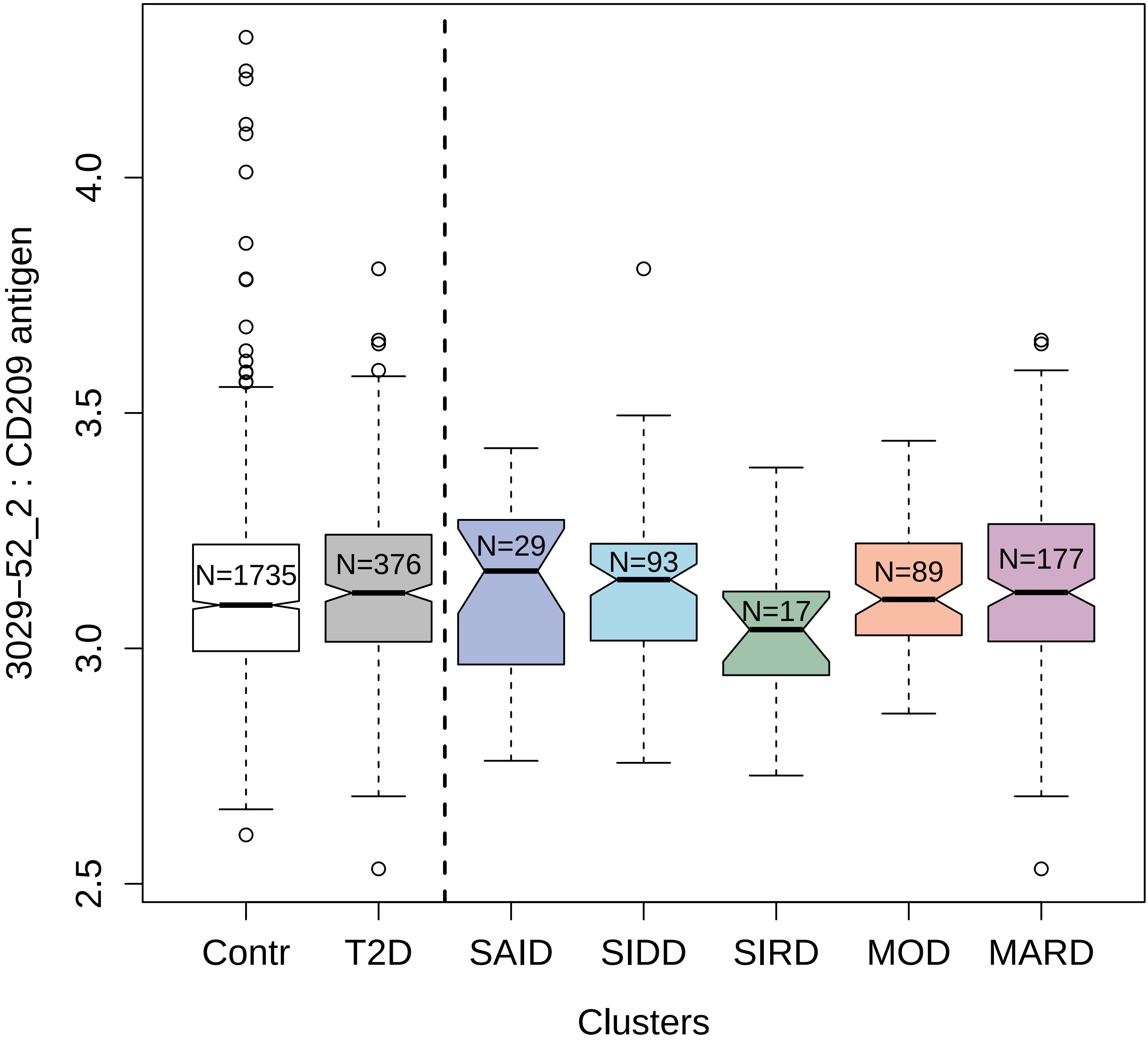

3029−52_2 : CD209 antigen
2.5
3.0
3.5
4.0
N=1735
N=376
N=29
N=93
N=17
N=89
N=177
Contr
T2D
SAID
SIDD
SIRD
MOD
MARD
Clusters

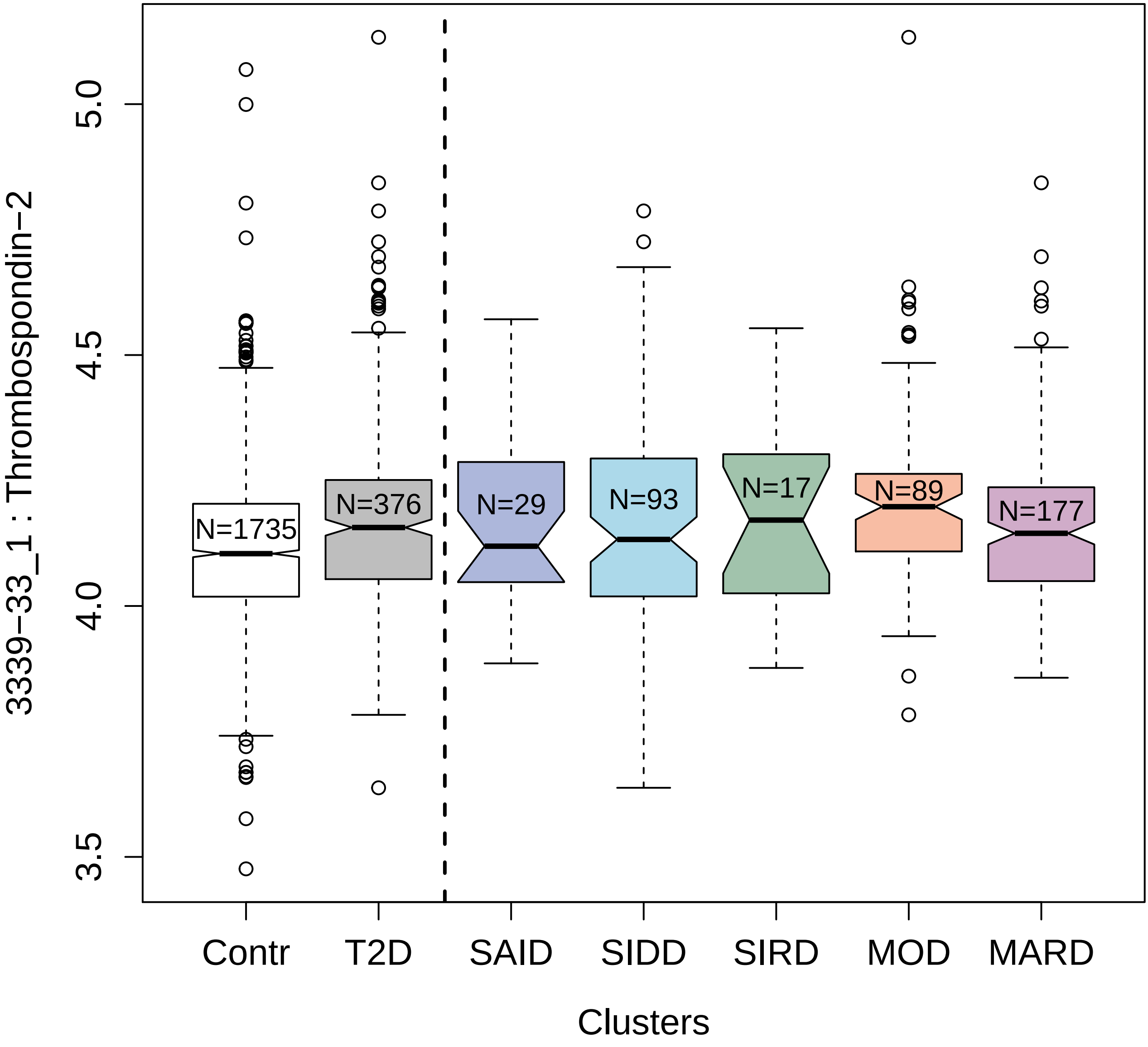
3339−33_1 : Thrombospondin−2
5.0
4.5
4.0
3.5
N=1735
N=376
N=29
N=93
N=17
N=89
N=177
Contr
T2D
SAID
SIDD
SIRD
MOD
MARD
Clusters

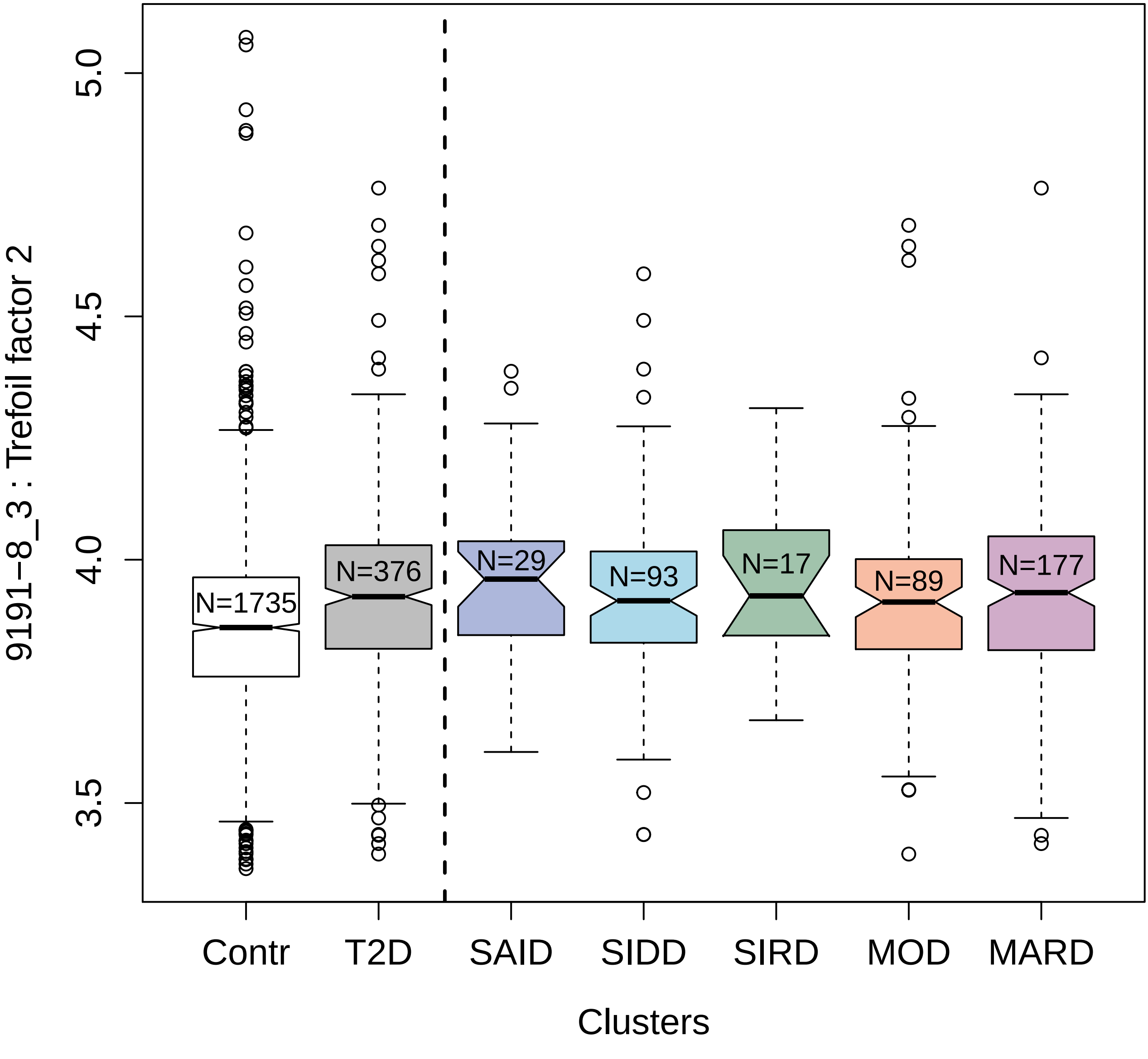
9191−8_3 : Trefoil factor 2
5.0
4.5
4.0
3.5
N=1735
N=376
N=29
N=93
N=17
N=89
N=177
Contr
T2D
SAID
SIDD
SIRD
MOD
MARD
Clusters

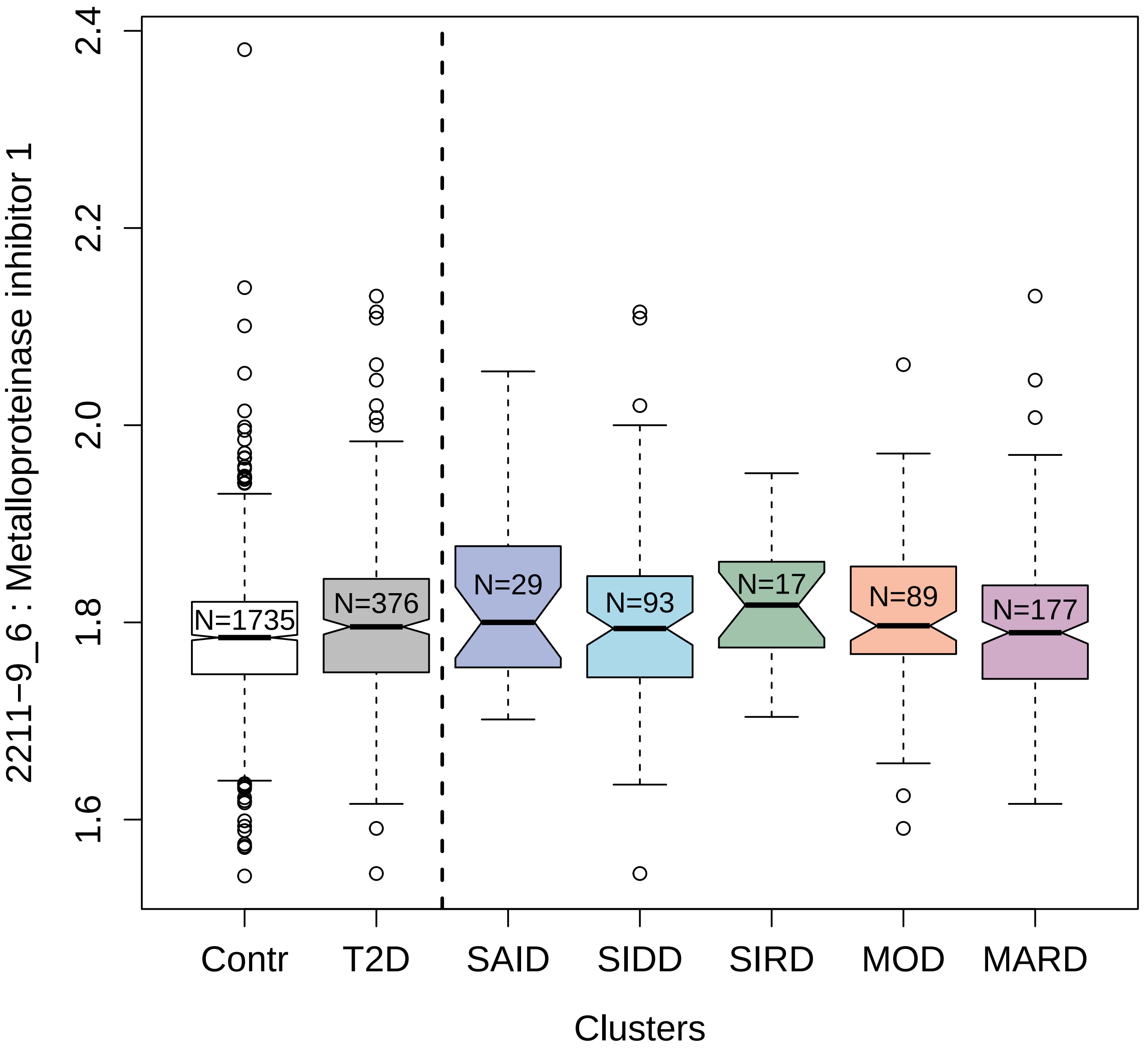

2211−9_6 : Metalloproteinase inhibitor 1
2.4
2.2
2.0
1.8
1.6
N=1735
N=376
N=29
N=93
N=17
N=89
N=177
Contr
T2D
SAID
SIDD
SIRD
MOD
MARD
Clusters

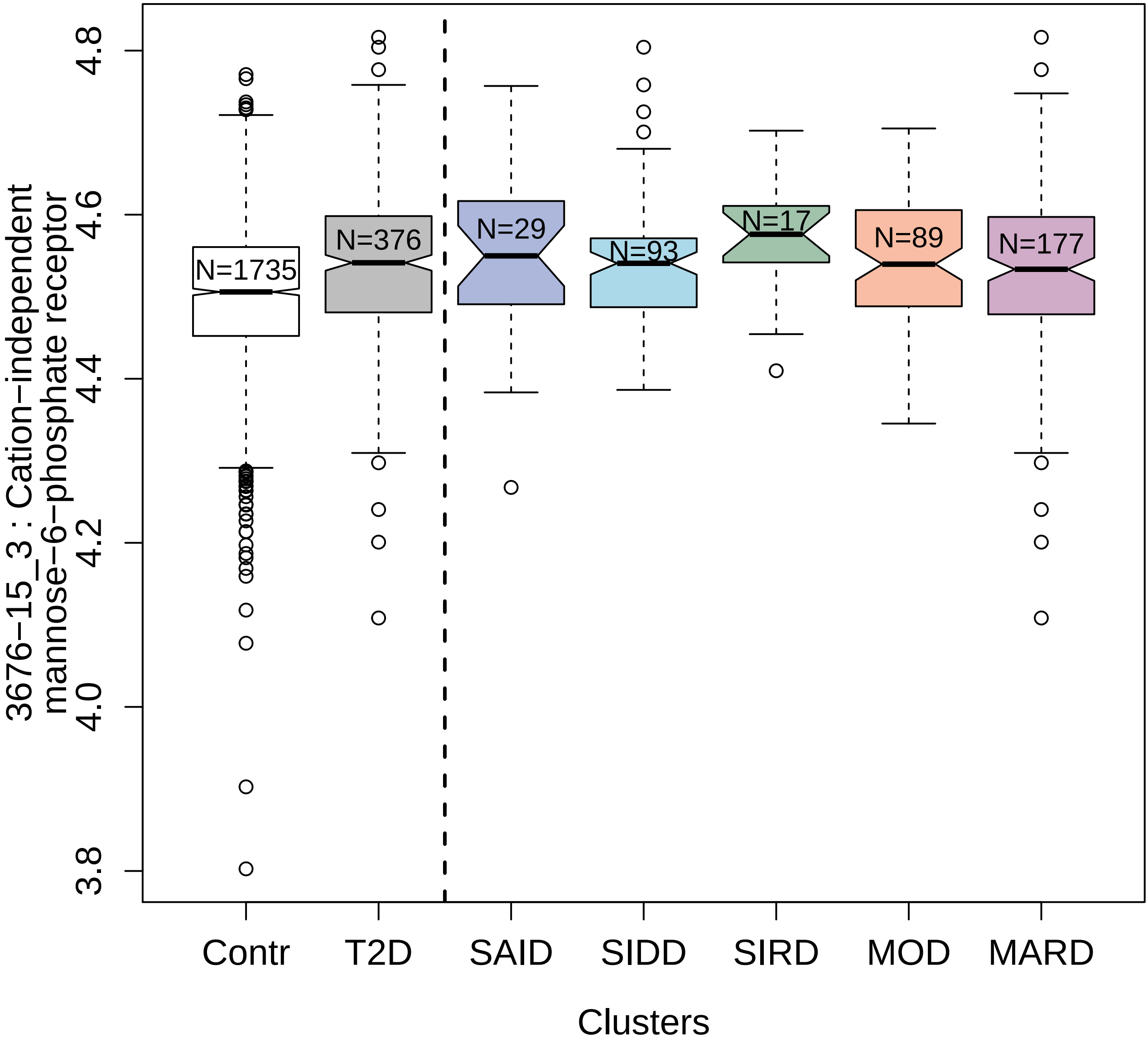

3676−15_3 : Cation−independent mannose−6−phosphate receptor
4.8
4.6
4.4
4.2
4.0
3.8
N=1735
N=376
N=29
N=93
N=17
N=89
N=177
Contr
T2D
SAID
SIDD
SIRD
MOD
MARD
Clusters

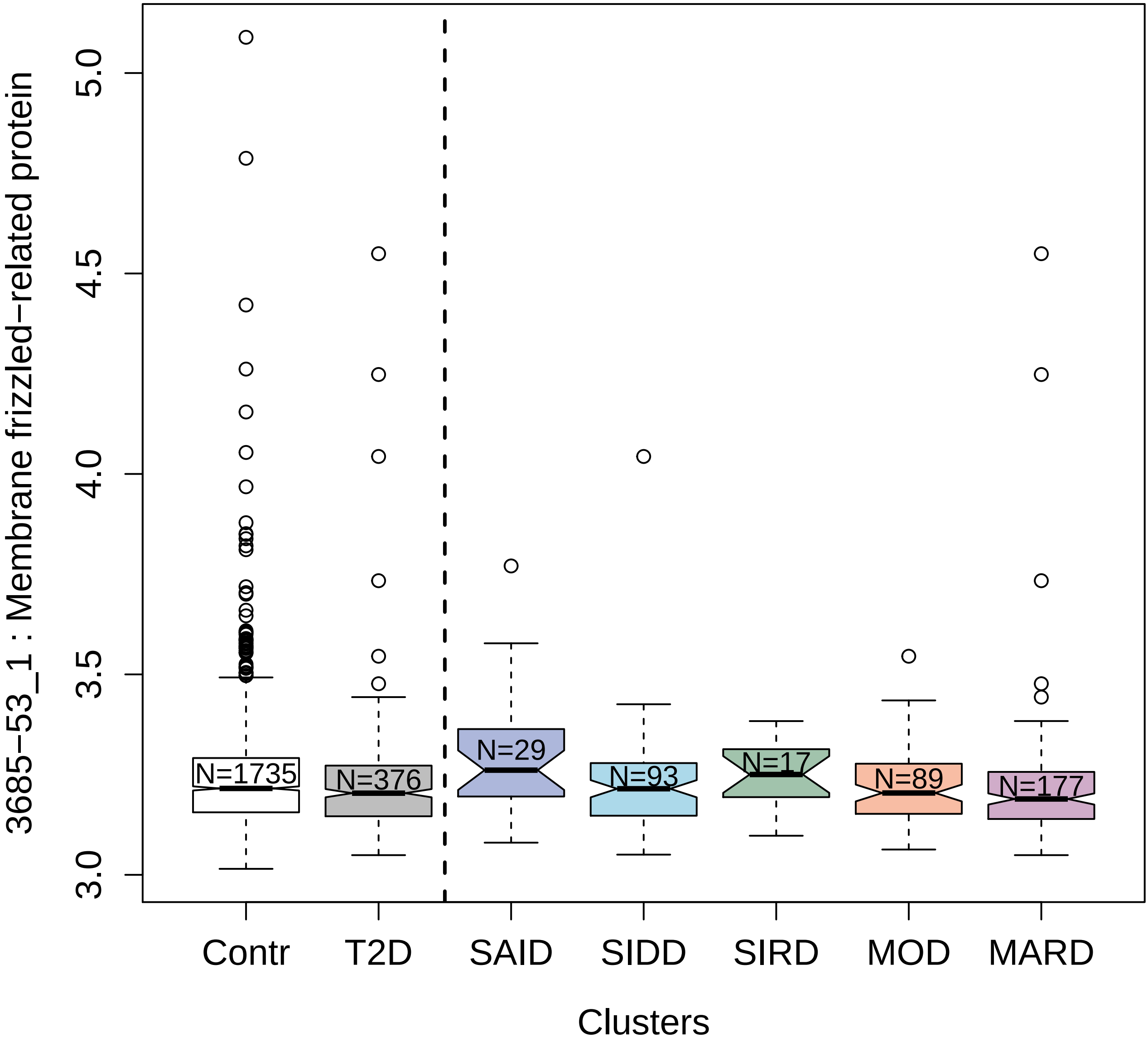

3685−53_1 : Membrane frizzled−related protein
5.0
4.5
4.0
3.5
3.0
N=1735
N=376
N=29
N=93
N=17
N=89
N=177
Contr
T2D
SAID
SIDD
SIRD
MOD
MARD
Clusters

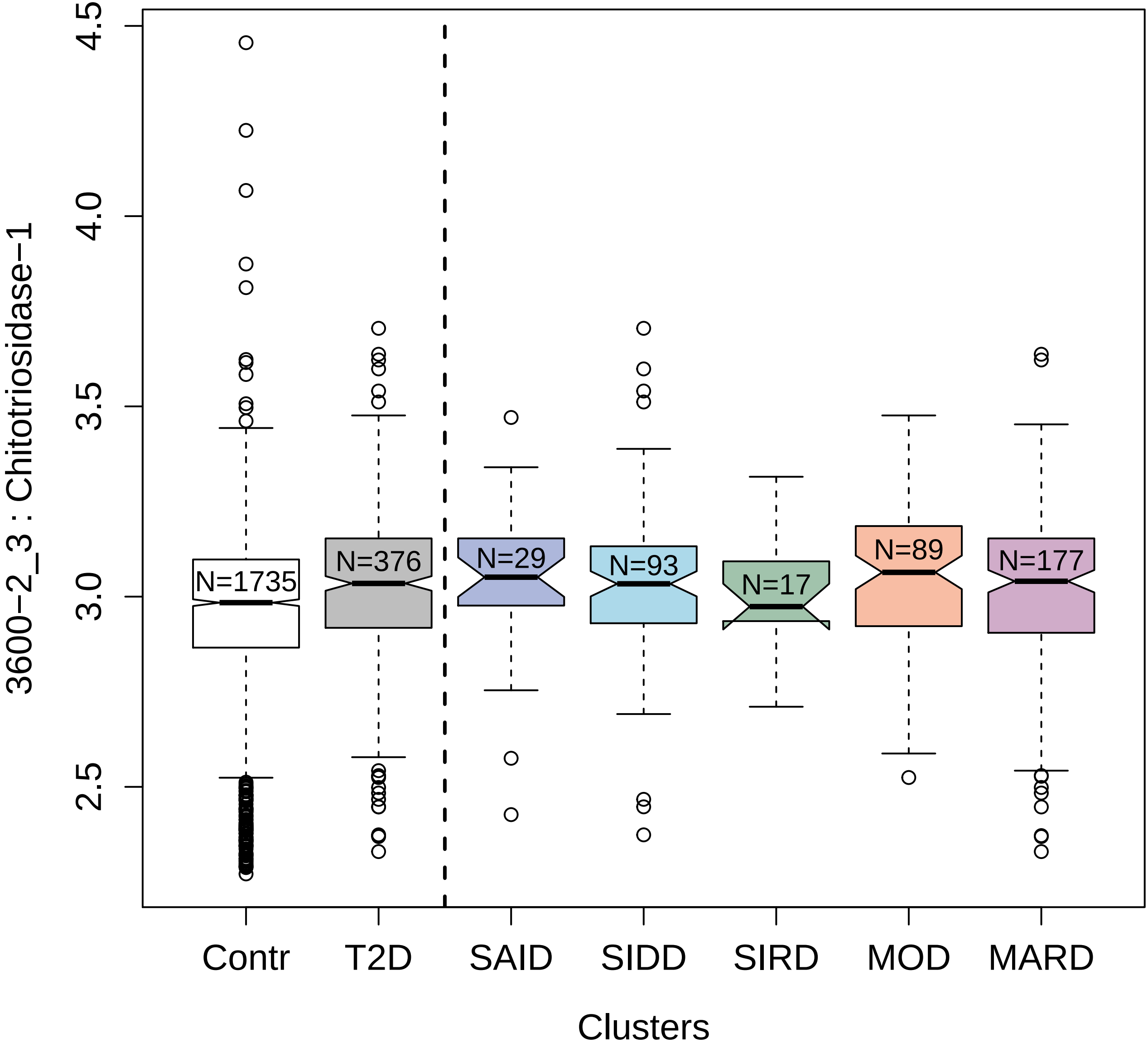
3600−2_3 : Chitotriosidase−1
4.5
4.0
3.5
3.0
2.5
N=1735
N=376
N=29
N=93
N=17
N=89
N=177
Contr
T2D
SAID
SIDD
SIRD
MOD
MARD
Clusters

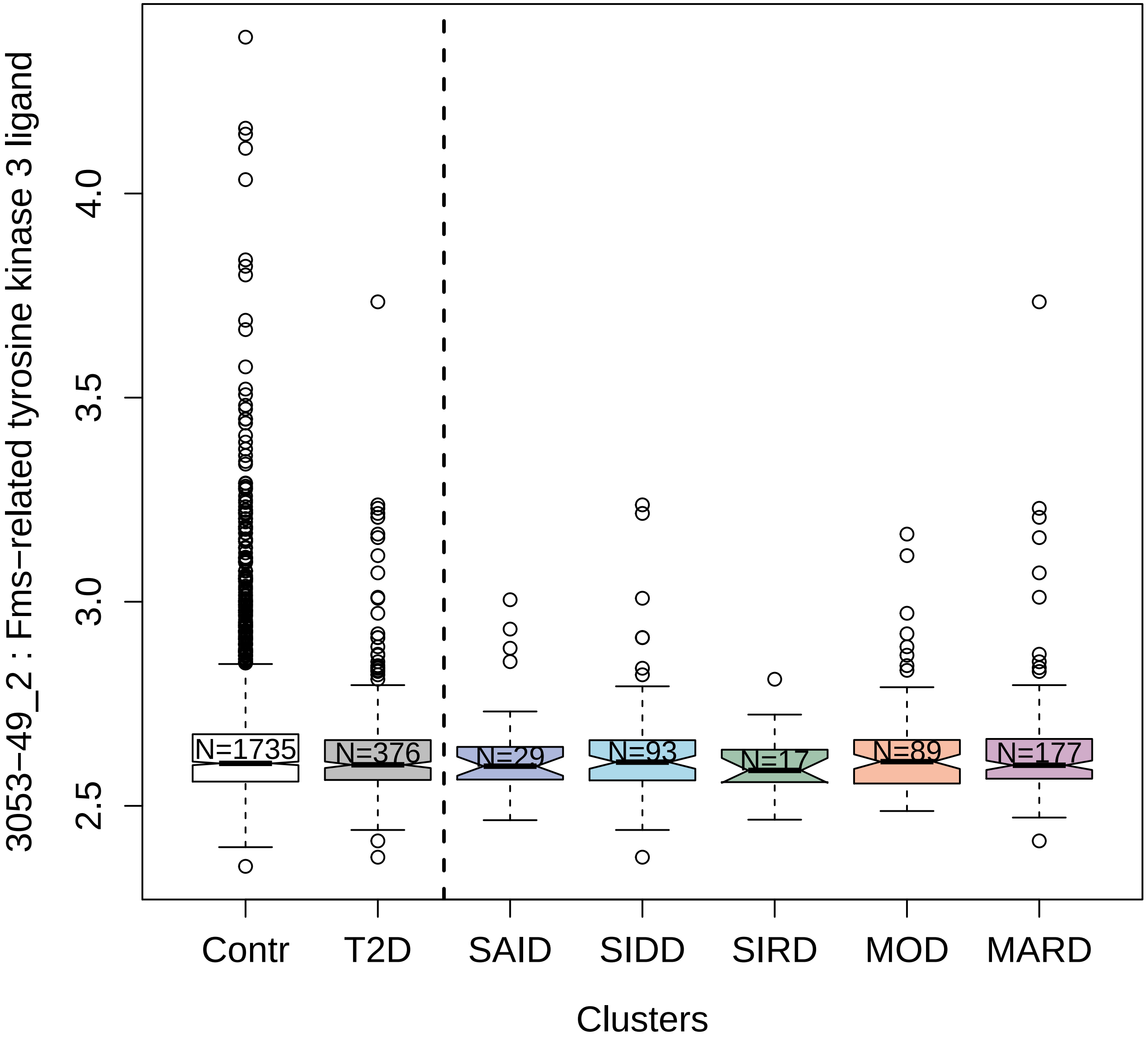
3053−49_2 : Fms−related tyrosine kinase 3 ligand
4.0
3.5
3.0
2.5
N=1735
N=376
N=29
N=93
N=17
N=89
N=177
Contr
T2D
SAID
SIDD
SIRD
MOD
MARD
Clusters

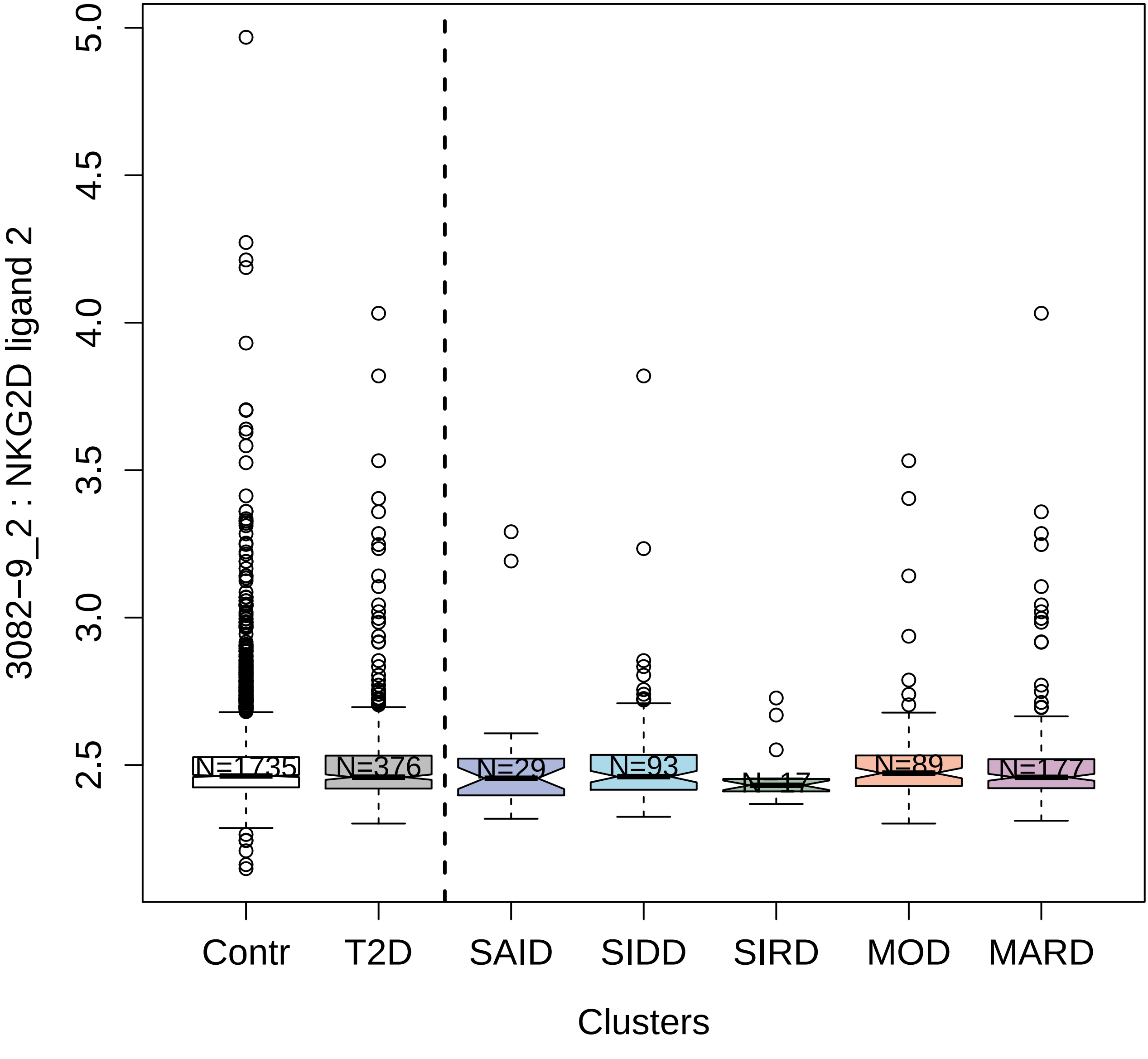

3082−9_2 : NKG2D ligand 2
5.0
4.5
4.0
3.5
3.0
2.5
N=1735
N=376
N=29
N=93
N=17
N=89
N=177
Contr
T2D
SAID
SIDD
SIRD
MOD
MARD
Clusters

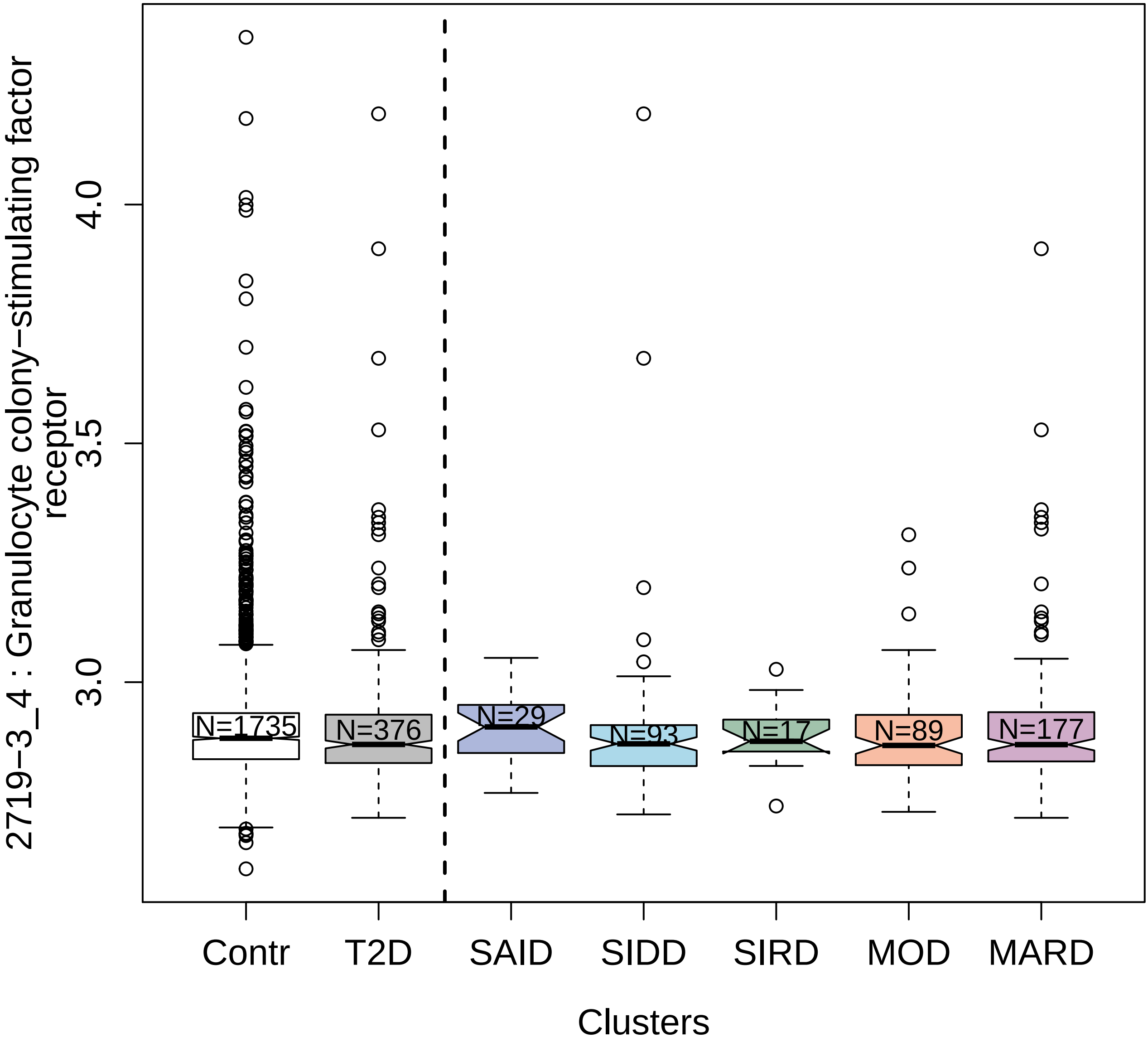

2719−3_4 : Granulocyte colony−stimulating factor receptor
3.0
3.5
4.0
N=1735
N=376
N=29
N=93
N=17
N=89
N=177
Contr
T2D
SAID
SIDD
SIRD
MOD
MARD
Clusters

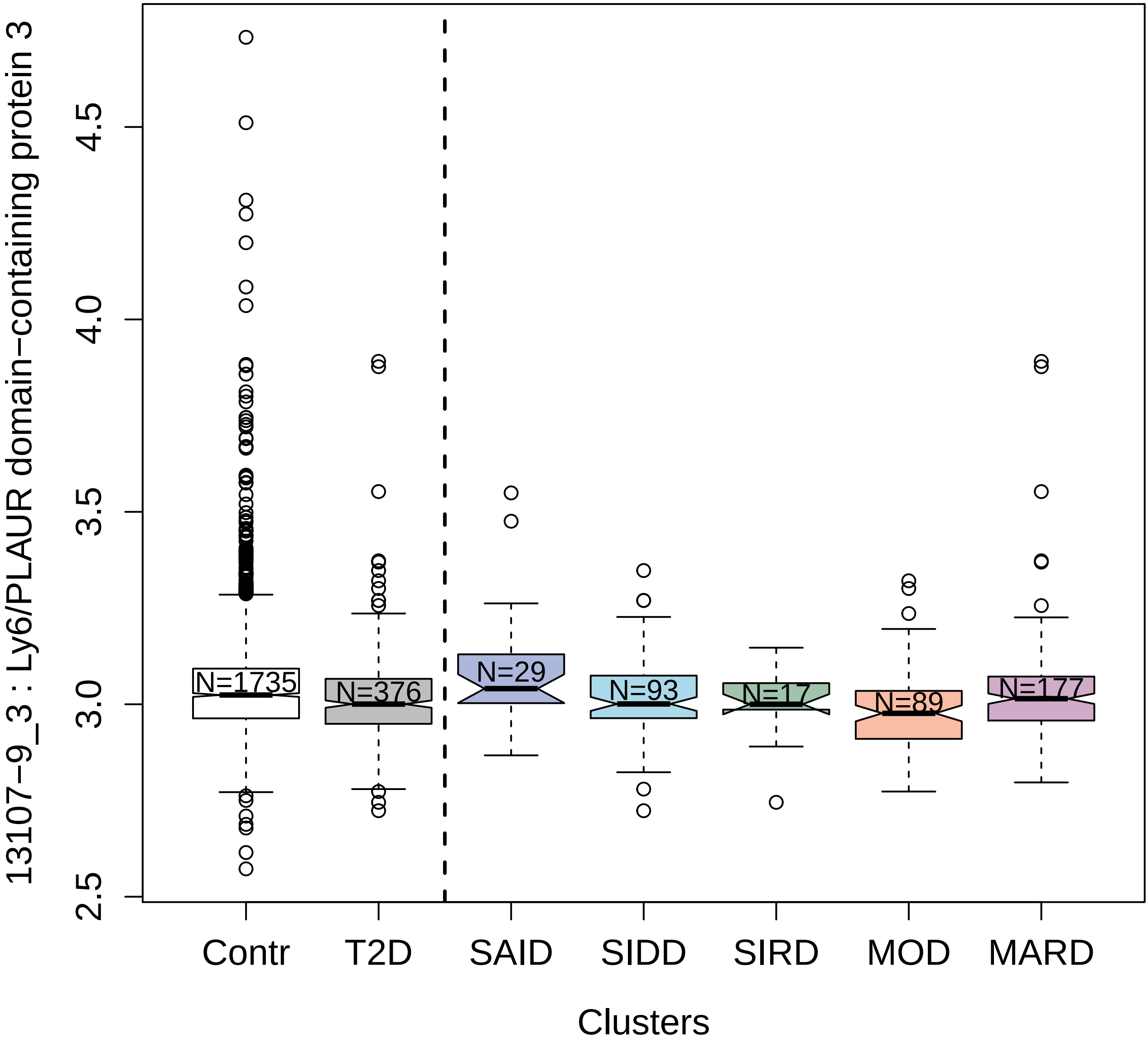

13107−9_3 : Ly6/PLAUR domain−containing protein 3
2.5
3.0
3.5
4.0
4.5
N=1735
N=376
N=29
N=93
N=17
N=89
N=177
Contr
T2D
SAID
SIDD
SIRD
MOD
MARD
Clusters

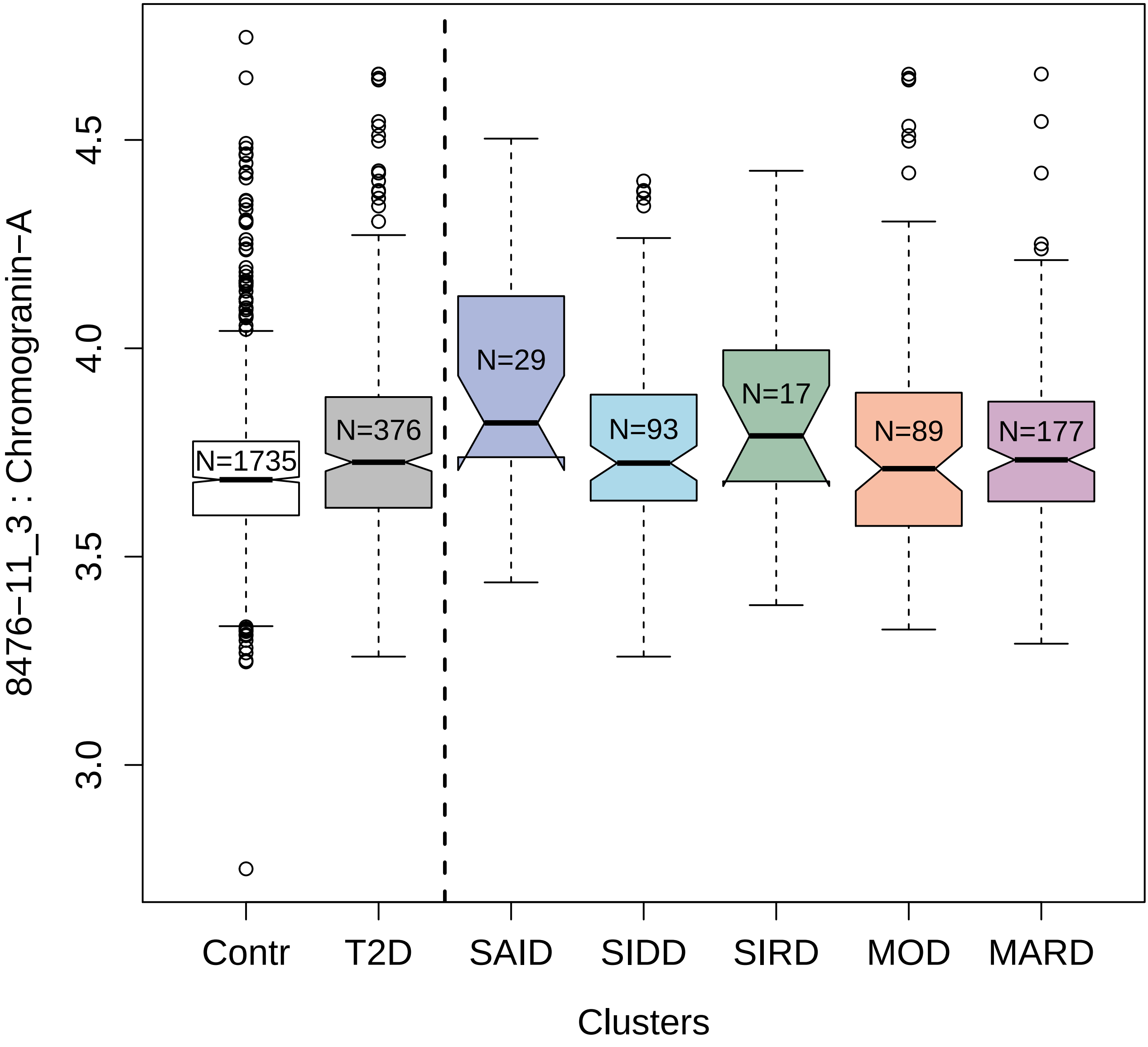

8476−11_3 : Chromogranin−A
3.0
3.5
4.0
4.5
N=1735
N=376
N=29
N=93
N=17
N=89
N=177
Contr
T2D
SAID
SIDD
SIRD
MOD
MARD
Clusters

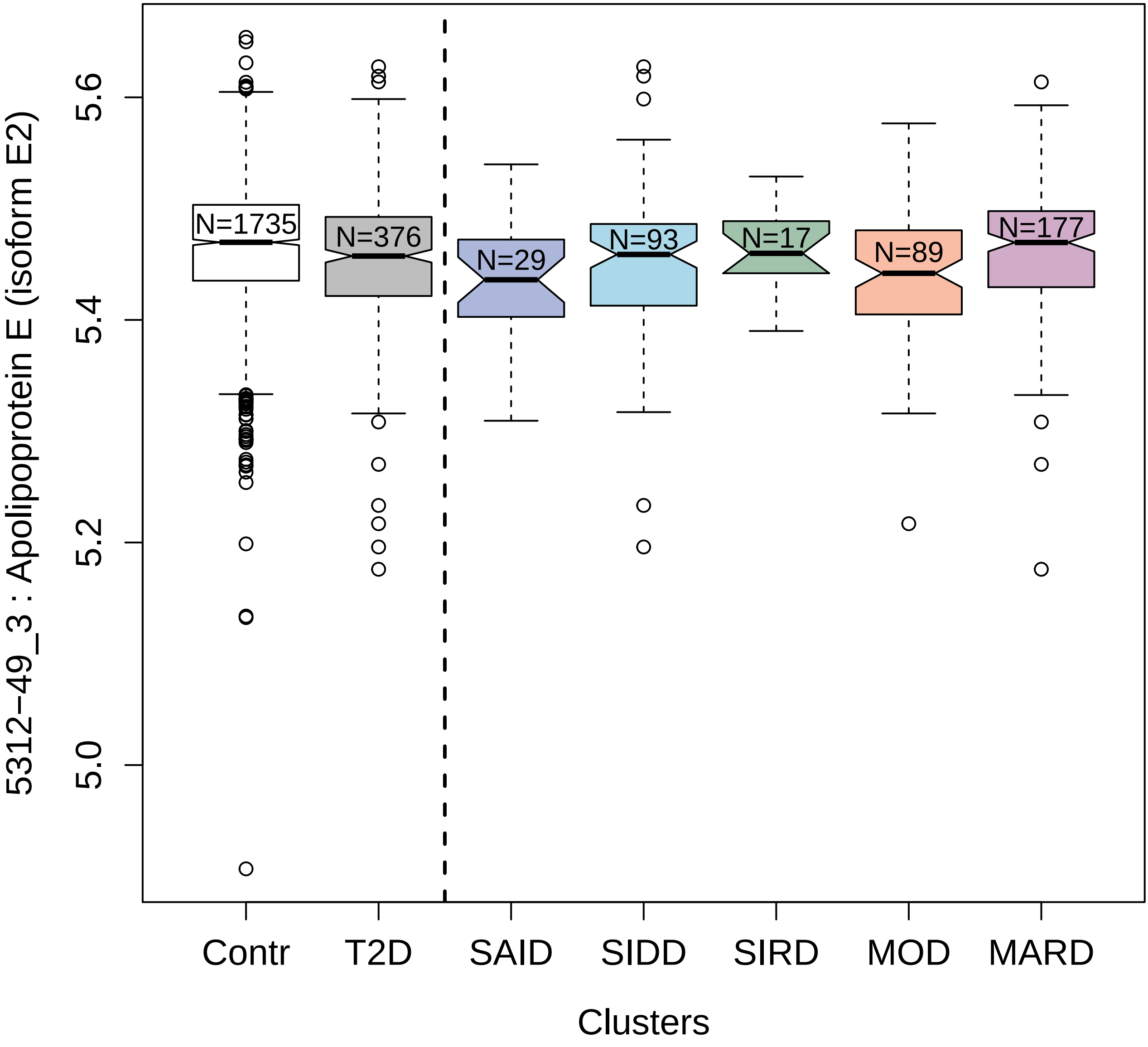

5312−49_3 : Apolipoprotein E (isoform E2)
5.0
5.2
5.4
5.6
N=1735
N=376
N=29
N=93
N=17
N=89
N=177
Contr
T2D
SAID
SIDD
SIRD
MOD
MARD
Clusters

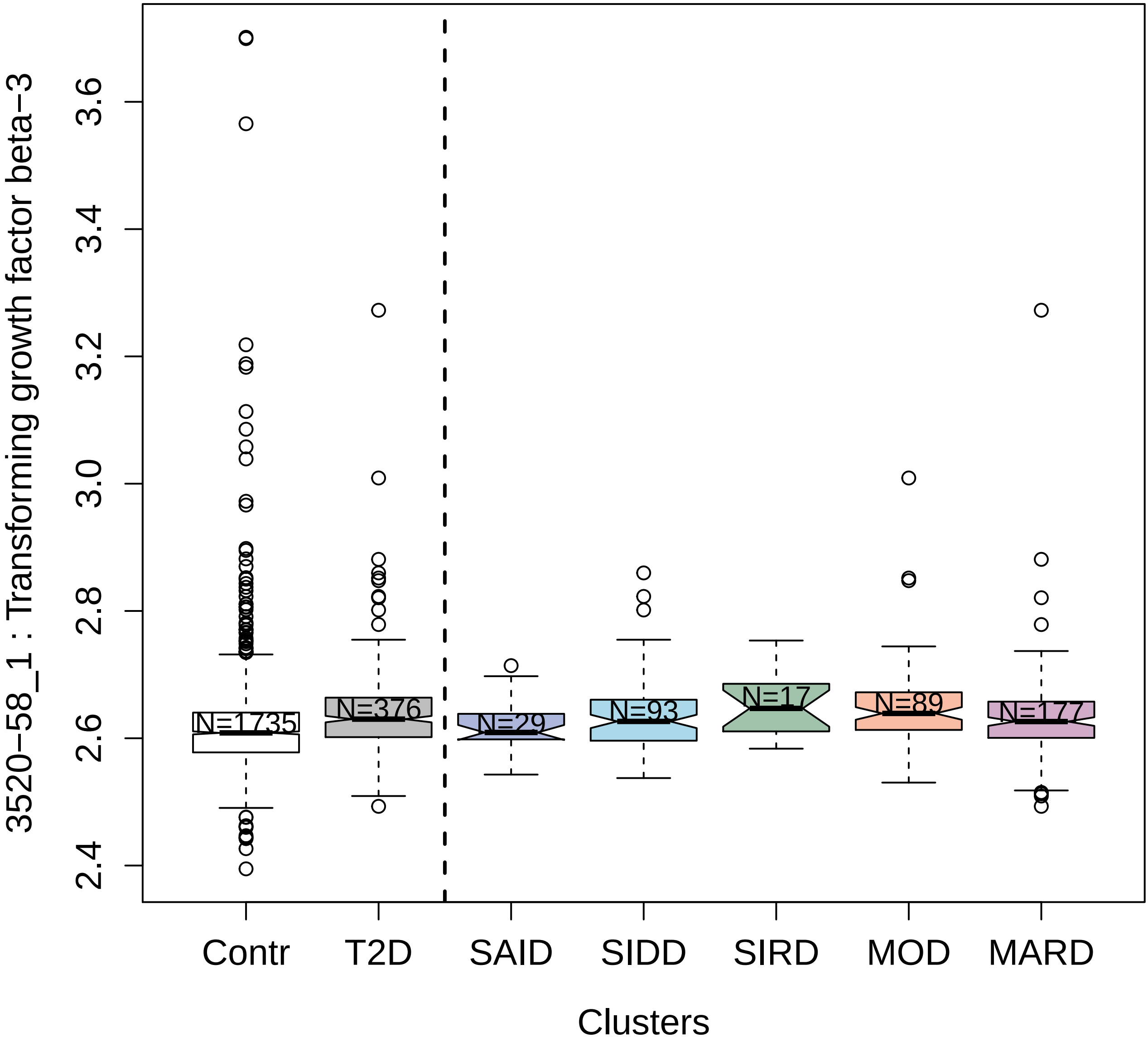

3520−58_1 : Transforming growth factor beta−3
2.4
2.6
2.8
3.0
3.2
3.4
3.6
N=1735
N=376
N=29
N=93
N=17
N=89
N=177
Contr
T2D
SAID
SIDD
SIRD
MOD
MARD
Clusters

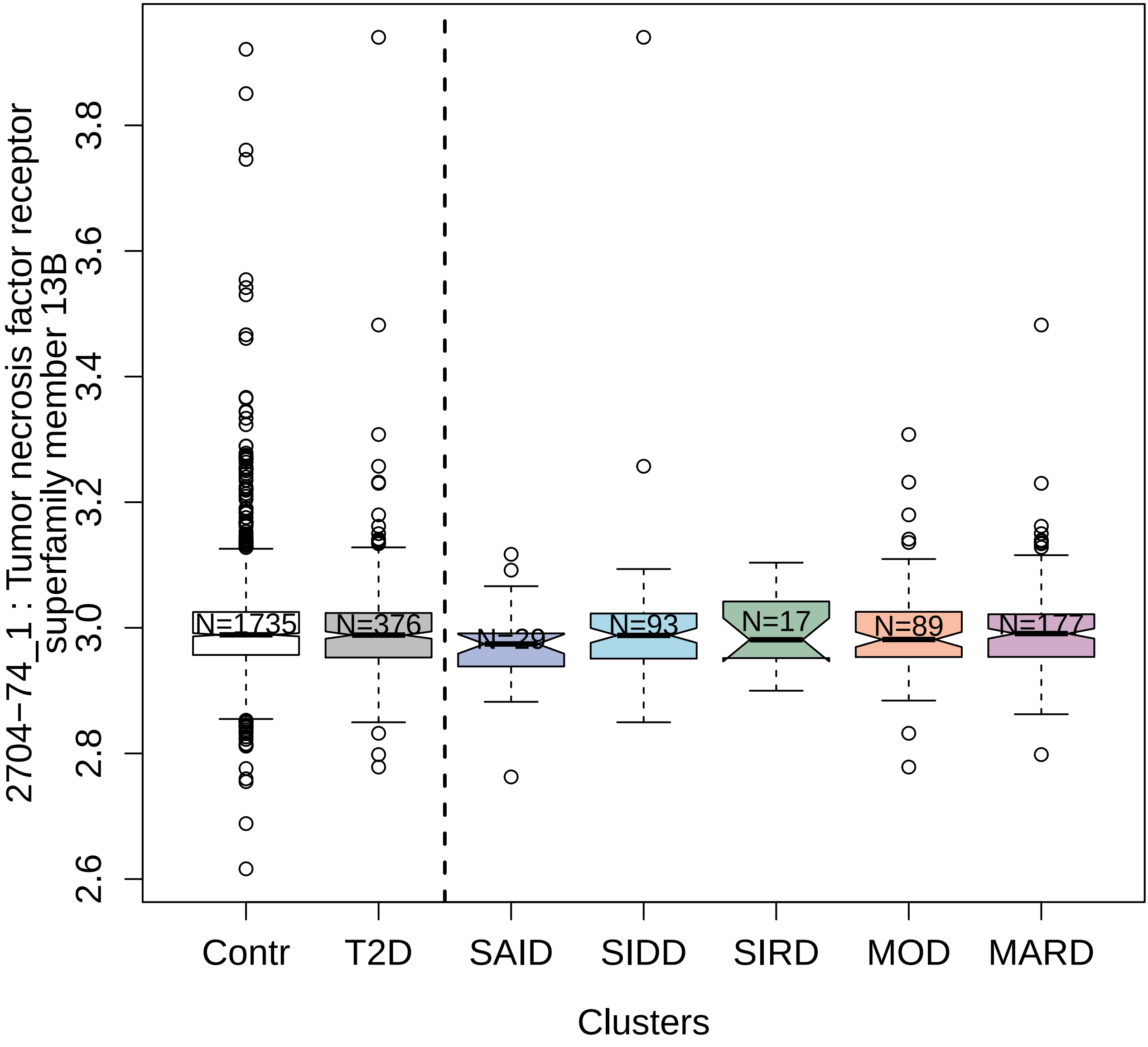
2704−74_1 : Tumor necrosis factor receptor superfamily member 13B
2.6
2.8
3.0
3.2
3.4
3.6
3.8
N=1735
N=376
N=29
N=93
N=17
N=89
N=177
Contr
T2D
SAID
SIDD
SIRD
MOD
MARD
Clusters

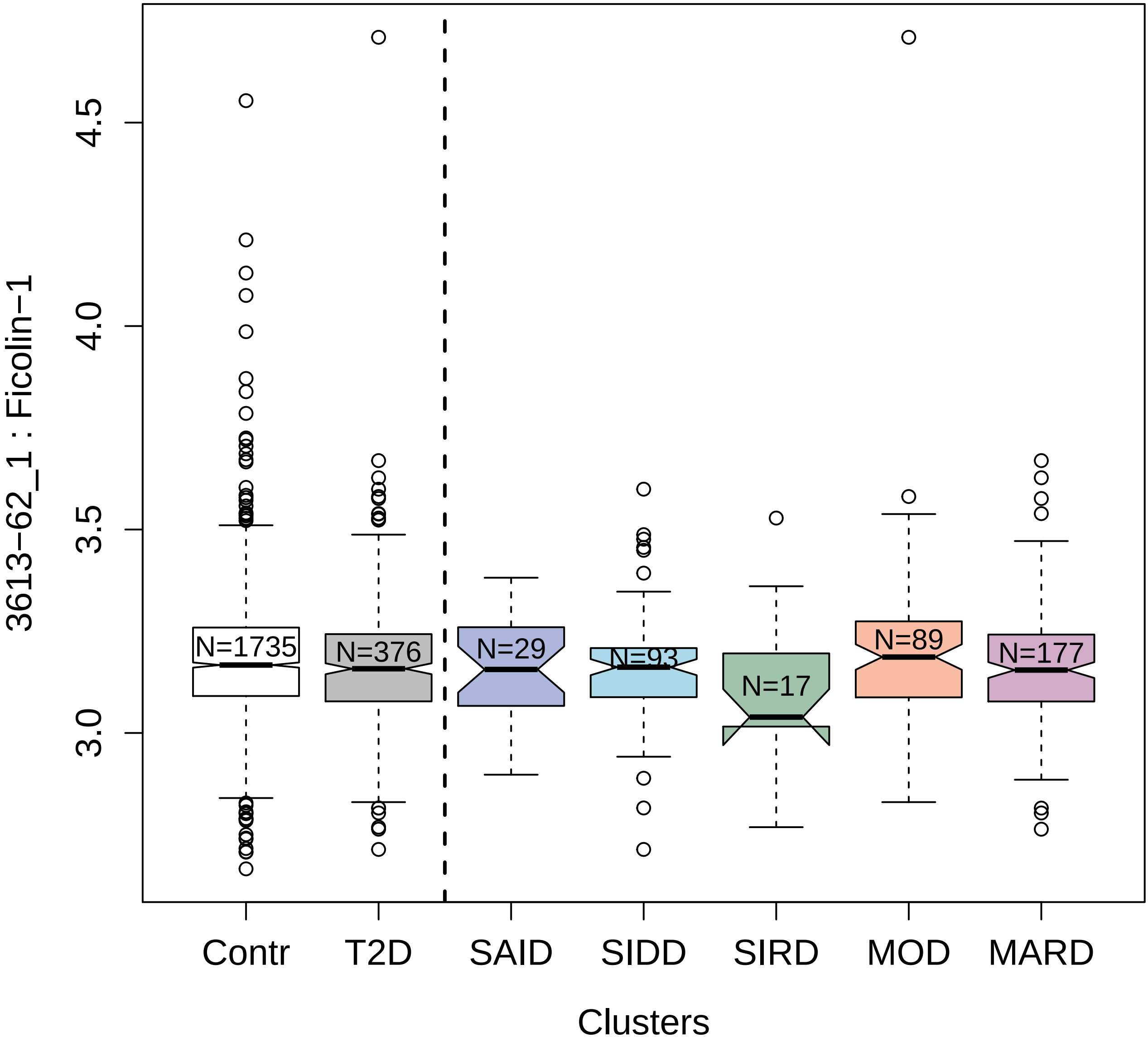

3613−62_1 : Ficolin−1
4.5
4.0
3.5
3.0
N=1735
N=376
N=29
N=93
N=17
N=89
N=177
Contr
T2D
SAID
SIDD
SIRD
MOD
MARD
Clusters

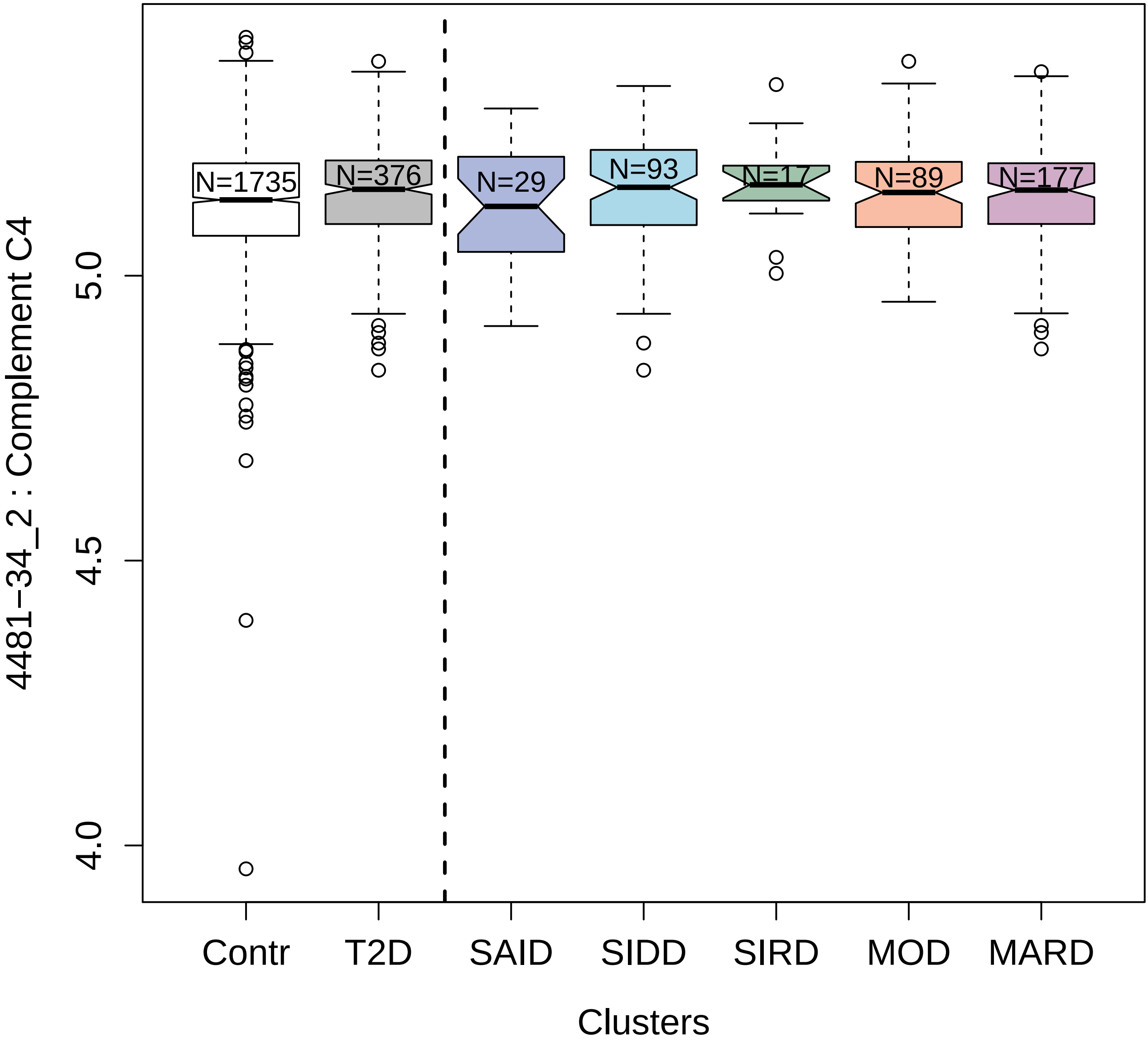

4481−34_2 : Complement C4
5.0
4.5
4.0
N=1735
N=376
N=29
N=93
N=17
N=89
N=177
Contr
T2D
SAID
SIDD
SIRD
MOD
MARD
Clusters

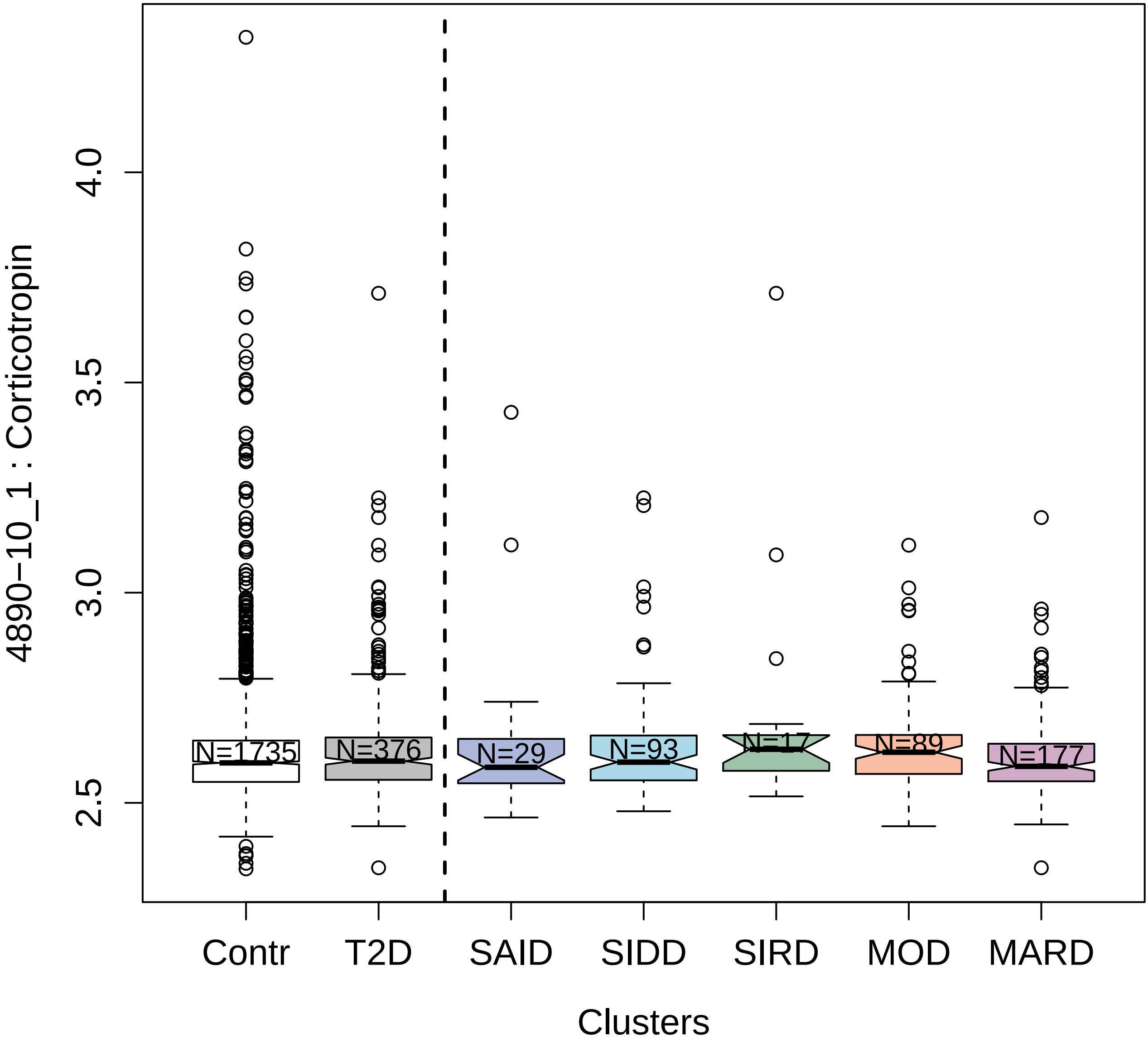

4890−10_1 : Corticotropin
4.0
3.5
3.0
2.5
N=1735
N=376
N=29
N=93
N=17
N=89
N=177
Contr
T2D
SAID
SIDD
SIRD
MOD
MARD
Clusters

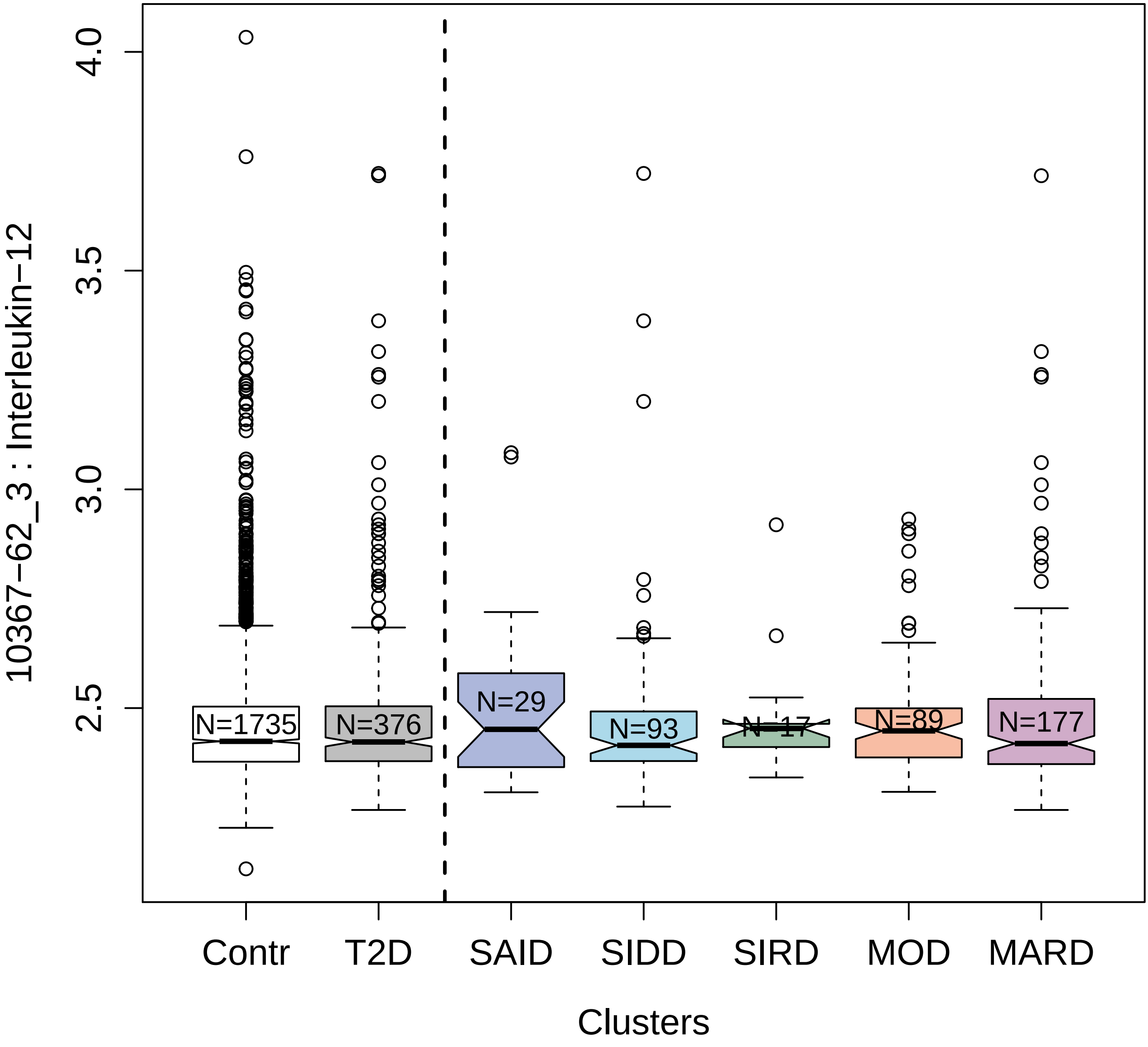

10367−62_3 : Interleukin−12
4.0
3.5
3.0
2.5
N=1735
N=376
N=29
N=93
N=17
N=89
N=177
Contr
T2D
SAID
SIDD
SIRD
MOD
MARD
Clusters

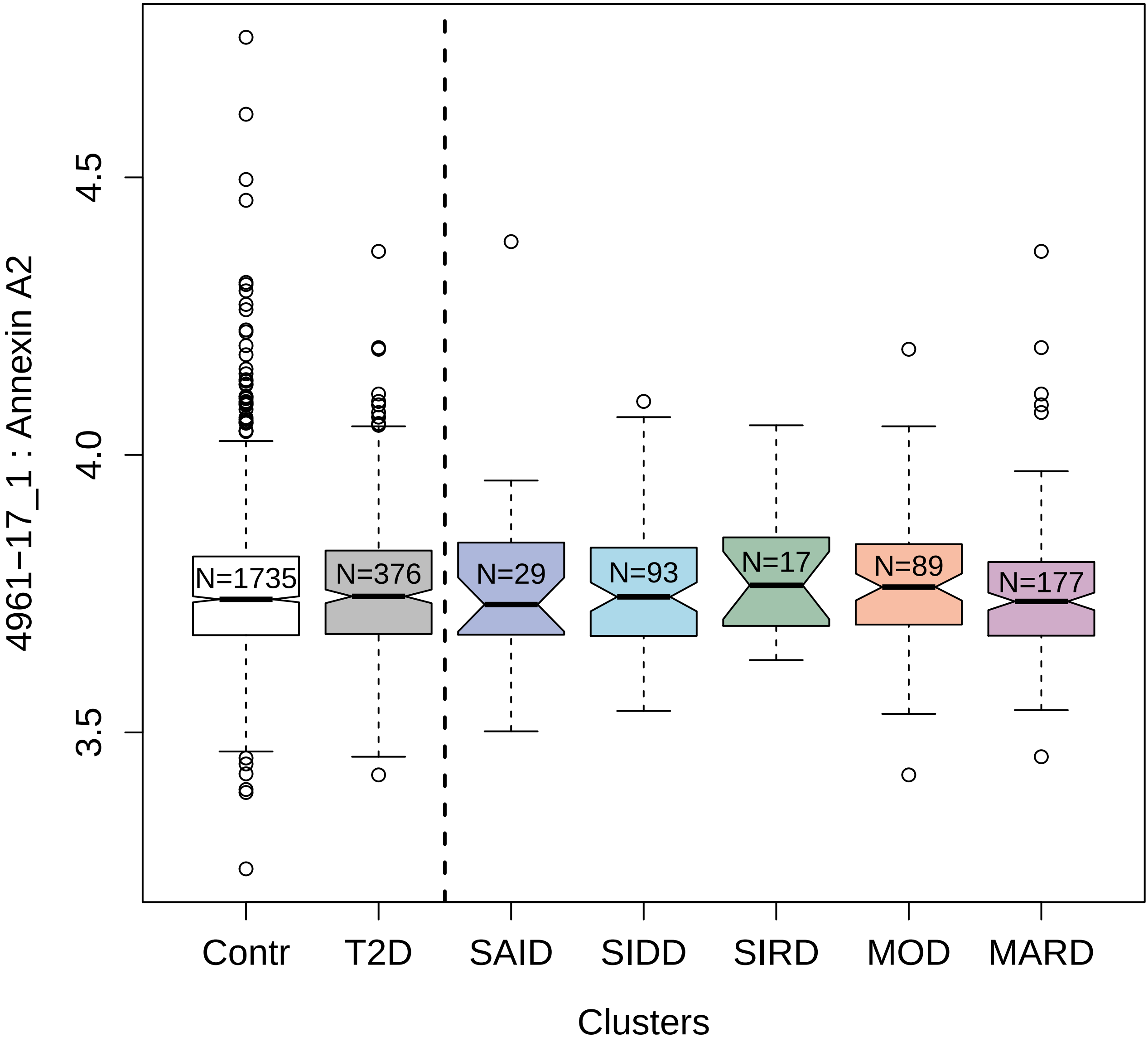

4961−17_1 : Annexin A2
4.5
4.0
3.5
N=1735
N=376
N=29
N=93
N=17
N=89
N=177
Contr
T2D
SAID
SIDD
SIRD
MOD
MARD
Clusters

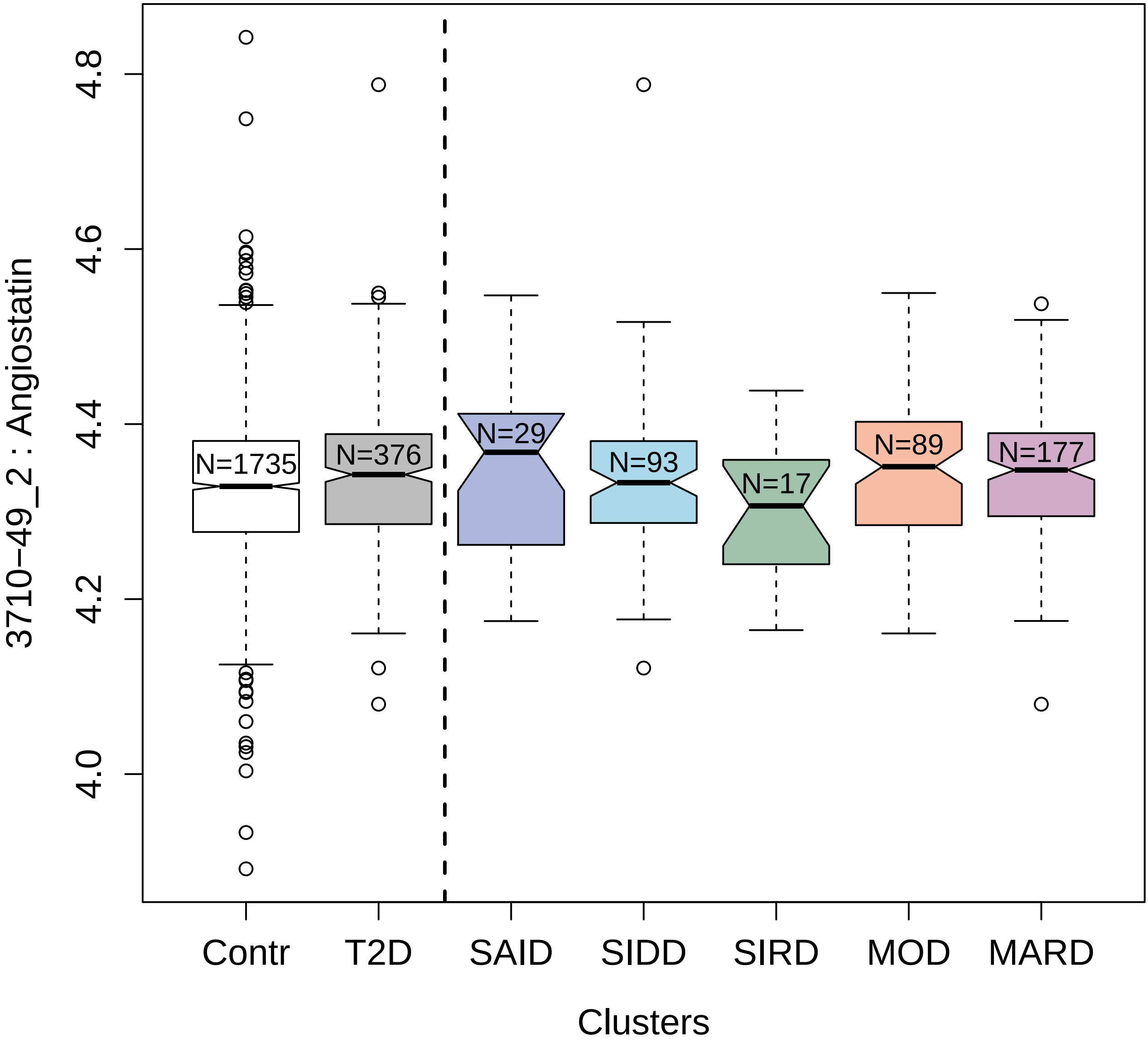
3710−49_2 : Angiostatin
4.0
4.2
4.4
4.6
4.8
N=1735
N=376
N=29
N=93
N=17
N=89
N=177
Contr
T2D
SAID
SIDD
SIRD
MOD
MARD
Clusters

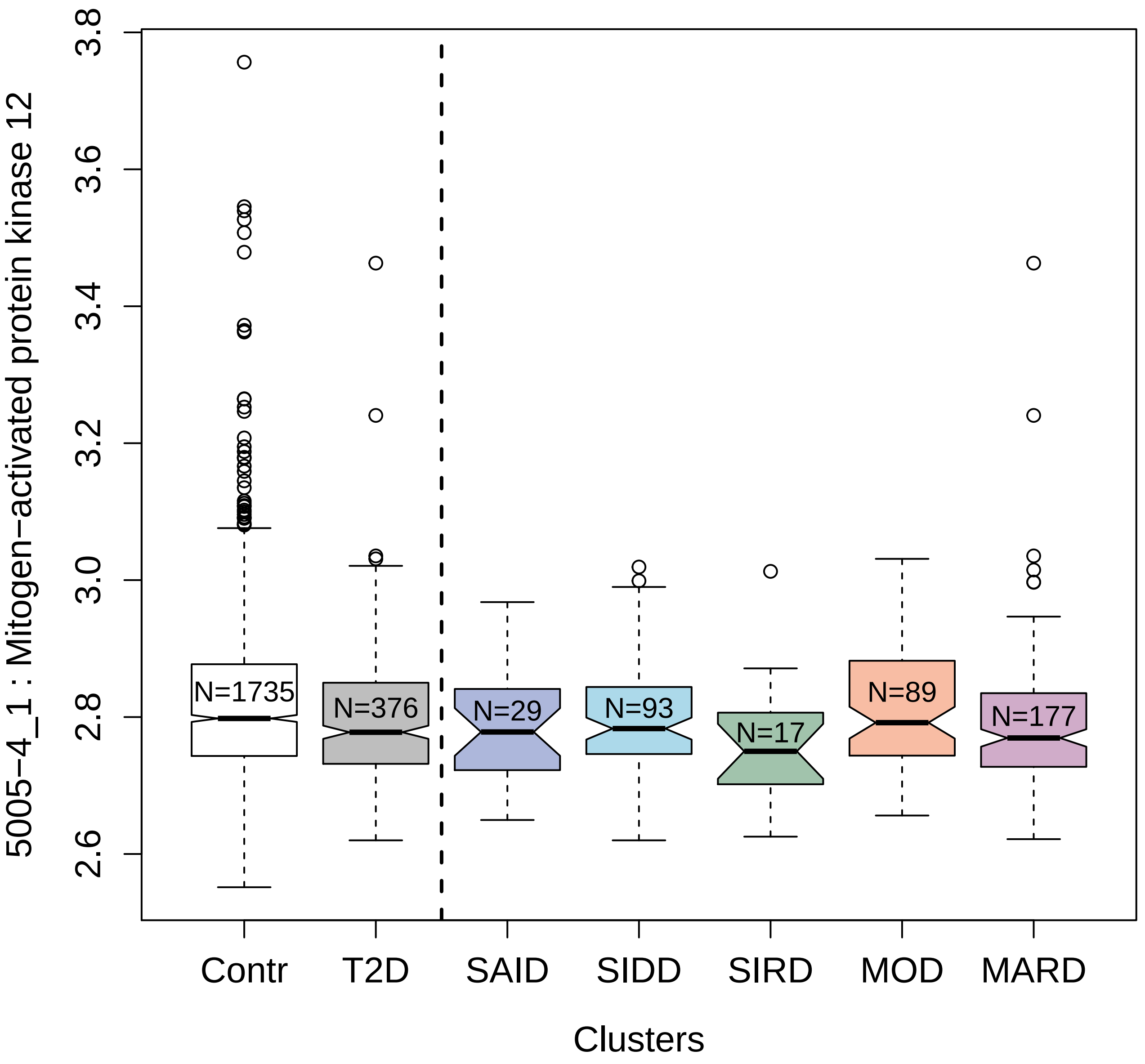

5005−4_1 : Mitogen−activated protein kinase 12
3.8
3.6
3.4
3.2
3.0
2.8
2.6
N=1735
N=376
N=29
N=93
N=17
N=89
N=177
Contr
T2D
SAID
SIDD
SIRD
MOD
MARD
Clusters

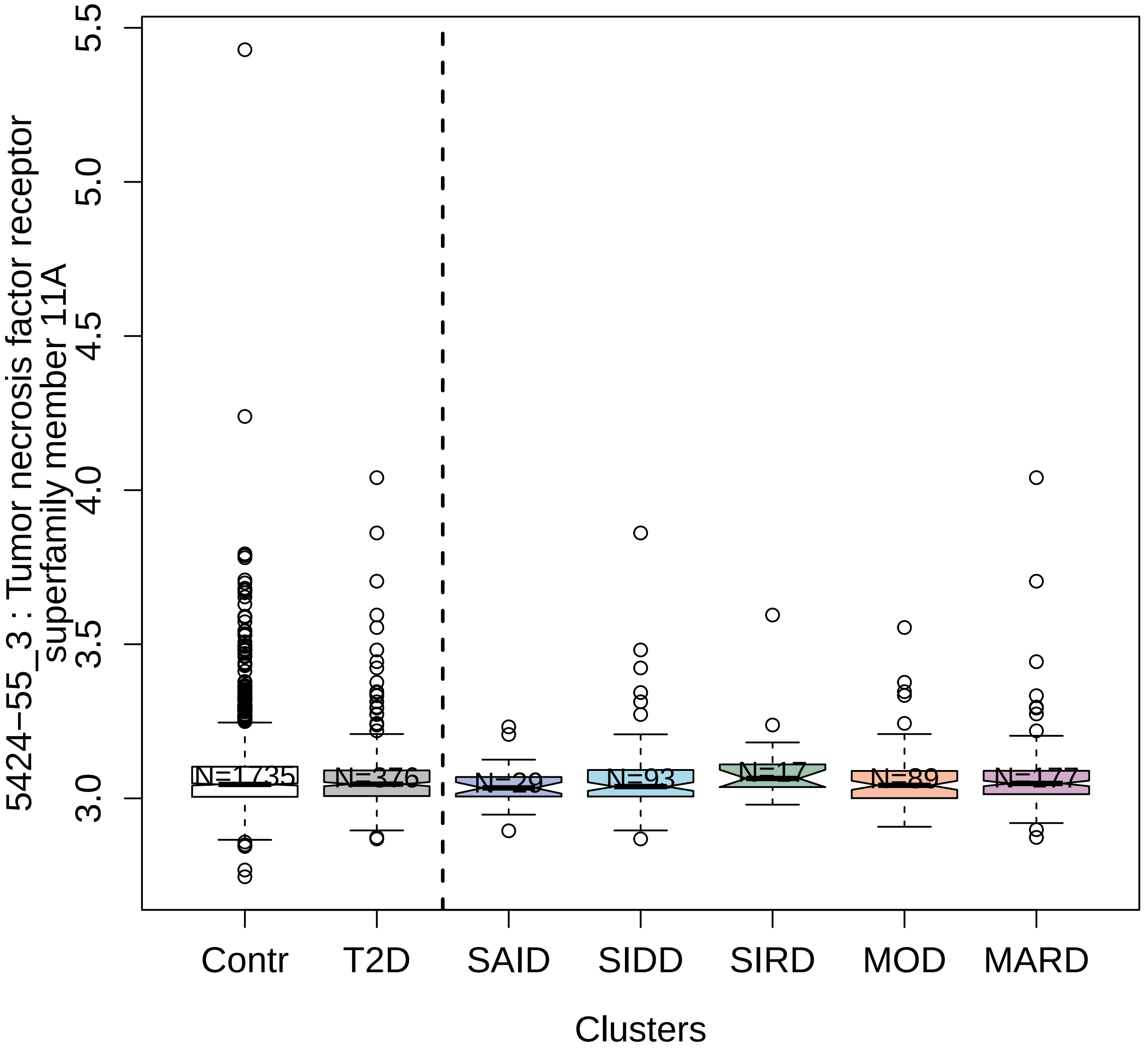
5424−55_3 : Tumor necrosis factor receptor
superfamily member 11A
3.0
3.5
4.0
4.5
5.0
5.5
N=1735
N=376
N=29
N=93
N=17
N=89
N=177
Contr
T2D
SAID
SIDD
SIRD
MOD
MARD
Clusters

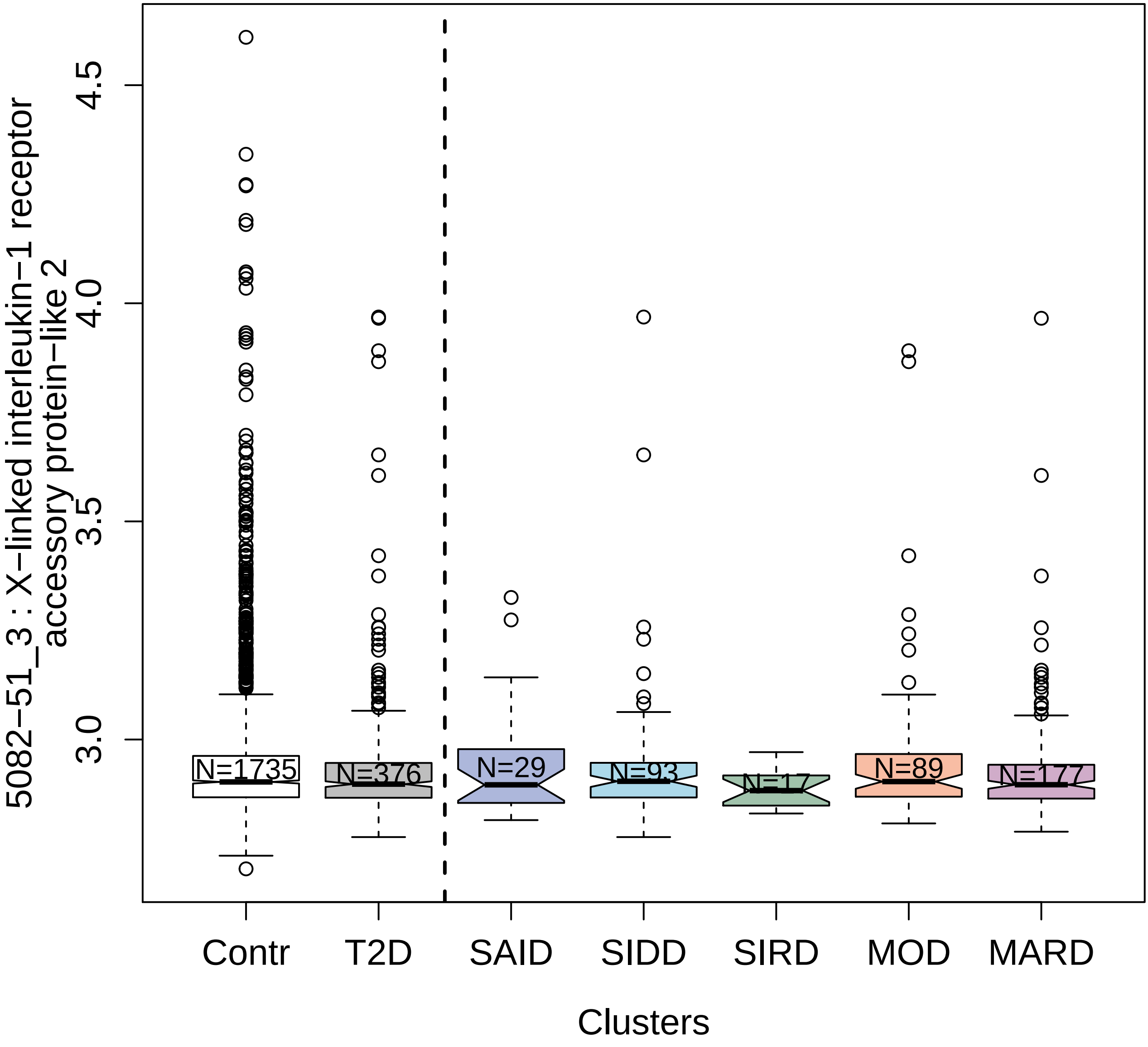

5082−51_3 : X−linked interleukin−1 receptor accessory protein−like 2
4.5
4.0
3.5
3.0
N=1735
N=376
N=29
N=93
N=17
N=89
N=177
Contr
T2D
SAID
SIDD
SIRD
MOD
MARD
Clusters

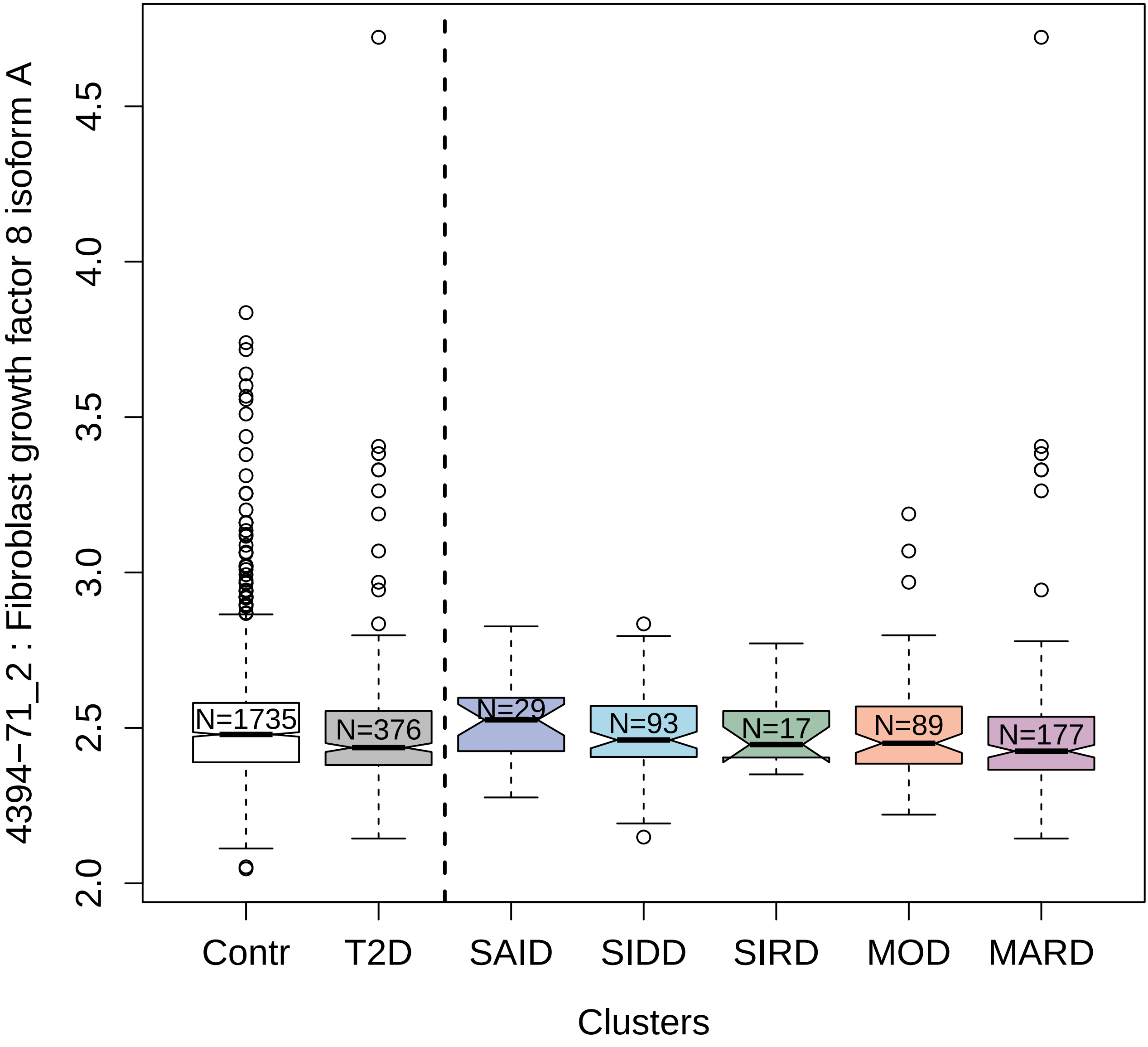
4394−71_2 : Fibroblast growth factor 8 isoform A
2.0
2.5
3.0
3.5
4.0
4.5
N=1735
N=376
N=29
N=93
N=17
N=89
N=177
Contr
T2D
SAID
SIDD
SIRD
MOD
MARD
Clusters

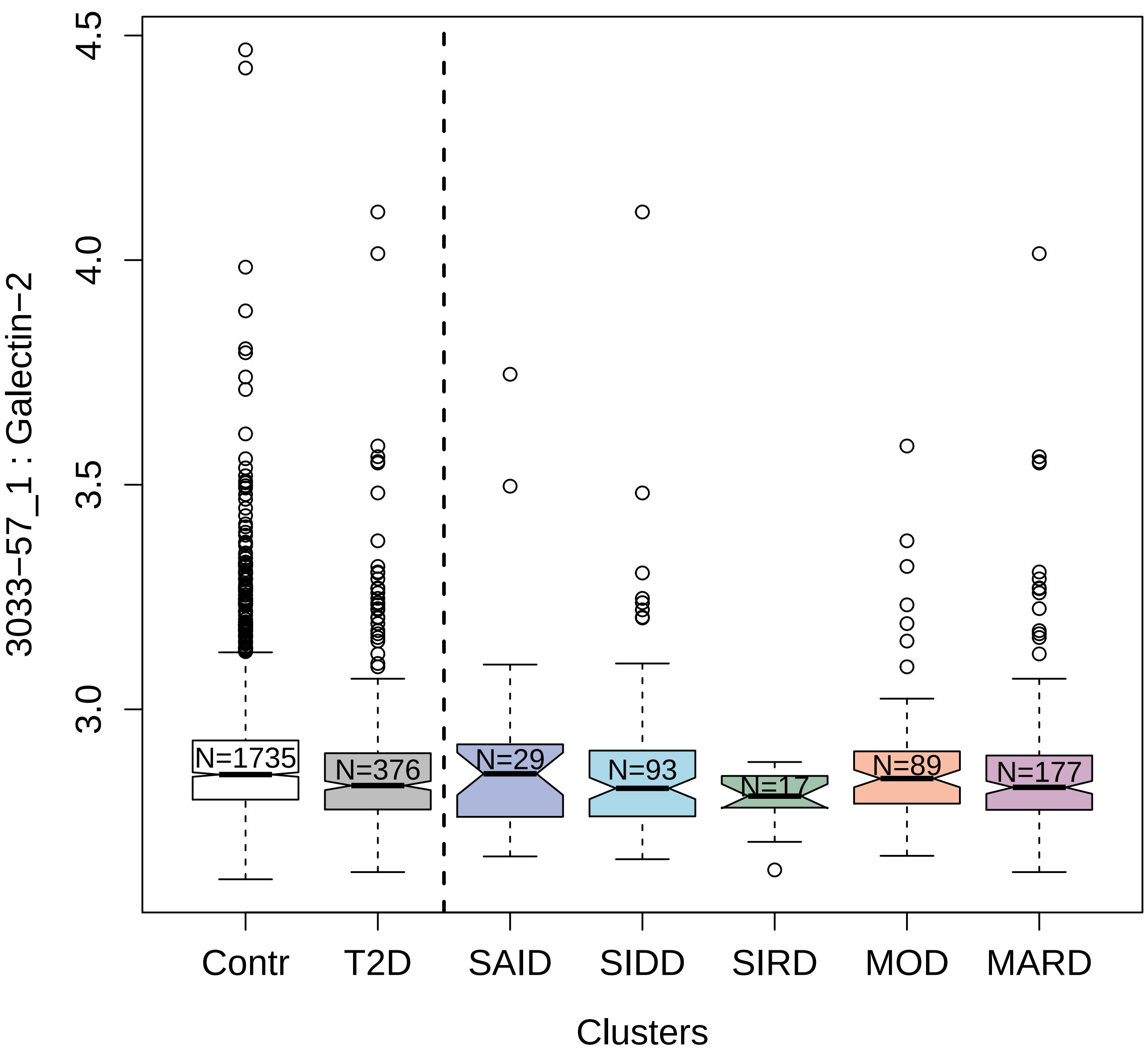

3033−57_1 : Galectin−2
4.5
4.0
3.5
3.0
N=1735
N=376
N=29
N=93
N=17
N=89
N=177
Contr
T2D
SAID
SIDD
SIRD
MOD
MARD
Clusters

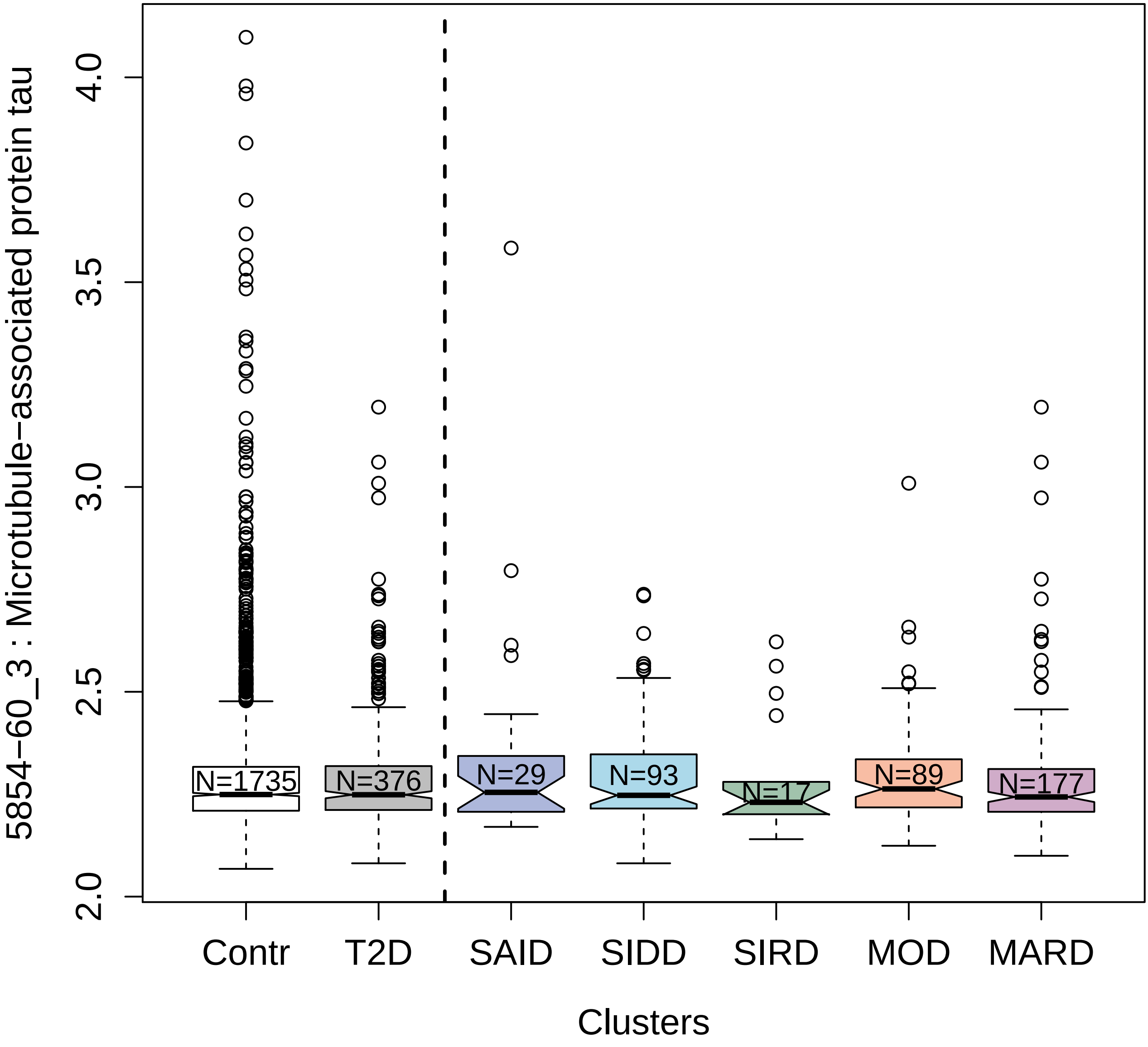

5854−60_3 : Microtubule−associated protein tau
2.0
2.5
3.0
3.5
4.0
N=1735
N=376
N=29
N=93
N=17
N=89
N=177
Contr
T2D
SAID
SIDD
SIRD
MOD
MARD
Clusters

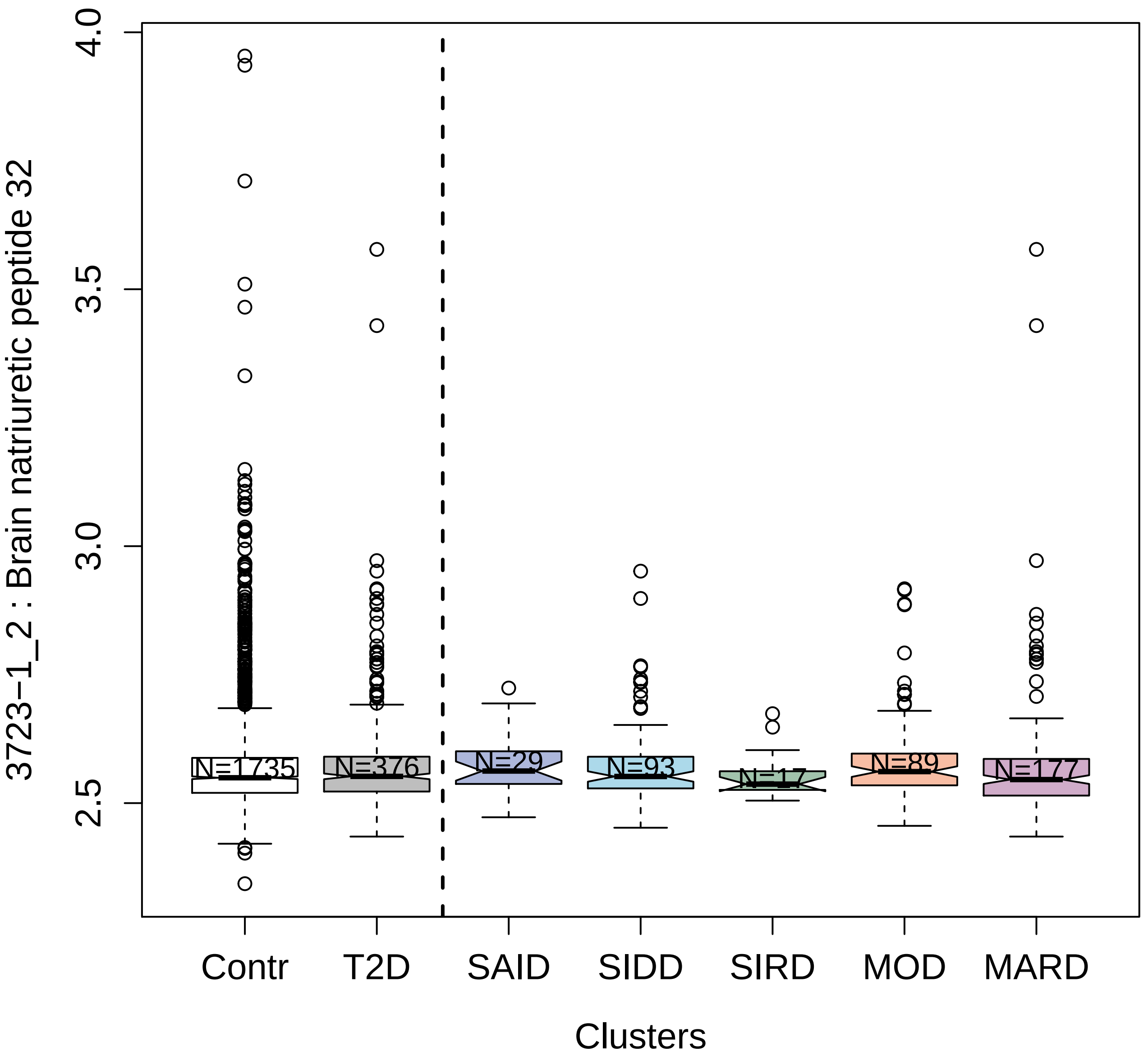

3723−1_2 : Brain natriuretic peptide 32
4.0
3.5
3.0
2.5
N=1735
N=376
N=29
N=93
N=17
N=89
N=177
Contr
T2D
SAID
SIDD
SIRD
MOD
MARD
Clusters

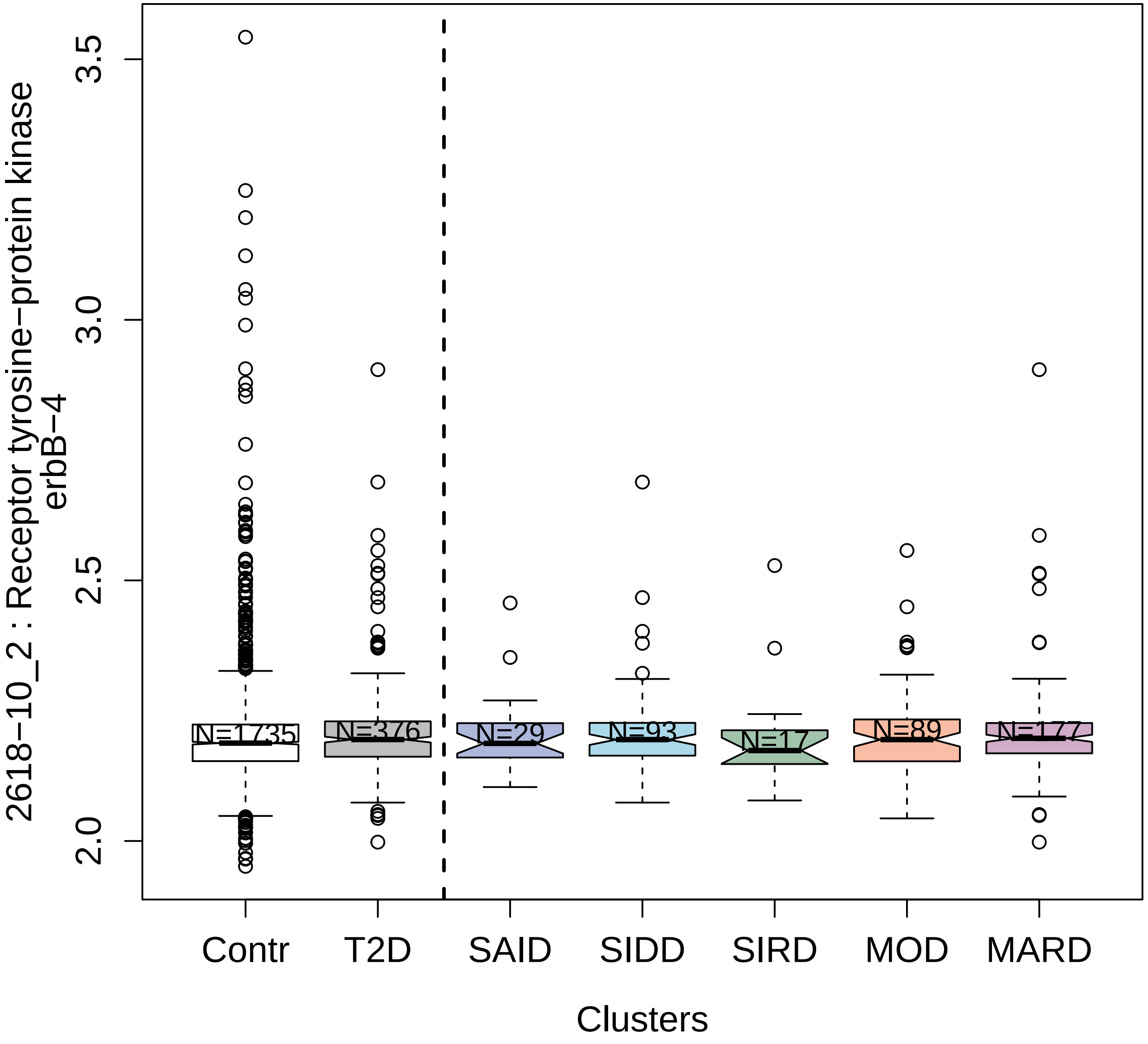

2618−10_2 : Receptor tyrosine−protein kinase erbB−4
3.5
3.0
2.5
2.0
N=1735
N=376
N=29
N=93
N=17
N=89
N=177
Contr
T2D
SAID
SIDD
SIRD
MOD
MARD
Clusters

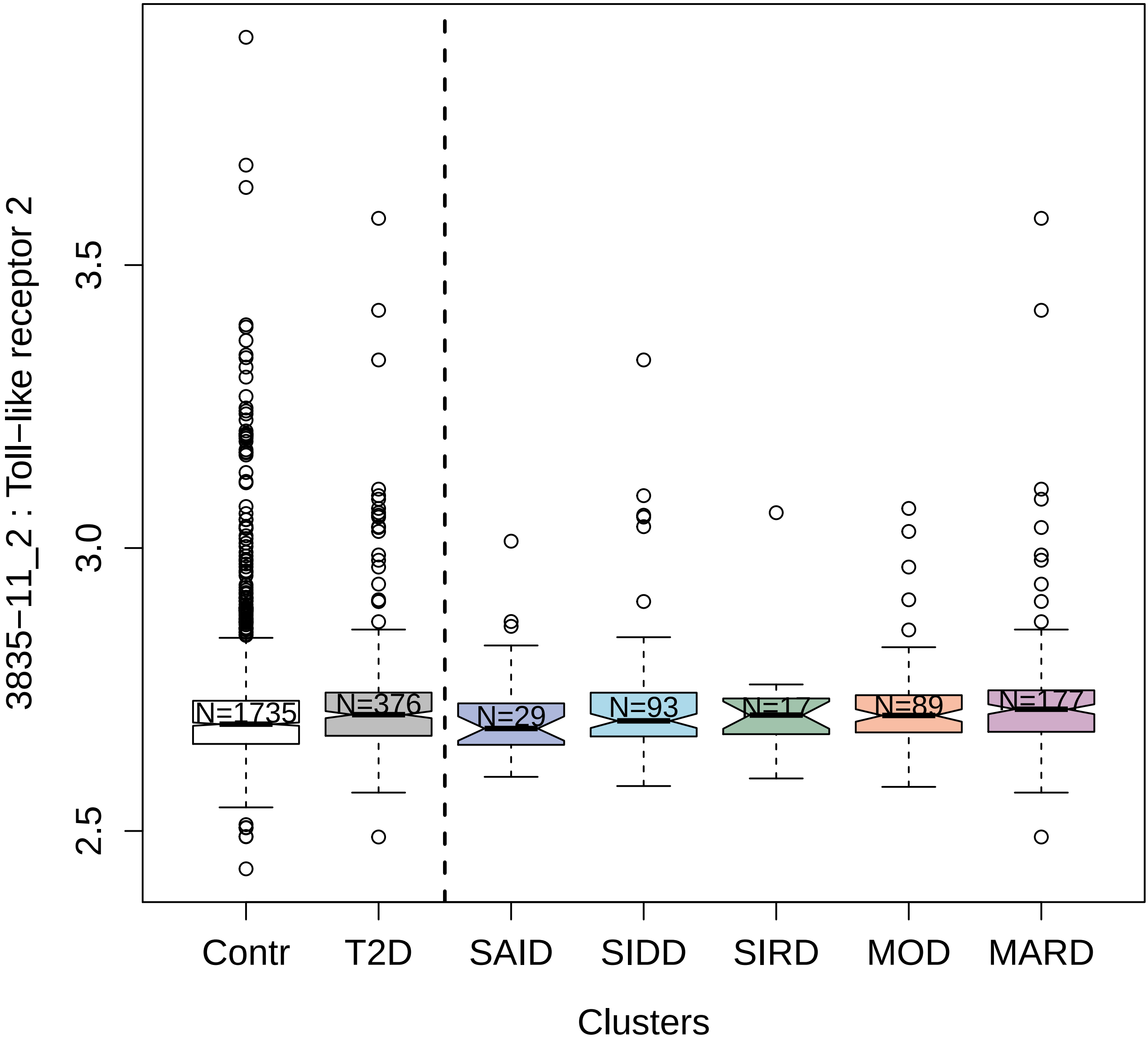

3835−11_2 : Toll−like receptor 2
3.5
3.0
2.5
N=1735
N=376
N=29
N=93
N=17
N=89
N=177
Contr
T2D
SAID
SIDD
SIRD
MOD
MARD
Clusters

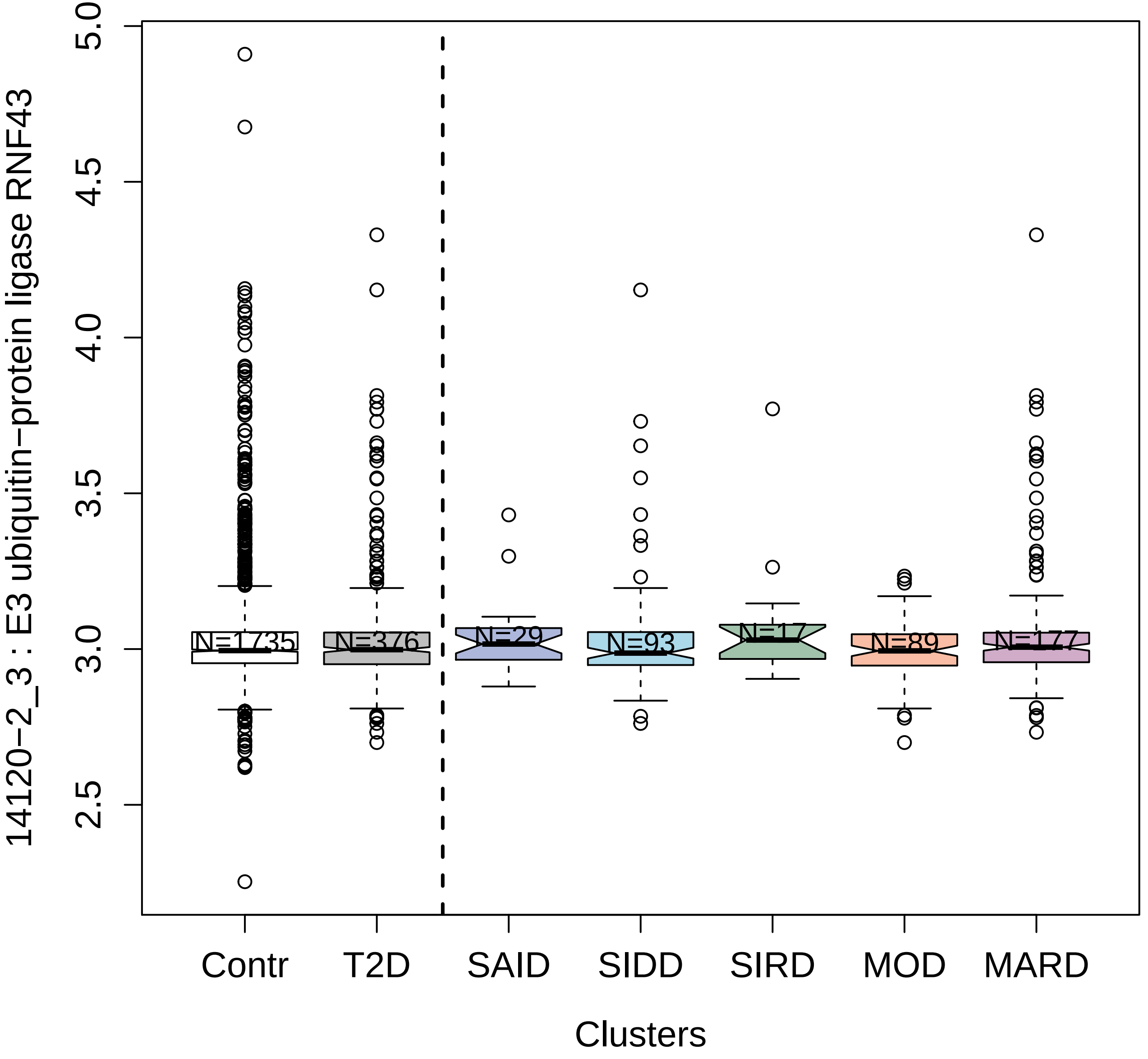

14120−2_3 : E3 ubiquitin−protein ligase RNF43
5.0
4.5
4.0
3.5
3.0
2.5
N=1735
N=376
N=29
N=93
N=17
N=89
N=177
Contr
T2D
SAID
SIDD
SIRD
MOD
MARD
Clusters

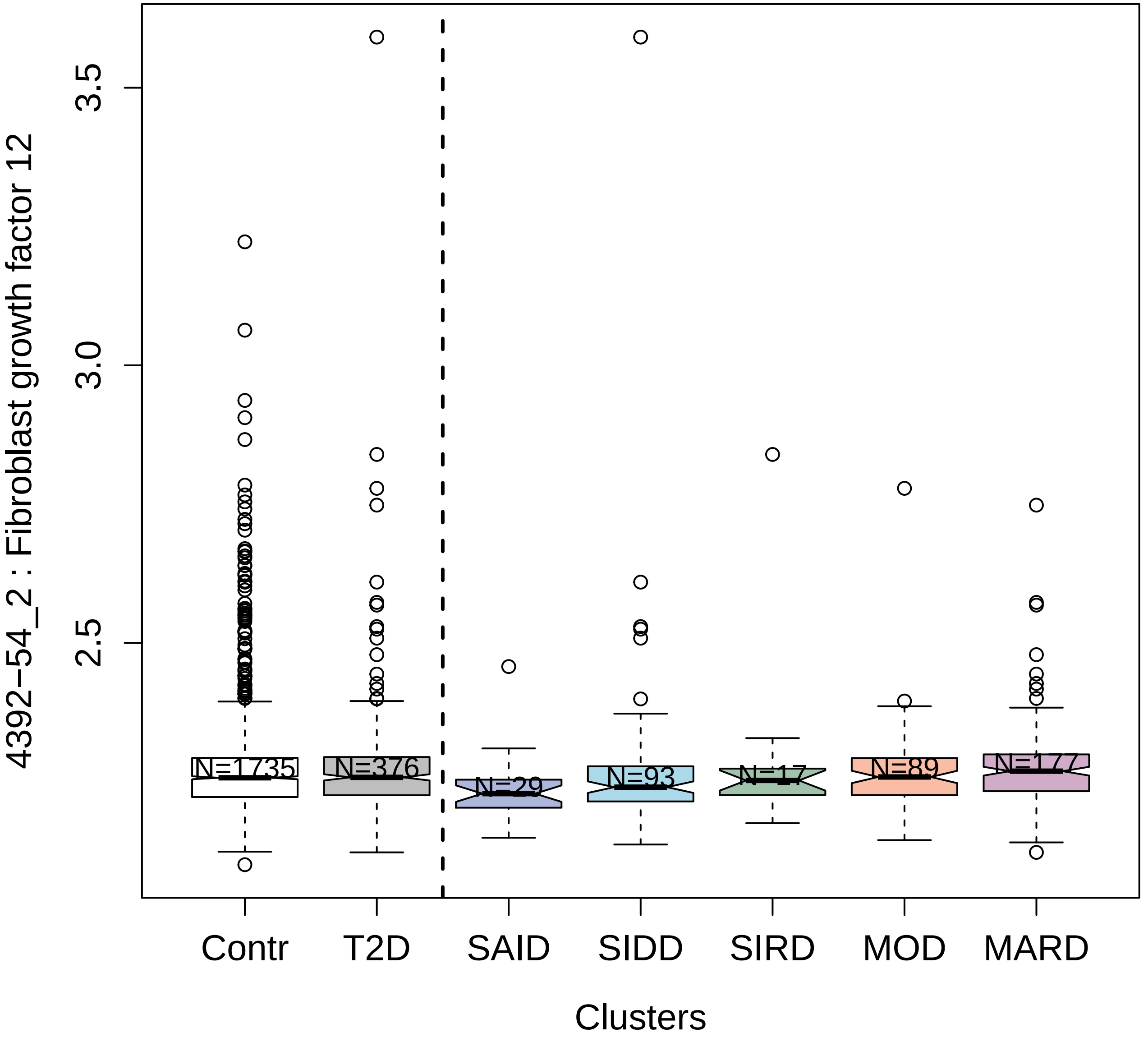

4392−54_2 : Fibroblast growth factor 12
3.5
3.0
2.5
N=1735
N=376
N=29
N=93
N=17
N=89
N=177
Contr
T2D
SAID
SIDD
SIRD
MOD
MARD
Clusters

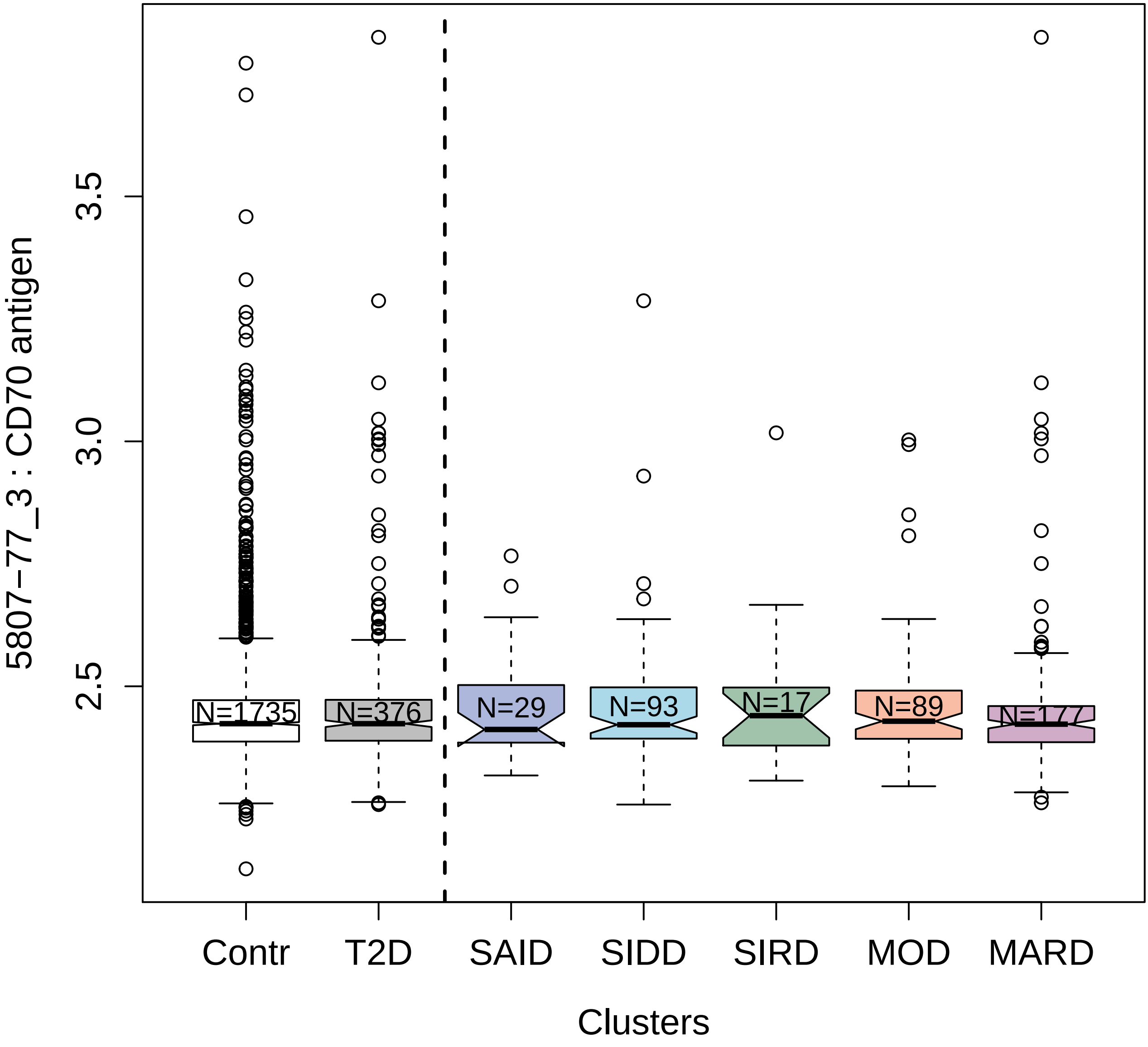

5807−77_3 : CD70 antigen
2.5
3.0
3.5
N=1735
N=376
N=29
N=93
N=17
N=89
N=177
Contr
T2D
SAID
SIDD
SIRD
MOD
MARD
Clusters

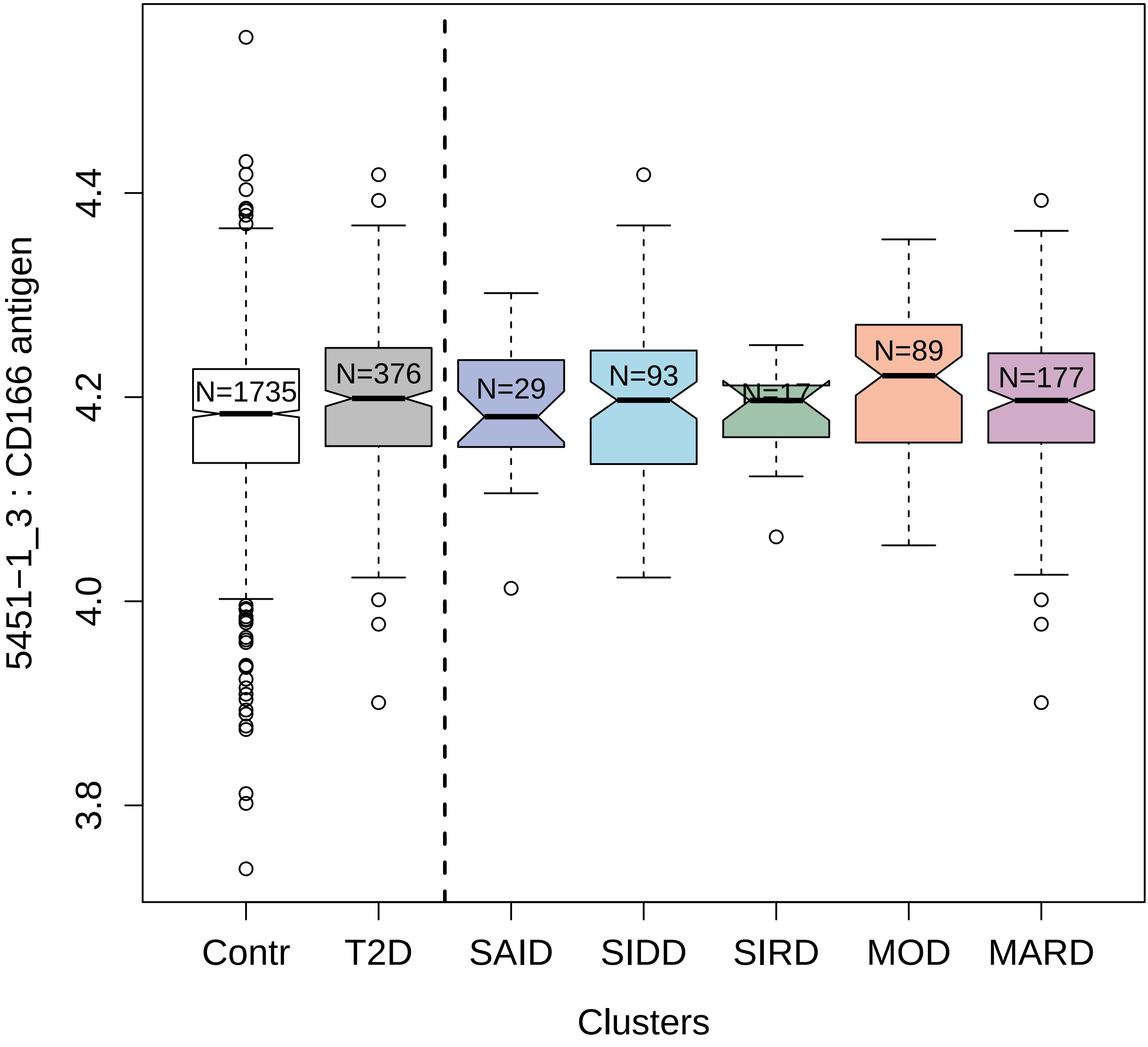

5451−1_3 : CD166 antigen
3.8
4.0
4.2
4.4
N=1735
N=376
N=29
N=93
N=17
N=89
N=177
Contr
T2D
SAID
SIDD
SIRD
MOD
MARD
Clusters

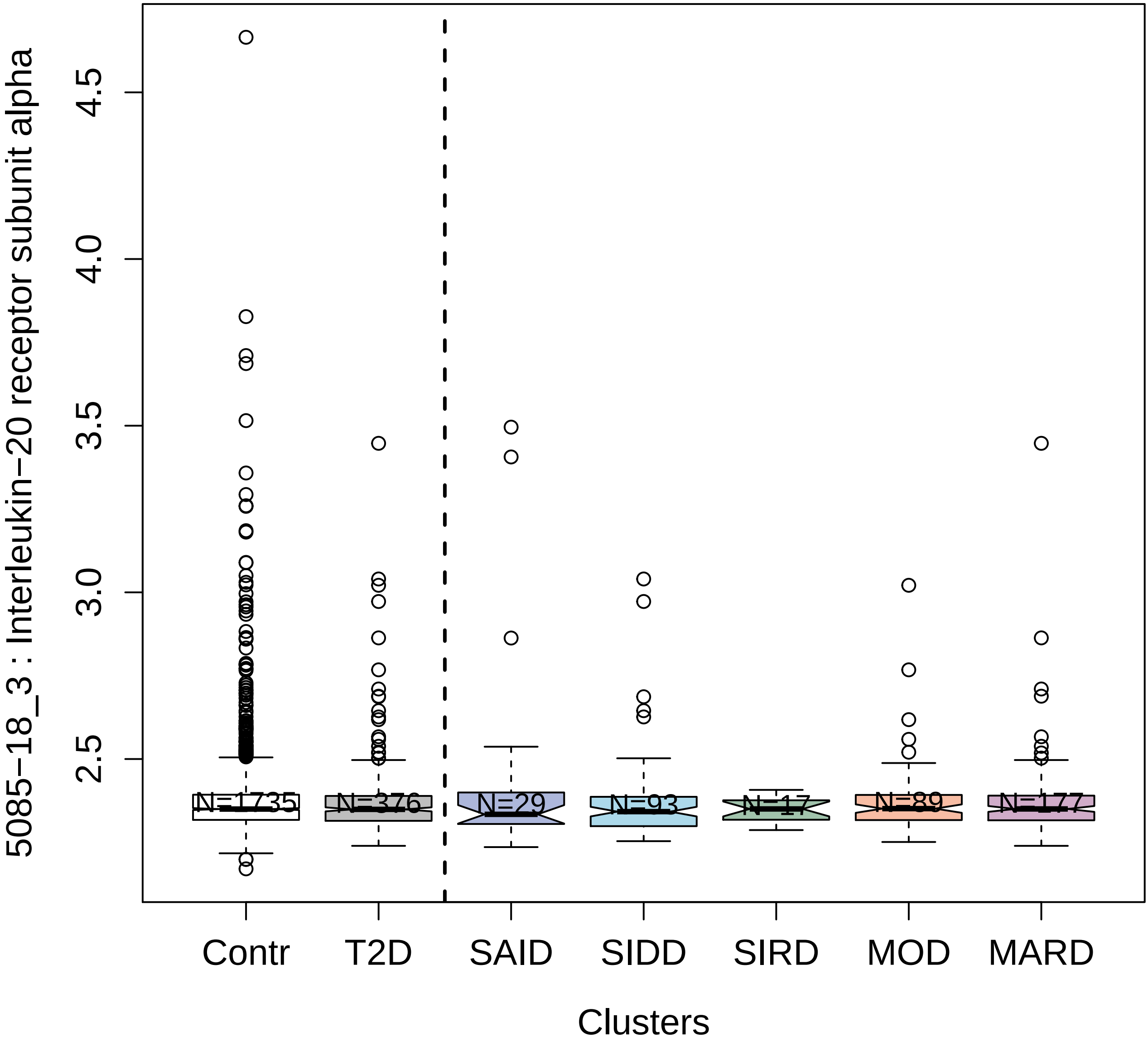
5085−18_3 : Interleukin−20 receptor subunit alpha
4.5
4.0
3.5
3.0
2.5
N=1735
N=376
N=29
N=93
N=17
N=89
N=177
Contr
T2D
SAID
SIDD
SIRD
MOD
MARD
Clusters

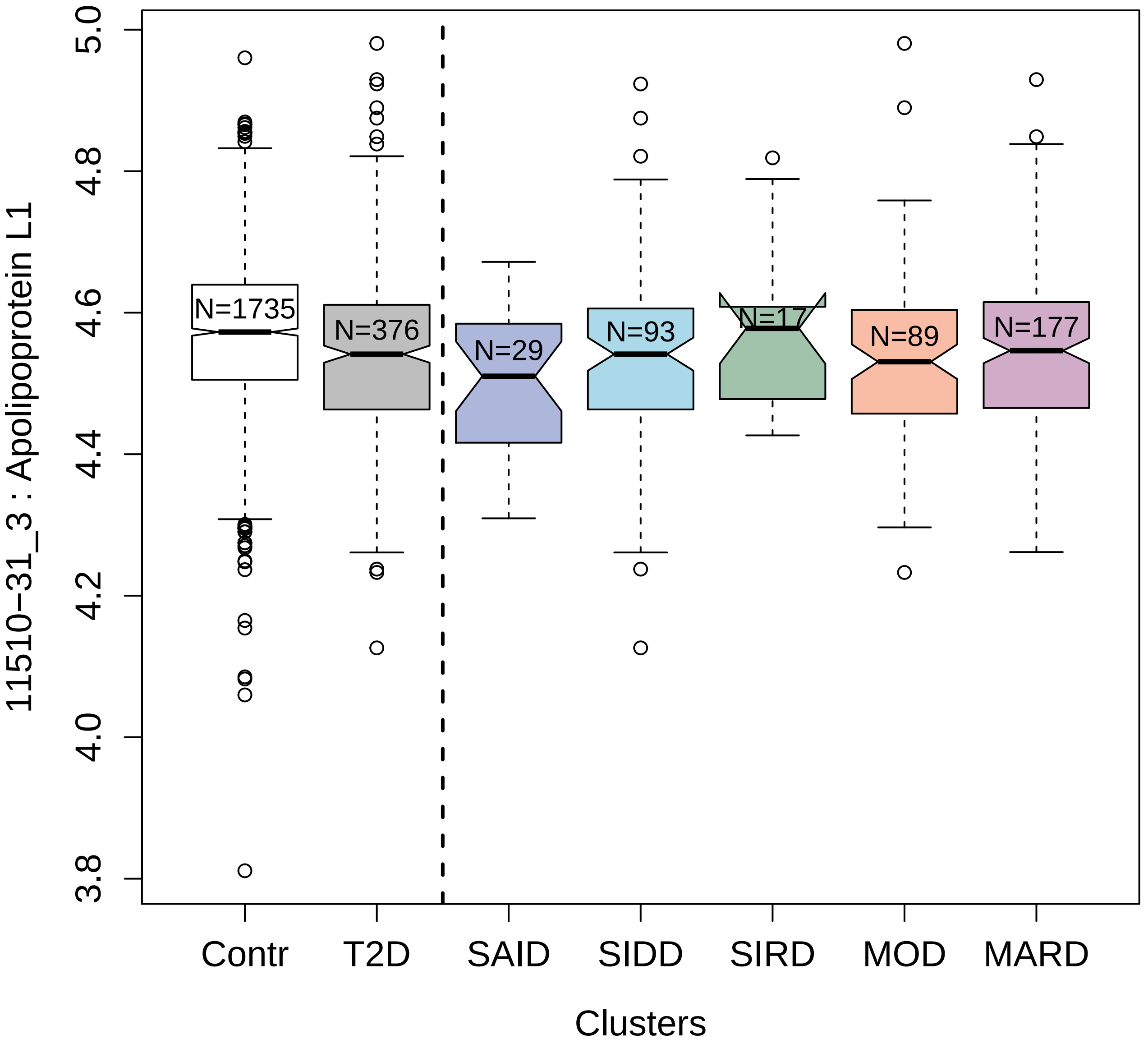

N=1735
N=376
N=29
N=93
N=17
N=89
N=177
5.0
4.8
4.6
4.4
4.2
4.0
3.8
11510−31_3 : Apolipoprotein L1
Contr
T2D
SAID
SIDD
SIRD
MOD
MARD
Clusters

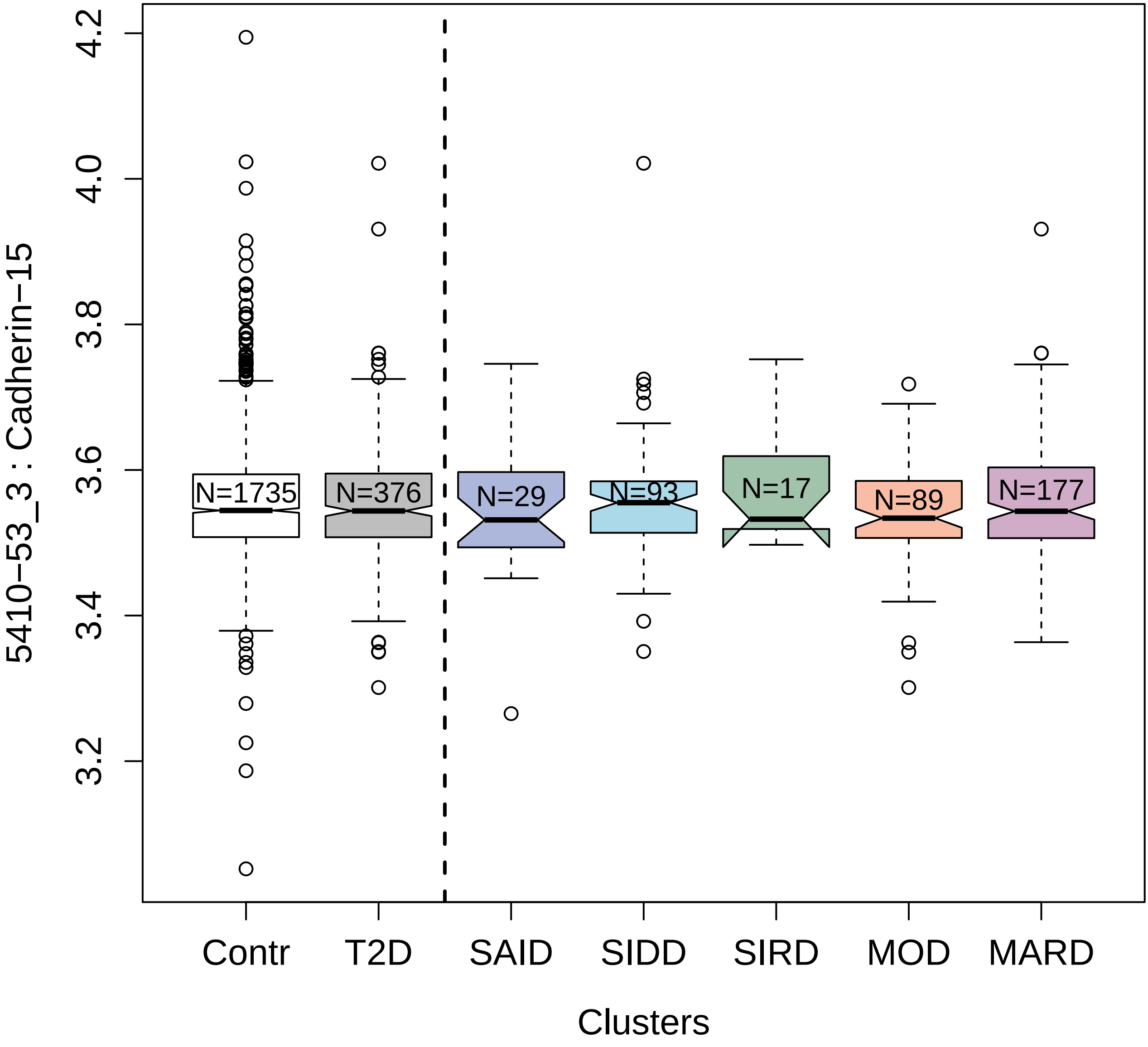

5410−53_3 : Cadherin−15
4.2
4.0
3.8
3.6
3.4
3.2
N=1735
N=376
N=29
N=93
N=17
N=89
N=177
Contr
T2D
SAID
SIDD
SIRD
MOD
MARD
Clusters

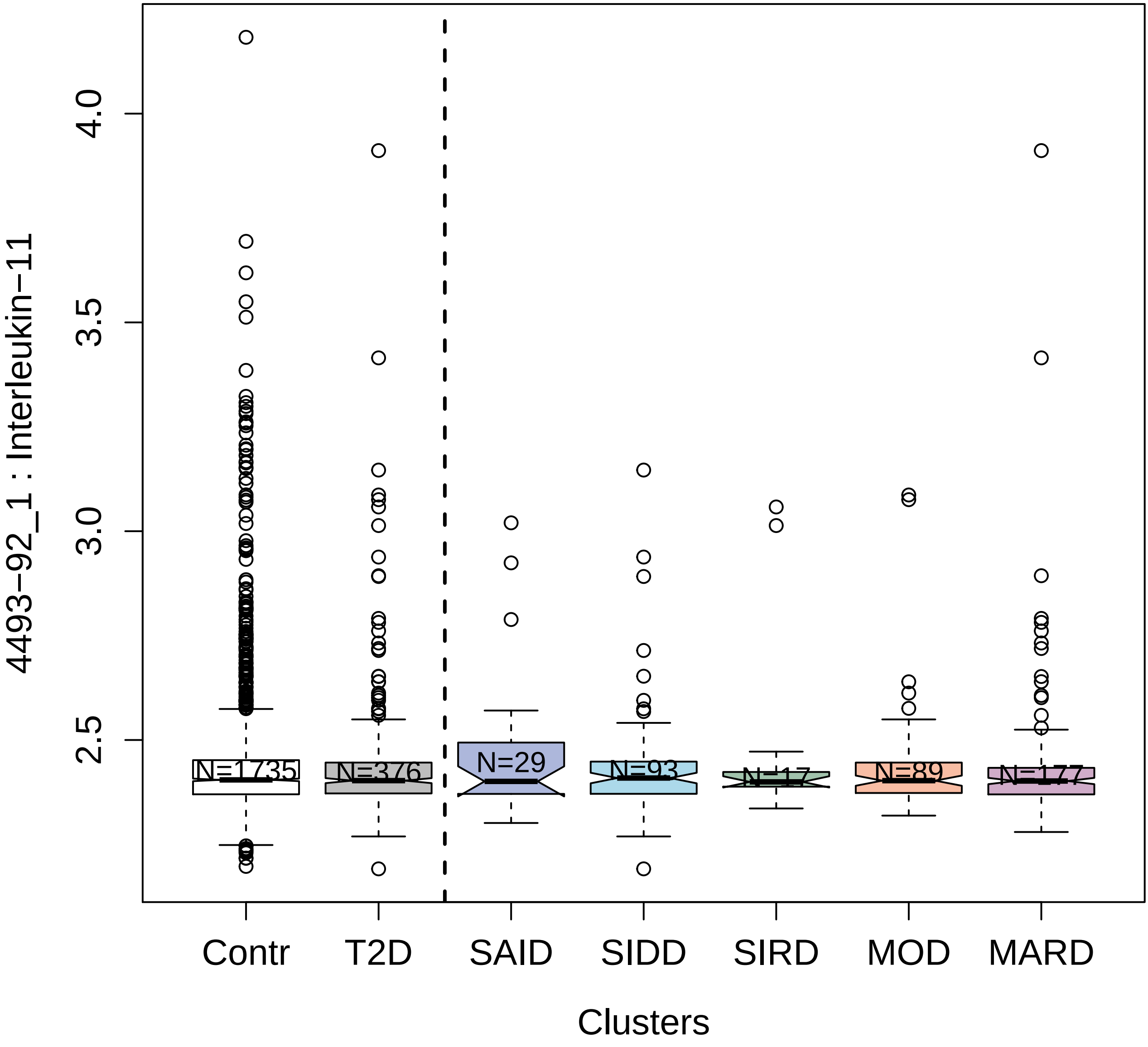

4493−92_1 : Interleukin−11
4.0
3.5
3.0
2.5
N=1735
N=376
N=29
N=93
N=17
N=89
N=177
Contr
T2D
SAID
SIDD
SIRD
MOD
MARD
Clusters

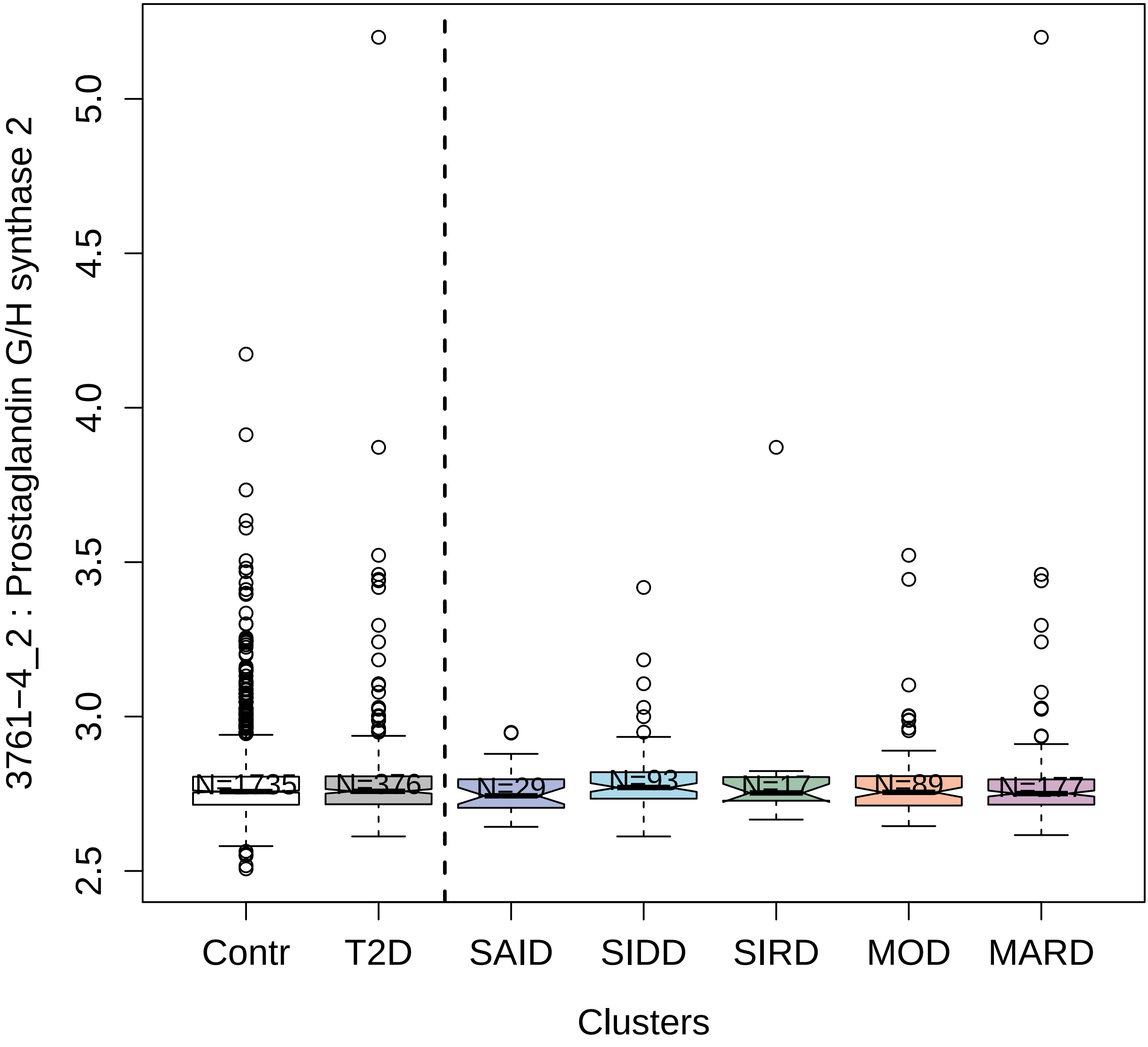

3761−4_2 : Prostaglandin G/H synthase 2
5.0
4.5
4.0
3.5
3.0
2.5
N=1735
N=376
N=29
N=93
N=17
N=89
N=177
Contr
T2D
SAID
SIDD
SIRD
MOD
MARD
Clusters

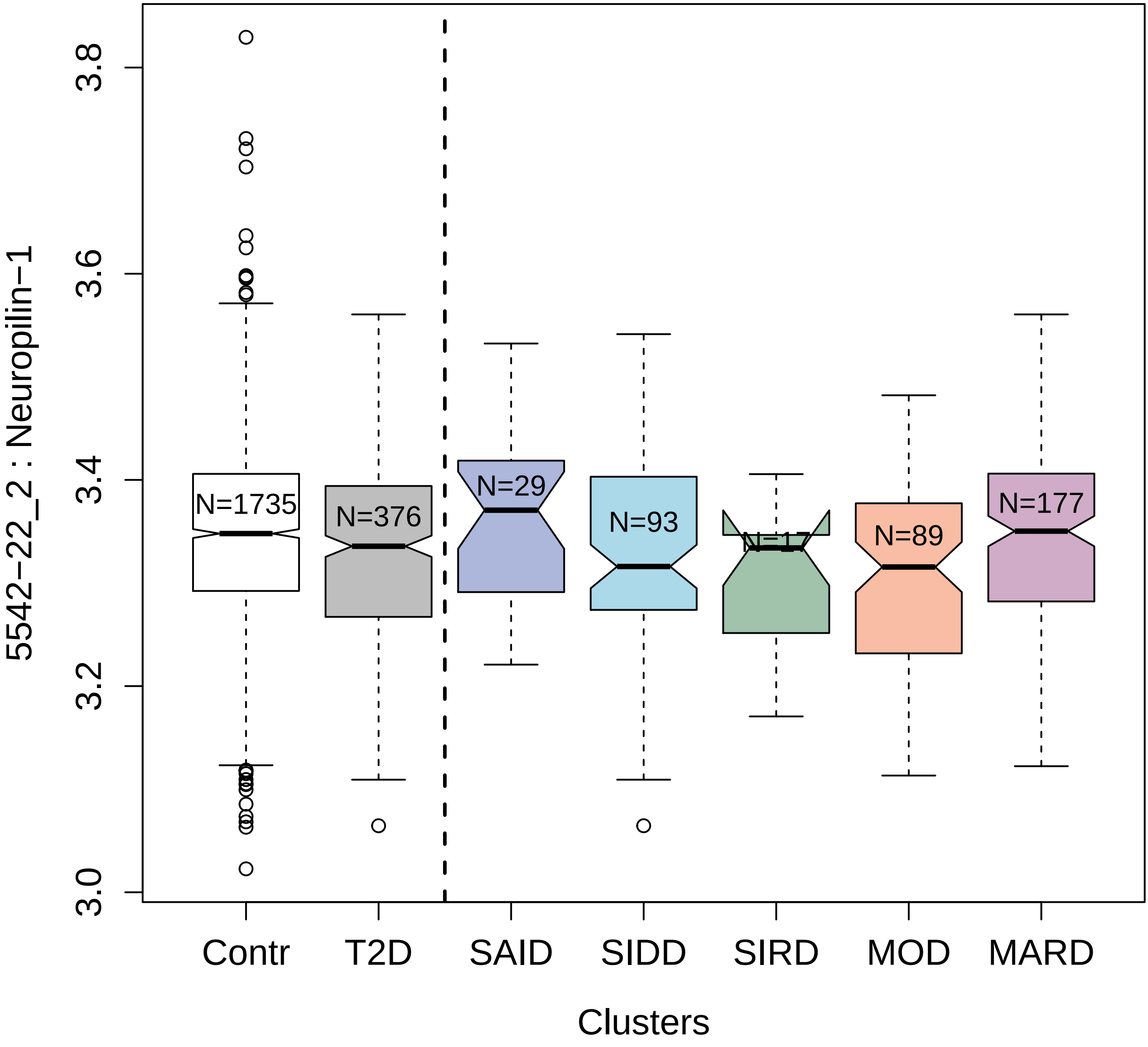

5542−22_2 : Neuropilin−1
3.0
3.2
3.4
3.6
3.8
N=1735
N=376
N=29
N=93
N=89
N=177
Contr
T2D
SAID
SIDD
SIRD
MOD
MARD
Clusters

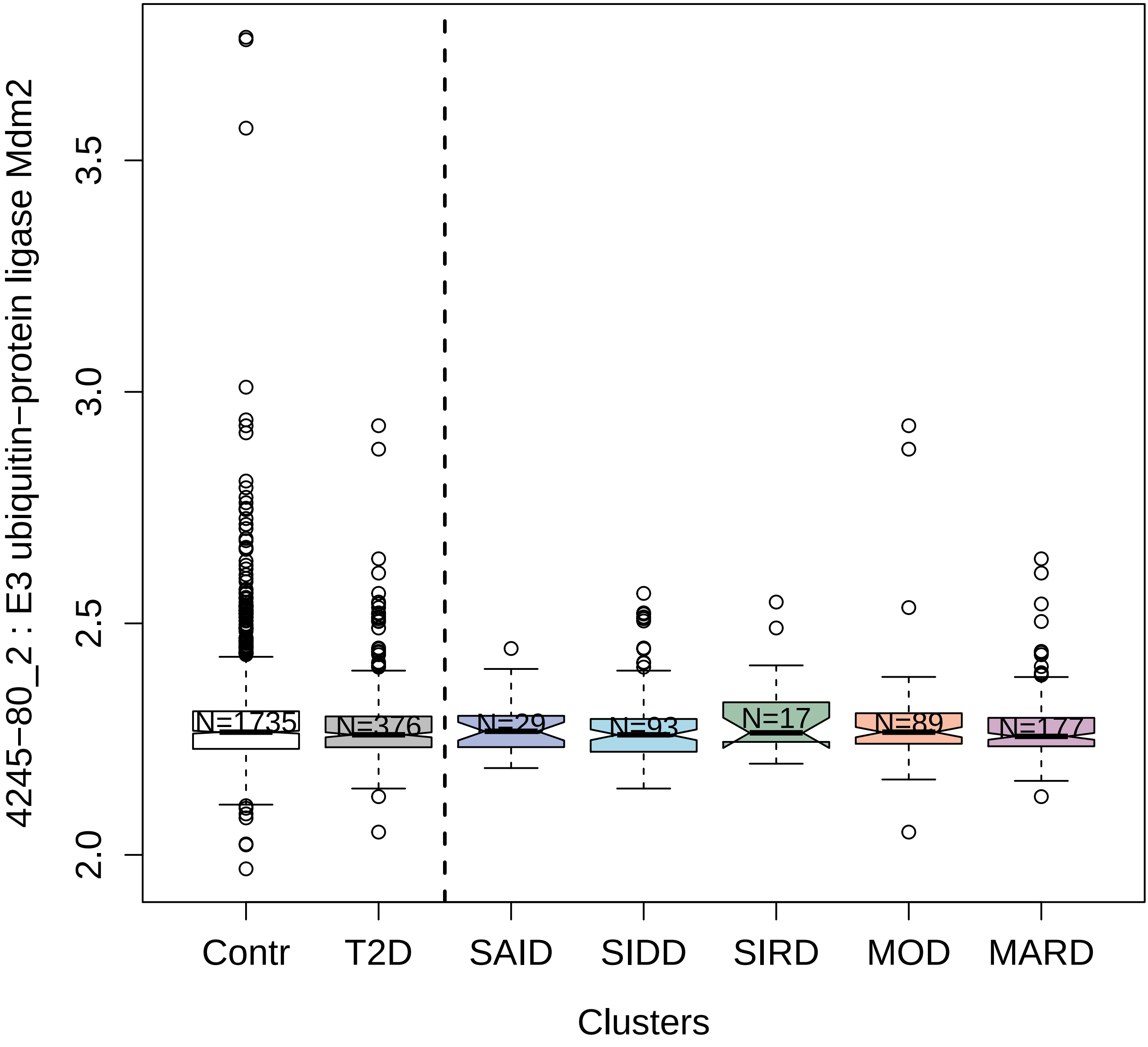

4245−80_2 : E3 ubiquitin−protein ligase Mdm2
2.0
2.5
3.0
3.5
N=1735
N=376
N=29
N=93
N=17
N=89
N=177
Contr
T2D
SAID
SIDD
SIRD
MOD
MARD
Clusters

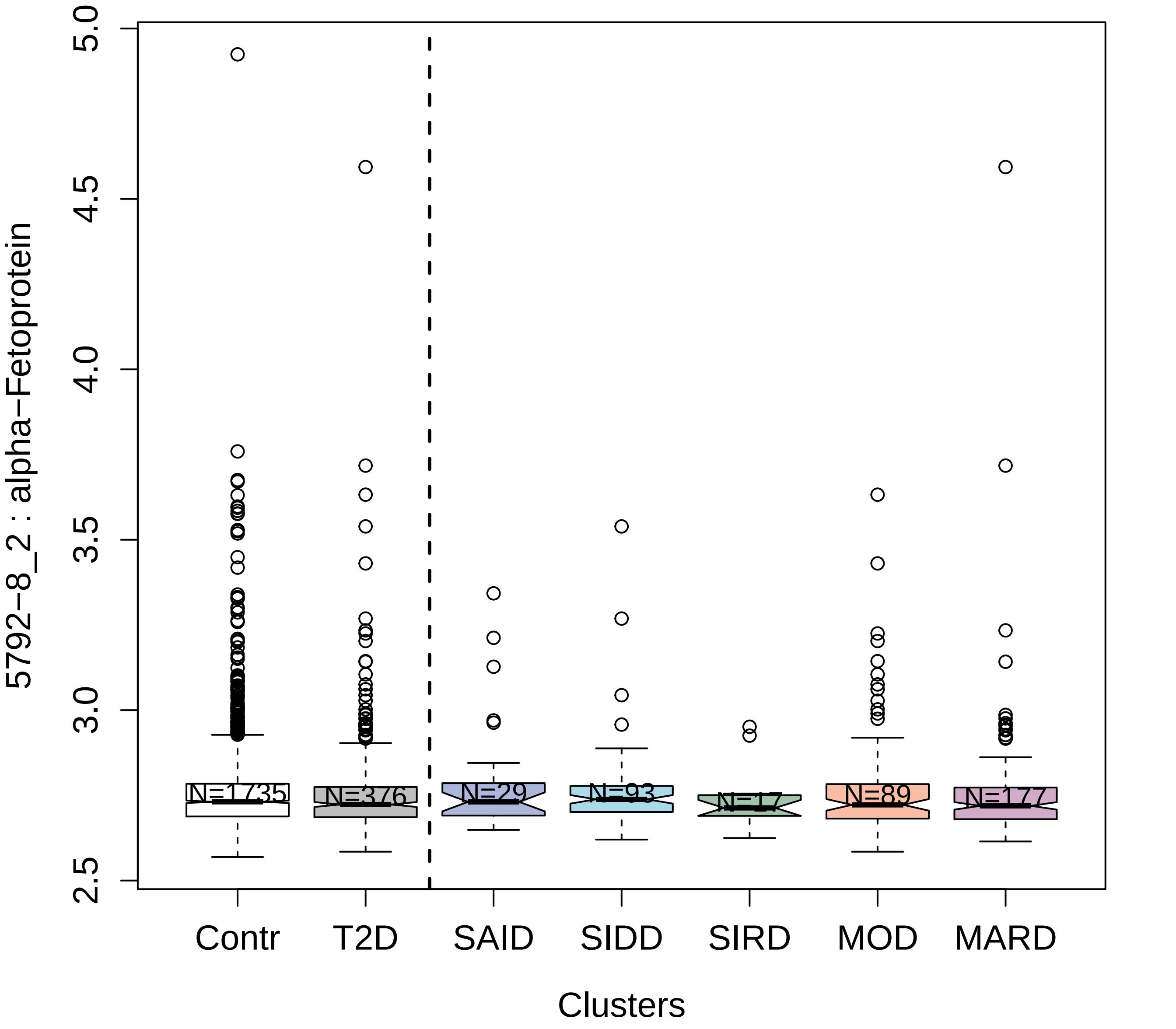

5792−8_2 : alpha−Fetoprotein
5.0
4.5
4.0
3.5
3.0
2.5
N=1735
N=376
N=29
N=93
N=17
N=89
N=177
Contr
T2D
SAID
SIDD
SIRD
MOD
MARD
Clusters

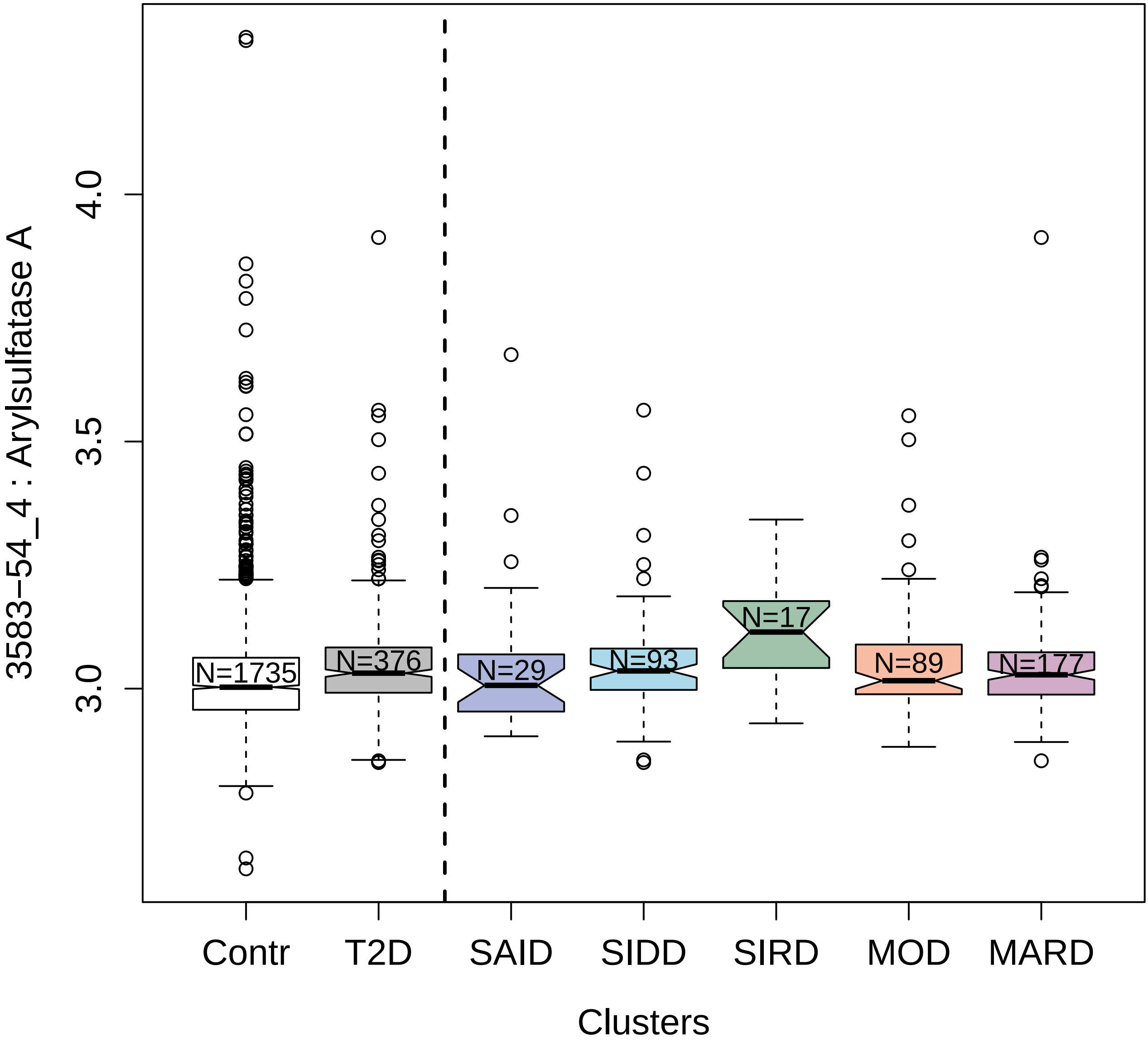

3583−54_4 : Arylsulfatase A
4.0
3.5
3.0
N=1735
N=376
N=29
N=93
N=17
N=89
N=177
Contr
T2D
SAID
SIDD
SIRD
MOD
MARD
Clusters

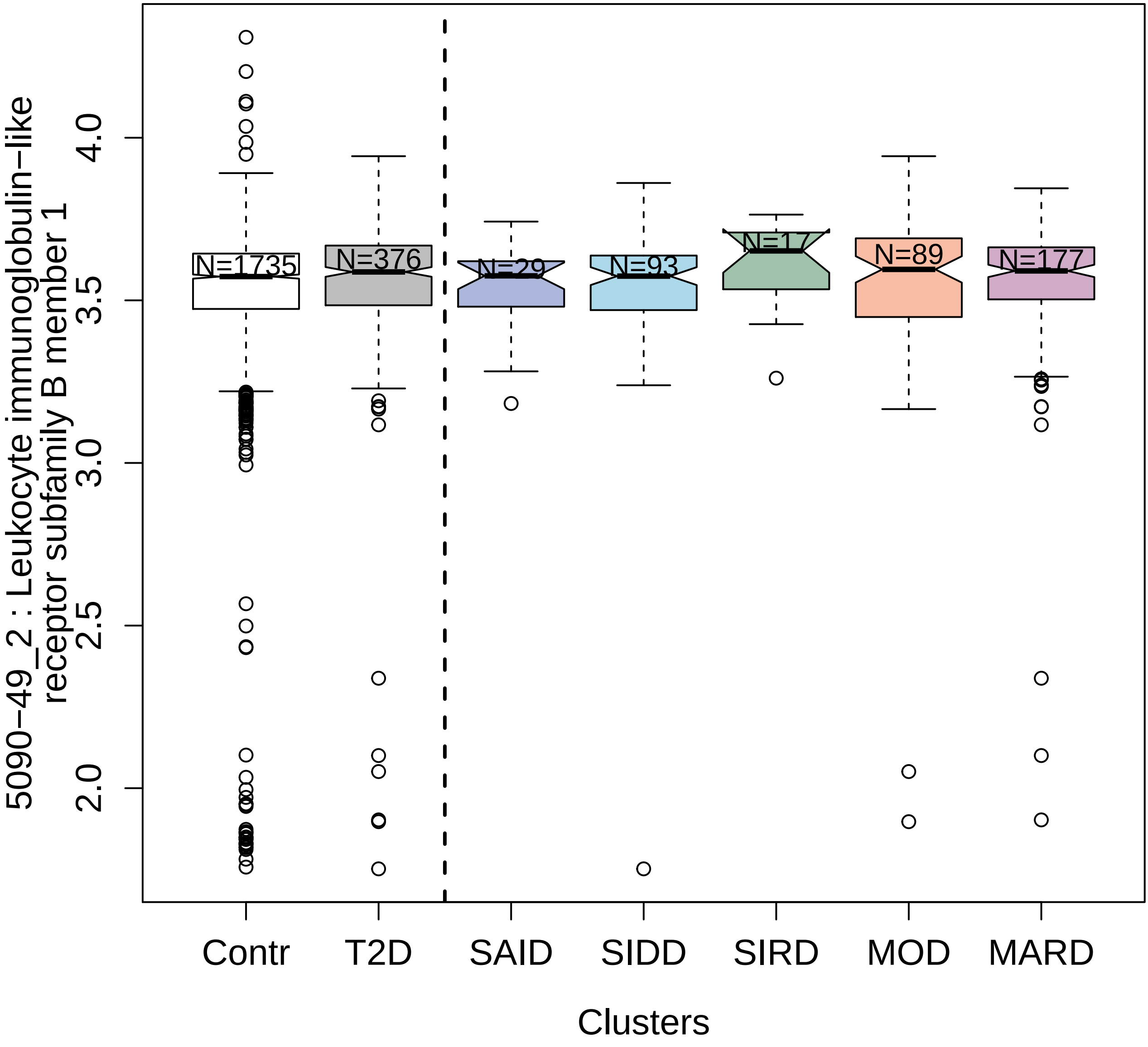

5090−49_2 : Leukocyte immunoglobulin−like
receptor subfamily B member 1
2.0
2.5
3.0
3.5
4.0
N=1735
N=376
N=29
N=93
N=17
N=89
N=177
Contr
T2D
SAID
SIDD
SIRD
MOD
MARD
Clusters

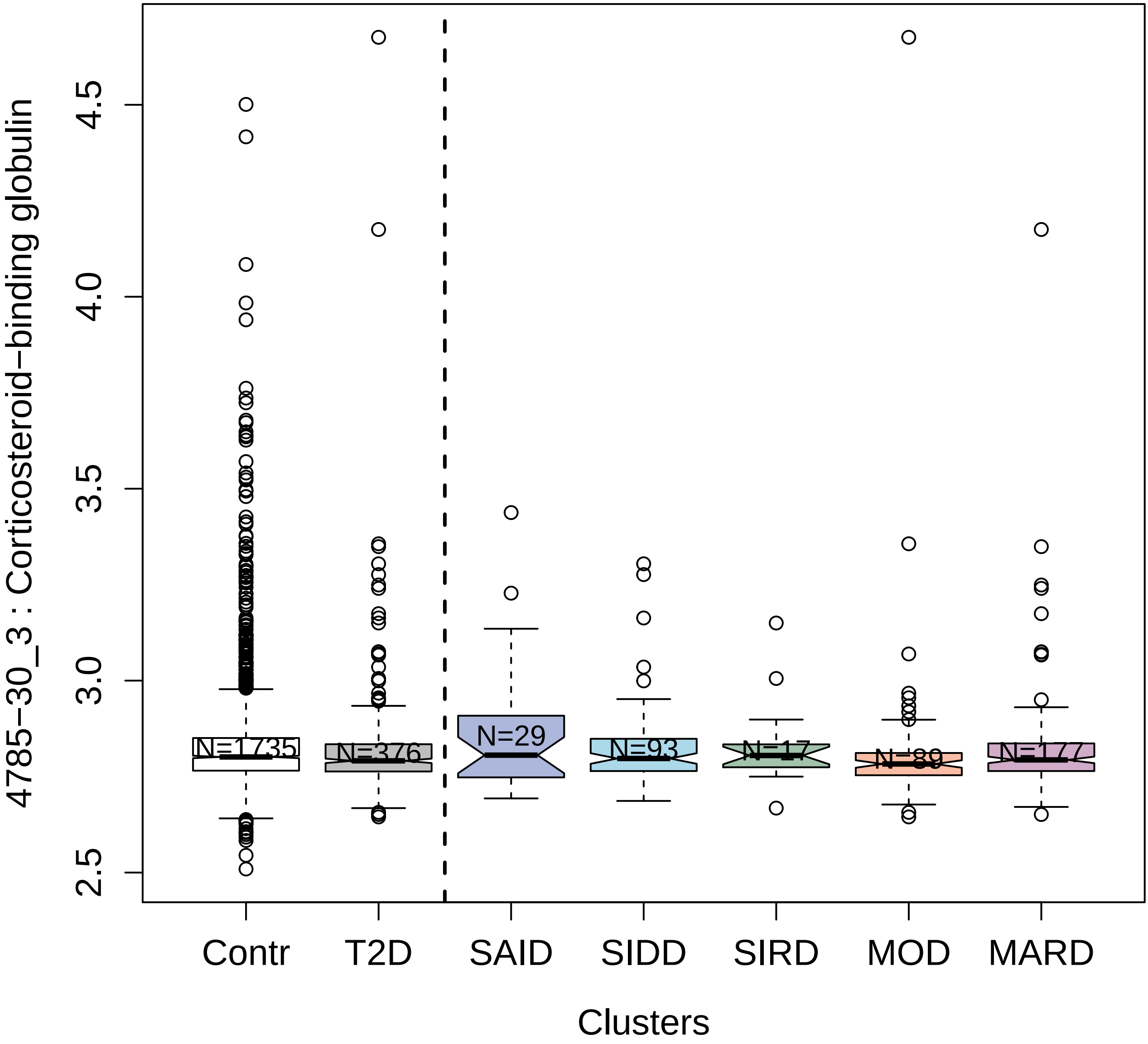

4785−30_3 : Corticosteroid−binding globulin
2.5
3.0
3.5
4.0
4.5
N=1735
N=376
N=29
N=93
N=17
N=89
N=177
Contr
T2D
SAID
SIDD
SIRD
MOD
MARD
Clusters

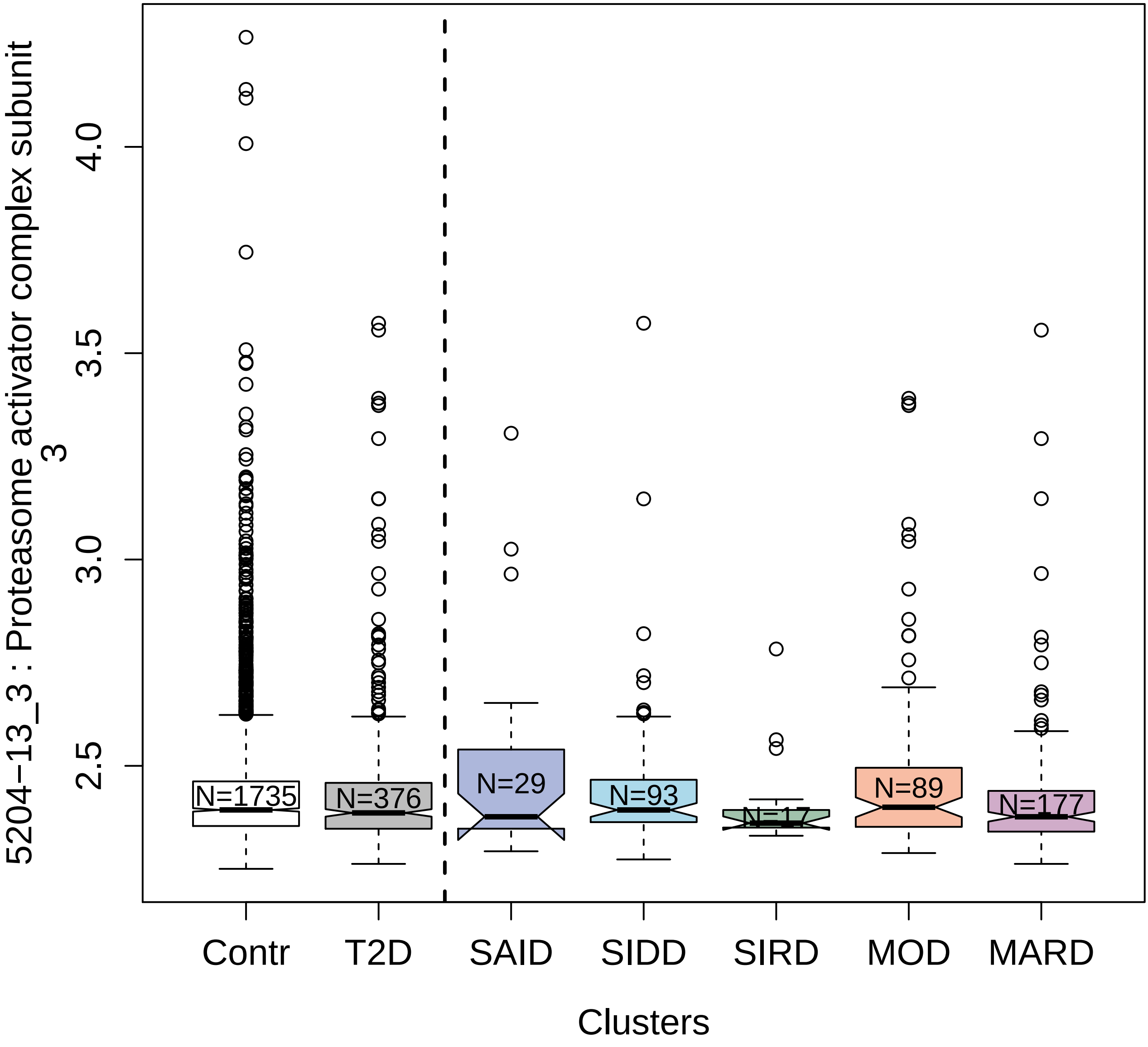
5204−13_3 : Proteasome activator complex subunit 3
Clusters
Contr
T2D
SAID
SIDD
SIRD
MOD
MARD
N=1735
N=376
N=29
N=93
N=17
N=89
N=177
2.5
3.0
3.5
4.0

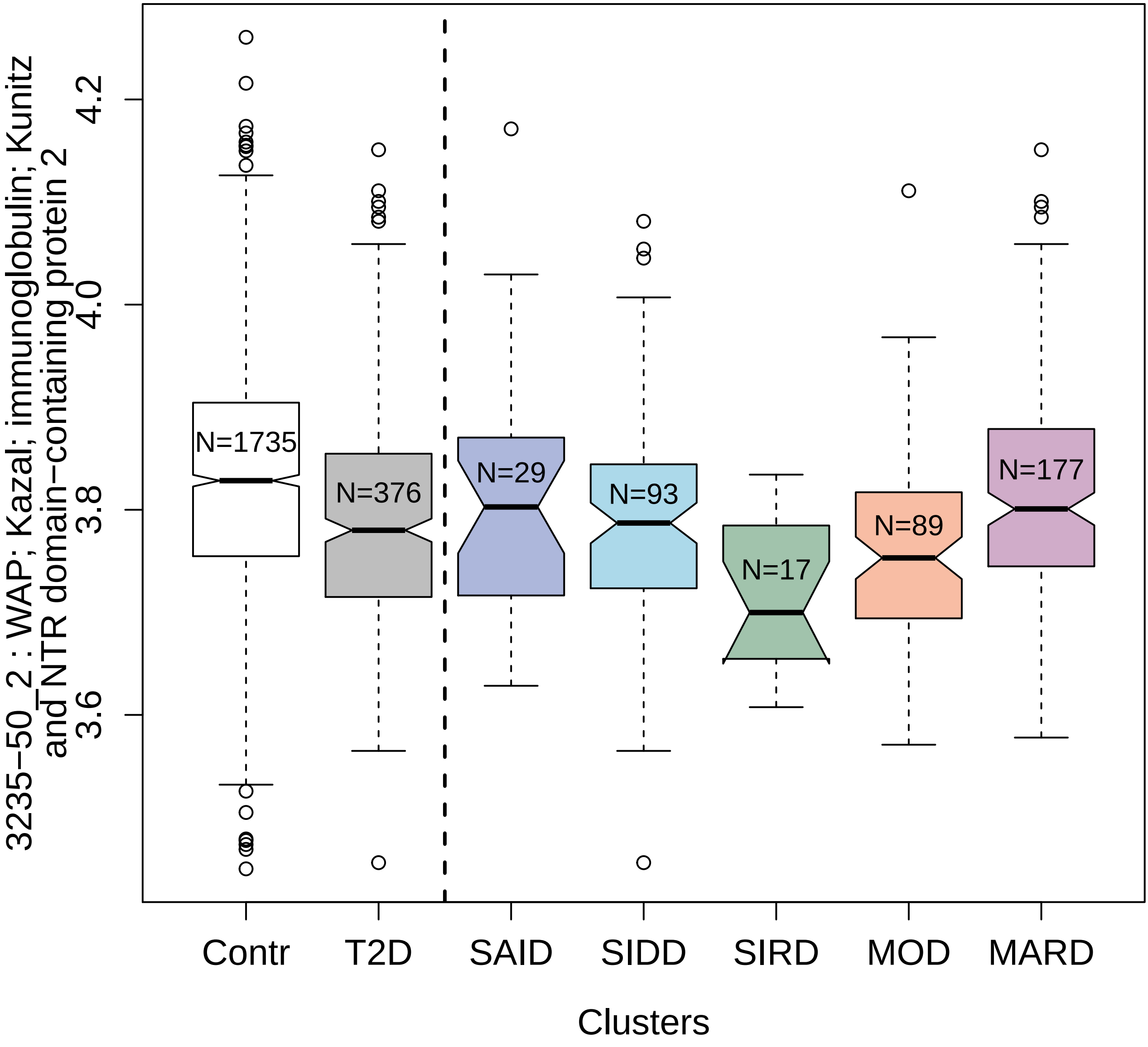

3235−50_2 : WAP, Kazal, immunoglobulin, Kunitz and NTR domain−containing protein 2
4.2
4.0
3.8
3.6
N=1735
N=376
N=29
N=93
N=17
N=89
N=177
Contr
T2D
SAID
SIDD
SIRD
MOD
MARD
Clusters

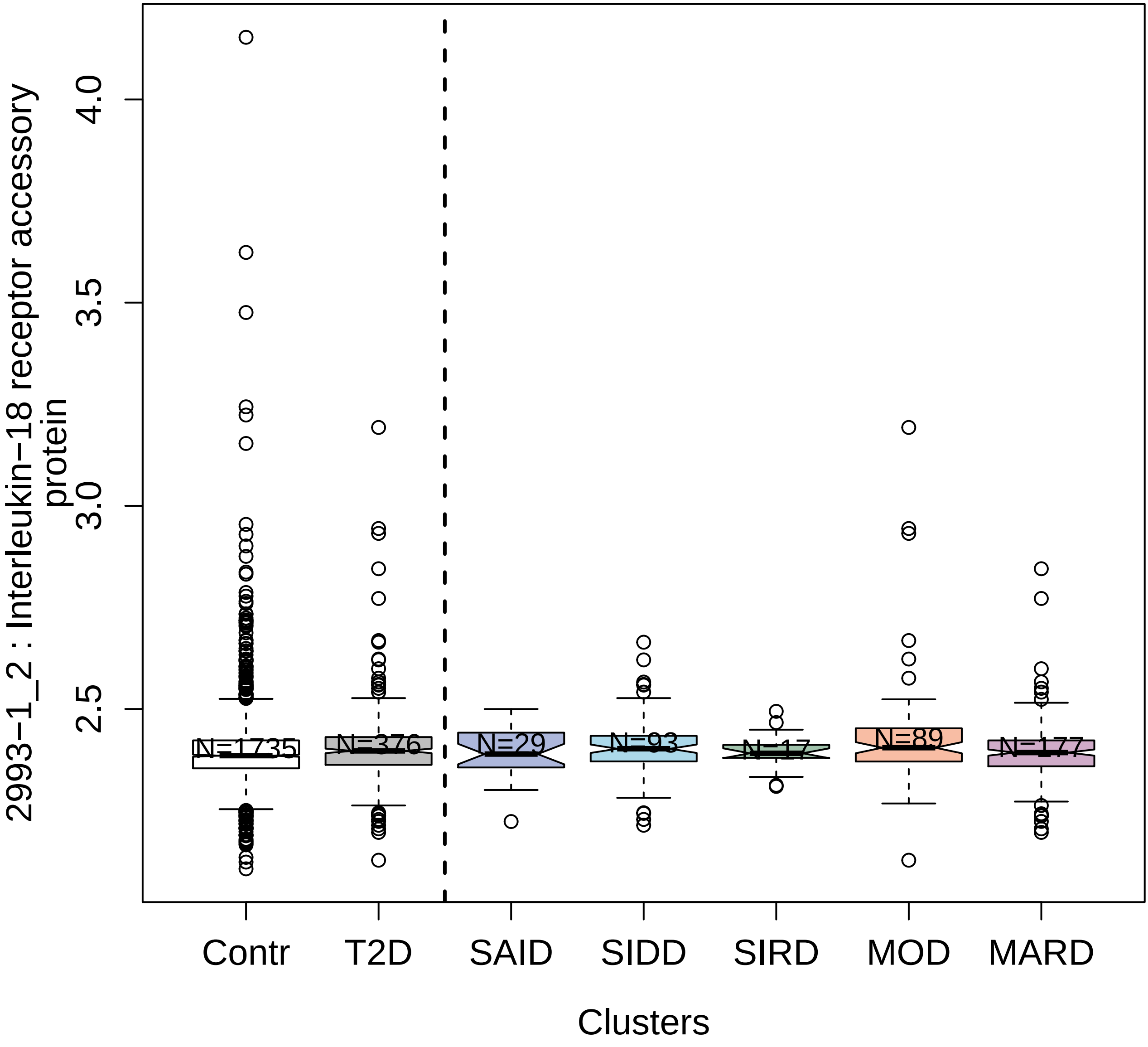

2993−1_2 : Interleukin−18 receptor accessory protein
4.0
3.5
3.0
2.5
N=1735
N=376
N=29
N=93
N=17
N=89
N=177
Contr
T2D
SAID
SIDD
SIRD
MOD
MARD
Clusters

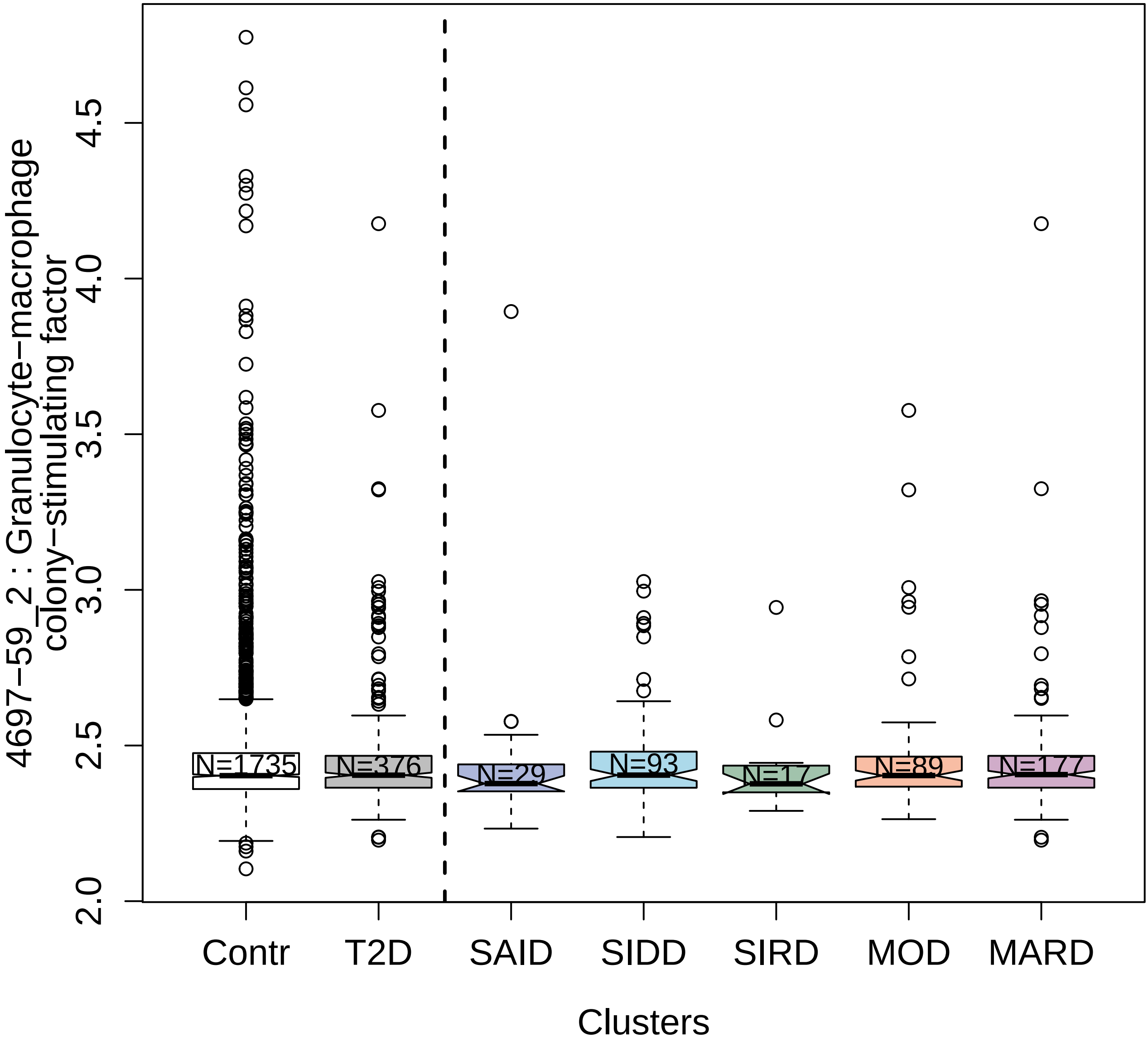

4697−59_2 : Granulocyte−macrophage
colony−stimulating factor
2.0
2.5
3.0
3.5
4.0
4.5
N=1735
N=376
N=29
N=93
N=17
N=89
N=177
Contr
T2D
SAID
SIDD
SIRD
MOD
MARD
Clusters

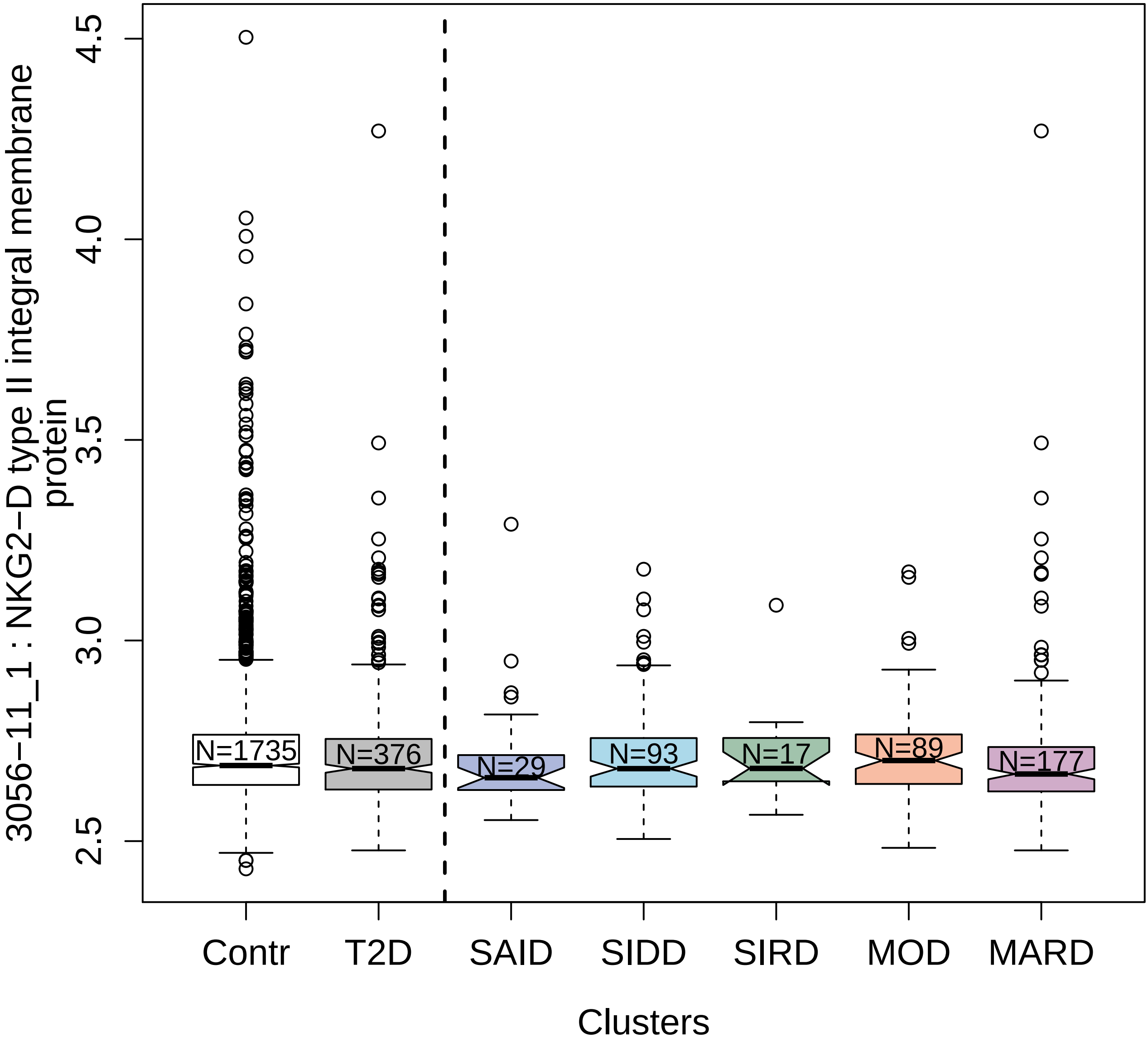

3056−11_1 : NKG2−D type II integral membrane protein
4.5
4.0
3.5
3.0
2.5
N=1735
N=376
N=29
N=93
N=17
N=89
N=177
Contr
T2D
SAID
SIDD
SIRD
MOD
MARD
Clusters

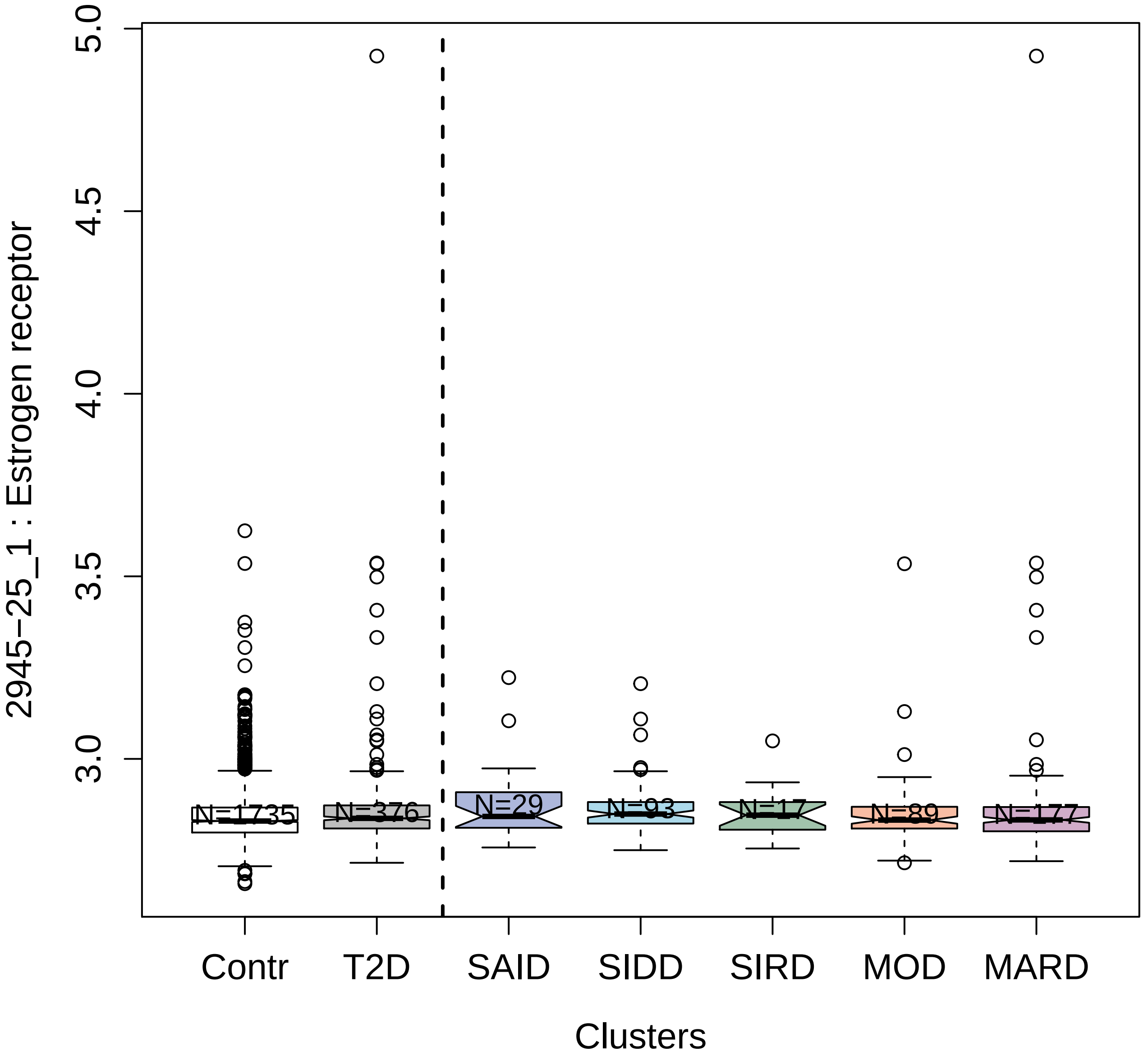

2945−25_1 : Estrogen receptor
5.0
4.5
4.0
3.5
3.0
N=1735
N=376
N=29
N=93
N=17
N=89
N=177
Contr
T2D
SAID
SIDD
SIRD
MOD
MARD
Clusters

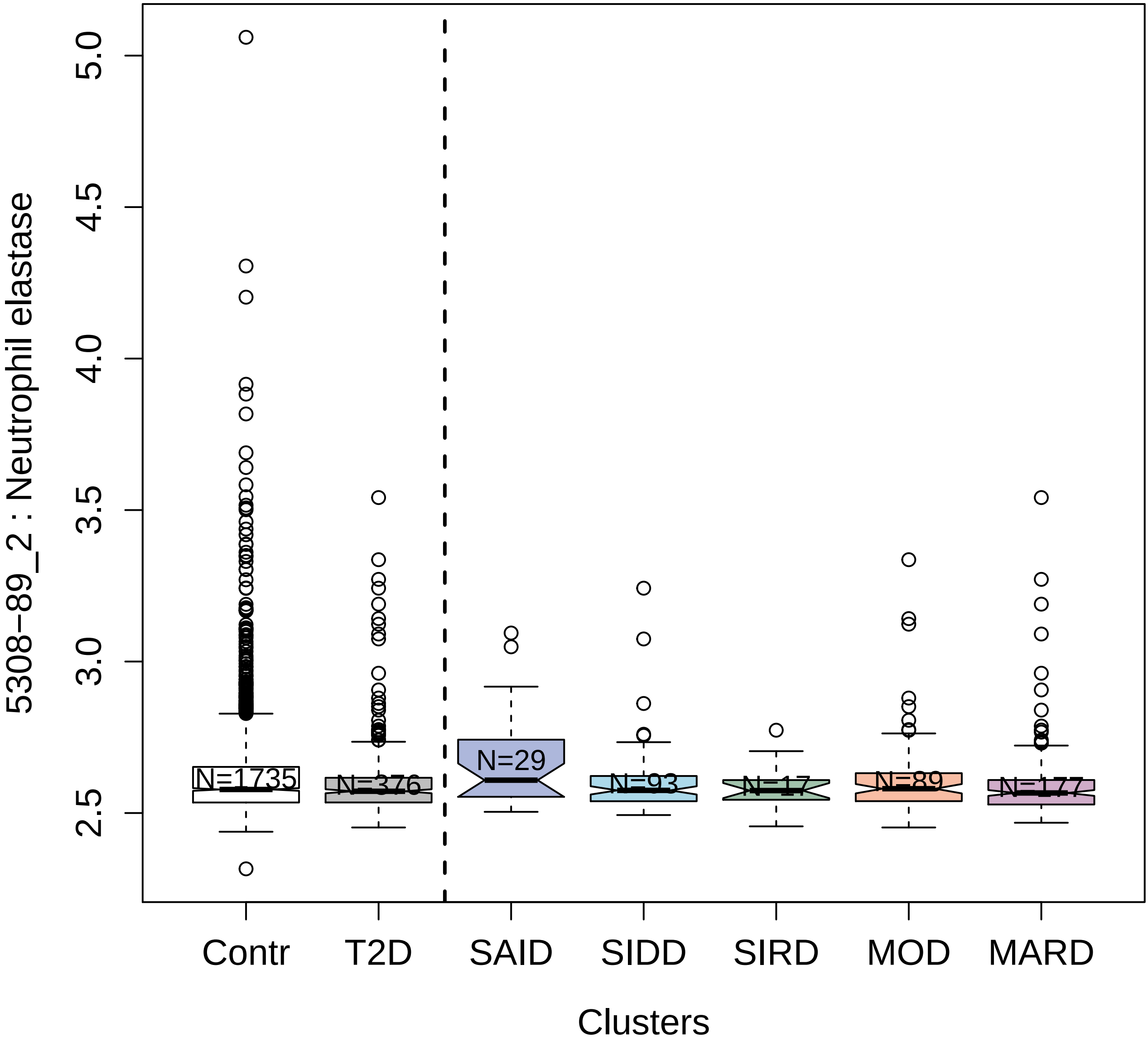

5308−89_2 : Neutrophil elastase
5.0
4.5
4.0
3.5
3.0
2.5
N=1735
N=376
N=29
N=93
N=17
N=89
N=177
Contr
T2D
SAID
SIDD
SIRD
MOD
MARD
Clusters

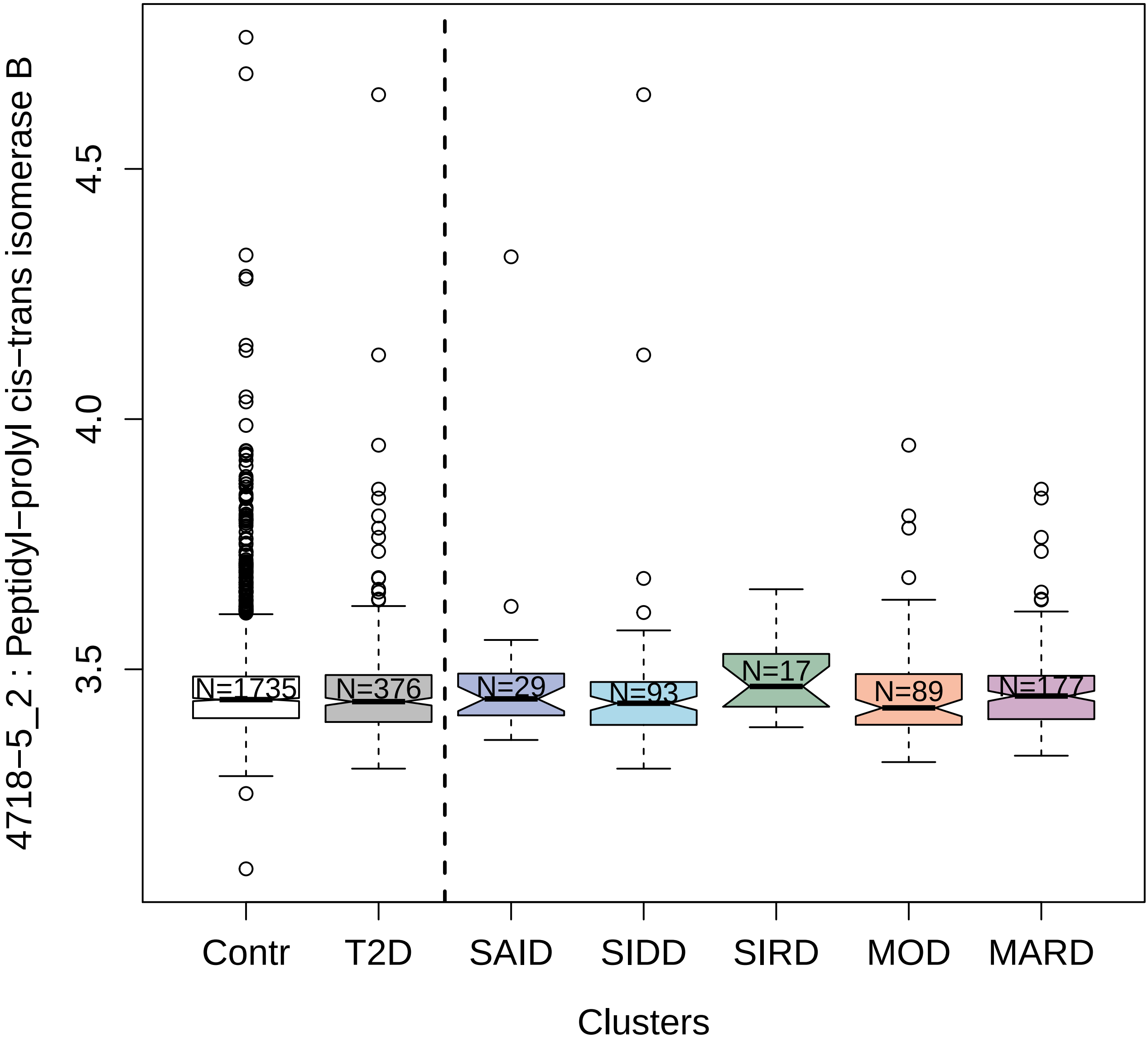

4718−5_2 : Peptidyl−prolyl cis−trans isomerase B
4.5
4.0
3.5
N=1735
N=376
N=29
N=93
N=17
N=89
N=177
Contr
T2D
SAID
SIDD
SIRD
MOD
MARD
Clusters

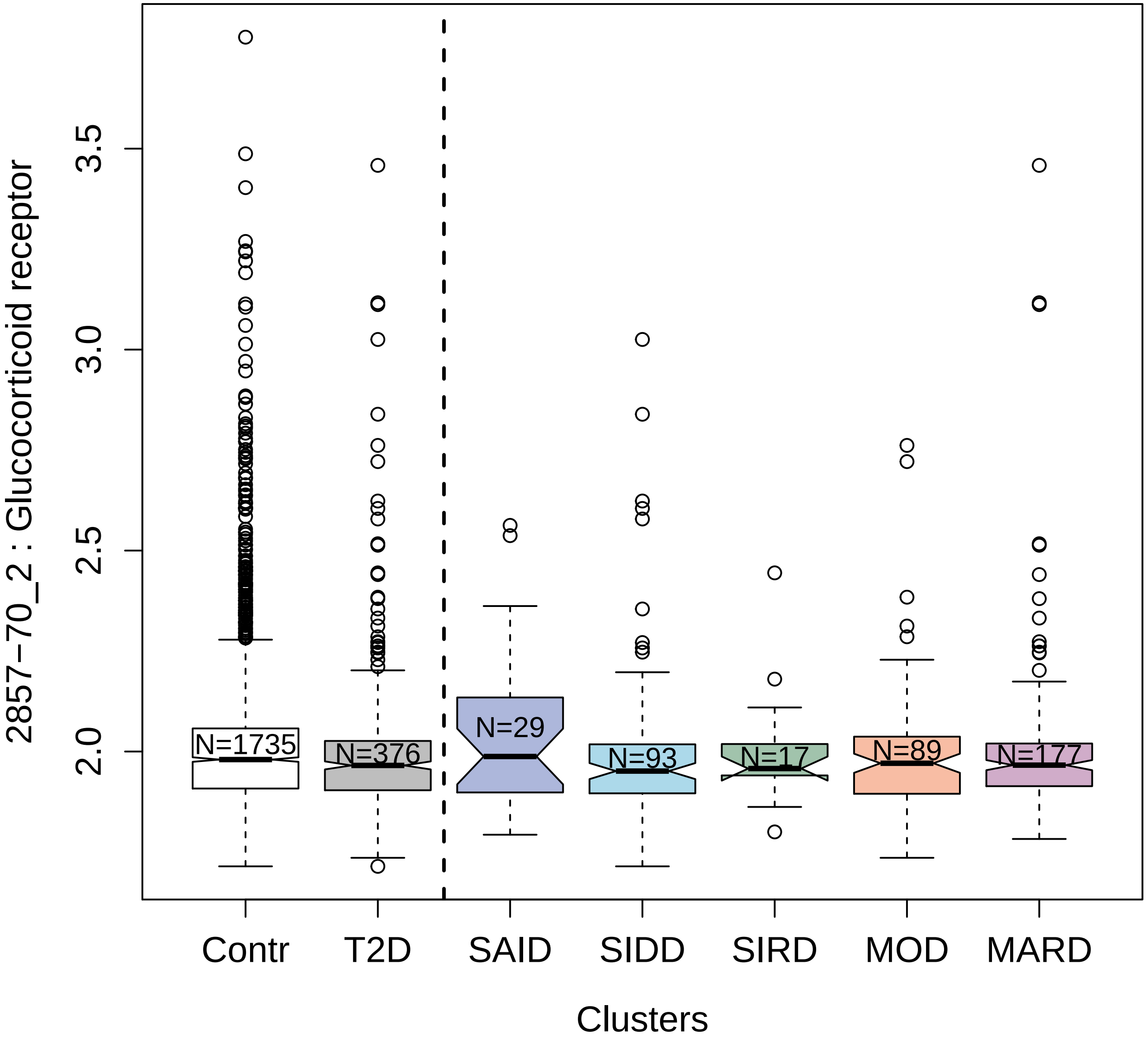

2857−70_2 : Glucocorticoid receptor
3.5
3.0
2.5
2.0
N=1735
N=376
N=29
N=93
N=17
N=89
N=177
Contr
T2D
SAID
SIDD
SIRD
MOD
MARD
Clusters

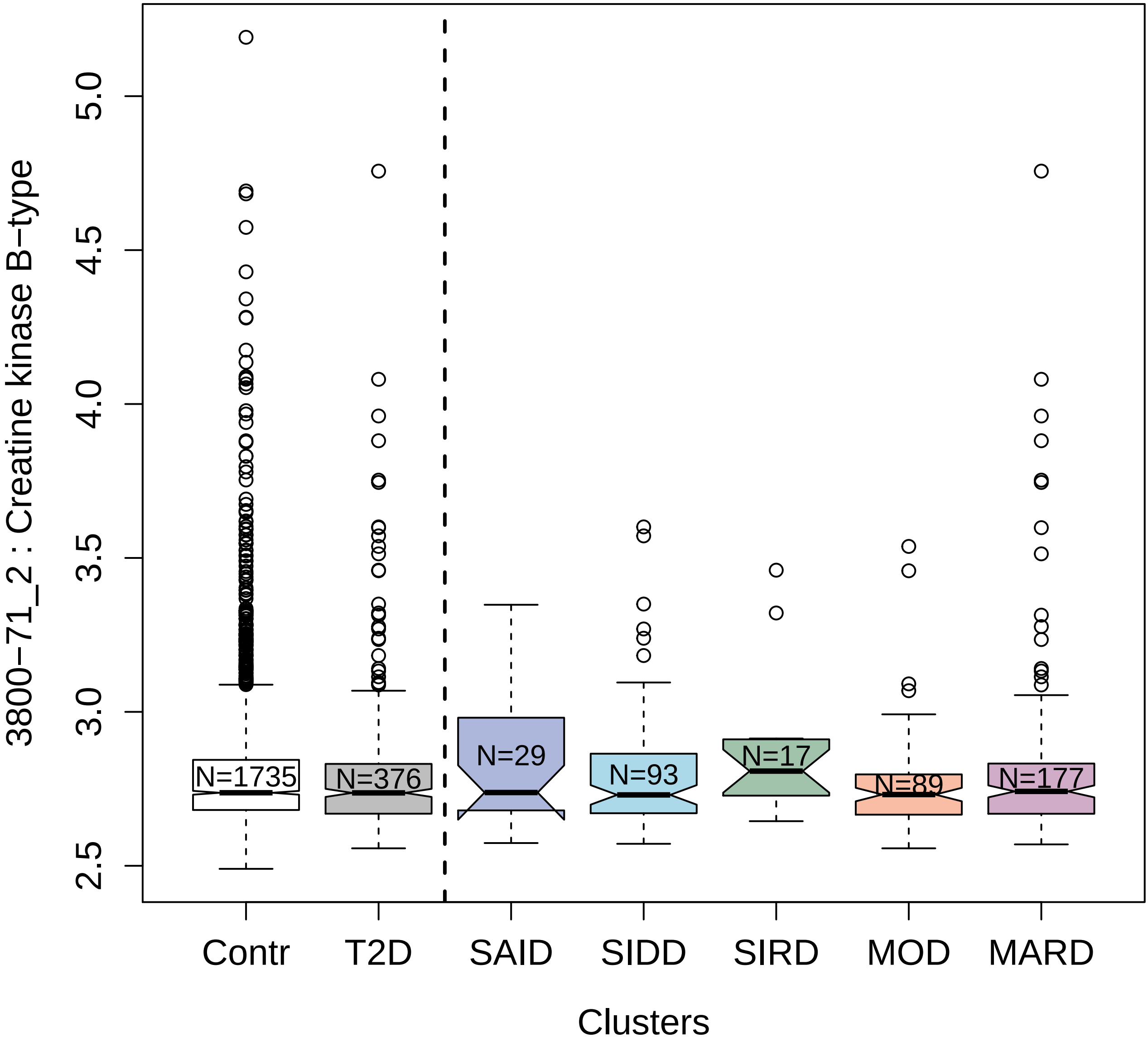
3800−71_2 : Creatine kinase B−type
5.0
4.5
4.0
3.5
3.0
2.5
N=1735
N=376
N=29
N=93
N=17
N=89
N=177
Contr
T2D
SAID
SIDD
SIRD
MOD
MARD
Clusters

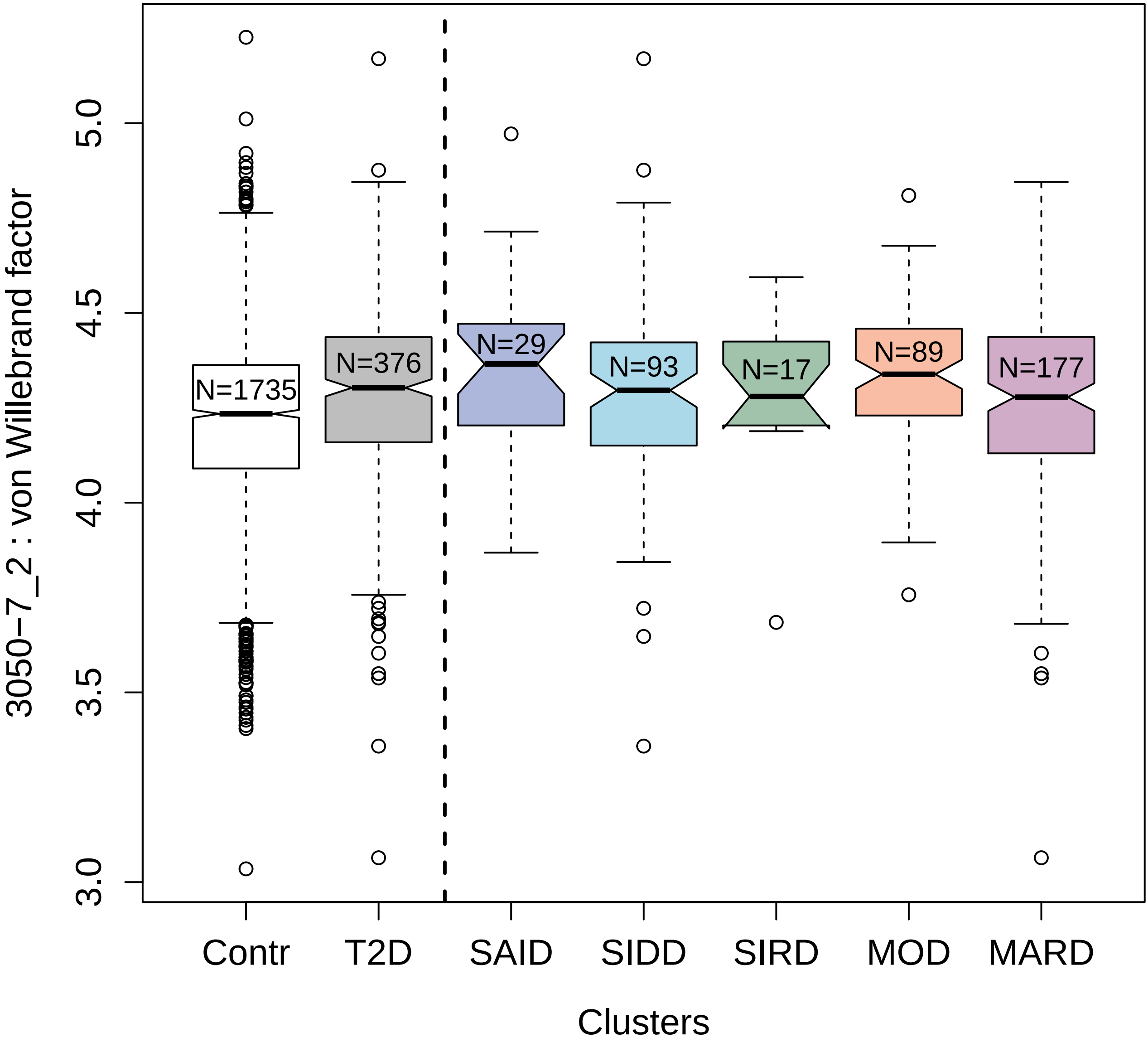
3050−7_2 : von Willebrand factor
3.0
3.5
4.0
4.5
5.0
N=1735
N=376
N=29
N=93
N=17
N=89
N=177
Contr
T2D
SAID
SIDD
SIRD
MOD
MARD
Clusters

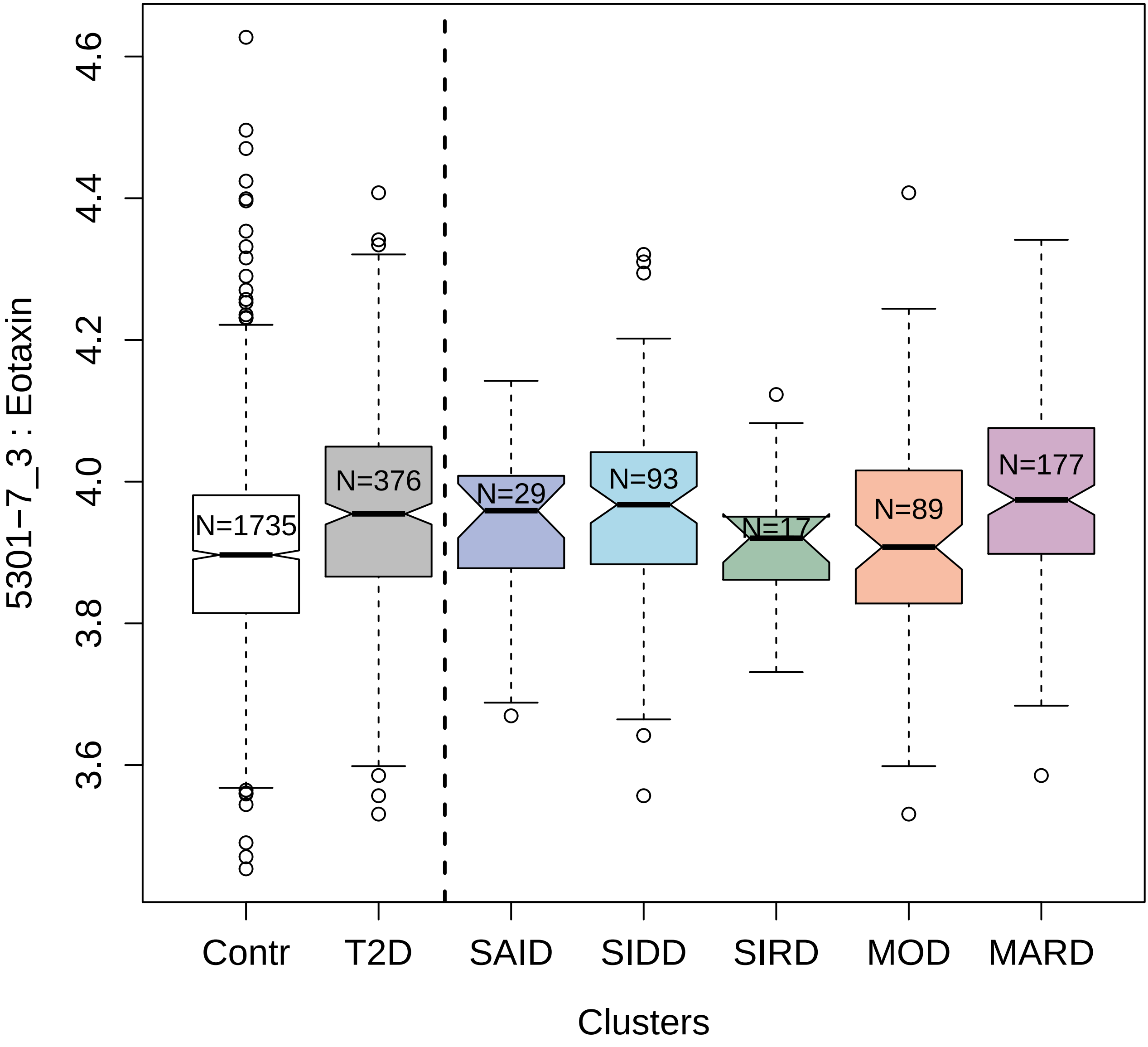

5301−7_3 : Eotaxin
4.6
4.4
4.2
4.0
3.8
3.6
N=1735
N=376
N=29
N=93
N=17
N=89
N=177
Contr
T2D
SAID
SIDD
SIRD
MOD
MARD
Clusters

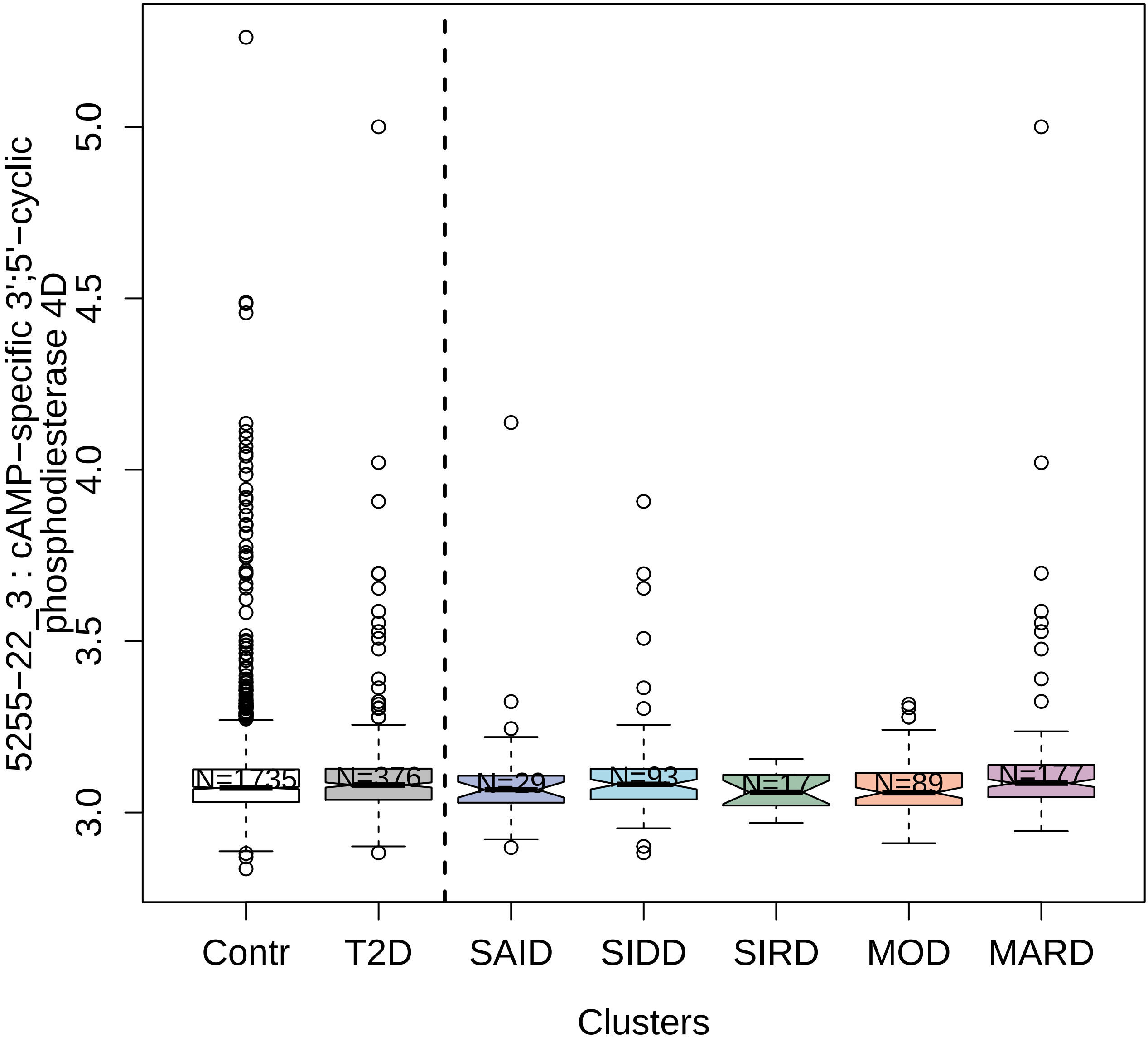
5255−22_3 : cAMP−specific 3',5'−cyclic phosphodiesterase 4D
5.0
4.5
4.0
3.5
3.0
N=1735
N=376
N=29
N=93
N=17
N=89
N=177
Contr
T2D
SAID
SIDD
SIRD
MOD
MARD
Clusters

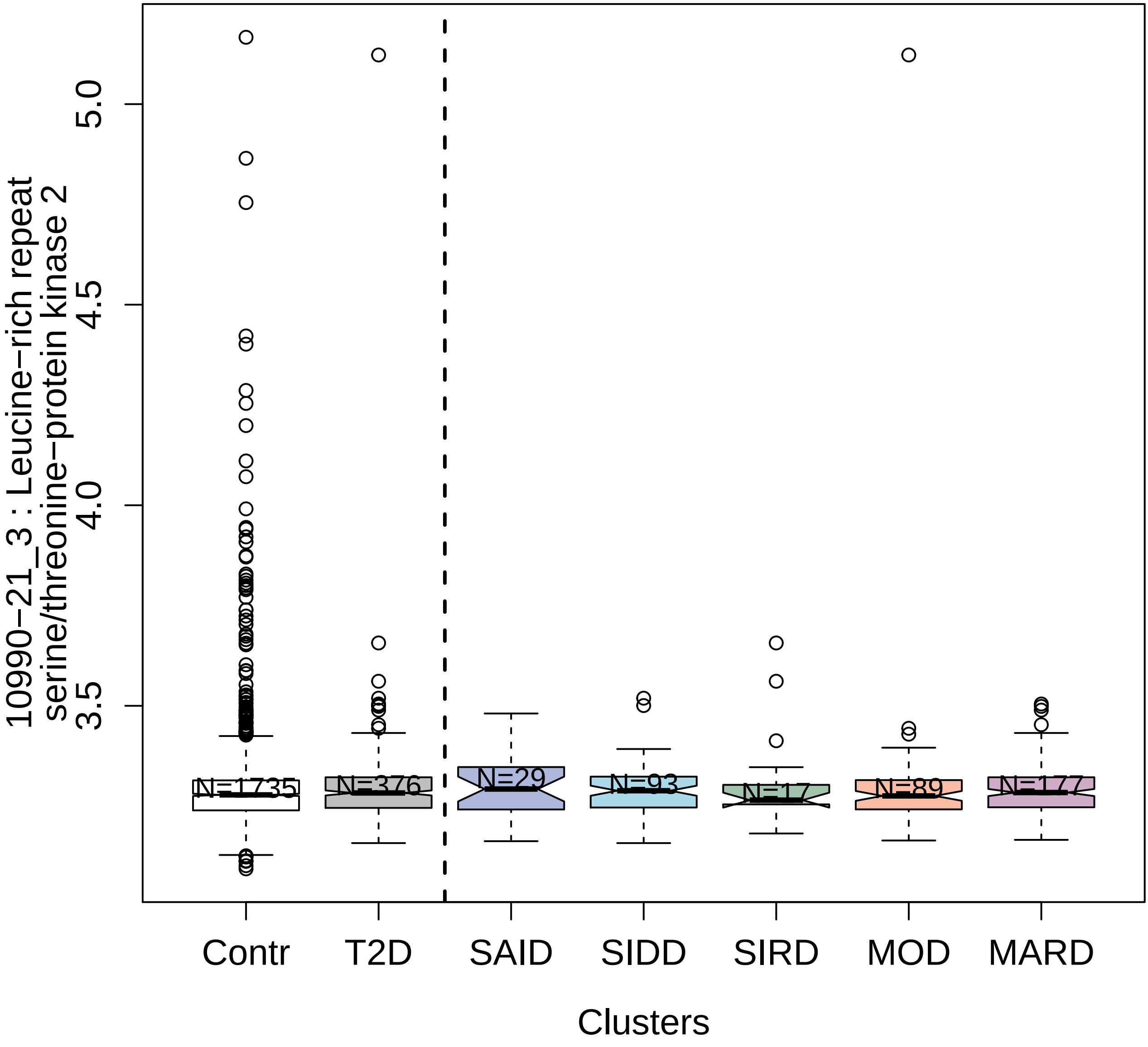
10990−21_3 : Leucine−rich repeat
serine/threonine−protein kinase 2
5.0
4.5
4.0
3.5
N=1735
N=376
N=29
N=93
N=17
N=89
N=177
Contr
T2D
SAID
SIDD
SIRD
MOD
MARD
Clusters

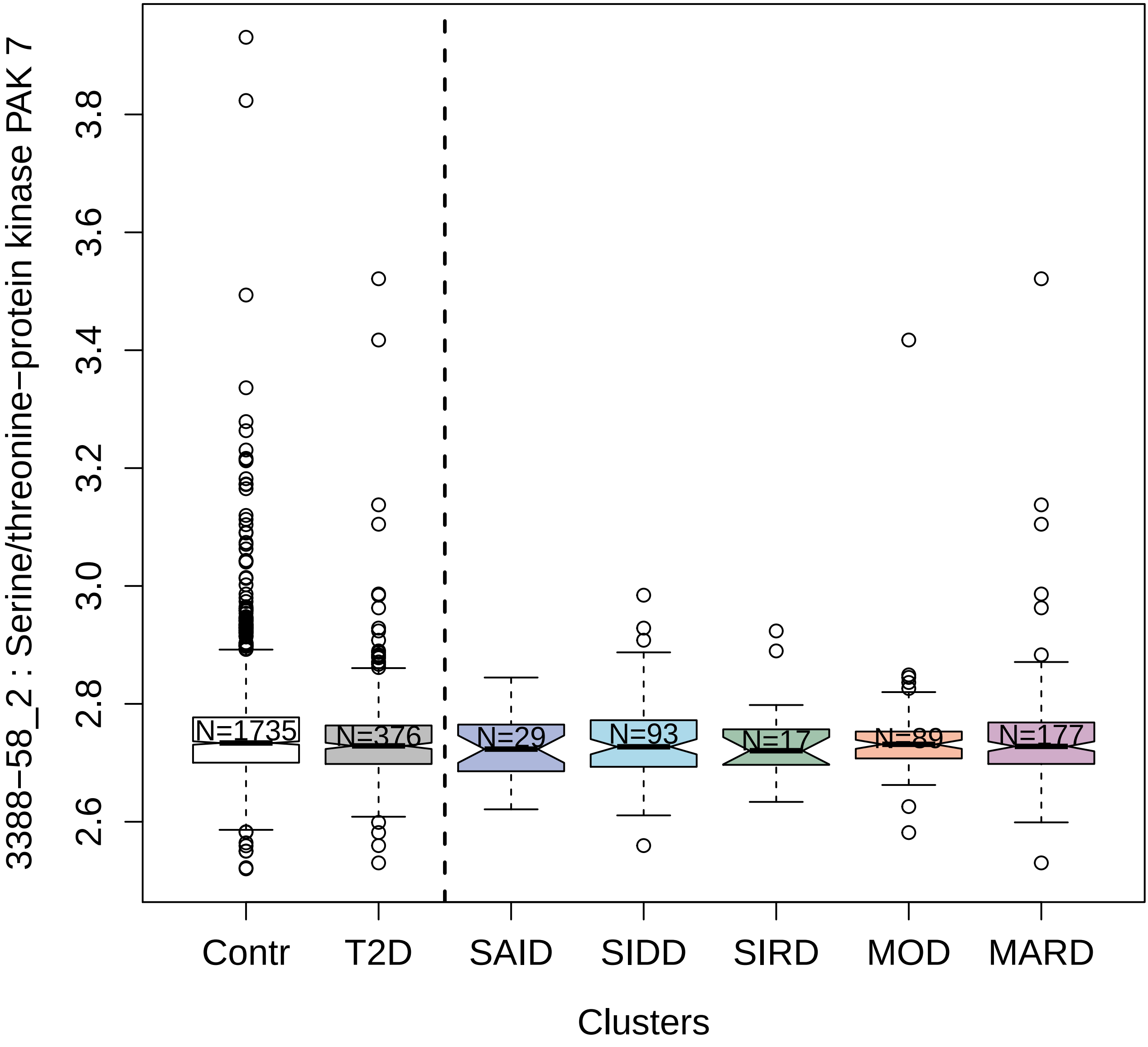
3388−58_2 : Serine/threonine−protein kinase PAK 7
2.6
2.8
3.0
3.2
3.4
3.6
3.8
N=1735
N=376
N=29
N=93
N=17
N=89
N=177
Contr
T2D
SAID
SIDD
SIRD
MOD
MARD
Clusters

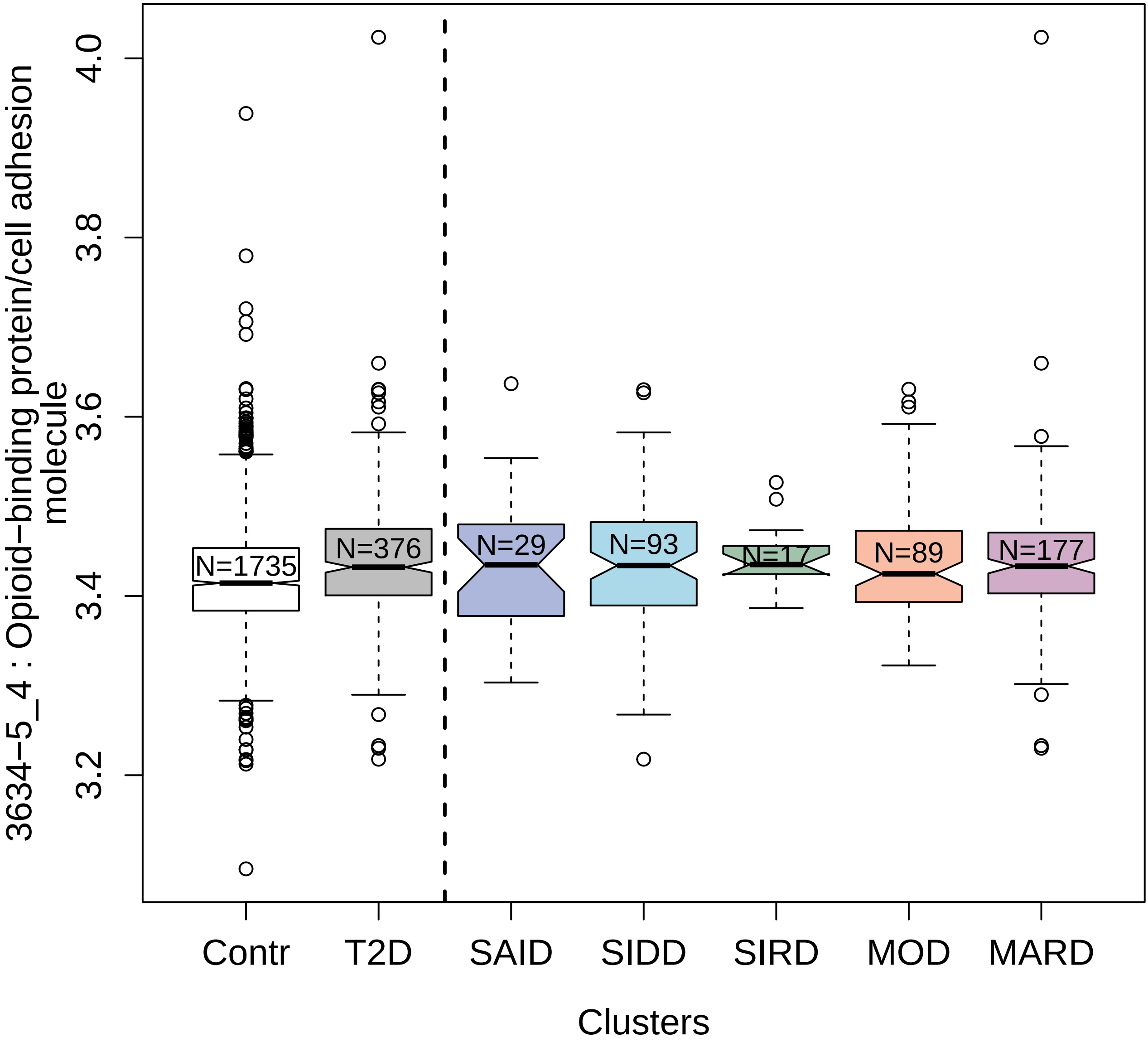
3634−5_4 : Opioid−binding protein/cell adhesion molecule
3.2
3.4
3.6
3.8
4.0
N=1735
N=376
N=29
N=93
N=17
N=89
N=177
Contr
T2D
SAID
SIDD
SIRD
MOD
MARD
Clusters

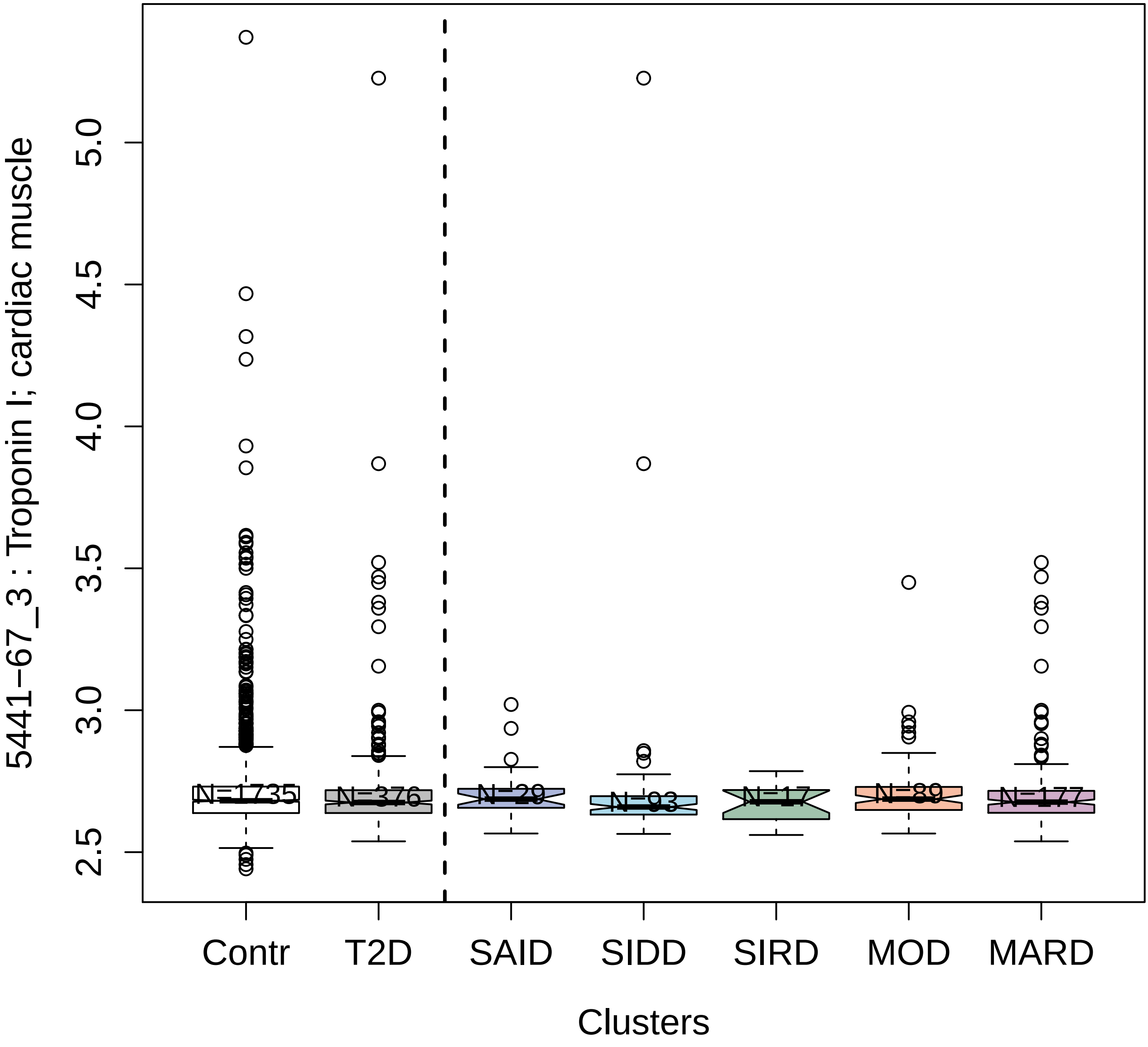

5441−67_3 : Troponin I; cardiac muscle
5.0
4.5
4.0
3.5
3.0
2.5
N=1735
N=376
N=29
N=93
N=17
N=89
N=177
Contr
T2D
SAID
SIDD
SIRD
MOD
MARD
Clusters

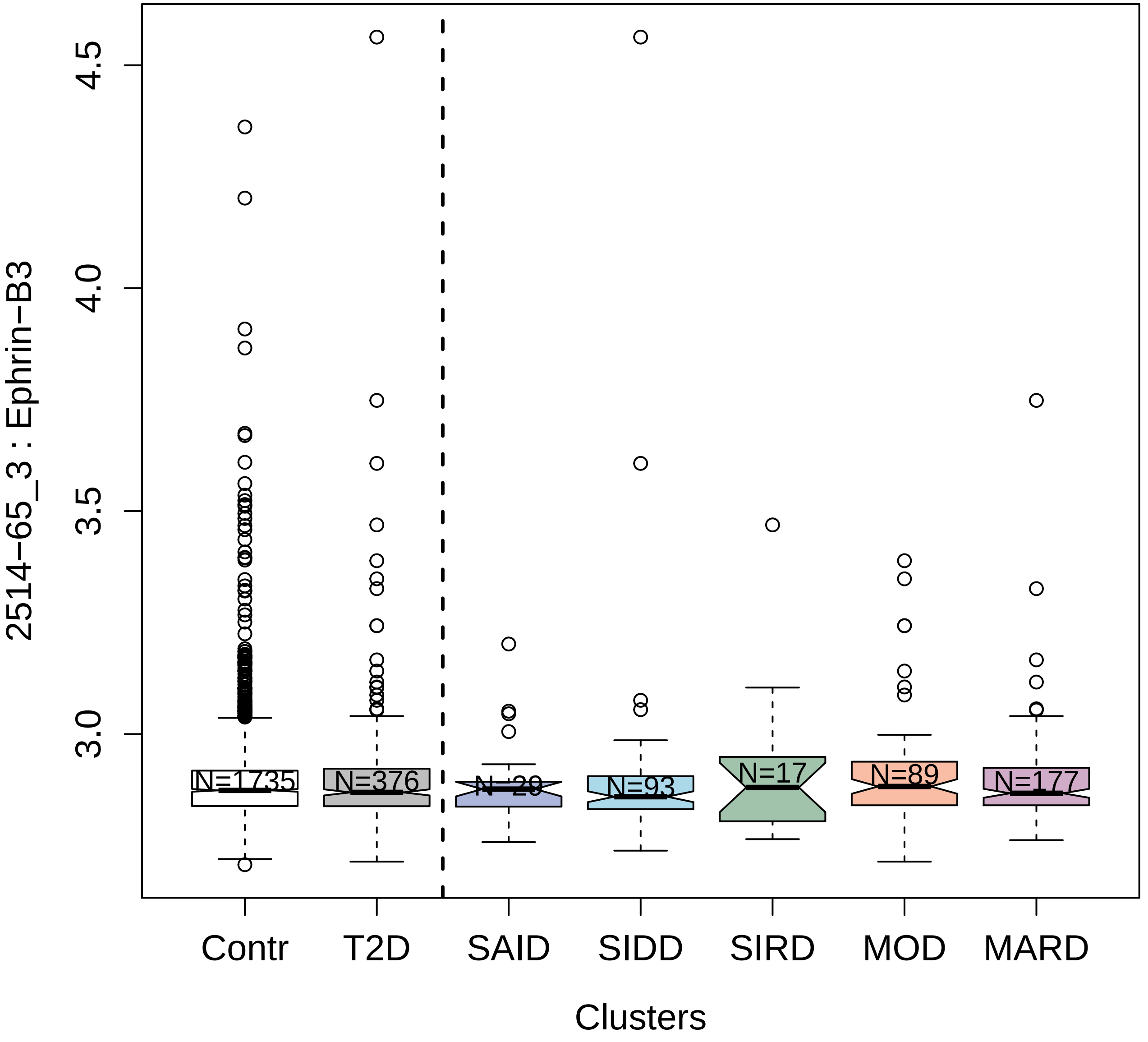

2514−65_3 : Ephrin−B3
4.5
4.0
3.5
3.0
N=1735
N=376
N=29
N=93
N=17
N=89
N=177
Contr
T2D
SAID
SIDD
SIRD
MOD
MARD
Clusters

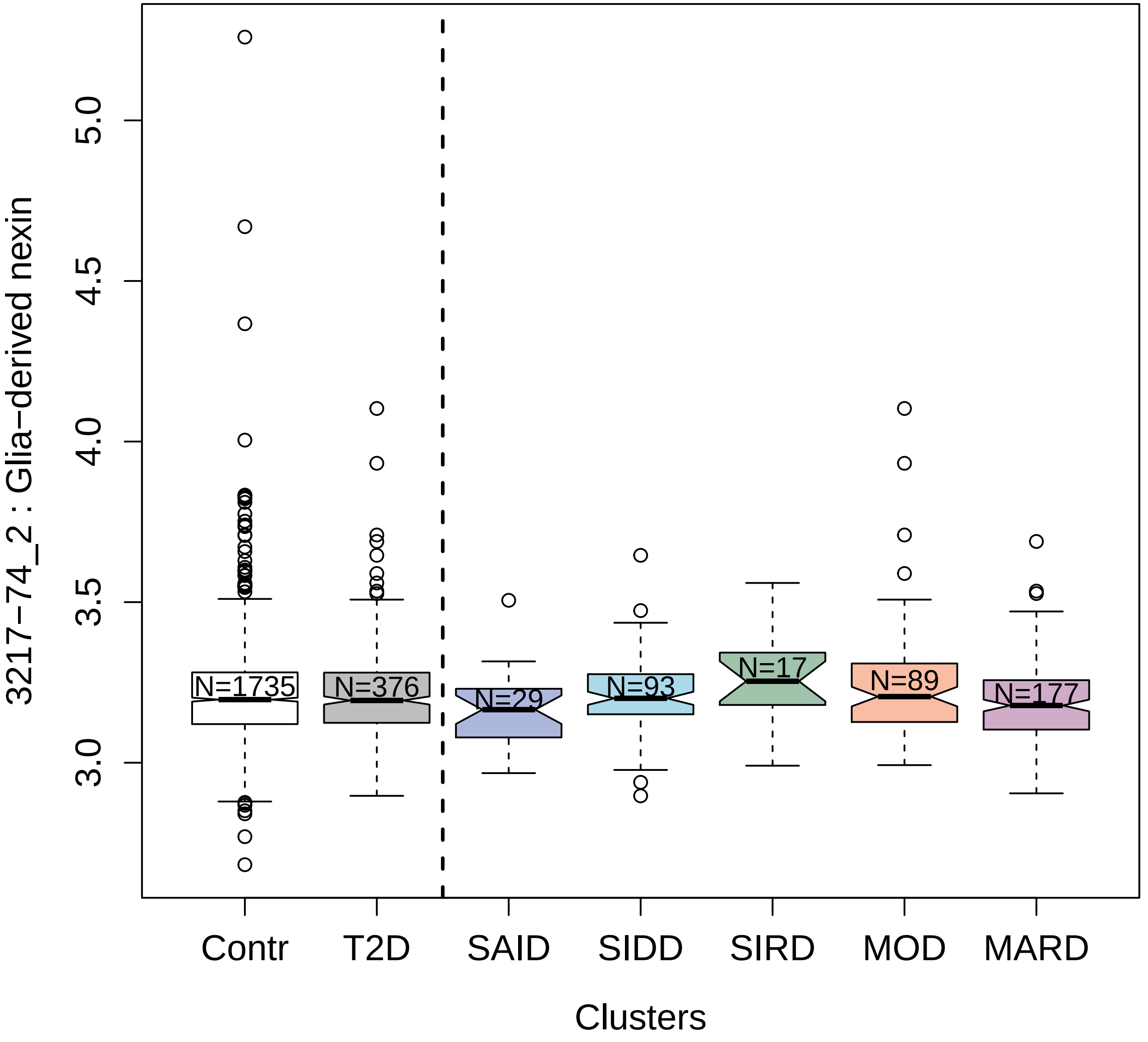

3217−74_2 : Glia−derived nexin
5.0
4.5
4.0
3.5
3.0
N=1735
N=376
N=29
N=93
N=17
N=89
N=177
Contr
T2D
SAID
SIDD
SIRD
MOD
MARD
Clusters

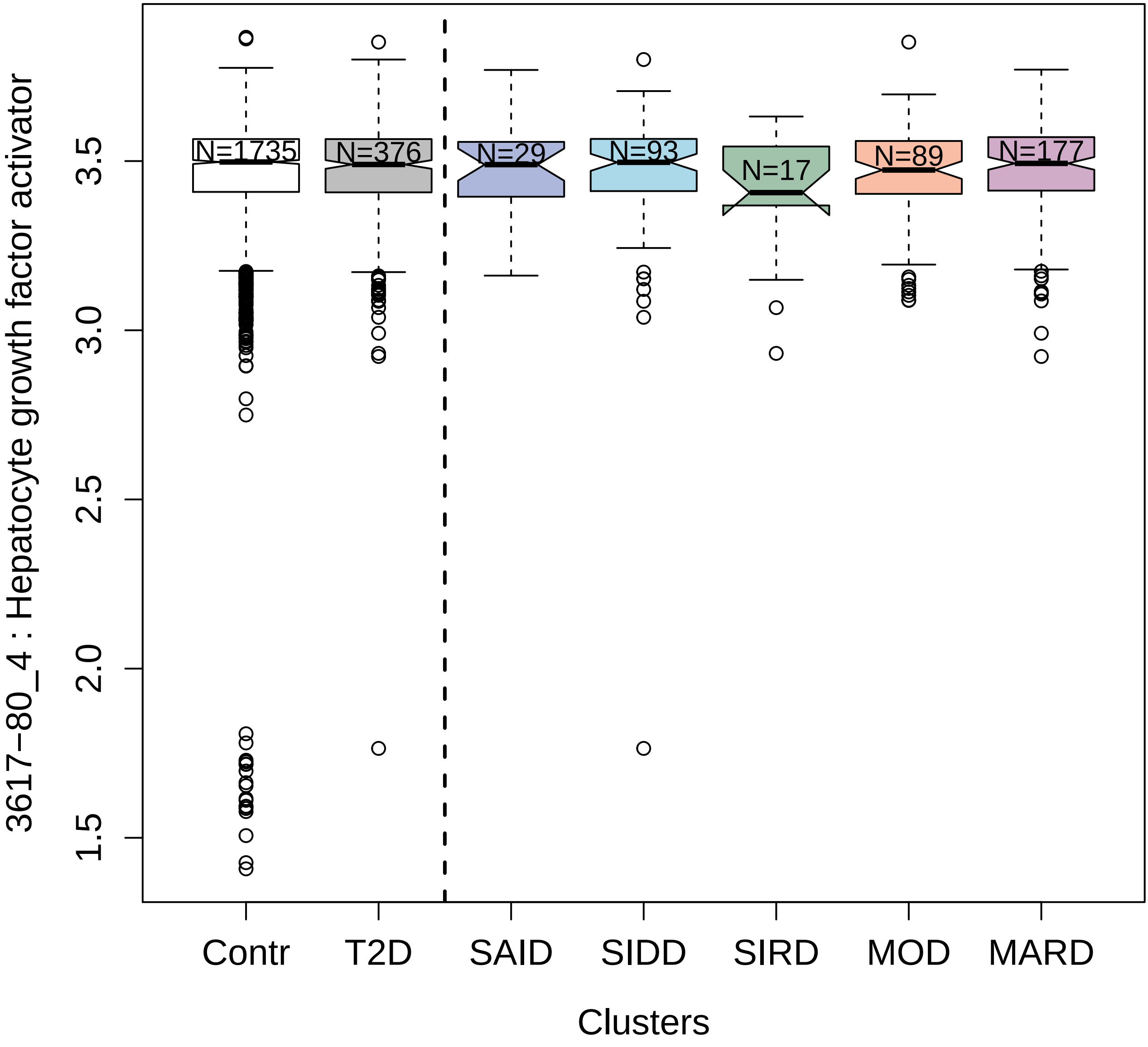

3617−80_4 : Hepatocyte growth factor activator
1.5
2.0
2.5
3.0
3.5
N=1735
N=376
N=29
N=93
N=17
N=89
N=177
Contr
T2D
SAID
SIDD
SIRD
MOD
MARD
Clusters

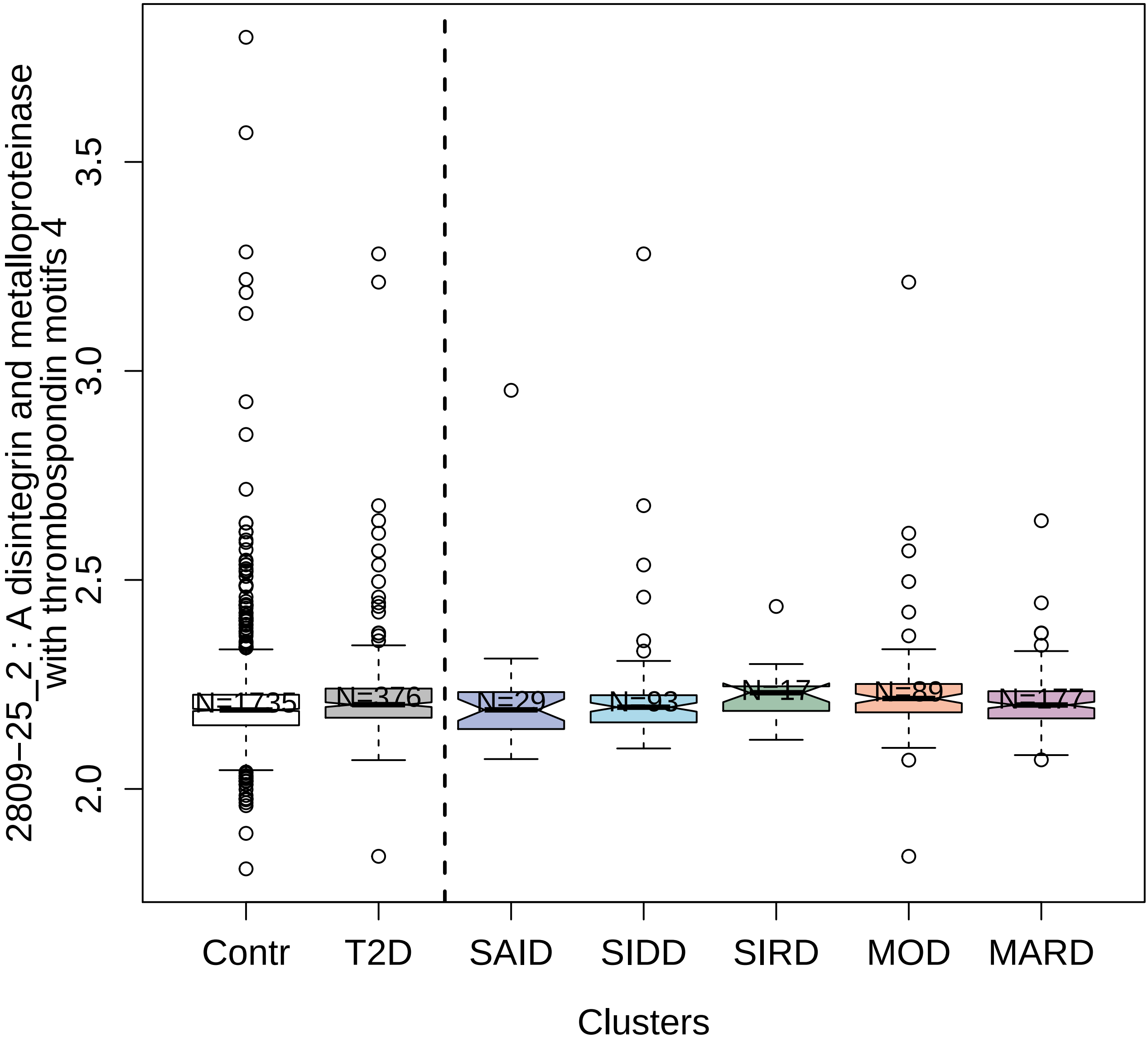
2809−25_2 : A disintegrin and metalloproteinase
_with thrombospondin motifs 4
2.0
2.5
3.0
3.5
N=1735
N=376
N=29
N=93
N=17
N=89
N=177
Contr
T2D
SAID
SIDD
SIRD
MOD
MARD
Clusters

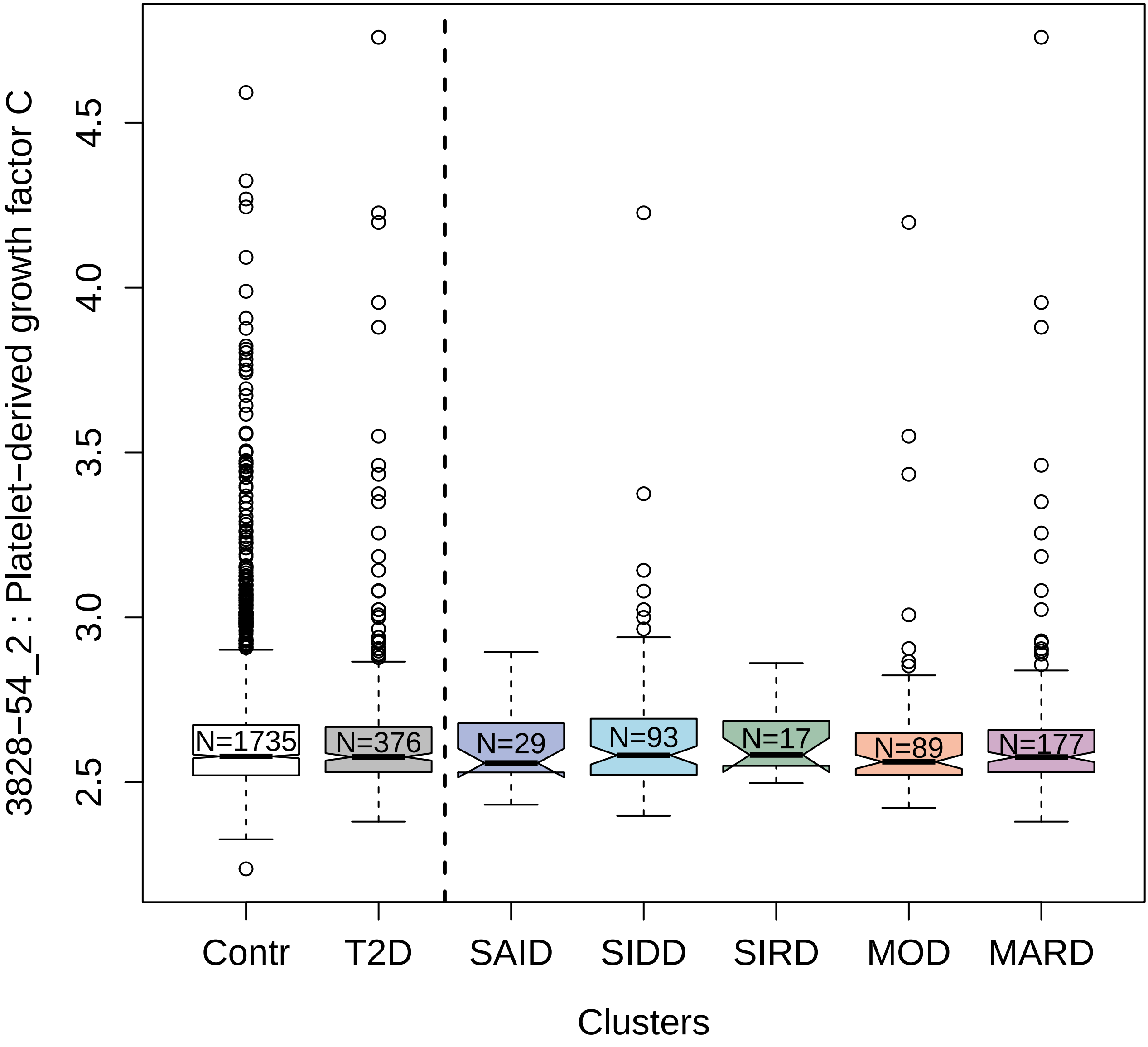

3828−54_2 : Platelet−derived growth factor C
2.5
3.0
3.5
4.0
4.5
N=1735
N=376
N=29
N=93
N=17
N=89
N=177
Contr
T2D
SAID
SIDD
SIRD
MOD
MARD
Clusters

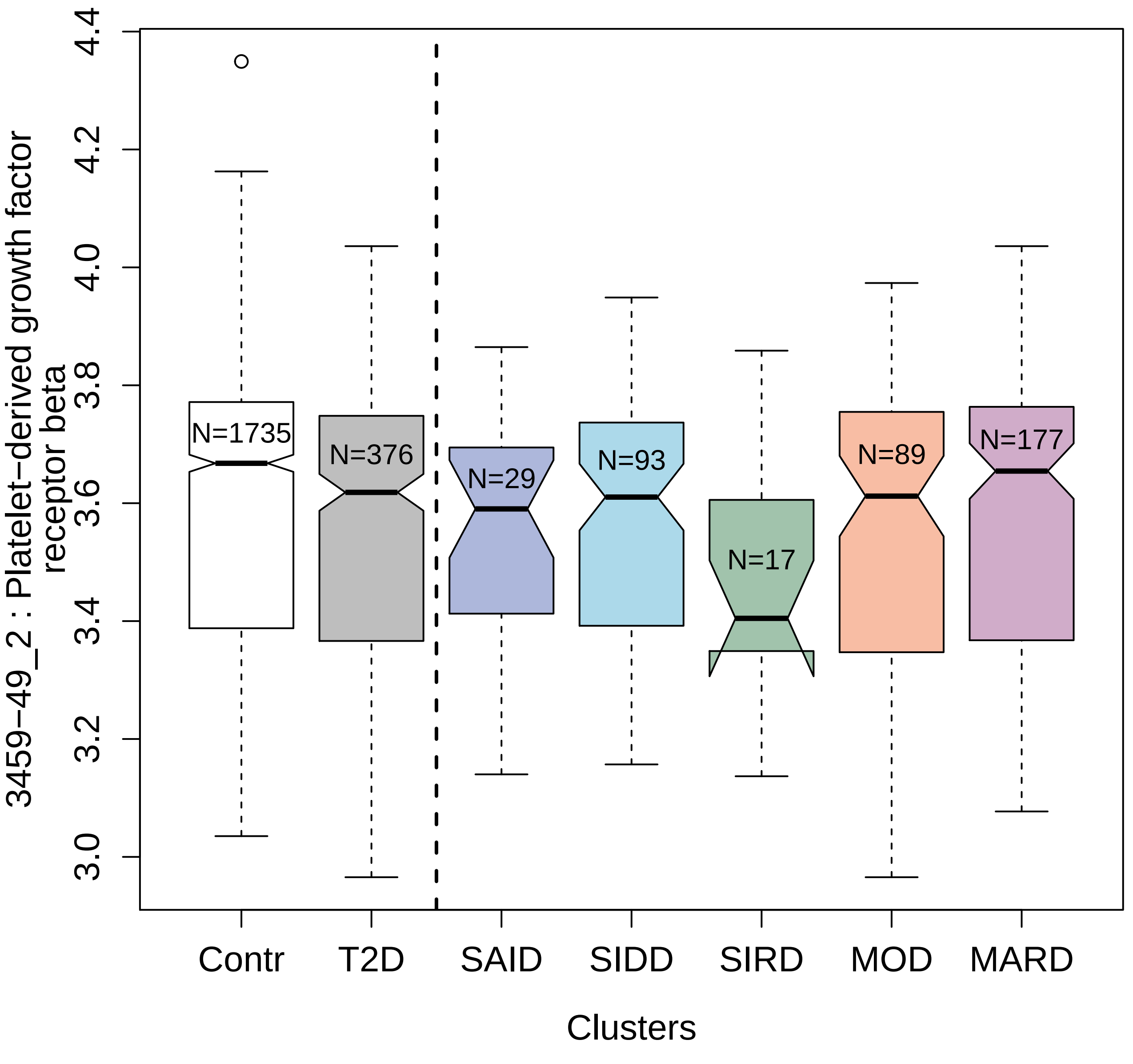

3459−49_2 : Platelet−derived growth factor receptor beta
4.4
4.2
4.0
3.8
3.6
3.4
3.2
3.0
N=1735
N=376
N=29
N=93
N=17
N=89
N=177
Contr
T2D
SAID
SIDD
SIRD
MOD
MARD
Clusters

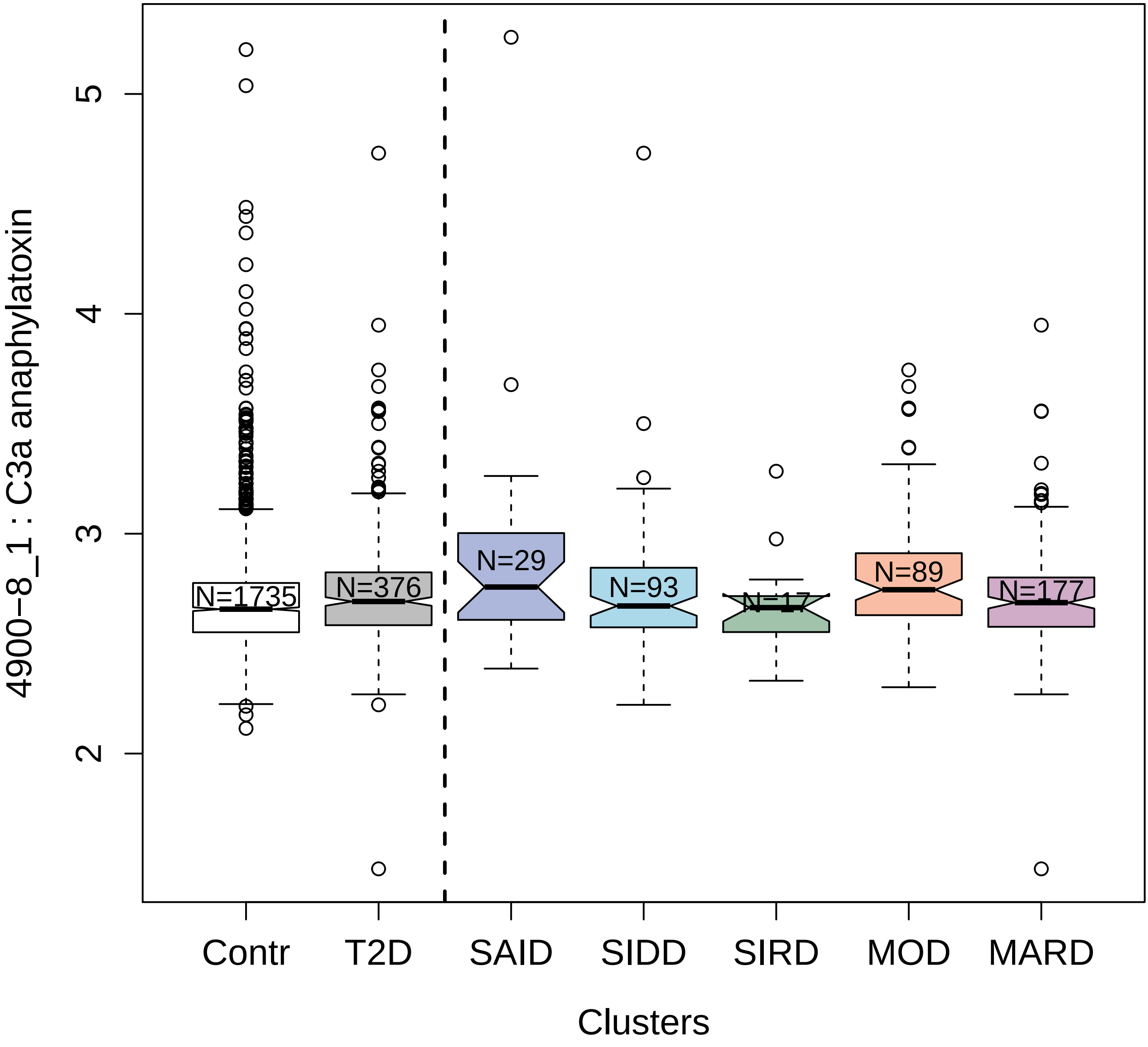

4900−8_1 : C3a anaphylatoxin
5
4
3
2
N=1735
N=376
N=29
N=93
N=17
N=89
N=177
Contr
T2D
SAID
SIDD
SIRD
MOD
MARD
Clusters

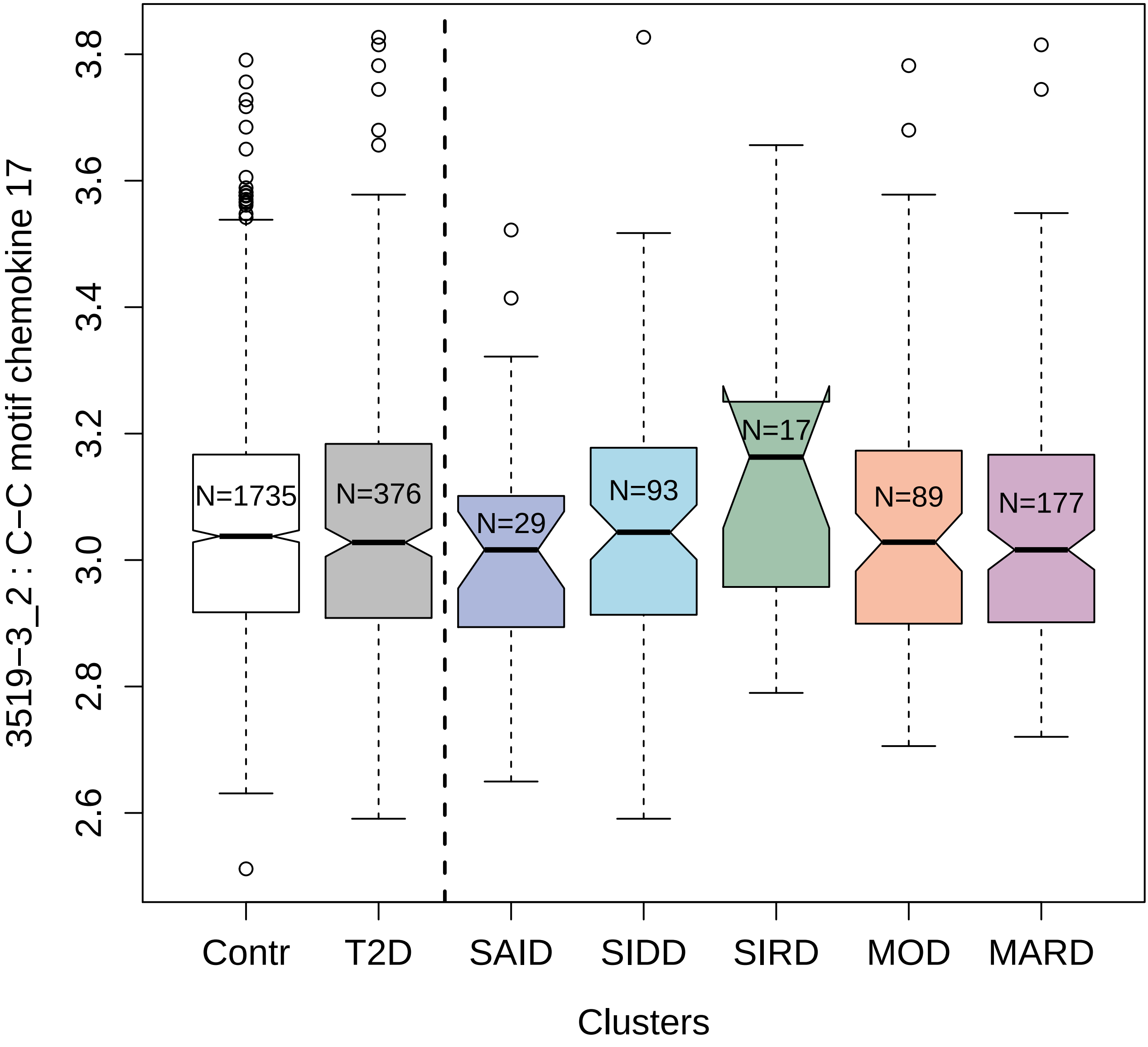

3519−3_2 : C−C motif chemokine 17
2.6
2.8
3.0
3.2
3.4
3.6
3.8
N=1735
N=376
N=29
N=93
N=17
N=89
N=177
Contr
T2D
SAID
SIDD
SIRD
MOD
MARD
Clusters

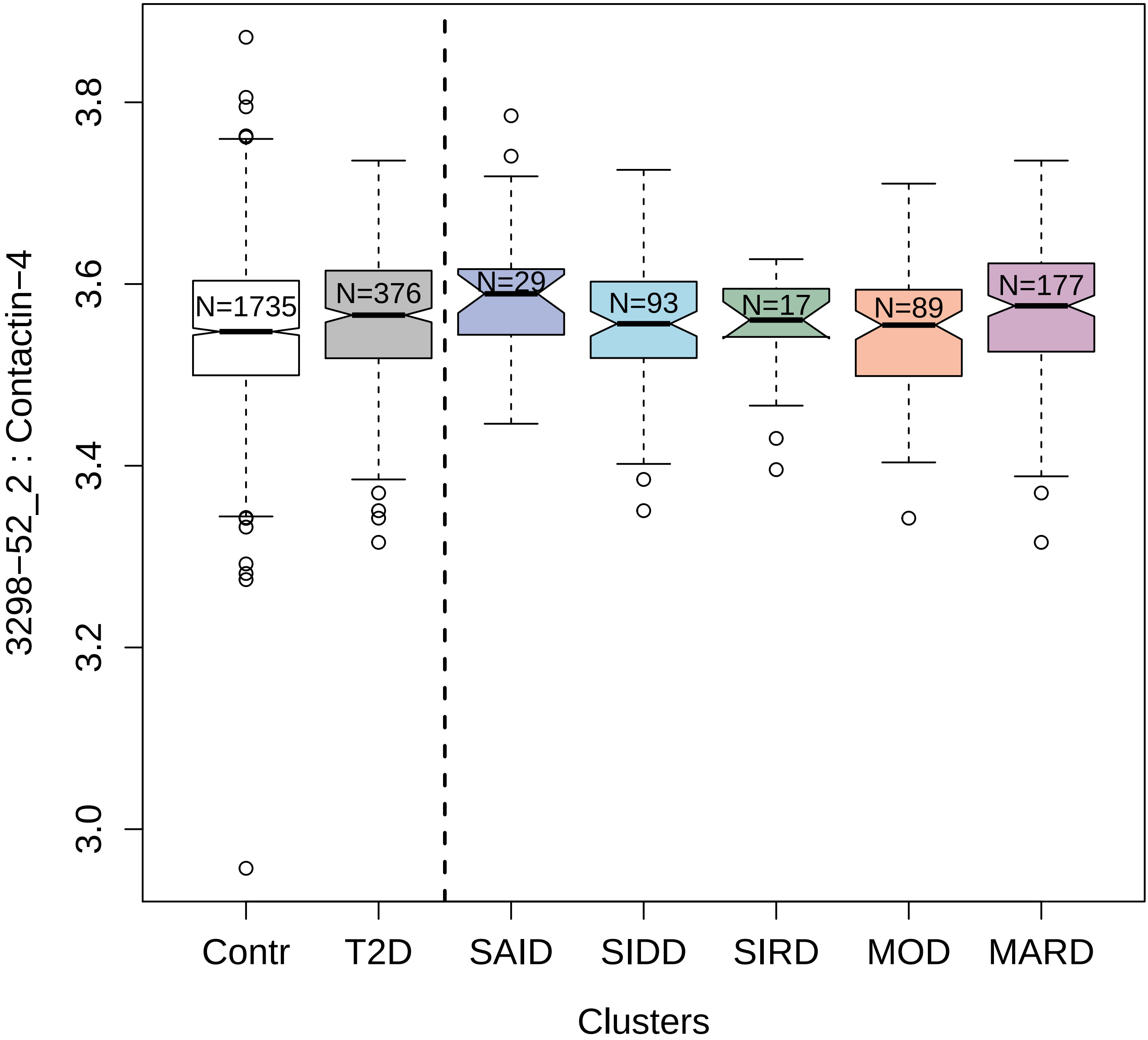

3298−52_2 : Contactin−4
3.0
3.2
3.4
3.6
3.8
N=1735
N=376
N=29
N=93
N=17
N=89
N=177
Contr
T2D
SAID
SIDD
SIRD
MOD
MARD
Clusters

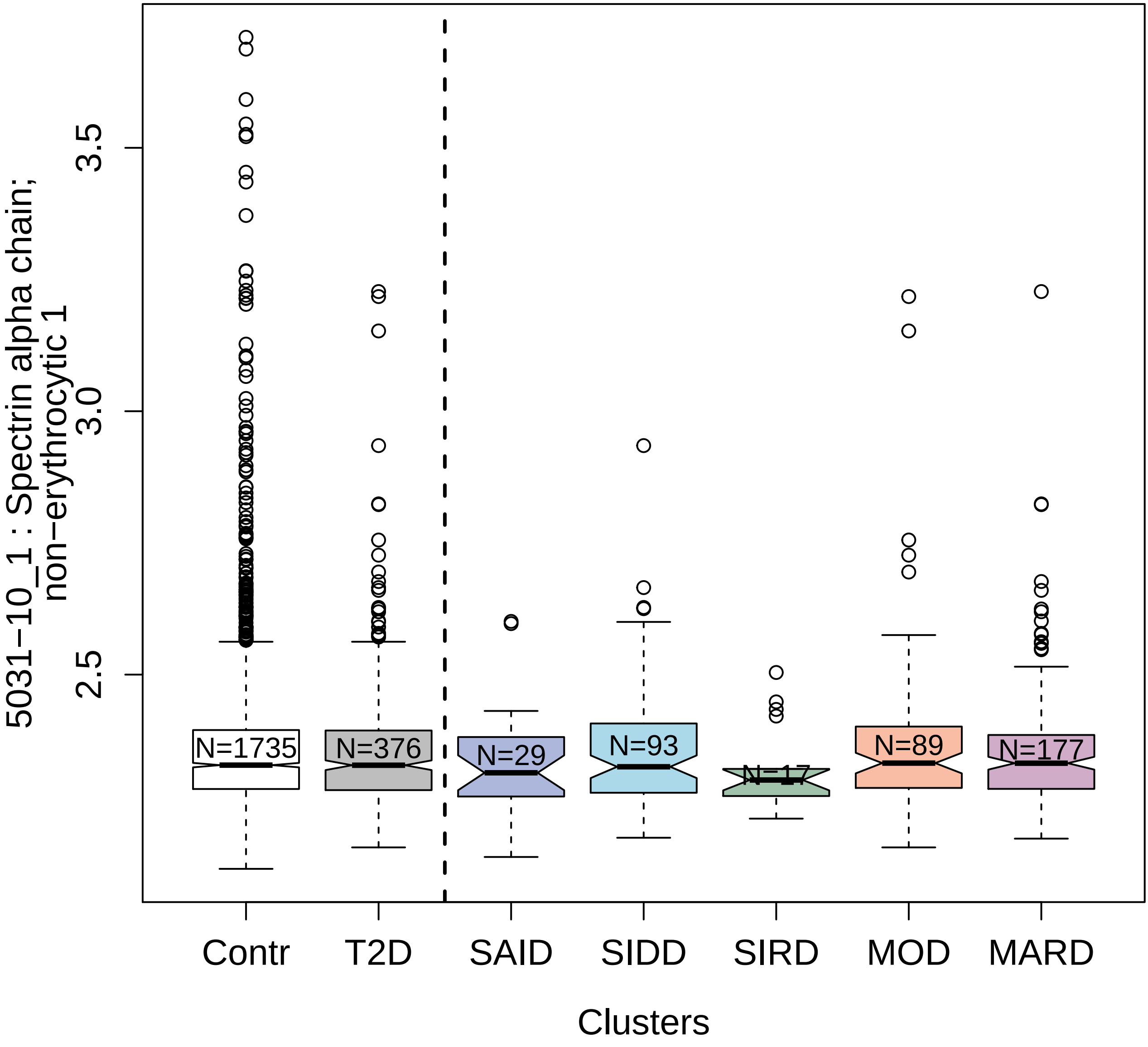

5031−10_1 : Spectrin alpha chain;
non−erythrocytic 1
3.5
3.0
2.5
N=1735
N=376
N=29
N=93
N=17
N=89
N=177
Contr
T2D
SAID
SIDD
SIRD
MOD
MARD
Clusters

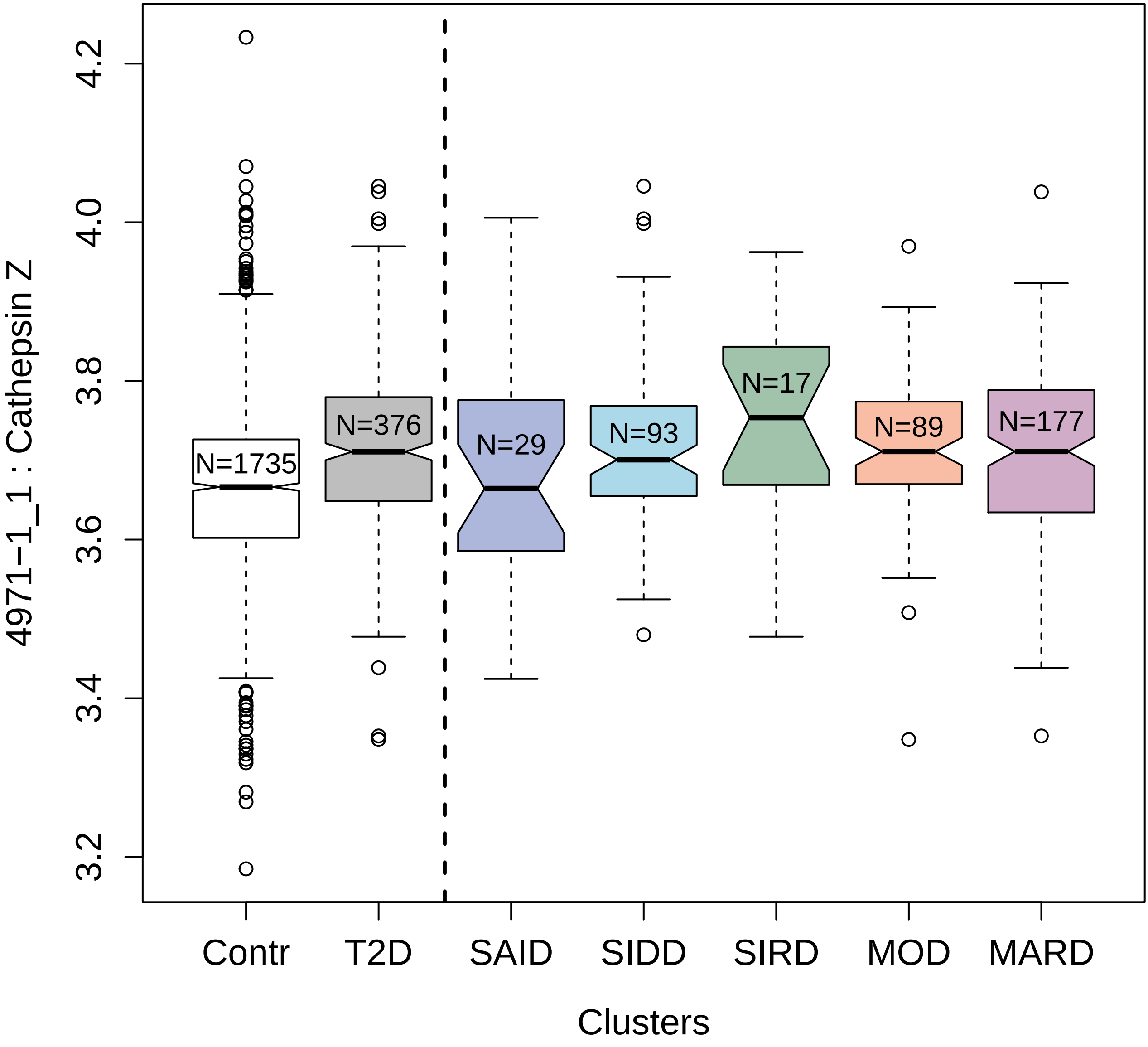

4971−1_1 : Cathepsin Z
4.2
4.0
3.8
3.6
3.4
3.2
N=1735
N=376
N=29
N=93
N=17
N=89
N=177
Contr
T2D
SAID
SIDD
SIRD
MOD
MARD
Clusters

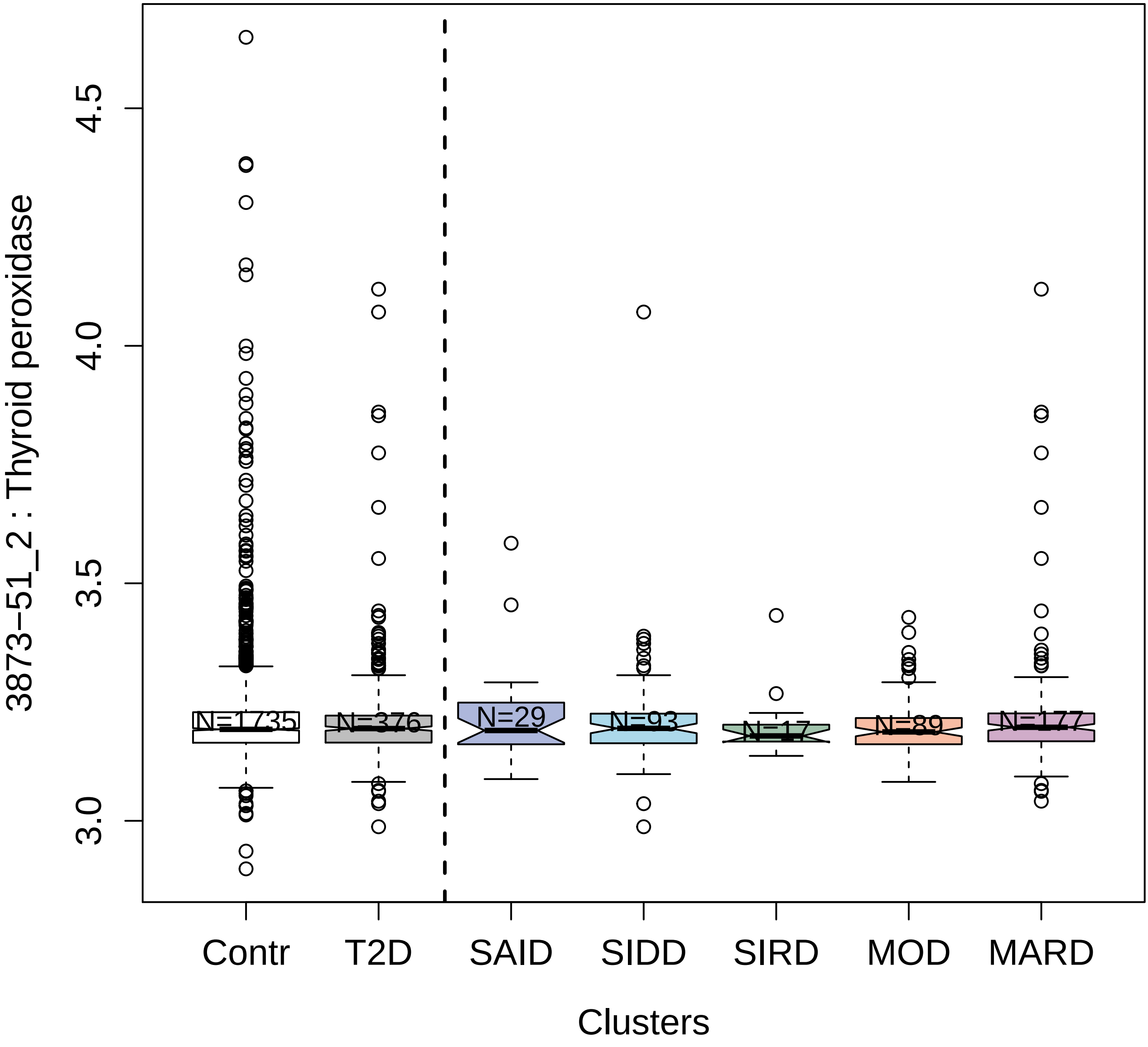

3873−51_2 : Thyroid peroxidase
3.0
3.5
4.0
4.5
N=1735
N=376
N=29
N=93
N=17
N=89
N=177
Contr
T2D
SAID
SIDD
SIRD
MOD
MARD
Clusters

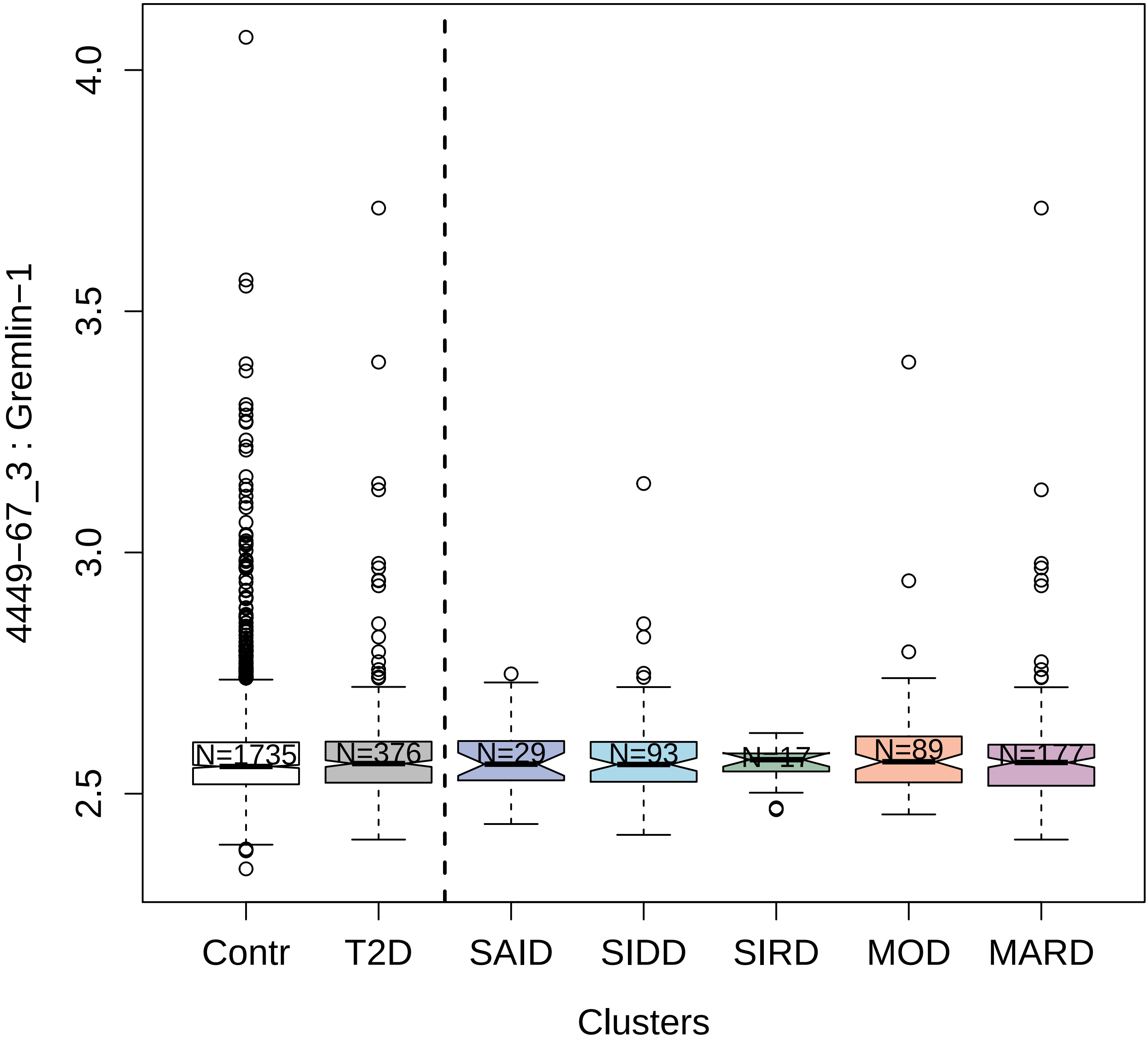

4449−67_3 : Gremlin−1
4.0
3.5
3.0
2.5
N=1735
N=376
N=29
N=93
N=17
N=89
N=177
Contr
T2D
SAID
SIDD
SIRD
MOD
MARD
Clusters

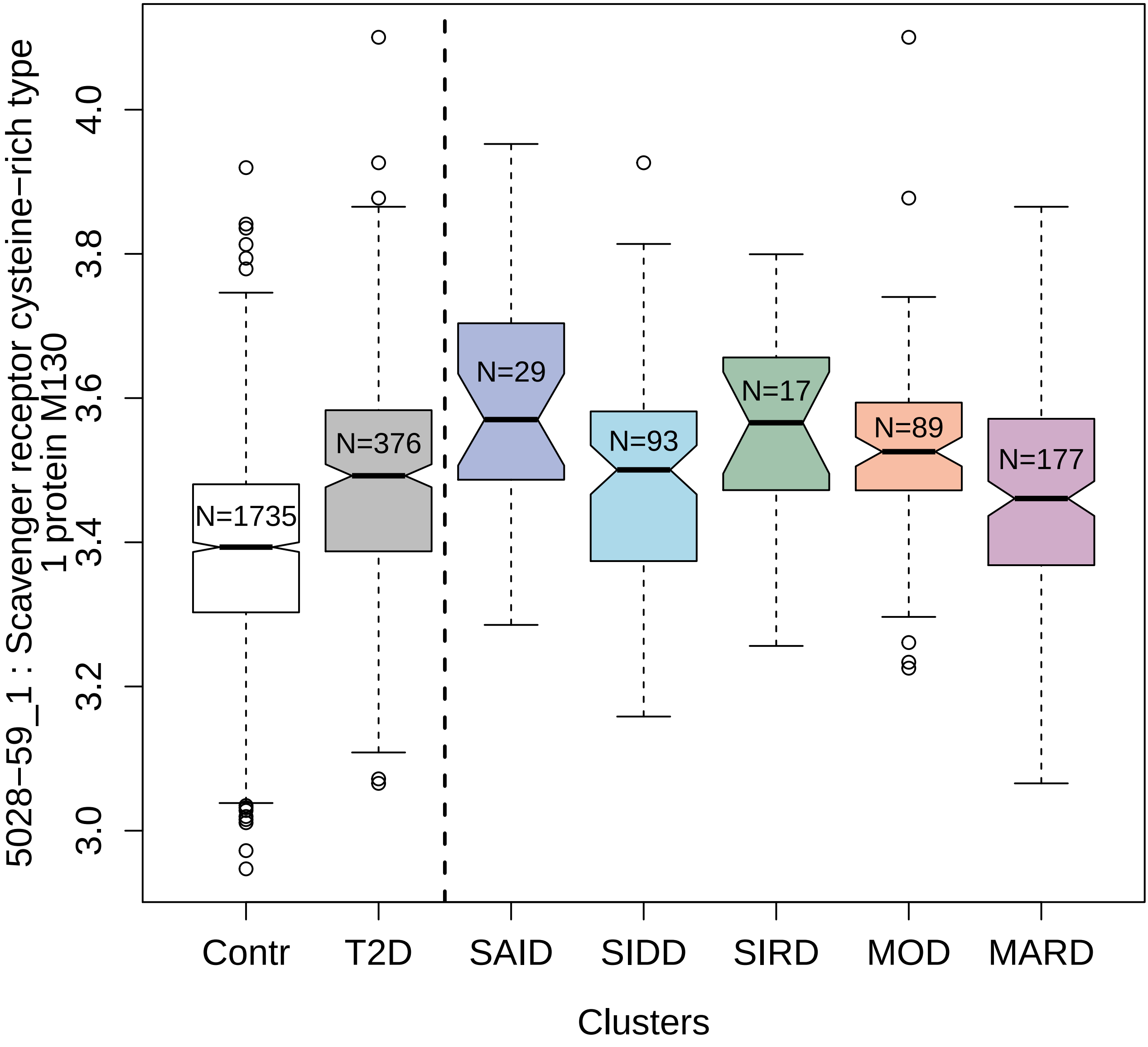

5028−59_1 : Scavenger receptor cysteine−rich type 1 protein M130
4.0
3.8
3.6
3.4
3.2
3.0
N=1735
N=376
N=29
N=93
N=17
N=89
N=177
Contr
T2D
SAID
SIDD
SIRD
MOD
MARD
Clusters

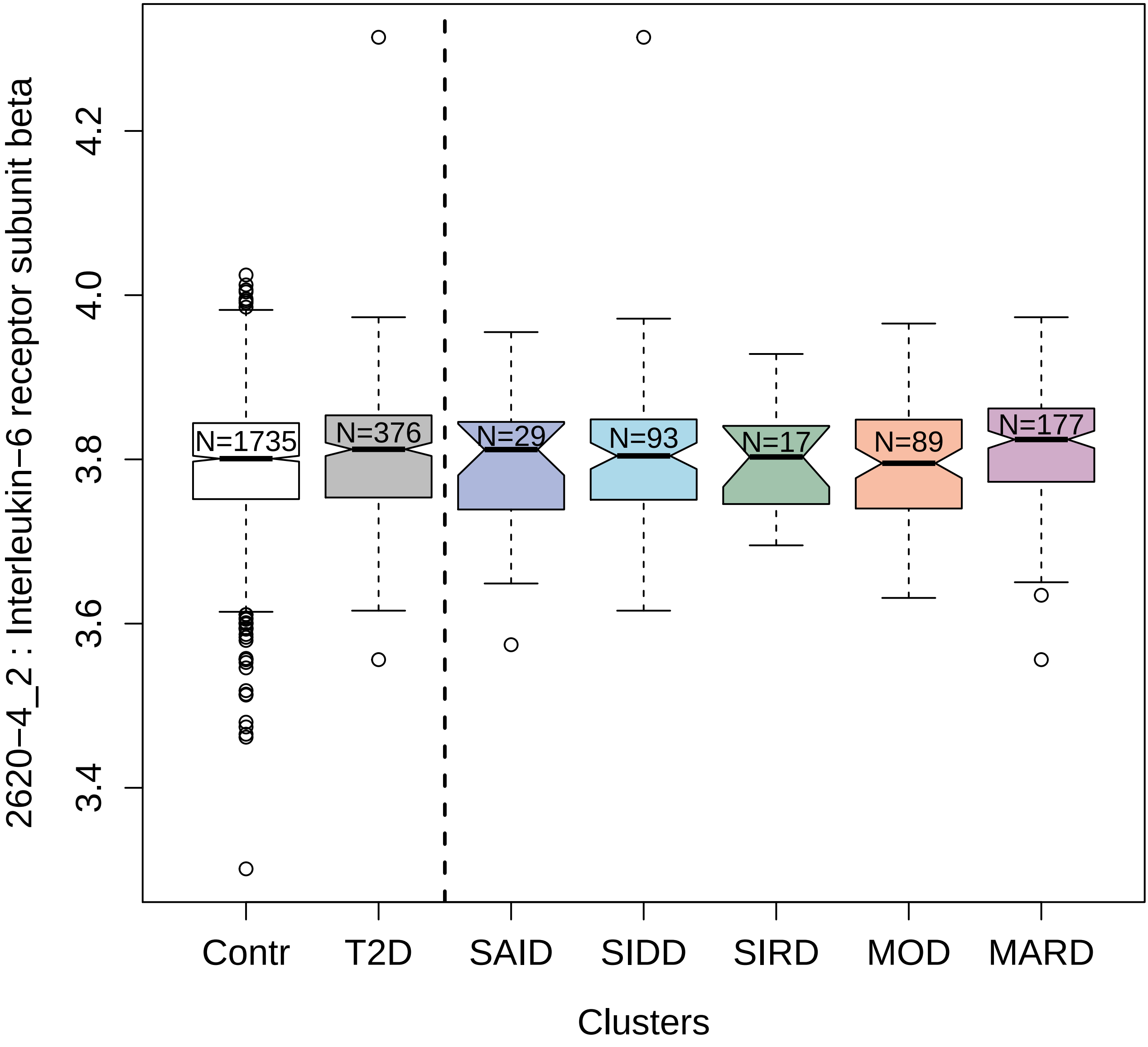

2620−4_2 : Interleukin−6 receptor subunit beta
3.4
3.6
3.8
4.0
4.2
N=1735
N=376
N=29
N=93
N=17
N=89
N=177
Contr
T2D
SAID
SIDD
SIRD
MOD
MARD
Clusters

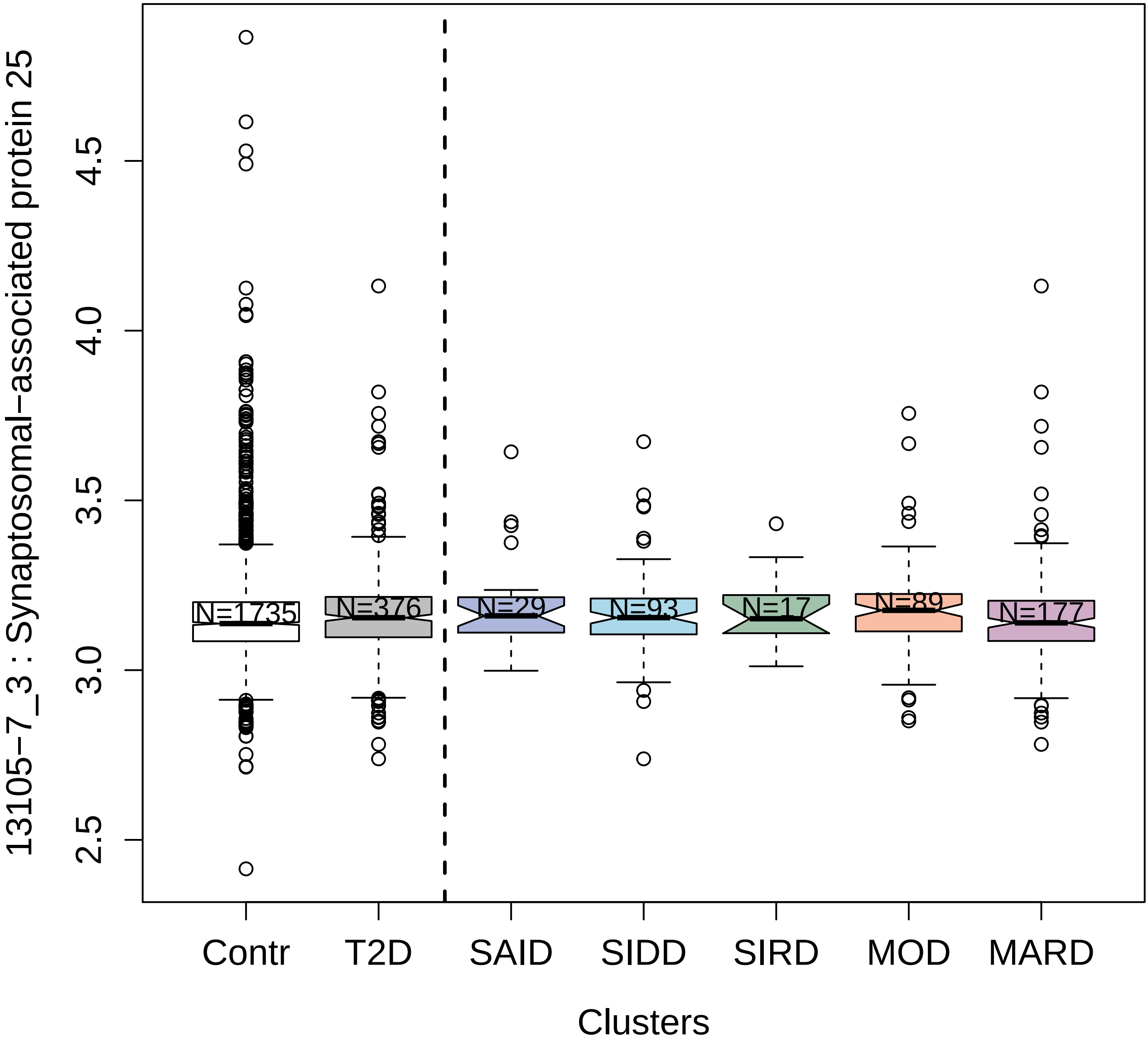

13105−7_3 : Synaptosomal−associated protein 25
2.5
3.0
3.5
4.0
4.5
N=1735
N=376
N=29
N=93
N=17
N=89
N=177
Contr
T2D
SAID
SIDD
SIRD
MOD
MARD
Clusters

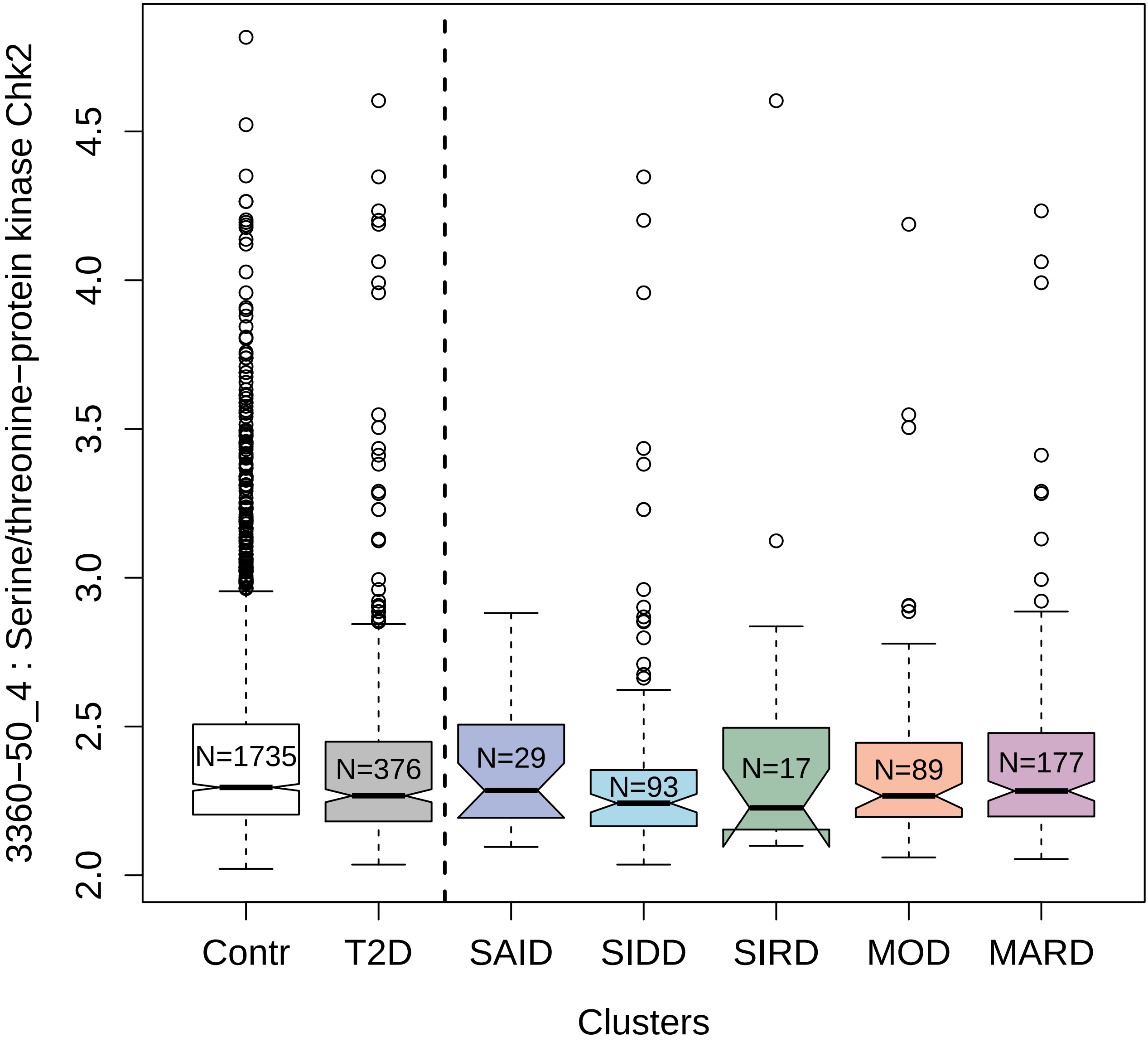

3360−50_4 : Serine/threonine−protein kinase Chk2
2.0
2.5
3.0
3.5
4.0
4.5
N=1735
N=376
N=29
N=93
N=17
N=89
N=177
Contr
T2D
SAID
SIDD
SIRD
MOD
MARD
Clusters

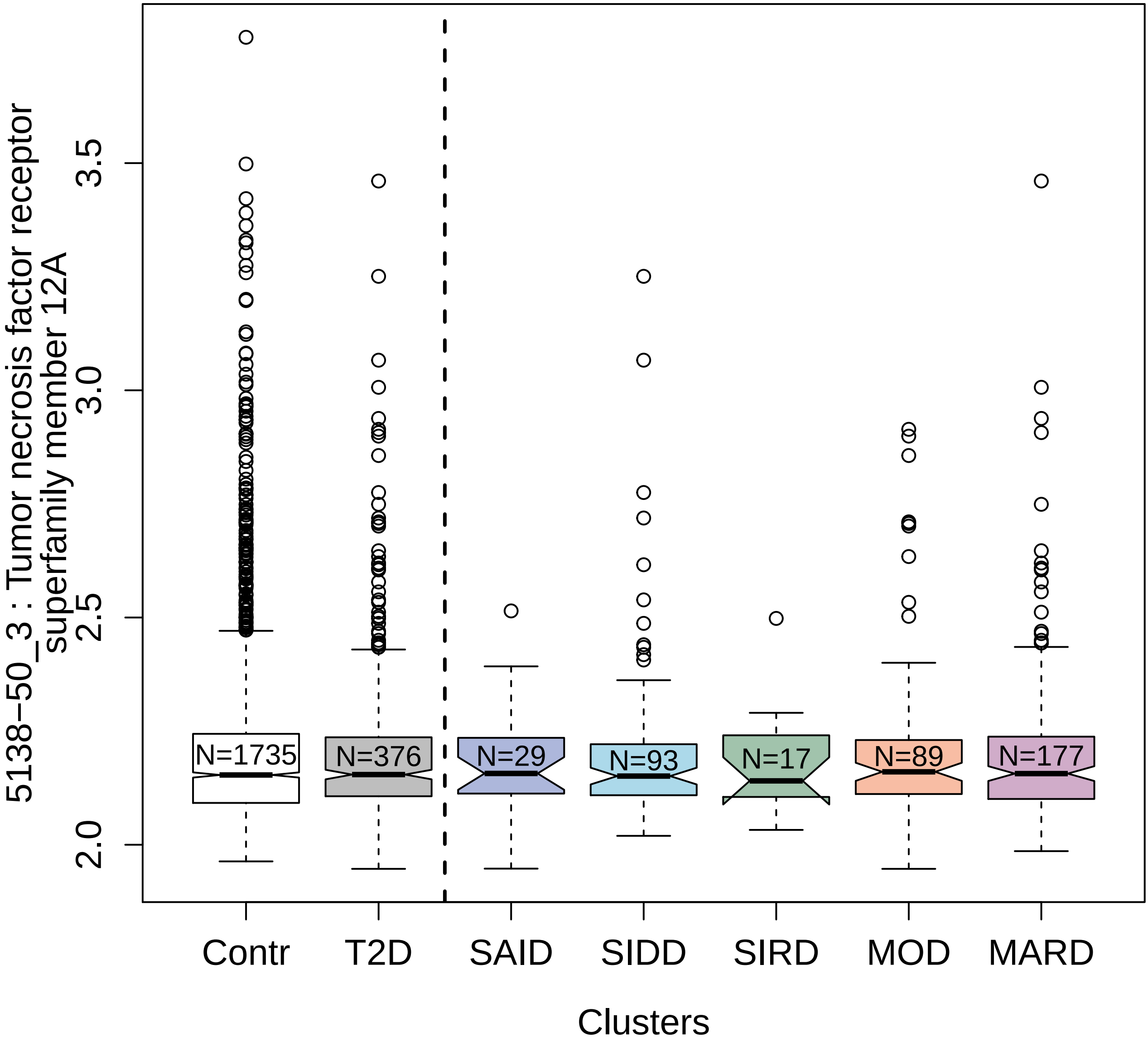
5138−50_3 : Tumor necrosis factor receptor
superfamily member 12A
2.0
2.5
3.0
3.5
N=1735
N=376
N=29
N=93
N=17
N=89
N=177
Contr
T2D
SAID
SIDD
SIRD
MOD
MARD
Clusters

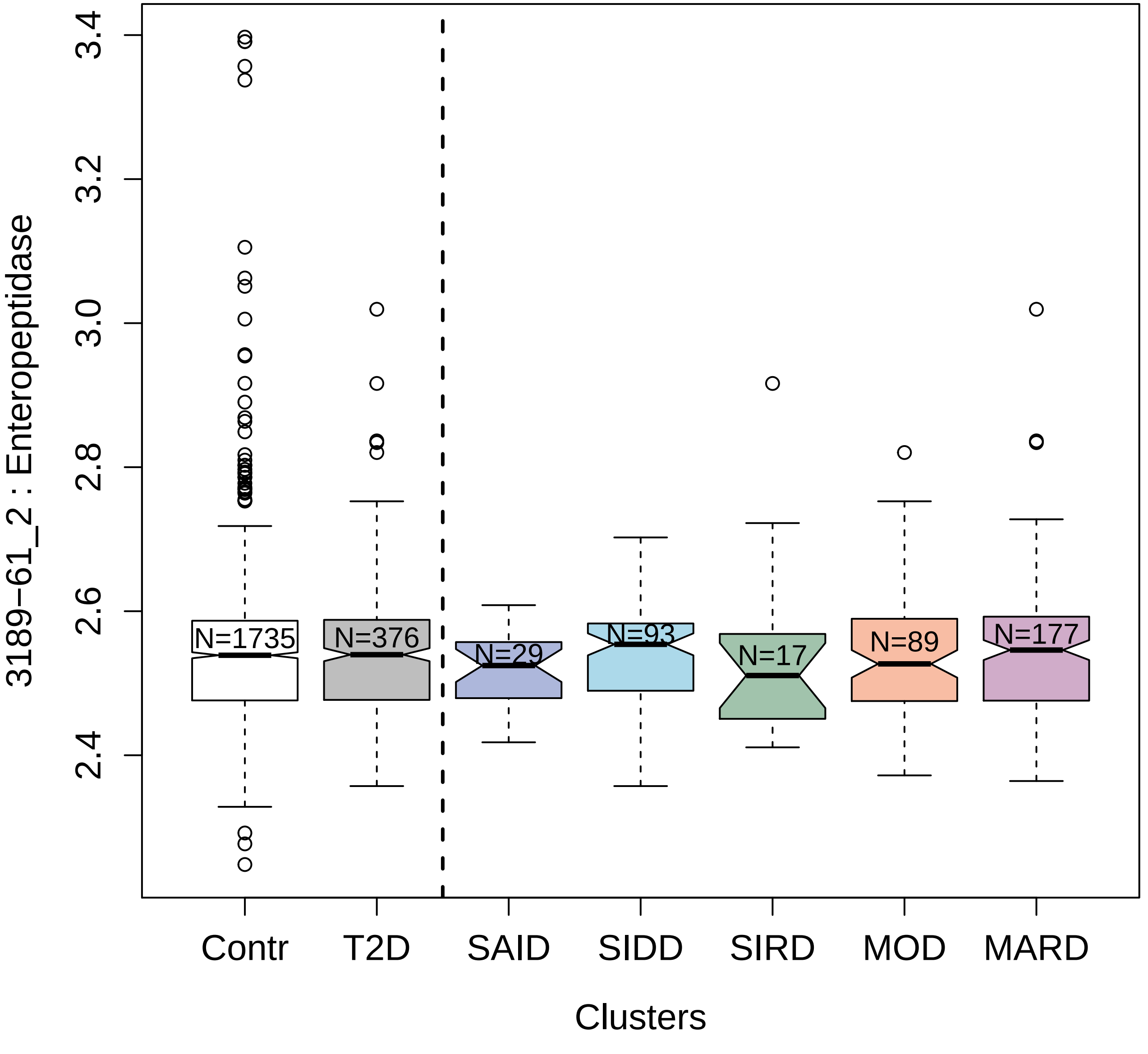
3189−61_2 : Enteropeptidase
3.4
3.2
3.0
2.8
2.6
2.4
N=1735
N=376
N=29
N=93
N=17
N=89
N=177
Contr
T2D
SAID
SIDD
SIRD
MOD
MARD
Clusters

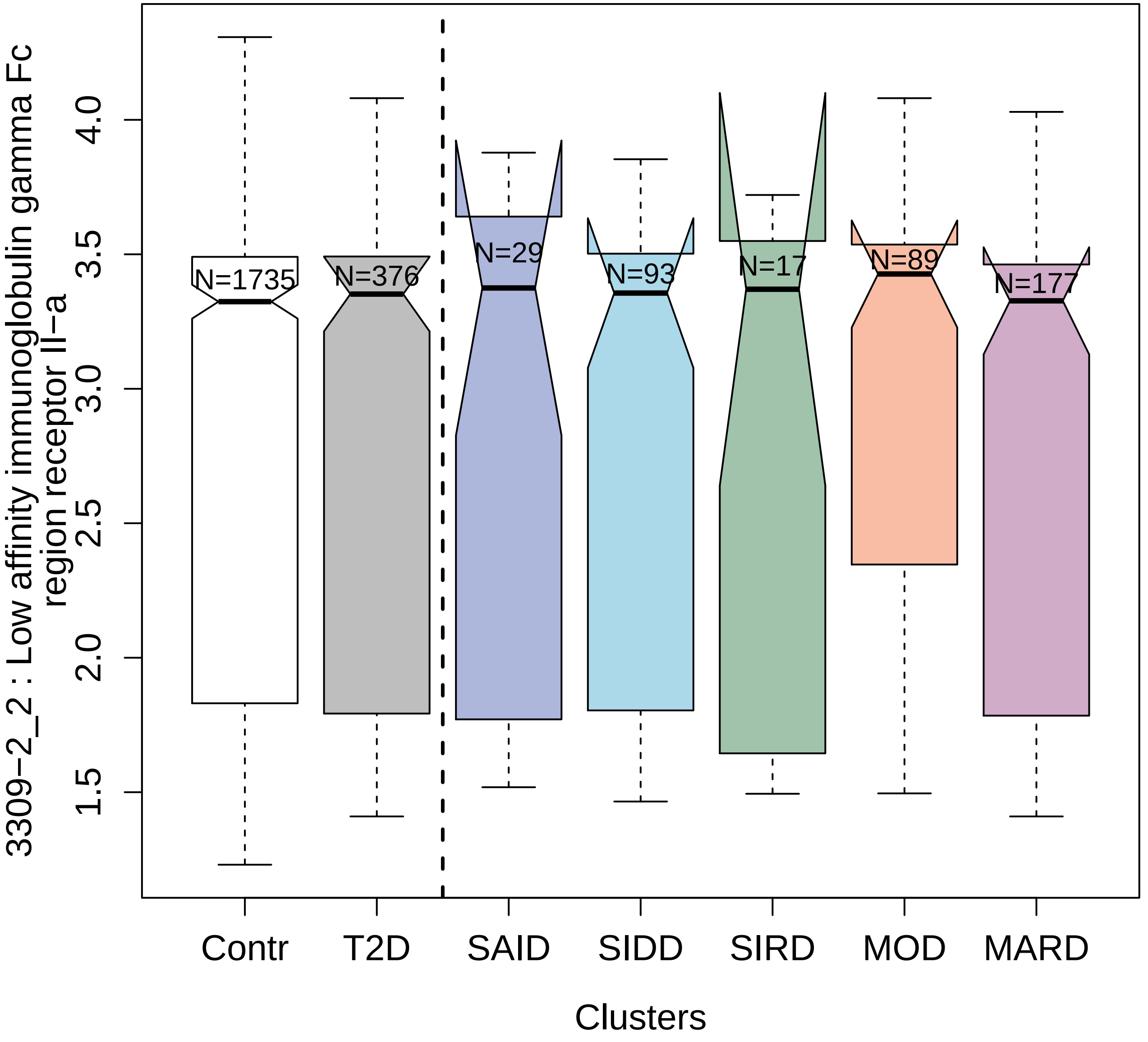

3309−2_2 : Low affinity immunoglobulin gamma Fc region receptor II−a
1.5
2.0
2.5
3.0
3.5
4.0
N=1735
N=376
N=29
N=93
N=17
N=89
N=177
Contr
T2D
SAID
SIDD
SIRD
MOD
MARD
Clusters

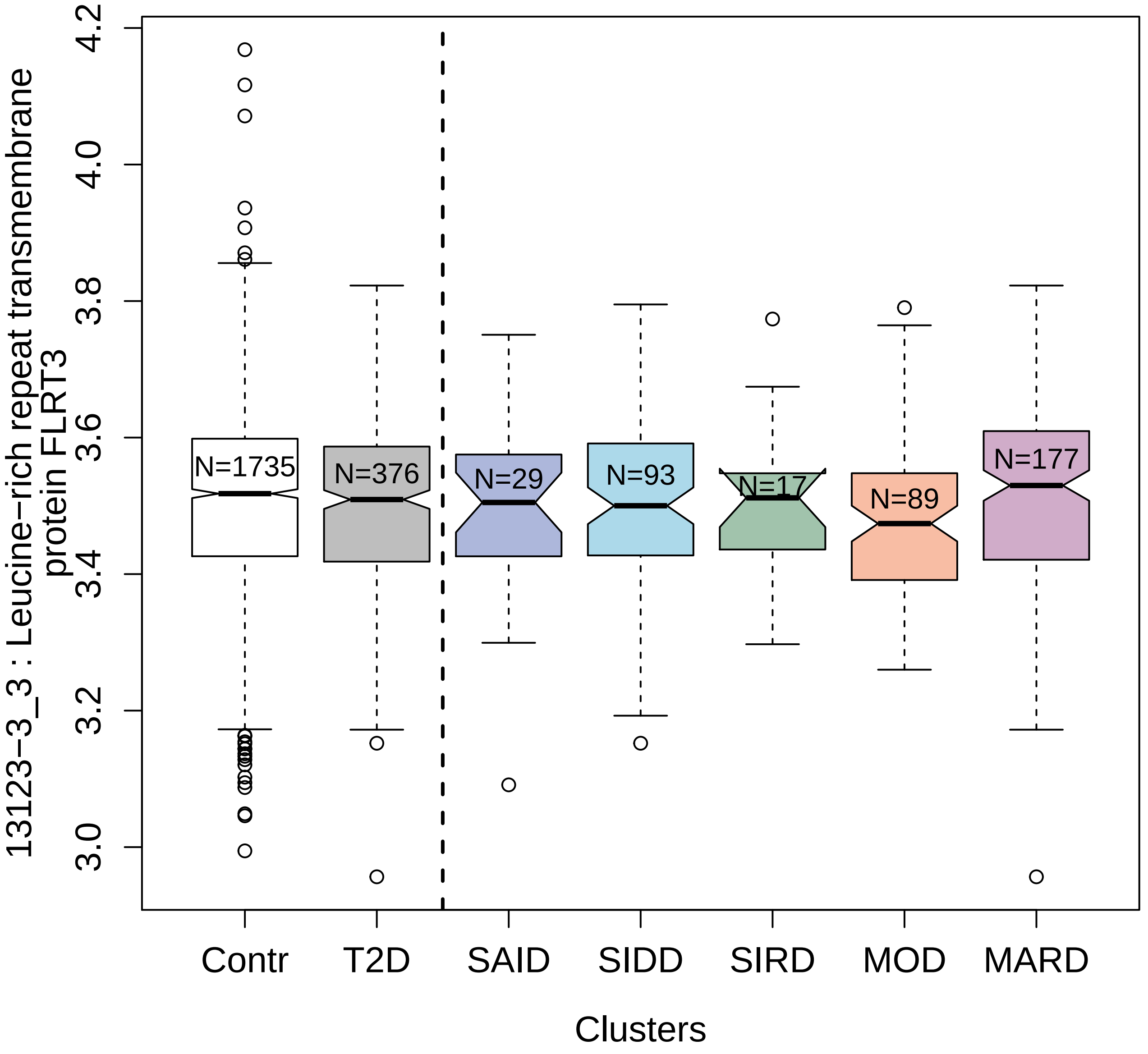

13123−3_3 : Leucine−rich repeat transmembrane
protein FLRT3
4.2
4.0
3.8
3.6
3.4
3.2
3.0
N=1735
N=376
N=29
N=93
N=17
N=89
N=177
Contr
T2D
SAID
SIDD
SIRD
MOD
MARD
Clusters

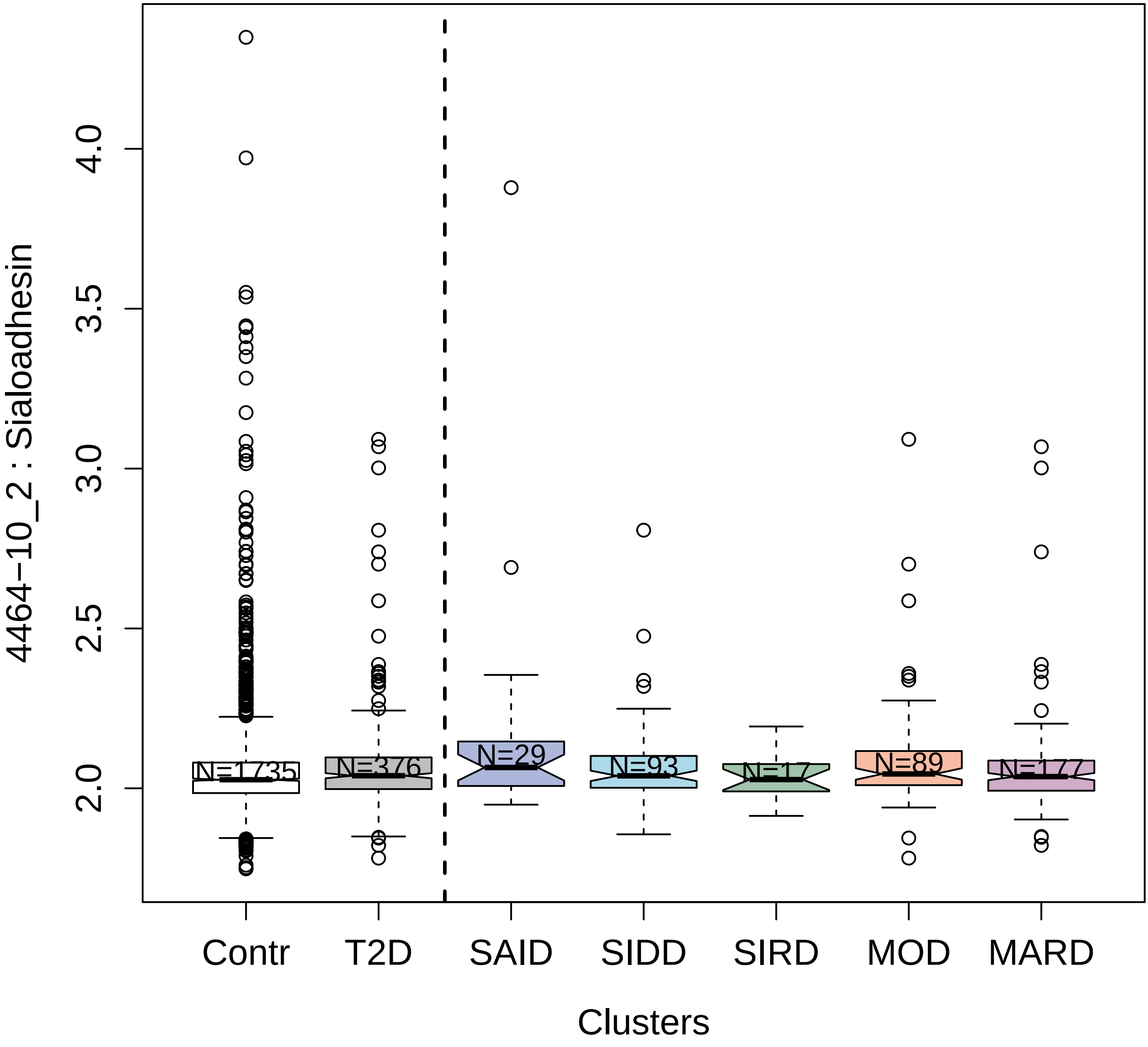
4464−10_2 : Sialoadhesin
4.0
3.5
3.0
2.5
2.0
N=1735
N=376
N=29
N=93
N=17
N=89
N=177
Contr
T2D
SAID
SIDD
SIRD
MOD
MARD
Clusters

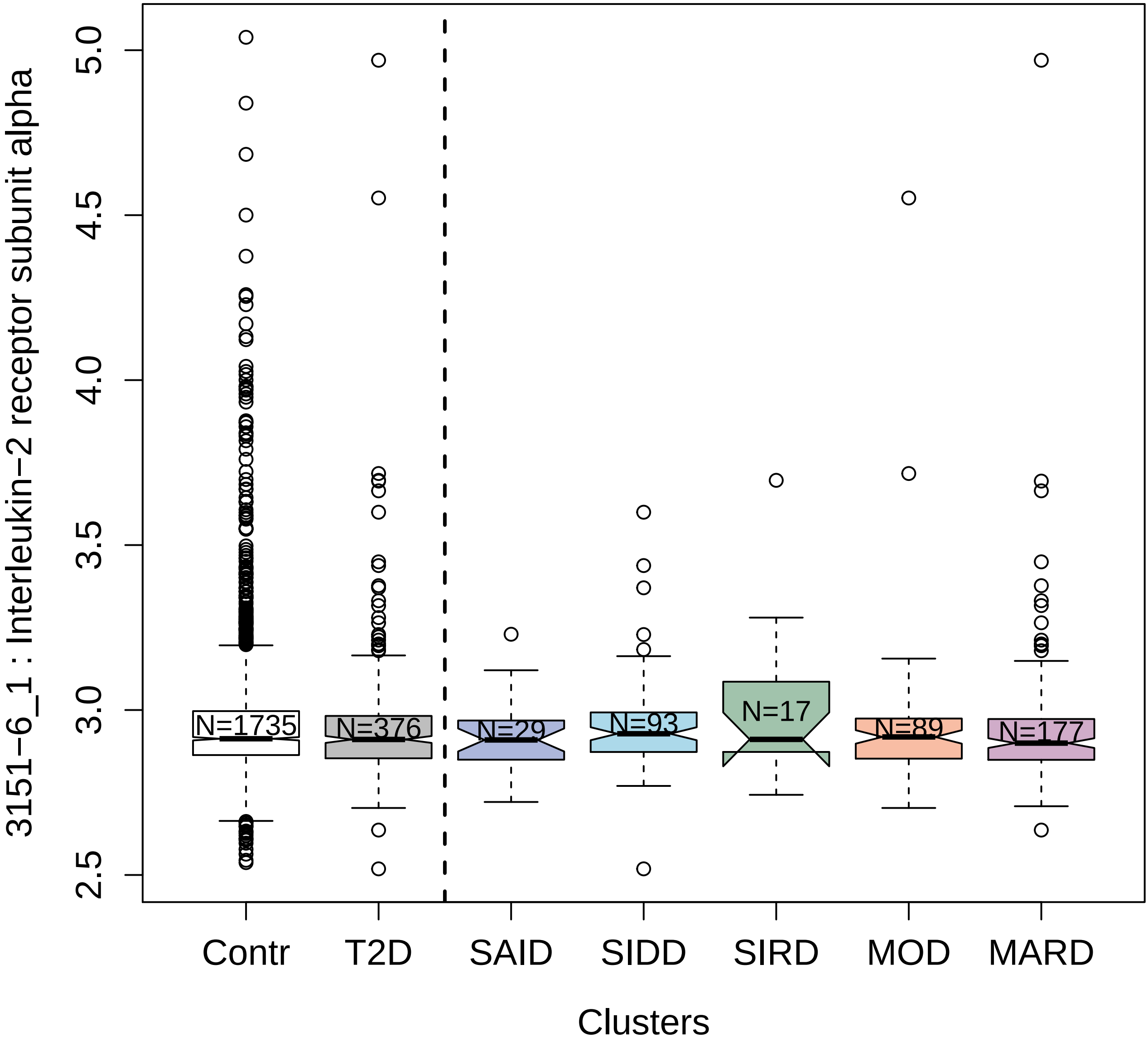

3151−6_1 : Interleukin−2 receptor subunit alpha
5.0
4.5
4.0
3.5
3.0
2.5
N=1735
N=376
N=29
N=93
N=17
N=89
N=177
Contr
T2D
SAID
SIDD
SIRD
MOD
MARD
Clusters

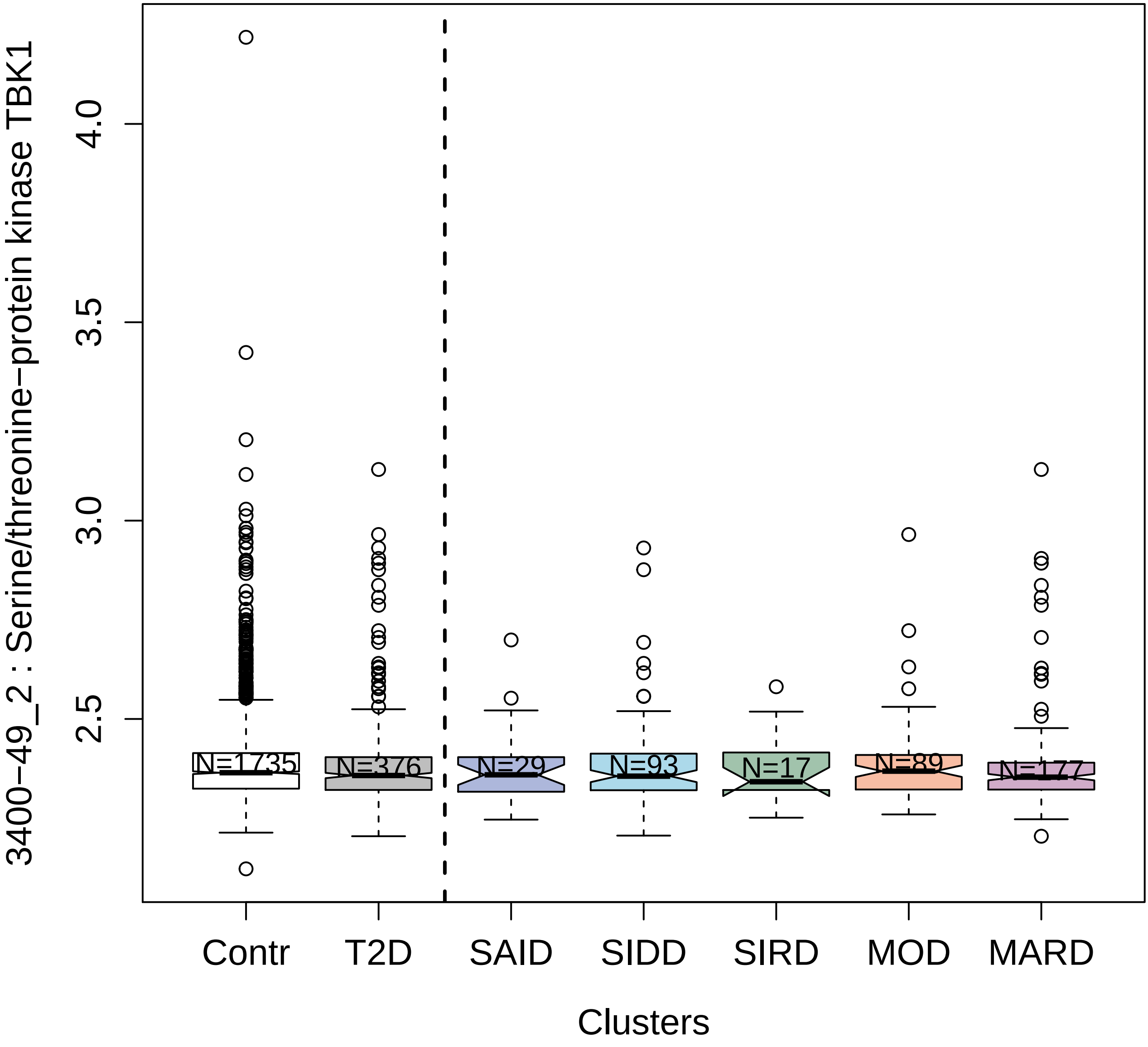

3400−49_2 : Serine/threonine−protein kinase TBK1
4.0
3.5
3.0
2.5
N=1735
N=376
N=29
N=93
N=17
N=89
N=177
Contr
T2D
SAID
SIDD
SIRD
MOD
MARD
Clusters

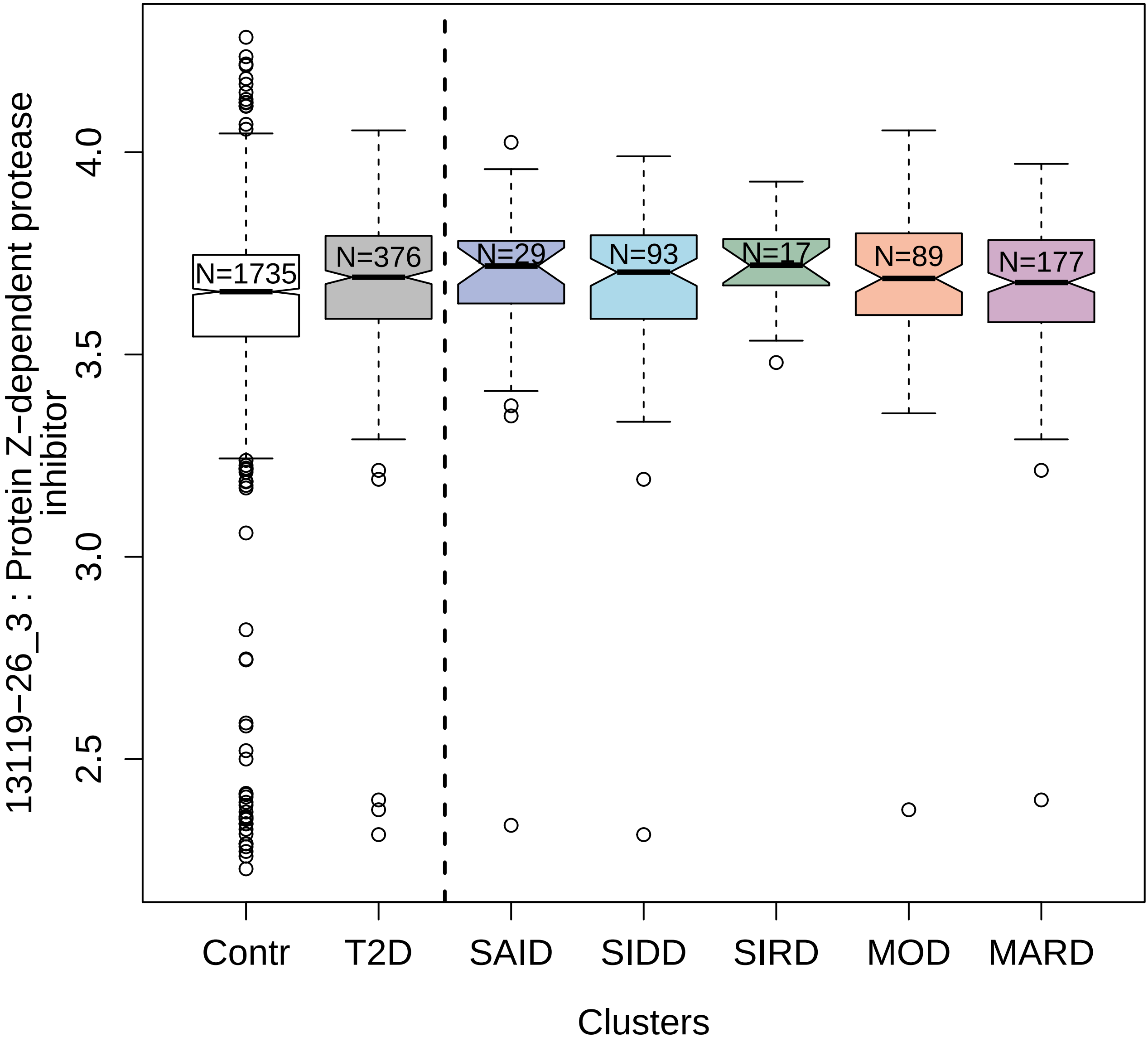

13119−26_3 : Protein Z−dependent protease inhibitor
4.0
3.5
3.0
2.5
N=1735
N=376
N=29
N=93
N=17
N=89
N=177
Contr
T2D
SAID
SIDD
SIRD
MOD
MARD
Clusters

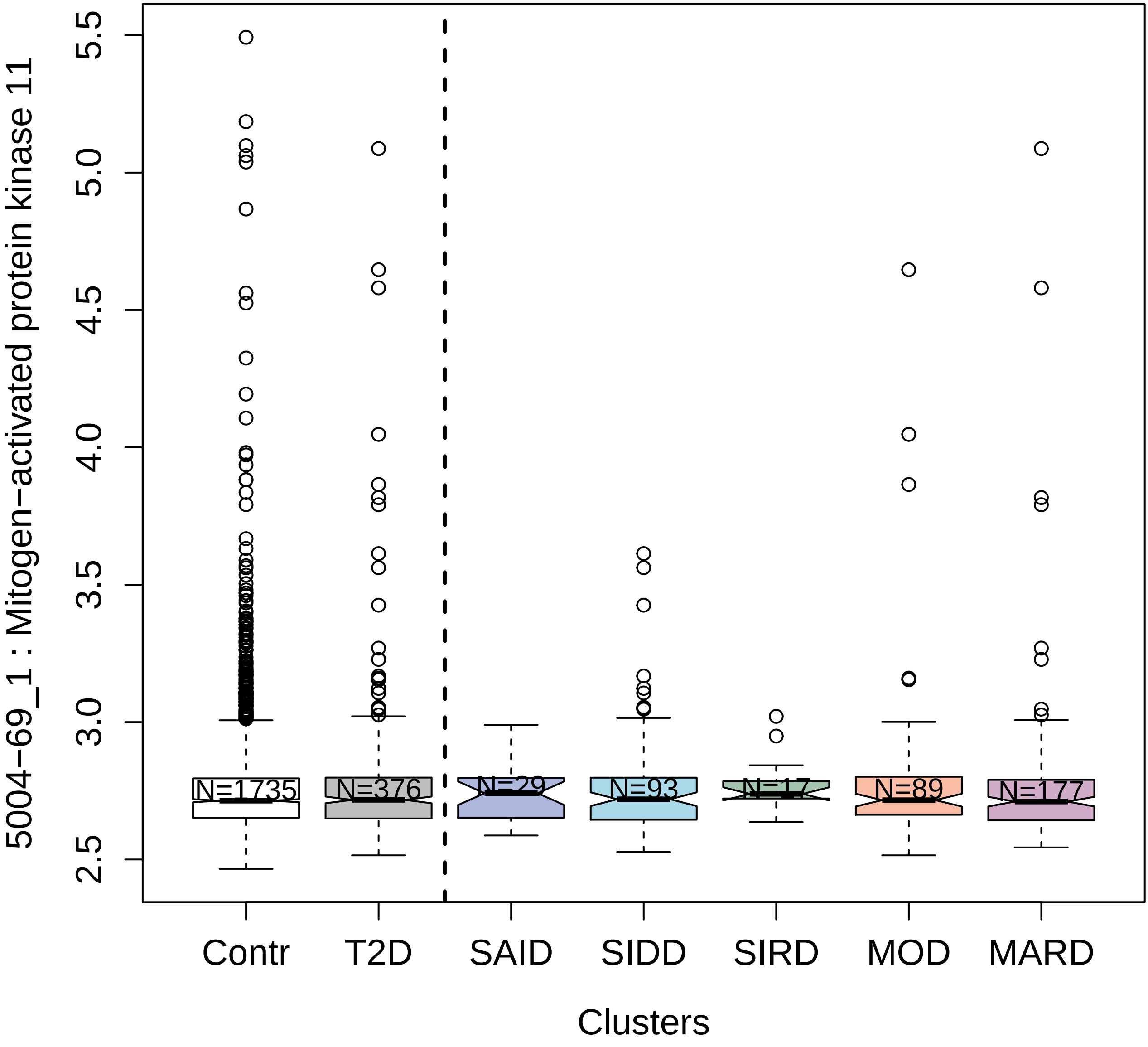

5004−69_1 : Mitogen−activated protein kinase 11
2.5
3.0
3.5
4.0
4.5
5.0
5.5
N=1735
N=376
N=29
N=93
N=17
N=89
N=177
Contr
T2D
SAID
SIDD
SIRD
MOD
MARD
Clusters

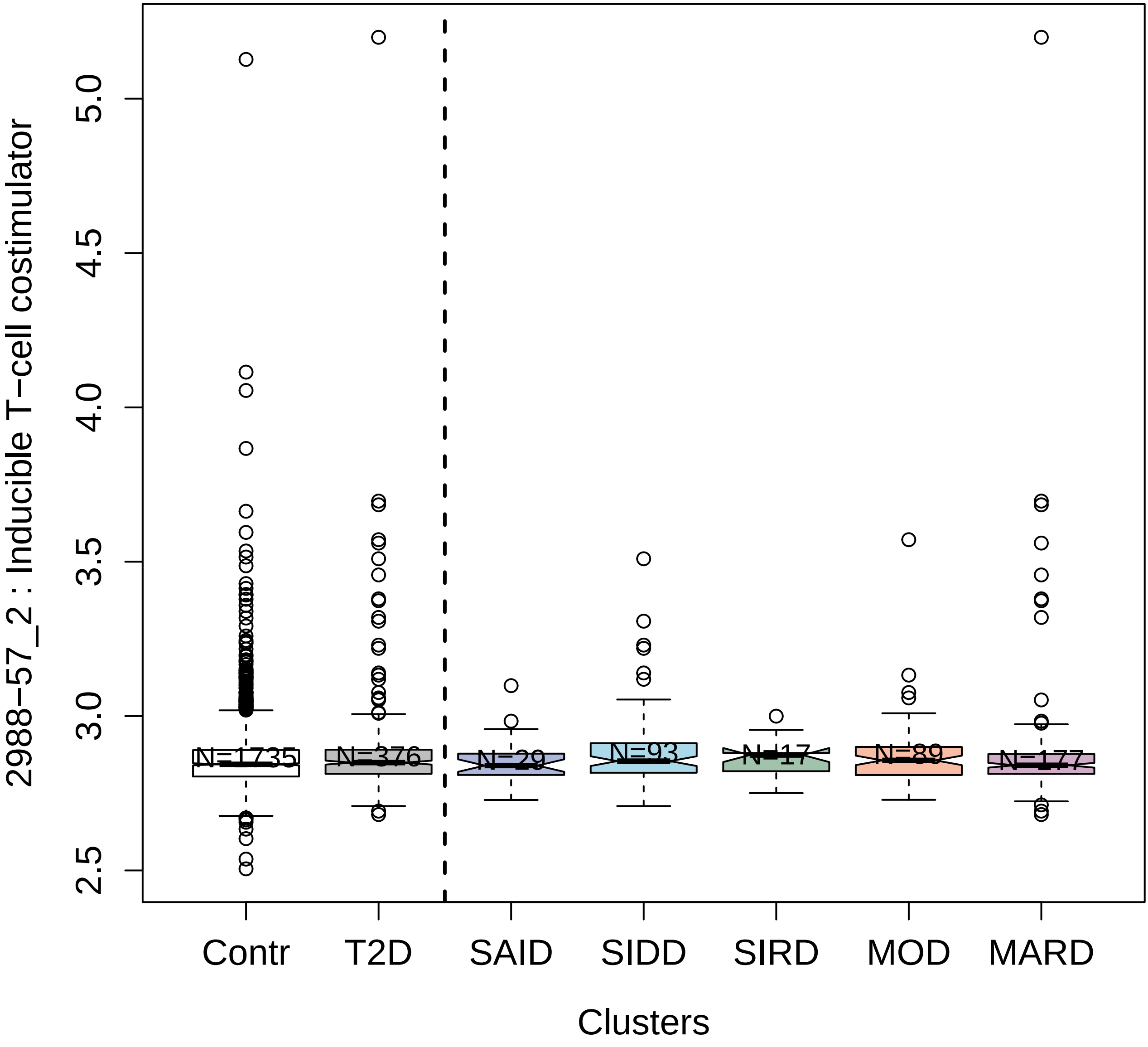

2988−57_2 : Inducible T−cell costimulator
2.5
3.0
3.5
4.0
4.5
5.0
N=1735
N=376
N=29
N=93
N=17
N=89
N=177
Contr
T2D
SAID
SIDD
SIRD
MOD
MARD
Clusters

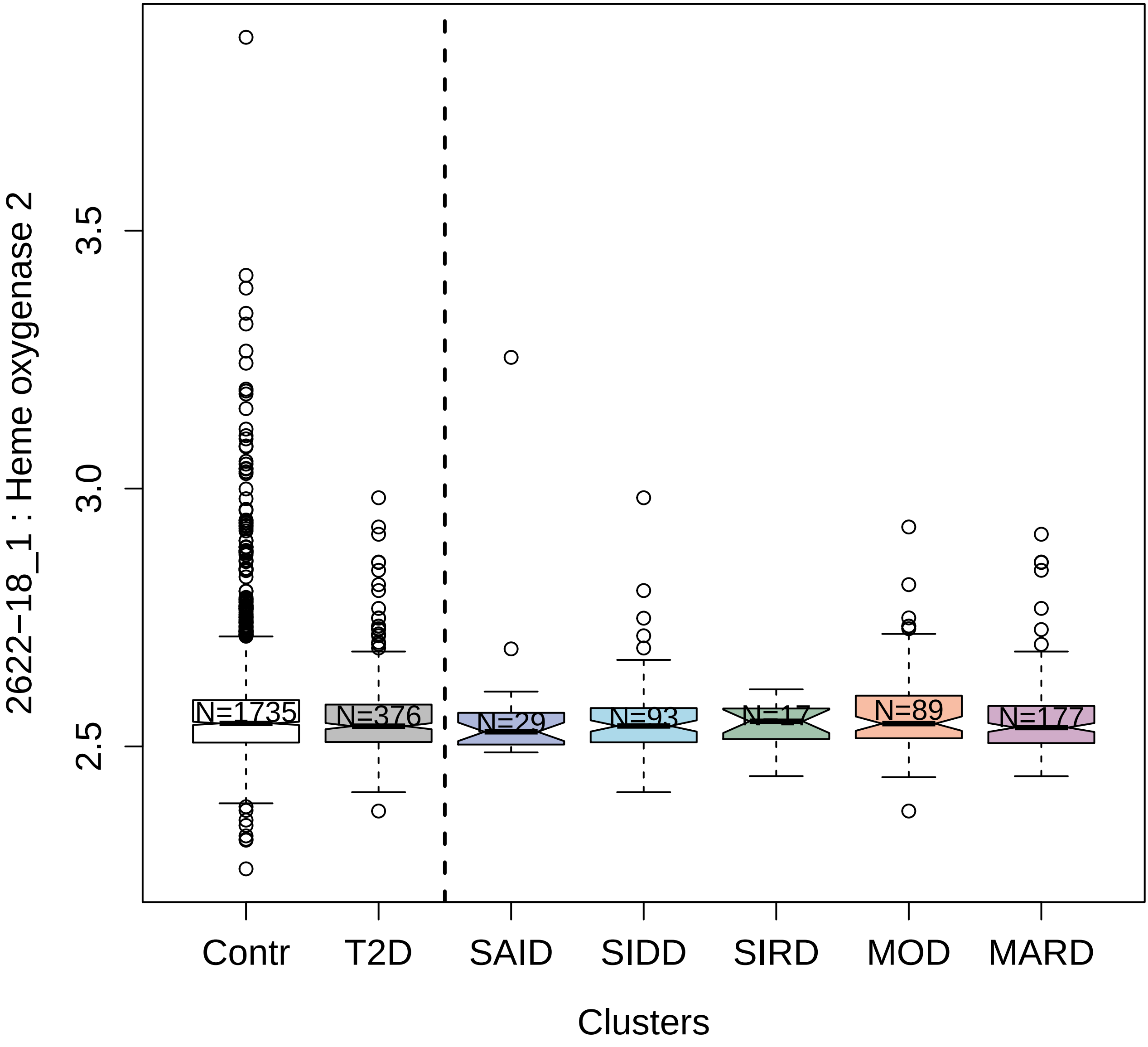
2622−18_1 : Heme oxygenase 2
3.5
3.0
2.5
N=1735
N=376
N=29
N=93
N=17
N=89
N=177
Contr
T2D
SAID
SIDD
SIRD
MOD
MARD
Clusters

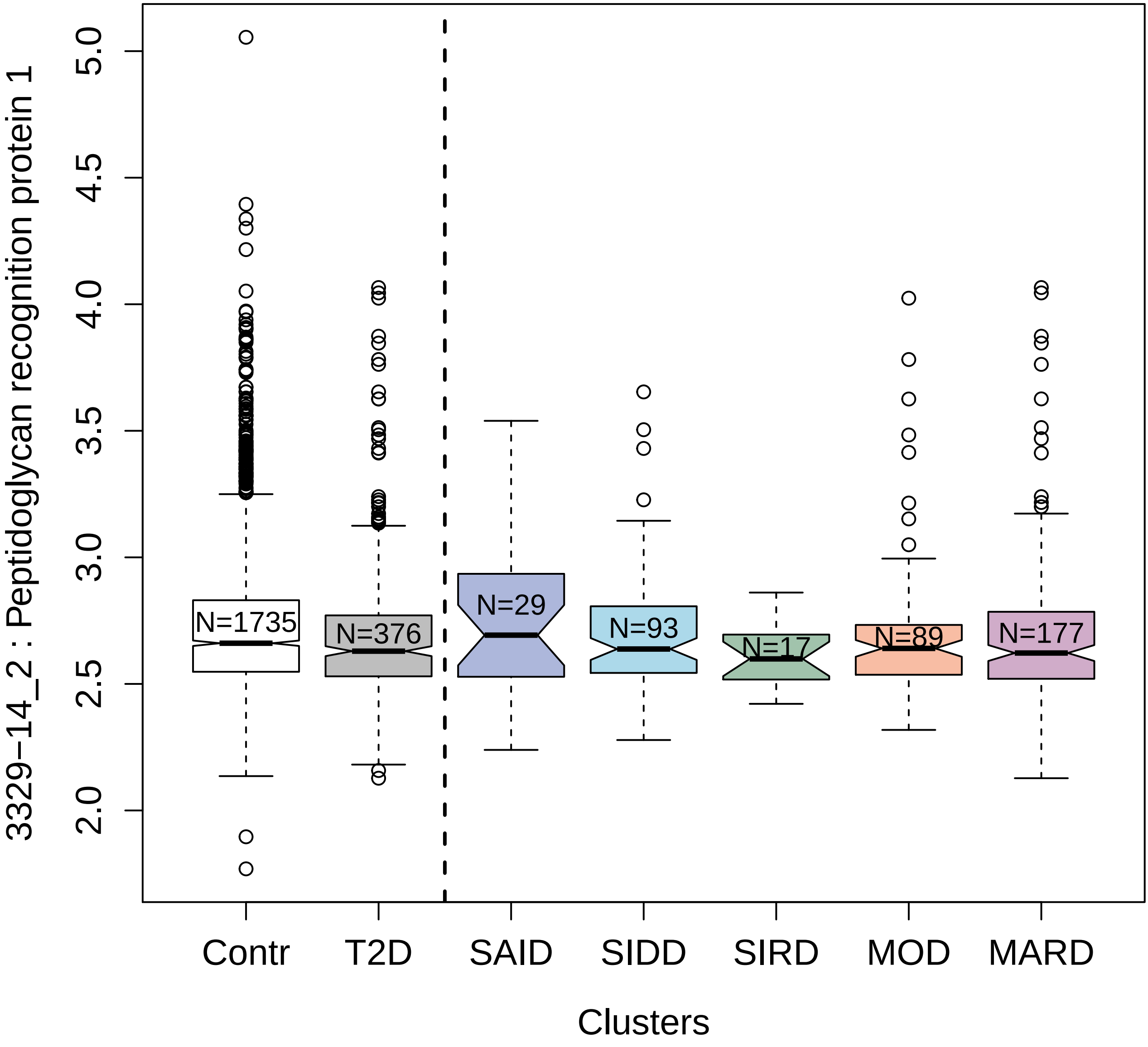
3329−14_2 : Peptidoglycan recognition protein 1
5.0
4.5
4.0
3.5
3.0
2.5
2.0
N=1735
N=376
N=29
N=93
N=17
N=89
N=177
Contr
T2D
SAID
SIDD
SIRD
MOD
MARD
Clusters

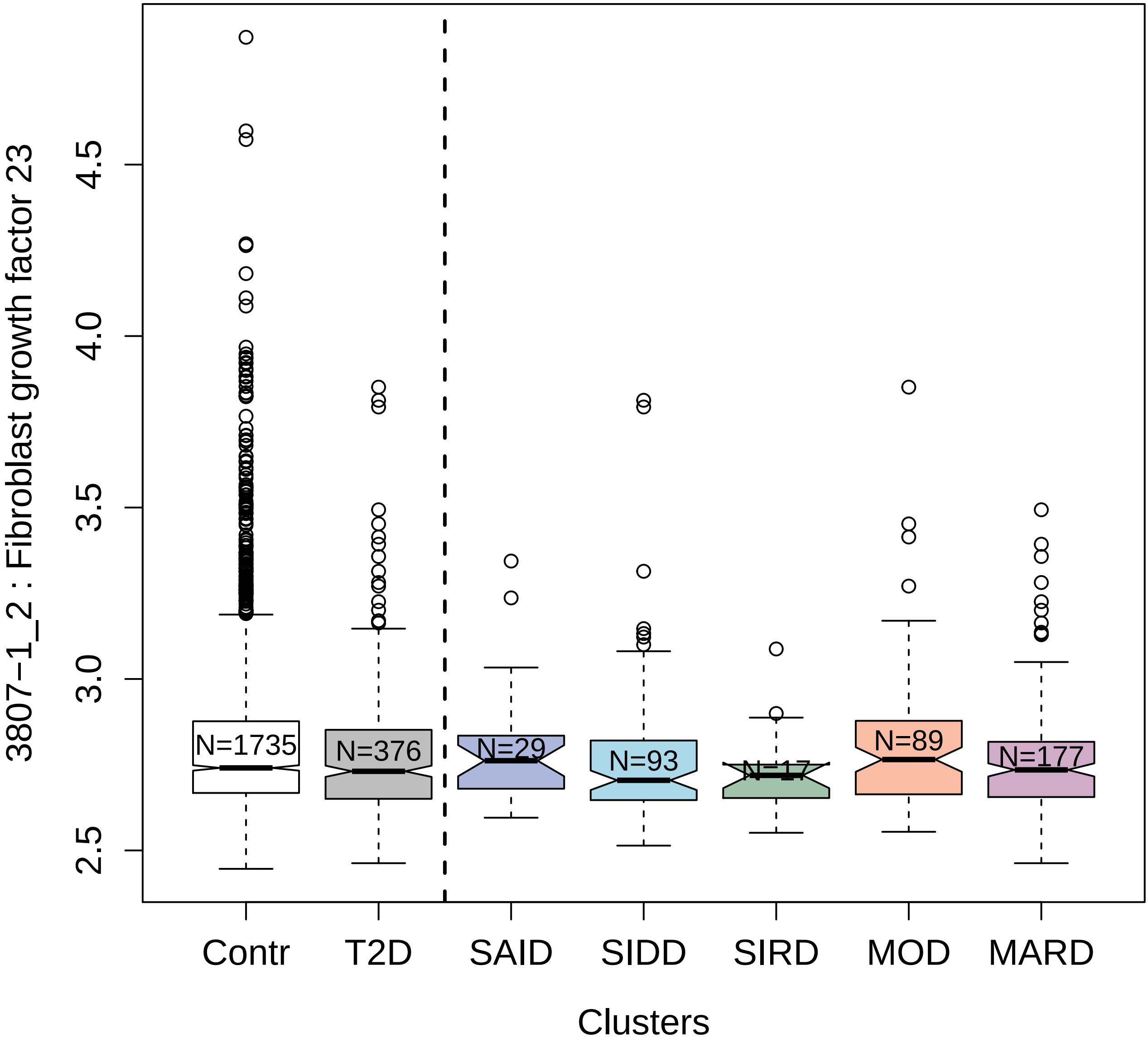

3807−1_2 : Fibroblast growth factor 23
2.5
3.0
3.5
4.0
4.5
N=1735
N=376
N=29
N=93
N=17
N=89
N=177
Contr
T2D
SAID
SIDD
SIRD
MOD
MARD
Clusters

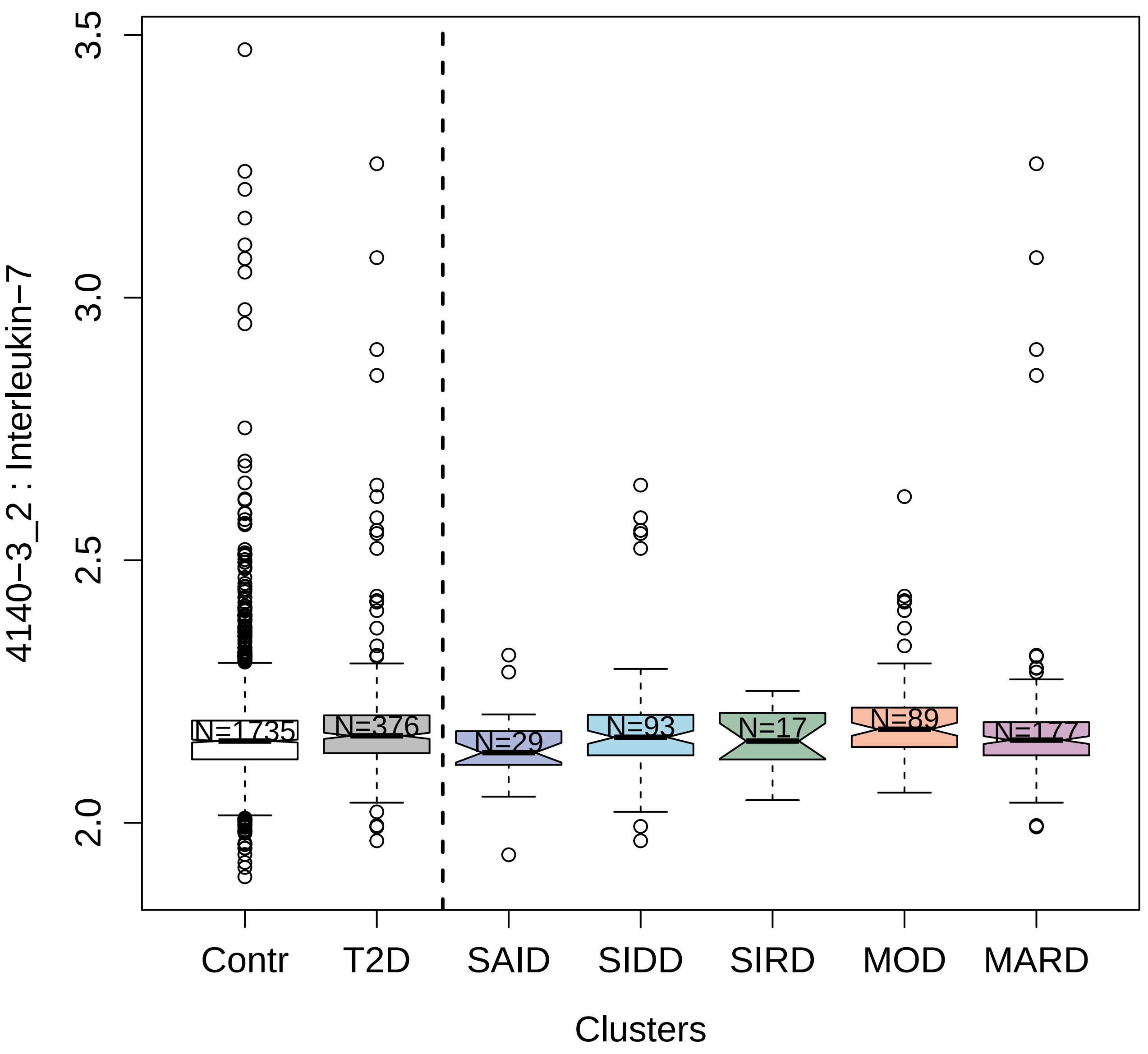

4140−3_2 : Interleukin−7
3.5
3.0
2.5
2.0
N=1735
N=376
N=29
N=93
N=17
N=89
N=177
Contr
T2D
SAID
SIDD
SIRD
MOD
MARD
Clusters

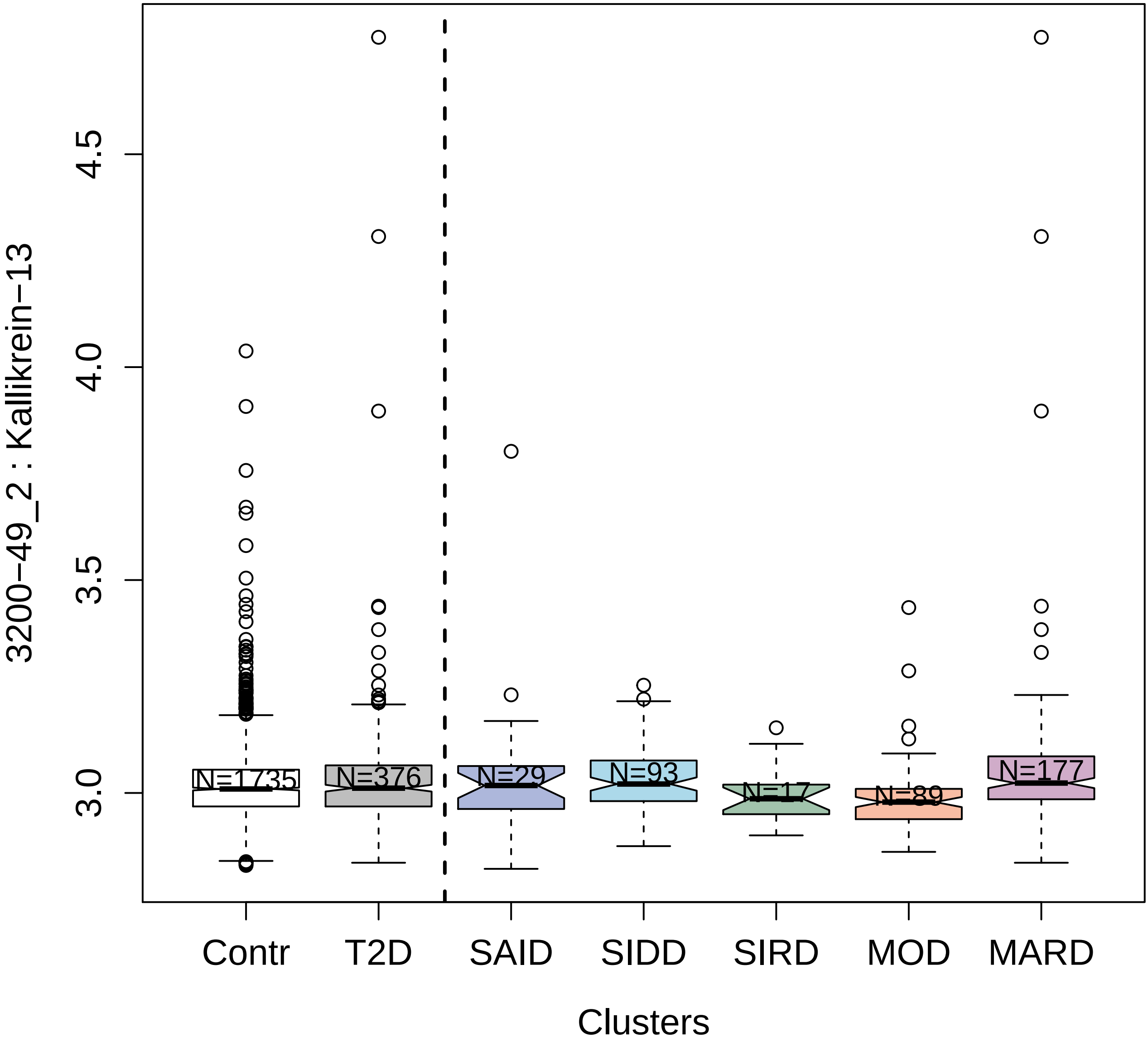

3200−49_2 : Kallikrein−13
3.0
3.5
4.0
4.5
N=1735
N=376
N=29
N=93
N=17
N=89
N=177
Contr
T2D
SAID
SIDD
SIRD
MOD
MARD
Clusters

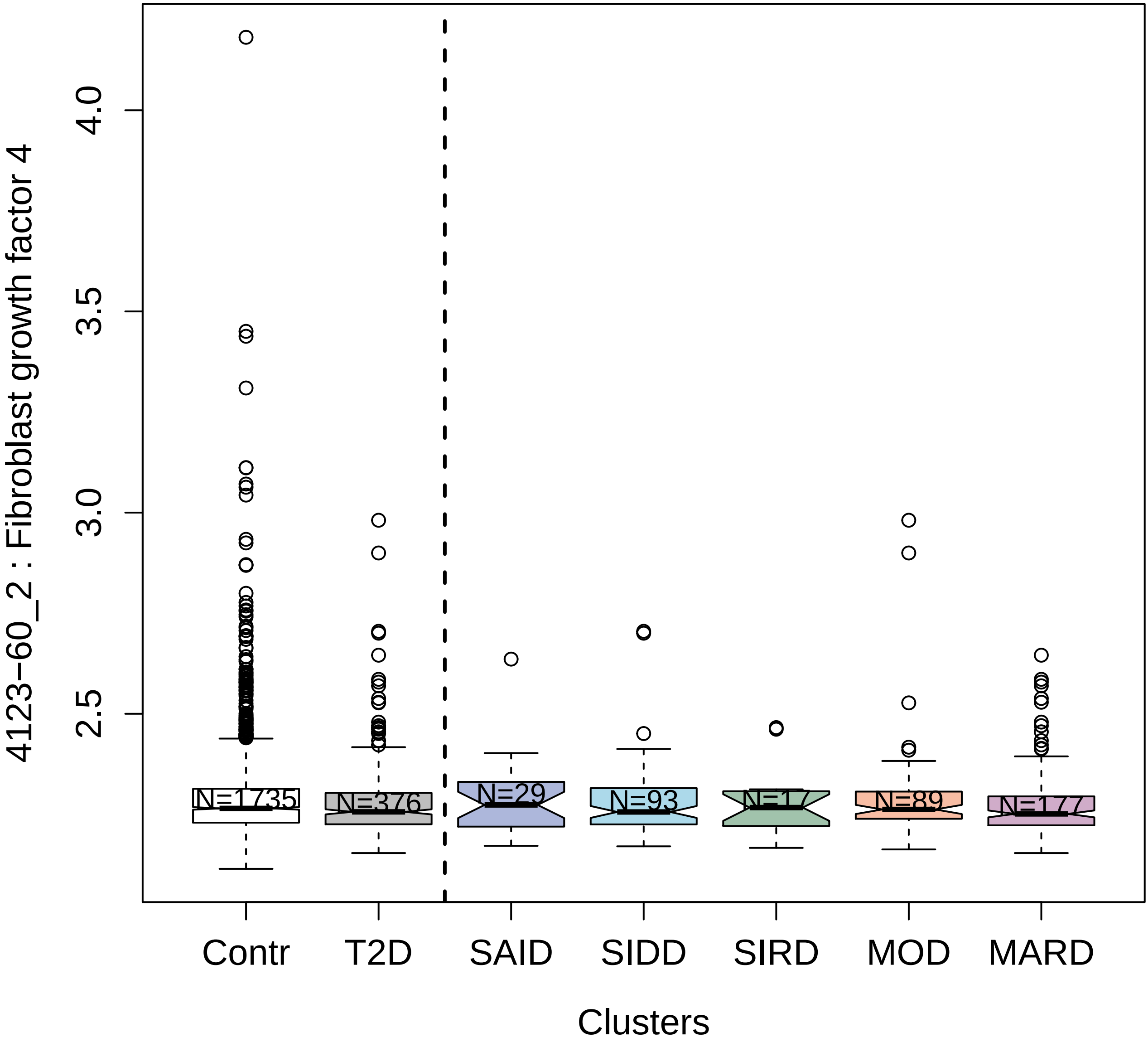
4123−60_2 : Fibroblast growth factor 4
4.0
3.5
3.0
2.5
N=1735
N=376
N=29
N=93
N=17
N=89
N=177
Contr
T2D
SAID
SIDD
SIRD
MOD
MARD
Clusters

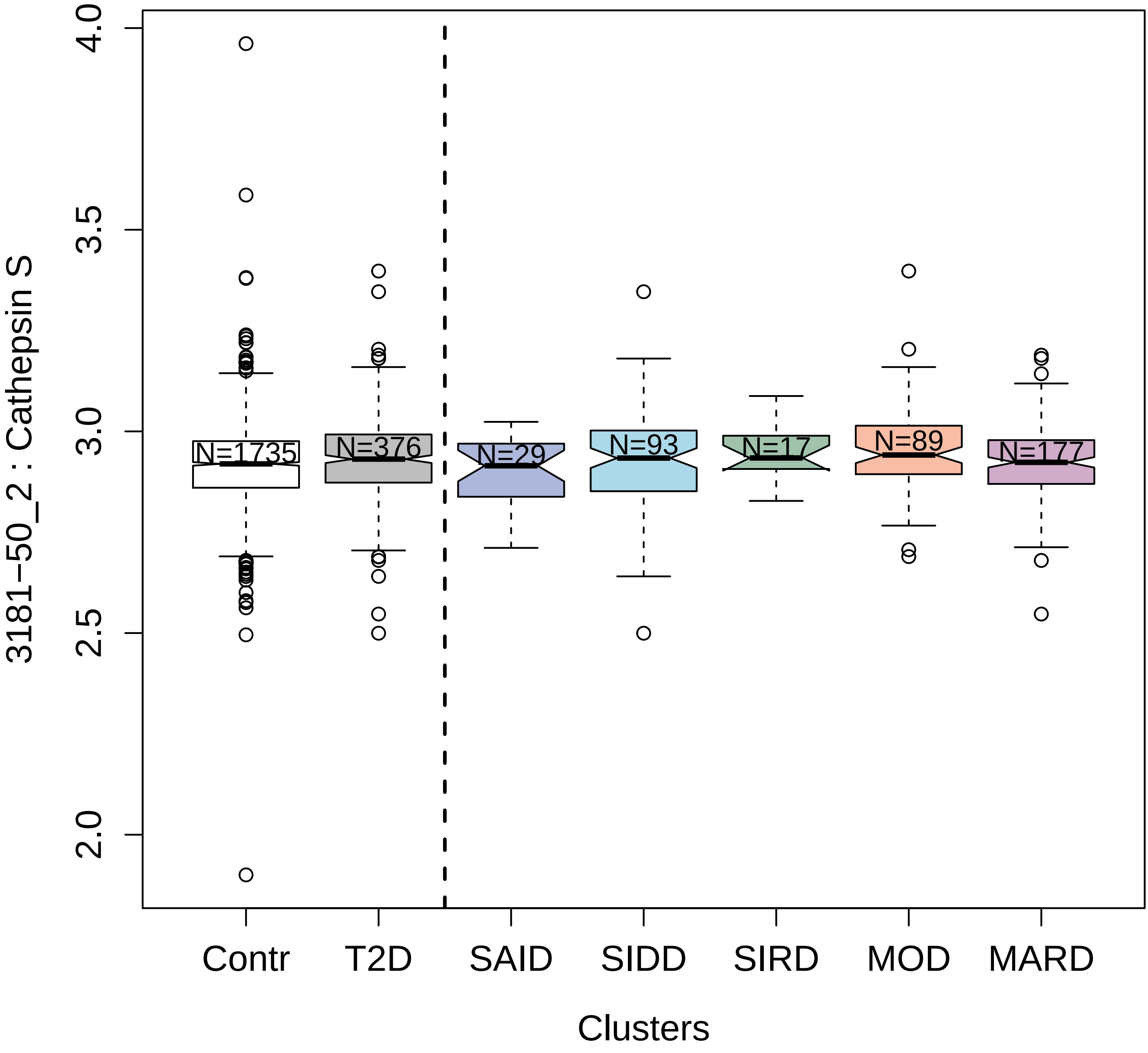

3181−50_2 : Cathepsin S
4.0
3.5
3.0
2.5
2.0
N=1735
N=376
N=29
N=93
N=17
N=89
N=177
Contr
T2D
SAID
SIDD
SIRD
MOD
MARD
Clusters

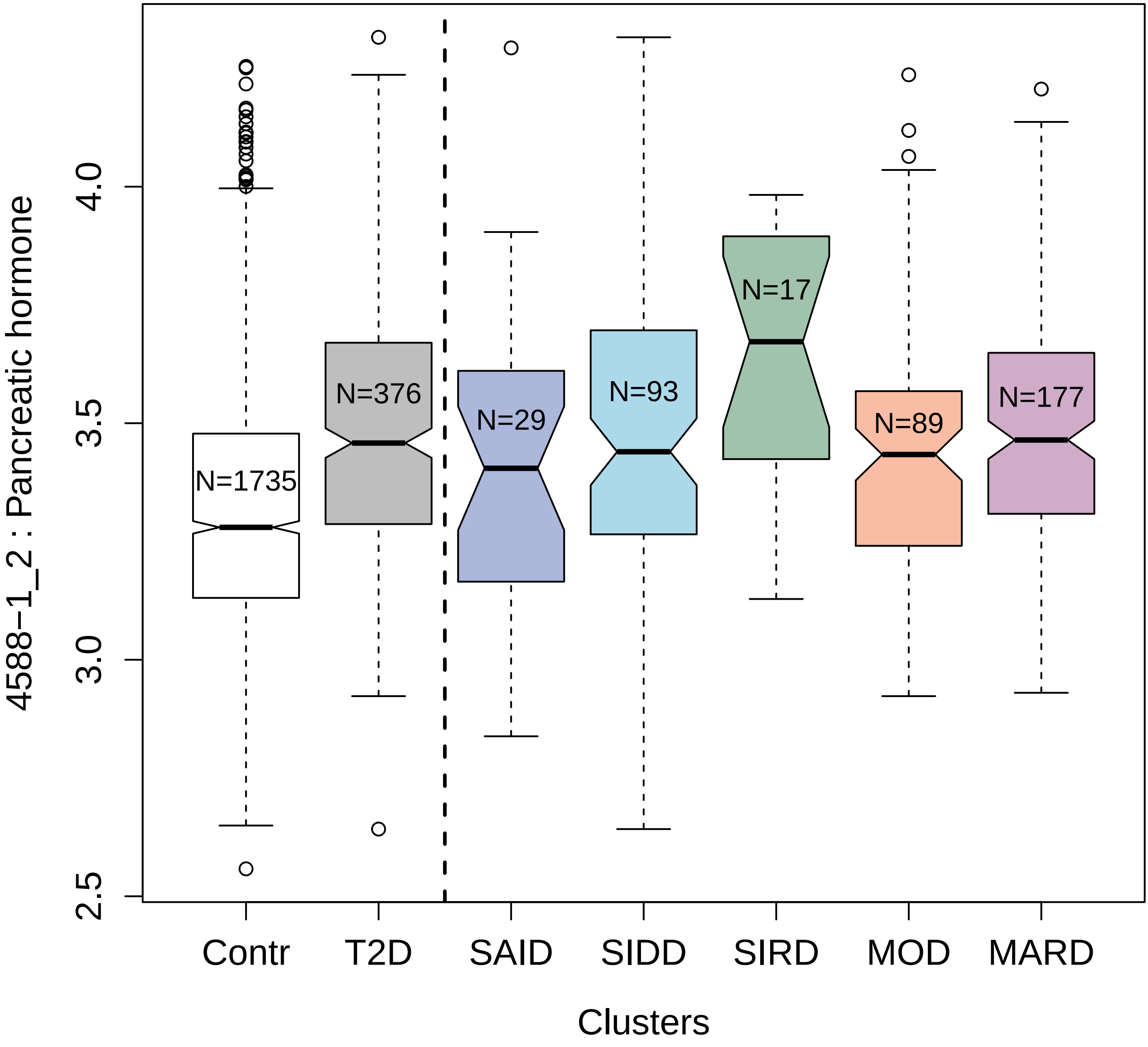

4588−1_2 : Pancreatic hormone
2.5
3.0
3.5
4.0
N=1735
N=376
N=29
N=93
N=17
N=89
N=177
Contr
T2D
SAID
SIDD
SIRD
MOD
MARD
Clusters

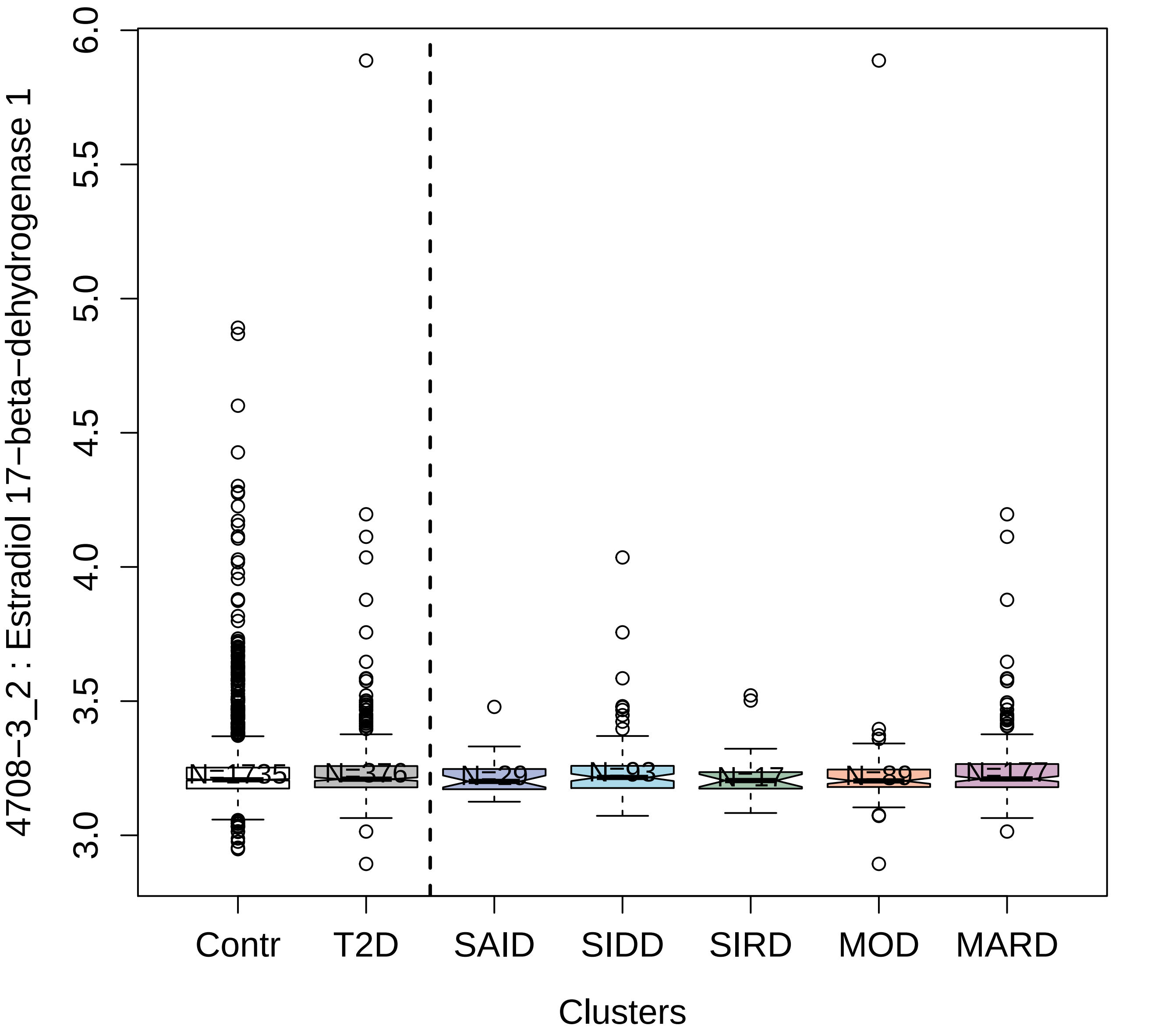

4708−3_2 : Estradiol 17−beta−dehydrogenase 1
6.0
5.5
5.0
4.5
4.0
3.5
3.0
N=1735
N=376
N=29
N=93
N=17
N=89
N=177
Contr
T2D
SAID
SIDD
SIRD
MOD
MARD
Clusters

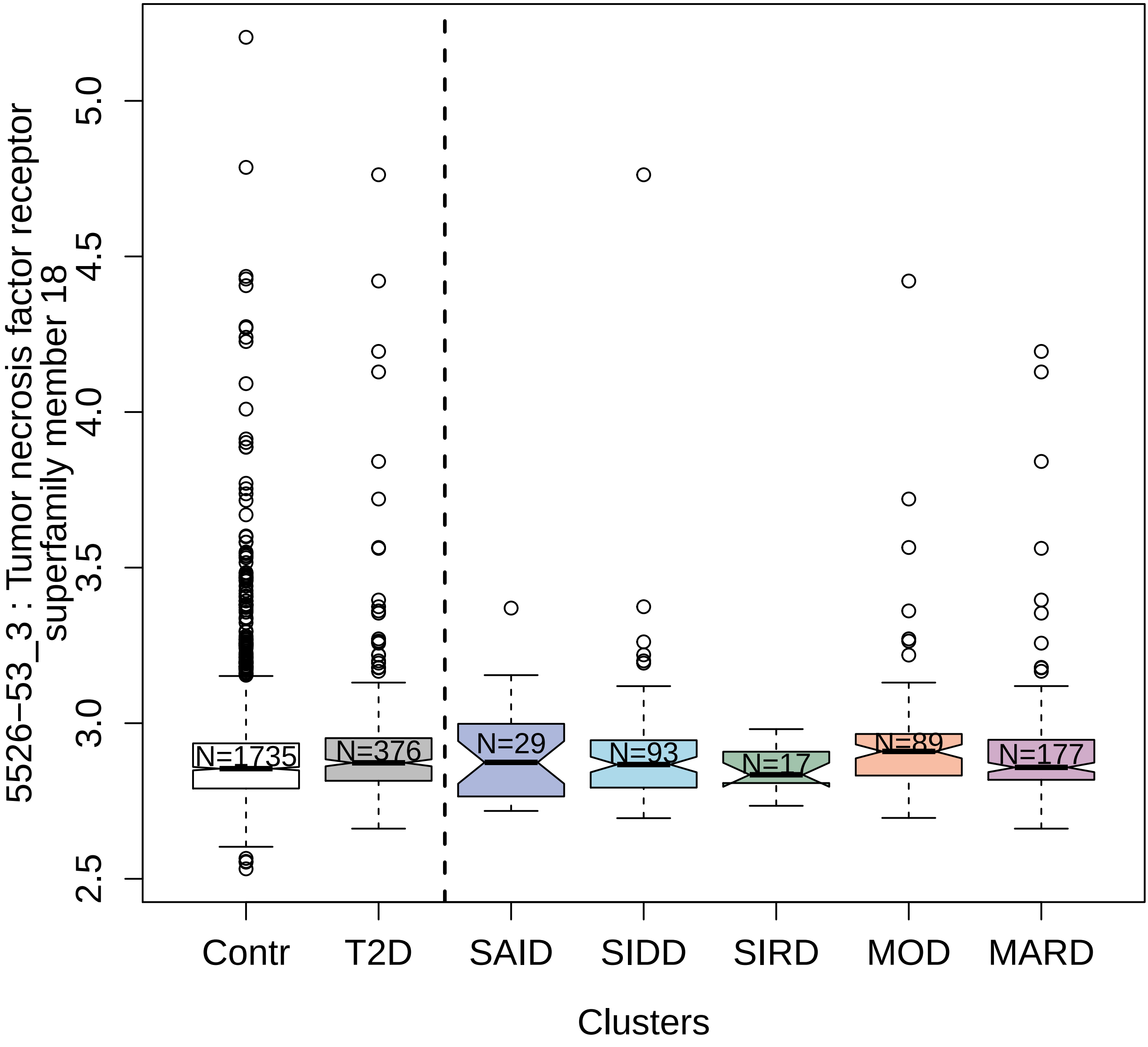
5526−53_3 : Tumor necrosis factor receptor
superfamily member 18
2.5
3.0
3.5
4.0
4.5
5.0
N=1735
N=376
N=29
N=93
N=17
N=89
N=177
Contr
T2D
SAID
SIDD
SIRD
MOD
MARD
Clusters

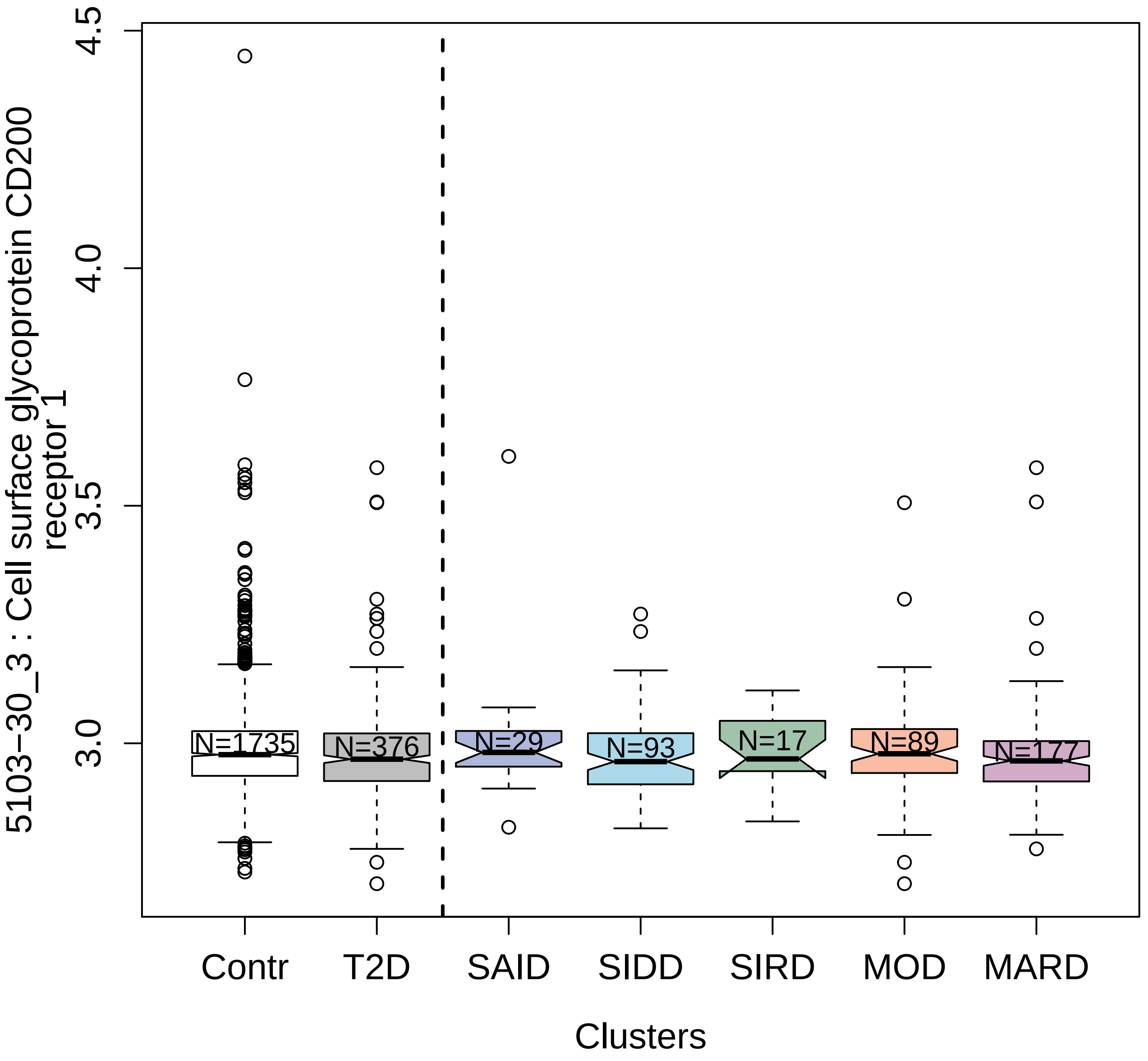

5103−30_3 : Cell surface glycoprotein CD200
receptor 1
4.5
4.0
3.5
3.0
N=1735
N=376
N=29
N=93
N=17
N=89
N=177
Contr
T2D
SAID
SIDD
SIRD
MOD
MARD
Clusters

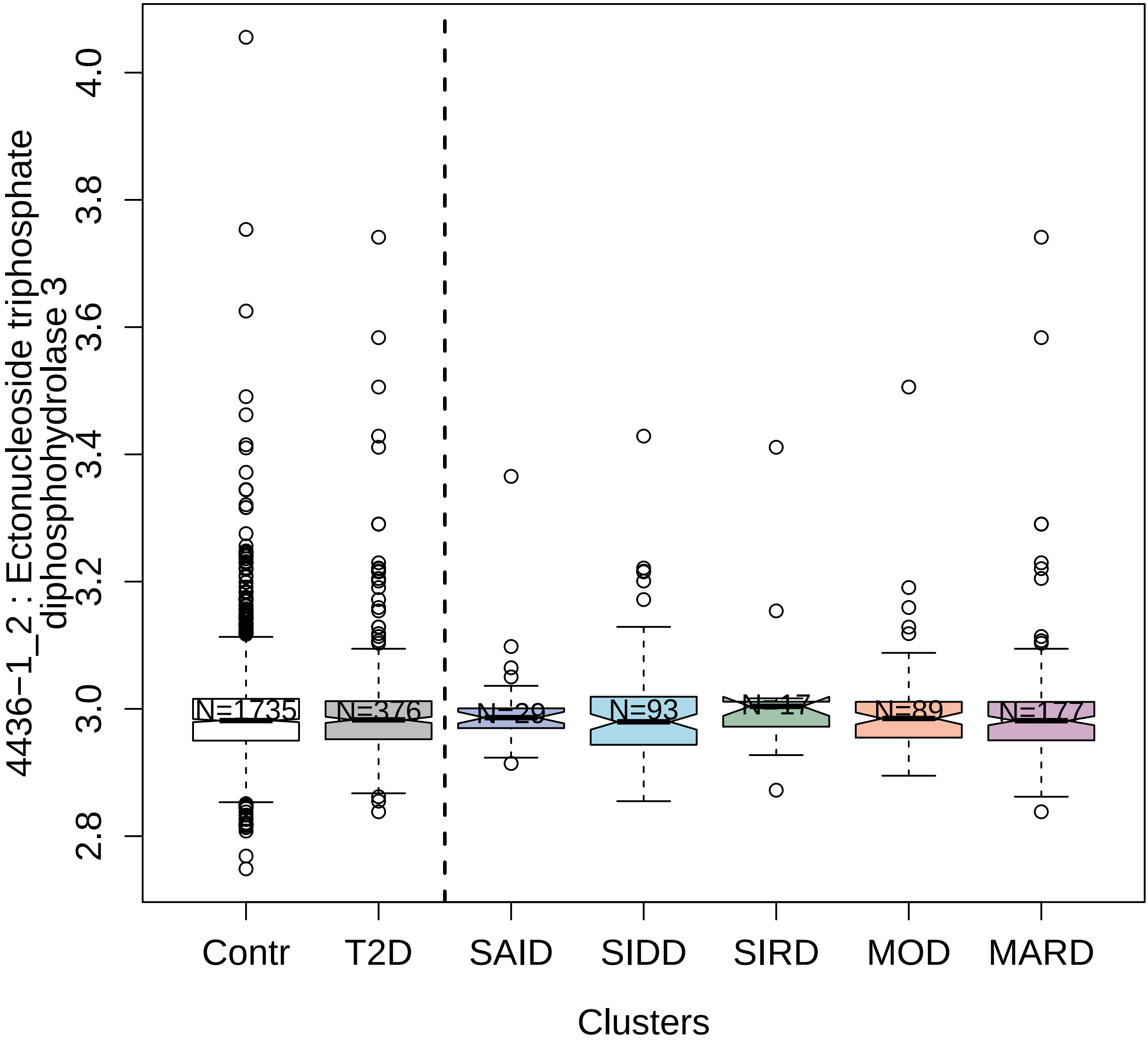
4436−1_2 : Ectonucleoside triphosphate diphosphohydrolase 3
4.0
3.8
3.6
3.4
3.2
3.0
2.8
N=1735
N=376
N=29
N=93
N=17
N=89
N=177
Contr
T2D
SAID
SIDD
SIRD
MOD
MARD
Clusters

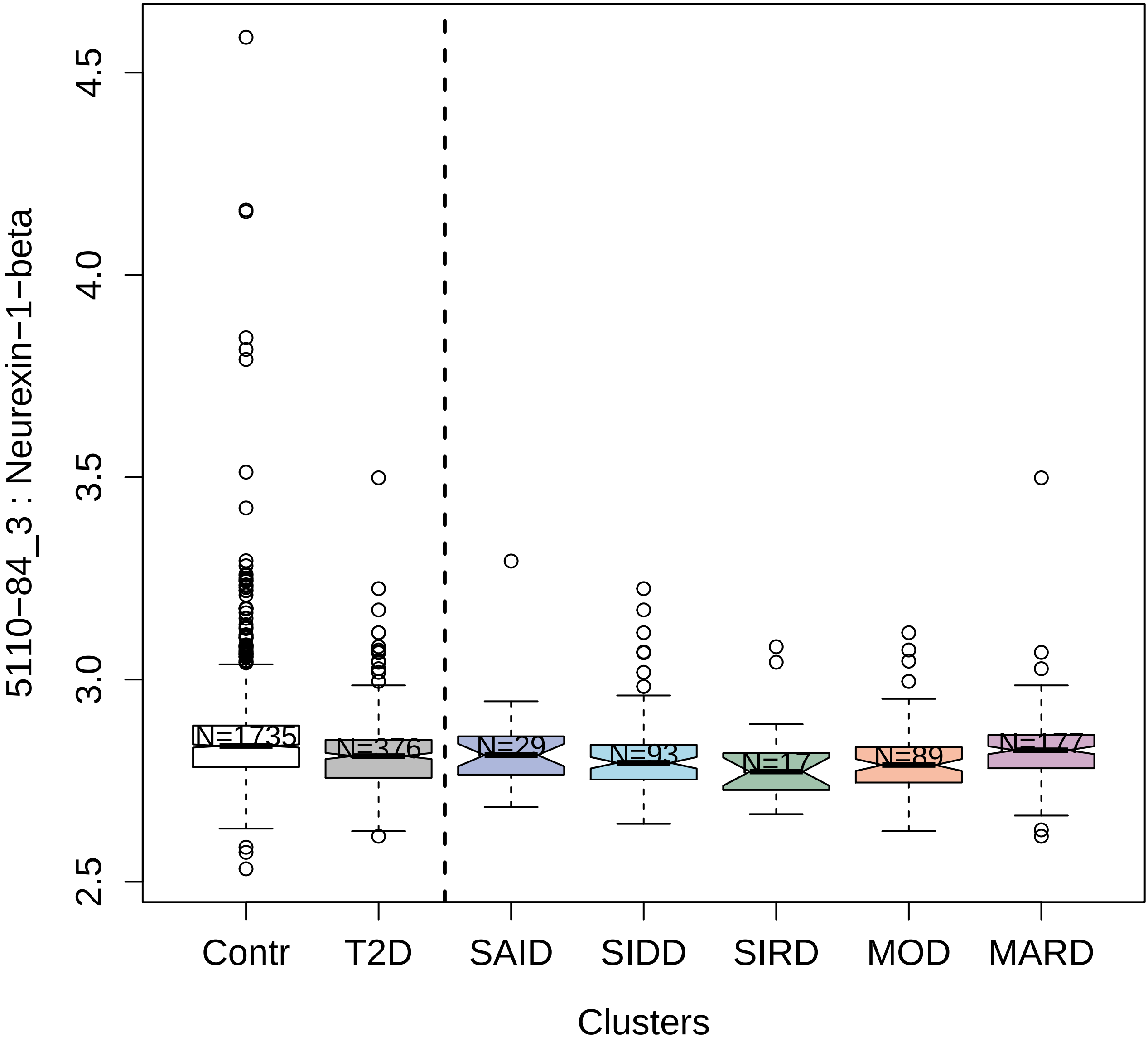

5110−84_3 : Neurexin−1−beta
4.5
4.0
3.5
3.0
2.5
N=1735
N=376
N=29
N=93
N=17
N=89
N=177
Contr
T2D
SAID
SIDD
SIRD
MOD
MARD
Clusters

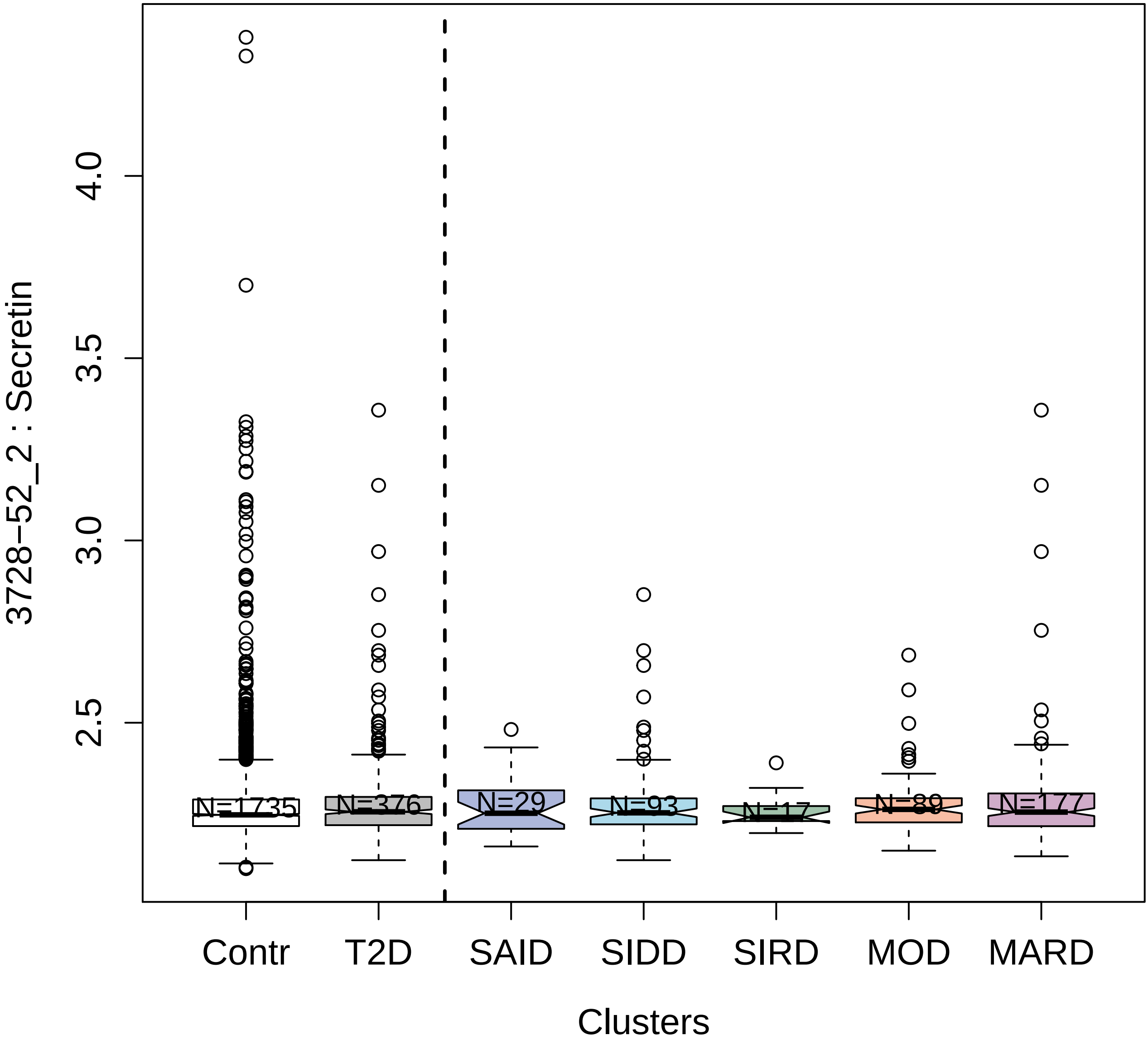
3728−52_2 : Secretin
4.0
3.5
3.0
2.5
N=1735
N=376
N=29
N=93
N=17
N=89
N=177
Contr
T2D
SAID
SIDD
SIRD
MOD
MARD
Clusters

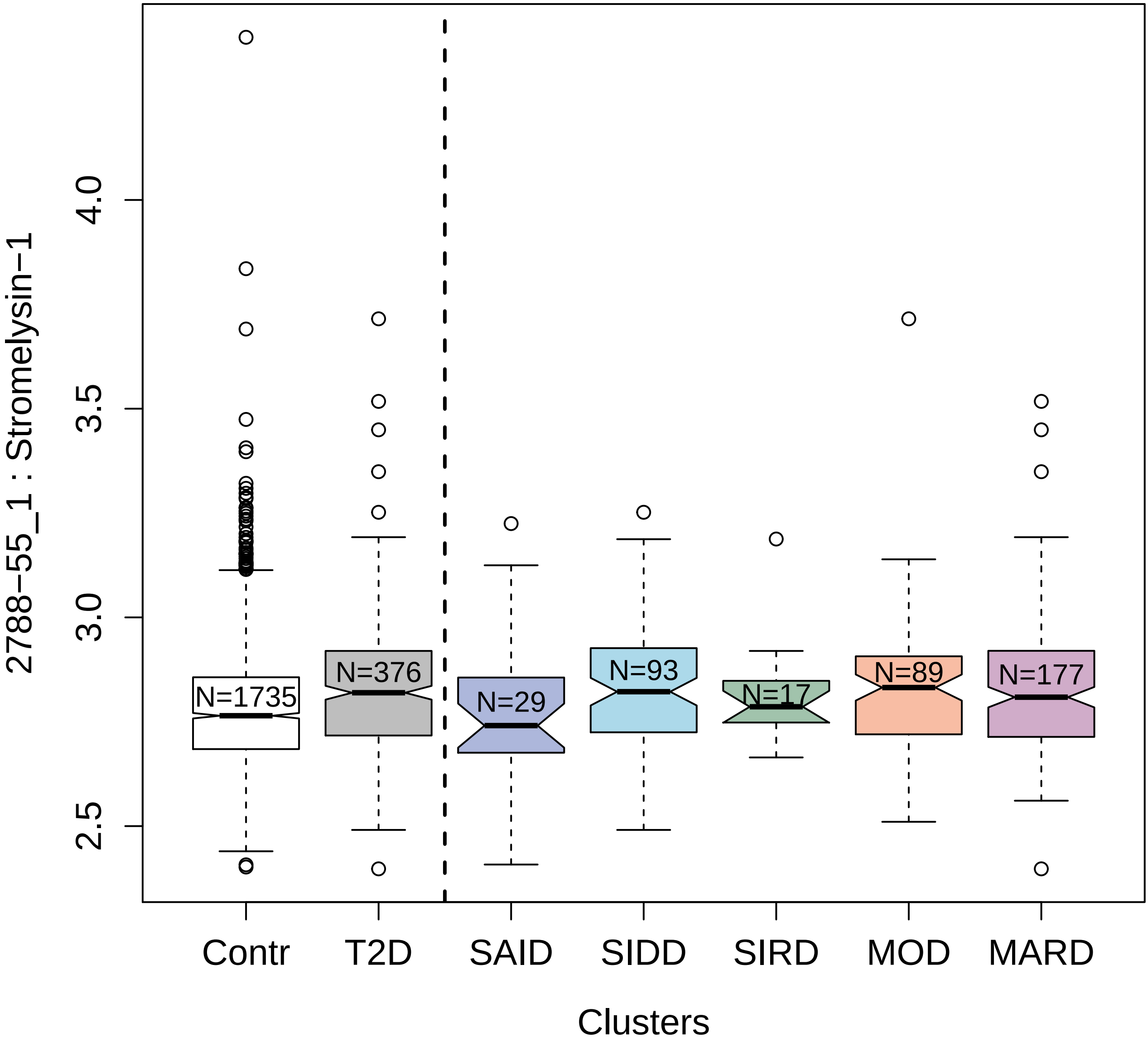
2788−55_1 : Stromelysin−1
4.0
3.5
3.0
2.5
N=1735
N=376
N=29
N=93
N=17
N=89
N=177
Contr
T2D
SAID
SIDD
SIRD
MOD
MARD
Clusters

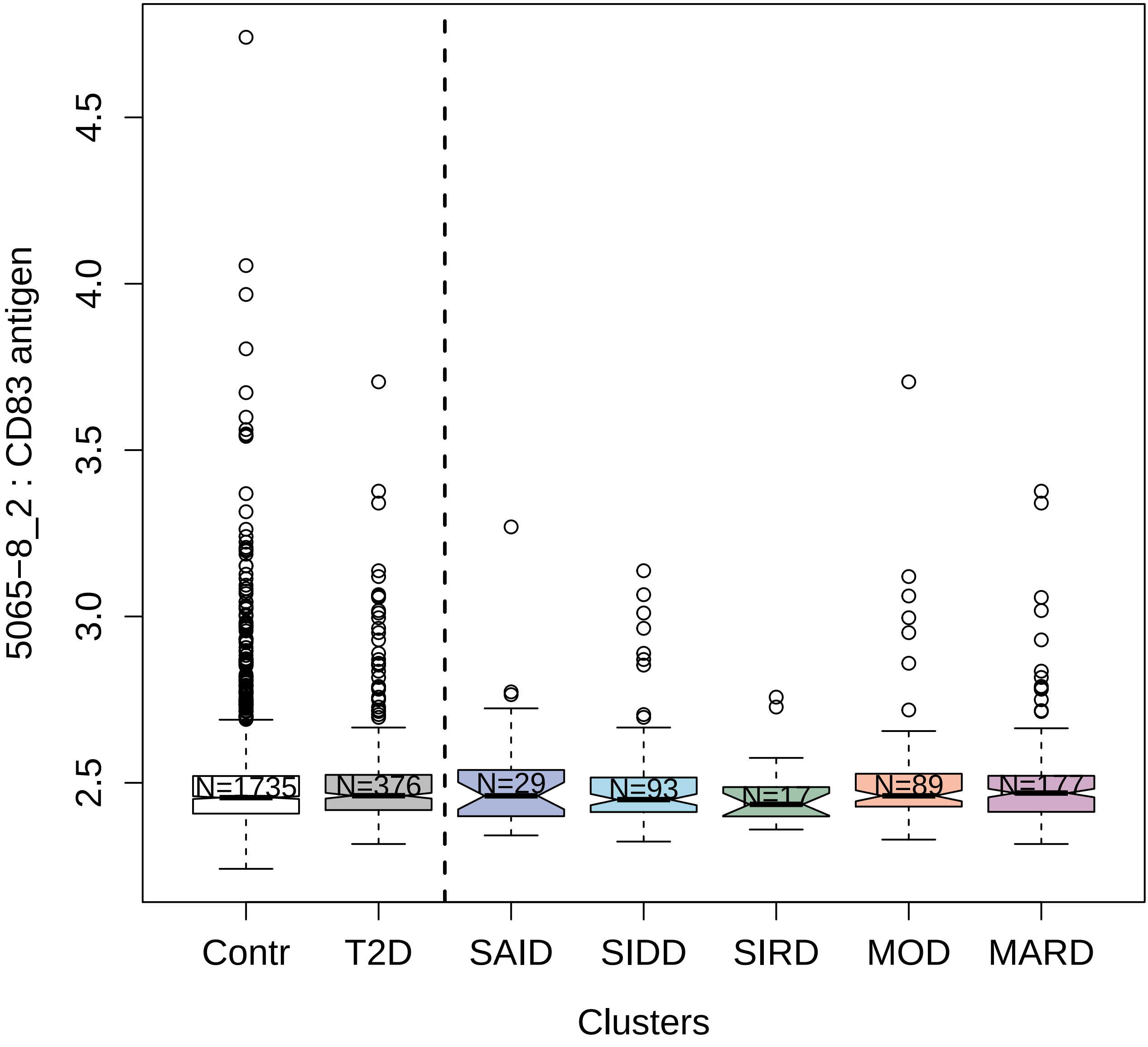

5065−8_2 : CD83 antigen
4.5
4.0
3.5
3.0
2.5
N=1735
N=376
N=29
N=93
N=17
N=89
N=177
Contr
T2D
SAID
SIDD
SIRD
MOD
MARD
Clusters

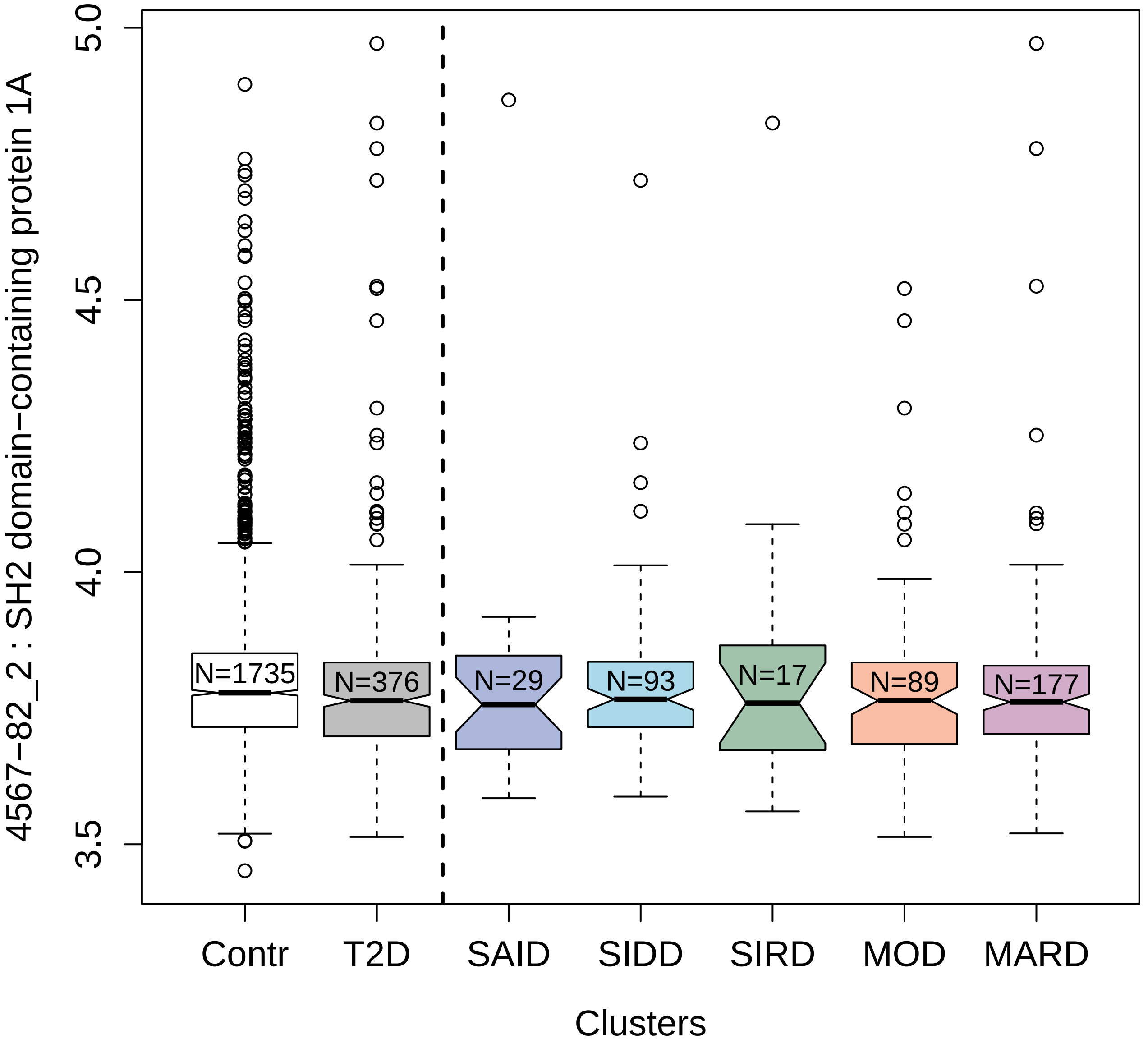

4567−82_2 : SH2 domain−containing protein 1A
5.0
4.5
4.0
3.5
N=1735
N=376
N=29
N=93
N=17
N=89
N=177
Contr
T2D
SAID
SIDD
SIRD
MOD
MARD
Clusters

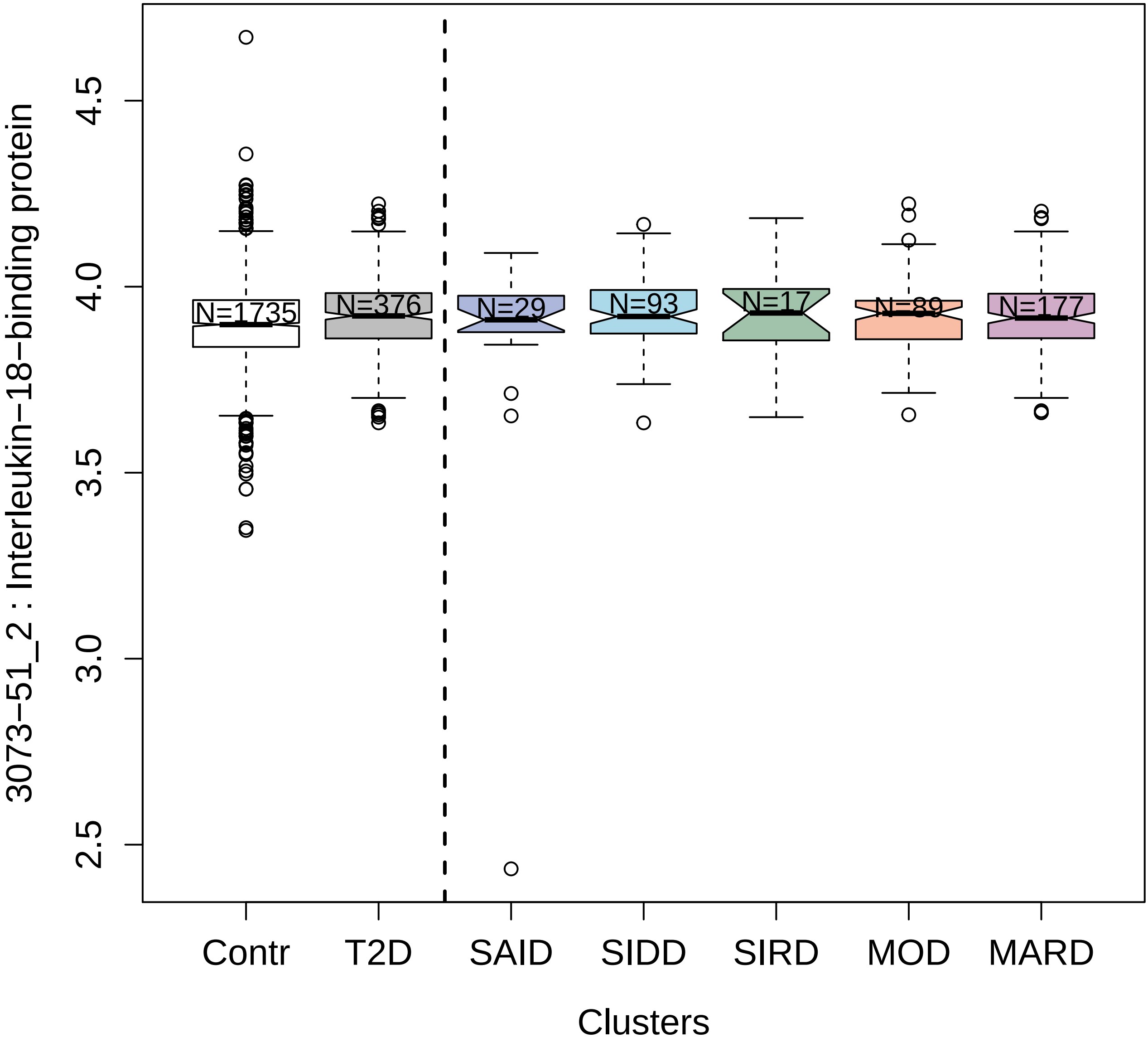

3073−51_2 : Interleukin−18−binding protein
4.5
4.0
3.5
3.0
2.5
N=1735
N=376
N=29
N=93
N=17
N=89
N=177
Contr
T2D
SAID
SIDD
SIRD
MOD
MARD
Clusters

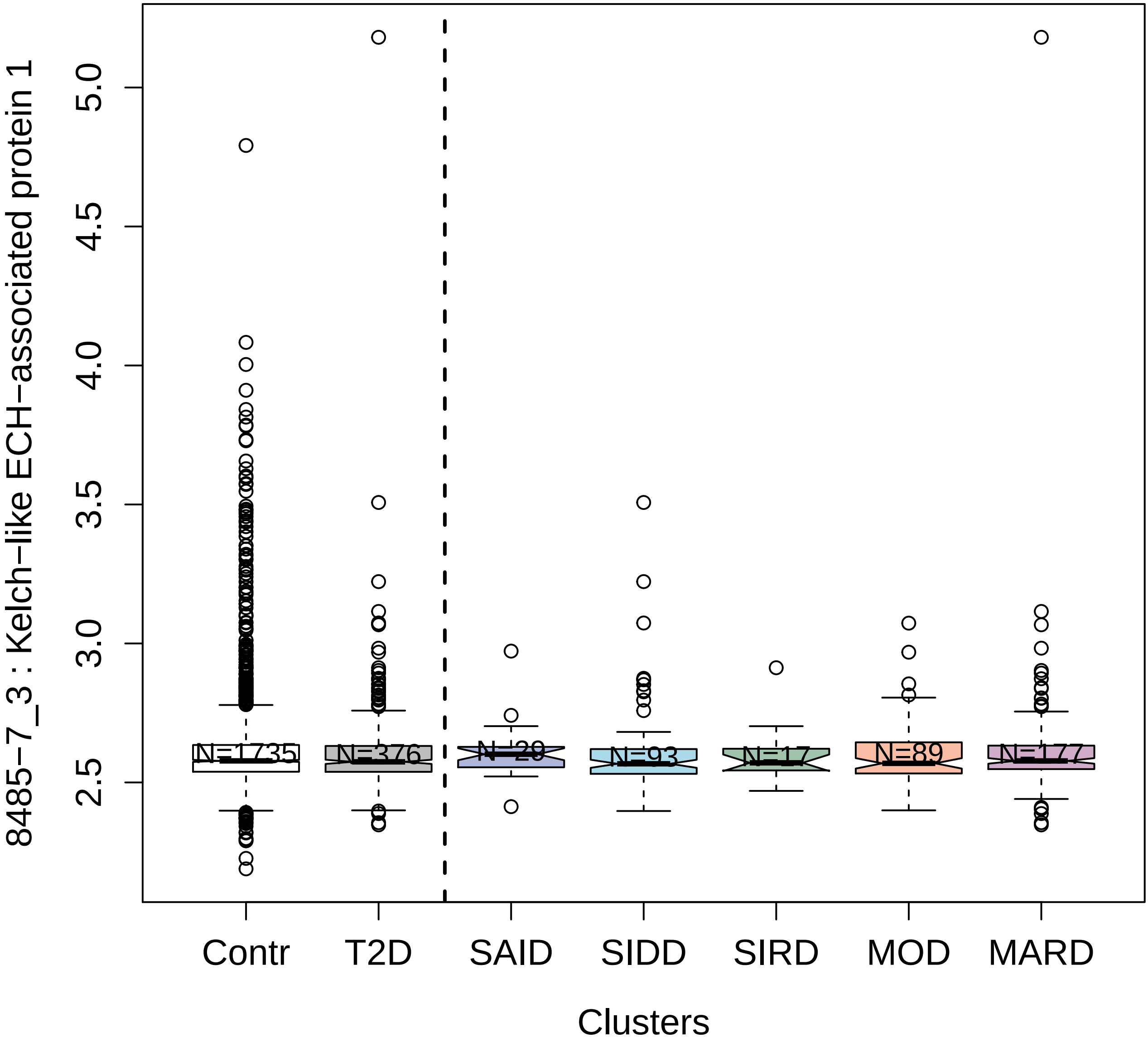

8485−7_3 : Kelch−like ECH−associated protein 1
5.0
4.5
4.0
3.5
3.0
2.5
N=1735
N=376
N=20
N=93
N=17
N=89
N=177
Contr
T2D
SAID
SIDD
SIRD
MOD
MARD
Clusters

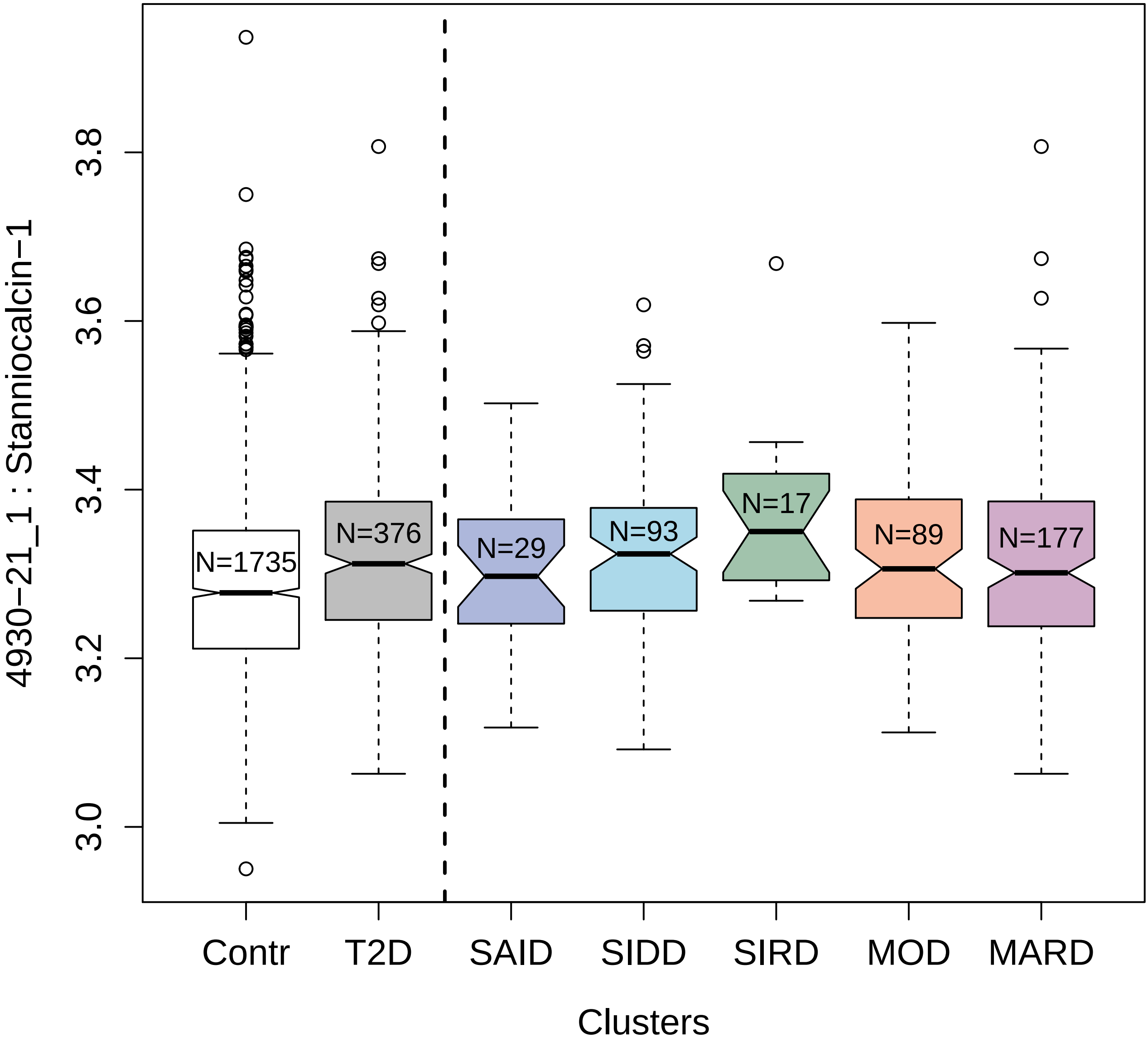

4930−21_1 : Stanniocalcin−1
3.0
3.2
3.4
3.6
3.8
N=1735
N=376
N=29
N=93
N=17
N=89
N=177
Contr
T2D
SAID
SIDD
SIRD
MOD
MARD
Clusters

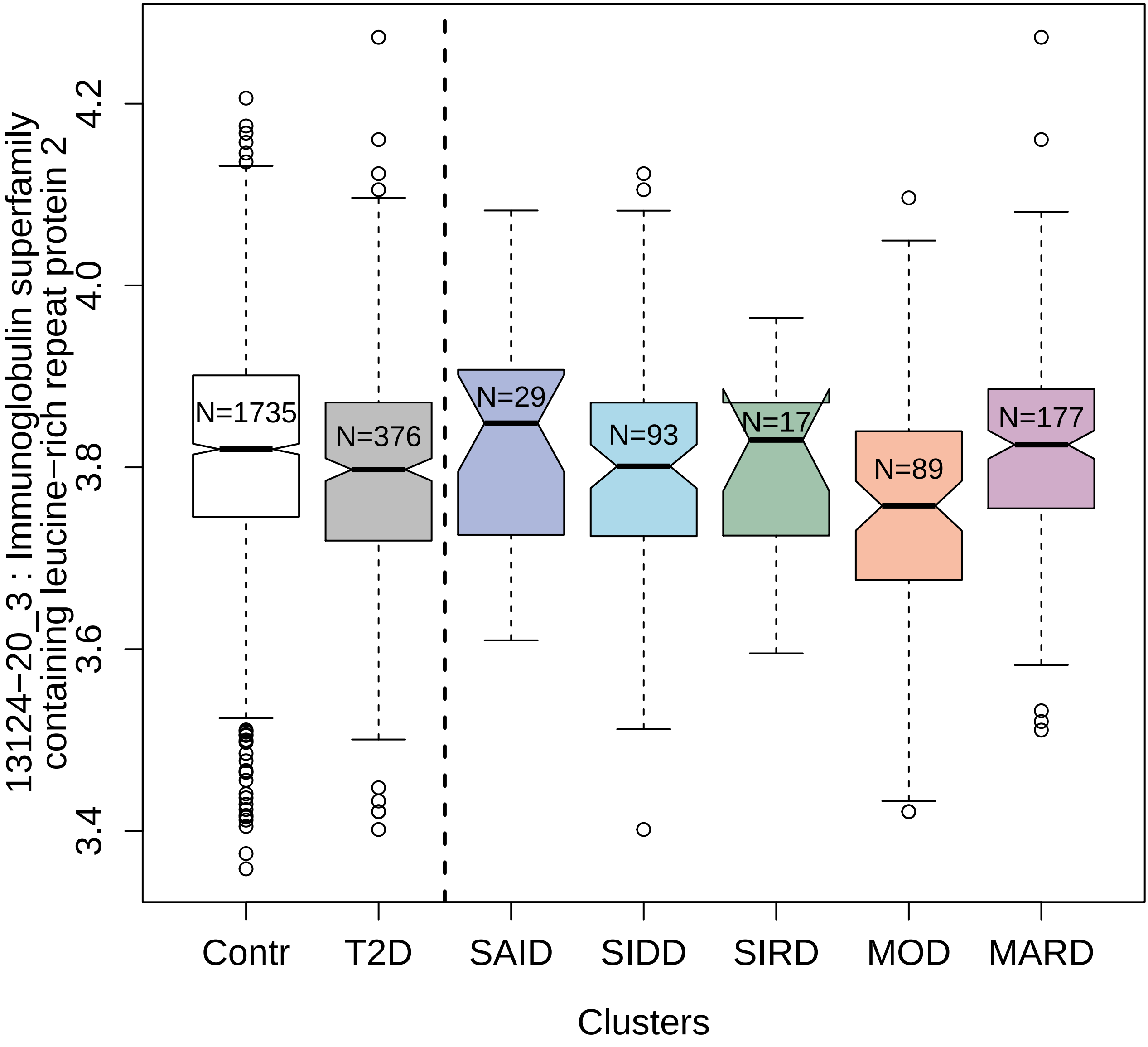

13124−20_3 : Immunoglobulin superfamily
containing leucine−rich repeat protein 2
3.4
3.6
3.8
4.0
4.2
N=1735
N=376
N=29
N=93
N=17
N=89
N=177
Contr
T2D
SAID
SIDD
SIRD
MOD
MARD
Clusters

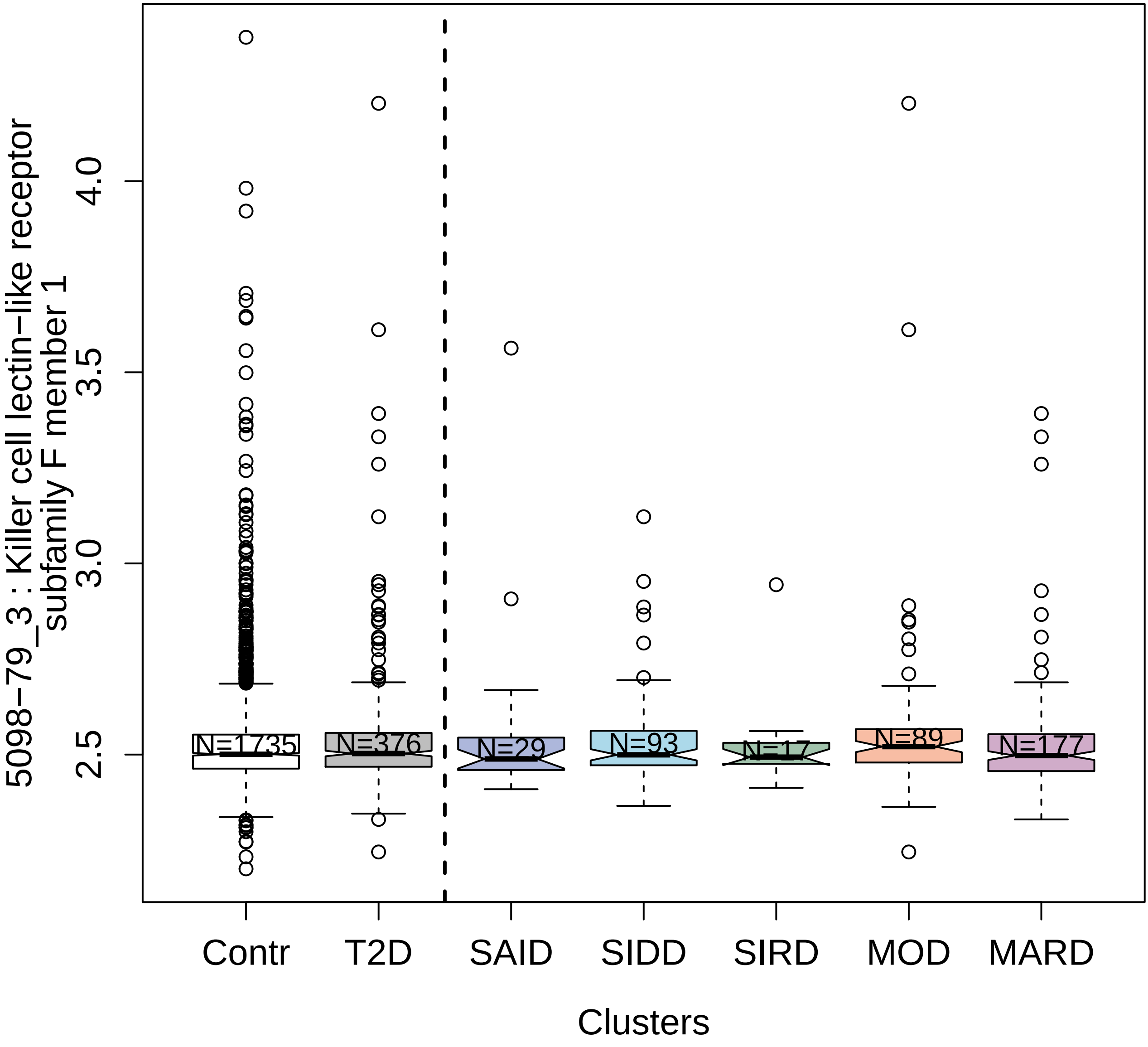

5098−79_3 : Killer cell lectin−like receptor subfamily F member 1
2.5
3.0
3.5
4.0
N=1735
N=376
N=29
N=93
N=17
N=89
N=177
Contr
T2D
SAID
SIDD
SIRD
MOD
MARD
Clusters

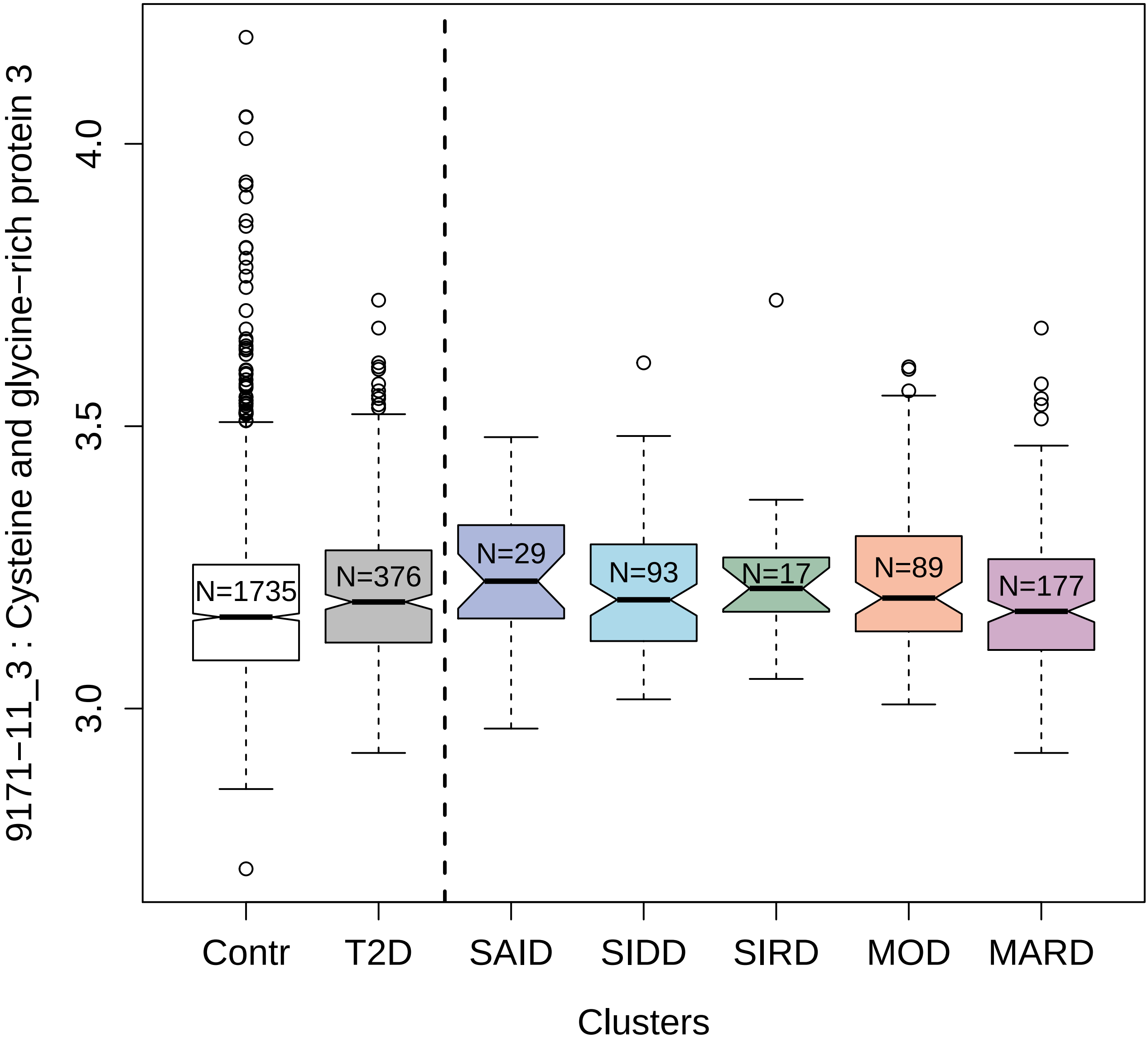
9171−11_3 : Cysteine and glycine−rich protein 3
4.0
3.5
3.0
N=1735
N=376
N=29
N=93
N=17
N=89
N=177
Contr
T2D
SAID
SIDD
SIRD
MOD
MARD
Clusters

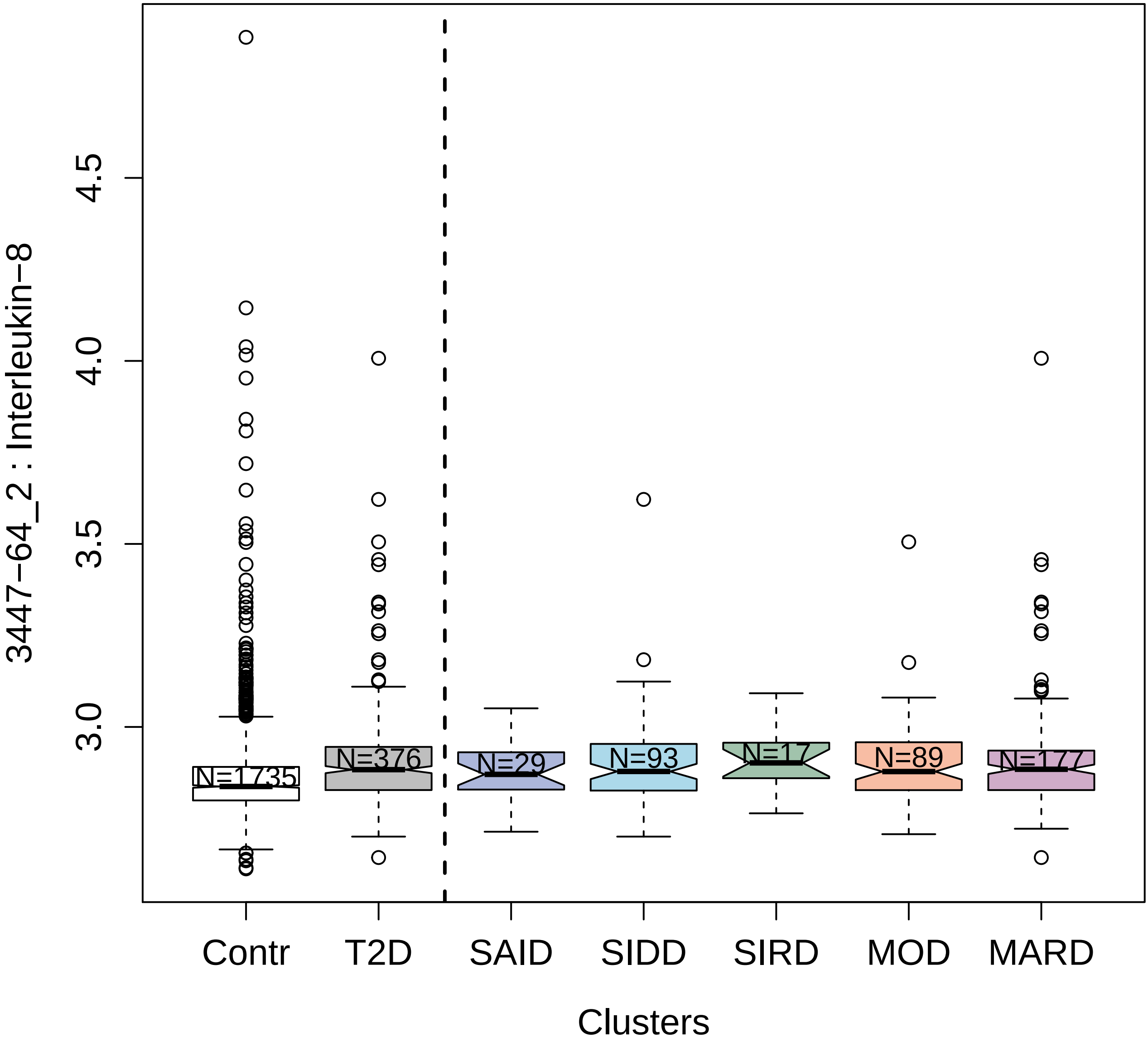

3447−64_2 : Interleukin−8
4.5
4.0
3.5
3.0
N=1735
N=376
N=29
N=93
N=17
N=89
N=177
Contr
T2D
SAID
SIDD
SIRD
MOD
MARD
Clusters

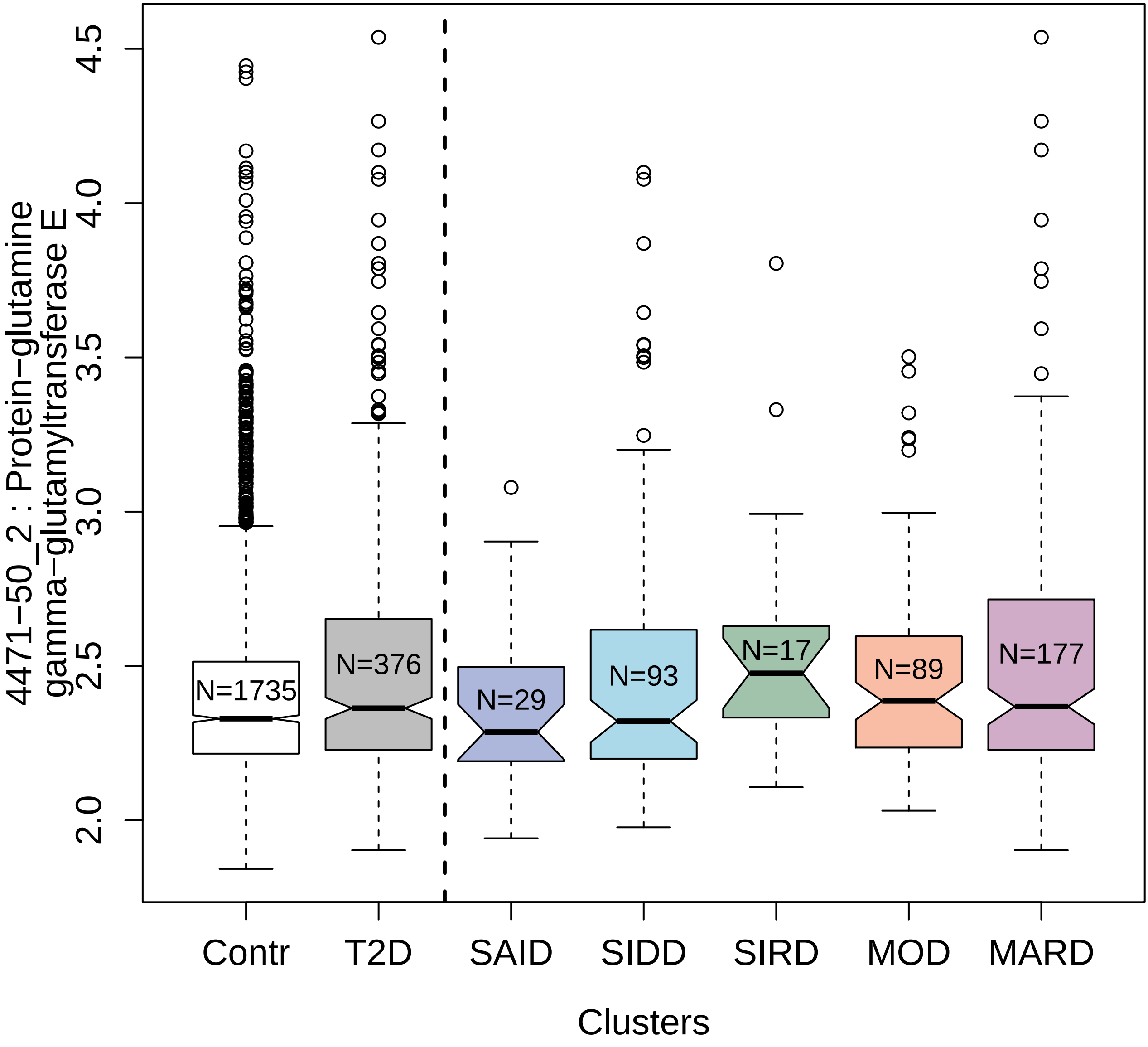

4471−50_2 : Protein−glutamine
gamma−glutamyltransferase E
2.0
2.5
3.0
3.5
4.0
4.5
N=1735
N=376
N=29
N=93
N=17
N=89
N=177
Contr
T2D
SAID
SIDD
SIRD
MOD
MARD
Clusters

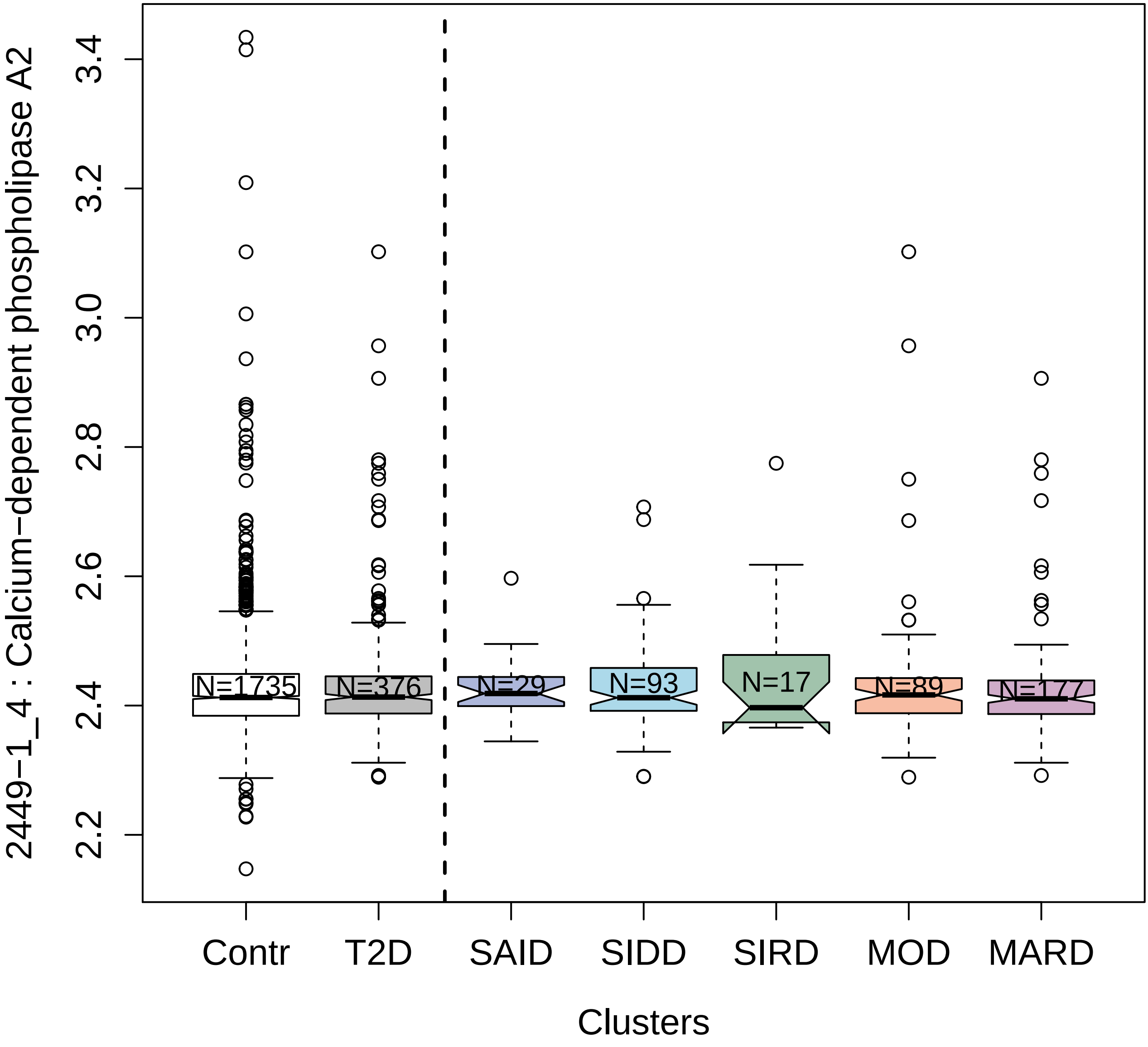

2449−1_4 : Calcium−dependent phospholipase A2
2.2
2.4
2.6
2.8
3.0
3.2
3.4
N=1735
N=376
N=29
N=93
N=17
N=89
N=177
Contr
T2D
SAID
SIDD
SIRD
MOD
MARD
Clusters

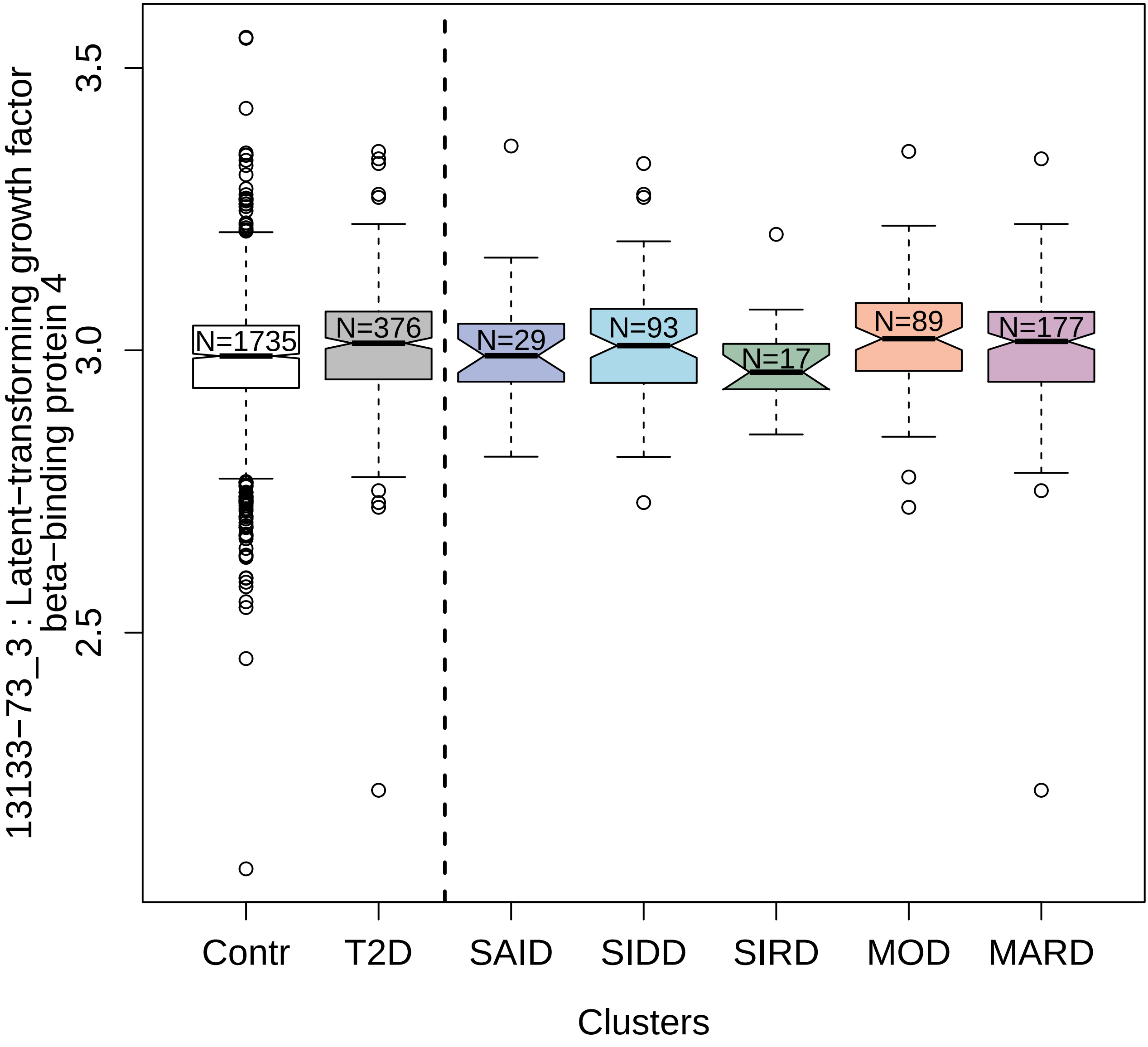
13133−73_3 : Latent−transforming growth factor
beta−binding protein 4
3.5
3.0
2.5
N=1735
N=376
N=29
N=93
N=17
N=89
N=177
Contr
T2D
SAID
SIDD
SIRD
MOD
MARD
Clusters

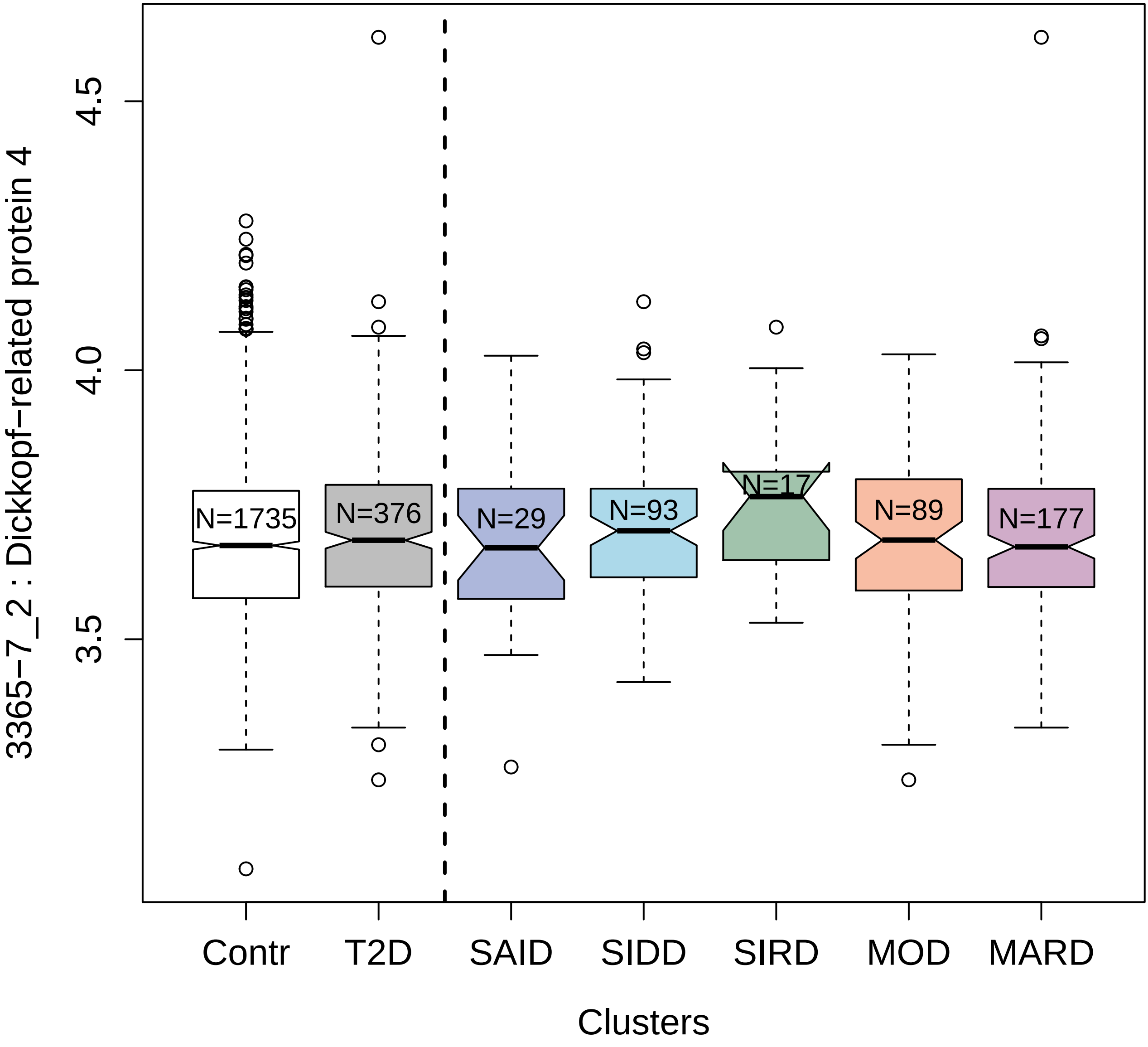
3365−7_2 : Dickkopf−related protein 4
4.5
4.0
3.5
N=1735
N=376
N=29
N=93
N=17
N=89
N=177
Contr
T2D
SAID
SIDD
SIRD
MOD
MARD
Clusters

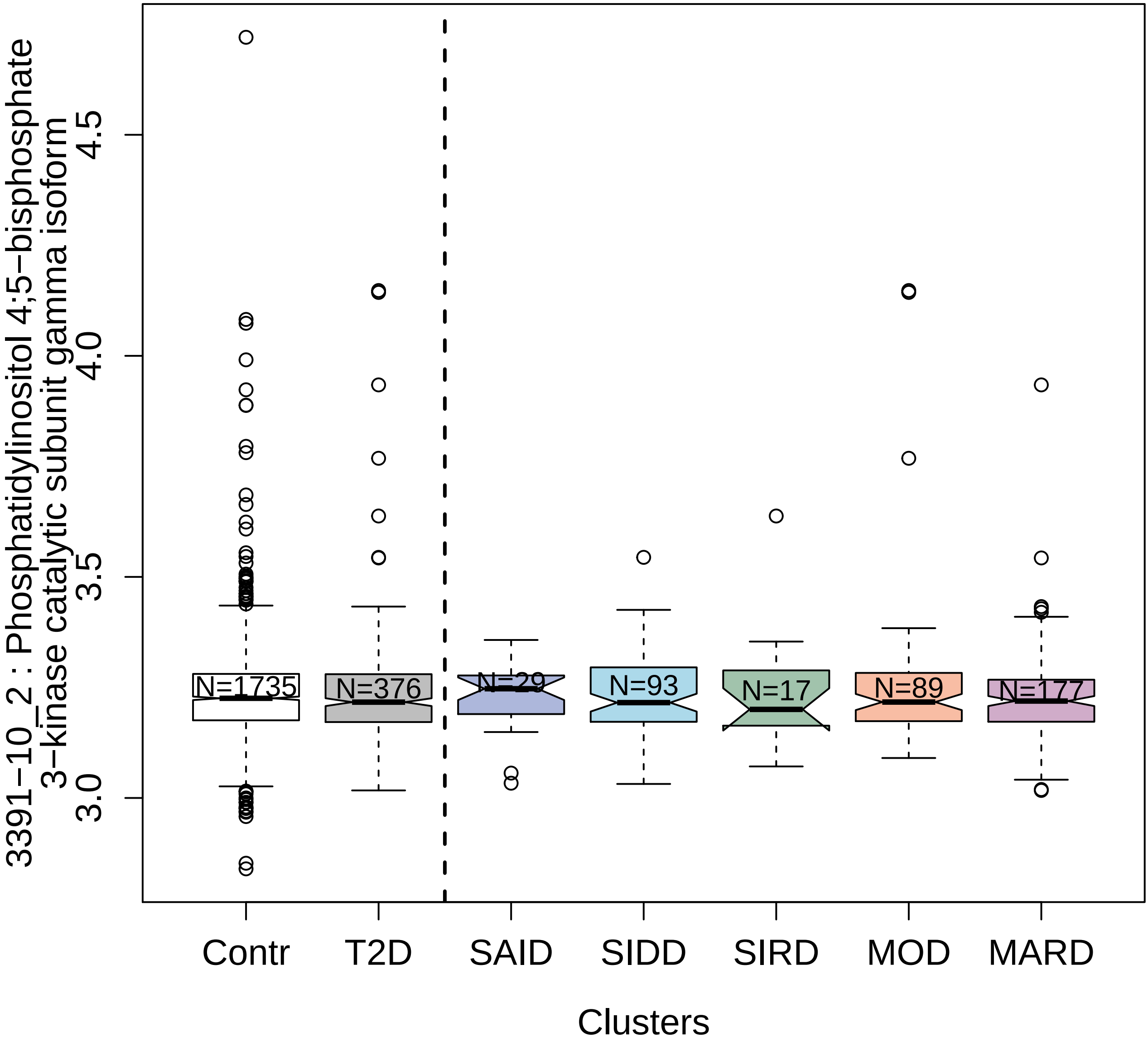
3391−10_2 : Phosphatidylinositol 4,5−bisphosphate
3−kinase catalytic subunit gamma isoform
4.5
4.0
3.5
3.0
N=1735
N=376
N=29
N=93
N=17
N=89
N=177
Contr
T2D
SAID
SIDD
SIRD
MOD
MARD
Clusters

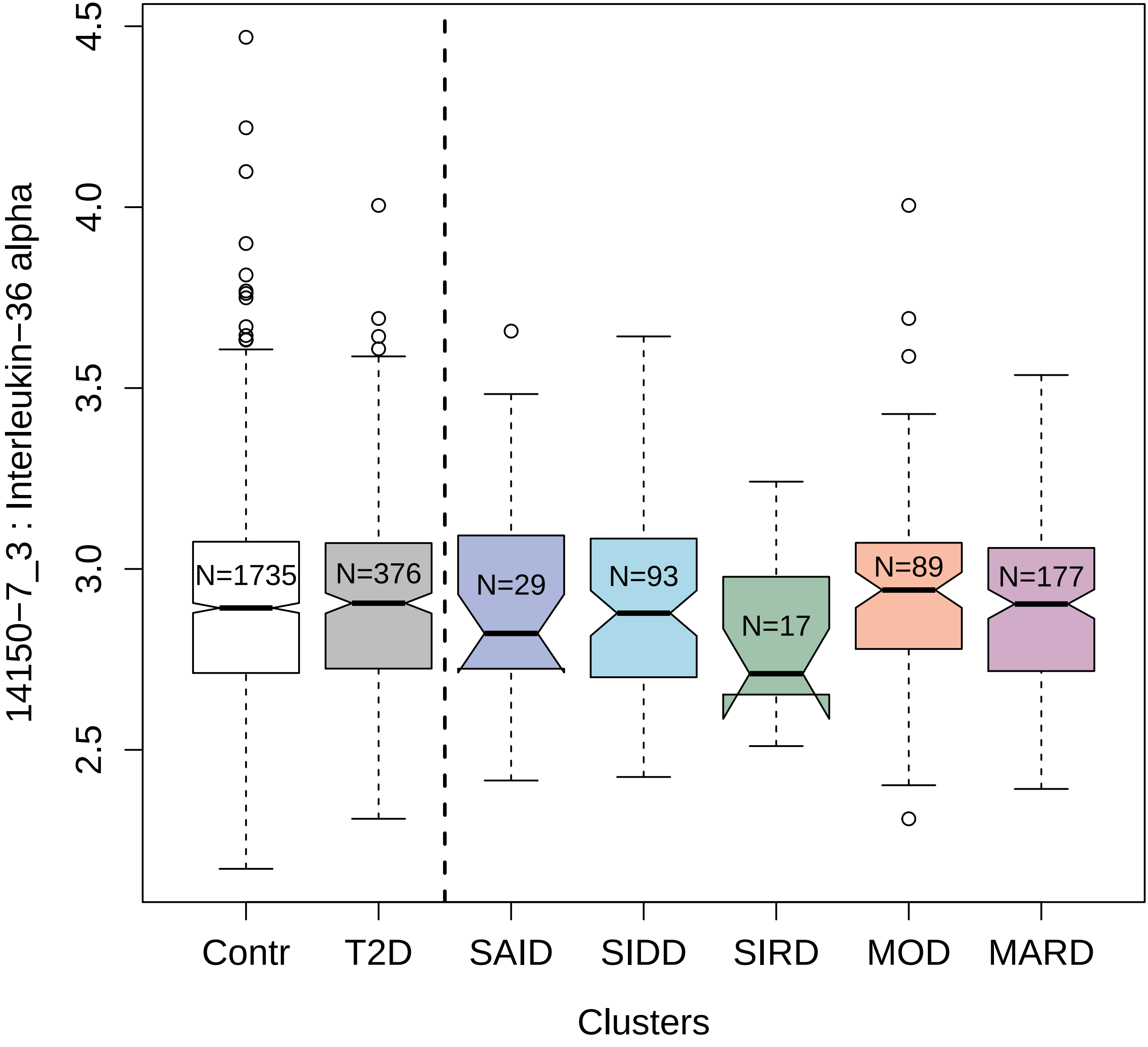

14150−7_3 : Interleukin−36 alpha
4.5
4.0
3.5
3.0
2.5
N=1735
N=376
N=29
N=93
N=17
N=89
N=177
Contr
T2D
SAID
SIDD
SIRD
MOD
MARD
Clusters
