## Supplementary Figure 8 for "Metabolic and proteomic signatures of type 2 diabetes subtypes in an Arab population"

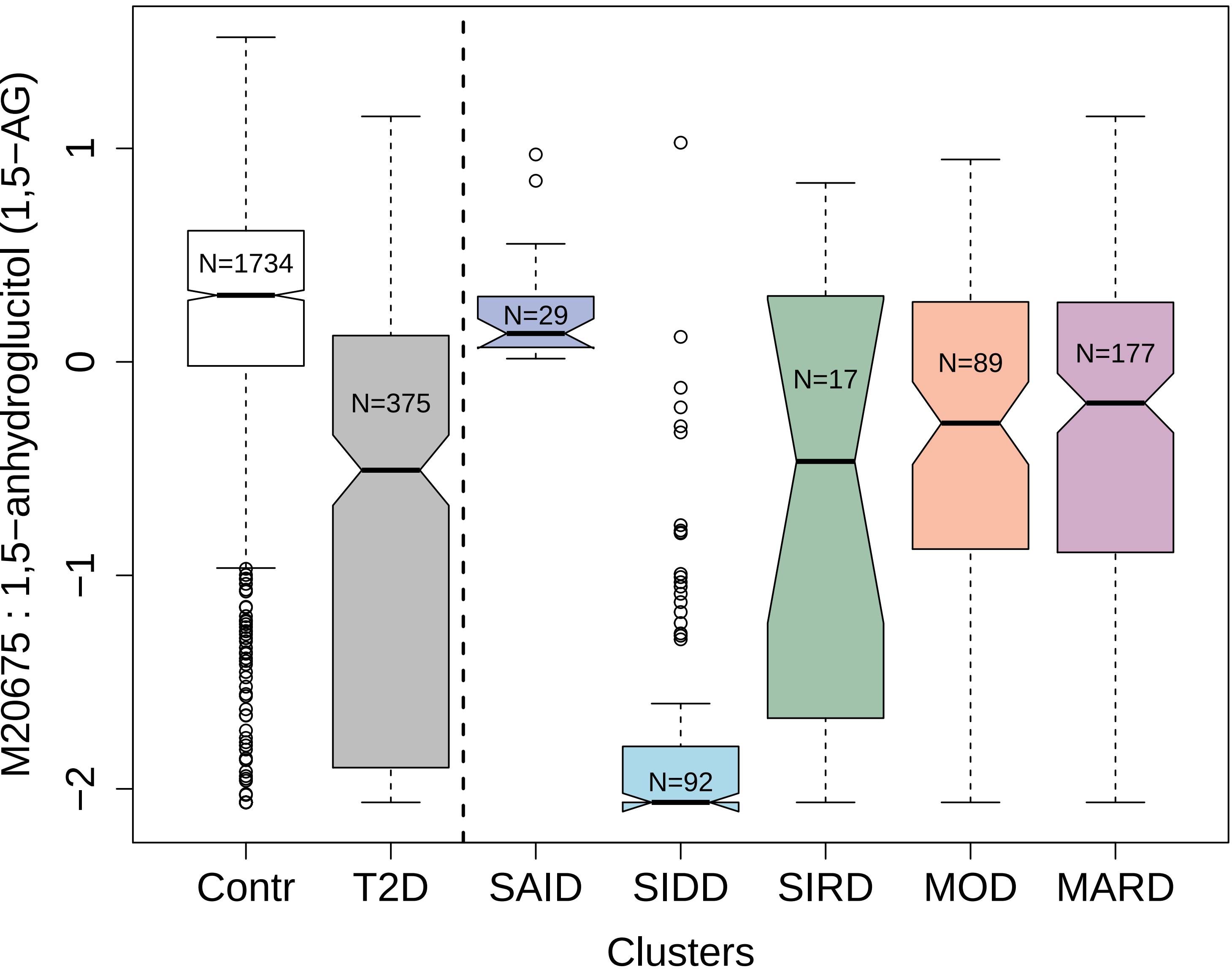
M20675 : 1,5−anhydroglucitol (1,5−AG)
1
0
−1
−2
N=1734
N=375
N=29
N=92
N=17
N=89
N=177
Contr
T2D
SAID
SIDD
SIRD
MOD
MARD
Clusters

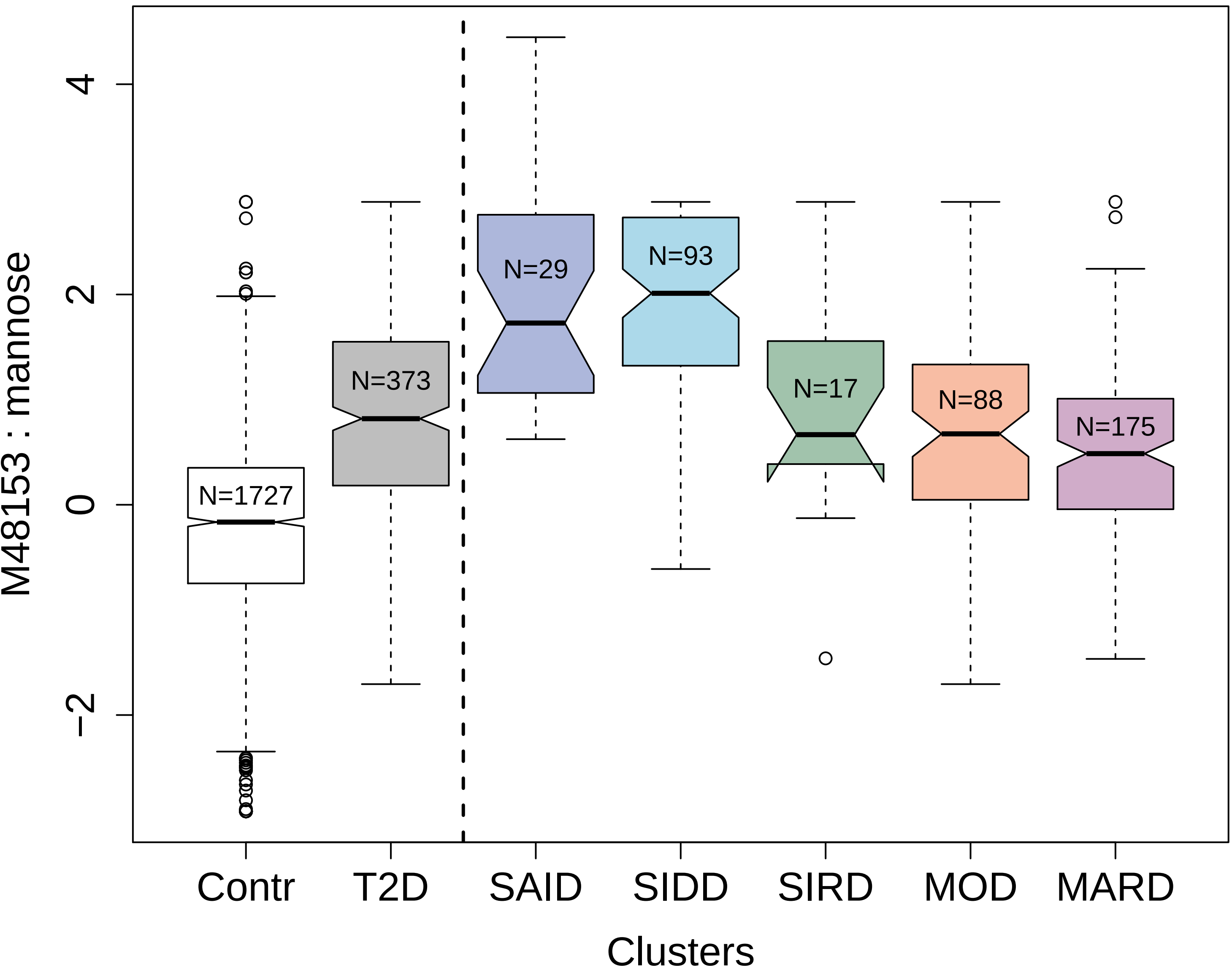
M48153 : mannose
4
2
0
−2
N=1727
N=373
N=29
N=93
N=17
N=88
N=175
Contr
T2D
SAID
SIDD
SIRD
MOD
MARD
Clusters

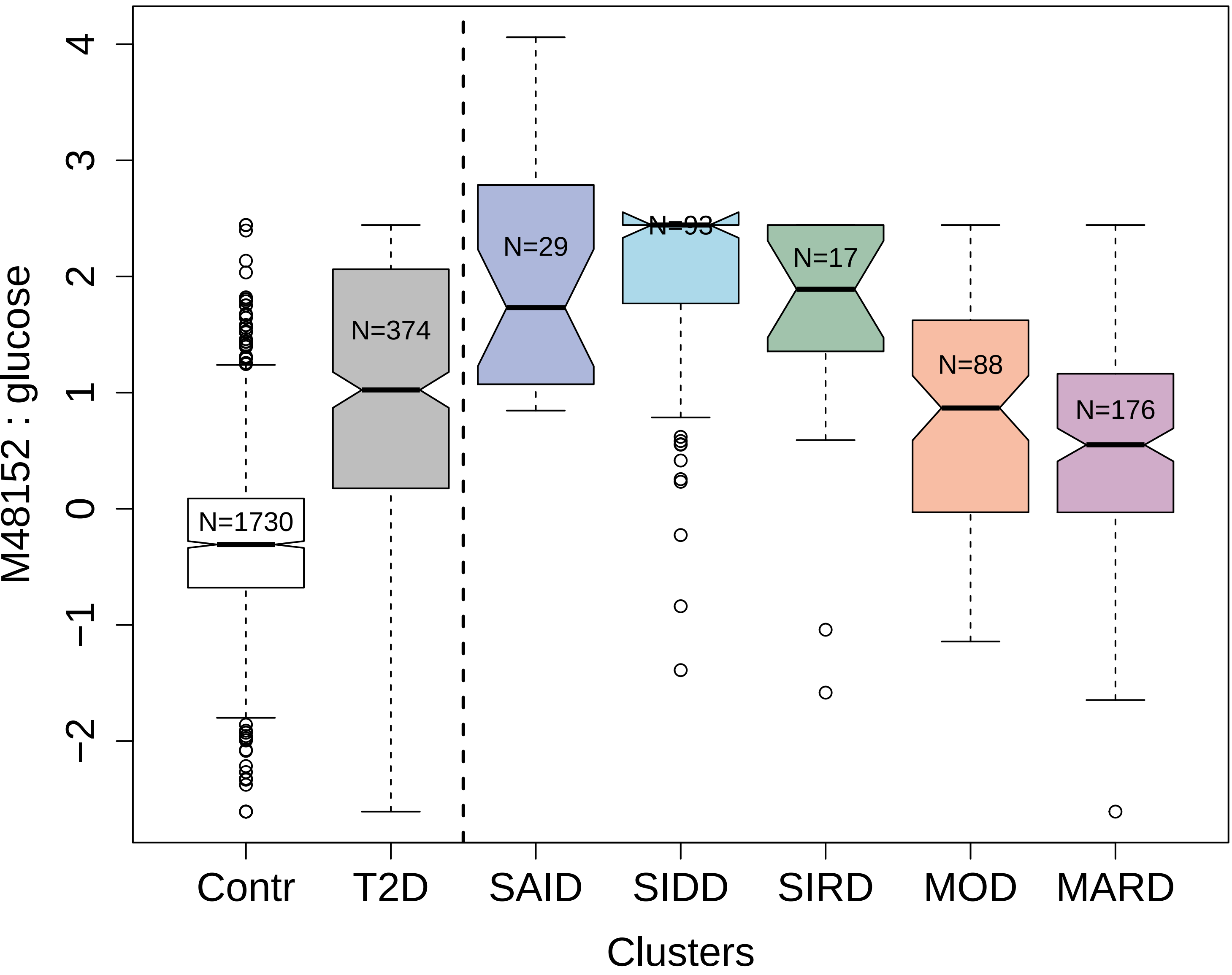

M48152 : glucose
4
3
2
1
0
−1
−2
N=1730
N=374
N=29
N=93
N=17
N=88
N=176
Contr
T2D
SAID
SIDD
SIRD
MOD
MARD
Clusters

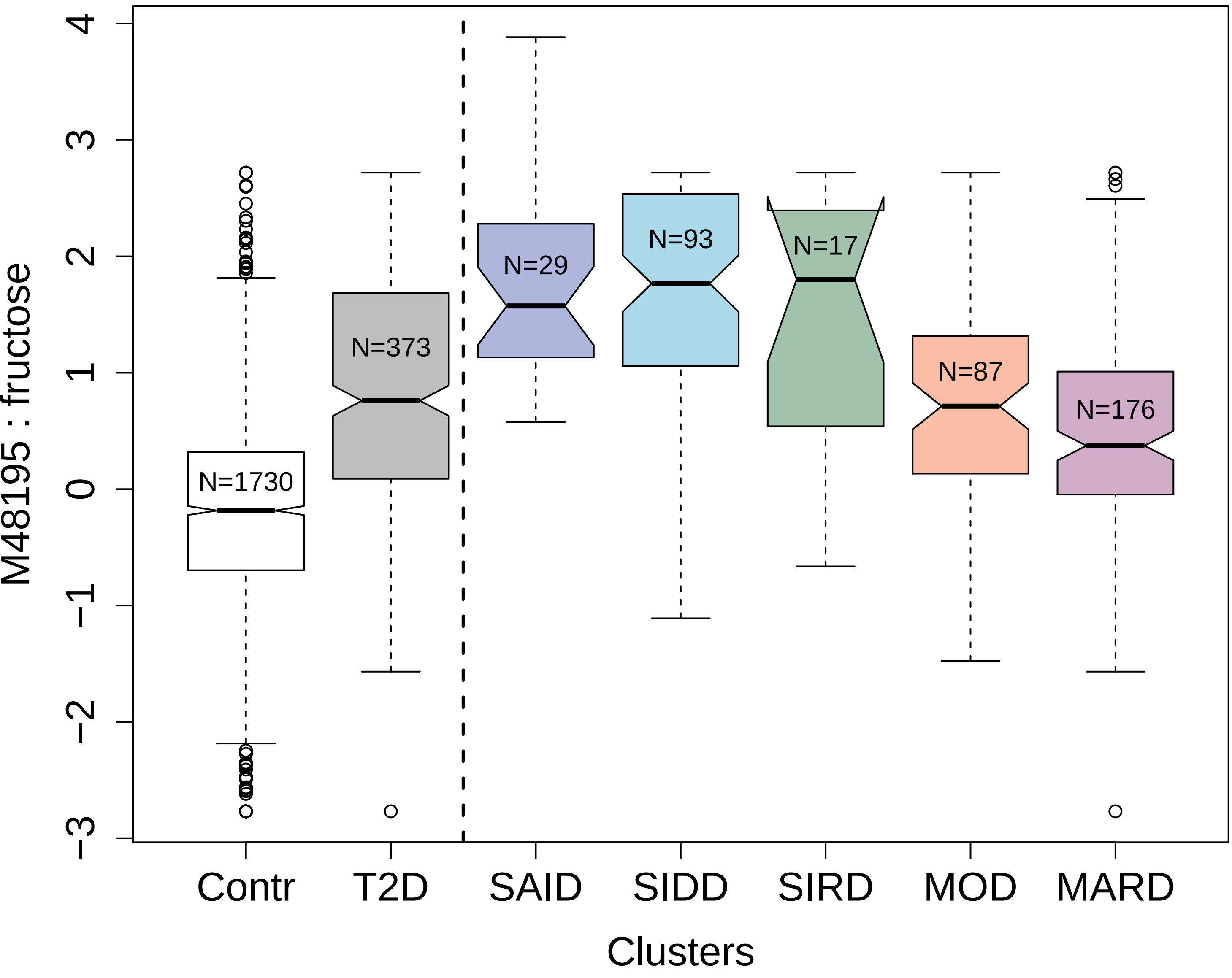

M48195 : fructose
4
3
2
1
0
−1
−2
−3
N=1730
N=373
N=29
N=93
N=17
N=87
N=176
Contr
T2D
SAID
SIDD
SIRD
MOD
MARD
Clusters

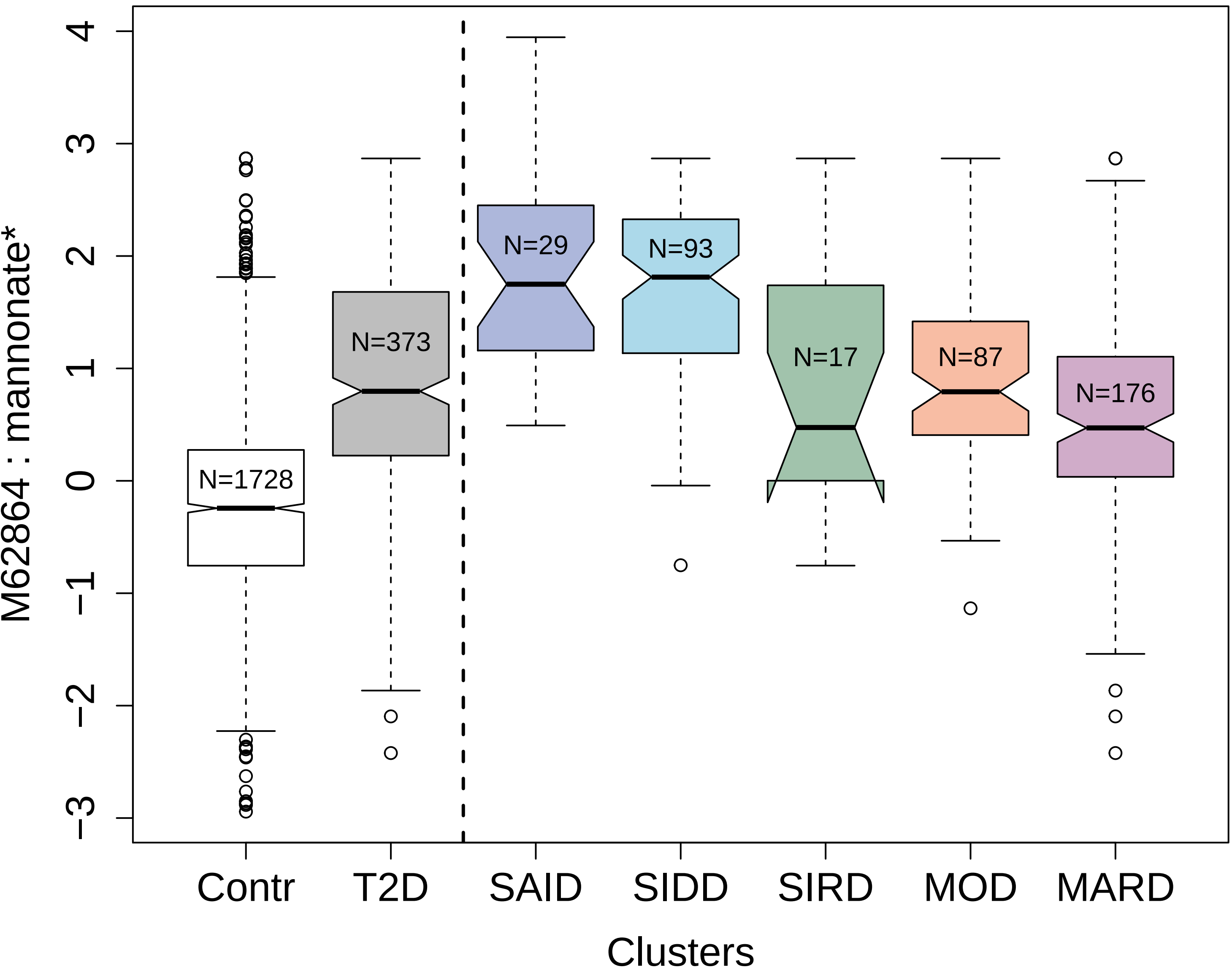
M62864 : mannonate*
4
3
2
1
0
−1
−2
−3
N=1728
N=373
N=29
N=93
N=17
N=87
N=176
Contr
T2D
SAID
SIDD
SIRD
MOD
MARD
Clusters

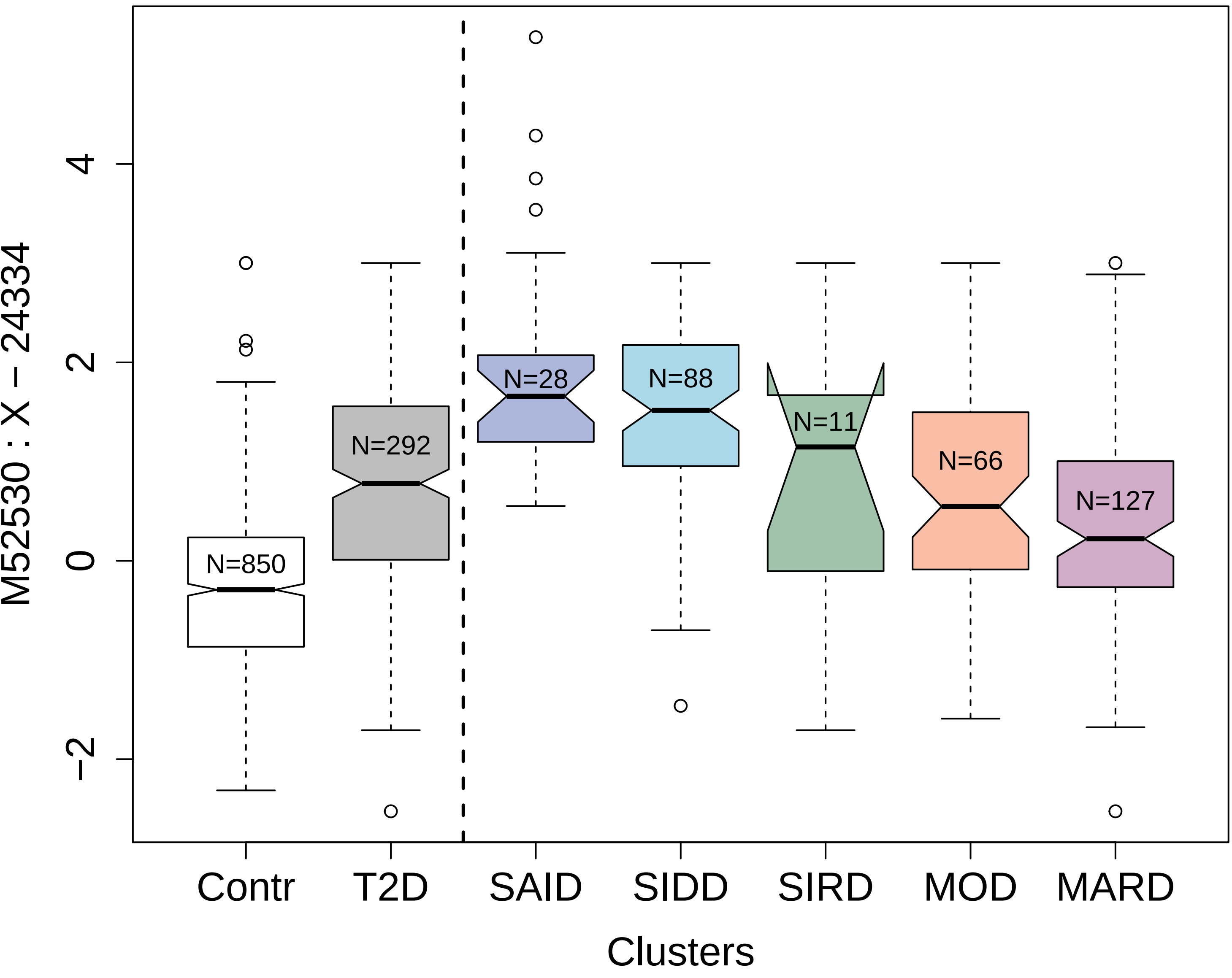

M52530 : X − 24334
−2
0
2
4
N=850
N=292
N=28
N=88
N=11
N=66
N=127
Contr
T2D
SAID
SIDD
SIRD
MOD
MARD
Clusters

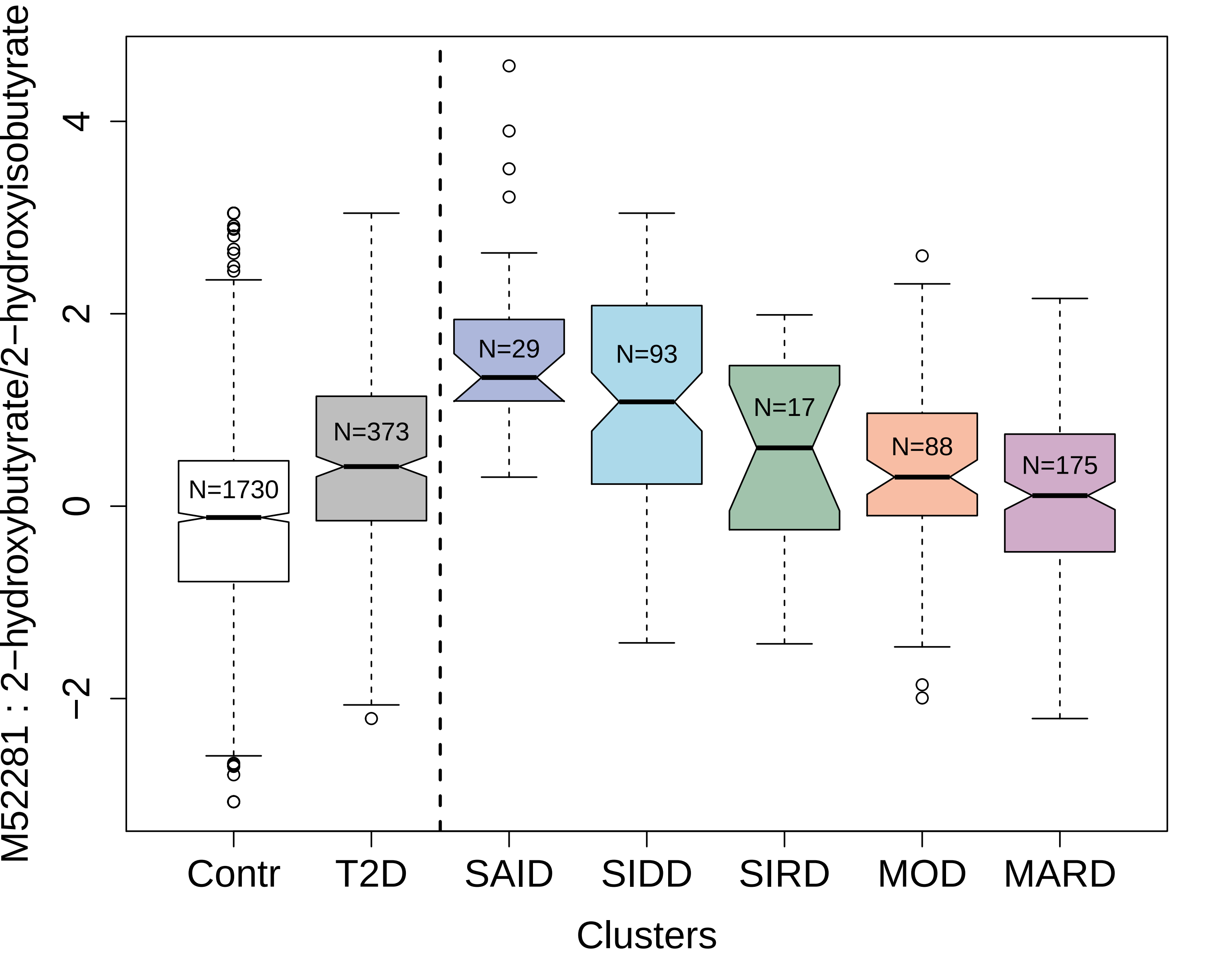
M52281 : 2−hydroxybutyrate/2−hydroxyisobutyrate
4
2
0
−2
N=1730
N=373
N=29
N=93
N=17
N=88
N=175
Contr
T2D
SAID
SIDD
SIRD
MOD
MARD
Clusters

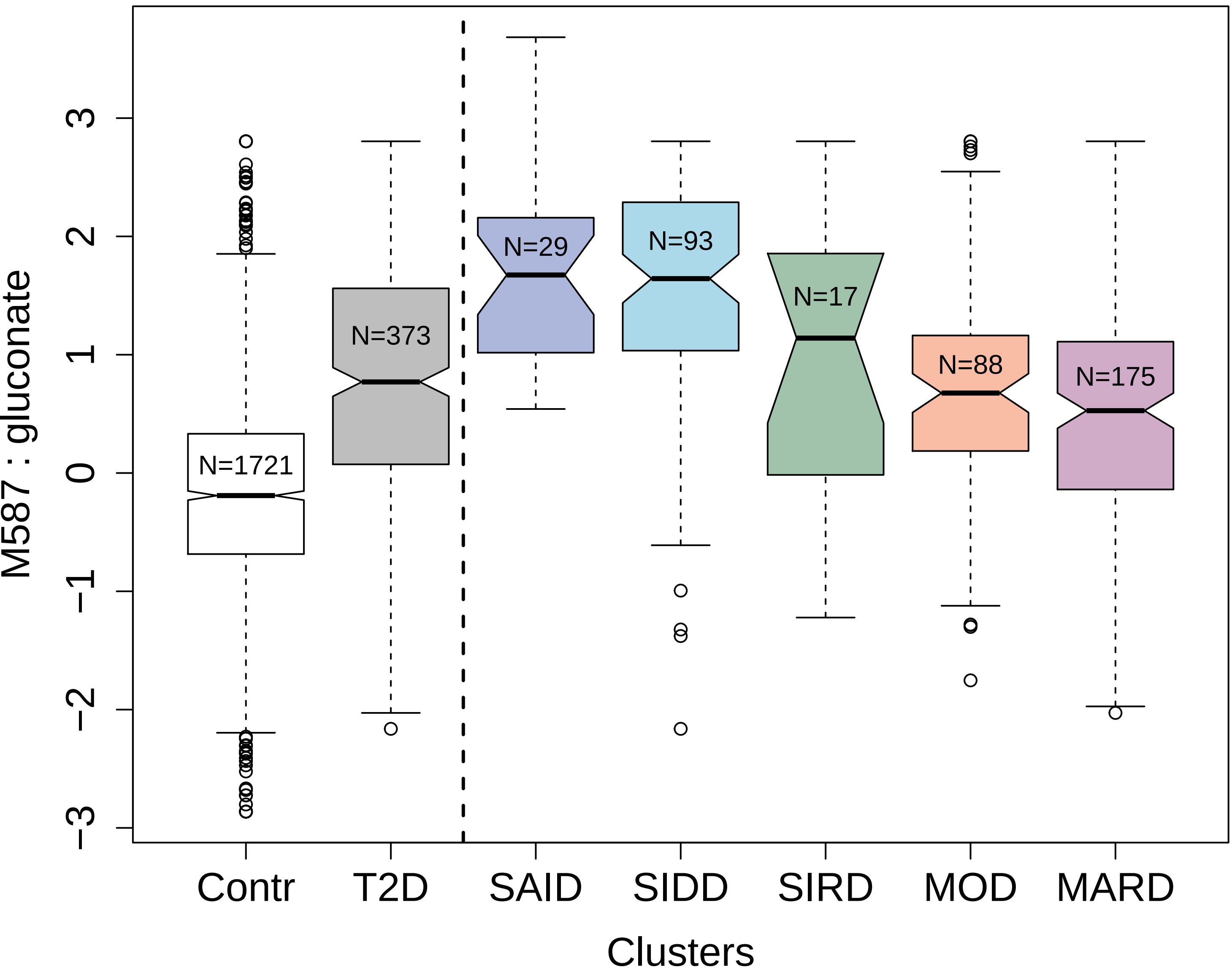

M587 : gluconate
3
2
1
0
−1
−2
−3
N=1721
N=373
N=29
N=93
N=17
N=88
N=175
Contr
T2D
SAID
SIDD
SIRD
MOD
MARD
Clusters

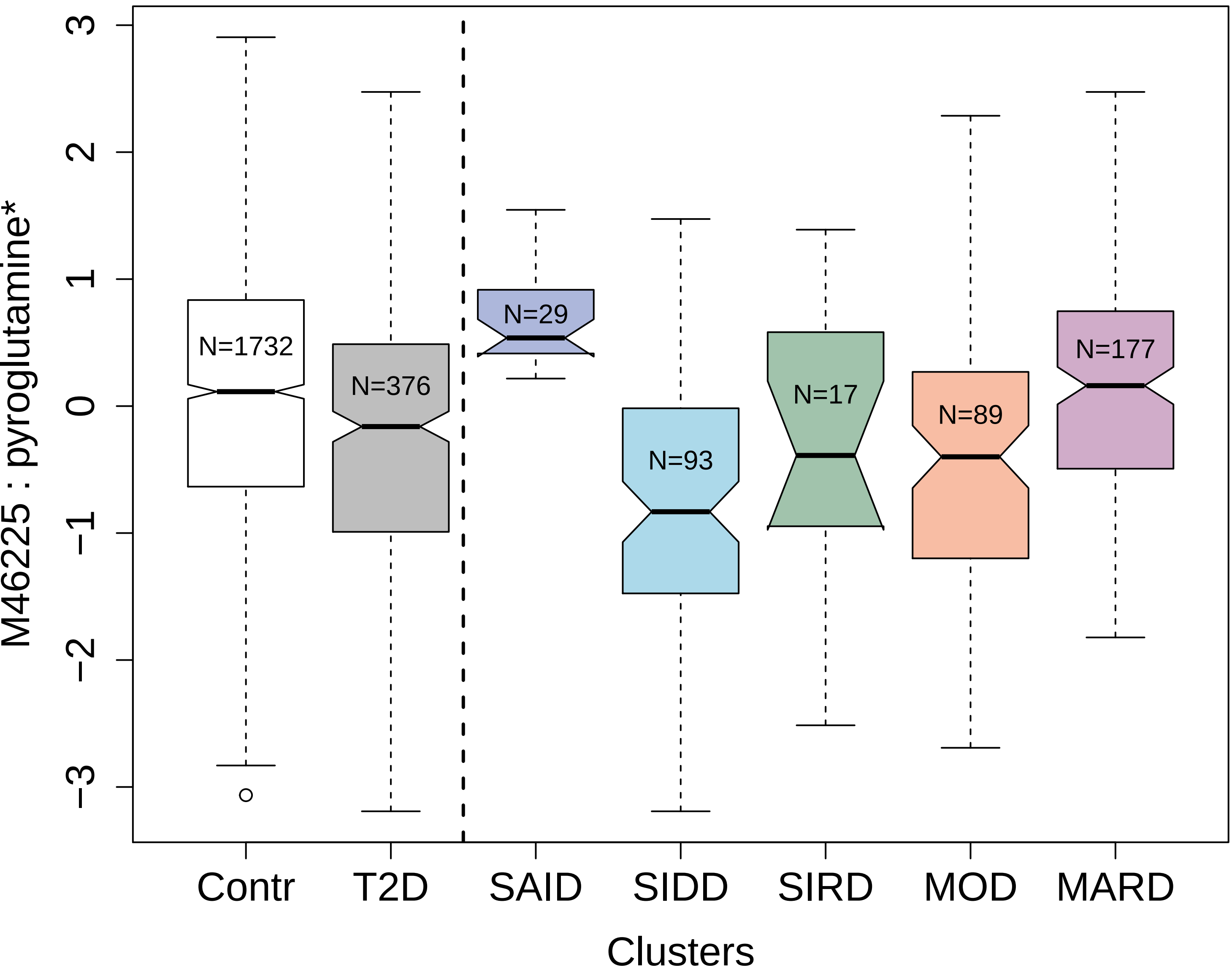
M46225 : pyroglutamine*
3
2
1
0
−1
−2
−3
N=1732
N=376
N=29
N=93
N=17
N=89
N=177
Contr
T2D
SAID
SIDD
SIRD
MOD
MARD
Clusters

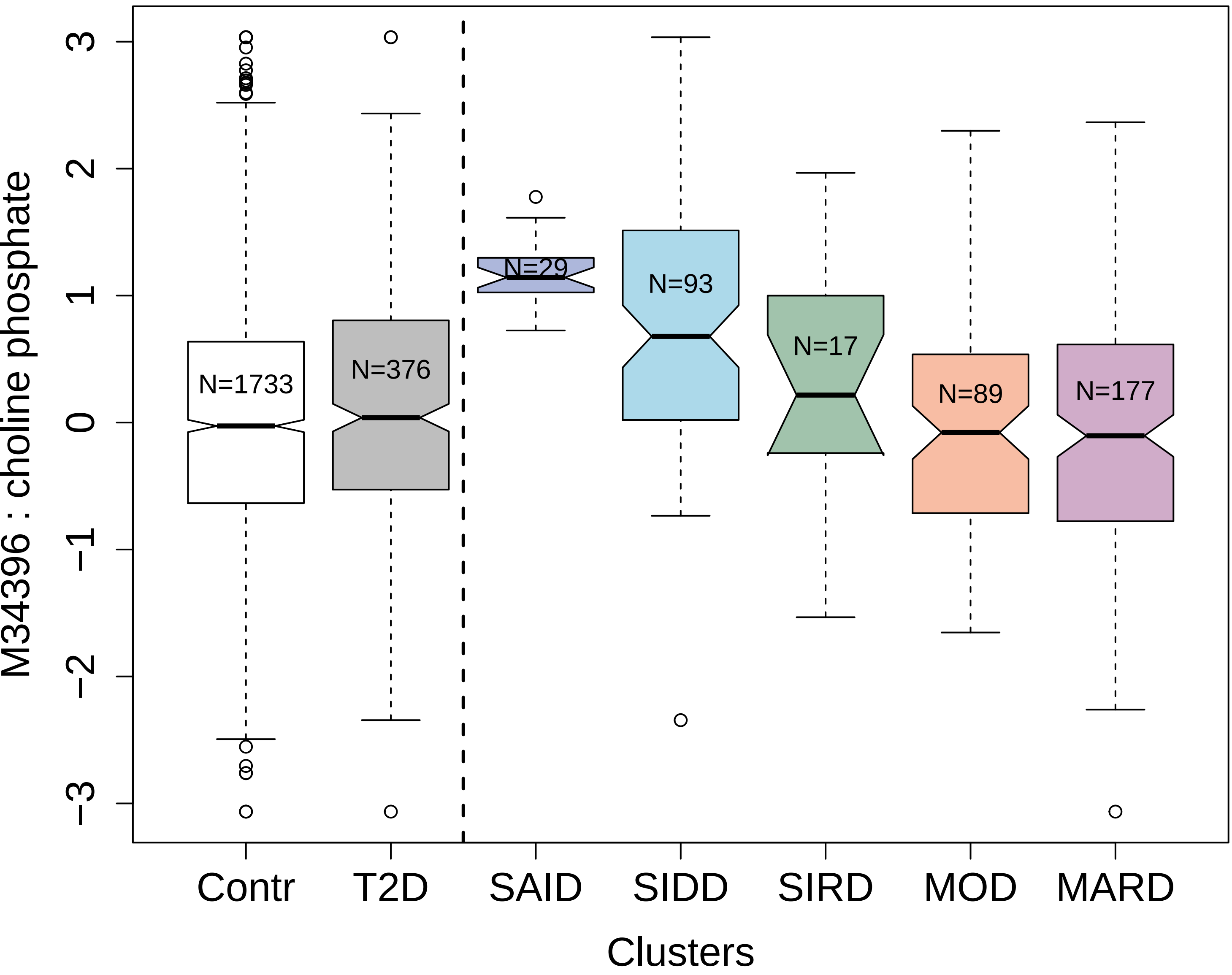

M34396 : choline phosphate
3
2
1
0
−1
−2
−3
N=1733
N=376
N=29
N=93
N=17
N=89
N=177
Contr
T2D
SAID
SIDD
SIRD
MOD
MARD
Clusters

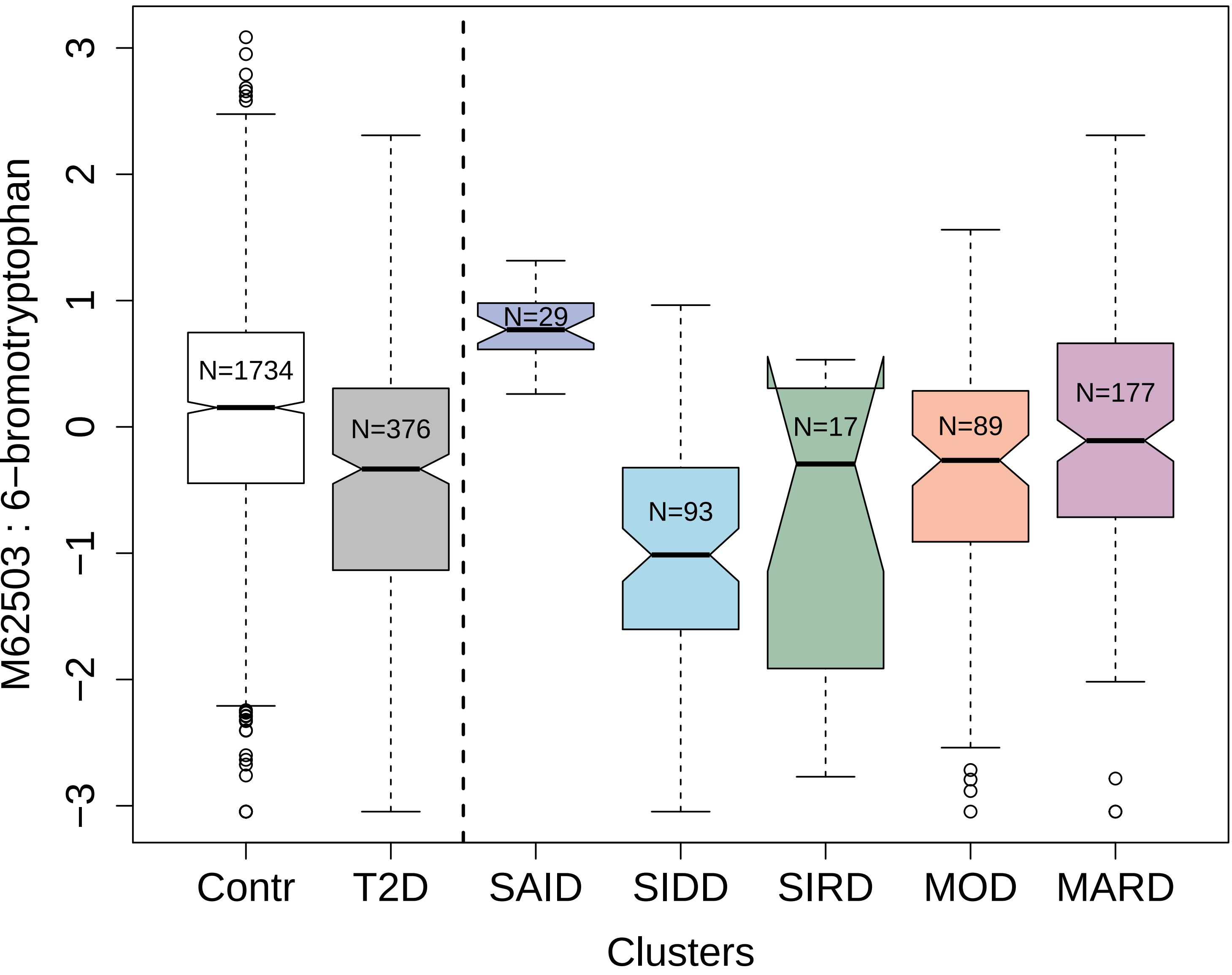

M62503 : 6−bromotryptophan
Clusters
Contr
T2D
SAID
SIDD
SIRD
MOD
MARD
N=1734
N=376
N=29
N=93
N=17
N=89
N=177
3
2
1
0
−1
−2
−3

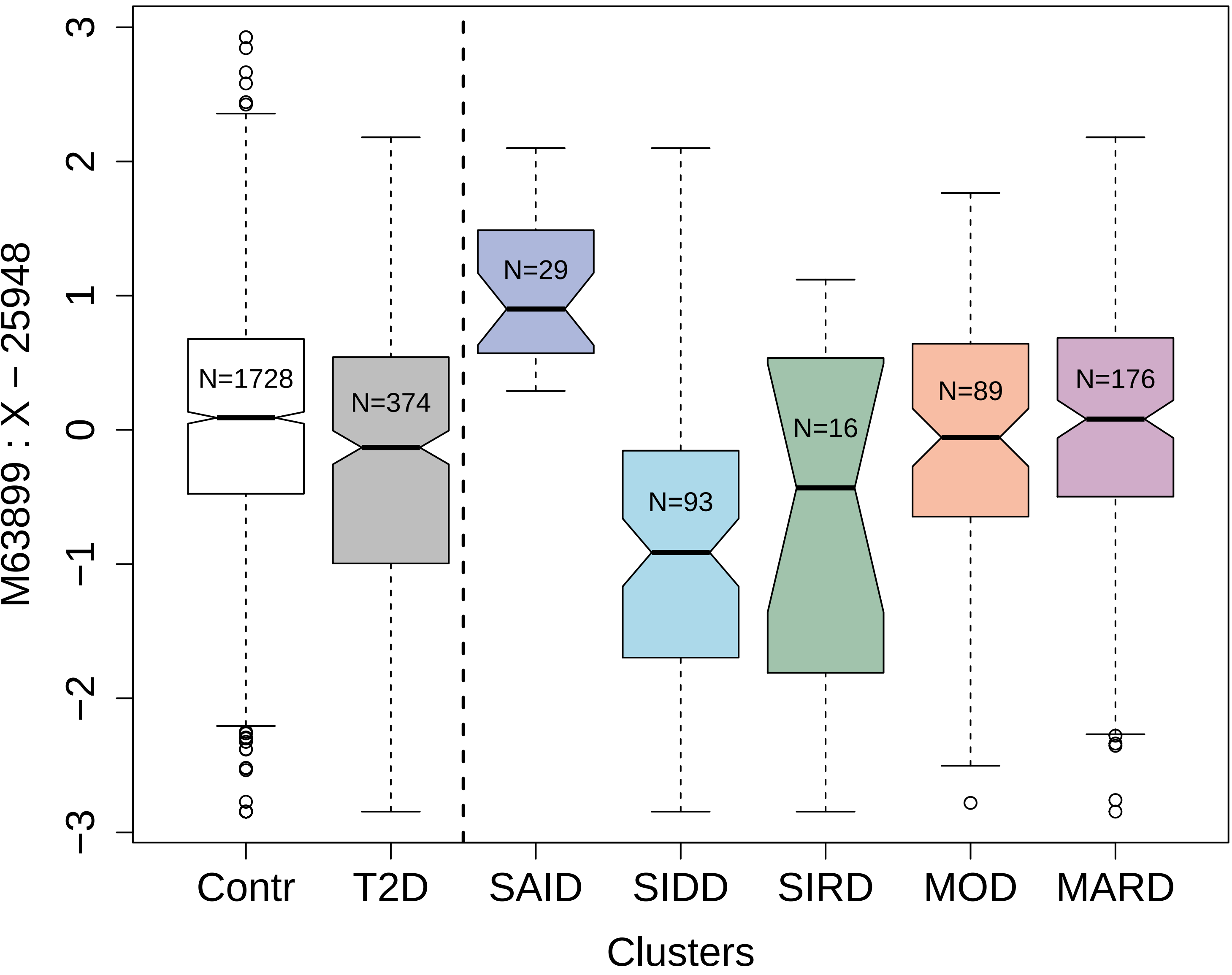

M63899 : X − 25948
3
2
1
0
−1
−2
−3
N=1728
N=374
N=29
N=93
N=16
N=89
N=176
Contr
T2D
SAID
SIDD
SIRD
MOD
MARD
Clusters

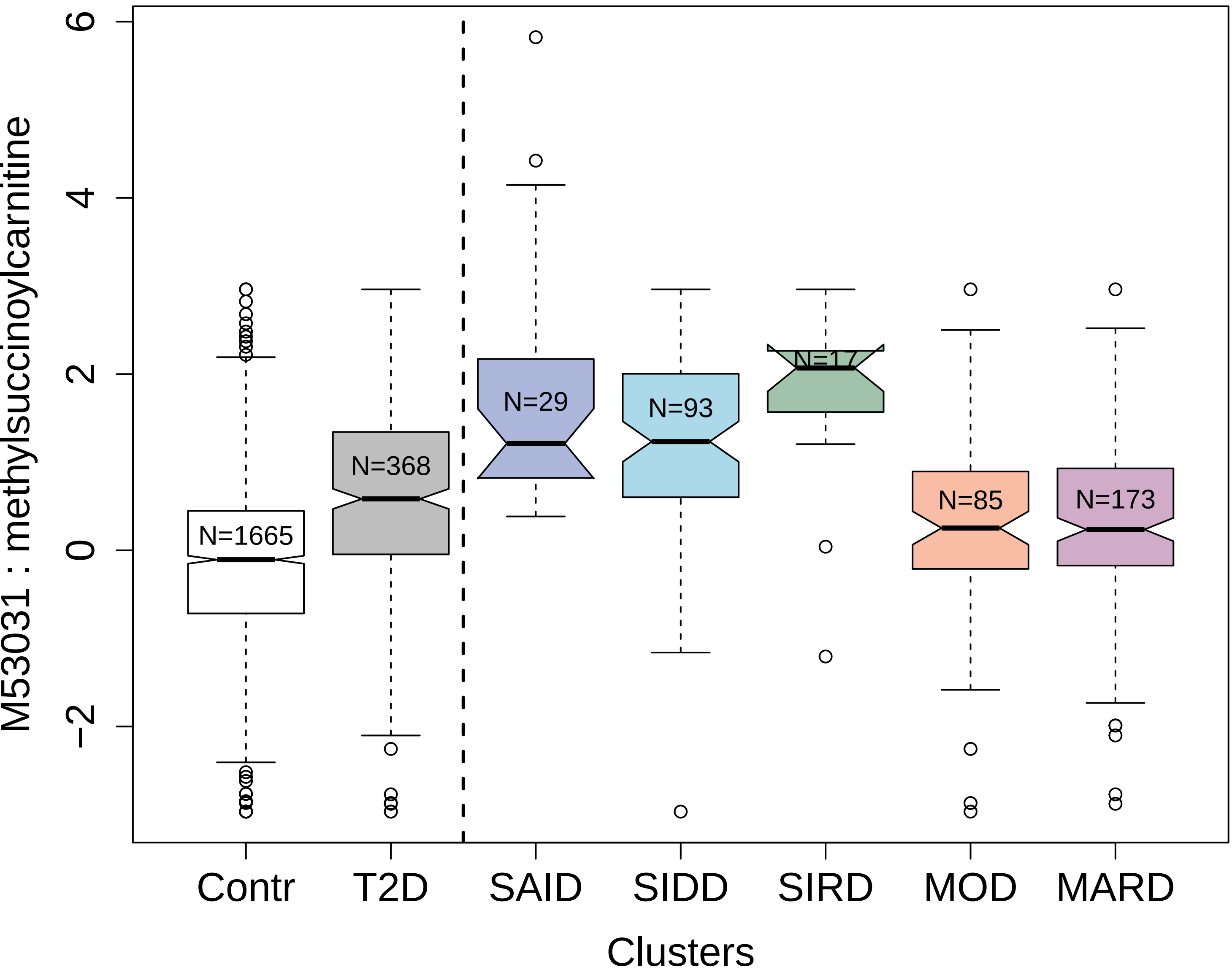

M53031 : methylsuccinoylcarnitine
Clusters
Contr
T2D
SAID
SIDD
SIRD
MOD
MARD
N=1665
N=368
N=29
N=93
N=17
N=85
N=173
6
4
2
0
−2

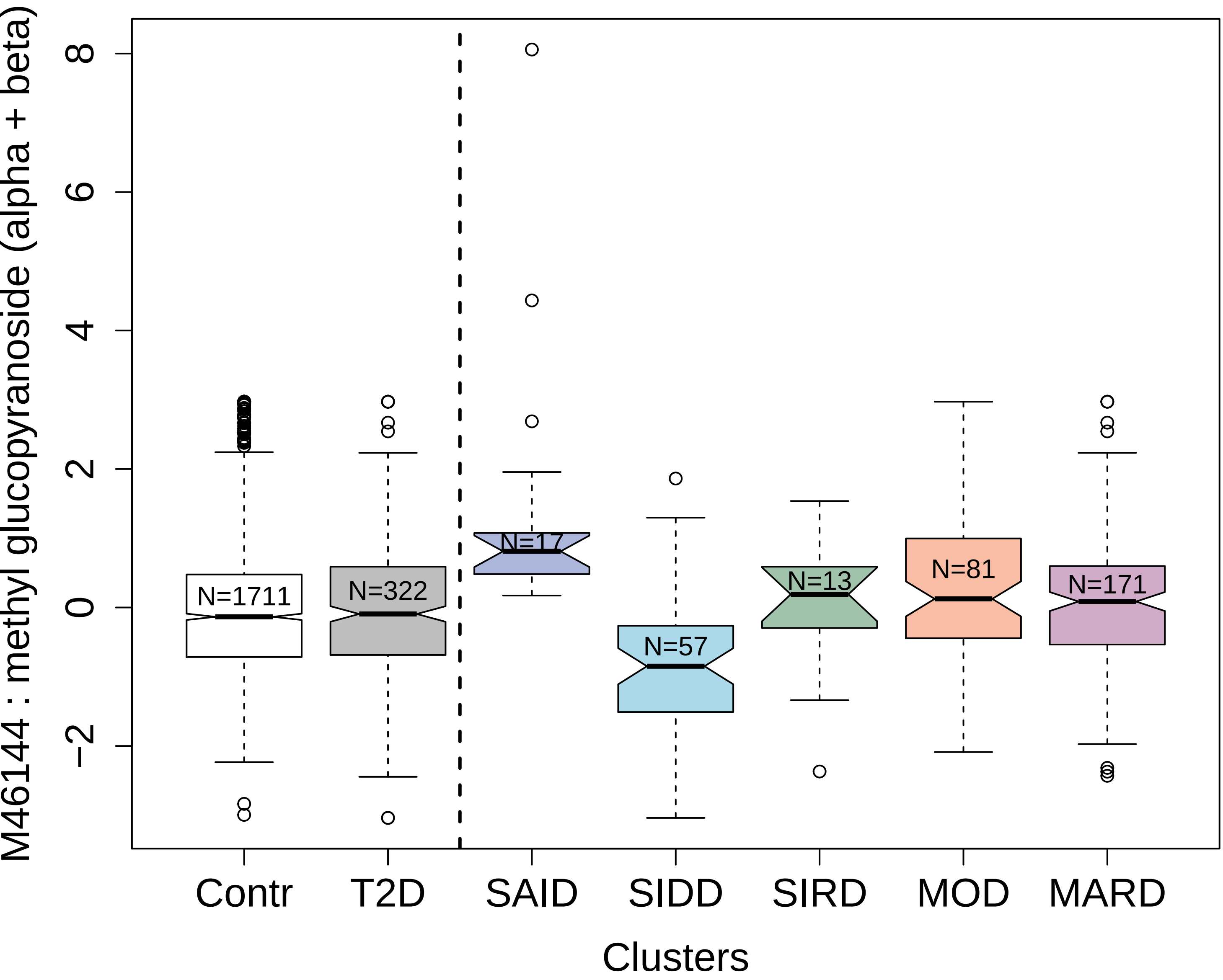

M46144 : methyl glucopyranoside (alpha + beta)
8
6
4
2
0
−2
N=1711
N=322
N=17
N=57
N=13
N=81
N=171
Contr
T2D
SAID
SIDD
SIRD
MOD
MARD
Clusters

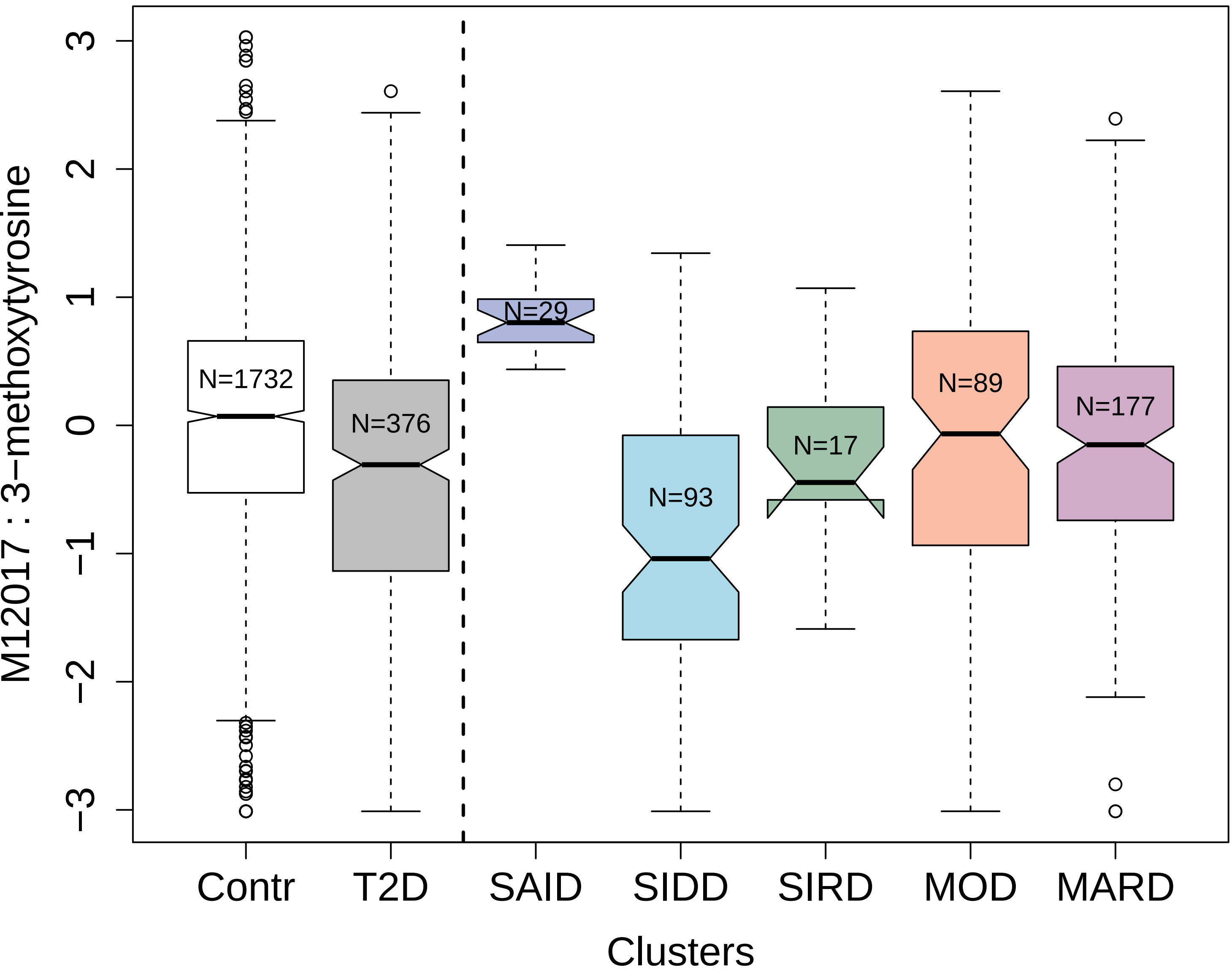
M12017 : 3−methoxytyrosine
3
2
1
0
−1
−2
−3
N=1732
N=376
N=29
N=93
N=17
N=89
N=177
Contr
T2D
SAID
SIDD
SIRD
MOD
MARD
Clusters

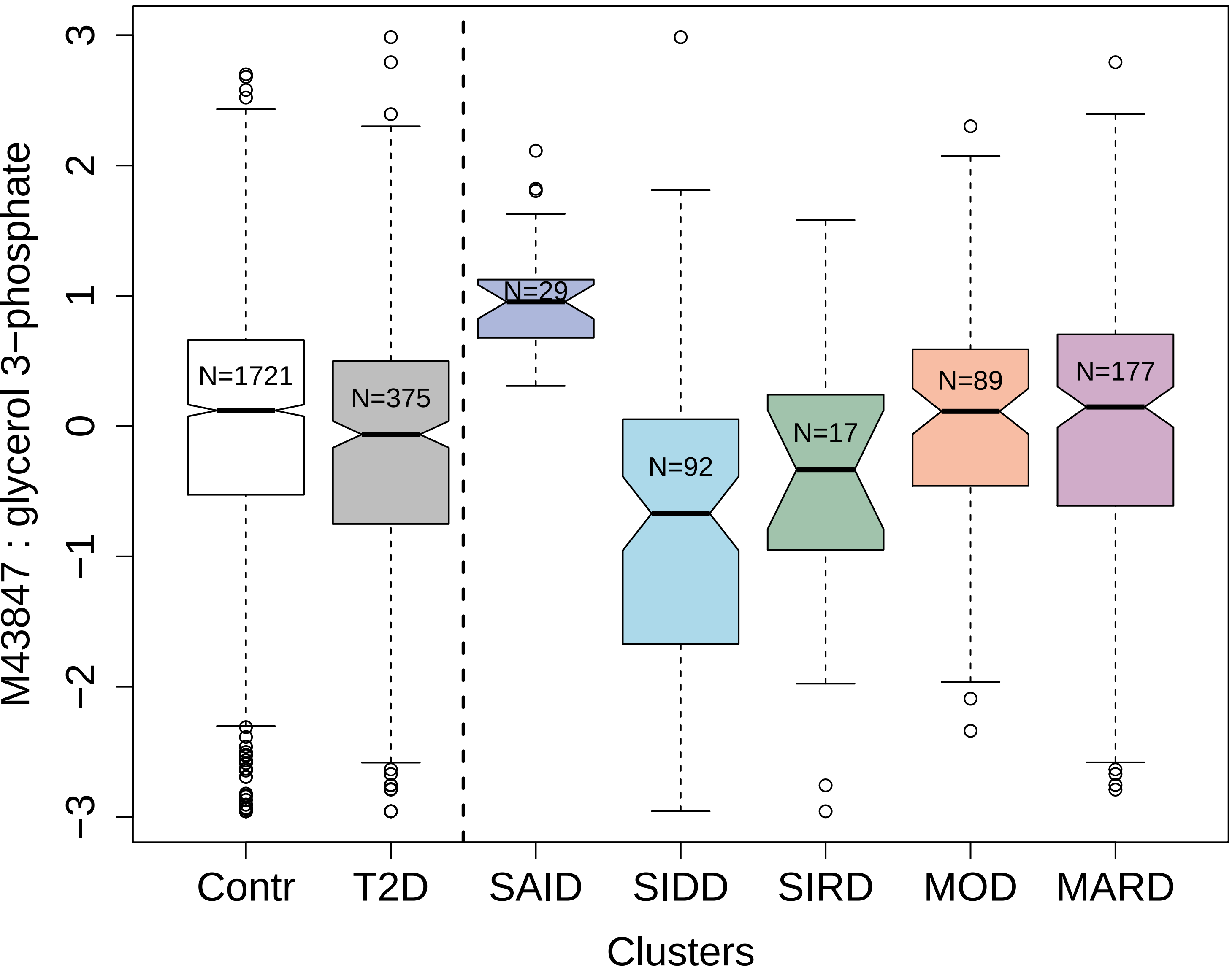
M43847 : glycerol 3−phosphate
3
2
1
0
−1
−2
−3
N=1721
N=375
N=29
N=92
N=17
N=89
N=177
Contr
T2D
SAID
SIDD
SIRD
MOD
MARD
Clusters

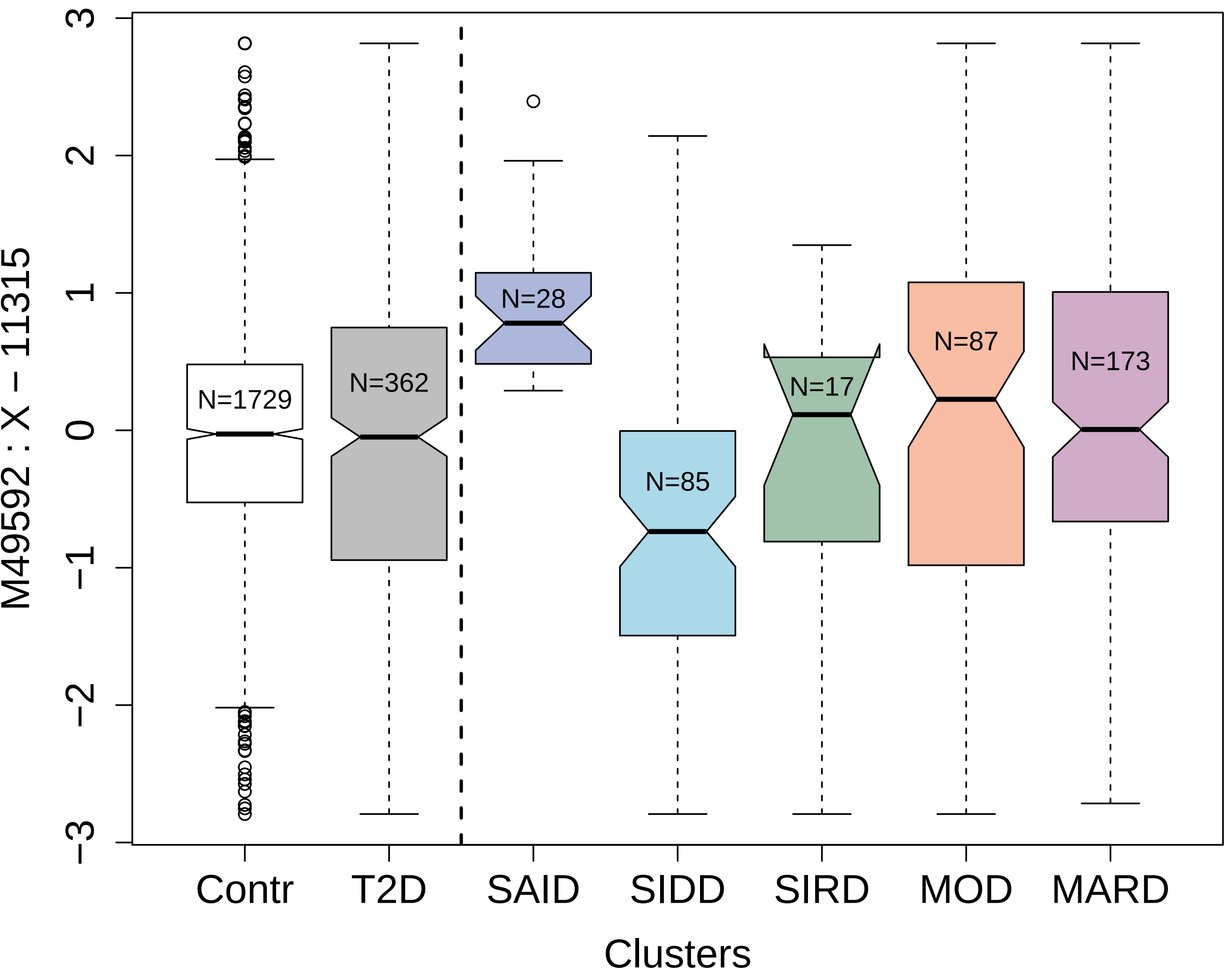

M49592 : X − 11315
3
2
1
0
−1
−2
−3
N=1729
N=362
N=28
N=85
N=17
N=87
N=173
Contr
T2D
SAID
SIDD
SIRD
MOD
MARD
Clusters

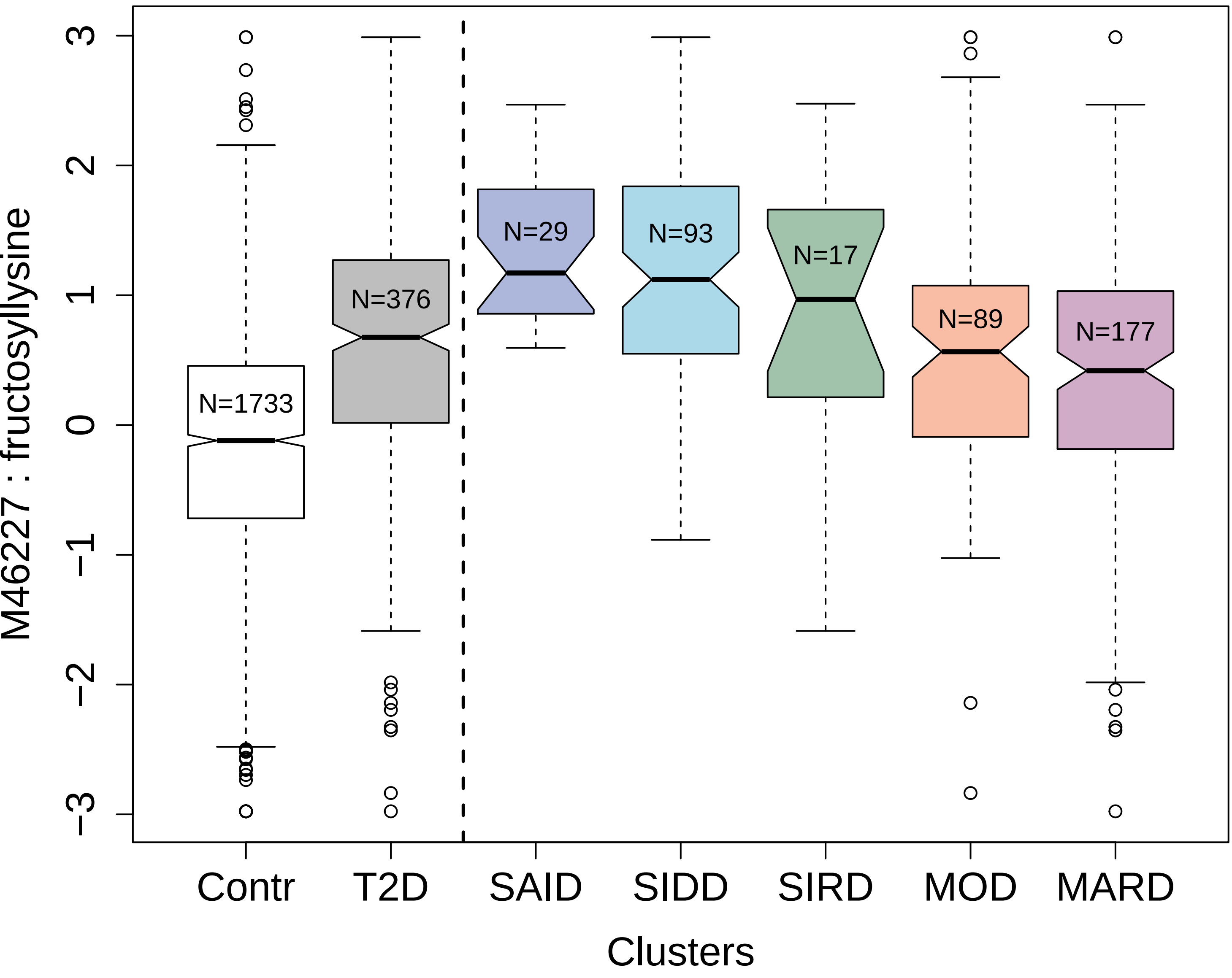

M46227 : fructosyllysine
3
2
1
0
−1
−2
−3
N=1733
N=376
N=29
N=93
N=17
N=89
N=177
Contr
T2D
SAID
SIDD
SIRD
MOD
MARD
Clusters

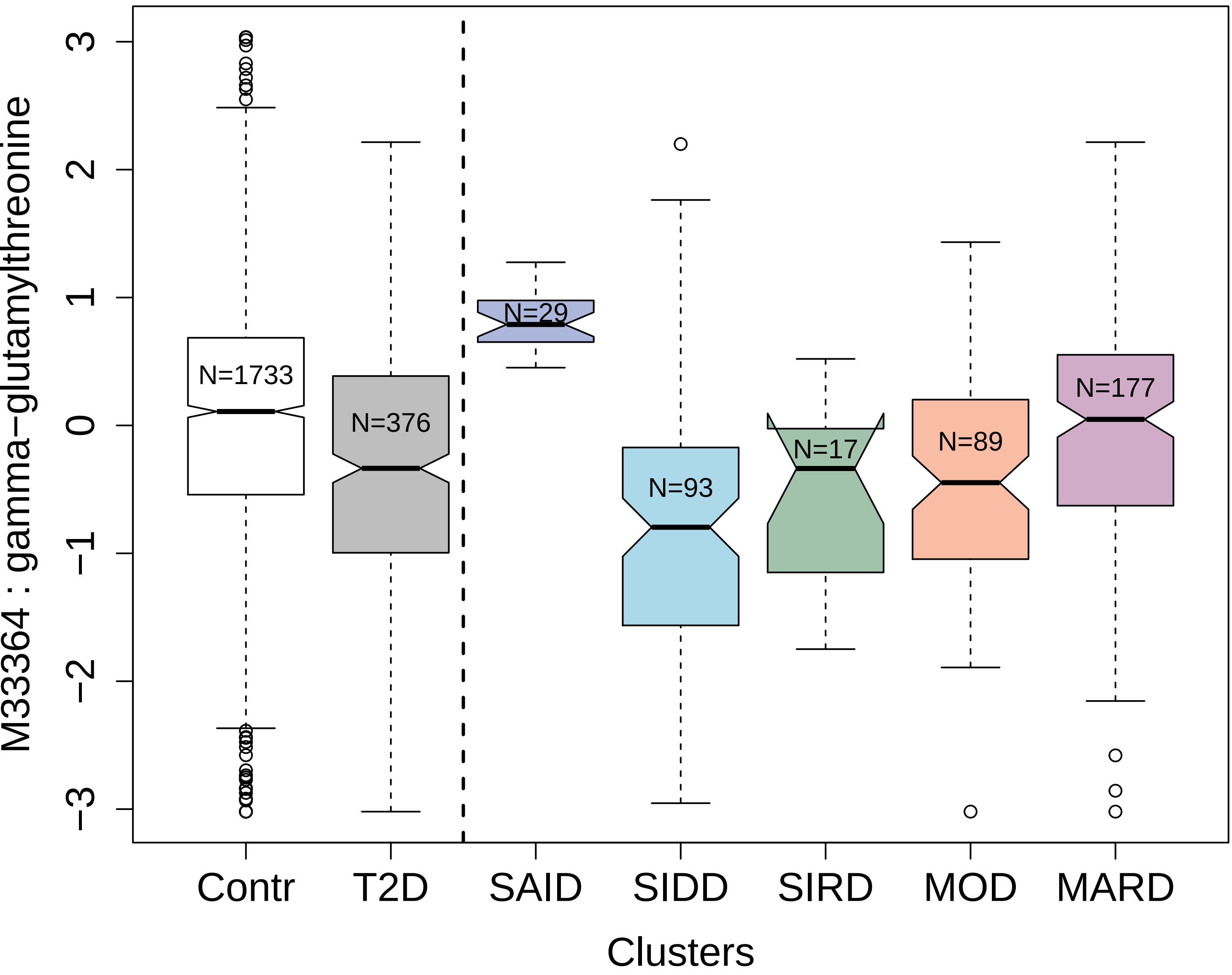

M33364 : gamma−glutamylthreonine
3
2
1
0
−1
−2
−3
N=1733
N=376
N=29
N=93
N=17
N=89
N=177
Contr
T2D
SAID
SIDD
SIRD
MOD
MARD
Clusters

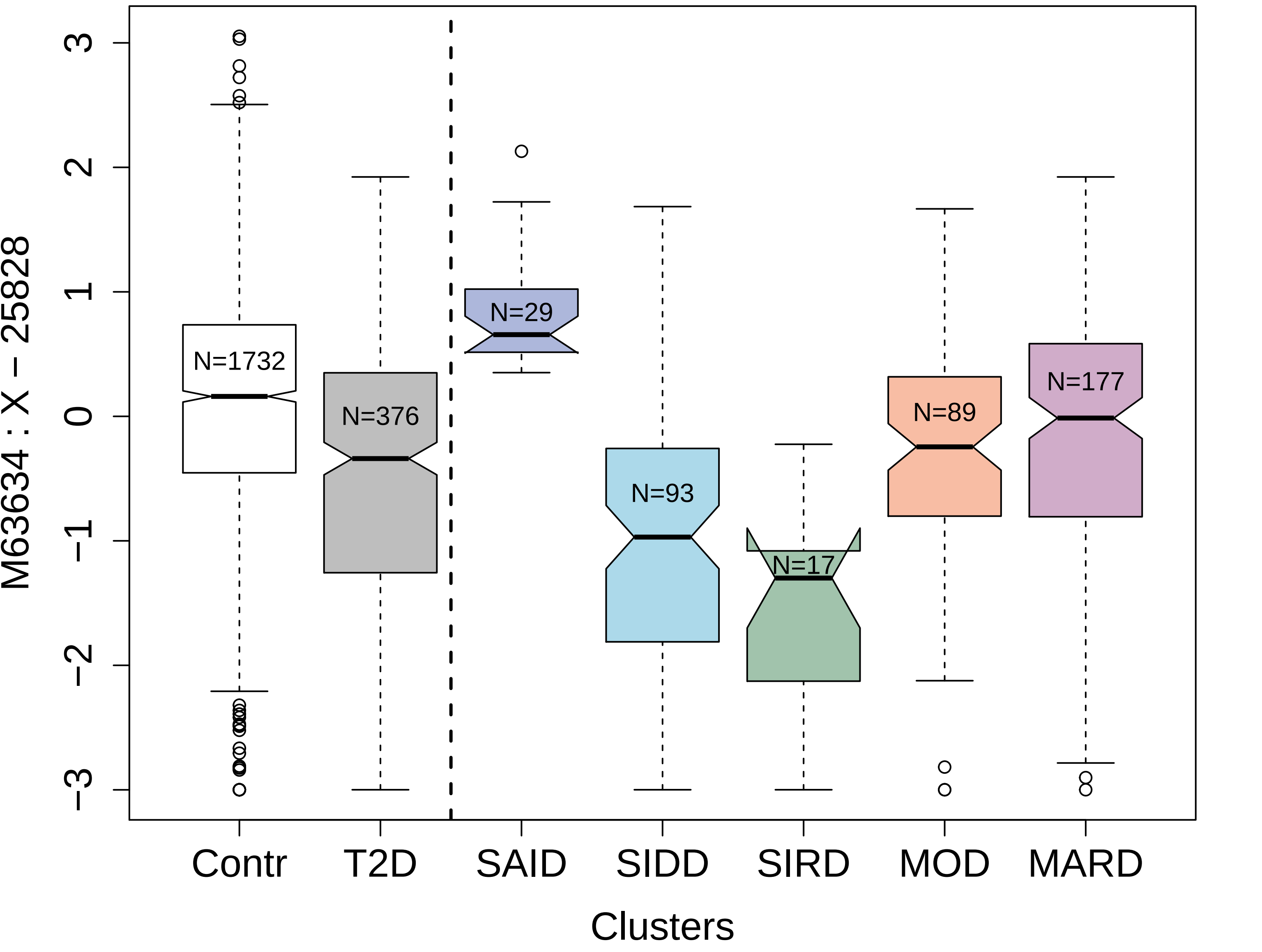

M63634 : X − 25828
3
2
1
0
−1
−2
−3
N=1732
N=376
N=29
N=93
N=17
N=89
N=177
Contr
T2D
SAID
SIDD
SIRD
MOD
MARD
Clusters

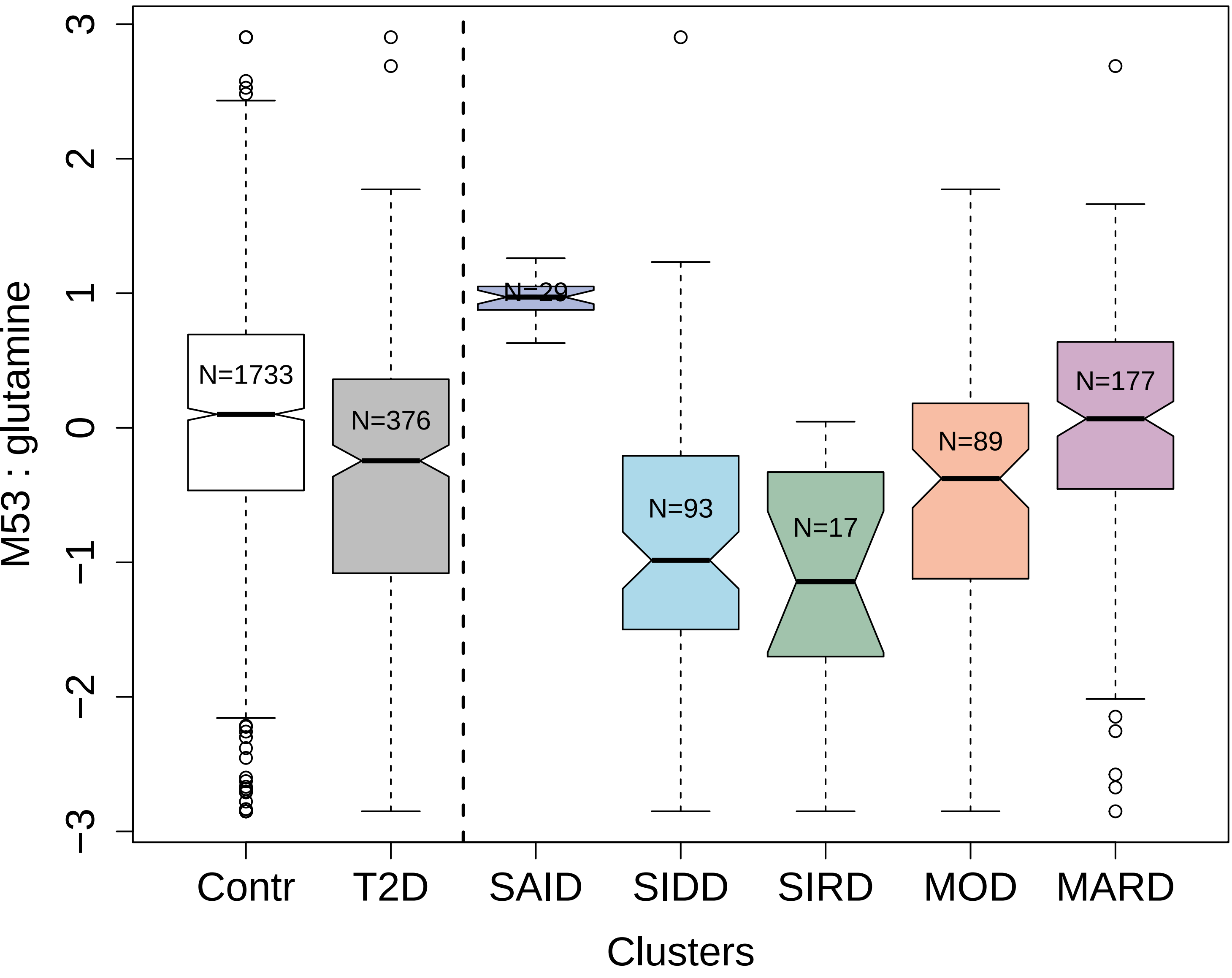
M53 : glutamine
3
2
1
0
−1
−2
−3
N=1733
N=376
N=29
N=93
N=17
N=89
N=177
Contr
T2D
SAID
SIDD
SIRD
MOD
MARD
Clusters

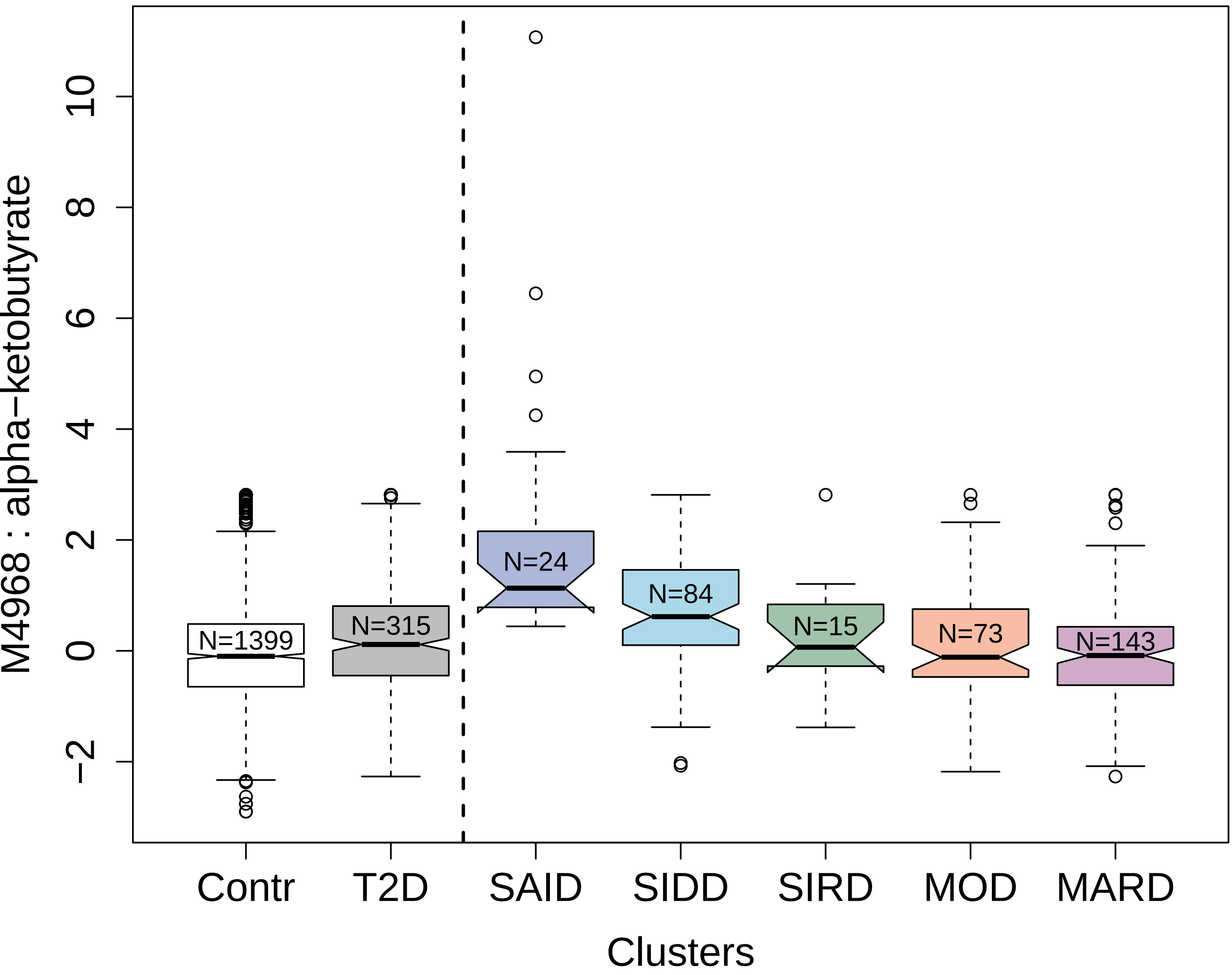
M4968 : alpha−ketobutyrate
10
8
6
4
2
0
−2
N=1399
N=315
N=24
N=84
N=15
N=73
N=143
Contr
T2D
SAID
SIDD
SIRD
MOD
MARD
Clusters

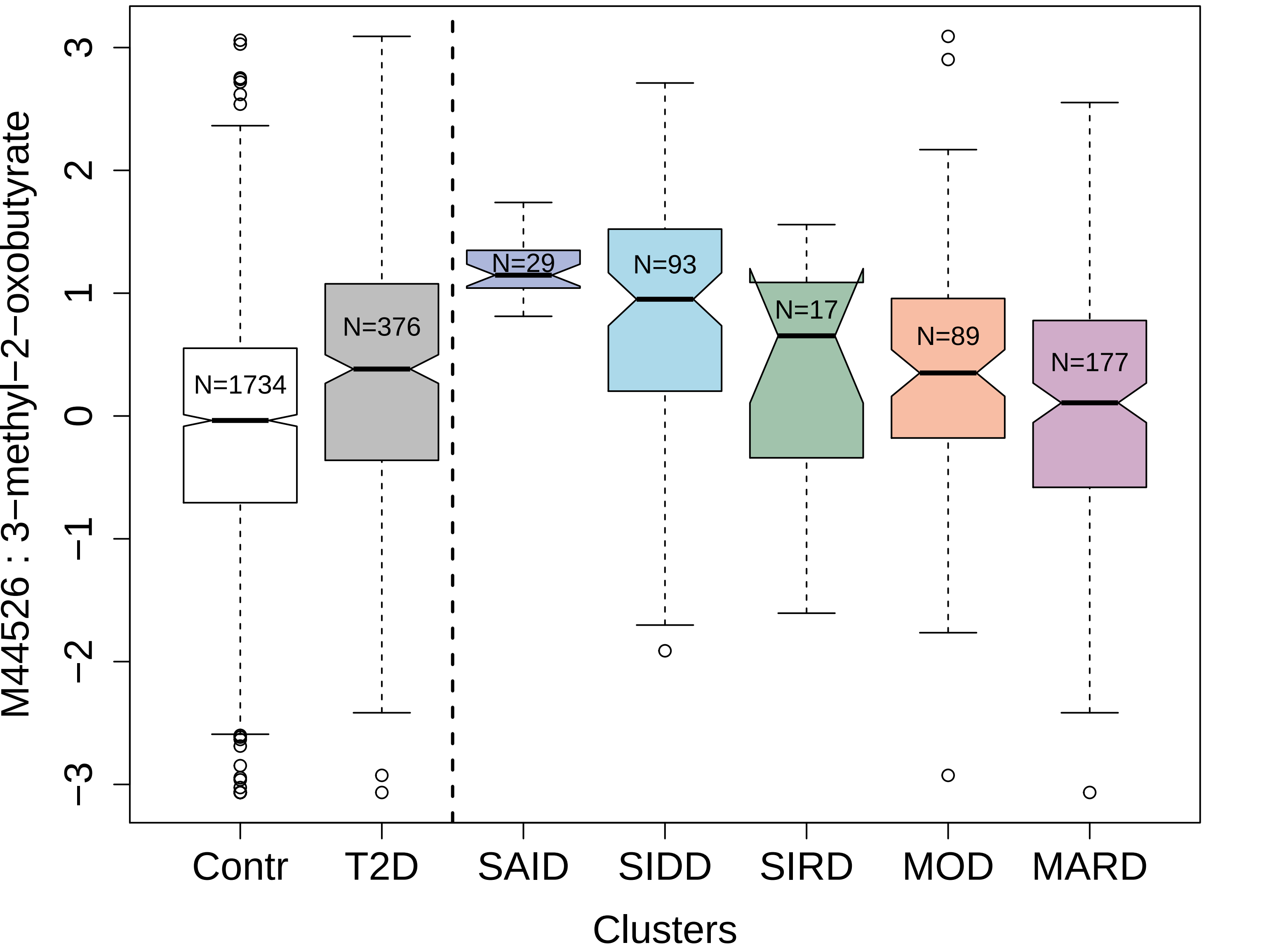

M44526 : 3−methyl−2−oxobutyrate
3
2
1
0
−1
−2
−3
N=1734
N=376
N=29
N=93
N=17
N=89
N=177
Contr
T2D
SAID
SIDD
SIRD
MOD
MARD
Clusters

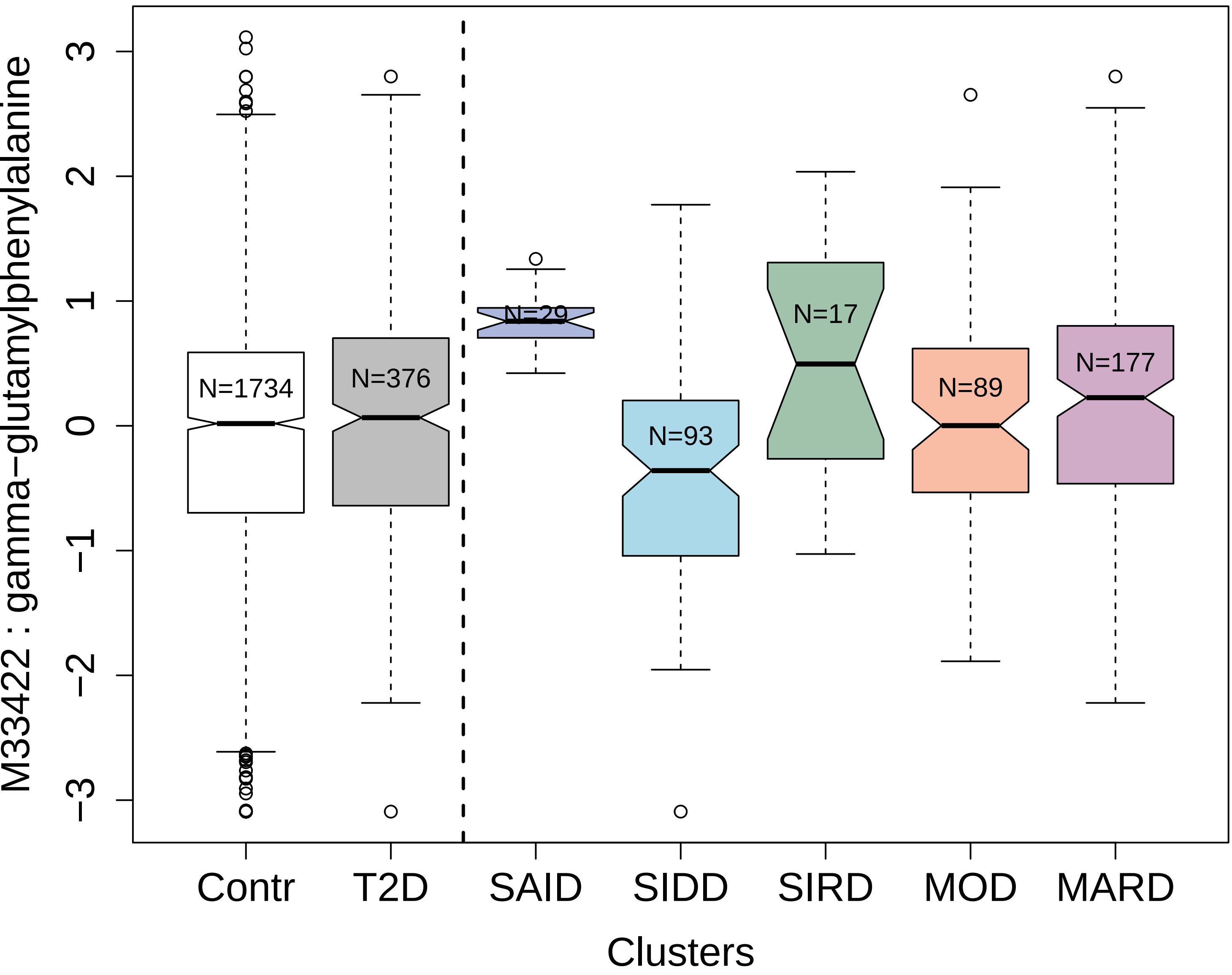

M33422 : gamma−glutamylphenylalanine
3
2
1
0
−1
−2
−3
N=1734
N=376
N=29
N=93
N=17
N=89
N=177
Contr
T2D
SAID
SIDD
SIRD
MOD
MARD
Clusters

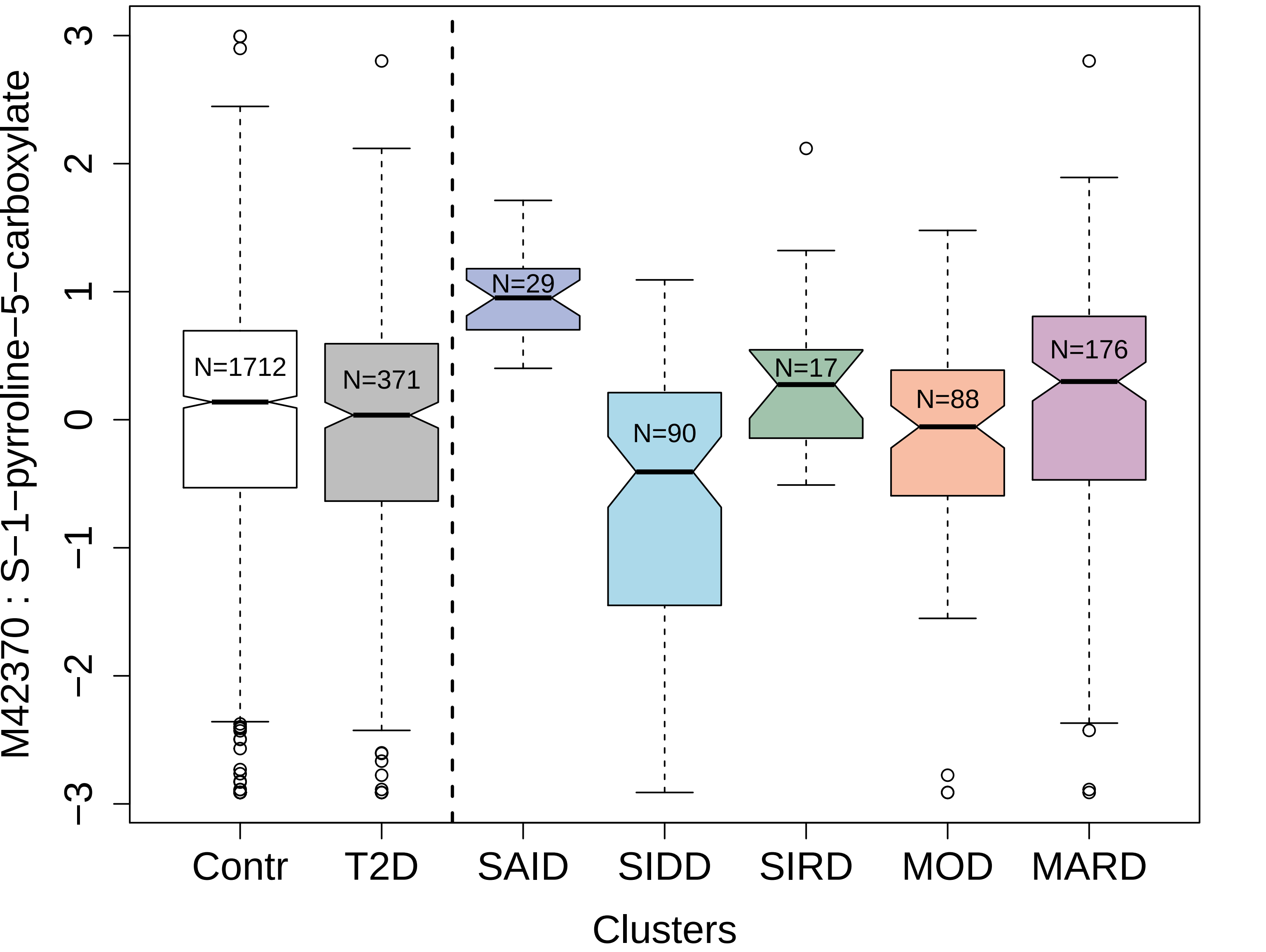
M42370 : S−1−pyrroline−5−carboxylate
3
2
1
0
−1
−2
−3
N=1712
N=371
N=29
N=90
N=17
N=88
N=176
Contr
T2D
SAID
SIDD
SIRD
MOD
MARD
Clusters

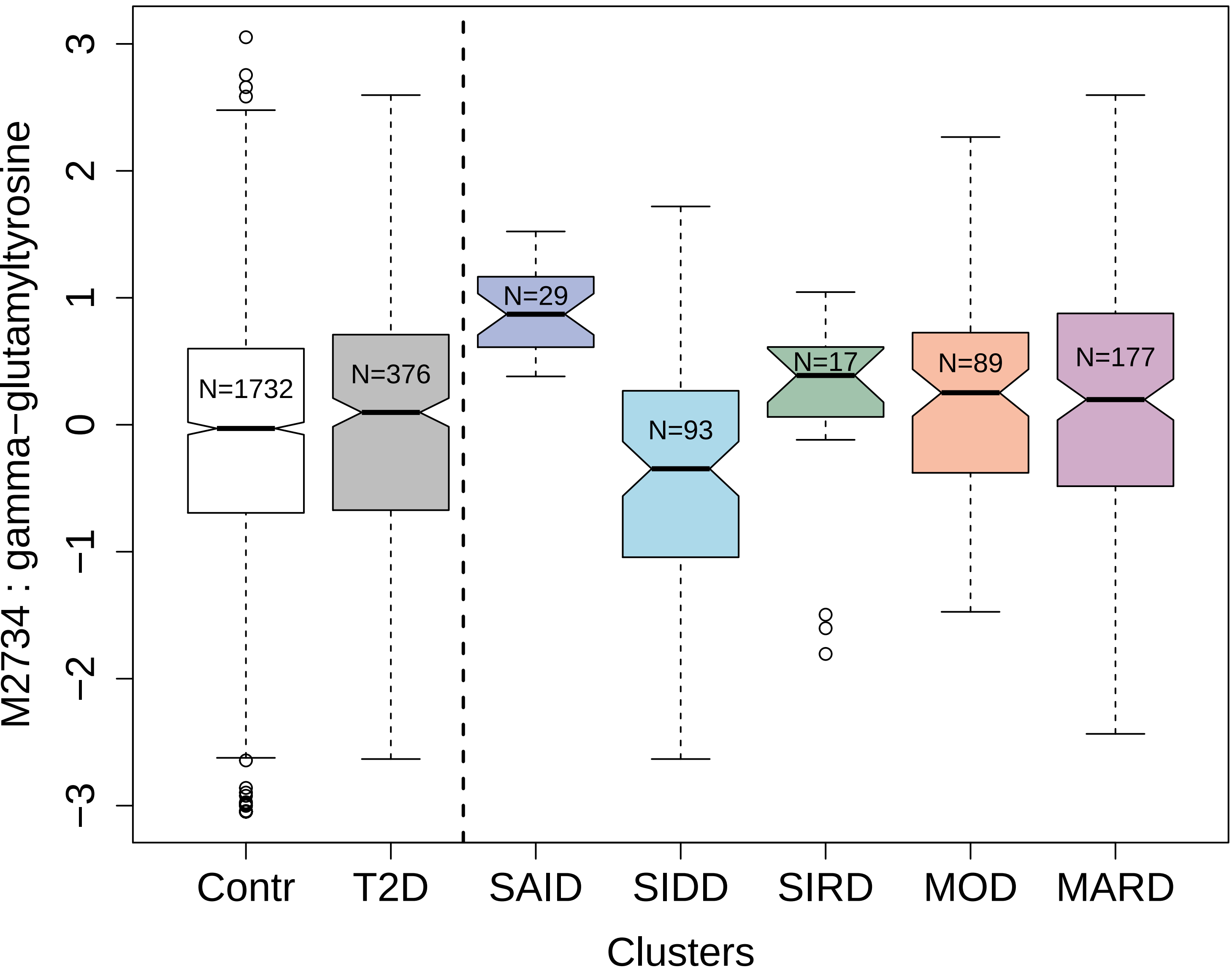
M2734 : gamma−glutamyltyrosine
3
2
1
0
−1
−2
−3
N=1732
N=376
N=29
N=93
N=17
N=89
N=177
Contr
T2D
SAID
SIDD
SIRD
MOD
MARD
Clusters

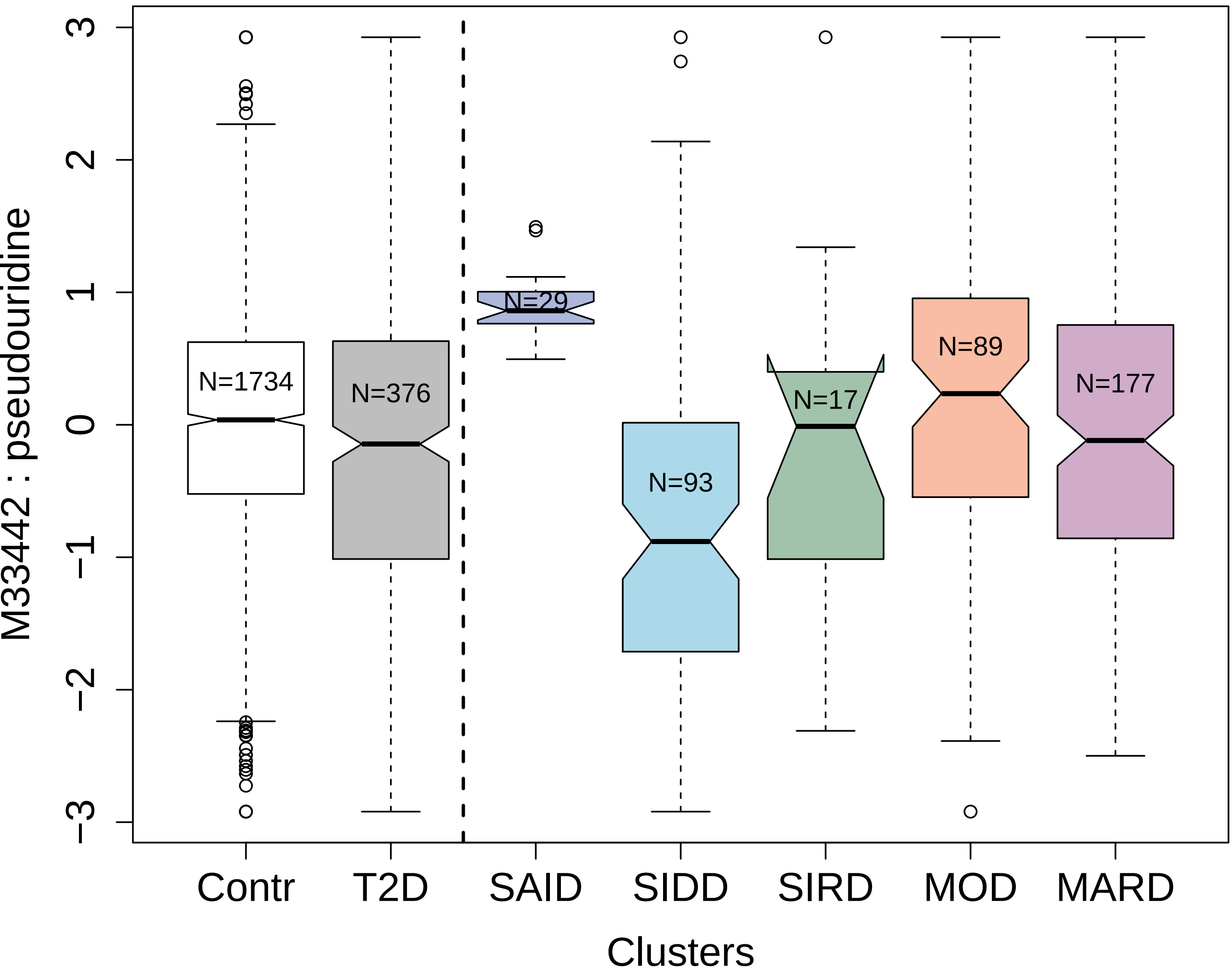

M33442 : pseudouridine
3
2
1
0
−1
−2
−3
N=1734
N=376
N=29
N=93
N=17
N=89
N=177
Contr
T2D
SAID
SIDD
SIRD
MOD
MARD
Clusters

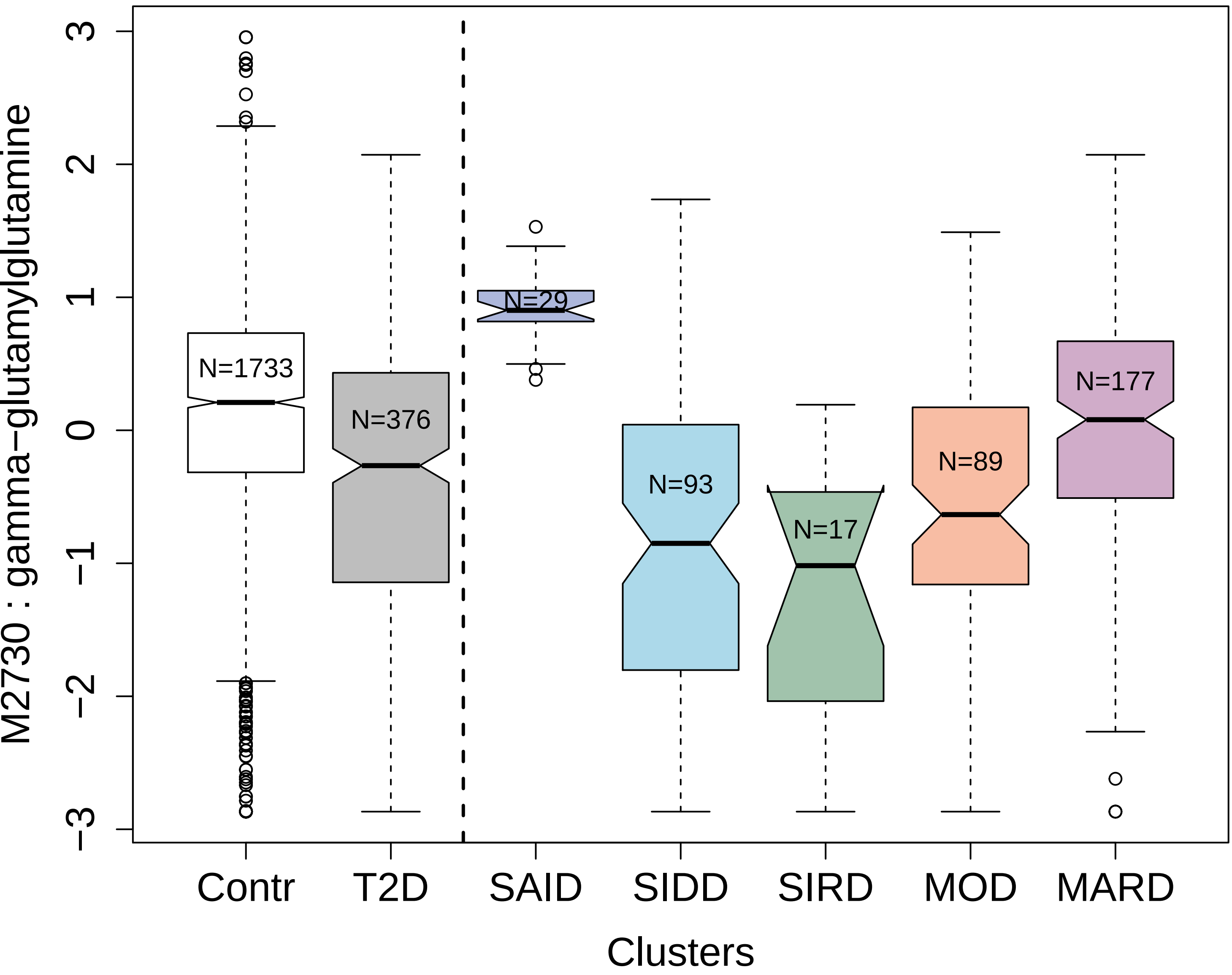

M2730 : gamma−glutamylglutamine
3
2
1
0
−1
−2
−3
N=1733
N=376
N=29
N=93
N=17
N=89
N=177
Contr
T2D
SAID
SIDD
SIRD
MOD
MARD
Clusters

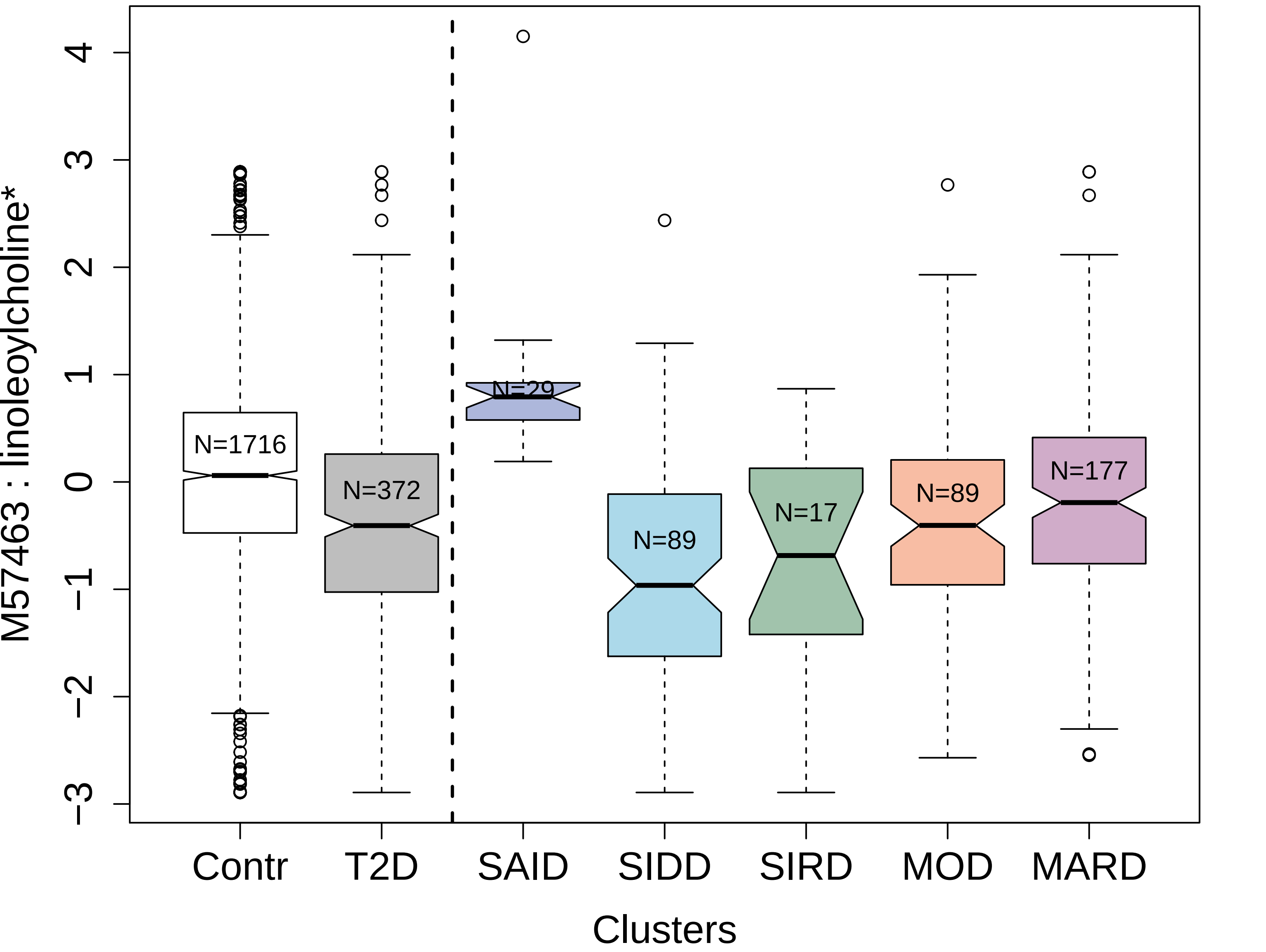
M57463 : linoleoylcholine*
Clusters
Contr
T2D
SAID
SIDD
SIRD
MOD
MARD
N=1716
N=372
N=29
N=89
N=17
N=89
N=177
4
3
2
1
0
−1
−2
−3

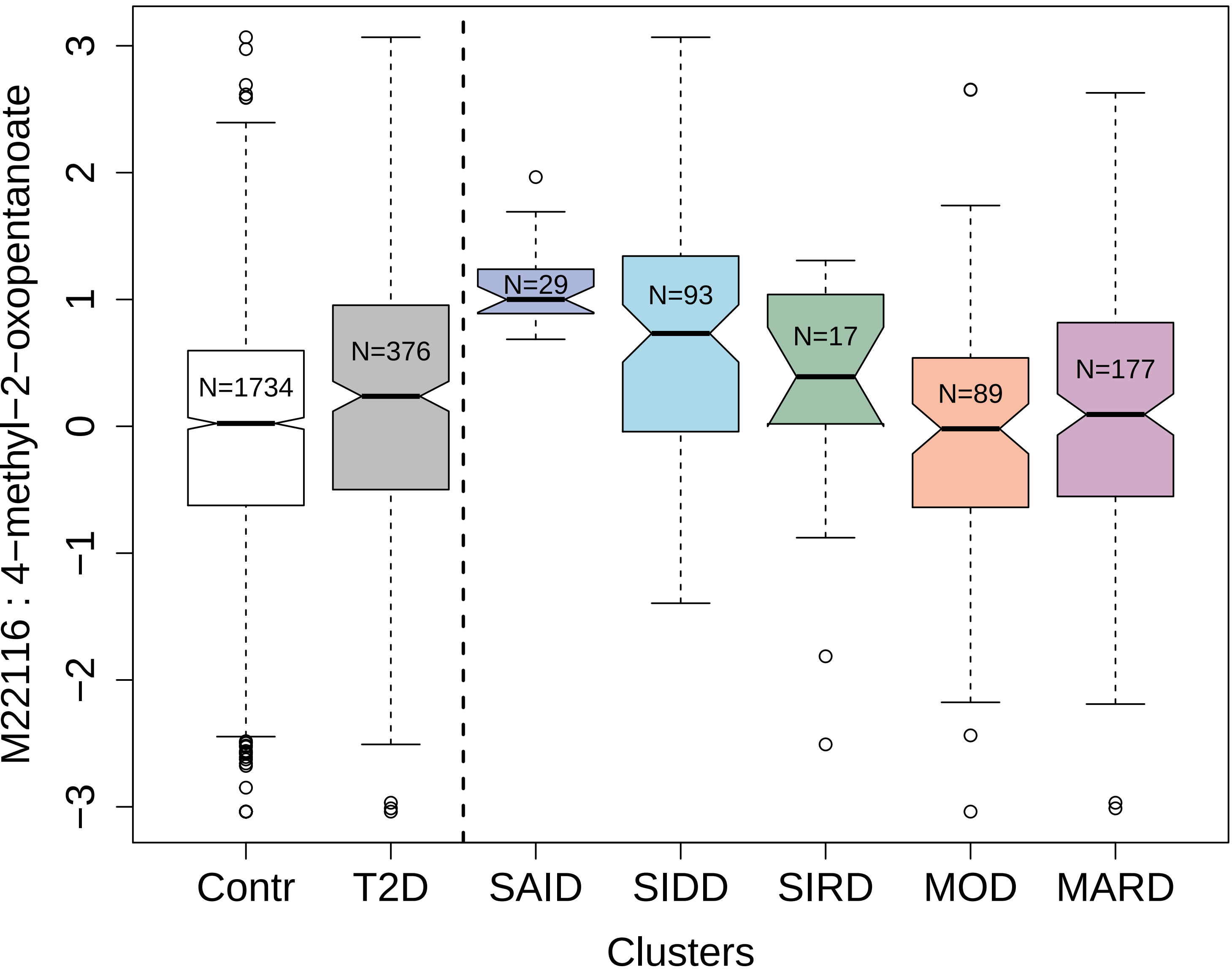
M22116 : 4−methyl−2−oxopentanoate
3
2
1
0
−1
−2
−3
N=1734
N=376
N=29
N=93
N=17
N=89
N=177
Contr
T2D
SAID
SIDD
SIRD
MOD
MARD
Clusters

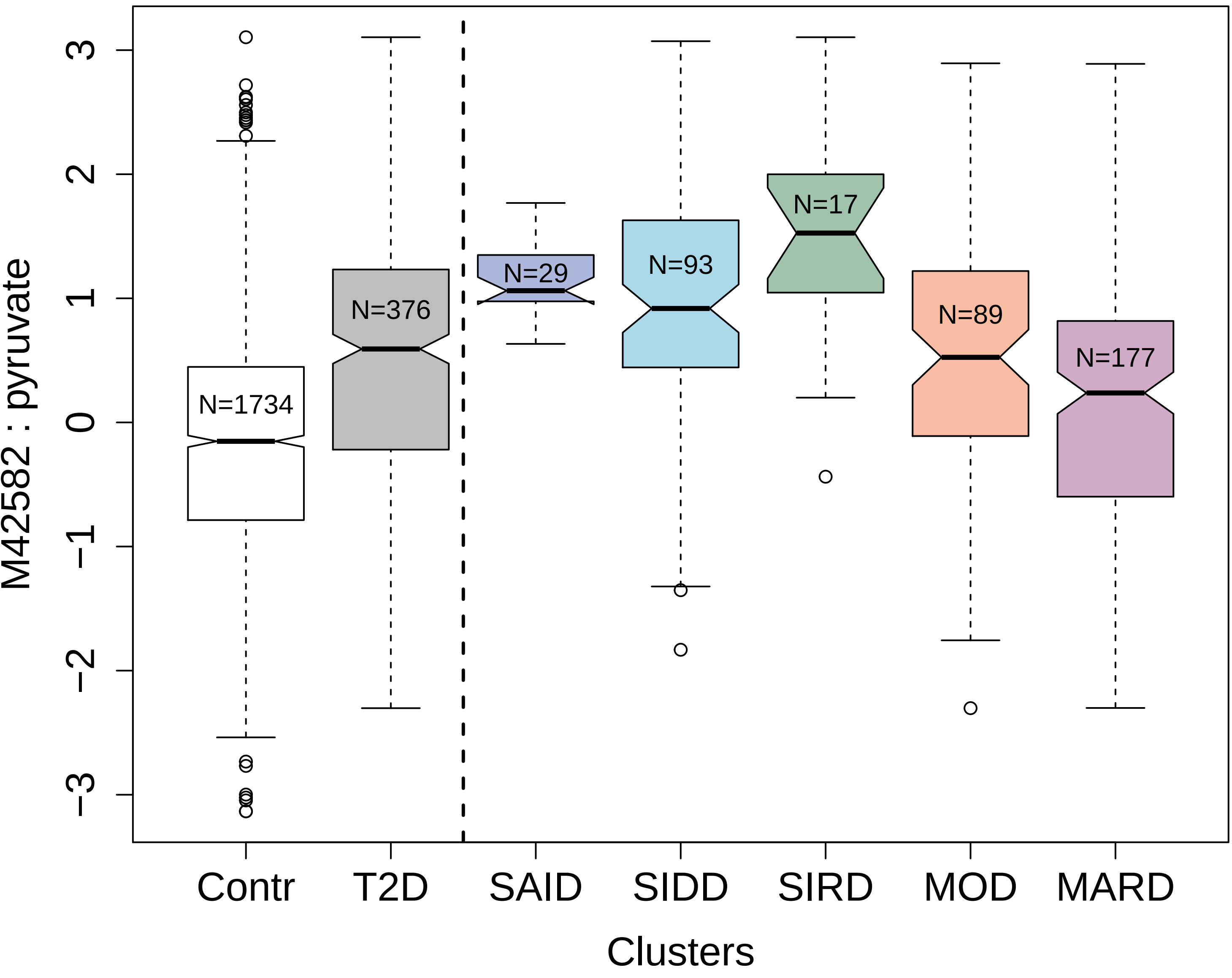

M42582 : pyruvate
3
2
1
0
−1
−2
−3
N=1734
N=376
N=29
N=93
N=17
N=89
N=177
Contr
T2D
SAID
SIDD
SIRD
MOD
MARD
Clusters

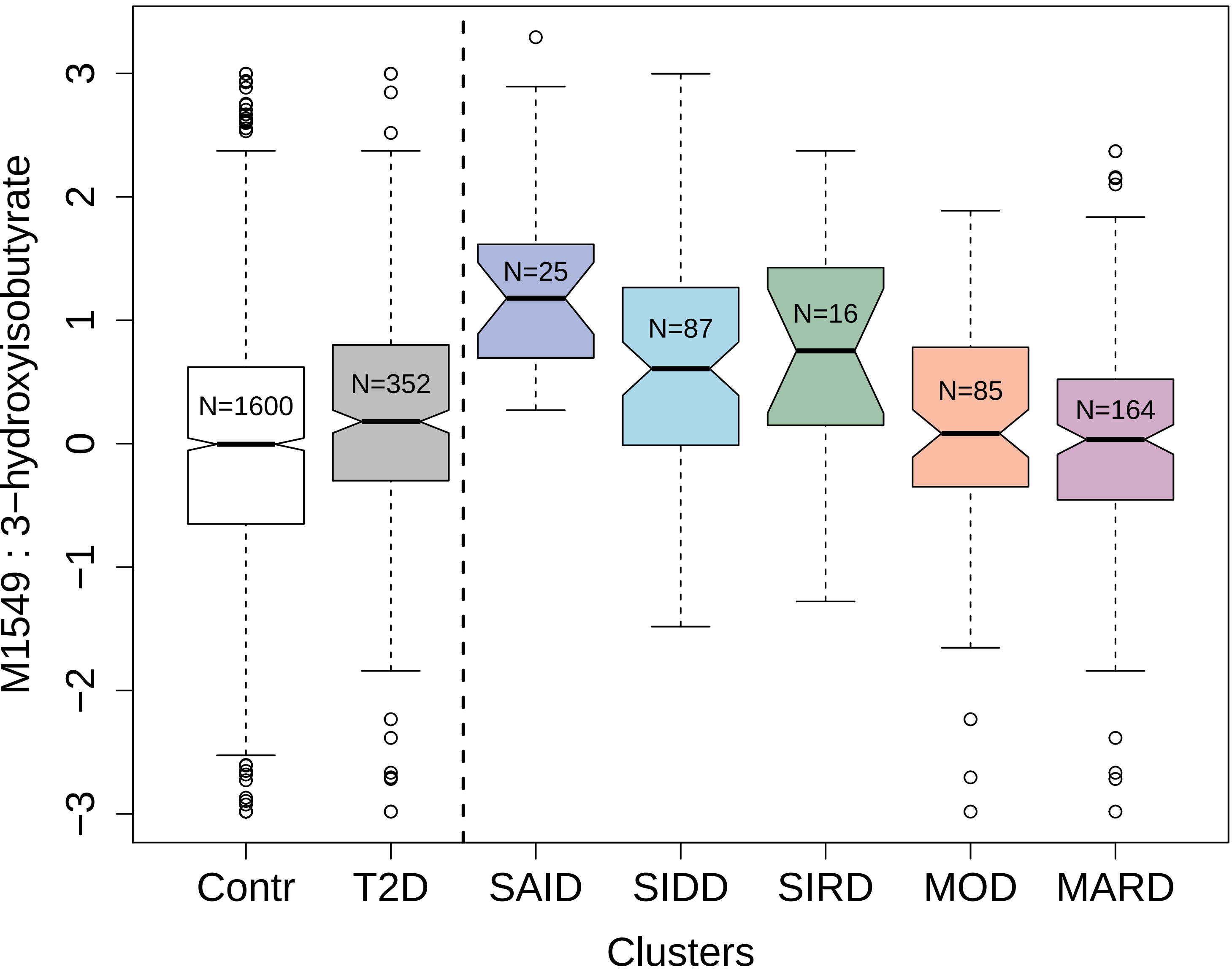

M1549 : 3−hydroxyisobutyrate
3
2
1
0
−1
−2
−3
N=1600
N=352
N=25
N=87
N=16
N=85
N=164
Contr
T2D
SAID
SIDD
SIRD
MOD
MARD
Clusters

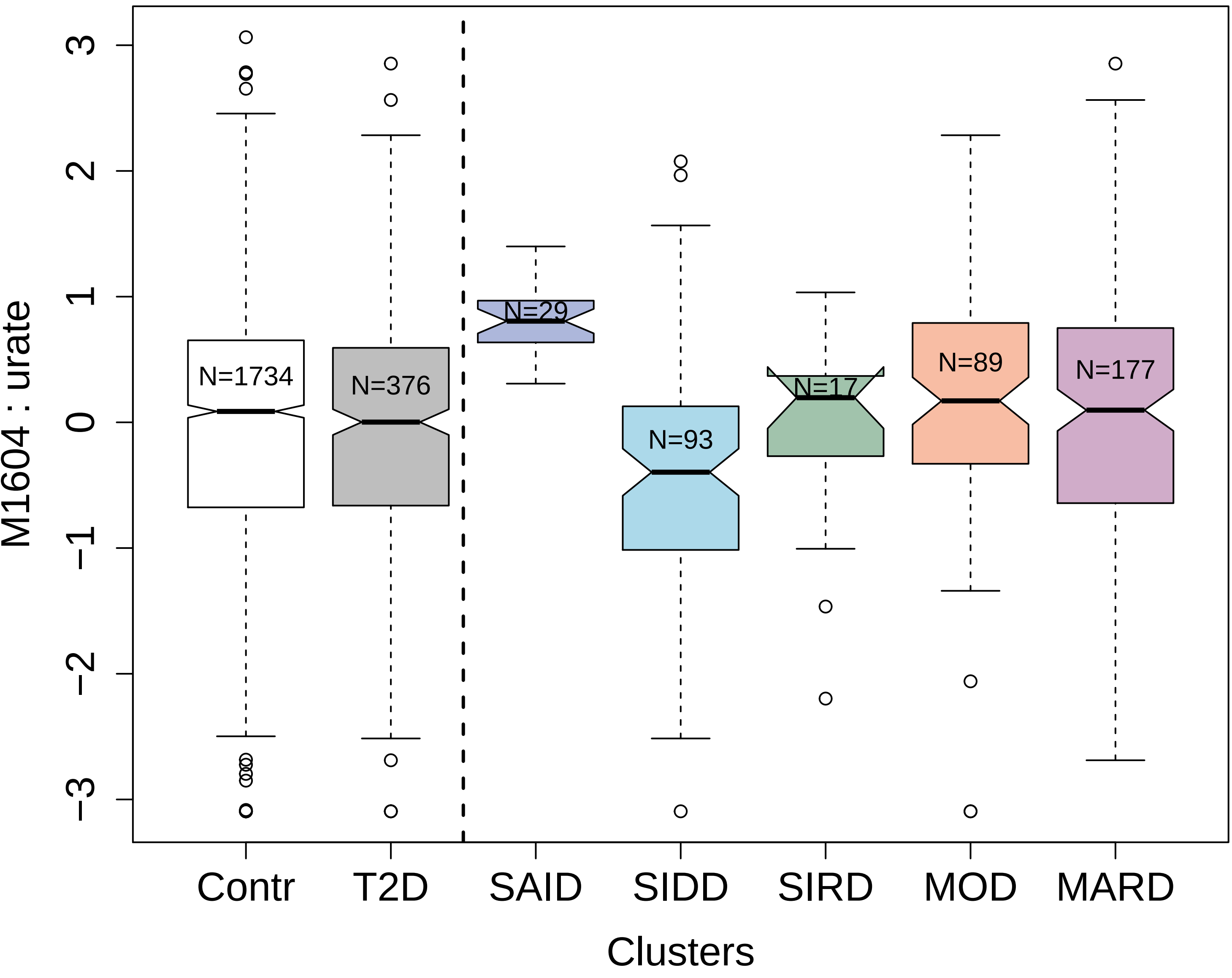

M1604 : urate
3
2
1
0
−1
−2
−3
N=1734
N=376
N=29
N=93
N=17
N=89
N=177
Contr
T2D
SAID
SIDD
SIRD
MOD
MARD
Clusters

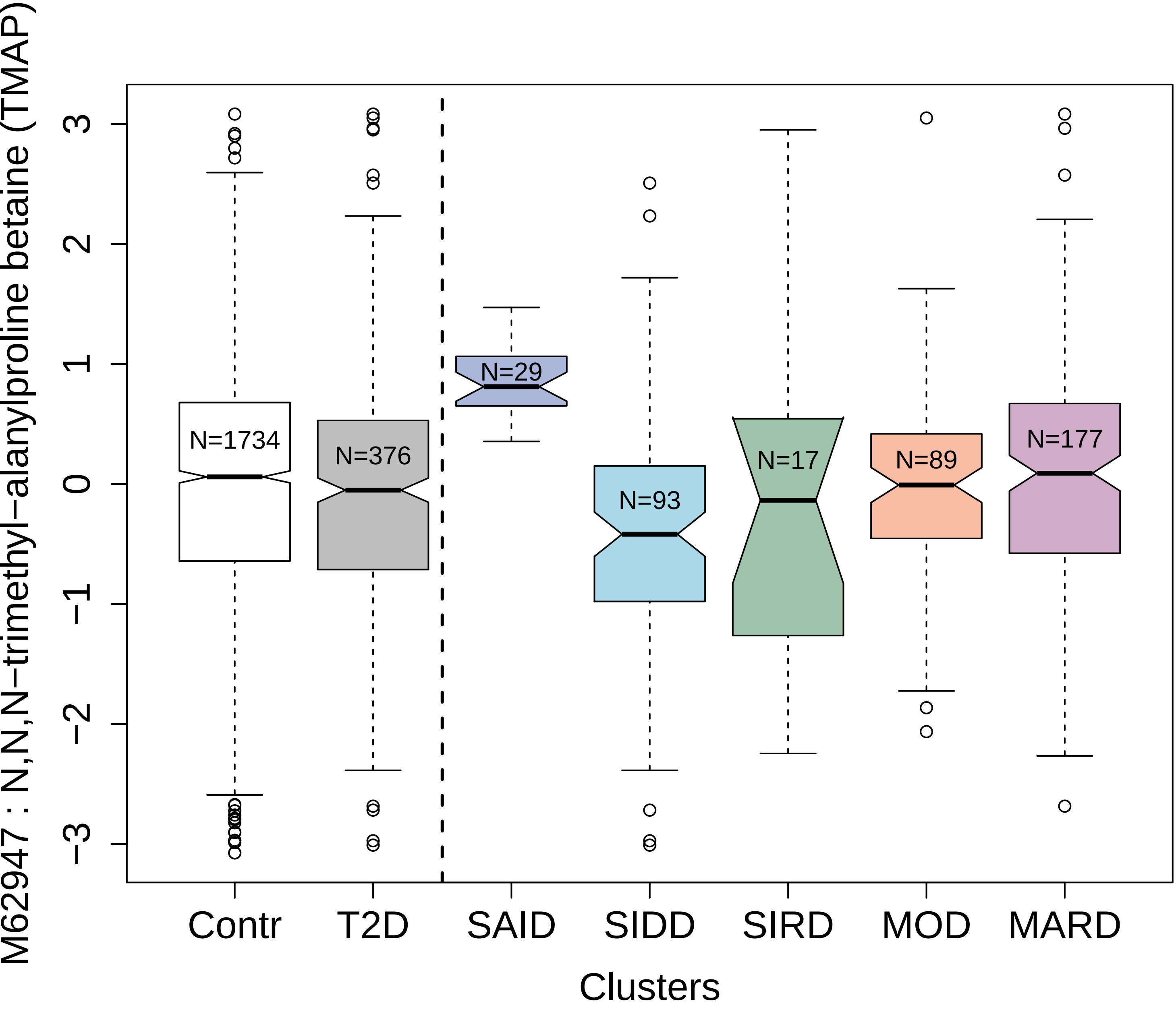

M62947 : N,N,N−trimethyl−alanylproline betaine (TMAP)
3
2
1
0
−1
−2
−3
N=1734
N=376
N=29
N=93
N=17
N=89
N=177
Contr
T2D
SAID
SIDD
SIRD
MOD
MARD
Clusters

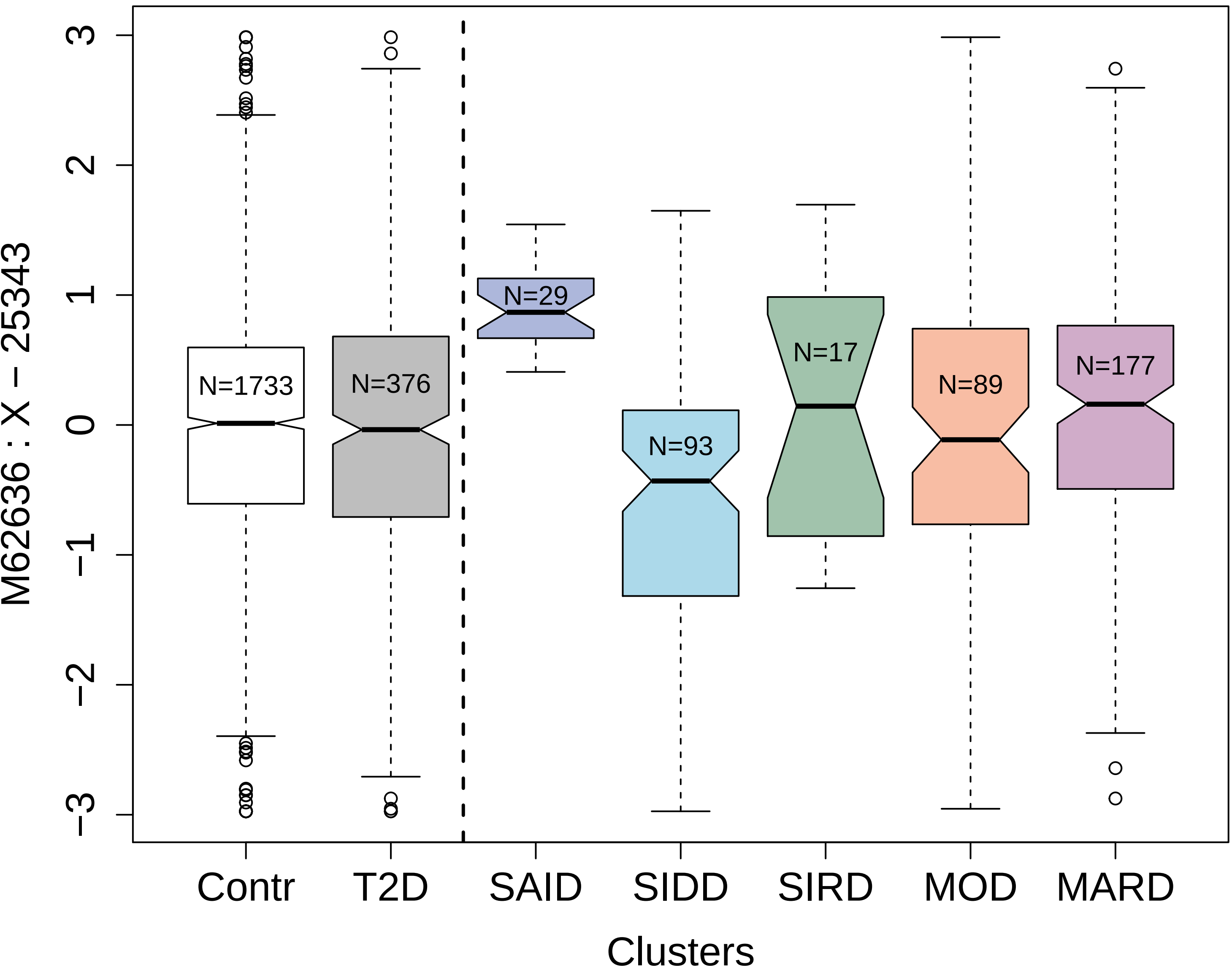

M62636 : X − 25343
3
2
1
0
−1
−2
−3
N=1733
N=376
N=29
N=93
N=17
N=89
N=177
Contr
T2D
SAID
SIDD
SIRD
MOD
MARD
Clusters

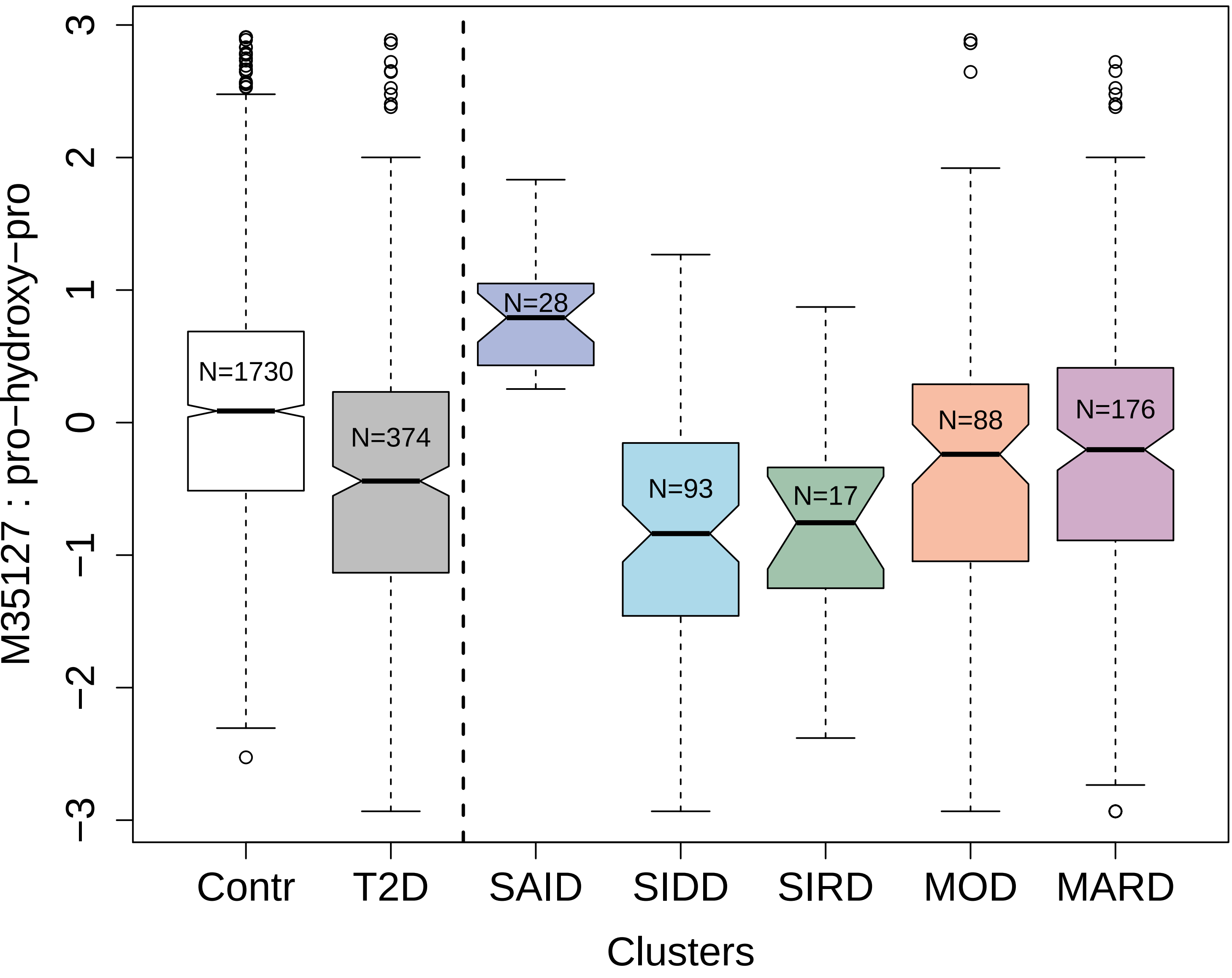

M35127 : pro−hydroxy−pro
3
2
1
0
−1
−2
−3
N=1730
N=374
N=28
N=93
N=17
N=88
N=176
Contr
T2D
SAID
SIDD
SIRD
MOD
MARD
Clusters

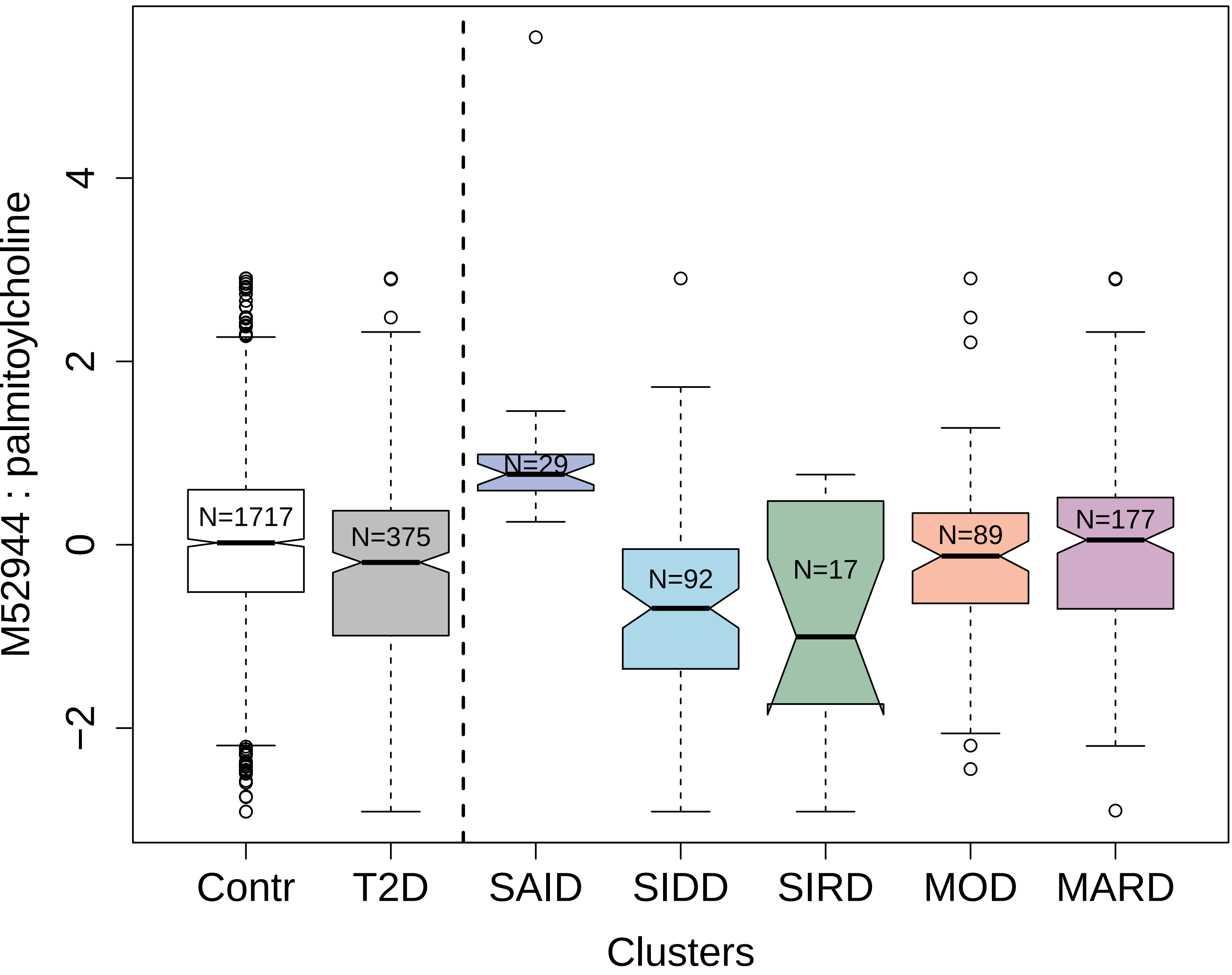

M52944 : palmitoylcholine
4
2
0
−2
N=1717
N=375
N=29
N=92
N=17
N=89
N=177
Contr
T2D
SAID
SIDD
SIRD
MOD
MARD
Clusters

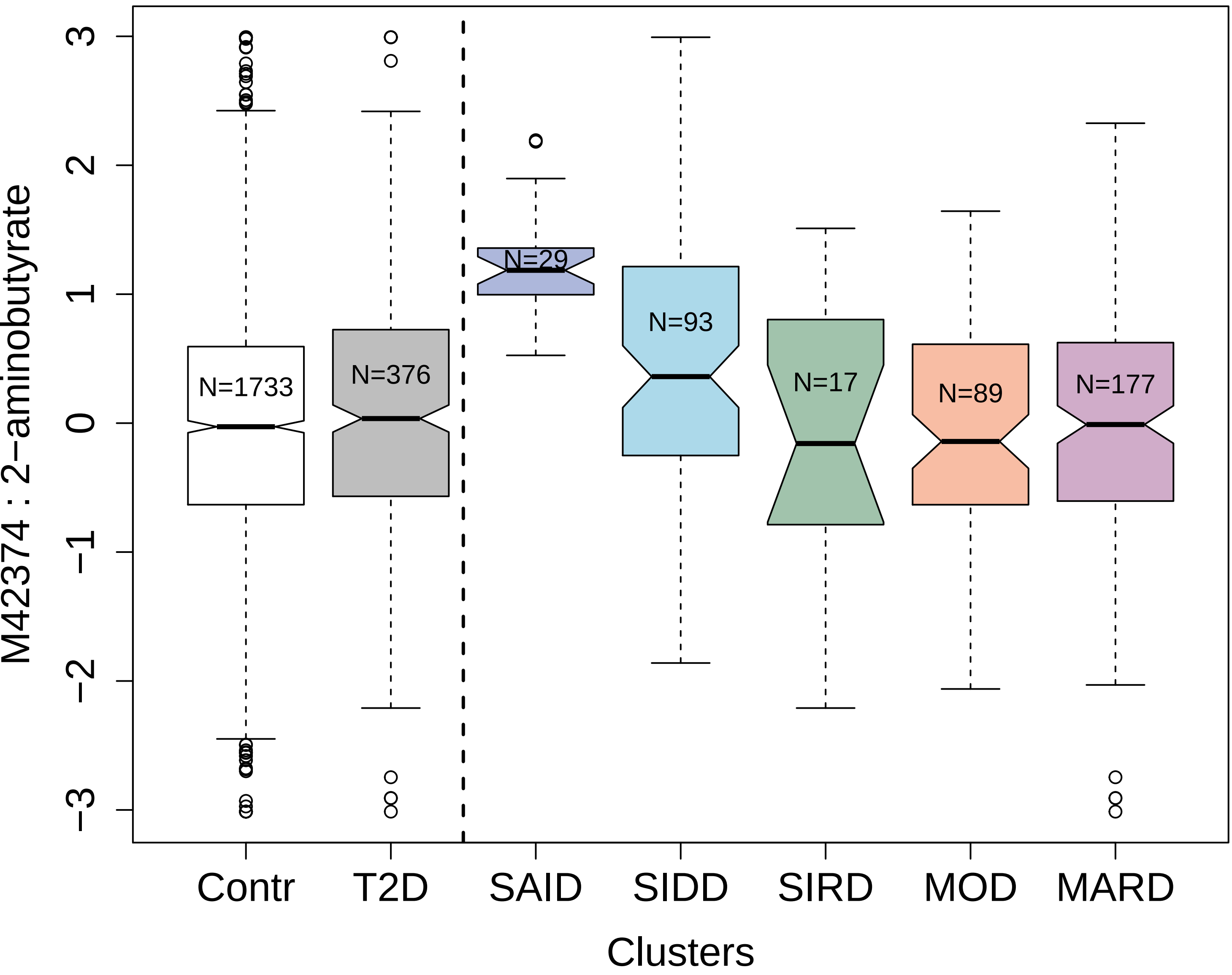
M42374 : 2−aminobutyrate
3
2
1
0
−1
−2
−3
N=1733
N=376
N=29
N=93
N=17
N=89
N=177
Contr
T2D
SAID
SIDD
SIRD
MOD
MARD
Clusters

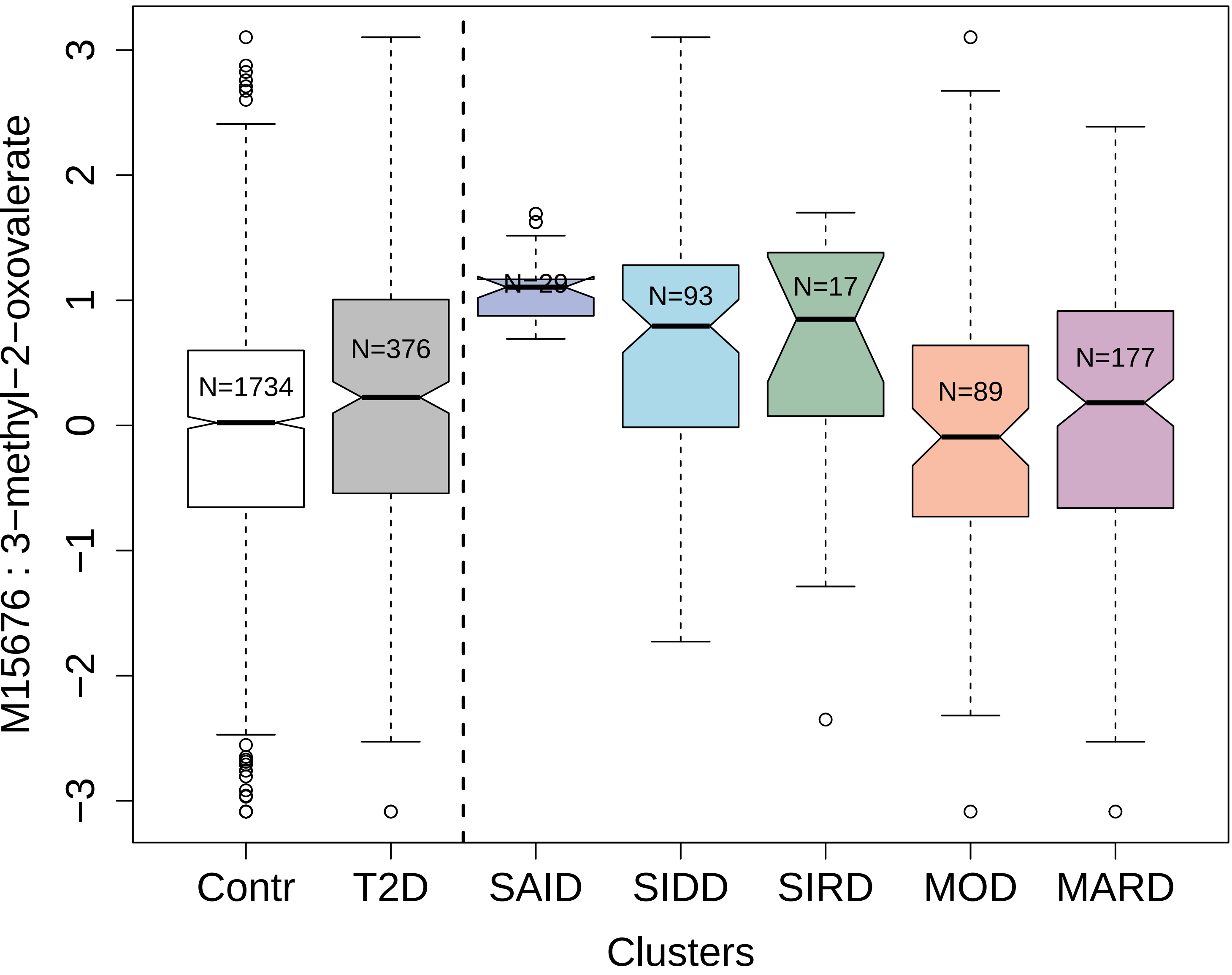

M15676 : 3−methyl−2−oxovalerate
3
2
1
0
−1
−2
−3
N=1734
N=376
N=29
N=93
N=17
N=89
N=177
Contr
T2D
SAID
SIDD
SIRD
MOD
MARD
Clusters

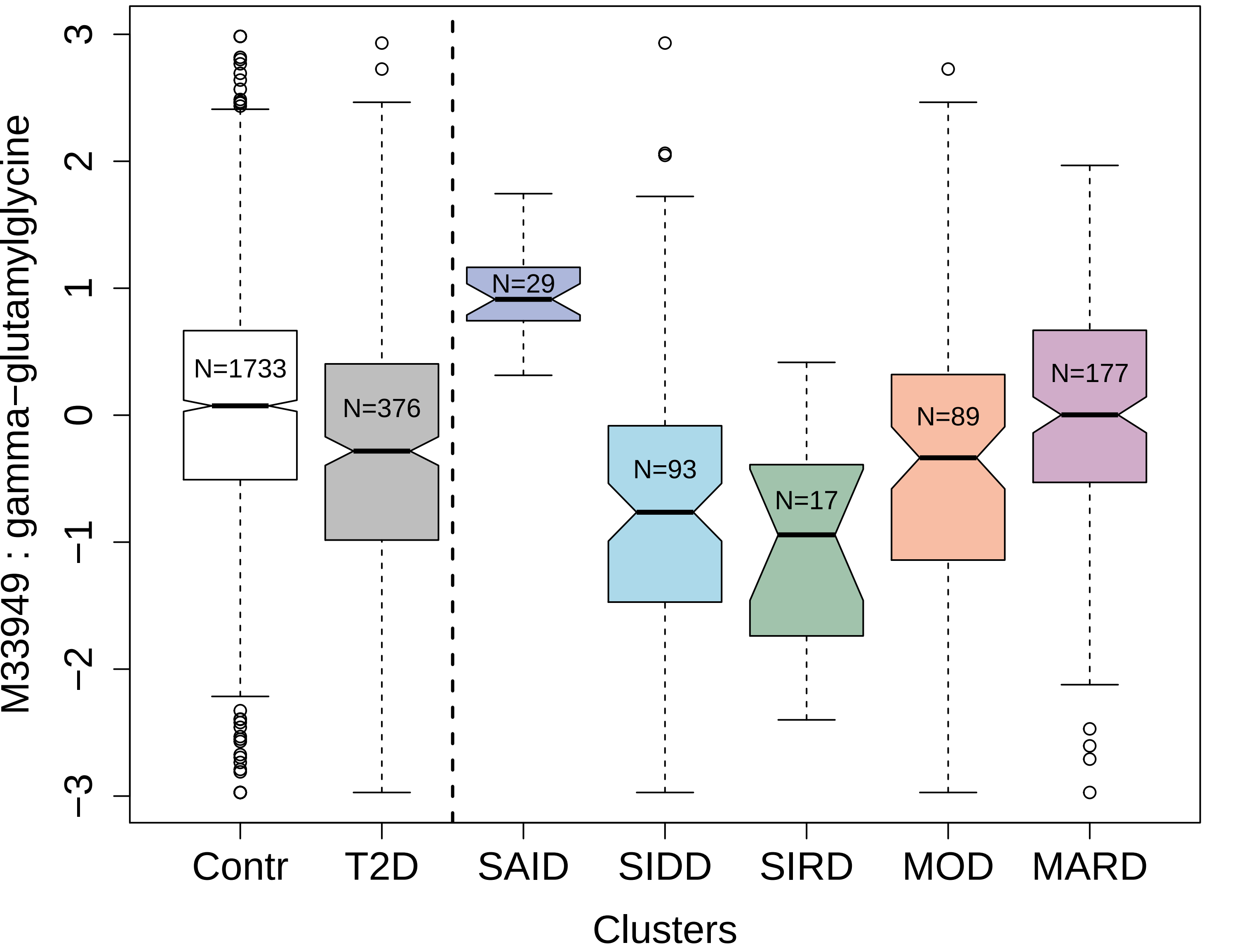

M33949 : gamma−glutamylglycine
3
2
1
0
−1
−2
−3
N=1733
N=376
N=29
N=93
N=17
N=89
N=177
Contr
T2D
SAID
SIDD
SIRD
MOD
MARD
Clusters

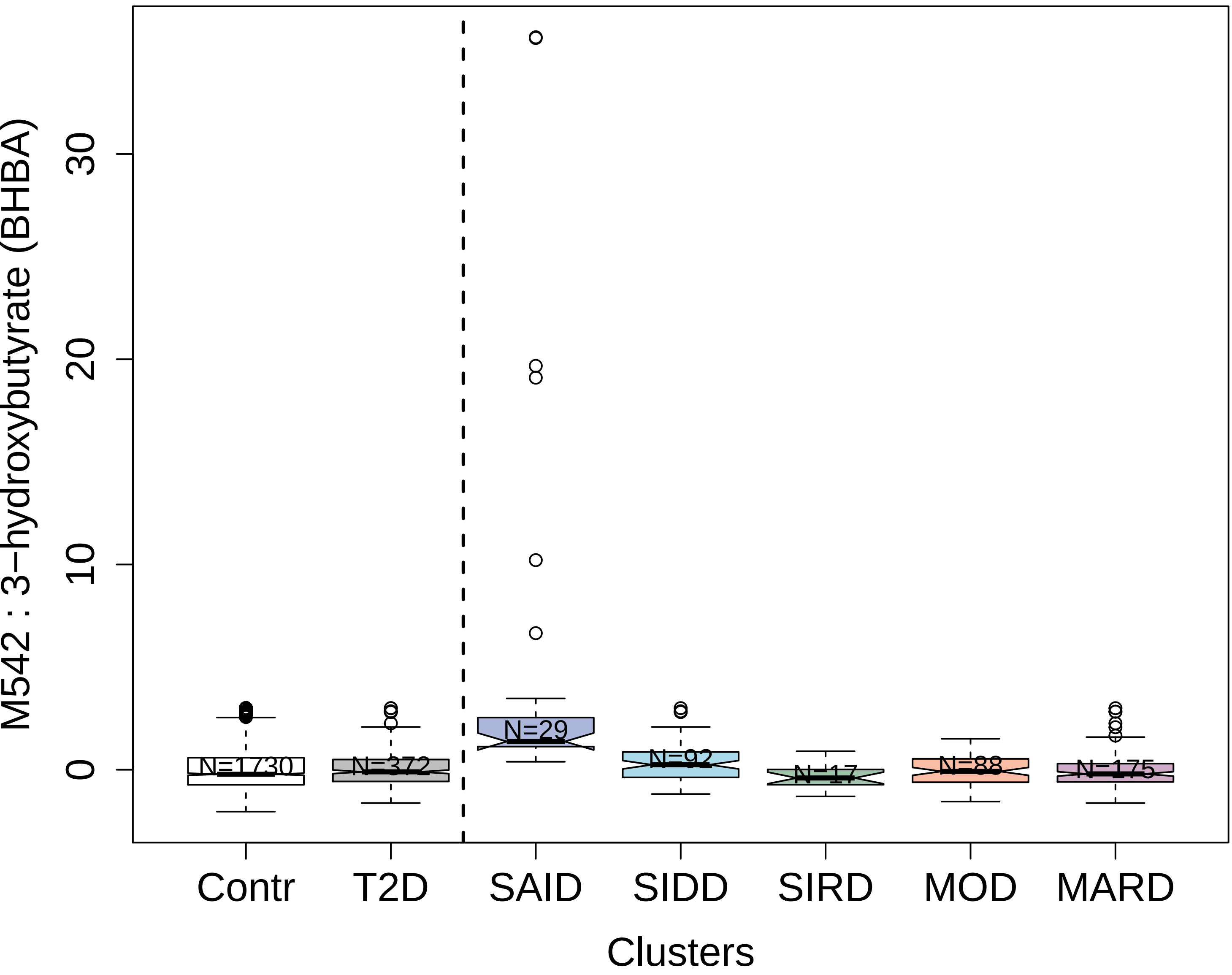

M542 : 3−hydroxybutyrate (BHBA)
0
10
20
30
N=1730
N=372
N=29
N=92
N=17
N=88
N=175
Contr
T2D
SAID
SIDD
SIRD
MOD
MARD
Clusters

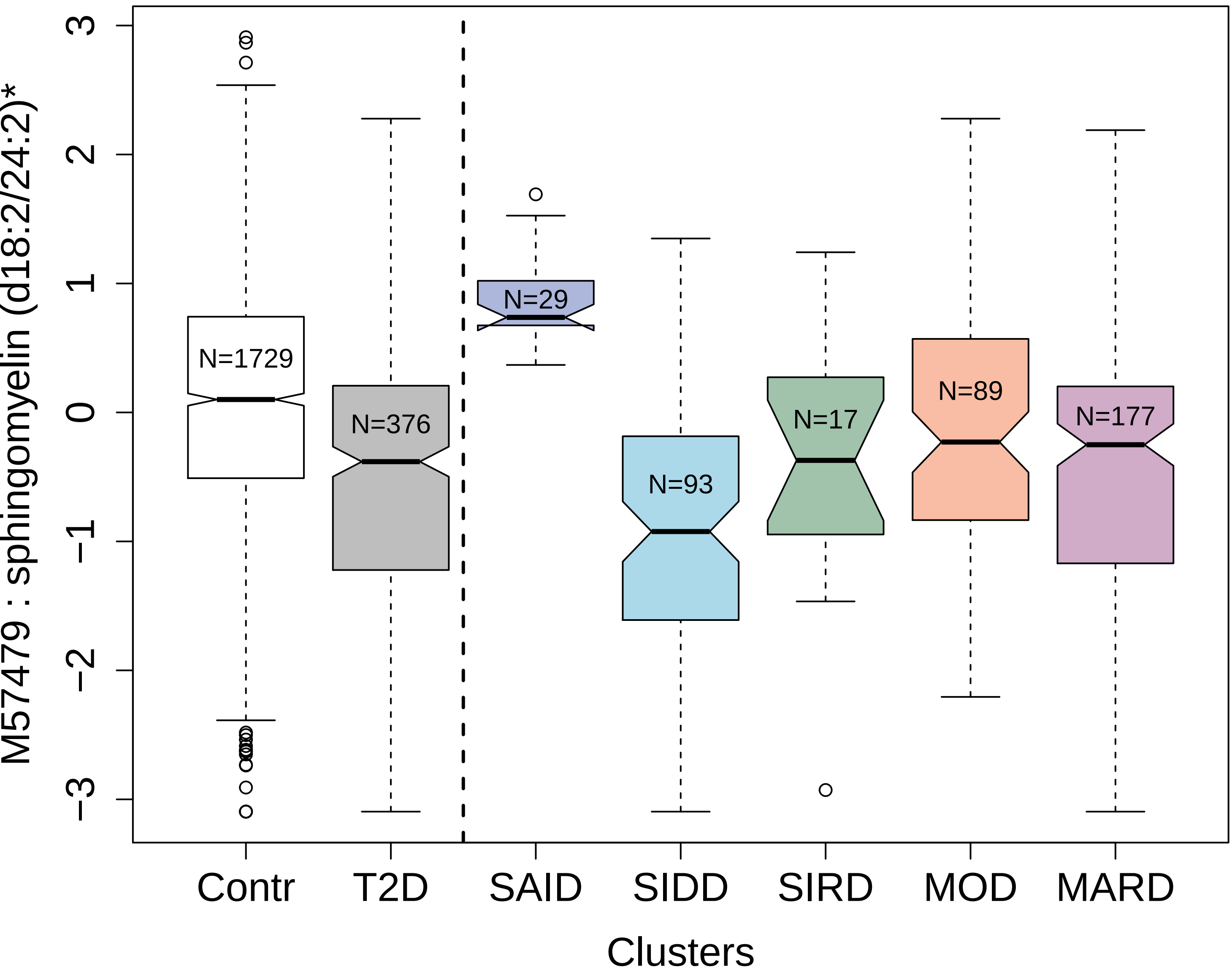

M57479 : sphingomyelin (d18:2/24:2)*
3
2
1
0
−1
−2
−3
N=1729
N=376
N=29
N=93
N=17
N=89
N=177
Contr
T2D
SAID
SIDD
SIRD
MOD
MARD
Clusters

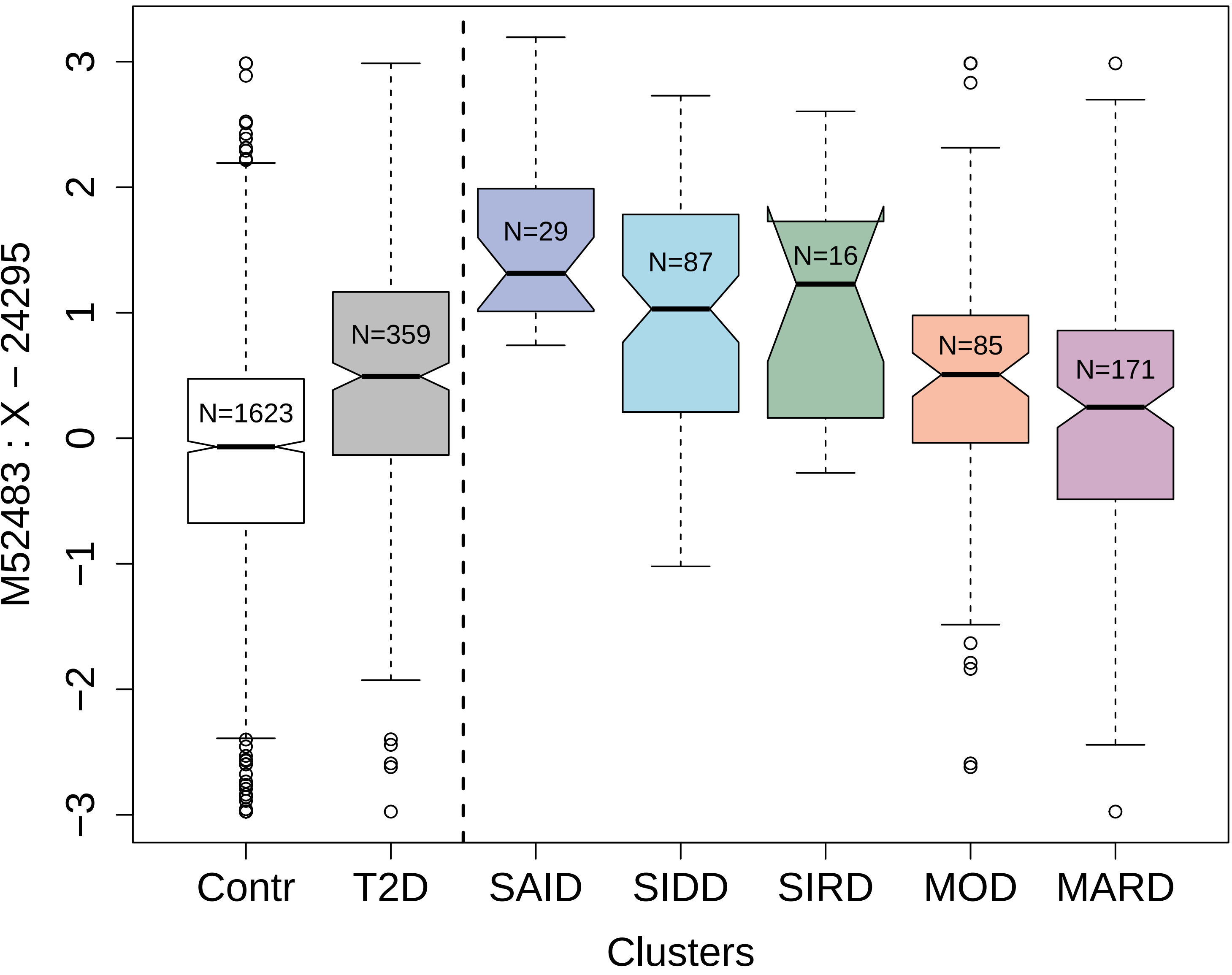
M52483 : X − 24295
3
2
1
0
−1
−2
−3
N=1623
N=359
N=29
N=87
N=16
N=85
N=171
Contr
T2D
SAID
SIDD
SIRD
MOD
MARD
Clusters

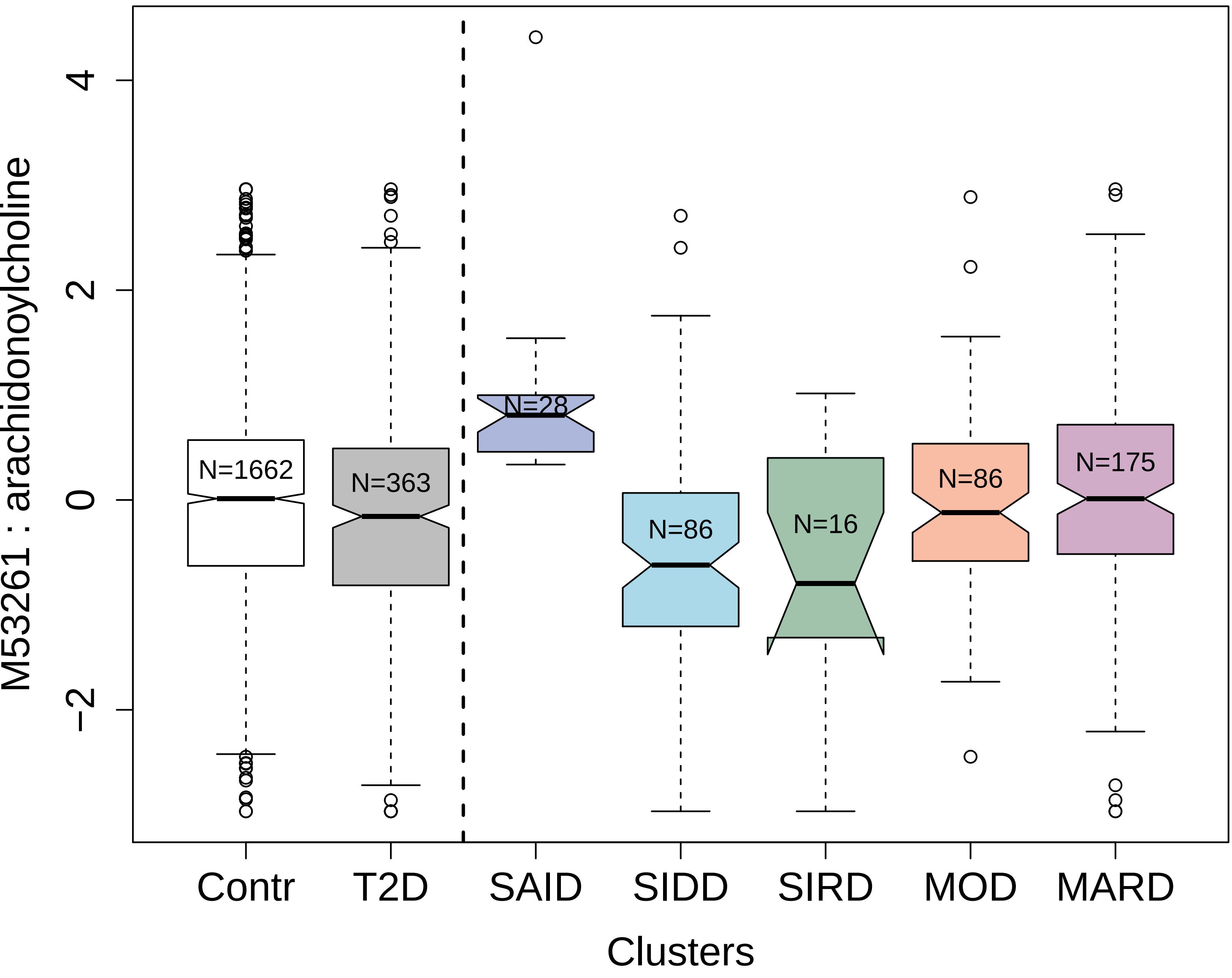
M53261 : arachidonoylcholine
4
2
0
−2
N=1662
N=363
N=28
N=86
N=16
N=86
N=175
Contr
T2D
SAID
SIDD
SIRD
MOD
MARD
Clusters

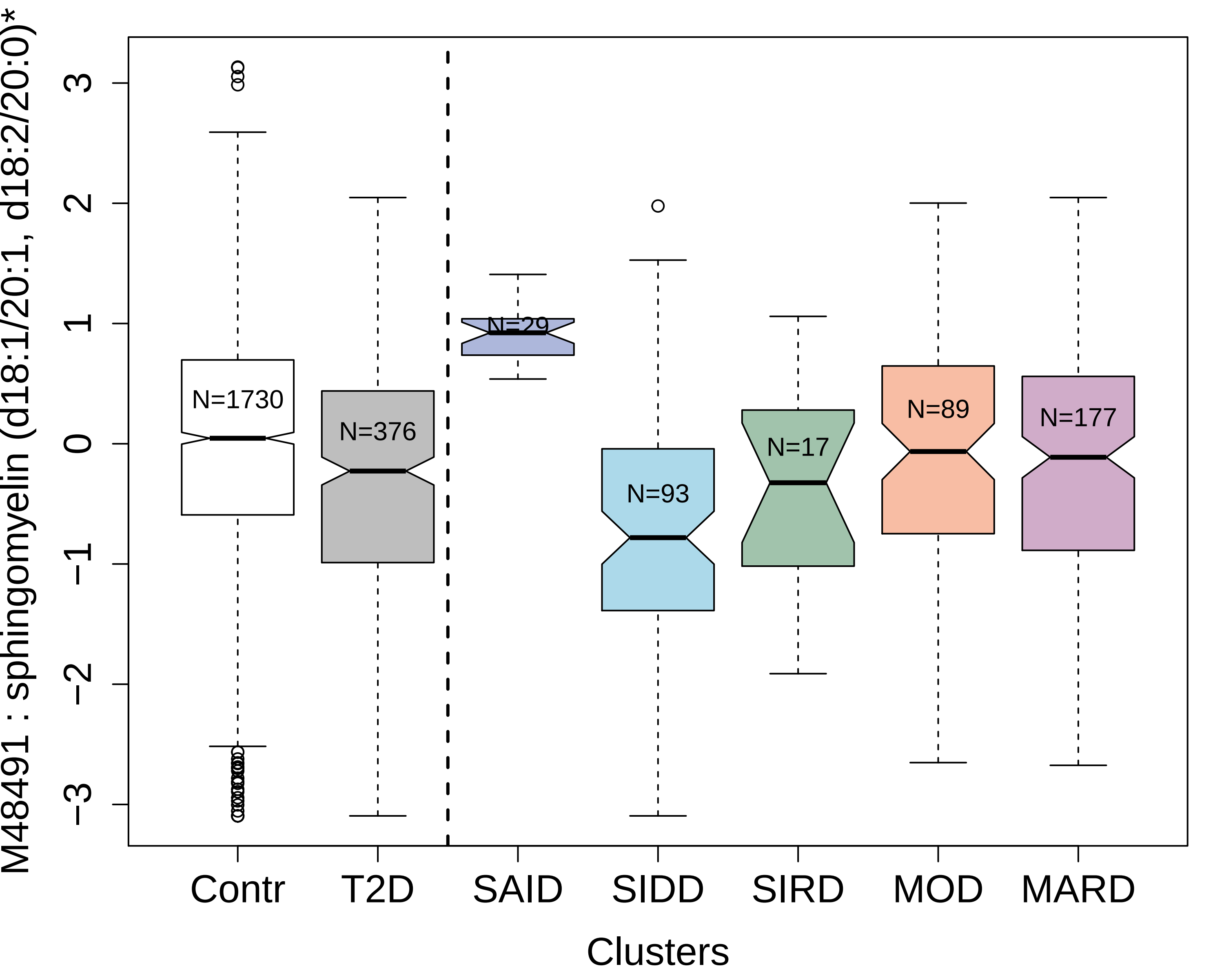

M48491 : sphingomyelin (d18:1/20:1, d18:2/20:0)*
3
2
1
0
−1
−2
−3
N=1730
N=376
N=29
N=93
N=17
N=89
N=177
Contr
T2D
SAID
SIDD
SIRD
MOD
MARD
Clusters

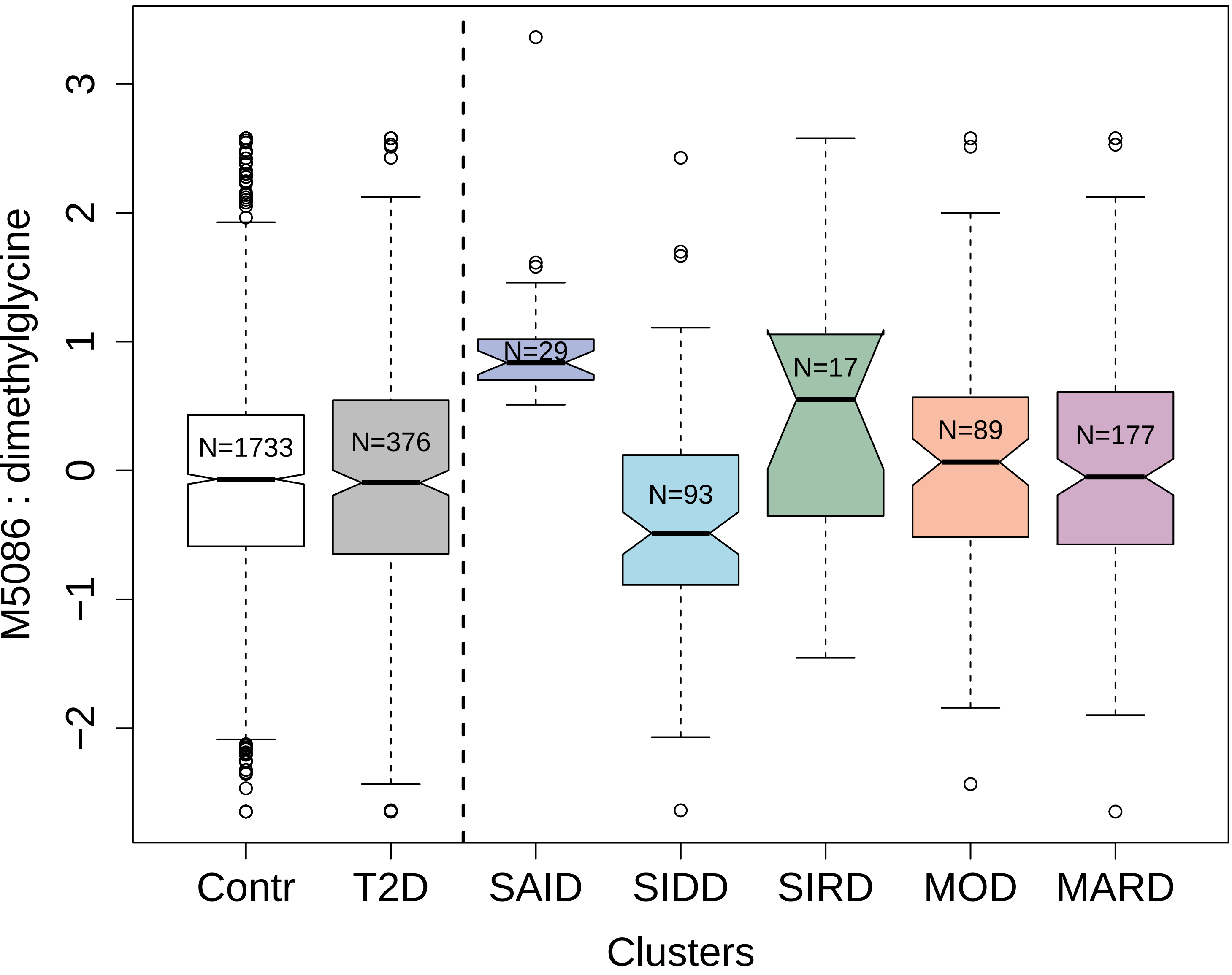

M5086 : dimethylglycine
3
2
1
0
−1
−2
N=1733
N=376
N=29
N=93
N=17
N=89
N=177
Contr
T2D
SAID
SIDD
SIRD
MOD
MARD
Clusters

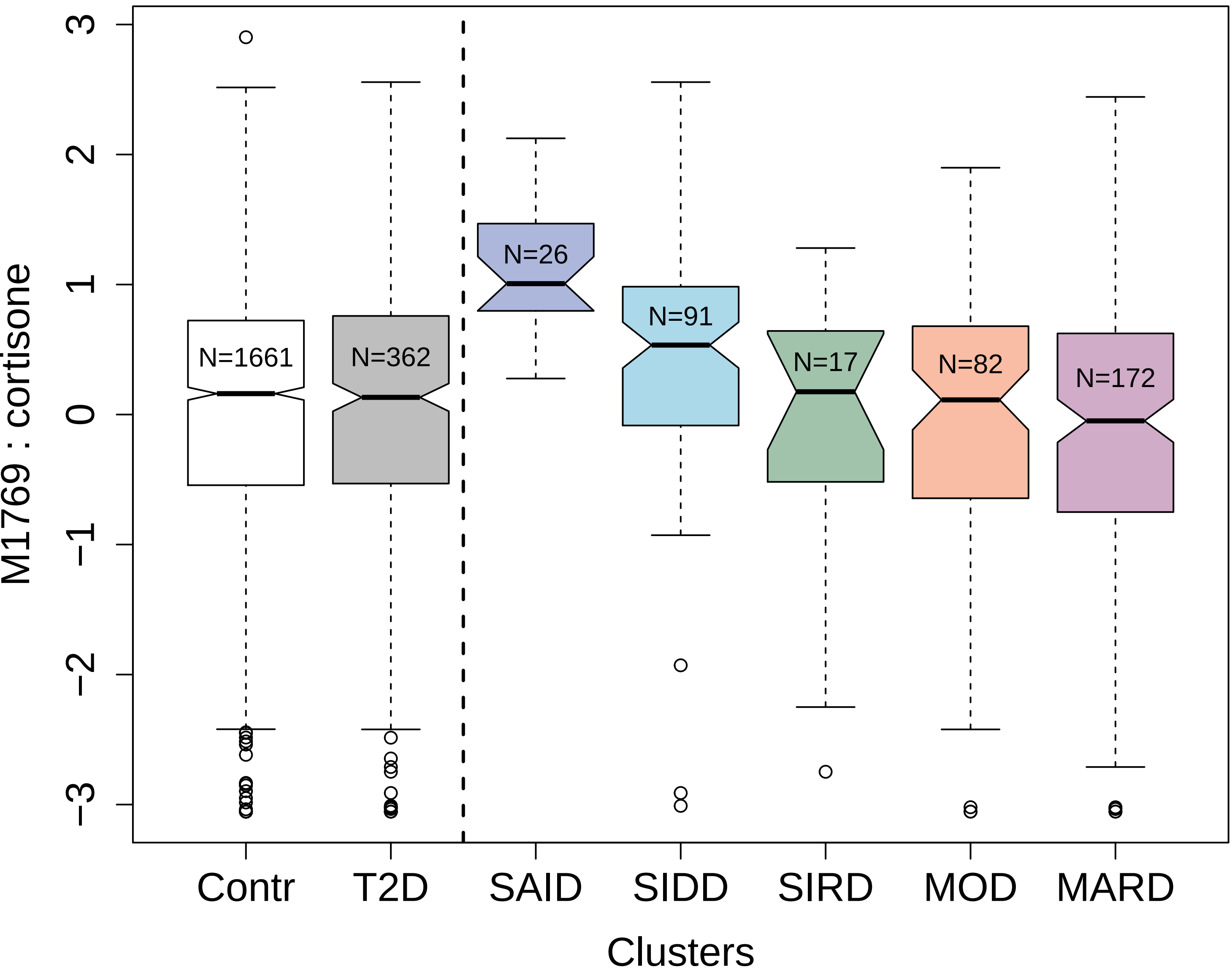
M1769 : cortisone
3
2
1
0
−1
−2
−3
N=1661
N=362
N=26
N=91
N=17
N=82
N=172
Contr
T2D
SAID
SIDD
SIRD
MOD
MARD
Clusters

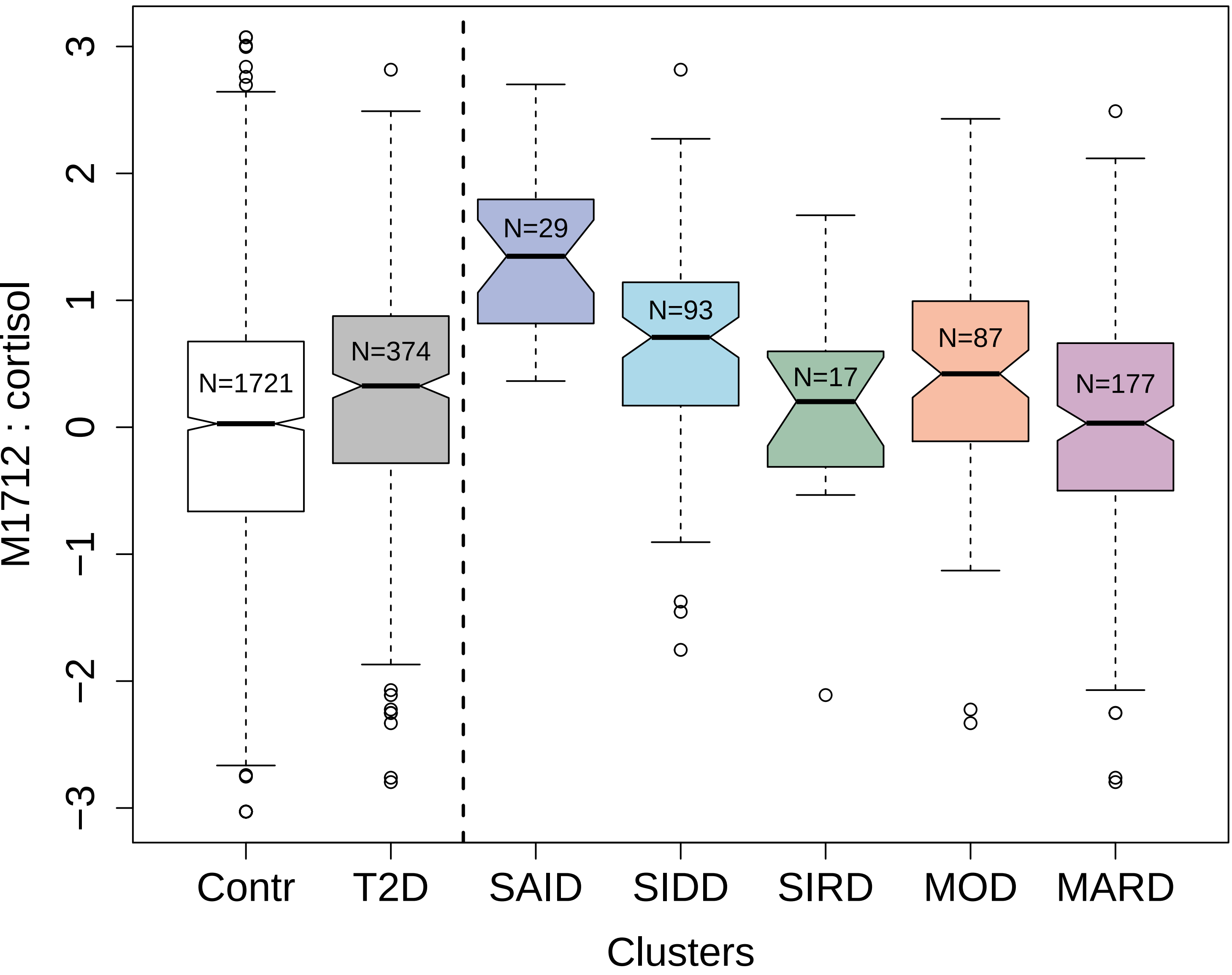
M1712 : cortisol
3
2
1
0
−1
−2
−3
N=1721
N=374
N=29
N=93
N=17
N=87
N=177
Contr
T2D
SAID
SIDD
SIRD
MOD
MARD
Clusters

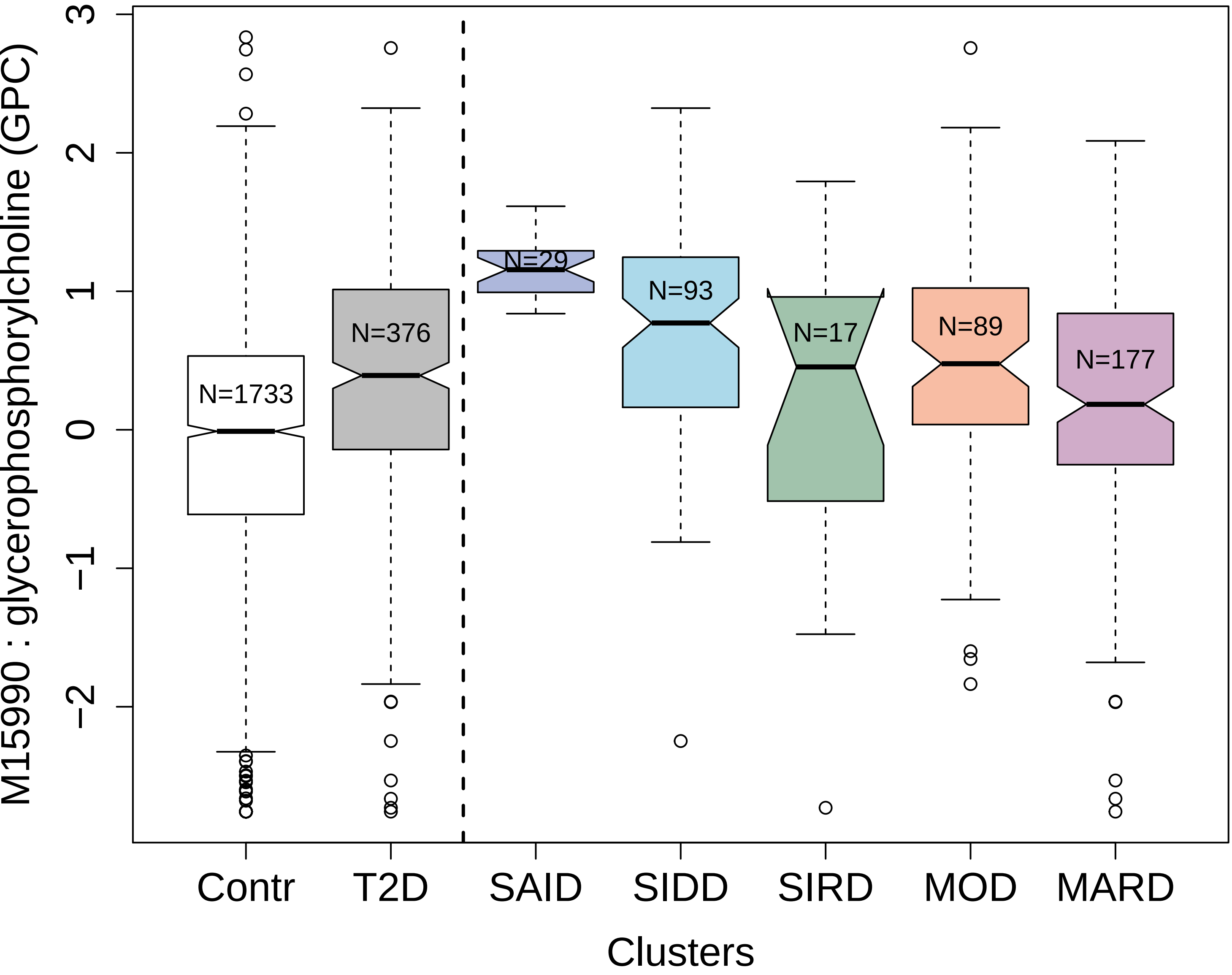

M15990 : glycerophosphorylcholine (GPC)
3
2
1
0
−1
−2
N=1733
N=376
N=29
N=93
N=17
N=89
N=177
Contr
T2D
SAID
SIDD
SIRD
MOD
MARD
Clusters

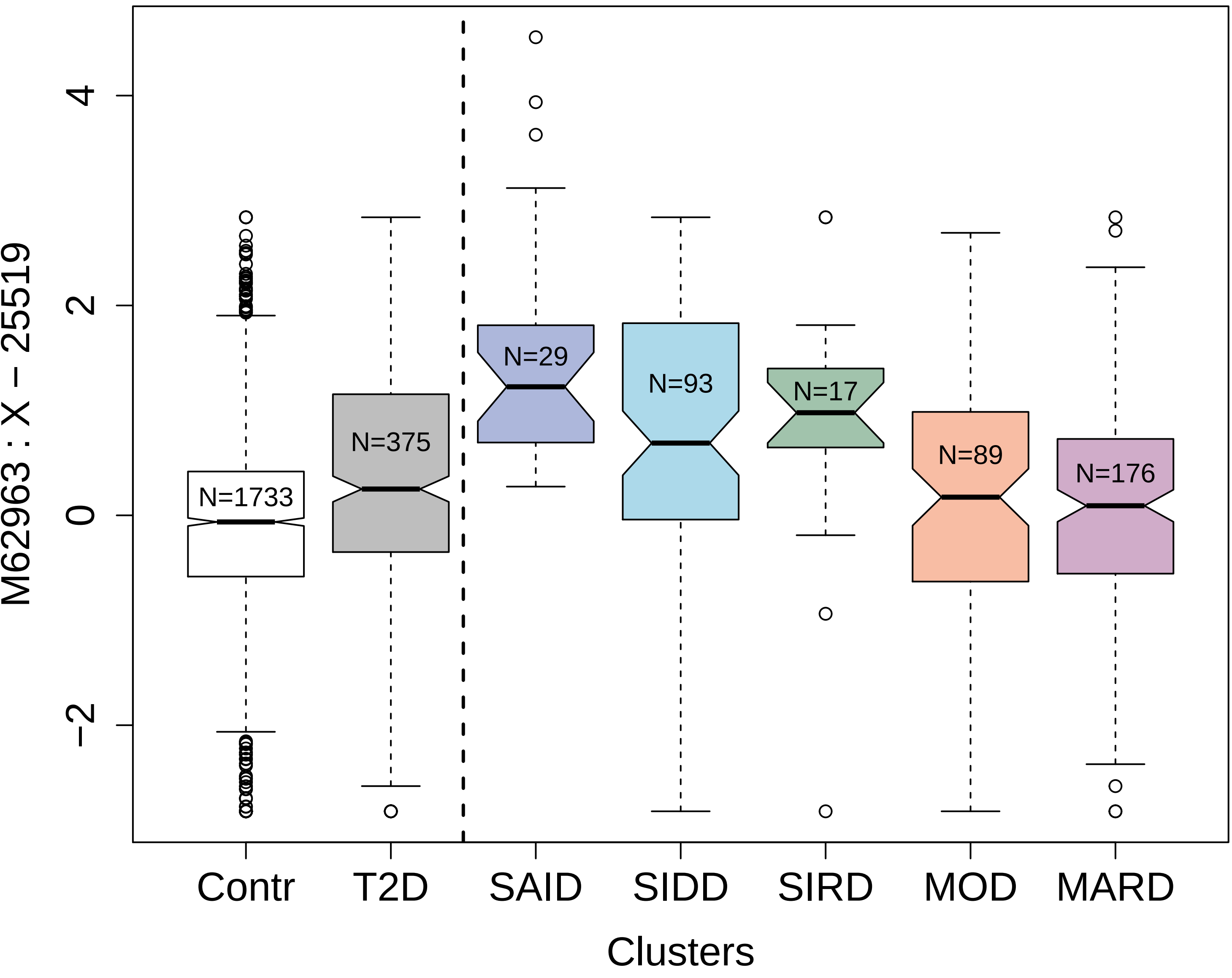
M62963 : X − 25519
4
2
0
−2
N=1733
N=375
N=29
N=93
N=17
N=89
N=176
Contr
T2D
SAID
SIDD
SIRD
MOD
MARD
Clusters

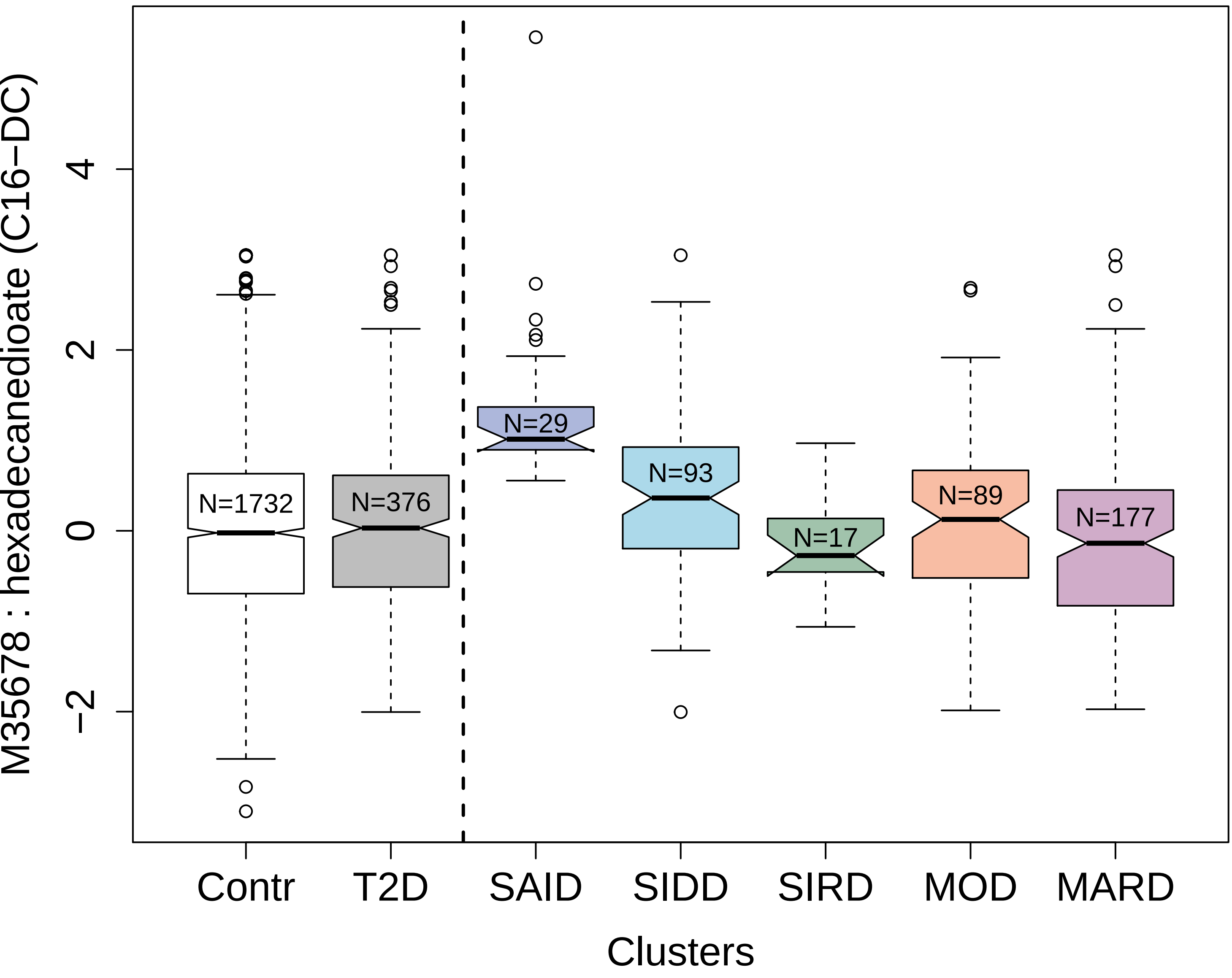
M35678 : hexadecanediodate (C16−DC)
Clusters
Contr
T2D
SAID
SIDD
SIRD
MOD
MARD
N=1732
N=376
N=29
N=93
N=17
N=89
N=177
−2
0
2
4

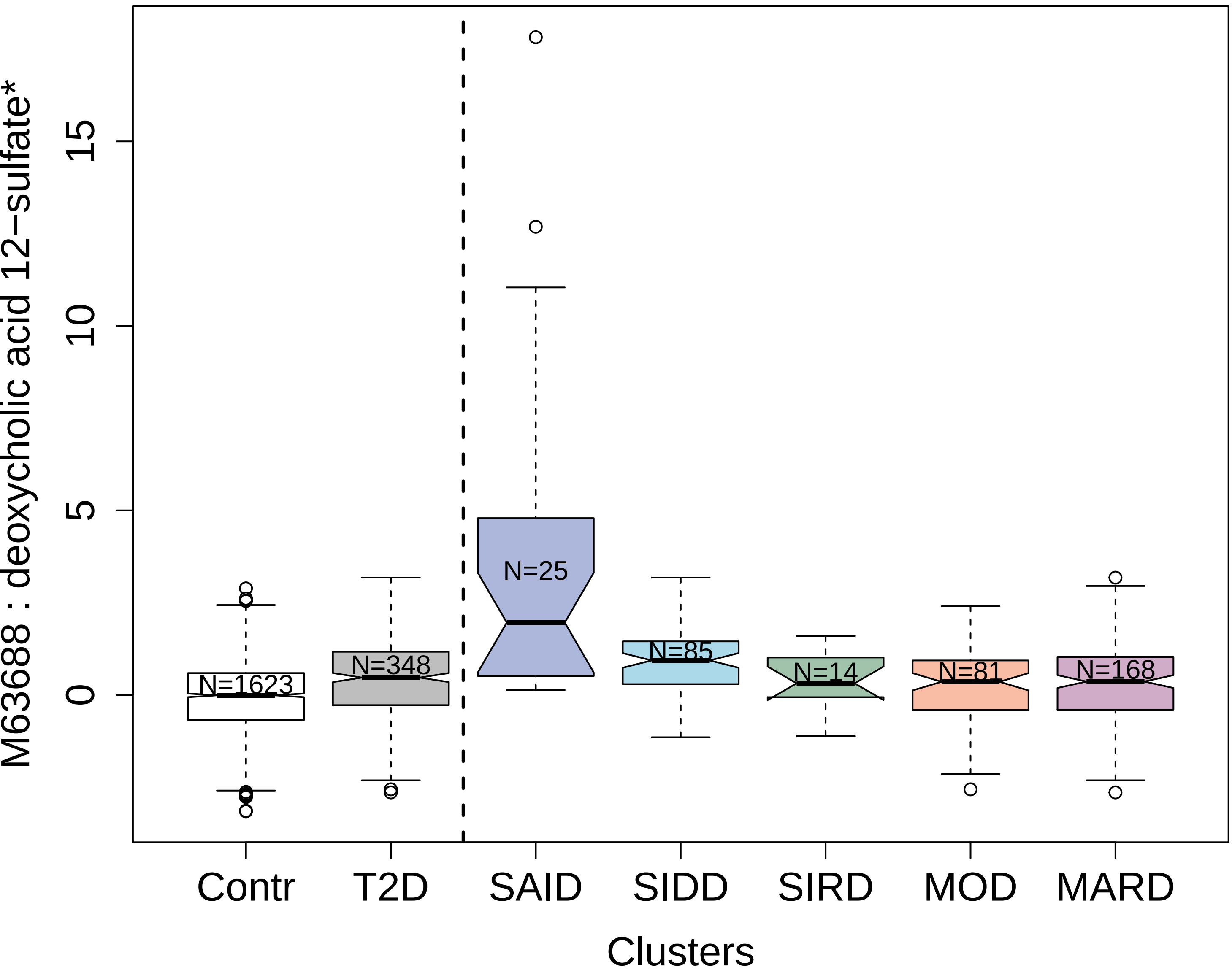
M63688 : deoxycholic acid 12−sulfate*
0
5
10
15
N=1623
N=348
N=25
N=85
N=14
N=81
N=168
Contr
T2D
SAID
SIDD
SIRD
MOD
MARD
Clusters

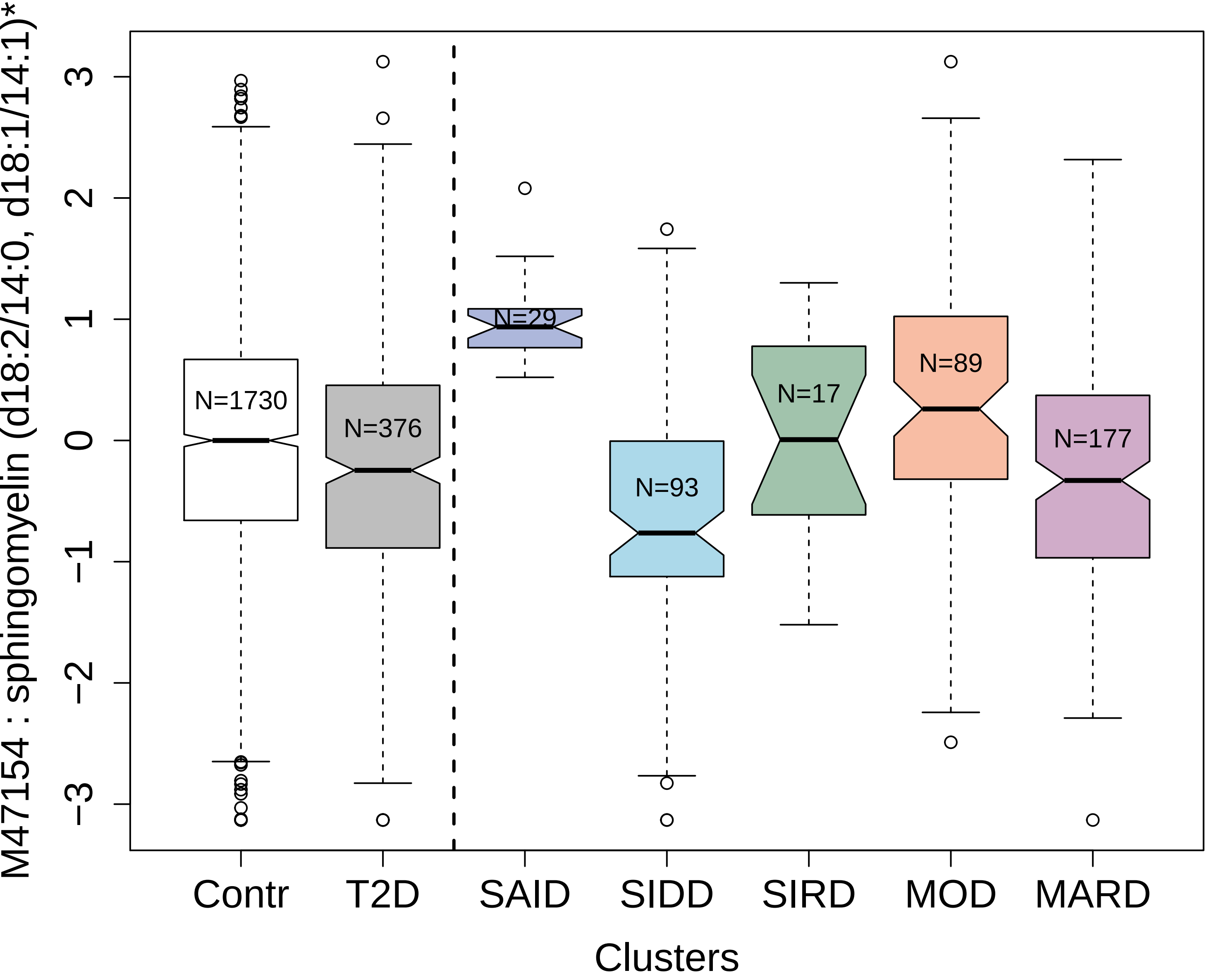

M47154 : sphingomyelin (d18:2/14:0, d18:1/14:1)*
3
2
1
0
−1
−2
−3
N=1730
N=376
N=29
N=93
N=17
N=89
N=177
Contr
T2D
SAID
SIDD
SIRD
MOD
MARD
Clusters

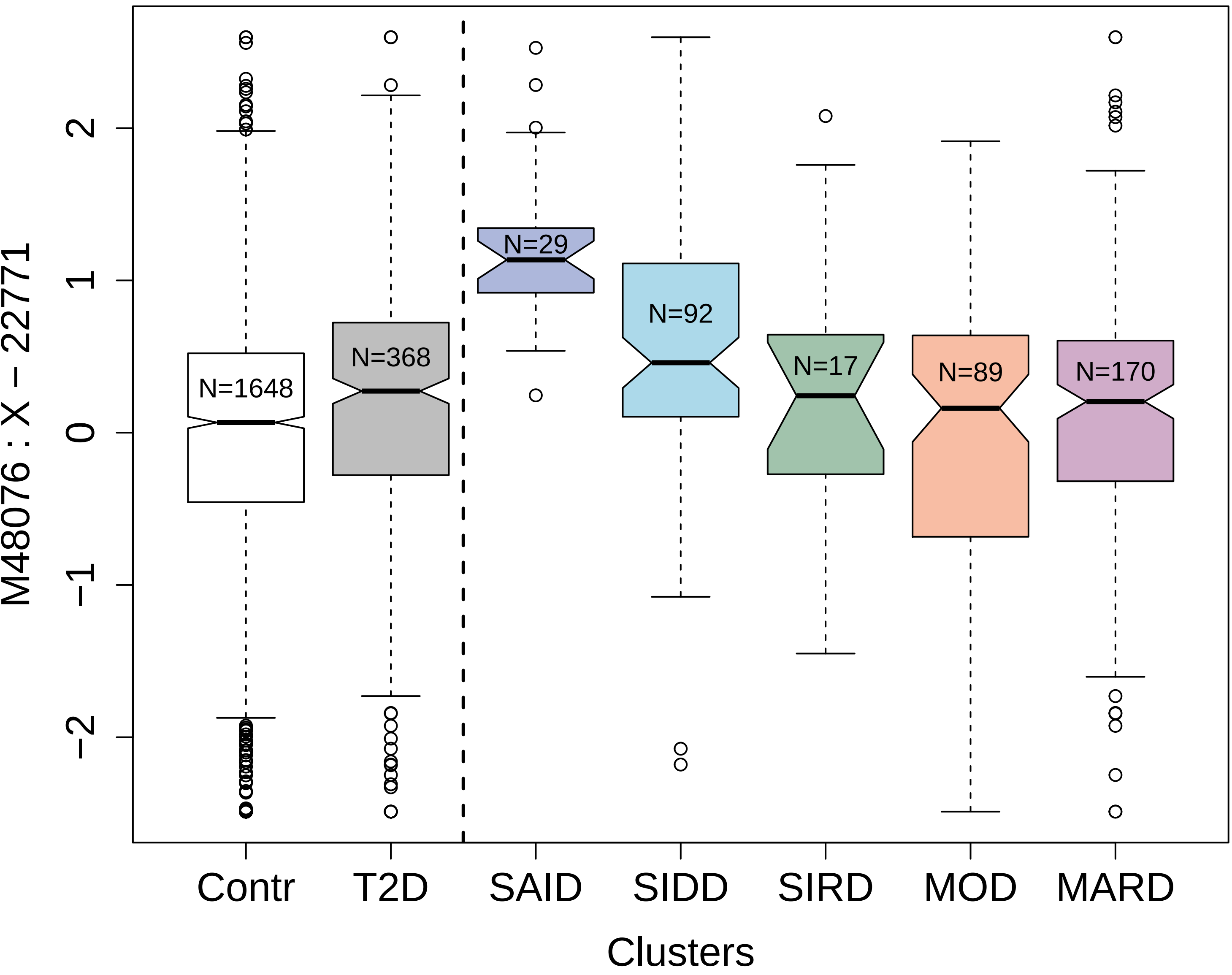
M48076 : X − 22771
2
1
0
−1
−2
N=1648
N=368
N=29
N=92
N=17
N=89
N=170
Contr
T2D
SAID
SIDD
SIRD
MOD
MARD
Clusters

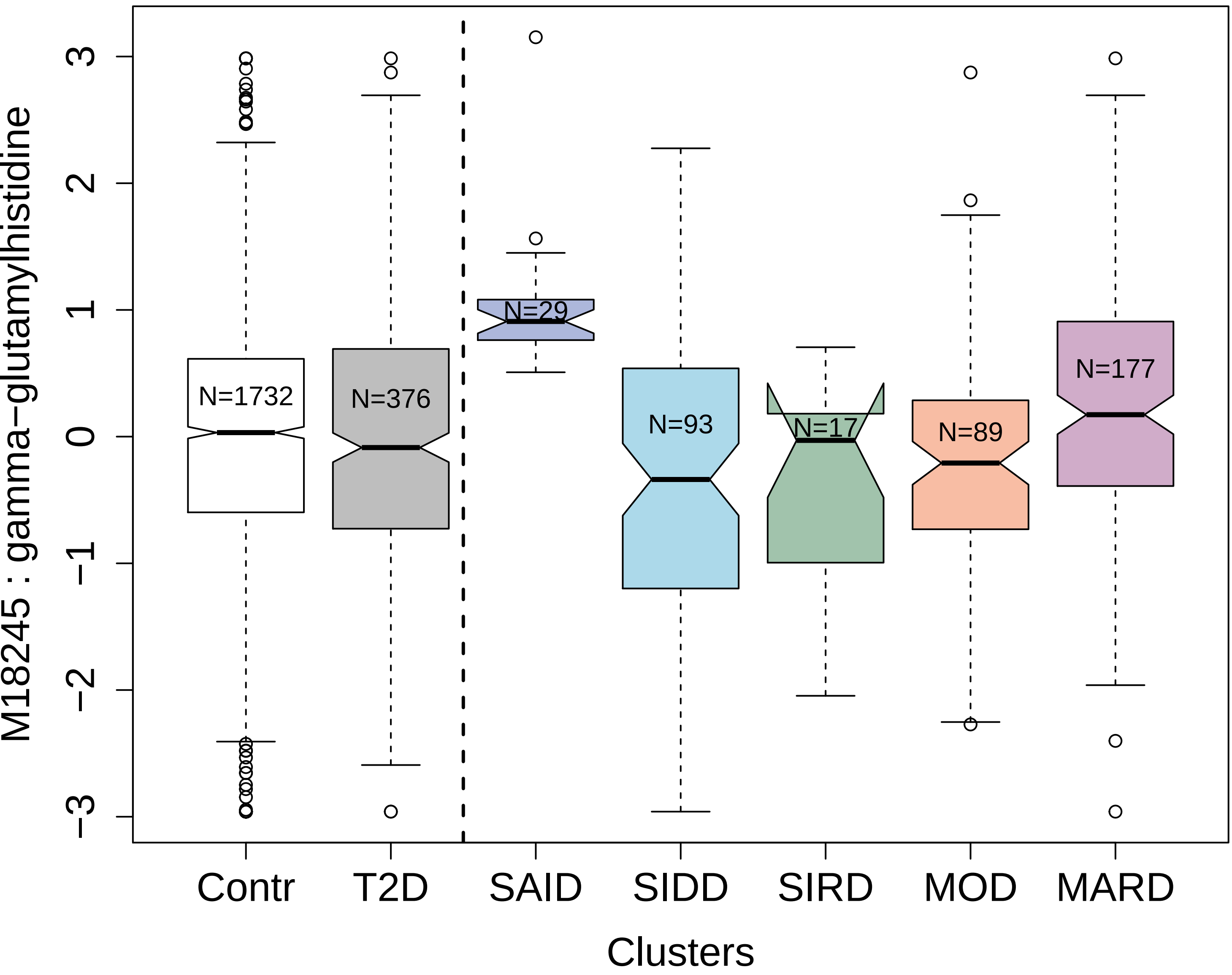

M18245 : gamma−glutamylhistidine
3
2
1
0
−1
−2
−3
N=1732
N=376
N=29
N=93
N=17
N=89
N=177
Contr
T2D
SAID
SIDD
SIRD
MOD
MARD
Clusters

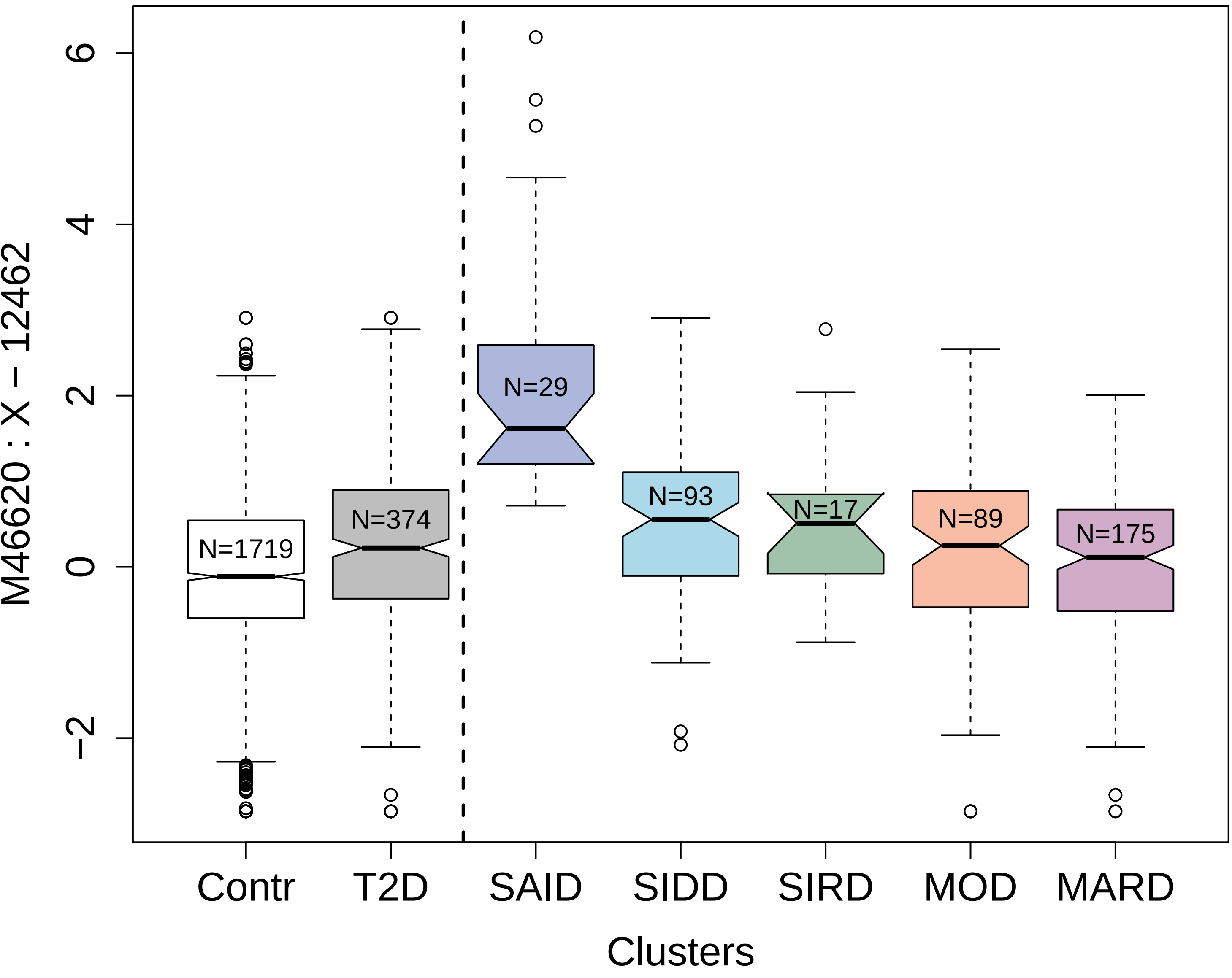

M46620 : X − 12462
6
4
2
0
−2
N=1719
N=374
N=29
N=93
N=17
N=89
N=175
Contr
T2D
SAID
SIDD
SIRD
MOD
MARD
Clusters

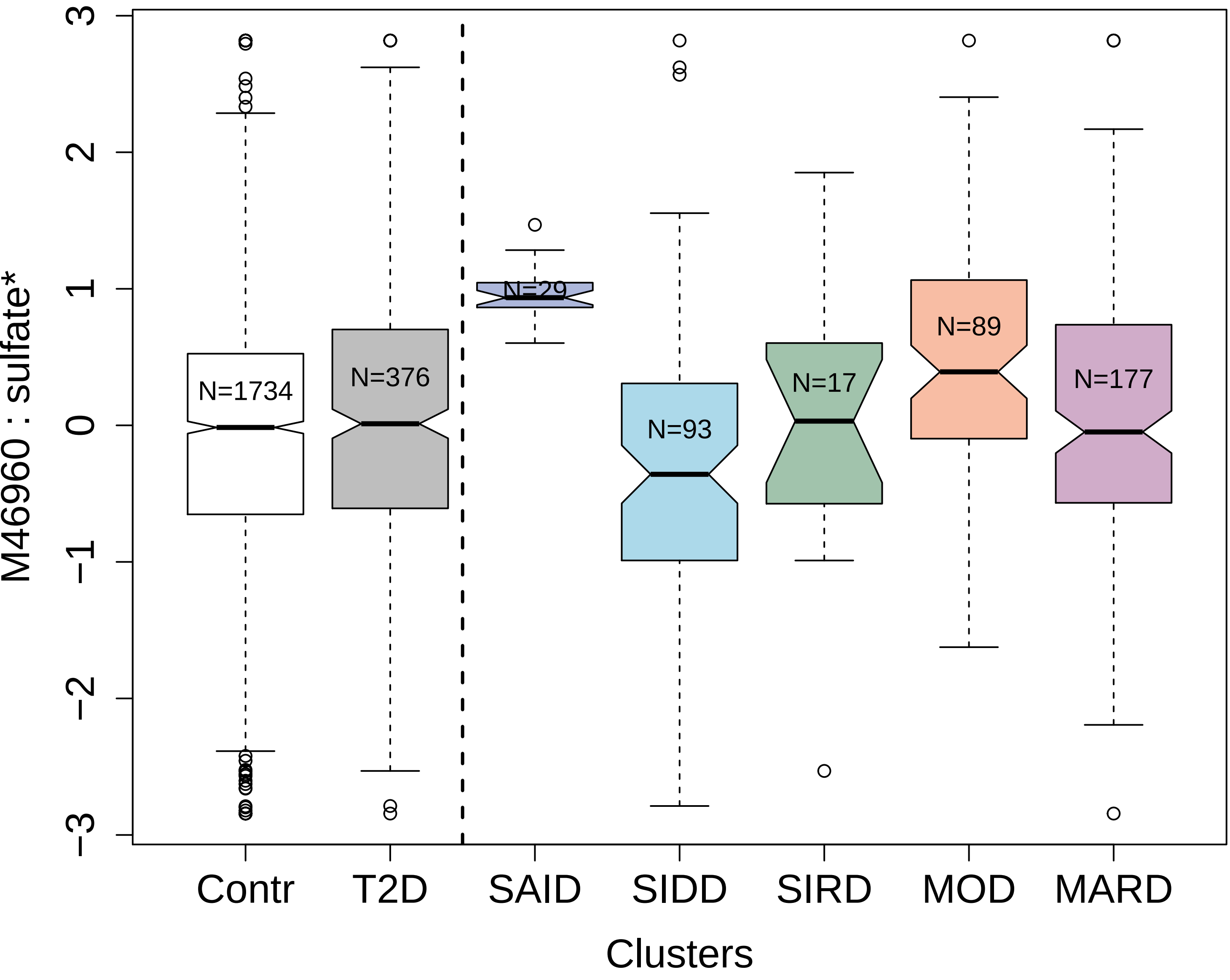

M46960 : sulfate*
3
2
1
0
−1
−2
−3
N=1734
N=376
N=29
N=93
N=17
N=89
N=177
Contr
T2D
SAID
SIDD
SIRD
MOD
MARD
Clusters

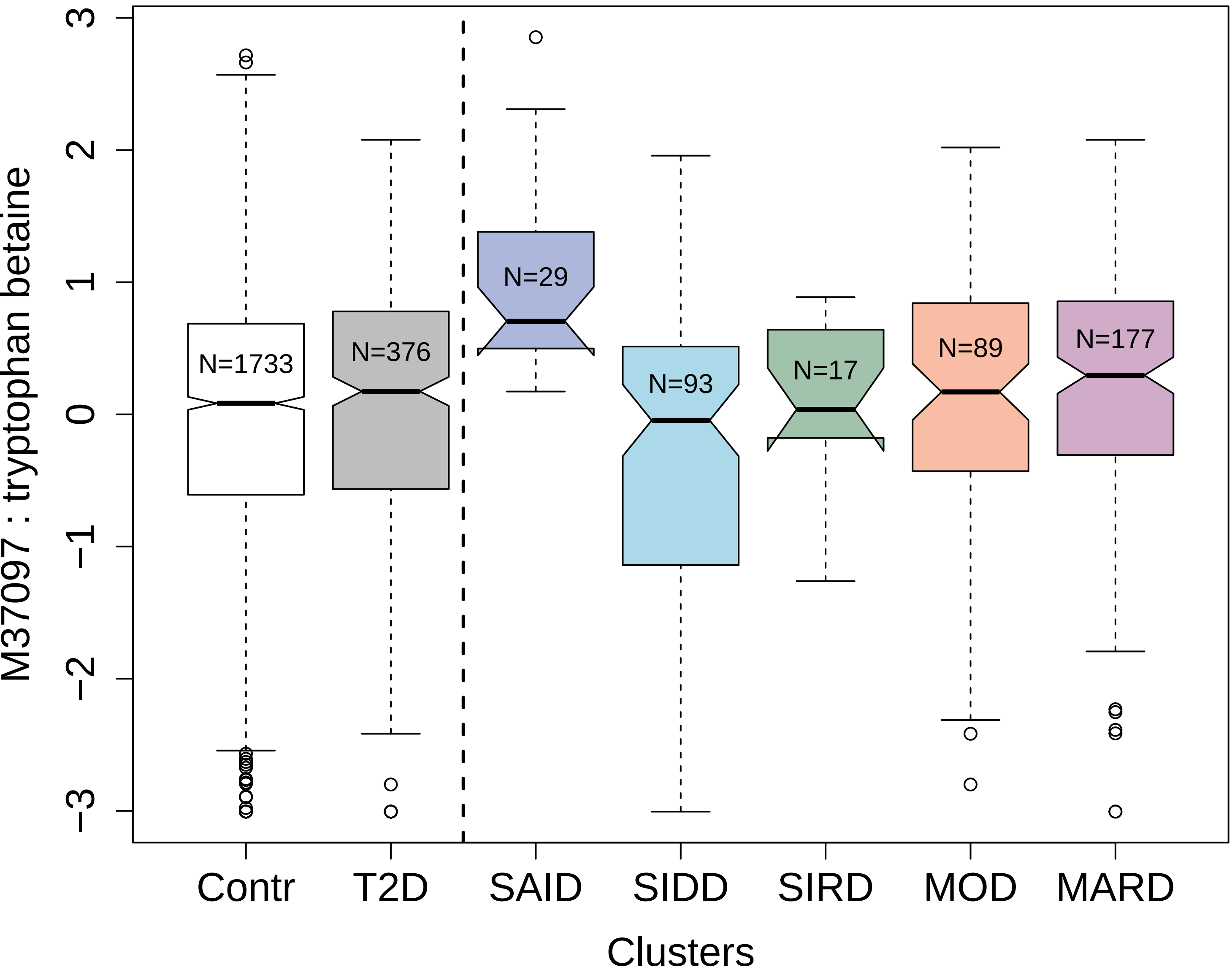

M37097 : tryptophan betaine
3
2
1
0
−1
−2
−3
N=1733
N=376
N=29
N=93
N=17
N=89
N=177
Contr
T2D
SAID
SIDD
SIRD
MOD
MARD
Clusters

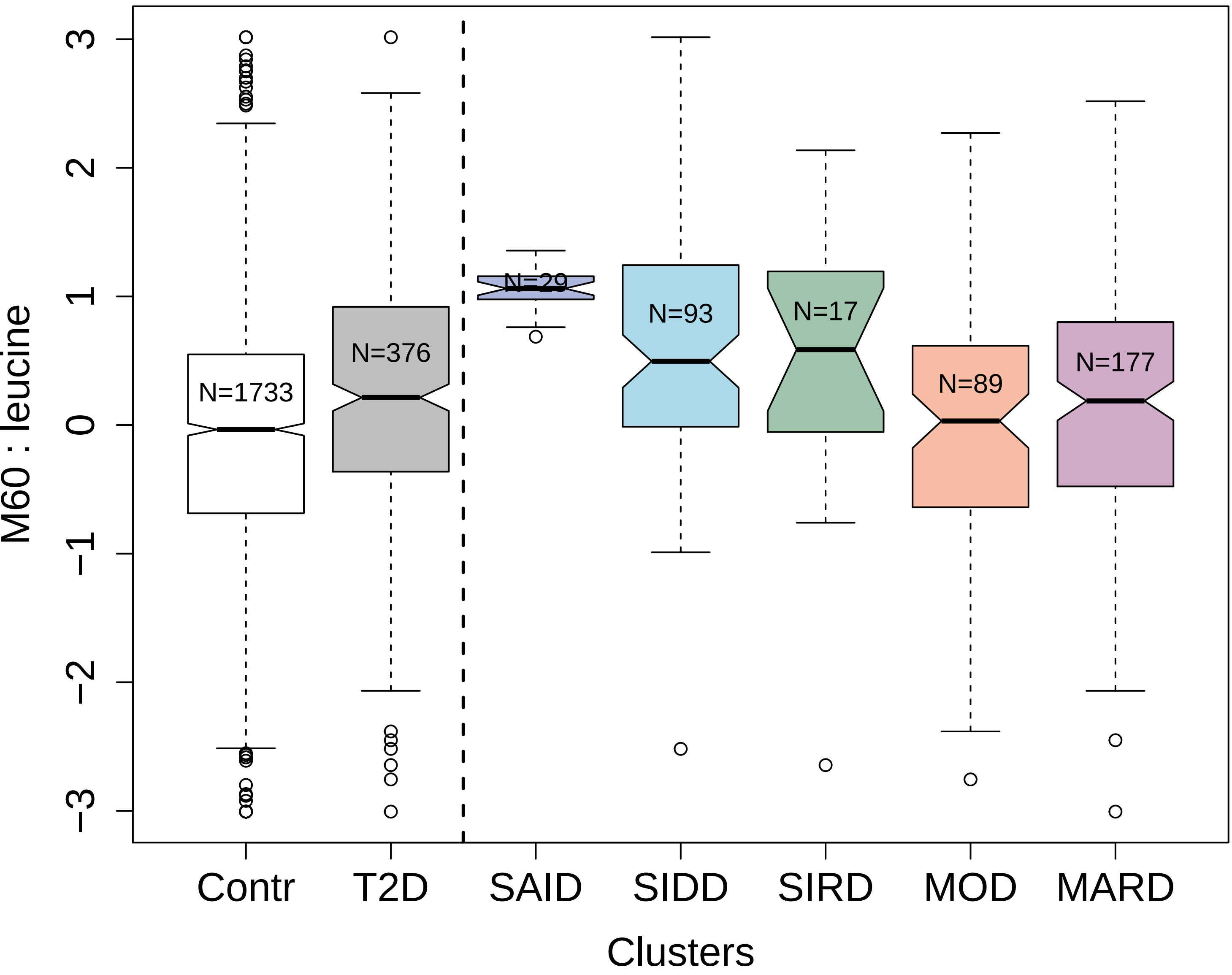

M60 : leucine
3
2
1
0
−1
−2
−3
N=1733
N=376
N=29
N=93
N=17
N=89
N=177
Contr
T2D
SAID
SIDD
SIRD
MOD
MARD
Clusters

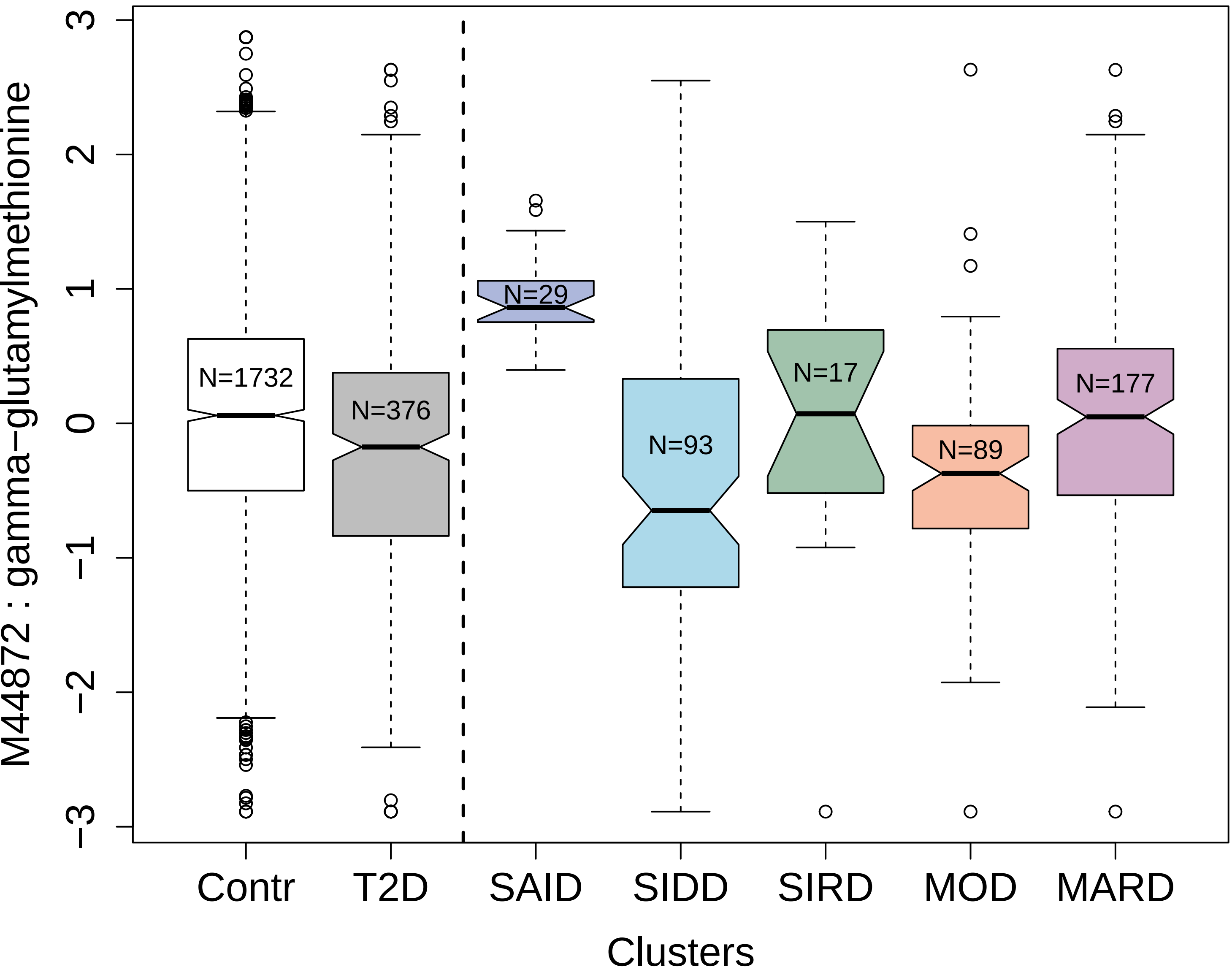

M44872 : gamma−glutamylmethionine
3
2
1
0
−1
−2
−3
N=1732
N=376
N=29
N=93
N=17
N=89
N=177
Contr
T2D
SAID
SIDD
SIRD
MOD
MARD
Clusters

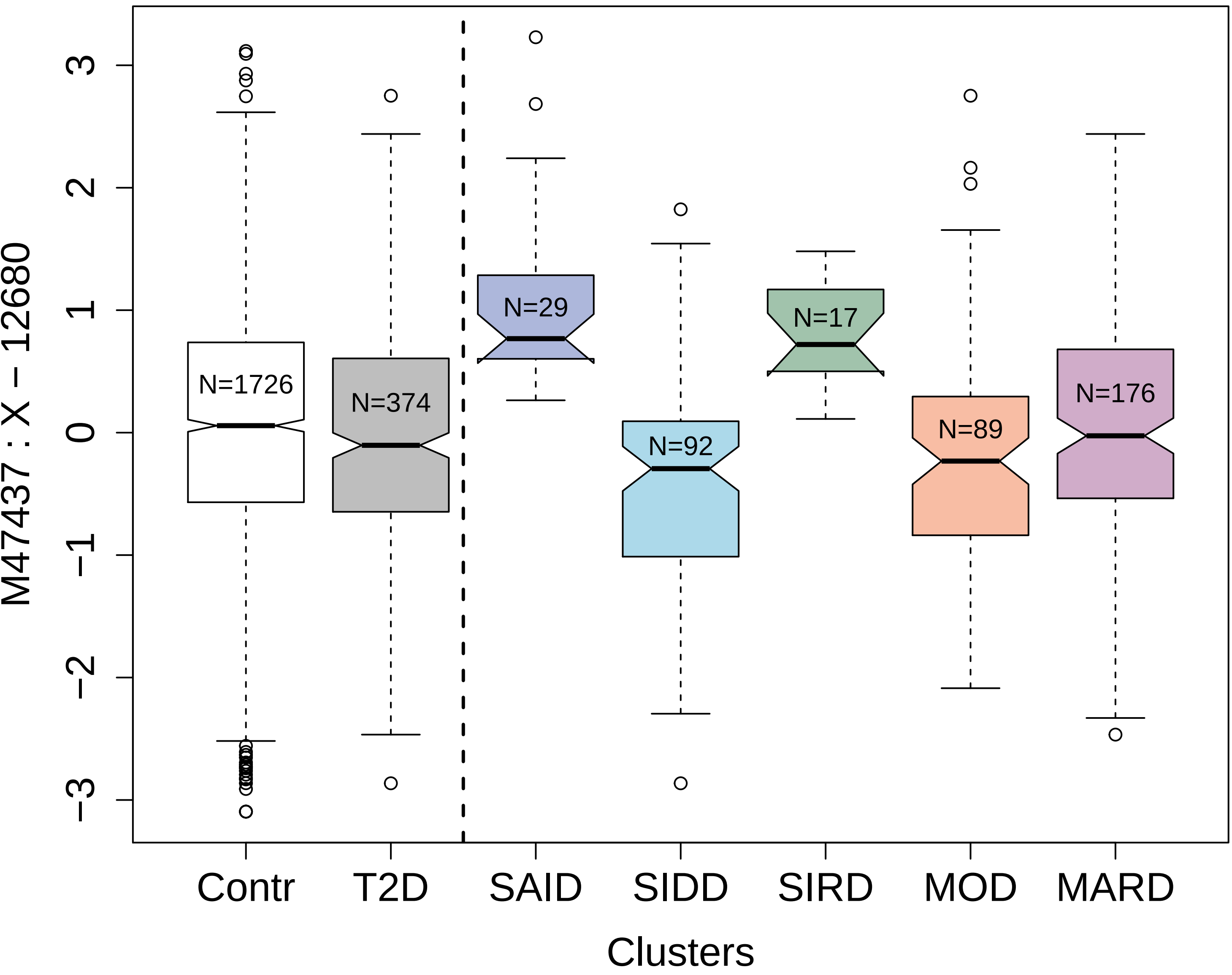

M47437 : X − 12680
3
2
1
0
−1
−2
−3
N=1726
N=374
N=29
N=92
N=17
N=89
N=176
Contr
T2D
SAID
SIDD
SIRD
MOD
MARD
Clusters

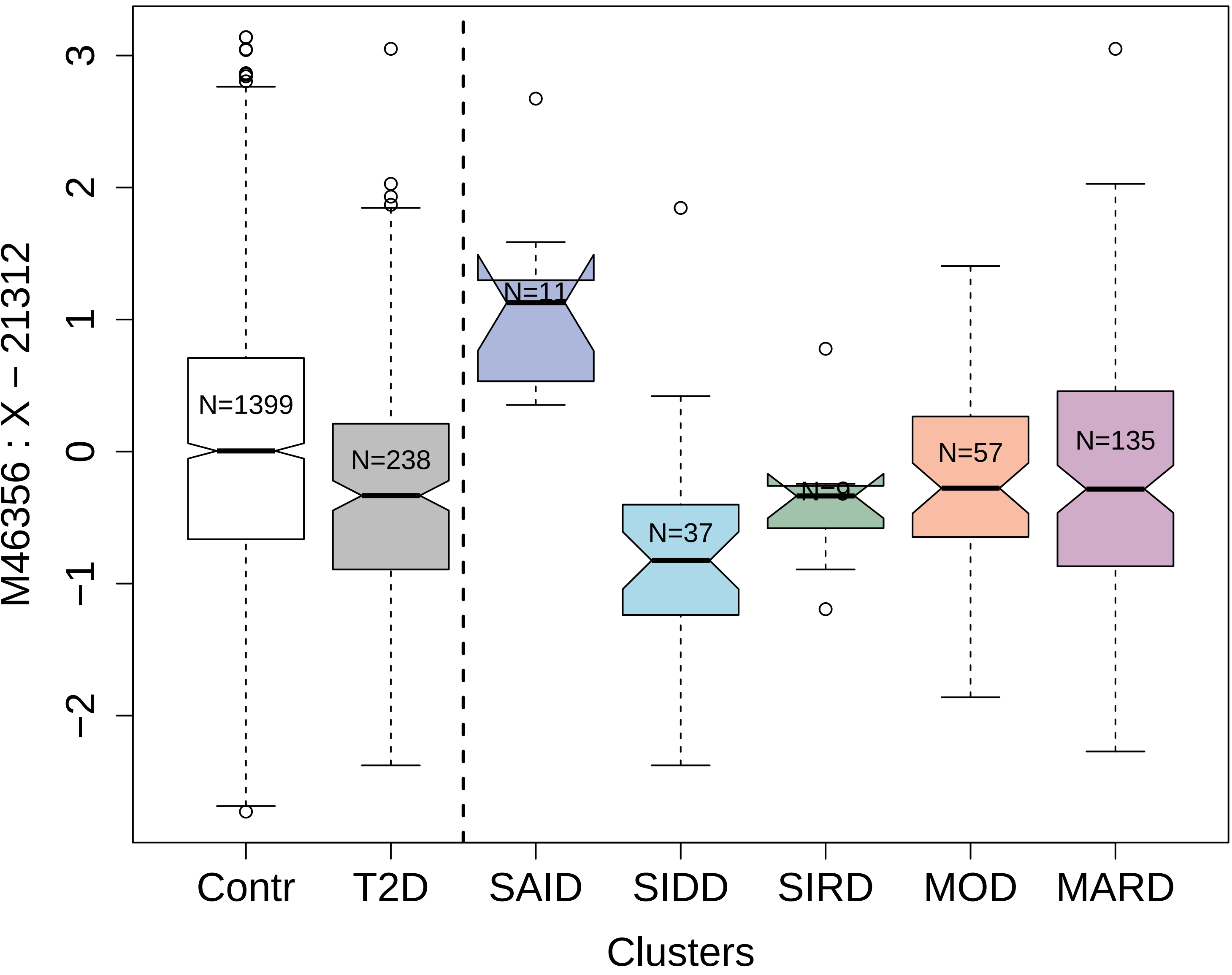

M46356 : X − 21312
Clusters
Contr
T2D
SAID
SIDD
SIRD
MOD
MARD
N=1399
N=238
N=11
N=37
N=9
N=57
N=135
3
2
1
0
−1
−2

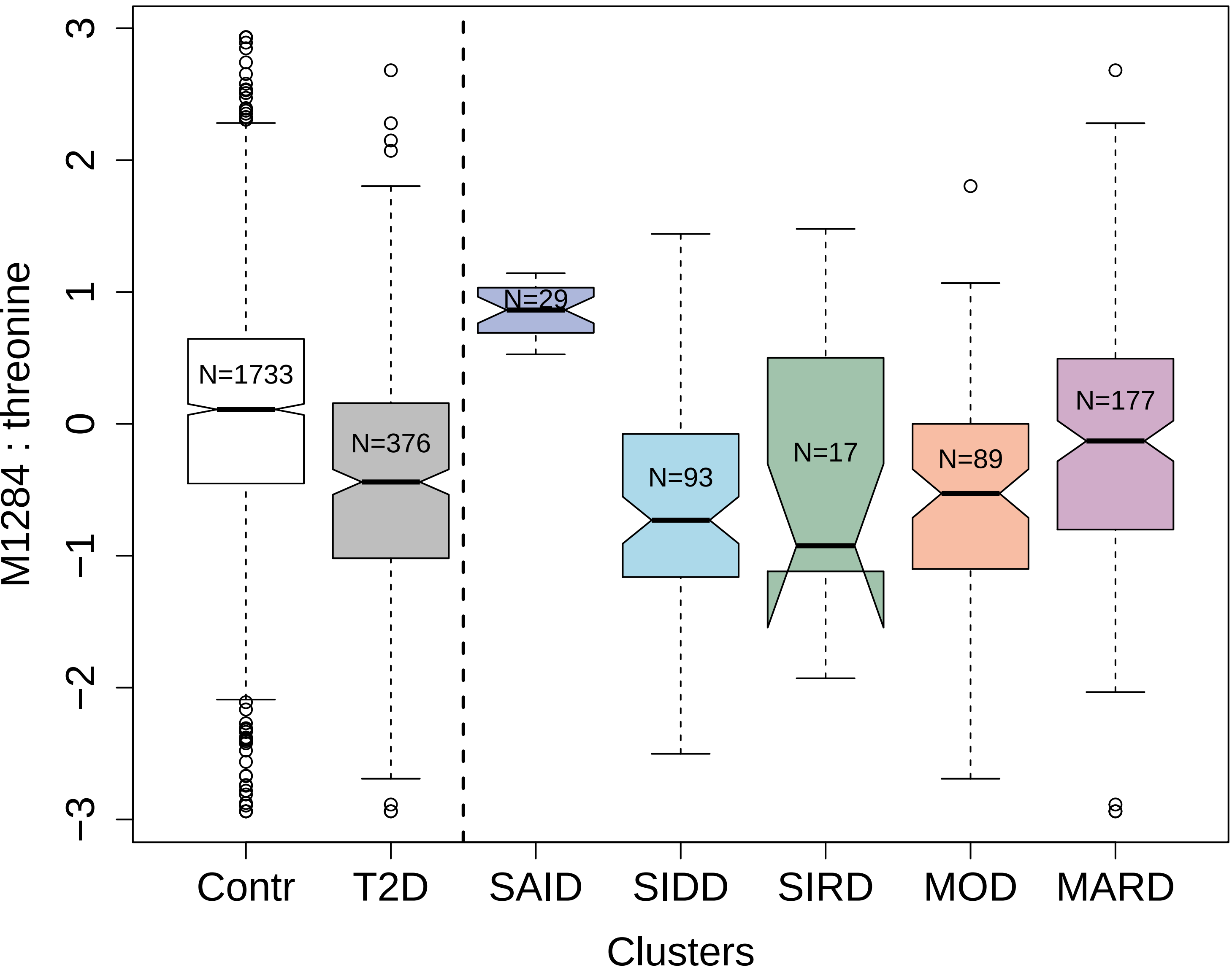

M1284 : threonine
3
2
1
0
−1
−2
−3
N=1733
N=376
N=29
N=93
N=17
N=89
N=177
Contr
T2D
SAID
SIDD
SIRD
MOD
MARD
Clusters

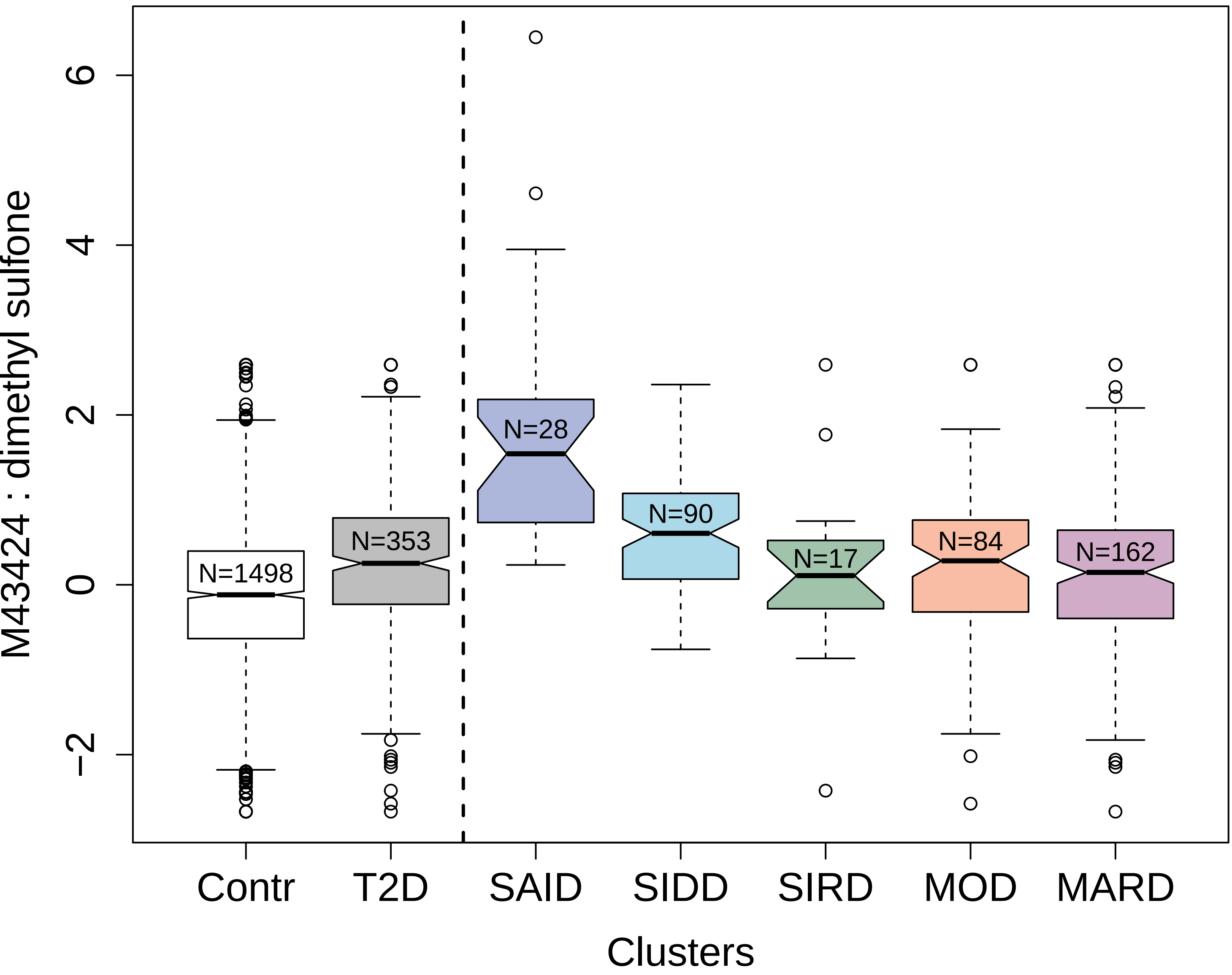

M43424 : dimethyl sulfone
6
4
2
0
−2
N=1498
N=353
N=28
N=90
N=17
N=84
N=162
Contr
T2D
SAID
SIDD
SIRD
MOD
MARD
Clusters

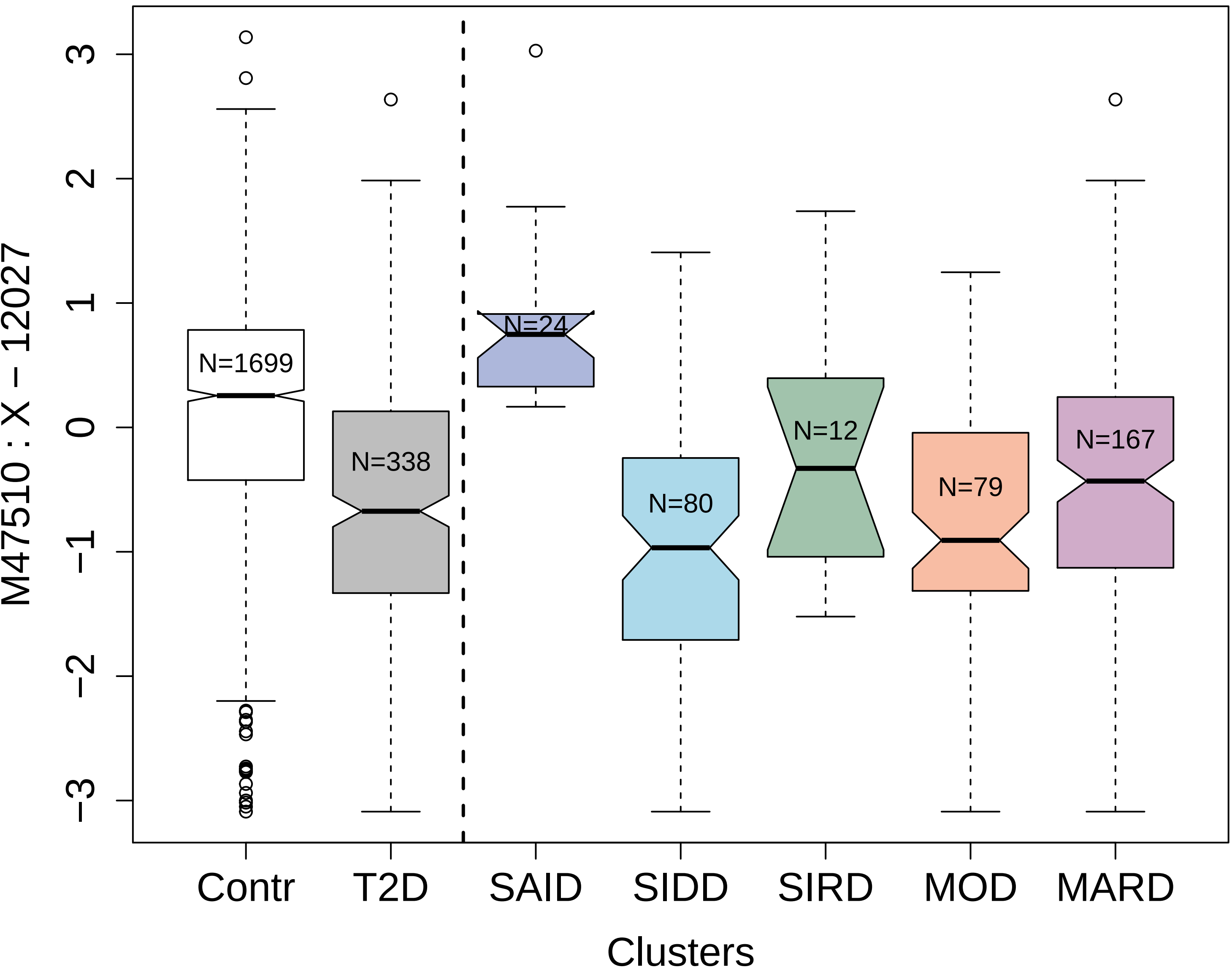

M47510 : X − 12027
3
2
1
0
−1
−2
−3
N=1699
N=338
N=24
N=80
N=12
N=79
N=167
Contr
T2D
SAID
SIDD
SIRD
MOD
MARD
Clusters

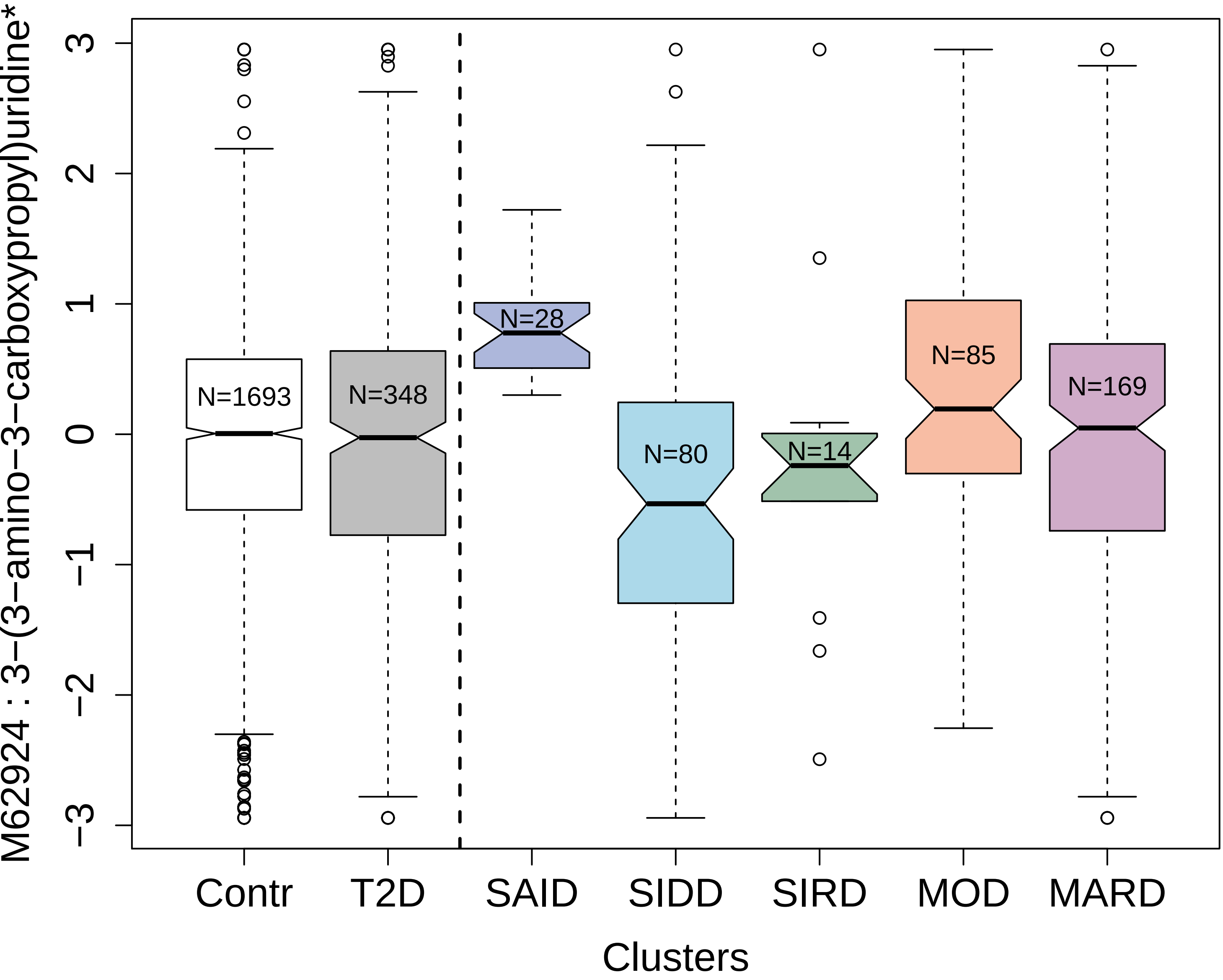

M62924 : 3−(3−amino−3−carboxypropyl)uridine*
3
2
1
0
−1
−2
−3
N=1693
N=348
N=28
N=80
N=14
N=85
N=169
Contr
T2D
SAID
SIDD
SIRD
MOD
MARD
Clusters

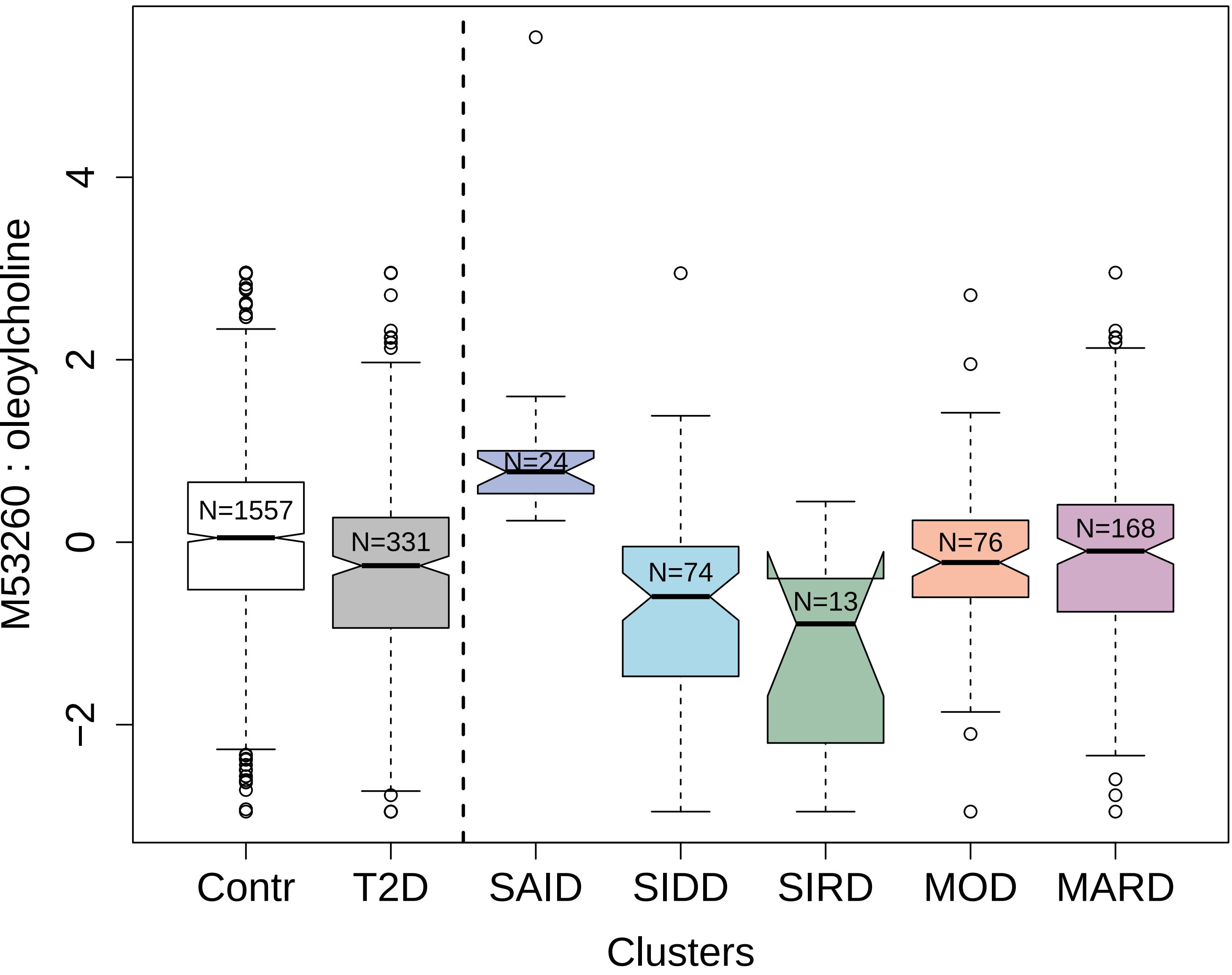
M53260 : oleoylcholine
4
2
0
−2
N=1557
N=331
N=24
N=74
N=13
N=76
N=168
Contr
T2D
SAID
SIDD
SIRD
MOD
MARD
Clusters

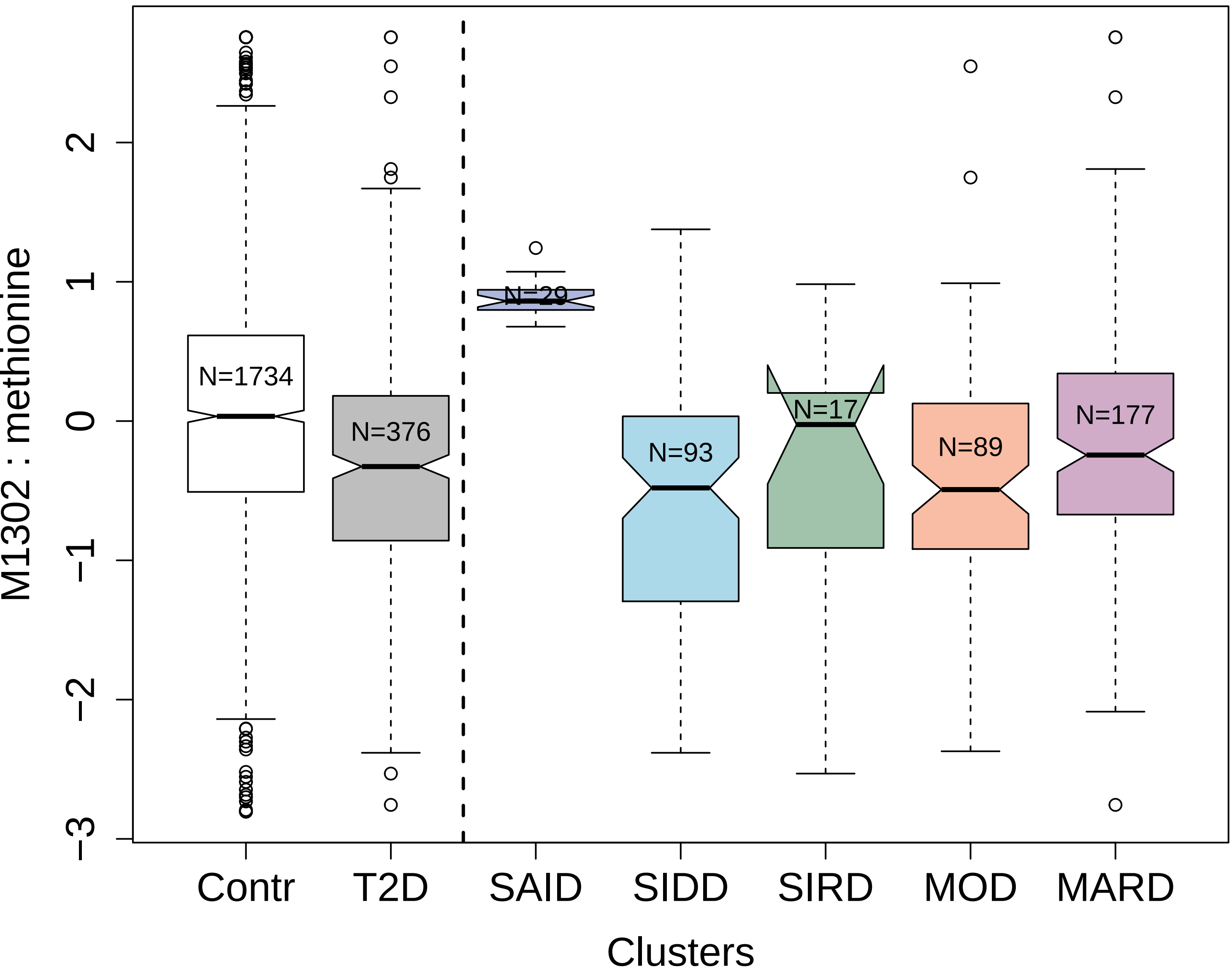

M1302 : methionine
−3
−2
−1
0
1
2
N=1734
N=376
N=29
N=93
N=17
N=89
N=177
Contr
T2D
SAID
SIDD
SIRD
MOD
MARD
Clusters

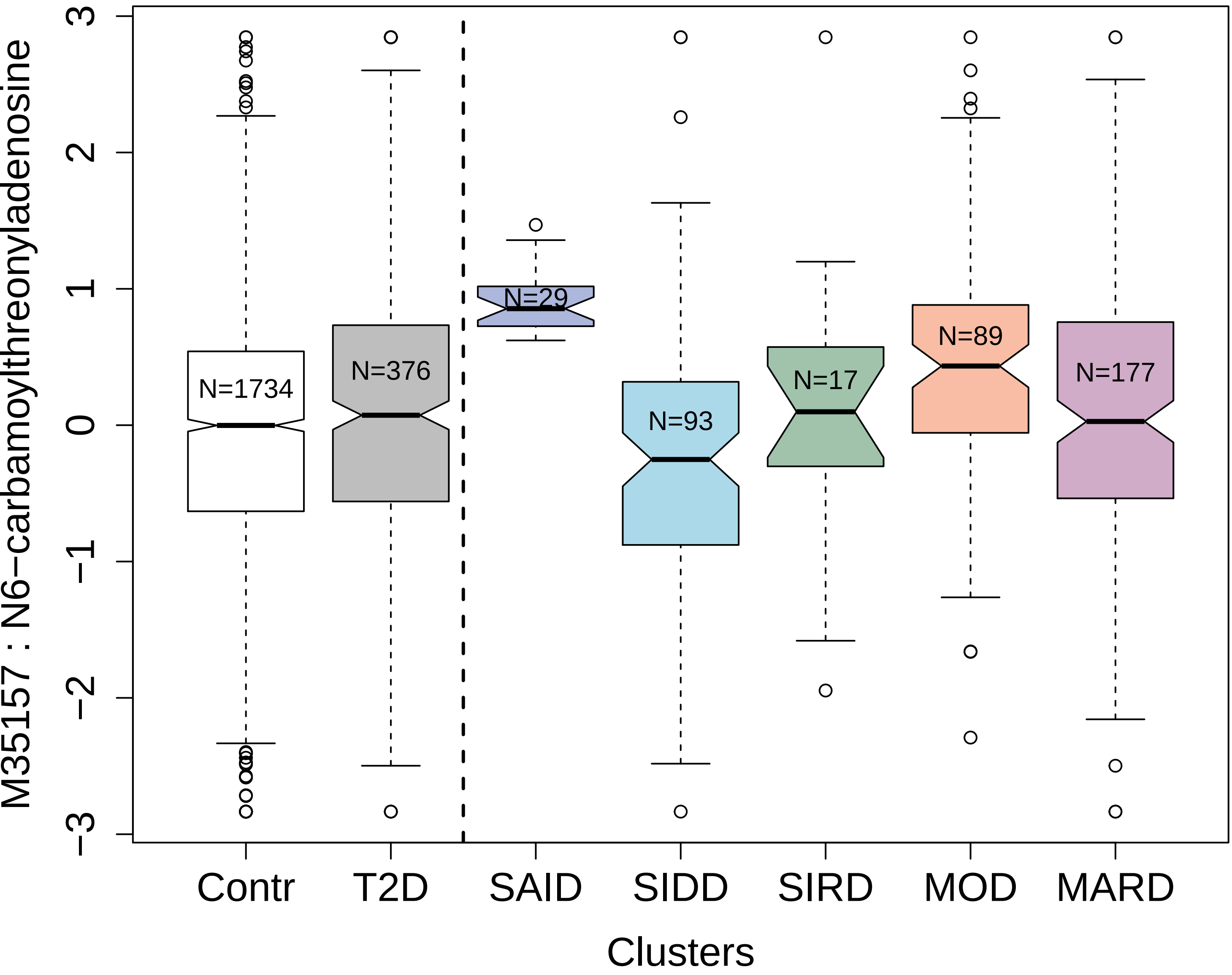
M35157 : N6−carbamoylthreonyladenosine
3
2
1
0
−1
−2
−3
N=1734
N=376
N=29
N=93
N=17
N=89
N=177
Contr
T2D
SAID
SIDD
SIRD
MOD
MARD
Clusters

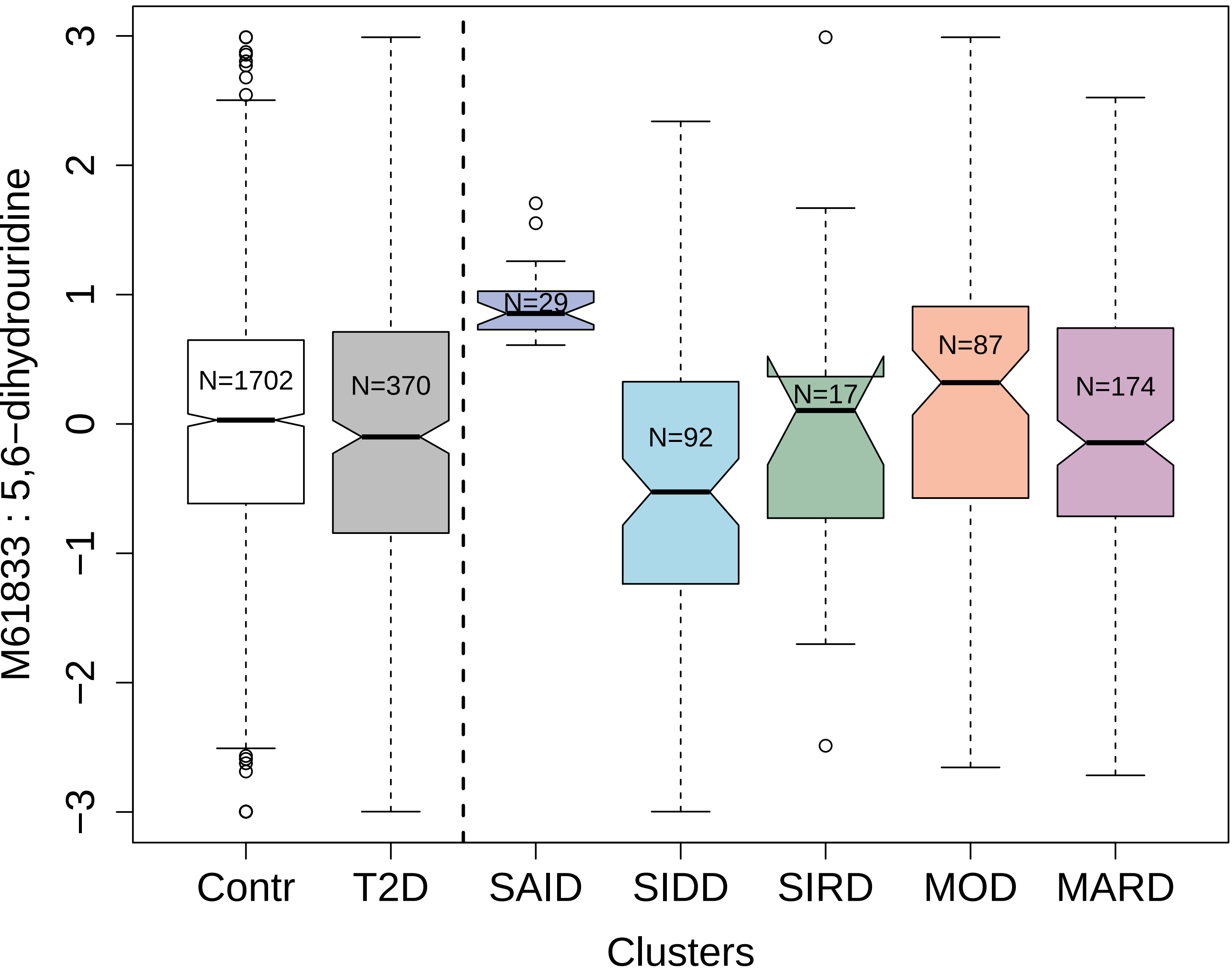
M61833 : 5,6−dihydrouridine
3
2
1
0
−1
−2
−3
N=1702
N=370
N=29
N=92
N=17
N=87
N=174
Contr
T2D
SAID
SIDD
SIRD
MOD
MARD
Clusters

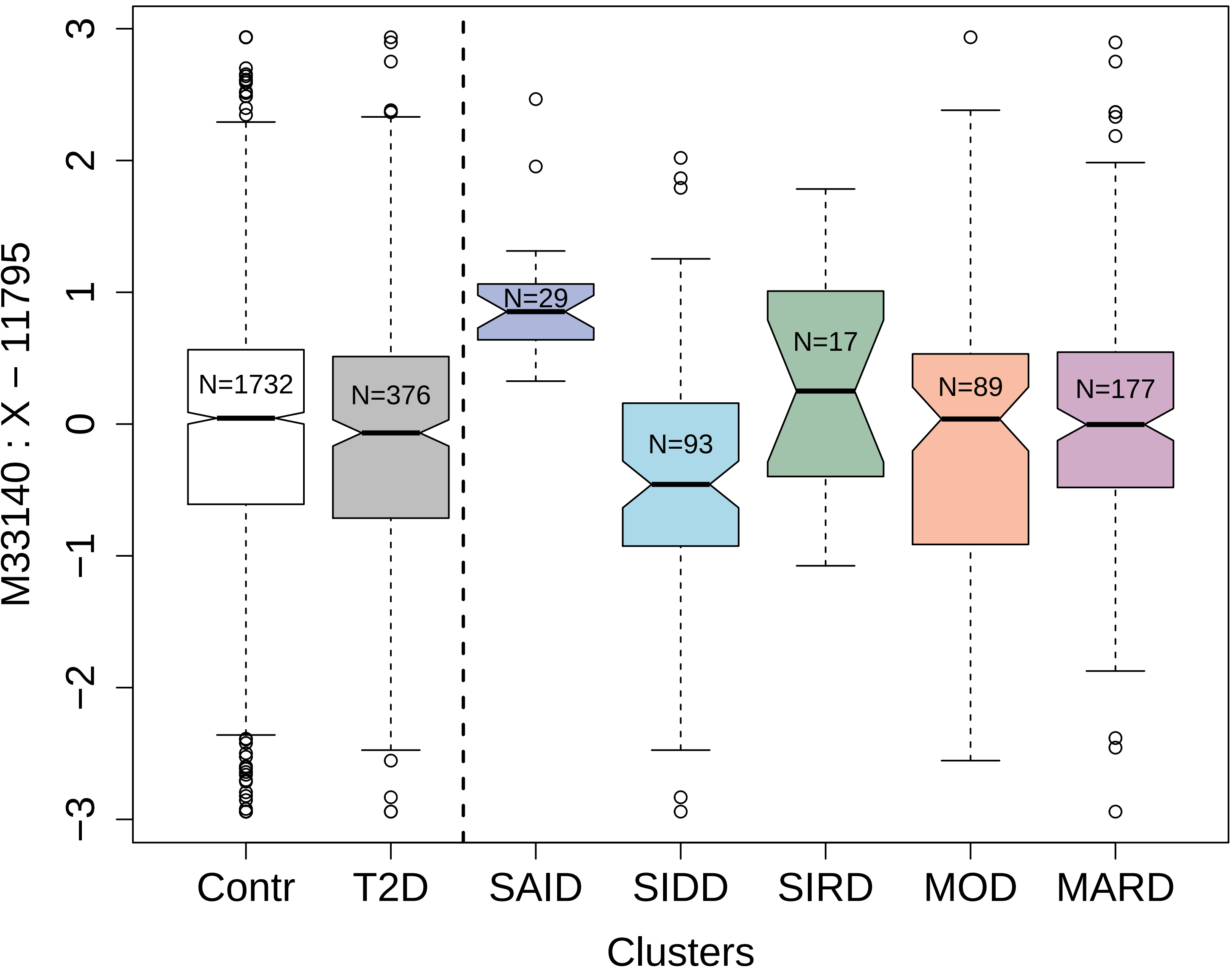

M33140 : X − 11795
3
2
1
0
−1
−2
−3
N=1732
N=376
N=29
N=93
N=17
N=89
N=177
Contr
T2D
SAID
SIDD
SIRD
MOD
MARD
Clusters

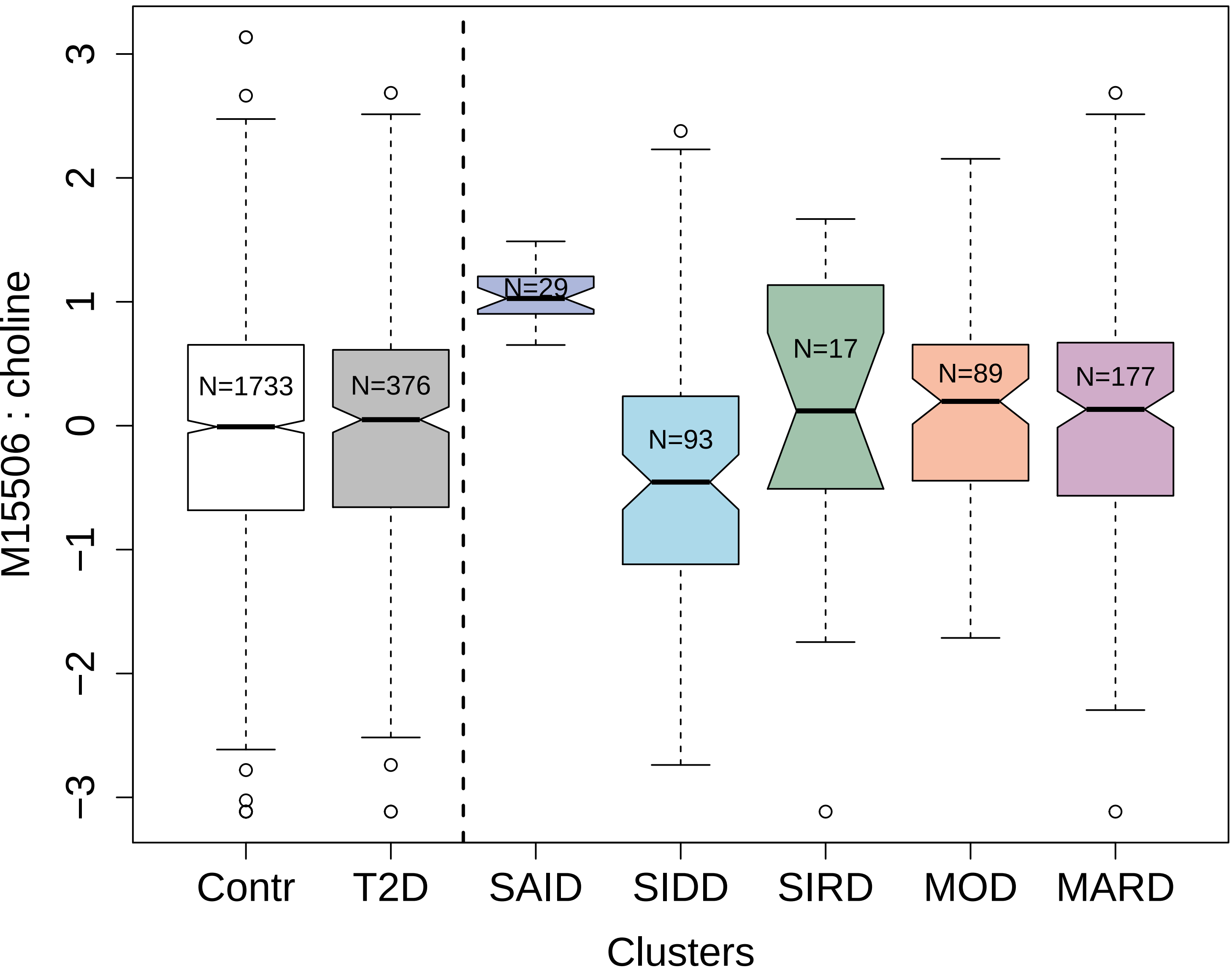

M15506 : choline
3
2
1
0
−1
−2
−3
N=1733
N=376
N=29
N=93
N=17
N=89
N=177
Contr
T2D
SAID
SIDD
SIRD
MOD
MARD
Clusters

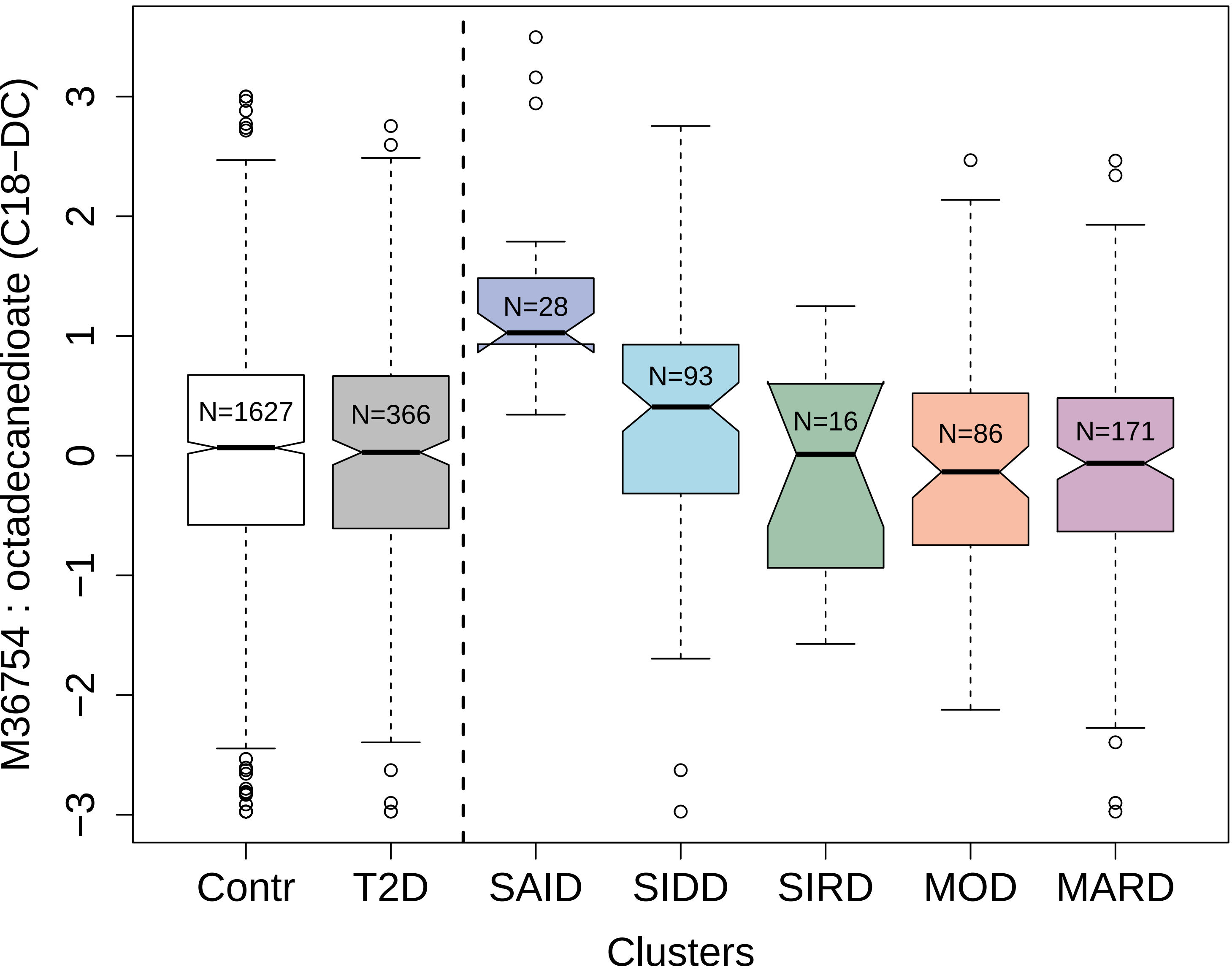
M36754 : octadecanedioate (C18−DC)
3
2
1
0
−1
−2
−3
N=1627
N=366
N=28
N=93
N=16
N=86
N=171
Contr
T2D
SAID
SIDD
SIRD
MOD
MARD
Clusters

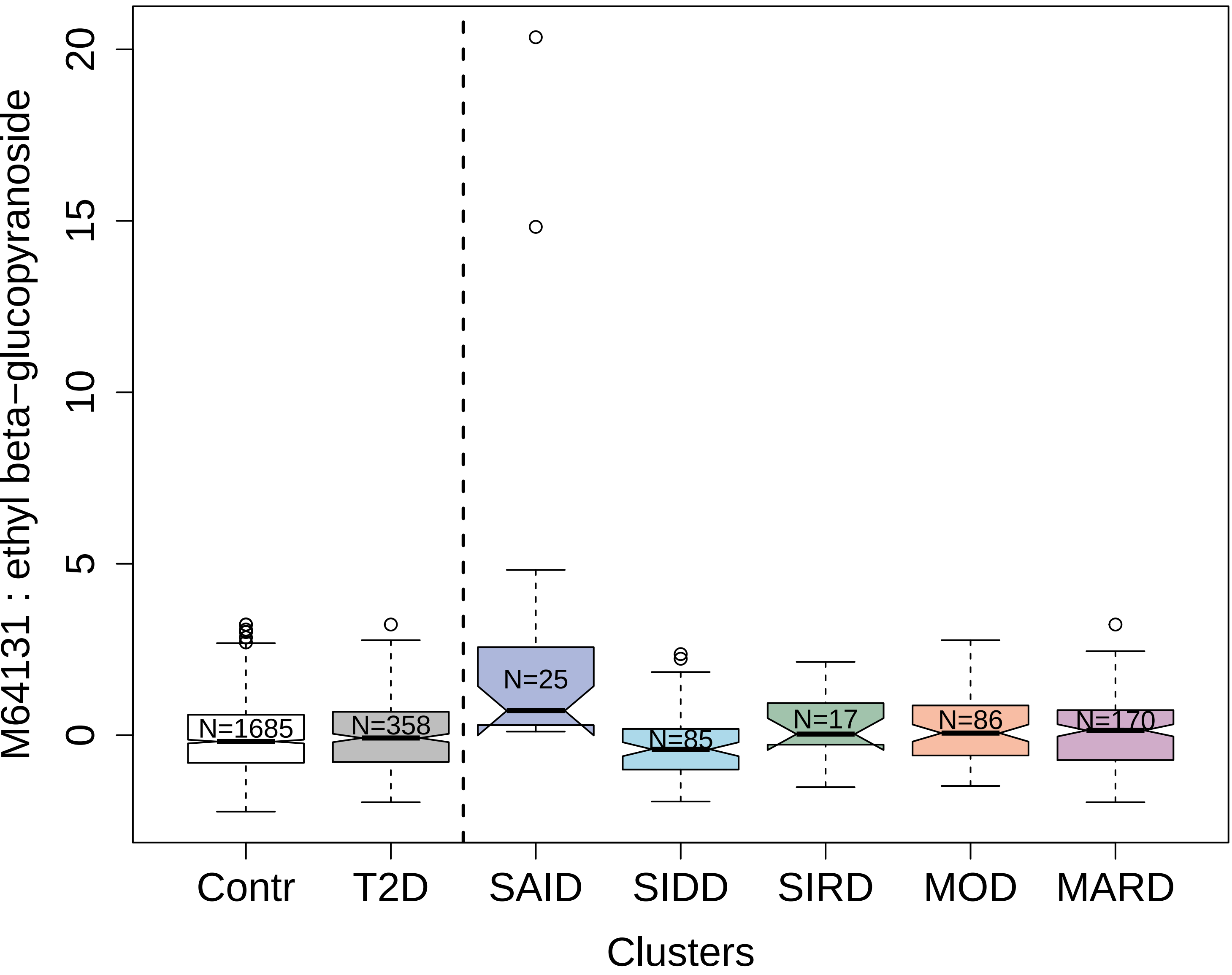

M64131 : ethyl beta−glucopyranoside
0
5
10
15
20
N=1685
N=358
N=25
N=85
N=17
N=86
N=170
Contr
T2D
SAID
SIDD
SIRD
MOD
MARD
Clusters

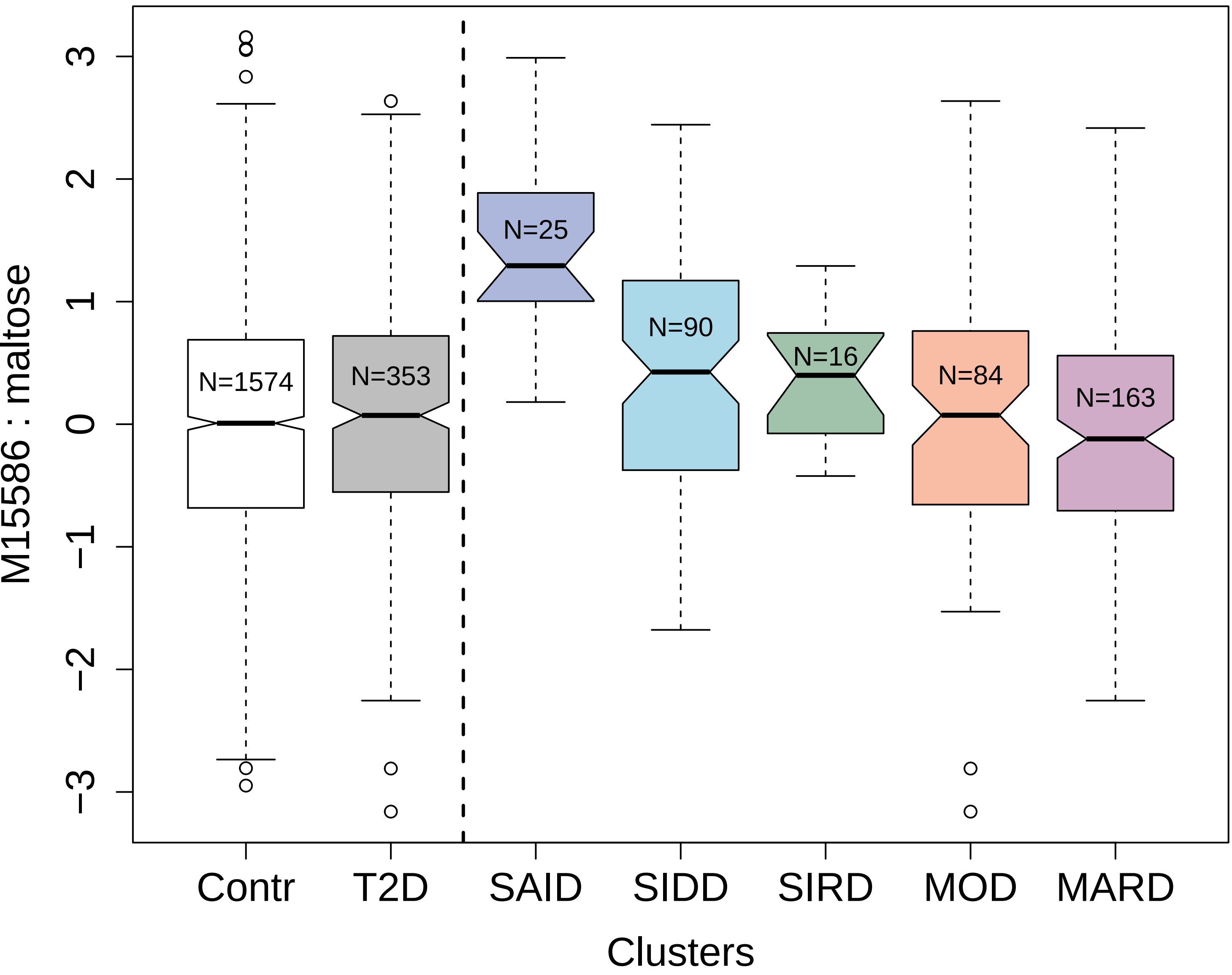

M15586 : maltose
3
2
1
0
−1
−2
−3
N=1574
N=353
N=25
N=90
N=16
N=84
N=163
Contr
T2D
SAID
SIDD
SIRD
MOD
MARD
Clusters

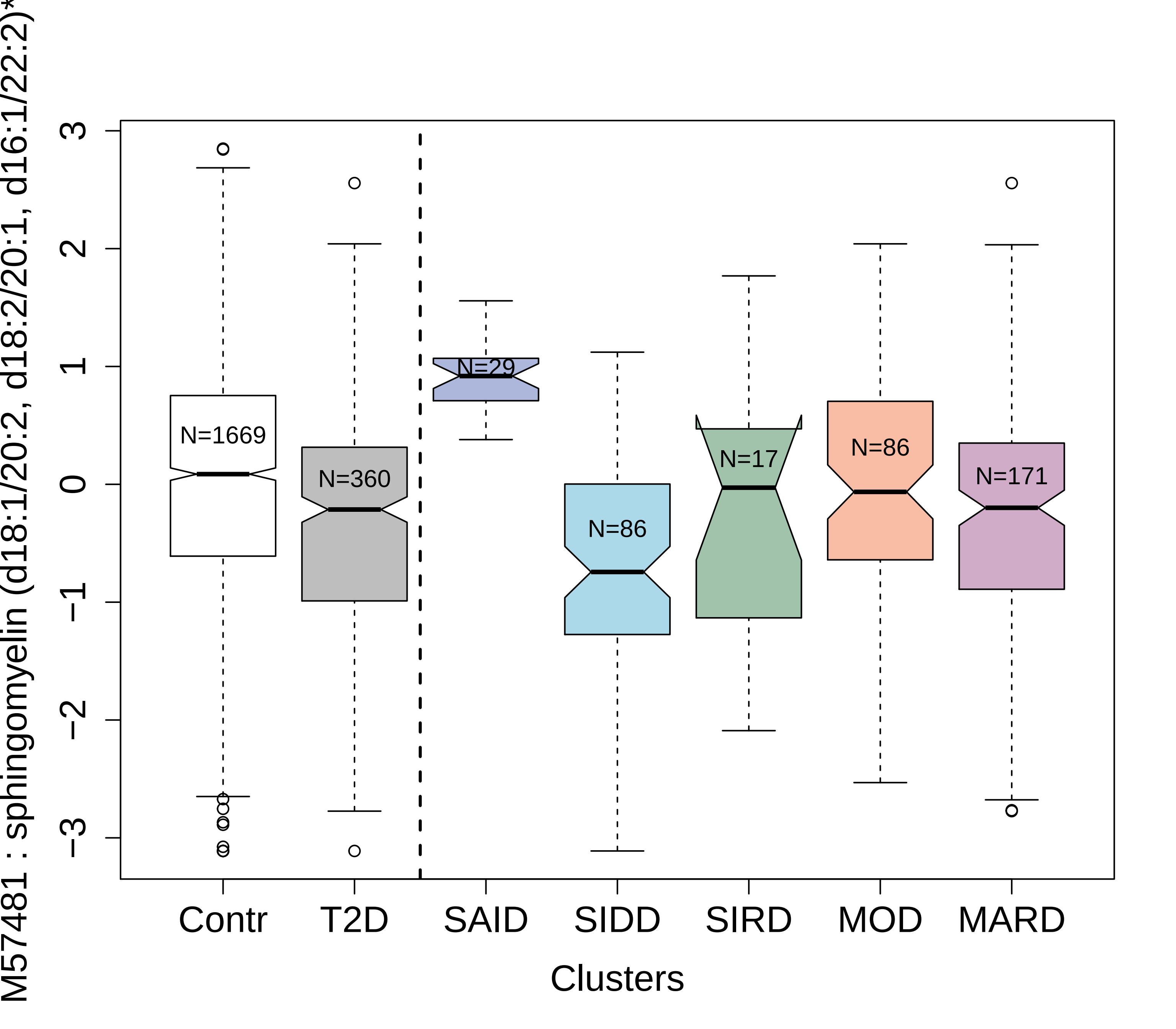

M57481 : sphingomyelin (d18:1/20:2, d18:2/20:1, d16:1/22:2)*
3
2
1
0
−1
−2
−3
N=1669
N=360
N=29
N=86
N=17
N=86
N=171
Contr
T2D
SAID
SIDD
SIRD
MOD
MARD
Clusters

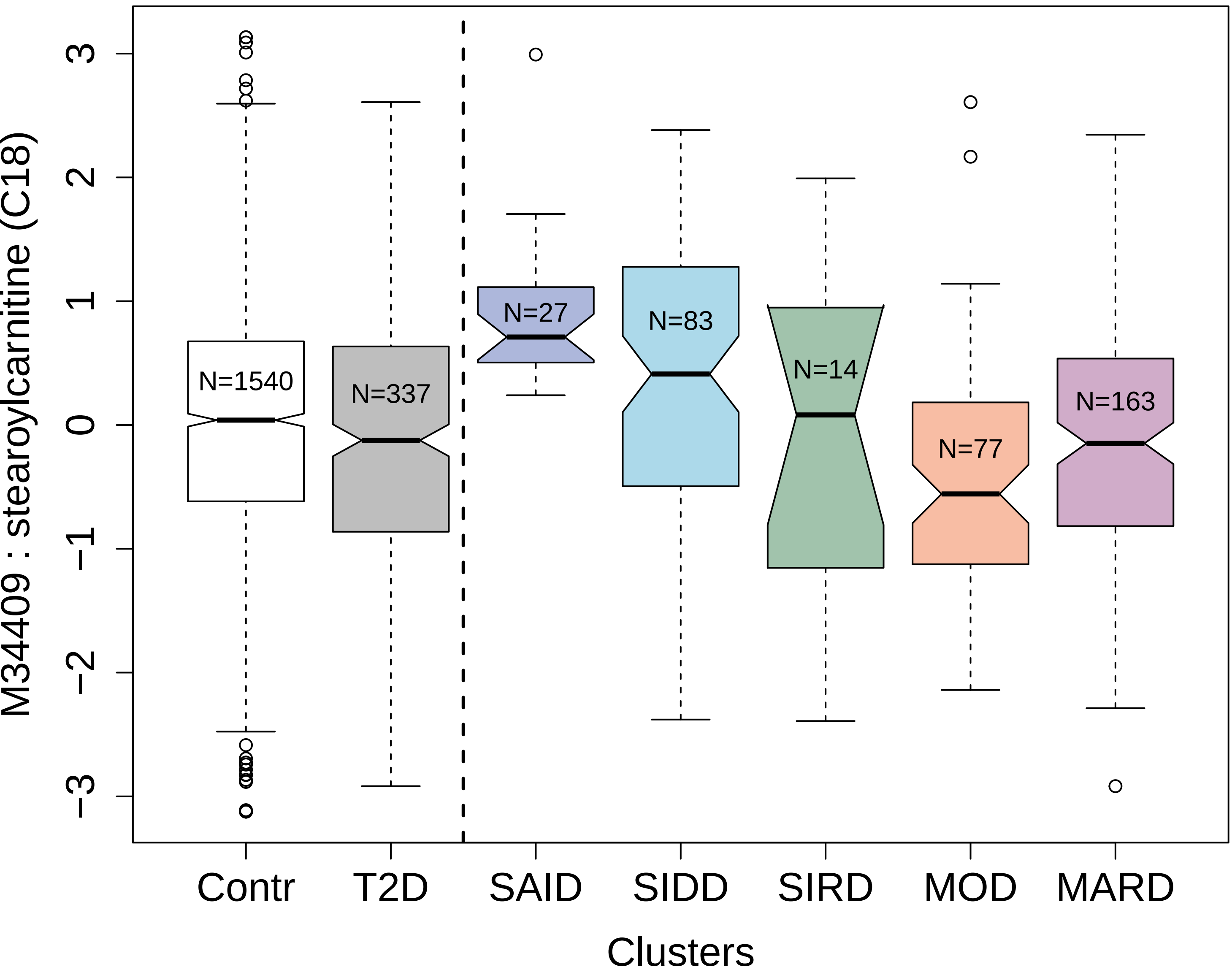

M34409 : stearoylcarnitine (C18)
3
2
1
0
−1
−2
−3
N=1540
N=337
N=27
N=83
N=14
N=77
N=163
Contr
T2D
SAID
SIDD
SIRD
MOD
MARD
Clusters

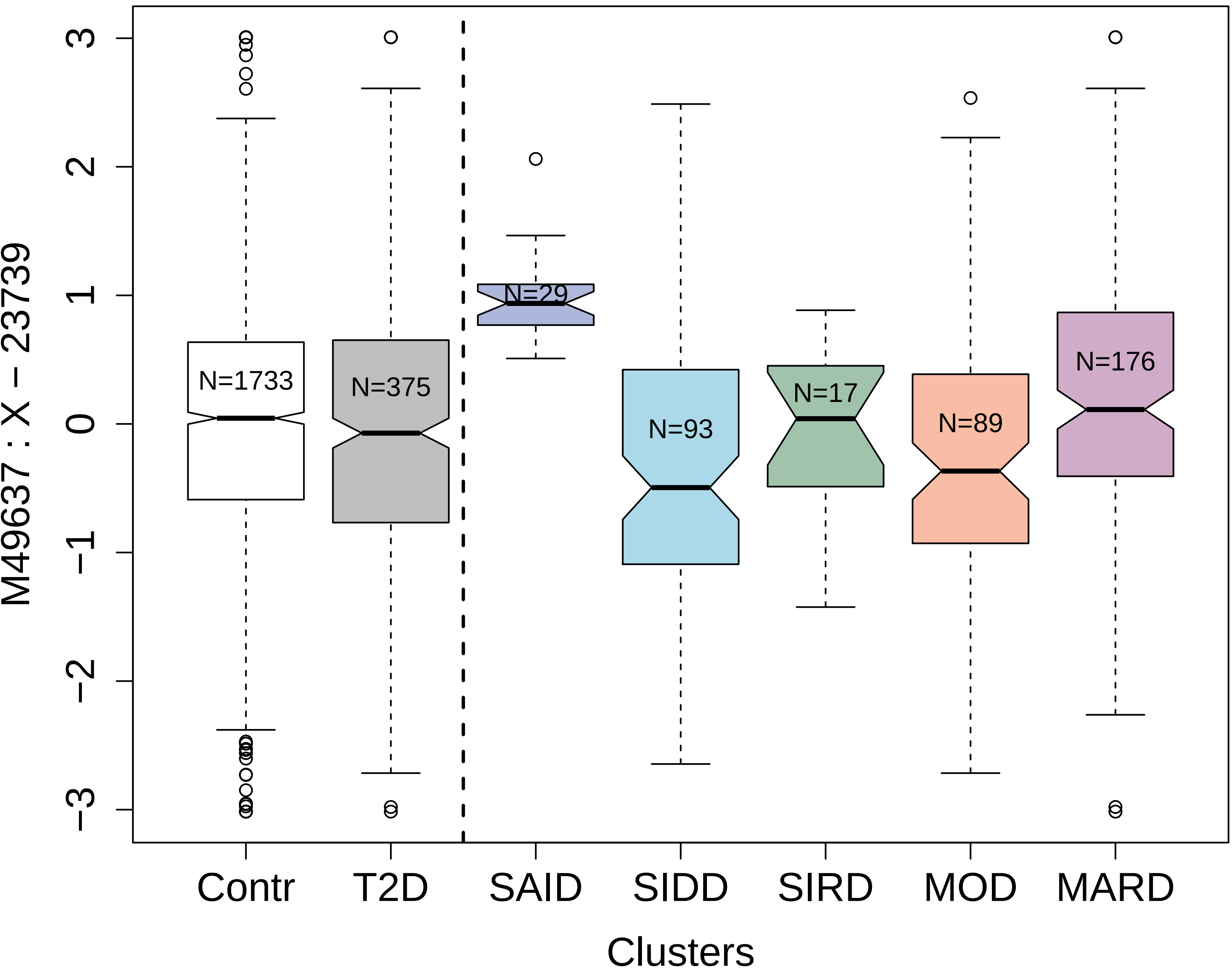

M49637 : X − 23739
3
2
1
0
−1
−2
−3
N=1733
N=375
N=29
N=93
N=17
N=89
N=176
Contr
T2D
SAID
SIDD
SIRD
MOD
MARD
Clusters

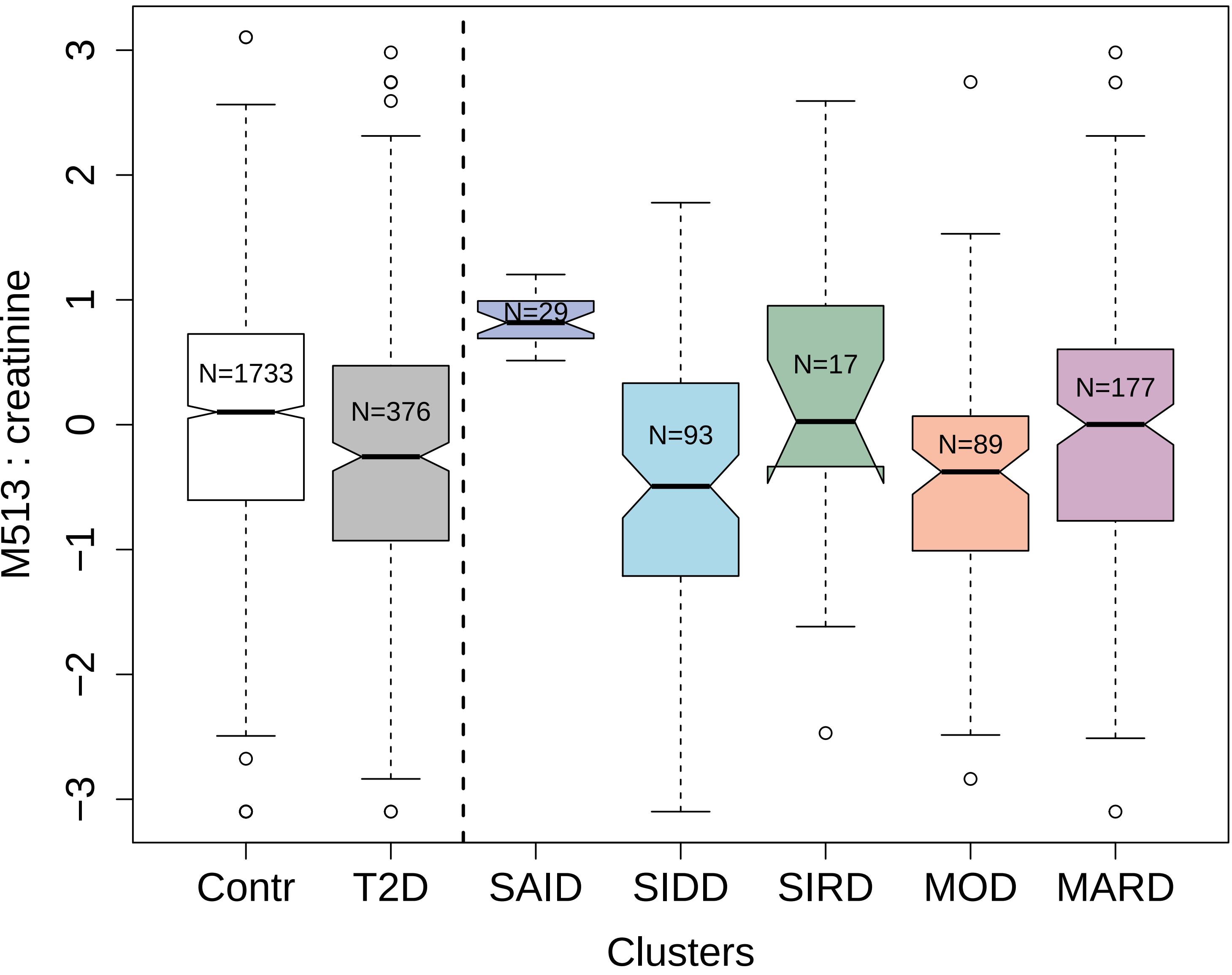
M513 : creatinine
Clusters
Contr
T2D
SAID
SIDD
SIRD
MOD
MARD
N=1733
N=376
N=29
N=93
N=17
N=89
N=177
3
2
1
0
−1
−2
−3

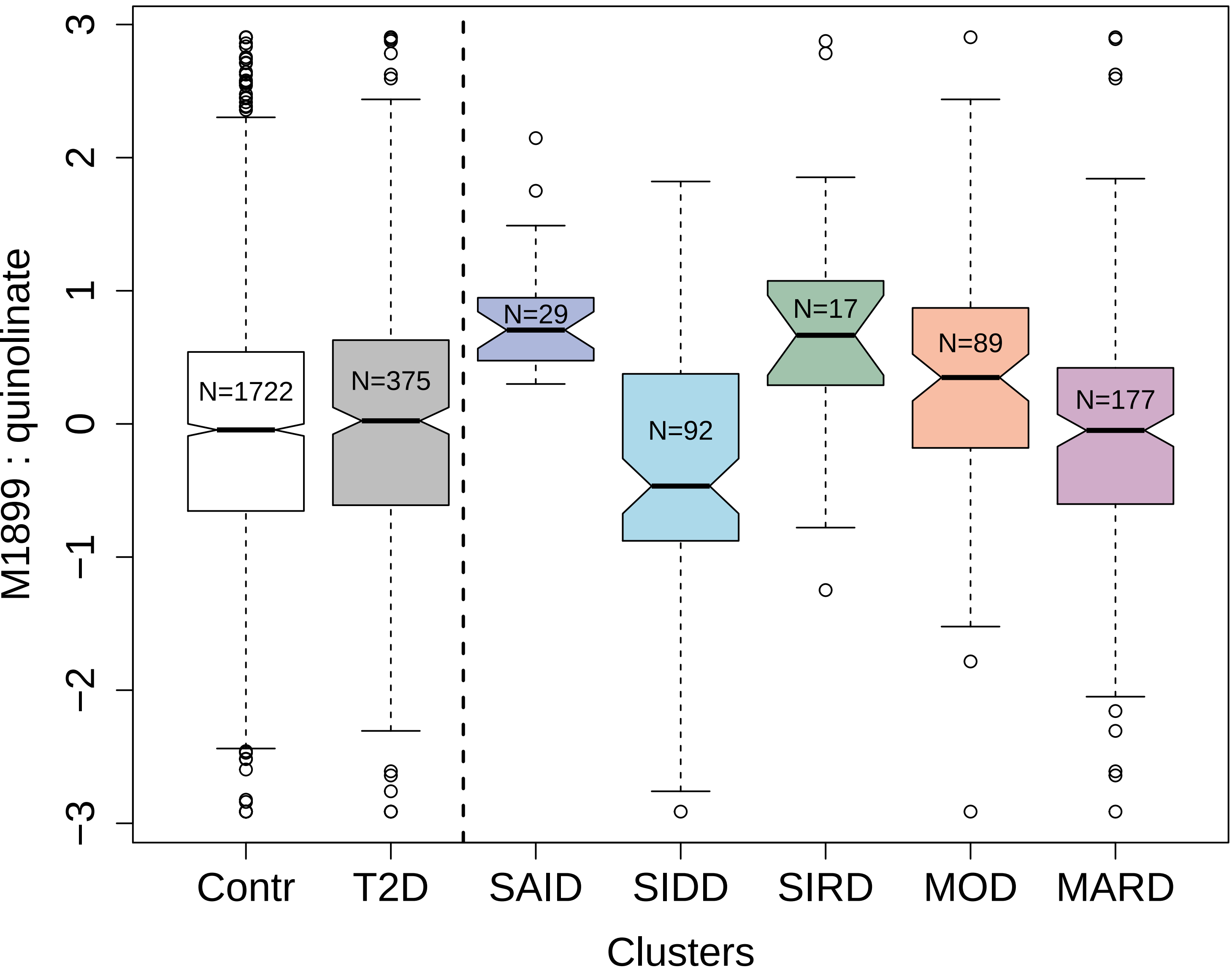

M1899 : quinolinate
3
2
1
0
−1
−2
−3
N=1722
N=375
N=29
N=92
N=17
N=89
N=177
Contr
T2D
SAID
SIDD
SIRD
MOD
MARD
Clusters

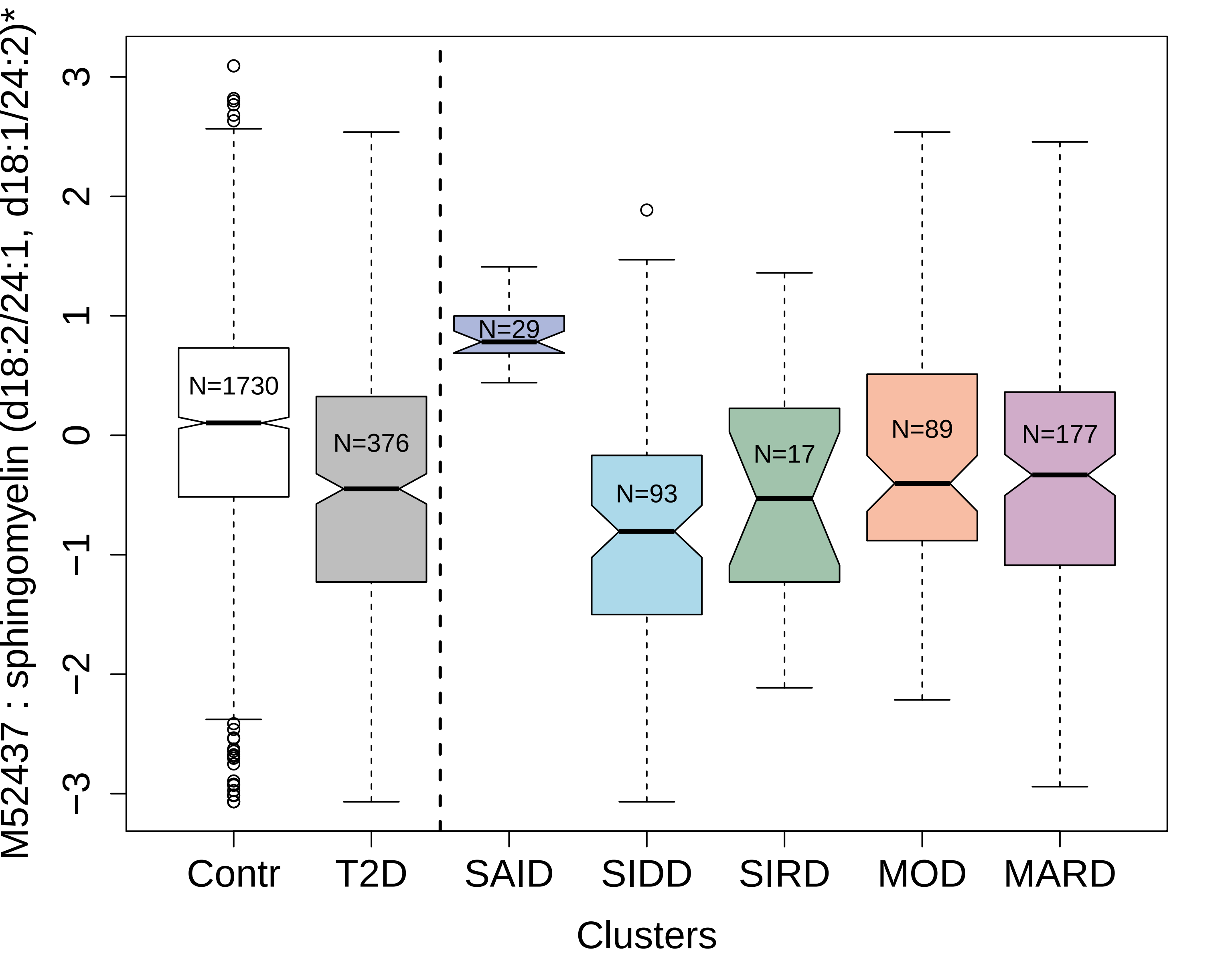

M52437 : sphingomyelin (d18:2/24:1, d18:1/24:2)*
3
2
1
0
−1
−2
−3
N=1730
N=376
N=29
N=93
N=17
N=89
N=177
Contr
T2D
SAID
SIDD
SIRD
MOD
MARD
Clusters

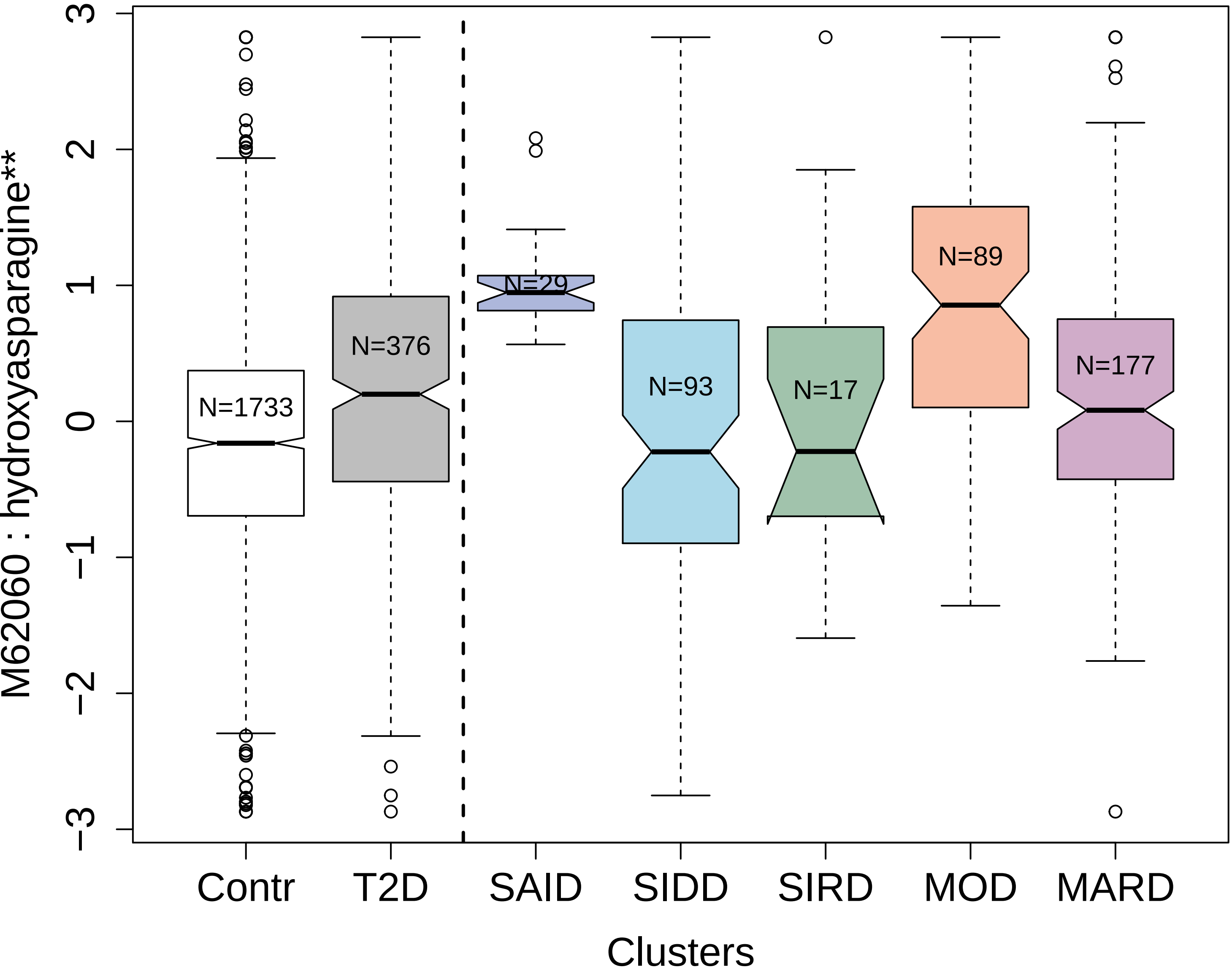

M62060 : hydroxyasparagine**
3
2
1
0
−1
−2
−3
N=1733
N=376
N=29
N=93
N=17
N=89
N=177
Contr
T2D
SAID
SIDD
SIRD
MOD
MARD
Clusters

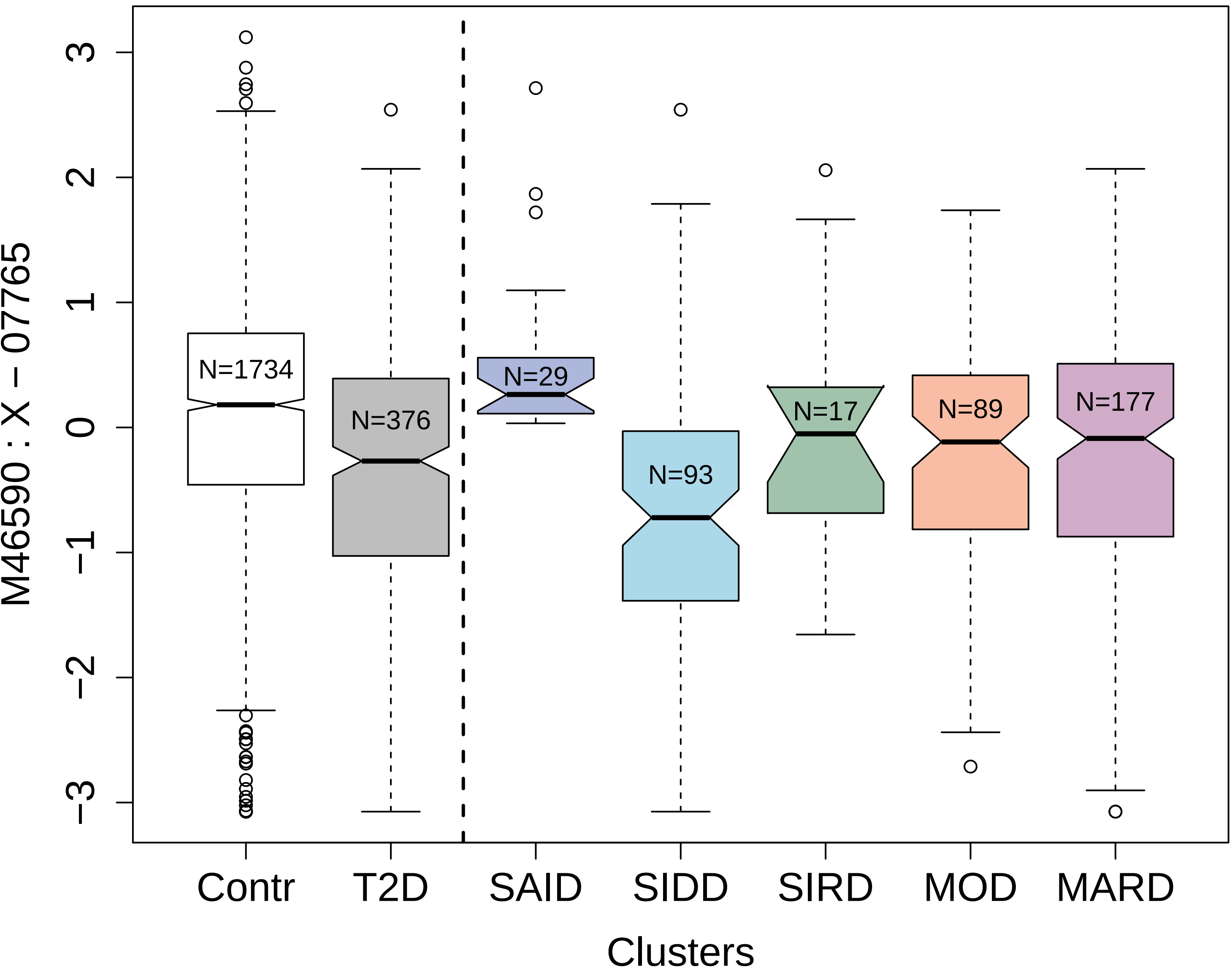

M46590 : X − 07765
3
2
1
0
−1
−2
−3
N=1734
N=376
N=29
N=93
N=17
N=89
N=177
Contr
T2D
SAID
SIDD
SIRD
MOD
MARD
Clusters

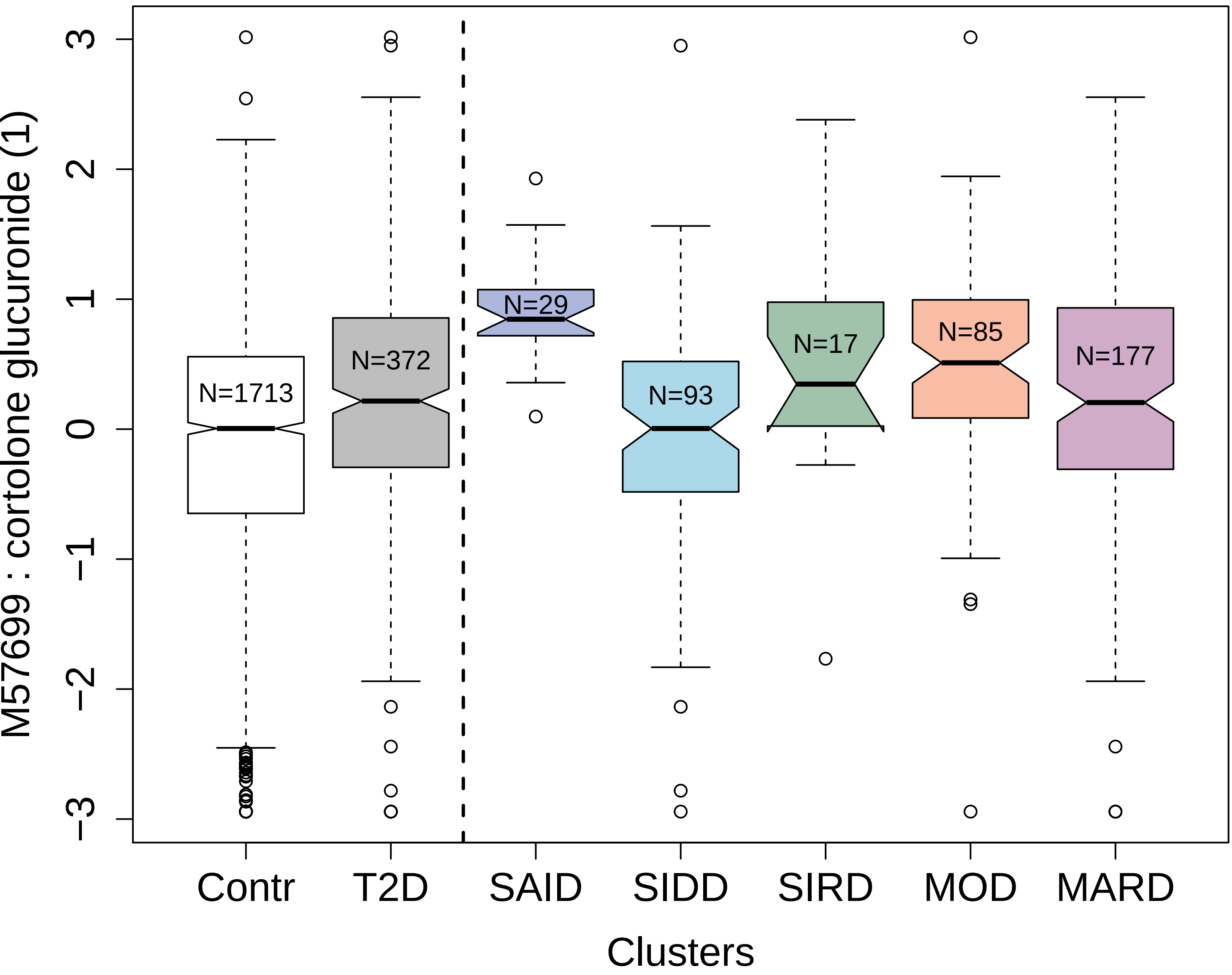

M57699 : cortolone glucuronide (1)
3
2
1
0
−1
−2
−3
N=1713
N=372
N=29
N=93
N=17
N=85
N=177
Contr
T2D
SAID
SIDD
SIRD
MOD
MARD
Clusters

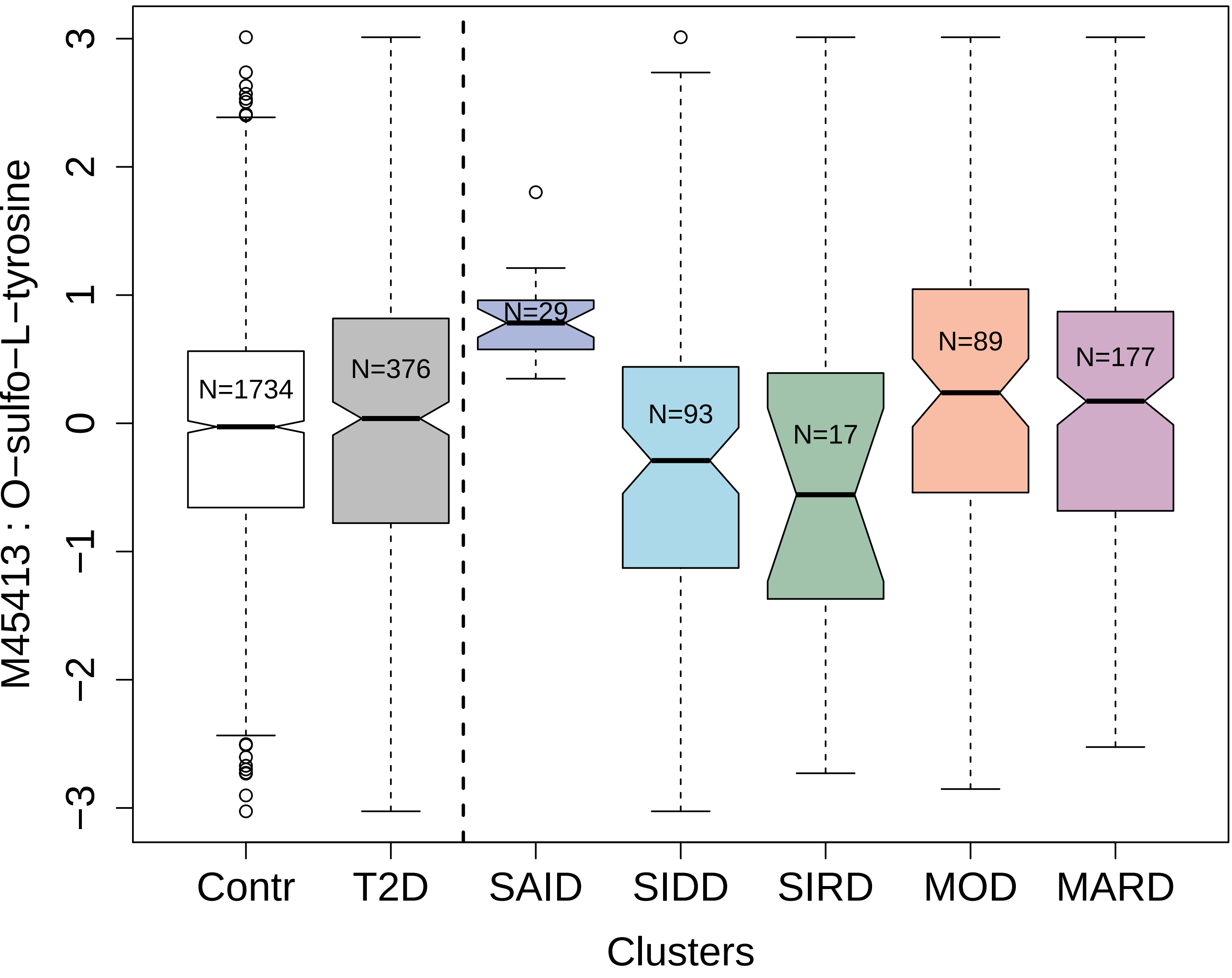

M45413 : O−sulfo−L−tyrosine
3
2
1
0
−1
−2
−3
N=1734
N=376
N=29
N=93
N=17
N=89
N=177
Contr
T2D
SAID
SIDD
SIRD
MOD
MARD
Clusters

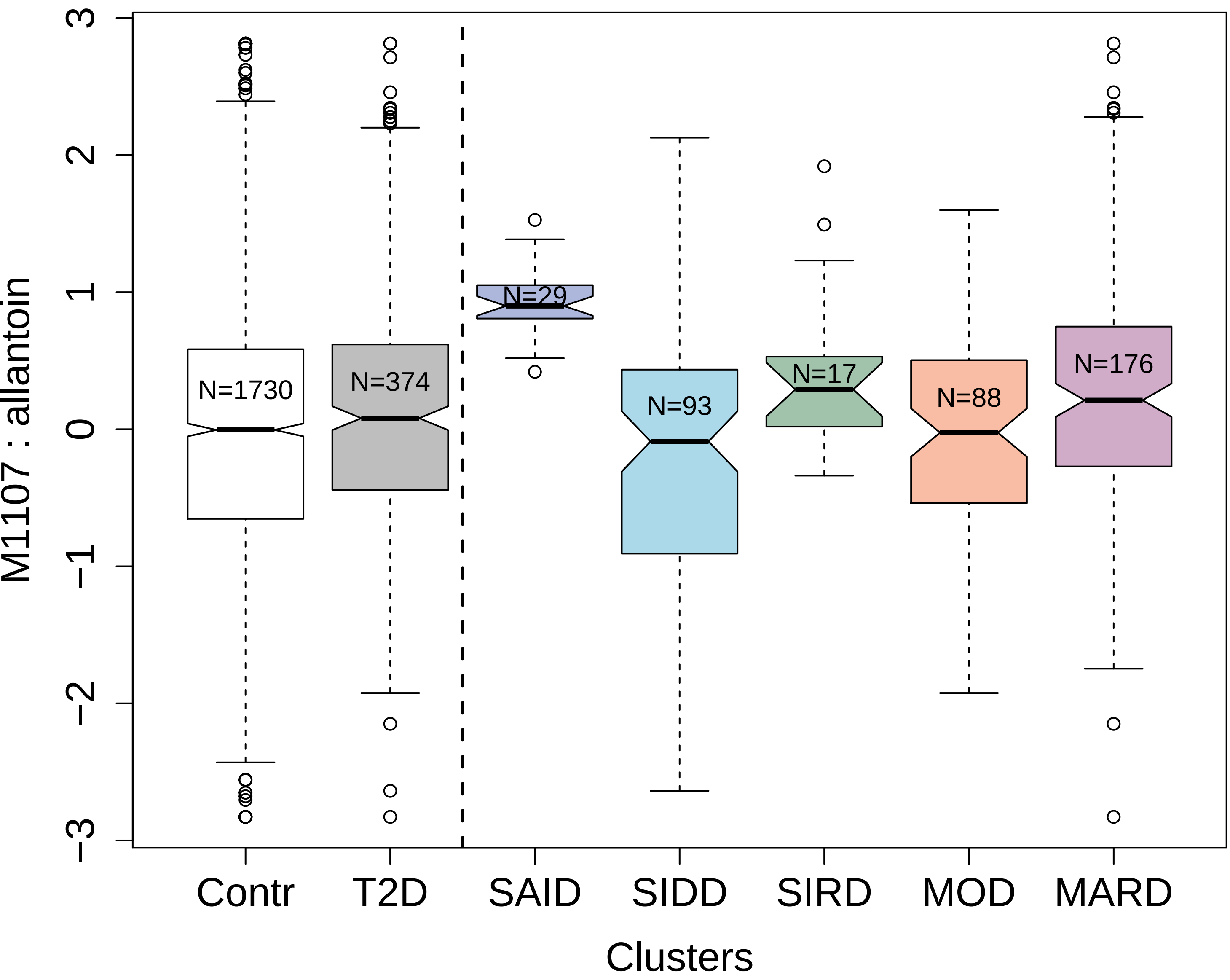

M1107 : allantoin
3
2
1
0
−1
−2
−3
N=1730
N=374
N=29
N=93
N=17
N=88
N=176
Contr
T2D
SAID
SIDD
SIRD
MOD
MARD
Clusters

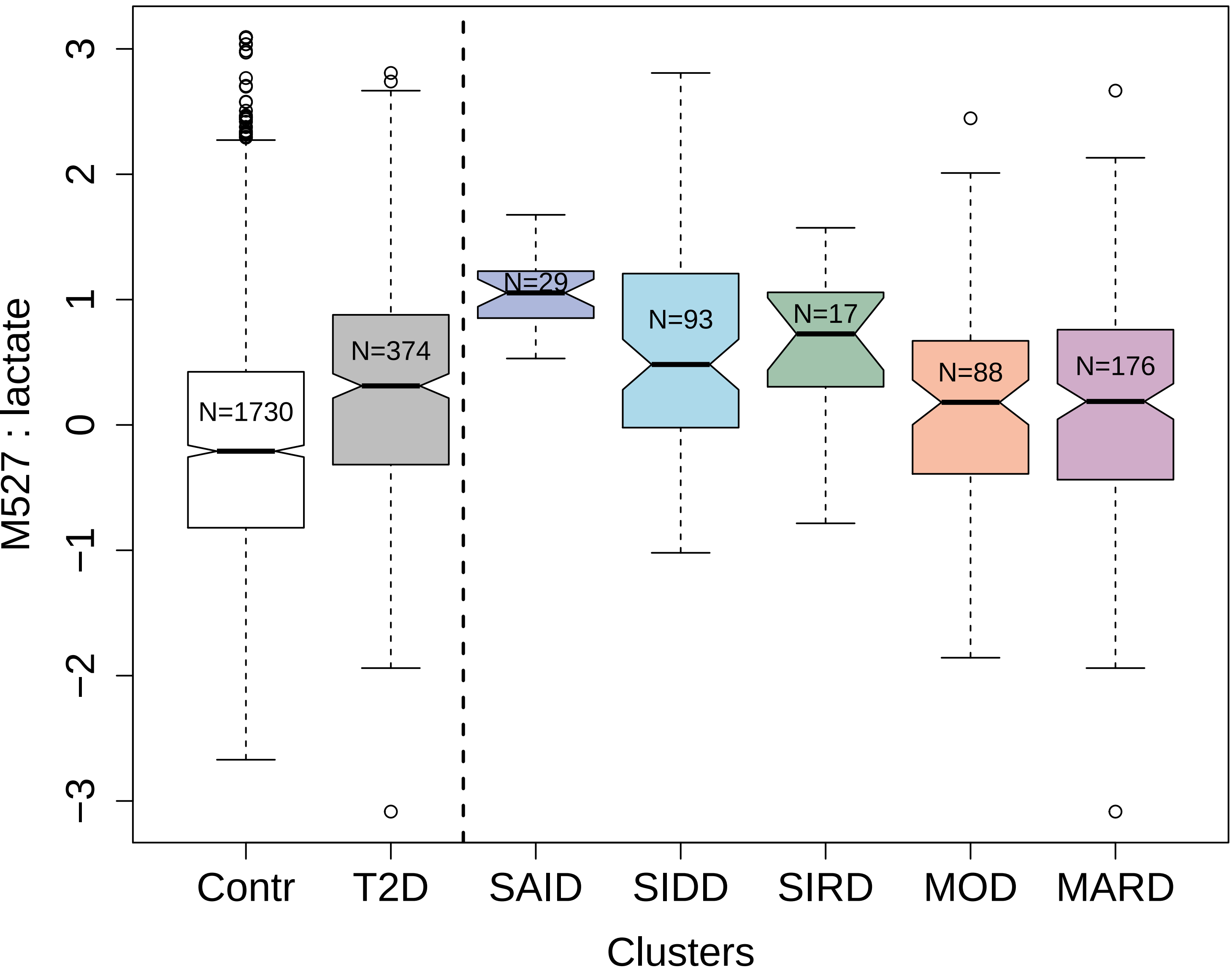

M527 : lactate
3
2
1
0
−1
−2
−3
N=1730
N=374
N=29
N=93
N=17
N=88
N=176
Contr
T2D
SAID
SIDD
SIRD
MOD
MARD
Clusters

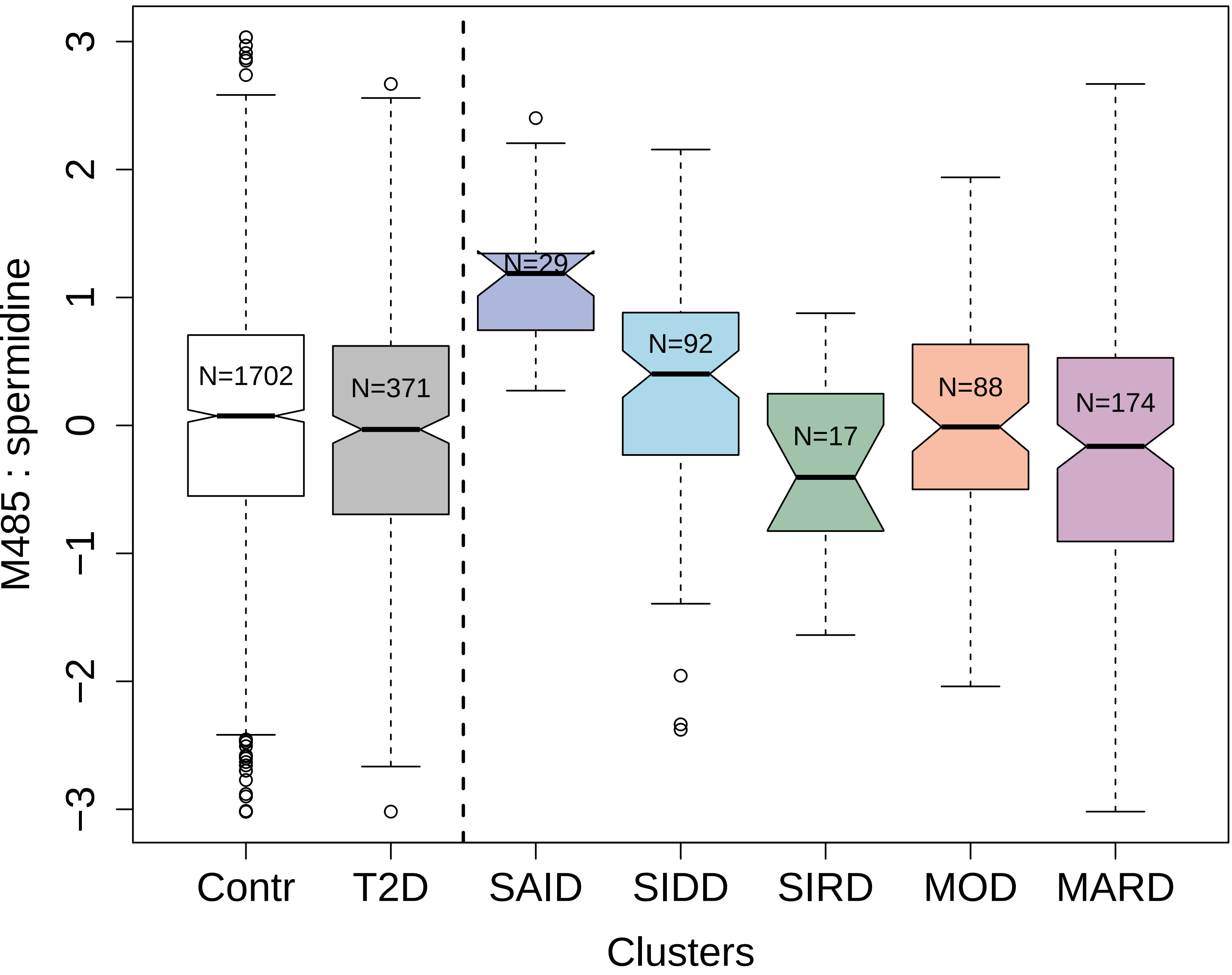
M485 : spermidine
Clusters
Contr
T2D
SAID
SIDD
SIRD
MOD
MARD
N=1702
N=371
N=29
N=92
N=17
N=88
N=174
3
2
1
0
−1
−2
−3

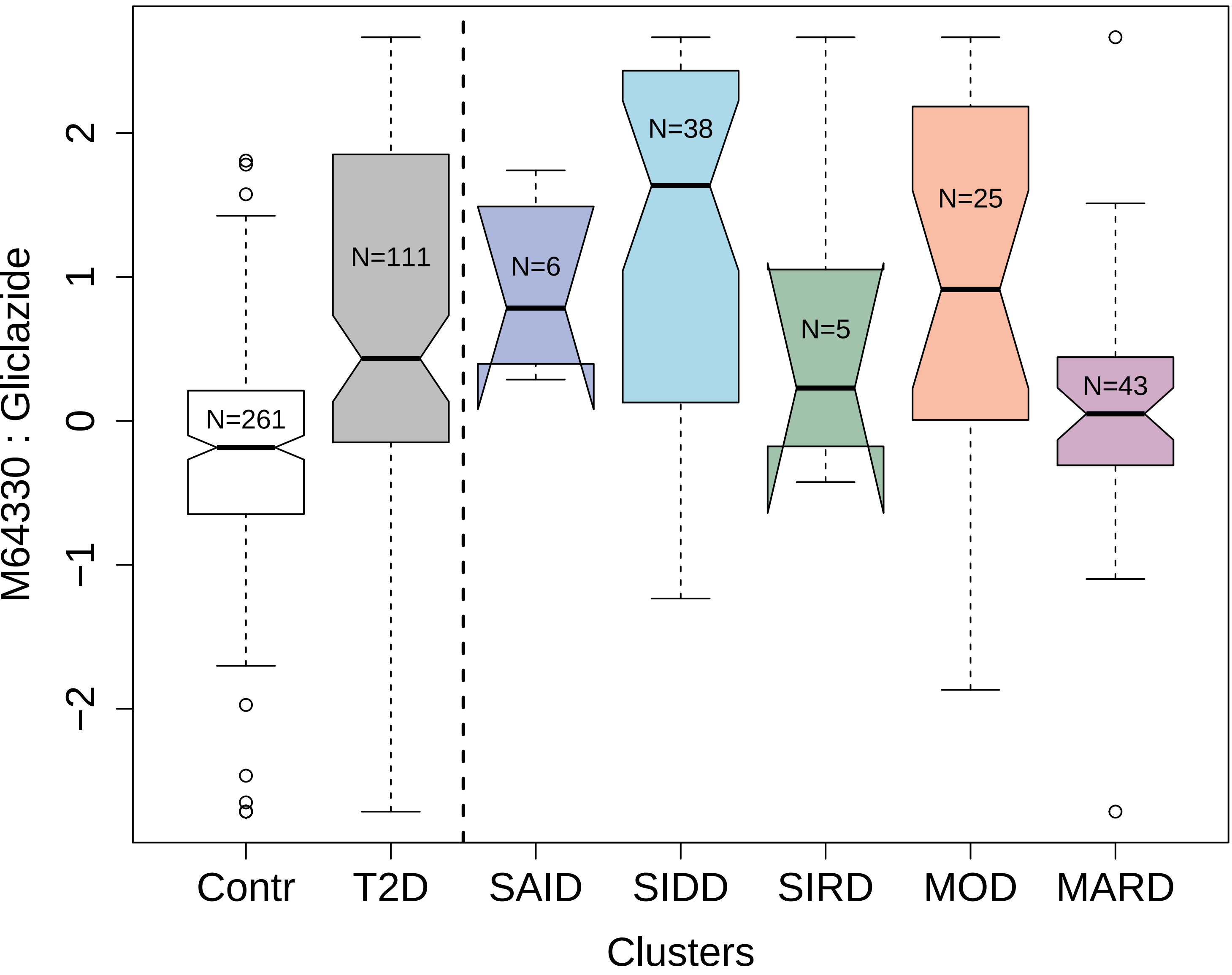

M64330 : Gliclazide
Clusters
Contr
T2D
SAID
SIDD
SIRD
MOD
MARD
N=261
N=111
N=6
N=38
N=5
N=25
N=43
−2
−1
0
1
2

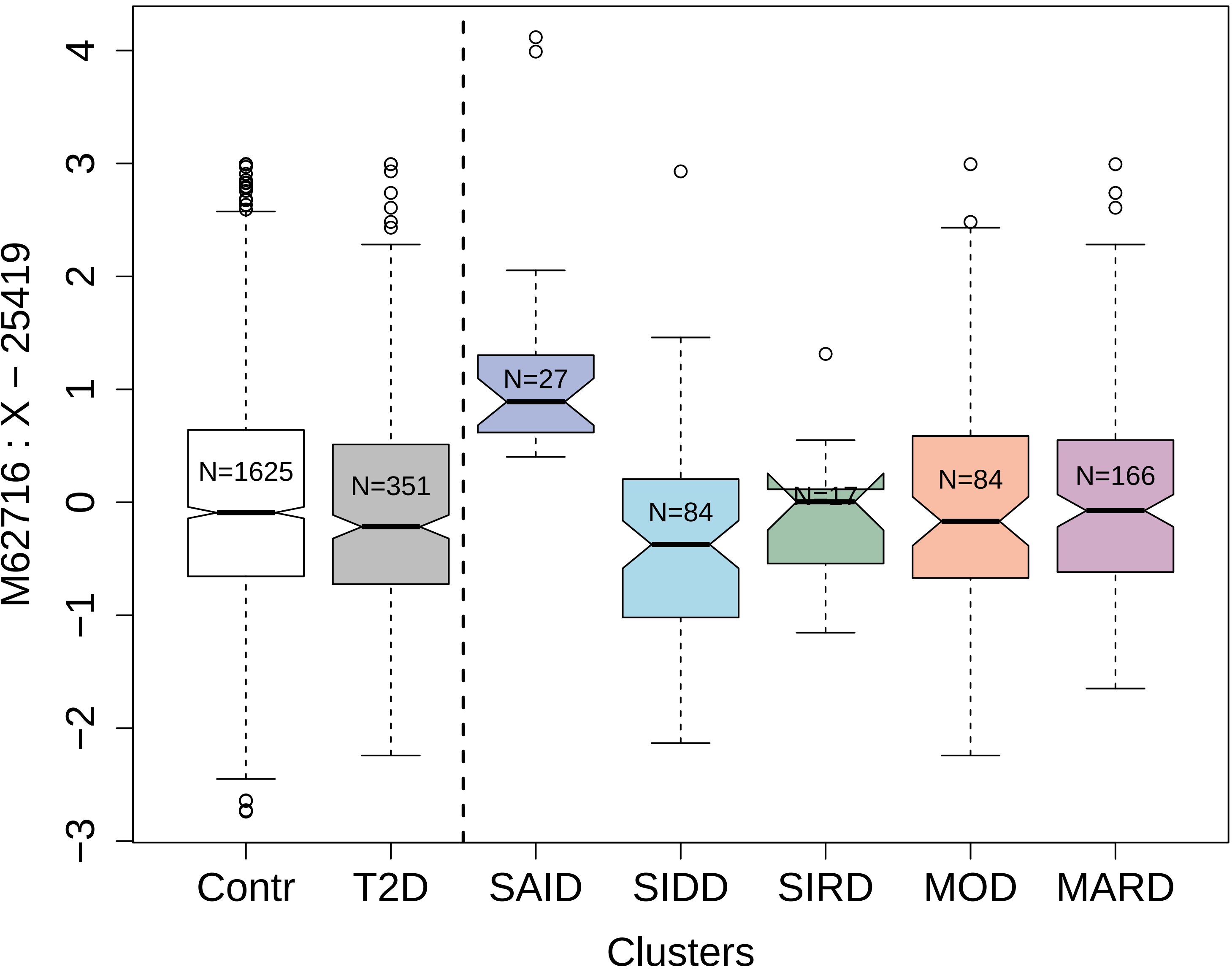
M62716 : X − 25419
4
3
2
1
0
−1
−2
−3
N=1625
N=351
N=27
N=84
N=17
N=84
N=166
Contr
T2D
SAID
SIDD
SIRD
MOD
MARD
Clusters

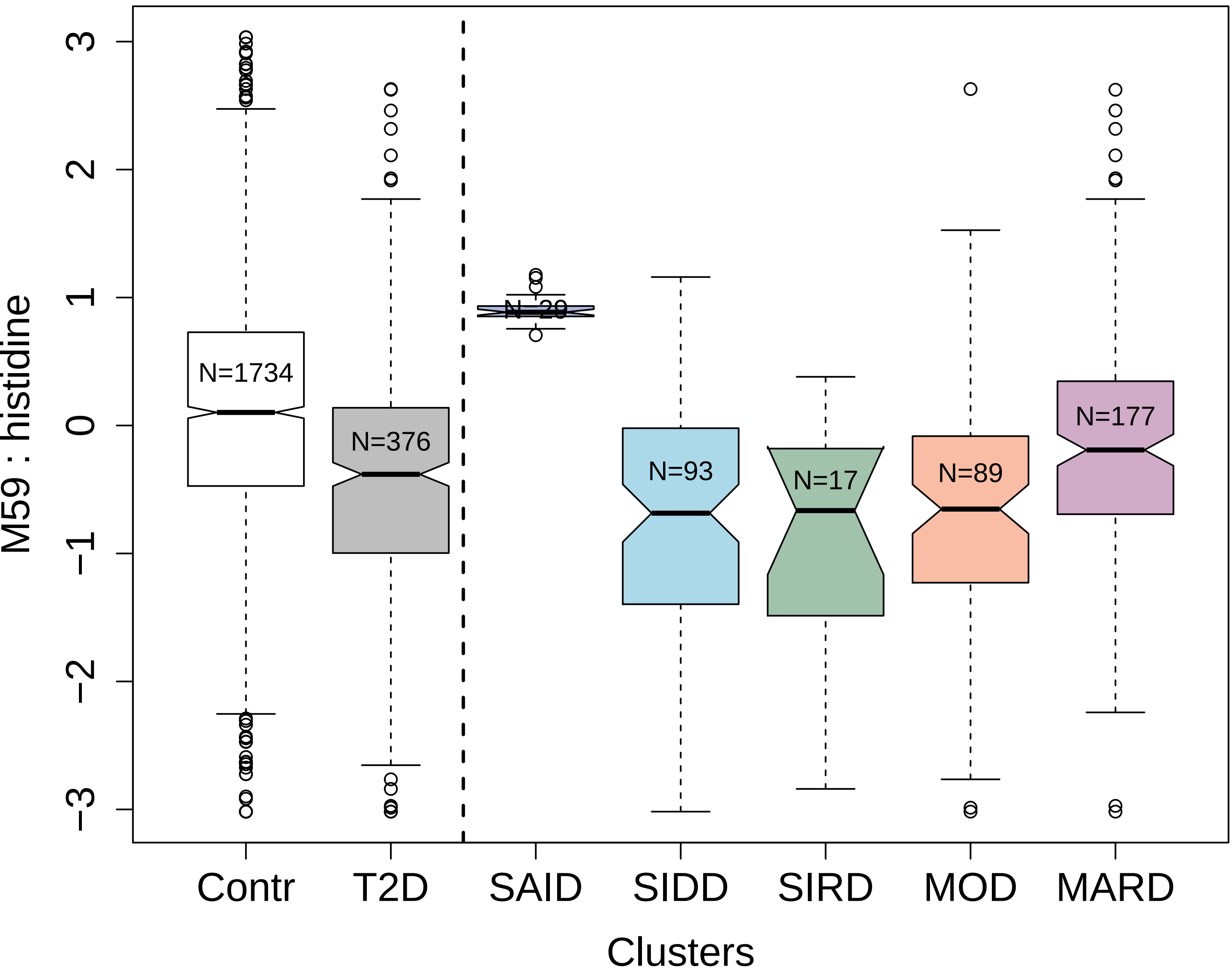

M59 : histidine
3
2
1
0
−1
−2
−3
N=1734
N=376
N=29
N=93
N=17
N=89
N=177
Contr
T2D
SAID
SIDD
SIRD
MOD
MARD
Clusters

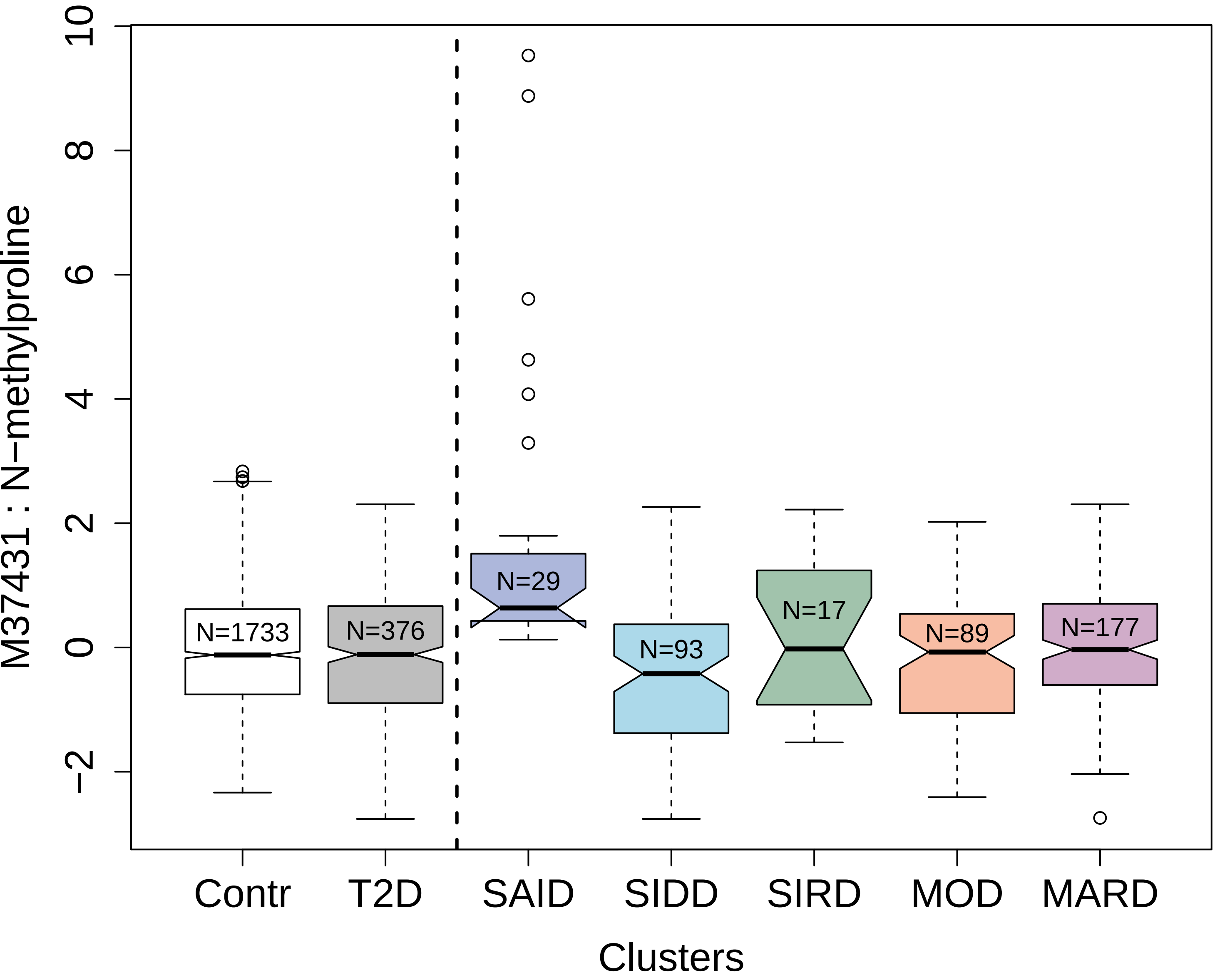
M37431 : N−methylproline
Clusters
Contr
T2D
SAID
SIDD
SIRD
MOD
MARD
N=1733
N=376
N=29
N=93
N=17
N=89
N=177
10
8
6
4
2
0
−2

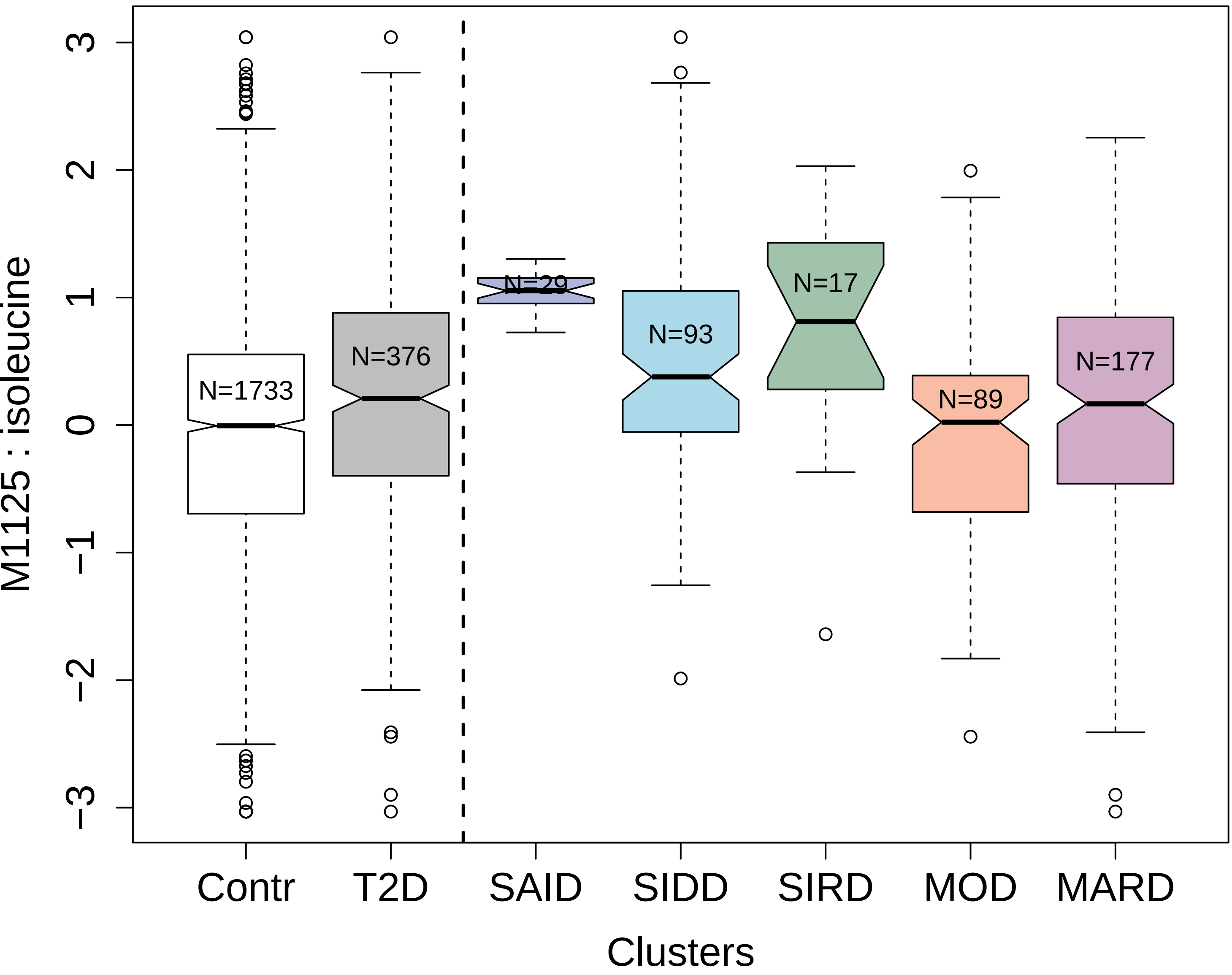
M1125 : isoleucine
3
2
1
0
−1
−2
−3
N=1733
N=376
N=29
N=93
N=17
N=89
N=177
Contr
T2D
SAID
SIDD
SIRD
MOD
MARD
Clusters

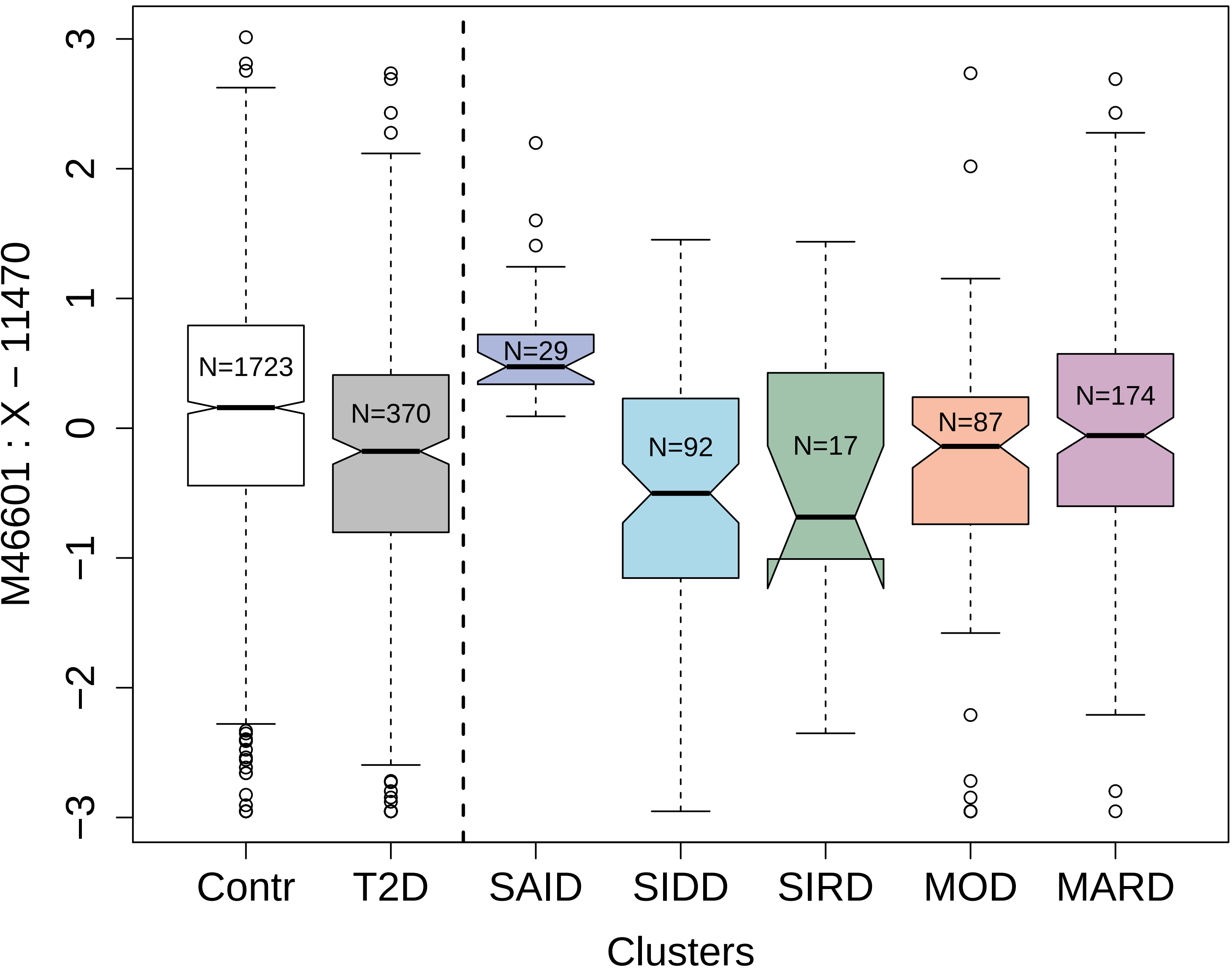

M46601 : X − 11470
3
2
1
0
−1
−2
−3
N=1723
N=370
N=29
N=92
N=17
N=87
N=174
Contr
T2D
SAID
SIDD
SIRD
MOD
MARD
Clusters

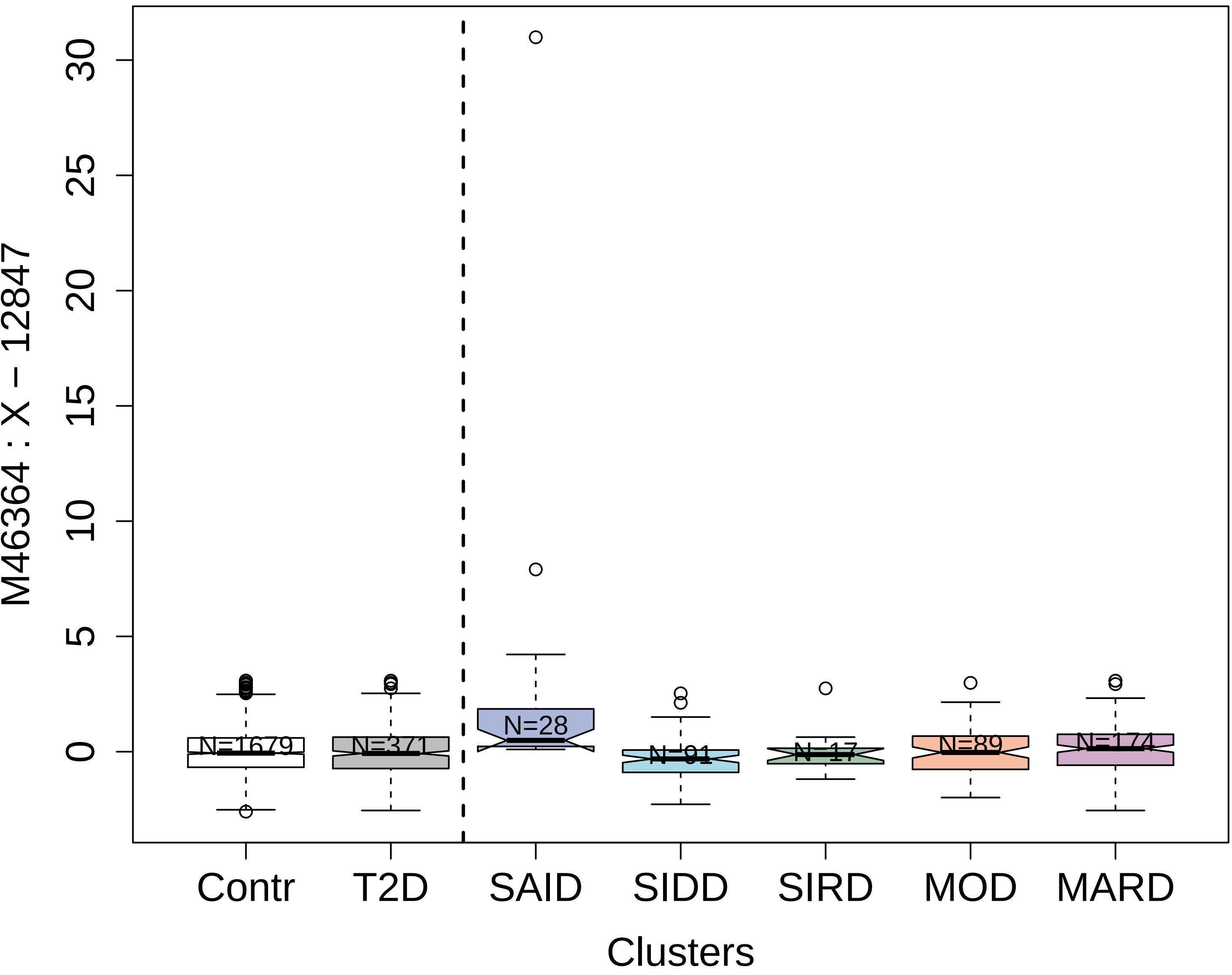

M46364 : X − 12847
0
5
10
15
20
25
30
N=1679
N=371
N=28
N=91
N=17
N=89
N=174
Contr
T2D
SAID
SIDD
SIRD
MOD
MARD
Clusters

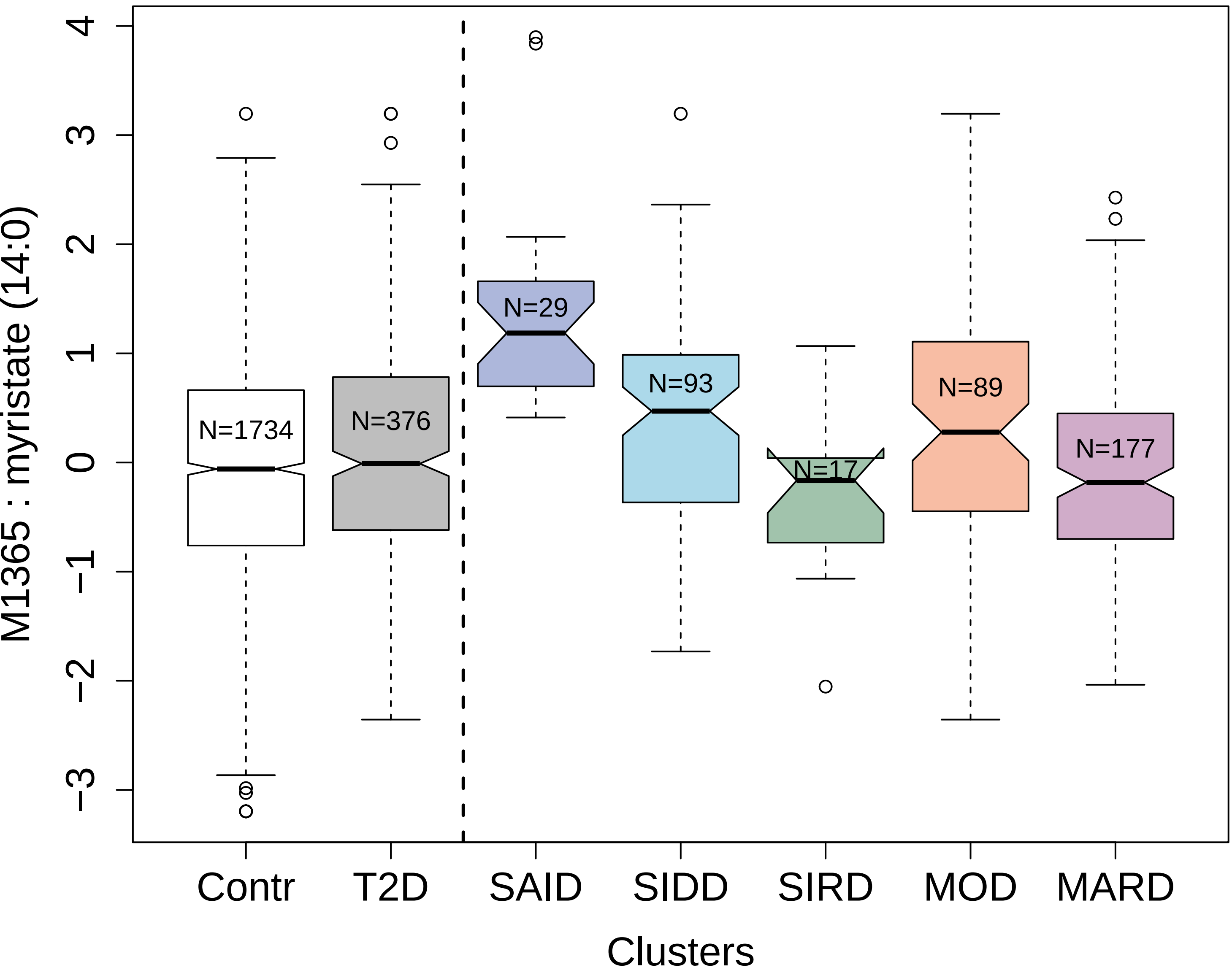

M1365 : myristate (14:0)
4
3
2
1
0
−1
−2
−3
N=1734
N=376
N=29
N=93
N=17
N=89
N=177
Contr
T2D
SAID
SIDD
SIRD
MOD
MARD
Clusters

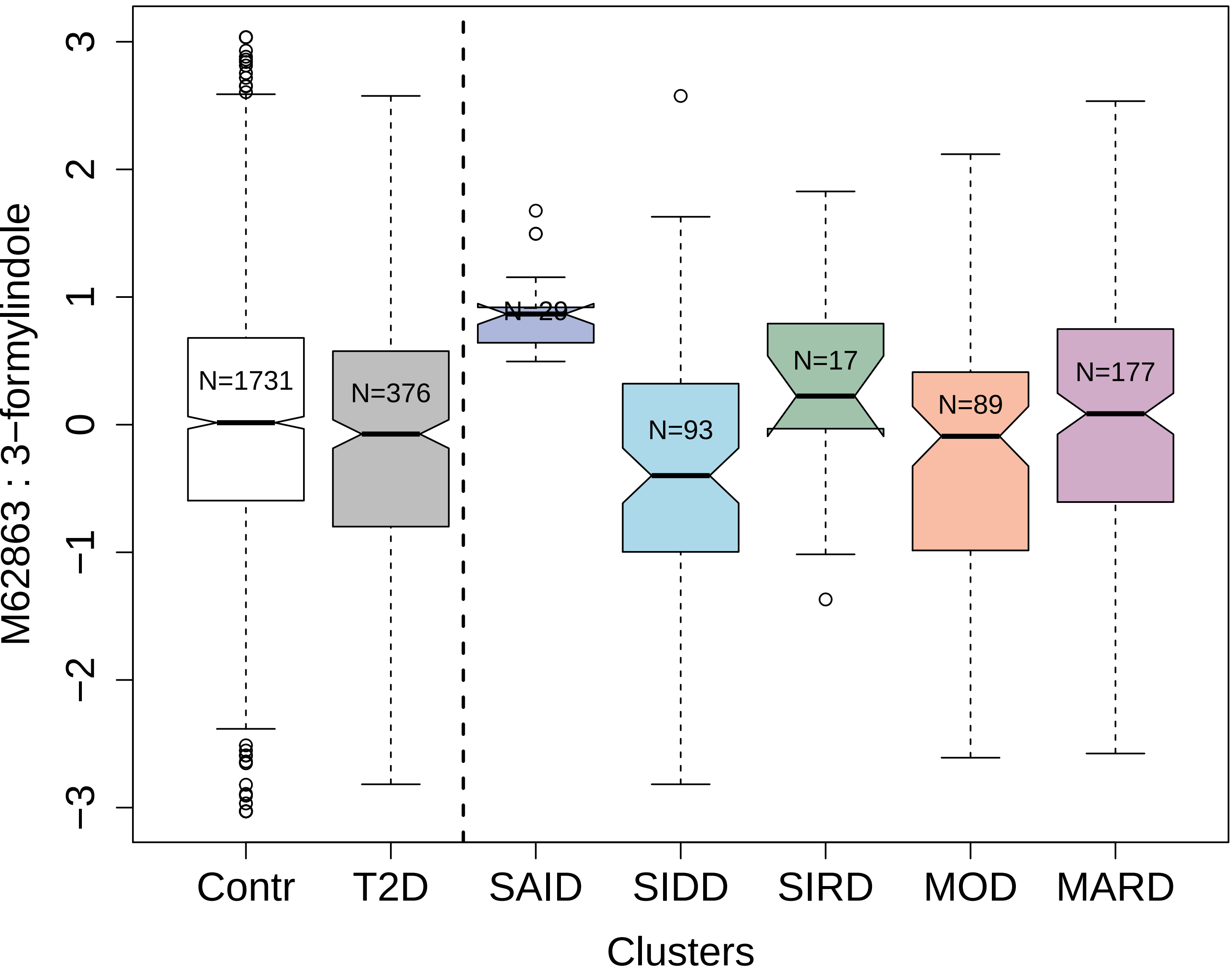
M62863 : 3−formylindole
Clusters
Contr
T2D
SAID
SIDD
SIRD
MOD
MARD
N=1731
N=376
N=29
N=93
N=17
N=89
N=177
3
2
1
0
−1
−2
−3

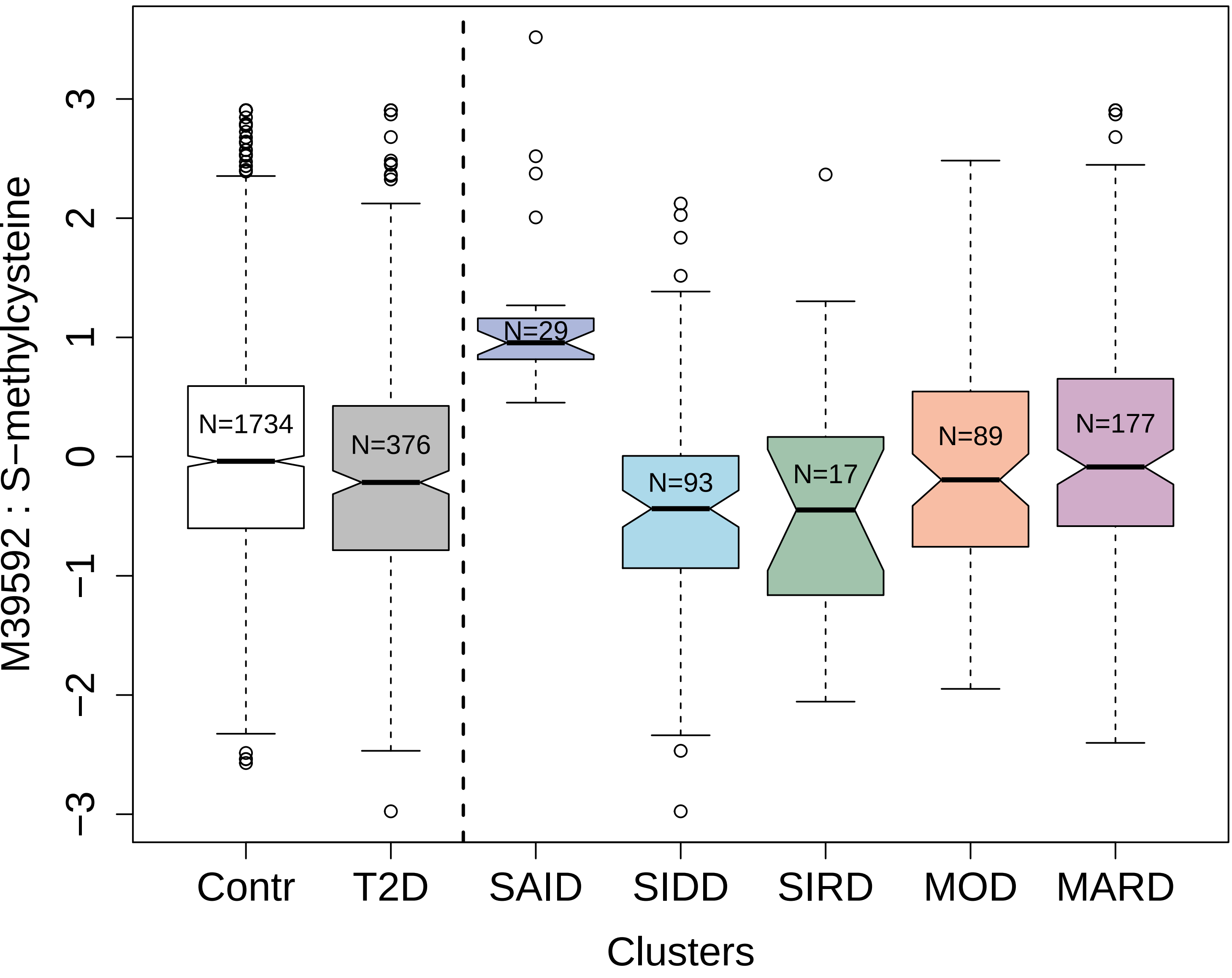
M39592 : S−methylcysteine
3
2
1
0
−1
−2
−3
N=1734
N=376
N=29
N=93
N=17
N=89
N=177
Contr
T2D
SAID
SIDD
SIRD
MOD
MARD
Clusters

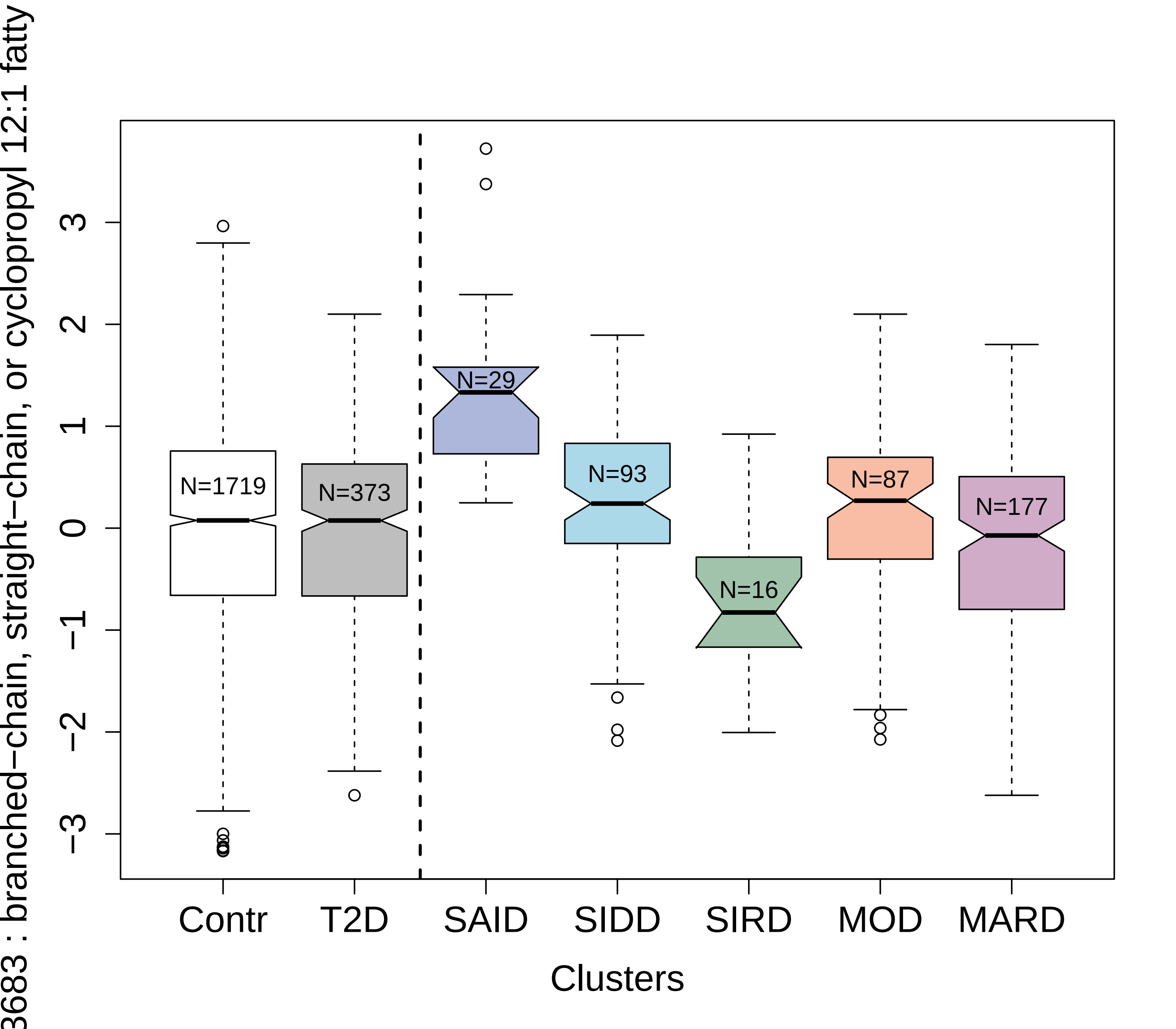
3683 : branched−chain, straight−chain, or cyclopropyl 12:1 fatty
3
2
1
0
−1
−2
−3
N=1719
N=373
N=29
N=93
N=16
N=87
N=177
Contr
T2D
SAID
SIDD
SIRD
MOD
MARD
Clusters

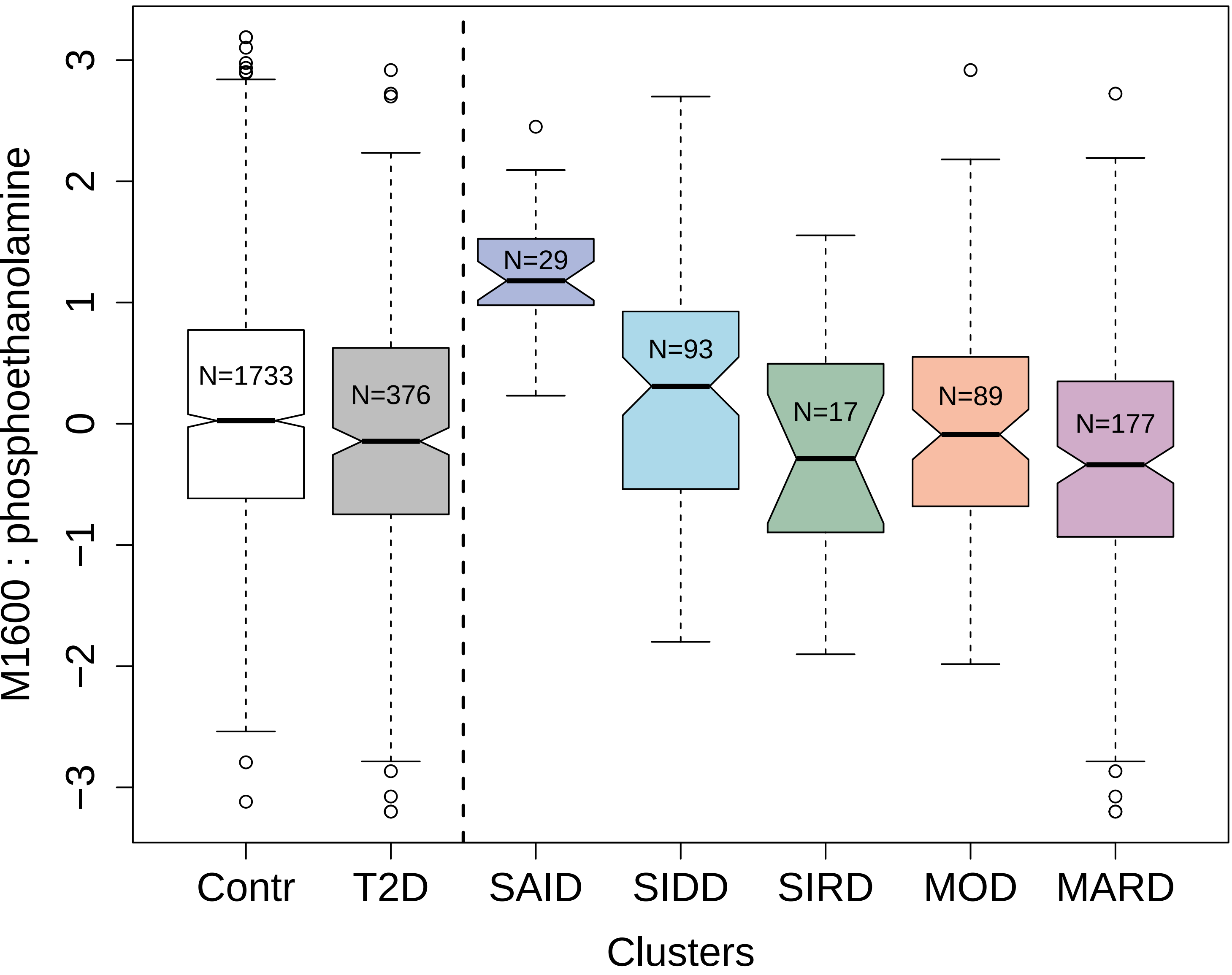

M1600 : phosphoethanolamine
3
2
1
0
−1
−2
−3
N=1733
N=376
N=29
N=93
N=17
N=89
N=177
Contr
T2D
SAID
SIDD
SIRD
MOD
MARD
Clusters

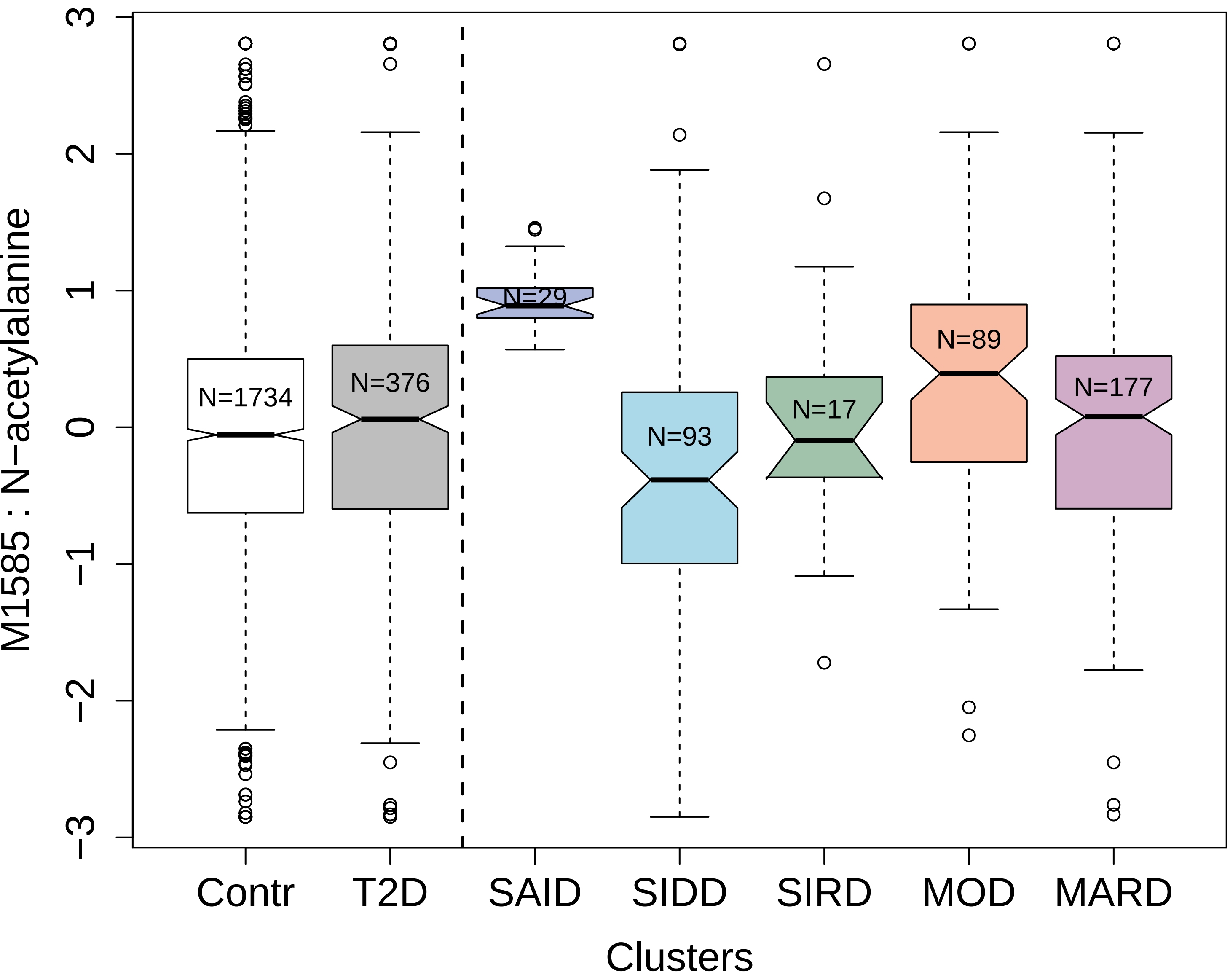

M1585 : N−acetylalanine
3
2
1
0
−1
−2
−3
N=1734
N=376
N=29
N=93
N=17
N=89
N=177
Contr
T2D
SAID
SIDD
SIRD
MOD
MARD
Clusters

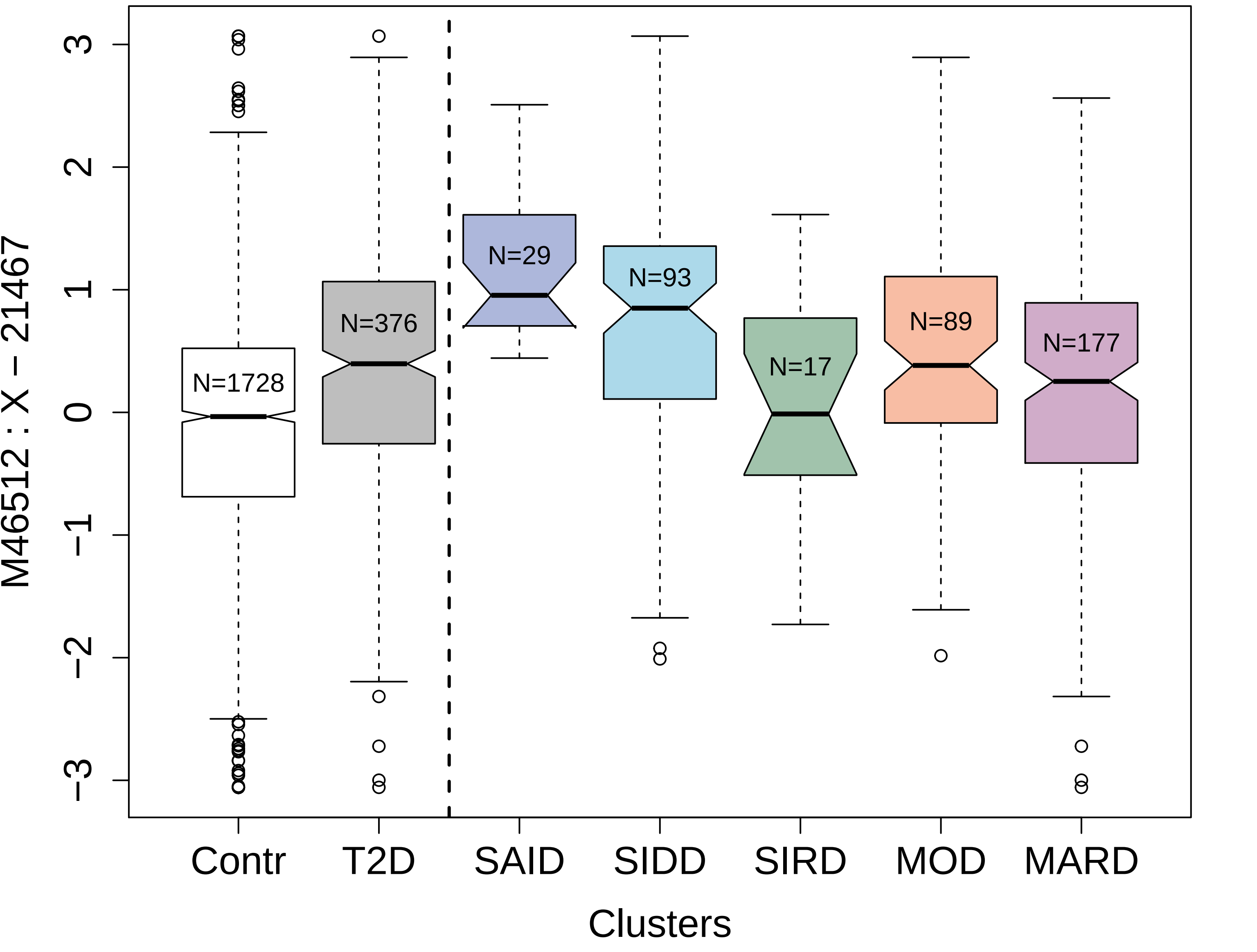

M46512 : X − 21467
3
2
1
0
−1
−2
−3
N=1728
N=376
N=29
N=93
N=17
N=89
N=177
Contr
T2D
SAID
SIDD
SIRD
MOD
MARD
Clusters

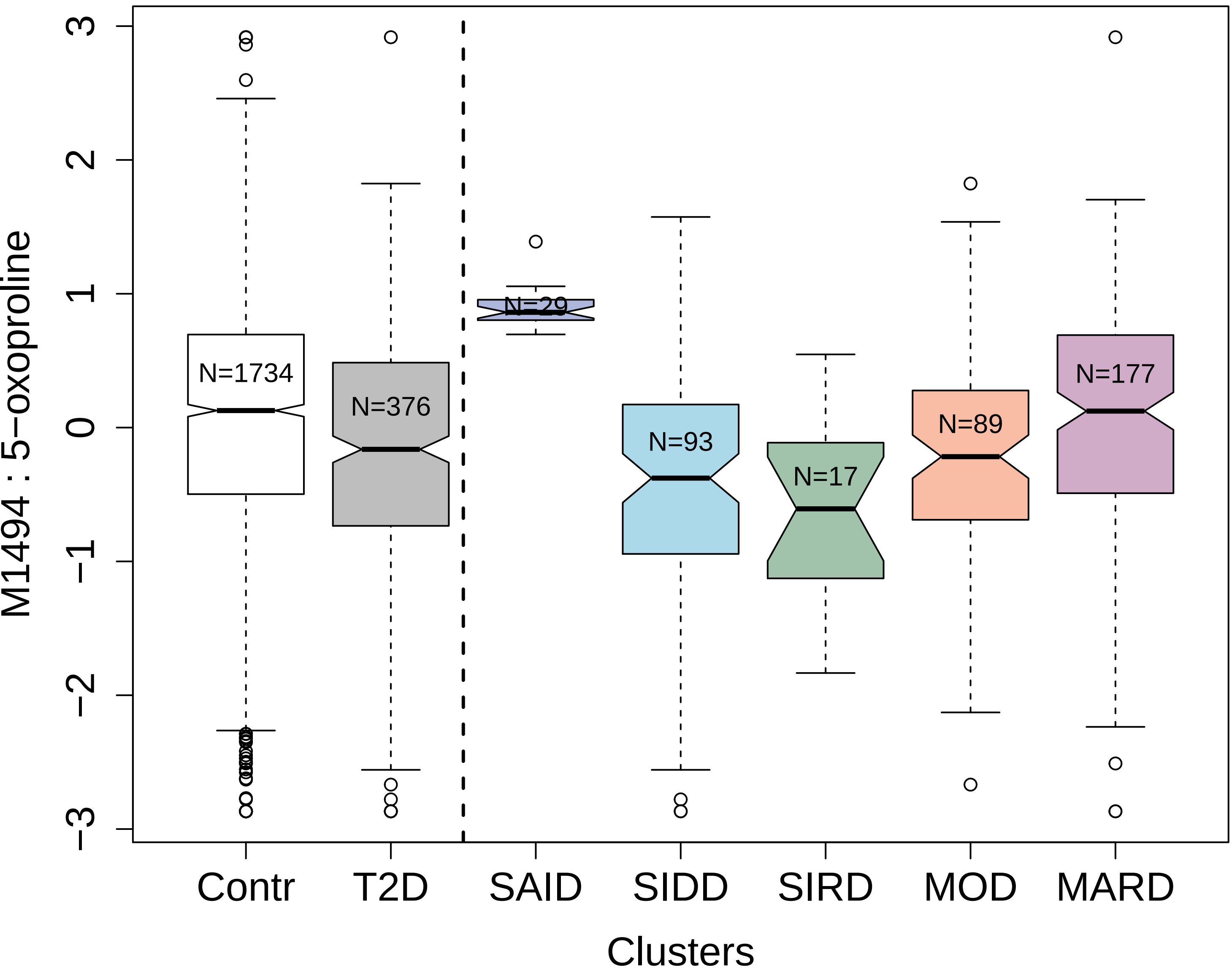

M1494 : 5−oxoproline
3
2
1
0
−1
−2
−3
N=1734
N=376
N=29
N=93
N=17
N=89
N=177
Contr
T2D
SAID
SIDD
SIRD
MOD
MARD
Clusters

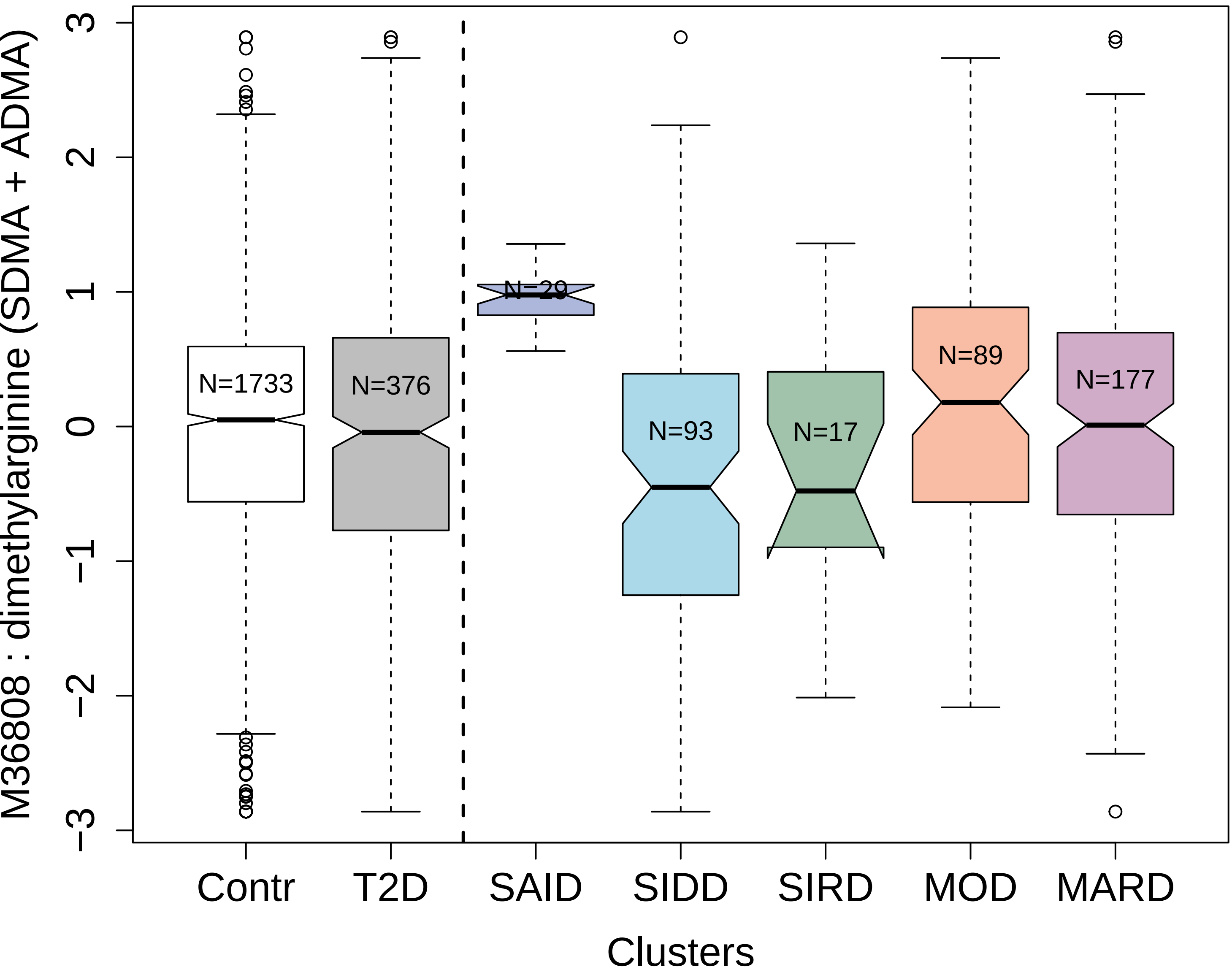
M36808 : dimethylarginine (SDMA + ADMA)
3
2
1
0
−1
−2
−3
N=1733
N=376
N=29
N=93
N=17
N=89
N=177
Contr
T2D
SAID
SIDD
SIRD
MOD
MARD
Clusters

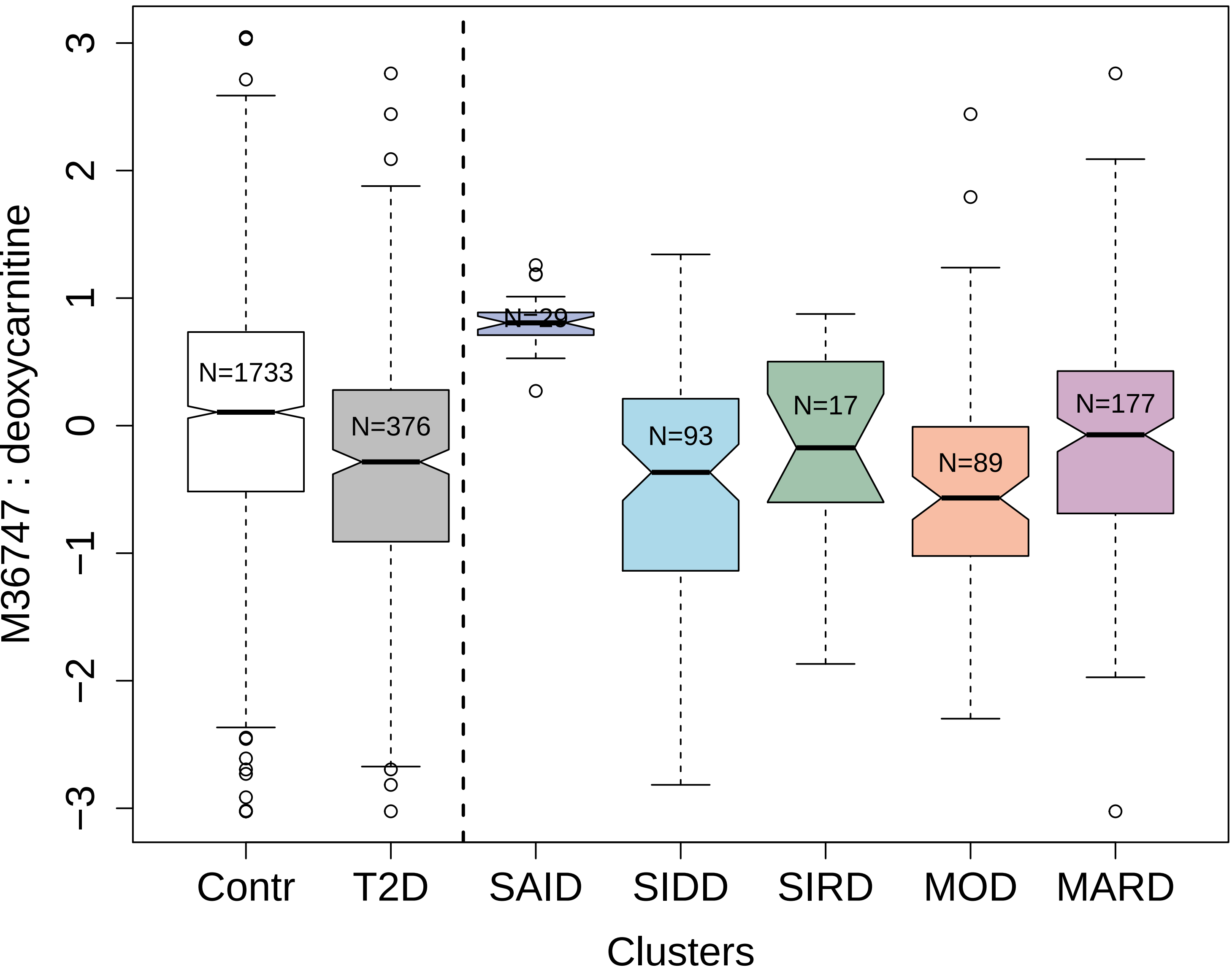

M36747 : deoxycarnitine
3
2
1
0
−1
−2
−3
N=1733
N=376
N=29
N=93
N=17
N=89
N=177
Contr
T2D
SAID
SIDD
SIRD
MOD
MARD
Clusters

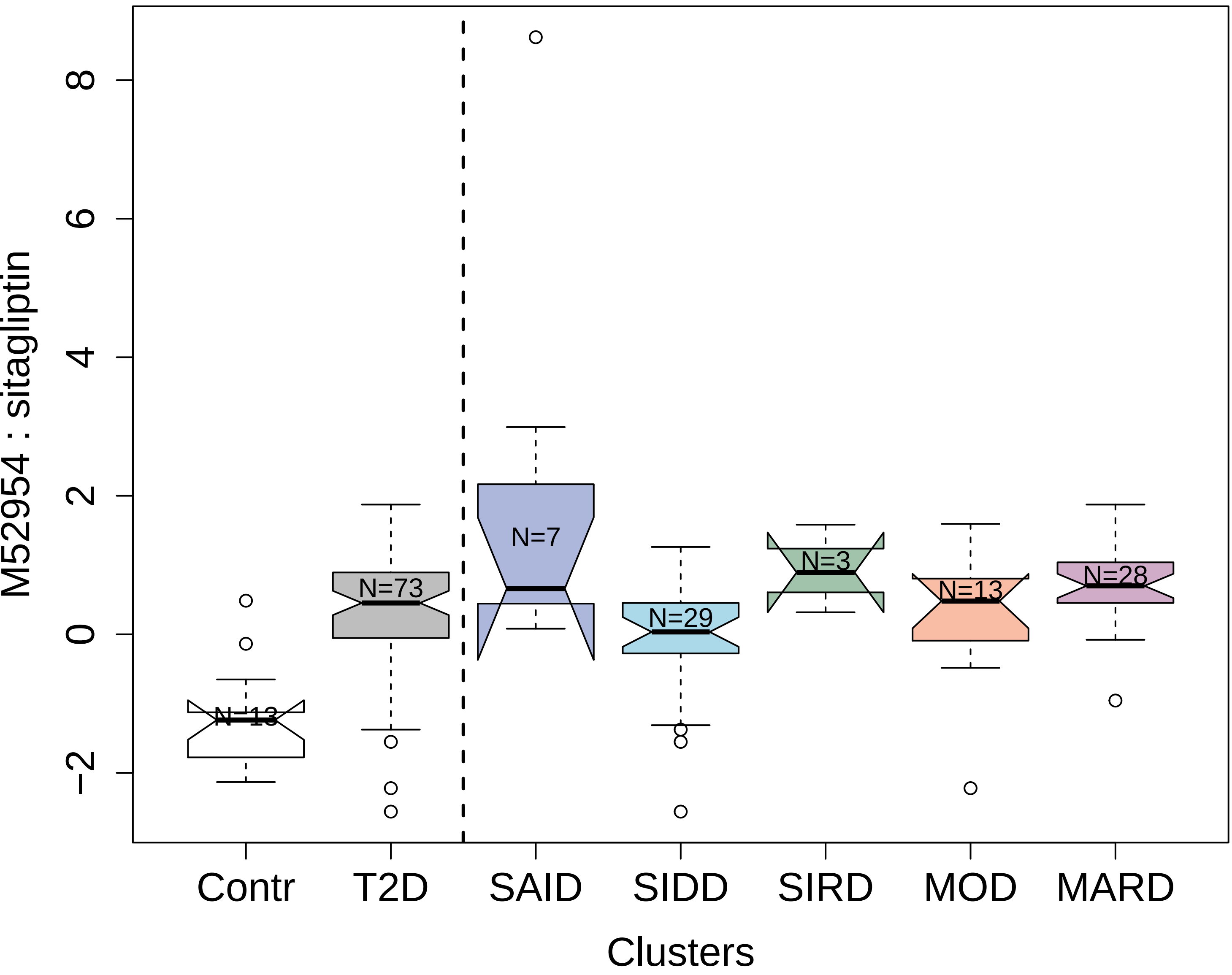

M52954 : sitagliptin
Clusters
Contr
T2D
SAID
SIDD
SIRD
MOD
MARD
N=13
N=73
N=7
N=29
N=3
N=13
N=28
−2
0
2
4
6
8

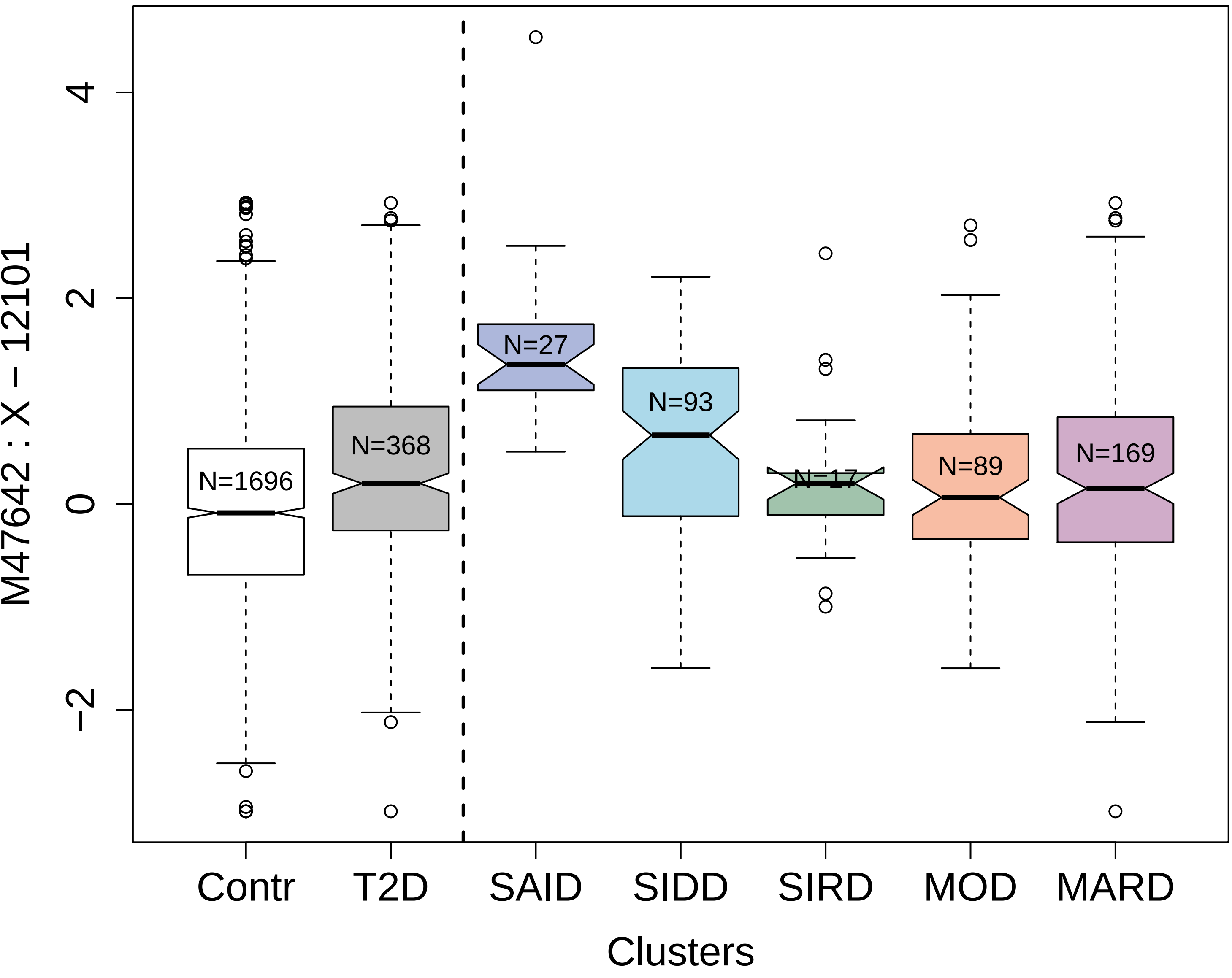

M47642 : X − 12101
4
2
0
−2
N=1696
N=368
N=27
N=93
N=17
N=89
N=169
Contr
T2D
SAID
SIDD
SIRD
MOD
MARD
Clusters

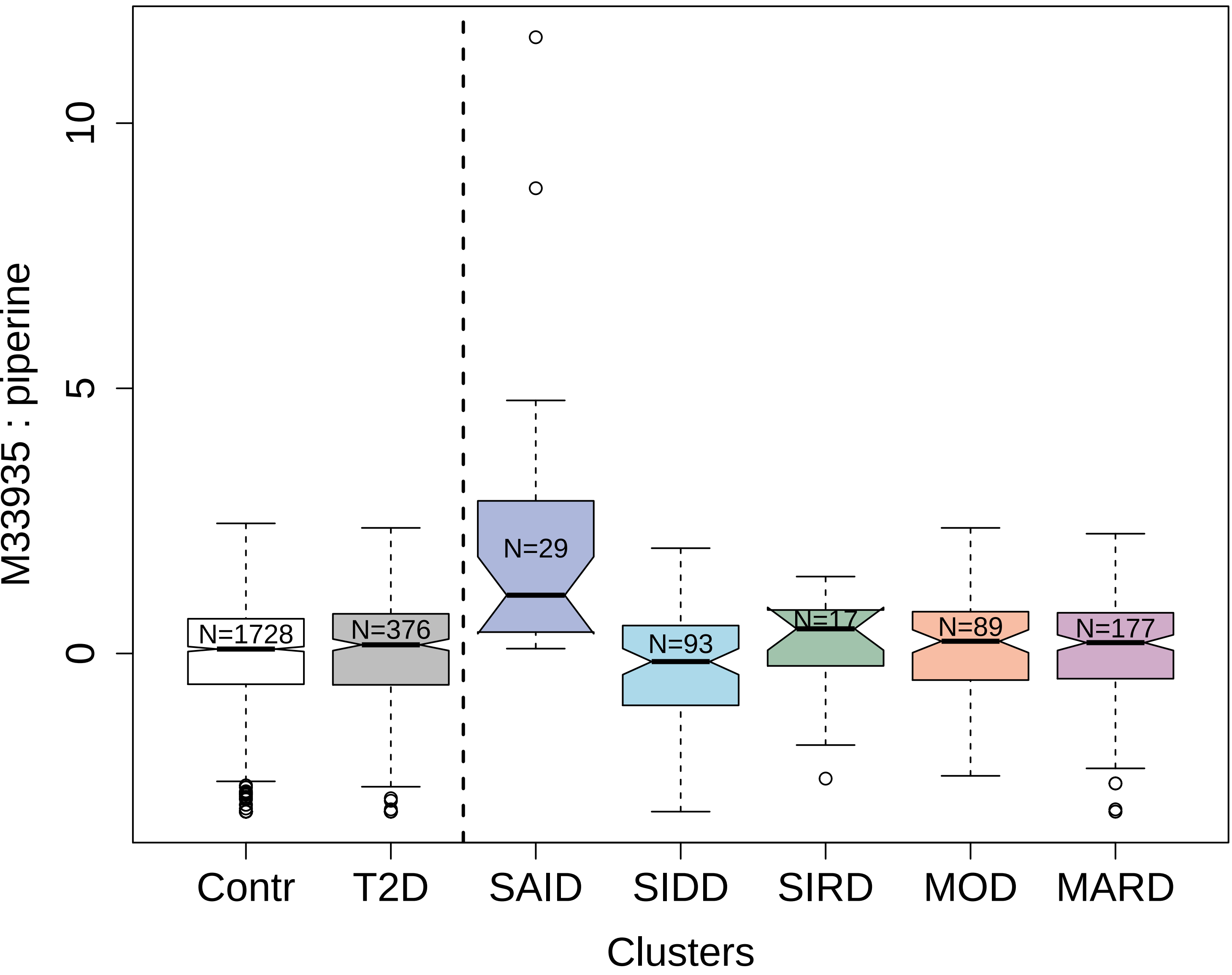

M33935 : piperine
0
5
10
N=1728
N=376
N=29
N=93
N=17
N=89
N=177
Contr
T2D
SAID
SIDD
SIRD
MOD
MARD
Clusters

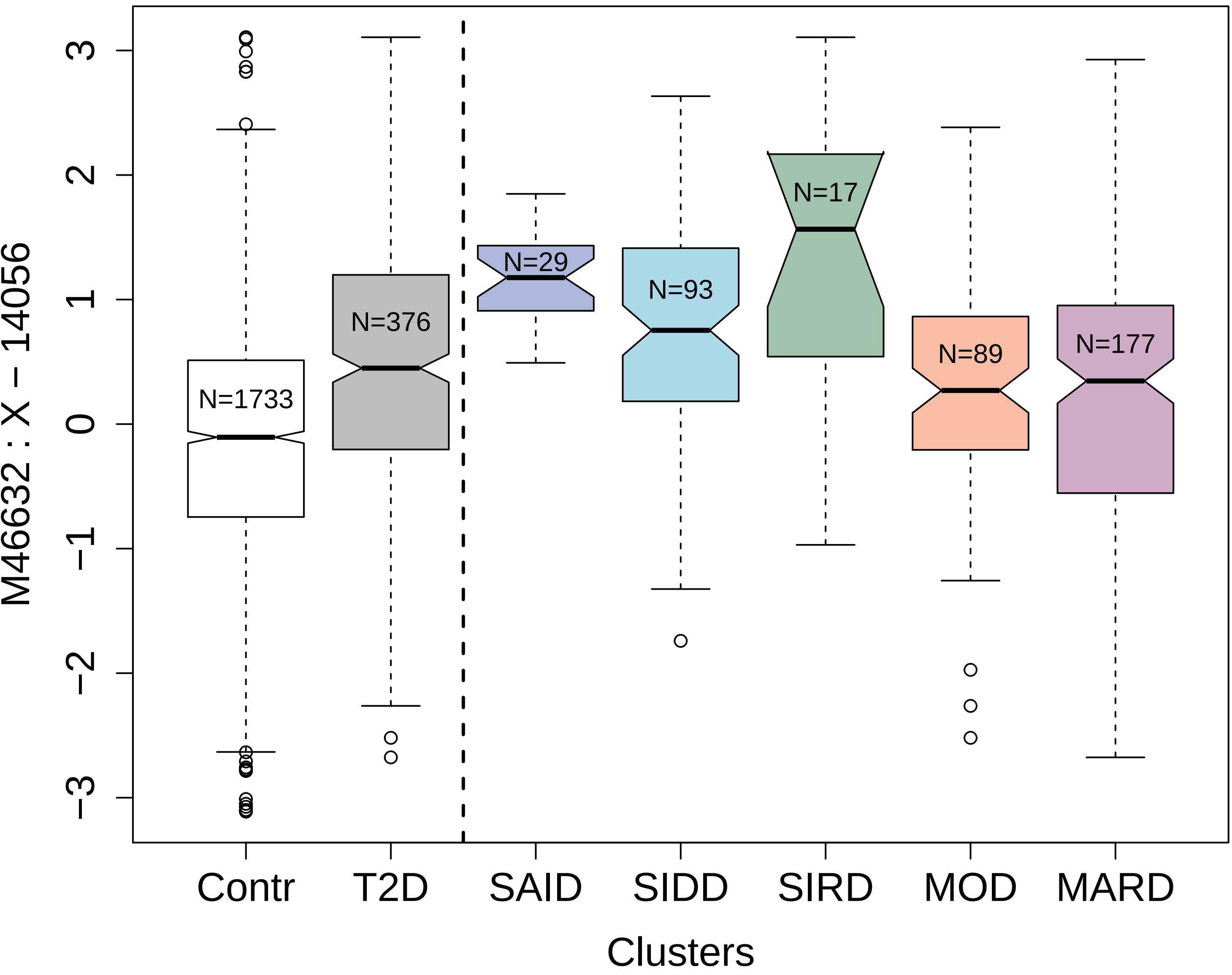

M46632 : X − 14056
3
2
1
0
−1
−2
−3
N=1733
N=376
N=29
N=93
N=17
N=89
N=177
Contr
T2D
SAID
SIDD
SIRD
MOD
MARD
Clusters

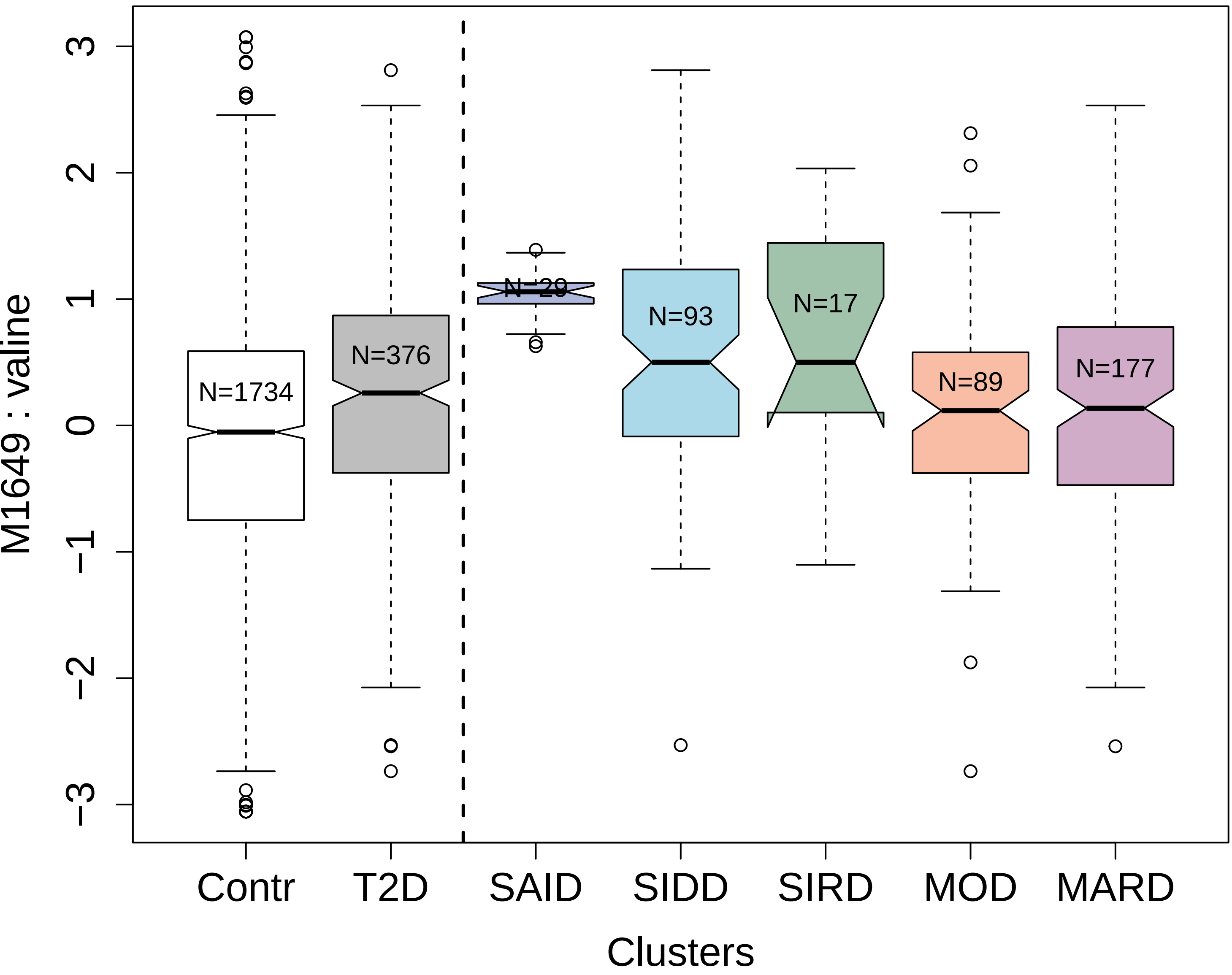

M1649 : valine
3
2
1
0
−1
−2
−3
N=1734
N=376
N=29
N=93
N=17
N=89
N=177
Contr
T2D
SAID
SIDD
SIRD
MOD
MARD
Clusters

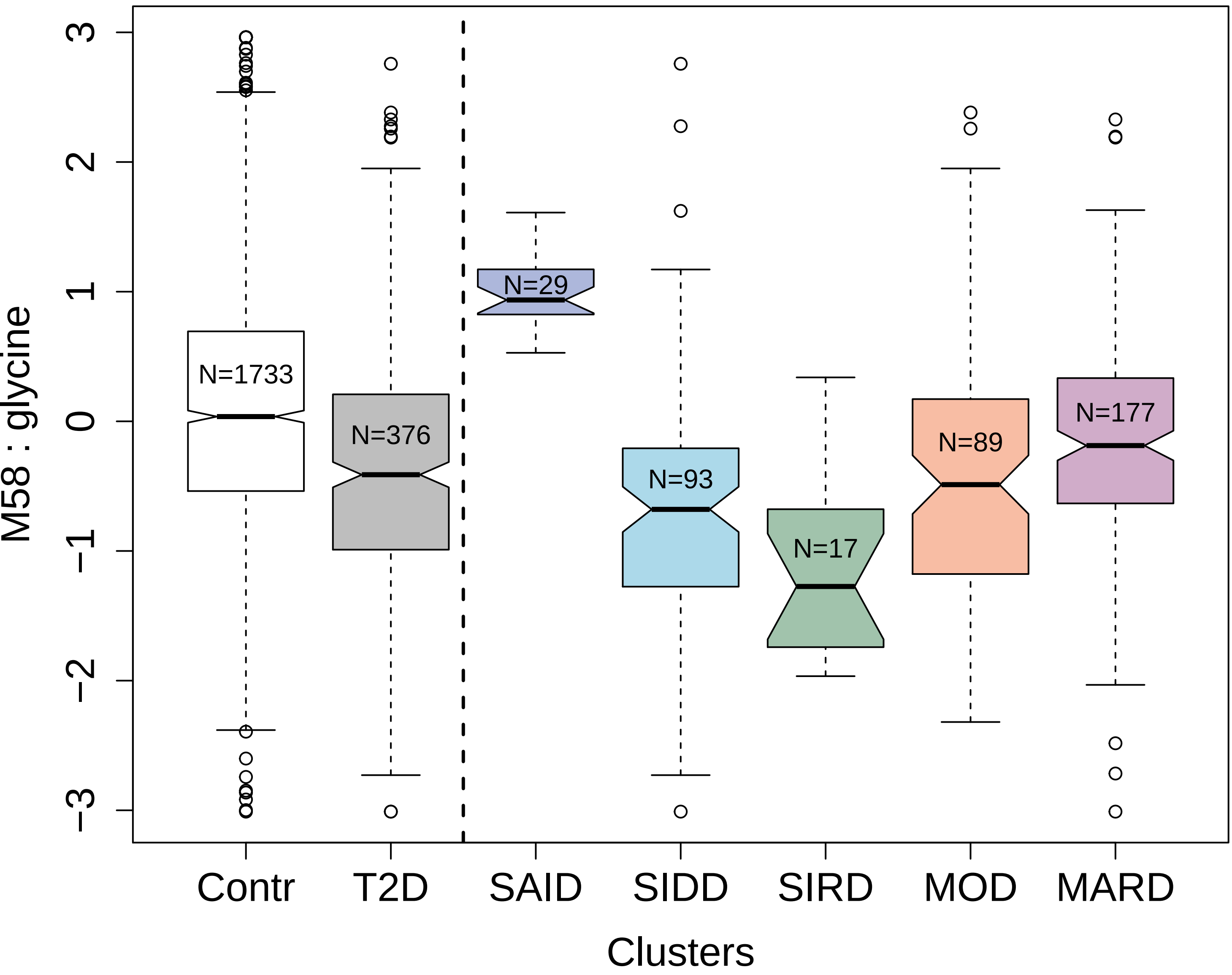

M58 : glycine
3
2
1
0
−1
−2
−3
N=1733
N=376
N=29
N=93
N=17
N=89
N=177
Contr
T2D
SAID
SIDD
SIRD
MOD
MARD
Clusters

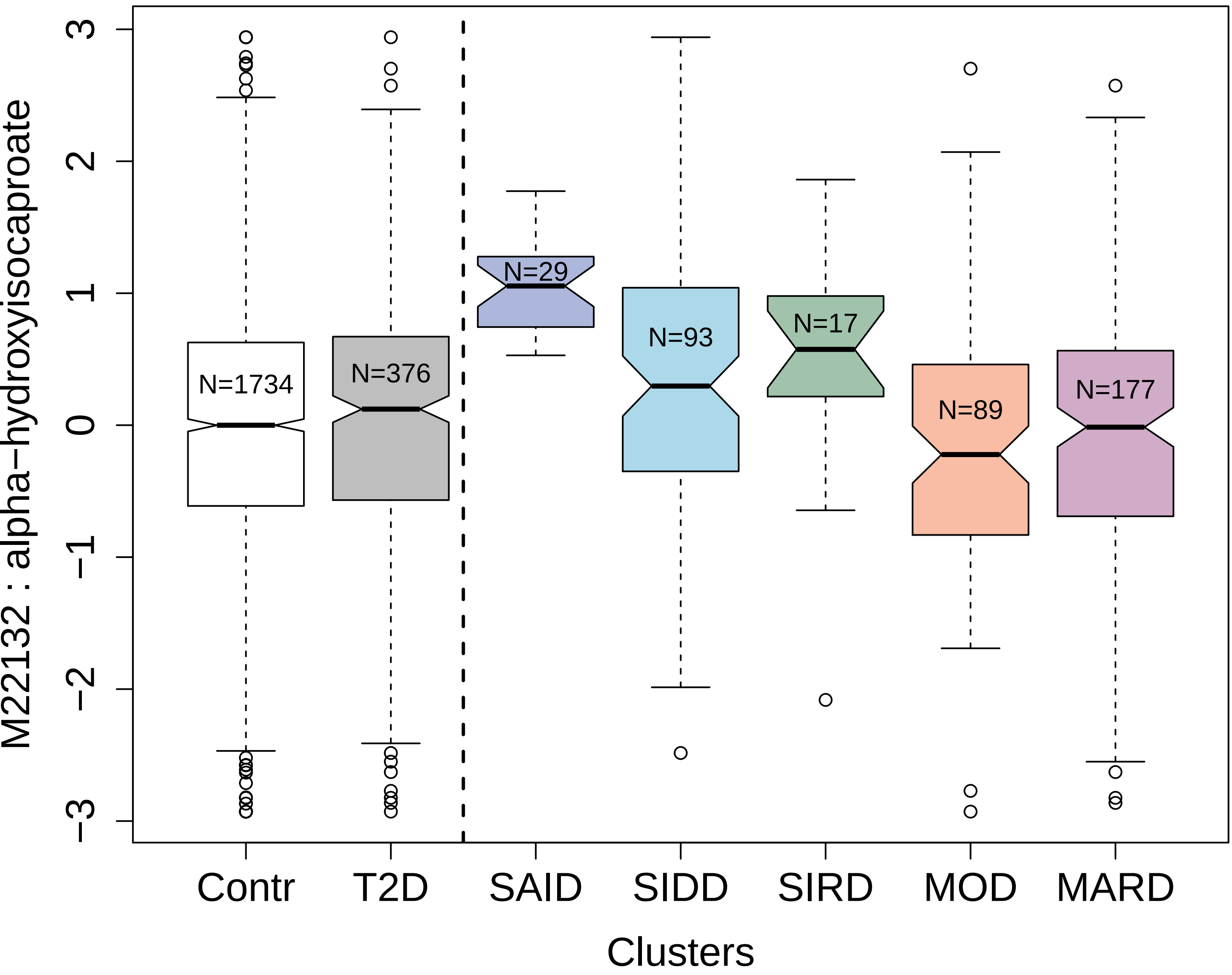

M22132 : alpha−hydroxyisocaproate
3
2
1
0
−1
−2
−3
N=1734
N=376
N=29
N=93
N=17
N=89
N=177
Contr
T2D
SAID
SIDD
SIRD
MOD
MARD
Clusters

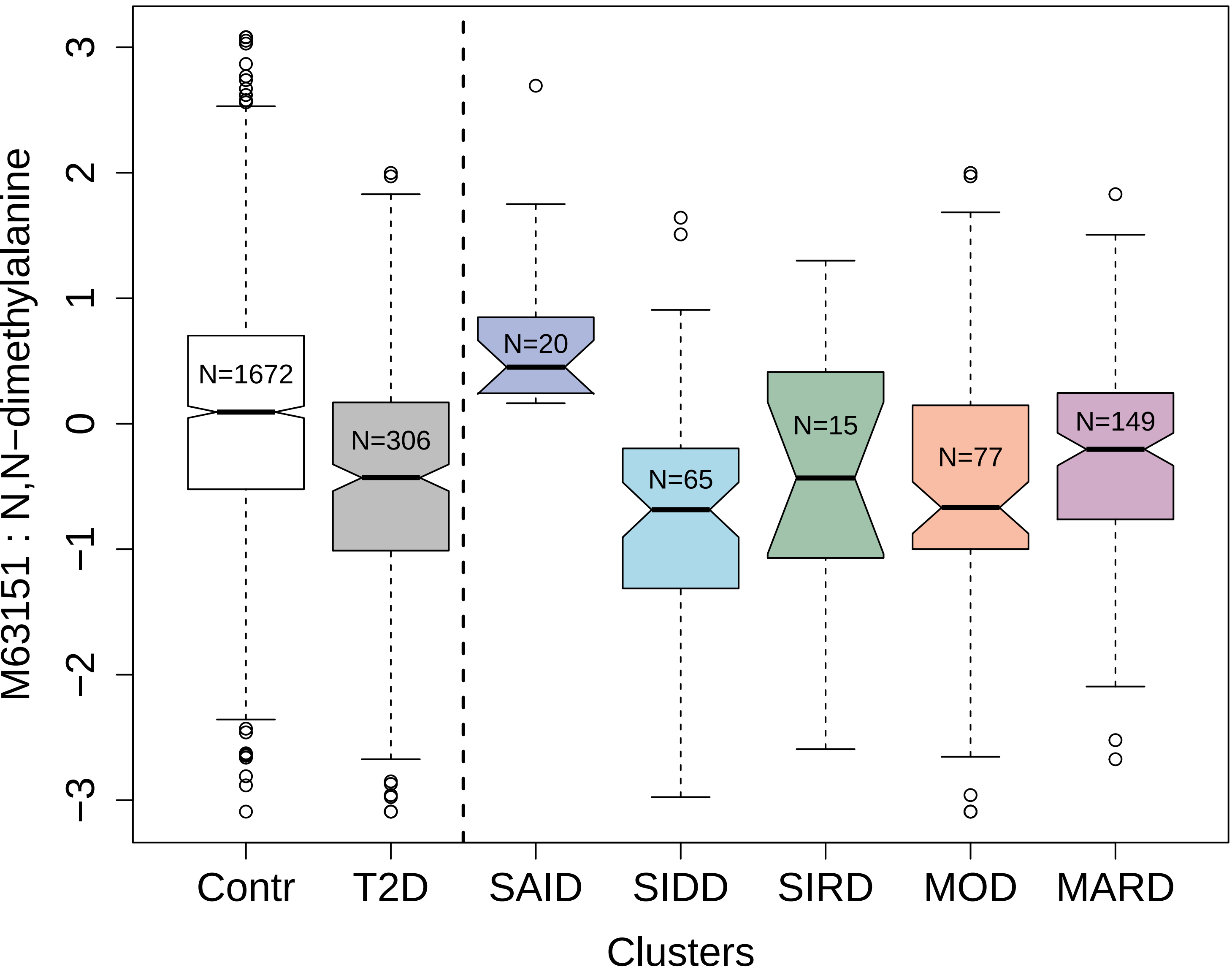

M63151 : N,N−dimethylalanine
3
2
1
0
−1
−2
−3
N=1672
N=306
N=20
N=65
N=15
N=77
N=149
Contr
T2D
SAID
SIDD
SIRD
MOD
MARD
Clusters

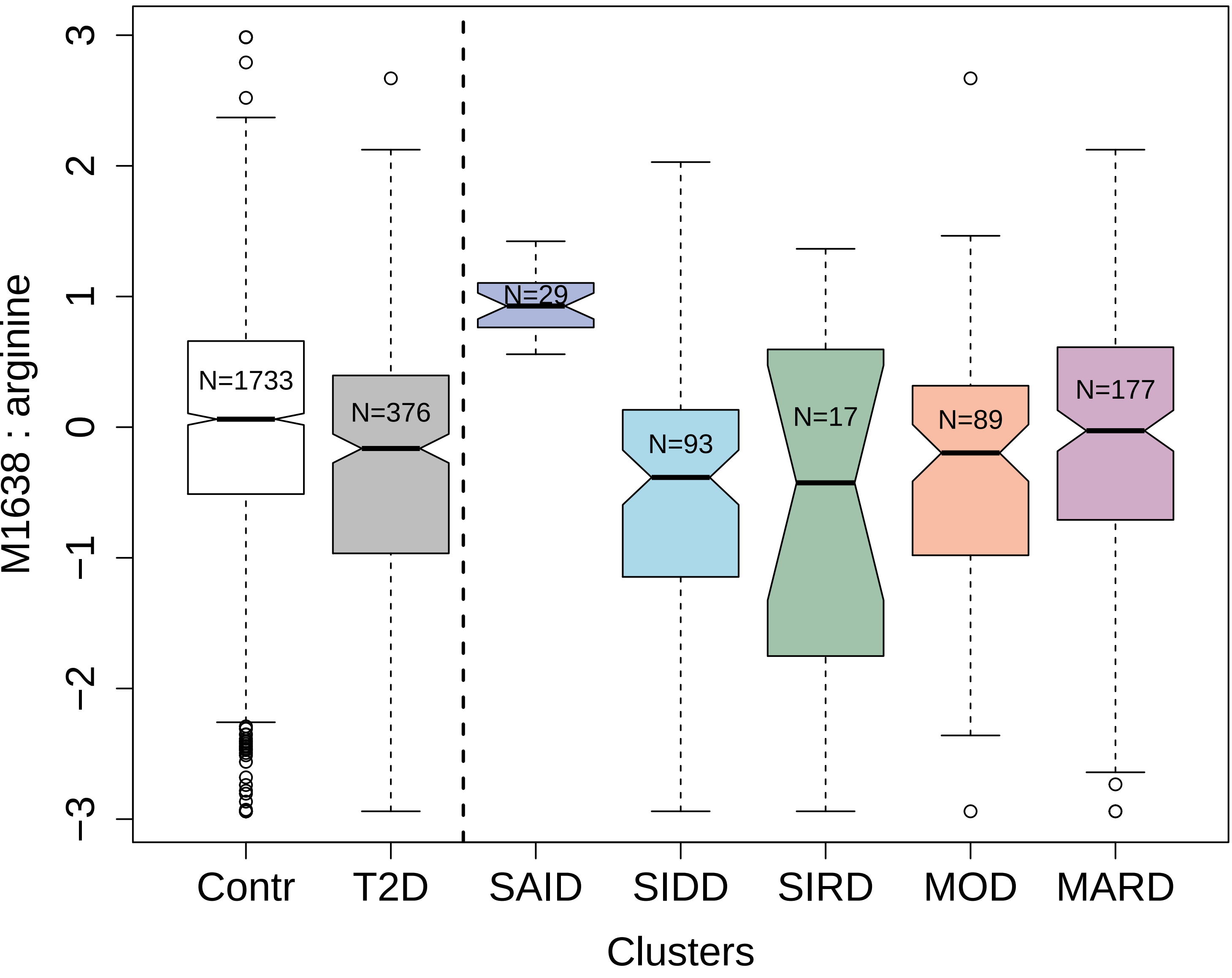

M1638 : arginine
3
2
1
0
−1
−2
−3
N=1733
N=376
N=29
N=93
N=17
N=89
N=177
Contr
T2D
SAID
SIDD
SIRD
MOD
MARD
Clusters

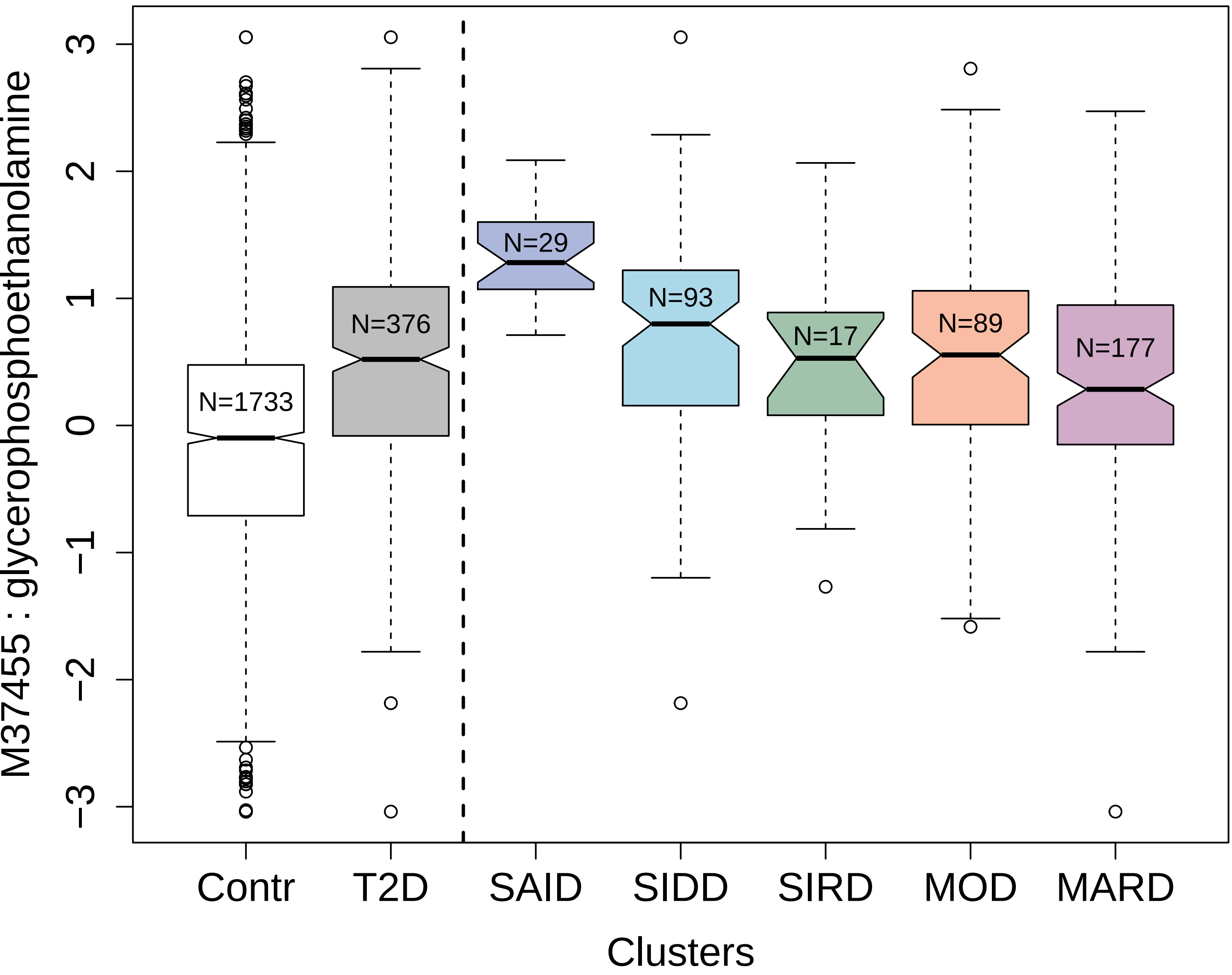

M37455 : glycerophosphoethanolamine
3
2
1
0
−1
−2
−3
N=1733
N=376
N=29
N=93
N=17
N=89
N=177
Contr
T2D
SAID
SIDD
SIRD
MOD
MARD
Clusters

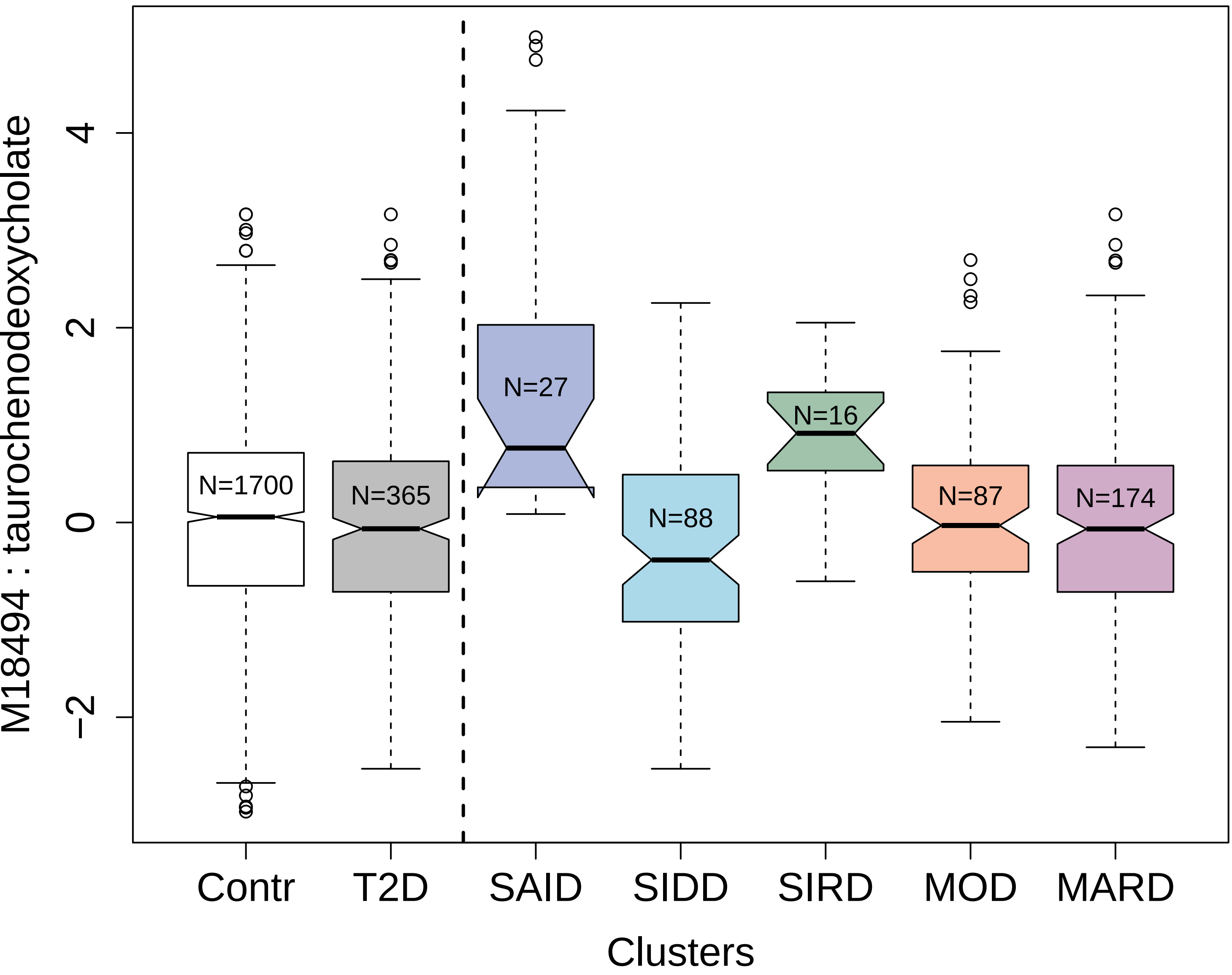

M18494 : taurochenodeoxycholate
4
2
0
−2
N=1700
N=365
N=27
N=88
N=16
N=87
N=174
Contr
T2D
SAID
SIDD
SIRD
MOD
MARD
Clusters

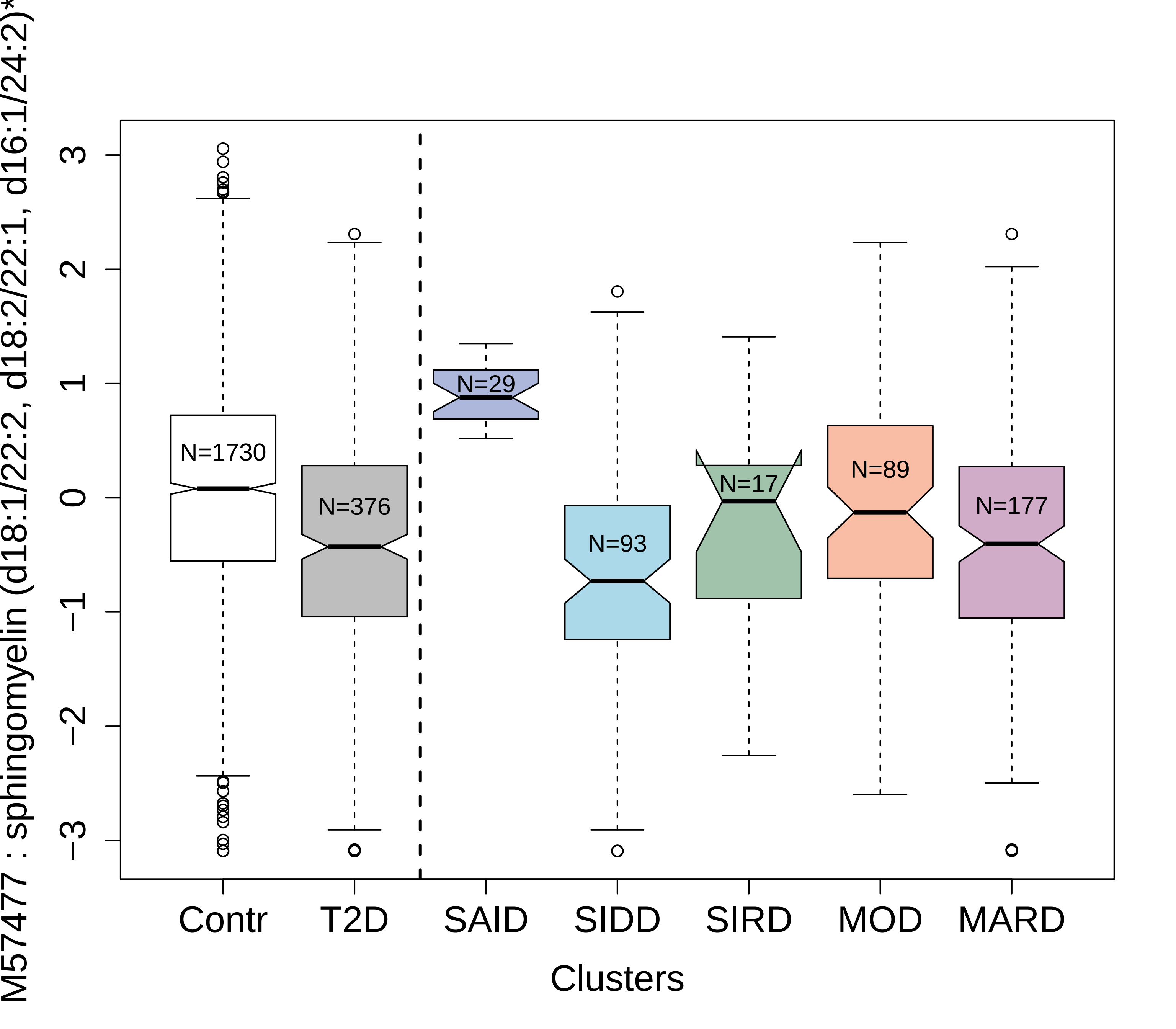

M57477 : sphingomyelin (d18:1/22:2, d18:2/22:1, d16:1/24:2)*
3
2
1
0
−1
−2
−3
N=1730
N=376
N=29
N=93
N=17
N=89
N=177
Contr
T2D
SAID
SIDD
SIRD
MOD
MARD
Clusters

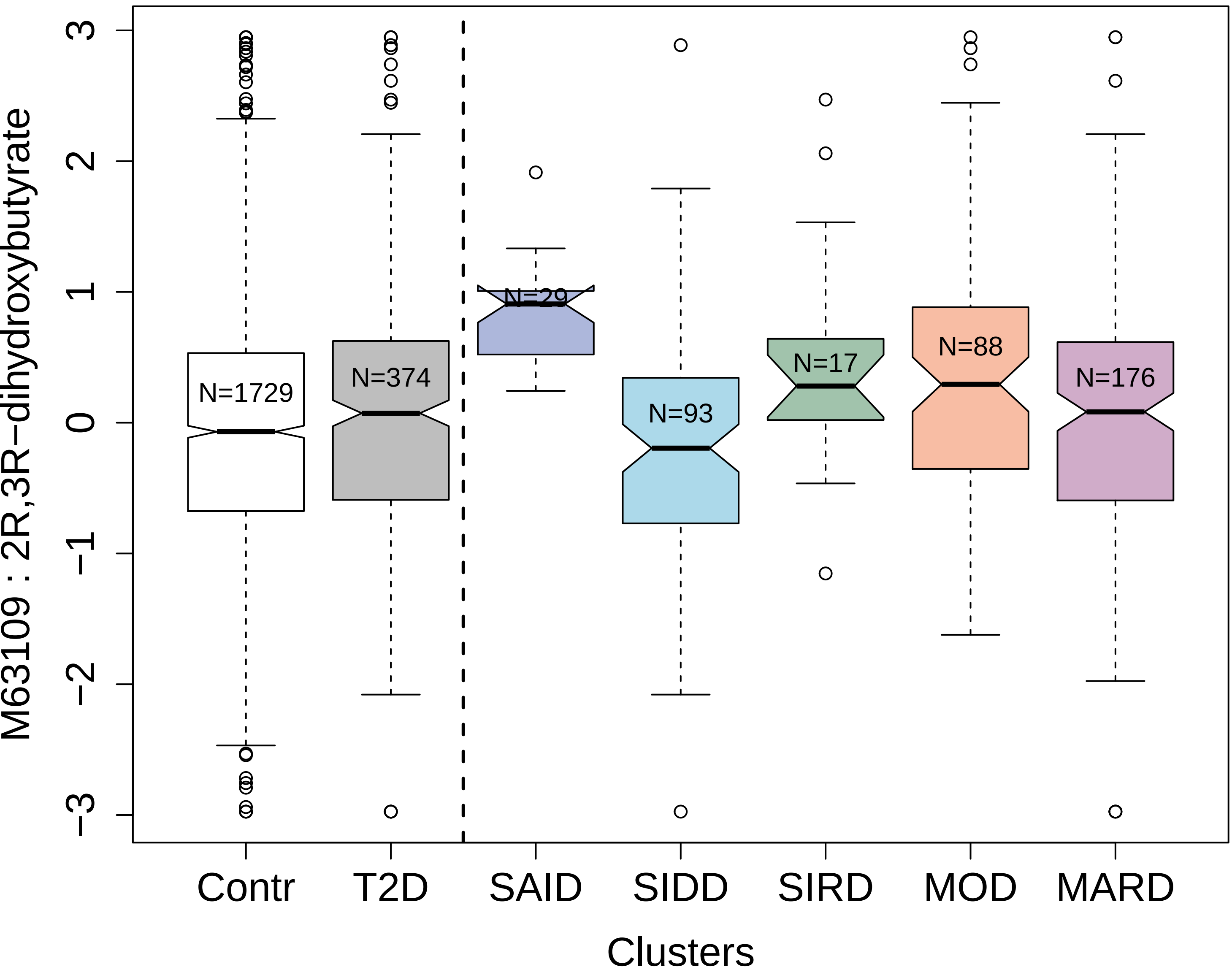
M63109 : 2R,3R−dihydroxybutyrate
Clusters
Contr
T2D
SAID
SIDD
SIRD
MOD
MARD
N=1729
N=374
N=29
N=93
N=17
N=88
N=176
3
2
1
0
−1
−2
−3

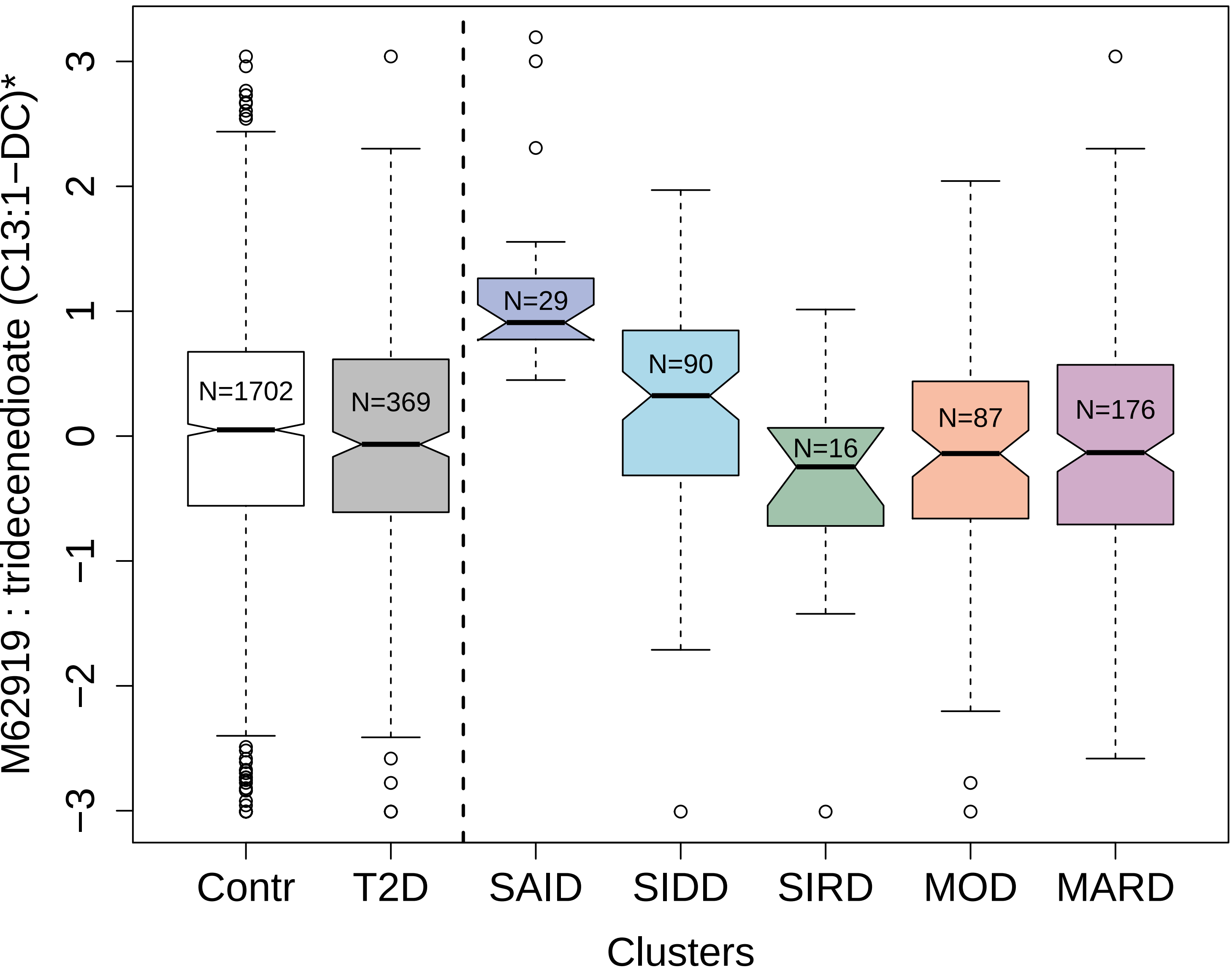
M62919 : tridecenedioate (C13:1−DC)*
3
2
1
0
−1
−2
−3
N=1702
N=369
N=29
N=90
N=16
N=87
N=176
Contr
T2D
SAID
SIDD
SIRD
MOD
MARD
Clusters

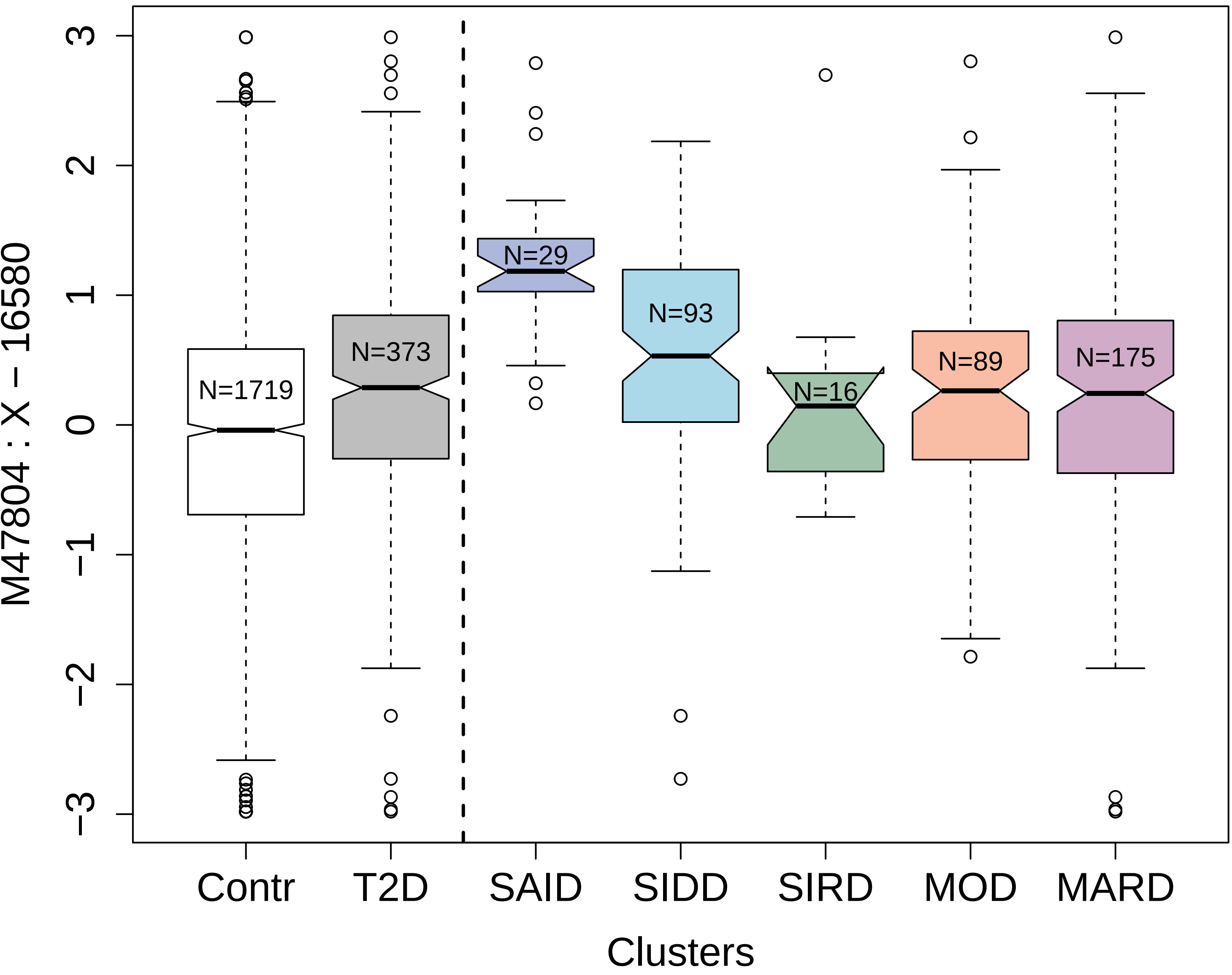
M47804 : X − 16580
3
2
1
0
−1
−2
−3
N=1719
N=373
N=29
N=93
N=16
N=89
N=175
Contr
T2D
SAID
SIDD
SIRD
MOD
MARD
Clusters

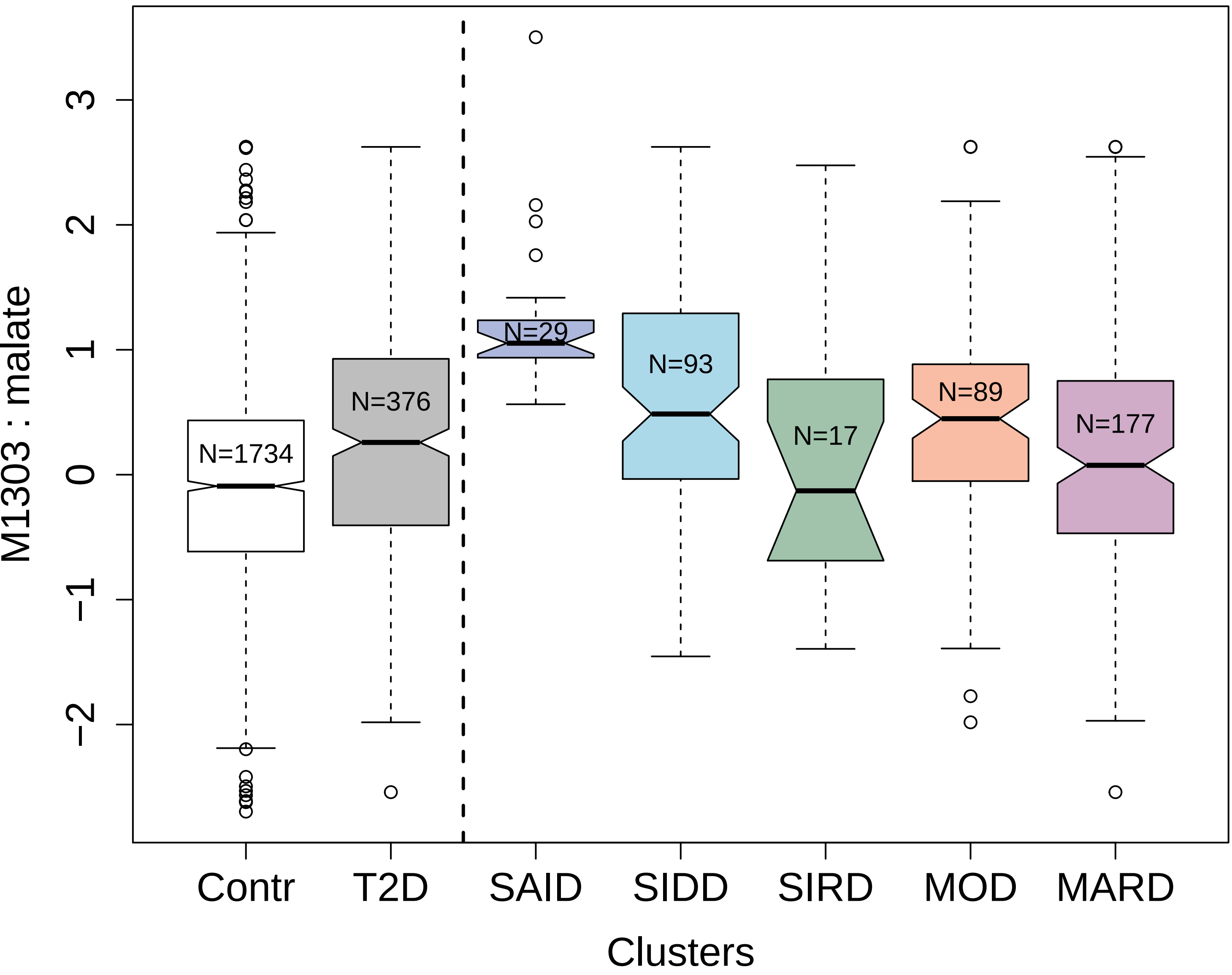
M1303 : malate
3
2
1
0
−1
−2
N=1734
N=376
N=29
N=93
N=17
N=89
N=177
Contr
T2D
SAID
SIDD
SIRD
MOD
MARD
Clusters

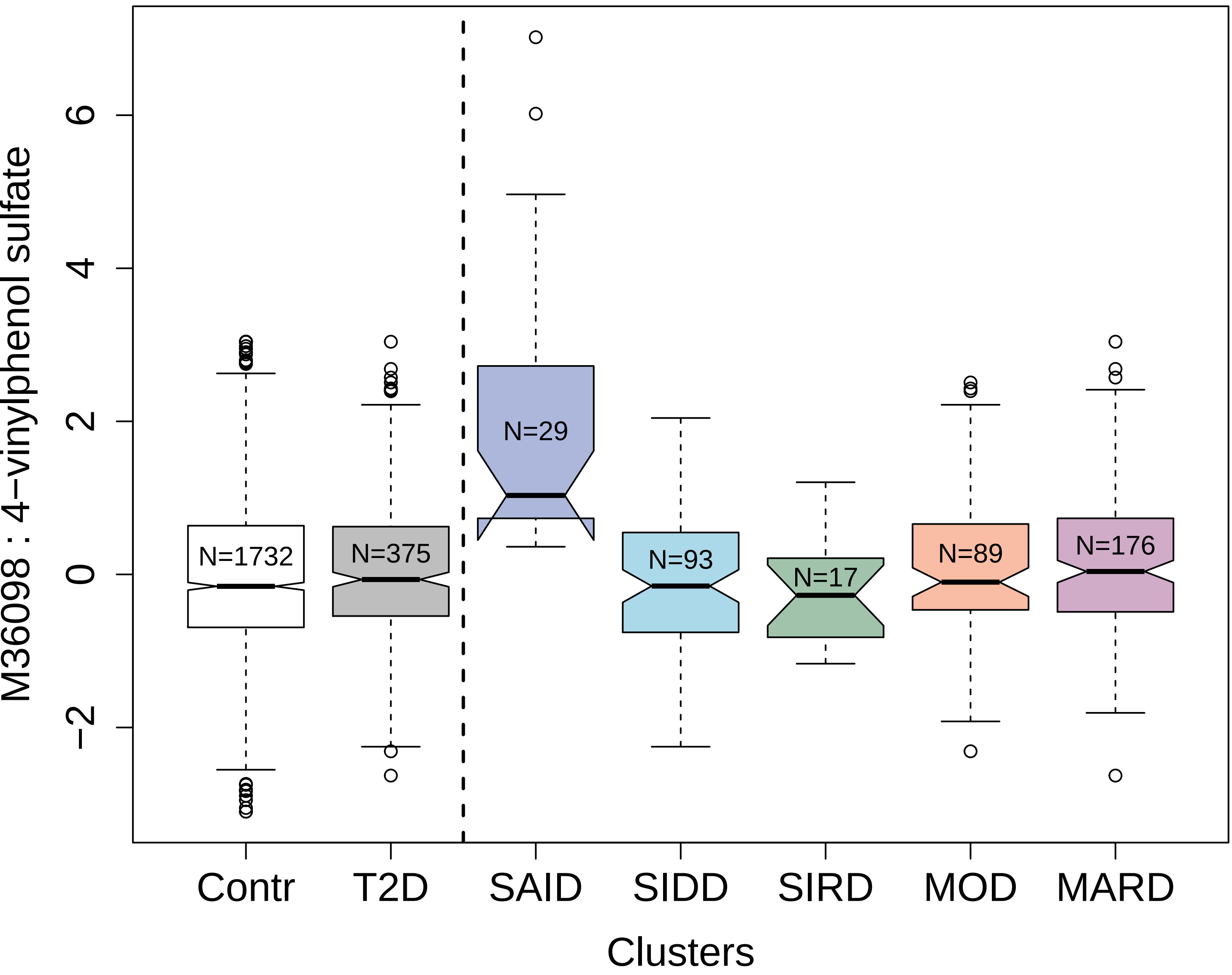
M36098 : 4−vinylphenol sulfate
−2
0
2
4
6
N=1732
N=375
N=29
N=93
N=17
N=89
N=176
Contr
T2D
SAID
SIDD
SIRD
MOD
MARD
Clusters

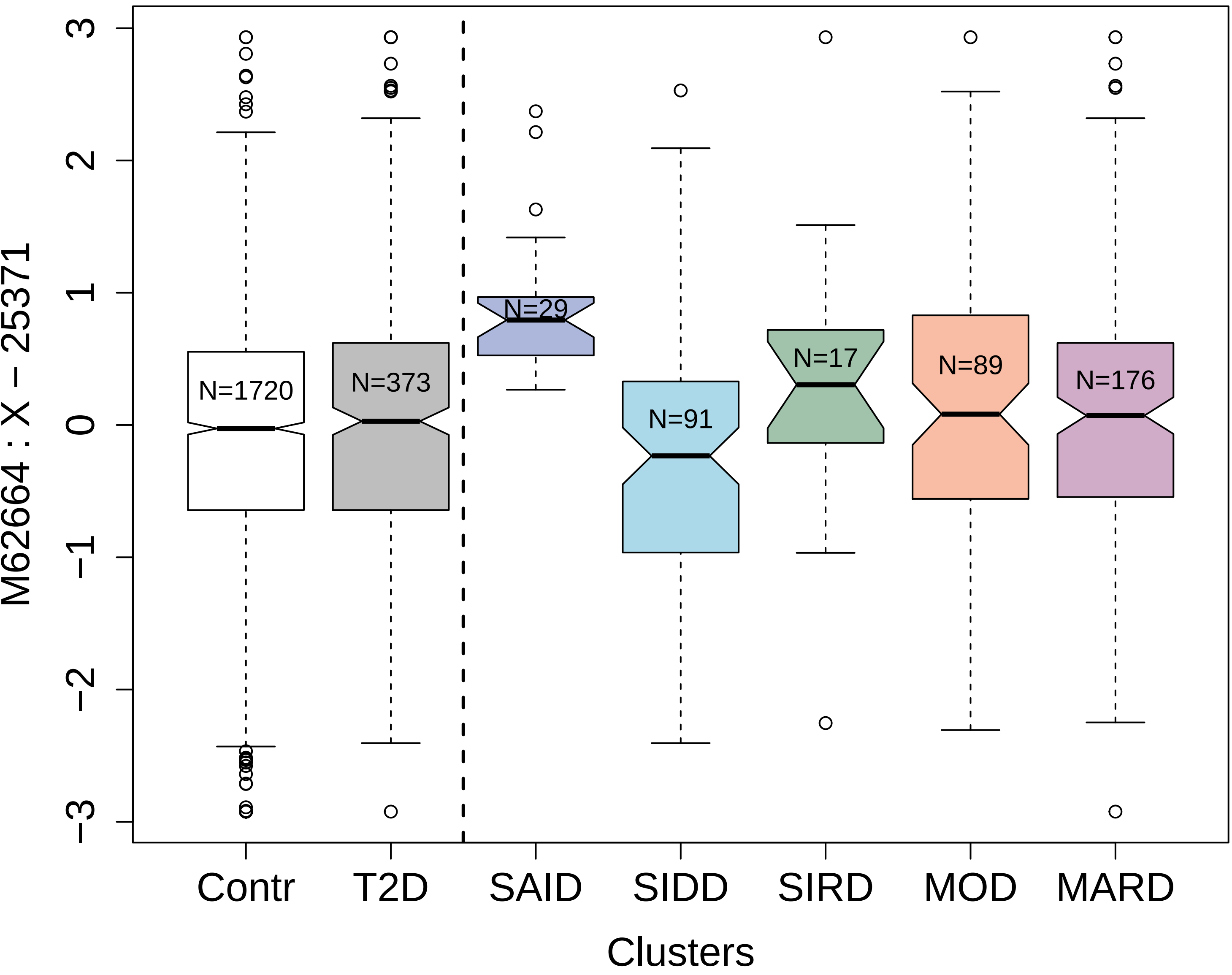

M62664 : X − 25371
3
2
1
0
−1
−2
−3
N=1720
N=373
N=29
N=91
N=17
N=89
N=176
Contr
T2D
SAID
SIDD
SIRD
MOD
MARD
Clusters

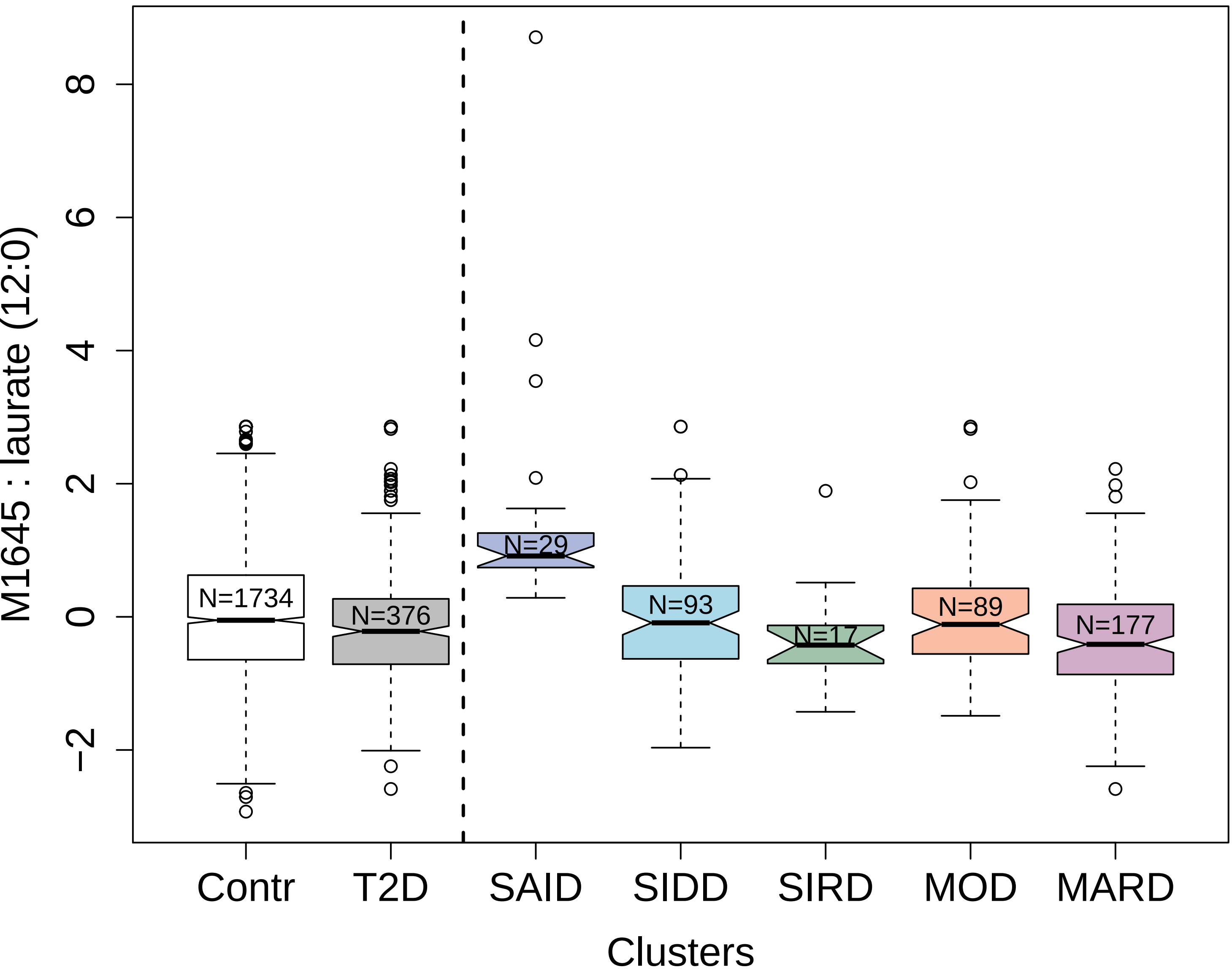
M1645 : laurate (12:0)
8
6
4
2
0
−2
N=1734
N=376
N=29
N=93
N=17
N=89
N=177
Contr
T2D
SAID
SIDD
SIRD
MOD
MARD
Clusters

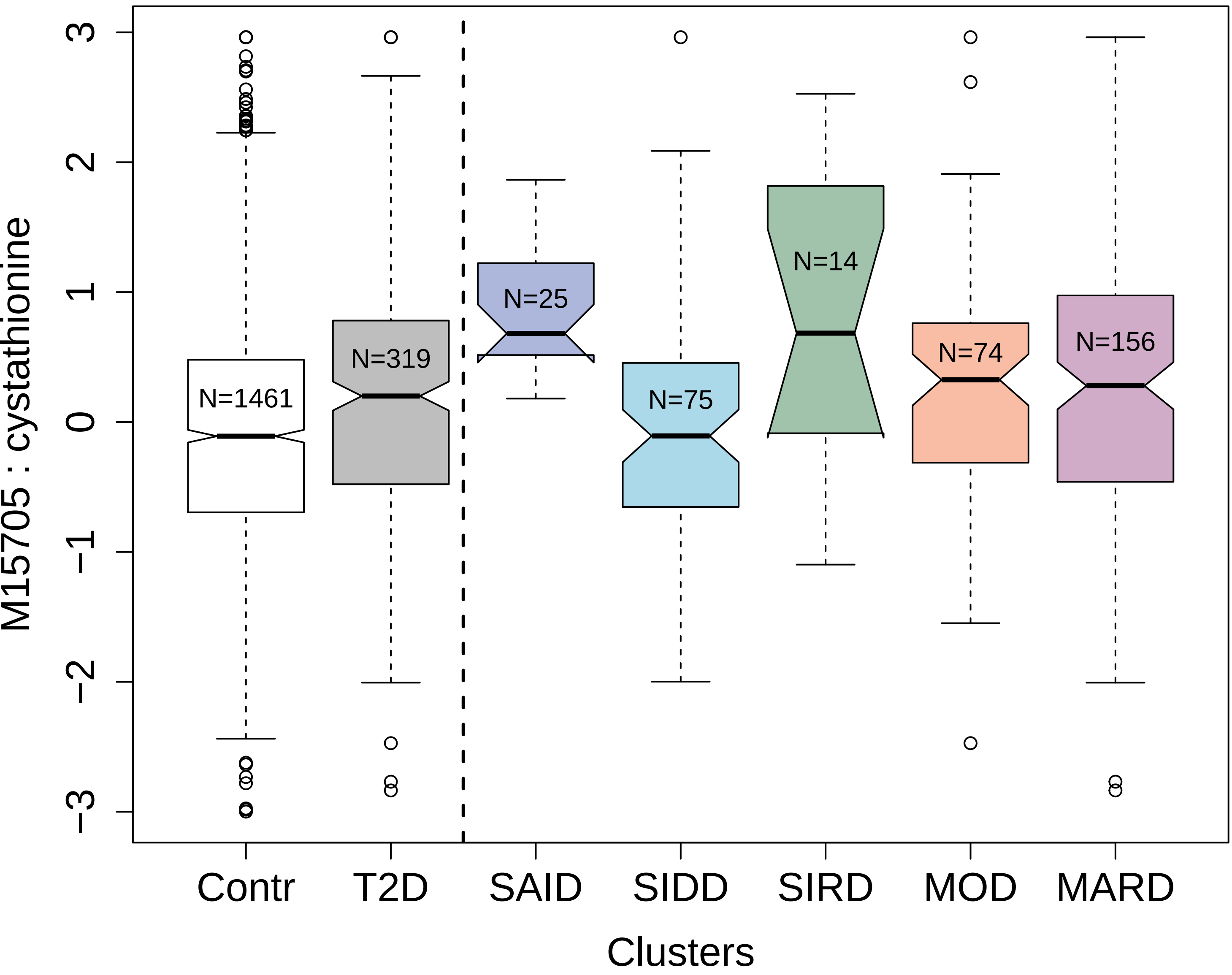

M15705 : cystathionine
3
2
1
0
−1
−2
−3
N=1461
N=319
N=25
N=75
N=14
N=74
N=156
Contr
T2D
SAID
SIDD
SIRD
MOD
MARD
Clusters

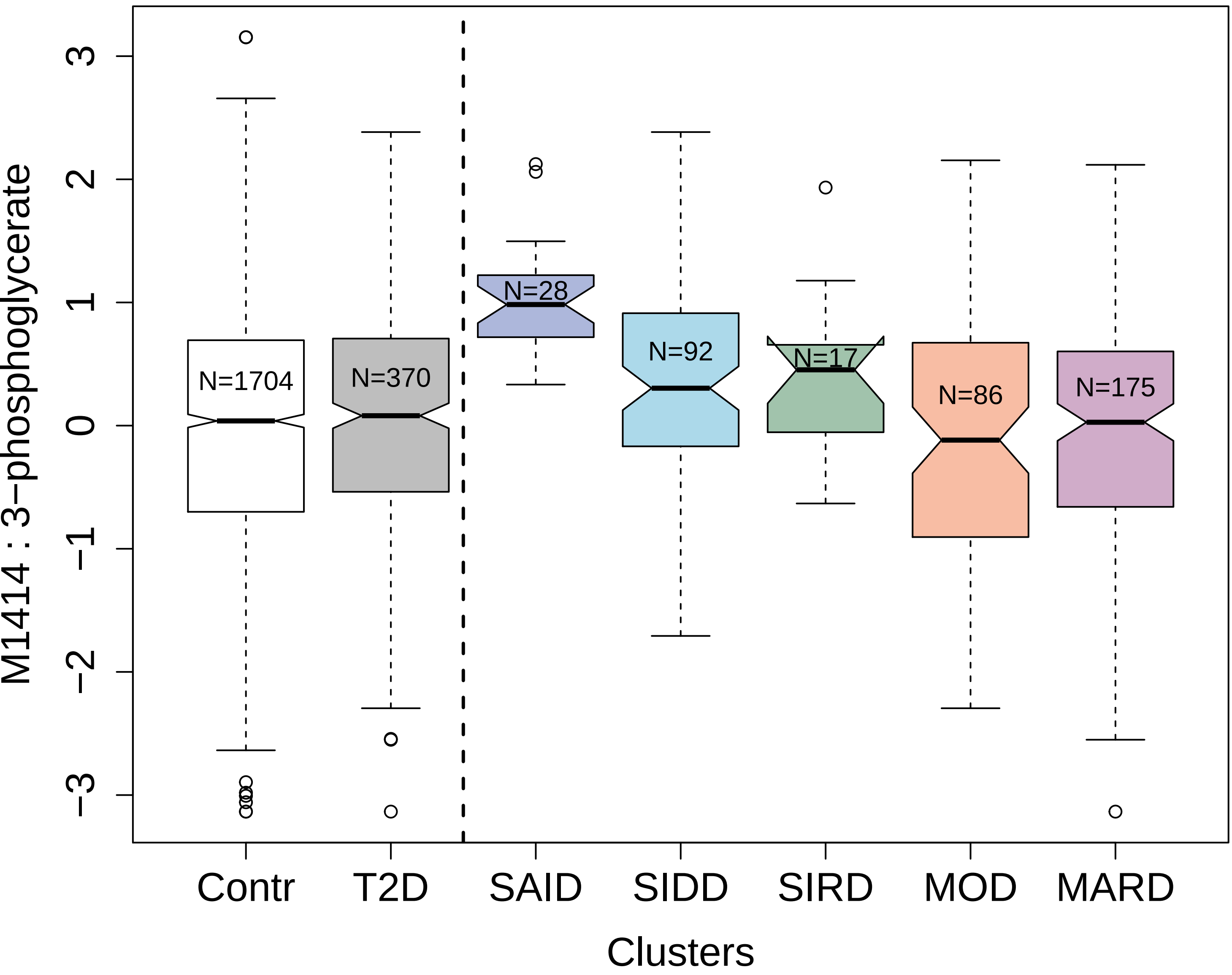

M1414 : 3−phosphoglycerate
3
2
1
0
−1
−2
−3
N=1704
N=370
N=28
N=92
N=17
N=86
N=175
Contr
T2D
SAID
SIDD
SIRD
MOD
MARD
Clusters

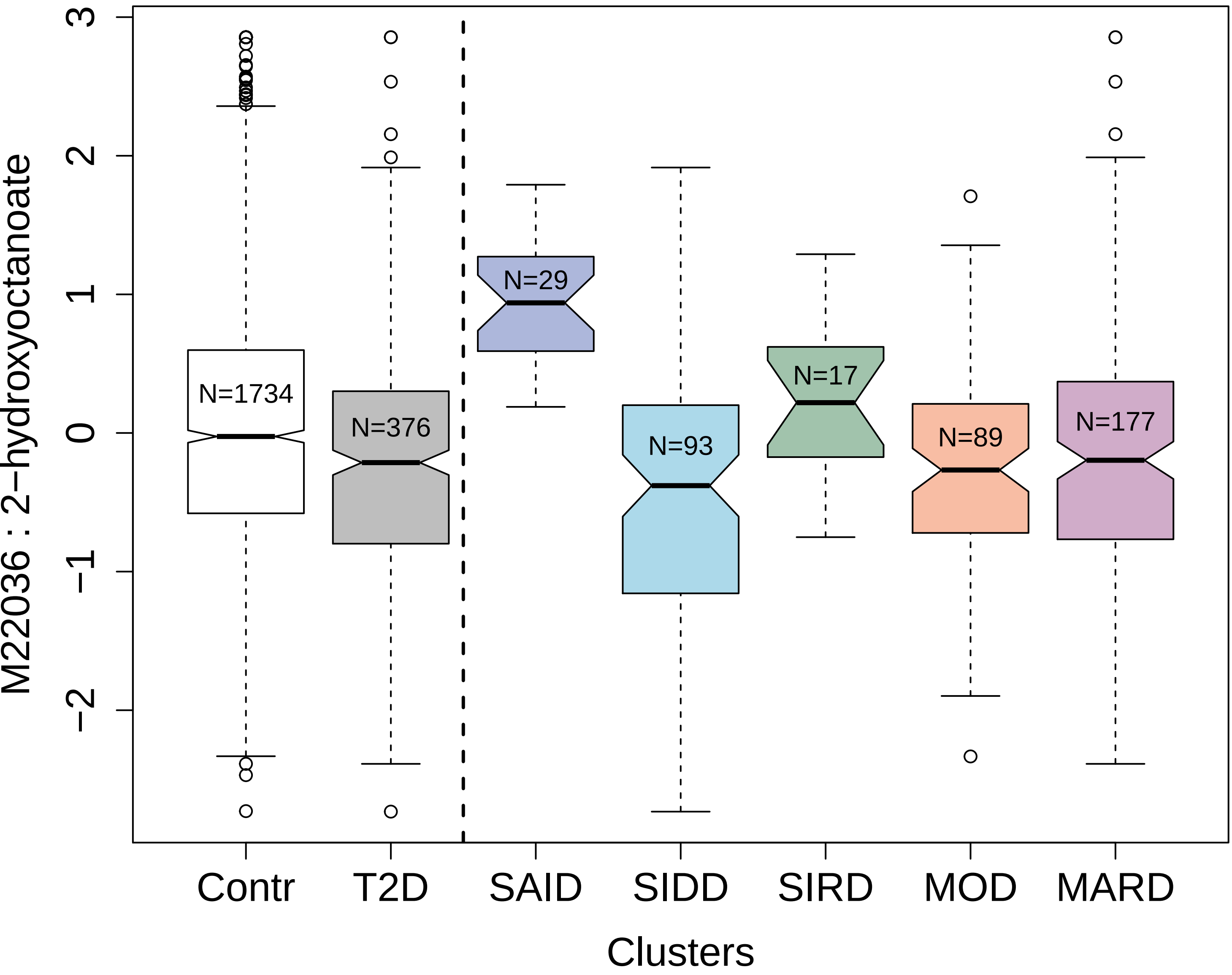
M22036 : 2−hydroxyoctanoate
3
2
1
0
−1
−2
N=1734
N=376
N=29
N=93
N=17
N=89
N=177
Contr
T2D
SAID
SIDD
SIRD
MOD
MARD
Clusters

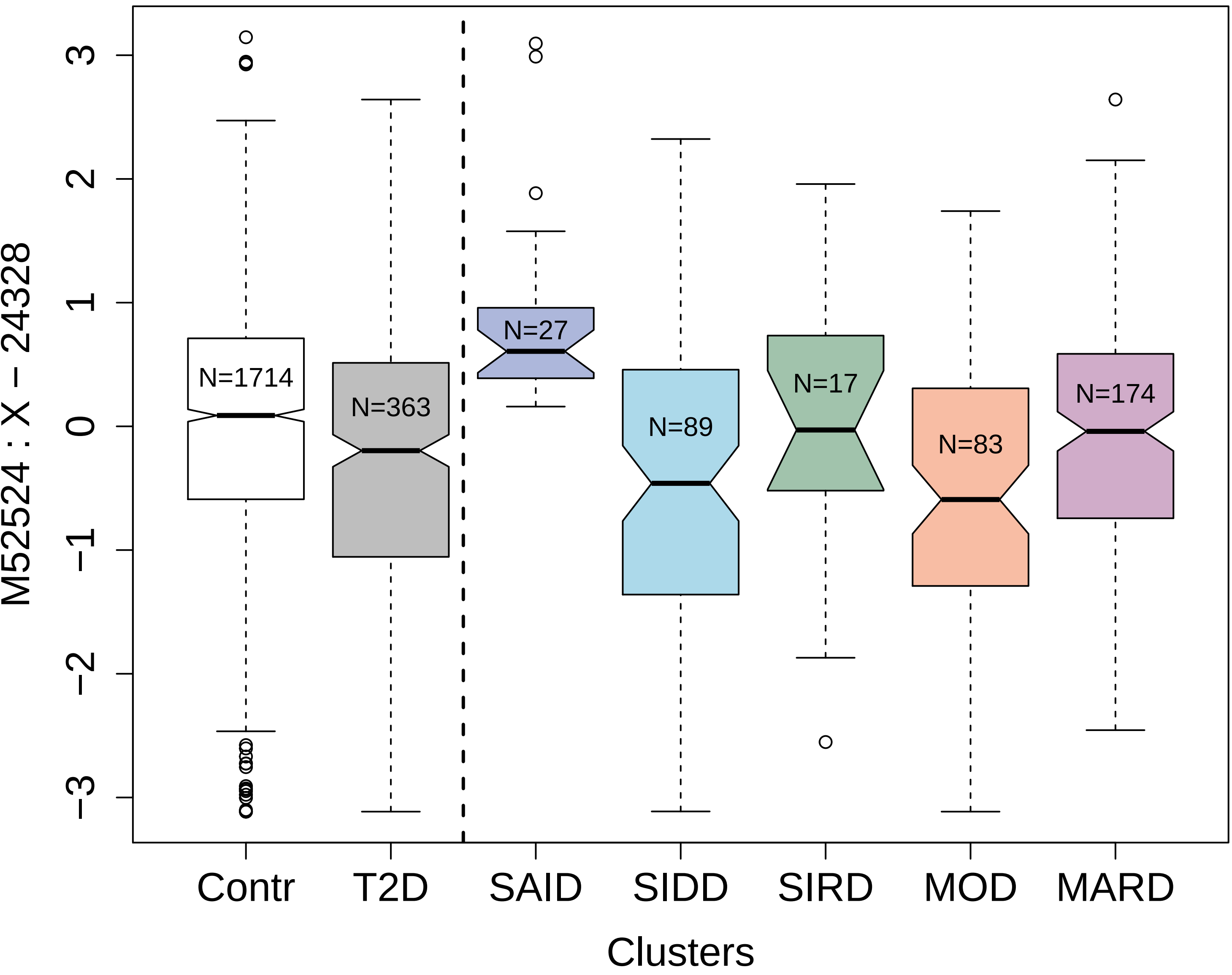
M52524 : X − 24328
3
2
1
0
−1
−2
−3
N=1714
N=363
N=27
N=89
N=17
N=83
N=174
Contr
T2D
SAID
SIDD
SIRD
MOD
MARD
Clusters

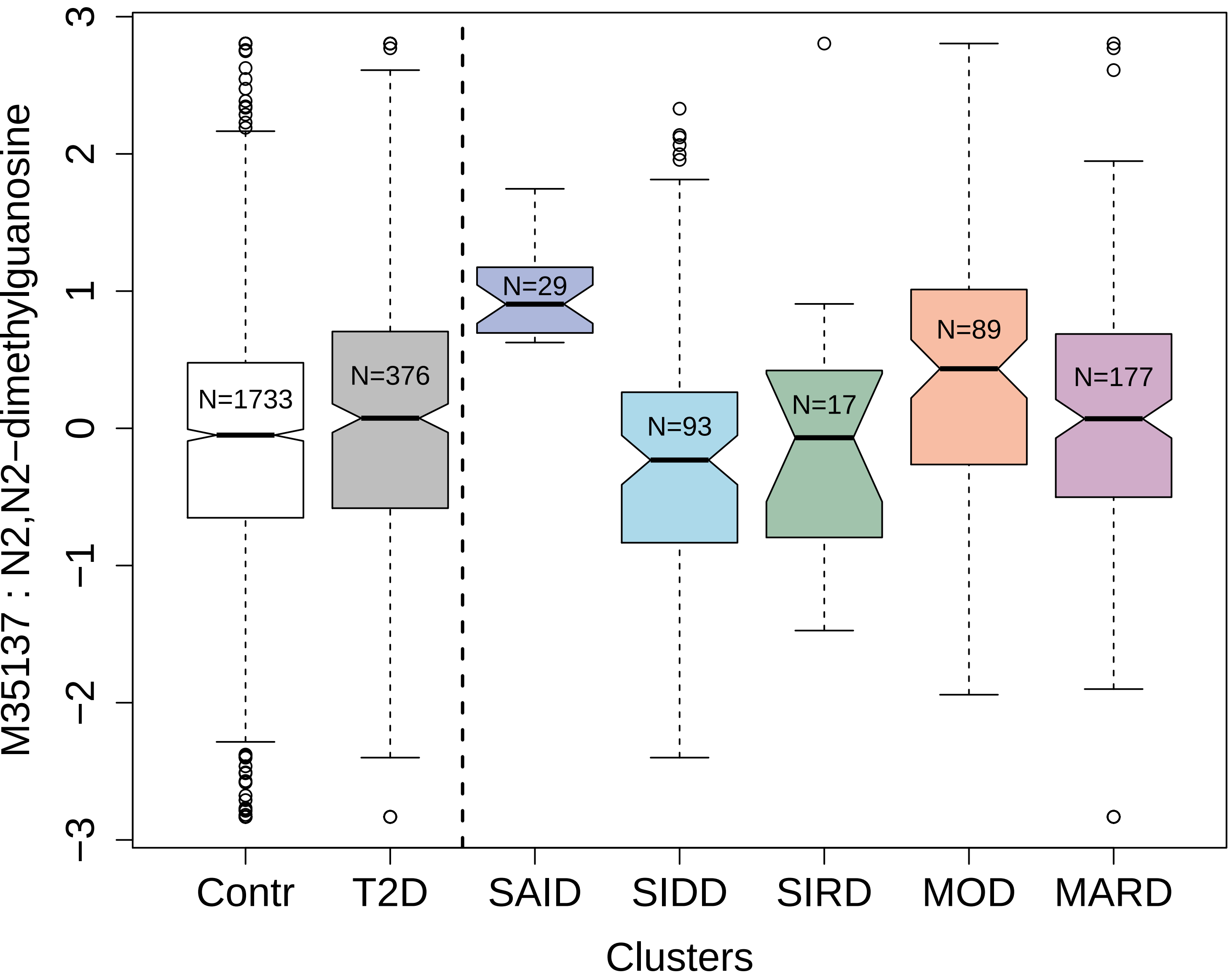

M35137 : N2,N2−dimethylguanosine
3
2
1
0
−1
−2
−3
N=1733
N=376
N=29
N=93
N=17
N=89
N=177
Contr
T2D
SAID
SIDD
SIRD
MOD
MARD
Clusters

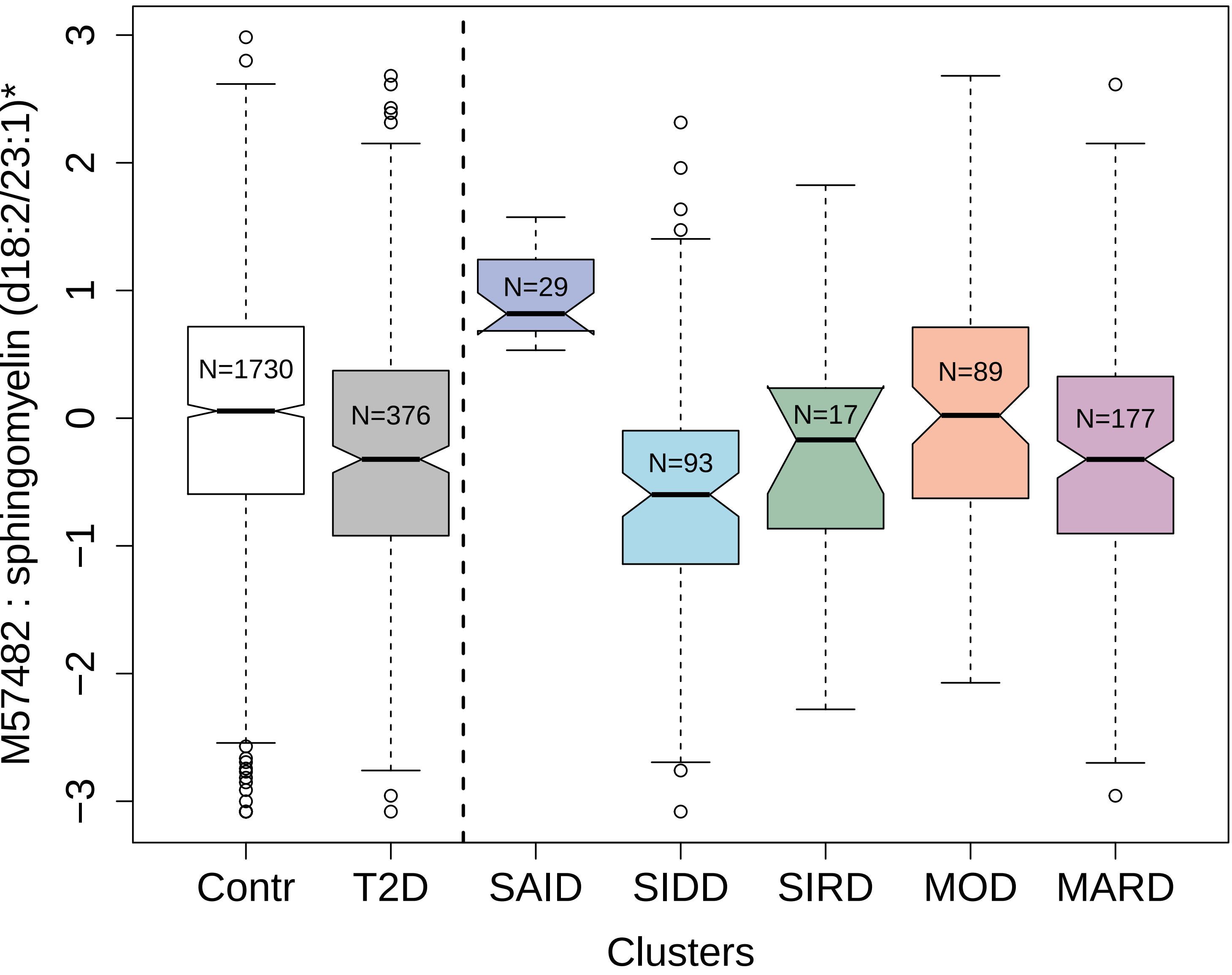

M57482 : sphingomyelin (d18:2/23:1)*
3
2
1
0
−1
−2
−3
N=1730
N=376
N=29
N=93
N=17
N=89
N=177
Contr
T2D
SAID
SIDD
SIRD
MOD
MARD
Clusters

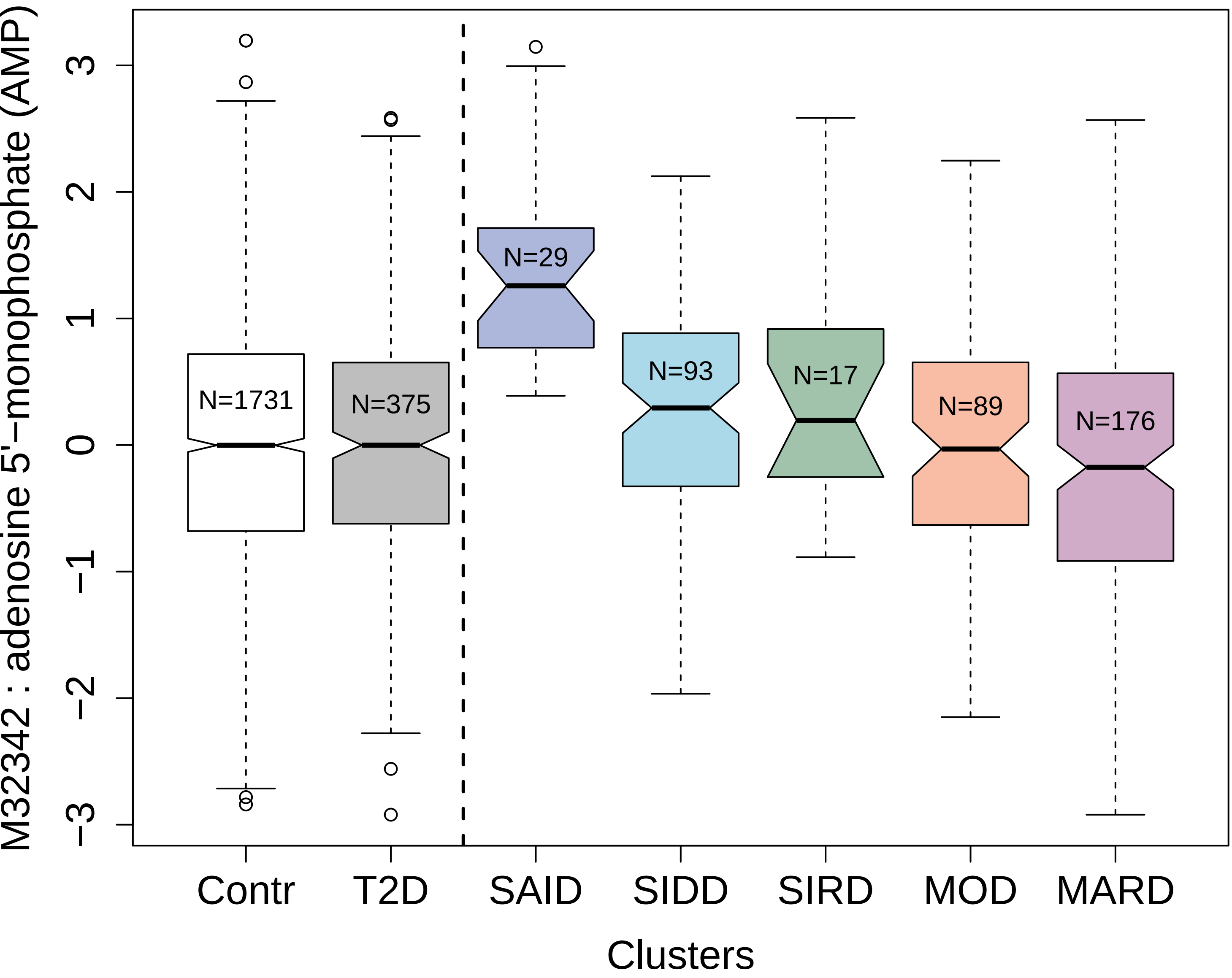
M32342 : adenosine 5'−monophosphate (AMP)
−3
−2
−1
0
1
2
3
N=1731
N=375
N=29
N=93
N=17
N=89
N=176
Contr
T2D
SAID
SIDD
SIRD
MOD
MARD
Clusters

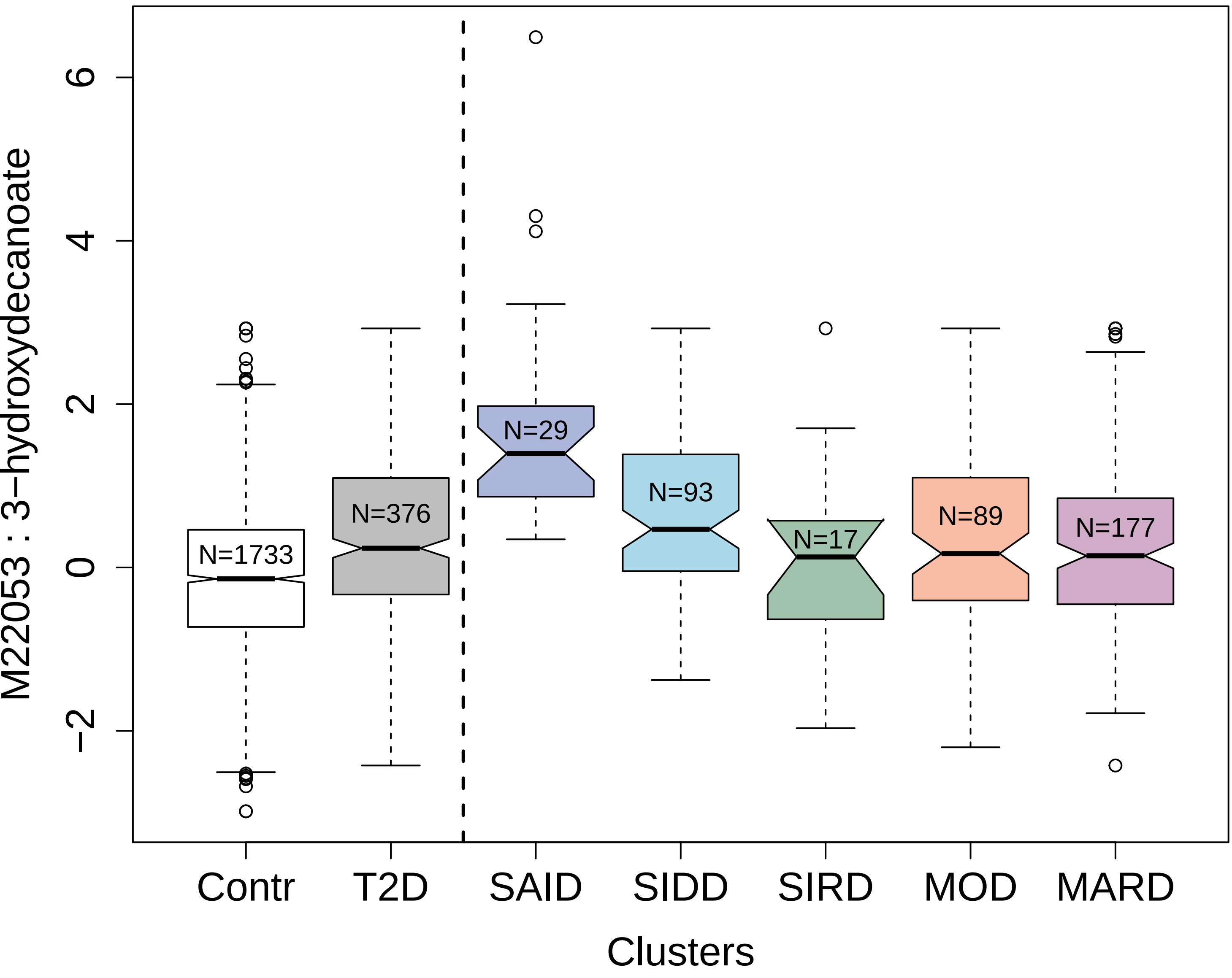
M22053 : 3−hydroxydecanoate
6
4
2
0
−2
N=1733
N=376
N=29
N=93
N=17
N=89
N=177
Contr
T2D
SAID
SIDD
SIRD
MOD
MARD
Clusters

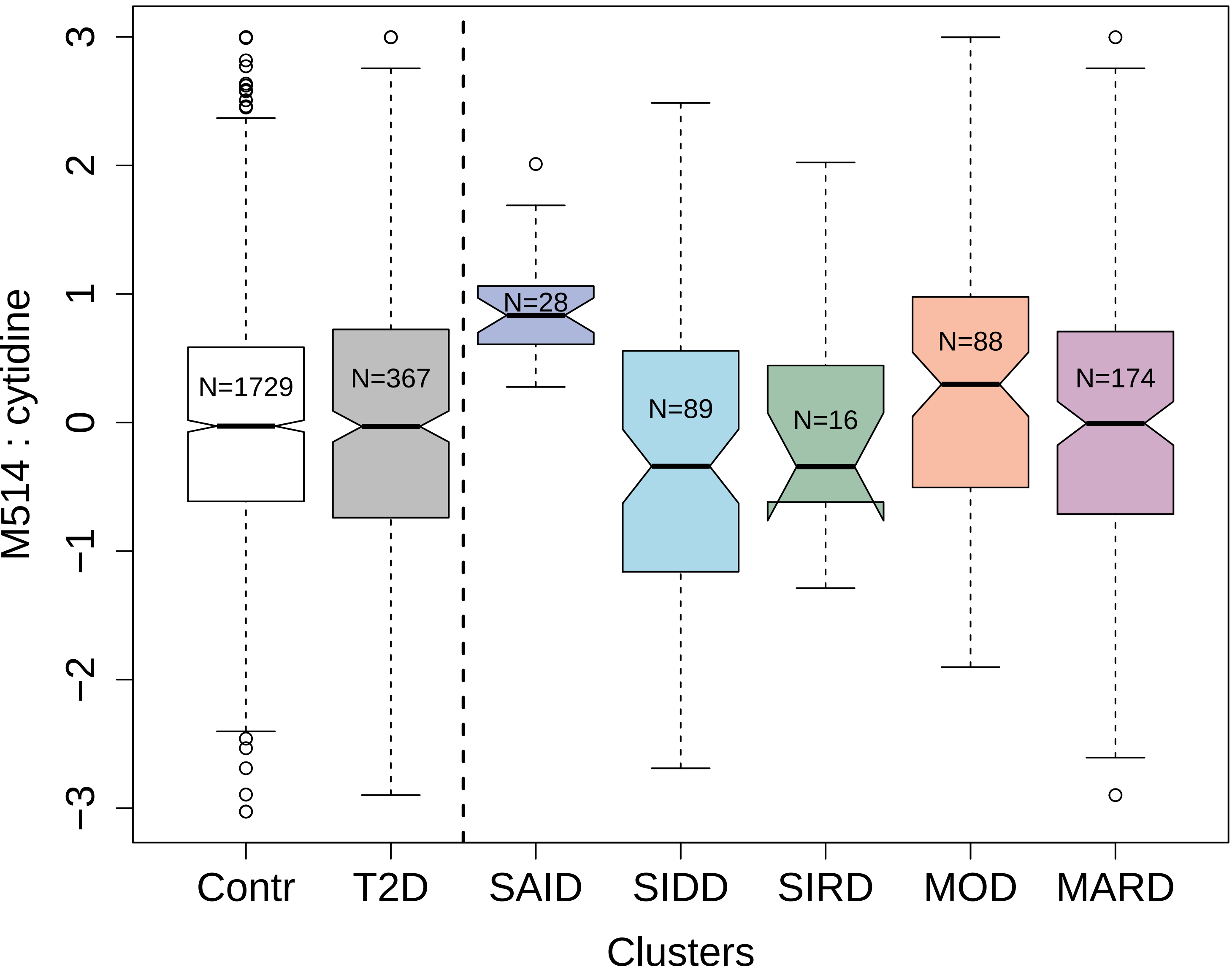
M514 : cytidine
3
2
1
0
−1
−2
−3
N=1729
N=367
N=28
N=89
N=16
N=88
N=174
Contr
T2D
SAID
SIDD
SIRD
MOD
MARD
Clusters

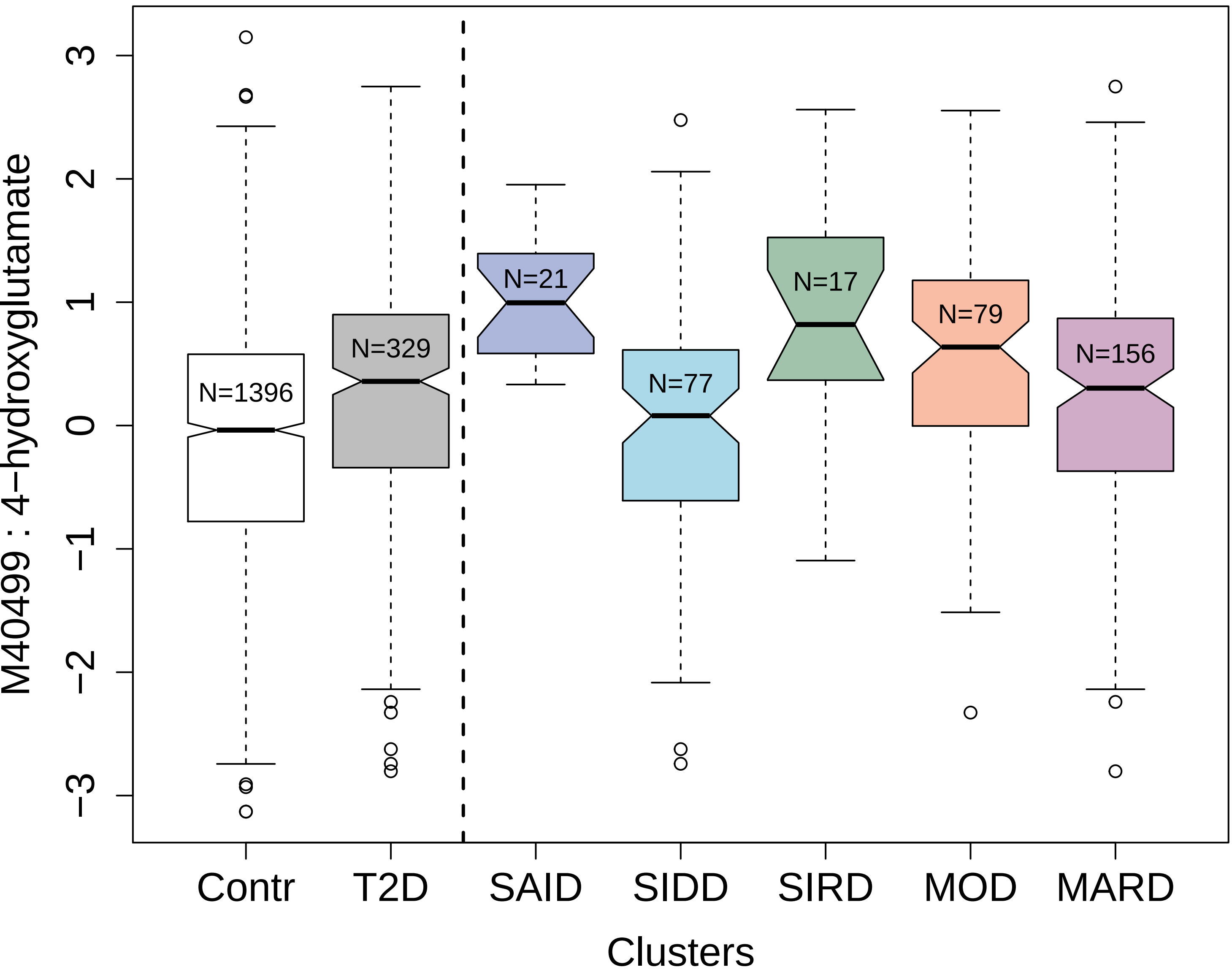

M40499 : 4−hydroxyglutamate
3
2
1
0
−1
−2
−3
N=1396
N=329
N=21
N=77
N=17
N=79
N=156
Contr
T2D
SAID
SIDD
SIRD
MOD
MARD
Clusters

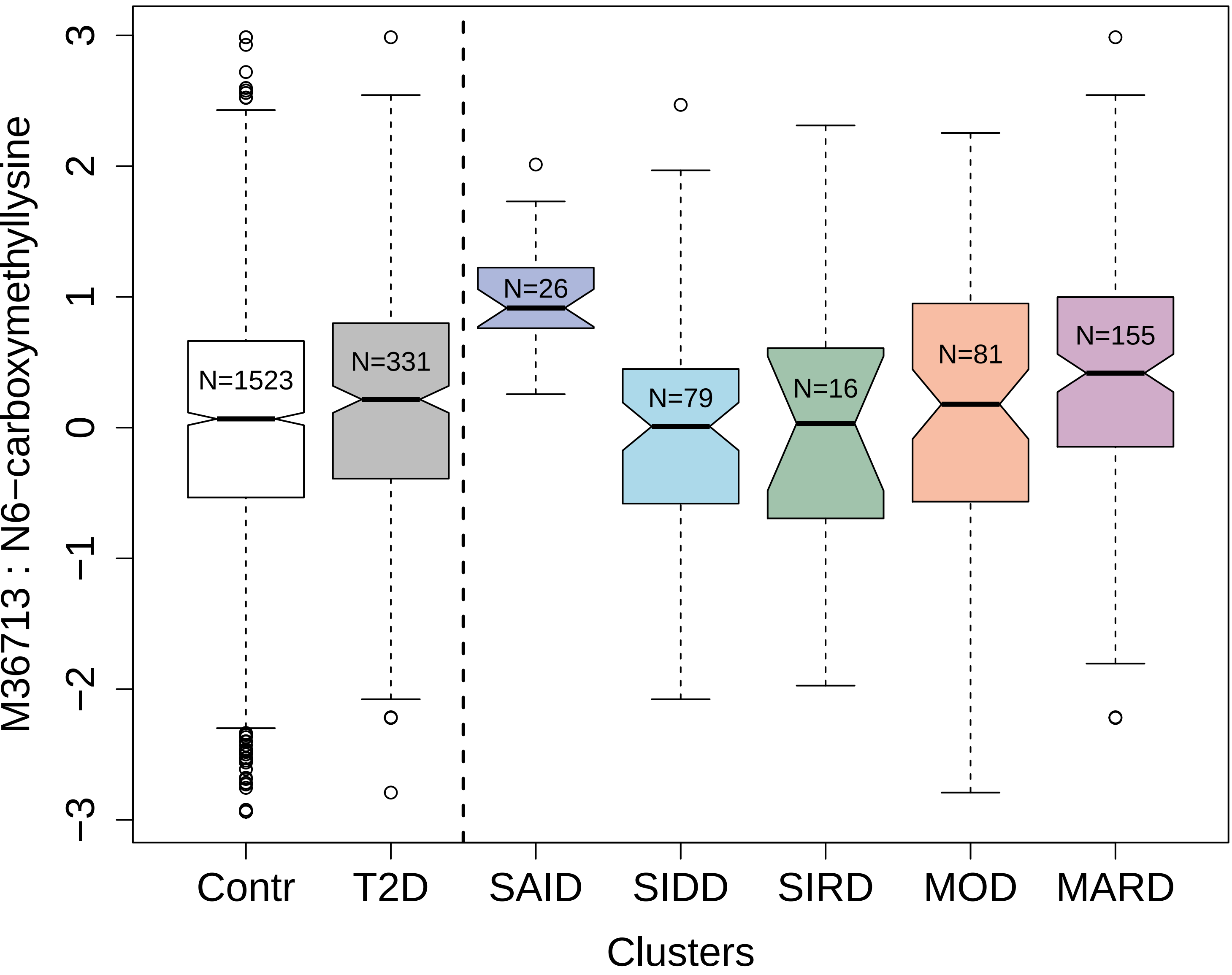

M36713 : N6−carboxymethyllysine
3
2
1
0
−1
−2
−3
N=1523
N=331
N=26
N=79
N=16
N=81
N=155
Contr
T2D
SAID
SIDD
SIRD
MOD
MARD
Clusters

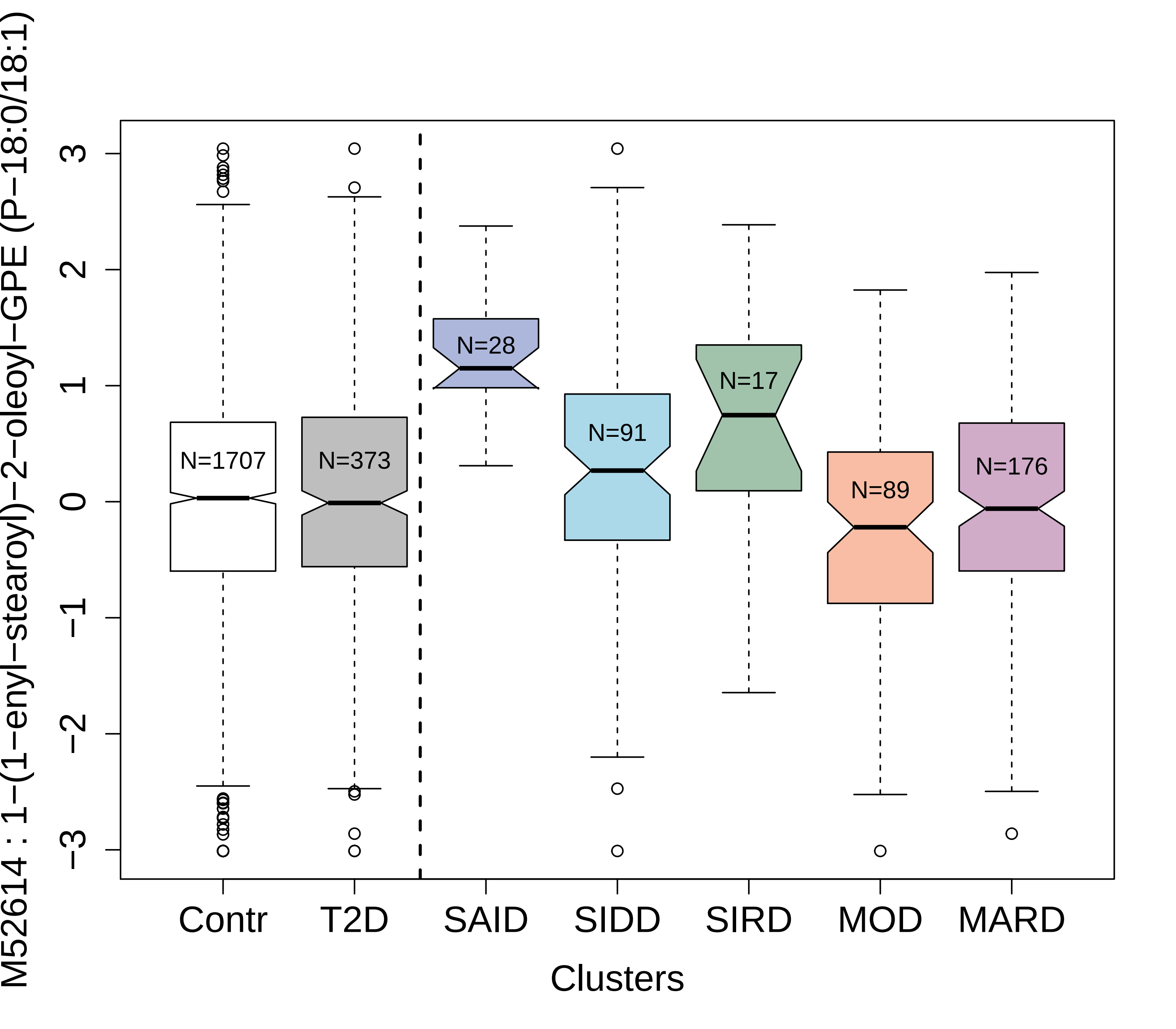
M52614 : 1−(1−enyl−stearoyl)−2−oleoyl−GPE (P−18:0/18:1)
3
2
1
0
−1
−2
−3
N=1707
N=373
N=28
N=91
N=17
N=89
N=176
Contr
T2D
SAID
SIDD
SIRD
MOD
MARD
Clusters

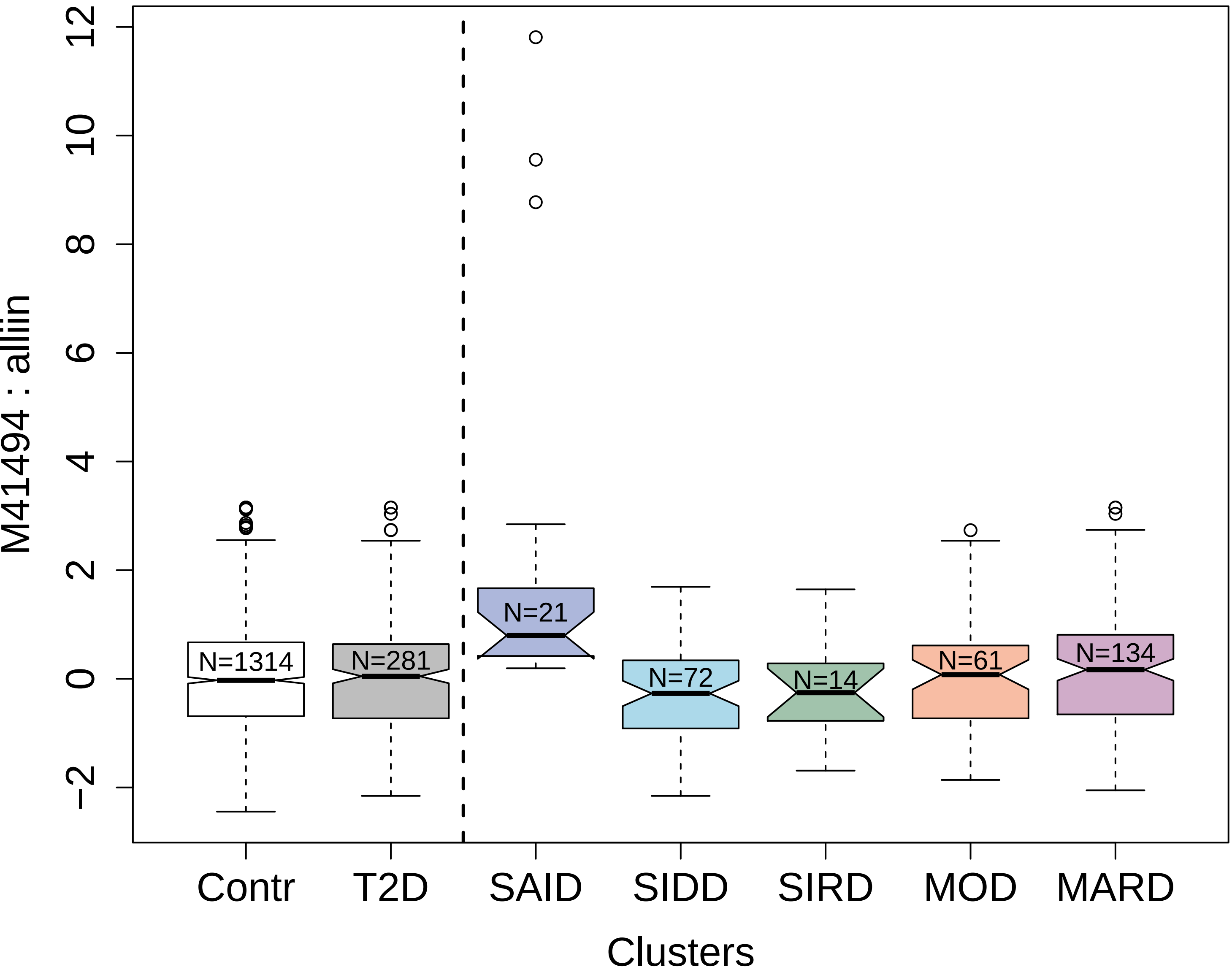

M41494 : alliin
12
10
8
6
4
2
0
−2
N=1314
N=281
N=21
N=72
N=14
N=61
N=134
Contr
T2D
SAID
SIDD
SIRD
MOD
MARD
Clusters

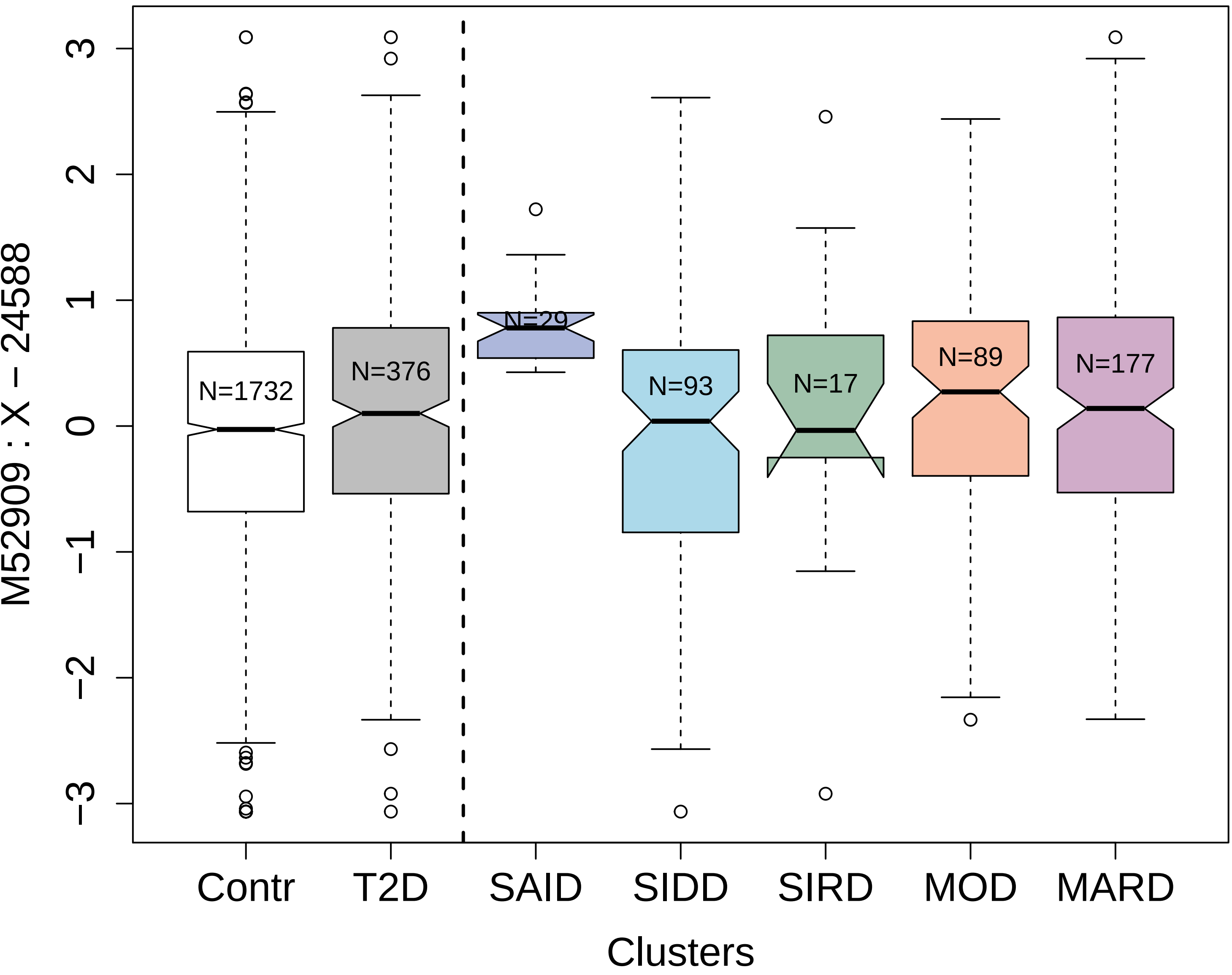
M52909 : X − 24588
3
2
1
0
−1
−2
−3
N=1732
N=376
N=29
N=93
N=17
N=89
N=177
Contr
T2D
SAID
SIDD
SIRD
MOD
MARD
Clusters

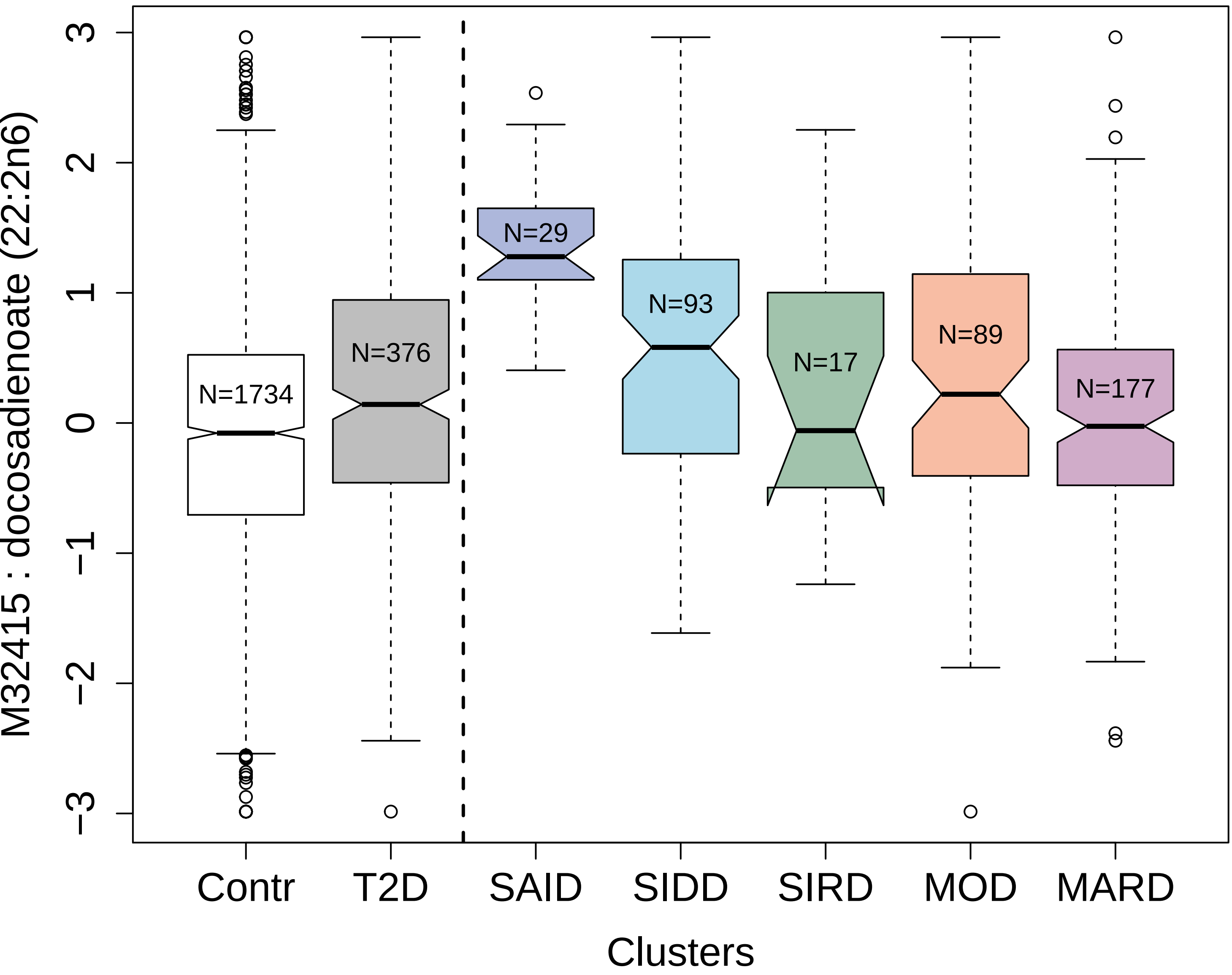

M32415 : docosadienoate (22:2n6)
3
2
1
0
−1
−2
−3
N=1734
N=376
N=29
N=93
N=17
N=89
N=177
Contr
T2D
SAID
SIDD
SIRD
MOD
MARD
Clusters

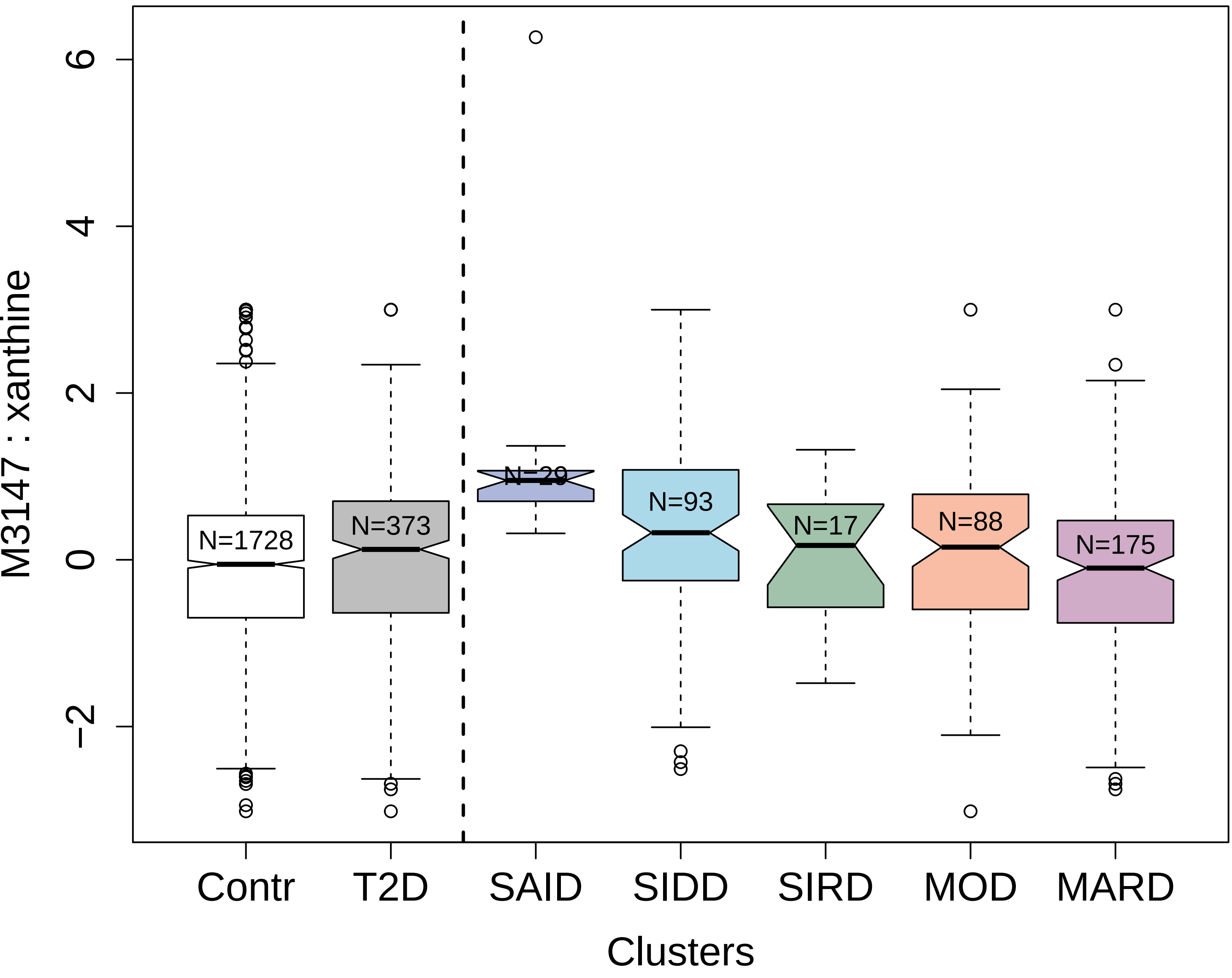
M3147 : xanthine
Clusters
Contr
T2D
SAID
SIDD
SIRD
MOD
MARD
N=1728
N=373
N=29
N=93
N=17
N=88
N=175
6
4
2
0
−2

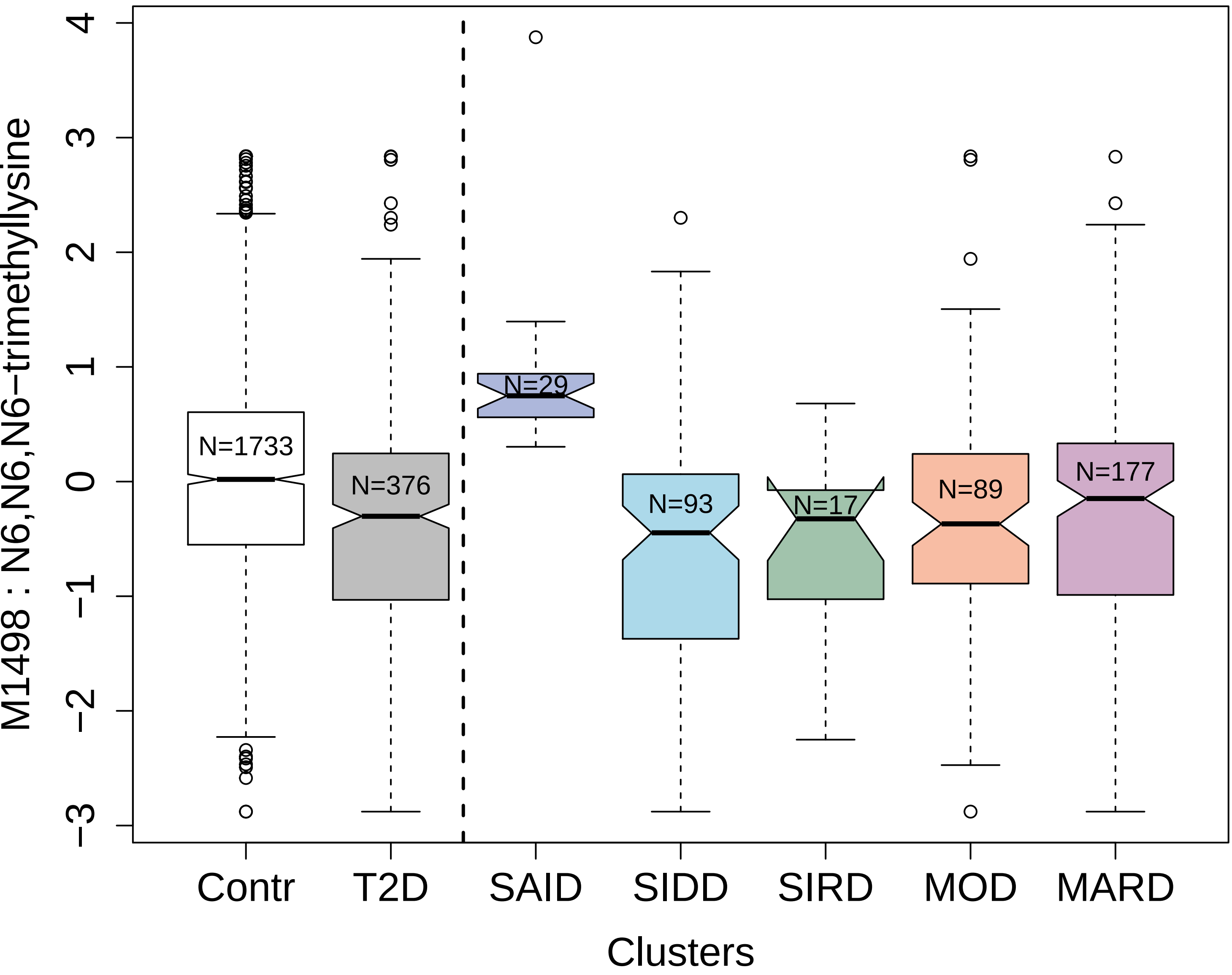
M1498 : N6,N6,N6−trimethyllysine
4
3
2
1
0
−1
−2
−3
N=1733
N=376
N=29
N=93
N=17
N=89
N=177
Contr
T2D
SAID
SIDD
SIRD
MOD
MARD
Clusters

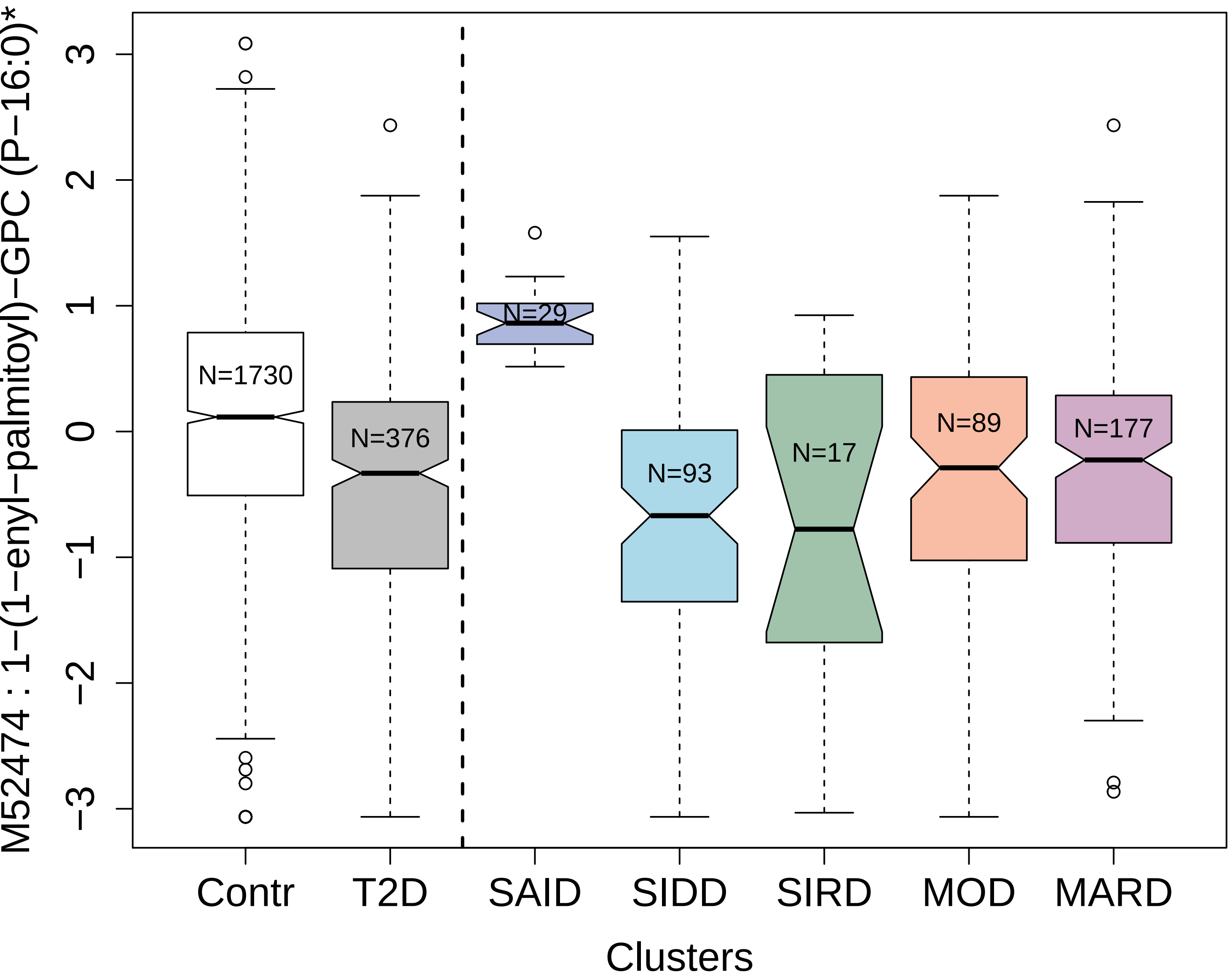

M52474 : 1−(1−enyl−palmitoyl)−GPC (P−16:0)*
3
2
1
0
−1
−2
−3
N=1730
N=376
N=29
N=93
N=17
N=89
N=177
Contr
T2D
SAID
SIDD
SIRD
MOD
MARD
Clusters

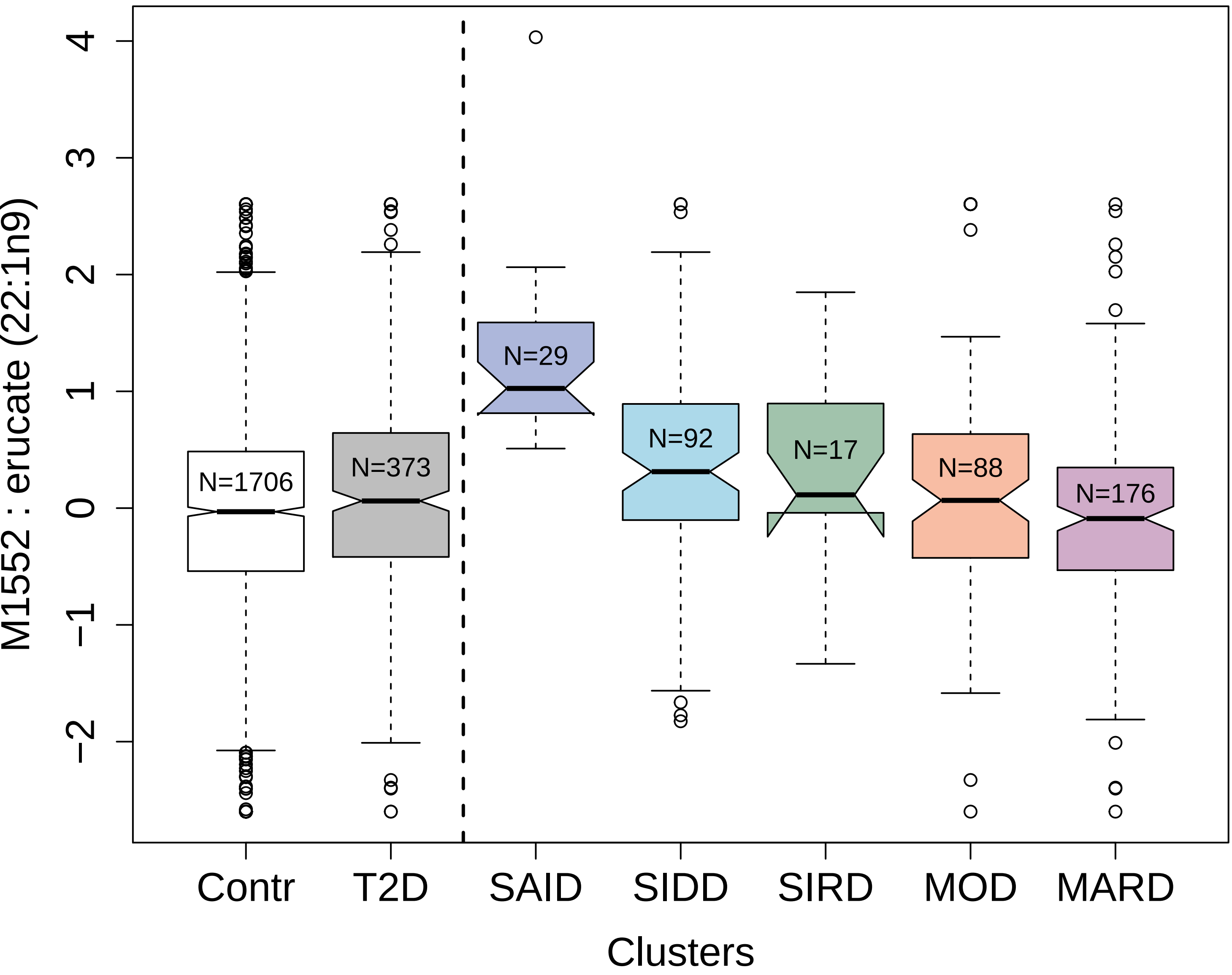
M1552 : erucate (22:1n9)
4
3
2
1
0
−1
−2
N=1706
N=373
N=29
N=92
N=17
N=88
N=176
Contr
T2D
SAID
SIDD
SIRD
MOD
MARD
Clusters

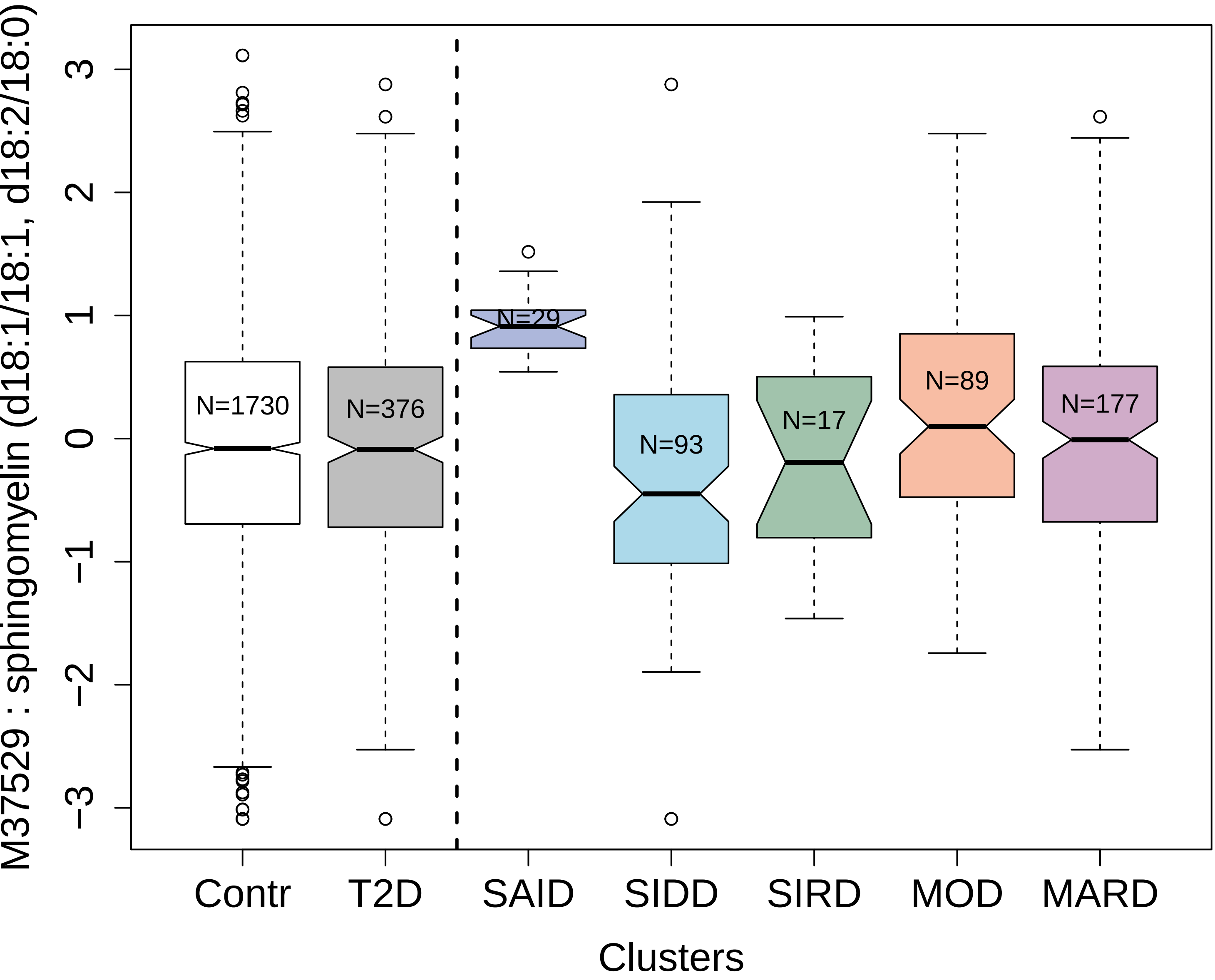

M37529 : sphingomyelin (d18:1/18:1, d18:2/18:0)
3
2
1
0
−1
−2
−3
N=1730
N=376
N=29
N=93
N=17
N=89
N=177
Contr
T2D
SAID
SIDD
SIRD
MOD
MARD
Clusters

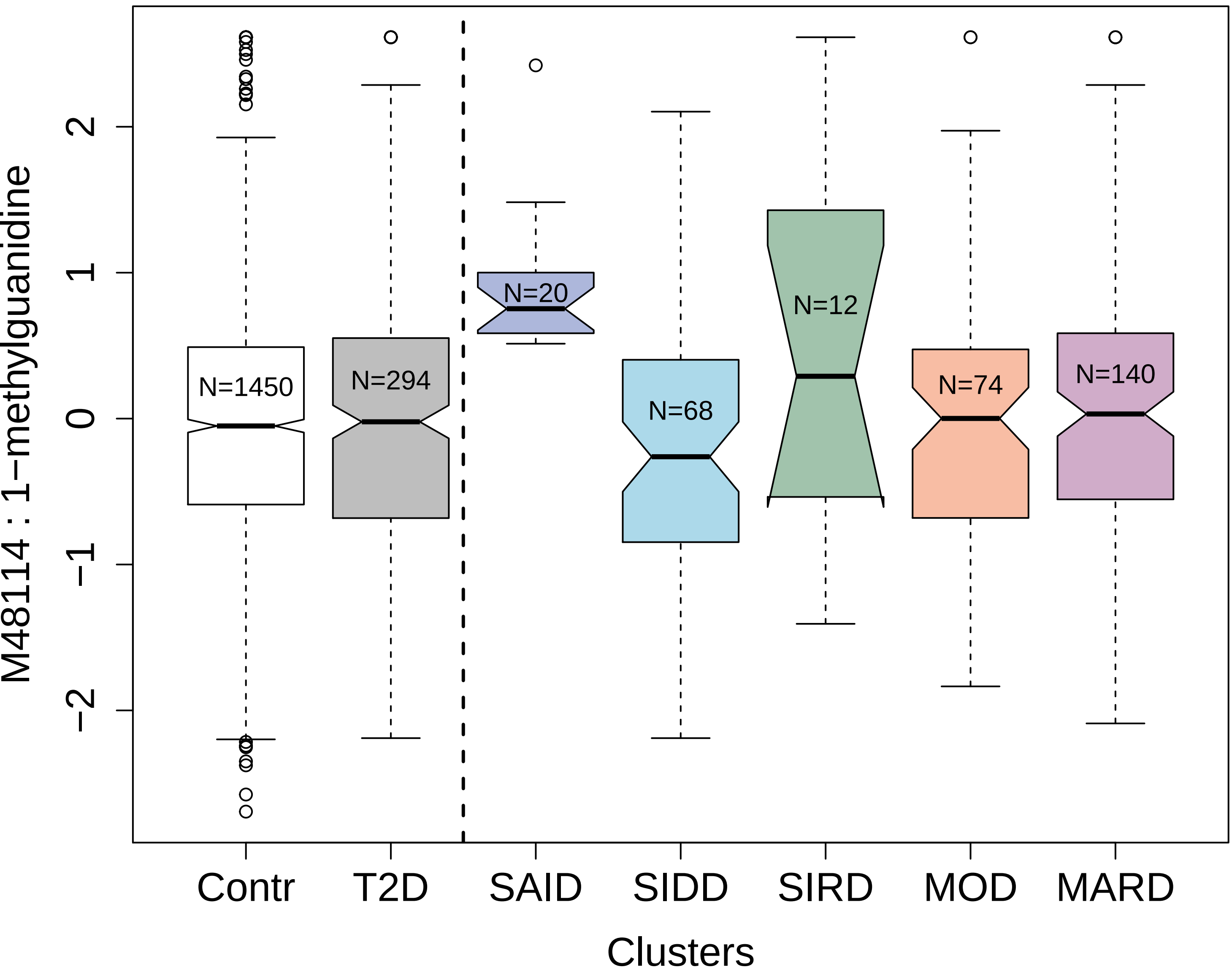

M48114 : 1−methylguanidine
2
1
0
−1
−2
N=1450
N=294
N=20
N=68
N=12
N=74
N=140
Contr
T2D
SAID
SIDD
SIRD
MOD
MARD
Clusters

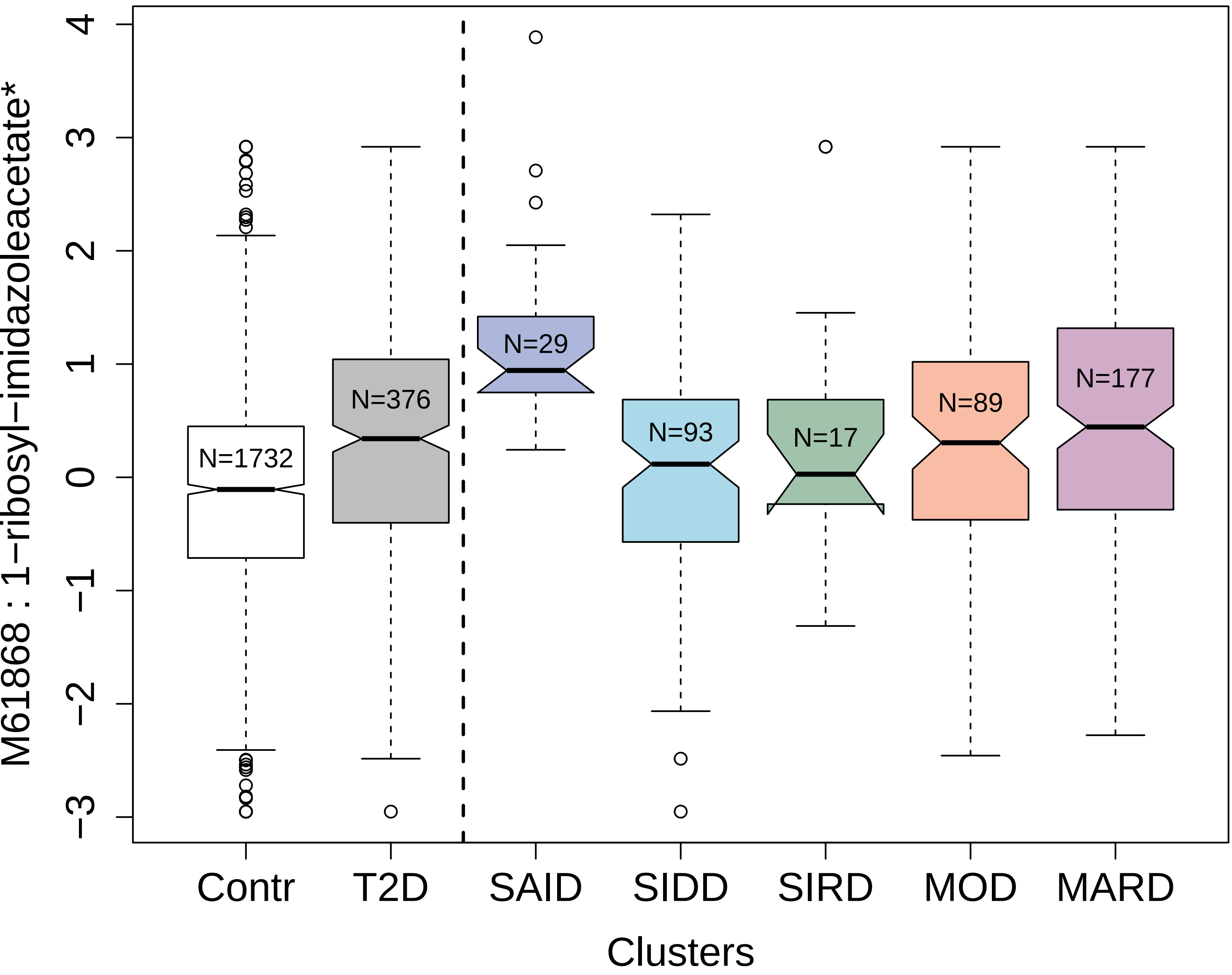
M61868 : 1−ribosyl−imidazoleacetate*
4
3
2
1
0
−1
−2
−3
N=1732
N=376
N=29
N=93
N=17
N=89
N=177
Contr
T2D
SAID
SIDD
SIRD
MOD
MARD
Clusters

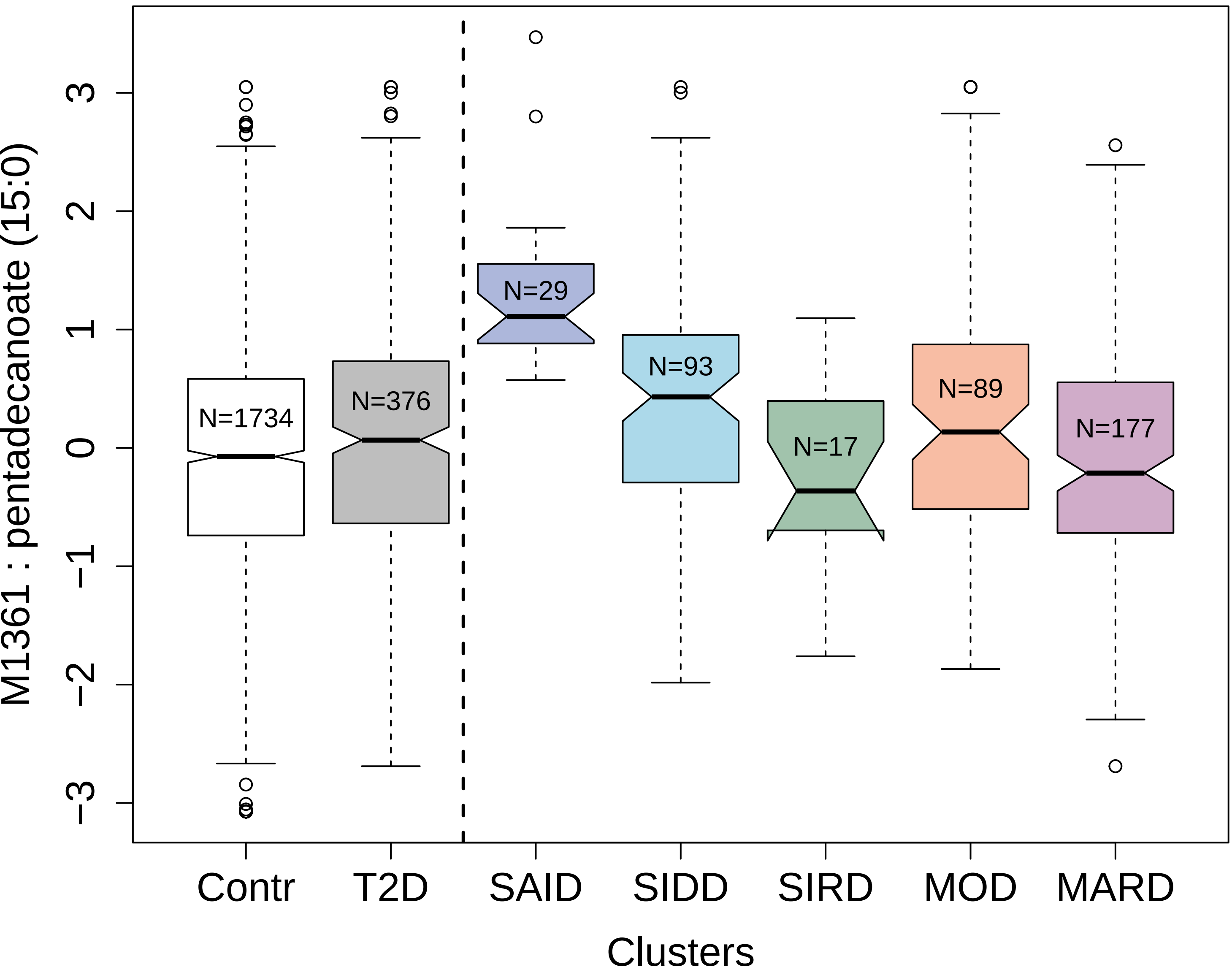
M1361 : pentadecanoate (15:0)
3
2
1
0
−1
−2
−3
N=1734
N=376
N=29
N=93
N=17
N=89
N=177
Contr
T2D
SAID
SIDD
SIRD
MOD
MARD
Clusters

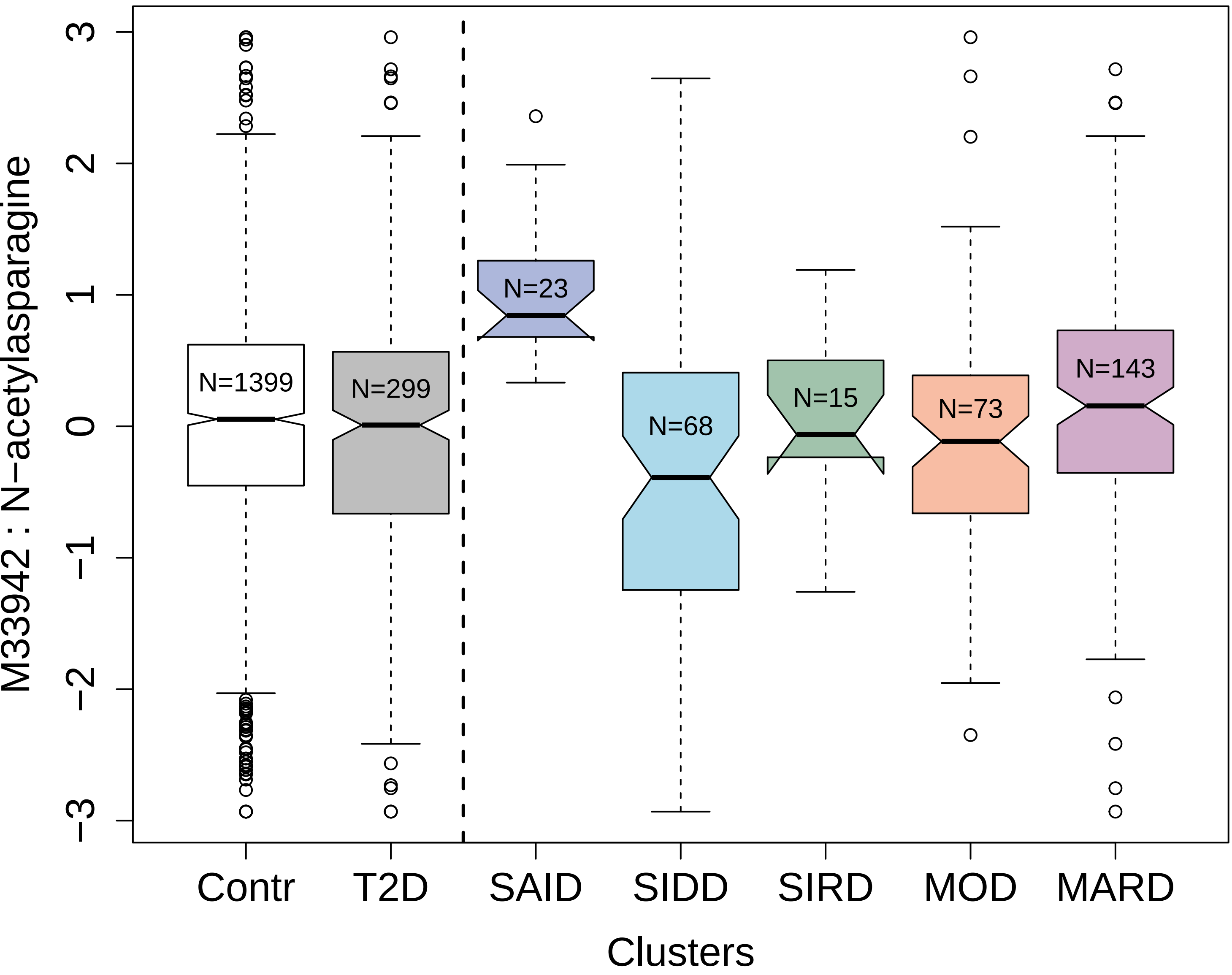

M33942 : N−acetylasparagine
3
2
1
0
−1
−2
−3
N=1399
N=299
N=23
N=68
N=15
N=73
N=143
Contr
T2D
SAID
SIDD
SIRD
MOD
MARD
Clusters

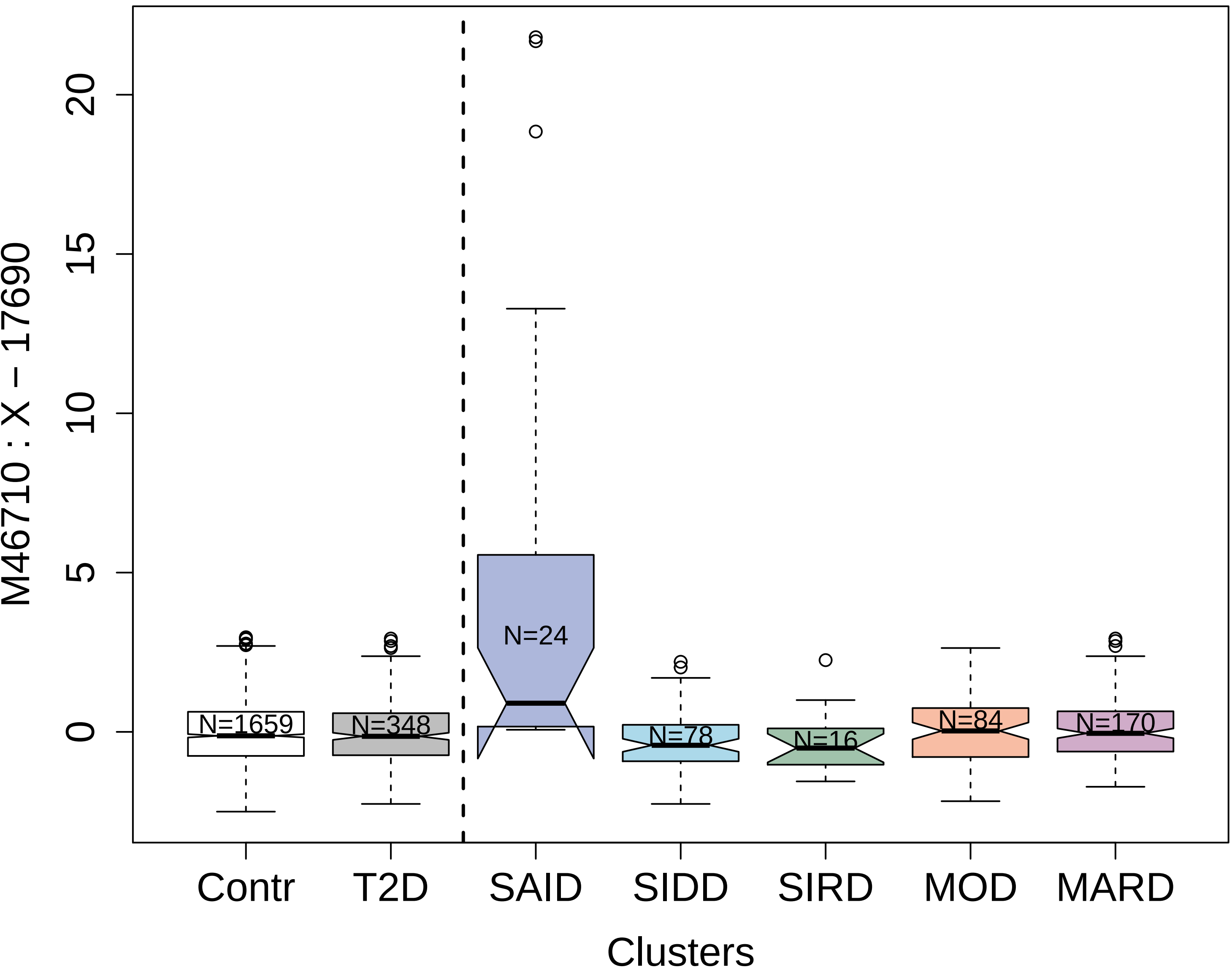

M46710 : X − 17690
0
5
10
15
20
N=1659
N=348
N=24
N=78
N=16
N=84
N=170
Contr
T2D
SAID
SIDD
SIRD
MOD
MARD
Clusters

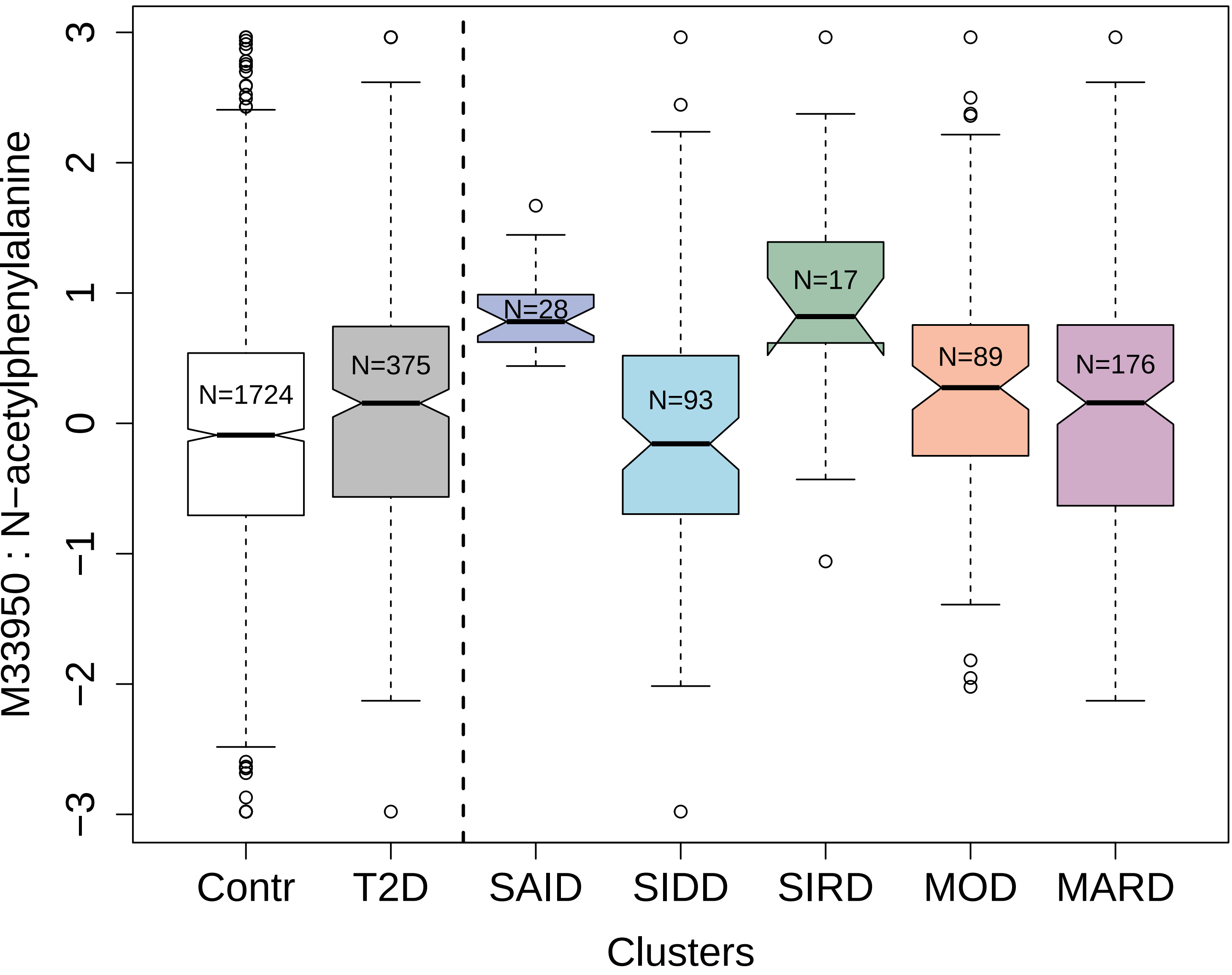

M33950 : N−acetylphenylalanine
3
2
1
0
−1
−2
−3
N=1724
N=375
N=28
N=93
N=17
N=89
N=176
Contr
T2D
SAID
SIDD
SIRD
MOD
MARD
Clusters

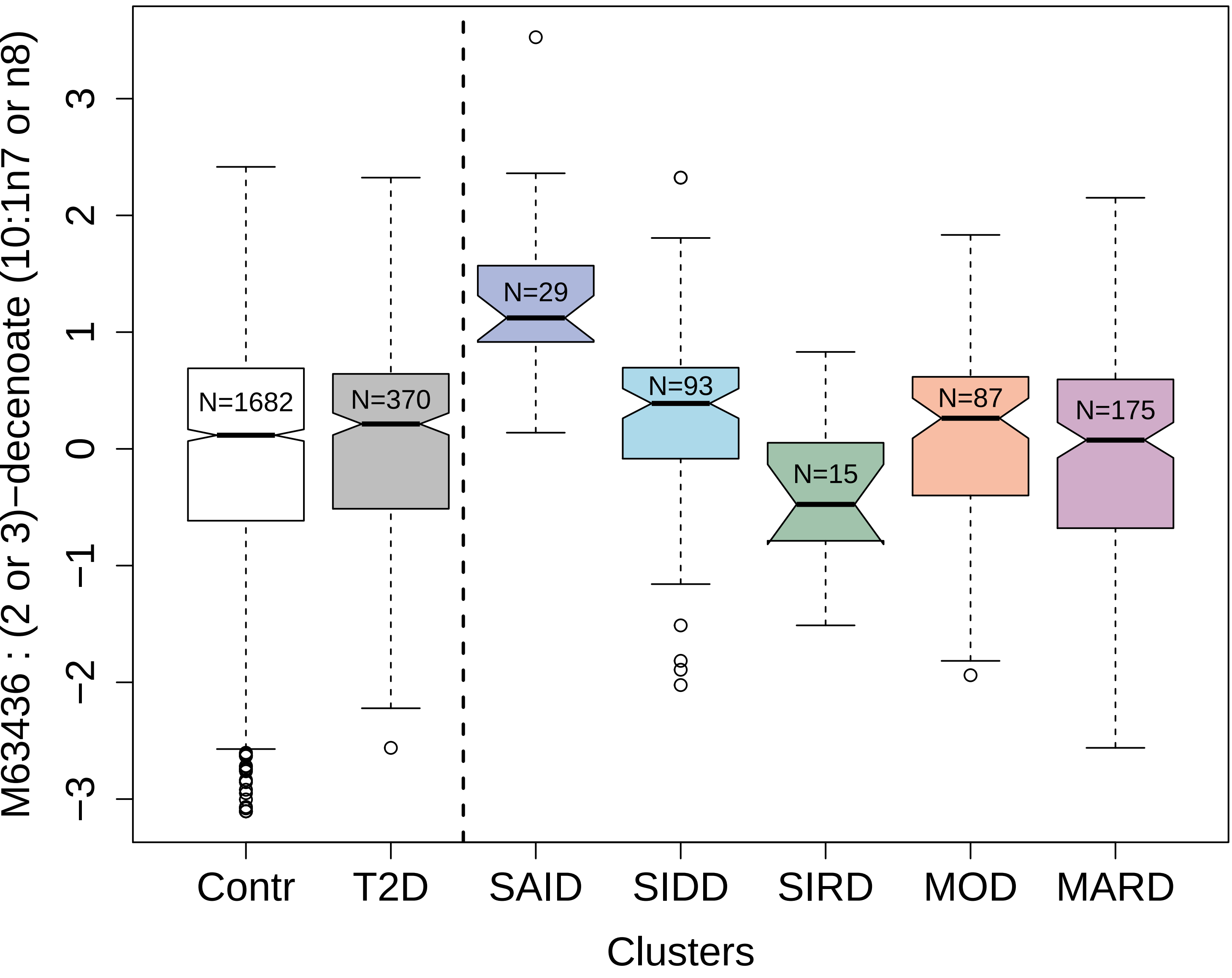

M63436 : (2 or 3)−decenoate (10:1n7 or n8)
3
2
1
0
−1
−2
−3
N=1682
N=370
N=29
N=93
N=15
N=87
N=175
Contr
T2D
SAID
SIDD
SIRD
MOD
MARD
Clusters

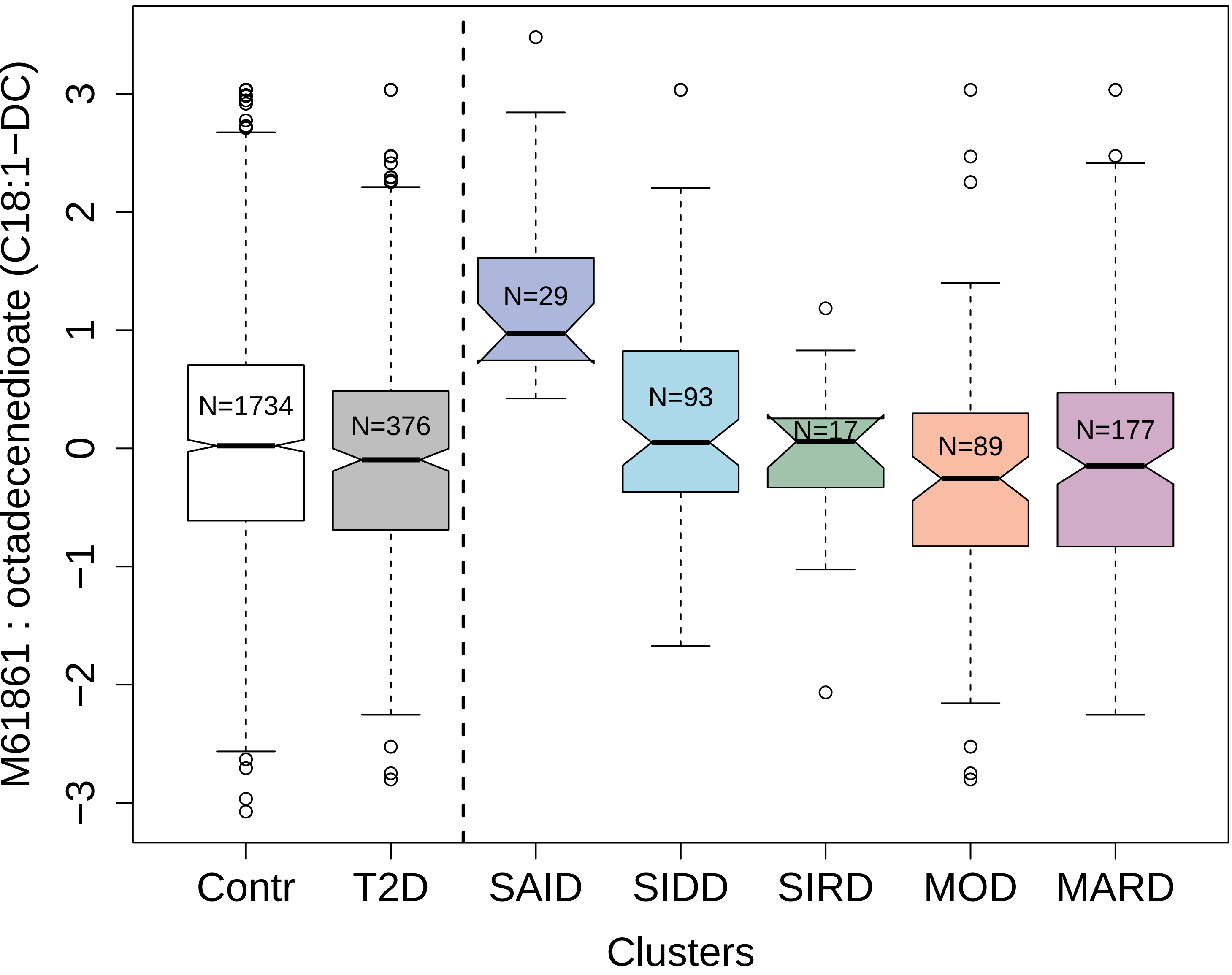
M61861 : octadecenedioate (C18:1−DC)
3
2
1
0
−1
−2
−3
N=1734
N=376
N=29
N=93
N=17
N=89
N=177
Contr
T2D
SAID
SIDD
SIRD
MOD
MARD
Clusters

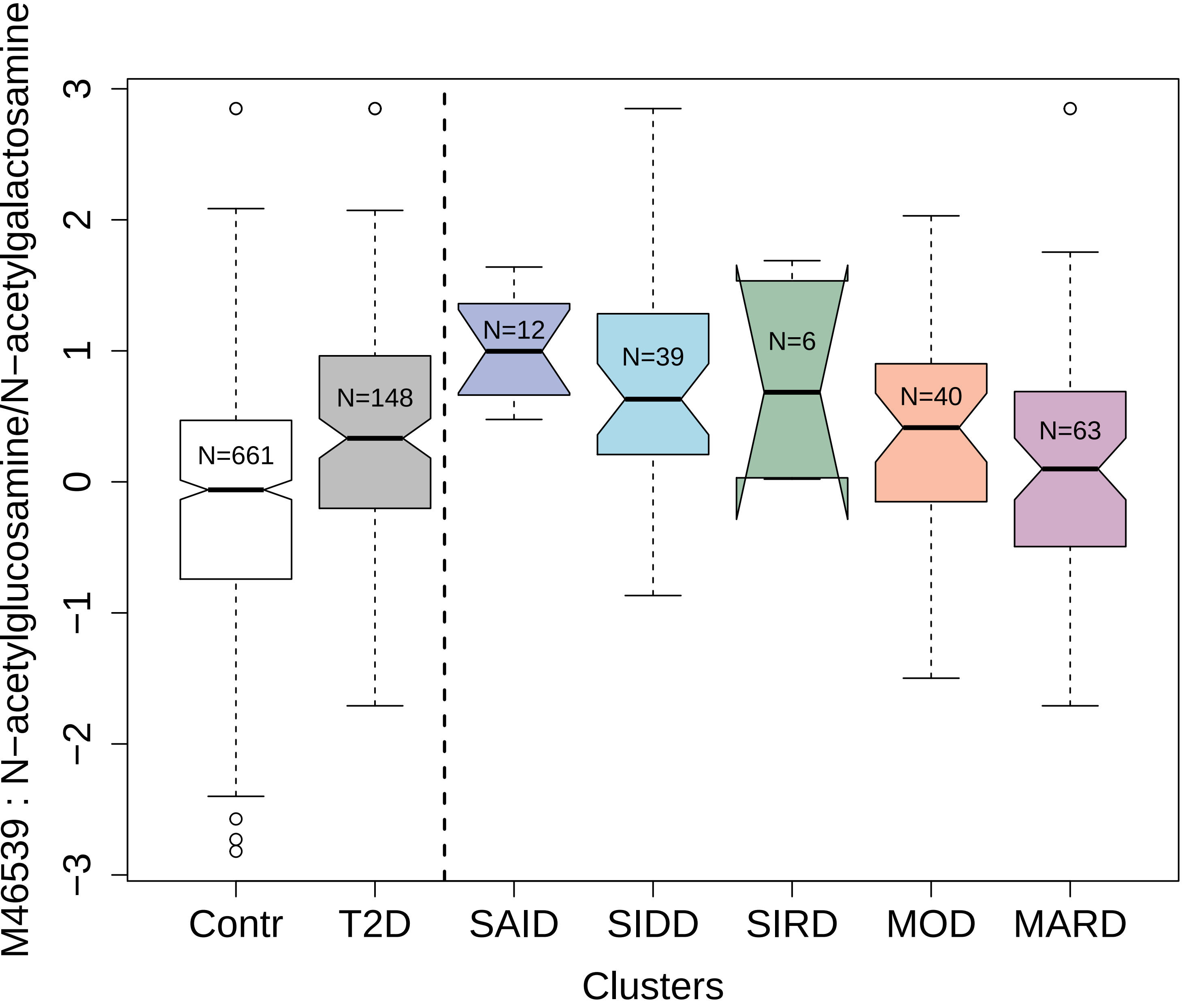

M46539 : N−acetylglucosamine/N−acetylgalactosamine
3
2
1
0
−1
−2
−3
N=661
N=148
N=12
N=39
N=6
N=40
N=63
Contr
T2D
SAID
SIDD
SIRD
MOD
MARD
Clusters

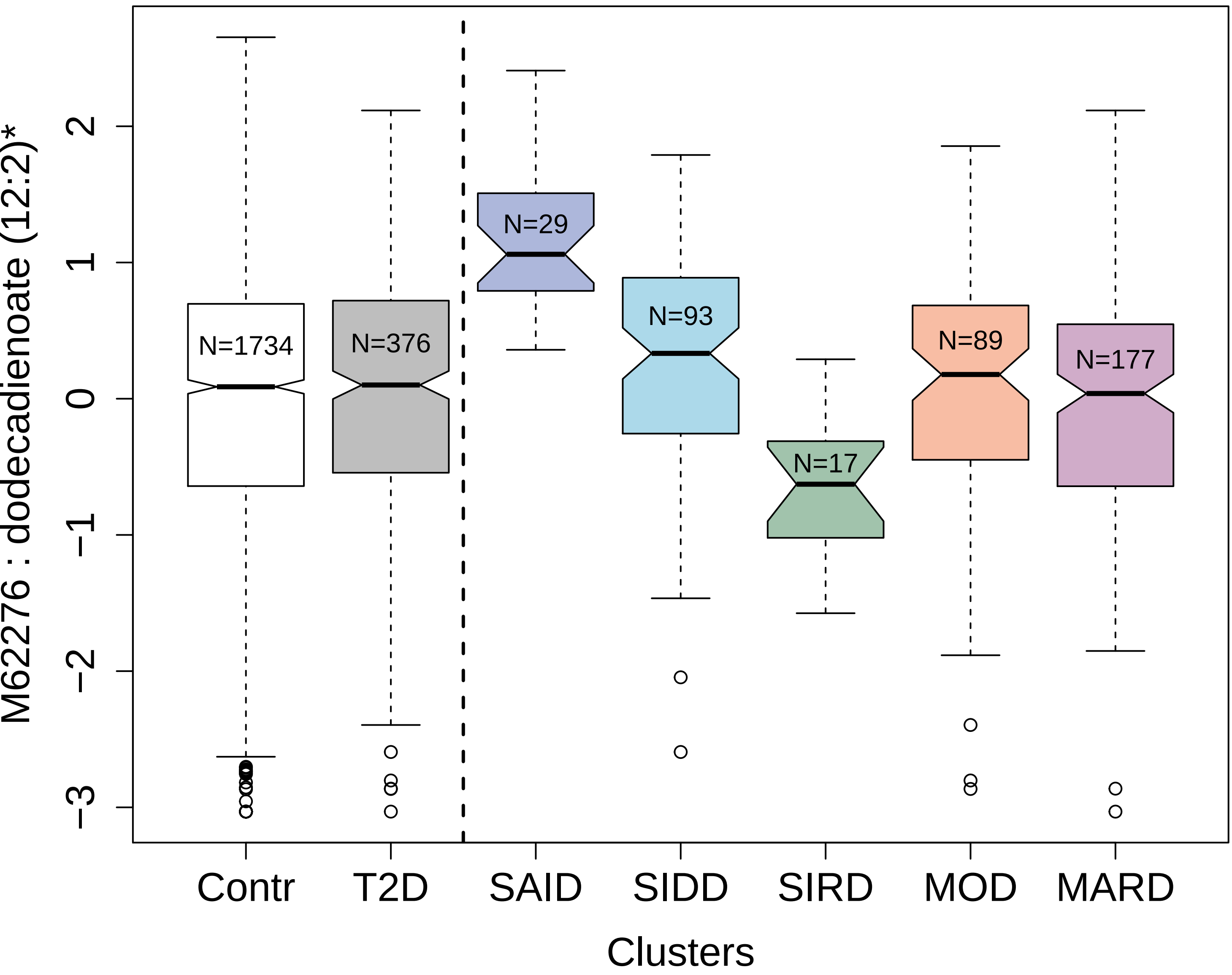

M62276 : dodecadienoate (12:2)*
2
1
0
−1
−2
−3
N=1734
N=376
N=29
N=93
N=17
N=89
N=177
Contr
T2D
SAID
SIDD
SIRD
MOD
MARD
Clusters

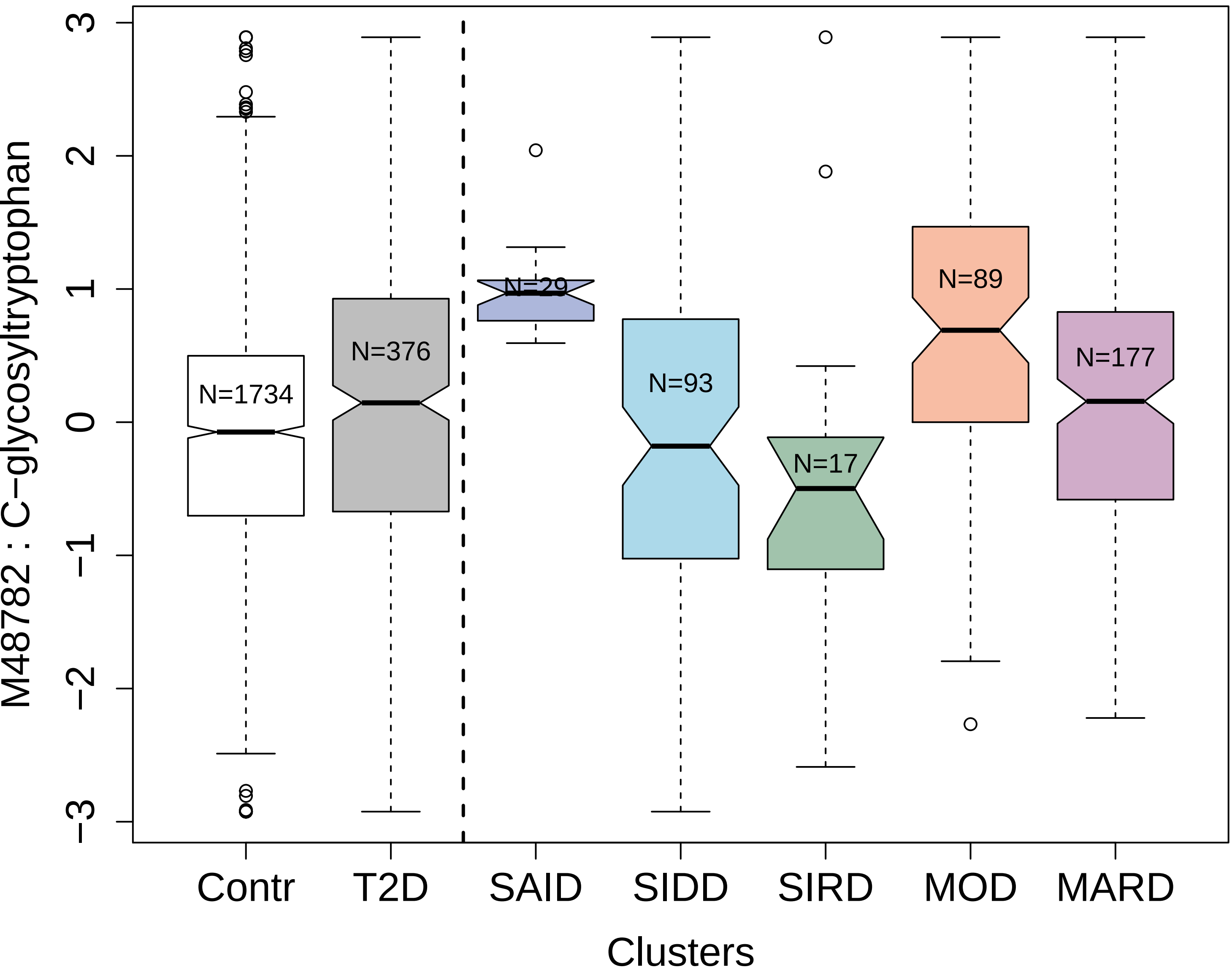
M48782 : C−glycosyltryptophan
3
2
1
0
−1
−2
−3
N=1734
N=376
N=29
N=93
N=17
N=89
N=177
Contr
T2D
SAID
SIDD
SIRD
MOD
MARD
Clusters

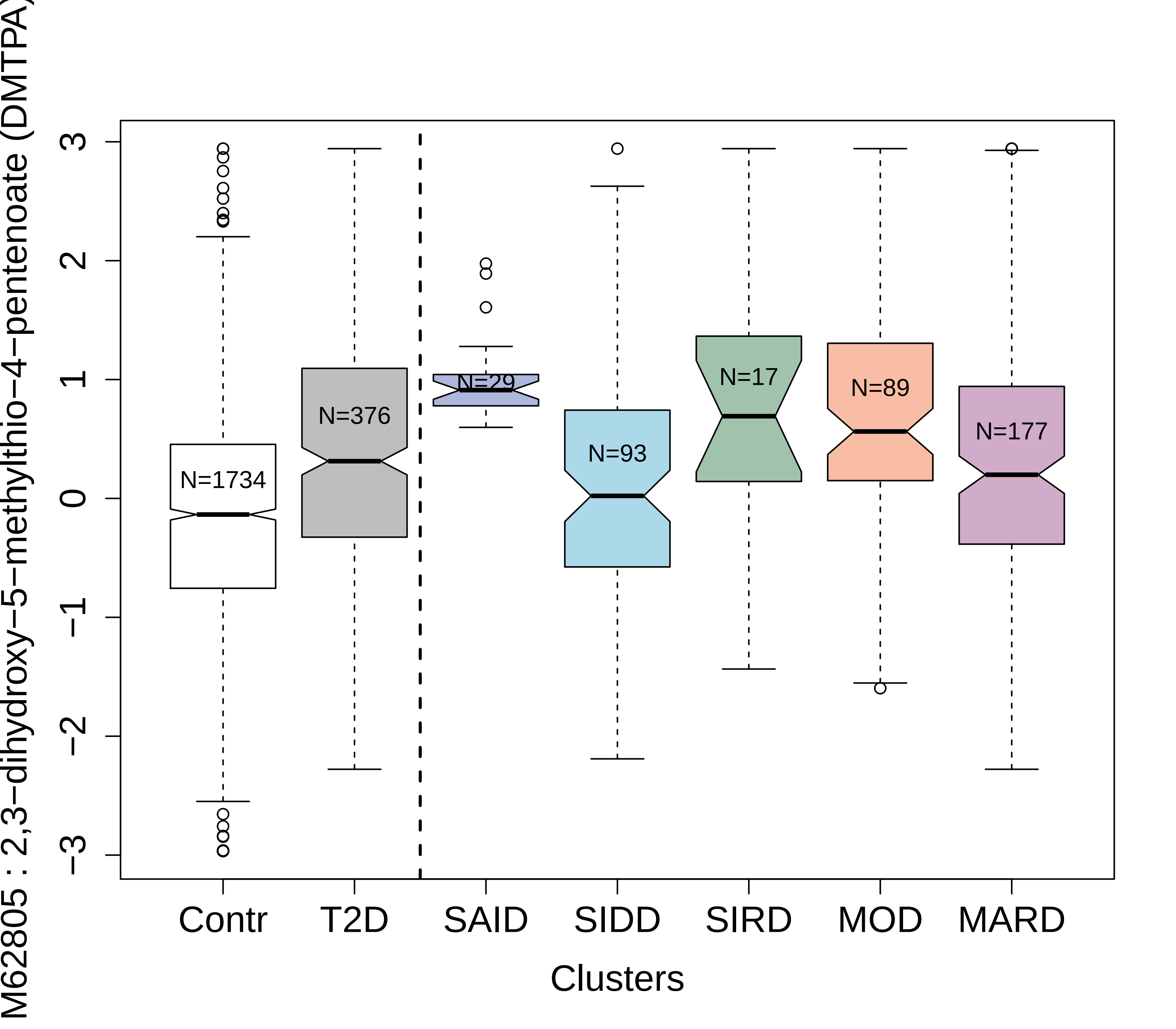
M62805 : 2,3−dihydroxy−5−methylthio−4−pentenoate (DMTPA)
3
2
1
0
−1
−2
−3
N=1734
N=376
N=29
N=93
N=17
N=89
N=177
Contr
T2D
SAID
SIDD
SIRD
MOD
MARD
Clusters

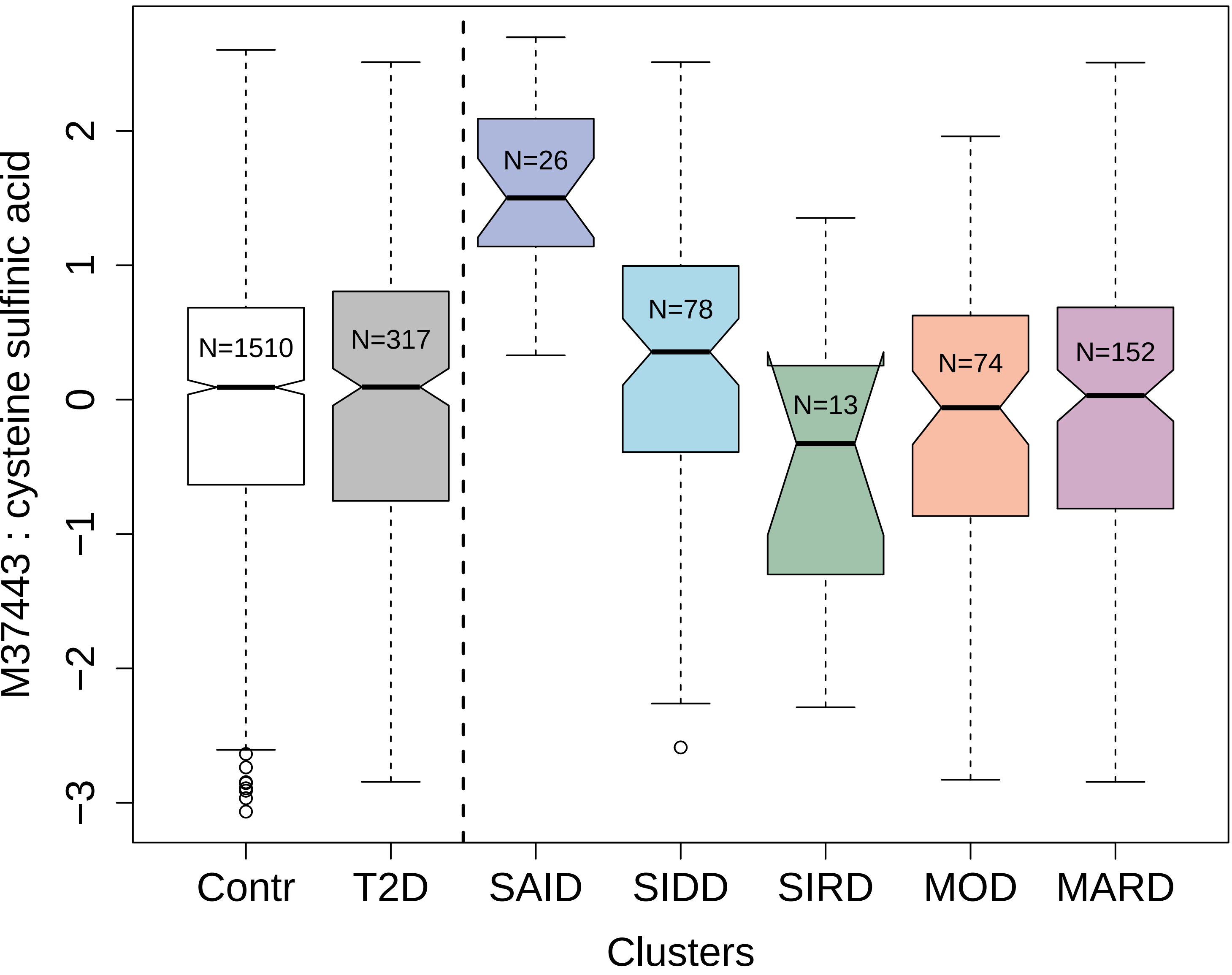
M37443 : cysteine sulfinic acid
Clusters
Contr
T2D
SAID
SIDD
SIRD
MOD
MARD
N=1510
N=317
N=26
N=78
N=13
N=74
N=152
2
1
0
−1
−2
−3

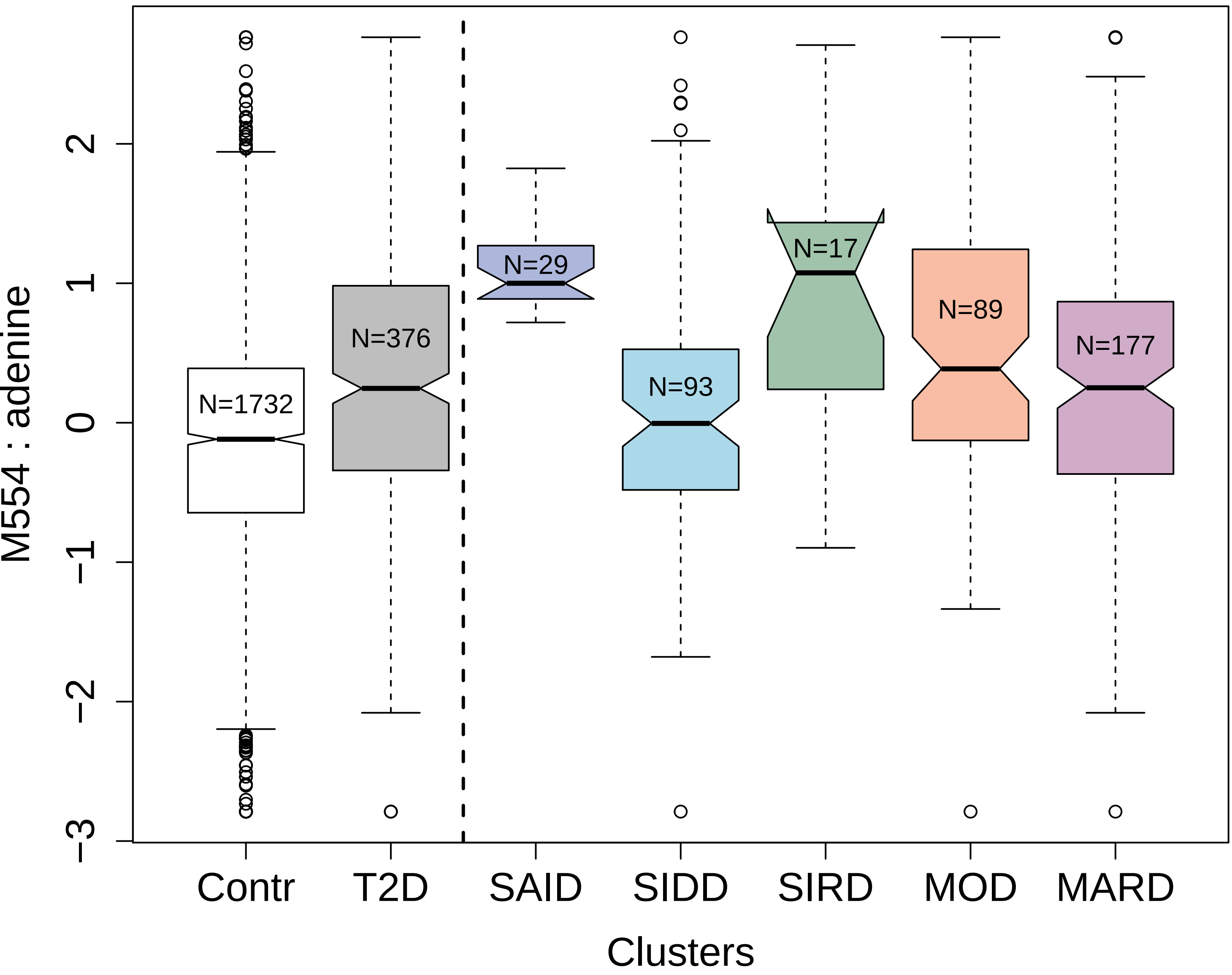
M554 : adenine
Clusters
Contr
T2D
SAID
SIDD
SIRD
MOD
MARD
N=1732
N=376
N=29
N=93
N=17
N=89
N=177
−3
−2
−1
0
1
2

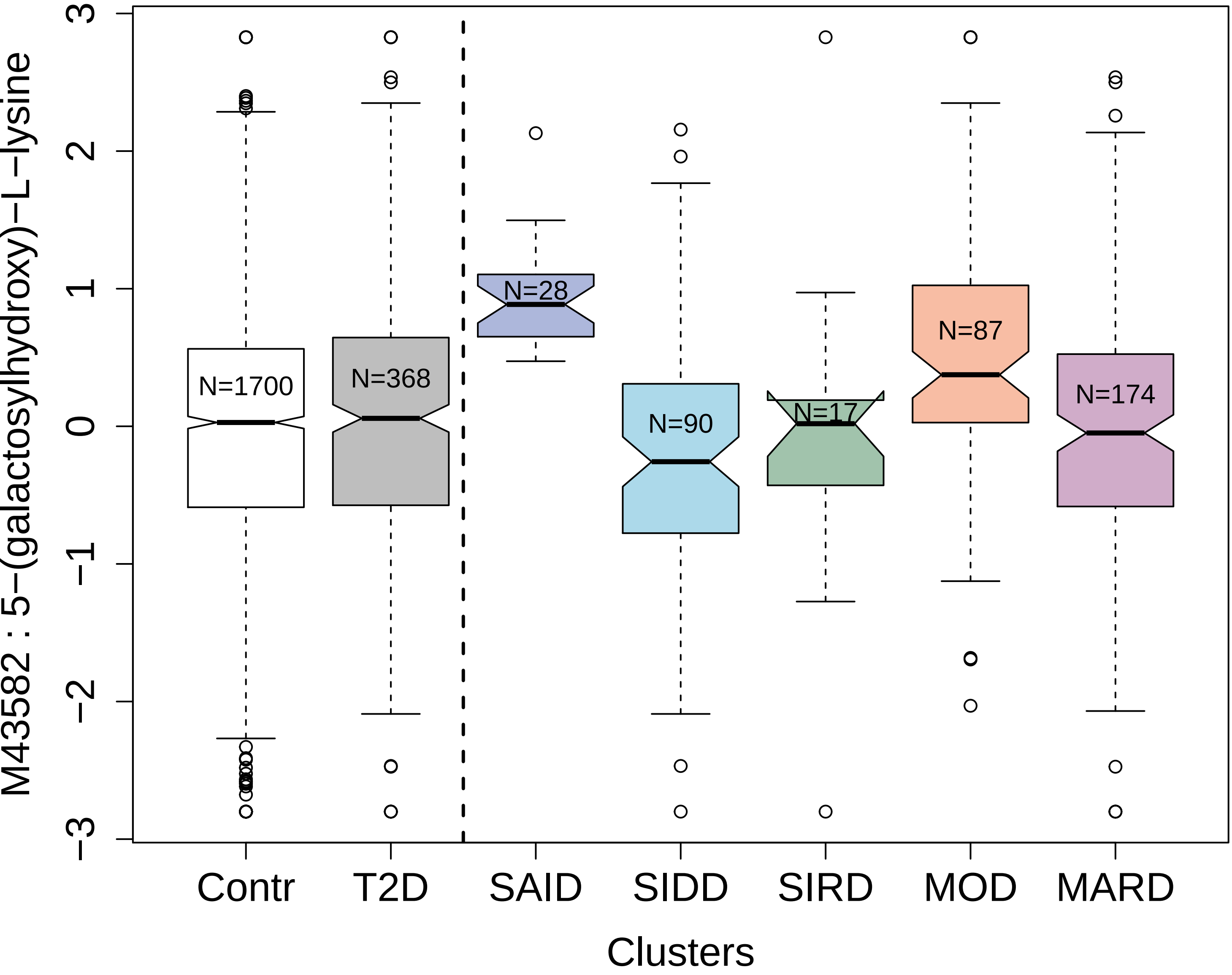

M43582 : 5−(galactosylhydroxy)−L−lysine
3
2
1
0
−1
−2
−3
N=1700
N=368
N=28
N=90
N=17
N=87
N=174
Contr
T2D
SAID
SIDD
SIRD
MOD
MARD
Clusters

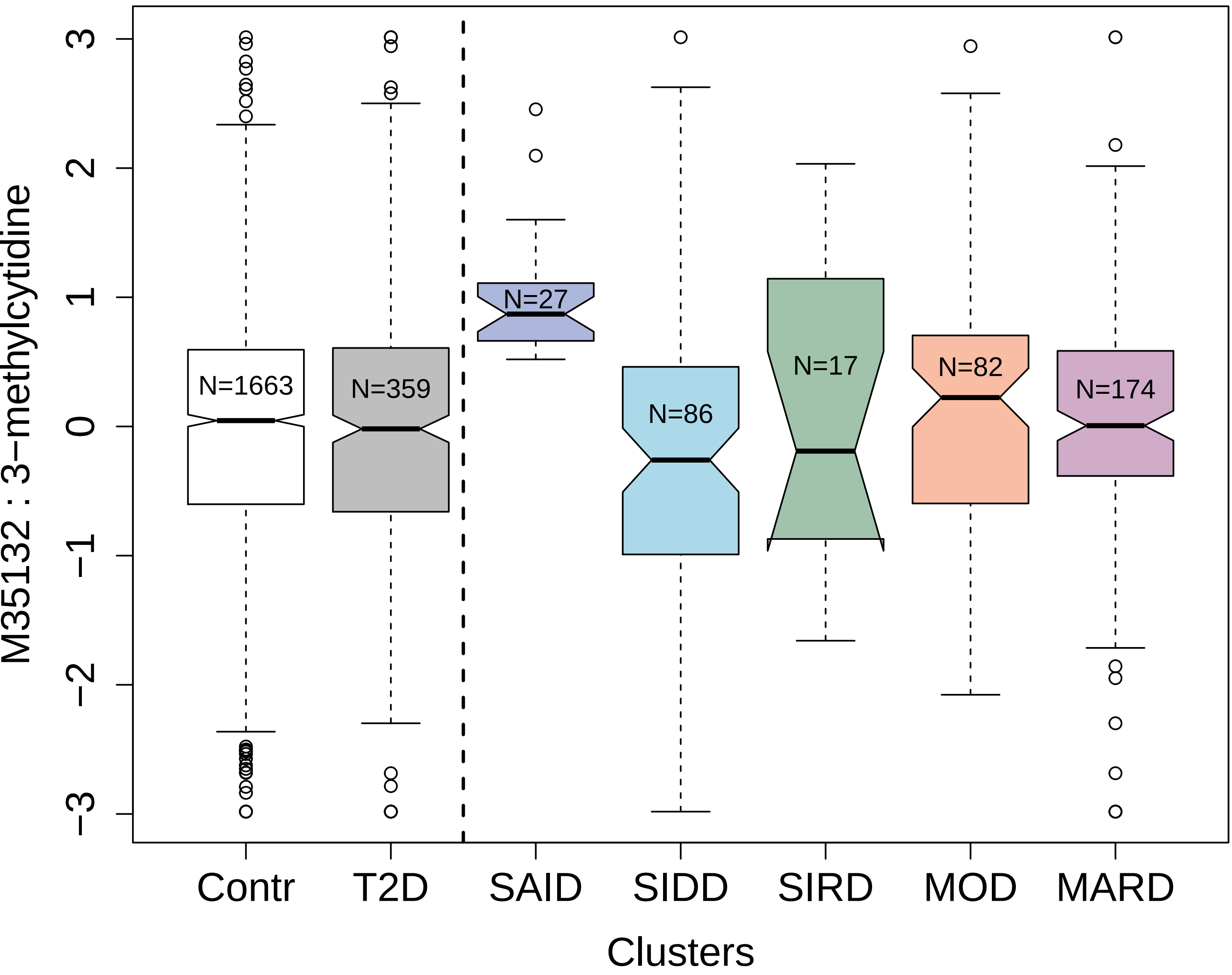
M35132 : 3−methylcytidine
3
2
1
0
−1
−2
−3
N=1663
N=359
N=27
N=86
N=17
N=82
N=174
Contr
T2D
SAID
SIDD
SIRD
MOD
MARD
Clusters

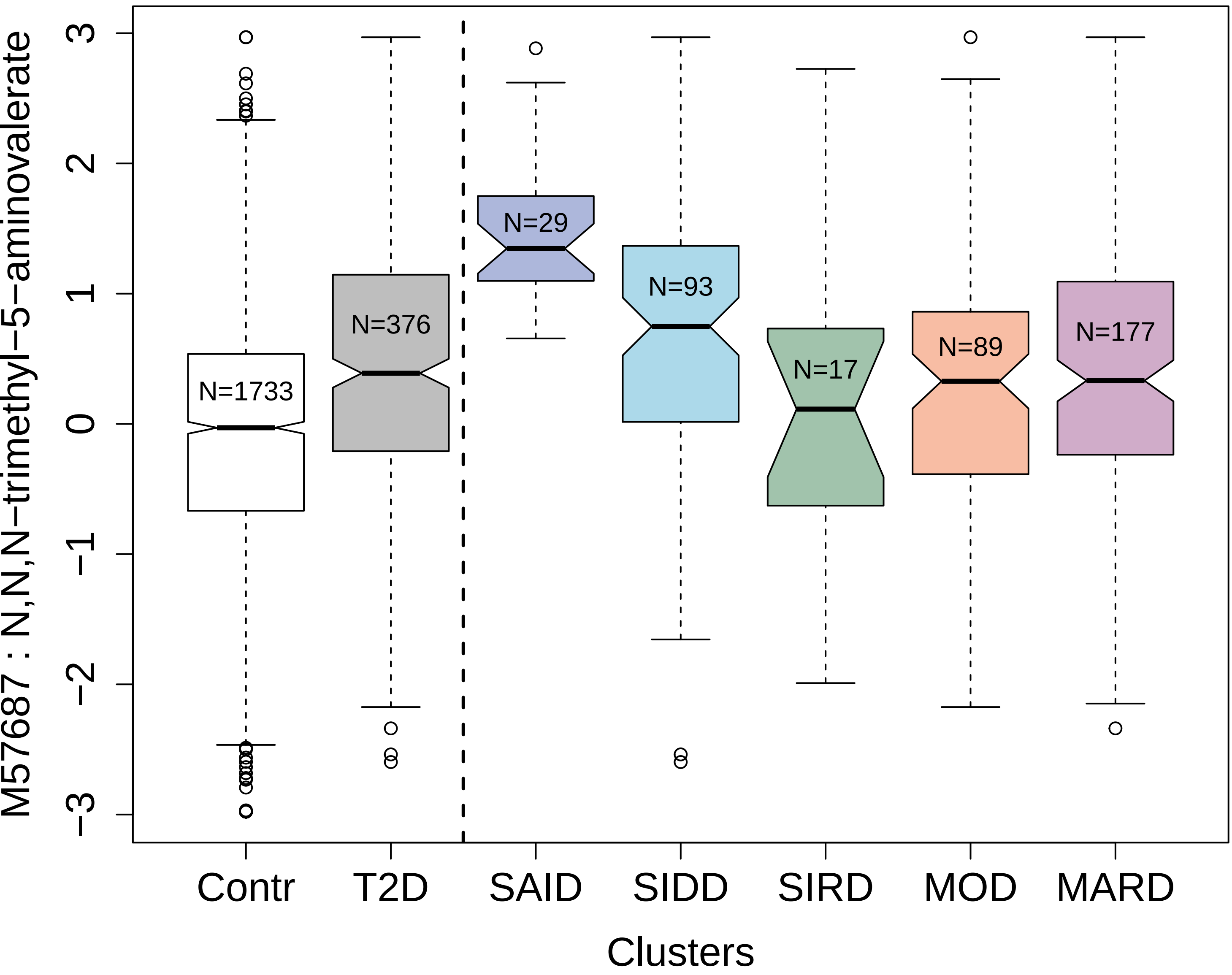

M57687 : N,N,N−trimethyl−5−aminovalerate
3
2
1
0
−1
−2
−3
N=1733
N=376
N=29
N=93
N=17
N=89
N=177
Contr
T2D
SAID
SIDD
SIRD
MOD
MARD
Clusters

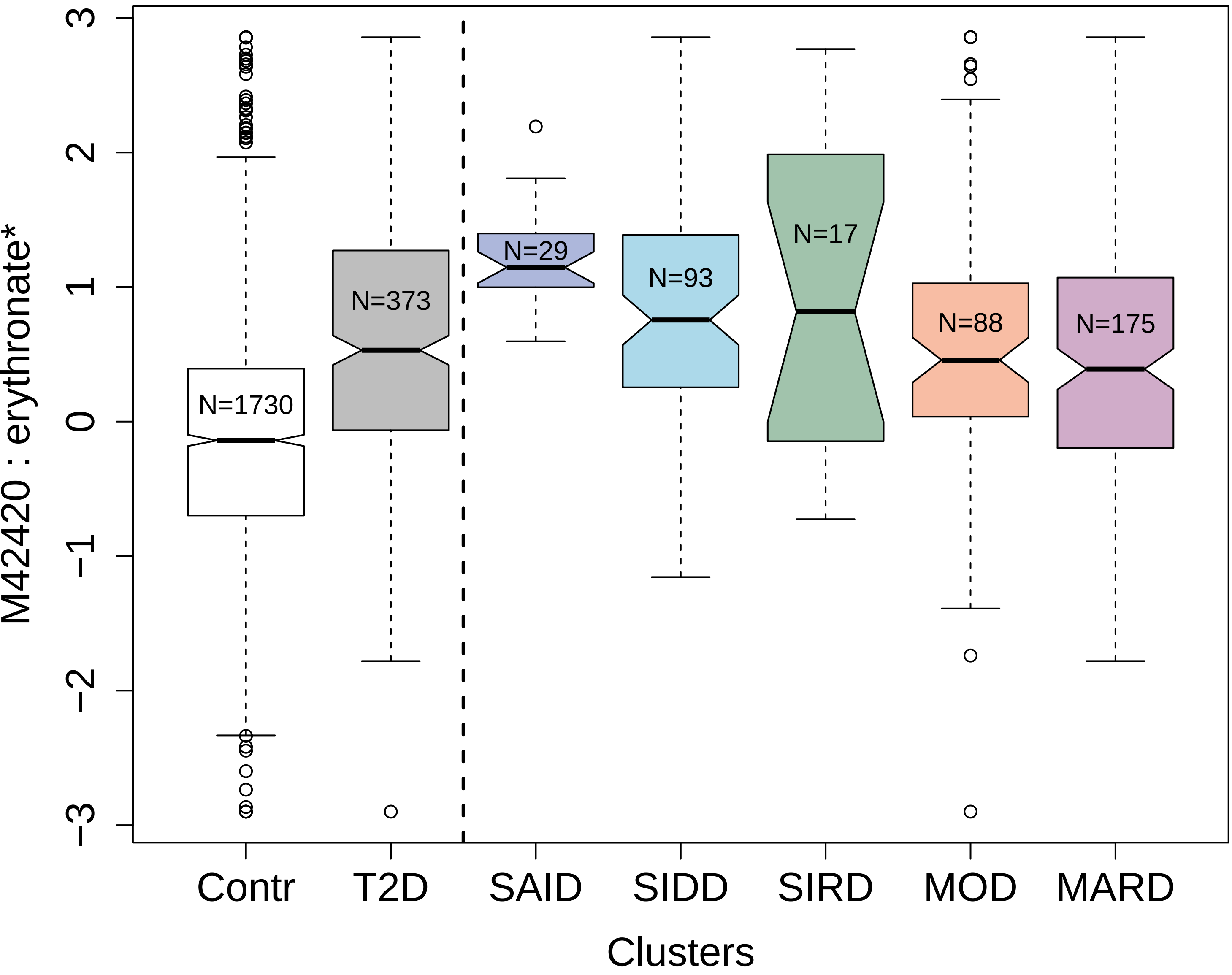
M42420 : erythronate*
3
2
1
0
−1
−2
−3
N=1730
N=373
N=29
N=93
N=17
N=88
N=175
Contr
T2D
SAID
SIDD
SIRD
MOD
MARD
Clusters

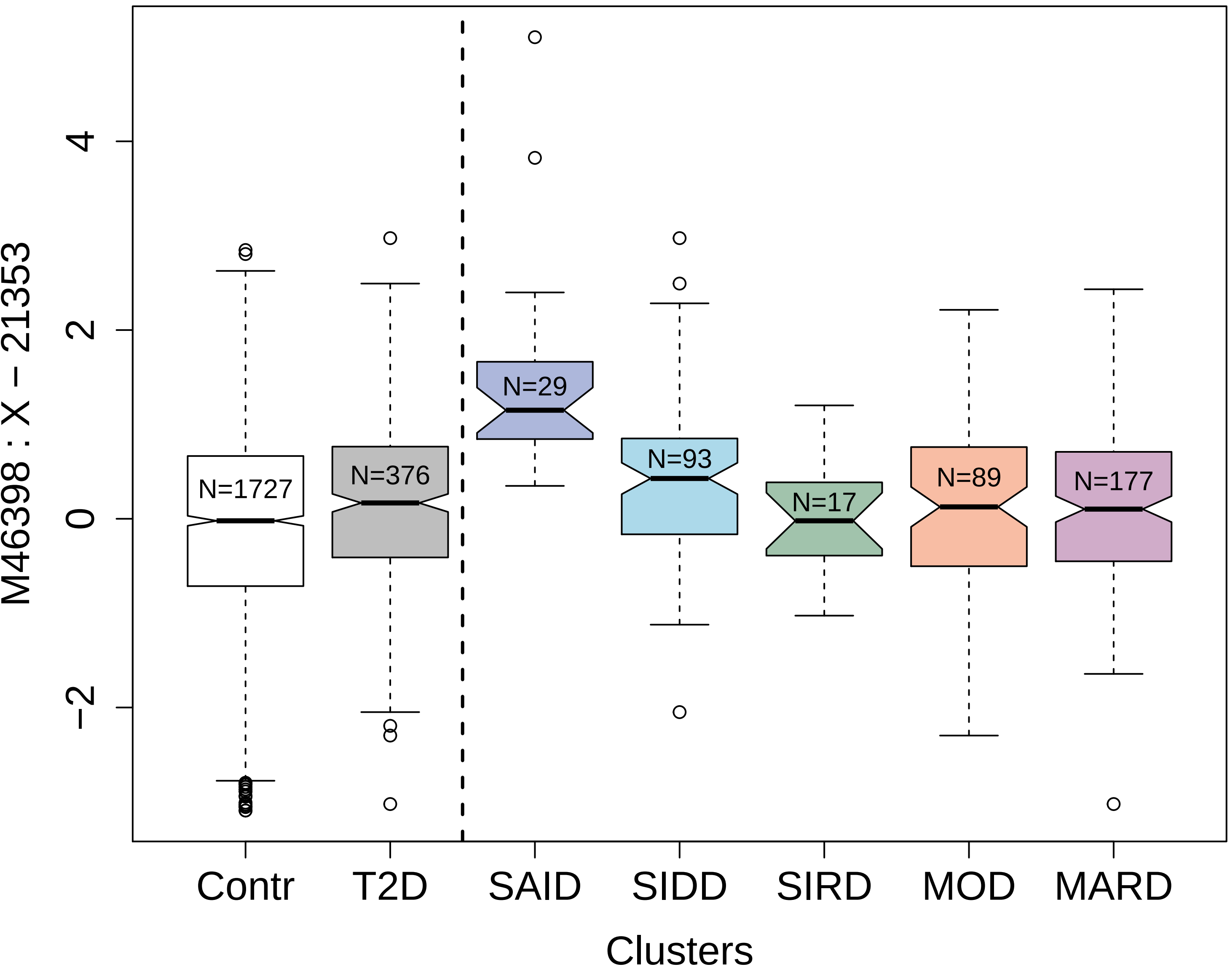

M46398 : X − 21353
4
2
0
−2
N=1727
N=376
N=29
N=93
N=17
N=89
N=177
Contr
T2D
SAID
SIDD
SIRD
MOD
MARD
Clusters

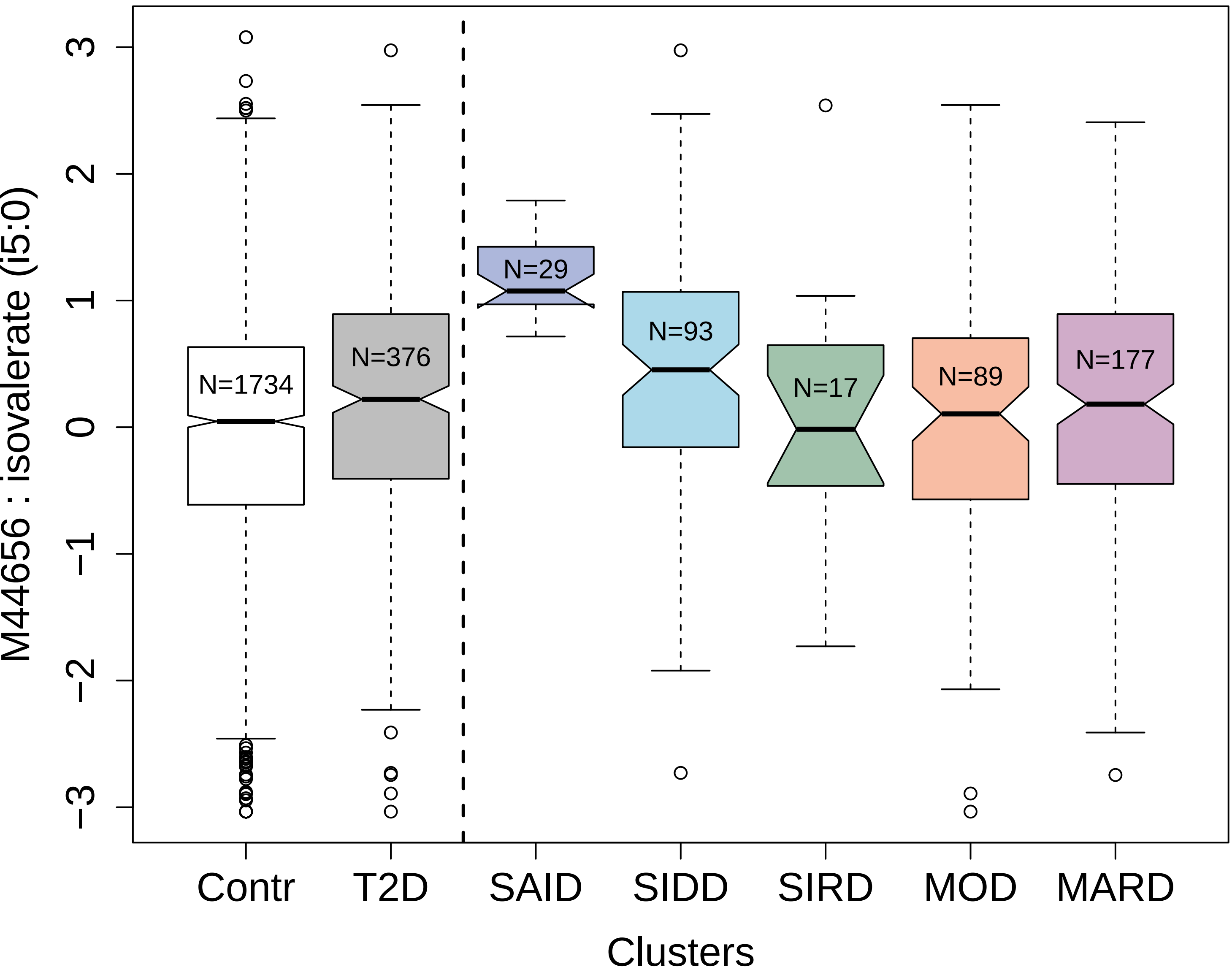

M44656 : isovalerate (i5:0)
3
2
1
0
−1
−2
−3
N=1734
N=376
N=29
N=93
N=17
N=89
N=177
Contr
T2D
SAID
SIDD
SIRD
MOD
MARD
Clusters

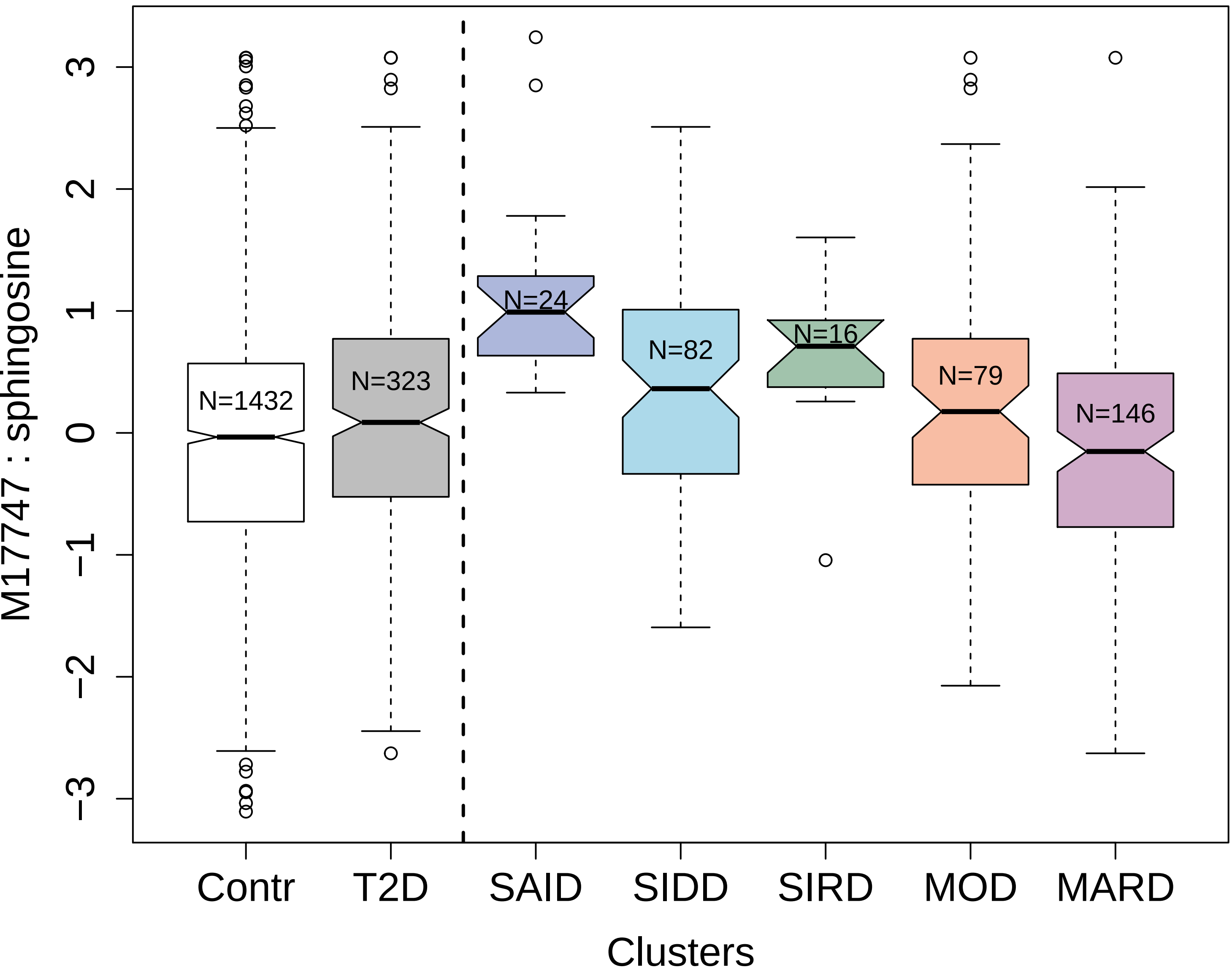
M17747 : sphingosine
3
2
1
0
−1
−2
−3
N=1432
N=323
N=24
N=82
N=16
N=79
N=146
Contr
T2D
SAID
SIDD
SIRD
MOD
MARD
Clusters

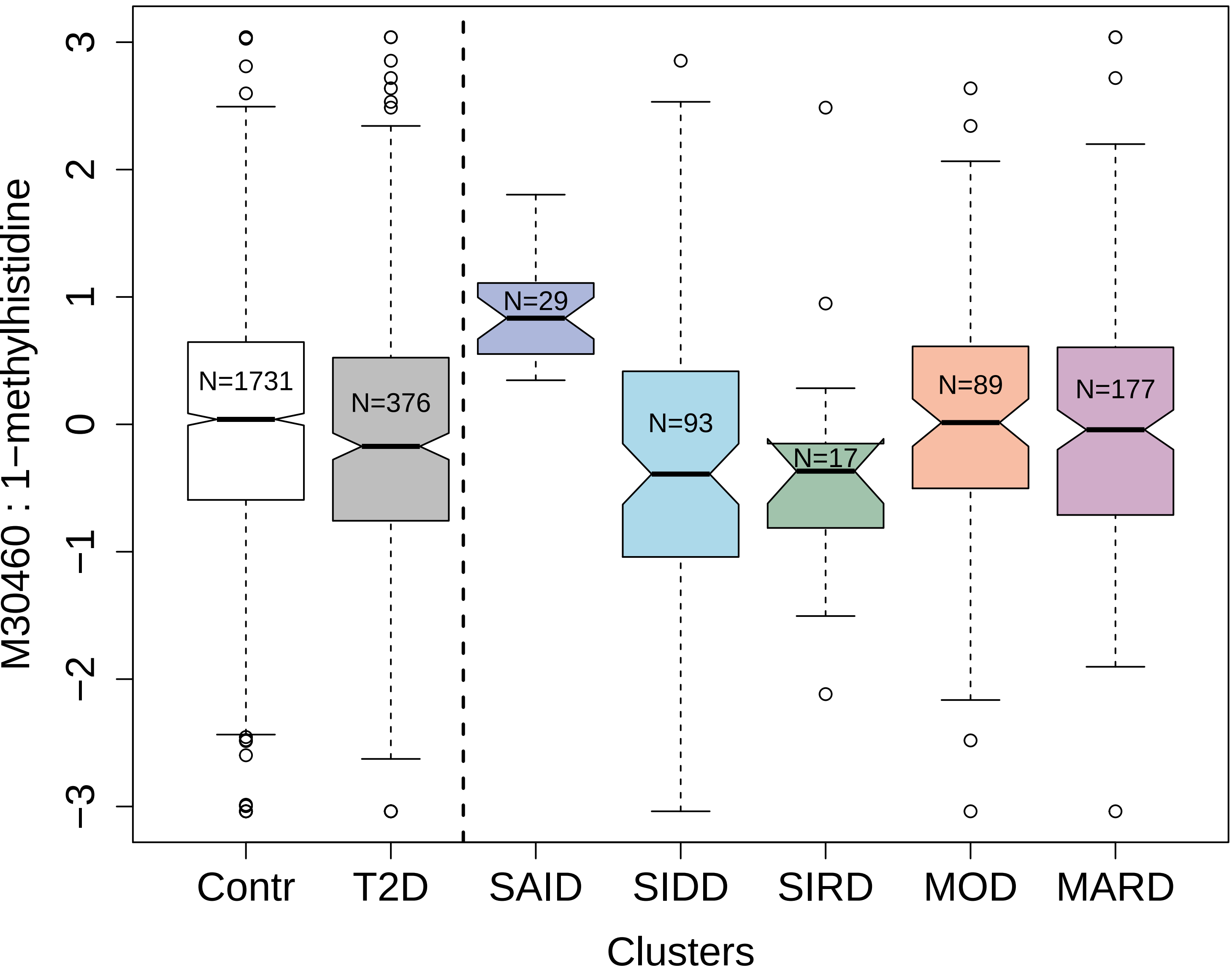
M30460 : 1−methylhistidine
3
2
1
0
−1
−2
−3
N=1731
N=376
N=29
N=93
N=17
N=89
N=177
Contr
T2D
SAID
SIDD
SIRD
MOD
MARD
Clusters

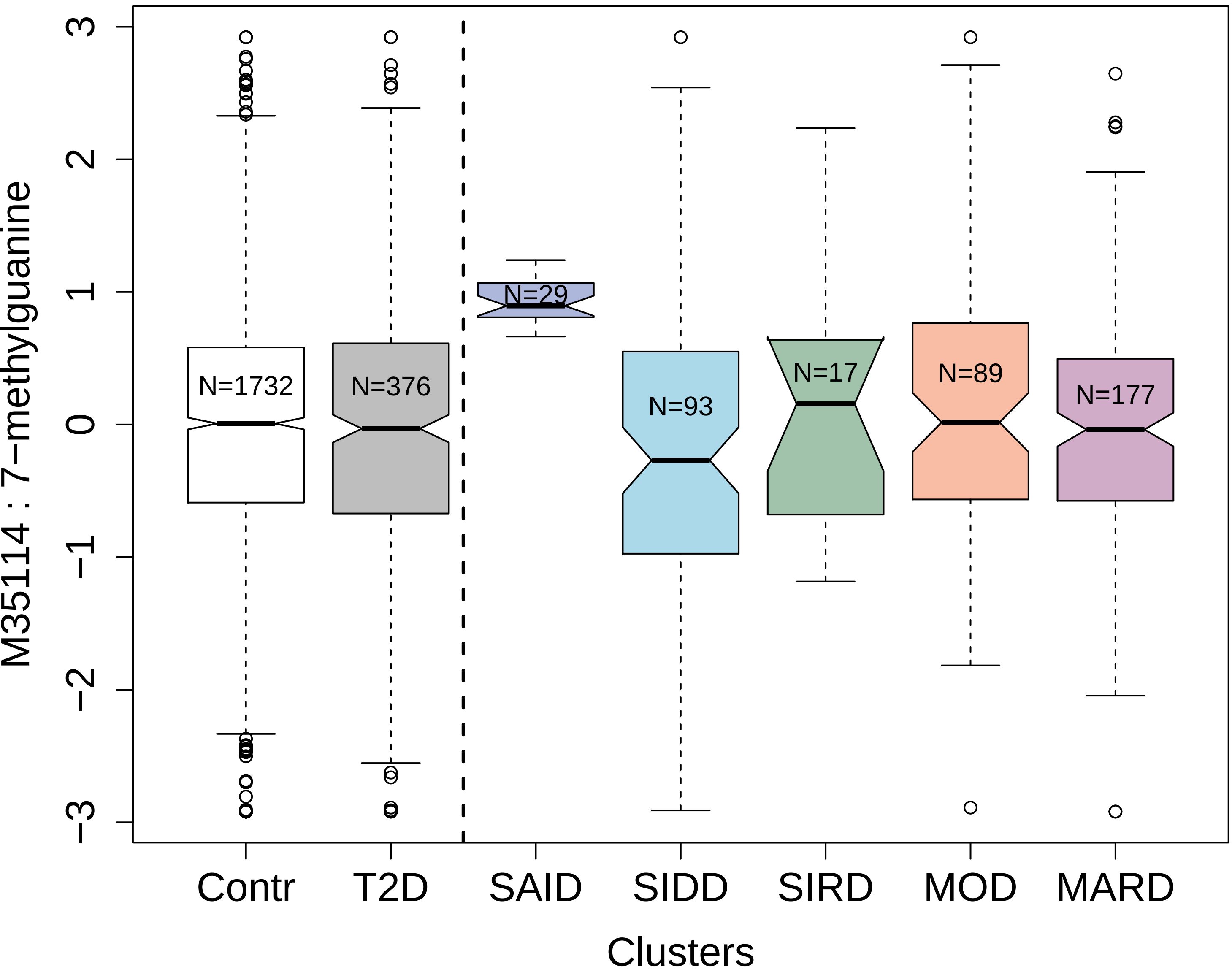
M35114 : 7−methylguanine
3
2
1
0
−1
−2
−3
N=1732
N=376
N=29
N=93
N=17
N=89
N=177
Contr
T2D
SAID
SIDD
SIRD
MOD
MARD
Clusters

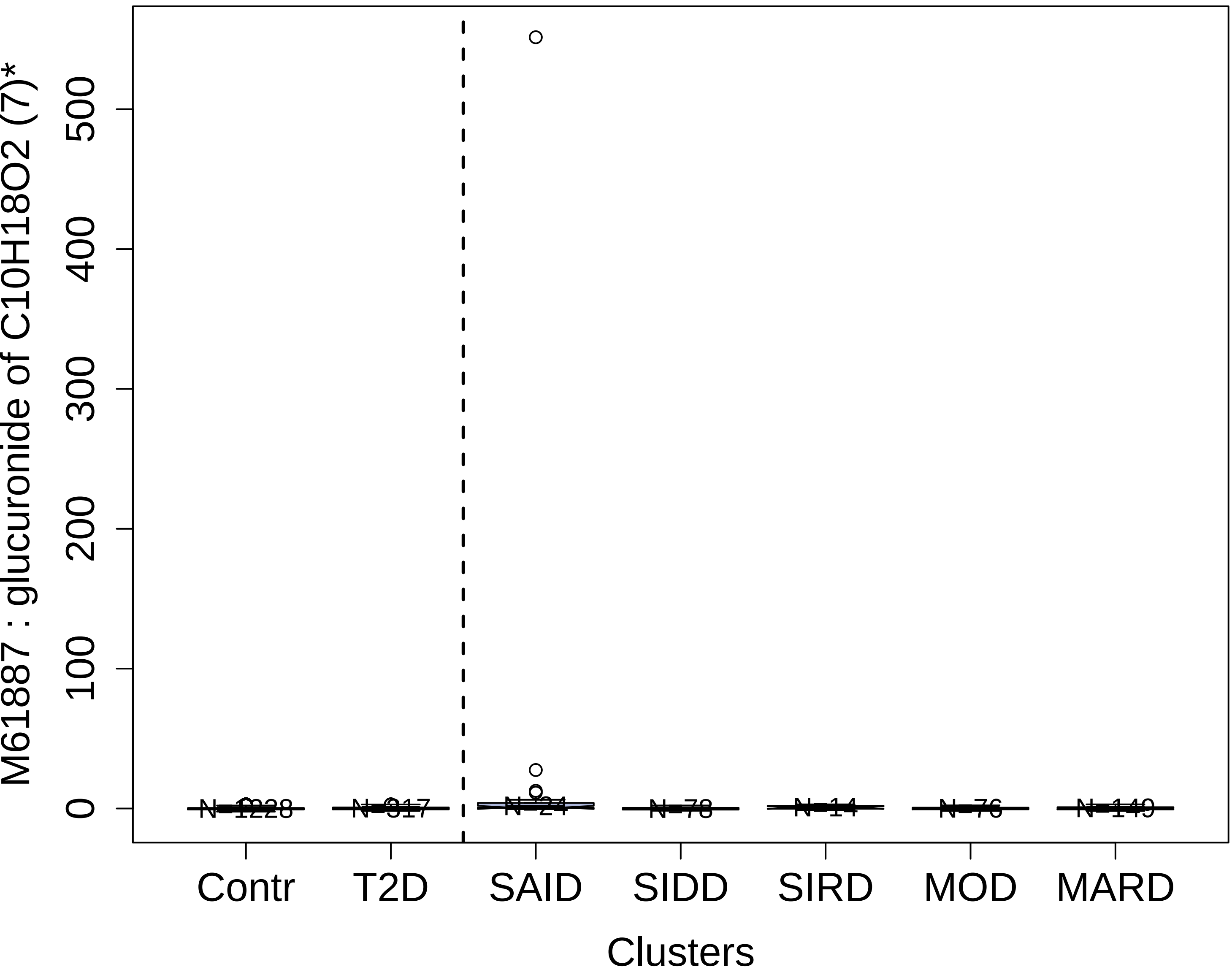
M61887 : glucuronide of C10H18O2 (7)*
0
100
200
300
400
500
N=1228
N=317
N=24
N=78
N=14
N=76
N=149
Contr
T2D
SAID
SIDD
SIRD
MOD
MARD
Clusters

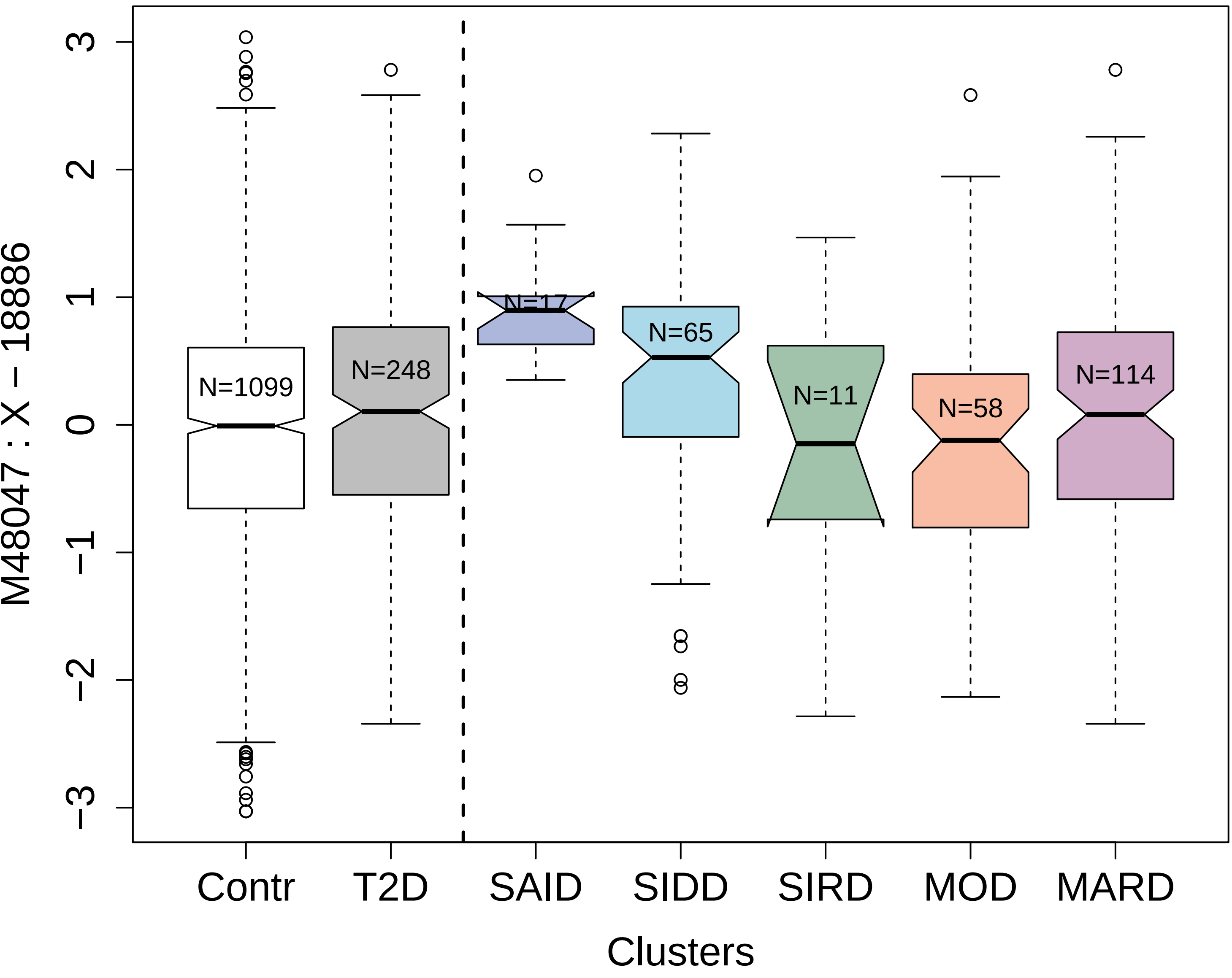
M48047 : X − 18886
3
2
1
0
−1
−2
−3
N=1099
N=248
N=17
N=65
N=11
N=58
N=114
Contr
T2D
SAID
SIDD
SIRD
MOD
MARD
Clusters

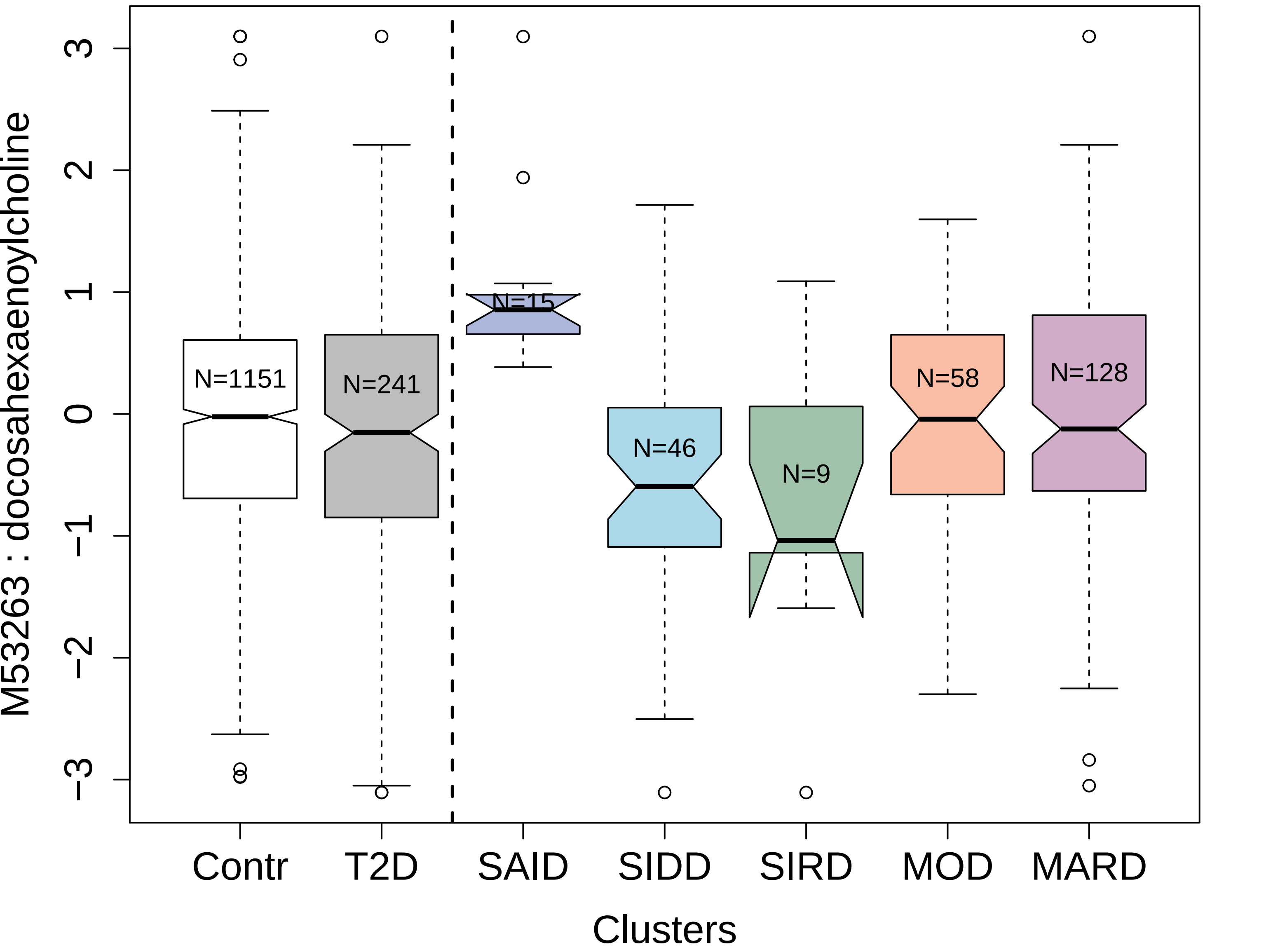
M53263 : docosahexaenoylcholine
3
2
1
0
−1
−2
−3
N=1151
N=241
N=15
N=46
N=9
N=58
N=128
Contr
T2D
SAID
SIDD
SIRD
MOD
MARD
Clusters

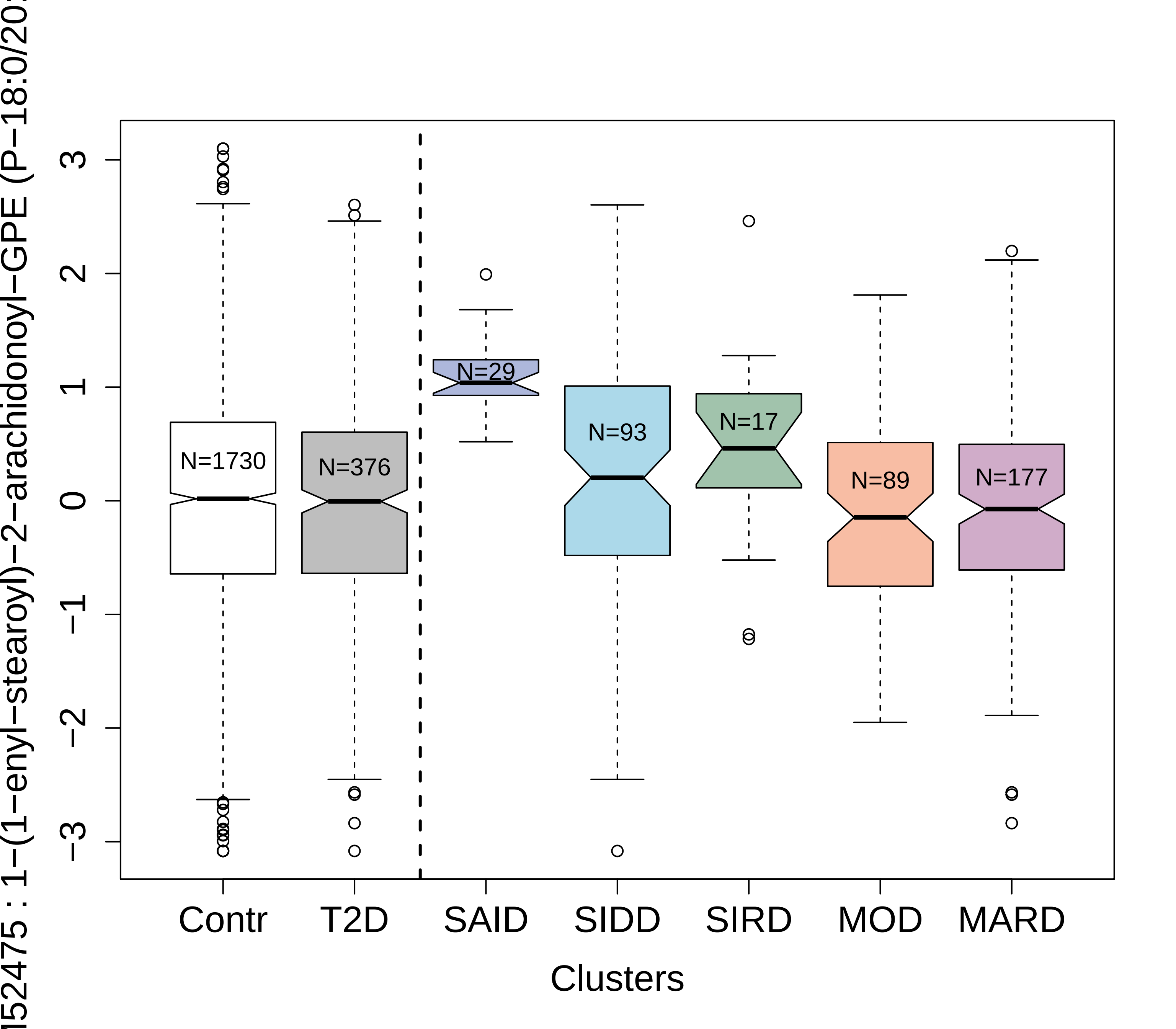
152475 : 1−(1−enyl−stearoyl)−2−arachidonoyl−GPE (P−18:0/20:
3
2
1
0
−1
−2
−3
N=1730
N=376
N=29
N=93
N=17
N=89
N=177
Contr
T2D
SAID
SIDD
SIRD
MOD
MARD
Clusters

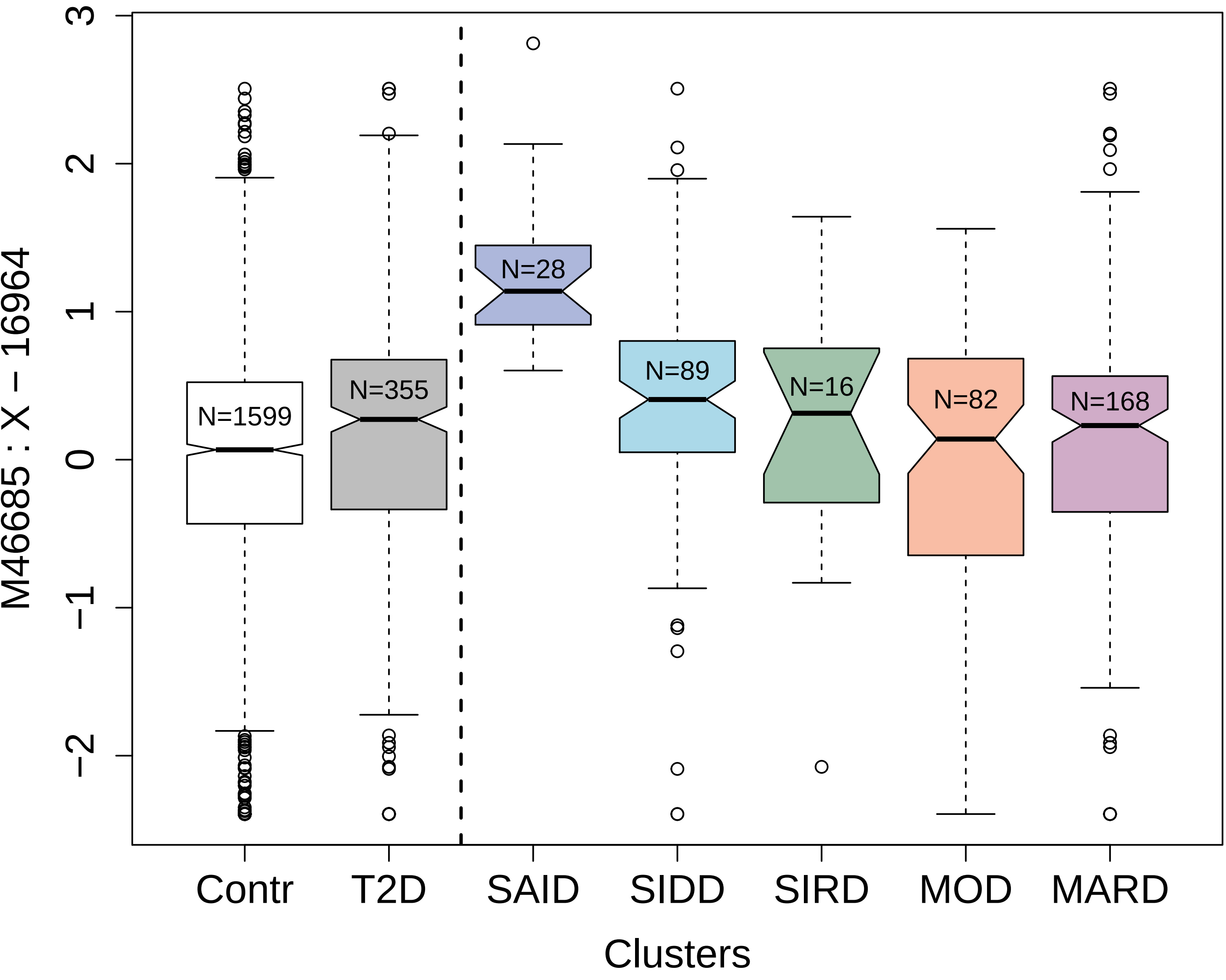
M46685 : X − 16964
3
2
1
0
−1
−2
N=1599
N=355
N=28
N=89
N=16
N=82
N=168
Contr
T2D
SAID
SIDD
SIRD
MOD
MARD
Clusters

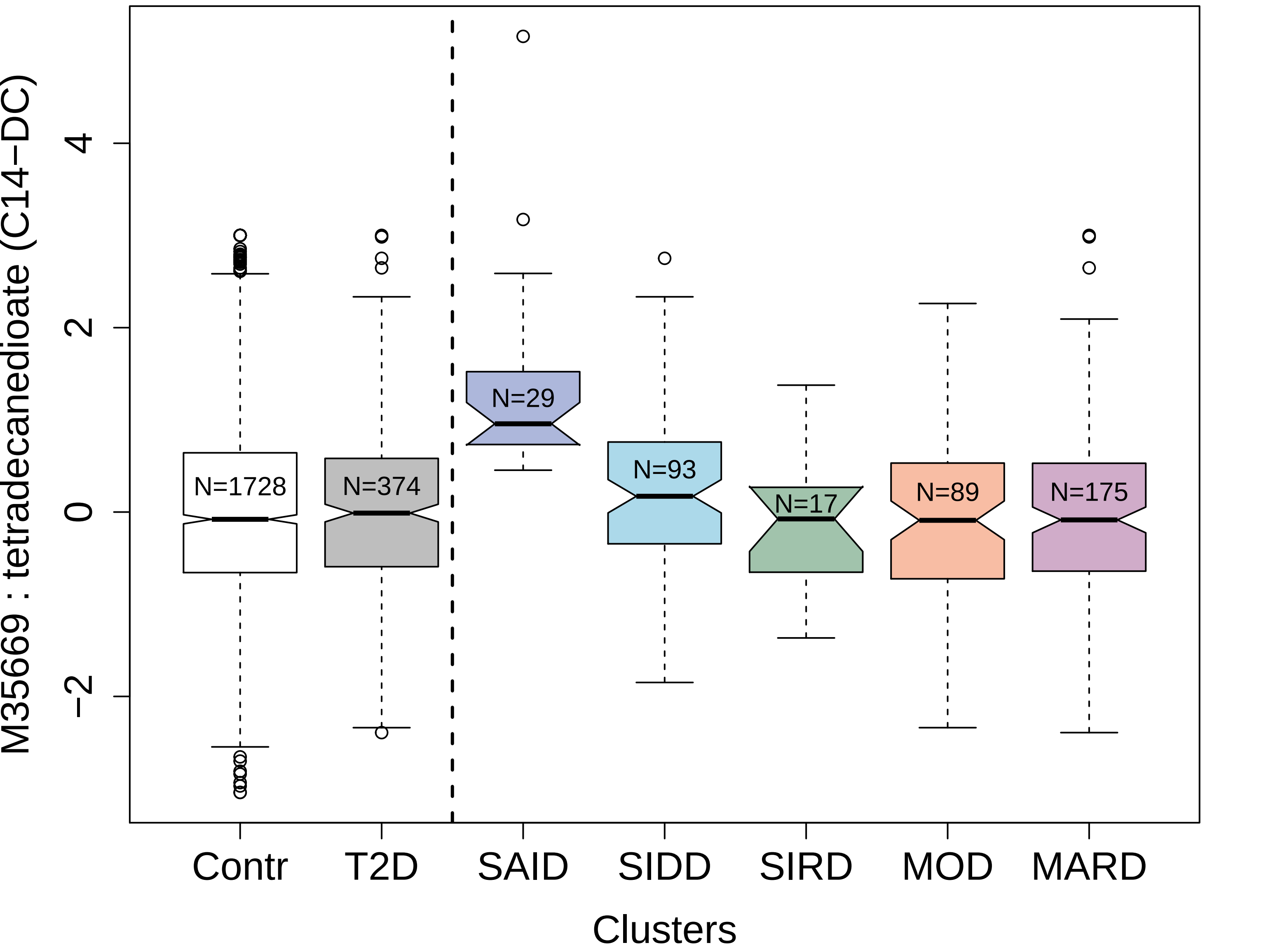

M35669 : tetradecanedioate (C14−DC)
4
2
0
−2
N=1728
N=374
N=29
N=93
N=17
N=89
N=175
Contr
T2D
SAID
SIDD
SIRD
MOD
MARD
Clusters

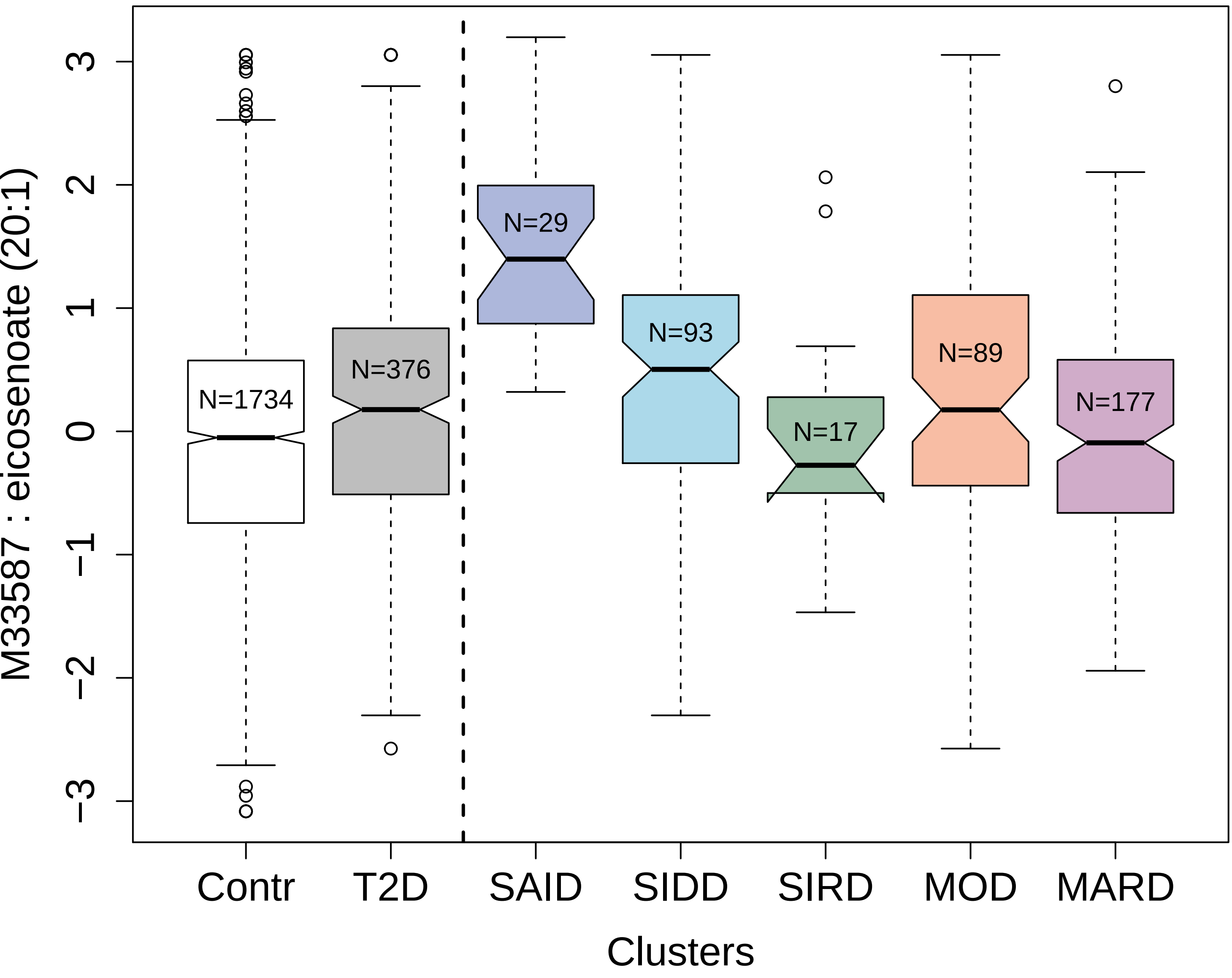

M33587 : eicosenoate (20:1)
3
2
1
0
−1
−2
−3
N=1734
N=376
N=29
N=93
N=17
N=89
N=177
Contr
T2D
SAID
SIDD
SIRD
MOD
MARD
Clusters

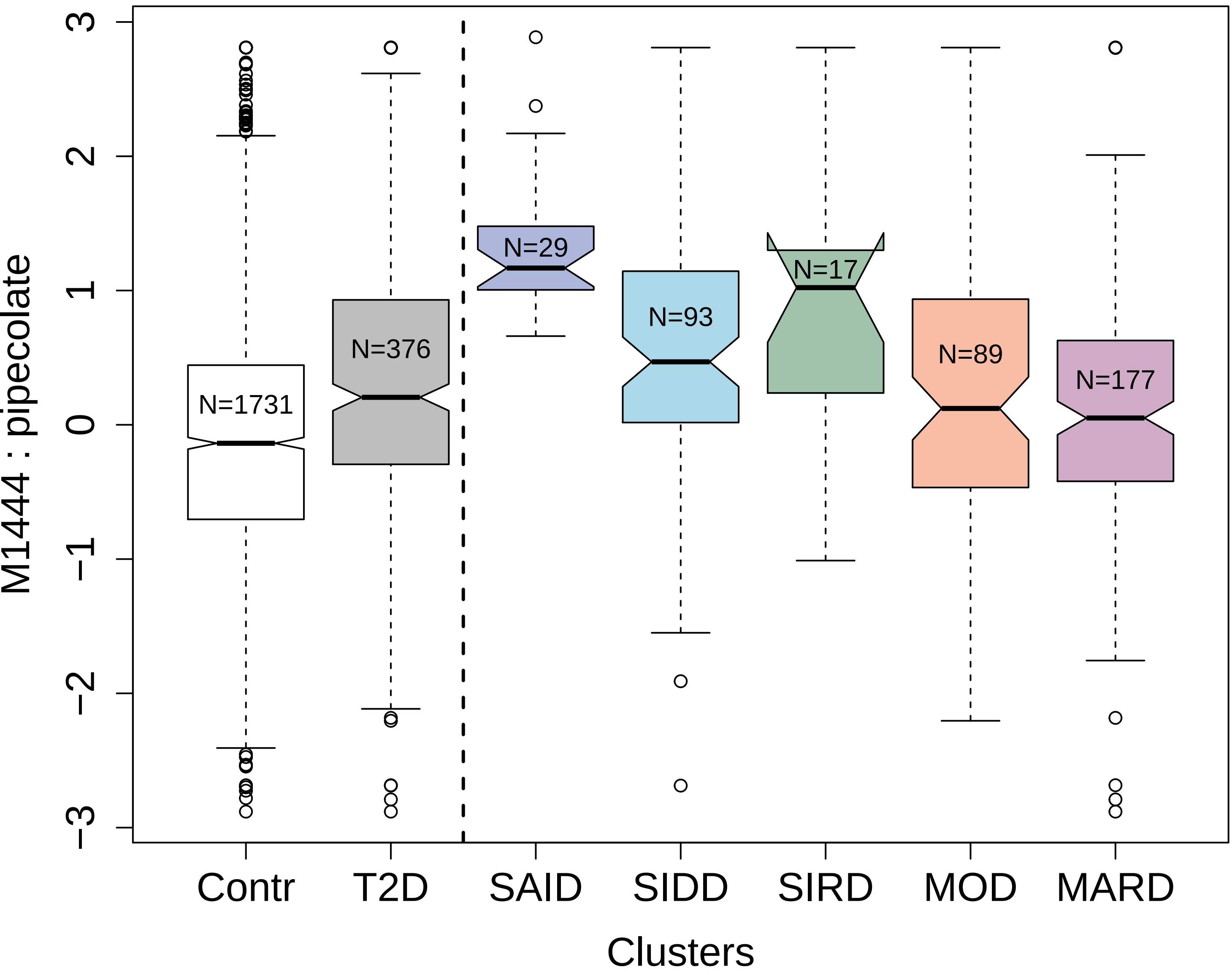
M1444 : pipecolate
3
2
1
0
−1
−2
−3
N=1731
N=376
N=29
N=93
N=17
N=89
N=177
Contr
T2D
SAID
SIDD
SIRD
MOD
MARD
Clusters

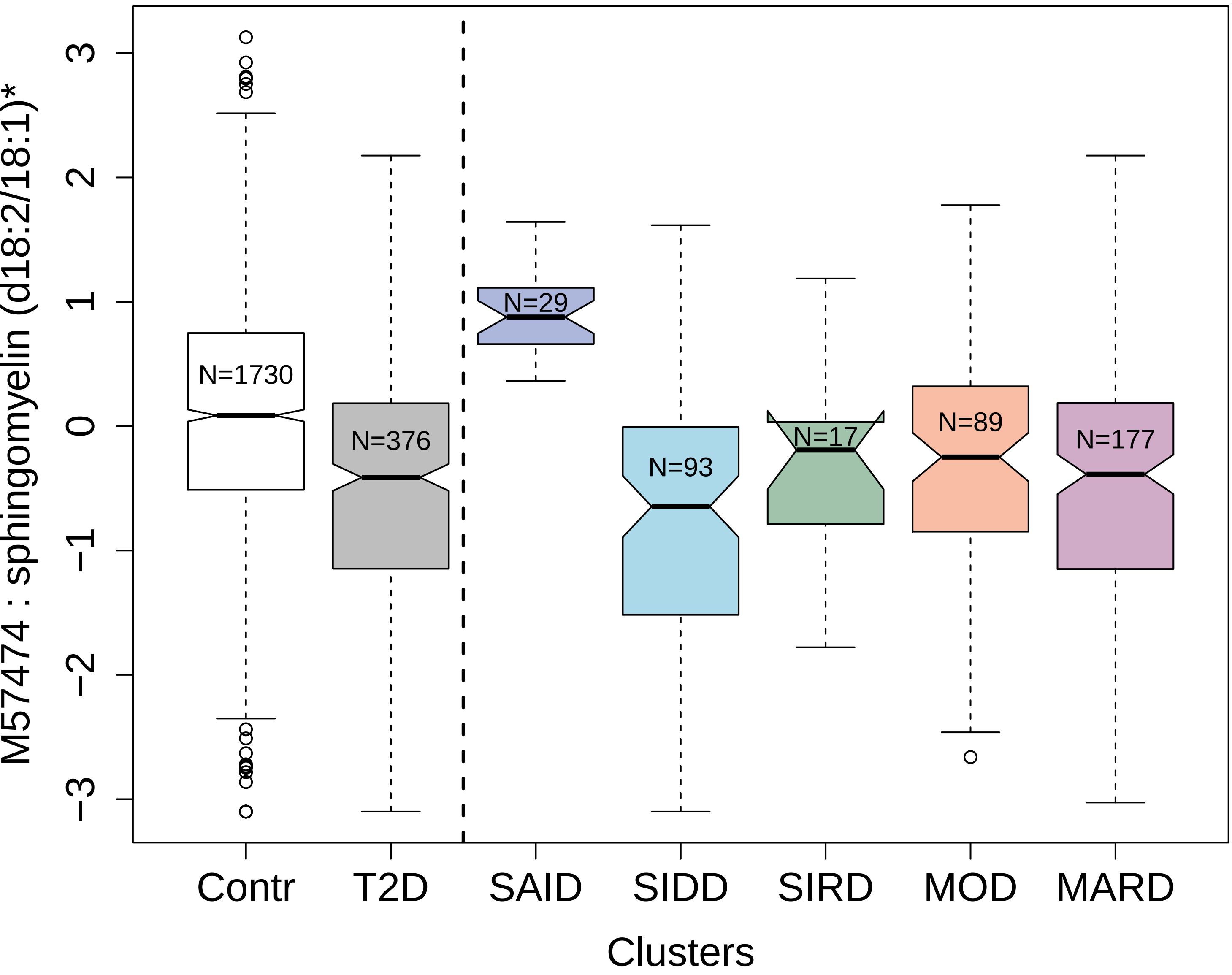

M57474 : sphingomyelin (d18:2/18:1)*
3
2
1
0
−1
−2
−3
N=1730
N=376
N=29
N=93
N=17
N=89
N=177
Contr
T2D
SAID
SIDD
SIRD
MOD
MARD
Clusters

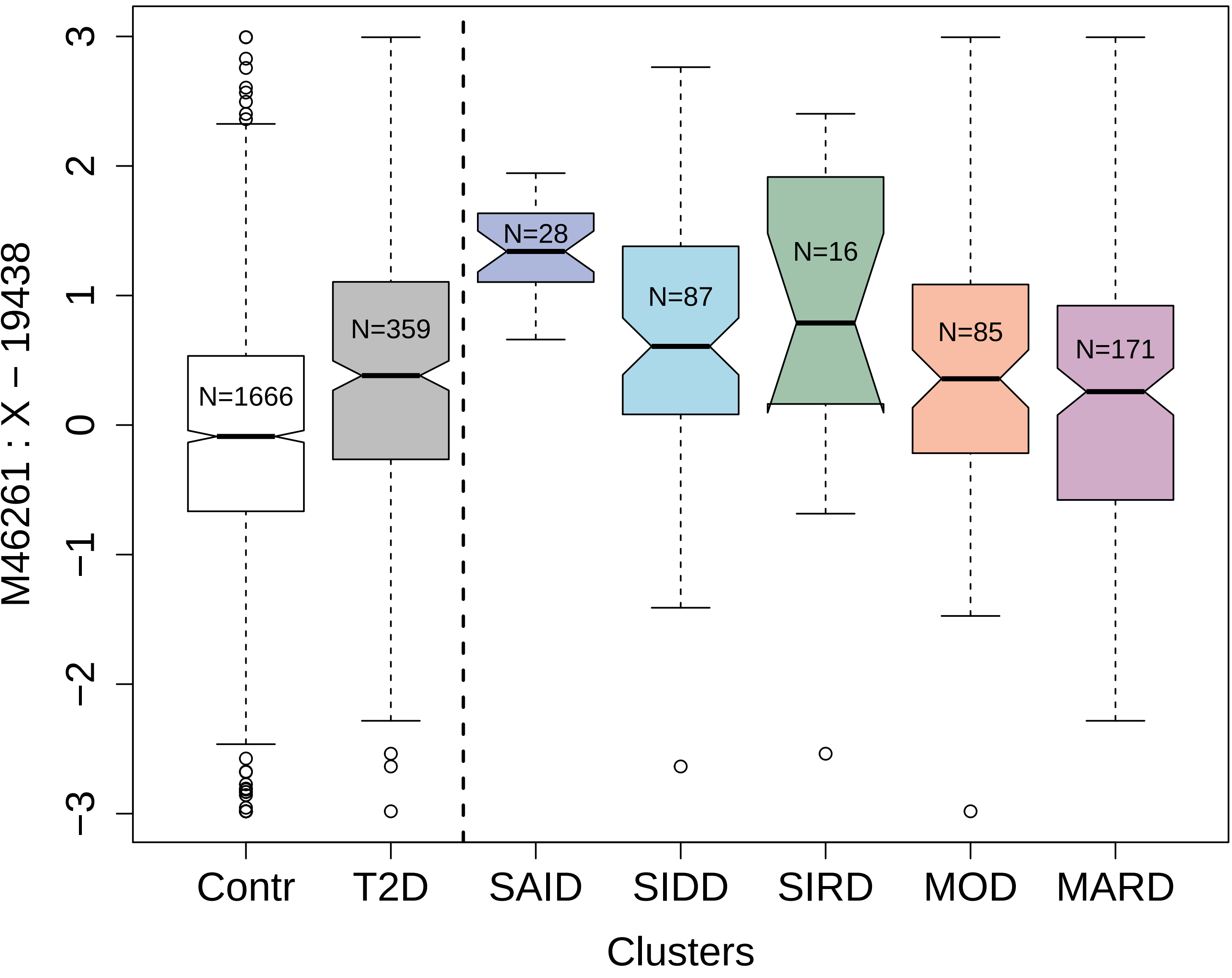

M46261 : X − 19438
3
2
1
0
−1
−2
−3
N=1666
N=359
N=28
N=87
N=16
N=85
N=171
Contr
T2D
SAID
SIDD
SIRD
MOD
MARD
Clusters

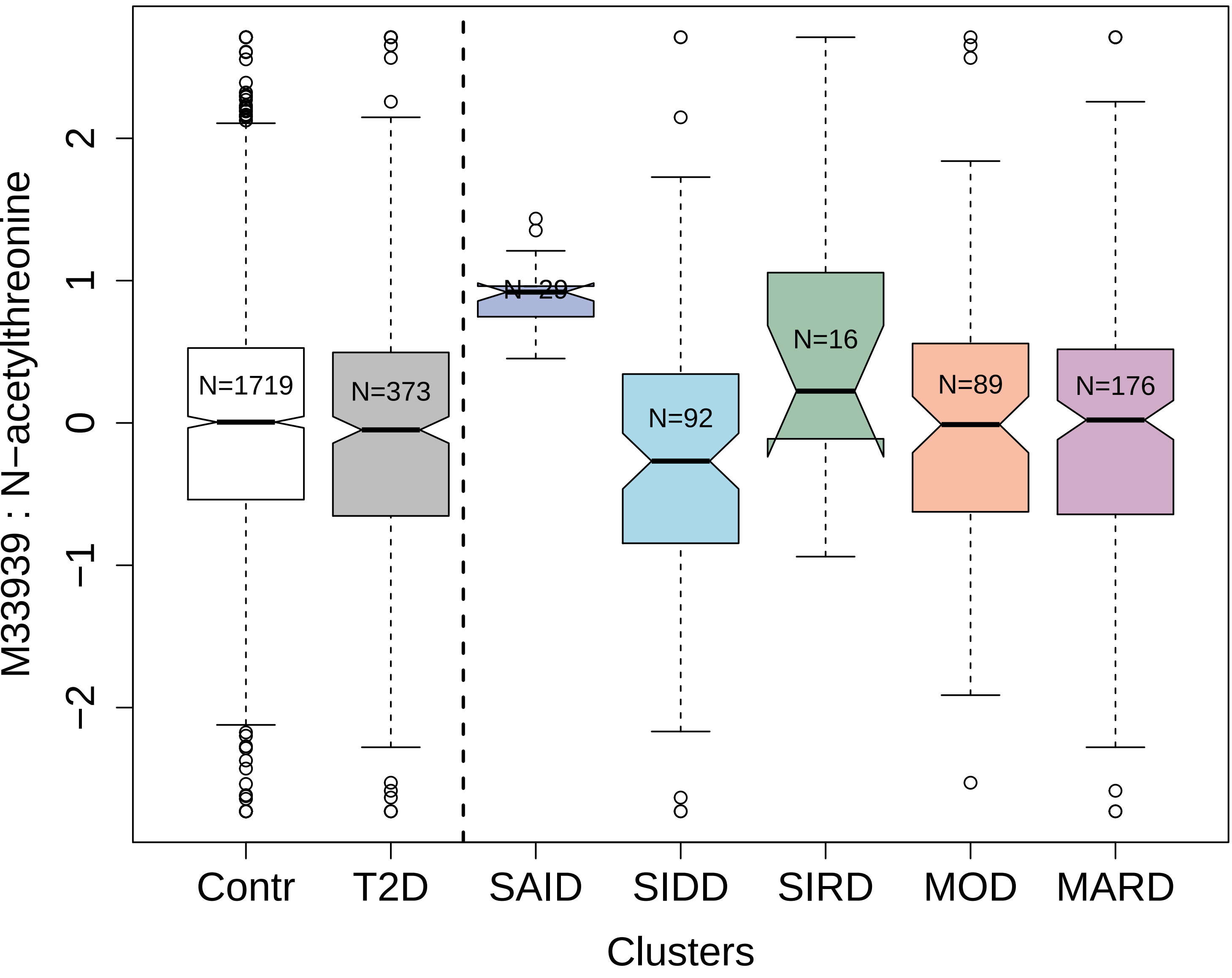

M33939 : N−acetylthreonine
2
1
0
−1
−2
N=1719
N=373
N=29
N=92
N=16
N=89
N=176
Contr
T2D
SAID
SIDD
SIRD
MOD
MARD
Clusters

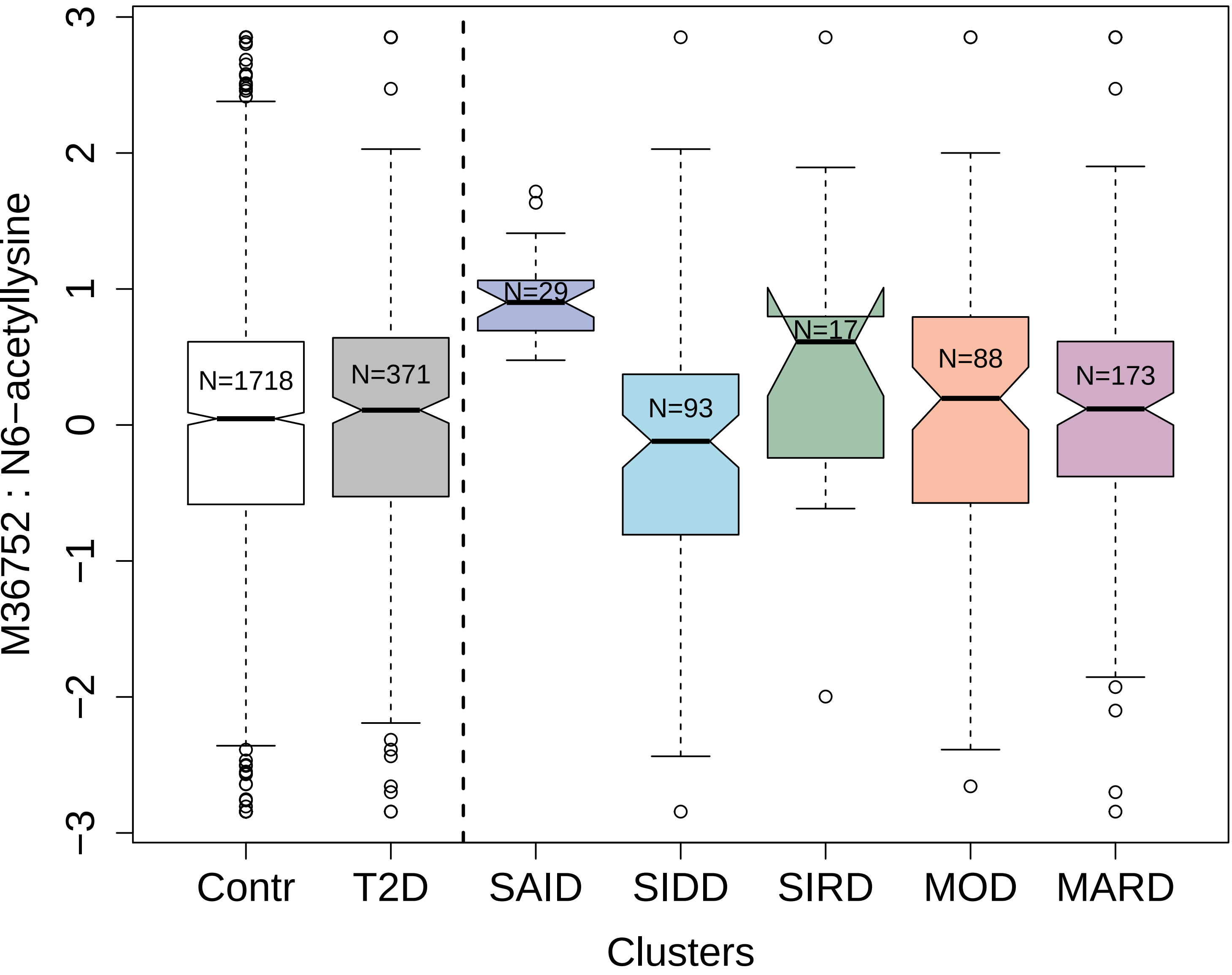
M36752 : N6−acetyllysine
3
2
1
0
−1
−2
−3
N=1718
N=371
N=29
N=93
N=17
N=88
N=173
Contr
T2D
SAID
SIDD
SIRD
MOD
MARD
Clusters

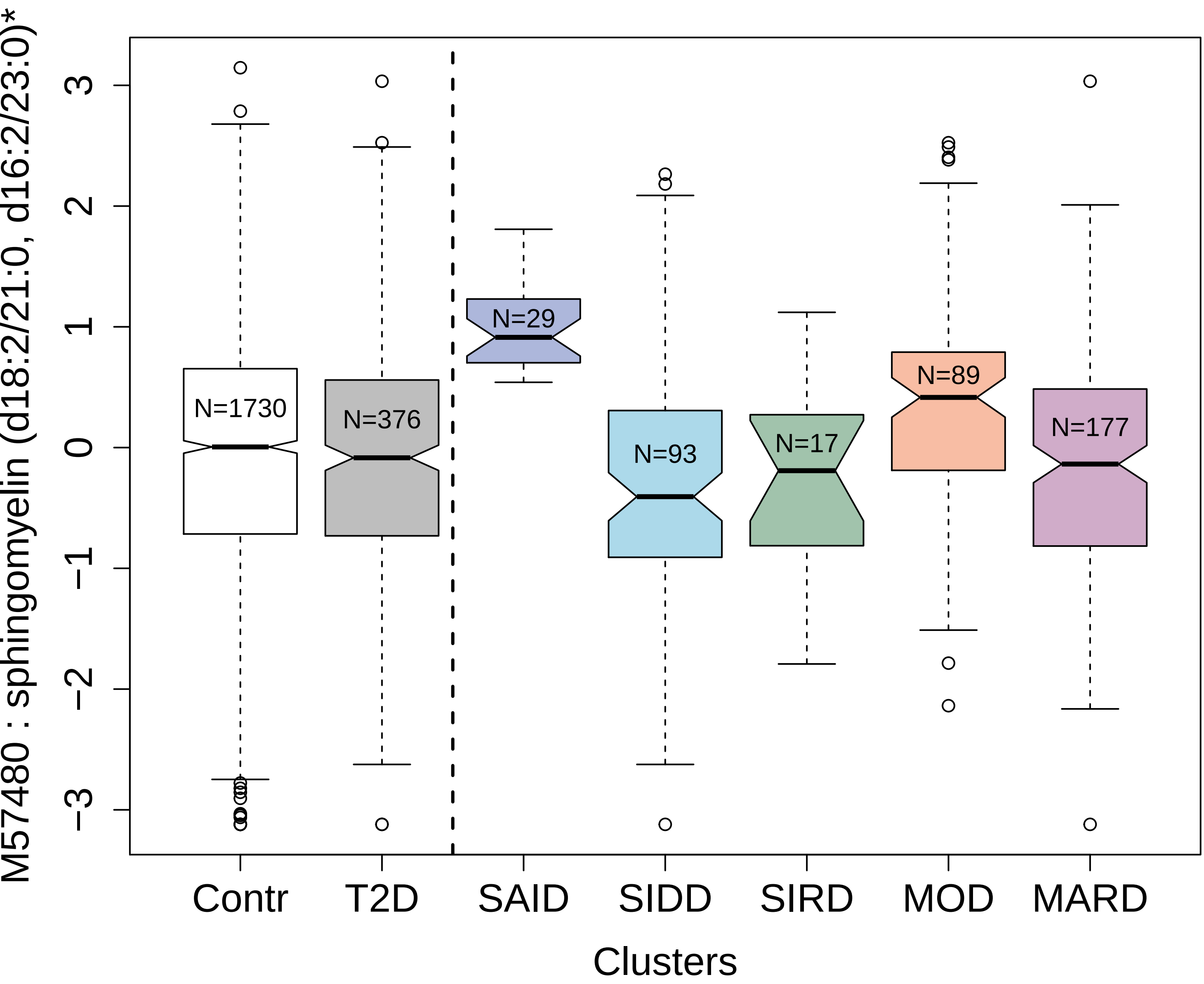
M57480 : sphingomyelin (d18:2/21:0, d16:2/23:0)*
3
2
1
0
−1
−2
−3
N=1730
N=376
N=29
N=93
N=17
N=89
N=177
Contr
T2D
SAID
SIDD
SIRD
MOD
MARD
Clusters

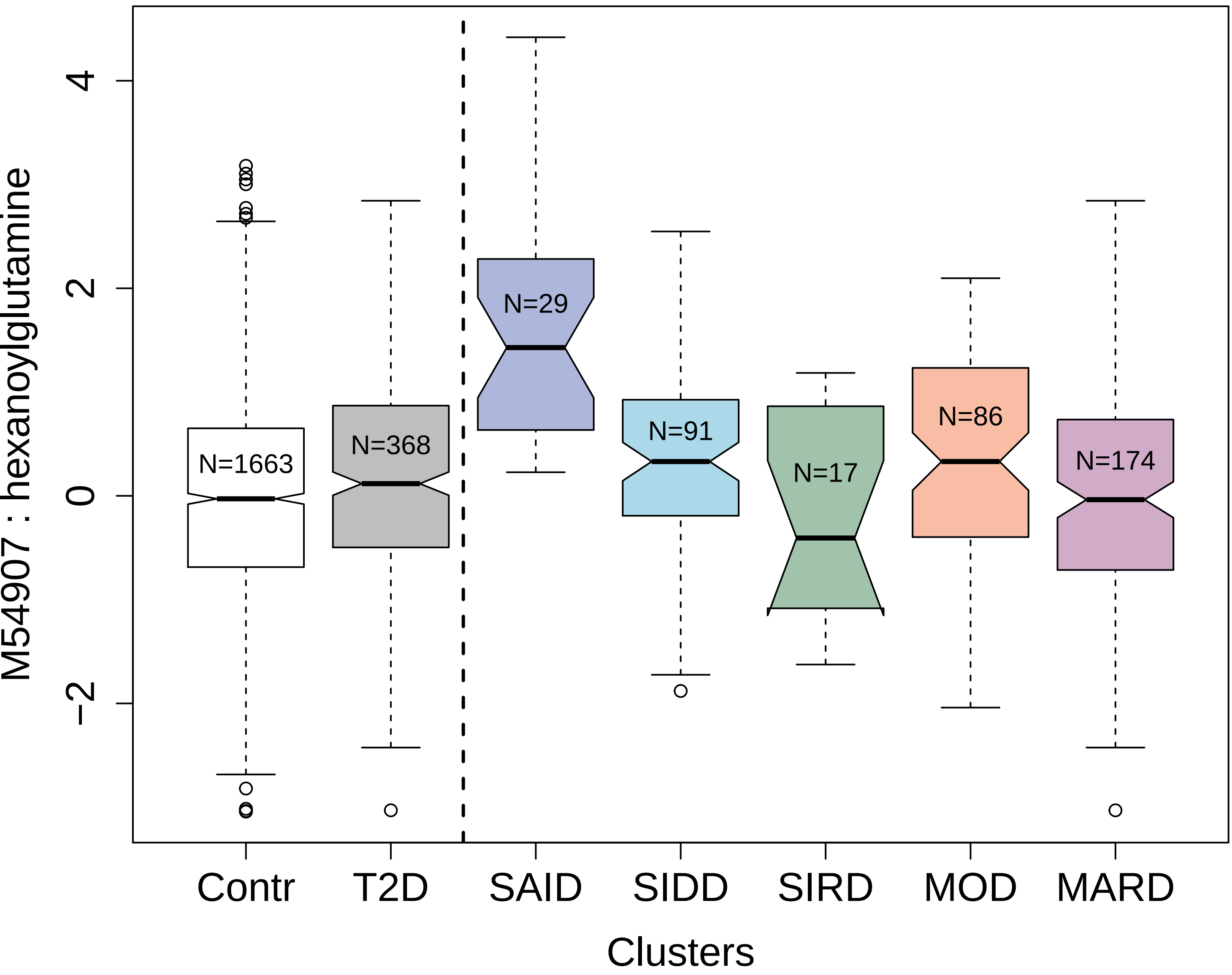

M54907 : hexanoylglutamine
4
2
0
−2
N=1663
N=368
N=29
N=91
N=17
N=86
N=174
Contr
T2D
SAID
SIDD
SIRD
MOD
MARD
Clusters

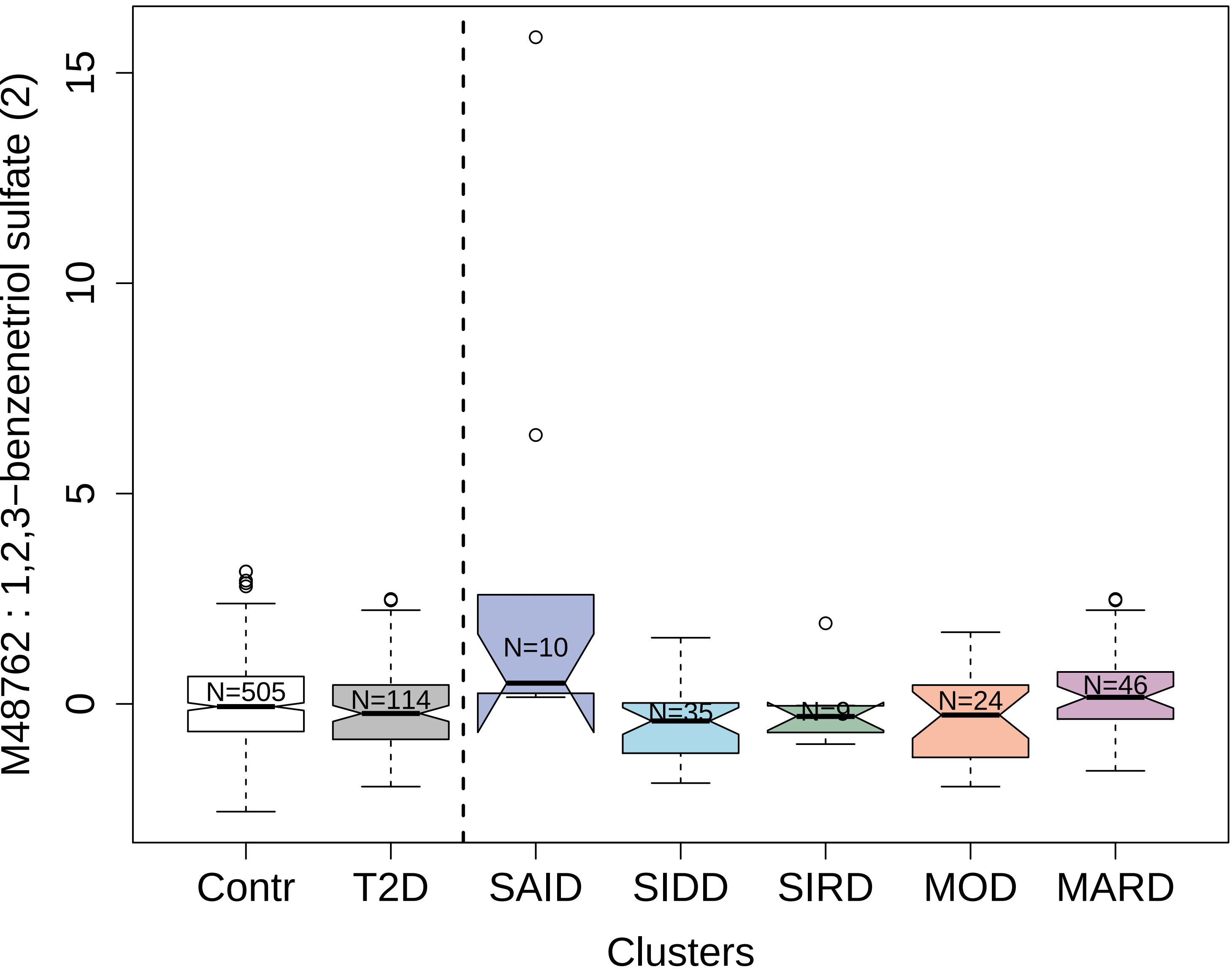

M48762 : 1,2,3−benzenetriol sulfate (2)
15
10
5
0
N=505
N=114
N=10
N=35
N=9
N=24
N=46
Contr
T2D
SAID
SIDD
SIRD
MOD
MARD
Clusters

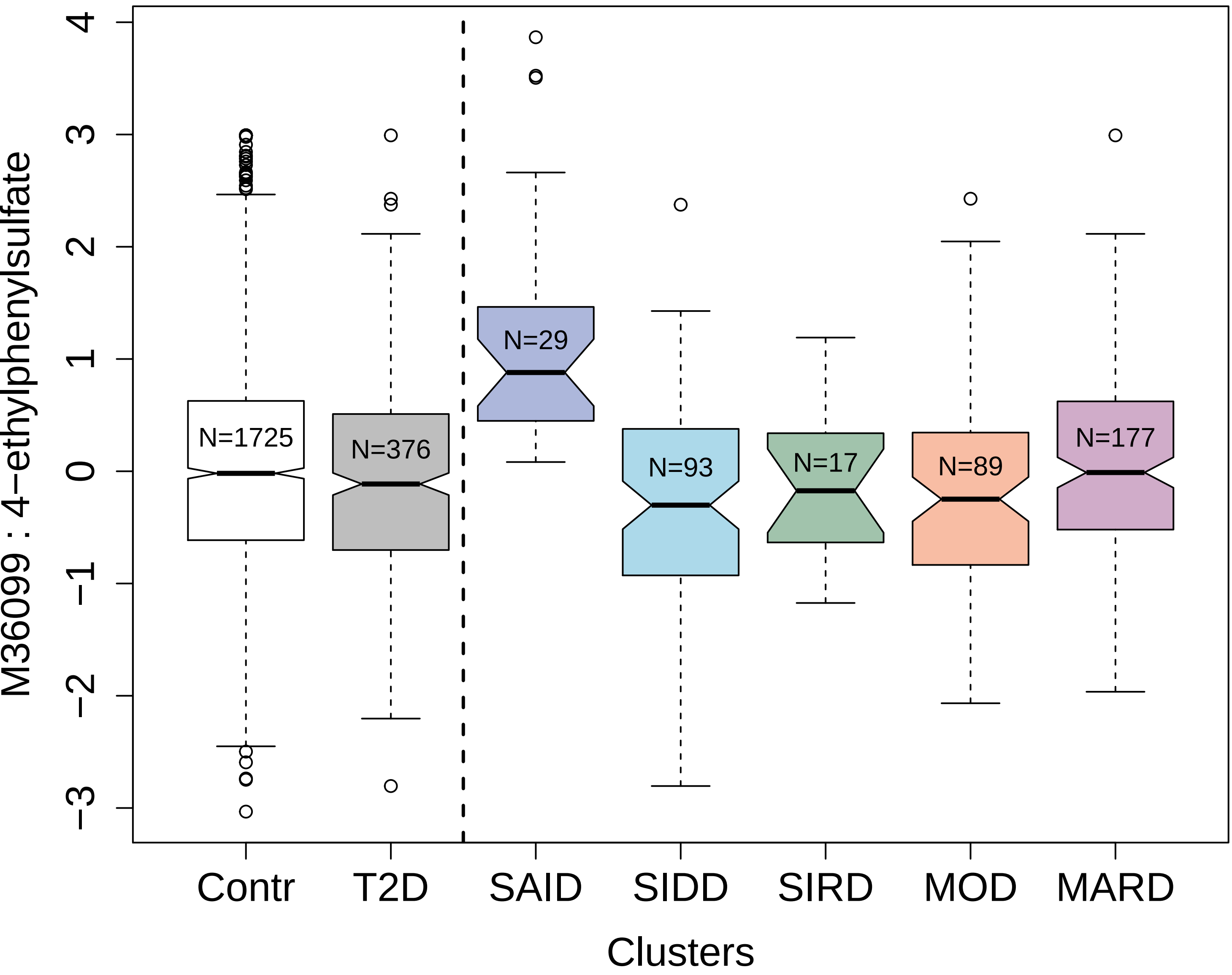

M36099 : 4−ethylphenylsulfate
4
3
2
1
0
−1
−2
−3
N=1725
N=376
N=29
N=93
N=17
N=89
N=177
Contr
T2D
SAID
SIDD
SIRD
MOD
MARD
Clusters

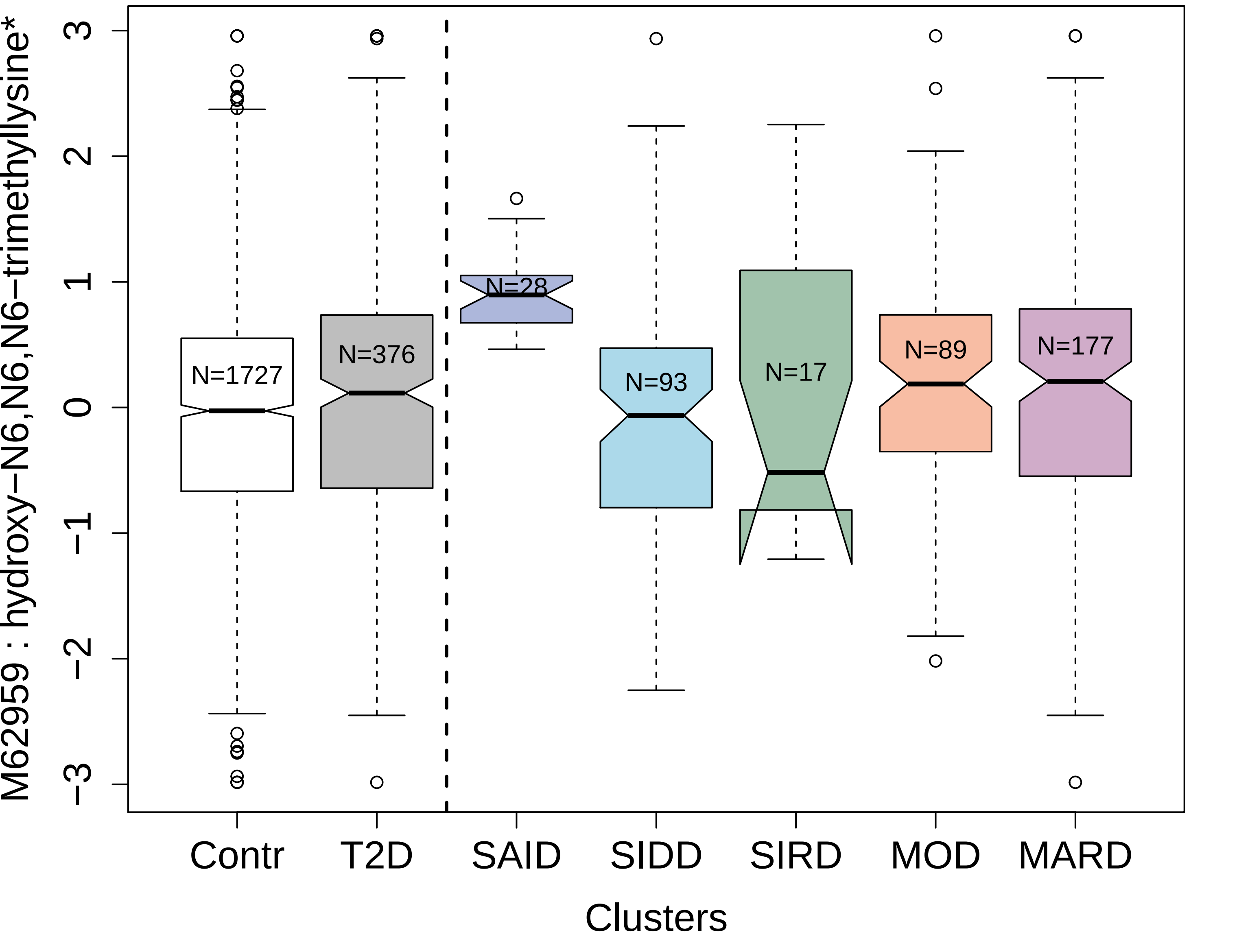

M62959 : hydroxy−N6,N6,N6−trimethyllysine*
3
2
1
0
−1
−2
−3
N=1727
N=376
N=28
N=93
N=17
N=89
N=177
Contr
T2D
SAID
SIDD
SIRD
MOD
MARD
Clusters

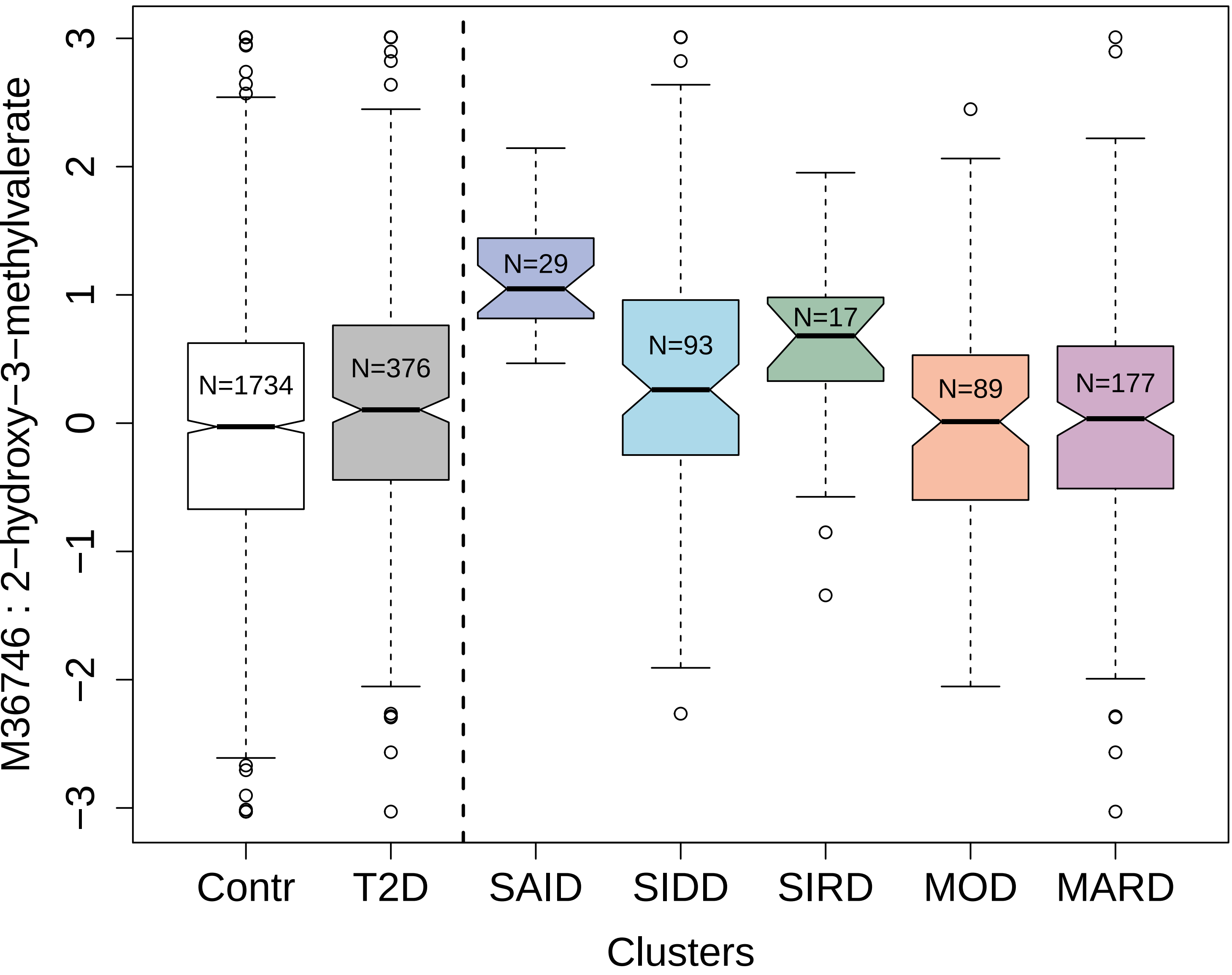

M36746 : 2−hydroxy−3−methylvalerate
3
2
1
0
−1
−2
−3
N=1734
N=376
N=29
N=93
N=17
N=89
N=177
Contr
T2D
SAID
SIDD
SIRD
MOD
MARD
Clusters

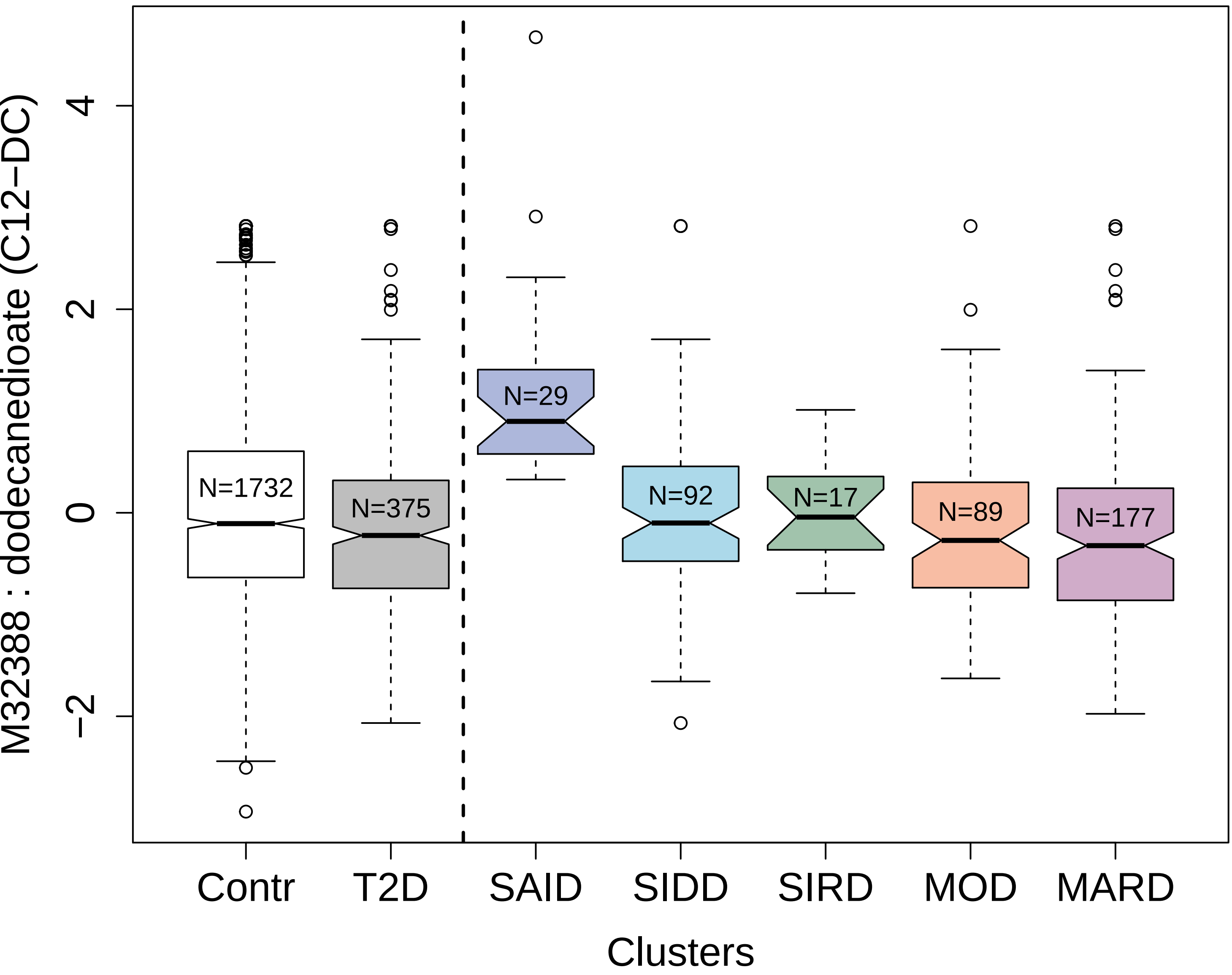
M32388 : dodecanedioate (C12−DC)
4
2
0
−2
N=1732
N=375
N=29
N=92
N=17
N=89
N=177
Contr
T2D
SAID
SIDD
SIRD
MOD
MARD
Clusters

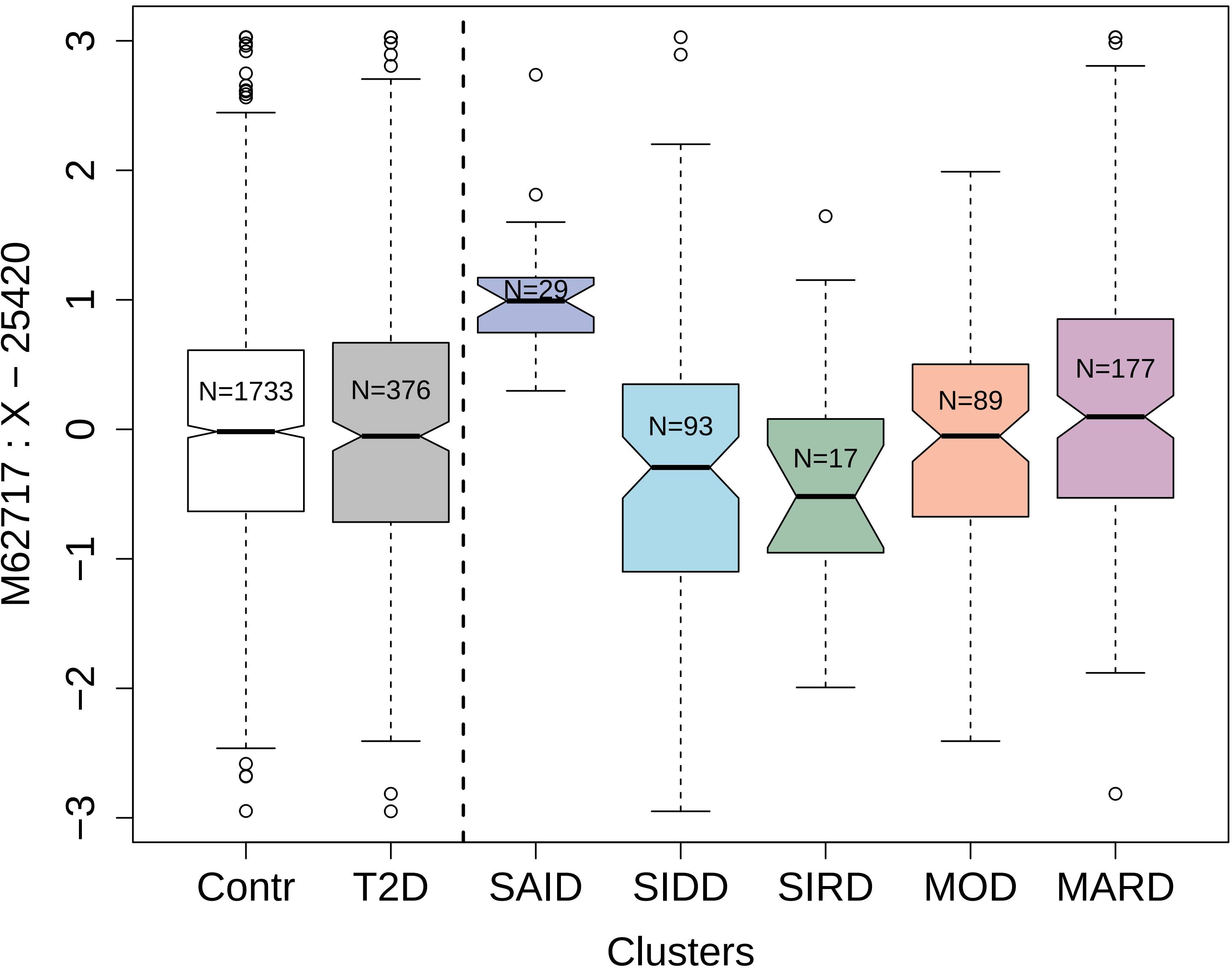
M62717 : X − 25420
3
2
1
0
−1
−2
−3
N=1733
N=376
N=29
N=93
N=17
N=89
N=177
Contr
T2D
SAID
SIDD
SIRD
MOD
MARD
Clusters

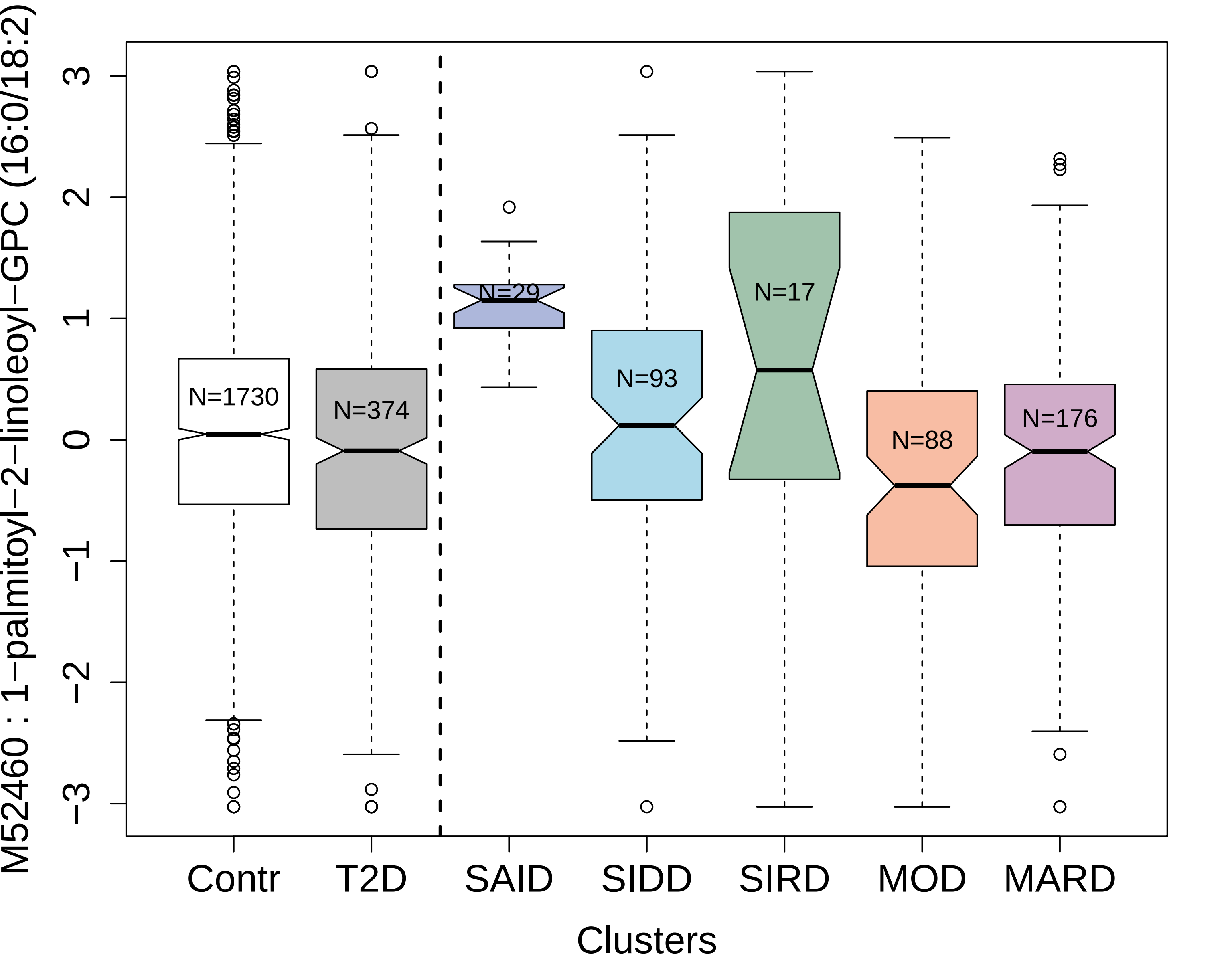
M52460 : 1−palmitoyl−2−linoleoyl−GPC (16:0/18:2)
−3
−2
−1
0
1
2
3
N=1730
N=374
N=29
N=93
N=17
N=88
N=176
Contr
T2D
SAID
SIDD
SIRD
MOD
MARD
Clusters

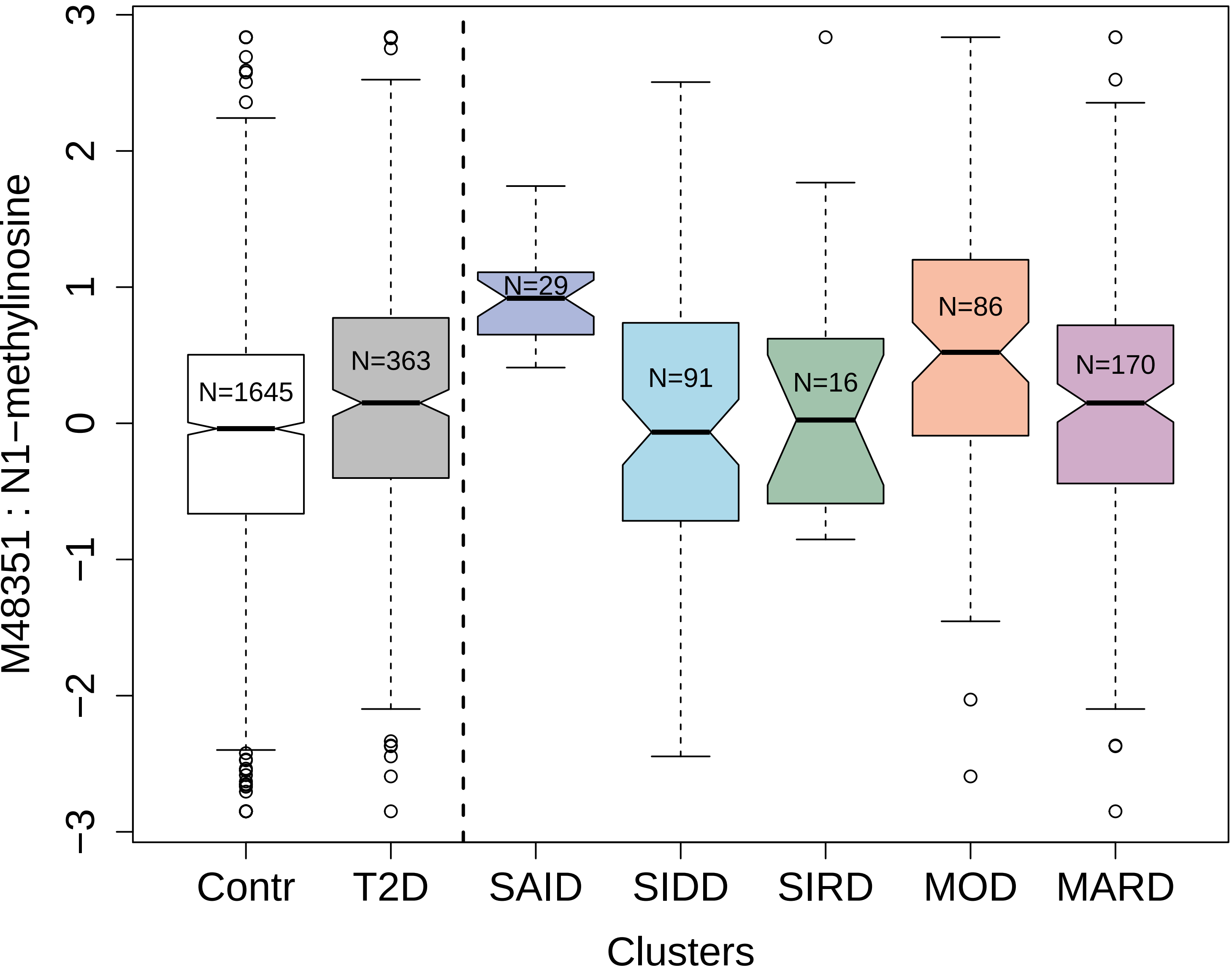

M48351 : N1−methylinosine
3
2
1
0
−1
−2
−3
N=1645
N=363
N=29
N=91
N=16
N=86
N=170
Contr
T2D
SAID
SIDD
SIRD
MOD
MARD
Clusters

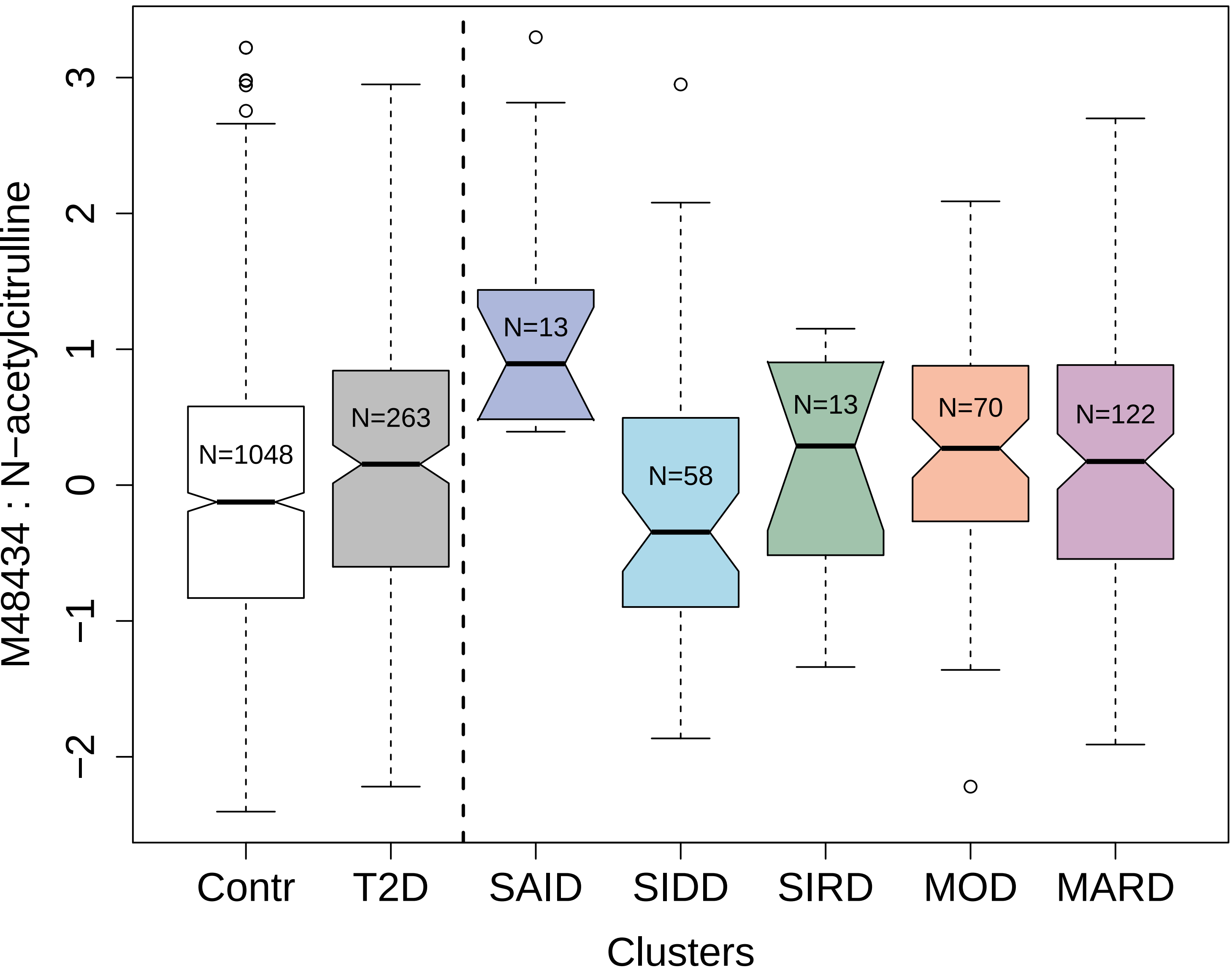

M48434 : N−acetylcitrulline
3
2
1
0
−1
−2
N=1048
N=263
N=13
N=58
N=13
N=70
N=122
Contr
T2D
SAID
SIDD
SIRD
MOD
MARD
Clusters

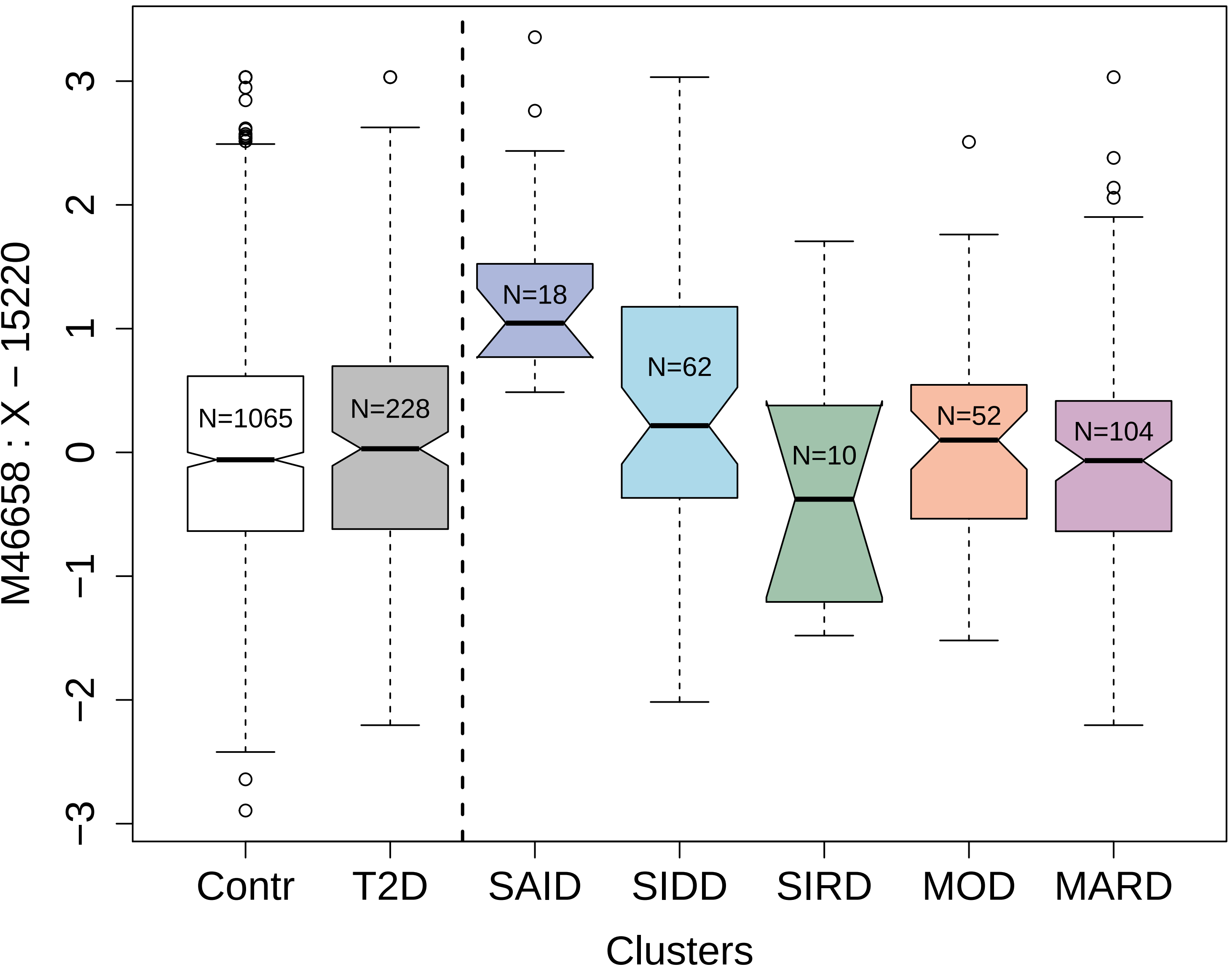

M46658 : X − 15220
3
2
1
0
−1
−2
−3
N=1065
N=228
N=18
N=62
N=10
N=52
N=104
Contr
T2D
SAID
SIDD
SIRD
MOD
MARD
Clusters

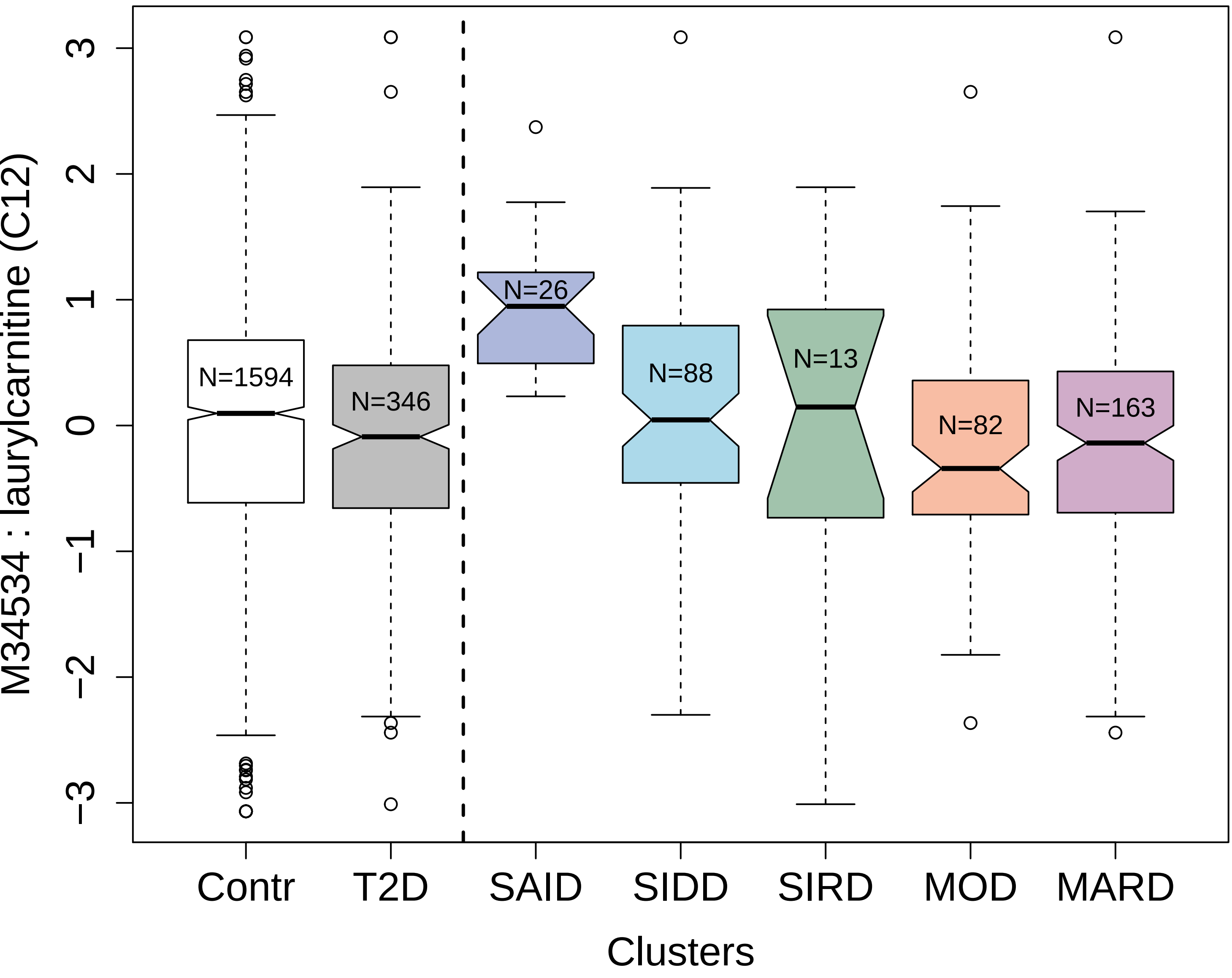
M34534 : laurylcarnitine (C12)
3
2
1
0
−1
−2
−3
N=1594
N=346
N=26
N=88
N=13
N=82
N=163
Contr
T2D
SAID
SIDD
SIRD
MOD
MARD
Clusters

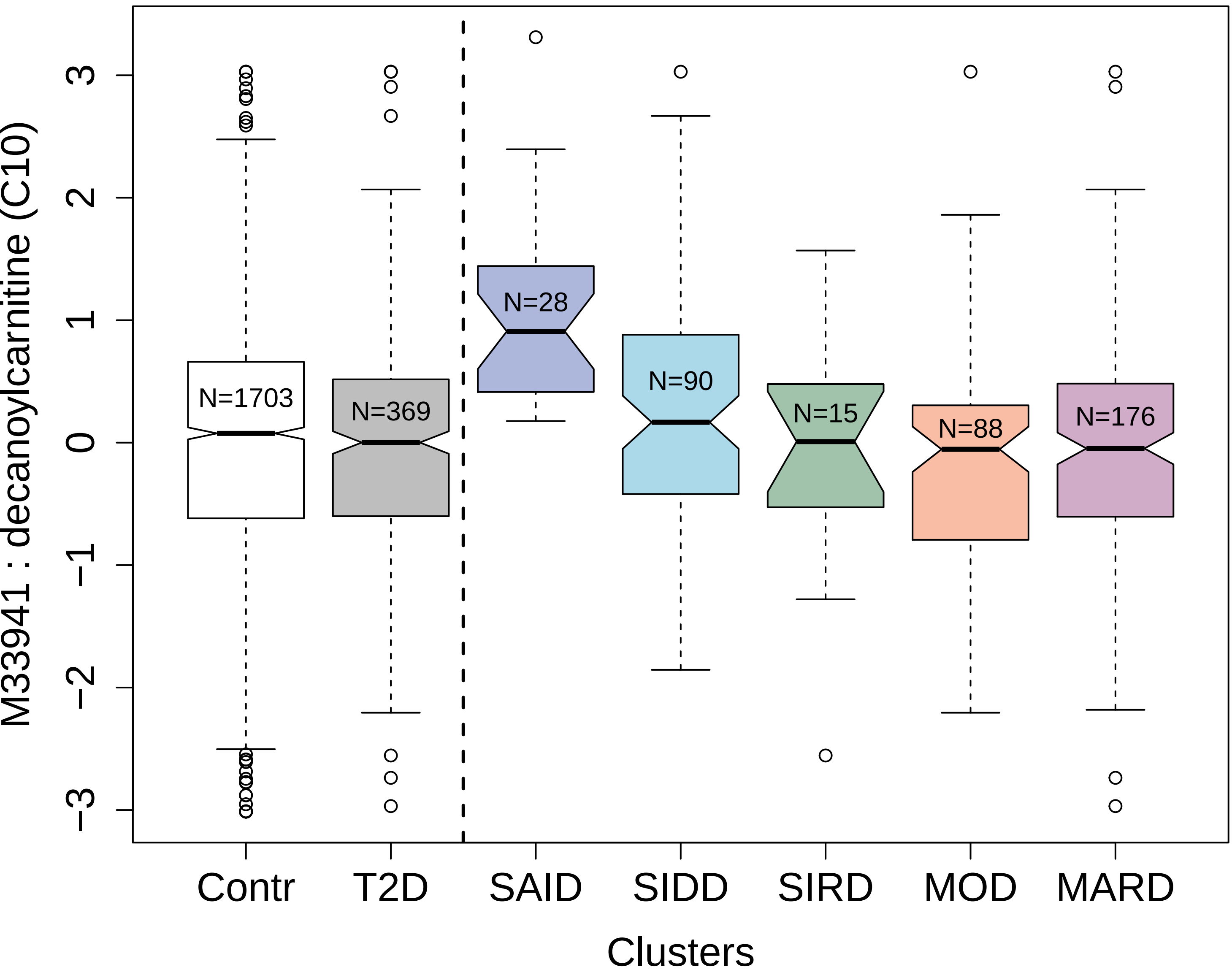
M33941 : decanoylcarnitine (C10)
3
2
1
0
−1
−2
−3
N=1703
N=369
N=28
N=90
N=15
N=88
N=176
Contr
T2D
SAID
SIDD
SIRD
MOD
MARD
Clusters

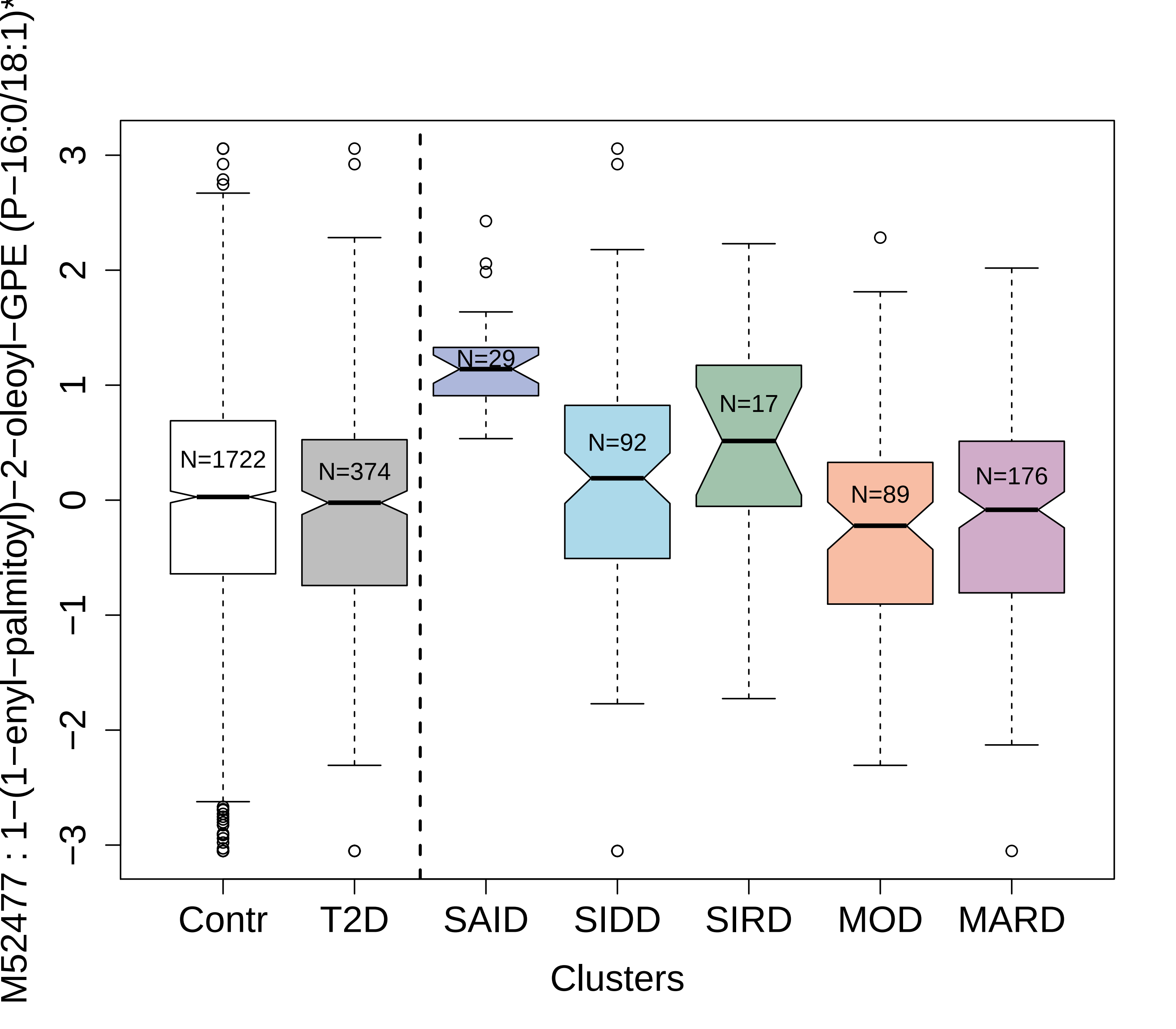
M52477 : 1−(1−enyl−palmitoyl)−2−oleoyl−GPE (P−16:0/18:1)*
3
2
1
0
−1
−2
−3
N=1722
N=374
N=29
N=92
N=17
N=89
N=176
Contr
T2D
SAID
SIDD
SIRD
MOD
MARD
Clusters

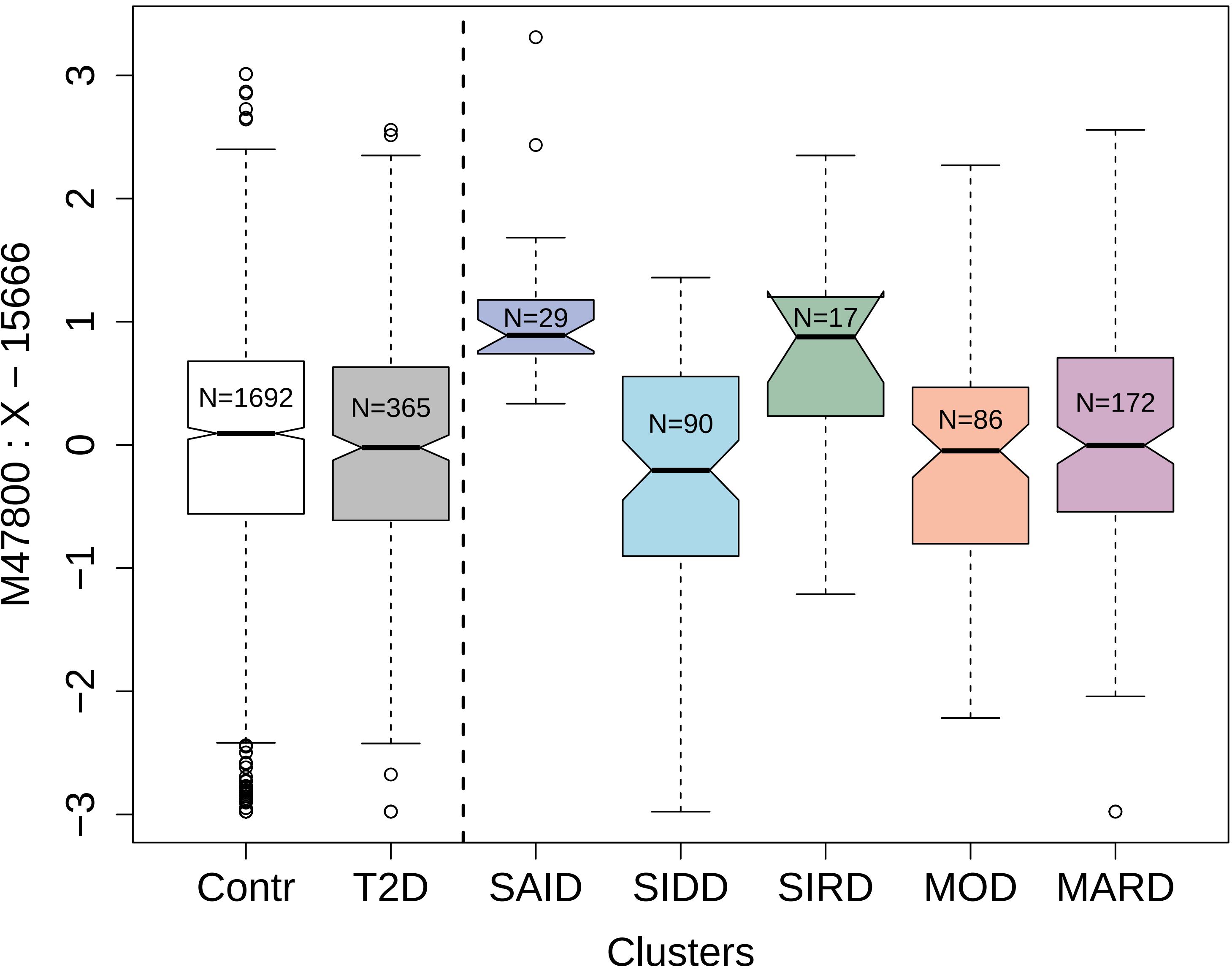
M47800 : X − 15666
3
2
1
0
−1
−2
−3
N=1692
N=365
N=29
N=90
N=17
N=86
N=172
Contr
T2D
SAID
SIDD
SIRD
MOD
MARD
Clusters

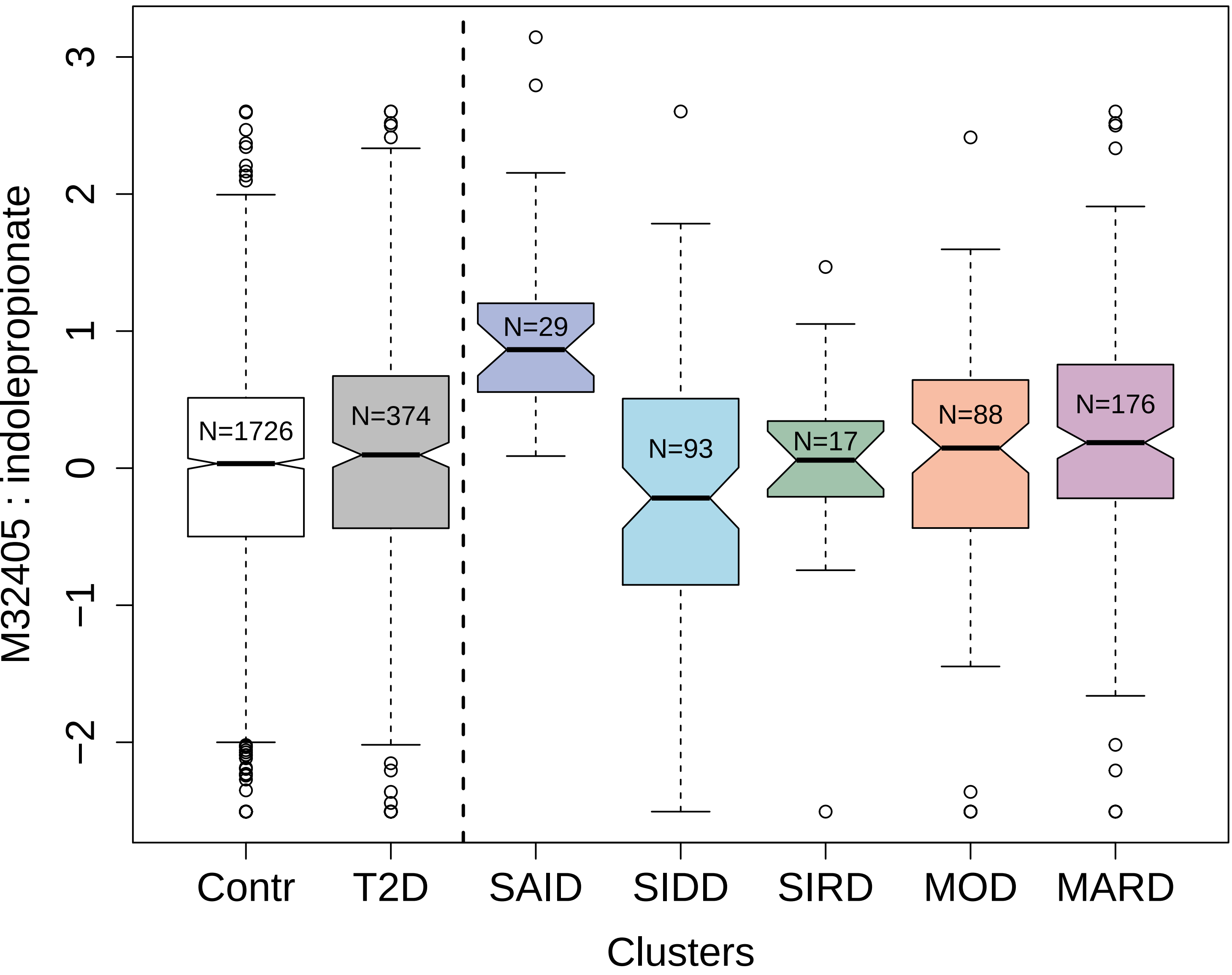

M32405 : indolepropionate
3
2
1
0
−1
−2
N=1726
N=374
N=29
N=93
N=17
N=88
N=176
Contr
T2D
SAID
SIDD
SIRD
MOD
MARD
Clusters

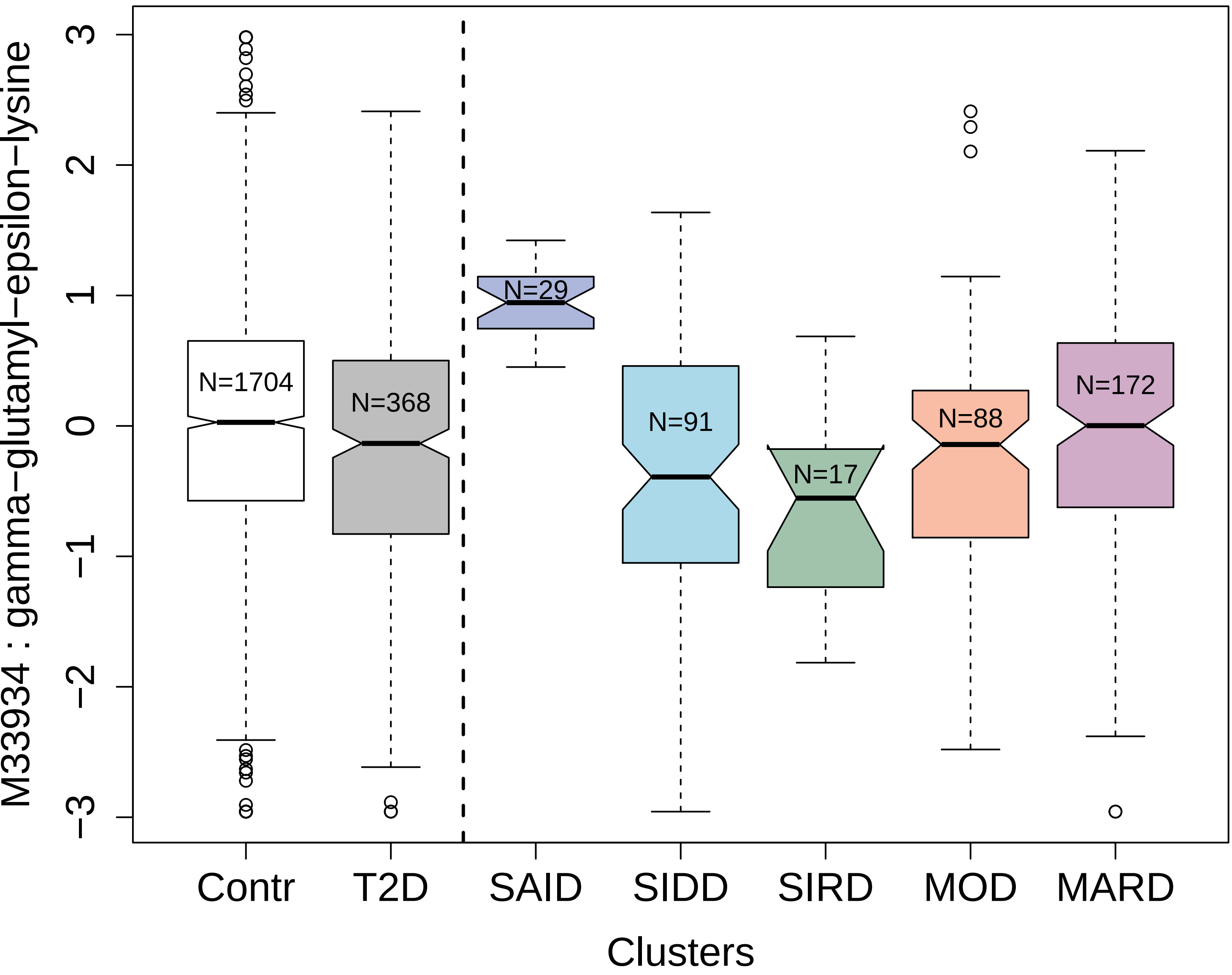
M33934 : gamma−glutamyl−epsilon−lysine
3
2
1
0
−1
−2
−3
N=1704
N=368
N=29
N=91
N=17
N=88
N=172
Contr
T2D
SAID
SIDD
SIRD
MOD
MARD
Clusters

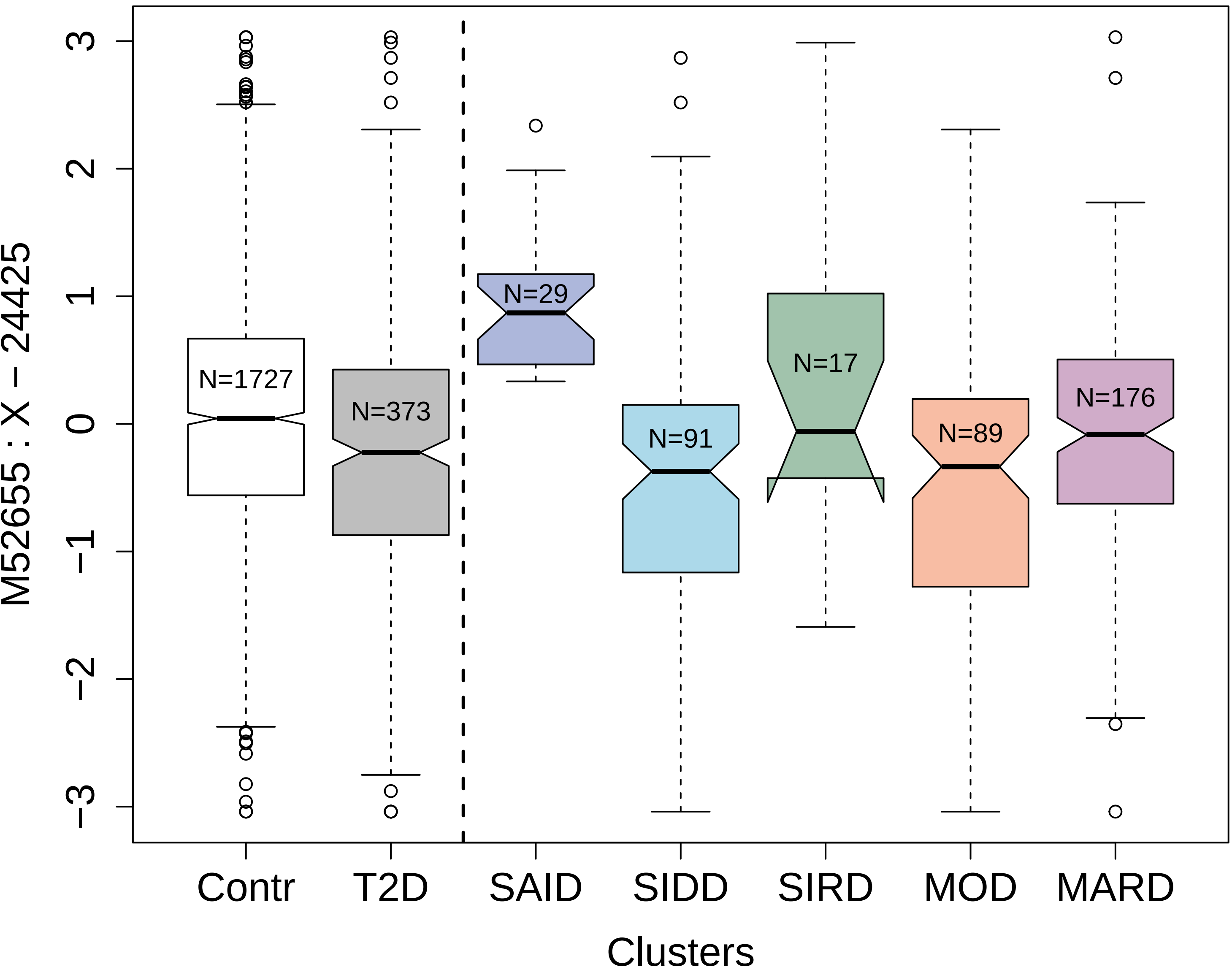

M52655 : X − 24425
3
2
1
0
−1
−2
−3
N=1727
N=373
N=29
N=91
N=17
N=89
N=176
Contr
T2D
SAID
SIDD
SIRD
MOD
MARD
Clusters

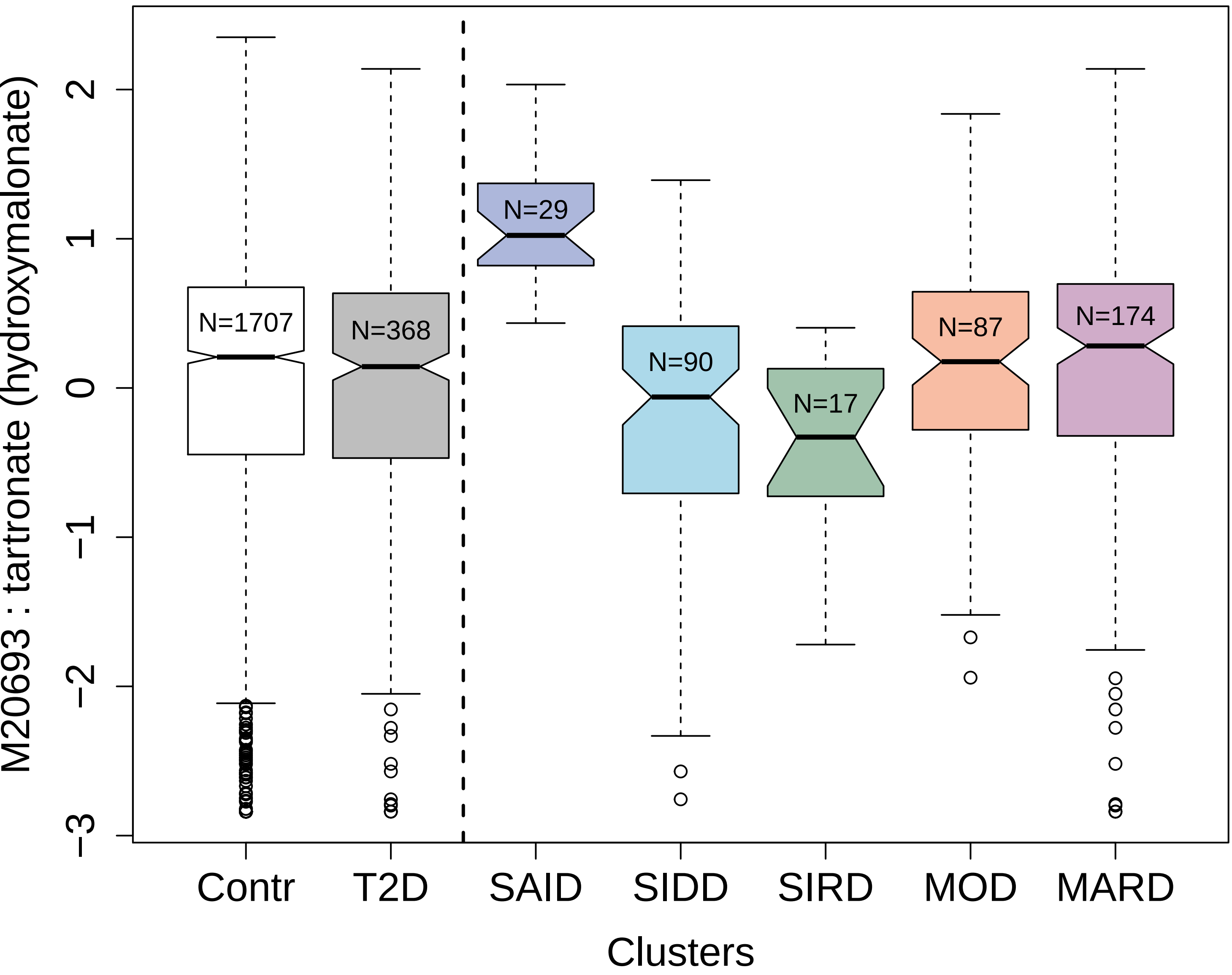

M20693 : tartronate (hydroxymalonate)
2
1
0
−1
−2
−3
N=1707
N=368
N=29
N=90
N=17
N=87
N=174
Contr
T2D
SAID
SIDD
SIRD
MOD
MARD
Clusters

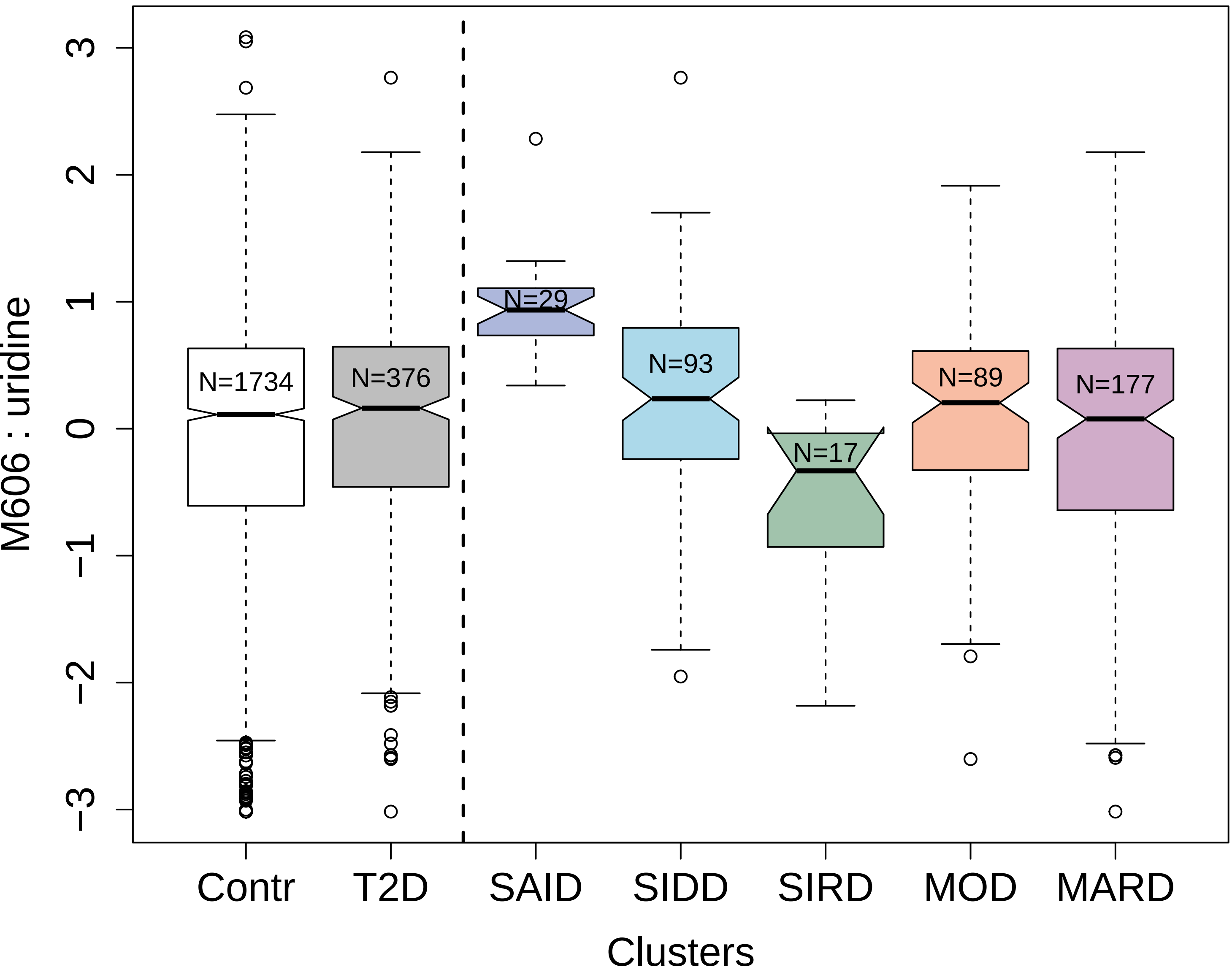
M606 : uridine
3
2
1
0
−1
−2
−3
N=1734
N=376
N=29
N=93
N=17
N=89
N=177
Contr
T2D
SAID
SIDD
SIRD
MOD
MARD
Clusters

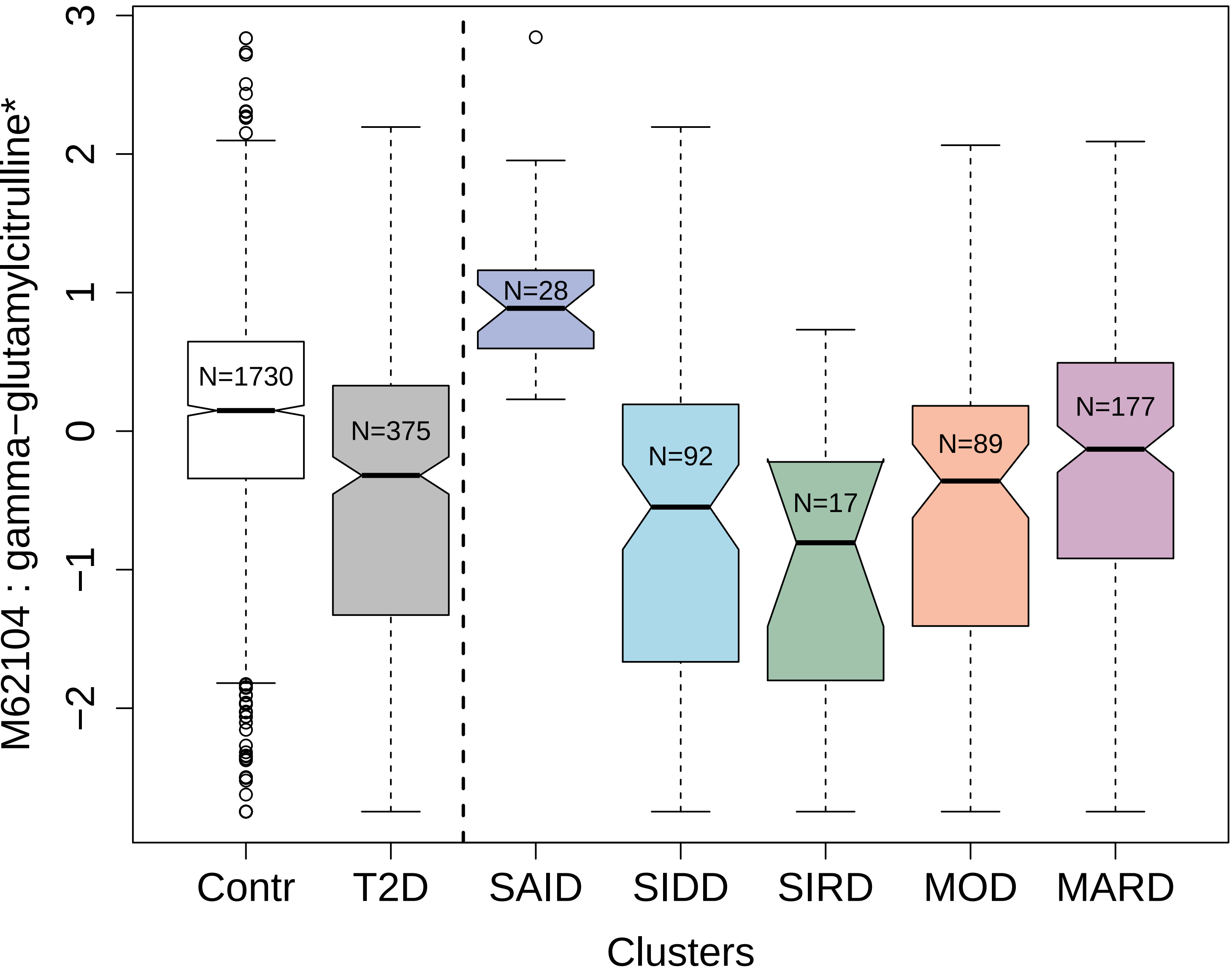
M62104 : gamma−glutamylcitrulline*
3
2
1
0
−1
−2
N=1730
N=375
N=28
N=92
N=17
N=89
N=177
Contr
T2D
SAID
SIDD
SIRD
MOD
MARD
Clusters

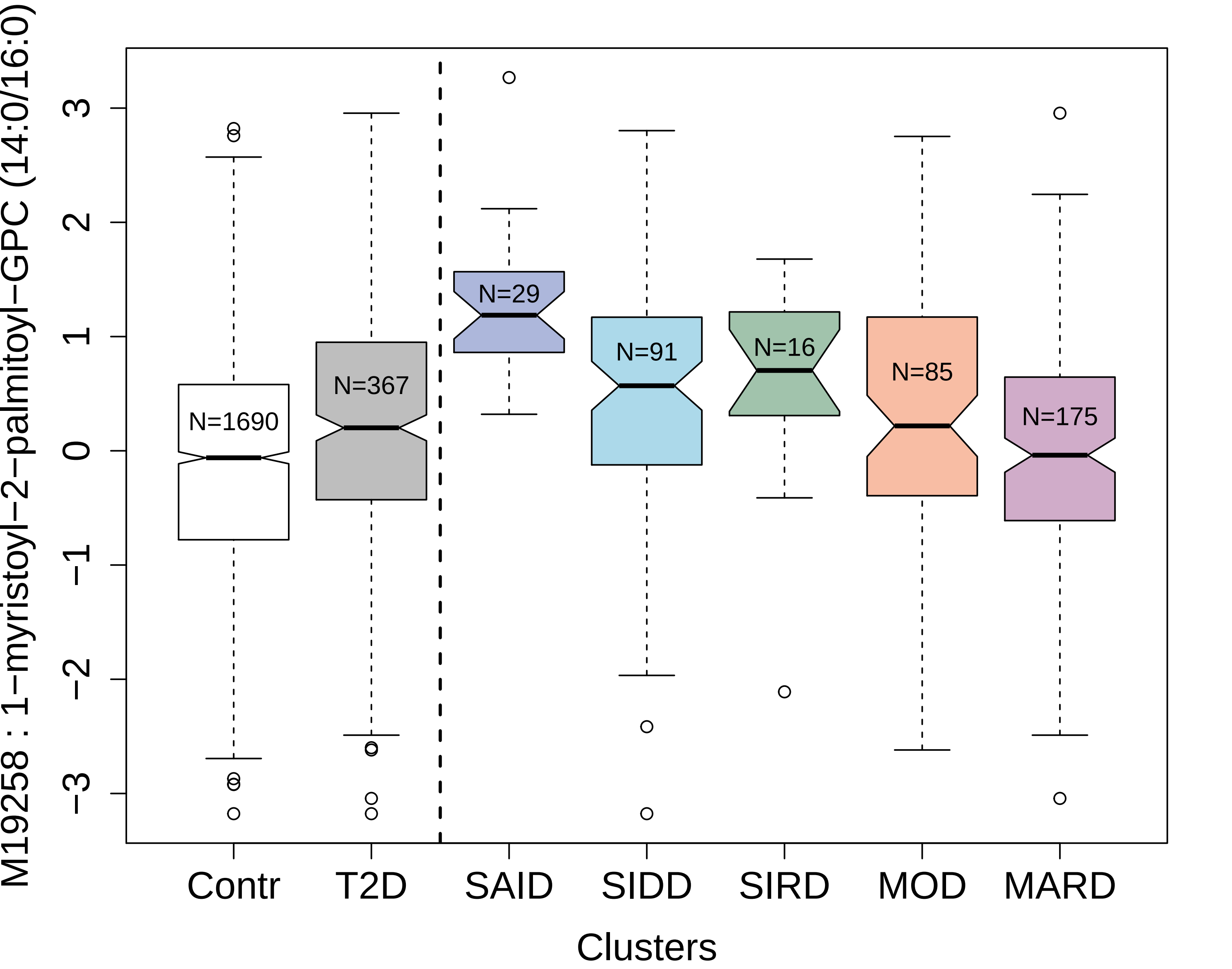

M19258 : 1−myristoyl−2−palmitoyl−GPC (14:0/16:0)
3
2
1
0
−1
−2
−3
N=1690
N=367
N=29
N=91
N=16
N=85
N=175
Contr
T2D
SAID
SIDD
SIRD
MOD
MARD
Clusters

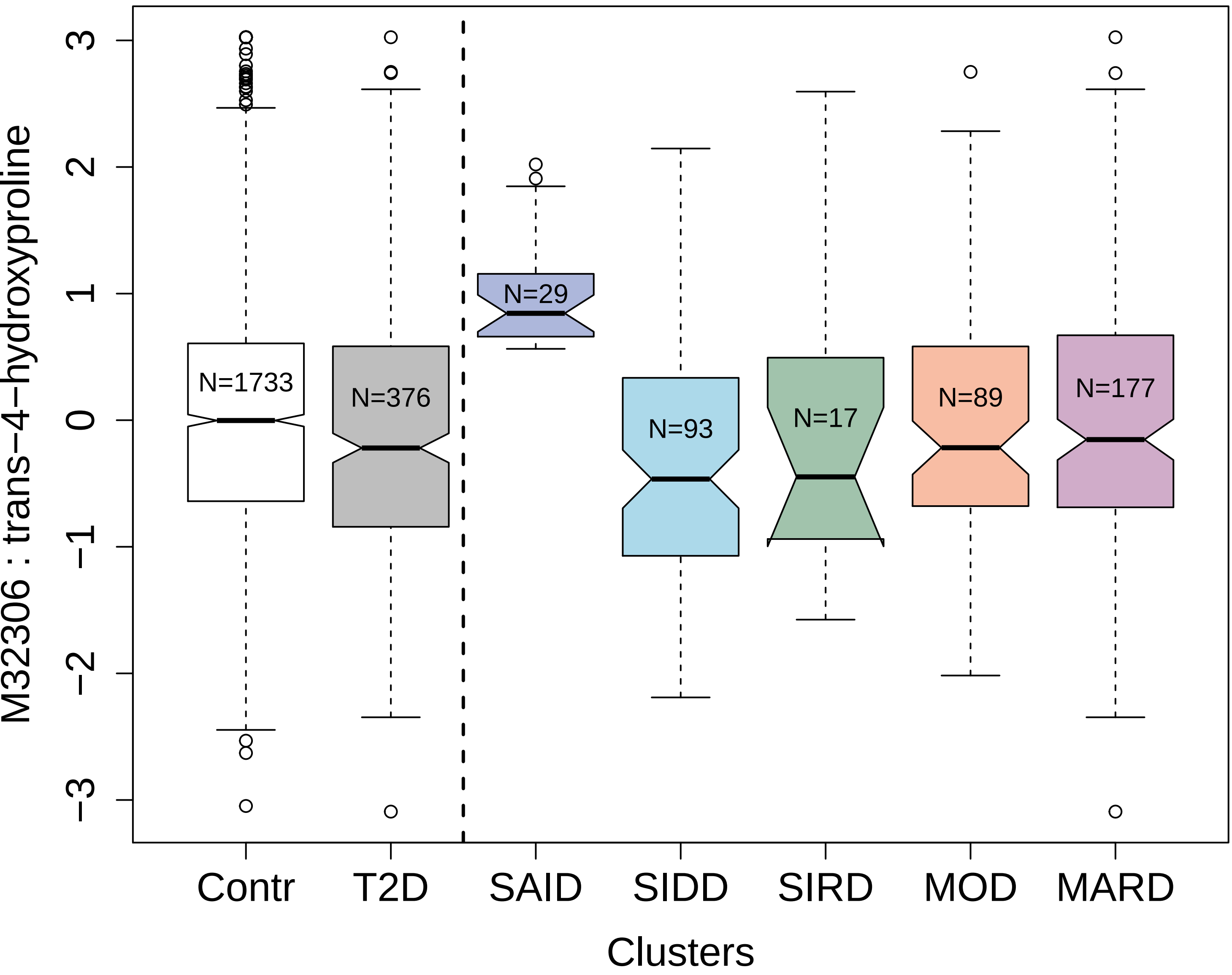

M32306 : trans−4−hydroxyproline
3
2
1
0
−1
−2
−3
N=1733
N=376
N=29
N=93
N=17
N=89
N=177
Contr
T2D
SAID
SIDD
SIRD
MOD
MARD
Clusters

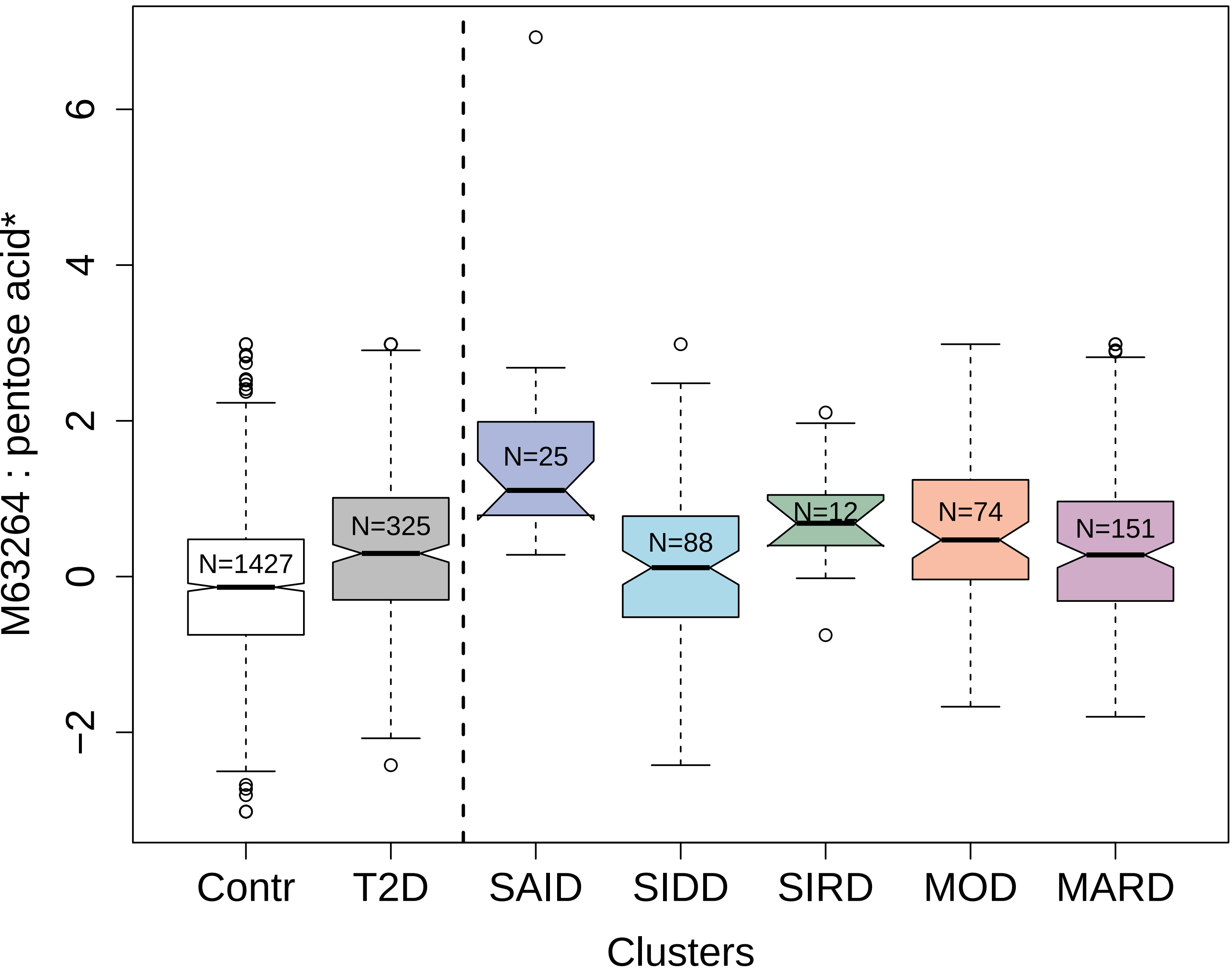

M63264 : pentose acid*
6
4
2
0
−2
N=1427
N=325
N=25
N=88
N=12
N=74
N=151
Contr
T2D
SAID
SIDD
SIRD
MOD
MARD
Clusters

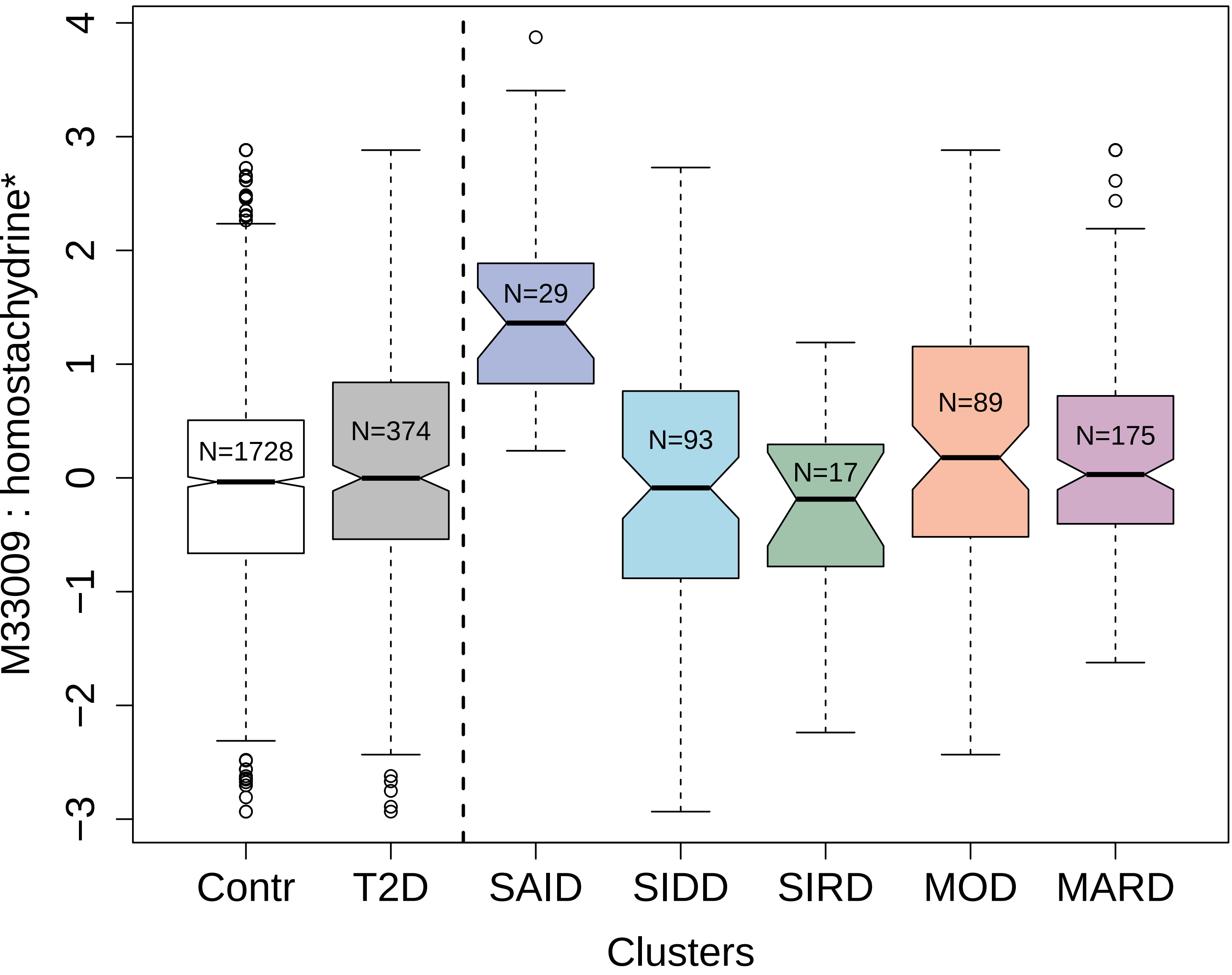
M33009 : homostachydrine*
4
3
2
1
0
−1
−2
−3
N=1728
N=374
N=29
N=93
N=17
N=89
N=175
Contr
T2D
SAID
SIDD
SIRD
MOD
MARD
Clusters

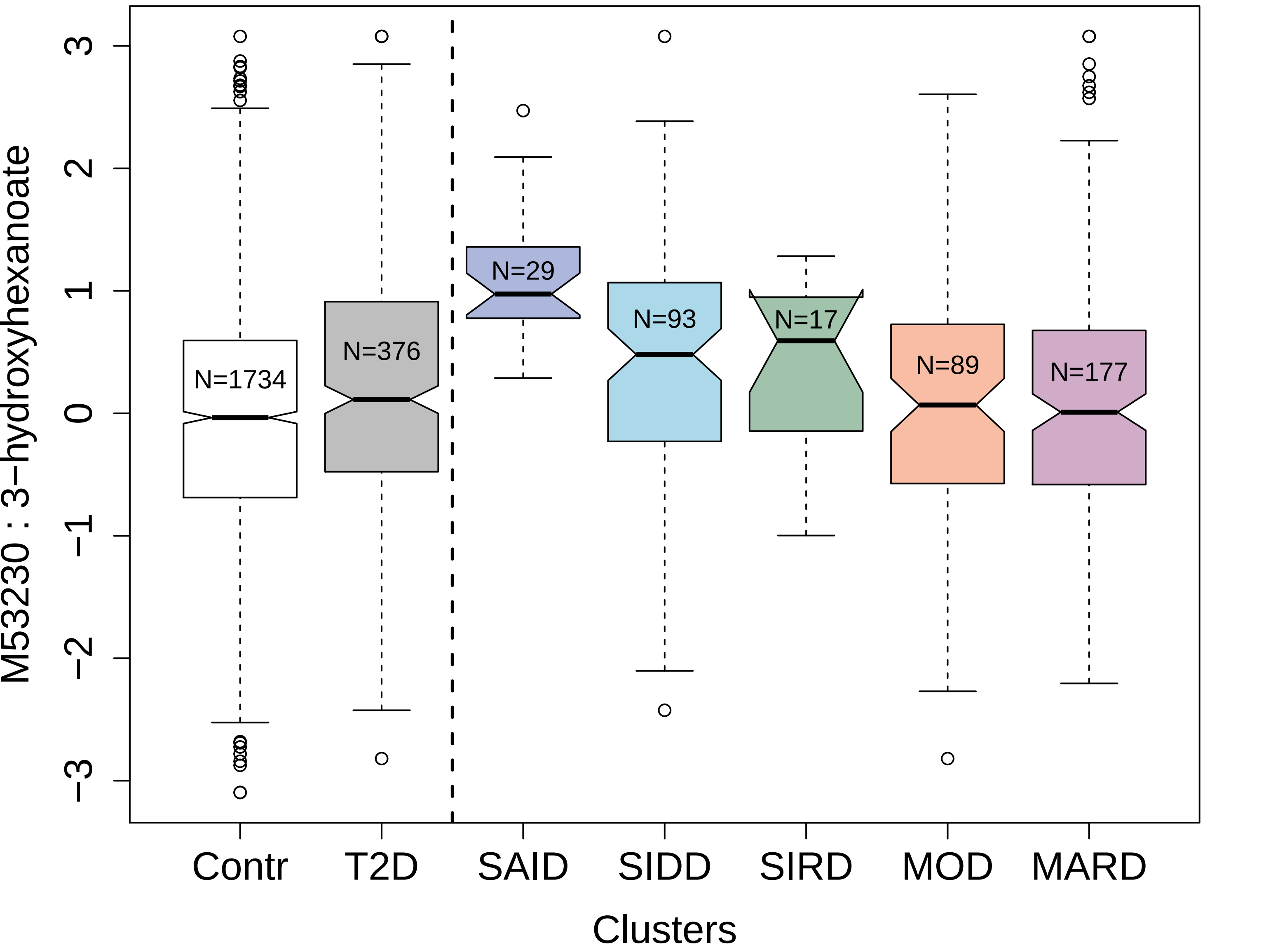
M53230 : 3−hydroxyhexanoate
Clusters
Contr
T2D
SAID
SIDD
SIRD
MOD
MARD
N=1734
N=376
N=29
N=93
N=17
N=89
N=177
3
2
1
0
−1
−2
−3

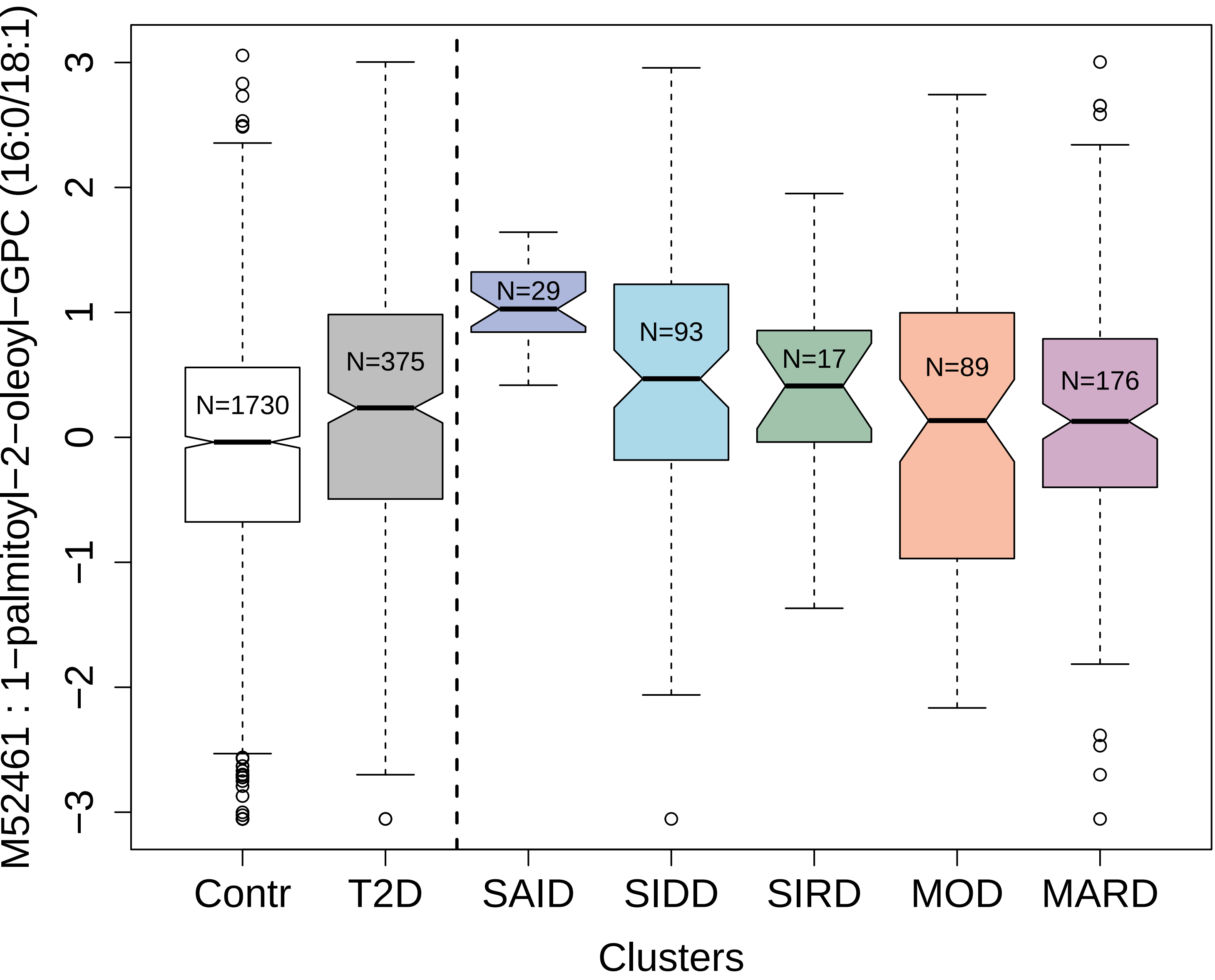

M52461 : 1−palmitoyl−2−oleoyl−GPC (16:0/18:1)
3
2
1
0
−1
−2
−3
N=1730
N=375
N=29
N=93
N=17
N=89
N=176
Contr
T2D
SAID
SIDD
SIRD
MOD
MARD
Clusters

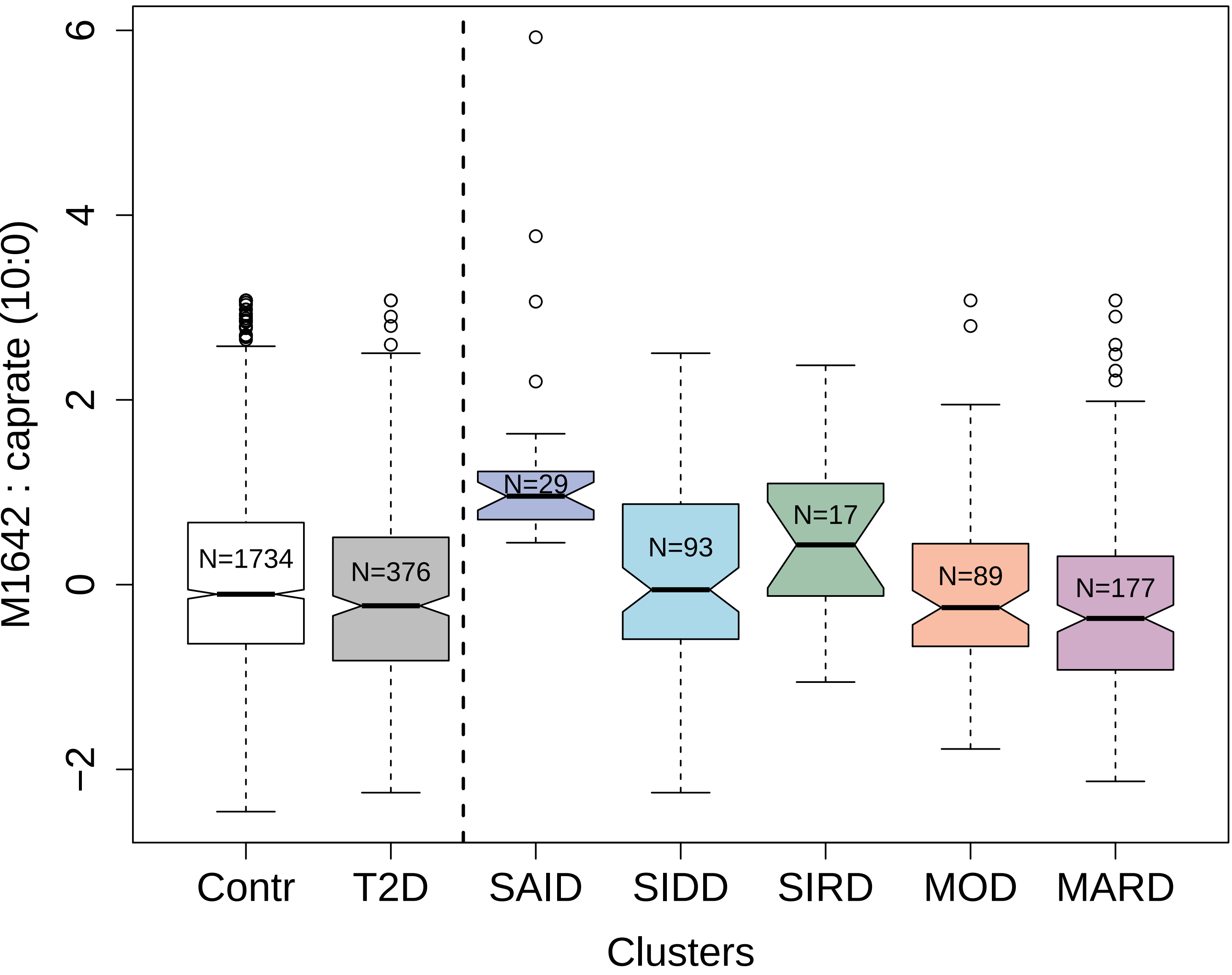
M1642 : caprate (10:0)
6
4
2
0
−2
N=1734
N=376
N=29
N=93
N=17
N=89
N=177
Contr
T2D
SAID
SIDD
SIRD
MOD
MARD
Clusters

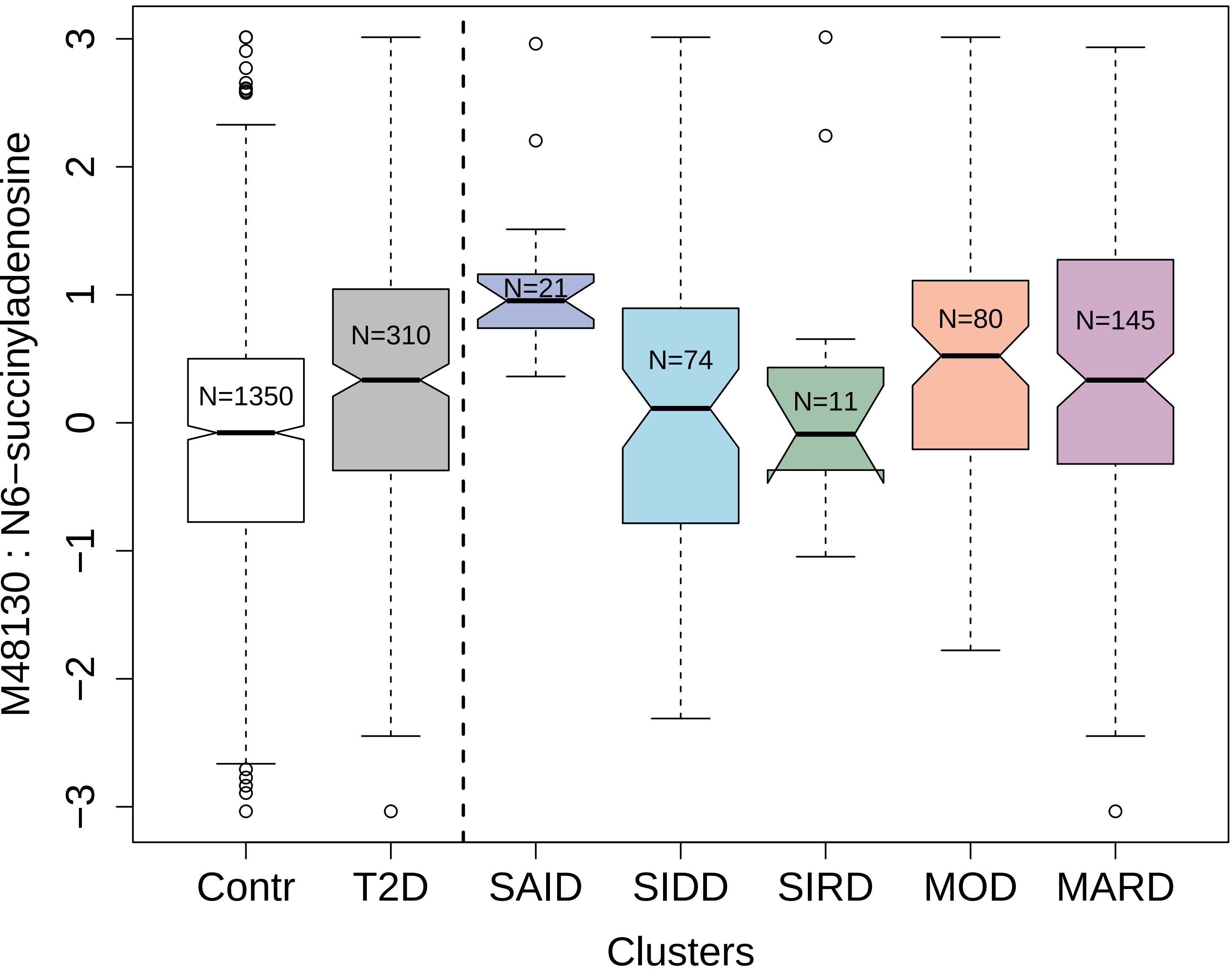

M48130 : N6−succinyladenosine
3
2
1
0
−1
−2
−3
N=1350
N=310
N=21
N=74
N=11
N=80
N=145
Contr
T2D
SAID
SIDD
SIRD
MOD
MARD
Clusters

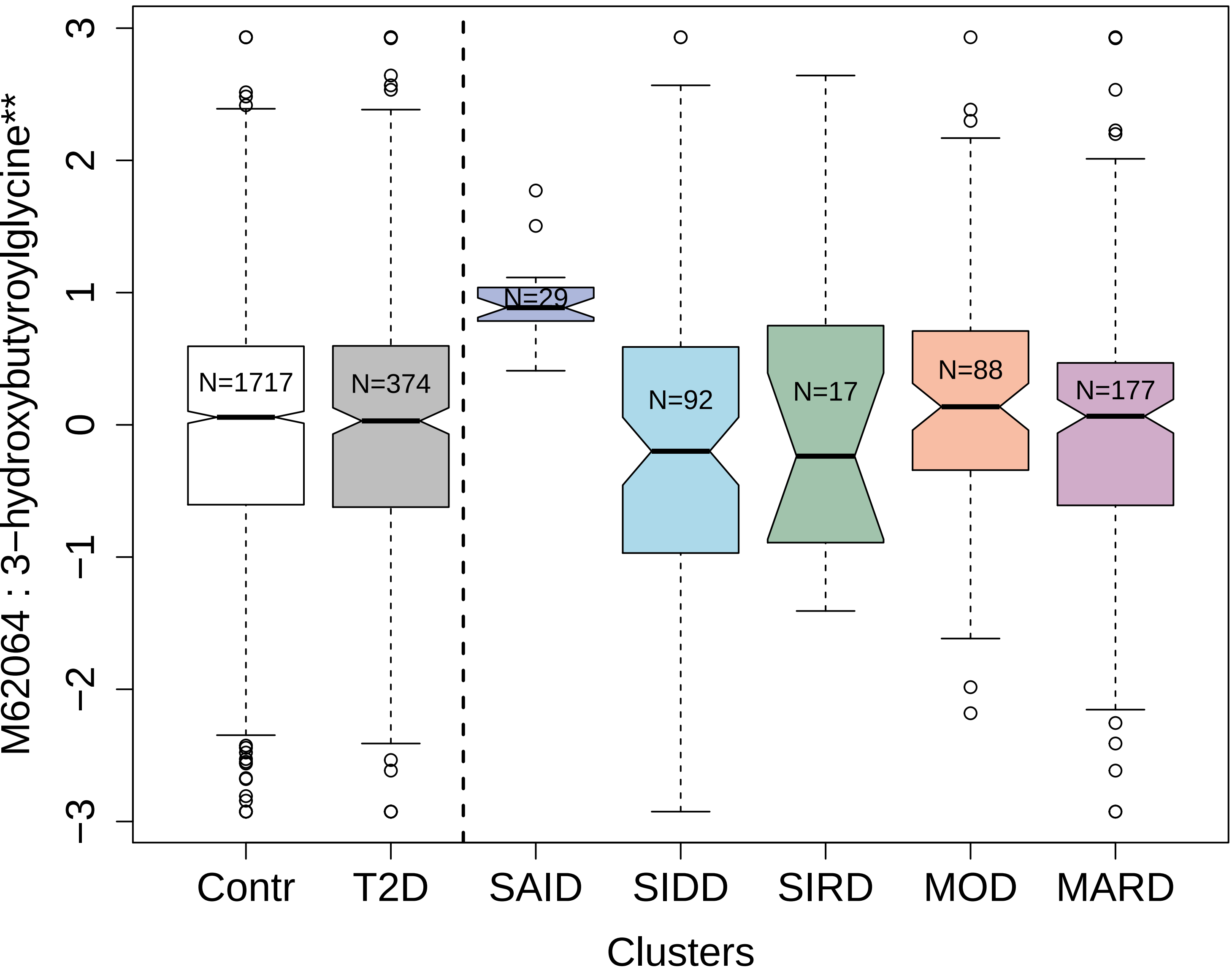

M62064 : 3−hydroxybutyroylglycine**
3
2
1
0
−1
−2
−3
N=1717
N=374
N=29
N=92
N=17
N=88
N=177
Contr
T2D
SAID
SIDD
SIRD
MOD
MARD
Clusters

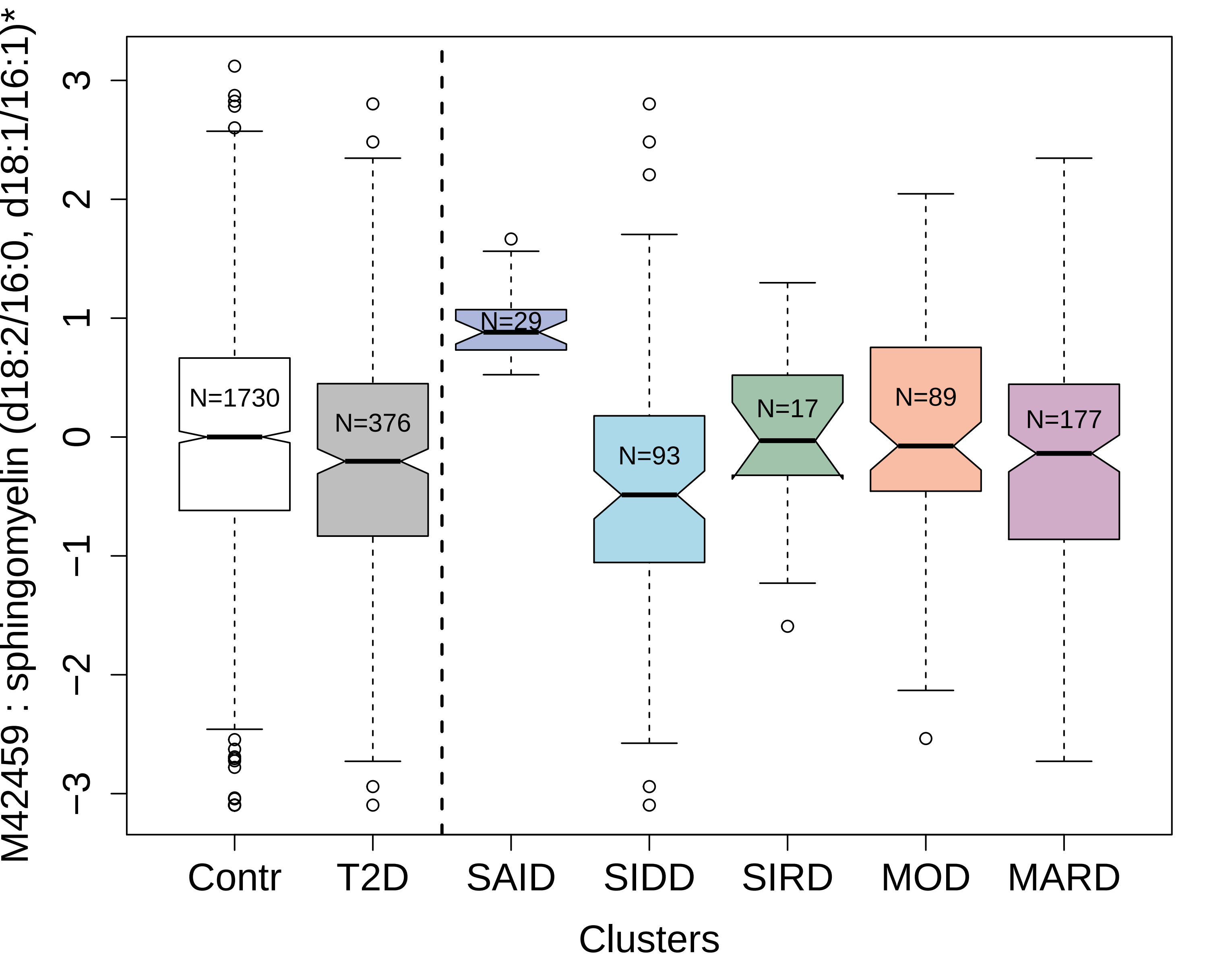
M42459 : sphingomyelin (d18:2/16:0, d18:1/16:1)*
3
2
1
0
−1
−2
−3
N=1730
N=376
N=29
N=93
N=17
N=89
N=177
Contr
T2D
SAID
SIDD
SIRD
MOD
MARD
Clusters

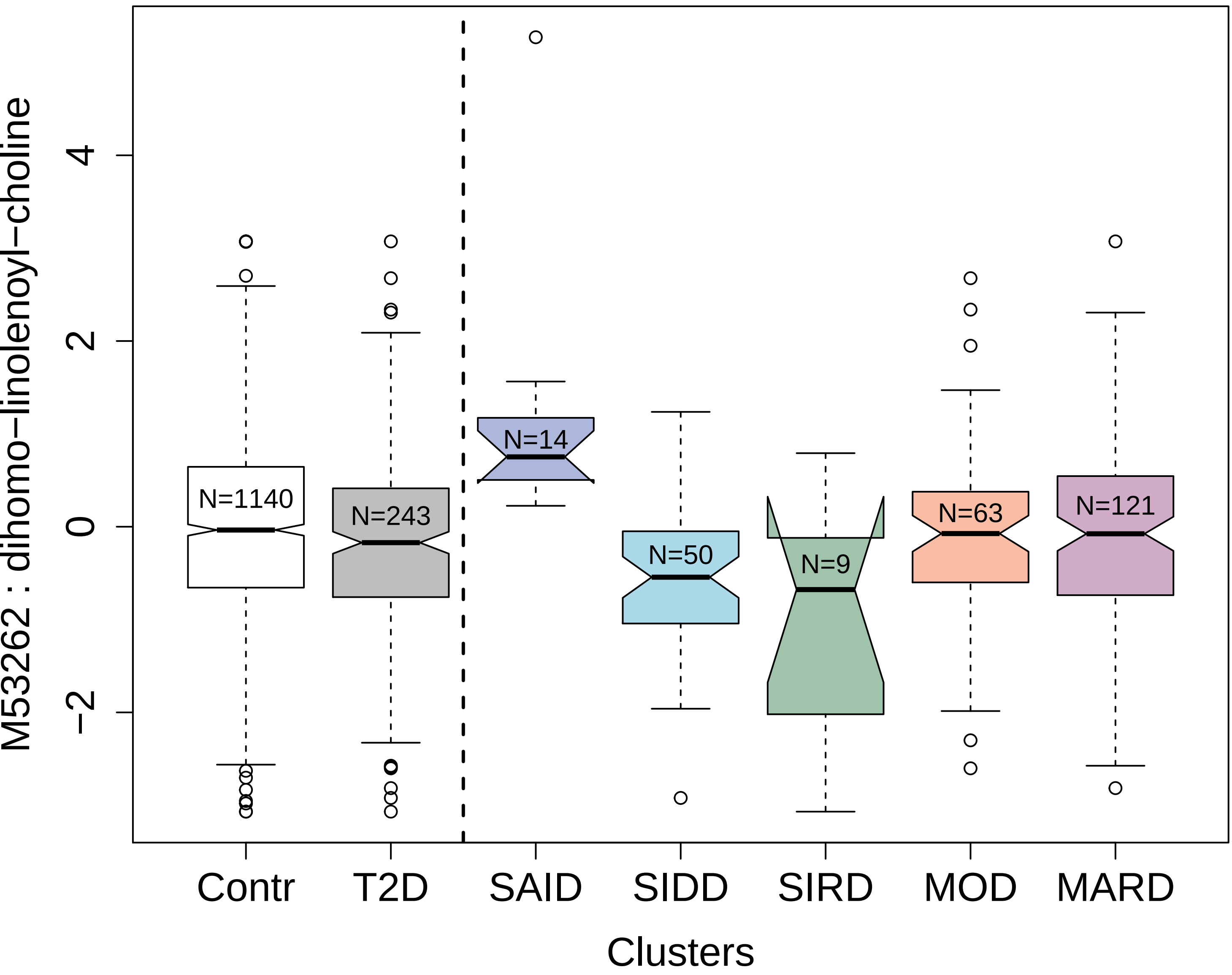
M53262 : dihomo−linolenoyl−choline
4
2
0
−2
N=1140
N=243
N=14
N=50
N=9
N=63
N=121
Contr
T2D
SAID
SIDD
SIRD
MOD
MARD
Clusters

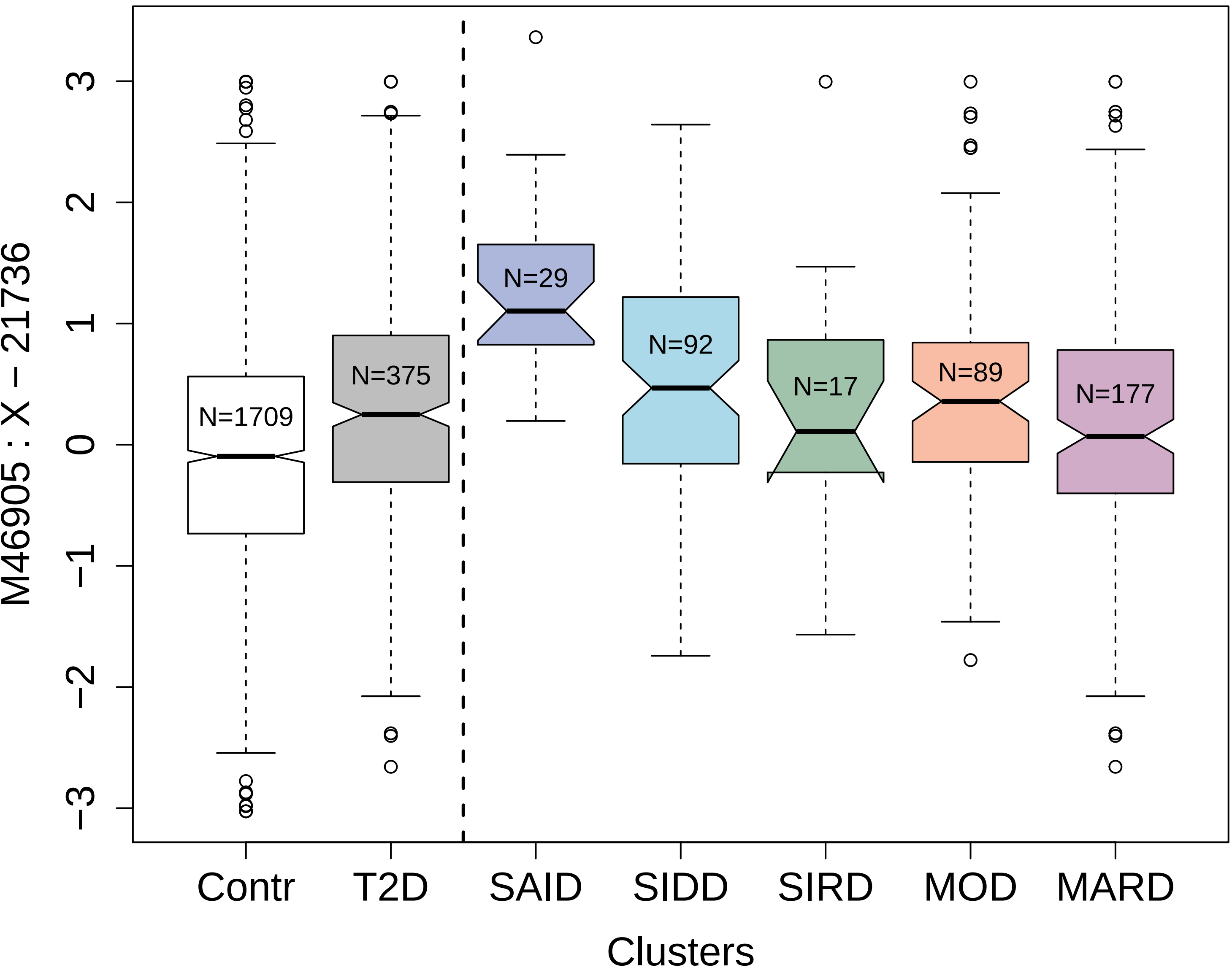

M46905 : X − 21736
3
2
1
0
−1
−2
−3
N=1709
N=375
N=29
N=92
N=17
N=89
N=177
Contr
T2D
SAID
SIDD
SIRD
MOD
MARD
Clusters

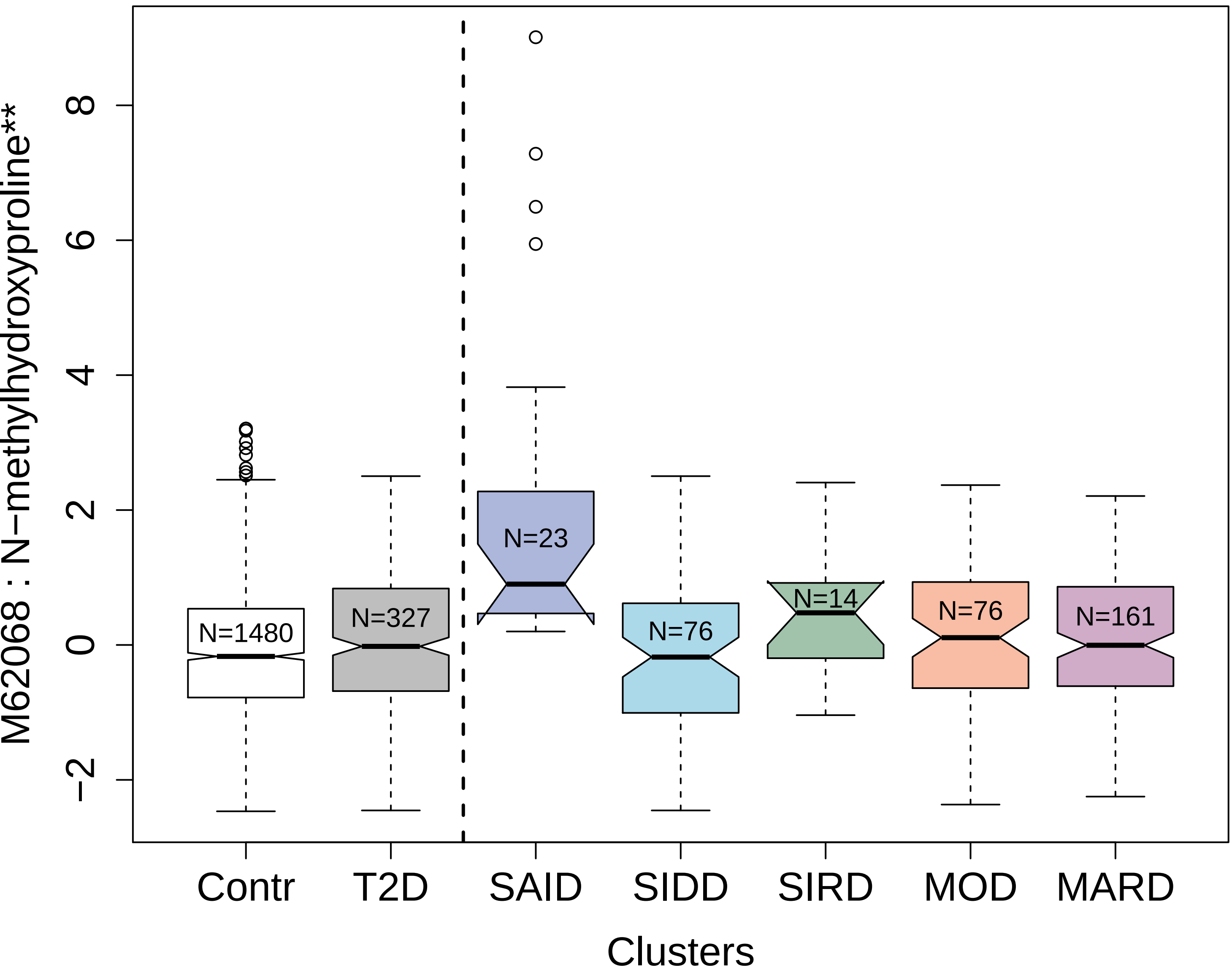

M62068 : N−methylhydroxyproline**
−2
0
2
4
6
8
N=1480
N=327
N=23
N=76
N=14
N=76
N=161
Contr
T2D
SAID
SIDD
SIRD
MOD
MARD
Clusters

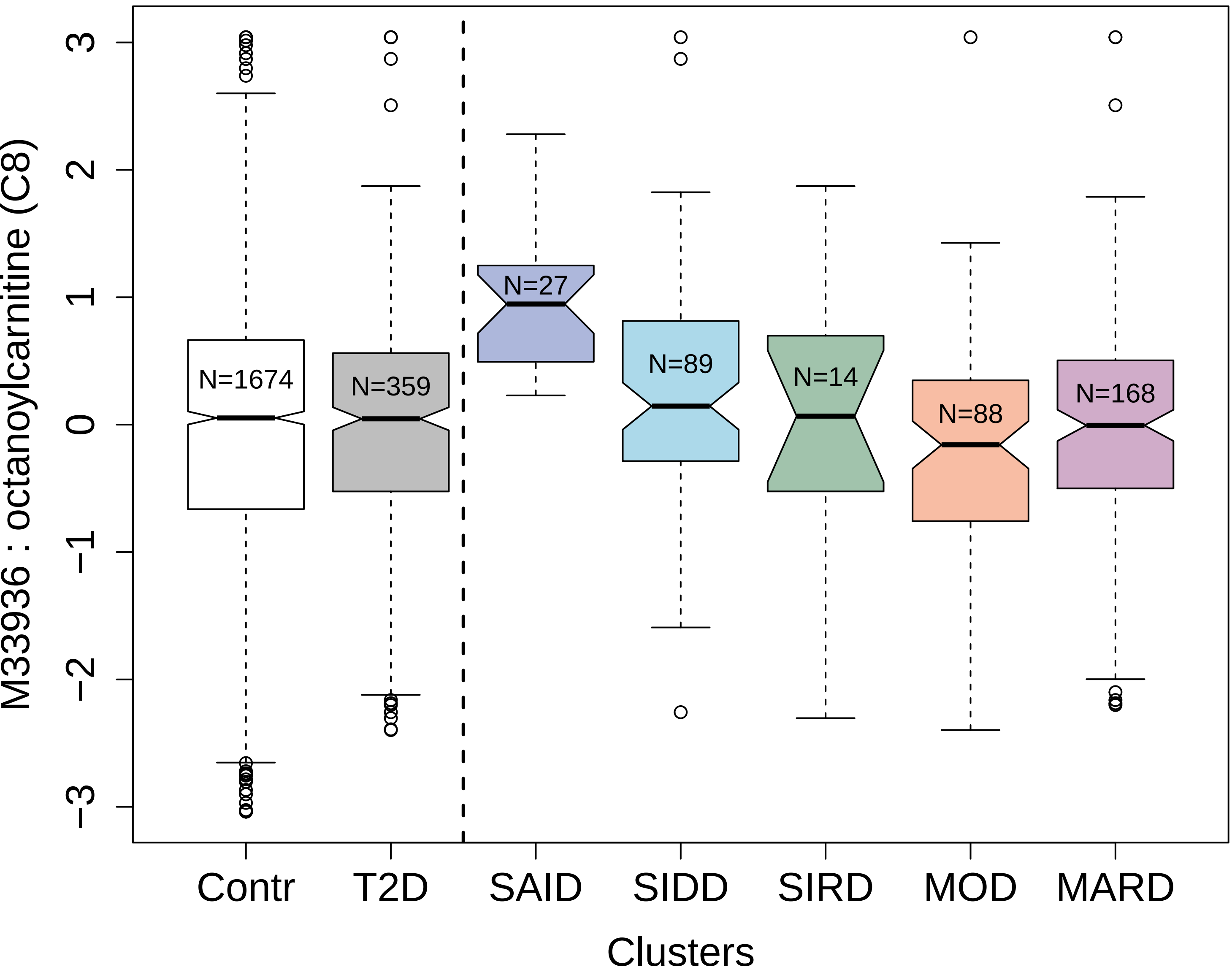

M33936 : octanoylcarnitine (C8)
3
2
1
0
−1
−2
−3
N=1674
N=359
N=27
N=89
N=14
N=88
N=168
Contr
T2D
SAID
SIDD
SIRD
MOD
MARD
Clusters

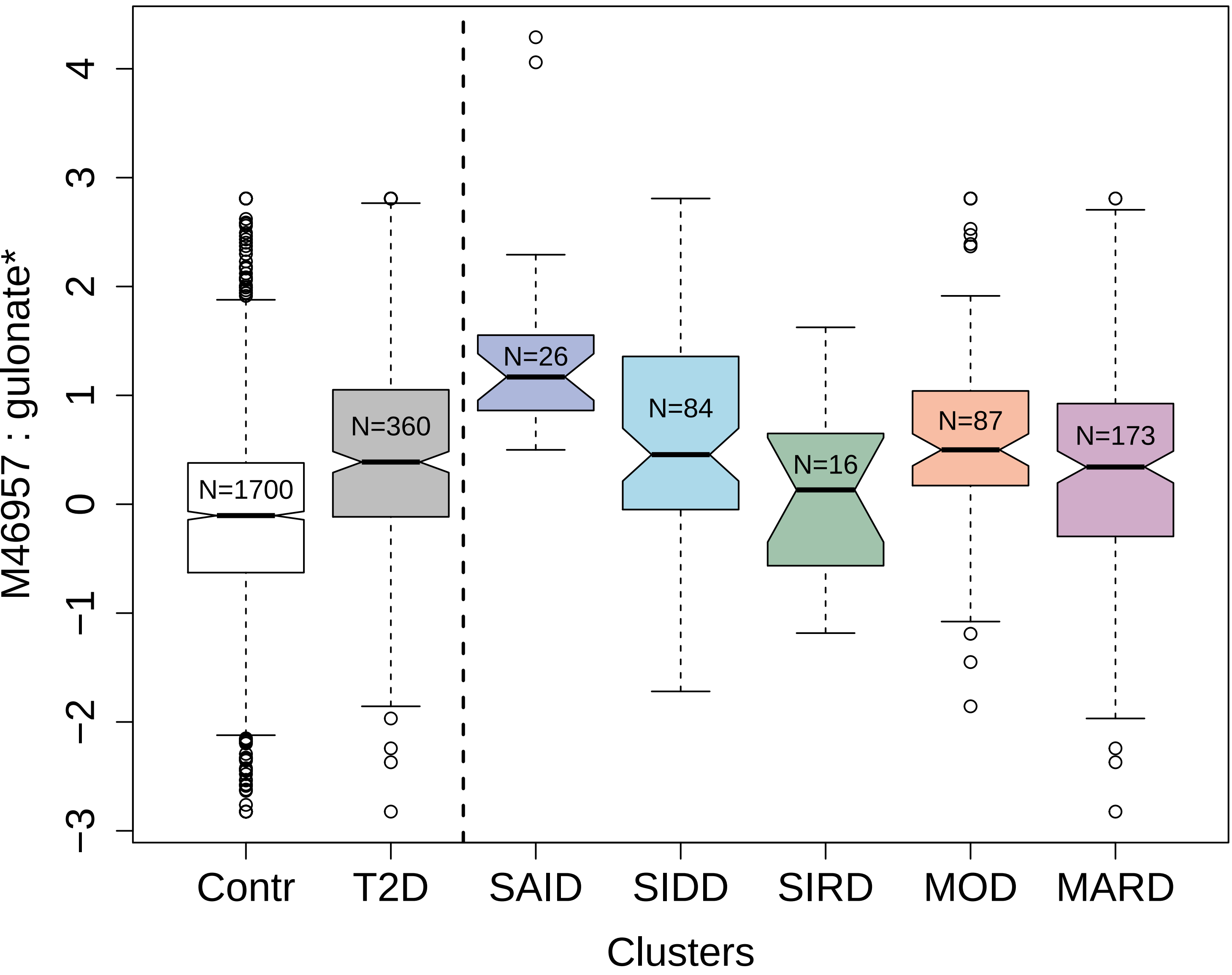

M46957 : gulonate*
4
3
2
1
0
−1
−2
−3
N=1700
N=360
N=26
N=84
N=16
N=87
N=173
Contr
T2D
SAID
SIDD
SIRD
MOD
MARD
Clusters

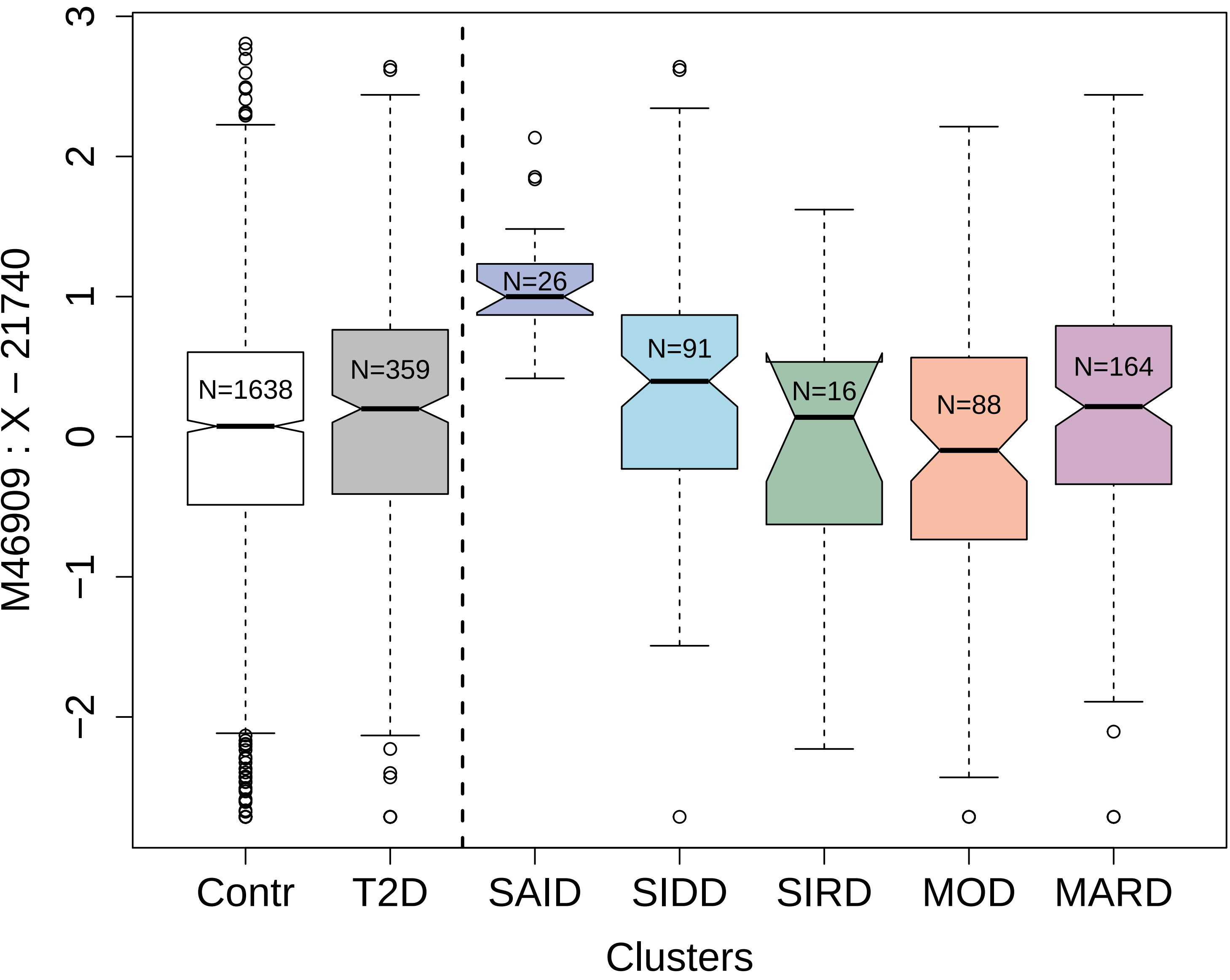
3
2
1
0
−1
−2
M46909 : X − 21740
N=1638
N=359
N=26
N=91
N=16
N=88
N=164
Contr
T2D
SAID
SIDD
SIRD
MOD
MARD
Clusters

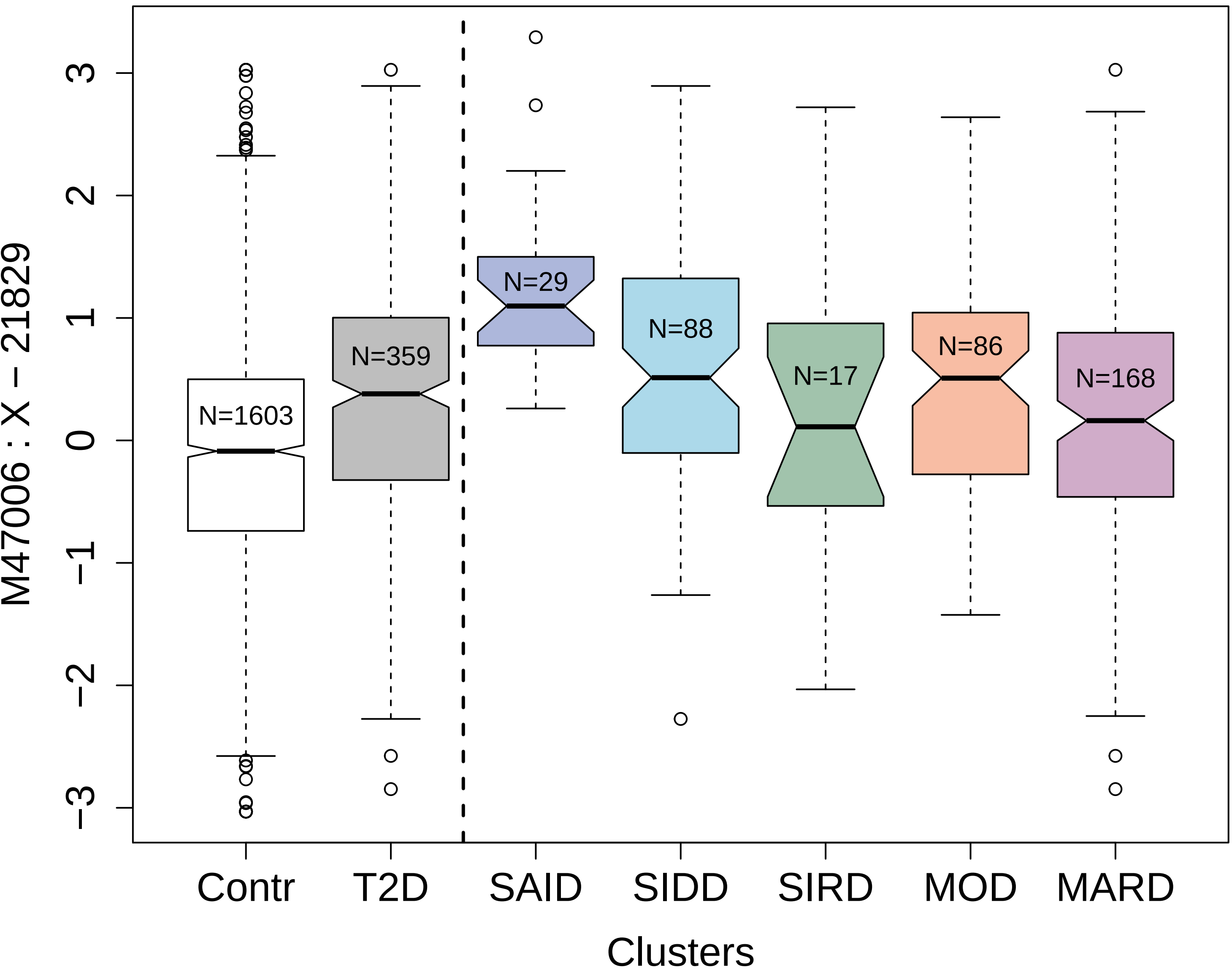

M47006 : X − 21829
3
2
1
0
−1
−2
−3
N=1603
N=359
N=29
N=88
N=17
N=86
N=168
Contr
T2D
SAID
SIDD
SIRD
MOD
MARD
Clusters

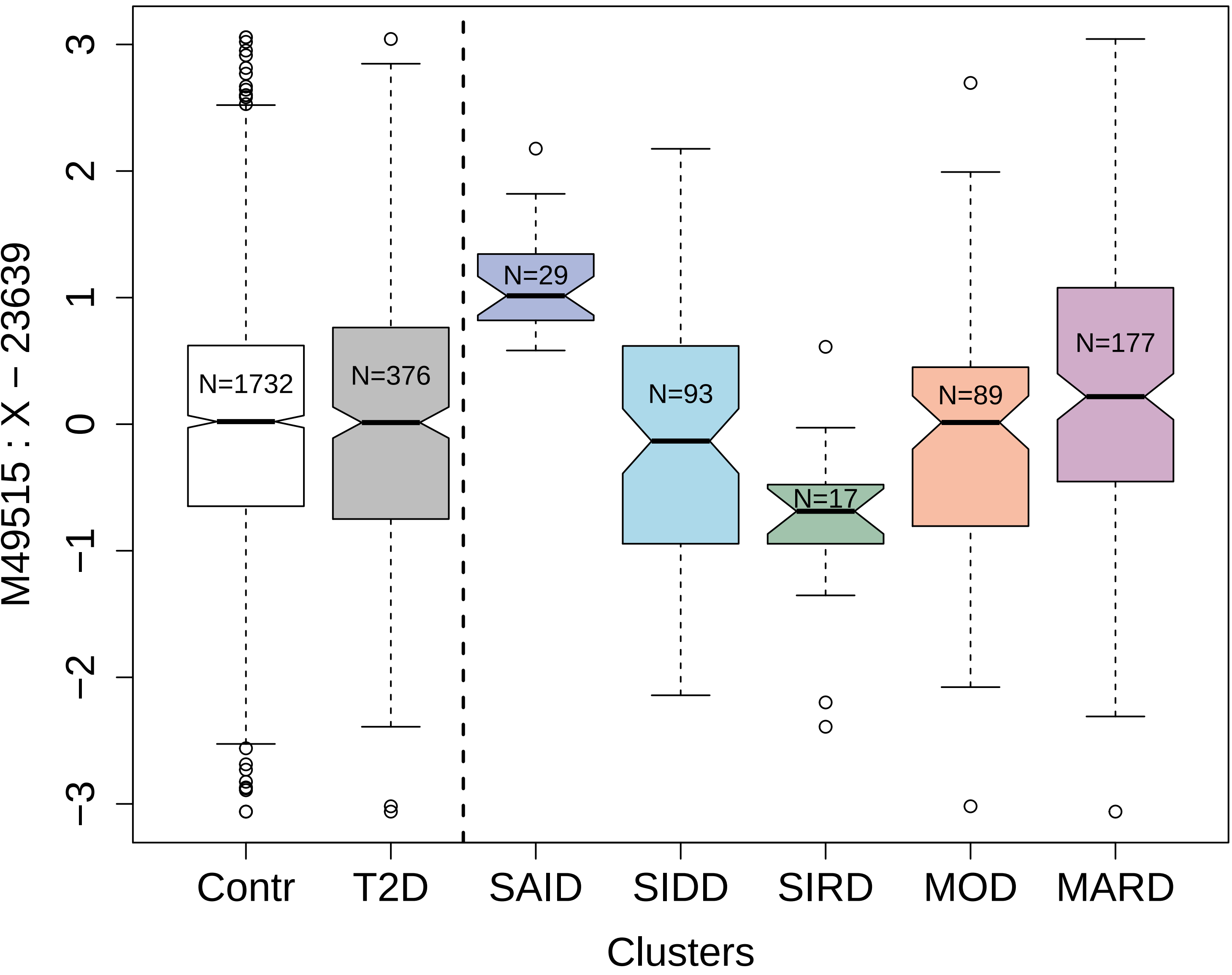
M49515 : X − 23639
3
2
1
0
−1
−2
−3
N=1732
N=376
N=29
N=93
N=17
N=89
N=177
Contr
T2D
SAID
SIDD
SIRD
MOD
MARD
Clusters

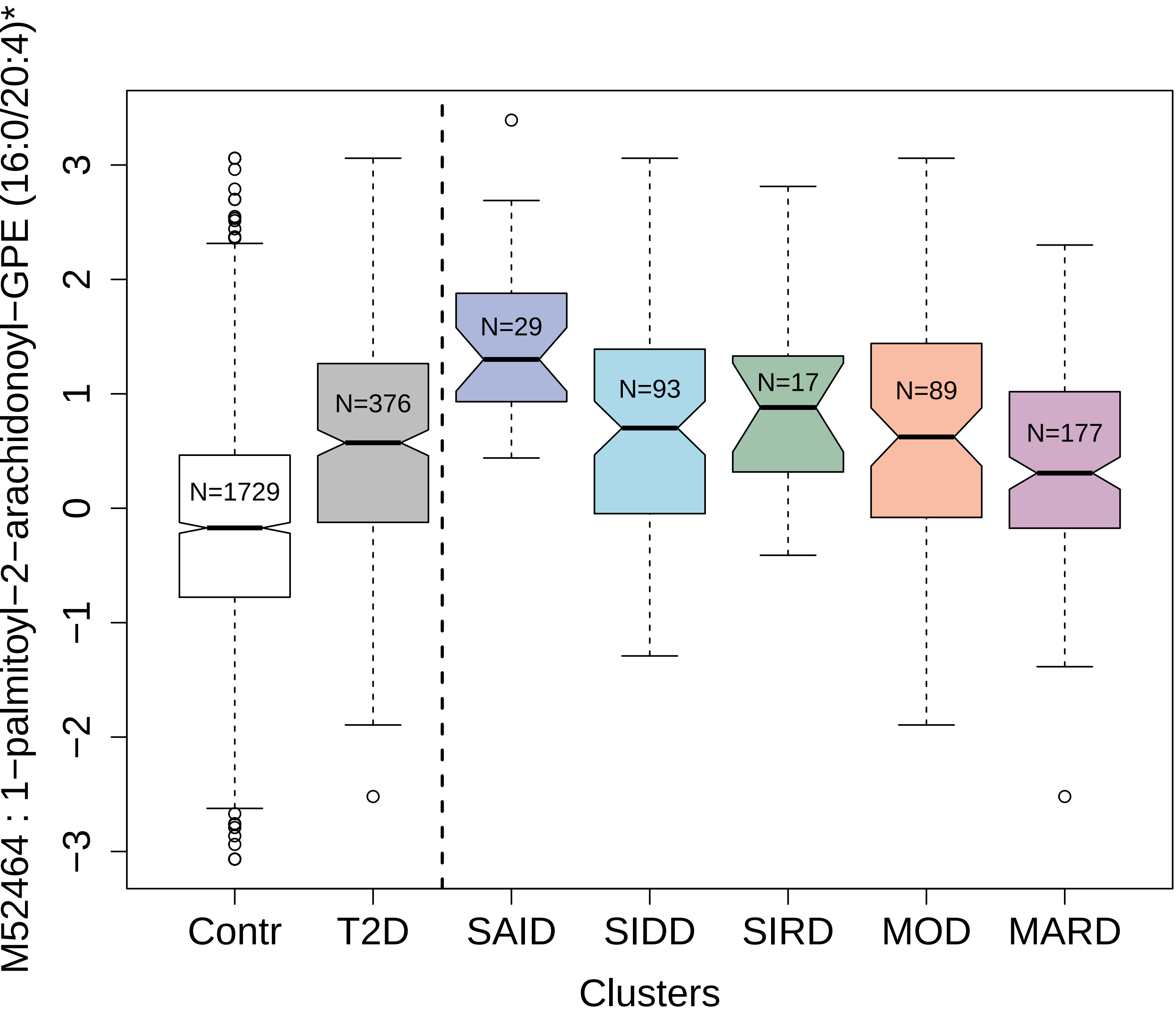
M52464 : 1−palmitoyl−2−arachidonoyl−GPE (16:0/20:4)*
3
2
1
0
−1
−2
−3
N=1729
N=376
N=29
N=93
N=17
N=89
N=177
Contr
T2D
SAID
SIDD
SIRD
MOD
MARD
Clusters

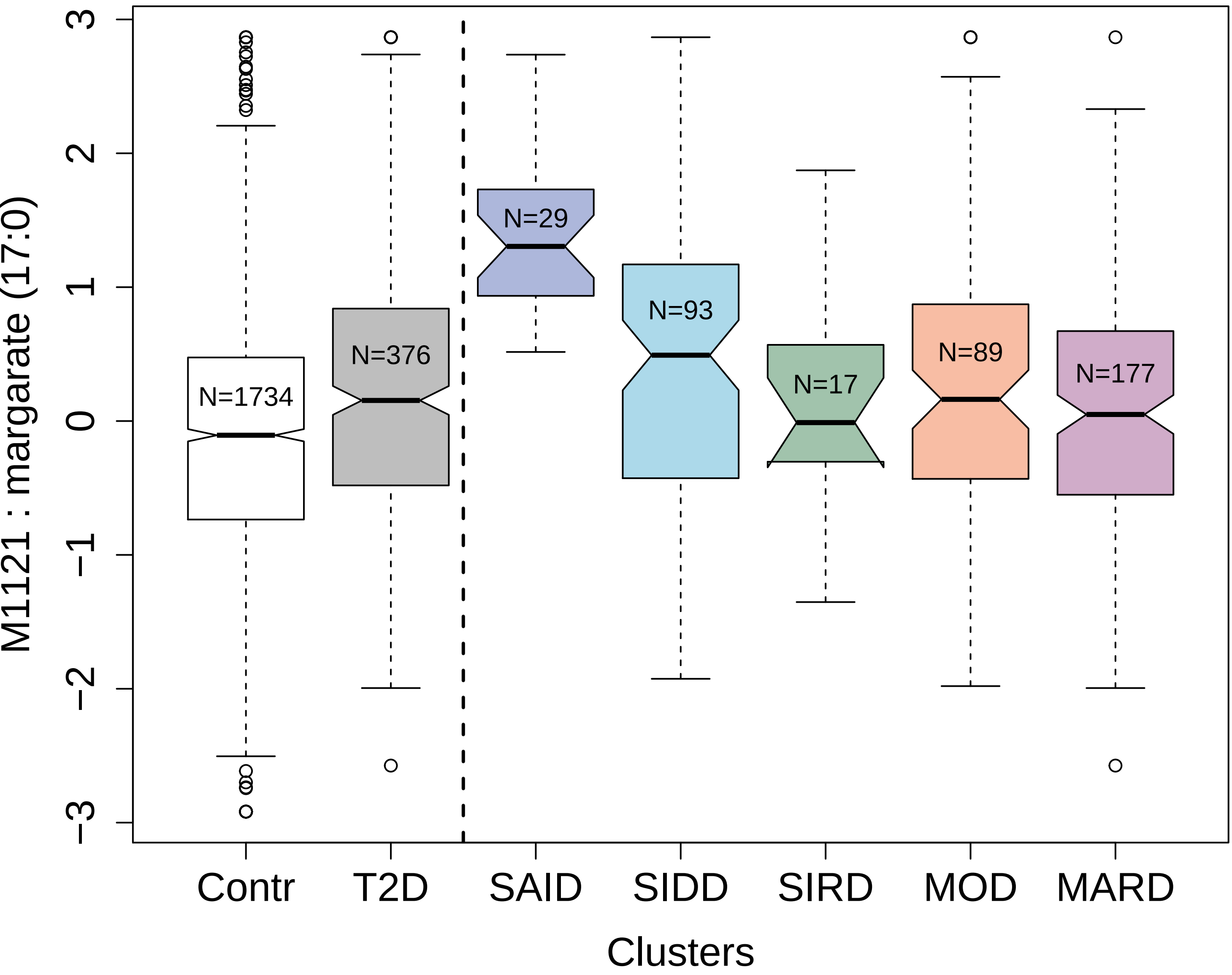
M1121 : margarate (17:0)
3
2
1
0
−1
−2
−3
N=1734
N=376
N=29
N=93
N=17
N=89
N=177
Contr
T2D
SAID
SIDD
SIRD
MOD
MARD
Clusters

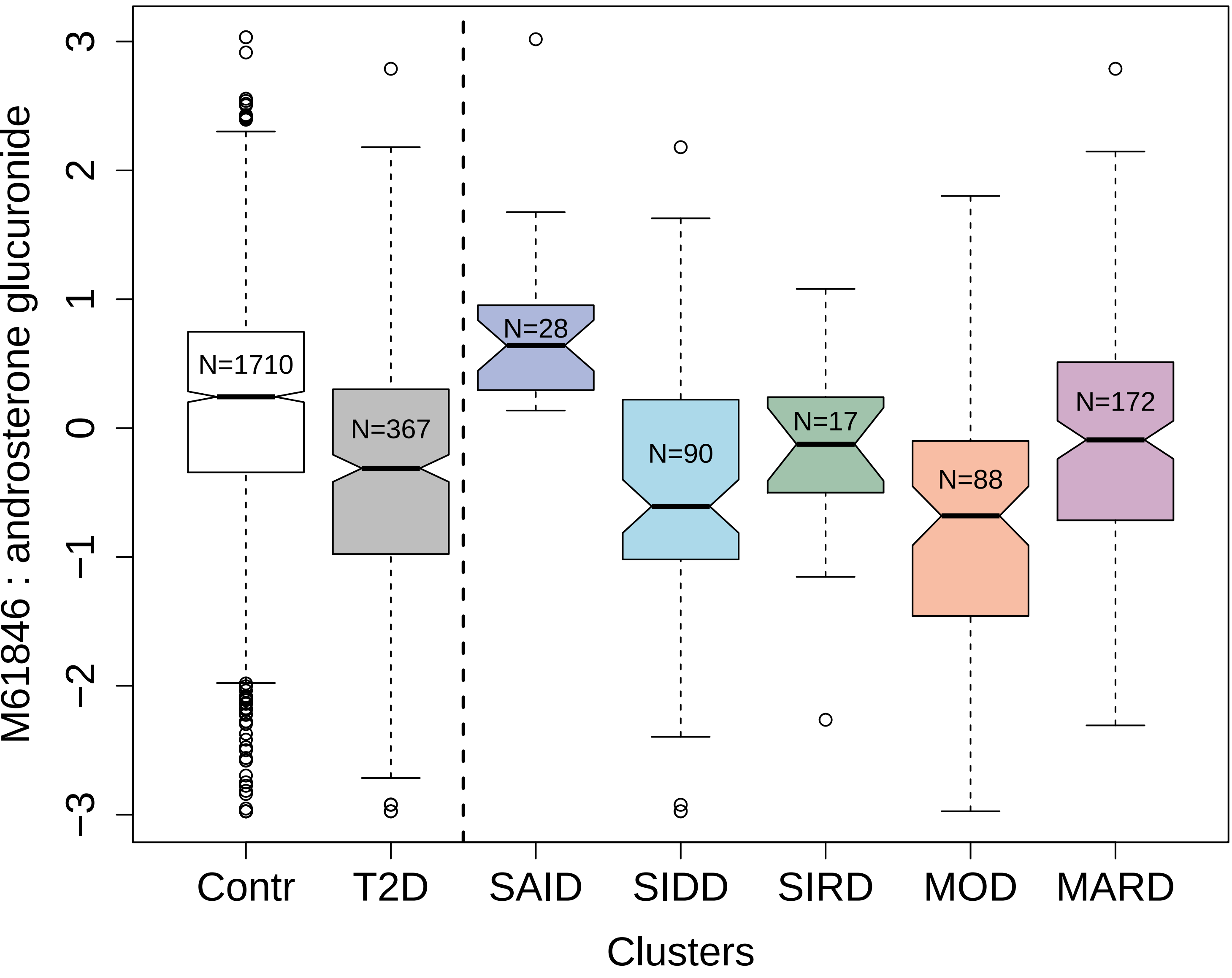
M61846 : androsterone glucuronide
3
2
1
0
−1
−2
−3
N=1710
N=367
N=28
N=90
N=17
N=88
N=172
Contr
T2D
SAID
SIDD
SIRD
MOD
MARD
Clusters

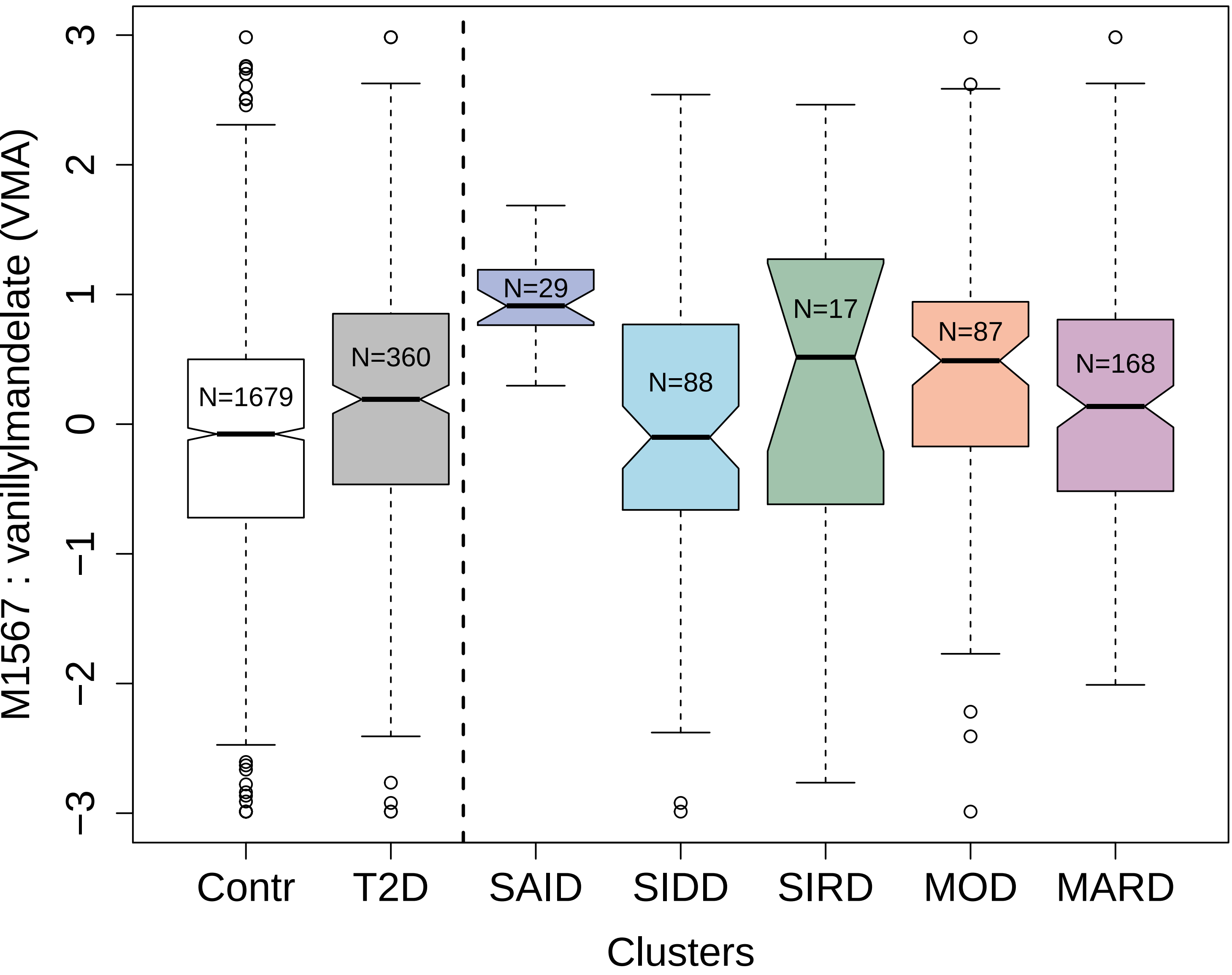
M1567 : vanillylmandelate (VMA)
3
2
1
0
−1
−2
−3
N=1679
N=360
N=29
N=88
N=17
N=87
N=168
Contr
T2D
SAID
SIDD
SIRD
MOD
MARD
Clusters

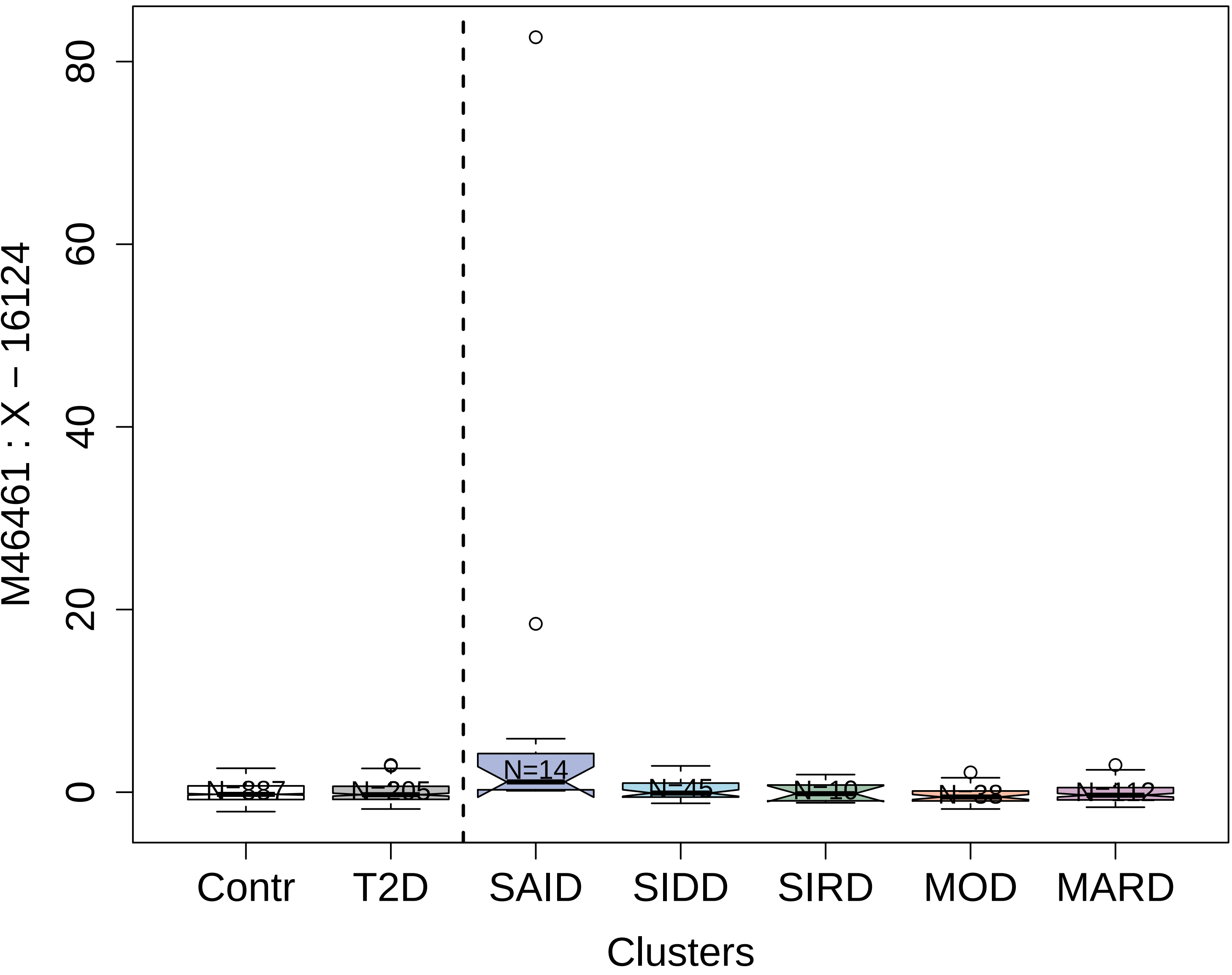

M46461 : X − 16124
0
20
40
60
80
N=887
N=205
N=14
N=45
N=10
N=38
N=112
Contr
T2D
SAID
SIDD
SIRD
MOD
MARD
Clusters

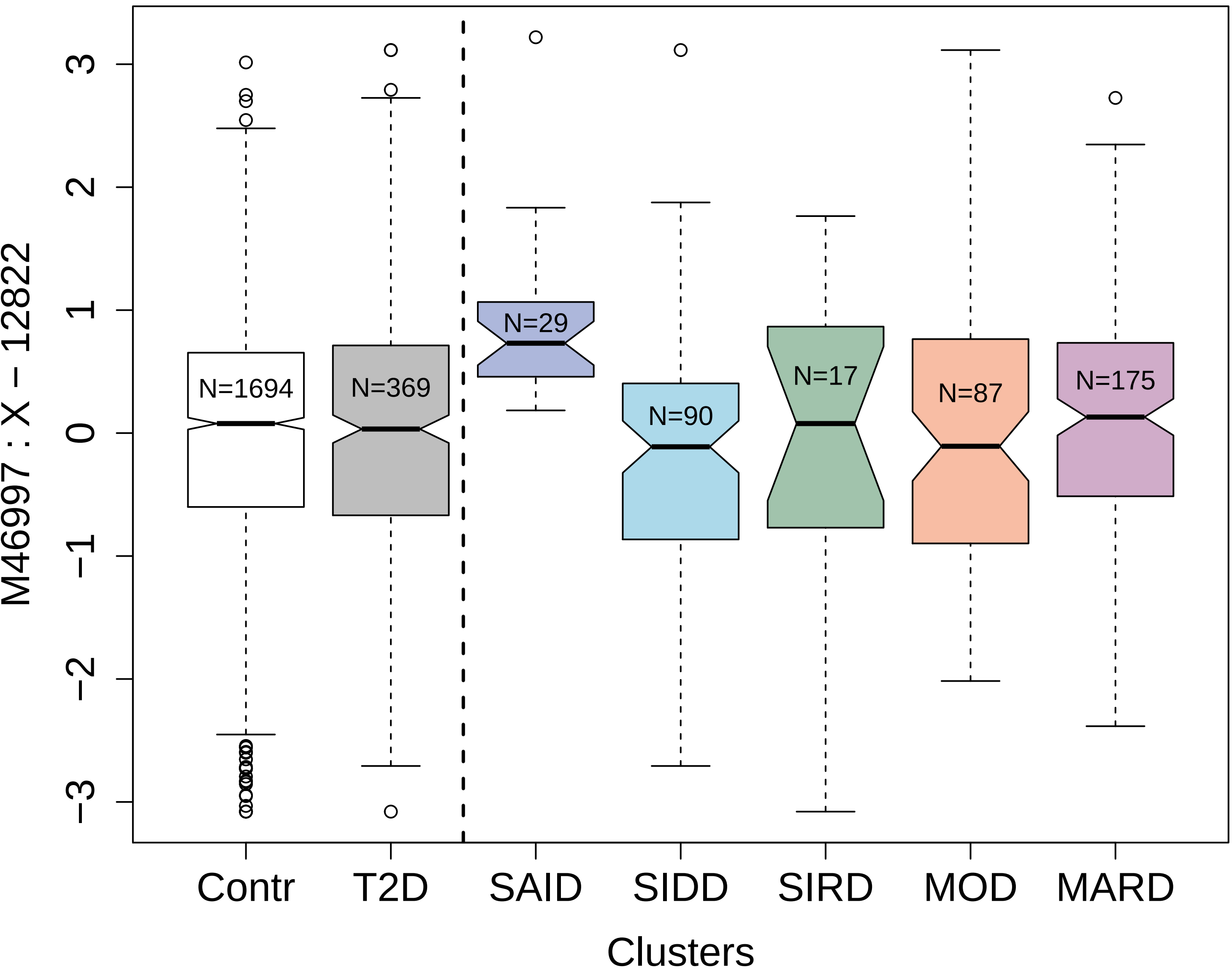

M46997 : X − 12822
3
2
1
0
−1
−2
−3
N=1694
N=369
N=29
N=90
N=17
N=87
N=175
Contr
T2D
SAID
SIDD
SIRD
MOD
MARD
Clusters

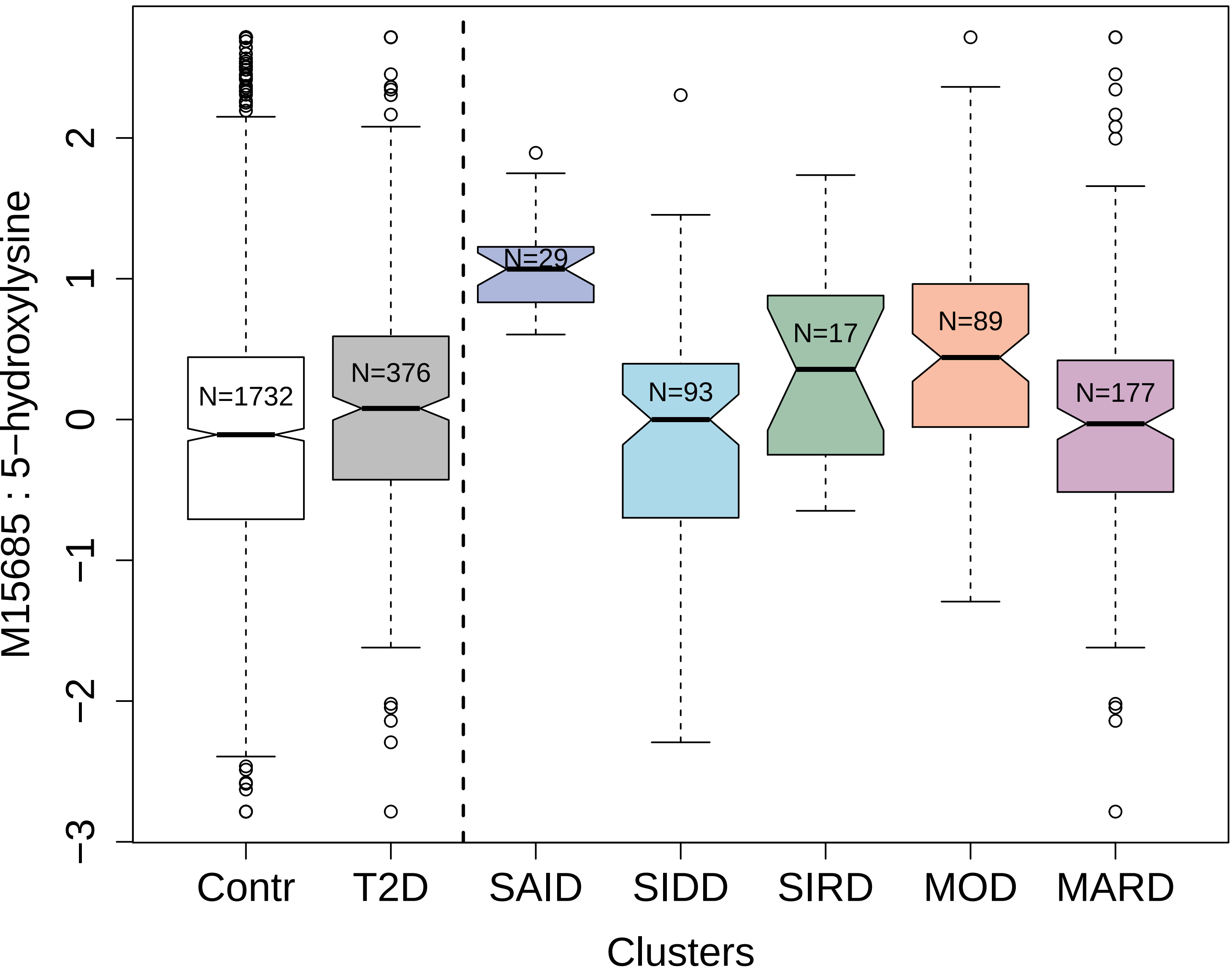

M15685 : 5−hydroxylysine
2
1
0
−1
−2
−3
N=1732
N=376
N=29
N=93
N=17
N=89
N=177
Contr
T2D
SAID
SIDD
SIRD
MOD
MARD
Clusters

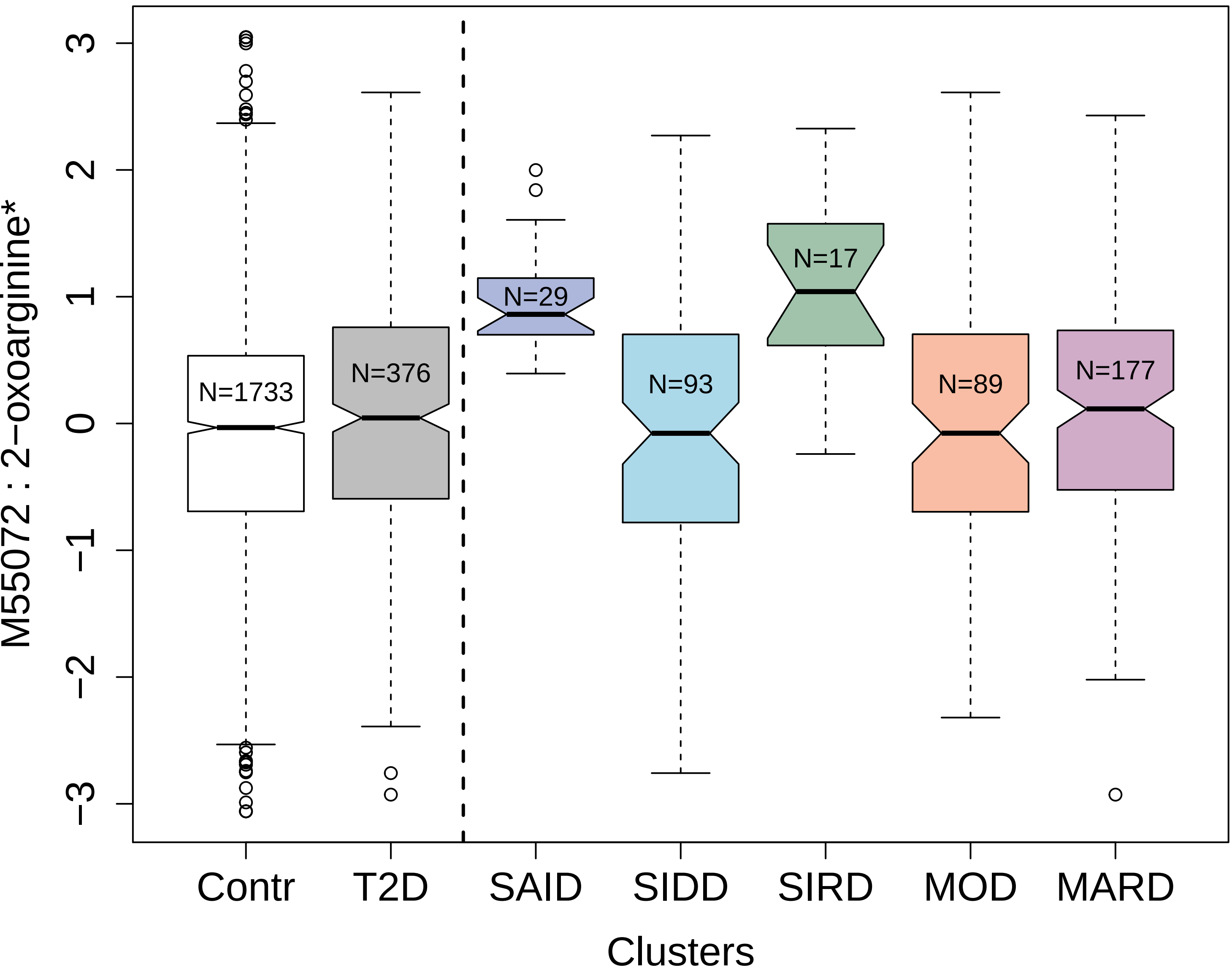
M55072 : 2−oxoarginine*
Clusters
Contr
T2D
SAID
SIDD
SIRD
MOD
MARD
N=1733
N=376
N=29
N=93
N=17
N=89
N=177
3
2
1
0
−1
−2
−3

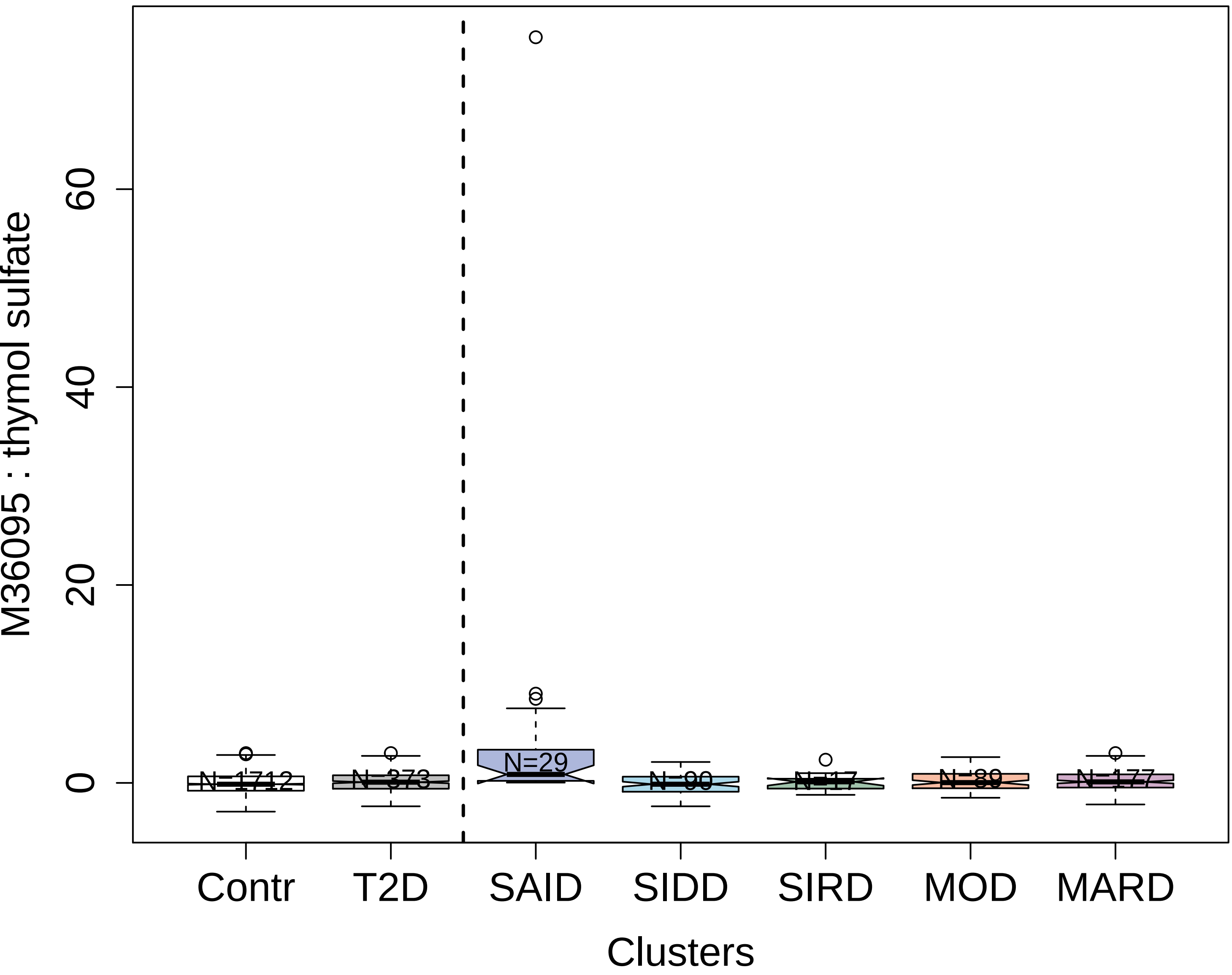
M36095 : thymol sulfate
0
20
40
60
N=1712
N=373
N=29
N=90
N=17
N=89
N=177
Contr
T2D
SAID
SIDD
SIRD
MOD
MARD
Clusters

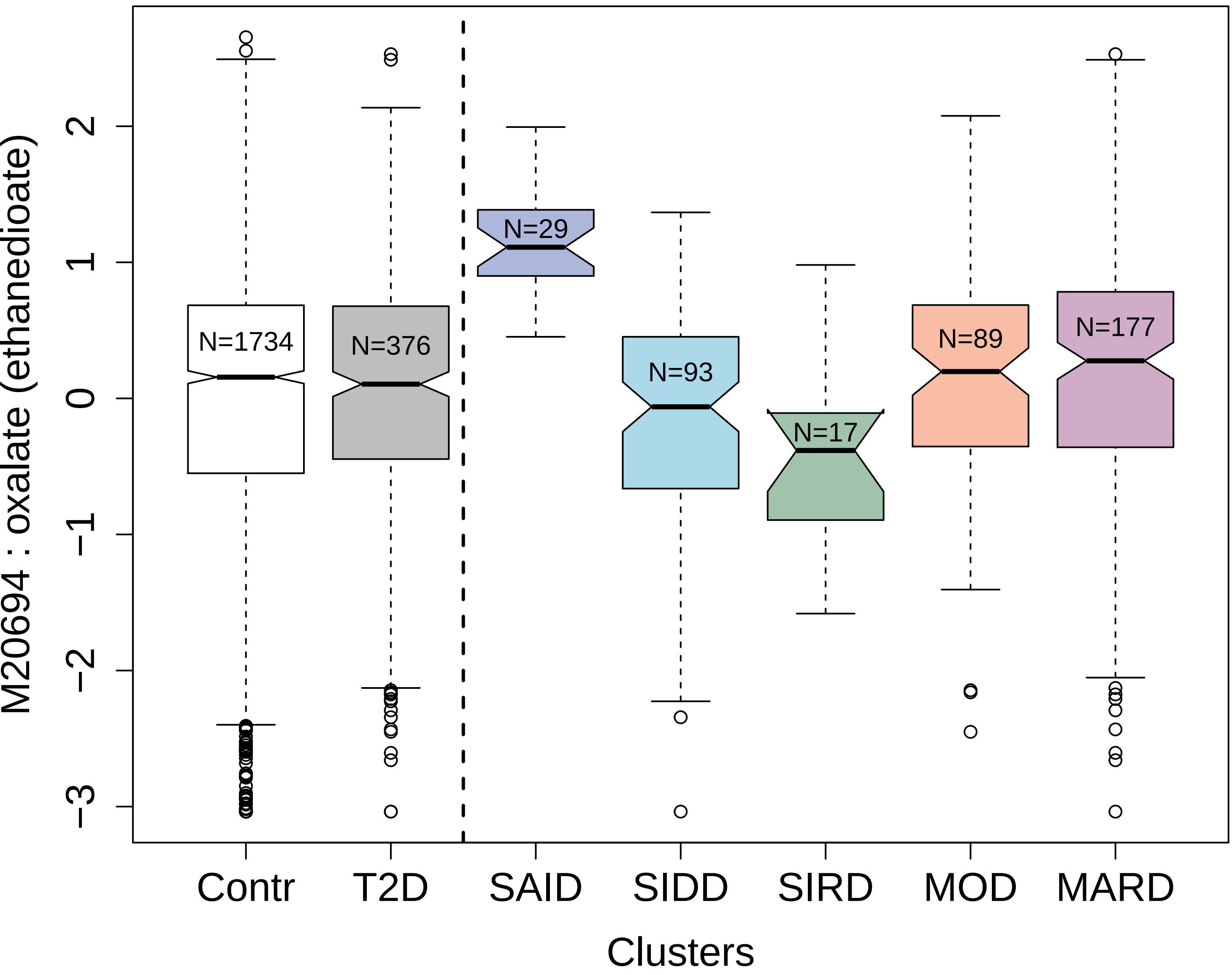

M20694 : oxalate (ethanedioate)
2
1
0
−1
−2
−3
N=1734
N=376
N=29
N=93
N=17
N=89
N=177
Contr
T2D
SAID
SIDD
SIRD
MOD
MARD
Clusters

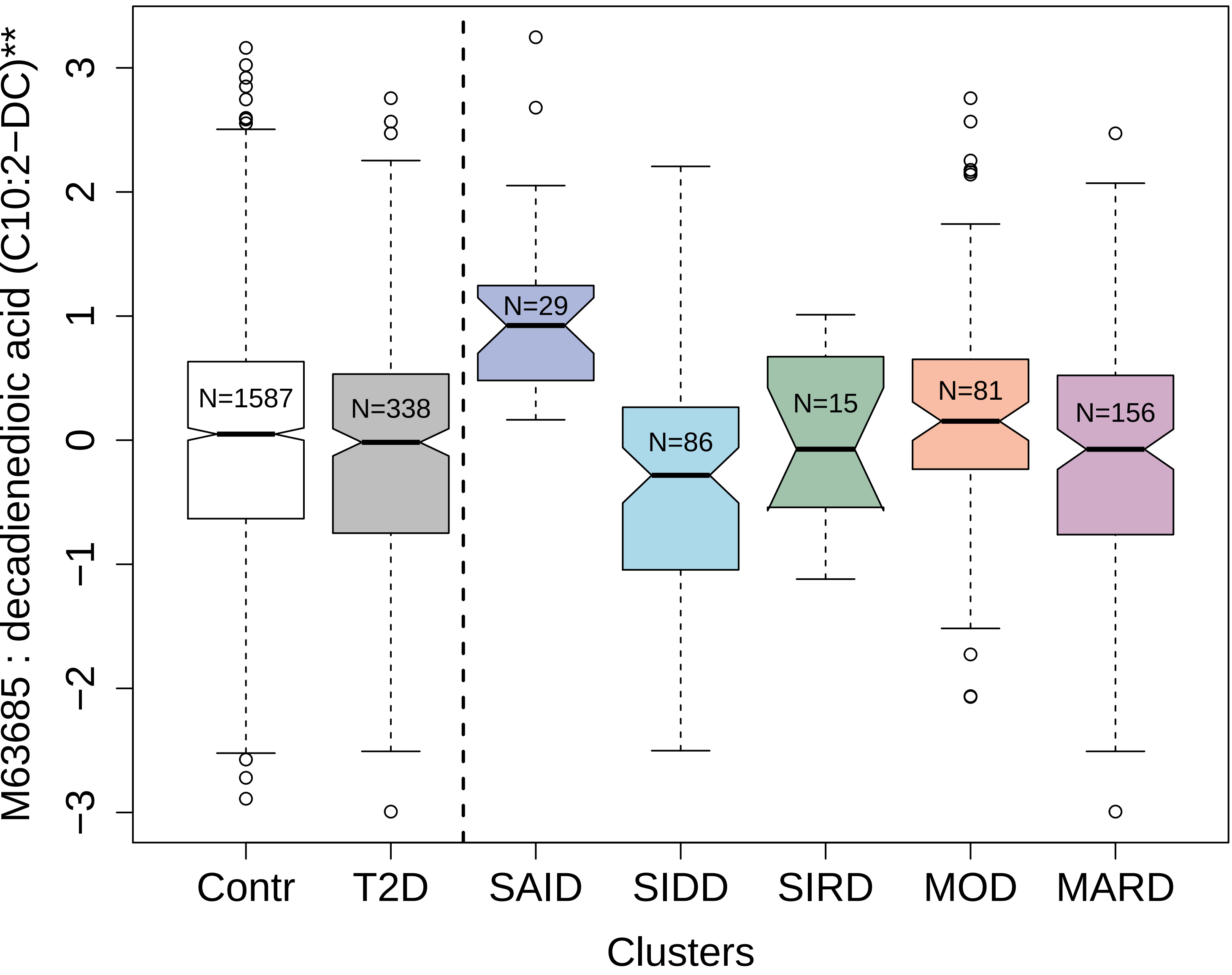

M63685 : decadienedioic acid (C10:2−DC)**
3
2
1
0
−1
−2
−3
N=1587
N=338
N=29
N=86
N=15
N=81
N=156
Contr
T2D
SAID
SIDD
SIRD
MOD
MARD
Clusters

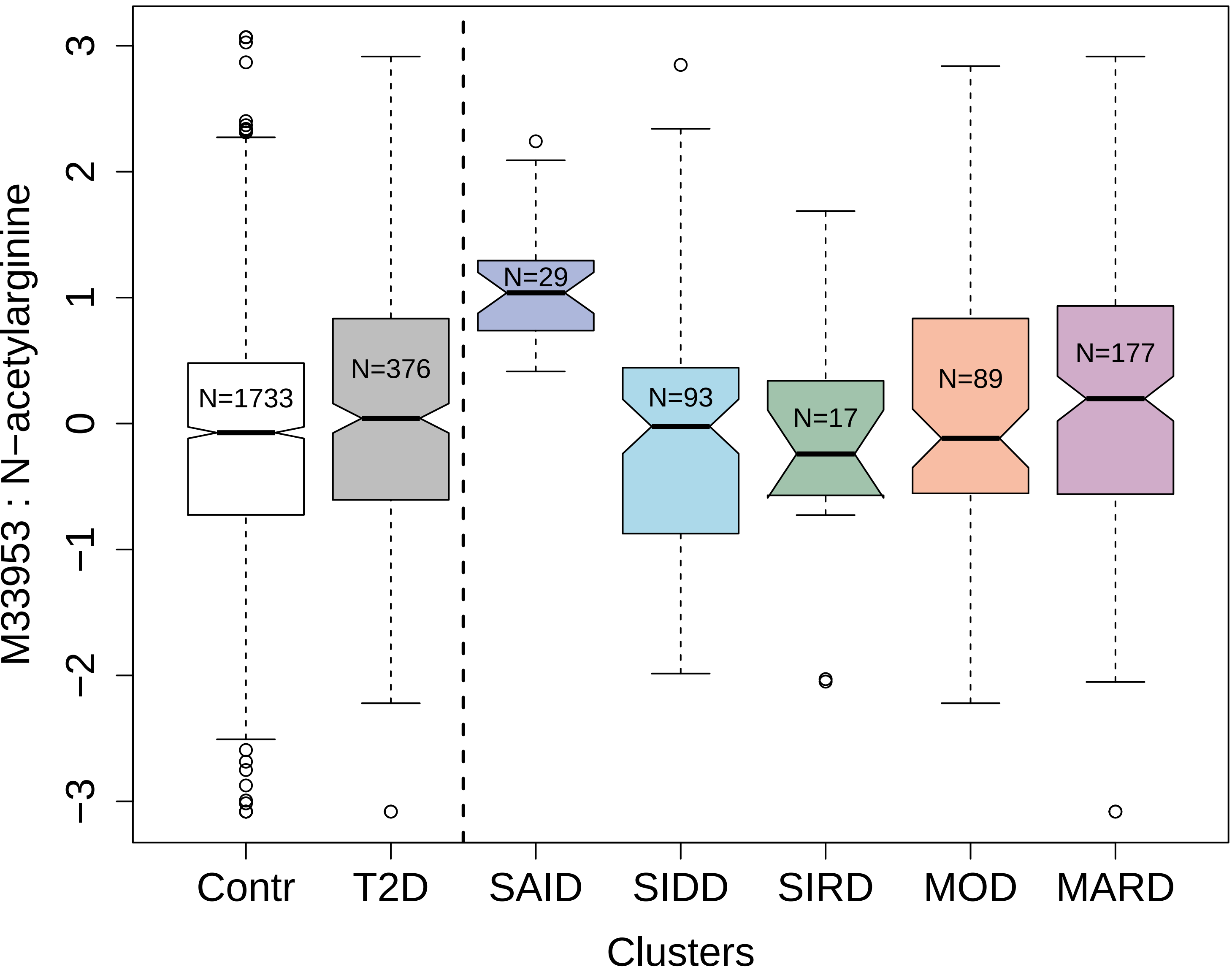

M33953 : N−acetylarginine
3
2
1
0
−1
−2
−3
N=1733
N=376
N=29
N=93
N=17
N=89
N=177
Contr
T2D
SAID
SIDD
SIRD
MOD
MARD
Clusters

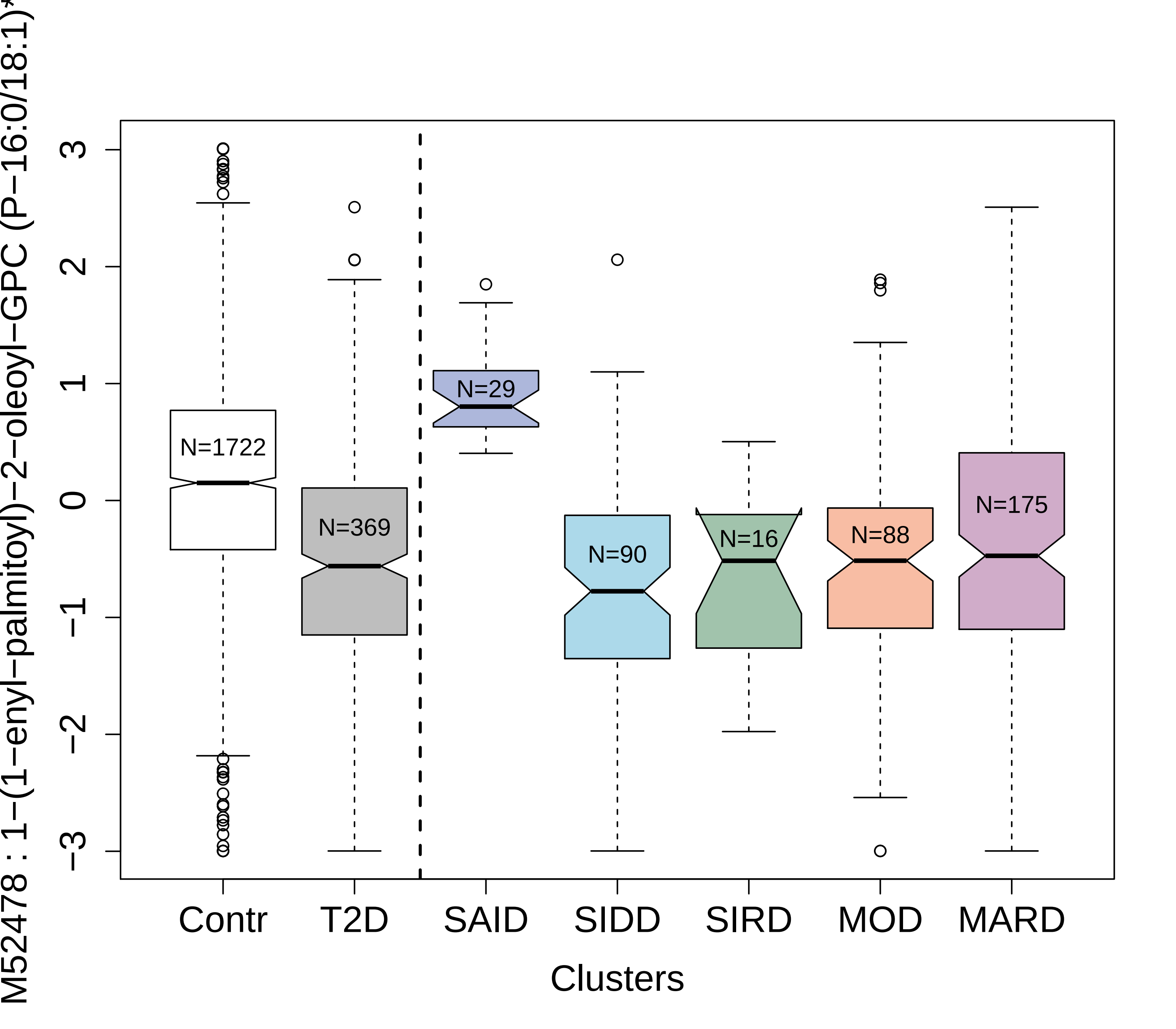
M52478 : 1−(1−enyl−palmitoyl)−2−oleoyl−GPC (P−16:0/18:1)*
3
2
1
0
−1
−2
−3
N=1722
N=369
N=29
N=90
N=16
N=88
N=175
Contr
T2D
SAID
SIDD
SIRD
MOD
MARD
Clusters

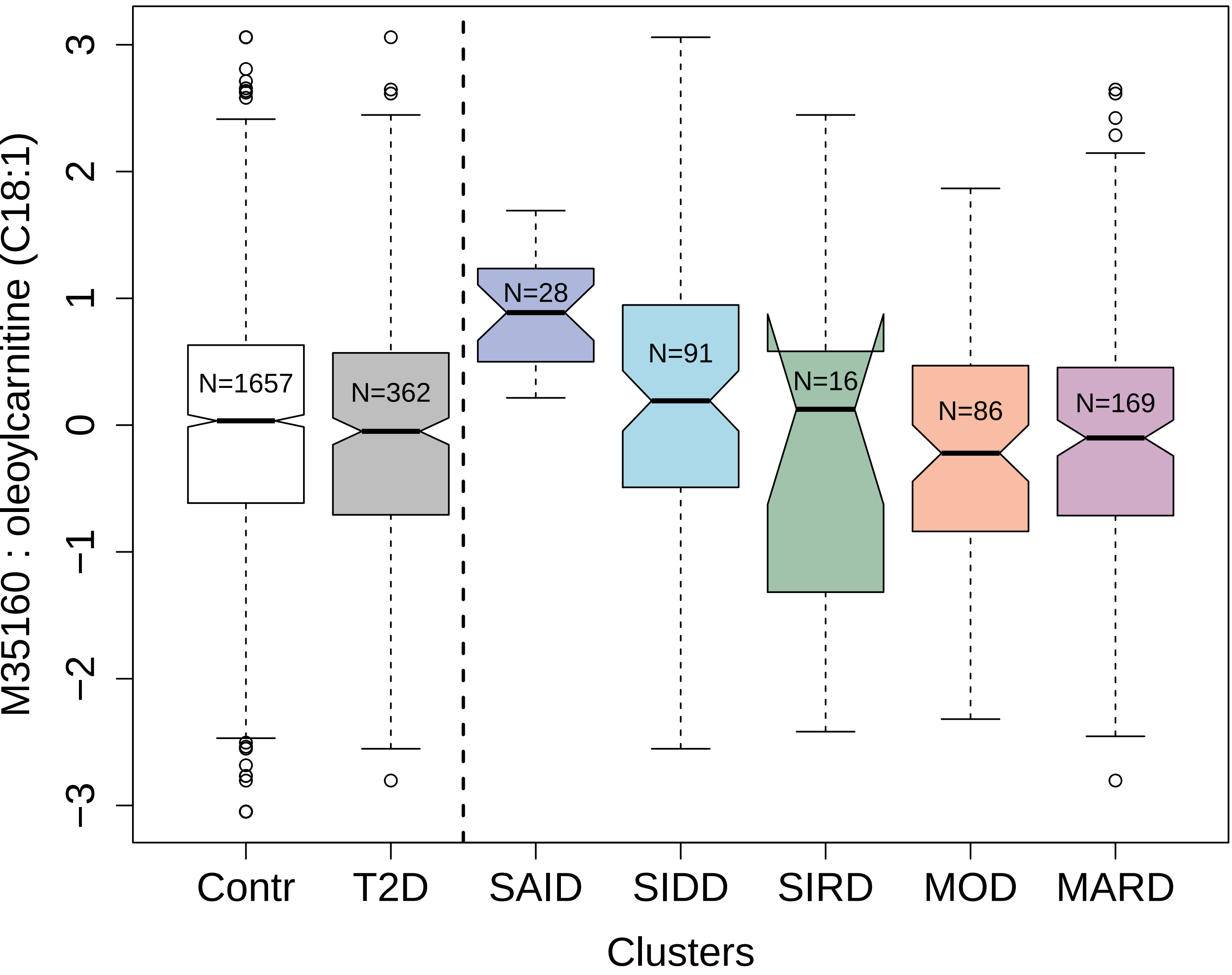

M35160 : oleoylcarnitine (C18:1)
3
2
1
0
−1
−2
−3
N=1657
N=362
N=28
N=91
N=16
N=86
N=169
Contr
T2D
SAID
SIDD
SIRD
MOD
MARD
Clusters

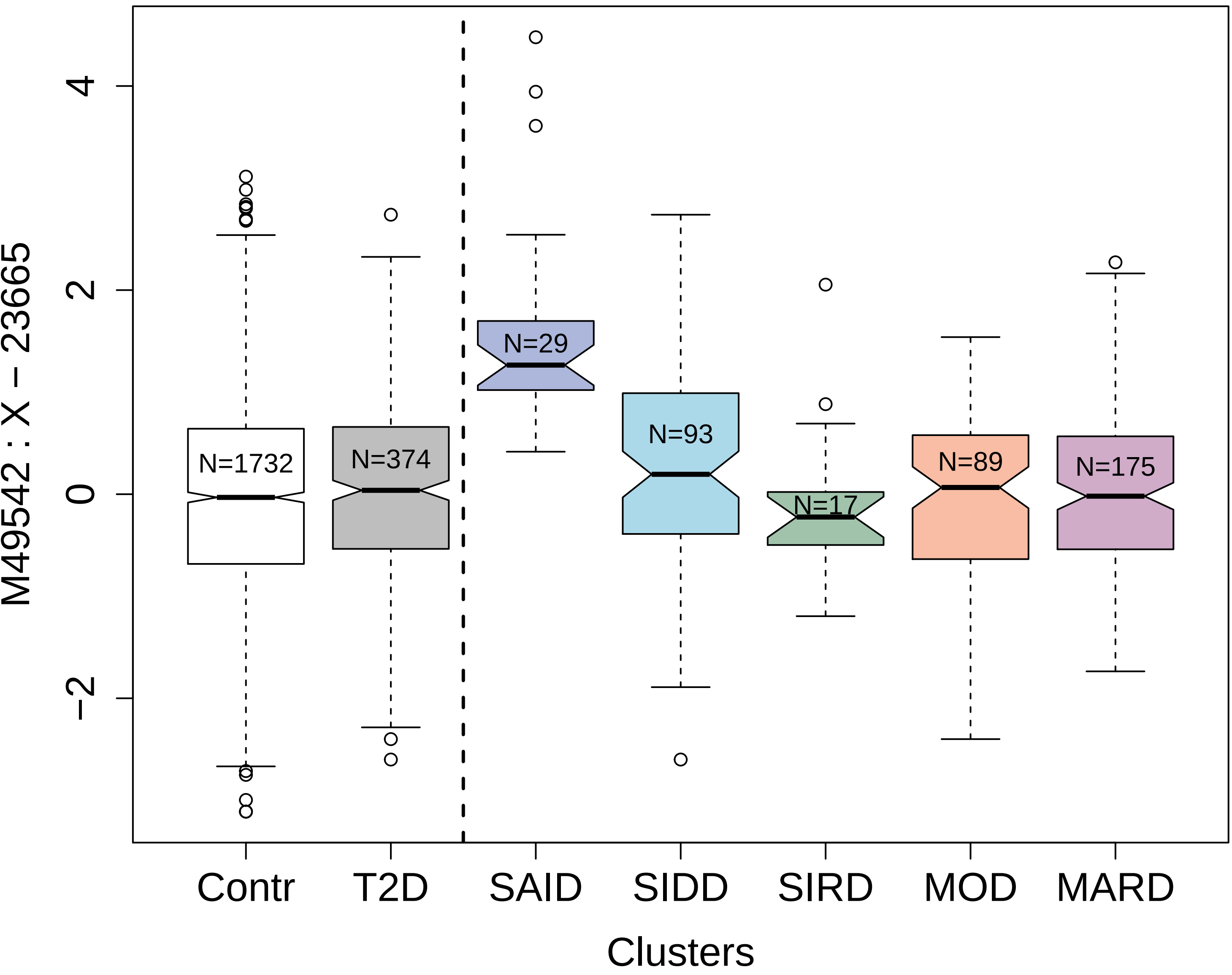

M49542 : X − 23665
4
2
0
−2
N=1732
N=374
N=29
N=93
N=17
N=89
N=175
Contr
T2D
SAID
SIDD
SIRD
MOD
MARD
Clusters

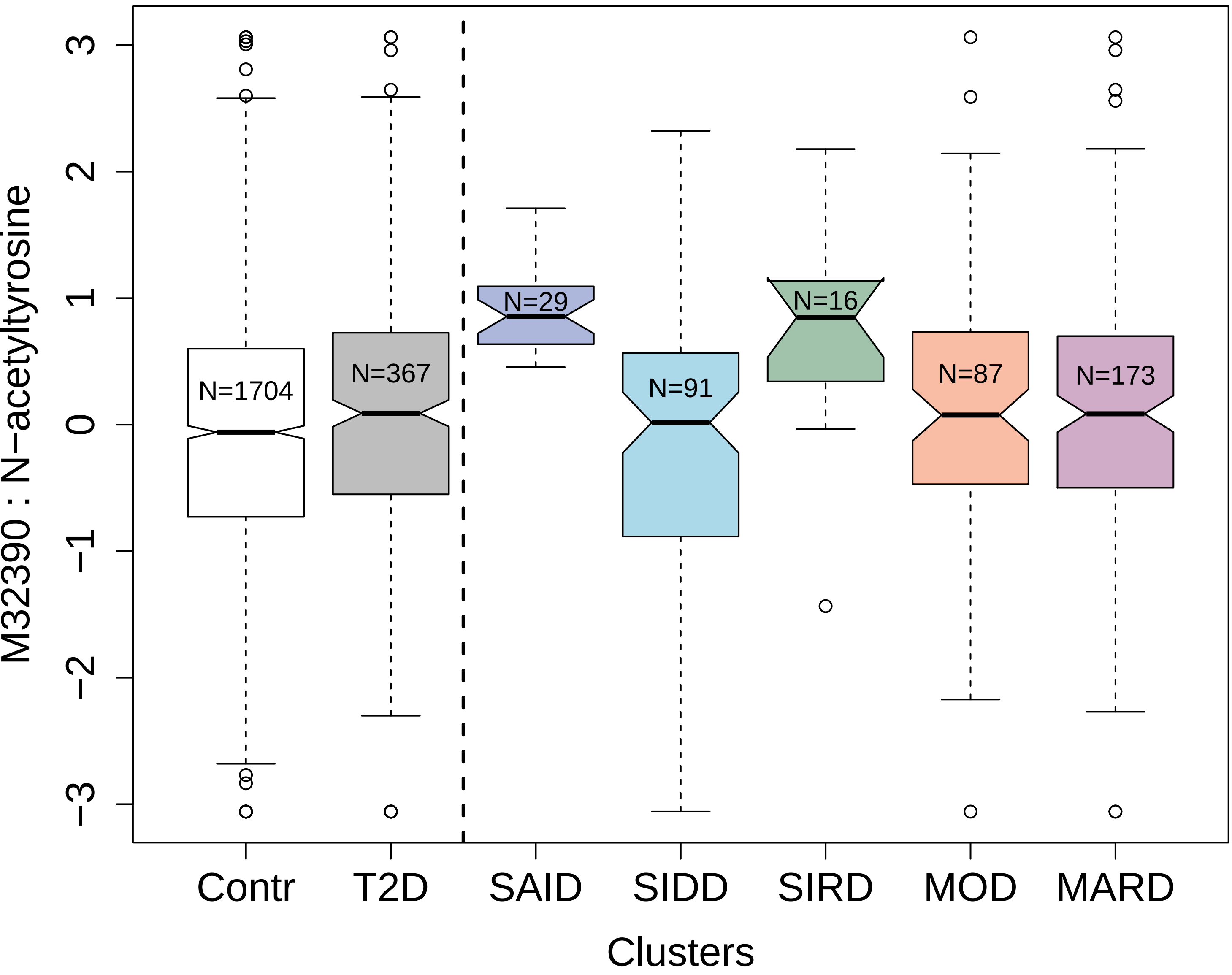
M32390 : N−acetyltyrosine
3
2
1
0
−1
−2
−3
N=1704
N=367
N=29
N=91
N=16
N=87
N=173
Contr
T2D
SAID
SIDD
SIRD
MOD
MARD
Clusters

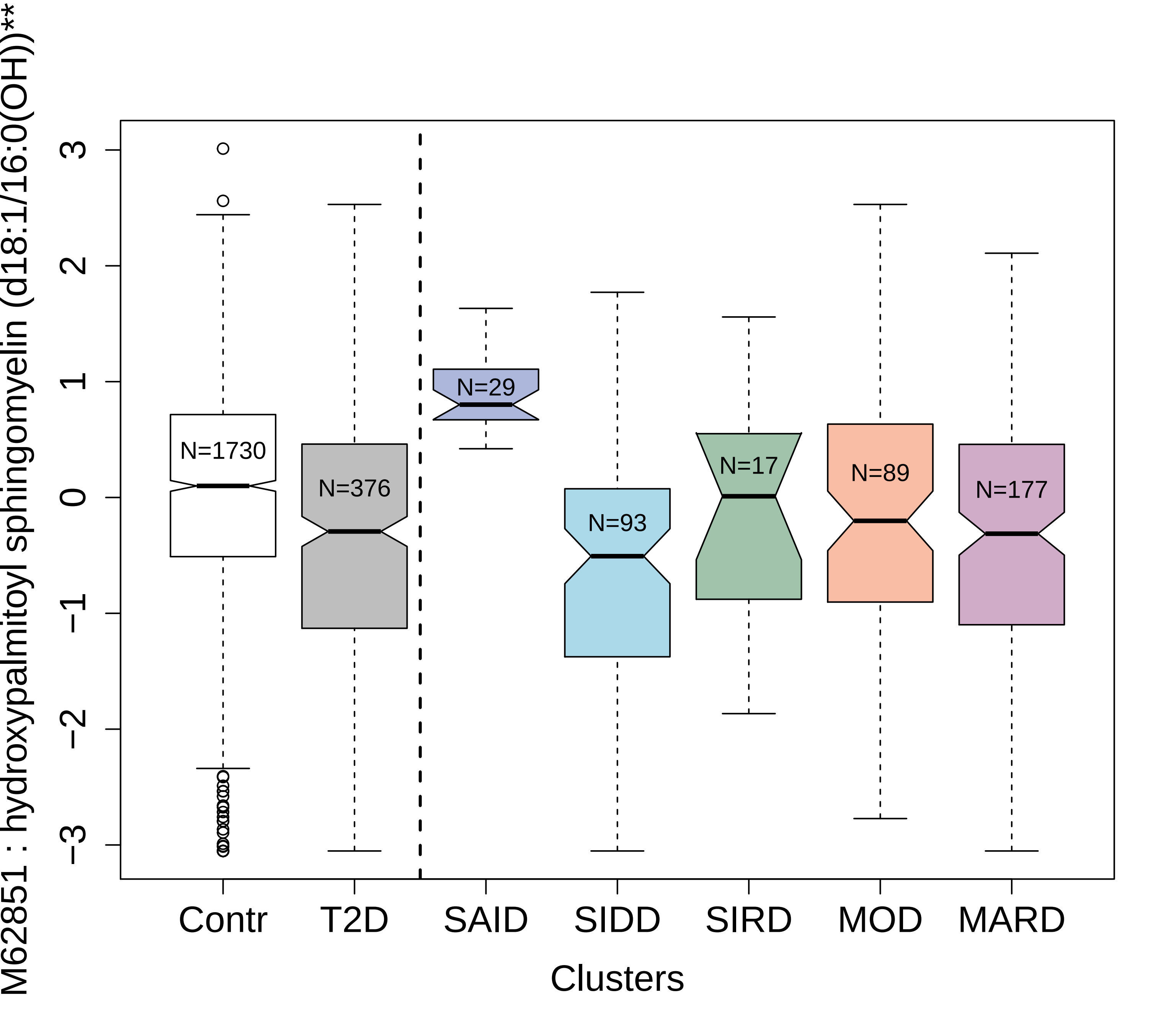
M62851 : hydroxypalmitoyl sphingomyelin (d18:1/16:0(OH))**
3
2
1
0
−1
−2
−3
N=1730
N=376
N=29
N=93
N=17
N=89
N=177
Contr
T2D
SAID
SIDD
SIRD
MOD
MARD
Clusters

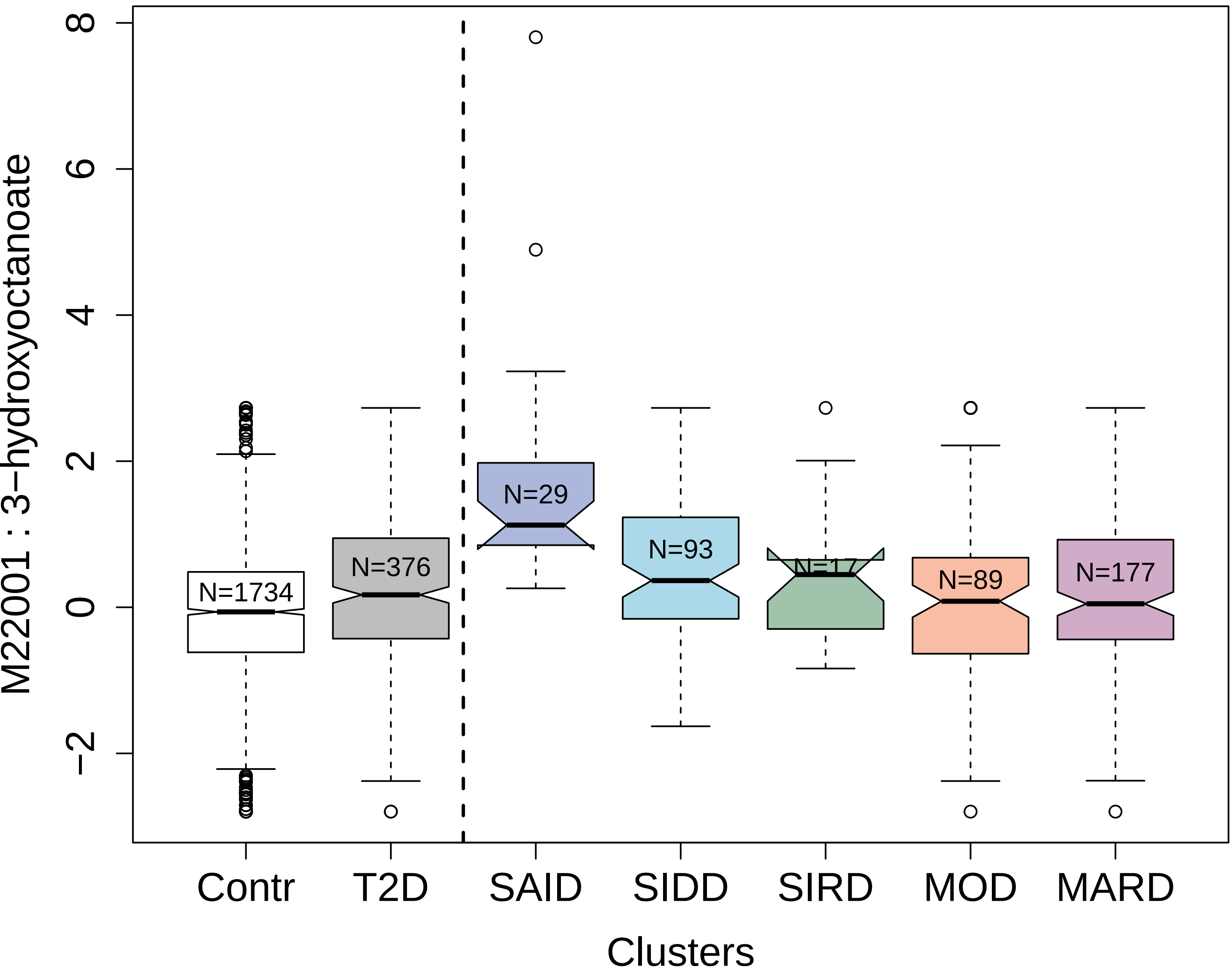

M22001 : 3−hydroxyoctanoate
8
6
4
2
0
−2
N=1734
N=376
N=29
N=93
N=17
N=89
N=177
Contr
T2D
SAID
SIDD
SIRD
MOD
MARD
Clusters

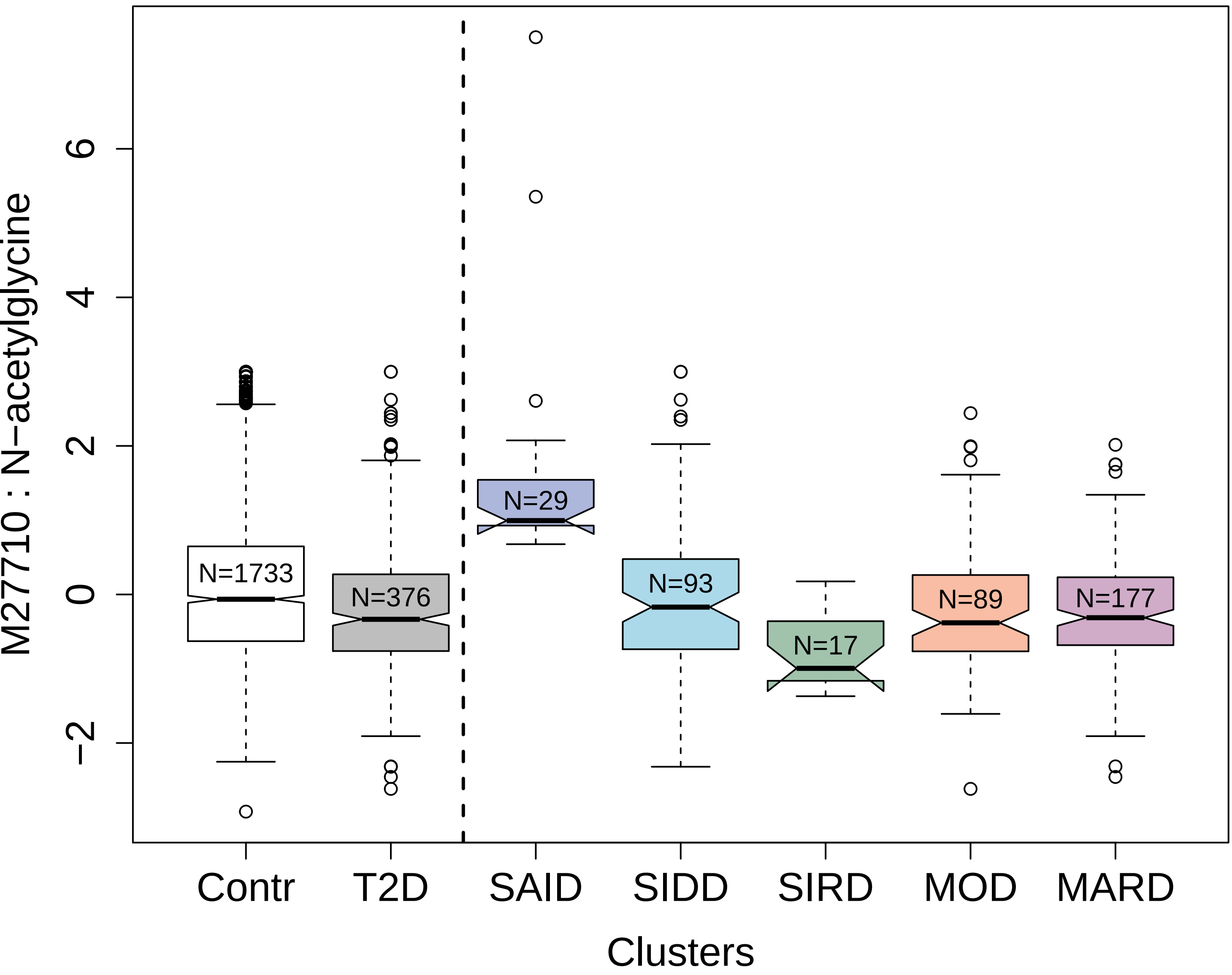
M27710 : N−acetylglycine
6
4
2
0
−2
N=1733
N=376
N=29
N=93
N=17
N=89
N=177
Contr
T2D
SAID
SIDD
SIRD
MOD
MARD
Clusters

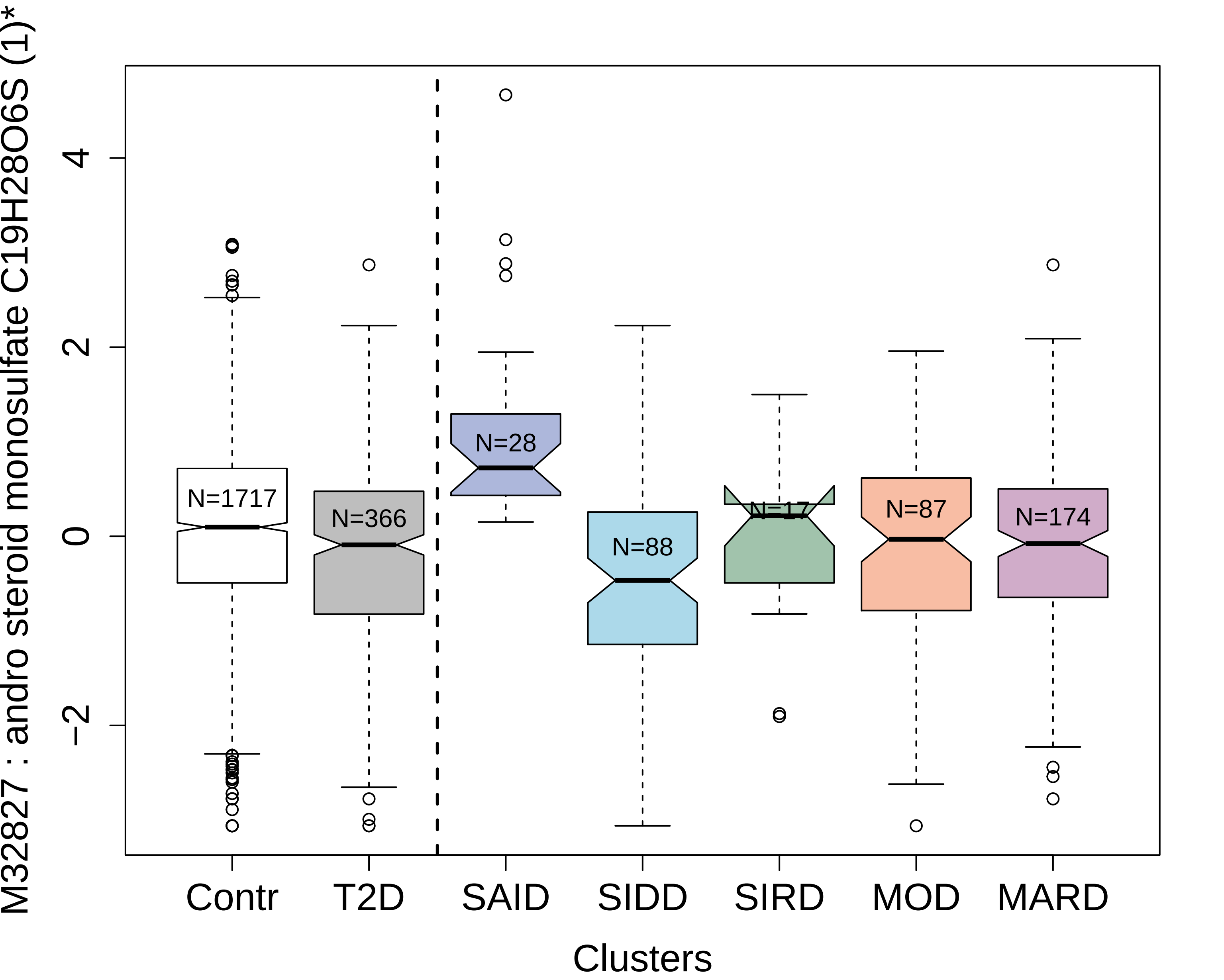

M32827 : andro steroid monosulfate C19H28O6S (1)*
4
2
0
−2
N=1717
N=366
N=28
N=88
N=17
N=87
N=174
Contr
T2D
SAID
SIDD
SIRD
MOD
MARD
Clusters

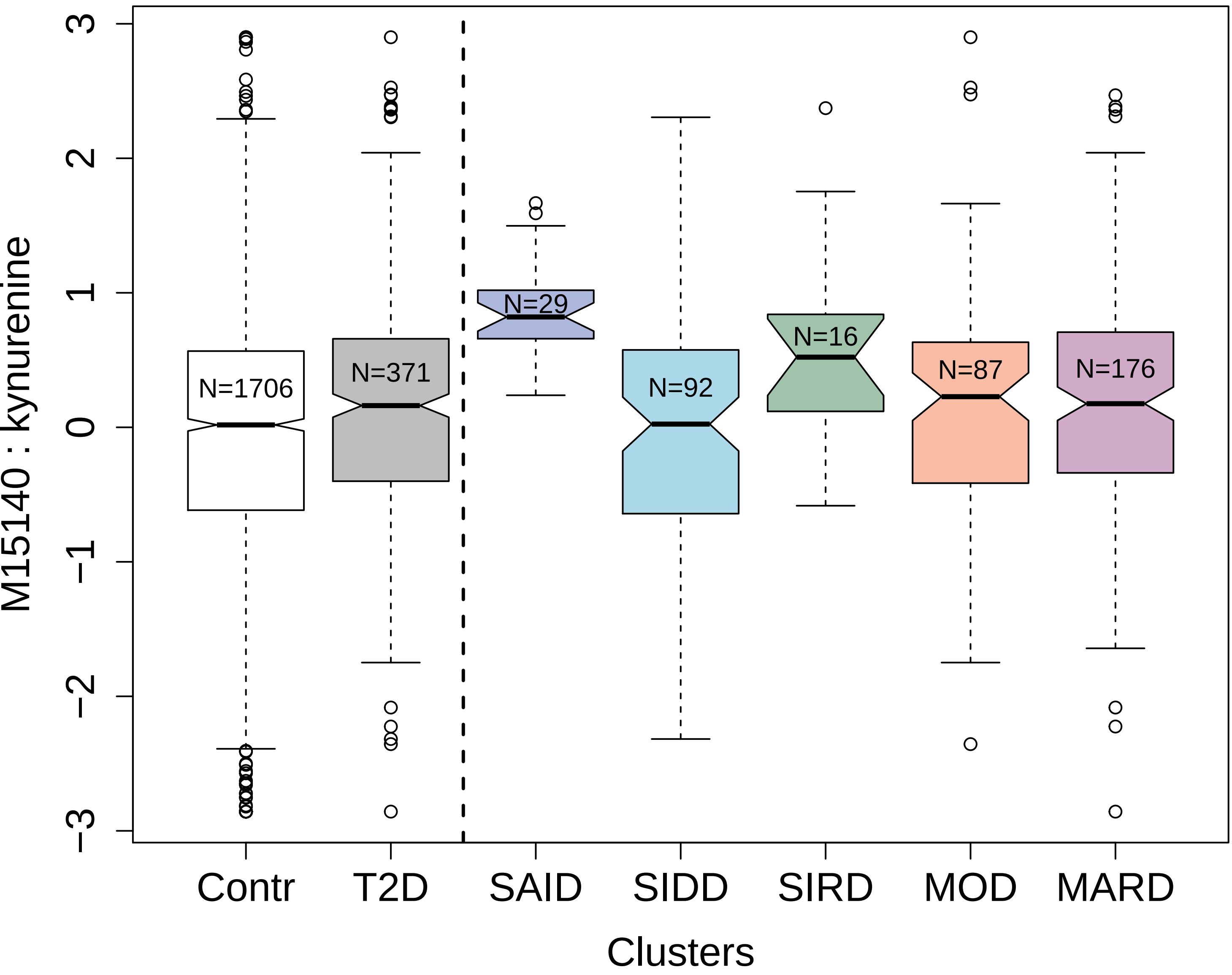
M15140 : kynurenine
3
2
1
0
−1
−2
−3
N=1706
N=371
N=29
N=92
N=16
N=87
N=176
Contr
T2D
SAID
SIDD
SIRD
MOD
MARD
Clusters

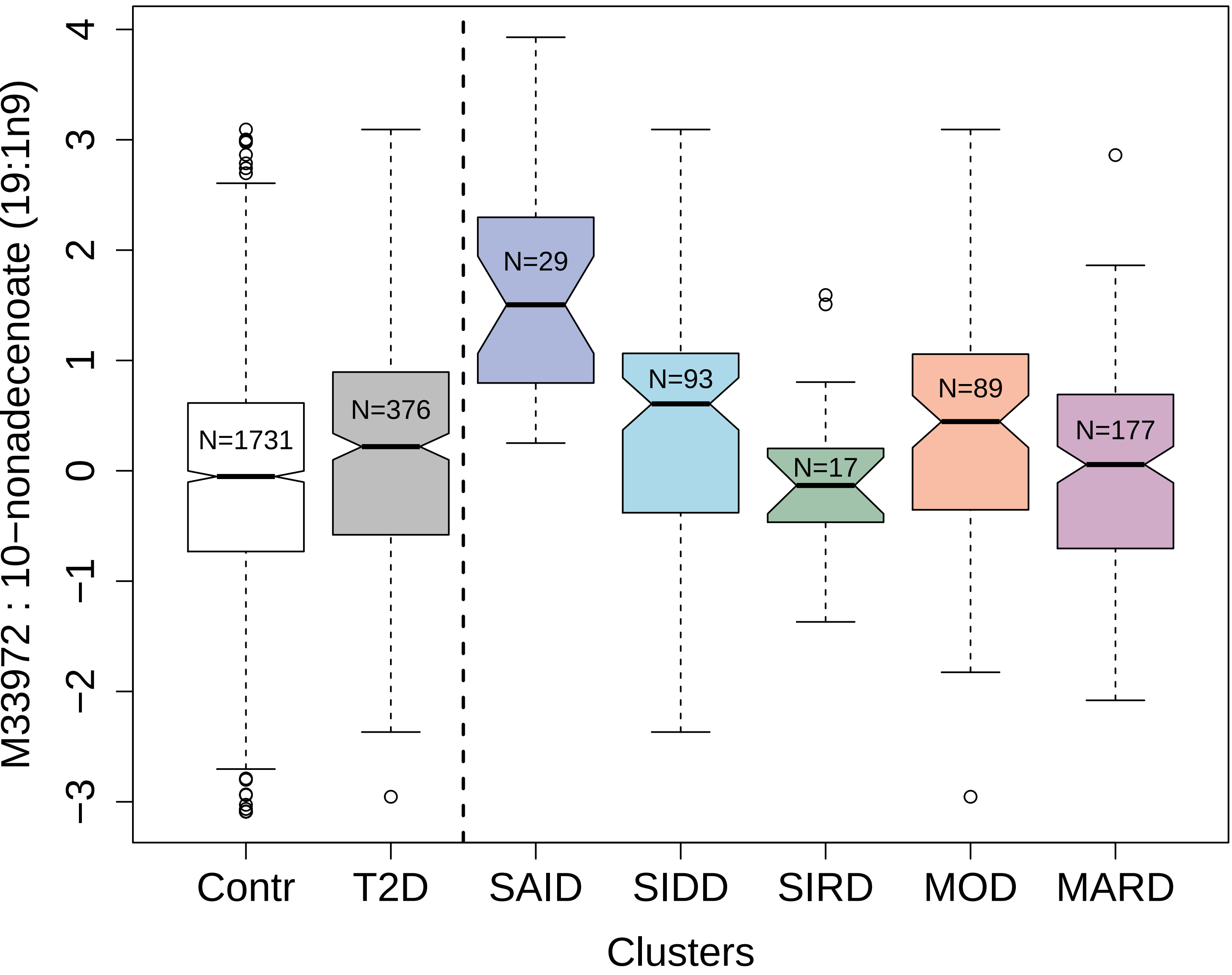
M33972 : 10−nonadecenoate (19:1n9)
Clusters
Contr
T2D
SAID
SIDD
SIRD
MOD
MARD
N=1731
N=376
N=29
N=93
N=17
N=89
N=177
4
3
2
1
0
−1
−2
−3

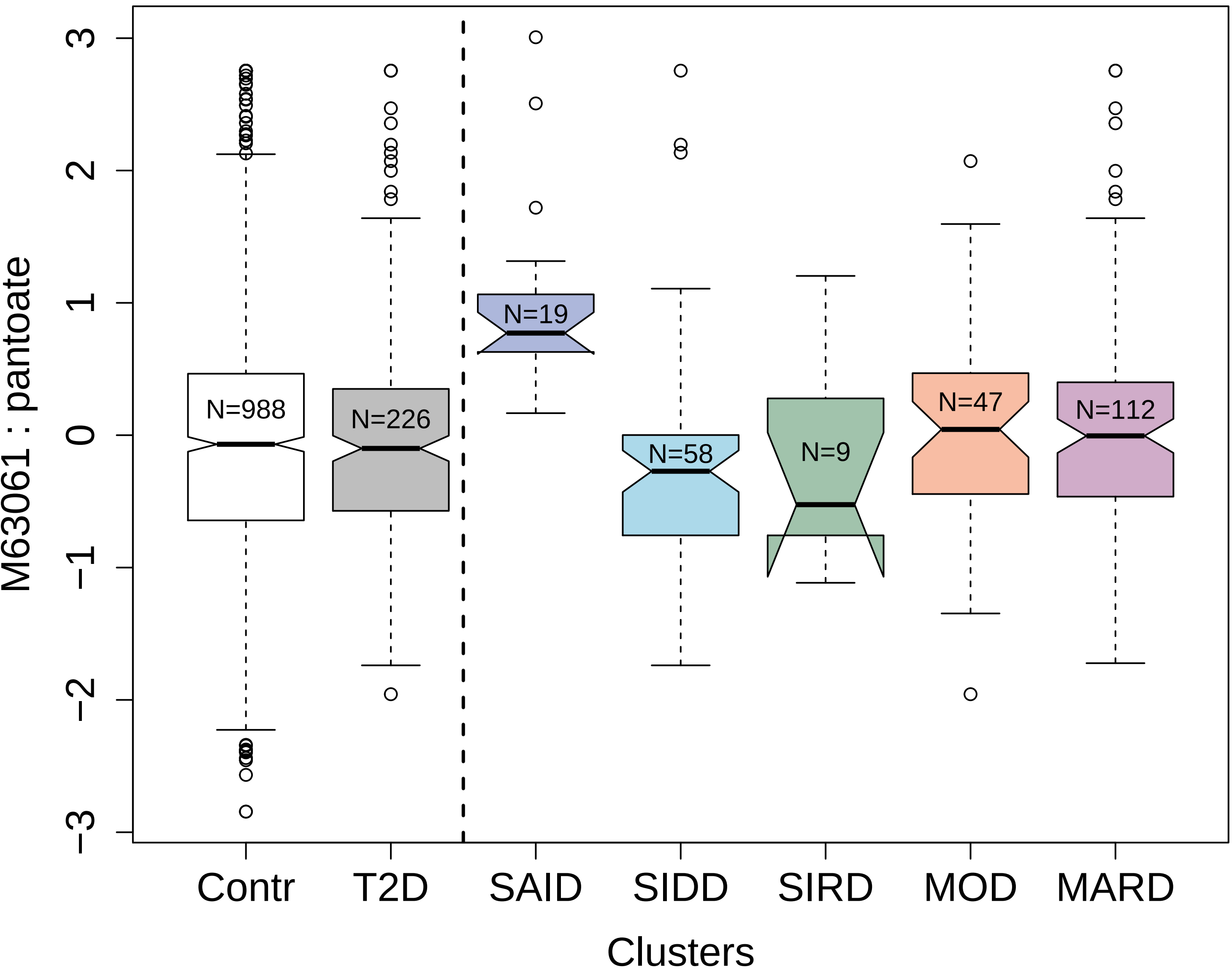

M63061 : pantoate
3
2
1
0
−1
−2
−3
N=988
N=226
N=19
N=58
N=9
N=47
N=112
Contr
T2D
SAID
SIDD
SIRD
MOD
MARD
Clusters

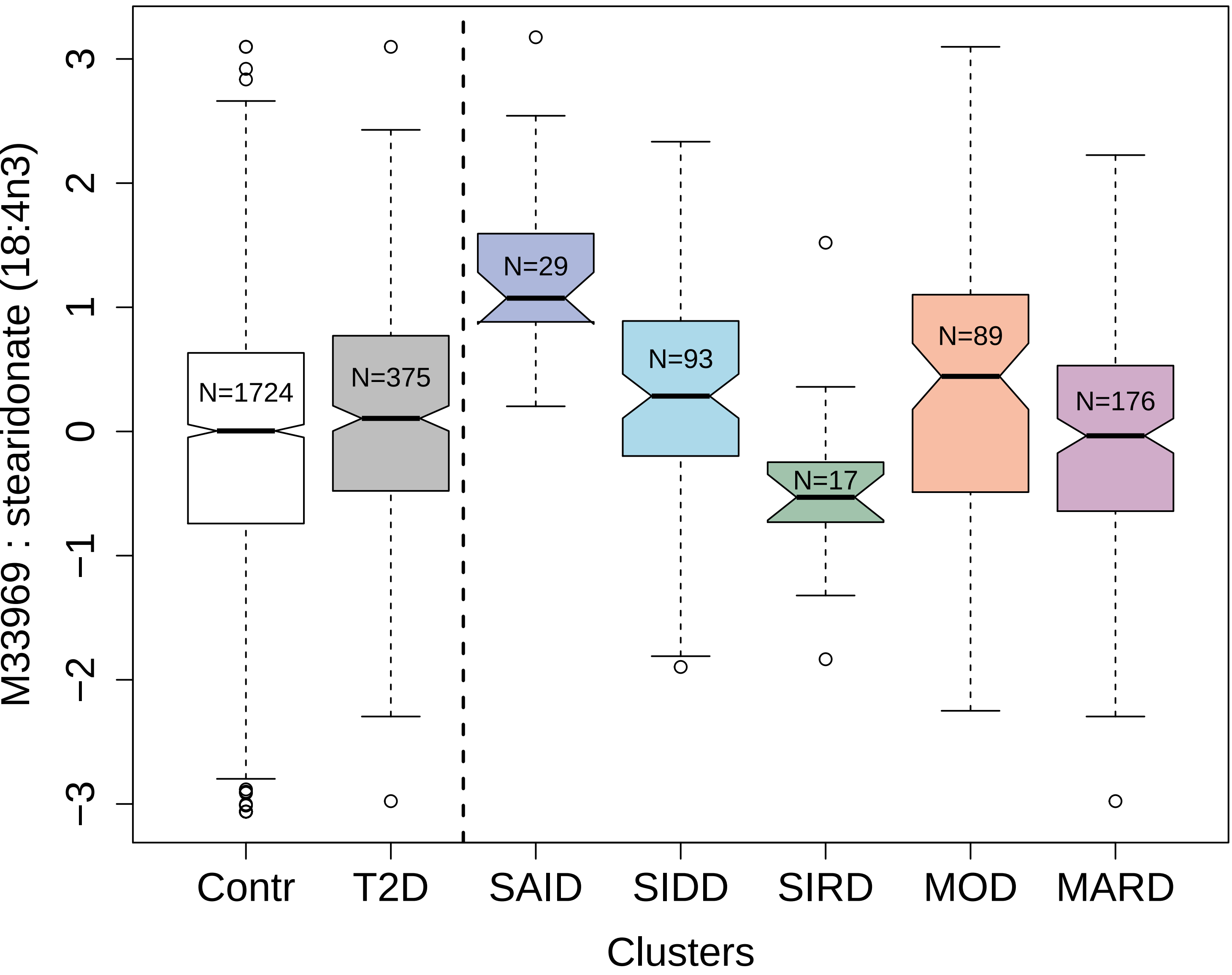

M33969 : stearidonate (18:4n3)
3
2
1
0
−1
−2
−3
N=1724
N=375
N=29
N=93
N=17
N=89
N=176
Contr
T2D
SAID
SIDD
SIRD
MOD
MARD
Clusters

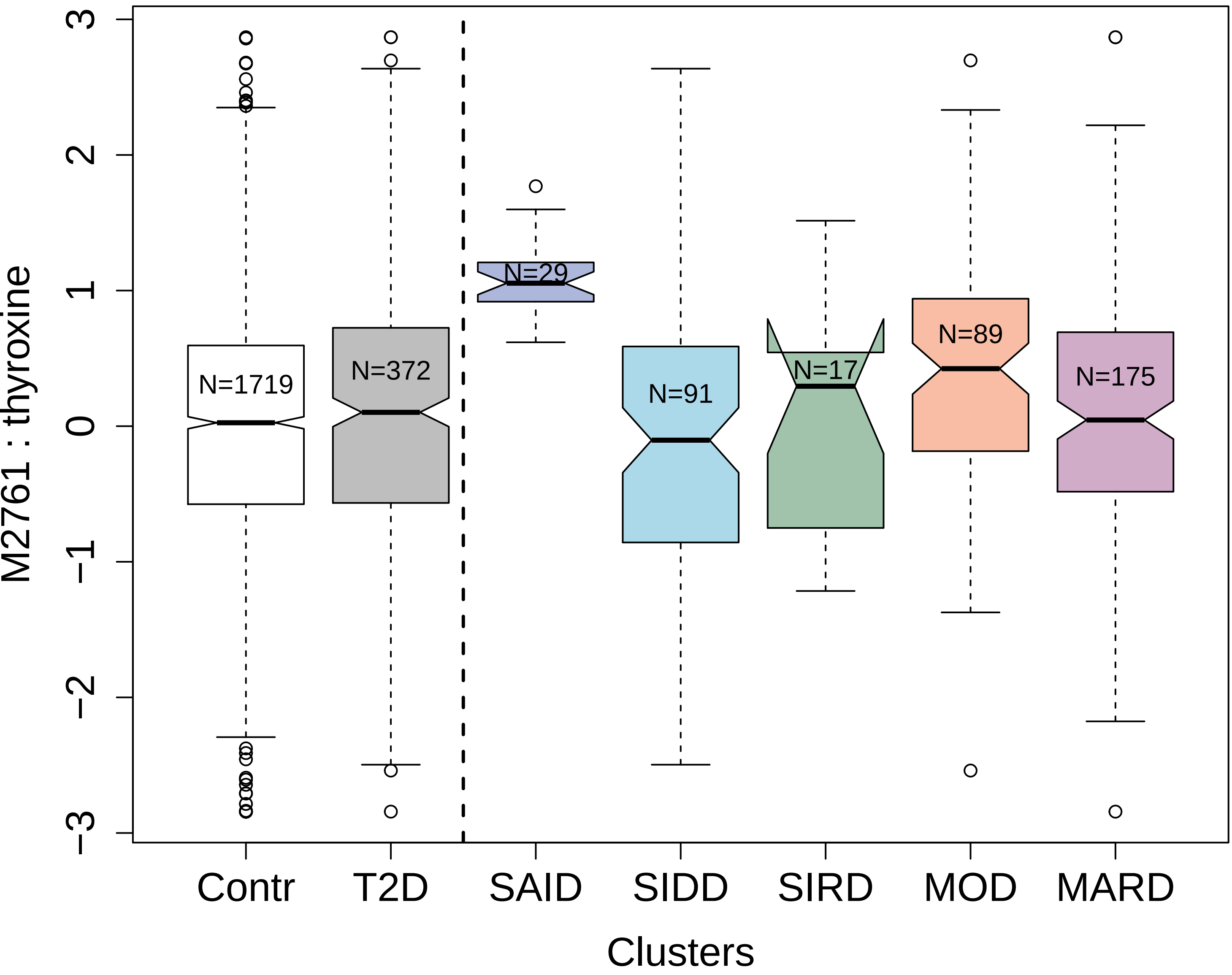
M2761 : thyroxine
3
2
1
0
−1
−2
−3
N=1719
N=372
N=29
N=91
N=17
N=89
N=175
Contr
T2D
SAID
SIDD
SIRD
MOD
MARD
Clusters

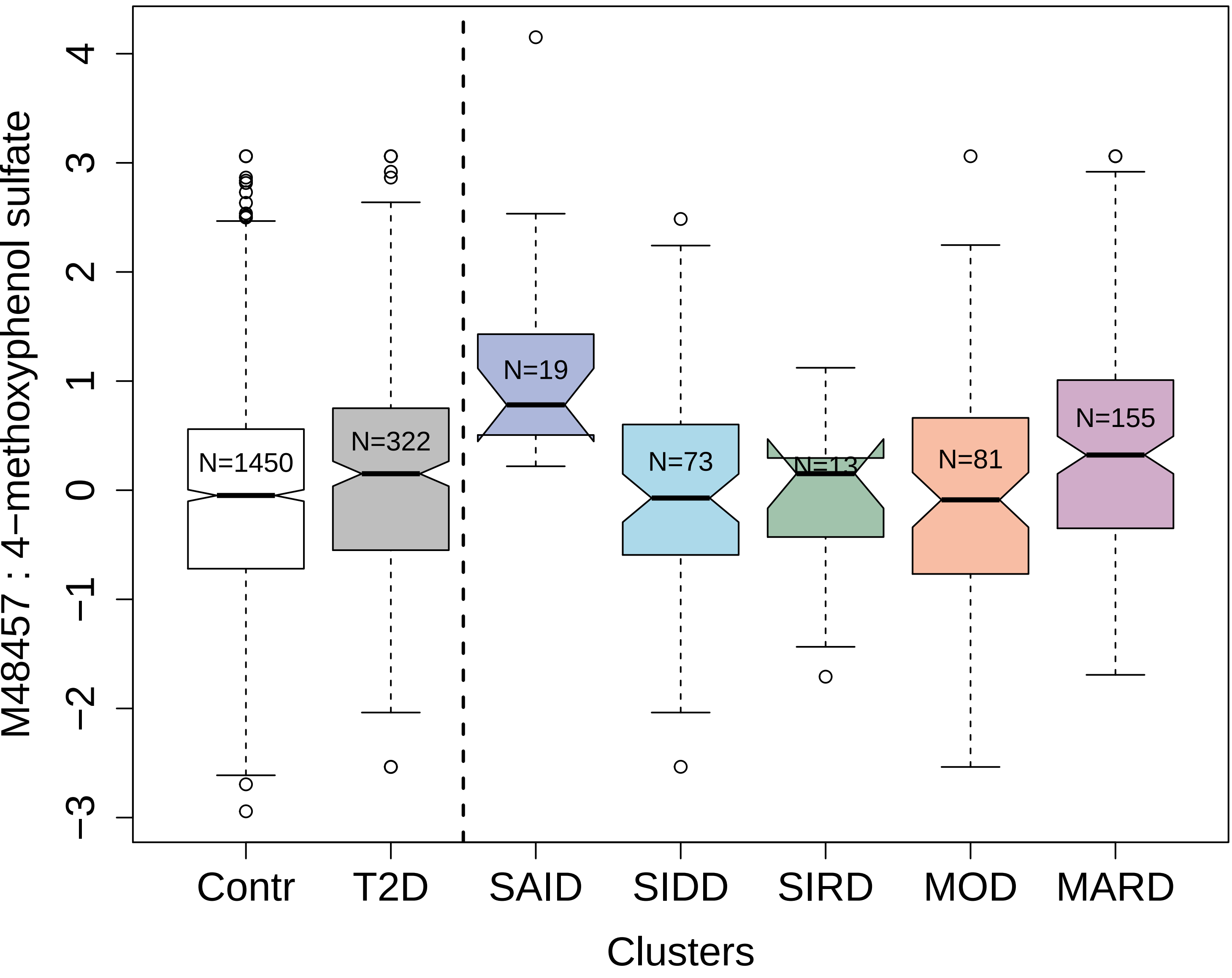

M48457 : 4−methoxyphenol sulfate
4
3
2
1
0
−1
−2
−3
N=1450
N=322
N=19
N=73
N=13
N=81
N=155
Contr
T2D
SAID
SIDD
SIRD
MOD
MARD
Clusters

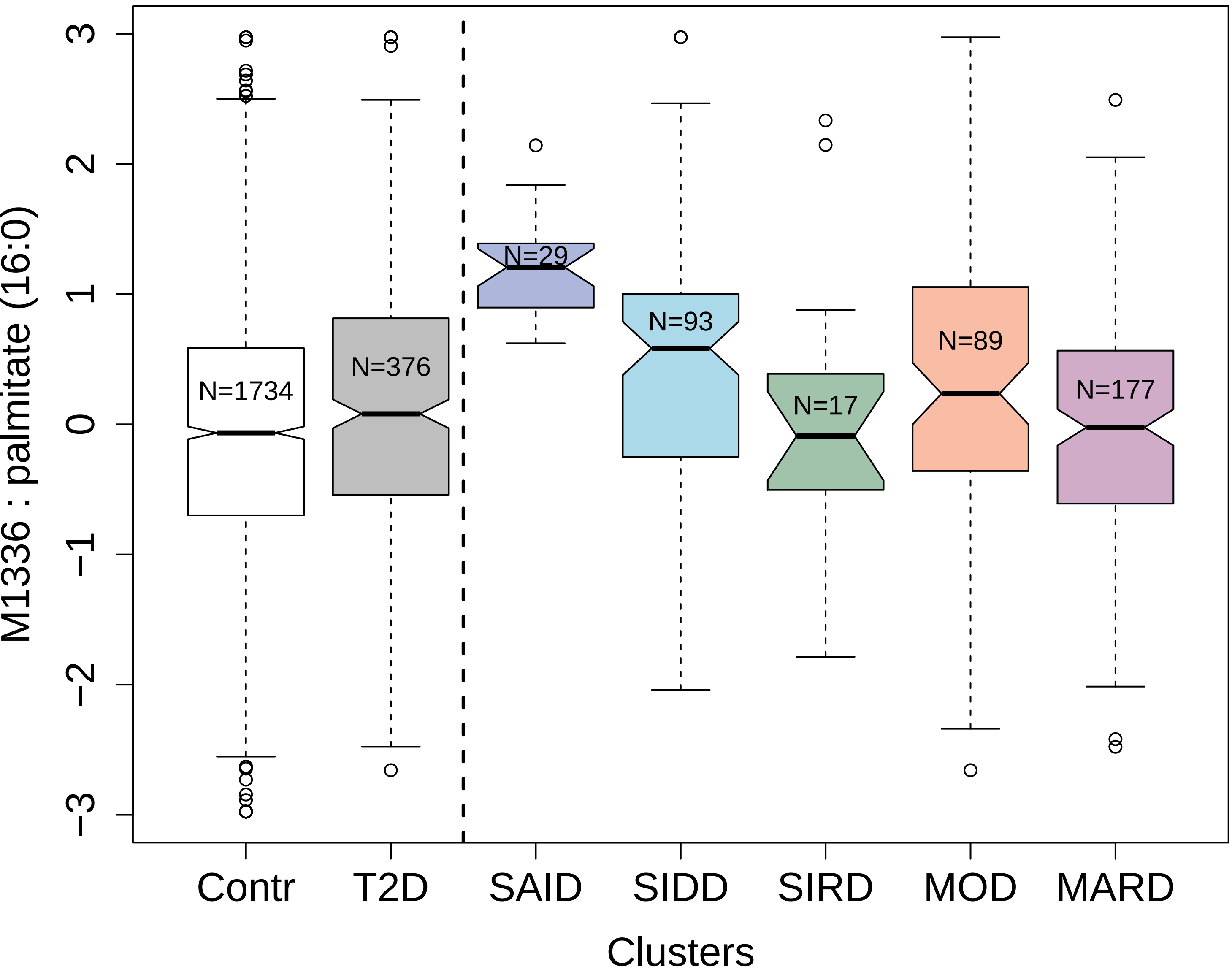

M1336 : palmitate (16:0)
3
2
1
0
−1
−2
−3
N=1734
N=376
N=29
N=93
N=17
N=89
N=177
Contr
T2D
SAID
SIDD
SIRD
MOD
MARD
Clusters

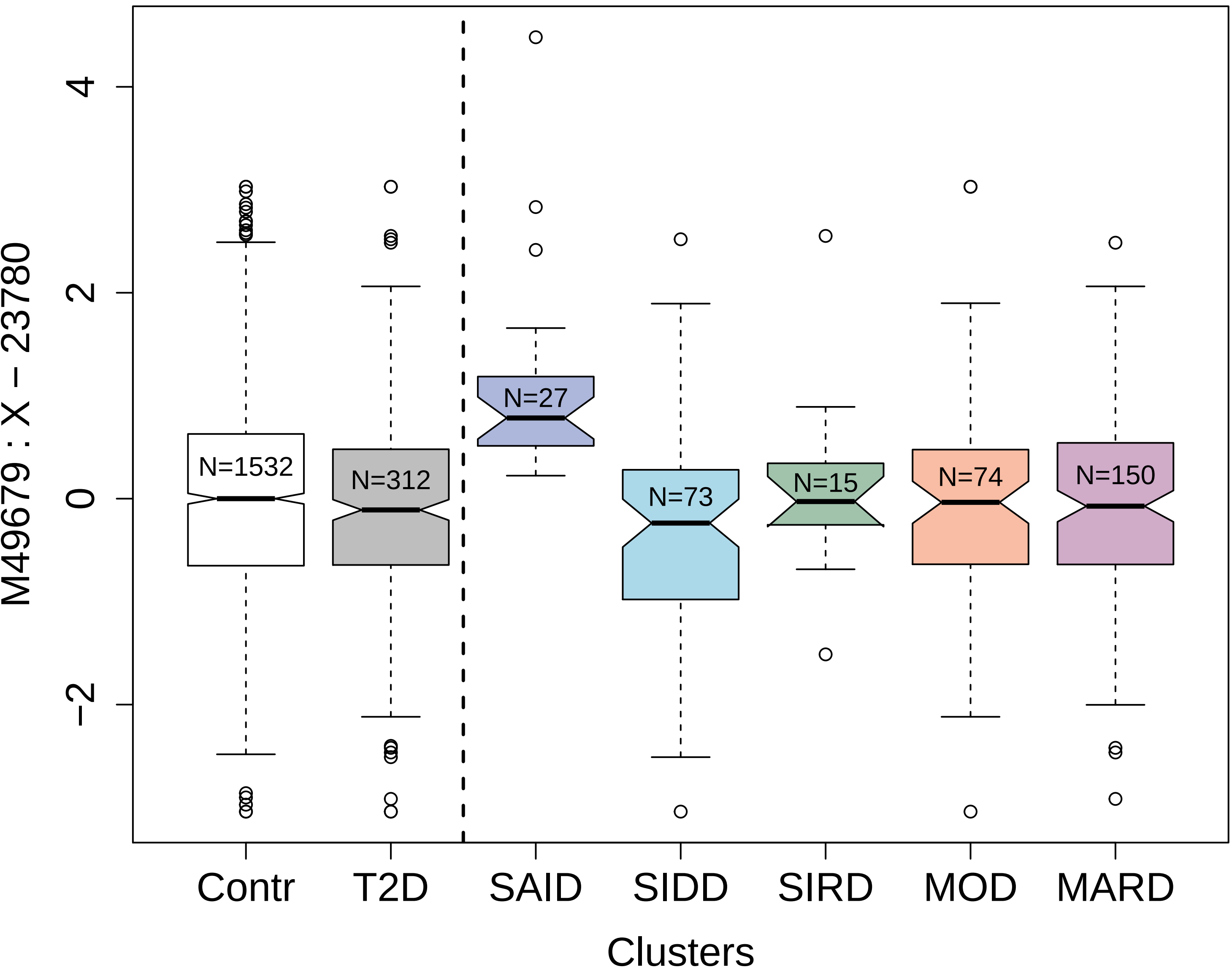

M49679 : X − 23780
4
2
0
−2
N=1532
N=312
N=27
N=73
N=15
N=74
N=150
Contr
T2D
SAID
SIDD
SIRD
MOD
MARD
Clusters

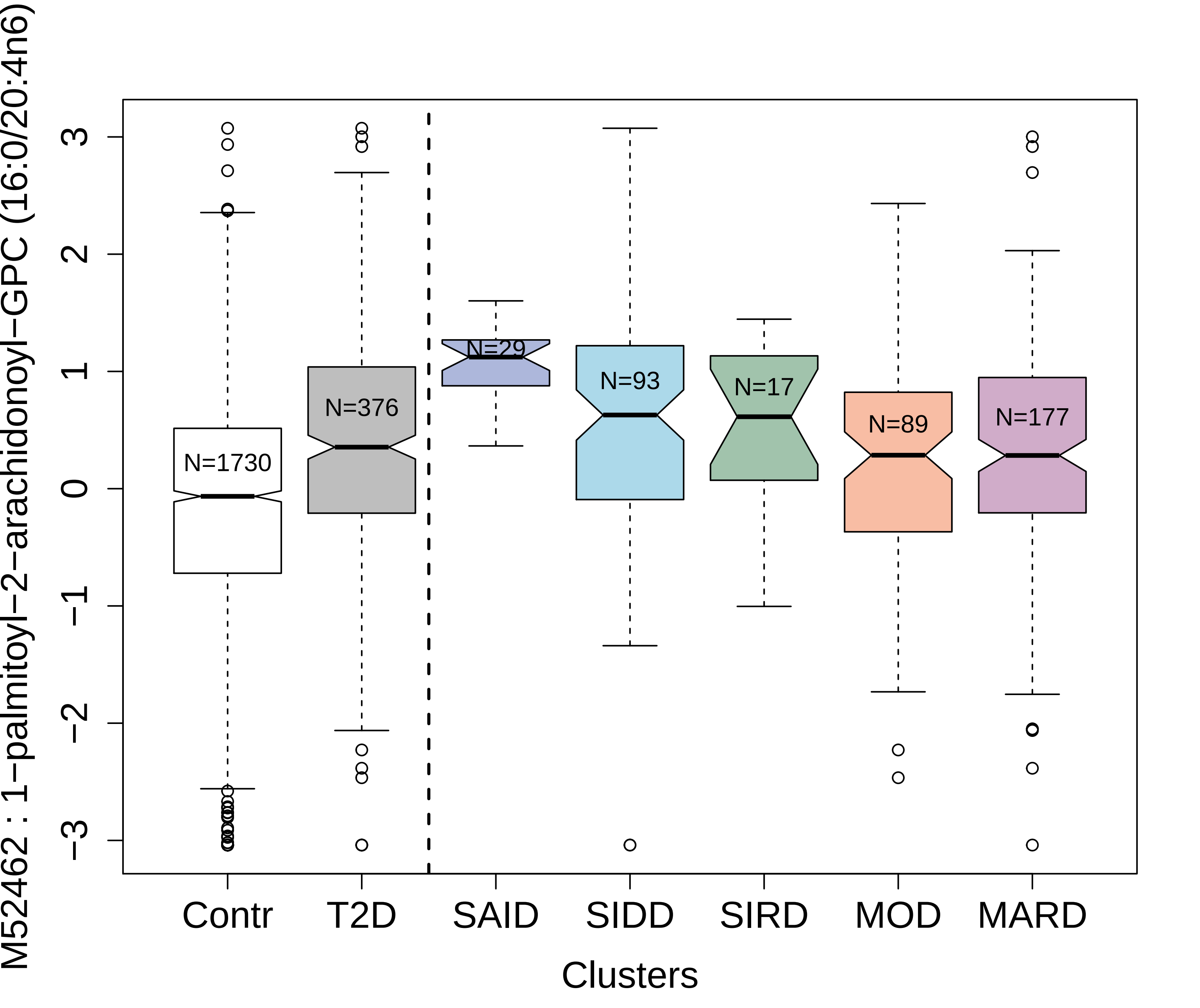
M52462 : 1−palmitoyl−2−arachidonoyl−GPC (16:0/20:4n6)
3
2
1
0
−1
−2
−3
N=1730
N=376
N=29
N=93
N=17
N=89
N=177
Contr
T2D
SAID
SIDD
SIRD
MOD
MARD
Clusters

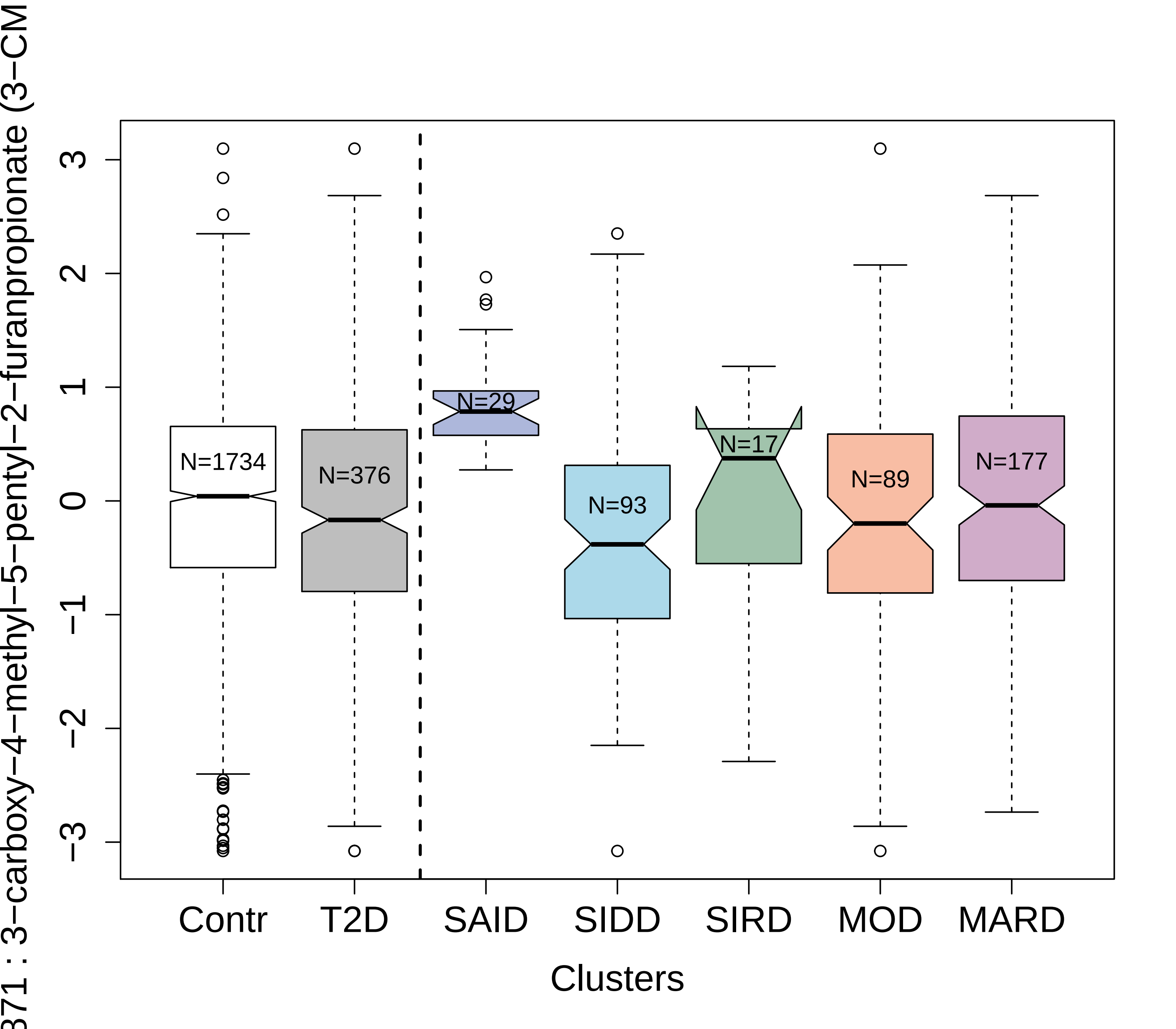
371 : 3−carboxy−4−methyl−5−pentyl−2−furanpropionate (3−CM
3
2
1
0
−1
−2
−3
N=1734
N=376
N=29
N=93
N=17
N=89
N=177
Contr
T2D
SAID
SIDD
SIRD
MOD
MARD
Clusters

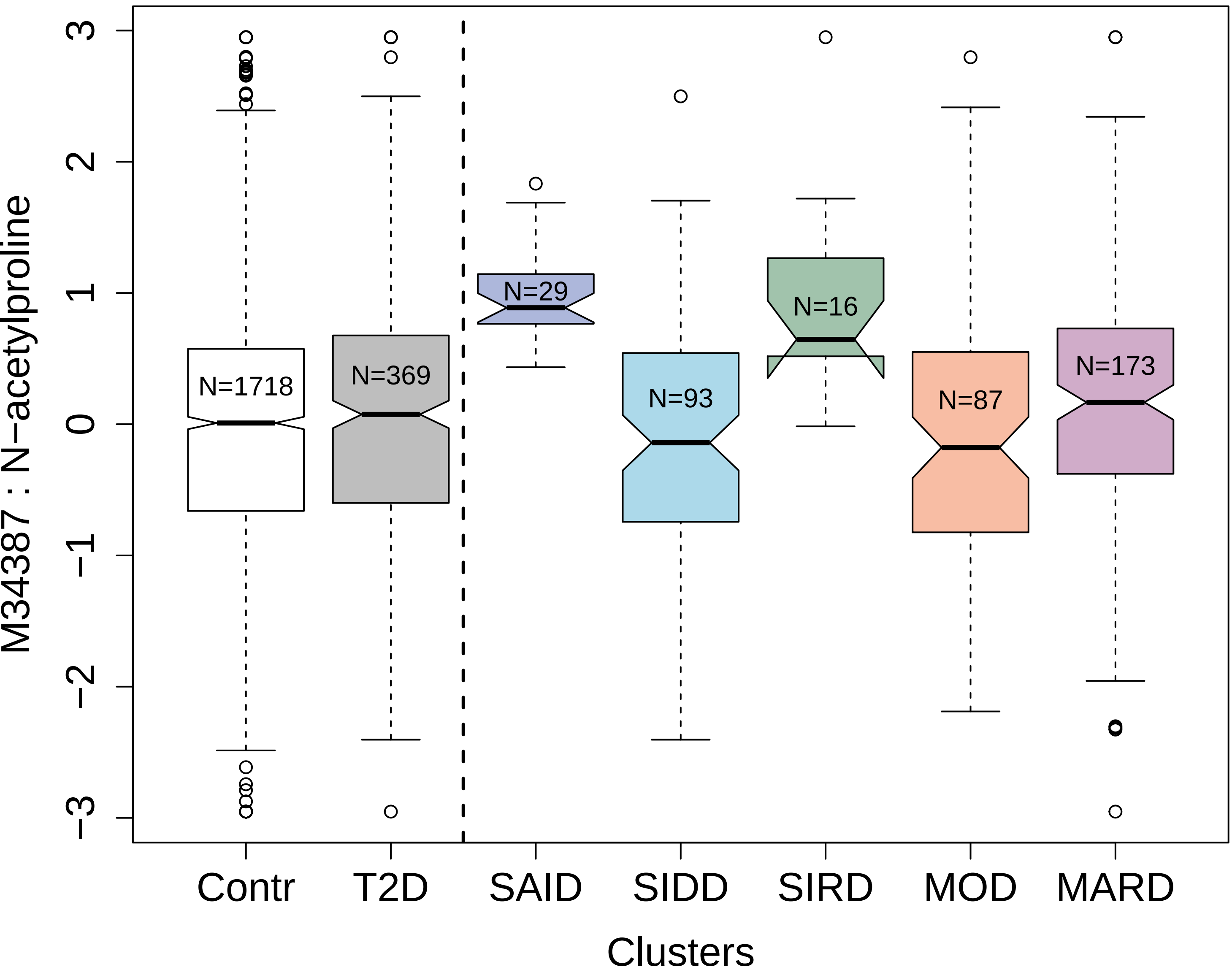

M34387 : N−acetylproline
3
2
1
0
−1
−2
−3
N=1718
N=369
N=29
N=93
N=16
N=87
N=173
Contr
T2D
SAID
SIDD
SIRD
MOD
MARD
Clusters

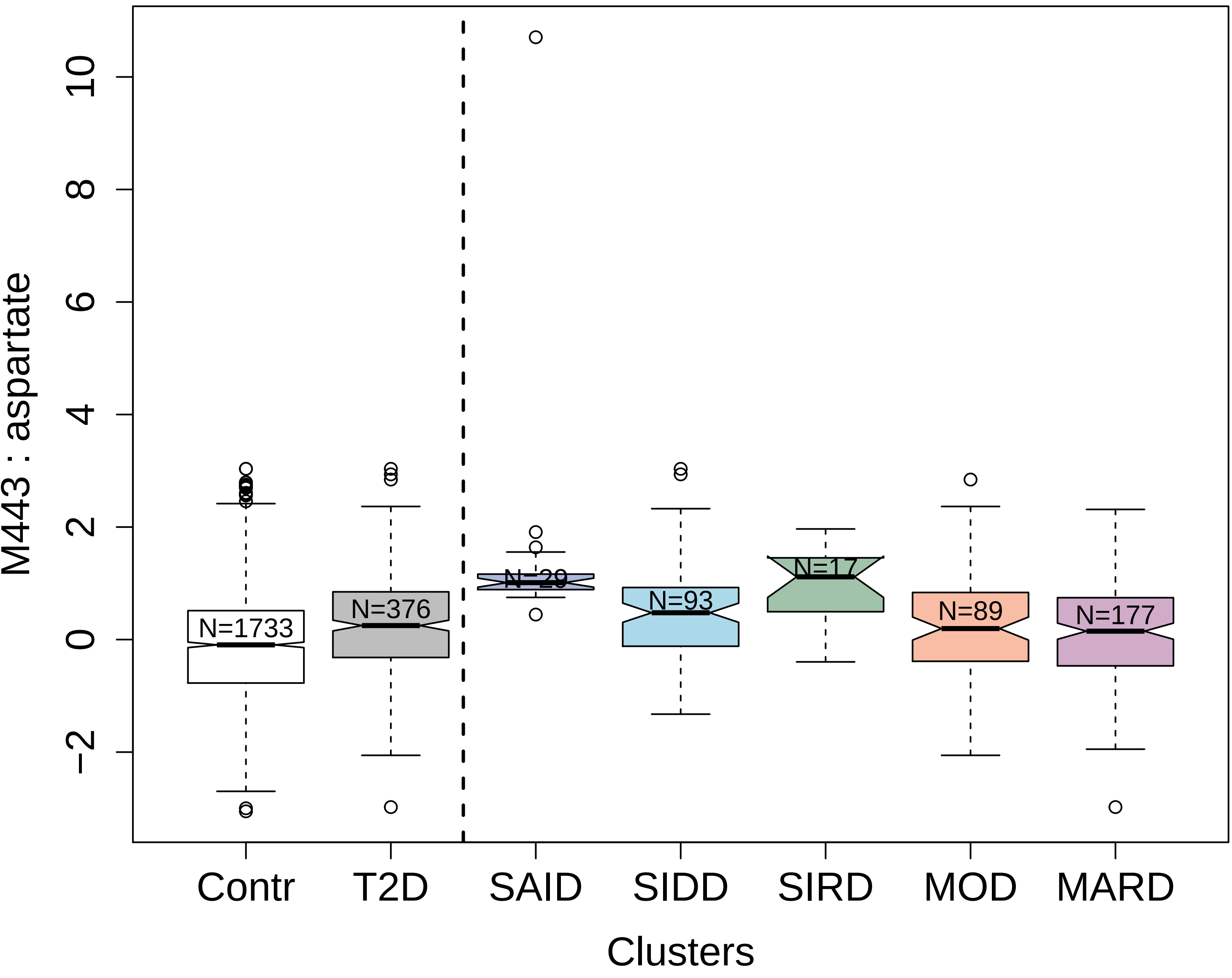

M443 : aspartate
10
8
6
4
2
0
−2
N=1733
N=376
N=29
N=93
N=17
N=89
N=177
Contr
T2D
SAID
SIDD
SIRD
MOD
MARD
Clusters

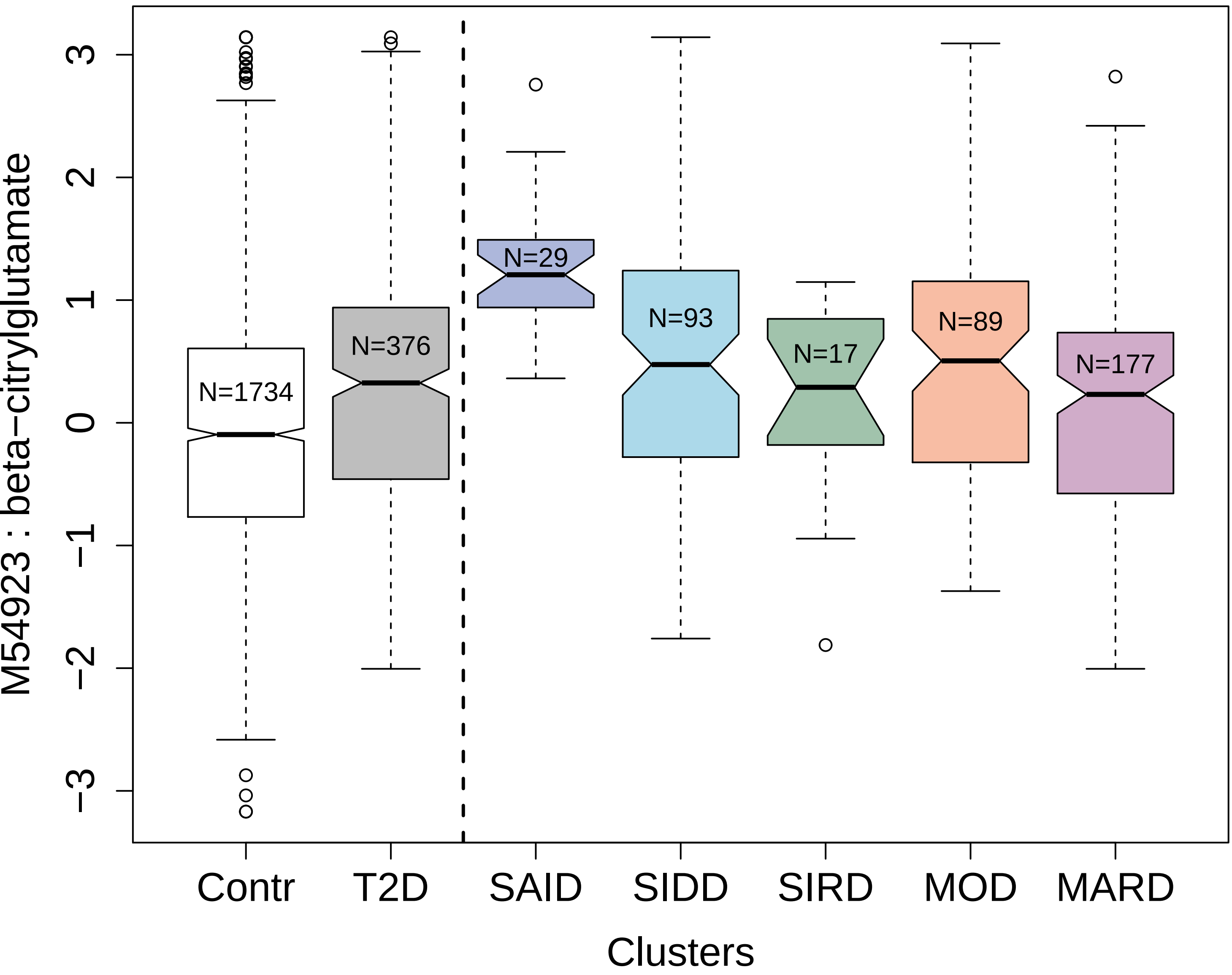

M54923 : beta−citrylglutamate
3
2
1
0
−1
−2
−3
N=1734
N=376
N=29
N=93
N=17
N=89
N=177
Contr
T2D
SAID
SIDD
SIRD
MOD
MARD
Clusters

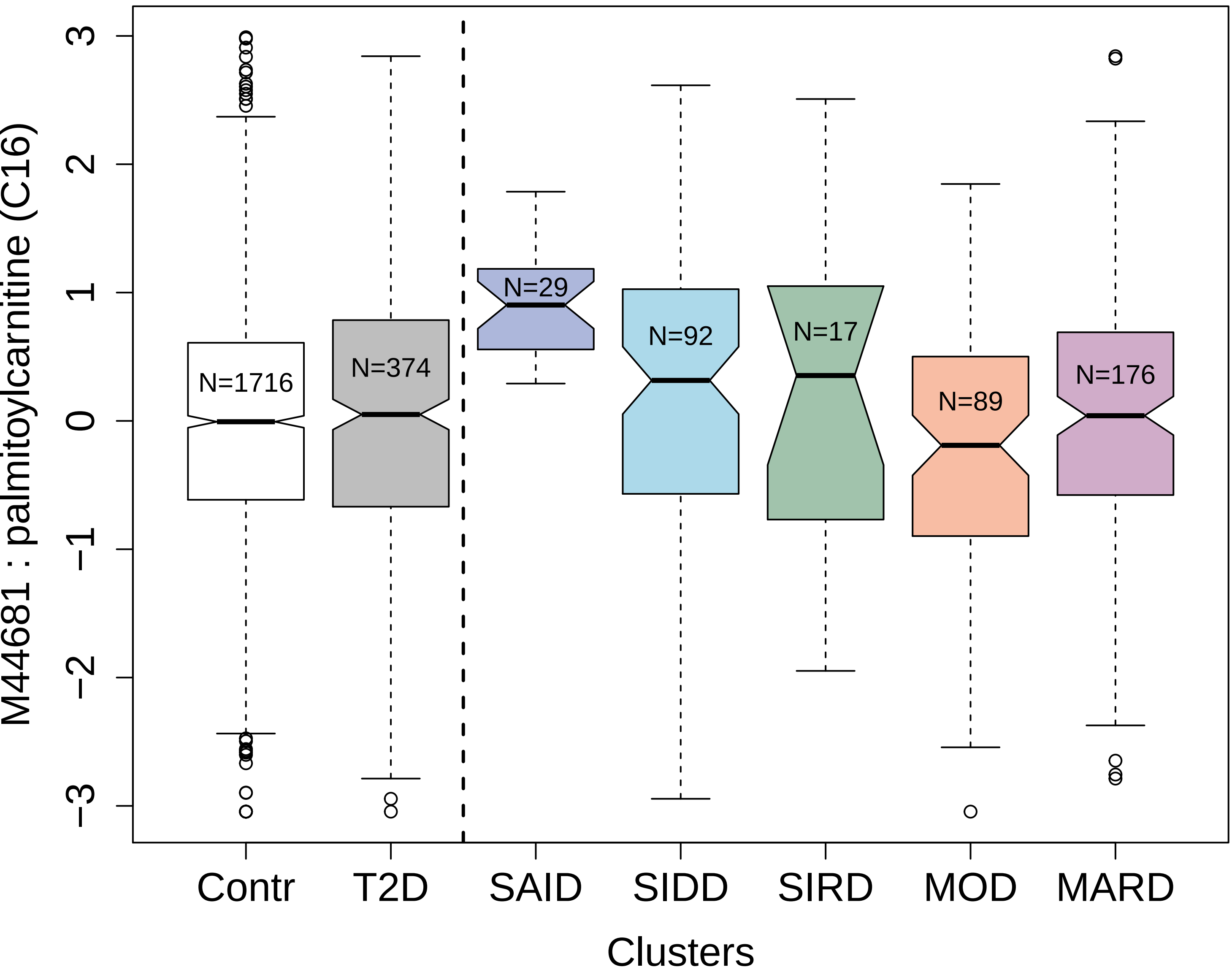
M44681 : palmitoylcarnitine (C16)
3
2
1
0
−1
−2
−3
N=1716
N=374
N=29
N=92
N=17
N=89
N=176
Contr
T2D
SAID
SIDD
SIRD
MOD
MARD
Clusters

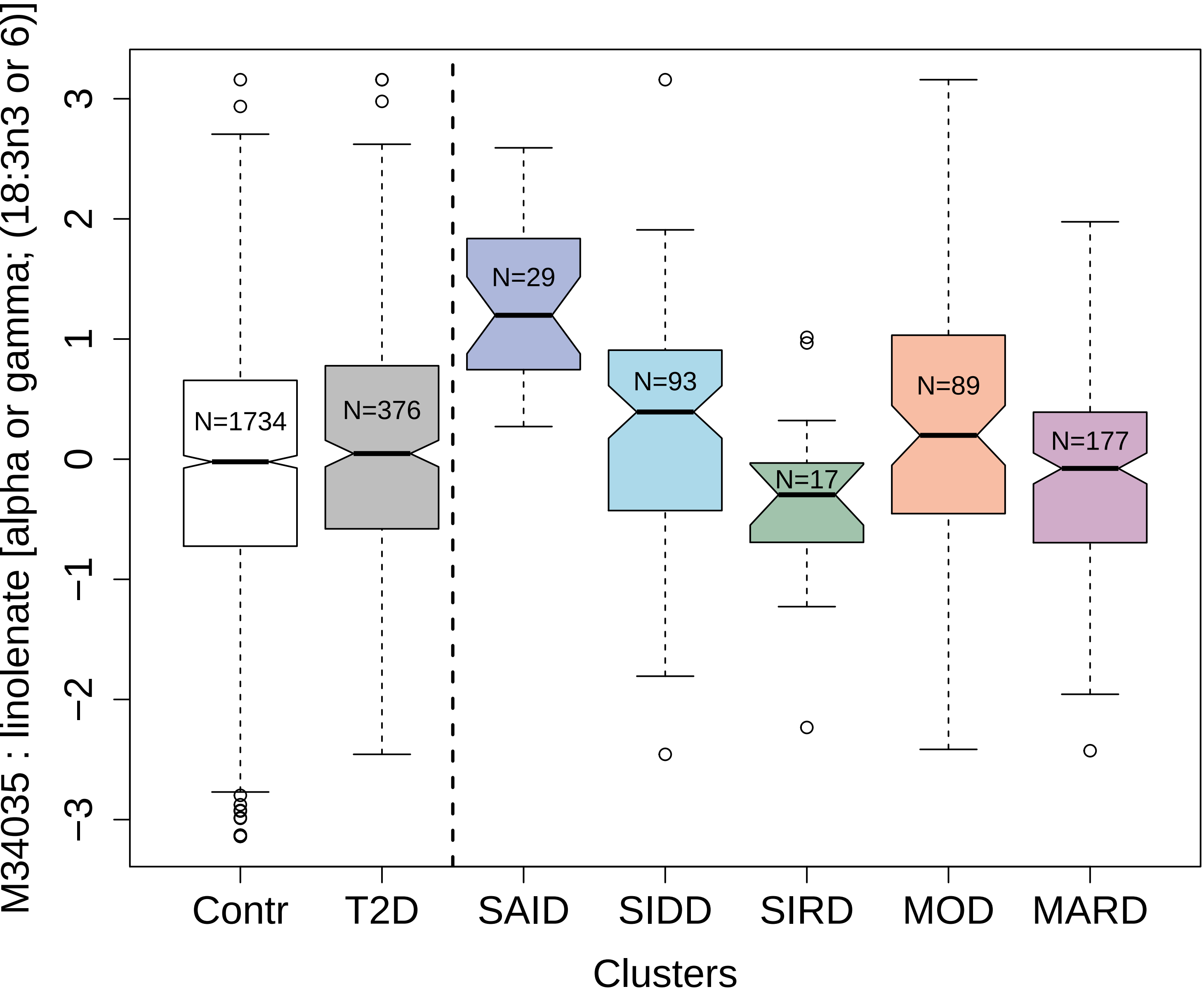

M34035 : linolenate [alpha or gamma; (18:3n3 or 6)]
3
2
1
0
−1
−2
−3
N=1734
N=376
N=29
N=93
N=17
N=89
N=177
Contr
T2D
SAID
SIDD
SIRD
MOD
MARD
Clusters

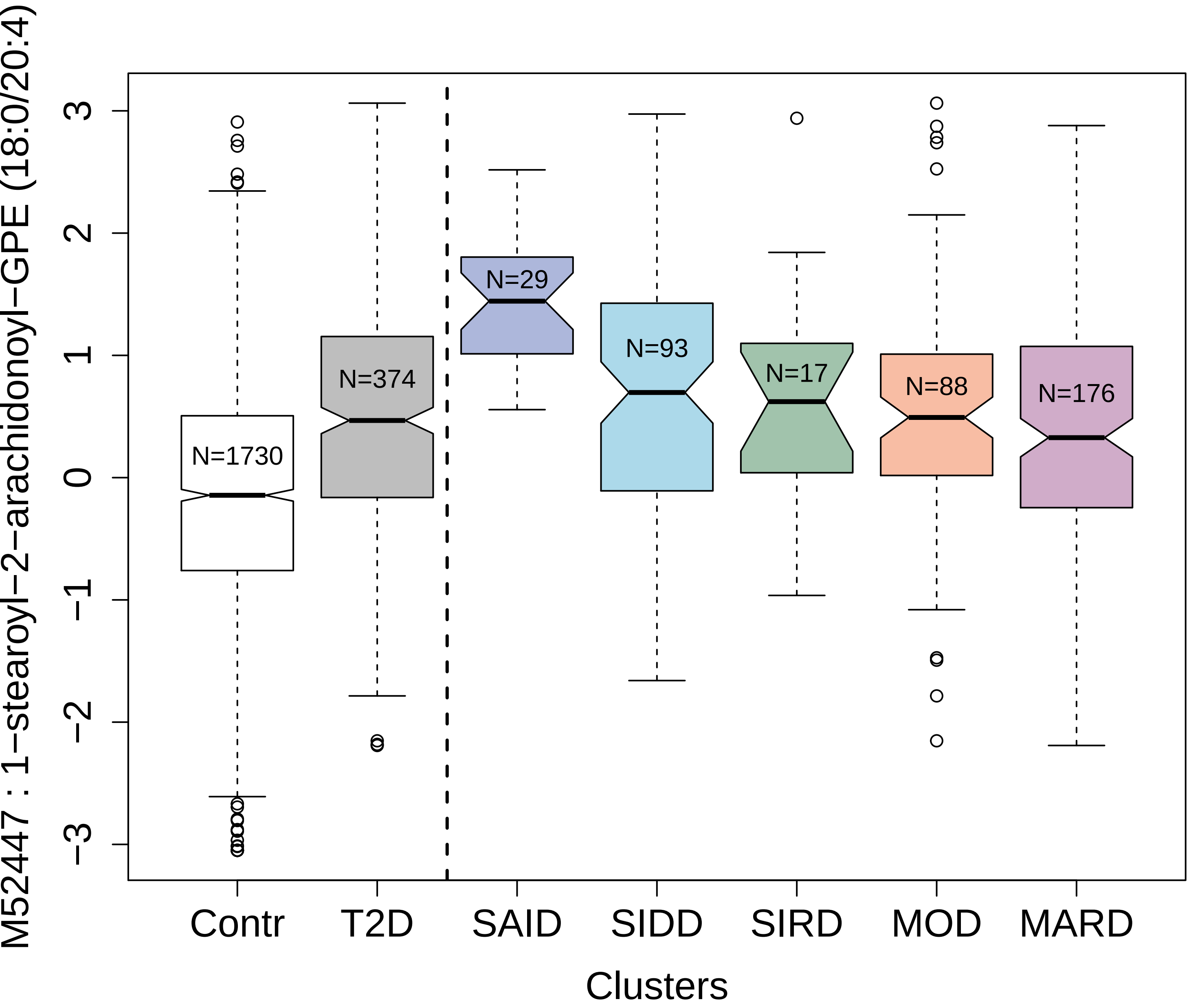

M52447 : 1−stearoyl−2−arachidonoyl−GPE (18:0/20:4)
3
2
1
0
−1
−2
−3
N=1730
N=374
N=29
N=93
N=17
N=88
N=176
Contr
T2D
SAID
SIDD
SIRD
MOD
MARD
Clusters

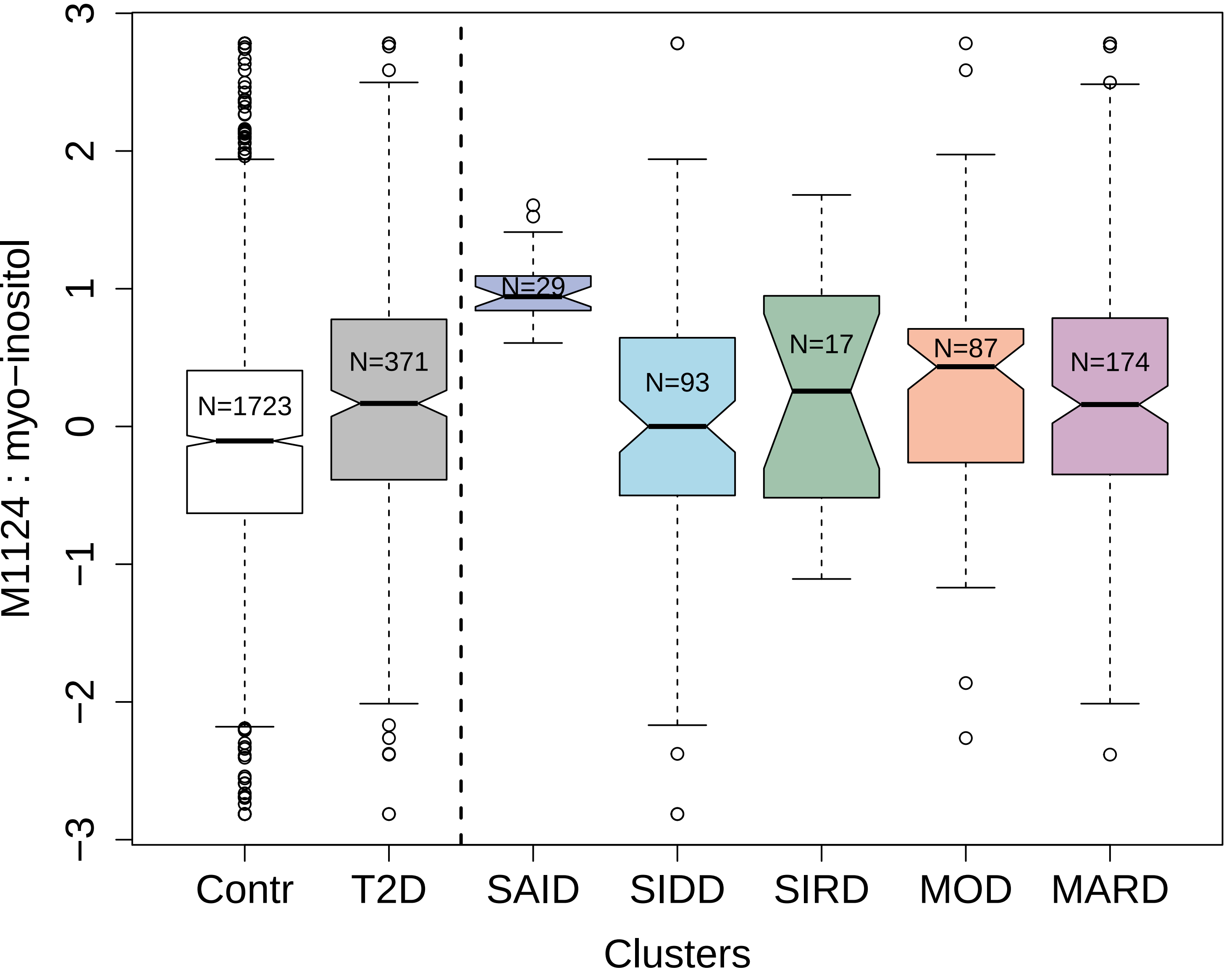
M1124 : myo−inositol
3
2
1
0
−1
−2
−3
N=1723
N=371
N=29
N=93
N=17
N=87
N=174
Contr
T2D
SAID
SIDD
SIRD
MOD
MARD
Clusters

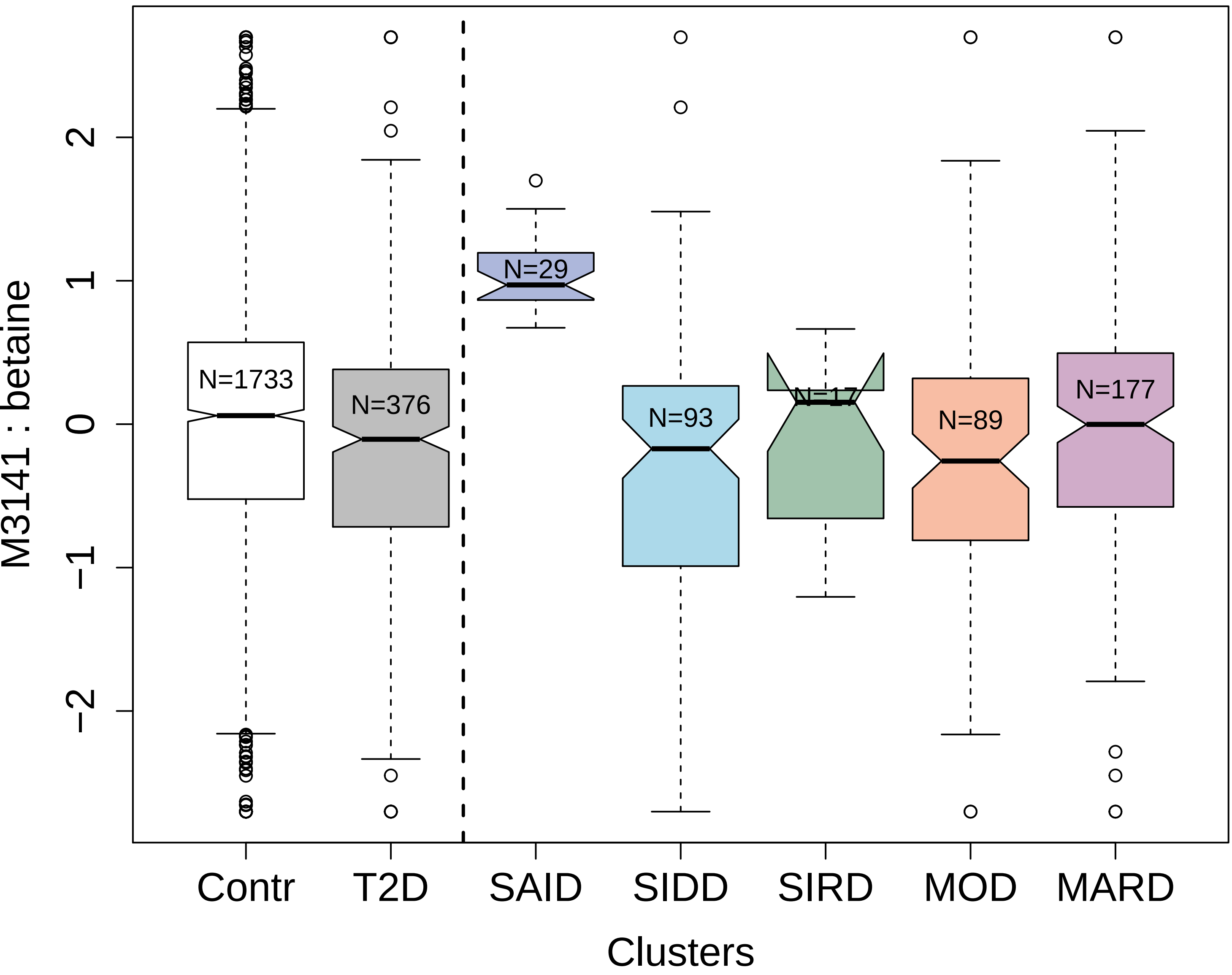
M3141 : betaine
2
1
0
−1
−2
N=1733
N=376
N=29
N=93
N=17
N=89
N=177
Contr
T2D
SAID
SIDD
SIRD
MOD
MARD
Clusters

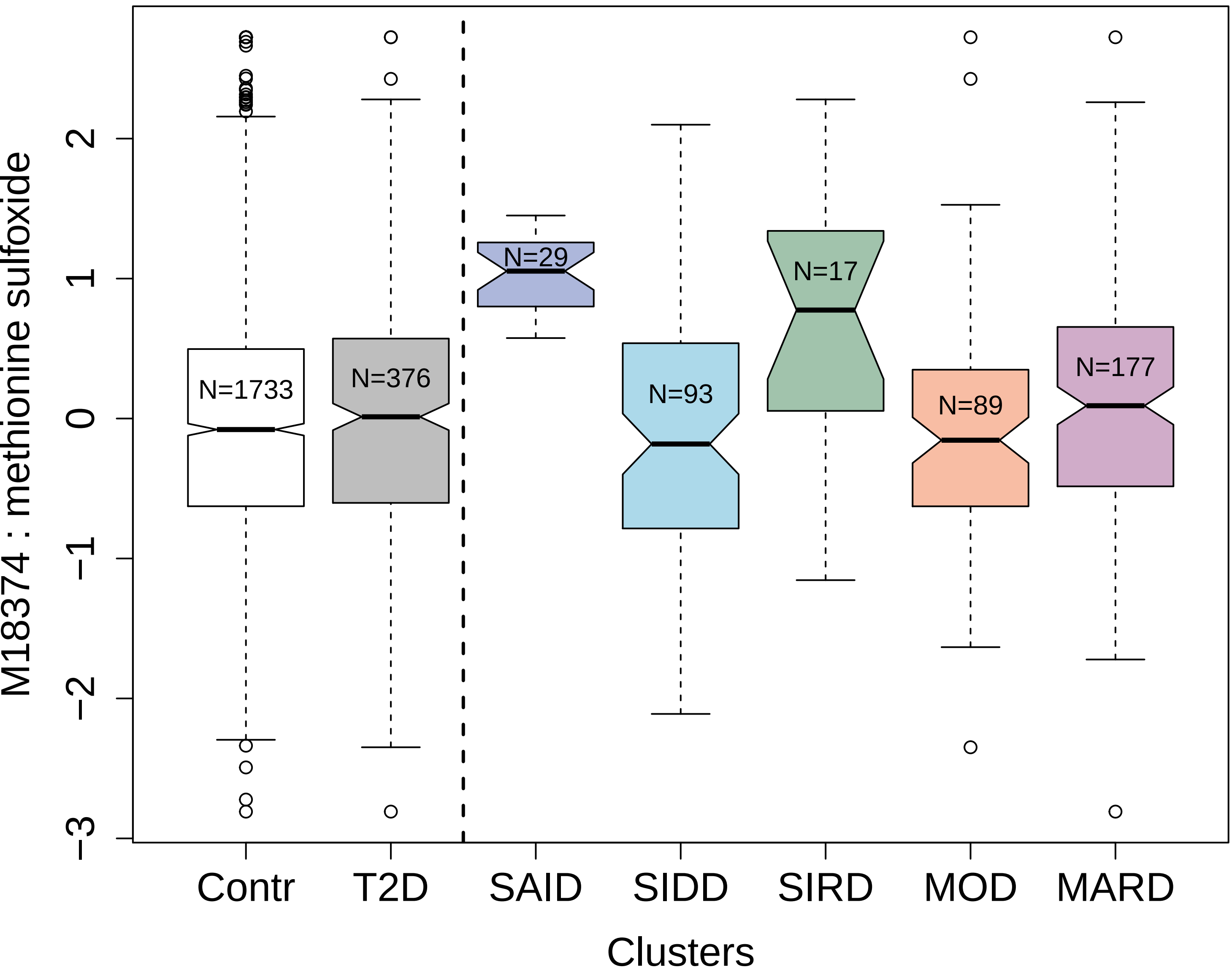

M18374 : methionine sulfoxide
Clusters
Contr
T2D
SAID
SIDD
SIRD
MOD
MARD
N=1733
N=376
N=29
N=93
N=17
N=89
N=177
2
1
0
−1
−2
−3

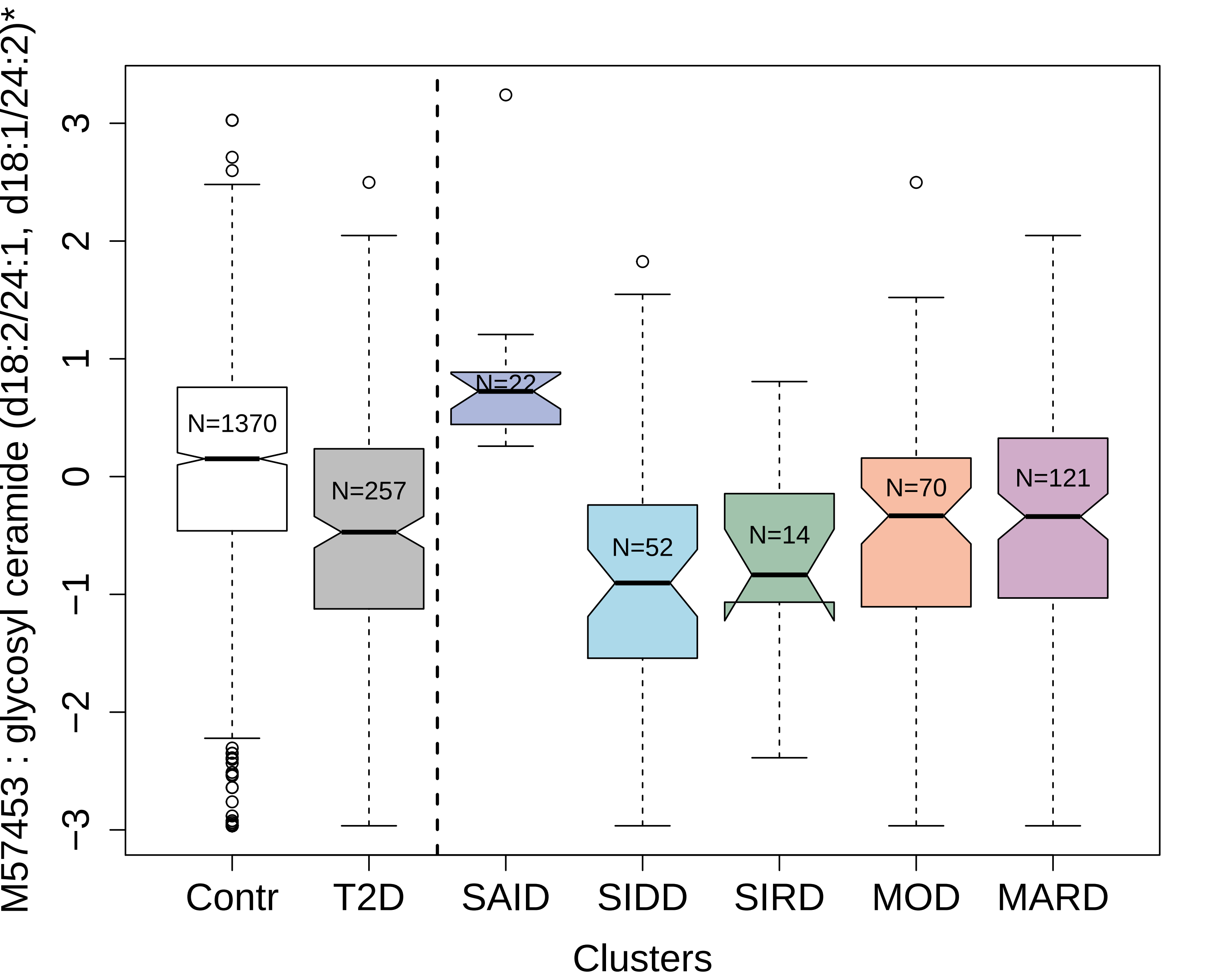
M57453 : glycosyl ceramide (d18:2/24:1, d18:1/24:2)*
3
2
1
0
−1
−2
−3
N=1370
N=257
N=22
N=52
N=14
N=70
N=121
Contr
T2D
SAID
SIDD
SIRD
MOD
MARD
Clusters

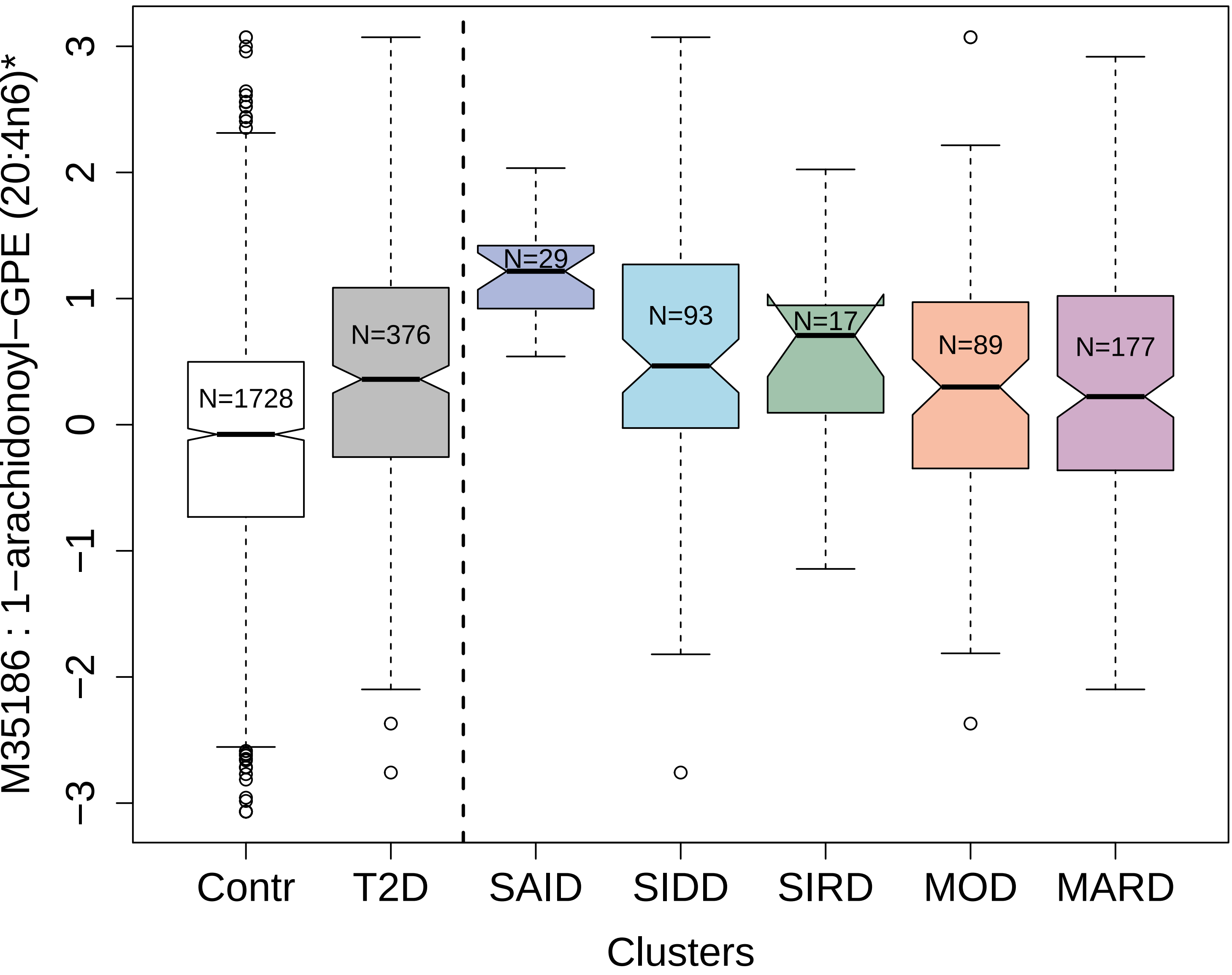
M35186 : 1−arachidonoyl−GPE (20:4n6)*
3
2
1
0
−1
−2
−3
N=1728
N=376
N=29
N=93
N=17
N=89
N=177
Contr
T2D
SAID
SIDD
SIRD
MOD
MARD
Clusters

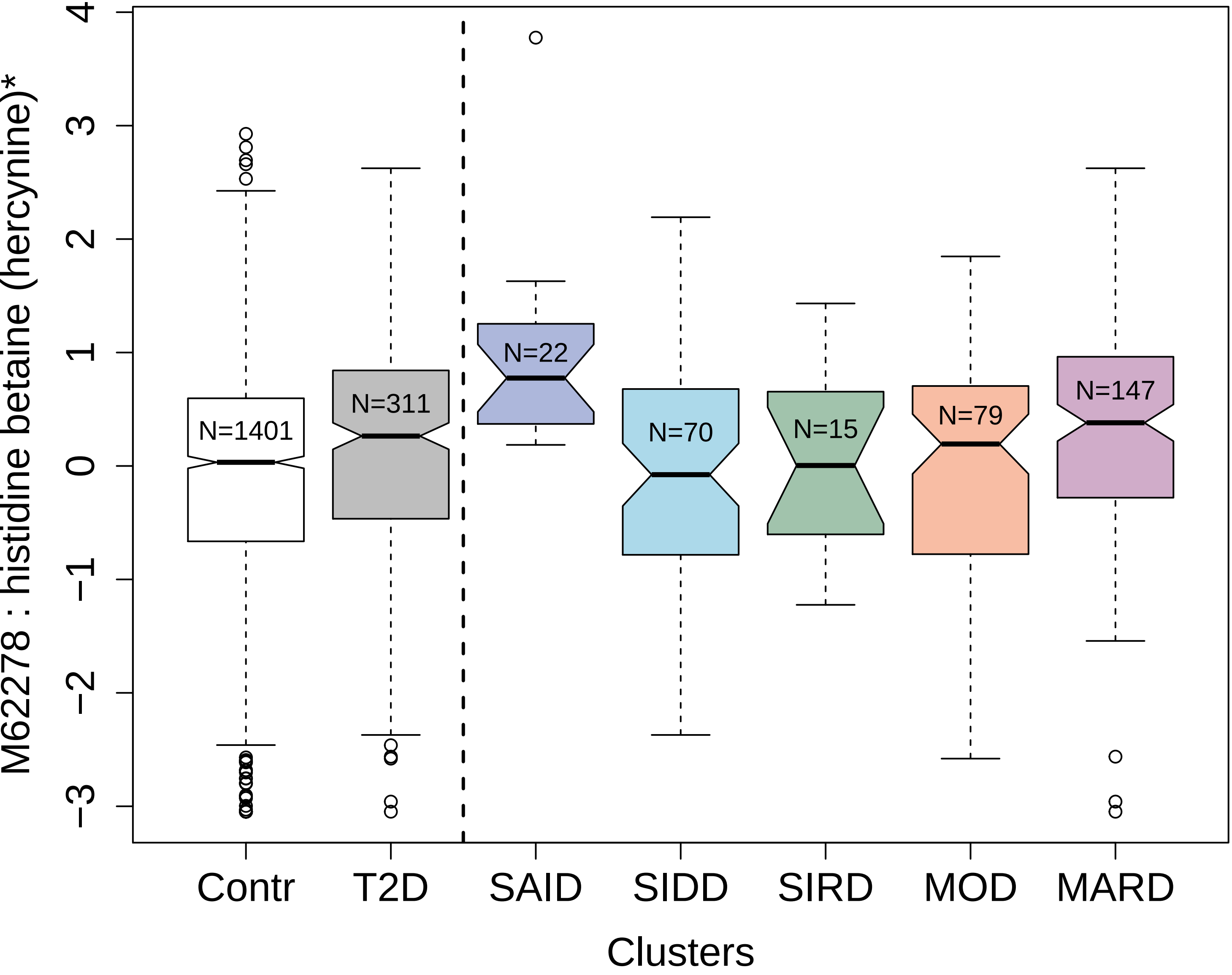
M62278 : histidine betaine (hercynine)*
4
3
2
1
0
−1
−2
−3
N=1401
N=311
N=22
N=70
N=15
N=79
N=147
Contr
T2D
SAID
SIDD
SIRD
MOD
MARD
Clusters

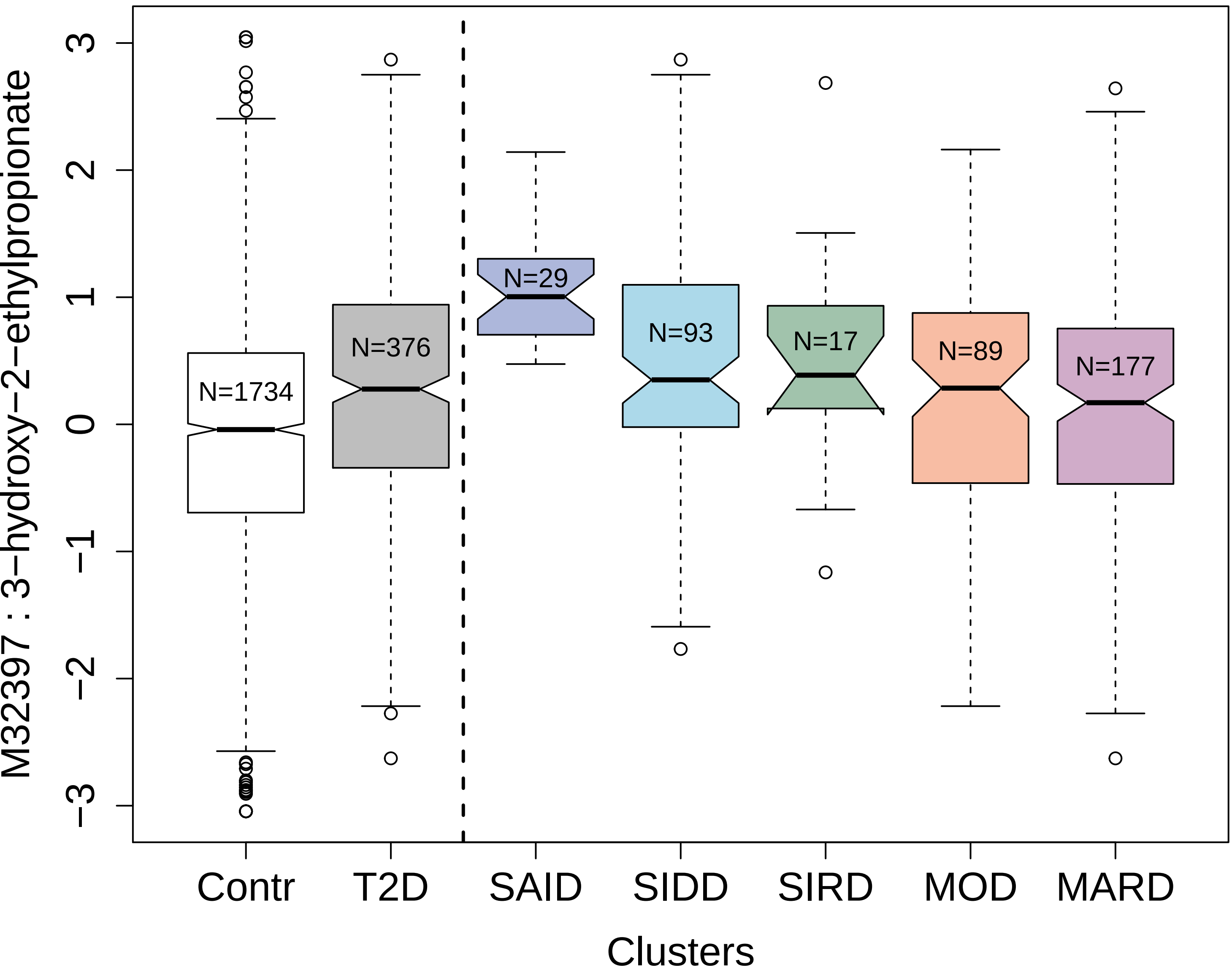
M32397 : 3−hydroxy−2−ethylpropionate
3
2
1
0
−1
−2
−3
N=1734
N=376
N=29
N=93
N=17
N=89
N=177
Contr
T2D
SAID
SIDD
SIRD
MOD
MARD
Clusters

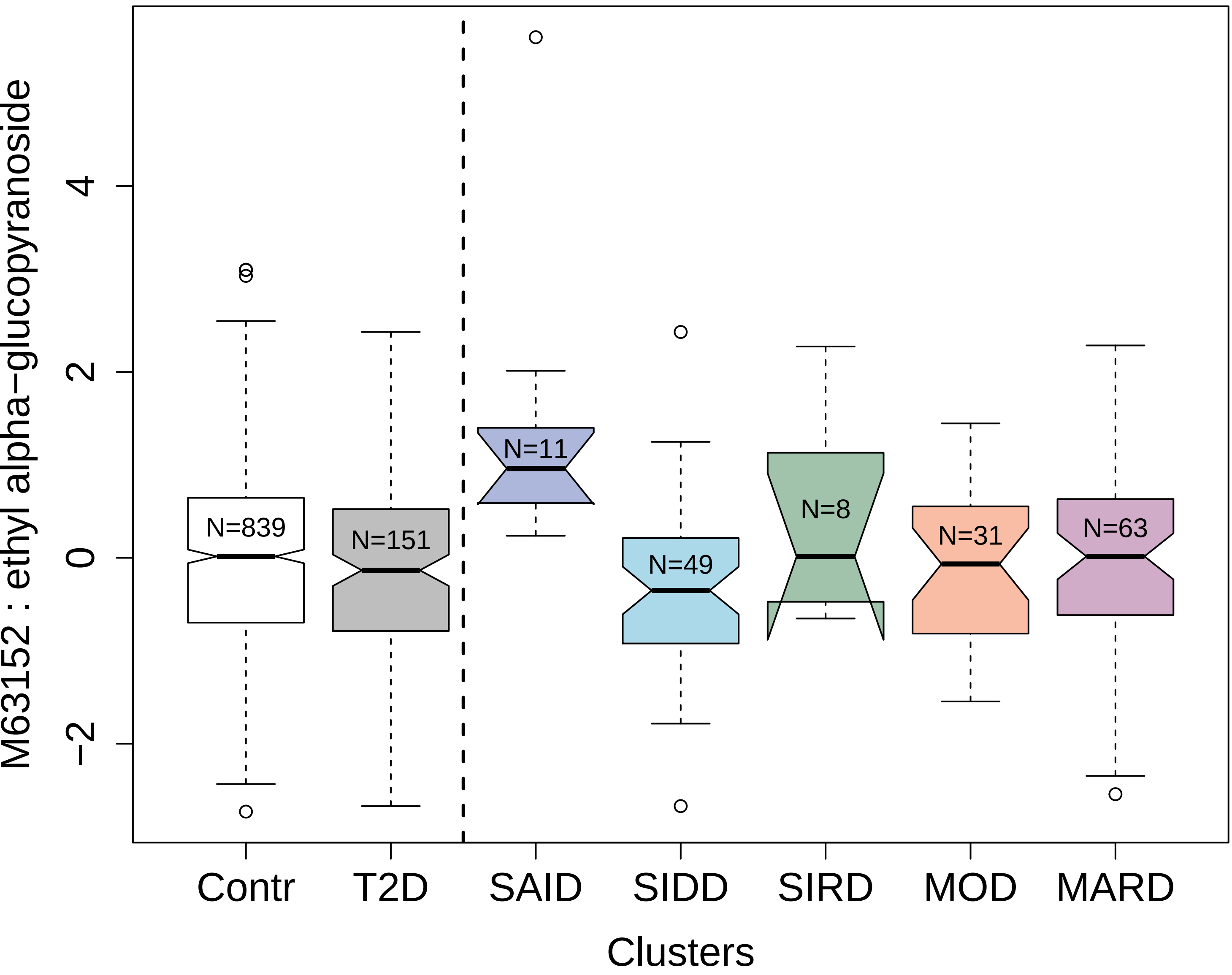

M63152 : ethyl alpha−glucopyranoside
−2
0
2
4
N=839
N=151
N=11
N=49
N=8
N=31
N=63
Contr
T2D
SAID
SIDD
SIRD
MOD
MARD
Clusters

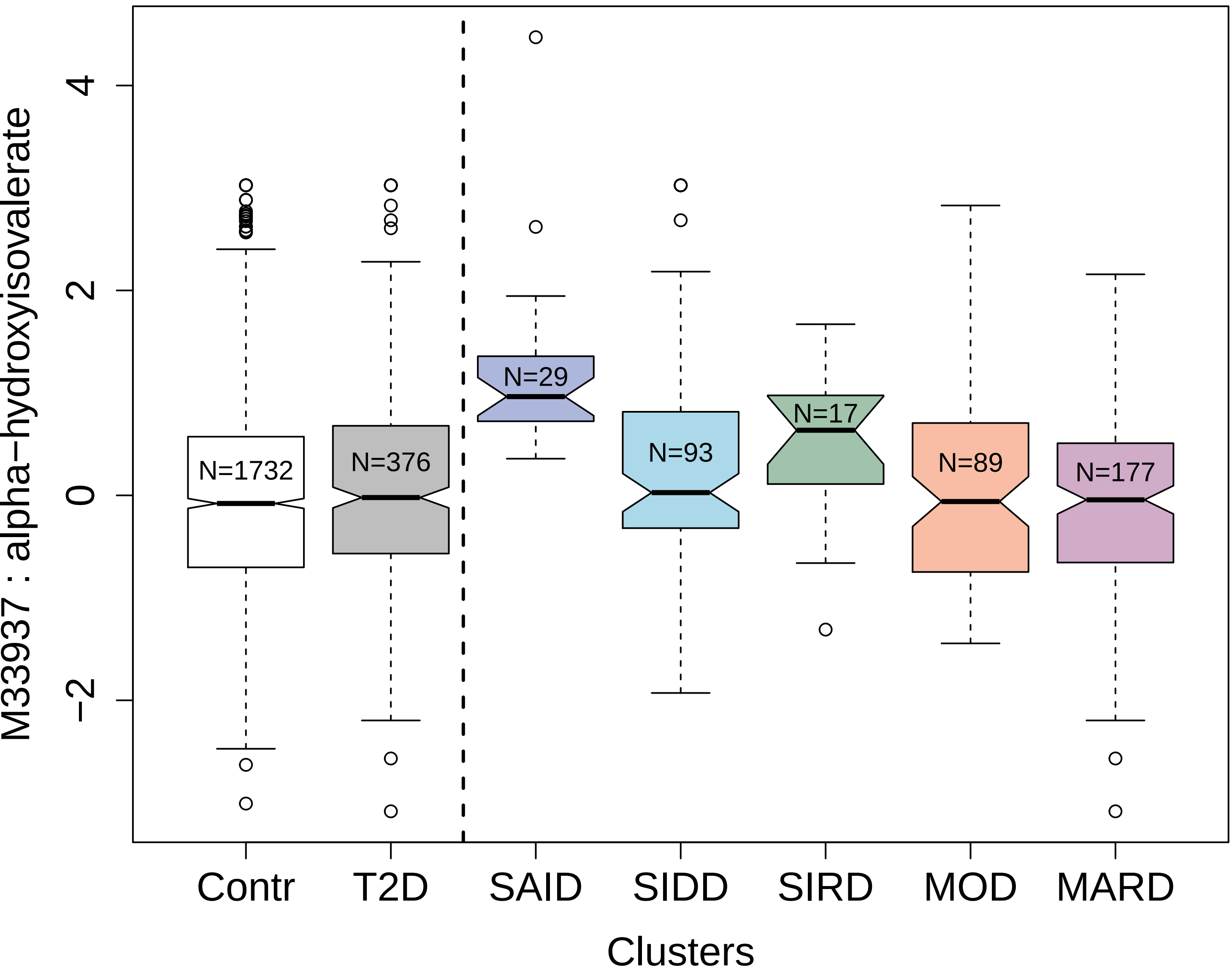

M33937 : alpha−hydroxyisovalerate
4
2
0
−2
N=1732
N=376
N=29
N=93
N=17
N=89
N=177
Contr
T2D
SAID
SIDD
SIRD
MOD
MARD
Clusters

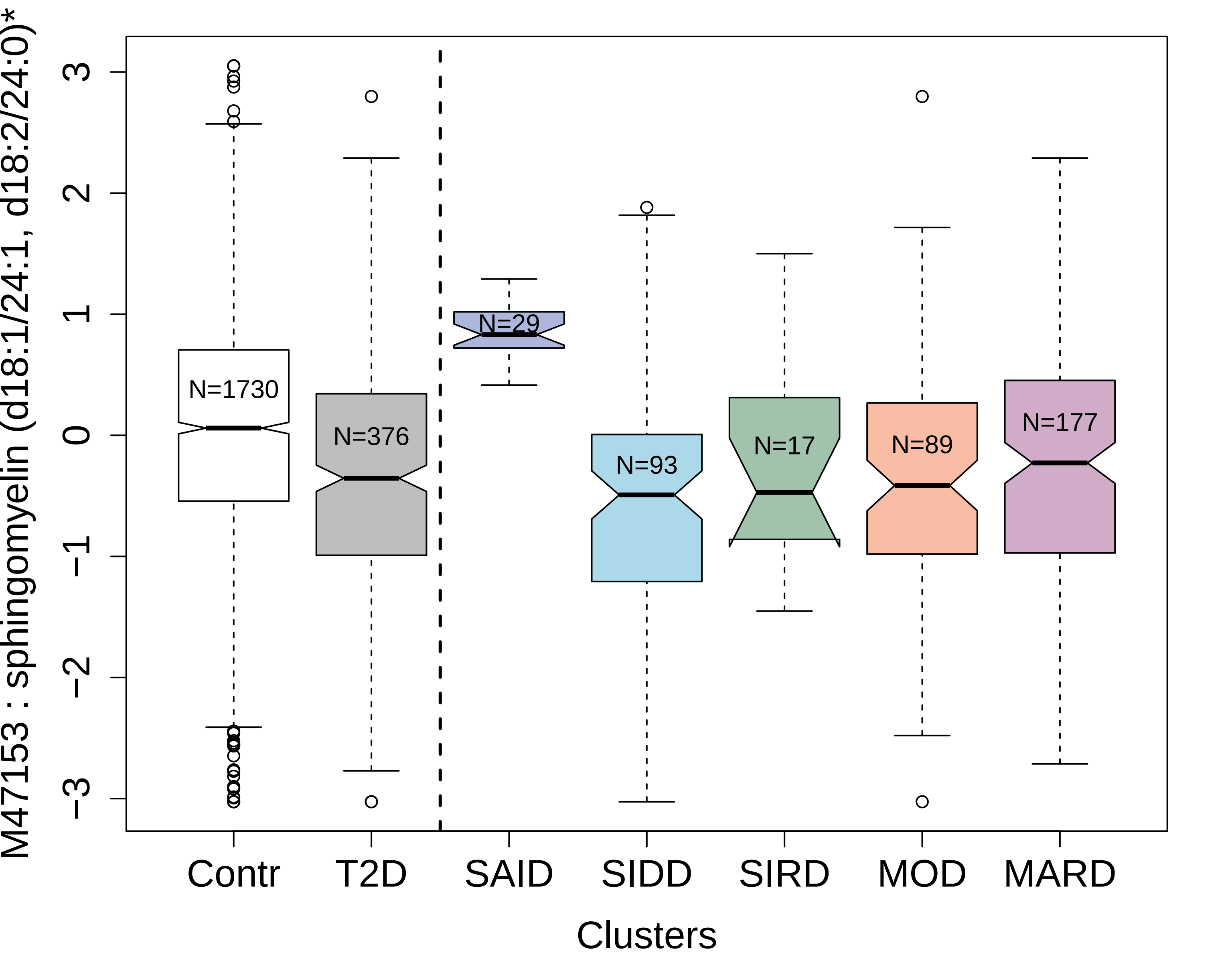
M47153 : sphingomyelin (d18:1/24:1, d18:2/24:0)*
3
2
1
0
−1
−2
−3
N=1730
N=376
N=29
N=93
N=17
N=89
N=177
Contr
T2D
SAID
SIDD
SIRD
MOD
MARD
Clusters

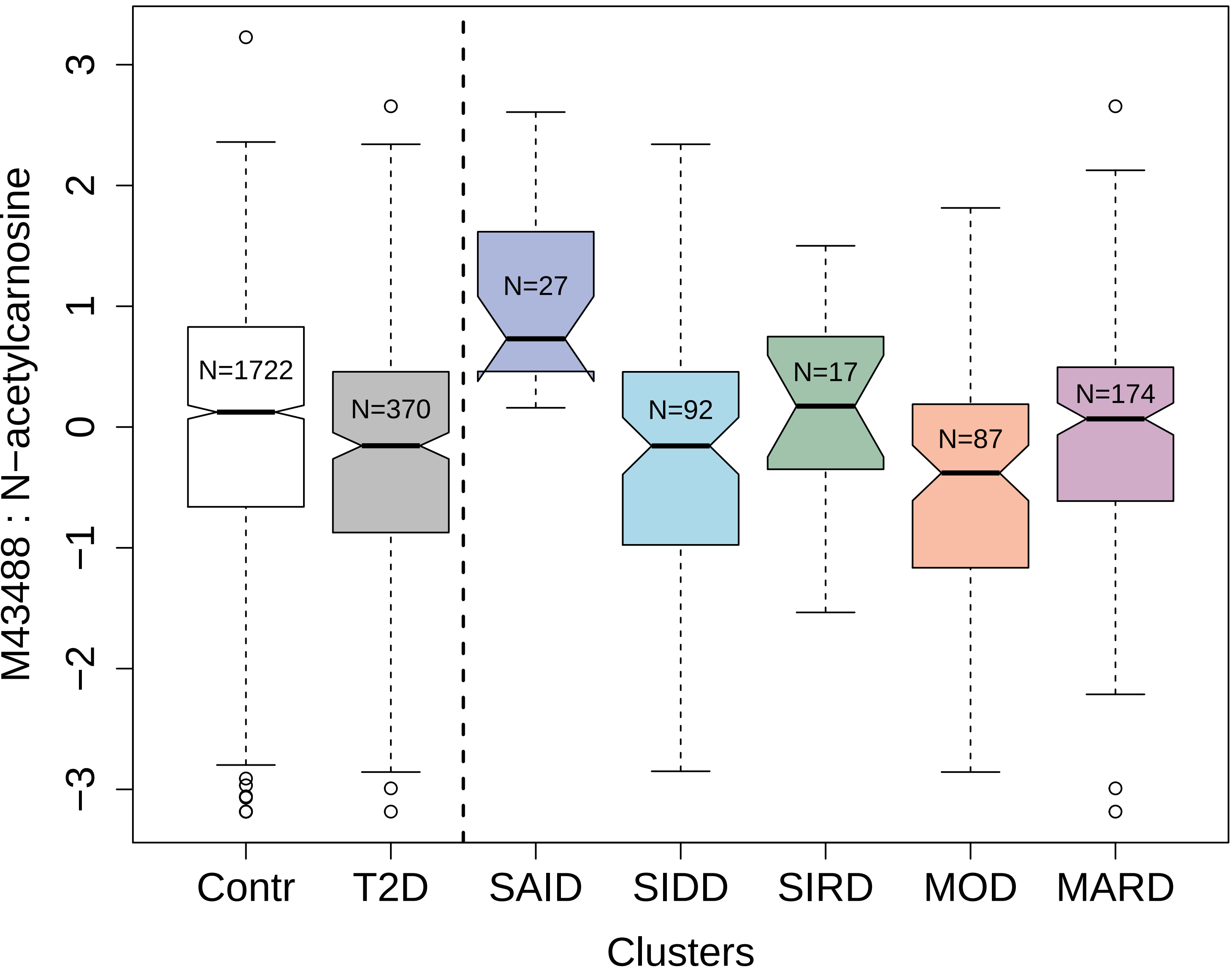

M43488 : N−acetylcarnosine
3
2
1
0
−1
−2
−3
N=1722
N=370
N=27
N=92
N=17
N=87
N=174
Contr
T2D
SAID
SIDD
SIRD
MOD
MARD
Clusters

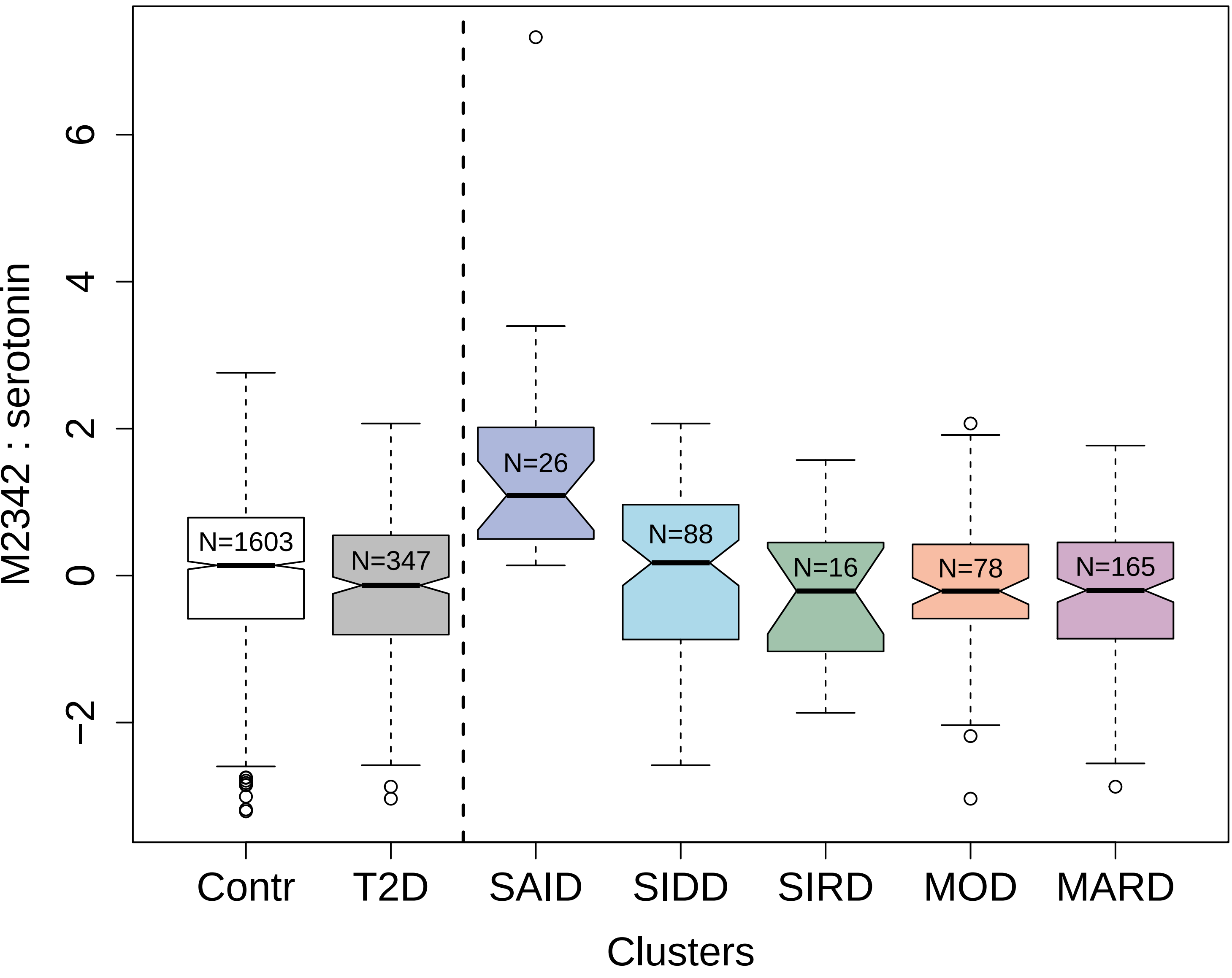

M2342 : serotonin
6
4
2
0
−2
N=1603
N=347
N=26
N=88
N=16
N=78
N=165
Contr
T2D
SAID
SIDD
SIRD
MOD
MARD
Clusters

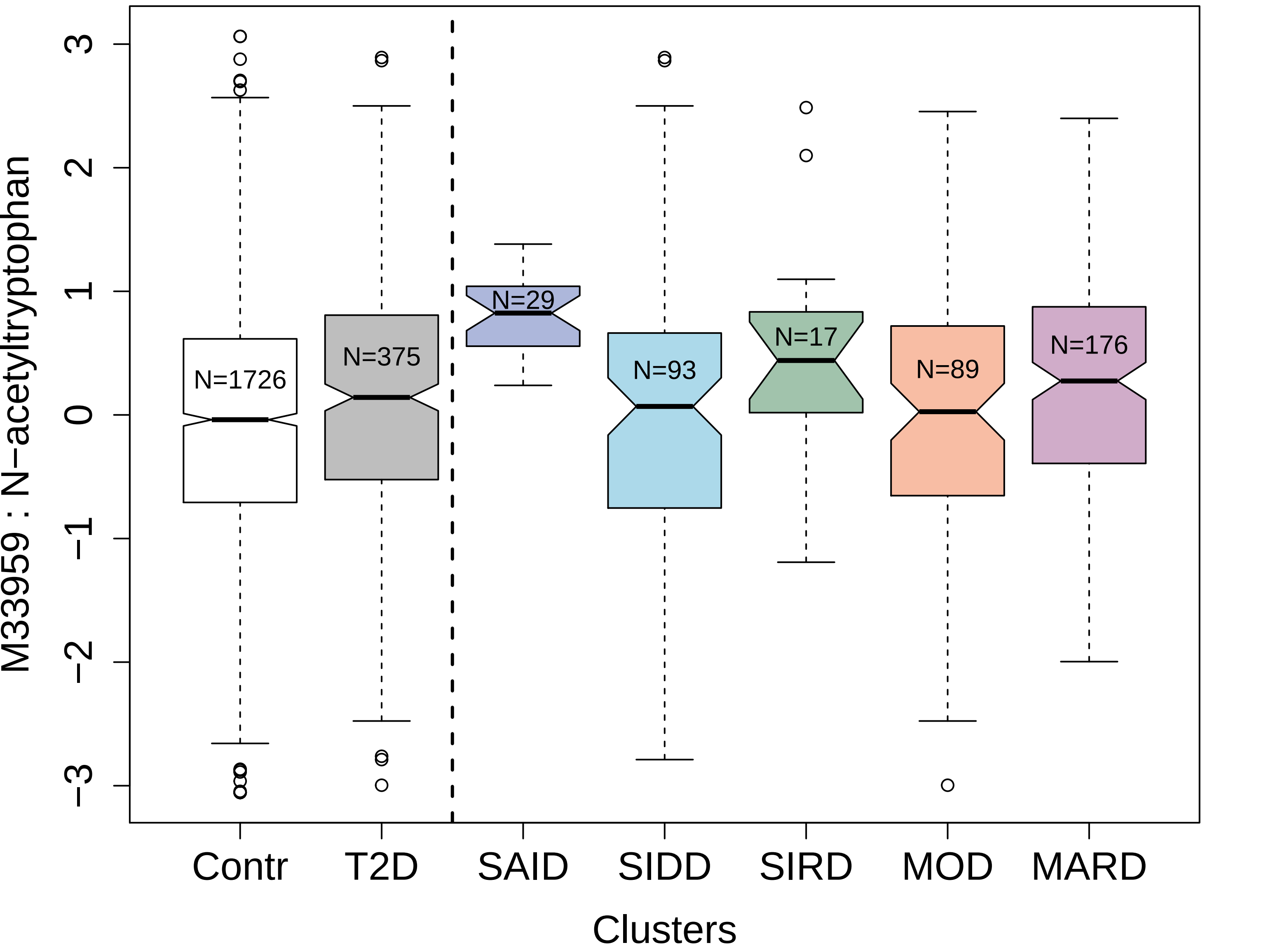

M33959 : N−acetyltryptophan
3
2
1
0
−1
−2
−3
N=1726
N=375
N=29
N=93
N=17
N=89
N=176
Contr
T2D
SAID
SIDD
SIRD
MOD
MARD
Clusters

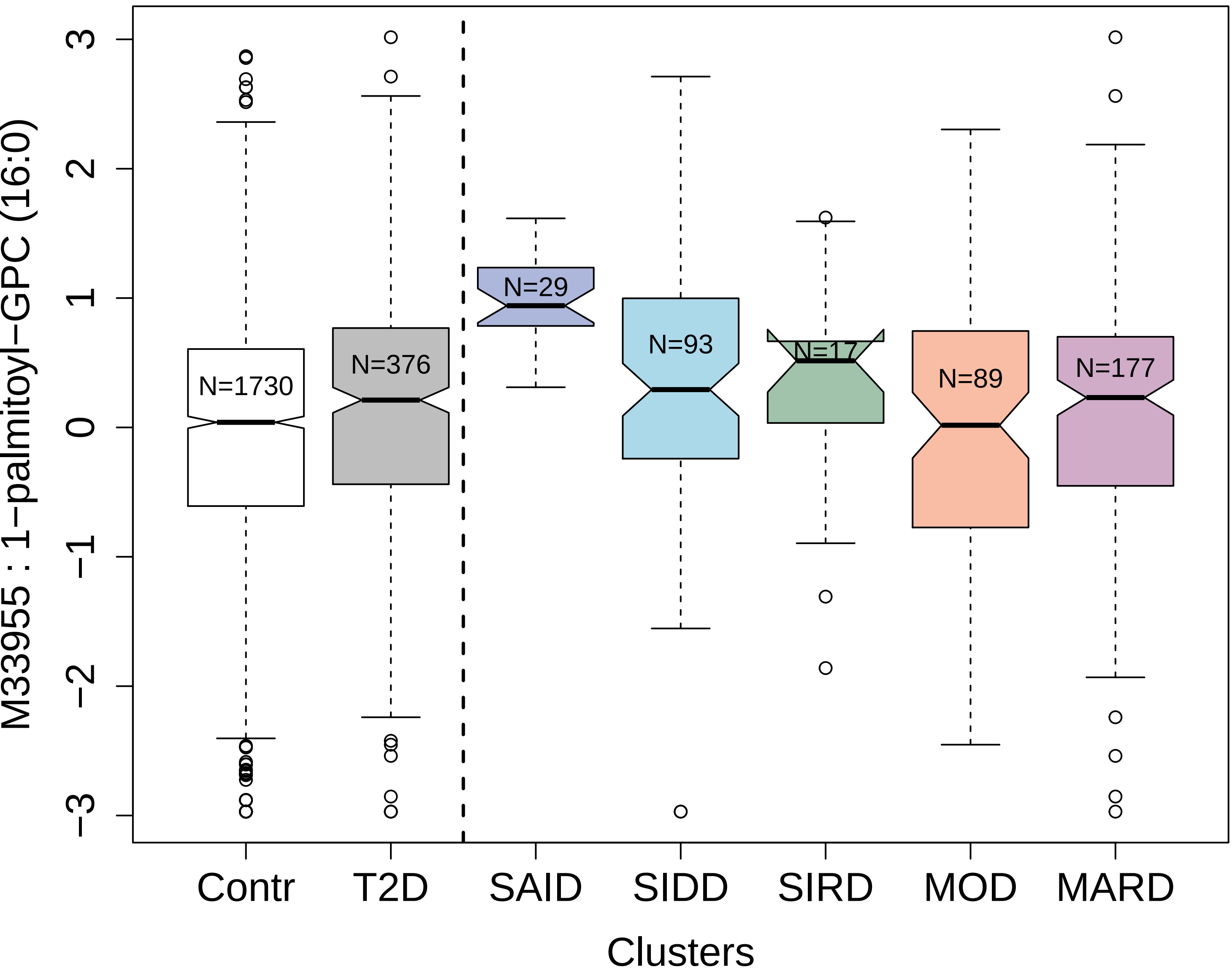

M33955 : 1−palmitoyl−GPC (16:0)
3
2
1
0
−1
−2
−3
N=1730
N=376
N=29
N=93
N=17
N=89
N=177
Contr
T2D
SAID
SIDD
SIRD
MOD
MARD
Clusters

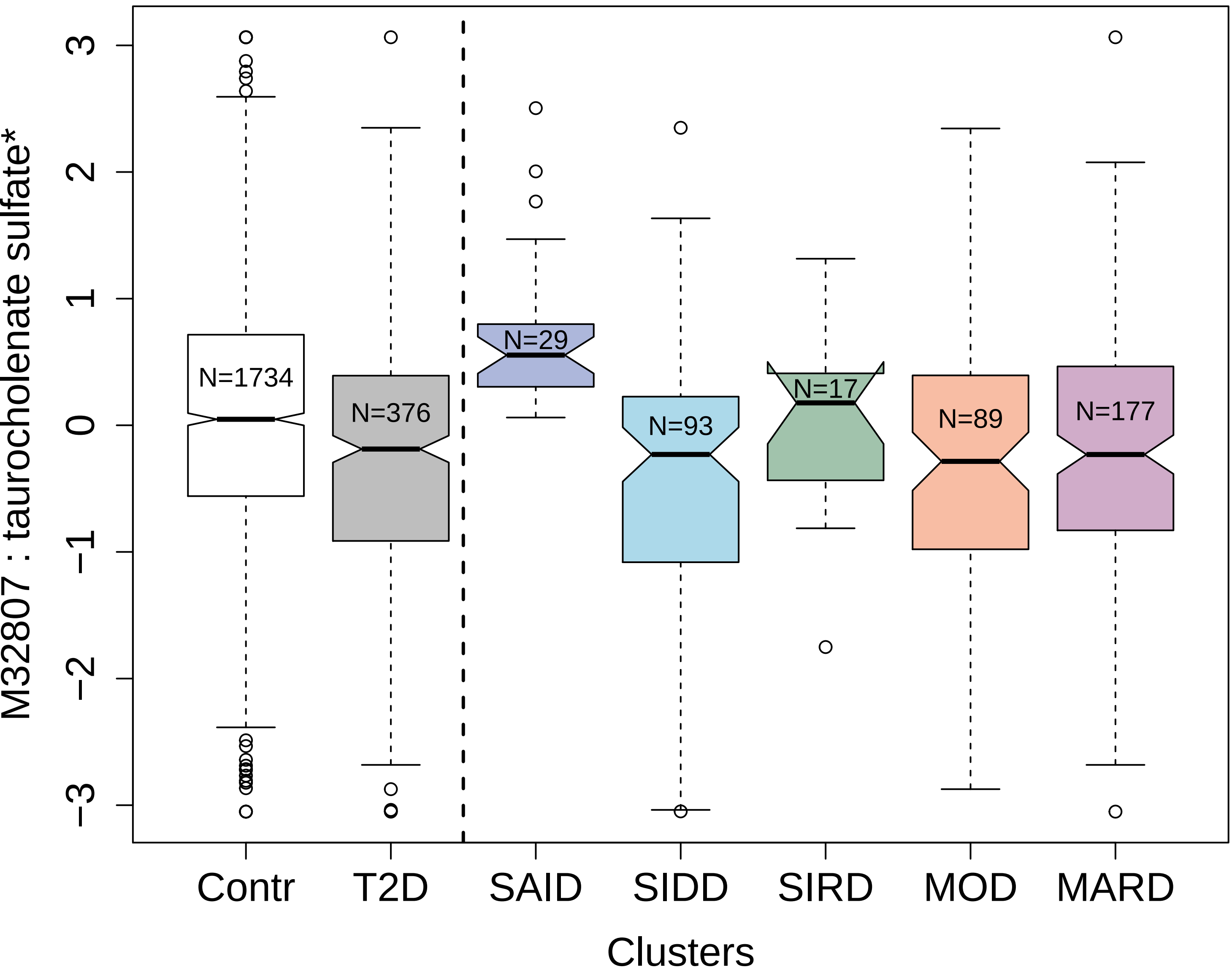

M32807 : taurocholenate sulfate*
3
2
1
0
−1
−2
−3
N=1734
N=376
N=29
N=93
N=17
N=89
N=177
Contr
T2D
SAID
SIDD
SIRD
MOD
MARD
Clusters

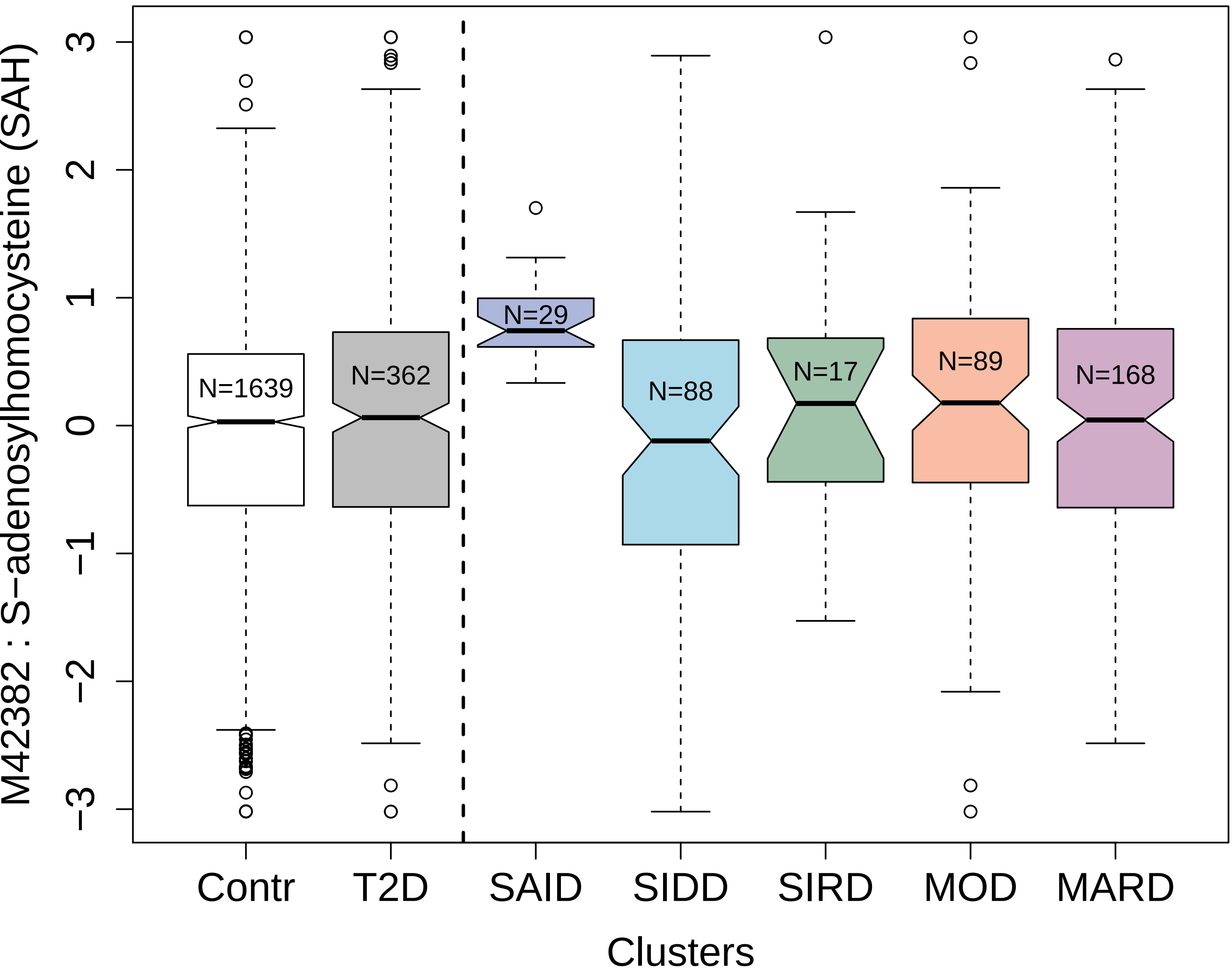
M42382 : S−adenosylhomocysteine (SAH)
3
2
1
0
−1
−2
−3
N=1639
N=362
N=29
N=88
N=17
N=89
N=168
Contr
T2D
SAID
SIDD
SIRD
MOD
MARD
Clusters

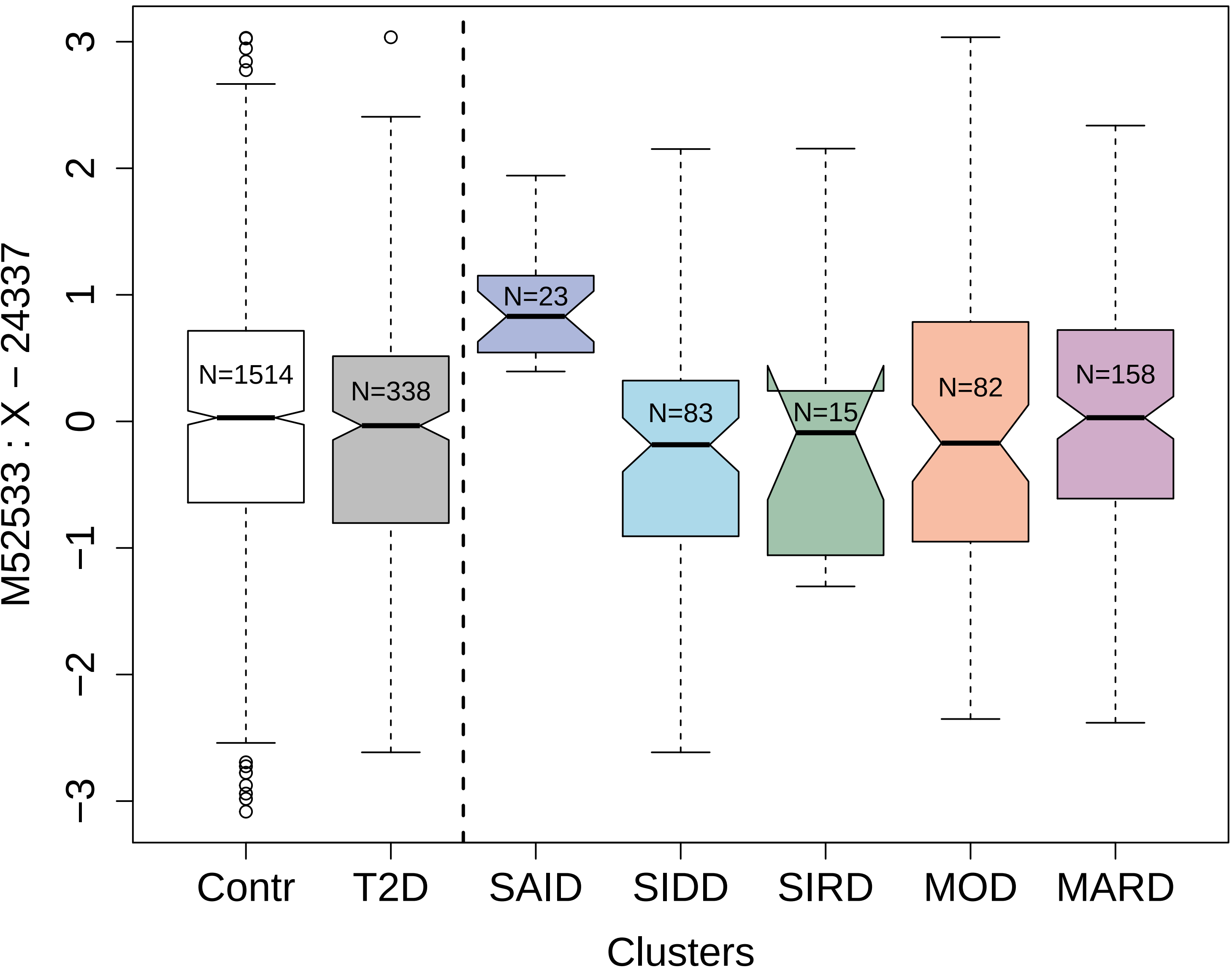

M52533 : X − 24337
3
2
1
0
−1
−2
−3
N=1514
N=338
N=23
N=83
N=15
N=82
N=158
Contr
T2D
SAID
SIDD
SIRD
MOD
MARD
Clusters

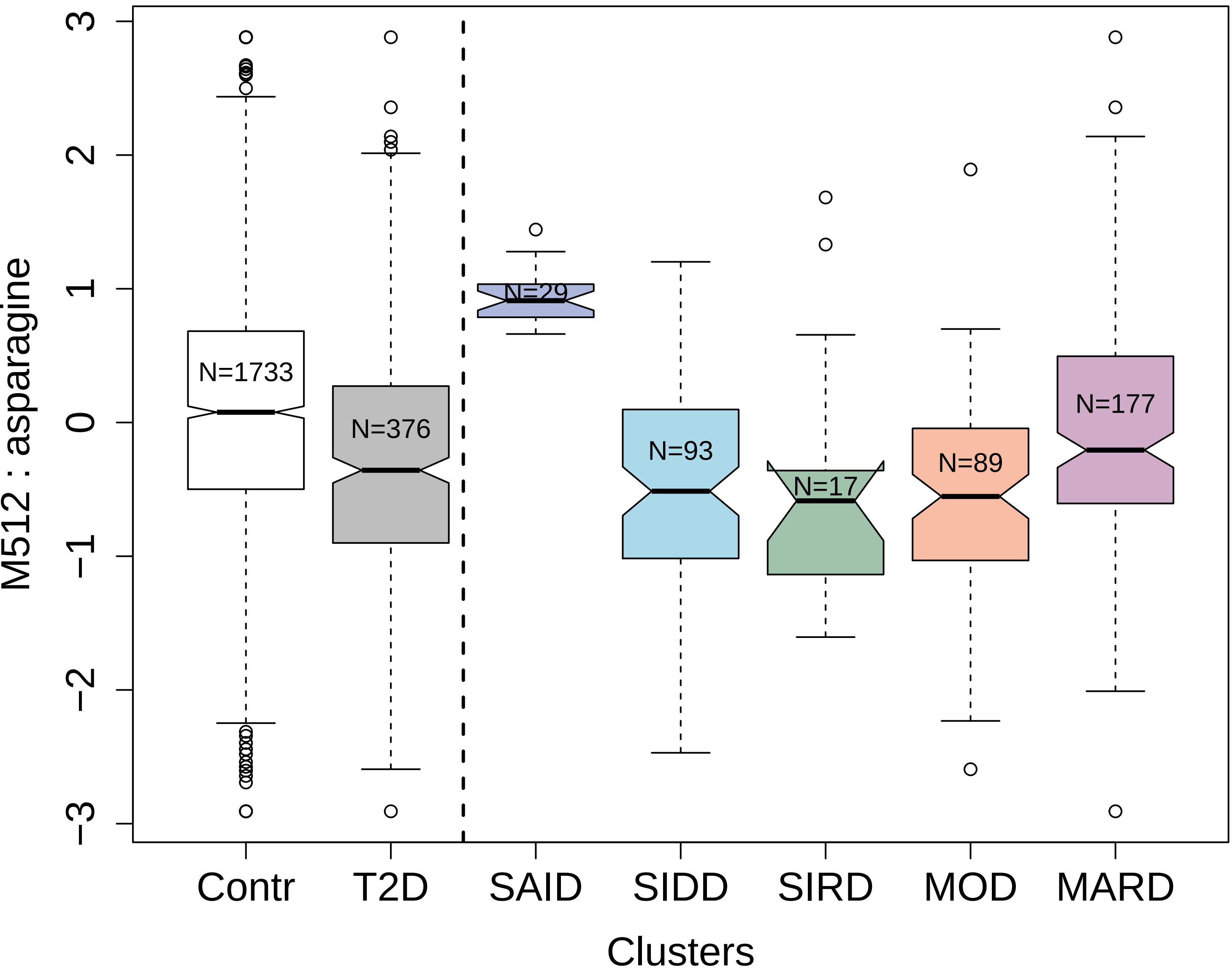

M512 : asparagine
3
2
1
0
−1
−2
−3
N=1733
N=376
N=29
N=93
N=17
N=89
N=177
Contr
T2D
SAID
SIDD
SIRD
MOD
MARD
Clusters

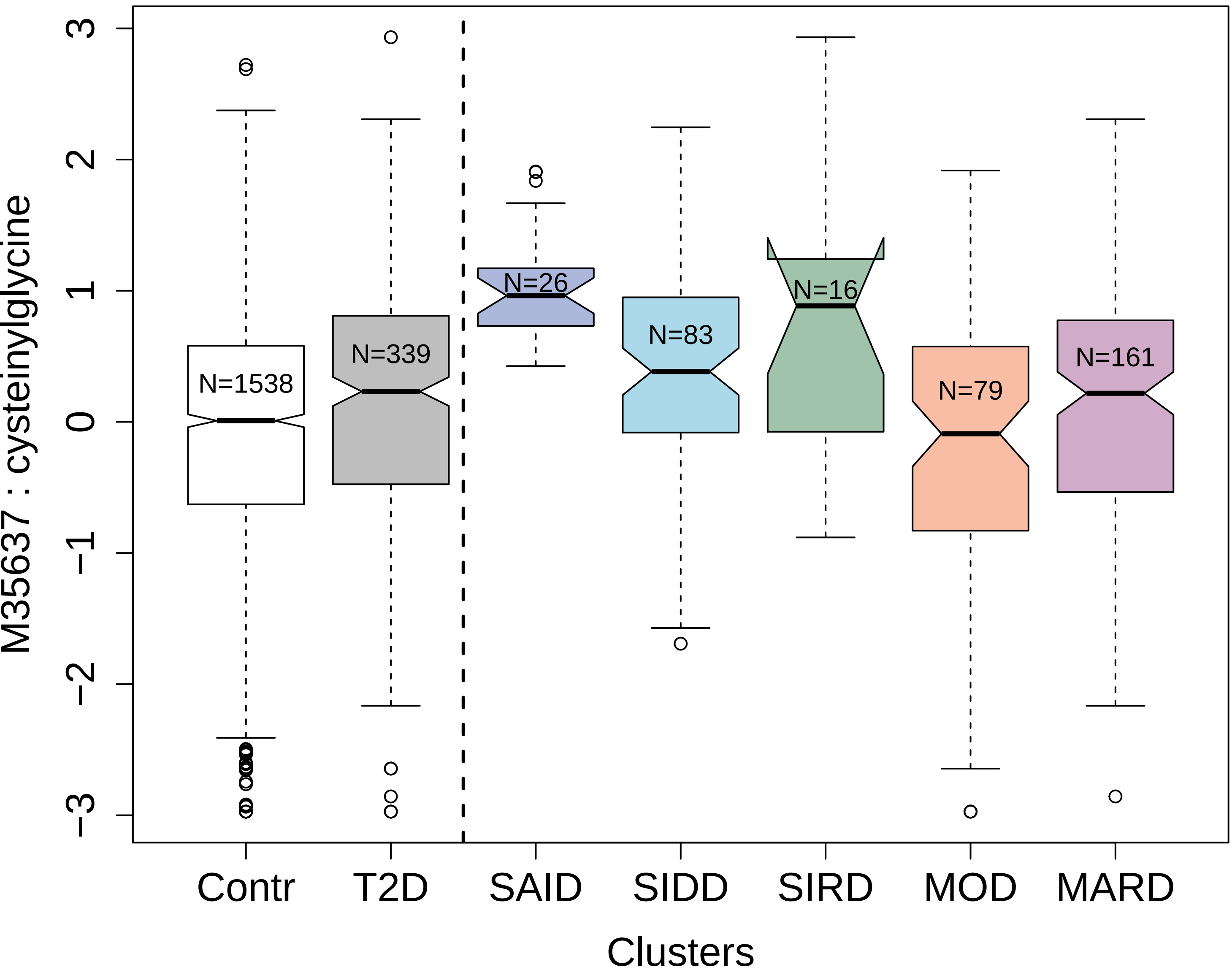

M35637 : cysteinylglycine
3
2
1
0
−1
−2
−3
N=1538
N=339
N=26
N=83
N=16
N=79
N=161
Contr
T2D
SAID
SIDD
SIRD
MOD
MARD
Clusters

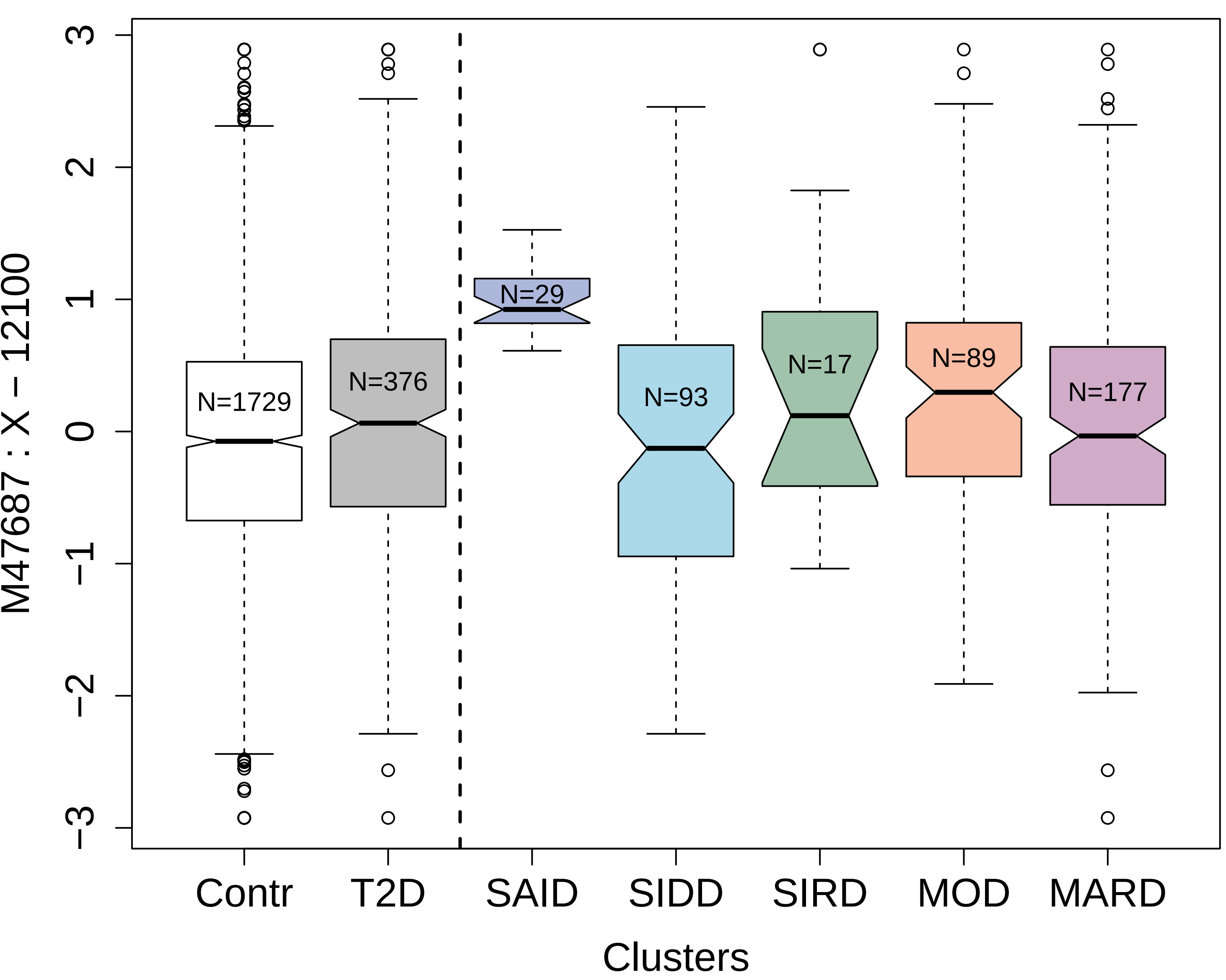

M47687 : X − 12100
3
2
1
0
−1
−2
−3
N=1729
N=376
N=29
N=93
N=17
N=89
N=177
Contr
T2D
SAID
SIDD
SIRD
MOD
MARD
Clusters

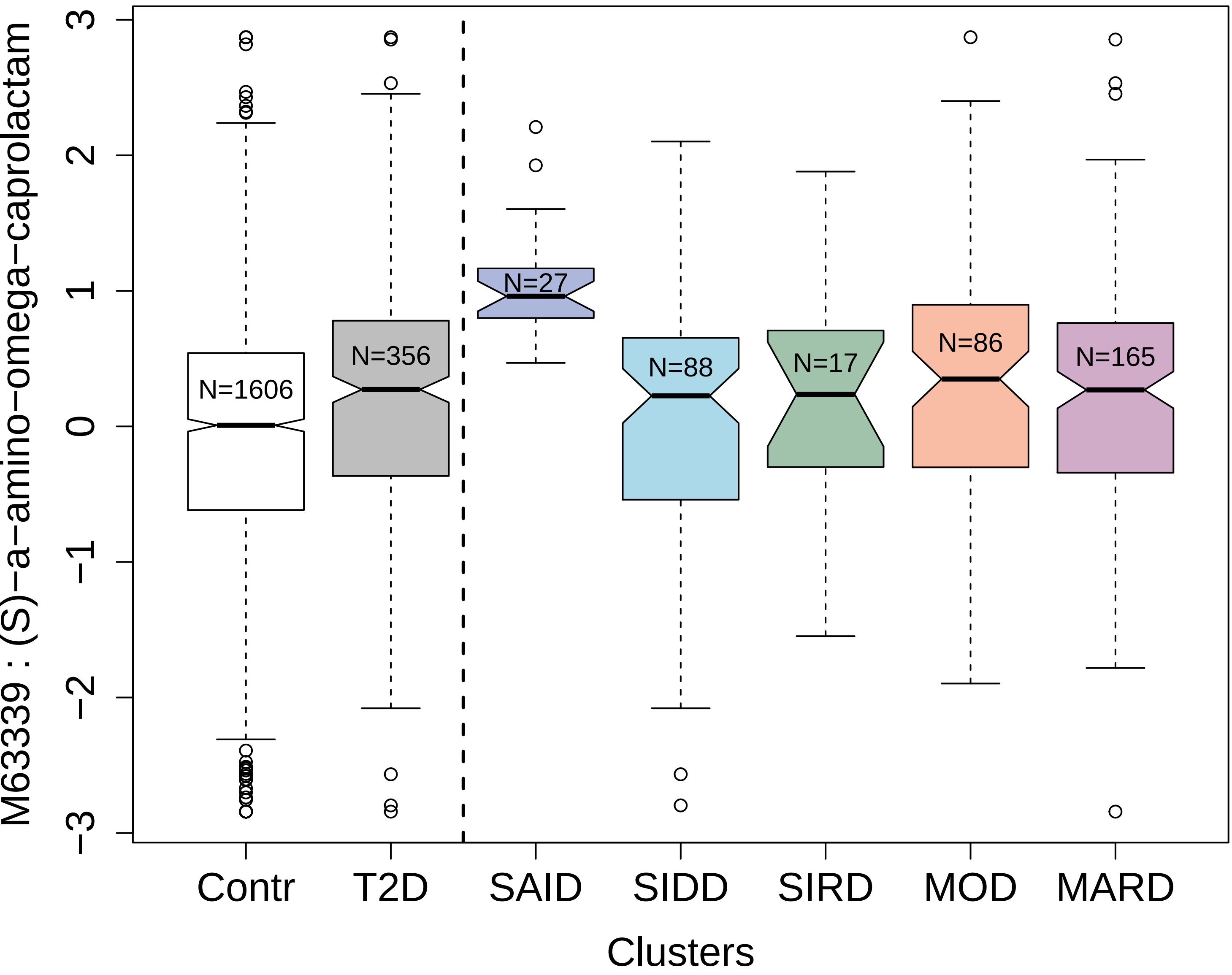
M6339 : (S)−a−amino−omega−caprolactam
3
2
1
0
−1
−2
−3
N=1606
N=356
N=27
N=88
N=17
N=86
N=165
Contr
T2D
SAID
SIDD
SIRD
MOD
MARD
Clusters

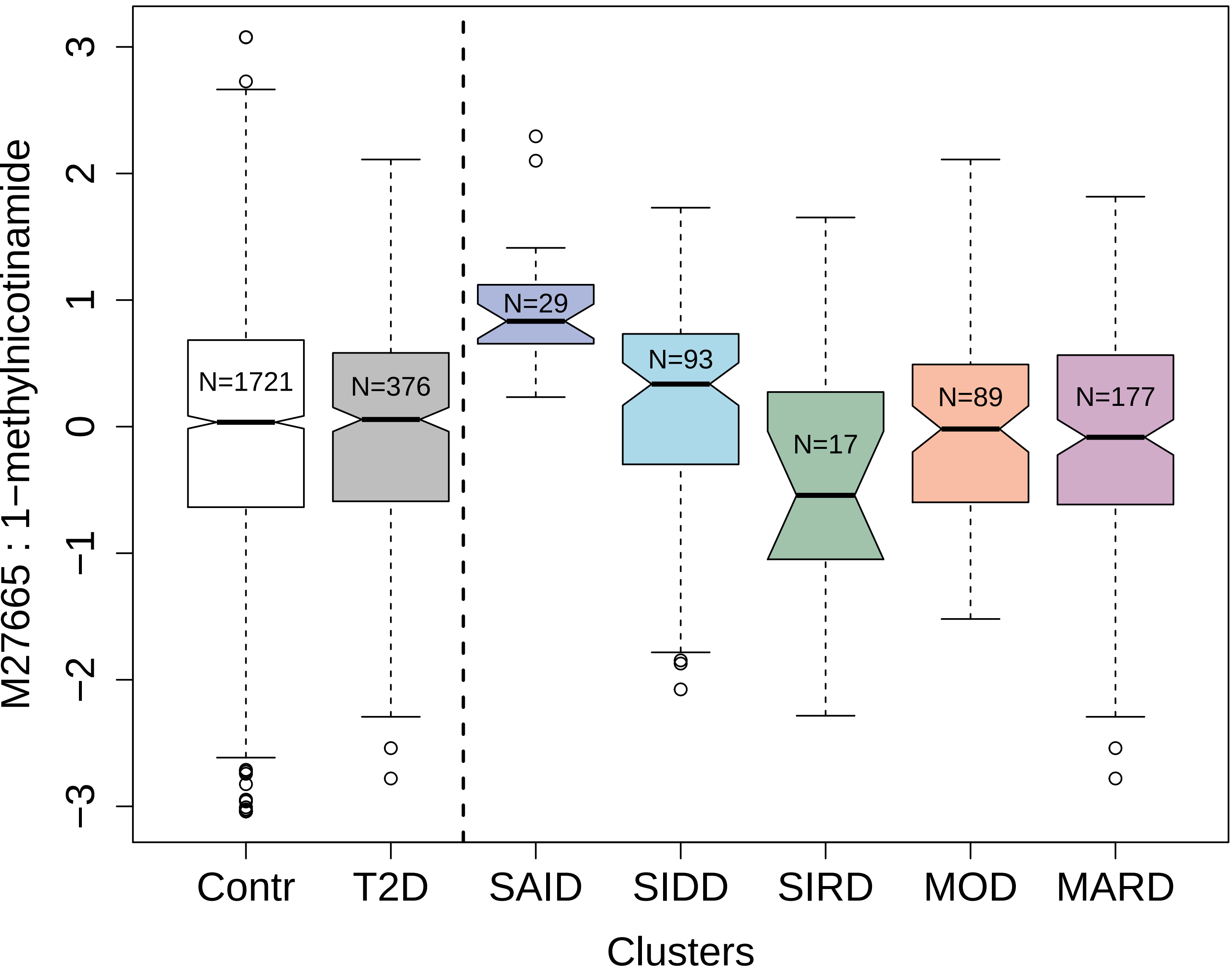

M27665 : 1−methylnicotinamide
3
2
1
0
−1
−2
−3
N=1721
N=376
N=29
N=93
N=17
N=89
N=177
Contr
T2D
SAID
SIDD
SIRD
MOD
MARD
Clusters

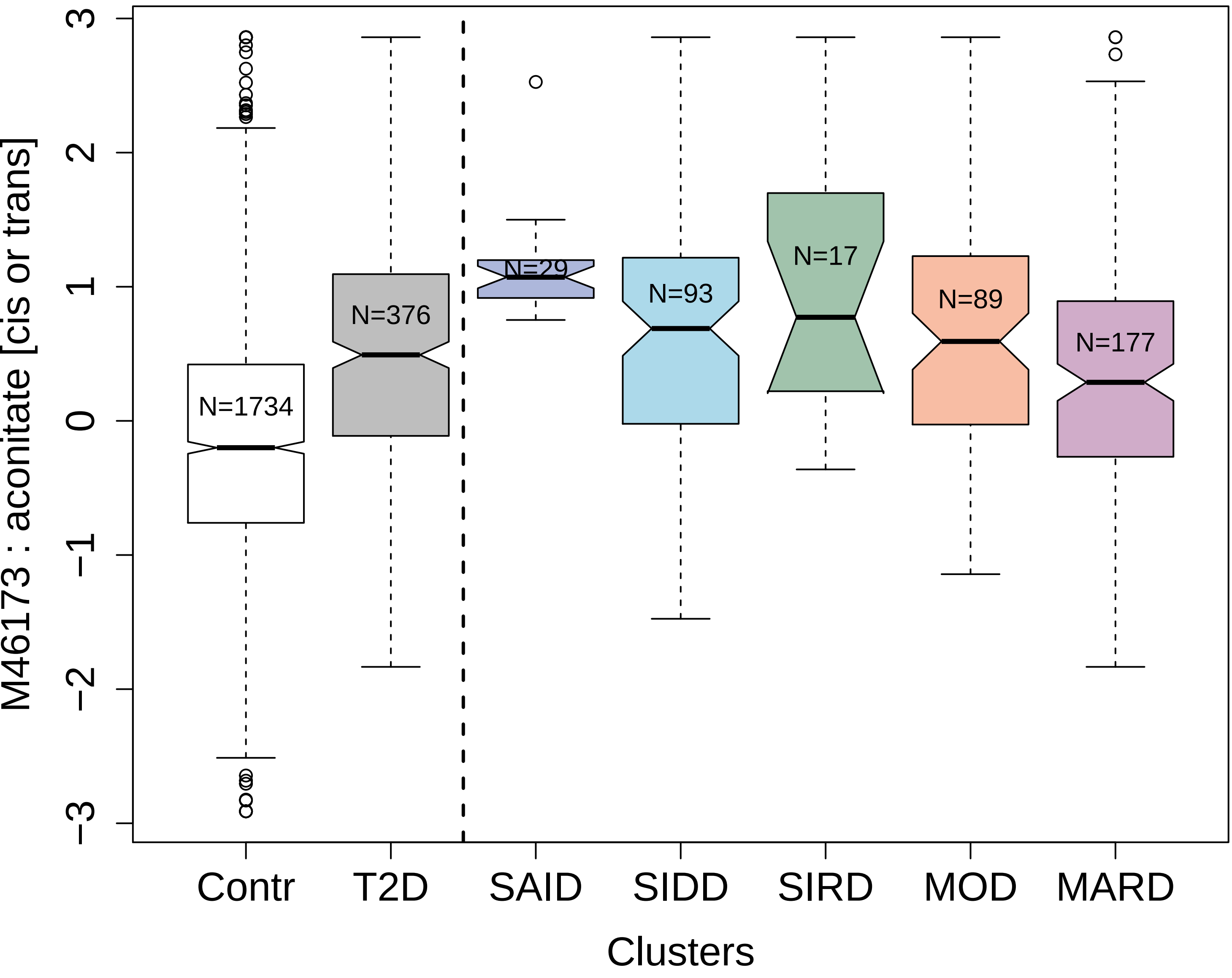

M46173 : aconitate [cis or trans]
3
2
1
0
−1
−2
−3
N=1734
N=376
N=29
N=93
N=17
N=89
N=177
Contr
T2D
SAID
SIDD
SIRD
MOD
MARD
Clusters

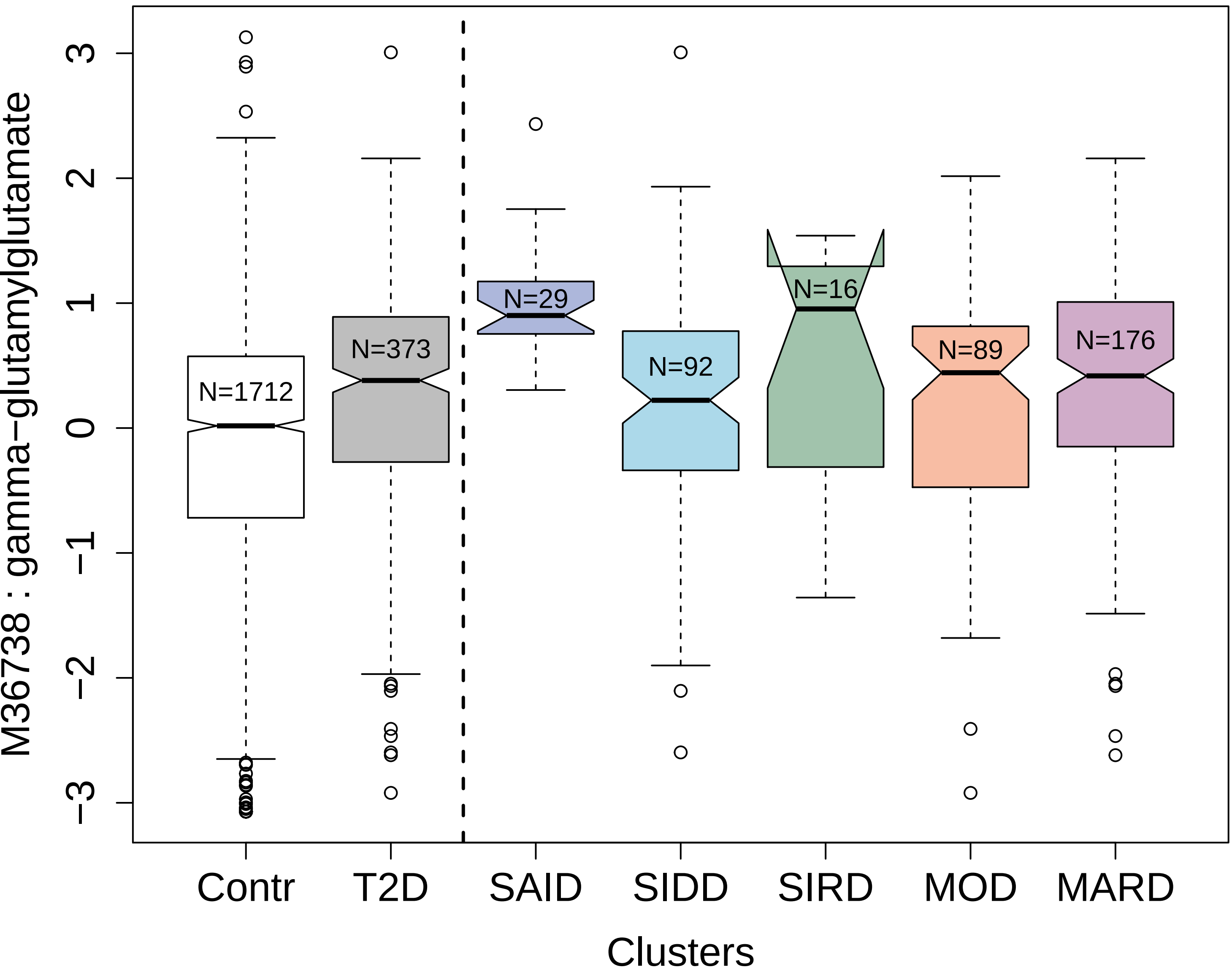

M36738 : gamma−glutamylglutamate
3
2
1
0
−1
−2
−3
N=1712
N=373
N=29
N=92
N=16
N=89
N=176
Contr
T2D
SAID
SIDD
SIRD
MOD
MARD
Clusters

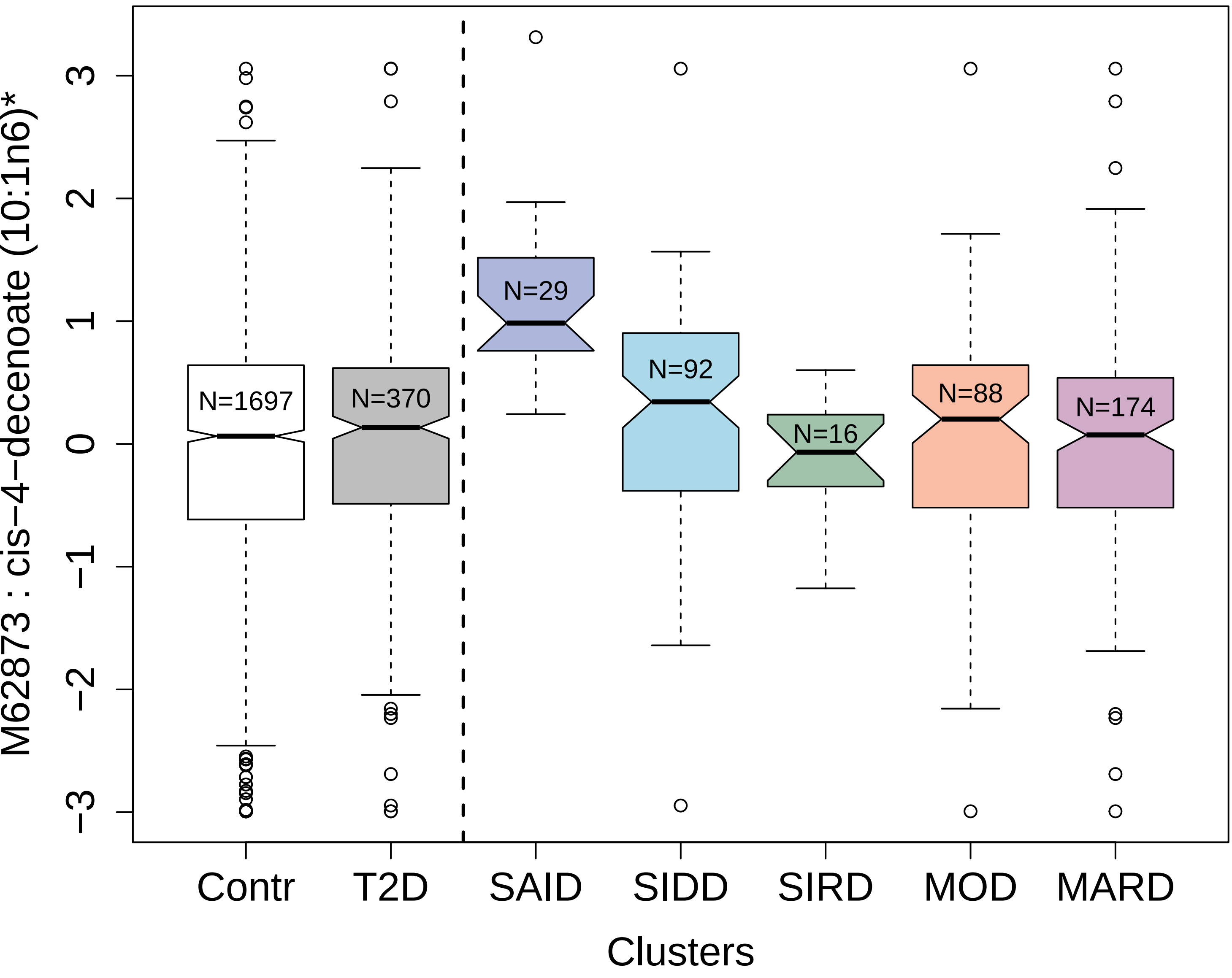

M62873 : cis−4−decenoate (10:1n6)*
3
2
1
0
−1
−2
−3
N=1697
N=370
N=29
N=92
N=16
N=88
N=174
Contr
T2D
SAID
SIDD
SIRD
MOD
MARD
Clusters

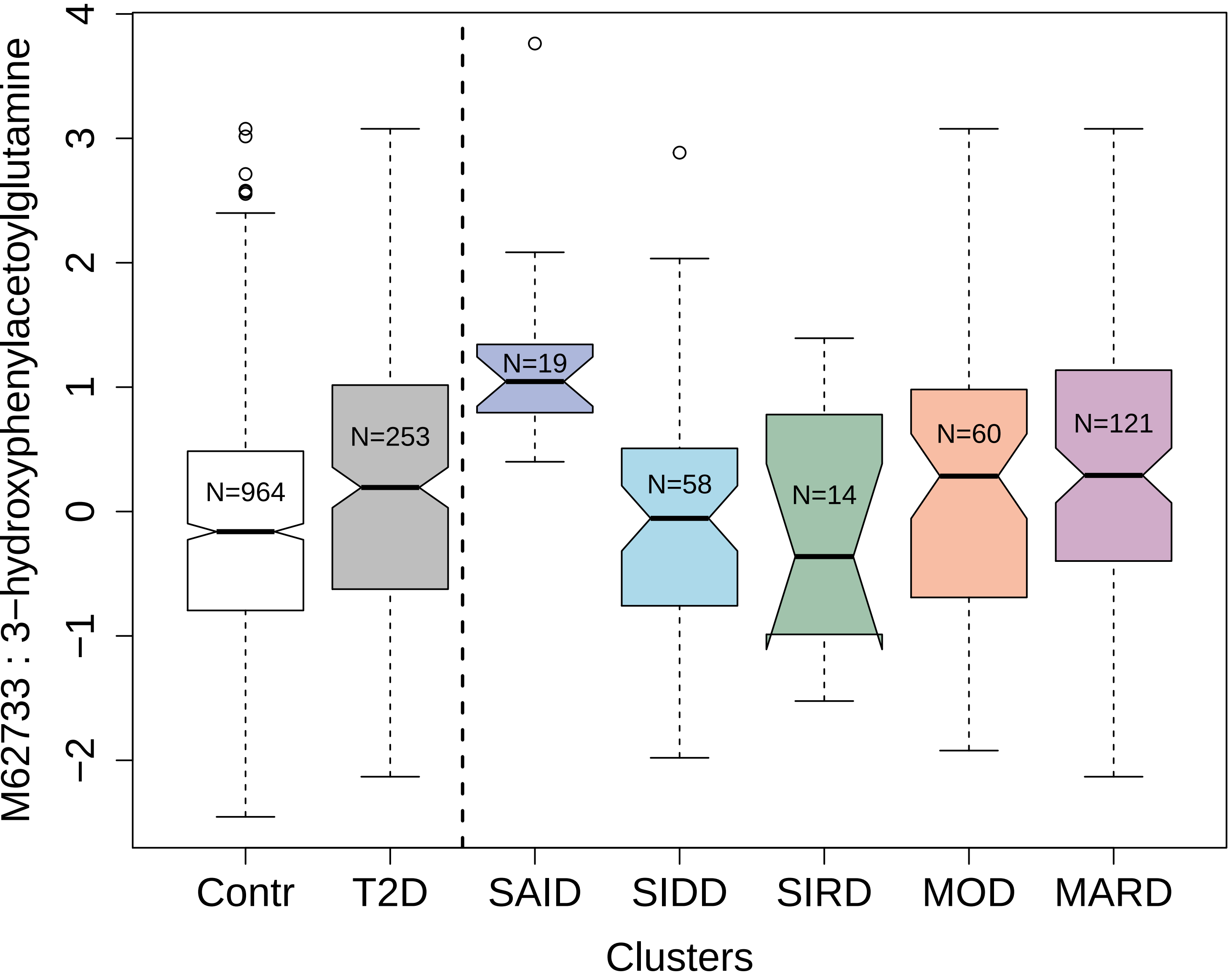
M62733 : 3−hydroxyphenylacetoylglutamine
4
3
2
1
0
−1
−2
N=964
N=253
N=19
N=58
N=14
N=60
N=121
Contr
T2D
SAID
SIDD
SIRD
MOD
MARD
Clusters

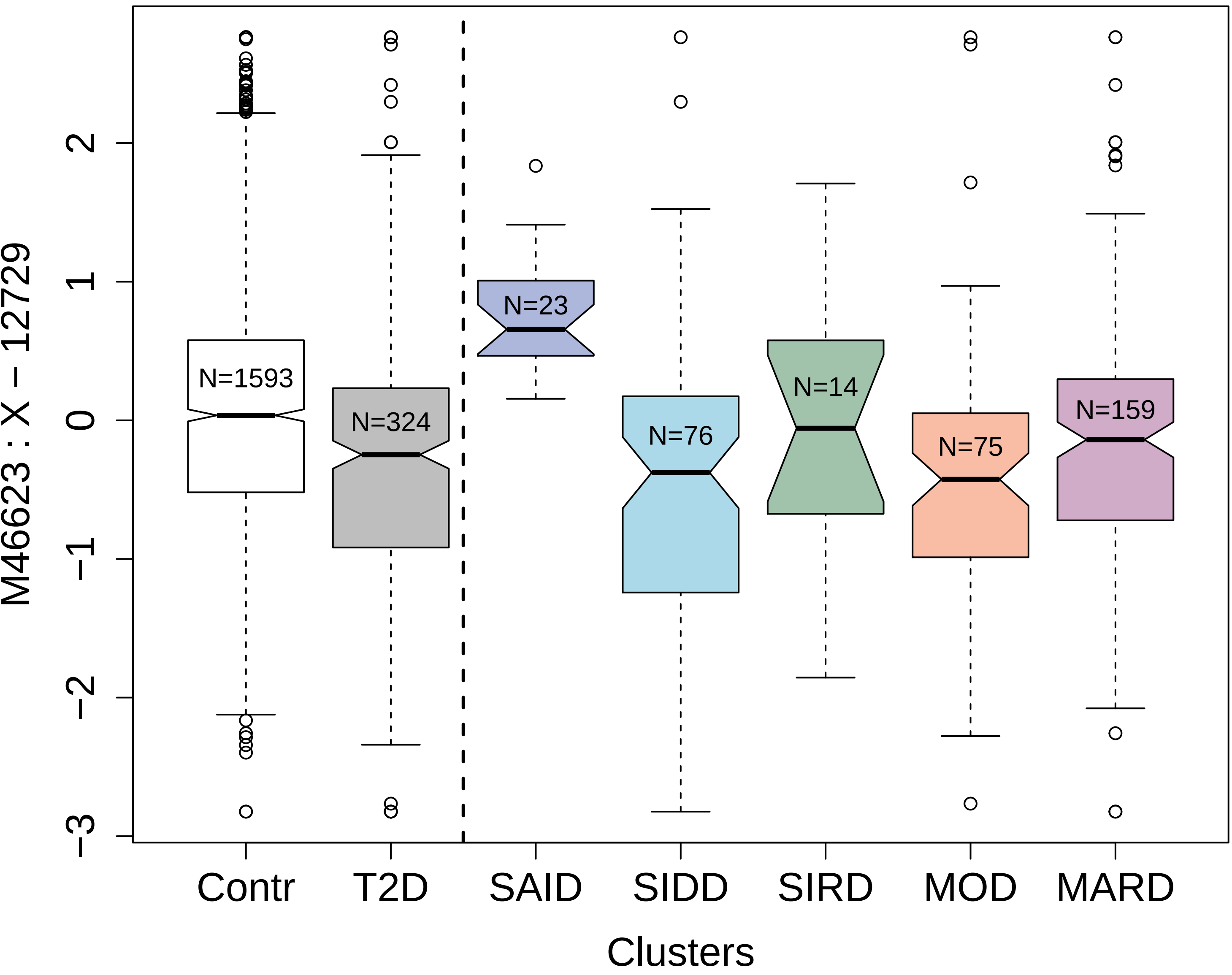
M46623 : X − 12729
2
1
0
−1
−2
−3
N=1593
N=324
N=23
N=76
N=14
N=75
N=159
Contr
T2D
SAID
SIDD
SIRD
MOD
MARD
Clusters

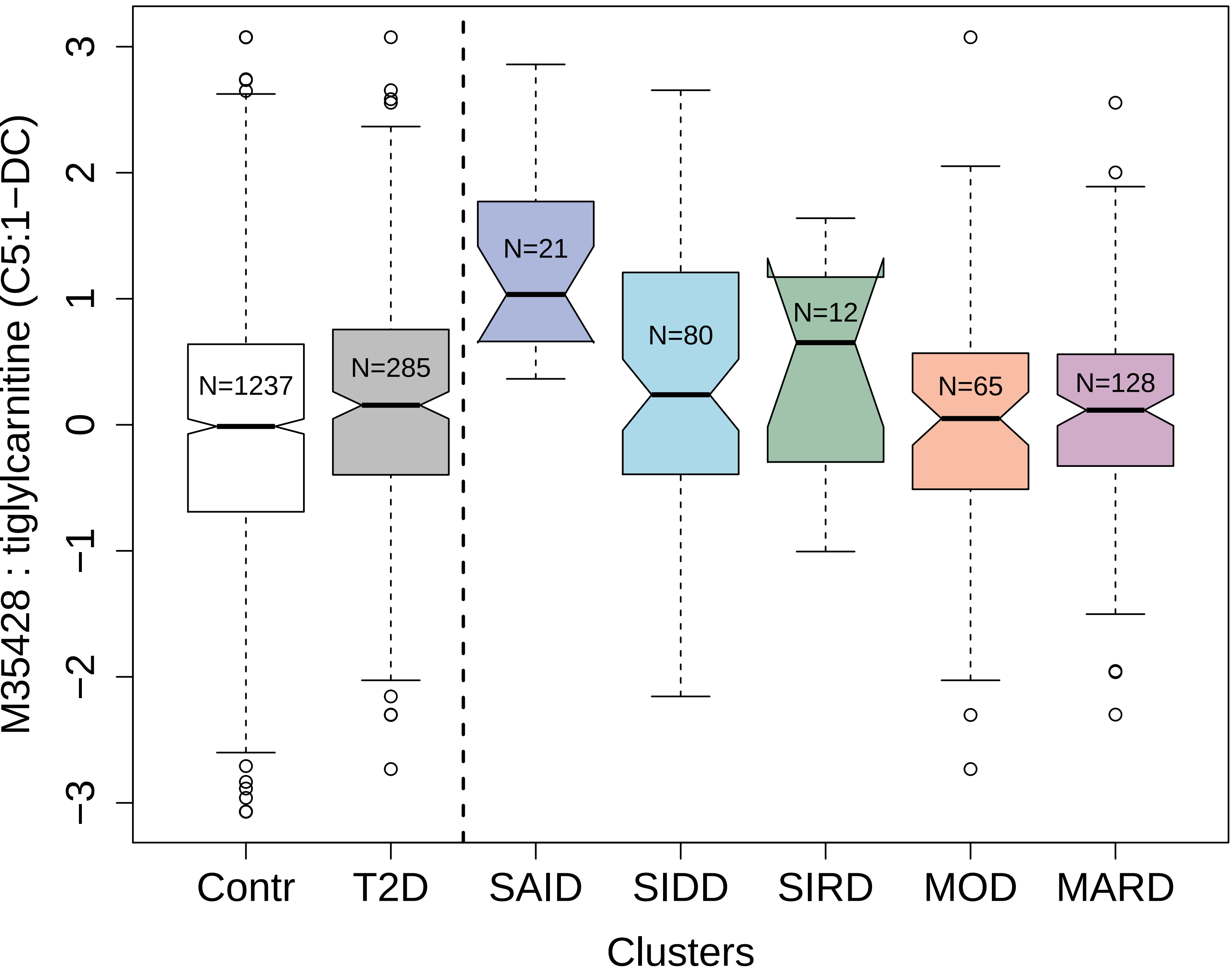

M35428 : tiglylcarnitine (C5:1−DC)
3
2
1
0
−1
−2
−3
N=1237
N=285
N=21
N=80
N=12
N=65
N=128
Contr
T2D
SAID
SIDD
SIRD
MOD
MARD
Clusters

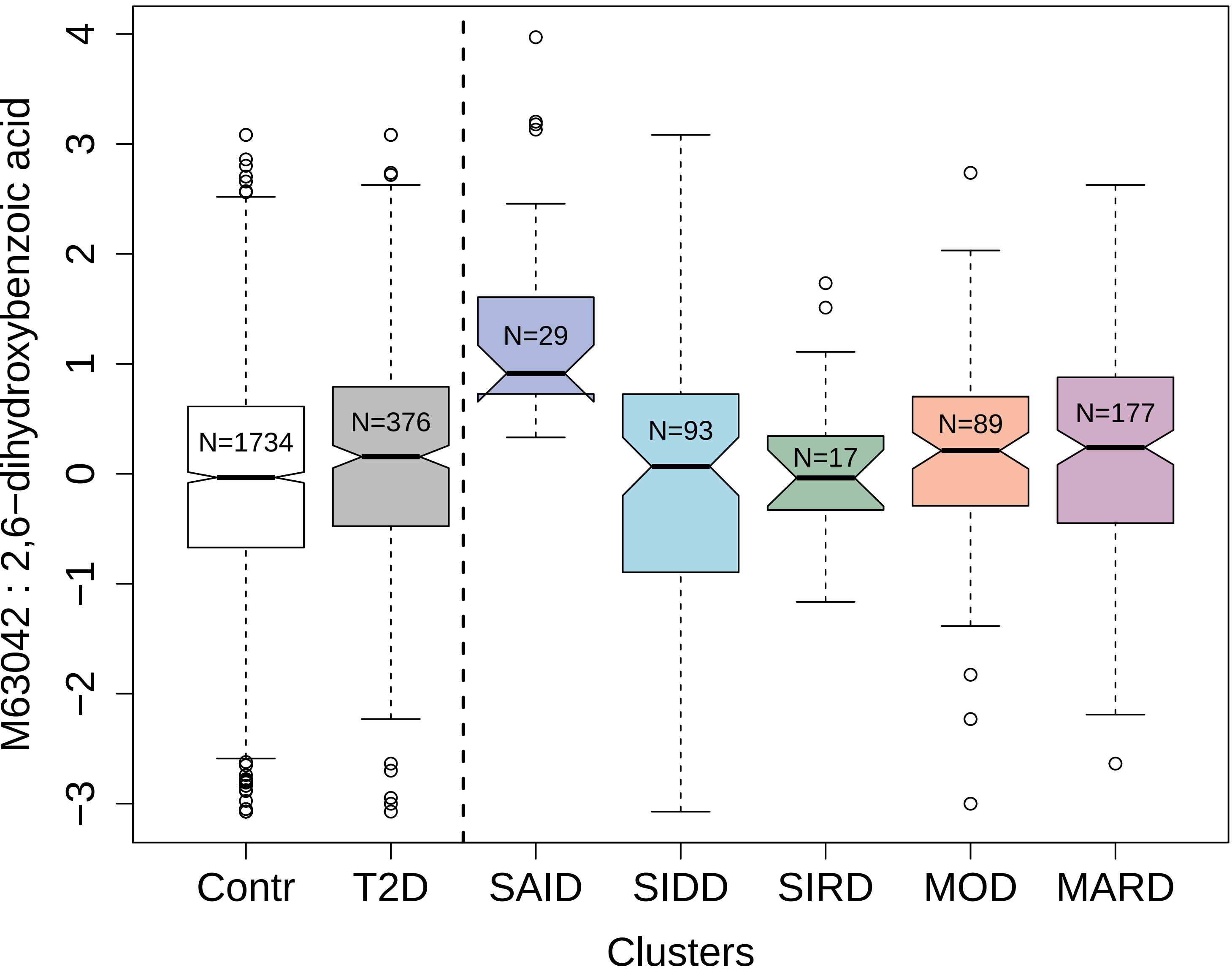

M63042 : 2,6−dihydroxybenzoic acid
4
3
2
1
0
−1
−2
−3
N=1734
N=376
N=29
N=93
N=17
N=89
N=177
Contr
T2D
SAID
SIDD
SIRD
MOD
MARD
Clusters

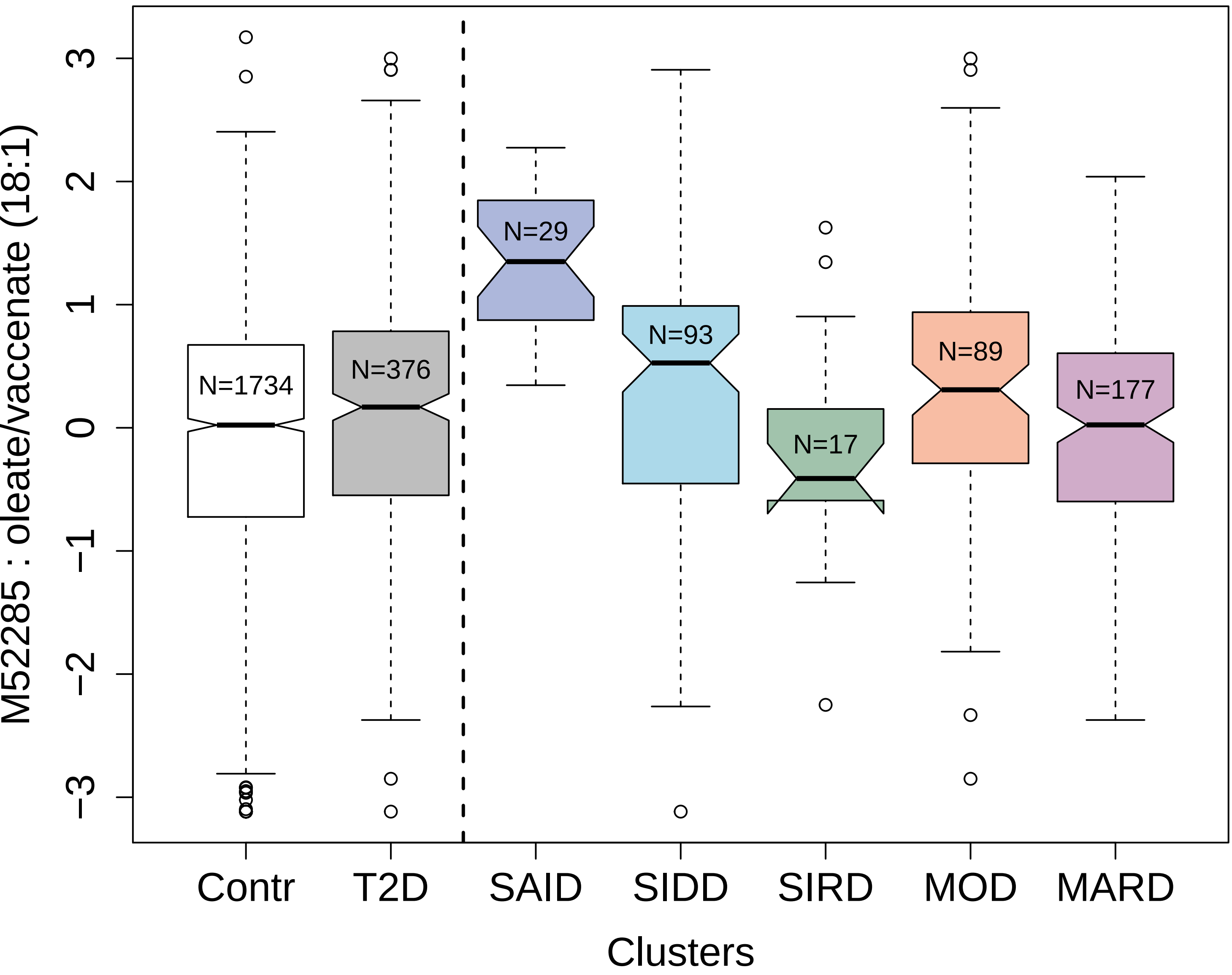
M52285 : oleate/vaccenate (18:1)
3
2
1
0
−1
−2
−3
N=1734
N=376
N=29
N=93
N=17
N=89
N=177
Contr
T2D
SAID
SIDD
SIRD
MOD
MARD
Clusters

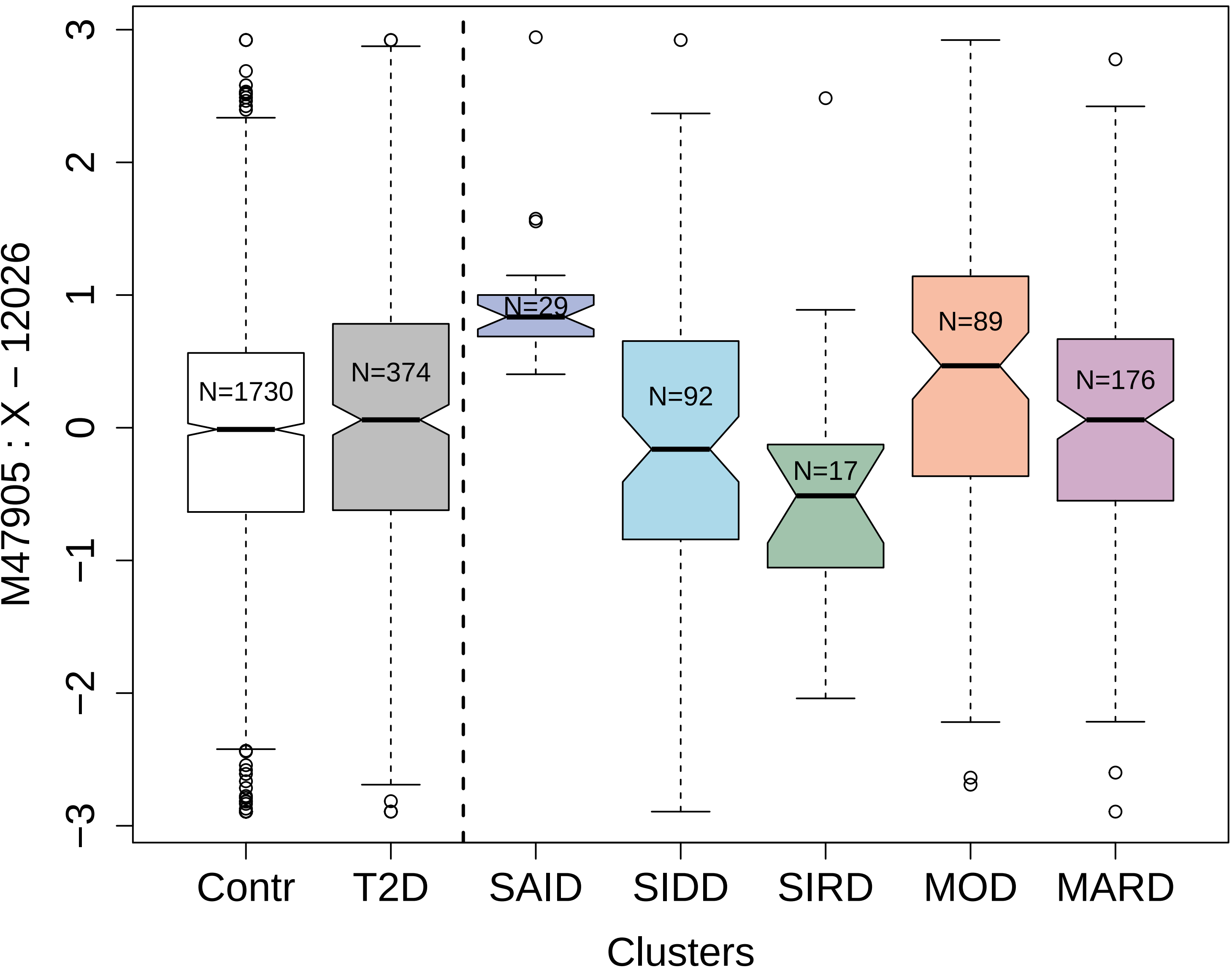
M47905 : X − 12026
3
2
1
0
−1
−2
−3
N=1730
N=374
N=29
N=92
N=17
N=89
N=176
Contr
T2D
SAID
SIDD
SIRD
MOD
MARD
Clusters

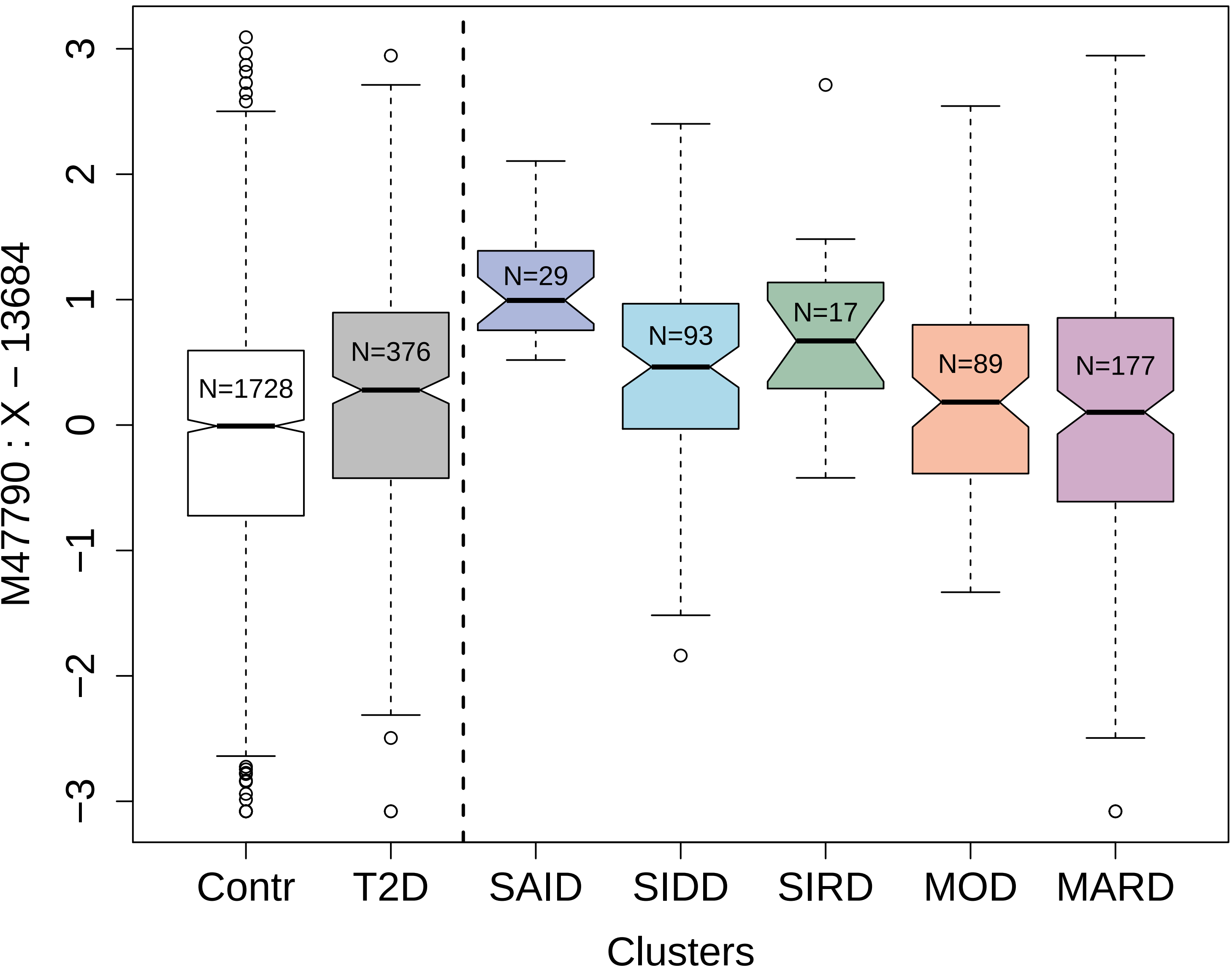

M47790 : X − 13684
3
2
1
0
−1
−2
−3
N=1728
N=376
N=29
N=93
N=17
N=89
N=177
Contr
T2D
SAID
SIDD
SIRD
MOD
MARD
Clusters

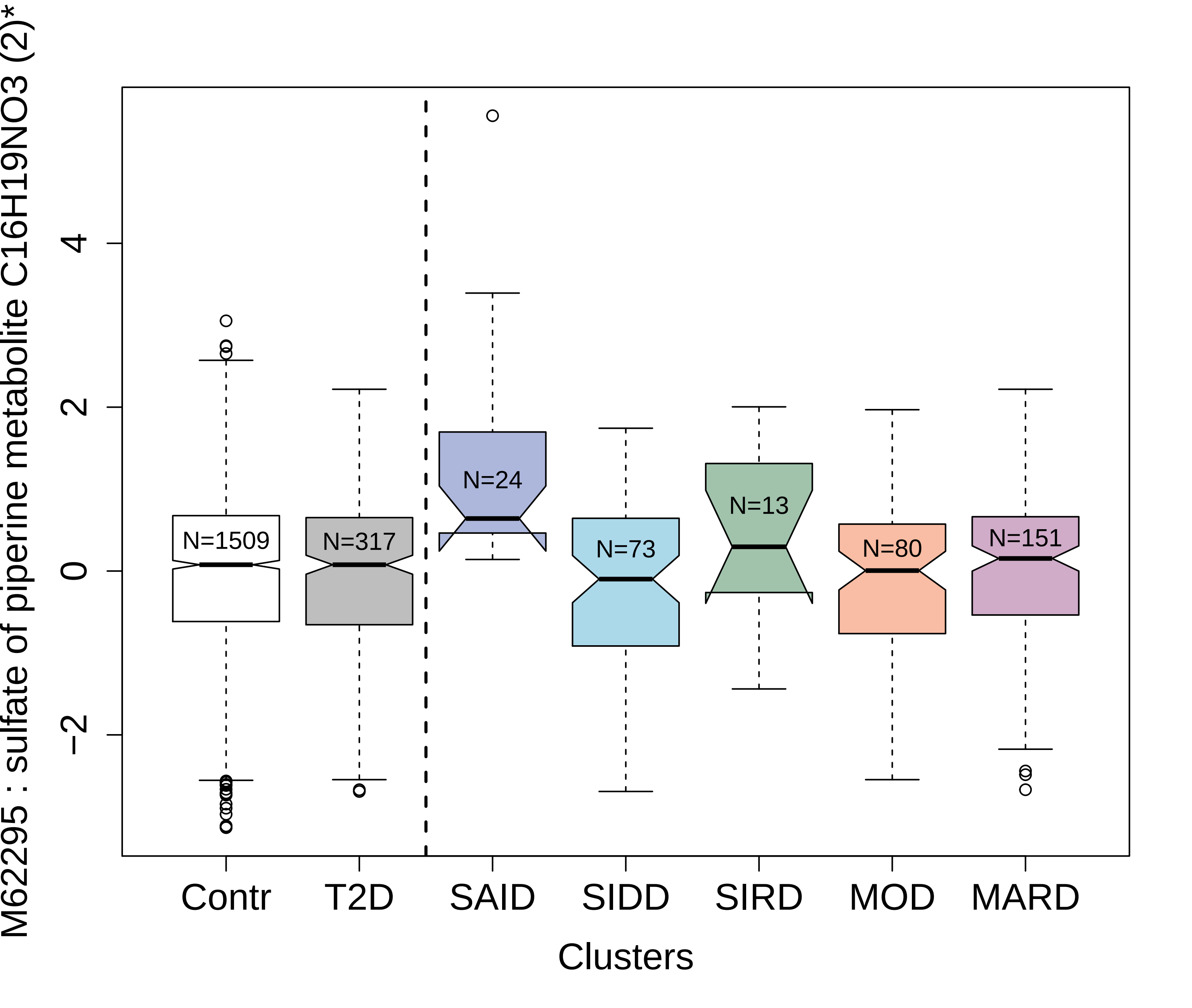

M62295 : sulfate of piperine metabolite C16H19NO3 (2)*
4
2
0
−2
N=1509
N=317
N=24
N=73
N=13
N=80
N=151
Contr
T2D
SAID
SIDD
SIRD
MOD
MARD
Clusters

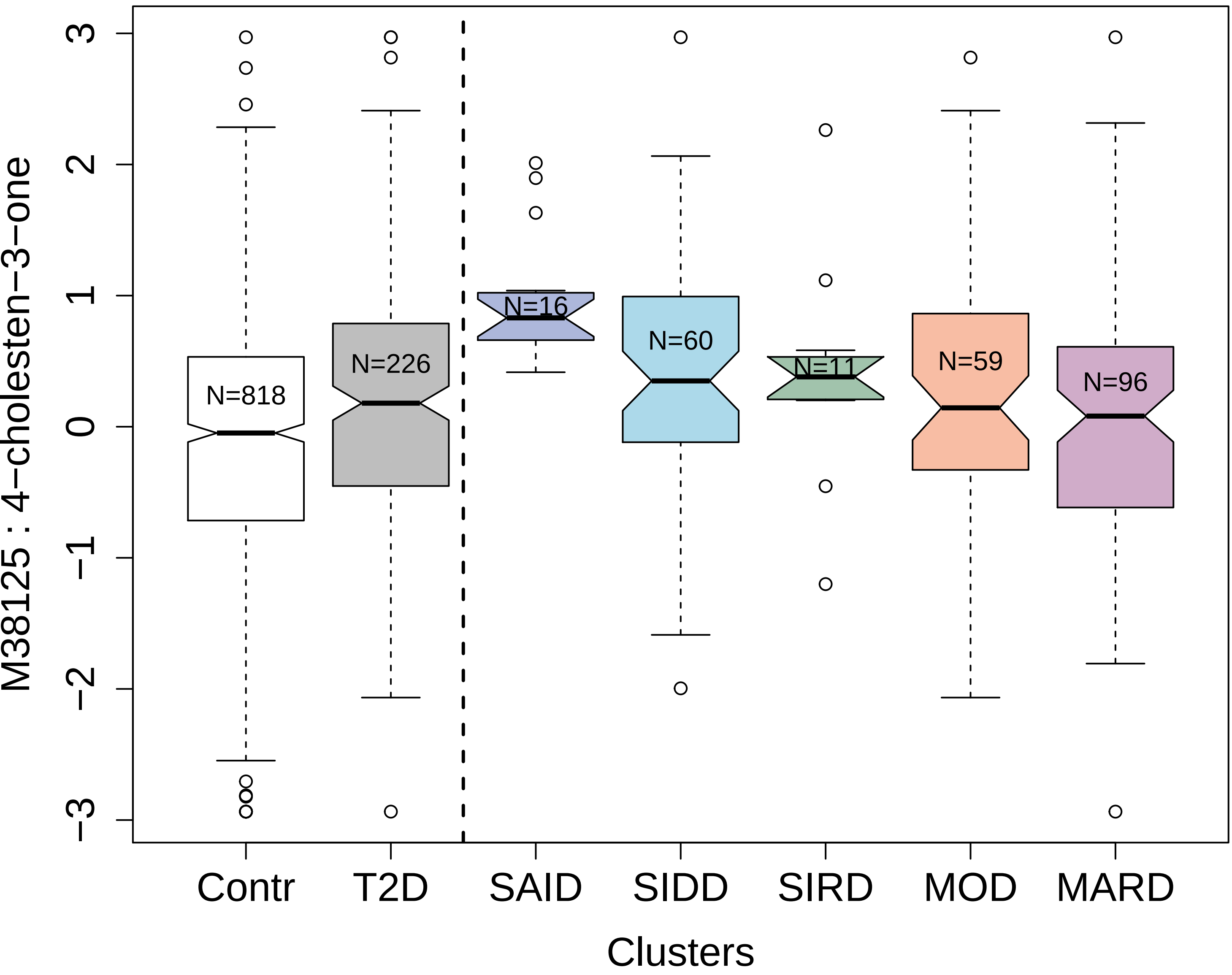

M38125 : 4−cholesten−3−one
3
2
1
0
−1
−2
−3
N=818
N=226
N=16
N=60
N=11
N=59
N=96
Contr
T2D
SAID
SIDD
SIRD
MOD
MARD
Clusters

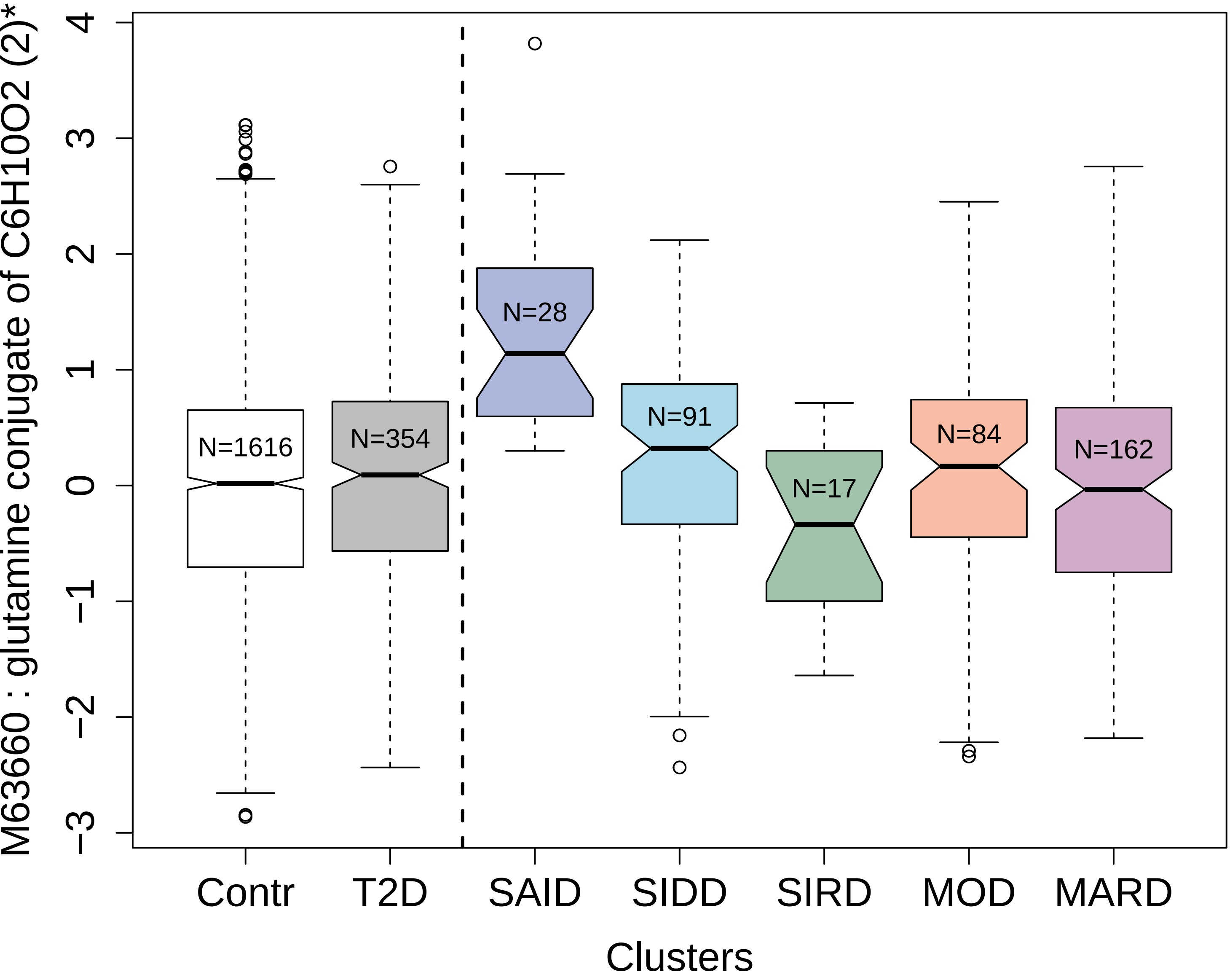
M63660 : glutamine conjugate of C6H10O2 (2)*
4
3
2
1
0
−1
−2
−3
N=1616
N=354
N=28
N=91
N=17
N=84
N=162
Contr
T2D
SAID
SIDD
SIRD
MOD
MARD
Clusters

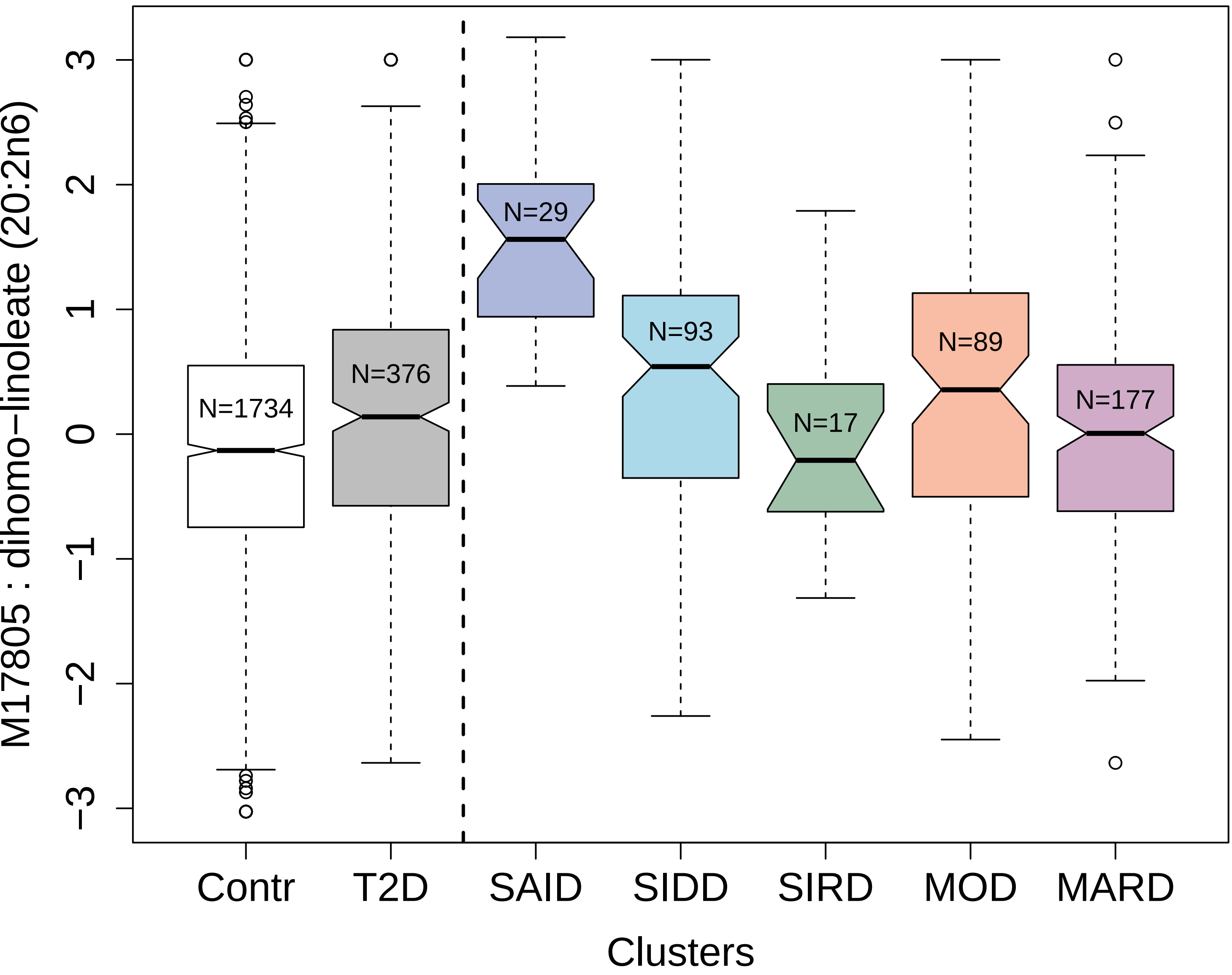
M17805 : dihomo−linoleate (20:2n6)
3
2
1
0
−1
−2
−3
N=1734
N=376
N=29
N=93
N=17
N=89
N=177
Contr
T2D
SAID
SIDD
SIRD
MOD
MARD
Clusters

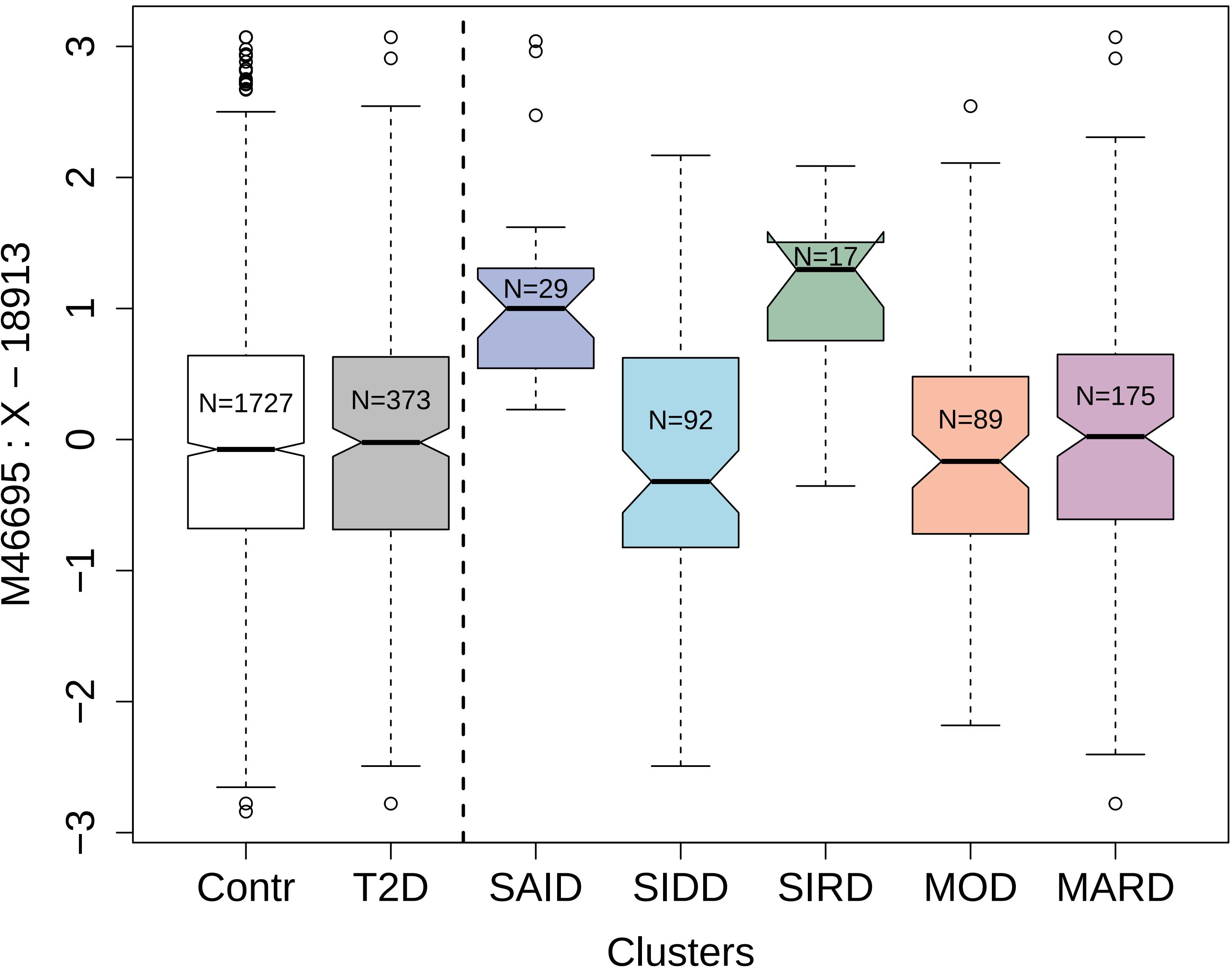

M46695 : X − 18913
3
2
1
0
−1
−2
−3
N=1727
N=373
N=29
N=92
N=17
N=89
N=175
Contr
T2D
SAID
SIDD
SIRD
MOD
MARD
Clusters

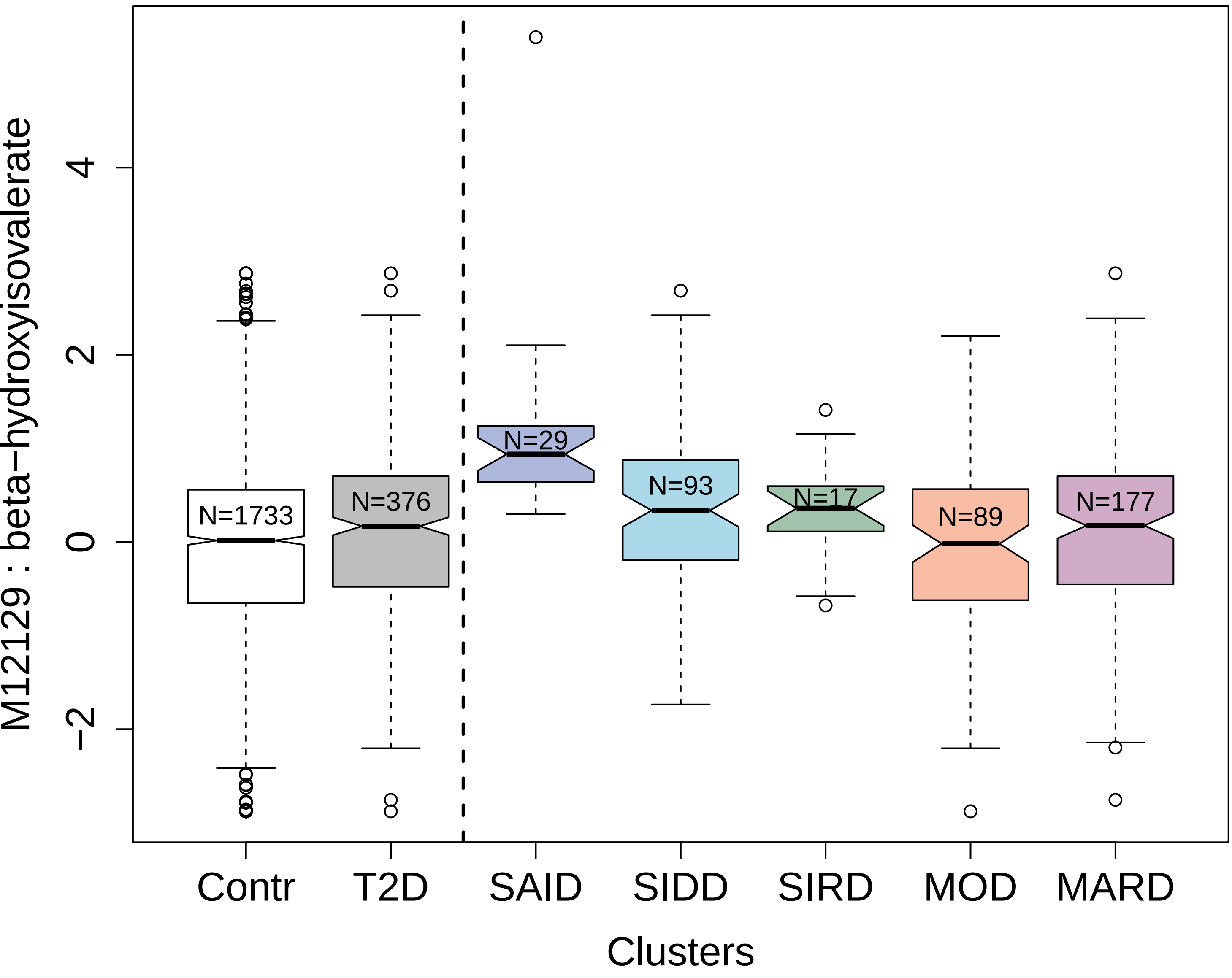

M12129 : beta−hydroxyisovalerate
4
2
0
−2
N=1733
N=376
N=29
N=93
N=17
N=89
N=177
Contr
T2D
SAID
SIDD
SIRD
MOD
MARD
Clusters

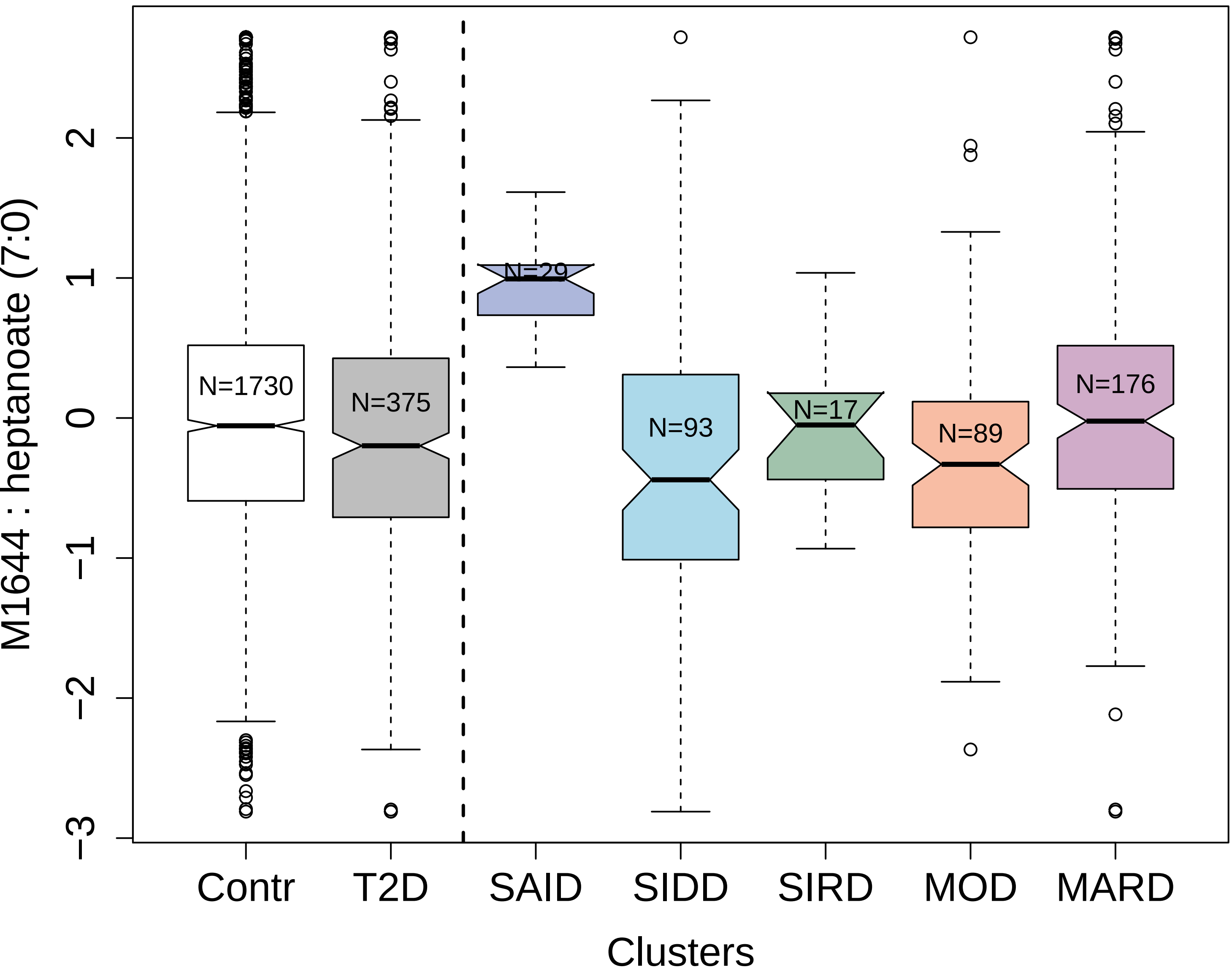
M1644 : heptanoate (7:0)
2
1
0
−1
−2
−3
N=1730
N=375
N=29
N=93
N=17
N=89
N=176
Contr
T2D
SAID
SIDD
SIRD
MOD
MARD
Clusters

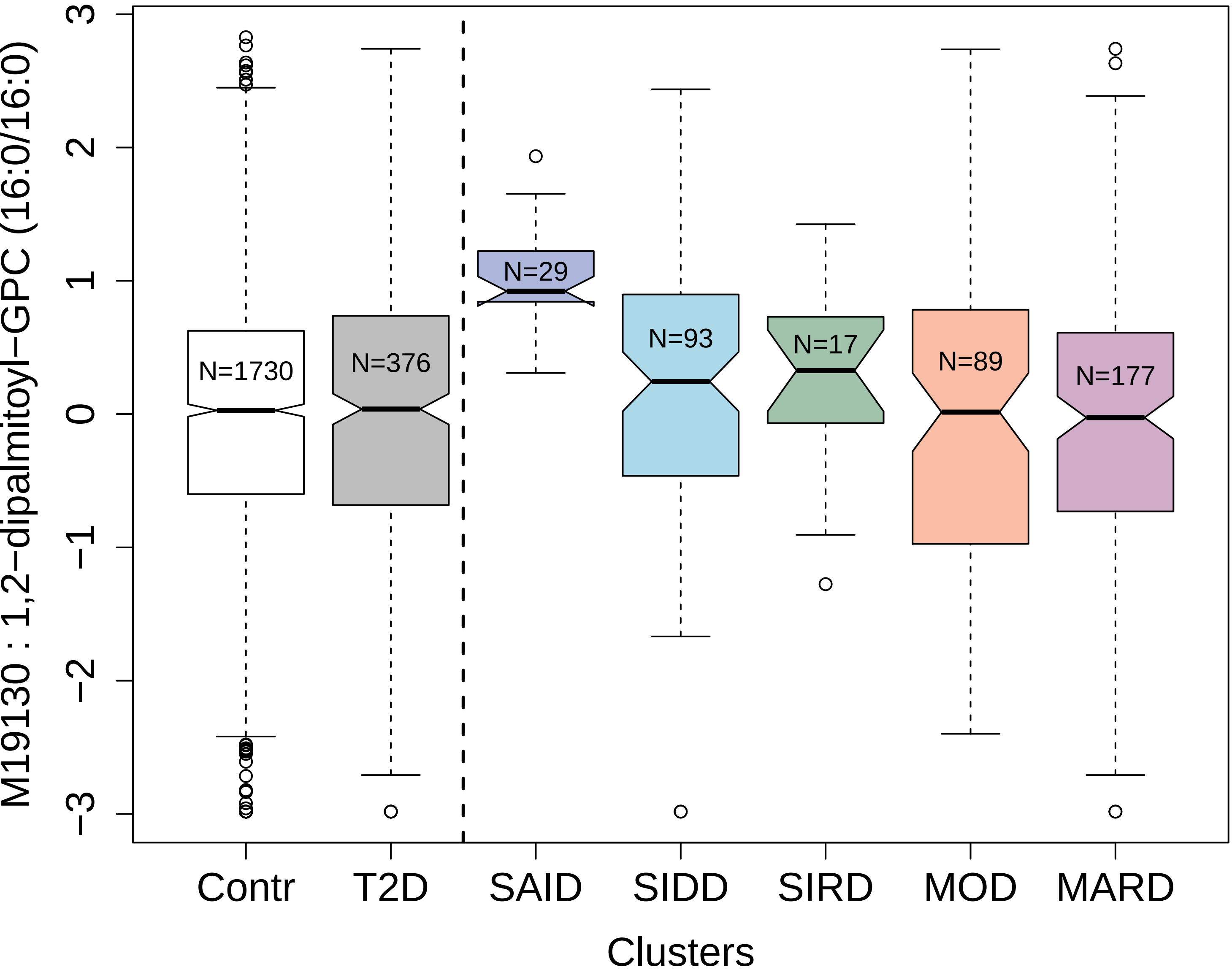

M19130 : 1,2−dipalmitoyl−GPC (16:0/16:0)
3
2
1
0
−1
−2
−3
N=1730
N=376
N=29
N=93
N=17
N=89
N=177
Contr
T2D
SAID
SIDD
SIRD
MOD
MARD
Clusters

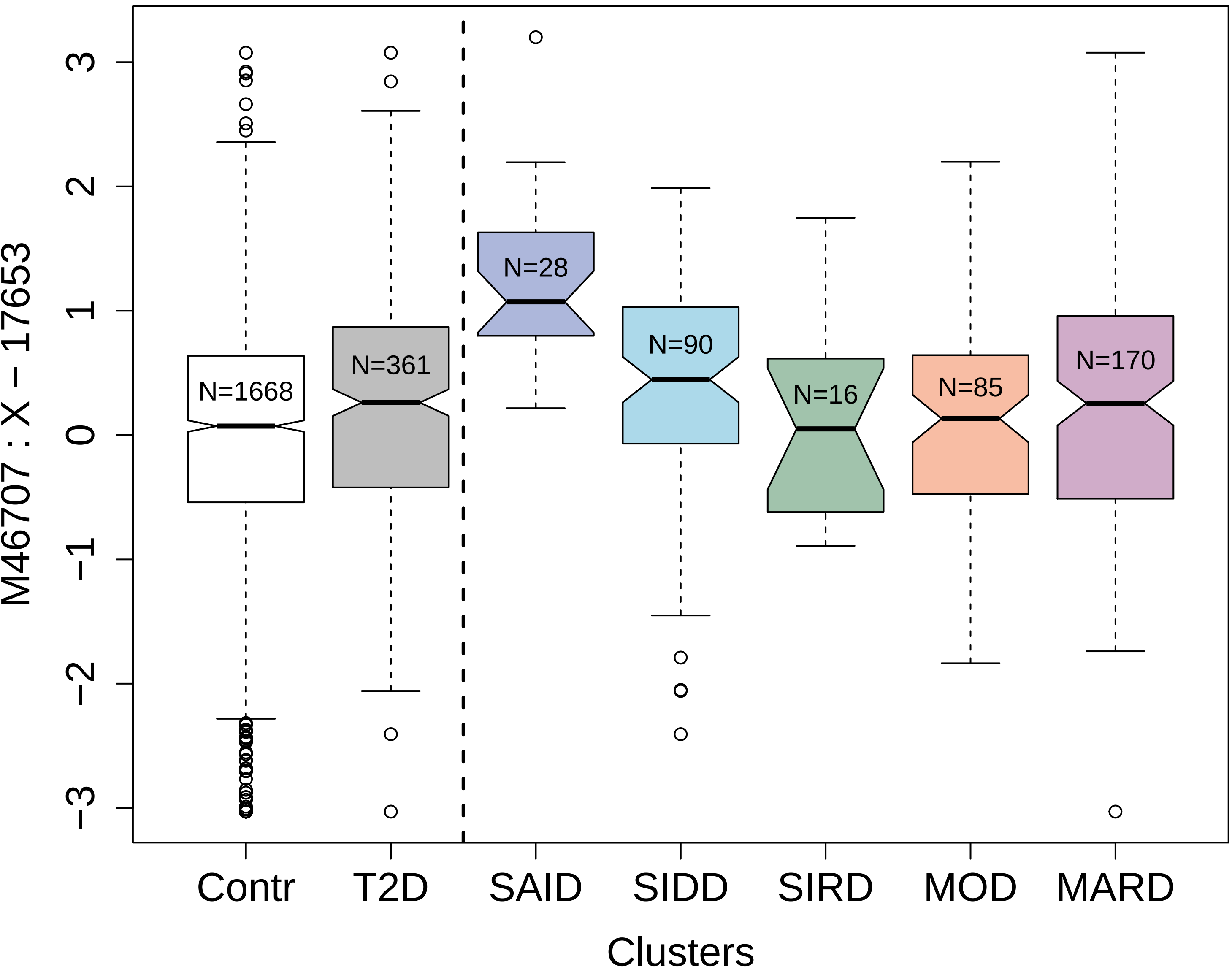

M46707 : X − 17653
3
2
1
0
−1
−2
−3
N=1668
N=361
N=28
N=90
N=16
N=85
N=170
Contr
T2D
SAID
SIDD
SIRD
MOD
MARD
Clusters

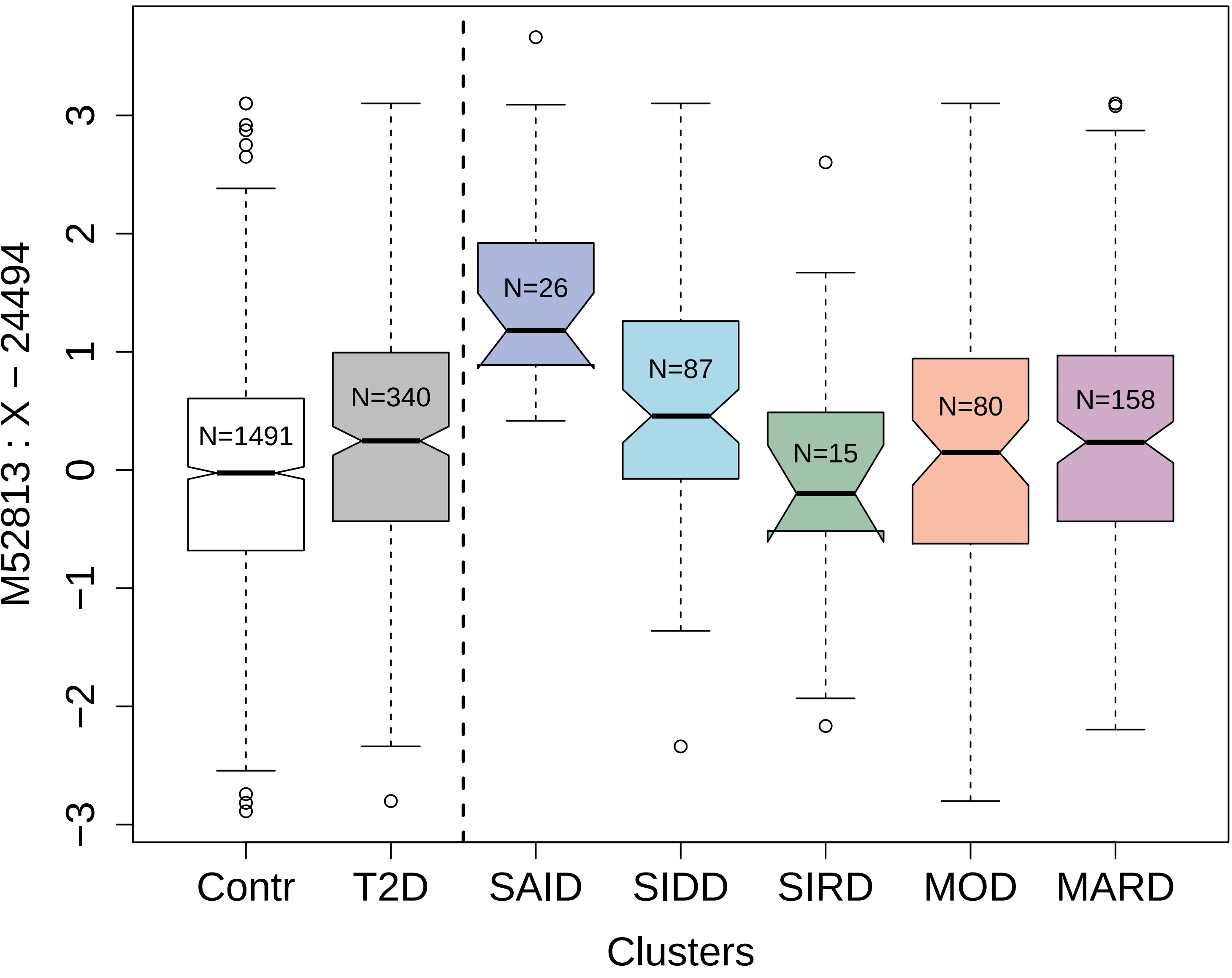

M52813 : X − 24494
−3
−2
−1
0
1
2
3
N=1491
N=340
N=26
N=87
N=15
N=80
N=158
Contr
T2D
SAID
SIDD
SIRD
MOD
MARD
Clusters

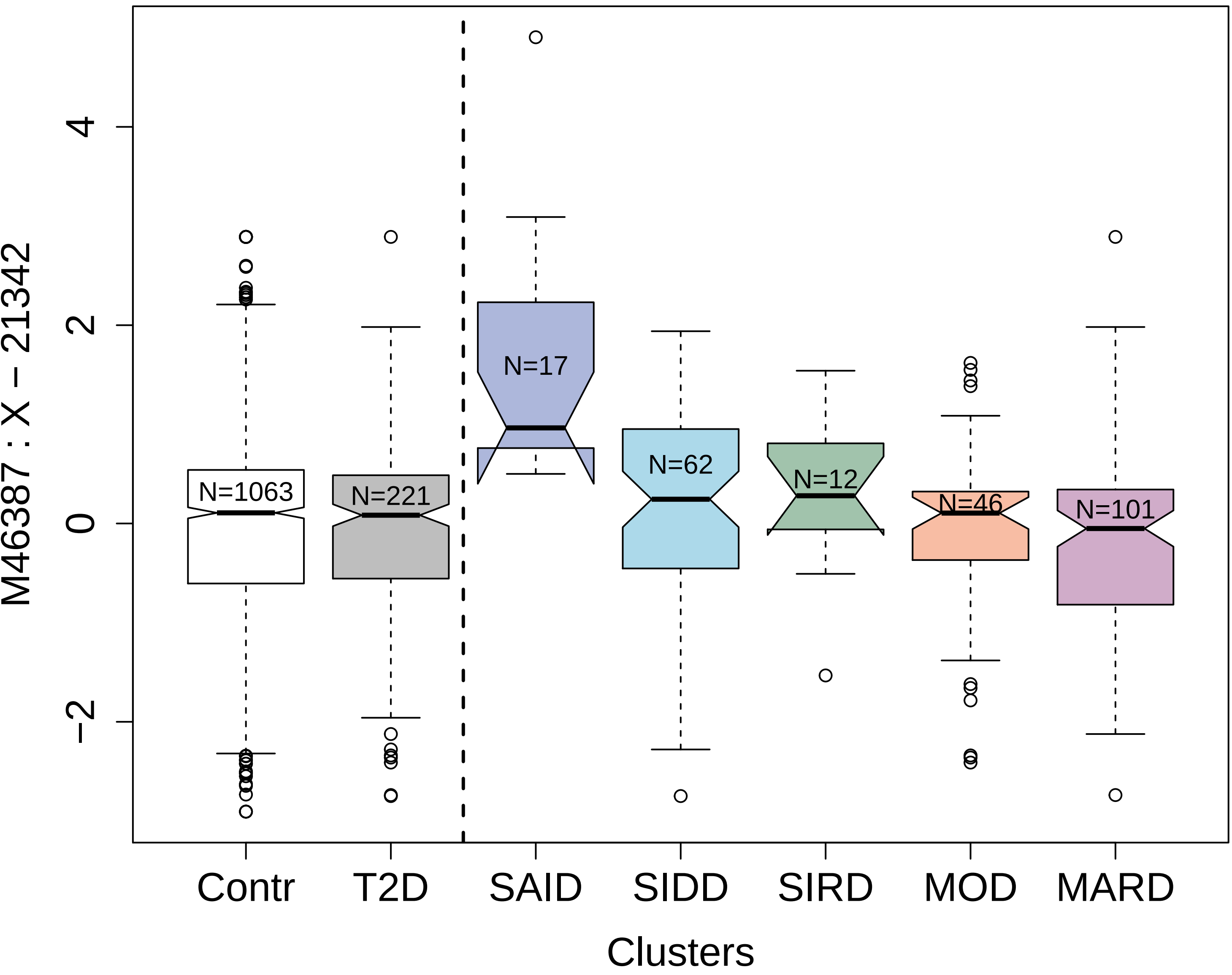

M46387 : X − 21342
4
2
0
−2
N=1063
N=221
N=17
N=62
N=12
N=46
N=101
Contr
T2D
SAID
SIDD
SIRD
MOD
MARD
Clusters

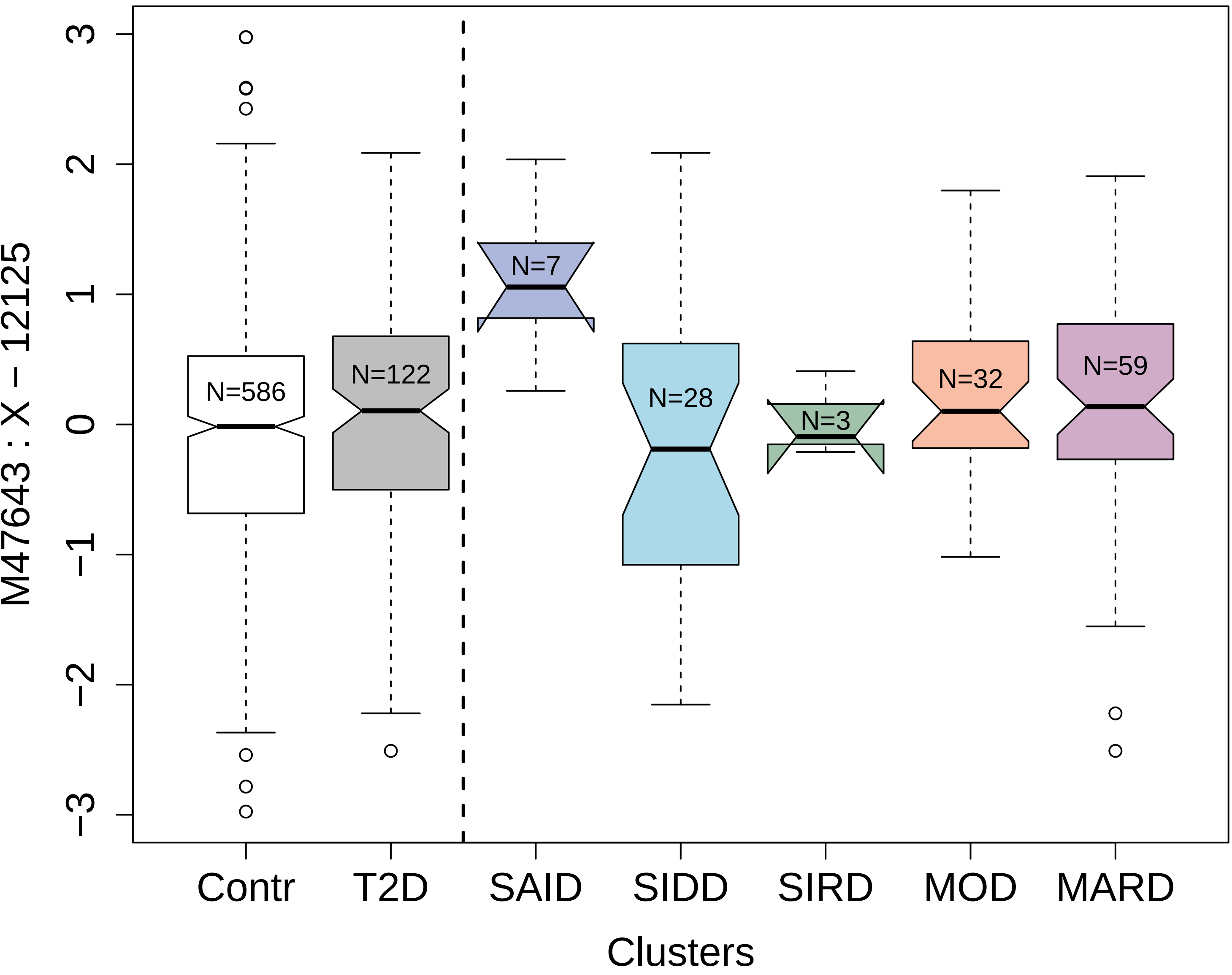

M47643 : X − 12125
3
2
1
0
−1
−2
−3
N=586
N=122
N=7
N=28
N=3
N=32
N=59
Contr
T2D
SAID
SIDD
SIRD
MOD
MARD
Clusters

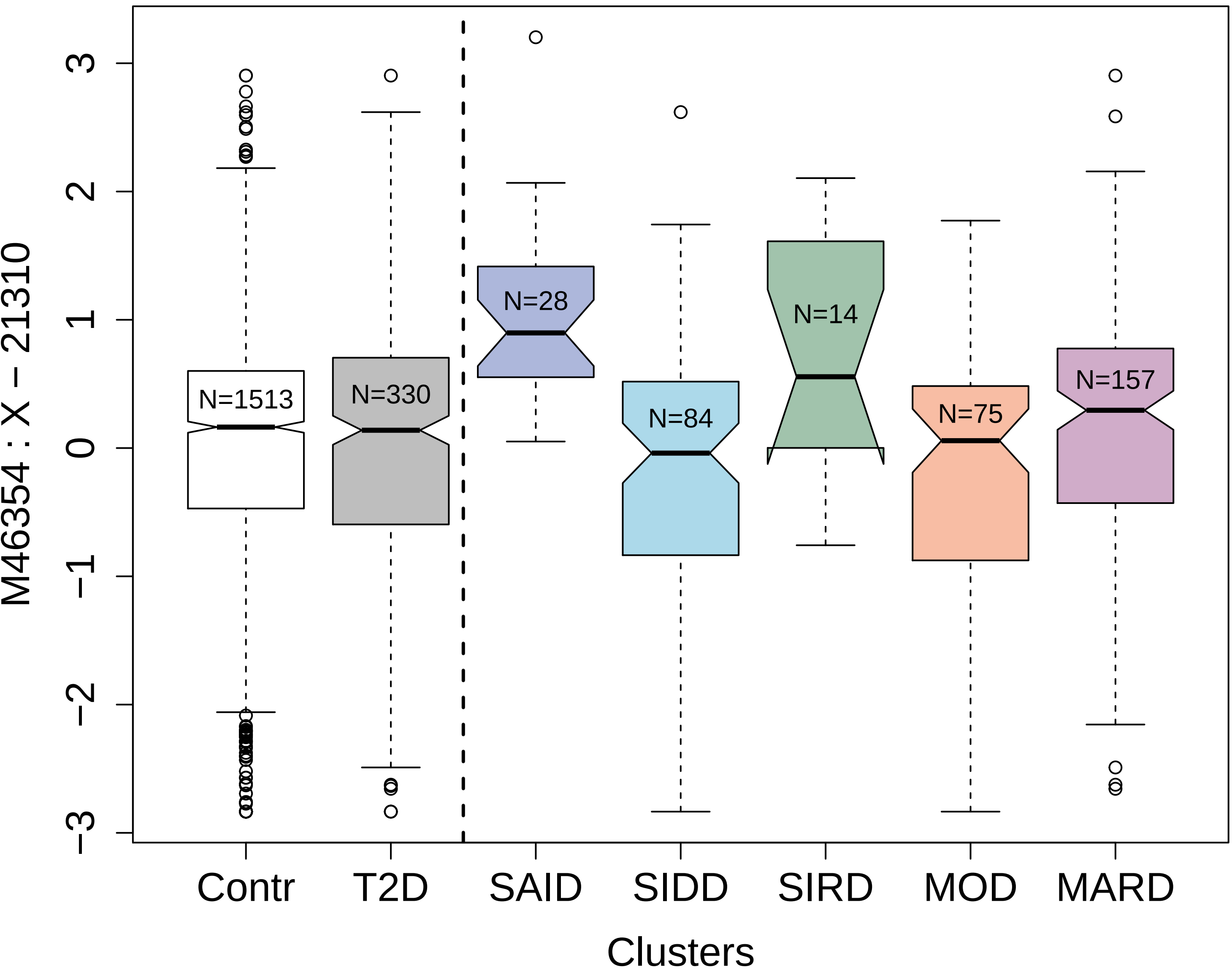
M46354 : X − 21310
3
2
1
0
−1
−2
−3
N=1513
N=330
N=28
N=84
N=14
N=75
N=157
Contr
T2D
SAID
SIDD
SIRD
MOD
MARD
Clusters

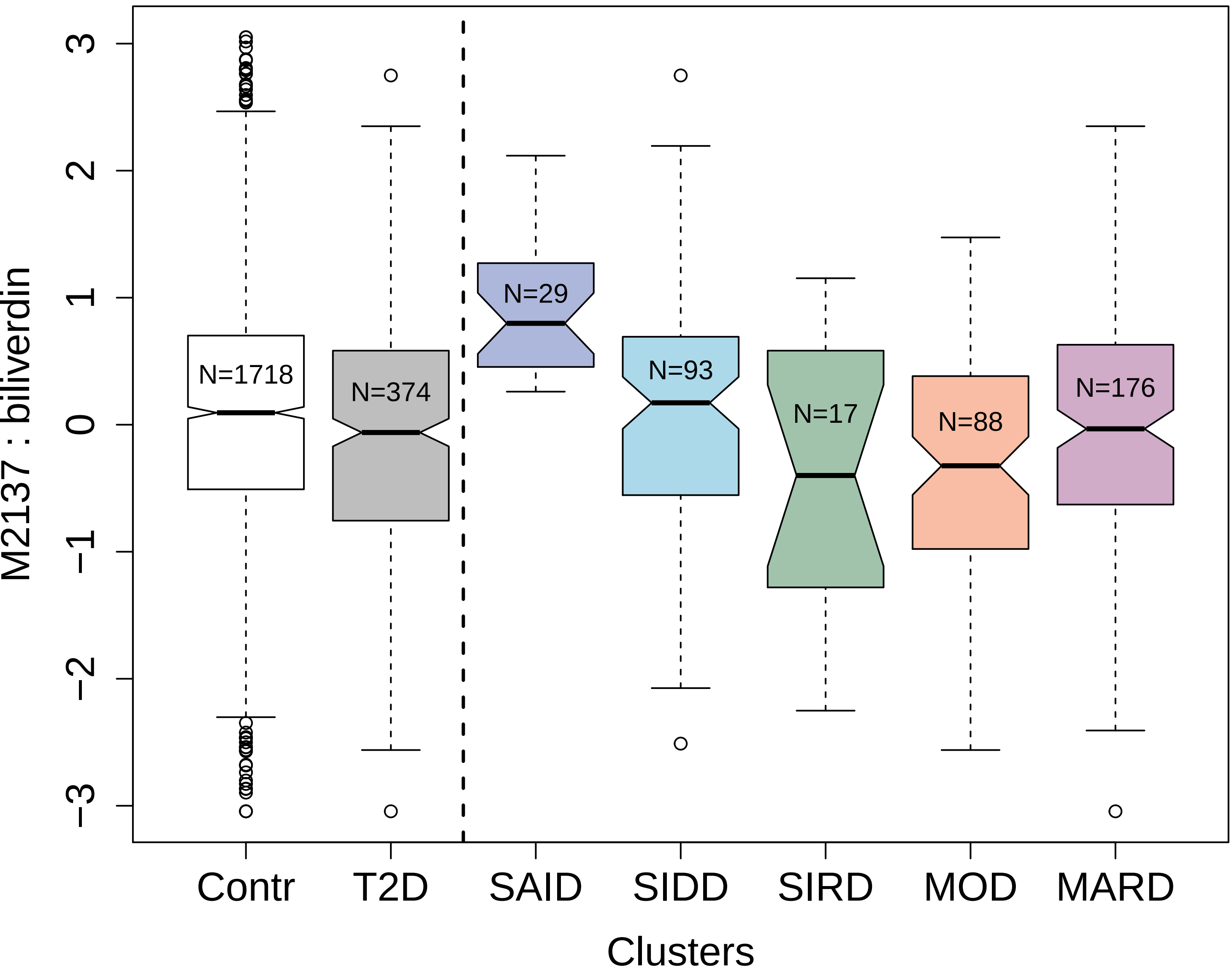

M2137 : biliverdin
3
2
1
0
−1
−2
−3
N=1718
N=374
N=29
N=93
N=17
N=88
N=176
Contr
T2D
SAID
SIDD
SIRD
MOD
MARD
Clusters

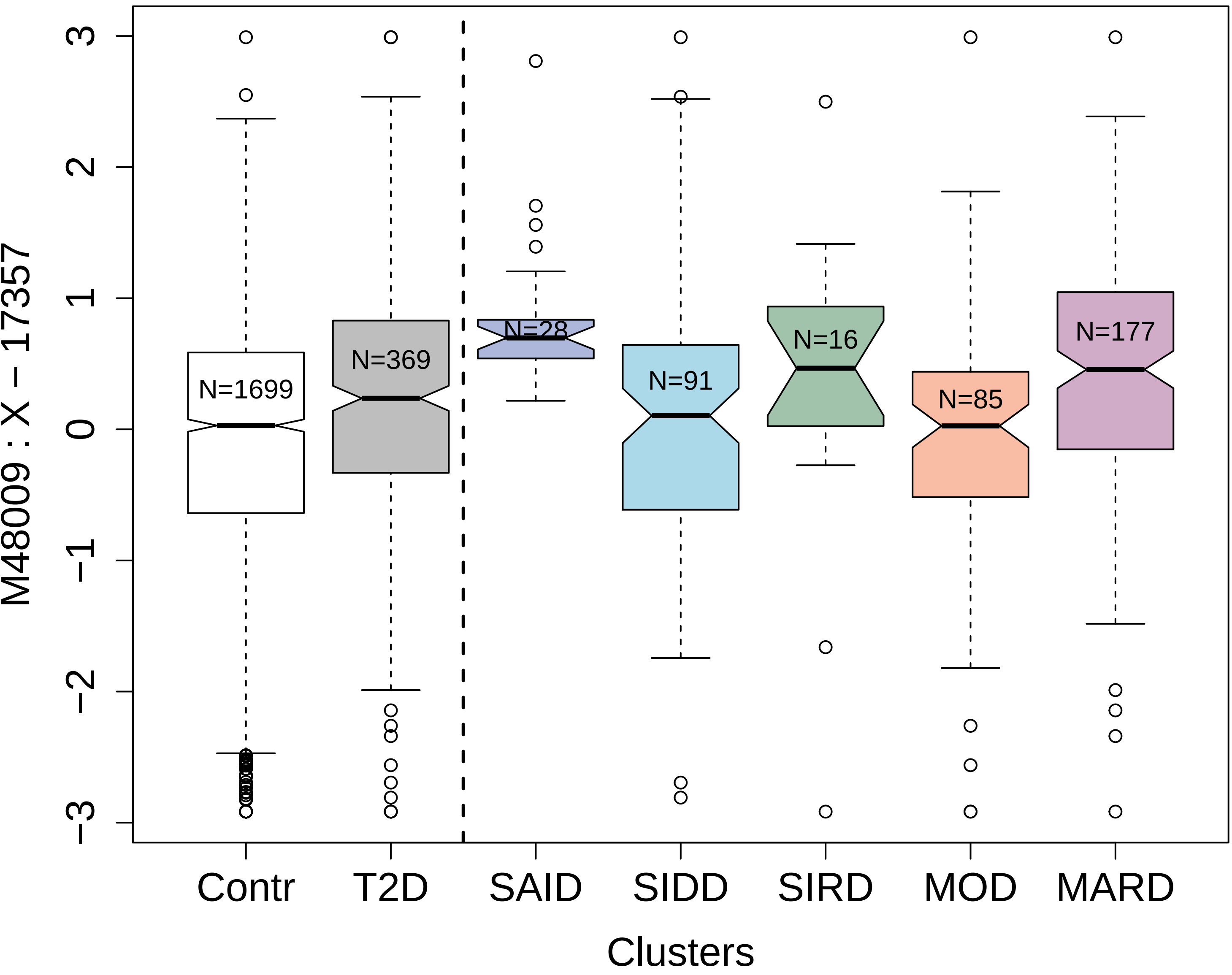

M48009 : X − 17357
3
2
1
0
−1
−2
−3
N=1699
N=369
N=28
N=91
N=16
N=85
N=177
Contr
T2D
SAID
SIDD
SIRD
MOD
MARD
Clusters

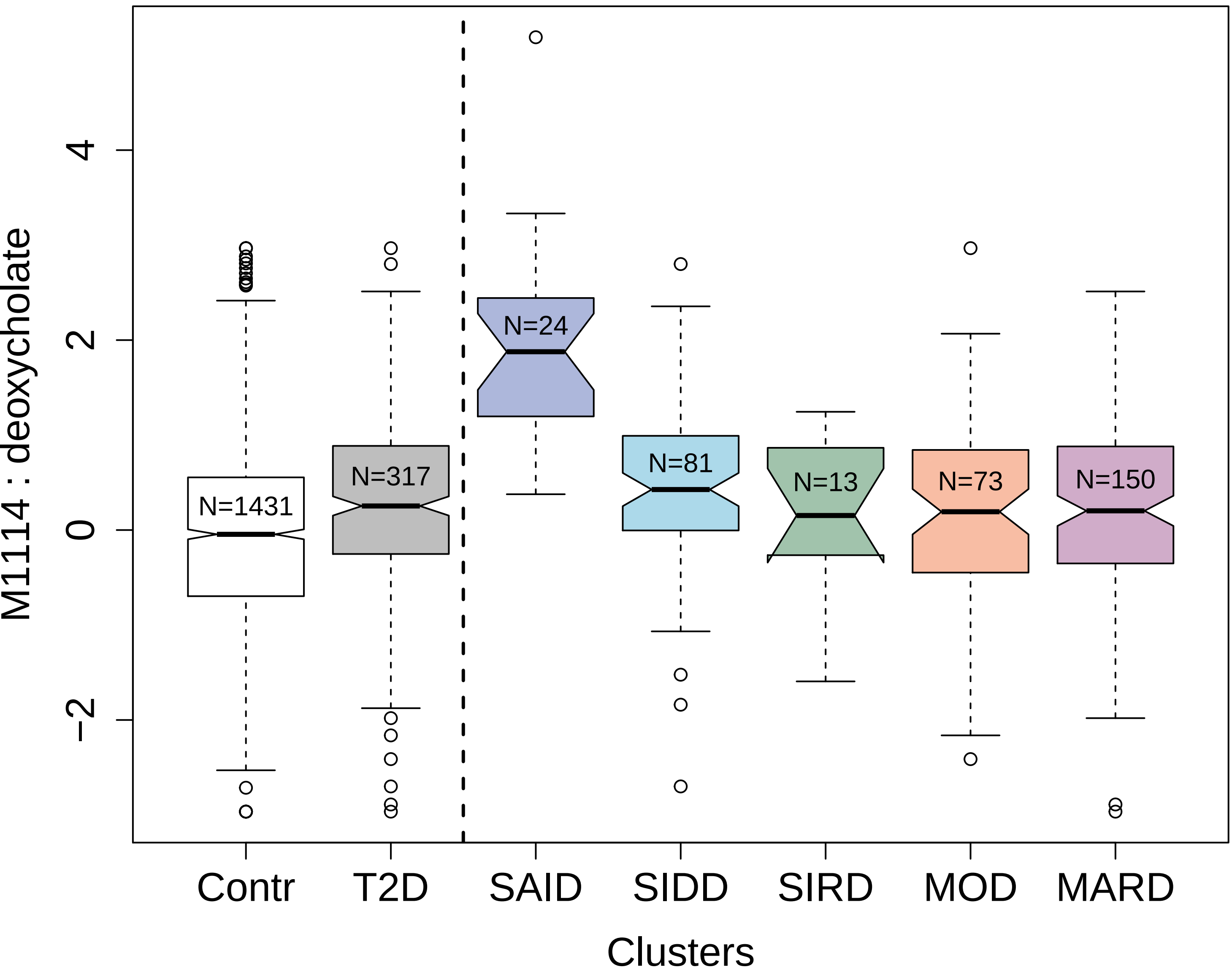
M1114 : deoxycholate
Clusters
Contr
T2D
SAID
SIDD
SIRD
MOD
MARD
N=1431
N=317
N=24
N=81
N=13
N=73
N=150
4
2
0
−2

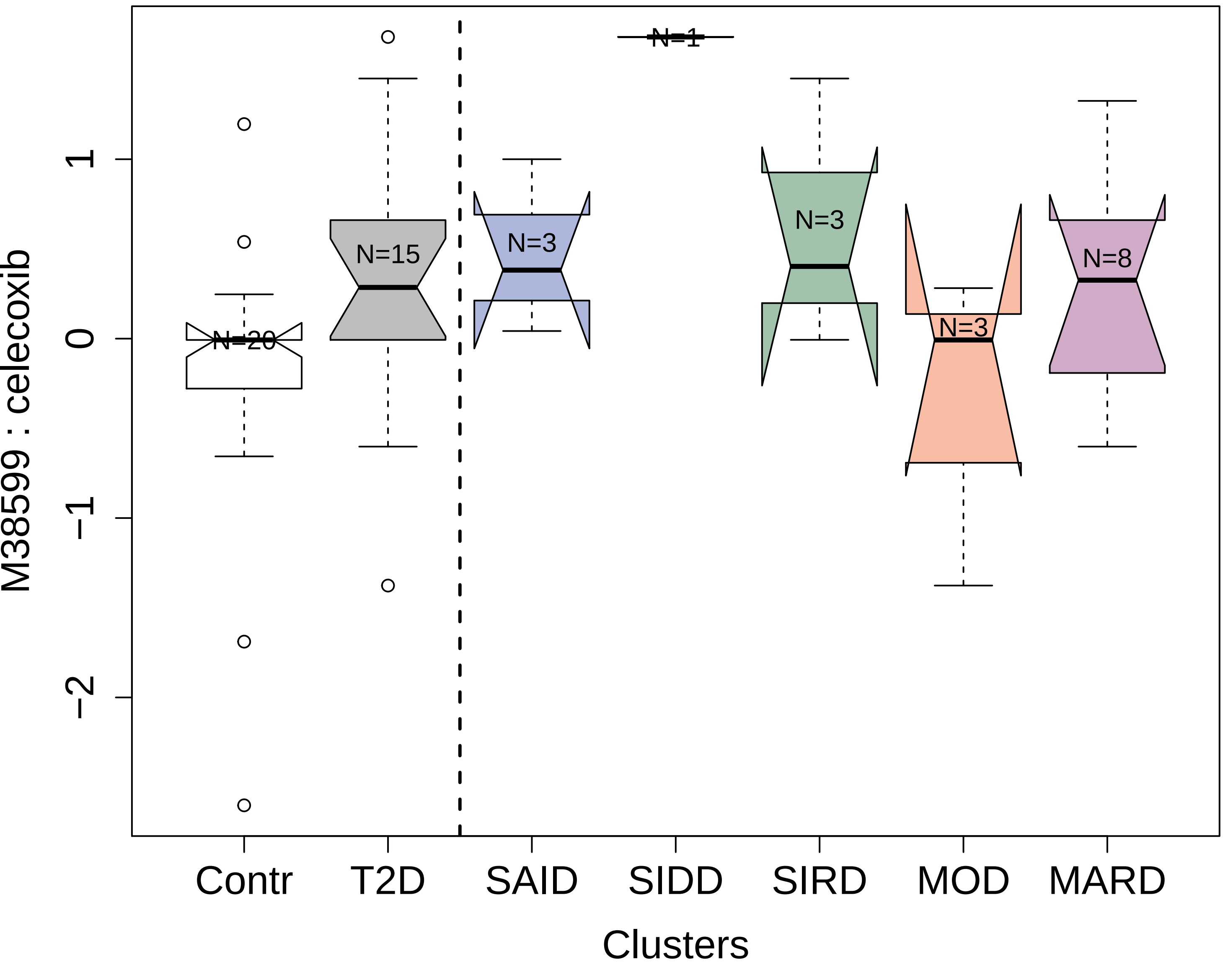
M38599 : celecoxib
1
0
−1
−2
N=20
N=15
N=3
N=1
N=3
N=3
N=8
Contr
T2D
SAID
SIDD
SIRD
MOD
MARD
Clusters

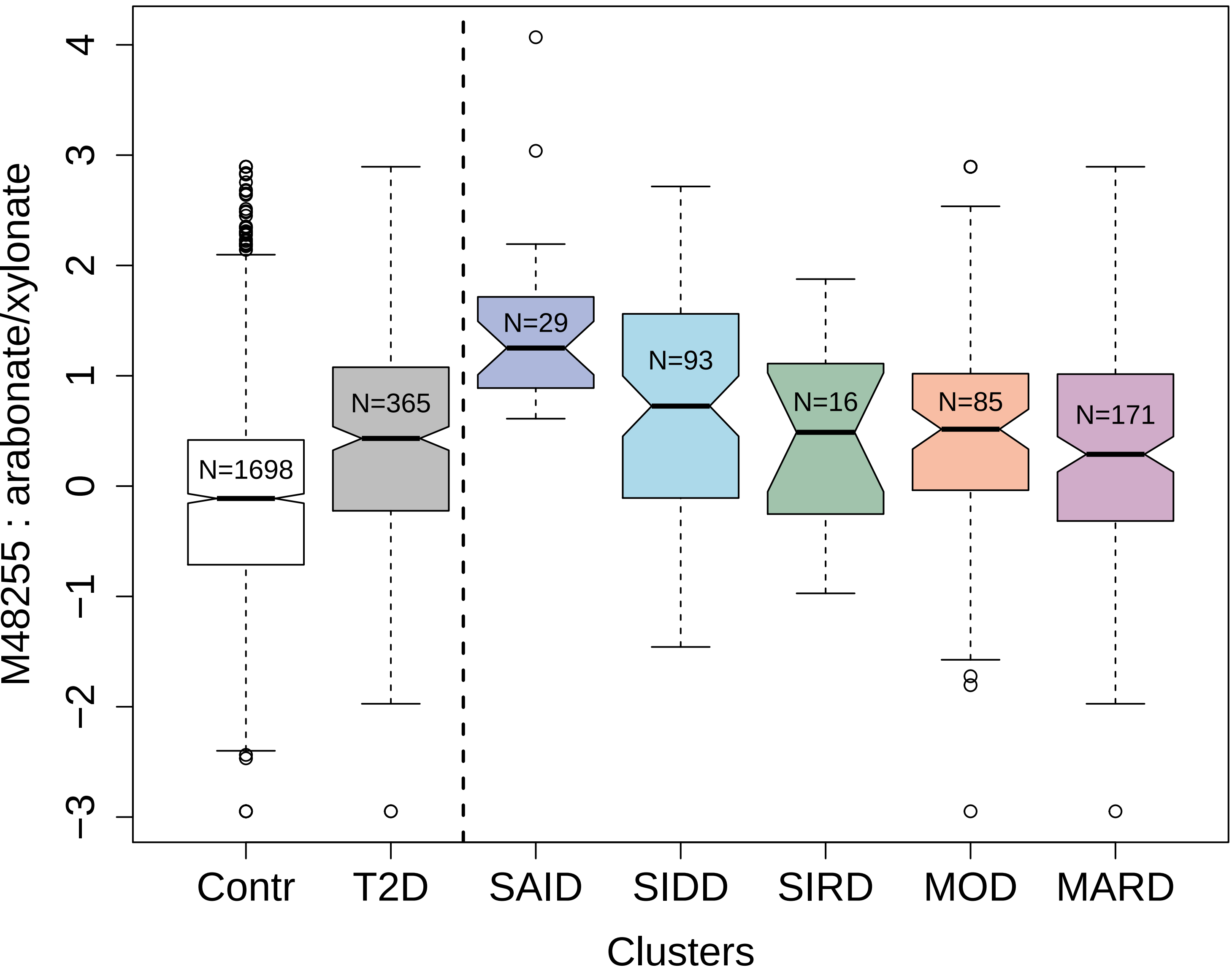

M48255 : arabonate/xylonate
4
3
2
1
0
−1
−2
−3
N=1698
N=365
N=29
N=93
N=16
N=85
N=171
Contr
T2D
SAID
SIDD
SIRD
MOD
MARD
Clusters

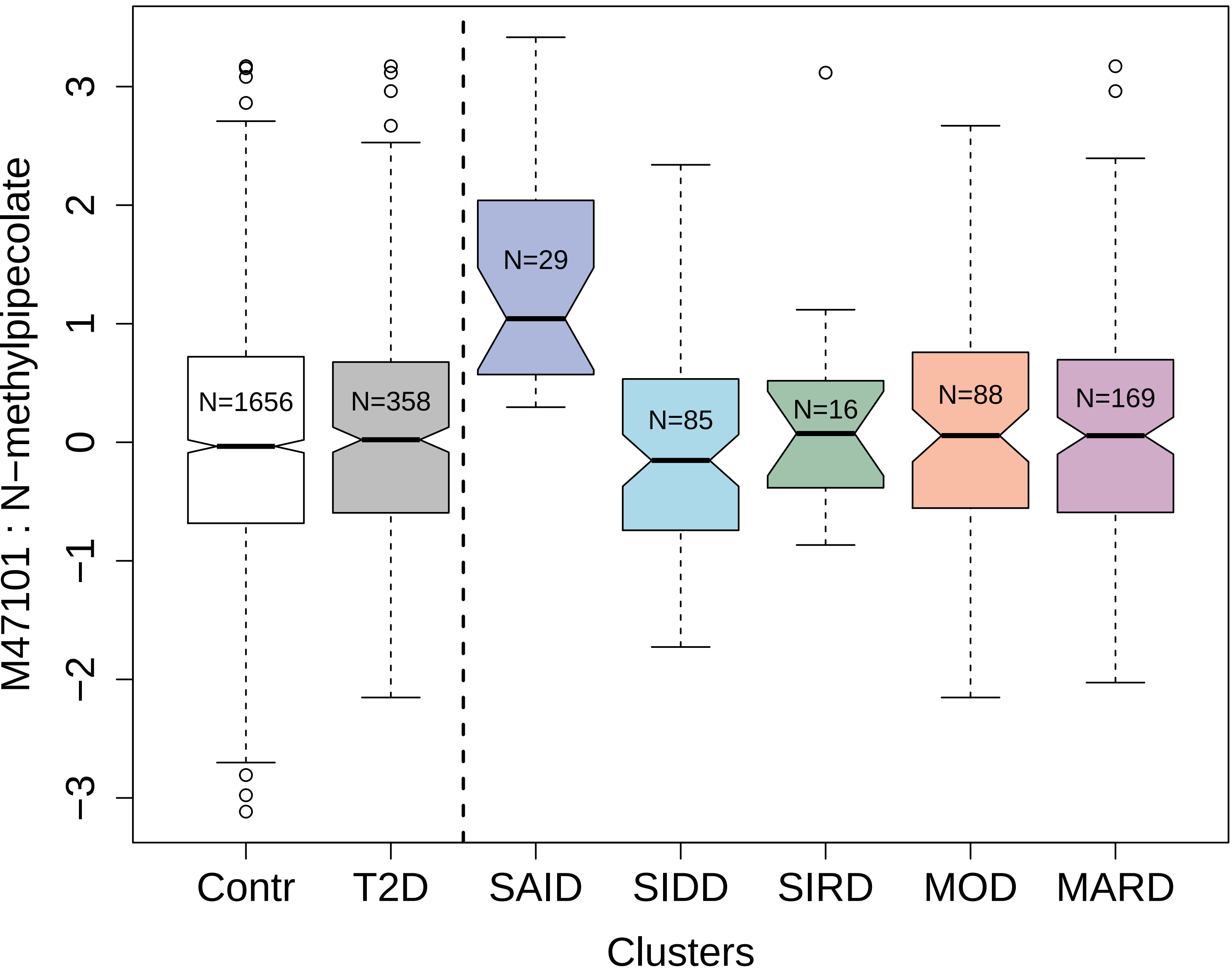

M47101 : N−methylpipecolate
3
2
1
0
−1
−2
−3
N=1656
N=358
N=29
N=85
N=16
N=88
N=169
Contr
T2D
SAID
SIDD
SIRD
MOD
MARD
Clusters

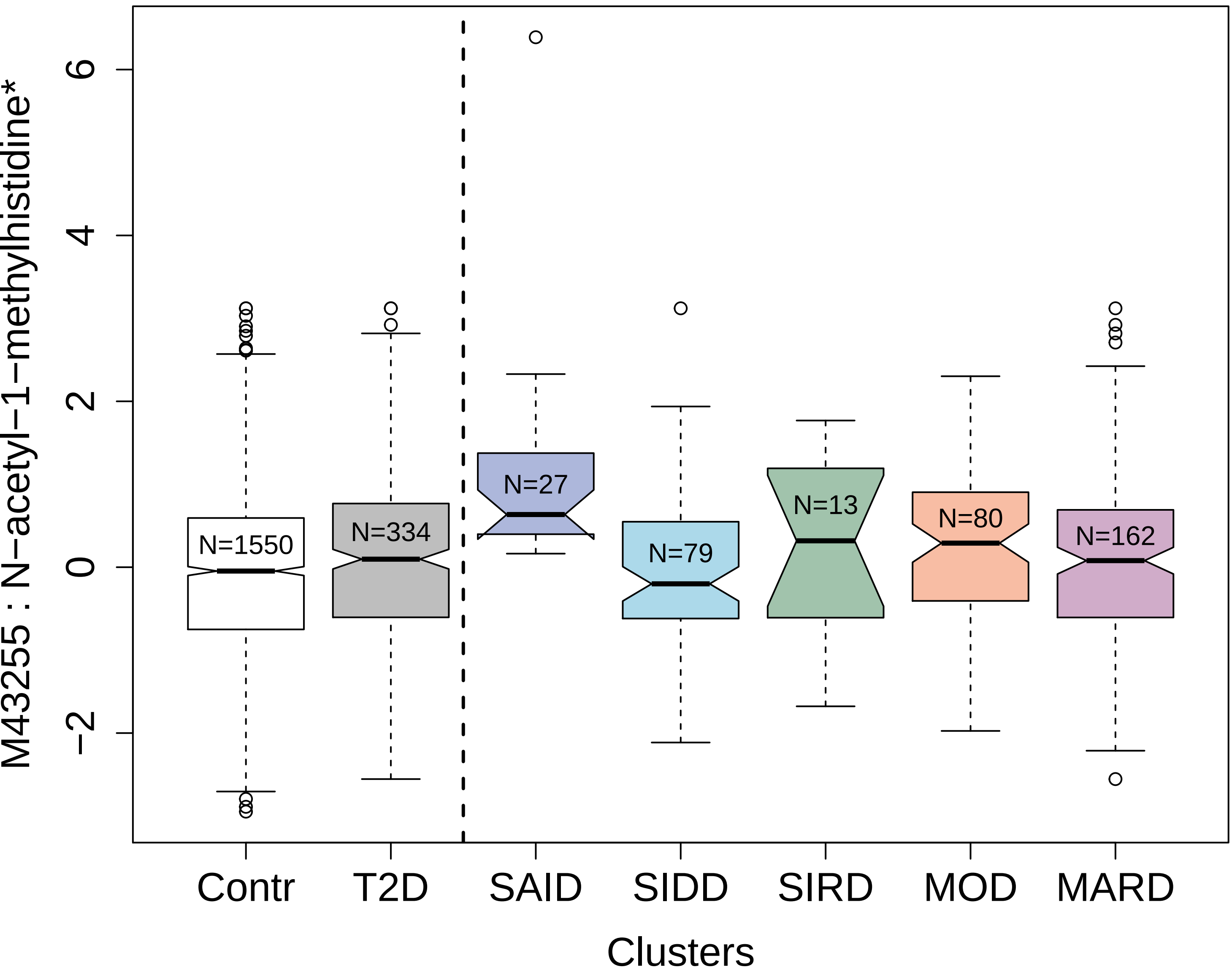

M43255 : N−acetyl−1−methylhistidine*
−2
0
2
4
6
N=1550
N=334
N=27
N=79
N=13
N=80
N=162
Contr
T2D
SAID
SIDD
SIRD
MOD
MARD
Clusters

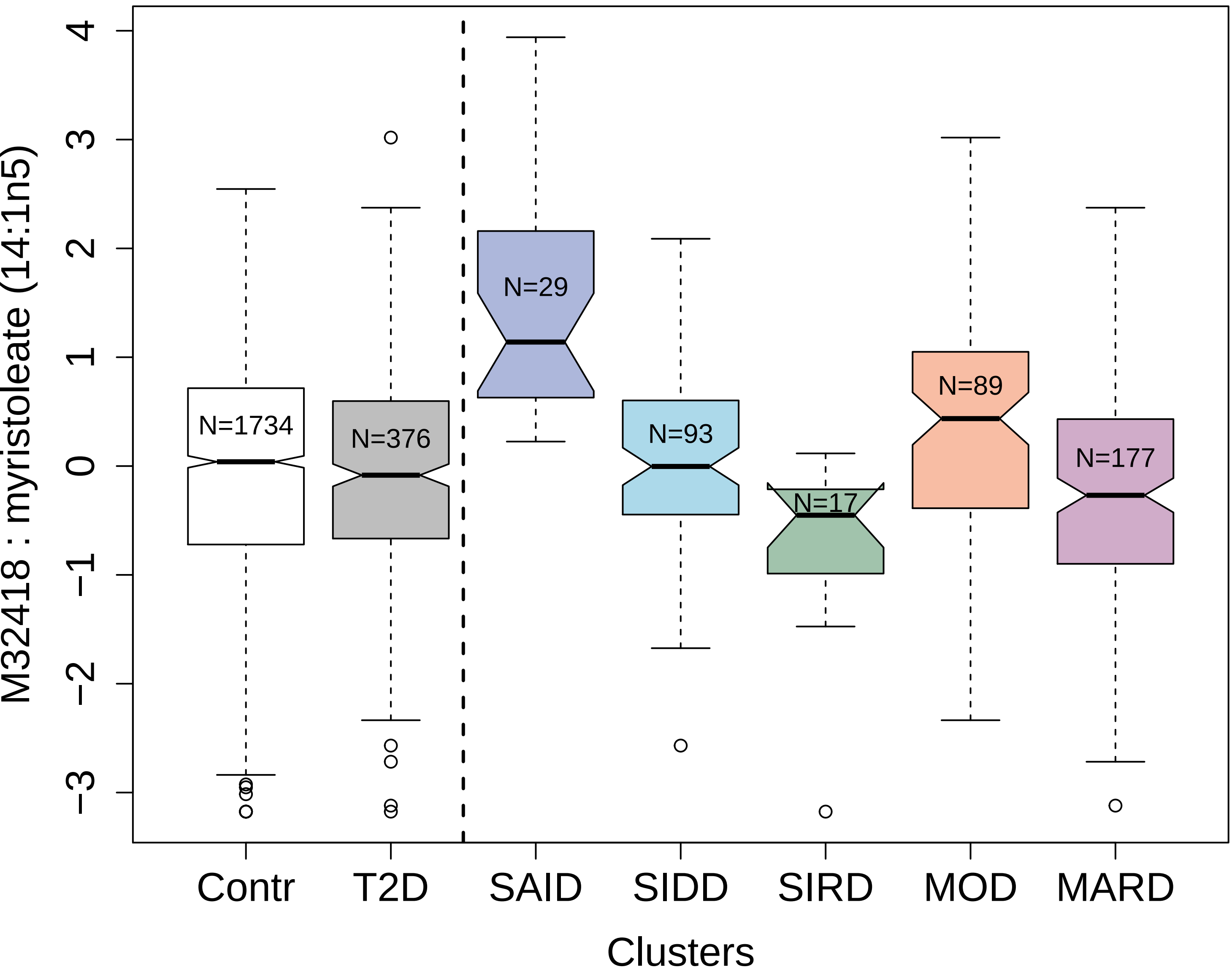

M32418 : myristoleate (14:1n5)
4
3
2
1
0
−1
−2
−3
N=1734
N=376
N=29
N=93
N=17
N=89
N=177
Contr
T2D
SAID
SIDD
SIRD
MOD
MARD
Clusters

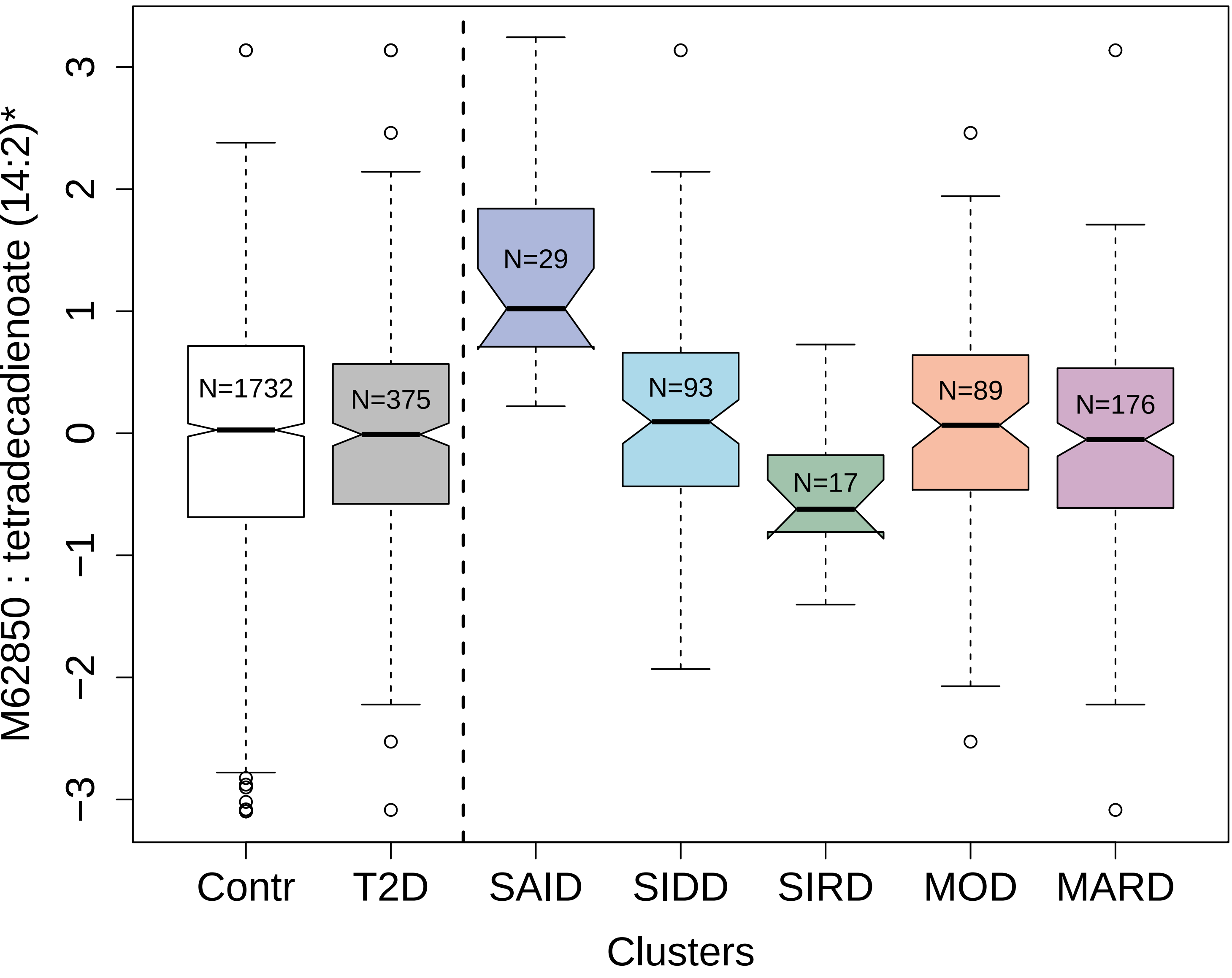
M62850 : tetradecadienoate (14:2)*
Clusters
Contr
T2D
SAID
SIDD
SIRD
MOD
MARD
N=1732
N=375
N=29
N=93
N=17
N=89
N=176
−3
−2
−1
0
1
2
3

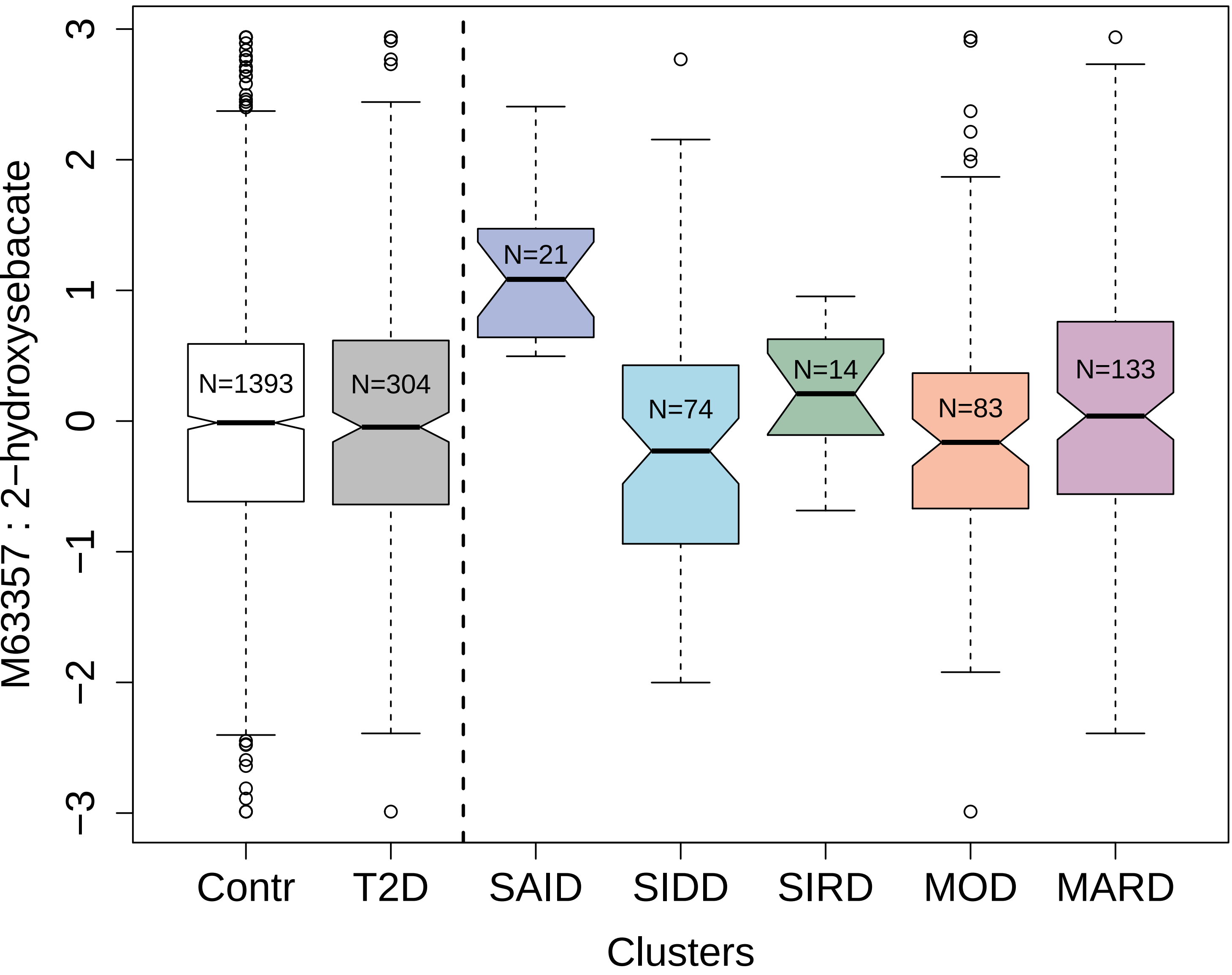

M63357 : 2−hydroxysebacate
3
2
1
0
−1
−2
−3
N=1393
N=304
N=21
N=74
N=14
N=83
N=133
Contr
T2D
SAID
SIDD
SIRD
MOD
MARD
Clusters

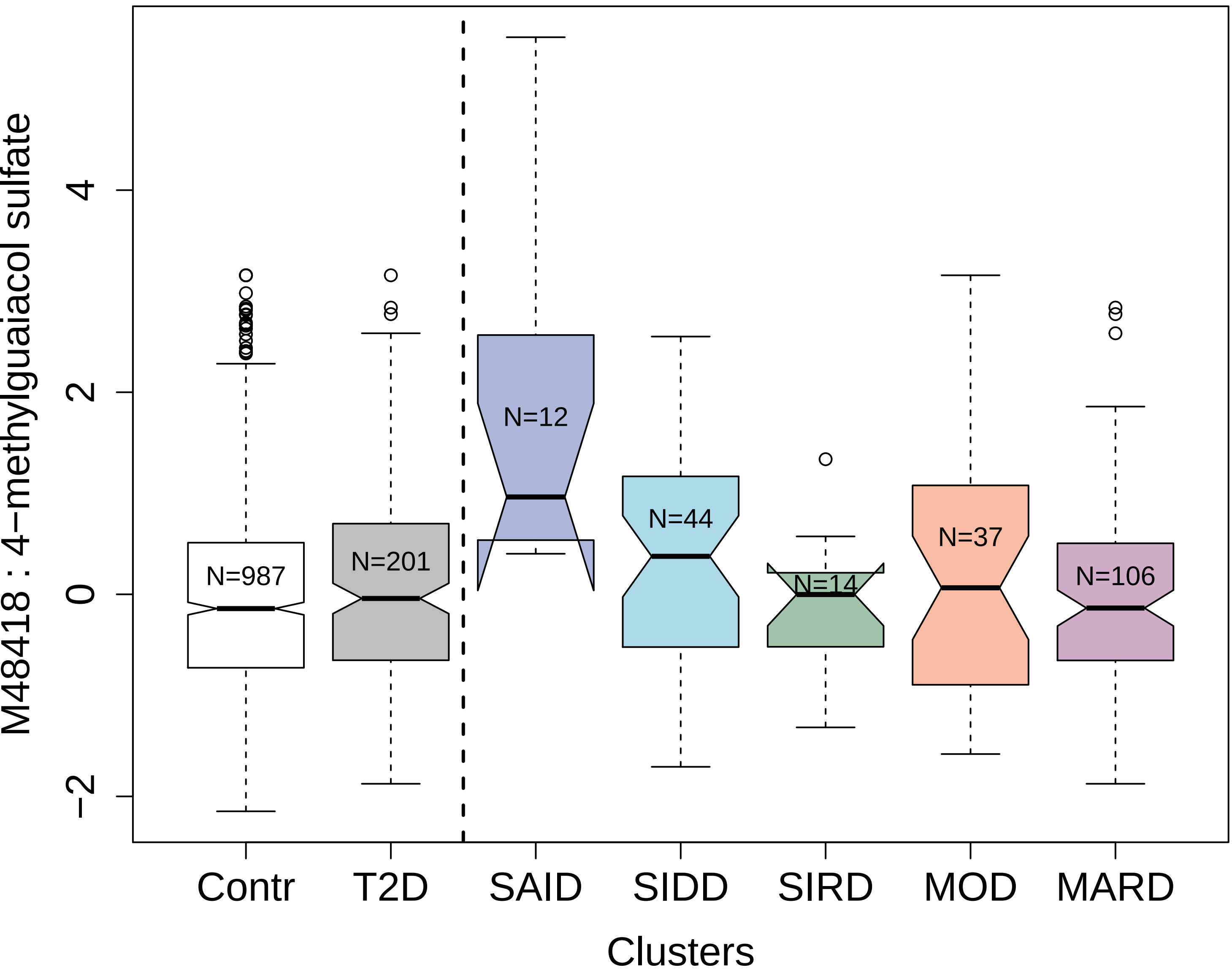
M48418 : 4−methylguaiacol sulfate
Clusters
Contr
T2D
SAID
SIDD
SIRD
MOD
MARD
N=987
N=201
N=12
N=44
N=14
N=37
N=106
−2
0
2
4

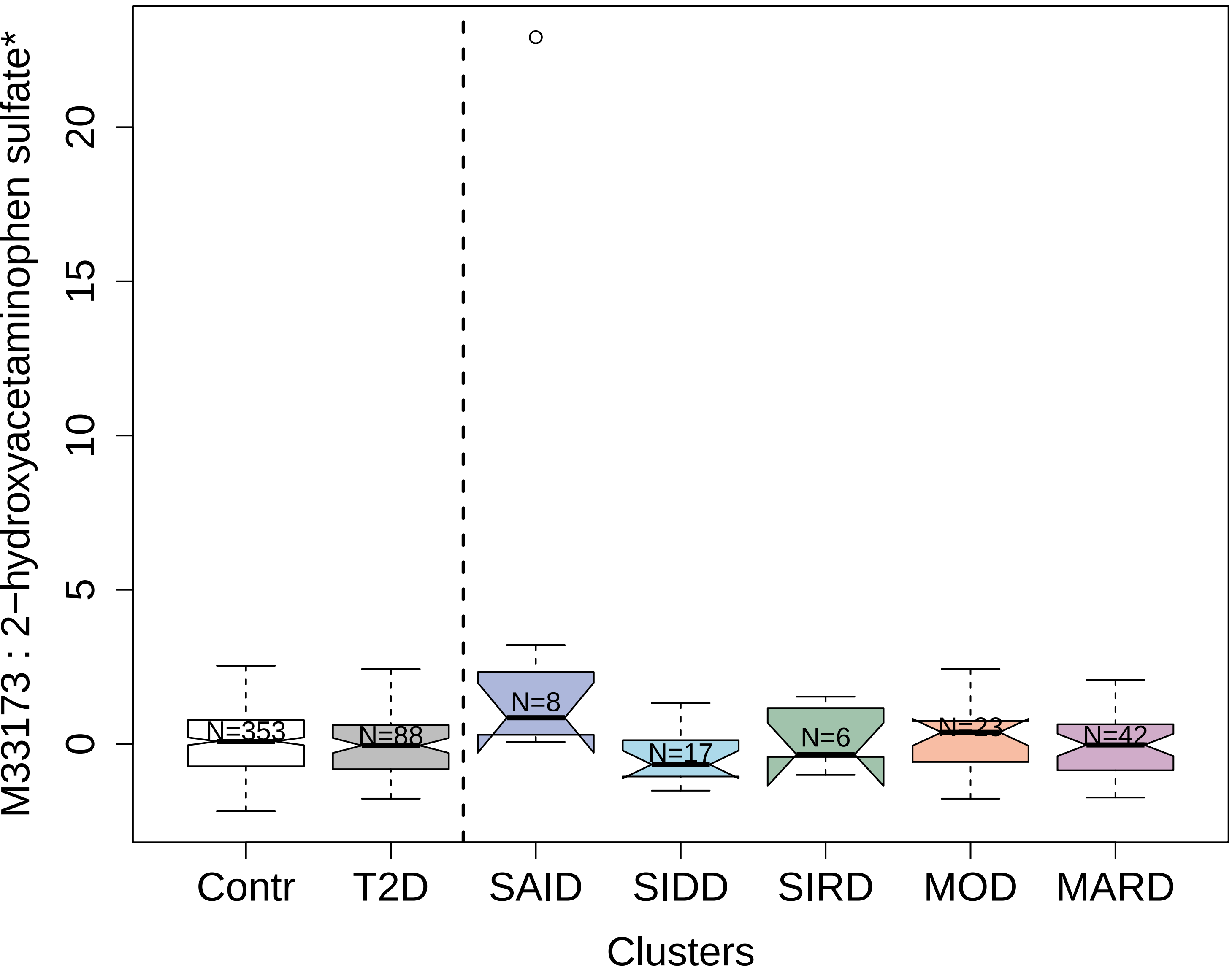

M33173 : 2−hydroxyacetaminophen sulfate*
0
5
10
15
20
N=353
N=88
N=8
N=17
N=6
N=23
N=42
Contr
T2D
SAID
SIDD
SIRD
MOD
MARD
Clusters

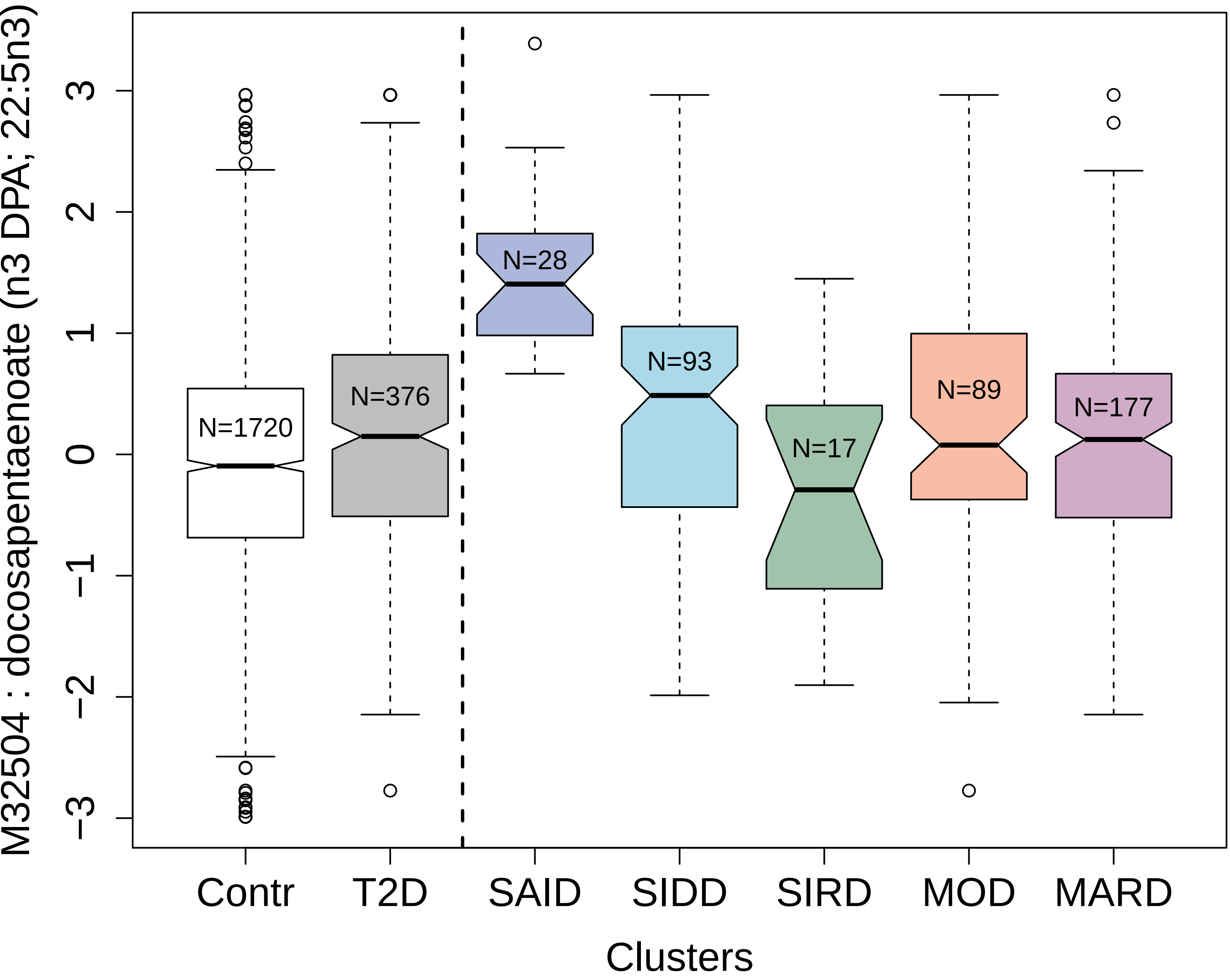

M32504 : docosapentaenoate (n3 DPA; 22:5n3)
3
2
1
0
−1
−2
−3
N=1720
N=376
N=28
N=93
N=17
N=89
N=177
Contr
T2D
SAID
SIDD
SIRD
MOD
MARD
Clusters

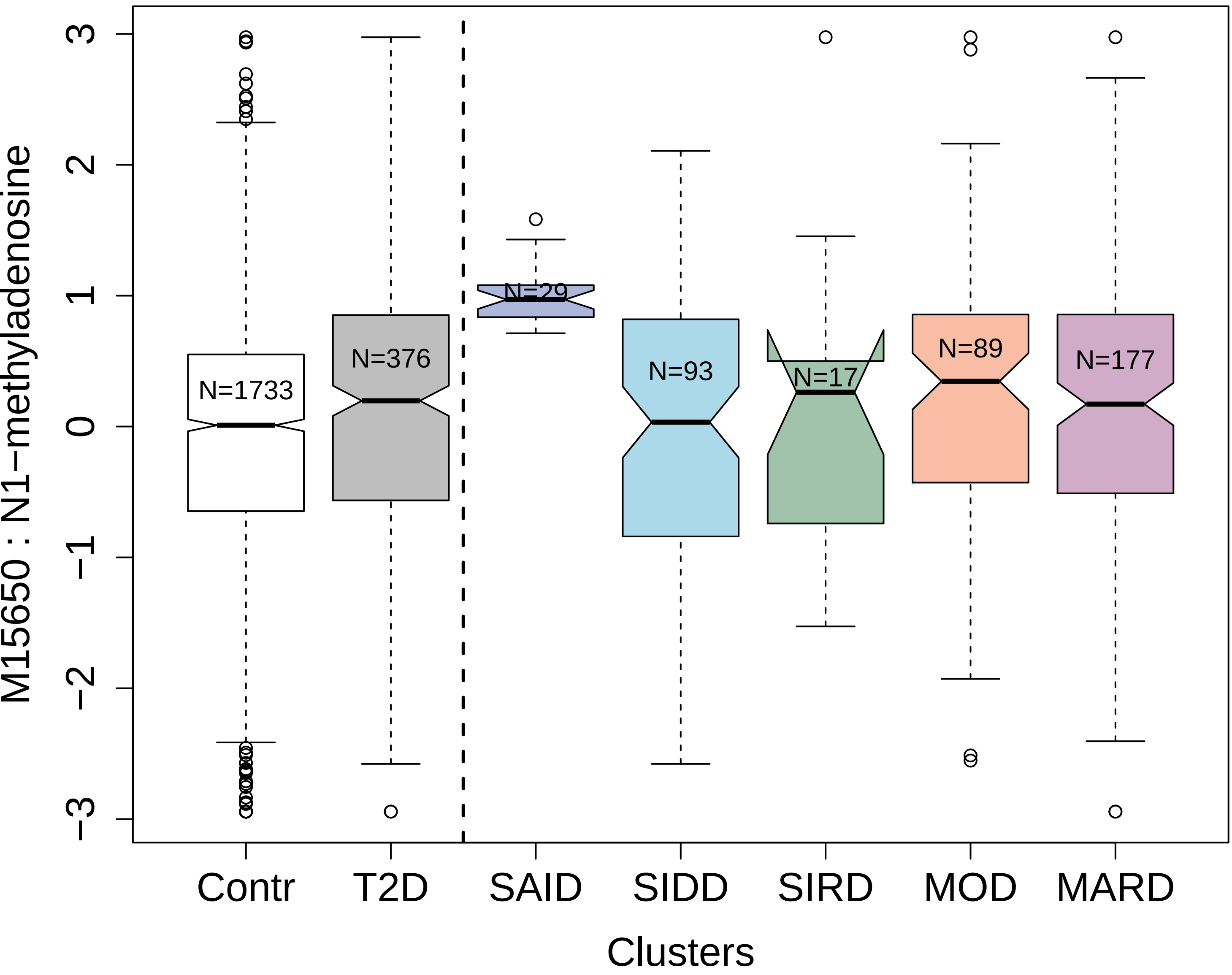
M15650 : N1−methyladenosine
3
2
1
0
−1
−2
−3
N=1733
N=376
N=29
N=93
N=17
N=89
N=177
Contr
T2D
SAID
SIDD
SIRD
MOD
MARD
Clusters

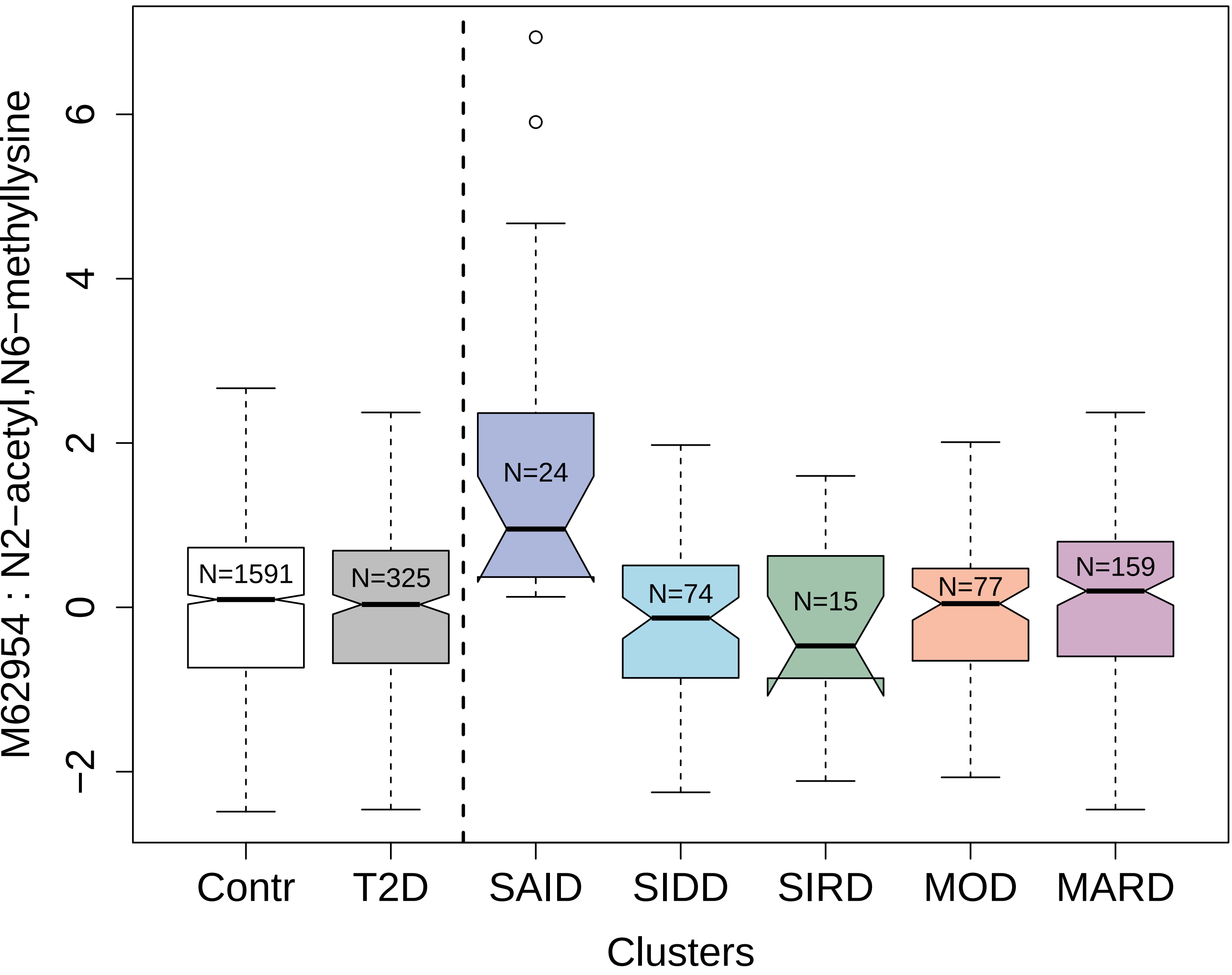

M62954 : N2−acetyl,N6−methyllysine
−2
0
2
4
6
N=1591
N=325
N=24
N=74
N=15
N=77
N=159
Contr
T2D
SAID
SIDD
SIRD
MOD
MARD
Clusters

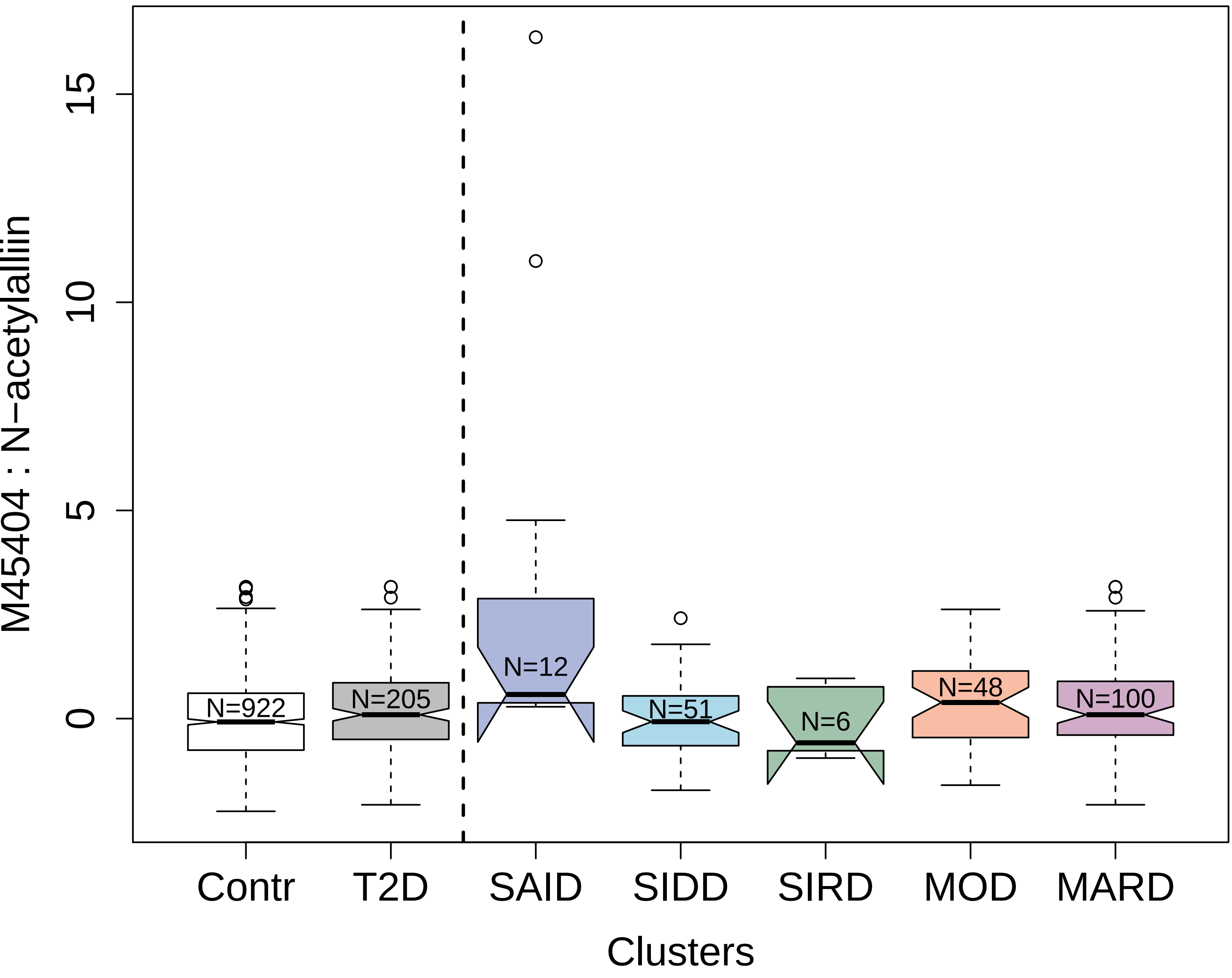

M45404 : N−acetylalliin
0
5
10
15
N=922
N=205
N=12
N=51
N=6
N=48
N=100
Contr
T2D
SAID
SIDD
SIRD
MOD
MARD
Clusters

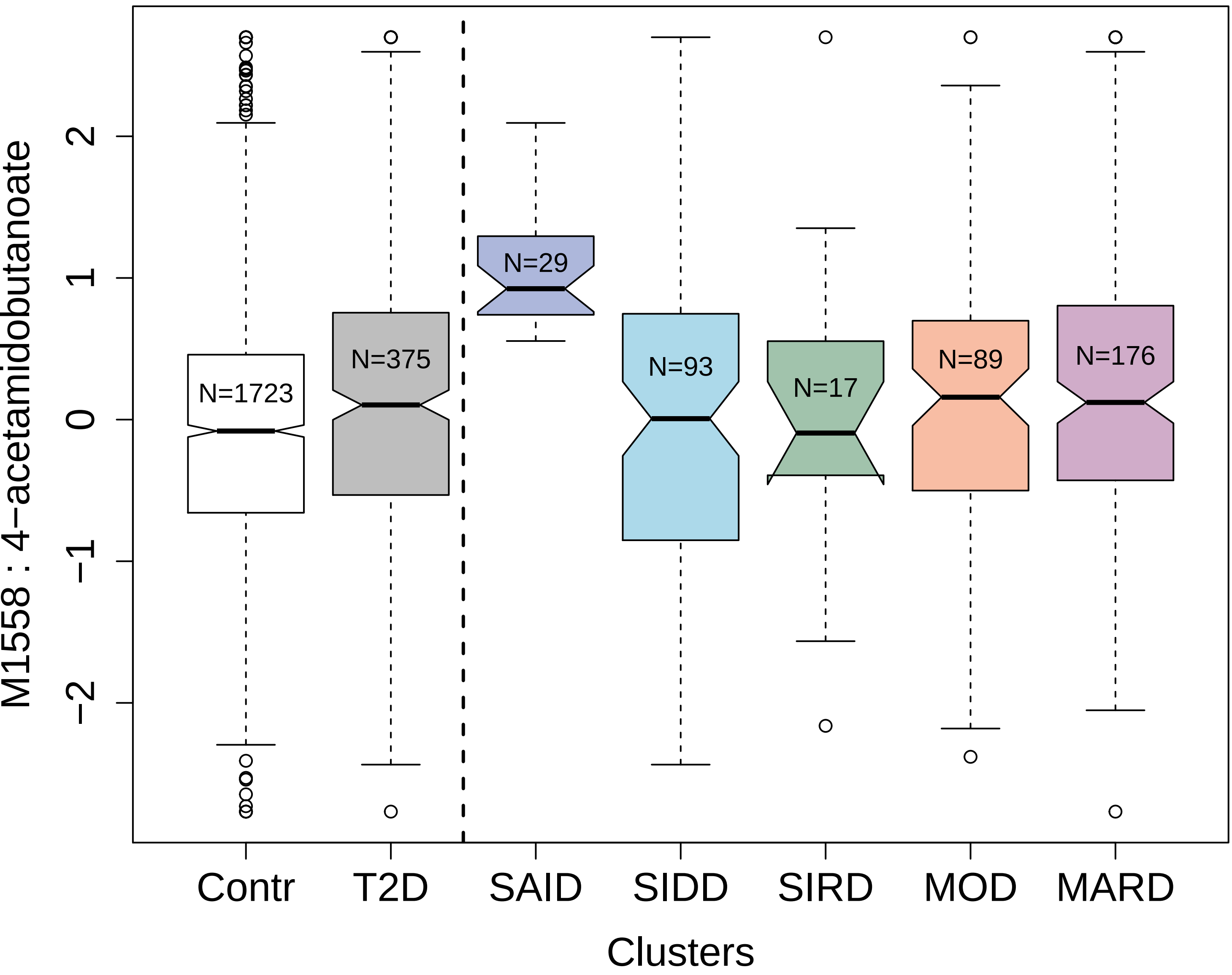
M1558 : 4−acetamidobutanoate
2
1
0
−1
−2
N=1723
N=375
N=29
N=93
N=17
N=89
N=176
Contr
T2D
SAID
SIDD
SIRD
MOD
MARD
Clusters

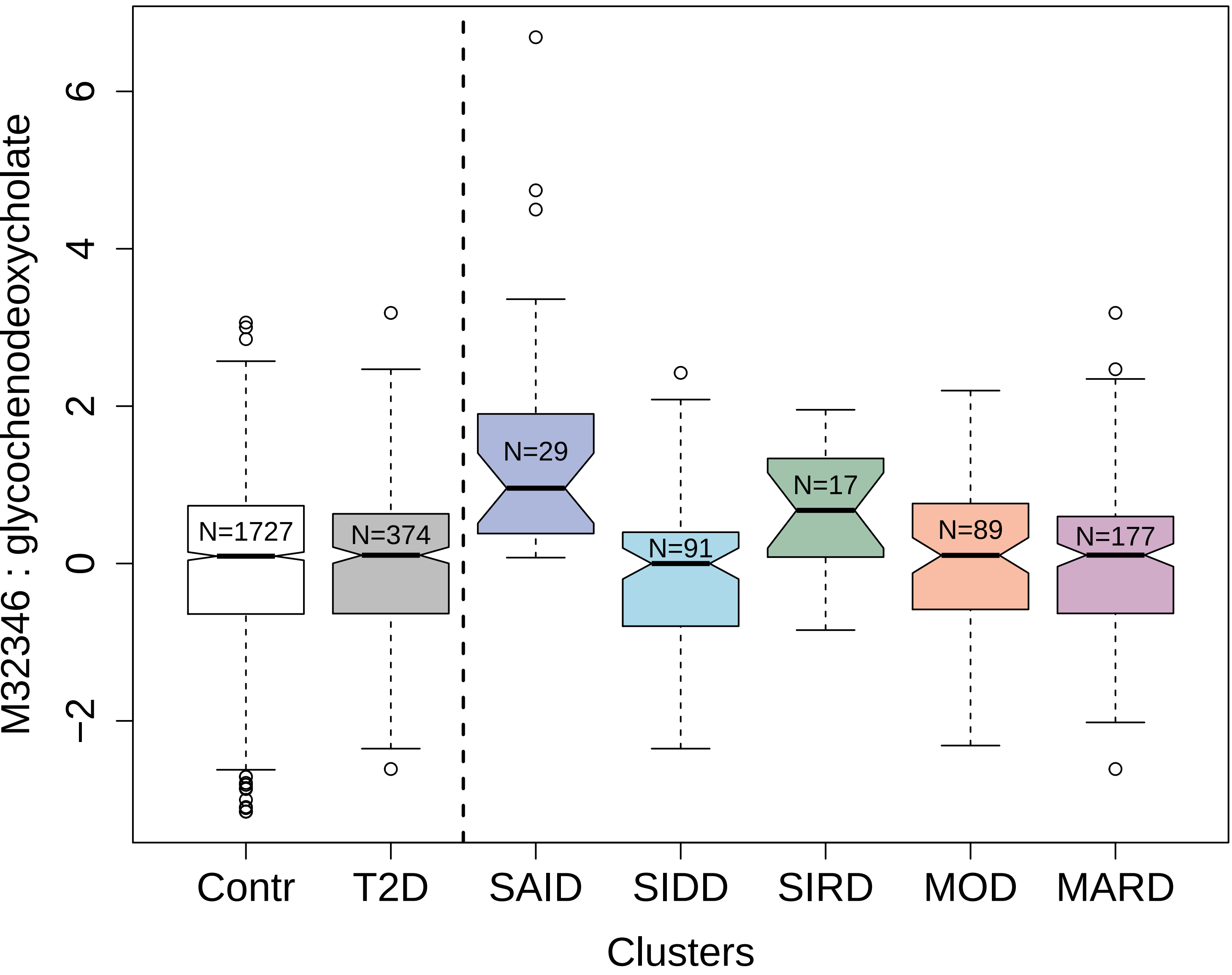
M32346 : glycochenodeoxycholate
Clusters
Contr
T2D
SAID
SIDD
SIRD
MOD
MARD
N=1727
N=374
N=29
N=91
N=17
N=89
N=177
6
4
2
0
−2

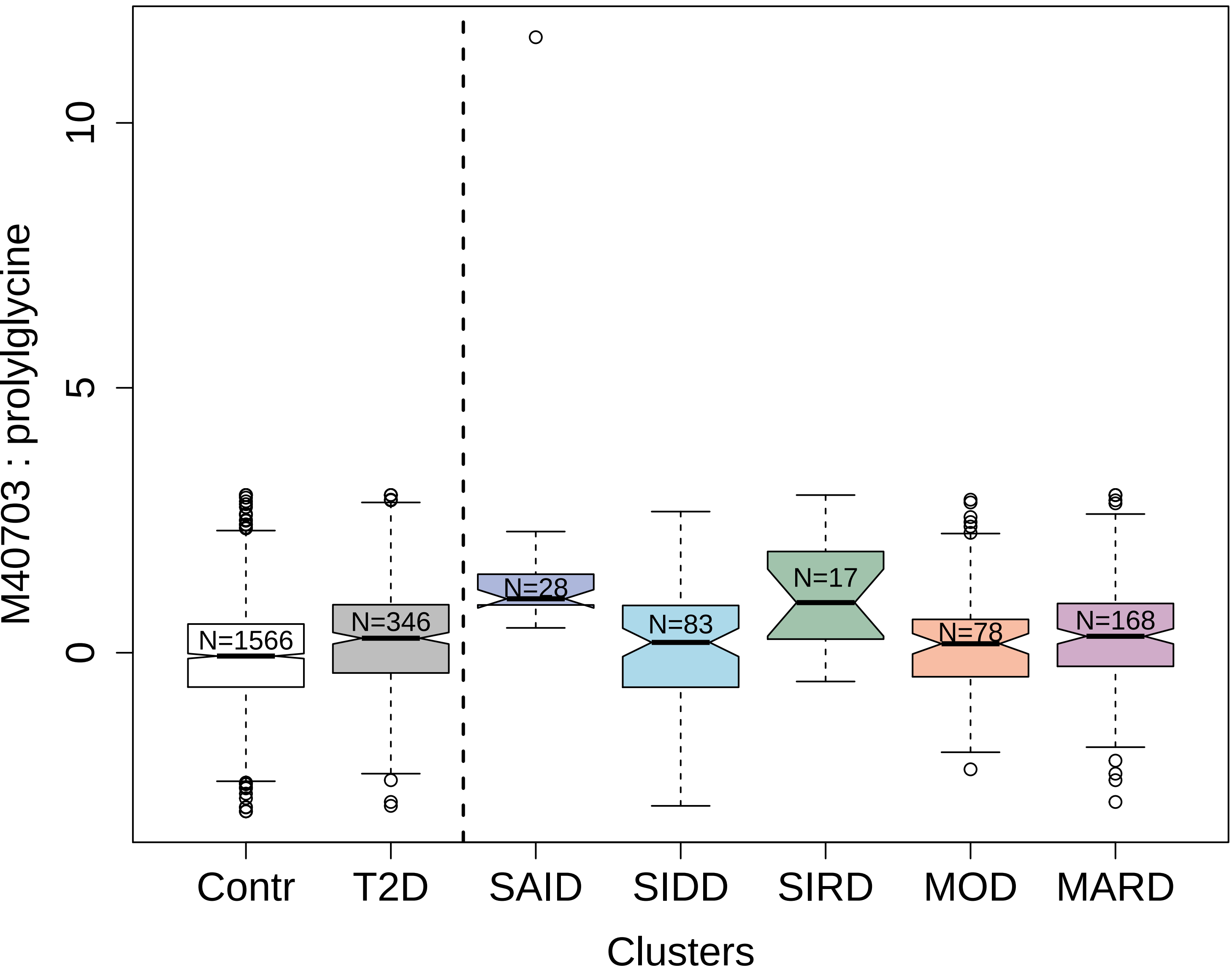

M40703 : prolylglycine
10
5
0
N=1566
N=346
N=28
N=83
N=17
N=78
N=168
Contr
T2D
SAID
SIDD
SIRD
MOD
MARD
Clusters

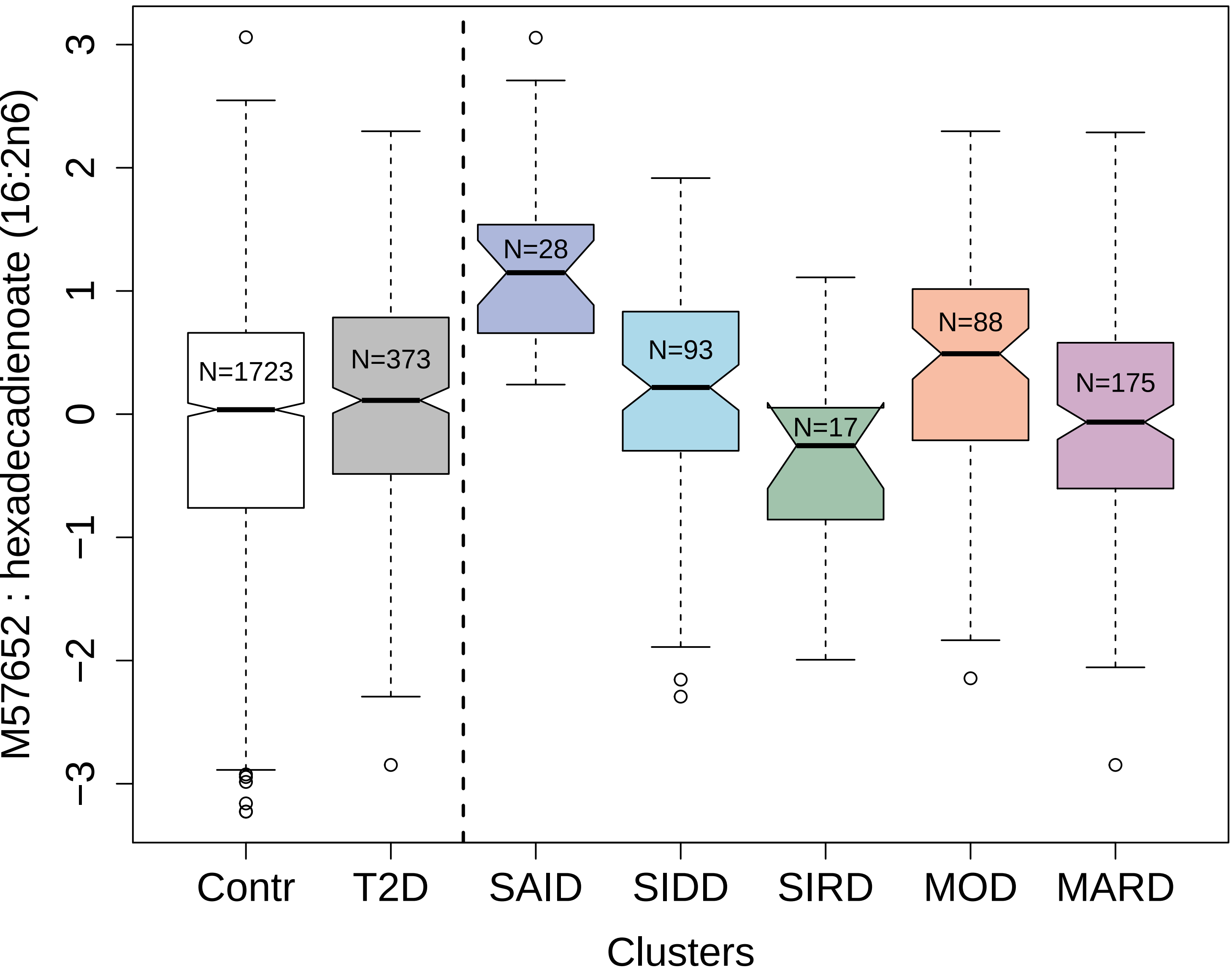

M57652 : hexadecadienoate (16:2n6)
3
2
1
0
−1
−2
−3
N=1723
N=373
N=28
N=93
N=17
N=88
N=175
Contr
T2D
SAID
SIDD
SIRD
MOD
MARD
Clusters

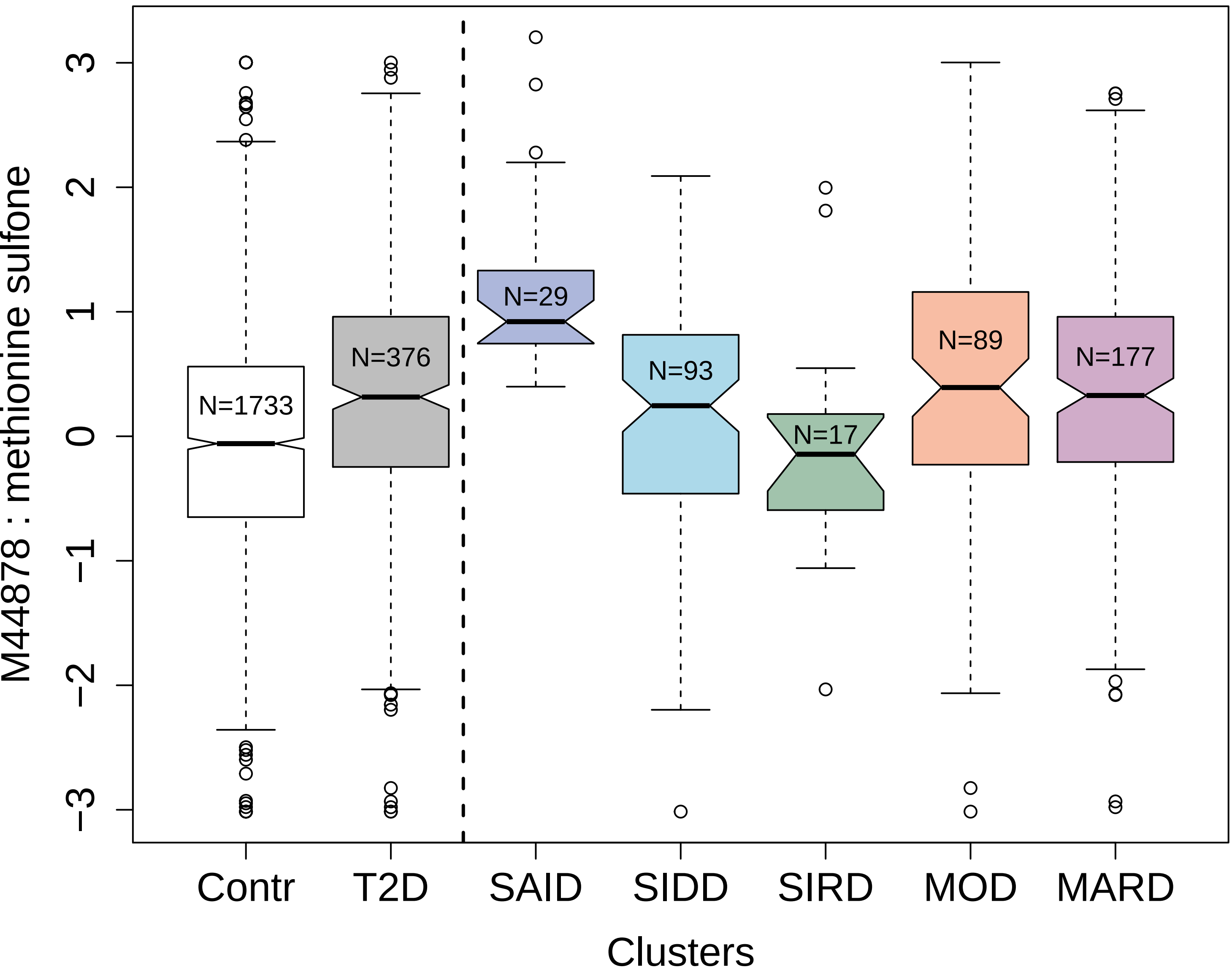

M44878 : methionine sulfone
3
2
1
0
−1
−2
−3
N=1733
N=376
N=29
N=93
N=17
N=89
N=177
Contr
T2D
SAID
SIDD
SIRD
MOD
MARD
Clusters

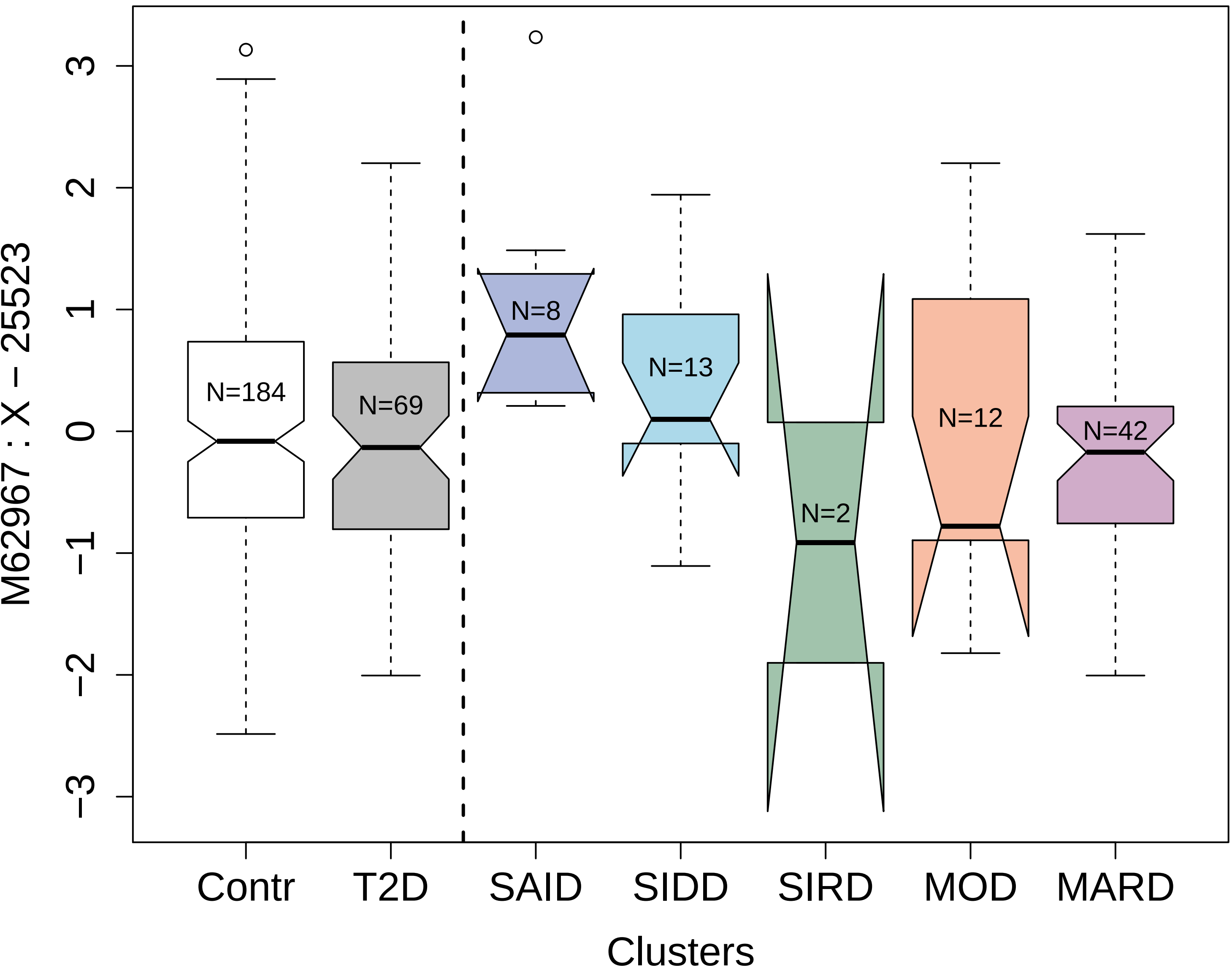

M62967 : X − 25523
3
2
1
0
−1
−2
−3
N=184
N=69
N=8
N=13
N=2
N=12
N=42
Contr
T2D
SAID
SIDD
SIRD
MOD
MARD
Clusters

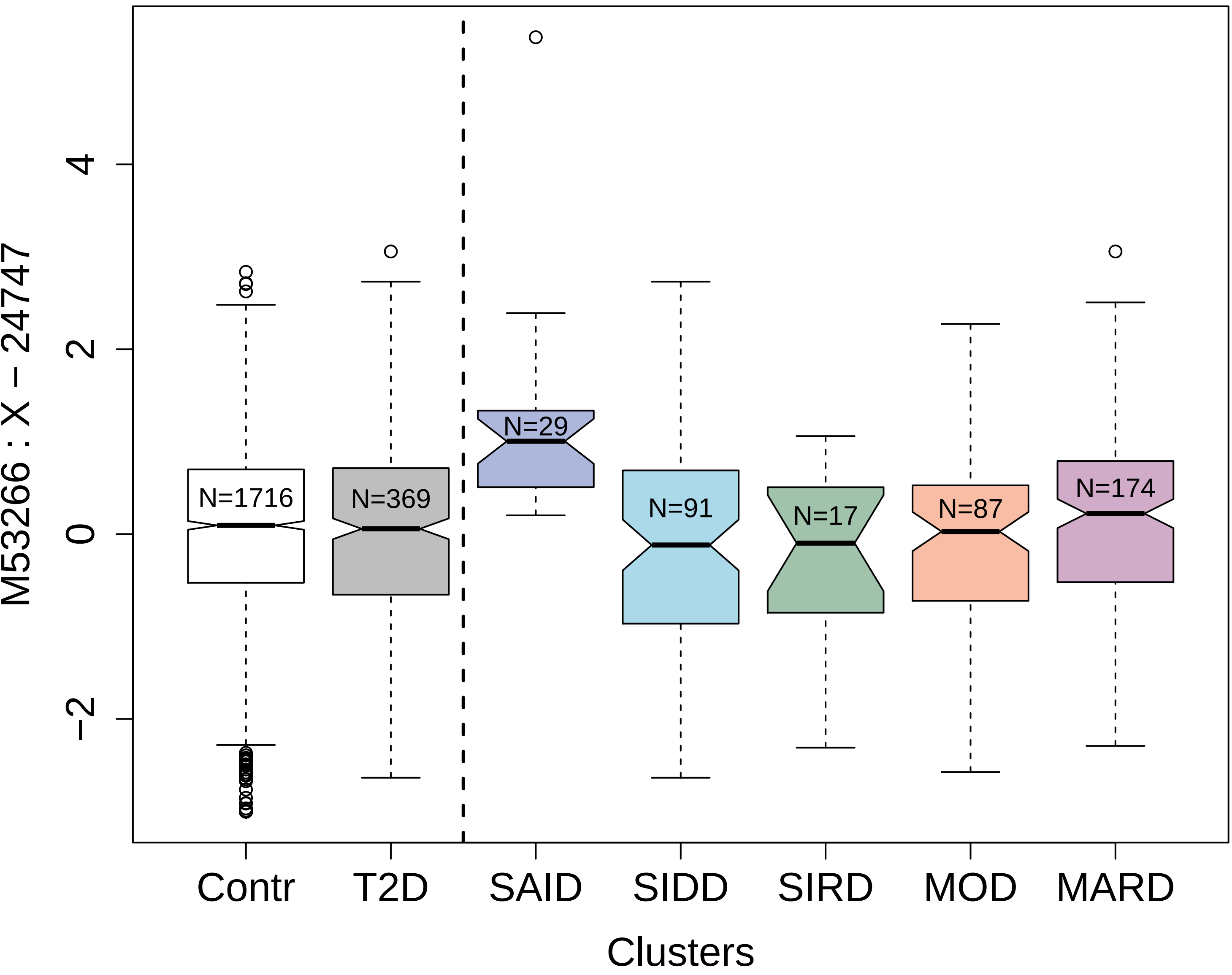
M53266 : X − 24747
−2
0
2
4
N=1716
N=369
N=29
N=91
N=17
N=87
N=174
Contr
T2D
SAID
SIDD
SIRD
MOD
MARD
Clusters

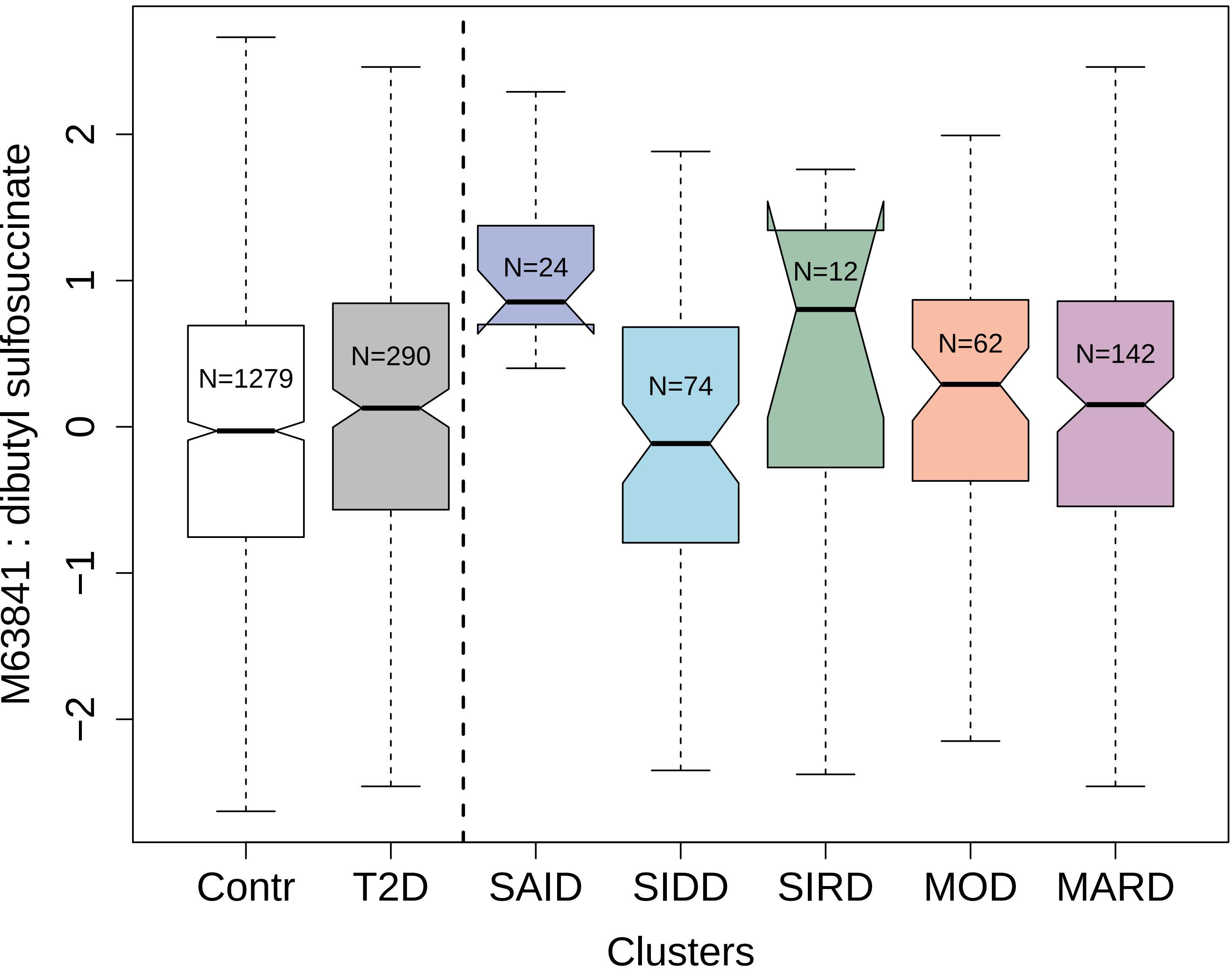

M63841 : dibutyl sulfosuccinate
2
1
0
−1
−2
N=1279
N=290
N=24
N=74
N=12
N=62
N=142
Contr
T2D
SAID
SIDD
SIRD
MOD
MARD
Clusters

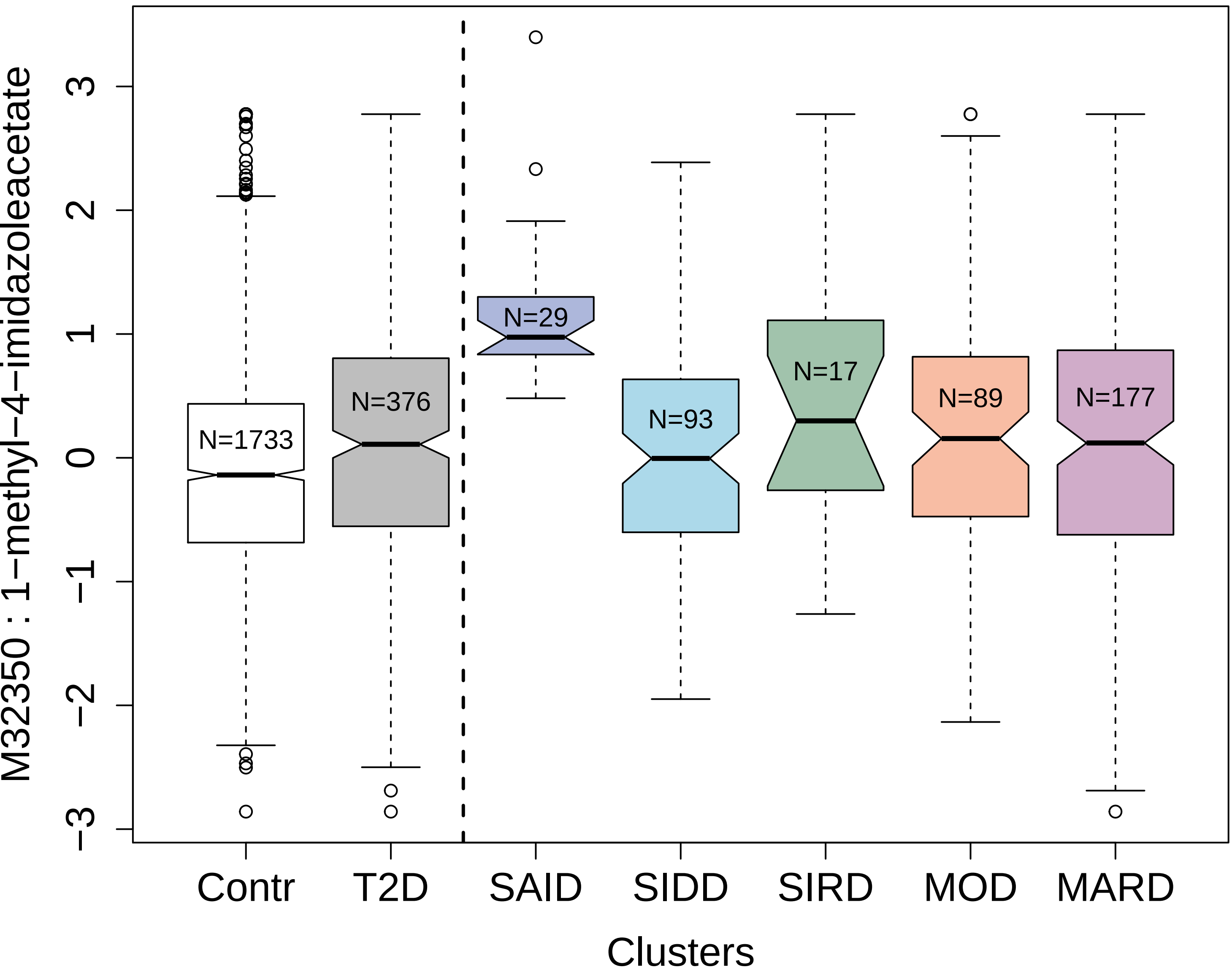
M32350 : 1−methyl−4−imidazoleacetate
3
2
1
0
−1
−2
−3
N=1733
N=376
N=29
N=93
N=17
N=89
N=177
Contr
T2D
SAID
SIDD
SIRD
MOD
MARD
Clusters

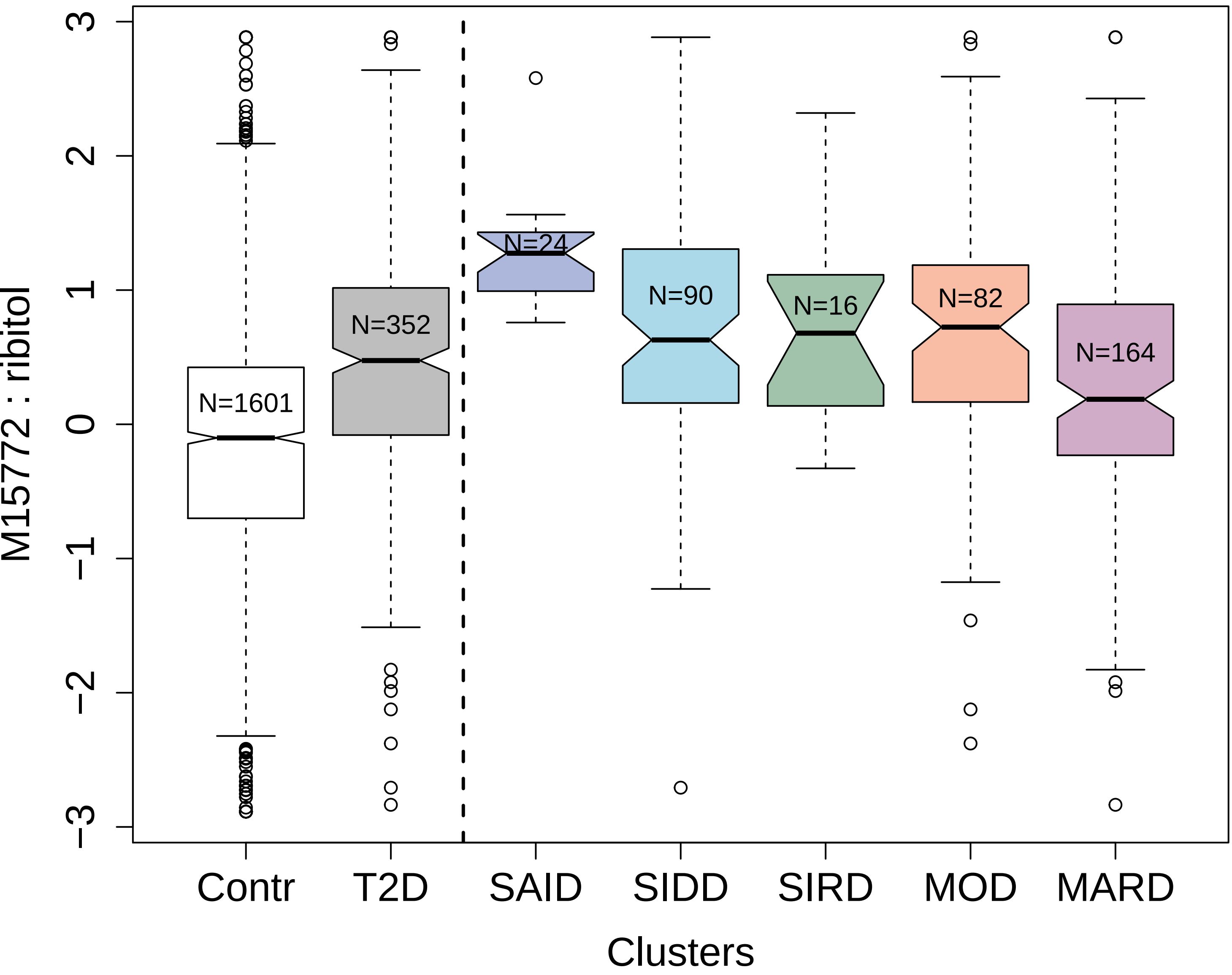
M15772 : ribitol
3
2
1
0
−1
−2
−3
N=1601
N=352
N=24
N=90
N=16
N=82
N=164
Contr
T2D
SAID
SIDD
SIRD
MOD
MARD
Clusters

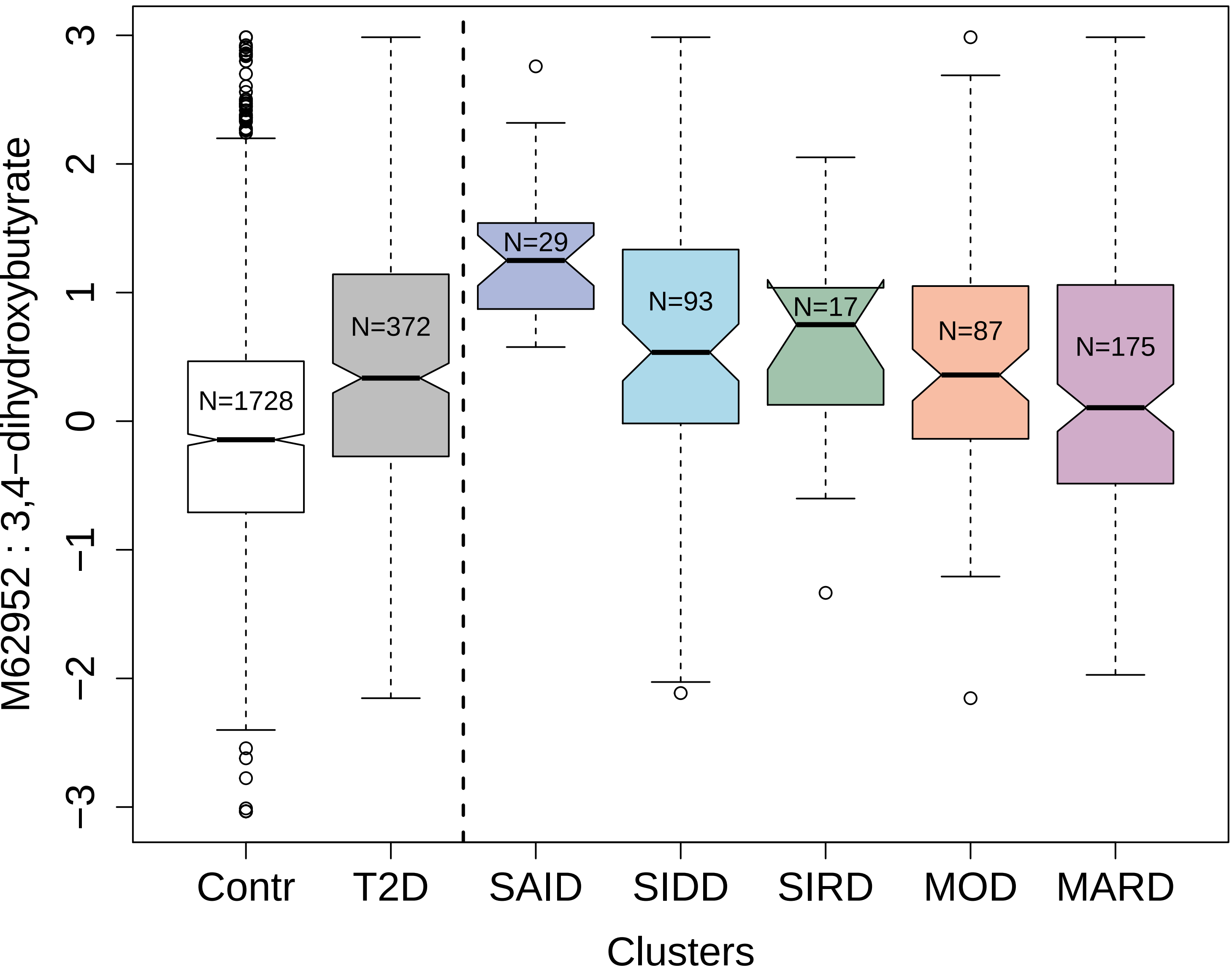

M62952 : 3,4−dihydroxybutyrate
3
2
1
0
−1
−2
−3
N=1728
N=372
N=29
N=93
N=17
N=87
N=175
Contr
T2D
SAID
SIDD
SIRD
MOD
MARD
Clusters

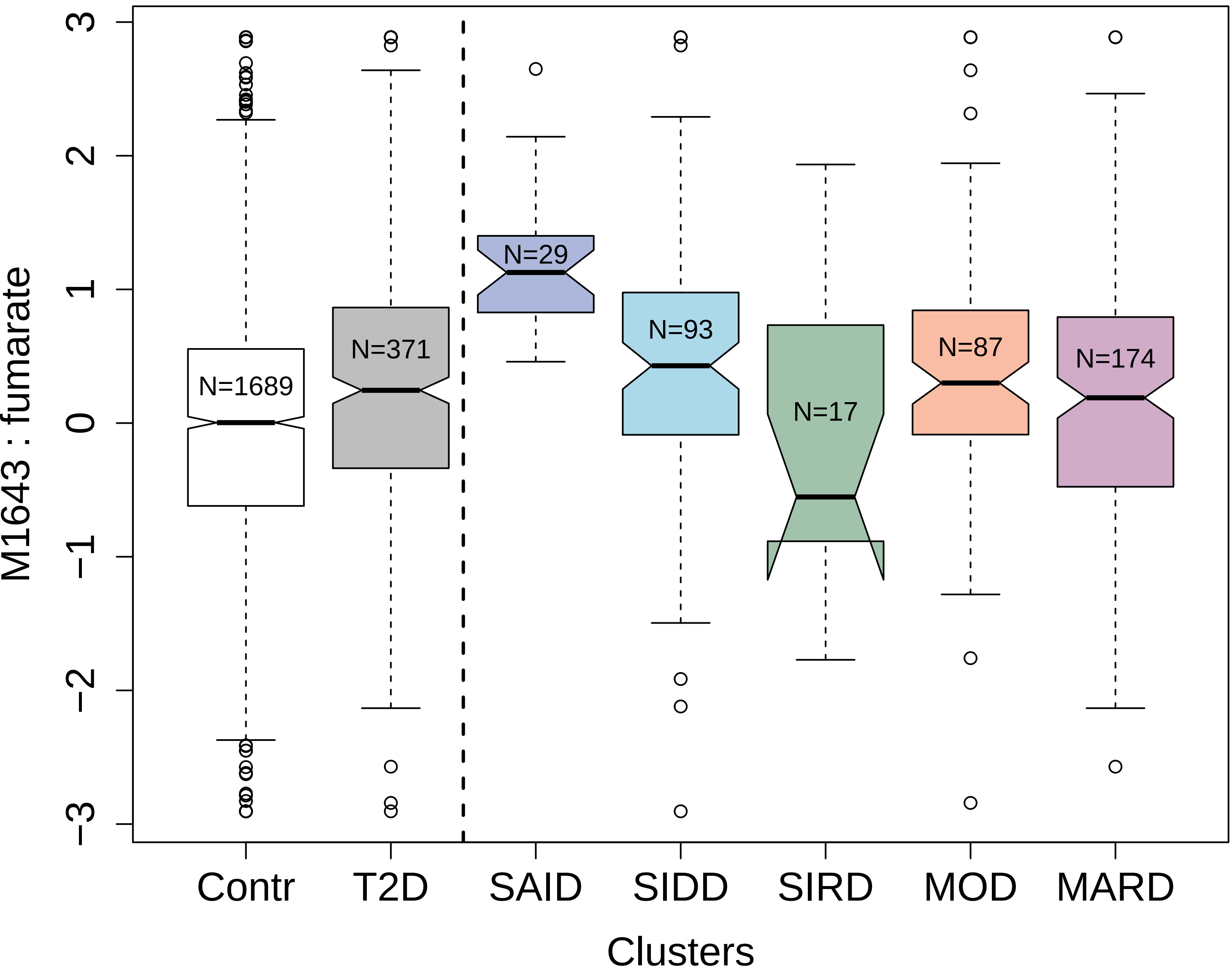

M1643 : fumarate
3
2
1
0
−1
−2
−3
N=1689
N=371
N=29
N=93
N=17
N=87
N=174
Contr
T2D
SAID
SIDD
SIRD
MOD
MARD
Clusters

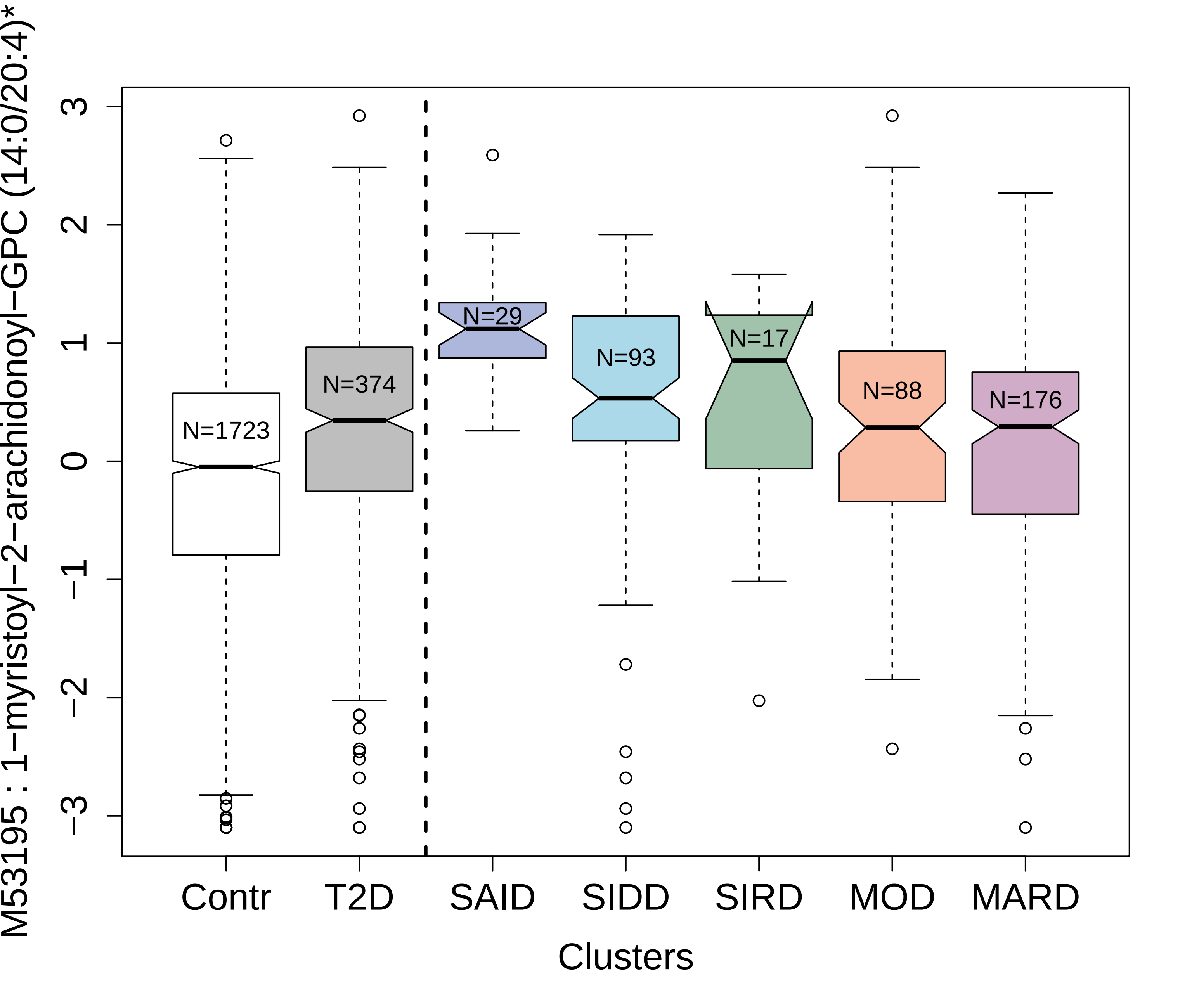
M53195 : 1−myristoyl−2−arachidonoyl−GPC (14:0/20:4)*
3
2
1
0
−1
−2
−3
N=1723
N=374
N=29
N=93
N=17
N=88
N=176
Contr
T2D
SAID
SIDD
SIRD
MOD
MARD
Clusters

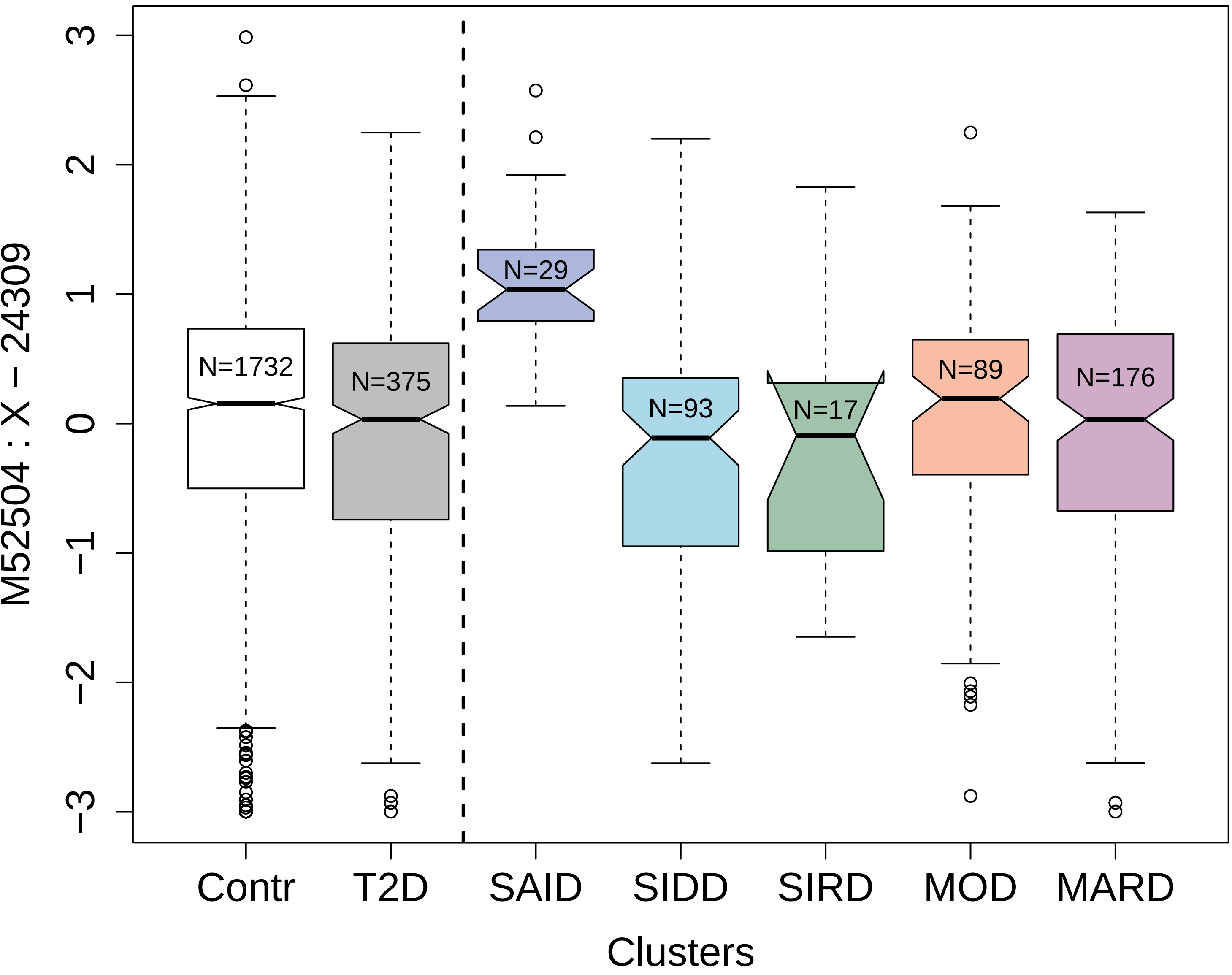

M52504 : X − 24309
3
2
1
0
−1
−2
−3
N=1732
N=375
N=29
N=93
N=17
N=89
N=176
Contr
T2D
SAID
SIDD
SIRD
MOD
MARD
Clusters

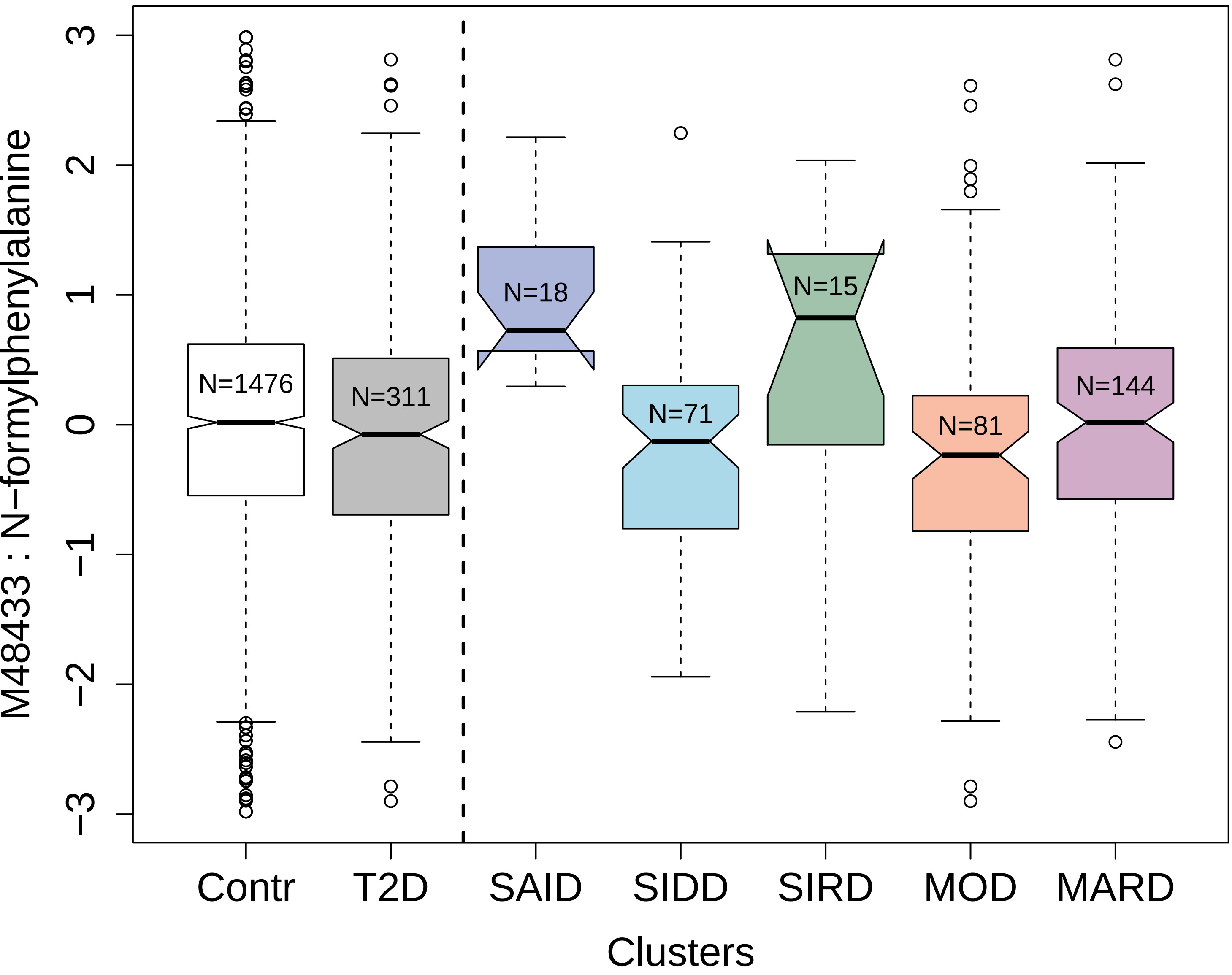

M48433 : N−formylphenylalanine
3
2
1
0
−1
−2
−3
N=1476
N=311
N=18
N=71
N=15
N=81
N=144
Contr
T2D
SAID
SIDD
SIRD
MOD
MARD
Clusters

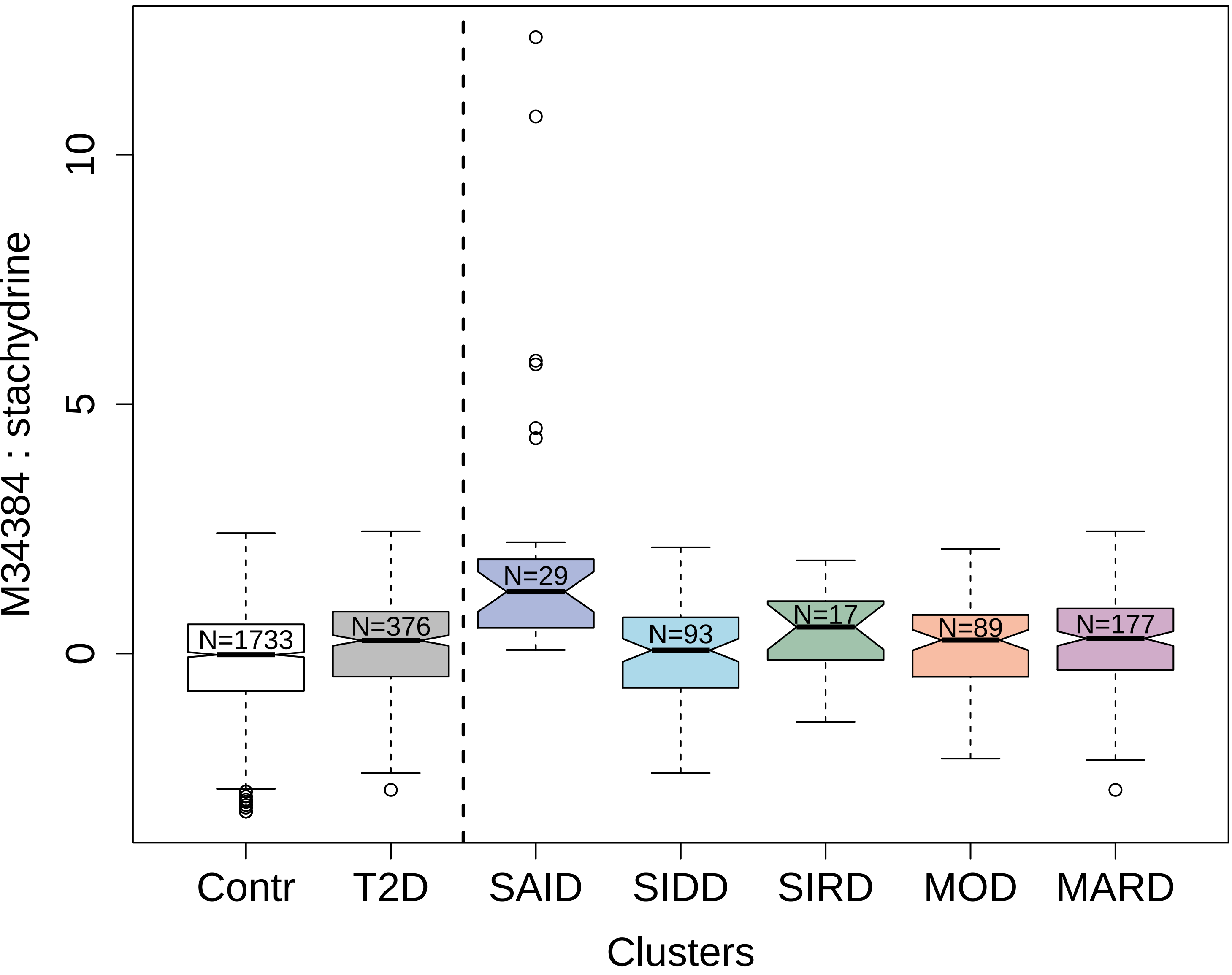

M34384 : stachydrine
0
5
10
N=1733
N=376
N=29
N=93
N=17
N=89
N=177
Contr
T2D
SAID
SIDD
SIRD
MOD
MARD
Clusters

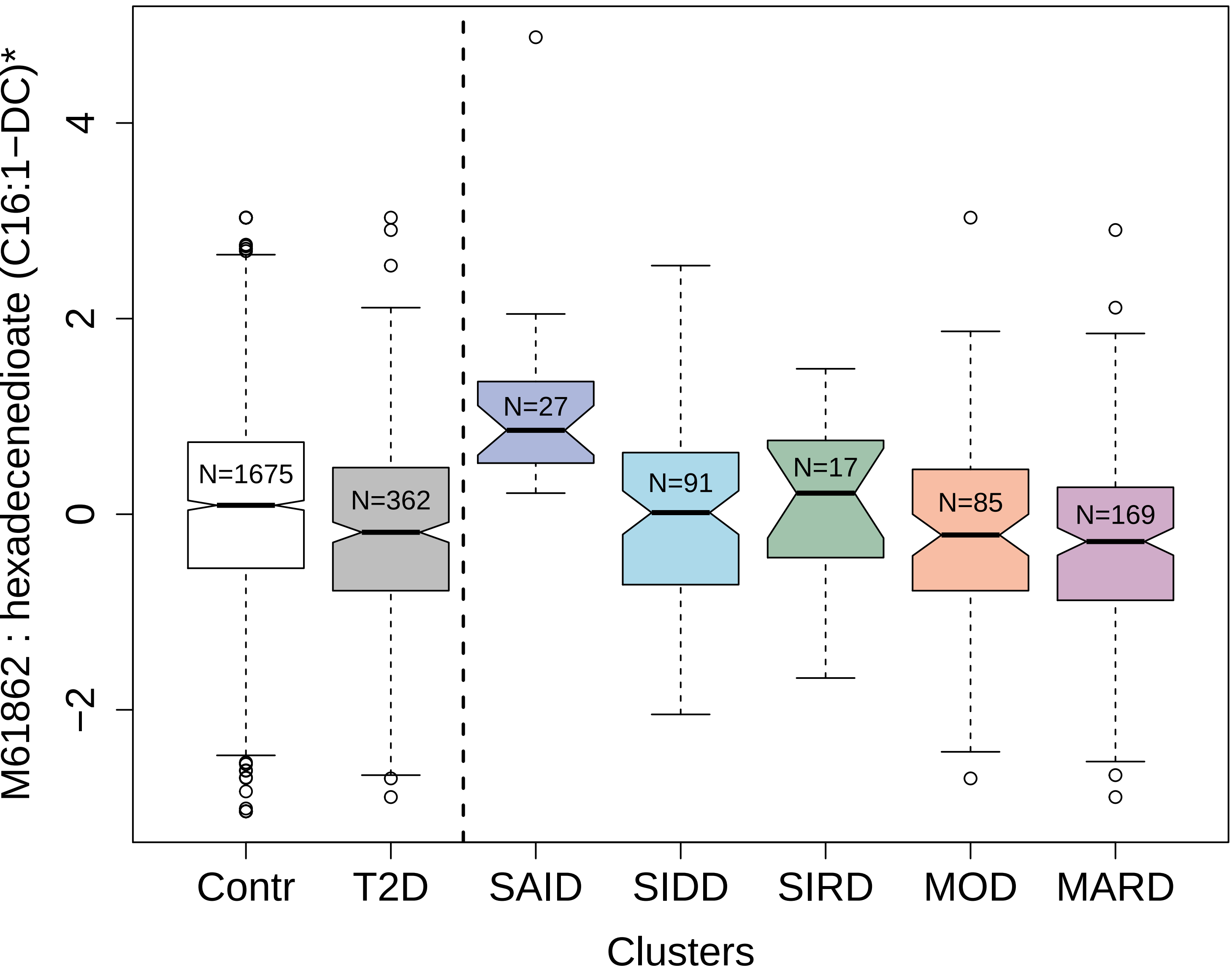

M61862 : hexadecenedioate (C16:1−DC)*
4
2
0
−2
N=1675
N=362
N=27
N=91
N=17
N=85
N=169
Contr
T2D
SAID
SIDD
SIRD
MOD
MARD
Clusters

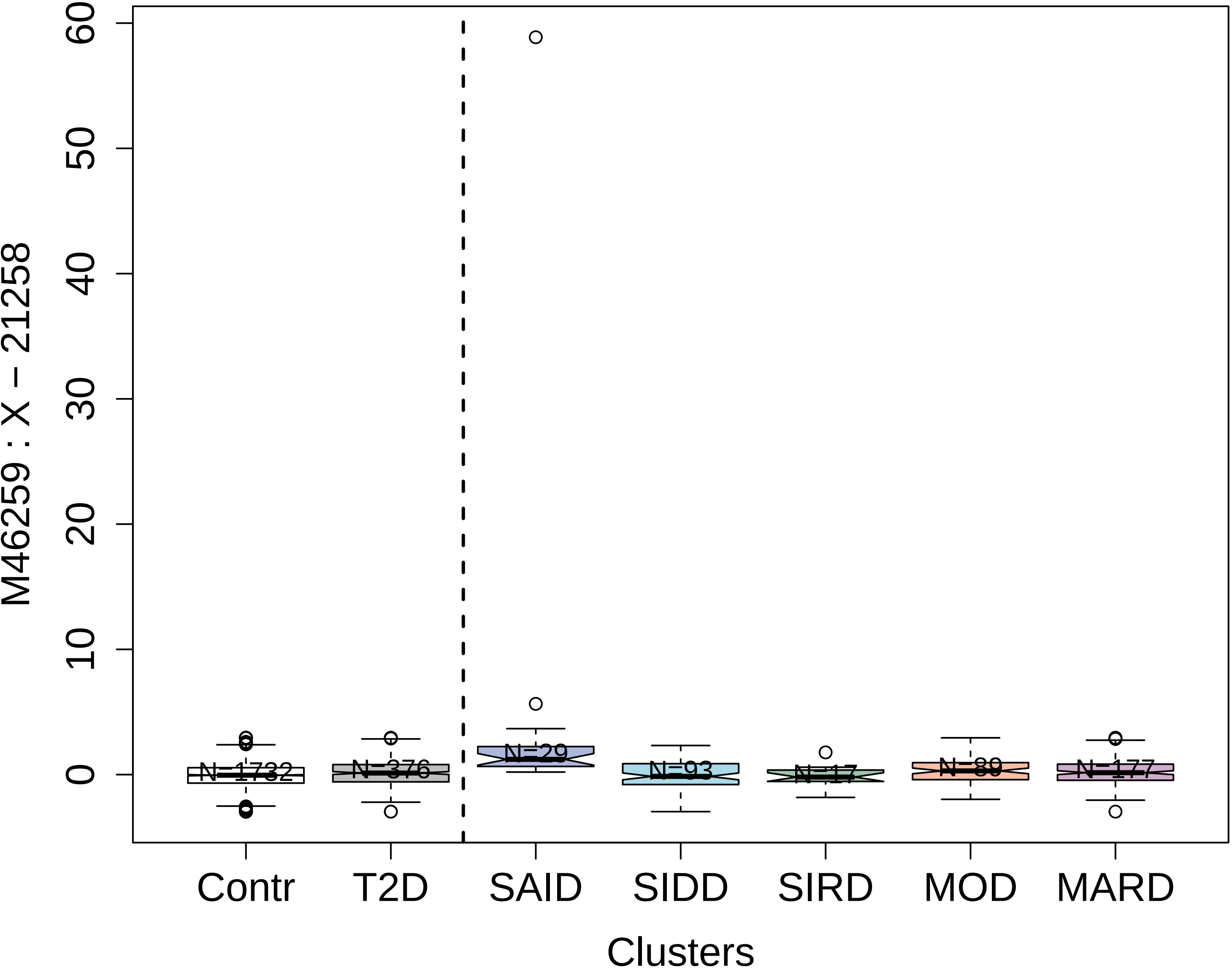

M46259 : X − 21258
0
10
20
30
40
50
60
N=1732
N=376
N=29
N=93
N=17
N=89
N=177
Contr
T2D
SAID
SIDD
SIRD
MOD
MARD
Clusters

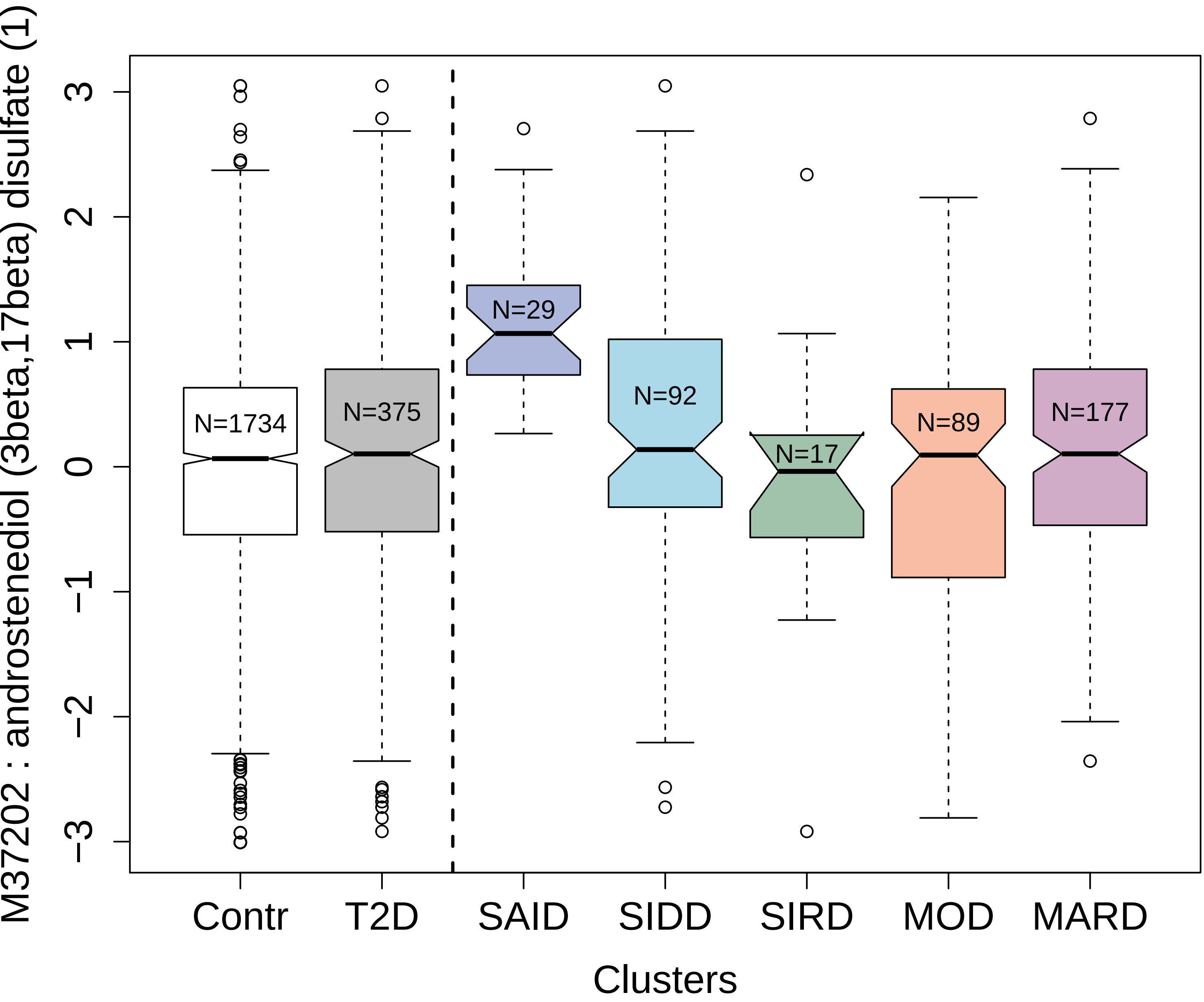

M37202 : androstenediol (3beta,17beta) disulfate (1)
3
2
1
0
−1
−2
−3
N=1734
N=375
N=29
N=92
N=17
N=89
N=177
Contr
T2D
SAID
SIDD
SIRD
MOD
MARD
Clusters

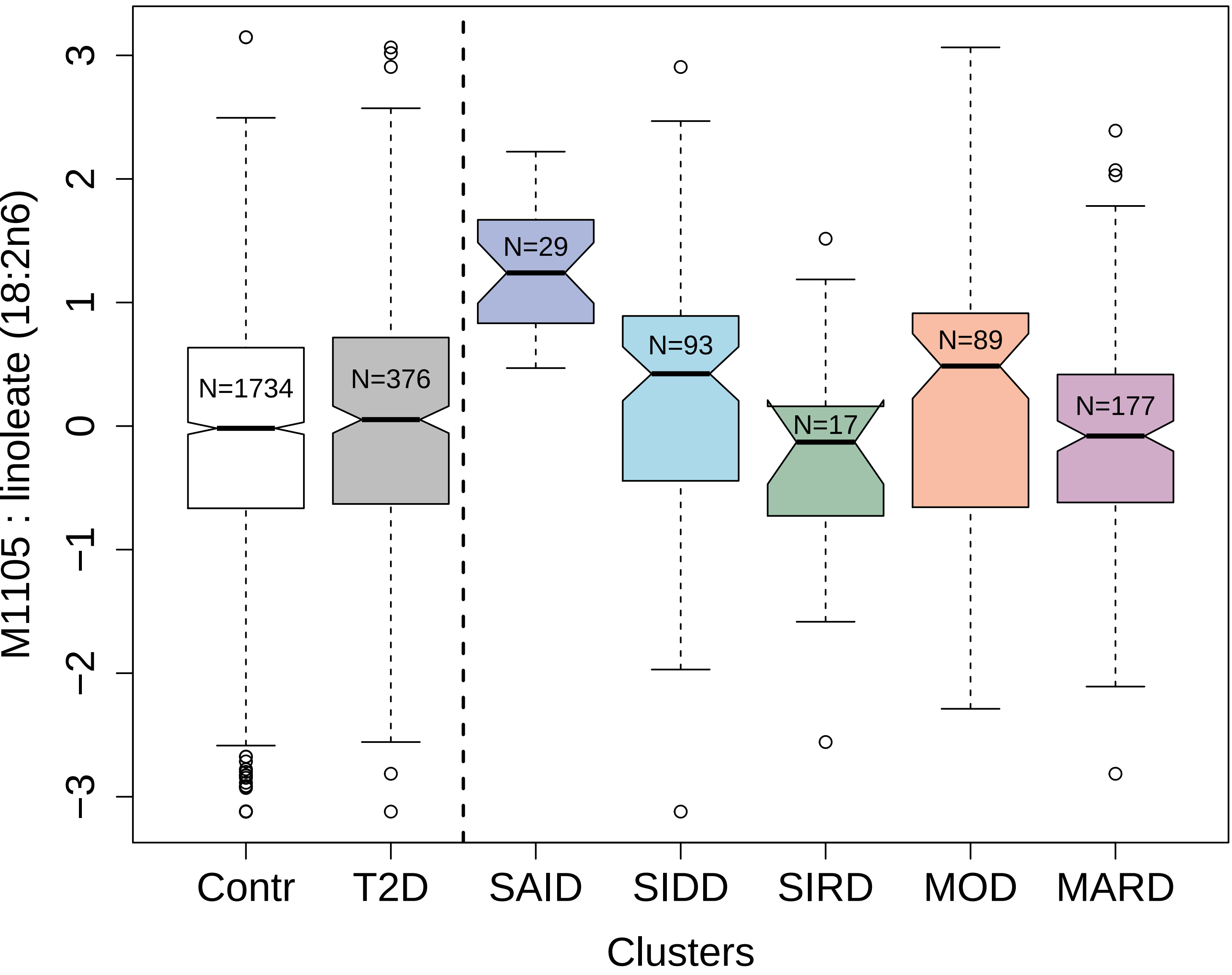

M1105 : linoleate (18:2n6)
3
2
1
0
−1
−2
−3
N=1734
N=376
N=29
N=93
N=17
N=89
N=177
Contr
T2D
SAID
SIDD
SIRD
MOD
MARD
Clusters

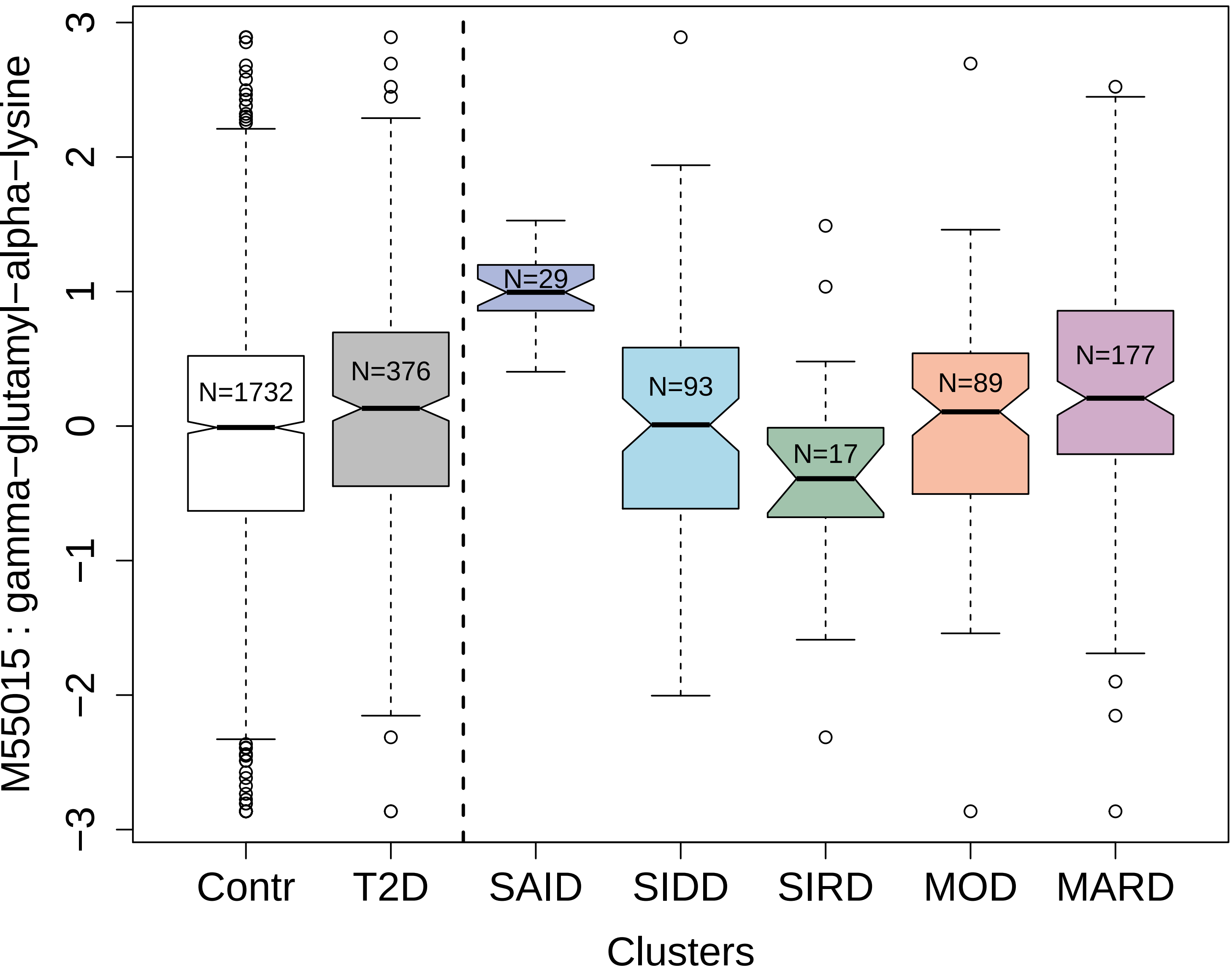

M55015 : gamma−glutamyl−alpha−lysine
3
2
1
0
−1
−2
−3
N=1732
N=376
N=29
N=93
N=17
N=89
N=177
Contr
T2D
SAID
SIDD
SIRD
MOD
MARD
Clusters

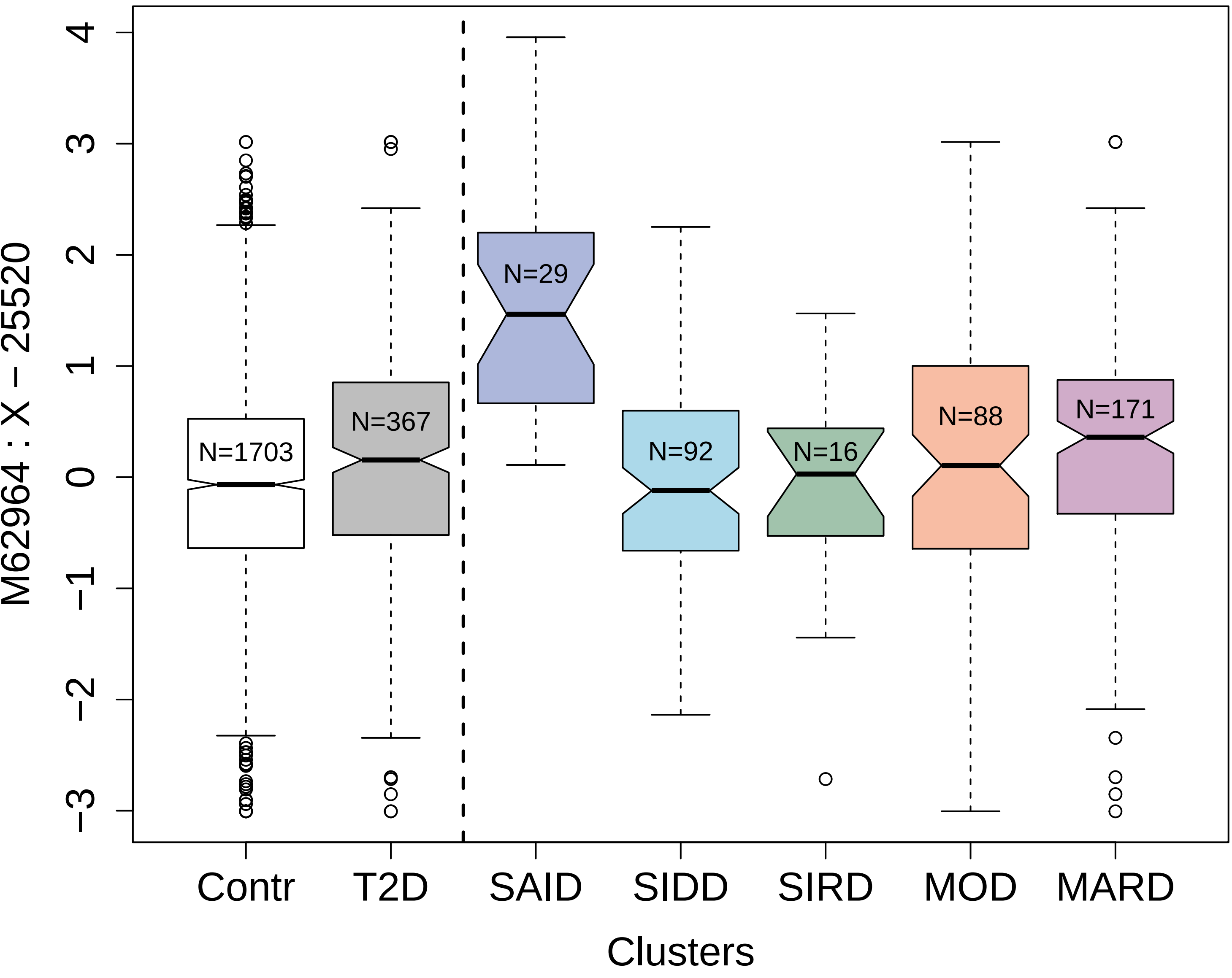

M62964 : X − 25520
4
3
2
1
0
−1
−2
−3
N=1703
N=367
N=29
N=92
N=16
N=88
N=171
Contr
T2D
SAID
SIDD
SIRD
MOD
MARD
Clusters

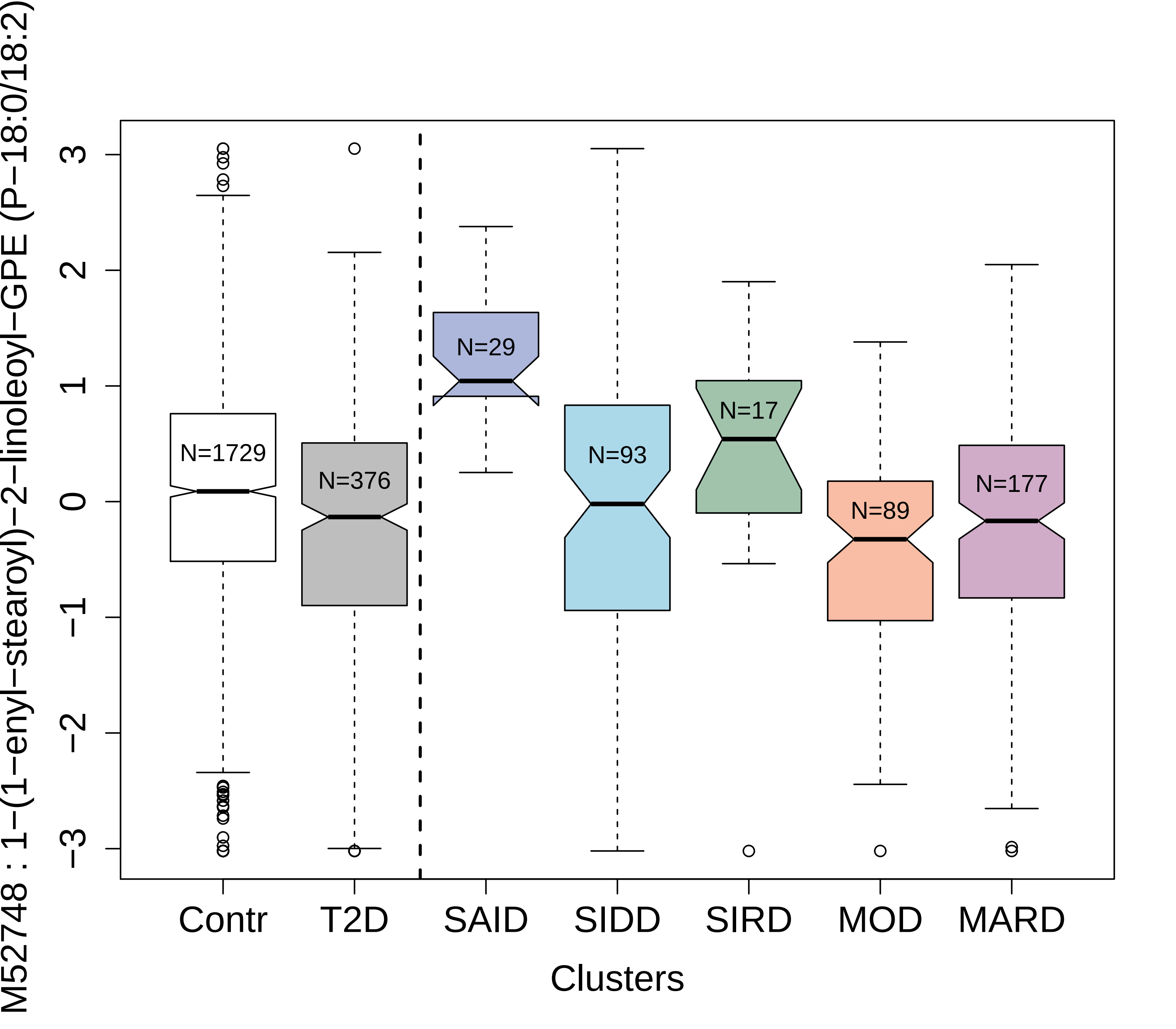
M52748 : 1−(1−enyl−stearoyl)−2−linoleoyl−GPE (P−18:0/18:2)
3
2
1
0
−1
−2
−3
N=1729
N=376
N=29
N=93
N=17
N=89
N=177
Contr
T2D
SAID
SIDD
SIRD
MOD
MARD
Clusters

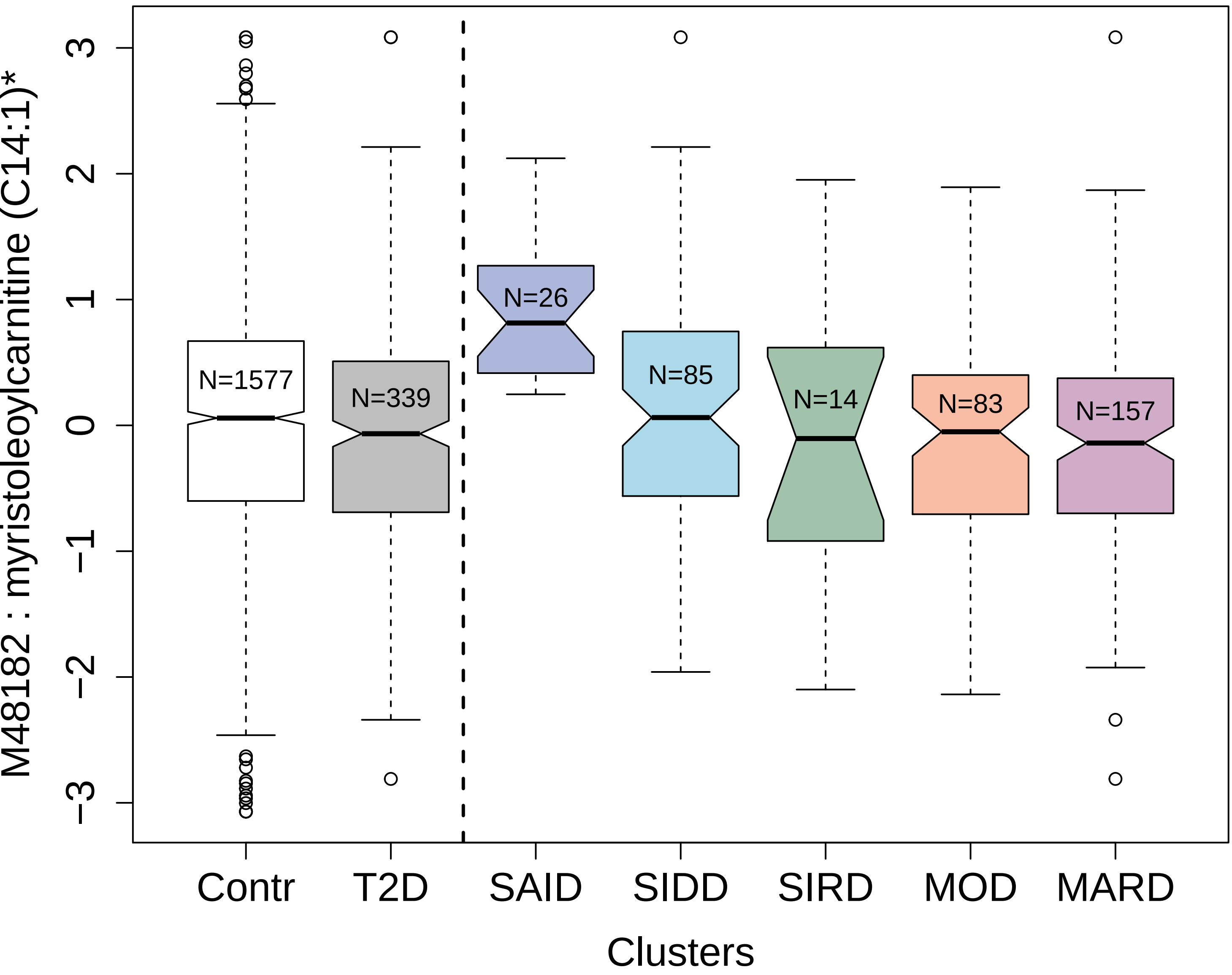

M48182 : myristoleoylcarnitine (C14:1)*
3
2
1
0
−1
−2
−3
N=1577
N=339
N=26
N=85
N=14
N=83
N=157
Contr
T2D
SAID
SIDD
SIRD
MOD
MARD
Clusters

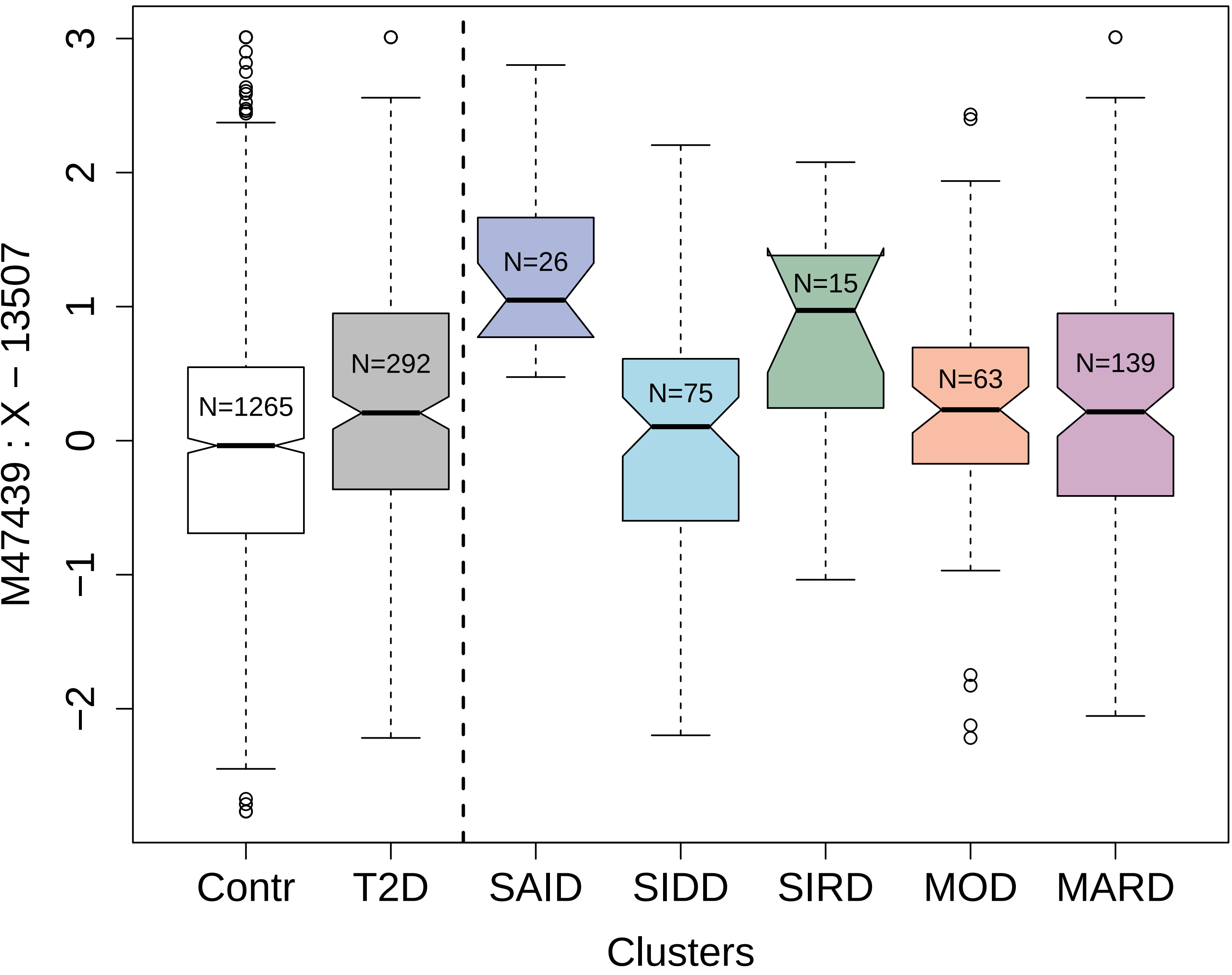
M47439 : X − 13507
3
2
1
0
−1
−2
N=1265
N=292
N=26
N=75
N=15
N=63
N=139
Contr
T2D
SAID
SIDD
SIRD
MOD
MARD
Clusters

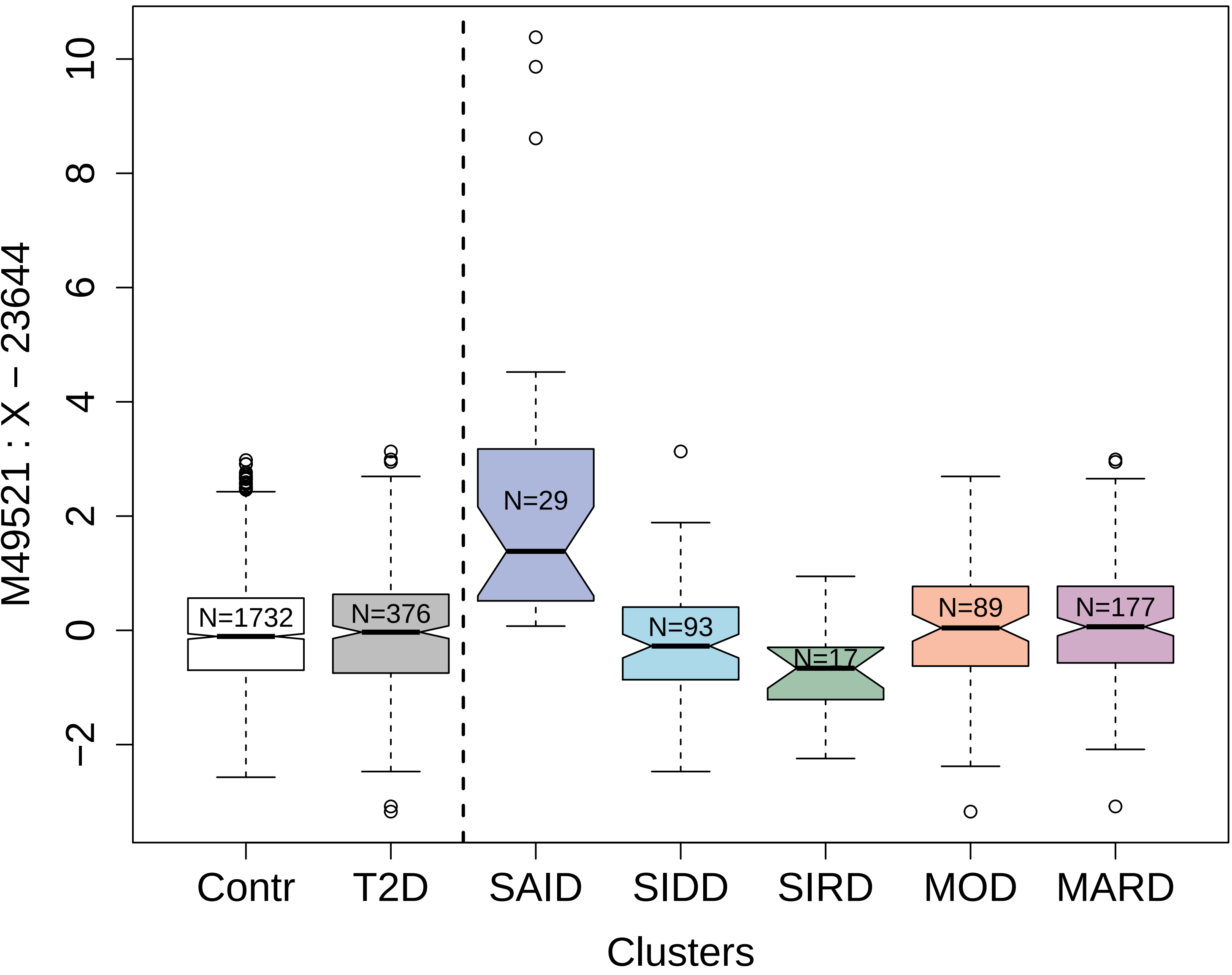

M49521 : X − 23644
10
8
6
4
2
0
−2
N=1732
N=376
N=29
N=93
N=17
N=89
N=177
Contr
T2D
SAID
SIDD
SIRD
MOD
MARD
Clusters

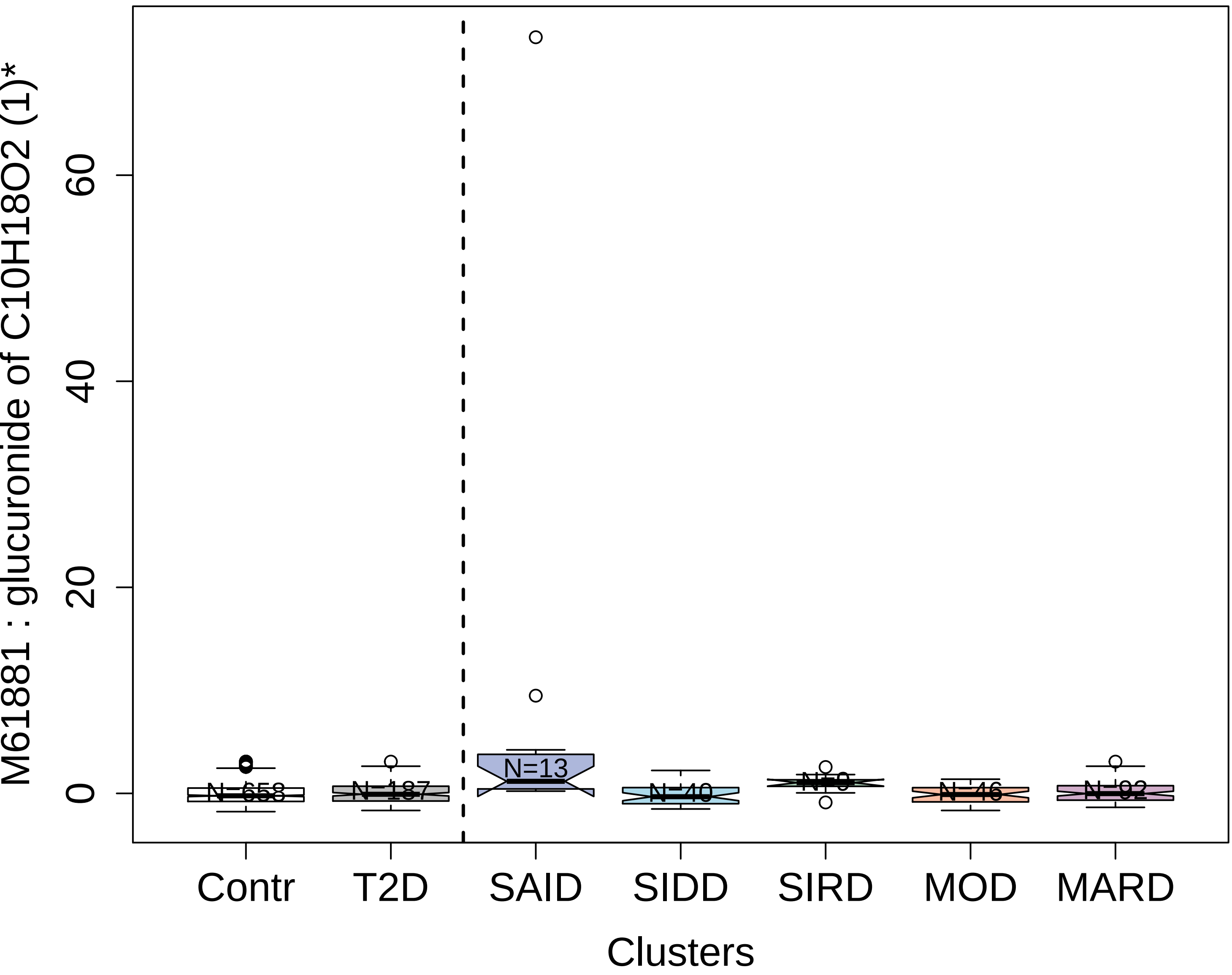
M61881 : glucuronide of C10H18O2 (1)*
0
20
40
60
N=658
N=187
N=13
N=40
N=9
N=46
N=92
Contr
T2D
SAID
SIDD
SIRD
MOD
MARD
Clusters

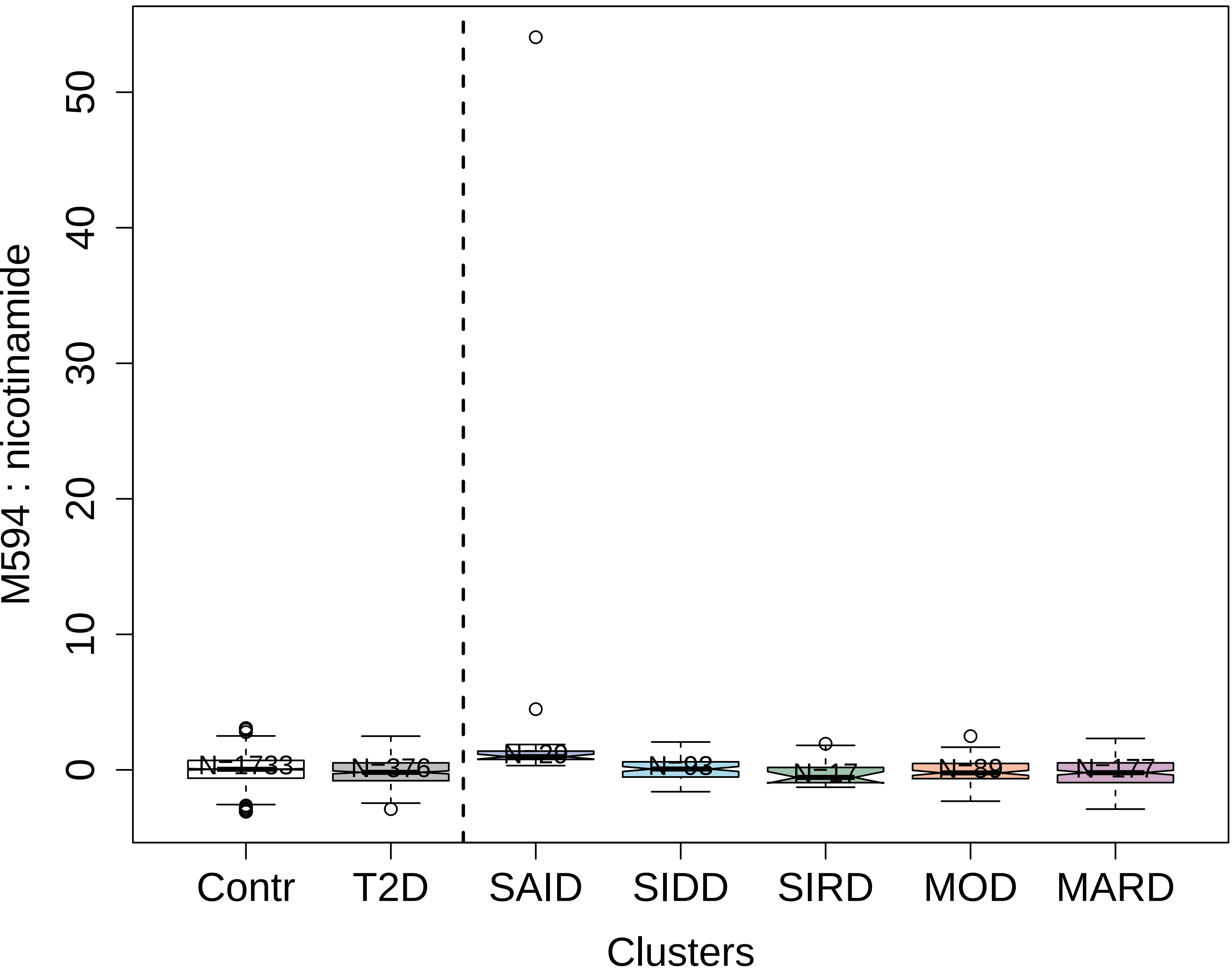

M594 : nicotinamide
0
10
20
30
40
50
N=1733
N=376
N=20
N=93
N=17
N=89
N=177
Contr
T2D
SAID
SIDD
SIRD
MOD
MARD
Clusters

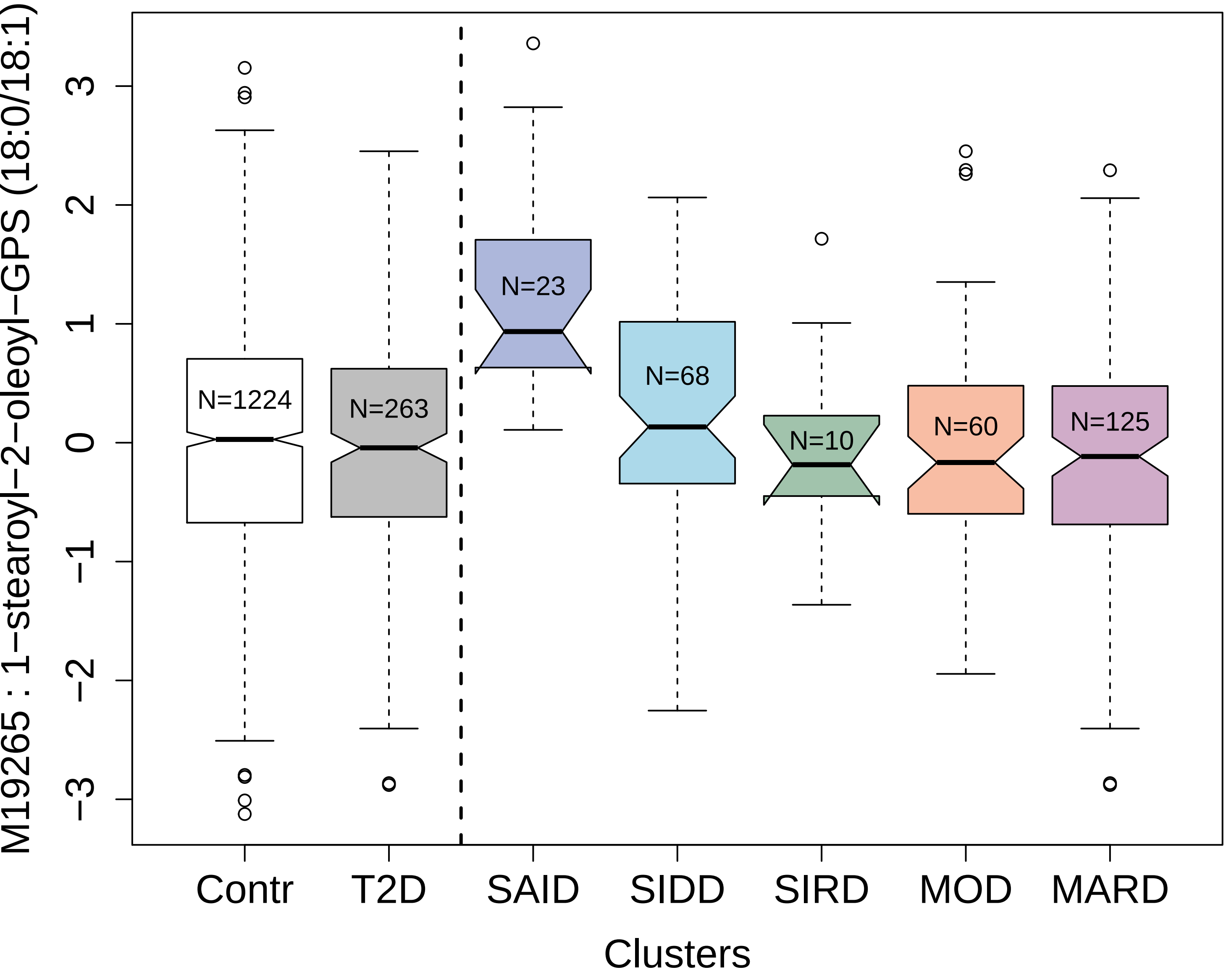

M19265 : 1−stearoyl−2−oleoyl−GPS (18:0/18:1)
3
2
1
0
−1
−2
−3
N=1224
N=263
N=23
N=68
N=10
N=60
N=125
Contr
T2D
SAID
SIDD
SIRD
MOD
MARD
Clusters

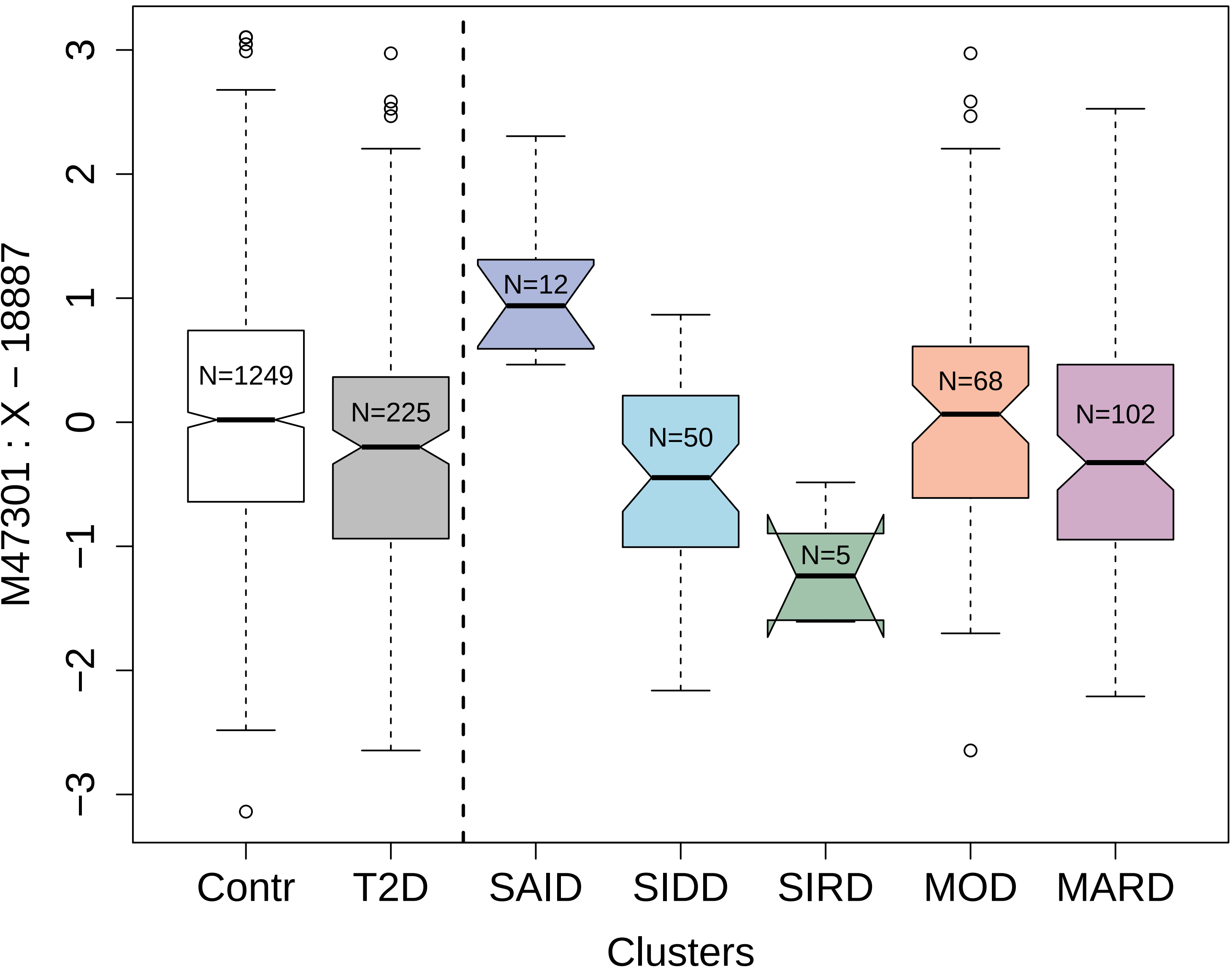

M47301 : X − 18887
3
2
1
0
−1
−2
−3
N=1249
N=225
N=12
N=50
N=5
N=68
N=102
Contr
T2D
SAID
SIDD
SIRD
MOD
MARD
Clusters

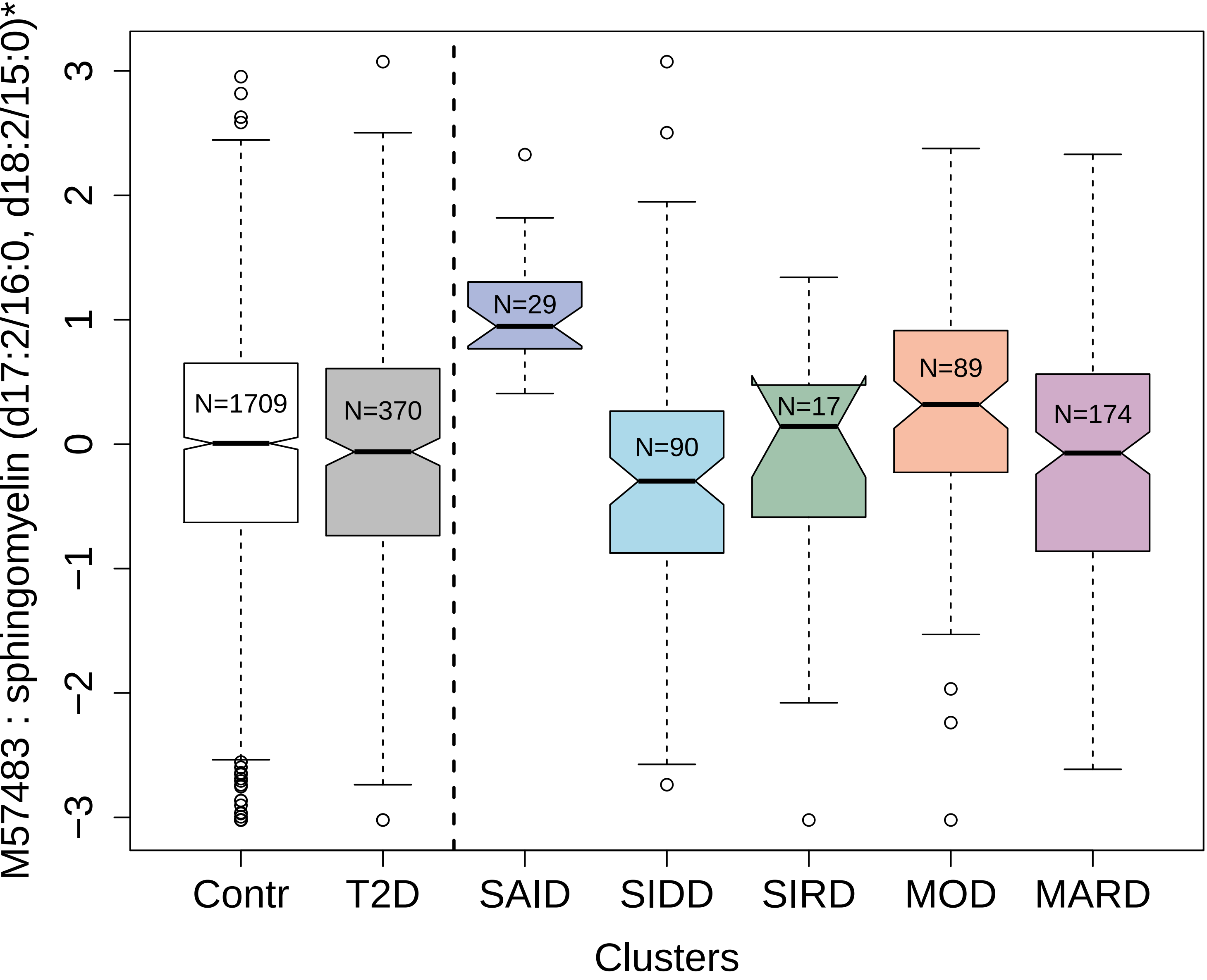

M57483 : sphingomyelin (d17:2/16:0, d18:2/15:0)*
3
2
1
0
−1
−2
−3
N=1709
N=370
N=29
N=90
N=17
N=89
N=174
Contr
T2D
SAID
SIDD
SIRD
MOD
MARD
Clusters

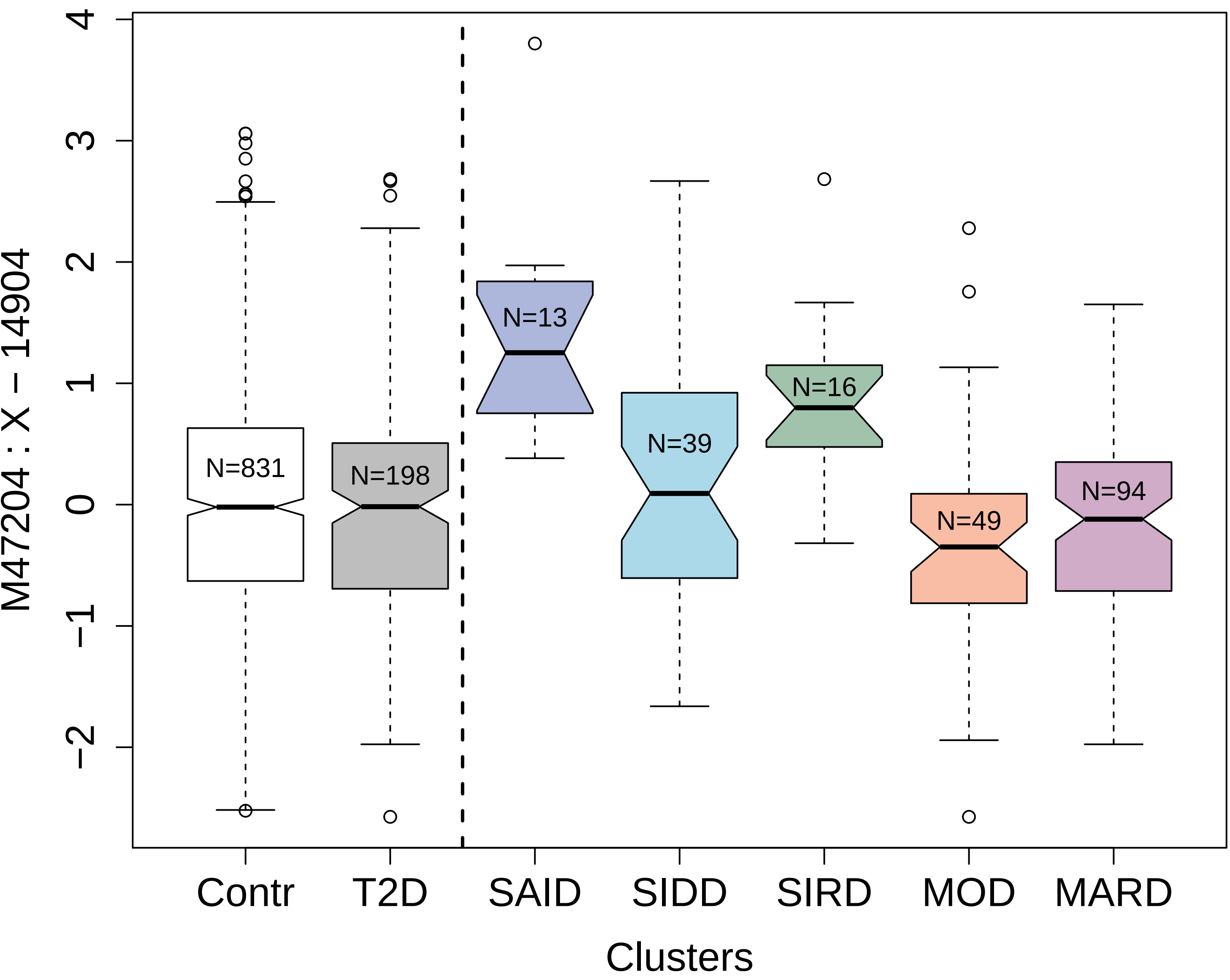

M47204 : X − 14904
4
3
2
1
0
−1
−2
N=831
N=198
N=13
N=39
N=16
N=49
N=94
Contr
T2D
SAID
SIDD
SIRD
MOD
MARD
Clusters

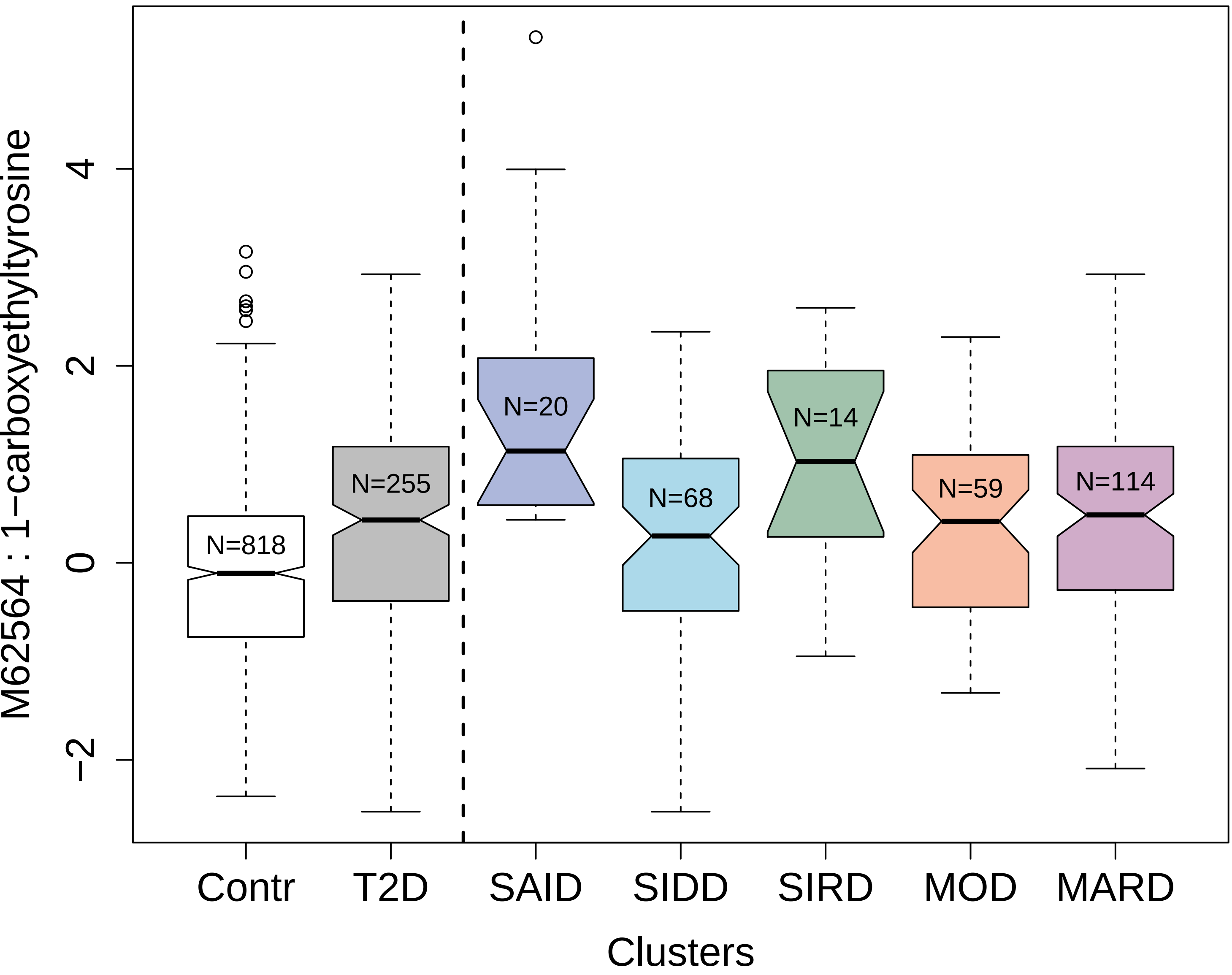
M62564 : 1−carboxyethyltyrosine
4
2
0
−2
N=818
N=255
N=20
N=68
N=14
N=59
N=114
Contr
T2D
SAID
SIDD
SIRD
MOD
MARD
Clusters

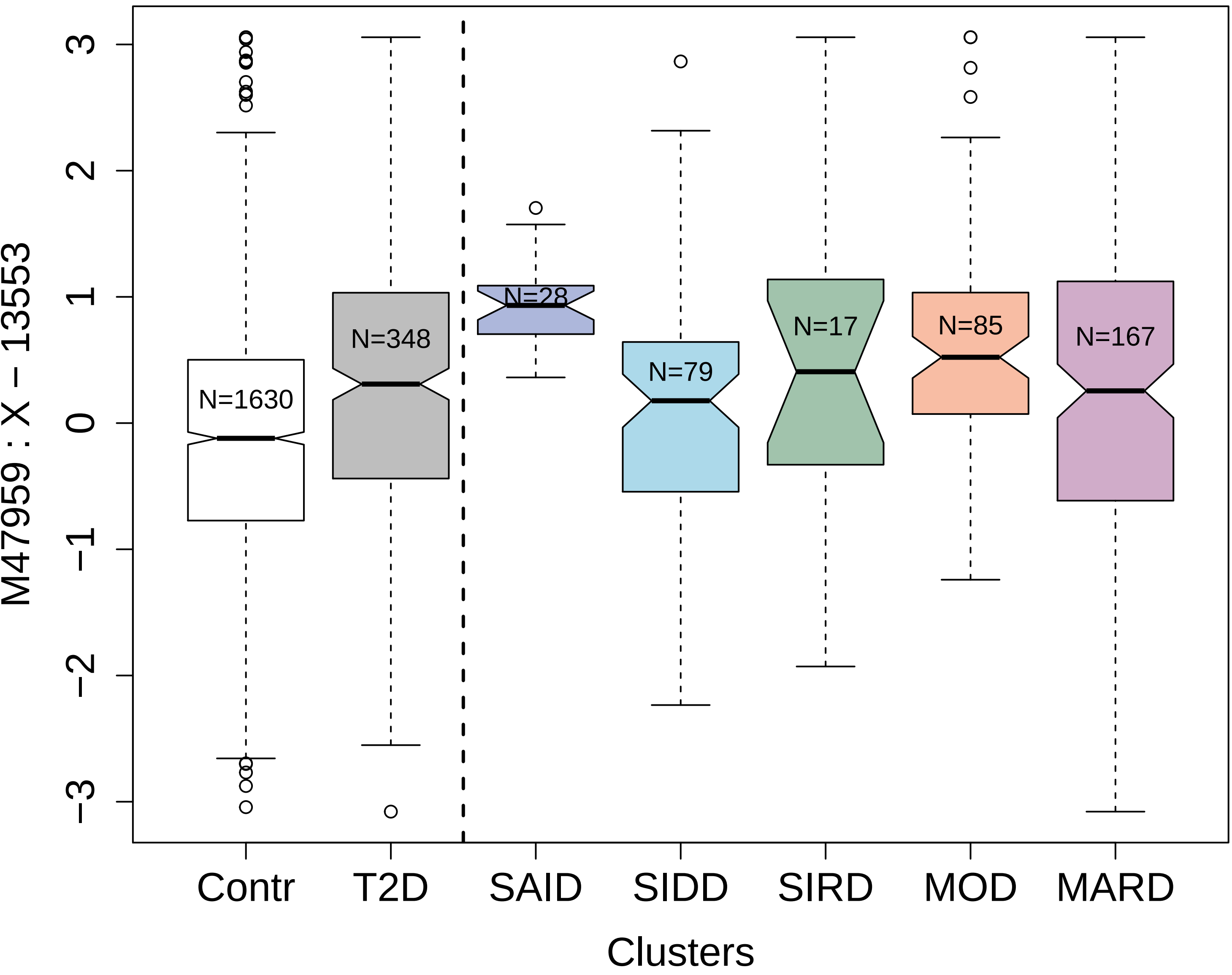
M47959 : X − 13553
3
2
1
0
−1
−2
−3
N=1630
N=348
N=28
N=79
N=17
N=85
N=167
Contr
T2D
SAID
SIDD
SIRD
MOD
MARD
Clusters

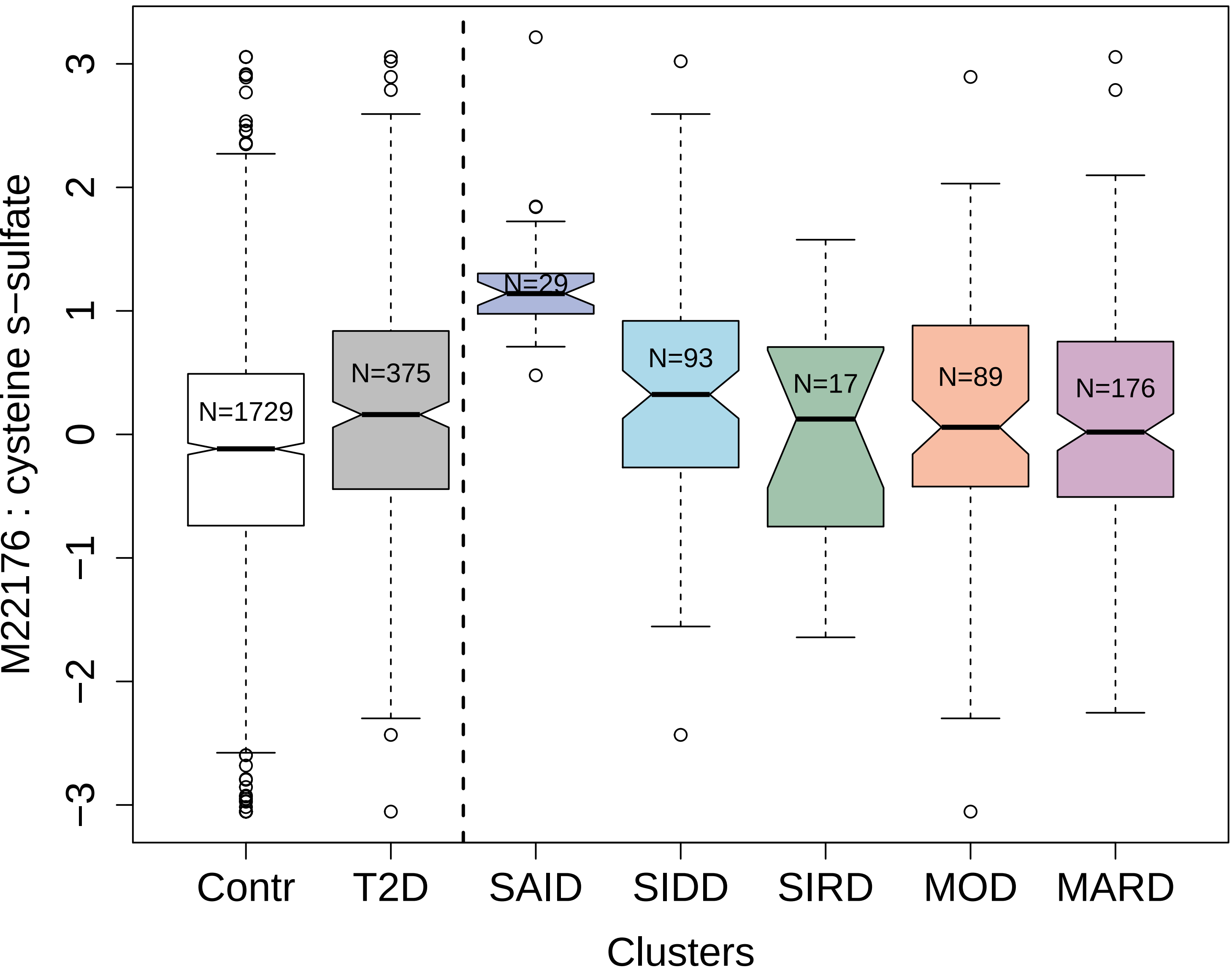
M22176 : cysteine s−sulfate
3
2
1
0
−1
−2
−3
N=1729
N=375
N=29
N=93
N=17
N=89
N=176
Contr
T2D
SAID
SIDD
SIRD
MOD
MARD
Clusters

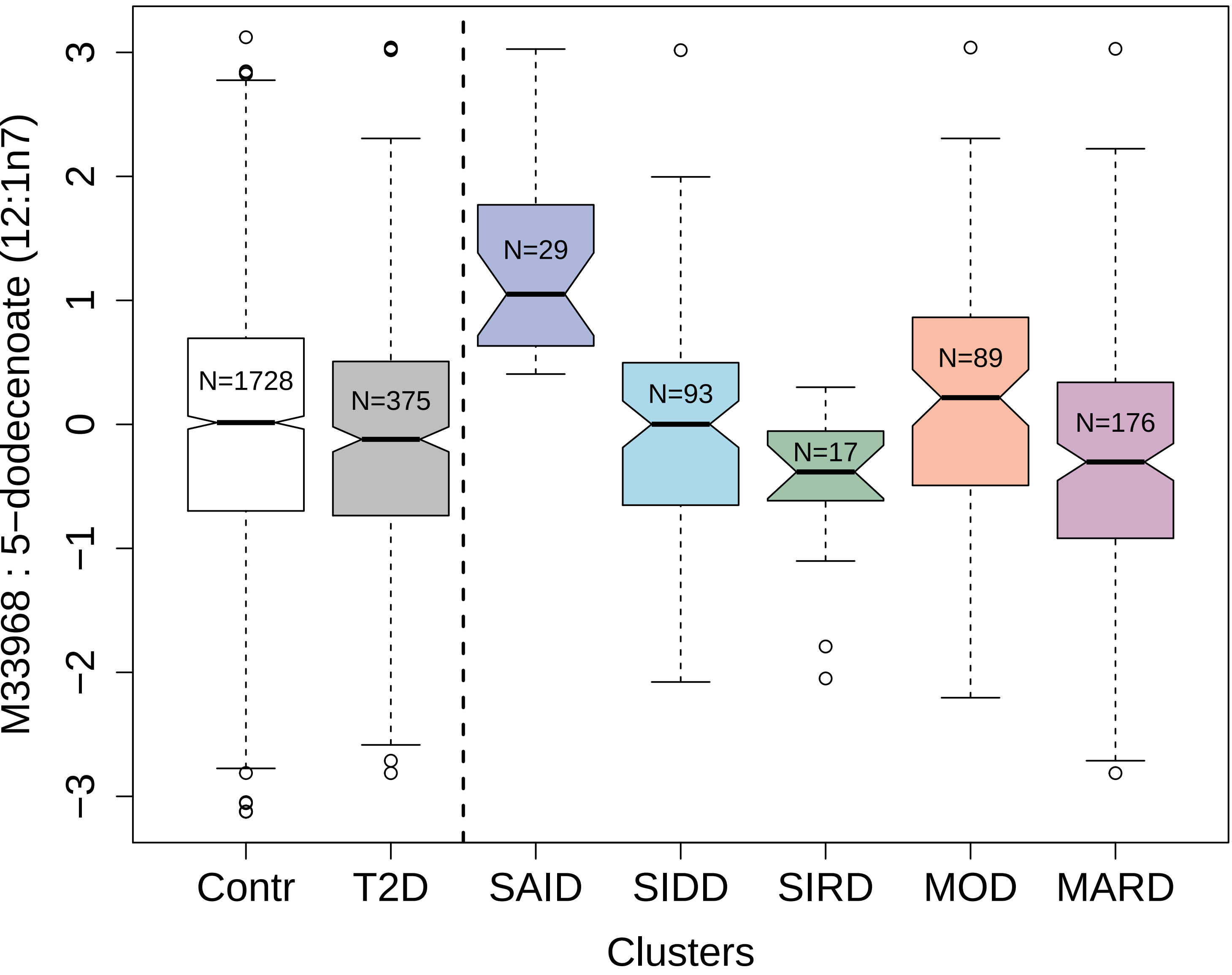
M33968 : 5−dodecenoate (12:1n7)
3
2
1
0
−1
−2
−3
N=1728
N=375
N=29
N=93
N=17
N=89
N=176
Contr
T2D
SAID
SIDD
SIRD
MOD
MARD
Clusters

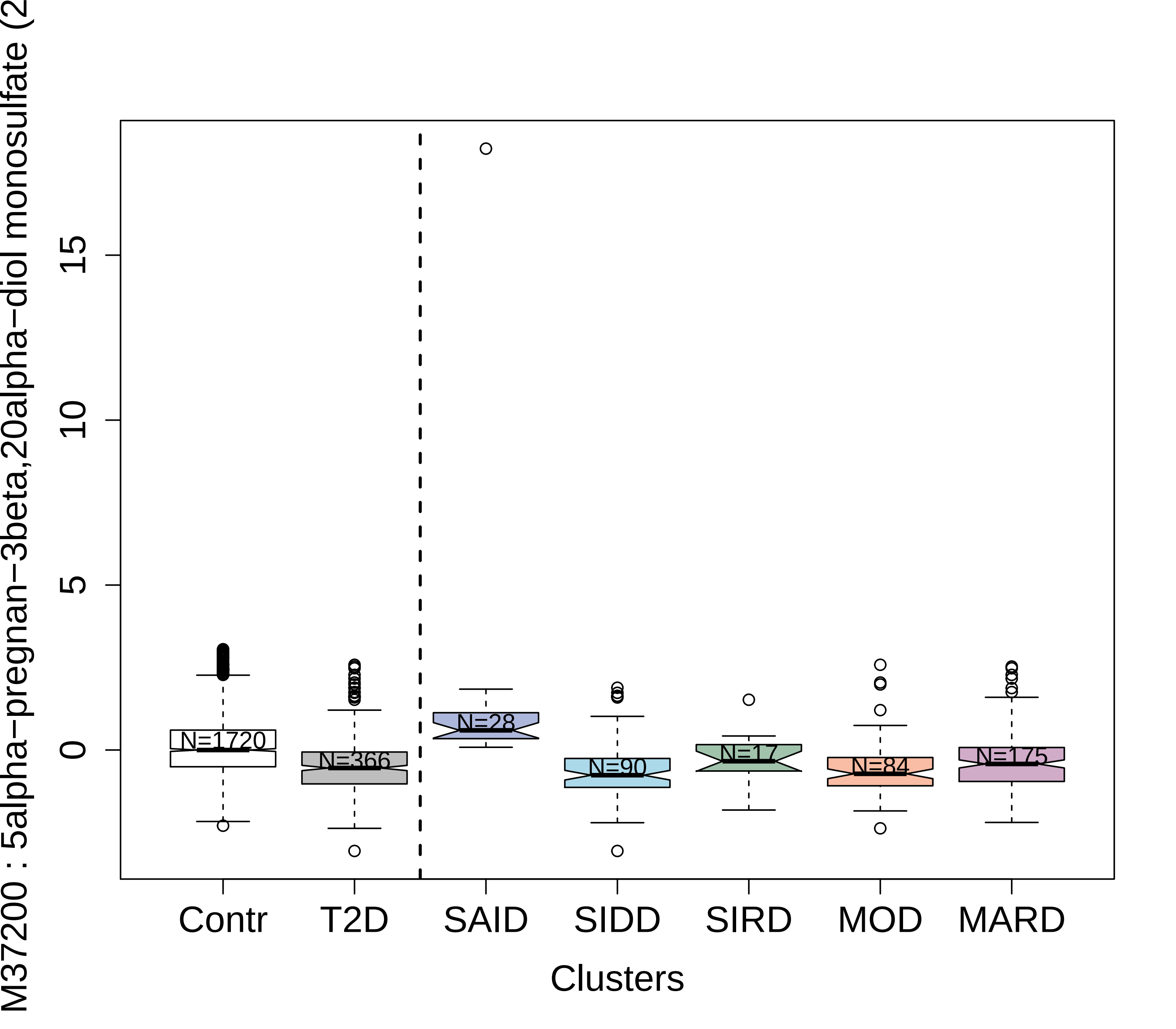

M37200 : 5alpha−pregnan−3beta,20alpha−diol monosulfate (2
0
5
10
15
N=1720
N=366
N=28
N=90
N=17
N=84
N=175
Contr
T2D
SAID
SIDD
SIRD
MOD
MARD
Clusters

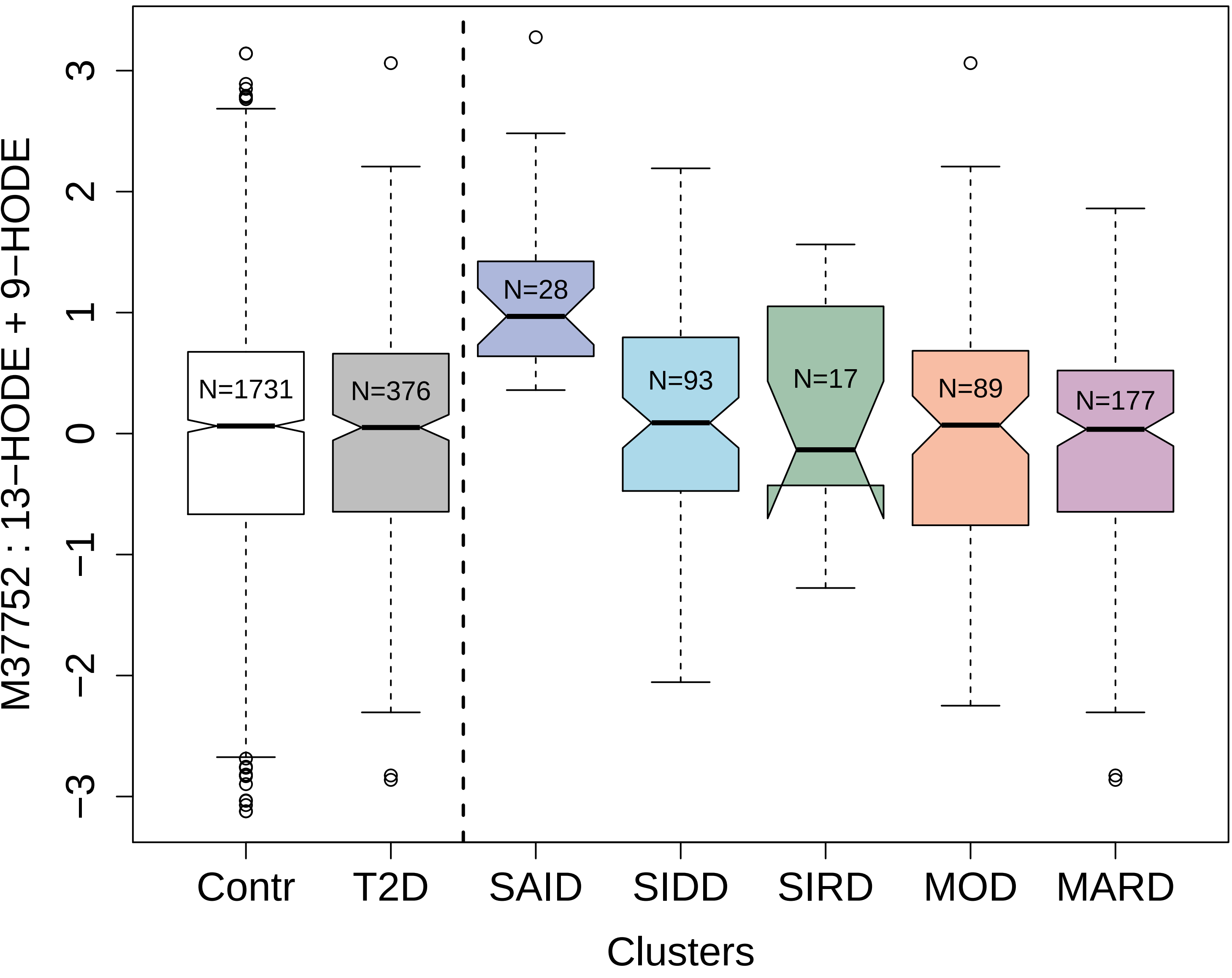

M37752 : 13−HODE + 9−HODE
−3
−2
−1
0
1
2
3
N=1731
N=376
N=28
N=93
N=17
N=89
N=177
Contr
T2D
SAID
SIDD
SIRD
MOD
MARD
Clusters

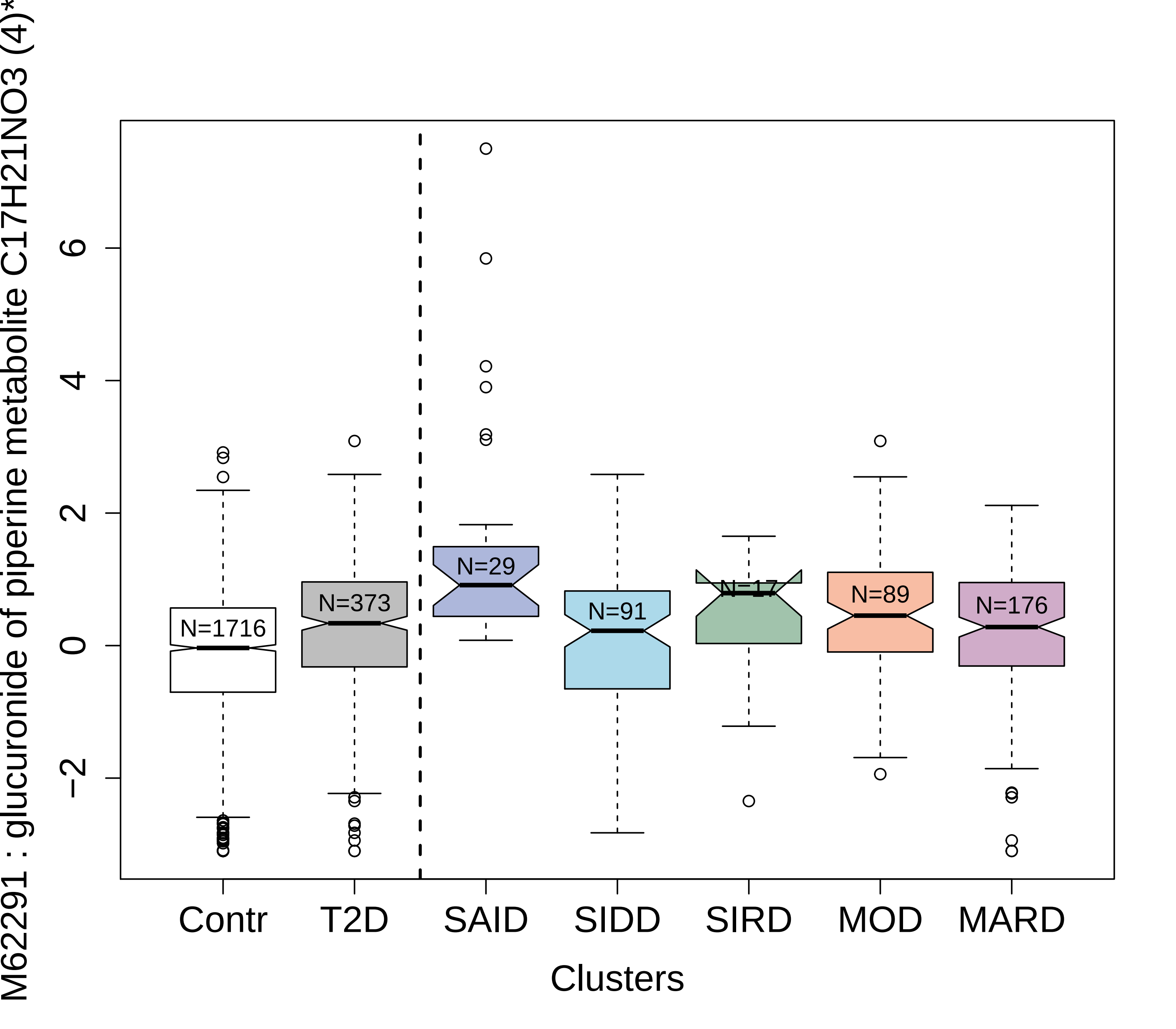
M62291 : glucuronide of piperine metabolite C17H21NO3 (4)*
6
4
2
0
−2
N=1716
N=373
N=29
N=91
N=17
N=89
N=176
Contr
T2D
SAID
SIDD
SIRD
MOD
MARD
Clusters

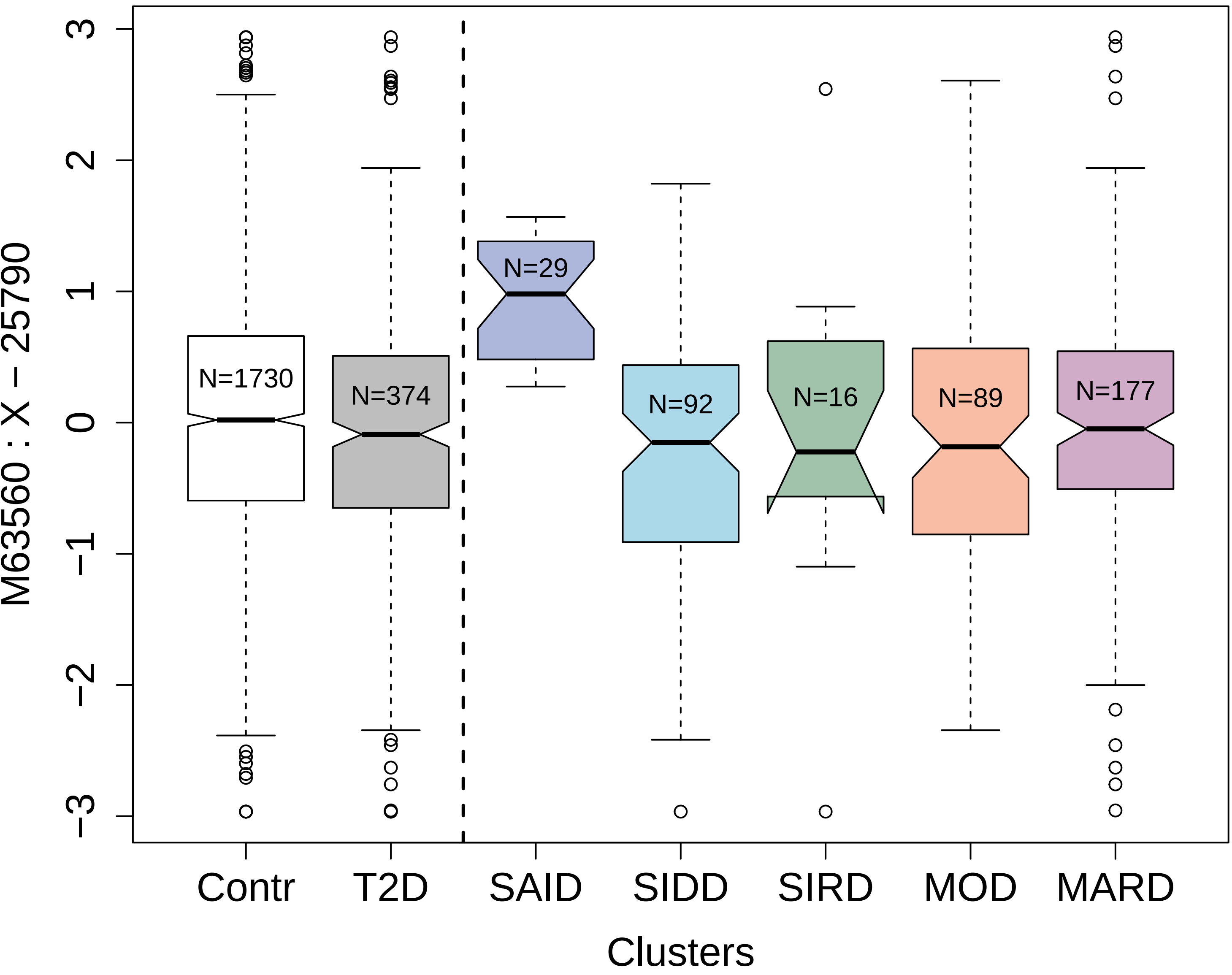

M63560 : X − 25790
3
2
1
0
−1
−2
−3
N=1730
N=374
N=29
N=92
N=16
N=89
N=177
Contr
T2D
SAID
SIDD
SIRD
MOD
MARD
Clusters

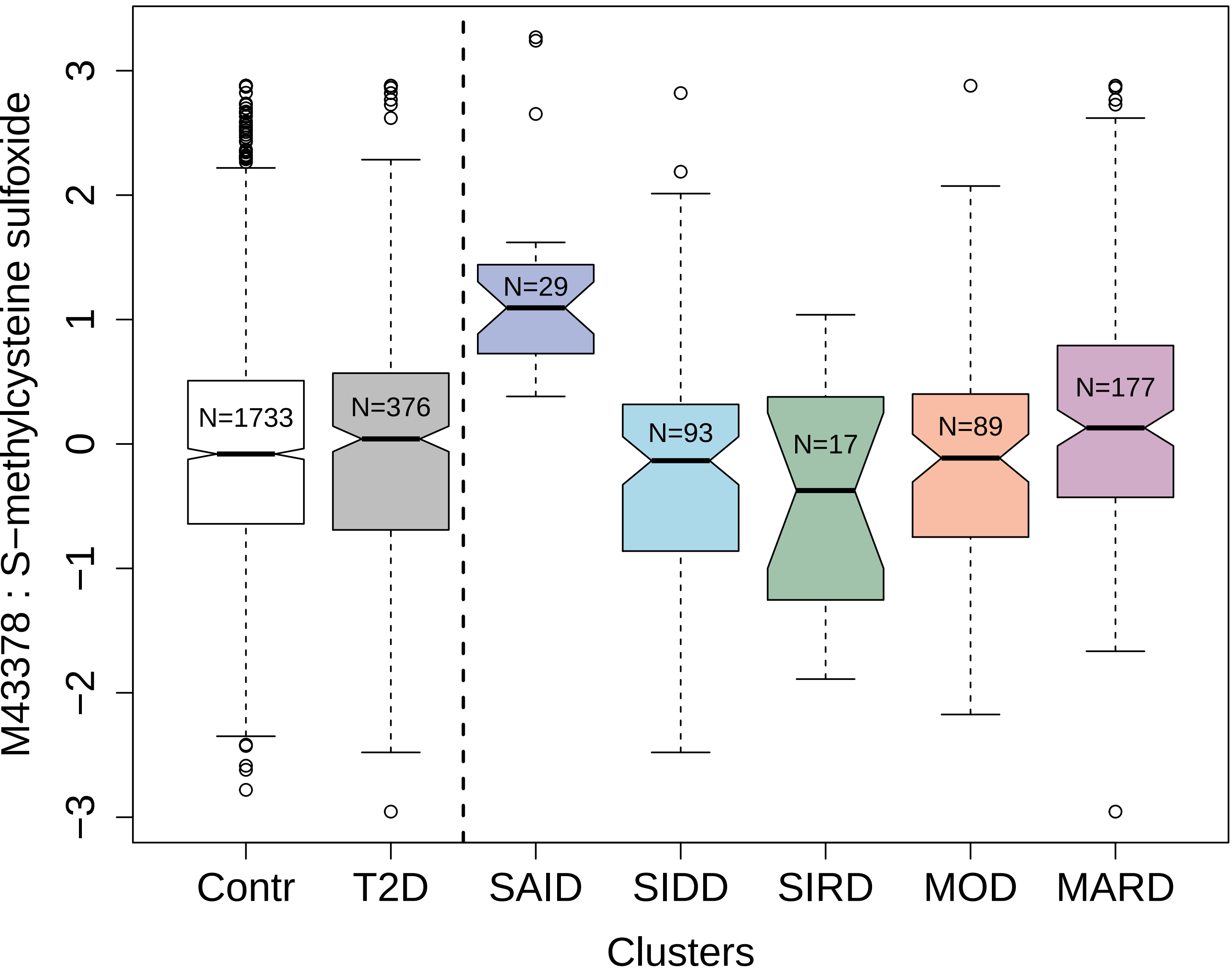

M43378 : S−methylcysteine sulfoxide
3
2
1
0
−1
−2
−3
N=1733
N=376
N=29
N=93
N=17
N=89
N=177
Contr
T2D
SAID
SIDD
SIRD
MOD
MARD
Clusters

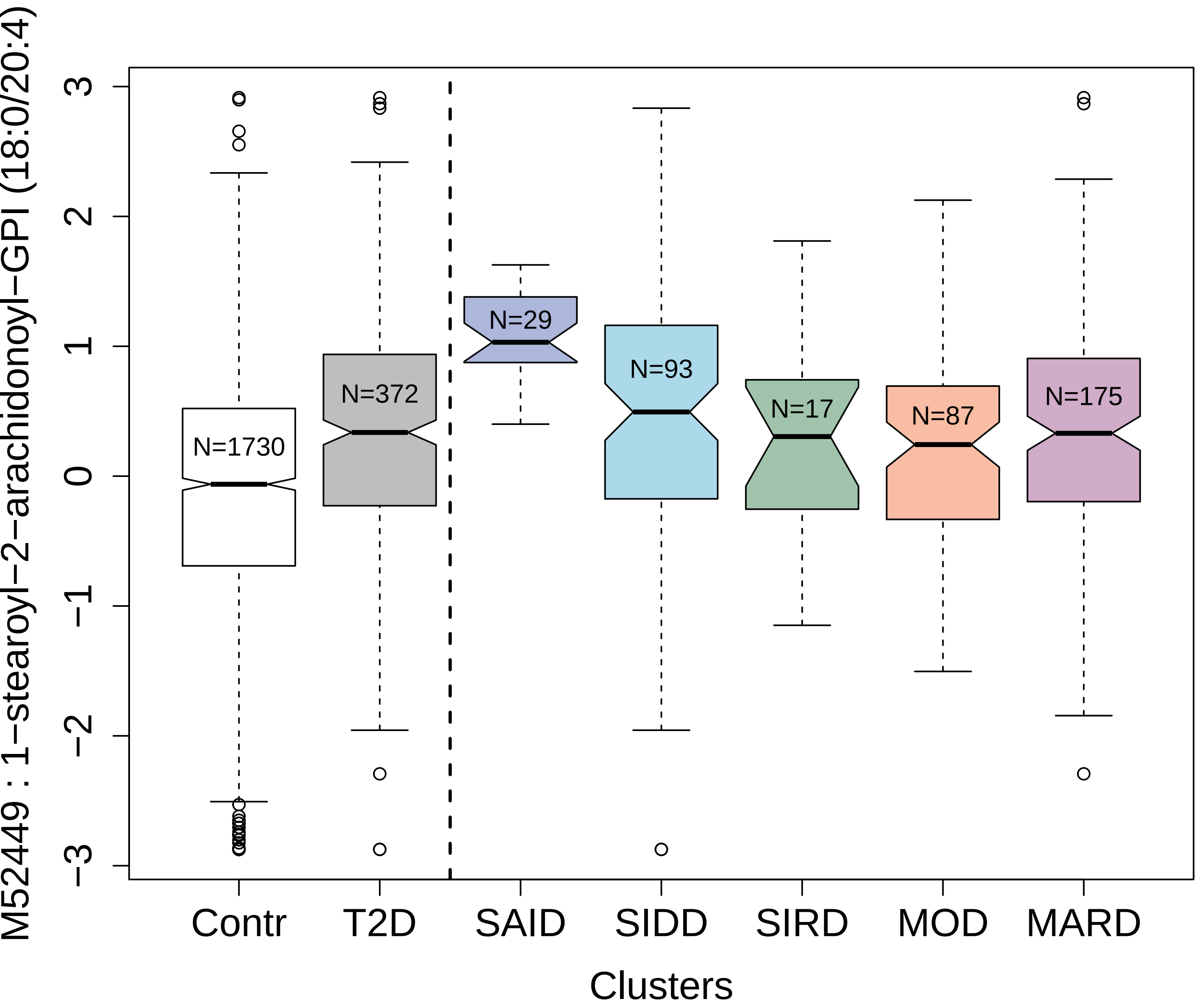

M52449 : 1−stearoyl−2−arachidonoyl−GPI (18:0/20:4)
3
2
1
0
−1
−2
−3
N=1730
N=372
N=29
N=93
N=17
N=87
N=175
Contr
T2D
SAID
SIDD
SIRD
MOD
MARD
Clusters

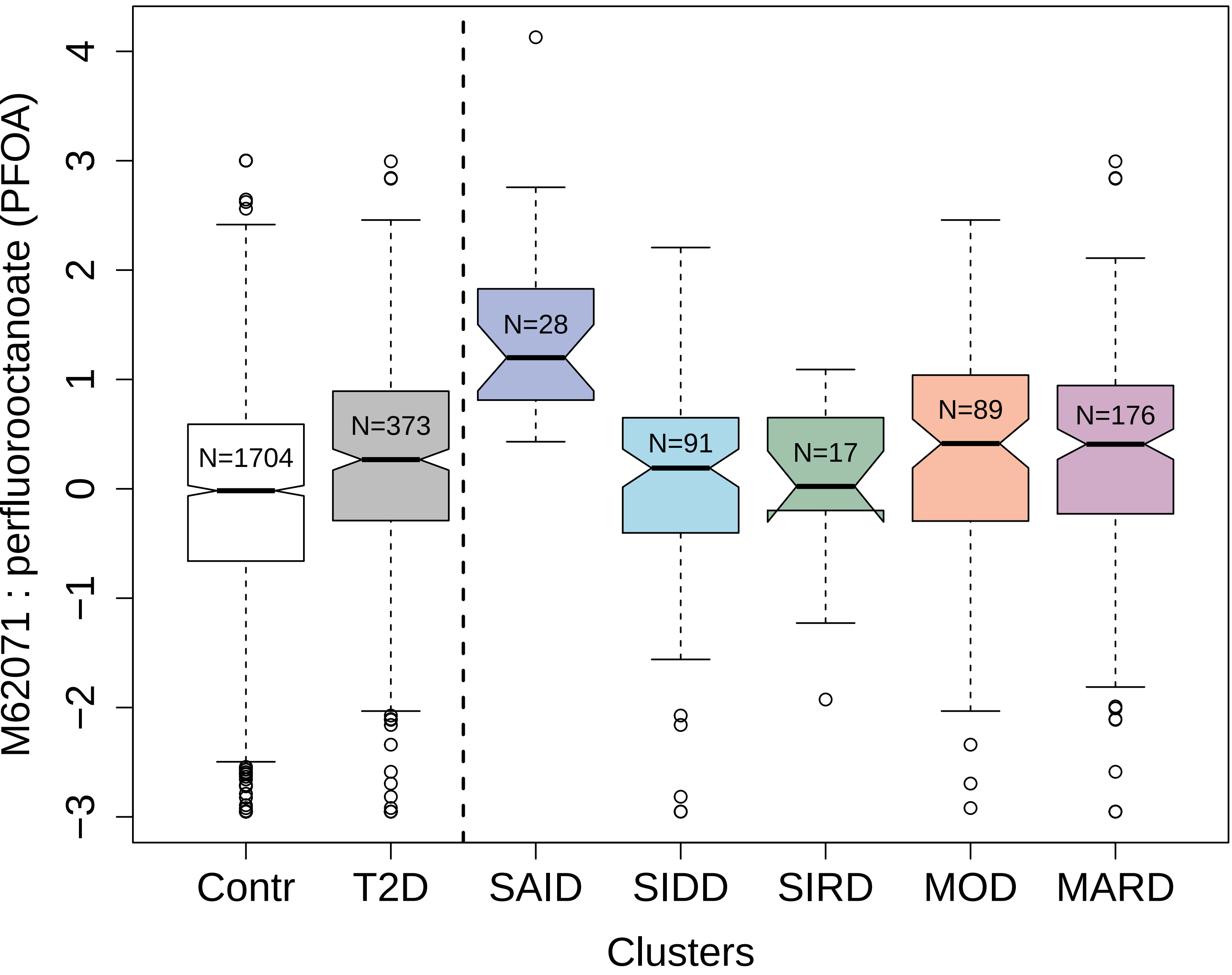

M62071 : perfluorooctanoate (PFOA)
4
3
2
1
0
−1
−2
−3
N=1704
N=373
N=28
N=91
N=17
N=89
N=176
Contr
T2D
SAID
SIDD
SIRD
MOD
MARD
Clusters

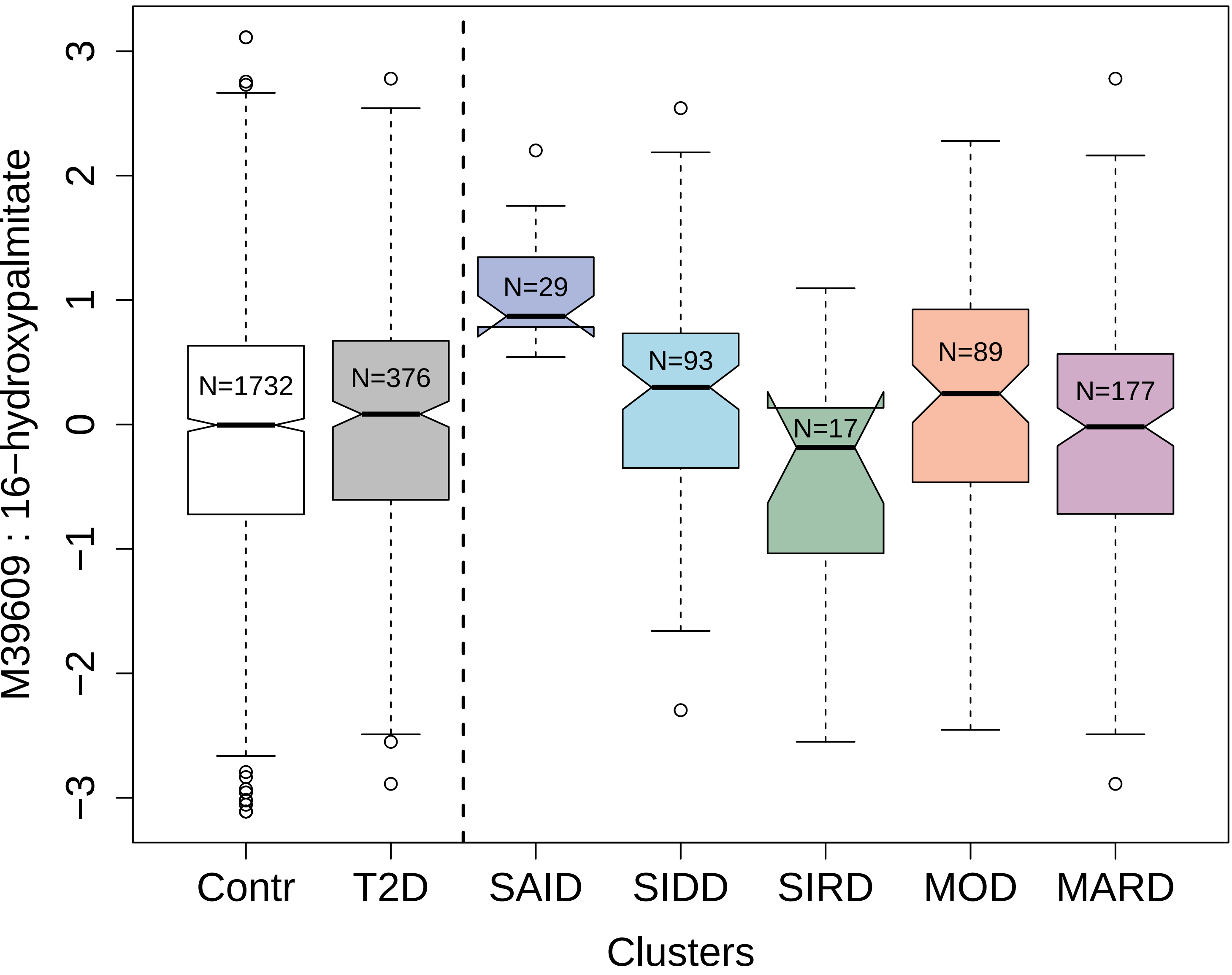

M39609 : 16−hydroxypalmitate
3
2
1
0
−1
−2
−3
N=1732
N=376
N=29
N=93
N=17
N=89
N=177
Contr
T2D
SAID
SIDD
SIRD
MOD
MARD
Clusters

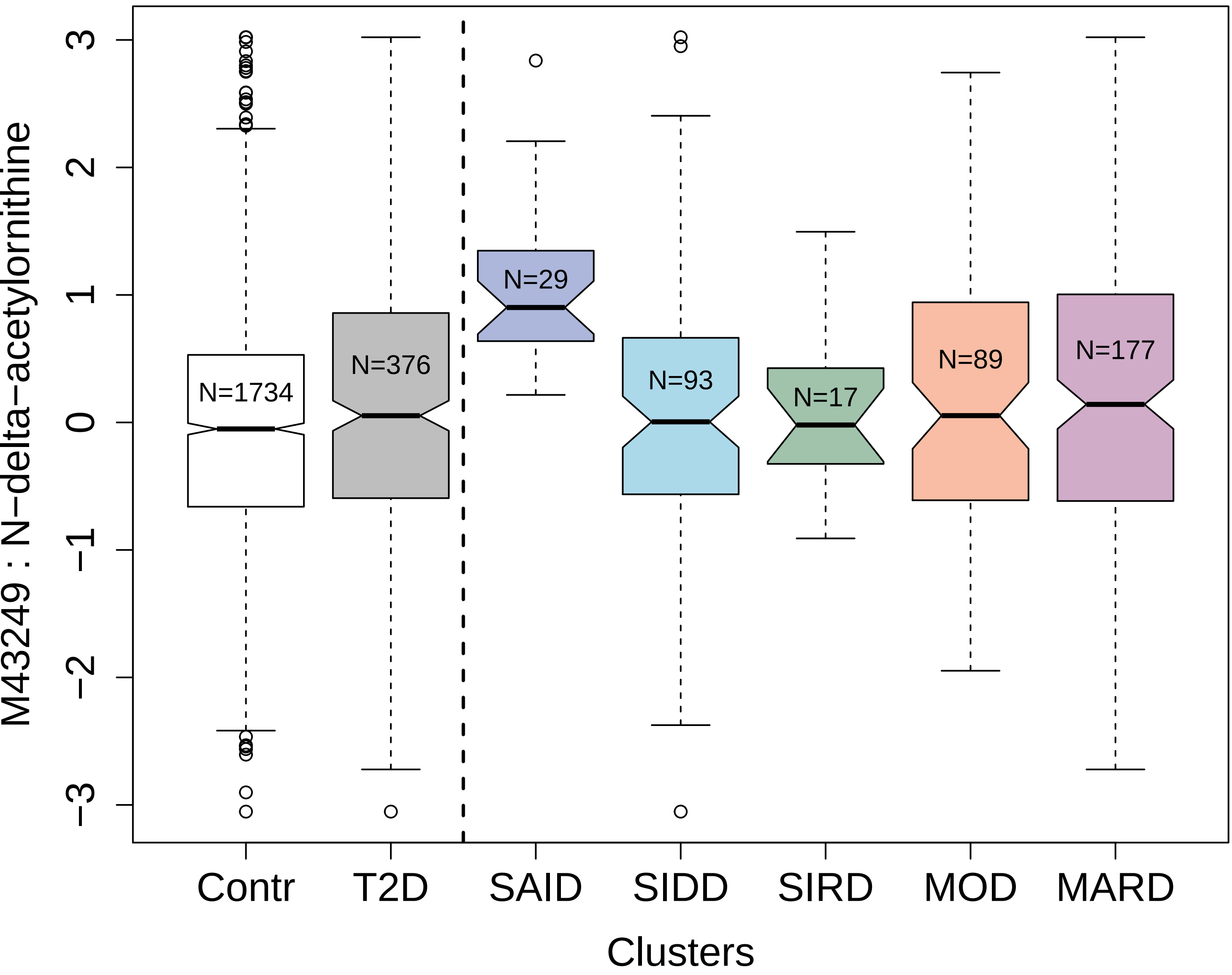

M43249 : N−delta−acetylornithine
3
2
1
0
−1
−2
−3
N=1734
N=376
N=29
N=93
N=17
N=89
N=177
Contr
T2D
SAID
SIDD
SIRD
MOD
MARD
Clusters

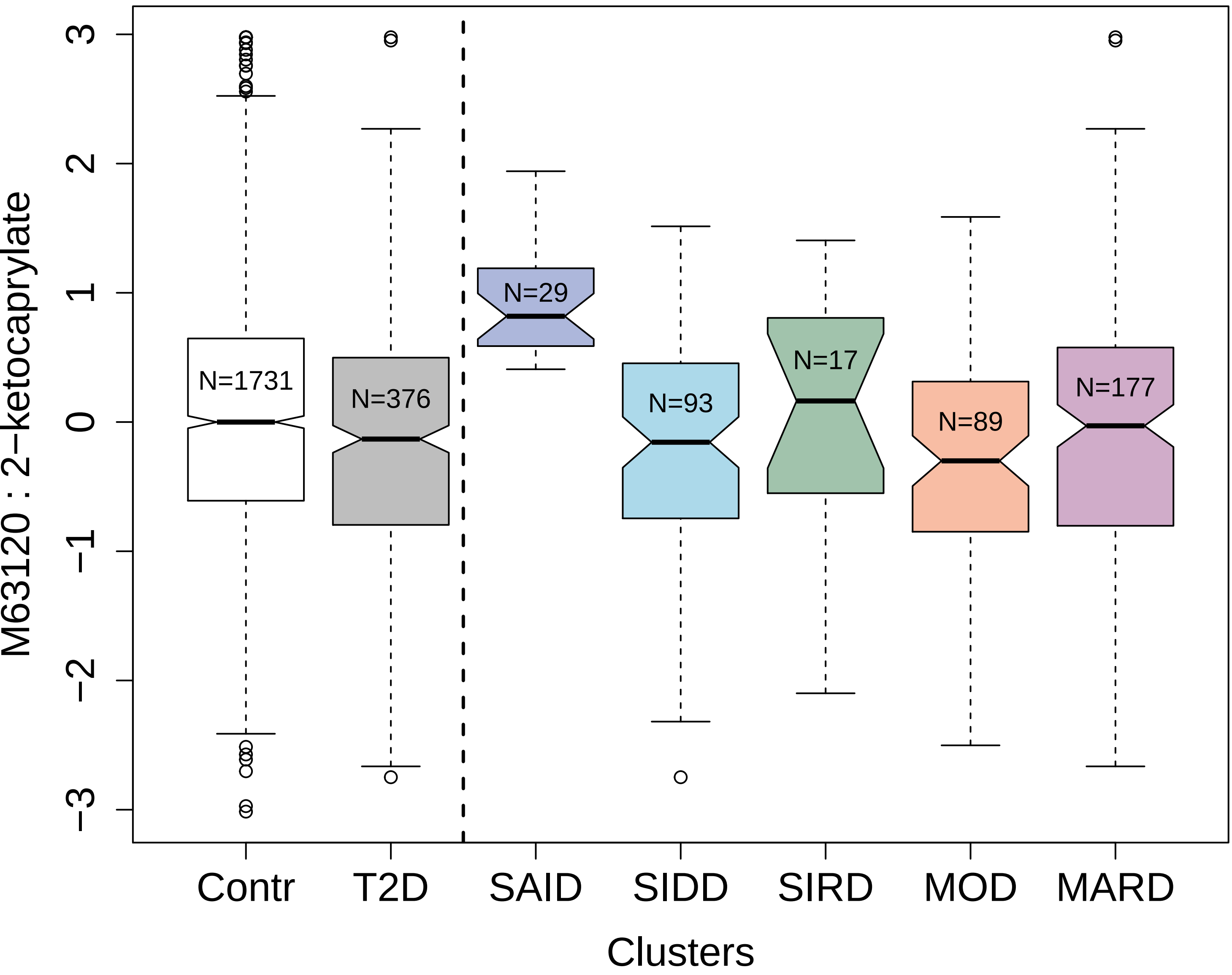

M63120 : 2−ketocaprylate
3
2
1
0
−1
−2
−3
N=1731
N=376
N=29
N=93
N=17
N=89
N=177
Contr
T2D
SAID
SIDD
SIRD
MOD
MARD
Clusters

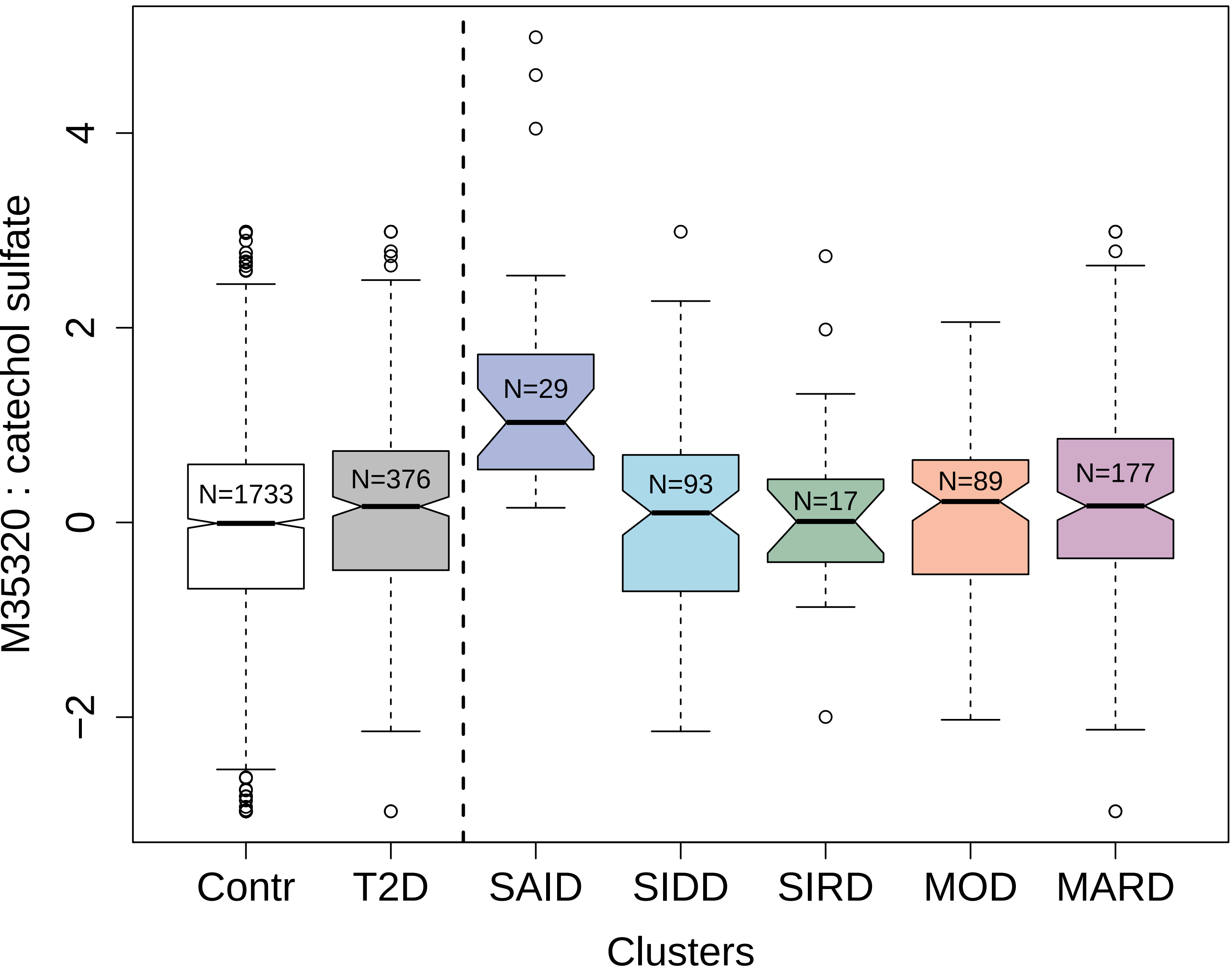

M35320 : catechol sulfate
4
2
0
−2
N=1733
N=376
N=29
N=93
N=17
N=89
N=177
Contr
T2D
SAID
SIDD
SIRD
MOD
MARD
Clusters

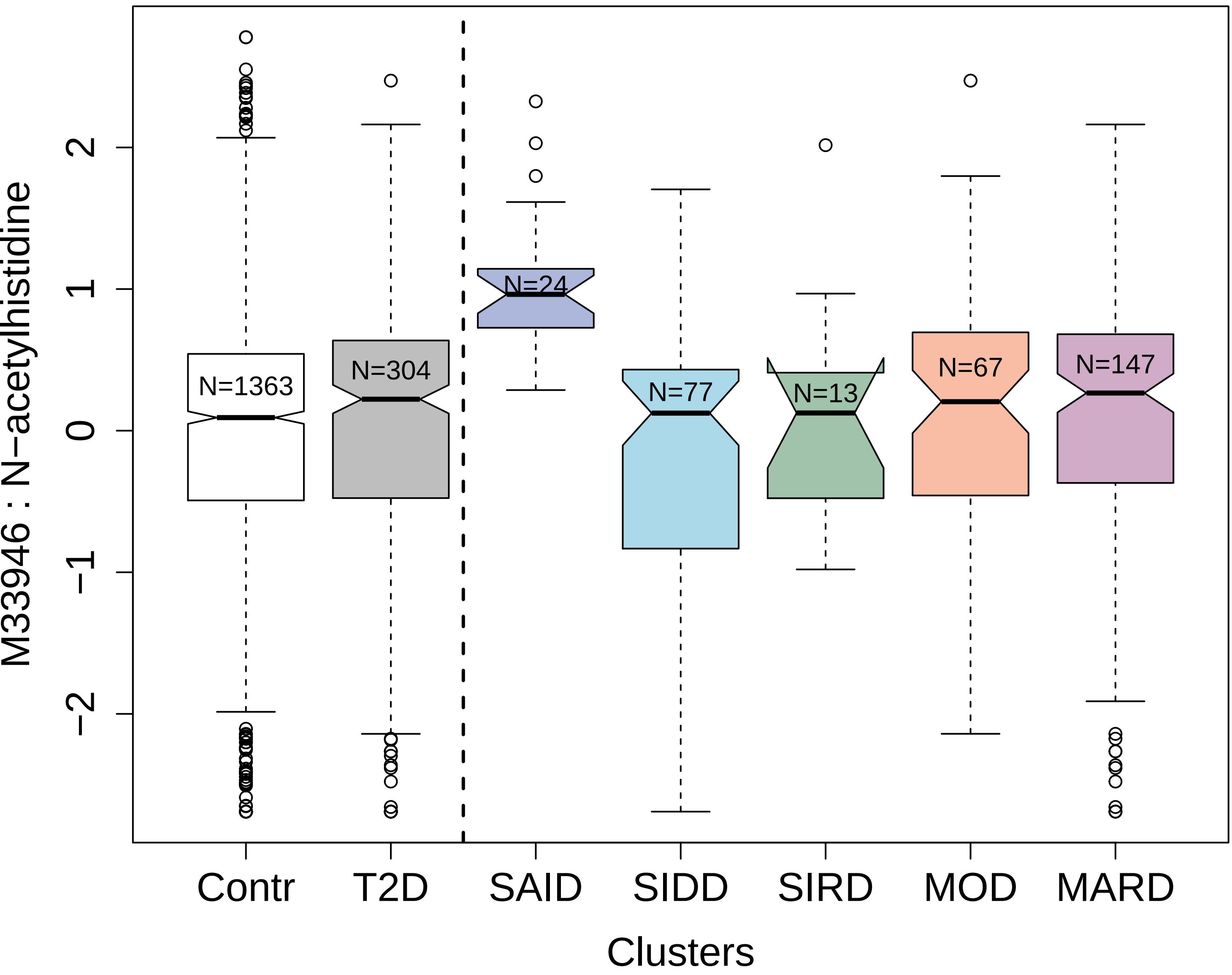
M33946 : N−acetylhistidine
N=1363
N=304
N=24
N=77
N=13
N=67
N=147
2
1
0
−1
−2
Contr
T2D
SAID
SIDD
SIRD
MOD
MARD
Clusters

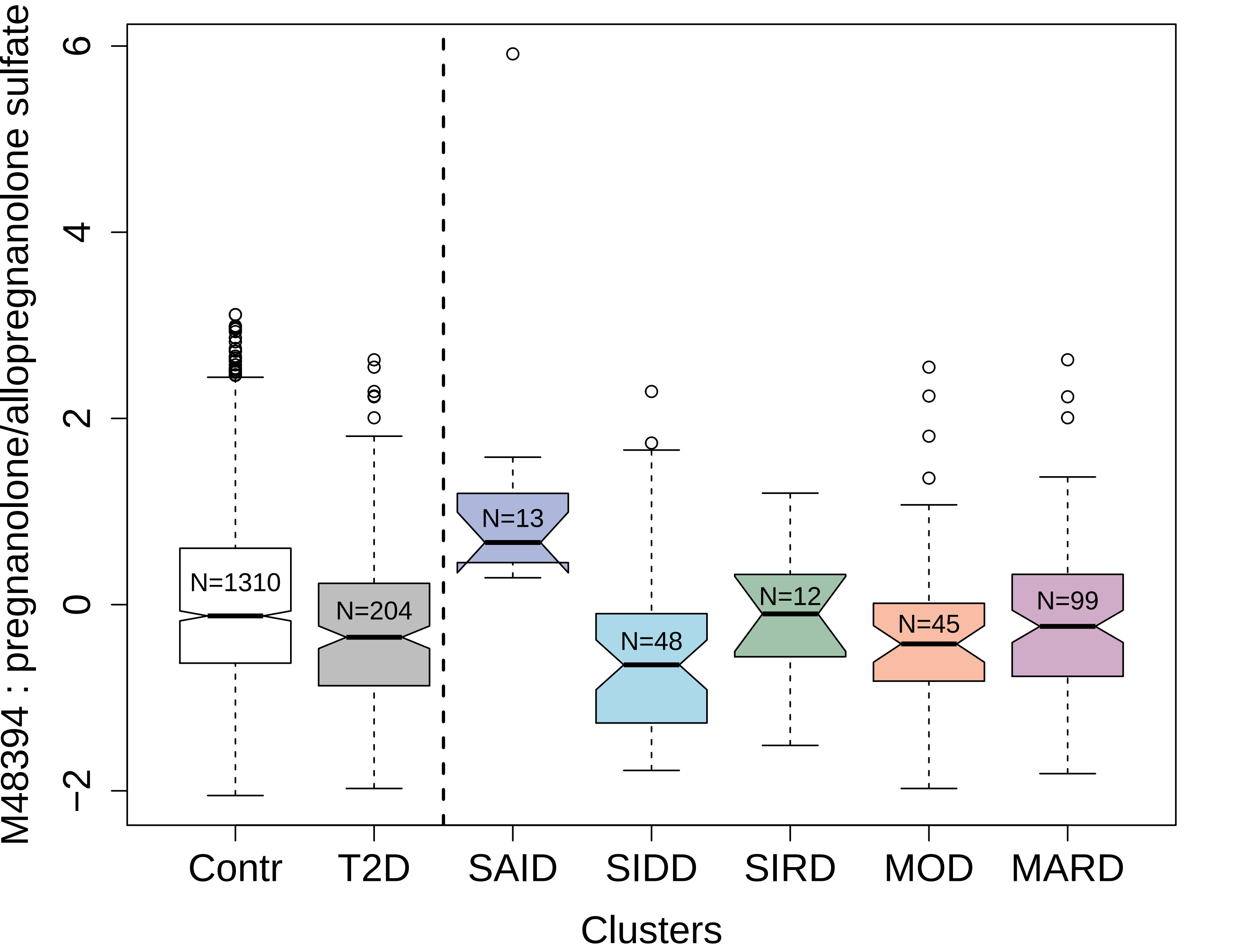
M48394 : pregnanolone/allopregnanolone sulfate
6
4
2
0
−2
N=1310
N=204
N=13
N=48
N=12
N=45
N=99
Contr
T2D
SAID
SIDD
SIRD
MOD
MARD
Clusters

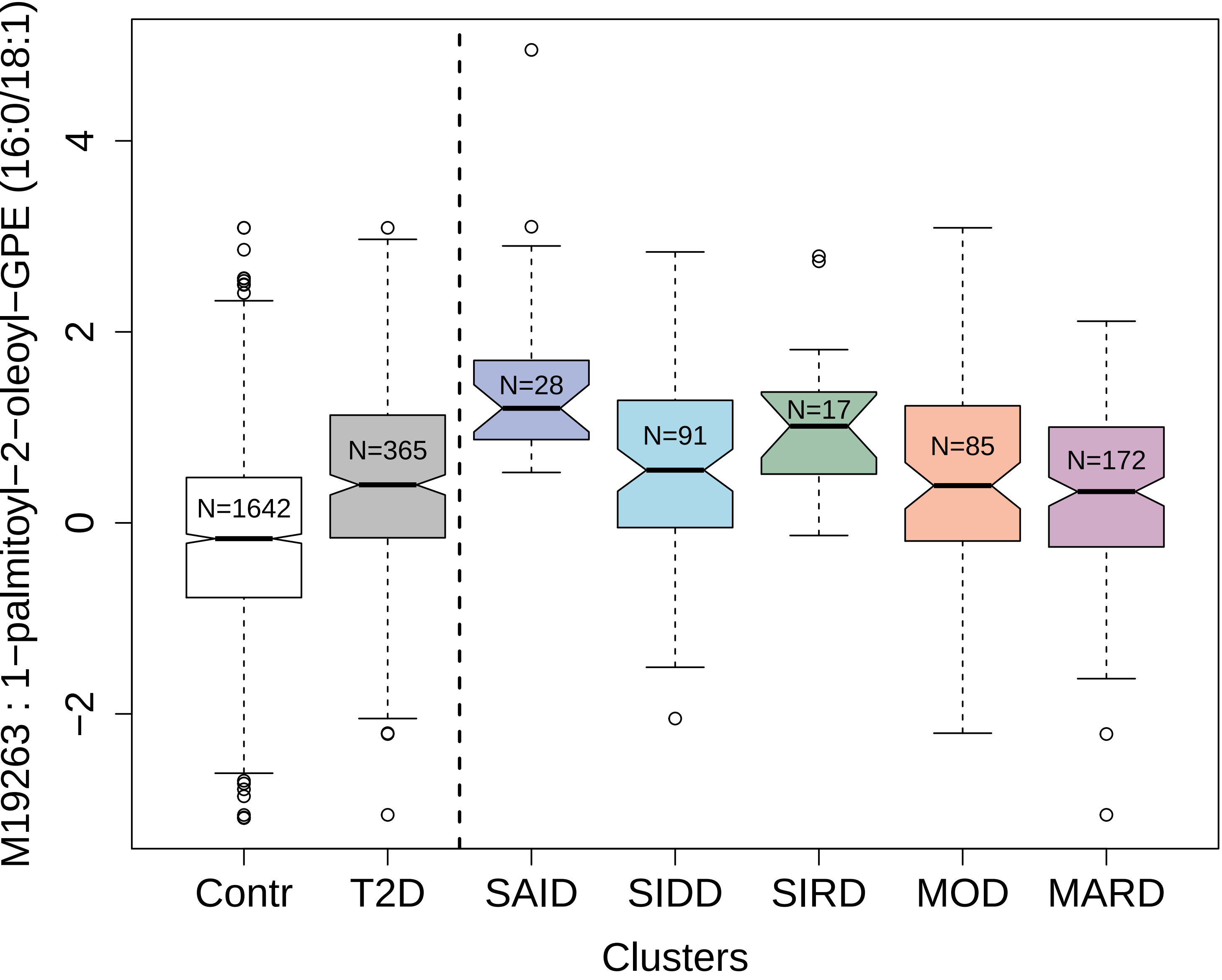

M19263 : 1−palmitoyl−2−oleoyl−GPE (16:0/18:1)
4
2
0
−2
N=1642
N=365
N=28
N=91
N=17
N=85
N=172
Contr
T2D
SAID
SIDD
SIRD
MOD
MARD
Clusters

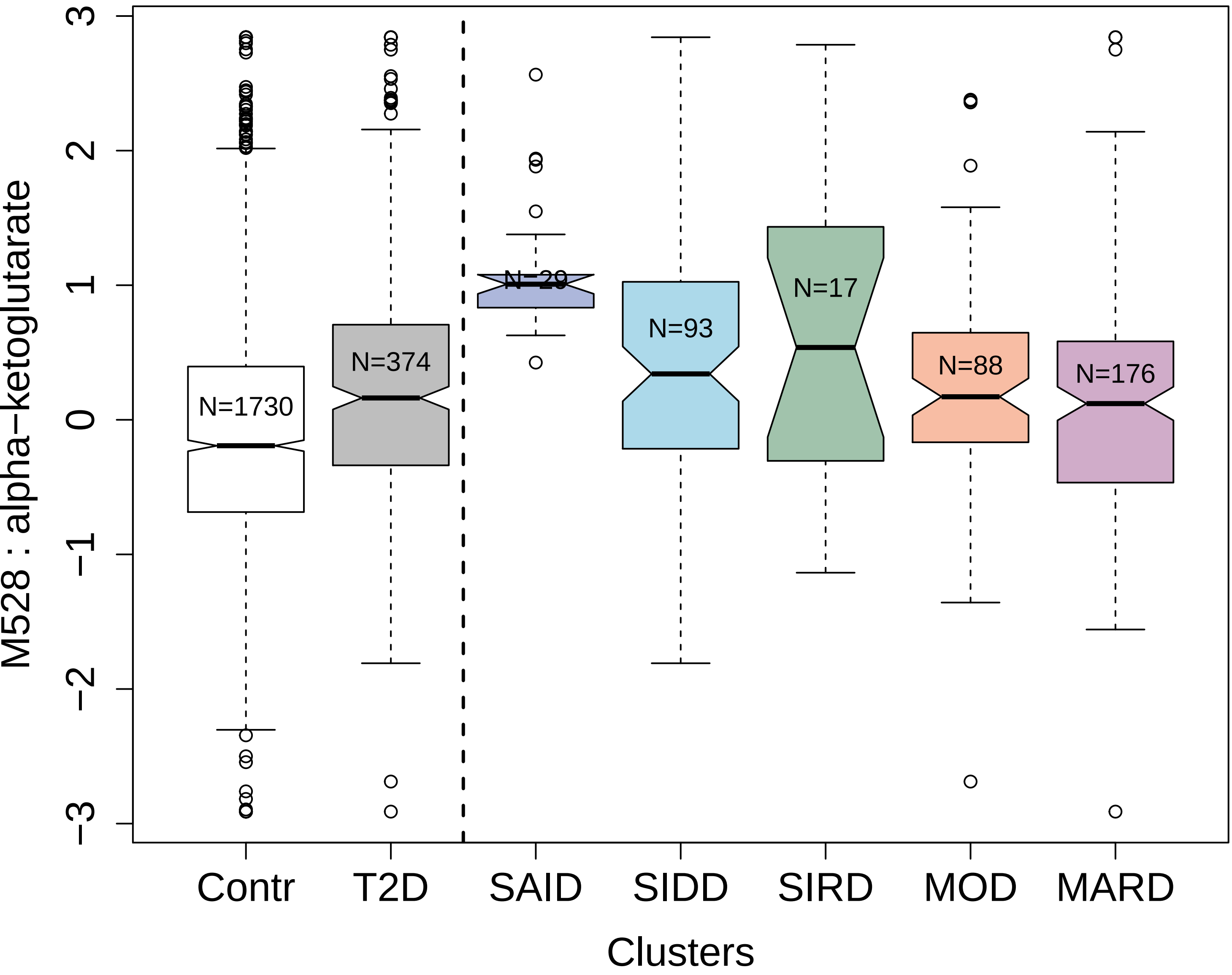

M528 : alpha−ketoglutarate
3
2
1
0
−1
−2
−3
N=1730
N=374
N=29
N=93
N=17
N=88
N=176
Contr
T2D
SAID
SIDD
SIRD
MOD
MARD
Clusters

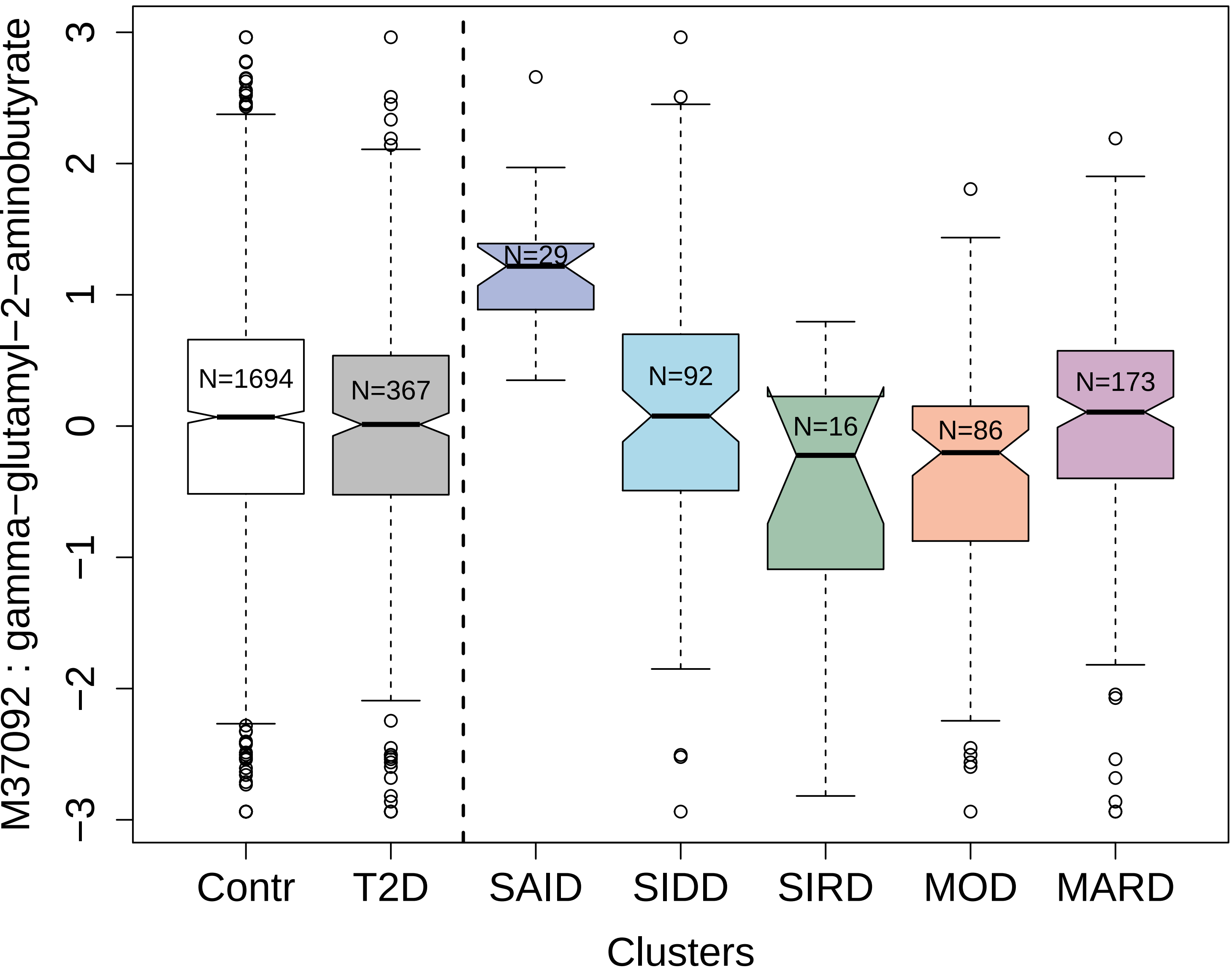
M37092 : gamma−glutamyl−2−aminobutyrate
3
2
1
0
−1
−2
−3
N=1694
N=367
N=29
N=92
N=16
N=86
N=173
Contr
T2D
SAID
SIDD
SIRD
MOD
MARD
Clusters

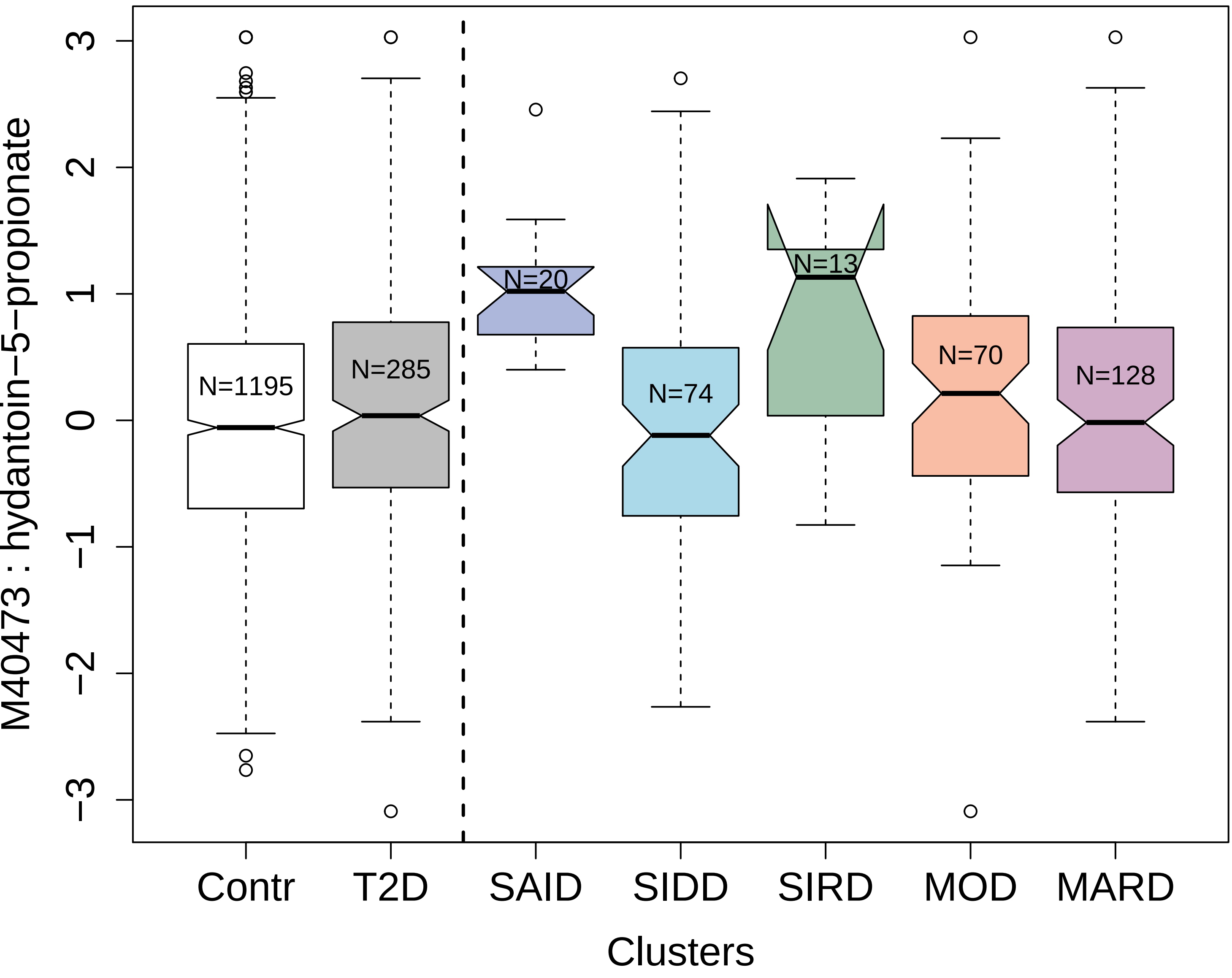

M40473 : hydantoin−5−propionate
3
2
1
0
−1
−2
−3
N=1195
N=285
N=20
N=74
N=13
N=70
N=128
Contr
T2D
SAID
SIDD
SIRD
MOD
MARD
Clusters

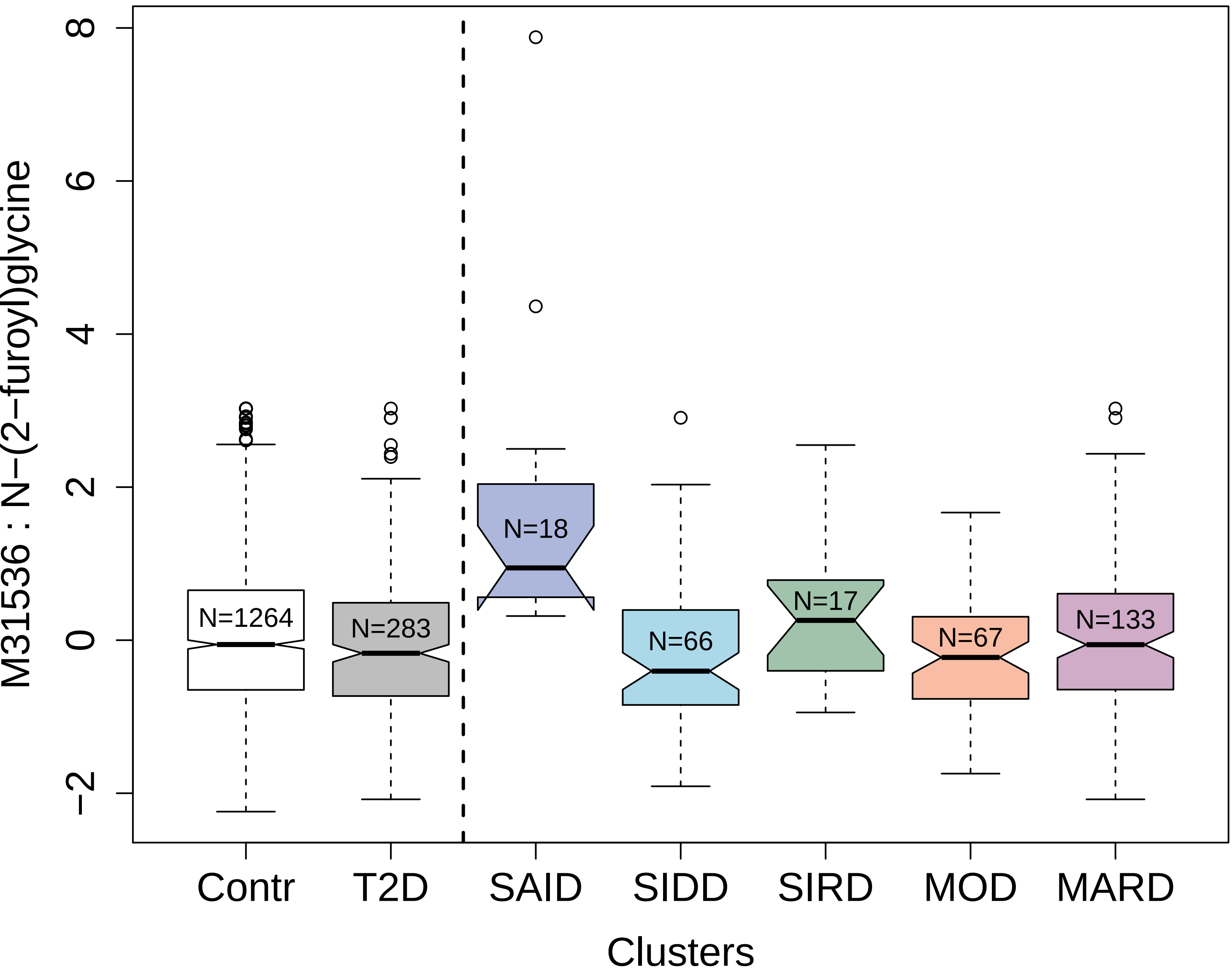
M31536 : N−(2−furoyl)glycine
8
6
4
2
0
−2
N=1264
N=283
N=18
N=66
N=17
N=67
N=133
Contr
T2D
SAID
SIDD
SIRD
MOD
MARD
Clusters

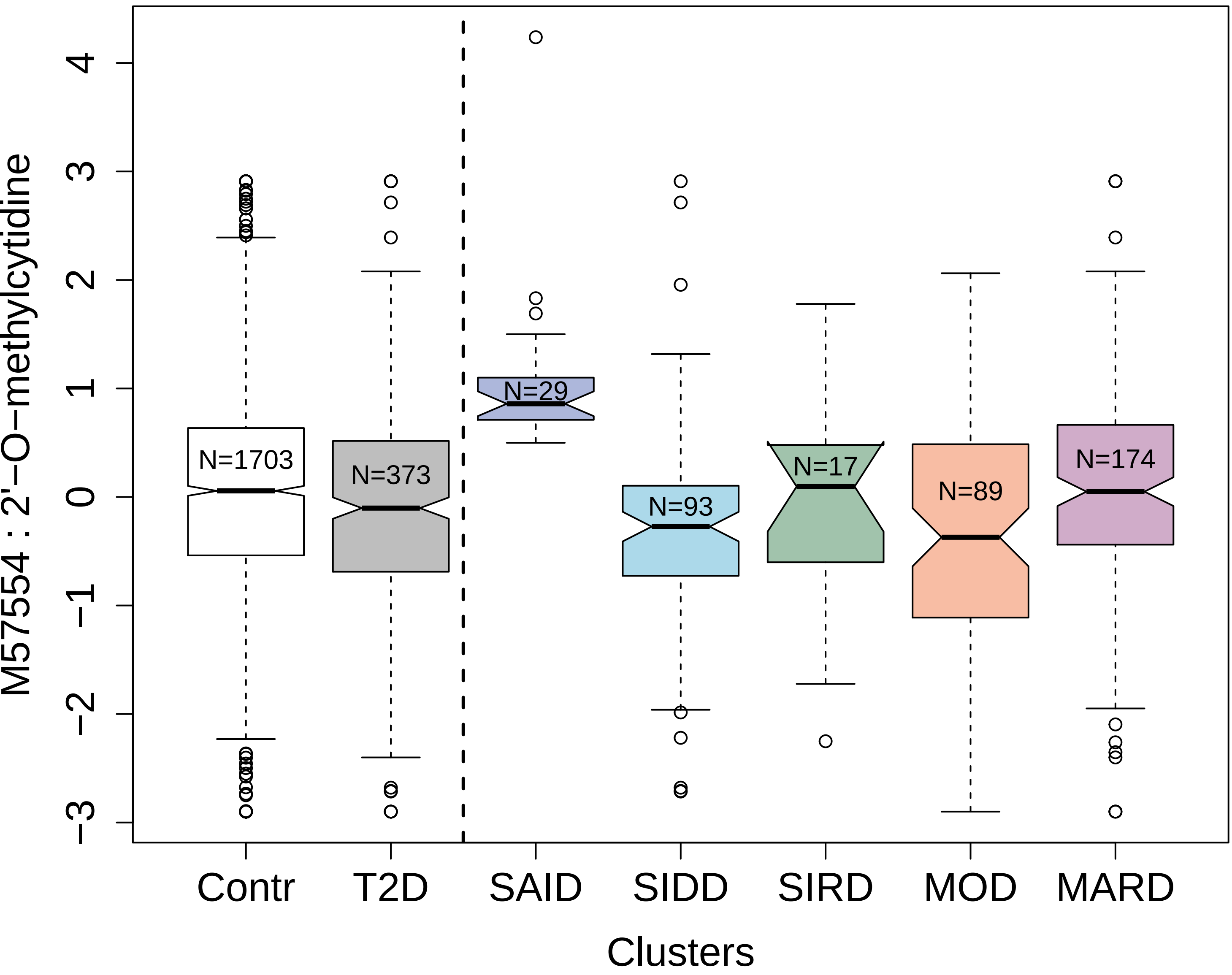
M57554 : 2'−O−methylcytidine
4
3
2
1
0
−1
−2
−3
N=1703
N=373
N=29
N=93
N=17
N=89
N=174
Contr
T2D
SAID
SIDD
SIRD
MOD
MARD
Clusters

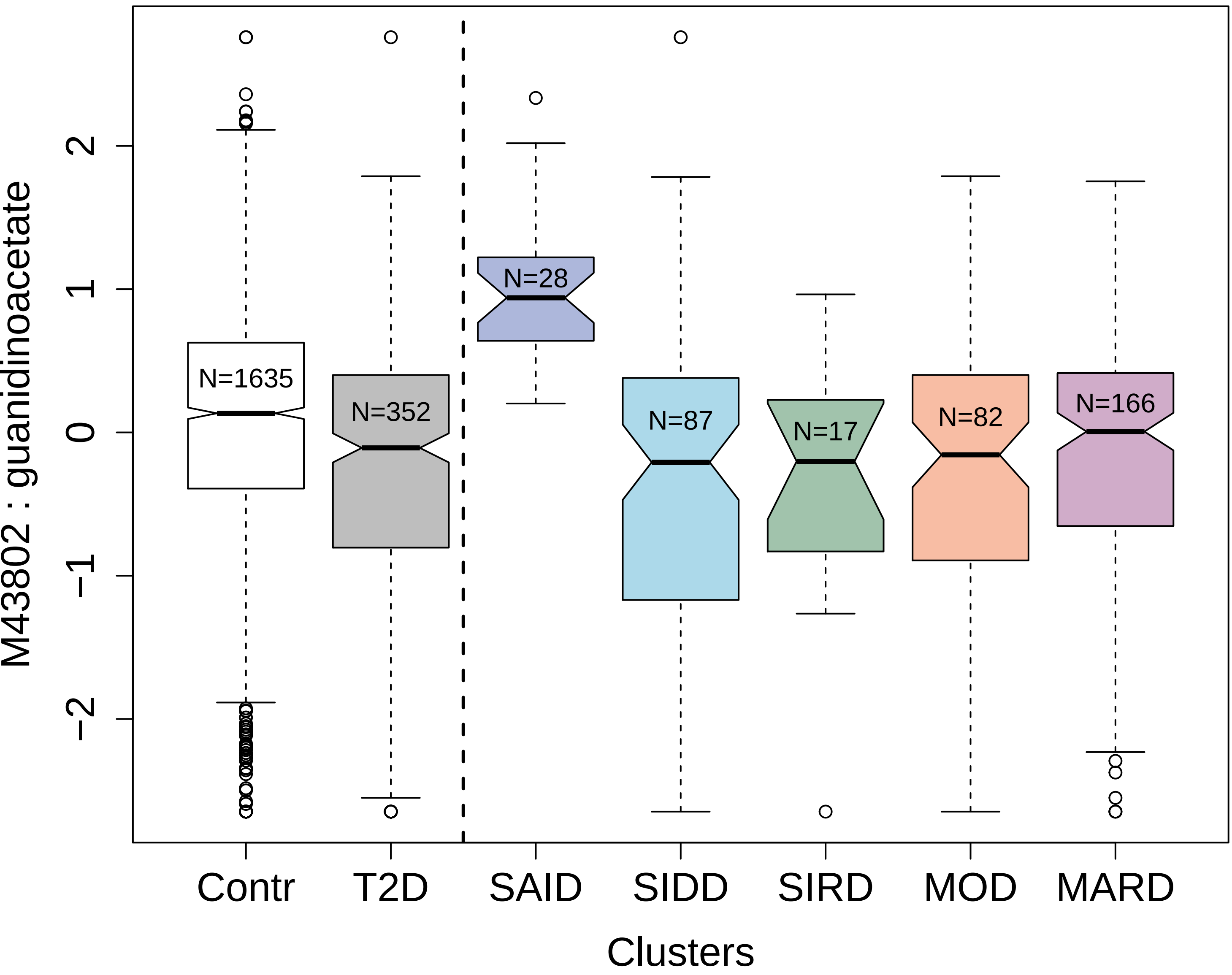

M43802 : guanidinoacetate
2
1
0
−1
−2
N=1635
N=352
N=28
N=87
N=17
N=82
N=166
Contr
T2D
SAID
SIDD
SIRD
MOD
MARD
Clusters

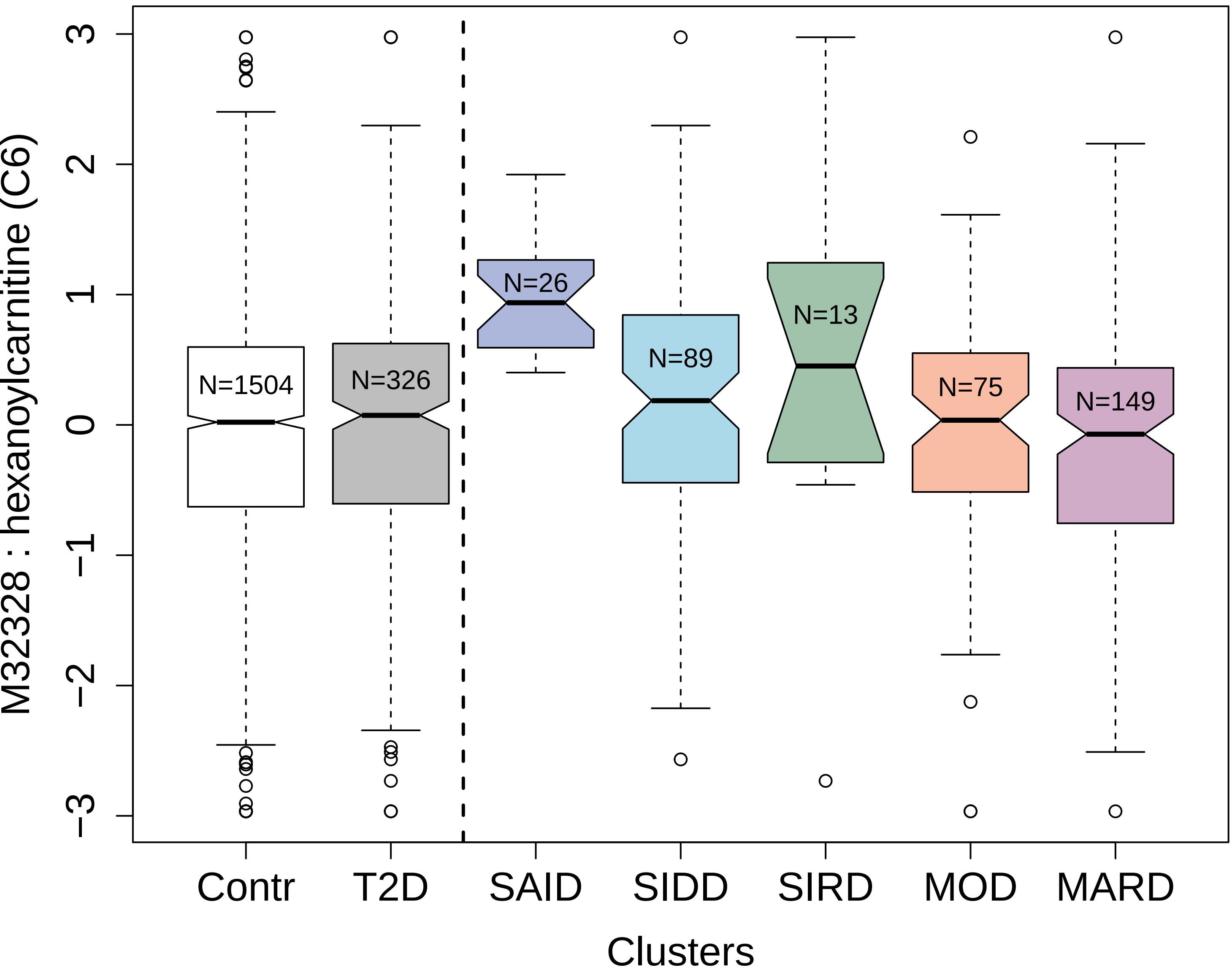
M32328 : hexanoylcarnitine (C6)
3
2
1
0
−1
−2
−3
N=1504
N=326
N=26
N=89
N=13
N=75
N=149
Contr
T2D
SAID
SIDD
SIRD
MOD
MARD
Clusters

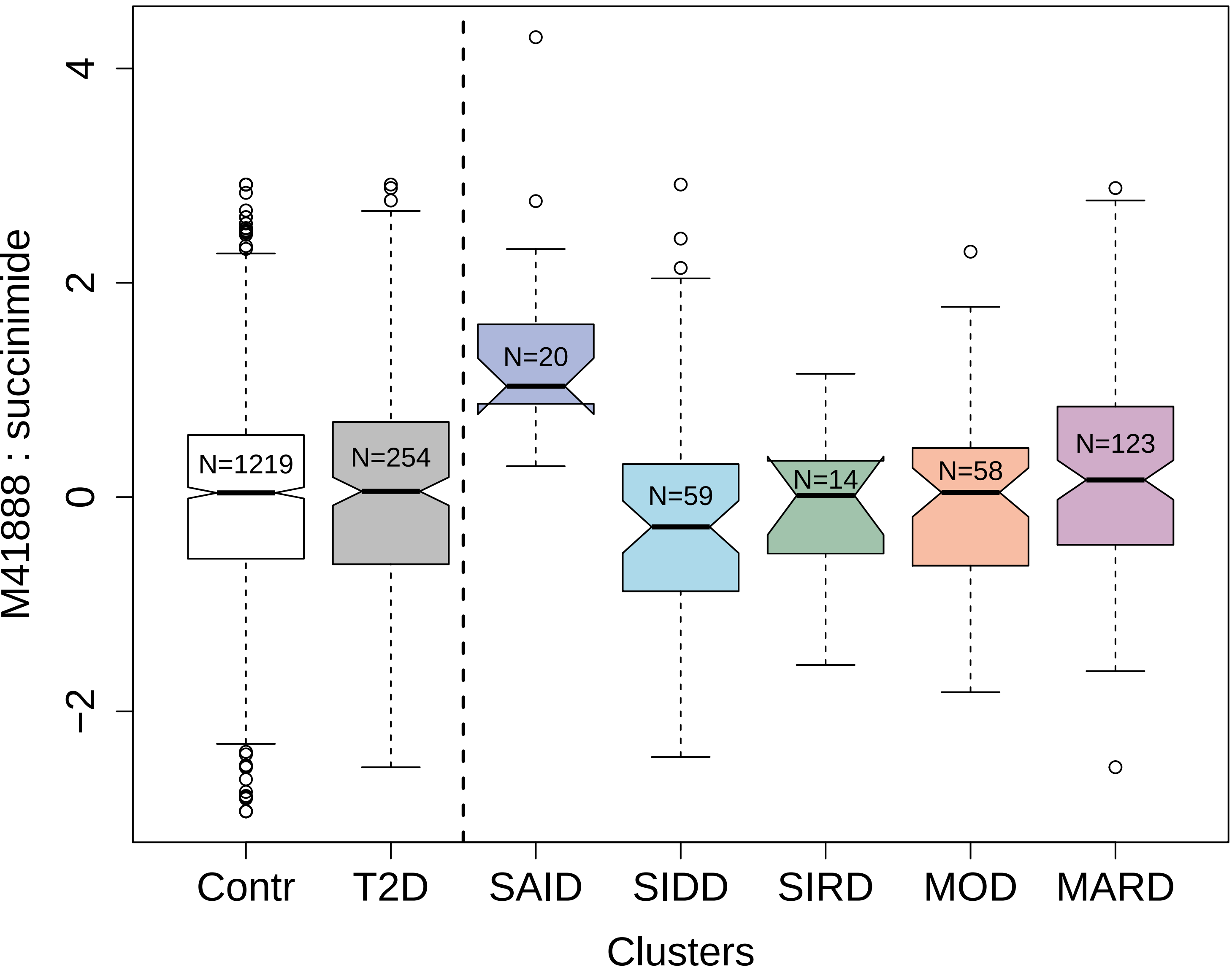

M41888 : succinimide
4
2
0
−2
N=1219
N=254
N=20
N=59
N=14
N=58
N=123
Contr
T2D
SAID
SIDD
SIRD
MOD
MARD
Clusters

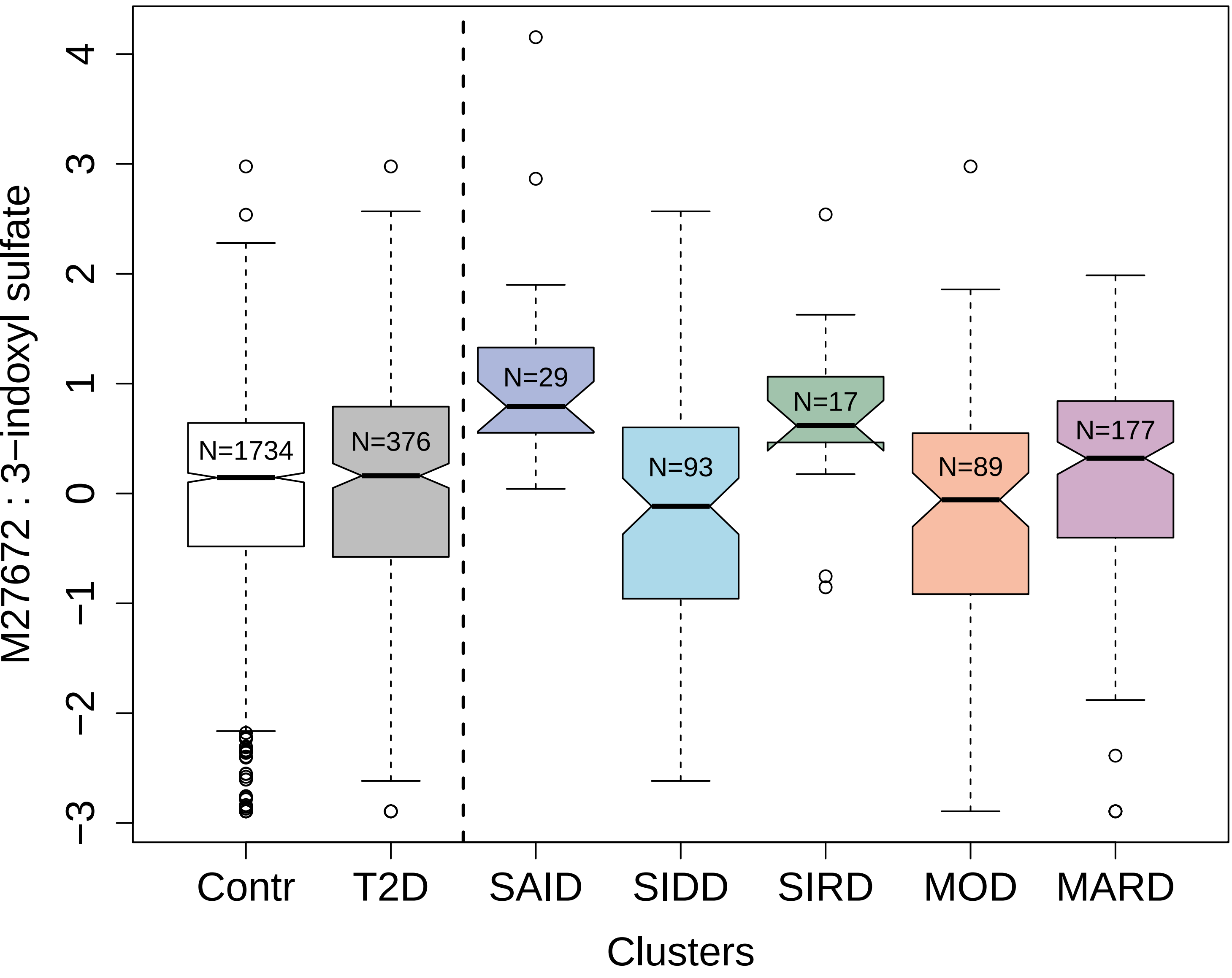

M27672 : 3−indoxyl sulfate
4
3
2
1
0
−1
−2
−3
N=1734
N=376
N=29
N=93
N=17
N=89
N=177
Contr
T2D
SAID
SIDD
SIRD
MOD
MARD
Clusters

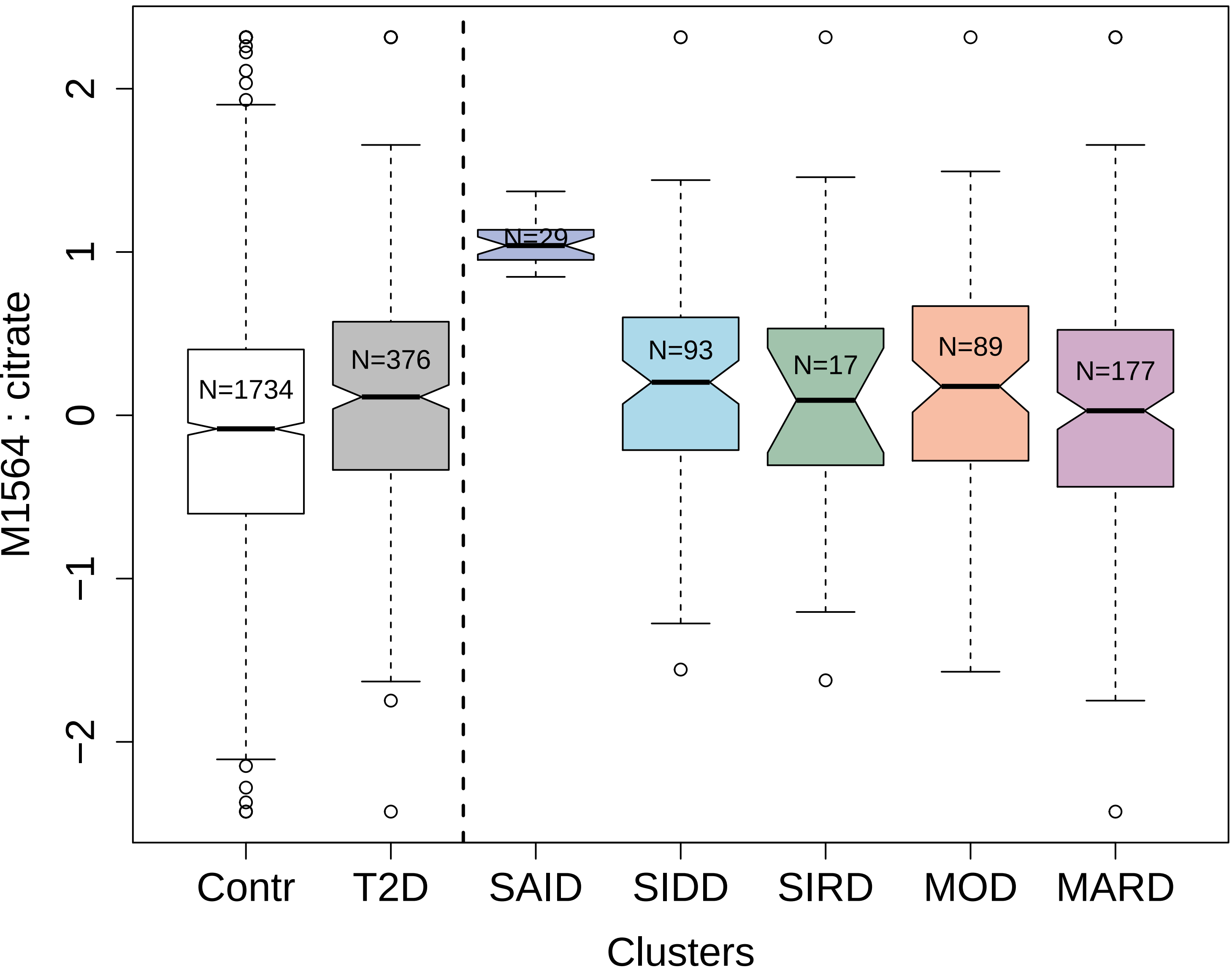

M1564 : citrate
2
1
0
−1
−2
N=1734
N=376
N=29
N=93
N=17
N=89
N=177
Contr
T2D
SAID
SIDD
SIRD
MOD
MARD
Clusters

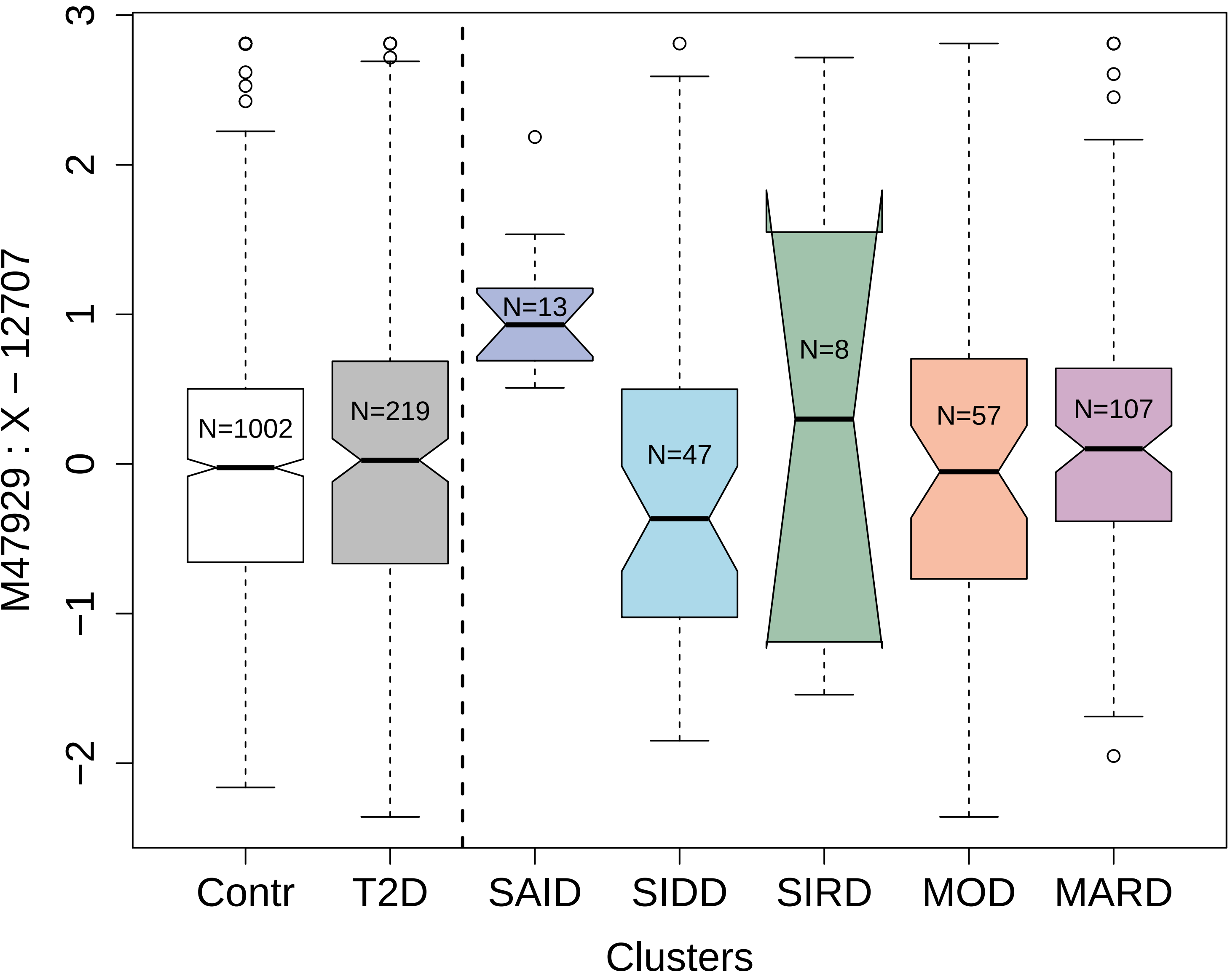

N=1002
N=219
N=13
N=47
N=8
N=57
N=107
Contr
T2D
SAID
SIDD
SIRD
MOD
MARD
Clusters
M47929 : X − 12707
3
2
1
0
−1
−2

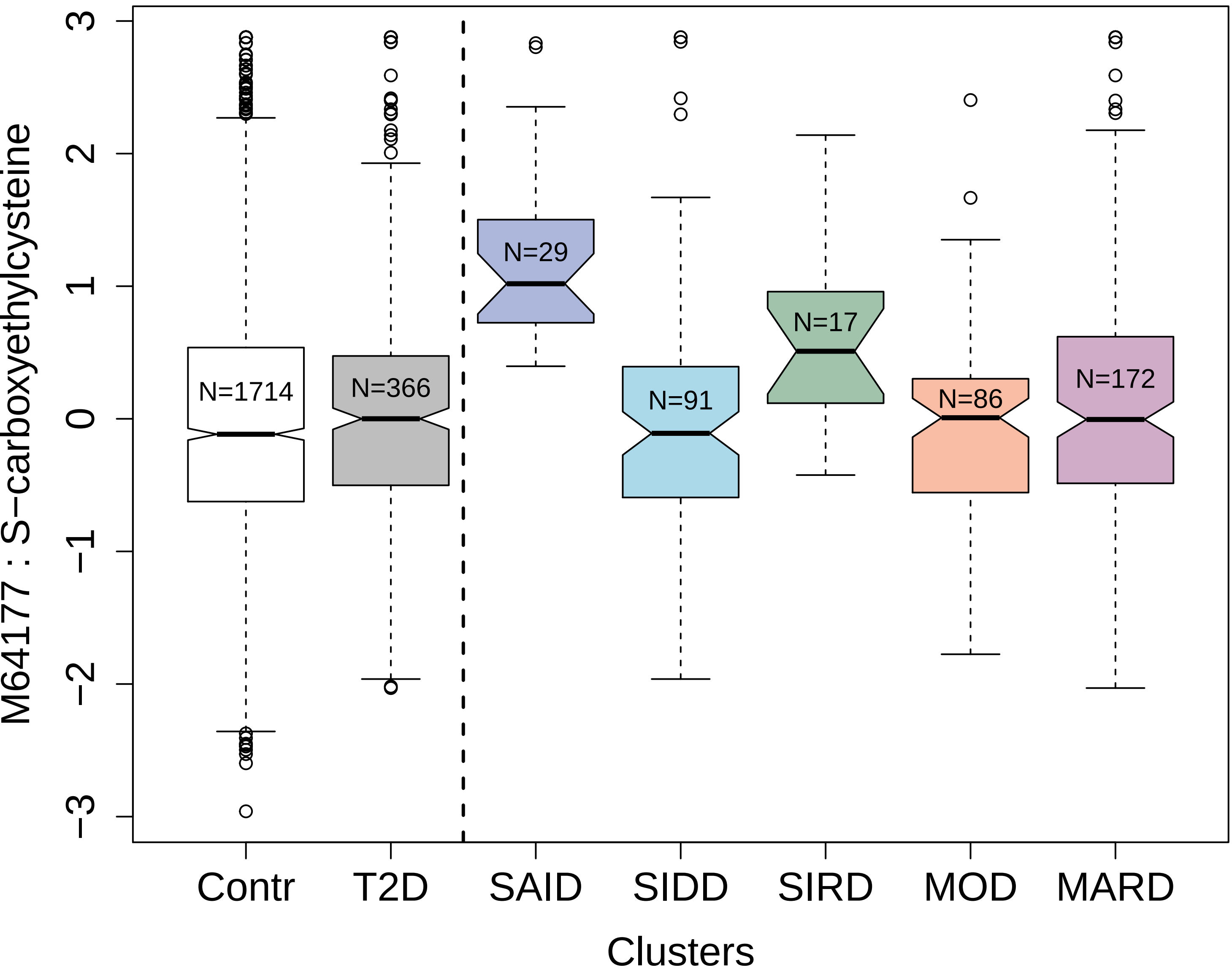

M64177 : S−carboxyethylcysteine
3
2
1
0
−1
−2
−3
N=1714
N=366
N=29
N=91
N=17
N=86
N=172
Contr
T2D
SAID
SIDD
SIRD
MOD
MARD
Clusters

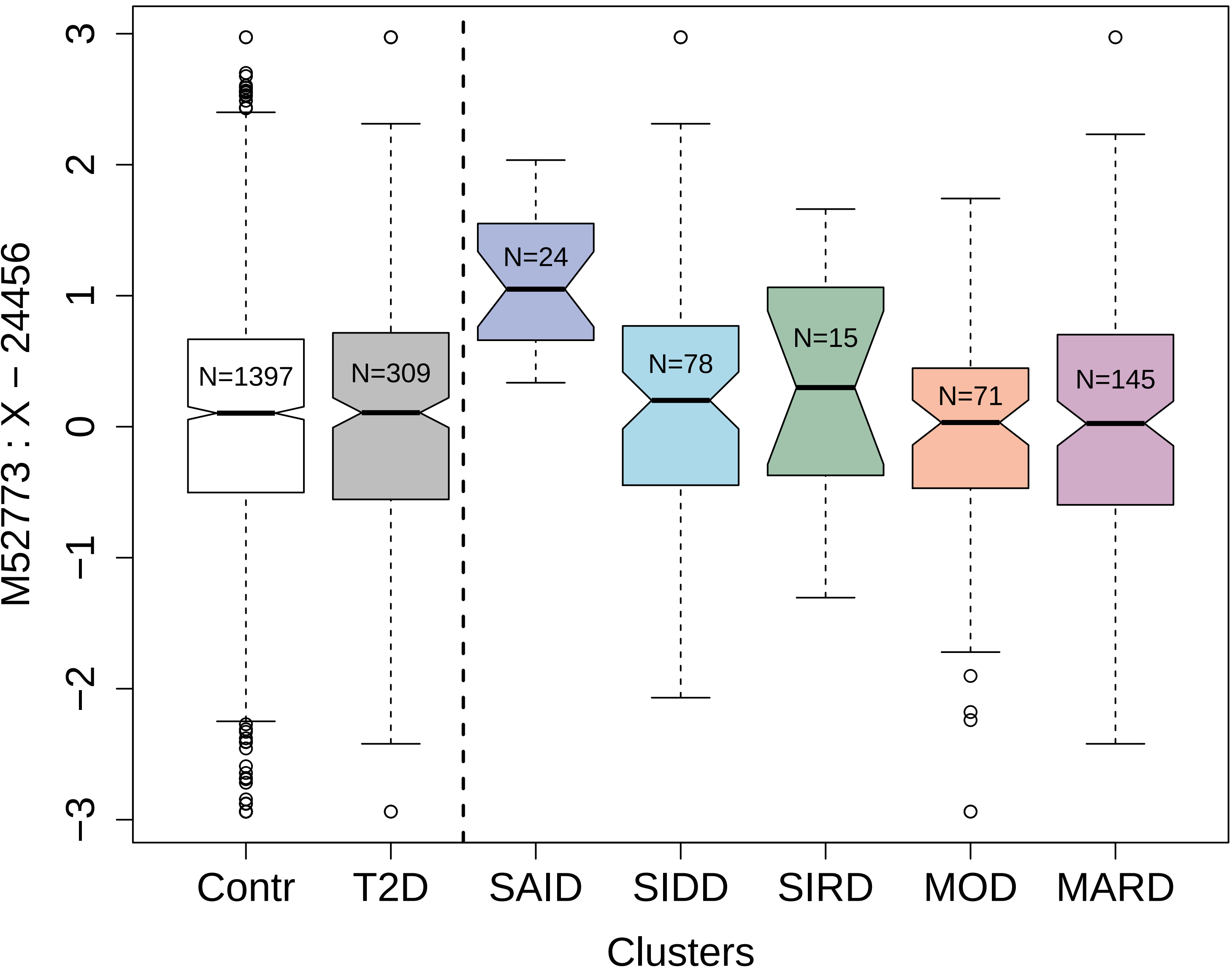
M52773 : X − 24456
3
2
1
0
−1
−2
−3
N=1397
N=309
N=24
N=78
N=15
N=71
N=145
Contr
T2D
SAID
SIDD
SIRD
MOD
MARD
Clusters

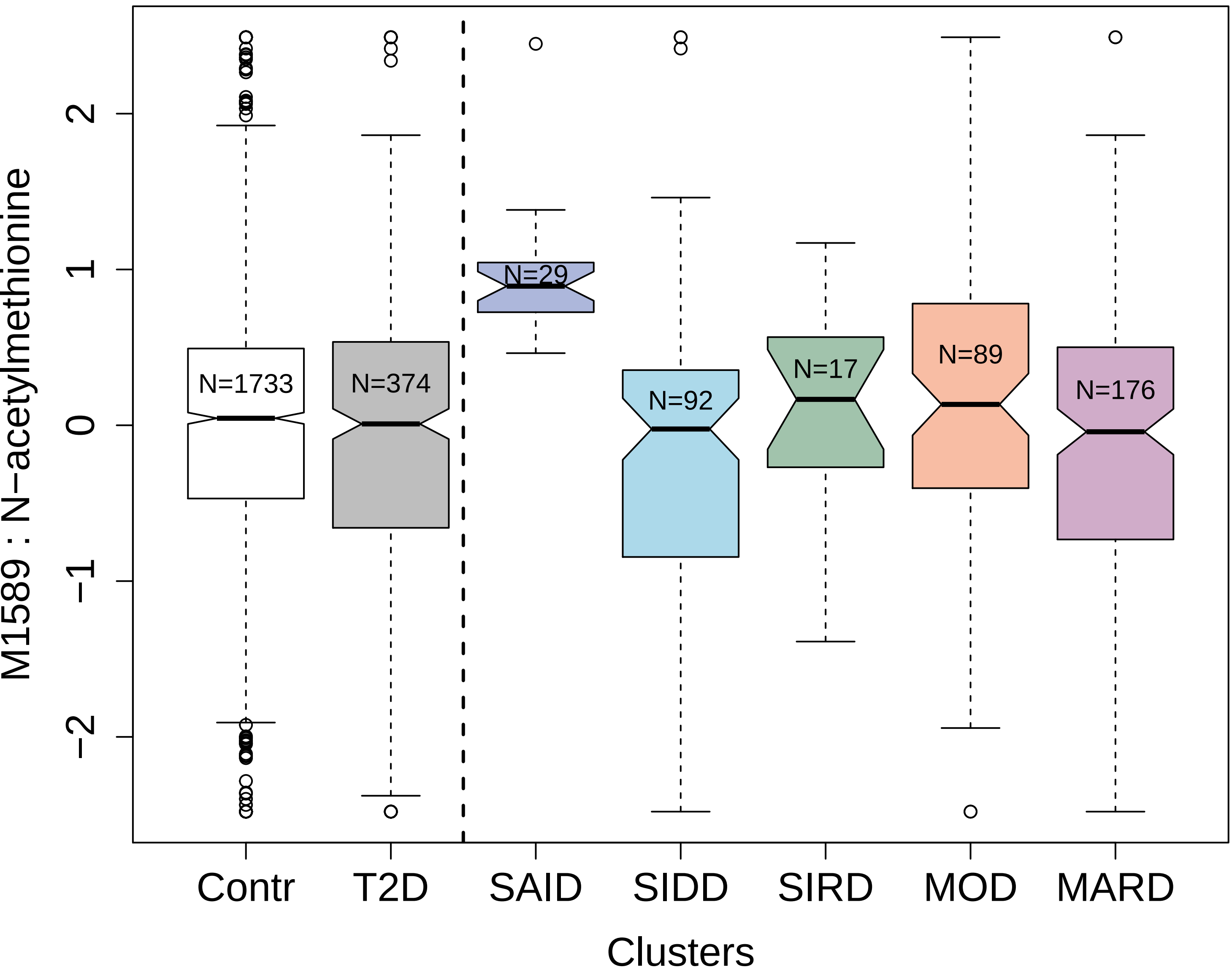

M1589 : N−acetylmethionine
Clusters
Contr
T2D
SAID
SIDD
SIRD
MOD
MARD
N=1733
N=374
N=29
N=92
N=17
N=89
N=176
−2
−1
0
1
2

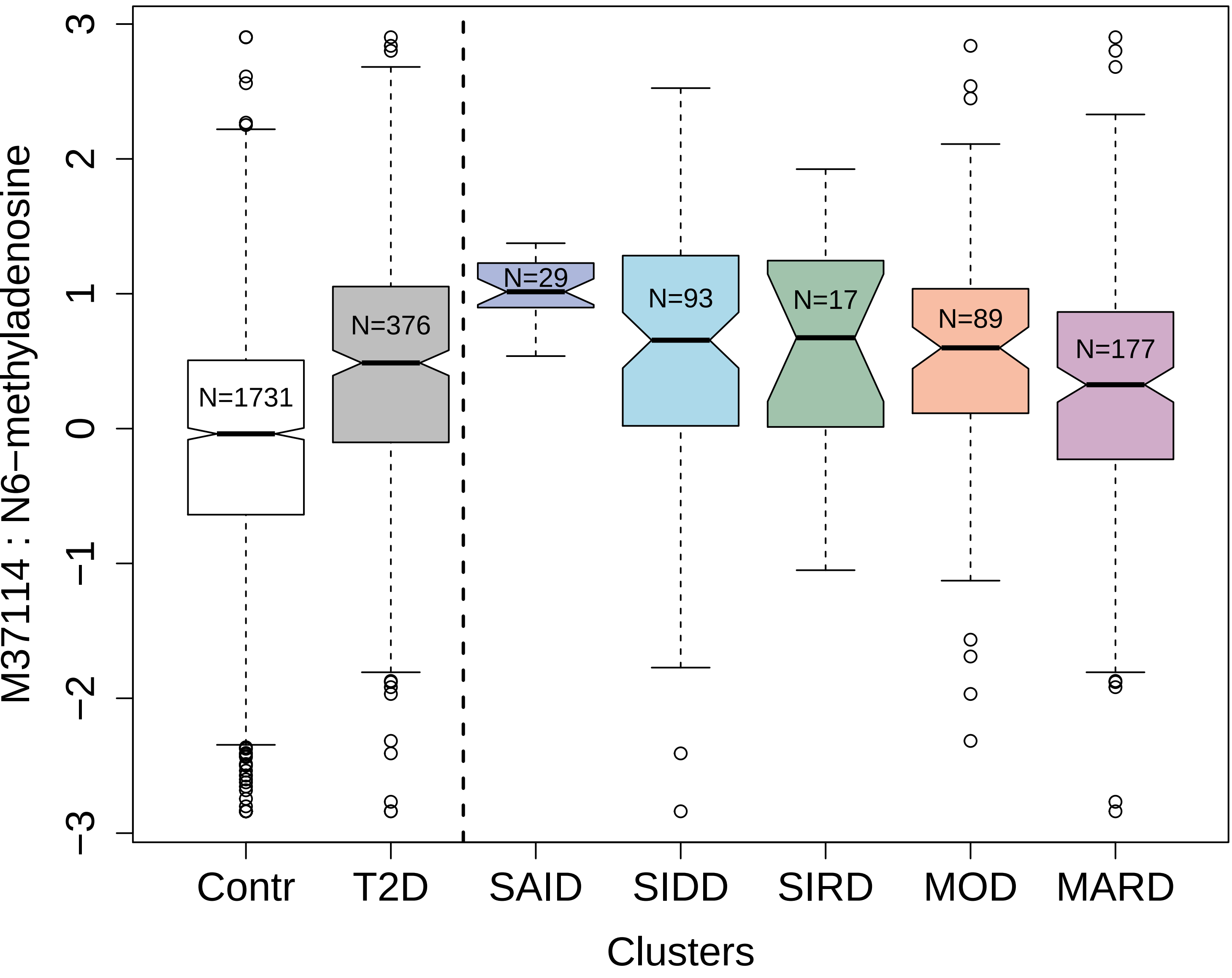
M37114 : N6−methyladenosine
Clusters
Contr
T2D
SAID
SIDD
SIRD
MOD
MARD
N=1731
N=376
N=29
N=93
N=17
N=89
N=177
3
2
1
0
−1
−2
−3

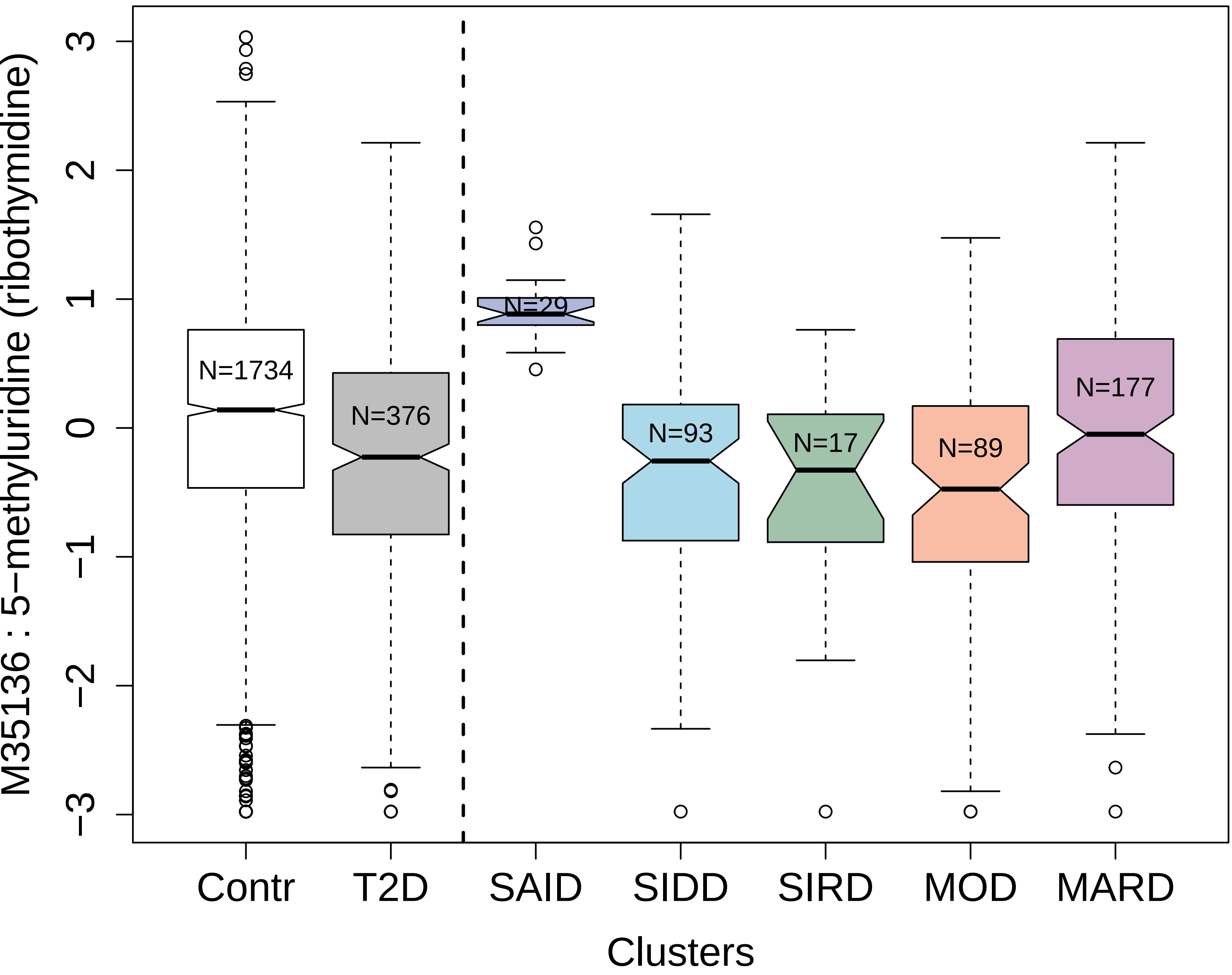
M35136 : 5−methyluridine (ribothymidine)
3
2
1
0
−1
−2
−3
N=1734
N=376
N=29
N=93
N=17
N=89
N=177
Contr
T2D
SAID
SIDD
SIRD
MOD
MARD
Clusters

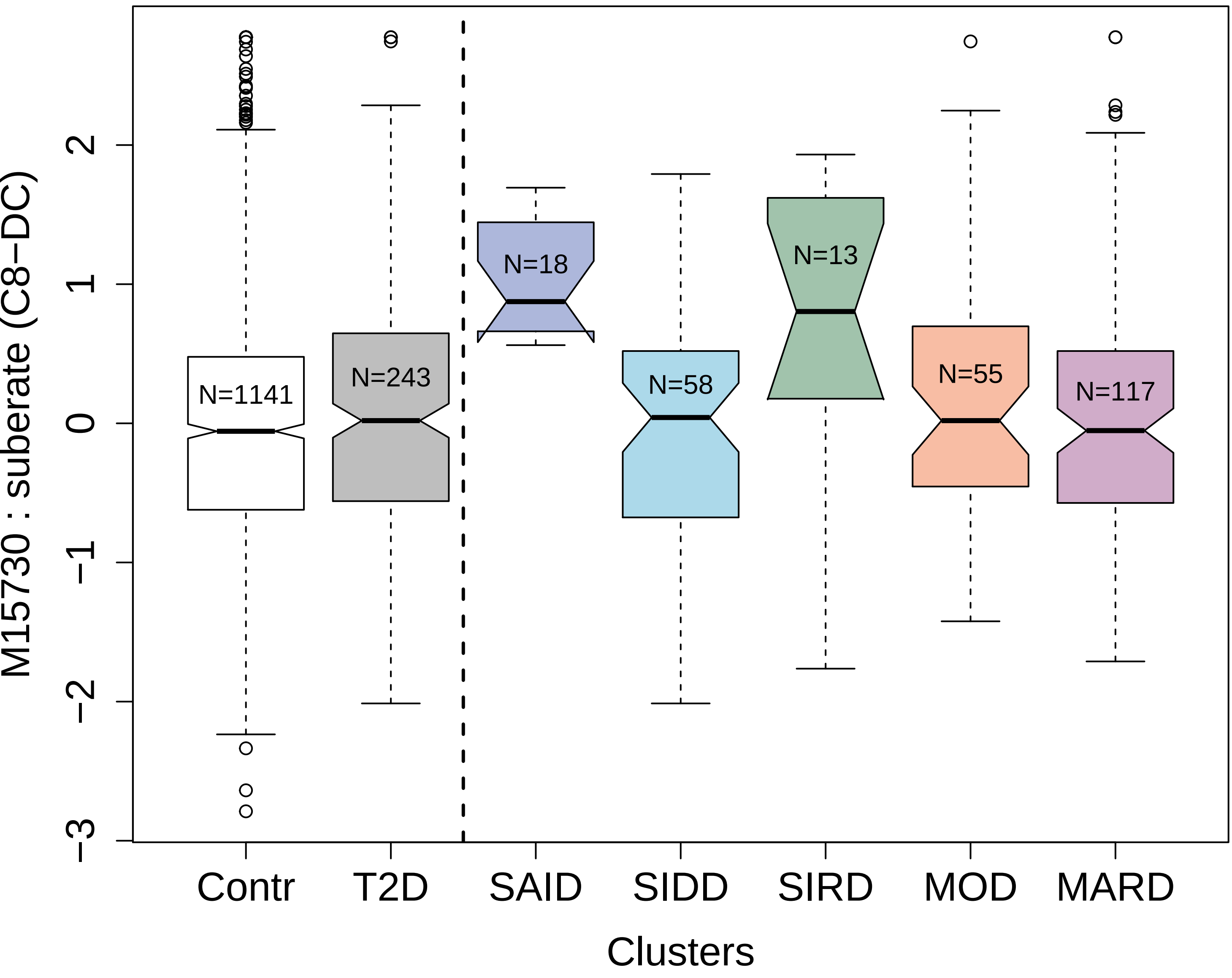

M15730 : suberate (C8−DC)
−3
−2
−1
0
1
2
N=1141
N=243
N=18
N=58
N=13
N=55
N=117
Contr
T2D
SAID
SIDD
SIRD
MOD
MARD
Clusters

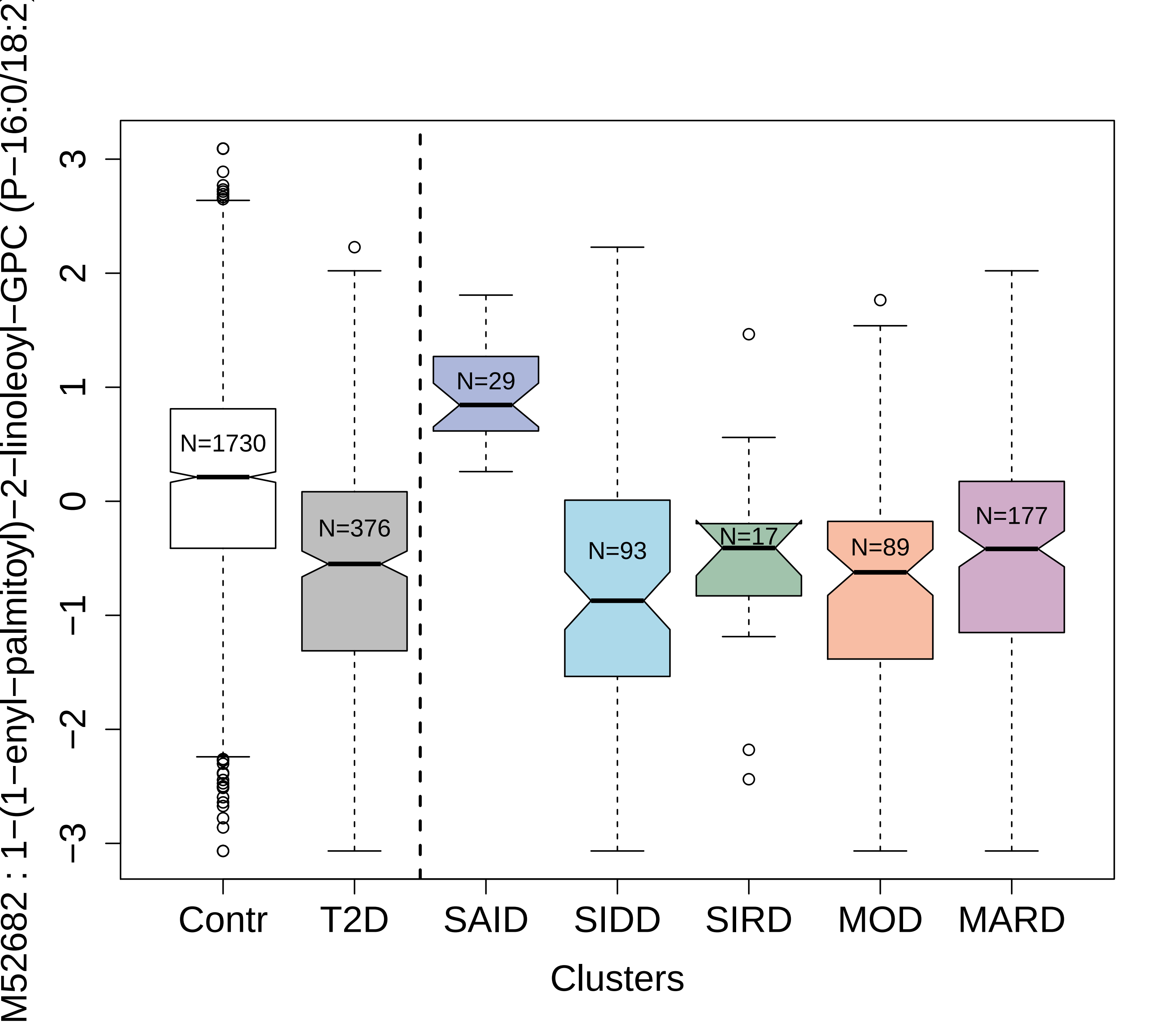
M52682 : 1−(1−enyl−palmitoyl)−2−linoleoyl−GPC (P−16:0/18:2)
3
2
1
0
−1
−2
−3
N=1730
N=376
N=29
N=93
N=17
N=89
N=177
Contr
T2D
SAID
SIDD
SIRD
MOD
MARD
Clusters

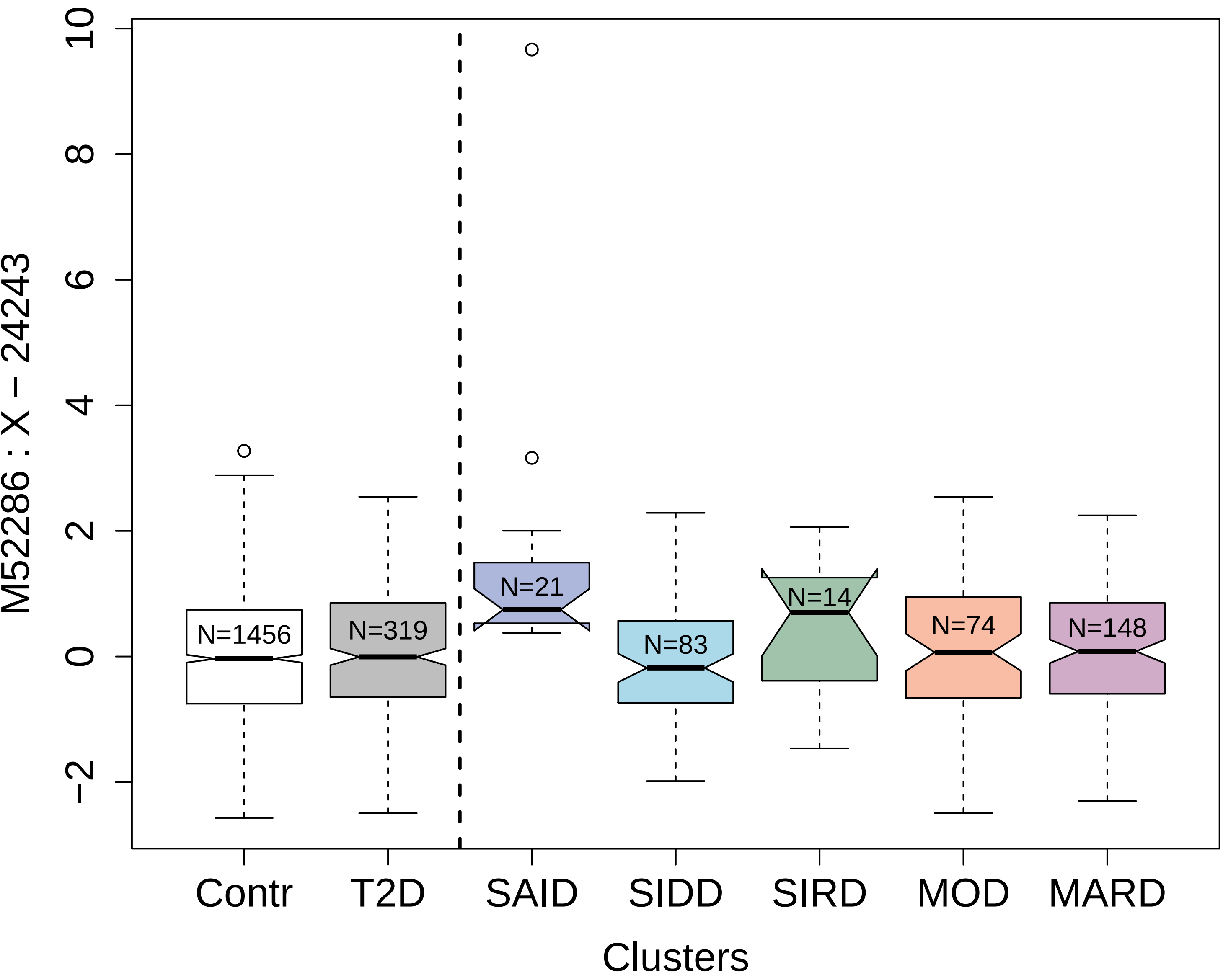

M52286 : X − 24243
10
8
6
4
2
0
−2
N=1456
N=319
N=21
N=83
N=14
N=74
N=148
Contr
T2D
SAID
SIDD
SIRD
MOD
MARD
Clusters

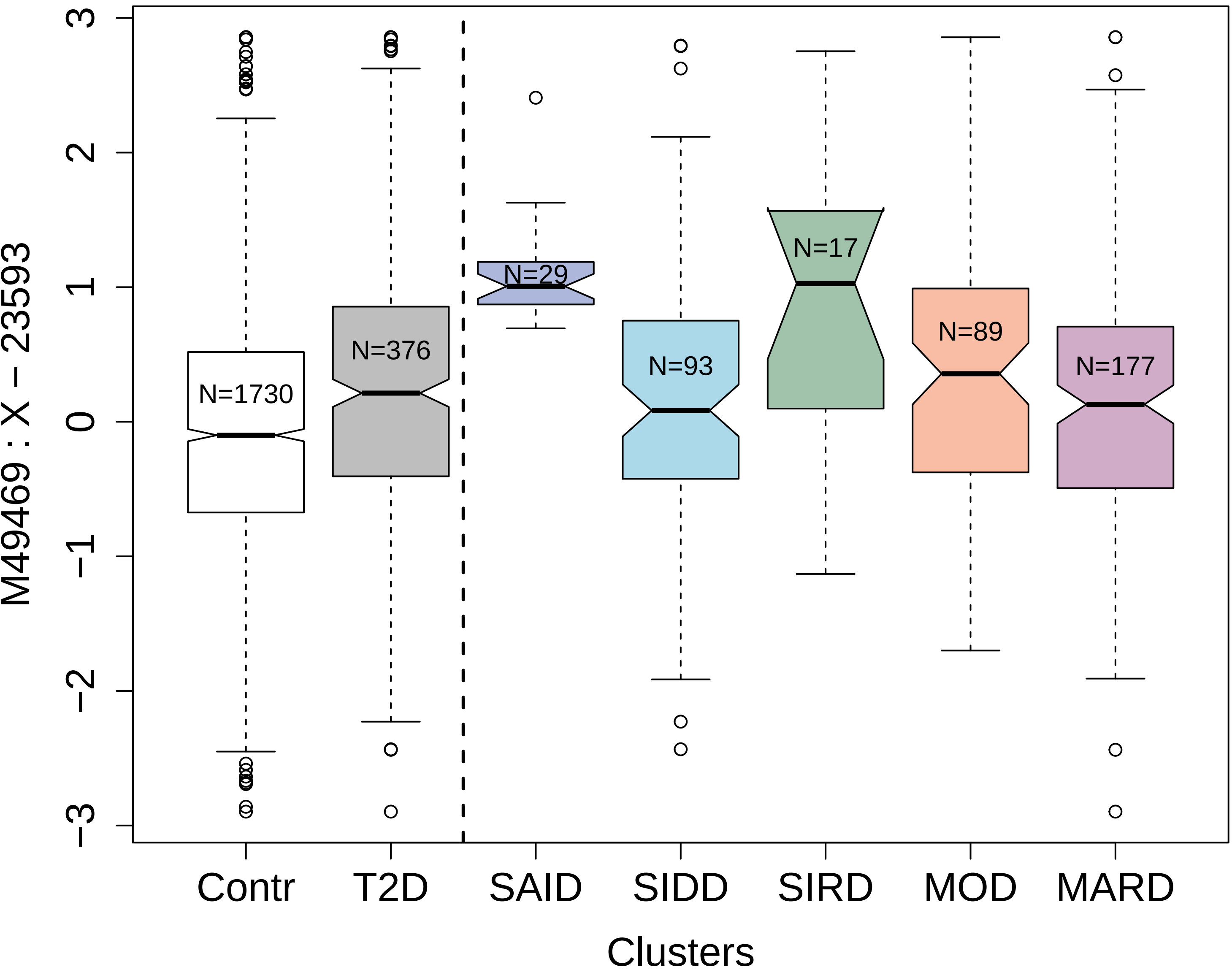

M49469 : X − 23593
3
2
1
0
−1
−2
−3
N=1730
N=376
N=29
N=93
N=17
N=89
N=177
Contr
T2D
SAID
SIDD
SIRD
MOD
MARD
Clusters

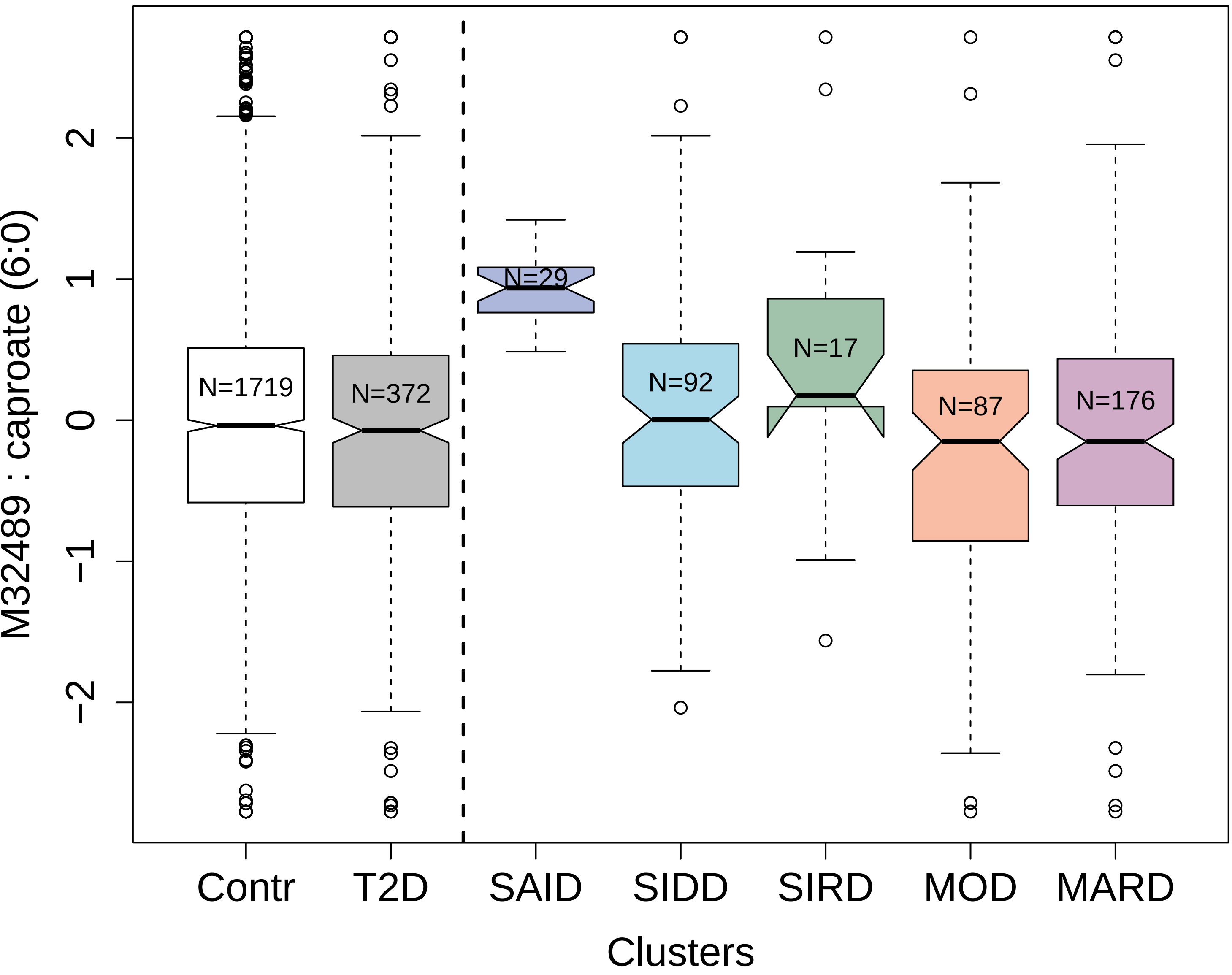

M32489 : caproate (6:0)
2
1
0
−1
−2
N=1719
N=372
N=29
N=92
N=17
N=87
N=176
Contr
T2D
SAID
SIDD
SIRD
MOD
MARD
Clusters

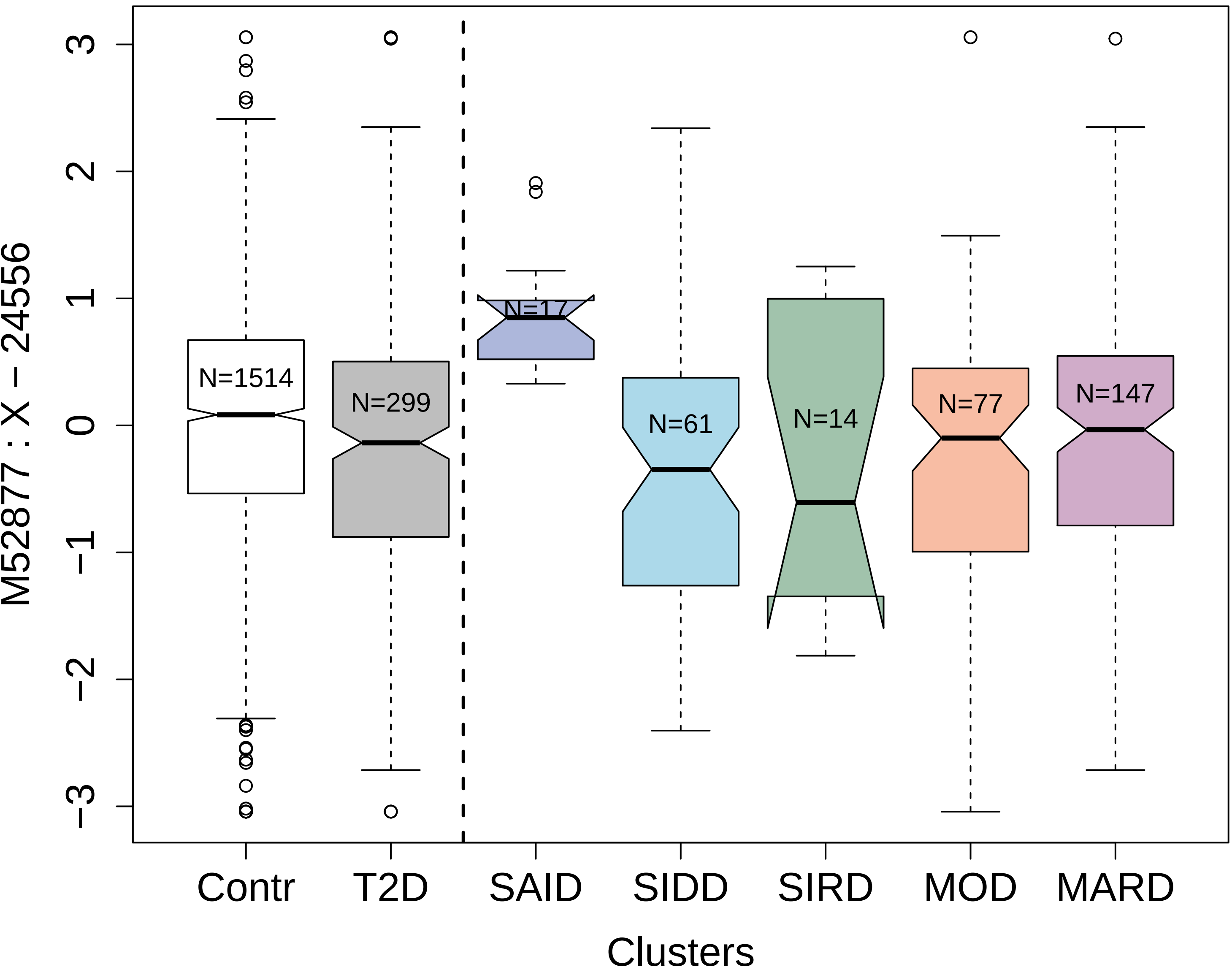

M52877 : X − 24556
3
2
1
0
−1
−2
−3
N=1514
N=299
N=17
N=61
N=14
N=77
N=147
Contr
T2D
SAID
SIDD
SIRD
MOD
MARD
Clusters

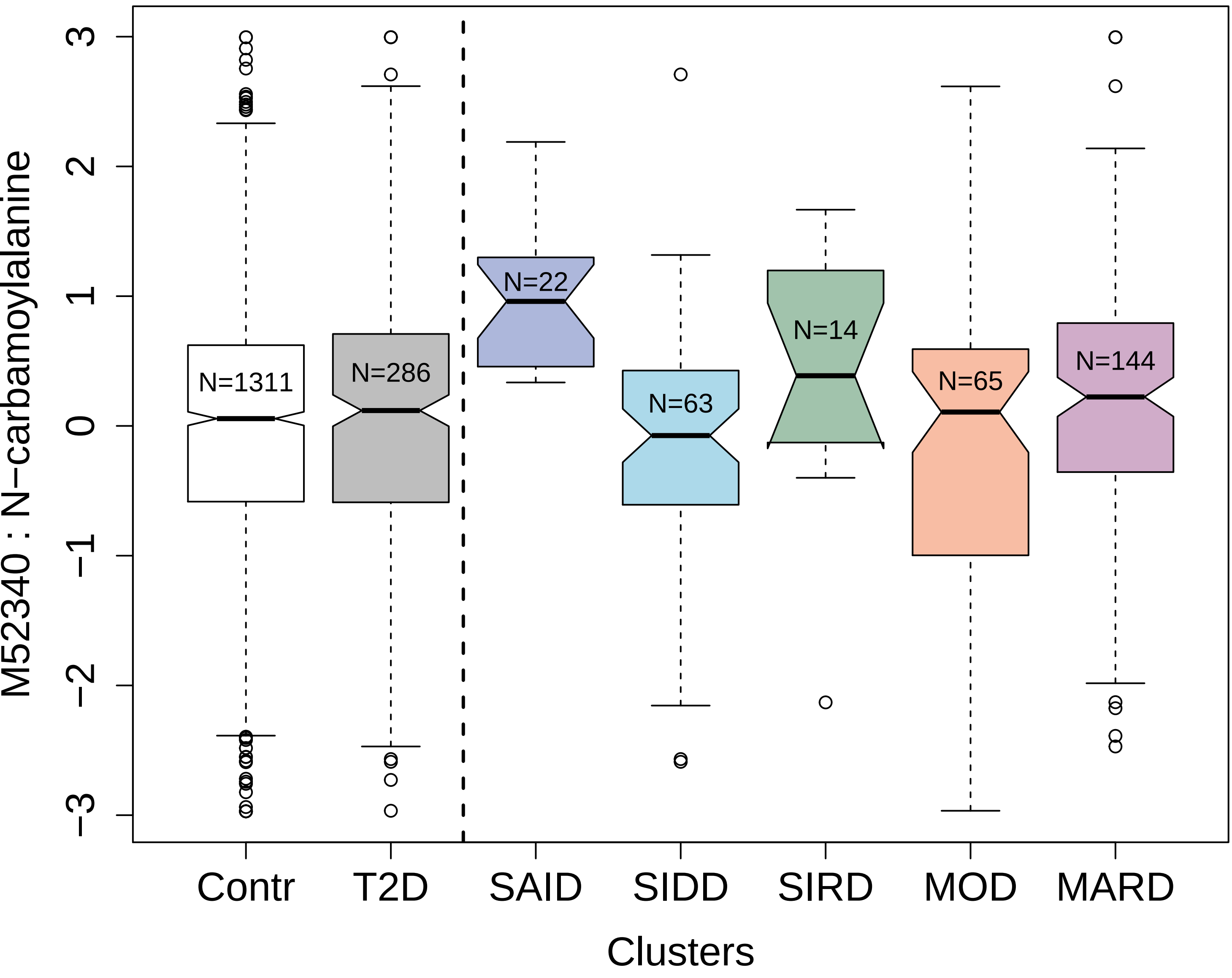
M52340 : N−carbamoylalanine
3
2
1
0
−1
−2
−3
N=1311
N=286
N=22
N=63
N=14
N=65
N=144
Contr
T2D
SAID
SIDD
SIRD
MOD
MARD
Clusters

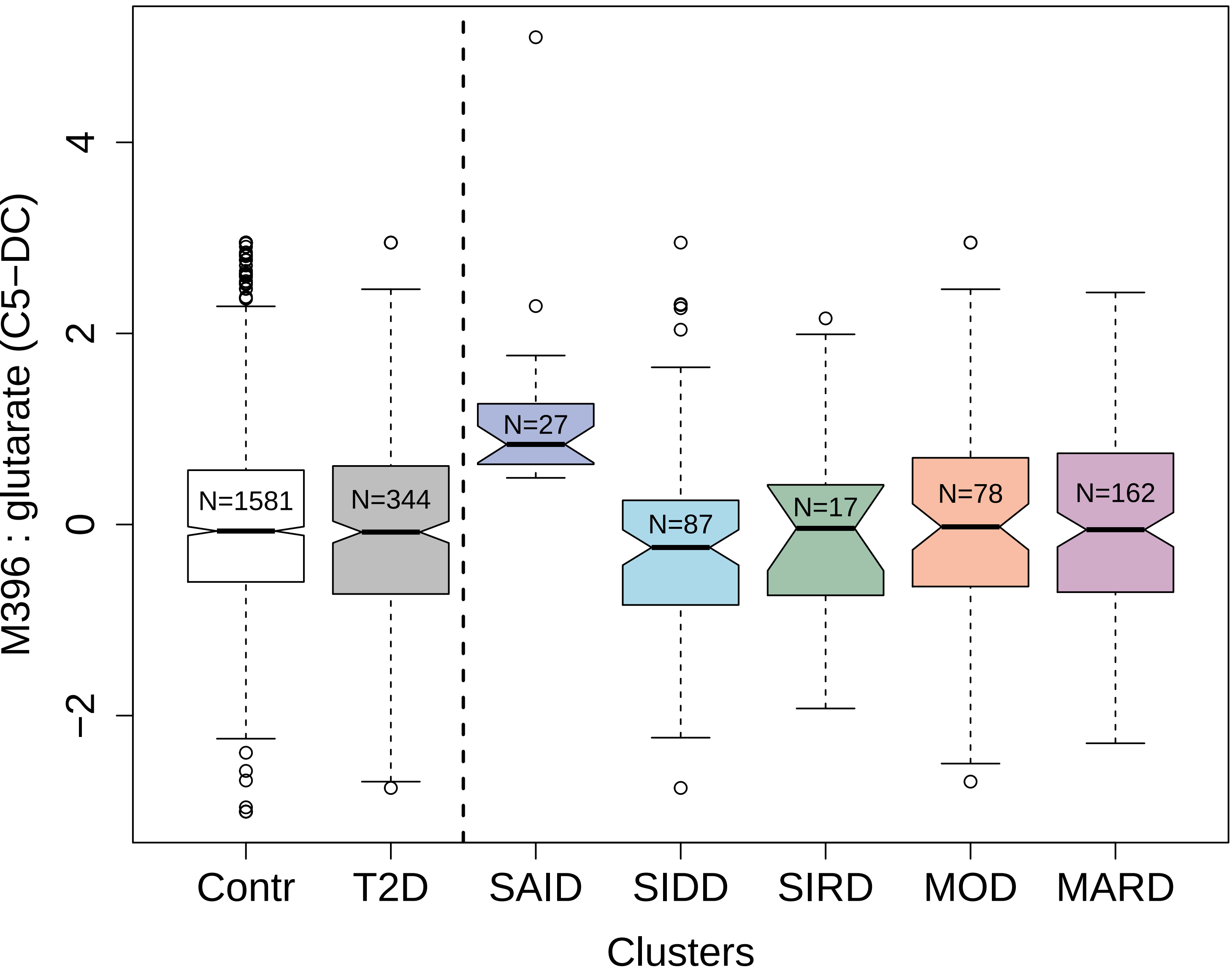
M396 : glutarate (C5−DC)
4
2
0
−2
N=1581
N=344
N=27
N=87
N=17
N=78
N=162
Contr
T2D
SAID
SIDD
SIRD
MOD
MARD
Clusters

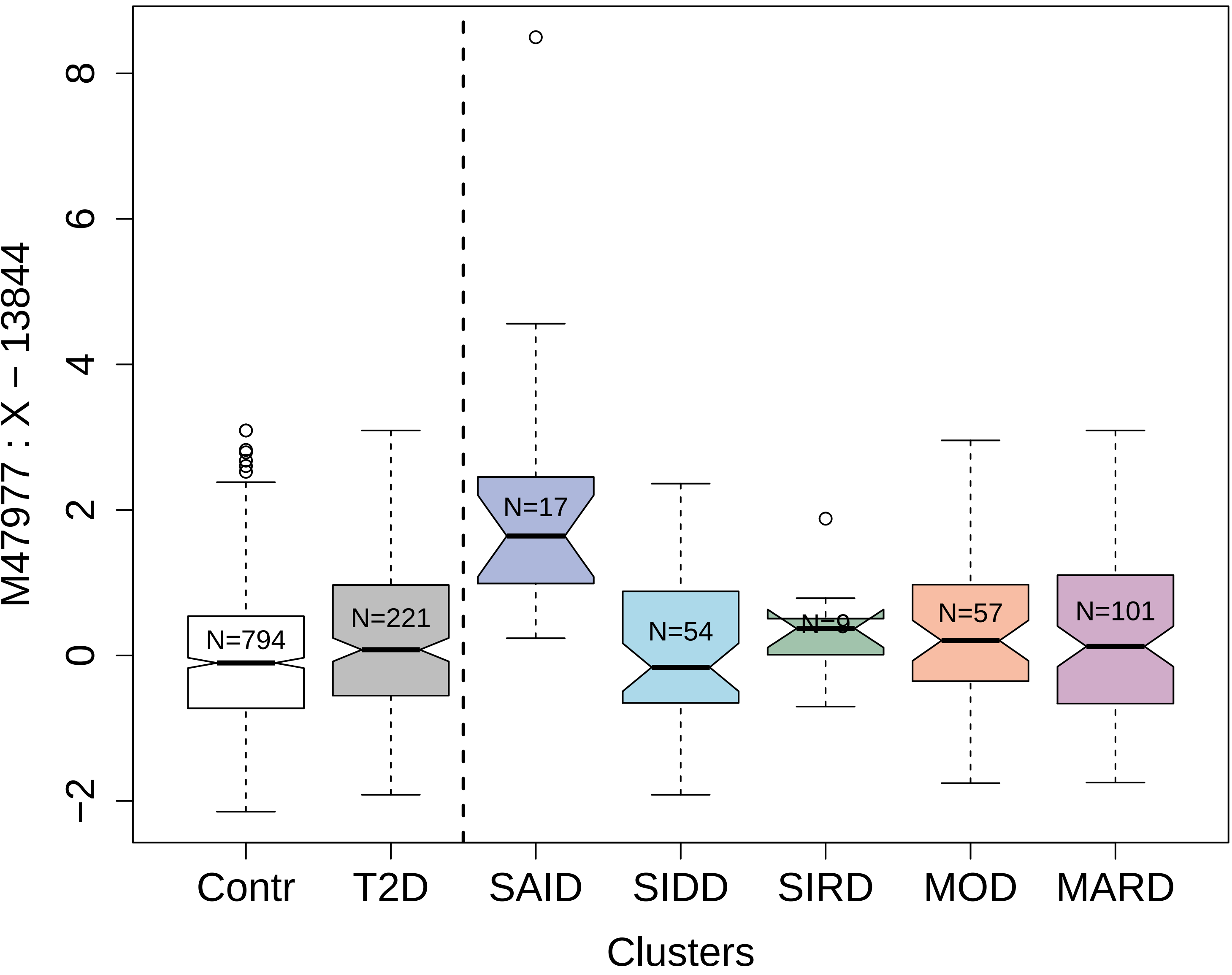

M47977 : X − 13844
−2
0
2
4
6
8
N=794
N=221
N=17
N=54
N=9
N=57
N=101
Contr
T2D
SAID
SIDD
SIRD
MOD
MARD
Clusters

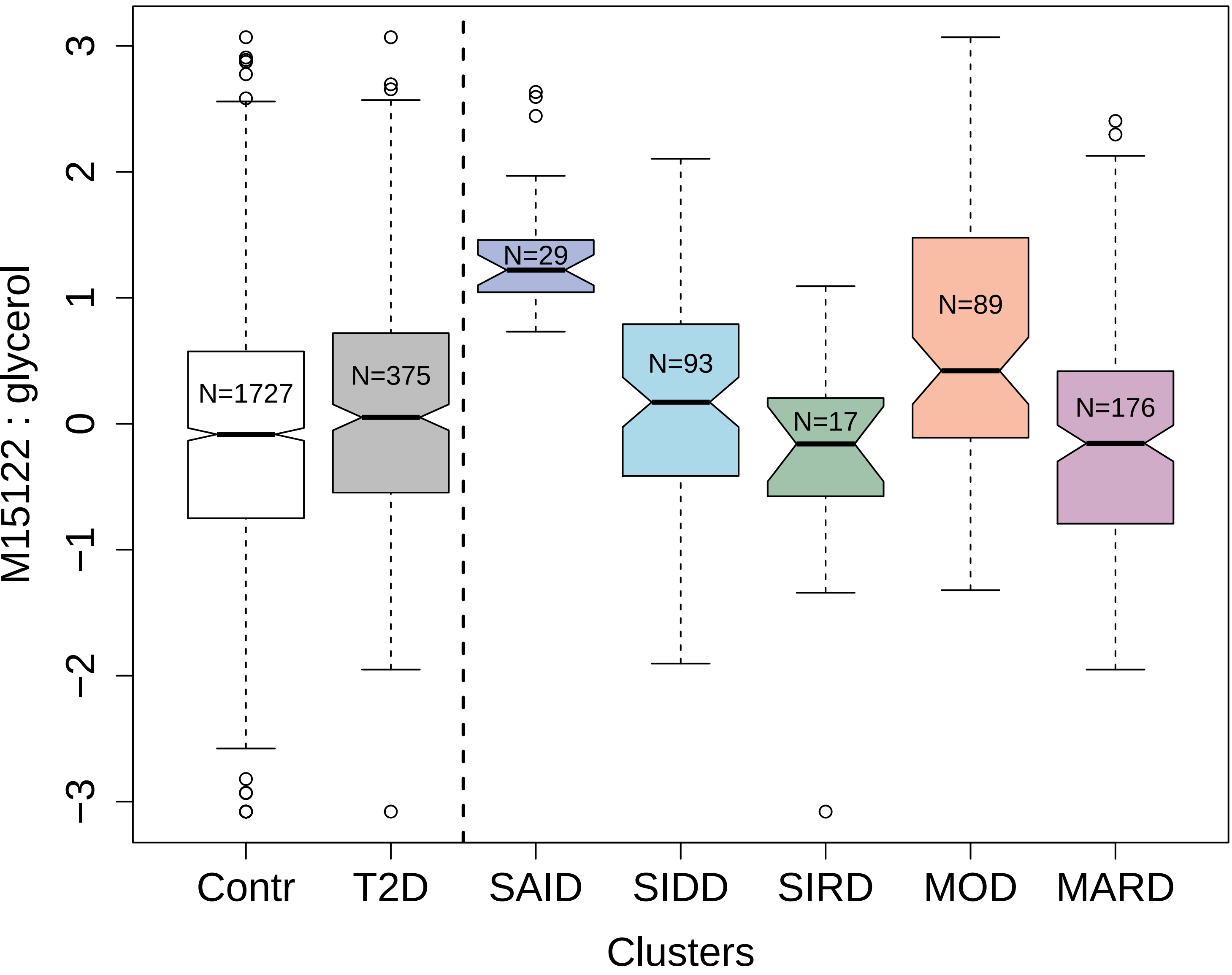

M15122 : glycerol
3
2
1
0
−1
−2
−3
N=1727
N=375
N=29
N=93
N=17
N=89
N=176
Contr
T2D
SAID
SIDD
SIRD
MOD
MARD
Clusters

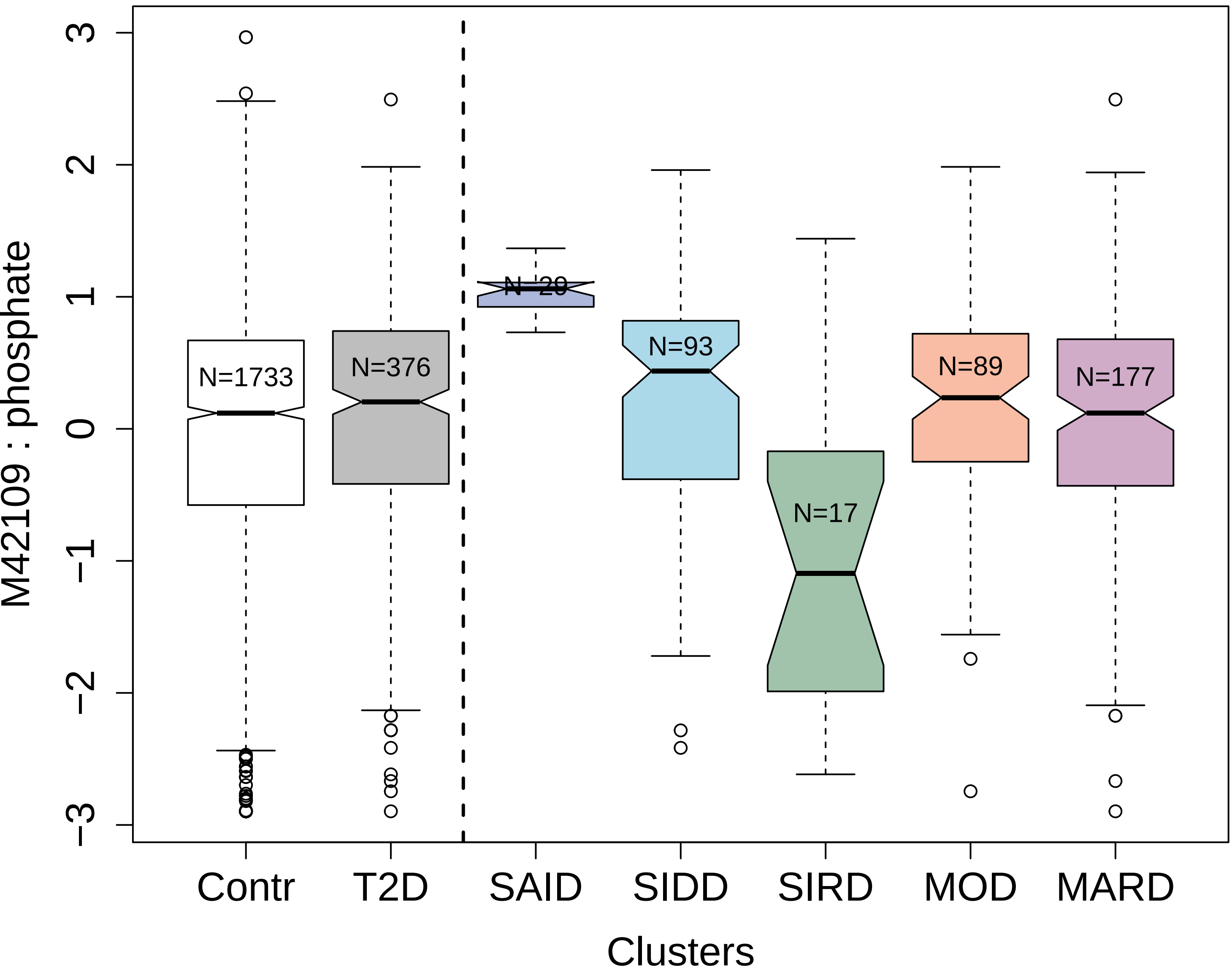

M42109 : phosphate
3
2
1
0
−1
−2
−3
N=1733
N=376
N=29
N=93
N=17
N=89
N=177
Contr
T2D
SAID
SIDD
SIRD
MOD
MARD
Clusters

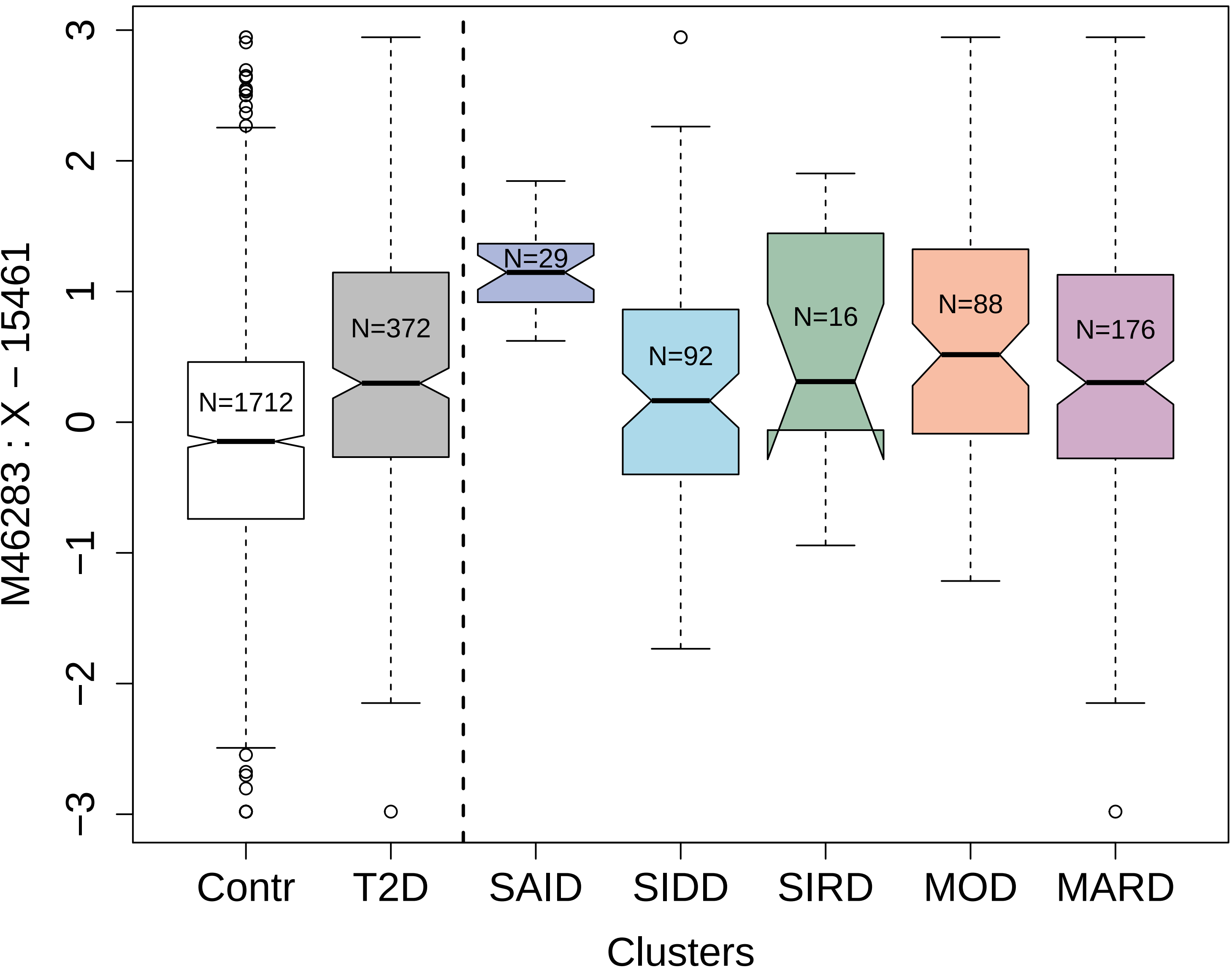

M46283 : X − 15461
Clusters
Contr
T2D
SAID
SIDD
SIRD
MOD
MARD
N=1712
N=372
N=29
N=92
N=16
N=88
N=176
3
2
1
0
−1
−2
−3

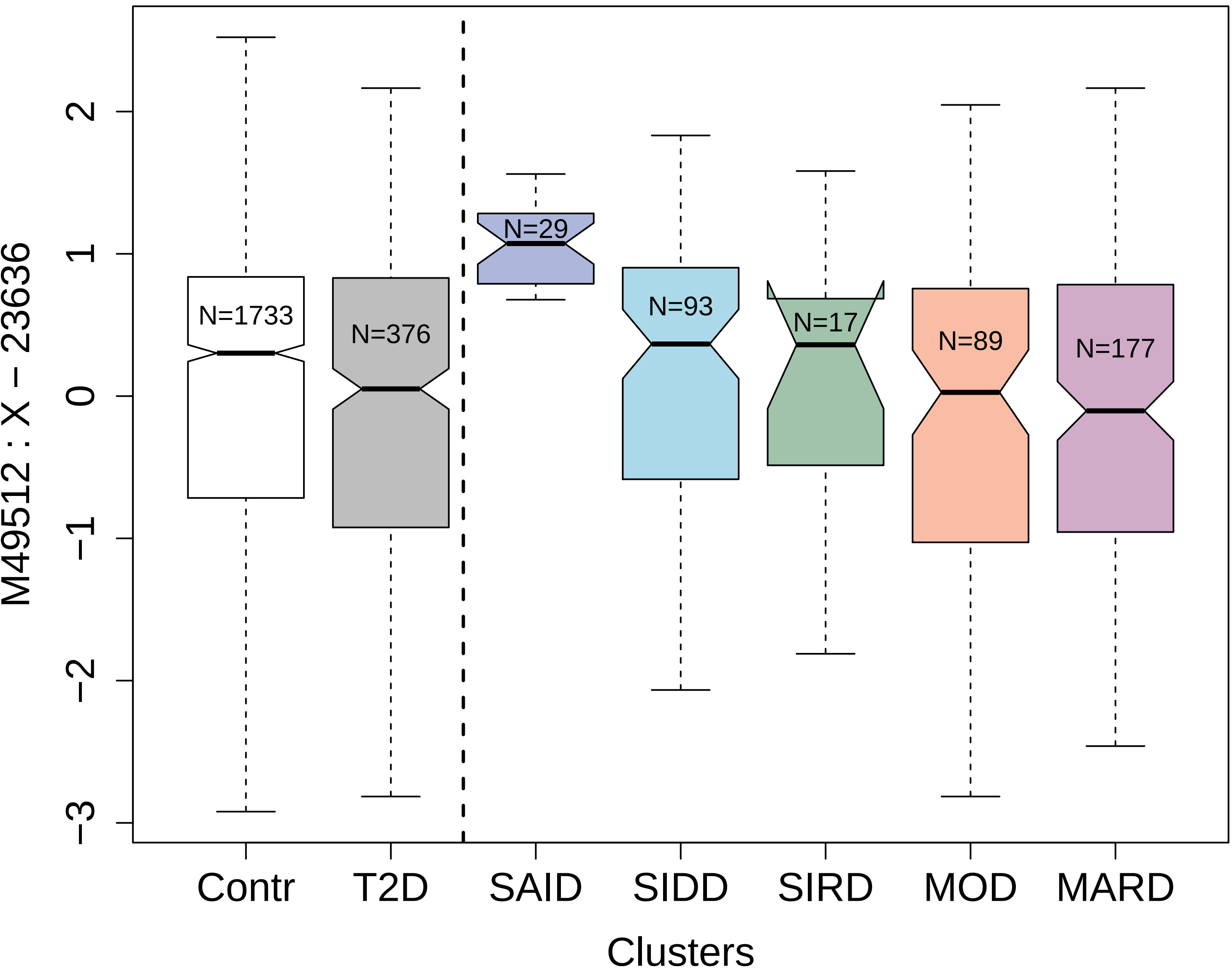

M49512 : X − 23636
2
1
0
−1
−2
−3
N=1733
N=376
N=29
N=93
N=17
N=89
N=177
Contr
T2D
SAID
SIDD
SIRD
MOD
MARD
Clusters

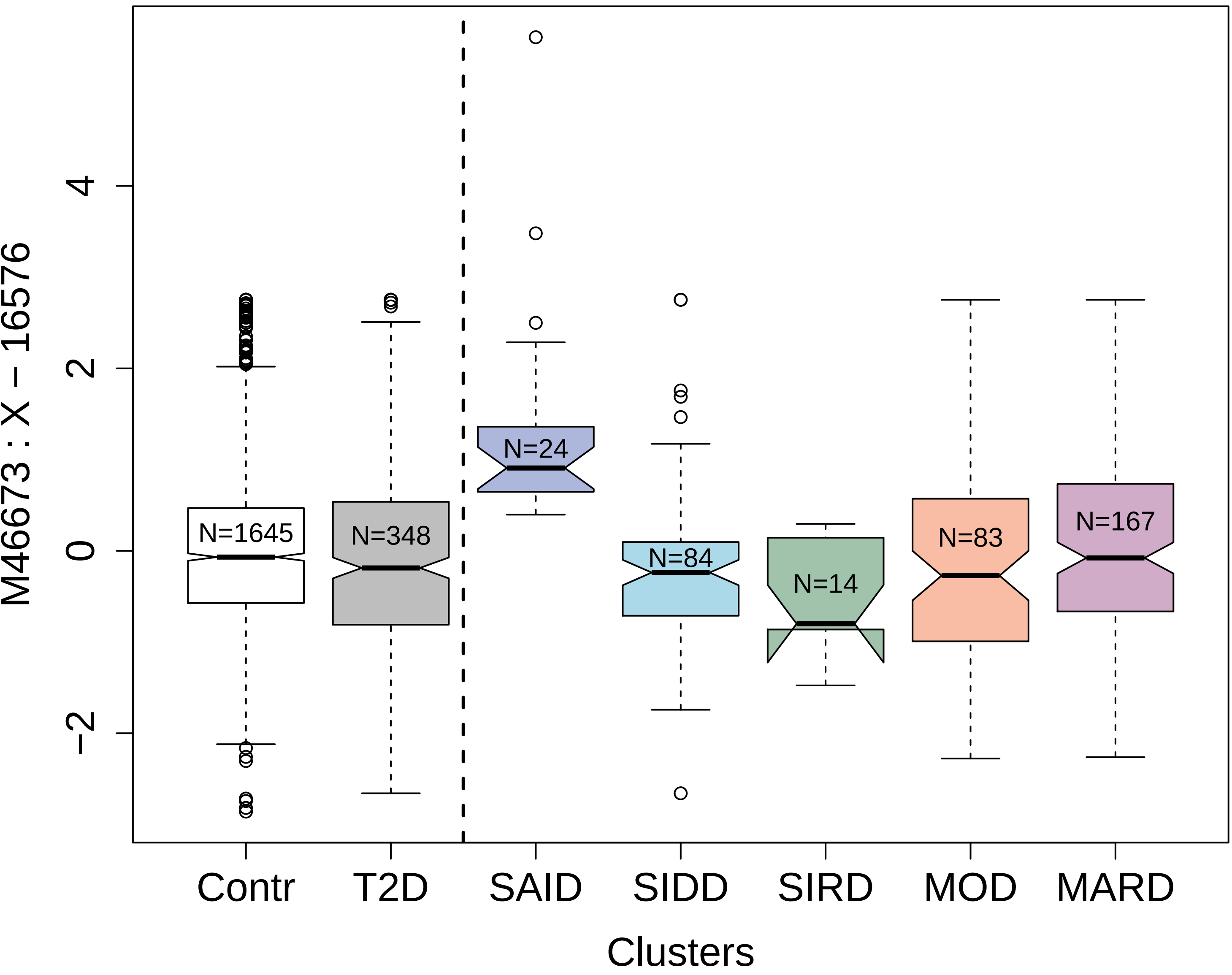

M46673 : X − 16576
4
2
0
−2
N=1645
N=348
N=24
N=84
N=14
N=83
N=167
Contr
T2D
SAID
SIDD
SIRD
MOD
MARD
Clusters

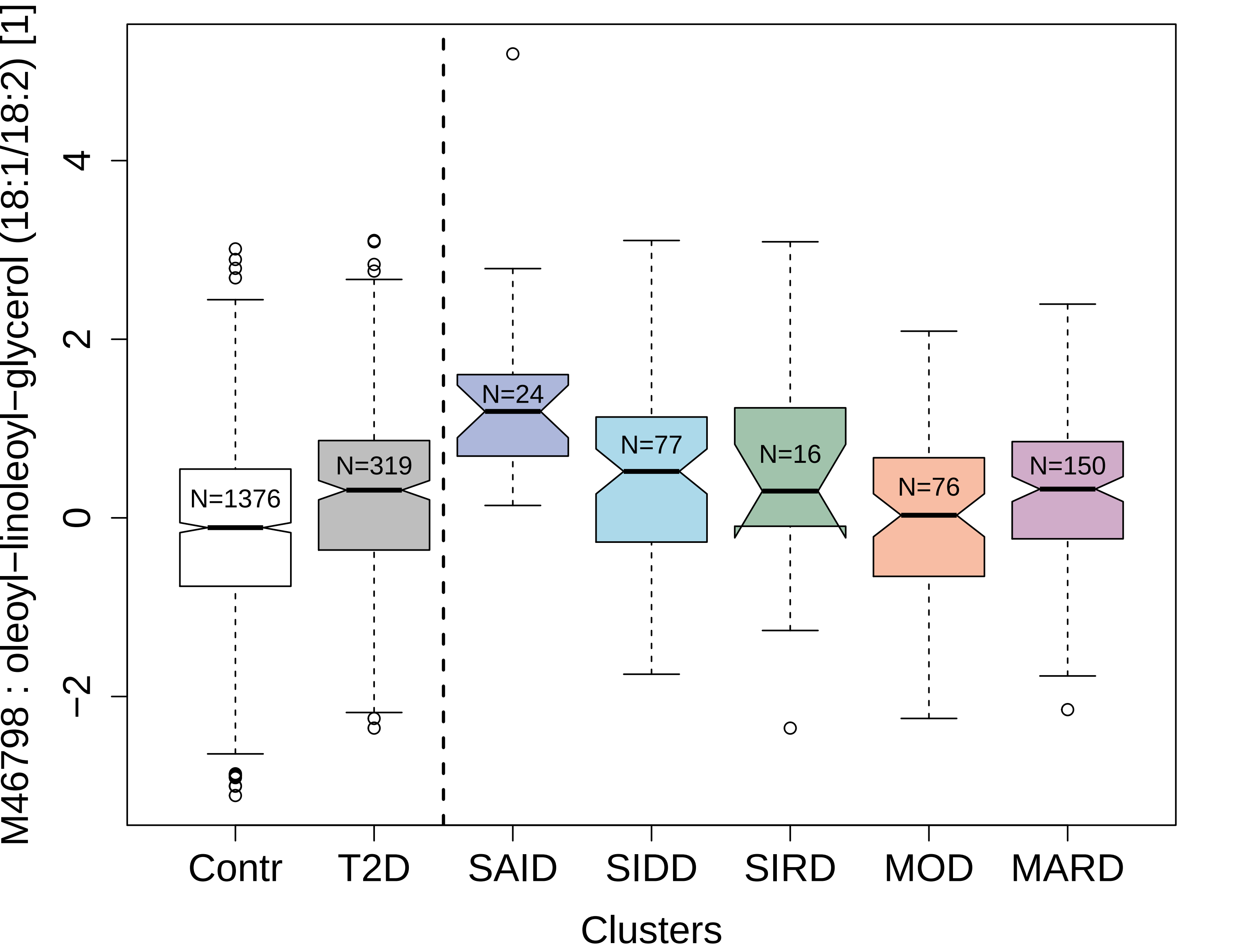
M46798 : oleoyl−linoleoyl−glycerol (18:1/18:2) [1]
4
2
0
−2
N=1376
N=319
N=24
N=77
N=16
N=76
N=150
Contr
T2D
SAID
SIDD
SIRD
MOD
MARD
Clusters

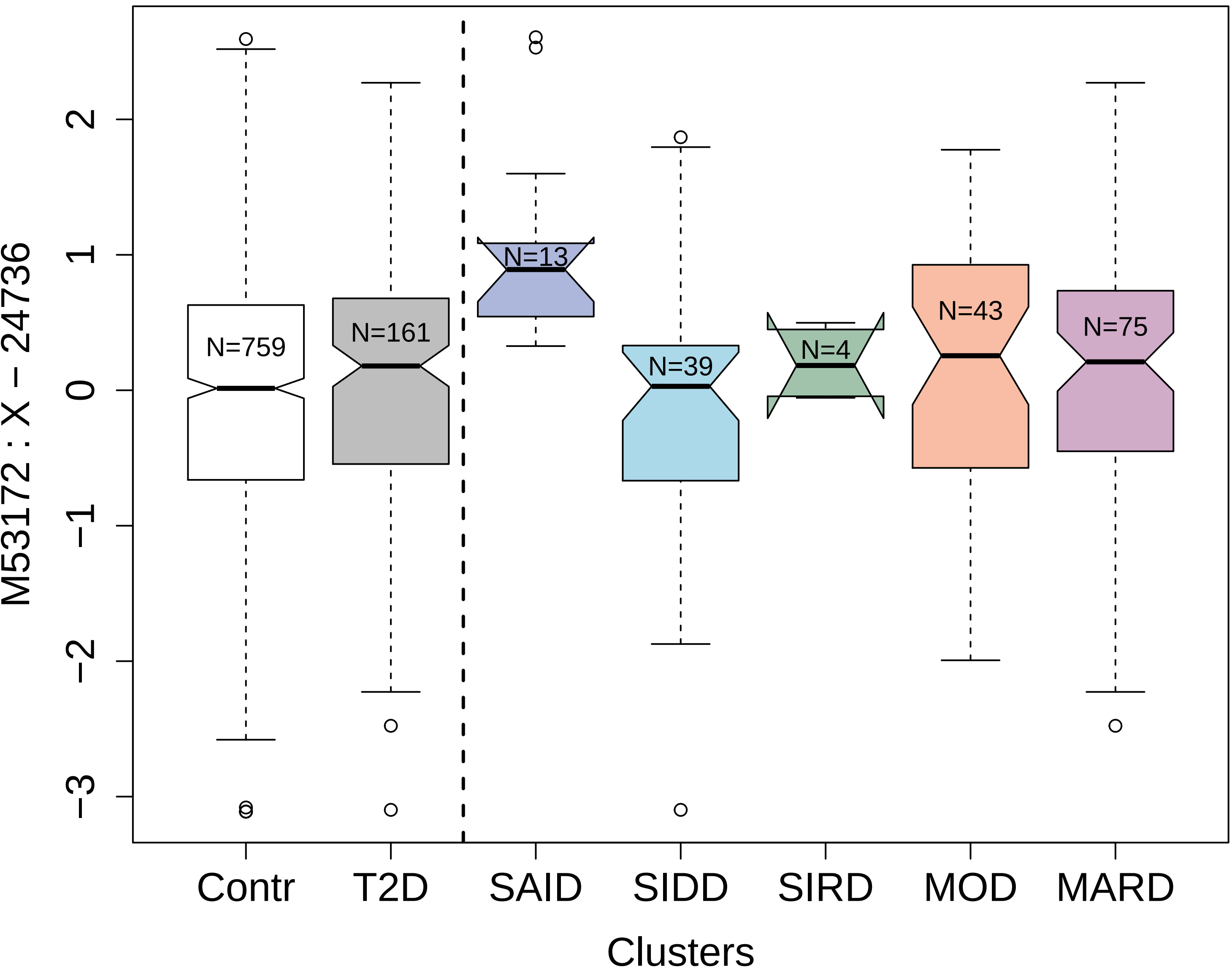

M53172 : X − 24736
Clusters
Contr
T2D
SAID
SIDD
SIRD
MOD
MARD
N=759
N=161
N=13
N=39
N=4
N=43
N=75
−3
−2
−1
0
1
2

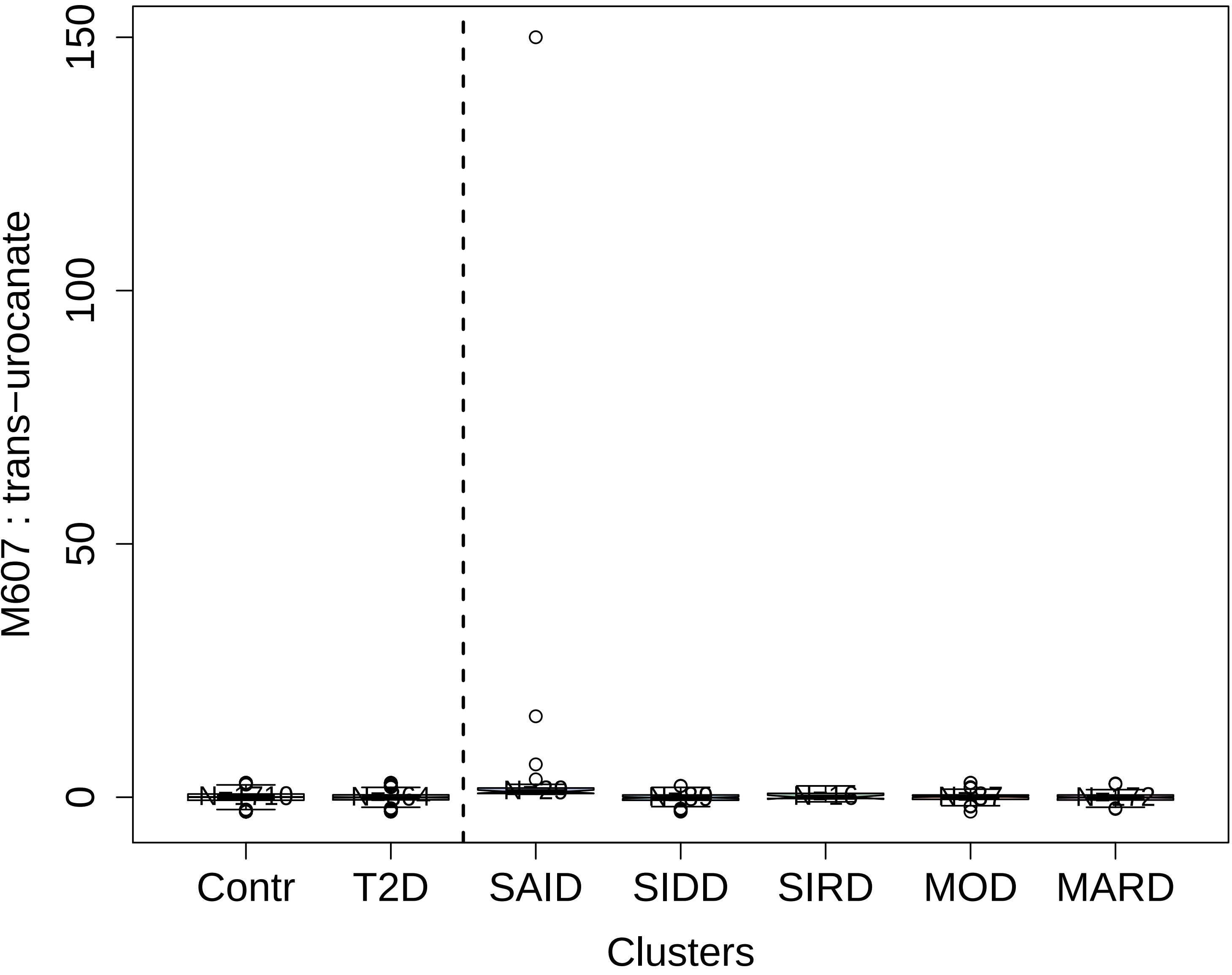

M607 : trans−urocanate
150
100
50
0
N=1710
N=364
N=20
N=80
N=16
N=87
N=172
Contr
T2D
SAID
SIDD
SIRD
MOD
MARD
Clusters

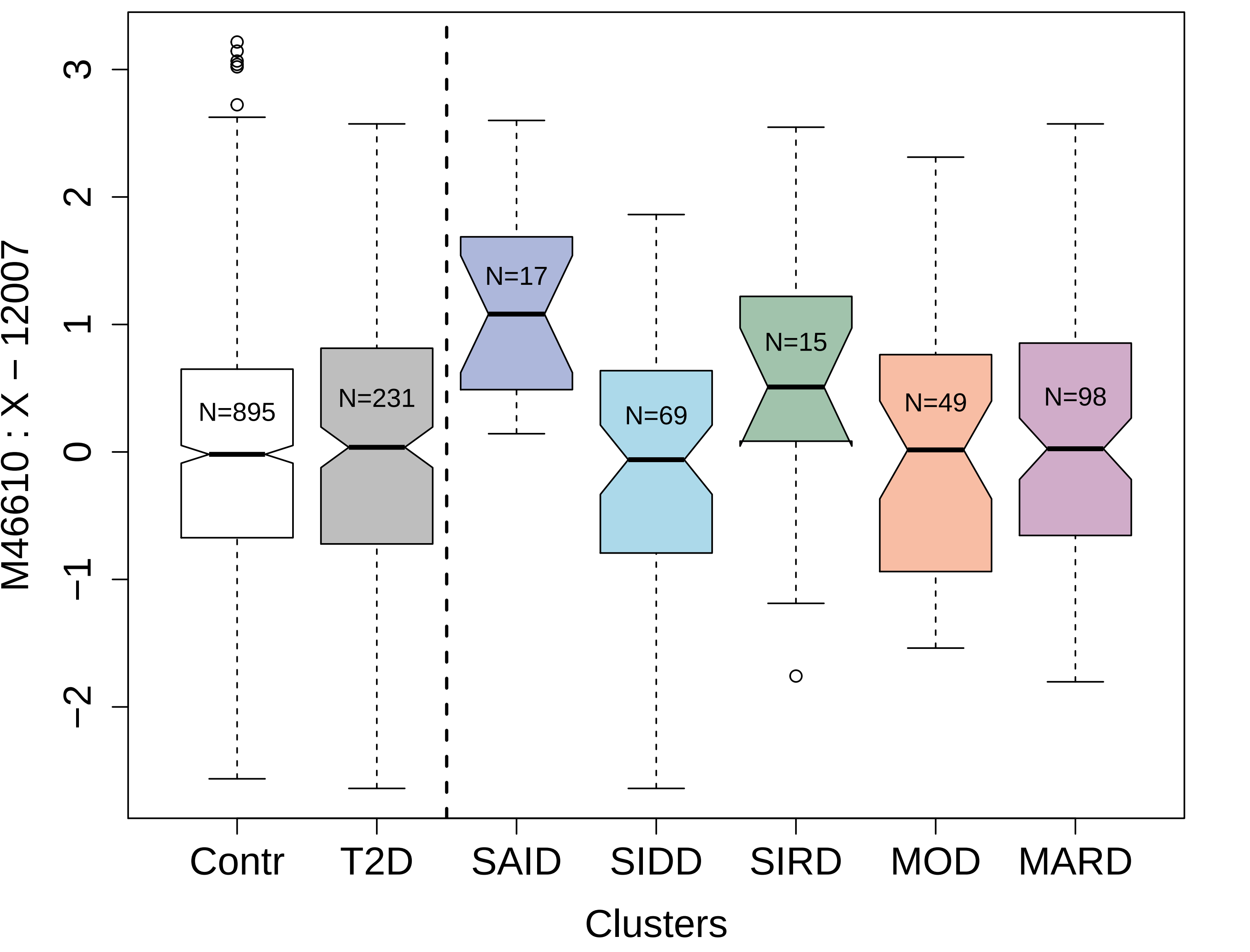

M46610 : X − 12007
3
2
1
0
−1
−2
N=895
N=231
N=17
N=69
N=15
N=49
N=98
Contr
T2D
SAID
SIDD
SIRD
MOD
MARD
Clusters

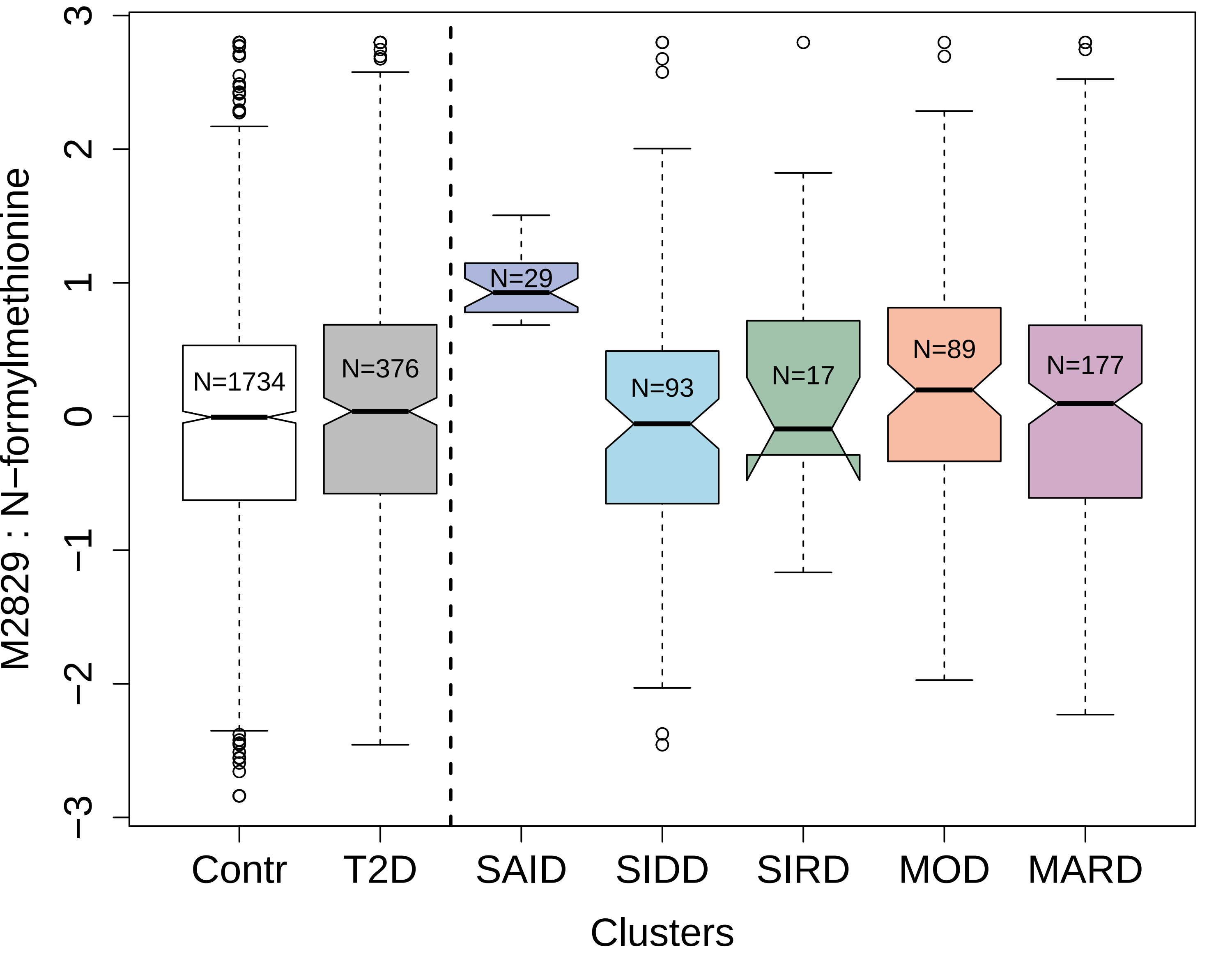

M2829 : N−formylmethionine
3
2
1
0
−1
−2
−3
N=1734
N=376
N=29
N=93
N=17
N=89
N=177
Contr
T2D
SAID
SIDD
SIRD
MOD
MARD
Clusters

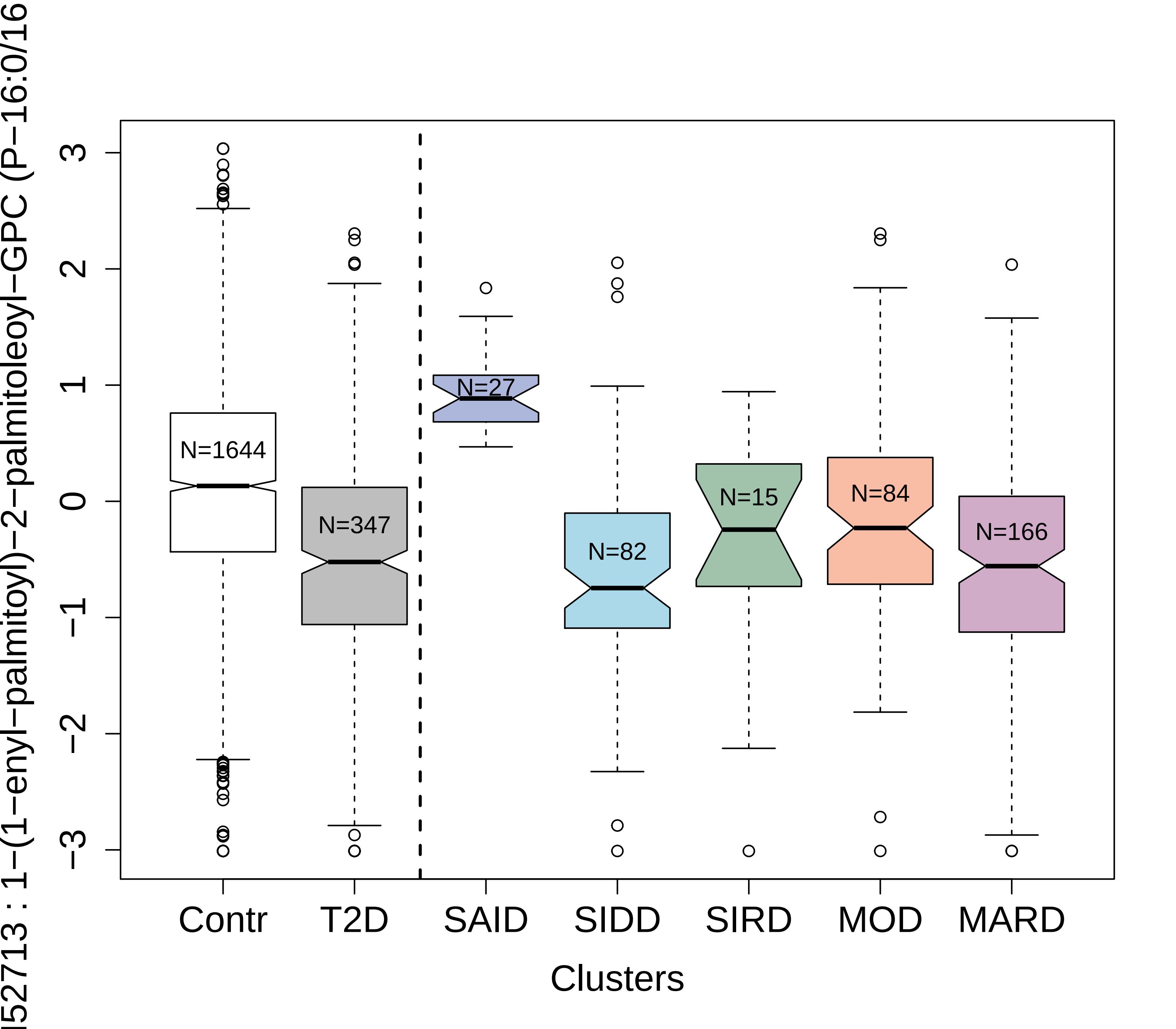

152713 : 1−(1−enyl−palmitoyl)−2−palmitoleoyl−GPC (P−16:0/16
3
2
1
0
−1
−2
−3
N=1644
N=347
N=27
N=82
N=15
N=84
N=166
Contr
T2D
SAID
SIDD
SIRD
MOD
MARD
Clusters

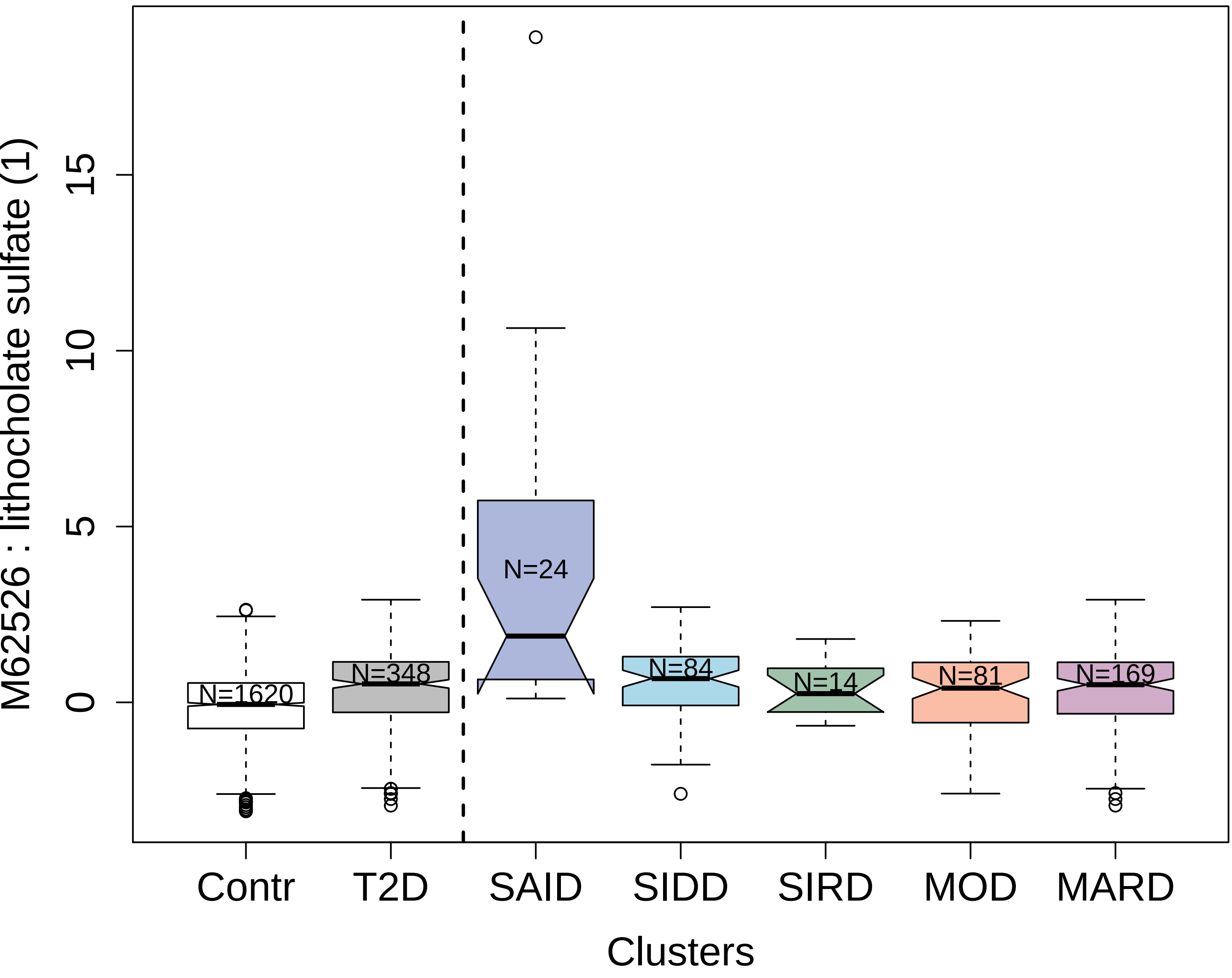

M62526 : lithocholate sulfate (1)
0
5
10
15
N=1620
N=348
N=24
N=84
N=14
N=81
N=169
Contr
T2D
SAID
SIDD
SIRD
MOD
MARD
Clusters

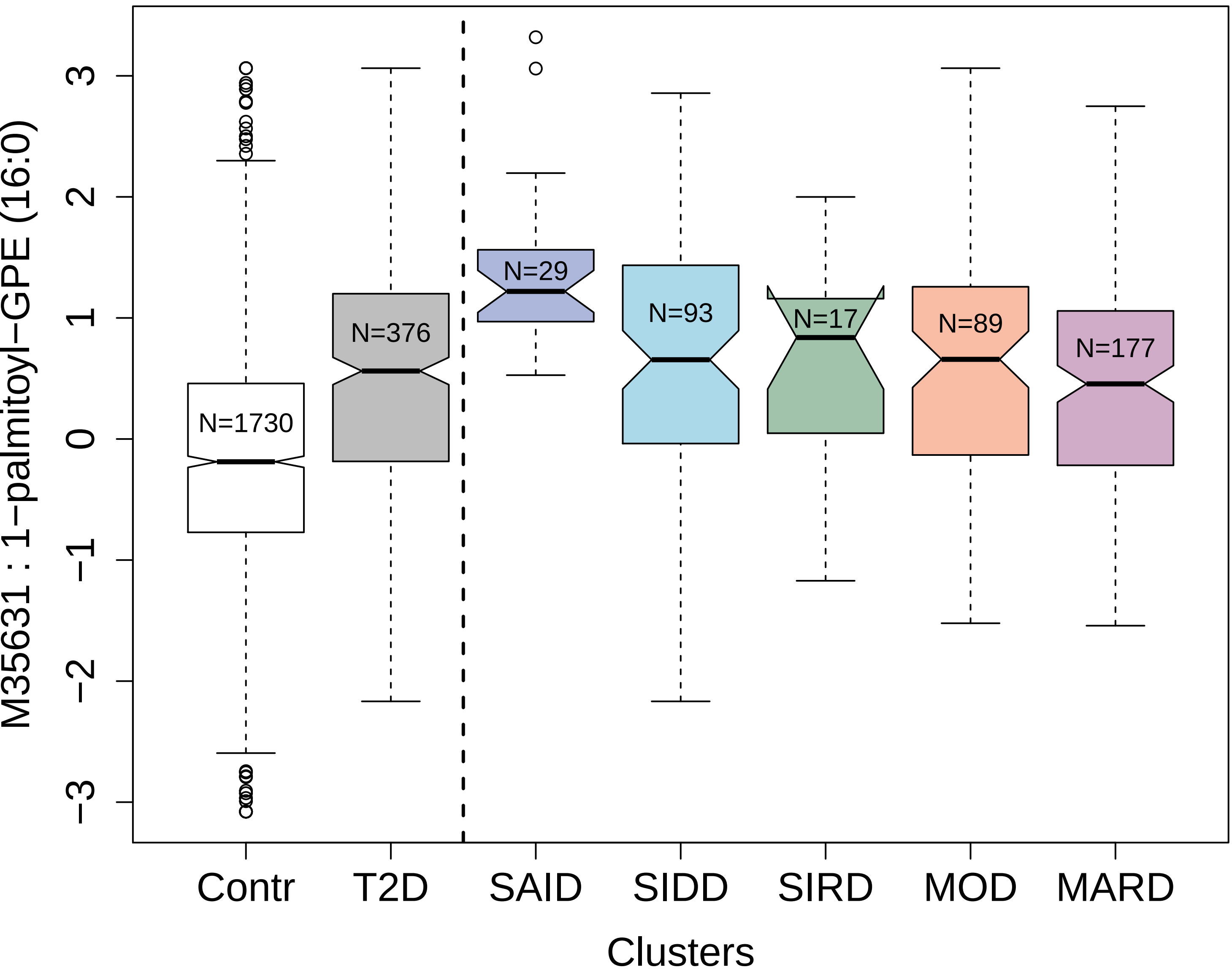
M35631 : 1−palmitoyl−GPE (16:0)
3
2
1
0
−1
−2
−3
N=1730
N=376
N=29
N=93
N=17
N=89
N=177
Contr
T2D
SAID
SIDD
SIRD
MOD
MARD
Clusters

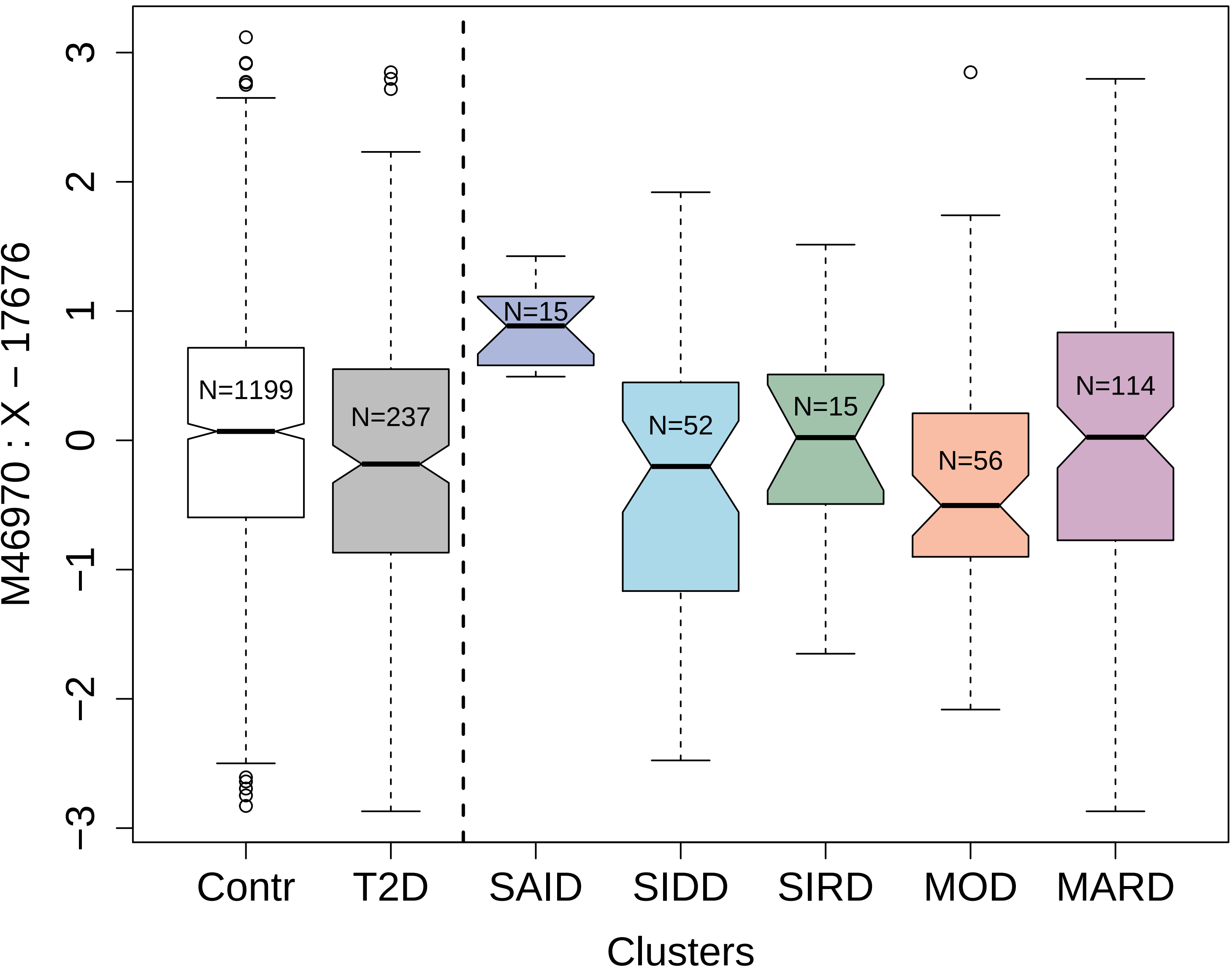

M46970 : X − 17676
3
2
1
0
−1
−2
−3
N=1199
N=237
N=15
N=52
N=15
N=56
N=114
Contr
T2D
SAID
SIDD
SIRD
MOD
MARD
Clusters

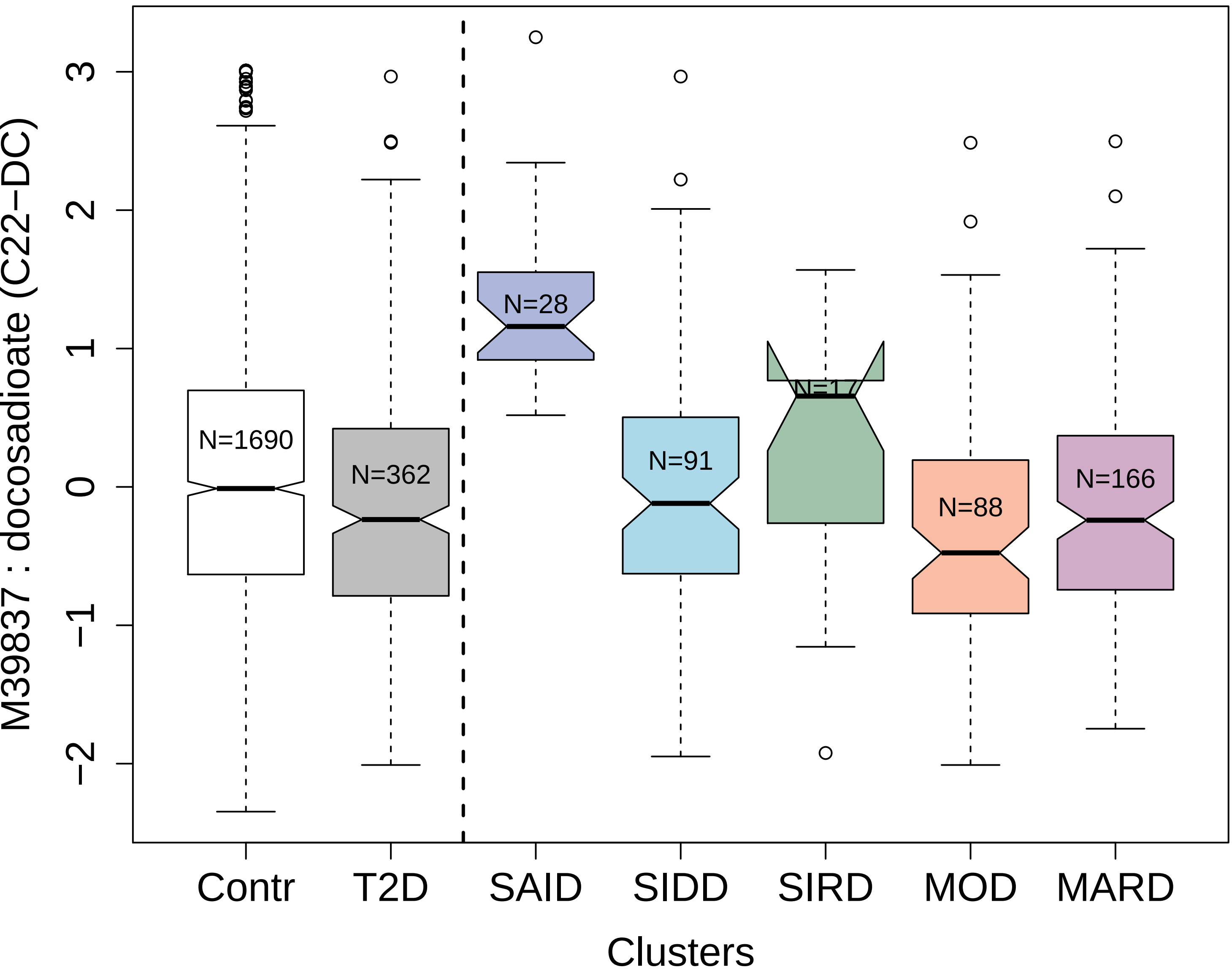

M39837 : docosadioate (C22−DC)
3
2
1
0
−1
−2
N=1690
N=362
N=28
N=91
N=17
N=88
N=166
Contr
T2D
SAID
SIDD
SIRD
MOD
MARD
Clusters

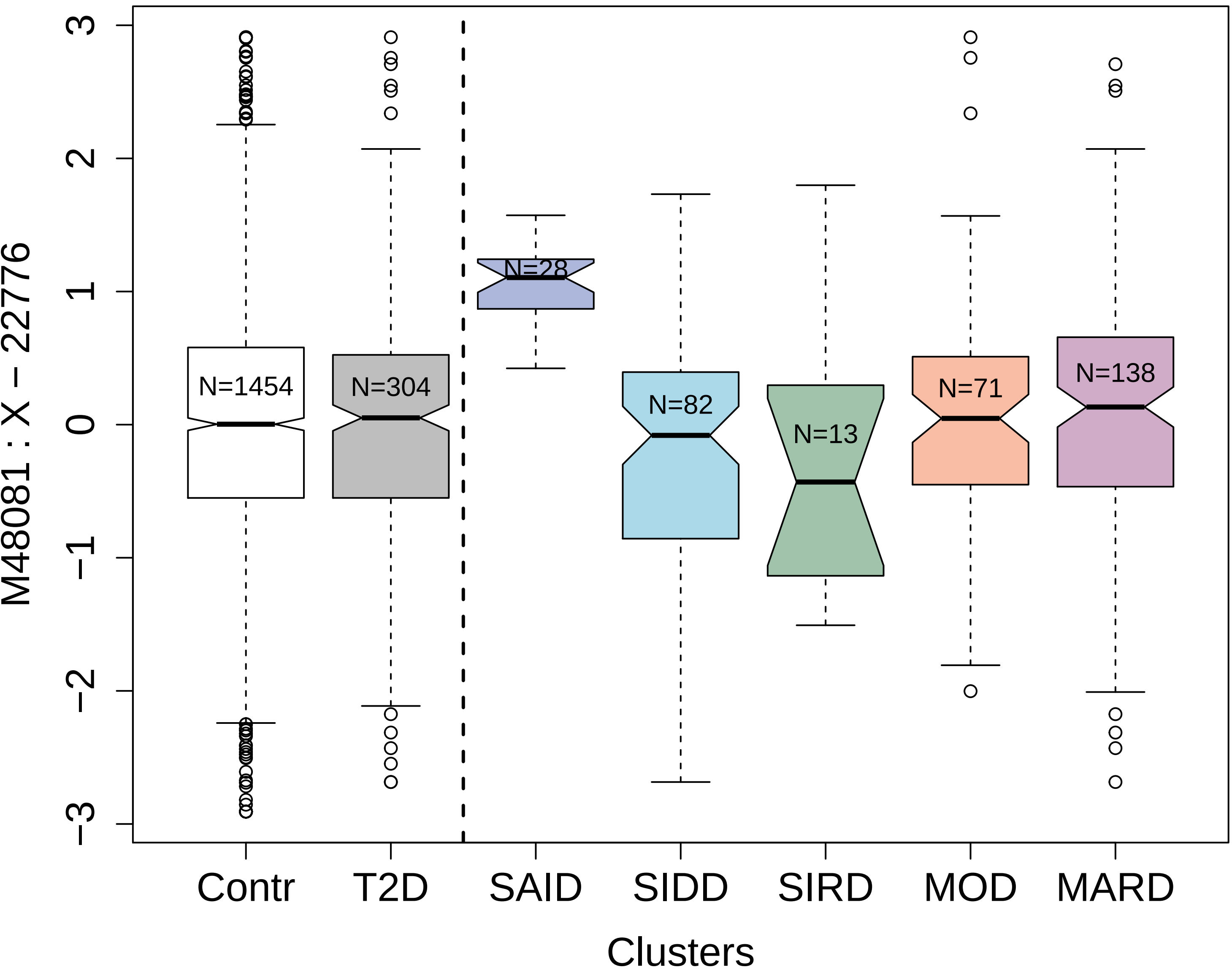

M48081 : X − 22776
3
2
1
0
−1
−2
−3
N=1454
N=304
N=28
N=82
N=13
N=71
N=138
Contr
T2D
SAID
SIDD
SIRD
MOD
MARD
Clusters

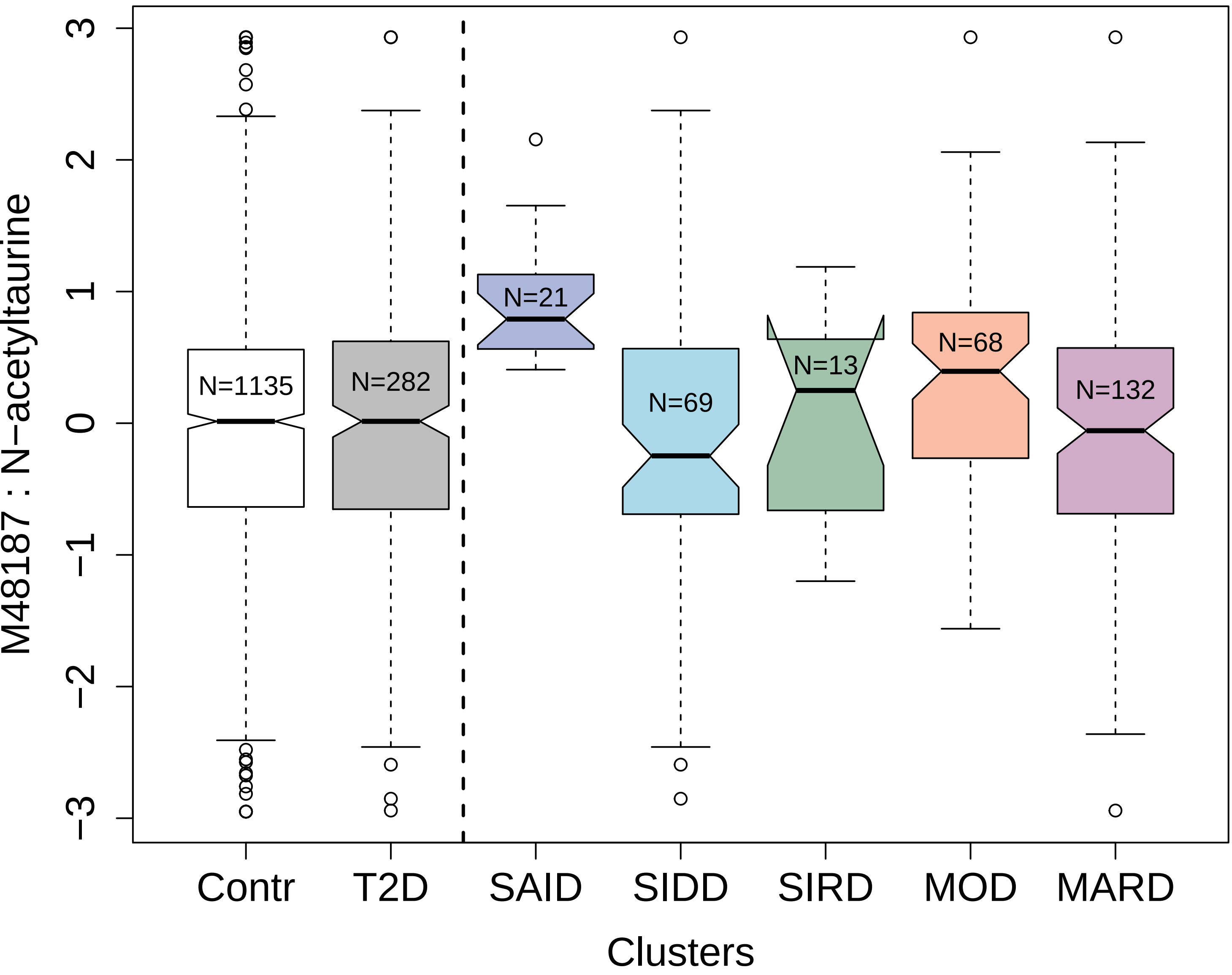

M48187 : N−acetyltaurine
3
2
1
0
−1
−2
−3
N=1135
N=282
N=21
N=69
N=13
N=68
N=132
Contr
T2D
SAID
SIDD
SIRD
MOD
MARD
Clusters

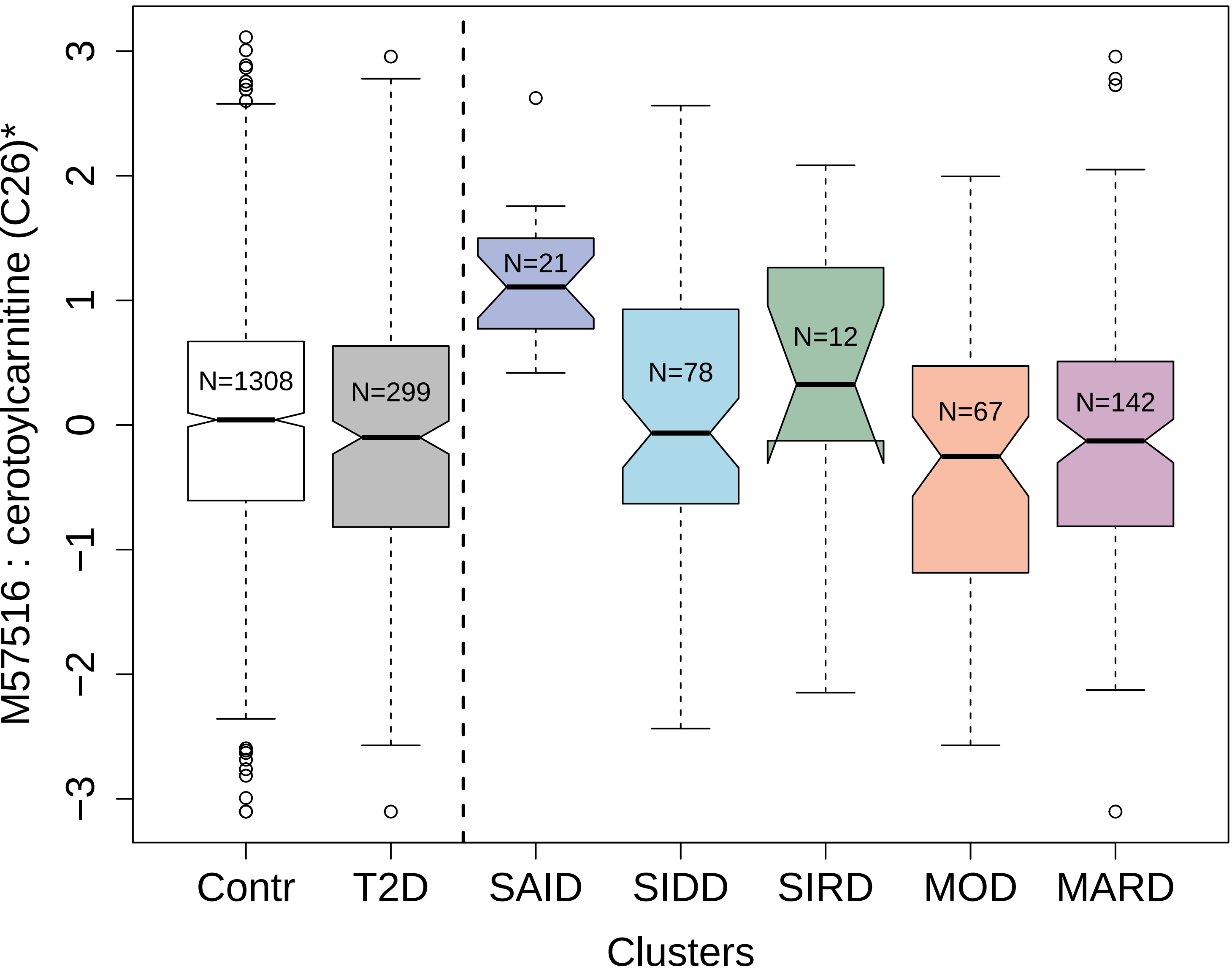

M57516 : cerotoylcarnitine (C26)*
3
2
1
0
−1
−2
−3
N=1308
N=299
N=21
N=78
N=12
N=67
N=142
Contr
T2D
SAID
SIDD
SIRD
MOD
MARD
Clusters

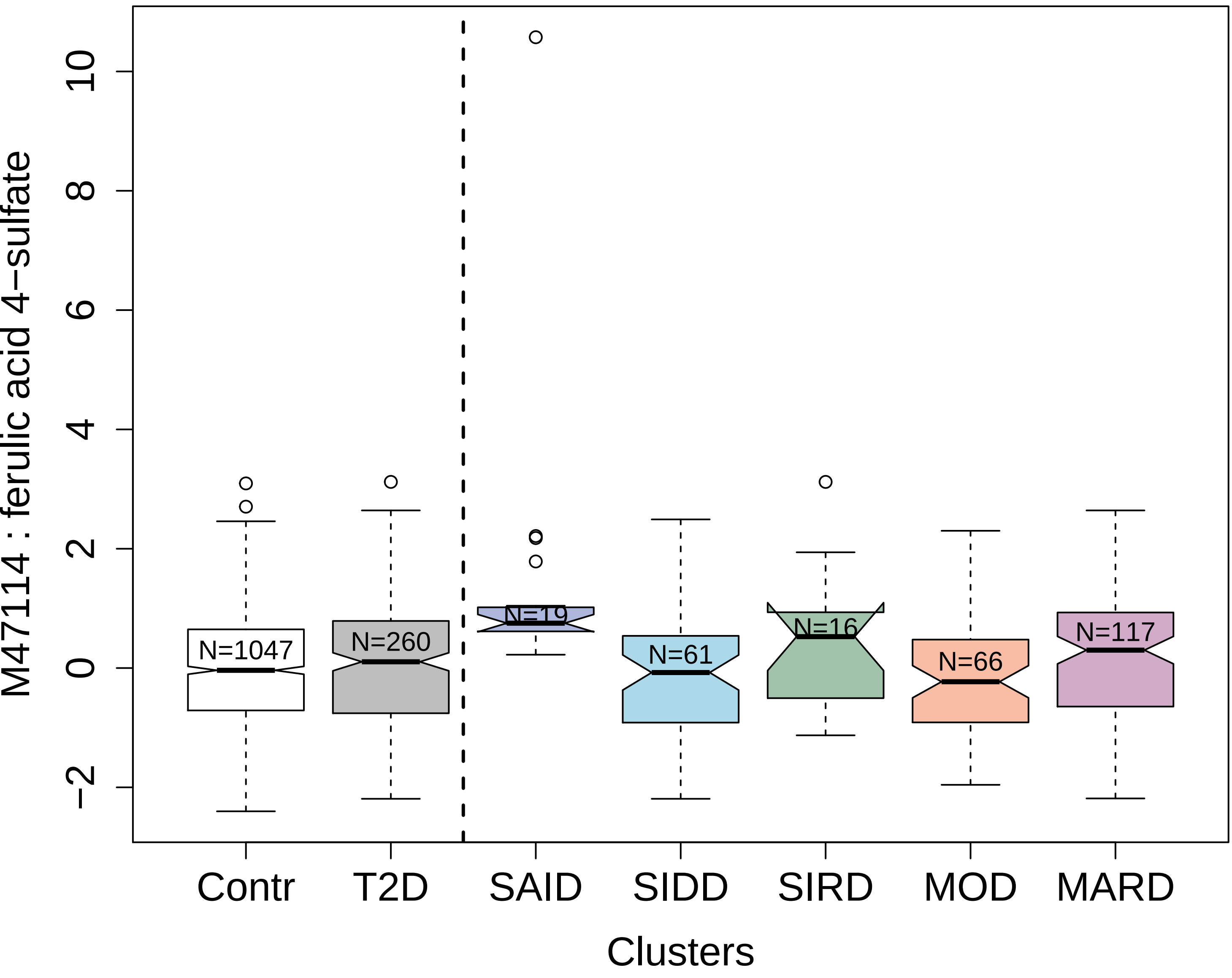

M47114 : ferulic acid 4−sulfate
−2
0
2
4
6
8
10
N=1047
N=260
N=19
N=61
N=16
N=66
N=117
Contr
T2D
SAID
SIDD
SIRD
MOD
MARD
Clusters

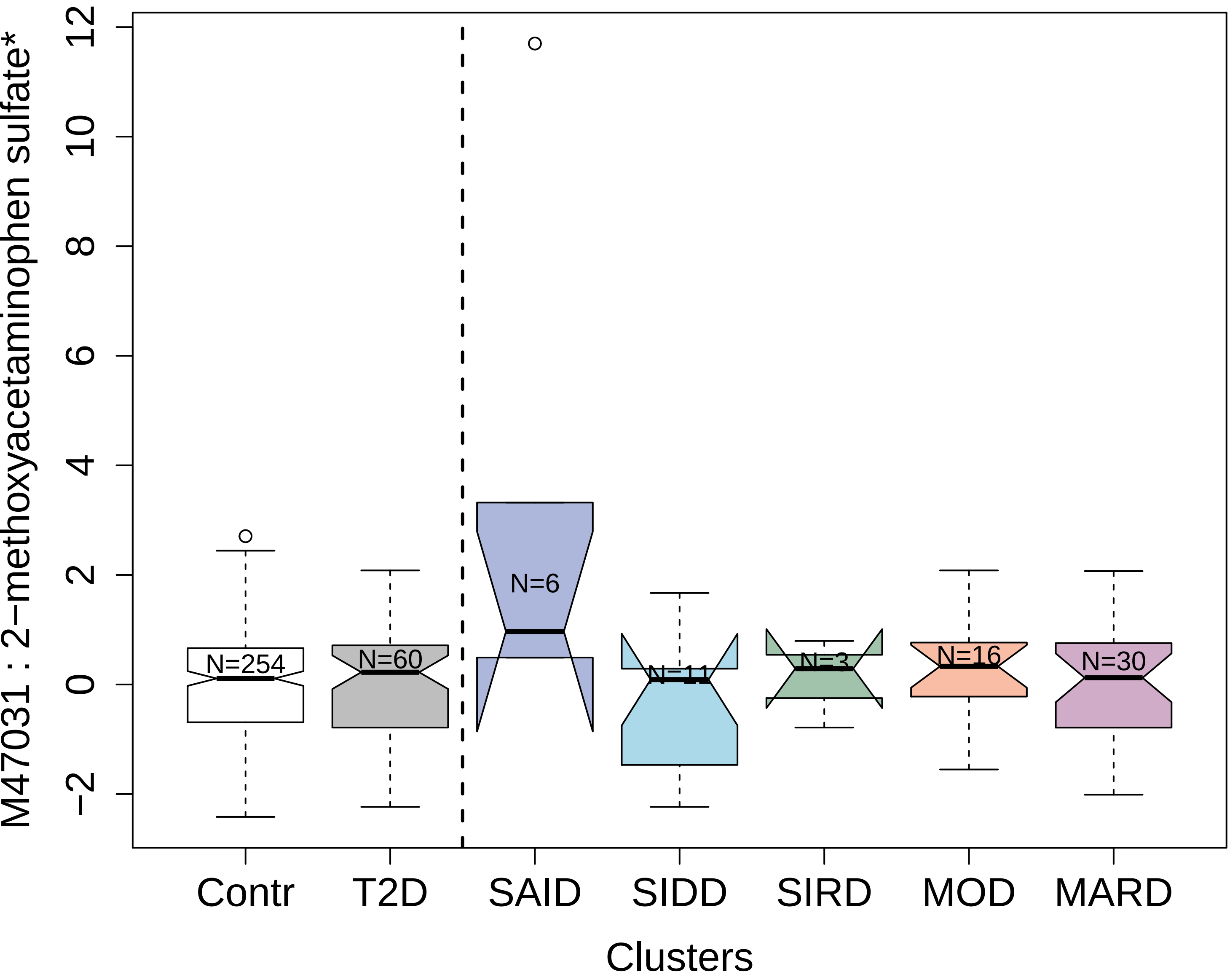

M47031 : 2−methoxyacetaminophen sulfate*
12
10
8
6
4
2
0
−2
N=254
N=60
N=6
N=11
N=3
N=16
N=30
Contr
T2D
SAID
SIDD
SIRD
MOD
MARD
Clusters

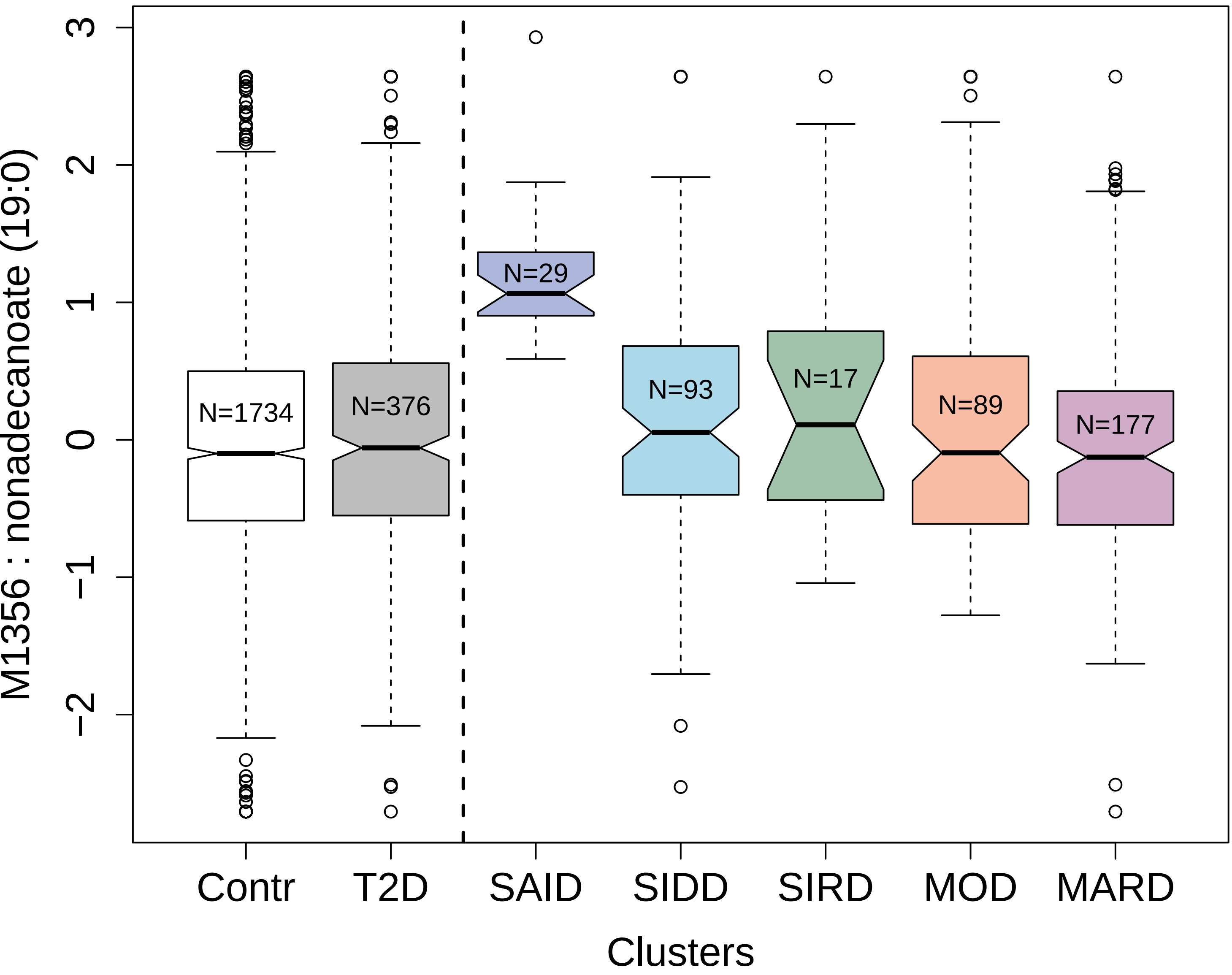
M1356 : nonadecanoate (19:0)
3
2
1
0
−1
−2
N=1734
N=376
N=29
N=93
N=17
N=89
N=177
Contr
T2D
SAID
SIDD
SIRD
MOD
MARD
Clusters

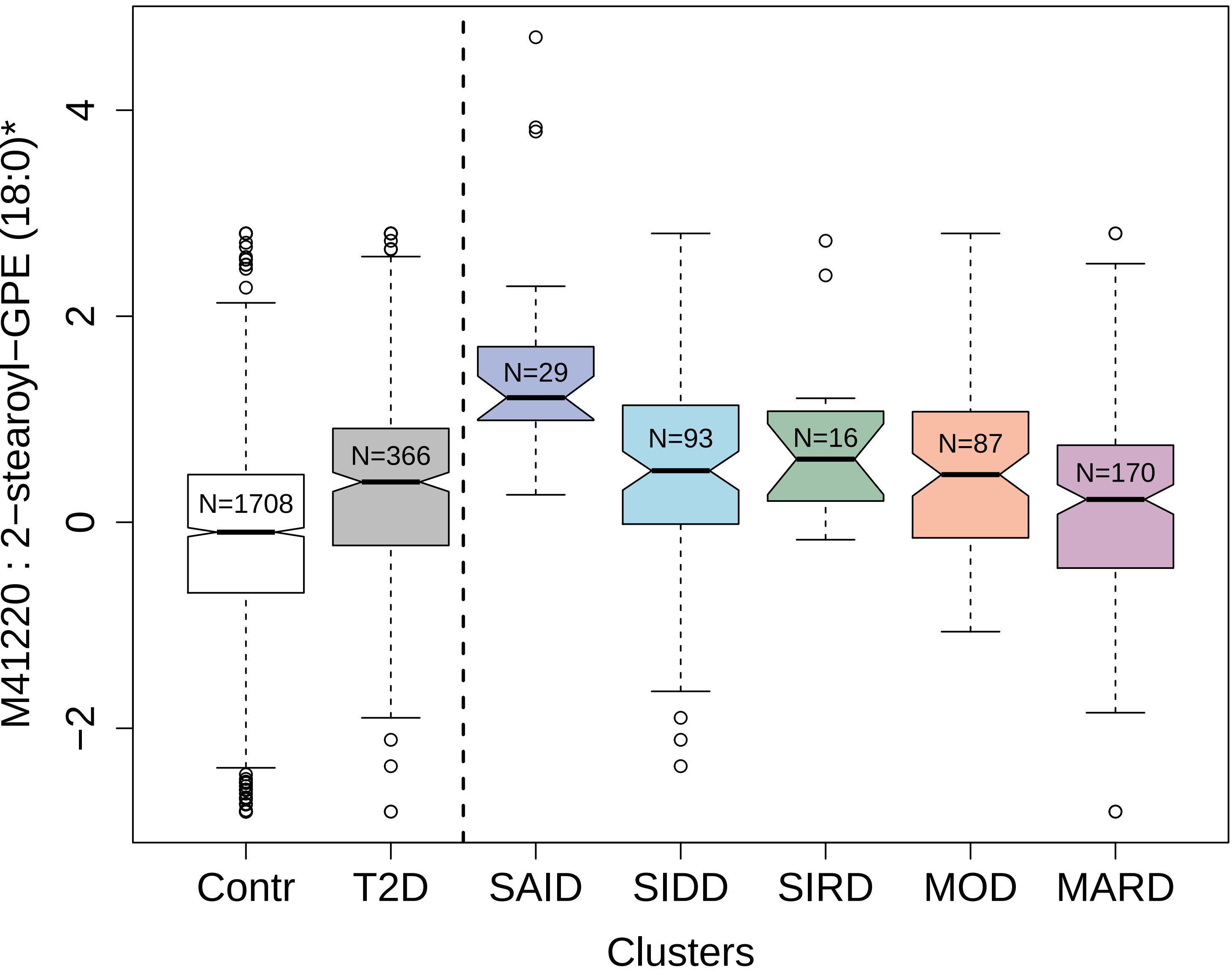

M41220 : 2−stearoyl−GPE (18:0)*
4
2
0
−2
N=1708
N=366
N=29
N=93
N=16
N=87
N=170
Contr
T2D
SAID
SIDD
SIRD
MOD
MARD
Clusters

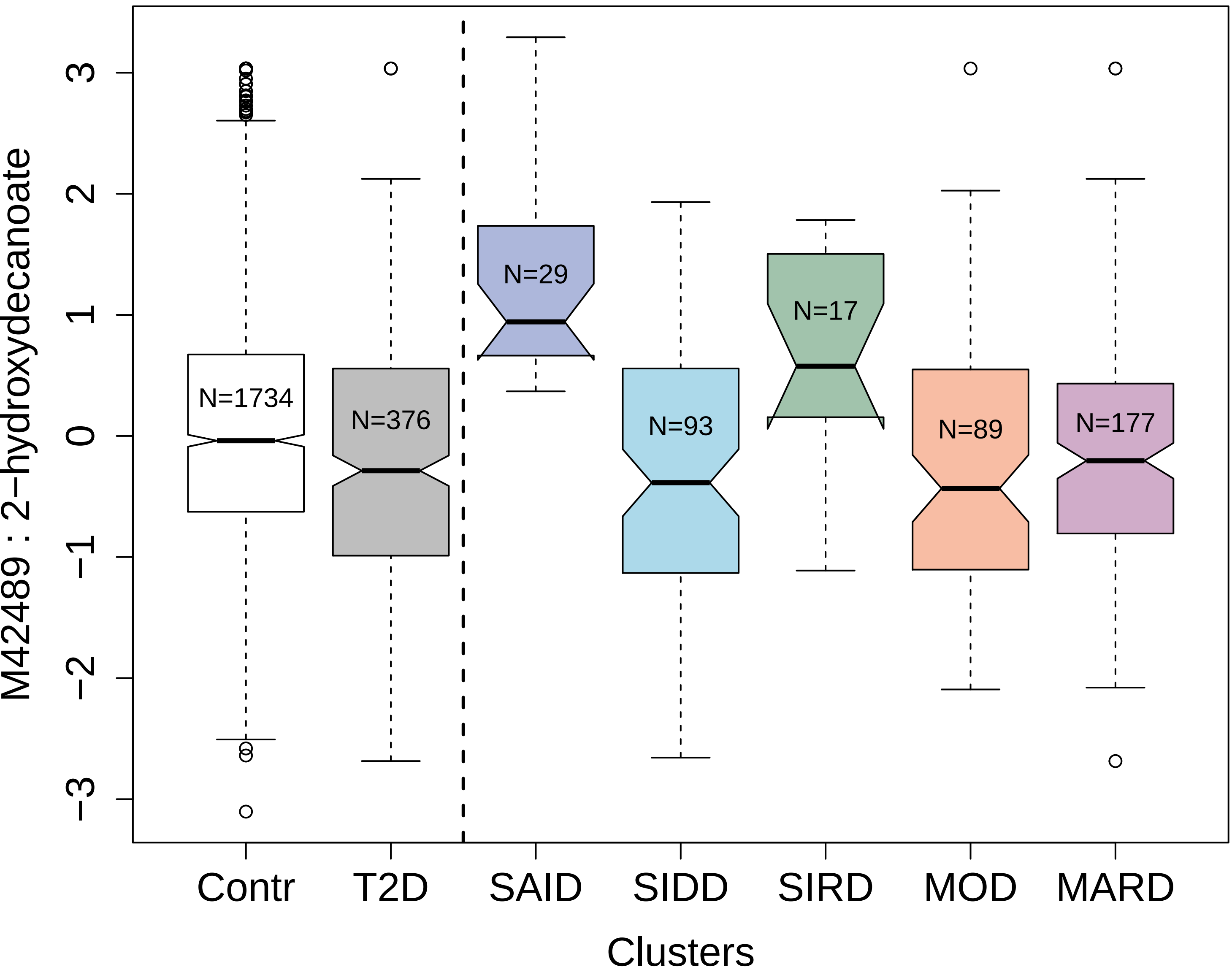

M42489 : 2−hydroxydecanoate
3
2
1
0
−1
−2
−3
N=1734
N=376
N=29
N=93
N=17
N=89
N=177
Contr
T2D
SAID
SIDD
SIRD
MOD
MARD
Clusters

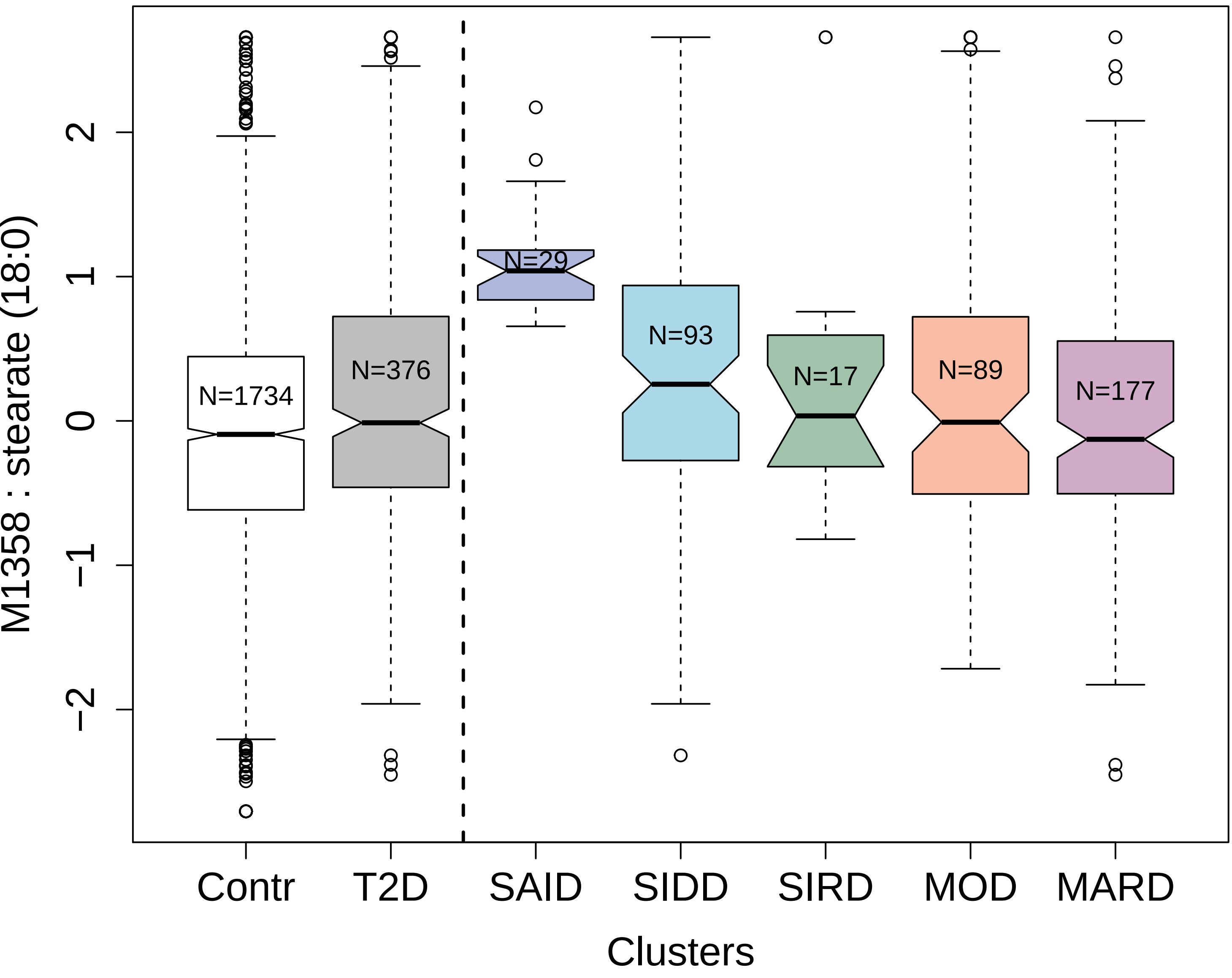

M1358 : stearate (18:0)
2
1
0
−1
−2
N=1734
N=376
N=29
N=93
N=17
N=89
N=177
Contr
T2D
SAID
SIDD
SIRD
MOD
MARD
Clusters

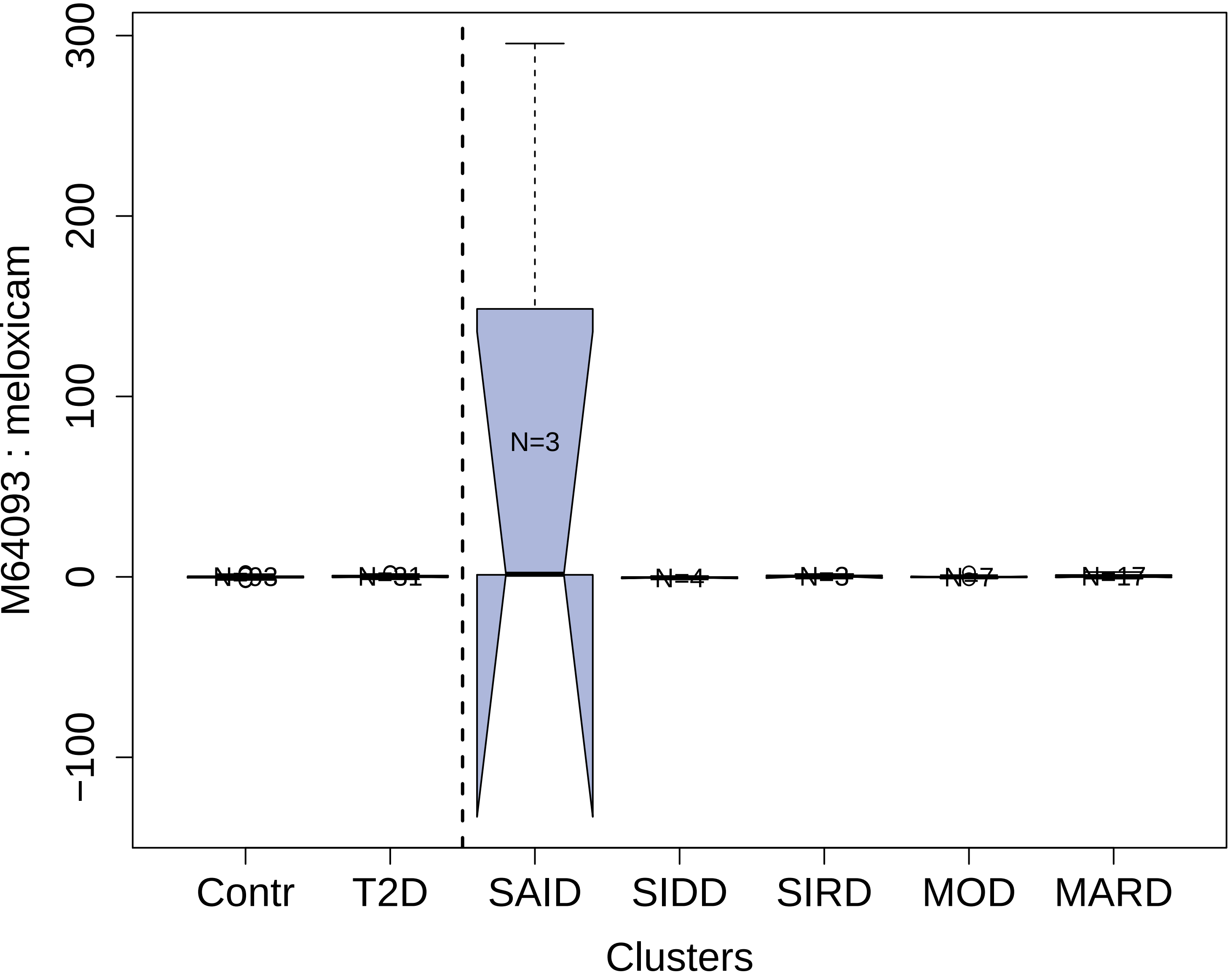

M64093 : meloxicam
300
200
100
0
−100
N=3
Contr
T2D
SAID
SIDD
SIRD
MOD
MARD
Clusters

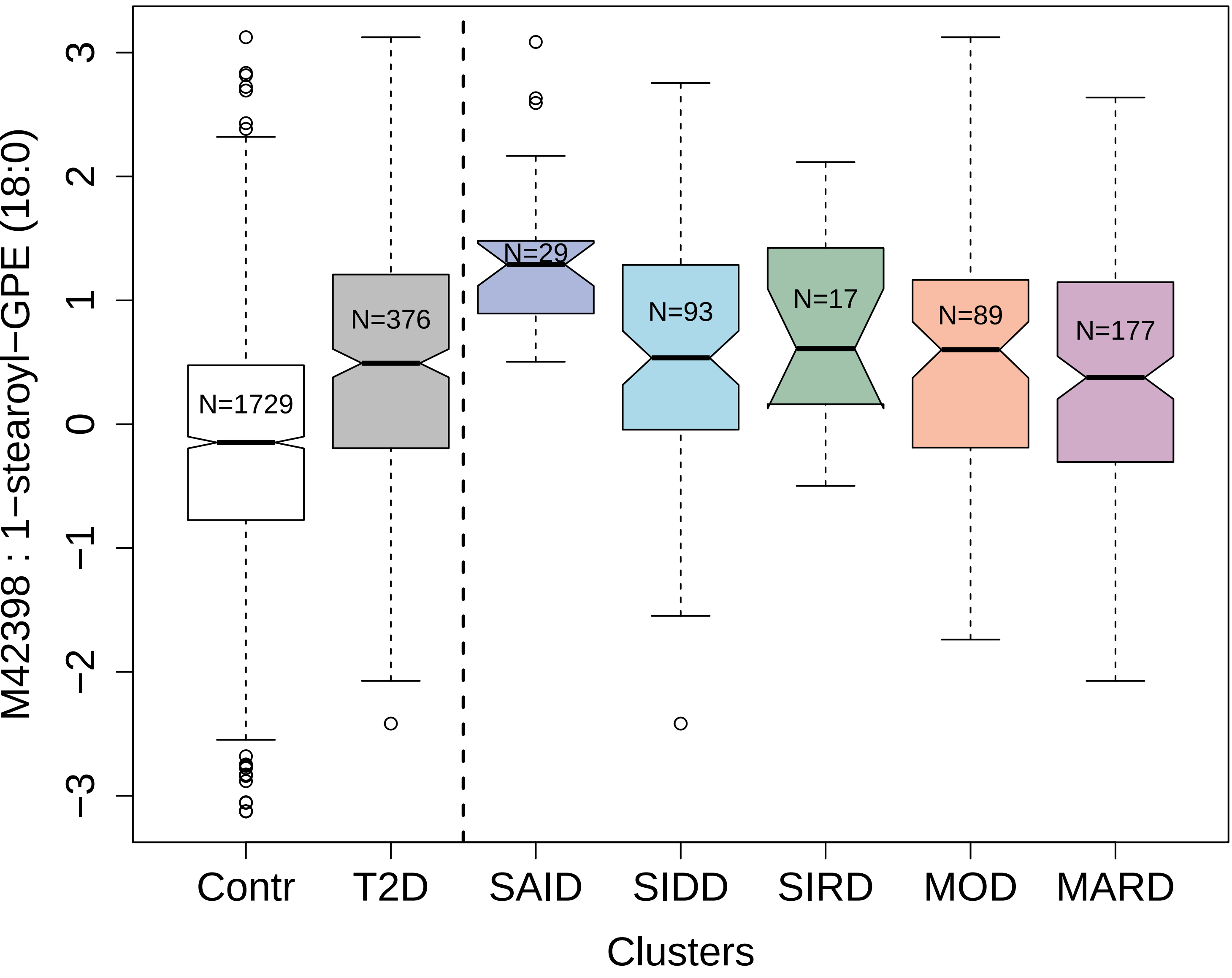

M42398 : 1−stearoyl−GPE (18:0)
3
2
1
0
−1
−2
−3
N=1729
N=376
N=29
N=93
N=17
N=89
N=177
Contr
T2D
SAID
SIDD
SIRD
MOD
MARD
Clusters

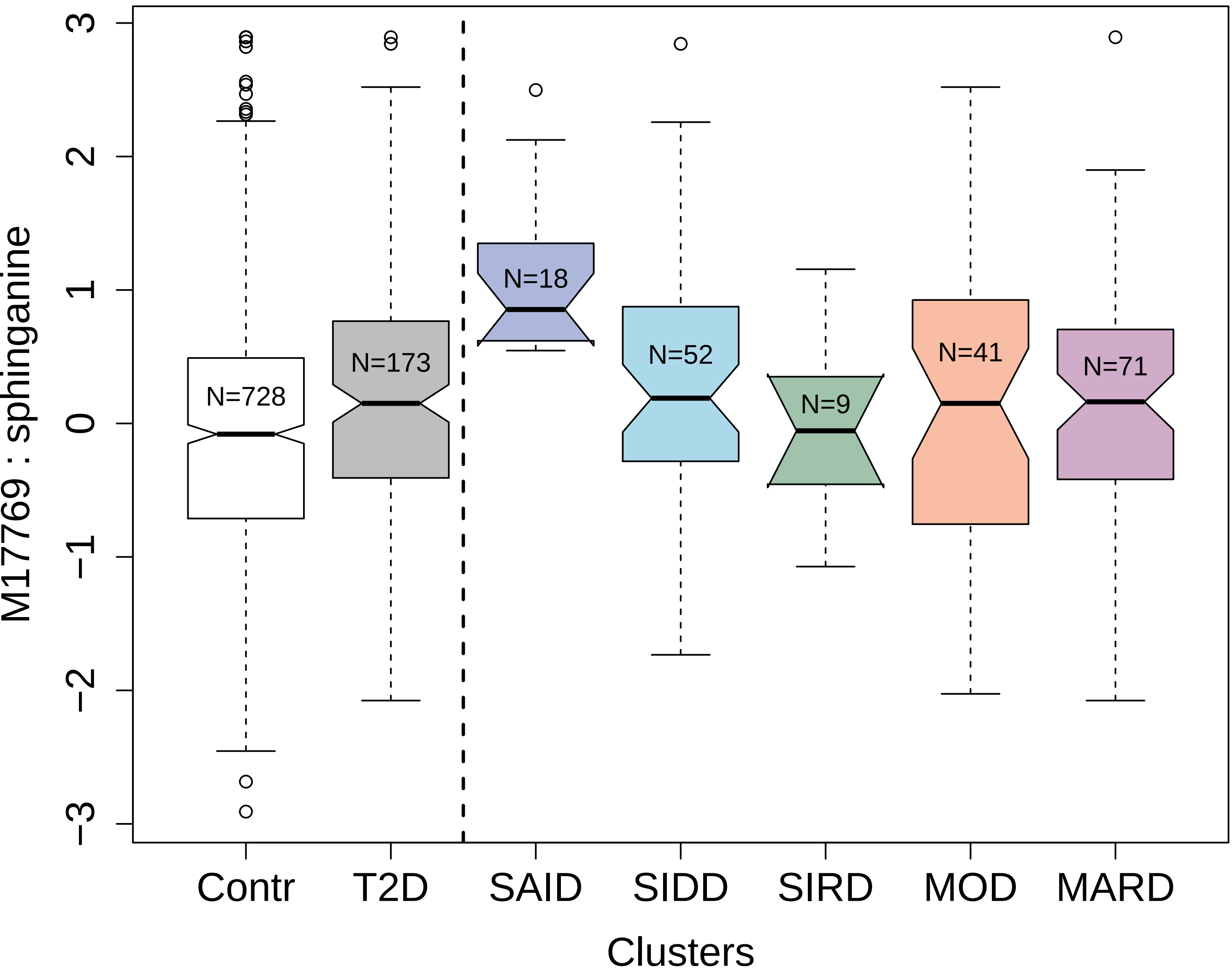

M17769 : sphinganine
3
2
1
0
−1
−2
−3
N=728
N=173
N=18
N=52
N=9
N=41
N=71
Contr
T2D
SAID
SIDD
SIRD
MOD
MARD
Clusters

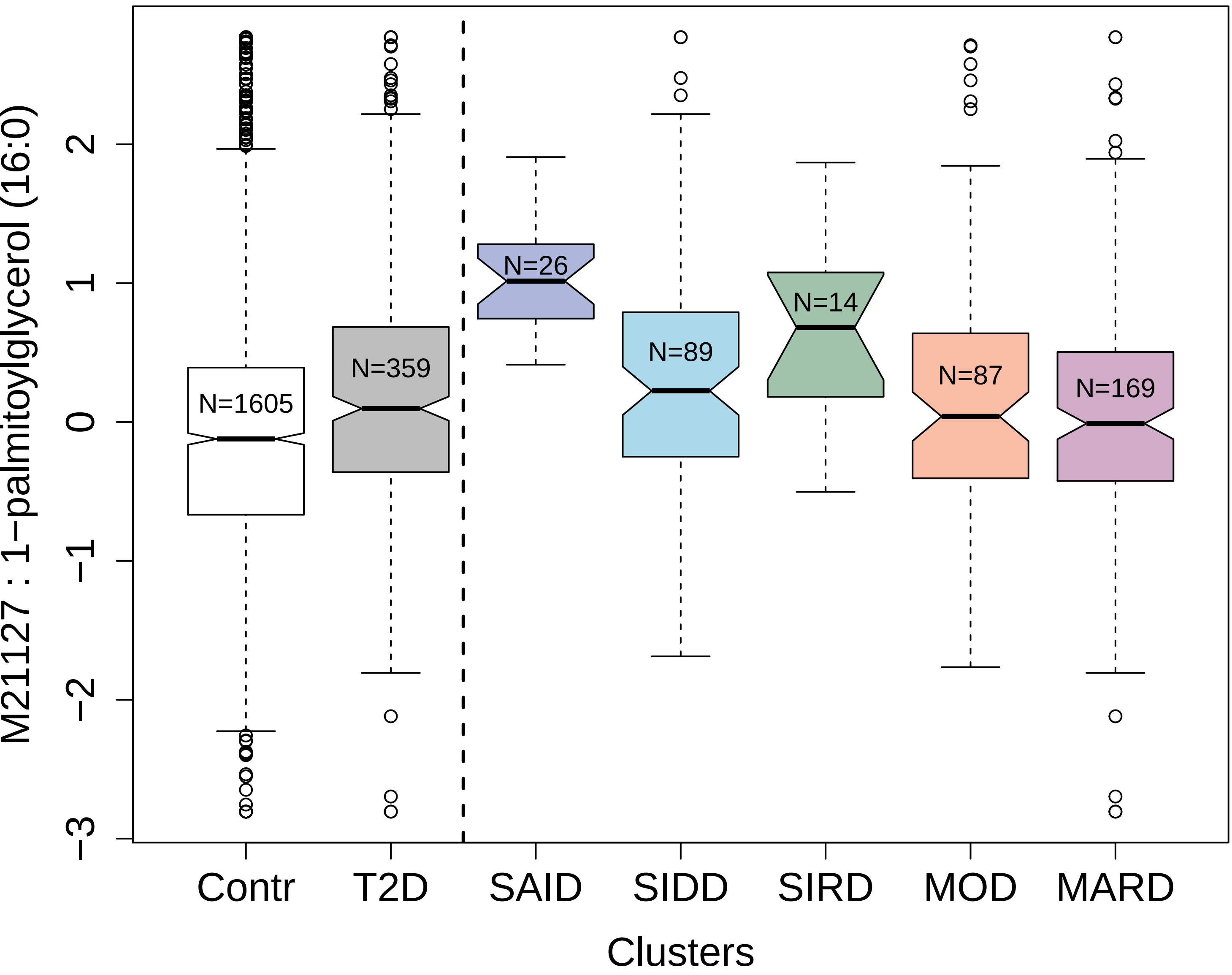
M21127 : 1−palmitoylglycerol (16:0)
−3
−2
−1
0
1
2
N=1605
N=359
N=26
N=89
N=14
N=87
N=169
Contr
T2D
SAID
SIDD
SIRD
MOD
MARD
Clusters

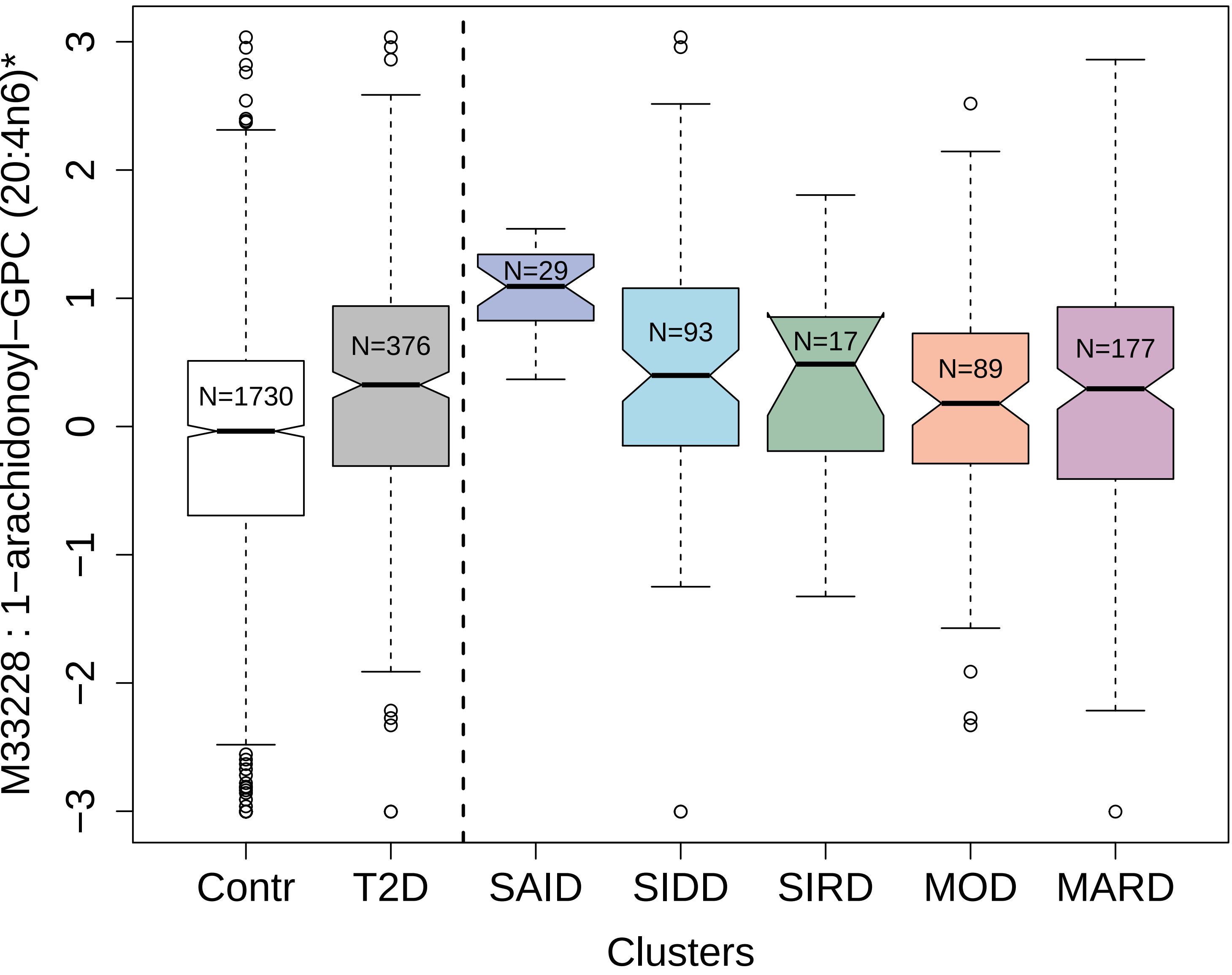
M33228 : 1−arachidonoyl−GPC (20:4n6)*
3
2
1
0
−1
−2
−3
N=1730
N=376
N=29
N=93
N=17
N=89
N=177
Contr
T2D
SAID
SIDD
SIRD
MOD
MARD
Clusters

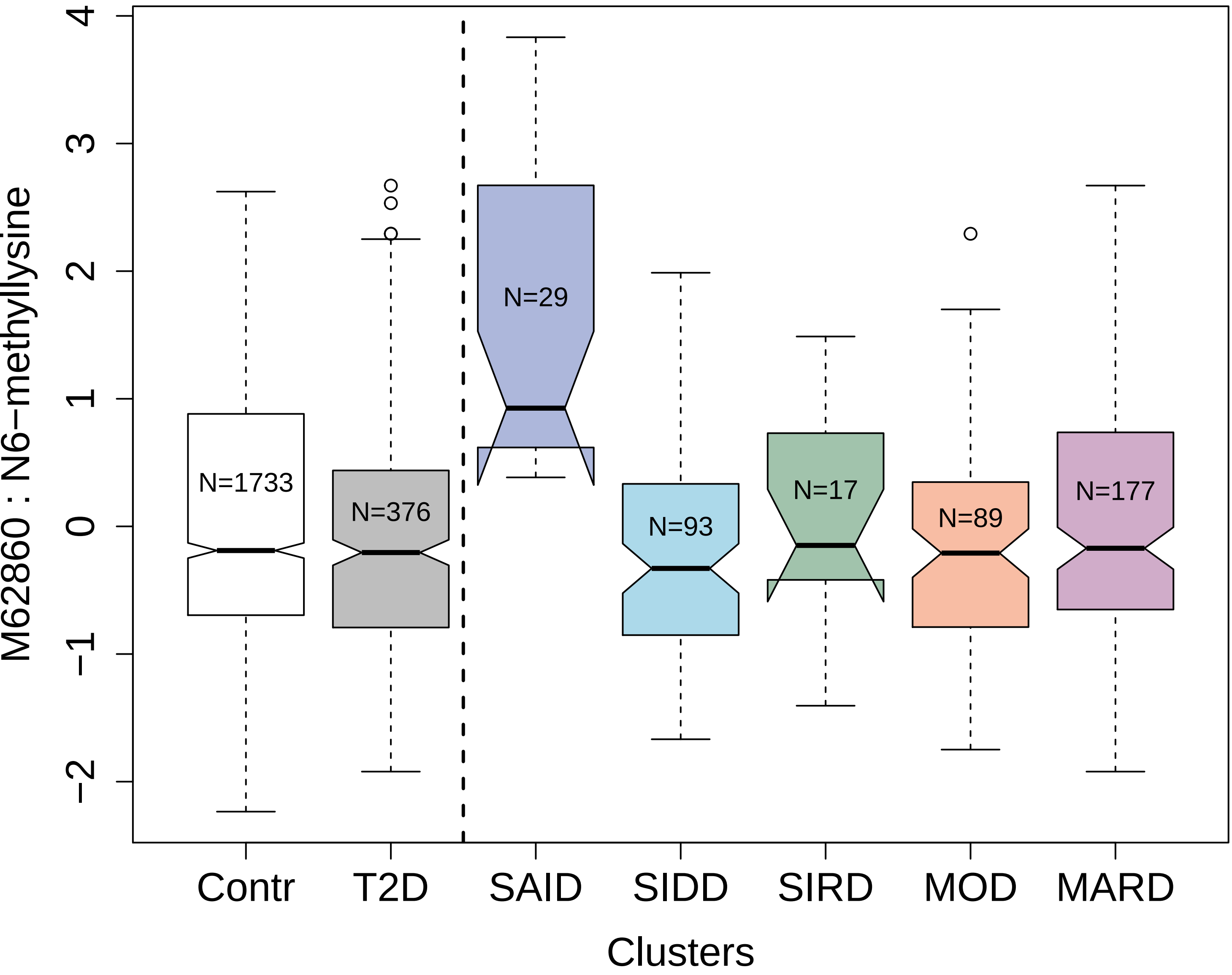

M62860 : N6−methyllysine
4
3
2
1
0
−1
−2
N=1733
N=376
N=29
N=93
N=17
N=89
N=177
Contr
T2D
SAID
SIDD
SIRD
MOD
MARD
Clusters

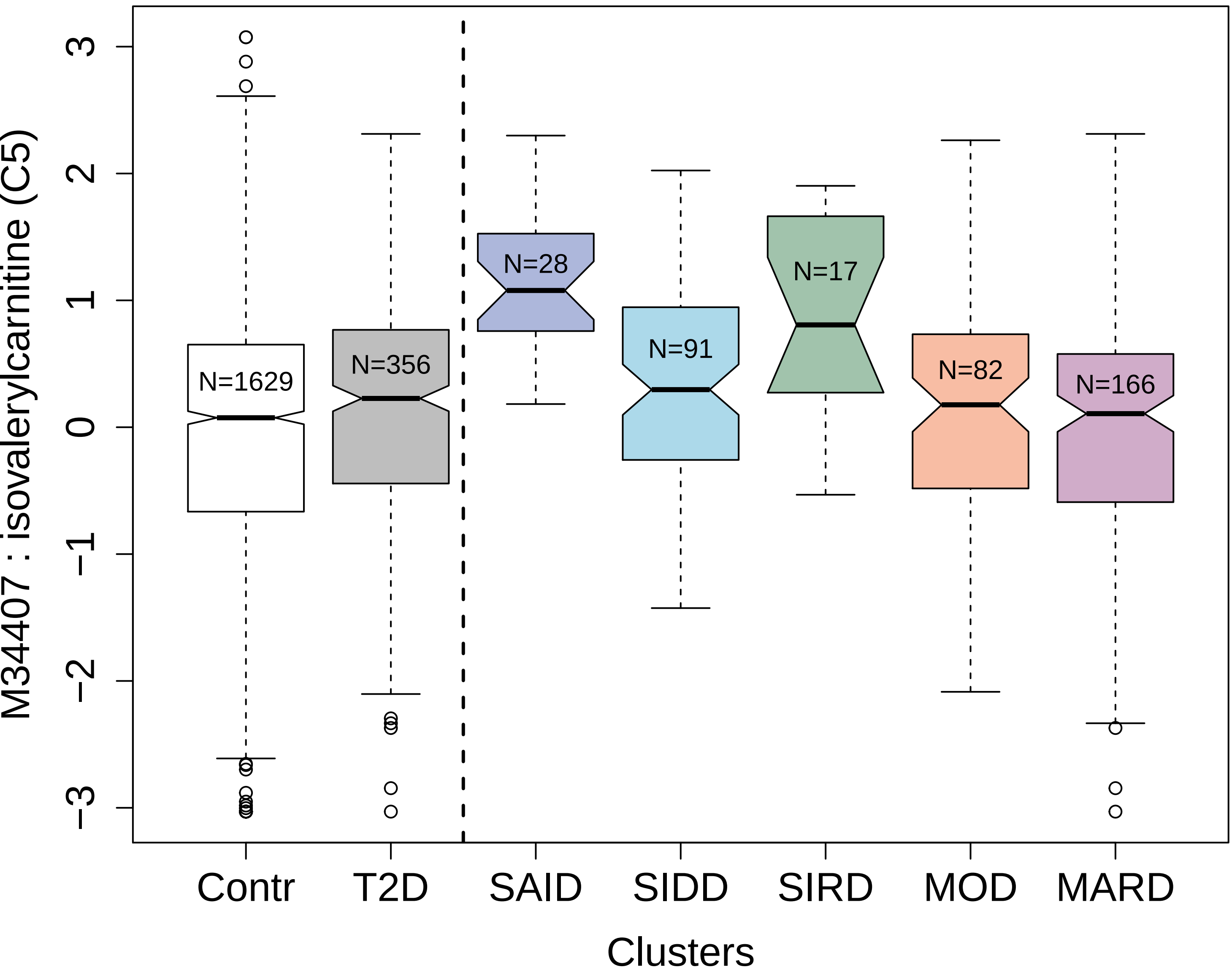
M34407 : isovalerylcarnitine (C5)
3
2
1
0
−1
−2
−3
N=1629
N=356
N=28
N=91
N=17
N=82
N=166
Contr
T2D
SAID
SIDD
SIRD
MOD
MARD
Clusters

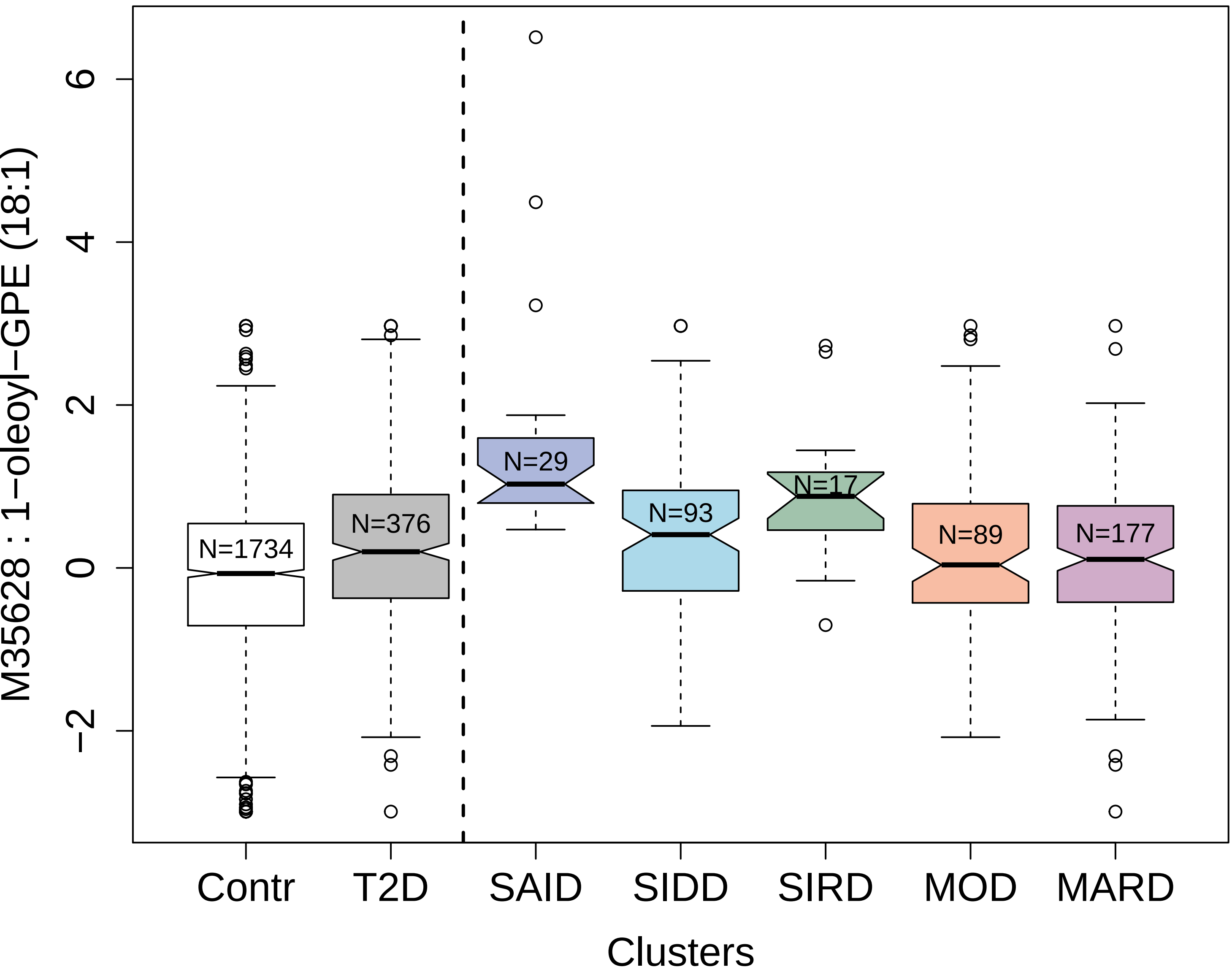

M35628 : 1−oleoyl−GPE (18:1)
6
4
2
0
−2
N=1734
N=376
N=29
N=93
N=17
N=89
N=177
Contr
T2D
SAID
SIDD
SIRD
MOD
MARD
Clusters

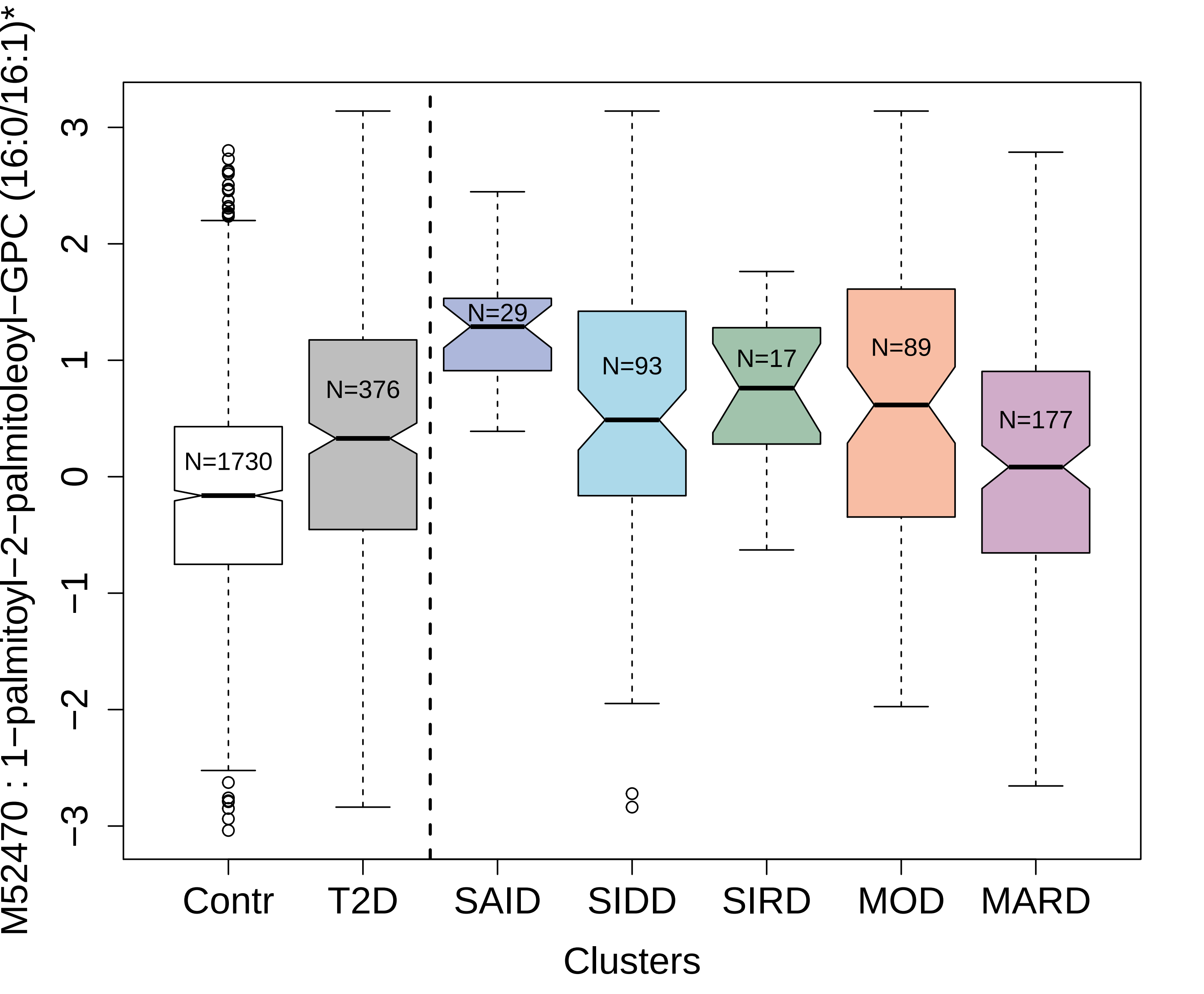

M52470 : 1−palmitoyl−2−palmitoleoyl−GPC (16:0/16:1)*
3
2
1
0
−1
−2
−3
N=1730
N=376
N=29
N=93
N=17
N=89
N=177
Contr
T2D
SAID
SIDD
SIRD
MOD
MARD
Clusters

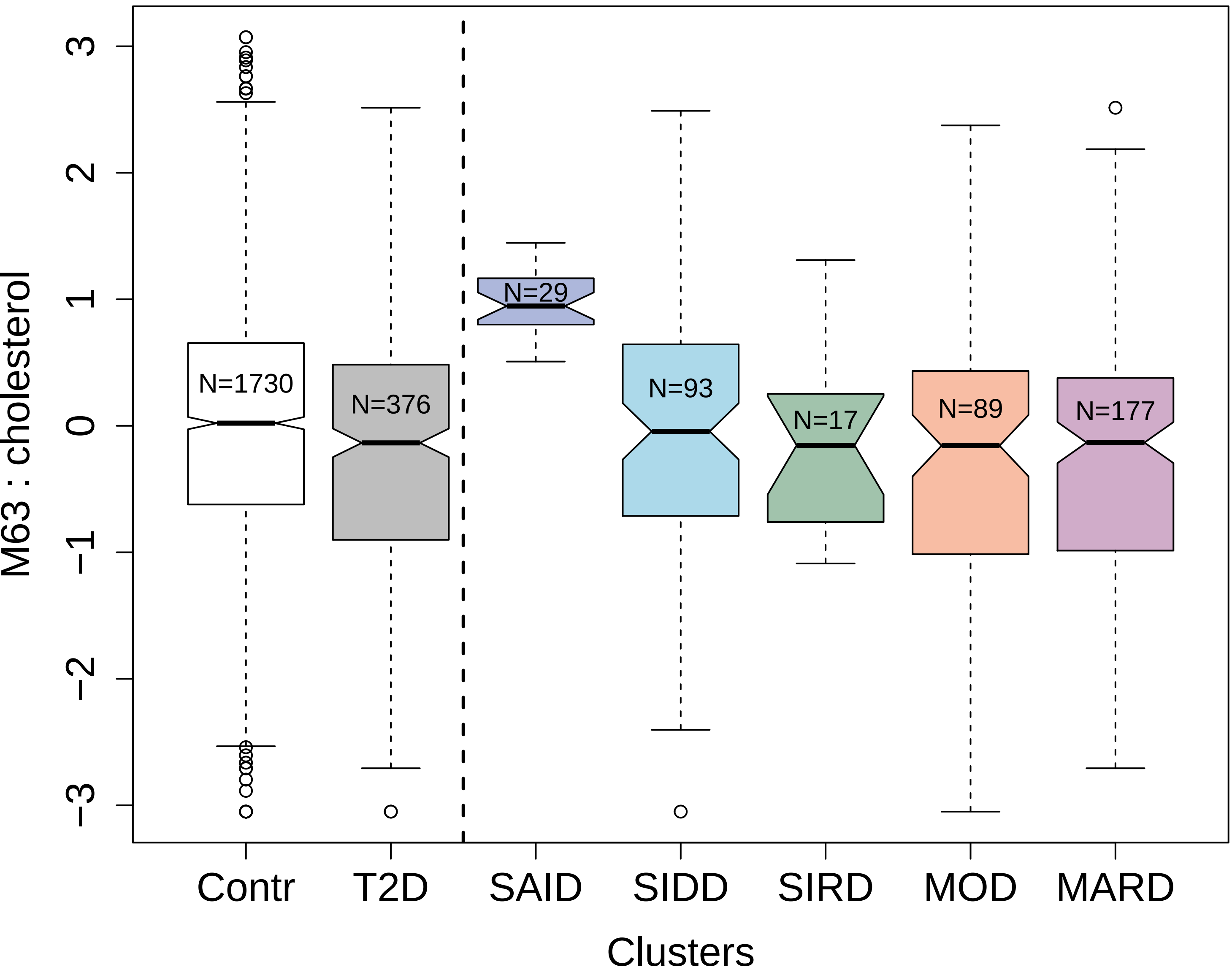
M63 : cholesterol
3
2
1
0
−1
−2
−3
N=1730
N=376
N=29
N=93
N=17
N=89
N=177
Contr
T2D
SAID
SIDD
SIRD
MOD
MARD
Clusters

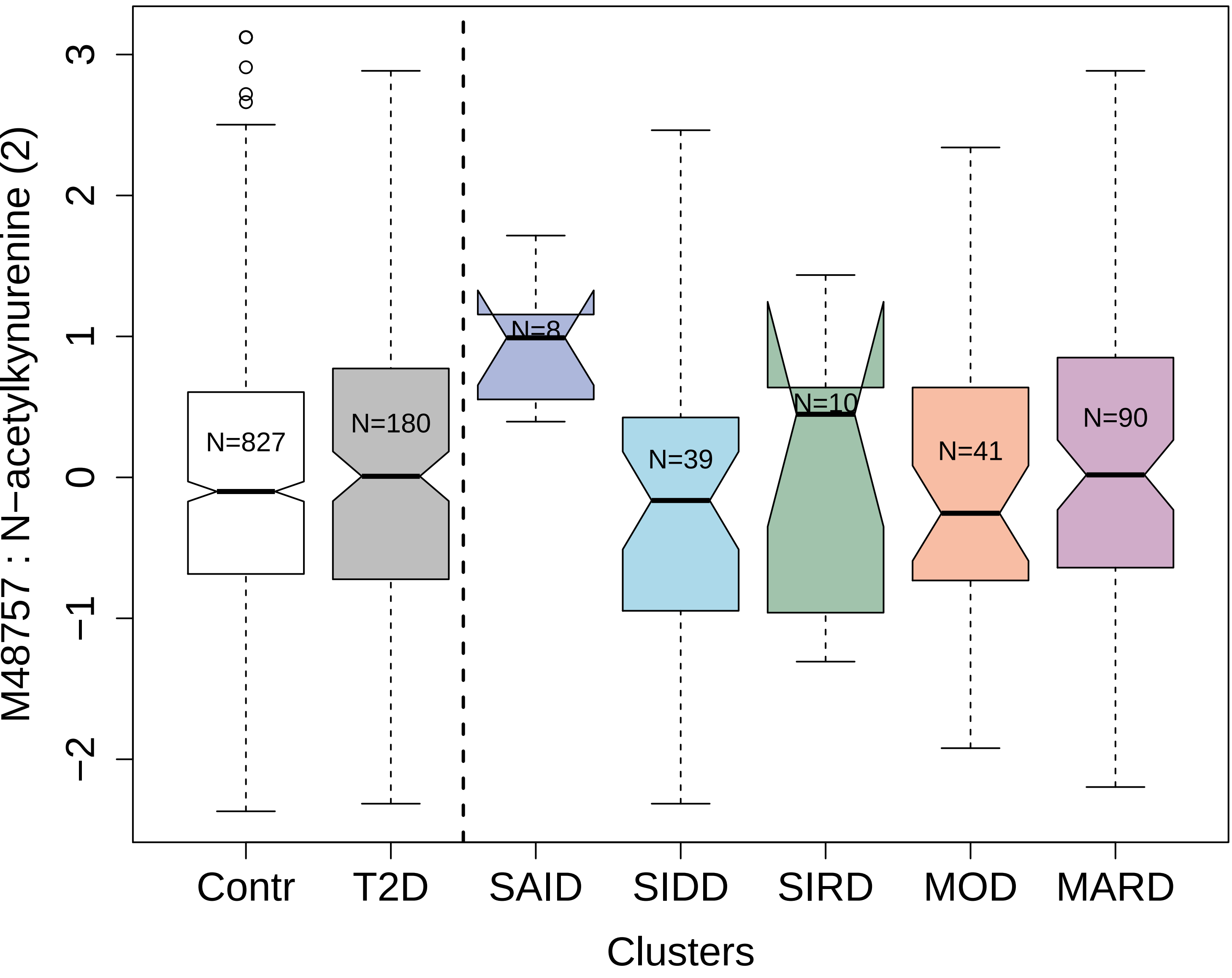

M48757 : N−acetylkynurenine (2)
3
2
1
0
−1
−2
N=827
N=180
N=8
N=39
N=10
N=41
N=90
Contr
T2D
SAID
SIDD
SIRD
MOD
MARD
Clusters

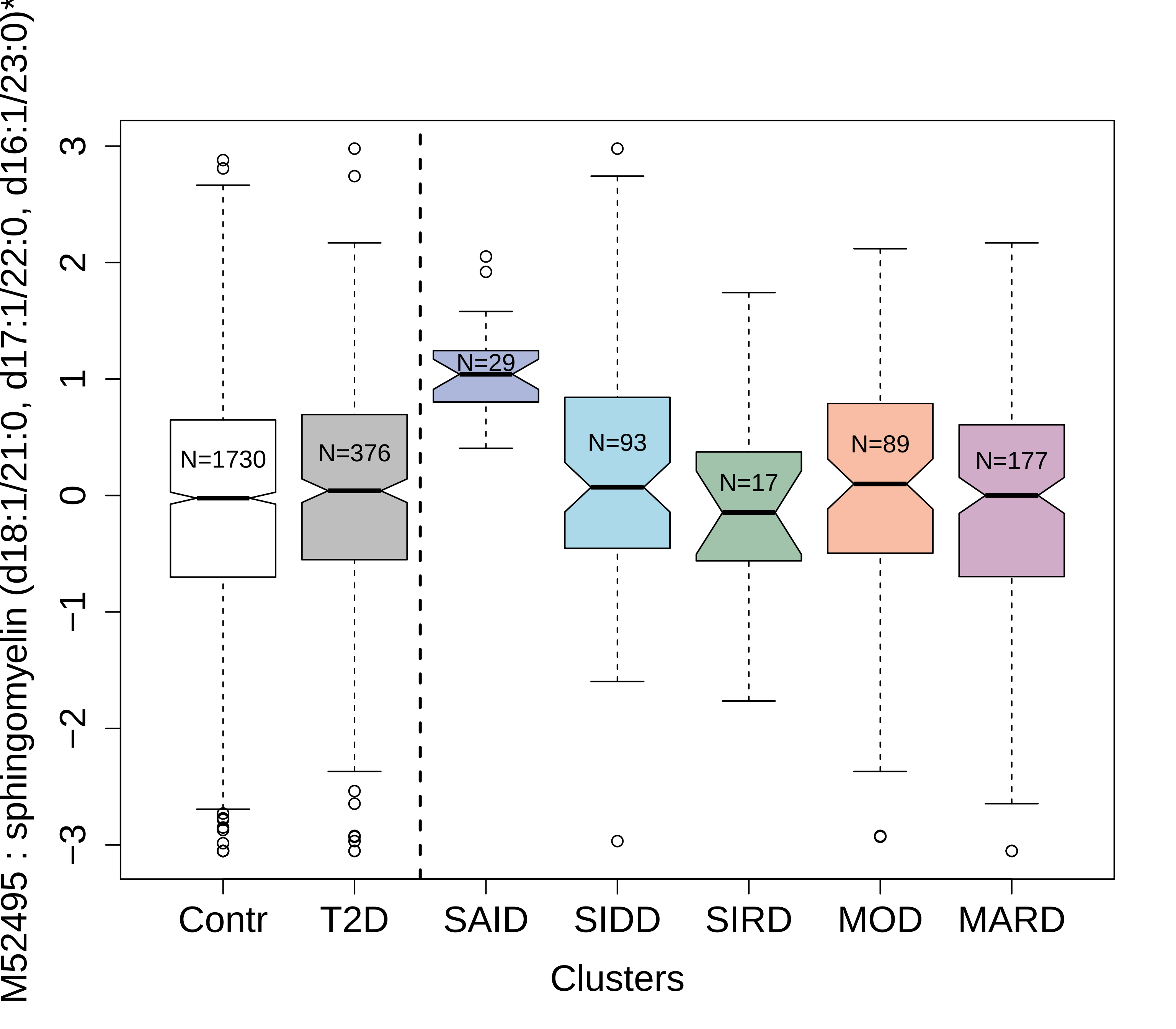
M52495 : sphingomyelin (d18:1/21:0, d17:1/22:0, d16:1/23:0)*
3
2
1
0
−1
−2
−3
N=1730
N=376
N=29
N=93
N=17
N=89
N=177
Contr
T2D
SAID
SIDD
SIRD
MOD
MARD
Clusters

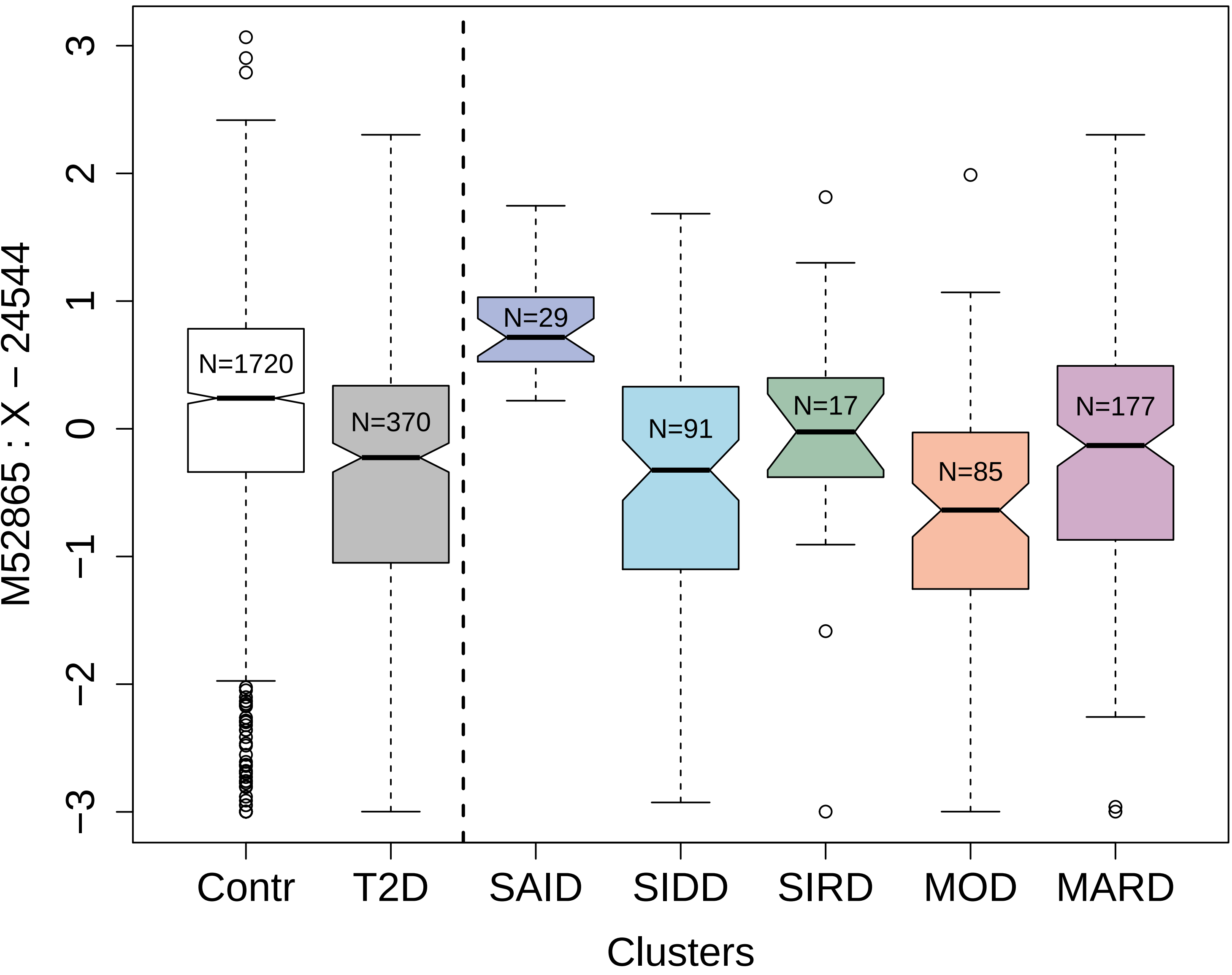
M52865 : X − 24544
3
2
1
0
−1
−2
−3
N=1720
N=370
N=29
N=91
N=17
N=85
N=177
Contr
T2D
SAID
SIDD
SIRD
MOD
MARD
Clusters

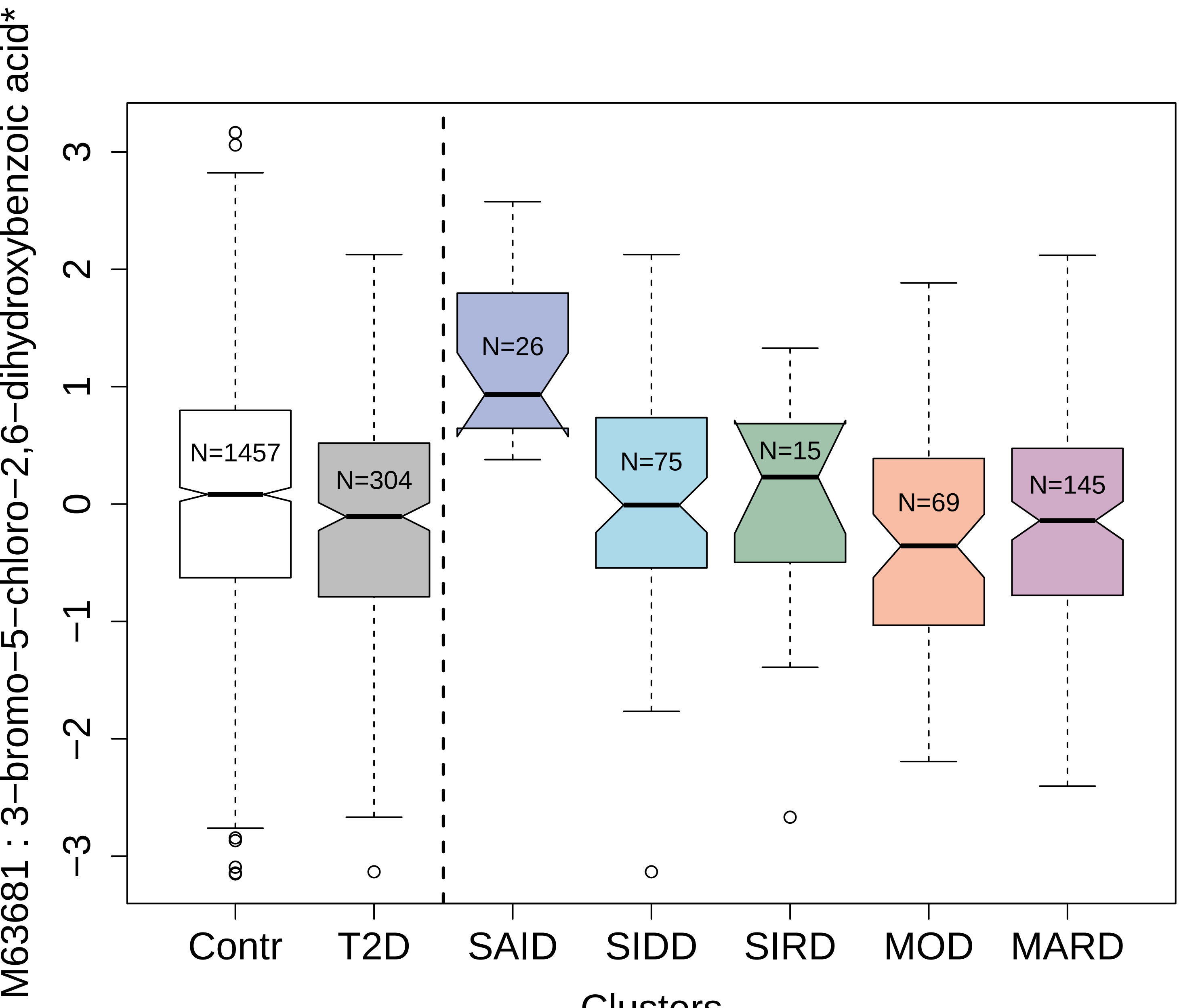
M63681 : 3−bromo−5−chloro−2,6−dihydroxybenzoic acid*
3
2
1
0
−1
−2
−3
N=1457
N=304
N=26
N=75
N=15
N=69
N=145
Contr
T2D
SAID
SIDD
SIRD
MOD
MARD
Clusters

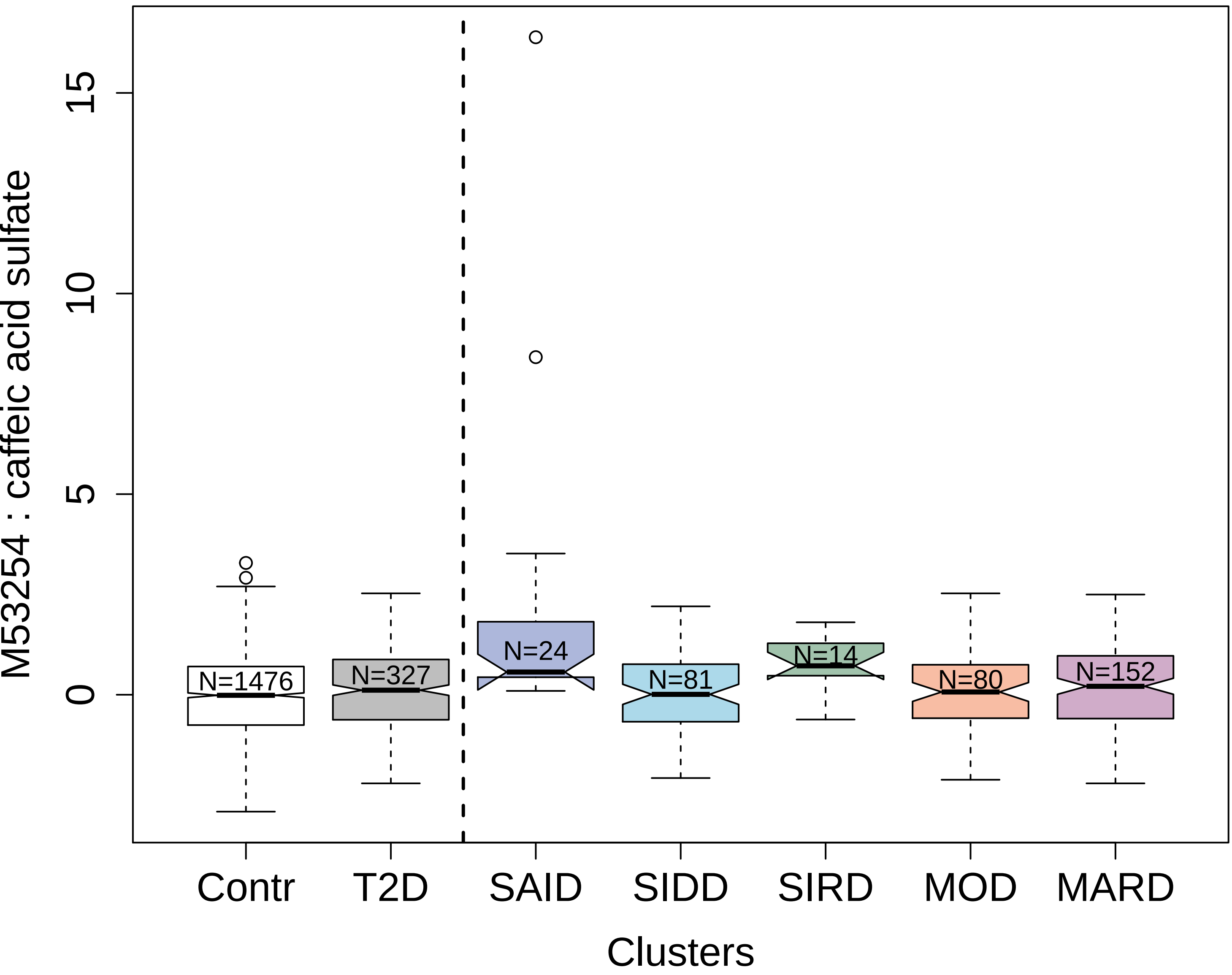

M53254 : caffeic acid sulfate
0
5
10
15
N=1476
N=327
N=24
N=81
N=14
N=80
N=152
Contr
T2D
SAID
SIDD
SIRD
MOD
MARD
Clusters

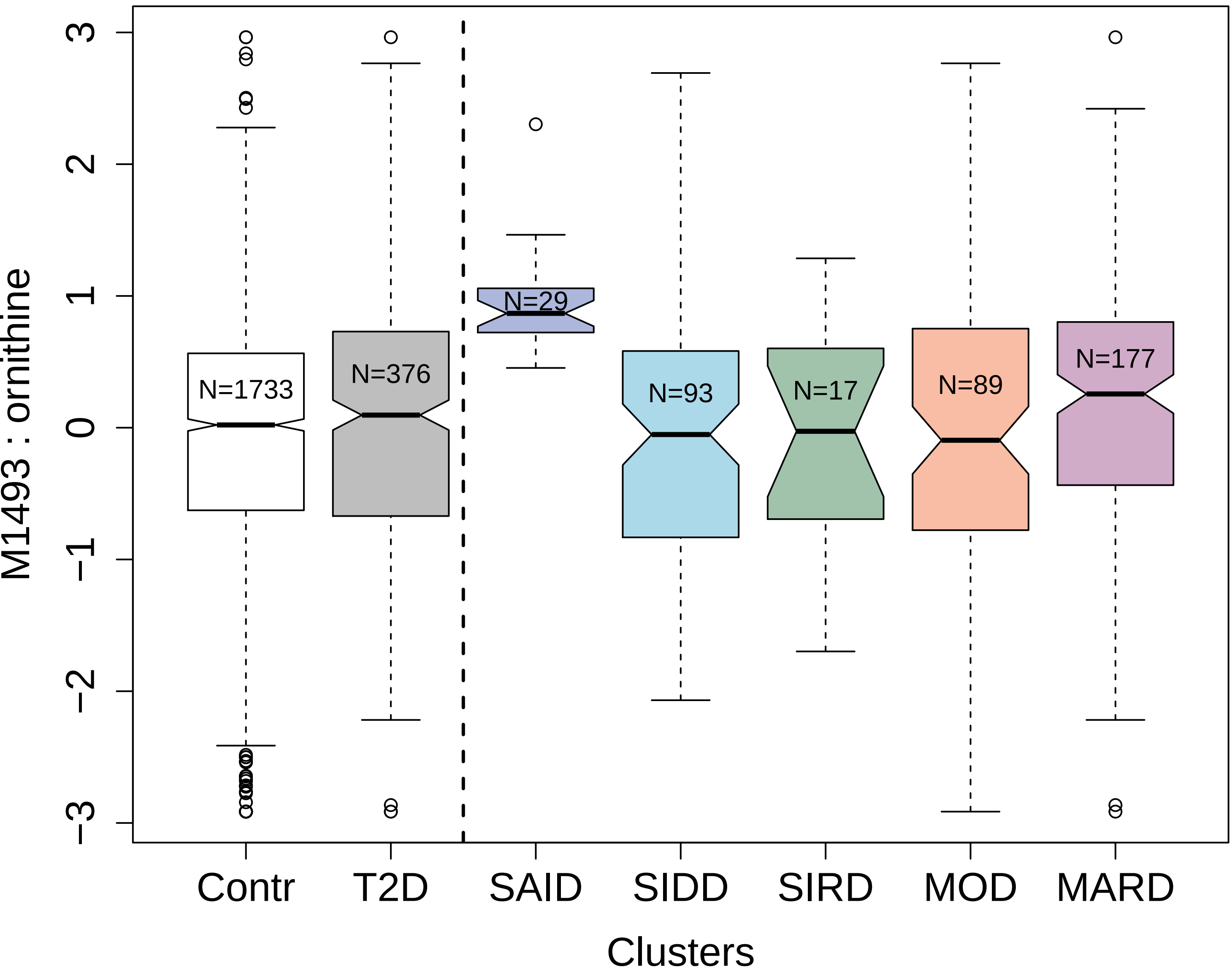

M1493 : ornithine
3
2
1
0
−1
−2
−3
N=1733
N=376
N=29
N=93
N=17
N=89
N=177
Contr
T2D
SAID
SIDD
SIRD
MOD
MARD
Clusters

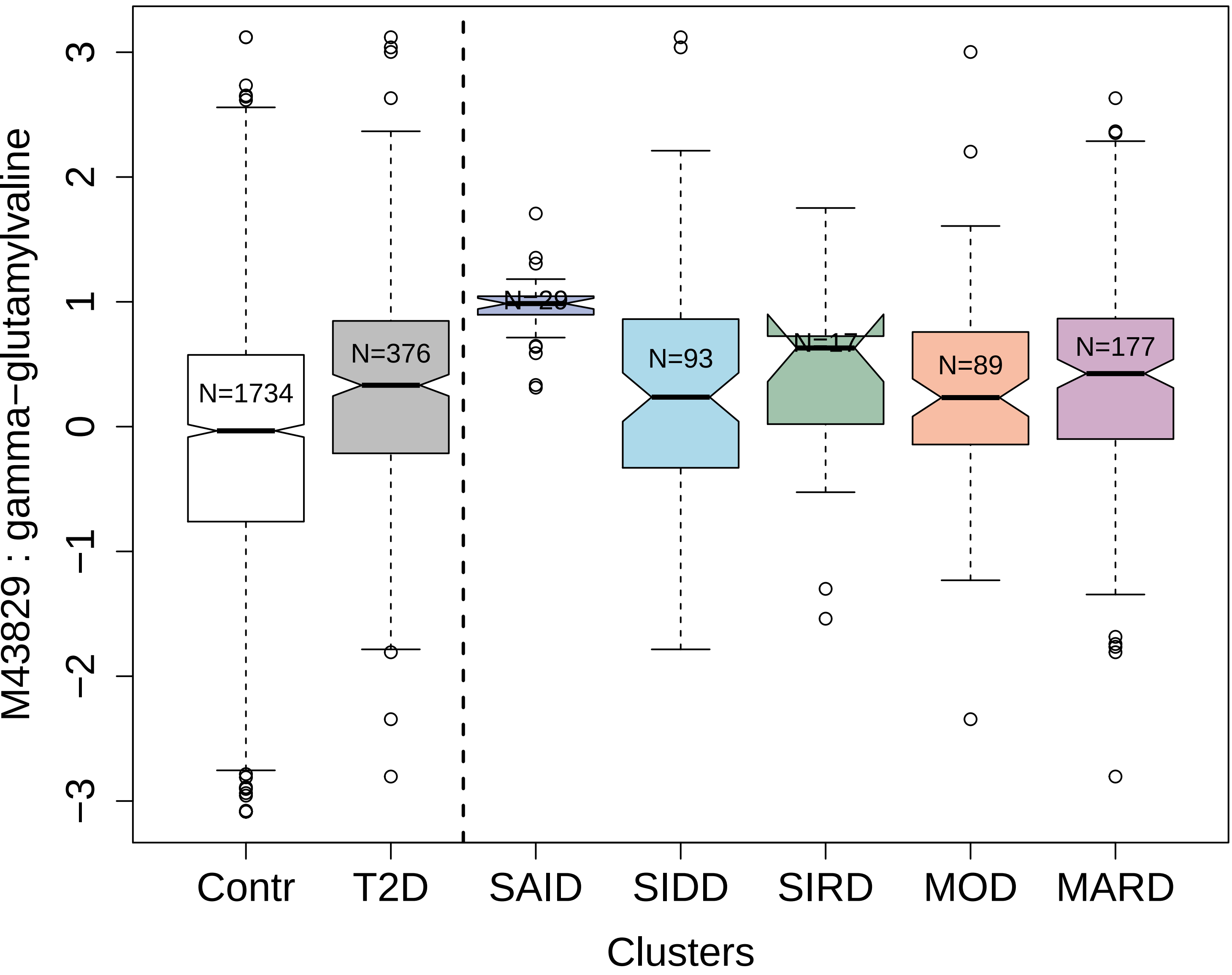
M43829 : gamma−glutamylvaline
Clusters
Contr
T2D
SAID
SIDD
SIRD
MOD
MARD
N=1734
N=376
N=29
N=93
N=17
N=89
N=177
3
2
1
0
−1
−2
−3

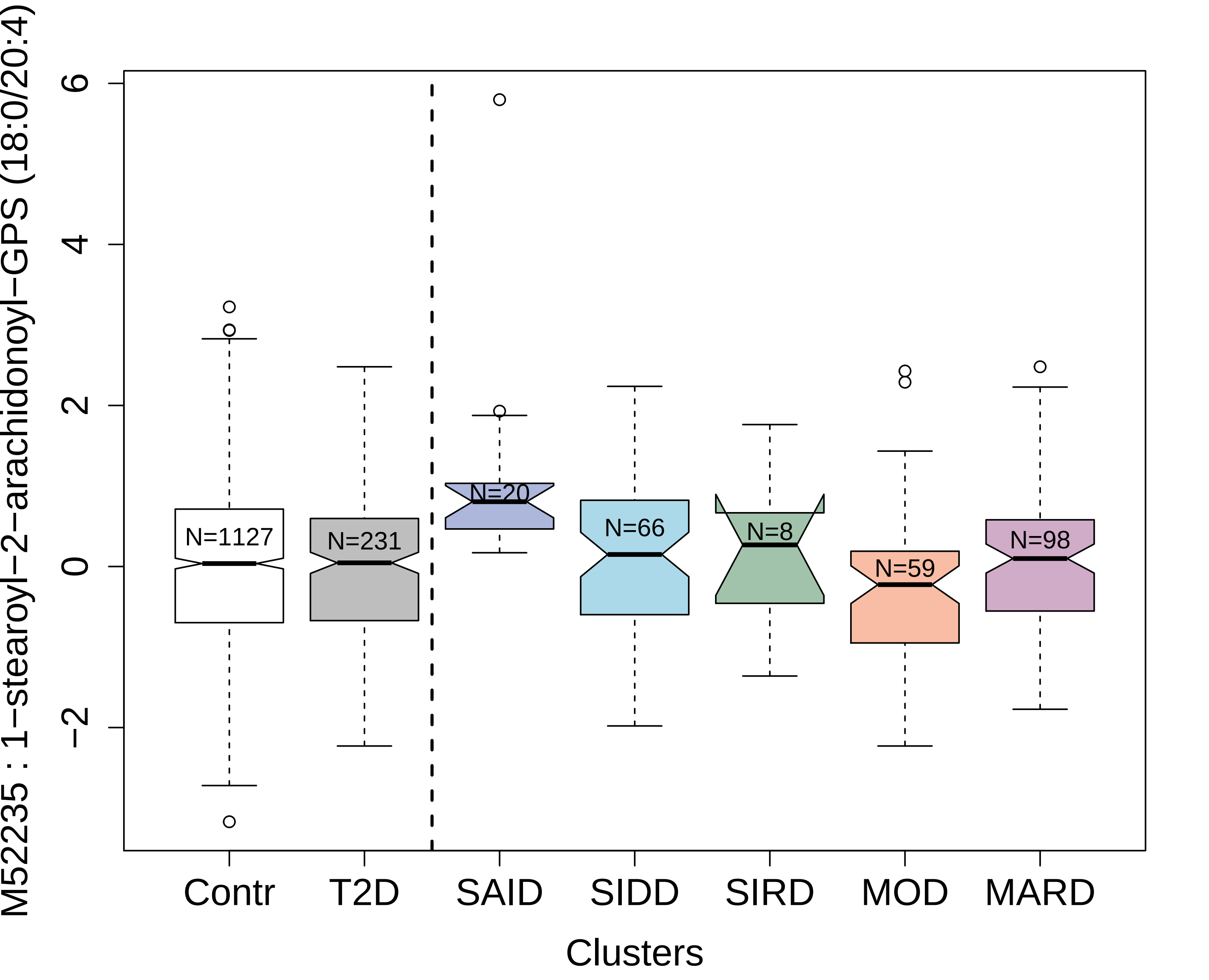

M52235 : 1−stearoyl−2−arachidonoyl−GPS (18:0/20:4)
6
4
2
0
−2
N=1127
N=231
N=20
N=66
N=8
N=59
N=98
Contr
T2D
SAID
SIDD
SIRD
MOD
MARD
Clusters

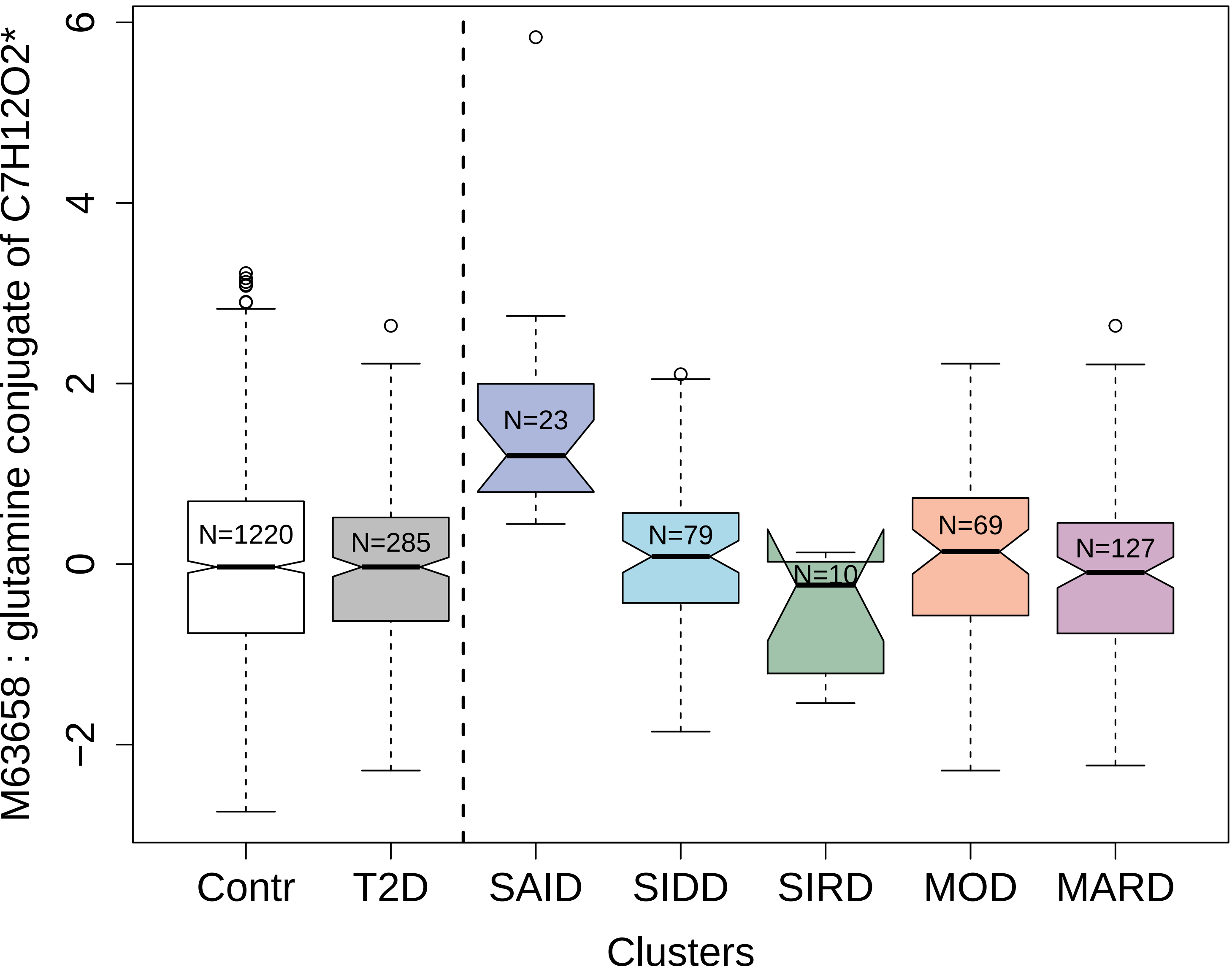
M63658 : glutamine conjugate of C7H12O2*
6
4
2
0
−2
N=1220
N=285
N=23
N=79
N=10
N=69
N=127
Contr
T2D
SAID
SIDD
SIRD
MOD
MARD
Clusters

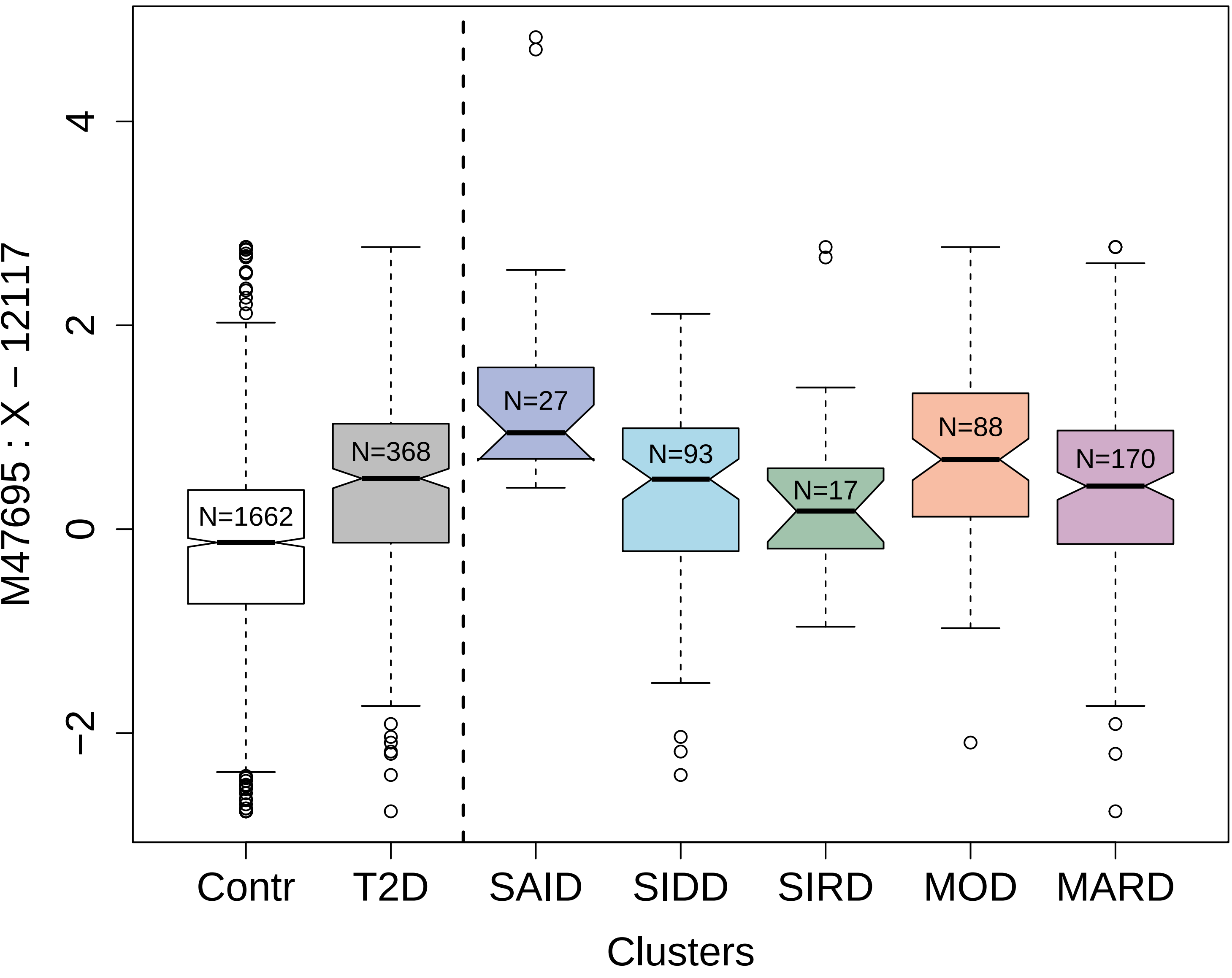

M47695 : X − 12117
4
2
0
−2
N=1662
N=368
N=27
N=93
N=17
N=88
N=170
Contr
T2D
SAID
SIDD
SIRD
MOD
MARD
Clusters

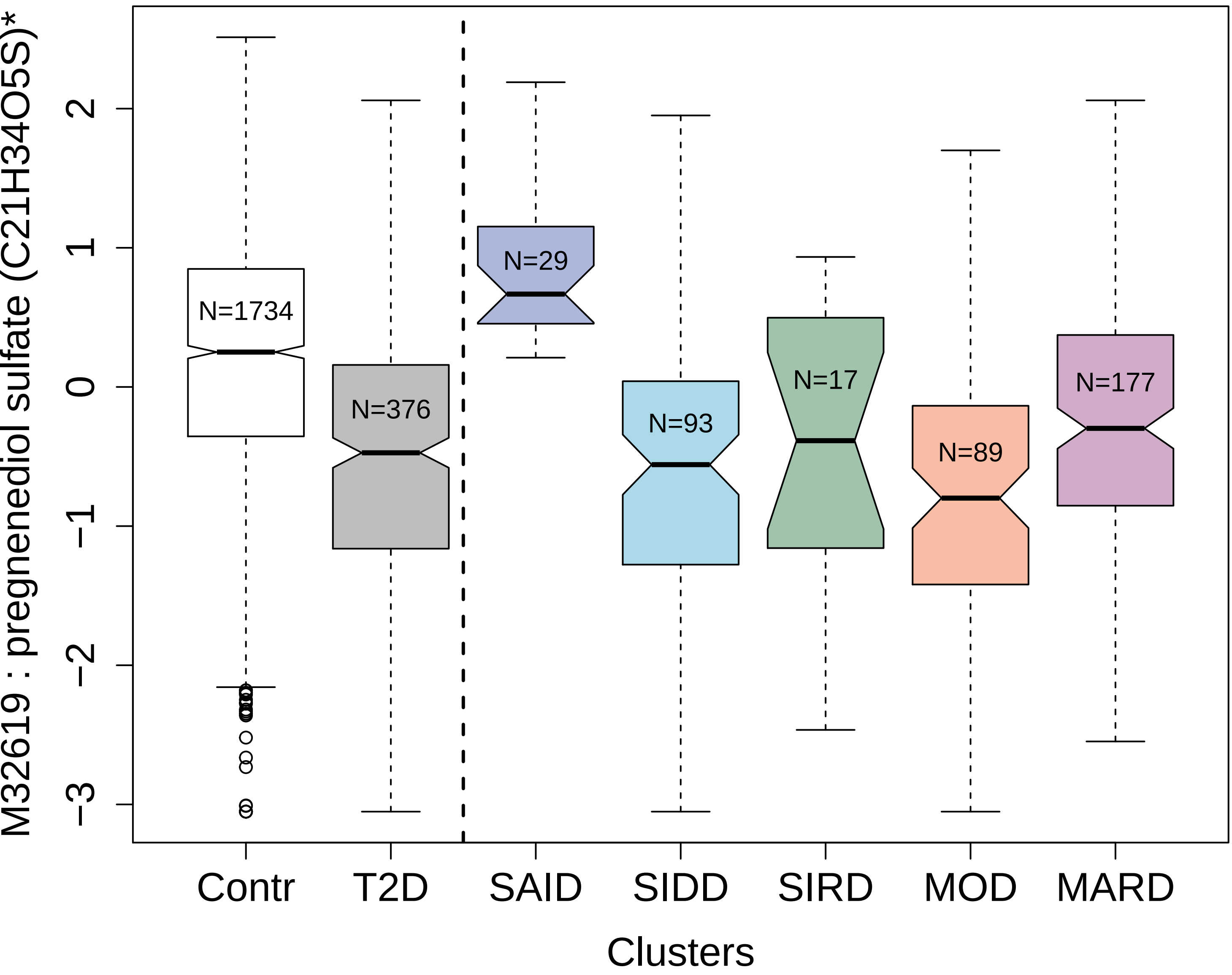

M32619 : pregnenediol sulfate (C21H34O5S)*
Clusters
Contr
T2D
SAID
SIDD
SIRD
MOD
MARD
N=1734
N=376
N=29
N=93
N=17
N=89
N=177
2
1
0
−1
−2
−3

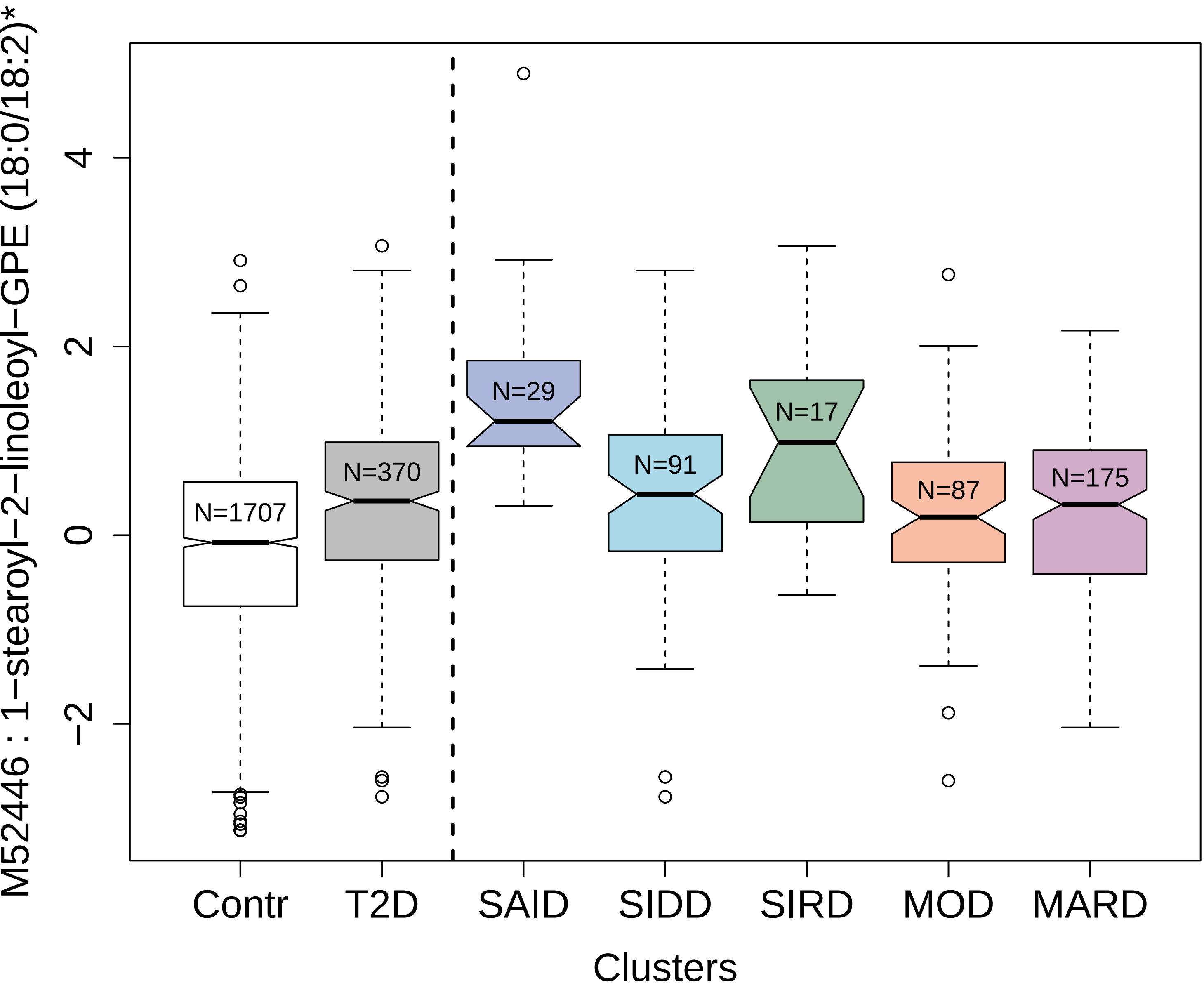
M52446 : 1−stearoyl−2−linoleoyl−GPE (18:0/18:2)*
4
2
0
−2
N=1707
N=370
N=29
N=91
N=17
N=87
N=175
Contr
T2D
SAID
SIDD
SIRD
MOD
MARD
Clusters

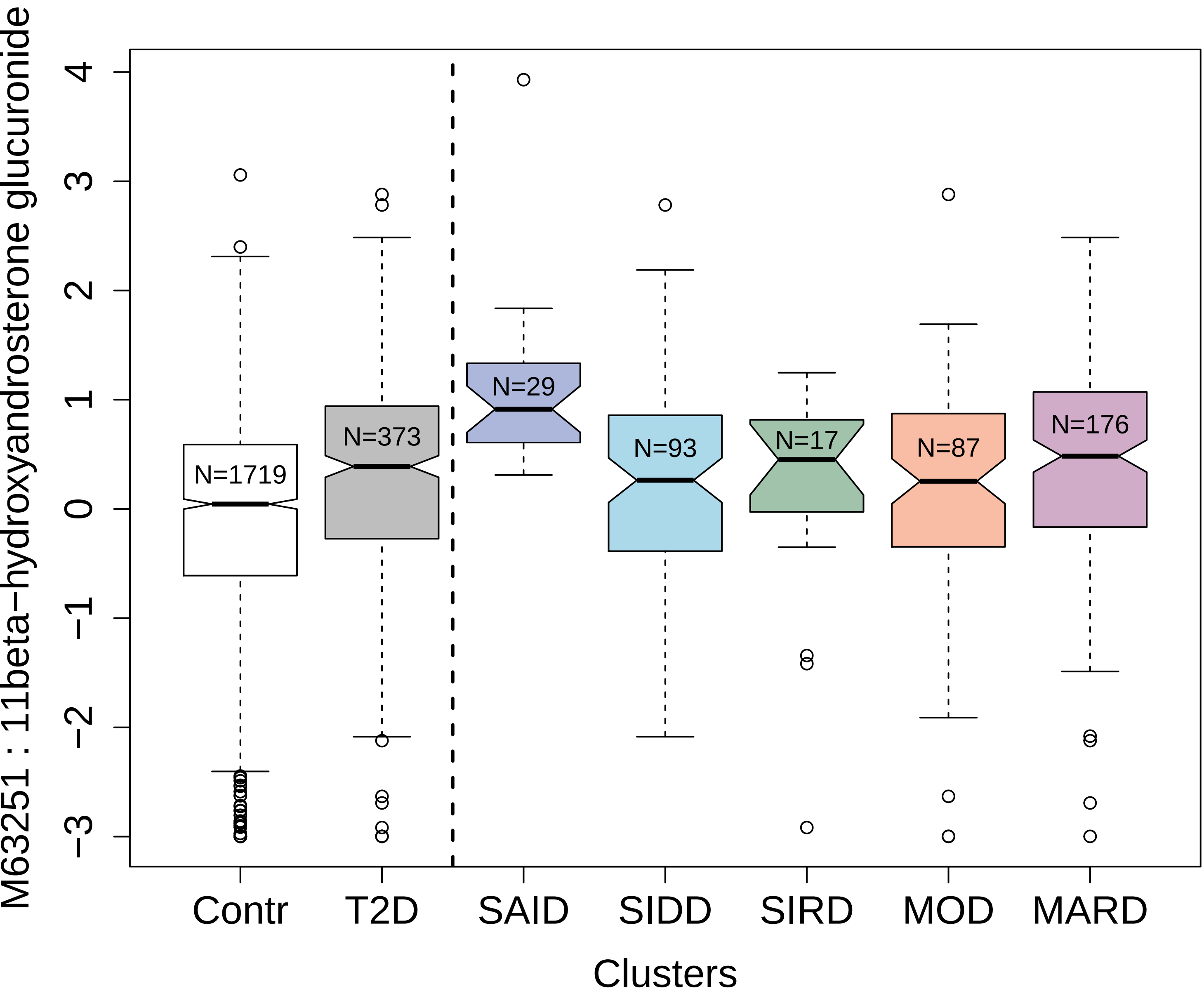

M63251 : 11beta−hydroxyandrosterone glucuronide
4
3
2
1
0
−1
−2
−3
N=1719
N=373
N=29
N=93
N=17
N=87
N=176
Contr
T2D
SAID
SIDD
SIRD
MOD
MARD
Clusters

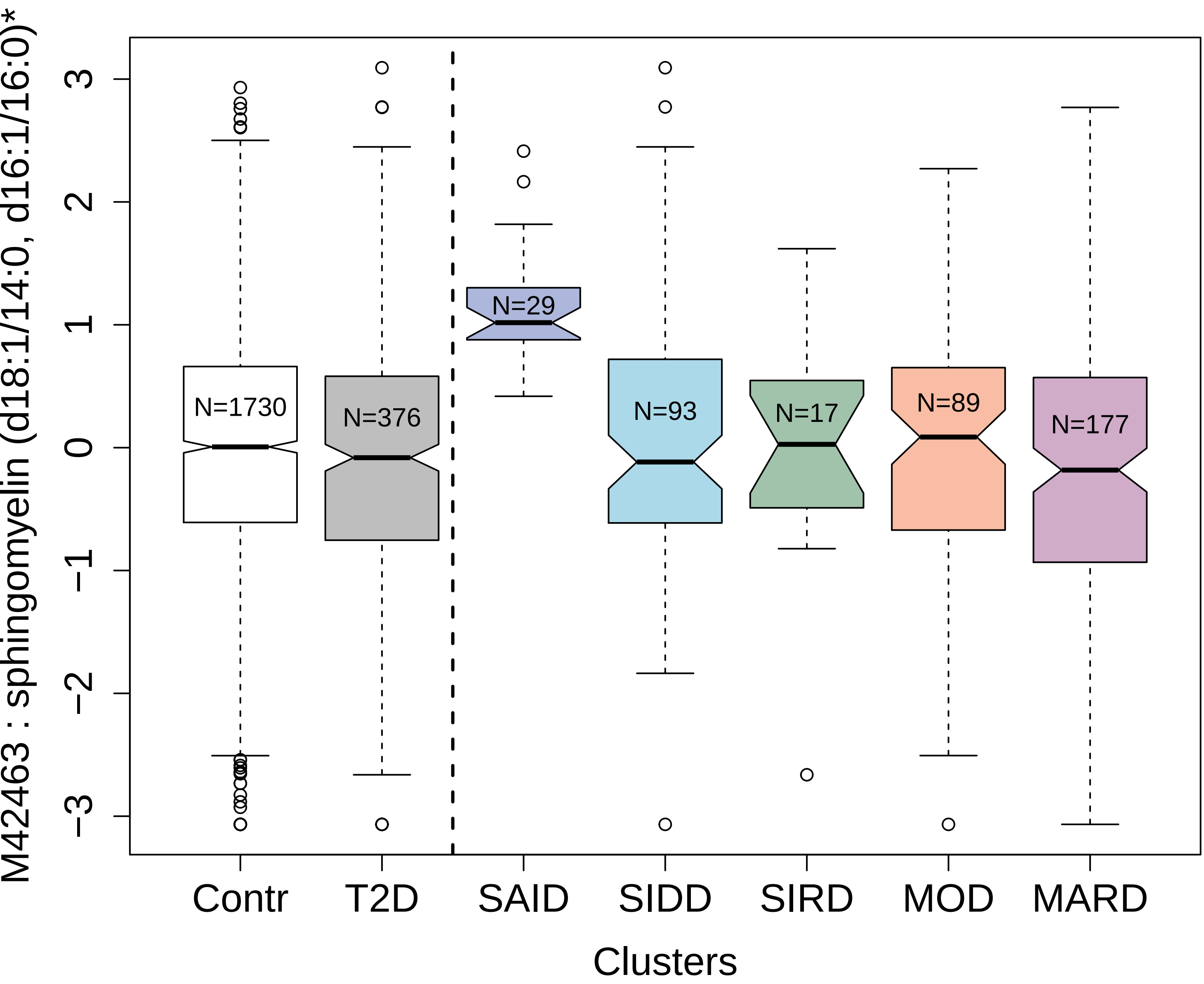
M42463 : sphingomyelin (d18:1/14:0, d16:1/16:0)*
3
2
1
0
−1
−2
−3
N=1730
N=376
N=29
N=93
N=17
N=89
N=177
Contr
T2D
SAID
SIDD
SIRD
MOD
MARD
Clusters

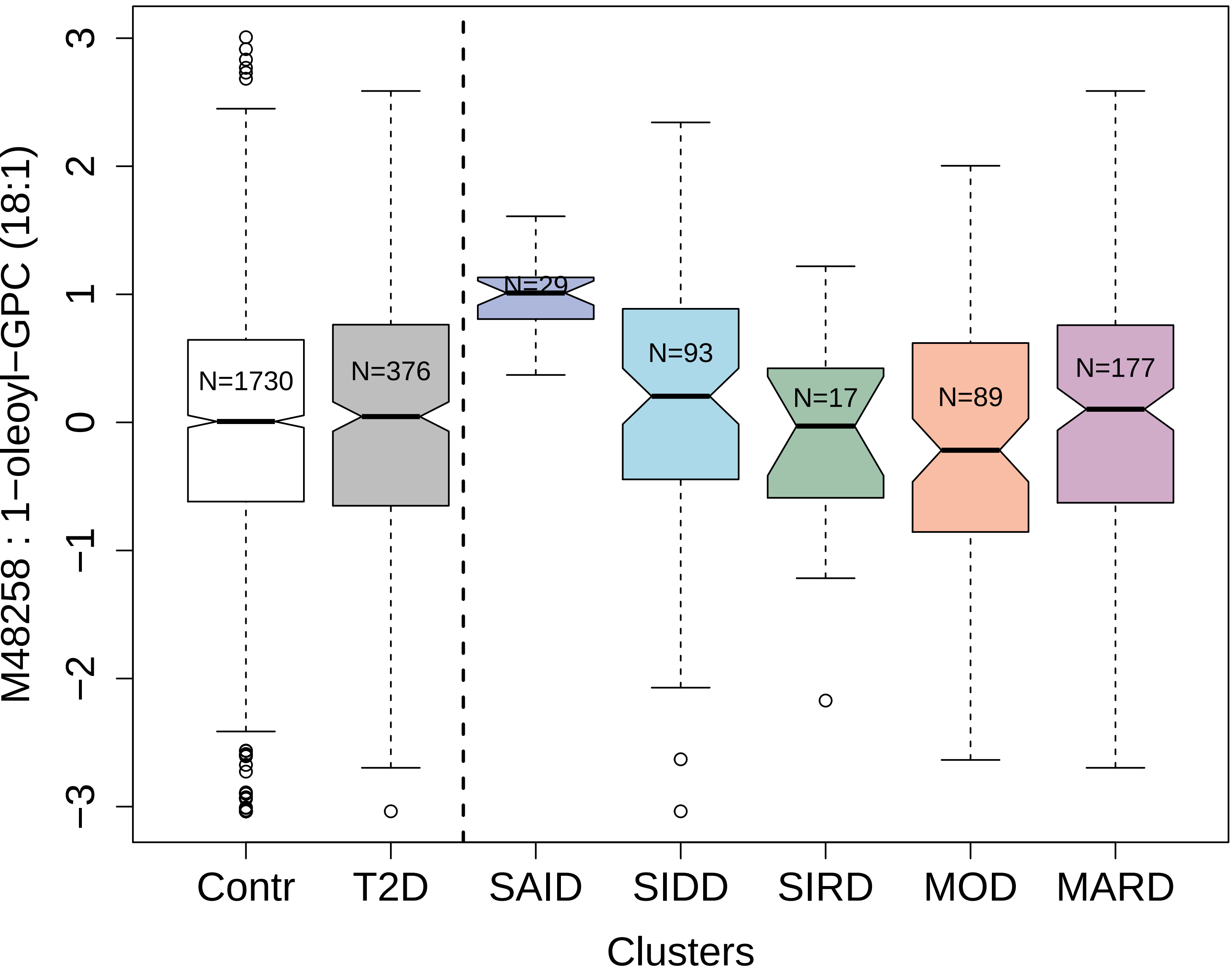

M48258 : 1−oleoyl−GPC (18:1)
3
2
1
0
−1
−2
−3
N=1730
N=376
N=29
N=93
N=17
N=89
N=177
Contr
T2D
SAID
SIDD
SIRD
MOD
MARD
Clusters

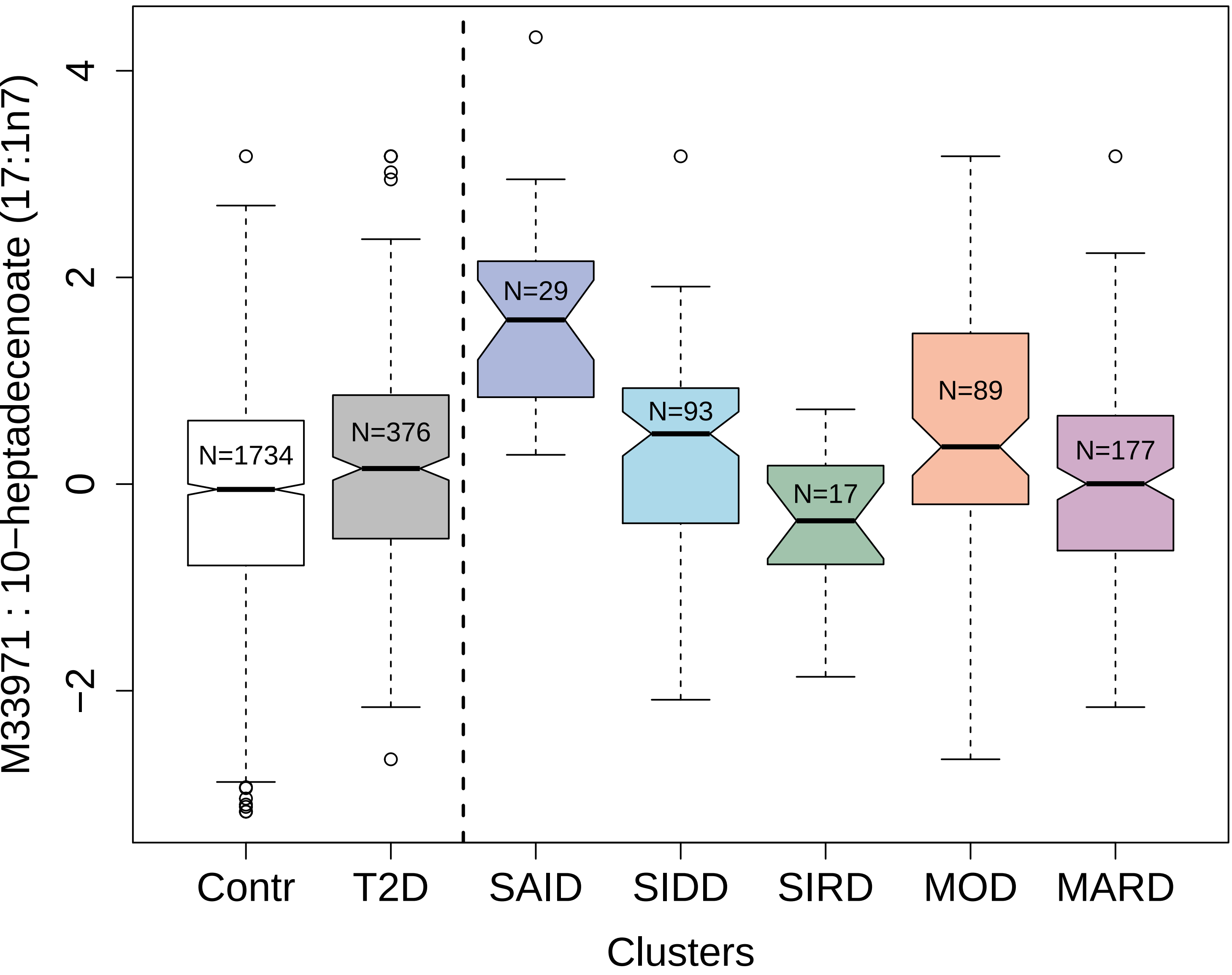

M33971 : 10−heptadecenoate (17:1n7)
4
2
0
−2
N=1734
N=376
N=29
N=93
N=17
N=89
N=177
Contr
T2D
SAID
SIDD
SIRD
MOD
MARD
Clusters

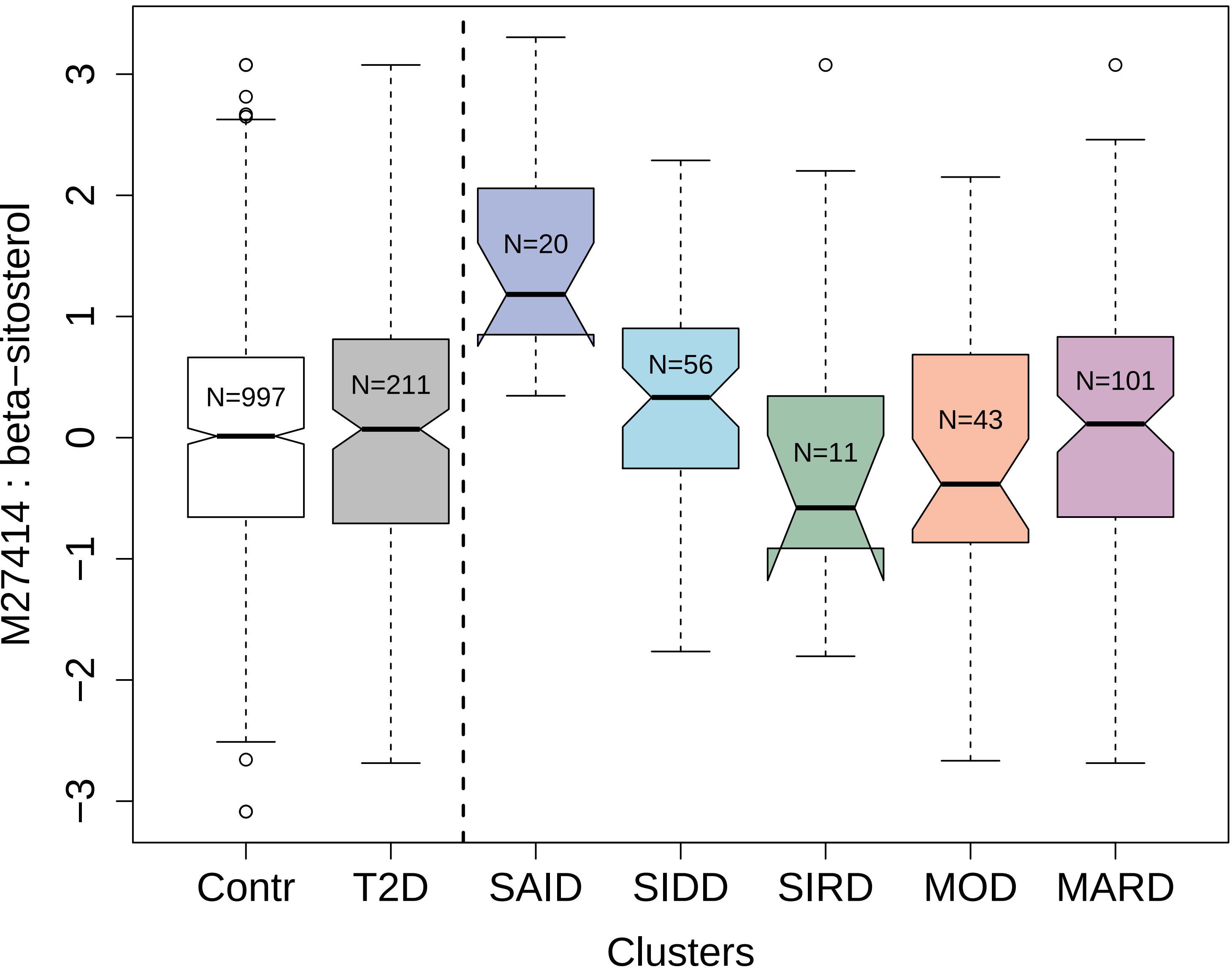
M27414 : beta−sitosterol
3
2
1
0
−1
−2
−3
N=997
N=211
N=20
N=56
N=11
N=43
N=101
Contr
T2D
SAID
SIDD
SIRD
MOD
MARD
Clusters

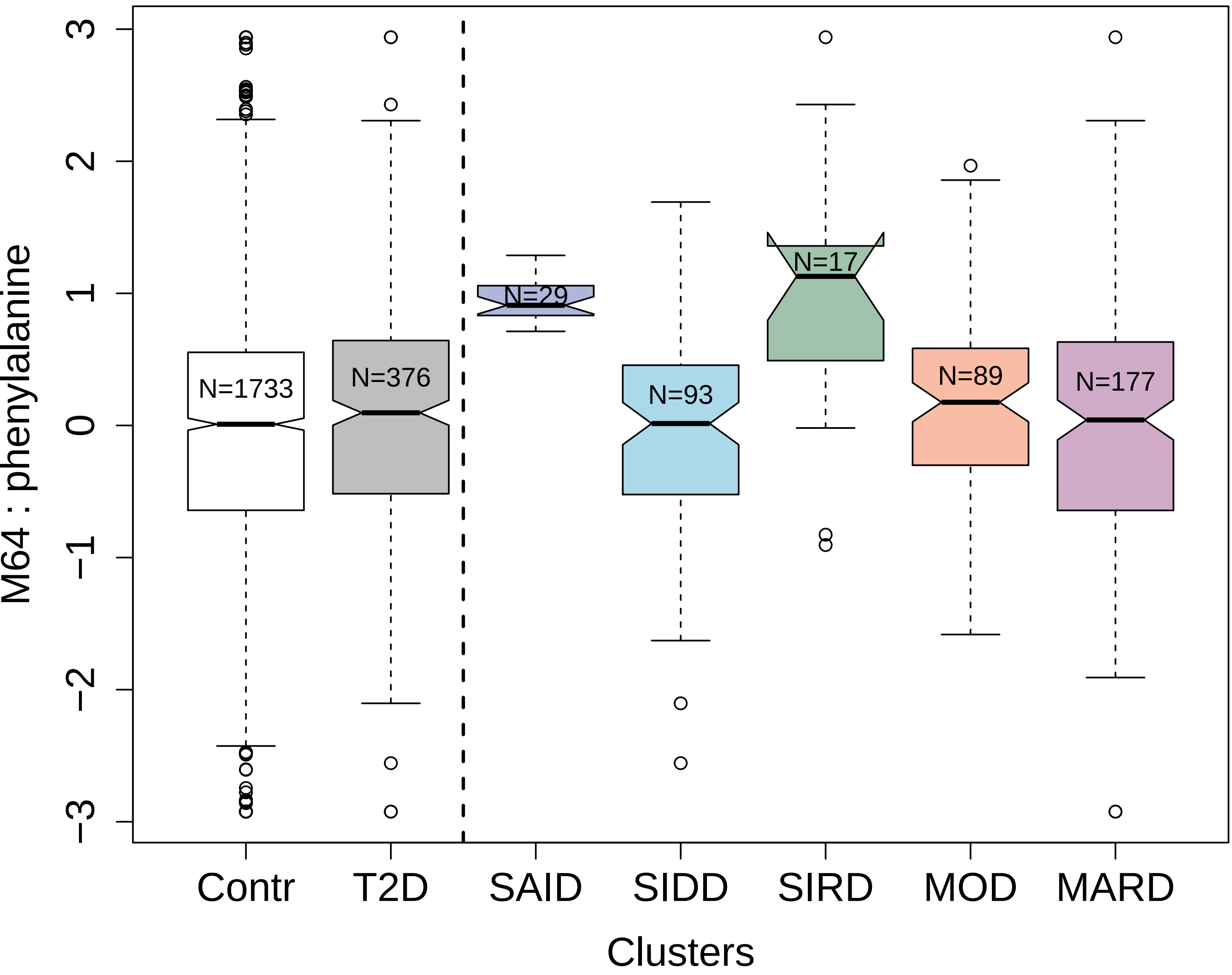

M64 : phenylalanine
3
2
1
0
−1
−2
−3
N=1733
N=376
N=29
N=93
N=17
N=89
N=177
Contr
T2D
SAID
SIDD
SIRD
MOD
MARD
Clusters

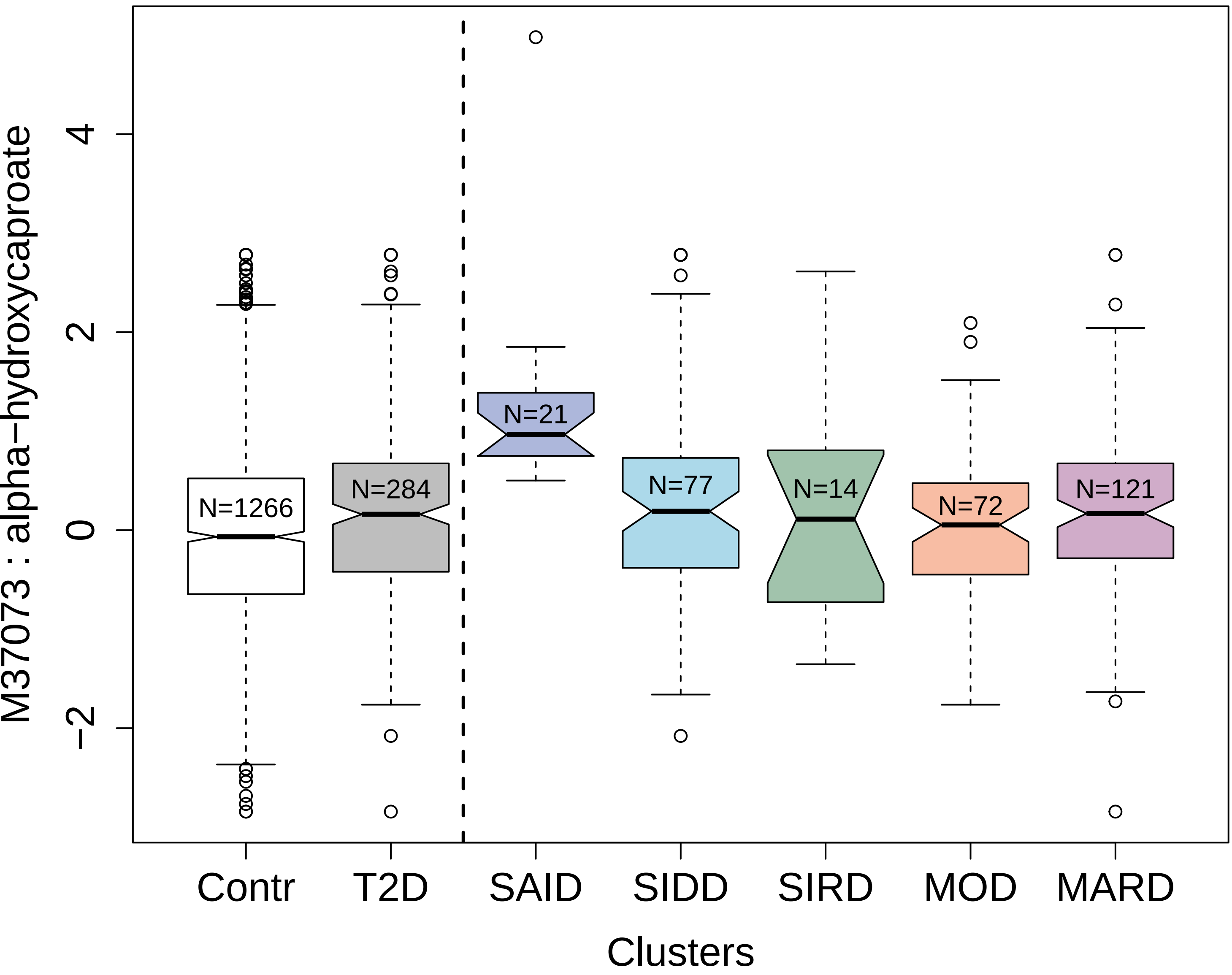
M37073 : alpha−hydroxycaproate
4
2
0
−2
N=1266
N=284
N=21
N=77
N=14
N=72
N=121
Contr
T2D
SAID
SIDD
SIRD
MOD
MARD
Clusters

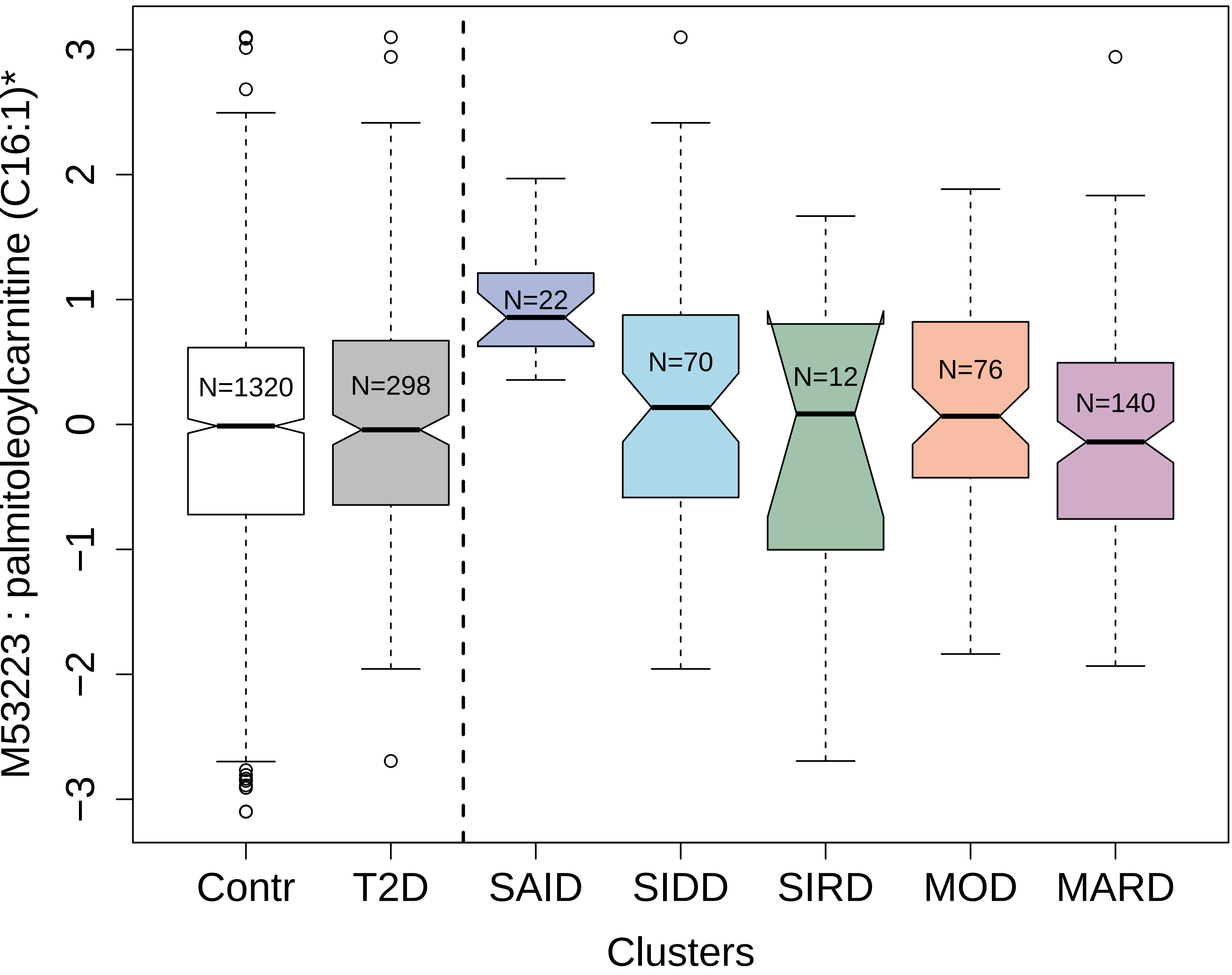
M53223 : palmitoleoylcarnitine (C16:1)*
3
2
1
0
−1
−2
−3
N=1320
N=298
N=22
N=70
N=12
N=76
N=140
Contr
T2D
SAID
SIDD
SIRD
MOD
MARD
Clusters

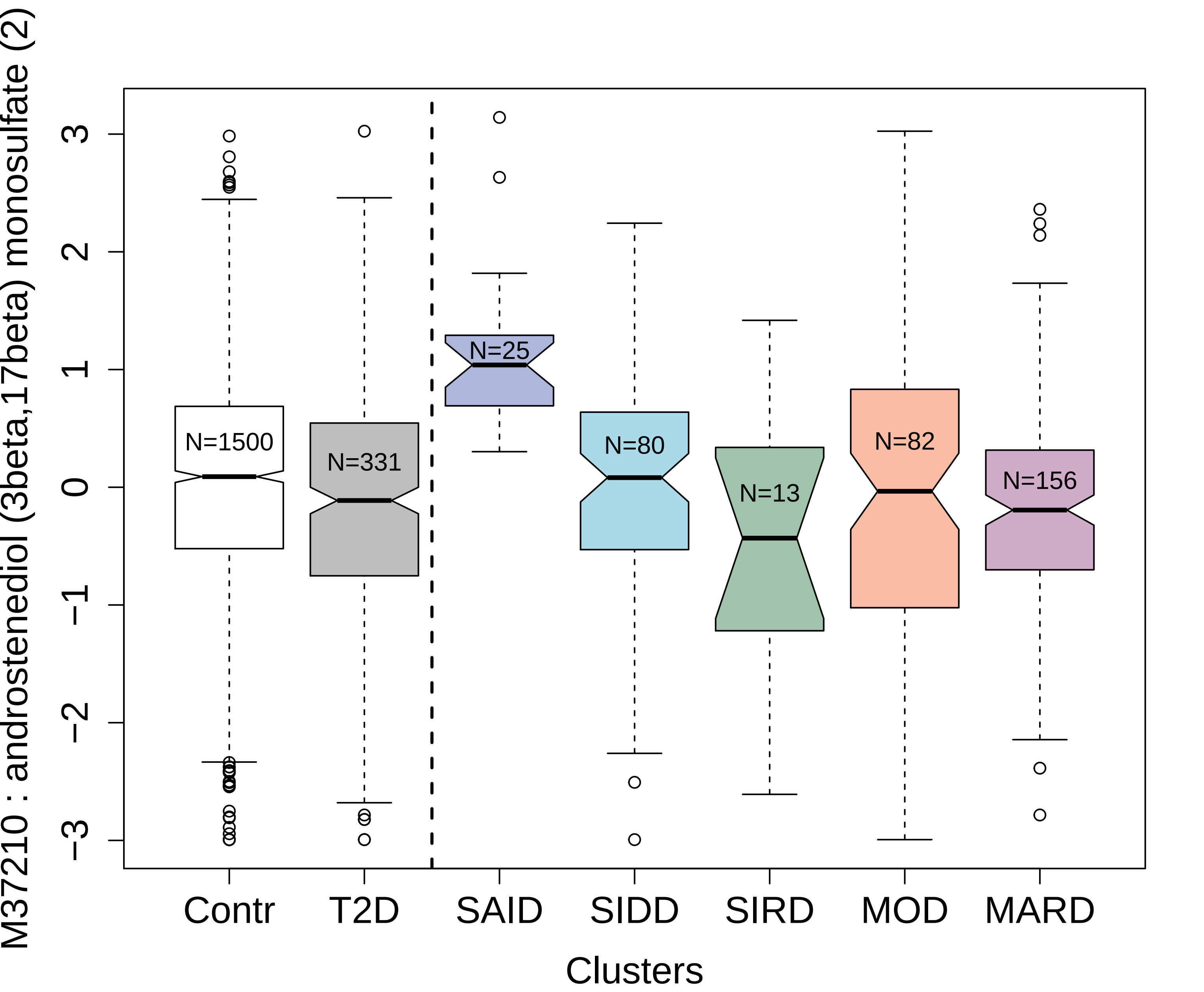
M37210 : androstenediol (3beta,17beta) monosulfate (2)
3
2
1
0
−1
−2
−3
N=1500
N=331
N=25
N=80
N=13
N=82
N=156
Contr
T2D
SAID
SIDD
SIRD
MOD
MARD
Clusters

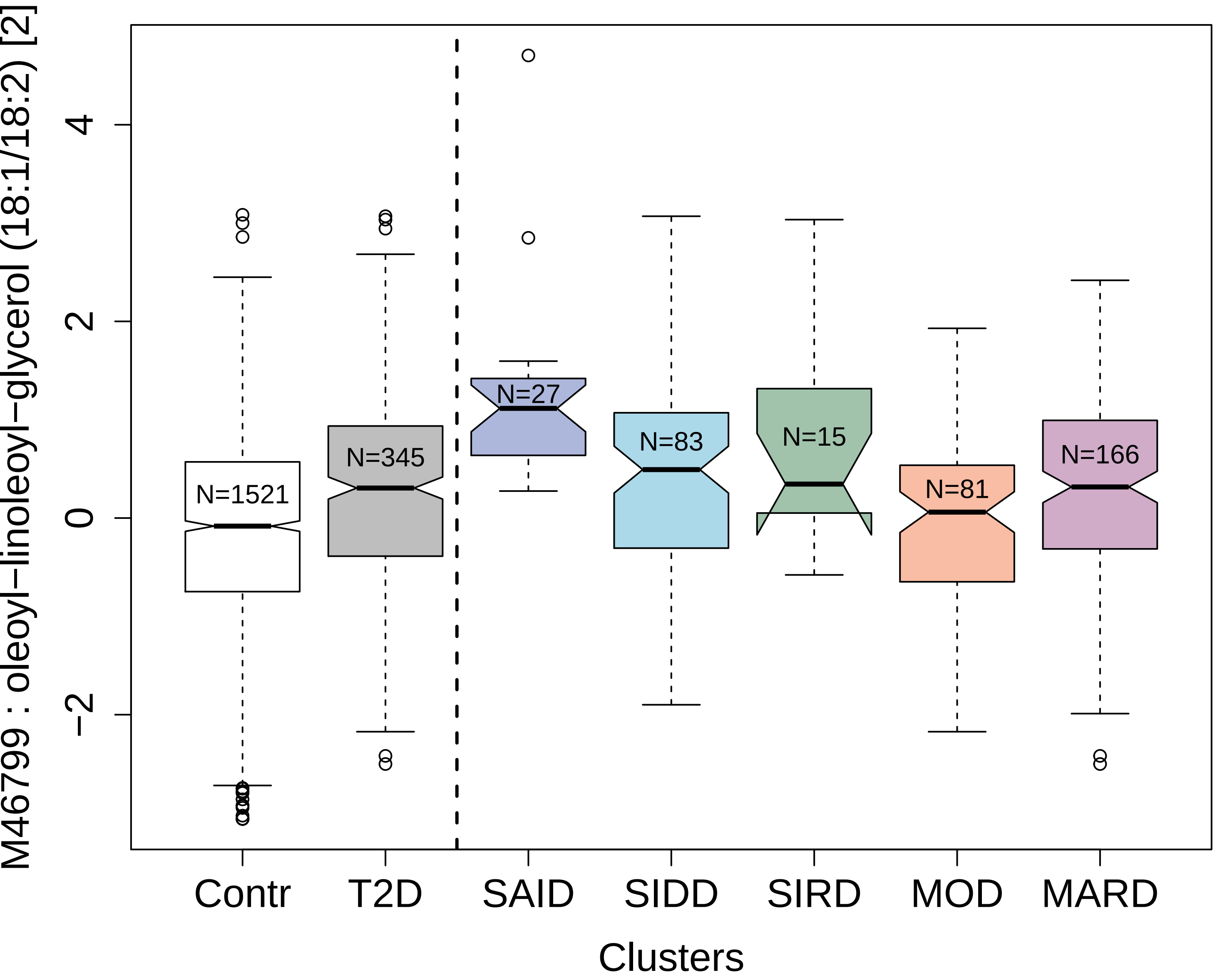
M46799 : oleoyl−linoleoyl−glycerol (18:1/18:2) [2]
4
2
0
−2
N=1521
N=345
N=27
N=83
N=15
N=81
N=166
Contr
T2D
SAID
SIDD
SIRD
MOD
MARD
Clusters

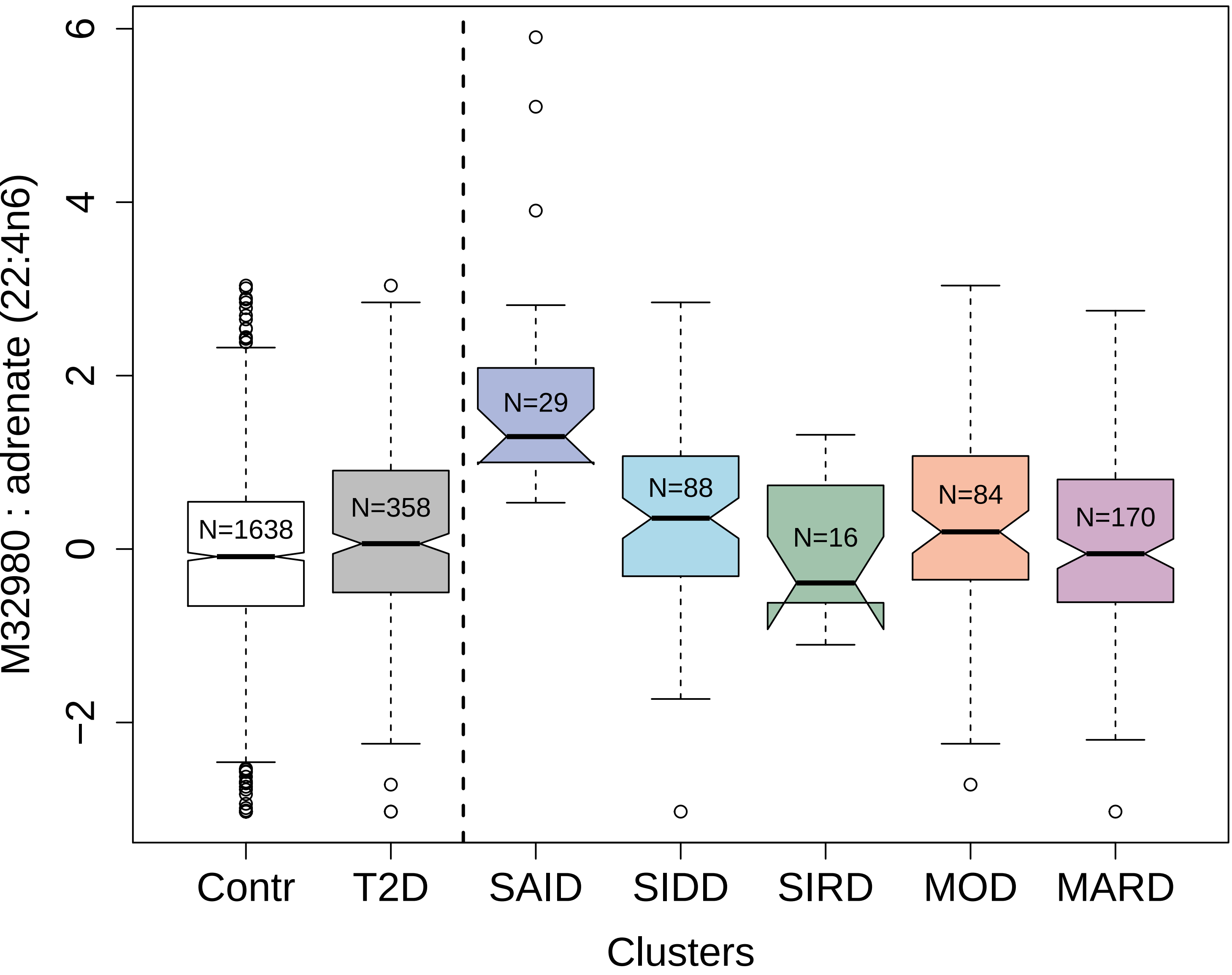

M32980 : adrenate (22:4n6)
6
4
2
0
−2
N=1638
N=358
N=29
N=88
N=16
N=84
N=170
Contr
T2D
SAID
SIDD
SIRD
MOD
MARD
Clusters

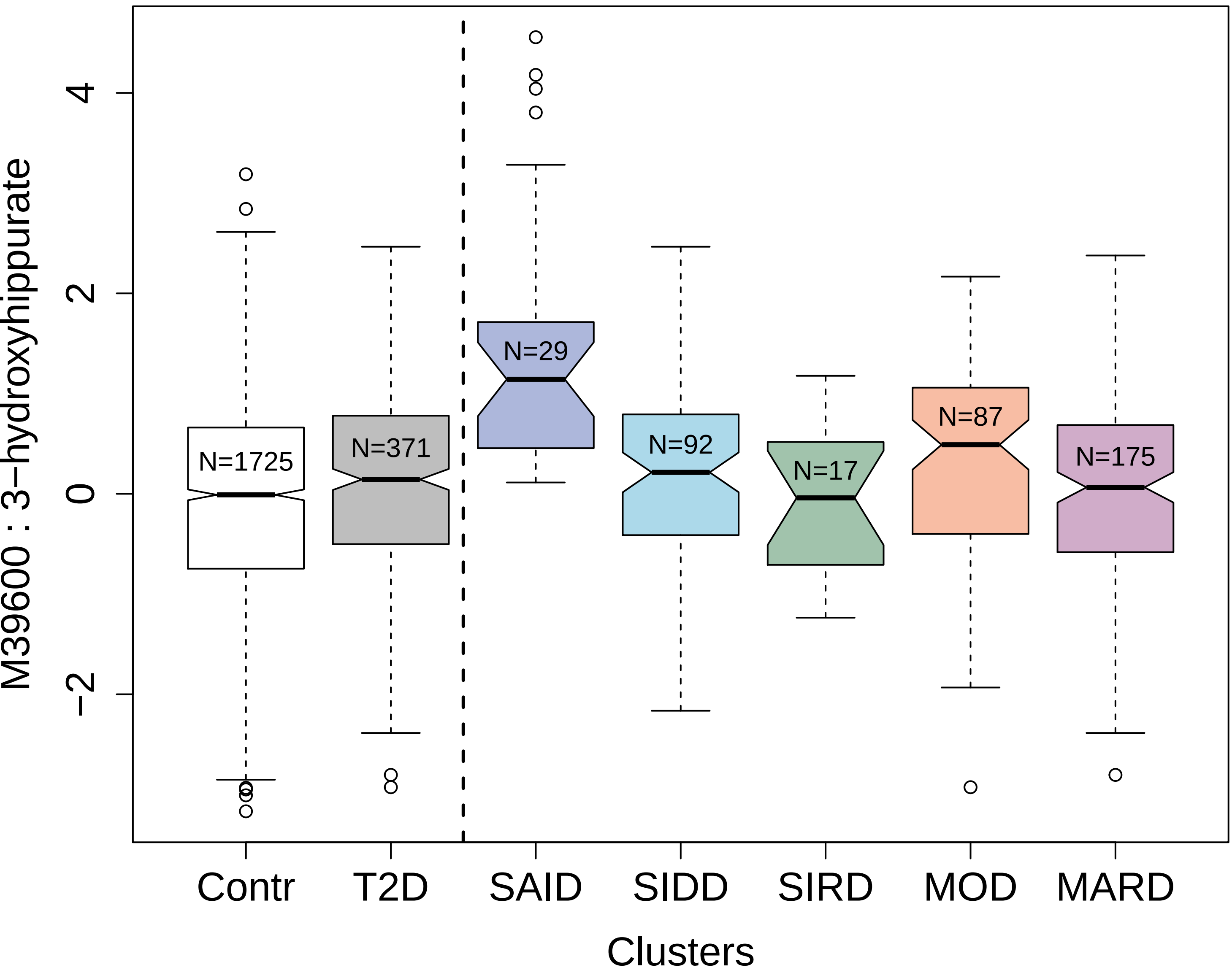

M39600 : 3−hydroxyhippurate
4
2
0
−2
N=1725
N=371
N=29
N=92
N=17
N=87
N=175
Contr
T2D
SAID
SIDD
SIRD
MOD
MARD
Clusters

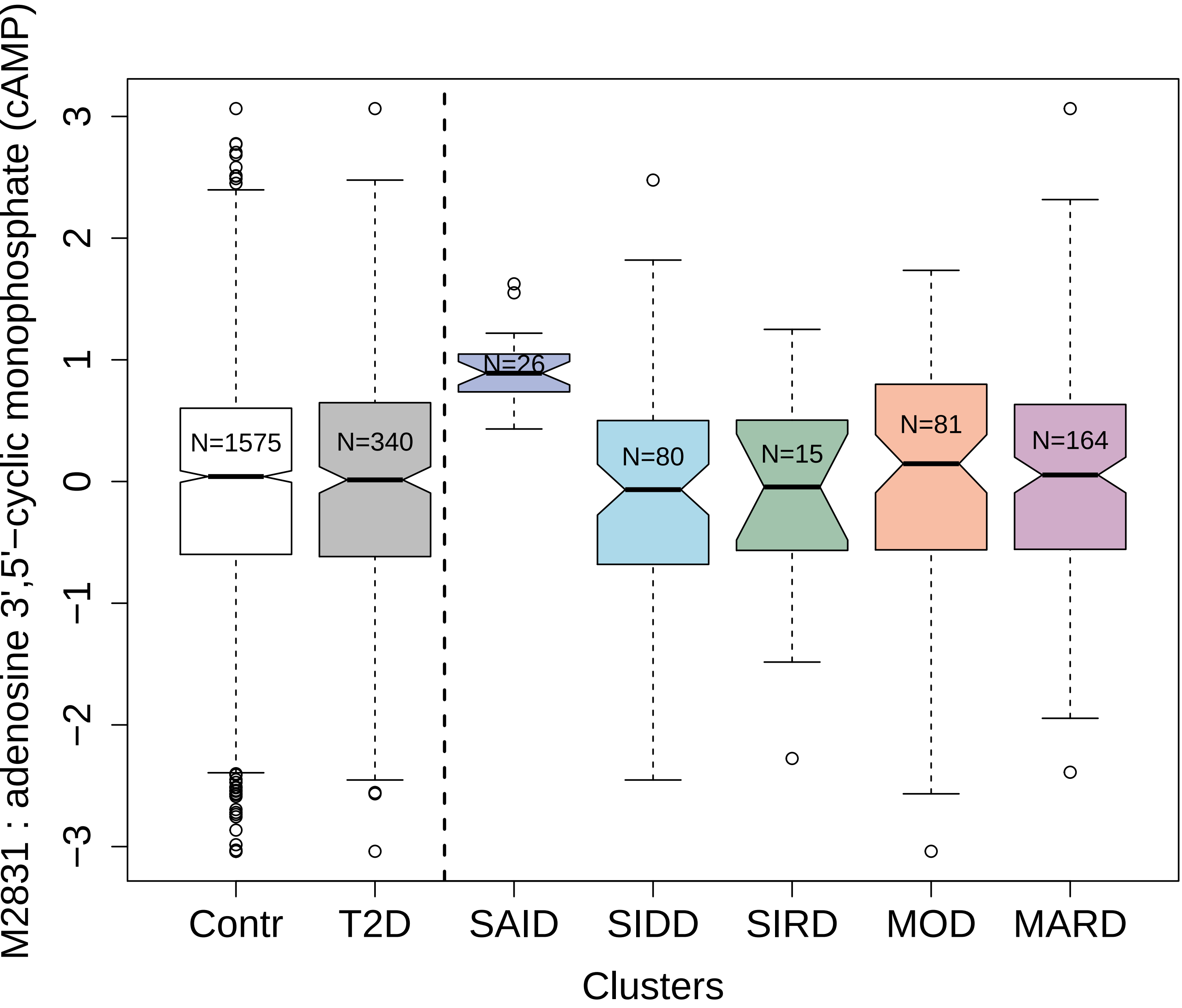
M2831 : adenosine 3',5'−cyclic monophosphate (cAMP)
3
2
1
0
−1
−2
−3
N=1575
N=340
N=26
N=80
N=15
N=81
N=164
Contr
T2D
SAID
SIDD
SIRD
MOD
MARD
Clusters

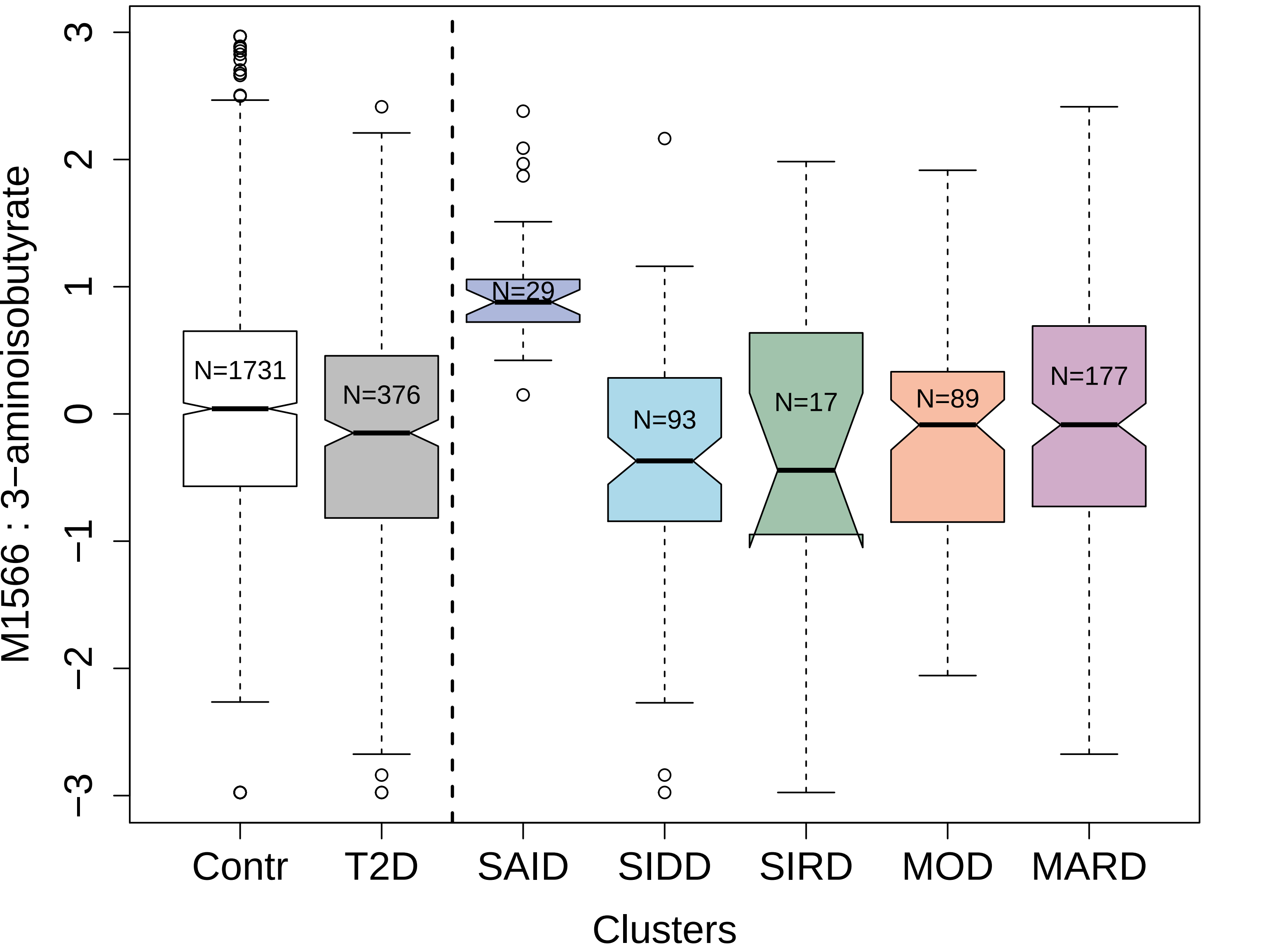

M1566 : 3−aminoisobutyrate
3
2
1
0
−1
−2
−3
N=1731
N=376
N=29
N=93
N=17
N=89
N=177
Contr
T2D
SAID
SIDD
SIRD
MOD
MARD
Clusters

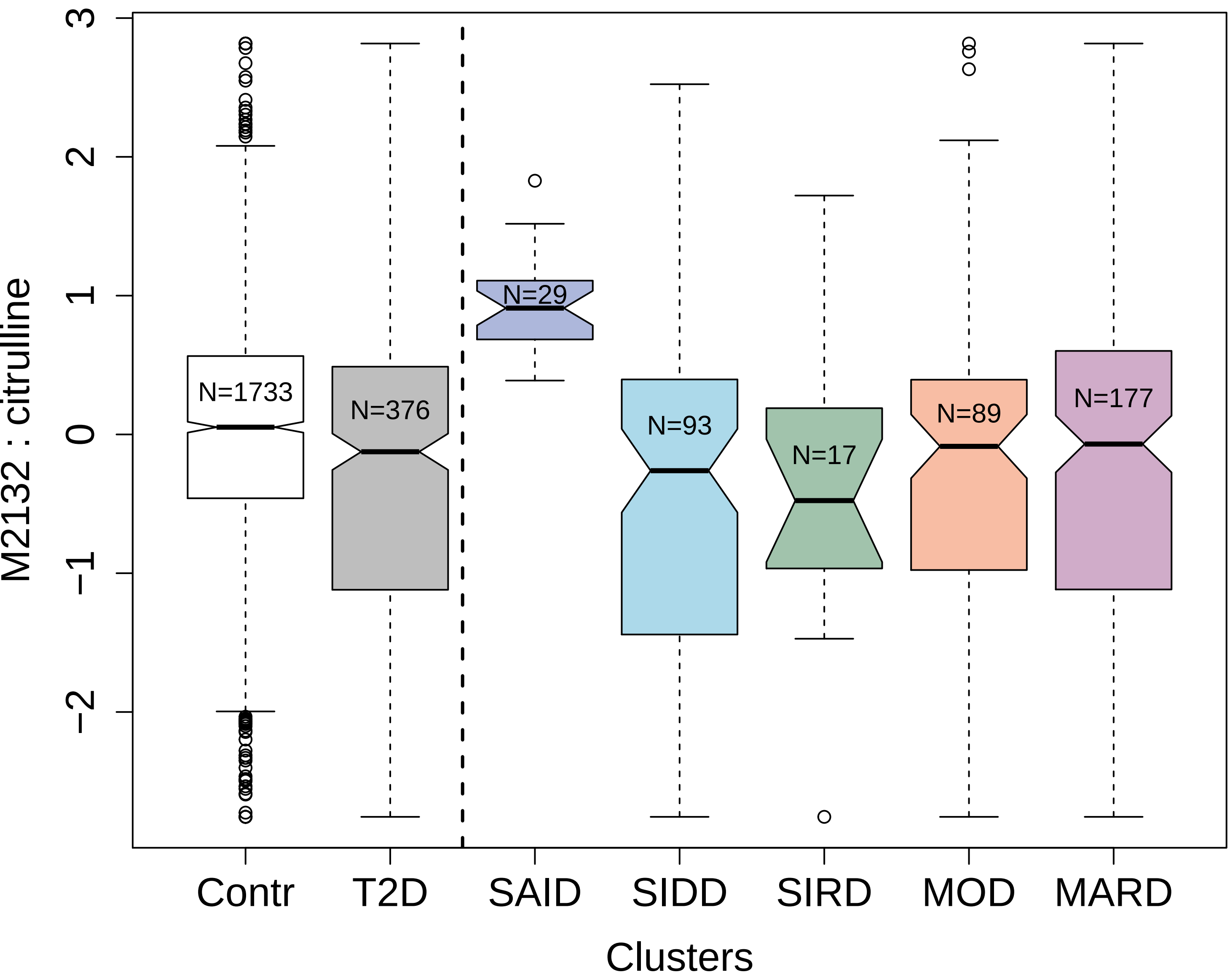
M2132 : citrulline
3
2
1
0
−1
−2
N=1733
N=376
N=29
N=93
N=17
N=89
N=177
Contr
T2D
SAID
SIDD
SIRD
MOD
MARD
Clusters

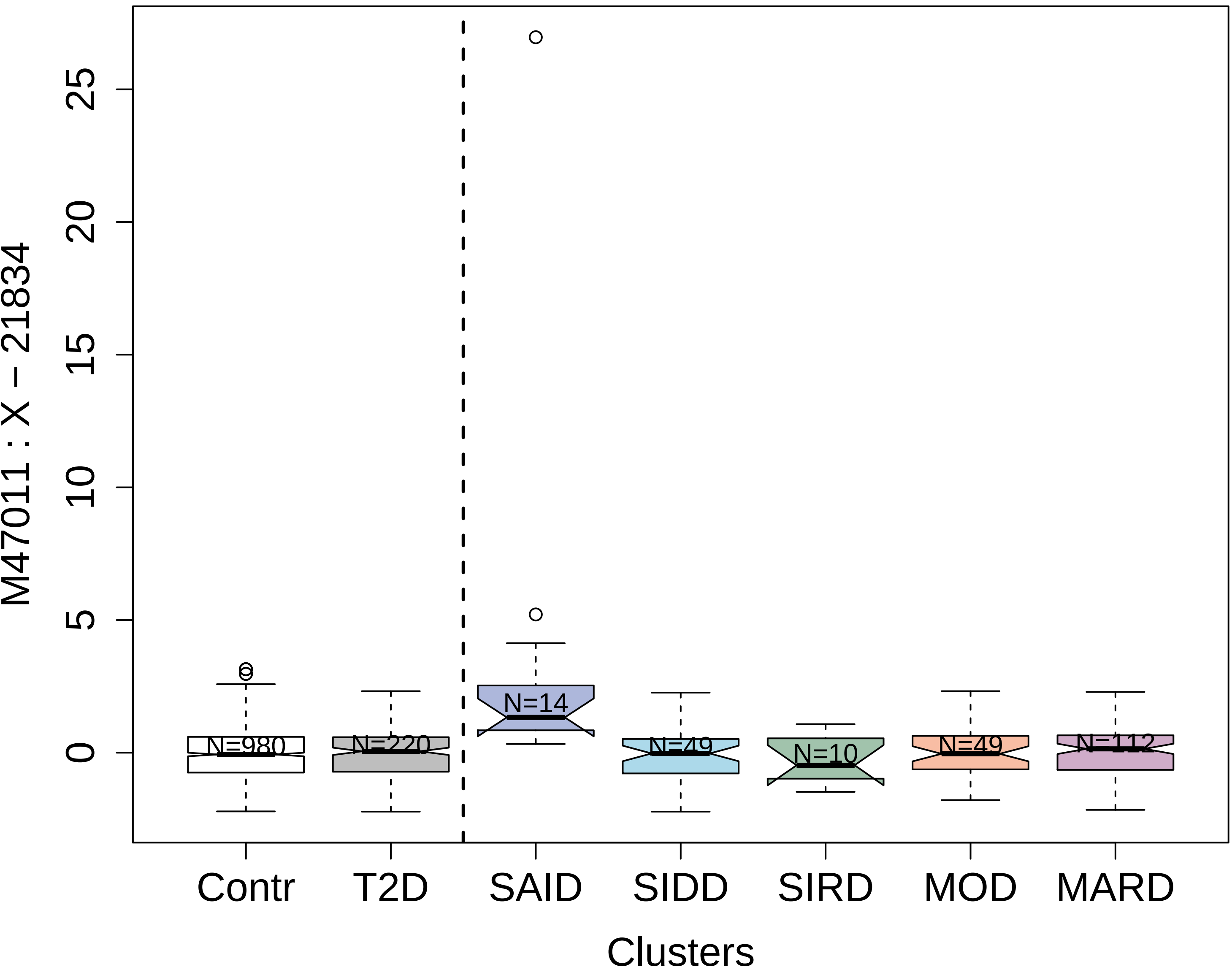
M47011 : X − 21834
0
5
10
15
20
25
N=980
N=220
N=14
N=49
N=10
N=49
N=112
Contr
T2D
SAID
SIDD
SIRD
MOD
MARD
Clusters

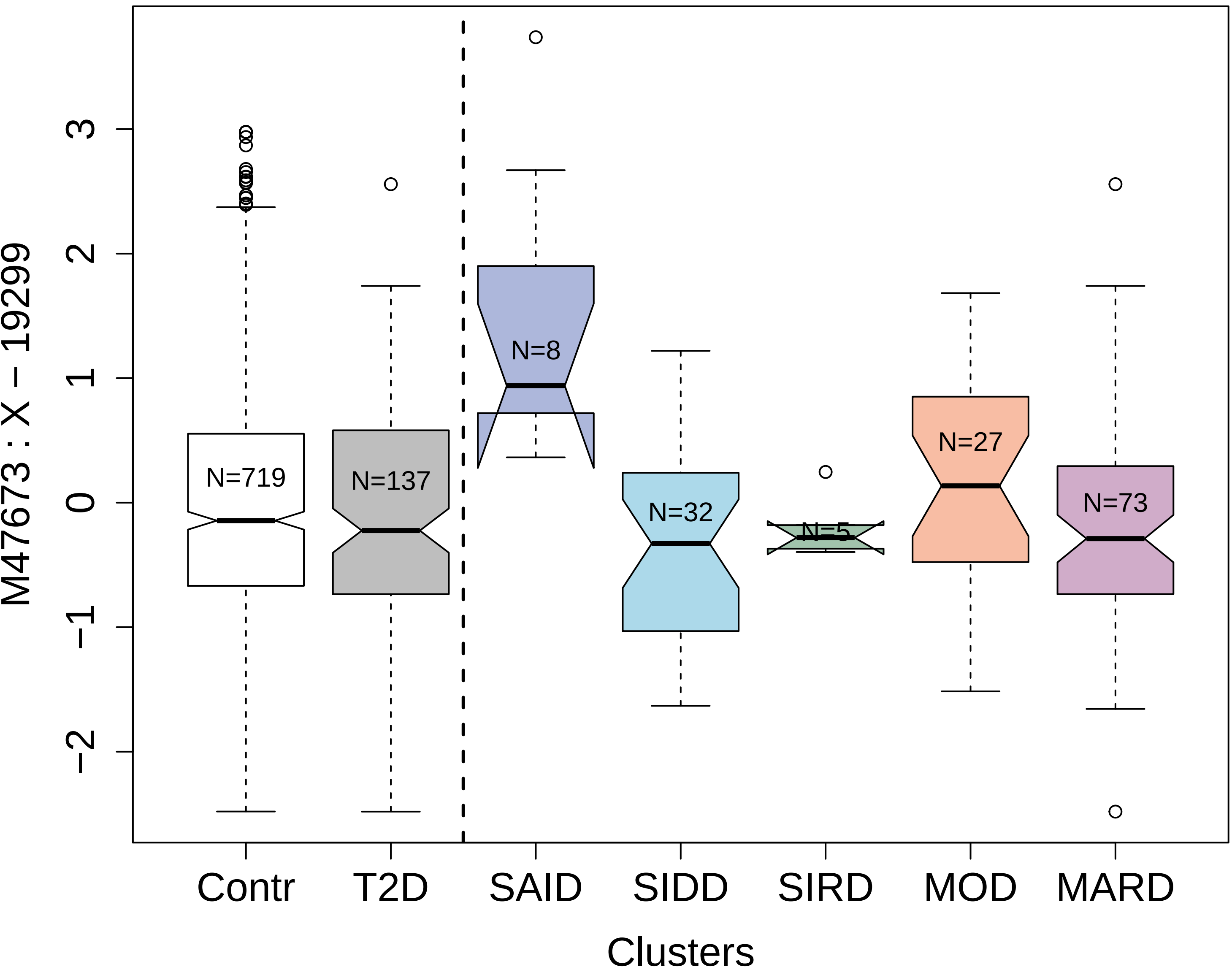

M47673 : X − 19299
3
2
1
0
−1
−2
N=719
N=137
N=8
N=32
N=5
N=27
N=73
Contr
T2D
SAID
SIDD
SIRD
MOD
MARD
Clusters

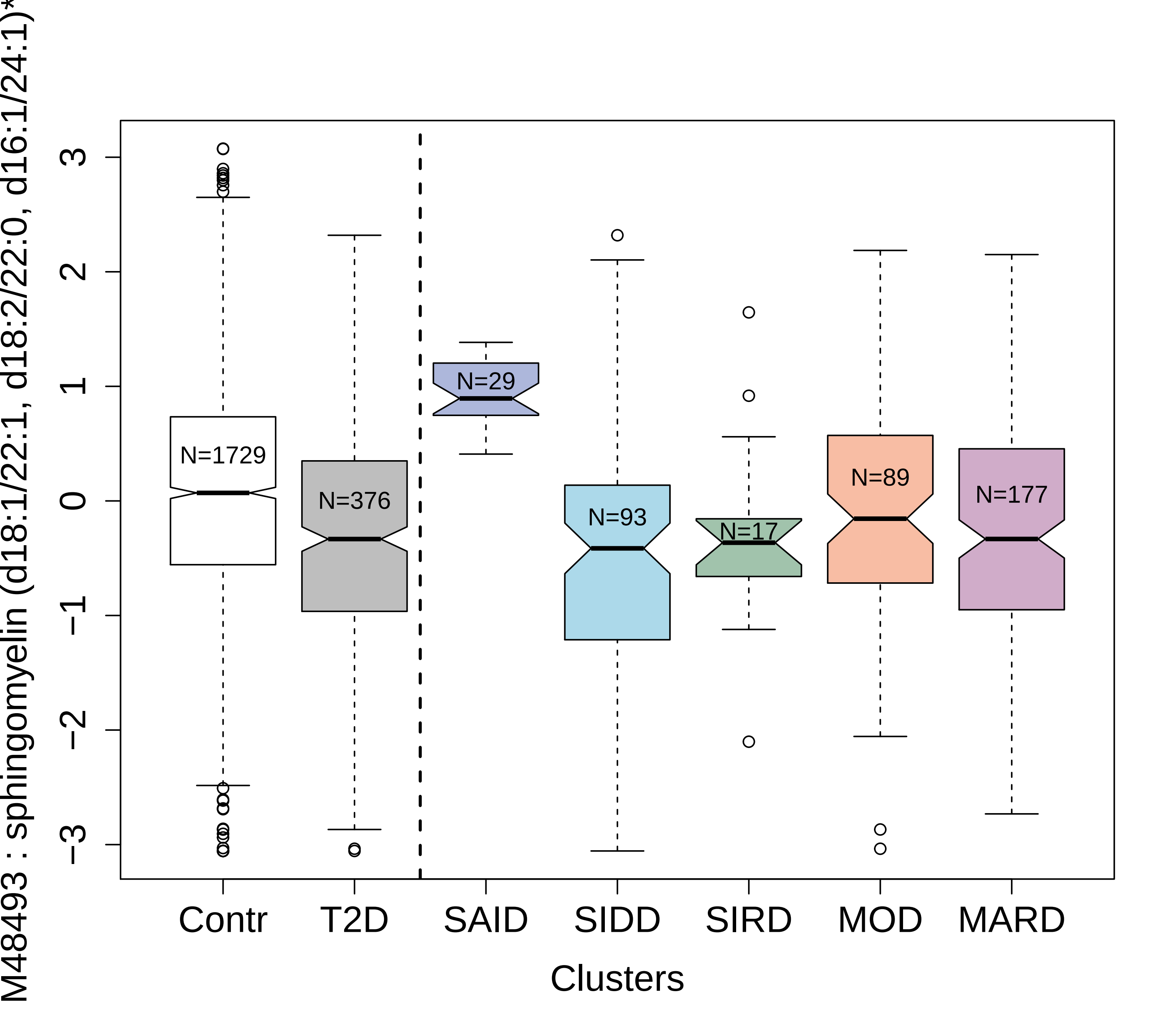
M48493 : sphingomyelin (d18:1/22:1, d18:2/22:0, d16:1/24:1)*
3
2
1
0
−1
−2
−3
N=1729
N=376
N=29
N=93
N=17
N=89
N=177
Contr
T2D
SAID
SIDD
SIRD
MOD
MARD
Clusters

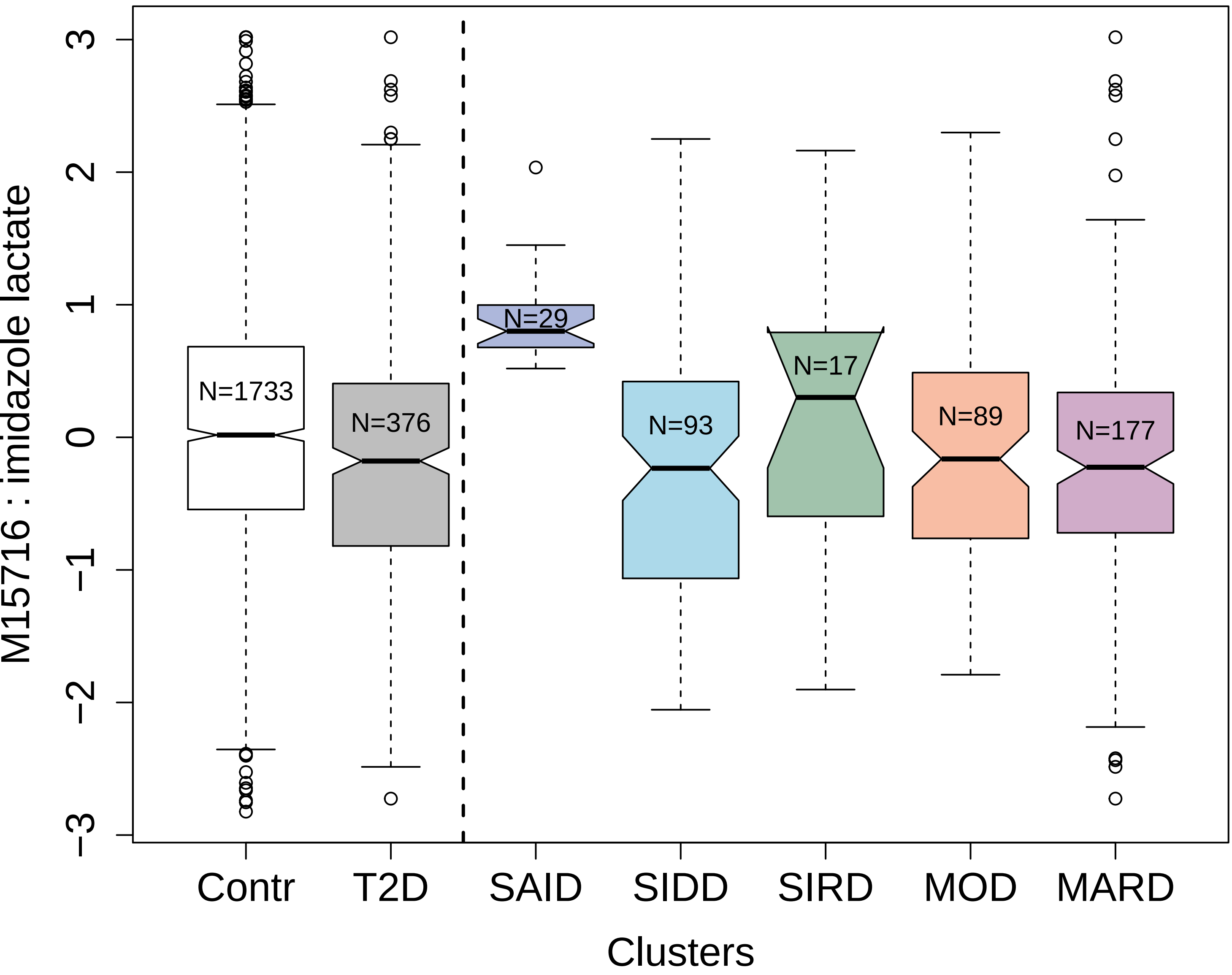
M15716 : imidazole lactate
3
2
1
0
−1
−2
−3
N=1733
N=376
N=29
N=93
N=17
N=89
N=177
Contr
T2D
SAID
SIDD
SIRD
MOD
MARD
Clusters

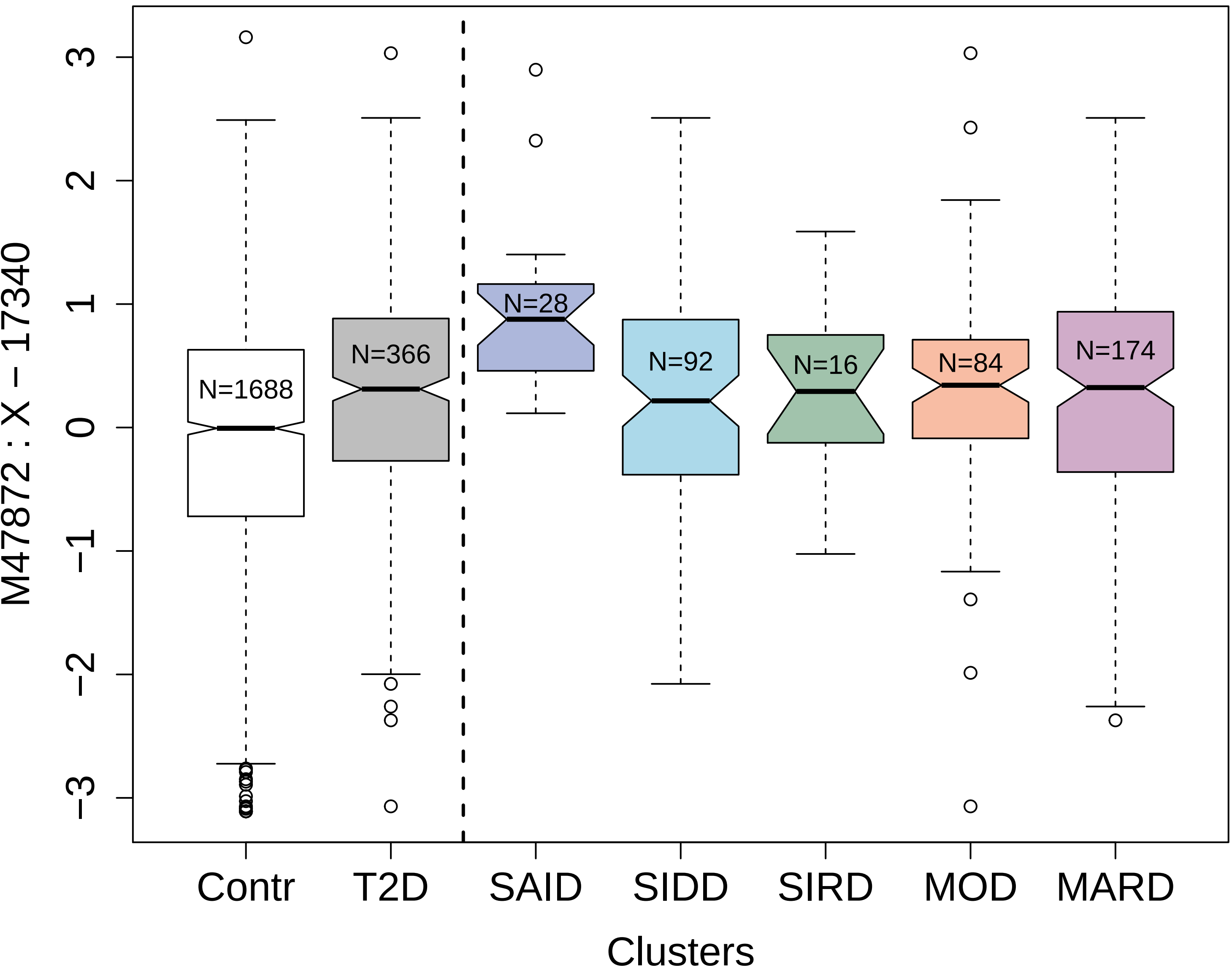

M47872 : X − 17340
3
2
1
0
−1
−2
−3
N=1688
N=366
N=28
N=92
N=16
N=84
N=174
Contr
T2D
SAID
SIDD
SIRD
MOD
MARD
Clusters

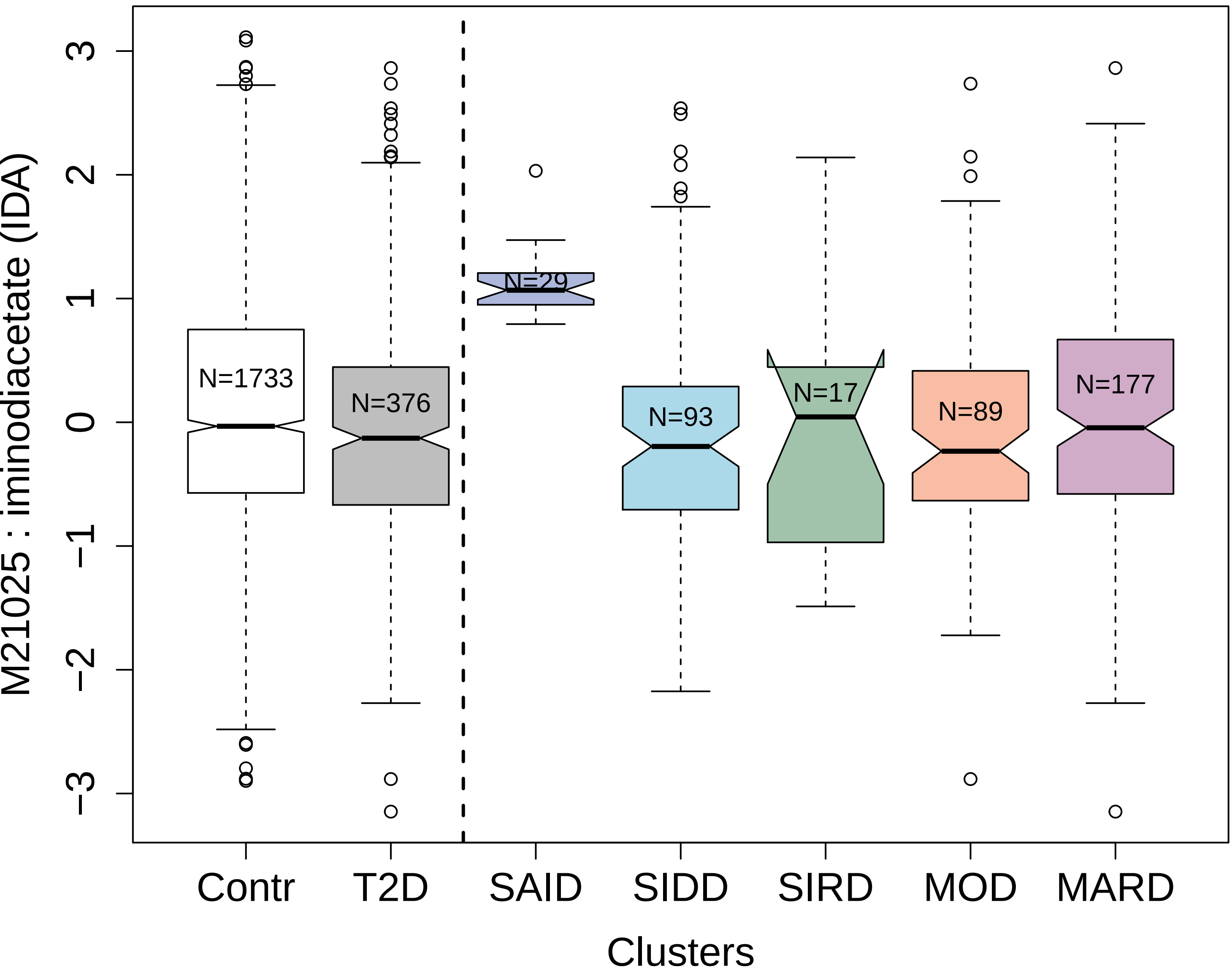

M21025 : iminodiacetate (IDA)
3
2
1
0
−1
−2
−3
N=1733
N=376
N=29
N=93
N=17
N=89
N=177
Contr
T2D
SAID
SIDD
SIRD
MOD
MARD
Clusters

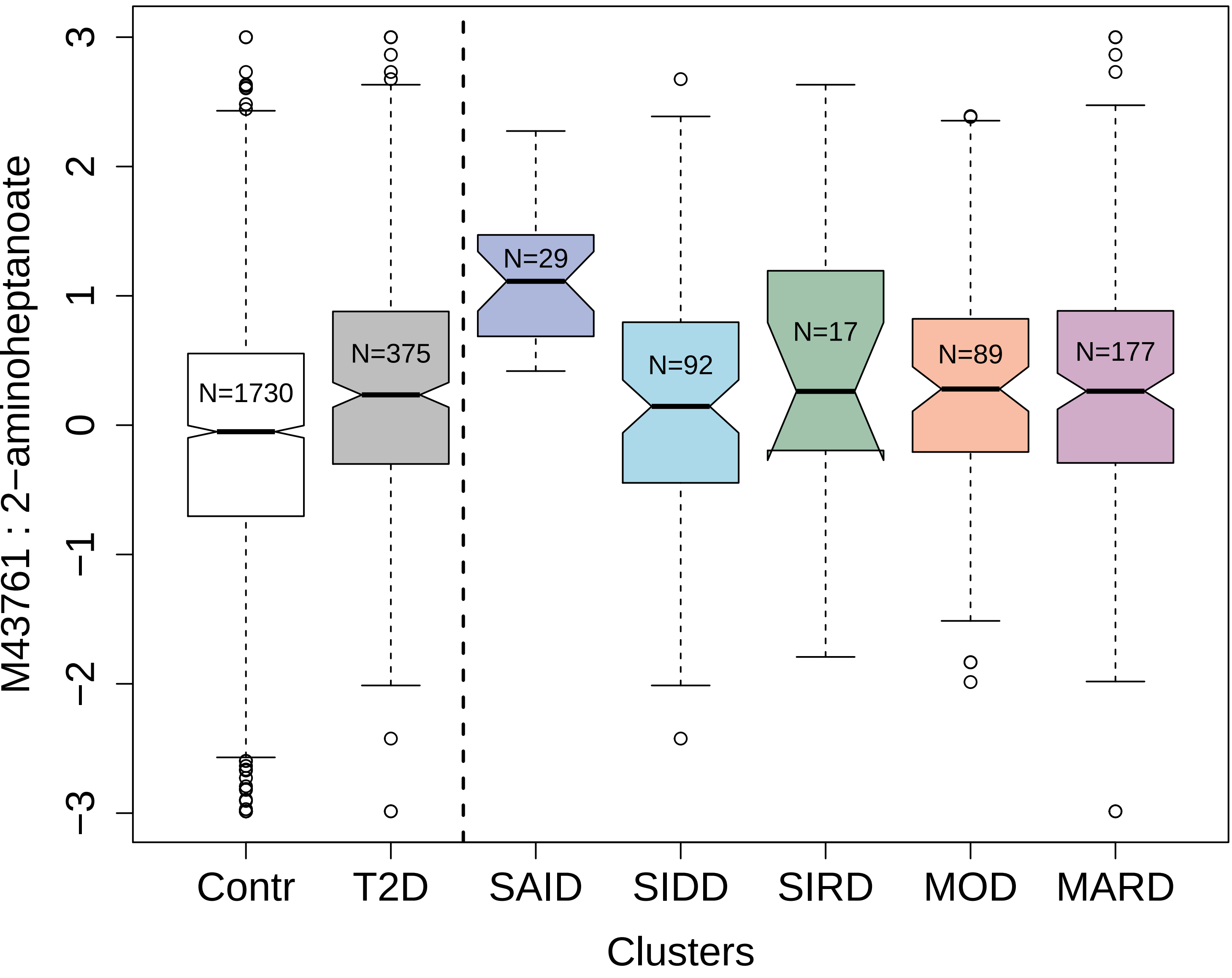

M43761 : 2−aminoheptanoate
3
2
1
0
−1
−2
−3
N=1730
N=375
N=29
N=92
N=17
N=89
N=177
Contr
T2D
SAID
SIDD
SIRD
MOD
MARD
Clusters

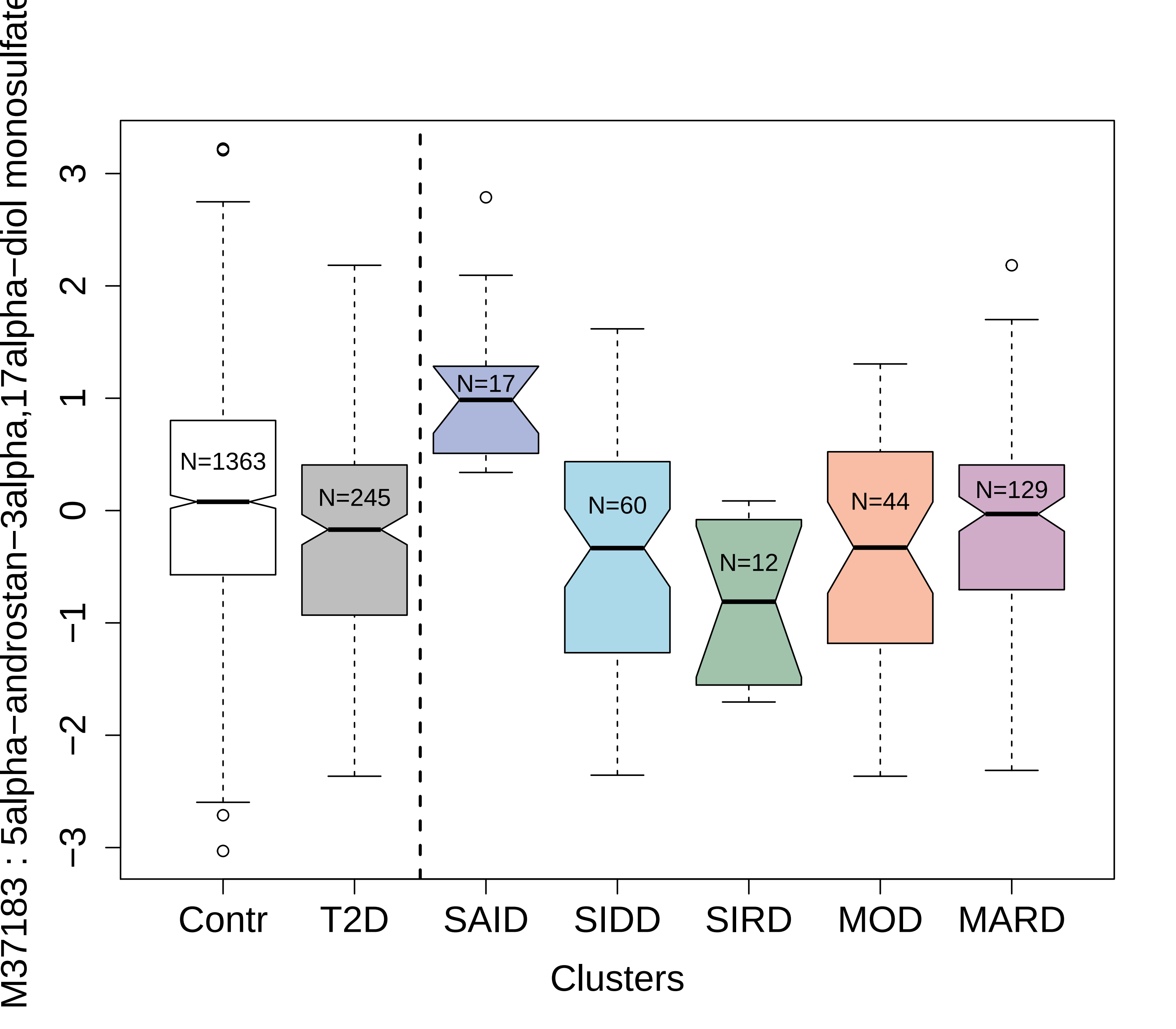
M37183 : 5alpha−androstan−3alpha,17alpha−diol monosulfate
3
2
1
0
−1
−2
−3
N=1363
N=245
N=17
N=60
N=12
N=44
N=129
Contr
T2D
SAID
SIDD
SIRD
MOD
MARD
Clusters

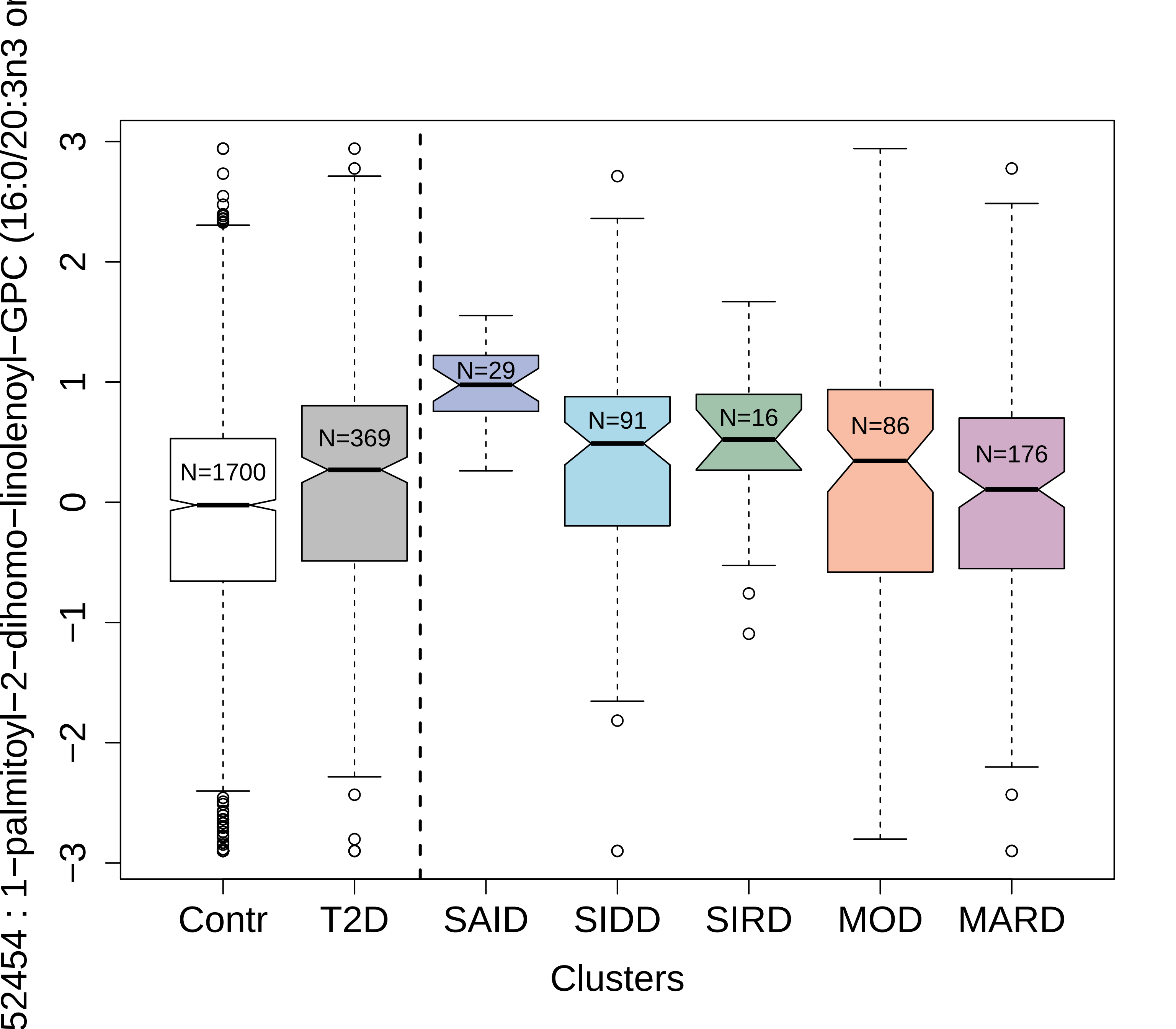

52454 : 1−palmitoyl−2−dihomo−linolenoyl−GPC (16:0/20:3n3 or
3
2
1
0
−1
−2
−3
N=1700
N=369
N=29
N=91
N=16
N=86
N=176
Contr
T2D
SAID
SIDD
SIRD
MOD
MARD
Clusters

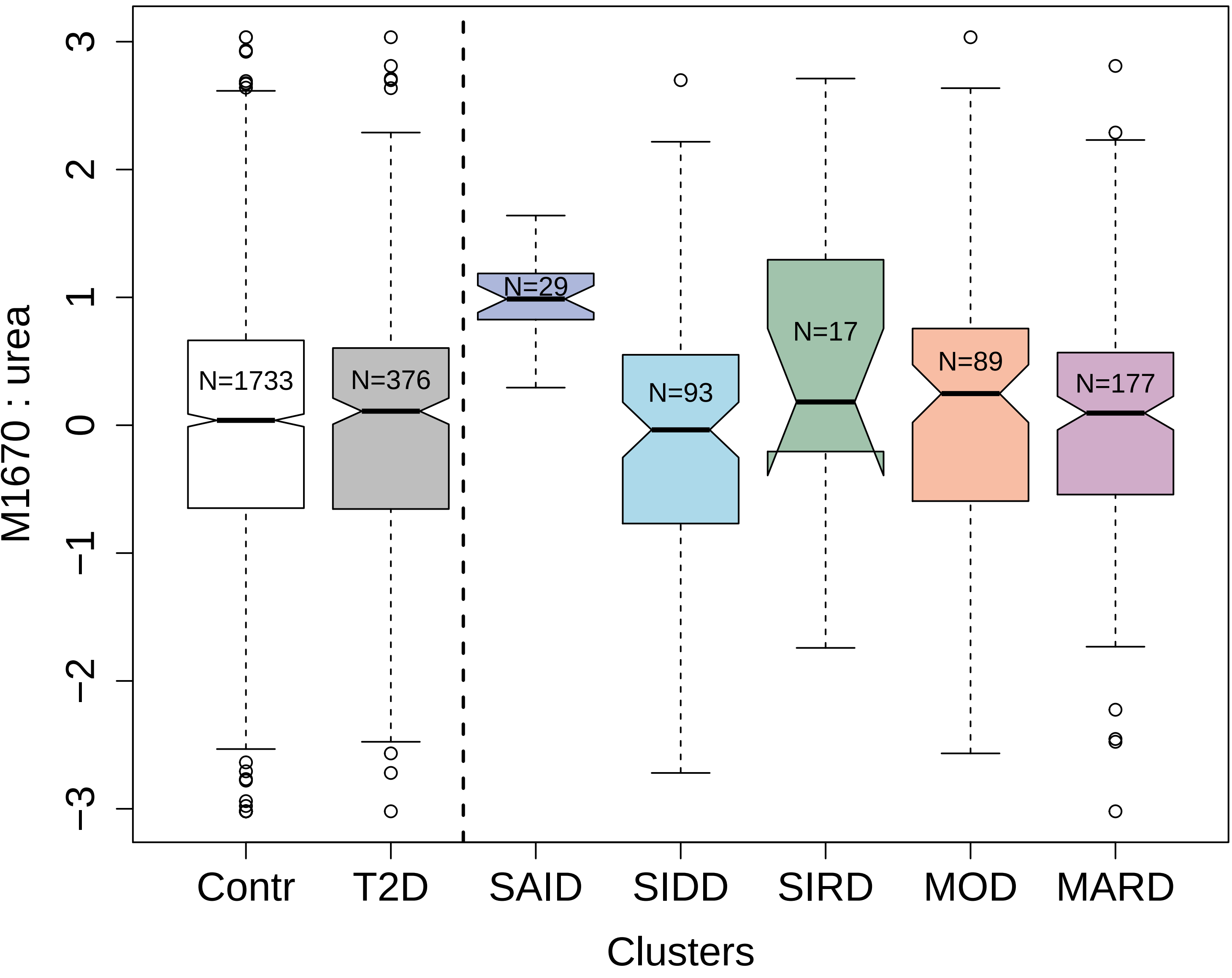

M1670 : urea
3
2
1
0
−1
−2
−3
N=1733
N=376
N=29
N=93
N=17
N=89
N=177
Contr
T2D
SAID
SIDD
SIRD
MOD
MARD
Clusters

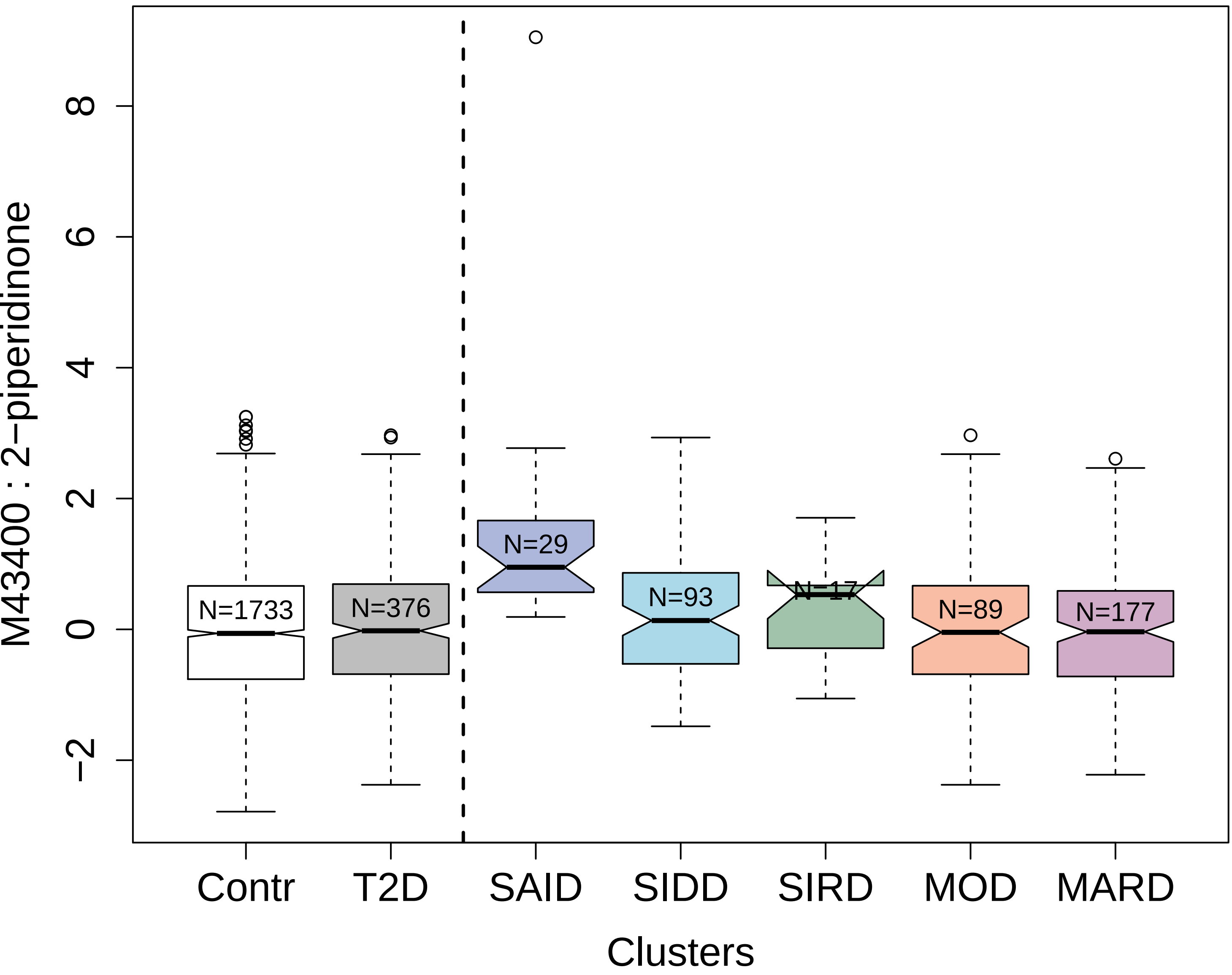

M43400 : 2−piperidinone
−2
0
2
4
6
8
N=1733
N=376
N=29
N=93
N=17
N=89
N=177
Contr
T2D
SAID
SIDD
SIRD
MOD
MARD
Clusters

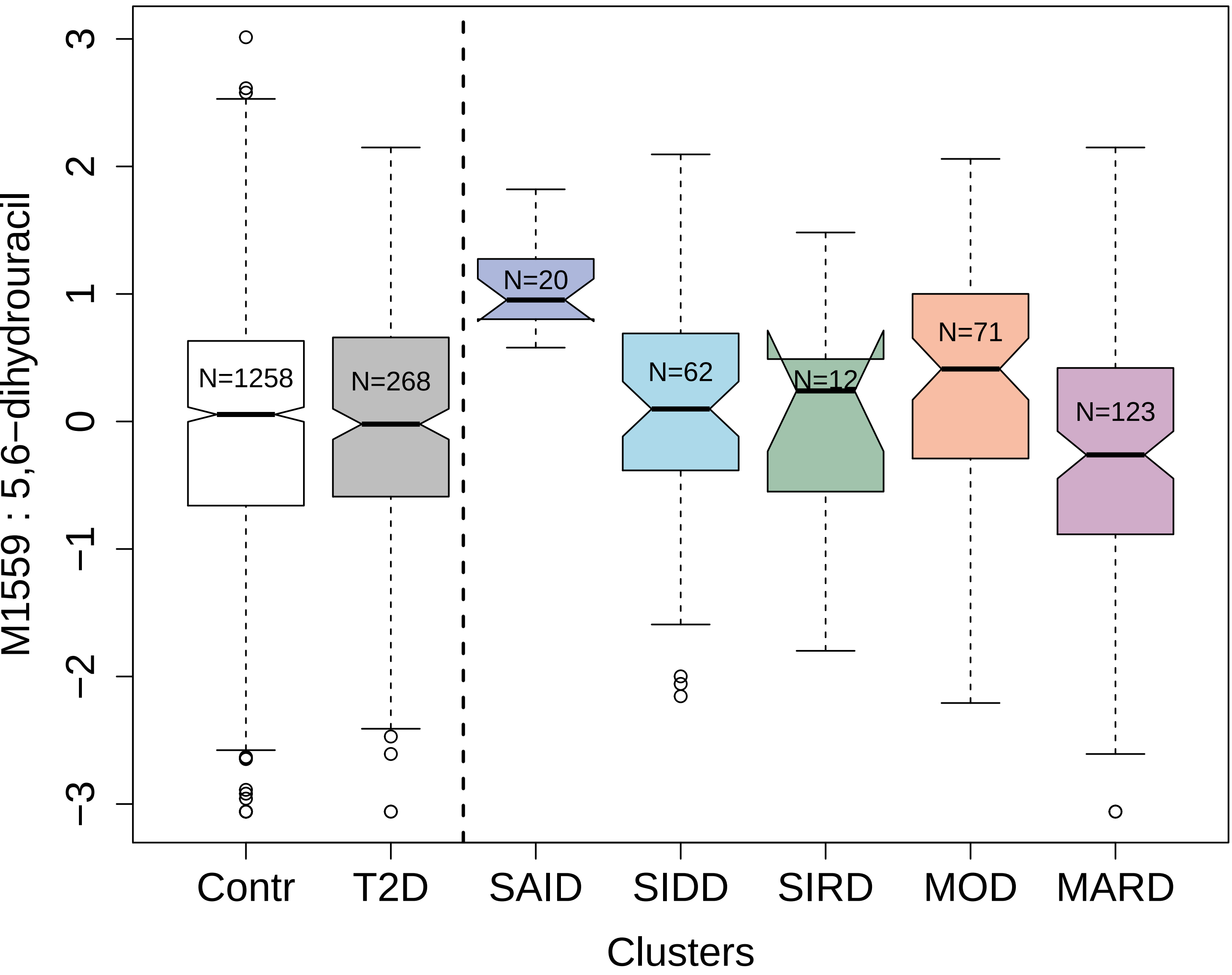

M1559 : 5,6−dihydrouracil
3
2
1
0
−1
−2
−3
N=1258
N=268
N=20
N=62
N=12
N=71
N=123
Contr
T2D
SAID
SIDD
SIRD
MOD
MARD
Clusters

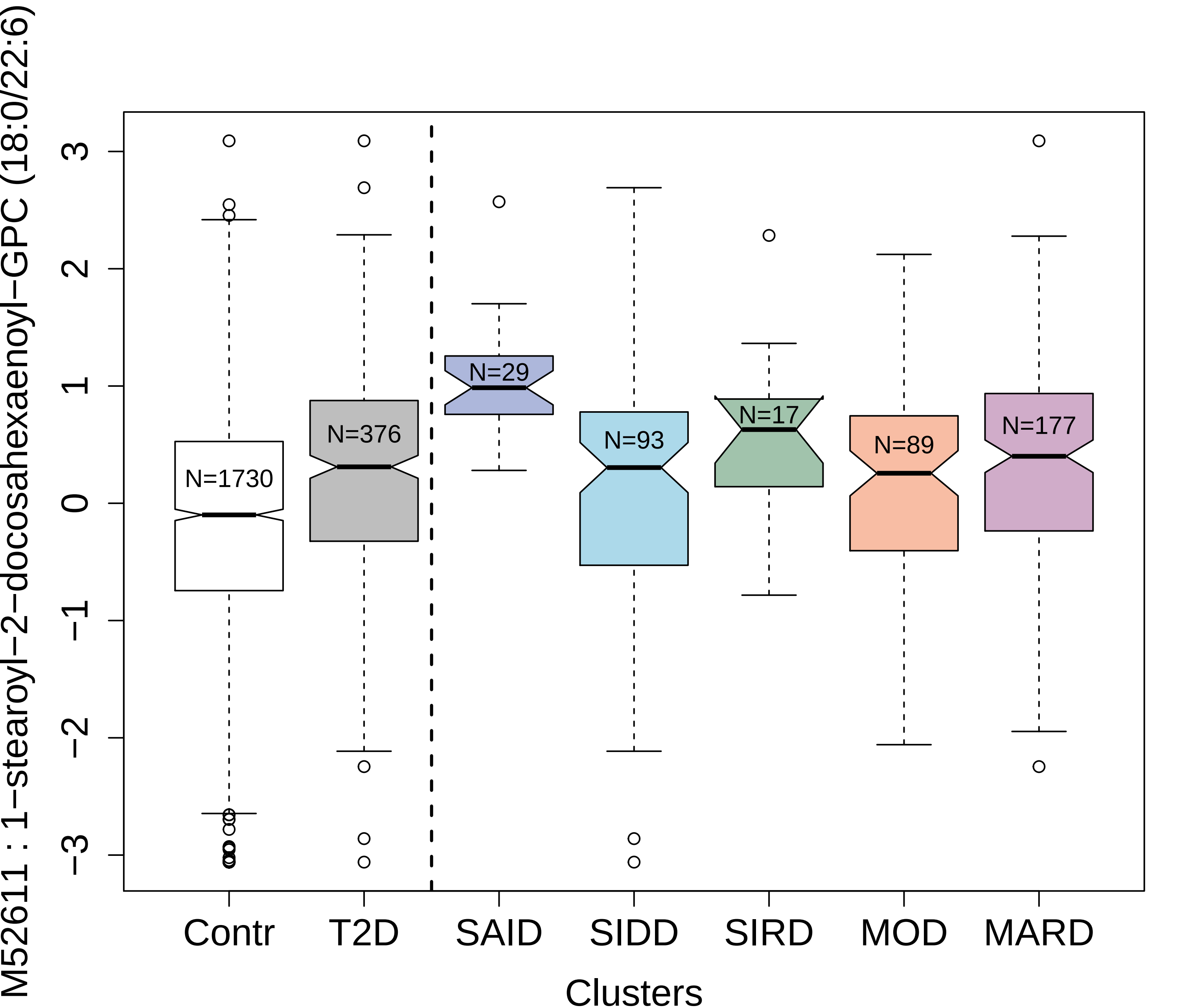
M52611 : 1−stearoyl−2−docosahexaenoyl−GPC (18:0/22:6)
3
2
1
0
−1
−2
−3
N=1730
N=376
N=29
N=93
N=17
N=89
N=177
Contr
T2D
SAID
SIDD
SIRD
MOD
MARD
Clusters

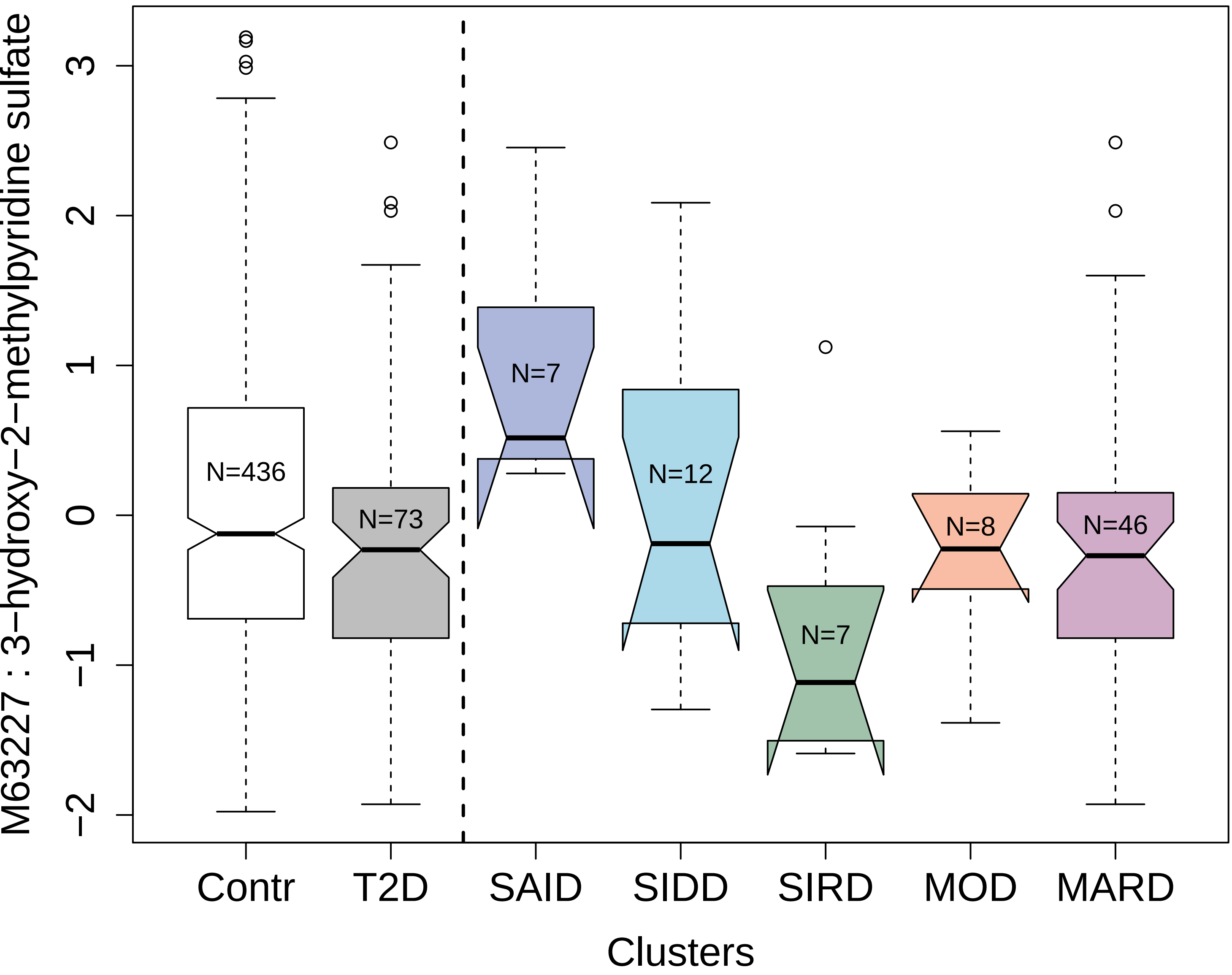

M63227 : 3−hydroxy−2−methylpyridine sulfate
−2
−1
0
1
2
3
N=436
N=73
N=7
N=12
N=7
N=8
N=46
Contr
T2D
SAID
SIDD
SIRD
MOD
MARD
Clusters

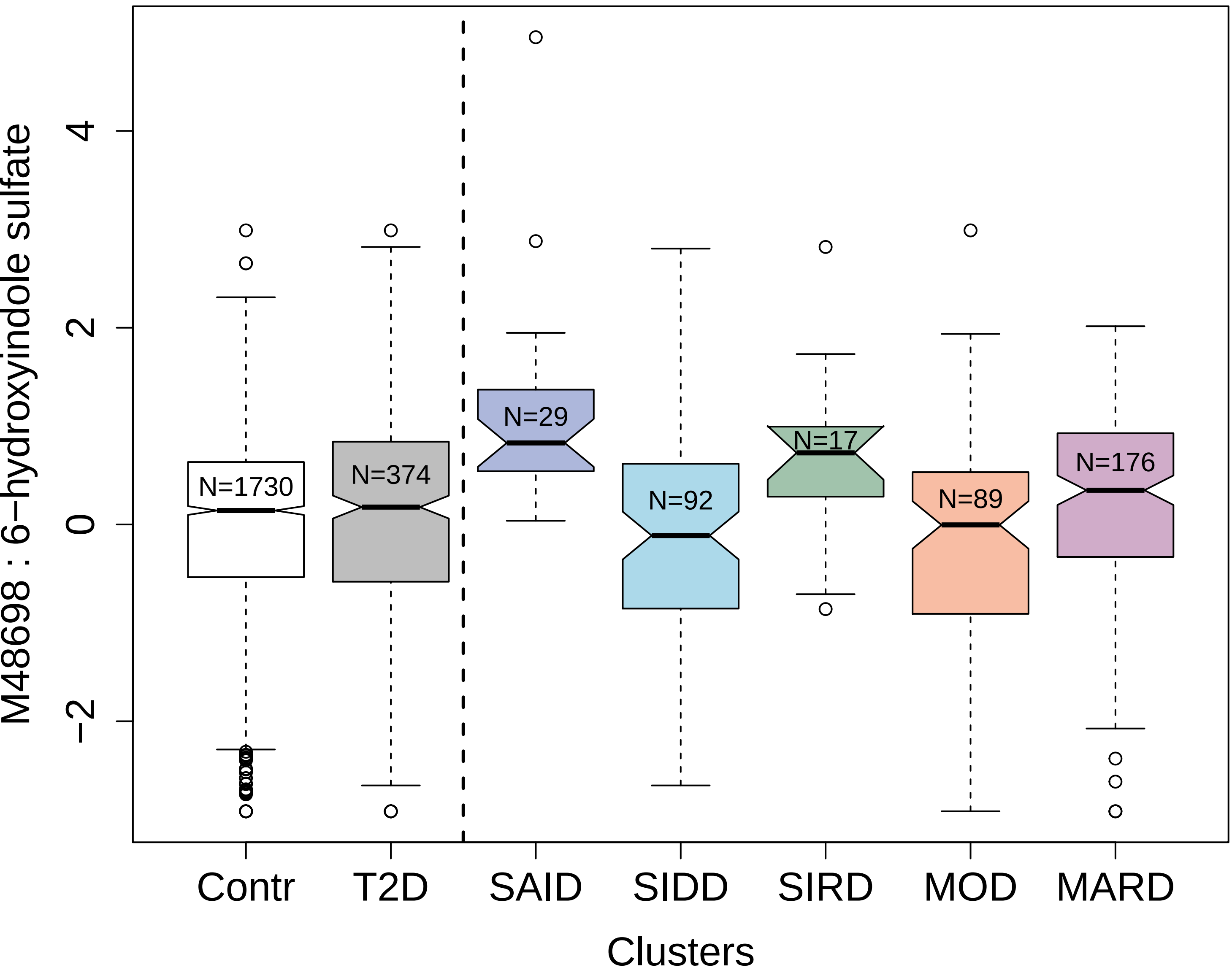
M48698 : 6−hydroxyindole sulfate
4
2
0
−2
N=1730
N=374
N=29
N=92
N=17
N=89
N=176
Contr
T2D
SAID
SIDD
SIRD
MOD
MARD
Clusters

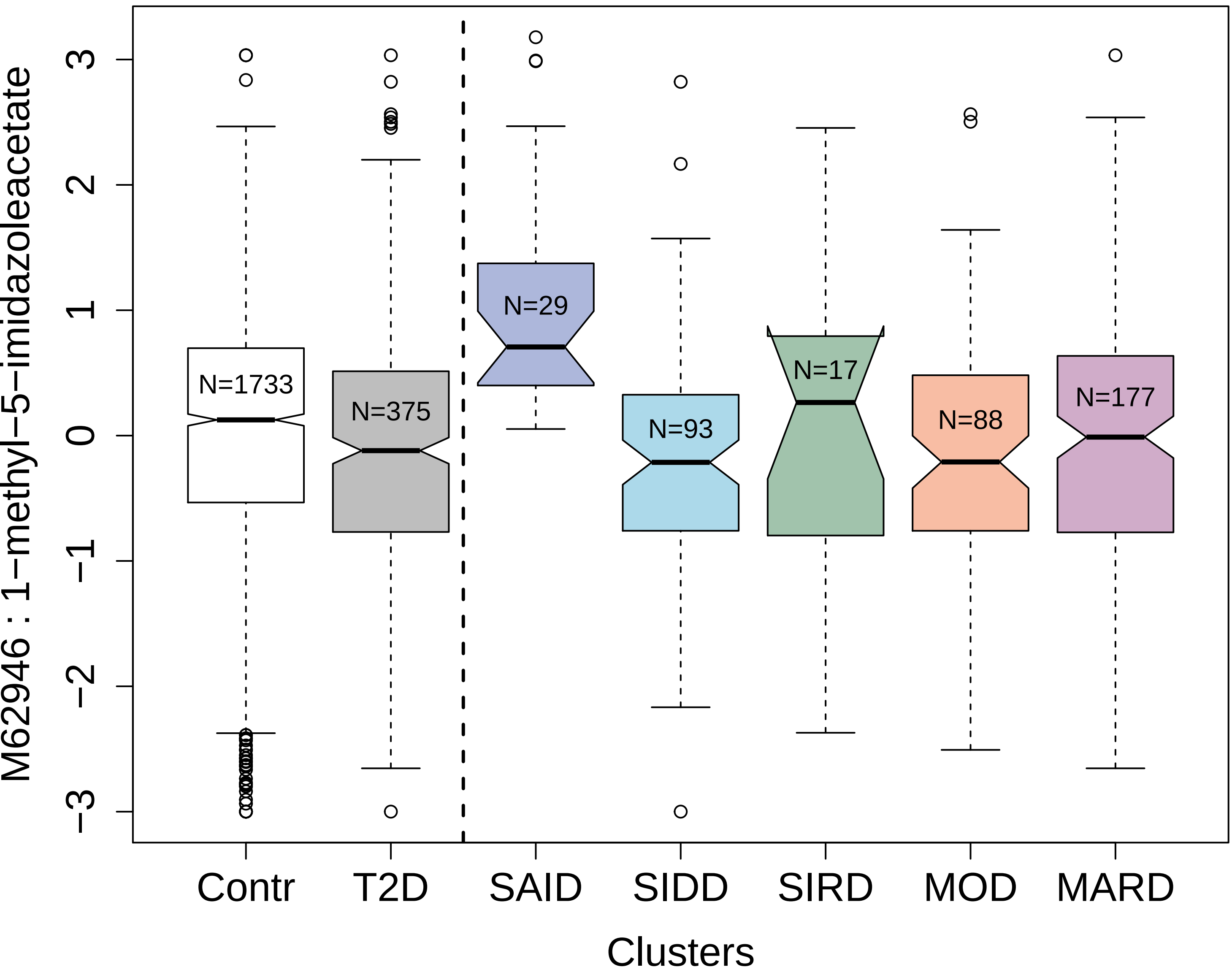

M62946 : 1−methyl−5−imidazoleacetate
−3
−2
−1
0
1
2
3
N=1733
N=375
N=29
N=93
N=17
N=88
N=177
Contr
T2D
SAID
SIDD
SIRD
MOD
MARD
Clusters

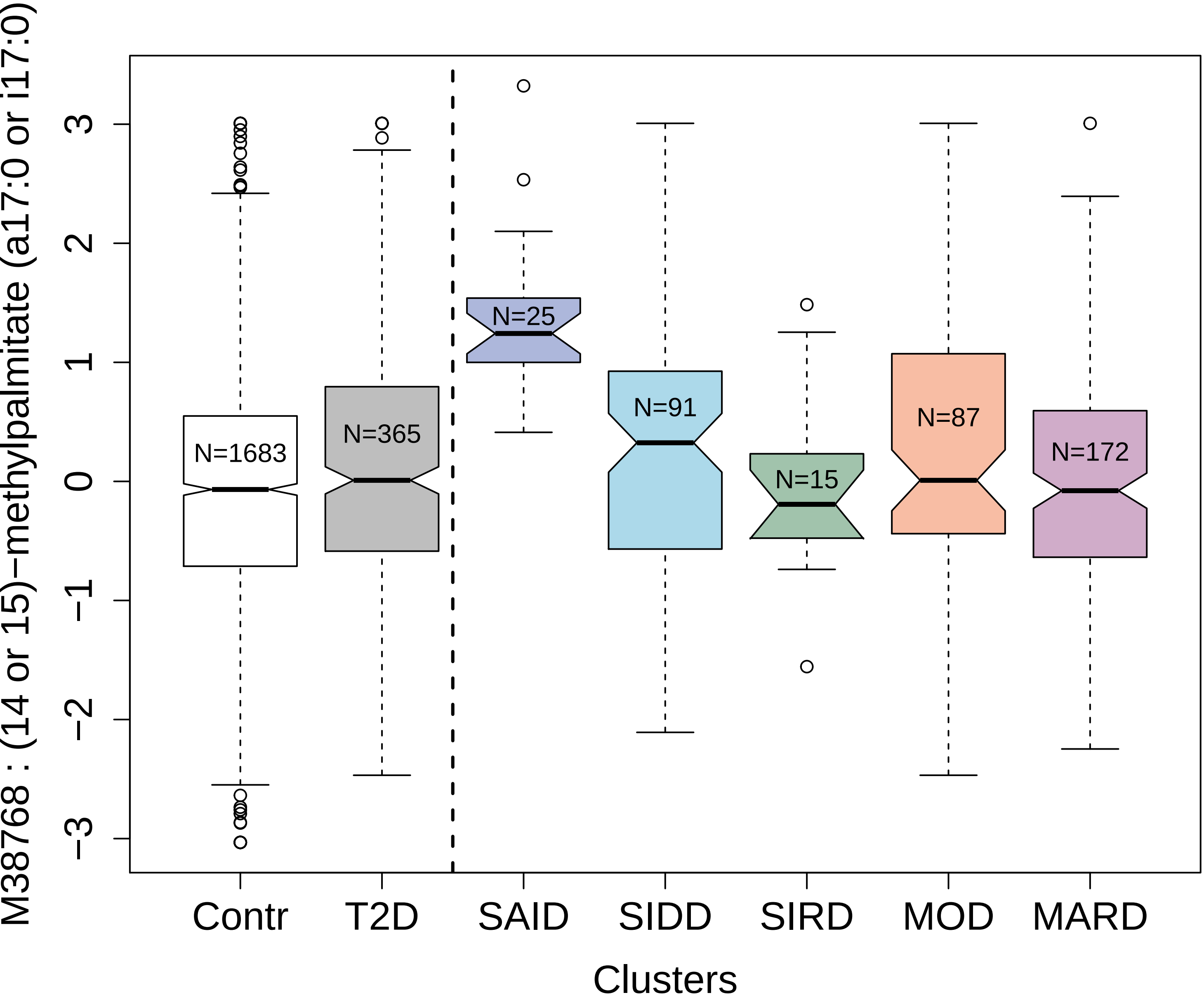

M38768 : (14 or 15)−methylpalmitate (a17:0 or i17:0)
3
2
1
0
−1
−2
−3
N=1683
N=365
N=25
N=91
N=15
N=87
N=172
Contr
T2D
SAID
SIDD
SIRD
MOD
MARD
Clusters

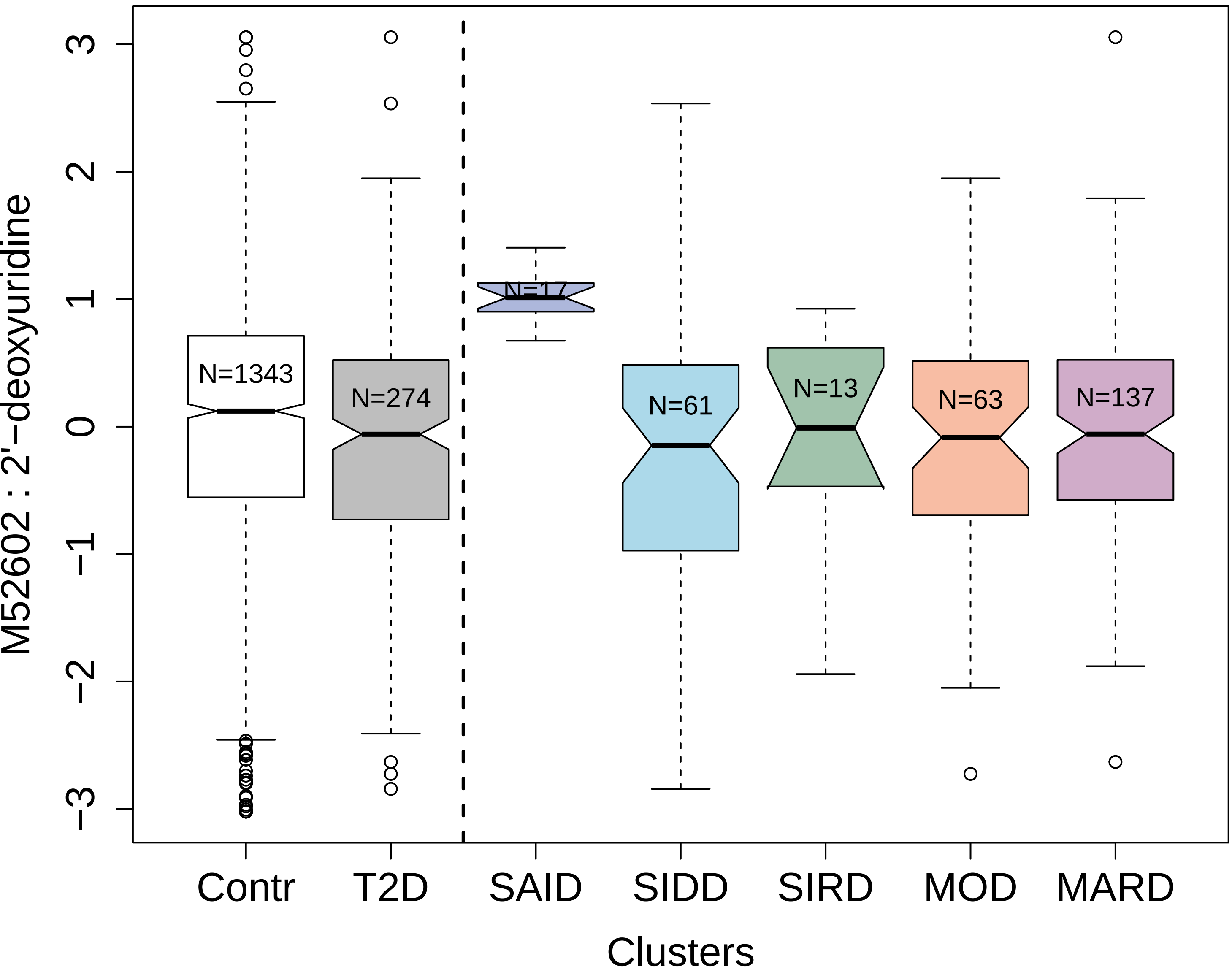
M52602 : 2'−deoxyuridine
3
2
1
0
−1
−2
−3
N=1343
N=274
N=17
N=61
N=13
N=63
N=137
Contr
T2D
SAID
SIDD
SIRD
MOD
MARD
Clusters

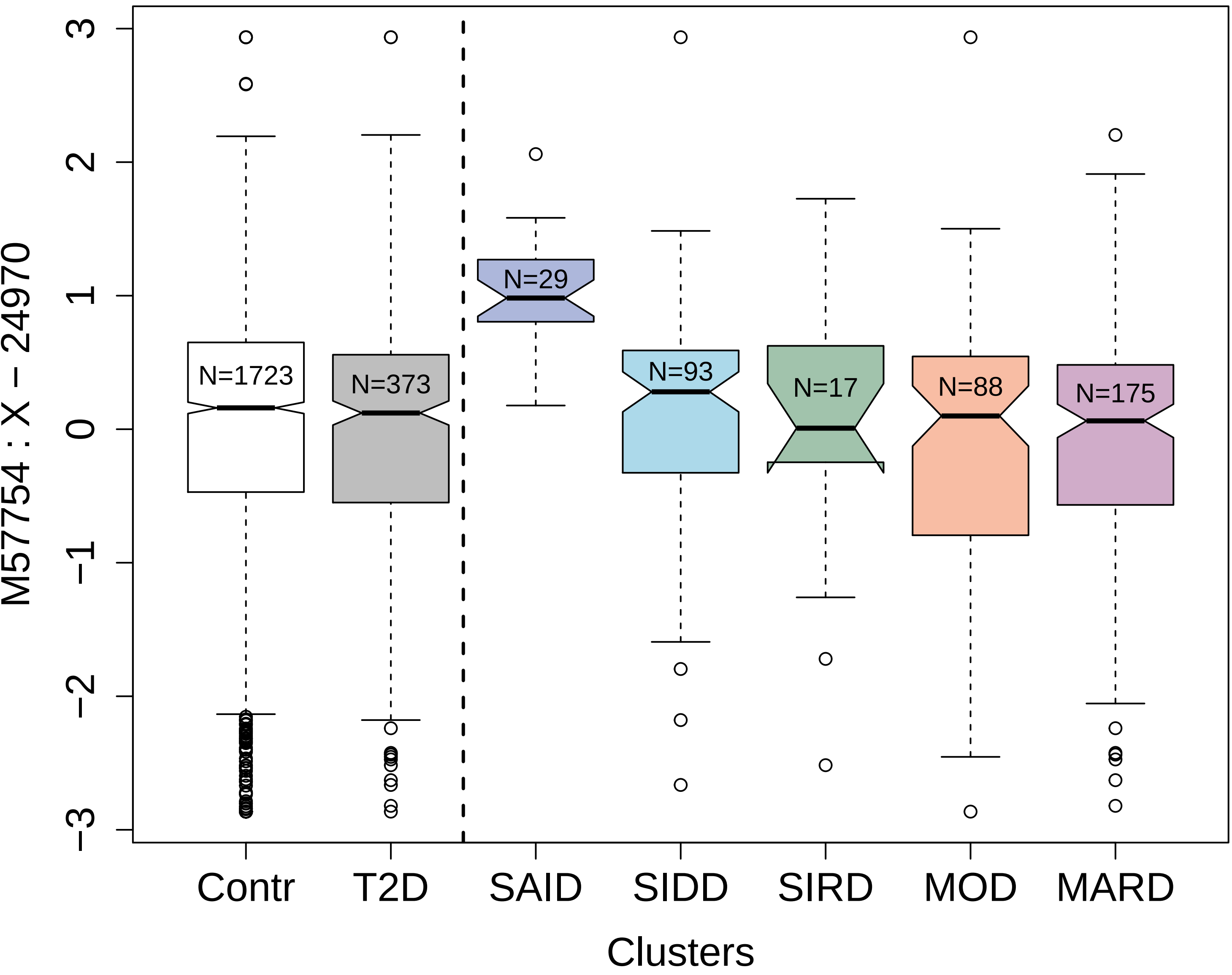
M57754 : X − 24970
3
2
1
0
−1
−2
−3
N=1723
N=373
N=29
N=93
N=17
N=88
N=175
Contr
T2D
SAID
SIDD
SIRD
MOD
MARD
Clusters

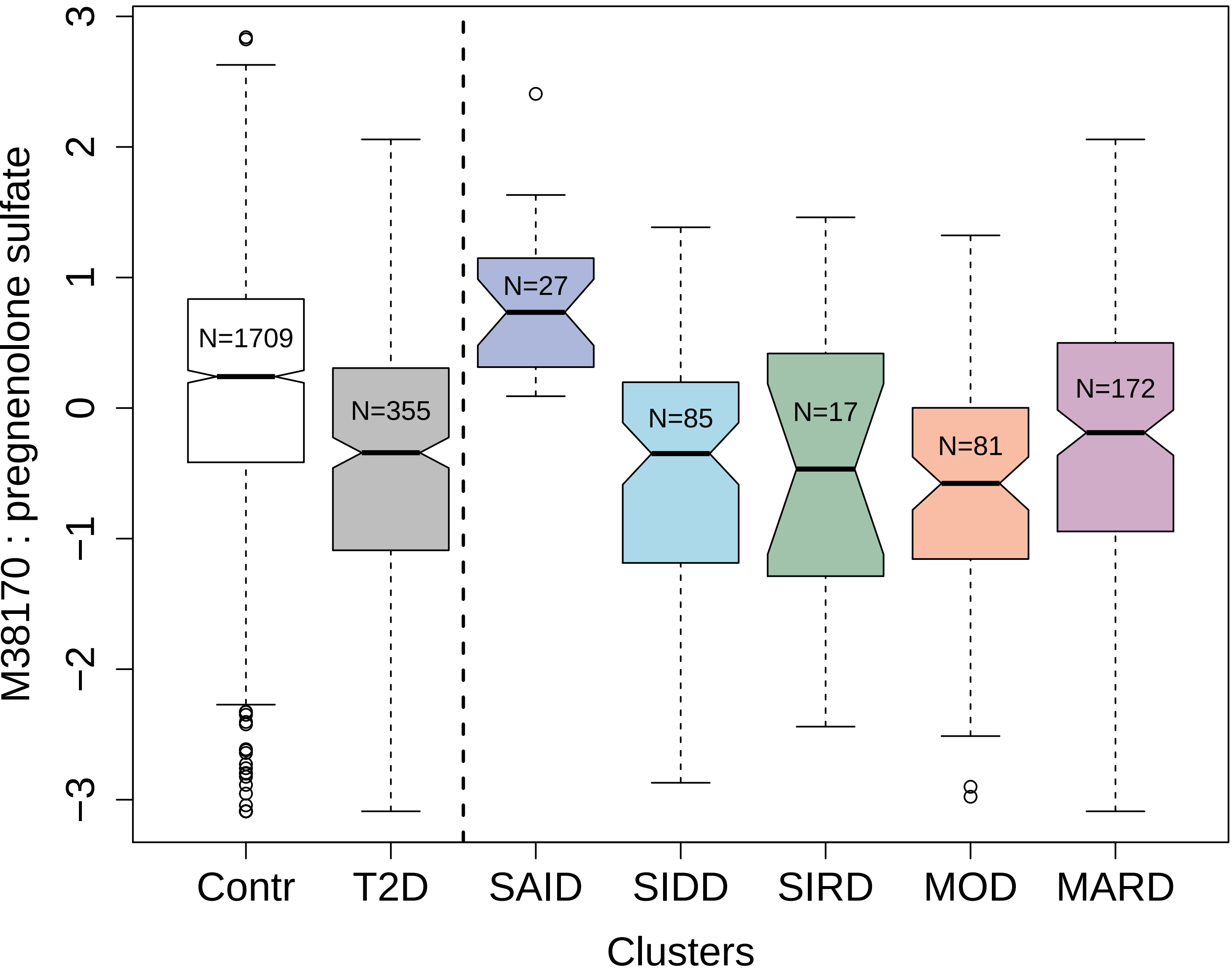
M38170 : pregnenolone sulfate
3
2
1
0
−1
−2
−3
N=1709
N=355
N=27
N=85
N=17
N=81
N=172
Contr
T2D
SAID
SIDD
SIRD
MOD
MARD
Clusters

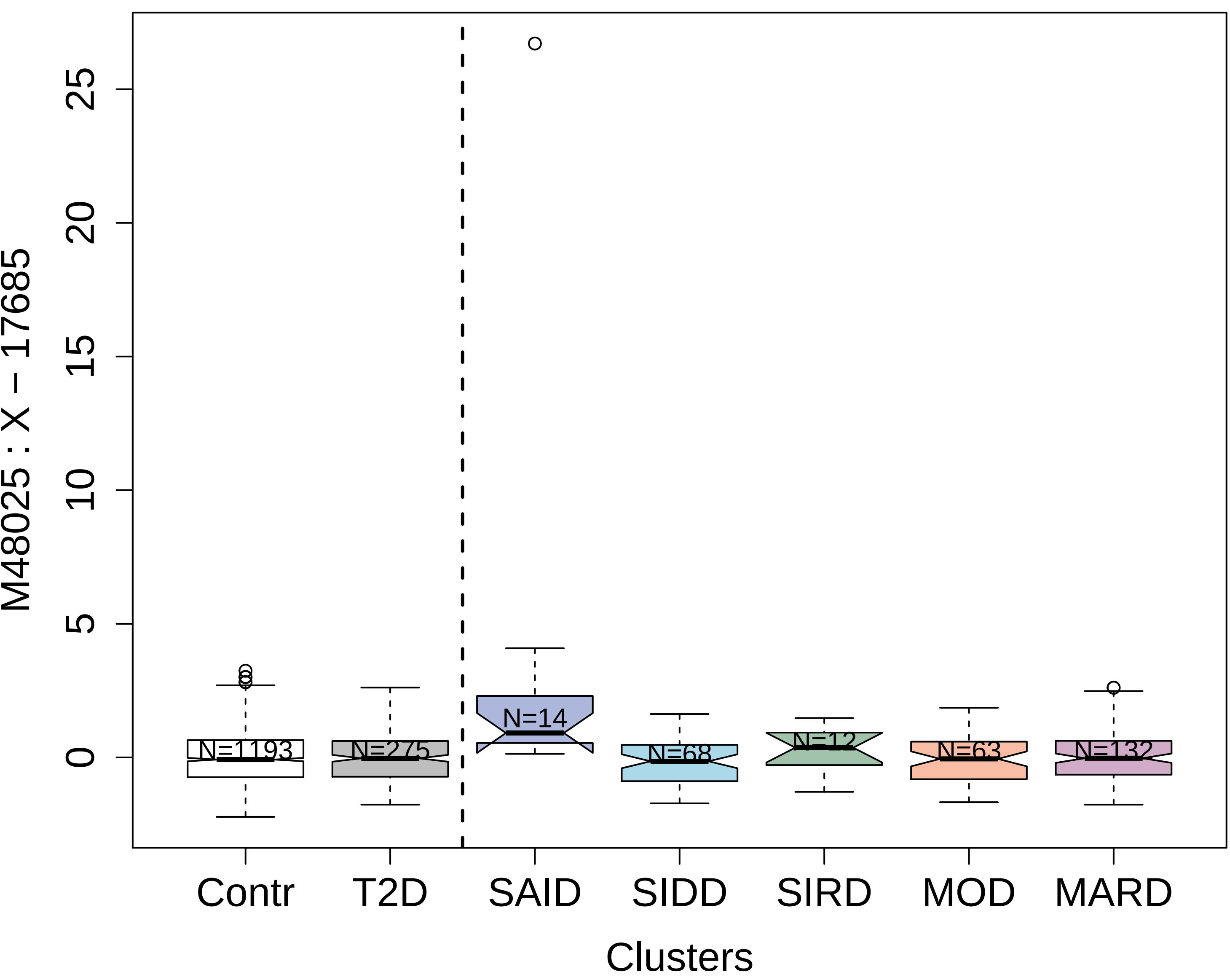
M48025 : X − 17685
0
5
10
15
20
25
N=1193
N=275
N=14
N=68
N=12
N=63
N=132
Contr
T2D
SAID
SIDD
SIRD
MOD
MARD
Clusters

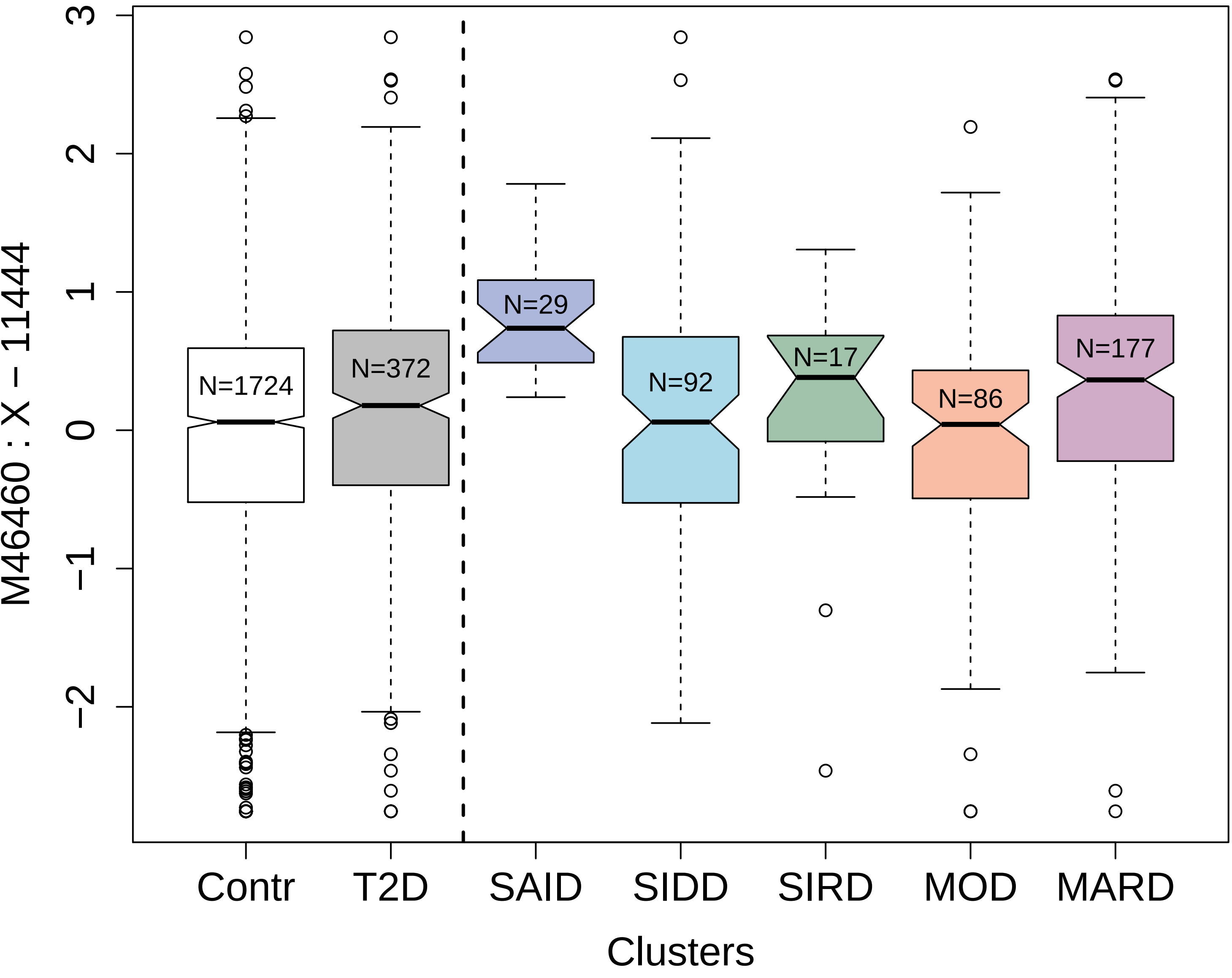

M46460 : X − 11444
3
2
1
0
−1
−2
N=1724
N=372
N=29
N=92
N=17
N=86
N=177
Contr
T2D
SAID
SIDD
SIRD
MOD
MARD
Clusters

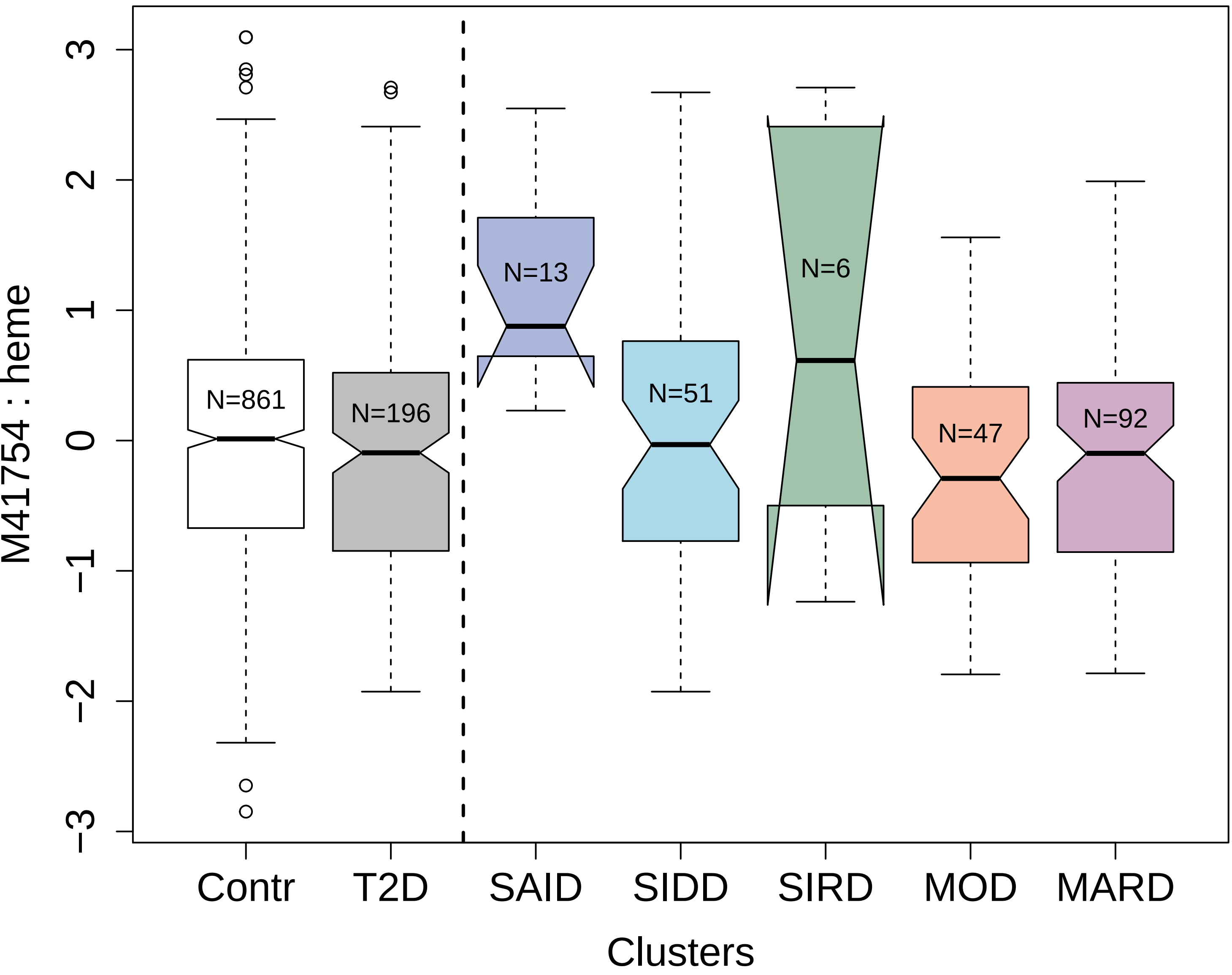

M41754 : heme
3
2
1
0
−1
−2
−3
N=861
N=196
N=13
N=51
N=6
N=47
N=92
Contr
T2D
SAID
SIDD
SIRD
MOD
MARD
Clusters

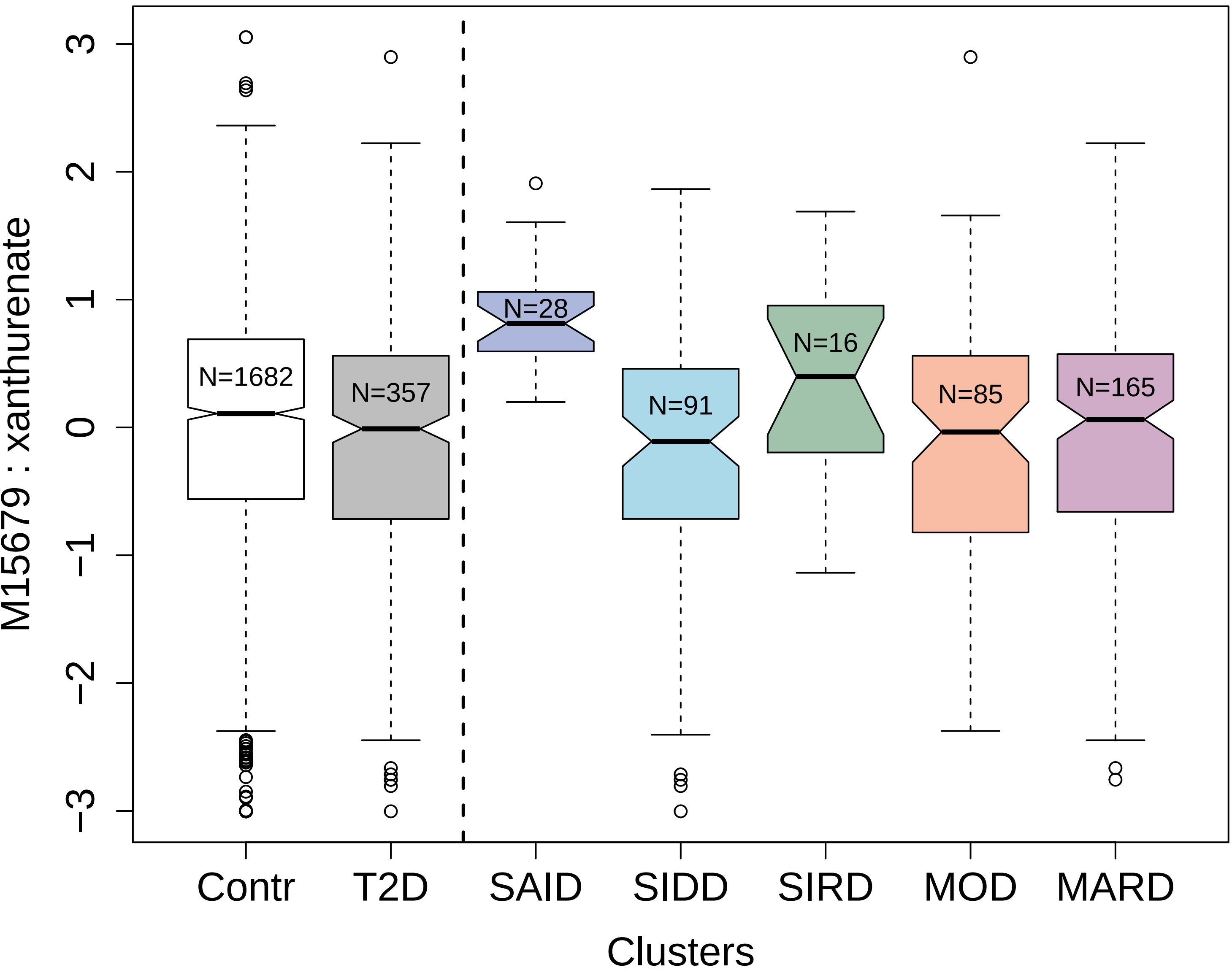

M15679 : xanthurenate
3
2
1
0
−1
−2
−3
N=1682
N=357
N=28
N=91
N=16
N=85
N=165
Contr
T2D
SAID
SIDD
SIRD
MOD
MARD
Clusters

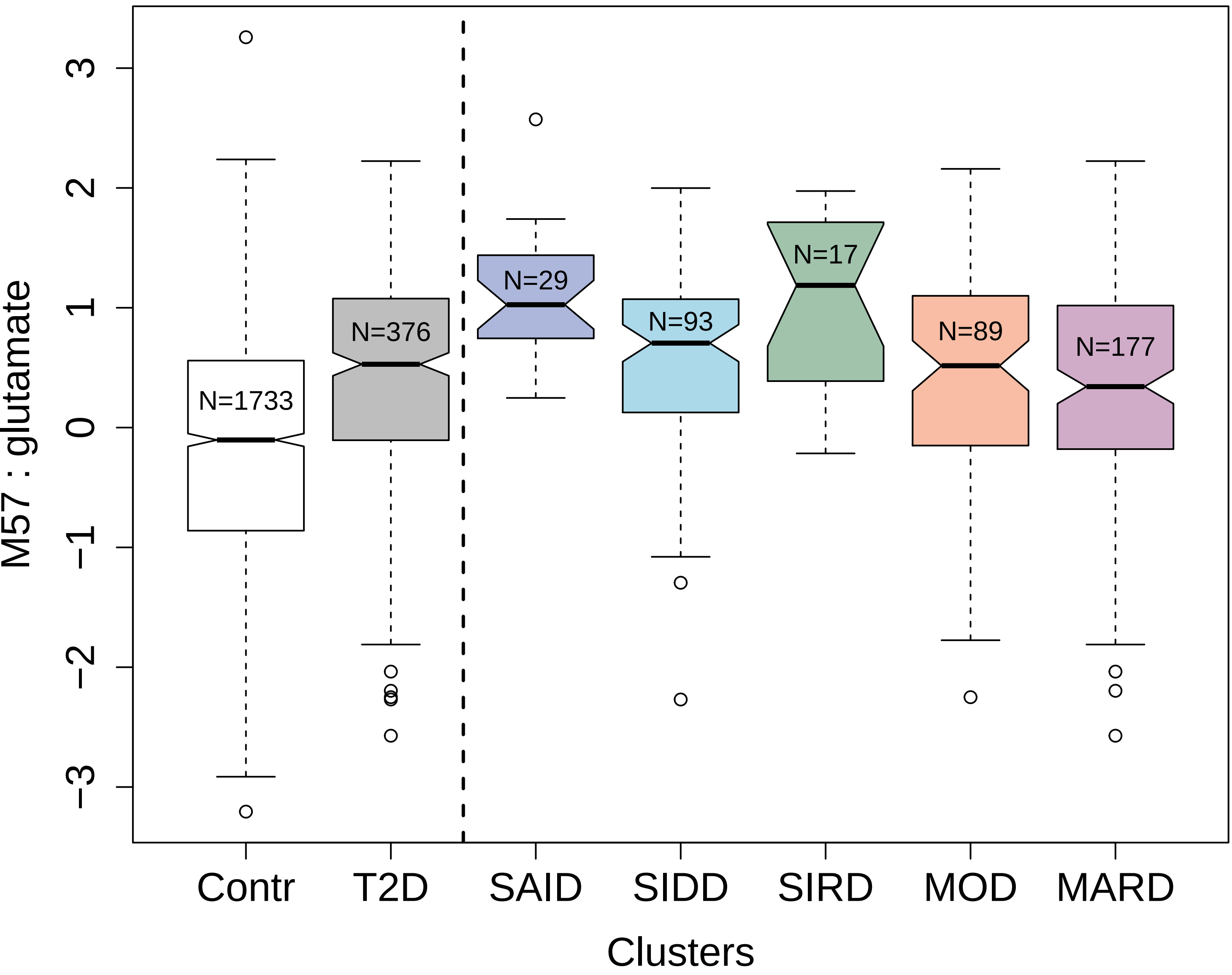

M57 : glutamate
3
2
1
0
−1
−2
−3
N=1733
N=376
N=29
N=93
N=17
N=89
N=177
Contr
T2D
SAID
SIDD
SIRD
MOD
MARD
Clusters

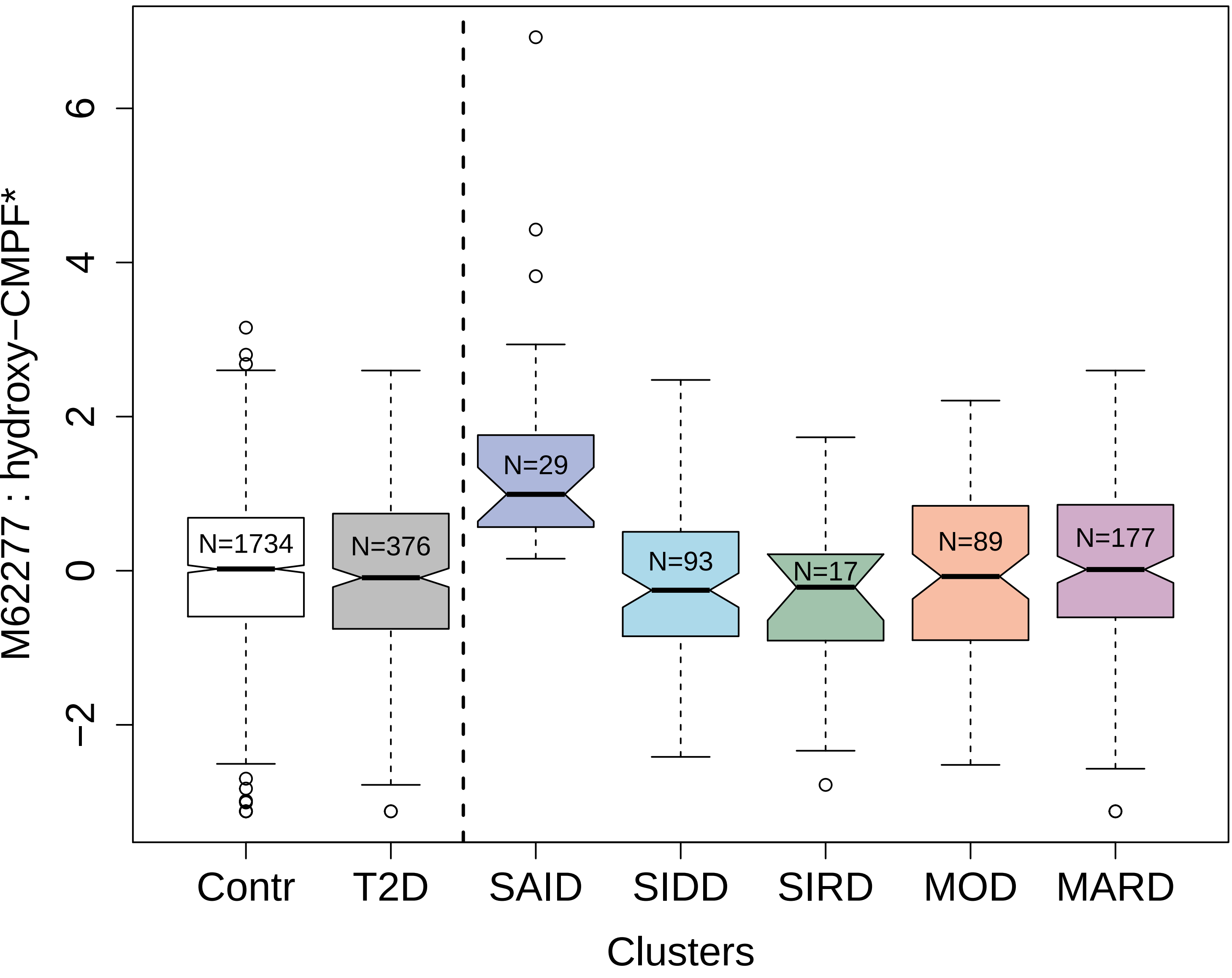

M62277 : hydroxy−CMPF*
6
4
2
0
−2
N=1734
N=376
N=29
N=93
N=17
N=89
N=177
Contr
T2D
SAID
SIDD
SIRD
MOD
MARD
Clusters

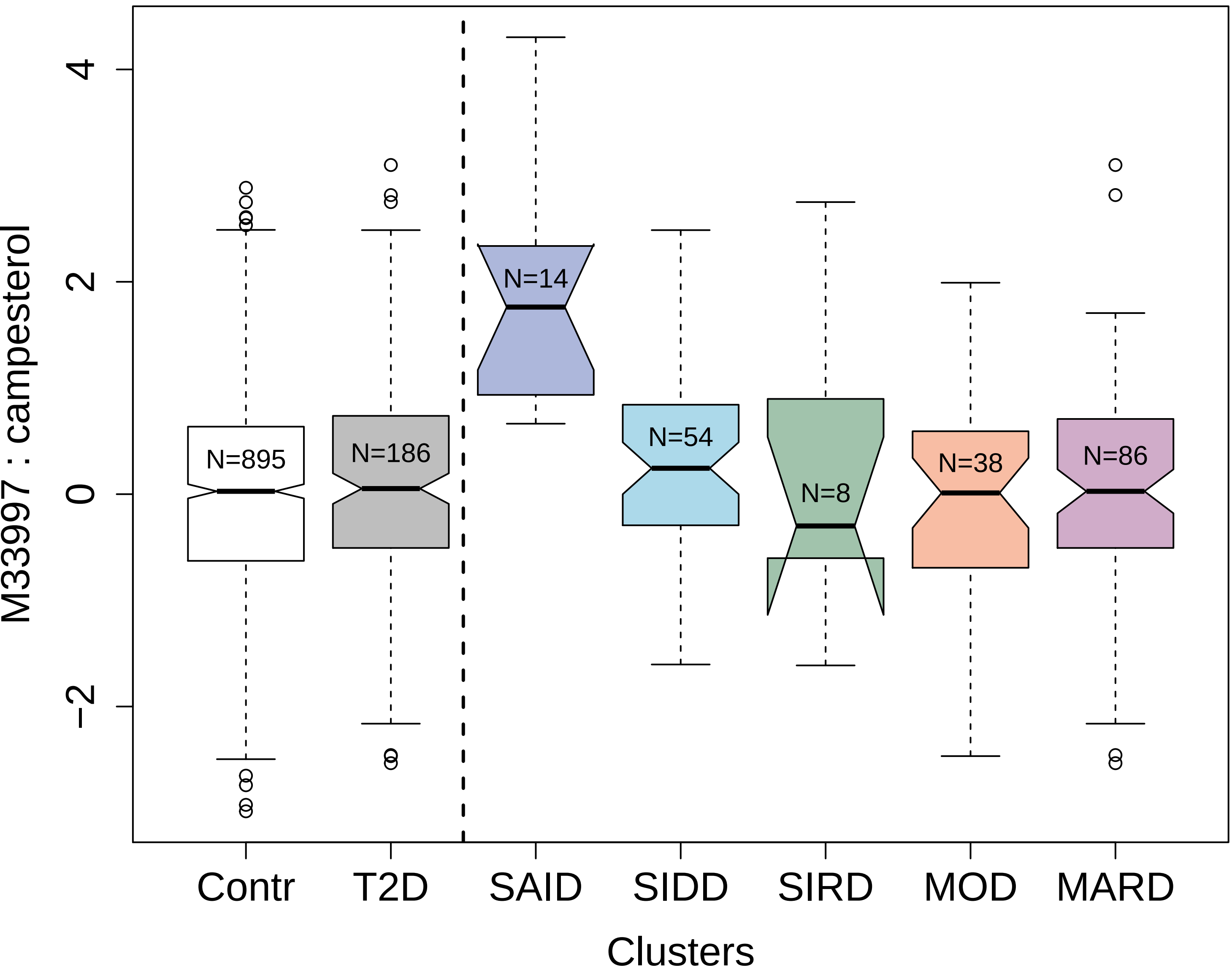

M33997 : campesterol
4
2
0
−2
N=895
N=186
N=14
N=54
N=8
N=38
N=86
Contr
T2D
SAID
SIDD
SIRD
MOD
MARD
Clusters

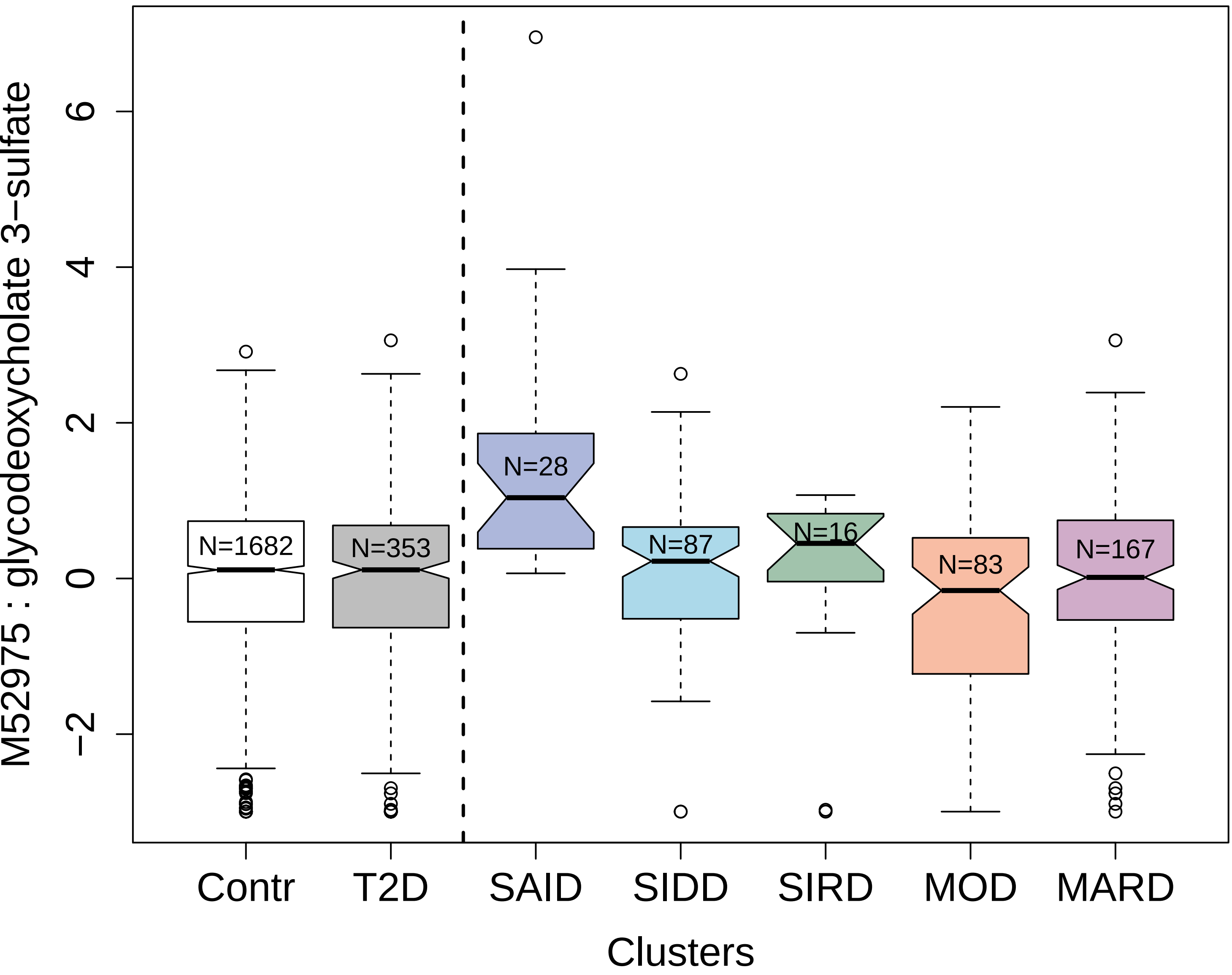
M52975 : glycodeoxycholate 3−sulfate
−2
0
2
4
6
N=1682
N=353
N=28
N=87
N=16
N=83
N=167
Contr
T2D
SAID
SIDD
SIRD
MOD
MARD
Clusters

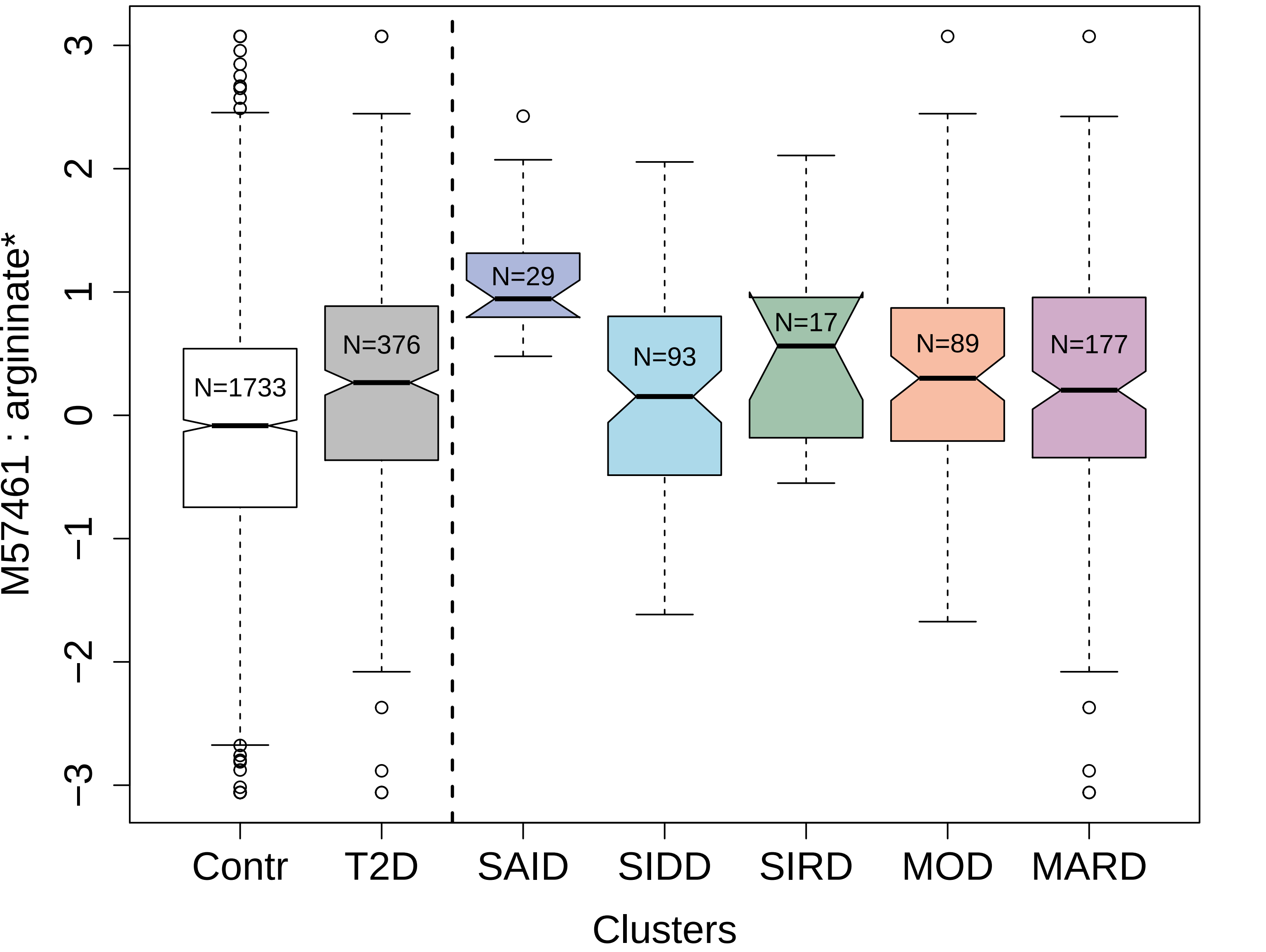

M57461 : argininate*
3
2
1
0
−1
−2
−3
N=1733
N=376
N=29
N=93
N=17
N=89
N=177
Contr
T2D
SAID
SIDD
SIRD
MOD
MARD
Clusters

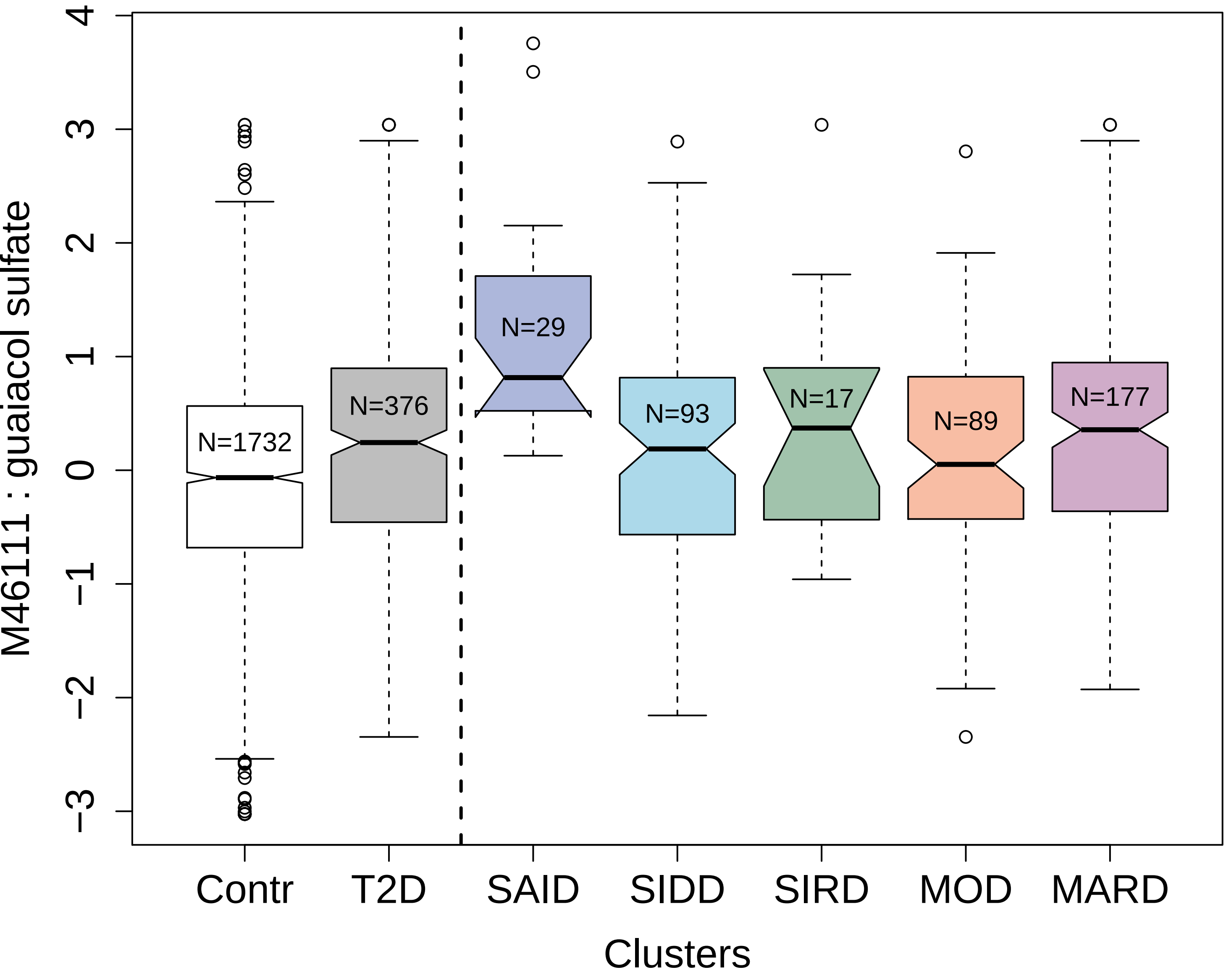

M46111 : guaiacol sulfate
4
3
2
1
0
−1
−2
−3
N=1732
N=376
N=29
N=93
N=17
N=89
N=177
Contr
T2D
SAID
SIDD
SIRD
MOD
MARD
Clusters

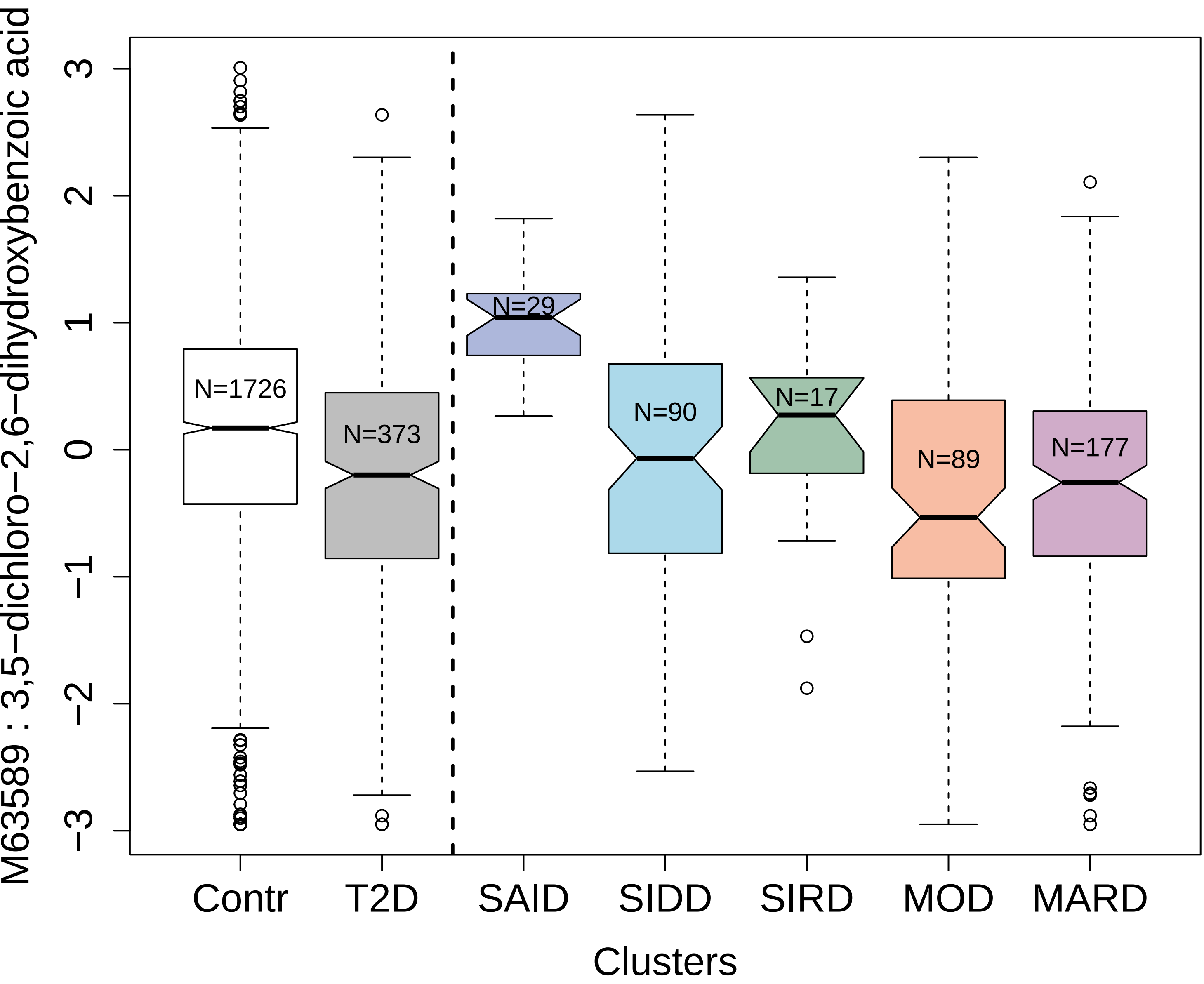
M63589 : 3,5−dichloro−2,6−dihydroxybenzoic acid
3
2
1
0
−1
−2
−3
N=1726
N=373
N=29
N=90
N=17
N=89
N=177
Contr
T2D
SAID
SIDD
SIRD
MOD
MARD
Clusters

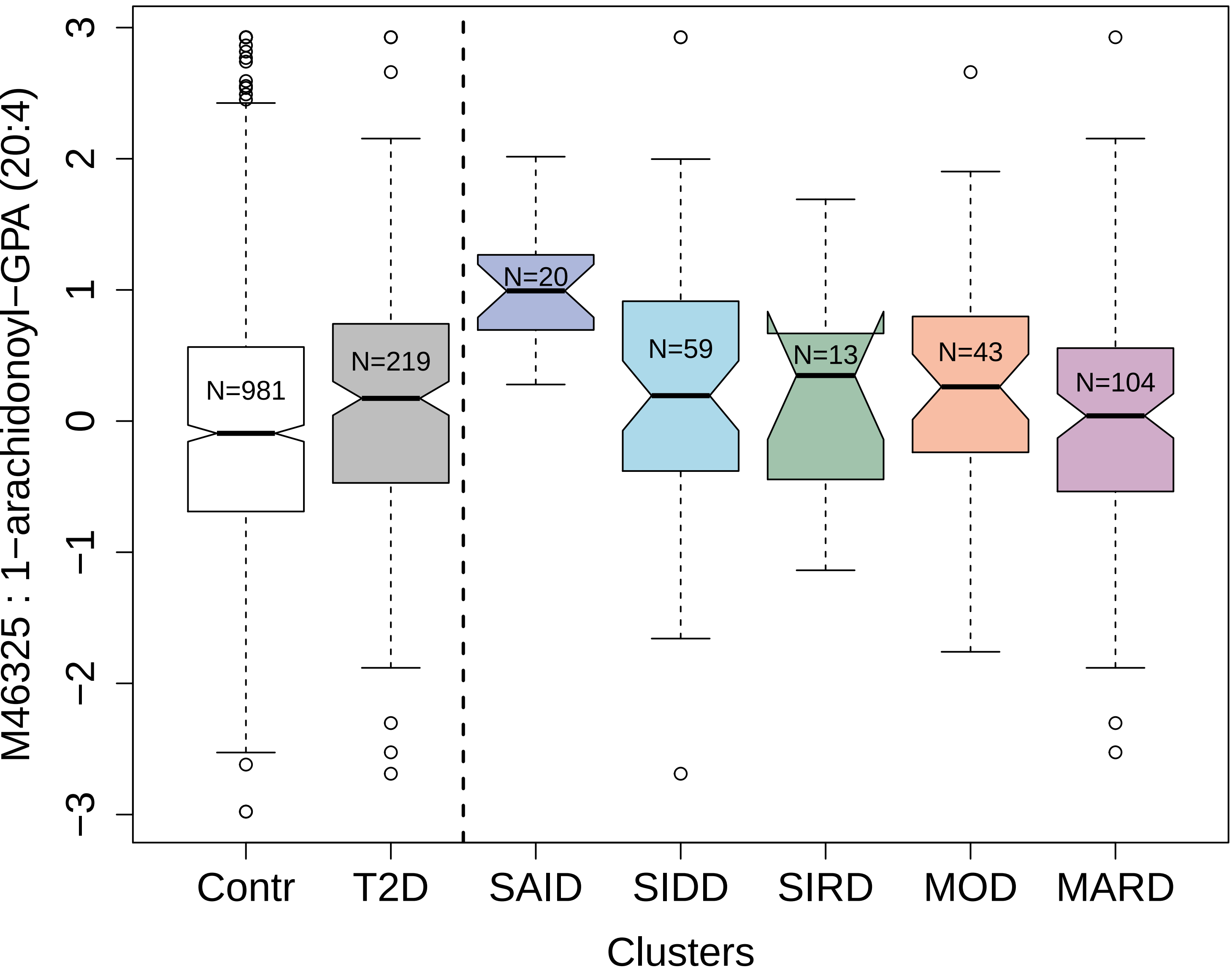

M46325 : 1−arachidonoyl−GPA (20:4)
3
2
1
0
−1
−2
−3
N=981
N=219
N=20
N=59
N=13
N=43
N=104
Contr
T2D
SAID
SIDD
SIRD
MOD
MARD
Clusters

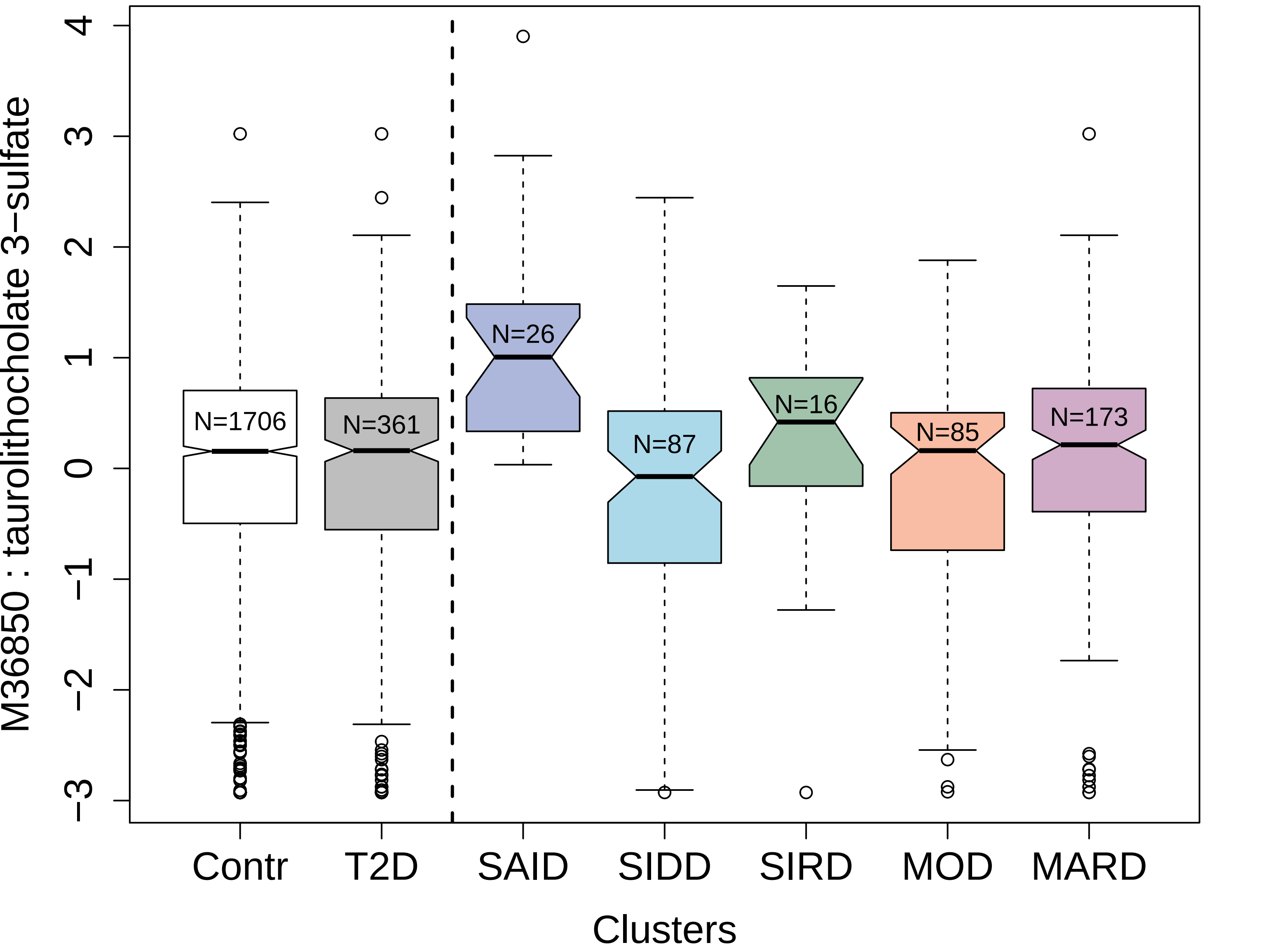

M36850 : taurolithocholate 3−sulfate
4
3
2
1
0
−1
−2
−3
N=1706
N=361
N=26
N=87
N=16
N=85
N=173
Contr
T2D
SAID
SIDD
SIRD
MOD
MARD
Clusters

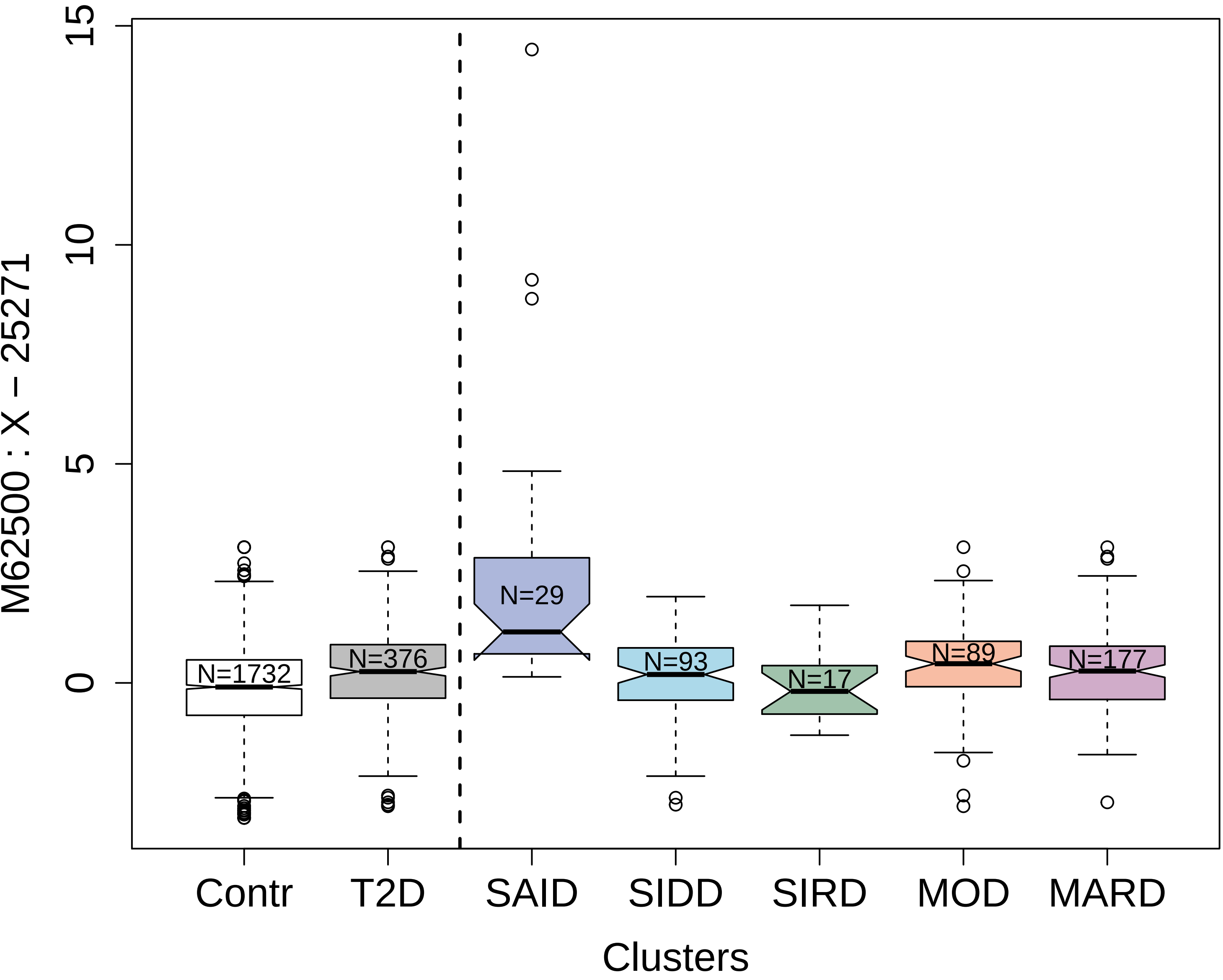
M62500 : X − 25271
15
10
5
0
N=1732
N=376
N=29
N=93
N=17
N=89
N=177
Contr
T2D
SAID
SIDD
SIRD
MOD
MARD
Clusters

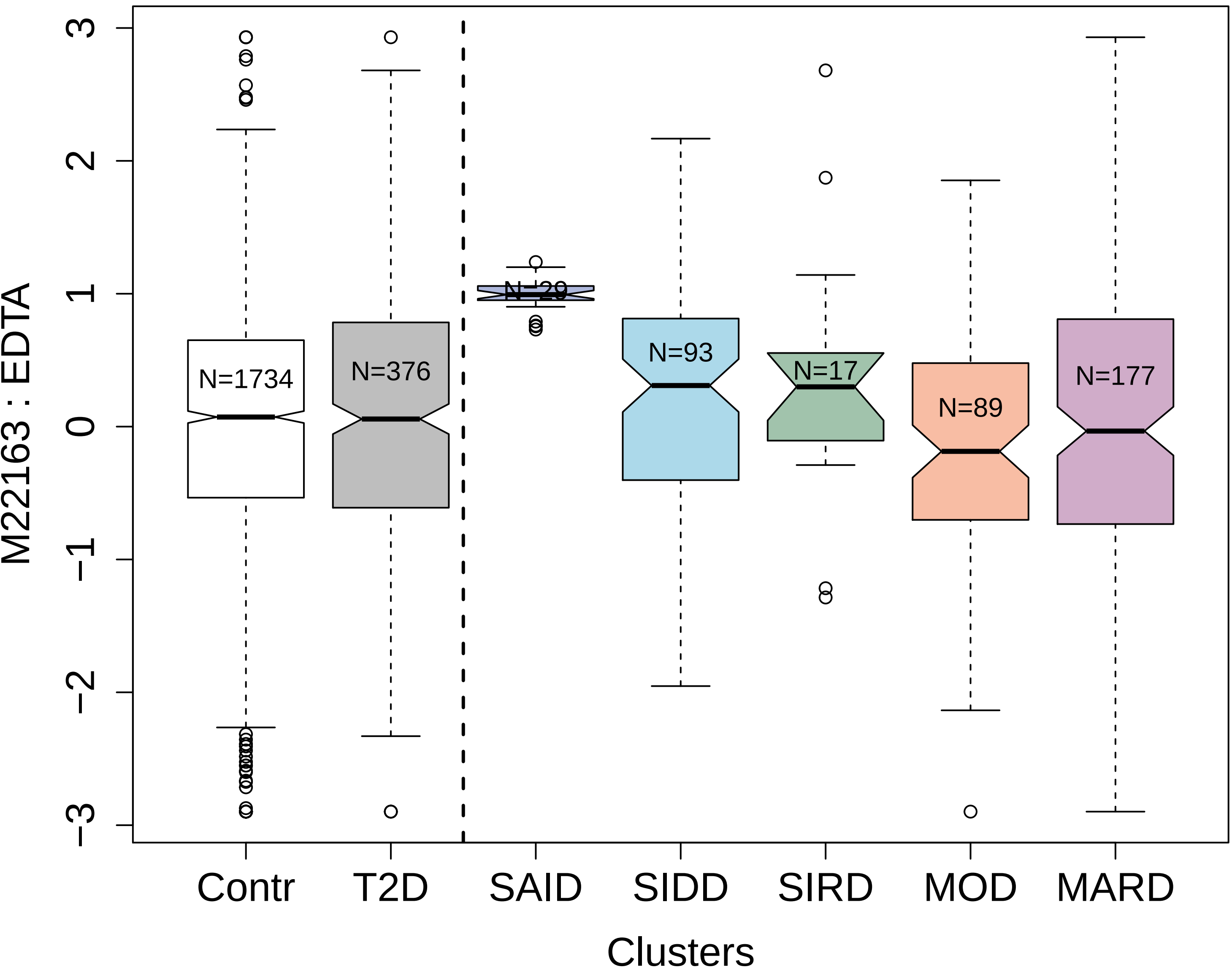

M22163 : EDTA
3
2
1
0
−1
−2
−3
N=1734
N=376
N=29
N=93
N=17
N=89
N=177
Contr
T2D
SAID
SIDD
SIRD
MOD
MARD
Clusters

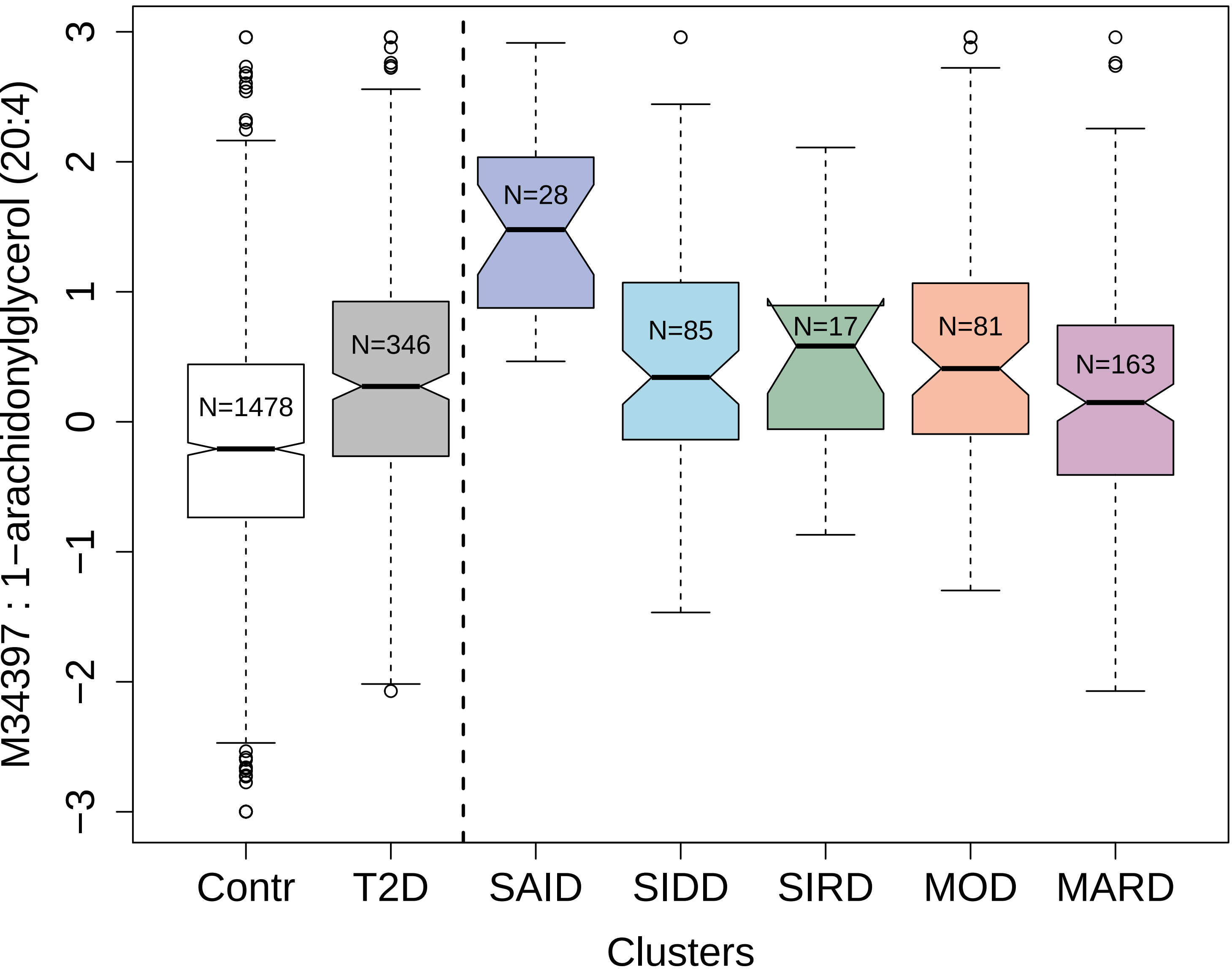

M34397 : 1−arachidonylglycerol (20:4)
3
2
1
0
−1
−2
−3
N=1478
N=346
N=28
N=85
N=17
N=81
N=163
Contr
T2D
SAID
SIDD
SIRD
MOD
MARD
Clusters

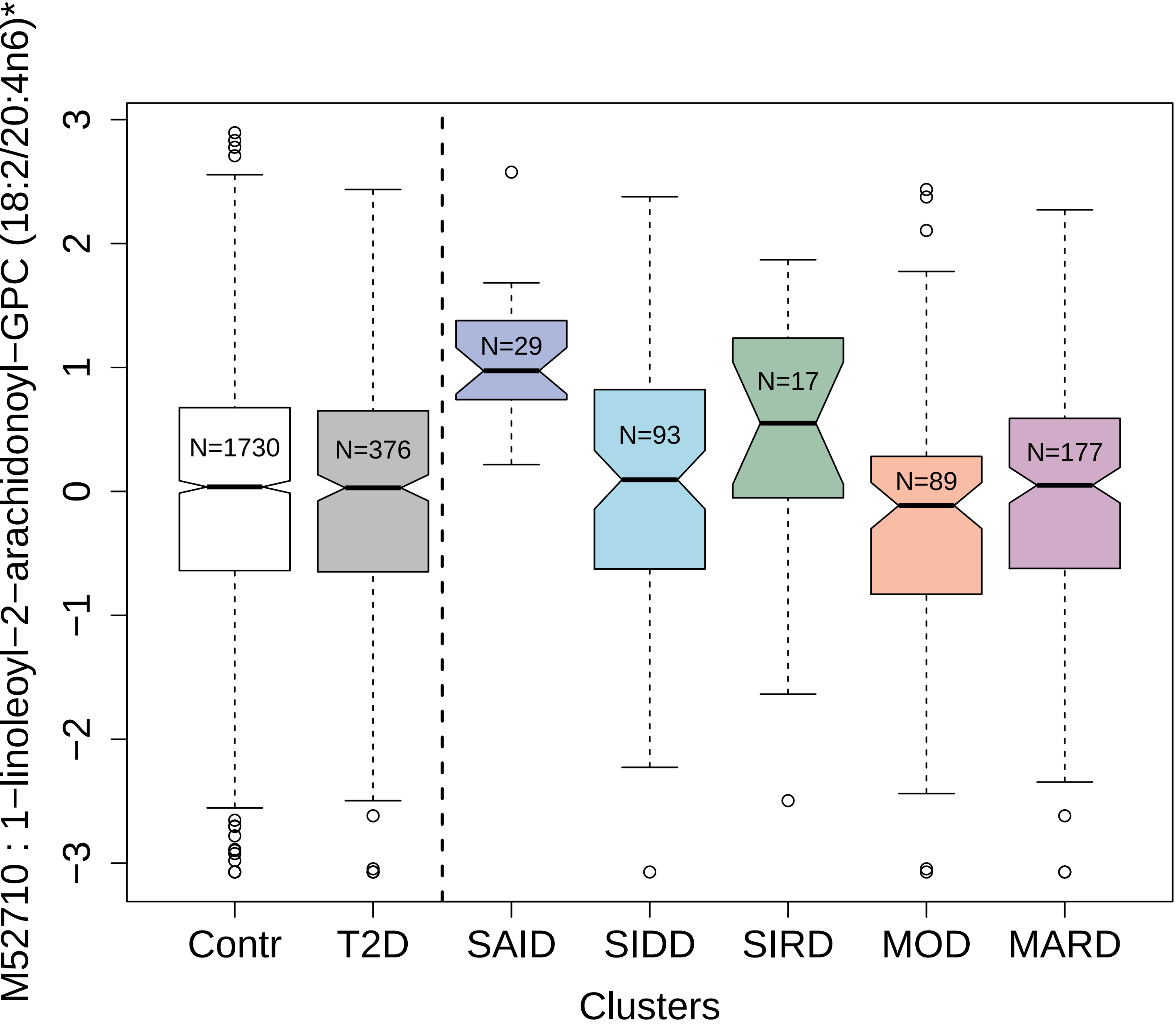
M52710 : 1−linoleoyl−2−arachidonoyl−GPC (18:2/20:4n6)*
3
2
1
0
−1
−2
−3
N=1730
N=376
N=29
N=93
N=17
N=89
N=177
Contr
T2D
SAID
SIDD
SIRD
MOD
MARD
Clusters

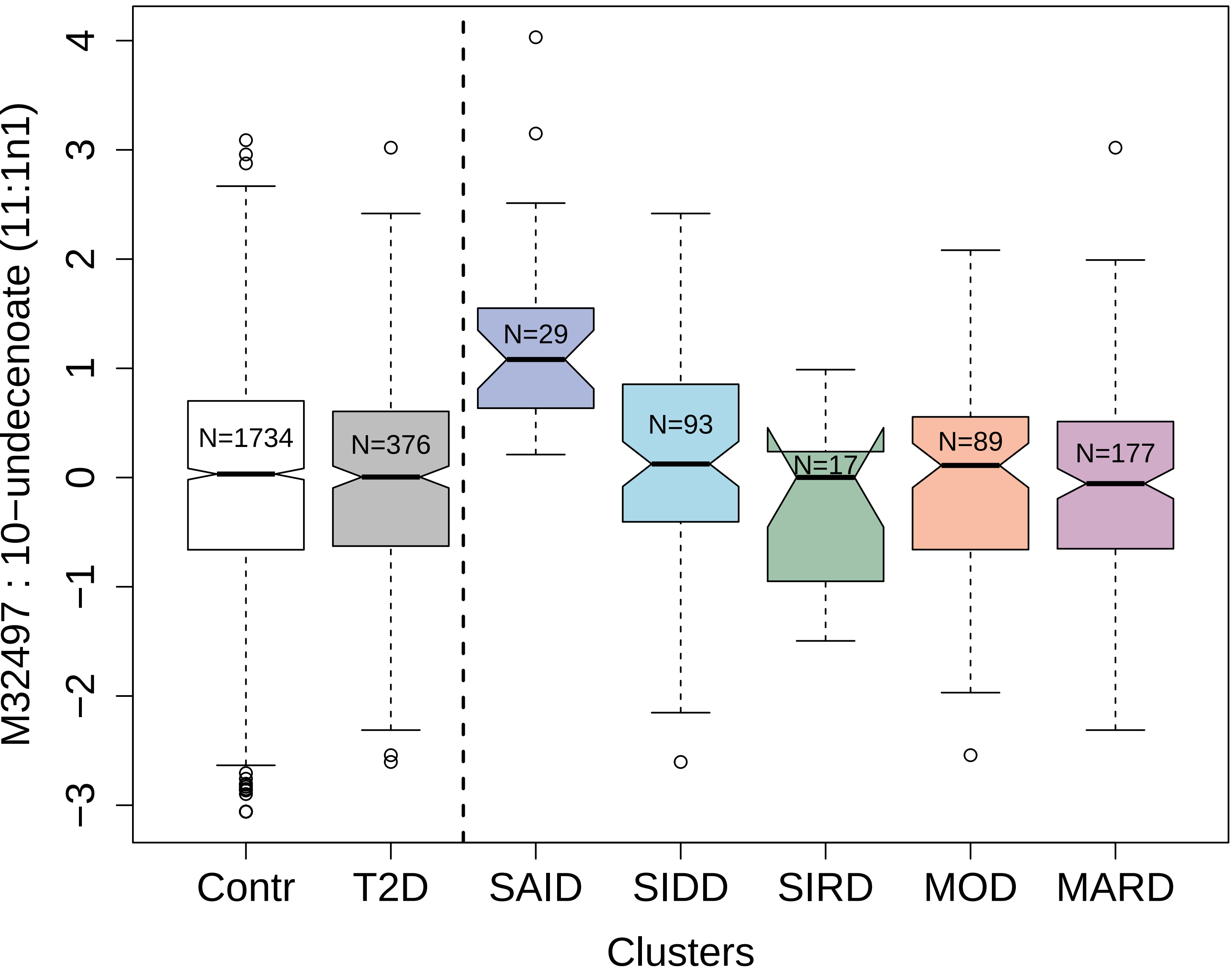

M32497 : 10−undecenoate (11:1n1)
4
3
2
1
0
−1
−2
−3
N=1734
N=376
N=29
N=93
N=17
N=89
N=177
Contr
T2D
SAID
SIDD
SIRD
MOD
MARD
Clusters

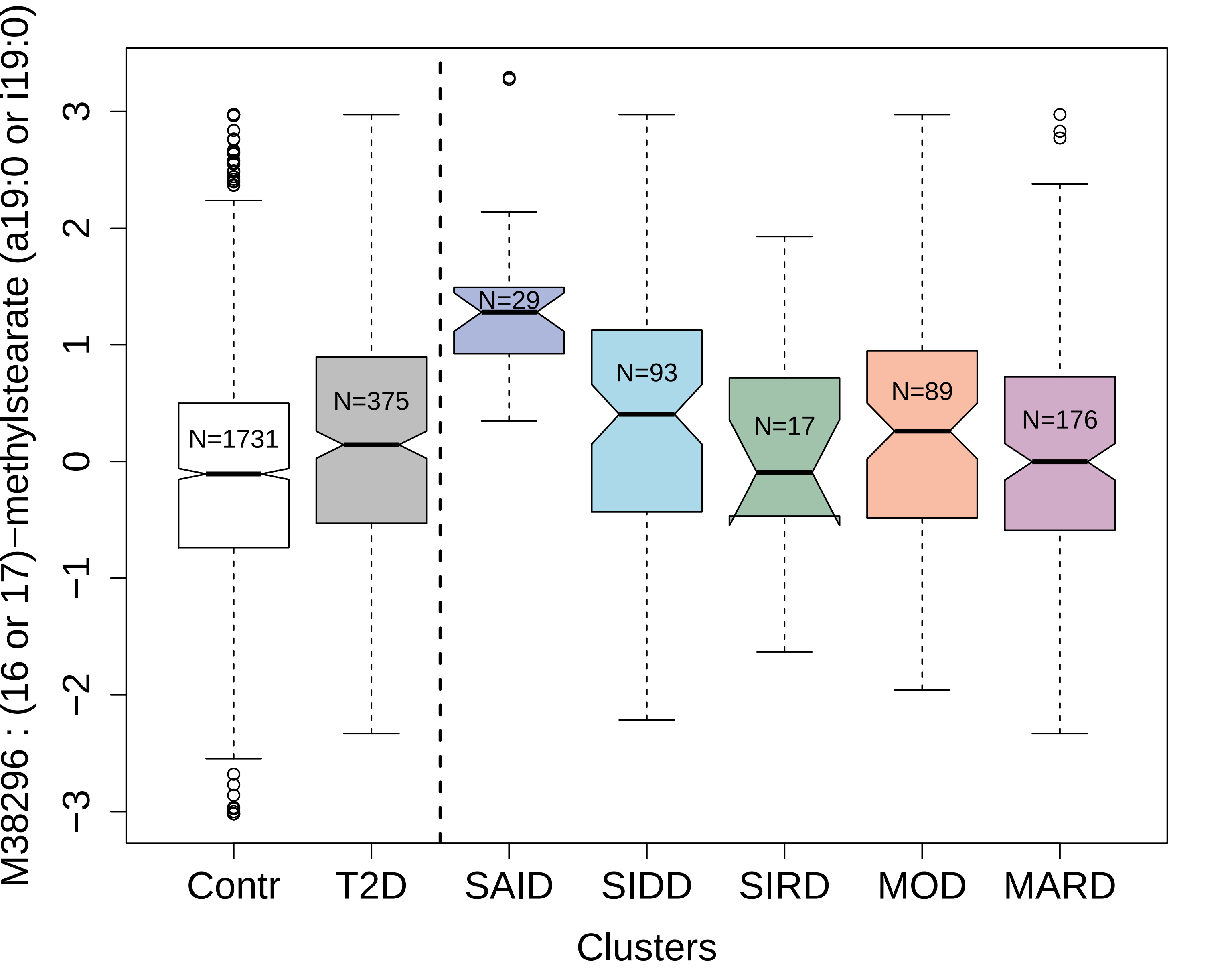
M38296 : (16 or 17)−methylstearate (a19:0 or i19:0)
3
2
1
0
−1
−2
−3
N=1731
N=375
N=29
N=93
N=17
N=89
N=176
Contr
T2D
SAID
SIDD
SIRD
MOD
MARD
Clusters

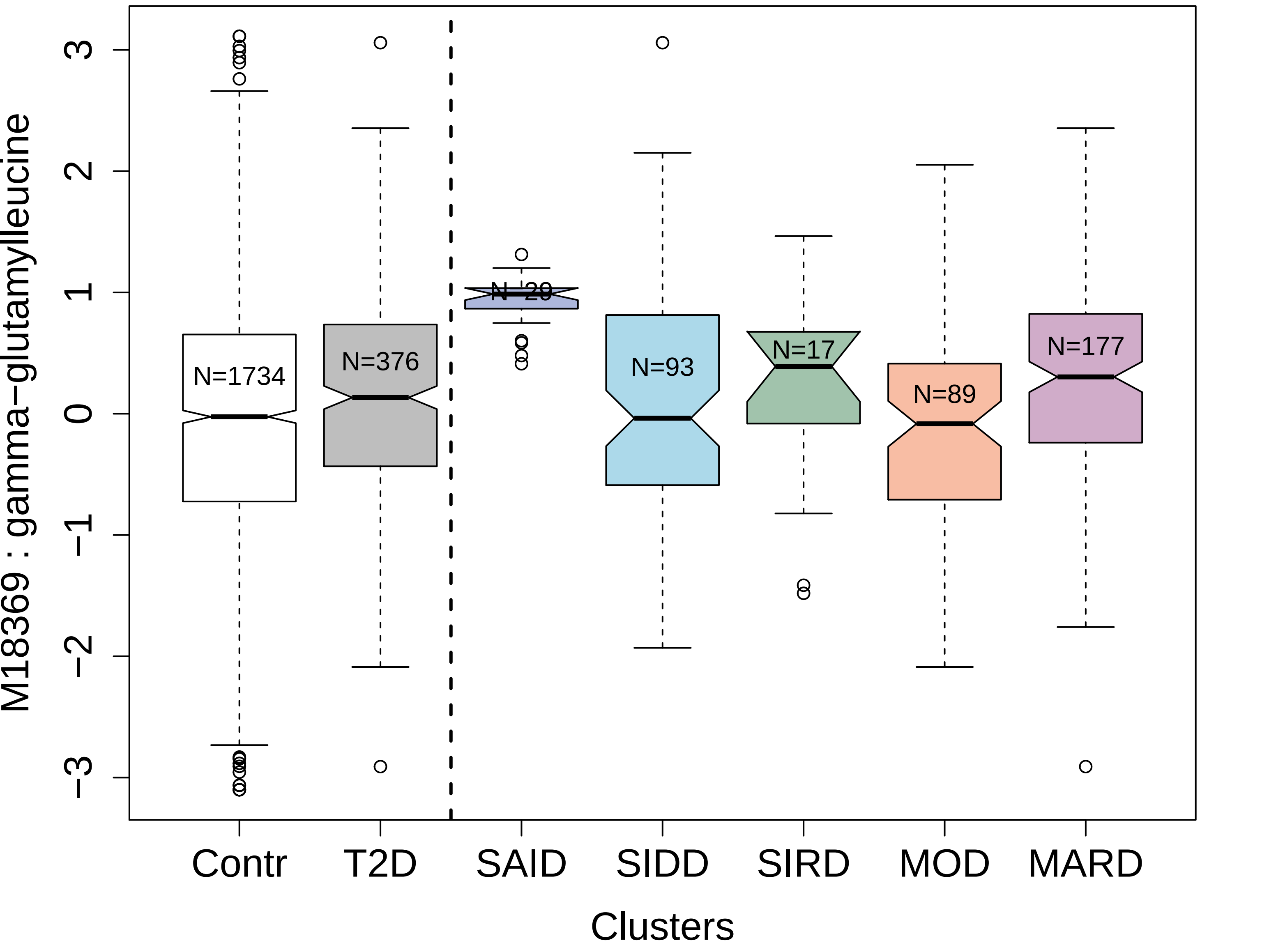

M18369 : gamma−glutamylleucine
3
2
1
0
−1
−2
−3
N=1734
N=376
N=93
N=17
N=89
N=177
Contr
T2D
SAID
SIDD
SIRD
MOD
MARD
Clusters

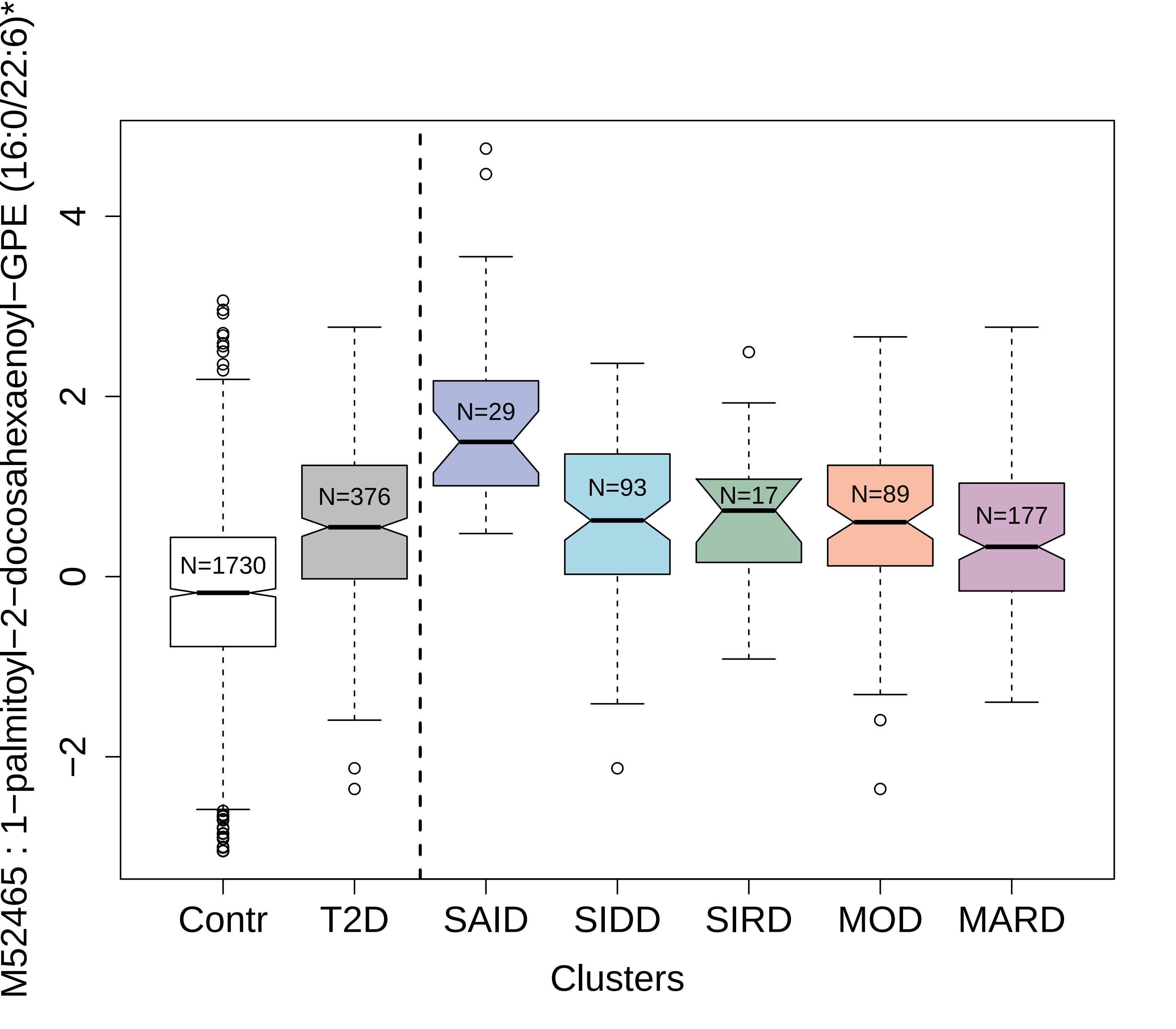

M52465 : 1−palmitoyl−2−docosahexaenoyl−GPE (16:0/22:6)*
4
2
0
−2
N=1730
N=376
N=29
N=93
N=17
N=89
N=177
Contr
T2D
SAID
SIDD
SIRD
MOD
MARD
Clusters

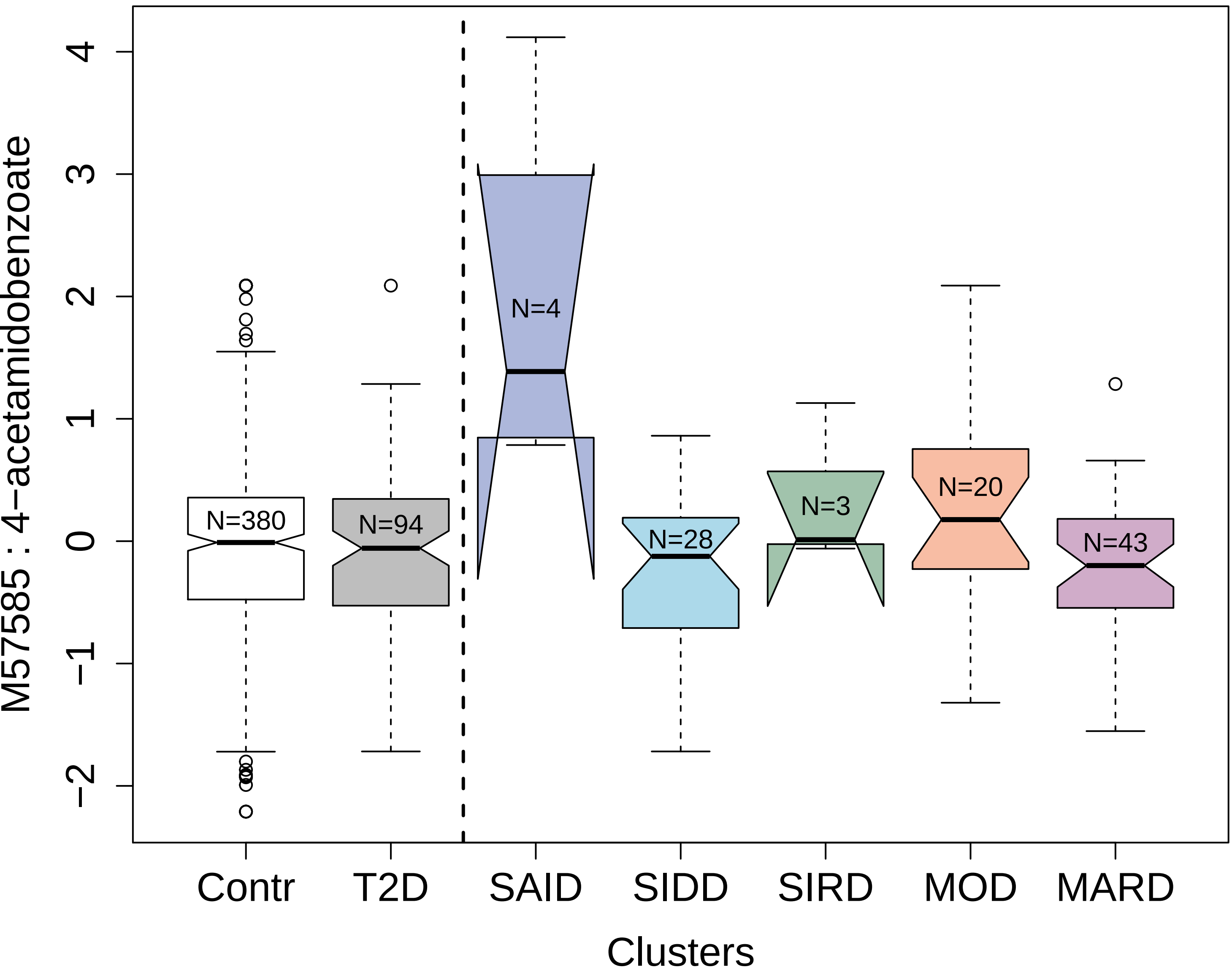

M57585 : 4−acetamidobenzoate
4
3
2
1
0
−1
−2
N=380
N=94
N=4
N=28
N=3
N=20
N=43
Contr
T2D
SAID
SIDD
SIRD
MOD
MARD
Clusters

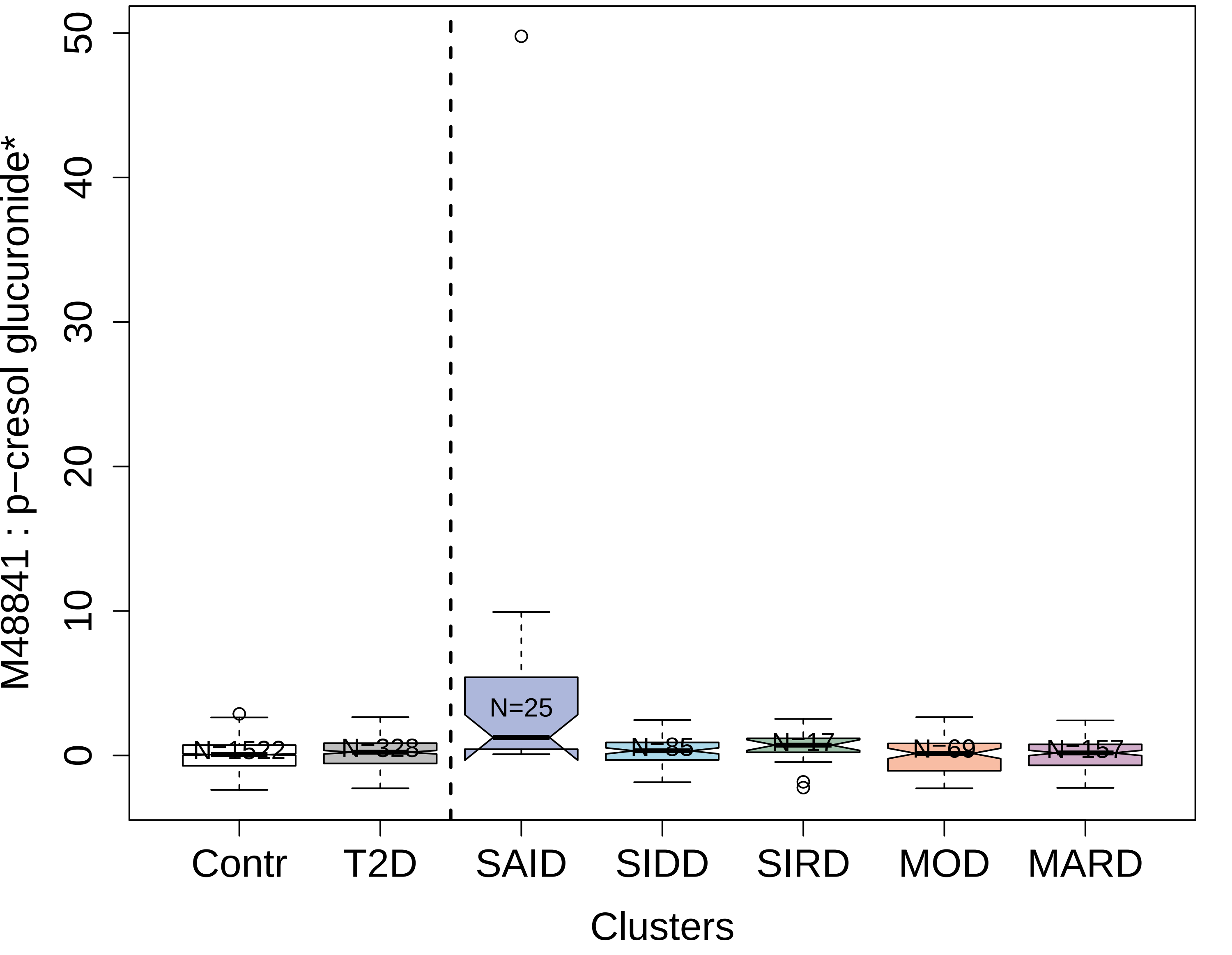

M48841 : p−cresol glucuronide*
0
10
20
30
40
50
N=1522
N=328
N=25
N=85
N=17
N=69
N=157
Contr
T2D
SAID
SIDD
SIRD
MOD
MARD
Clusters

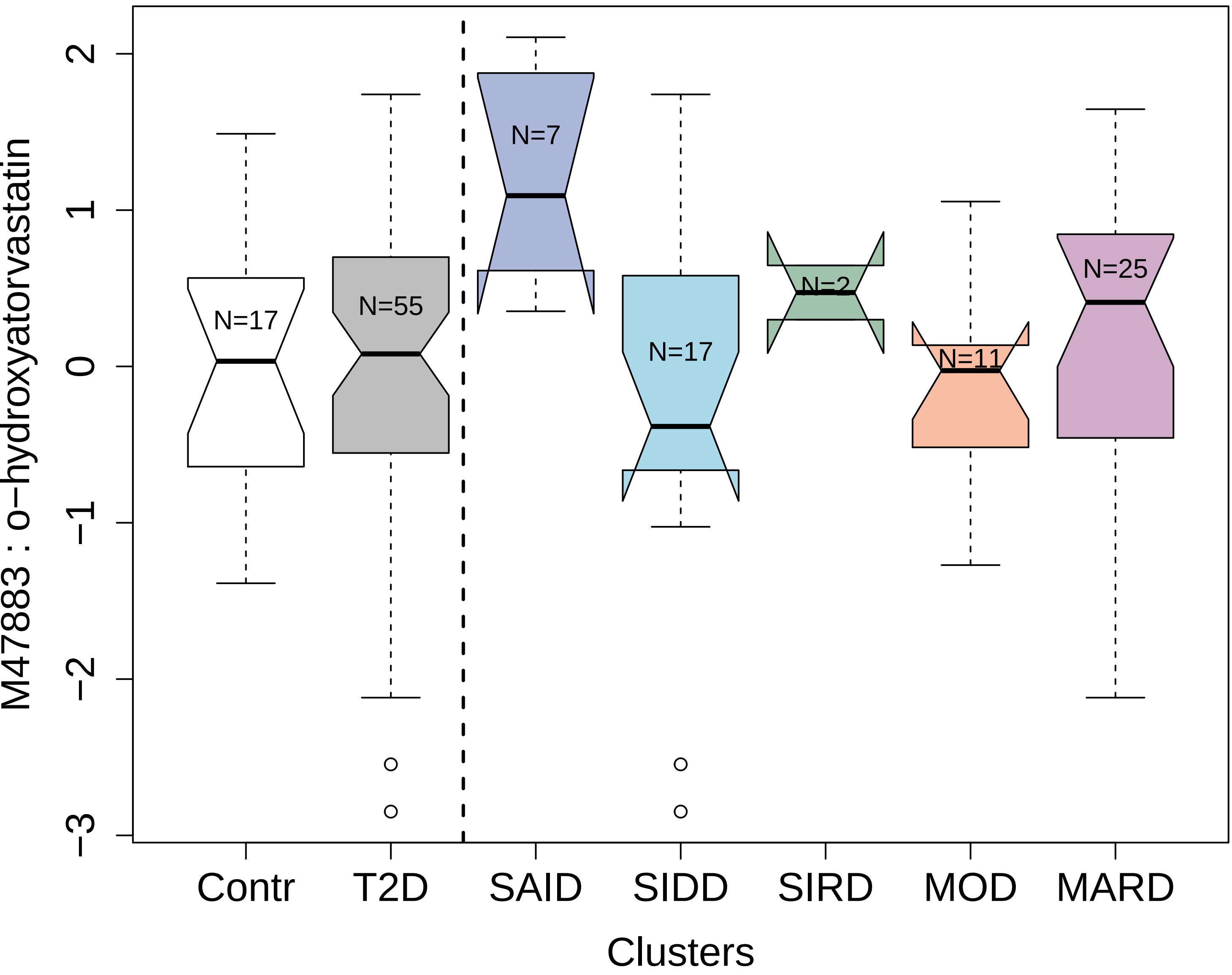
M47883 : o−hydroxyatorvastatin
Clusters
Contr
T2D
SAID
SIDD
SIRD
MOD
MARD
N=17
N=55
N=7
N=17
N=2
N=11
N=25
2
1
0
−1
−2
−3

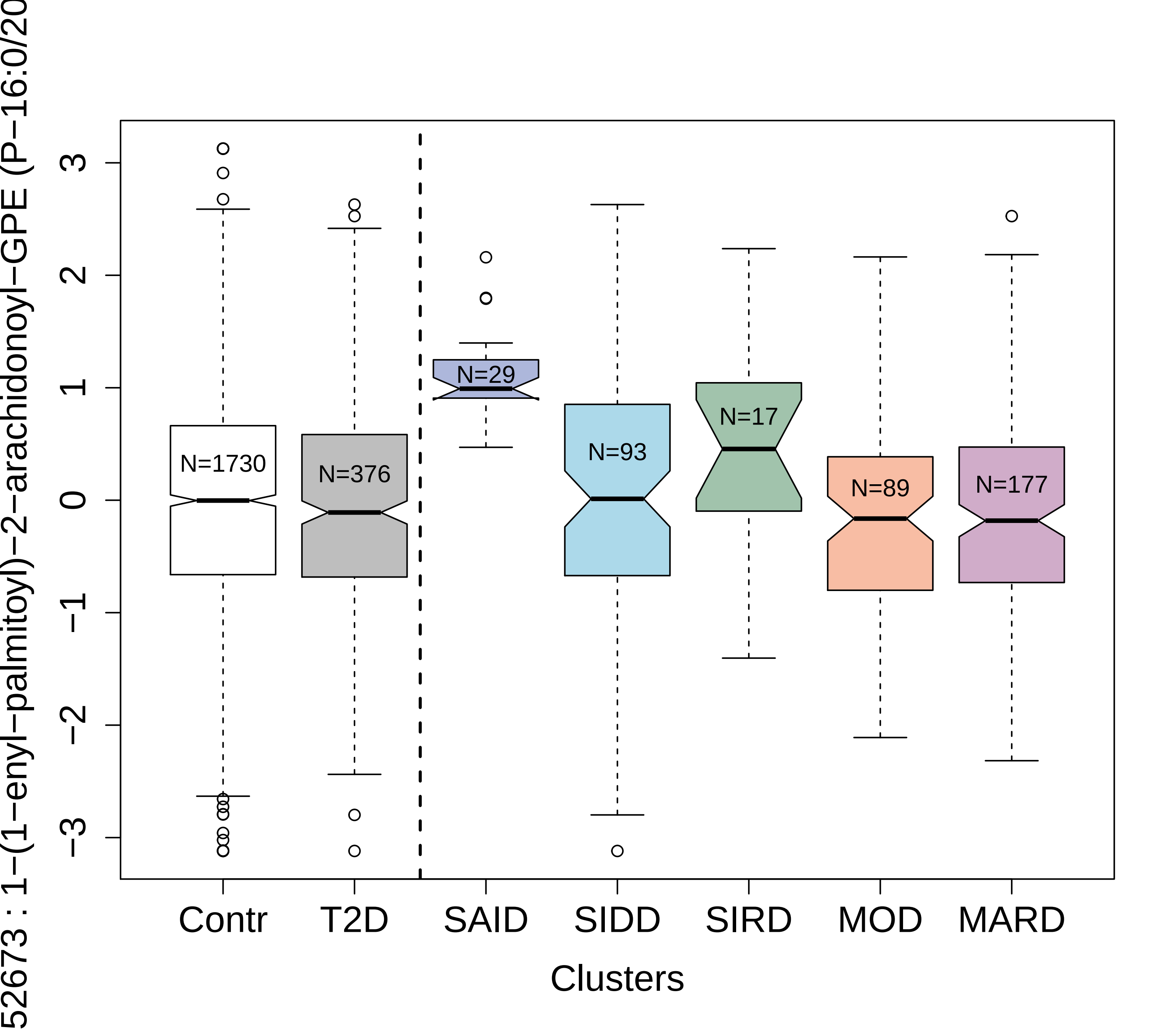

52673 : 1−(1−enyl−palmitoyl)−2−arachidonoyl−GPE (P−16:0/20
3
2
1
0
−1
−2
−3
N=1730
N=376
N=29
N=93
N=17
N=89
N=177
Contr
T2D
SAID
SIDD
SIRD
MOD
MARD
Clusters

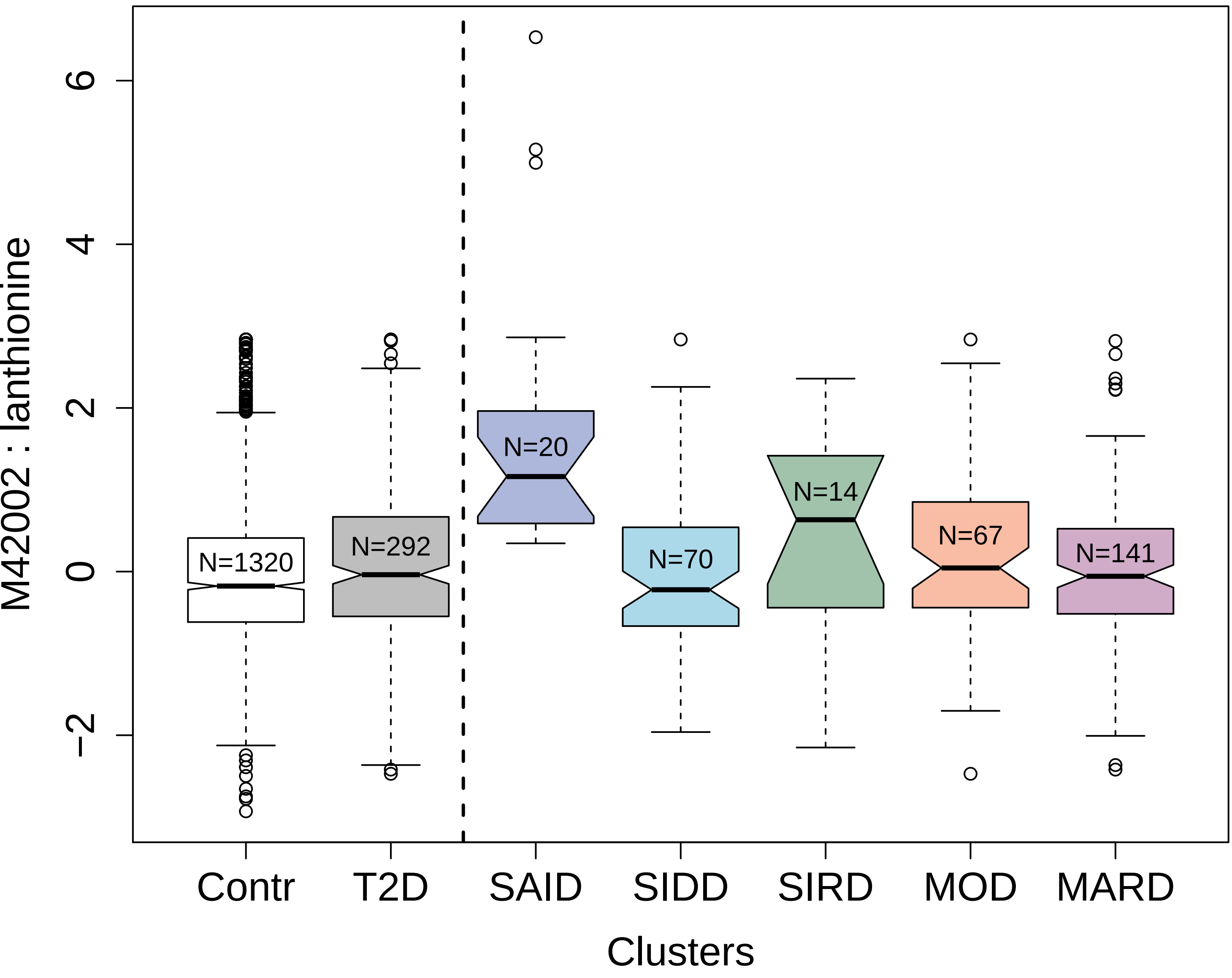
M42002 : lanthionine
6
4
2
0
−2
N=1320
N=292
N=20
N=70
N=14
N=67
N=141
Contr
T2D
SAID
SIDD
SIRD
MOD
MARD
Clusters

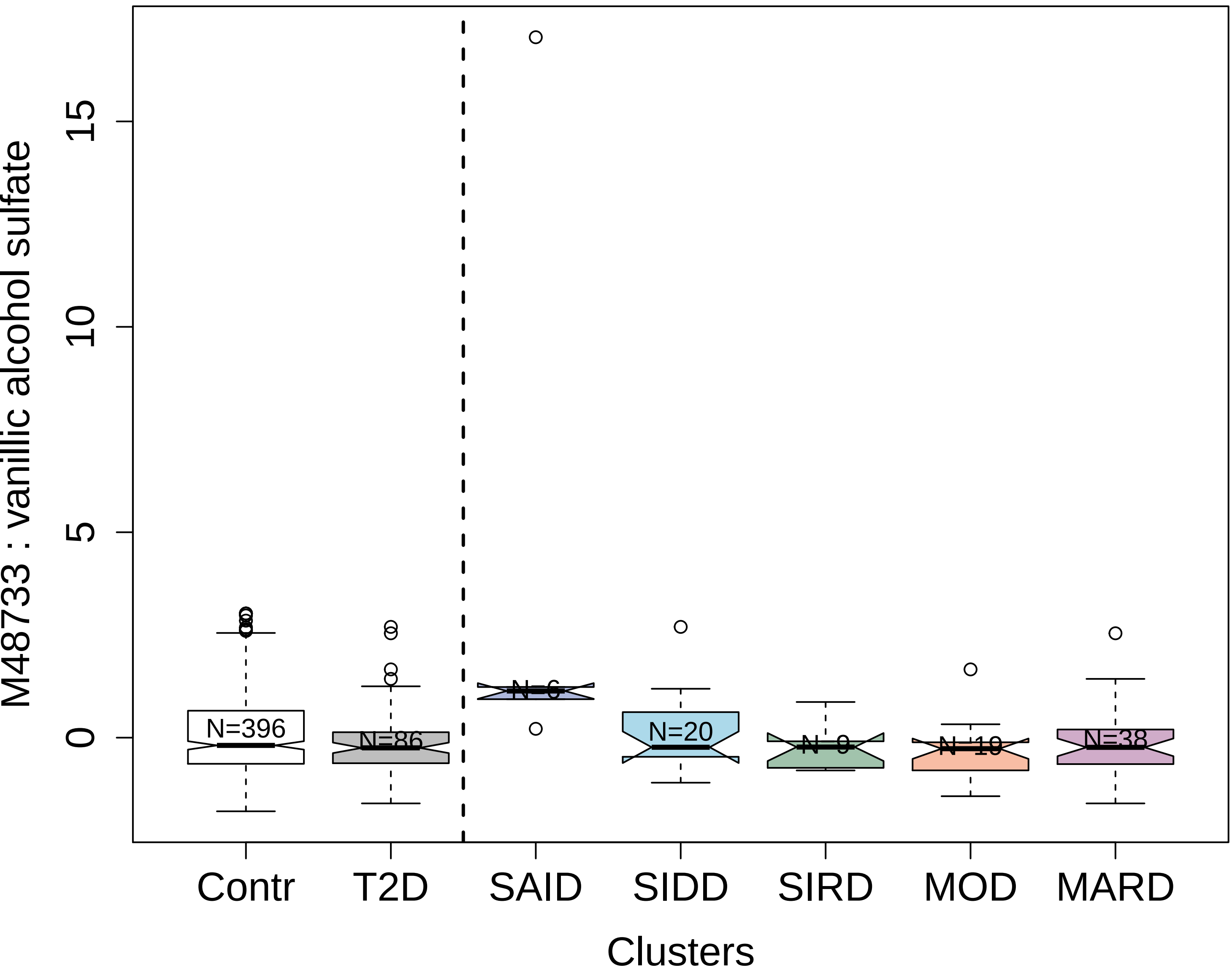
M48733 : vanillic alcohol sulfate
0
5
10
15
N=396
N=86
N=6
N=20
N=9
N=10
N=38
Contr
T2D
SAID
SIDD
SIRD
MOD
MARD
Clusters

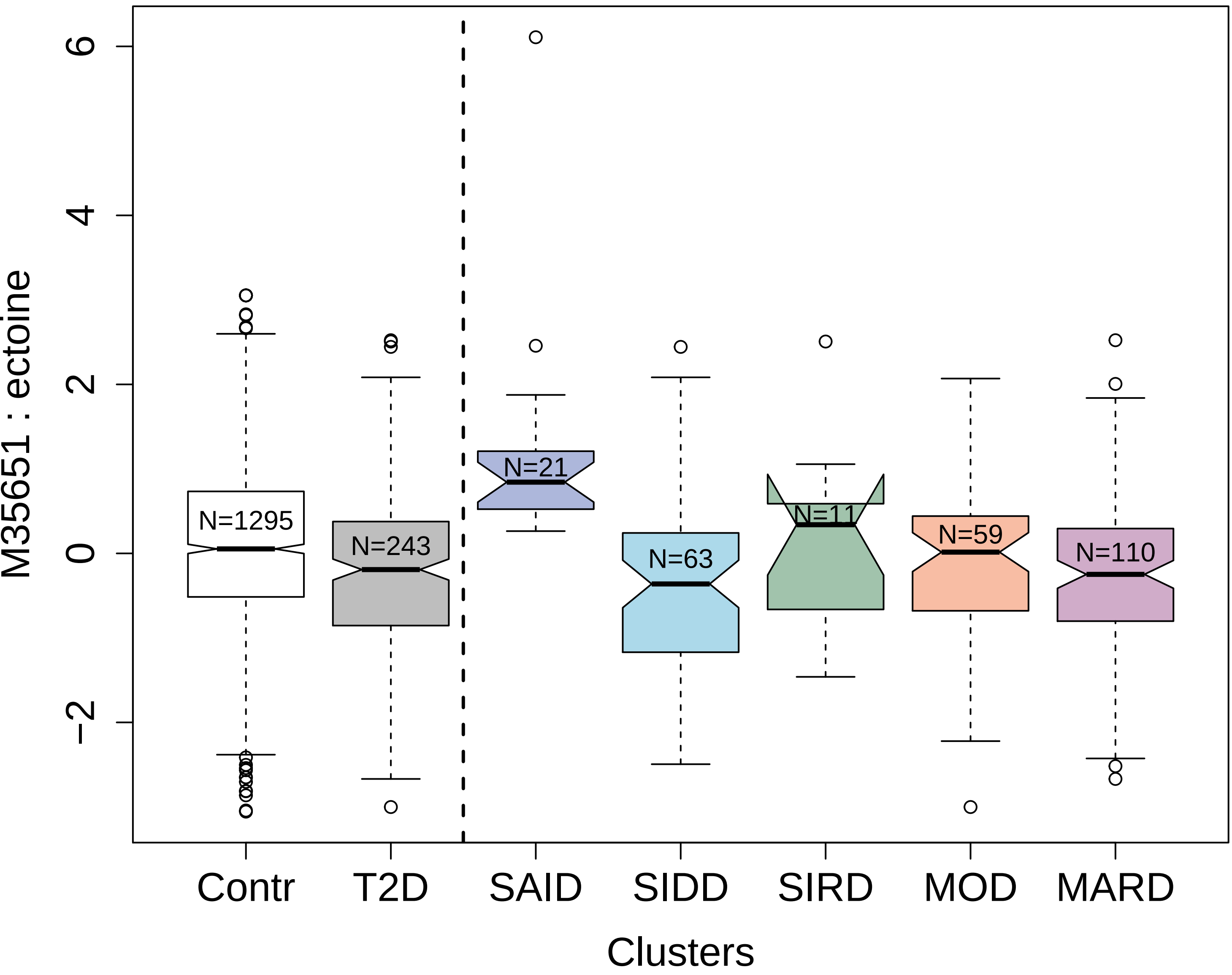

M35651 : ectoine
6
4
2
0
−2
N=1295
N=243
N=21
N=63
N=11
N=59
N=110
Contr
T2D
SAID
SIDD
SIRD
MOD
MARD
Clusters

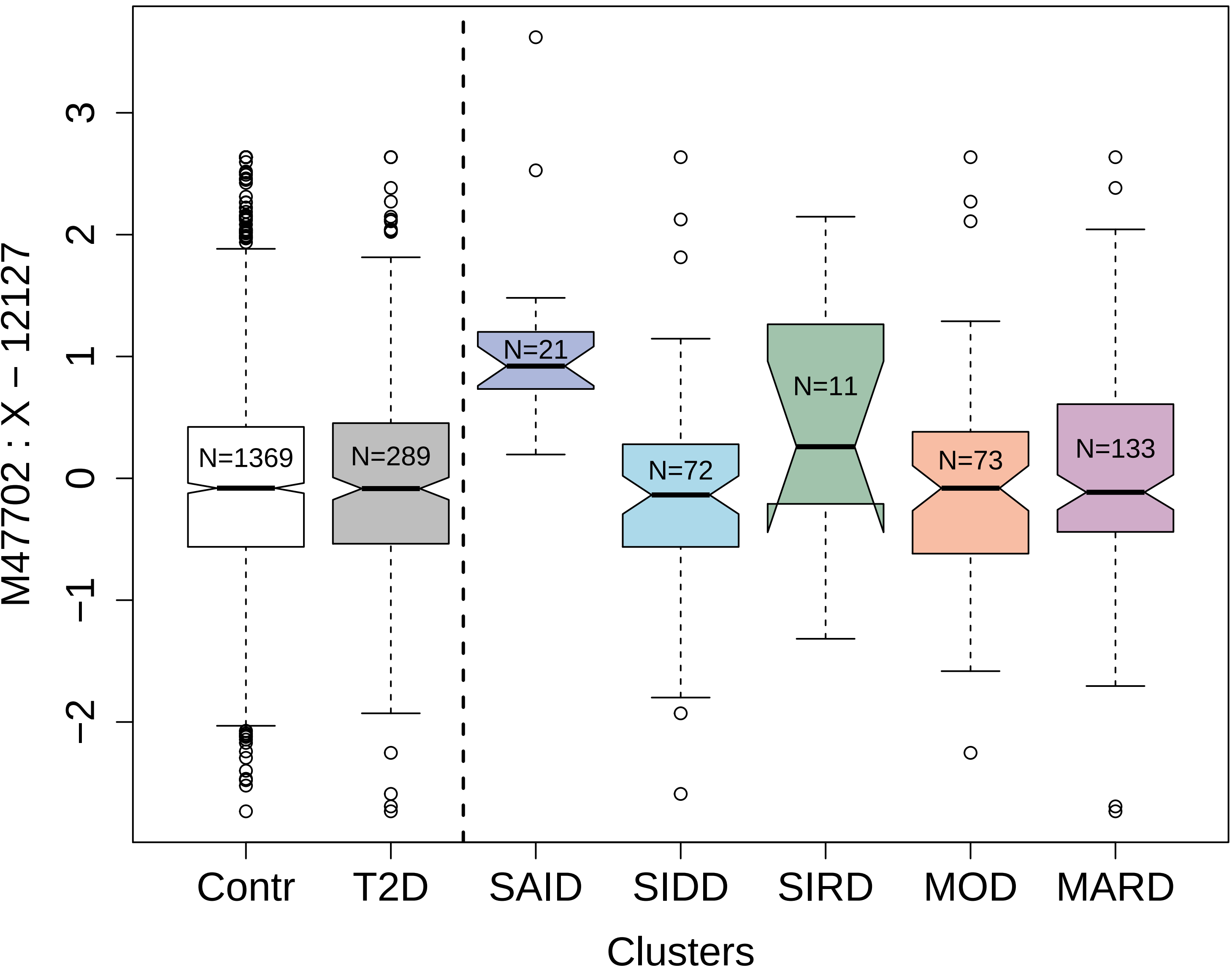

M47702 : X − 12127
3
2
1
0
−1
−2
N=1369
N=289
N=21
N=72
N=11
N=73
N=133
Contr
T2D
SAID
SIDD
SIRD
MOD
MARD
Clusters

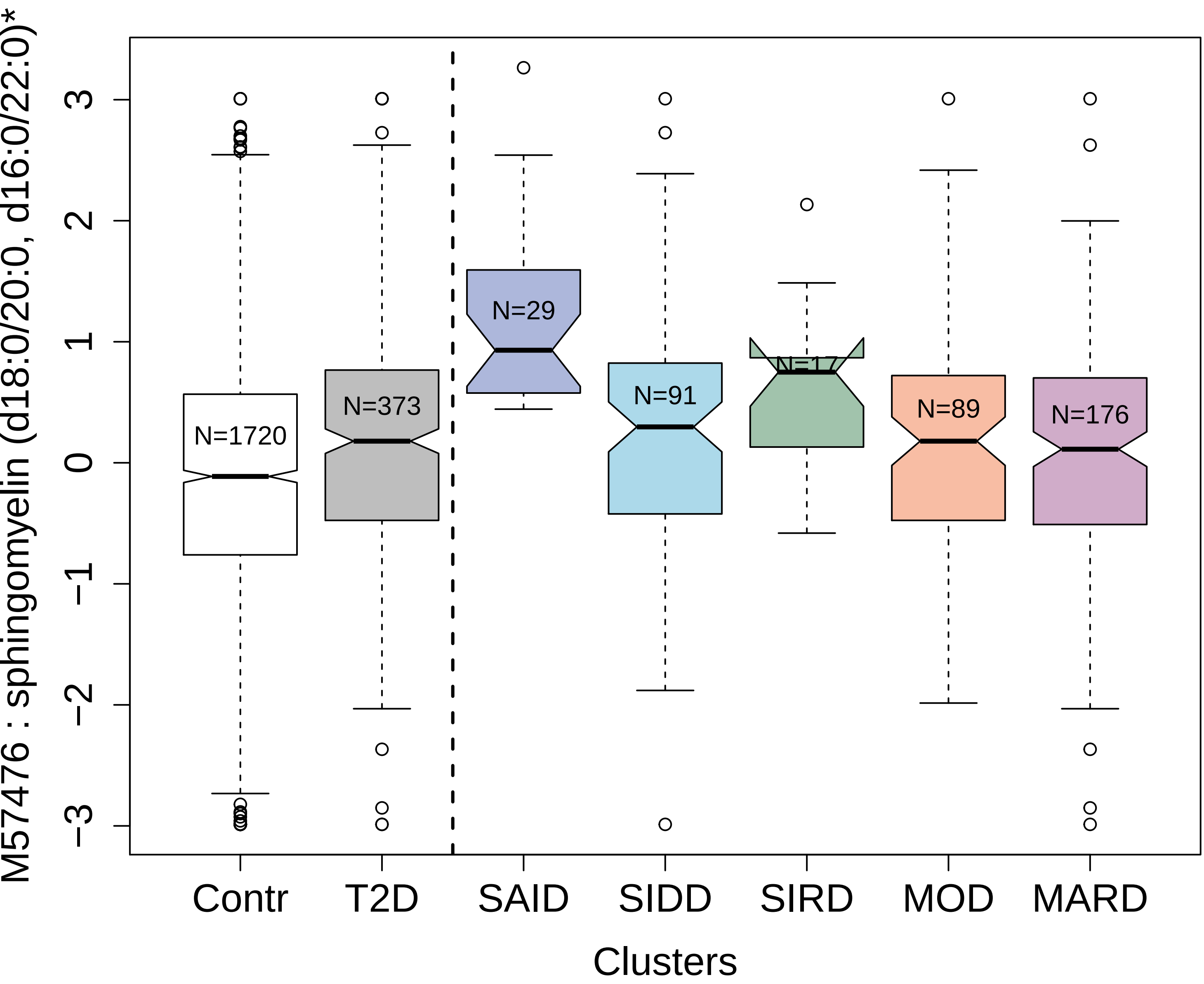

M57476 : sphingomyelin (d18:0/20:0, d16:0/22:0)*
3
2
1
0
−1
−2
−3
N=1720
N=373
N=29
N=91
N=17
N=89
N=176
Contr
T2D
SAID
SIDD
SIRD
MOD
MARD
Clusters

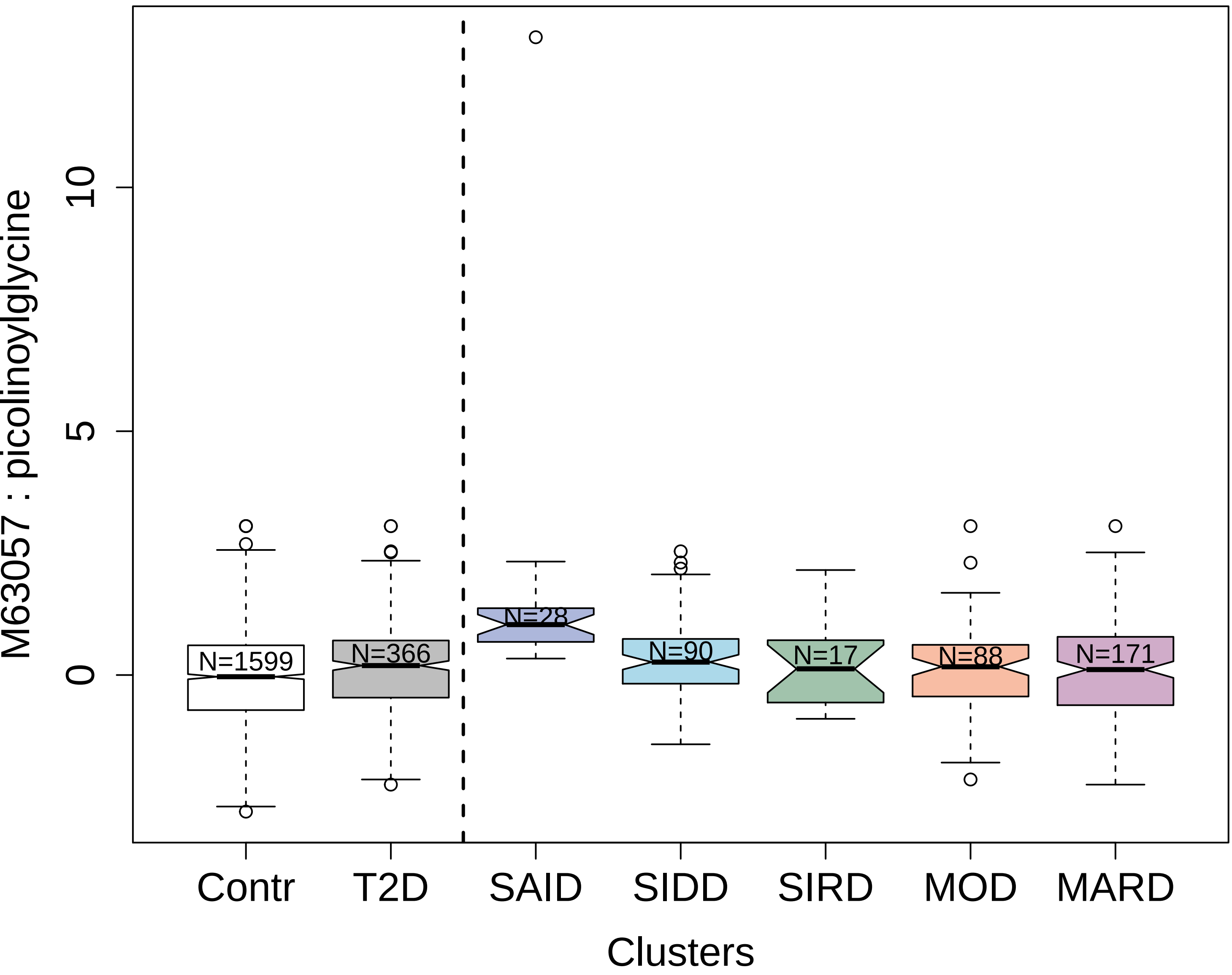
M63057 : picolinoylglycine
0
5
10
N=1599
N=366
N=28
N=90
N=17
N=88
N=171
Contr
T2D
SAID
SIDD
SIRD
MOD
MARD
Clusters

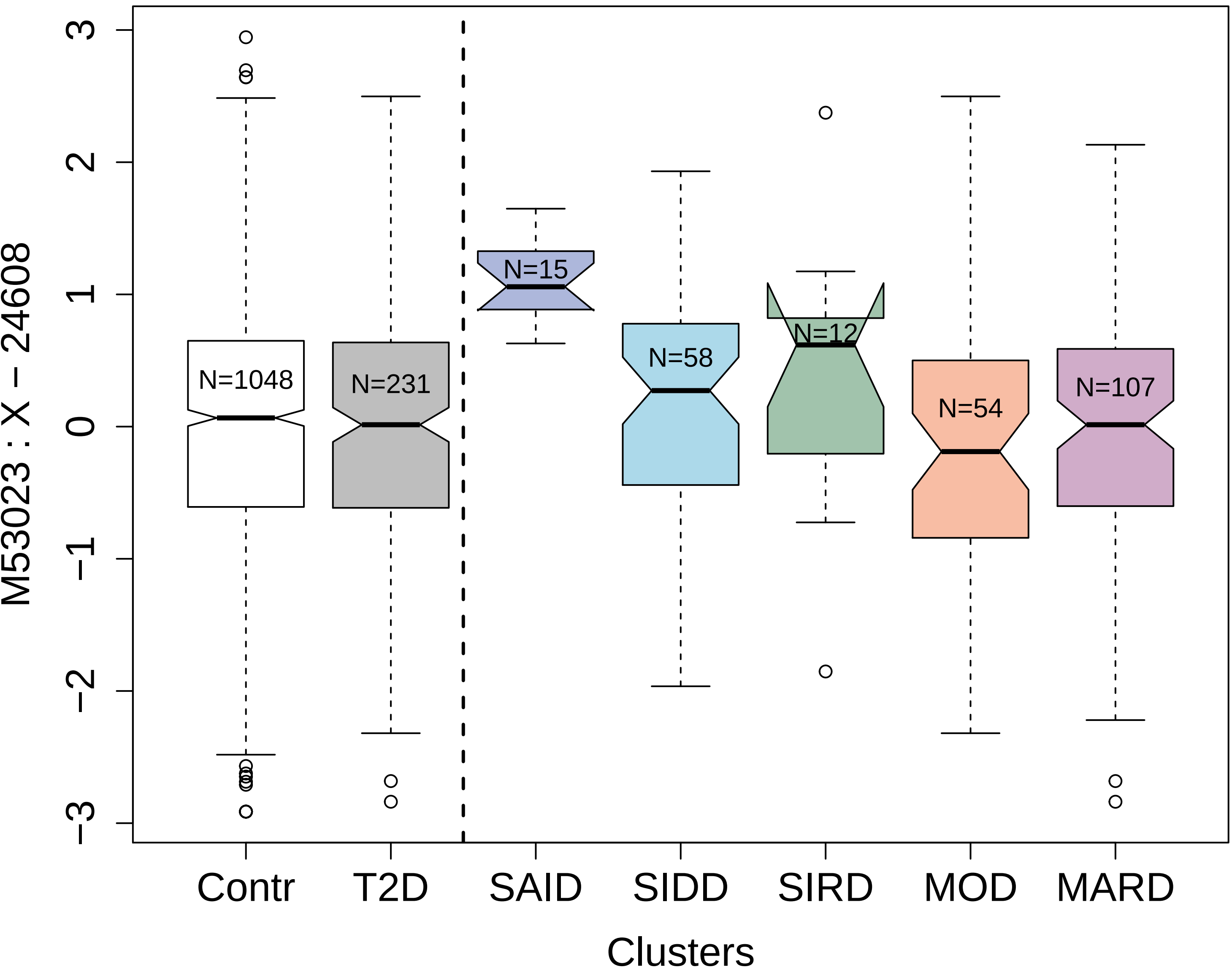

M53023 : X − 24608
3
2
1
0
−1
−2
−3
N=1048
N=231
N=15
N=58
N=12
N=54
N=107
Contr
T2D
SAID
SIDD
SIRD
MOD
MARD
Clusters

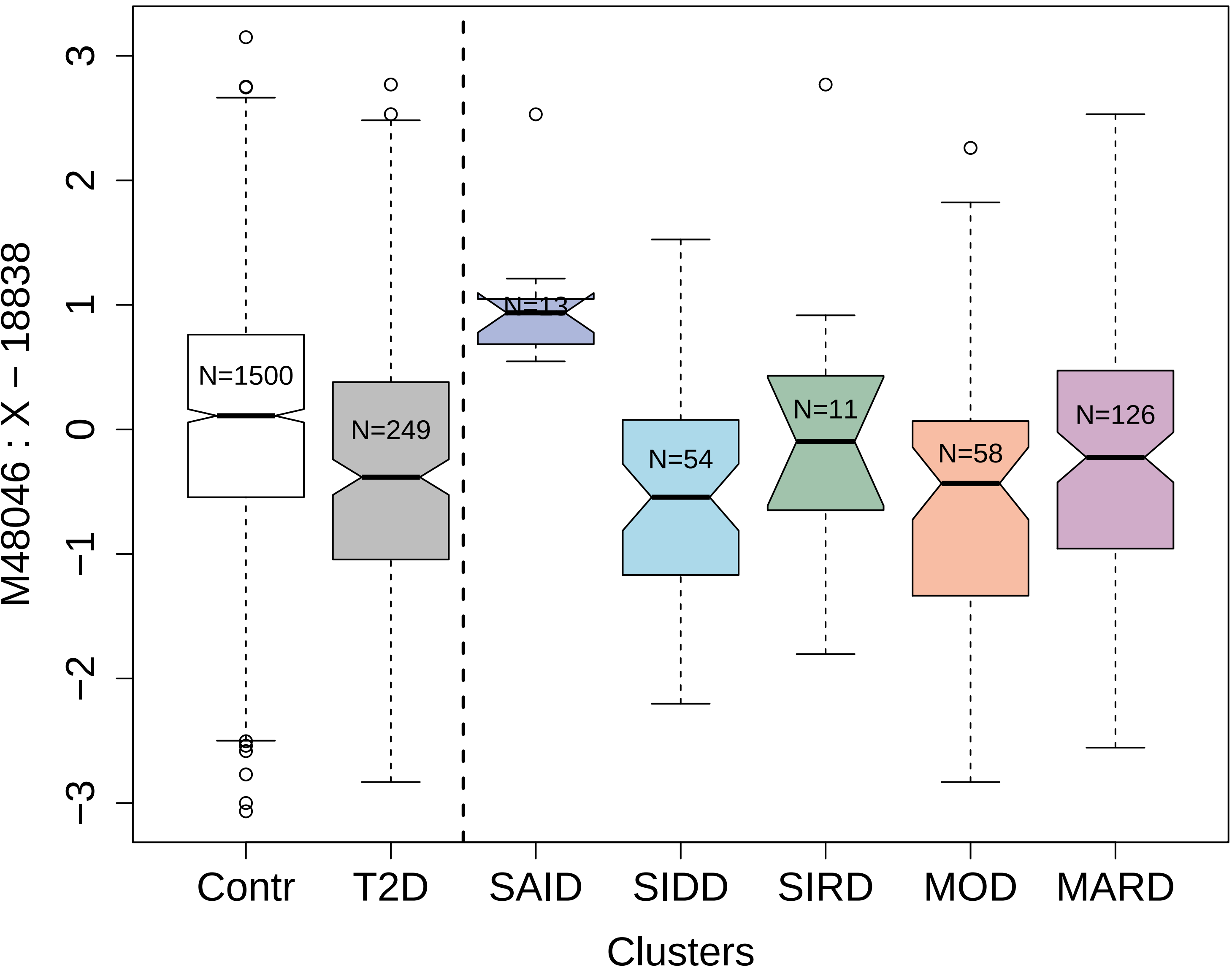

M48046 : X − 18838
3
2
1
0
−1
−2
−3
N=1500
N=249
N=13
N=54
N=11
N=58
N=126
Contr
T2D
SAID
SIDD
SIRD
MOD
MARD
Clusters

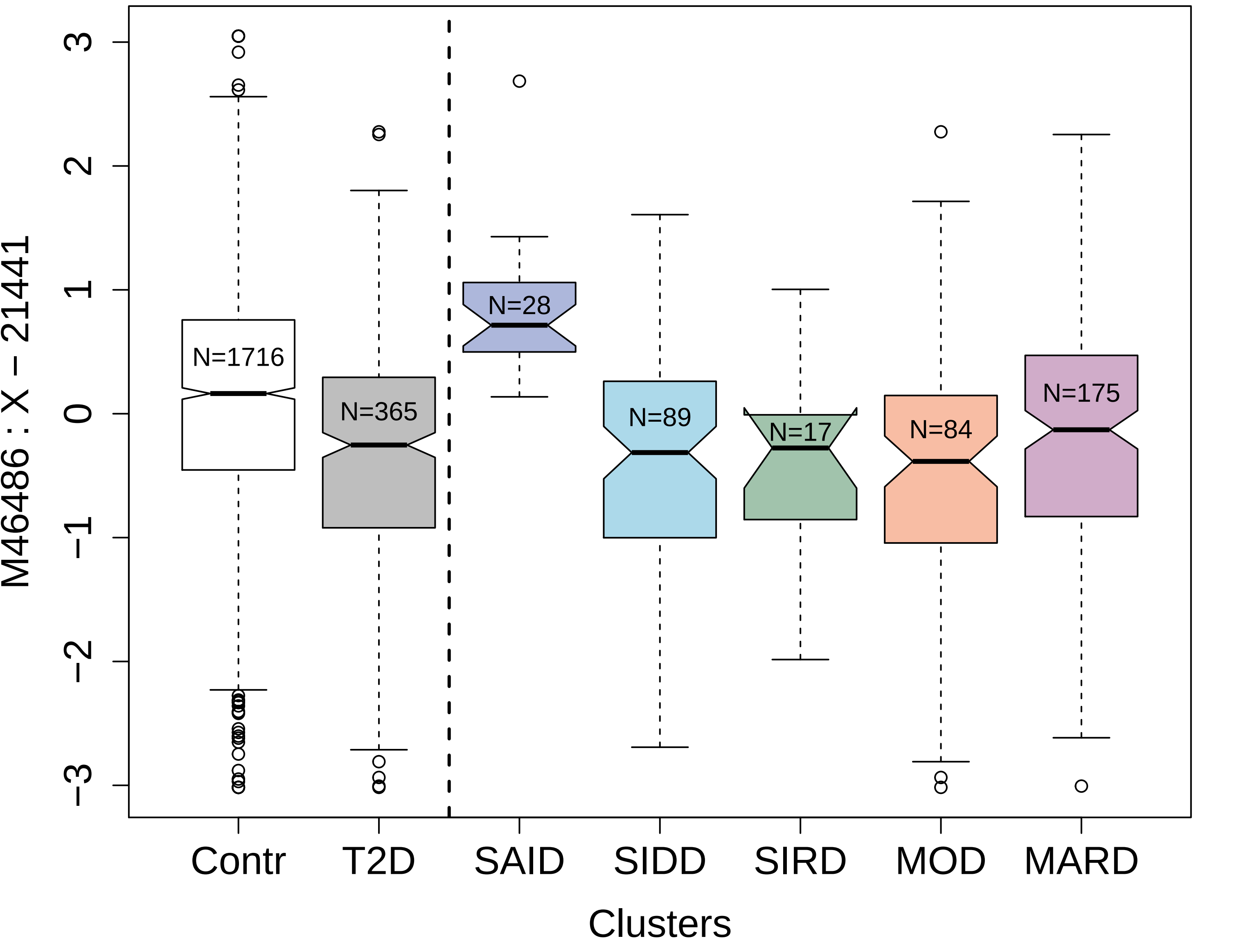

M46486 : X − 21441
3
2
1
0
−1
−2
−3
N=1716
N=365
N=28
N=89
N=17
N=84
N=175
Contr
T2D
SAID
SIDD
SIRD
MOD
MARD
Clusters

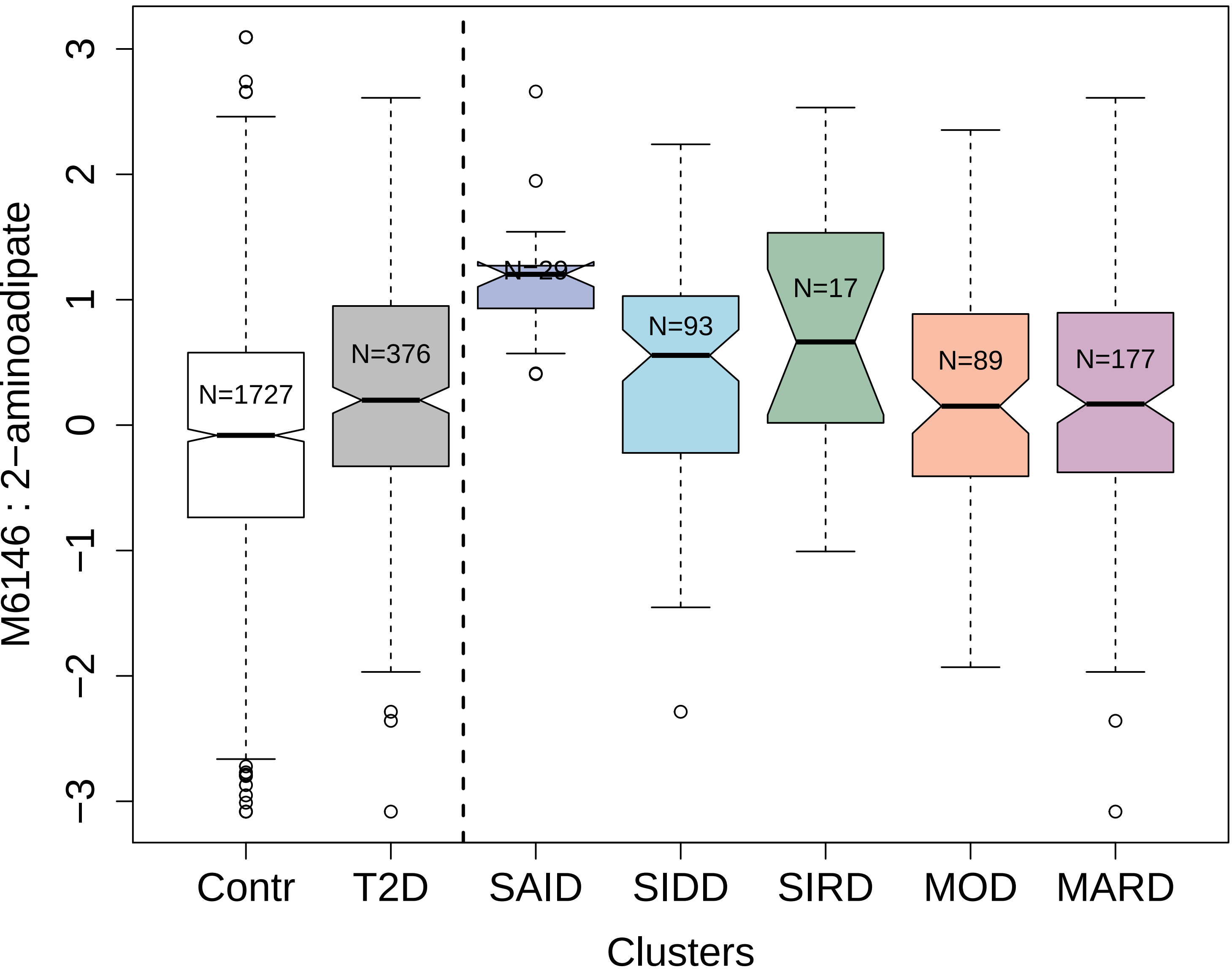
M6146 : 2−aminoadipate
Clusters
Contr
T2D
SAID
SIDD
SIRD
MOD
MARD
N=1727
N=376
N=29
N=93
N=17
N=89
N=177
3
2
1
0
−1
−2
−3

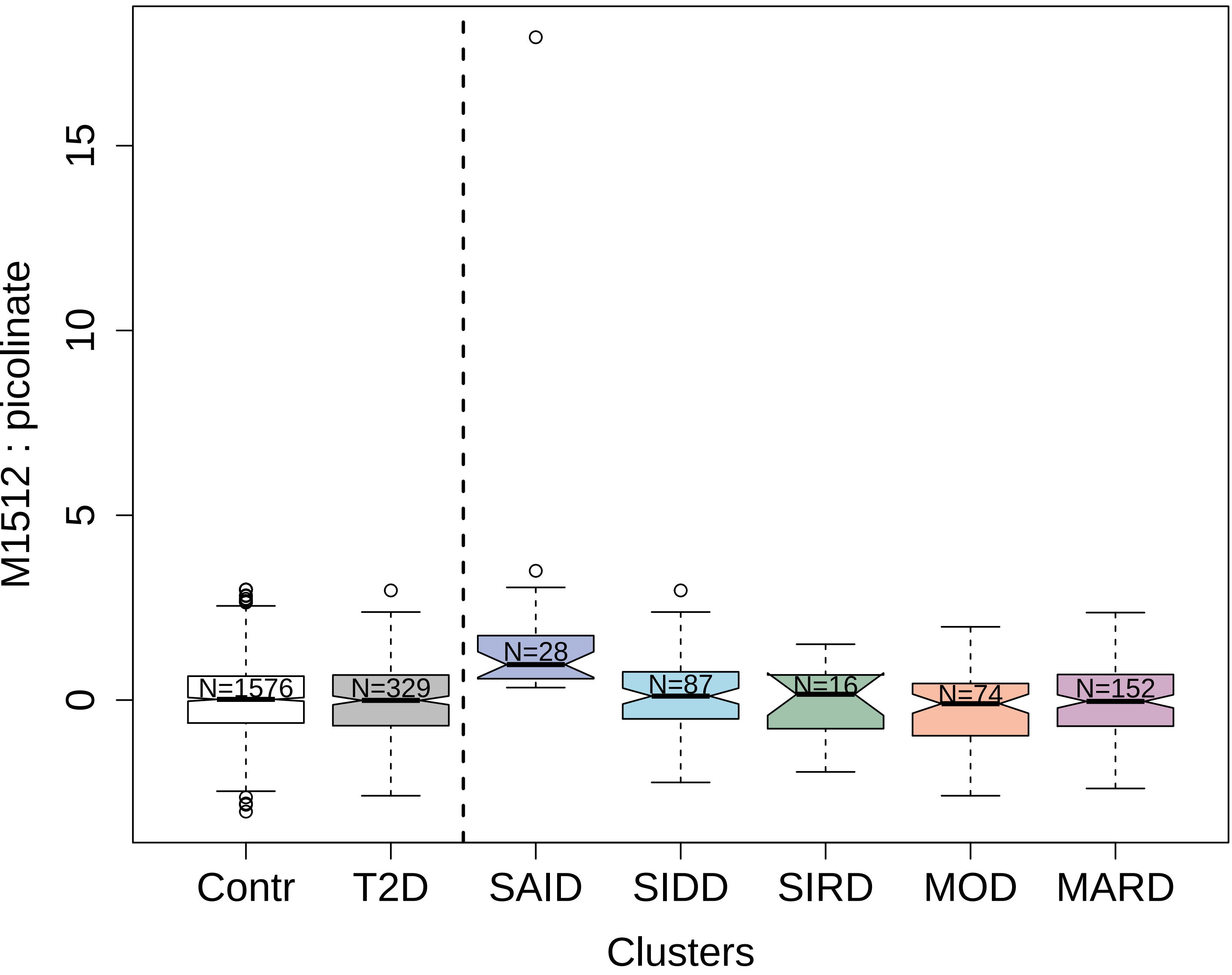

M1512 : picolinate
0
5
10
15
N=1576
N=329
N=28
N=87
N=16
N=74
N=152
Contr
T2D
SAID
SIDD
SIRD
MOD
MARD
Clusters

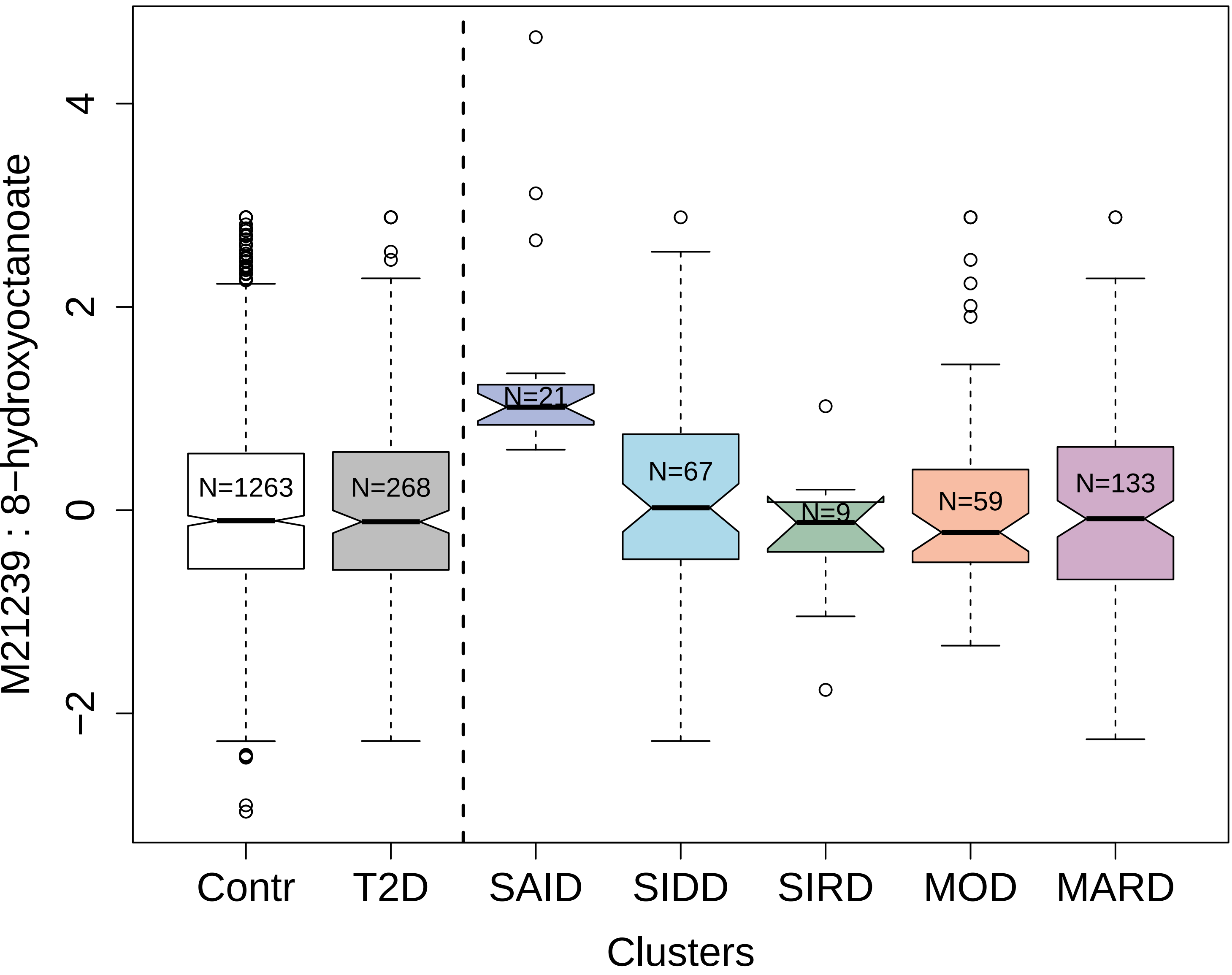

M21239 : 8−hydroxyoctanoate
4
2
0
−2
N=1263
N=268
N=21
N=67
N=9
N=59
N=133
Contr
T2D
SAID
SIDD
SIRD
MOD
MARD
Clusters

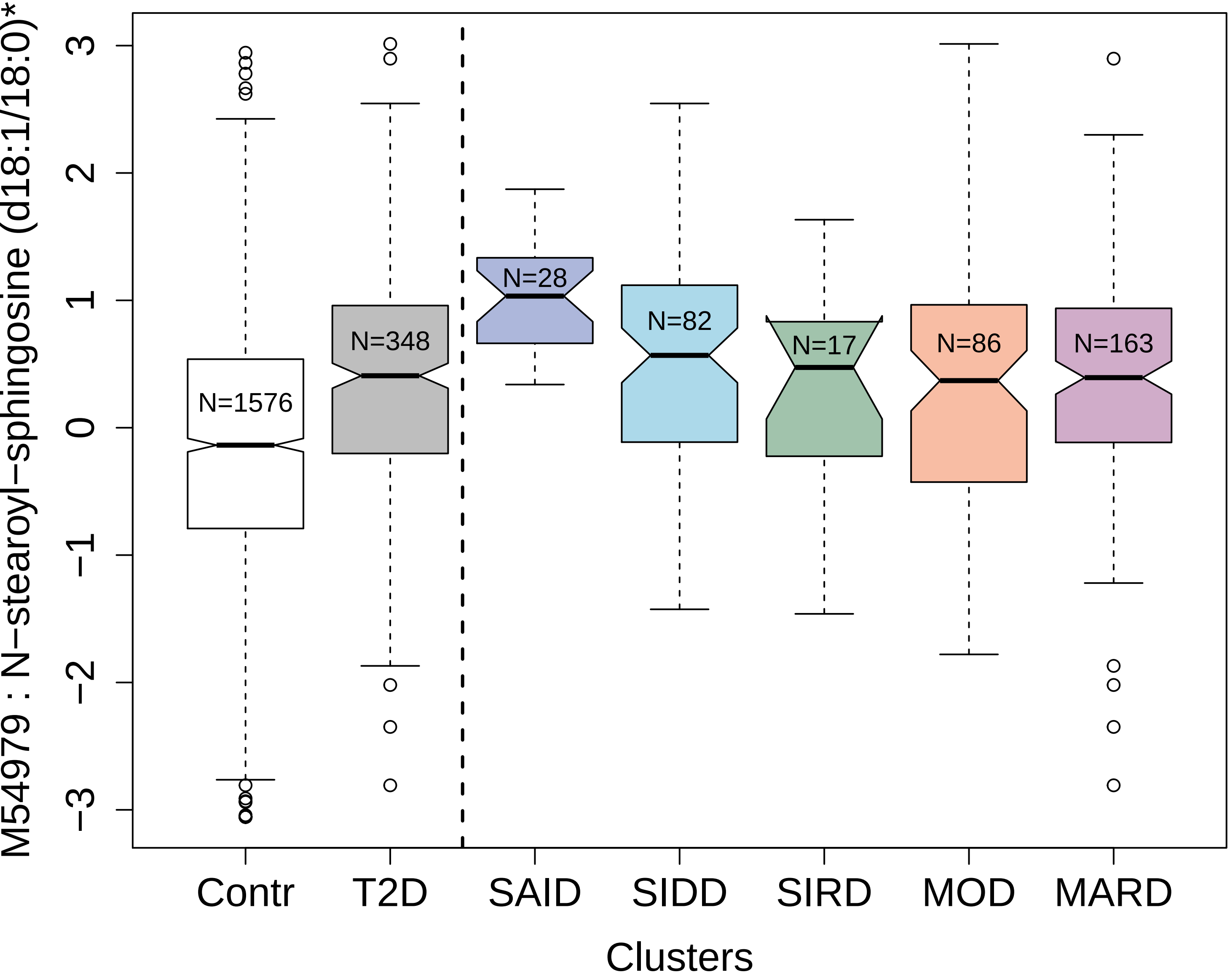

M54979 : N−stearoyl−sphingosine (d18:1/18:0)*
3
2
1
0
−1
−2
−3
N=1576
N=348
N=28
N=82
N=17
N=86
N=163
Contr
T2D
SAID
SIDD
SIRD
MOD
MARD
Clusters

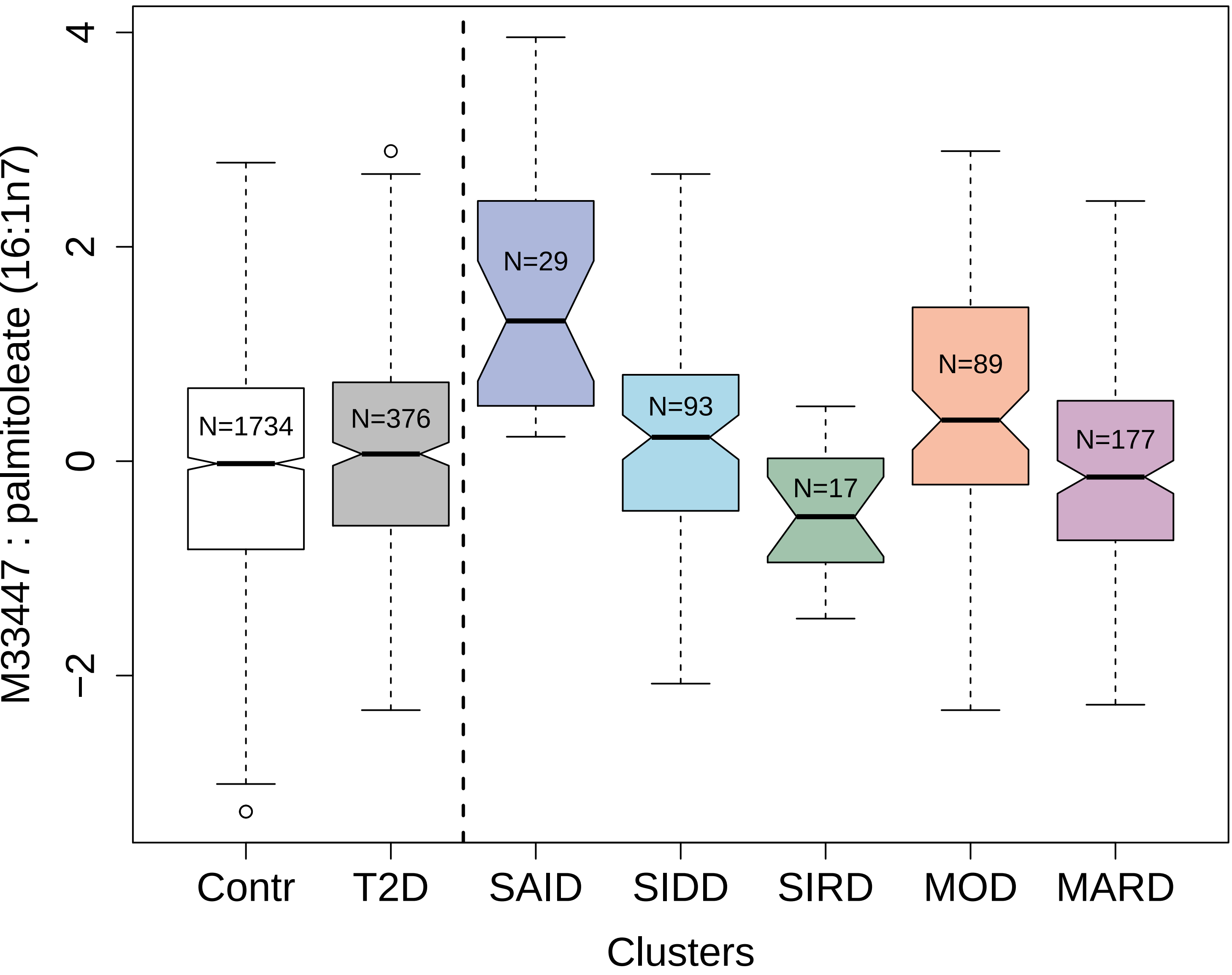

M33447 : palmitoleate (16:1n7)
4
2
0
−2
N=1734
N=376
N=29
N=93
N=17
N=89
N=177
Contr
T2D
SAID
SIDD
SIRD
MOD
MARD
Clusters

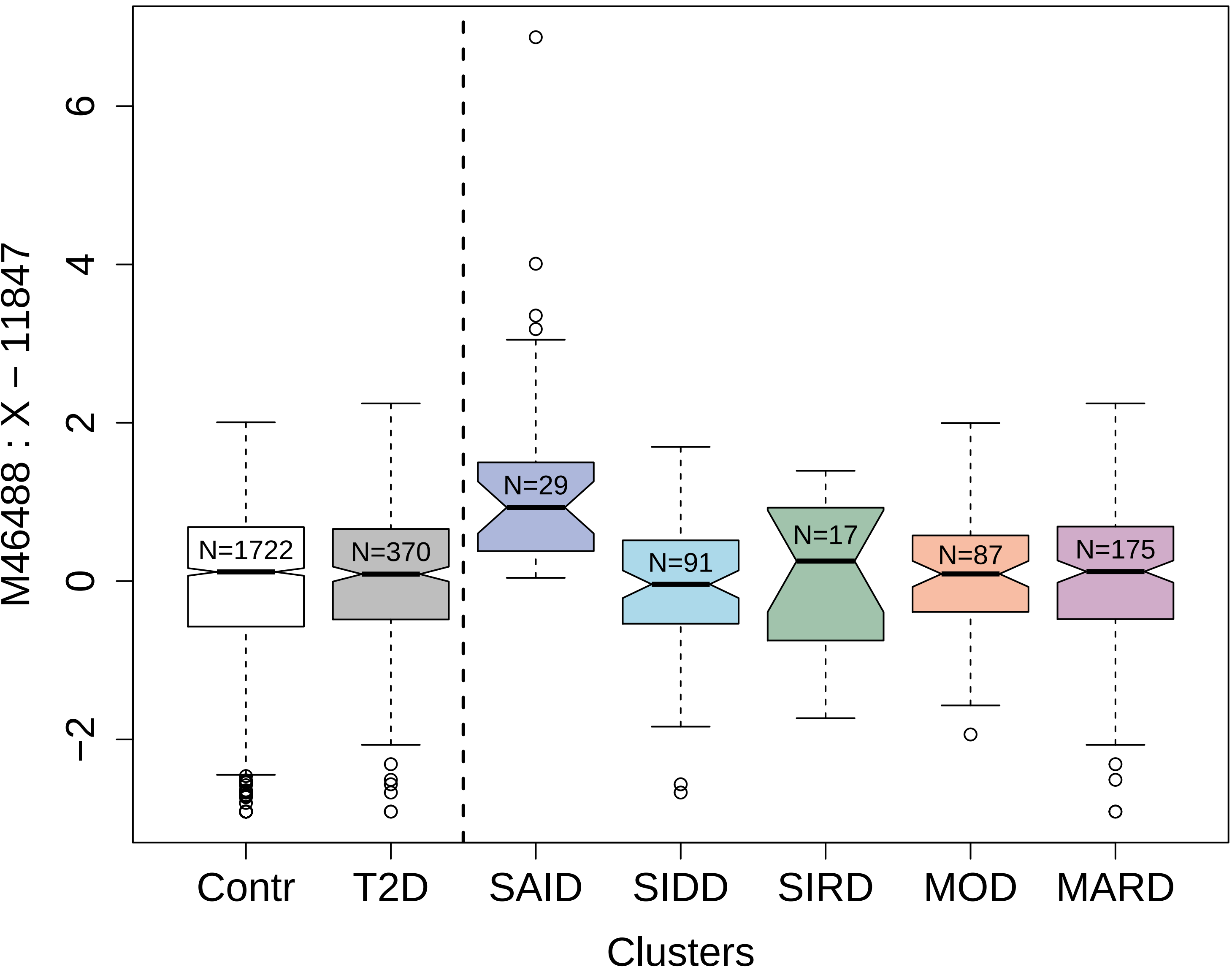

N=1722
N=370
N=29
N=91
N=17
N=87
N=175
6
4
2
0
−2
M46488 : X − 11847
Contr
T2D
SAID
SIDD
SIRD
MOD
MARD
Clusters

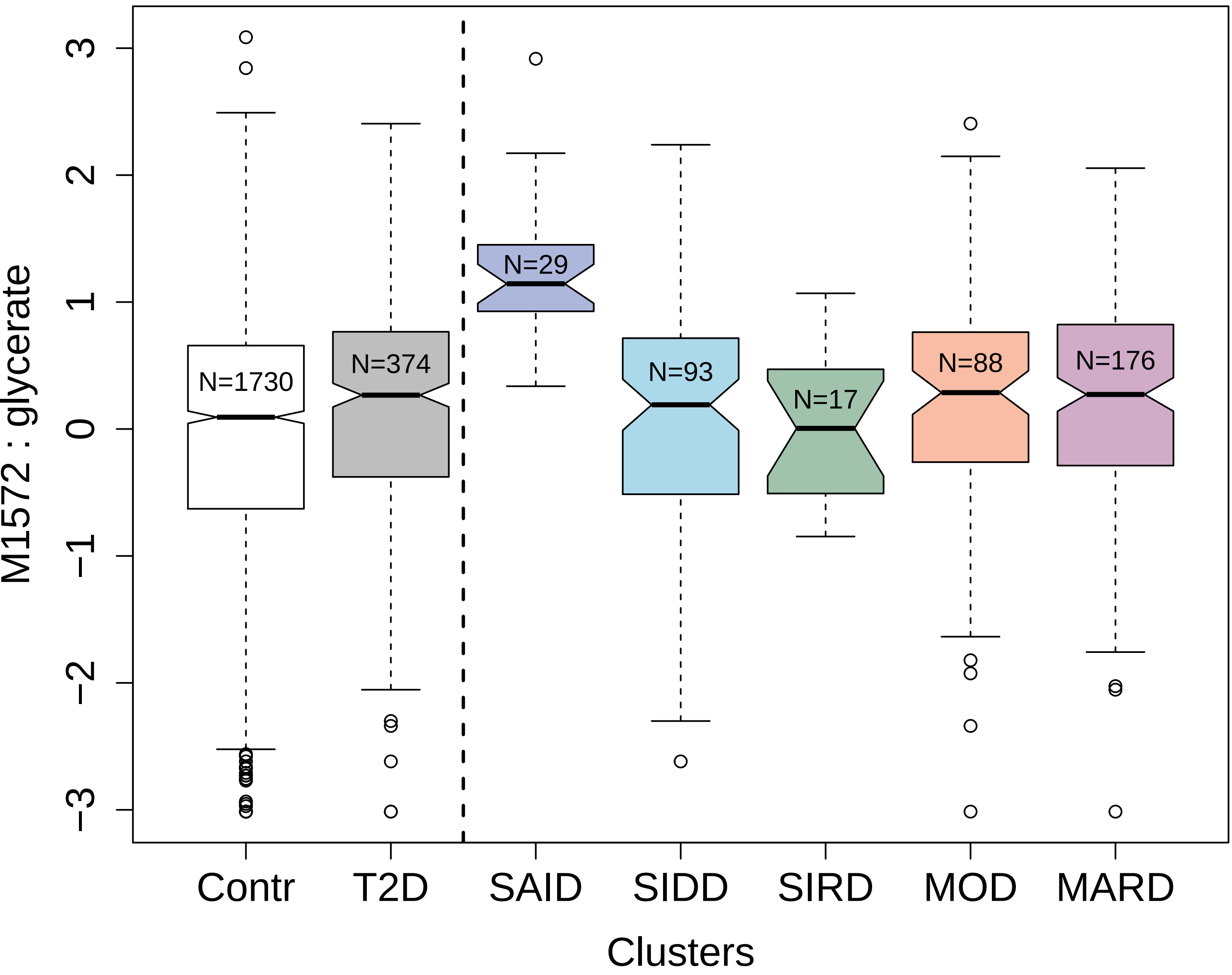
M1572 : glycerate
3
2
1
0
−1
−2
−3
N=1730
N=374
N=29
N=93
N=17
N=88
N=176
Contr
T2D
SAID
SIDD
SIRD
MOD
MARD
Clusters

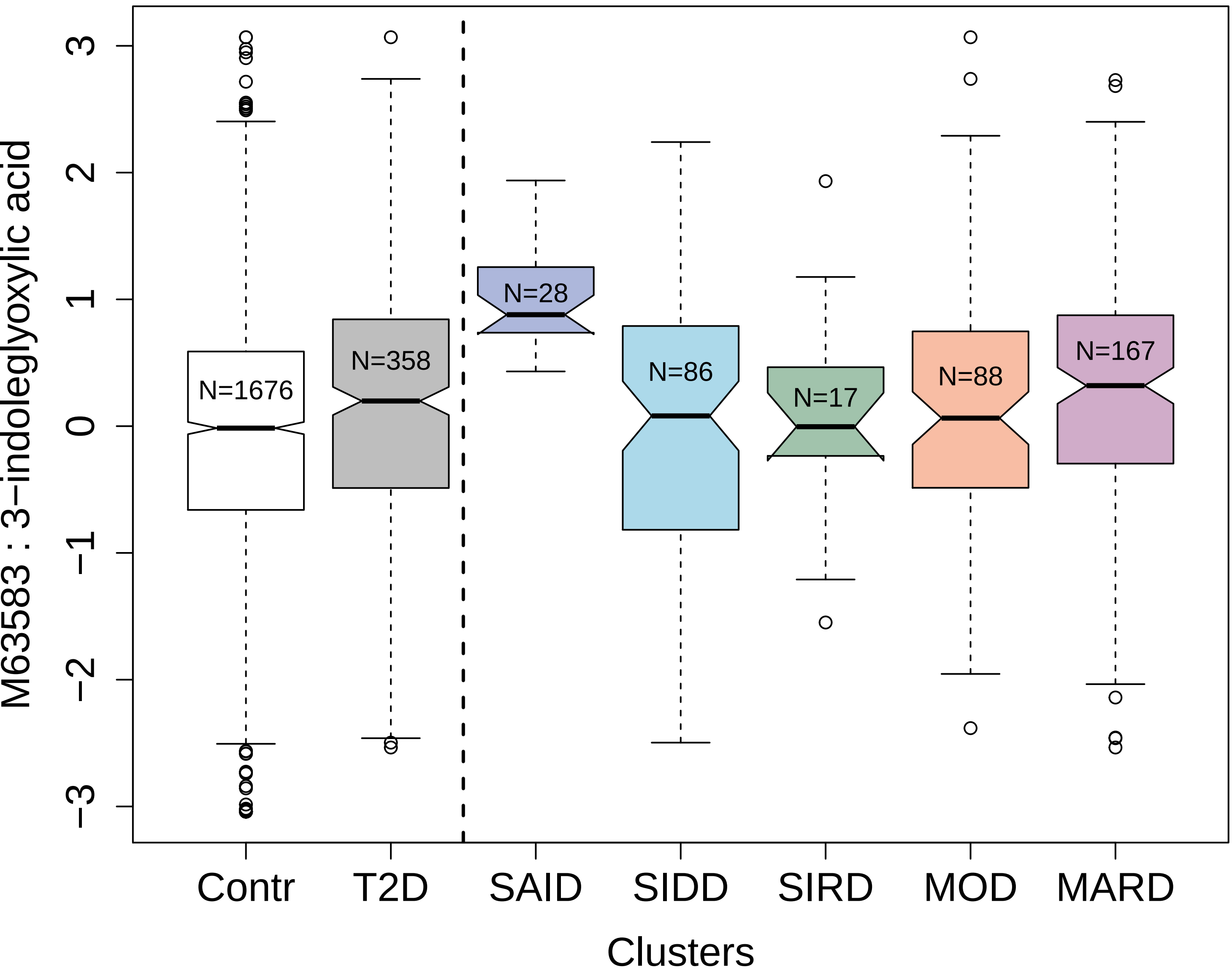

M63583 : 3−indoleglyoxylic acid
3
2
1
0
−1
−2
−3
N=1676
N=358
N=28
N=86
N=17
N=88
N=167
Contr
T2D
SAID
SIDD
SIRD
MOD
MARD
Clusters

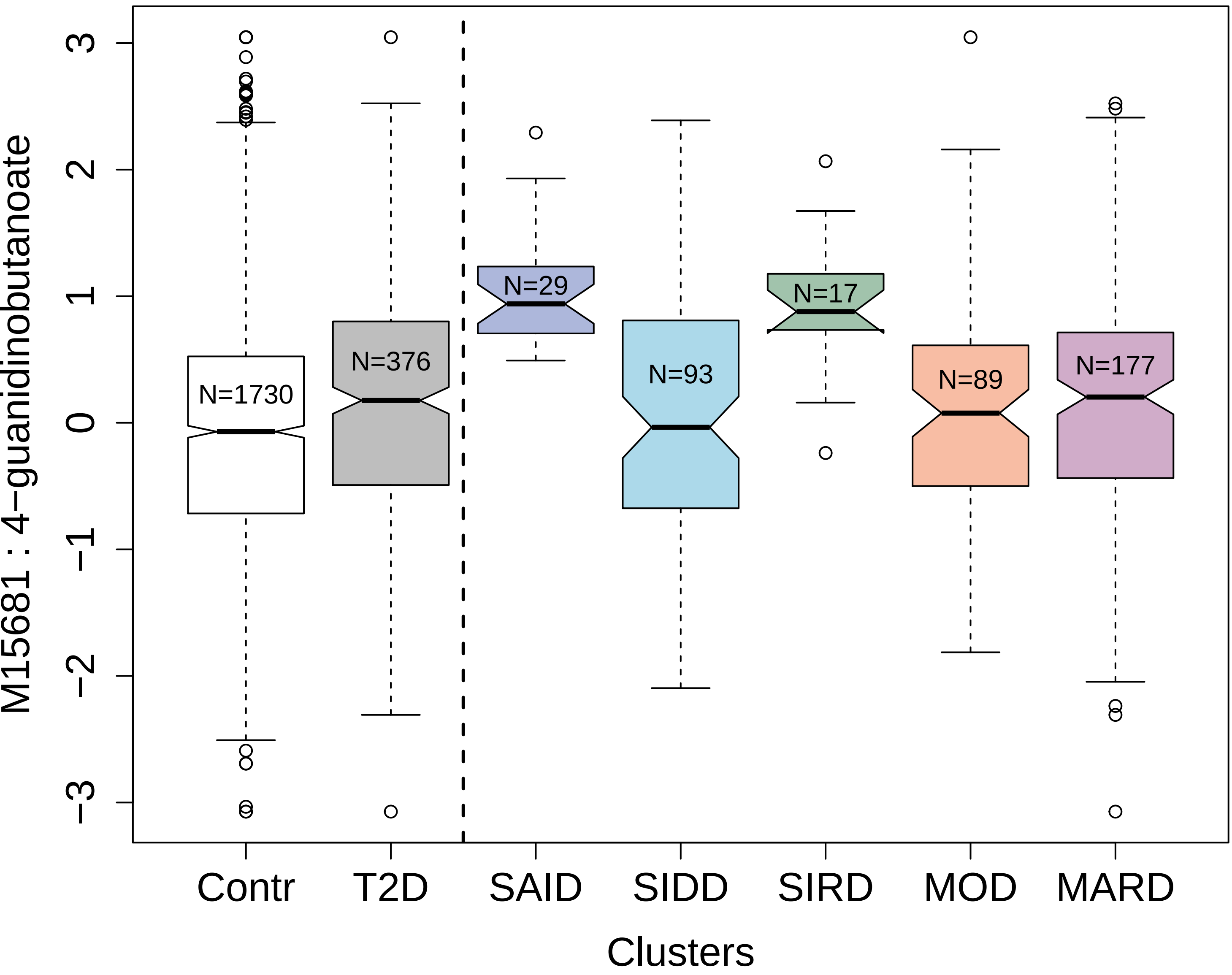

M15681 : 4−guanidinobutanoate
3
2
1
0
−1
−2
−3
N=1730
N=376
N=29
N=93
N=17
N=89
N=177
Contr
T2D
SAID
SIDD
SIRD
MOD
MARD
Clusters

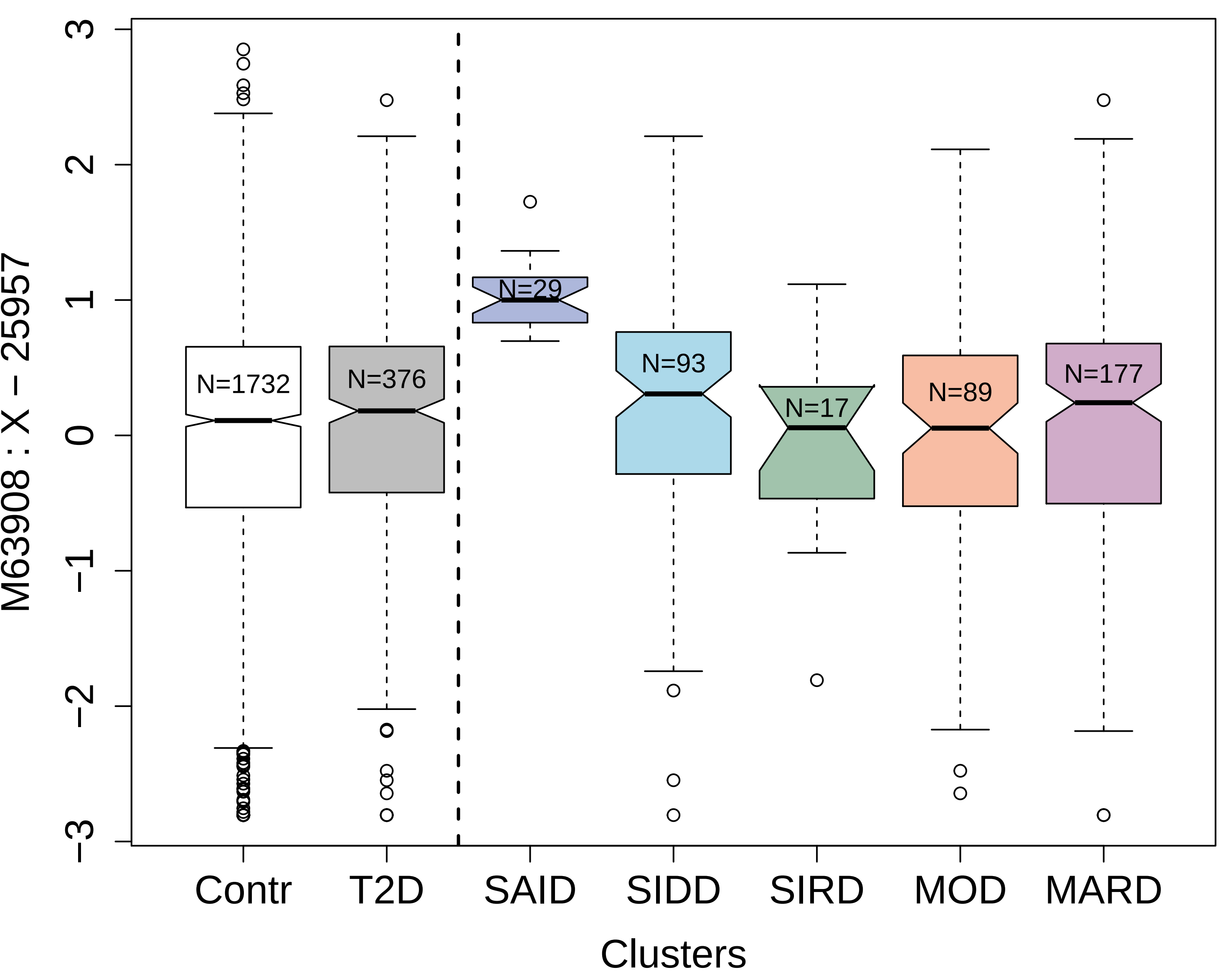

M63908 : X − 25957
3
2
1
0
−1
−2
−3
N=1732
N=376
N=29
N=93
N=17
N=89
N=177
Contr
T2D
SAID
SIDD
SIRD
MOD
MARD
Clusters

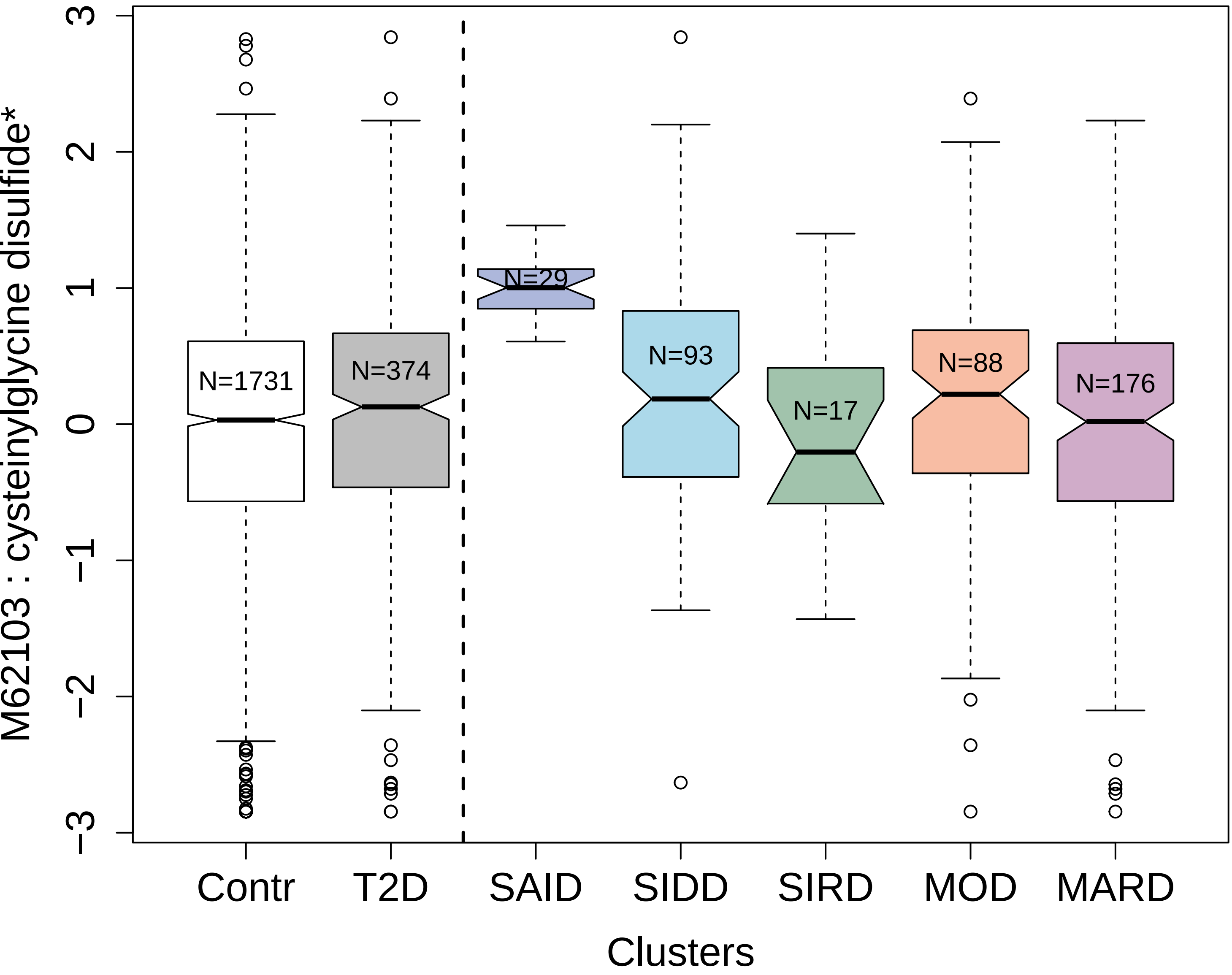

M62103 : cysteinylglycine disulfide*
3
2
1
0
−1
−2
−3
N=1731
N=374
N=29
N=93
N=17
N=88
N=176
Contr
T2D
SAID
SIDD
SIRD
MOD
MARD
Clusters

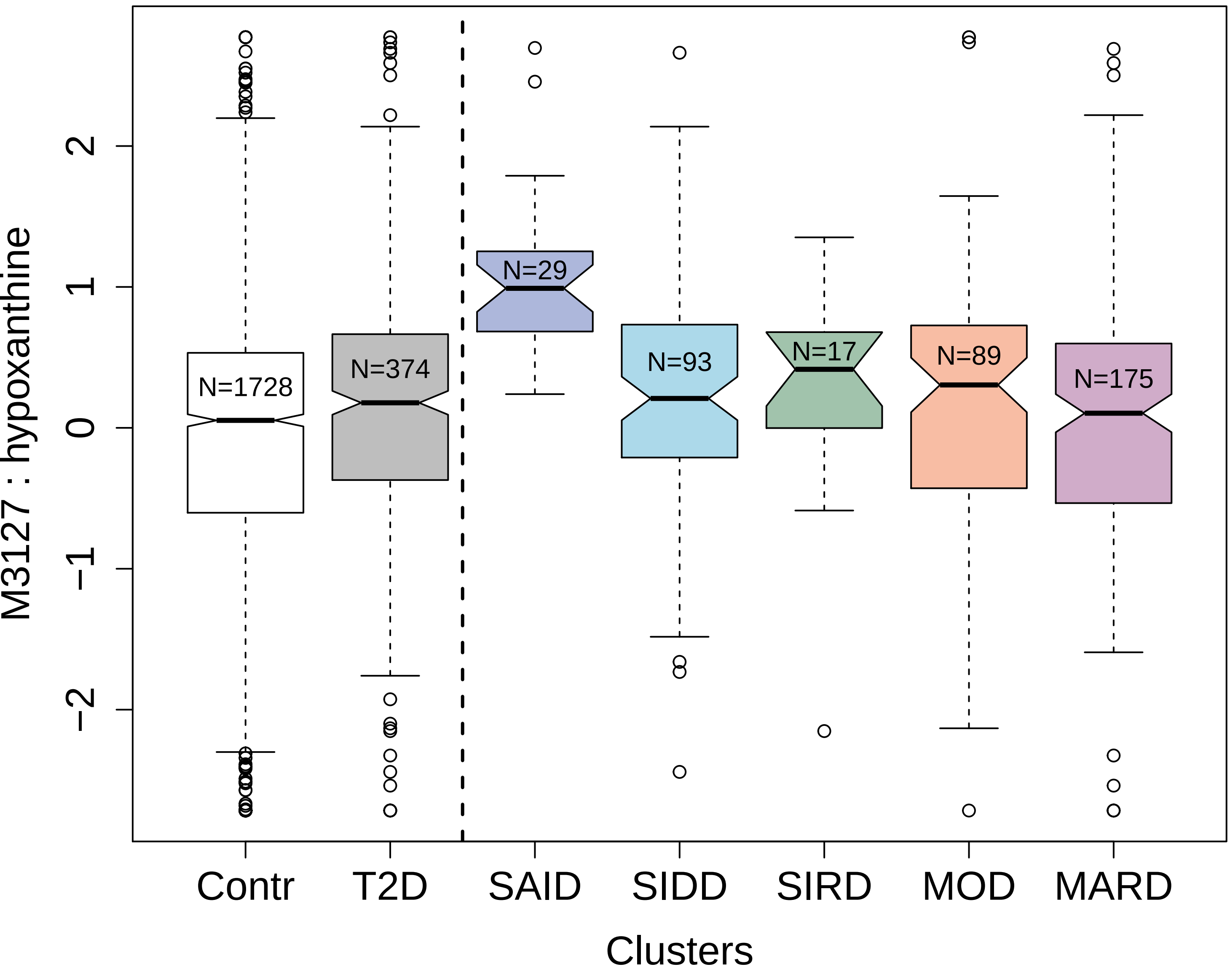
M3127 : hypoxanthine
Clusters
Contr
T2D
SAID
SIDD
SIRD
MOD
MARD
N=1728
N=374
N=29
N=93
N=17
N=89
N=175
2
1
0
−1
−2

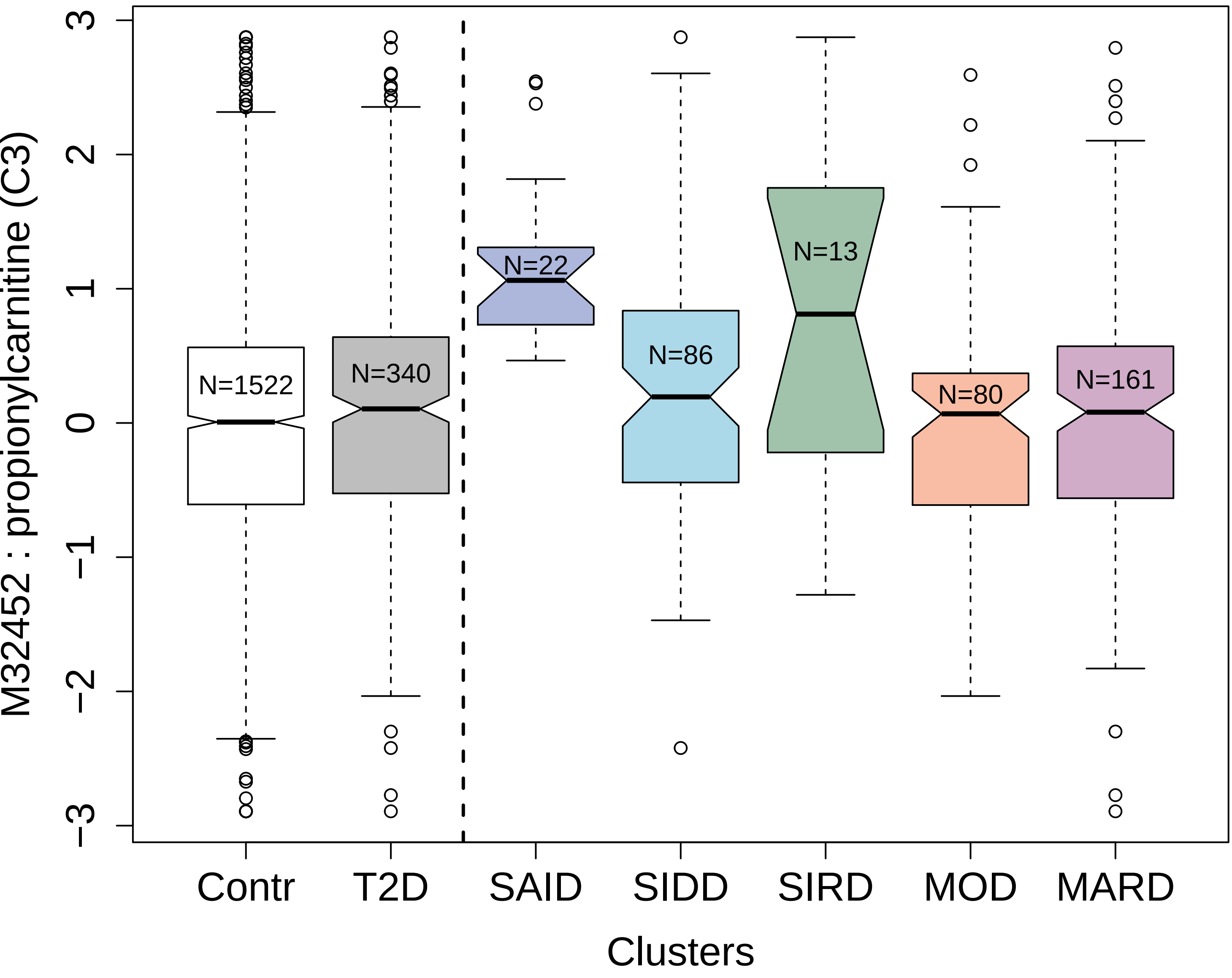

M32452 : propionylcarnitine (C3)
3
2
1
0
−1
−2
−3
N=1522
N=340
N=22
N=86
N=13
N=80
N=161
Contr
T2D
SAID
SIDD
SIRD
MOD
MARD
Clusters

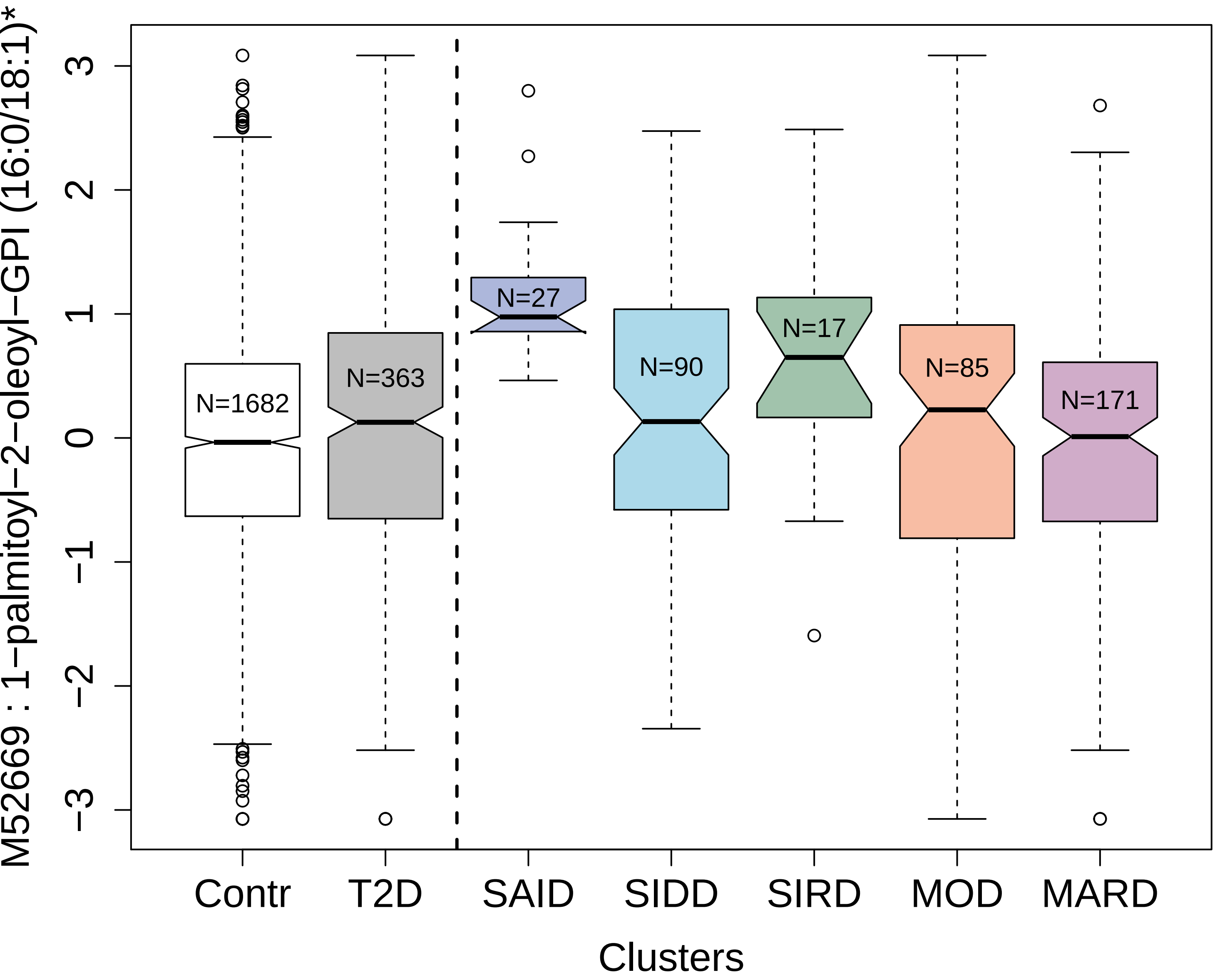
M52669 : 1−palmitoyl−2−oleoyl−GPI (16:0/18:1)*
3
2
1
0
−1
−2
−3
N=1682
N=363
N=27
N=90
N=17
N=85
N=171
Contr
T2D
SAID
SIDD
SIRD
MOD
MARD
Clusters

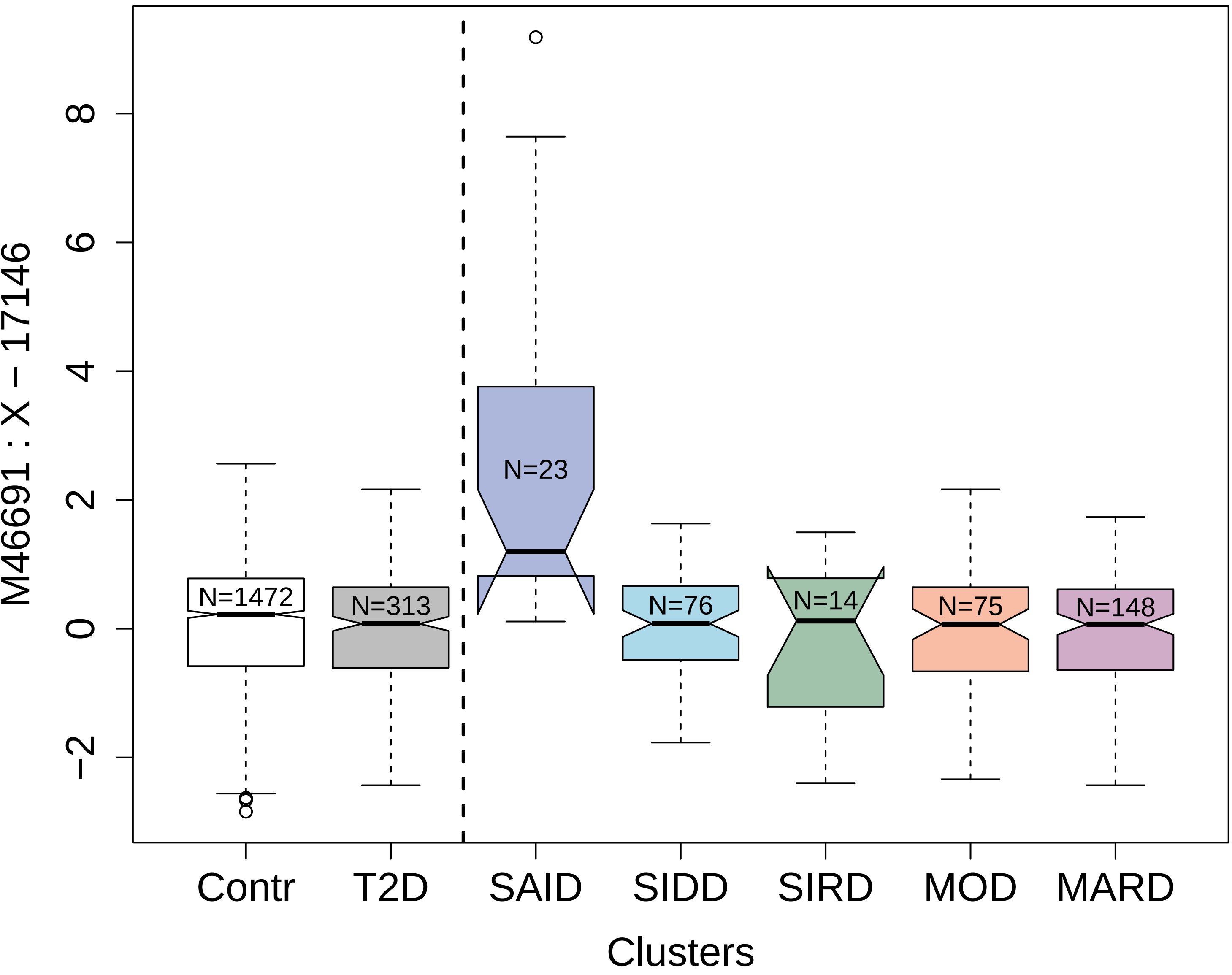

M46691 : X − 17146
−2
0
2
4
6
8
N=1472
N=313
N=23
N=76
N=14
N=75
N=148
Contr
T2D
SAID
SIDD
SIRD
MOD
MARD
Clusters

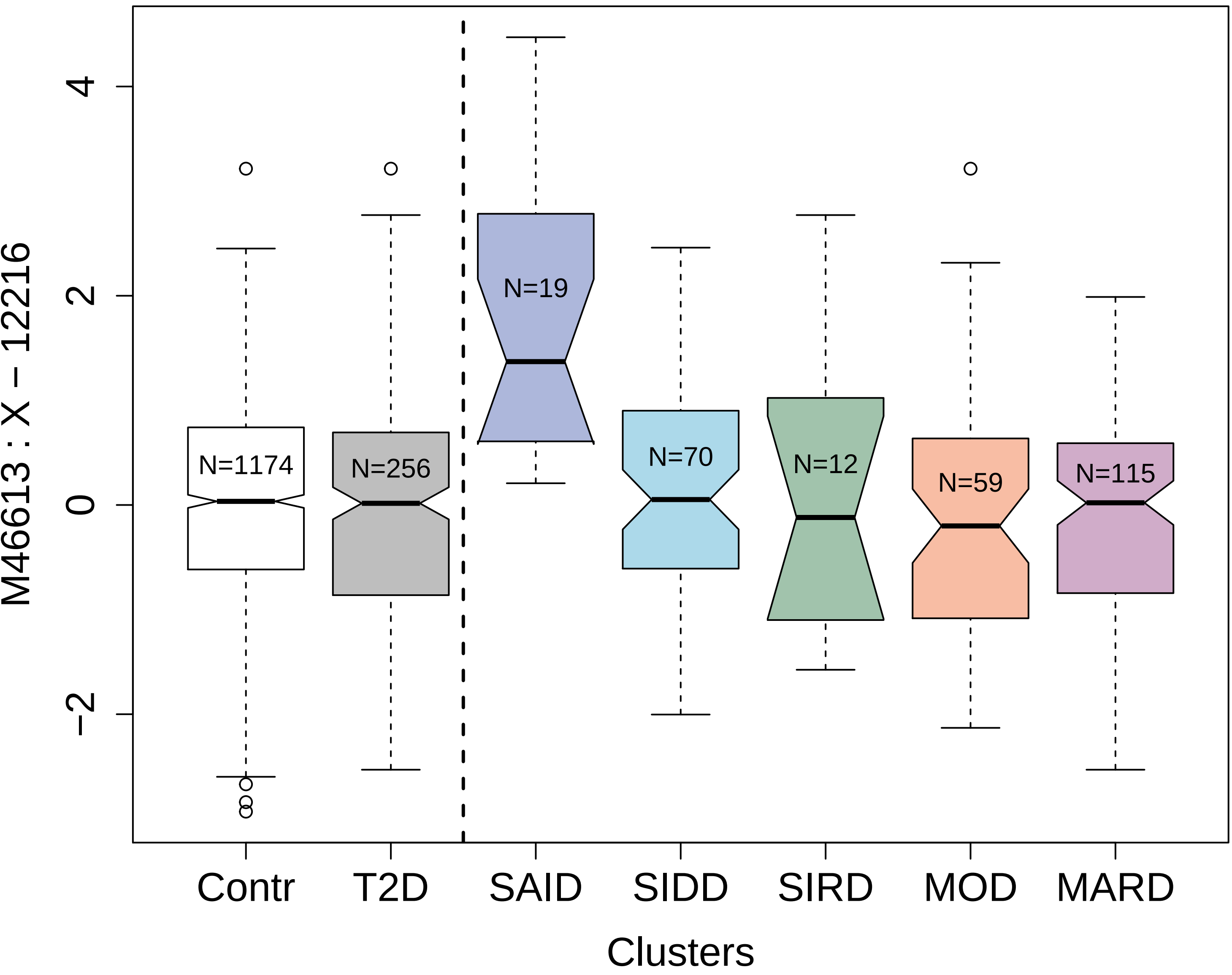
M46613 : X − 12216
4
2
0
−2
N=1174
N=256
N=19
N=70
N=12
N=59
N=115
Contr
T2D
SAID
SIDD
SIRD
MOD
MARD
Clusters

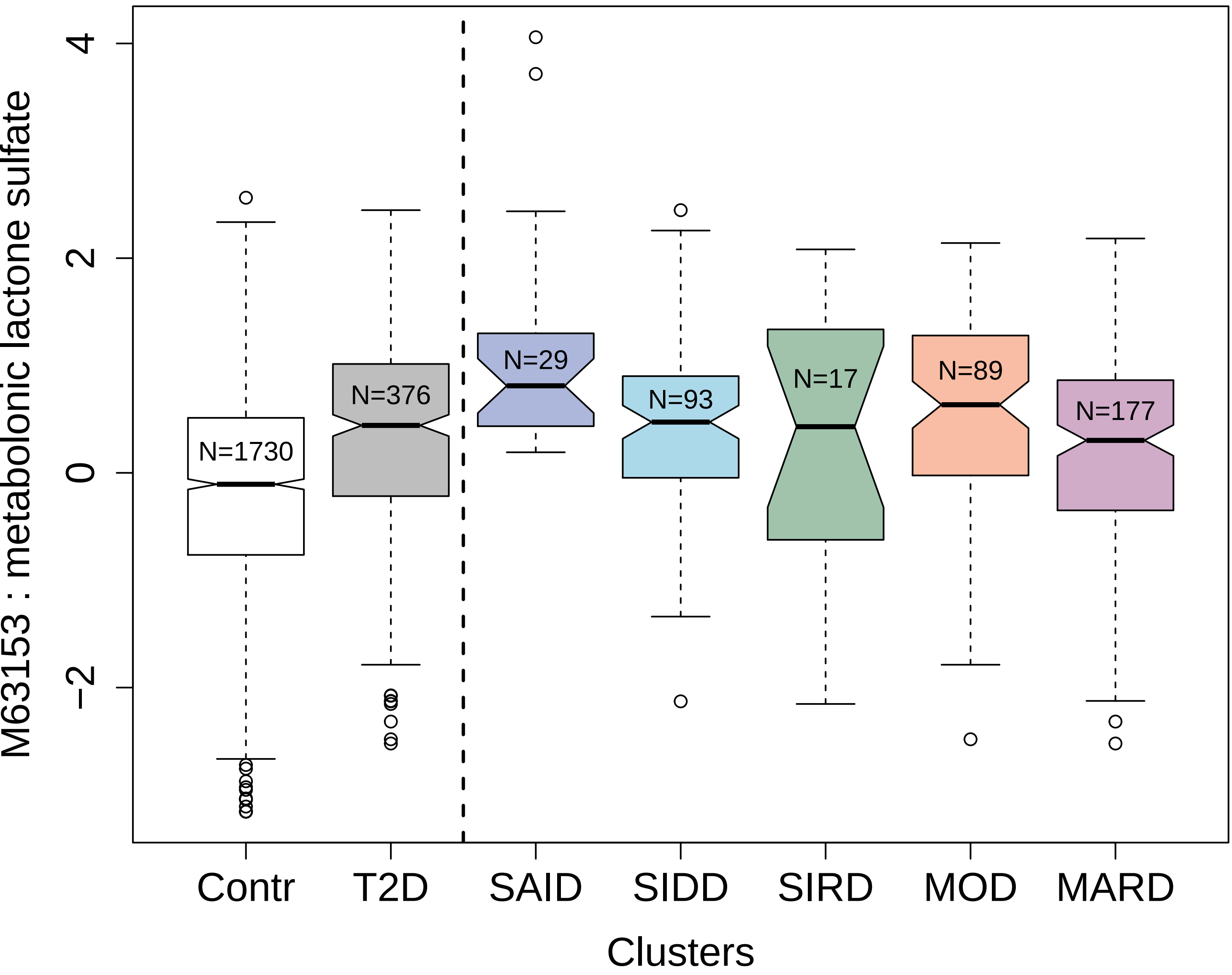

M63153 : metabolonic lactone sulfate
4
2
0
−2
N=1730
N=376
N=29
N=93
N=17
N=89
N=177
Contr
T2D
SAID
SIDD
SIRD
MOD
MARD
Clusters

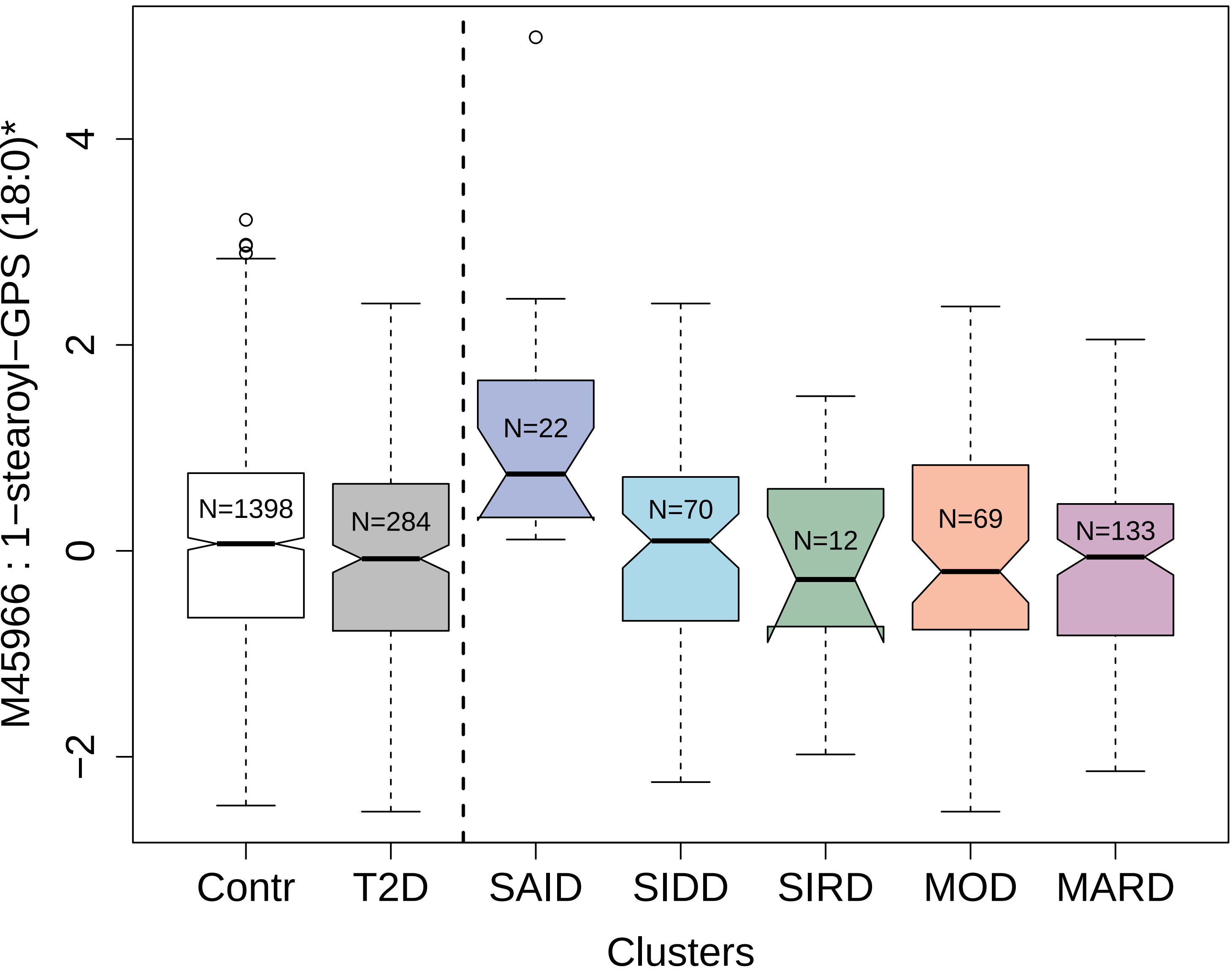
M45966 : 1−stearoyl−GPS (18:0)*
4
2
0
−2
N=1398
N=284
N=22
N=70
N=12
N=69
N=133
Contr
T2D
SAID
SIDD
SIRD
MOD
MARD
Clusters

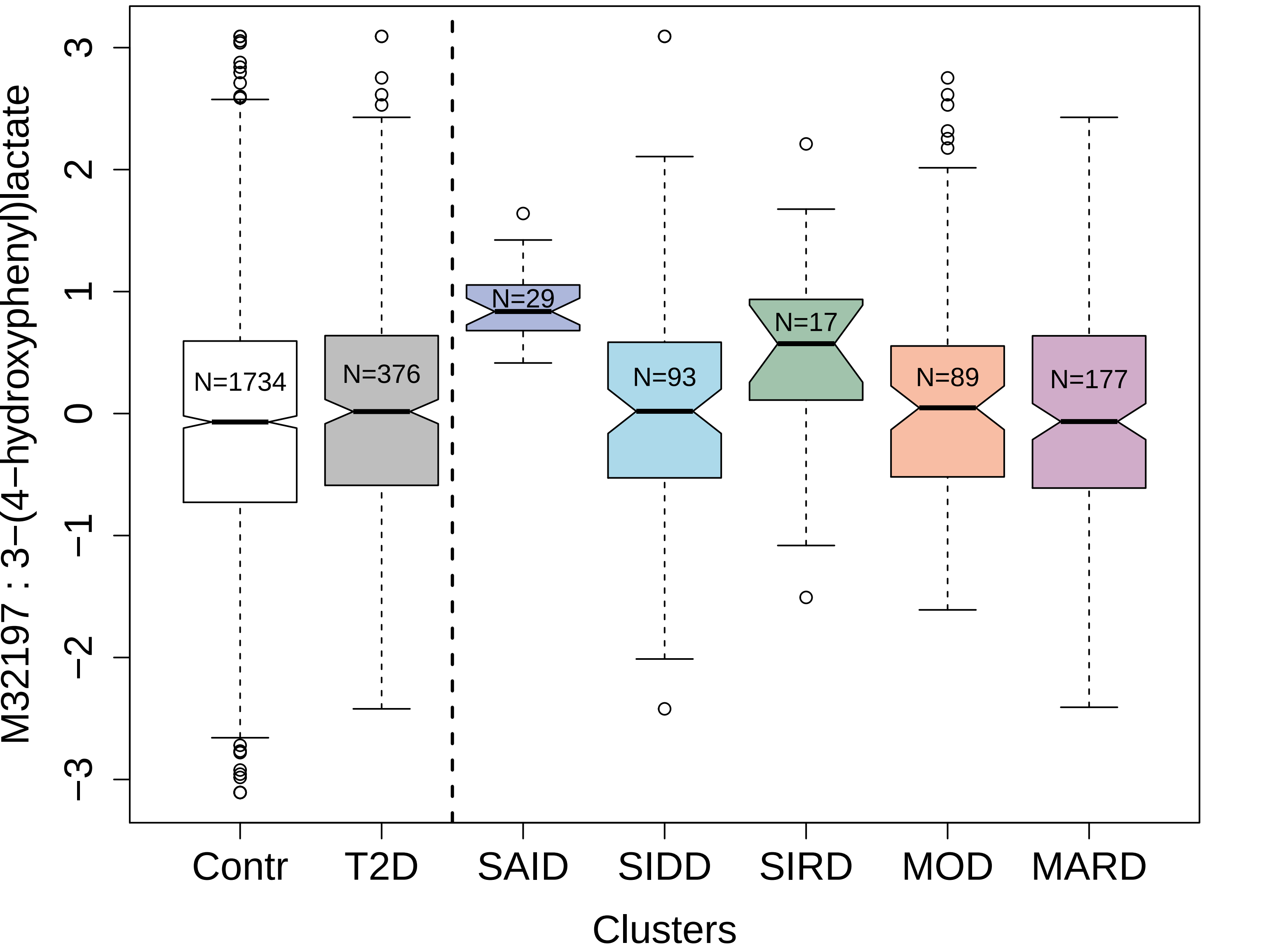

M32197 : 3−(4−hydroxyphenyl)lactate
3
2
1
0
−1
−2
−3
N=1734
N=376
N=29
N=93
N=17
N=89
N=177
Contr
T2D
SAID
SIDD
SIRD
MOD
MARD
Clusters

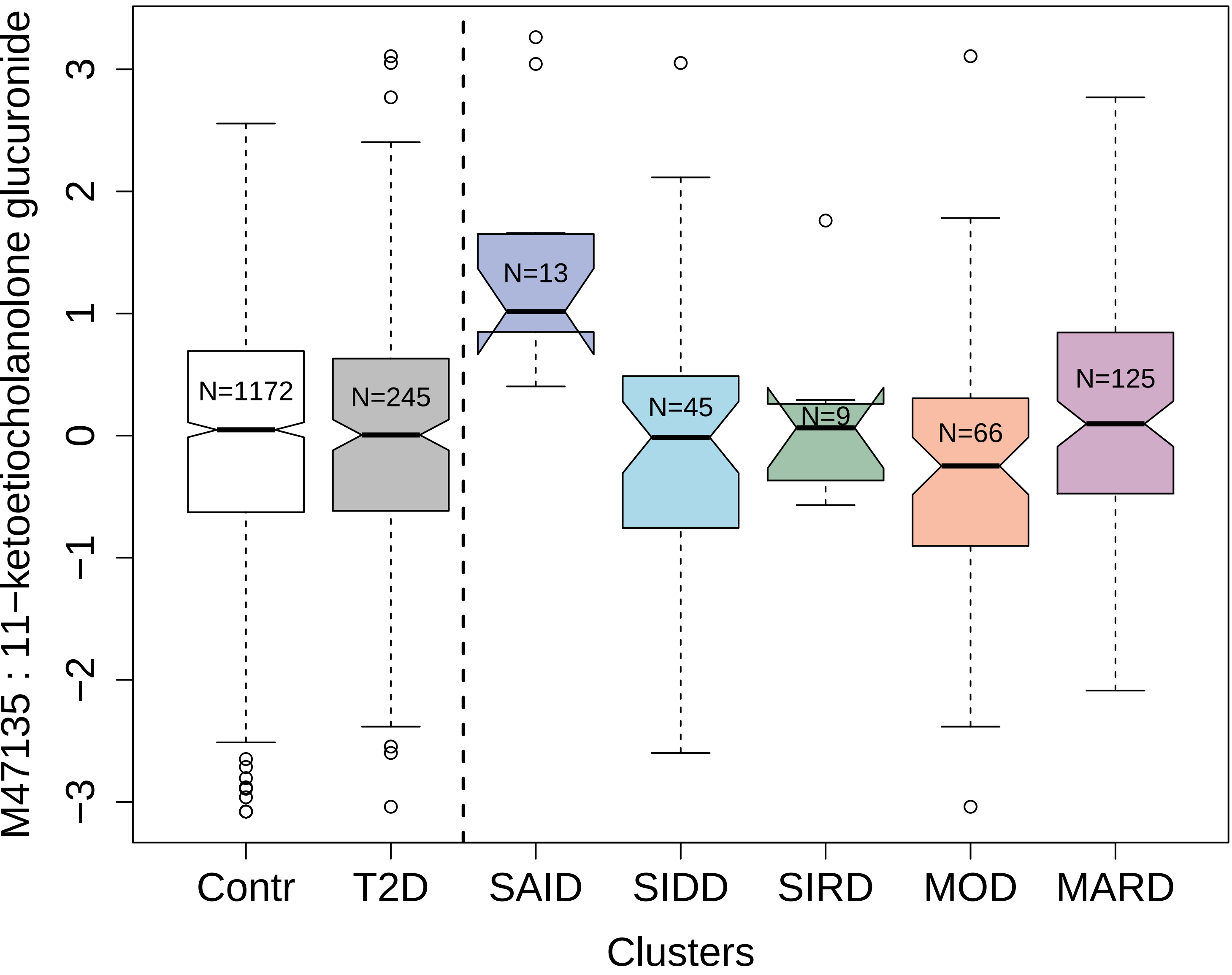
M47135 : 11−ketoetiocholanolone glucuronide
3
2
1
0
−1
−2
−3
N=1172
N=245
N=13
N=45
N=9
N=66
N=125
Contr
T2D
SAID
SIDD
SIRD
MOD
MARD
Clusters

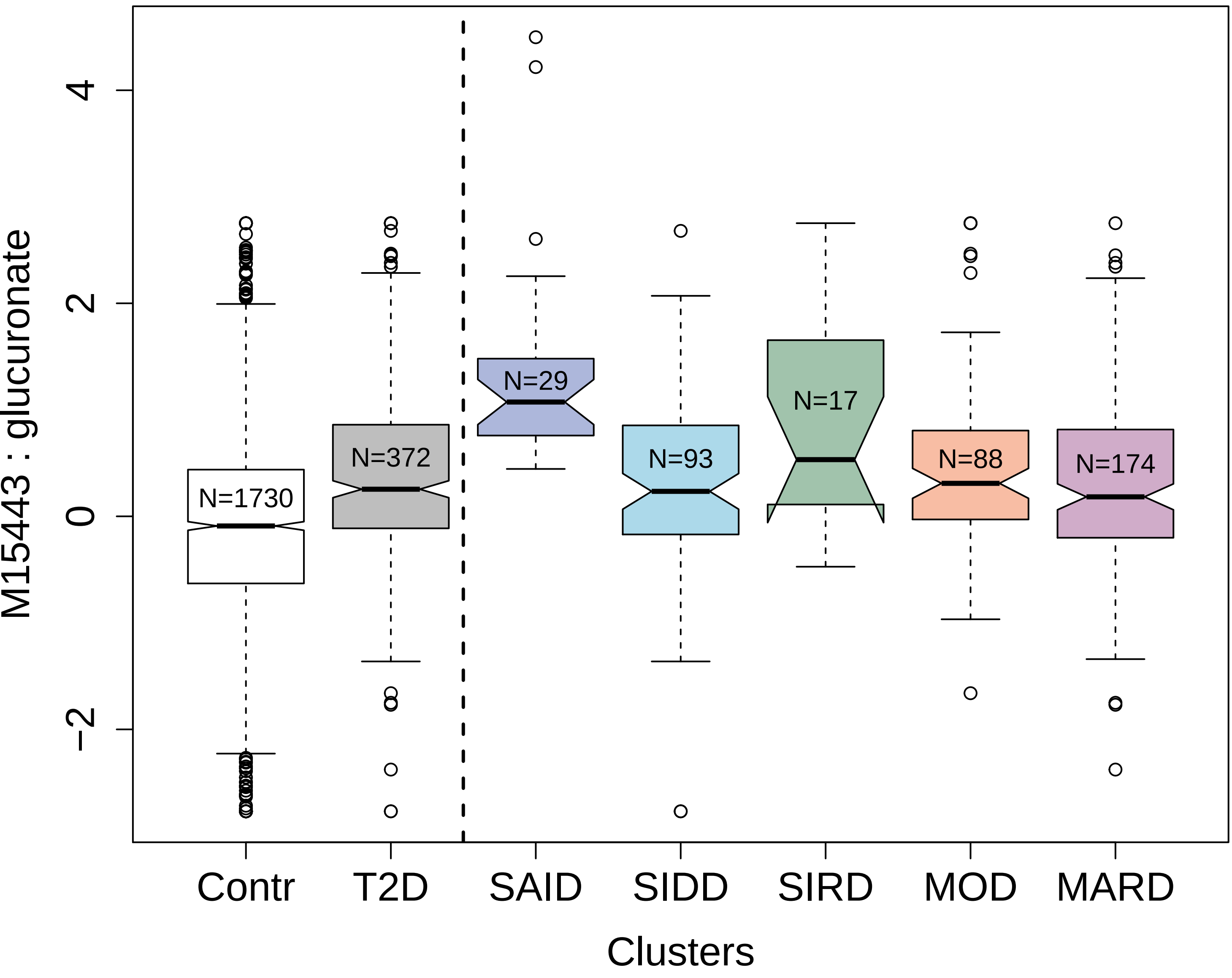

M15443 : glucuronate
−2
0
2
4
N=1730
N=372
N=29
N=93
N=17
N=88
N=174
Contr
T2D
SAID
SIDD
SIRD
MOD
MARD
Clusters

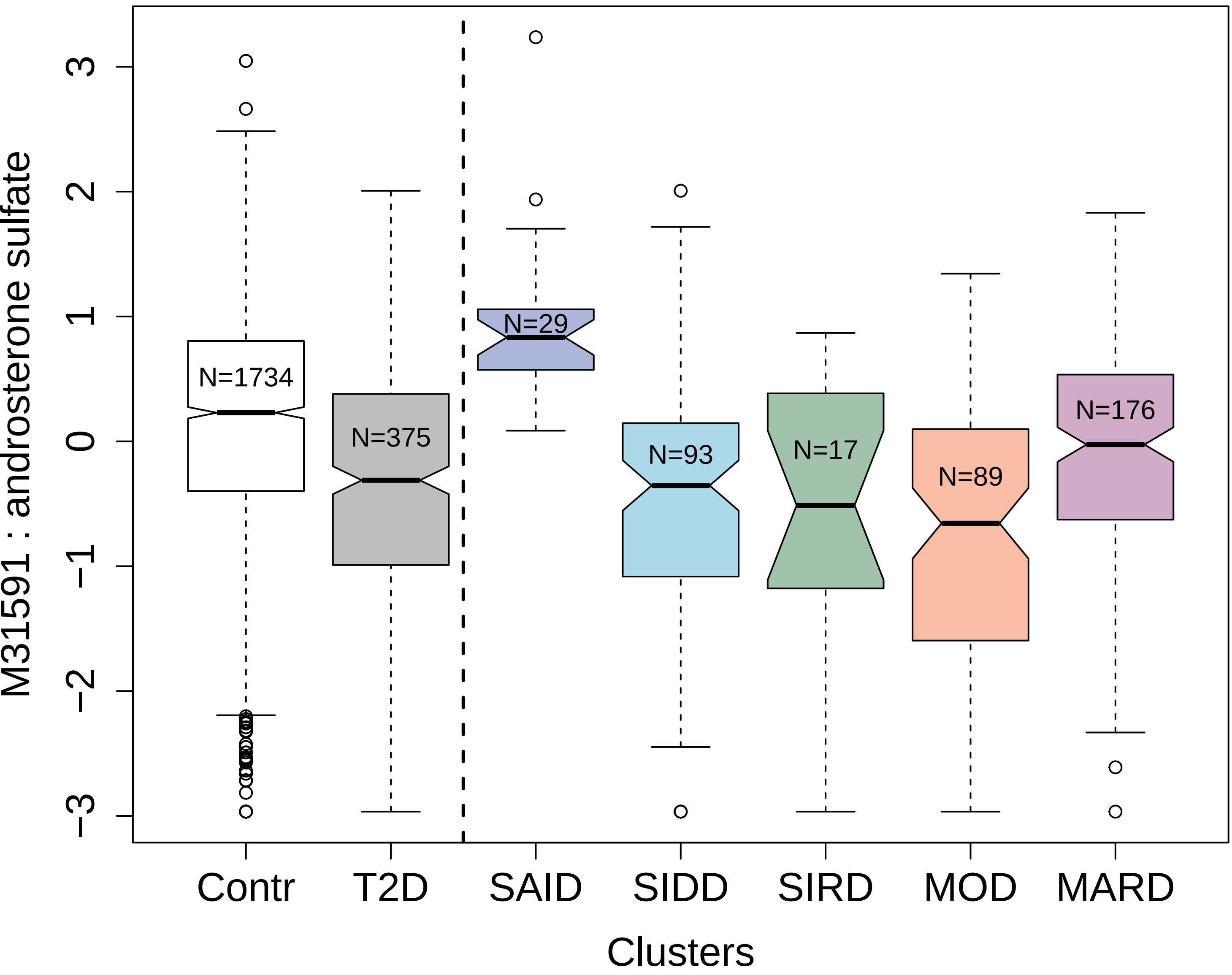
M31591 : androsterone sulfate
3
2
1
0
−1
−2
−3
N=1734
N=375
N=29
N=93
N=17
N=89
N=176
Contr
T2D
SAID
SIDD
SIRD
MOD
MARD
Clusters

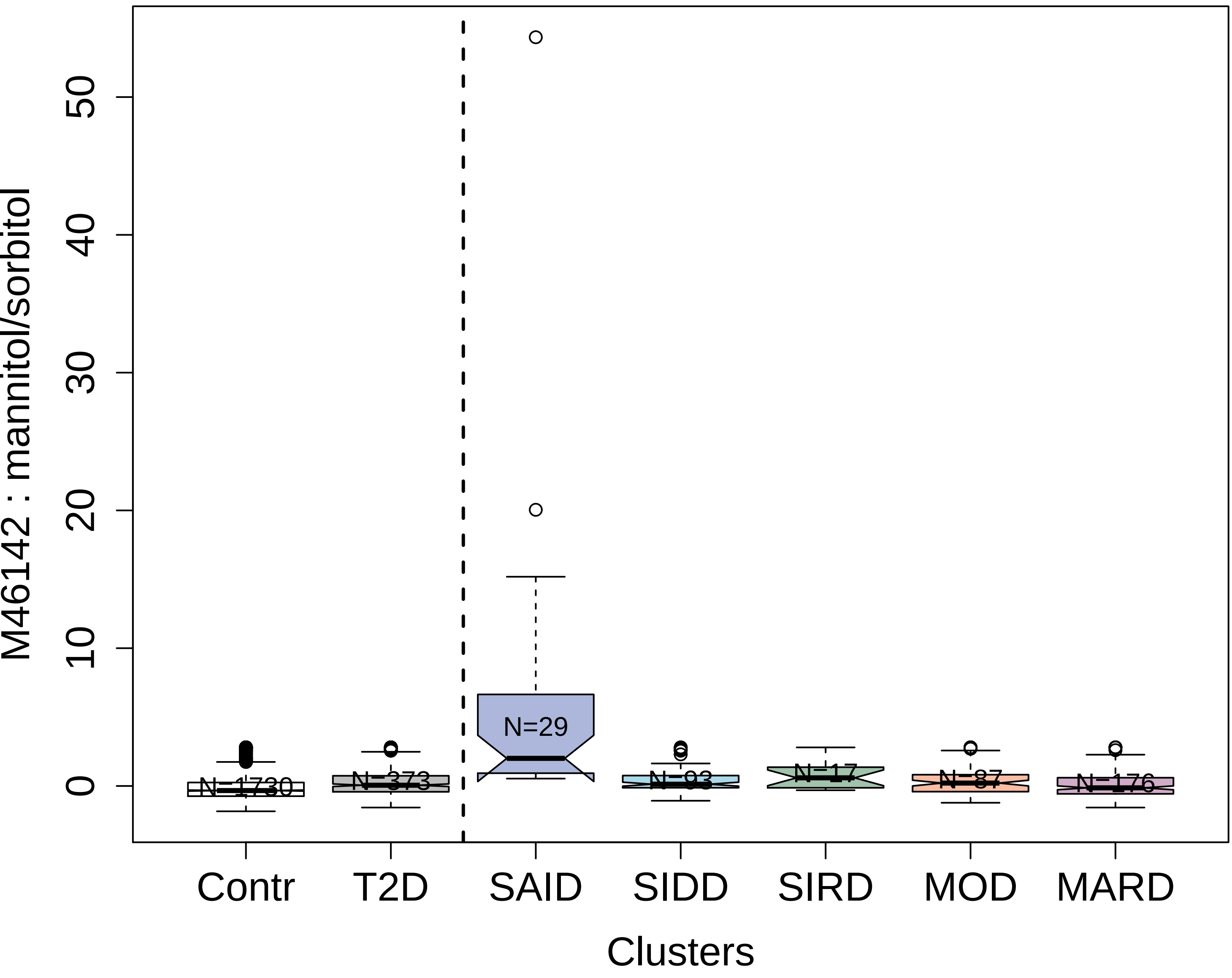
M46142 : mannitol/sorbitol
0
10
20
30
40
50
N=1730
N=373
N=29
N=93
N=17
N=87
N=176
Contr
T2D
SAID
SIDD
SIRD
MOD
MARD
Clusters

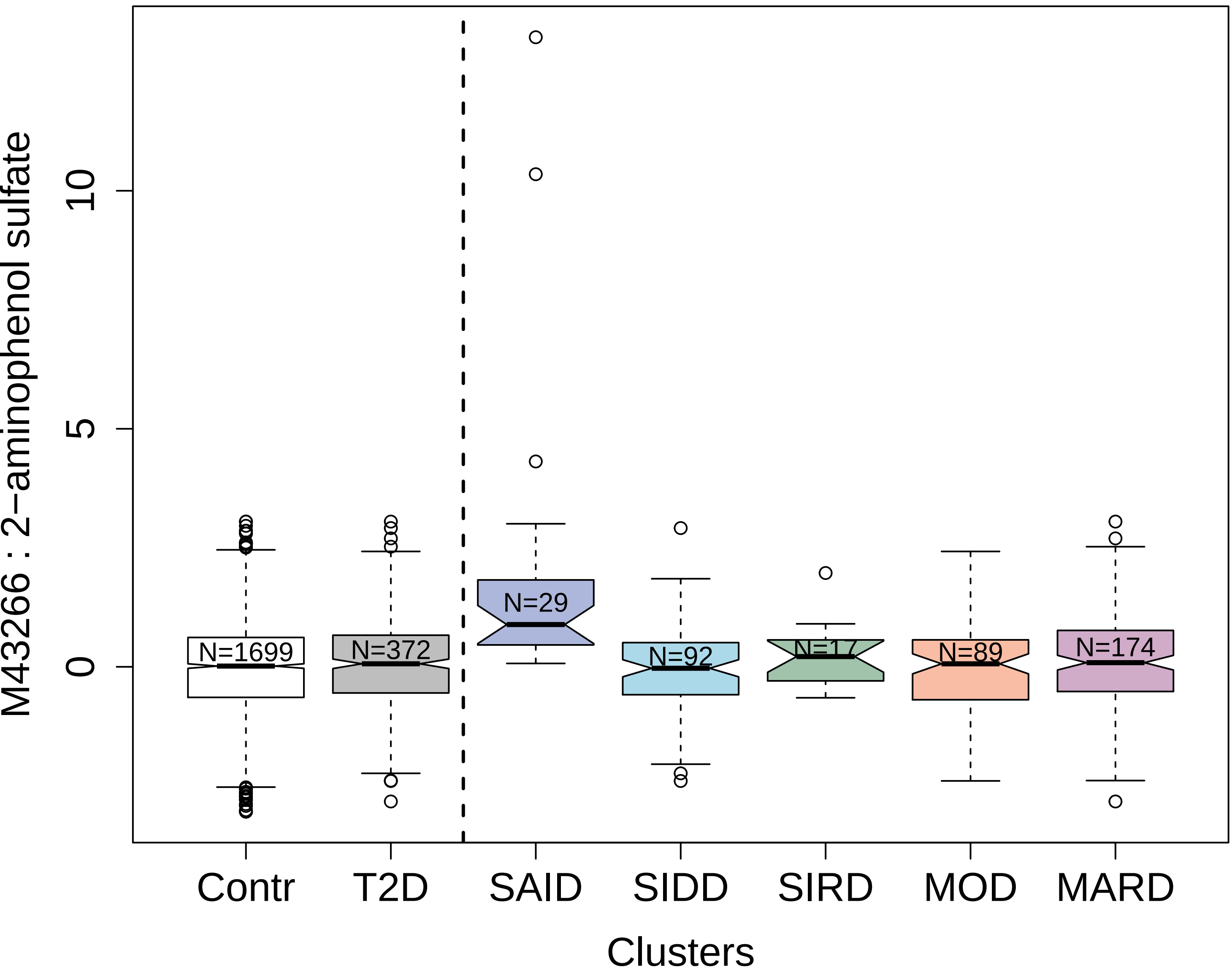

M43266 : 2−aminophenol sulfate
0
5
10
N=1699
N=372
N=29
N=92
N=17
N=89
N=174
Contr
T2D
SAID
SIDD
SIRD
MOD
MARD
Clusters

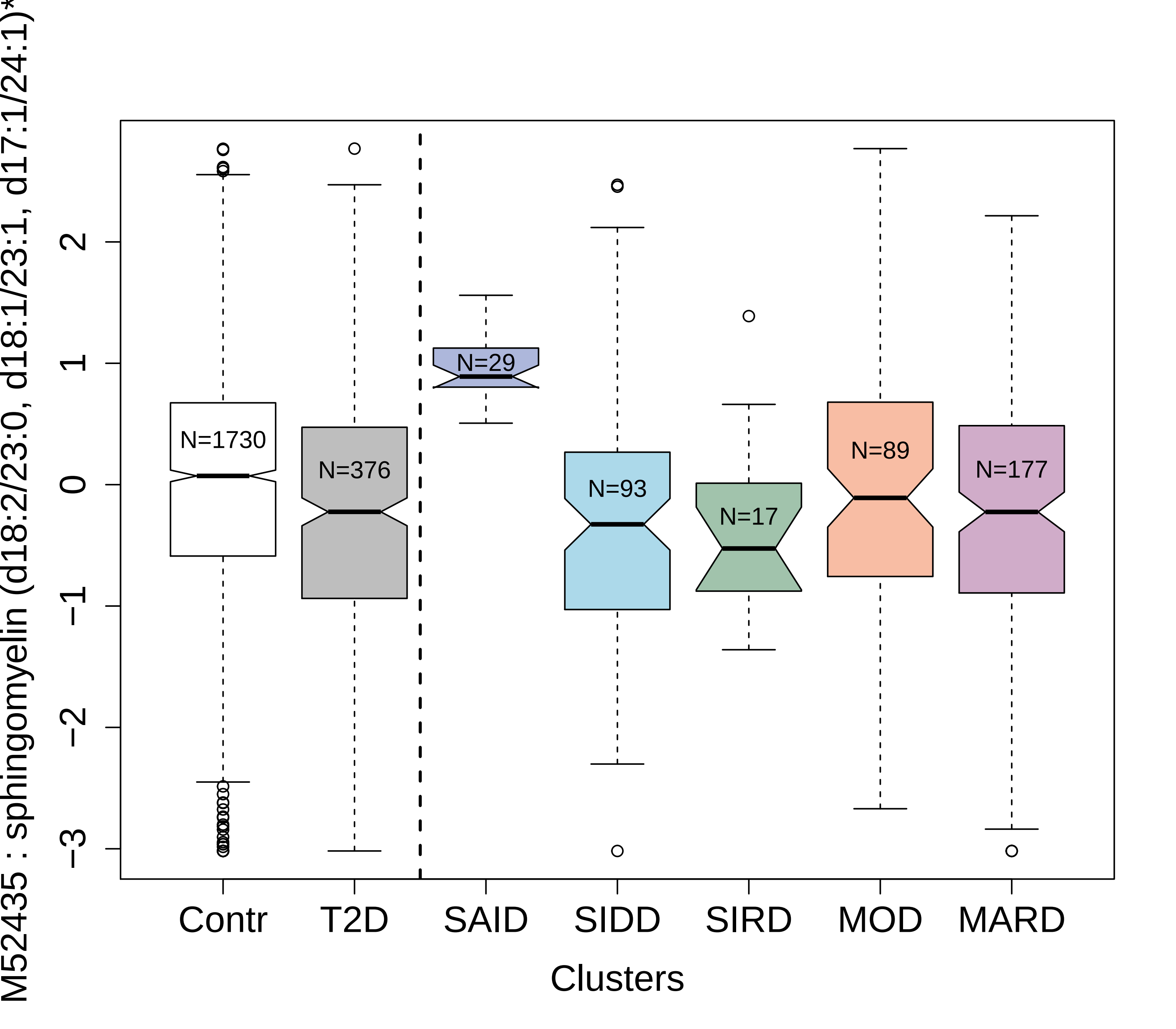

M52435 : sphingomyelin (d18:2/23:0, d18:1/23:1, d17:1/24:1)*
−3
−2
−1
0
1
2
N=1730
N=376
N=29
N=93
N=17
N=89
N=177
Contr
T2D
SAID
SIDD
SIRD
MOD
MARD
Clusters

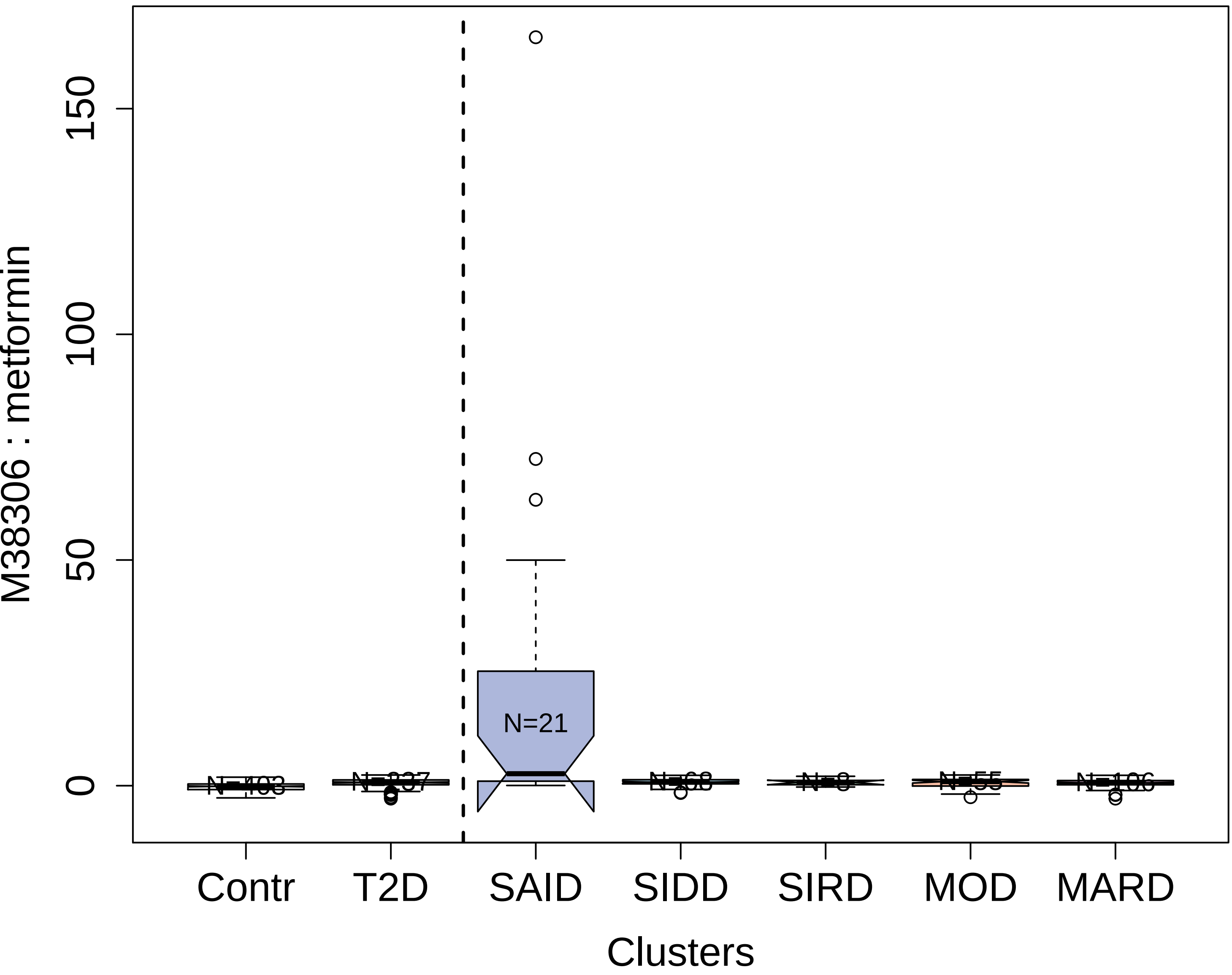
M38306 : metformin
0
50
100
150
N=21
Contr
T2D
SAID
SIDD
SIRD
MOD
MARD
Clusters

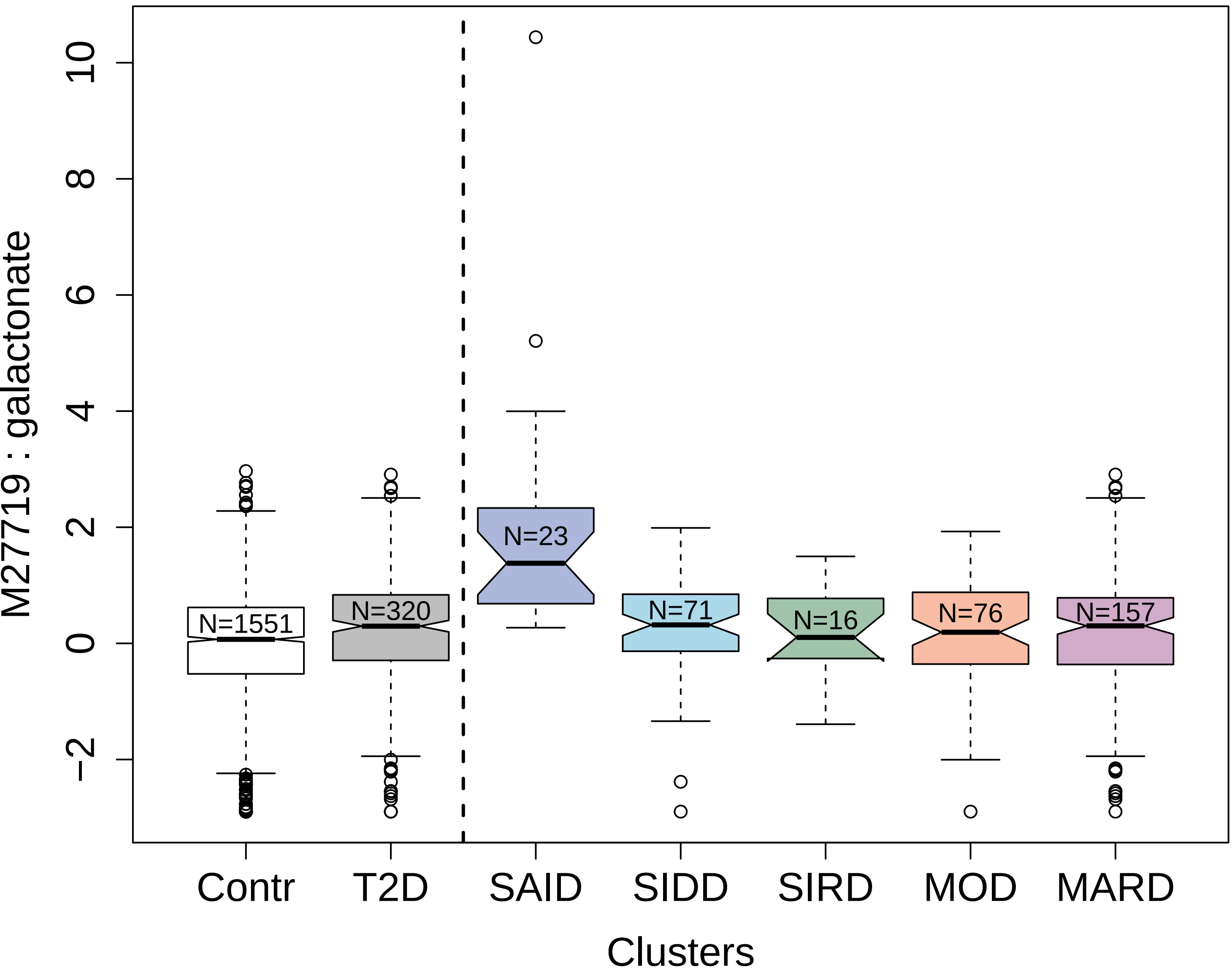
M27719 : galactonate
10
8
6
4
2
0
−2
N=1551
N=320
N=23
N=71
N=16
N=76
N=157
Contr
T2D
SAID
SIDD
SIRD
MOD
MARD
Clusters

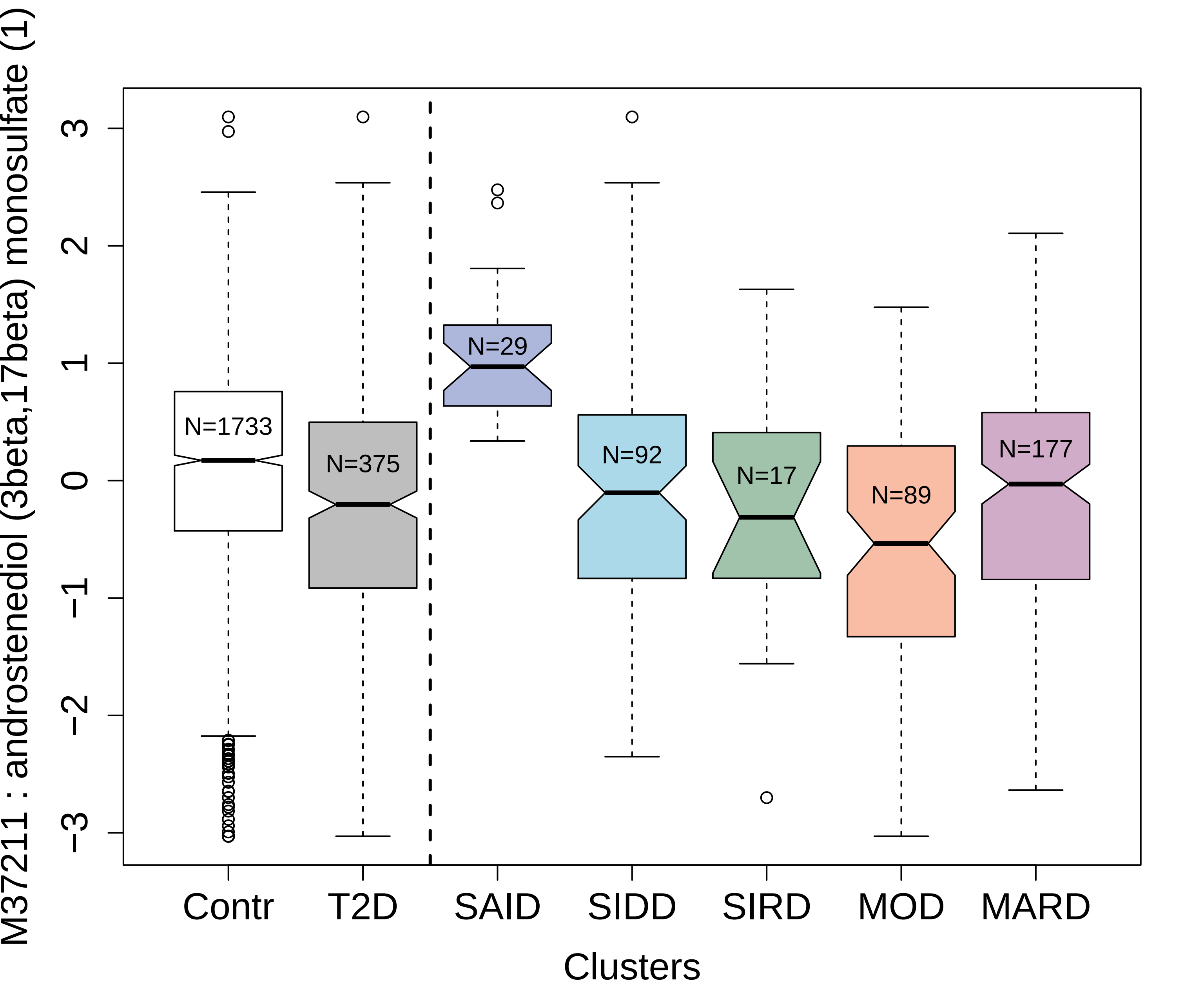
M37211 : androstenediol (3beta,17beta) monosulfate (1)
3
2
1
0
−1
−2
−3
N=1733
N=375
N=29
N=92
N=17
N=89
N=177
Contr
T2D
SAID
SIDD
SIRD
MOD
MARD
Clusters

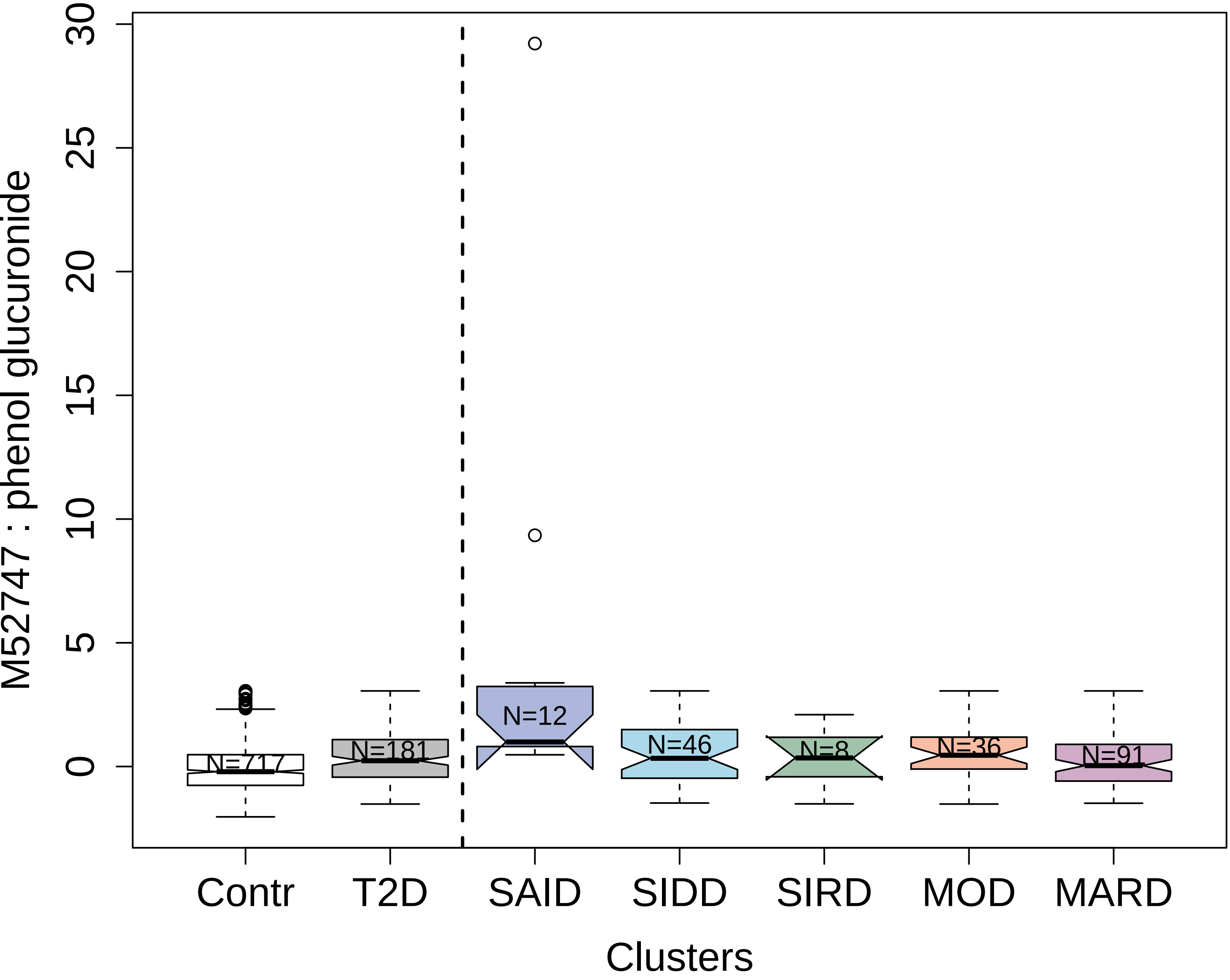

M52747 : phenol glucuronide
0
5
10
15
20
25
30
N=717
N=181
N=12
N=46
N=8
N=36
N=91
Contr
T2D
SAID
SIDD
SIRD
MOD
MARD
Clusters

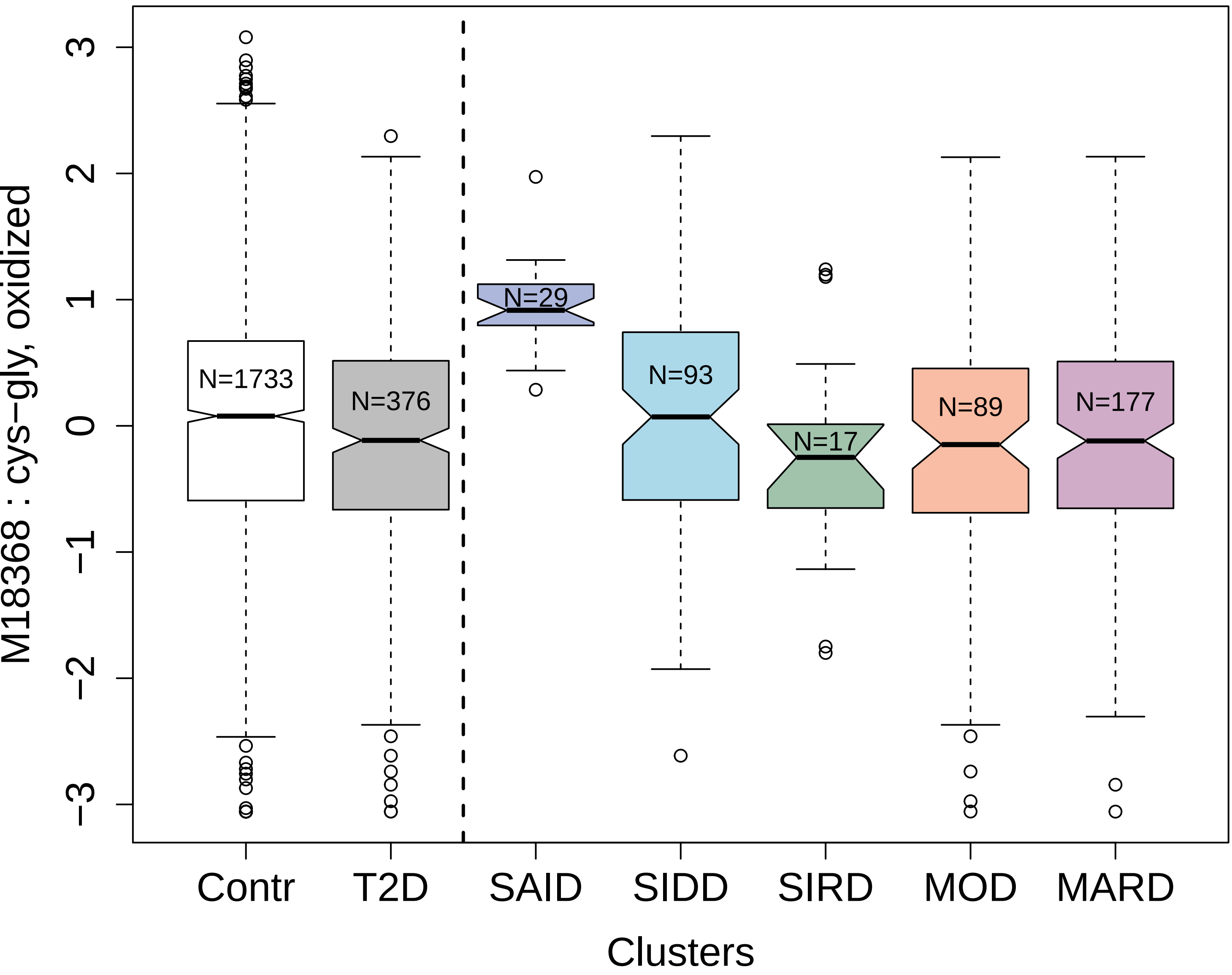
M18368 : cys−gly, oxidized
3
2
1
0
−1
−2
−3
N=1733
N=376
N=29
N=93
N=17
N=89
N=177
Contr
T2D
SAID
SIDD
SIRD
MOD
MARD
Clusters

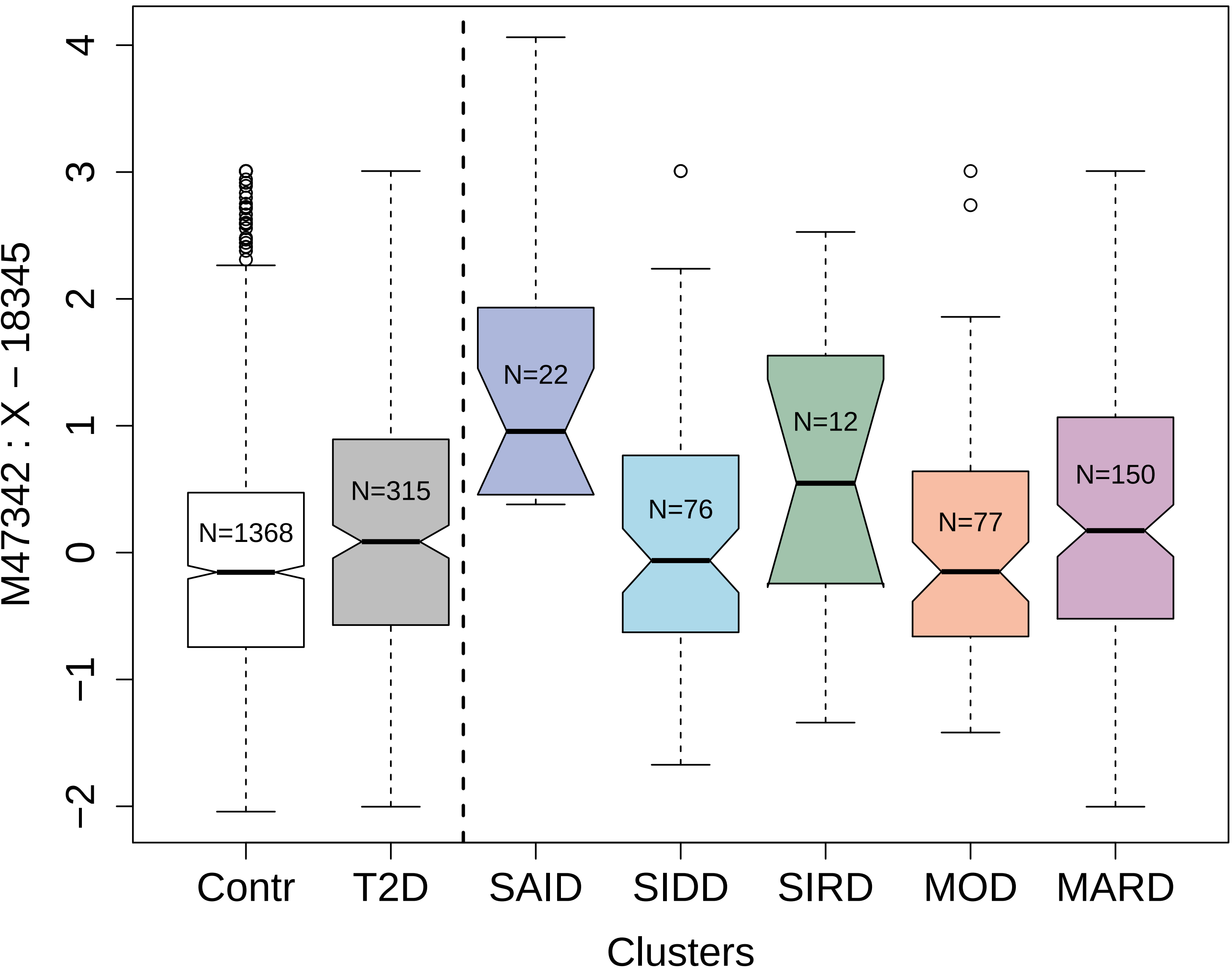
M47342 : X − 18345
4
3
2
1
0
−1
−2
N=1368
N=315
N=22
N=76
N=12
N=77
N=150
Contr
T2D
SAID
SIDD
SIRD
MOD
MARD
Clusters

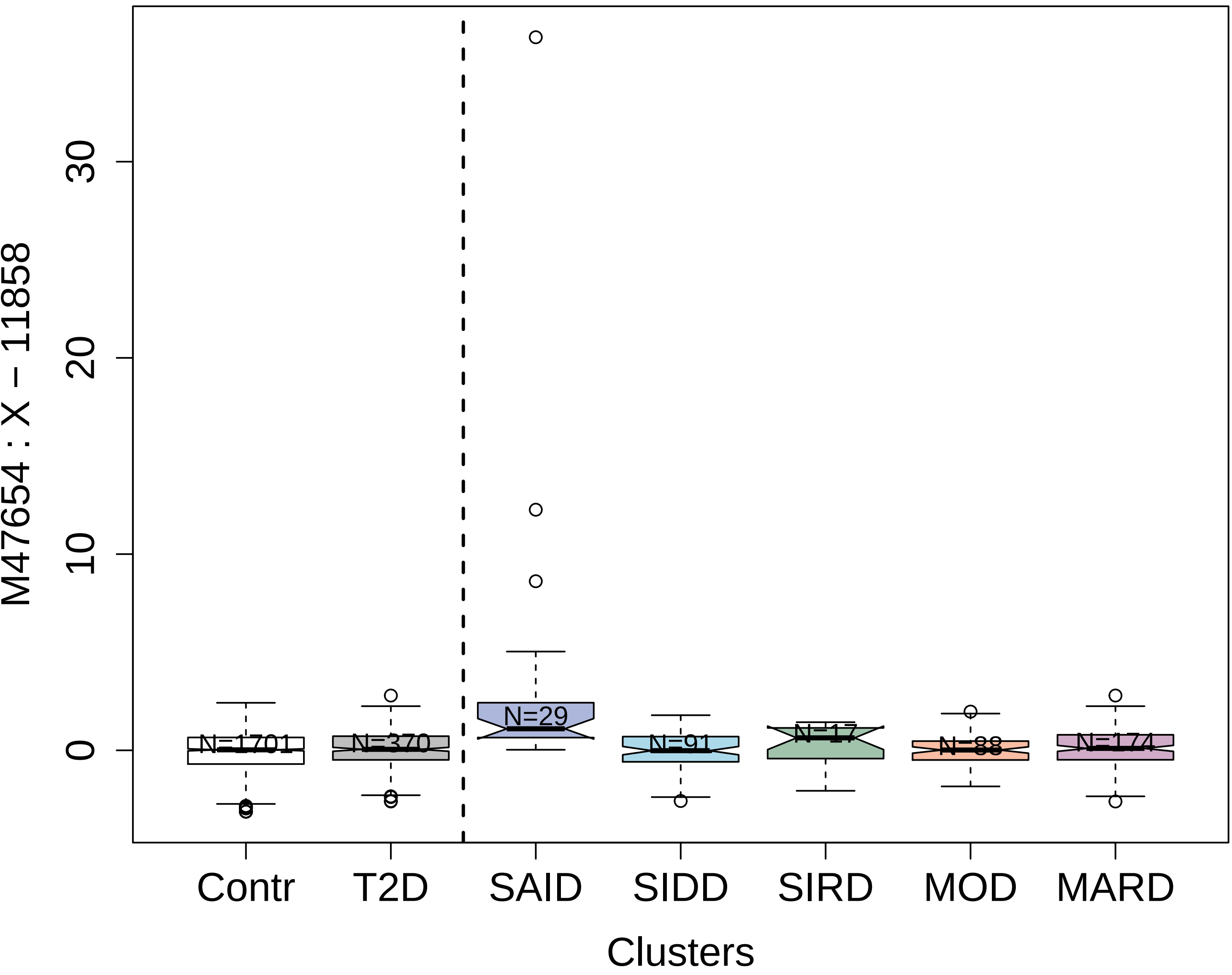

M47654 : X − 11858
0
10
20
30
N=1701
N=370
N=29
N=91
N=17
N=88
N=174
Contr
T2D
SAID
SIDD
SIRD
MOD
MARD
Clusters

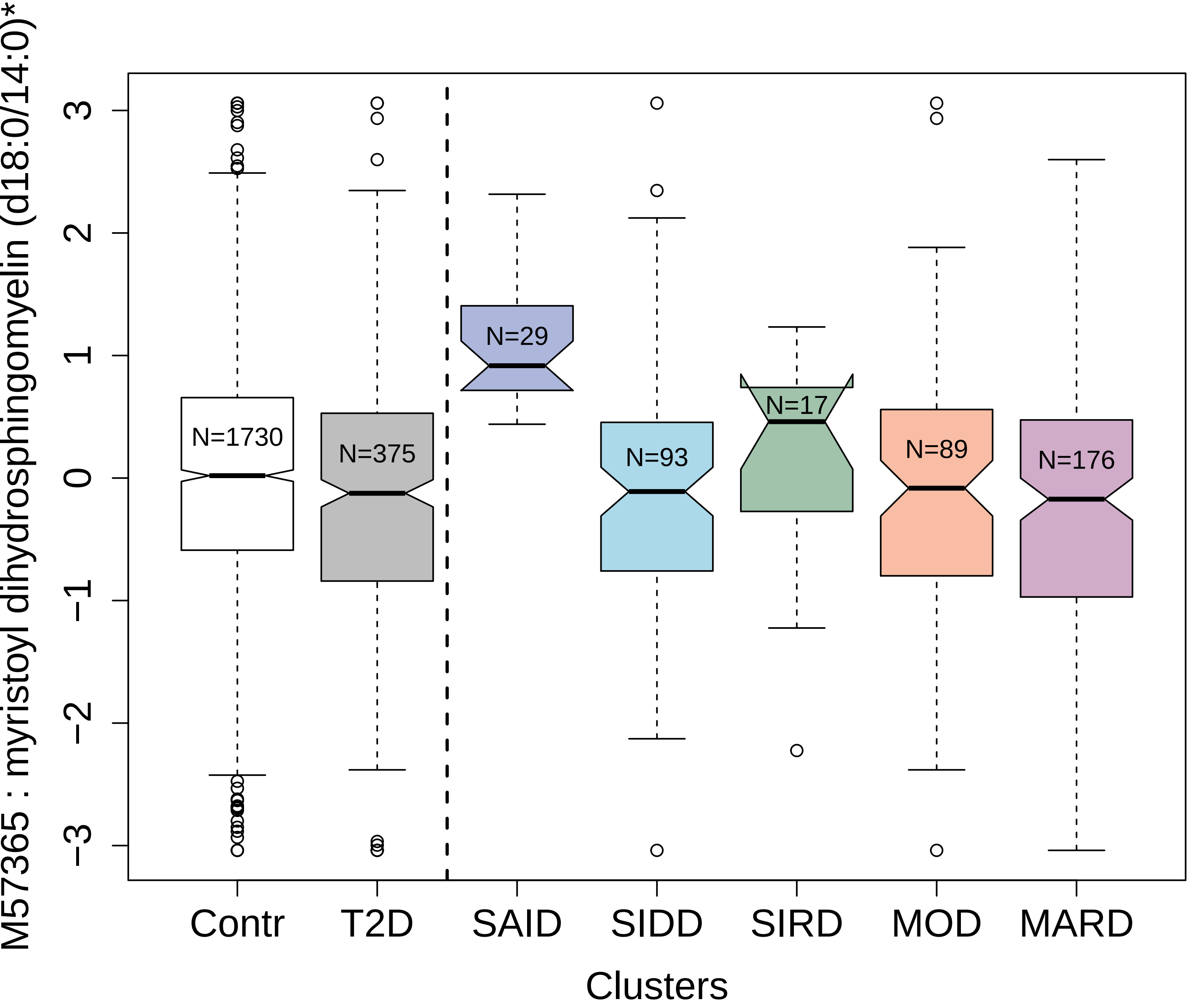

M57365 : myristoyl dihydrosphingomyelin (d18:0/14:0)*
3
2
1
0
−1
−2
−3
N=1730
N=375
N=29
N=93
N=17
N=89
N=176
Contr
T2D
SAID
SIDD
SIRD
MOD
MARD
Clusters

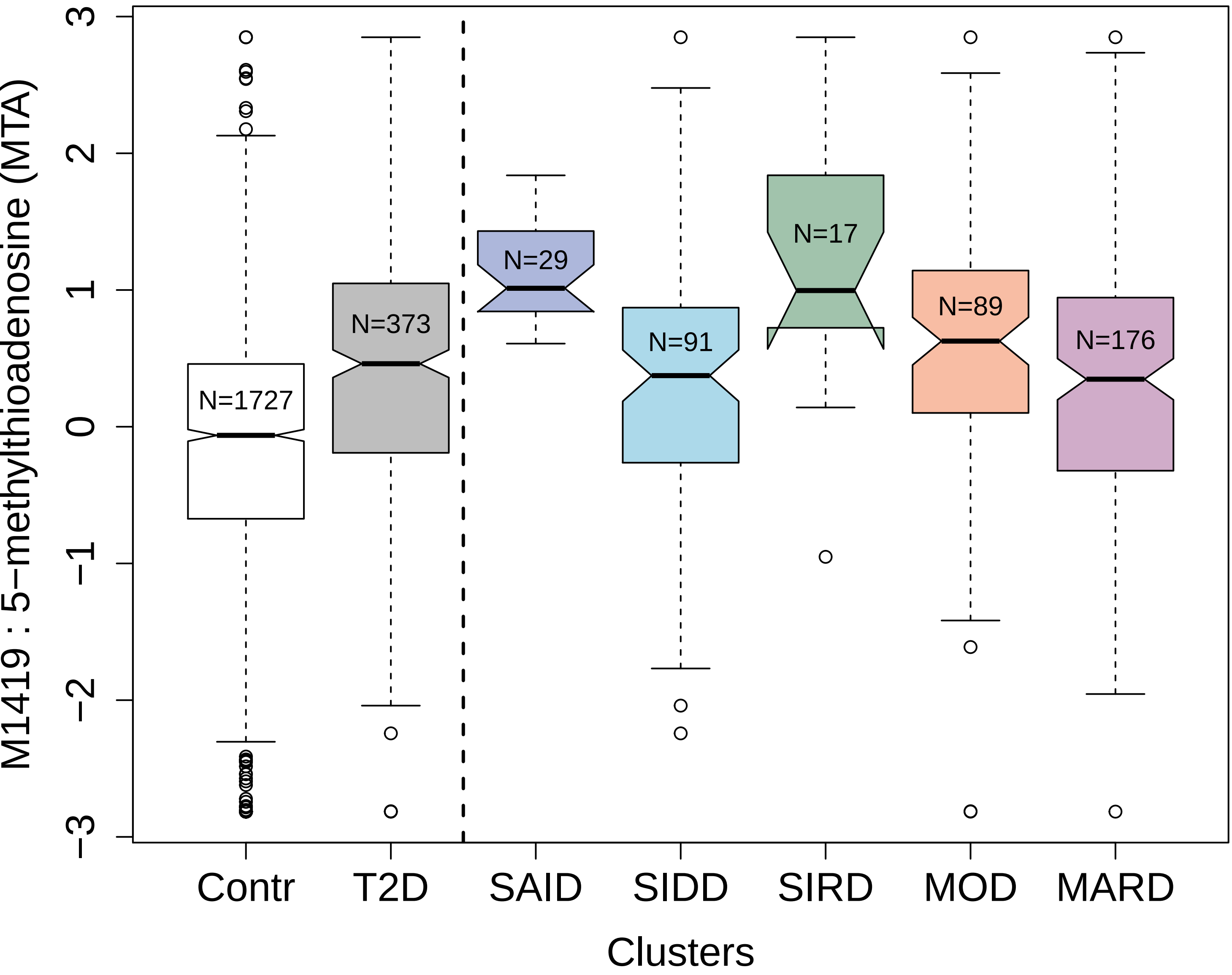

M1419 : 5−methylthioadenosine (MTA)
3
2
1
0
−1
−2
−3
N=1727
N=373
N=29
N=91
N=17
N=89
N=176
Contr
T2D
SAID
SIDD
SIRD
MOD
MARD
Clusters

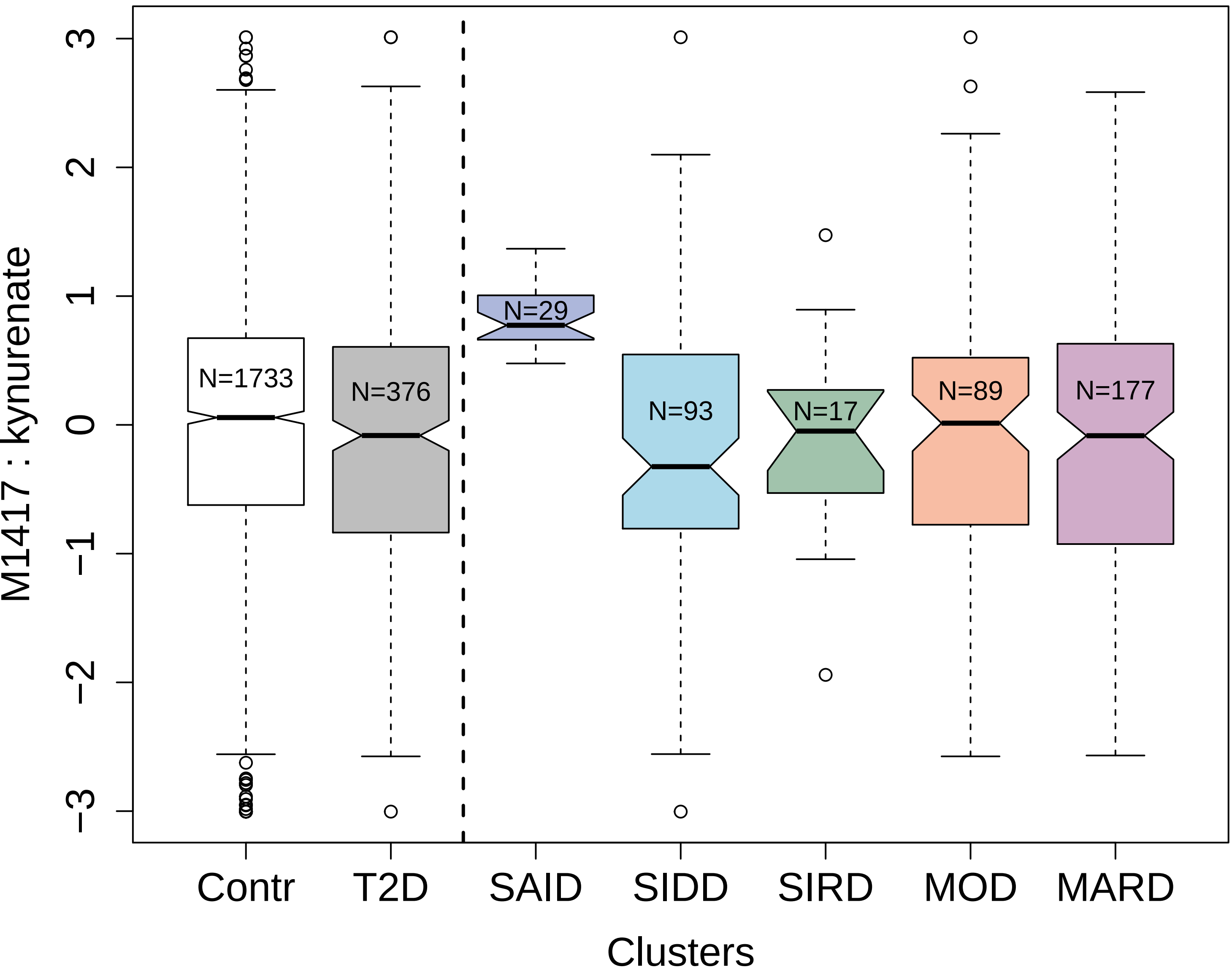
M1417 : kynurenate
3
2
1
0
−1
−2
−3
N=1733
N=376
N=29
N=93
N=17
N=89
N=177
Contr
T2D
SAID
SIDD
SIRD
MOD
MARD
Clusters

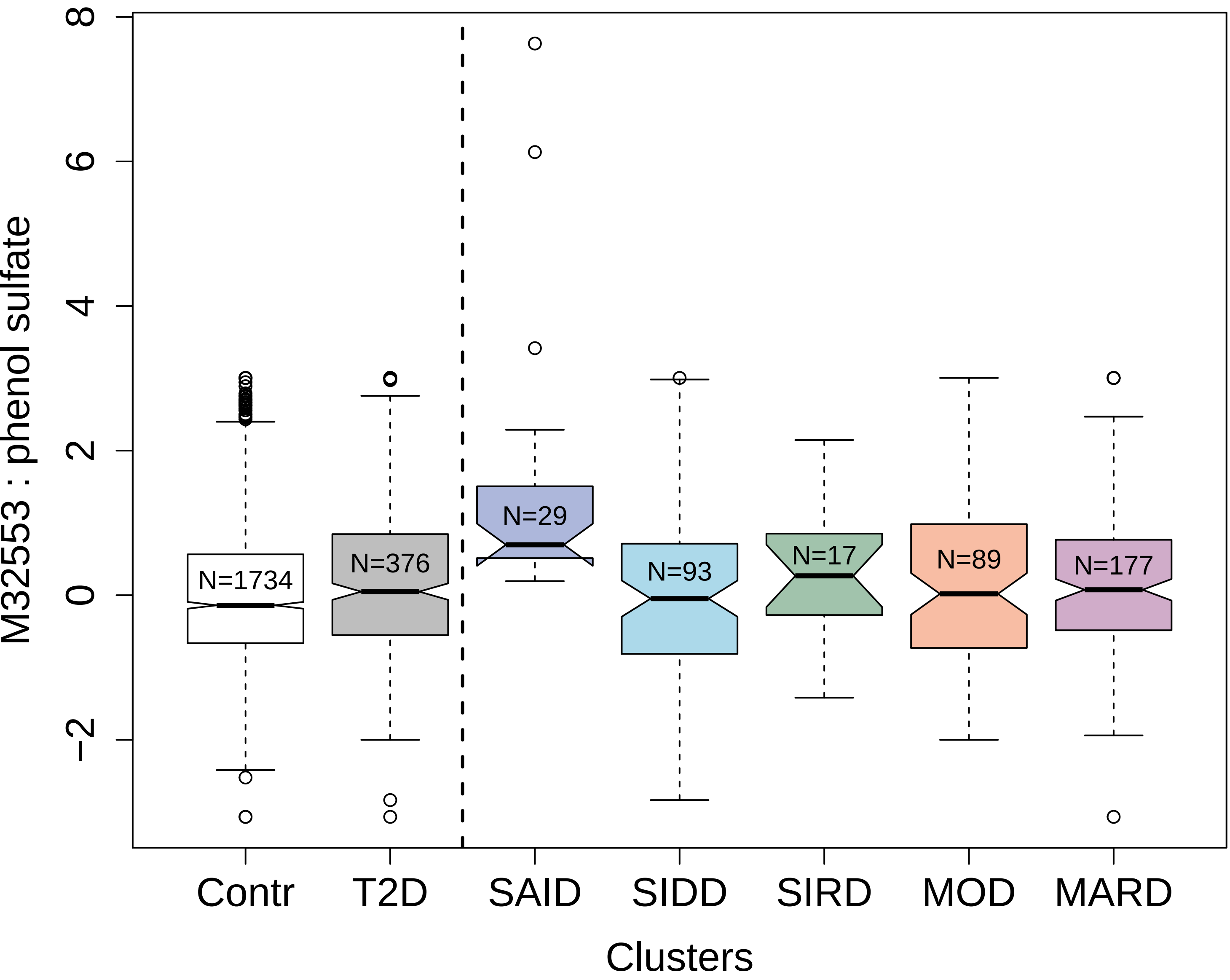
M32553 : phenol sulfate
8
6
4
2
0
−2
N=1734
N=376
N=29
N=93
N=17
N=89
N=177
Contr
T2D
SAID
SIDD
SIRD
MOD
MARD
Clusters

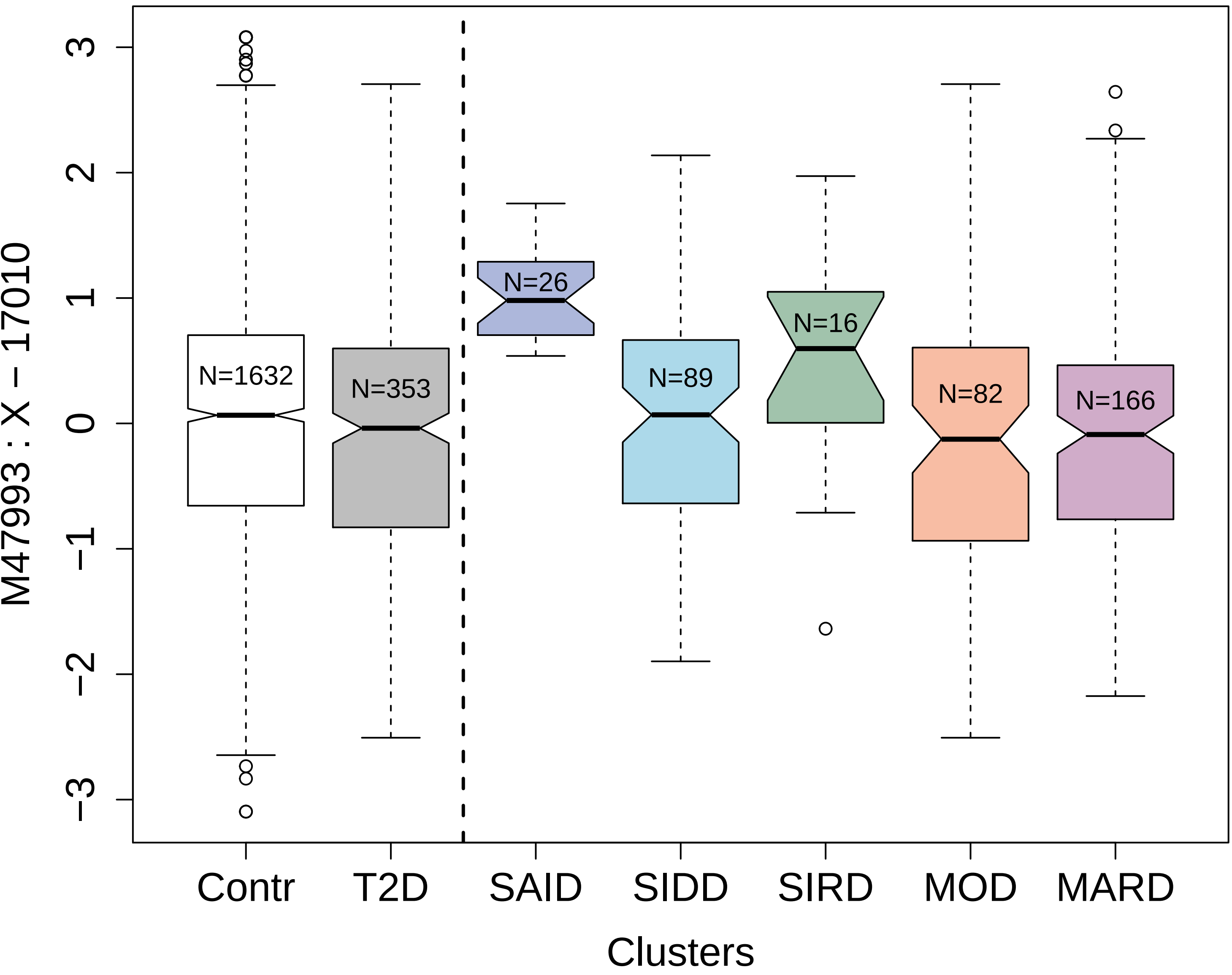

M47993 : X − 17010
3
2
1
0
−1
−2
−3
N=1632
N=353
N=26
N=89
N=16
N=82
N=166
Contr
T2D
SAID
SIDD
SIRD
MOD
MARD
Clusters

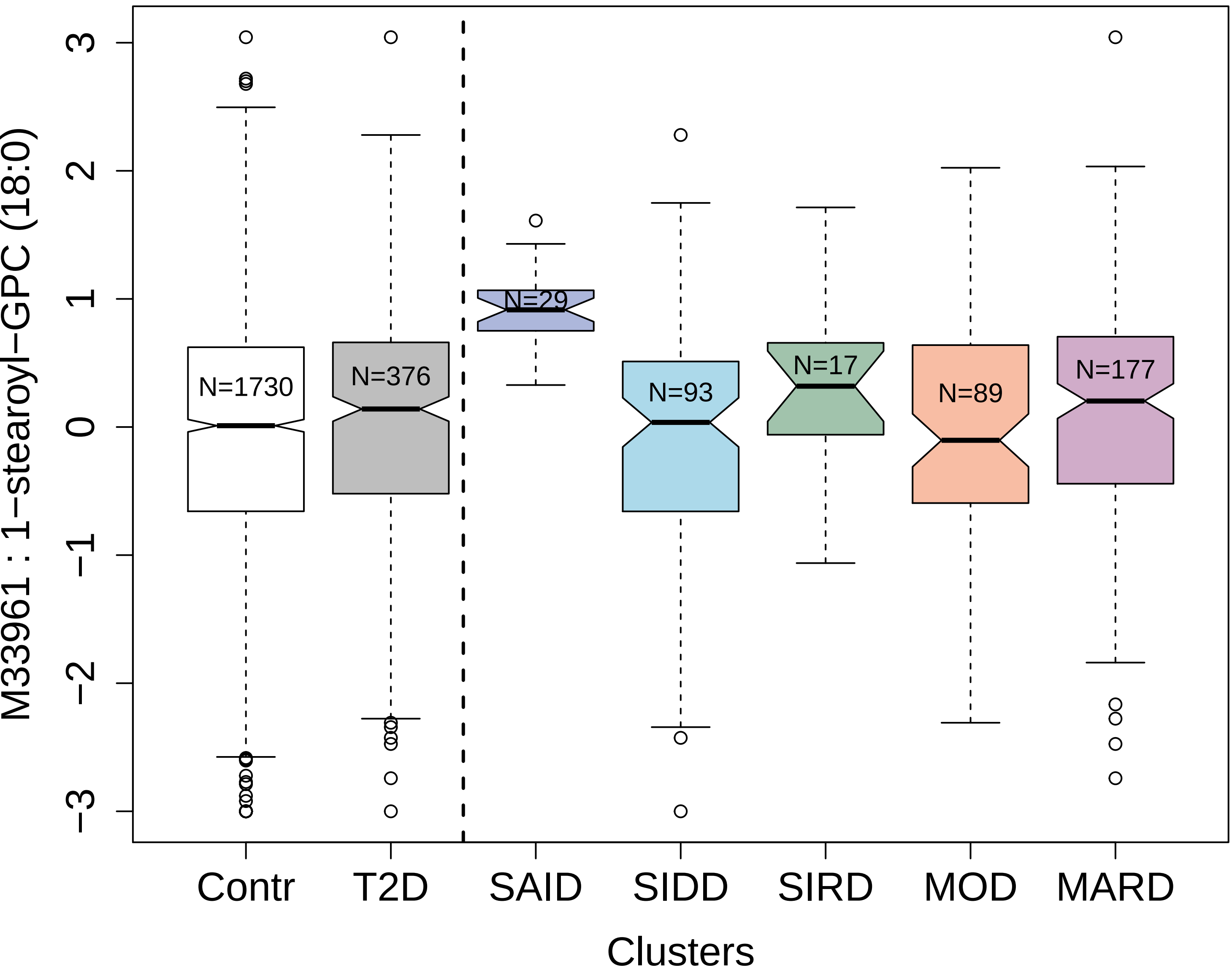

M33961 : 1−stearoyl−GPC (18:0)
3
2
1
0
−1
−2
−3
N=1730
N=376
N=29
N=93
N=17
N=89
N=177
Contr
T2D
SAID
SIDD
SIRD
MOD
MARD
Clusters

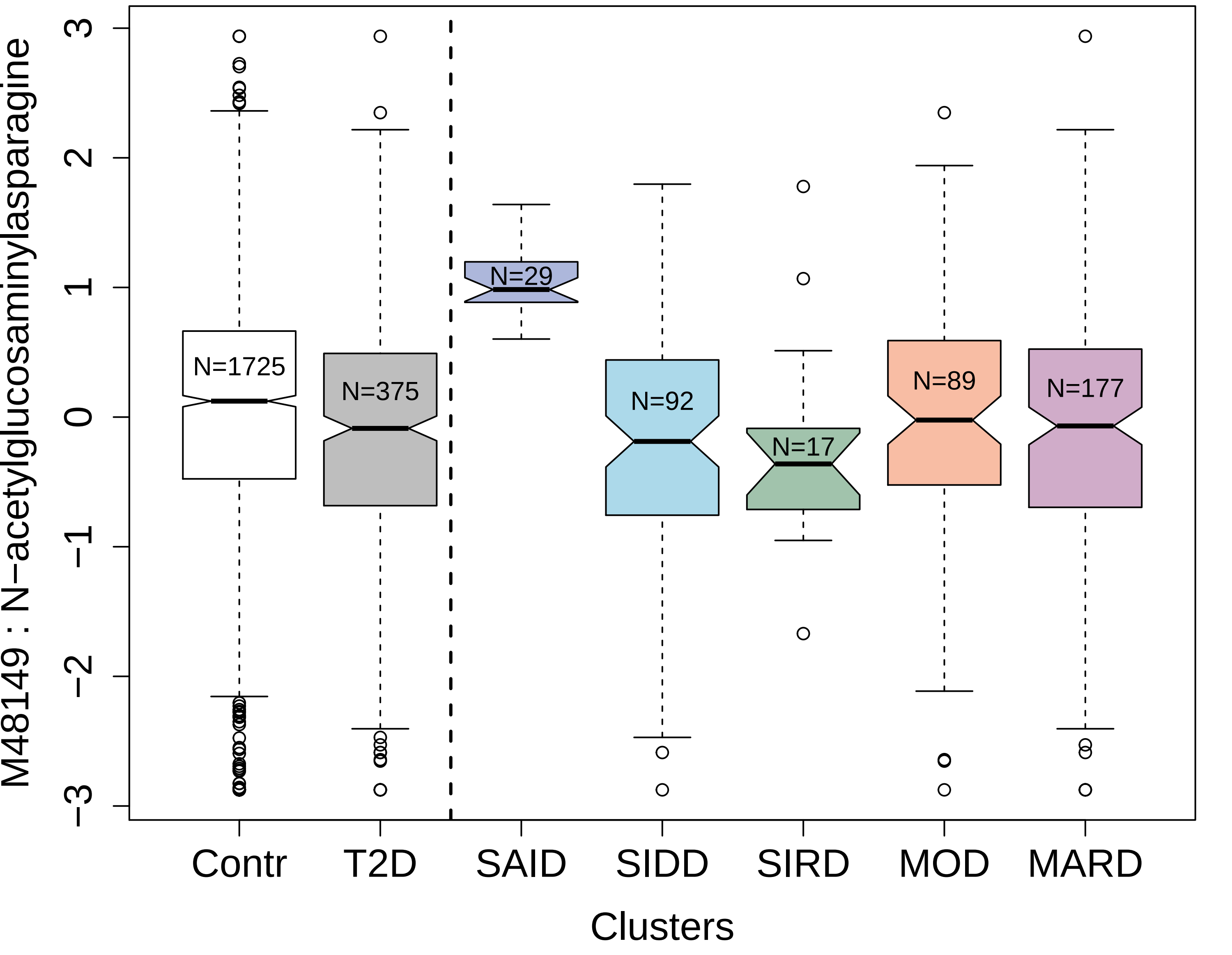

M48149 : N−acetylglucosaminylasparagine
3
2
1
0
−1
−2
−3
N=1725
N=375
N=29
N=92
N=17
N=89
N=177
Contr
T2D
SAID
SIDD
SIRD
MOD
MARD
Clusters

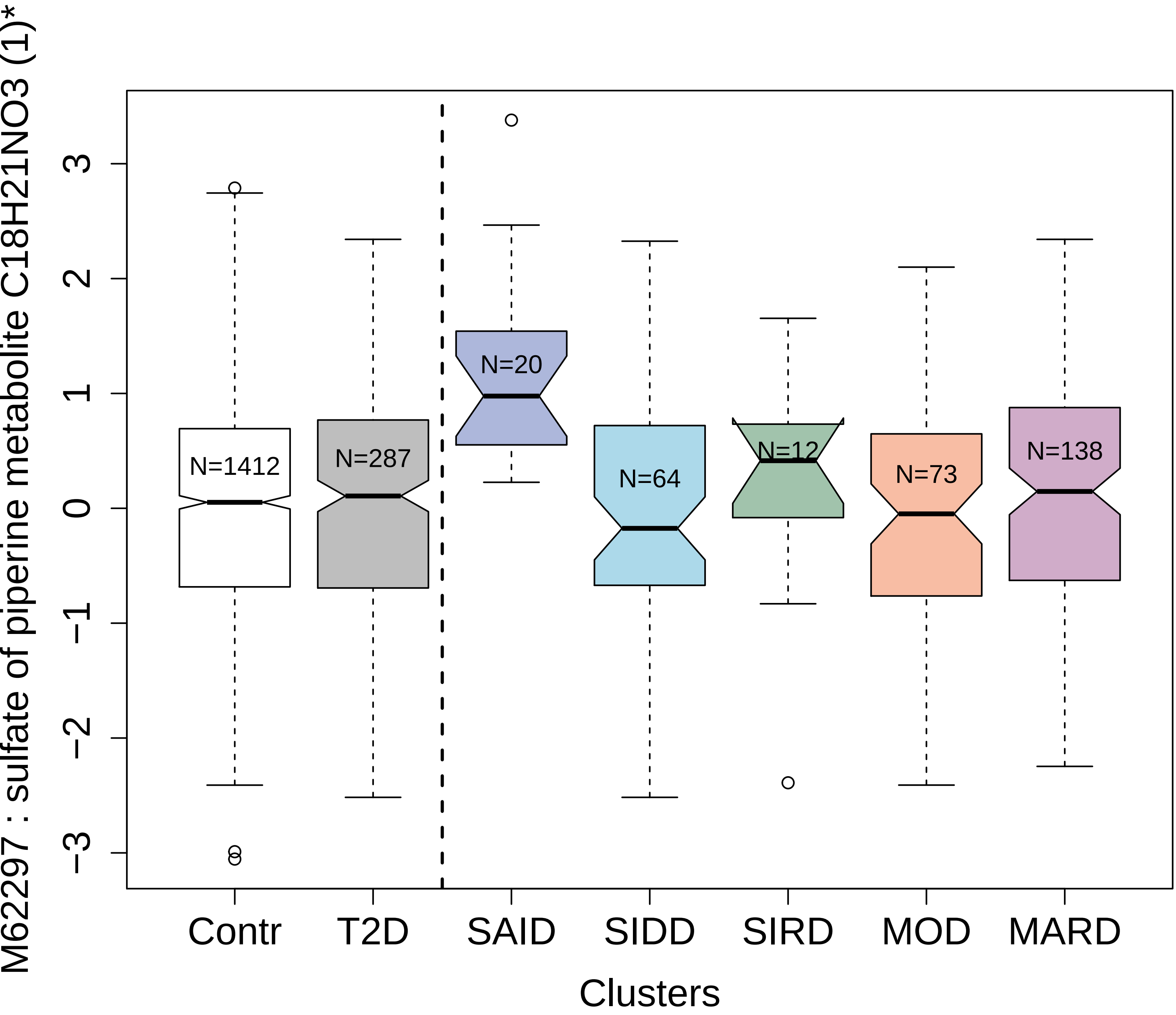
M62297 : sulfate of piperine metabolite C18H21NO3 (1)*
3
2
1
0
−1
−2
−3
N=1412
N=287
N=20
N=64
N=12
N=73
N=138
Contr
T2D
SAID
SIDD
SIRD
MOD
MARD
Clusters

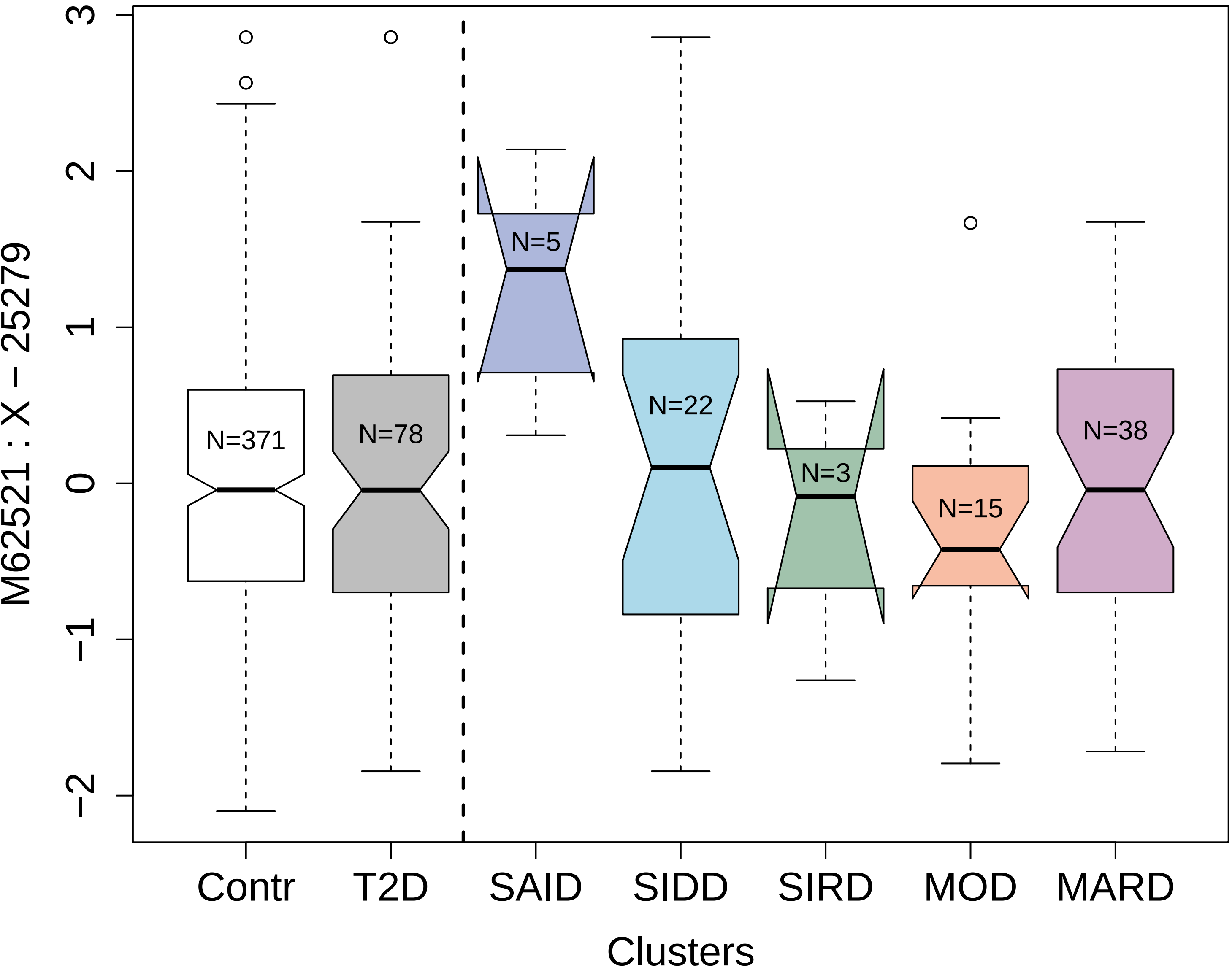

M62521 : X − 25279
3
2
1
0
−1
−2
N=371
N=78
N=5
N=22
N=3
N=15
N=38
Contr
T2D
SAID
SIDD
SIRD
MOD
MARD
Clusters

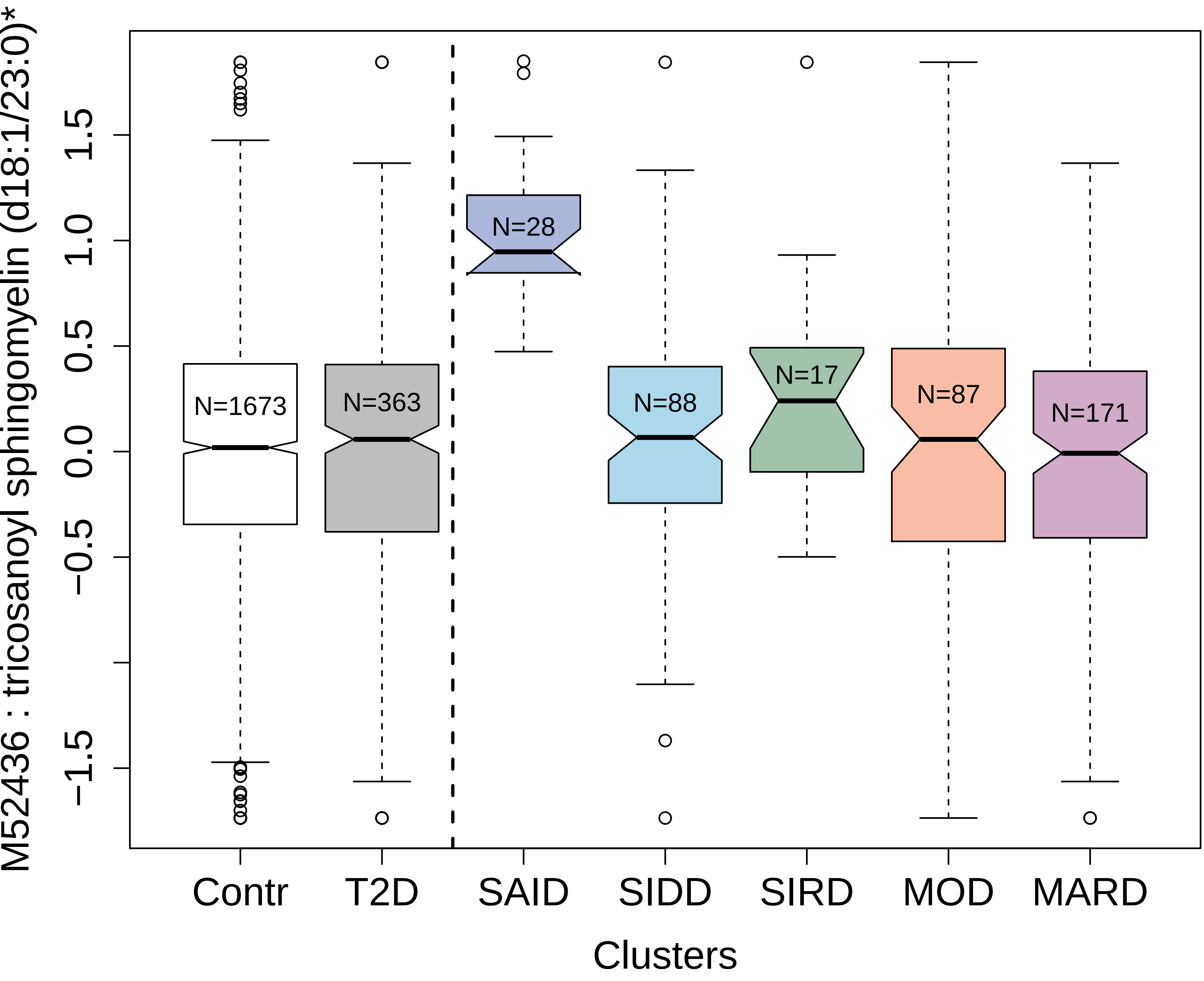
M52436 : tricosanoyl sphingomyelin (d18:1/23:0)*
1.5
1.0
0.5
0.0
−0.5
−1.5
N=1673
N=363
N=28
N=88
N=17
N=87
N=171
Contr
T2D
SAID
SIDD
SIRD
MOD
MARD
Clusters

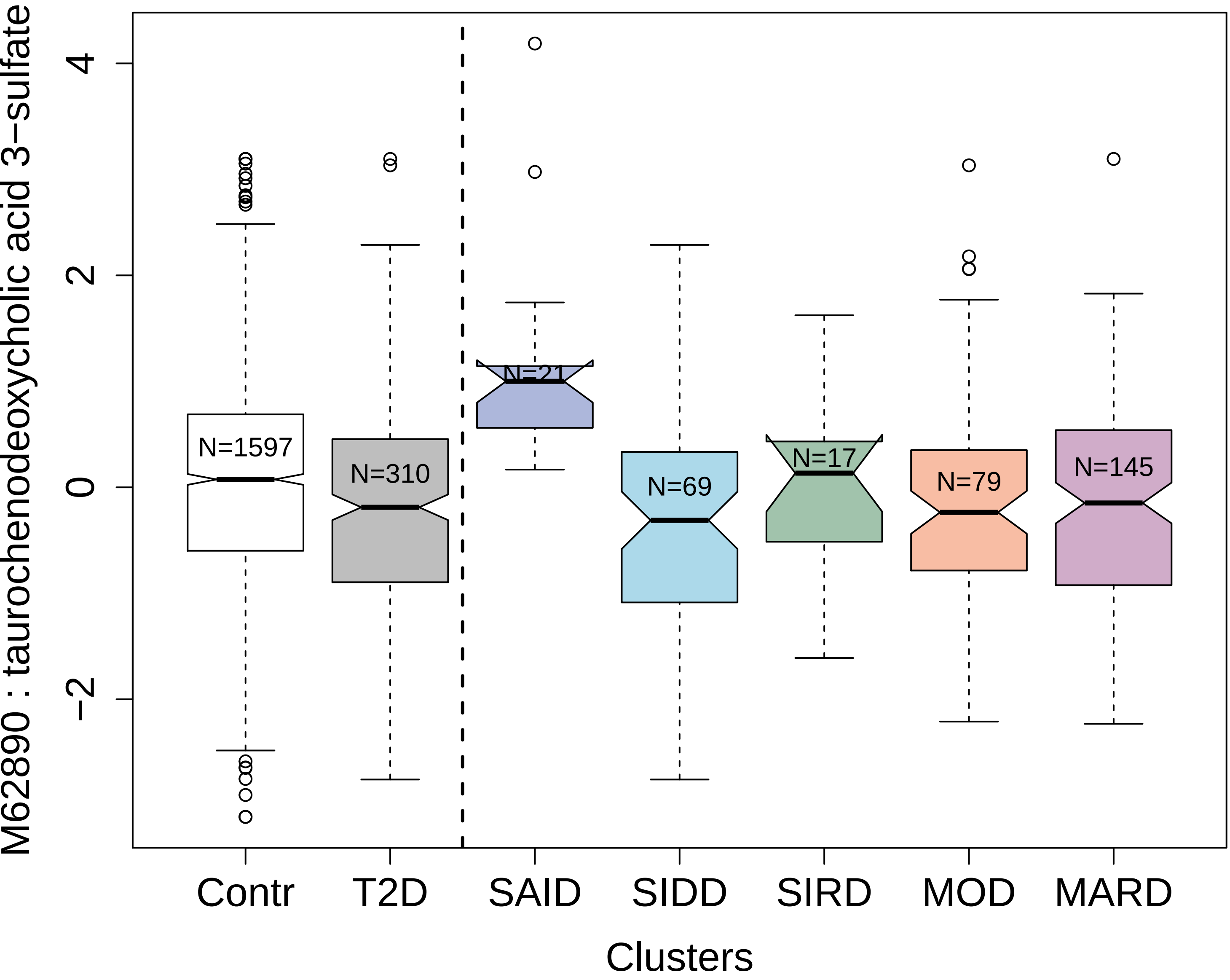
M62890 : taurochenodeoxycholic acid 3−sulfate
4
2
0
−2
N=1597
N=310
N=21
N=69
N=17
N=79
N=145
Contr
T2D
SAID
SIDD
SIRD
MOD
MARD
Clusters

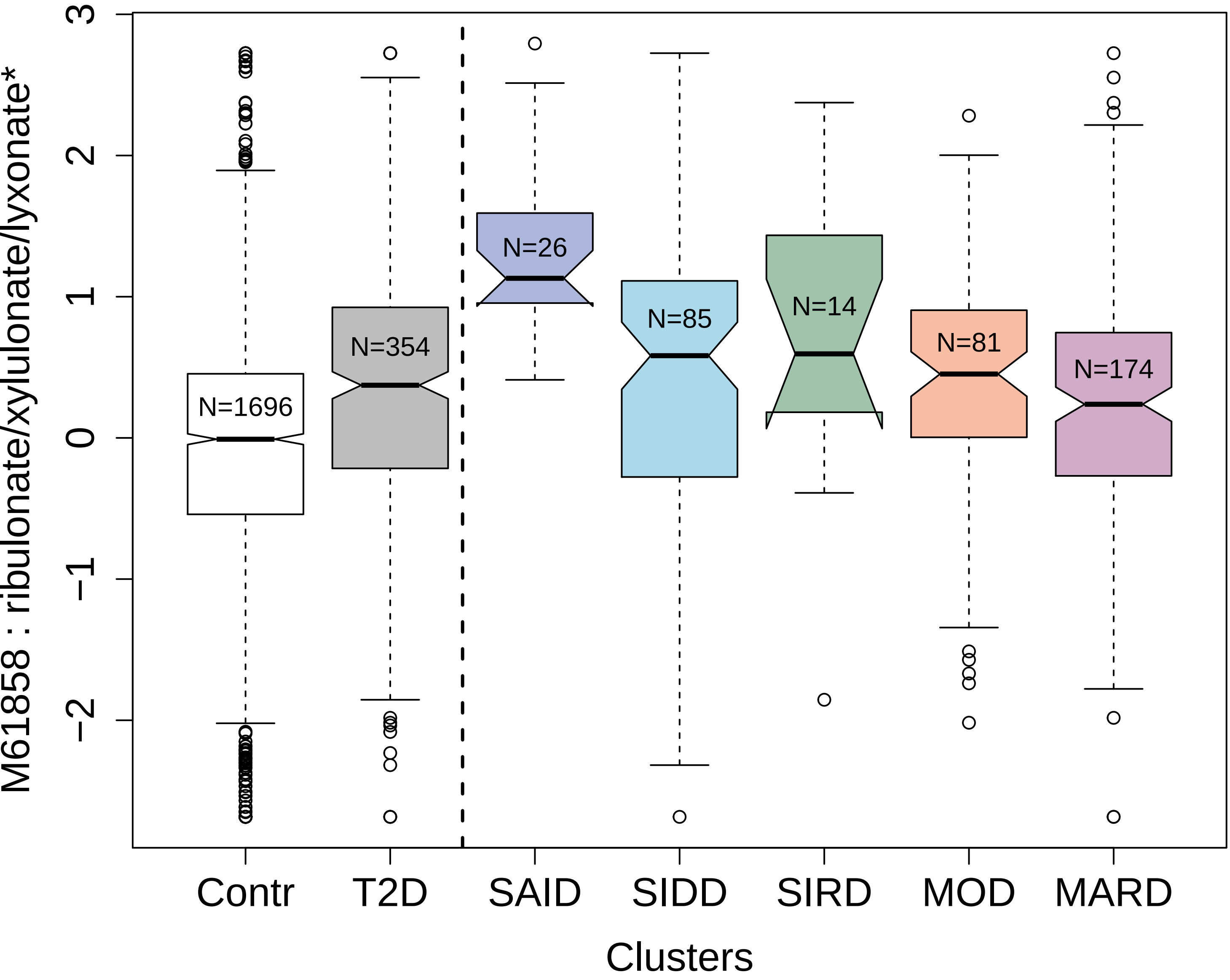
M61858 : ribulonate/xylulonate/lyxonate*
3
2
1
0
−1
−2
N=1696
N=354
N=26
N=85
N=14
N=81
N=174
Contr
T2D
SAID
SIDD
SIRD
MOD
MARD
Clusters

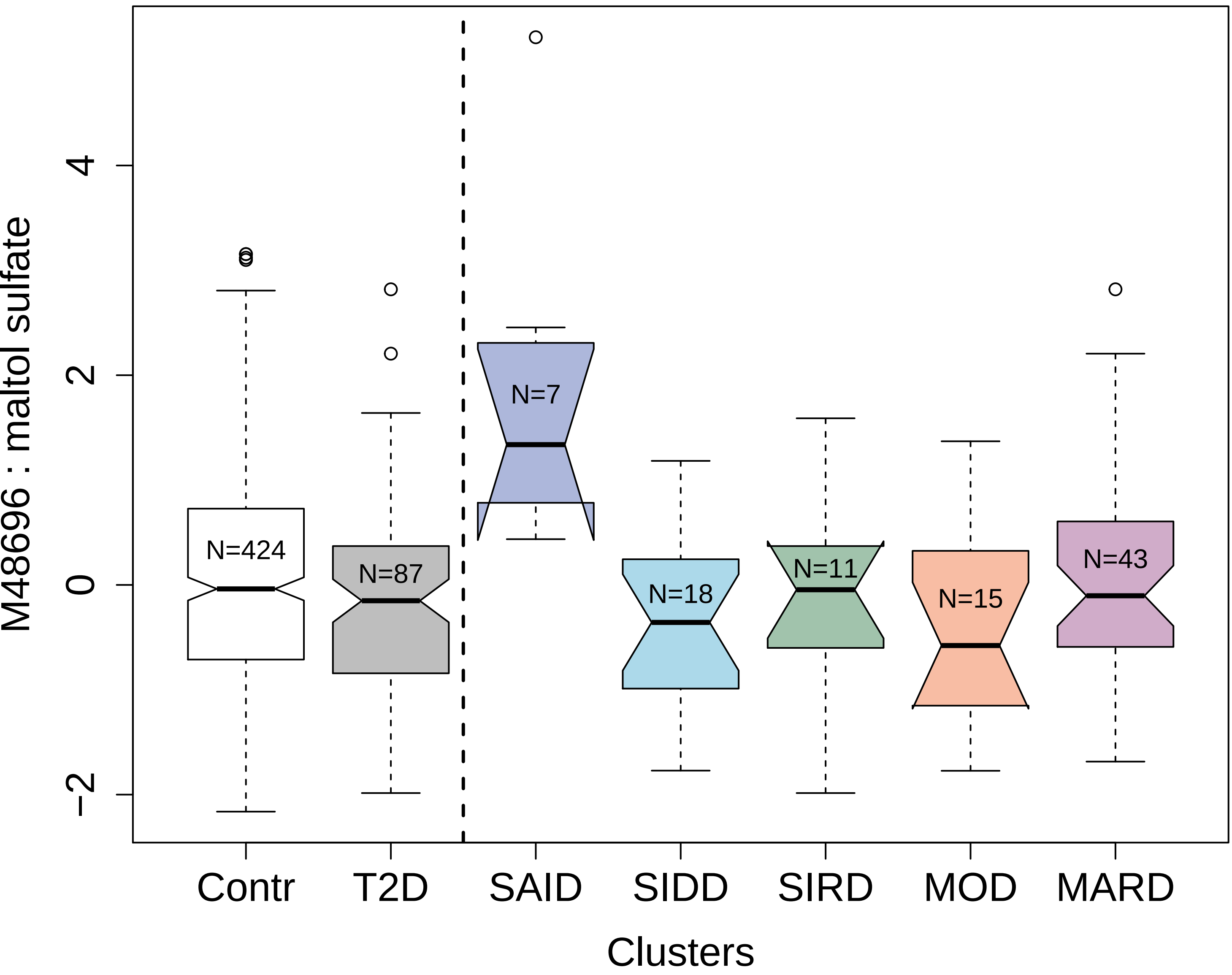
M48696 : maltol sulfate
4
2
0
−2
N=424
N=87
N=7
N=18
N=11
N=15
N=43
Contr
T2D
SAID
SIDD
SIRD
MOD
MARD
Clusters

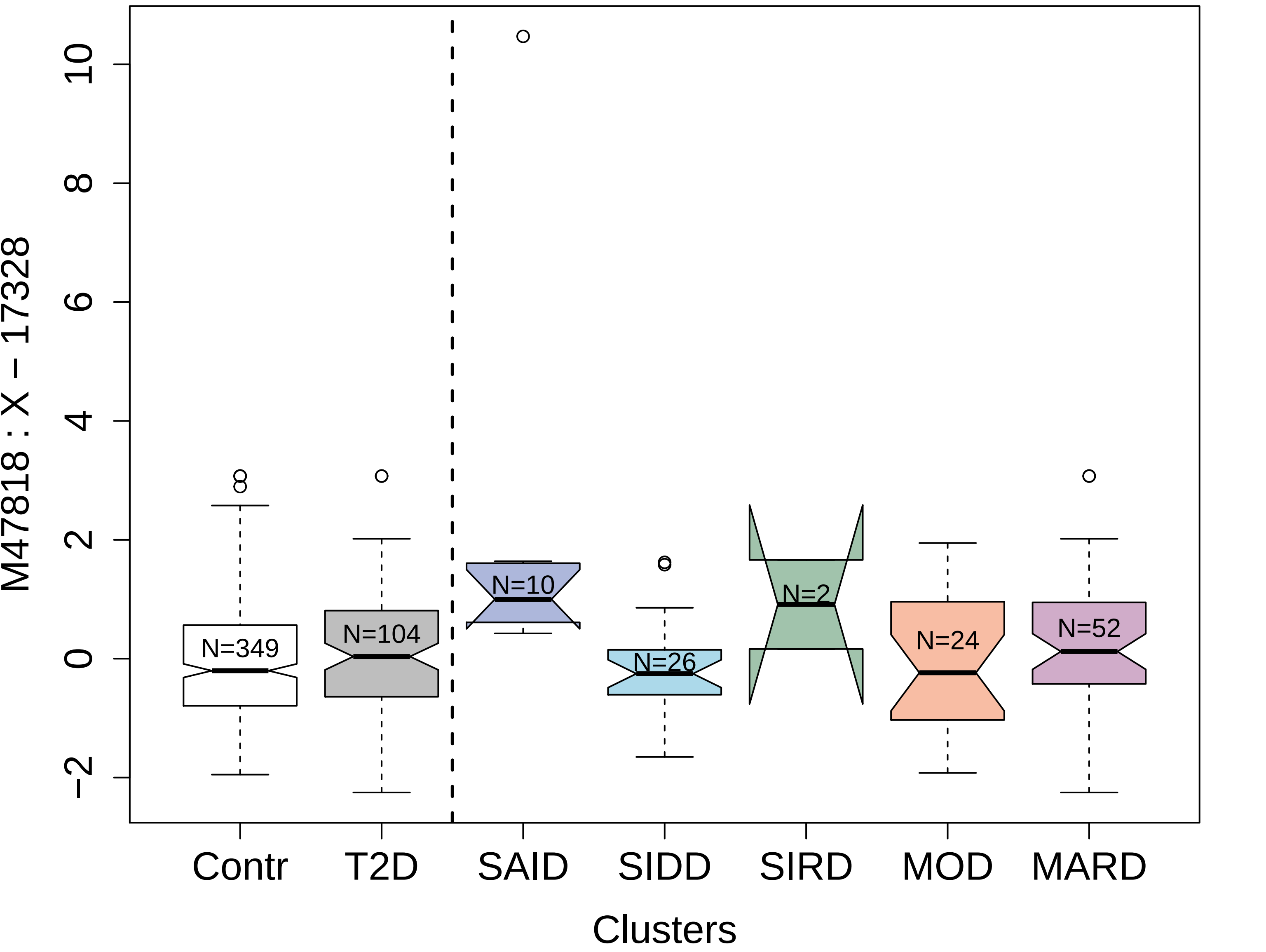

M47818 : X − 17328
10
8
6
4
2
0
−2
N=349
N=104
N=10
N=26
N=2
N=24
N=52
Contr
T2D
SAID
SIDD
SIRD
MOD
MARD
Clusters

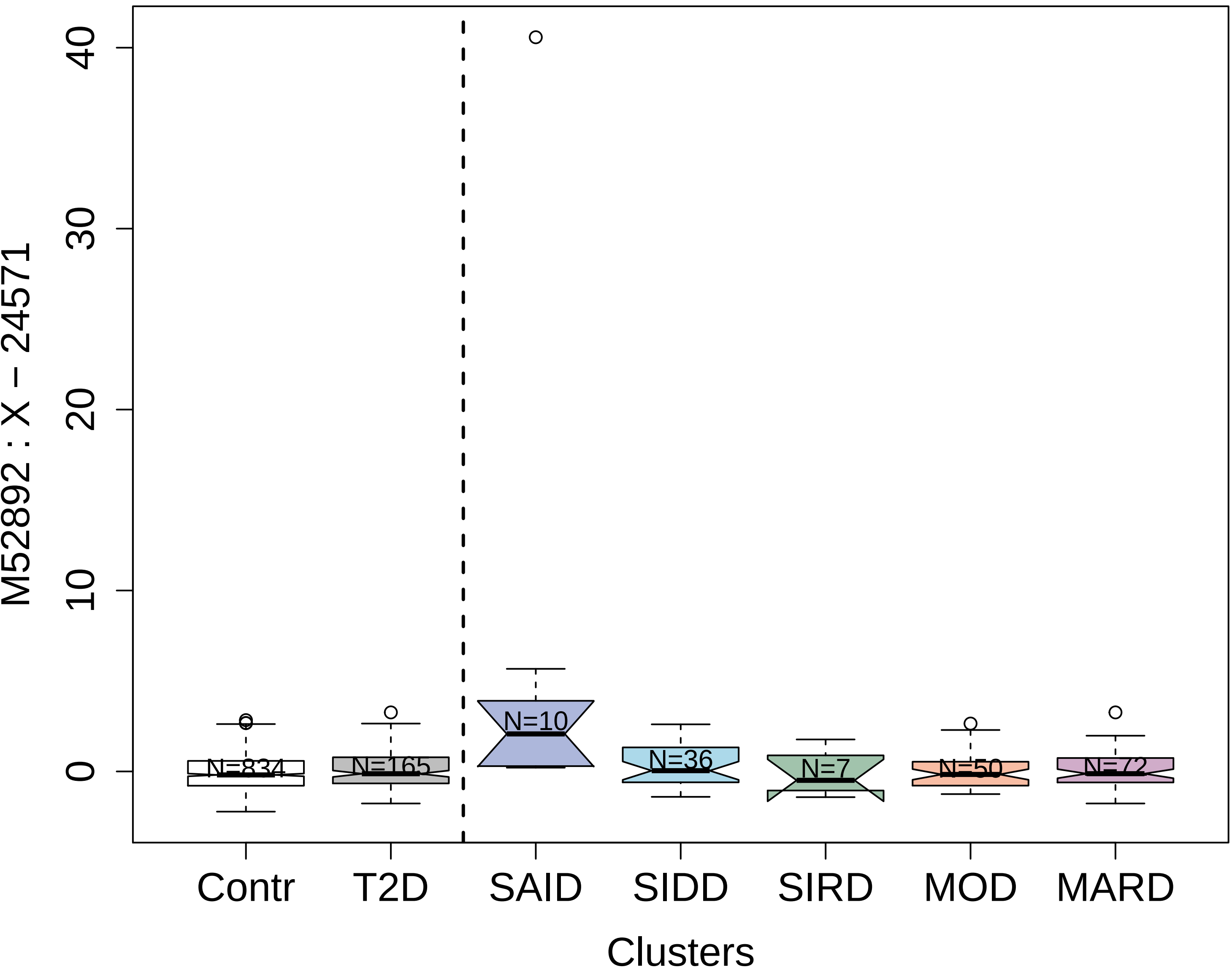

M52892 : X − 24571
0
10
20
30
40
N=834
N=165
N=10
N=36
N=7
N=50
N=72
Contr
T2D
SAID
SIDD
SIRD
MOD
MARD
Clusters

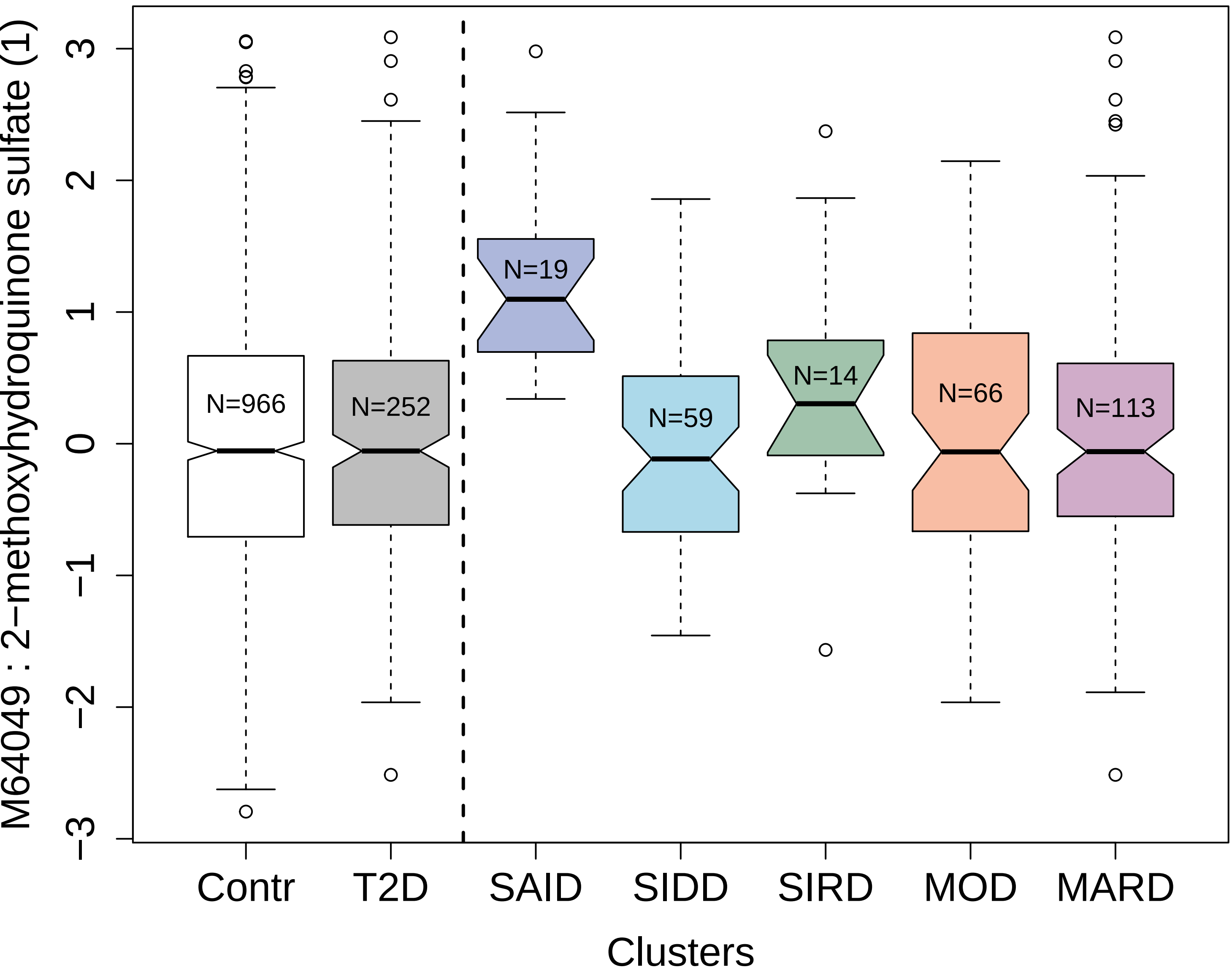

M64049 : 2−methoxyhydroquinone sulfate (1)
3
2
1
0
−1
−2
−3
N=966
N=252
N=19
N=59
N=14
N=66
N=113
Contr
T2D
SAID
SIDD
SIRD
MOD
MARD
Clusters

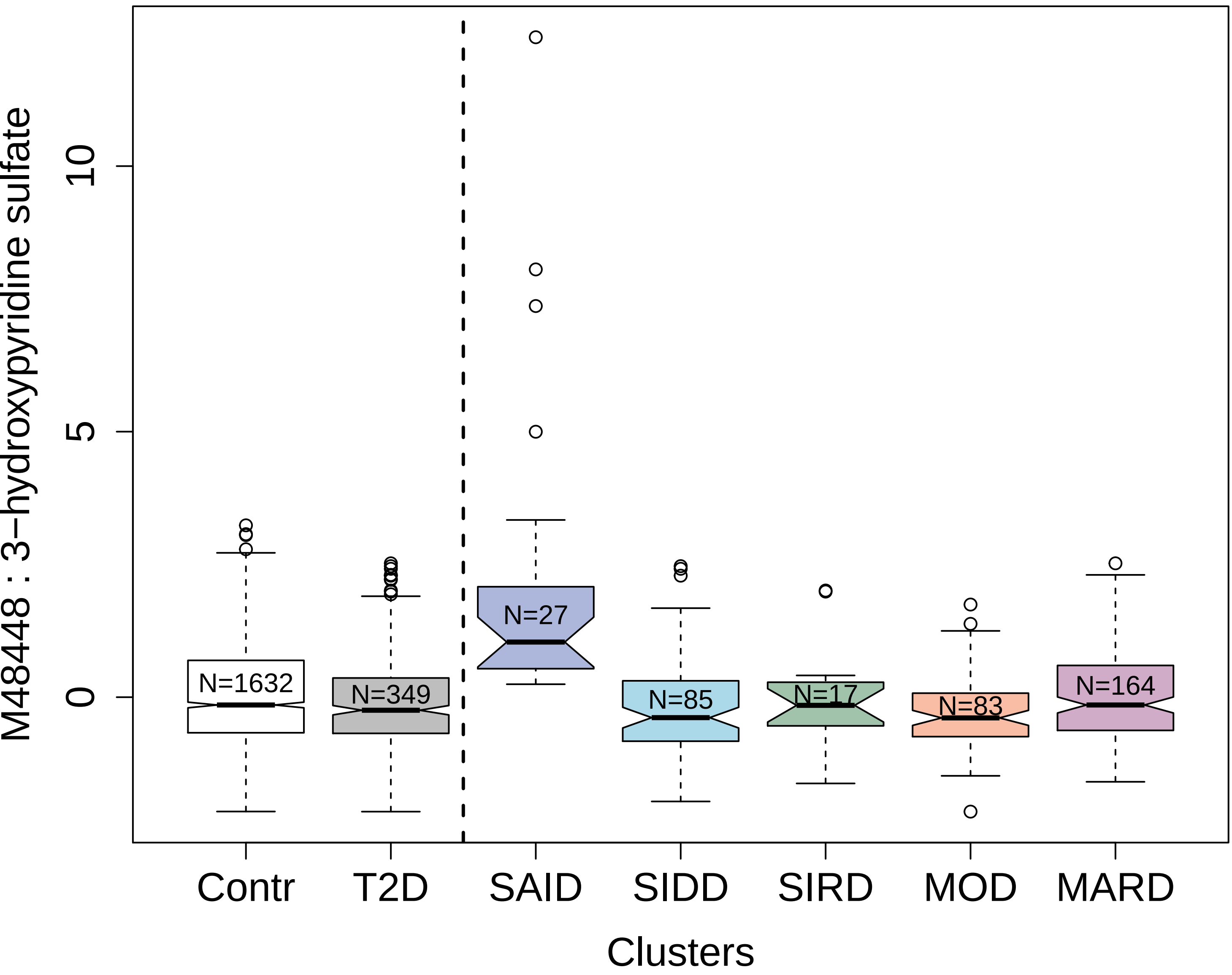
M48448 : 3−hydroxypyridine sulfate
0
5
10
N=1632
N=349
N=27
N=85
N=17
N=83
N=164
Contr
T2D
SAID
SIDD
SIRD
MOD
MARD
Clusters

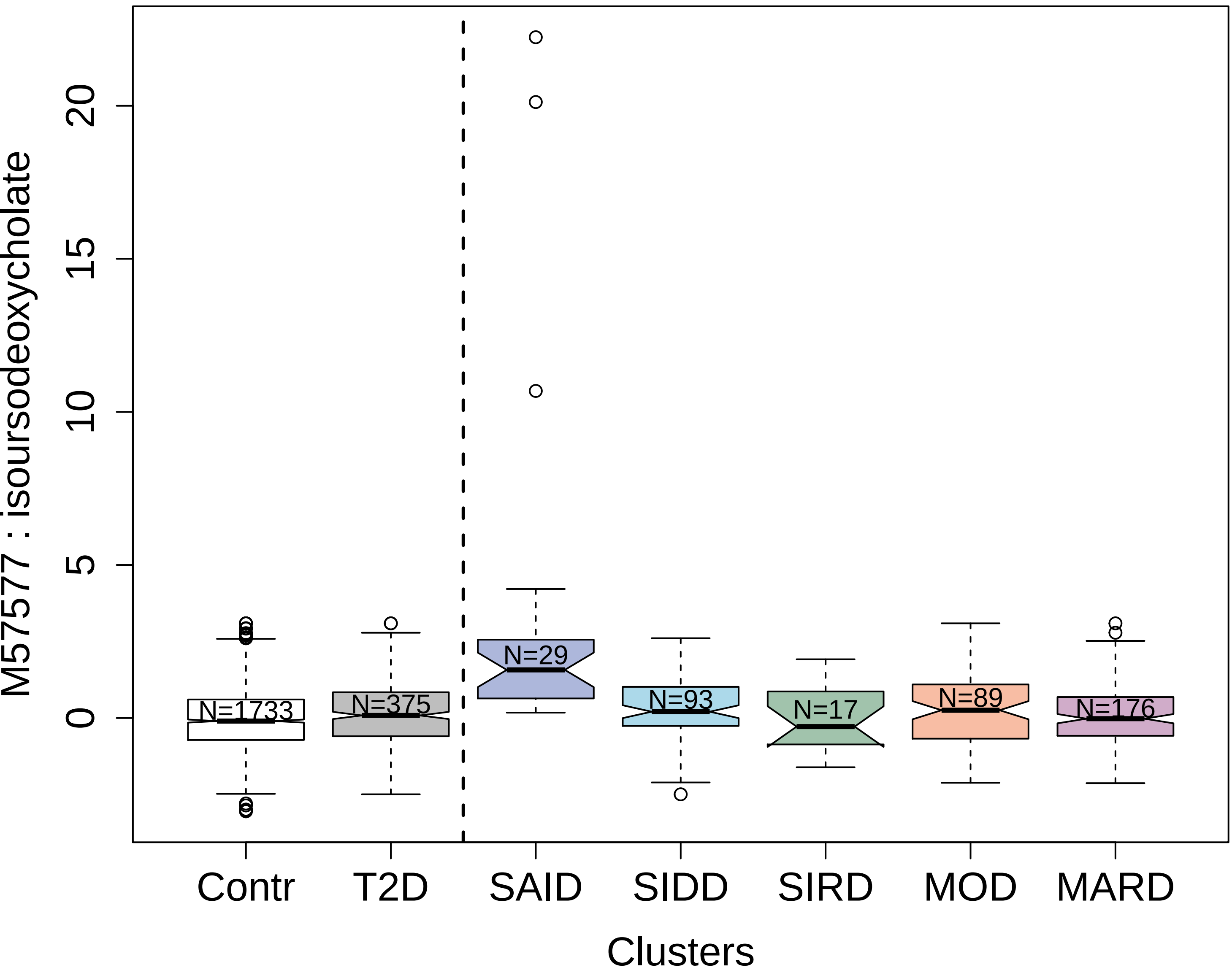
M57577 : isoursodeoxycholate
0
5
10
15
20
N=1733
N=375
N=29
N=93
N=17
N=89
N=176
Contr
T2D
SAID
SIDD
SIRD
MOD
MARD
Clusters

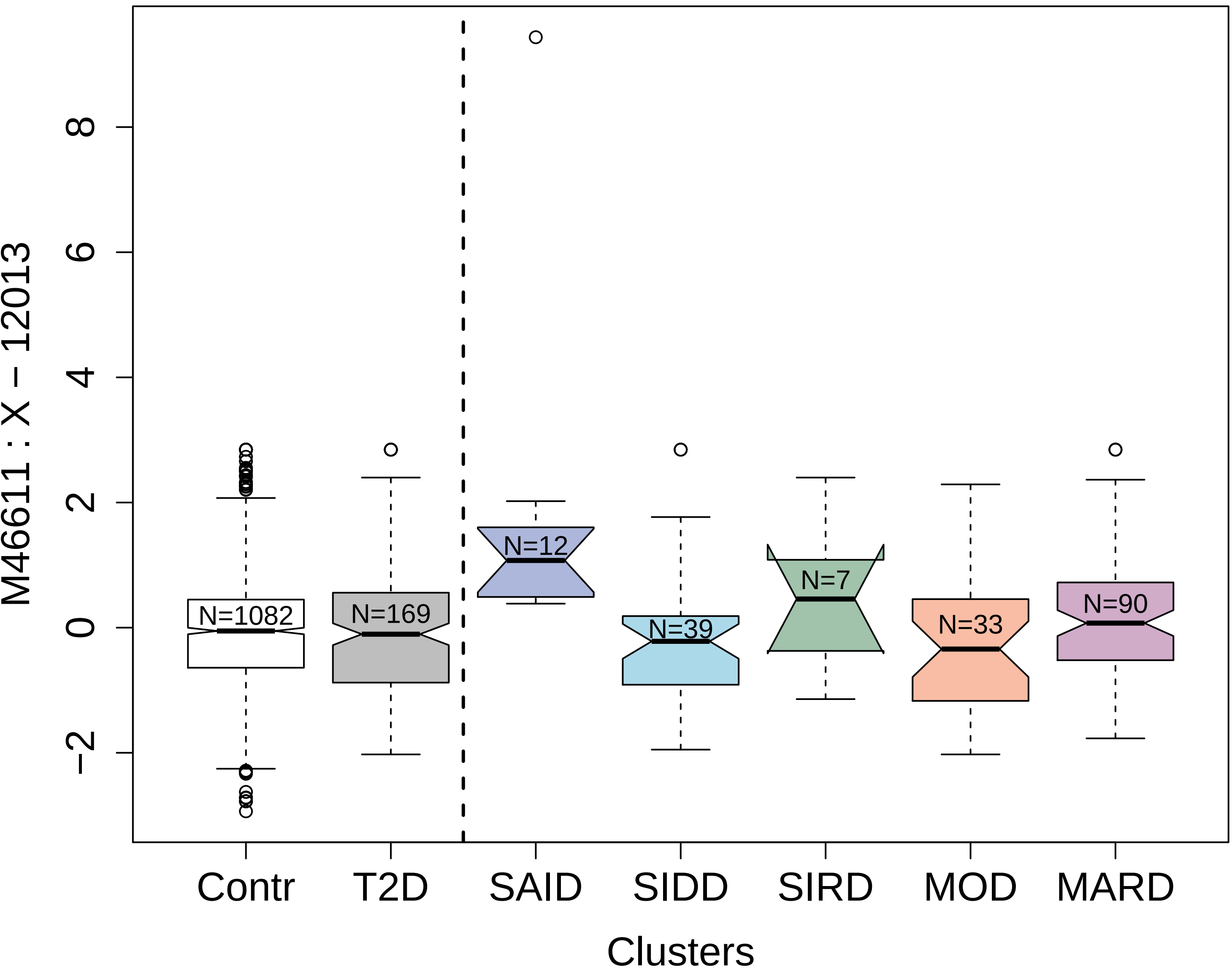

M46611 : X − 12013
8
6
4
2
0
−2
N=1082
N=169
N=12
N=39
N=7
N=33
N=90
Contr
T2D
SAID
SIDD
SIRD
MOD
MARD
Clusters

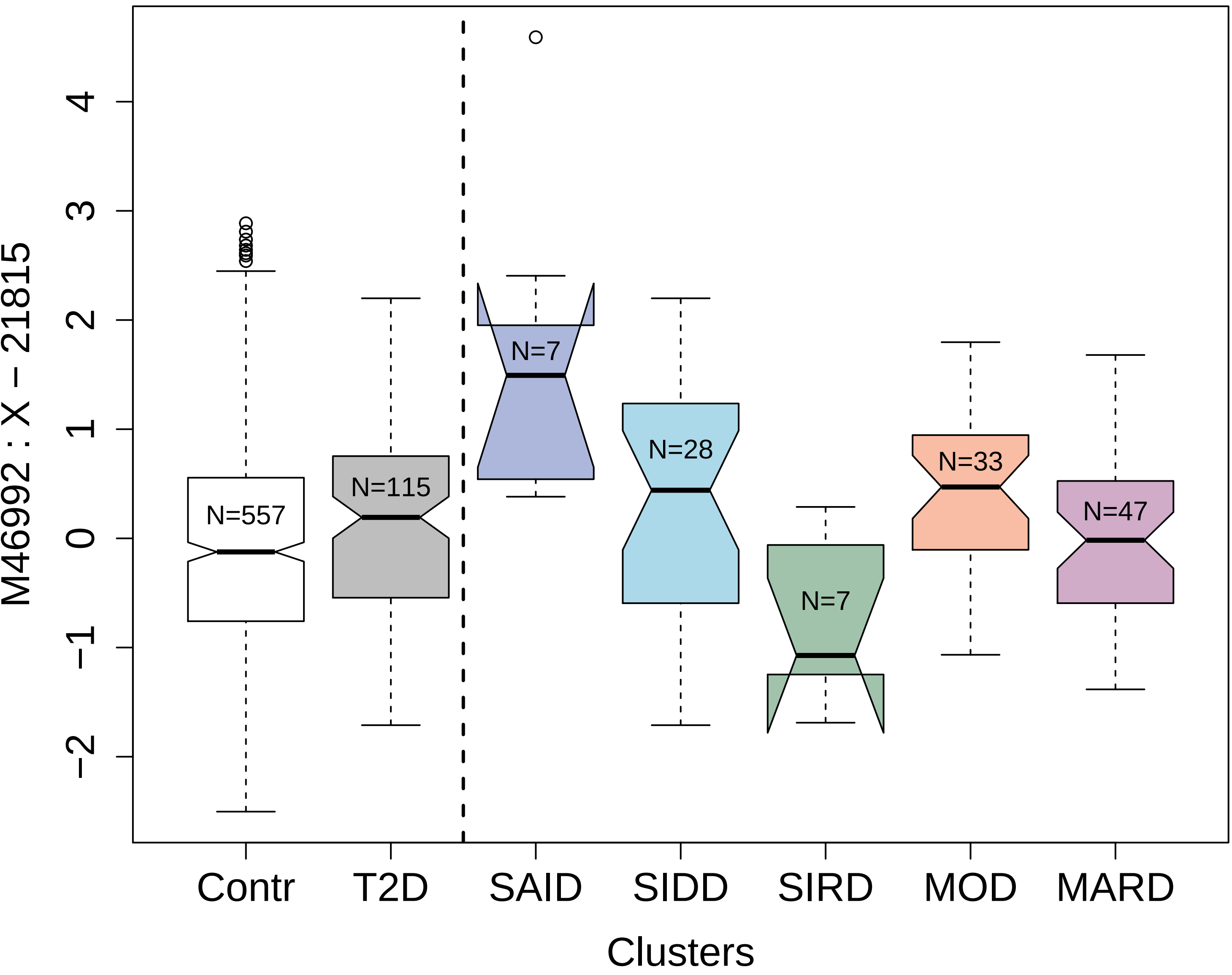

M46992 : X − 21815
4
3
2
1
0
−1
−2
N=557
N=115
N=7
N=28
N=7
N=33
N=47
Contr
T2D
SAID
SIDD
SIRD
MOD
MARD
Clusters

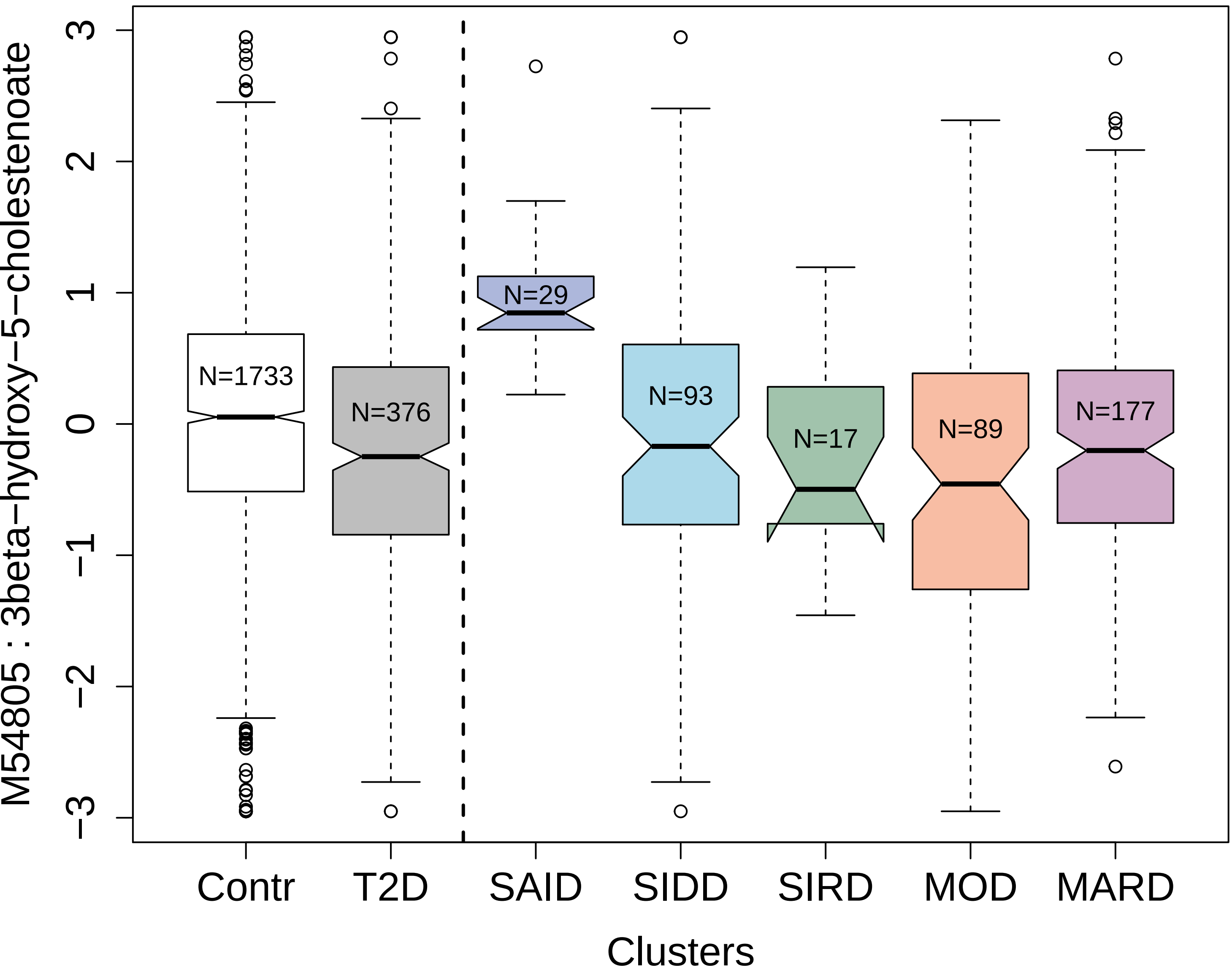
M54805 : 3beta−hydroxy−5−cholestenoate
3
2
1
0
−1
−2
−3
N=1733
N=376
N=29
N=93
N=17
N=89
N=177
Contr
T2D
SAID
SIDD
SIRD
MOD
MARD
Clusters

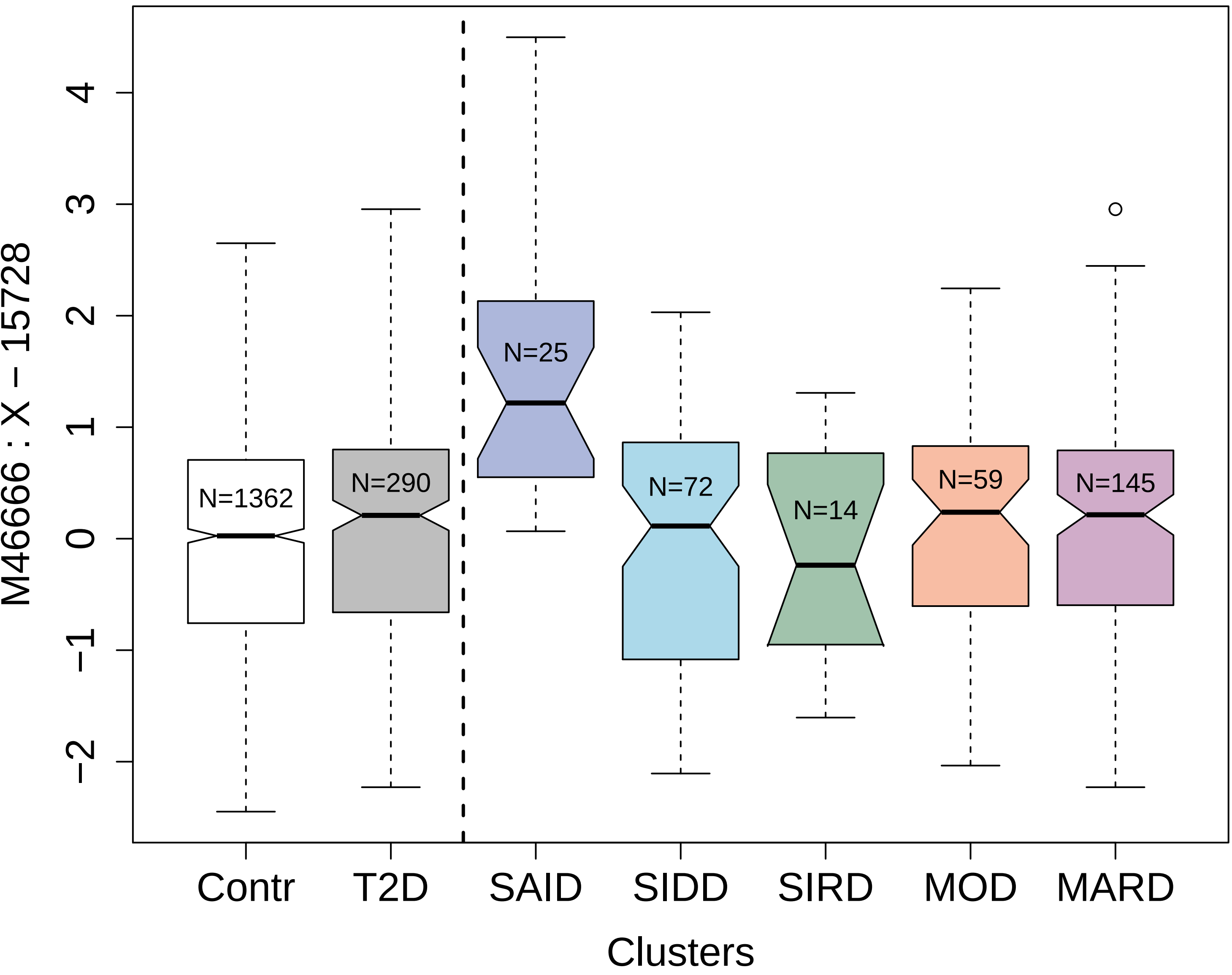

M46666 : X − 15728
4
3
2
1
0
−1
−2
N=1362
N=290
N=25
N=72
N=14
N=59
N=145
Contr
T2D
SAID
SIDD
SIRD
MOD
MARD
Clusters

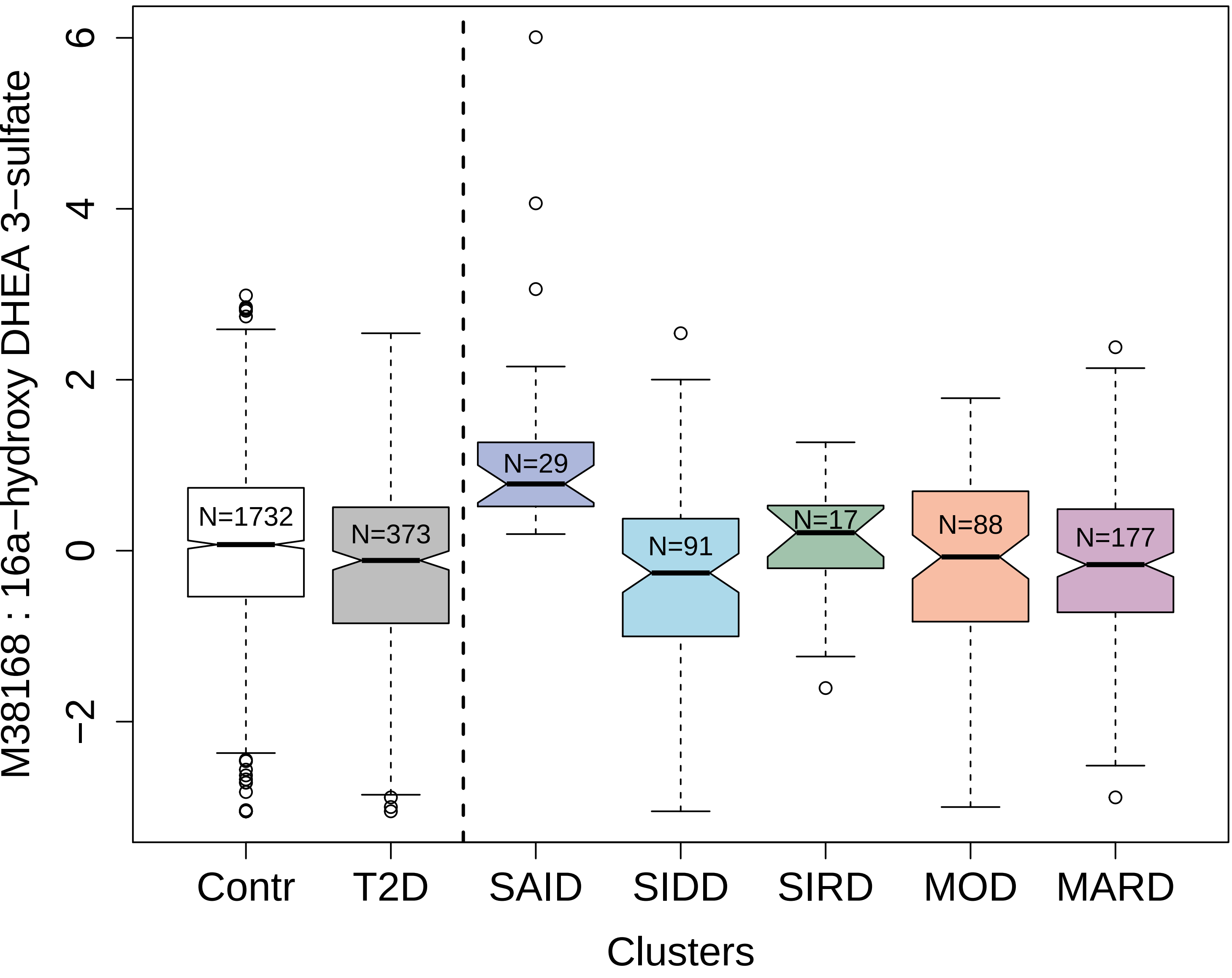

M38168 : 16a−hydroxy DHEA 3−sulfate
6
4
2
0
−2
N=1732
N=373
N=29
N=91
N=17
N=88
N=177
Contr
T2D
SAID
SIDD
SIRD
MOD
MARD
Clusters

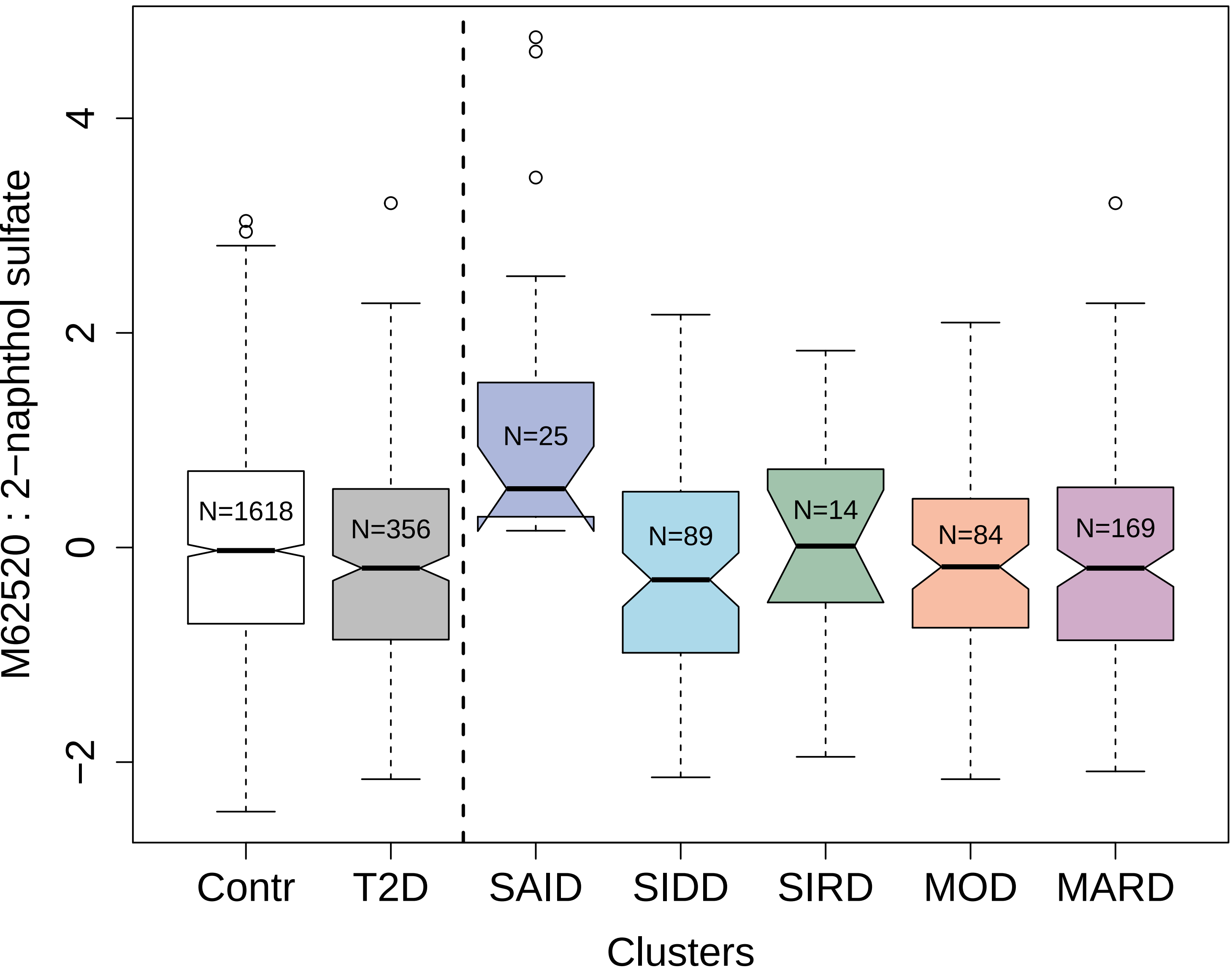
M62520 : 2−naphthol sulfate
−2
0
2
4
N=1618
N=356
N=25
N=89
N=14
N=84
N=169
Contr
T2D
SAID
SIDD
SIRD
MOD
MARD
Clusters

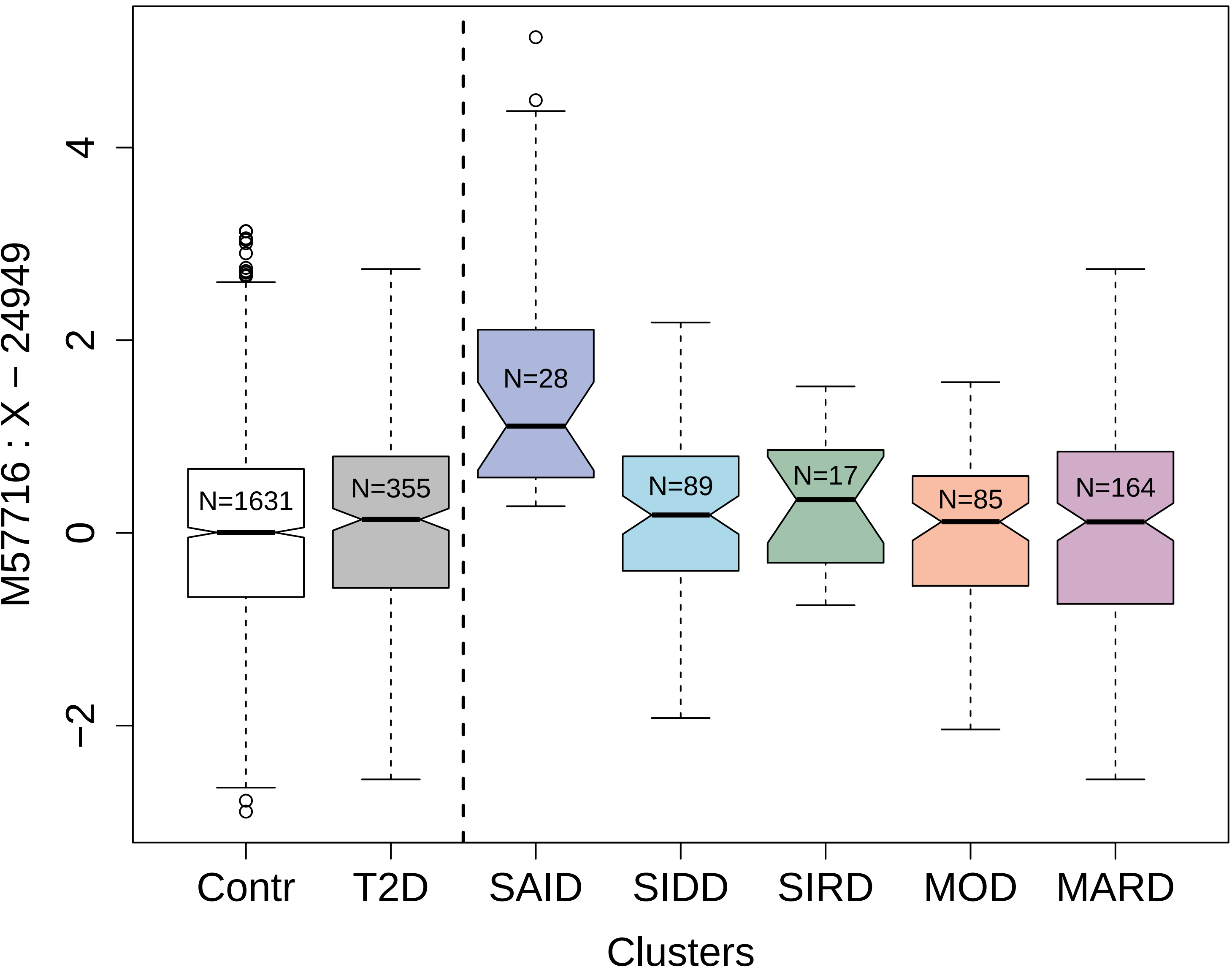

M57716 : X − 24949
4
2
0
−2
N=1631
N=355
N=28
N=89
N=17
N=85
N=164
Contr
T2D
SAID
SIDD
SIRD
MOD
MARD
Clusters

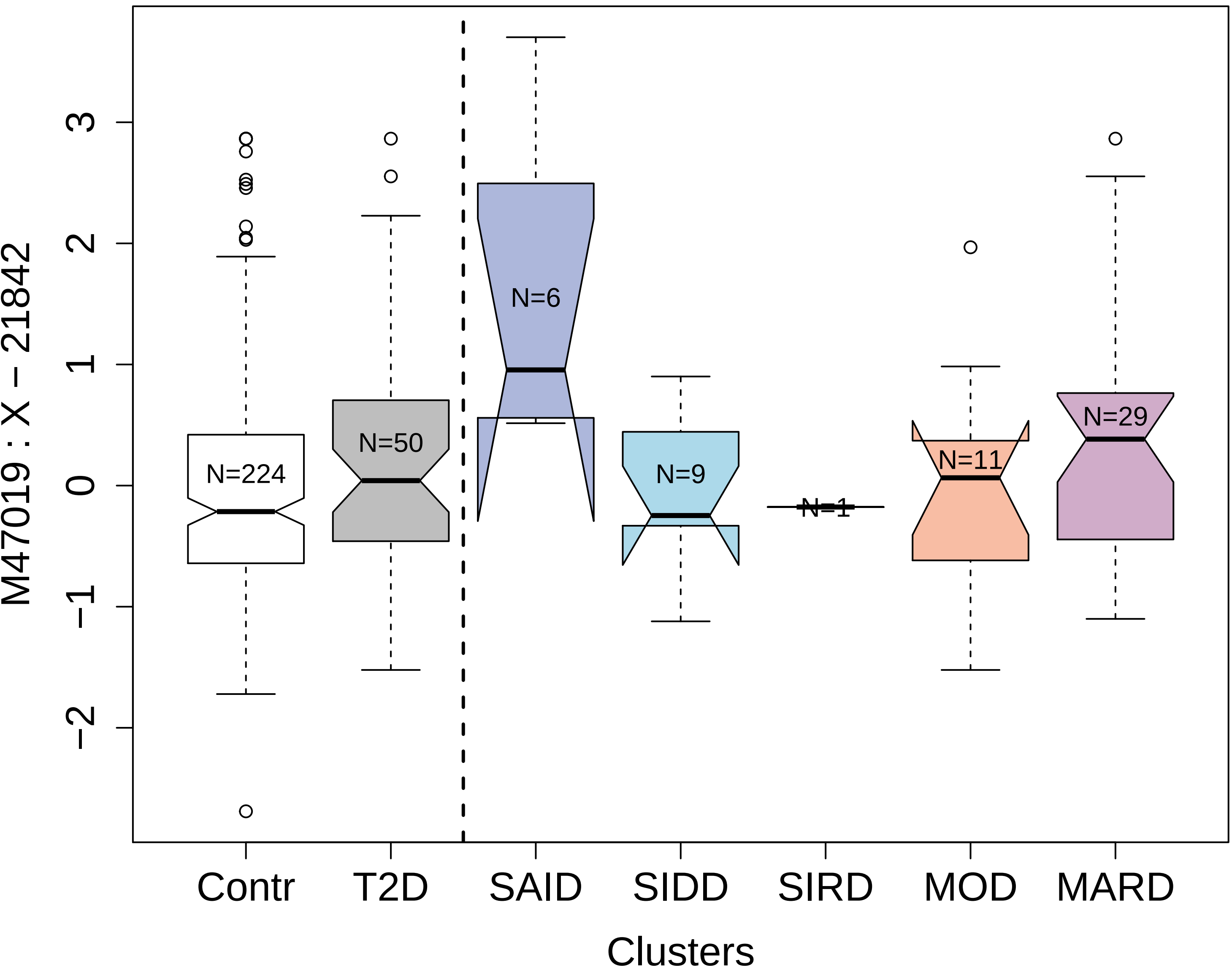

M47019 : X − 21842
3
2
1
0
−1
−2
N=224
N=50
N=6
N=9
N=1
N=11
N=29
Contr
T2D
SAID
SIDD
SIRD
MOD
MARD
Clusters

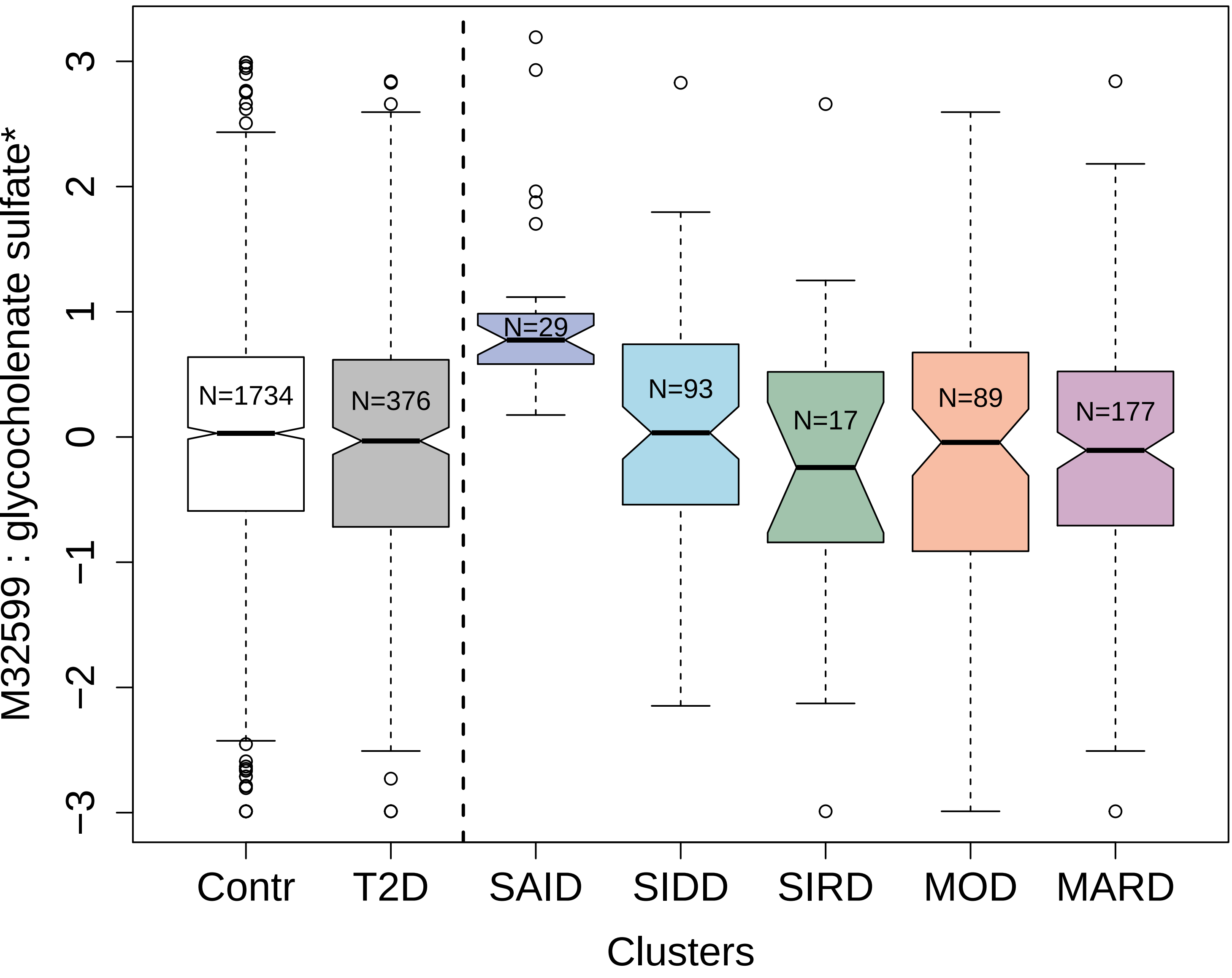

M32599 : glycocholenate sulfate*
3
2
1
0
−1
−2
−3
N=1734
N=376
N=29
N=93
N=17
N=89
N=177
Contr
T2D
SAID
SIDD
SIRD
MOD
MARD
Clusters

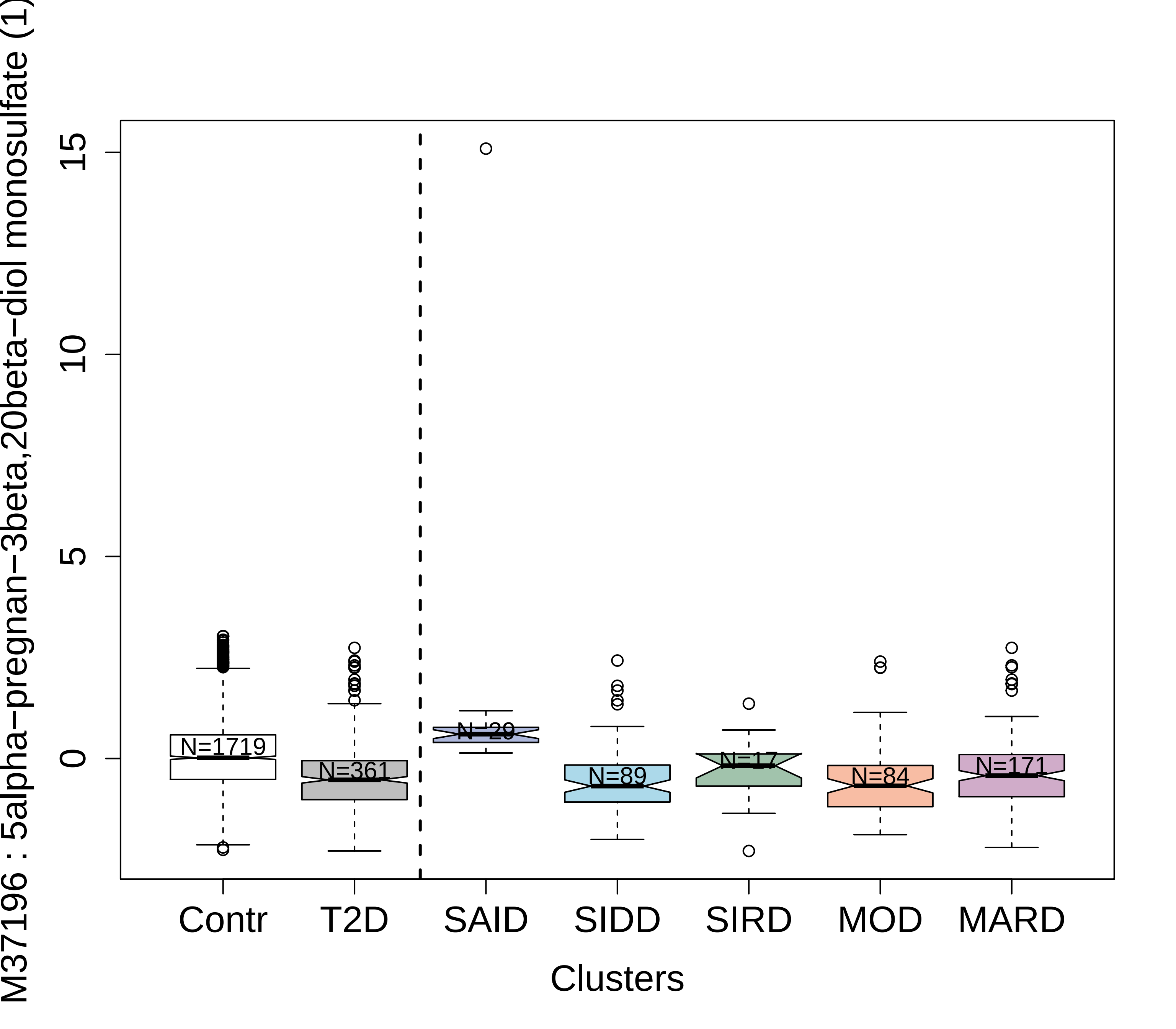
M37196 : 5alpha−pregnan−3beta,20beta−diol monosulfate (1)
15
10
5
0
N=1719
N=361
N=29
N=89
N=17
N=84
N=171
Contr
T2D
SAID
SIDD
SIRD
MOD
MARD
Clusters

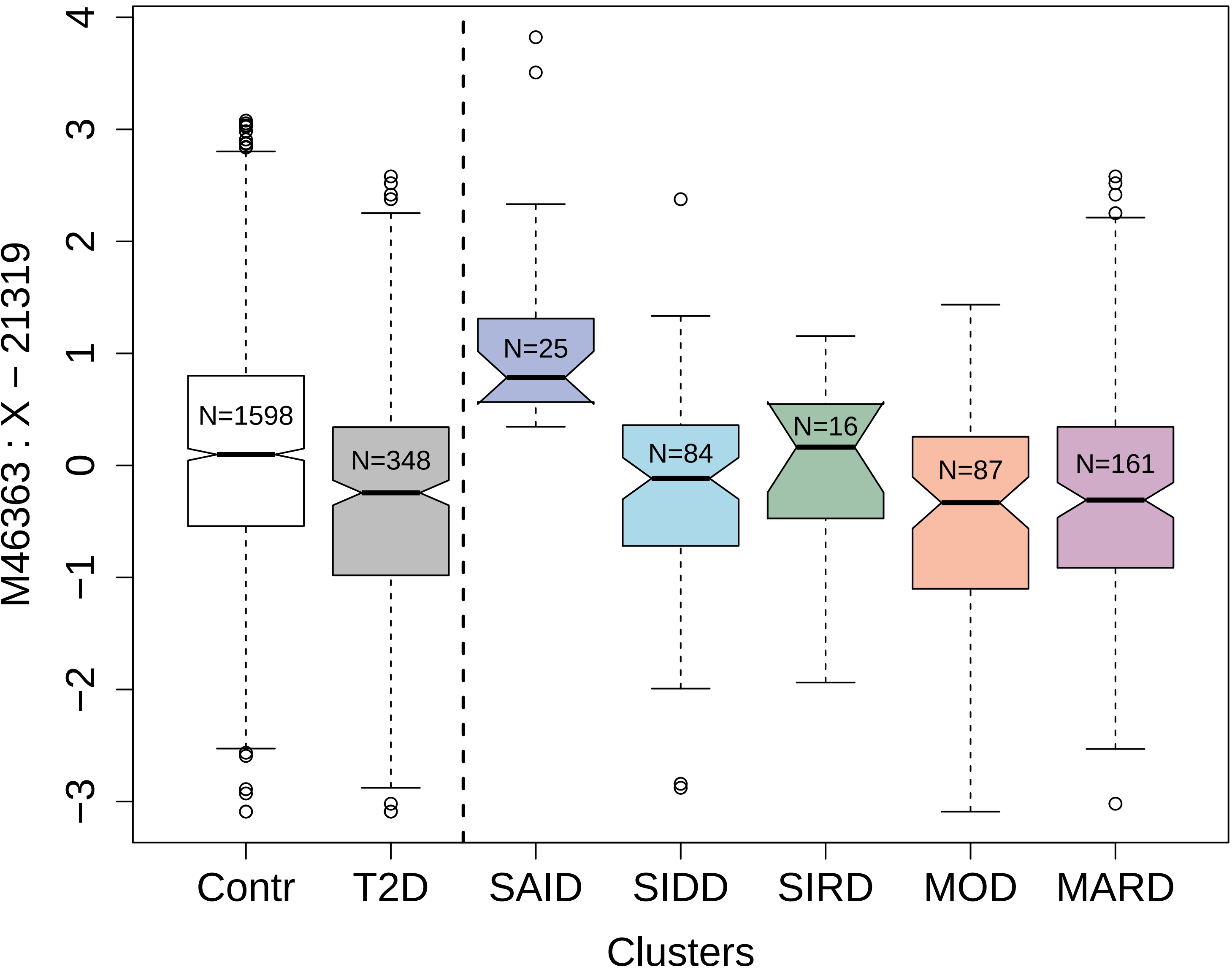

M46363 : X − 21319
4
3
2
1
0
−1
−2
−3
N=1598
N=348
N=25
N=84
N=16
N=87
N=161
Contr
T2D
SAID
SIDD
SIRD
MOD
MARD
Clusters

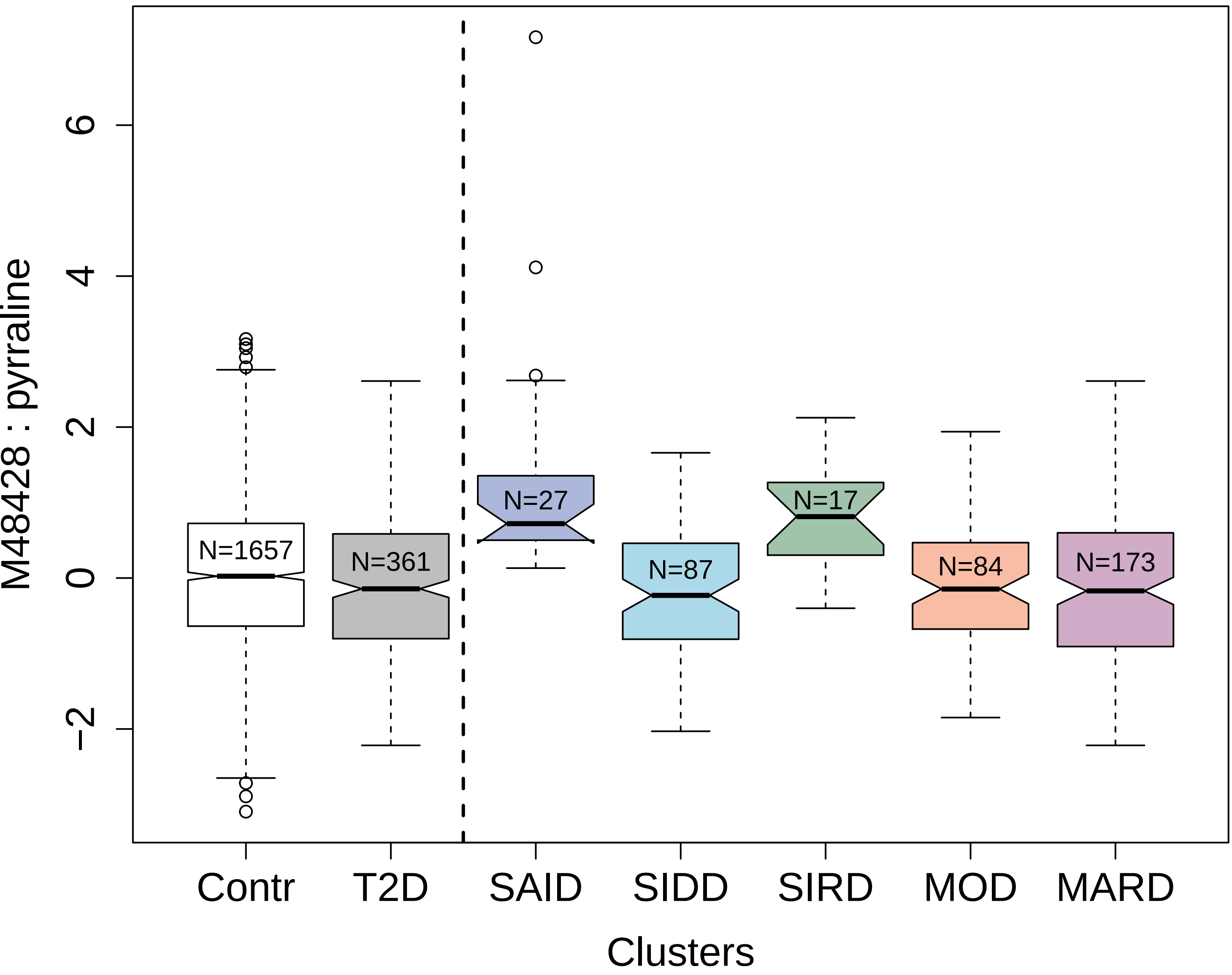

M48428 : pyrraline
Clusters
Contr
T2D
SAID
SIDD
SIRD
MOD
MARD
N=1657
N=361
N=27
N=87
N=17
N=84
N=173
6
4
2
0
−2

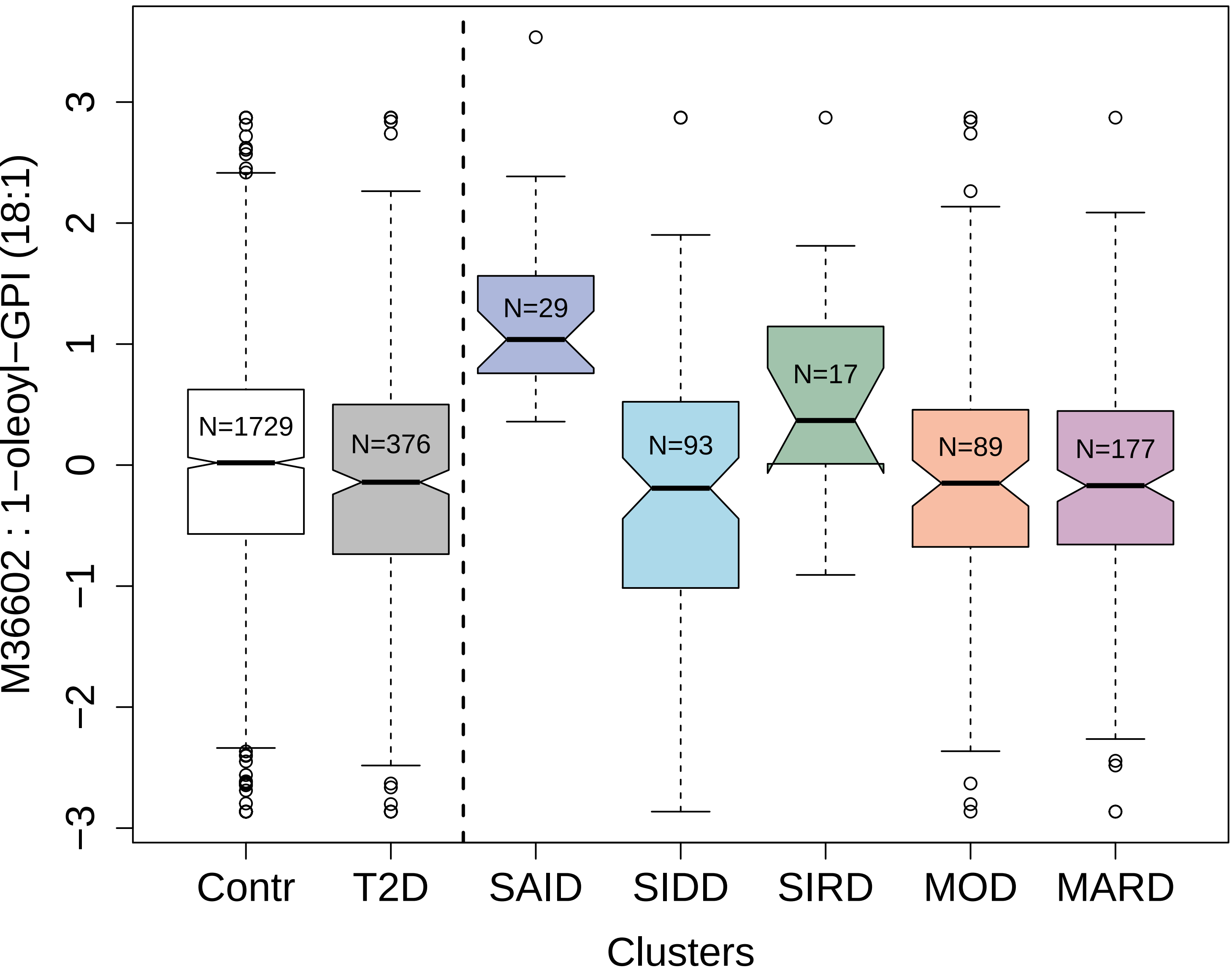

M36602 : 1−oleoyl−GPI (18:1)
3
2
1
0
−1
−2
−3
N=1729
N=376
N=29
N=93
N=17
N=89
N=177
Contr
T2D
SAID
SIDD
SIRD
MOD
MARD
Clusters

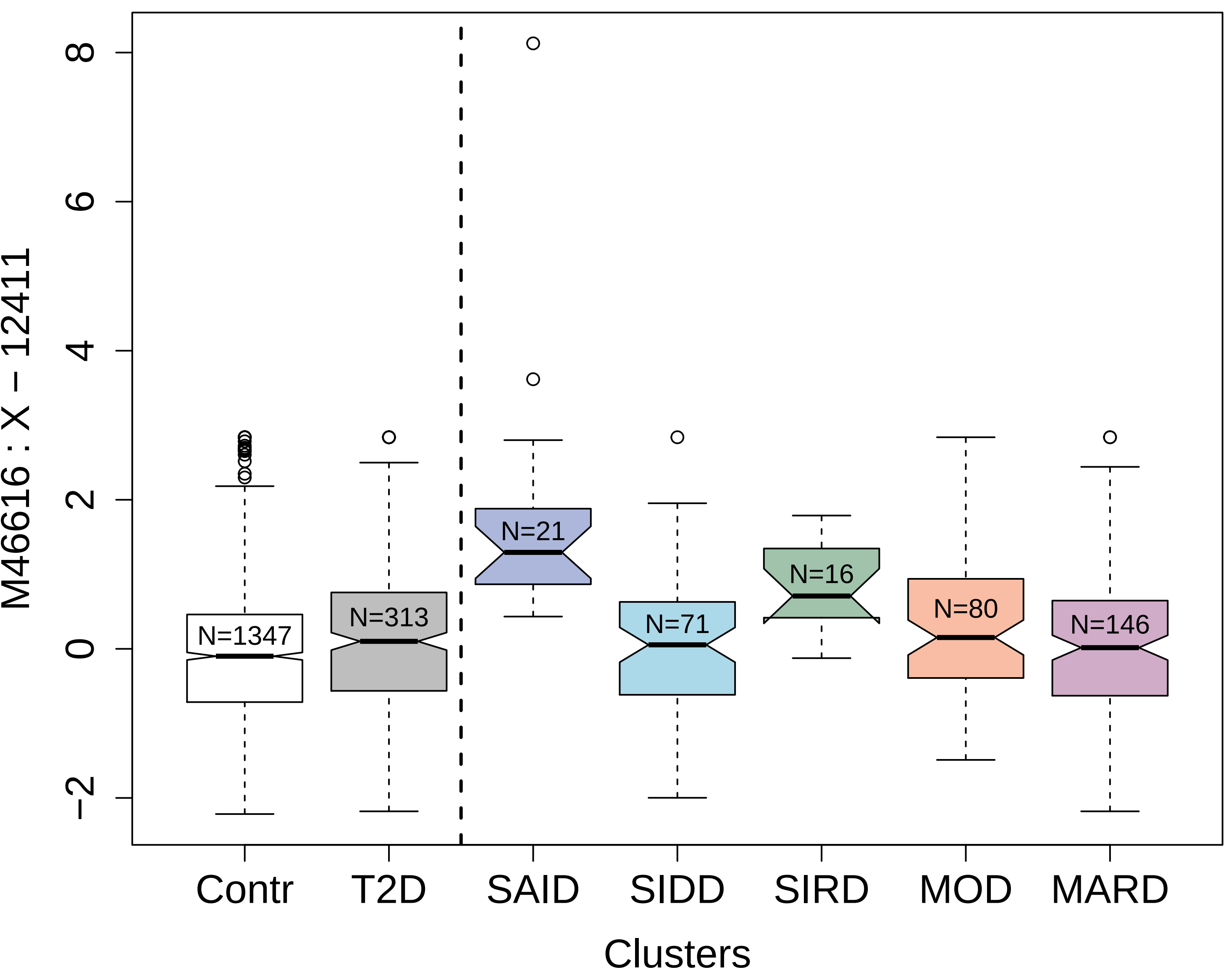

M46616 : X − 12411
8
6
4
2
0
−2
N=1347
N=313
N=21
N=71
N=16
N=80
N=146
Contr
T2D
SAID
SIDD
SIRD
MOD
MARD
Clusters

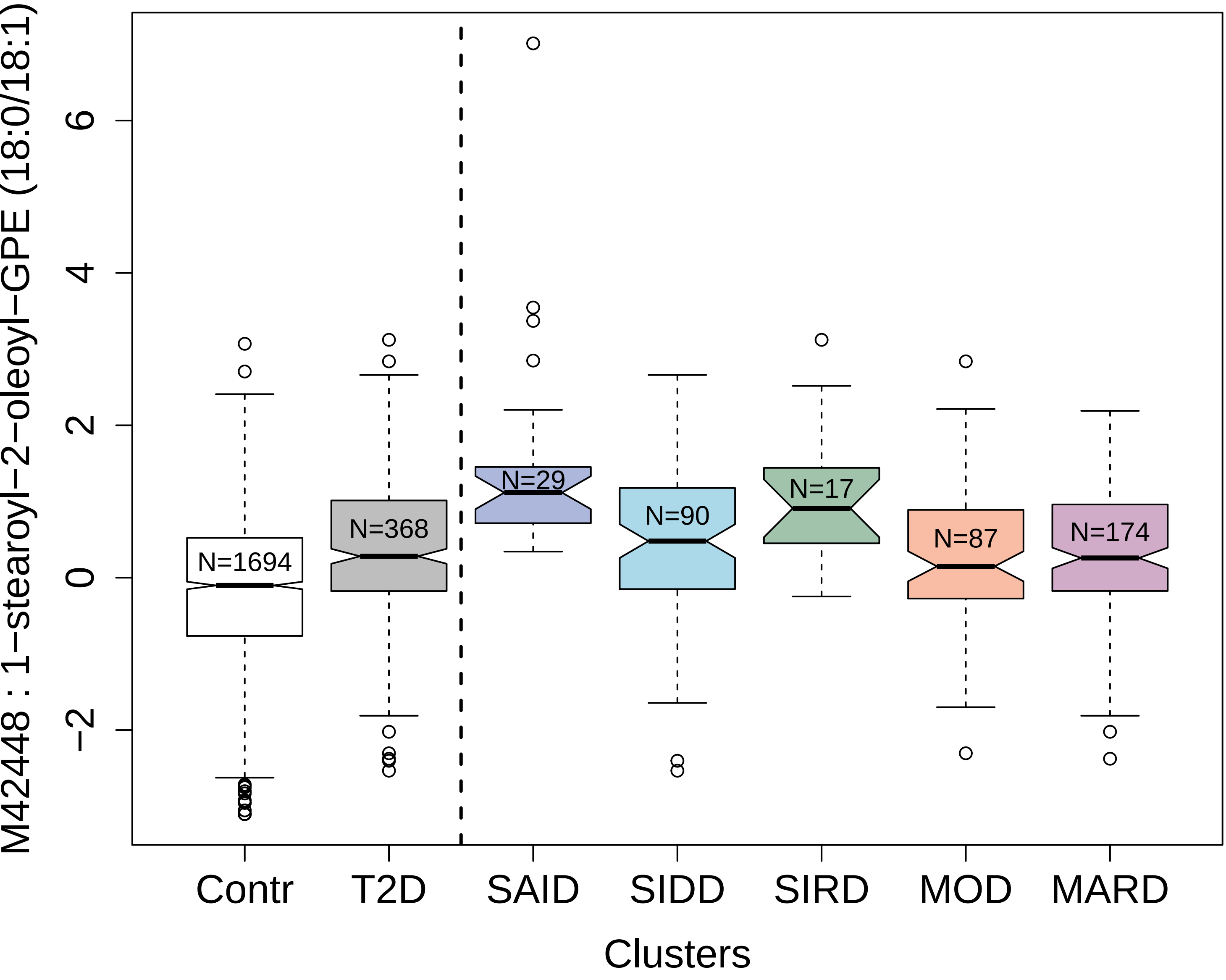
M42448 : 1−stearoyl−2−oleoyl−GPE (18:0/18:1)
6
4
2
0
−2
N=1694
N=368
N=29
N=90
N=17
N=87
N=174
Contr
T2D
SAID
SIDD
SIRD
MOD
MARD
Clusters

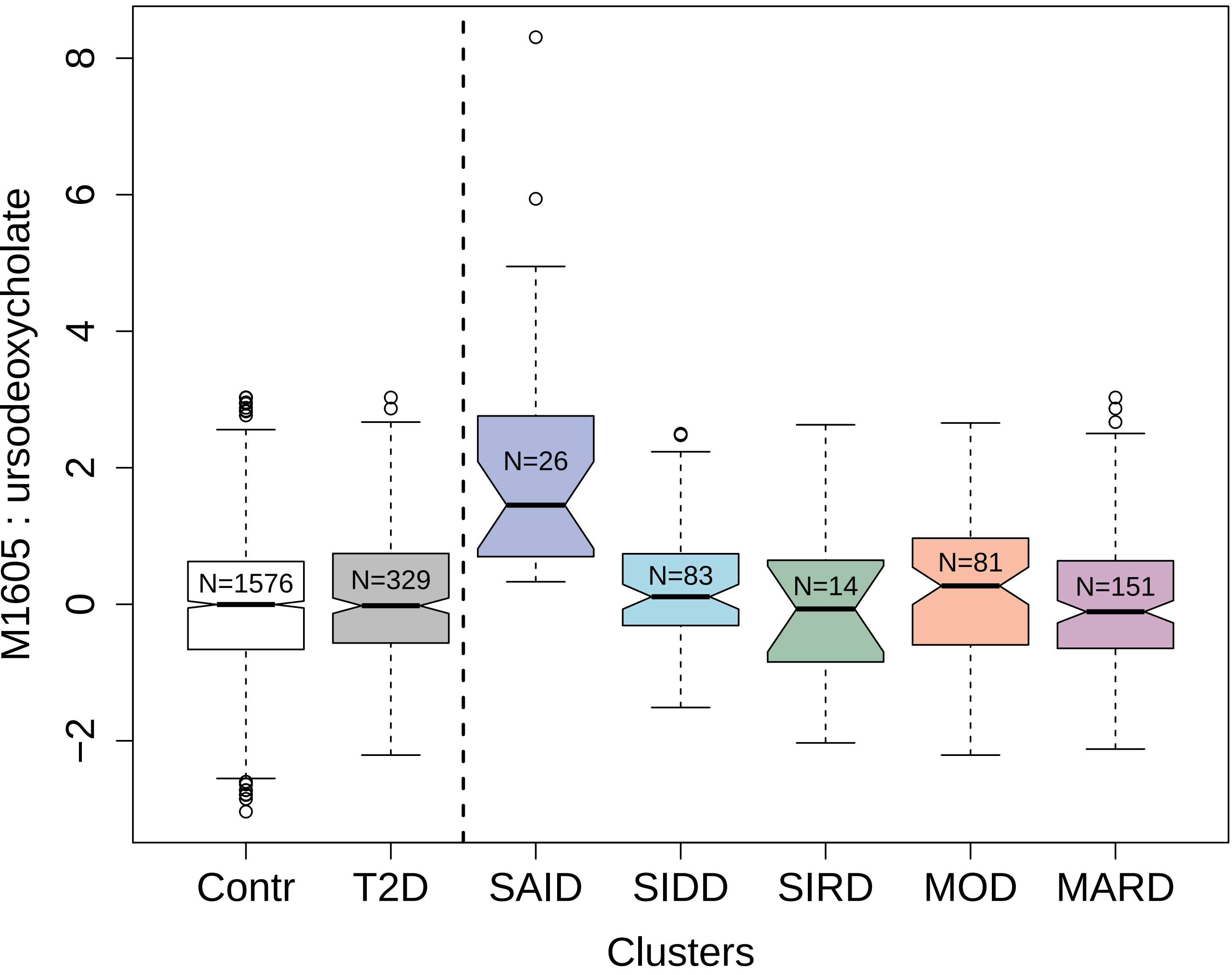

M1605 : ursodeoxycholate
Clusters
Contr
T2D
SAID
SIDD
SIRD
MOD
MARD
N=1576
N=329
N=26
N=83
N=14
N=81
N=151
−2
0
2
4
6
8

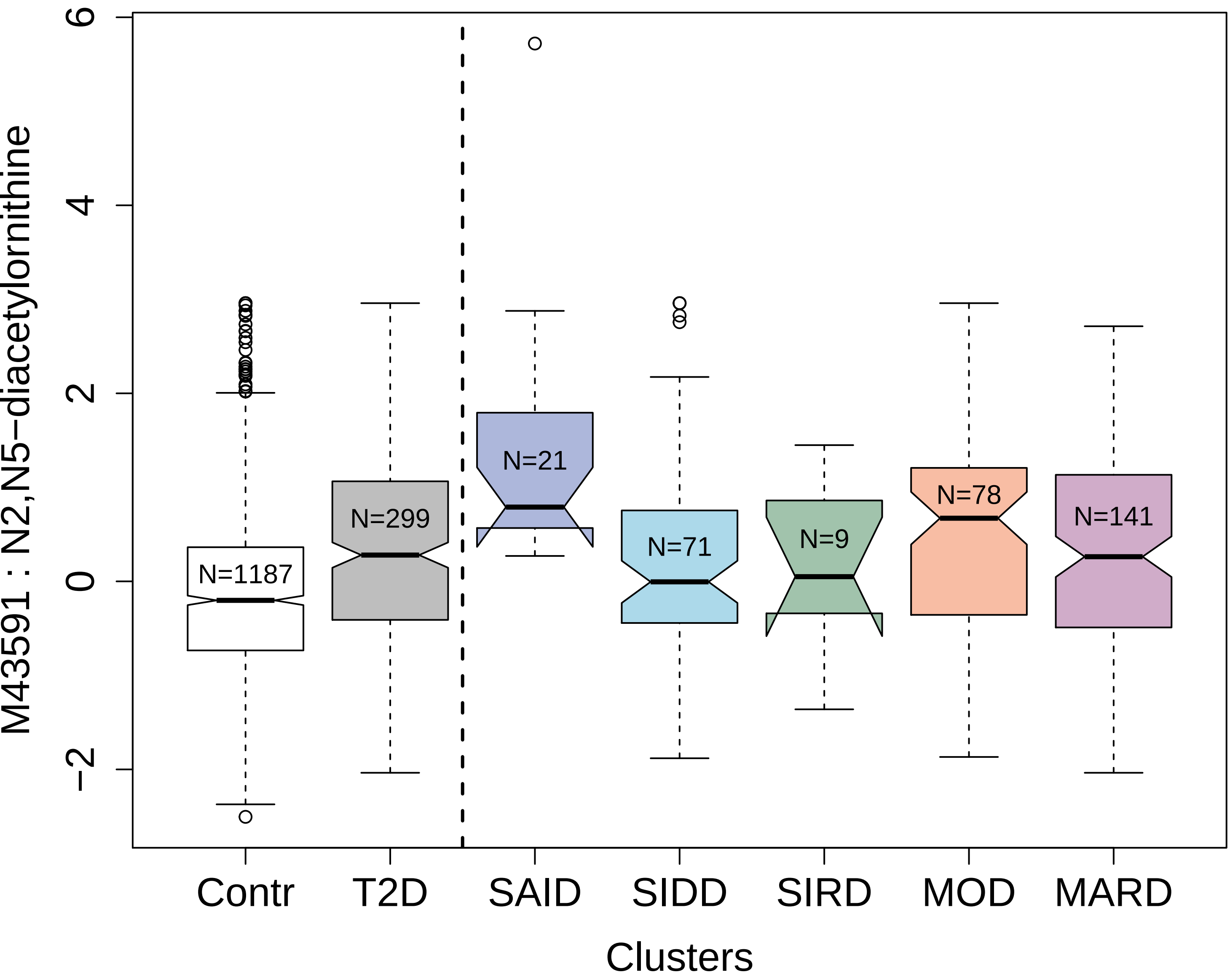

M43591 : N2,N5−diacetylornithine
6
4
2
0
−2
N=1187
N=299
N=21
N=71
N=9
N=78
N=141
Contr
T2D
SAID
SIDD
SIRD
MOD
MARD
Clusters

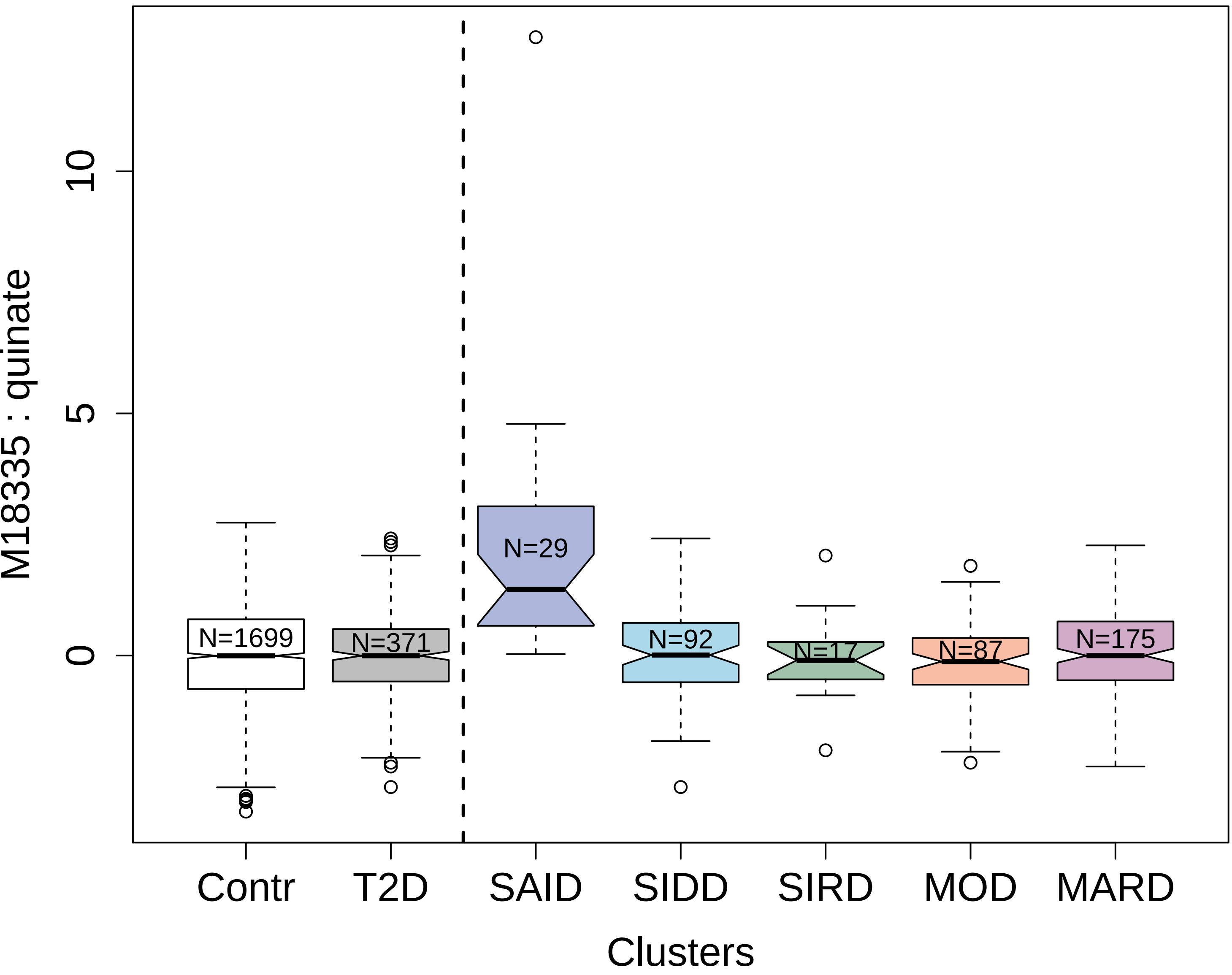

M18335 : quinate
0
5
10
N=1699
N=371
N=29
N=92
N=17
N=87
N=175
Contr
T2D
SAID
SIDD
SIRD
MOD
MARD
Clusters

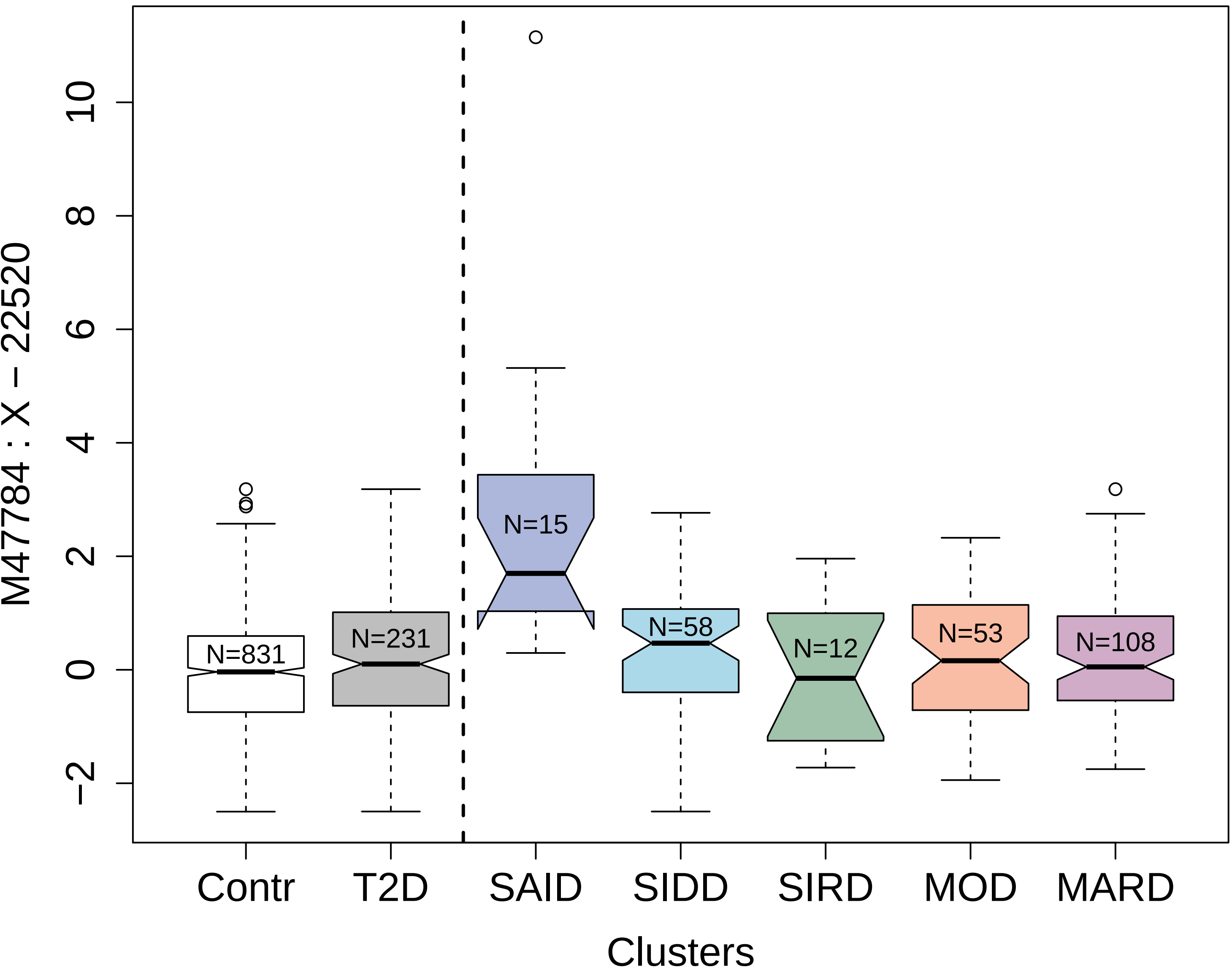
M47784 : X − 22520
10
8
6
4
2
0
−2
N=831
N=231
N=15
N=58
N=12
N=53
N=108
Contr
T2D
SAID
SIDD
SIRD
MOD
MARD
Clusters

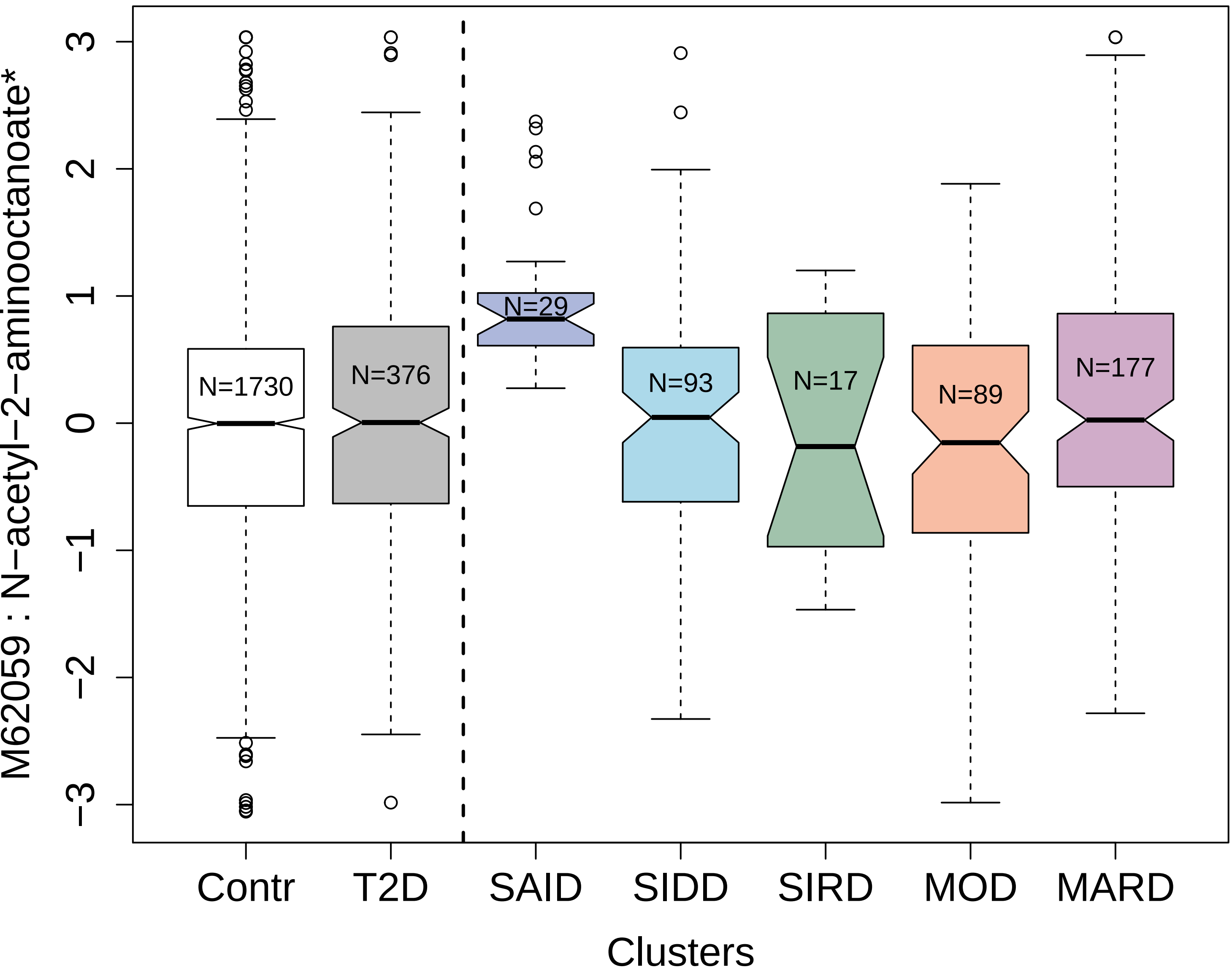

M62059 : N−acetyl−2−aminooctanoate*
3
2
1
0
−1
−2
−3
N=1730
N=376
N=29
N=93
N=17
N=89
N=177
Contr
T2D
SAID
SIDD
SIRD
MOD
MARD
Clusters

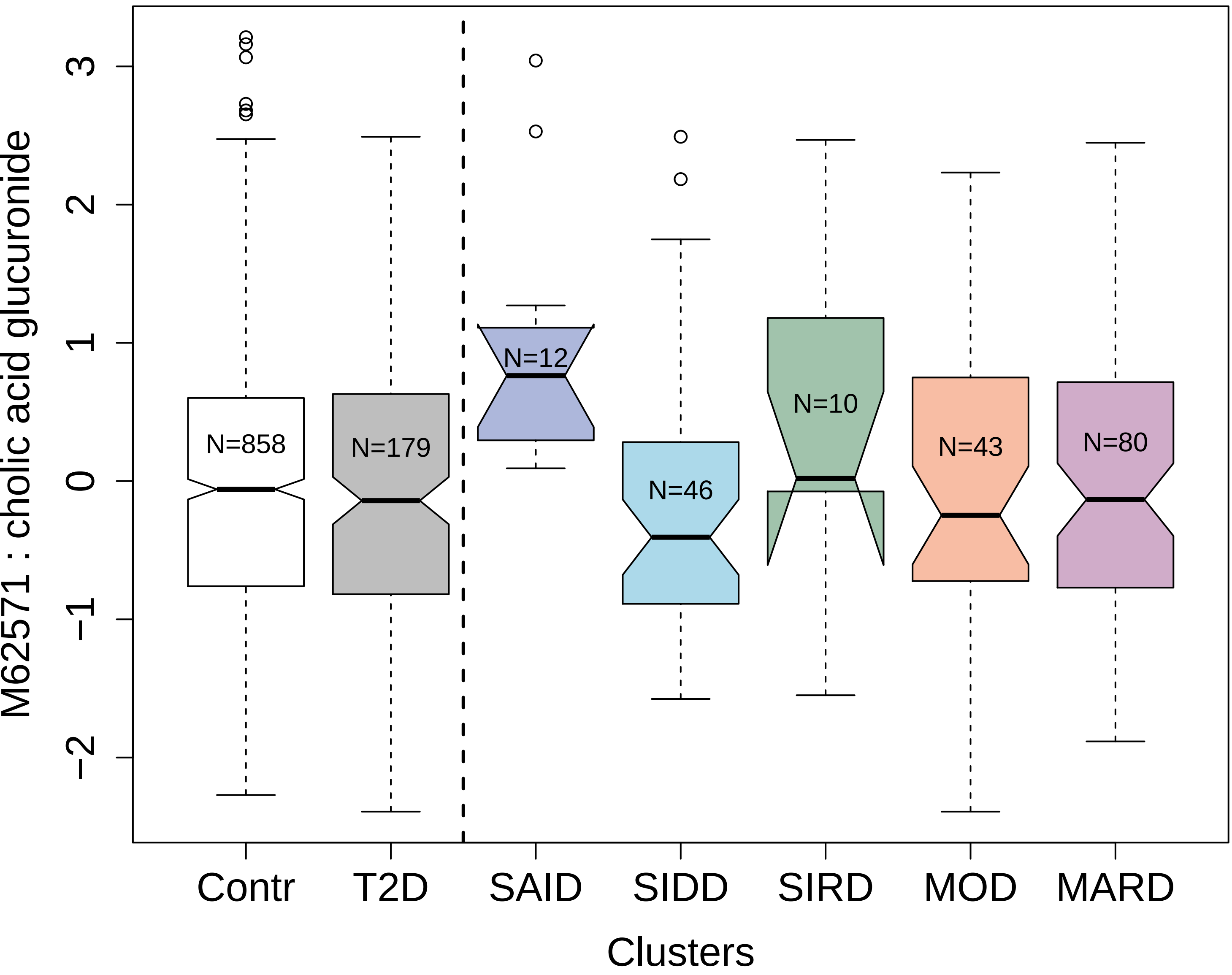

M62571 : cholic acid glucuronide
3
2
1
0
−1
−2
N=858
N=179
N=12
N=46
N=10
N=43
N=80
Contr
T2D
SAID
SIDD
SIRD
MOD
MARD
Clusters

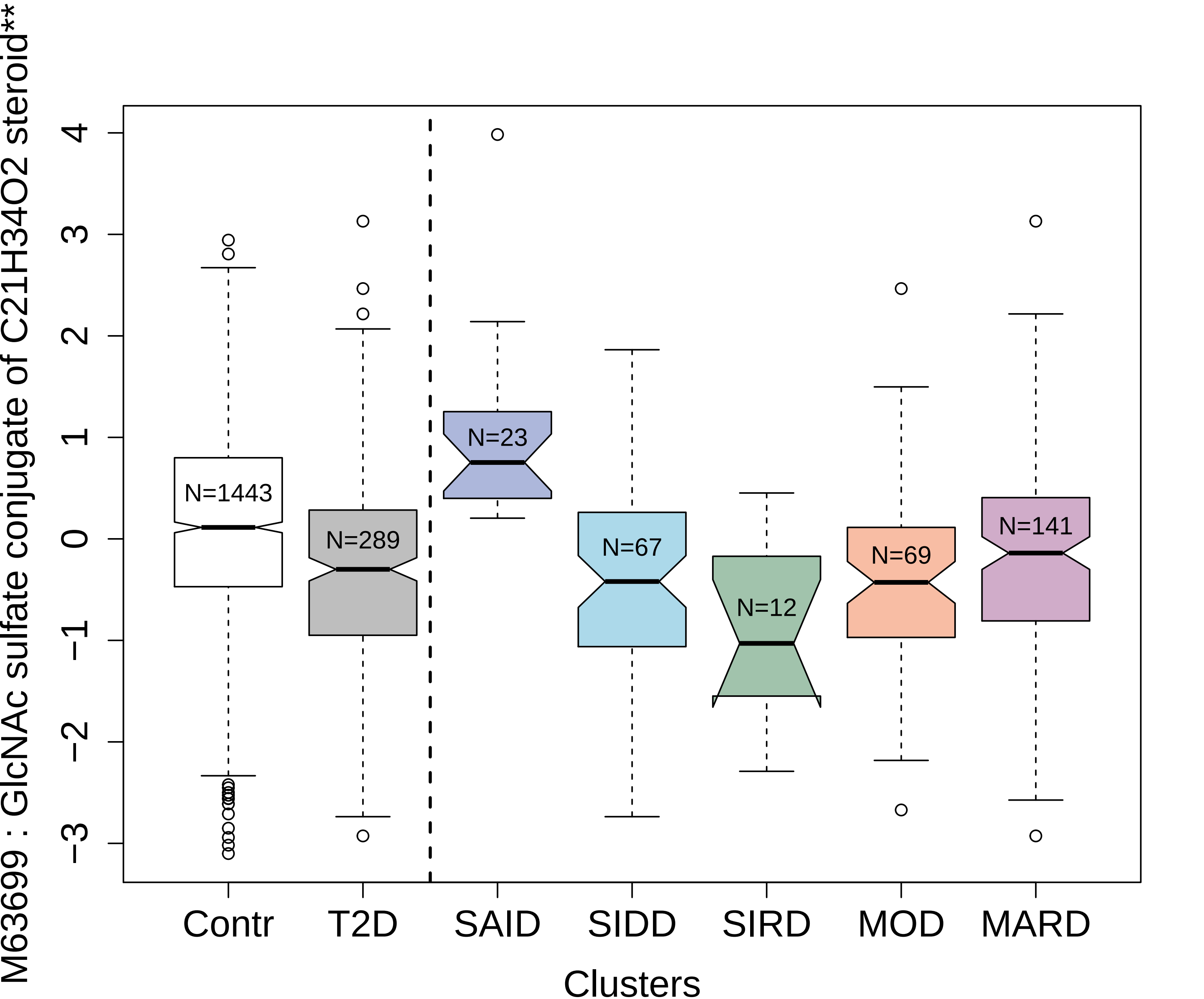

M63699 : GlcNAc sulfate conjugate of C21H34O2 steroid**
4
3
2
1
0
−1
−2
−3
N=1443
N=289
N=23
N=67
N=12
N=69
N=141
Contr
T2D
SAID
SIDD
SIRD
MOD
MARD
Clusters

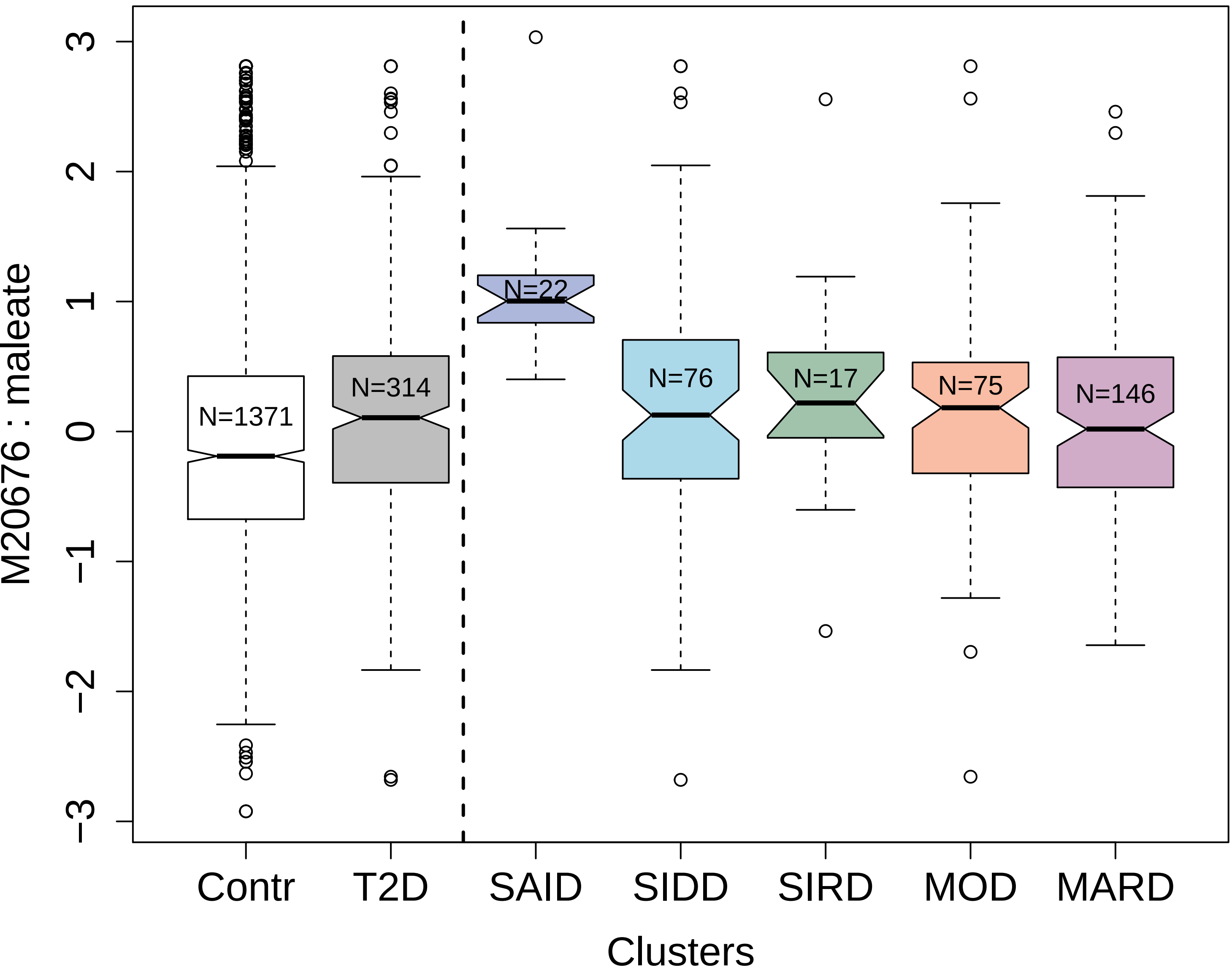

M20676 : maleate
3
2
1
0
−1
−2
−3
N=1371
N=314
N=22
N=76
N=17
N=75
N=146
Contr
T2D
SAID
SIDD
SIRD
MOD
MARD
Clusters

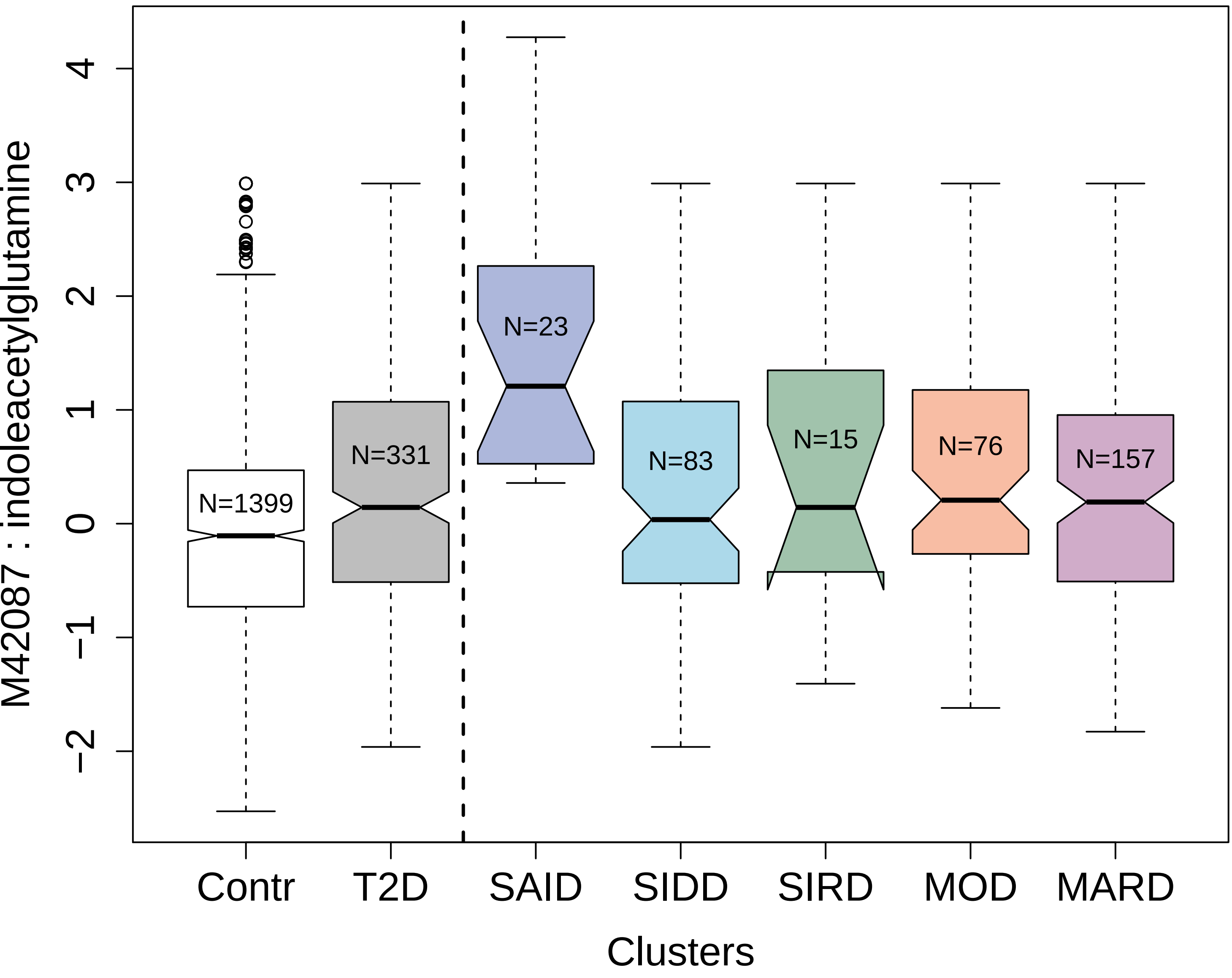

M42087 : indoleacetylglutamine
4
3
2
1
0
−1
−2
N=1399
N=331
N=23
N=83
N=15
N=76
N=157
Contr
T2D
SAID
SIDD
SIRD
MOD
MARD
Clusters

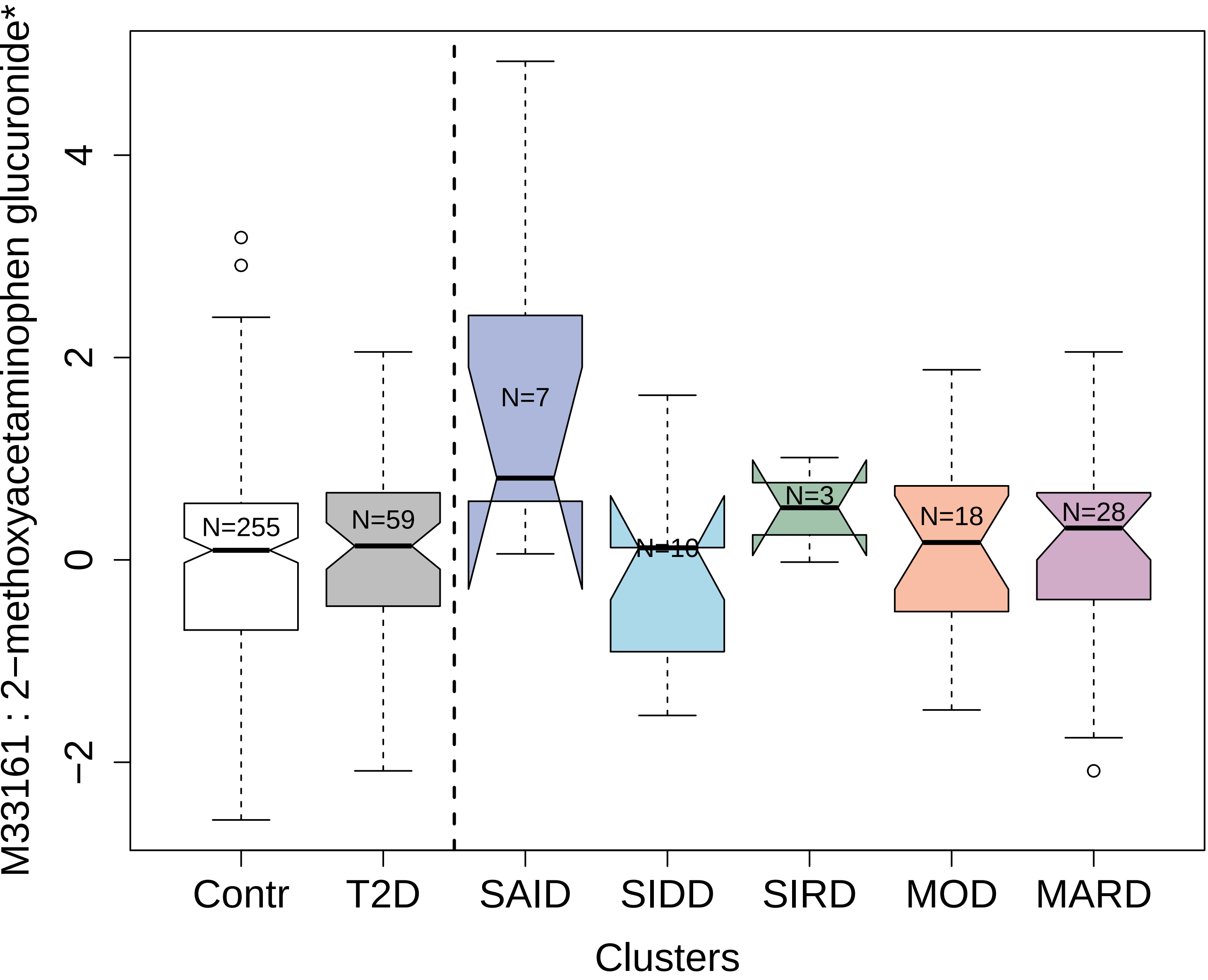
M33161 : 2−methoxyacetaminophen glucuronide*
−2
0
2
4
N=255
N=59
N=7
N=10
N=3
N=18
N=28
Contr
T2D
SAID
SIDD
SIRD
MOD
MARD
Clusters

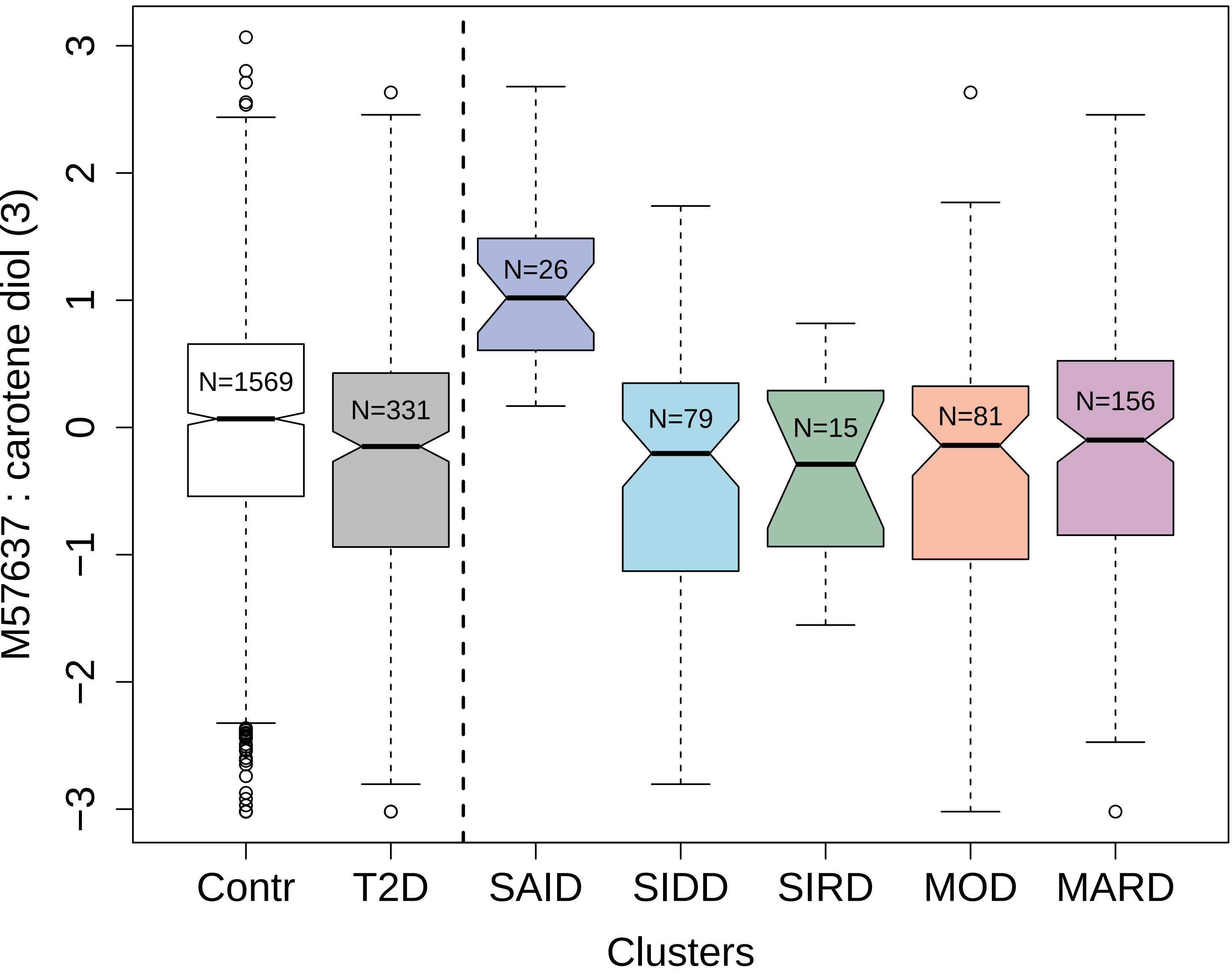
M57637 : carotene diol (3)
3
2
1
0
−1
−2
−3
N=1569
N=331
N=26
N=79
N=15
N=81
N=156
Contr
T2D
SAID
SIDD
SIRD
MOD
MARD
Clusters

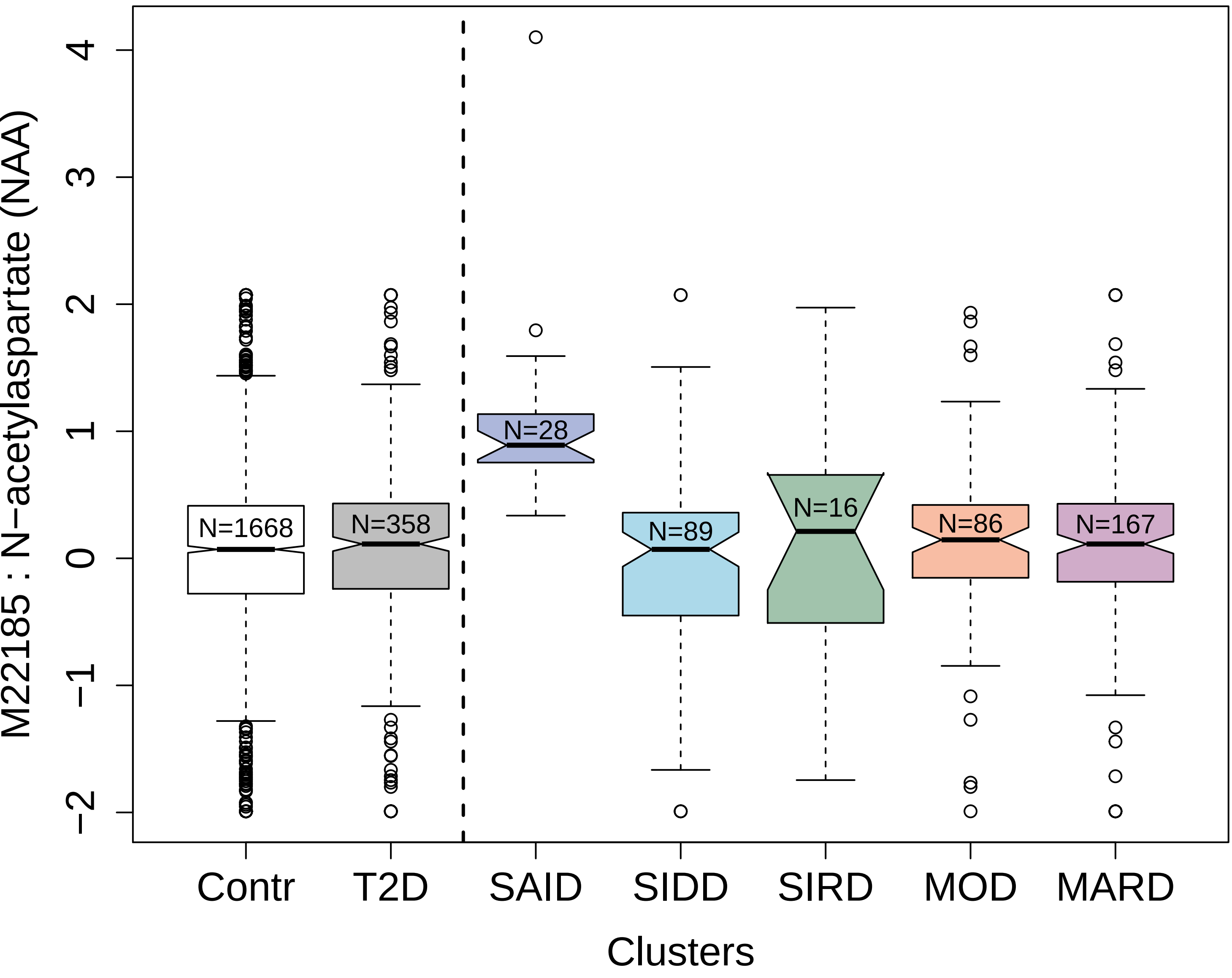

M22185 : N−acetylaspartate (NAA)
4
3
2
1
0
−1
−2
N=1668
N=358
N=28
N=89
N=16
N=86
N=167
Contr
T2D
SAID
SIDD
SIRD
MOD
MARD
Clusters

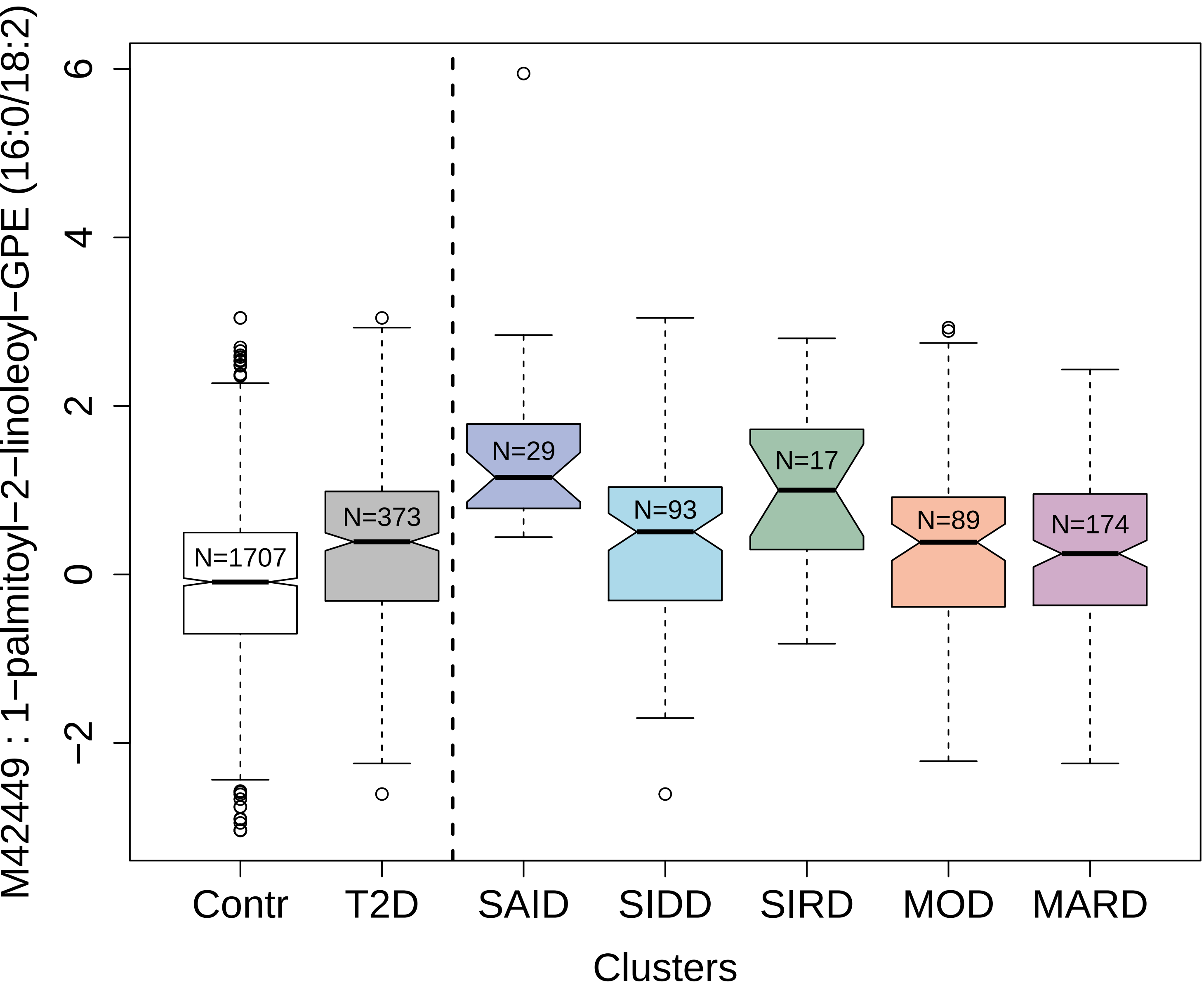
M42449 : 1−palmitoyl−2−linoleoyl−GPE (16:0/18:2)
6
4
2
0
−2
N=1707
N=373
N=29
N=93
N=17
N=89
N=174
Contr
T2D
SAID
SIDD
SIRD
MOD
MARD
Clusters

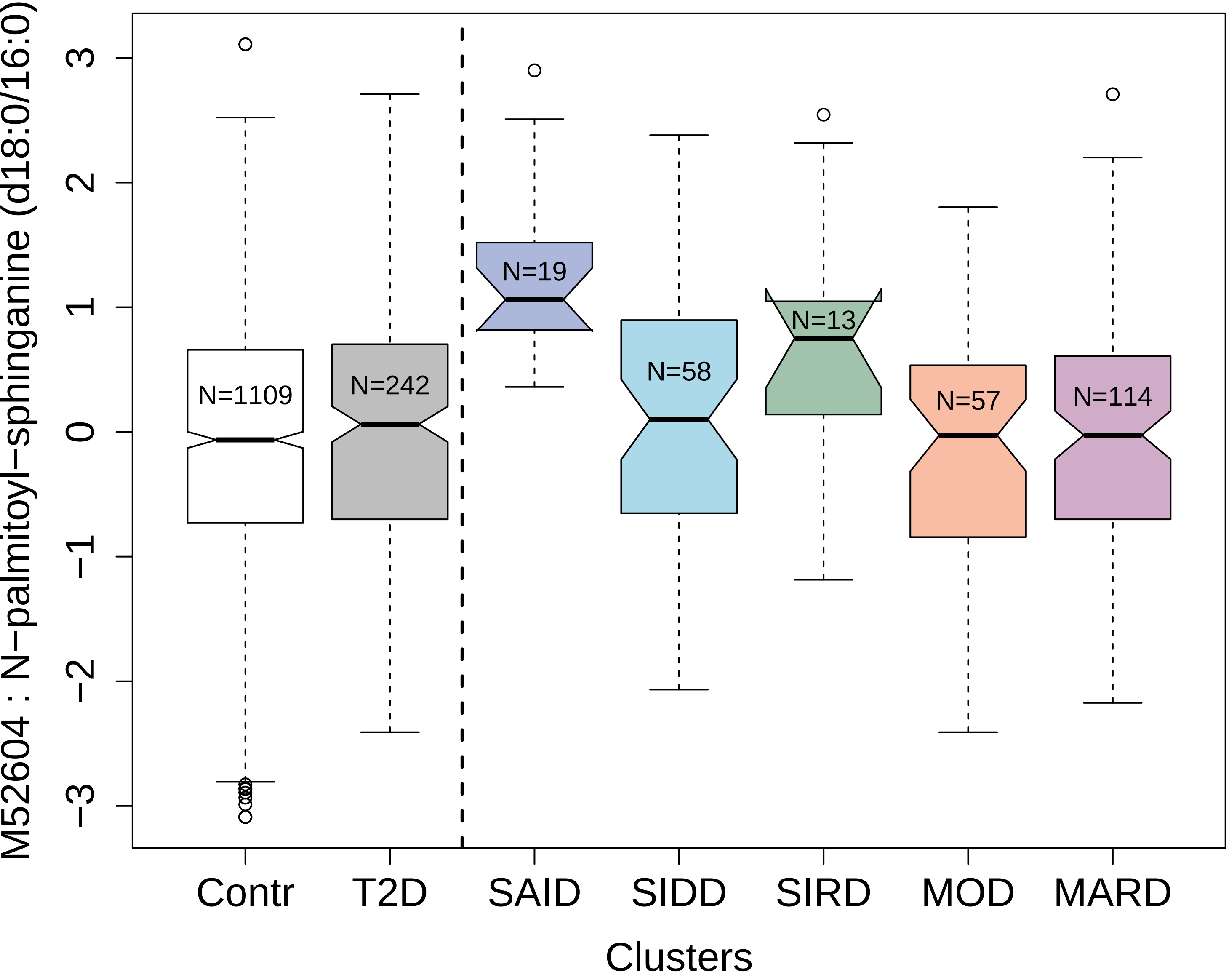

M52604 : N−palmitoyl−sphinganine (d18:0/16:0)
3
2
1
0
−1
−2
−3
N=1109
N=242
N=19
N=58
N=13
N=57
N=114
Contr
T2D
SAID
SIDD
SIRD
MOD
MARD
Clusters

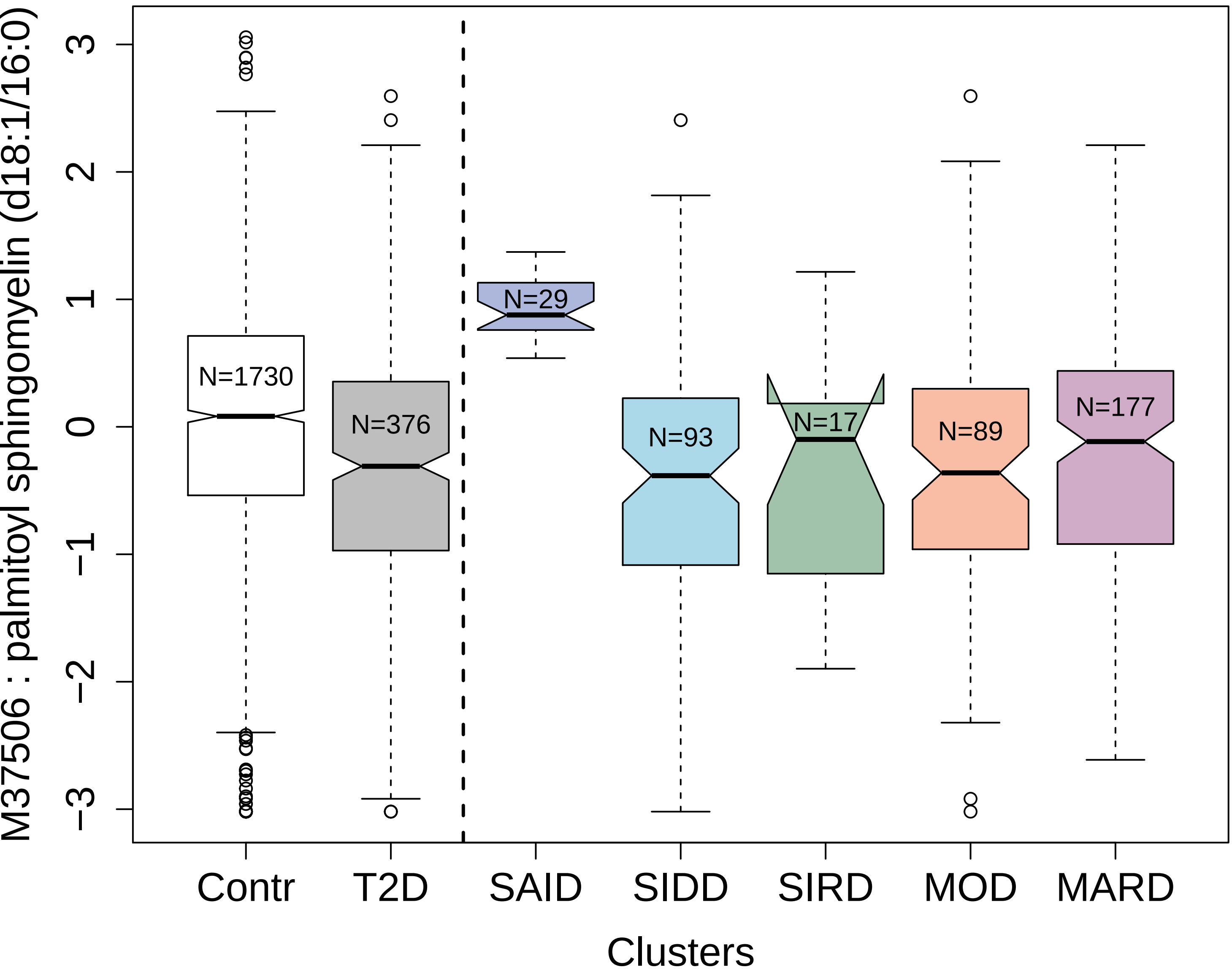
M37506 : palmitoyl sphingomyelin (d18:1/16:0)
3
2
1
0
−1
−2
−3
N=1730
N=376
N=29
N=93
N=17
N=89
N=177
Contr
T2D
SAID
SIDD
SIRD
MOD
MARD
Clusters

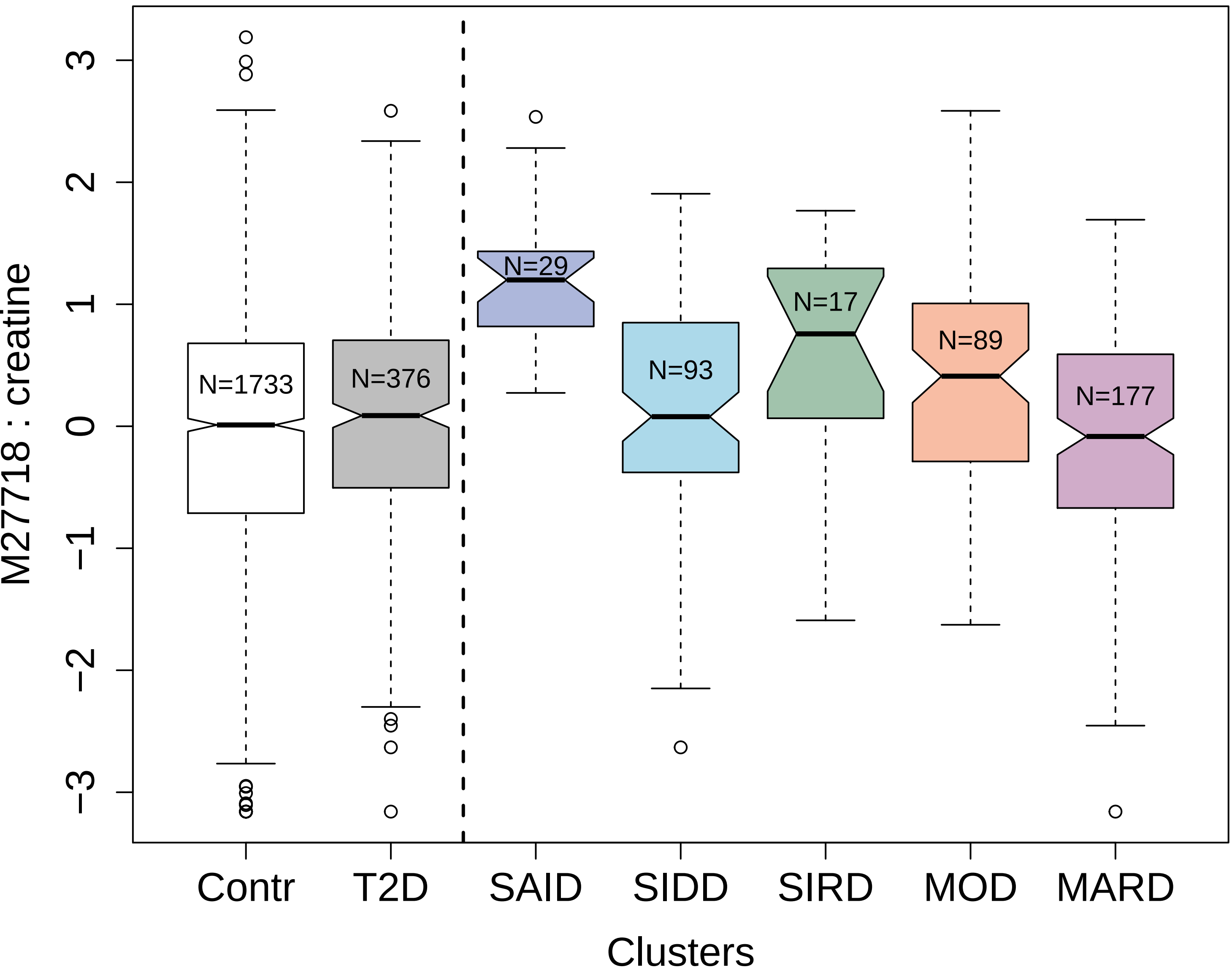
M27718 : creatine
3
2
1
0
−1
−2
−3
N=1733
N=376
N=29
N=93
N=17
N=89
N=177
Contr
T2D
SAID
SIDD
SIRD
MOD
MARD
Clusters

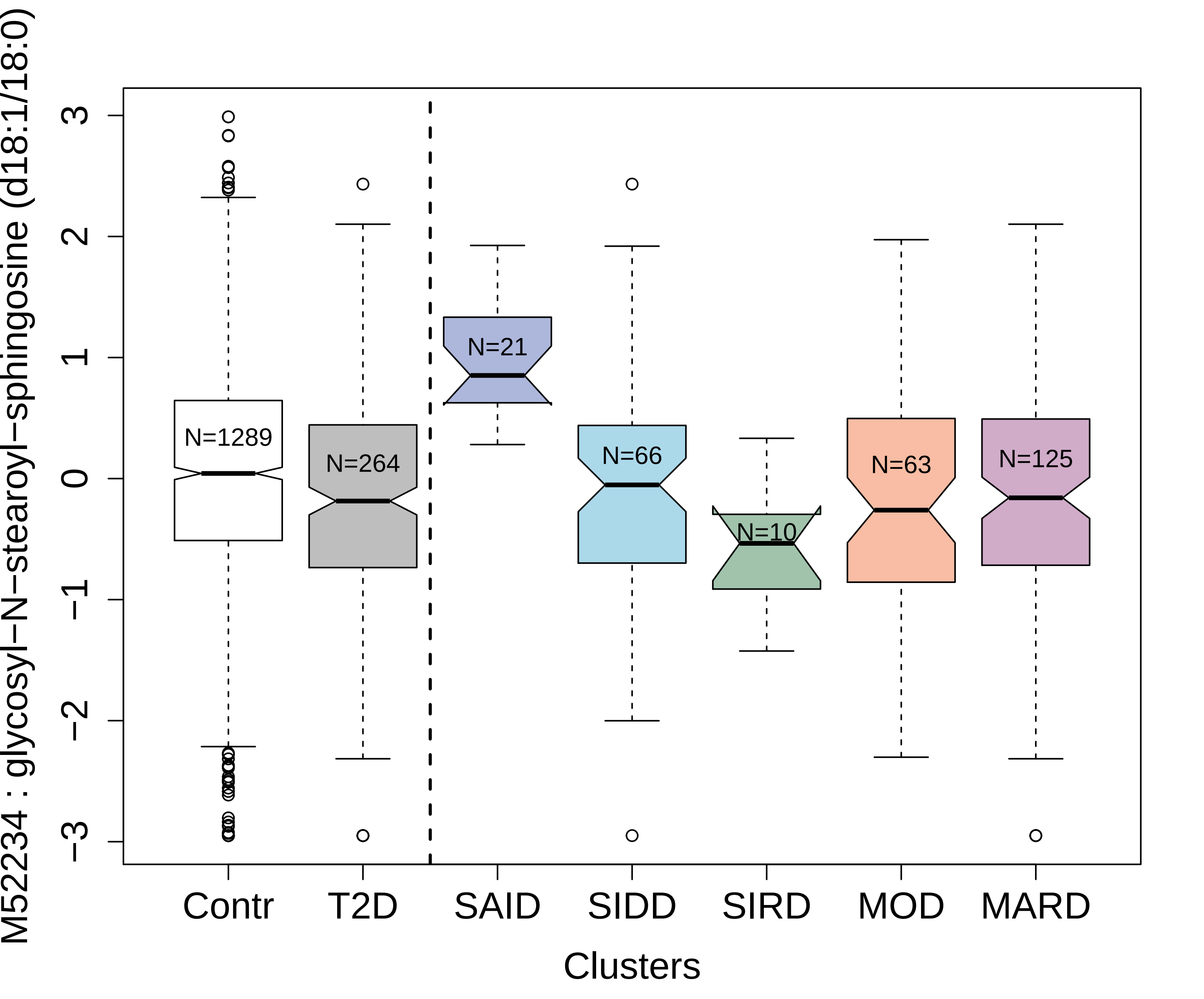

M52234 : glycosyl−N−stearoyl−sphingosine (d18:1/18:0)
3
2
1
0
−1
−2
−3
N=1289
N=264
N=21
N=66
N=10
N=63
N=125
Contr
T2D
SAID
SIDD
SIRD
MOD
MARD
Clusters

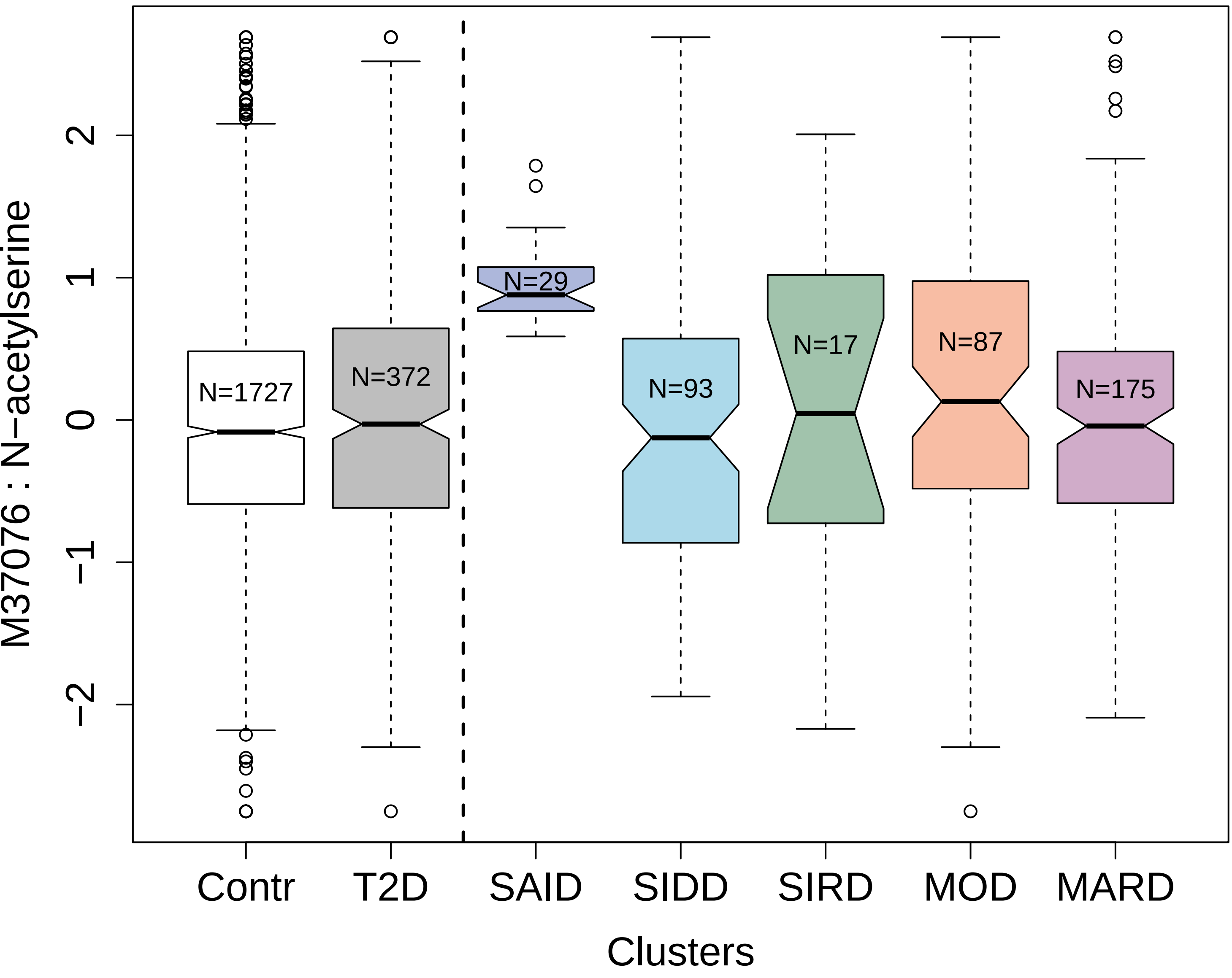
M37076 : N−acetylserine
N=1727
N=372
N=29
N=93
N=17
N=87
N=175
2
1
0
−1
−2
Contr
T2D
SAID
SIDD
SIRD
MOD
MARD
Clusters

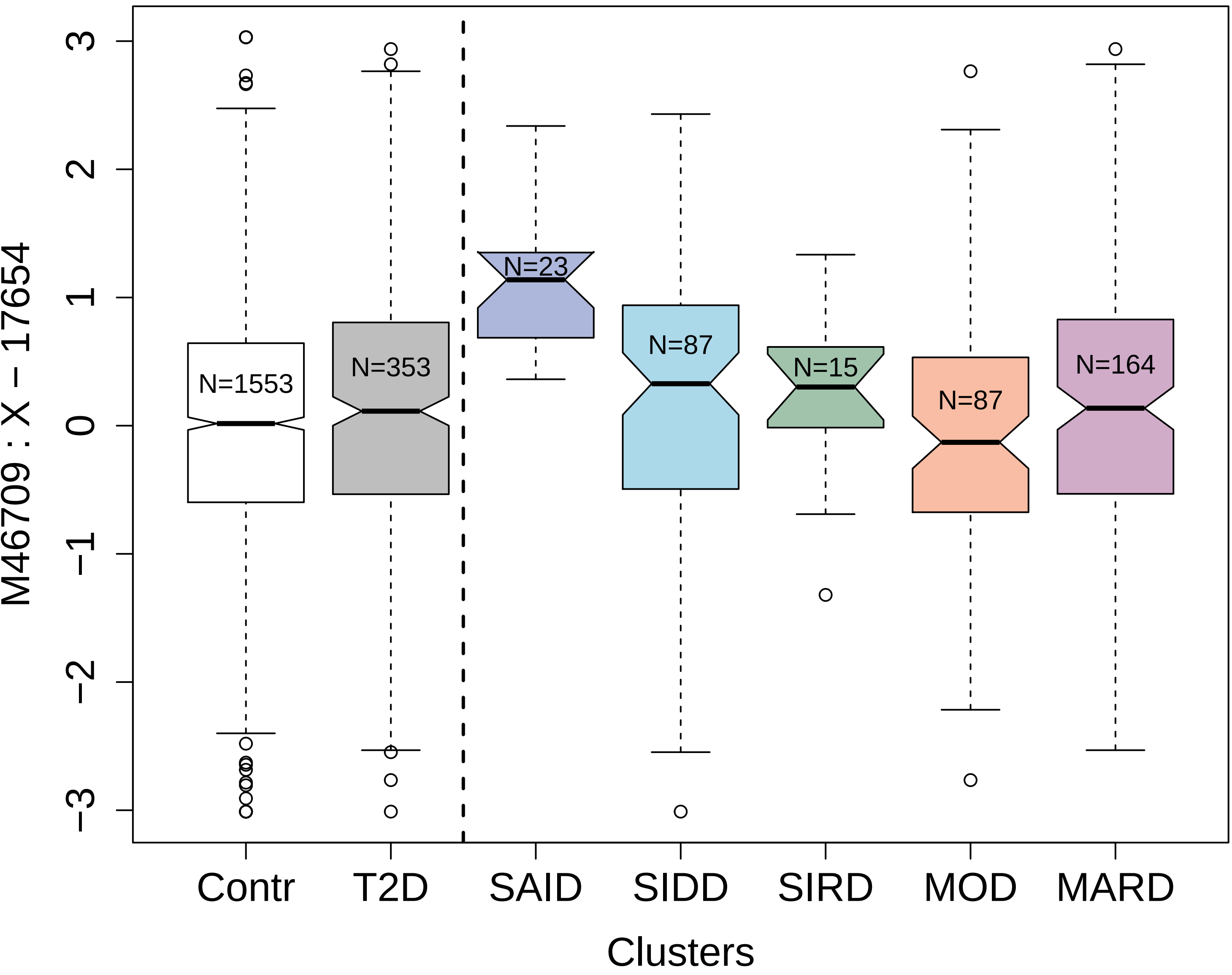

M46709 : X − 17654
3
2
1
0
−1
−2
−3
N=1553
N=353
N=23
N=87
N=15
N=87
N=164
Contr
T2D
SAID
SIDD
SIRD
MOD
MARD
Clusters

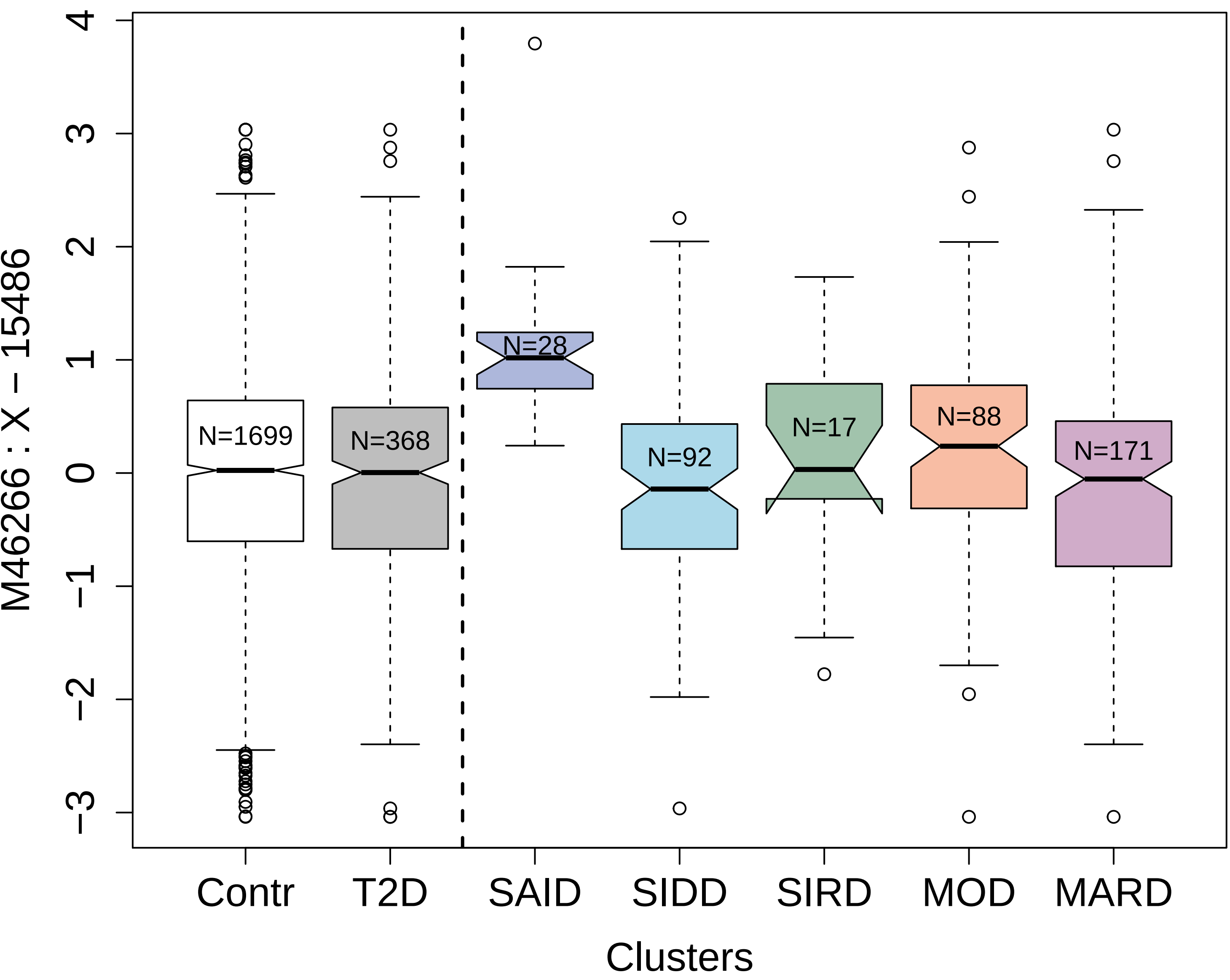

M46266 : X − 15486
4
3
2
1
0
−1
−2
−3
N=1699
N=368
N=28
N=92
N=17
N=88
N=171
Contr
T2D
SAID
SIDD
SIRD
MOD
MARD
Clusters

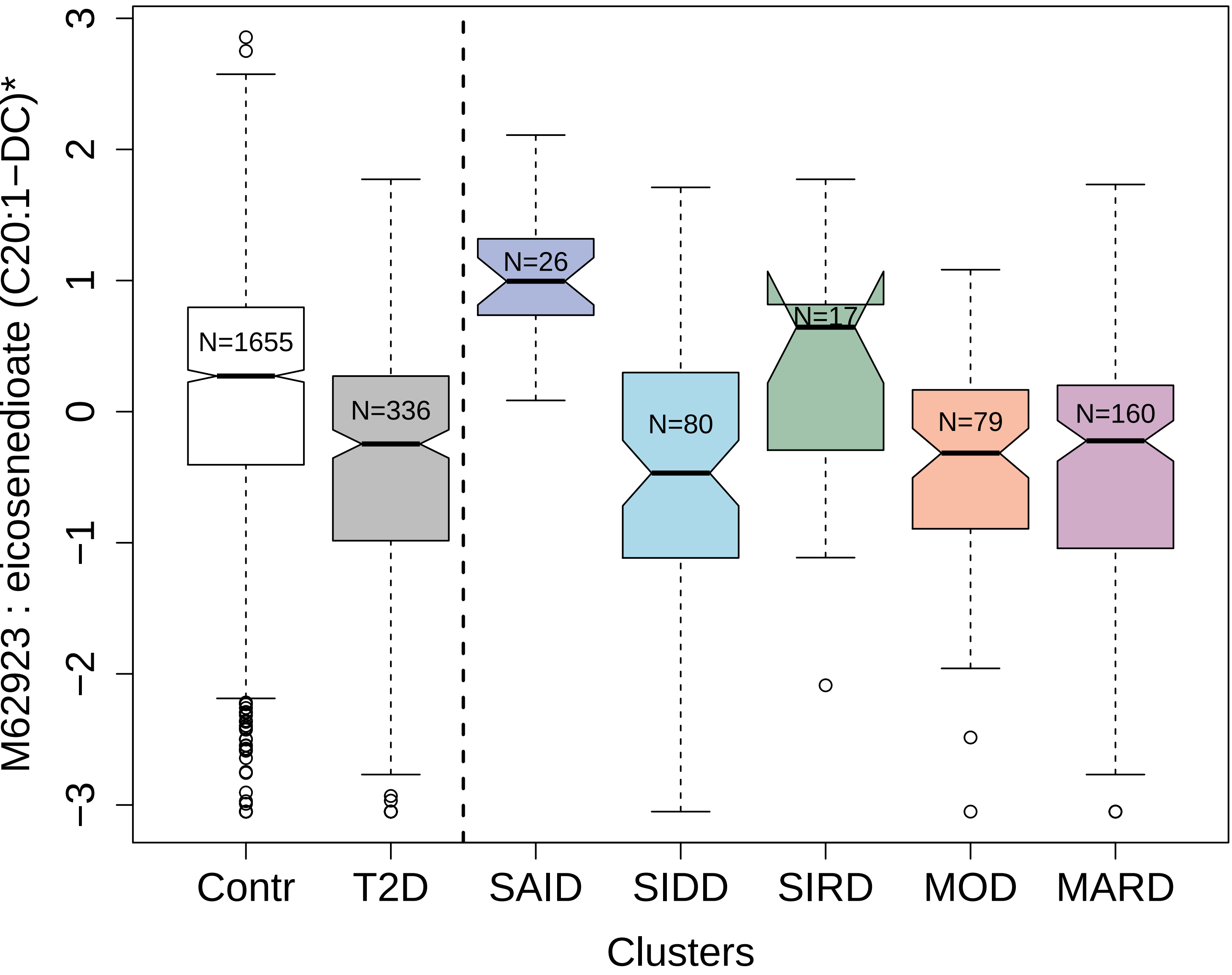

M62923 : eicosenedioate (C20:1−DC)*
3
2
1
0
−1
−2
−3
N=1655
N=336
N=26
N=80
N=17
N=79
N=160
Contr
T2D
SAID
SIDD
SIRD
MOD
MARD
Clusters

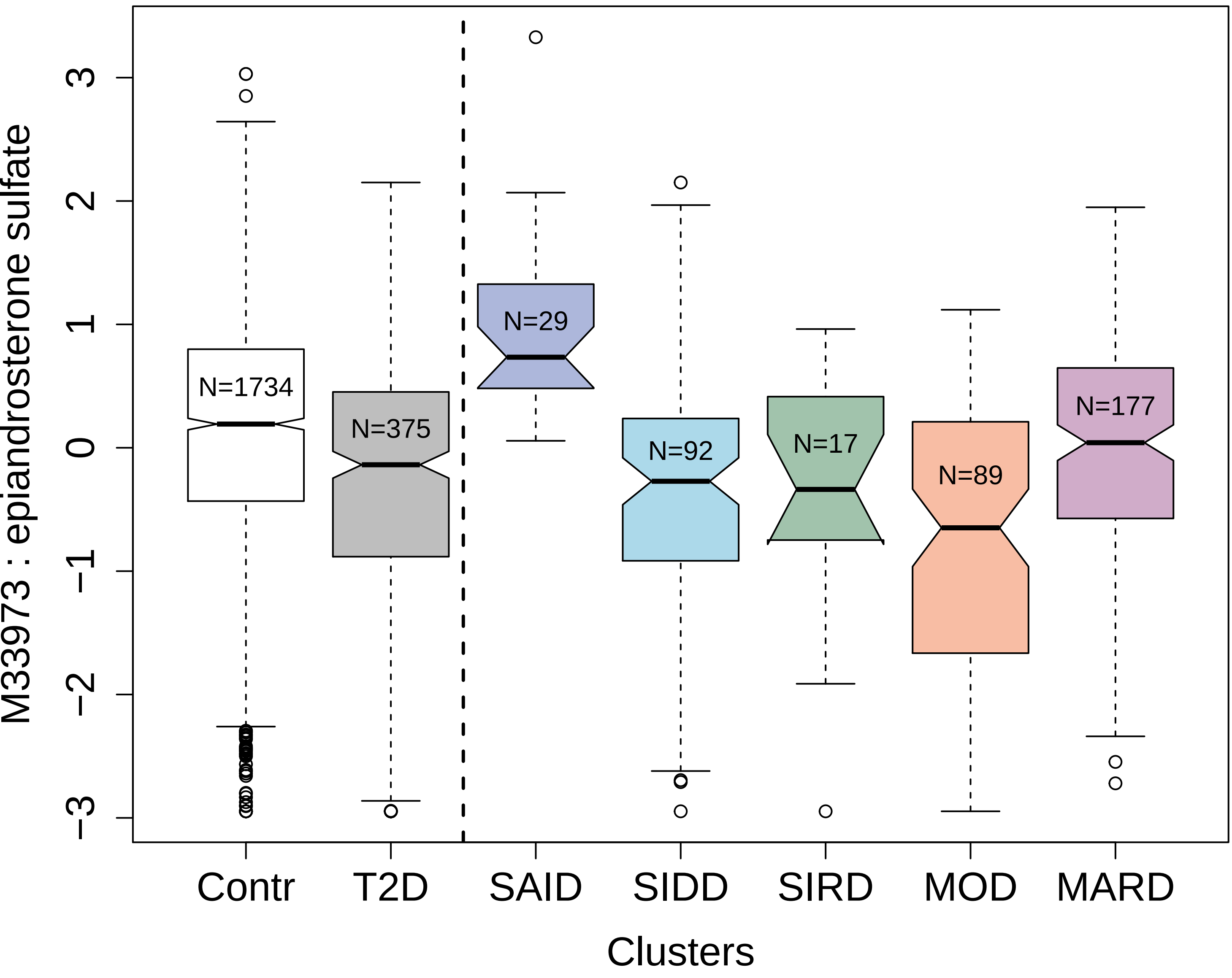

M33973 : epiandrosterone sulfate
3
2
1
0
−1
−2
−3
N=1734
N=375
N=29
N=92
N=17
N=89
N=177
Contr
T2D
SAID
SIDD
SIRD
MOD
MARD
Clusters

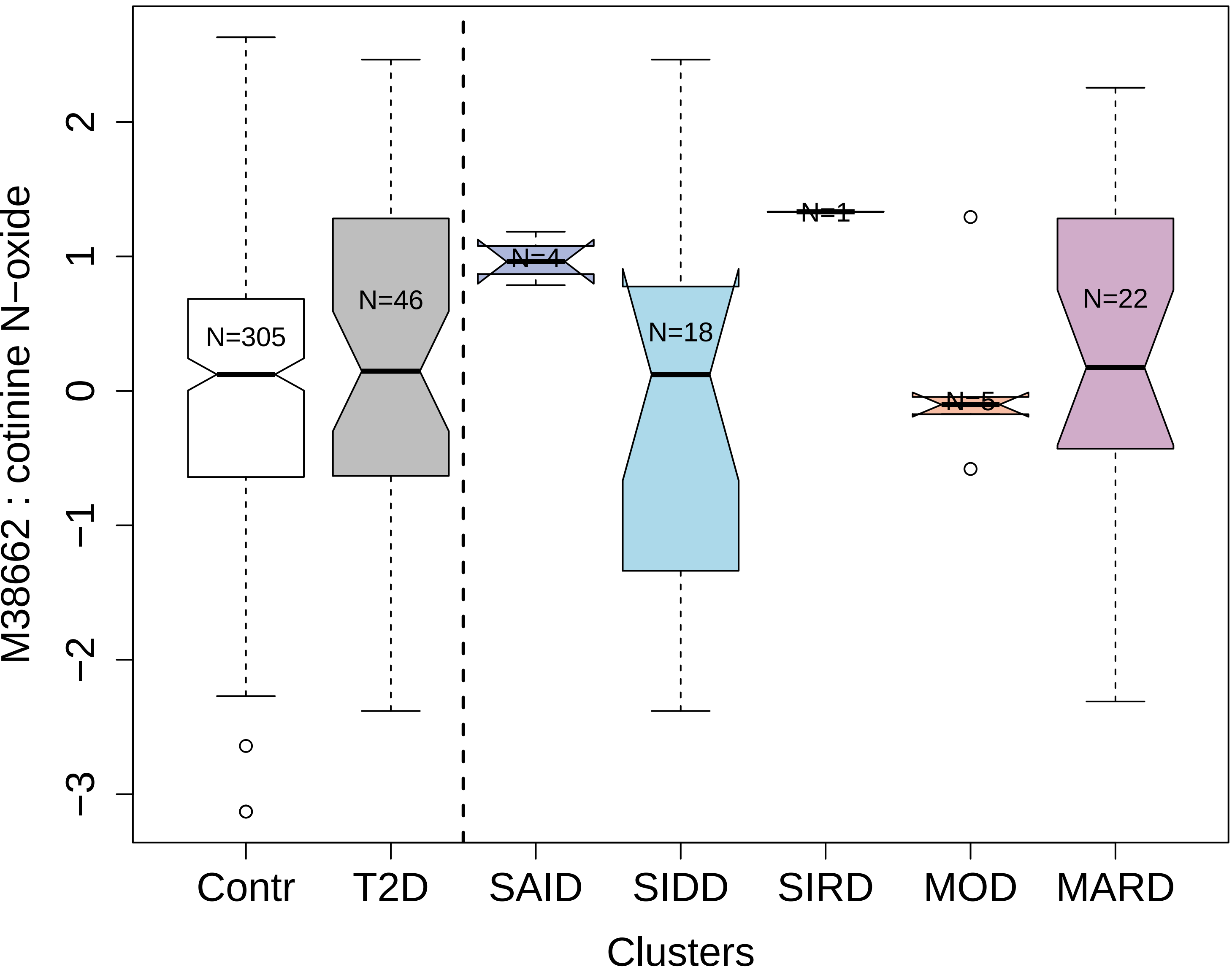

M38662 : cotinine N−oxide
−3
−2
−1
0
1
2
N=305
N=46
N=4
N=18
N=1
N=5
N=22
Contr
T2D
SAID
SIDD
SIRD
MOD
MARD
Clusters

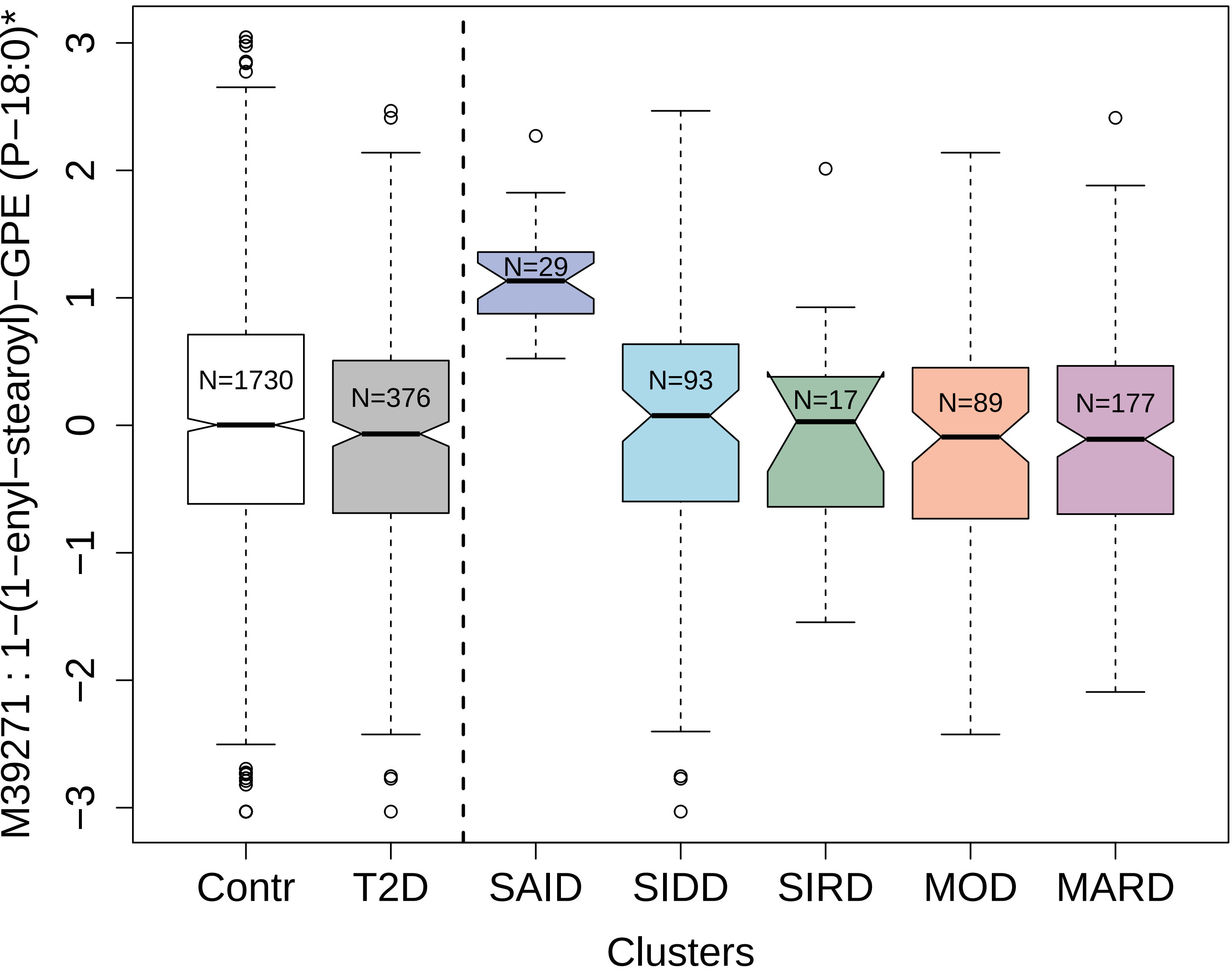
M39271 : 1−(1−enyl−stearoyl)−GPE (P−18:0)*
3
2
1
0
−1
−2
−3
N=1730
N=376
N=29
N=93
N=17
N=89
N=177
Contr
T2D
SAID
SIDD
SIRD
MOD
MARD
Clusters

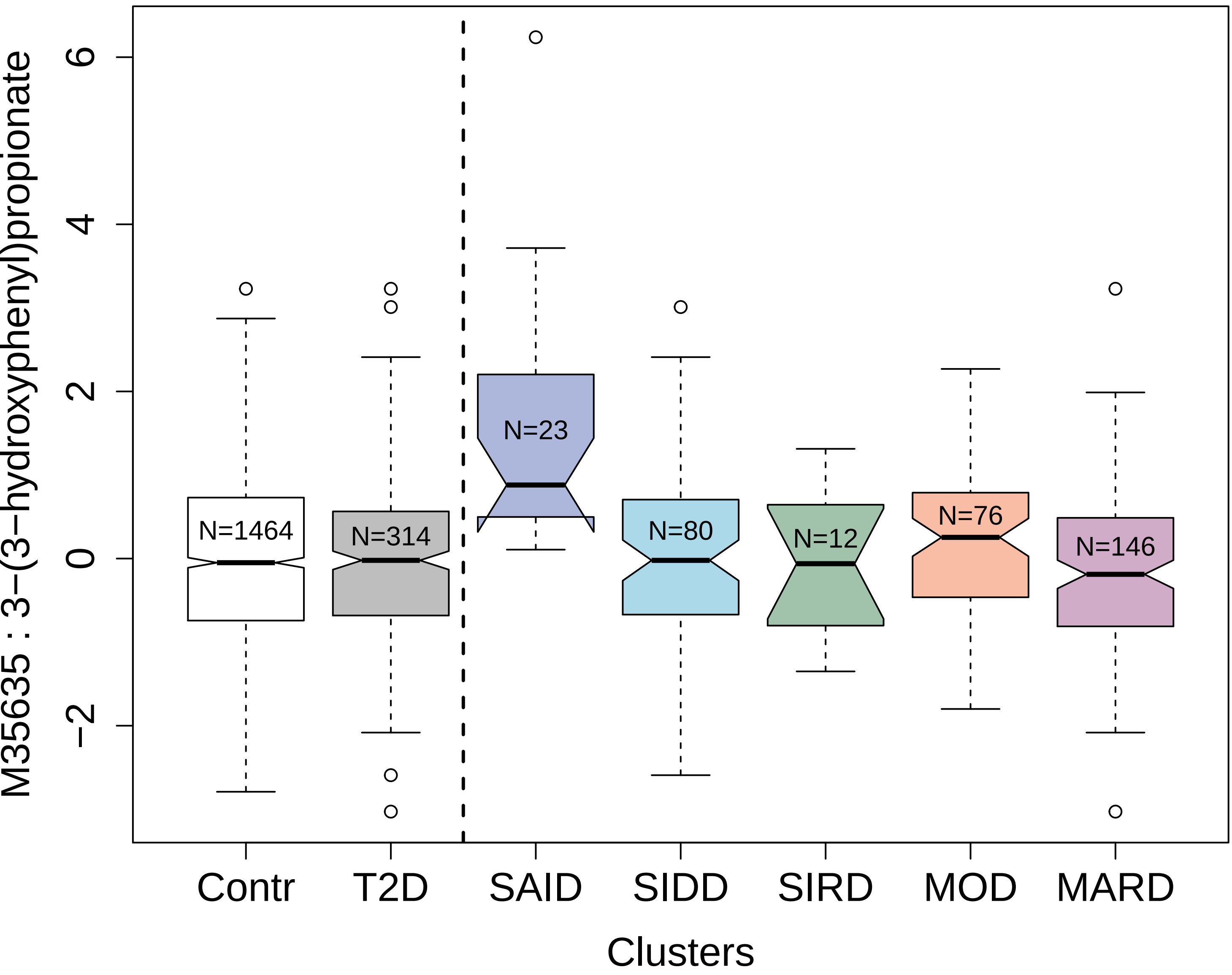

M35635 : 3−(3−hydroxyphenyl)propionate
−2
0
2
4
6
N=1464
N=314
N=23
N=80
N=12
N=76
N=146
Contr
T2D
SAID
SIDD
SIRD
MOD
MARD
Clusters

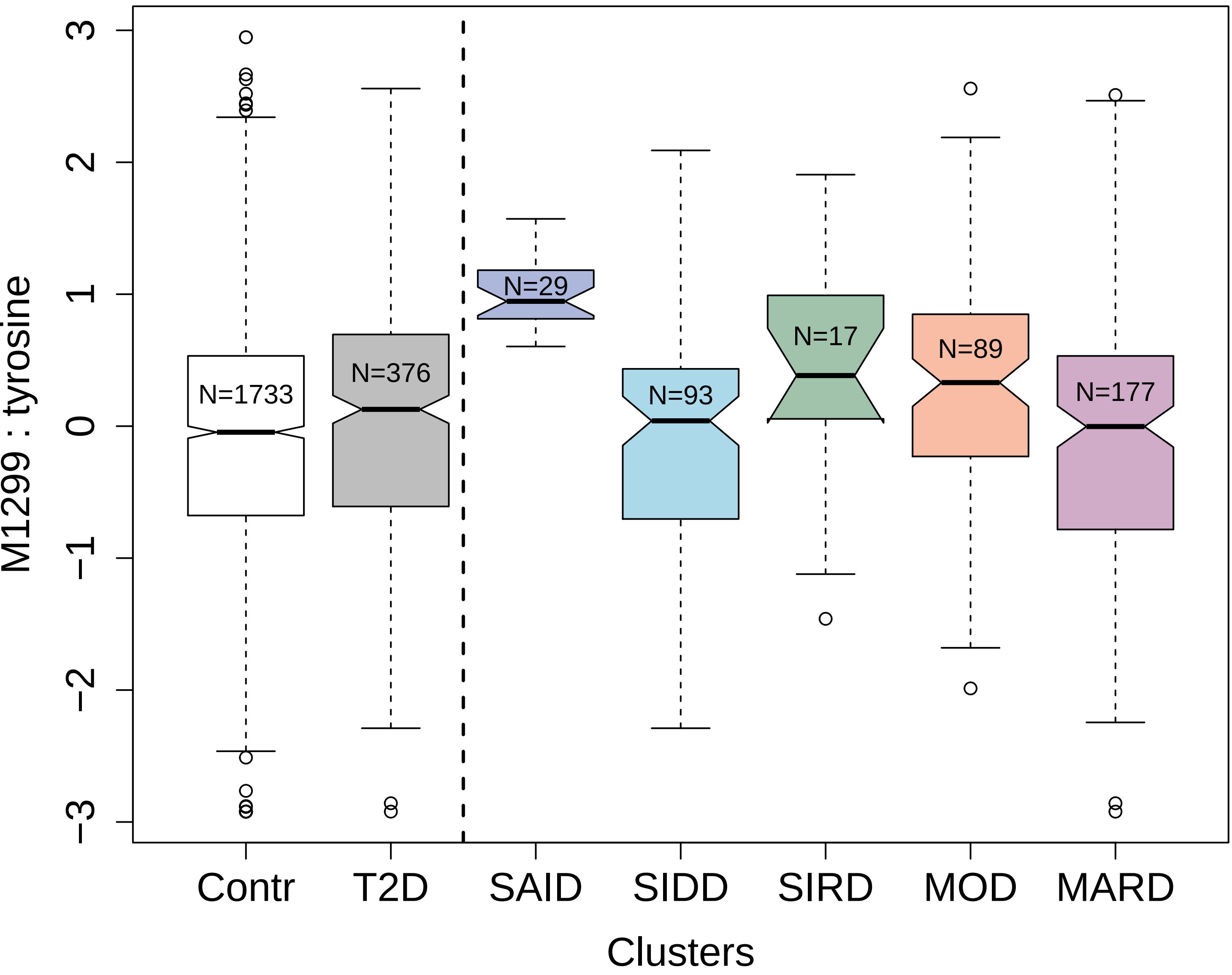
M1299 : tyrosine
3
2
1
0
−1
−2
−3
N=1733
N=376
N=29
N=93
N=17
N=89
N=177
Contr
T2D
SAID
SIDD
SIRD
MOD
MARD
Clusters

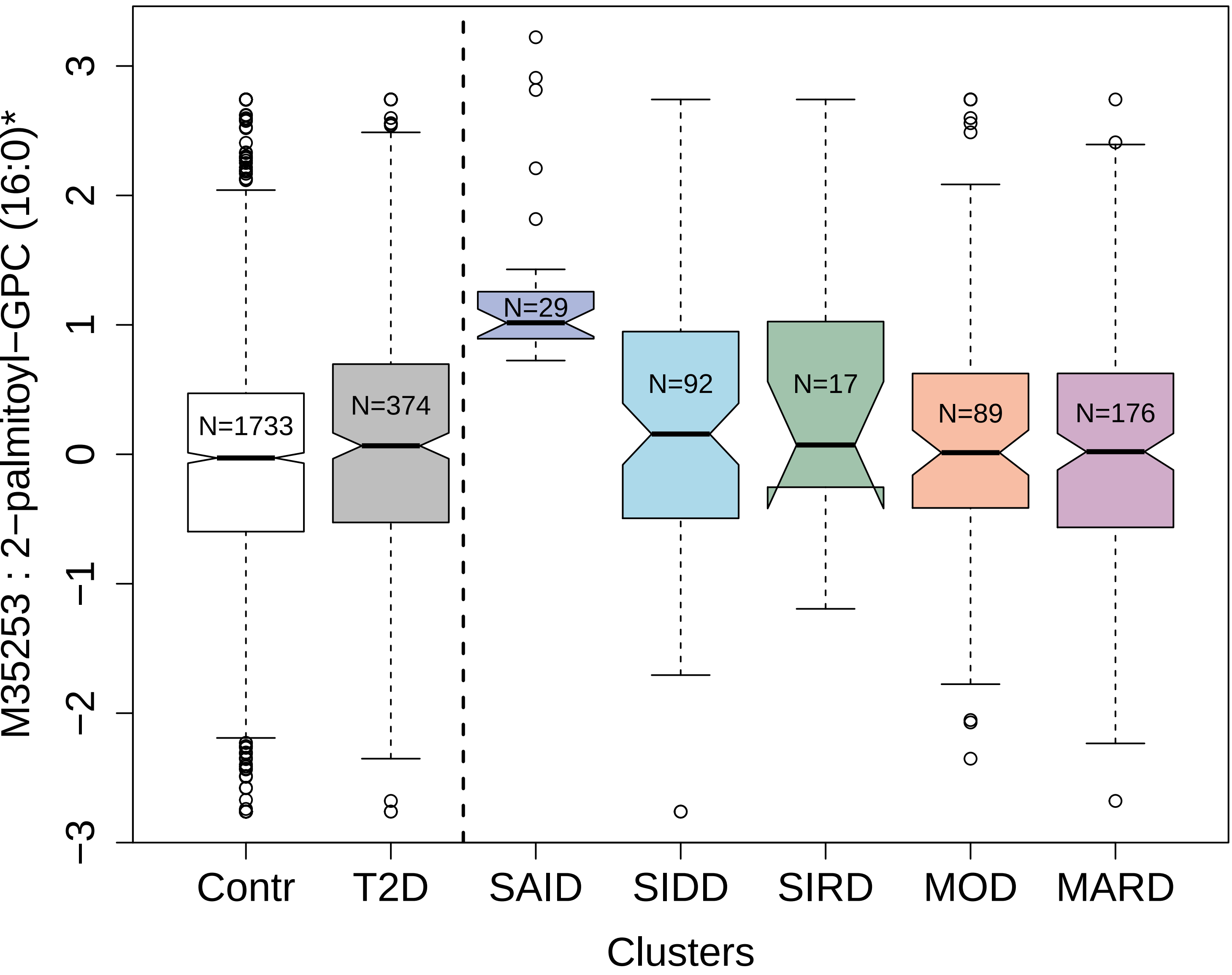

M35253 : 2−palmitoyl−GPC (16:0)*
3
2
1
0
−1
−2
−3
N=1733
N=374
N=29
N=92
N=17
N=89
N=176
Contr
T2D
SAID
SIDD
SIRD
MOD
MARD
Clusters

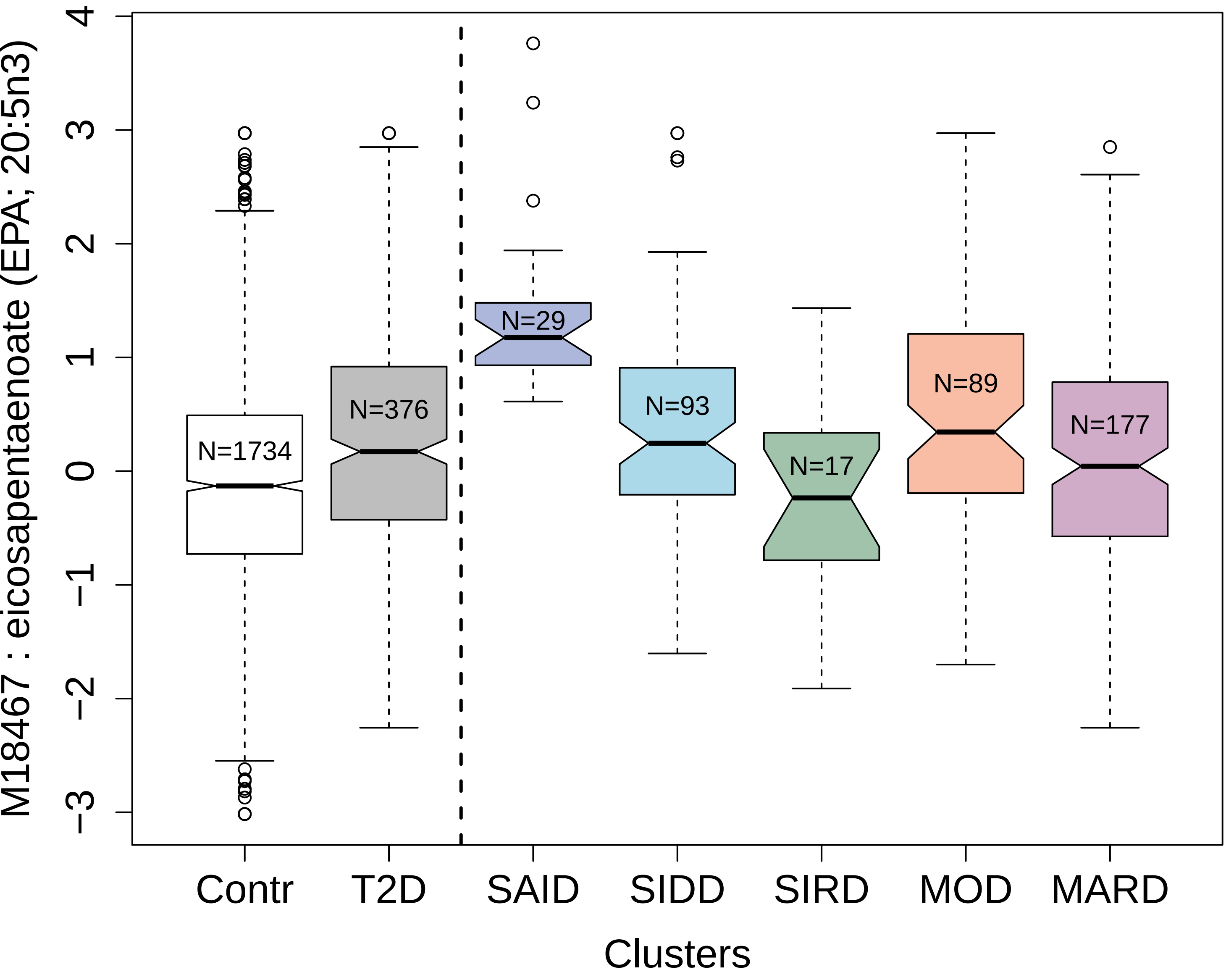

M18467 : eicosapentaenoate (EPA; 20:5n3)
4
3
2
1
0
−1
−2
−3
N=1734
N=376
N=29
N=93
N=17
N=89
N=177
Contr
T2D
SAID
SIDD
SIRD
MOD
MARD
Clusters

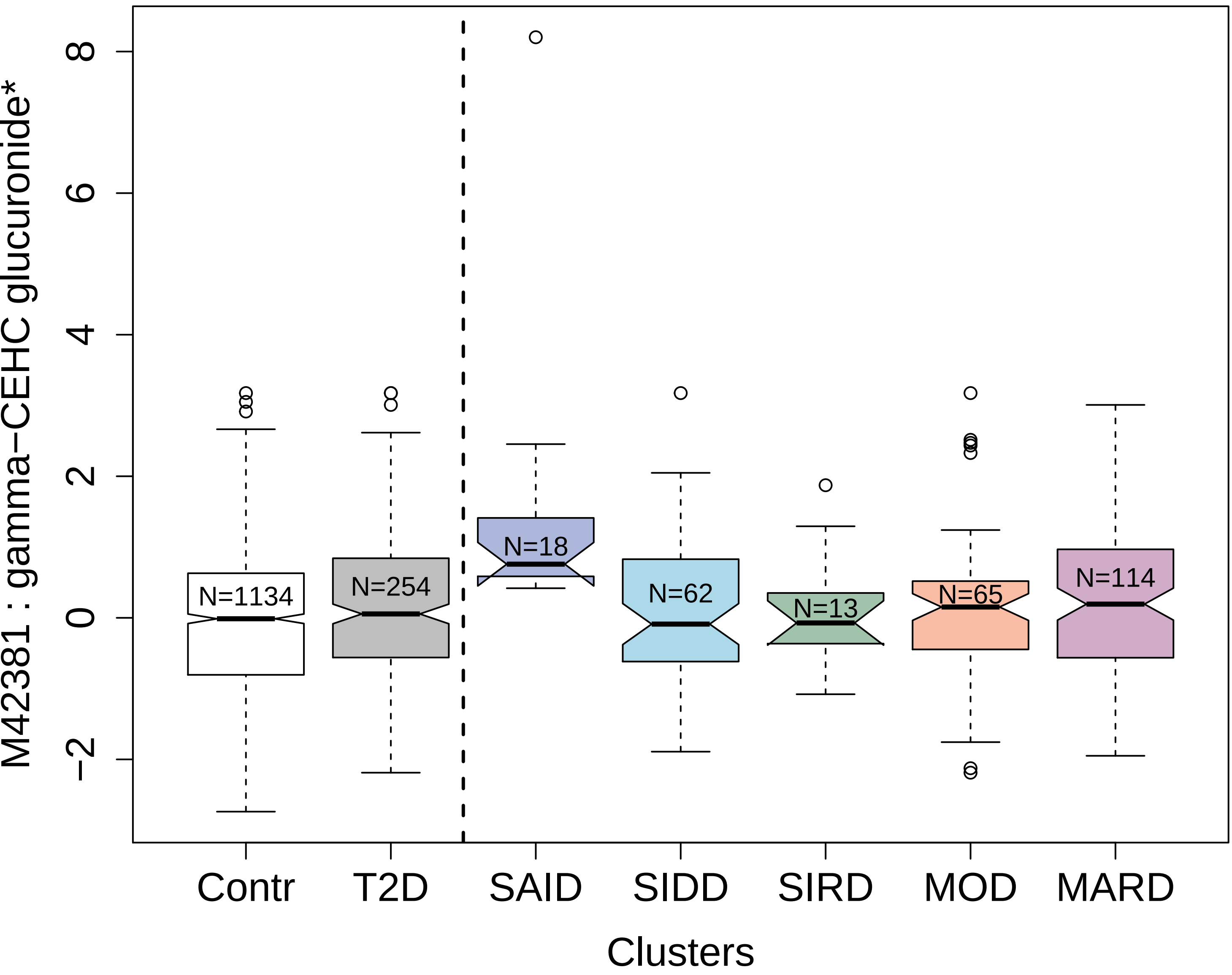

M42381 : gamma−CEHC glucuronide*
8
6
4
2
0
−2
N=1134
N=254
N=18
N=62
N=13
N=65
N=114
Contr
T2D
SAID
SIDD
SIRD
MOD
MARD
Clusters

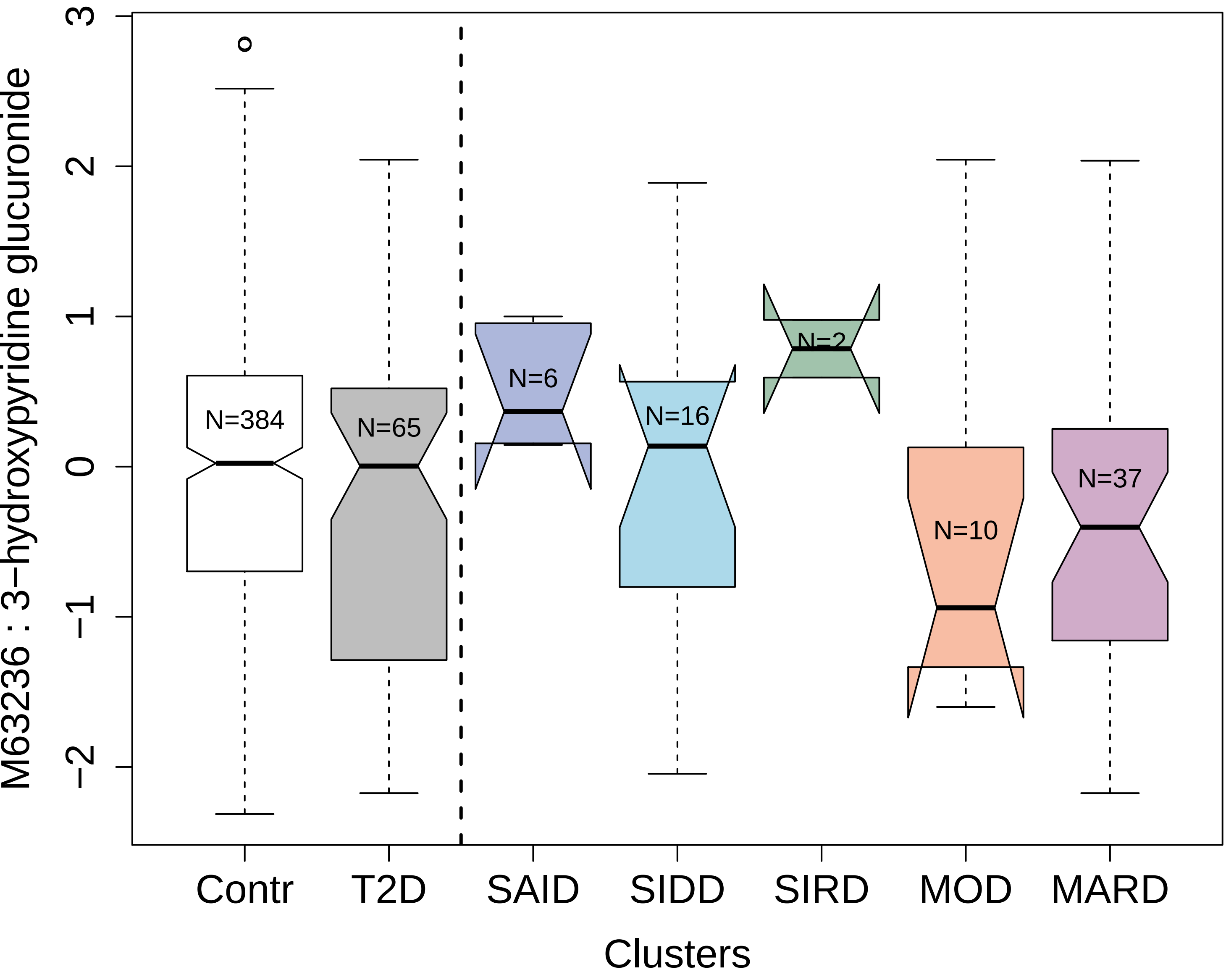

M63236 : 3−hydroxypyridine glucuronide
3
2
1
0
−1
−2
N=384
N=65
N=6
N=16
N=2
N=10
N=37
Contr
T2D
SAID
SIDD
SIRD
MOD
MARD
Clusters

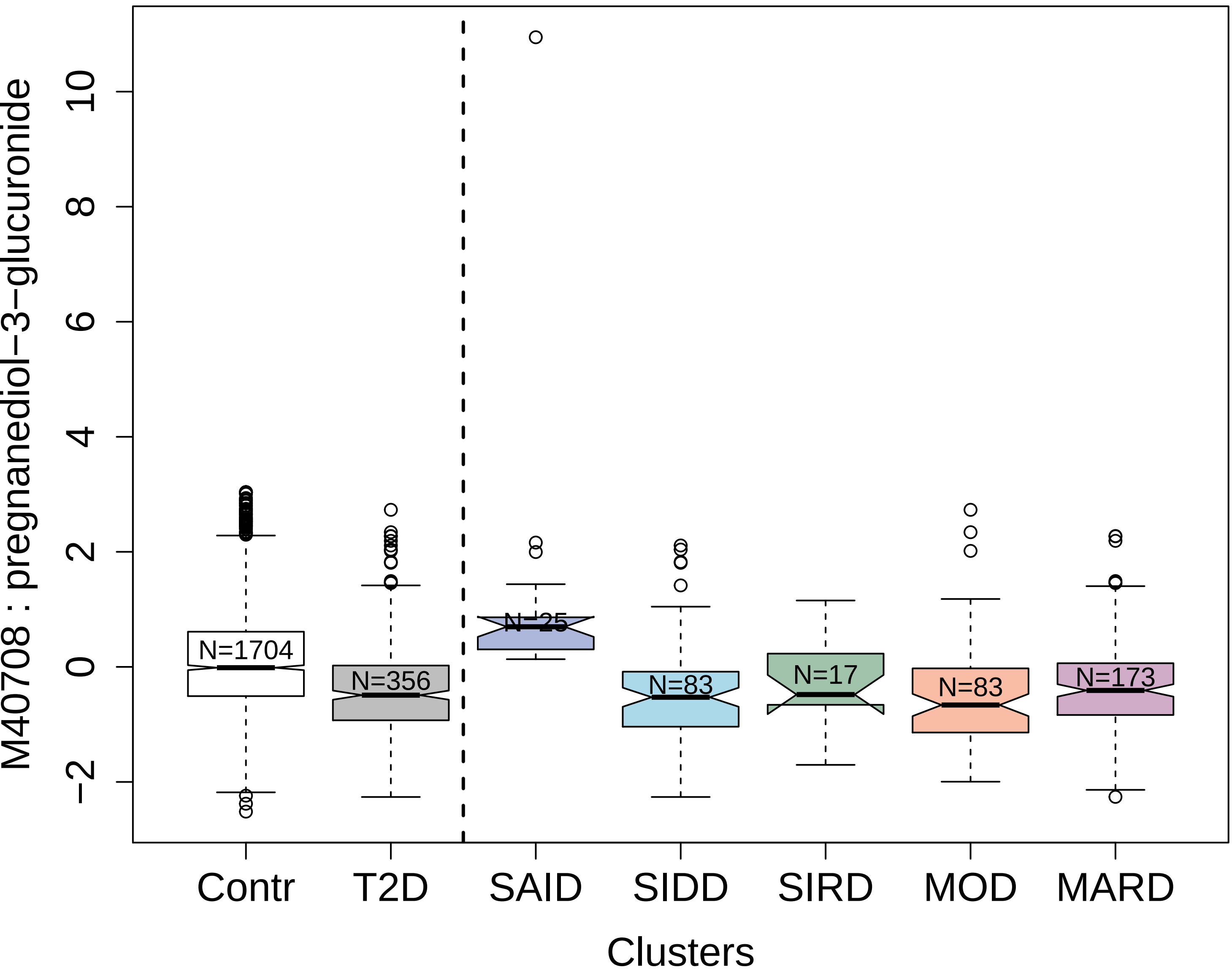

M40708 : pregnanediol−3−glucuronide
−2
0
2
4
6
8
10
N=1704
N=356
N=25
N=83
N=17
N=83
N=173
Contr
T2D
SAID
SIDD
SIRD
MOD
MARD
Clusters

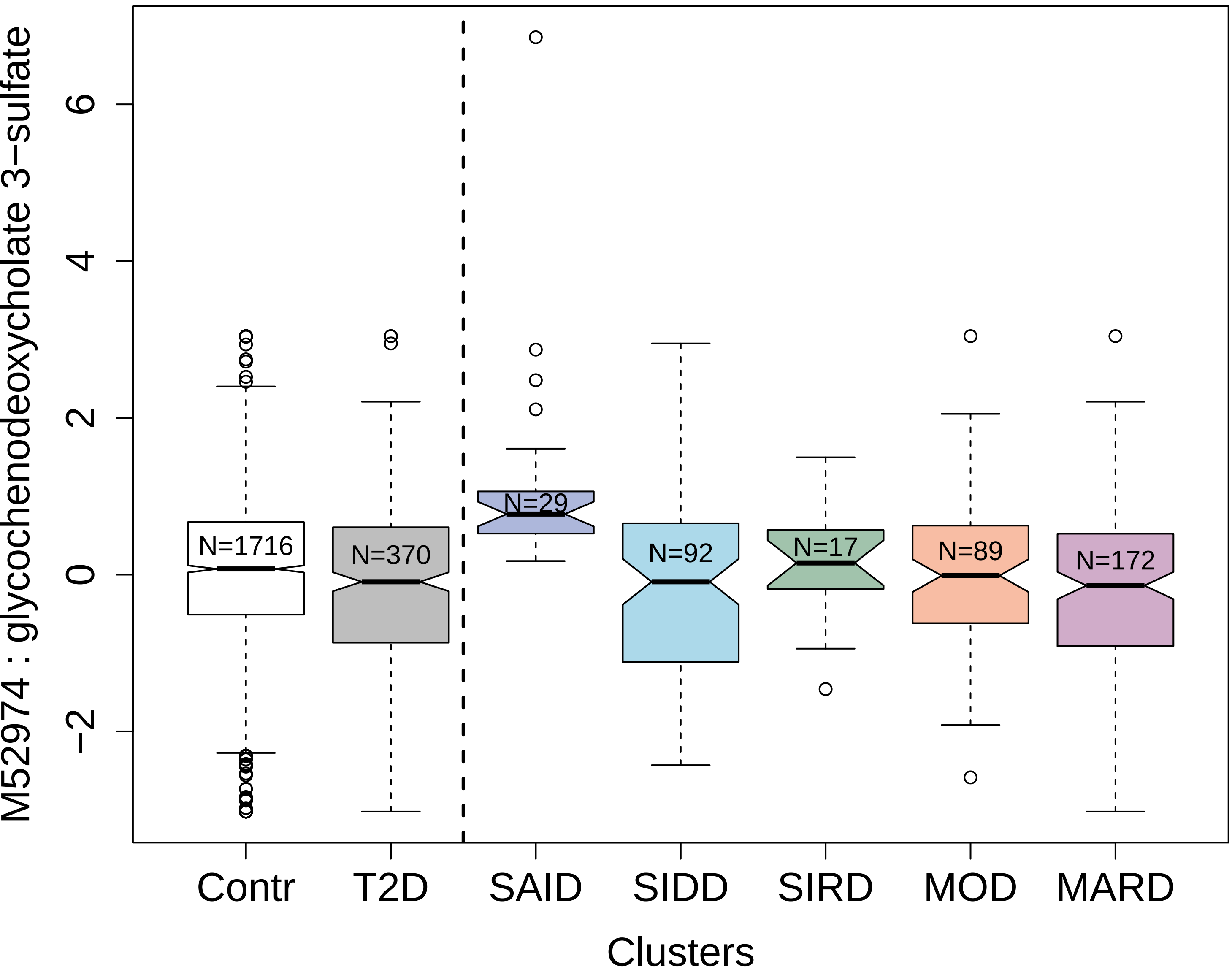
M52974 : glycochenodeoxycholate 3−sulfate
6
4
2
0
−2
N=1716
N=370
N=29
N=92
N=17
N=89
N=172
Contr
T2D
SAID
SIDD
SIRD
MOD
MARD
Clusters

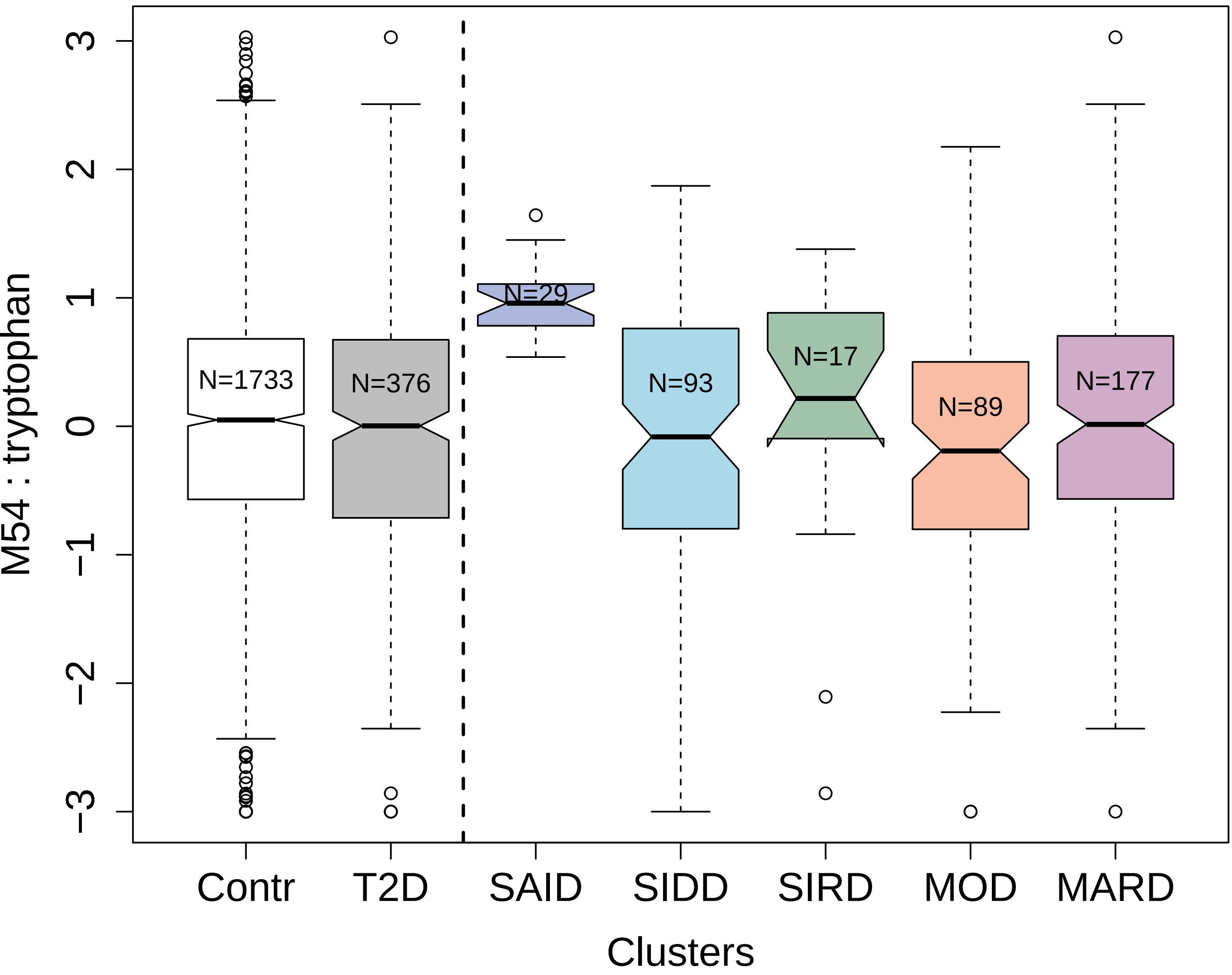

M54 : tryptophan
3
2
1
0
−1
−2
−3
N=1733
N=376
N=29
N=93
N=17
N=89
N=177
Contr
T2D
SAID
SIDD
SIRD
MOD
MARD
Clusters

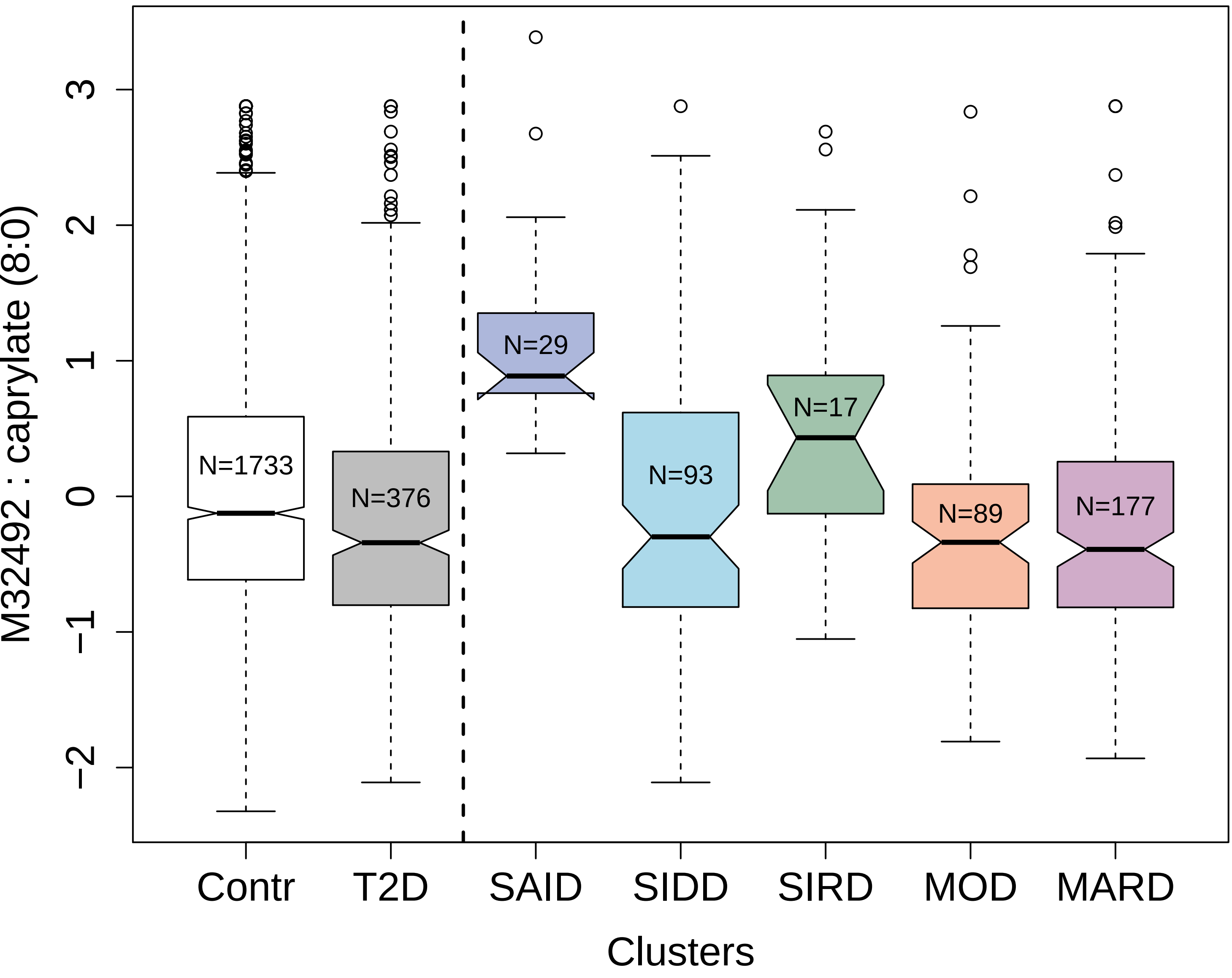

M32492 : caprylate (8:0)
3
2
1
0
−1
−2
N=1733
N=376
N=29
N=93
N=17
N=89
N=177
Contr
T2D
SAID
SIDD
SIRD
MOD
MARD
Clusters

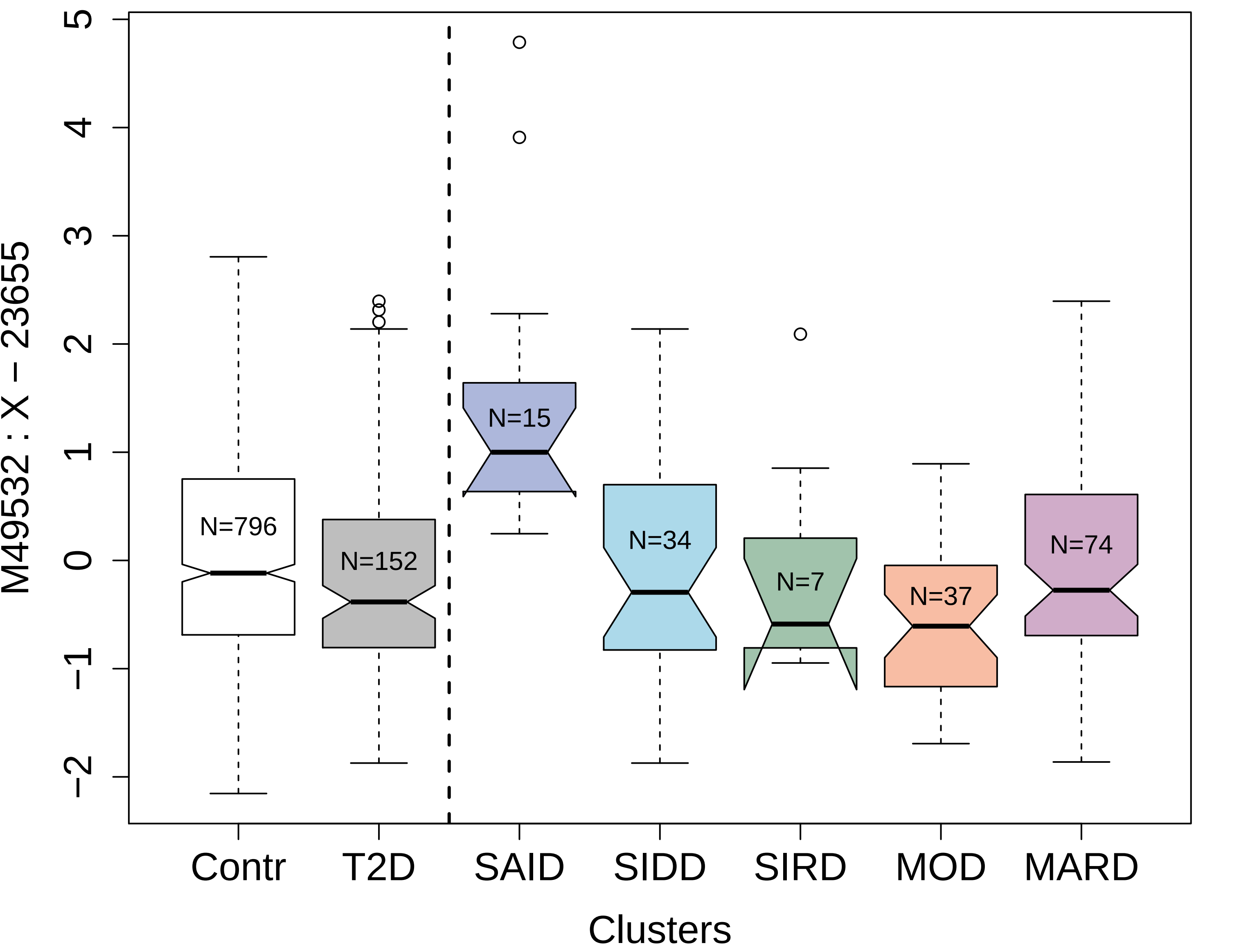
M49532 : X − 23655
5
4
3
2
1
0
−1
−2
N=796
N=152
N=15
N=34
N=7
N=37
N=74
Contr
T2D
SAID
SIDD
SIRD
MOD
MARD
Clusters

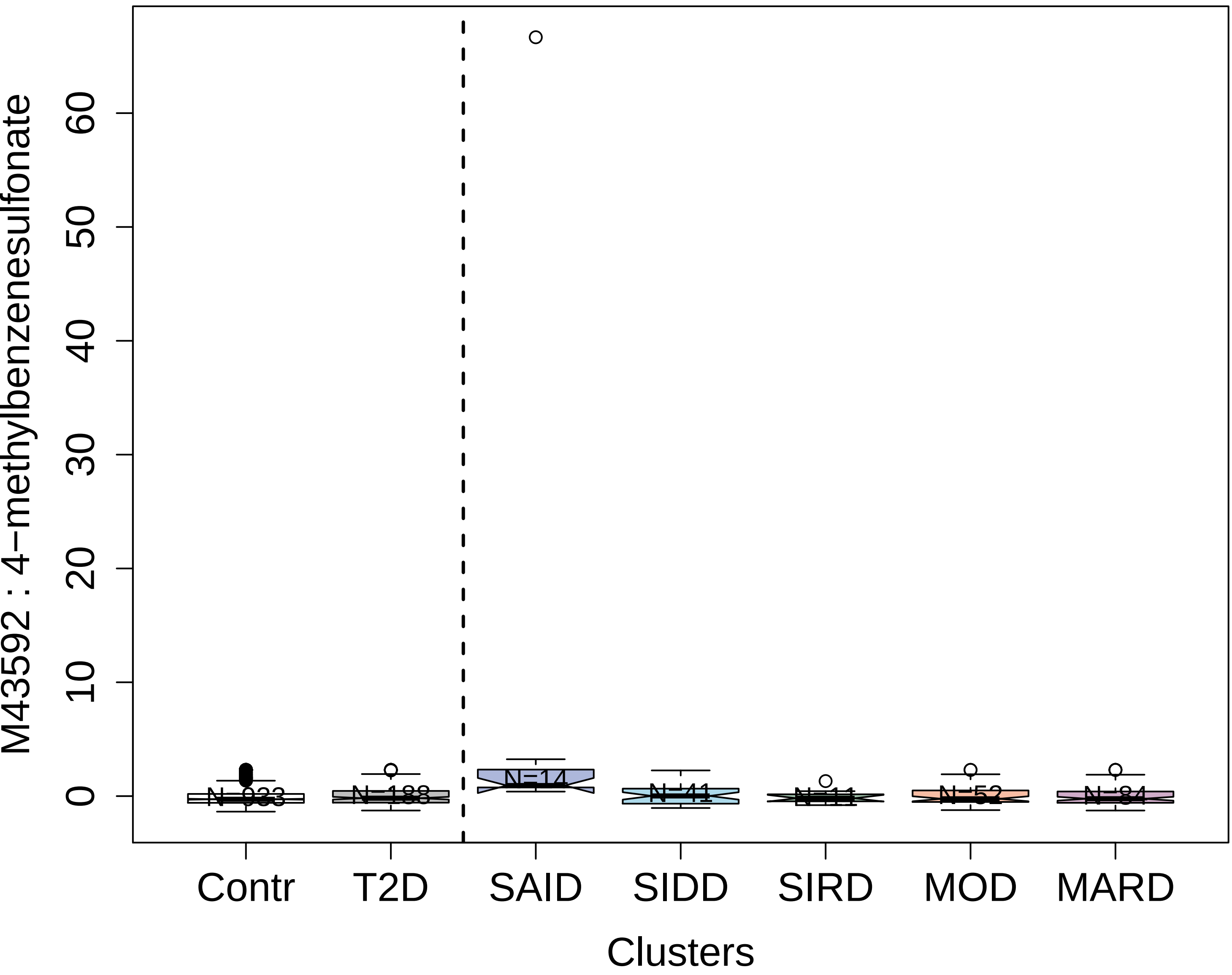
M43592 : 4−methylbenzenesulfonate
0
10
20
30
40
50
60
N=933
N=188
N=14
N=41
N=11
N=52
N=84
Contr
T2D
SAID
SIDD
SIRD
MOD
MARD
Clusters

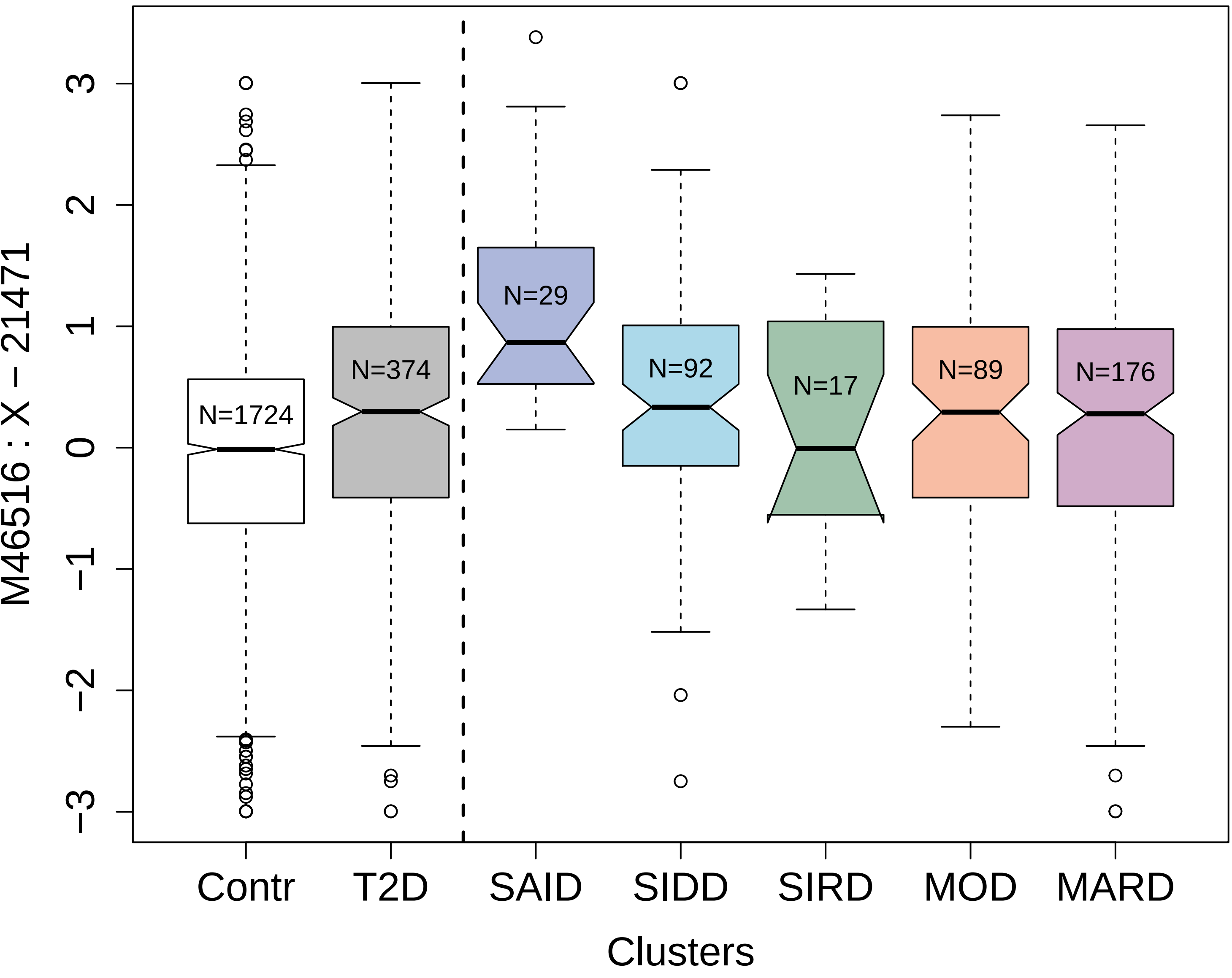

M46516 : X − 21471
3
2
1
0
−1
−2
−3
N=1724
N=374
N=29
N=92
N=17
N=89
N=176
Contr
T2D
SAID
SIDD
SIRD
MOD
MARD
Clusters

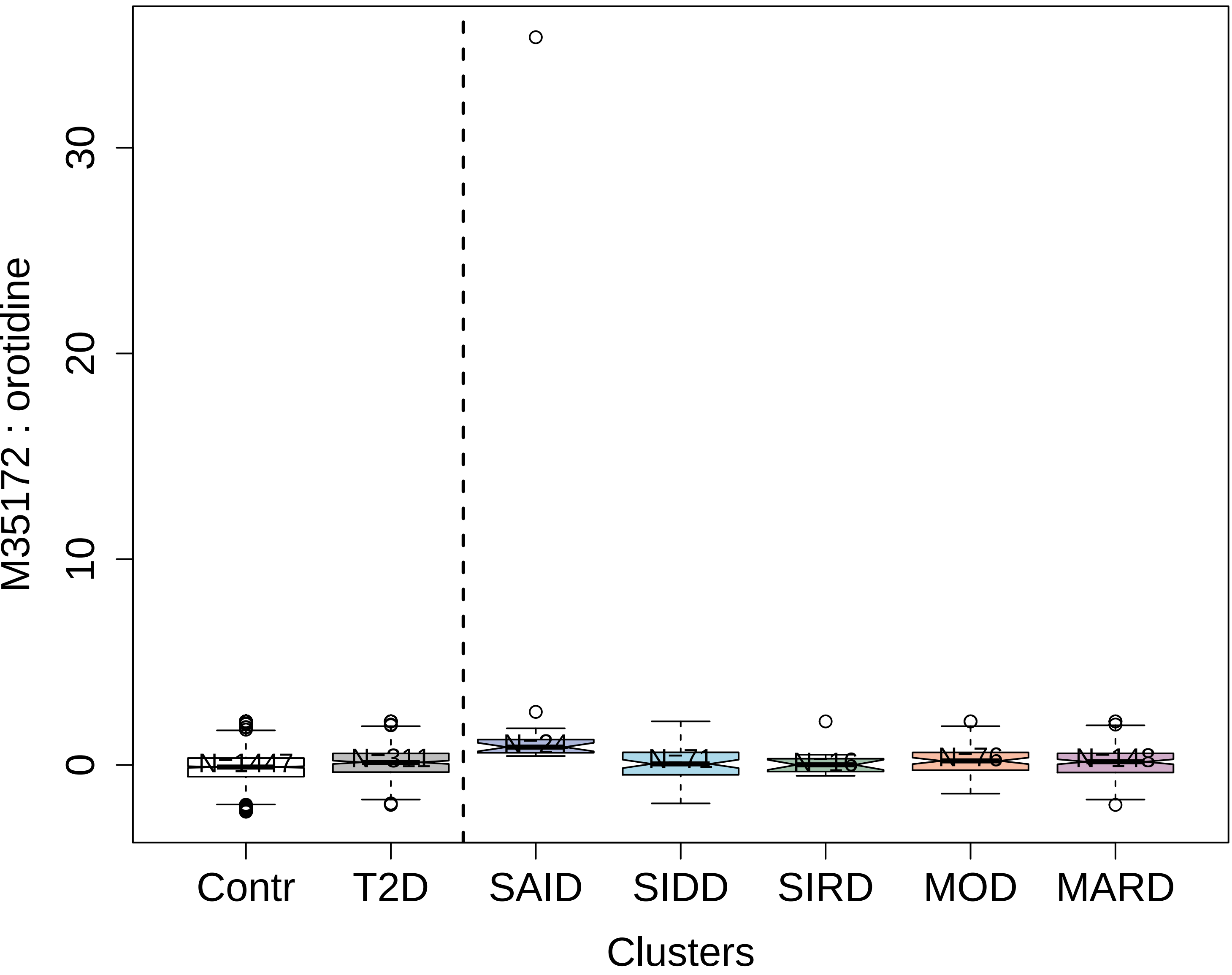

M35172 : orotidine
0
10
20
30
N=1447
N=311
N=24
N=71
N=16
N=76
N=148
Contr
T2D
SAID
SIDD
SIRD
MOD
MARD
Clusters

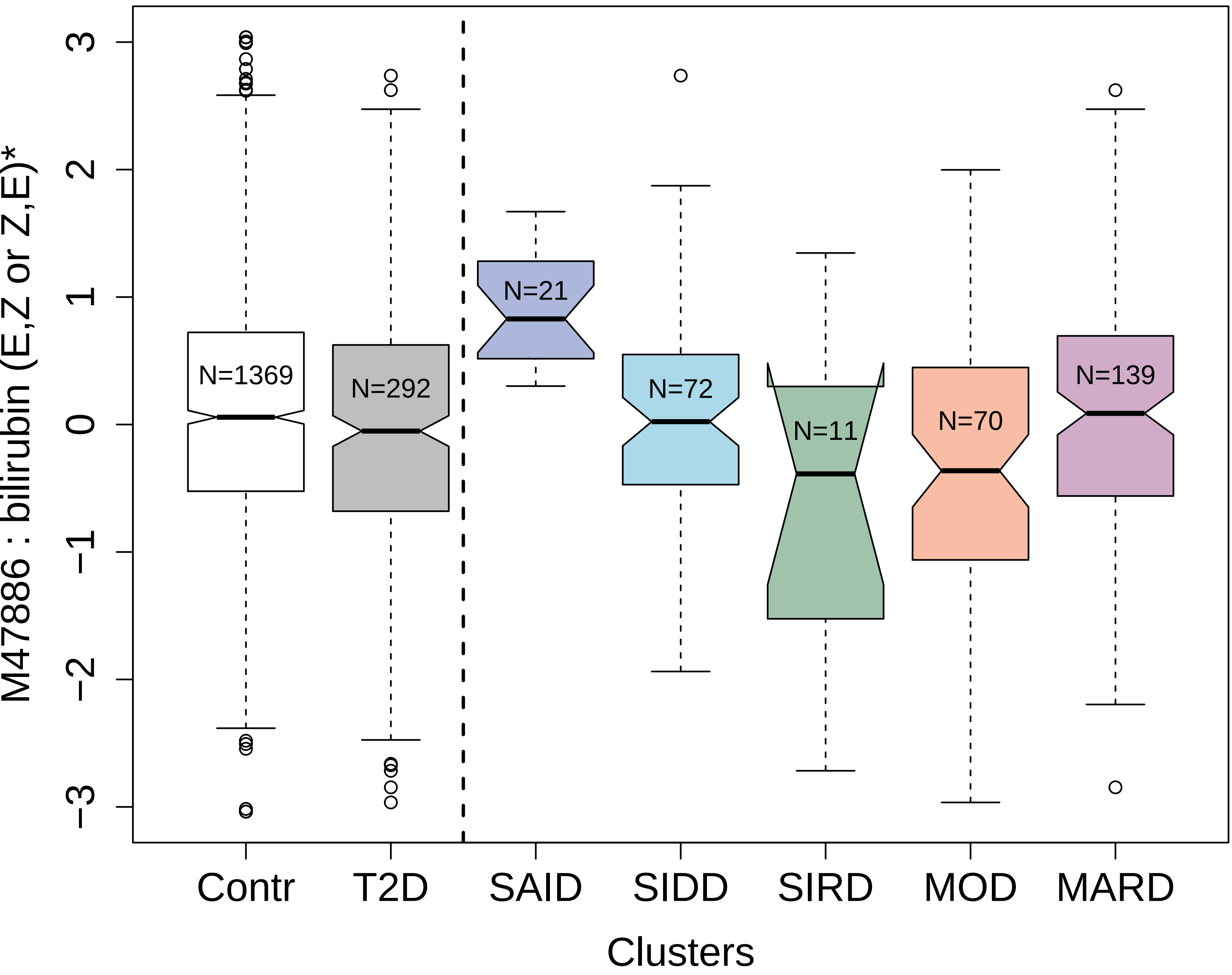
M47886 : bilirubin (E,Z or Z,E)*
3
2
1
0
−1
−2
−3
N=1369
N=292
N=21
N=72
N=11
N=70
N=139
Contr
T2D
SAID
SIDD
SIRD
MOD
MARD
Clusters

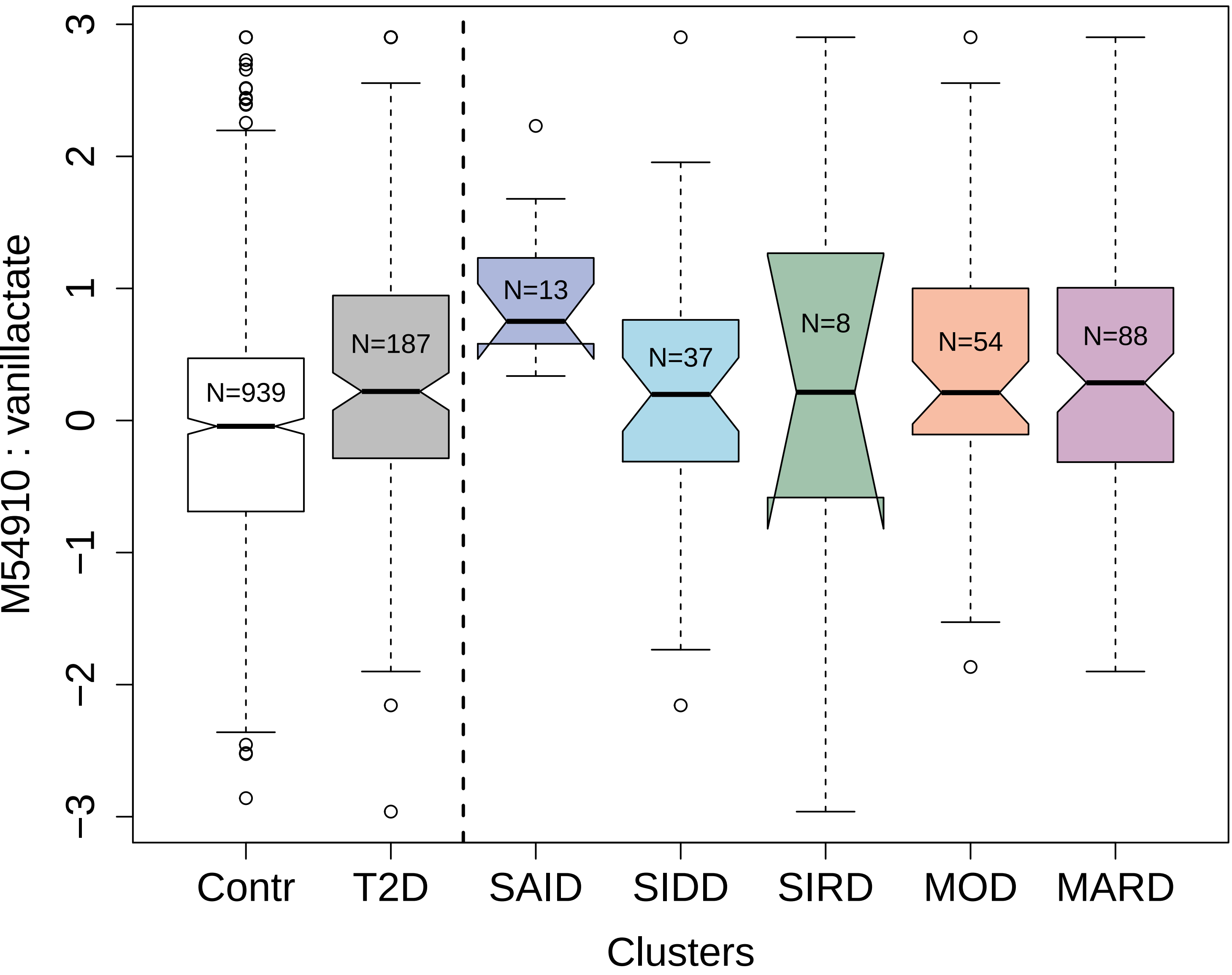
M54910 : vanillactate
3
2
1
0
−1
−2
−3
N=939
N=187
N=13
N=37
N=8
N=54
N=88
Contr
T2D
SAID
SIDD
SIRD
MOD
MARD
Clusters

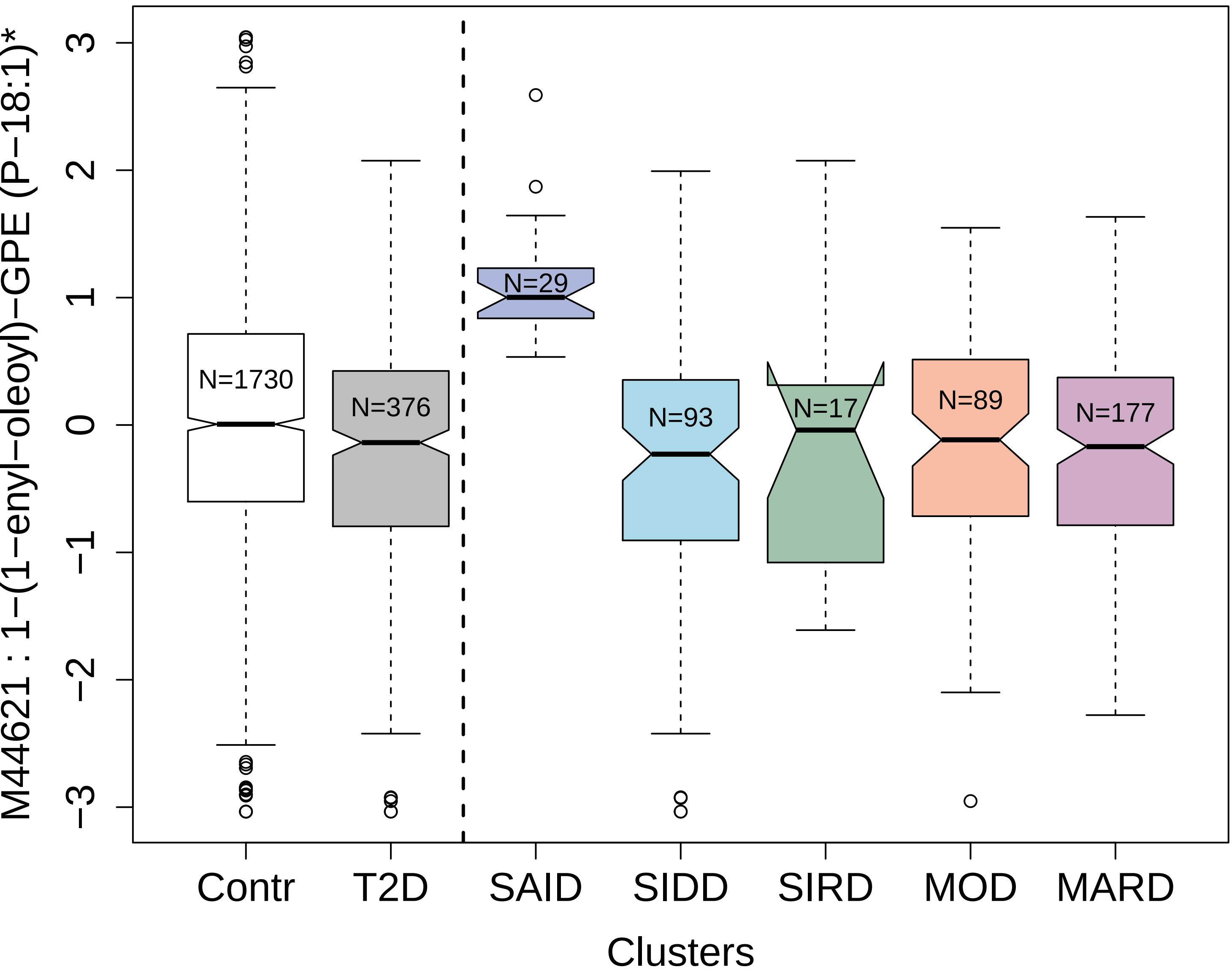

M44621 : 1−(1−enyl−oleoyl)−GPE (P−18:1)*
3
2
1
0
−1
−2
−3
N=1730
N=376
N=29
N=93
N=17
N=89
N=177
Contr
T2D
SAID
SIDD
SIRD
MOD
MARD
Clusters

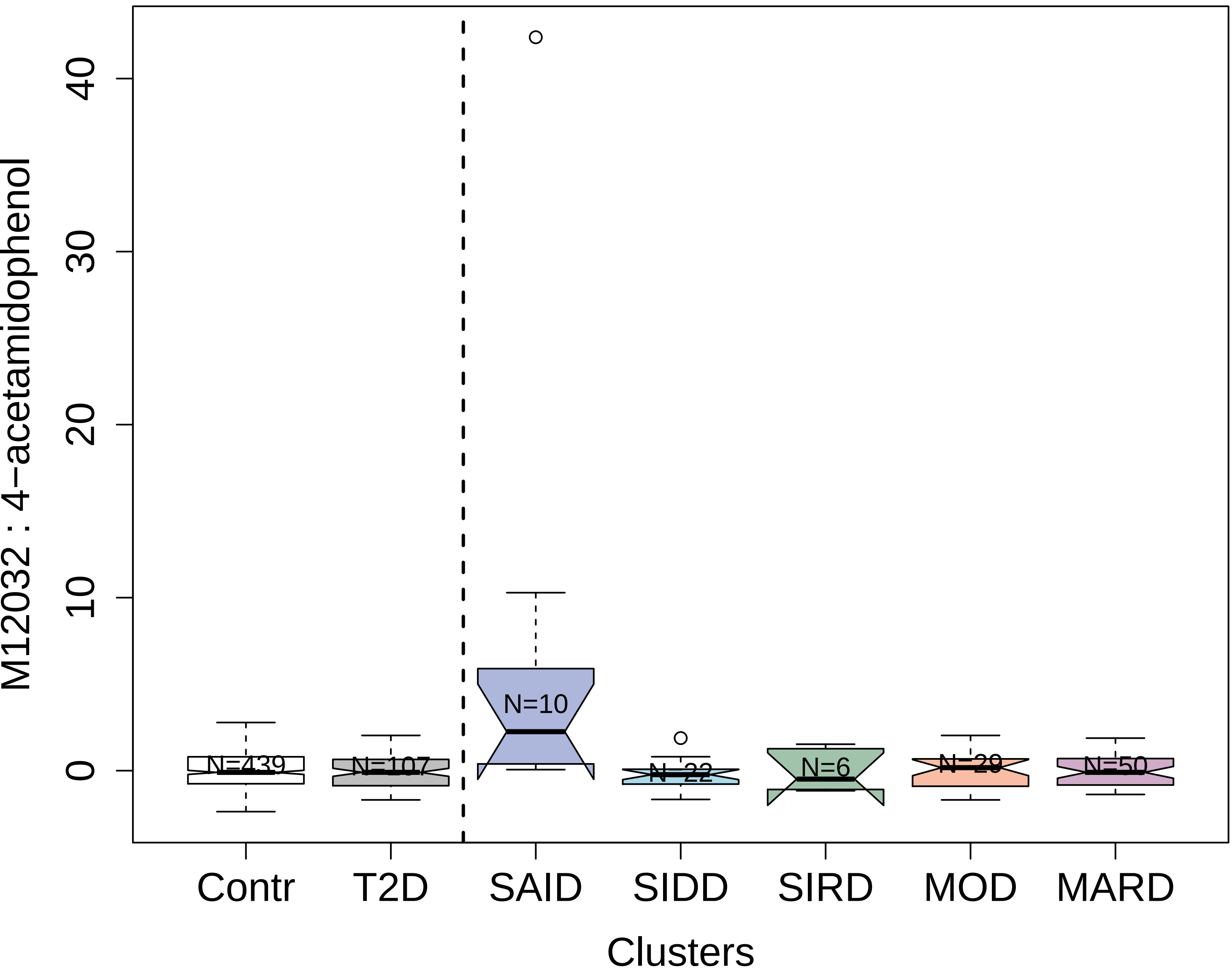

M12032 : 4−acetamidophenol
0
10
20
30
40
N=439
N=107
N=10
N=22
N=6
N=29
N=50
Contr
T2D
SAID
SIDD
SIRD
MOD
MARD
Clusters

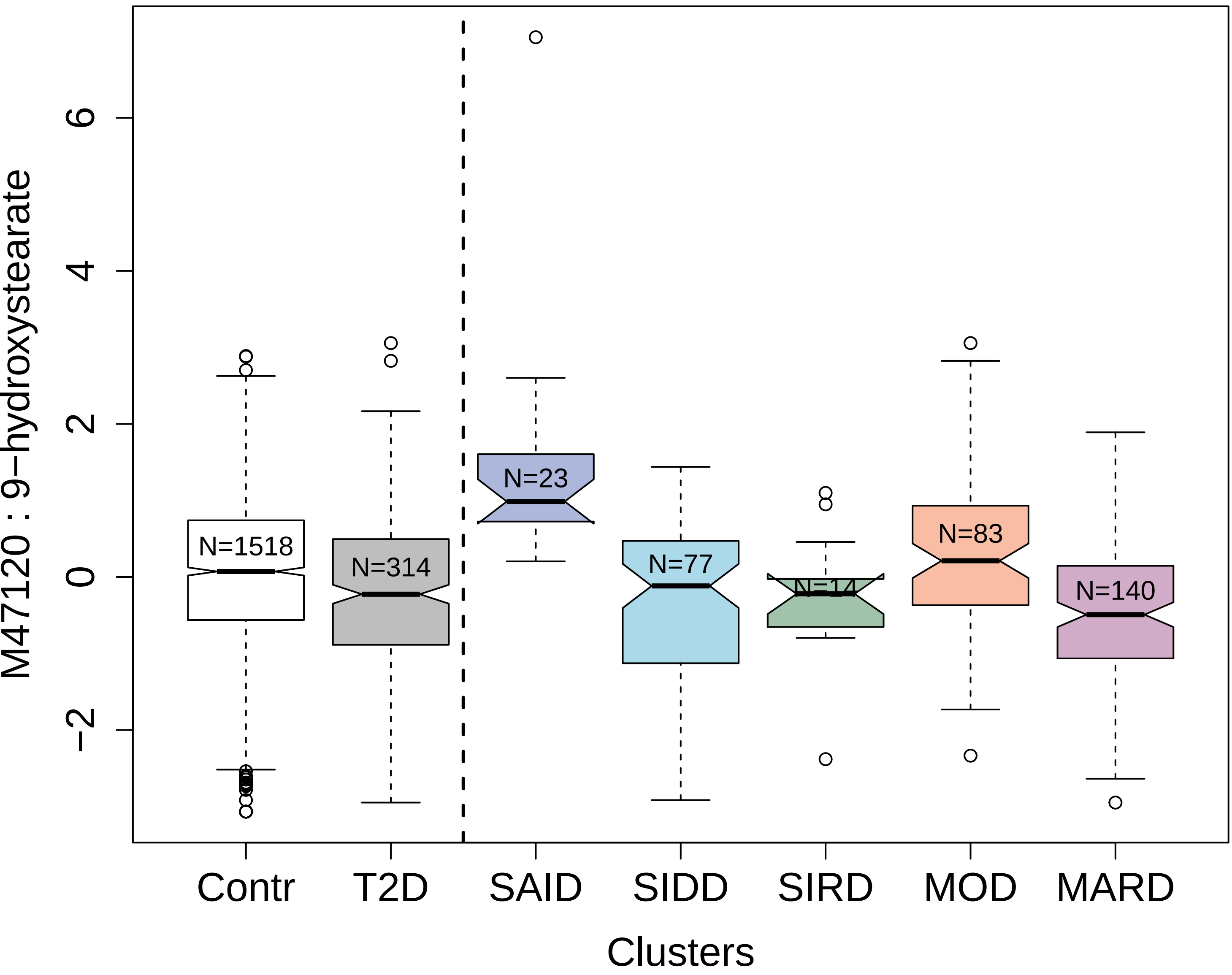

M47120 : 9−hydroxystearate
6
4
2
0
−2
N=1518
N=314
N=23
N=77
N=14
N=83
N=140
Contr
T2D
SAID
SIDD
SIRD
MOD
MARD
Clusters

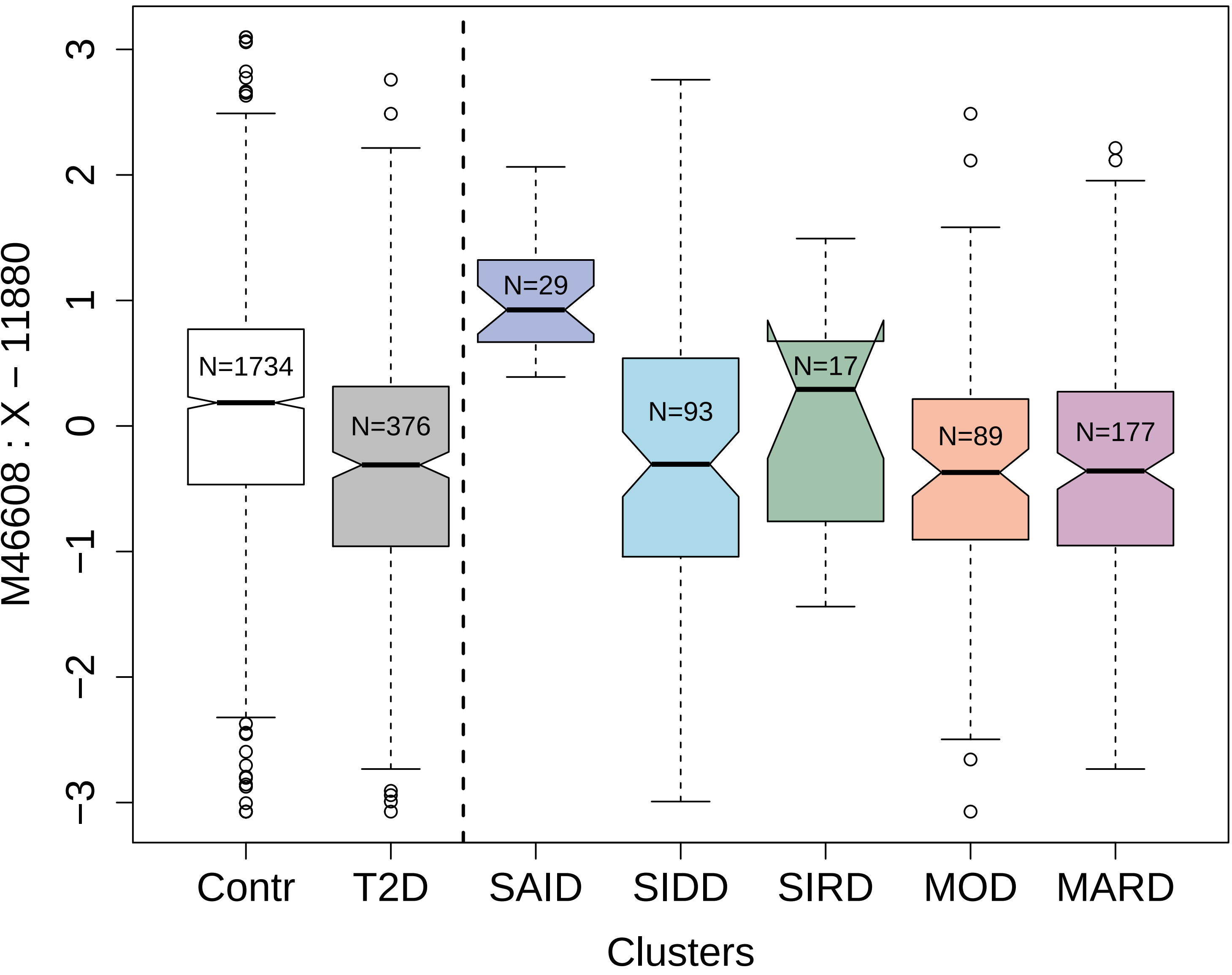
M46608 : X − 11880
Clusters
Contr
T2D
SAID
SIDD
SIRD
MOD
MARD
N=1734
N=376
N=29
N=93
N=17
N=89
N=177
3
2
1
0
−1
−2
−3

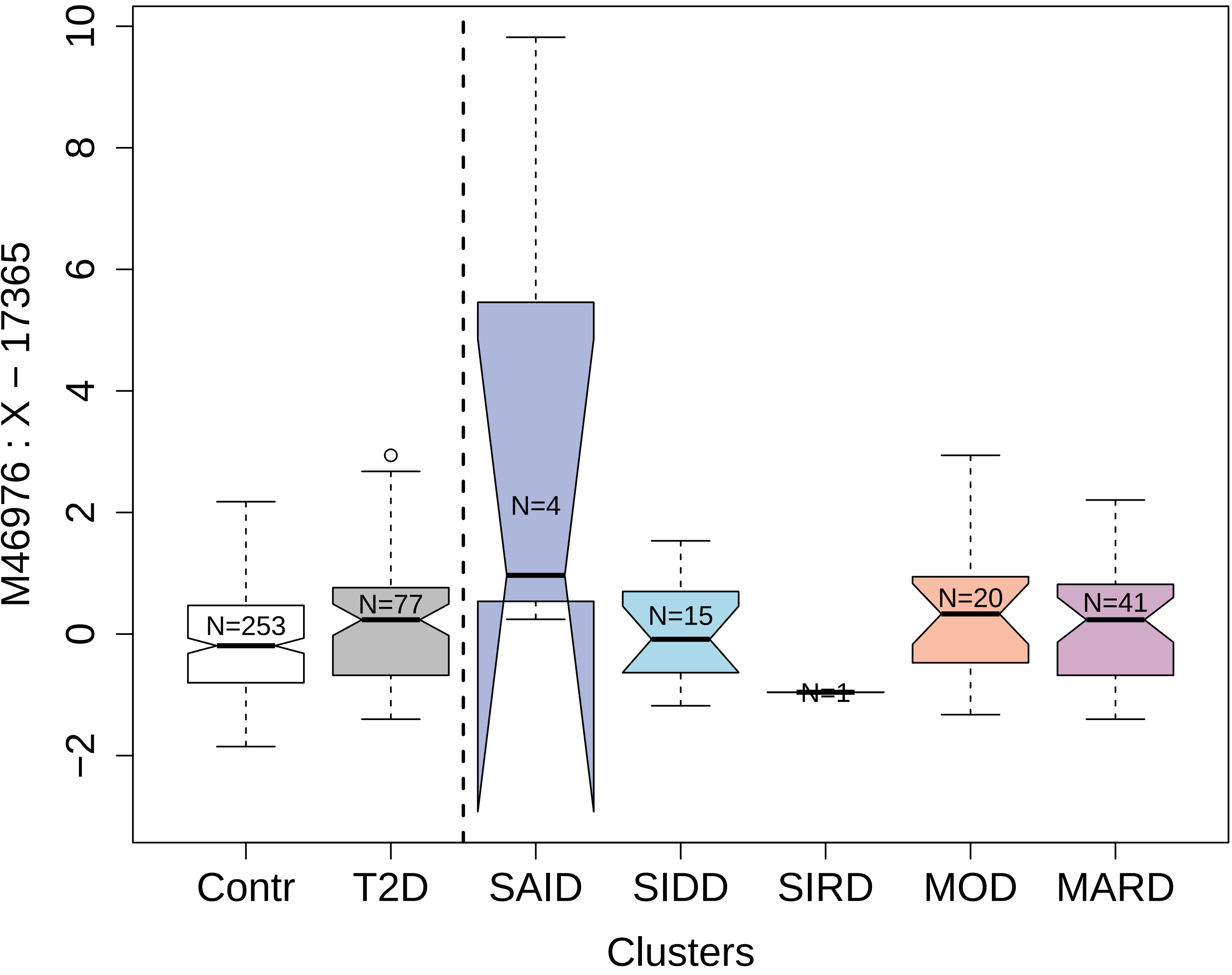
M46976 : X − 17365
10
8
6
4
2
0
−2
N=253
N=77
N=4
N=15
N=1
N=20
N=41
Contr
T2D
SAID
SIDD
SIRD
MOD
MARD
Clusters

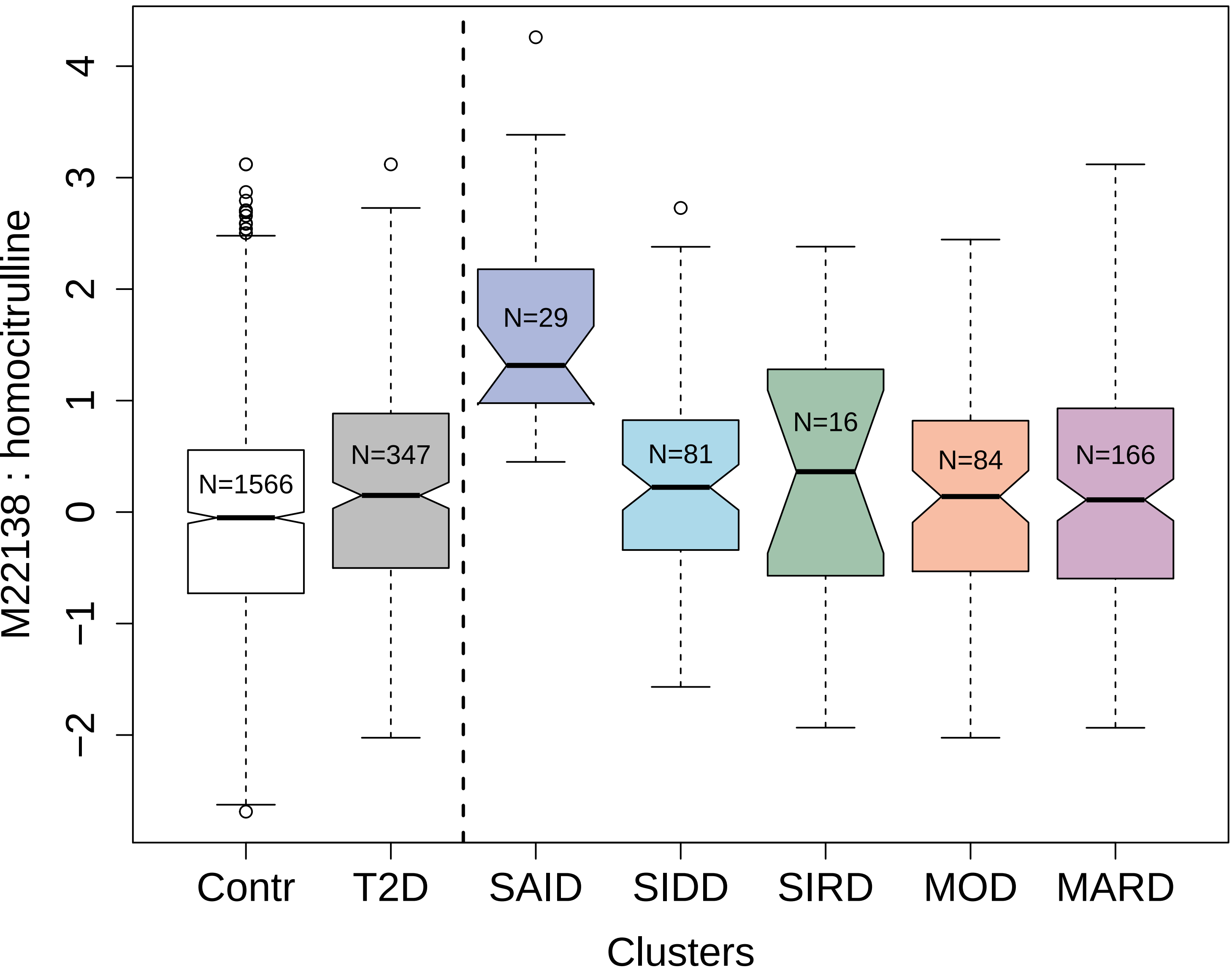
M22138 : homocitrulline
Clusters
Contr
T2D
SAID
SIDD
SIRD
MOD
MARD
N=1566
N=347
N=29
N=81
N=16
N=84
N=166
4
3
2
1
0
−1
−2

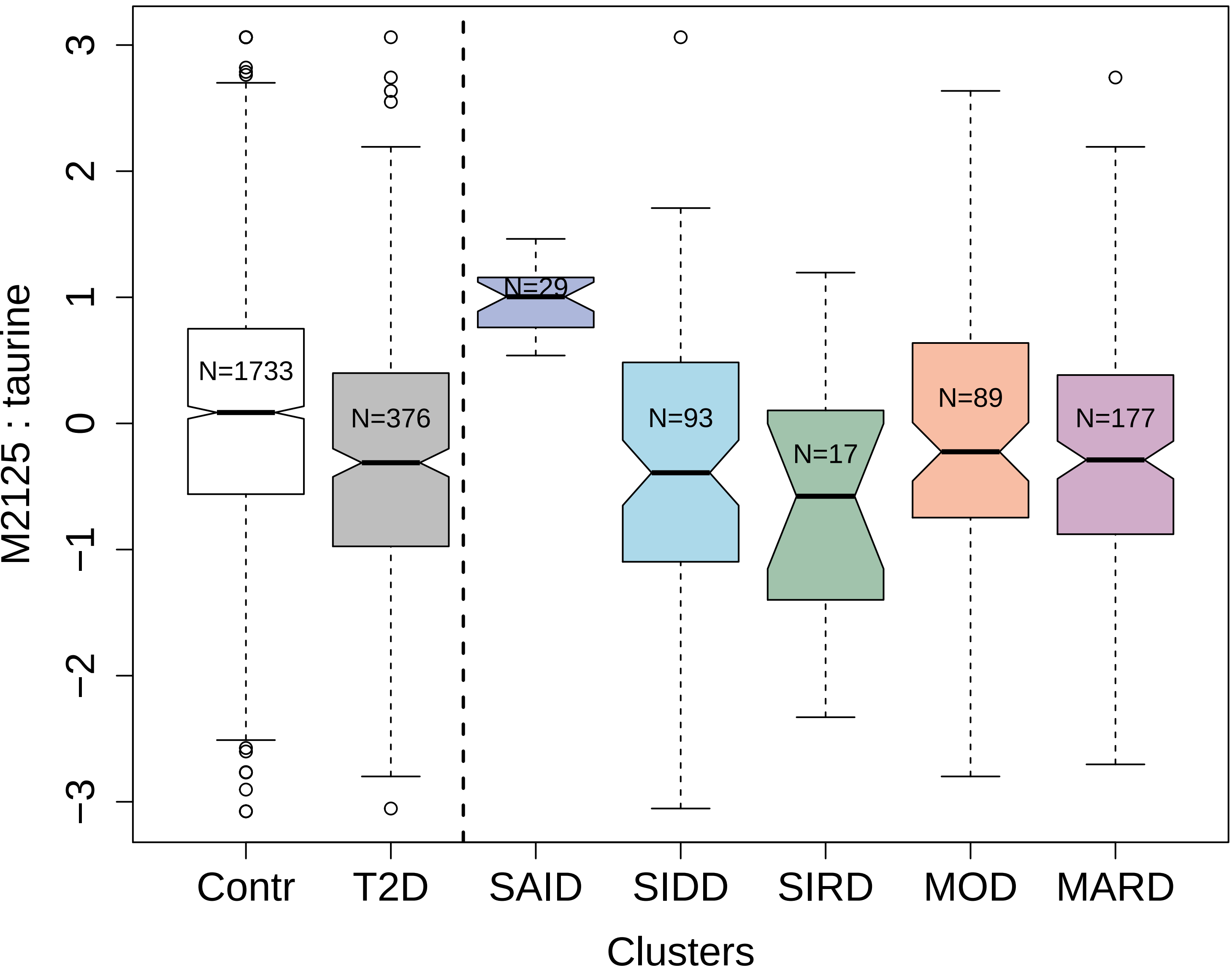
M2125 : taurine
Clusters
Contr
T2D
SAID
SIDD
SIRD
MOD
MARD
N=1733
N=376
N=29
N=93
N=17
N=89
N=177
3
2
1
0
−1
−2
−3

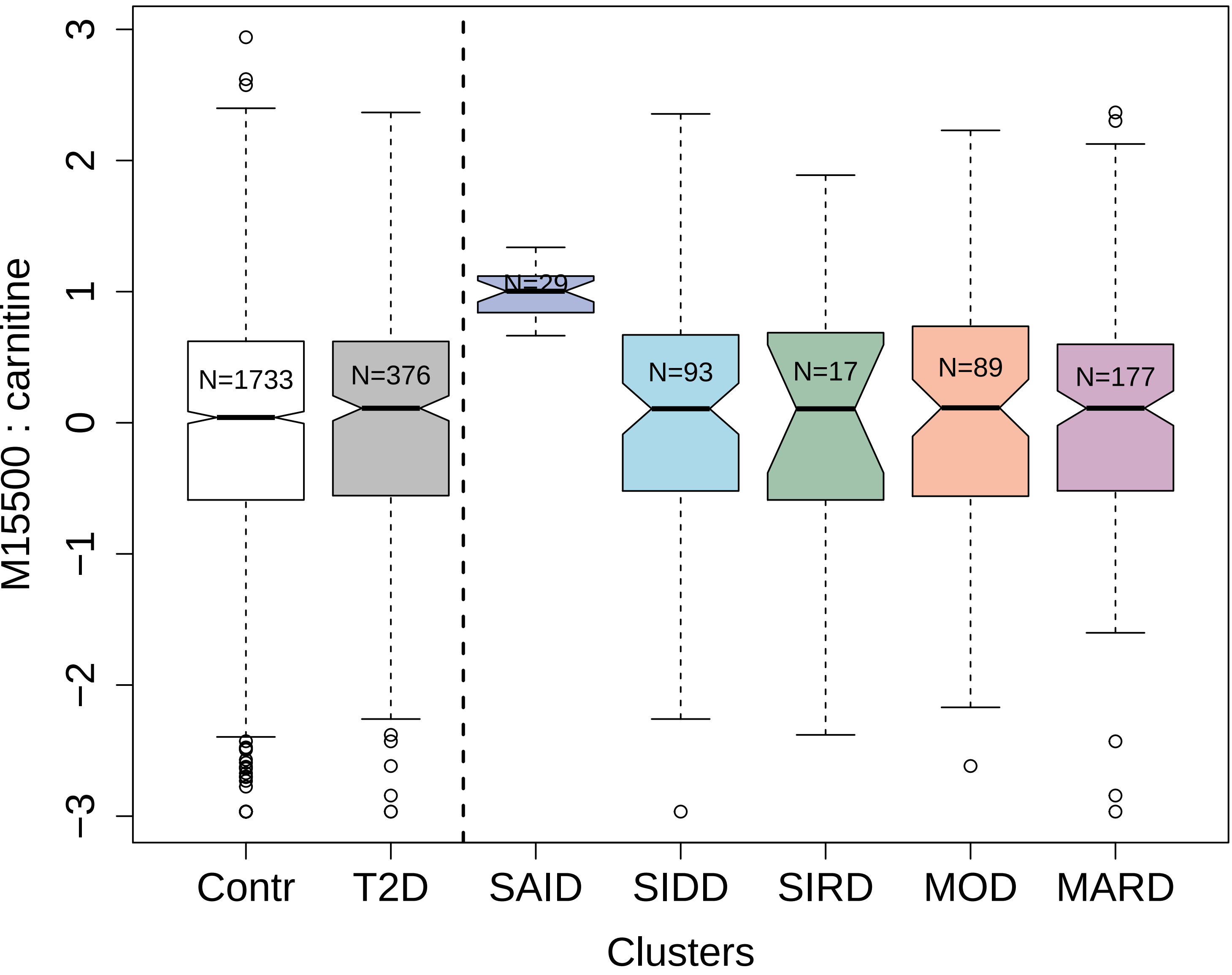

M15500 : carnitine
3
2
1
0
−1
−2
−3
N=1733
N=376
N=29
N=93
N=17
N=89
N=177
Contr
T2D
SAID
SIDD
SIRD
MOD
MARD
Clusters

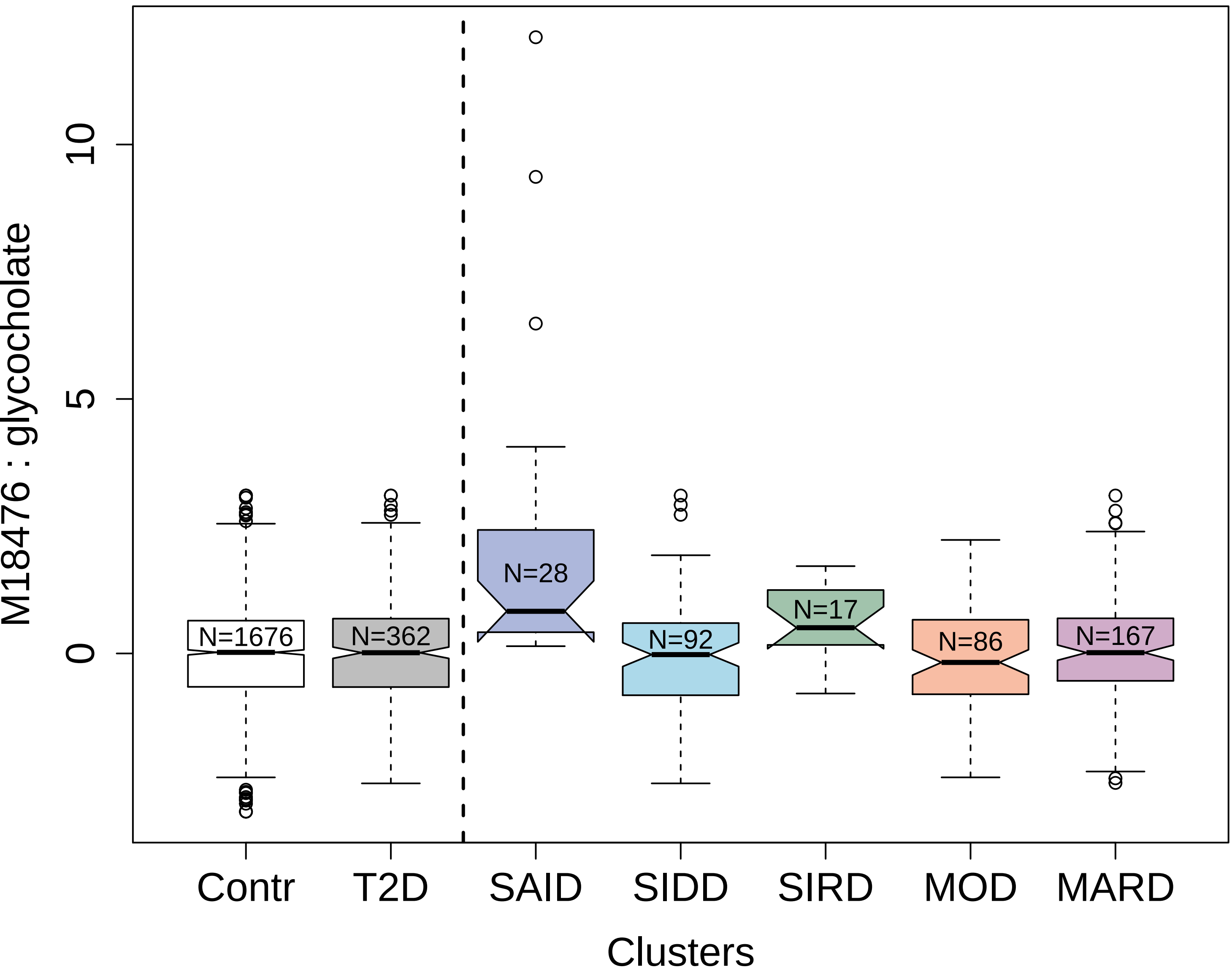

M18476 : glycocholate
0
5
10
N=1676
N=362
N=28
N=92
N=17
N=86
N=167
Contr
T2D
SAID
SIDD
SIRD
MOD
MARD
Clusters

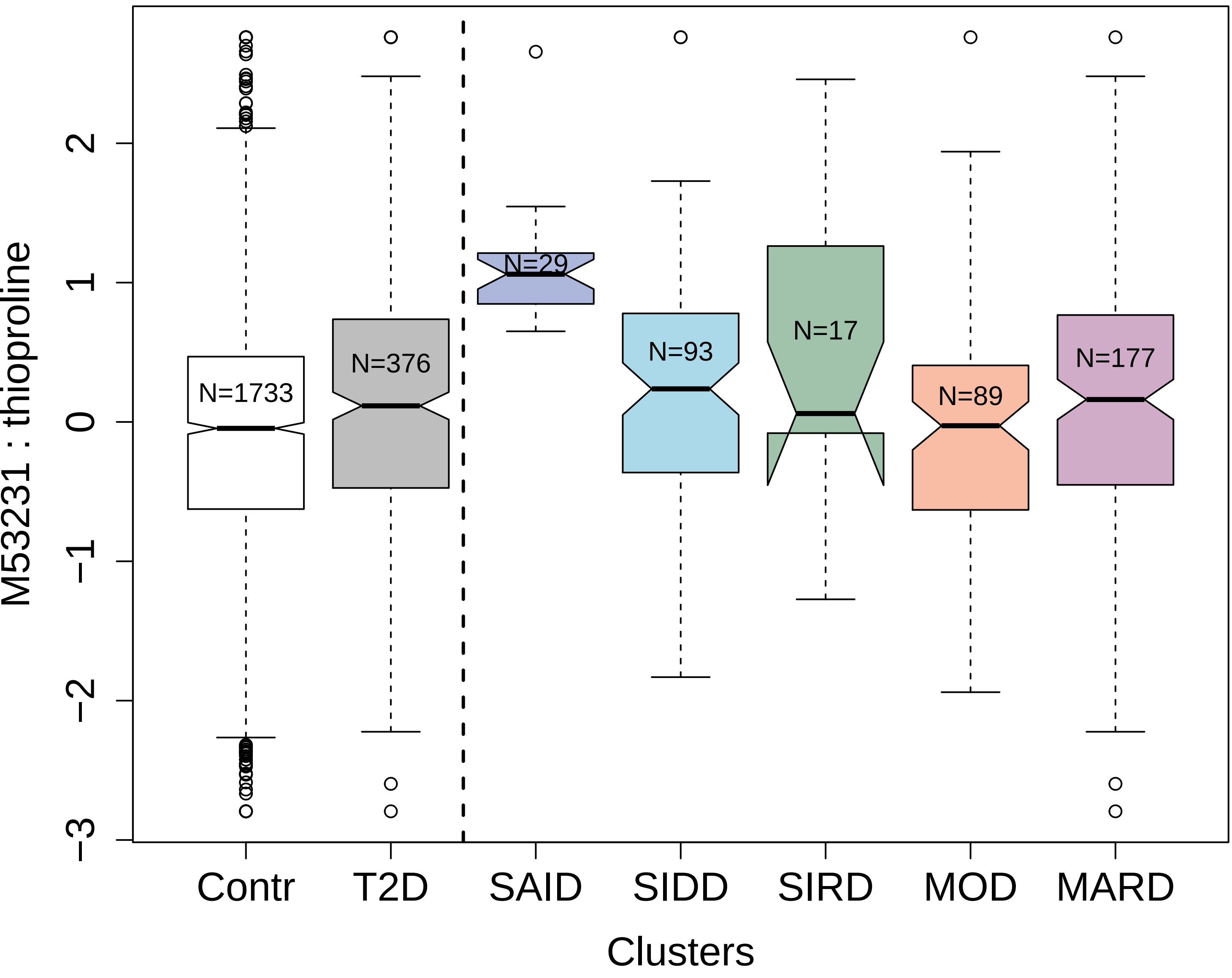

M53231 : thioproline
2
1
0
−1
−2
−3
N=1733
N=376
N=29
N=93
N=17
N=89
N=177
Contr
T2D
SAID
SIDD
SIRD
MOD
MARD
Clusters

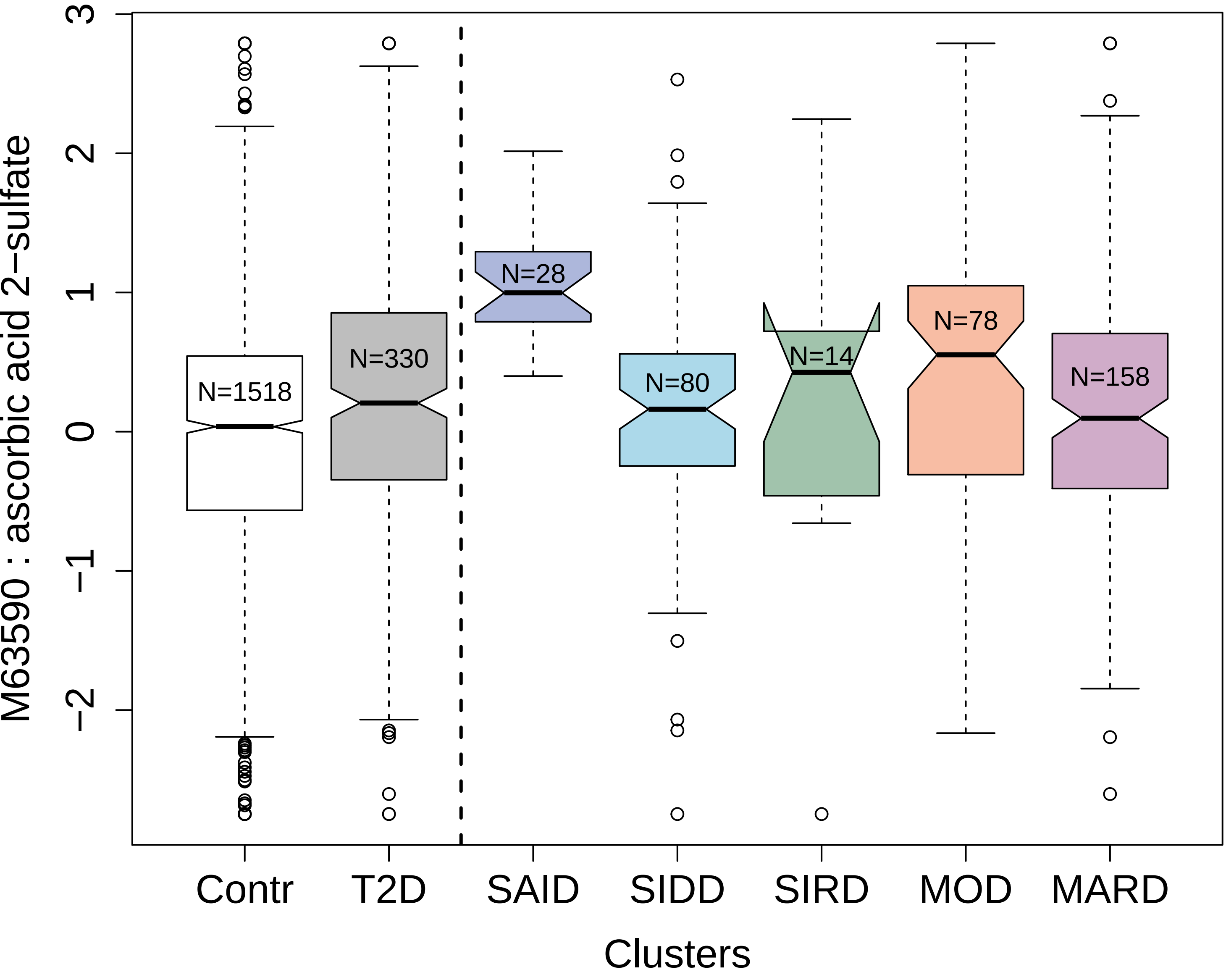

M63590 : ascorbic acid 2−sulfate
3
2
1
0
−1
−2
N=1518
N=330
N=28
N=80
N=14
N=78
N=158
Contr
T2D
SAID
SIDD
SIRD
MOD
MARD
Clusters

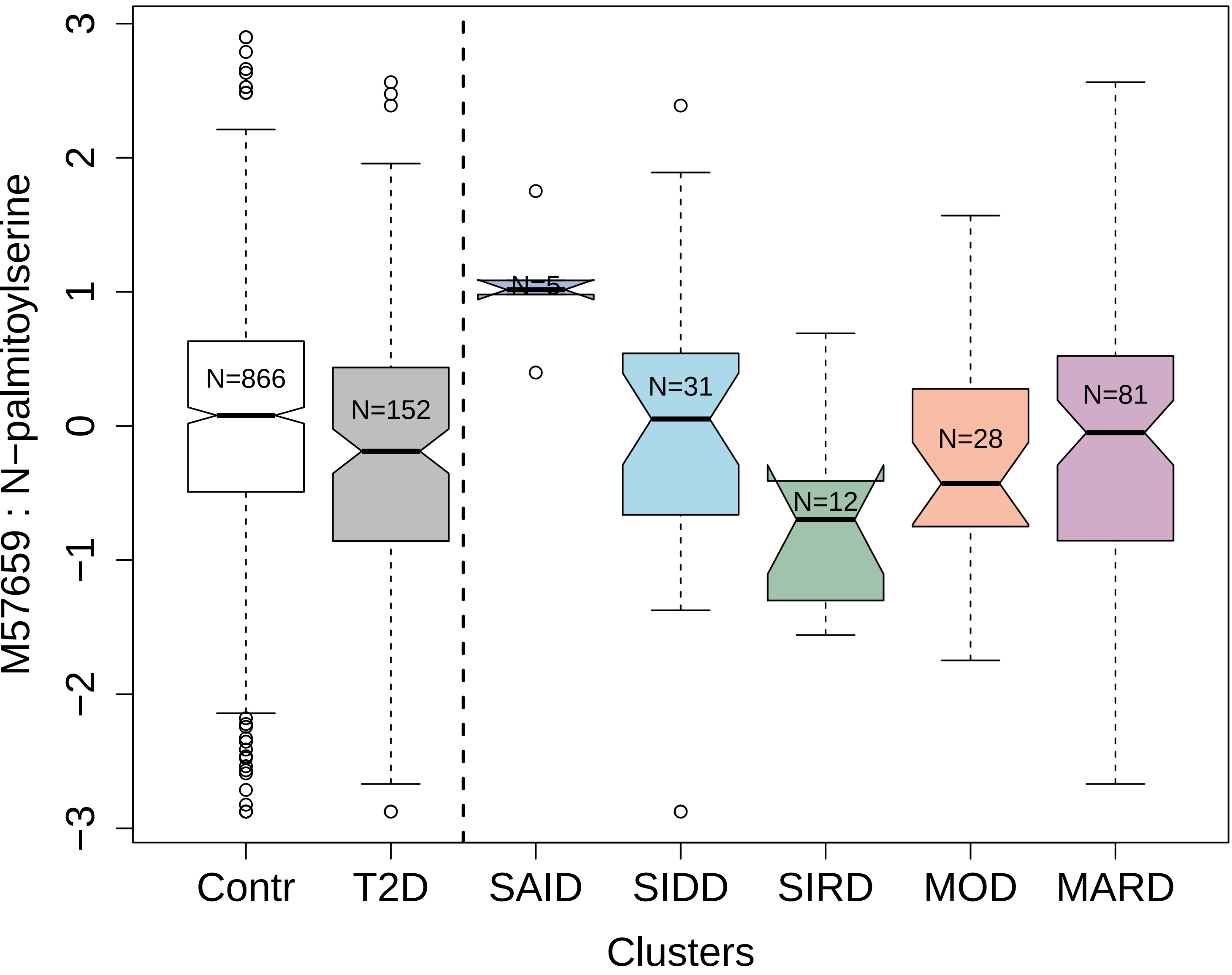
M57659 : N−palmitoylserine
3
2
1
0
−1
−2
−3
N=866
N=152
N=5
N=31
N=12
N=28
N=81
Contr
T2D
SAID
SIDD
SIRD
MOD
MARD
Clusters

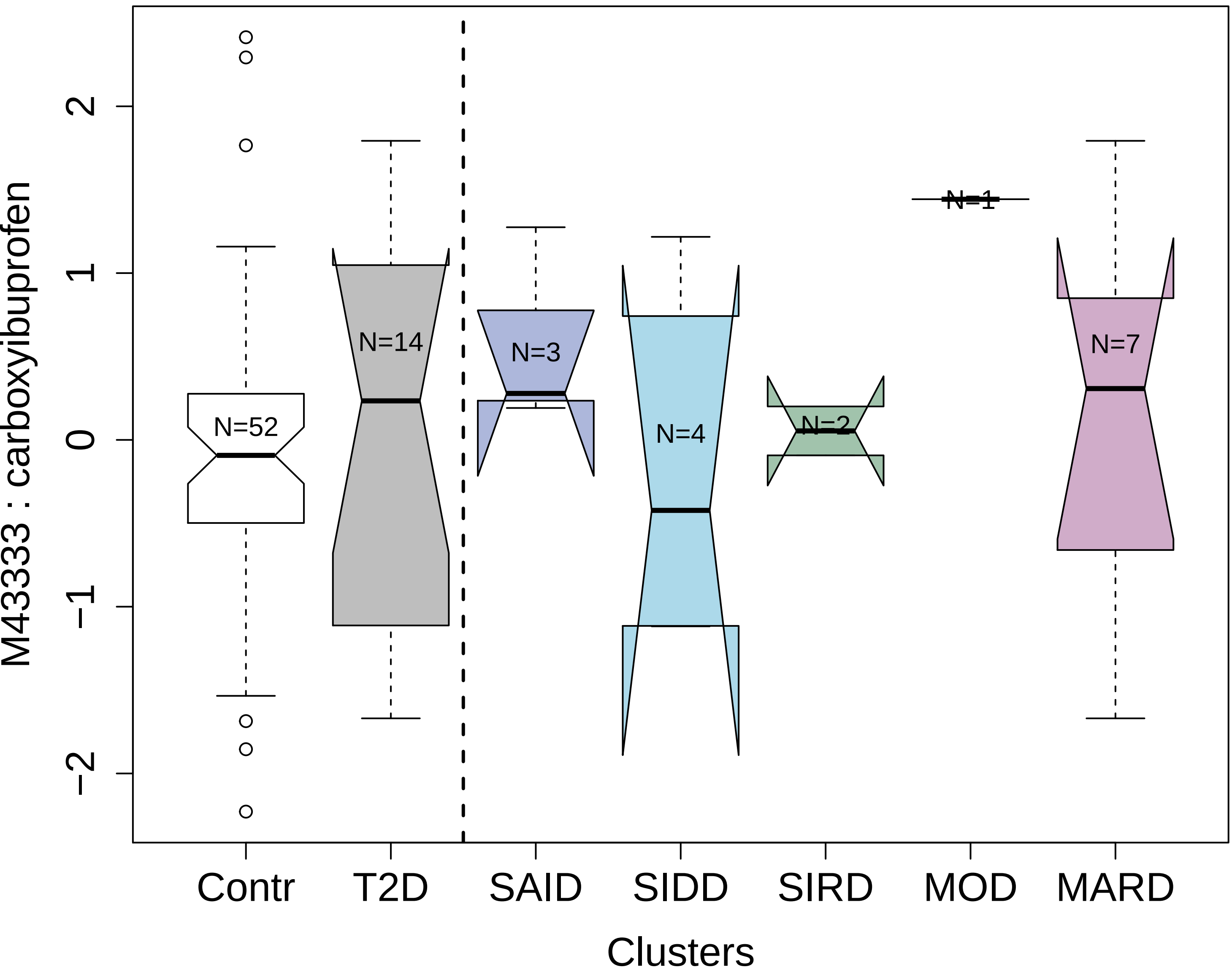

M43333 : carboxyibuprofen
2
1
0
−1
−2
N=52
N=14
N=3
N=4
N=2
N=1
N=7
Contr
T2D
SAID
SIDD
SIRD
MOD
MARD
Clusters

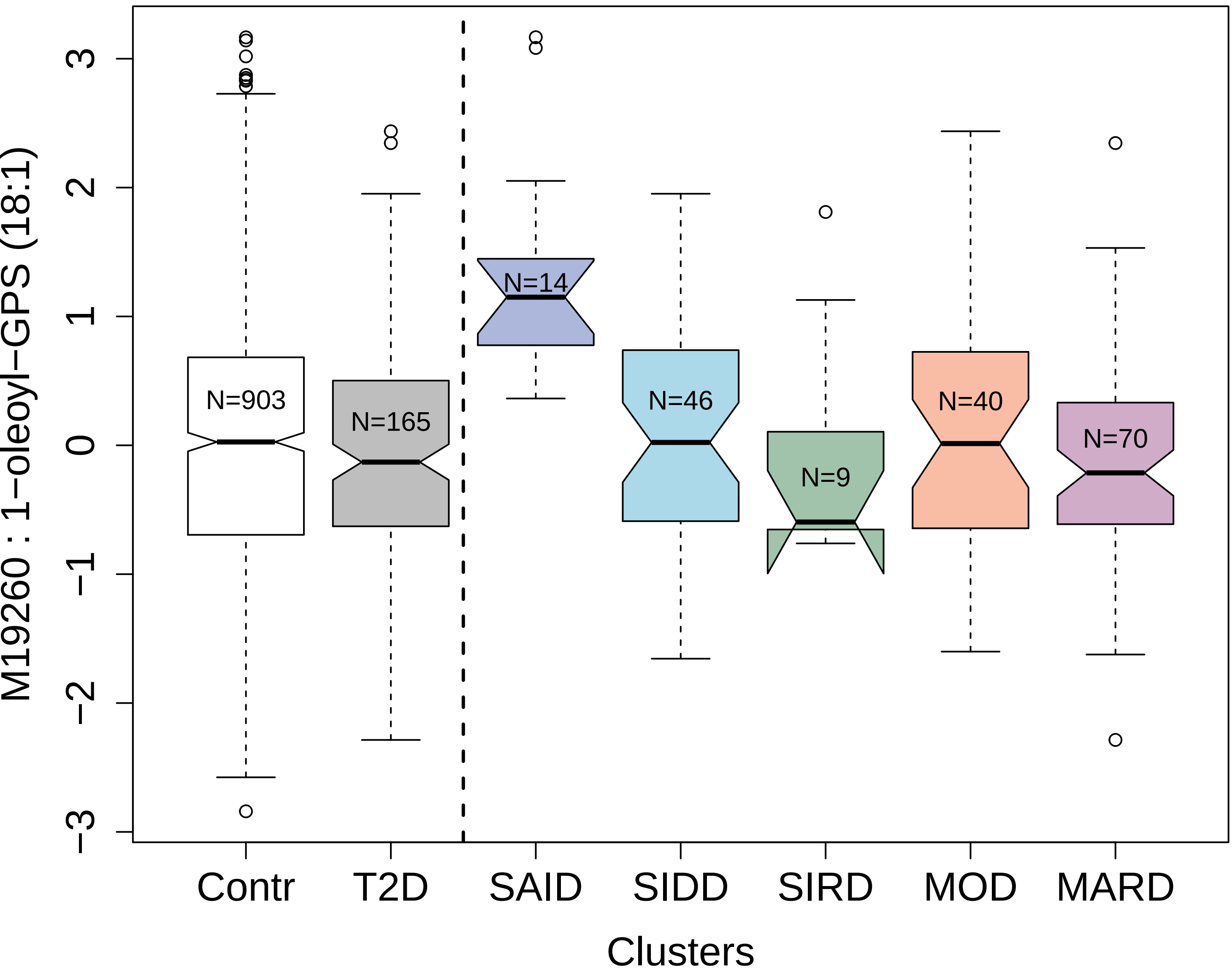

M19260 : 1−oleoyl−GPS (18:1)
3
2
1
0
−1
−2
−3
N=903
N=165
N=14
N=46
N=9
N=40
N=70
Contr
T2D
SAID
SIDD
SIRD
MOD
MARD
Clusters

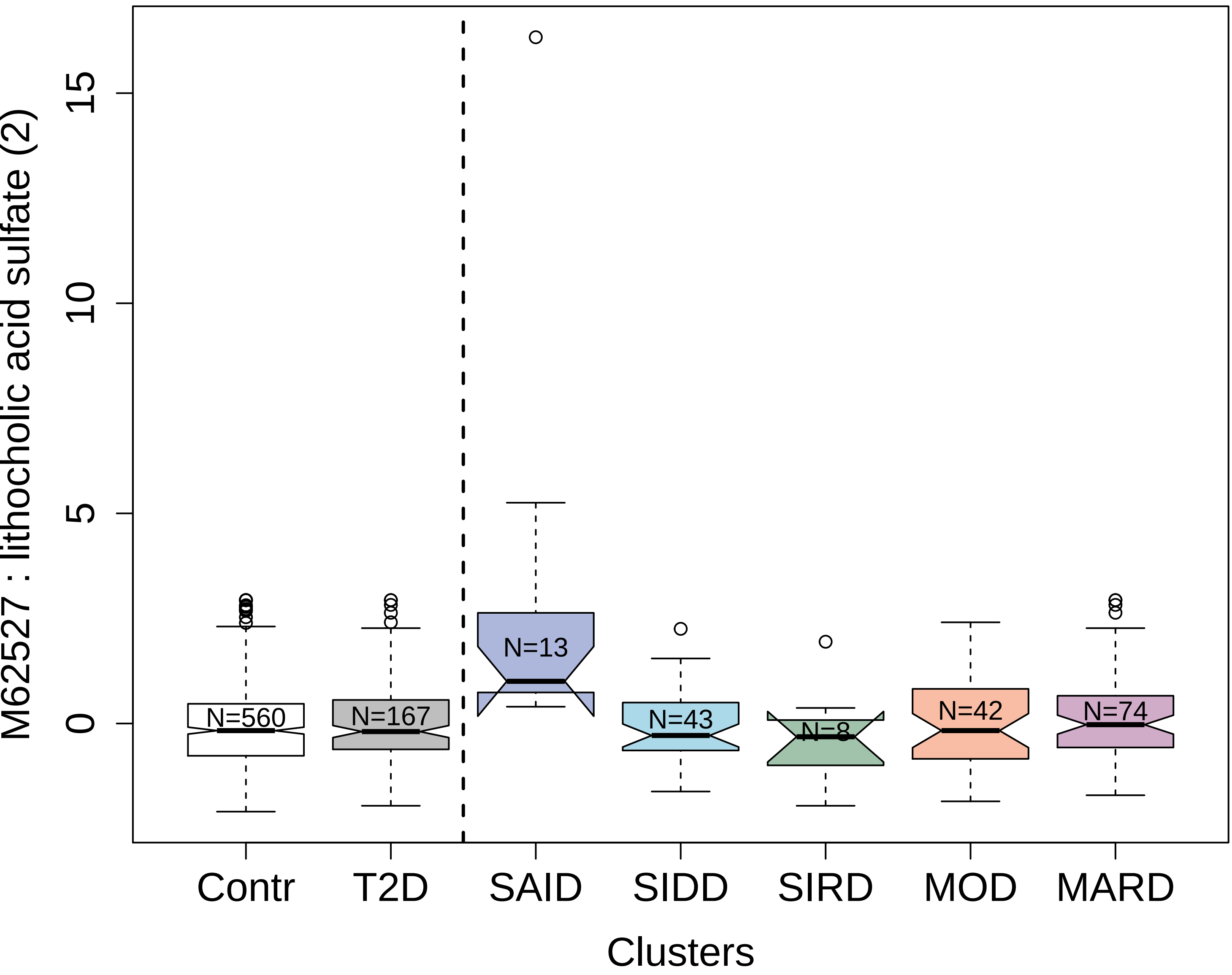

M62527 : lithocholic acid sulfate (2)
15
10
5
0
N=560
N=167
N=13
N=43
N=8
N=42
N=74
Contr
T2D
SAID
SIDD
SIRD
MOD
MARD
Clusters

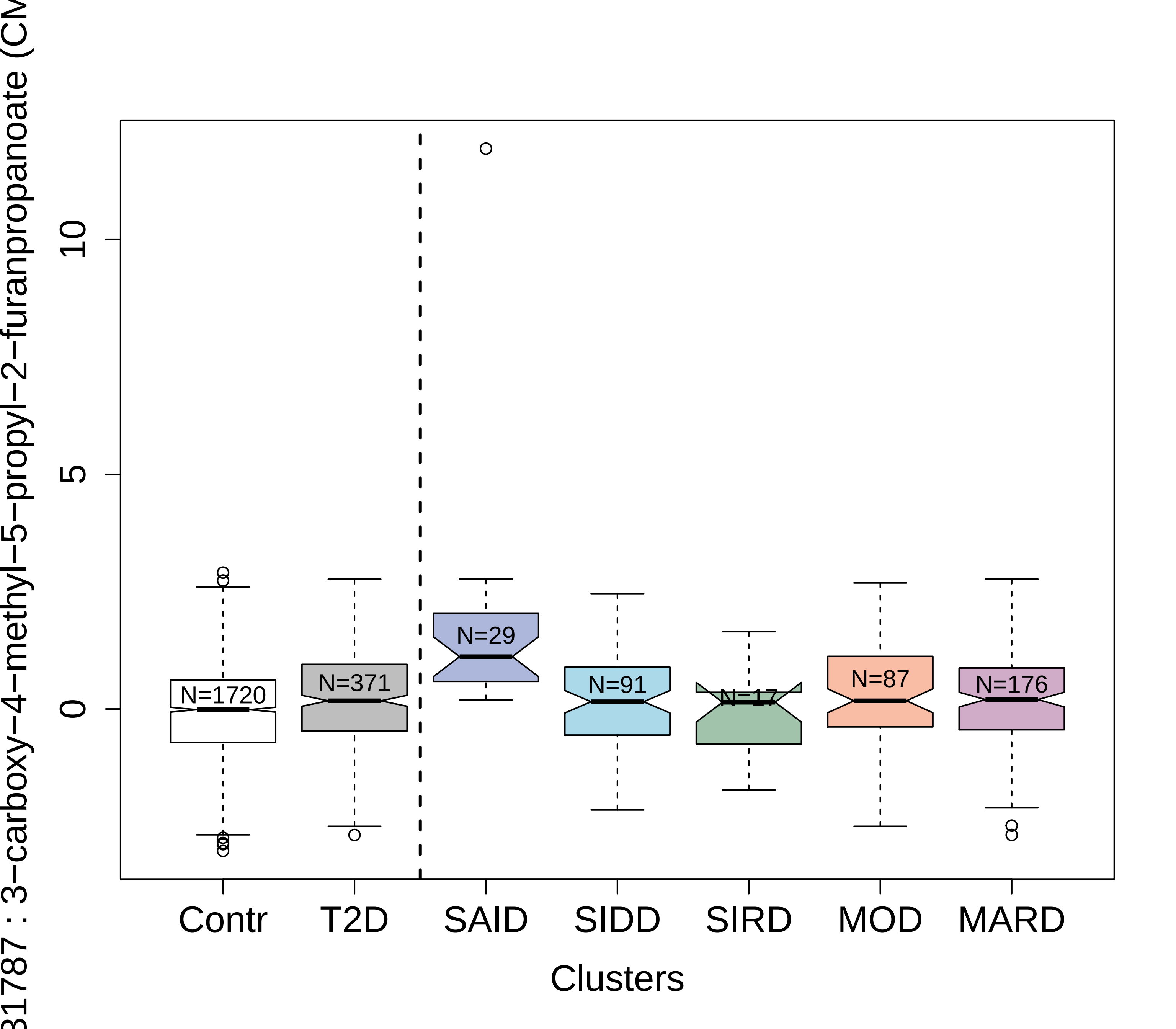

31787 : 3−carboxy−4−methyl−5−propyl−2−furanpropanoate (CM
0
5
10
N=1720
N=371
N=29
N=91
N=17
N=87
N=176
Contr
T2D
SAID
SIDD
SIRD
MOD
MARD
Clusters

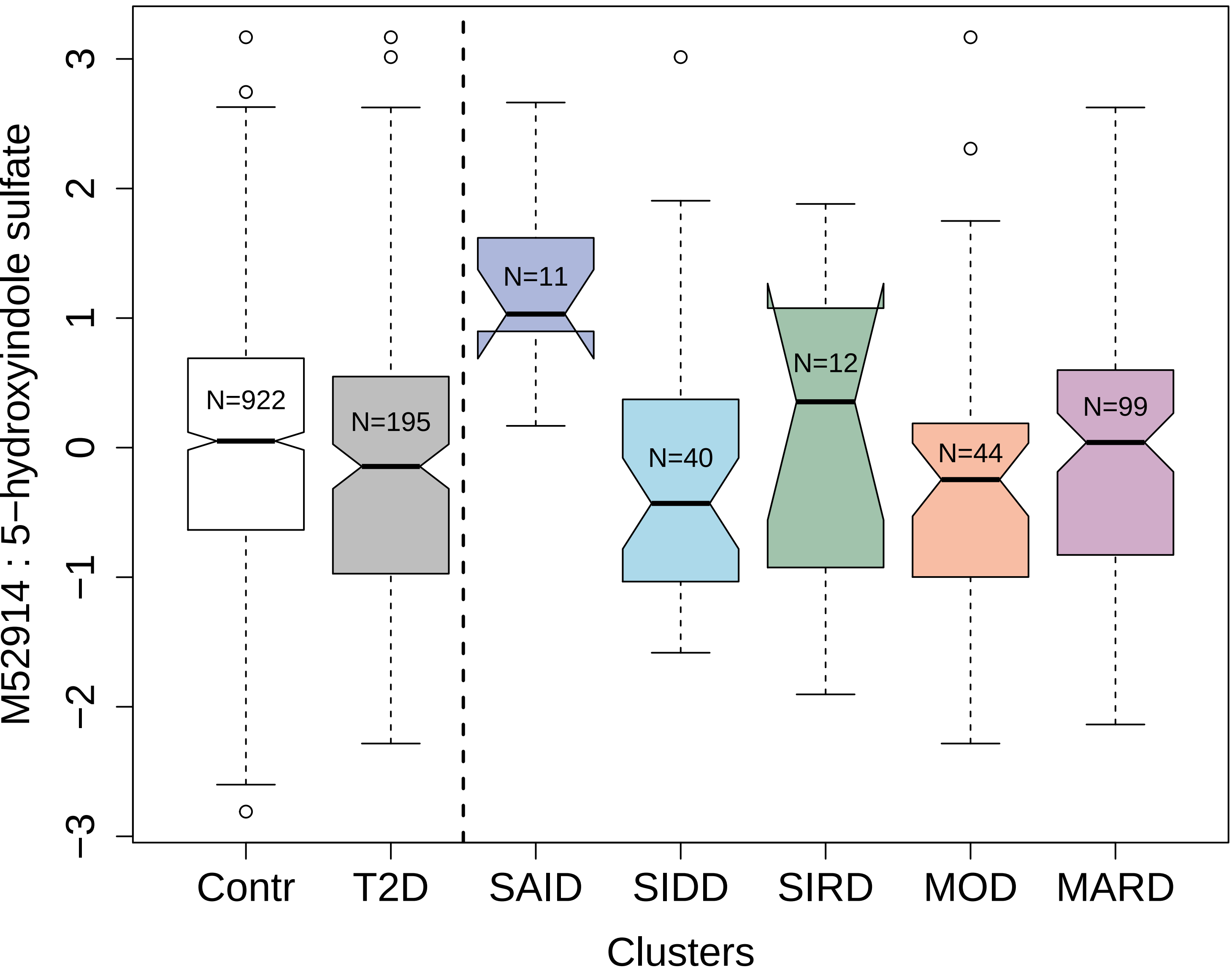

M52914 : 5−hydroxyindole sulfate
3
2
1
0
−1
−2
−3
N=922
N=195
N=11
N=40
N=12
N=44
N=99
Contr
T2D
SAID
SIDD
SIRD
MOD
MARD
Clusters

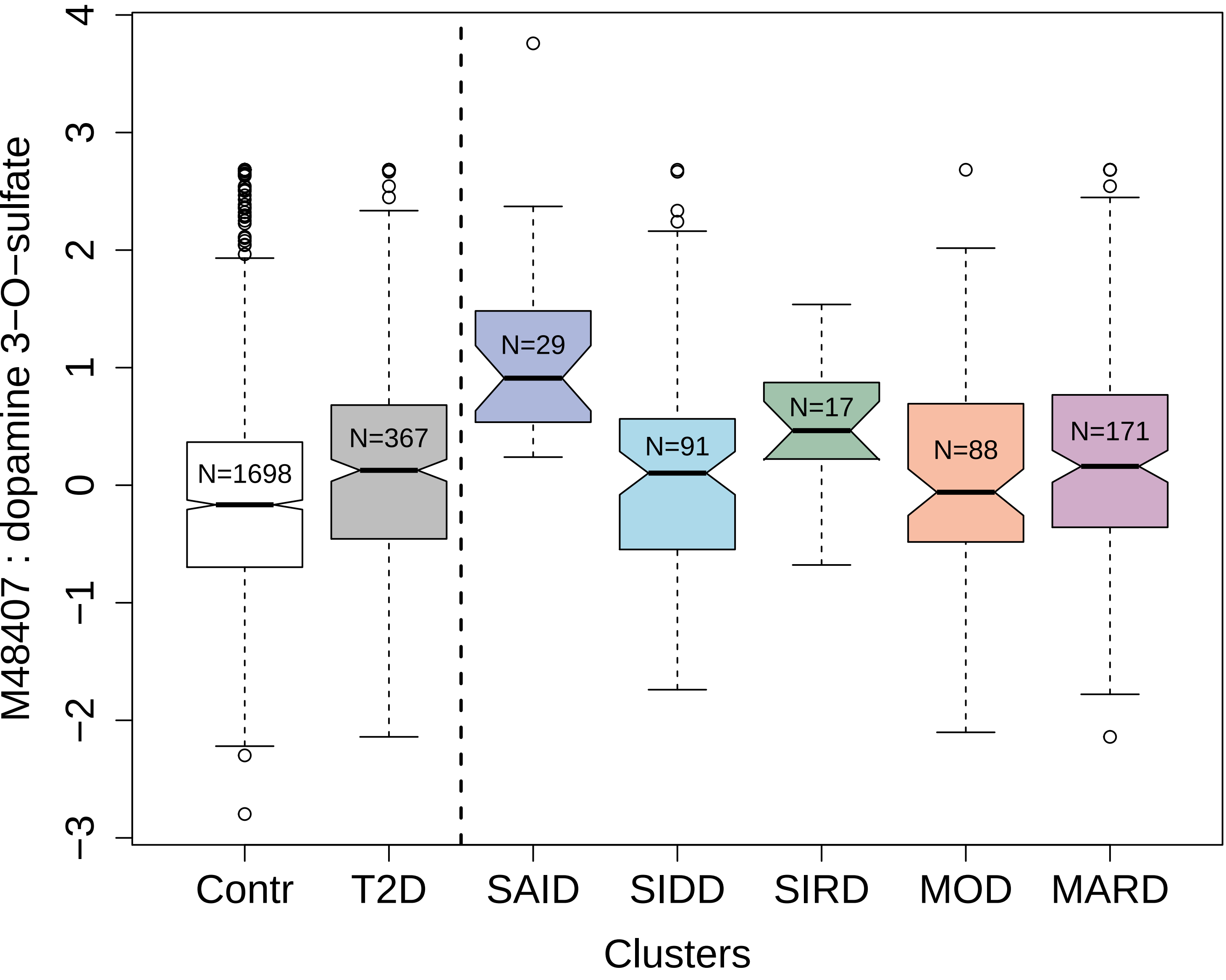

M48407 : dopamine 3−O−sulfate
4
3
2
1
0
−1
−2
−3
N=1698
N=367
N=29
N=91
N=17
N=88
N=171
Contr
T2D
SAID
SIDD
SIRD
MOD
MARD
Clusters

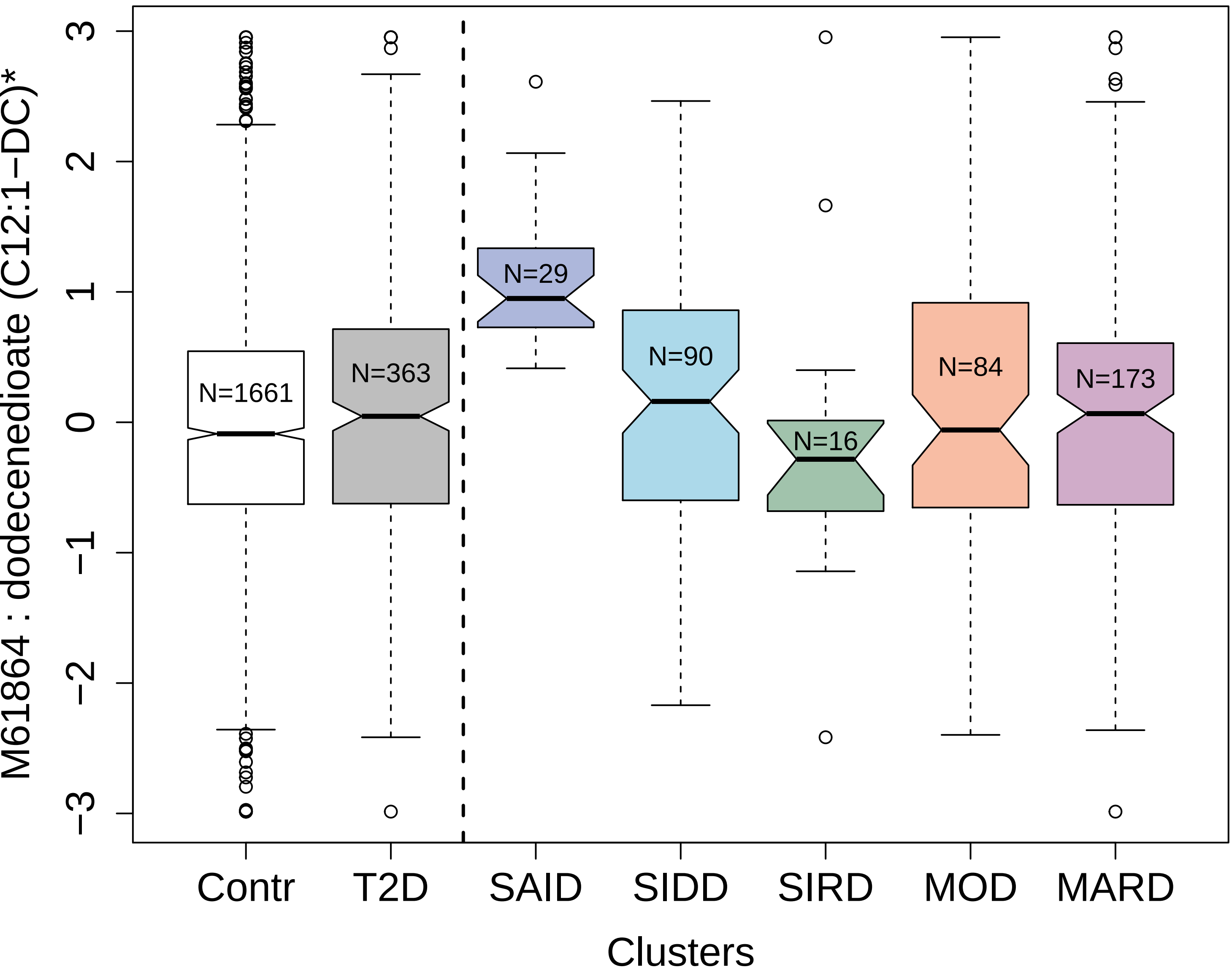

M61864 : dodecenedioate (C12:1−DC)*
3
2
1
0
−1
−2
−3
N=1661
N=363
N=29
N=90
N=16
N=84
N=173
Contr
T2D
SAID
SIDD
SIRD
MOD
MARD
Clusters

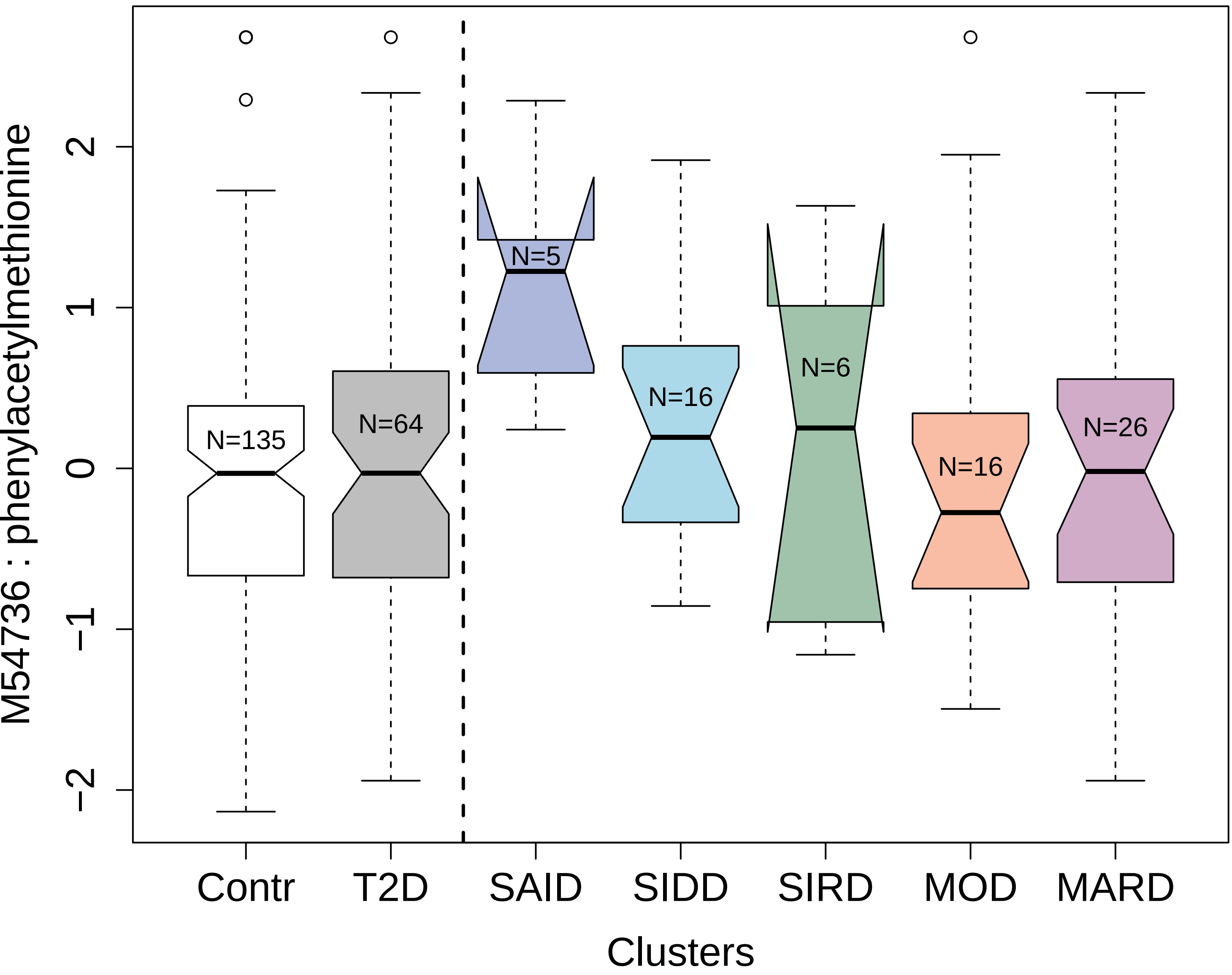
M54736 : phenylacetylmethionine
−2
−1
0
1
2
N=135
N=64
N=5
N=16
N=6
N=16
N=26
Contr
T2D
SAID
SIDD
SIRD
MOD
MARD
Clusters

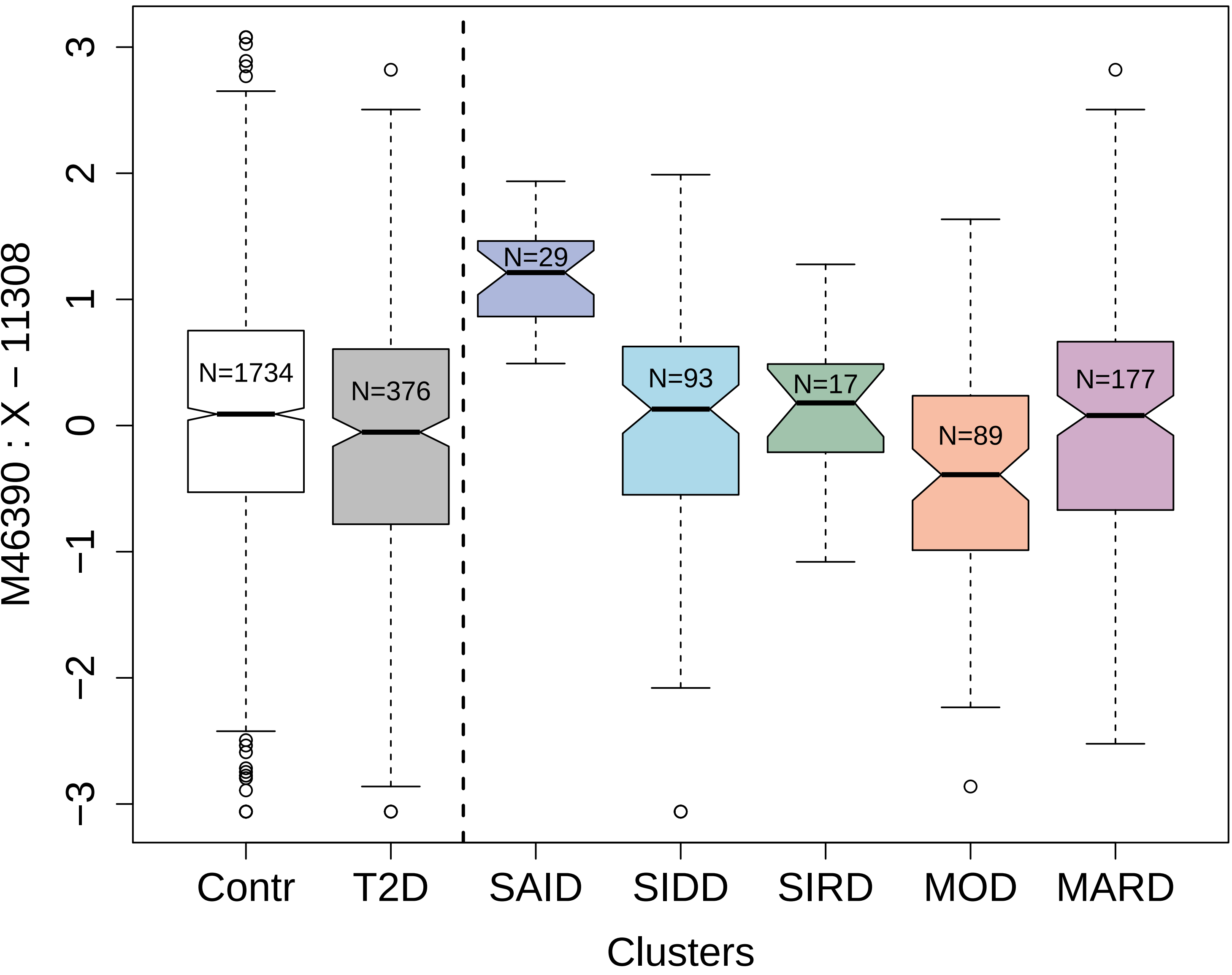

M46390 : X − 11308
3
2
1
0
−1
−2
−3
N=1734
N=376
N=29
N=93
N=17
N=89
N=177
Contr
T2D
SAID
SIDD
SIRD
MOD
MARD
Clusters

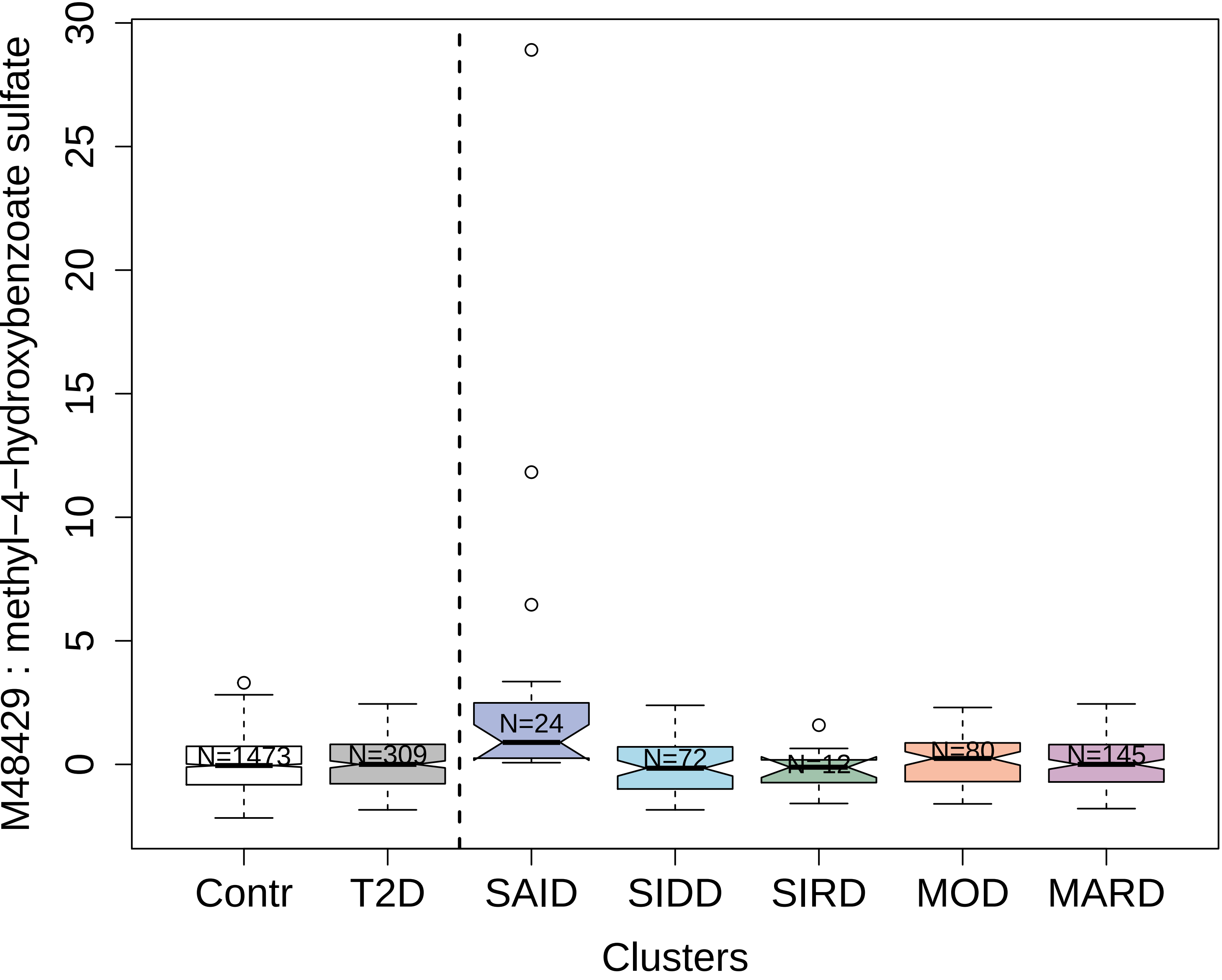

M48429 : methyl−4−hydroxybenzoate sulfate
30
25
20
15
10
5
0
N=1473
N=309
N=24
N=72
N=12
N=80
N=145
Contr
T2D
SAID
SIDD
SIRD
MOD
MARD
Clusters

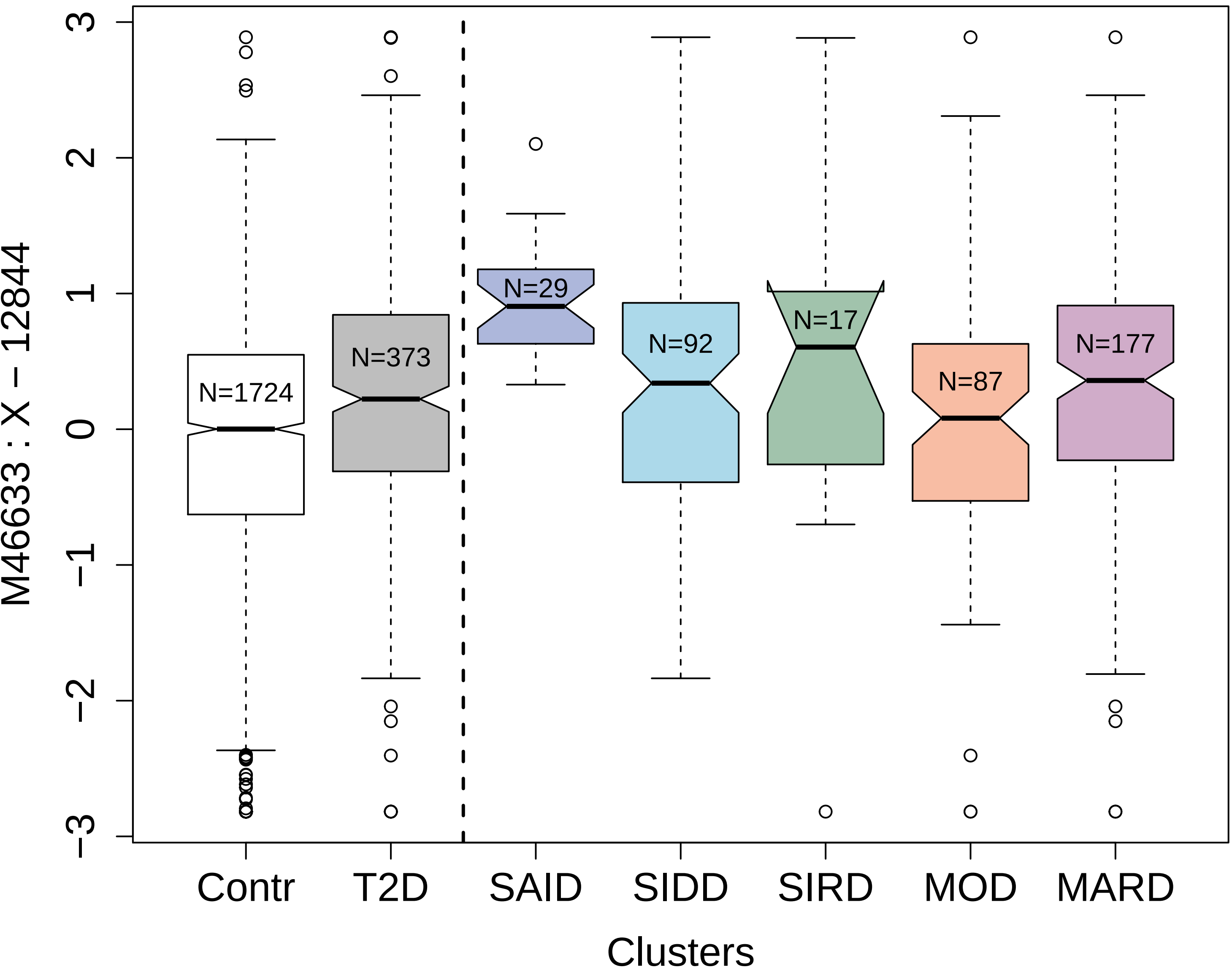

M46633 : X − 12844
3
2
1
0
−1
−2
−3
N=1724
N=373
N=29
N=92
N=17
N=87
N=177
Contr
T2D
SAID
SIDD
SIRD
MOD
MARD
Clusters

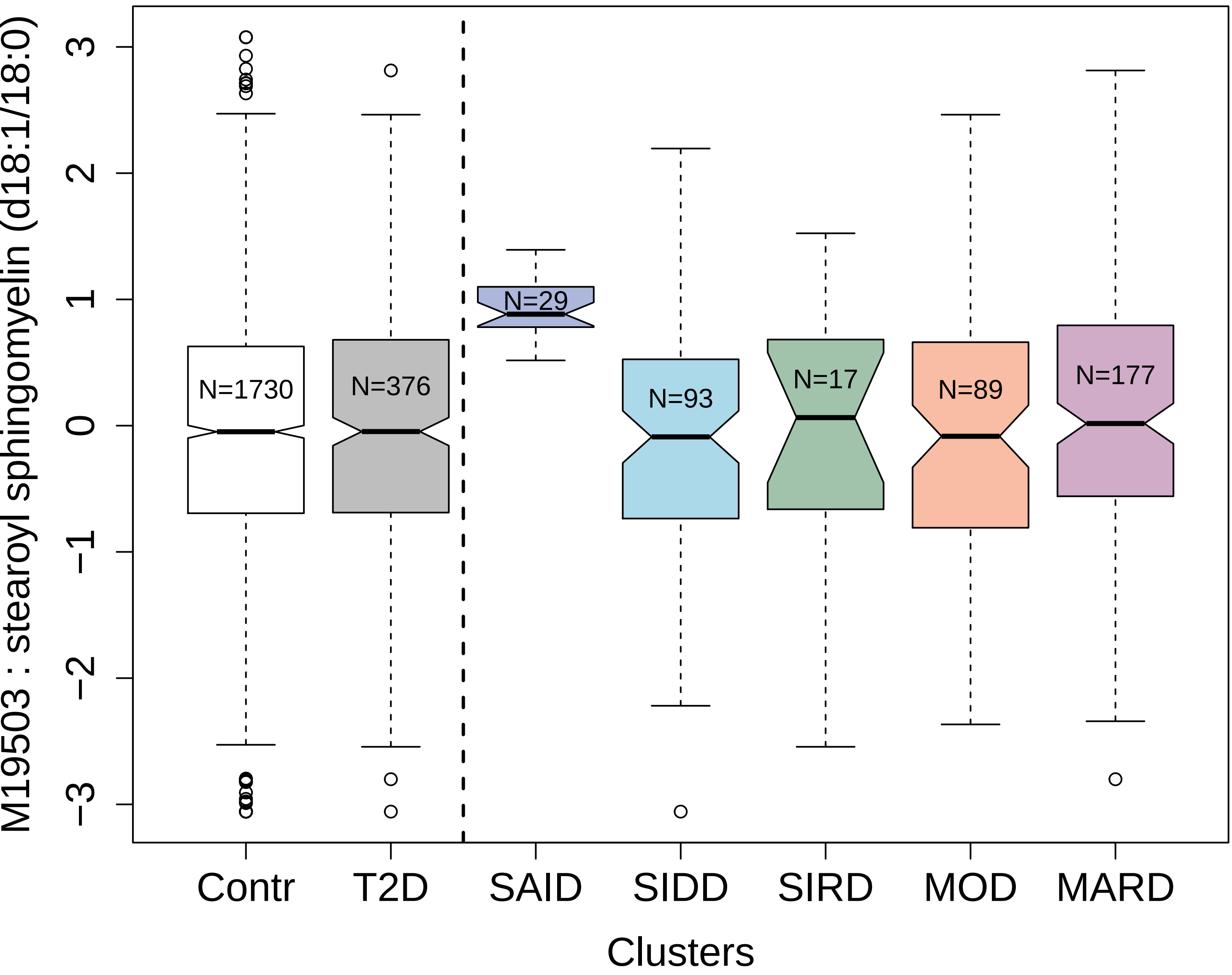
M19503 : stearoyl sphingomyelin (d18:1/18:0)
3
2
1
0
−1
−2
−3
N=1730
N=376
N=29
N=93
N=17
N=89
N=177
Contr
T2D
SAID
SIDD
SIRD
MOD
MARD
Clusters

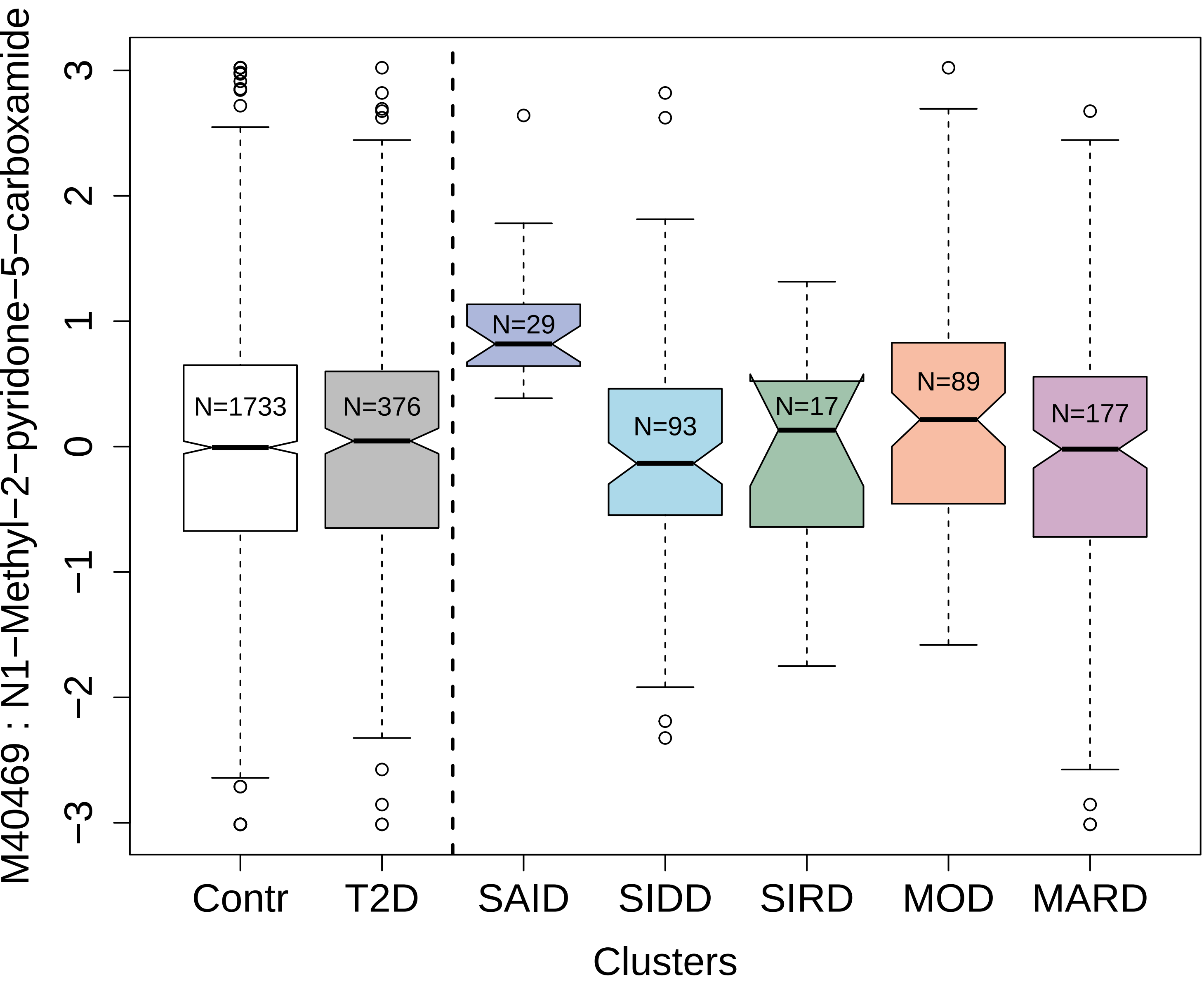
M40469 : N1−Methyl−2−pyridone−5−carboxamide
3
2
1
0
−1
−2
−3
N=1733
N=376
N=29
N=93
N=17
N=89
N=177
Contr
T2D
SAID
SIDD
SIRD
MOD
MARD
Clusters

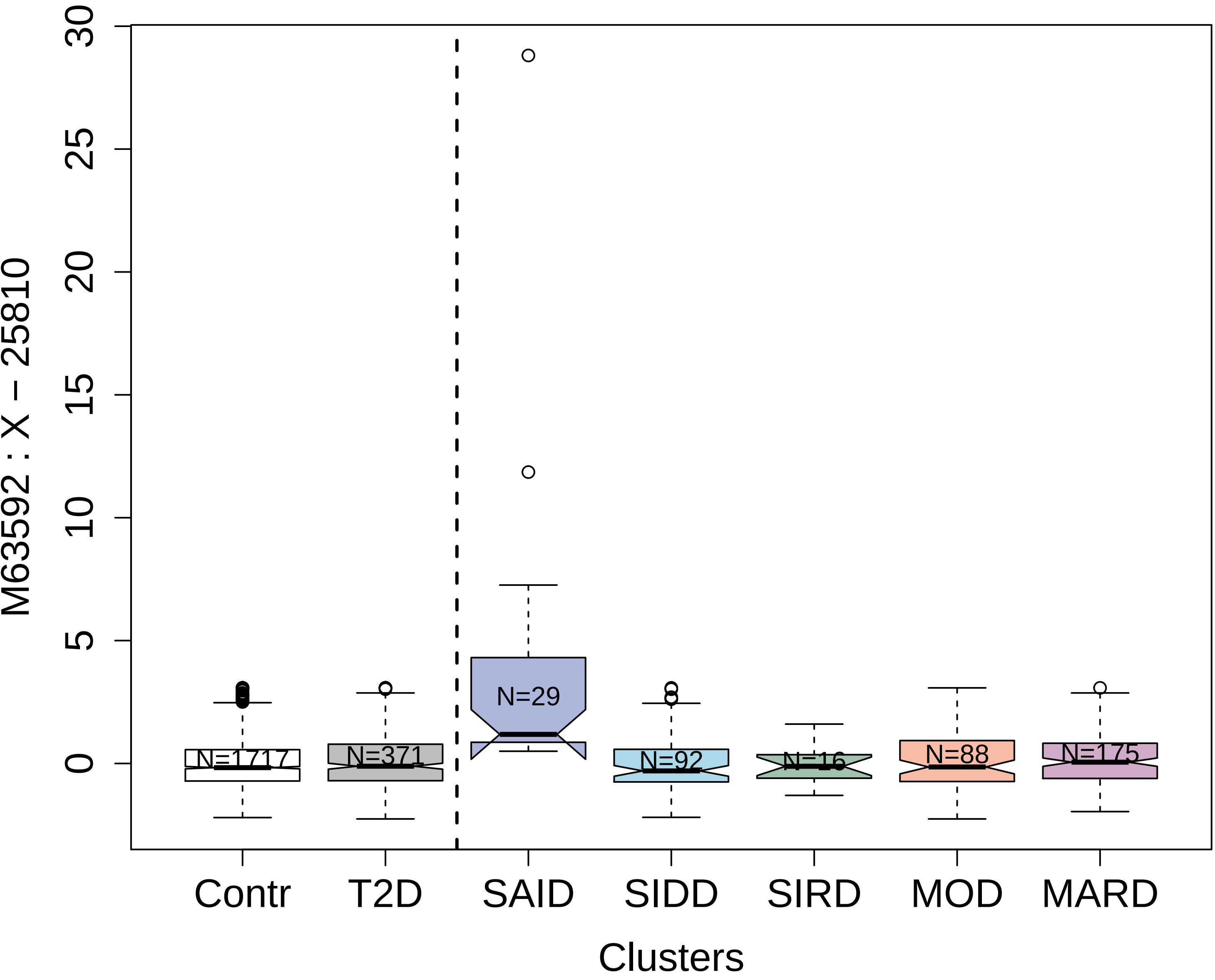

M63592 : X − 25810
30
25
20
15
10
5
0
N=1717
N=371
N=29
N=92
N=16
N=88
N=175
Contr
T2D
SAID
SIDD
SIRD
MOD
MARD
Clusters

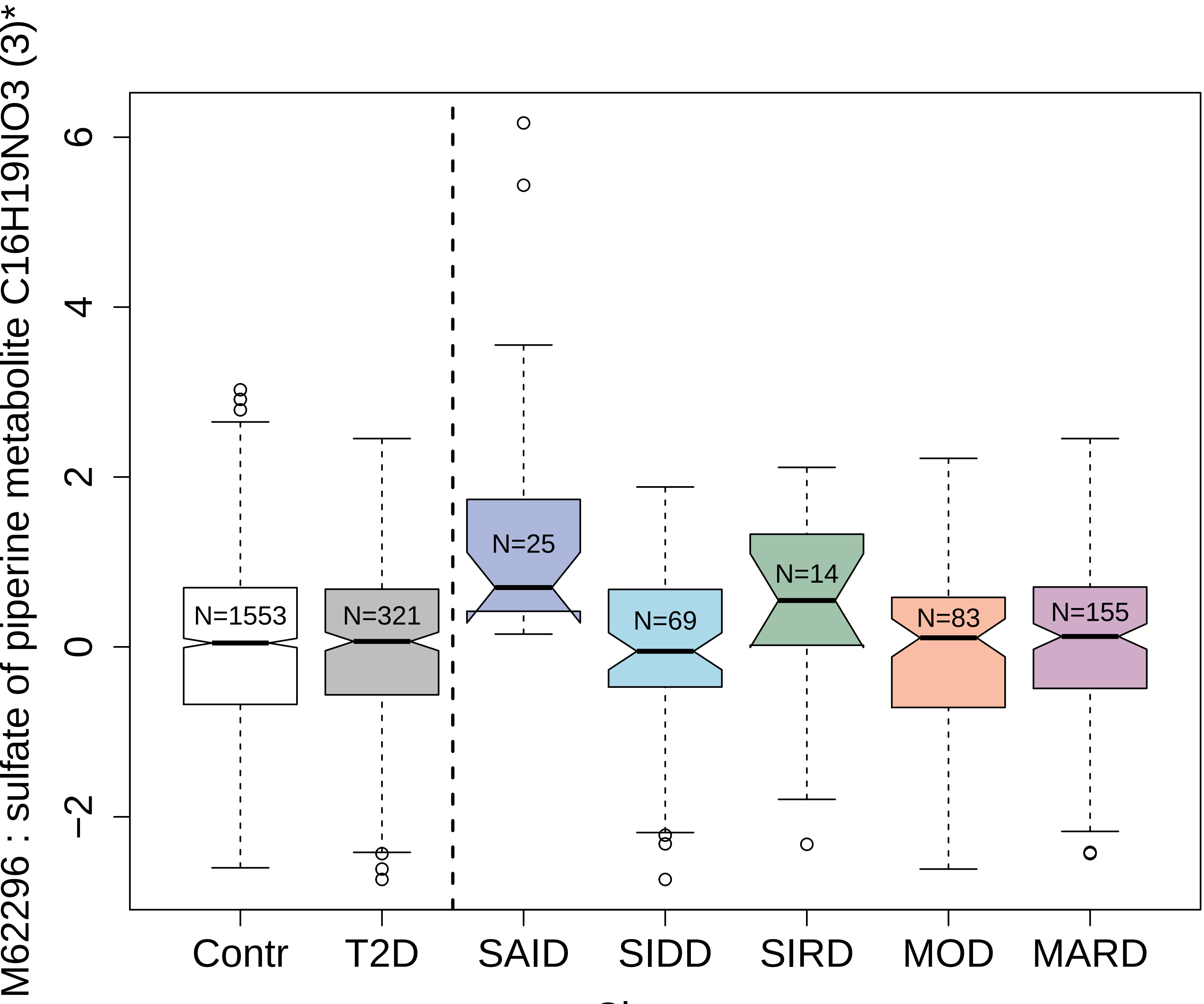

M62296 : sulfate of piperine metabolite C16H19NO3 (3)*
6
4
2
0
−2
N=1553
N=321
N=25
N=69
N=14
N=83
N=155
Contr
T2D
SAID
SIDD
SIRD
MOD
MARD
Clusters

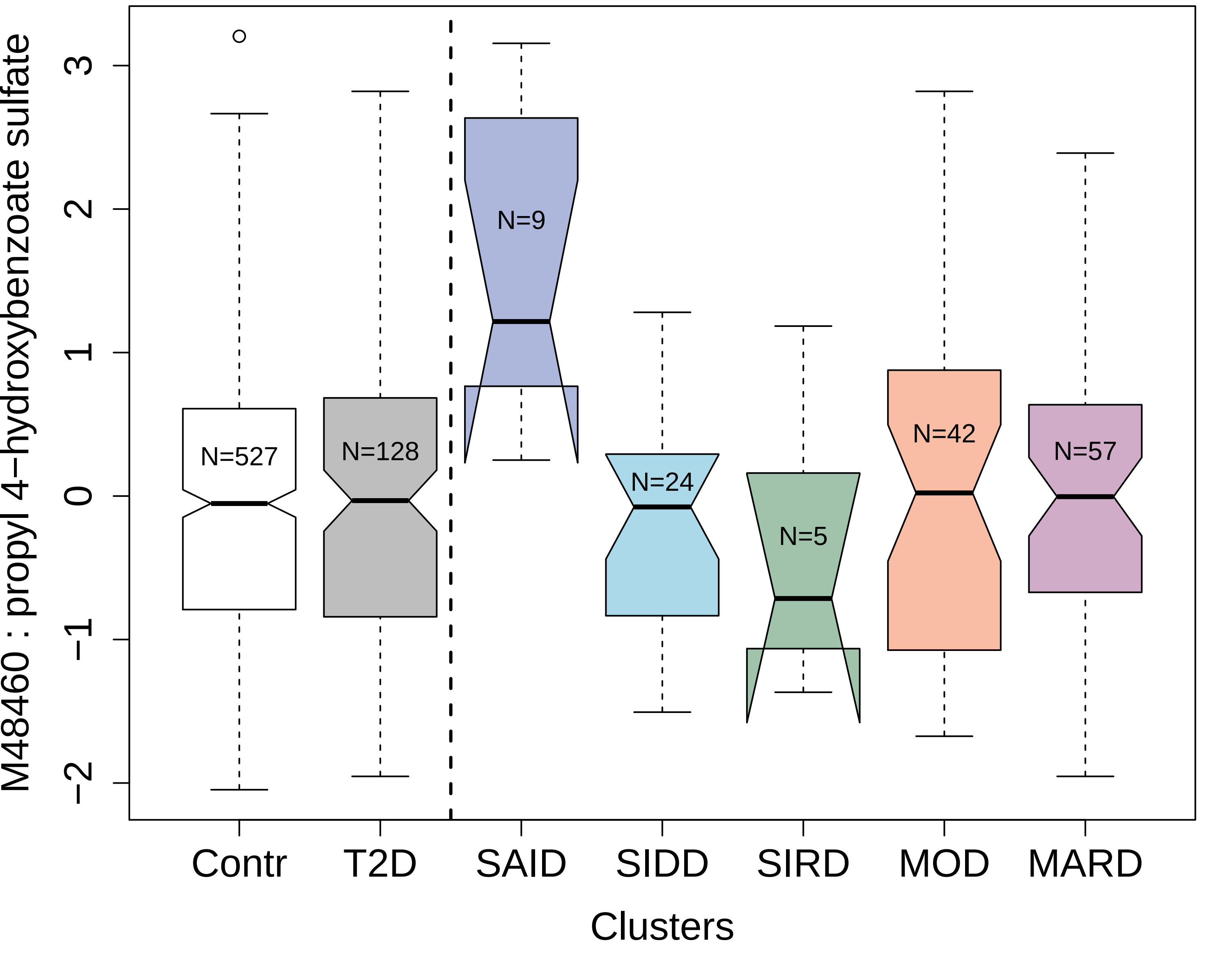
M48460 : propyl 4−hydroxybenzoate sulfate
−2
−1
0
1
2
3
N=527
N=128
N=9
N=24
N=5
N=42
N=57
Contr
T2D
SAID
SIDD
SIRD
MOD
MARD
Clusters

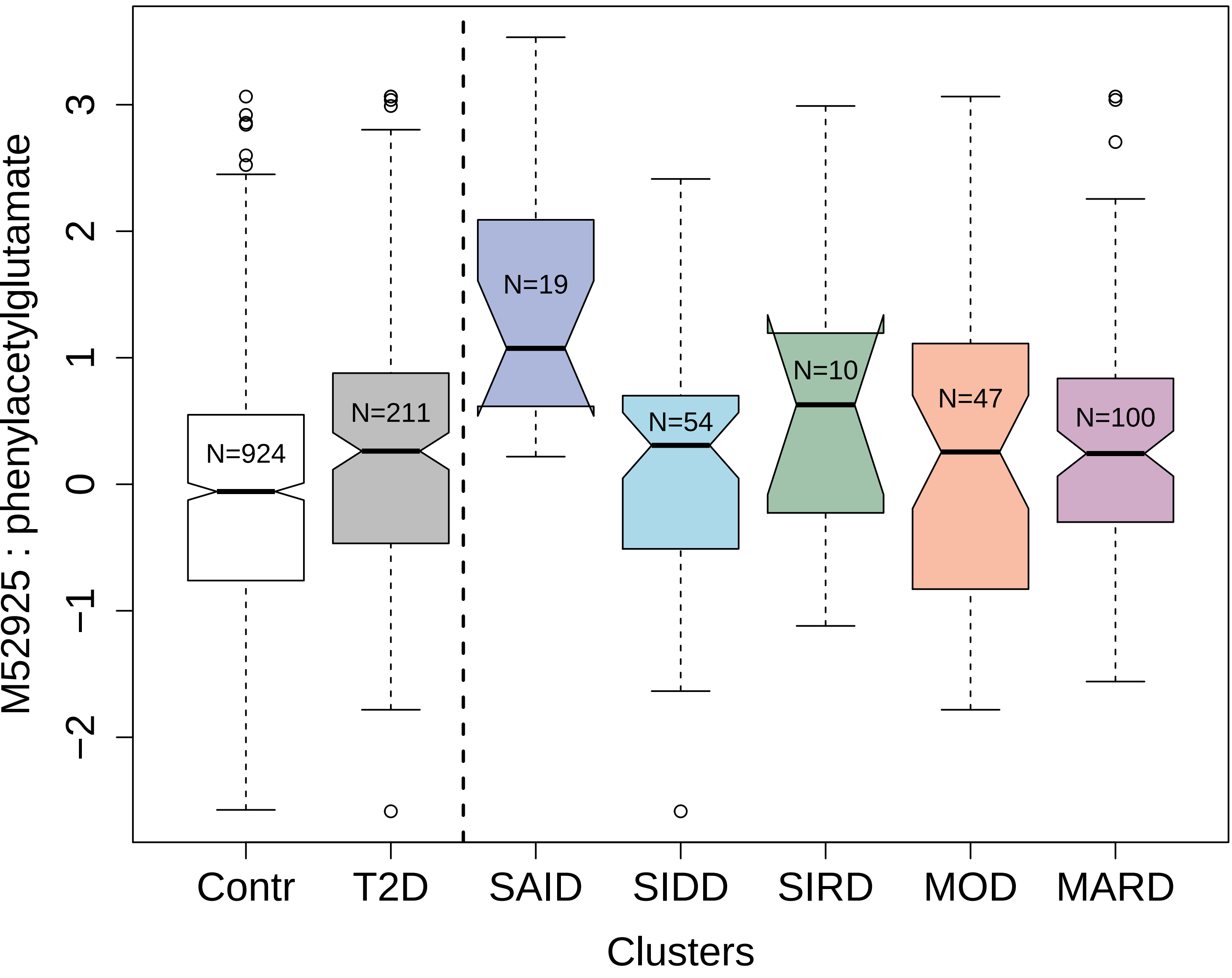

M52925 : phenylacetylglutamate
3
2
1
0
−1
−2
N=924
N=211
N=19
N=54
N=10
N=47
N=100
Contr
T2D
SAID
SIDD
SIRD
MOD
MARD
Clusters

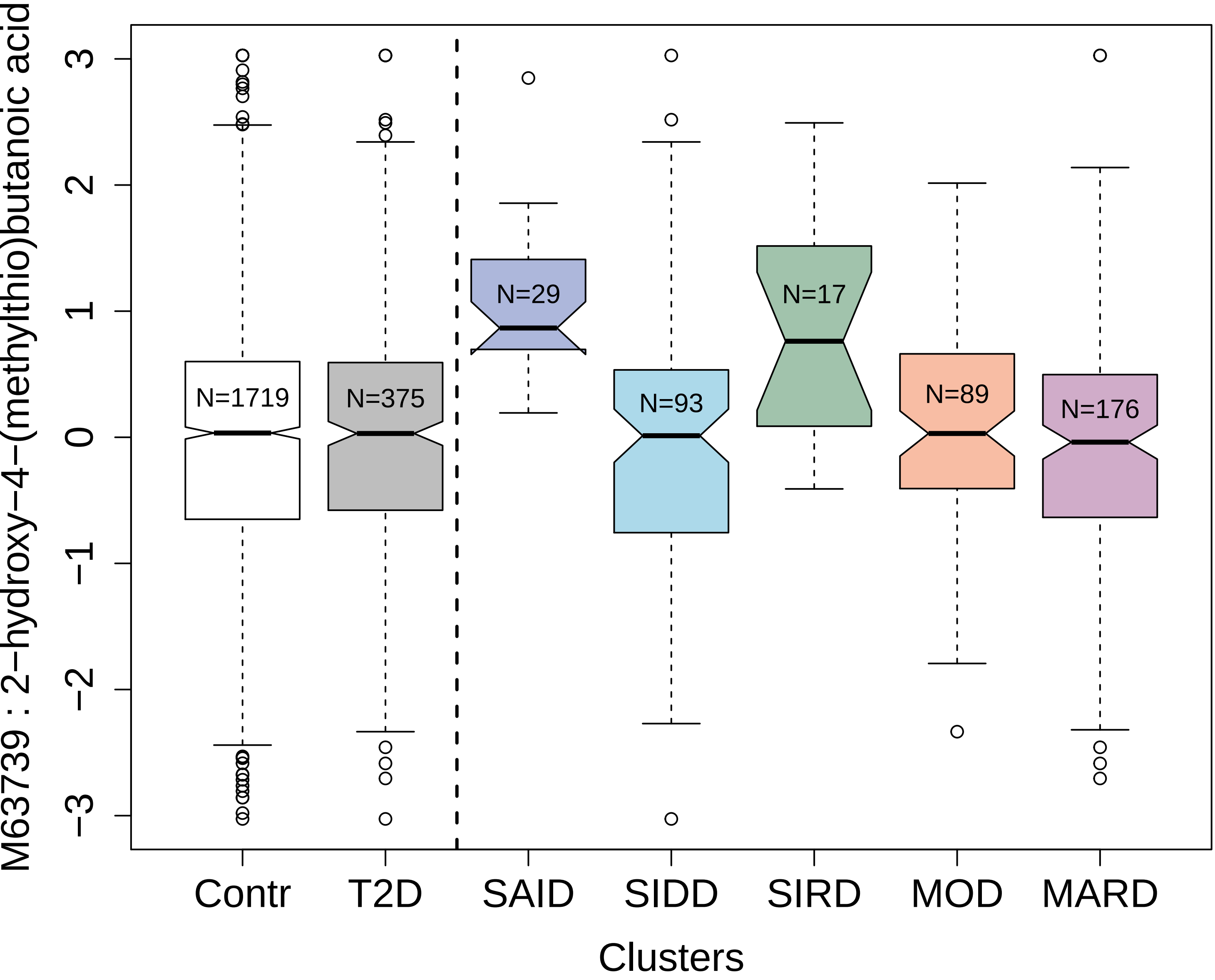

M63739 : 2−hydroxy−4−(methylthio)butanoic acid
3
2
1
0
−1
−2
−3
N=1719
N=375
N=29
N=93
N=17
N=89
N=176
Contr
T2D
SAID
SIDD
SIRD
MOD
MARD
Clusters

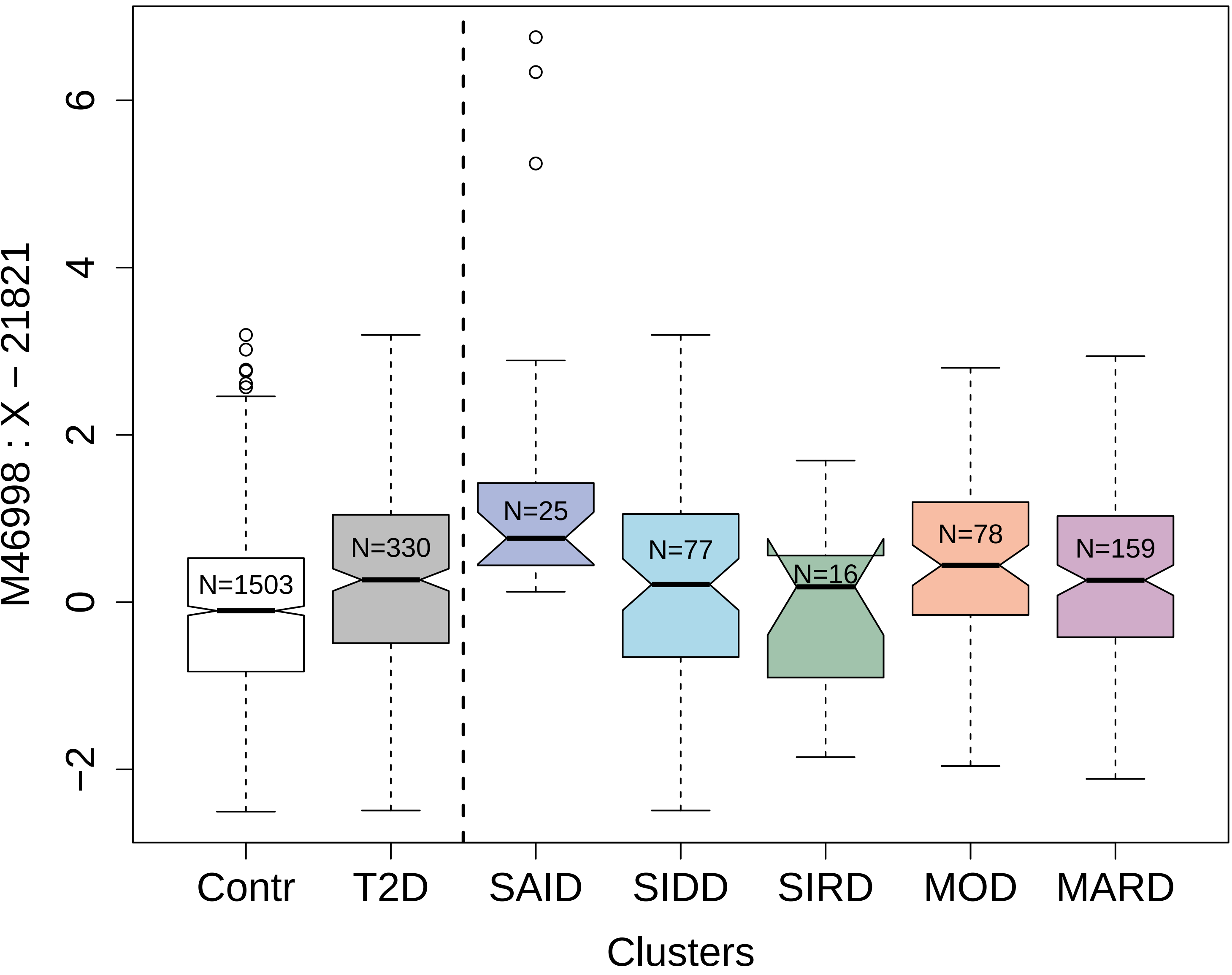

M46998 : X − 21821
6
4
2
0
−2
N=1503
N=330
N=25
N=77
N=16
N=78
N=159
Contr
T2D
SAID
SIDD
SIRD
MOD
MARD
Clusters

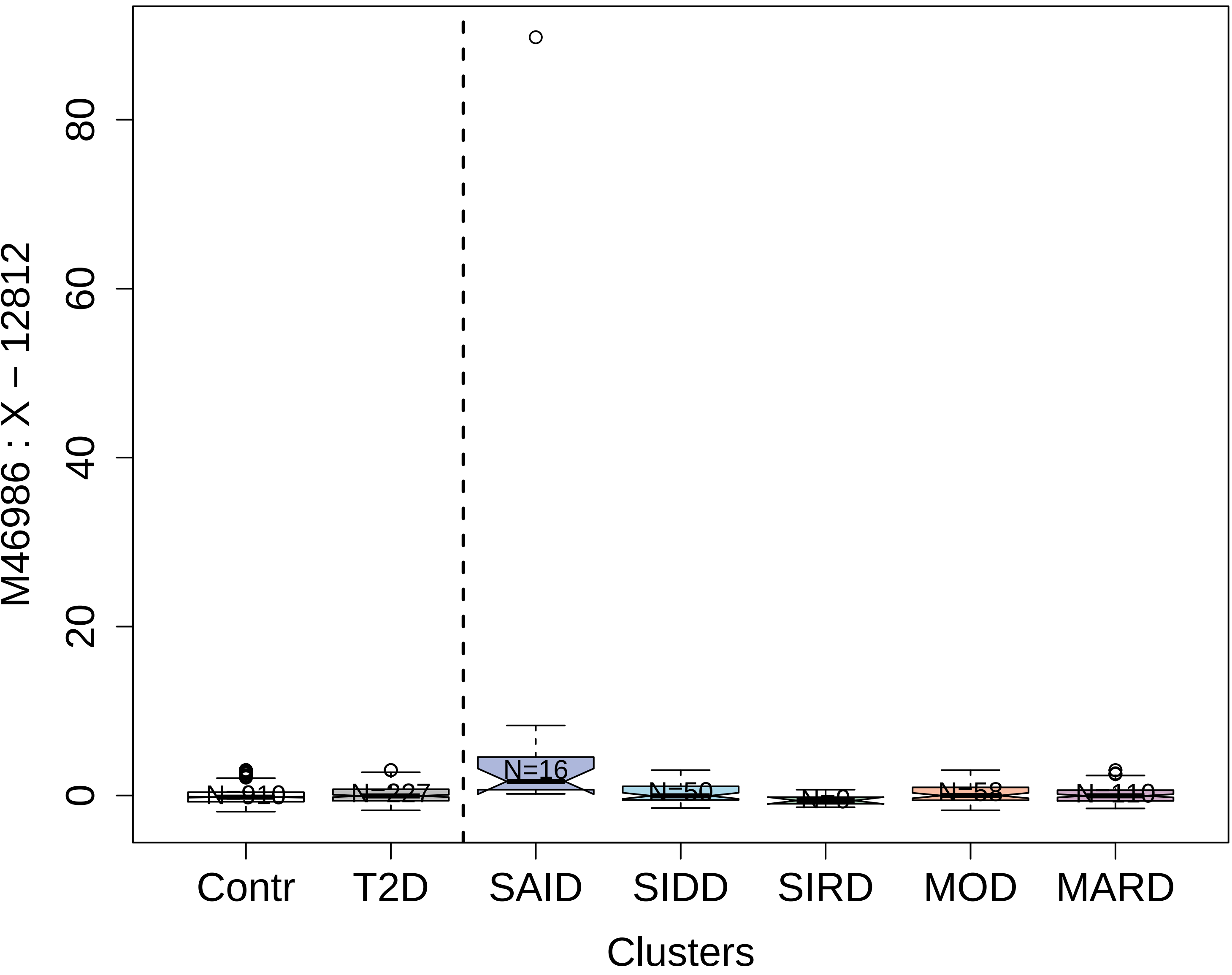

M46986 : X − 12812
0
20
40
60
80
N=910
N=227
N=16
N=50
N=0
N=58
N=110
Contr
T2D
SAID
SIDD
SIRD
MOD
MARD
Clusters

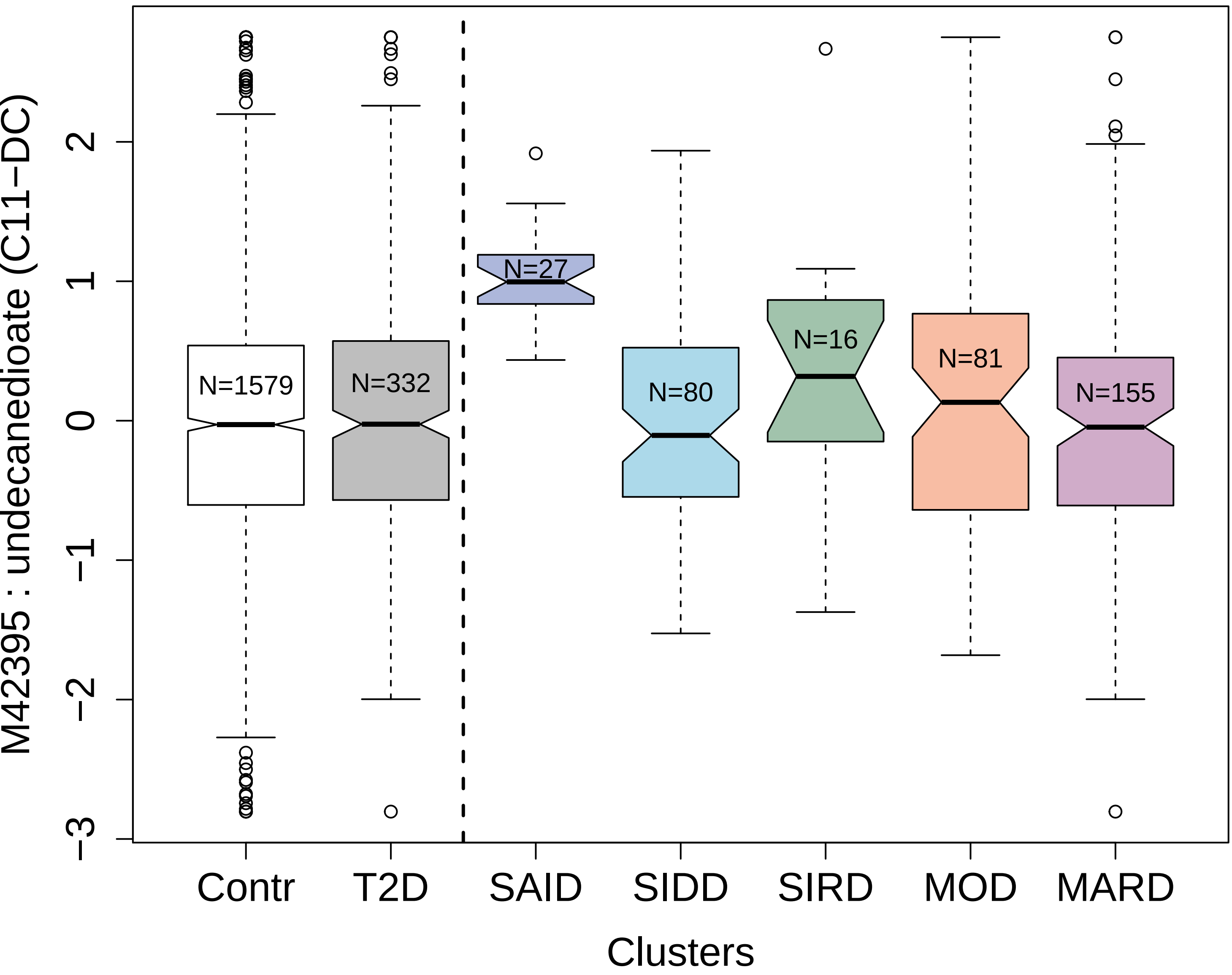
M42395 : undecanedioate (C11−DC)
−3
−2
−1
0
1
2
N=1579
N=332
N=27
N=80
N=16
N=81
N=155
Contr
T2D
SAID
SIDD
SIRD
MOD
MARD
Clusters

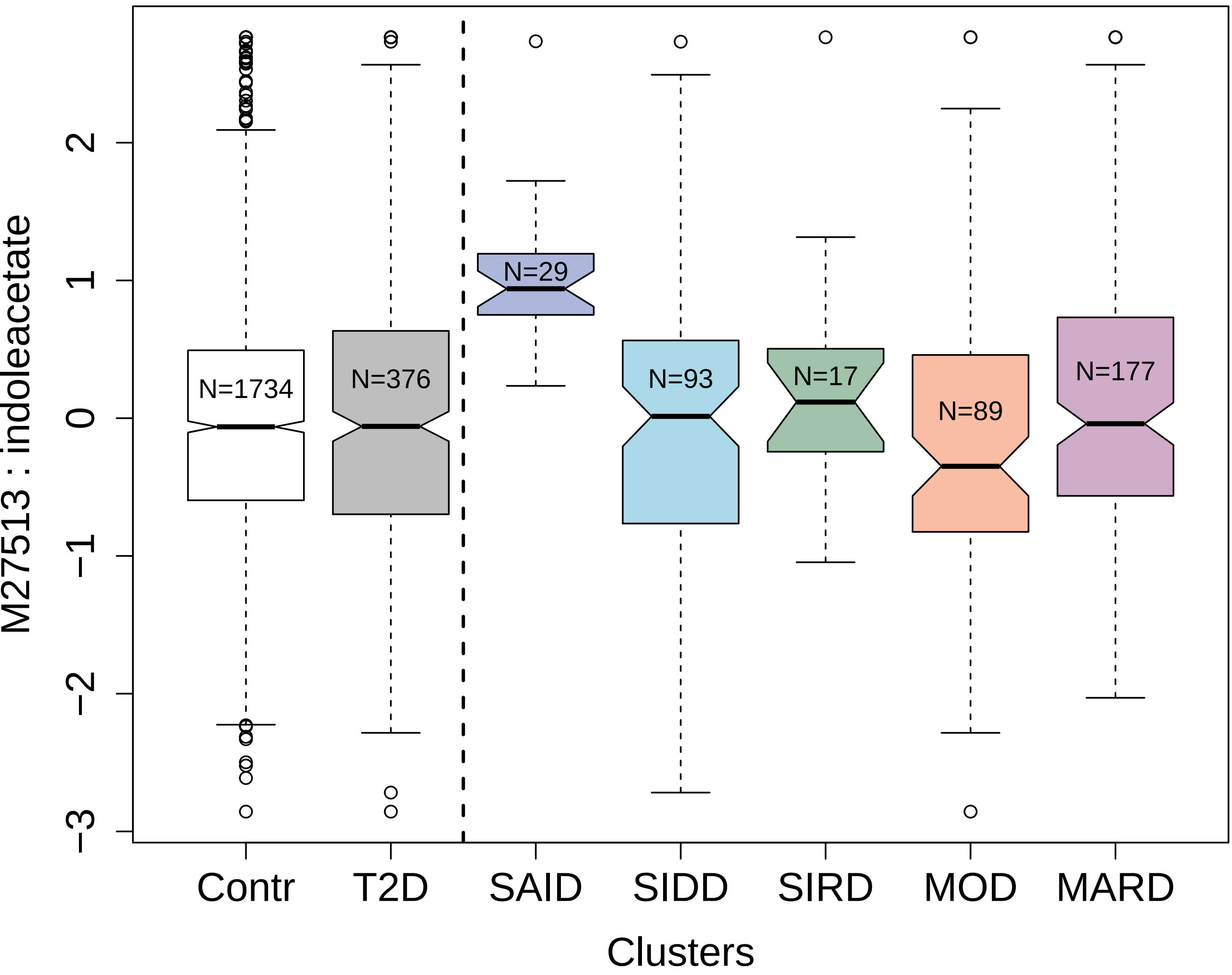

M27513 : indoleacetate
2
1
0
−1
−2
−3
N=1734
N=376
N=29
N=93
N=17
N=89
N=177
Contr
T2D
SAID
SIDD
SIRD
MOD
MARD
Clusters

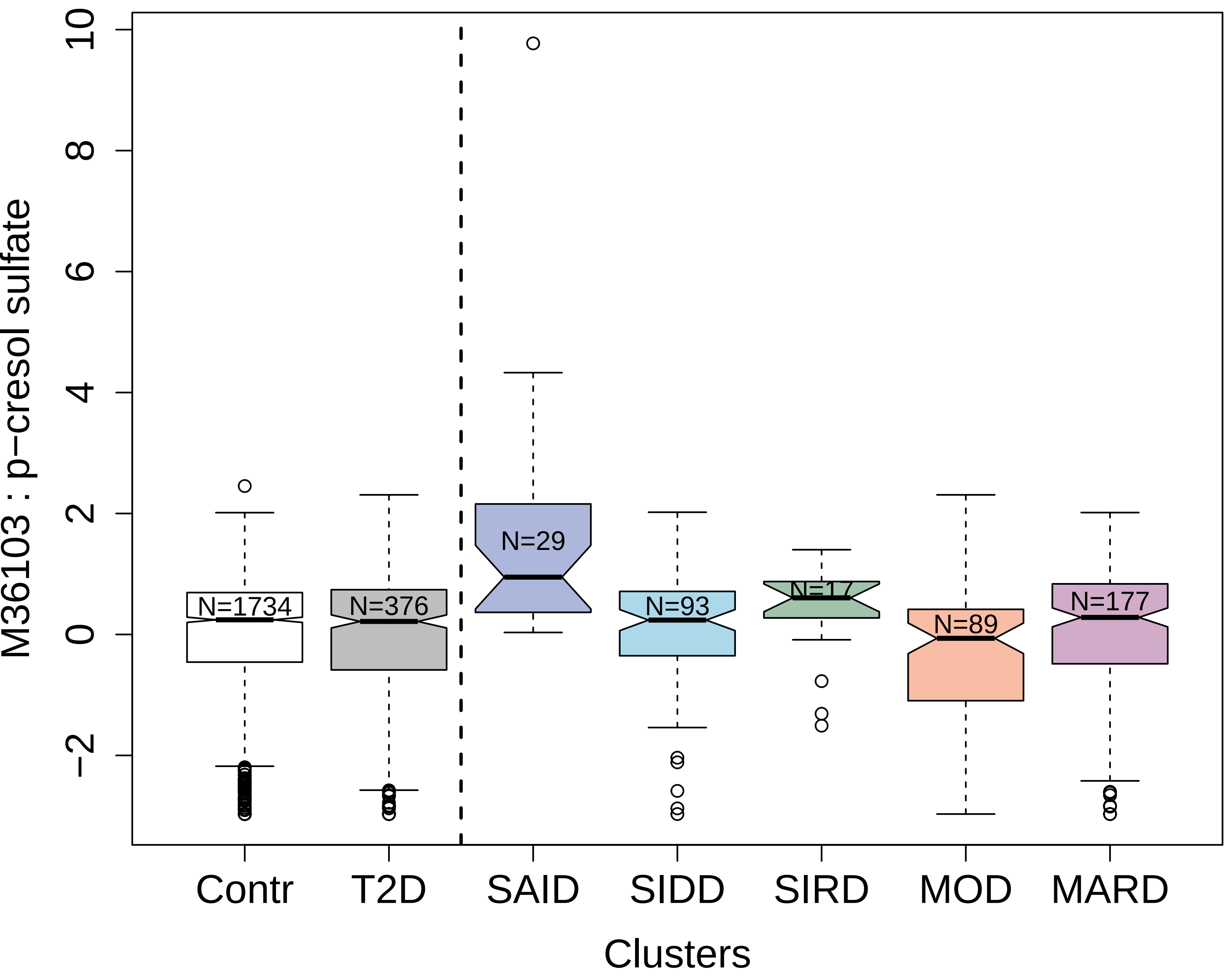
M36103 : p−cresol sulfate
10
8
6
4
2
0
−2
N=1734
N=376
N=29
N=93
N=17
N=89
N=177
Contr
T2D
SAID
SIDD
SIRD
MOD
MARD
Clusters

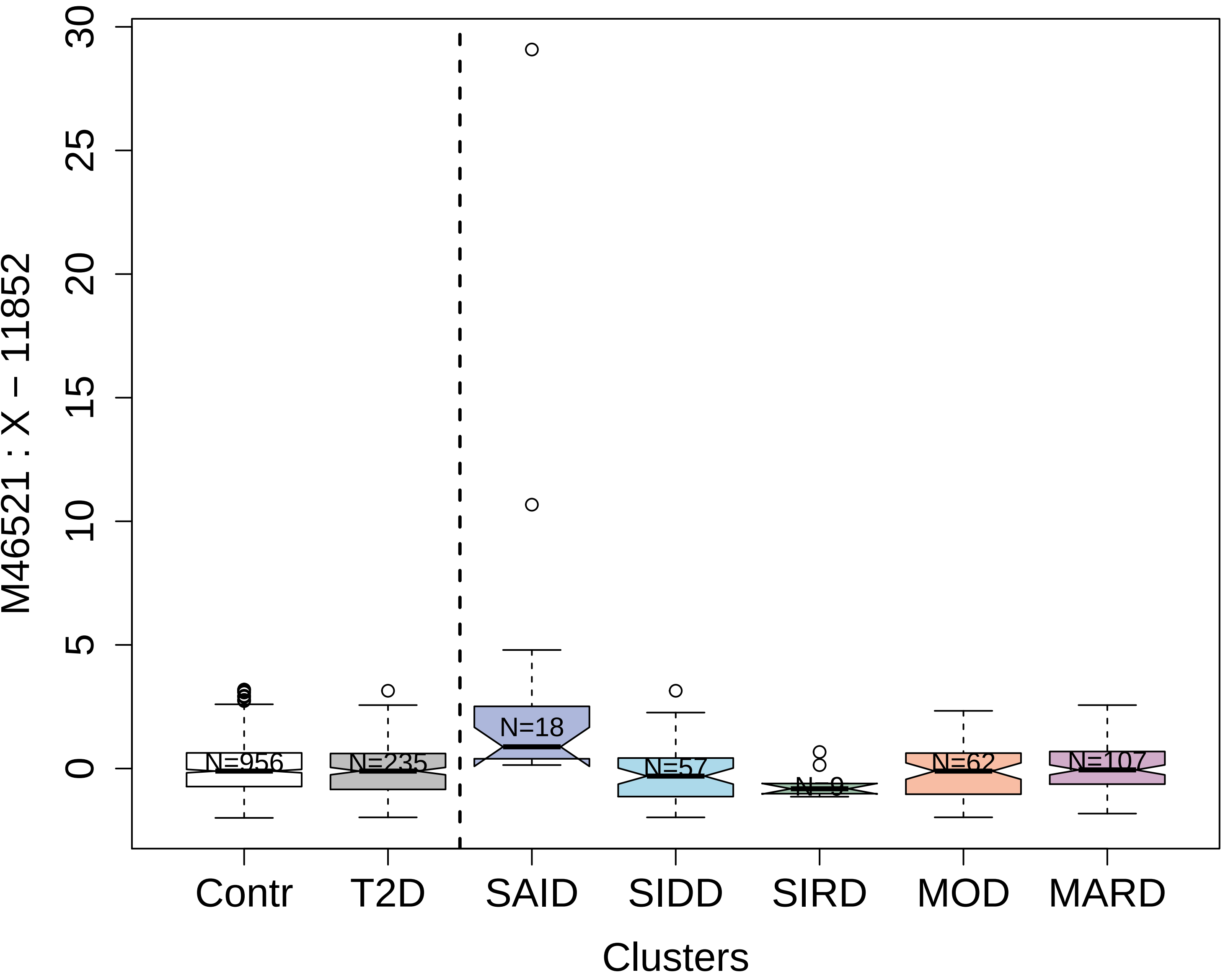
M46521 : X − 11852
30
25
20
15
10
5
0
N=956
N=235
N=18
N=57
N=9
N=62
N=107
Contr
T2D
SAID
SIDD
SIRD
MOD
MARD
Clusters

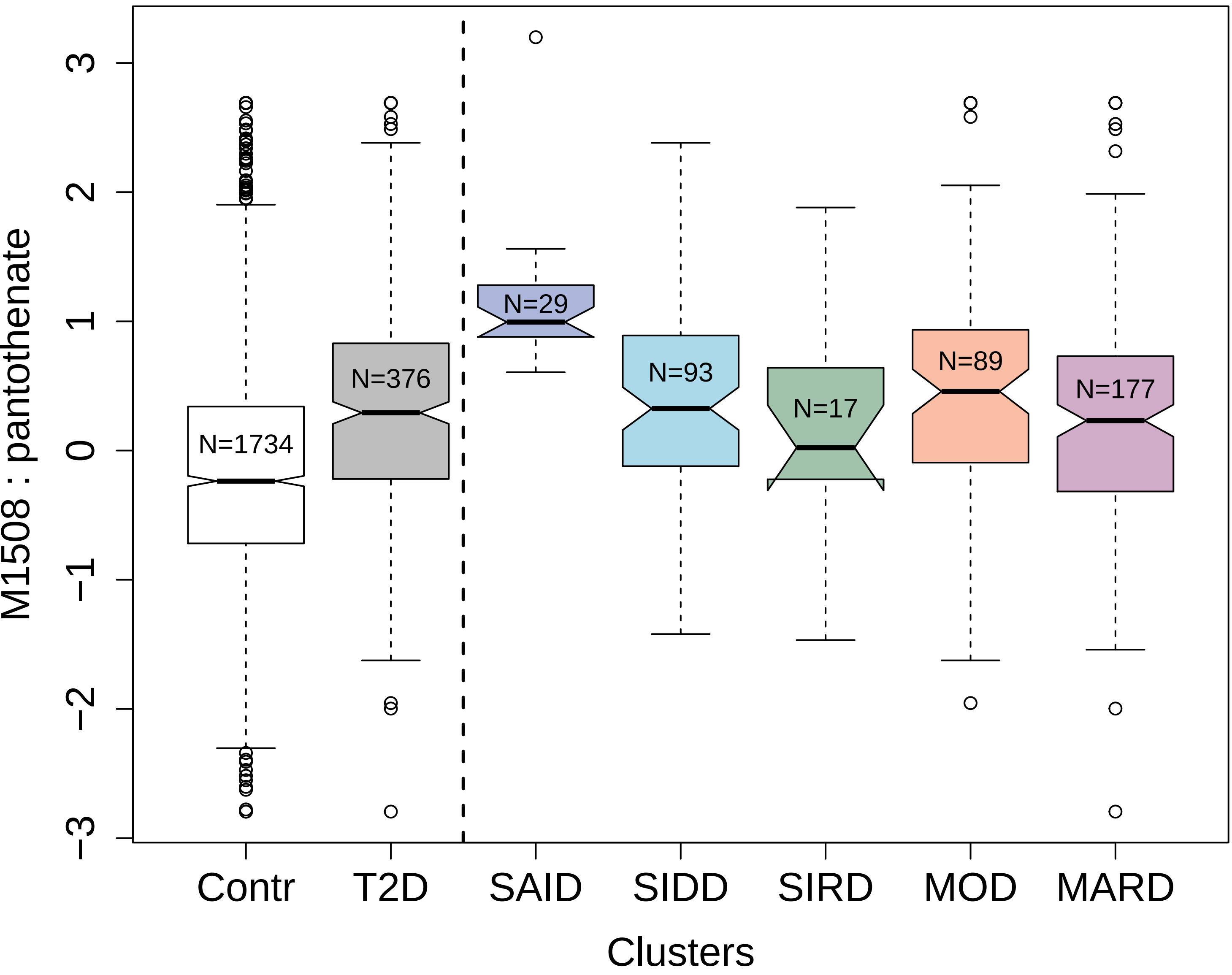

M1508 : pantothenate
3
2
1
0
−1
−2
−3
N=1734
N=376
N=29
N=93
N=17
N=89
N=177
Contr
T2D
SAID
SIDD
SIRD
MOD
MARD
Clusters

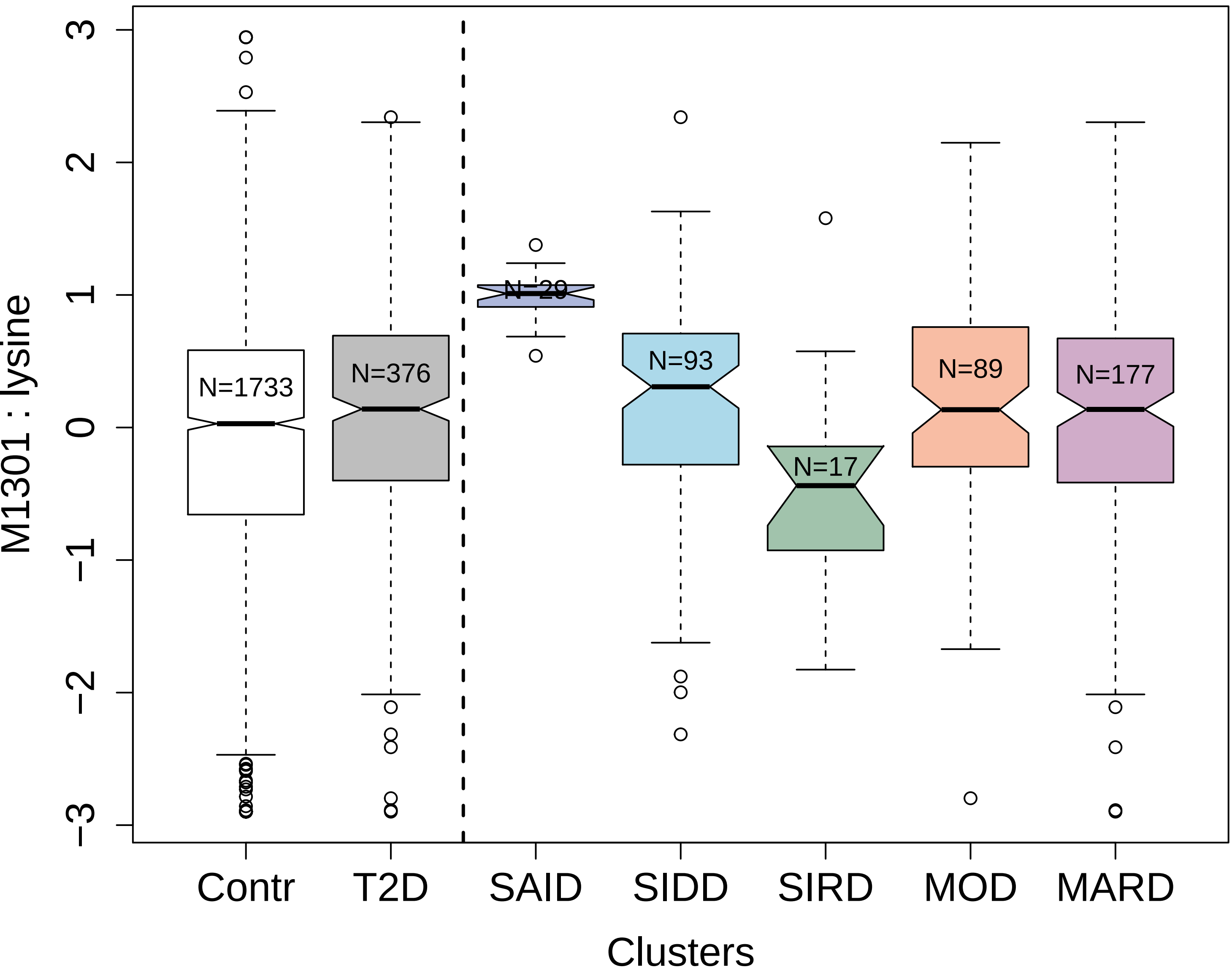

3
2
1
0
−1
−2
−3
M1301 : lysine
N=1733
N=376
N=29
N=93
N=17
N=89
N=177
Contr
T2D
SAID
SIDD
SIRD
MOD
MARD
Clusters

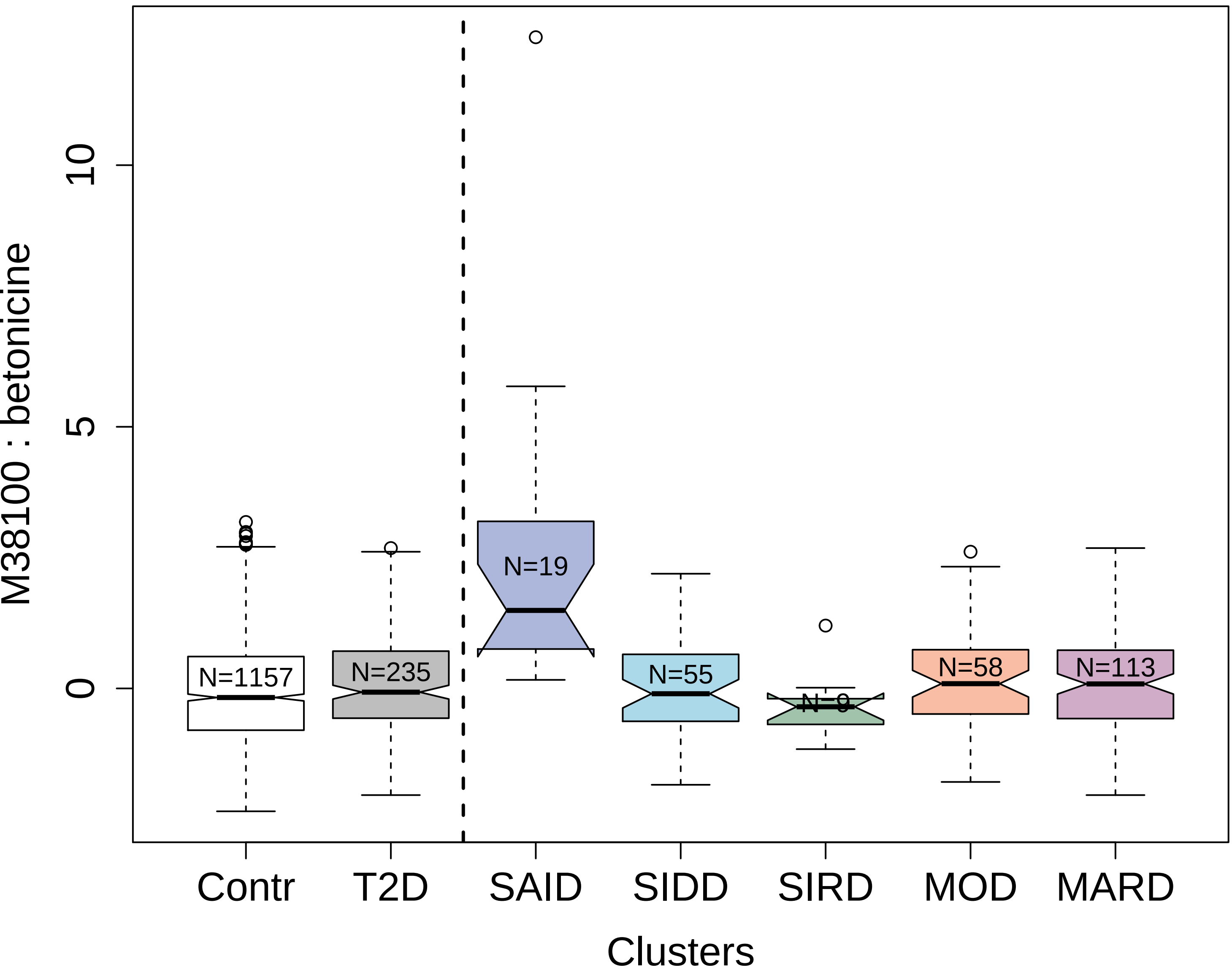
M38100 : betonicine
0
5
10
N=1157
N=235
N=19
N=55
N=9
N=58
N=113
Contr
T2D
SAID
SIDD
SIRD
MOD
MARD
Clusters

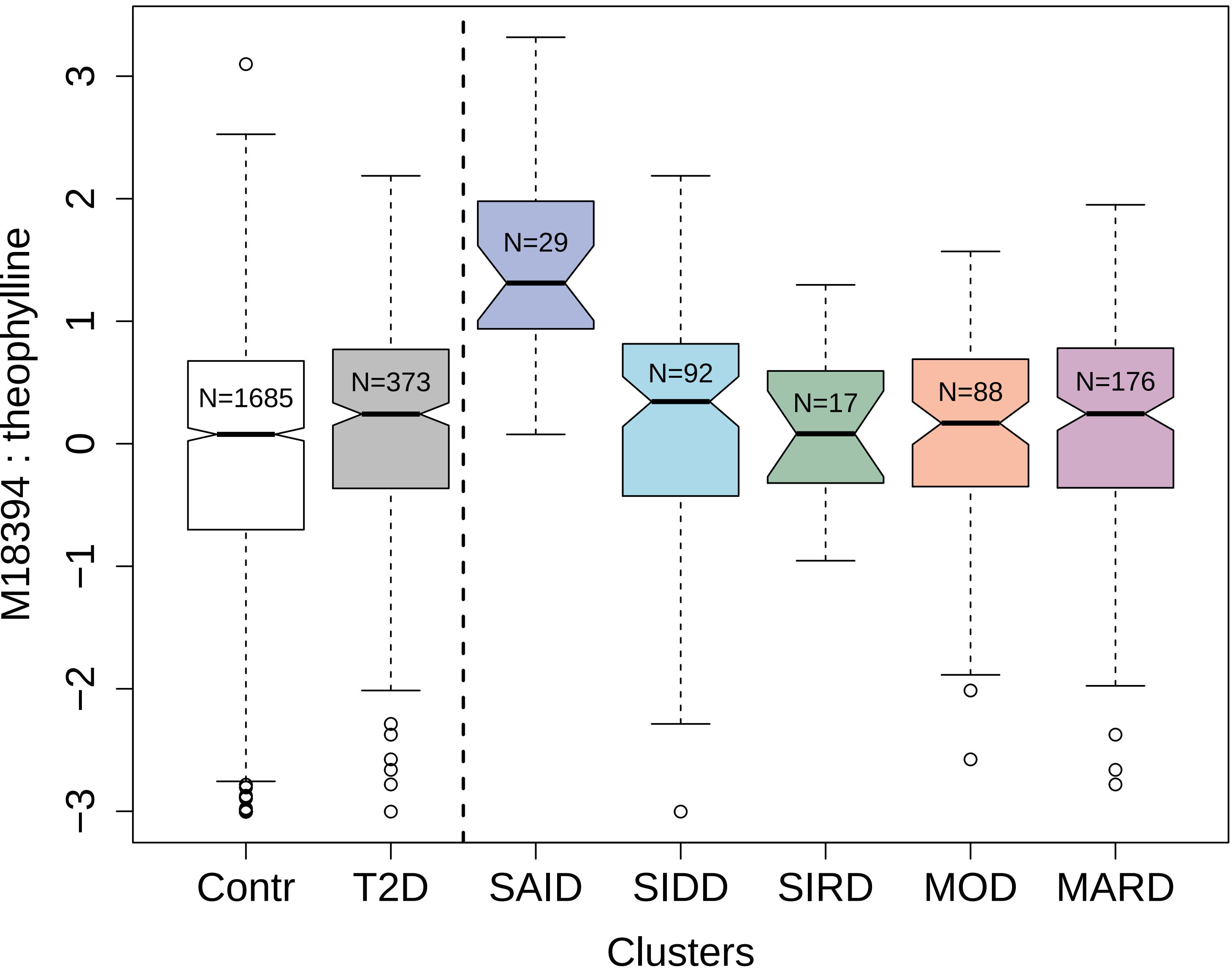

M18394 : theophylline
3
2
1
0
−1
−2
−3
N=1685
N=373
N=29
N=92
N=17
N=88
N=176
Contr
T2D
SAID
SIDD
SIRD
MOD
MARD
Clusters

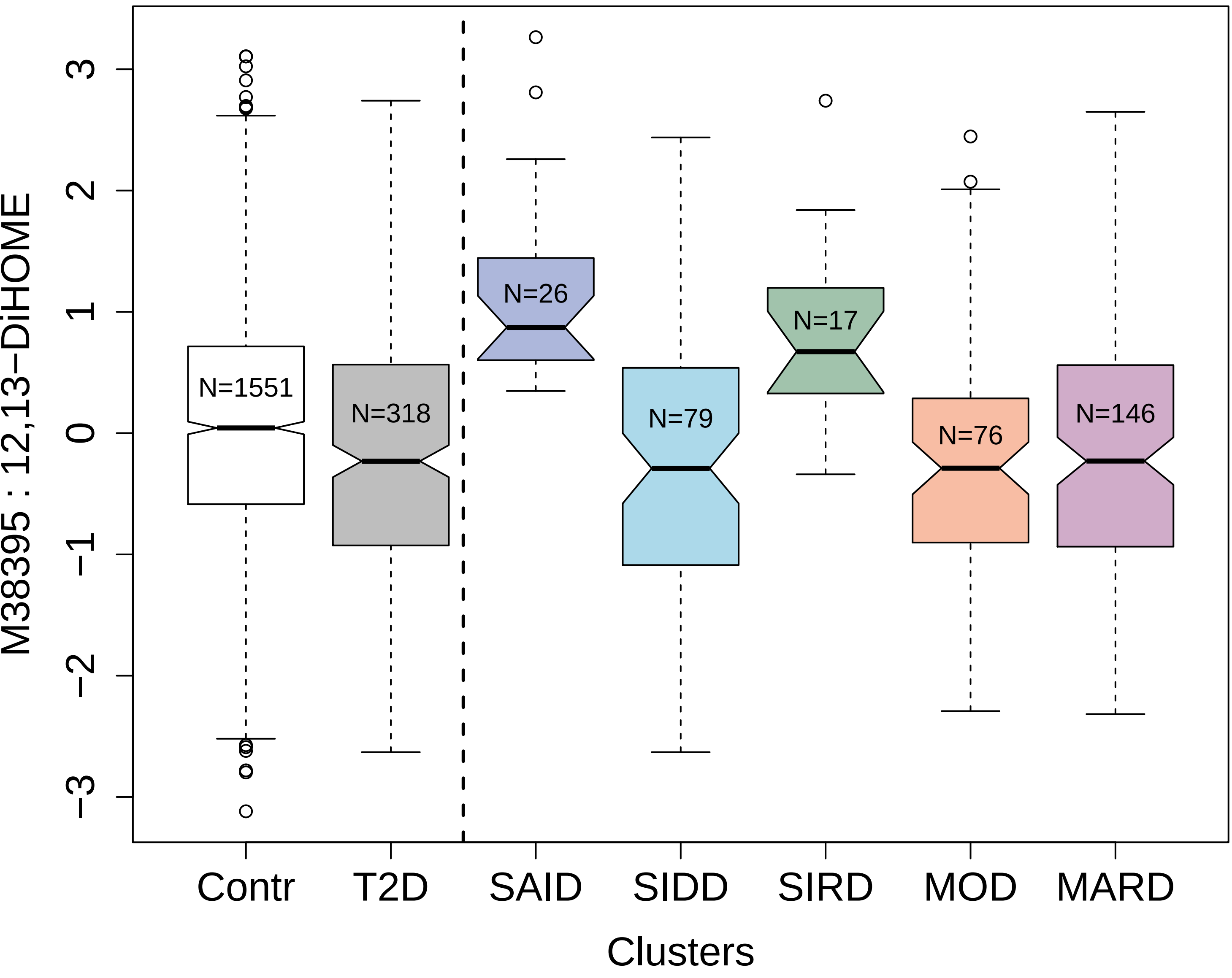

M38395 : 12,13−DiHOME
3
2
1
0
−1
−2
−3
N=1551
N=318
N=26
N=79
N=17
N=76
N=146
Contr
T2D
SAID
SIDD
SIRD
MOD
MARD
Clusters

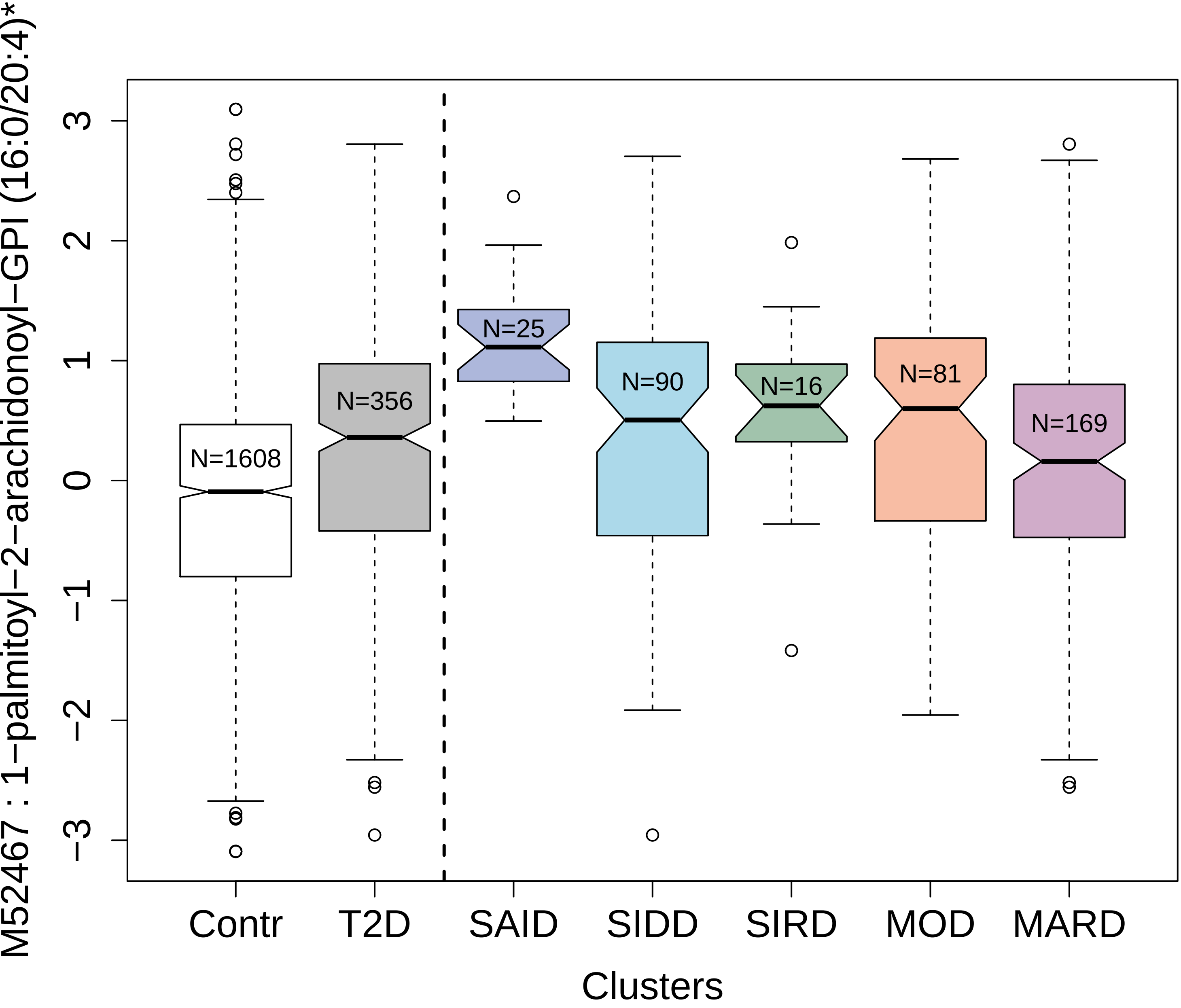

M52467 : 1−palmitoyl−2−arachidonoyl−GPI (16:0/20:4)*
3
2
1
0
−1
−2
−3
N=1608
N=356
N=25
N=90
N=16
N=81
N=169
Contr
T2D
SAID
SIDD
SIRD
MOD
MARD
Clusters

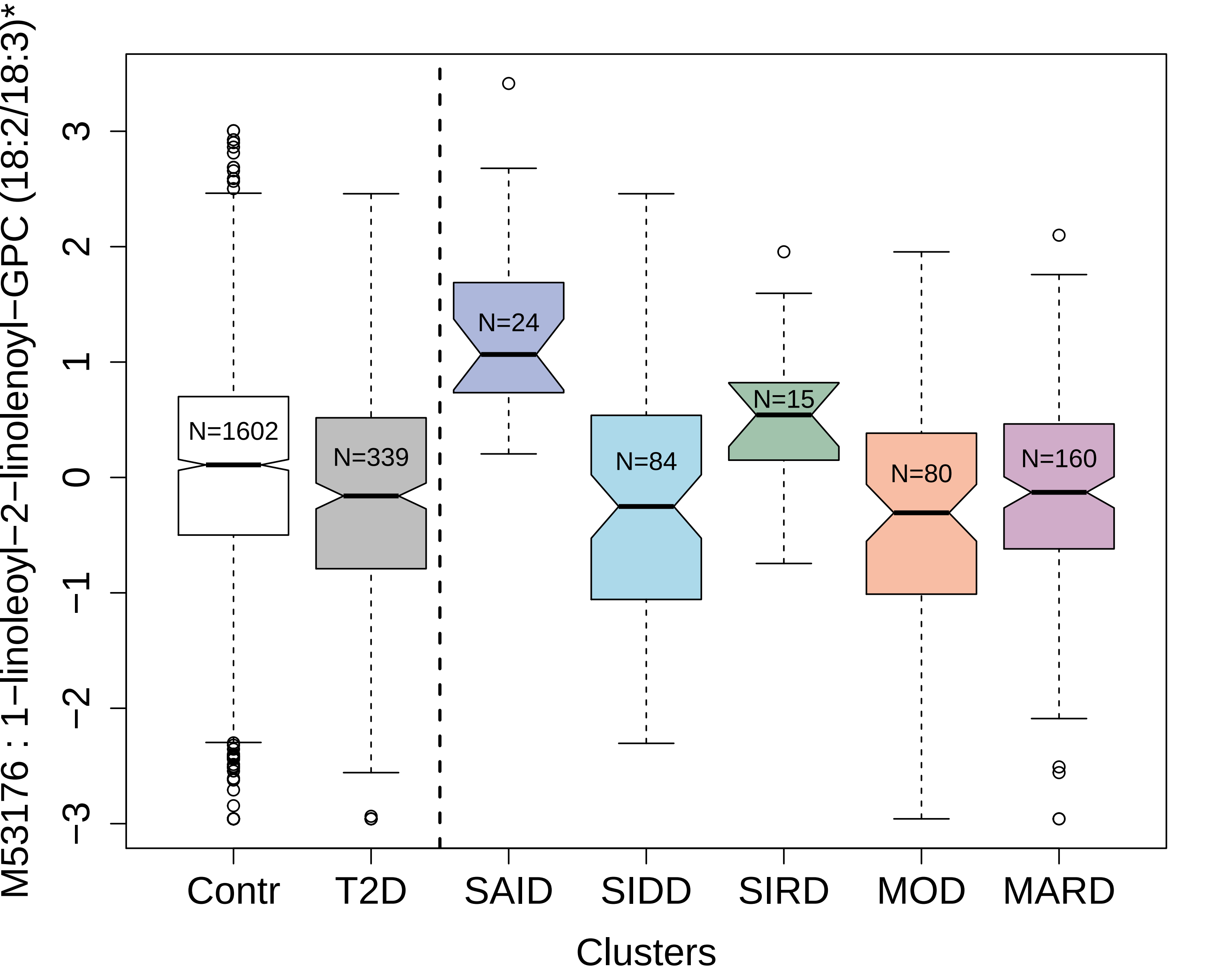

M53176 : 1−linoleoyl−2−linolenoyl−GPC (18:2/18:3)*
3
2
1
0
−1
−2
−3
N=1602
N=339
N=24
N=84
N=15
N=80
N=160
Contr
T2D
SAID
SIDD
SIRD
MOD
MARD
Clusters

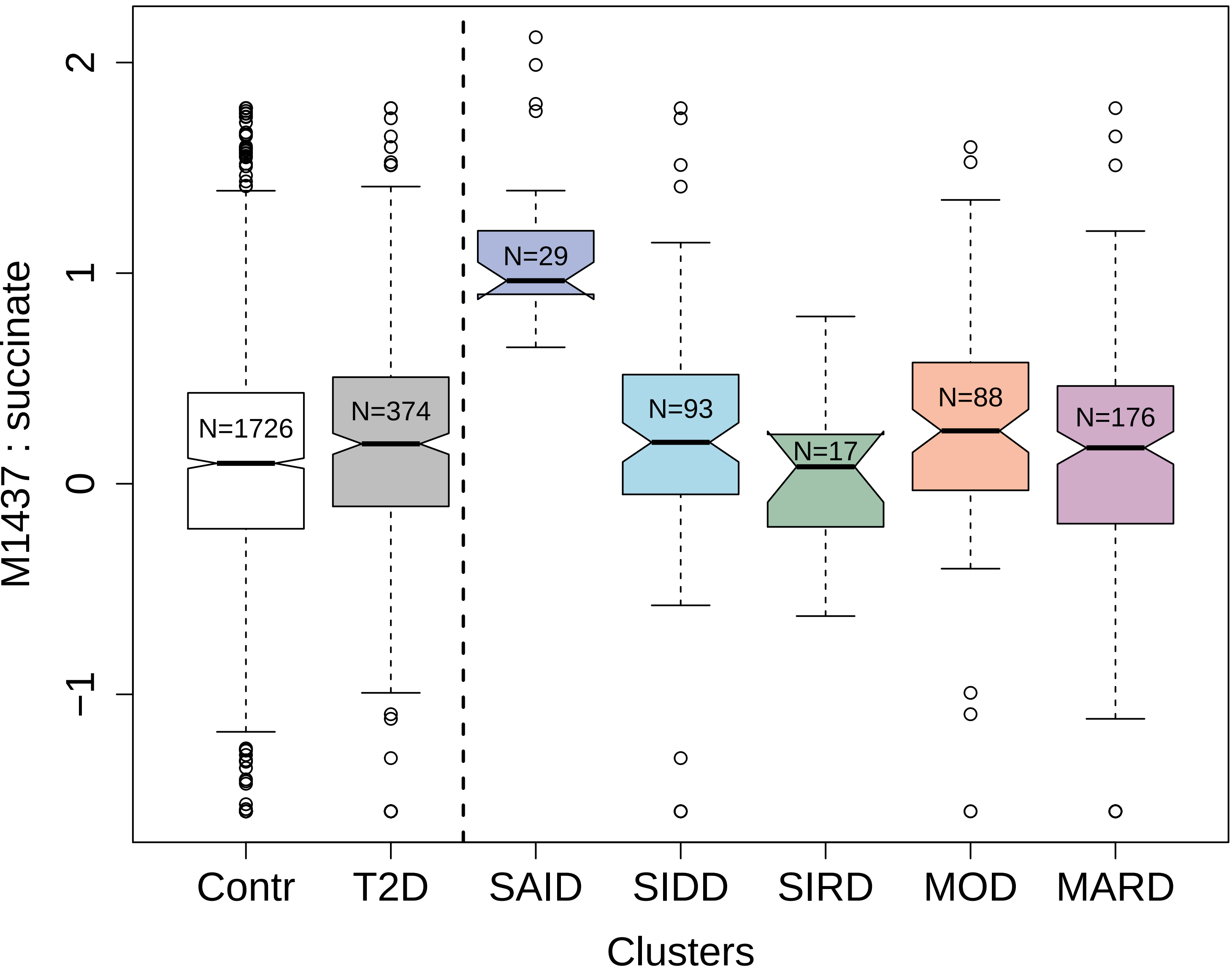
M1437 : succinate
2
1
0
−1
N=1726
N=374
N=29
N=93
N=17
N=88
N=176
Contr
T2D
SAID
SIDD
SIRD
MOD
MARD
Clusters

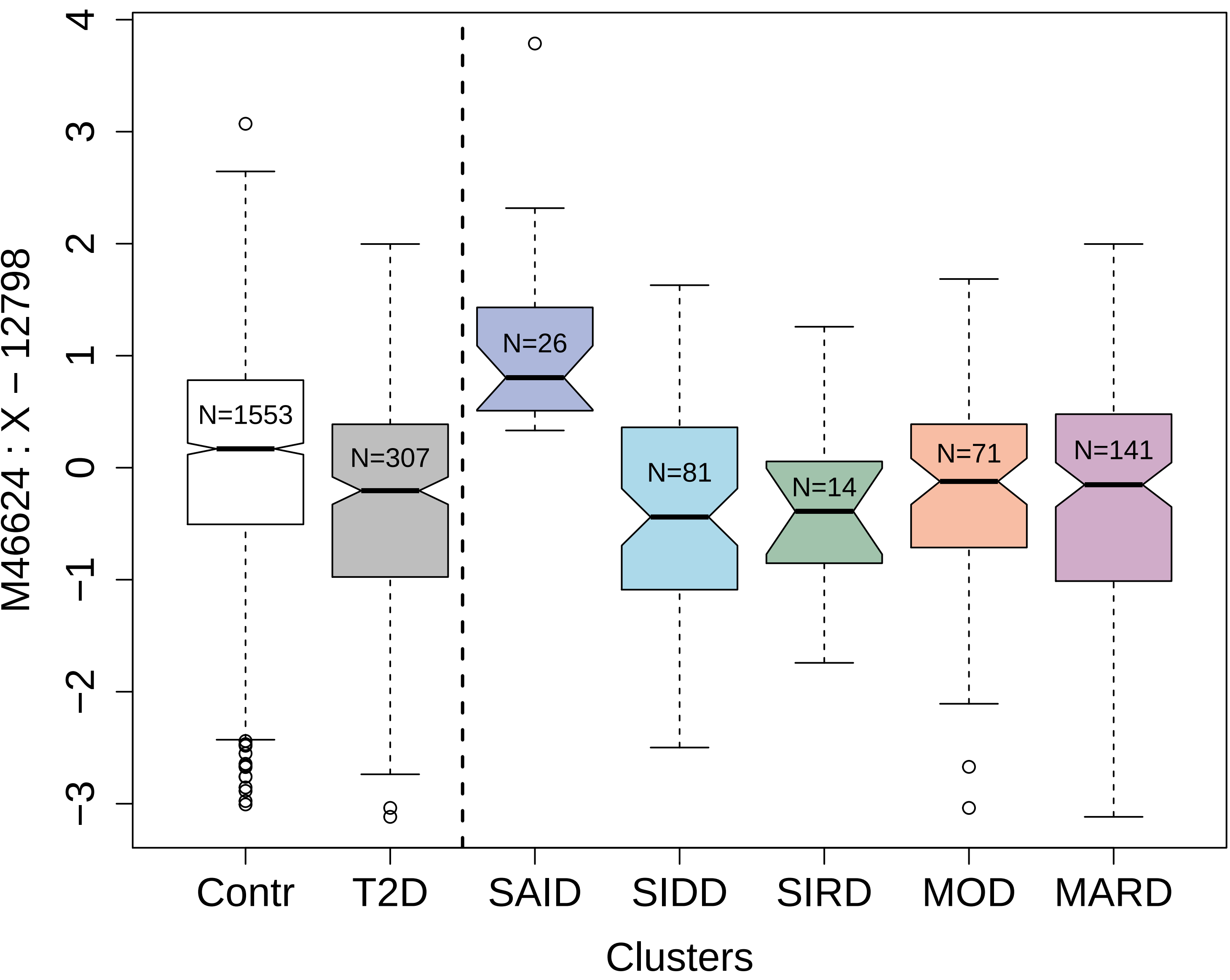

M46624 : X − 12798
4
3
2
1
0
−1
−2
−3
N=1553
N=307
N=26
N=81
N=14
N=71
N=141
Contr
T2D
SAID
SIDD
SIRD
MOD
MARD
Clusters

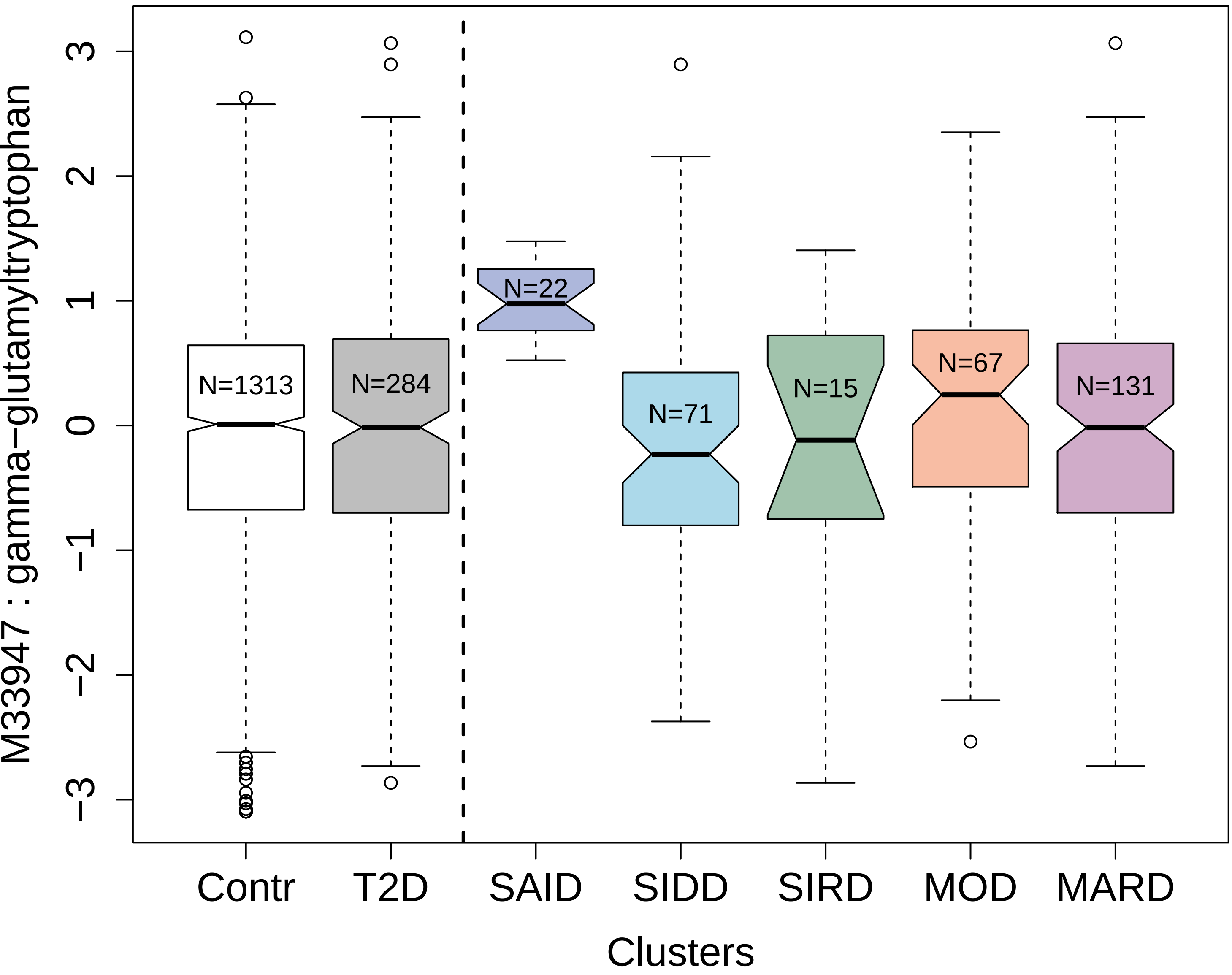

M33947 : gamma−glutamyltryptophan
3
2
1
0
−1
−2
−3
N=1313
N=284
N=22
N=71
N=15
N=67
N=131
Contr
T2D
SAID
SIDD
SIRD
MOD
MARD
Clusters

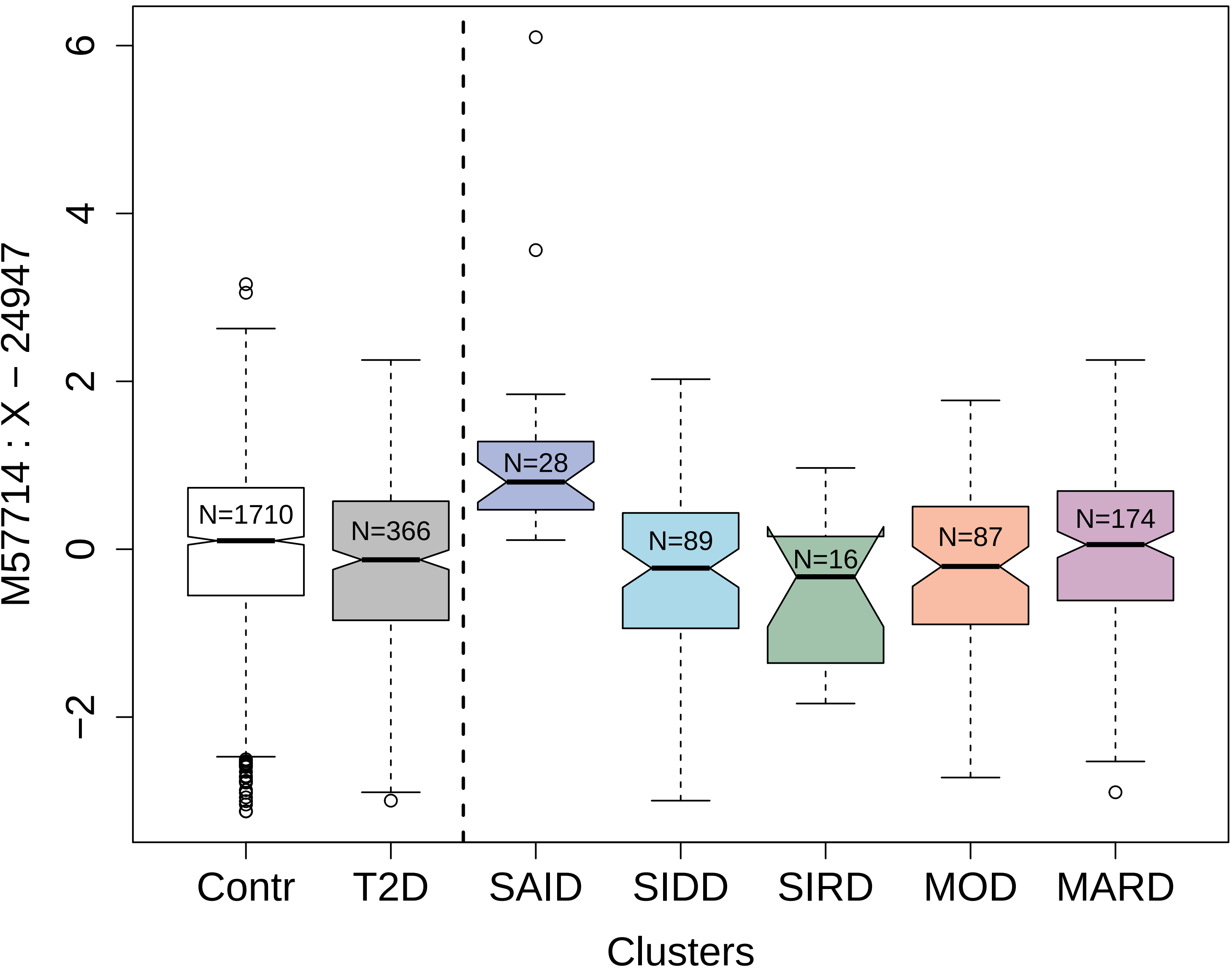

M57714 : X − 24947
6
4
2
0
−2
N=1710
N=366
N=28
N=89
N=16
N=87
N=174
Contr
T2D
SAID
SIDD
SIRD
MOD
MARD
Clusters

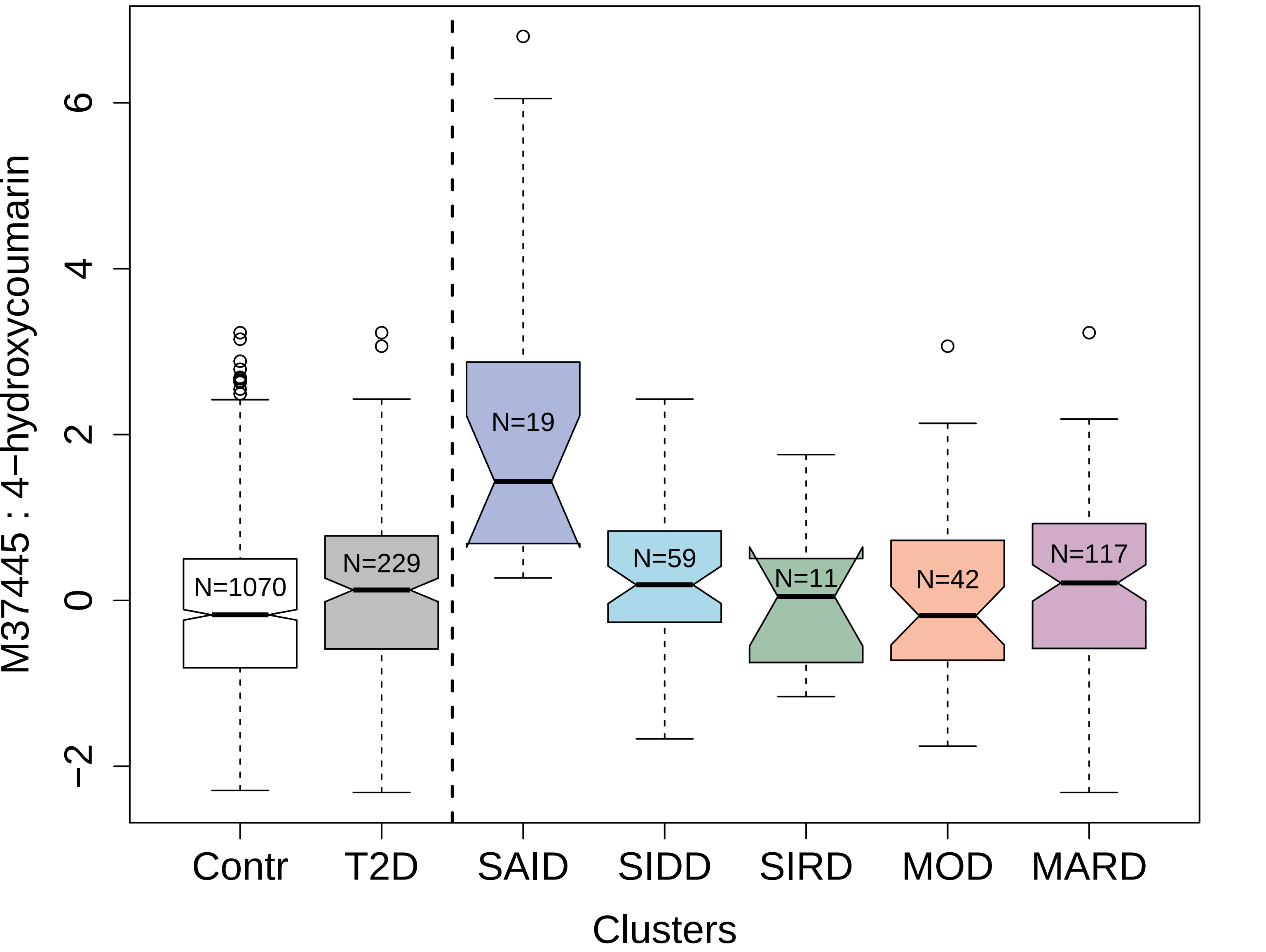

M37445 : 4−hydroxycoumarin
Clusters
Contr
T2D
SAID
SIDD
SIRD
MOD
MARD
N=1070
N=229
N=19
N=59
N=11
N=42
N=117
−2
0
2
4
6

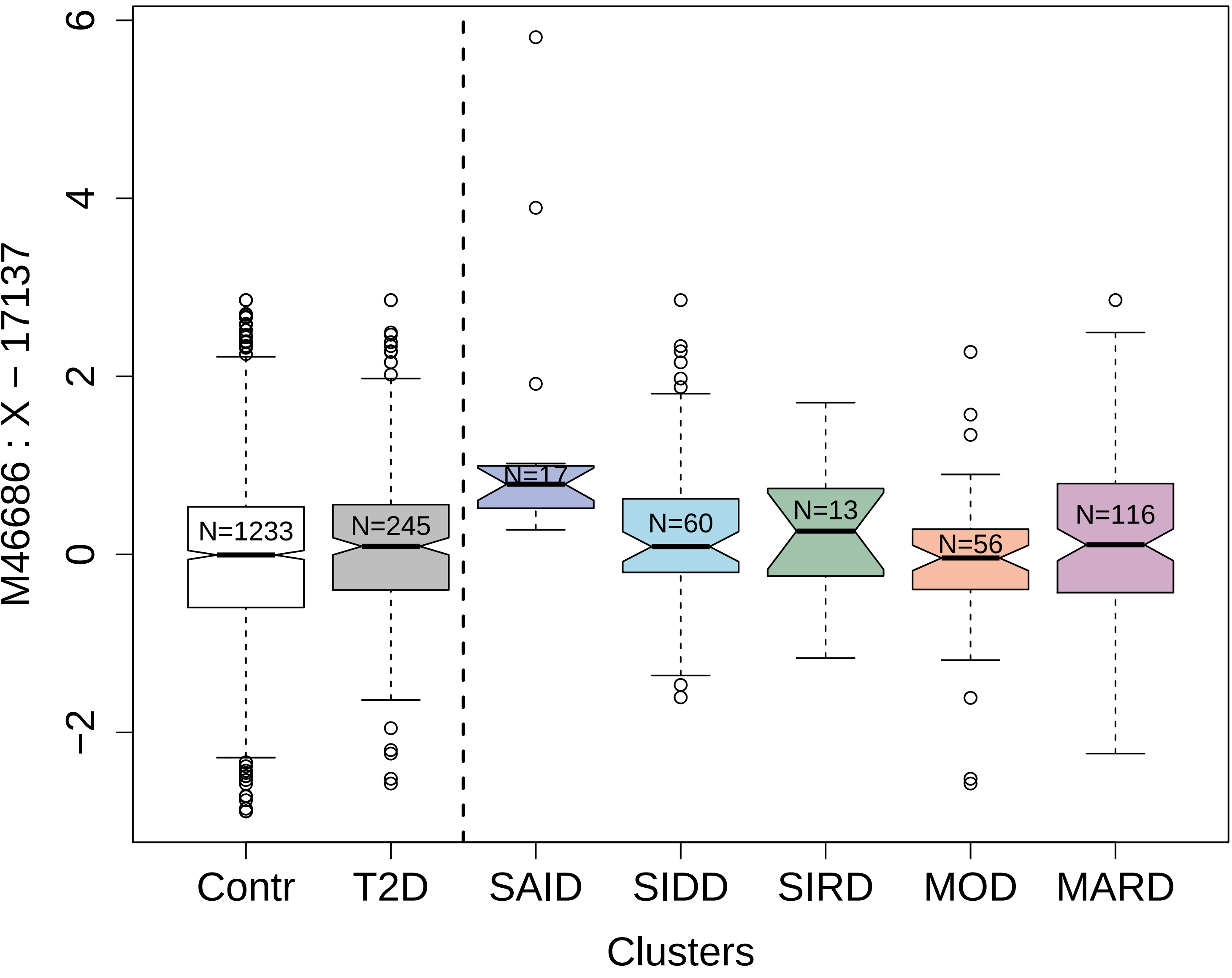

M46686 : X − 17137
6
4
2
0
−2
N=1233
N=245
N=17
N=60
N=13
N=56
N=116
Contr
T2D
SAID
SIDD
SIRD
MOD
MARD
Clusters

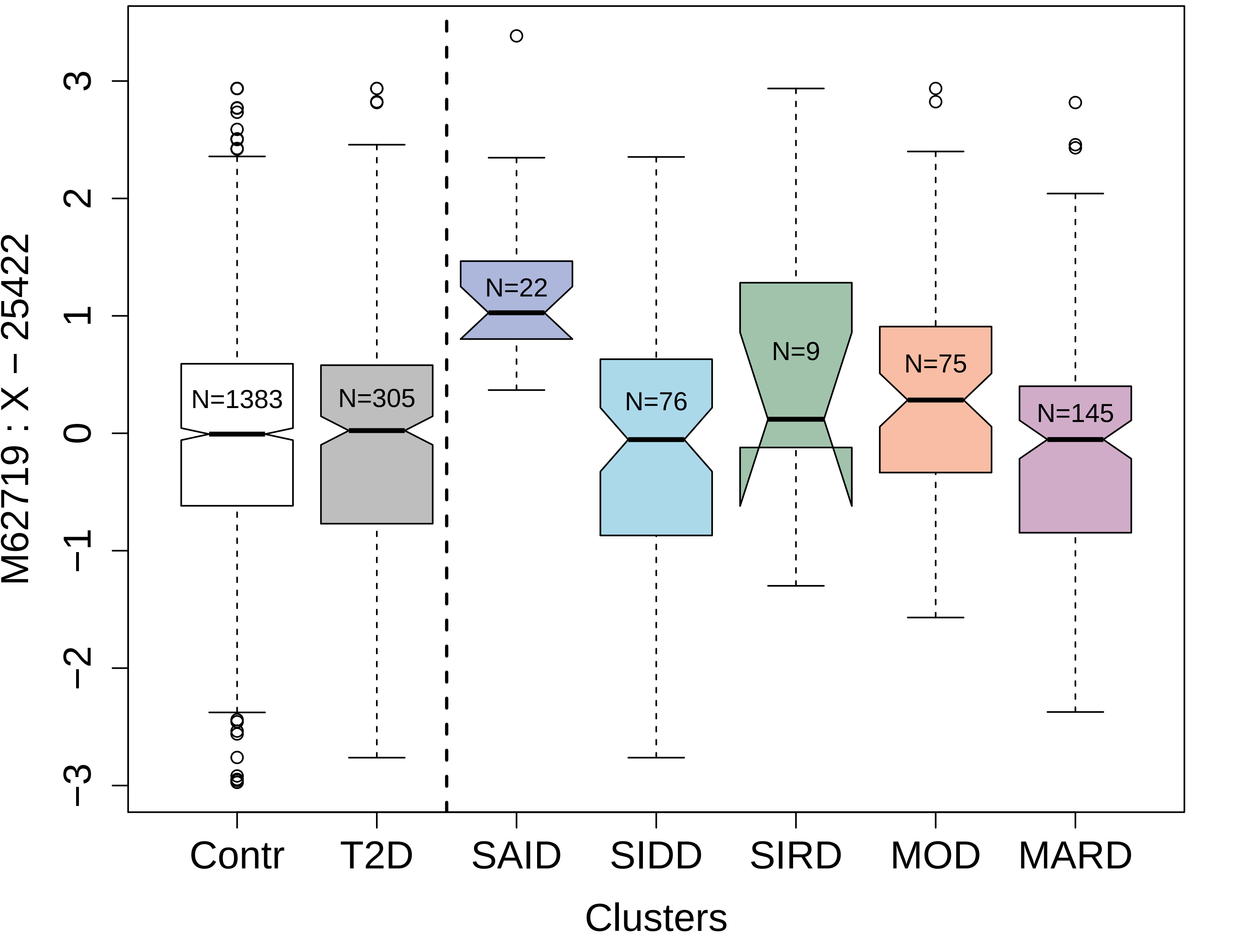
N=1383
N=305
N=22
N=76
N=9
N=75
N=145
Contr
T2D
SAID
SIDD
SIRD
MOD
MARD
Clusters
M62719 : X − 25422
3
2
1
0
−1
−2
−3

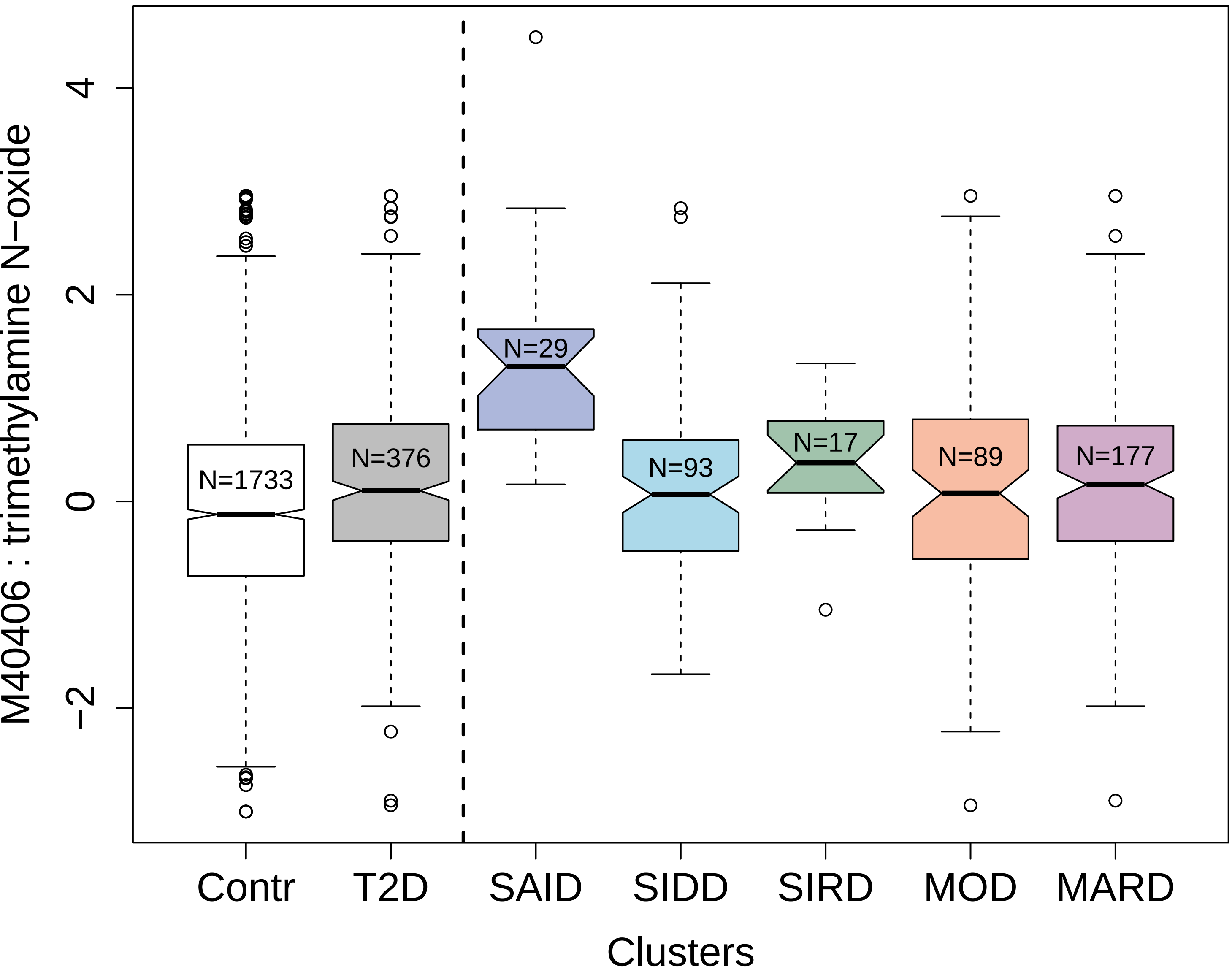

M40406 : trimethylamine N−oxide
4
2
0
−2
N=1733
N=376
N=29
N=93
N=17
N=89
N=177
Contr
T2D
SAID
SIDD
SIRD
MOD
MARD
Clusters

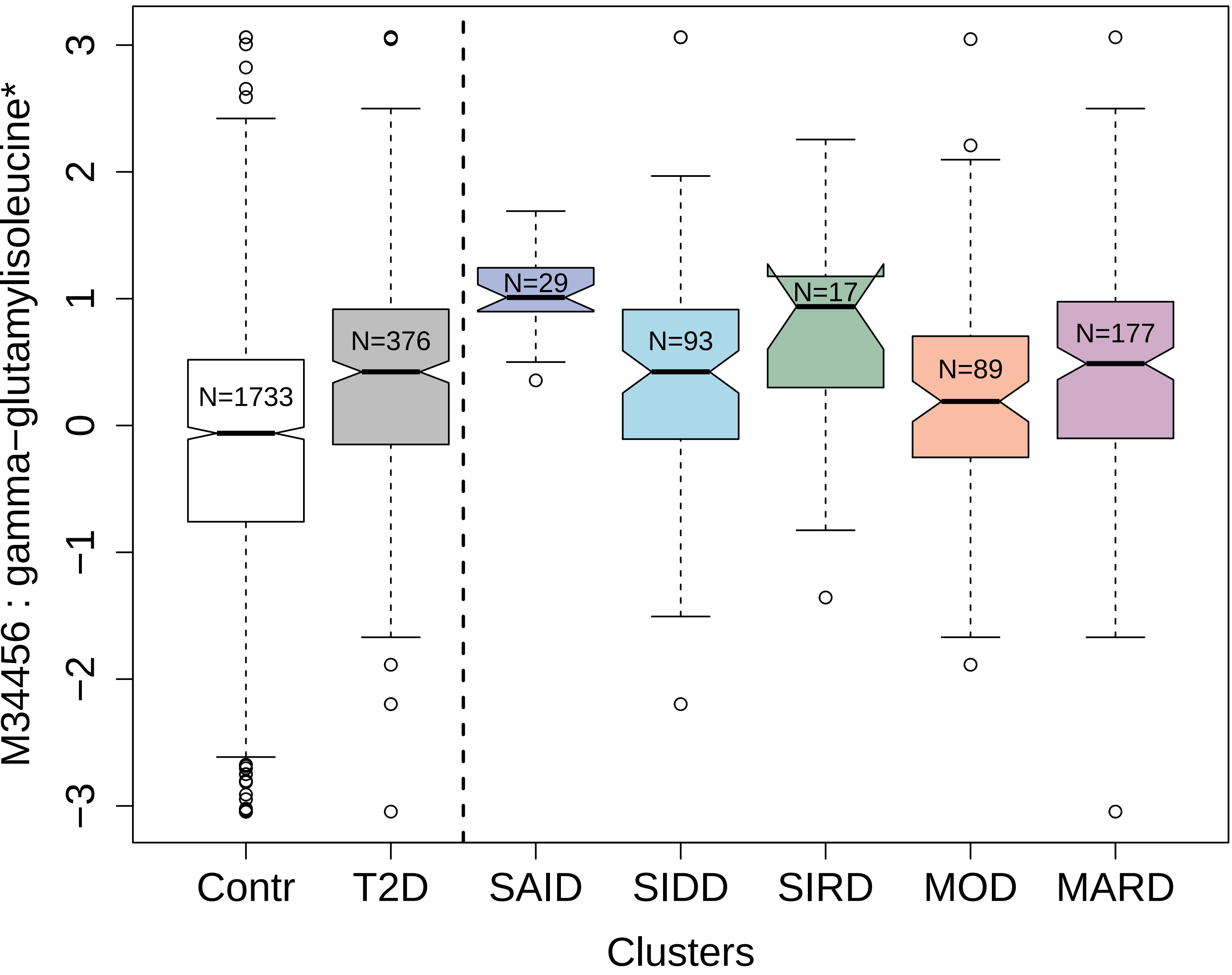
M34456 : gamma−glutamylisoleucine*
3
2
1
0
−1
−2
−3
N=1733
N=376
N=29
N=93
N=17
N=89
N=177
Contr
T2D
SAID
SIDD
SIRD
MOD
MARD
Clusters

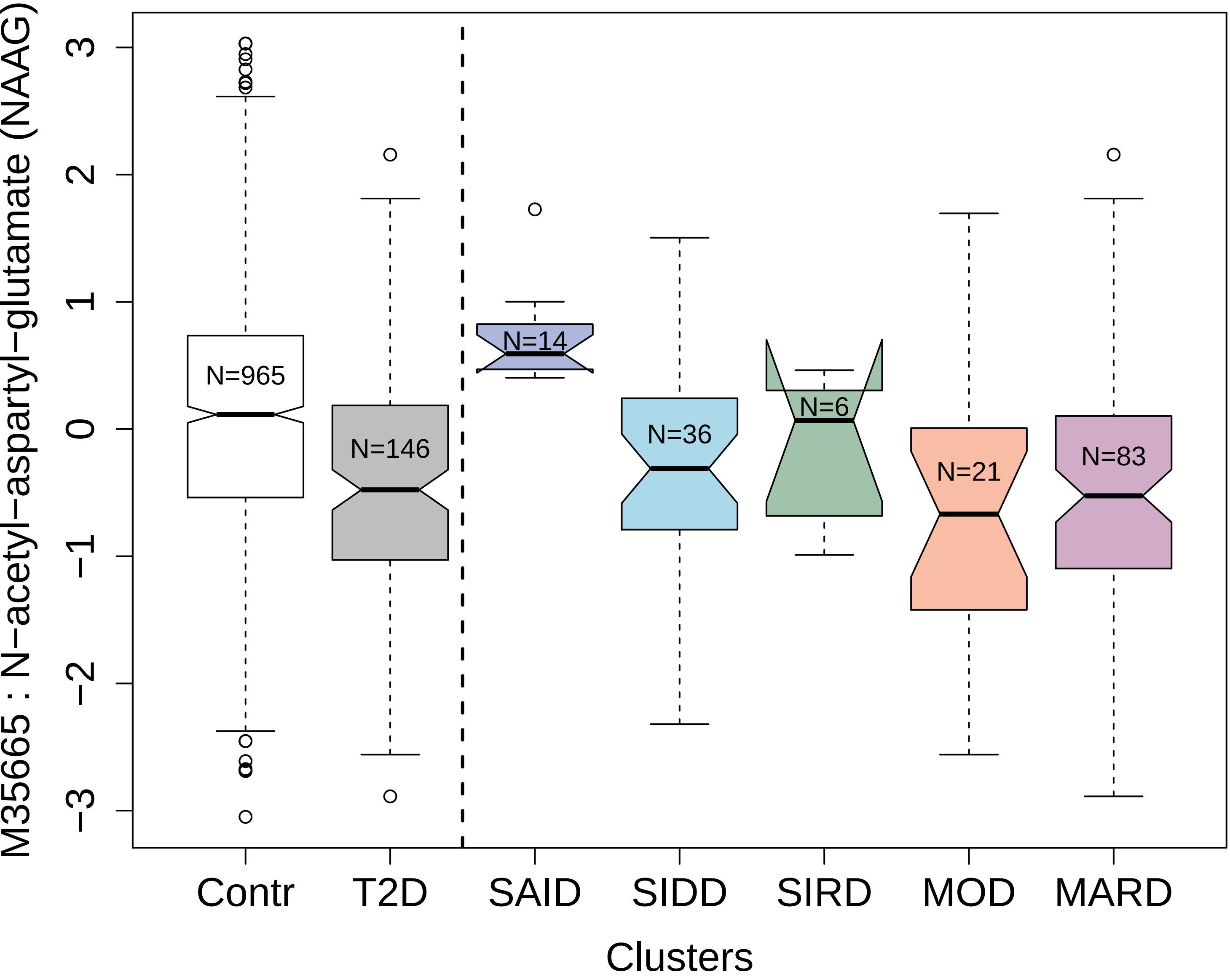
M35665 : N−acetyl−aspartyl−glutamate (NAAG)
3
2
1
0
−1
−2
−3
N=965
N=146
N=14
N=36
N=6
N=21
N=83
Contr
T2D
SAID
SIDD
SIRD
MOD
MARD
Clusters

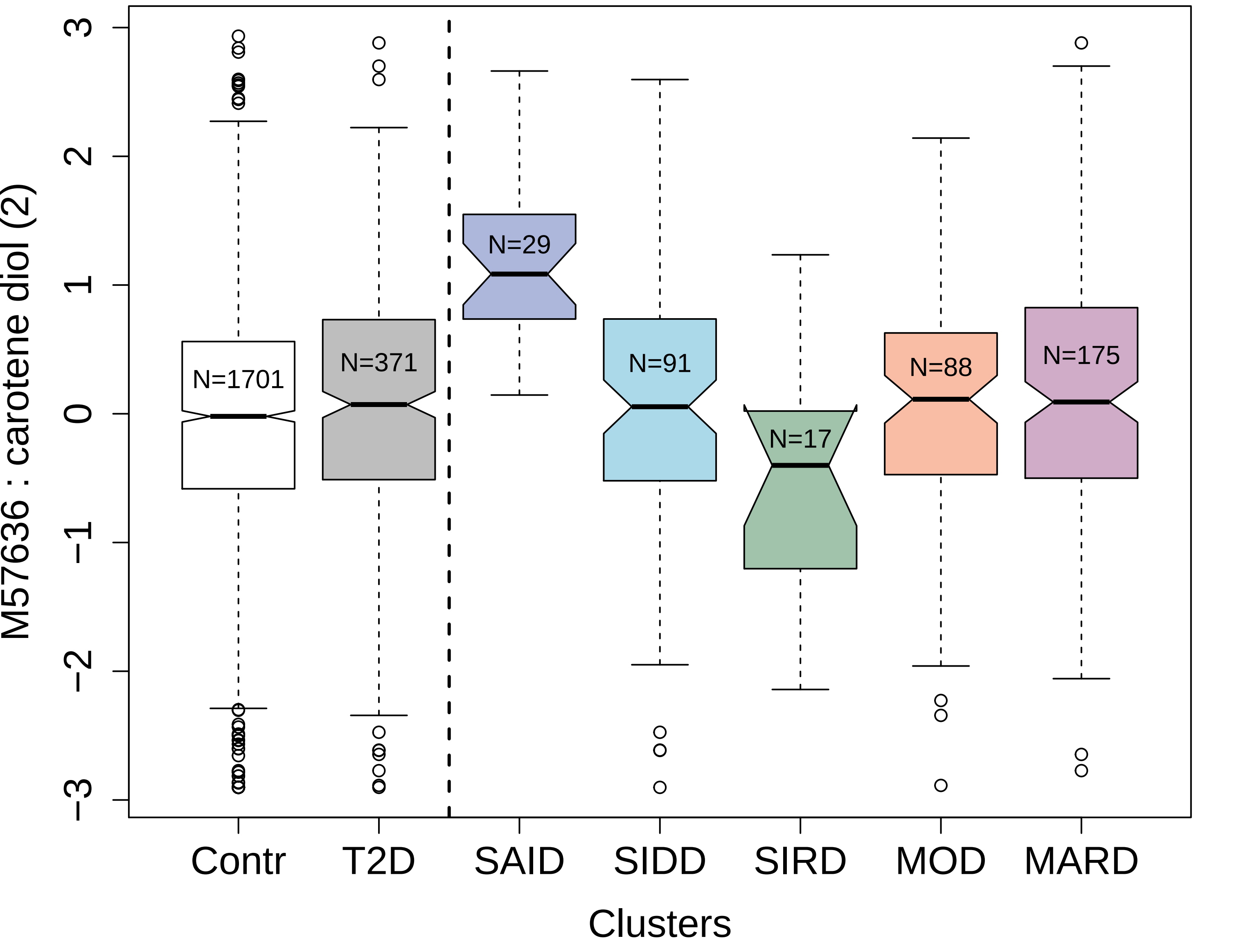

M57636 : carotene diol (2)
3
2
1
0
−1
−2
−3
N=1701
N=371
N=29
N=91
N=17
N=88
N=175
Contr
T2D
SAID
SIDD
SIRD
MOD
MARD
Clusters

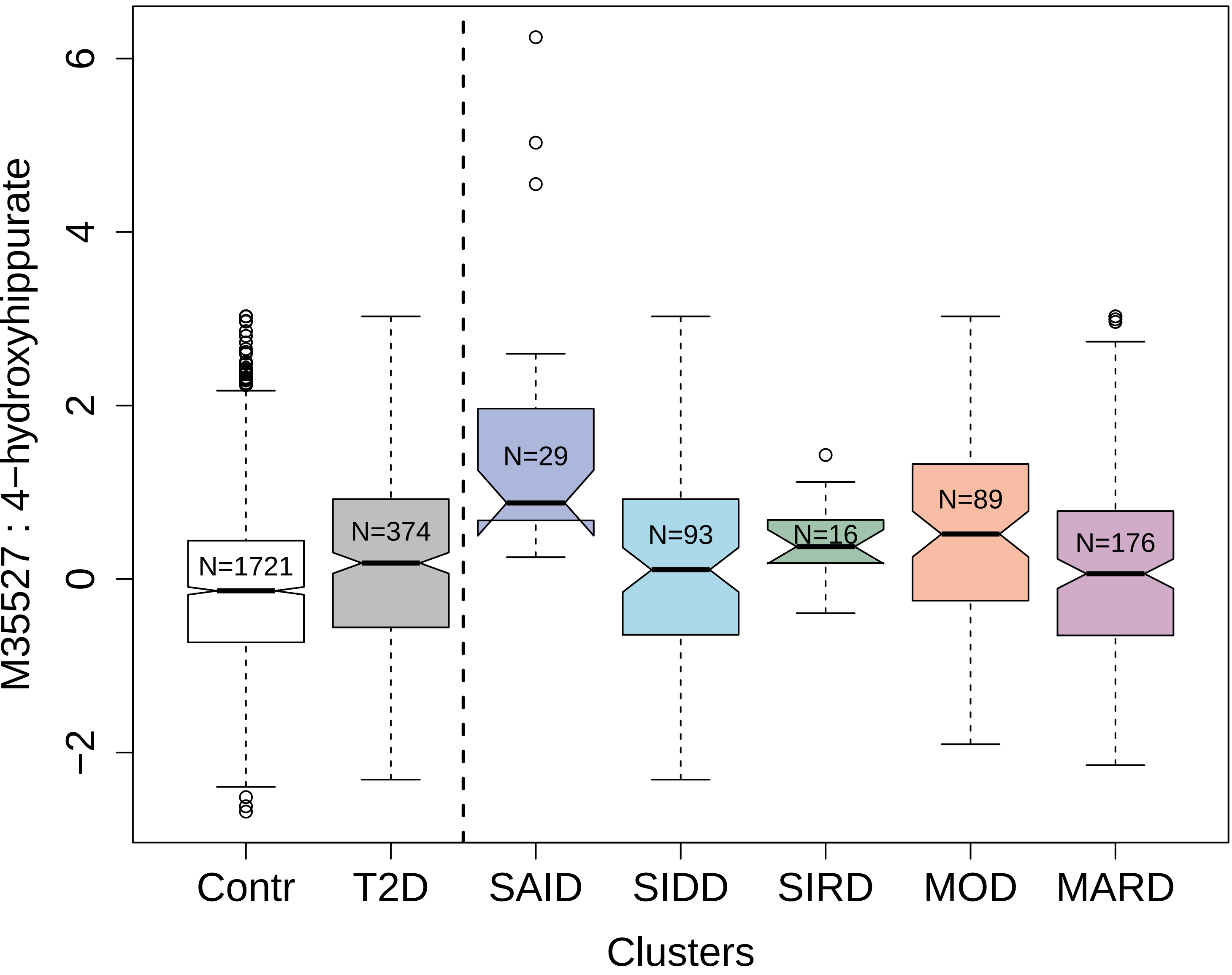

M35527 : 4−hydroxyhippurate
6
4
2
0
−2
N=1721
N=374
N=29
N=93
N=16
N=89
N=176
Contr
T2D
SAID
SIDD
SIRD
MOD
MARD
Clusters

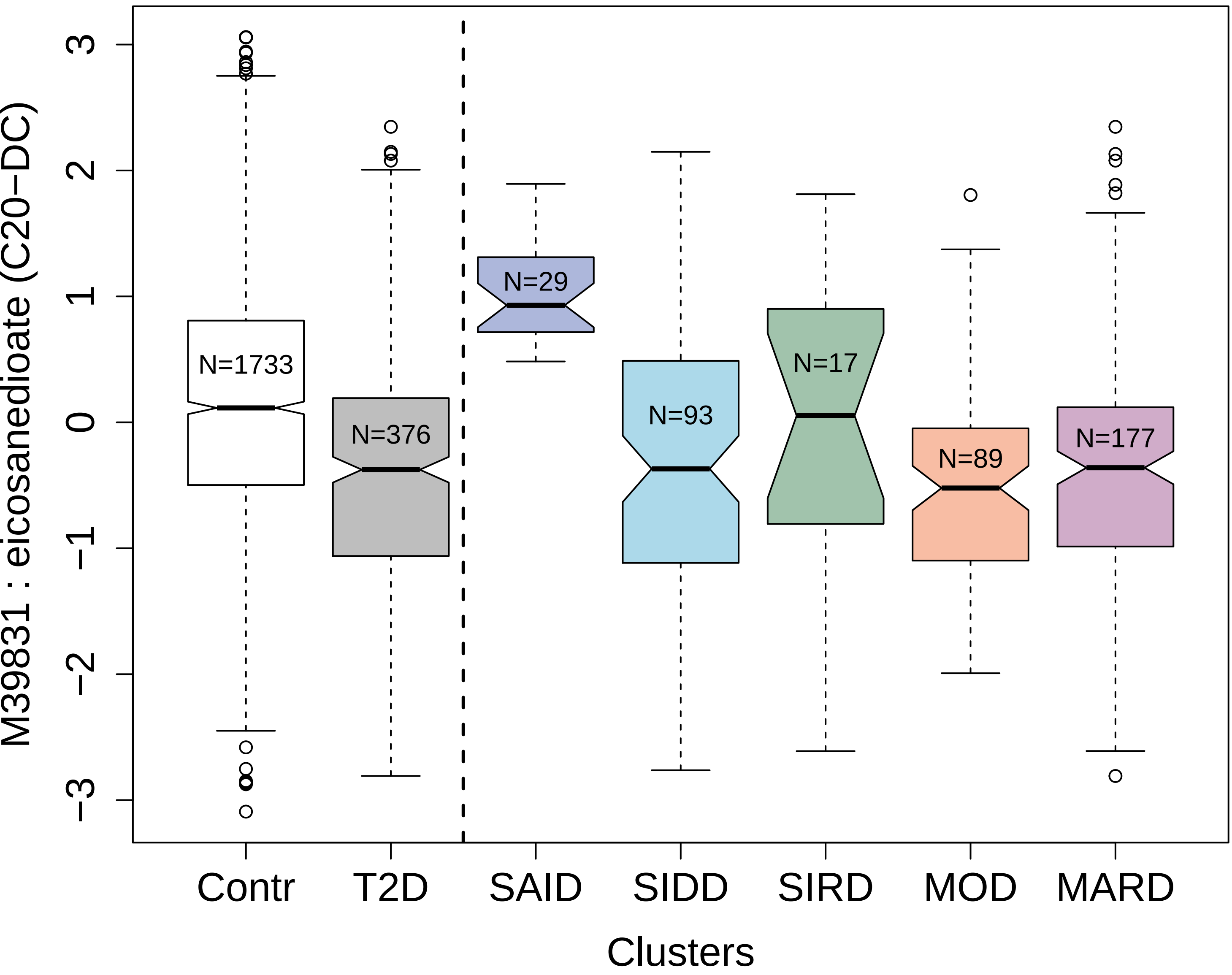
M39831 : eicosanedioate (C20−DC)
3
2
1
0
−1
−2
−3
N=1733
N=376
N=29
N=93
N=17
N=89
N=177
Contr
T2D
SAID
SIDD
SIRD
MOD
MARD
Clusters

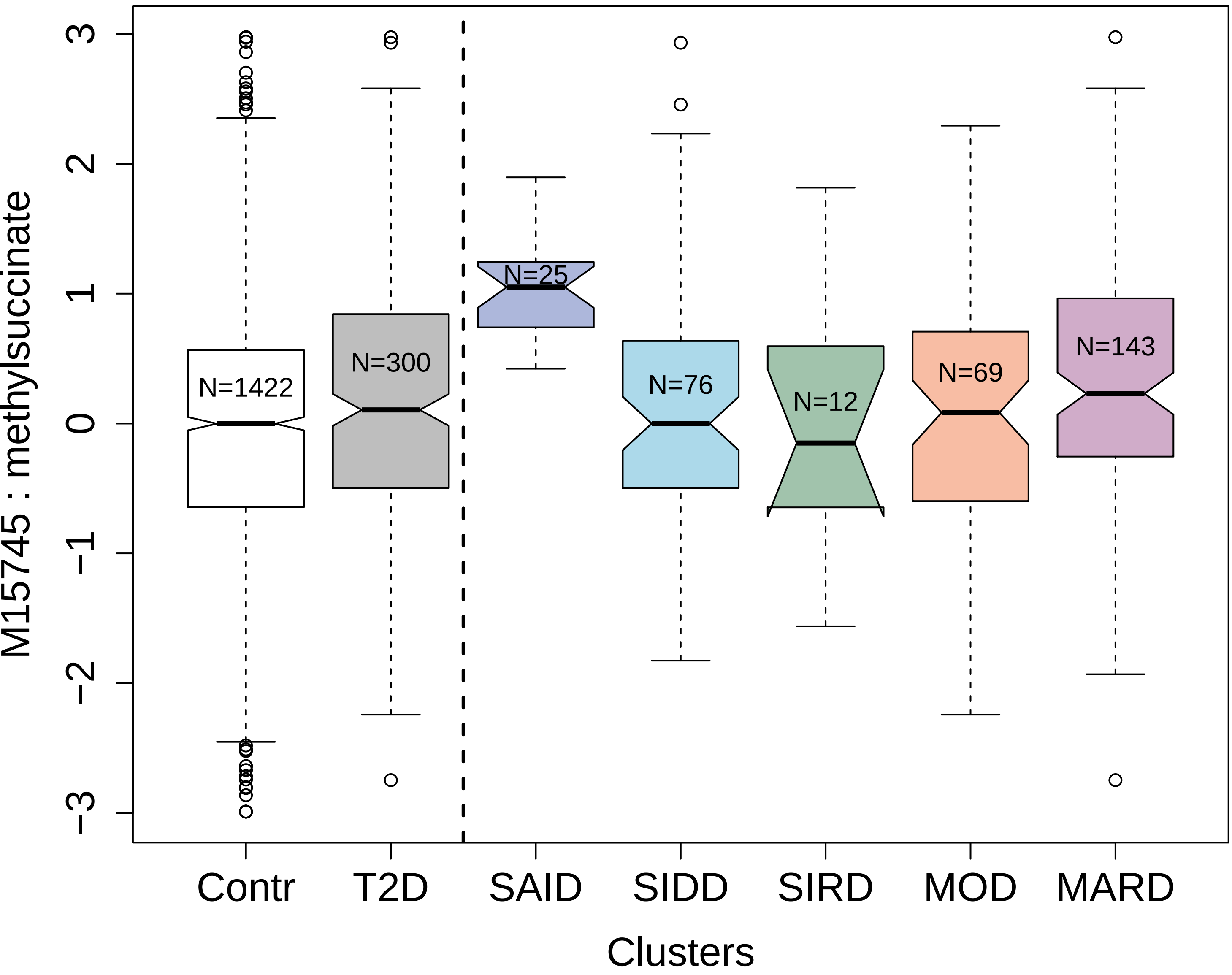

M15745 : methylsuccinate
3
2
1
0
−1
−2
−3
N=1422
N=300
N=25
N=76
N=12
N=69
N=143
Contr
T2D
SAID
SIDD
SIRD
MOD
MARD
Clusters

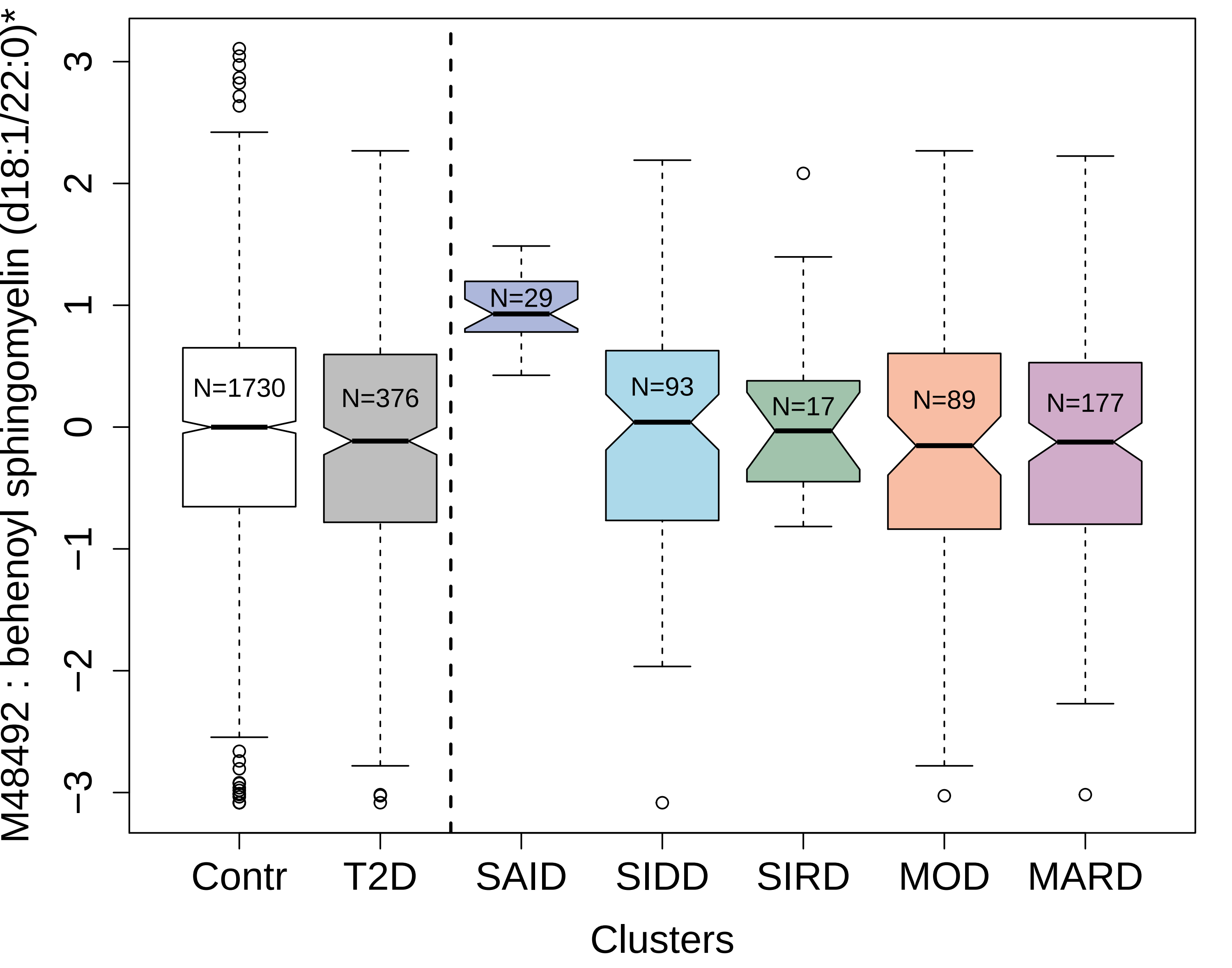
M48492 : behenoyl sphingomyelin (d18:1/22:0)*
3
2
1
0
−1
−2
−3
N=1730
N=376
N=29
N=93
N=17
N=89
N=177
Contr
T2D
SAID
SIDD
SIRD
MOD
MARD
Clusters

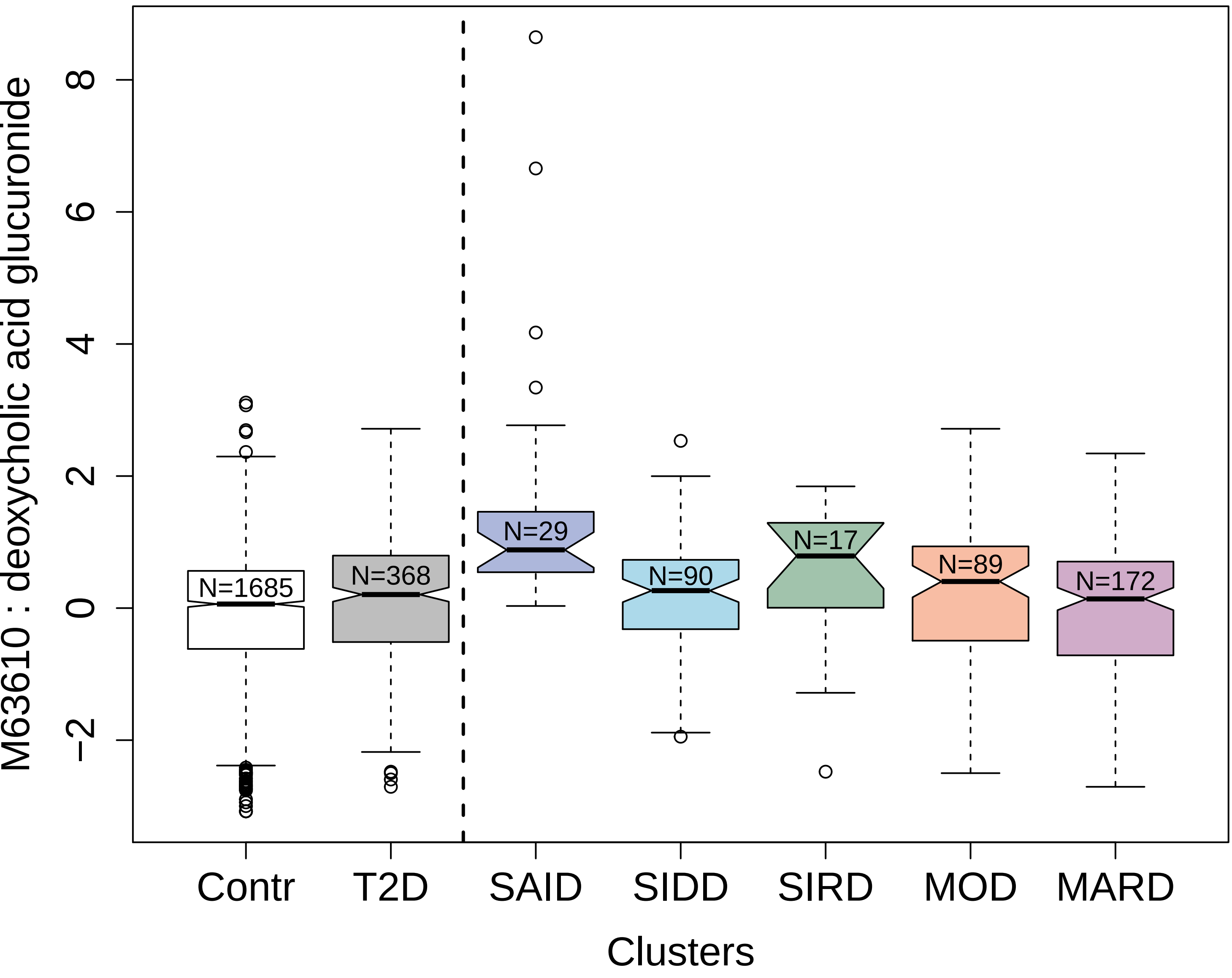
M63610 : deoxycholic acid glucuronide
8
6
4
2
0
−2
N=1685
N=368
N=29
N=90
N=17
N=89
N=172
Contr
T2D
SAID
SIDD
SIRD
MOD
MARD
Clusters

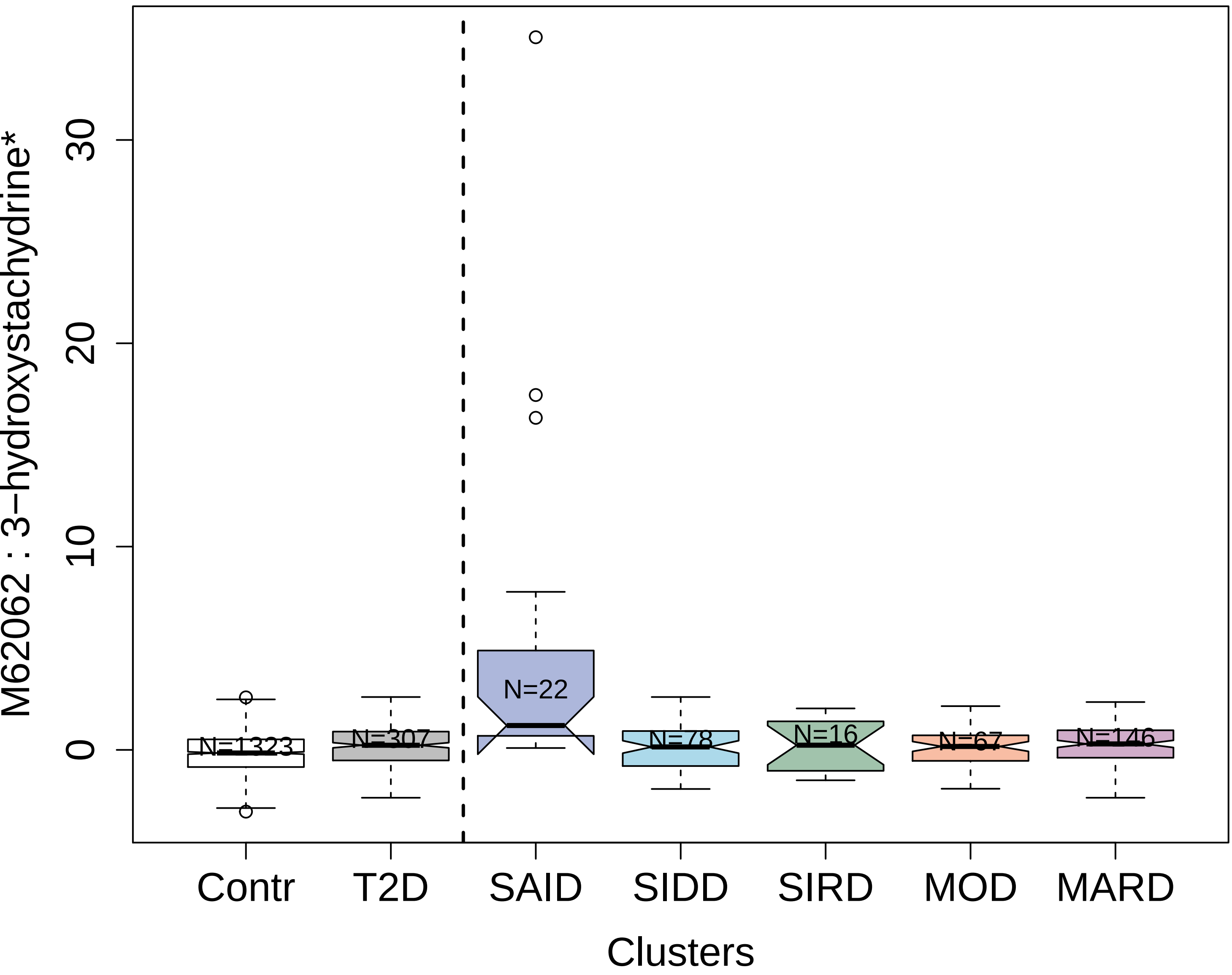
M62062 : 3−hydroxystachydrine*
0
10
20
30
N=1323
N=307
N=22
N=78
N=16
N=67
N=146
Contr
T2D
SAID
SIDD
SIRD
MOD
MARD
Clusters

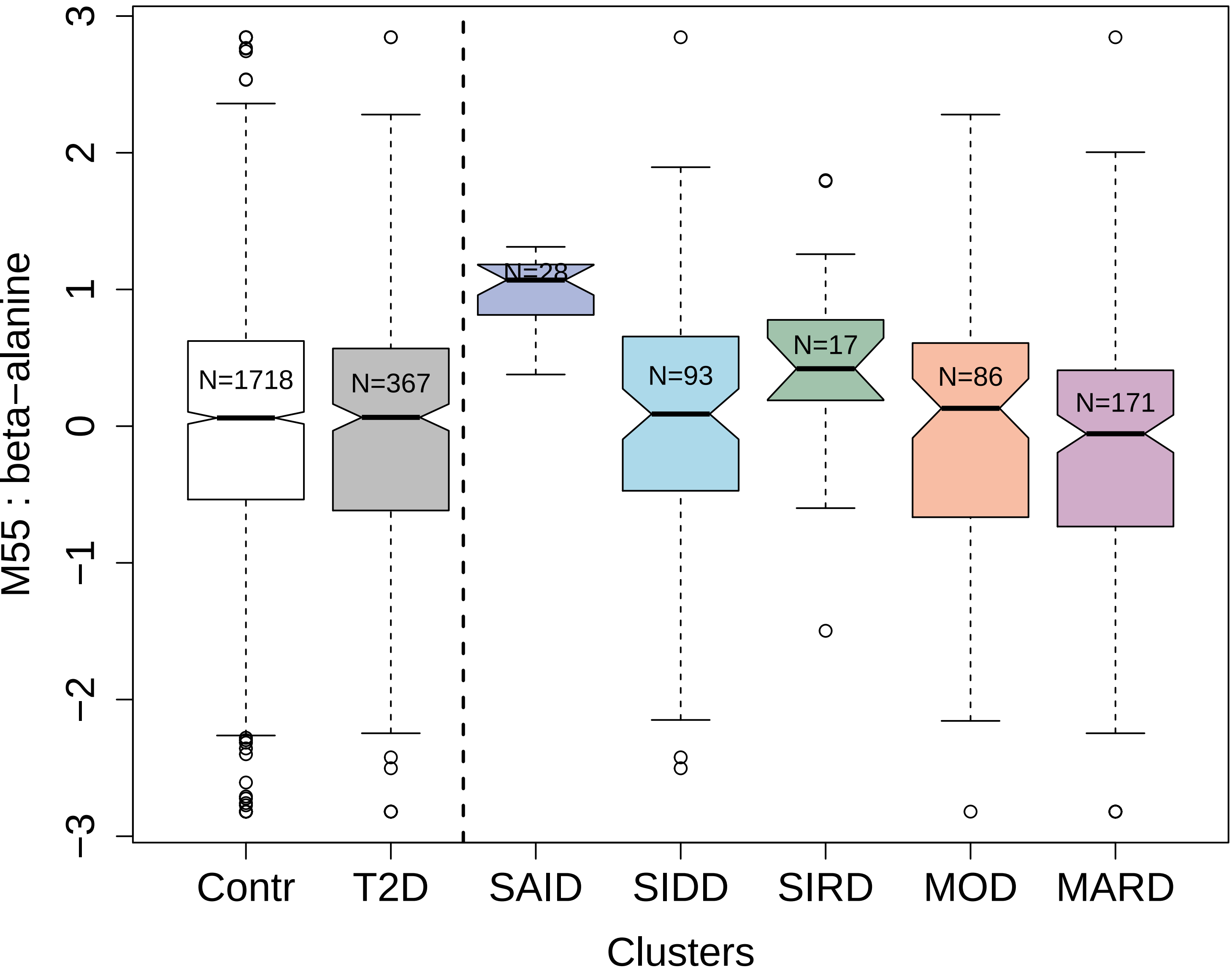

M55 : beta−alanine
3
2
1
0
−1
−2
−3
N=1718
N=367
N=28
N=93
N=17
N=86
N=171
Contr
T2D
SAID
SIDD
SIRD
MOD
MARD
Clusters

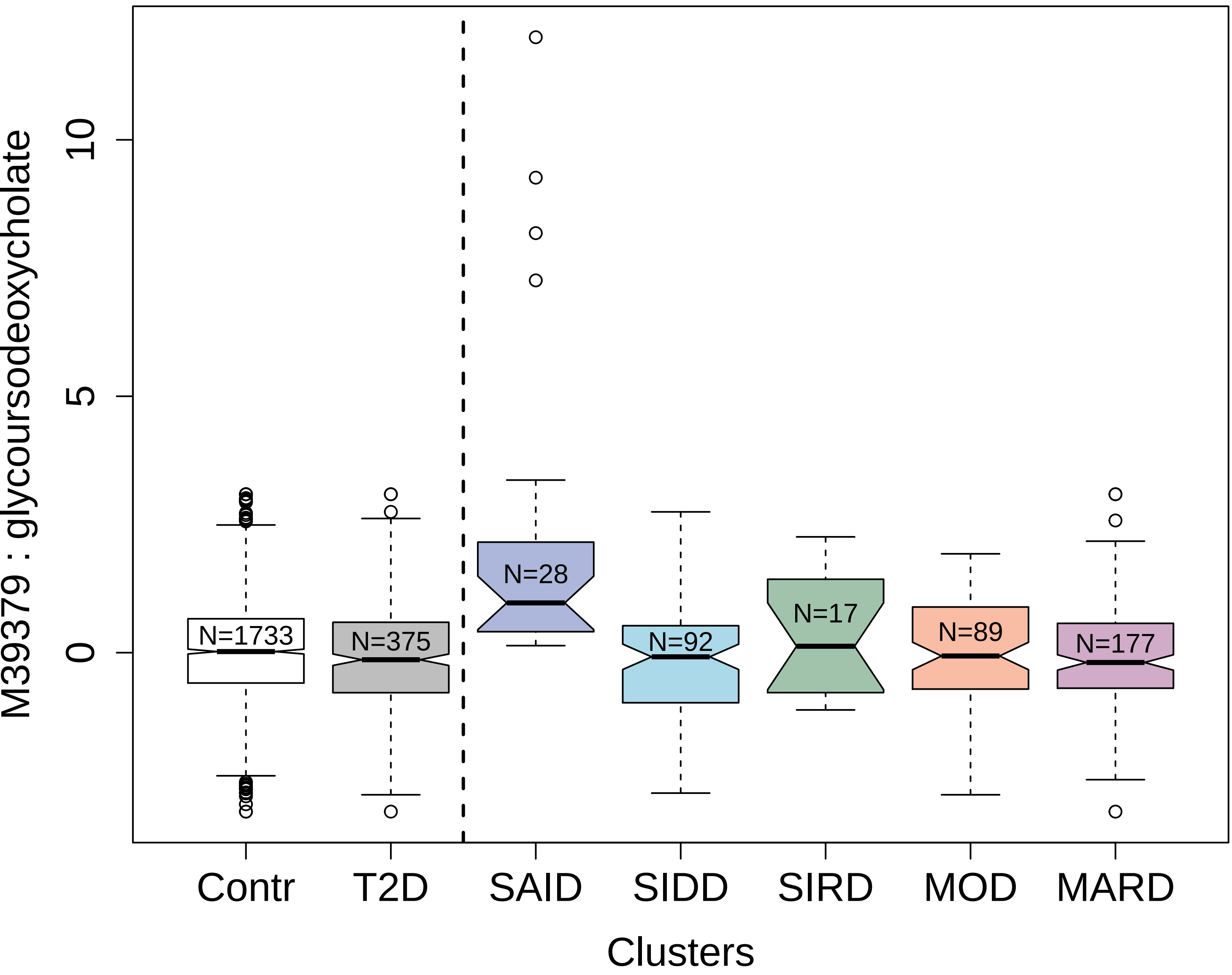

M39379 : glycoursodeoxycholate
0
5
10
N=1733
N=375
N=28
N=92
N=17
N=89
N=177
Contr
T2D
SAID
SIDD
SIRD
MOD
MARD
Clusters

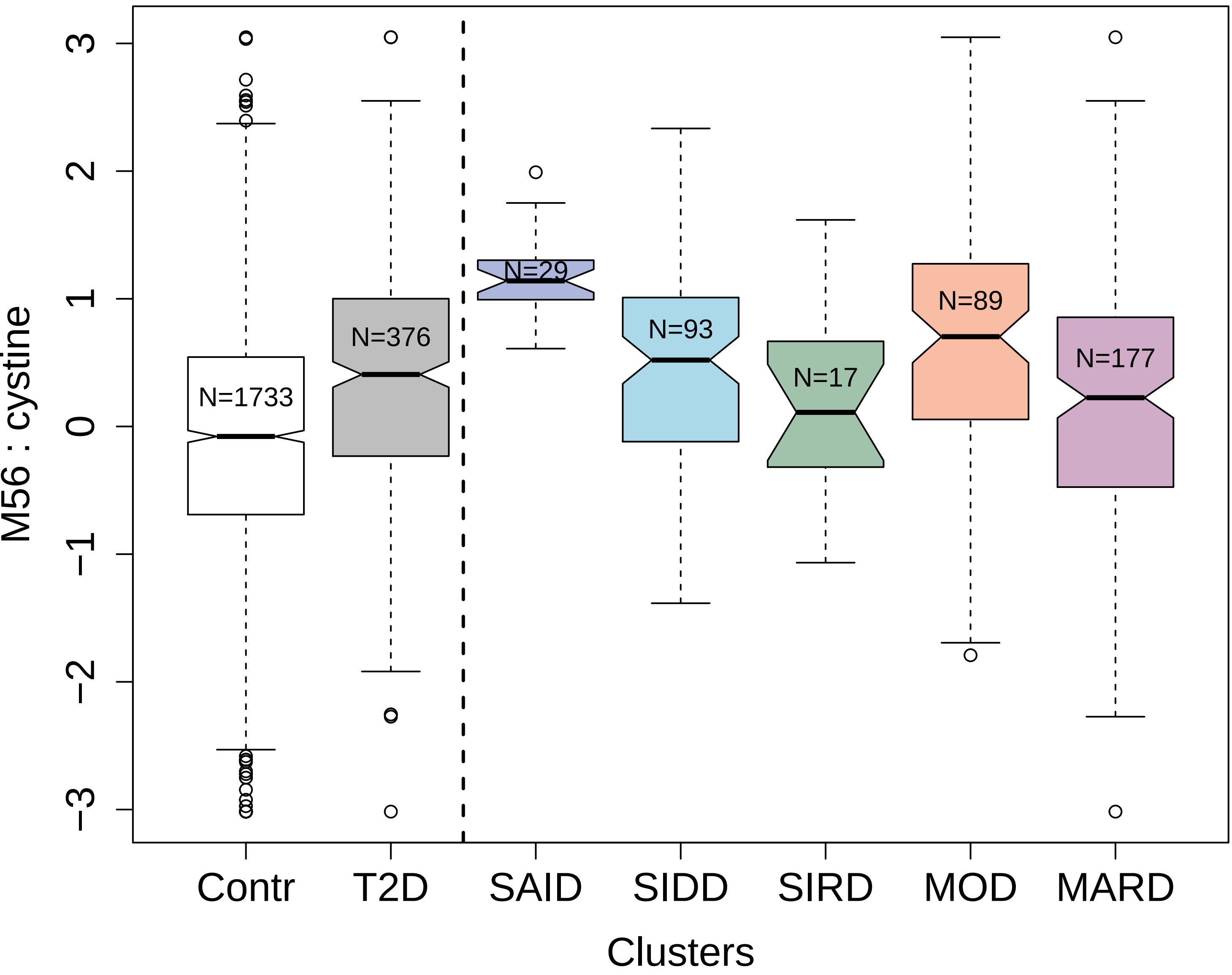

M56 : cystine
3
2
1
0
−1
−2
−3
N=1733
N=376
N=29
N=93
N=17
N=89
N=177
Contr
T2D
SAID
SIDD
SIRD
MOD
MARD
Clusters

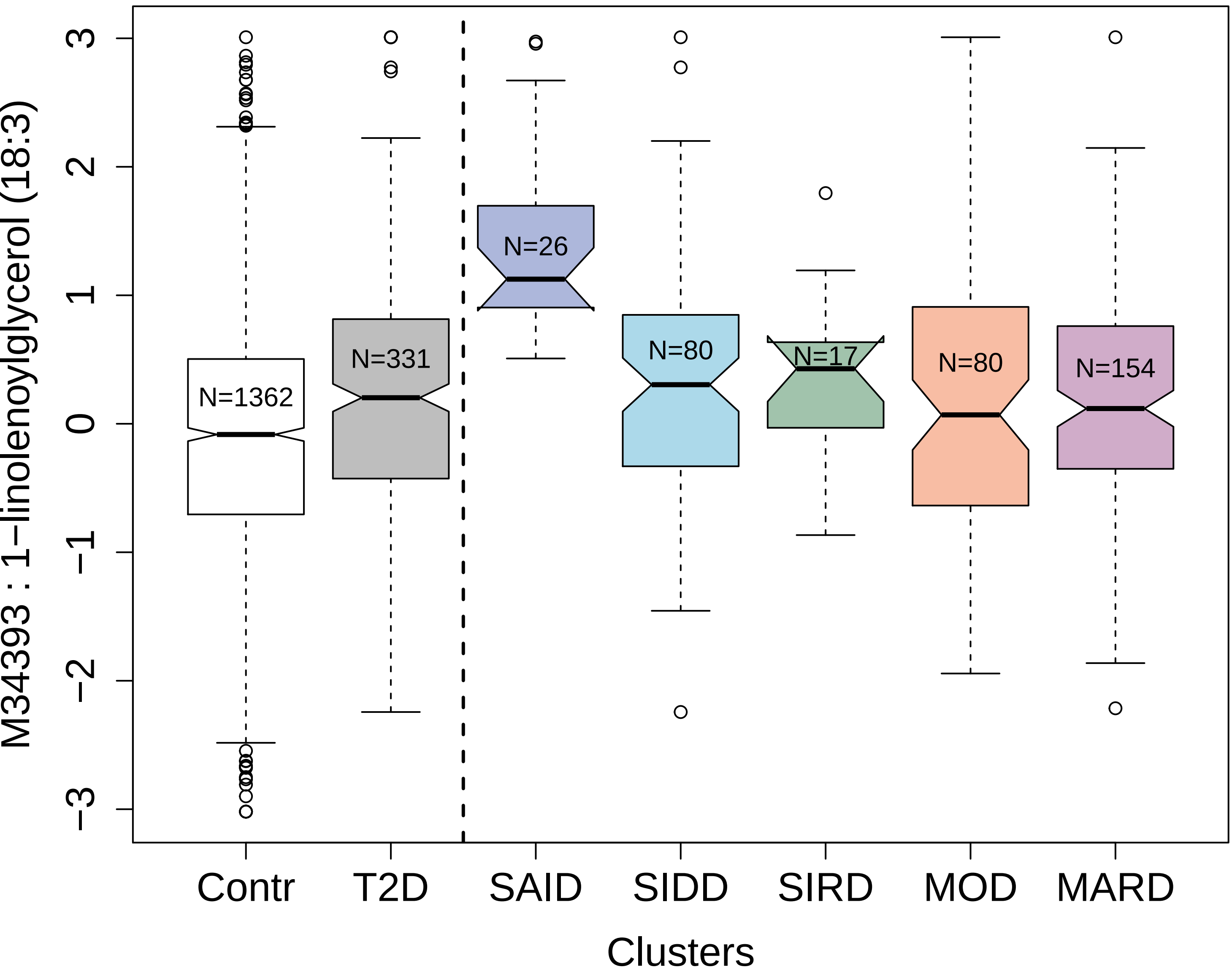

M34393 : 1−linolenoylglycerol (18:3)
3
2
1
0
−1
−2
−3
N=1362
N=331
N=26
N=80
N=17
N=80
N=154
Contr
T2D
SAID
SIDD
SIRD
MOD
MARD
Clusters

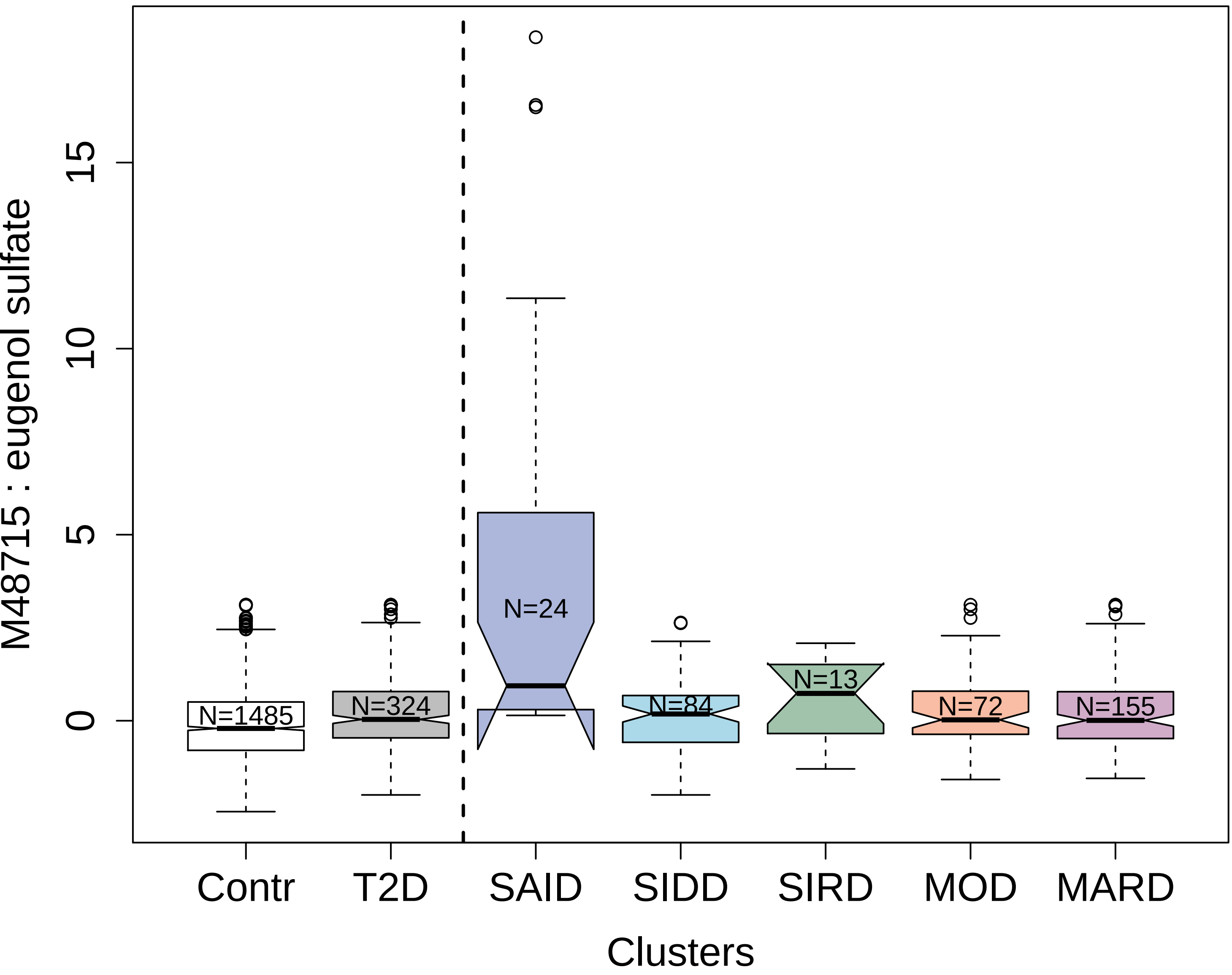
M48715 : eugenol sulfate
0
5
10
15
N=1485
N=324
N=24
N=84
N=13
N=72
N=155
Contr
T2D
SAID
SIDD
SIRD
MOD
MARD
Clusters

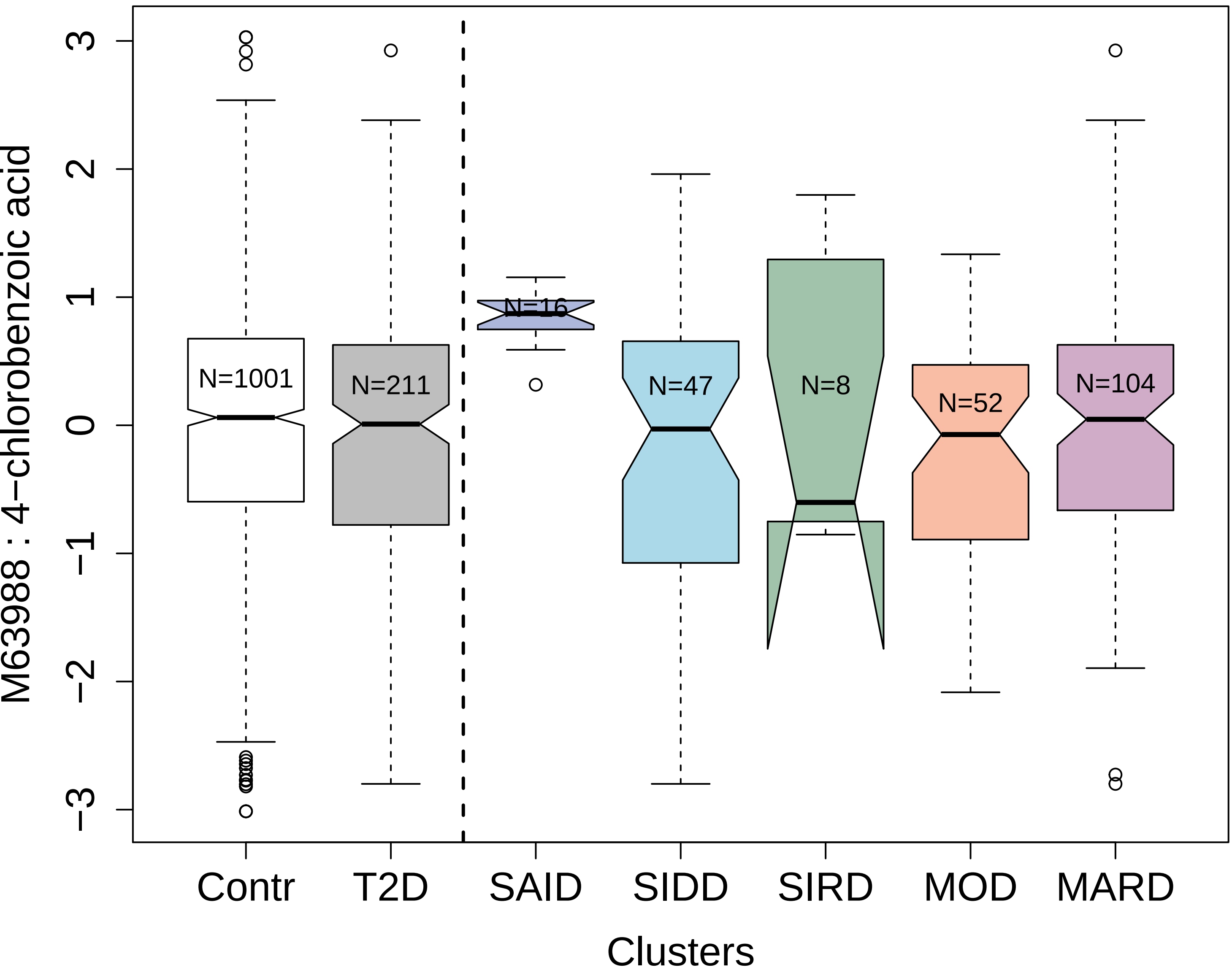
M63988 : 4−chlorobenzoic acid
3
2
1
0
−1
−2
−3
N=1001
N=211
N=16
N=47
N=8
N=52
N=104
Contr
T2D
SAID
SIDD
SIRD
MOD
MARD
Clusters

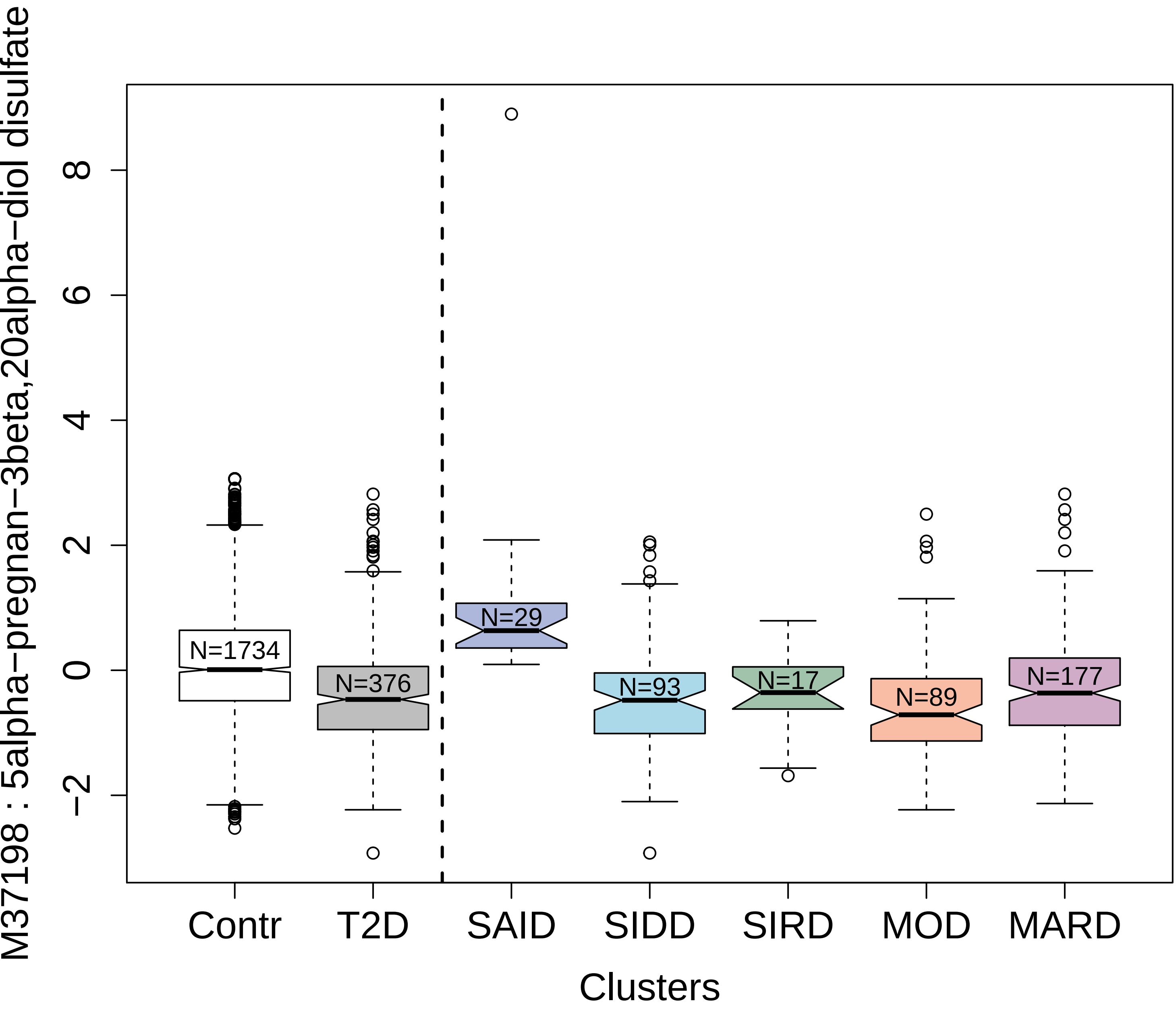
M37198 : 5alpha−pregnan−3beta,20alpha−diol disulfate
8
6
4
2
0
−2
N=1734
N=376
N=29
N=93
N=17
N=89
N=177
Contr
T2D
SAID
SIDD
SIRD
MOD
MARD
Clusters

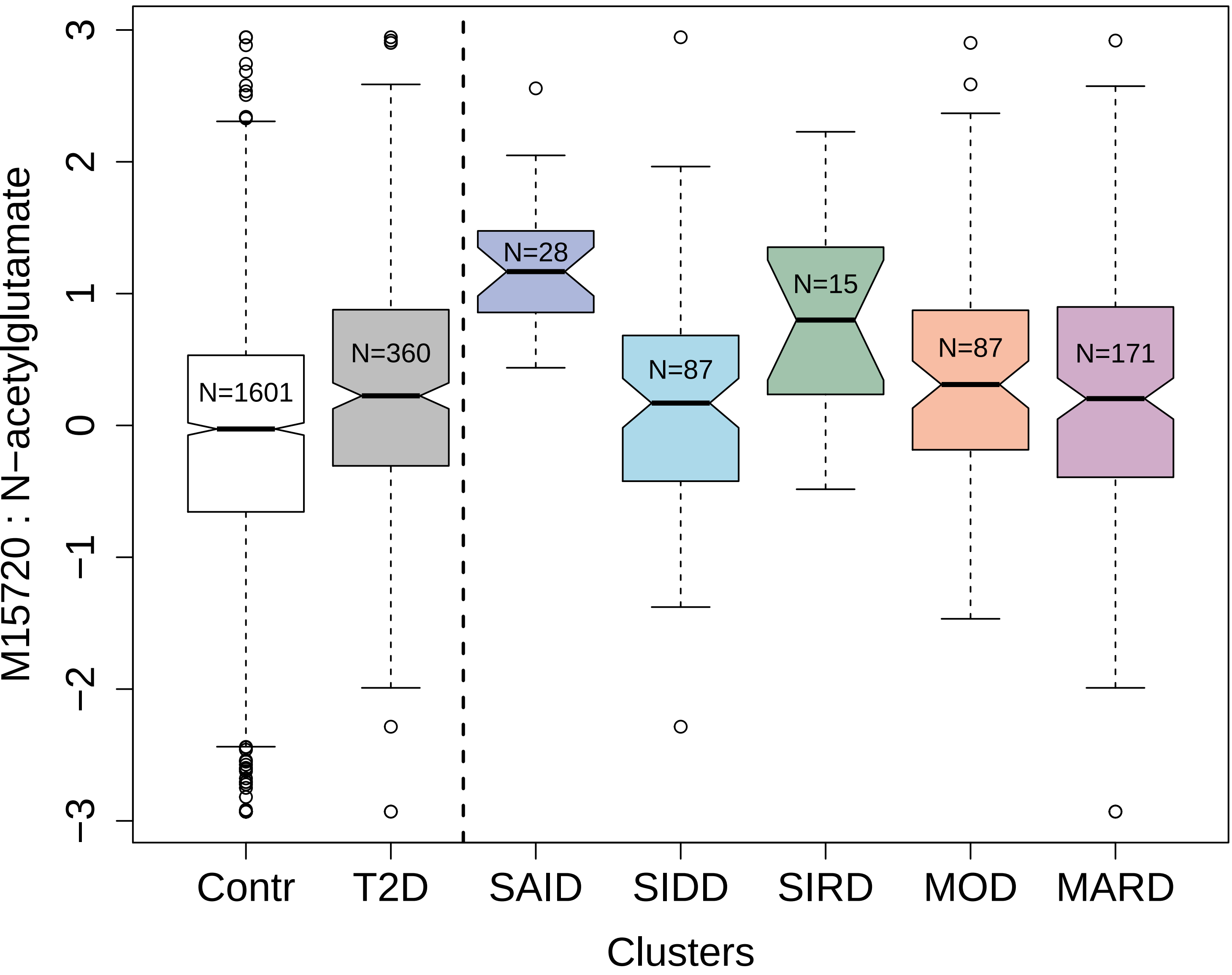

M15720 : N−acetylglutamate
3
2
1
0
−1
−2
−3
N=1601
N=360
N=28
N=87
N=15
N=87
N=171
Contr
T2D
SAID
SIDD
SIRD
MOD
MARD
Clusters

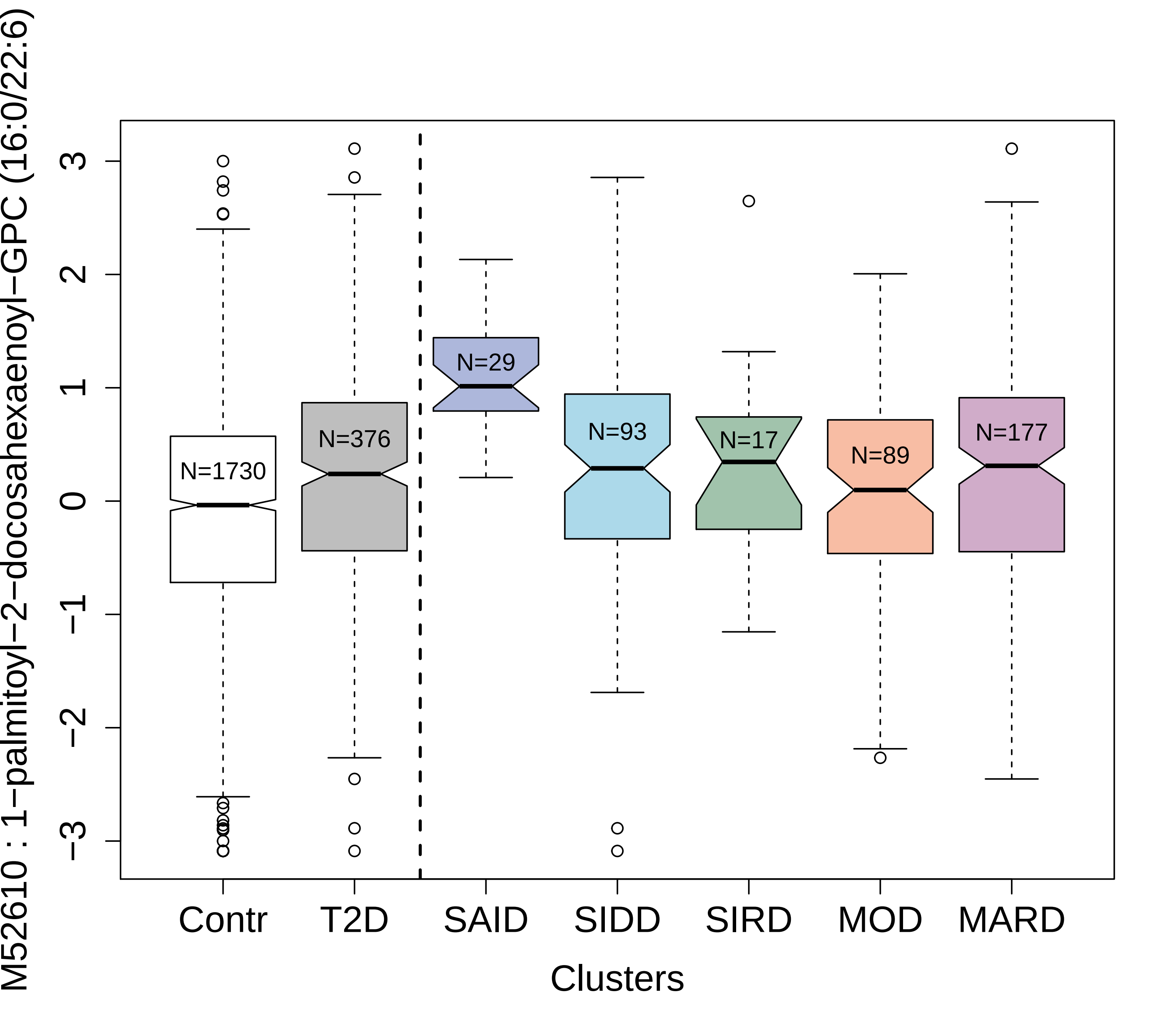
M52610 : 1−palmitoyl−2−docosahexaenoyl−GPC (16:0/22:6)
3
2
1
0
−1
−2
−3
N=1730
N=376
N=29
N=93
N=17
N=89
N=177
Contr
T2D
SAID
SIDD
SIRD
MOD
MARD
Clusters

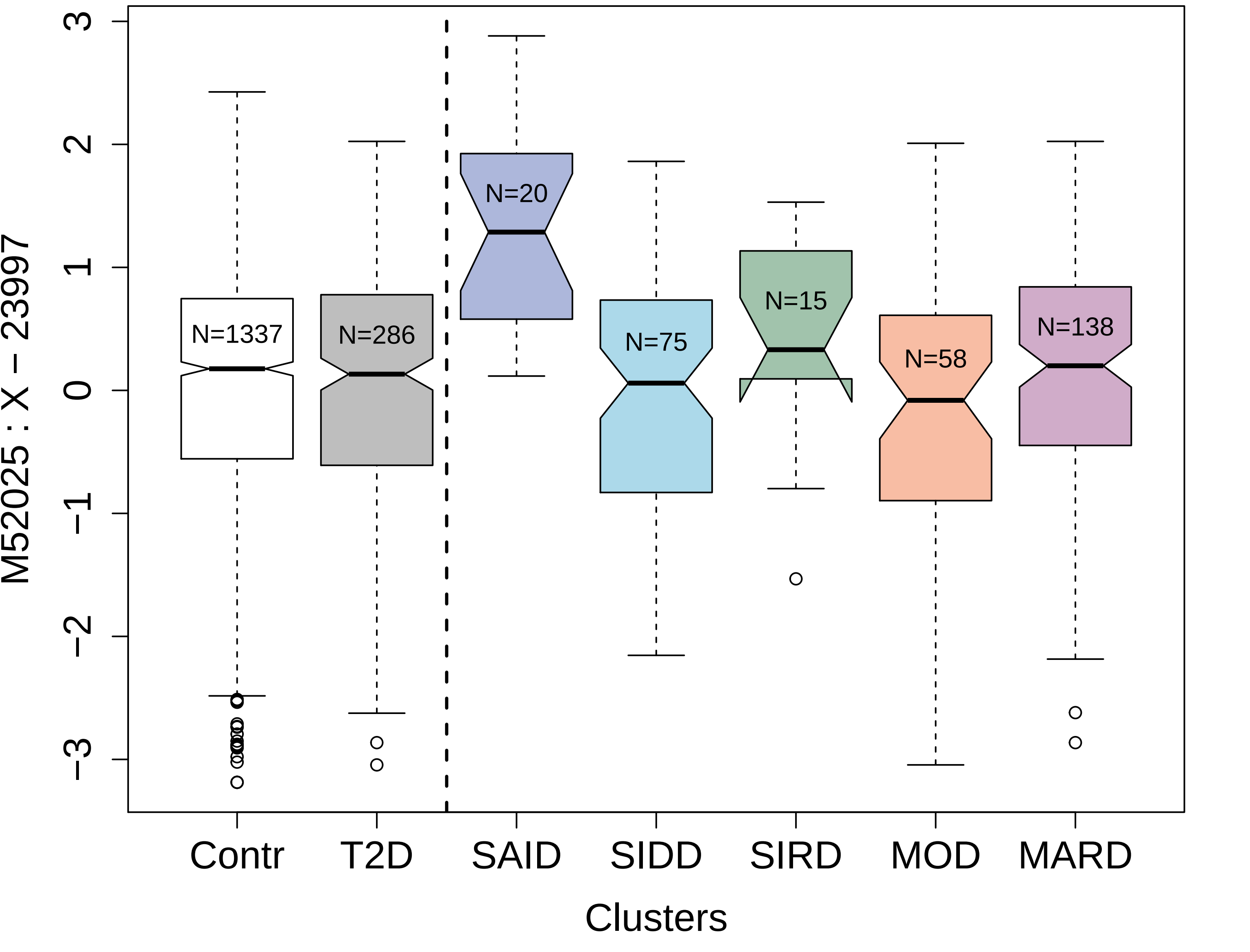

M52025 : X − 23997
3
2
1
0
−1
−2
−3
N=1337
N=286
N=20
N=75
N=15
N=58
N=138
Contr
T2D
SAID
SIDD
SIRD
MOD
MARD
Clusters

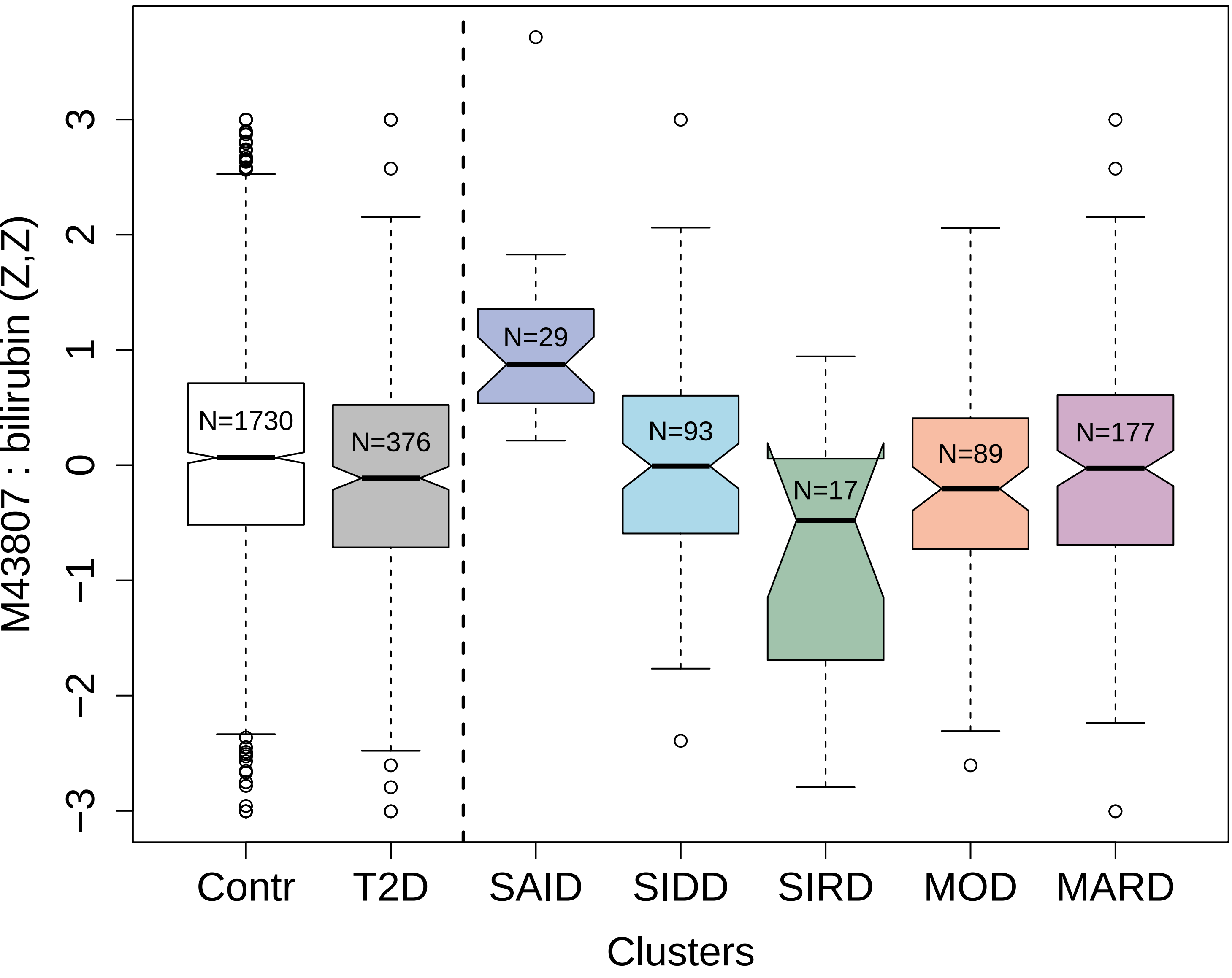

M43807 : bilirubin (Z,Z)
Clusters
Contr
T2D
SAID
SIDD
SIRD
MOD
MARD
N=1730
N=376
N=29
N=93
N=17
N=89
N=177
−3
−2
−1
0
1
2
3

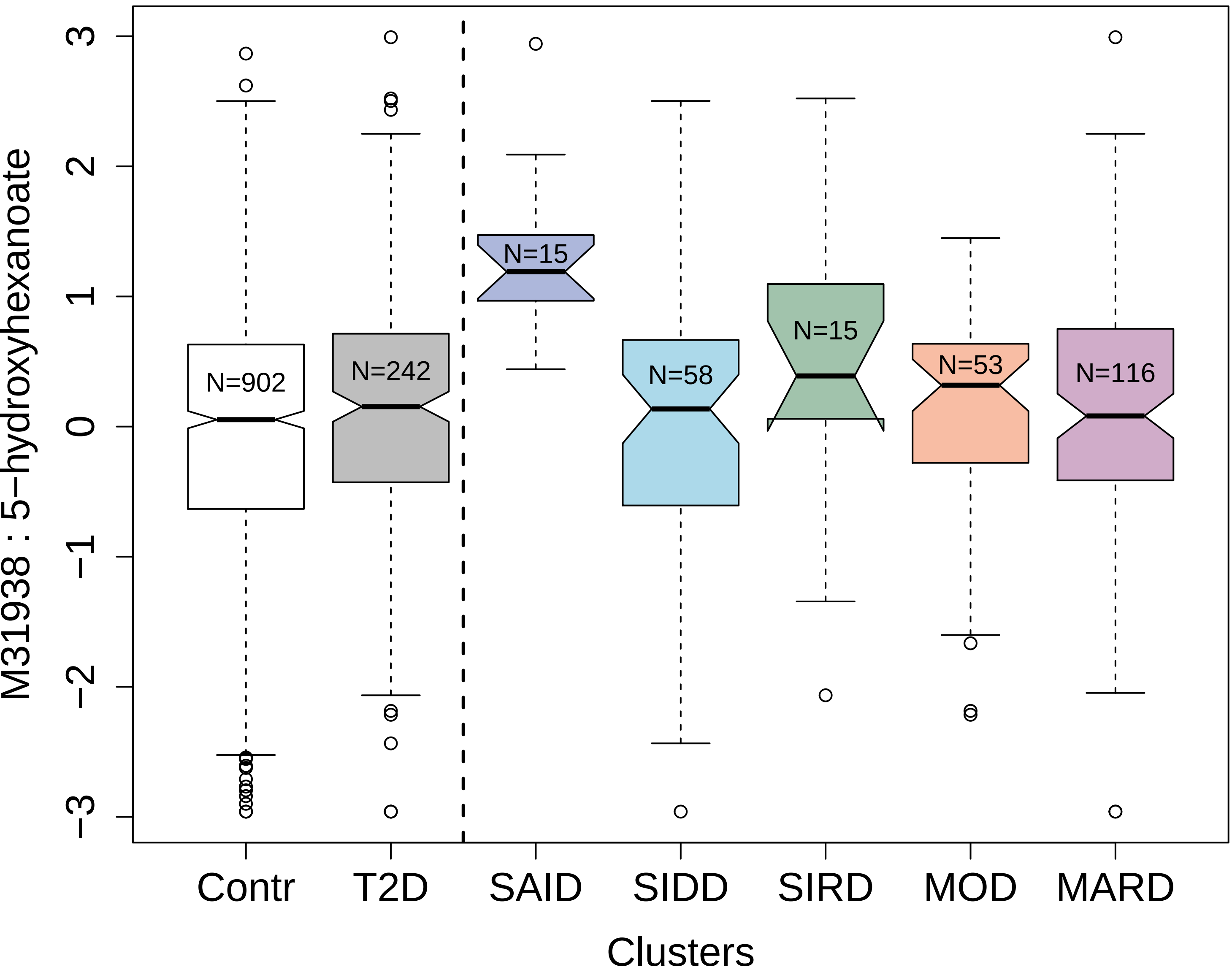
M31938 : 5−hydroxyhexanoate
3
2
1
0
−1
−2
−3
N=902
N=242
N=15
N=58
N=15
N=53
N=116
Contr
T2D
SAID
SIDD
SIRD
MOD
MARD
Clusters

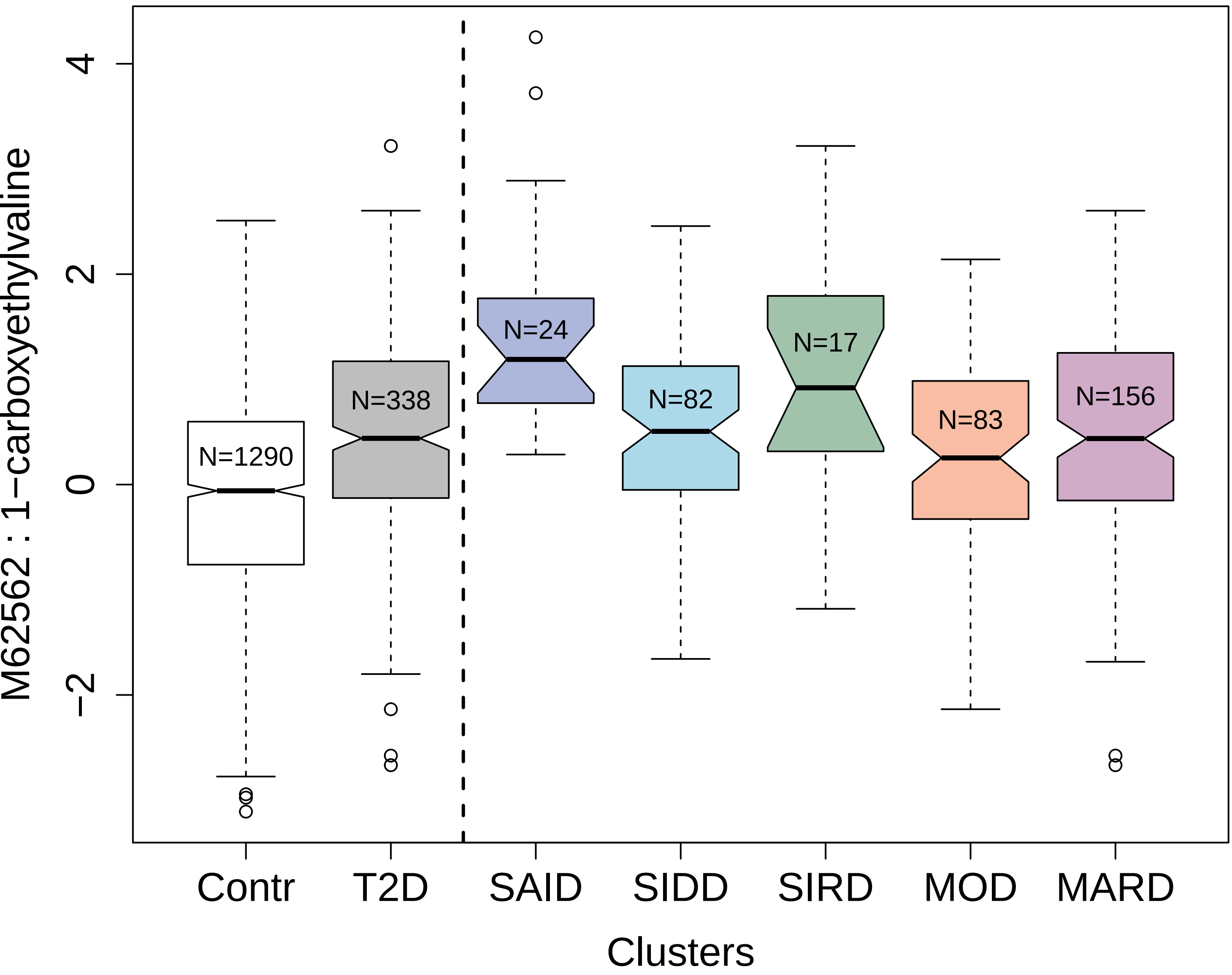
M62562 : 1−carboxyethylvaline
4
2
0
−2
N=1290
N=338
N=24
N=82
N=17
N=83
N=156
Contr
T2D
SAID
SIDD
SIRD
MOD
MARD
Clusters

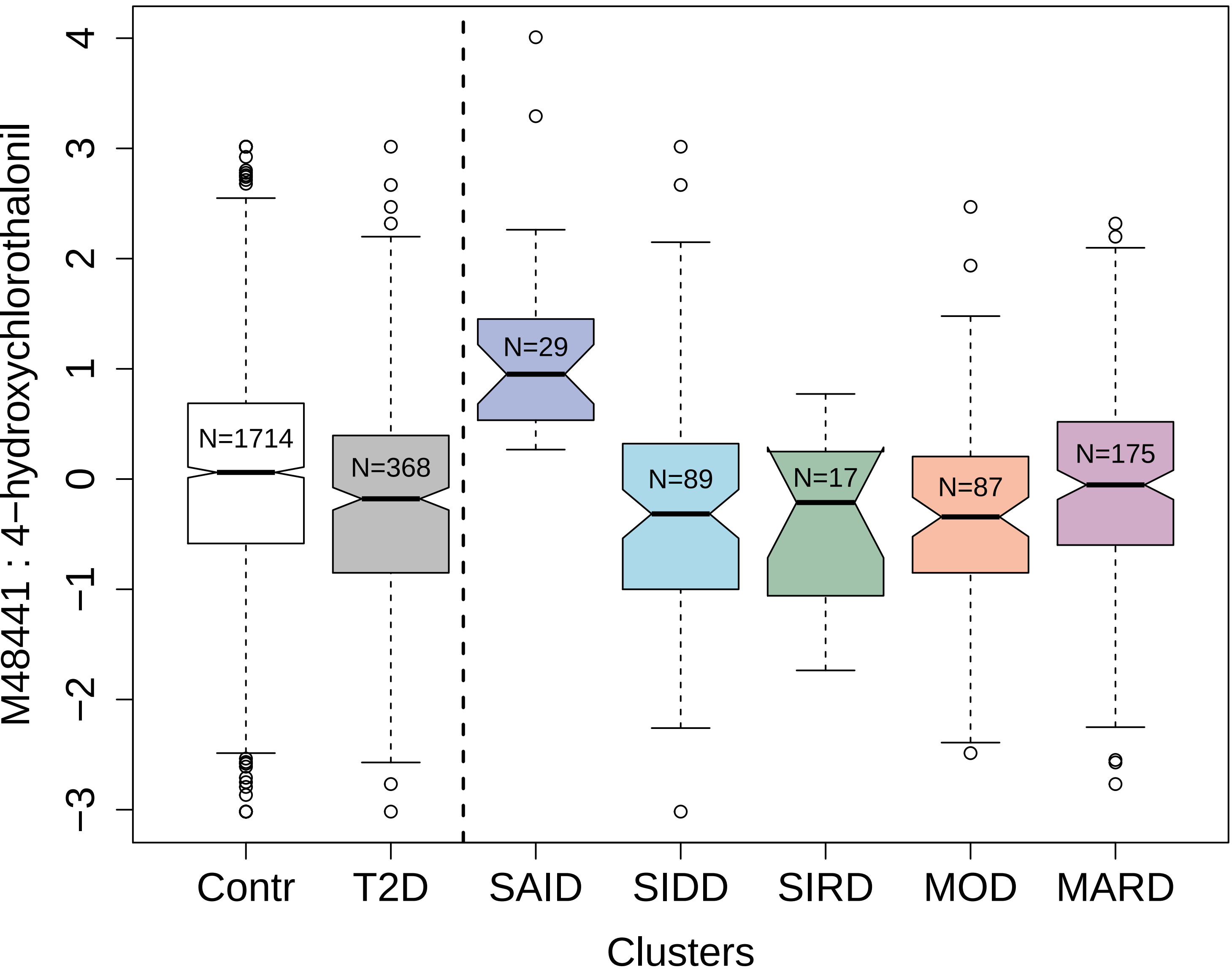

M48441 : 4−hydroxychlorothalonil
4
3
2
1
0
−1
−2
−3
N=1714
N=368
N=29
N=89
N=17
N=87
N=175
Contr
T2D
SAID
SIDD
SIRD
MOD
MARD
Clusters

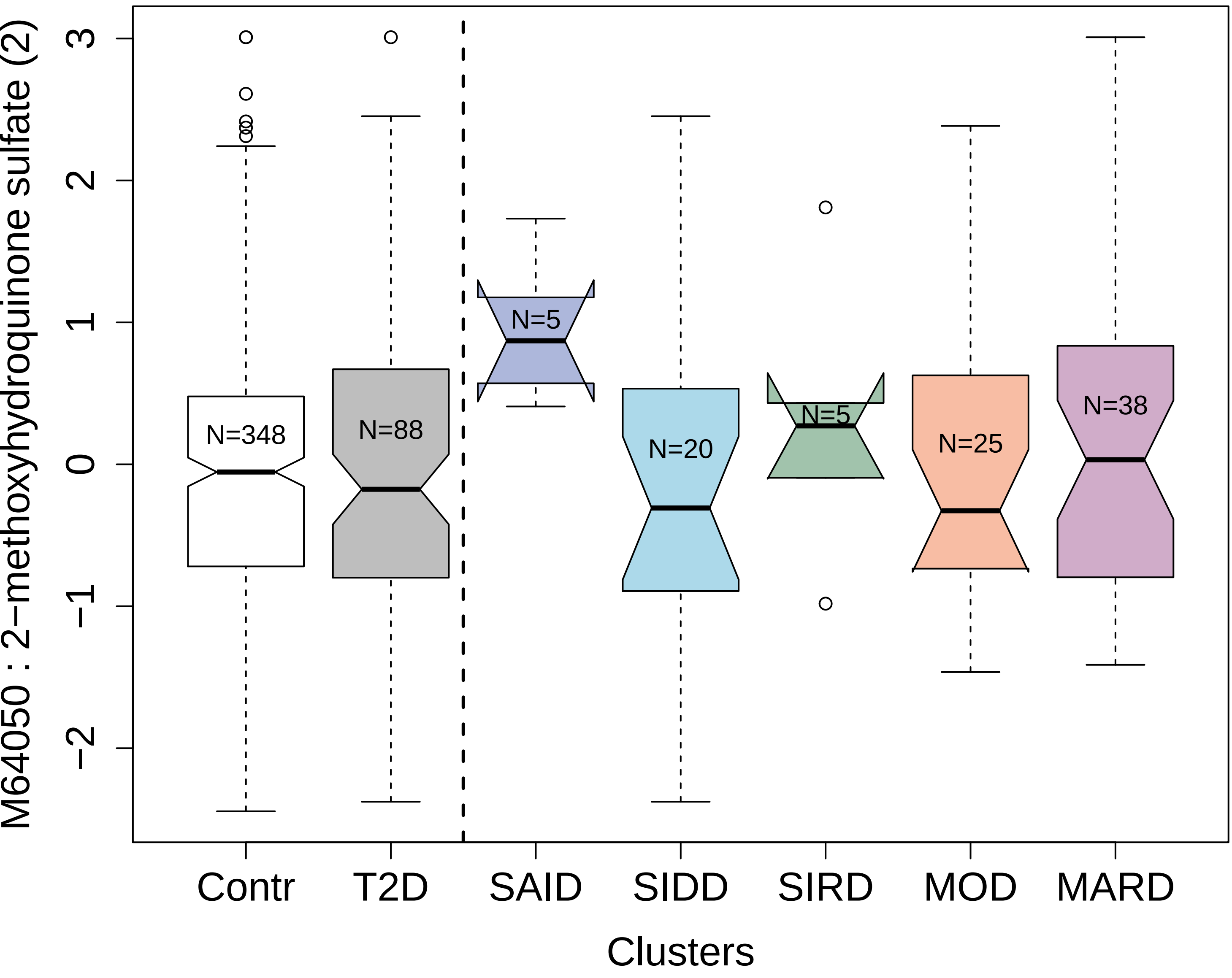

M64050 : 2−methoxyhydroquinone sulfate (2)
3
2
1
0
−1
−2
N=348
N=88
N=5
N=20
N=5
N=25
N=38
Contr
T2D
SAID
SIDD
SIRD
MOD
MARD
Clusters

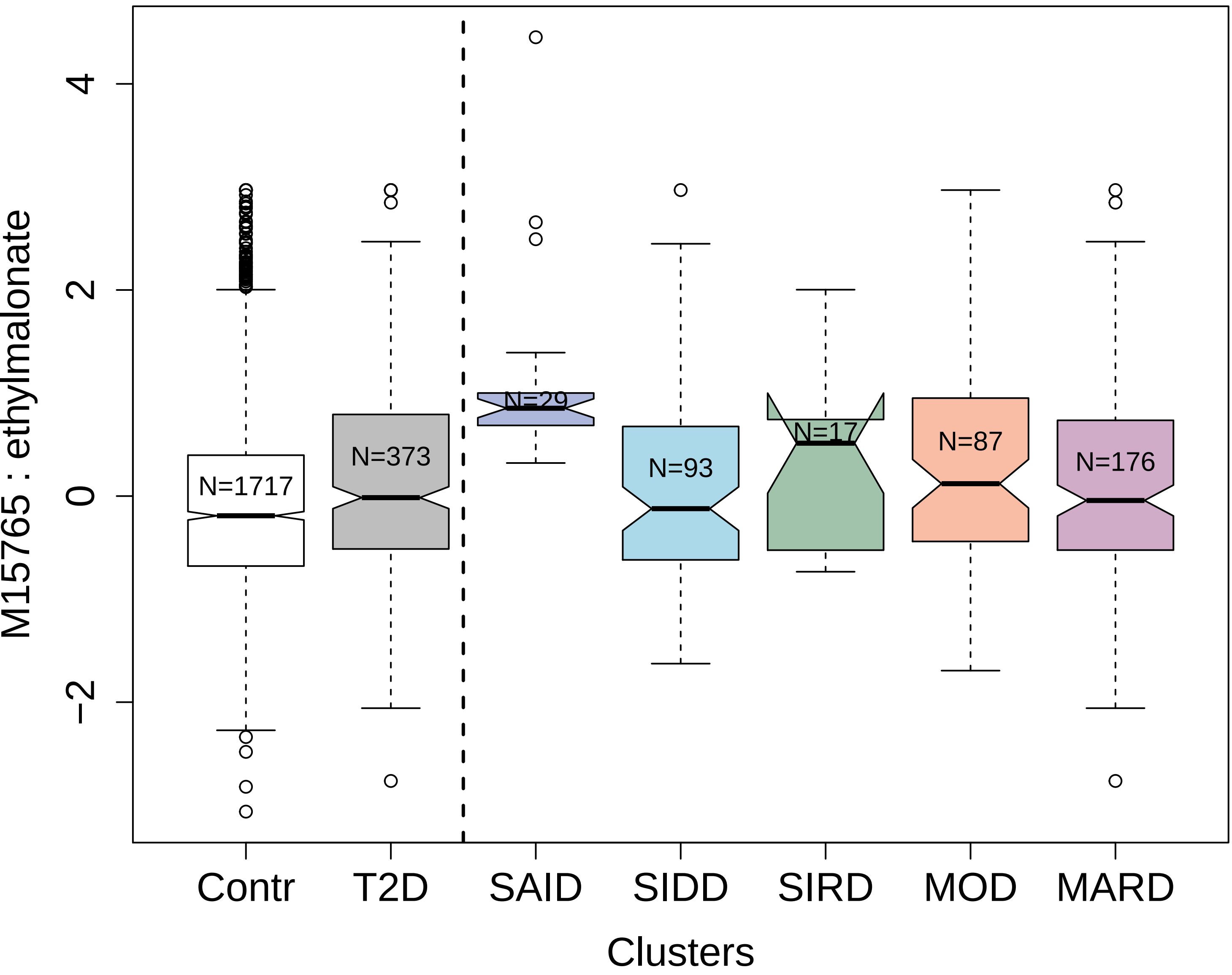

M15765 : ethylmalonate
4
2
0
−2
N=1717
N=373
N=29
N=93
N=17
N=87
N=176
Contr
T2D
SAID
SIDD
SIRD
MOD
MARD
Clusters

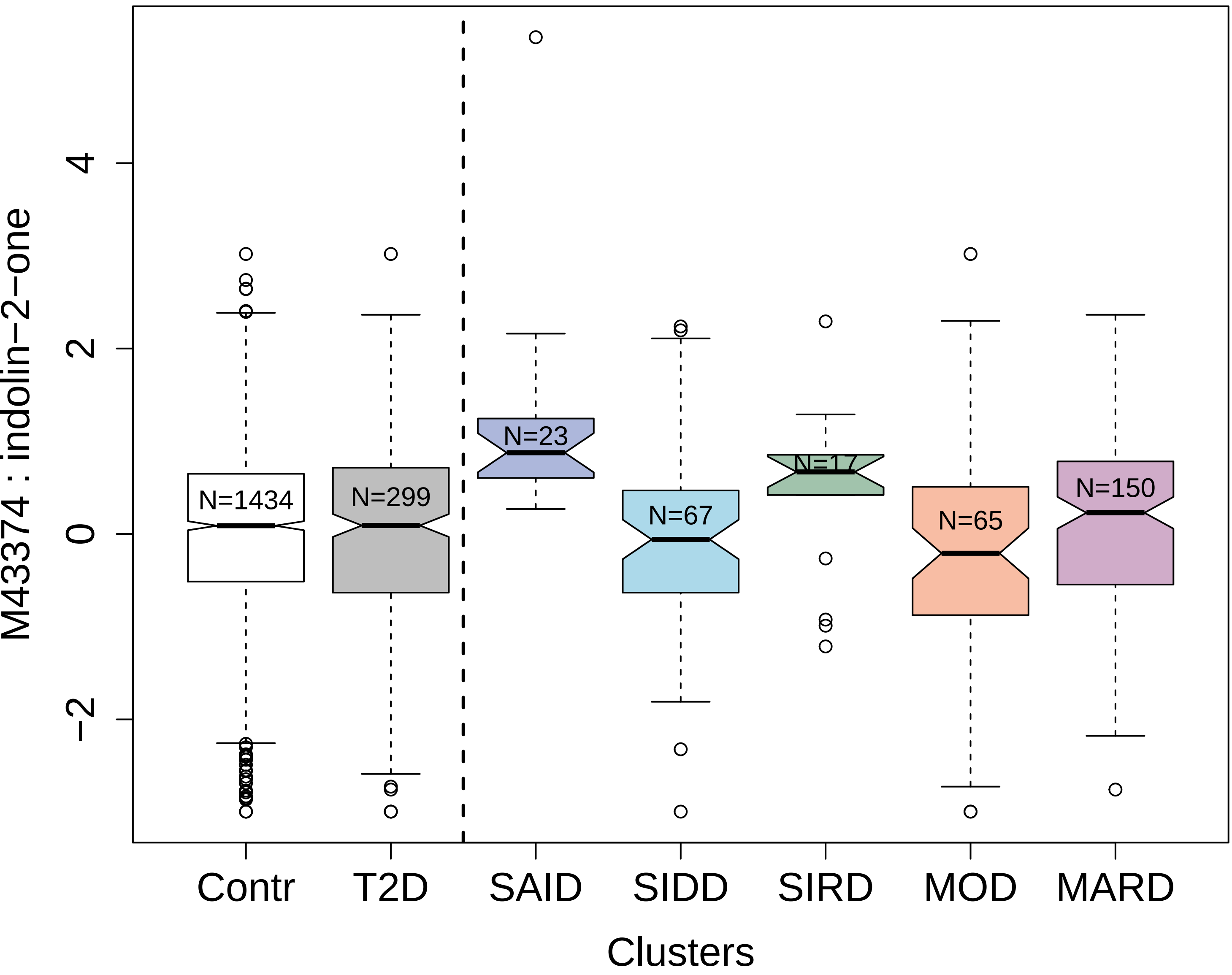

M43374 : indolin−2−one
4
2
0
−2
N=1434
N=299
N=23
N=67
N=17
N=65
N=150
Contr
T2D
SAID
SIDD
SIRD
MOD
MARD
Clusters

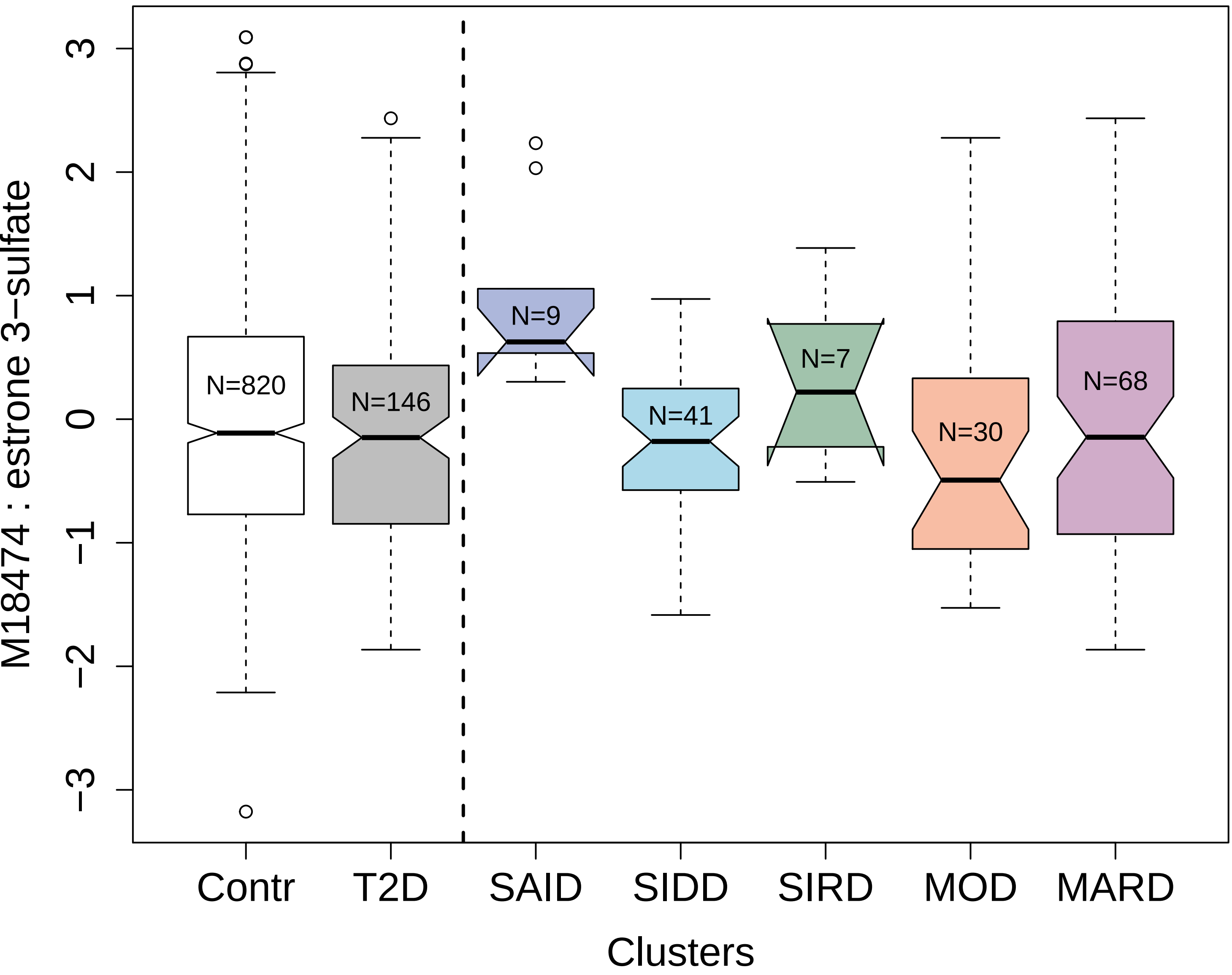

M18474 : estrone 3−sulfate
3
2
1
0
−1
−2
−3
N=820
N=146
N=9
N=41
N=7
N=30
N=68
Contr
T2D
SAID
SIDD
SIRD
MOD
MARD
Clusters

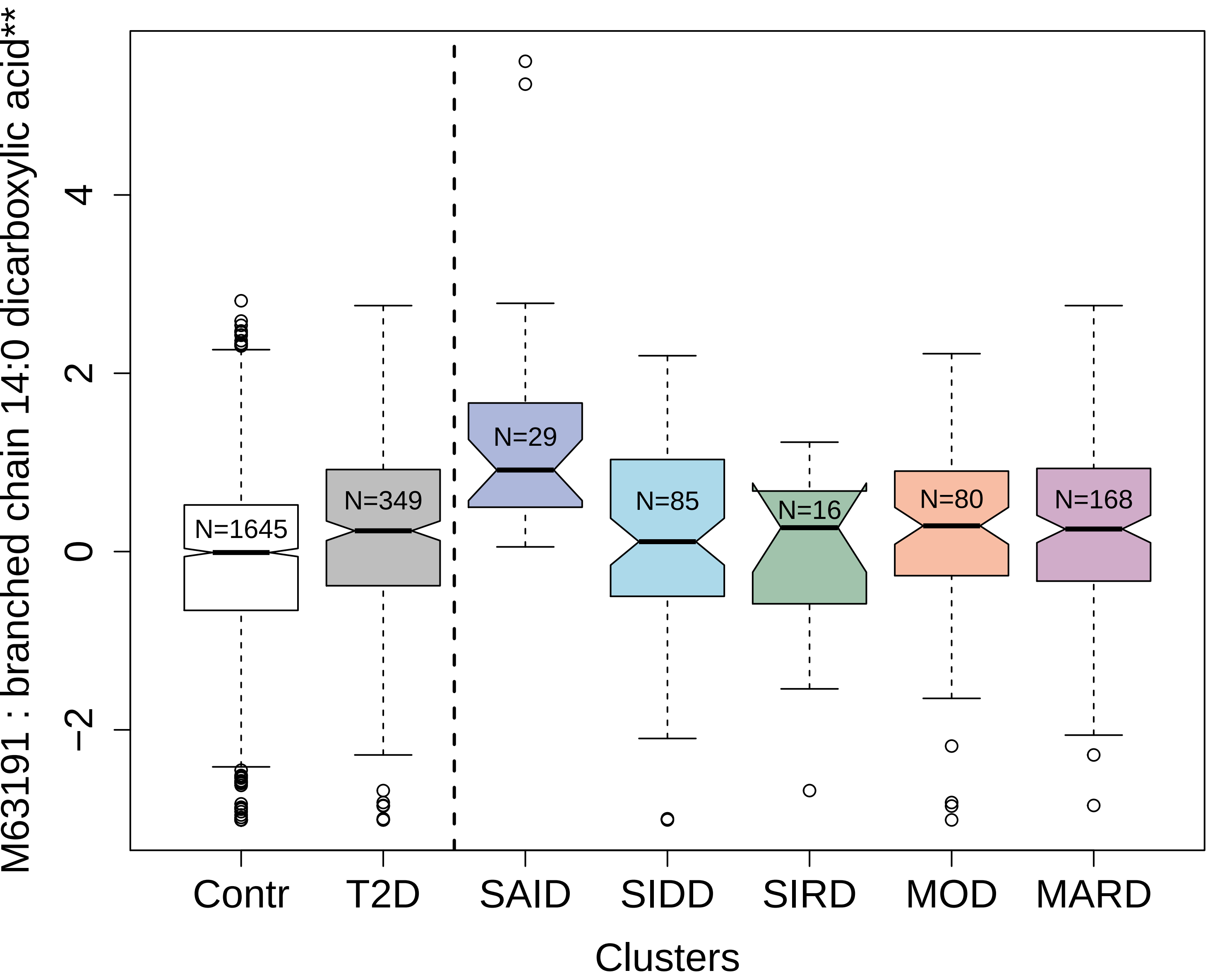

M63191 : branched chain 14:0 dicarboxylic acid**
4
2
0
−2
N=1645
N=349
N=29
N=85
N=16
N=80
N=168
Contr
T2D
SAID
SIDD
SIRD
MOD
MARD
Clusters

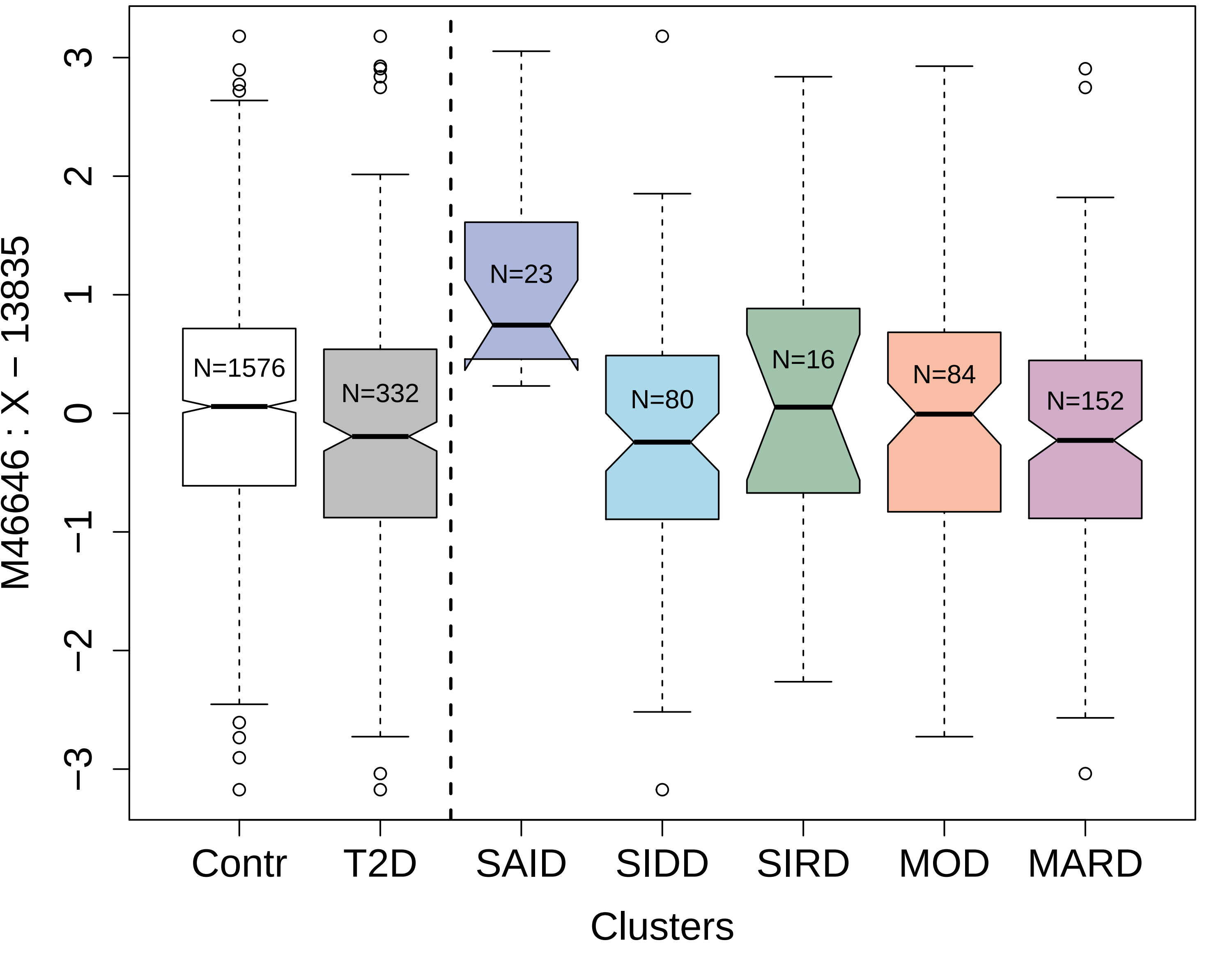
M46646 : X − 13835
3
2
1
0
−1
−2
−3
N=1576
N=332
N=23
N=80
N=16
N=84
N=152
Contr
T2D
SAID
SIDD
SIRD
MOD
MARD
Clusters

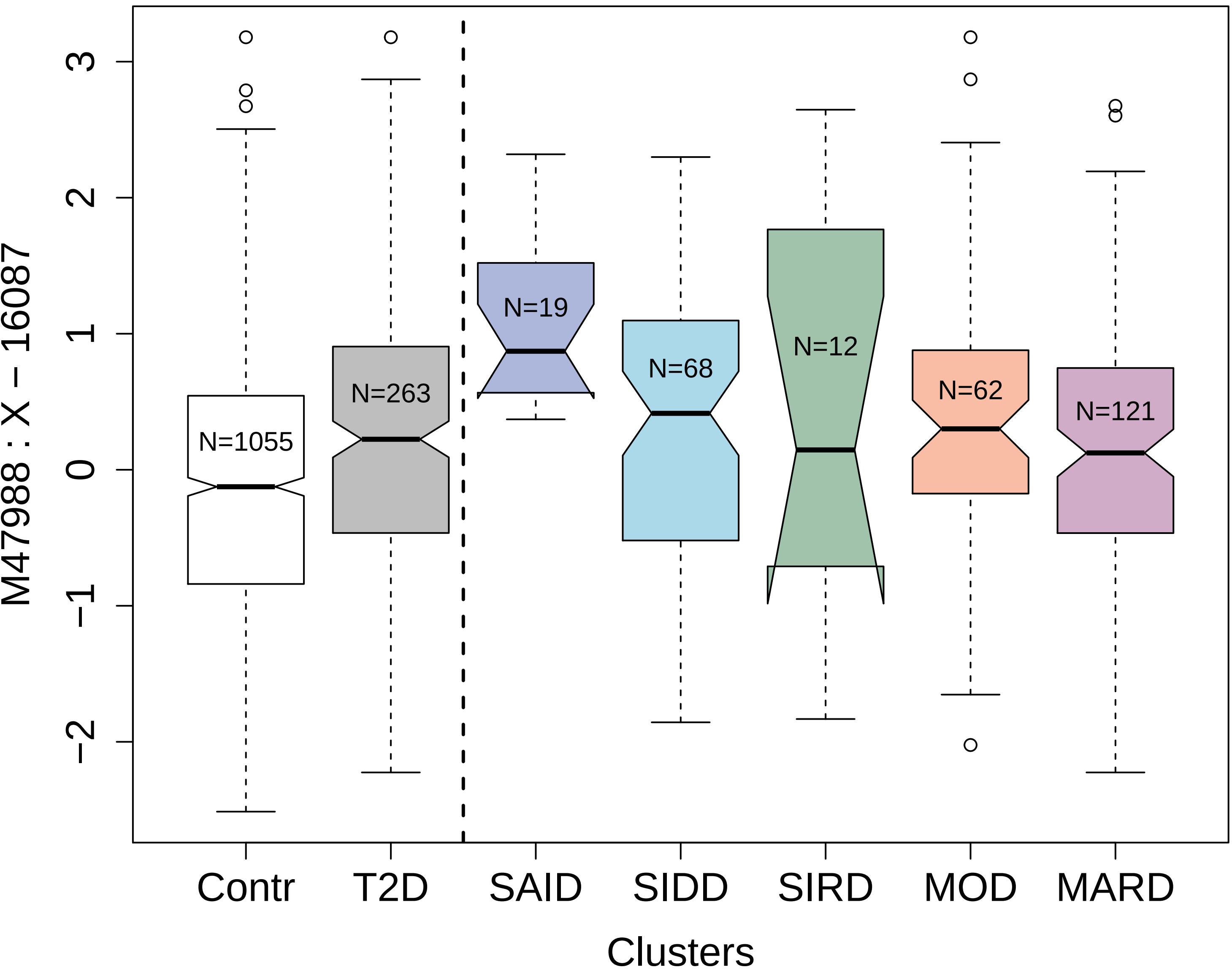

M47988 : X − 16087
3
2
1
0
−1
−2
N=1055
N=263
N=19
N=68
N=12
N=62
N=121
Contr
T2D
SAID
SIDD
SIRD
MOD
MARD
Clusters

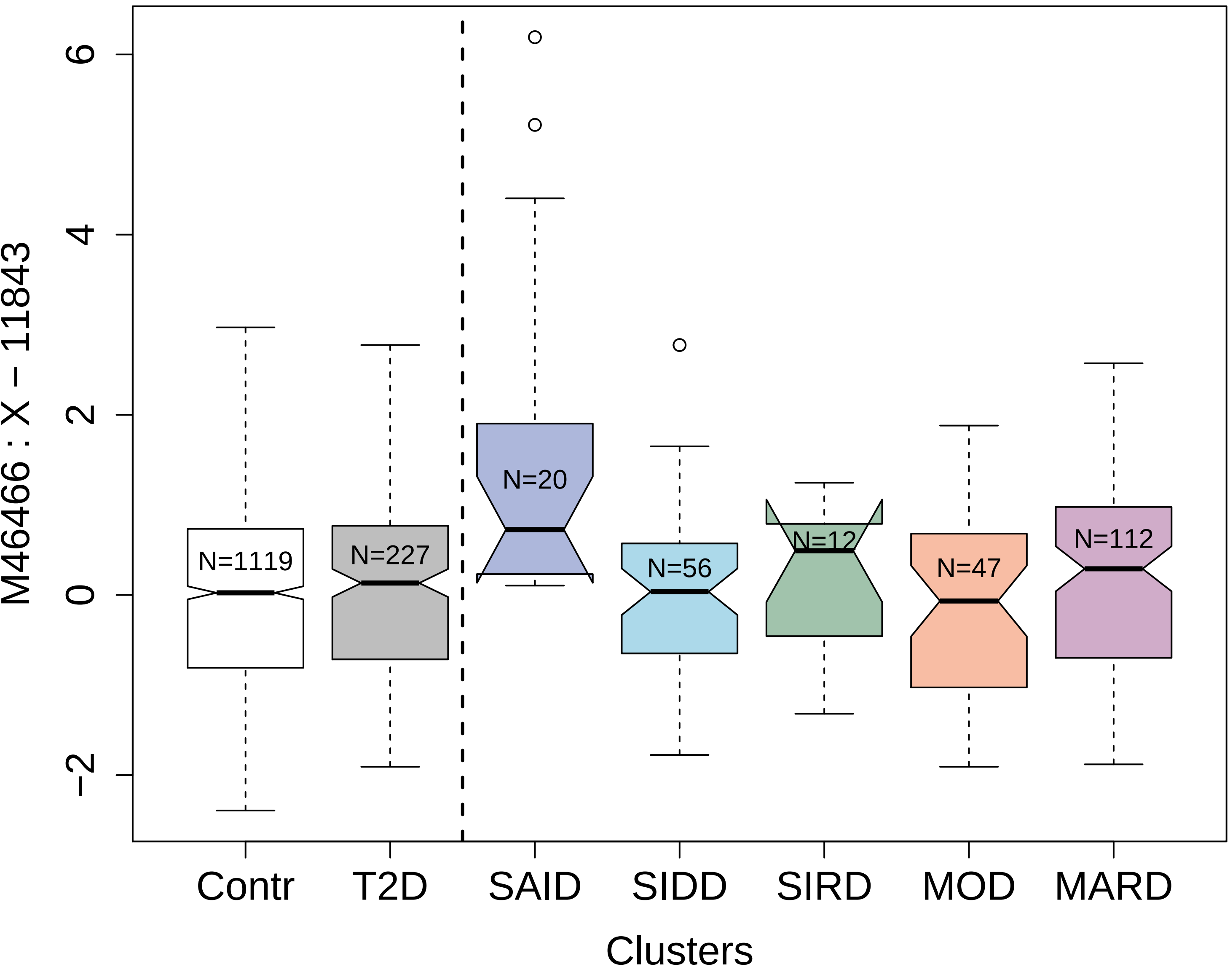

M46466 : X − 11843
−2
0
2
4
6
N=1119
N=227
N=20
N=56
N=12
N=47
N=112
Contr
T2D
SAID
SIDD
SIRD
MOD
MARD
Clusters

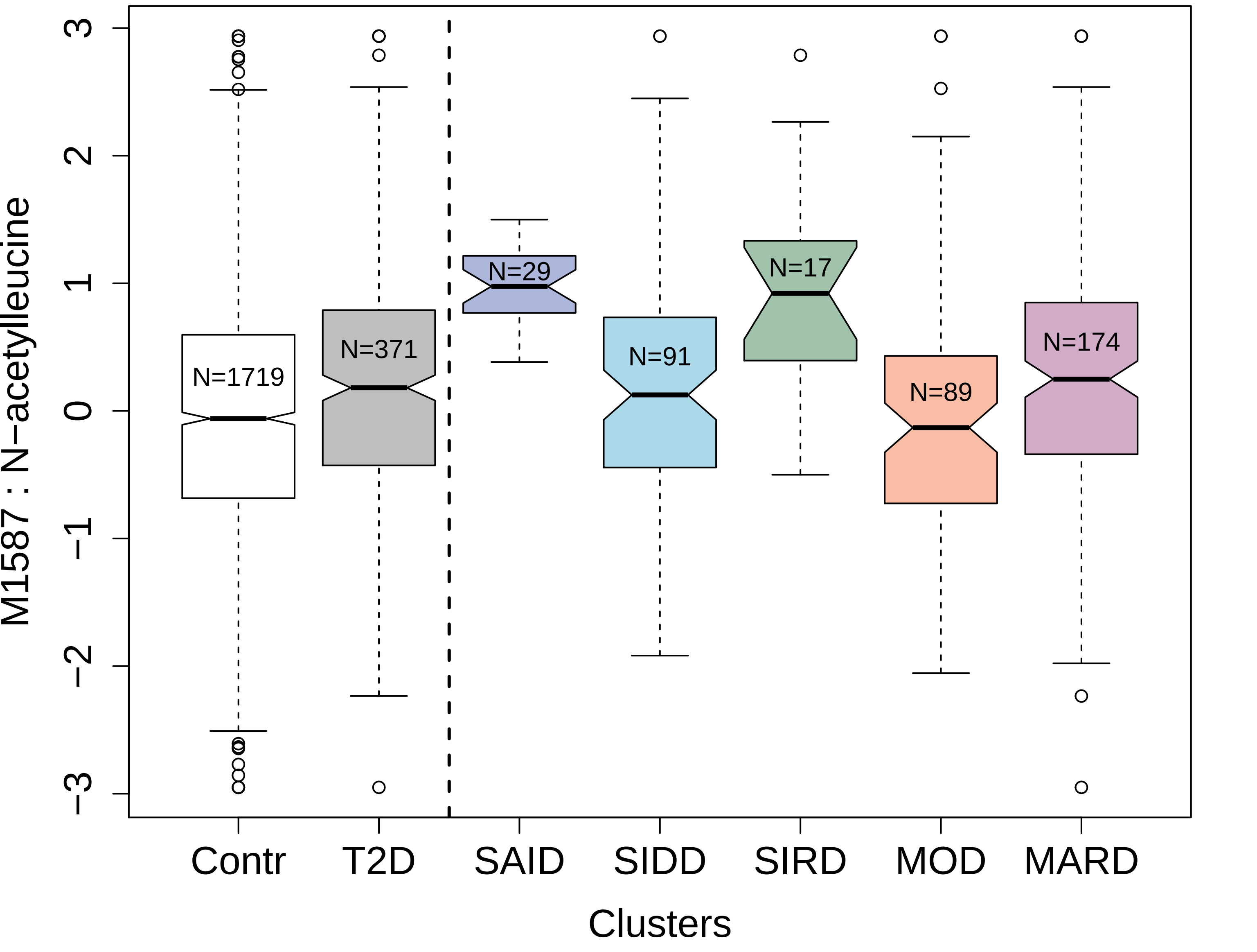

M1587 : N−acetylleucine
3
2
1
0
−1
−2
−3
N=1719
N=371
N=29
N=91
N=17
N=89
N=174
Contr
T2D
SAID
SIDD
SIRD
MOD
MARD
Clusters

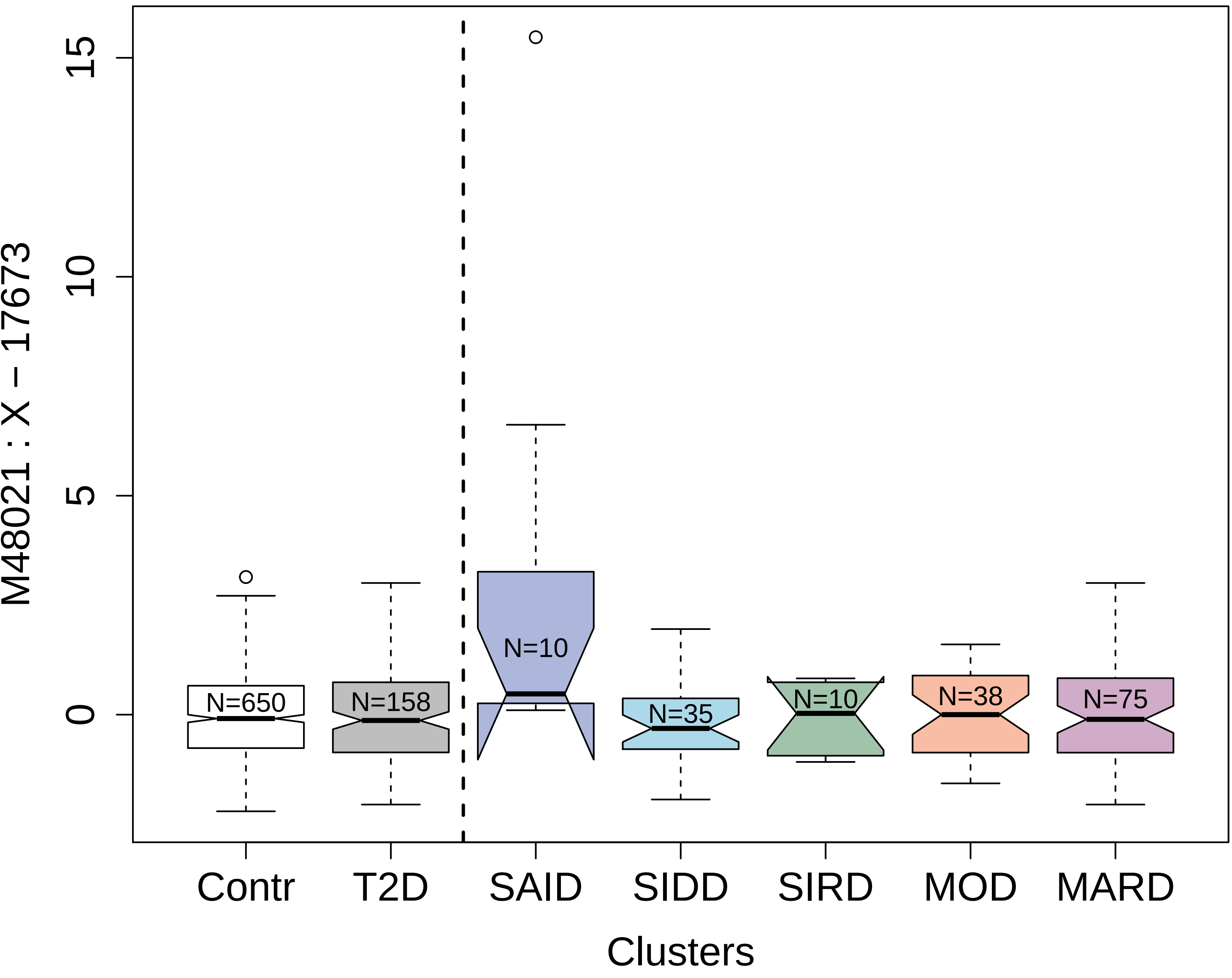

M48021 : X − 17673
0
5
10
15
N=650
N=158
N=10
N=35
N=10
N=38
N=75
Contr
T2D
SAID
SIDD
SIRD
MOD
MARD
Clusters

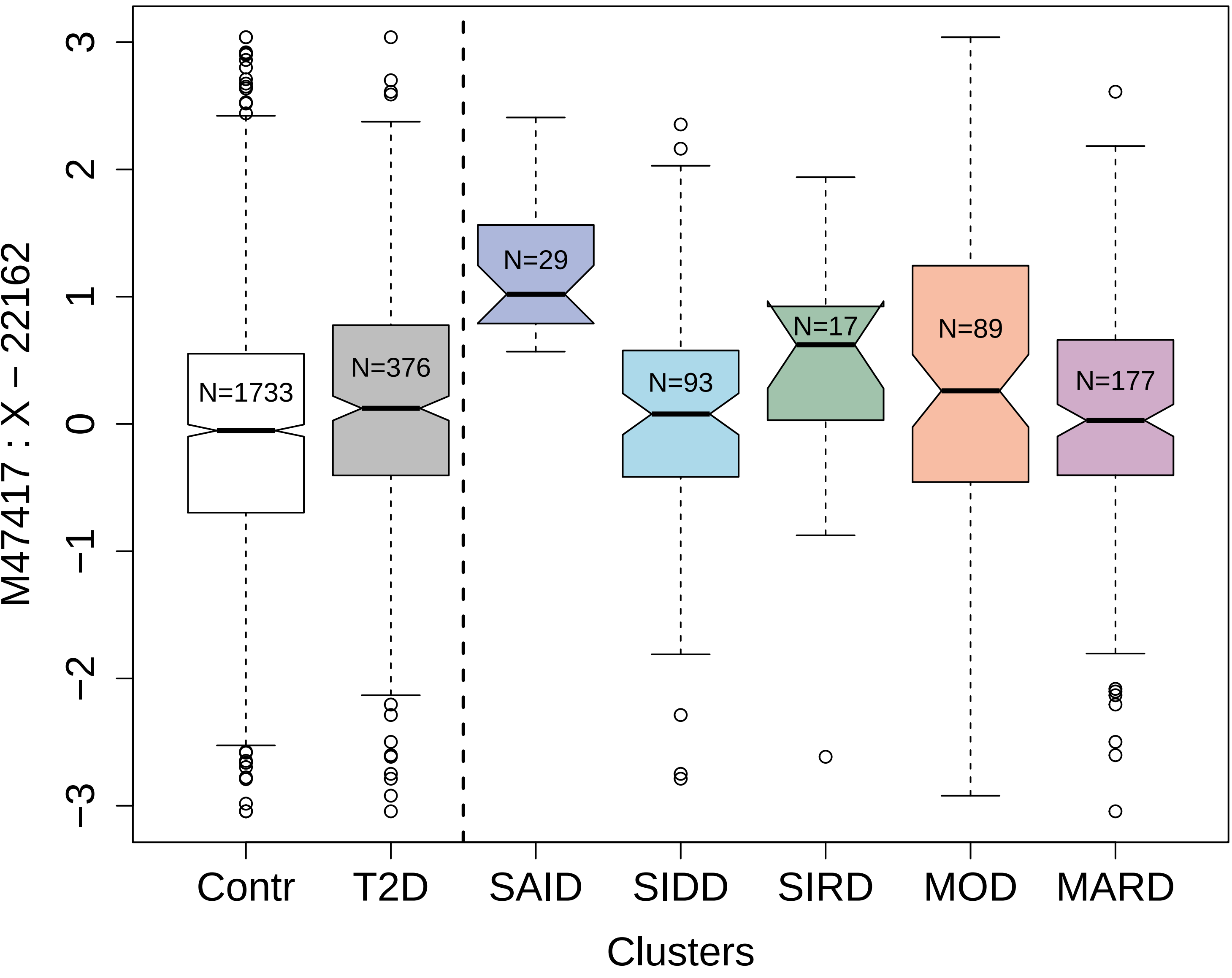

M47417 : X − 22162
3
2
1
0
−1
−2
−3
N=1733
N=376
N=29
N=93
N=17
N=89
N=177
Contr
T2D
SAID
SIDD
SIRD
MOD
MARD
Clusters

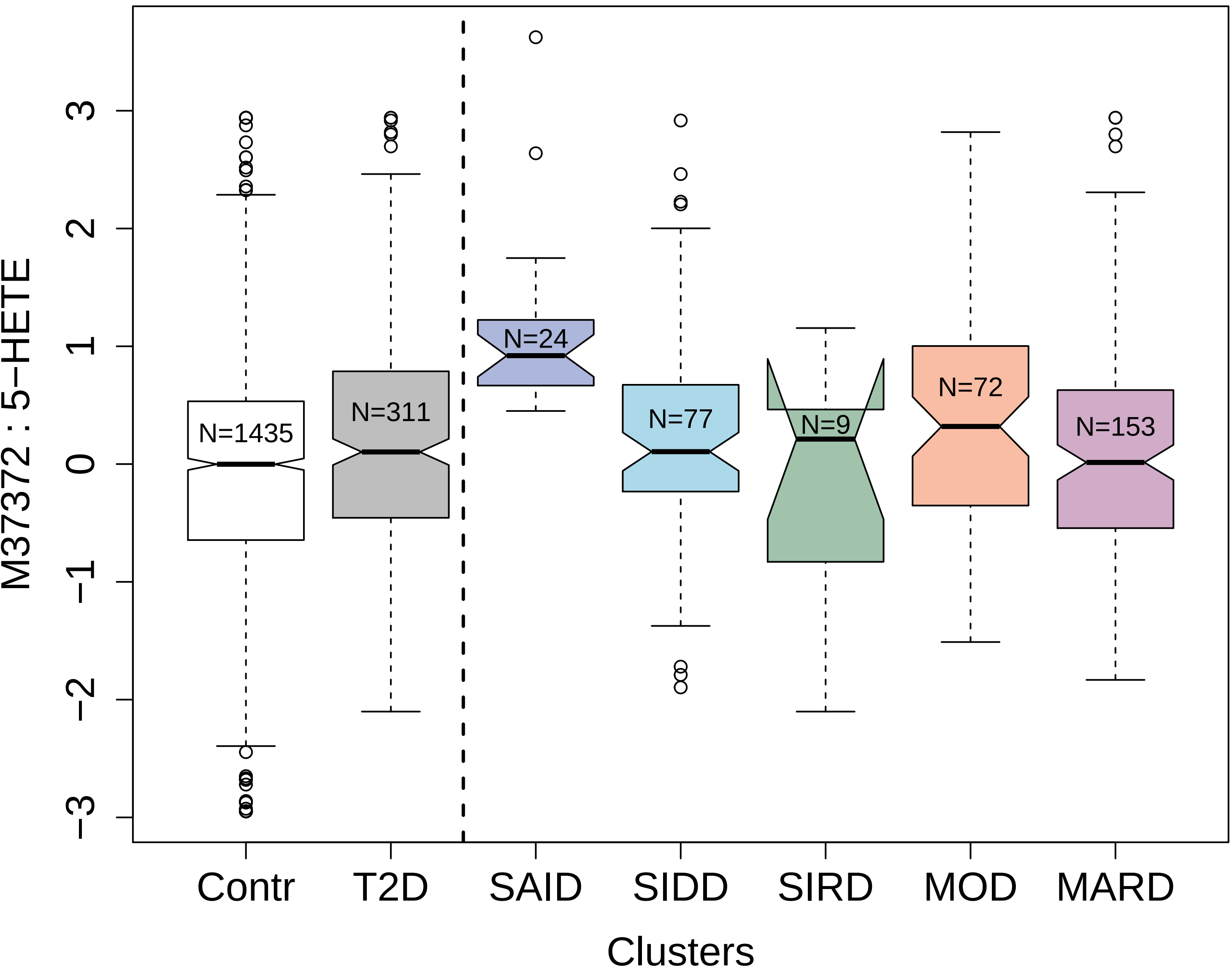
M37372 : 5−HETE
3
2
1
0
−1
−2
−3
N=1435
N=311
N=24
N=77
N=9
N=72
N=153
Contr
T2D
SAID
SIDD
SIRD
MOD
MARD
Clusters

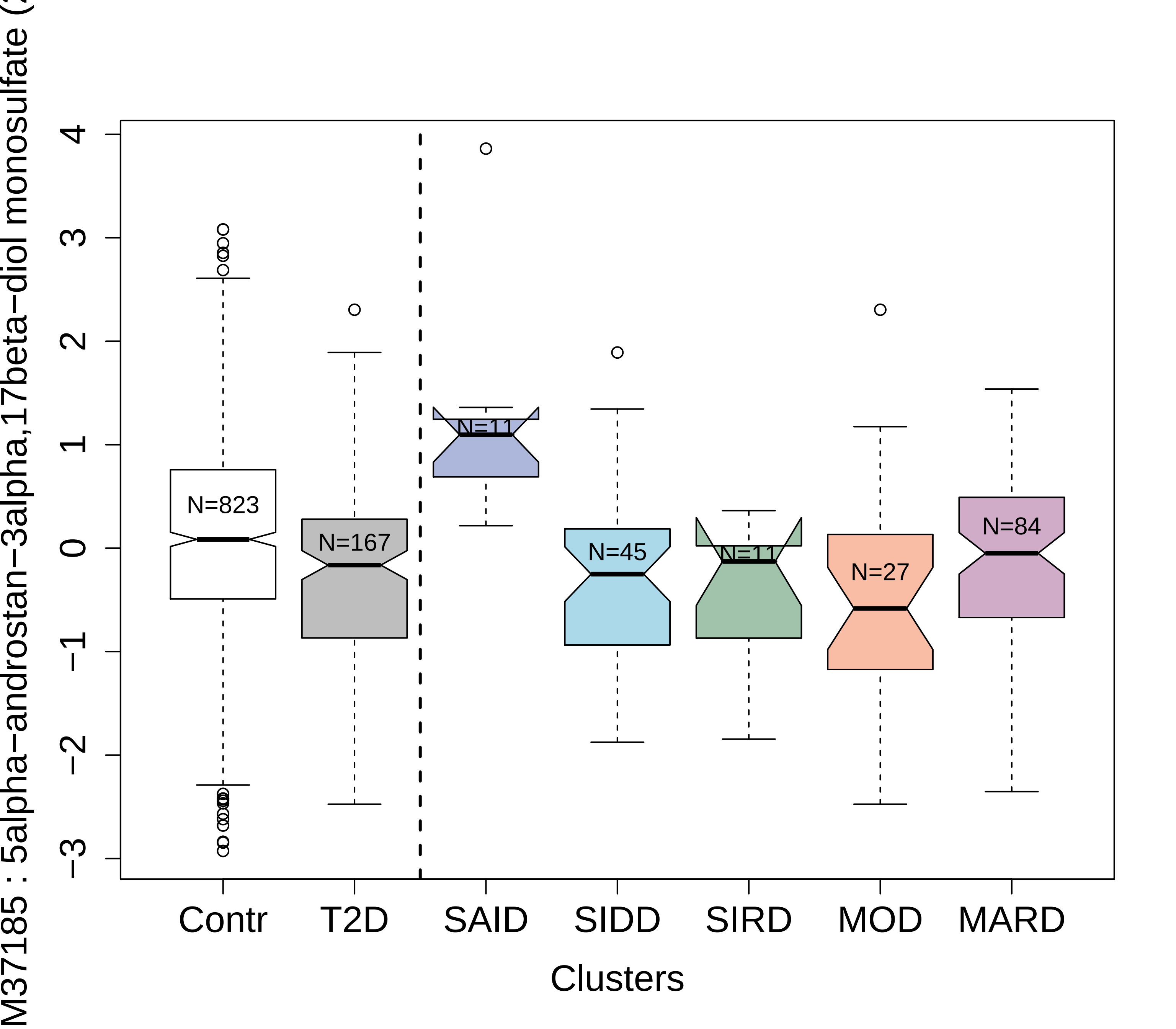

M37185 : 5alpha−androstan−3alpha,17beta−diol monosulfate (2)
4
3
2
1
0
−1
−2
−3
N=823
N=167
N=11
N=45
N=11
N=27
N=84
Contr
T2D
SAID
SIDD
SIRD
MOD
MARD
Clusters

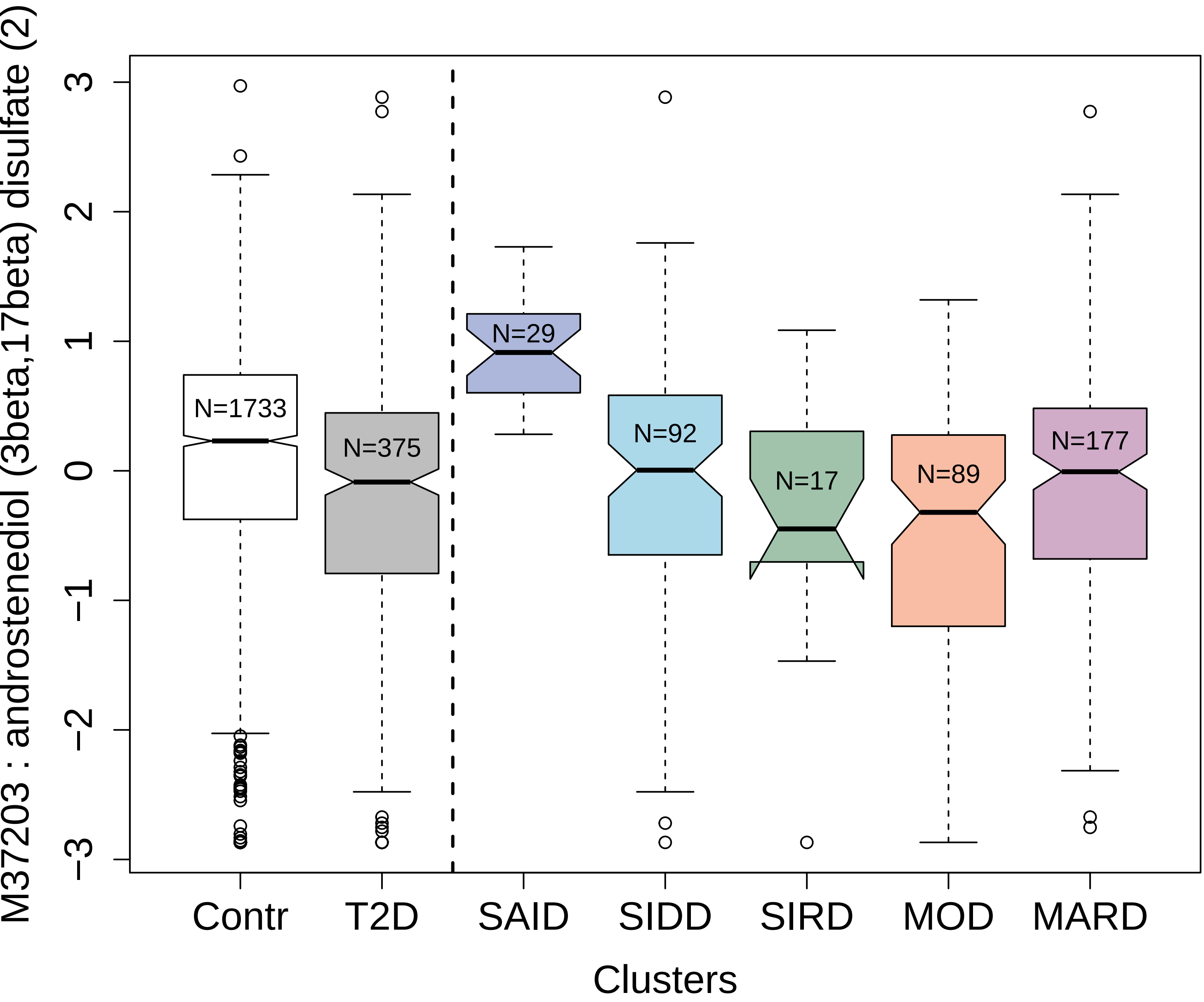

M37203 : androstenediol (3beta,17beta) disulfate (2)
3
2
1
0
−1
−2
−3
N=1733
N=375
N=29
N=92
N=17
N=89
N=177
Contr
T2D
SAID
SIDD
SIRD
MOD
MARD
Clusters

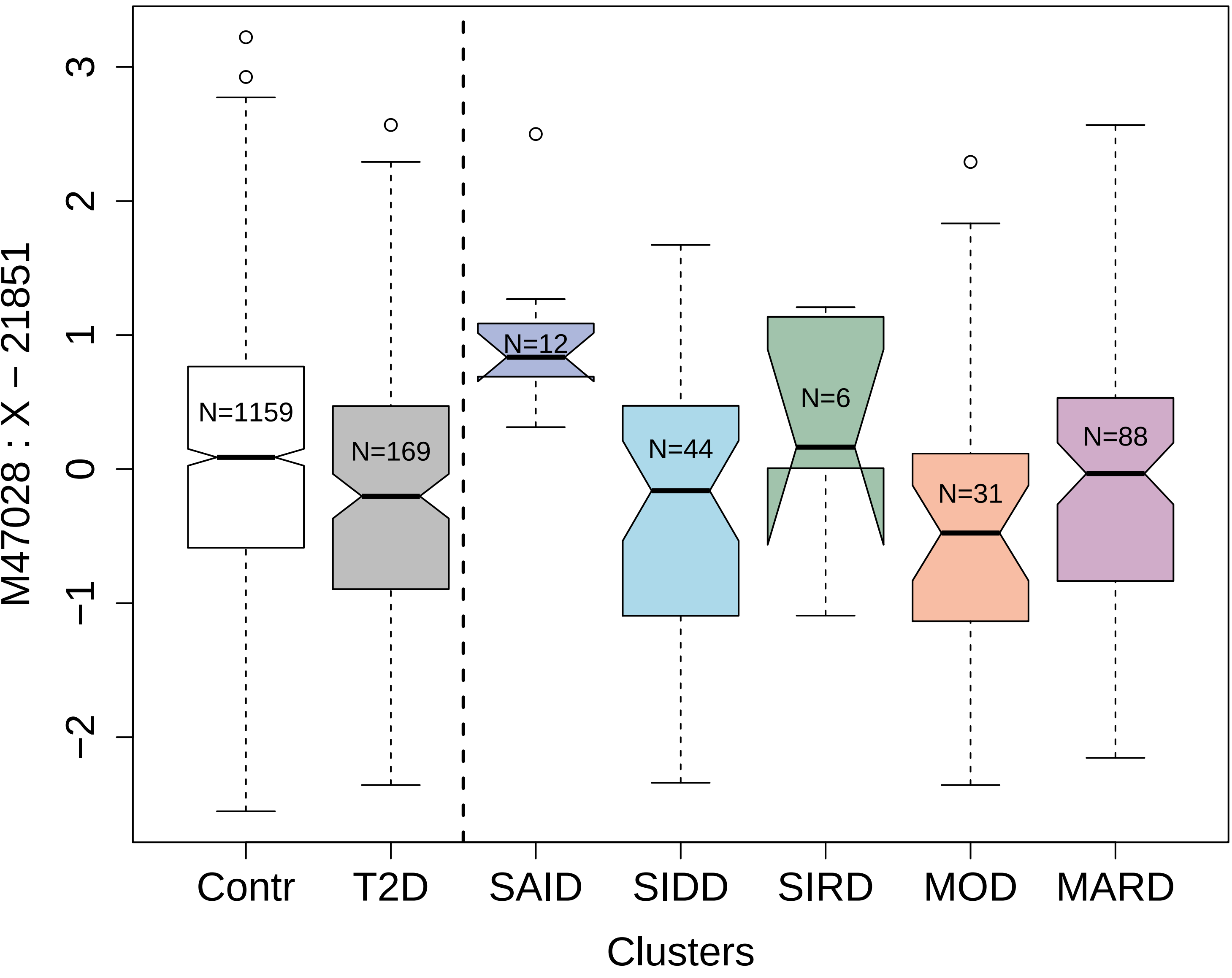

M47028 : X − 21851
3
2
1
0
−1
−2
N=1159
N=169
N=12
N=44
N=6
N=31
N=88
Contr
T2D
SAID
SIDD
SIRD
MOD
MARD
Clusters

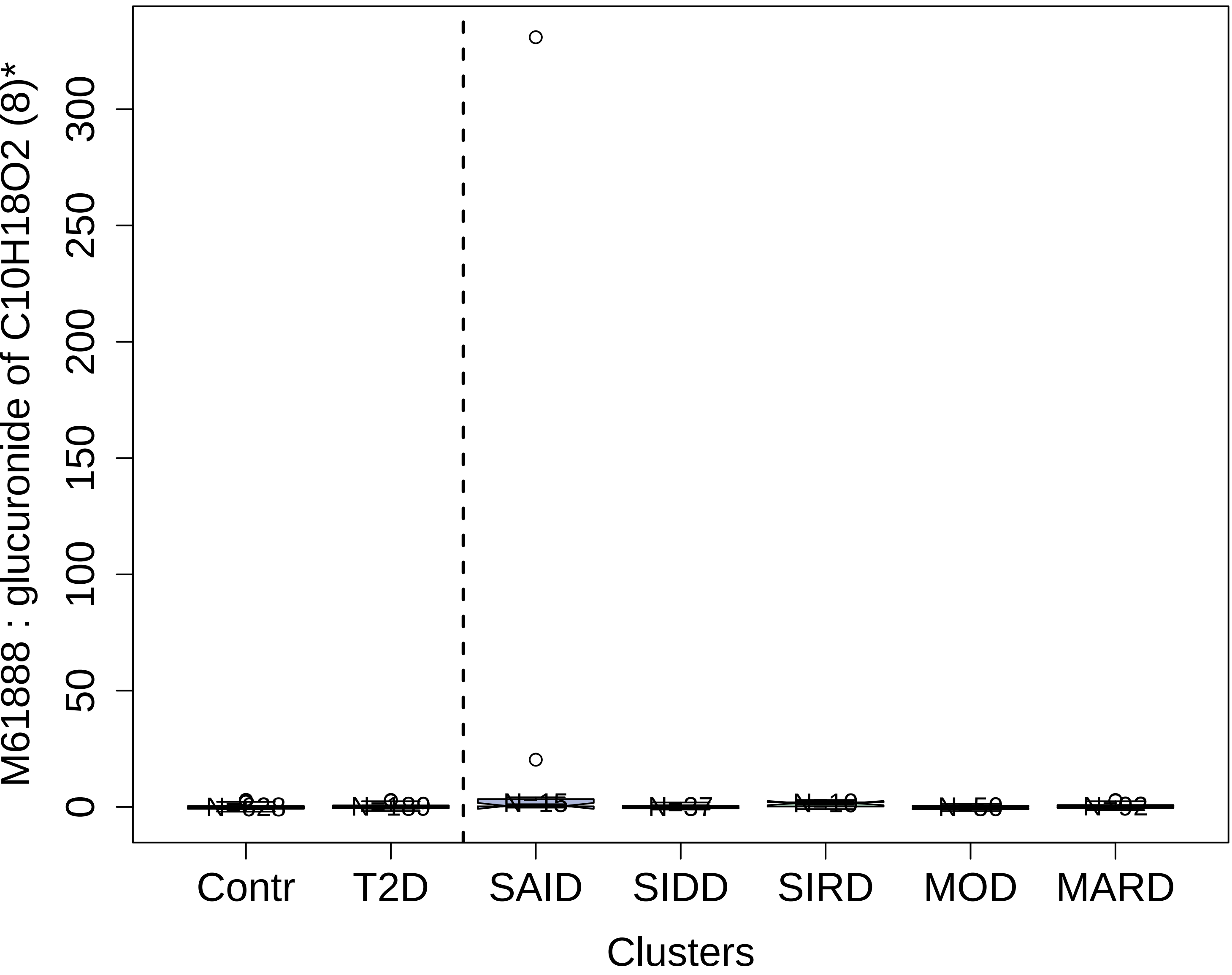
M61888 : glucuronide of C10H18O2 (8)*
0
50
100
150
200
250
300
Contr
T2D
SAID
SIDD
SIRD
MOD
MARD
Clusters

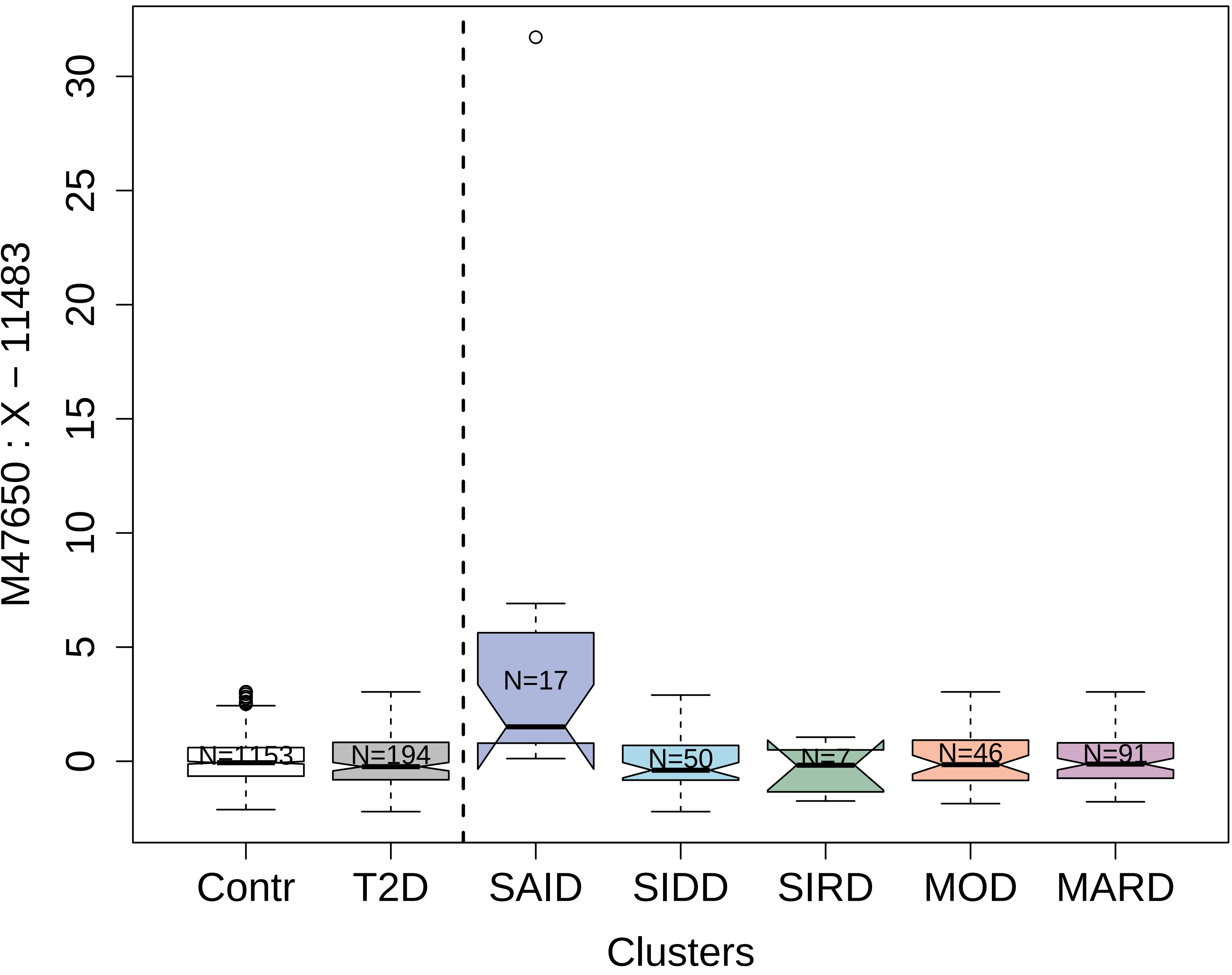
M47650 : X − 11483
0
5
10
15
20
25
30
N=1153
N=194
N=17
N=50
N=7
N=46
N=91
Contr
T2D
SAID
SIDD
SIRD
MOD
MARD
Clusters

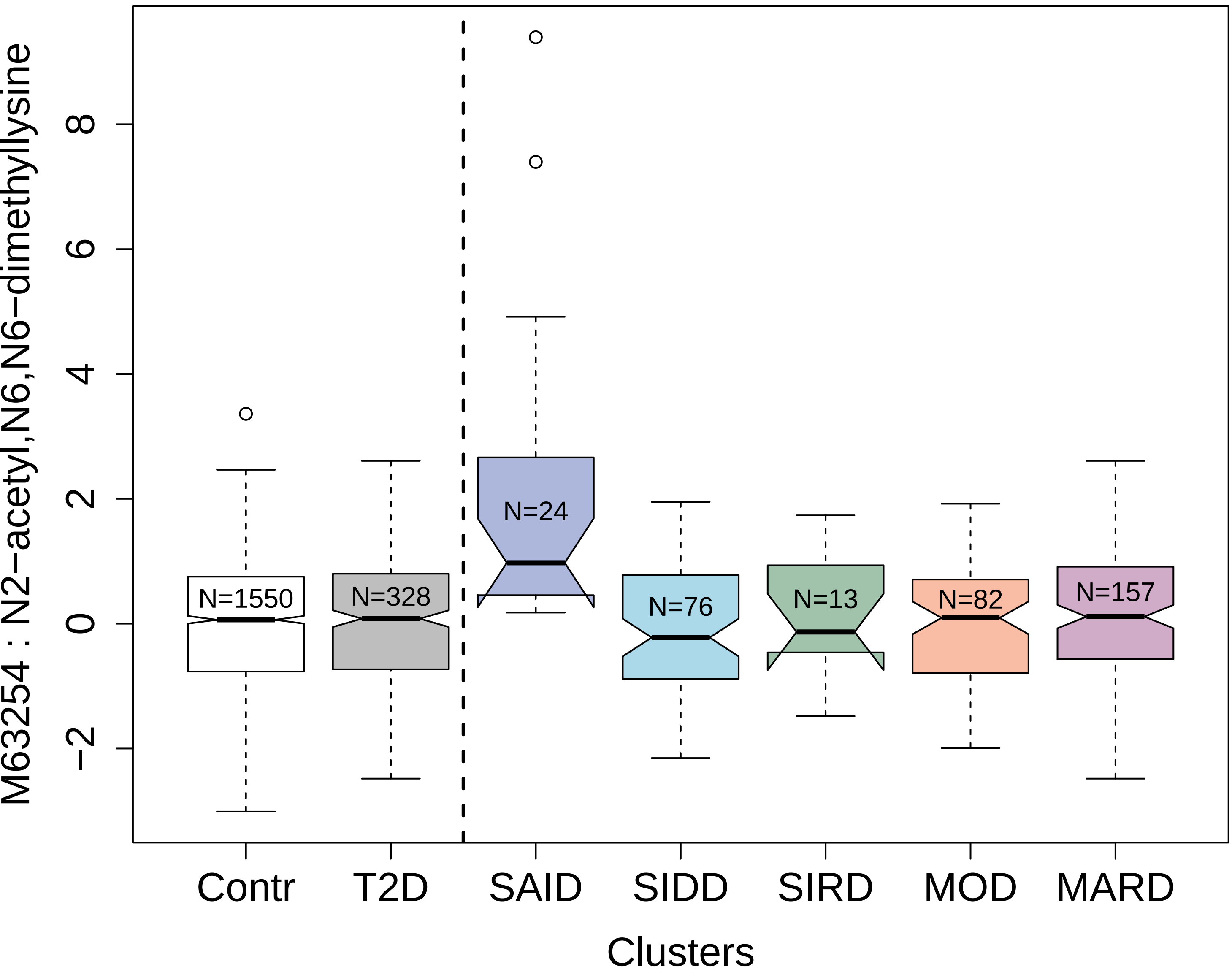
M63254 : N2−acetyl,N6,N6−dimethyllysine
−2
0
2
4
6
8
N=1550
N=328
N=24
N=76
N=13
N=82
N=157
Contr
T2D
SAID
SIDD
SIRD
MOD
MARD
Clusters

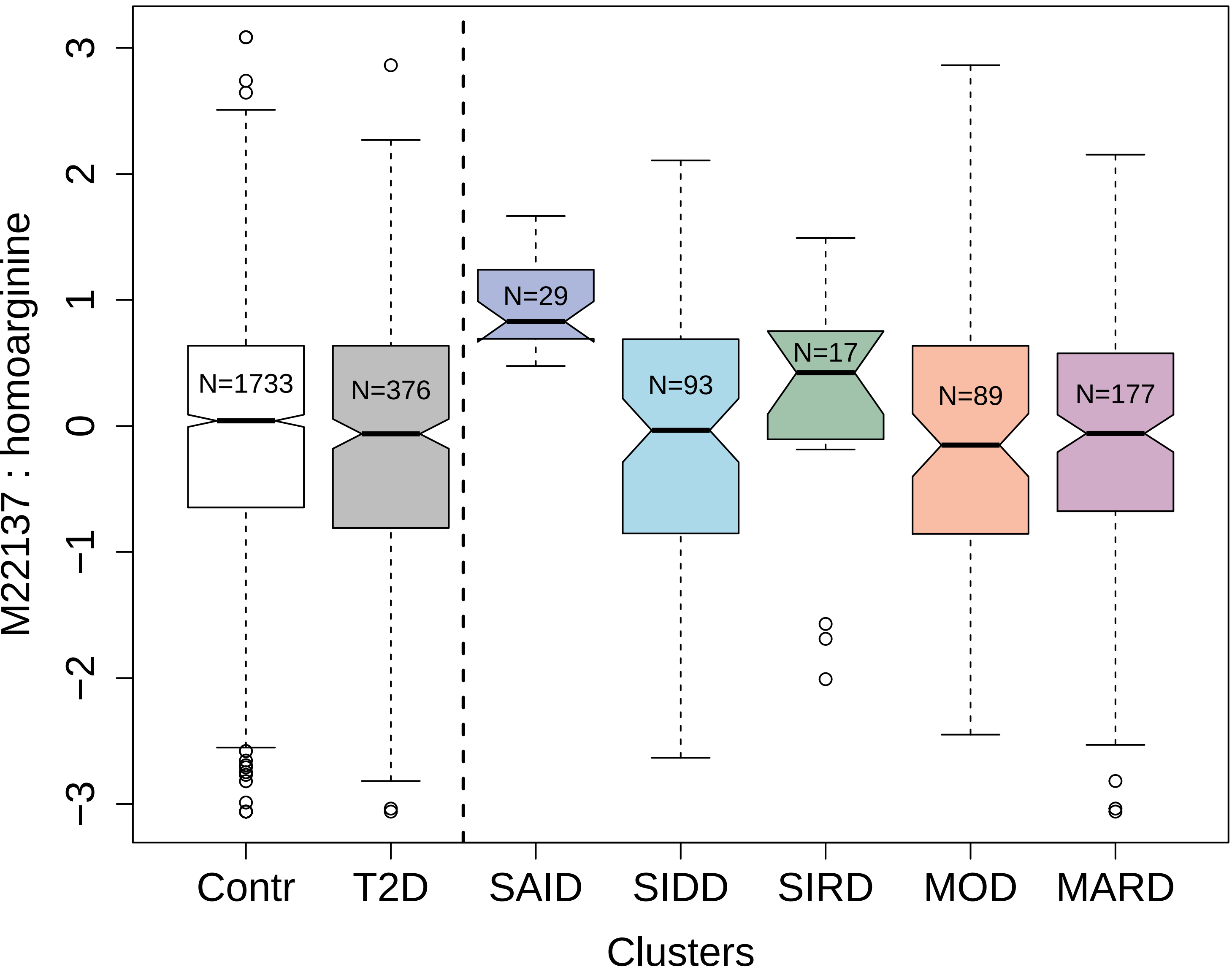

M22137 : homoarginine
3
2
1
0
−1
−2
−3
N=1733
N=376
N=29
N=93
N=17
N=89
N=177
Contr
T2D
SAID
SIDD
SIRD
MOD
MARD
Clusters

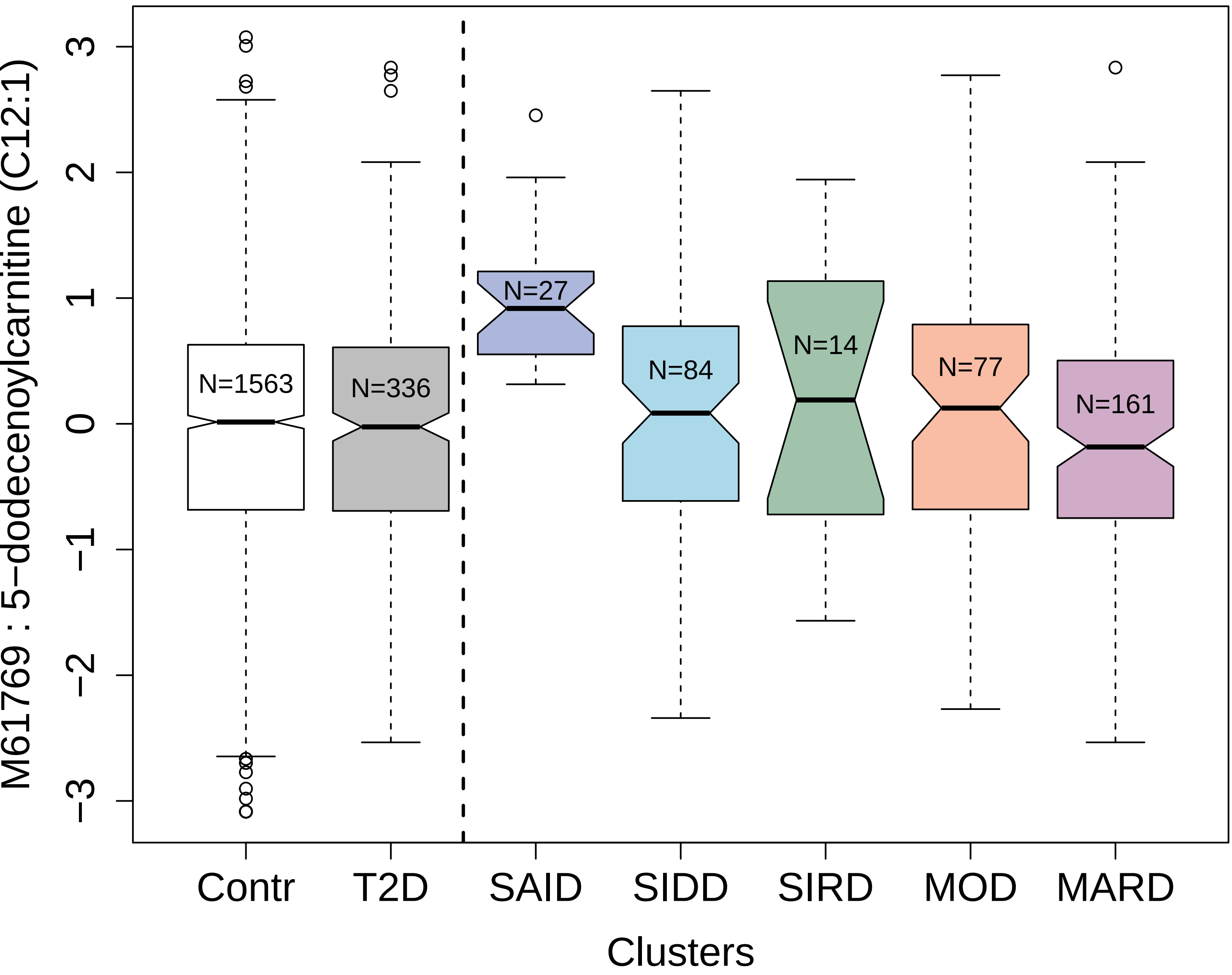
M61769 : 5−dodecenoylcarnitine (C12:1)
3
2
1
0
−1
−2
−3
N=1563
N=336
N=27
N=84
N=14
N=77
N=161
Contr
T2D
SAID
SIDD
SIRD
MOD
MARD
Clusters

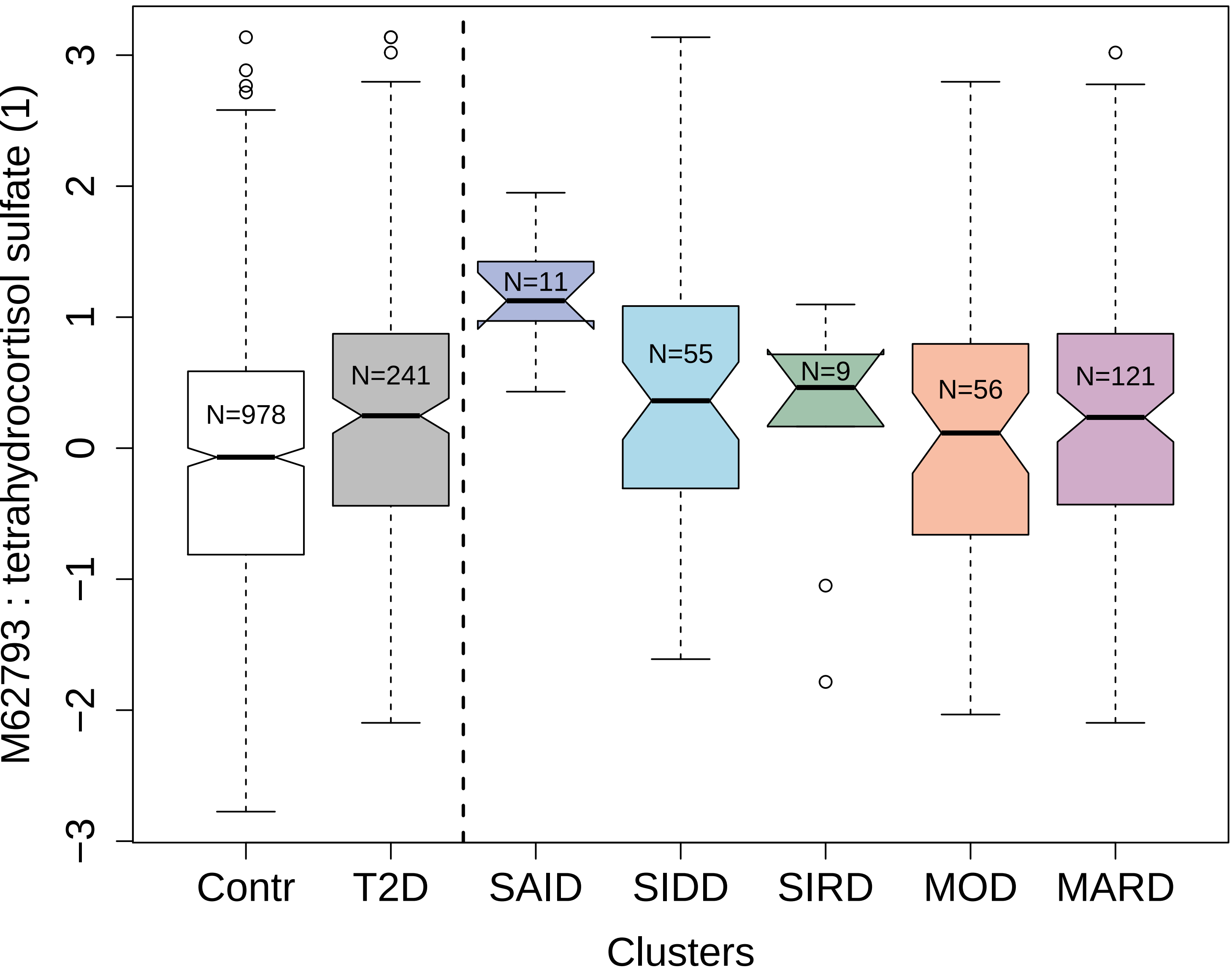
M62793 : tetrahydrocortisol sulfate (1)
3
2
1
0
−1
−2
−3
N=978
N=241
N=11
N=55
N=9
N=56
N=121
Contr
T2D
SAID
SIDD
SIRD
MOD
MARD
Clusters

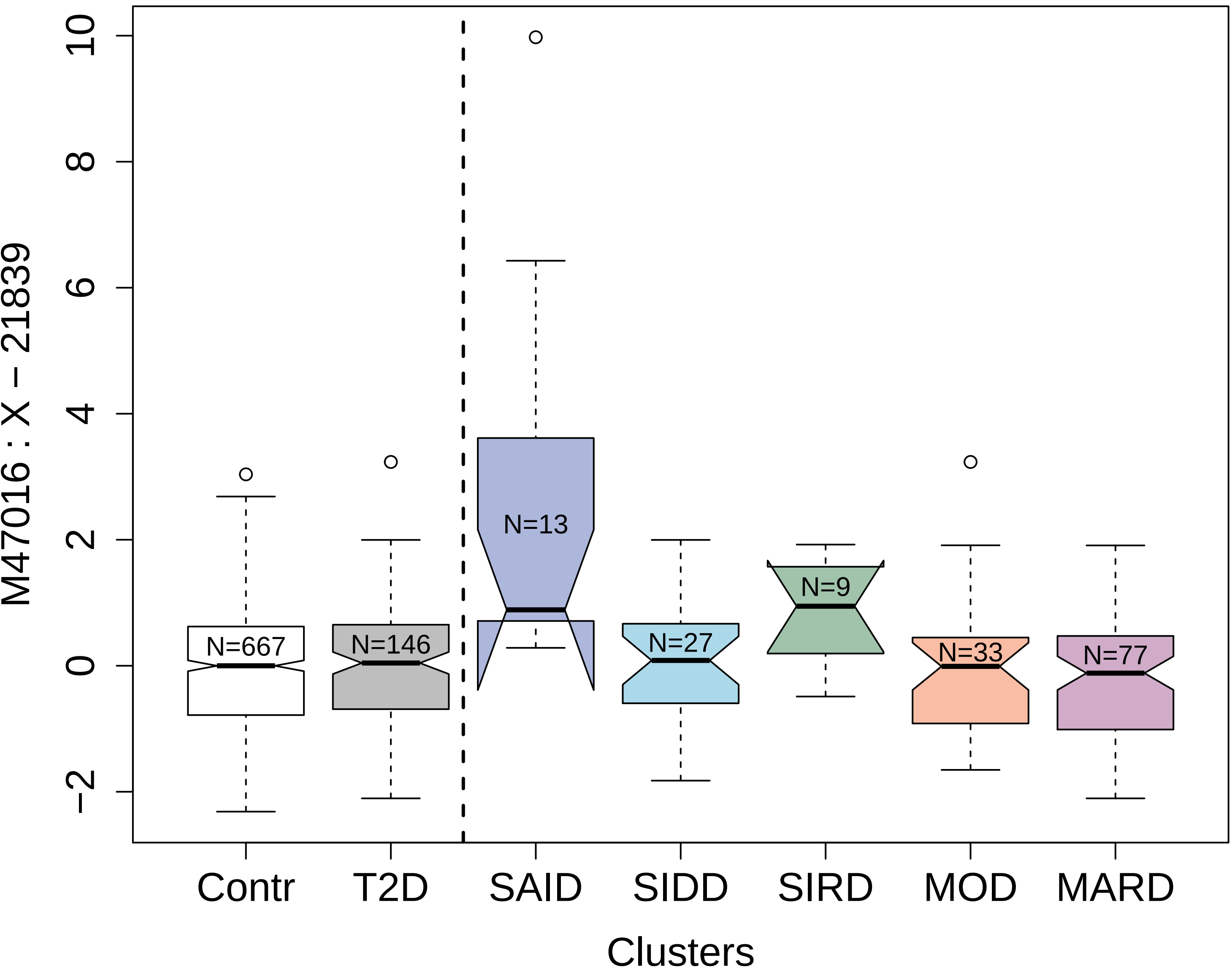

M47016 : X − 21839
10
8
6
4
2
0
−2
N=667
N=146
N=13
N=27
N=9
N=33
N=77
Contr
T2D
SAID
SIDD
SIRD
MOD
MARD
Clusters

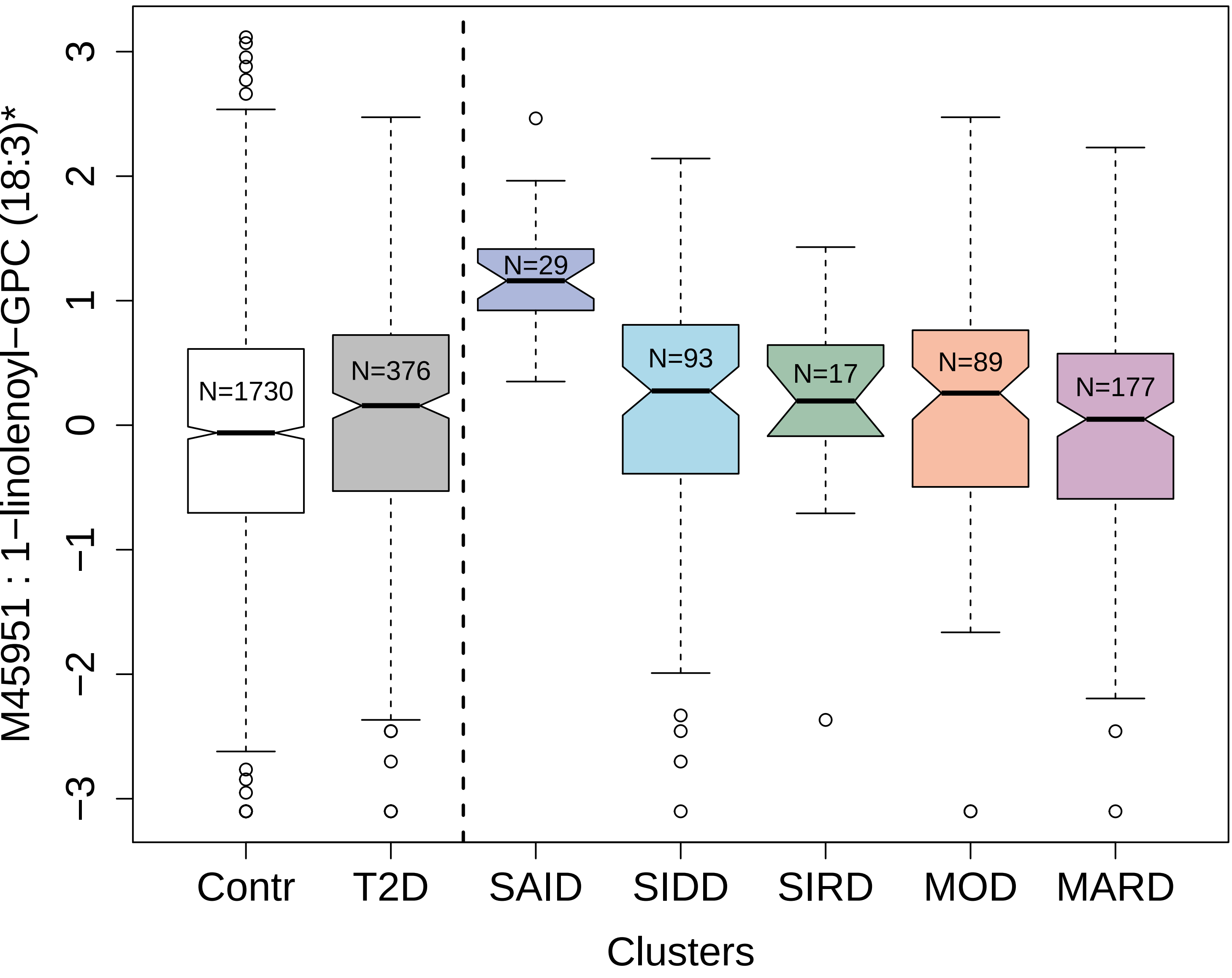

M45951 : 1−linolenoyl−GPC (18:3)*
3
2
1
0
−1
−2
−3
N=1730
N=376
N=29
N=93
N=17
N=89
N=177
Contr
T2D
SAID
SIDD
SIRD
MOD
MARD
Clusters

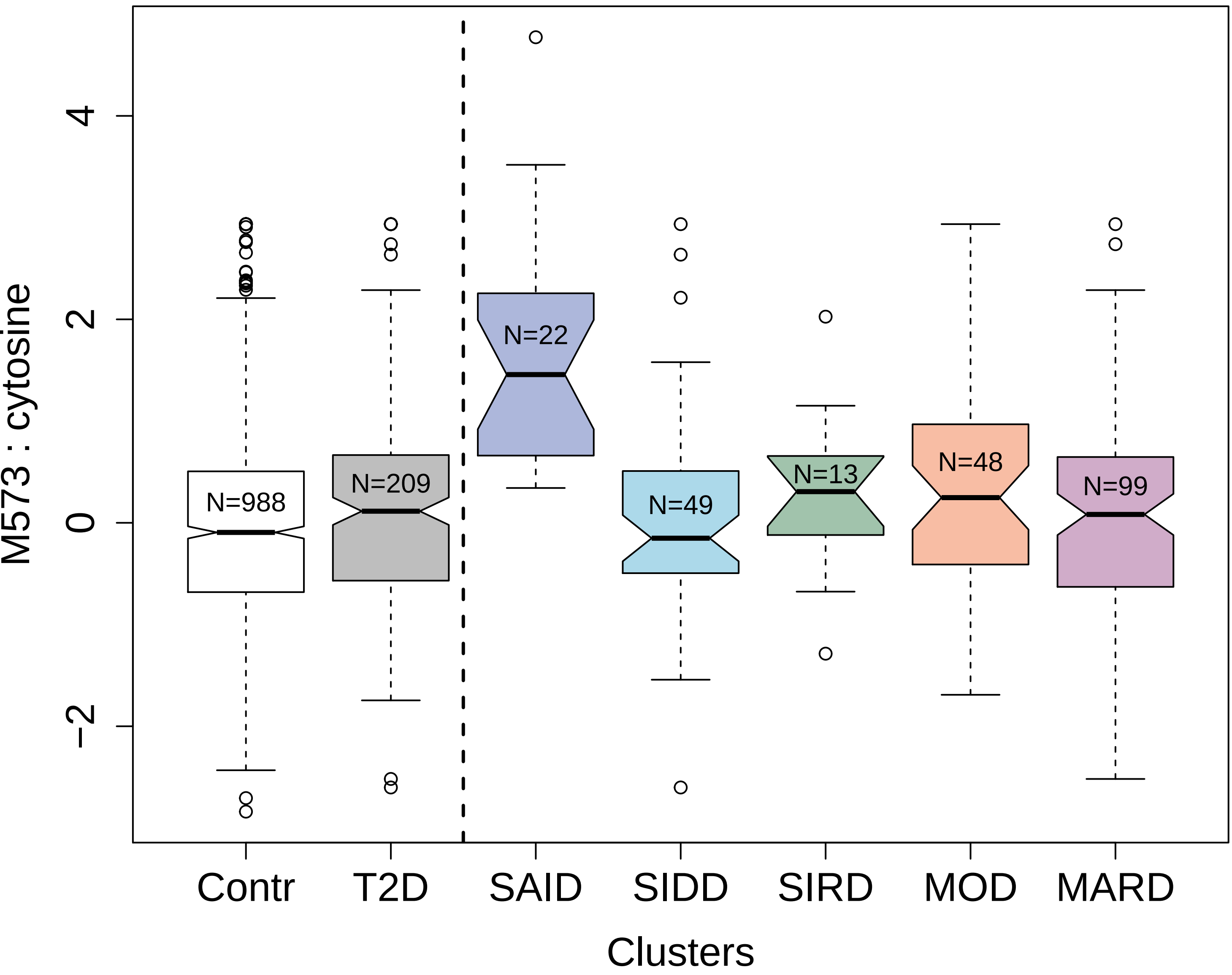

M573 : cytosine
4
2
0
−2
N=988
N=209
N=22
N=49
N=13
N=48
N=99
Contr
T2D
SAID
SIDD
SIRD
MOD
MARD
Clusters

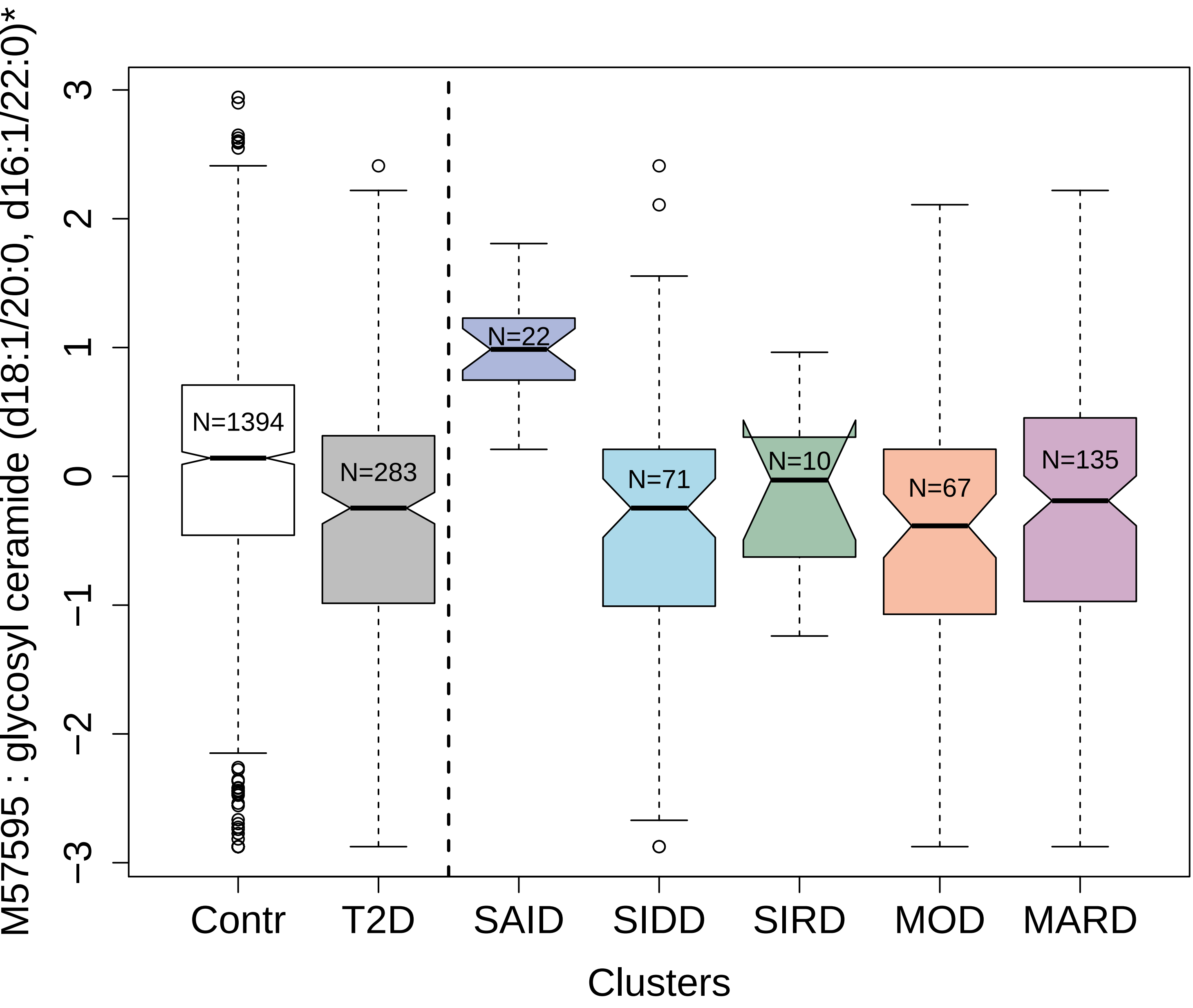
M57595 : glycosyl ceramide (d18:1/20:0, d16:1/22:0)*
3
2
1
0
−1
−2
−3
N=1394
N=283
N=22
N=71
N=10
N=67
N=135
Contr
T2D
SAID
SIDD
SIRD
MOD
MARD
Clusters

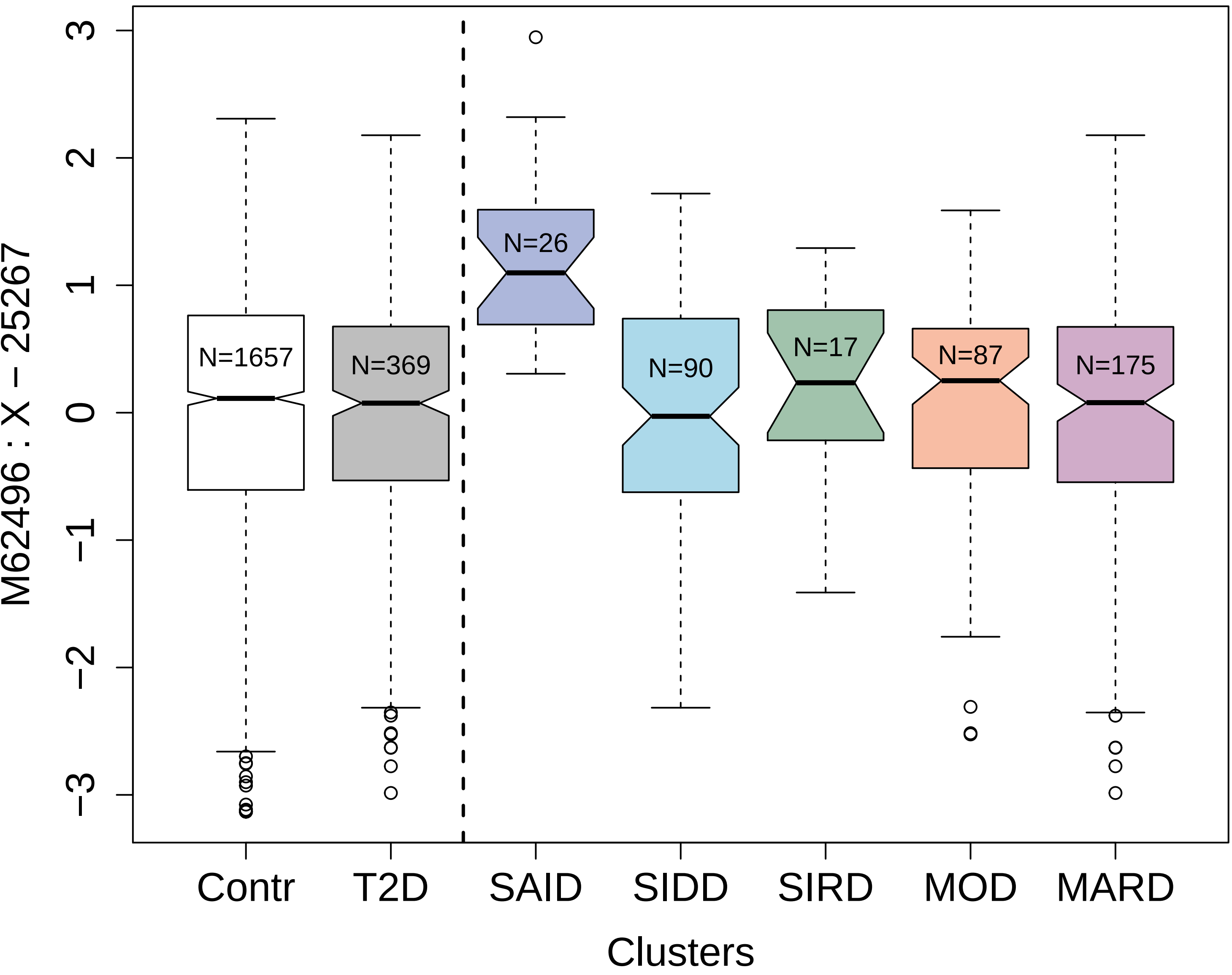
M62496 : X − 25267
3
2
1
0
−1
−2
−3
N=1657
N=369
N=26
N=90
N=17
N=87
N=175
Contr
T2D
SAID
SIDD
SIRD
MOD
MARD
Clusters

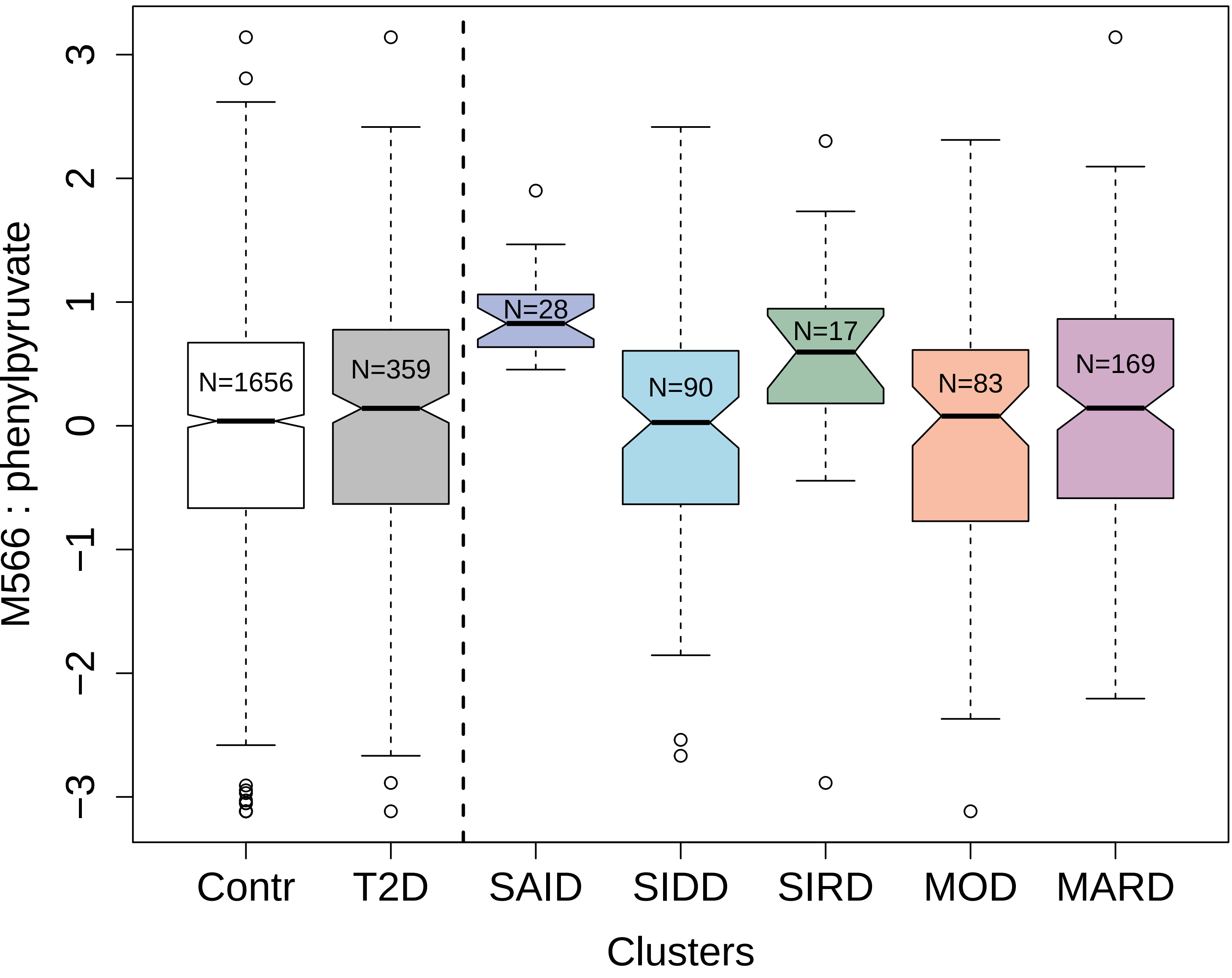
M566 : phenylpyruvate
3
2
1
0
−1
−2
−3
N=1656
N=359
N=28
N=90
N=17
N=83
N=169
Contr
T2D
SAID
SIDD
SIRD
MOD
MARD
Clusters

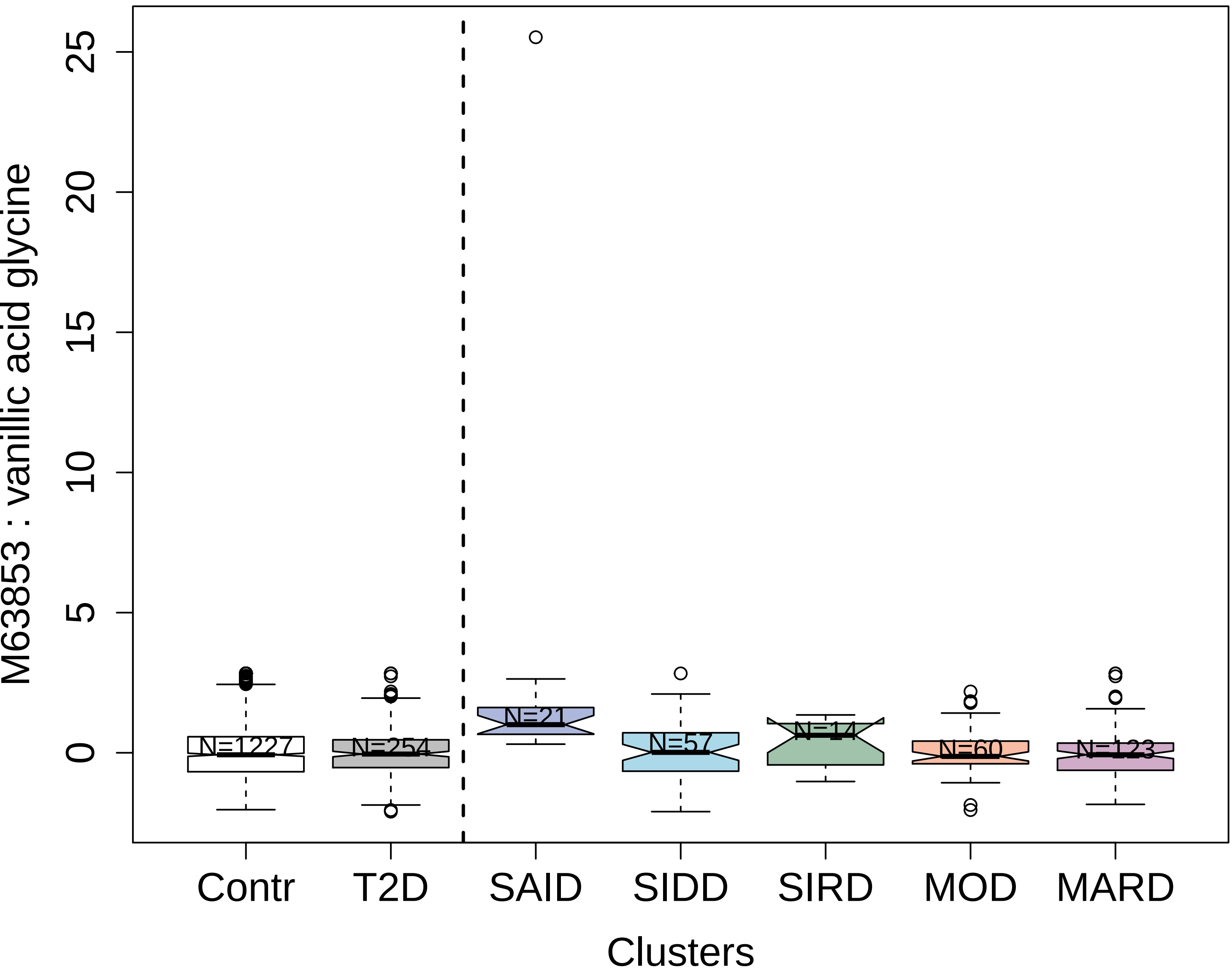

M63853 : vanillic acid glycine
0
5
10
15
20
25
N=1227
N=254
N=21
N=57
N=14
N=60
N=123
Contr
T2D
SAID
SIDD
SIRD
MOD
MARD
Clusters

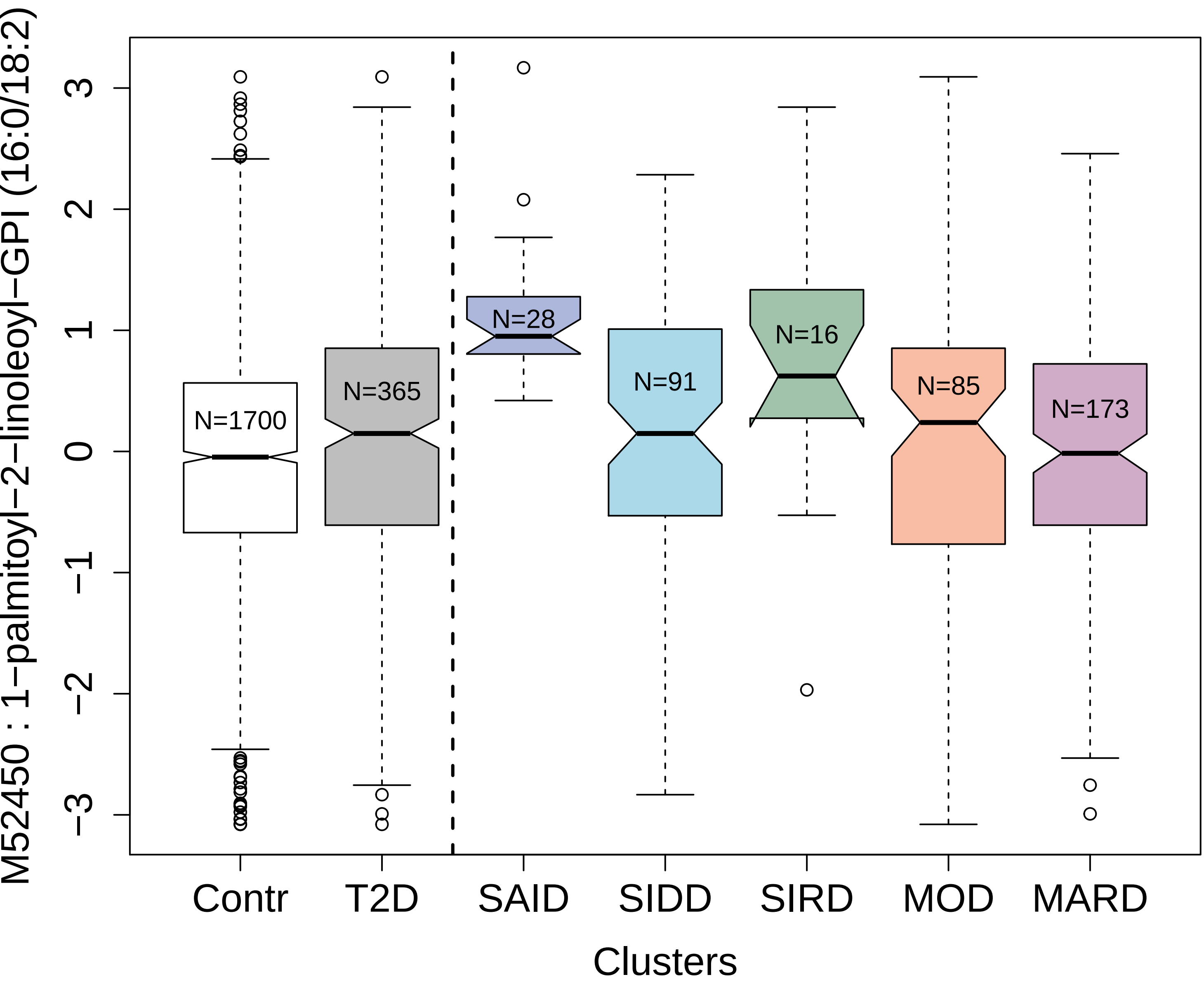
M52450 : 1−palmitoyl−2−linoleoyl−GPI (16:0/18:2)
3
2
1
0
−1
−2
−3
N=1700
N=365
N=28
N=91
N=16
N=85
N=173
Contr
T2D
SAID
SIDD
SIRD
MOD
MARD
Clusters

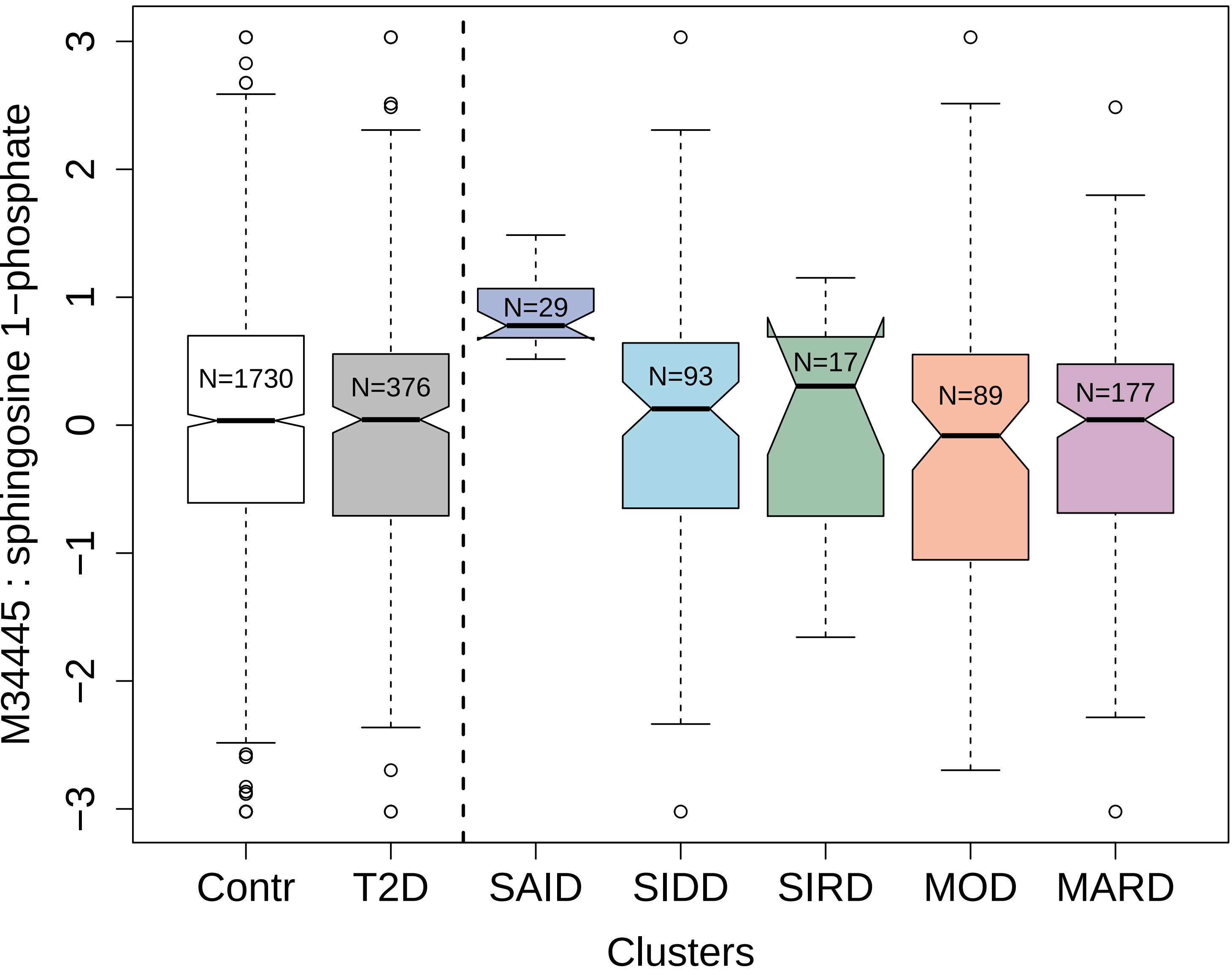
M34445 : sphingosine 1−phosphate
−3
−2
−1
0
1
2
3
N=1730
N=376
N=29
N=93
N=17
N=89
N=177
Contr
T2D
SAID
SIDD
SIRD
MOD
MARD
Clusters

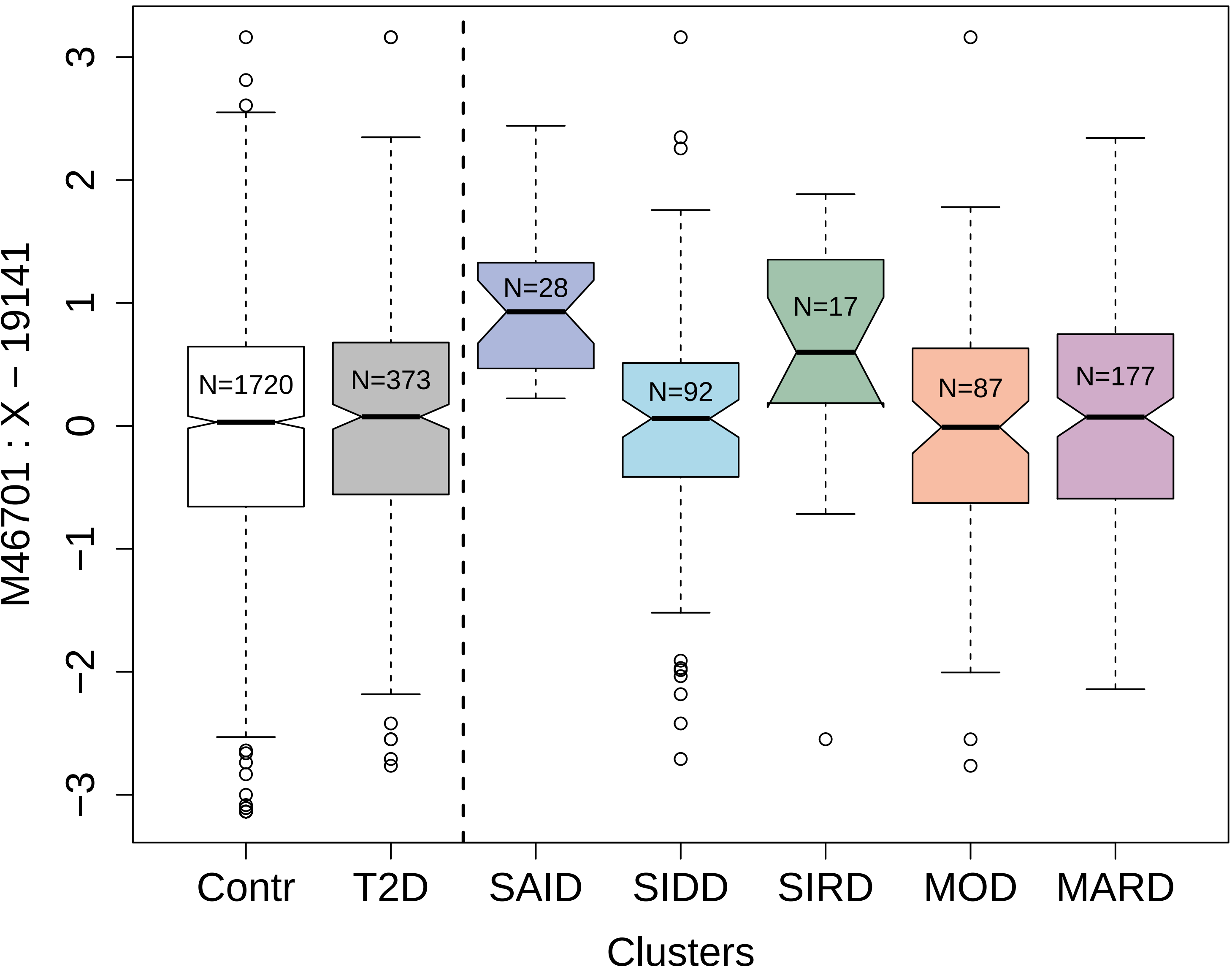
M46701 : X − 19141
3
2
1
0
−1
−2
−3
N=1720
N=373
N=28
N=92
N=17
N=87
N=177
Contr
T2D
SAID
SIDD
SIRD
MOD
MARD
Clusters

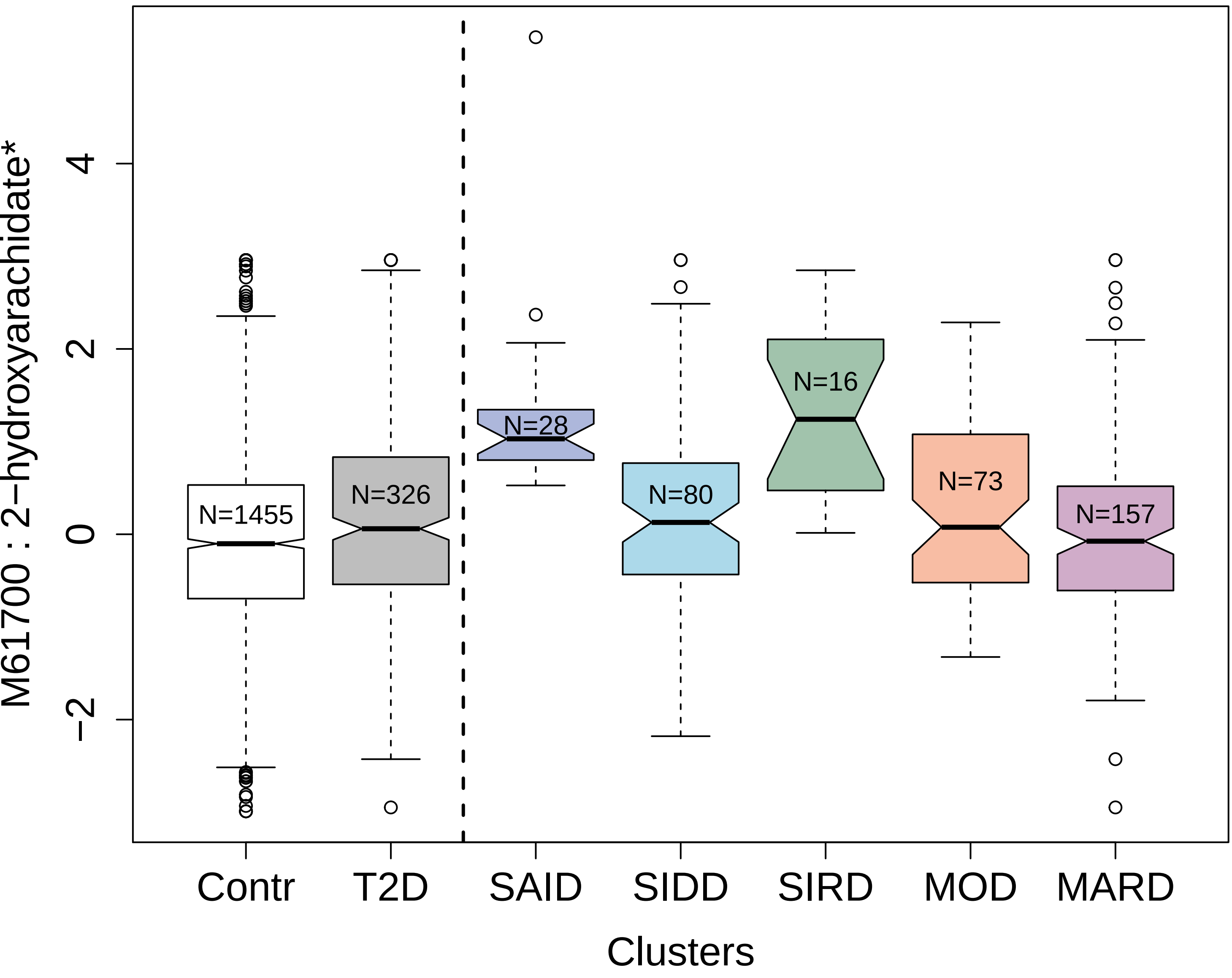

M61700 : 2−hydroxyarachidate*
Clusters
Contr
T2D
SAID
SIDD
SIRD
MOD
MARD
N=1455
N=326
N=28
N=80
N=16
N=73
N=157
4
2
0
−2

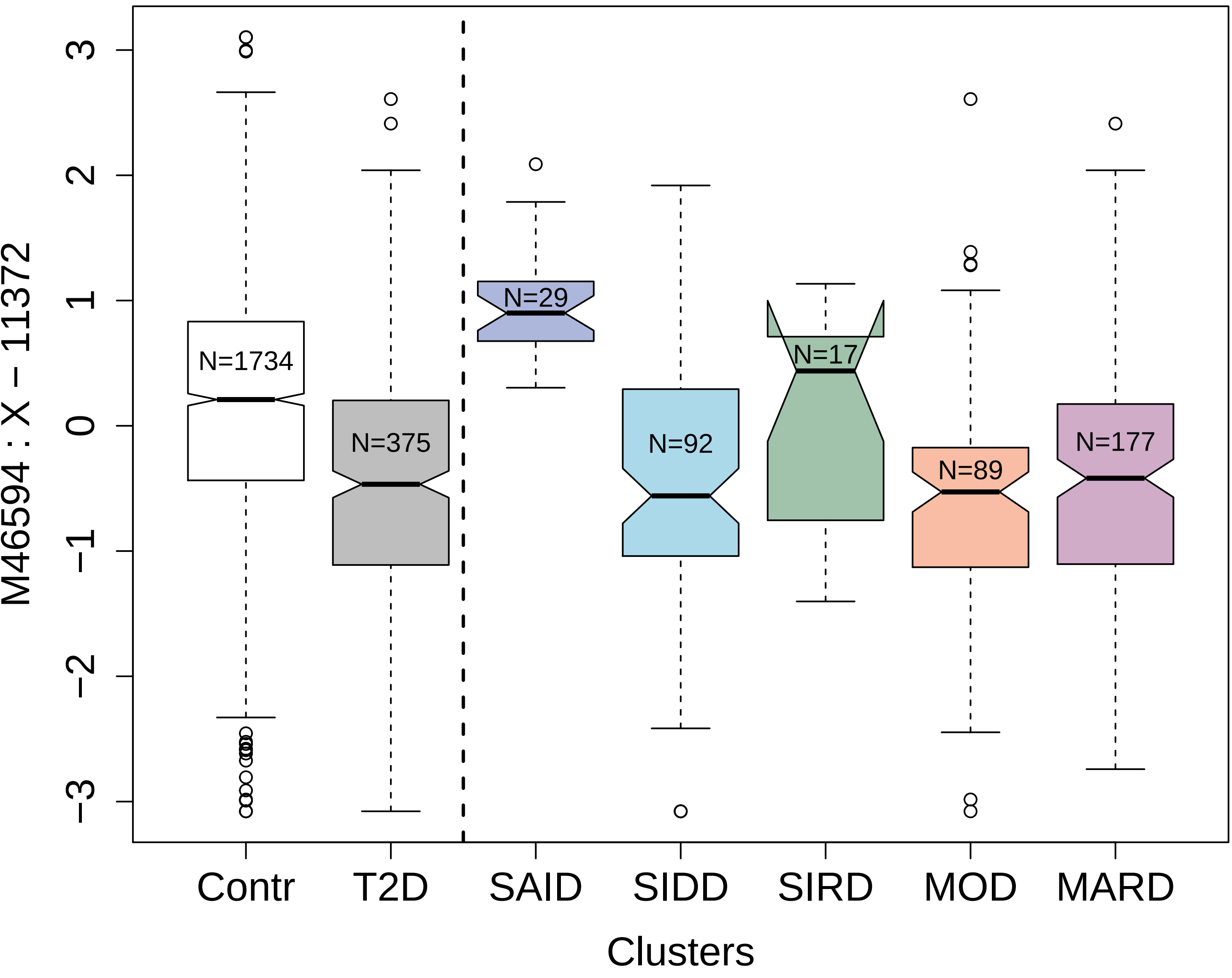
M46594 : X − 11372
3
2
1
0
−1
−2
−3
N=1734
N=375
N=29
N=92
N=17
N=89
N=177
Contr
T2D
SAID
SIDD
SIRD
MOD
MARD
Clusters

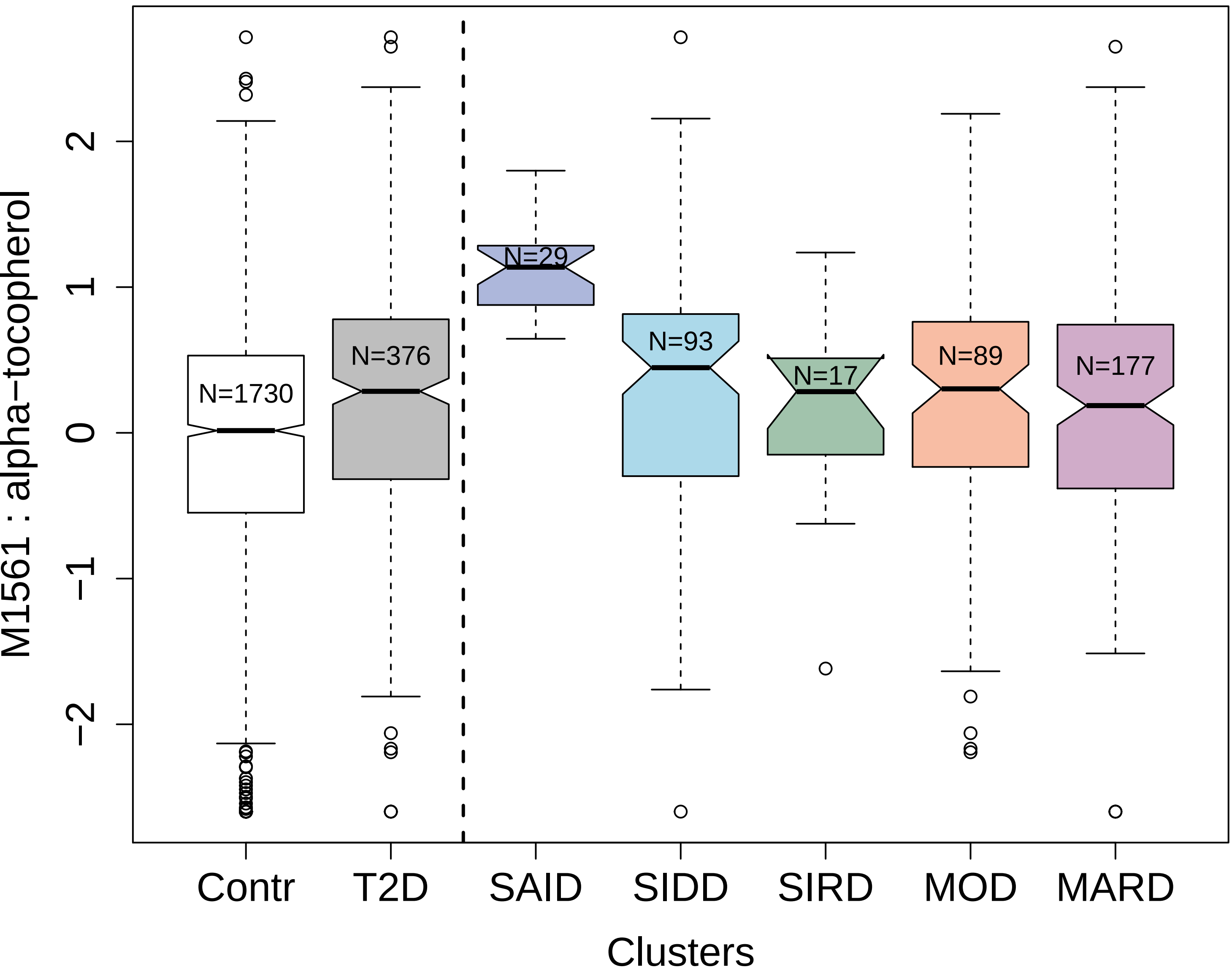

M1561 : alpha−tocopherol
2
1
0
−1
−2
N=1730
N=376
N=29
N=93
N=17
N=89
N=177
Contr
T2D
SAID
SIDD
SIRD
MOD
MARD
Clusters

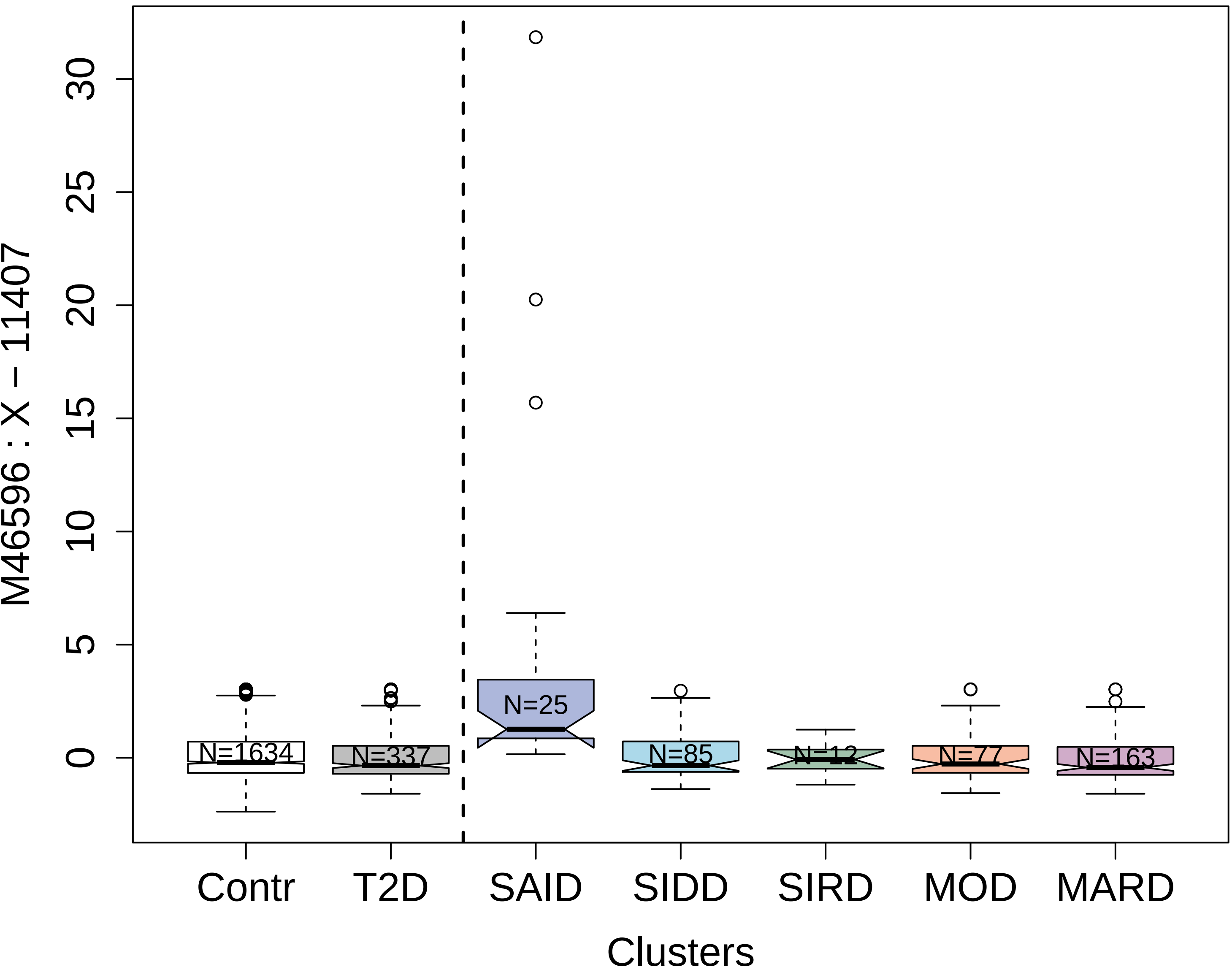

M46596 : X − 11407
0
5
10
15
20
25
30
N=1634
N=337
N=25
N=85
N=12
N=77
N=163
Contr
T2D
SAID
SIDD
SIRD
MOD
MARD
Clusters

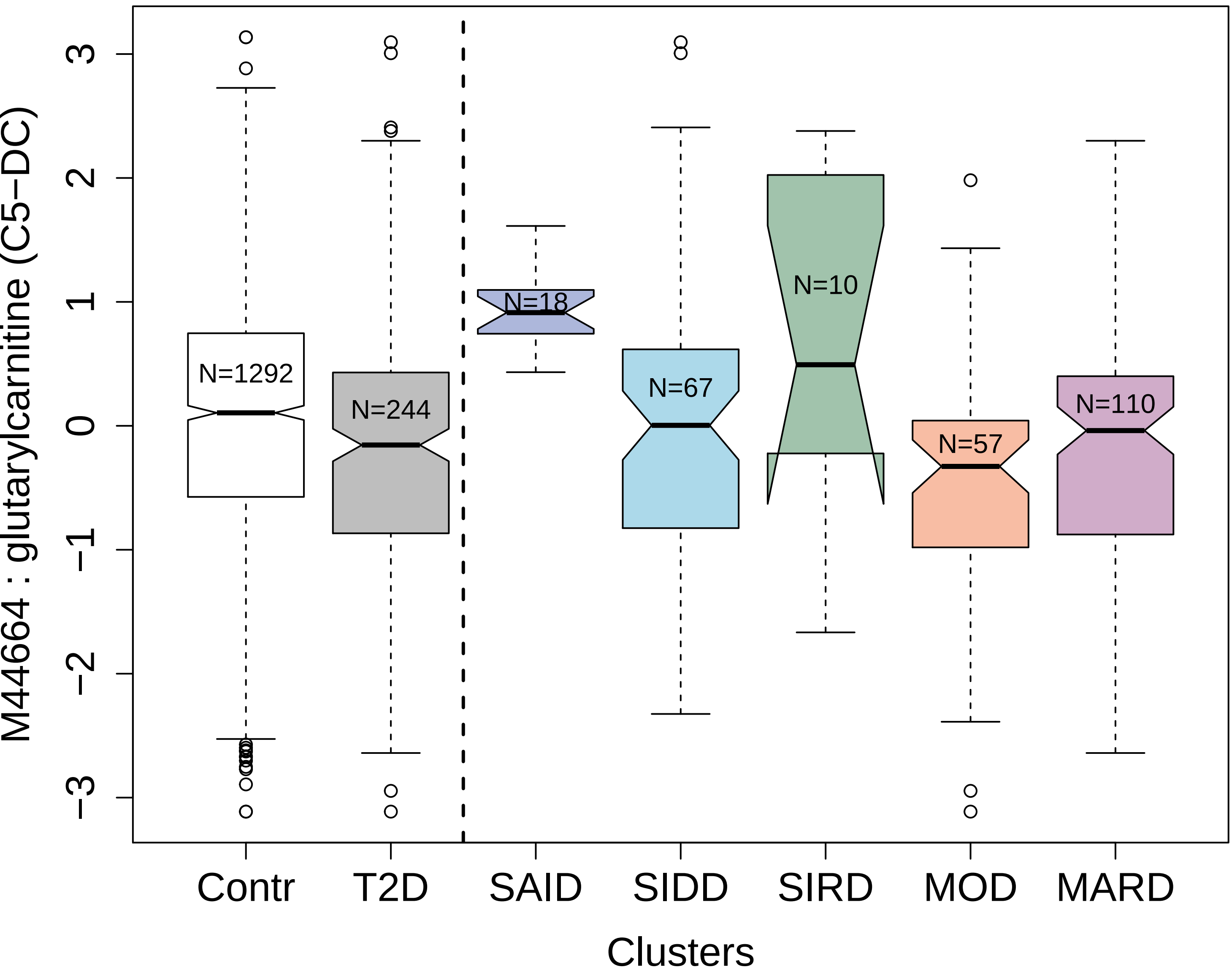

M44664 : glutarylcarnitine (C5−DC)
3
2
1
0
−1
−2
−3
N=1292
N=244
N=18
N=67
N=10
N=57
N=110
Contr
T2D
SAID
SIDD
SIRD
MOD
MARD
Clusters

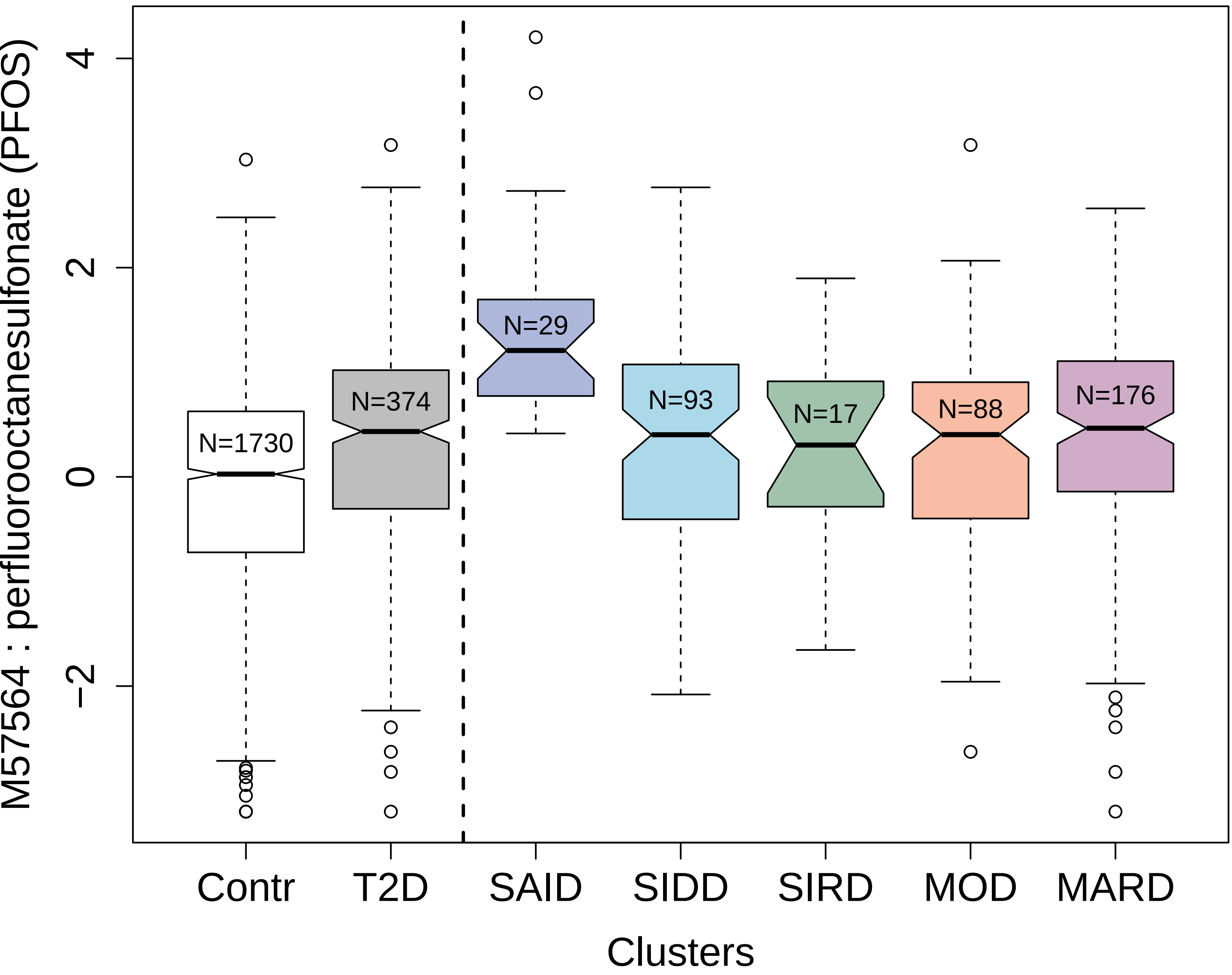

M57564 : perfluorooctanesulfonate (PFOS)
4
2
0
−2
N=1730
N=374
N=29
N=93
N=17
N=88
N=176
Contr
T2D
SAID
SIDD
SIRD
MOD
MARD
Clusters

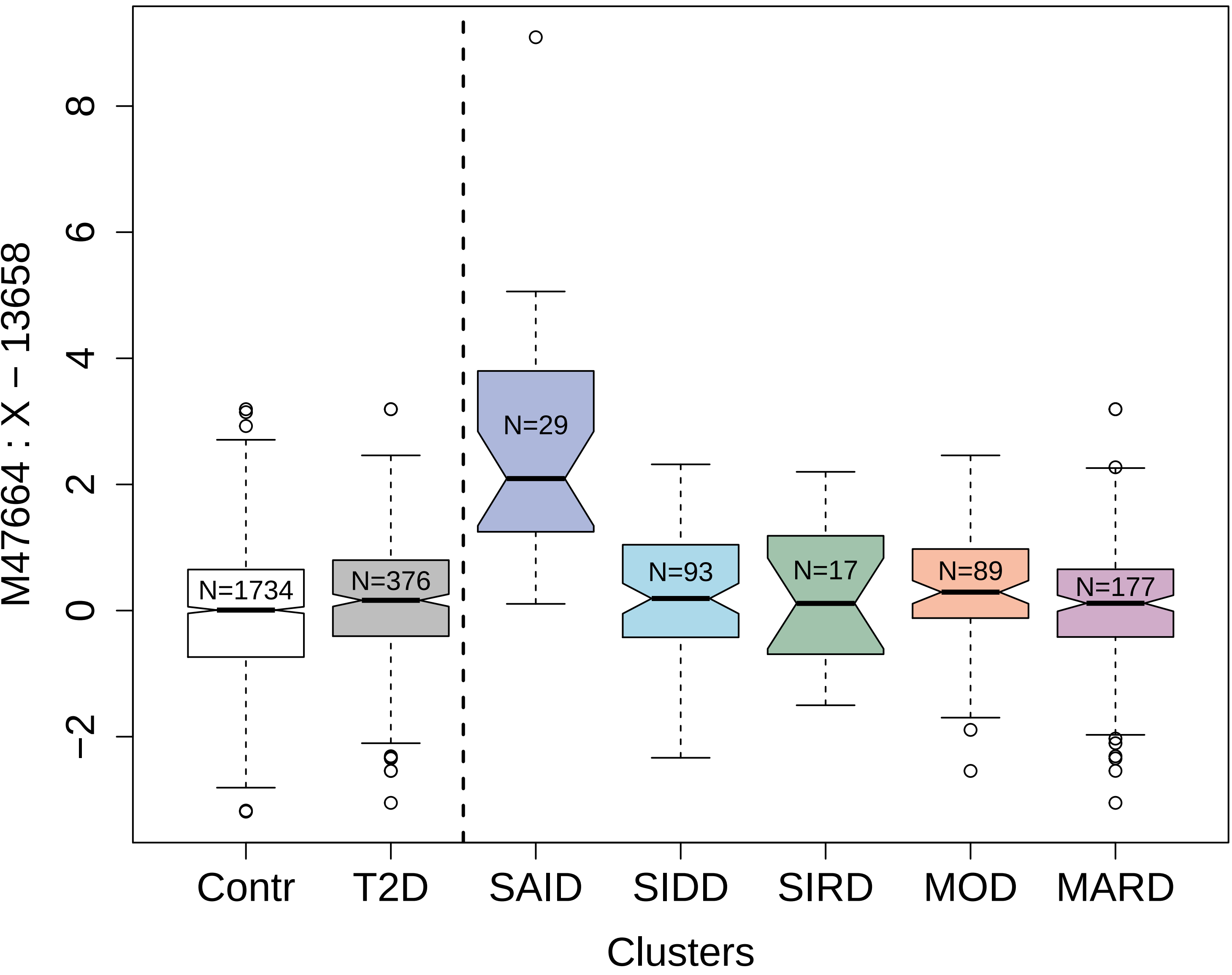
M47664 : X − 13658
8
6
4
2
0
−2
N=1734
N=376
N=29
N=93
N=17
N=89
N=177
Contr
T2D
SAID
SIDD
SIRD
MOD
MARD
Clusters

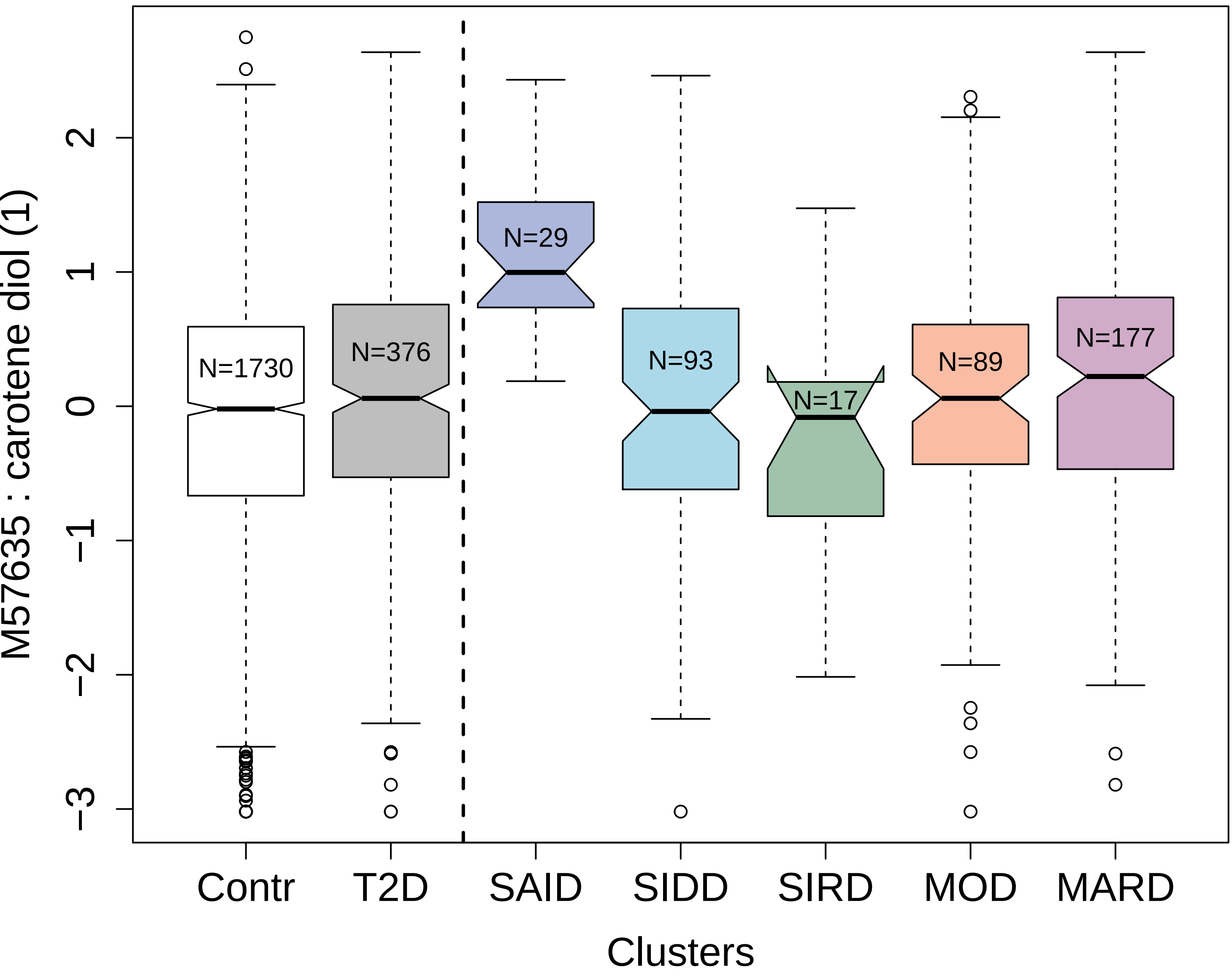

M57635 : carotene diol (1)
2
1
0
−1
−2
−3
N=1730
N=376
N=29
N=93
N=17
N=89
N=177
Contr
T2D
SAID
SIDD
SIRD
MOD
MARD
Clusters

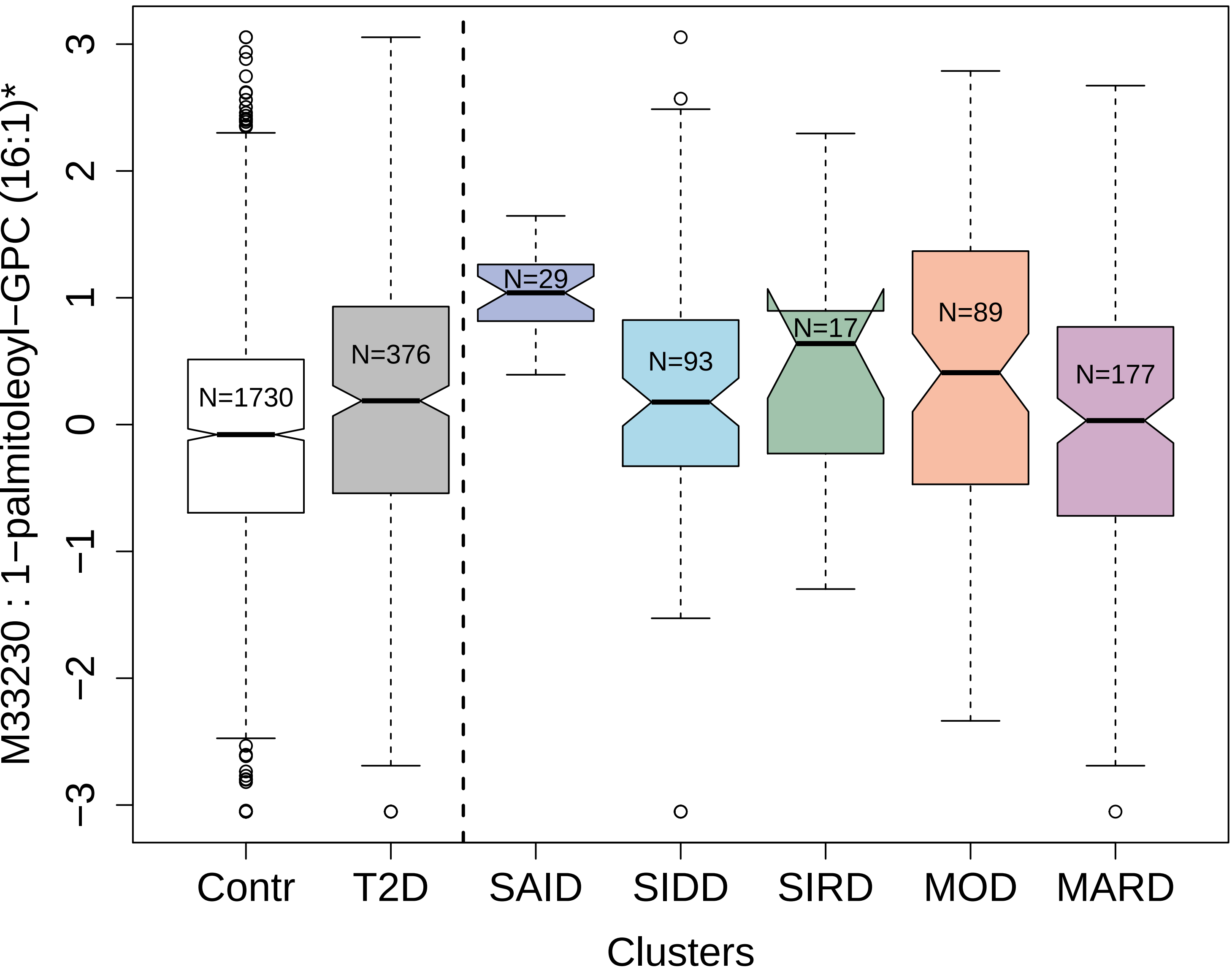
M33230 : 1−palmitoleoyl−GPC (16:1)*
3
2
1
0
−1
−2
−3
N=1730
N=376
N=29
N=93
N=17
N=89
N=177
Contr
T2D
SAID
SIDD
SIRD
MOD
MARD
Clusters

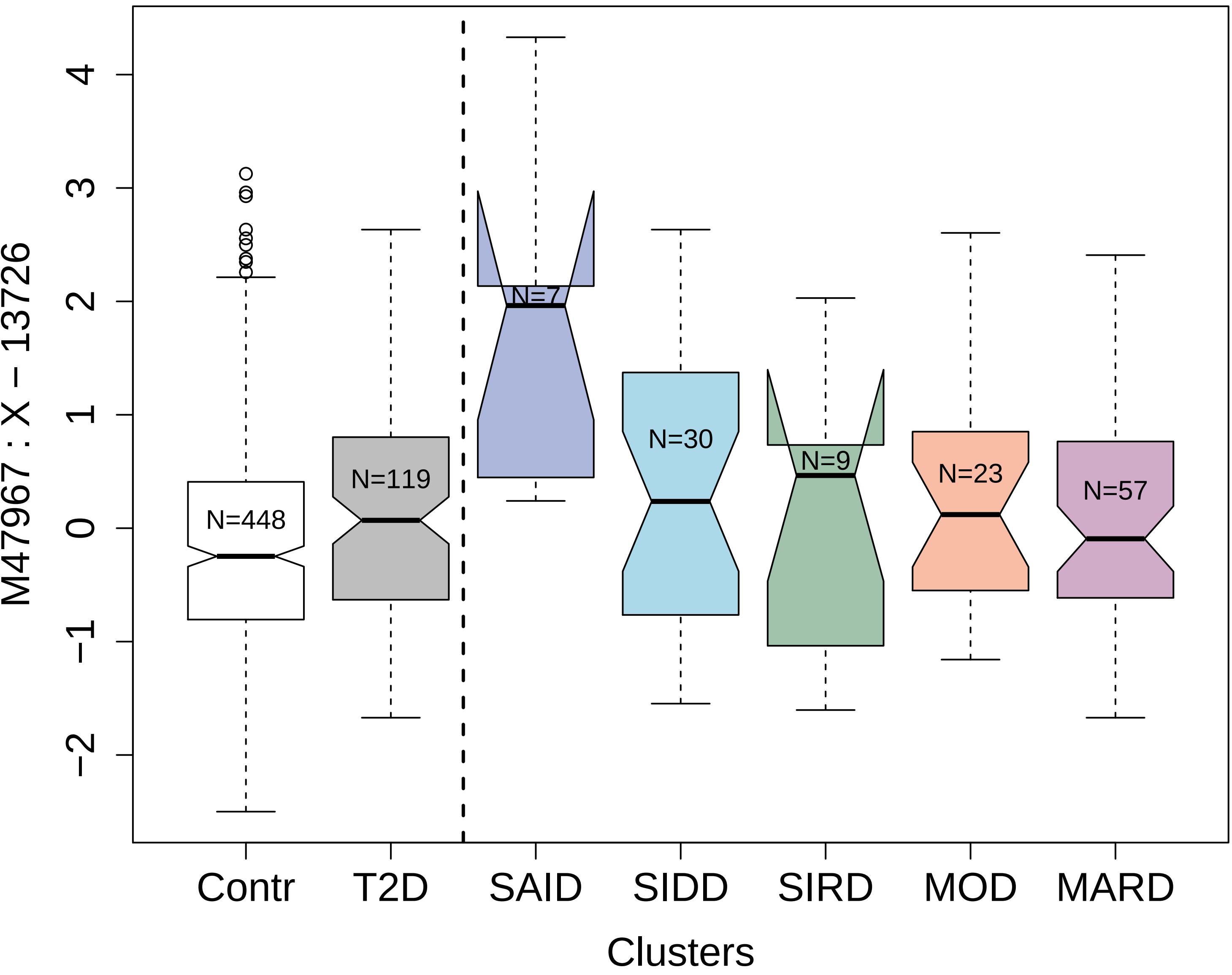

M47967 : X − 13726
4
3
2
1
0
−1
−2
N=448
N=119
N=7
N=30
N=9
N=23
N=57
Contr
T2D
SAID
SIDD
SIRD
MOD
MARD
Clusters

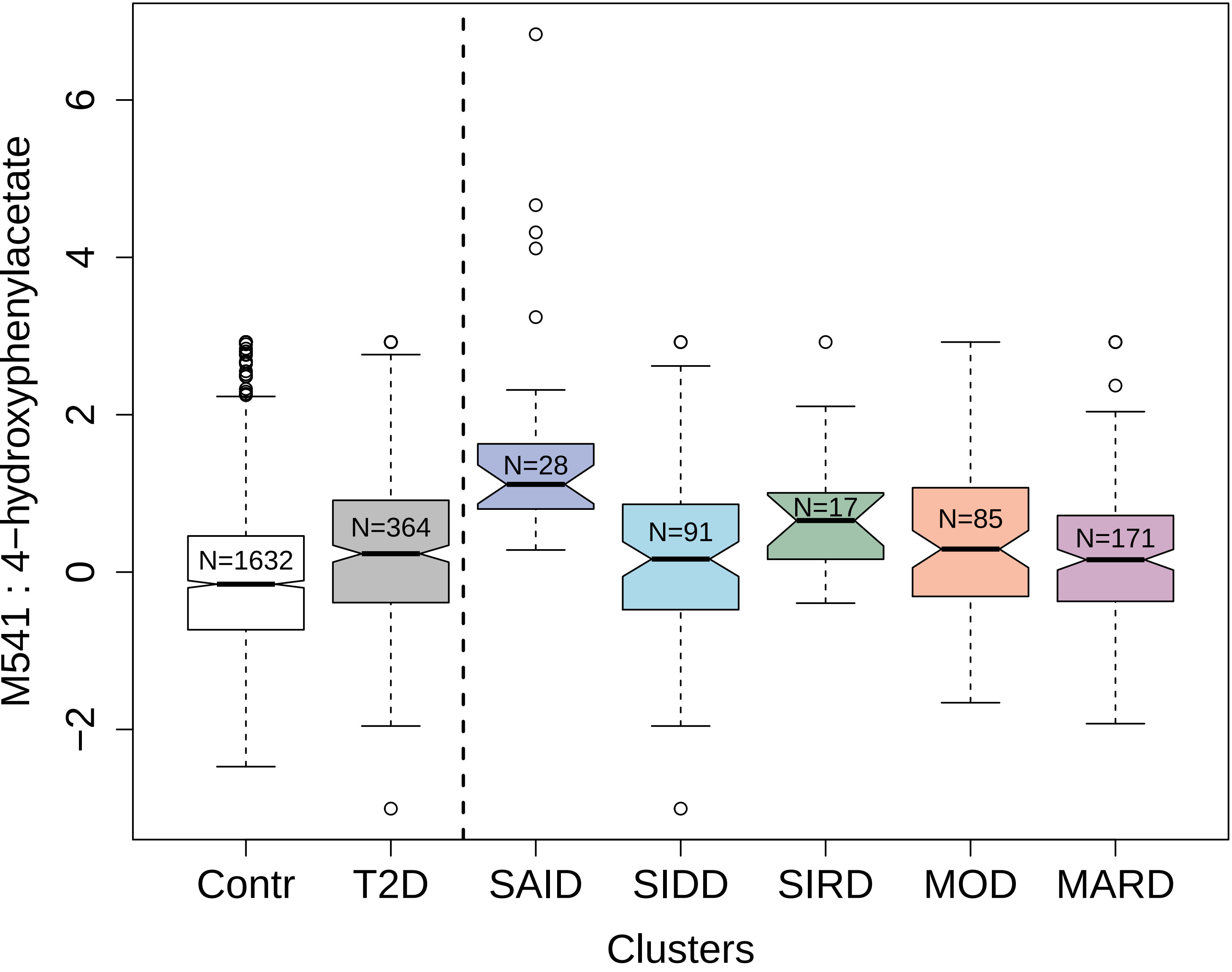

M541 : 4−hydroxyphenylacetate
6
4
2
0
−2
N=1632
N=364
N=28
N=91
N=17
N=85
N=171
Contr
T2D
SAID
SIDD
SIRD
MOD
MARD
Clusters

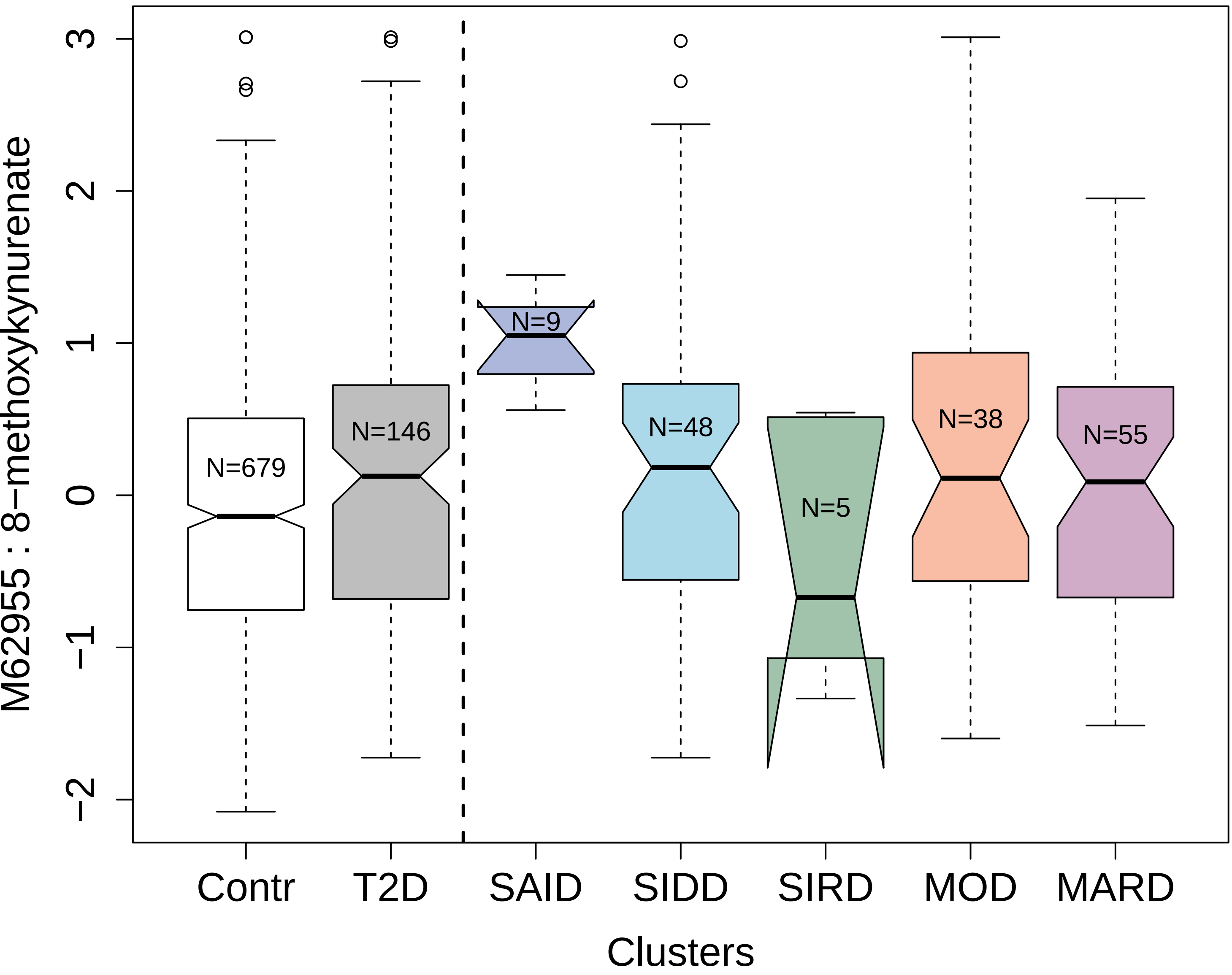

M62955 : 8−methoxykynurenate
3
2
1
0
−1
−2
N=679
N=146
N=9
N=48
N=5
N=38
N=55
Contr
T2D
SAID
SIDD
SIRD
MOD
MARD
Clusters

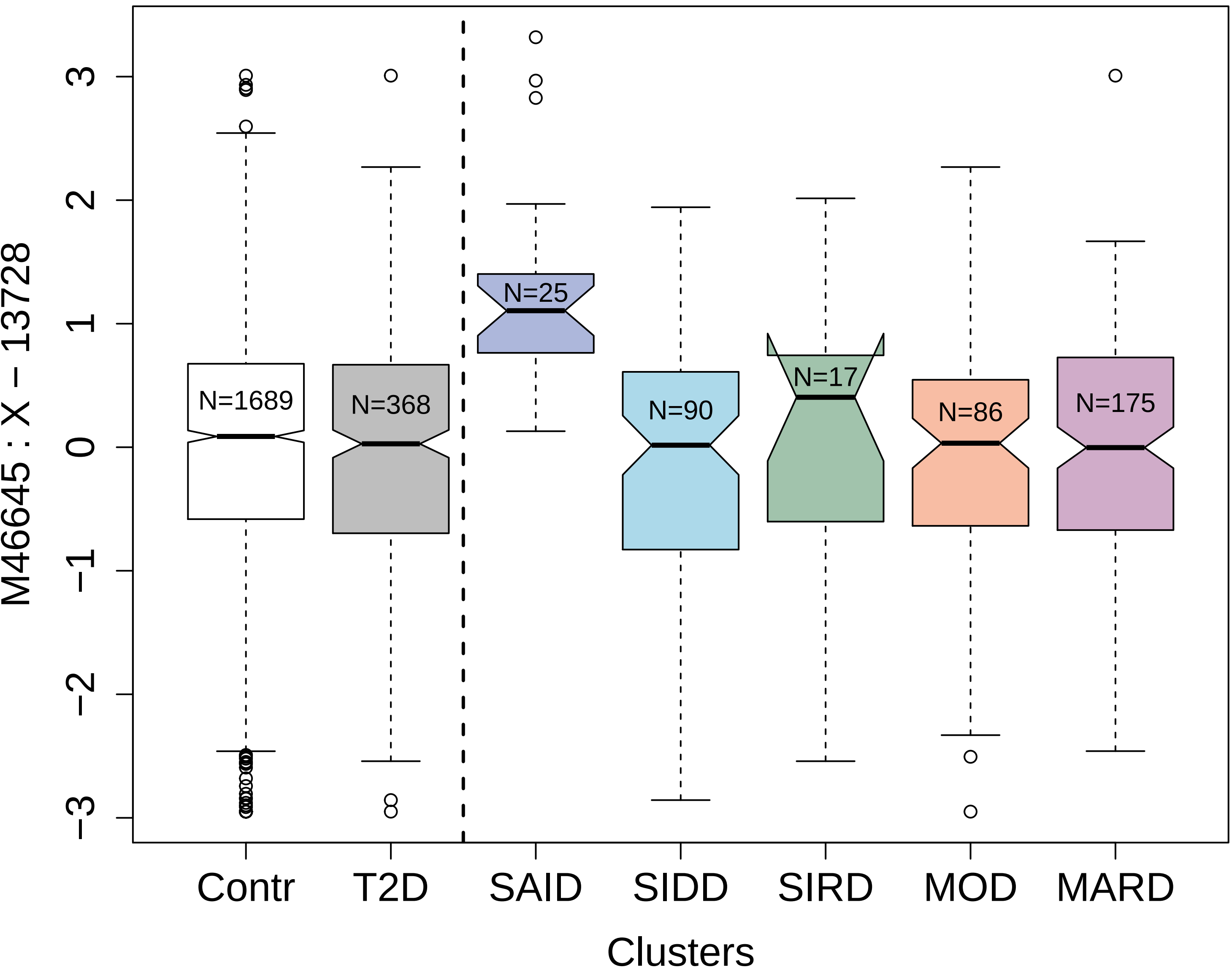

M46645 : X − 13728
3
2
1
0
−1
−2
−3
N=1689
N=368
N=25
N=90
N=17
N=86
N=175
Contr
T2D
SAID
SIDD
SIRD
MOD
MARD
Clusters

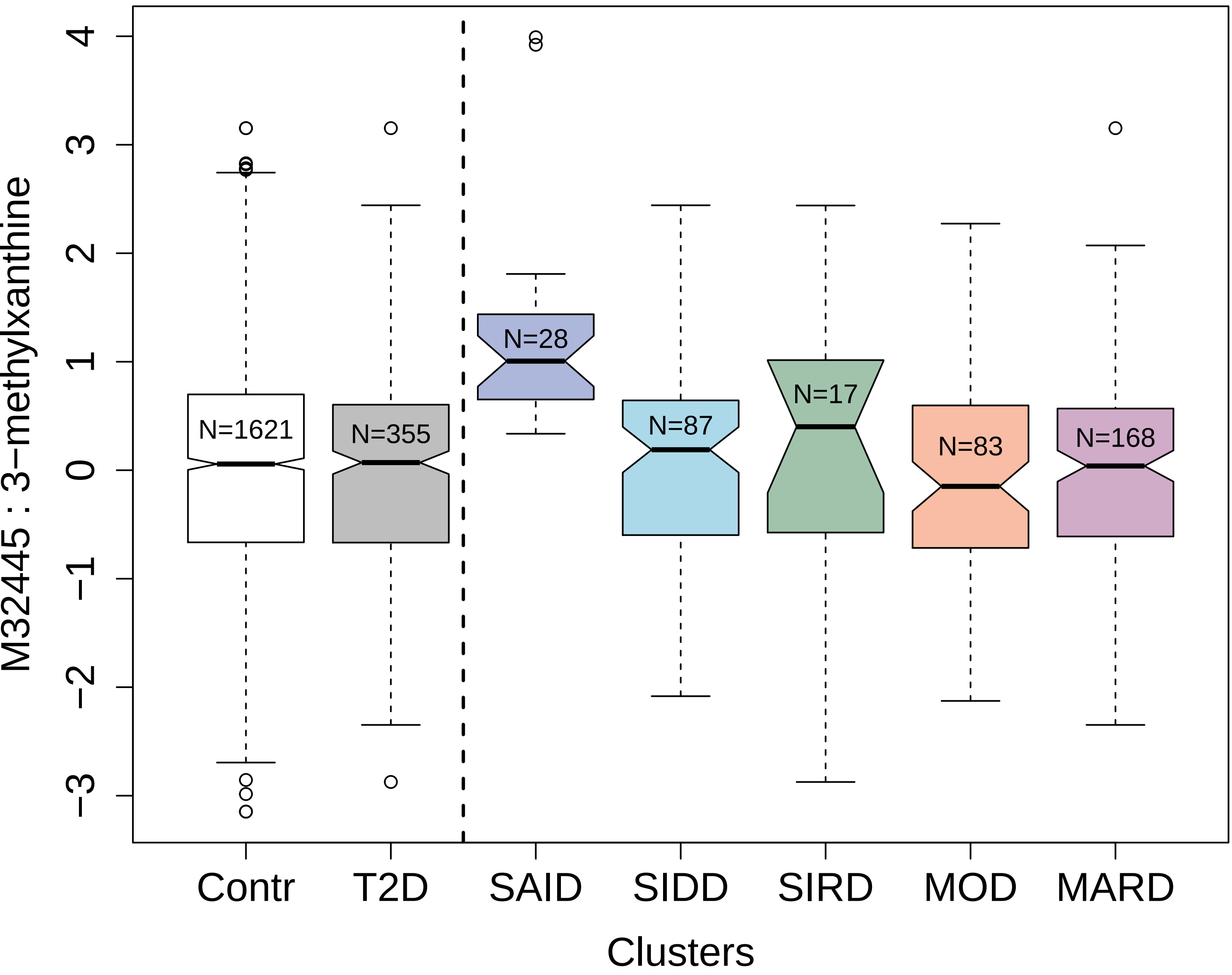
M32445 : 3−methylxanthine
4
3
2
1
0
−1
−2
−3
N=1621
N=355
N=28
N=87
N=17
N=83
N=168
Contr
T2D
SAID
SIDD
SIRD
MOD
MARD
Clusters

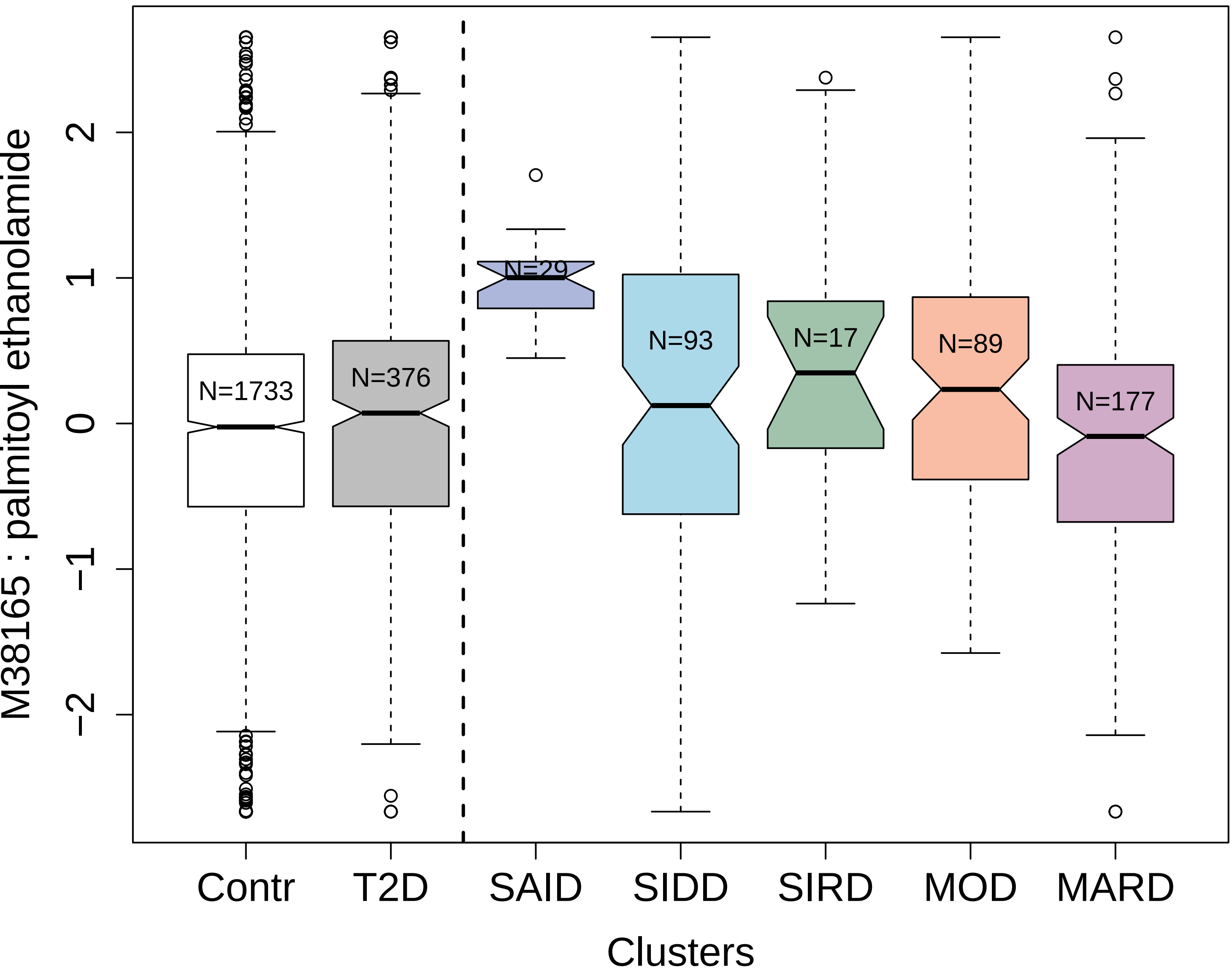

M38165 : palmitoyl ethanolamide
2
1
0
−1
−2
N=1733
N=376
N=29
N=93
N=17
N=89
N=177
Contr
T2D
SAID
SIDD
SIRD
MOD
MARD
Clusters

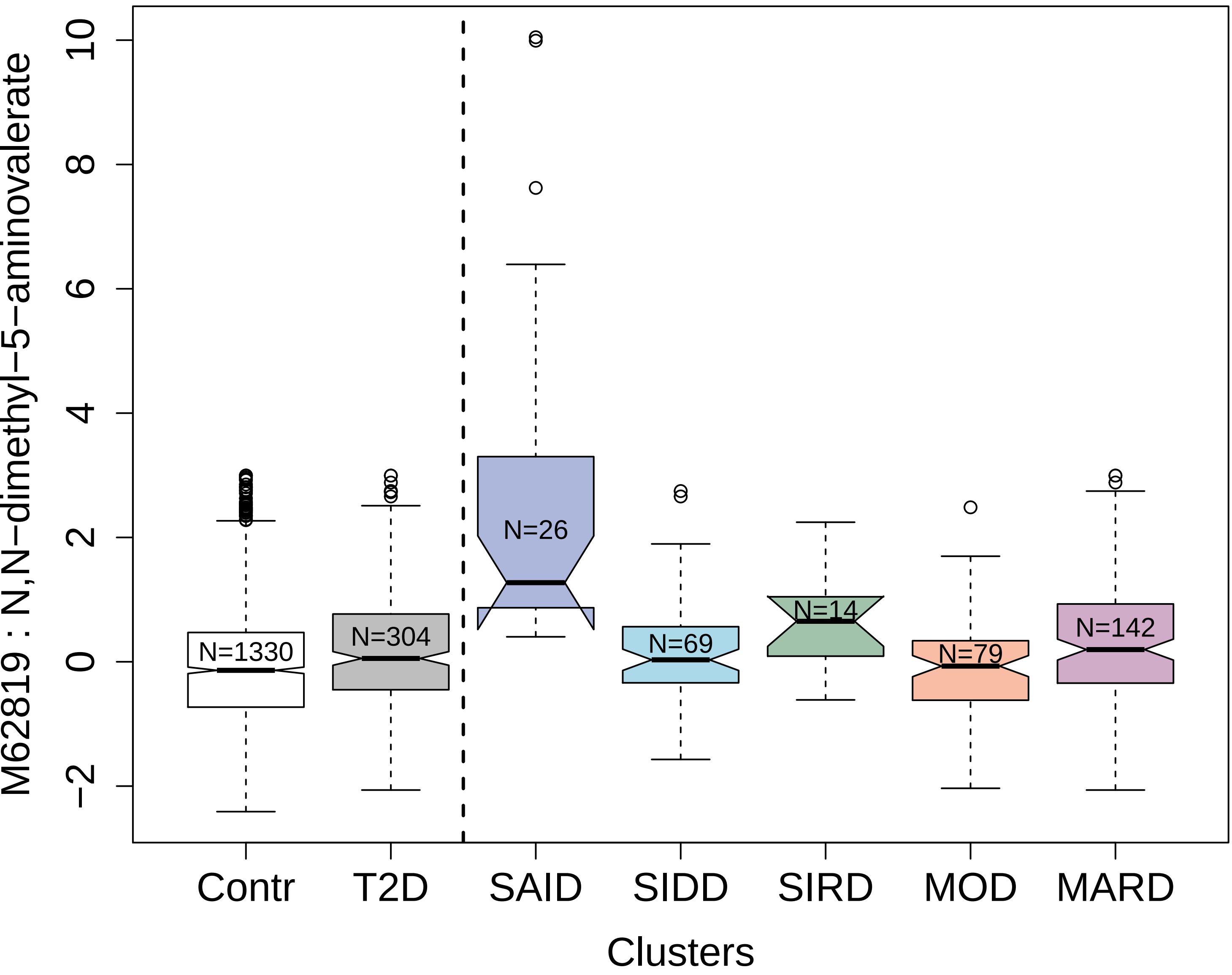

M62819 : N,N−dimethyl−5−aminovalerate
−2
0
2
4
6
8
10
N=1330
N=304
N=26
N=69
N=14
N=79
N=142
Contr
T2D
SAID
SIDD
SIRD
MOD
MARD
Clusters

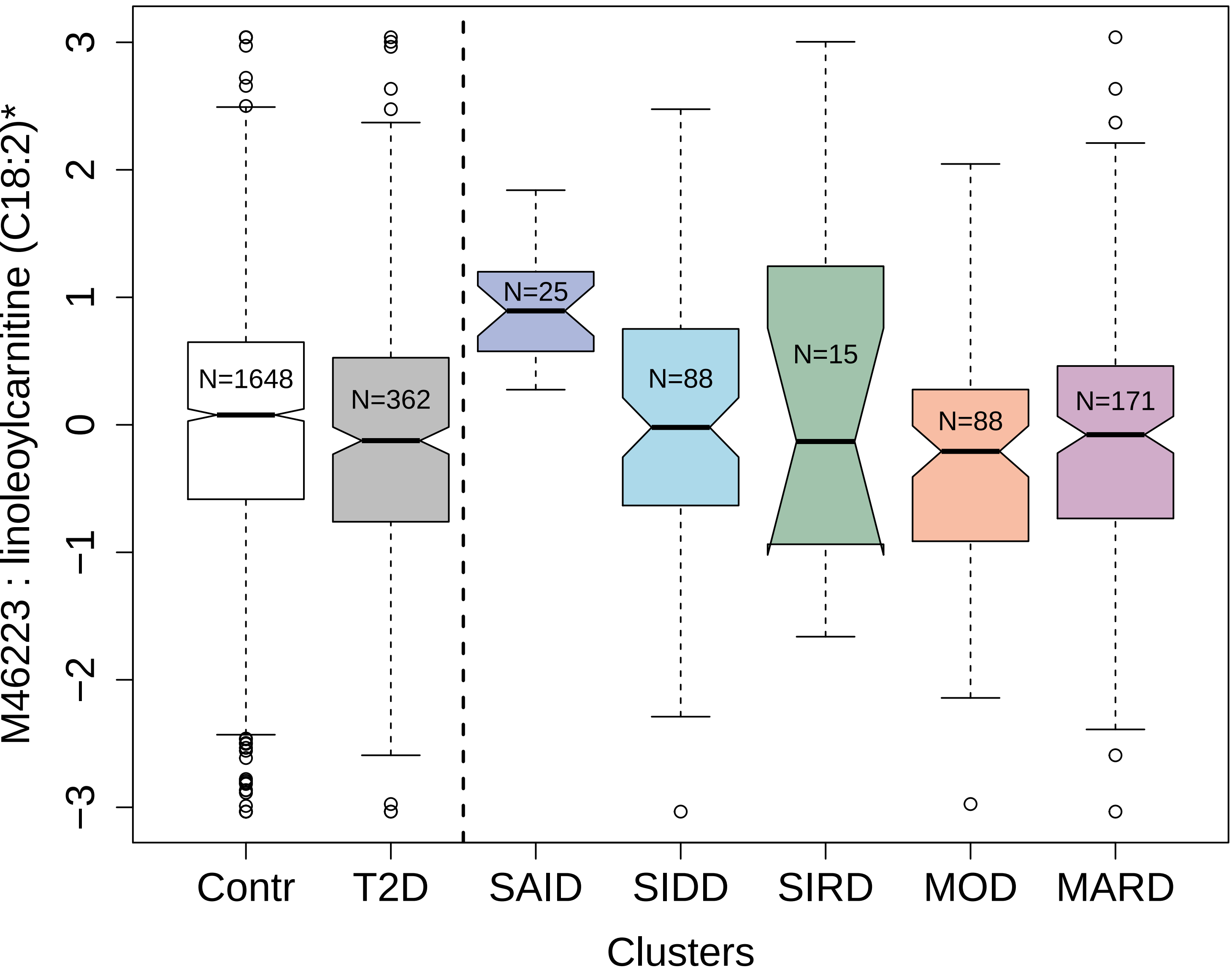

M46223 : linoleoylcarnitine (C18:2)*
3
2
1
0
−1
−2
−3
N=1648
N=362
N=25
N=88
N=15
N=88
N=171
Contr
T2D
SAID
SIDD
SIRD
MOD
MARD
Clusters

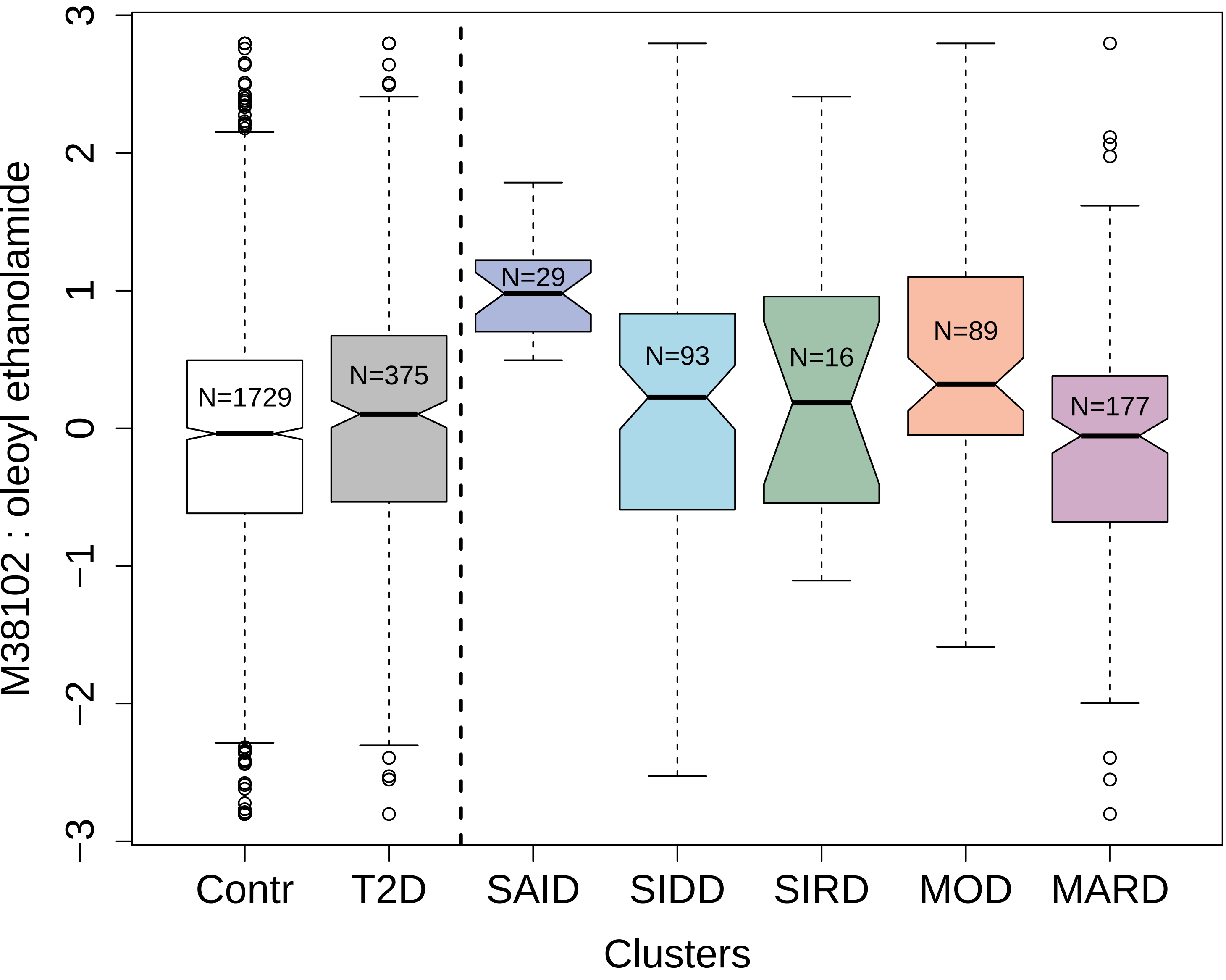

M38102 : oleoyl ethanolamide
3
2
1
0
−1
−2
−3
N=1729
N=375
N=29
N=93
N=16
N=89
N=177
Contr
T2D
SAID
SIDD
SIRD
MOD
MARD
Clusters

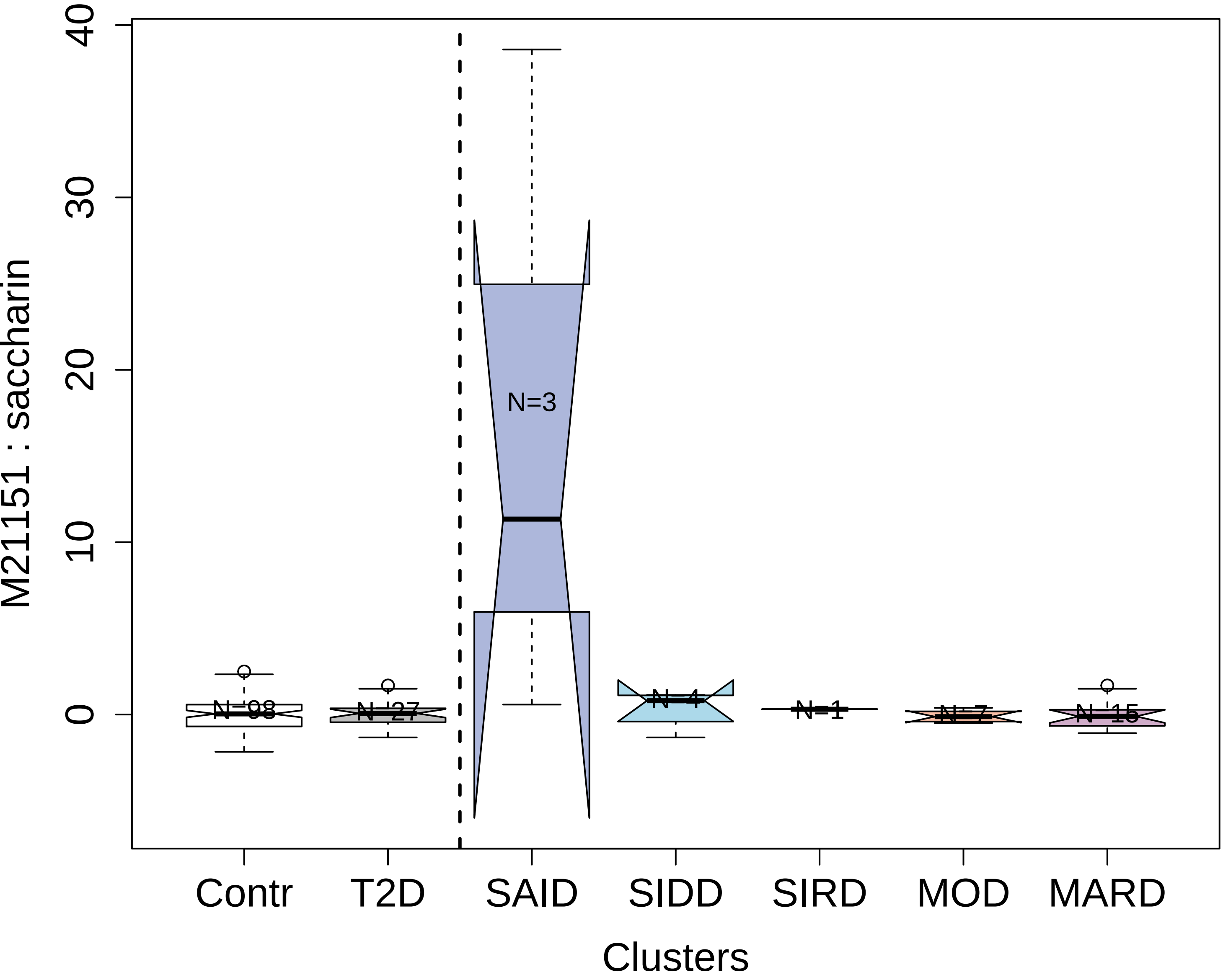

M21151 : saccharin
0
10
20
30
40
N=98
N=27
N=3
N=4
N=1
N=7
N=15
Contr
T2D
SAID
SIDD
SIRD
MOD
MARD
Clusters

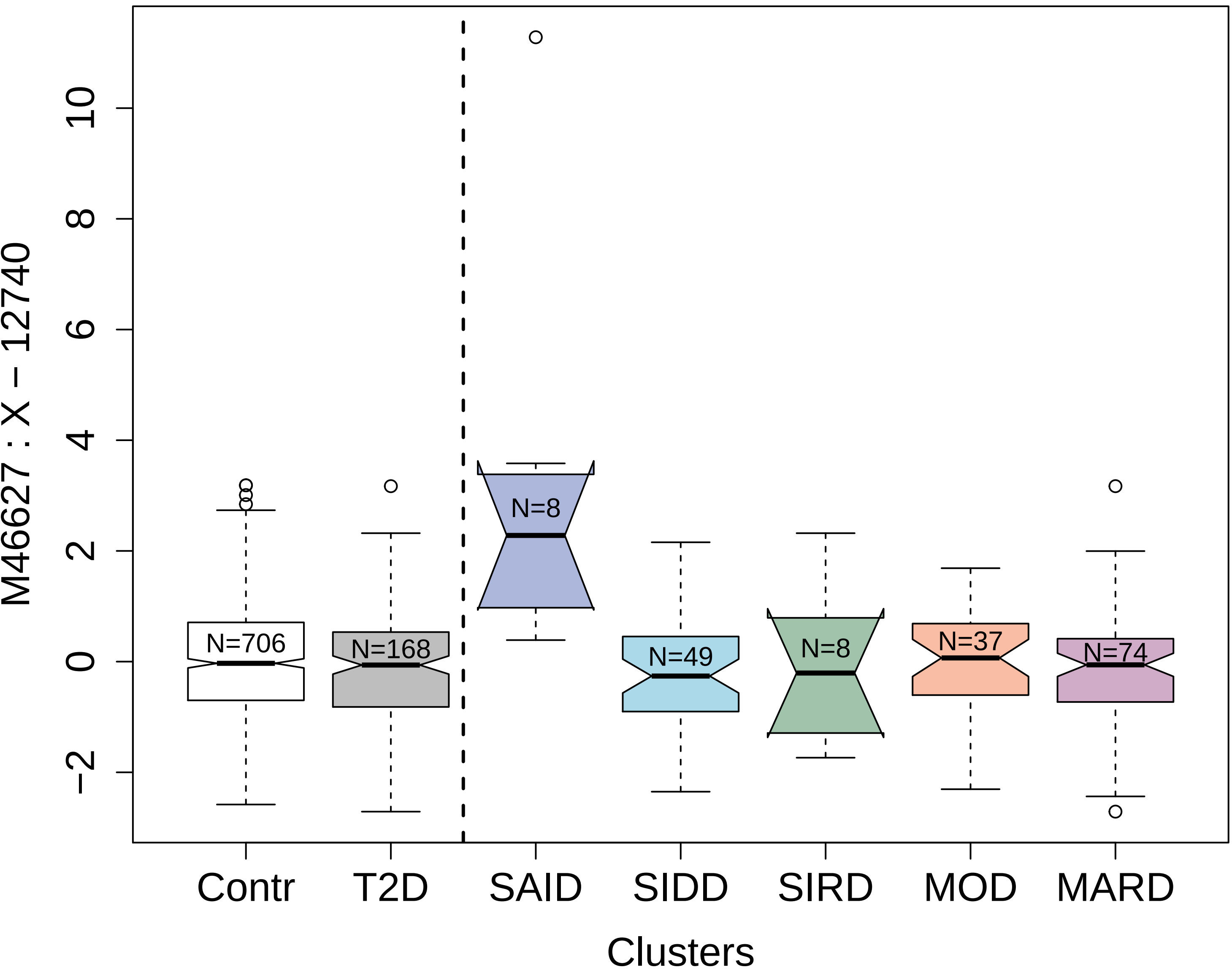
M46627 : X − 12740
10
8
6
4
2
0
−2
N=706
N=168
N=8
N=49
N=8
N=37
N=74
Contr
T2D
SAID
SIDD
SIRD
MOD
MARD
Clusters

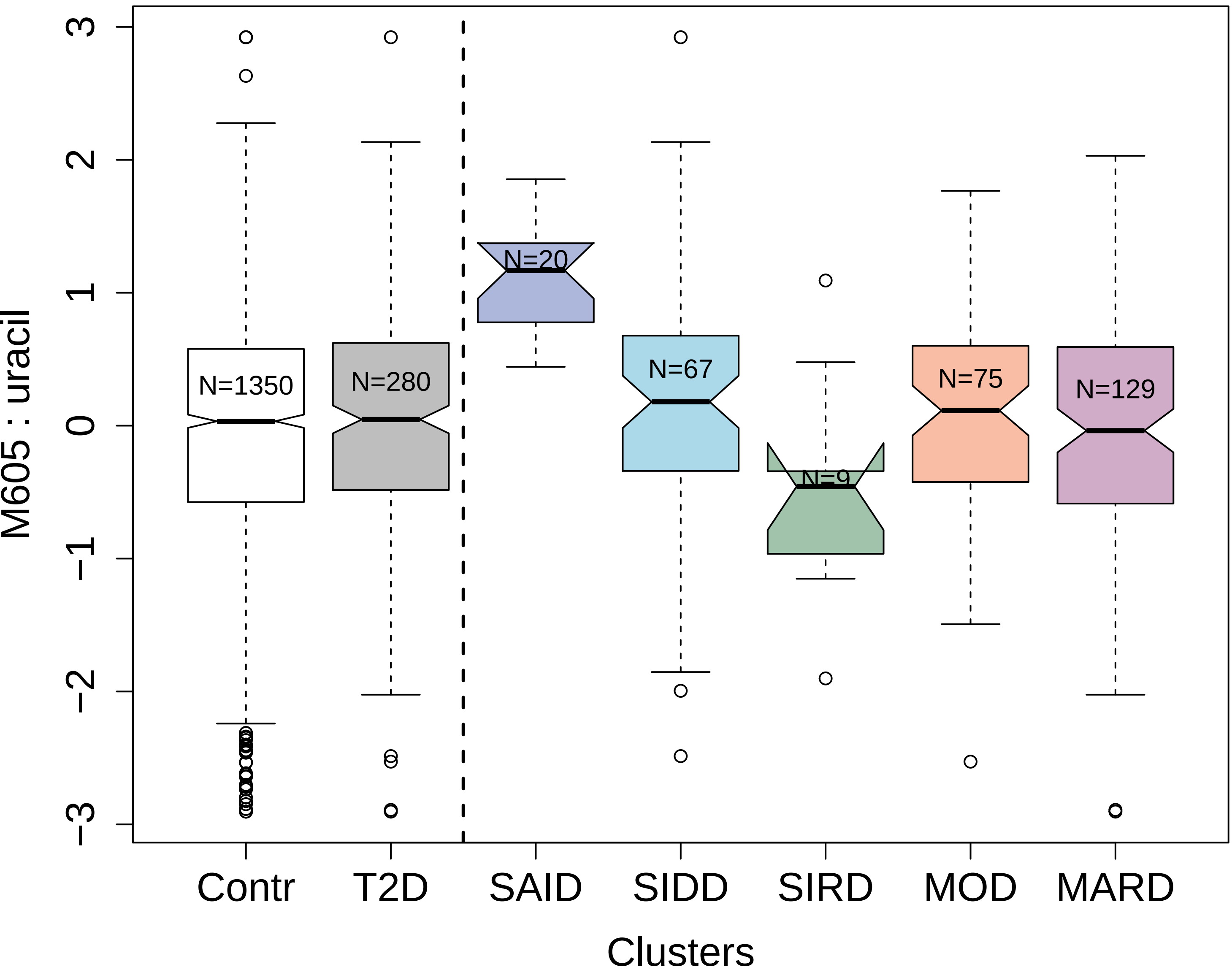
M605 : uracil
3
2
1
0
−1
−2
−3
N=1350
N=280
N=20
N=67
N=9
N=75
N=129
Contr
T2D
SAID
SIDD
SIRD
MOD
MARD
Clusters

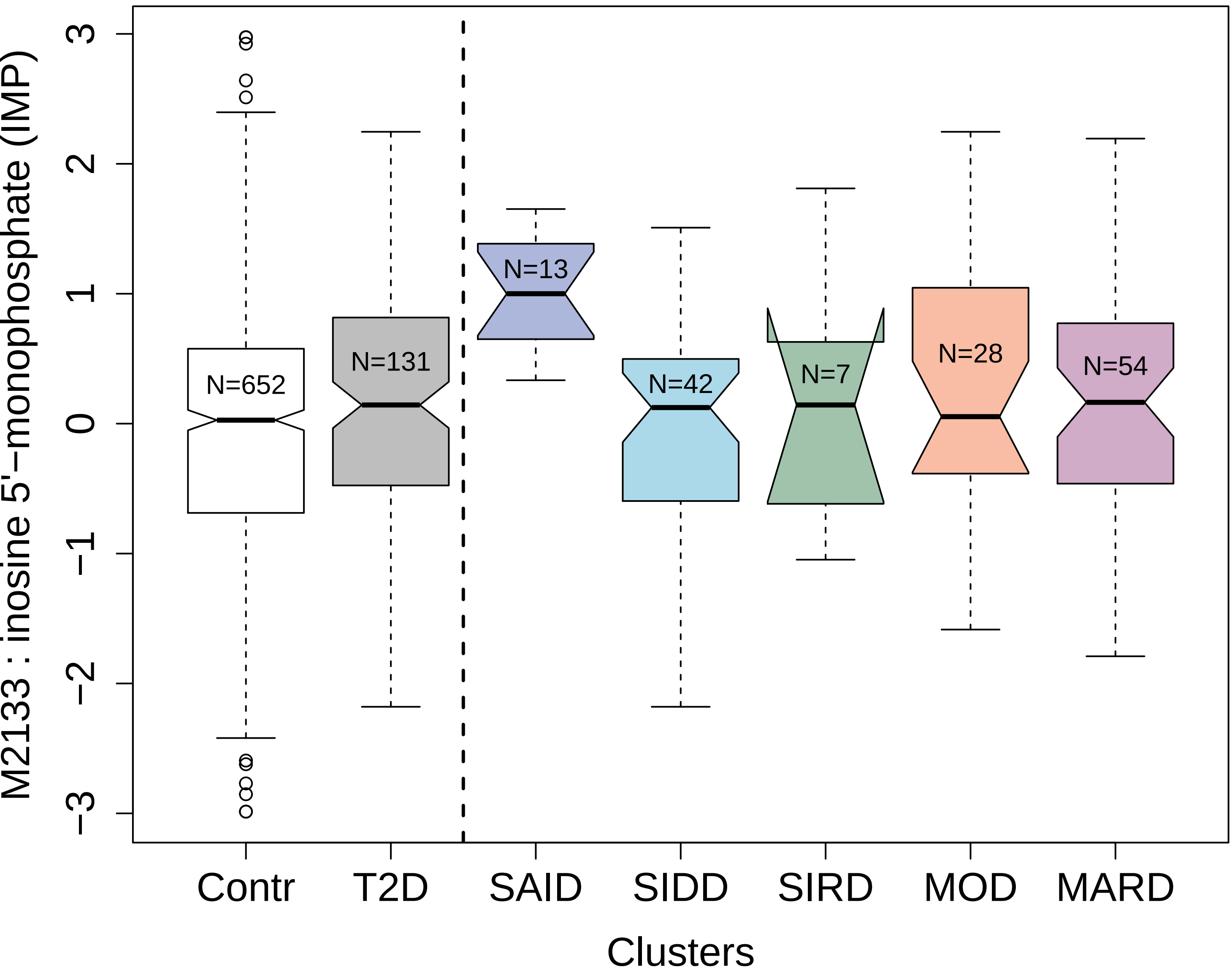
M2133 : inosine 5'−monophosphate (IMP)
3
2
1
0
−1
−2
−3
N=652
N=131
N=13
N=42
N=7
N=28
N=54
Contr
T2D
SAID
SIDD
SIRD
MOD
MARD
Clusters

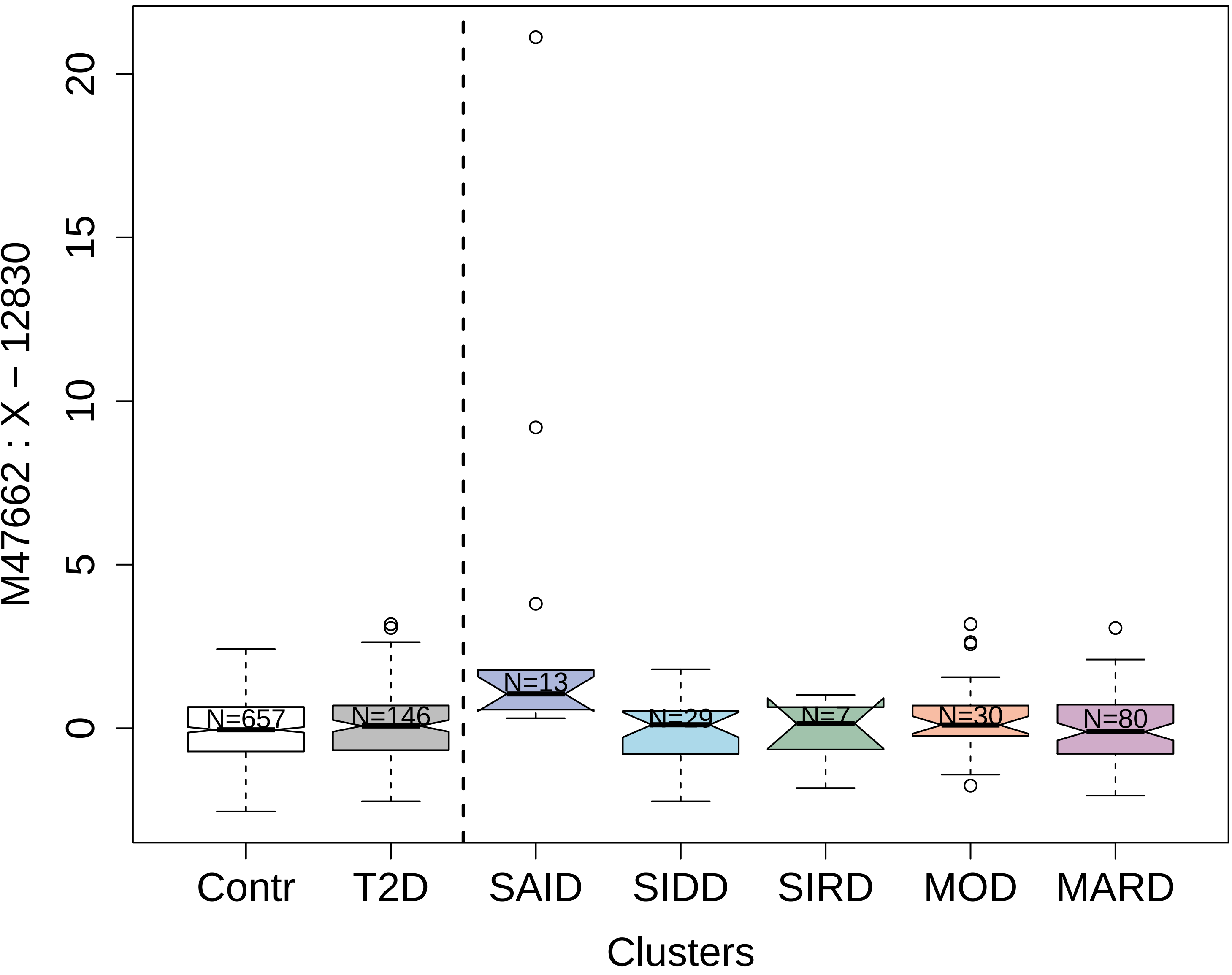
M47662 : X − 12830
0
5
10
15
20
N=657
N=146
N=13
N=29
N=7
N=30
N=80
Contr
T2D
SAID
SIDD
SIRD
MOD
MARD
Clusters

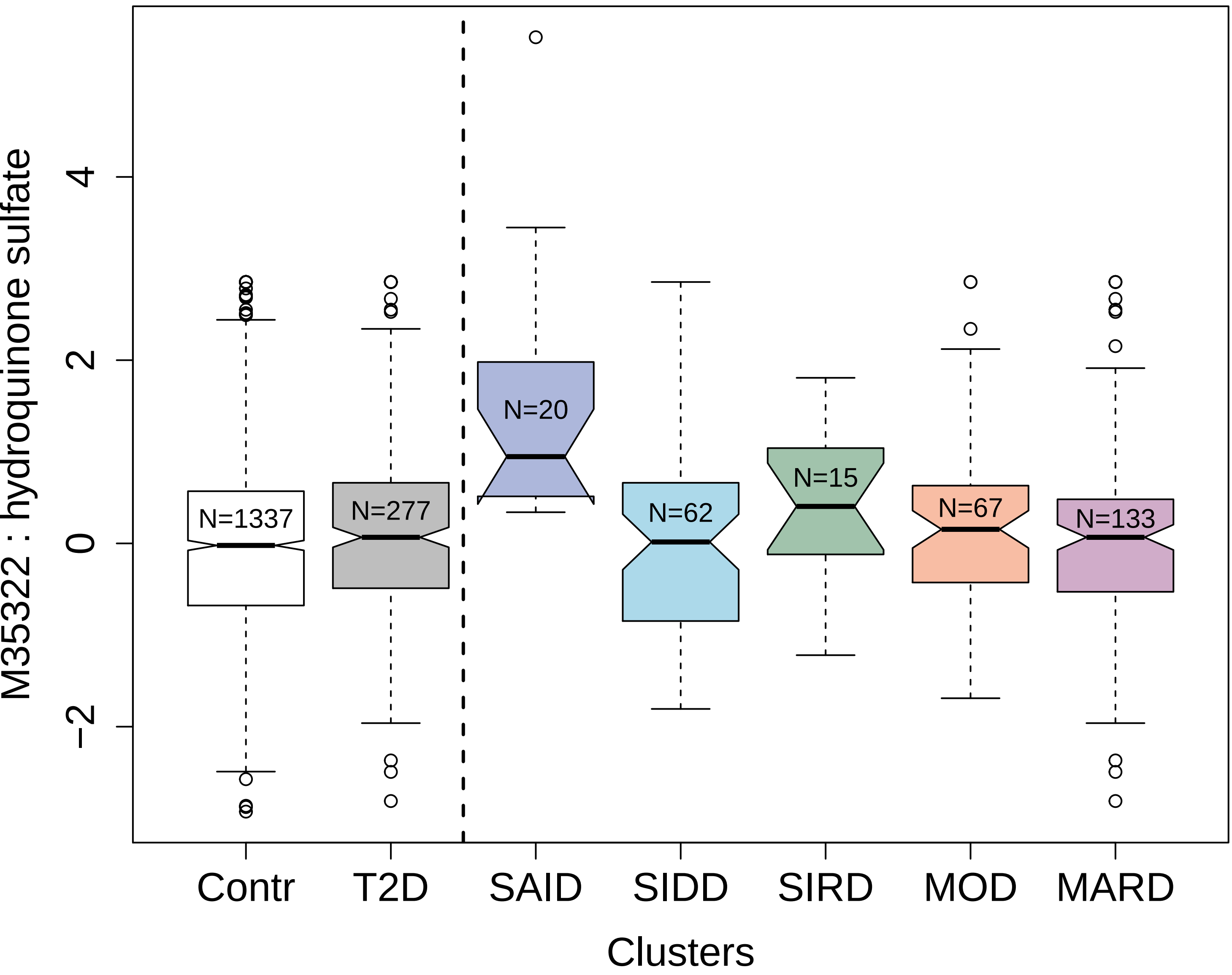

M35322 : hydroquinone sulfate
4
2
0
−2
N=1337
N=277
N=20
N=62
N=15
N=67
N=133
Contr
T2D
SAID
SIDD
SIRD
MOD
MARD
Clusters

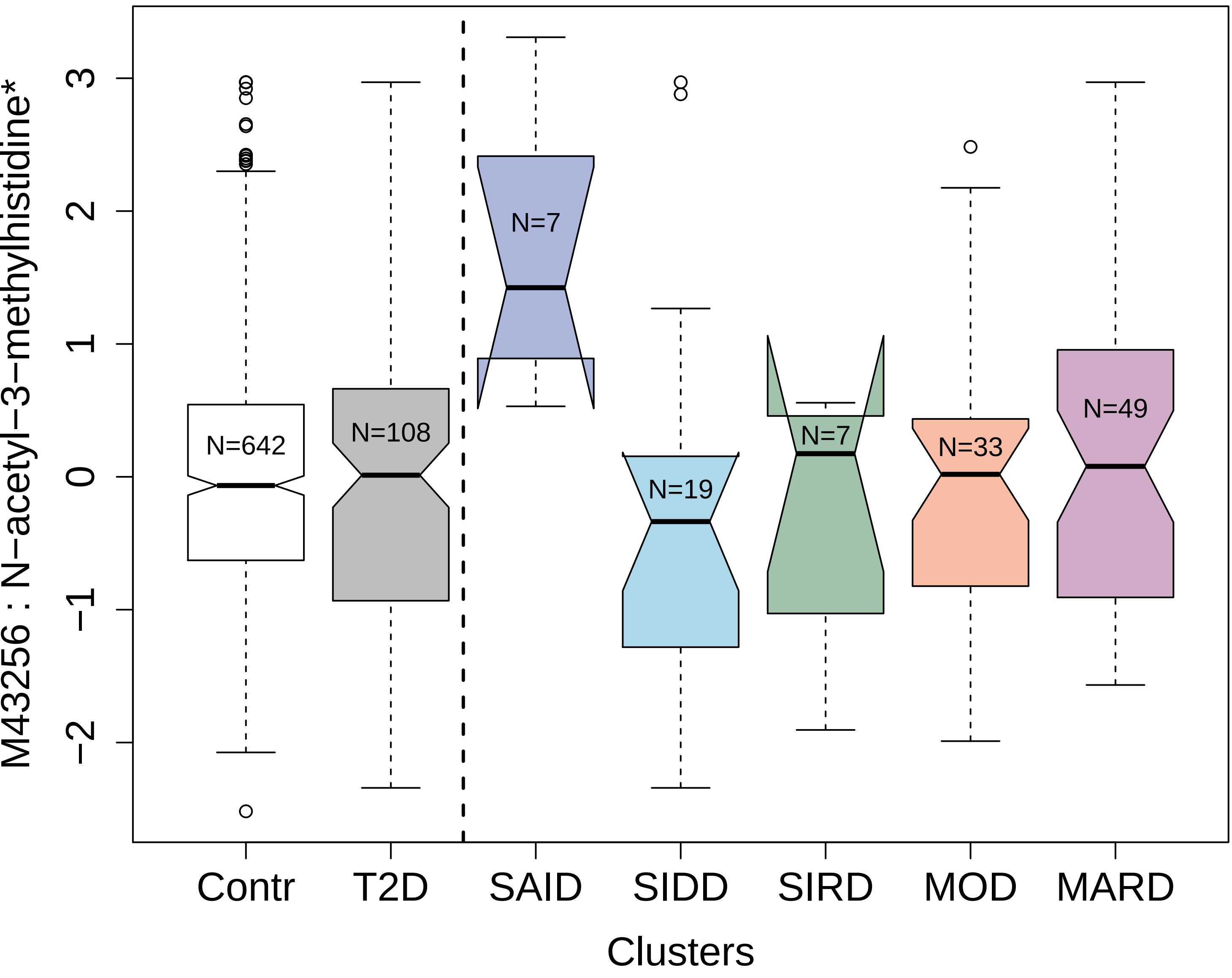
M43256 : N−acetyl−3−methylhistidine*
3
2
1
0
−1
−2
N=642
N=108
N=7
N=19
N=7
N=33
N=49
Contr
T2D
SAID
SIDD
SIRD
MOD
MARD
Clusters

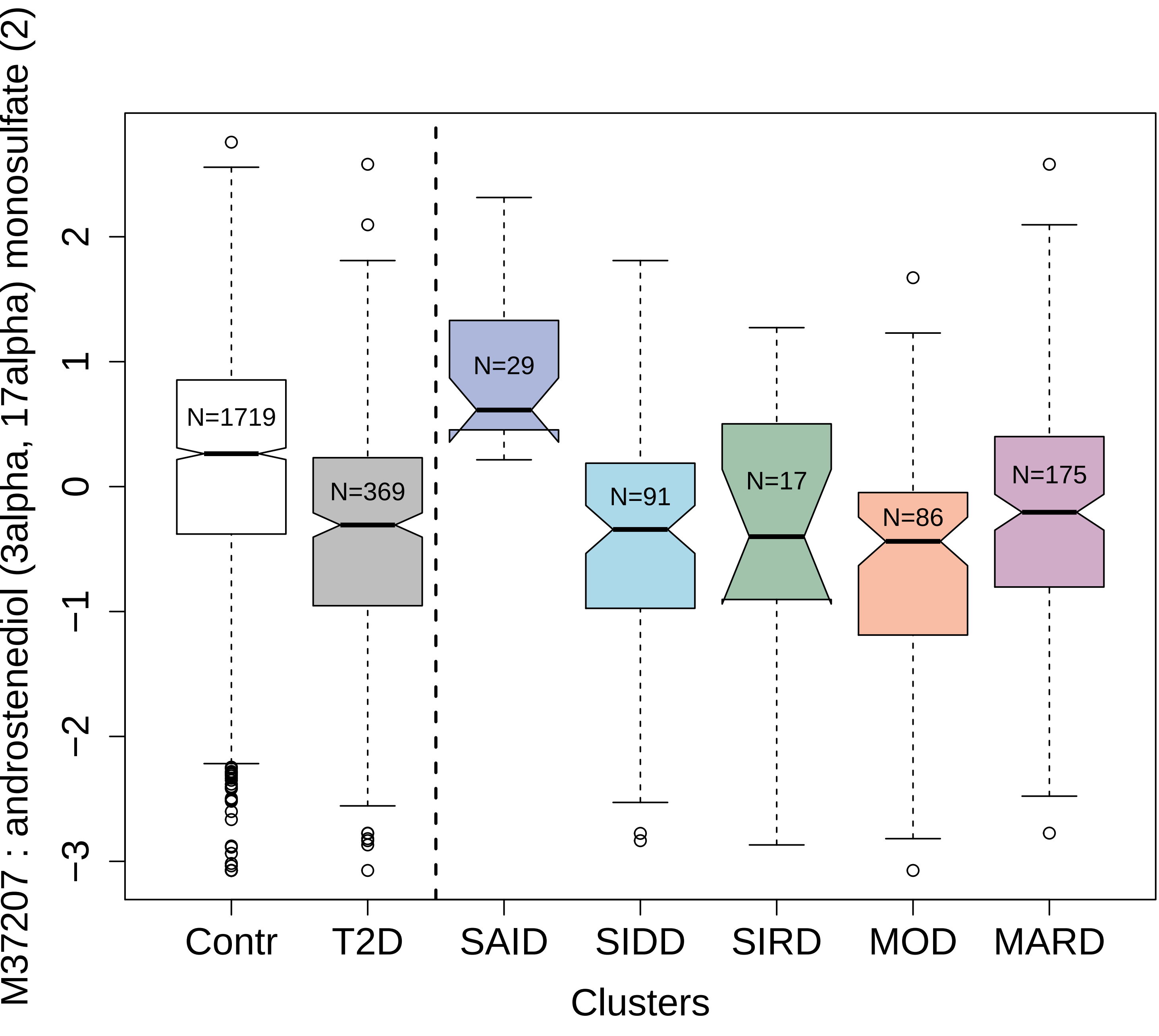
M37207 : androstenediol (3alpha, 17alpha) monosulfate (2)
−3
−2
−1
0
1
2
N=1719
N=369
N=29
N=91
N=17
N=86
N=175
Contr
T2D
SAID
SIDD
SIRD
MOD
MARD
Clusters

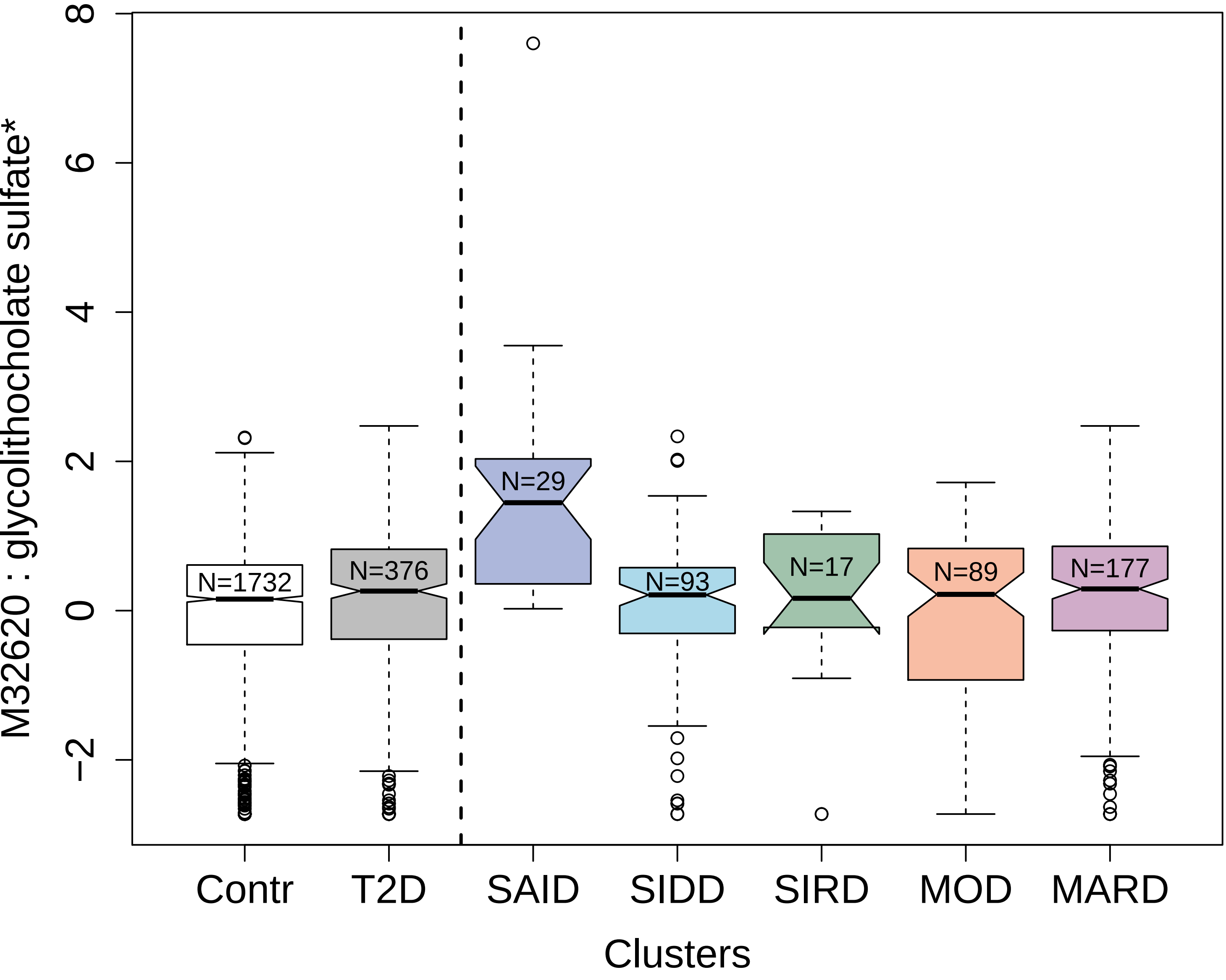

M32620 : glycolithocholate sulfate*
8
6
4
2
0
−2
N=1732
N=376
N=29
N=93
N=17
N=89
N=177
Contr
T2D
SAID
SIDD
SIRD
MOD
MARD
Clusters

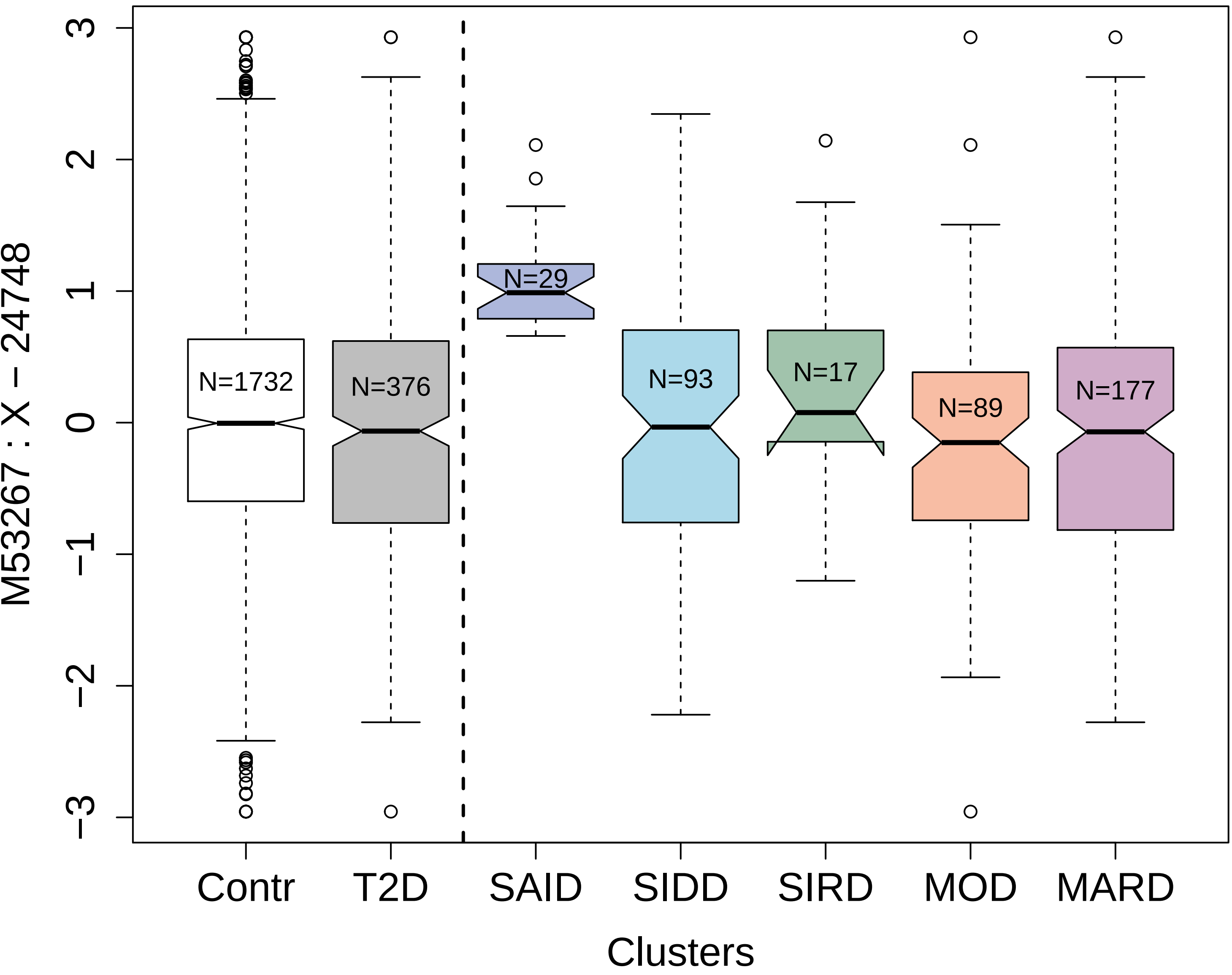
M53267 : X − 24748
3
2
1
0
−1
−2
−3
N=1732
N=376
N=29
N=93
N=17
N=89
N=177
Contr
T2D
SAID
SIDD
SIRD
MOD
MARD
Clusters

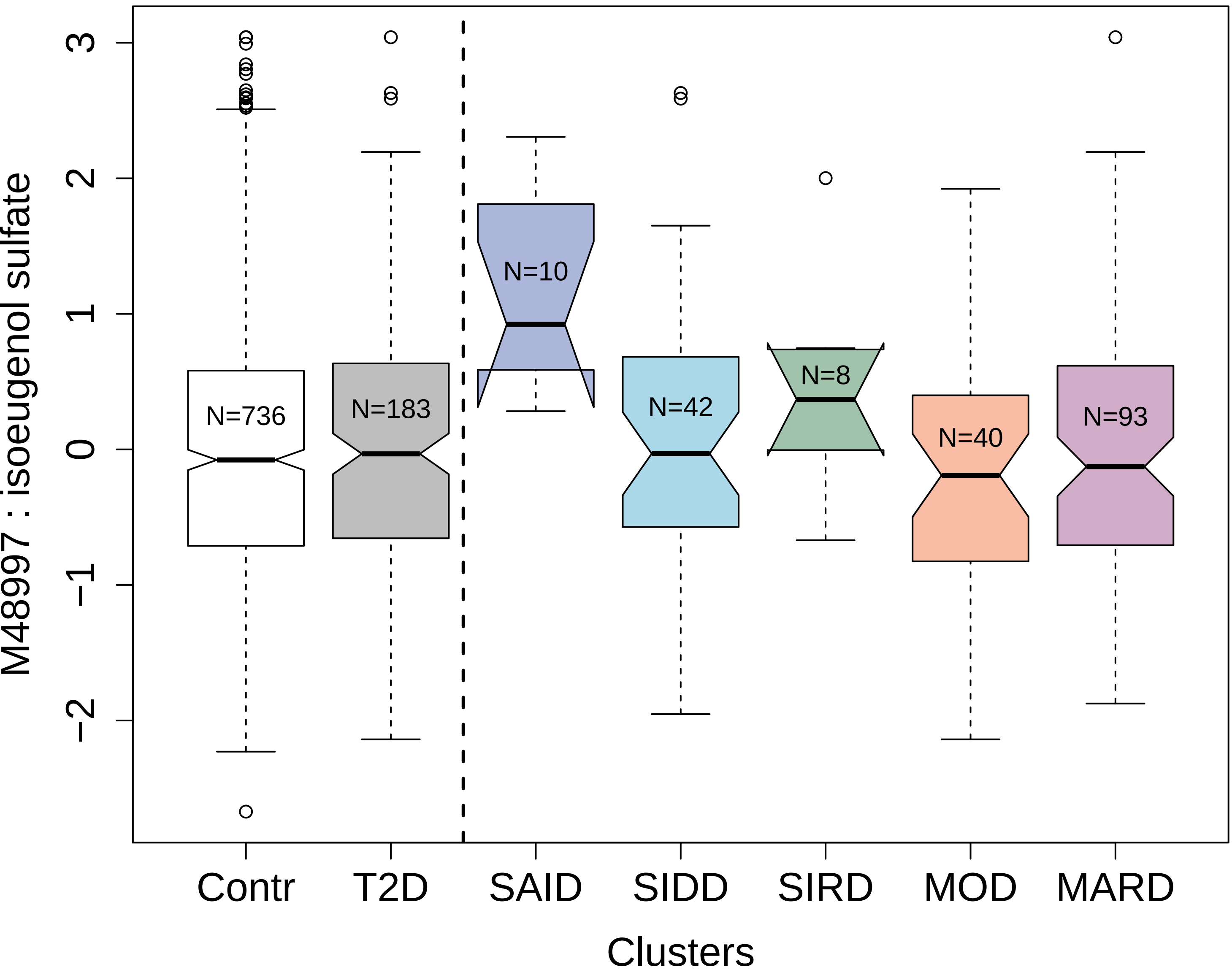
M48997 : isoeugenol sulfate
3
2
1
0
−1
−2
N=736
N=183
N=10
N=42
N=8
N=40
N=93
Contr
T2D
SAID
SIDD
SIRD
MOD
MARD
Clusters

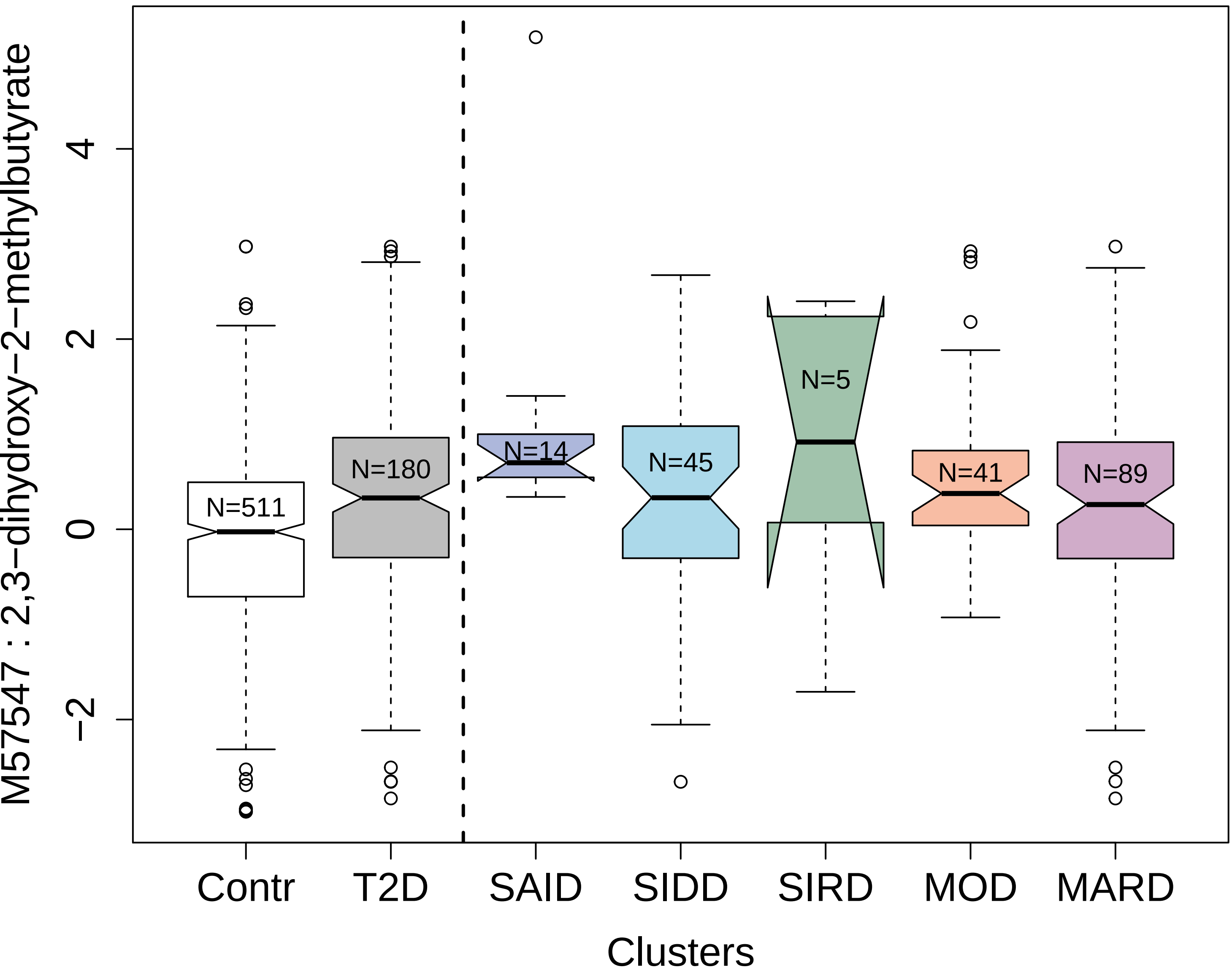

M57547 : 2,3−dihydroxy−2−methylbutyrate
Clusters
Contr
T2D
SAID
SIDD
SIRD
MOD
MARD
N=511
N=180
N=14
N=45
N=5
N=41
N=89
−2
0
2
4

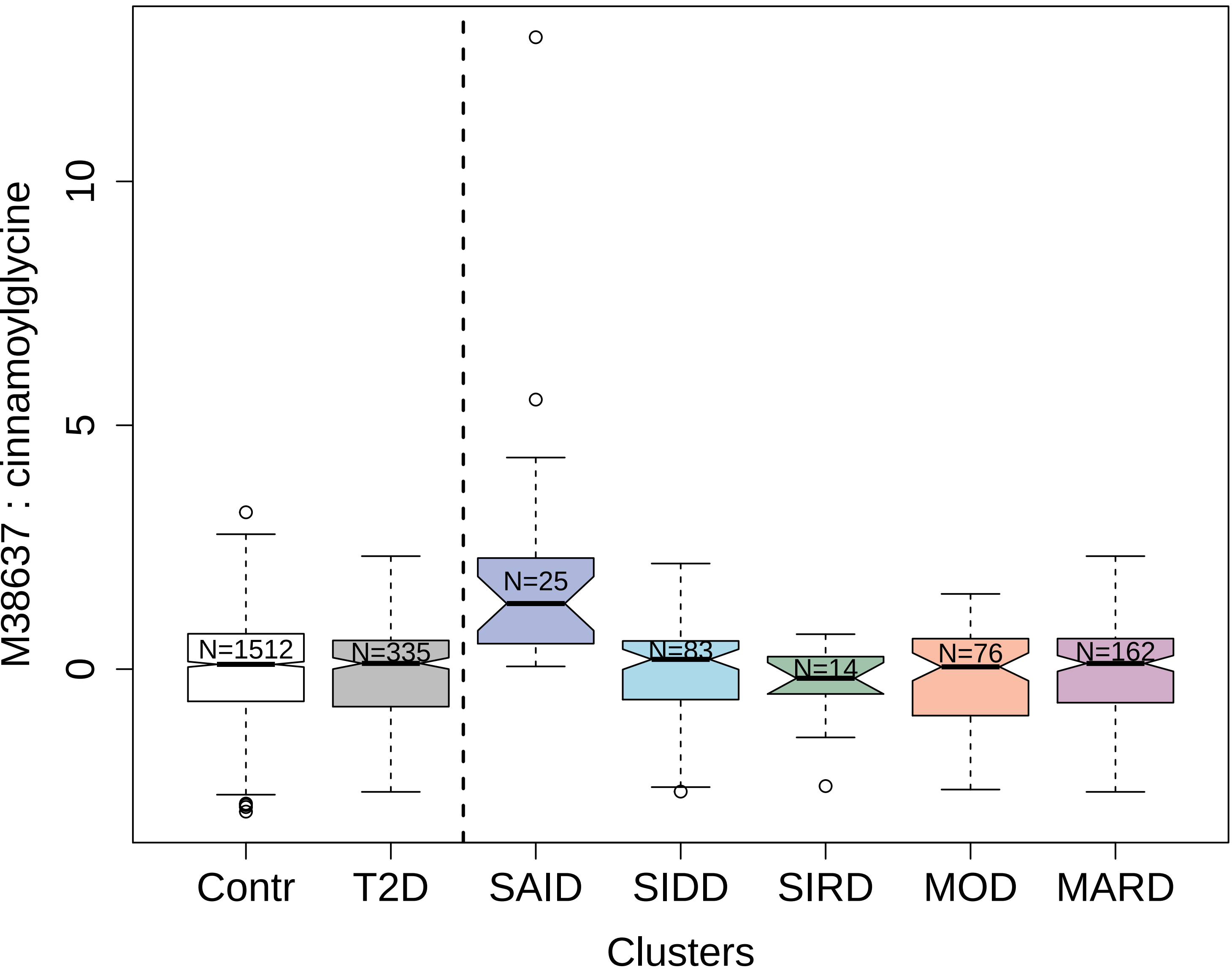
M38637 : cinnamoylglycine
0
5
10
N=1512
N=335
N=25
N=83
N=14
N=76
N=162
Contr
T2D
SAID
SIDD
SIRD
MOD
MARD
Clusters

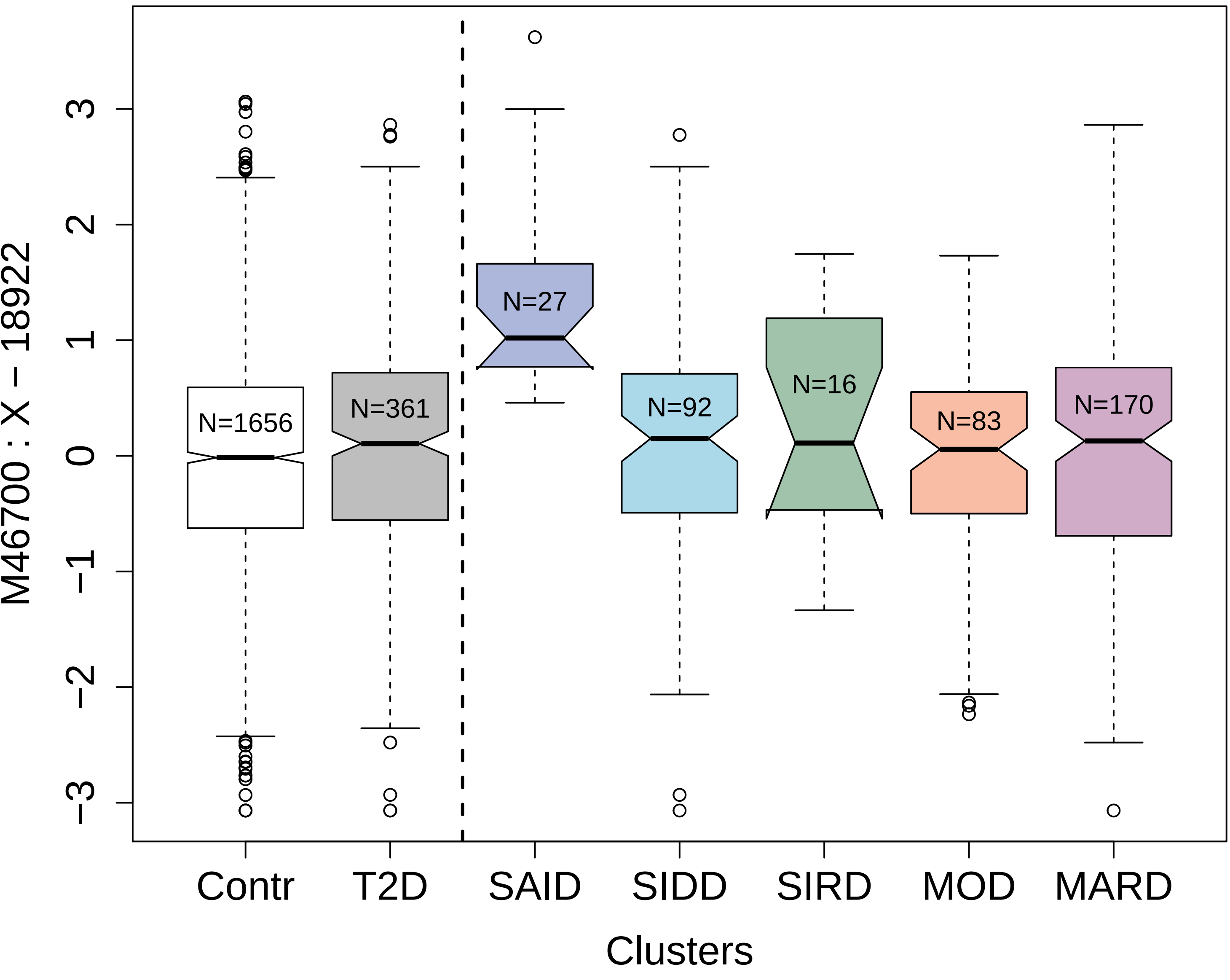

M46700 : X − 18922
3
2
1
0
−1
−2
−3
N=1656
N=361
N=27
N=92
N=16
N=83
N=170
Contr
T2D
SAID
SIDD
SIRD
MOD
MARD
Clusters

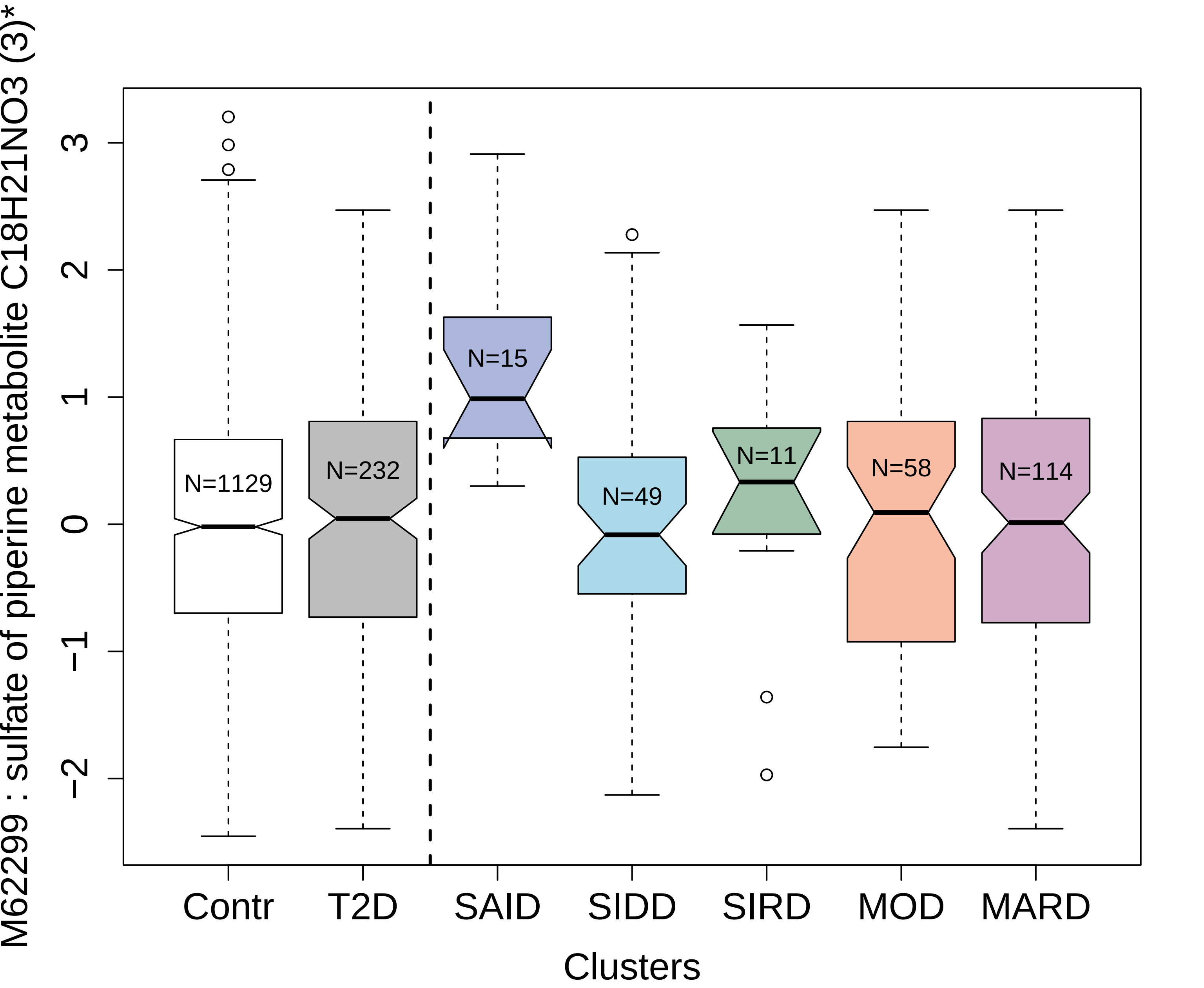
M62299 : sulfate of piperine metabolite C18H21NO3 (3)*
3
2
1
0
−1
−2
N=1129
N=232
N=15
N=49
N=11
N=58
N=114
Contr
T2D
SAID
SIDD
SIRD
MOD
MARD
Clusters

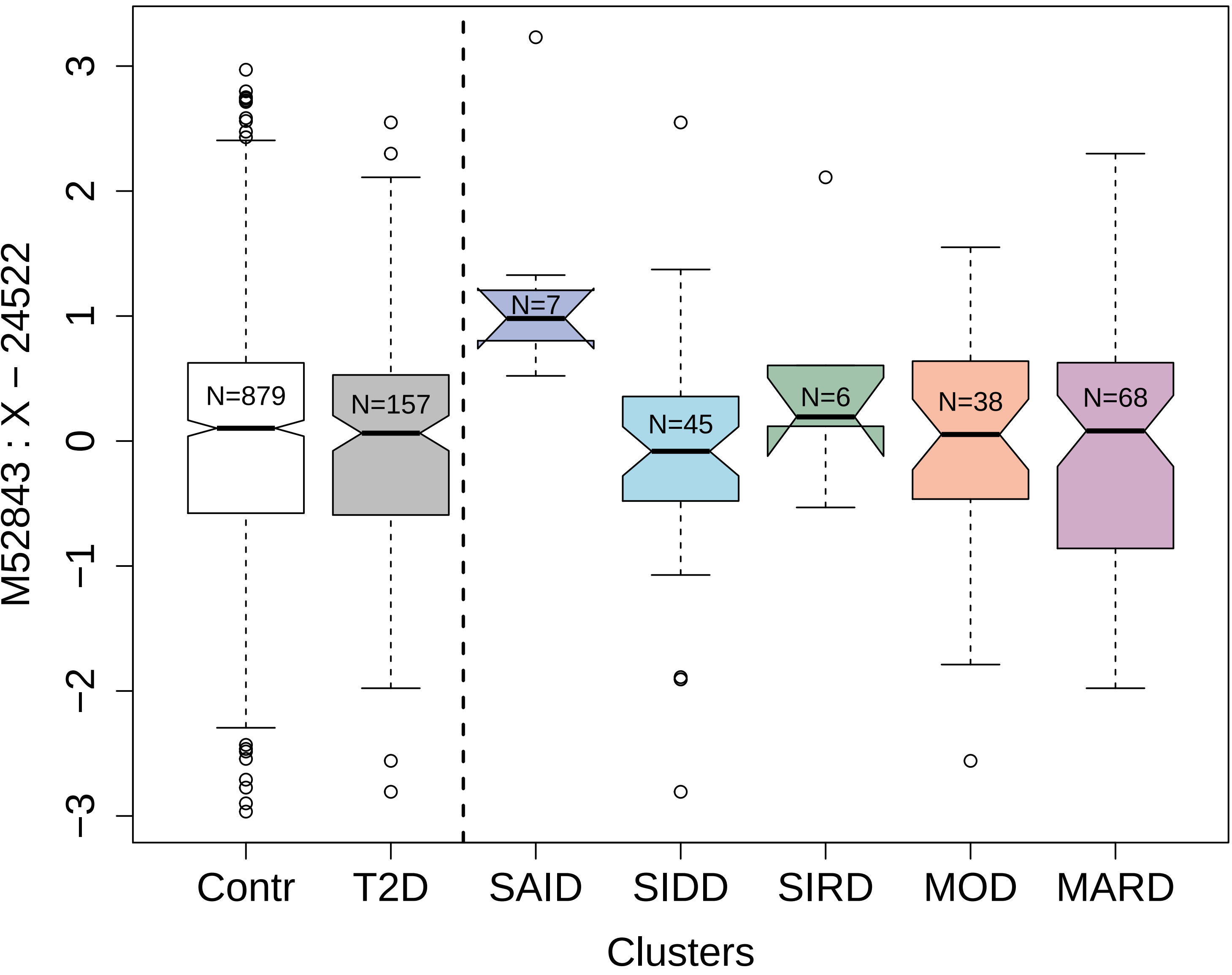

M52843 : X − 24522
3
2
1
0
−1
−2
−3
N=879
N=157
N=7
N=45
N=6
N=38
N=68
Contr
T2D
SAID
SIDD
SIRD
MOD
MARD
Clusters

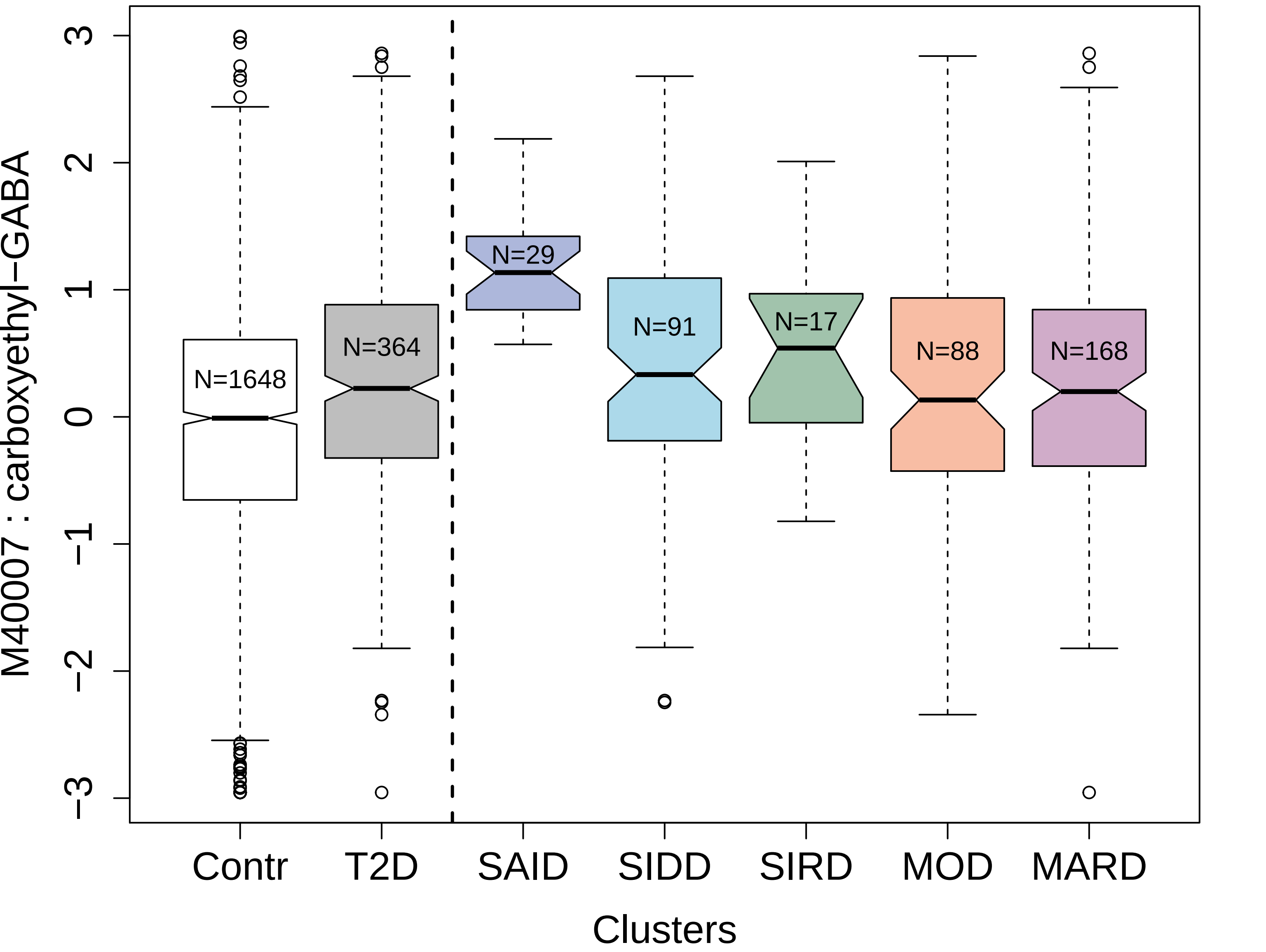
M40007 : carboxyethyl−GABA
3
2
1
0
−1
−2
−3
N=1648
N=364
N=29
N=91
N=17
N=88
N=168
Contr
T2D
SAID
SIDD
SIRD
MOD
MARD
Clusters

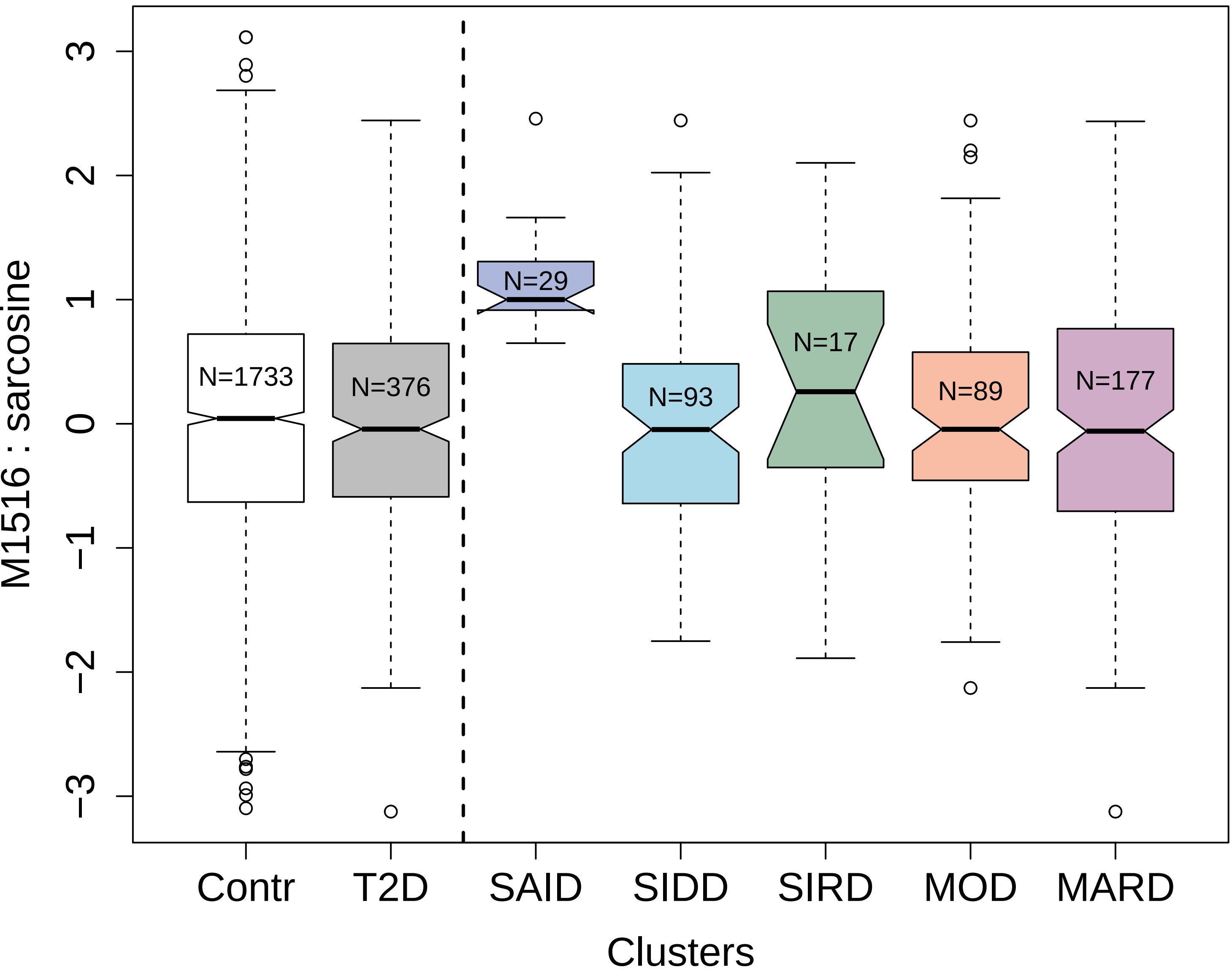

M1516 : sarcosine
3
2
1
0
−1
−2
−3
N=1733
N=376
N=29
N=93
N=17
N=89
N=177
Contr
T2D
SAID
SIDD
SIRD
MOD
MARD
Clusters

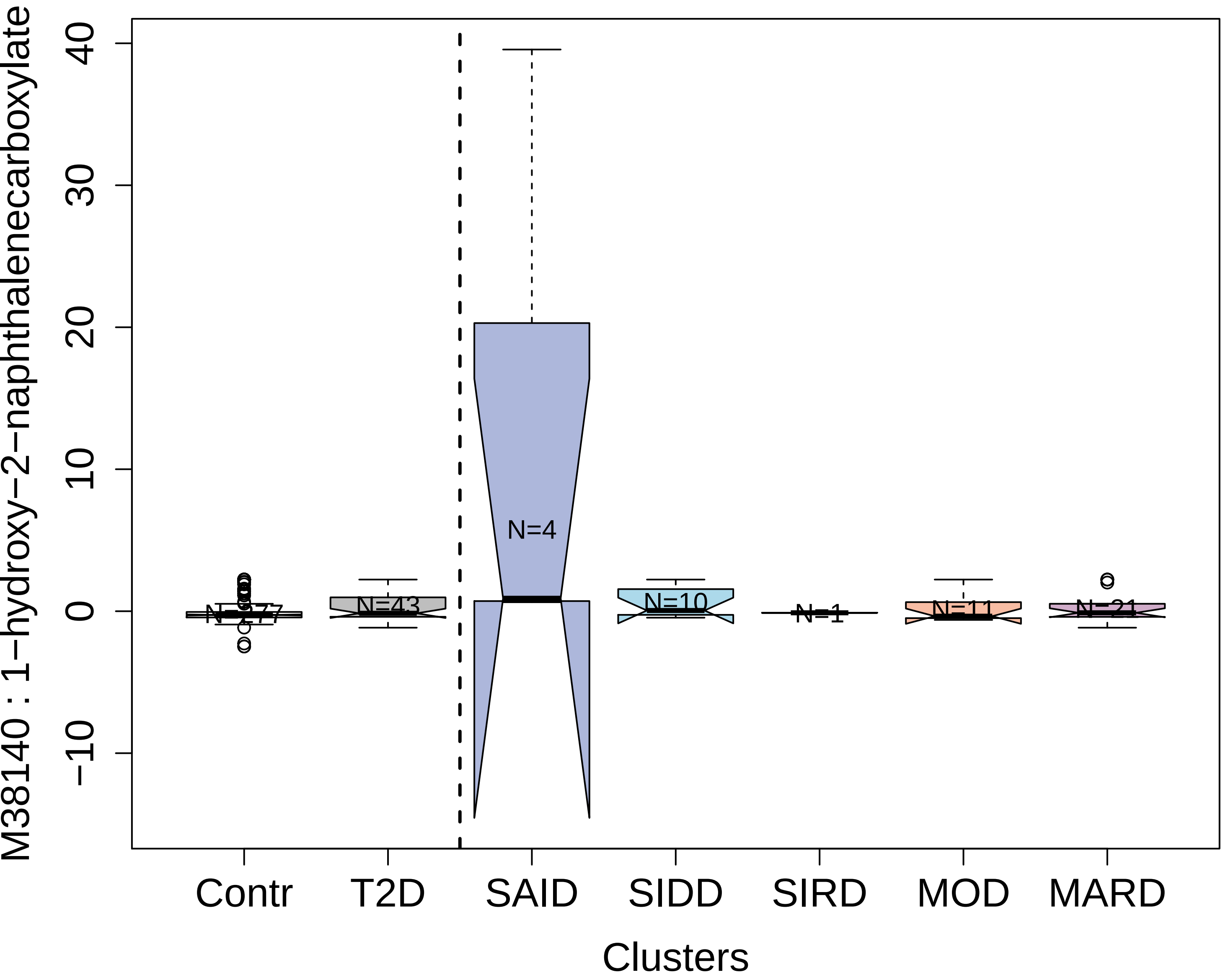
M38140 : 1−hydroxy−2−naphthalenecarboxylate
40
30
20
10
0
−10
N=177
N=43
N=4
N=10
N=1
N=11
N=21
Contr
T2D
SAID
SIDD
SIRD
MOD
MARD
Clusters

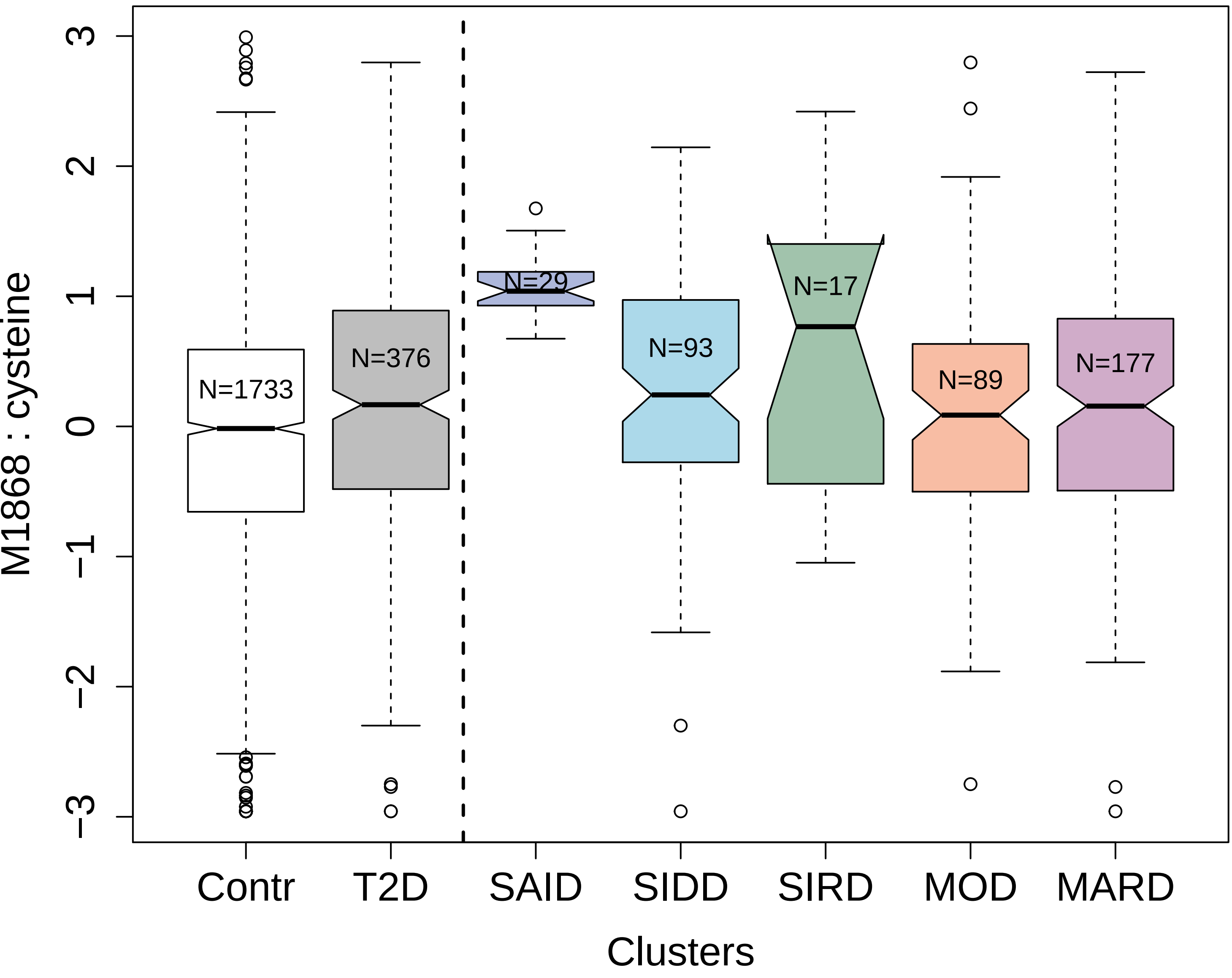

M1868 : cysteine
3
2
1
0
−1
−2
−3
N=1733
N=376
N=29
N=93
N=17
N=89
N=177
Contr
T2D
SAID
SIDD
SIRD
MOD
MARD
Clusters

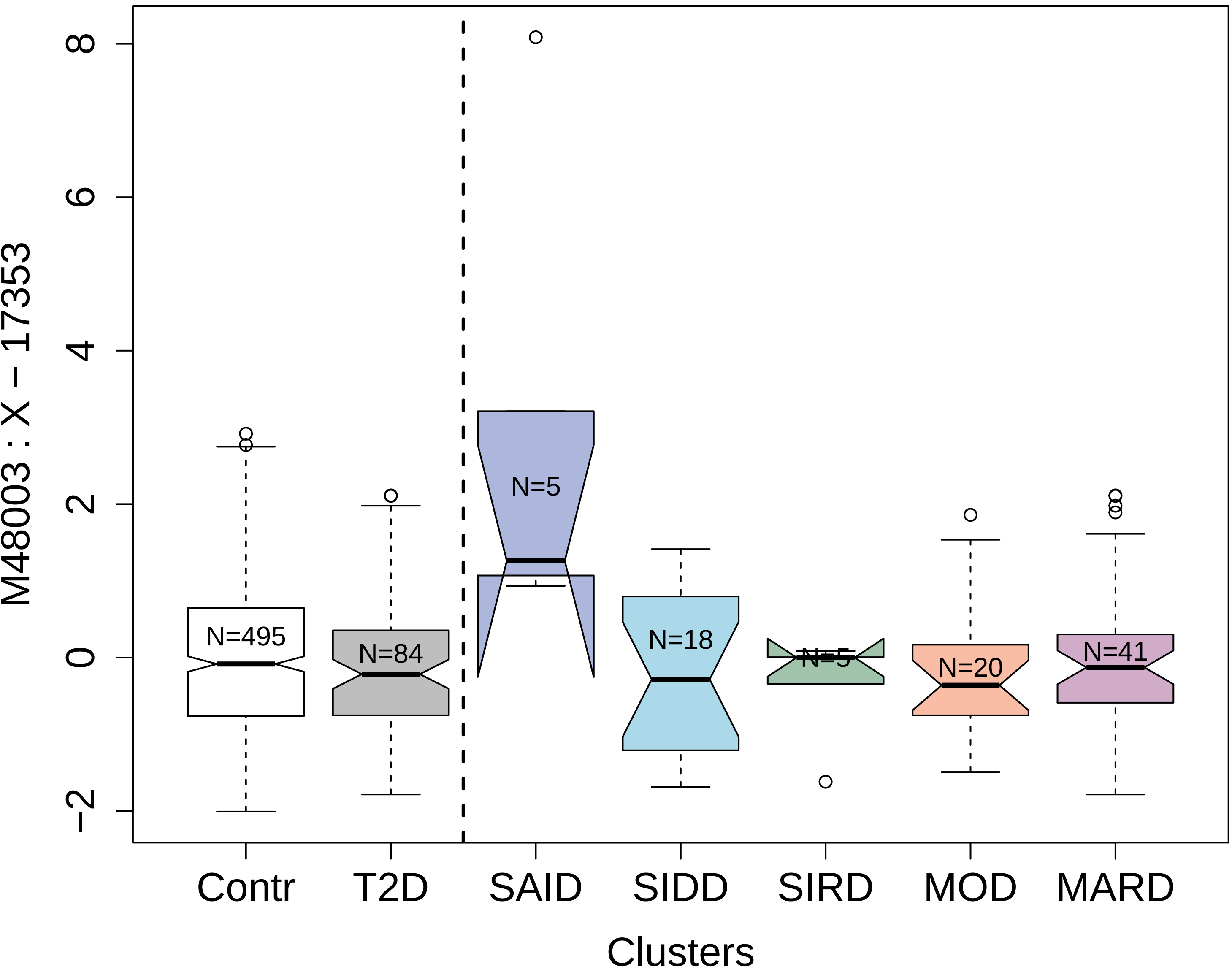
M48003 : X − 17353
8
6
4
2
0
−2
N=495
N=84
N=5
N=18
N=5
N=20
N=41
Contr
T2D
SAID
SIDD
SIRD
MOD
MARD
Clusters

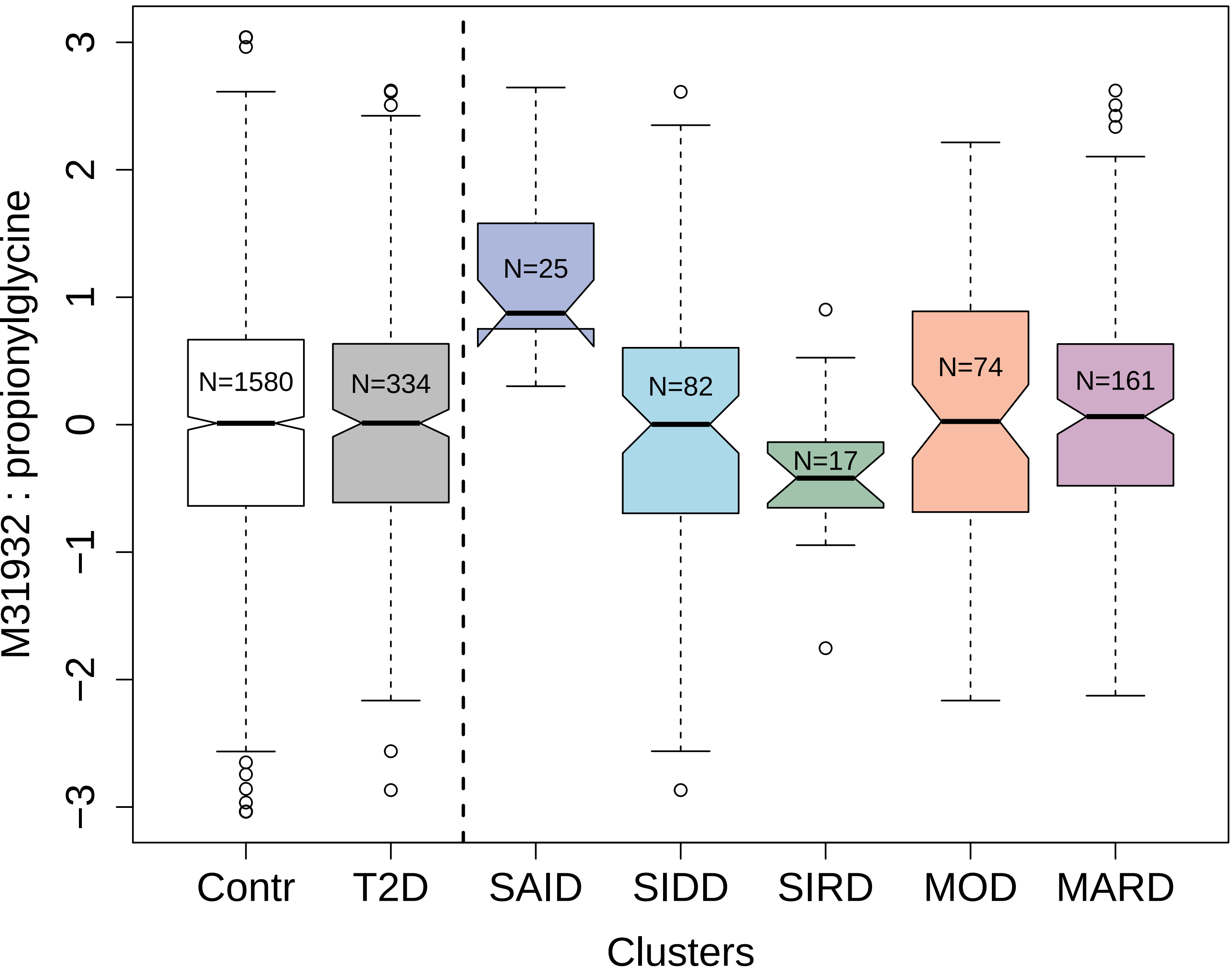

M31932 : propionylglycine
3
2
1
0
−1
−2
−3
N=1580
N=334
N=25
N=82
N=17
N=74
N=161
Contr
T2D
SAID
SIDD
SIRD
MOD
MARD
Clusters

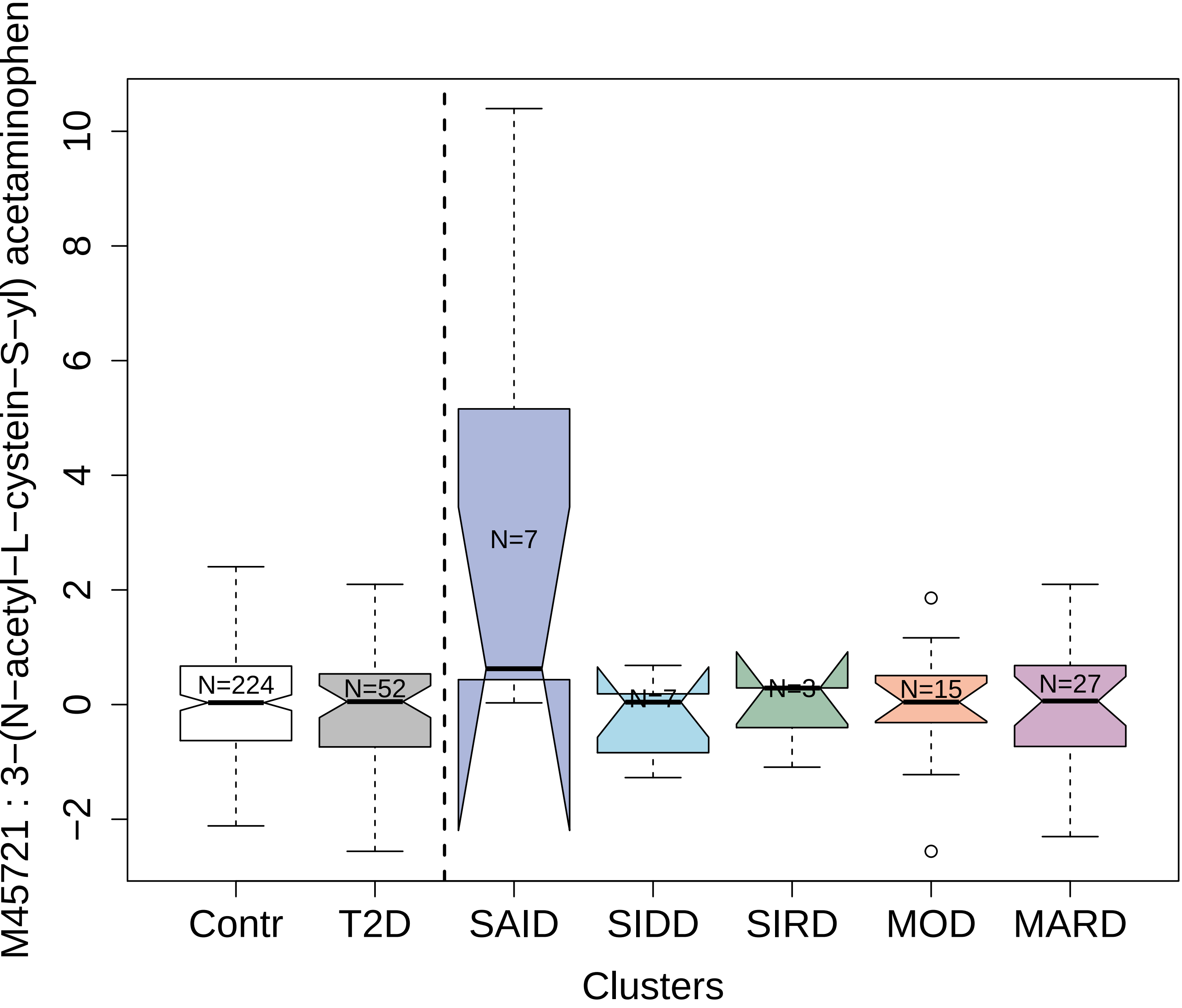
M45721 : 3−(N−acetyl−L−cystein−S−yl) acetaminophen
10
8
6
4
2
0
−2
N=224
N=52
N=7
N=7
N=3
N=15
N=27
Contr
T2D
SAID
SIDD
SIRD
MOD
MARD
Clusters

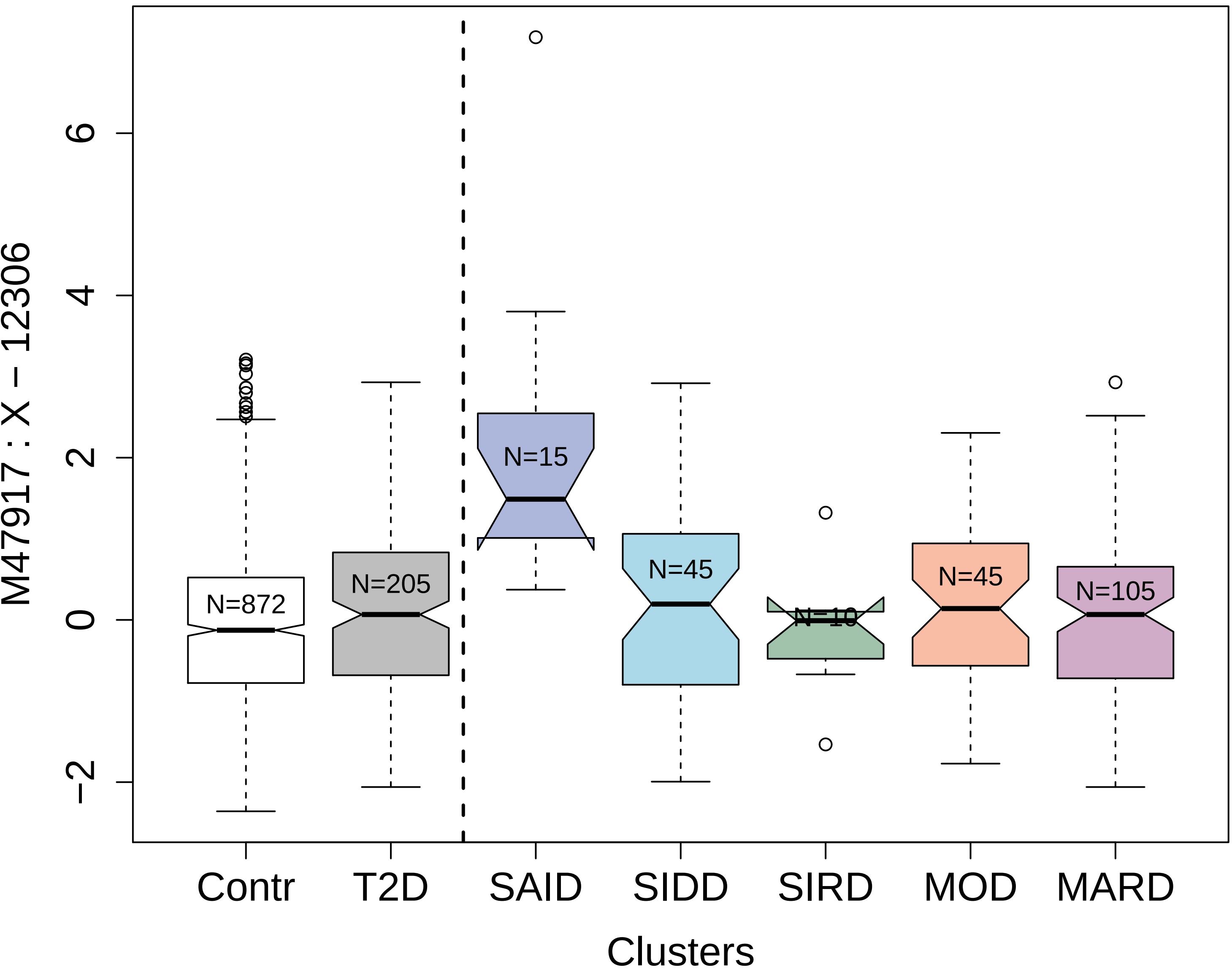

M47917 : X − 12306
6
4
2
0
−2
N=872
N=205
N=15
N=45
N=10
N=45
N=105
Contr
T2D
SAID
SIDD
SIRD
MOD
MARD
Clusters

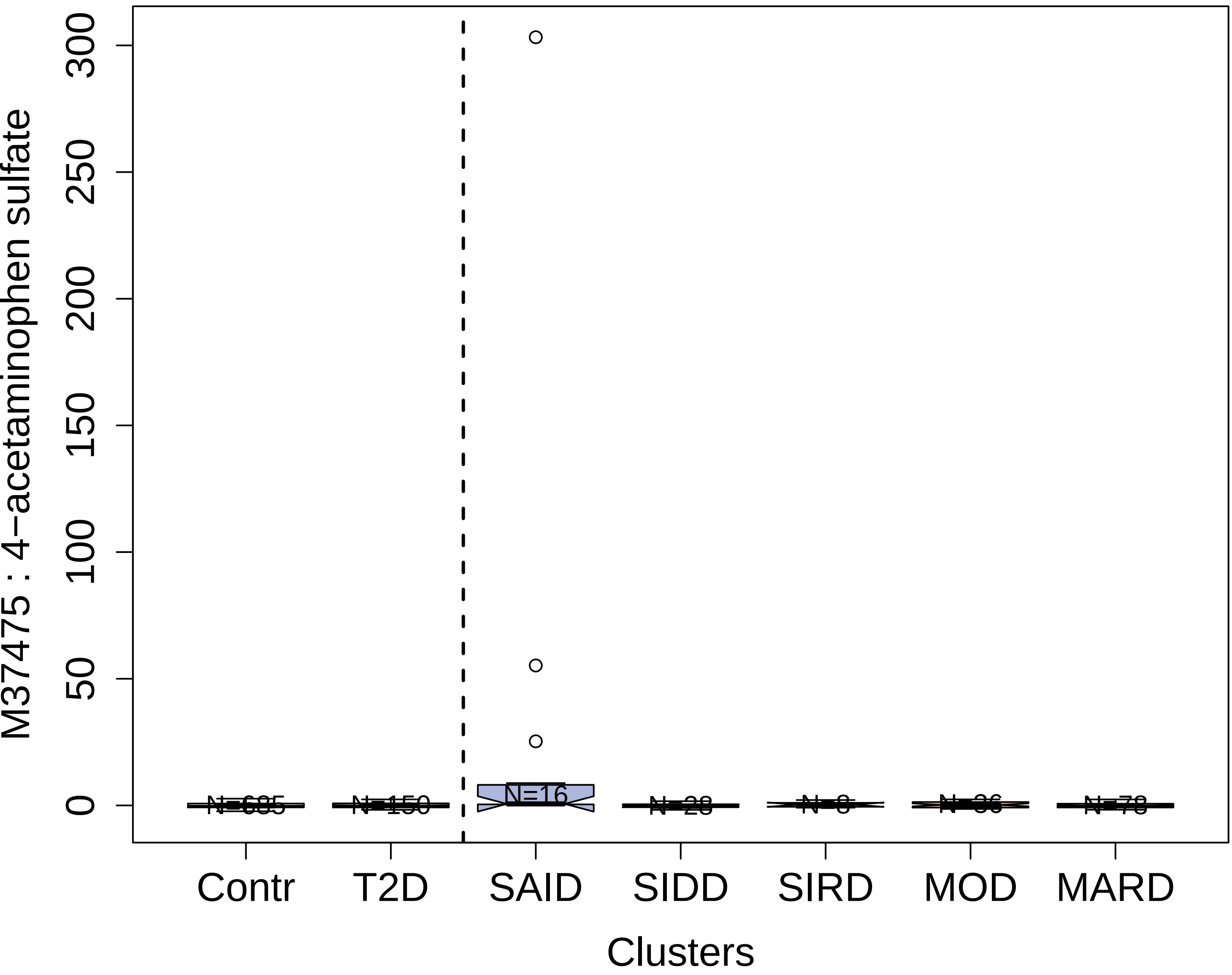
M37475 : 4−acetaminophen sulfate
Clusters
Contr
T2D
SAID
SIDD
SIRD
MOD
MARD
N=16
0
50
100
150
200
250
300

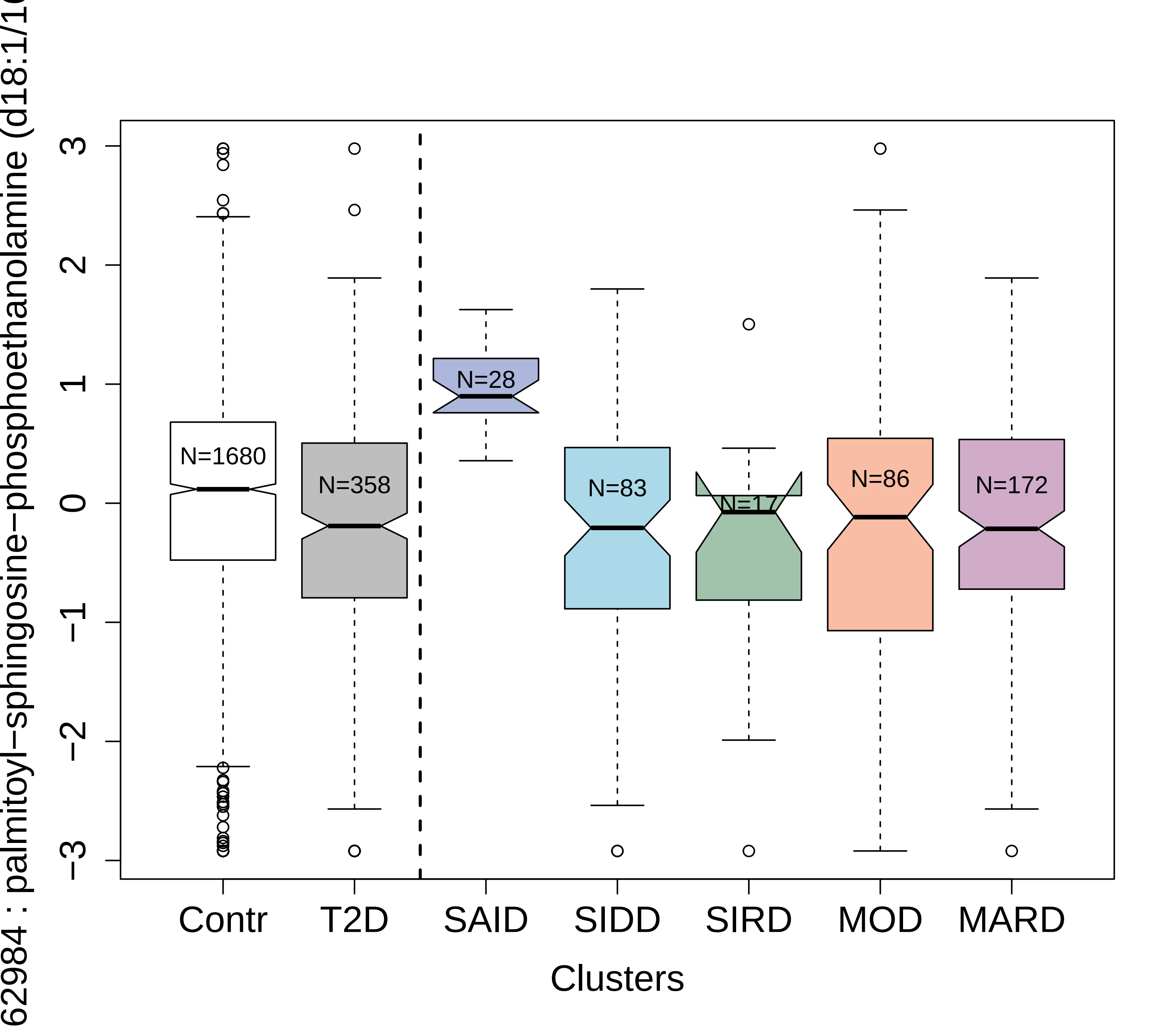

62984 : palmitoyl−sphingosine−phosphoethanolamine (d18:1/1
3
2
1
0
−1
−2
−3
N=1680
N=358
N=28
N=83
N=17
N=86
N=172
Contr
T2D
SAID
SIDD
SIRD
MOD
MARD
Clusters

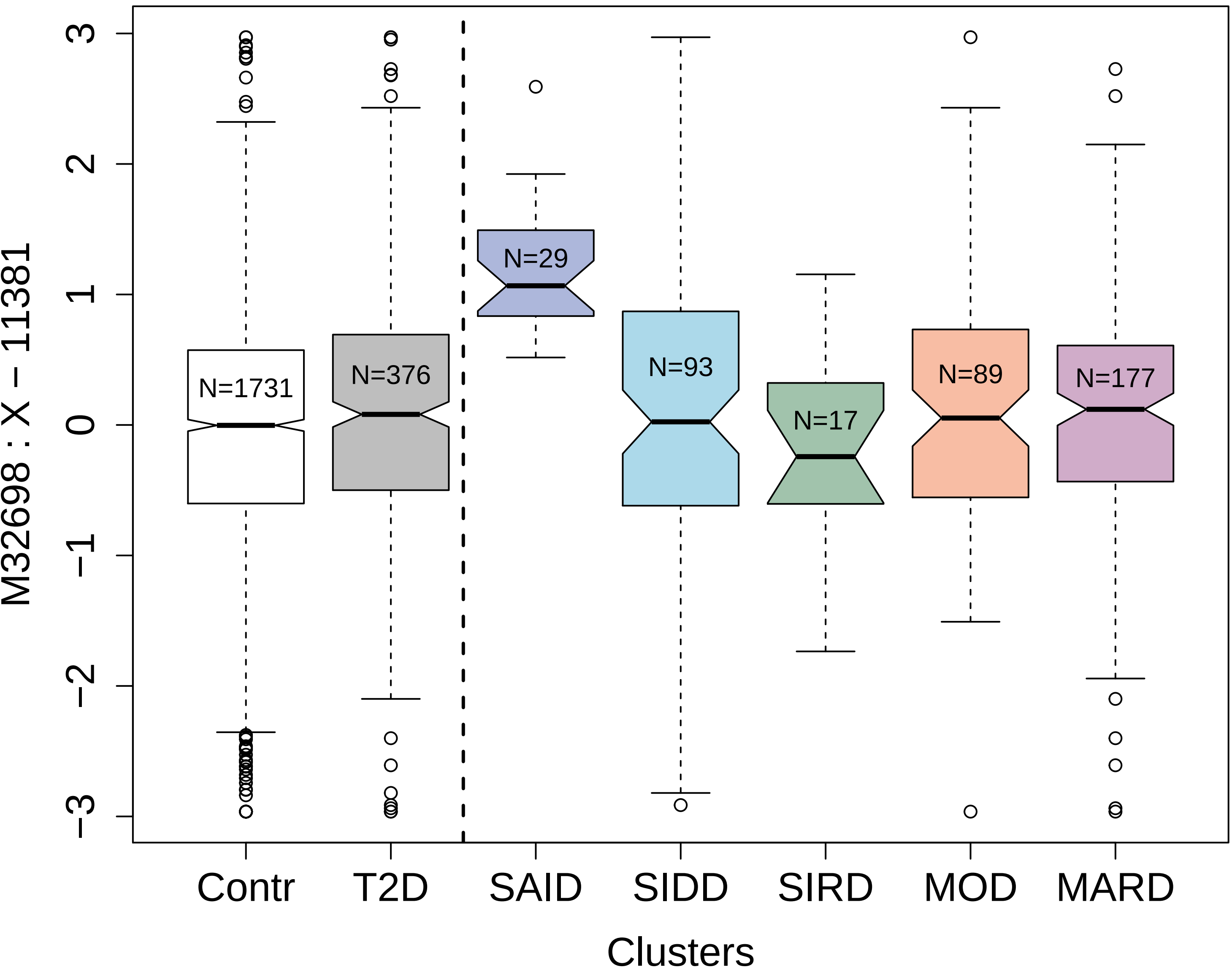

M32698 : X − 11381
3
2
1
0
−1
−2
−3
N=1731
N=376
N=29
N=93
N=17
N=89
N=177
Contr
T2D
SAID
SIDD
SIRD
MOD
MARD
Clusters

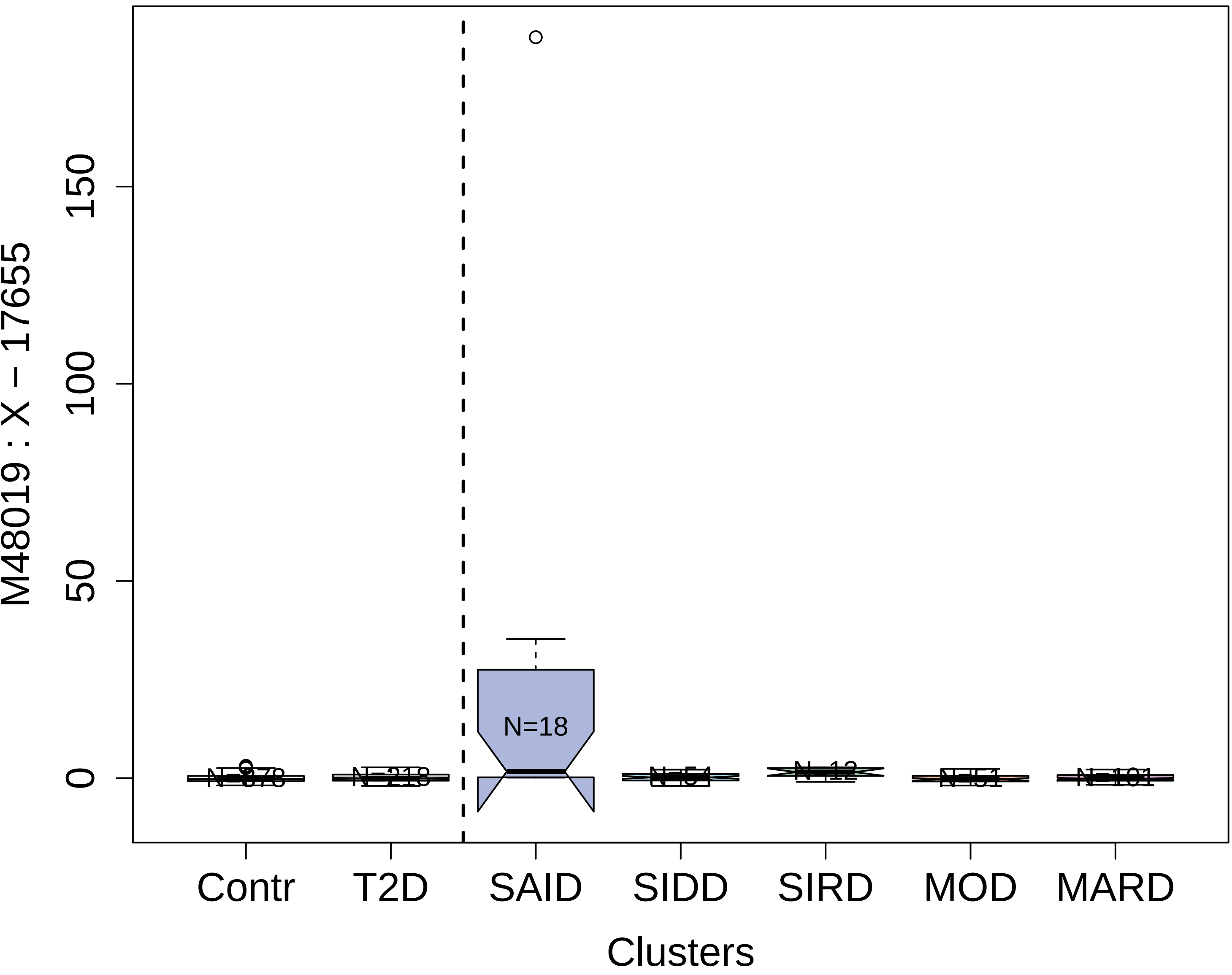

M48019 : X − 17655
0
50
100
150
N=18
Contr
T2D
SAID
SIDD
SIRD
MOD
MARD
Clusters

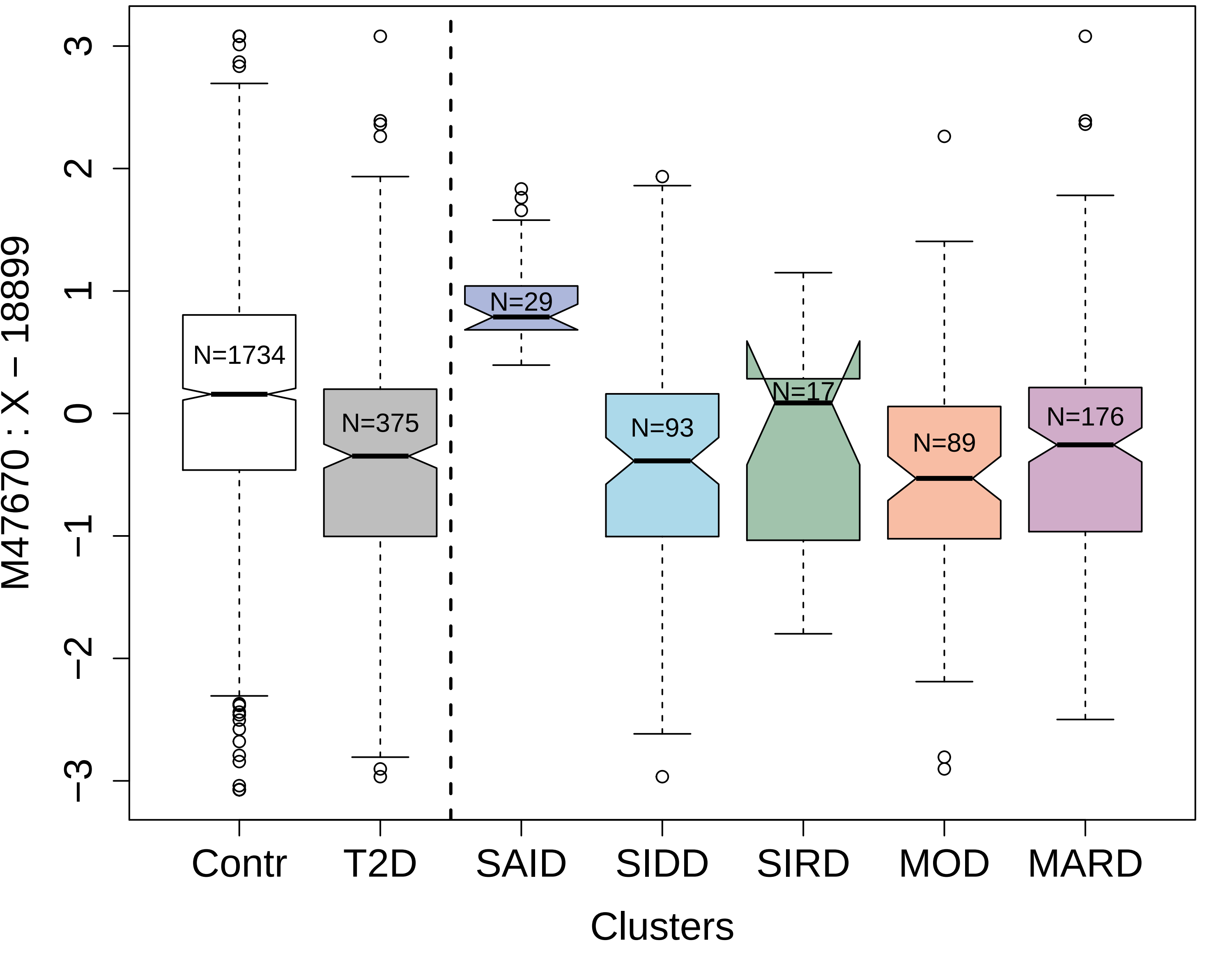

M47670 : X − 18899
Clusters
Contr
T2D
SAID
SIDD
SIRD
MOD
MARD
N=1734
N=375
N=29
N=93
N=17
N=89
N=176
3
2
1
0
−1
−2
−3

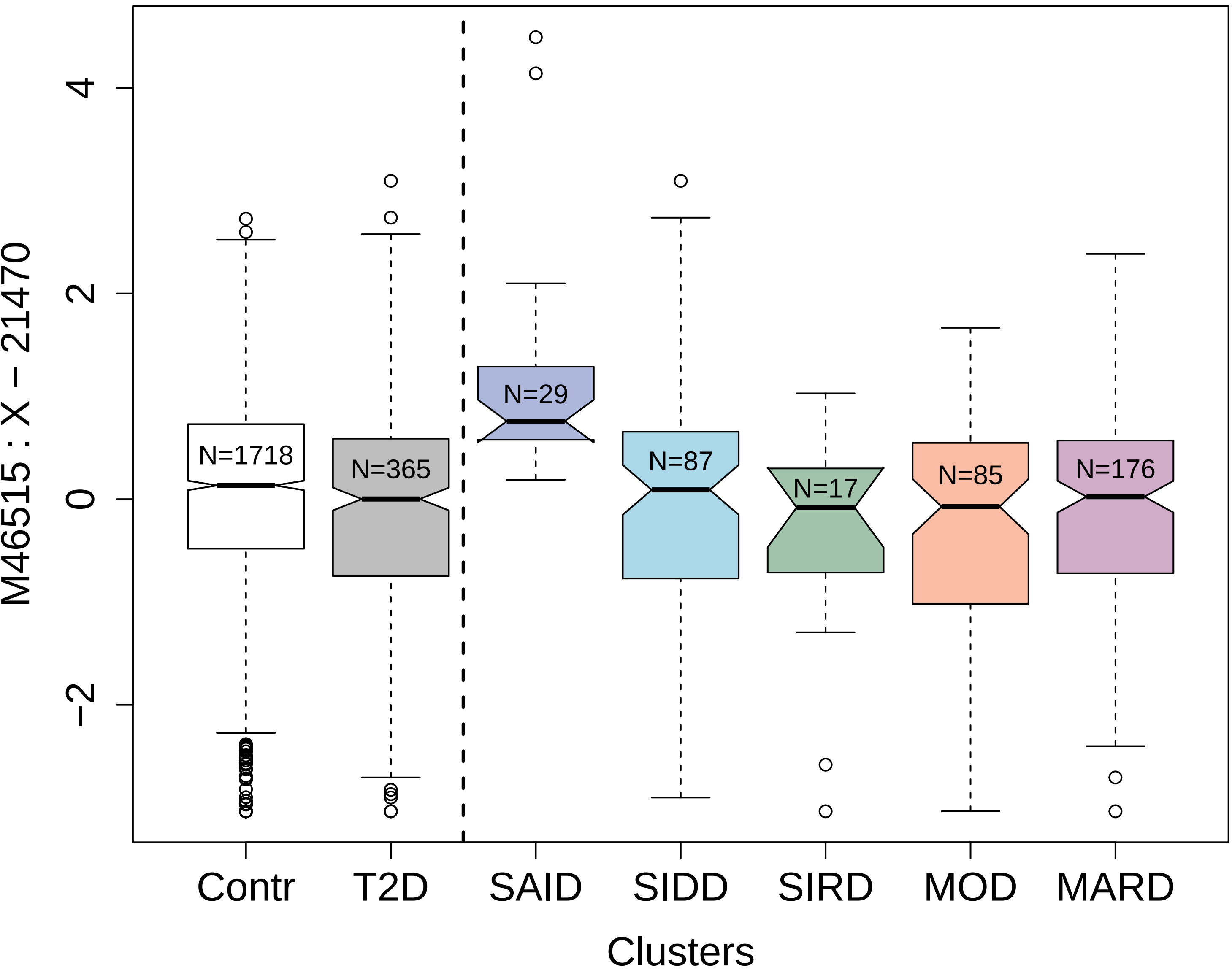

M46515 : X − 21470
4
2
0
−2
N=1718
N=365
N=29
N=87
N=17
N=85
N=176
Contr
T2D
SAID
SIDD
SIRD
MOD
MARD
Clusters

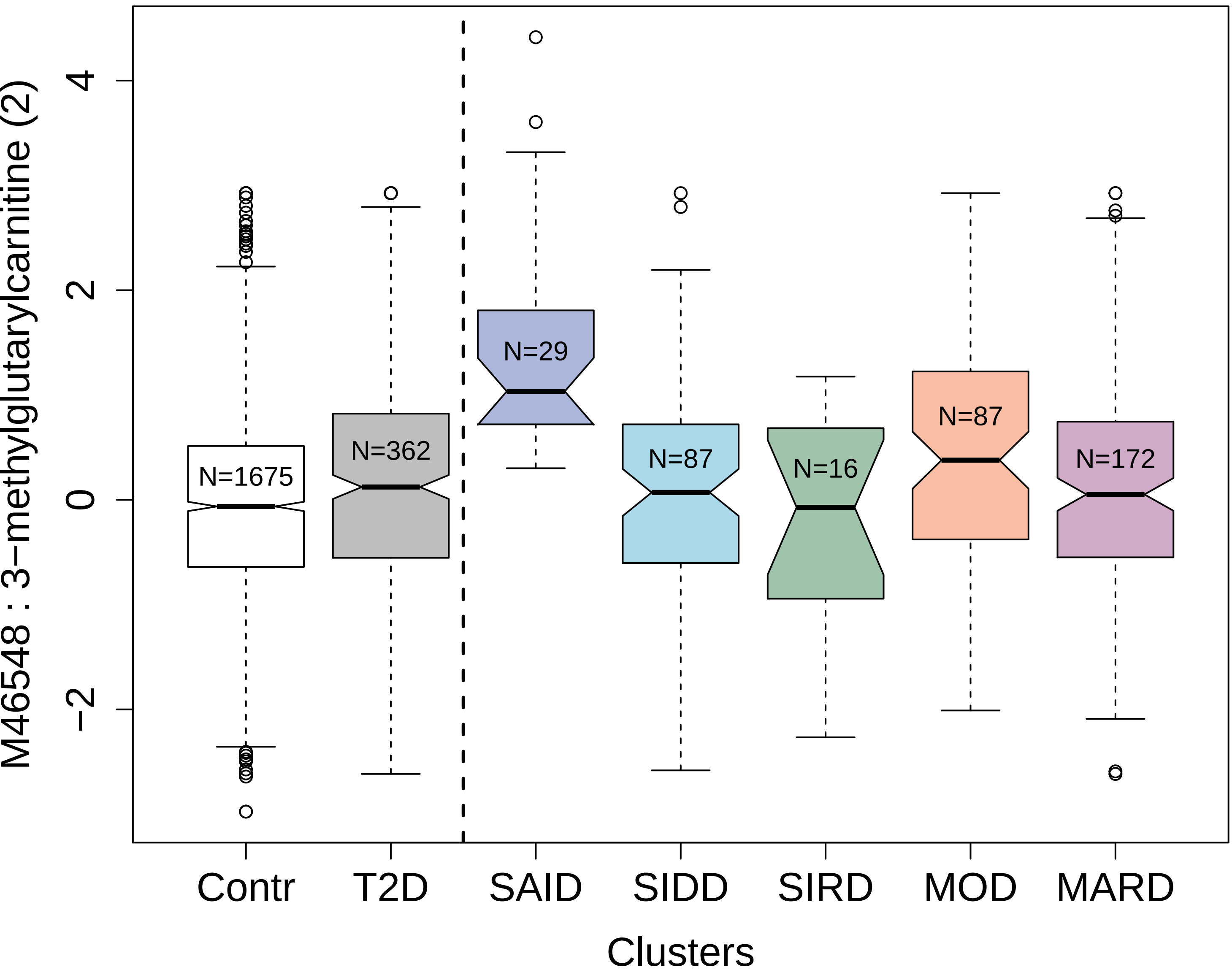

M46548 : 3−methylglutarylcarnitine (2)
4
2
0
−2
N=1675
N=362
N=29
N=87
N=16
N=87
N=172
Contr
T2D
SAID
SIDD
SIRD
MOD
MARD
Clusters

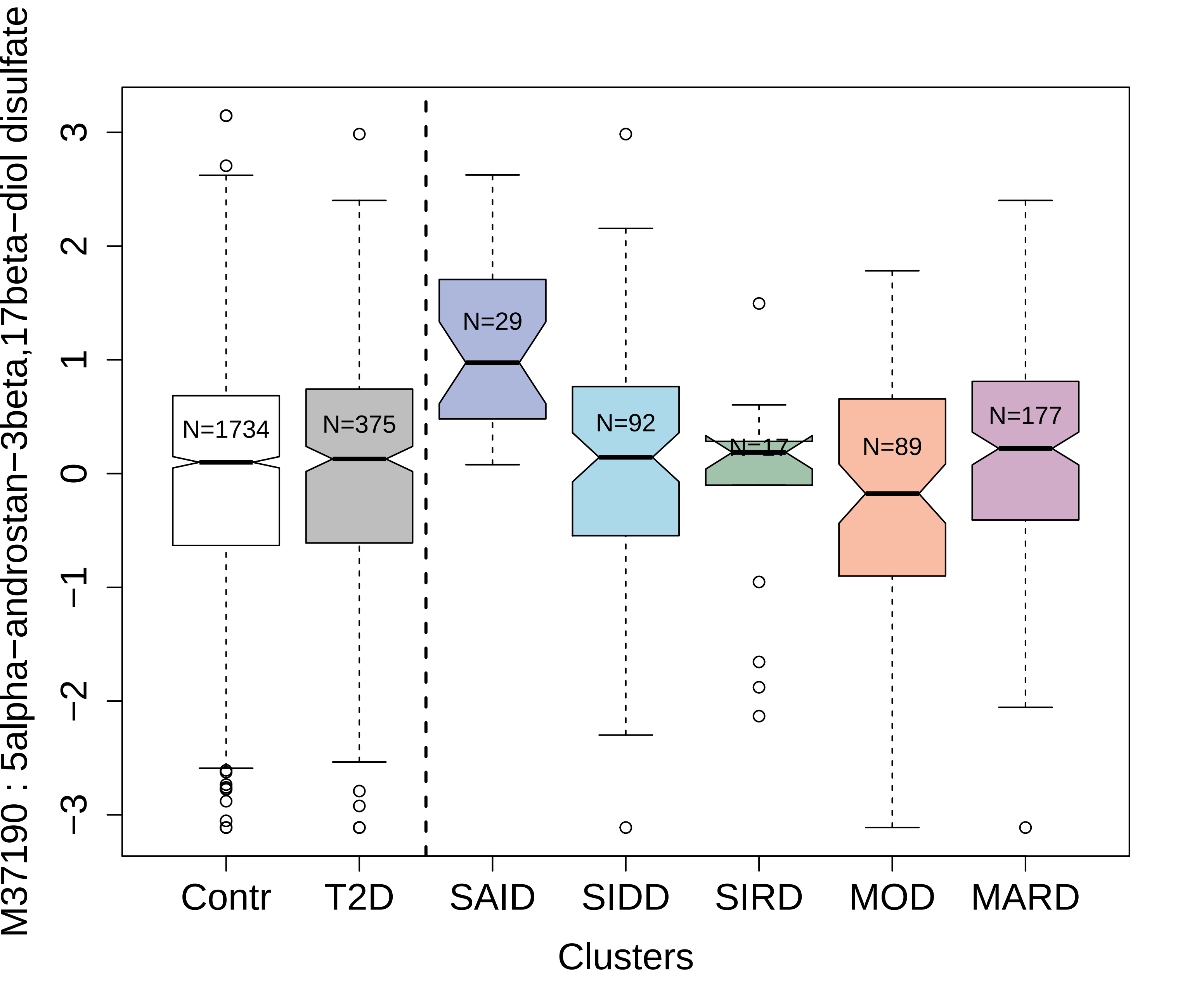

M37190 : 5alpha−androstan−3beta,17beta−diol disulfate
3
2
1
0
−1
−2
−3
N=1734
N=375
N=29
N=92
N=17
N=89
N=177
Contr
T2D
SAID
SIDD
SIRD
MOD
MARD
Clusters

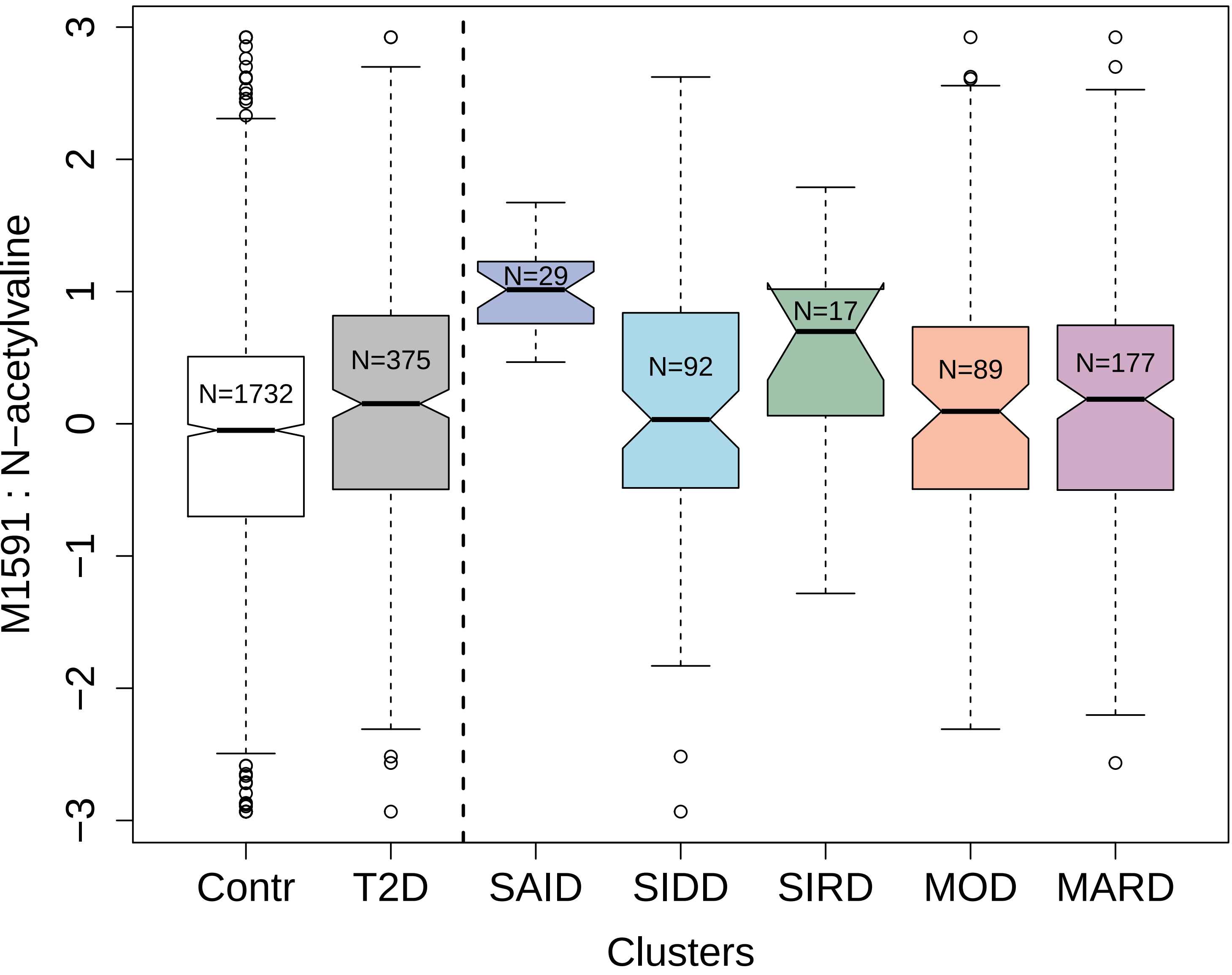
M1591 : N−acetylvaline
3
2
1
0
−1
−2
−3
N=1732
N=375
N=29
N=92
N=17
N=89
N=177
Contr
T2D
SAID
SIDD
SIRD
MOD
MARD
Clusters

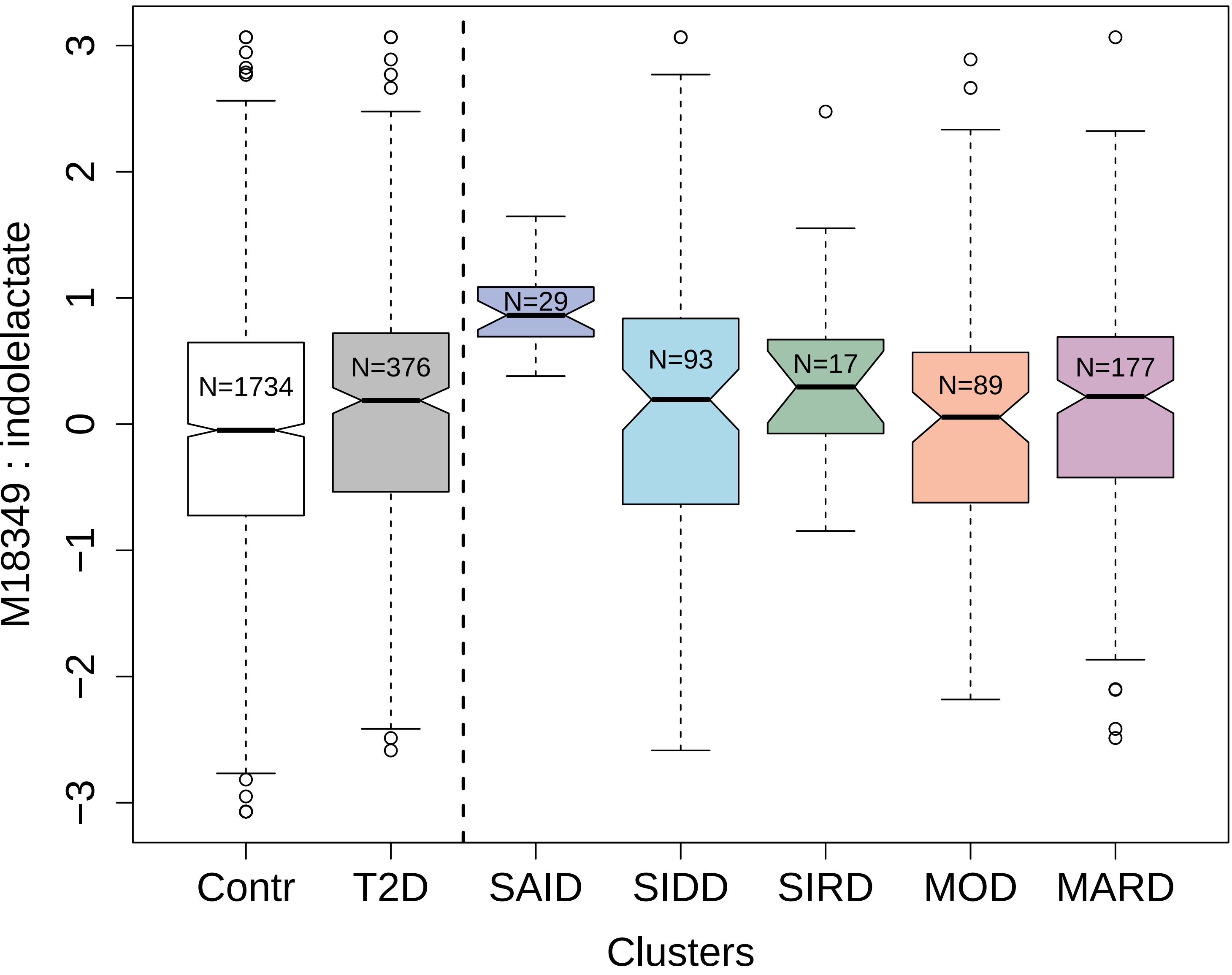
M18349 : indolelactate
3
2
1
0
−1
−2
−3
N=1734
N=376
N=29
N=93
N=17
N=89
N=177
Contr
T2D
SAID
SIDD
SIRD
MOD
MARD
Clusters

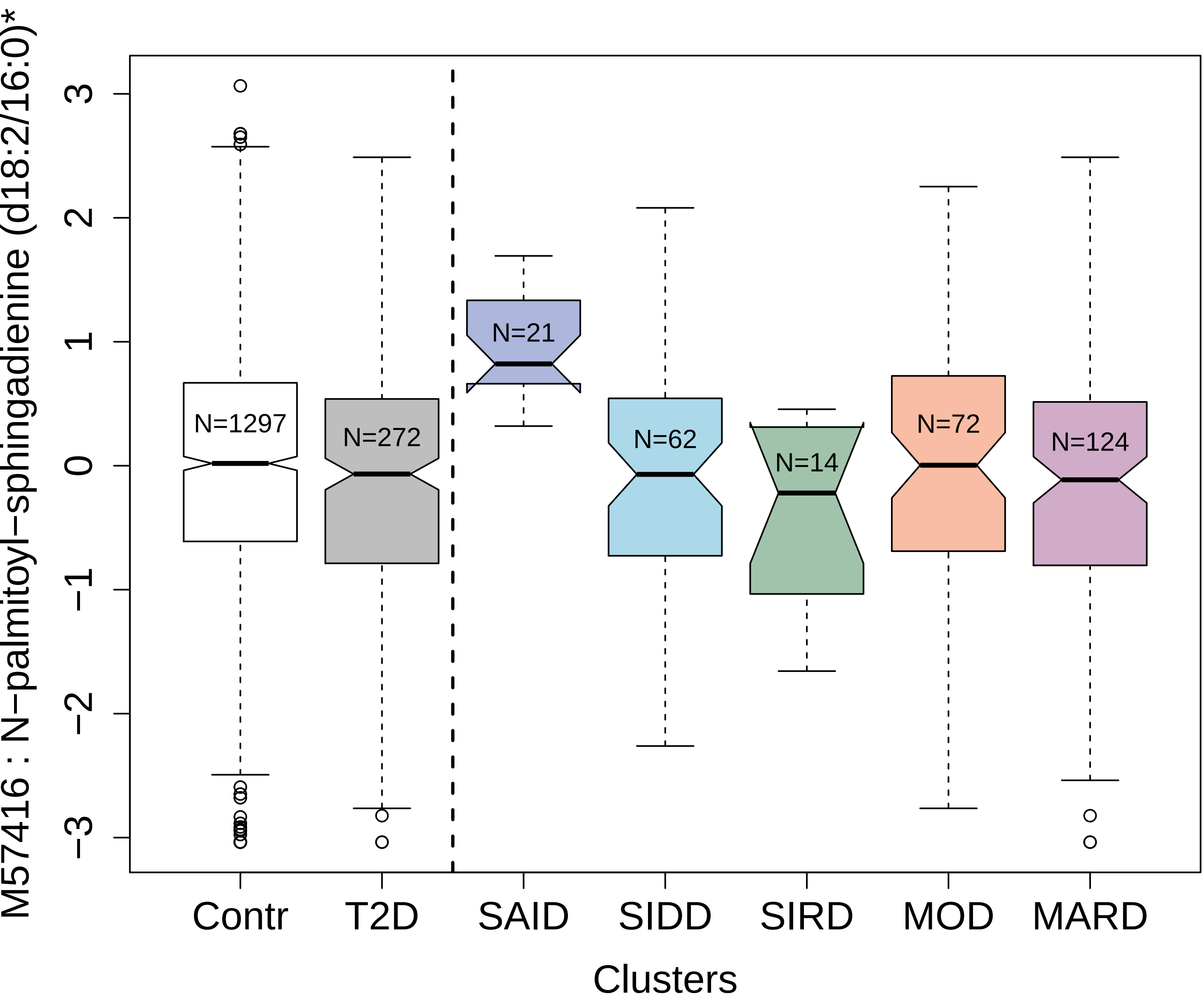

M57416 : N−palmitoyl−sphingadienine (d18:2/16:0)*
3
2
1
0
−1
−2
−3
N=1297
N=272
N=21
N=62
N=14
N=72
N=124
Contr
T2D
SAID
SIDD
SIRD
MOD
MARD
Clusters

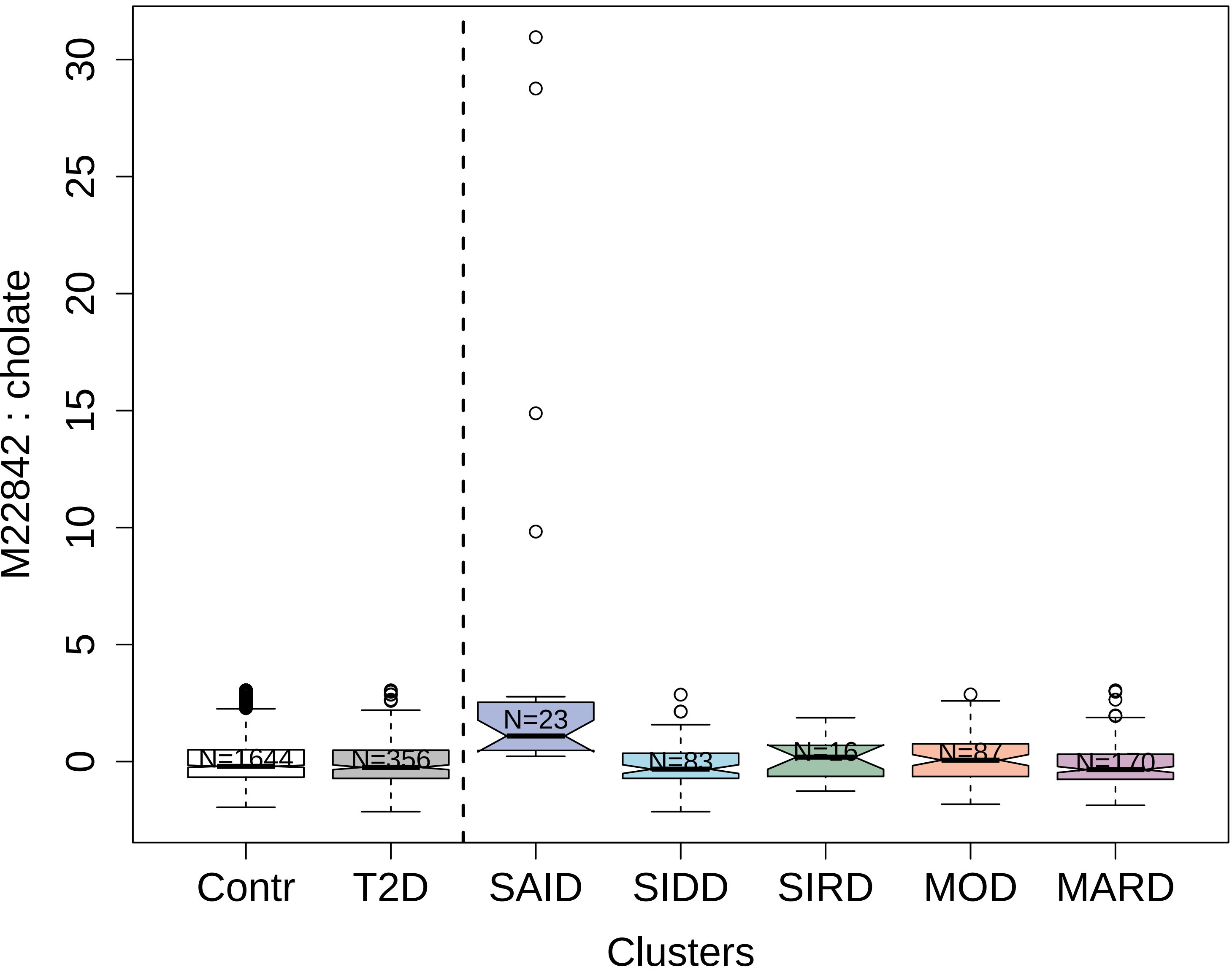
M22842 : cholate
0
5
10
15
20
25
30
N=1644
N=356
N=23
N=83
N=16
N=87
N=170
Contr
T2D
SAID
SIDD
SIRD
MOD
MARD
Clusters

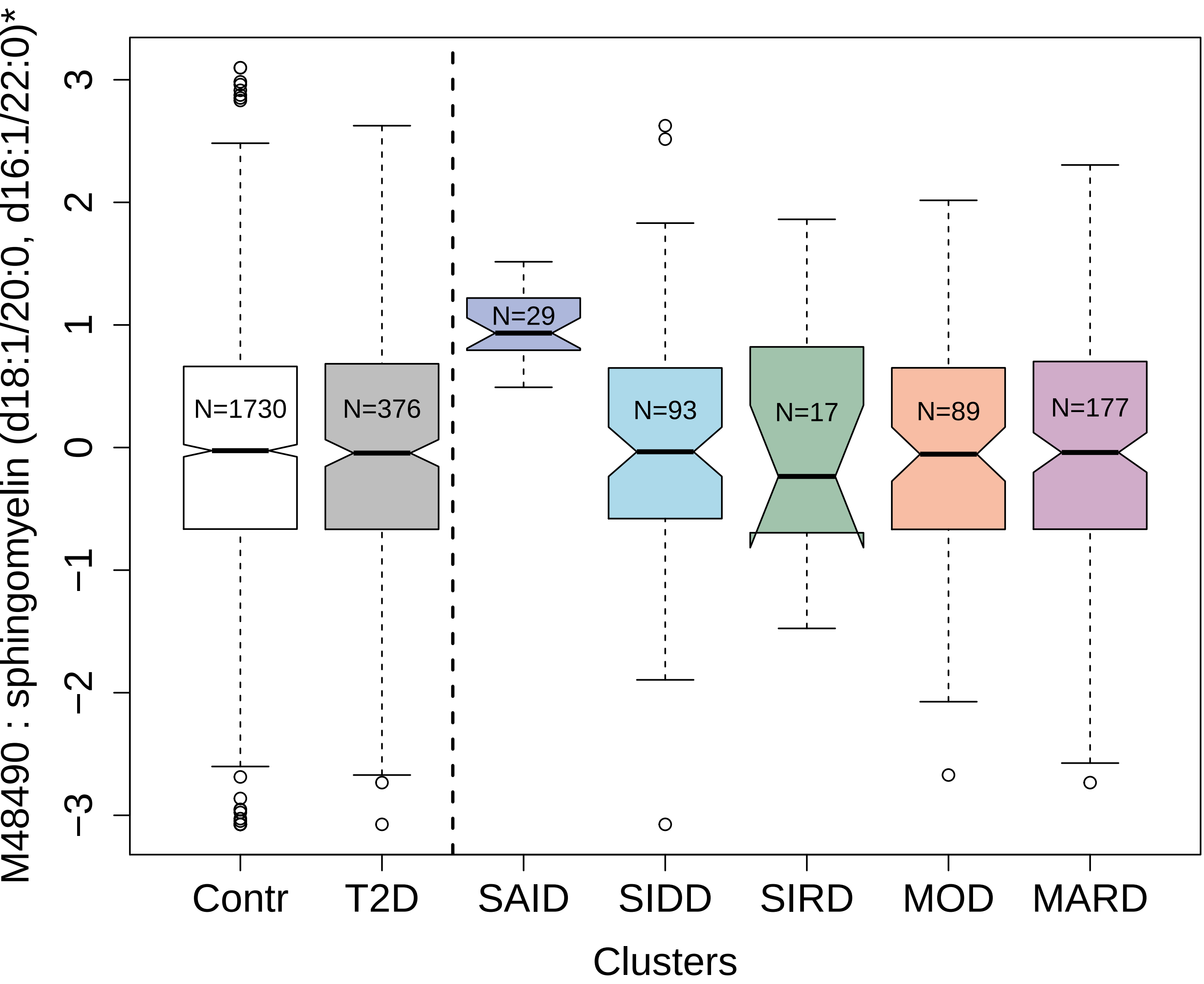
M48490 : sphingomyelin (d18:1/20:0, d16:1/22:0)*
3
2
1
0
−1
−2
−3
N=1730
N=376
N=29
N=93
N=17
N=89
N=177
Contr
T2D
SAID
SIDD
SIRD
MOD
MARD
Clusters

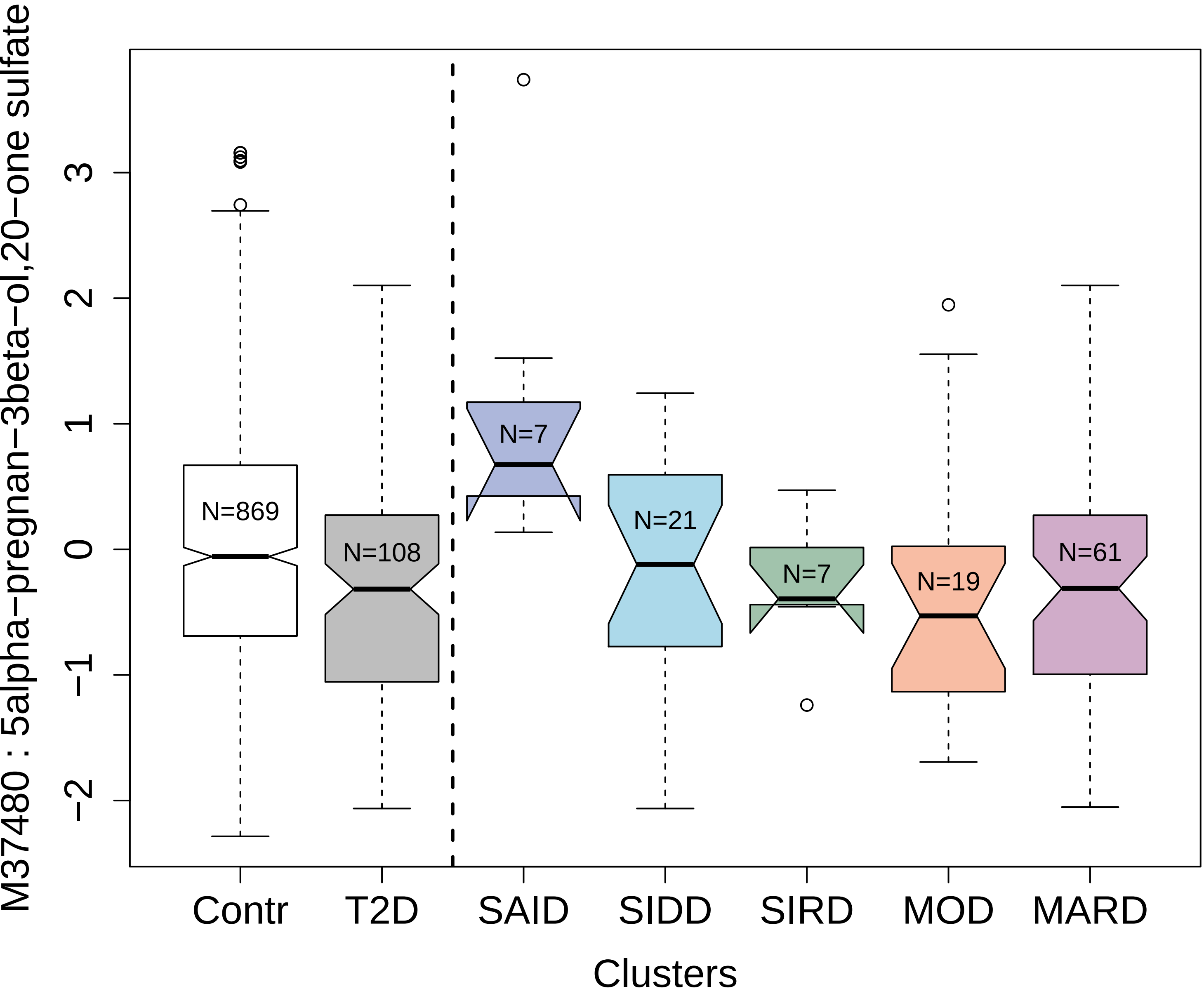

M37480 : 5alpha−pregnan−3beta−ol,20−one sulfate
3
2
1
0
−1
−2
N=869
N=108
N=7
N=21
N=7
N=19
N=61
Contr
T2D
SAID
SIDD
SIRD
MOD
MARD
Clusters

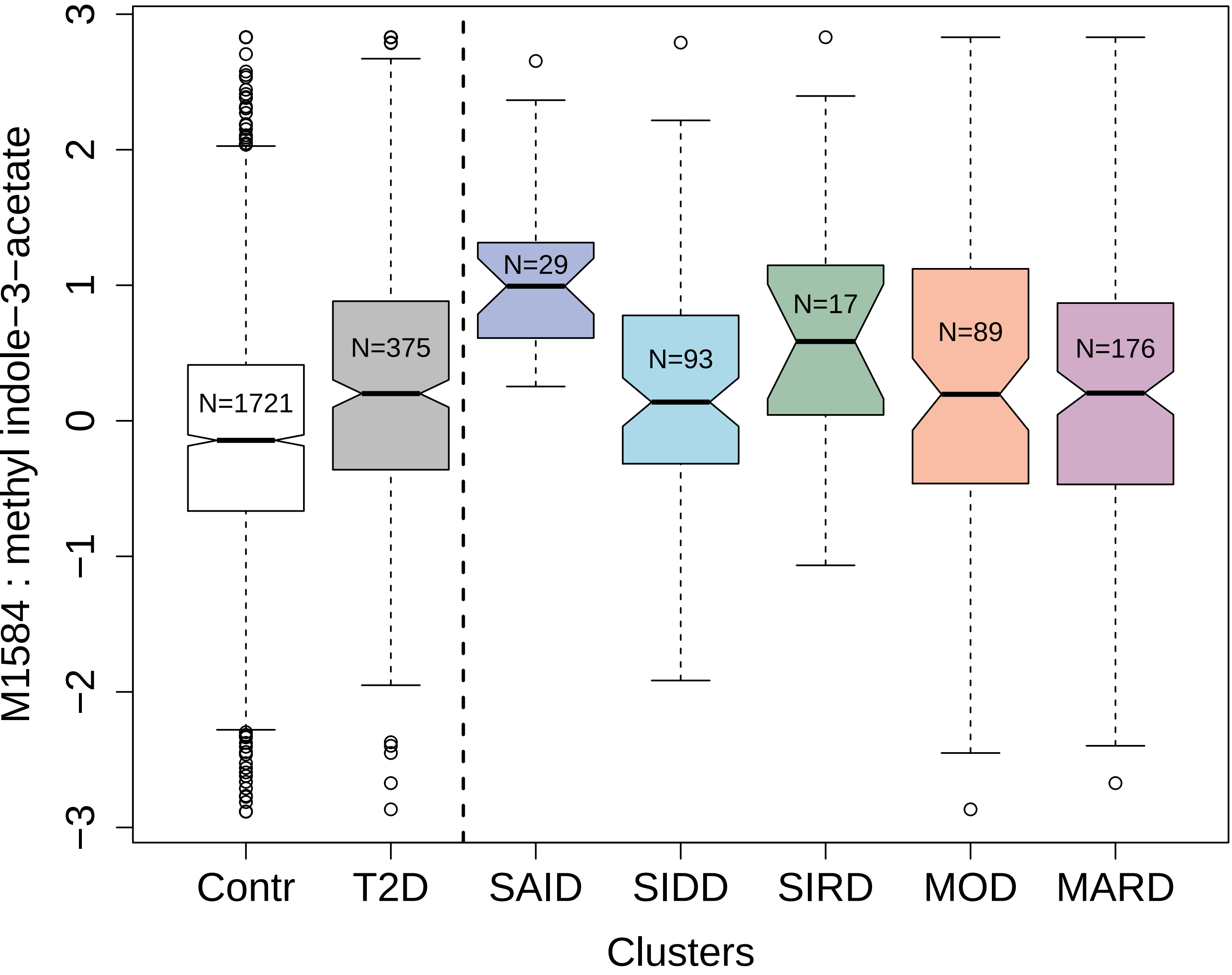

M1584 : methyl indole−3−acetate
3
2
1
0
−1
−2
−3
N=1721
N=375
N=29
N=93
N=17
N=89
N=176
Contr
T2D
SAID
SIDD
SIRD
MOD
MARD
Clusters

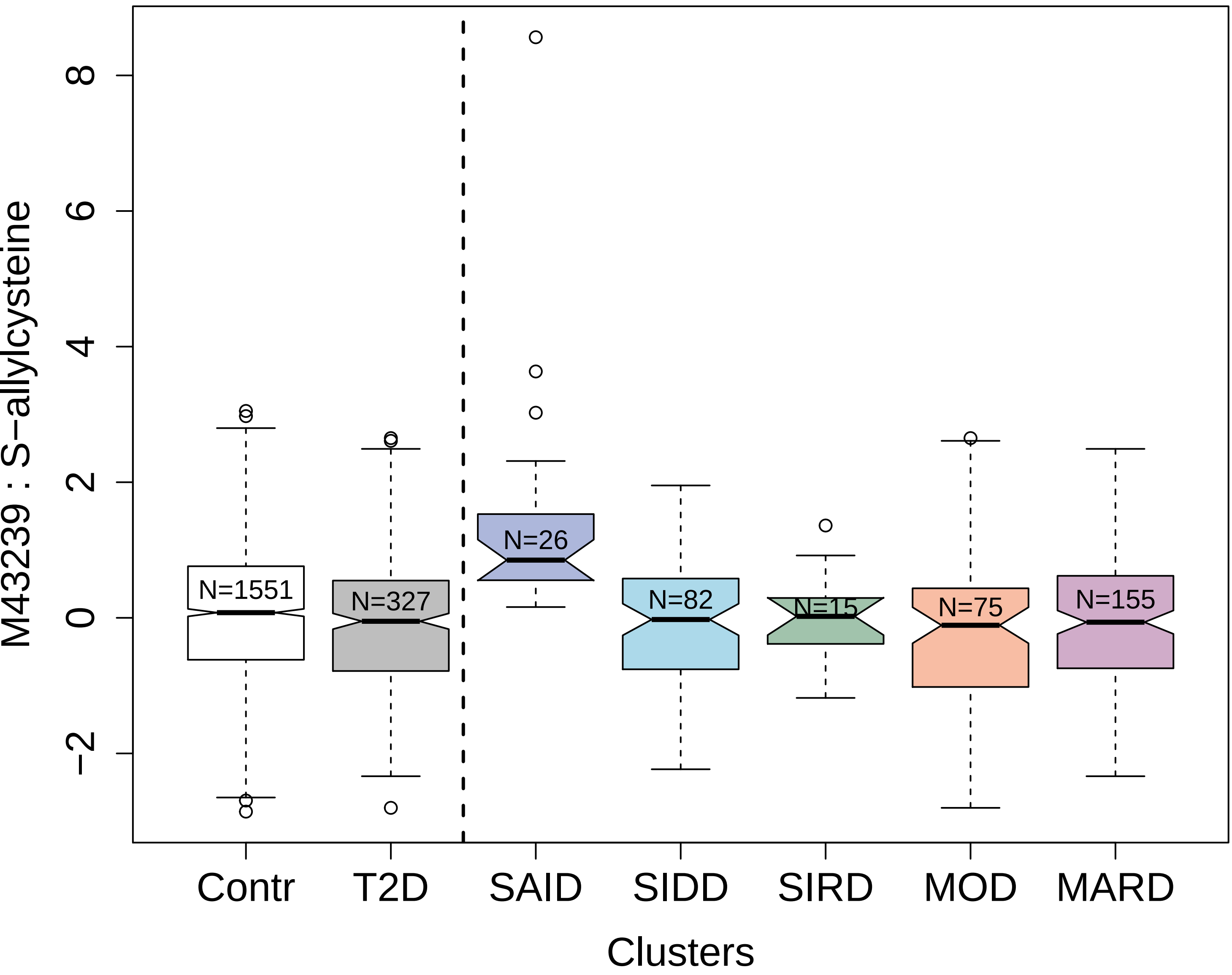
M43239 : S−allylcysteine
−2
0
2
4
6
8
N=1551
N=327
N=26
N=82
N=15
N=75
N=155
Contr
T2D
SAID
SIDD
SIRD
MOD
MARD
Clusters

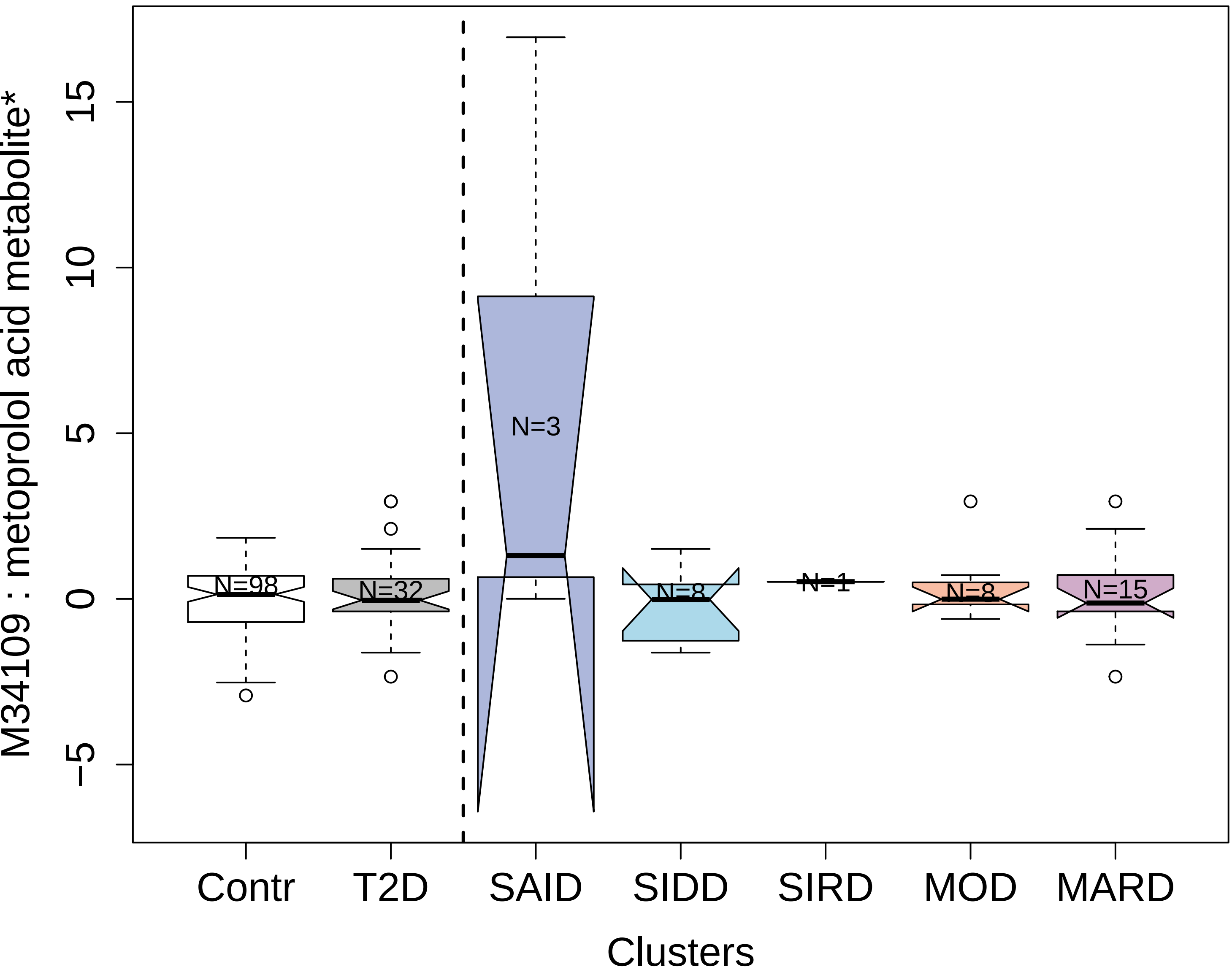

M34109 : metoprolol acid metabolite*
−5
0
5
10
15
N=98
N=32
N=3
N=8
N=1
N=8
N=15
Contr
T2D
SAID
SIDD
SIRD
MOD
MARD
Clusters

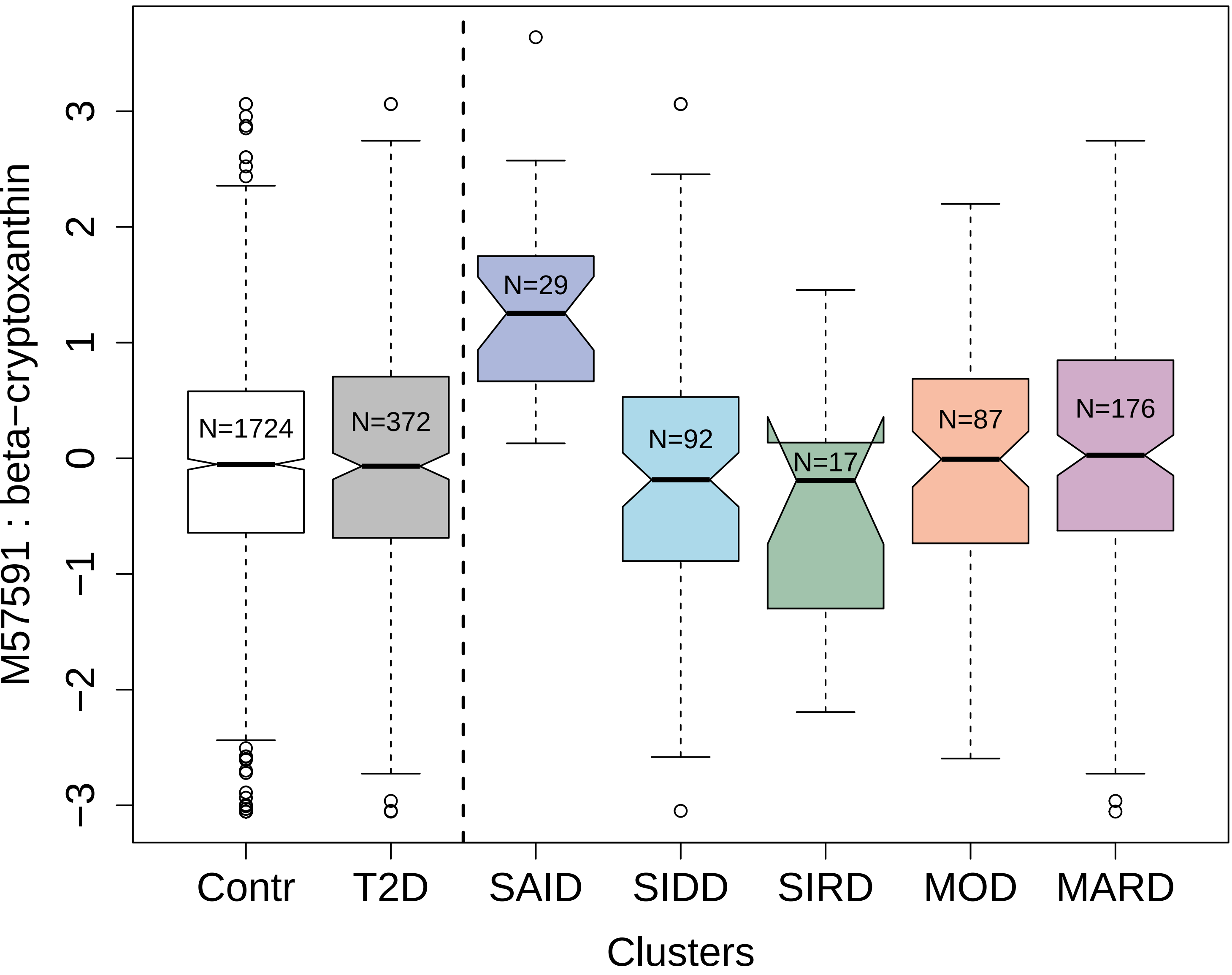

M57591 : beta−cryptoxanthin
3
2
1
0
−1
−2
−3
N=1724
N=372
N=29
N=92
N=17
N=87
N=176
Contr
T2D
SAID
SIDD
SIRD
MOD
MARD
Clusters

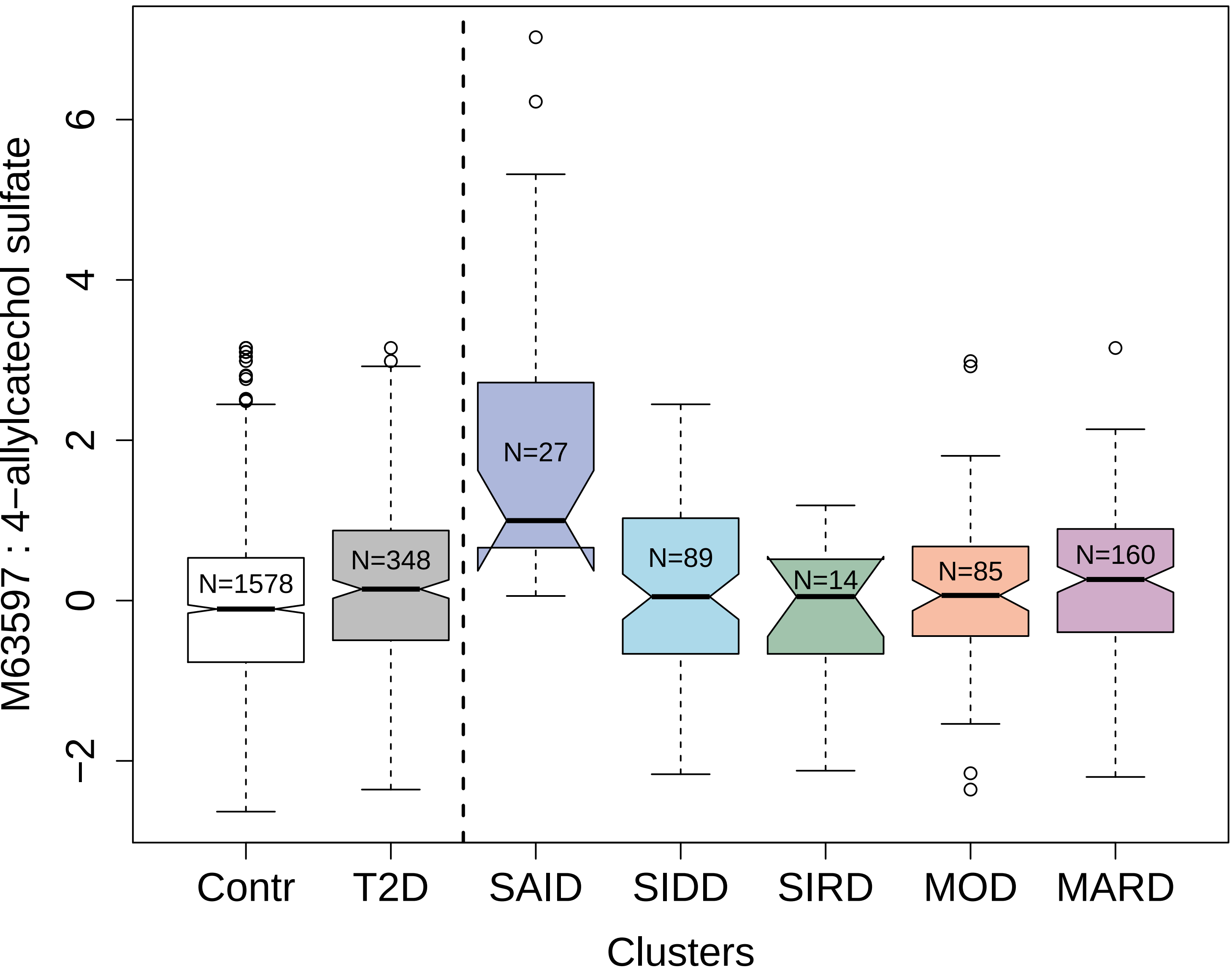
M63597 : 4−allylcatechol sulfate
6
4
2
0
−2
N=1578
N=348
N=27
N=89
N=14
N=85
N=160
Contr
T2D
SAID
SIDD
SIRD
MOD
MARD
Clusters

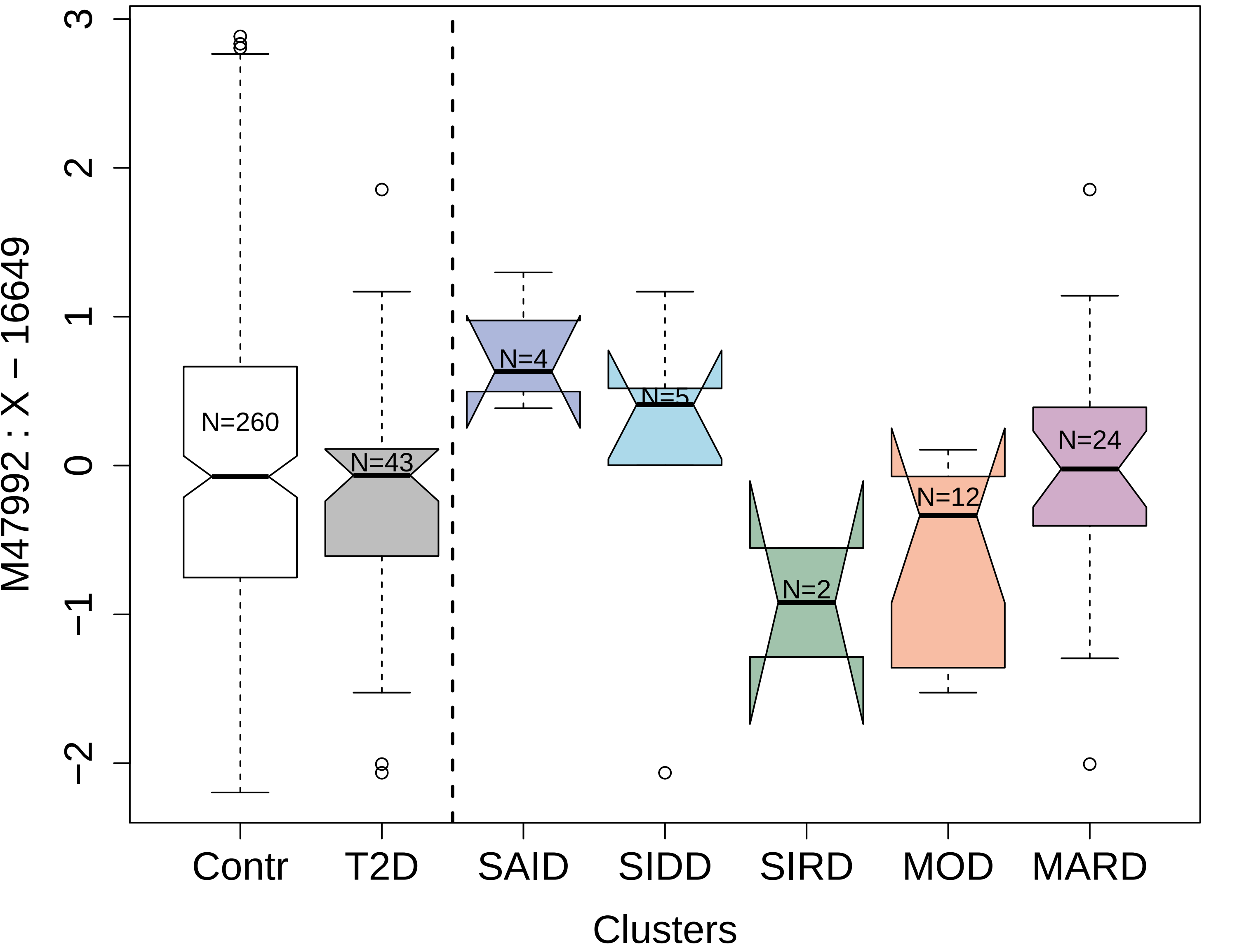

M47992 : X − 16649
3
2
1
0
−1
−2
N=260
N=43
N=4
N=5
N=2
N=12
N=24
Contr
T2D
SAID
SIDD
SIRD
MOD
MARD
Clusters

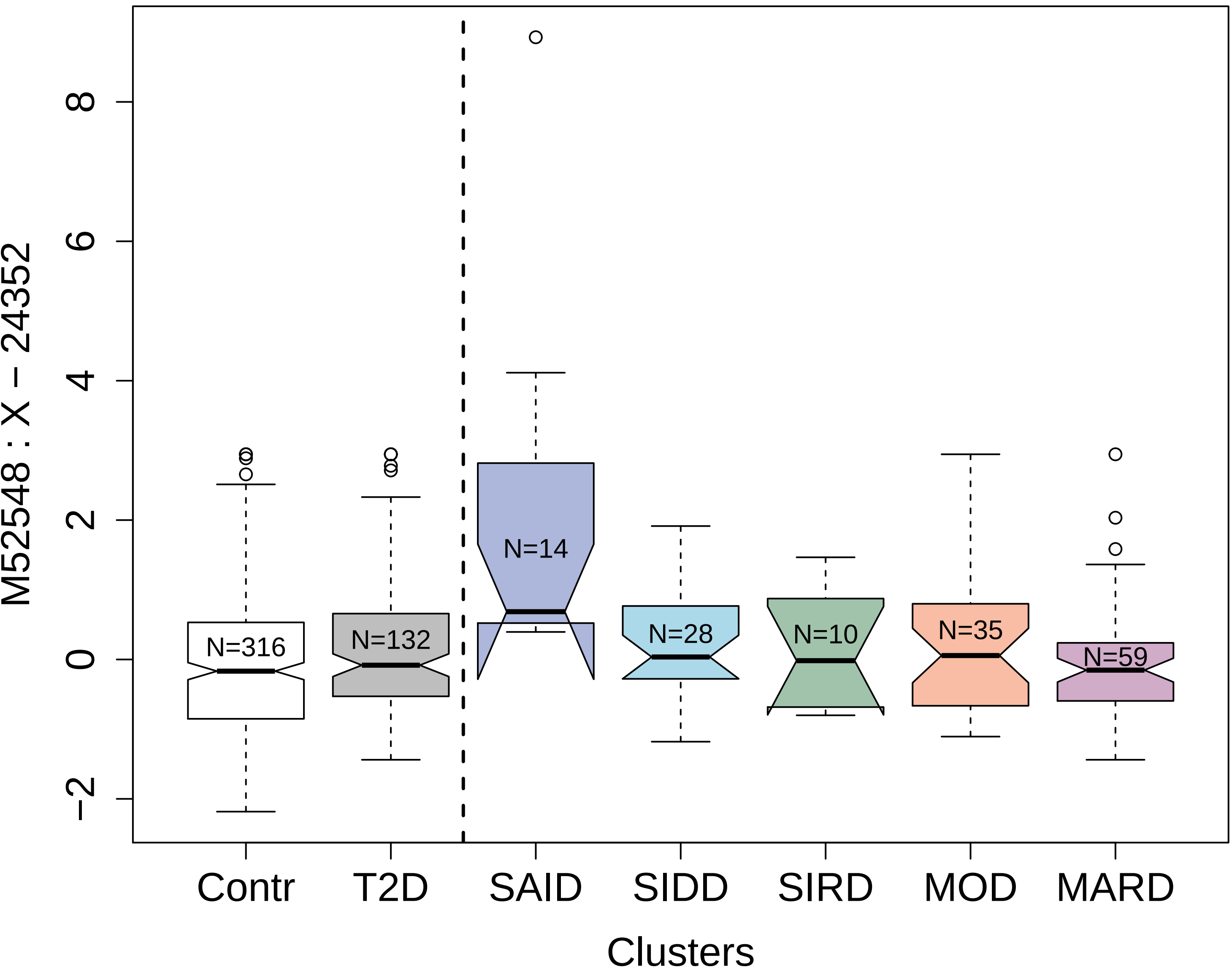
M52548 : X − 24352
−2
0
2
4
6
8
N=316
N=132
N=14
N=28
N=10
N=35
N=59
Contr
T2D
SAID
SIDD
SIRD
MOD
MARD
Clusters

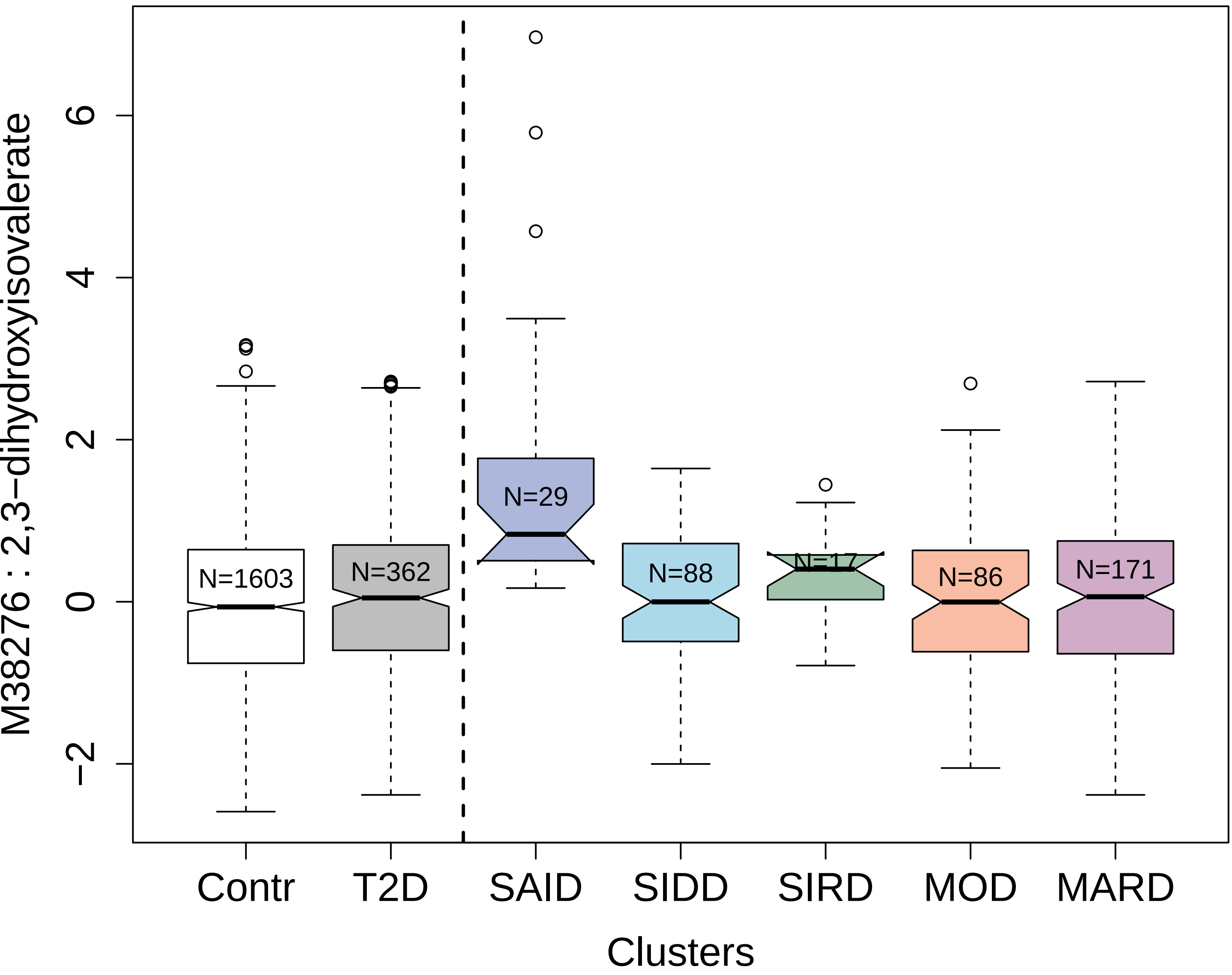
M38276 : 2,3−dihydroxyisovalerate
6
4
2
0
−2
N=1603
N=362
N=29
N=88
N=17
N=86
N=171
Contr
T2D
SAID
SIDD
SIRD
MOD
MARD
Clusters

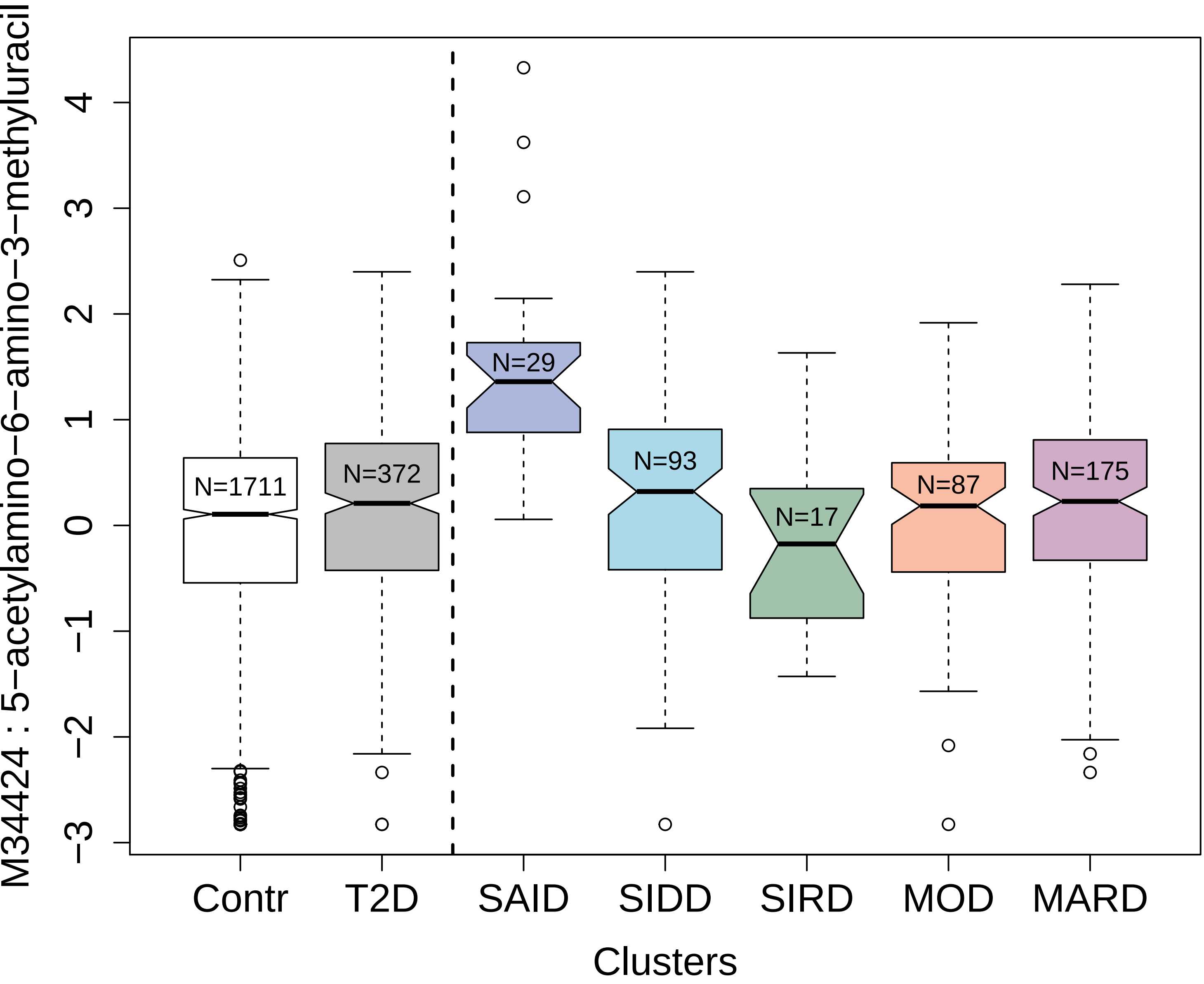

M34424 : 5−acetylamino−6−amino−3−methyluracil
4
3
2
1
0
−1
−2
−3
N=1711
N=372
N=29
N=93
N=17
N=87
N=175
Contr
T2D
SAID
SIDD
SIRD
MOD
MARD
Clusters

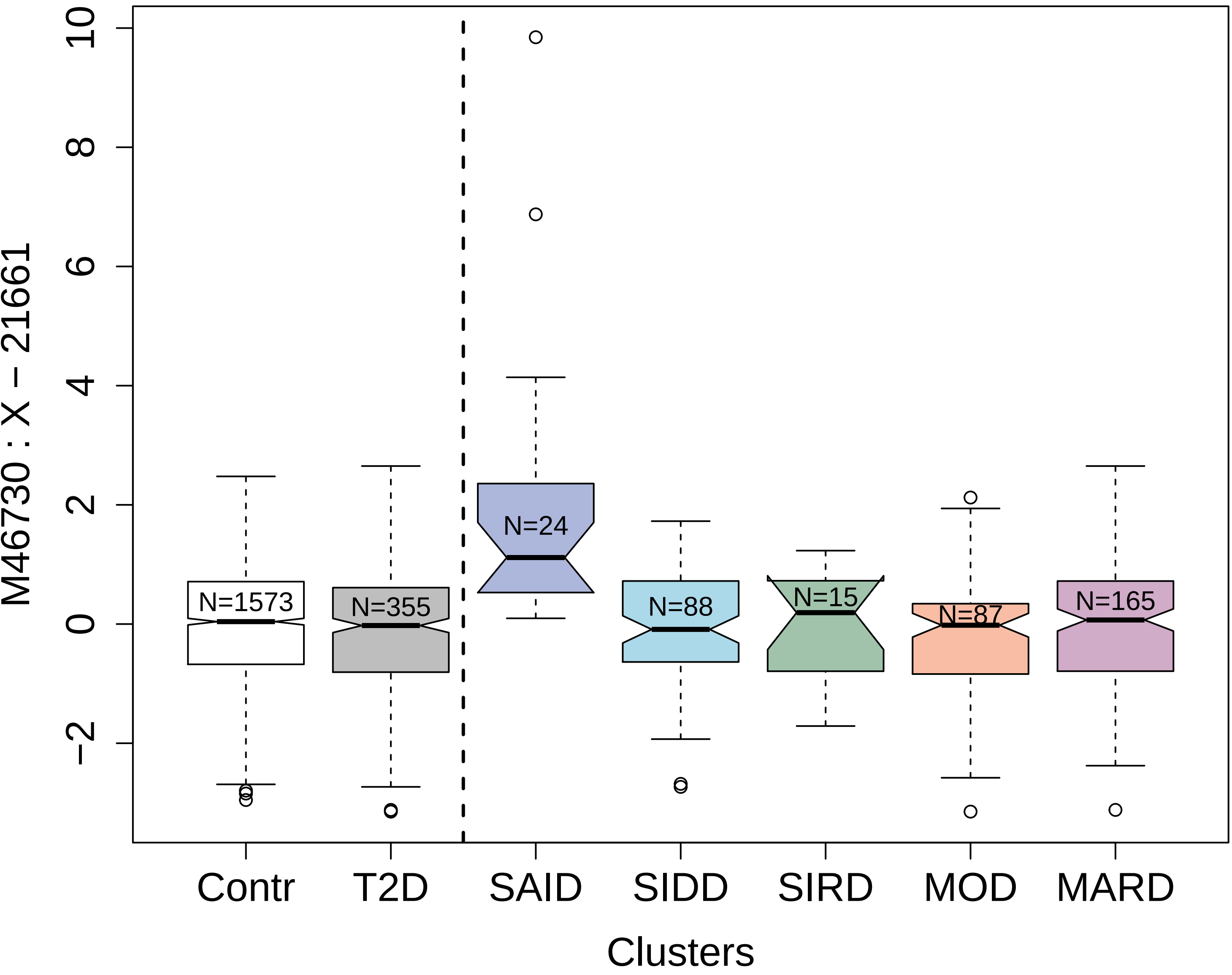
M46730 : X − 21661
10
8
6
4
2
0
−2
N=1573
N=355
N=24
N=88
N=15
N=87
N=165
Contr
T2D
SAID
SIDD
SIRD
MOD
MARD
Clusters

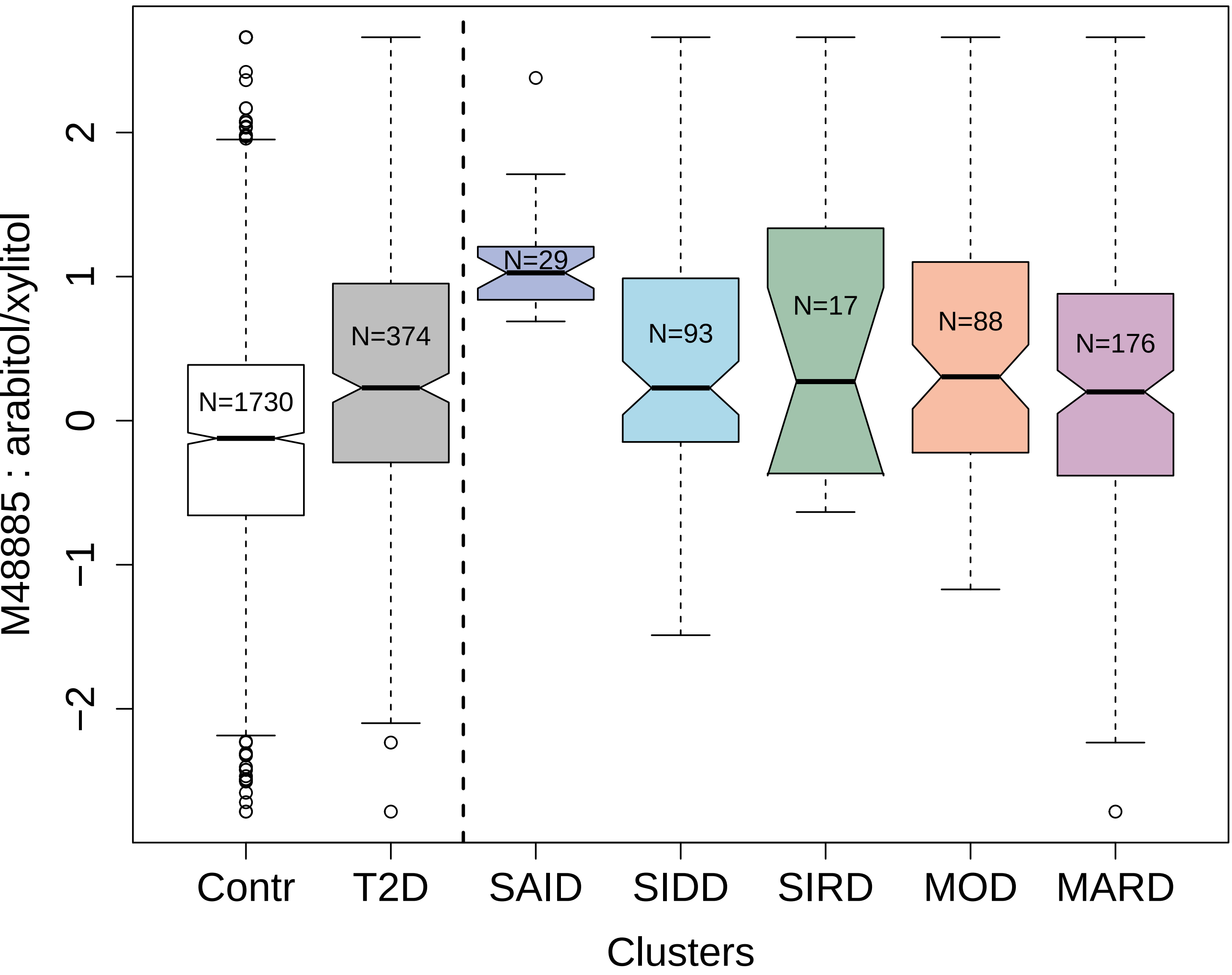

M48885 : arabitol/xylitol
2
1
0
−1
−2
N=1730
N=374
N=29
N=93
N=17
N=88
N=176
Contr
T2D
SAID
SIDD
SIRD
MOD
MARD
Clusters

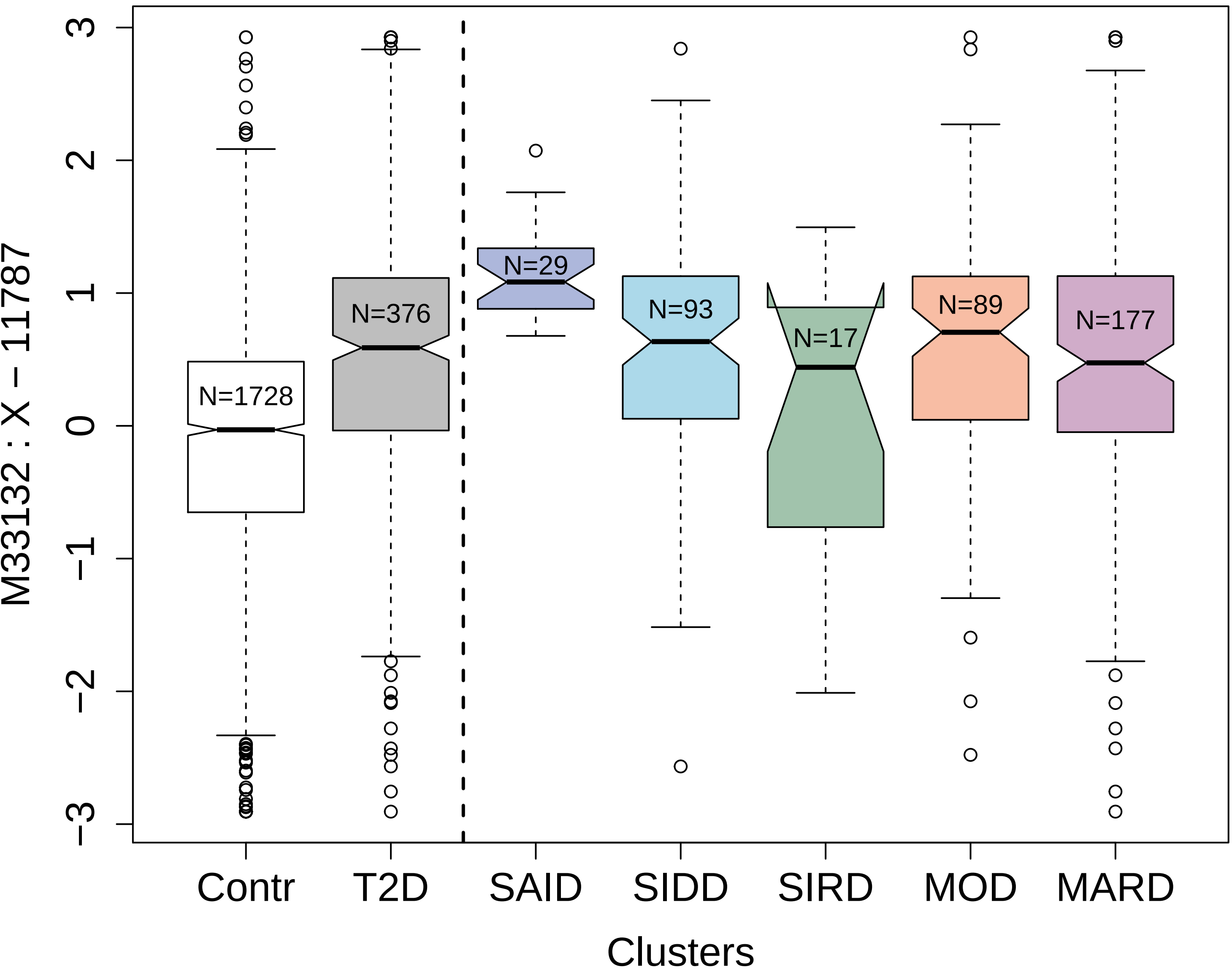

M33132 : X − 11787
3
2
1
0
−1
−2
−3
N=1728
N=376
N=29
N=93
N=17
N=89
N=177
Contr
T2D
SAID
SIDD
SIRD
MOD
MARD
Clusters

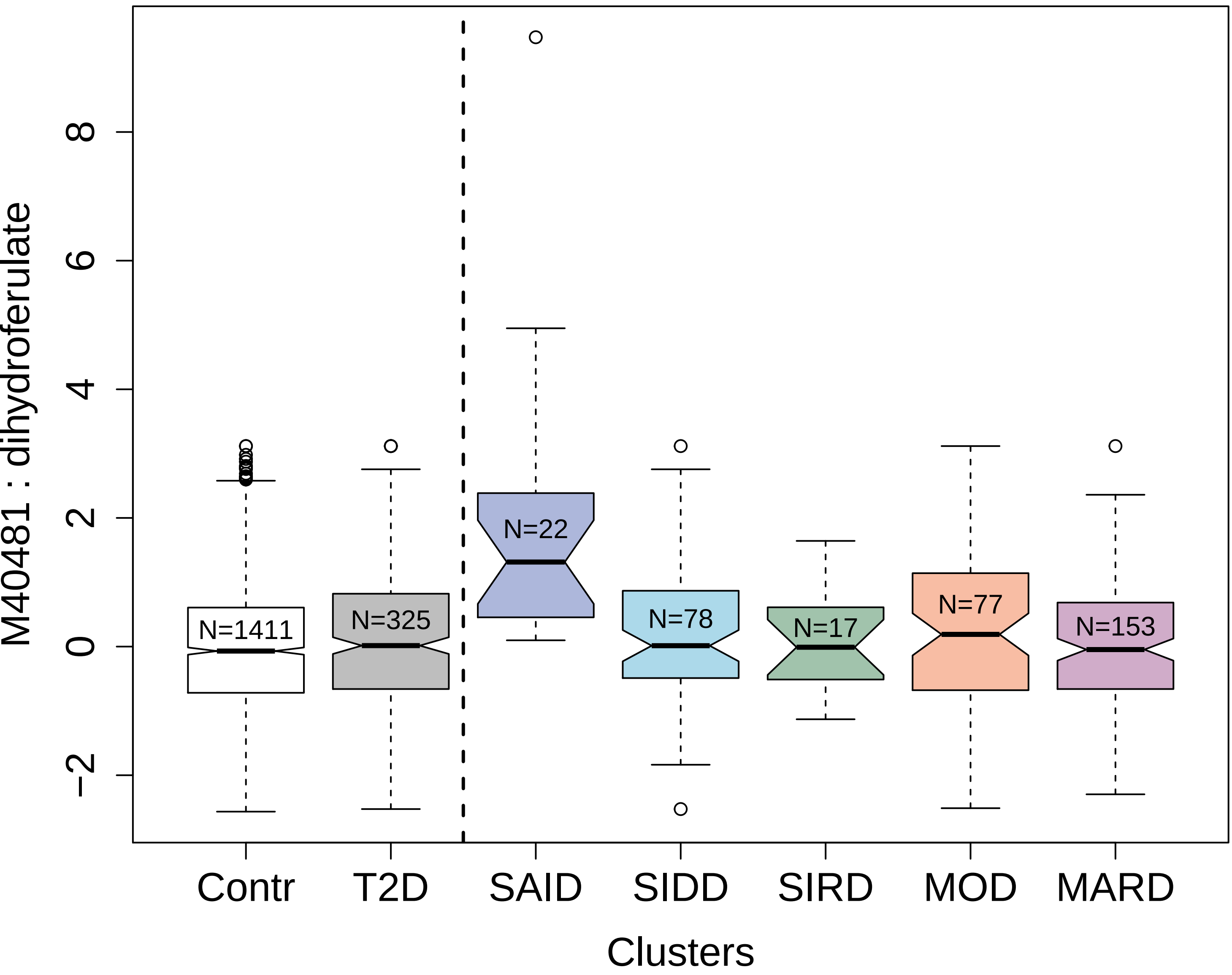
M40481 : dihydroferulate
−2
0
2
4
6
8
N=1411
N=325
N=22
N=78
N=17
N=77
N=153
Contr
T2D
SAID
SIDD
SIRD
MOD
MARD
Clusters

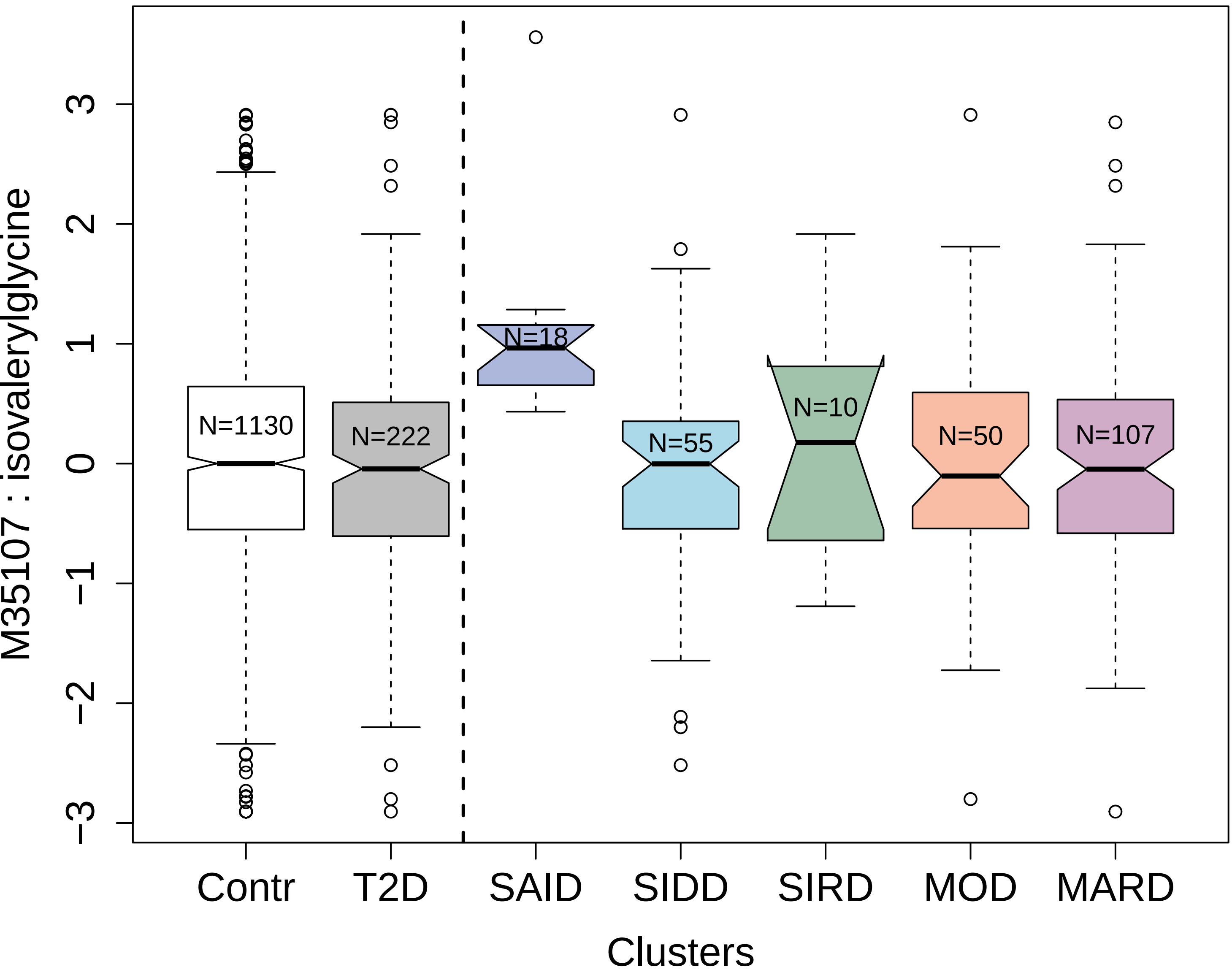
M35107 : isovalerylglycine
3
2
1
0
−1
−2
−3
N=1130
N=222
N=18
N=55
N=10
N=50
N=107
Contr
T2D
SAID
SIDD
SIRD
MOD
MARD
Clusters

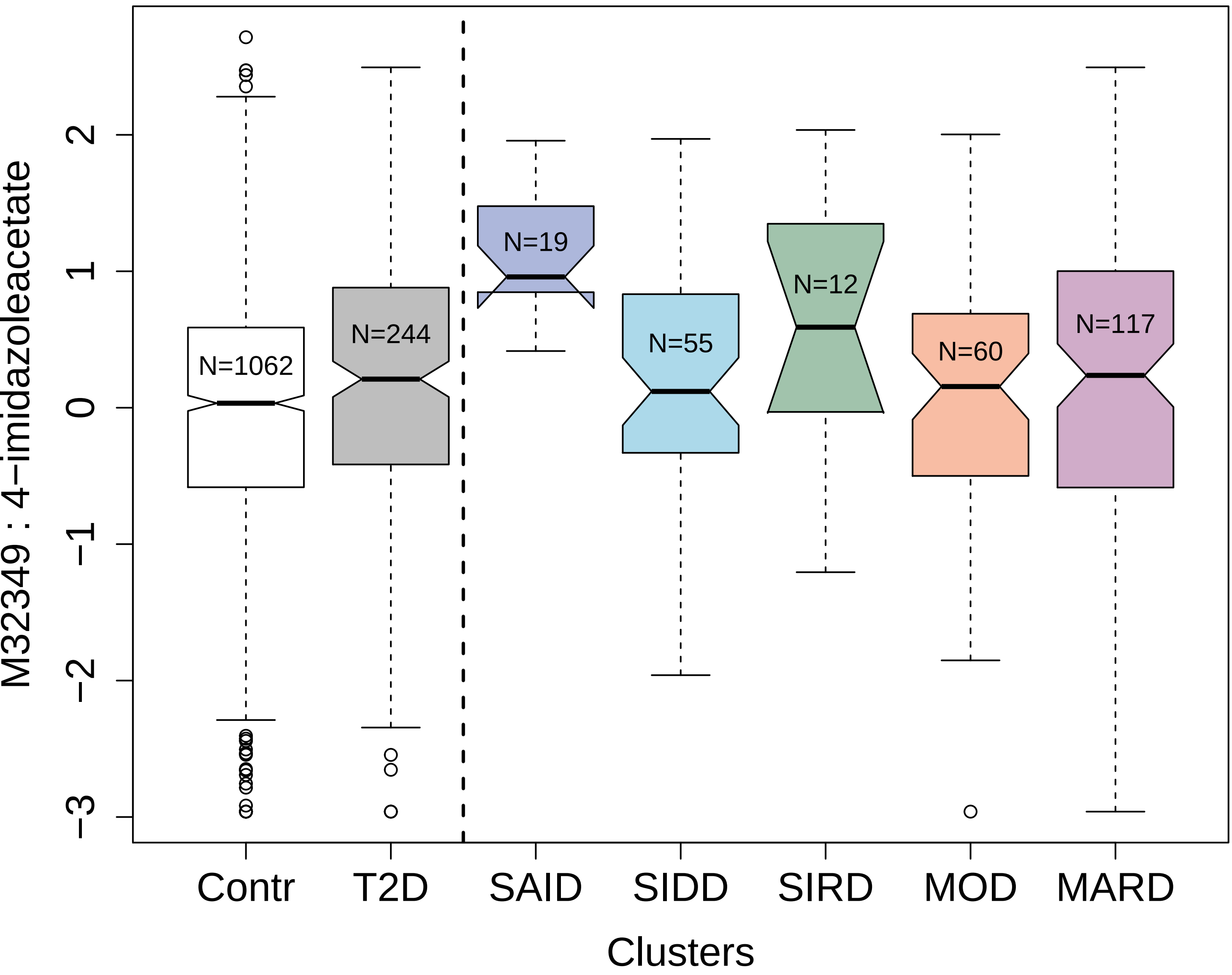
M32349 : 4−imidazoleacetate
2
1
0
−1
−2
−3
N=1062
N=244
N=19
N=55
N=12
N=60
N=117
Contr
T2D
SAID
SIDD
SIRD
MOD
MARD
Clusters

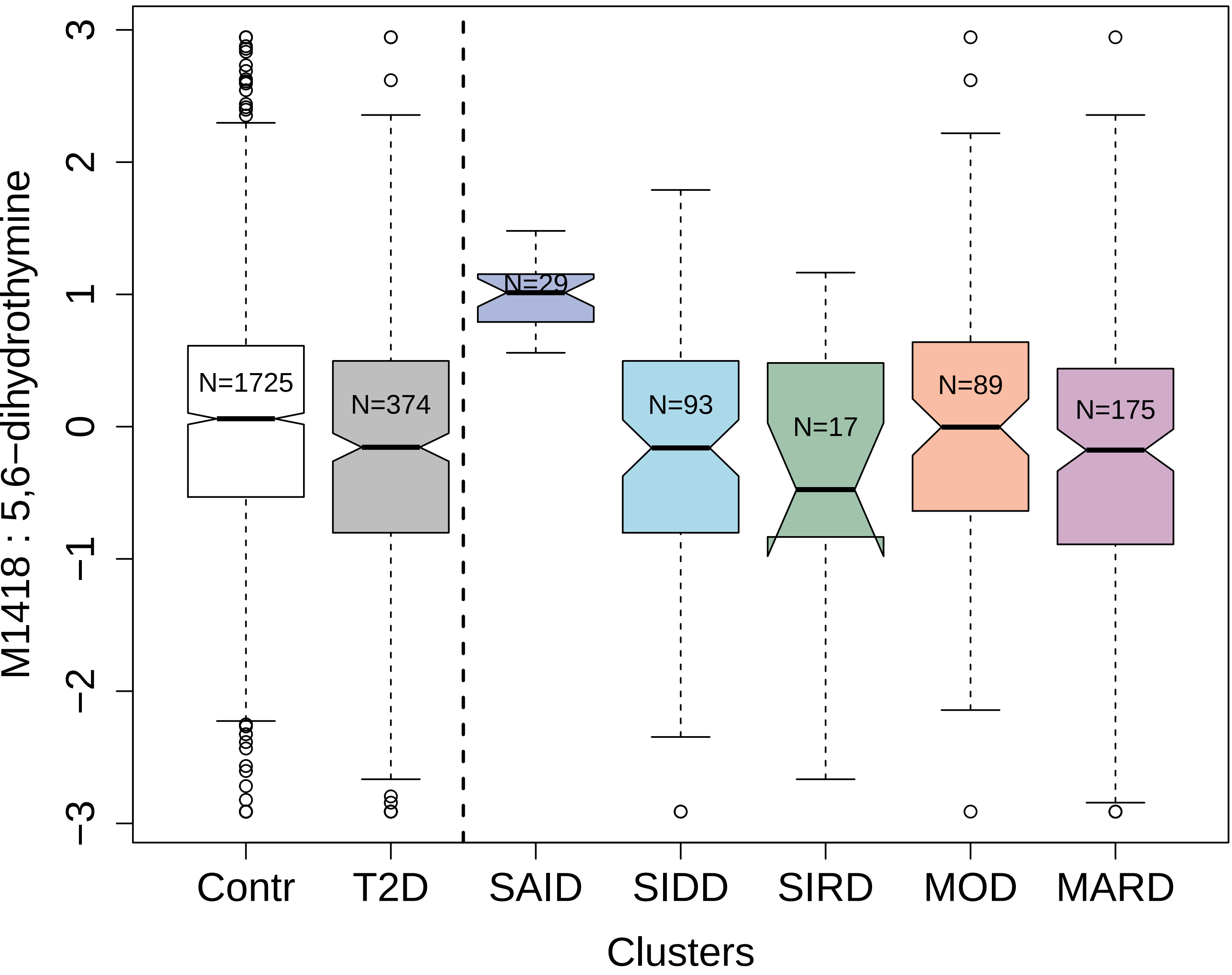

M1418 : 5,6−dihydrothymine
3
2
1
0
−1
−2
−3
N=1725
N=374
N=29
N=93
N=17
N=89
N=175
Contr
T2D
SAID
SIDD
SIRD
MOD
MARD
Clusters

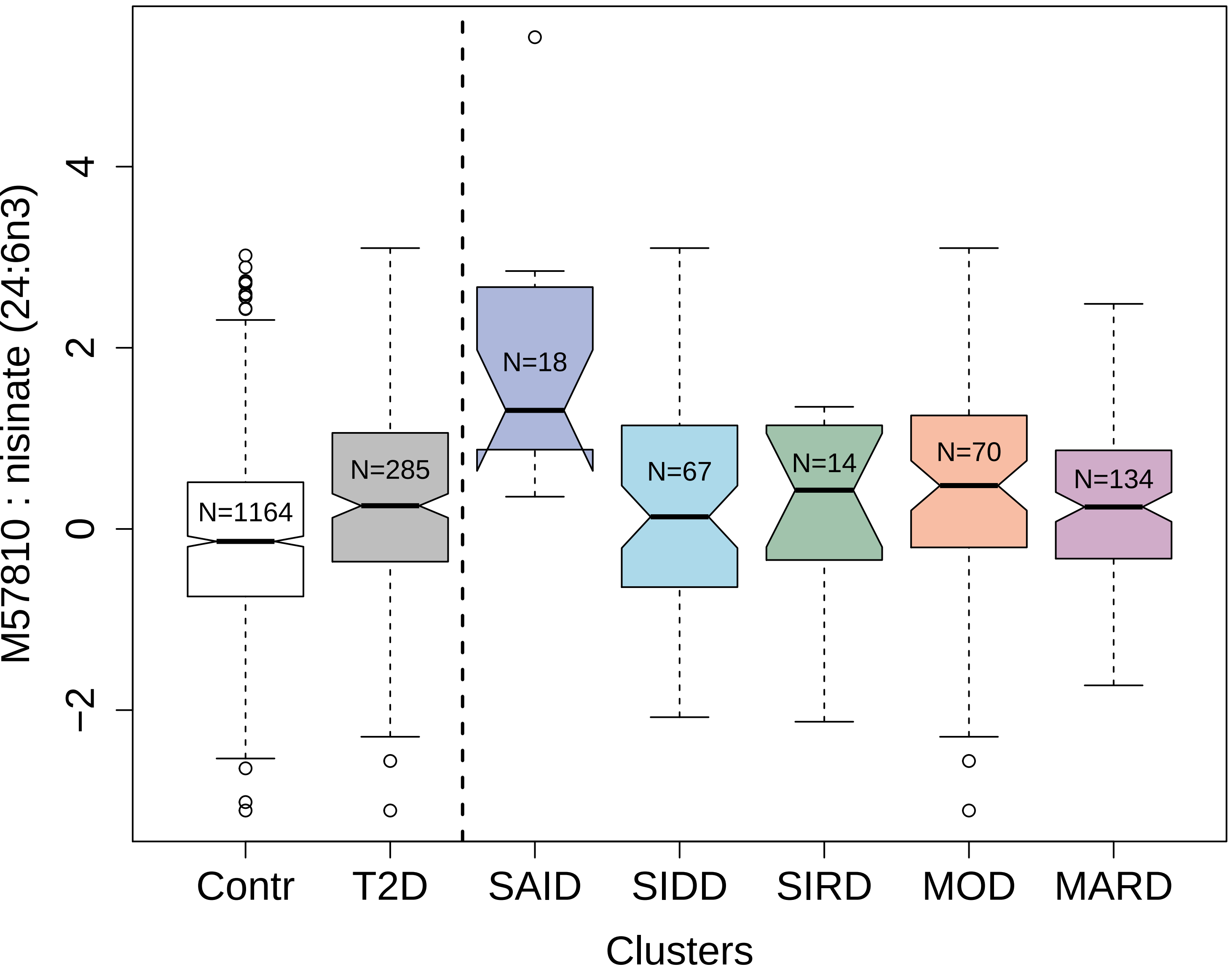
M57810 : nisinate (24:6n3)
4
2
0
−2
N=1164
N=285
N=18
N=67
N=14
N=70
N=134
Contr
T2D
SAID
SIDD
SIRD
MOD
MARD
Clusters

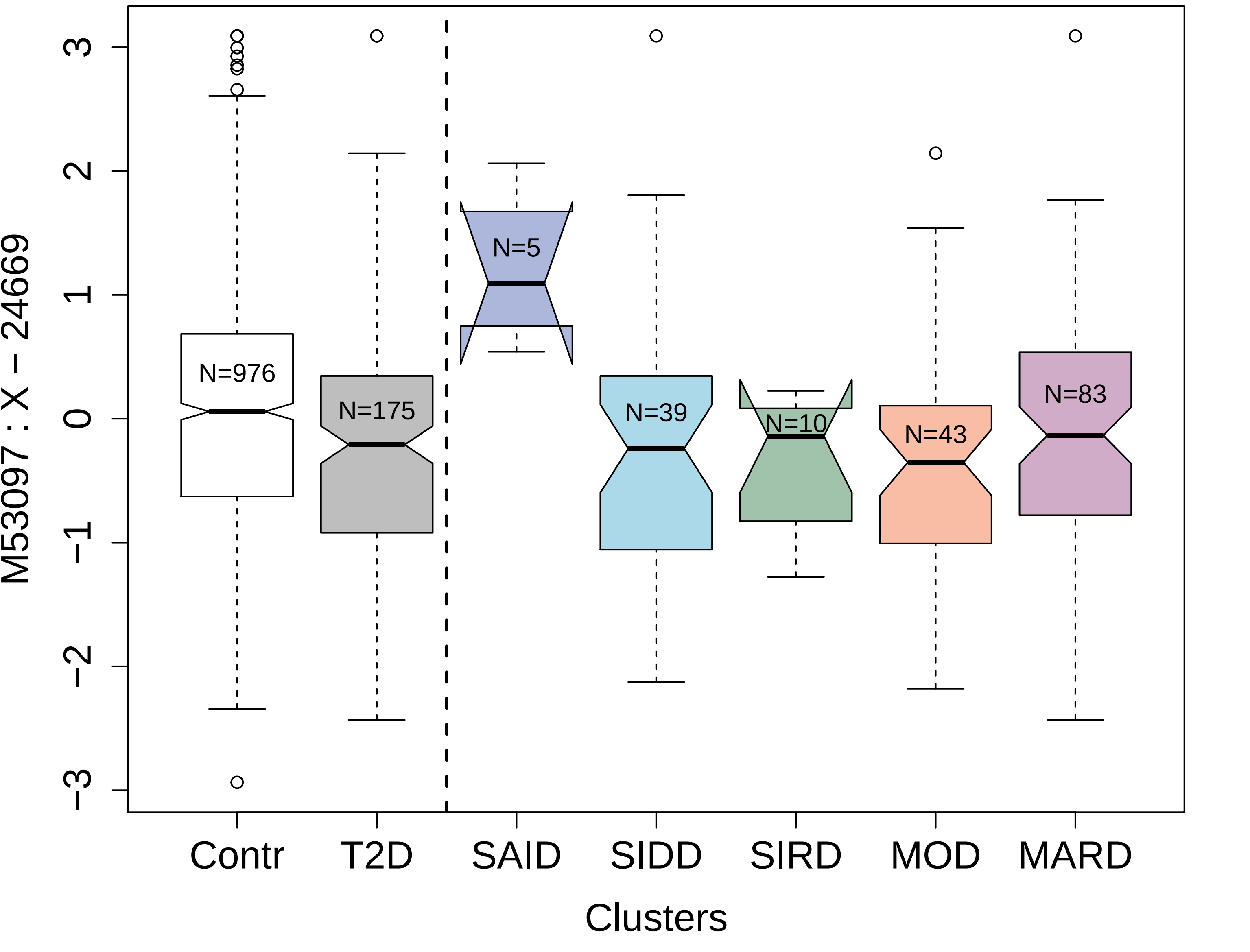

M53097 : X − 24669
Clusters
Contr
T2D
SAID
SIDD
SIRD
MOD
MARD
N=976
N=175
N=5
N=39
N=10
N=43
N=83
−3
−2
−1
0
1
2
3

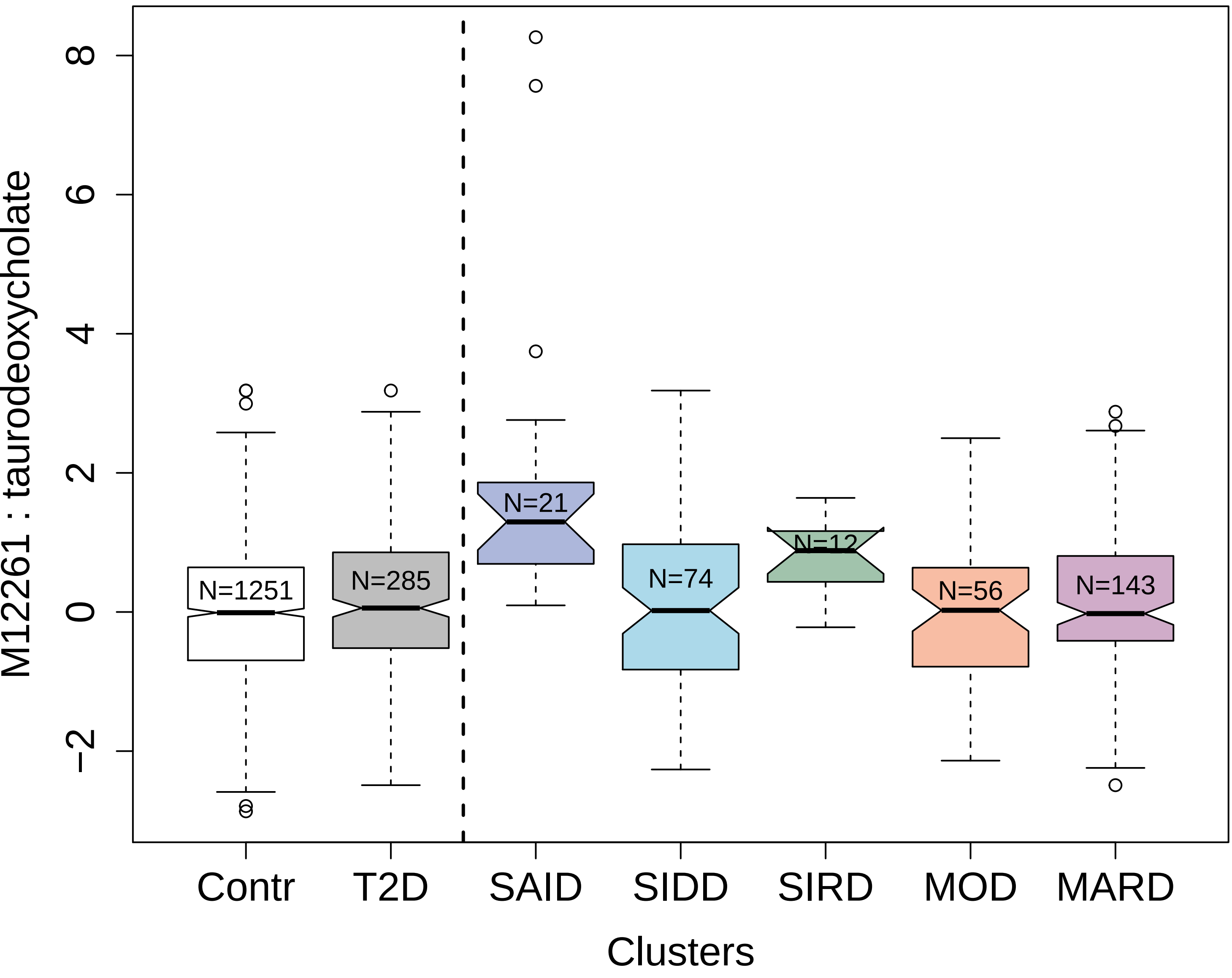

M12261 : taurodeoxycholate
8
6
4
2
0
−2
N=1251
N=285
N=21
N=74
N=12
N=56
N=143
Contr
T2D
SAID
SIDD
SIRD
MOD
MARD
Clusters

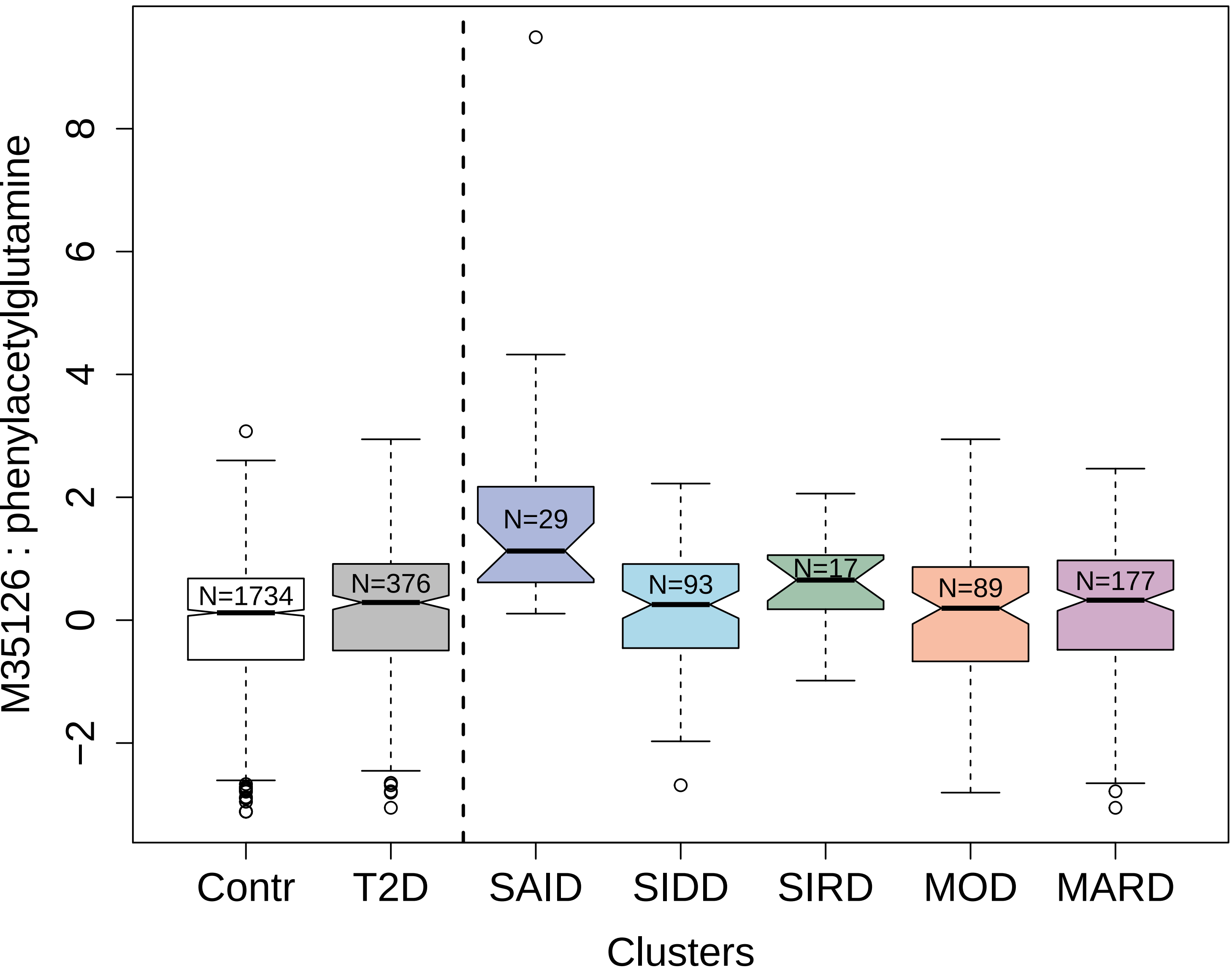
M35126 : phenylacetylglutamine
Clusters
Contr
T2D
SAID
SIDD
SIRD
MOD
MARD
N=1734
N=376
N=29
N=93
N=17
N=89
N=177
−2
0
2
4
6
8

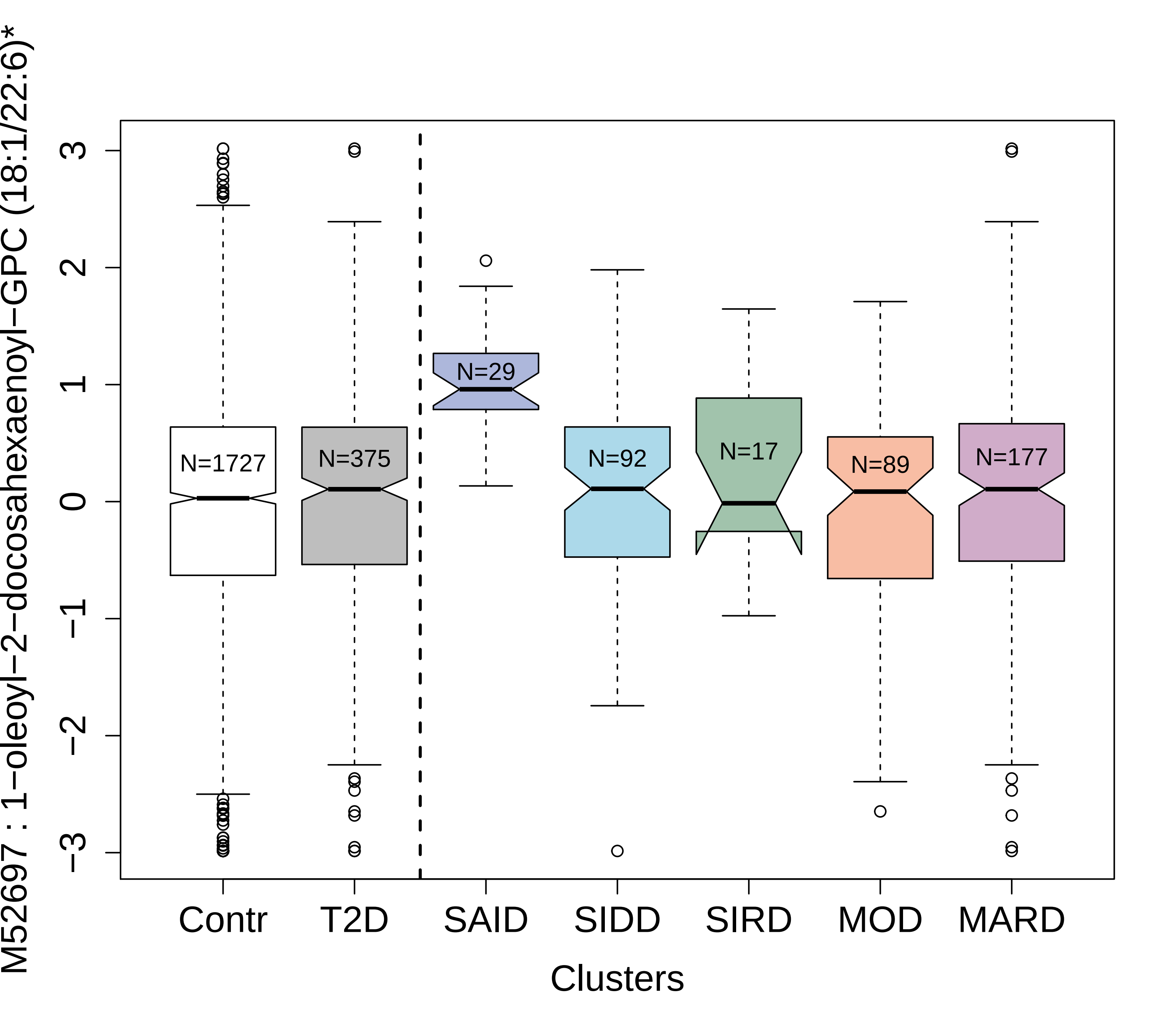
M52697 : 1−oleoyl−2−docosahexaenoyl−GPC (18:1/22:6)*
3
2
1
0
−1
−2
−3
N=1727
N=375
N=29
N=92
N=17
N=89
N=177
Contr
T2D
SAID
SIDD
SIRD
MOD
MARD
Clusters

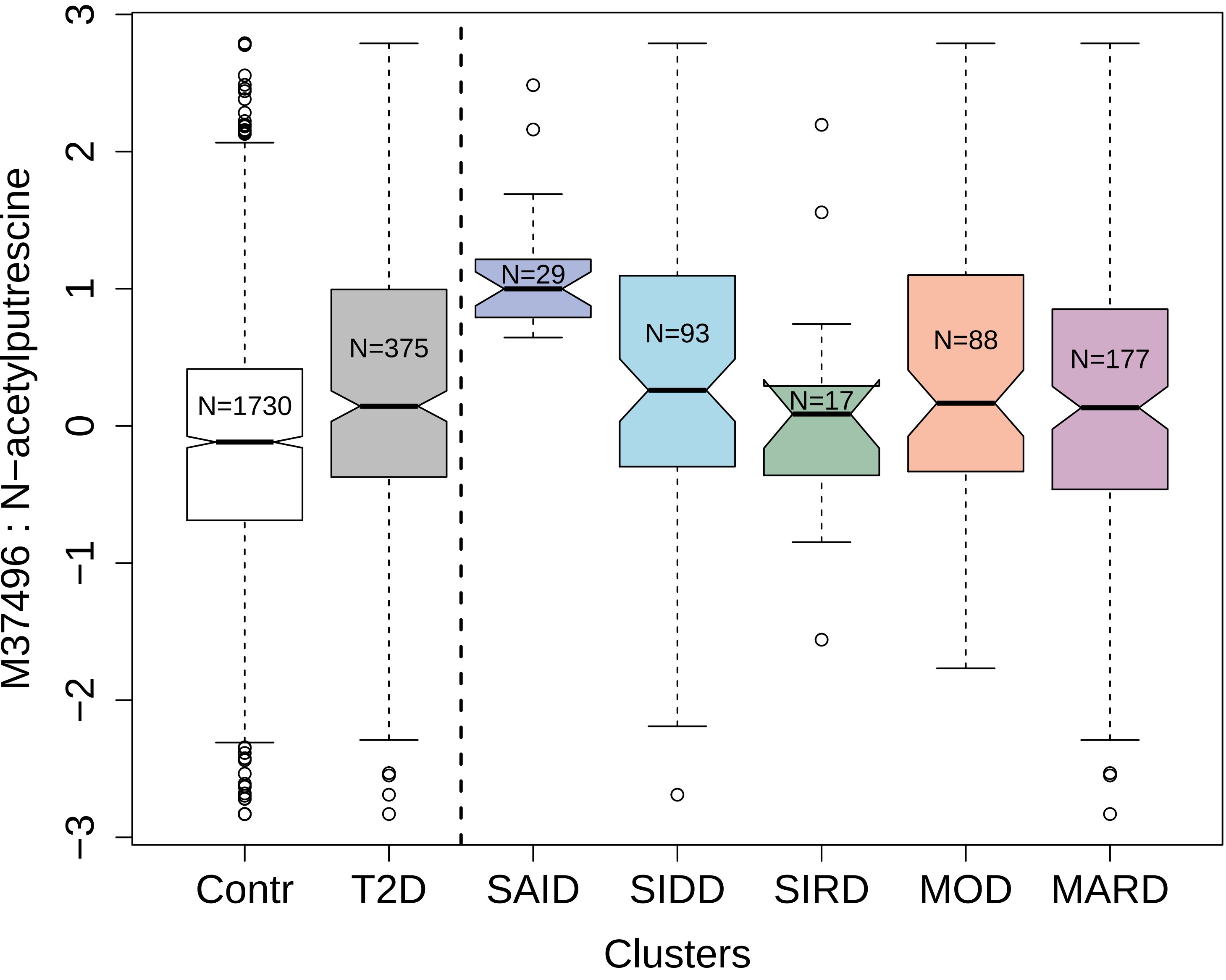
M37496 : N−acetylputrescine
Clusters
Contr
T2D
SAID
SIDD
SIRD
MOD
MARD
N=1730
N=375
N=29
N=93
N=17
N=88
N=177
3
2
1
0
−1
−2
−3

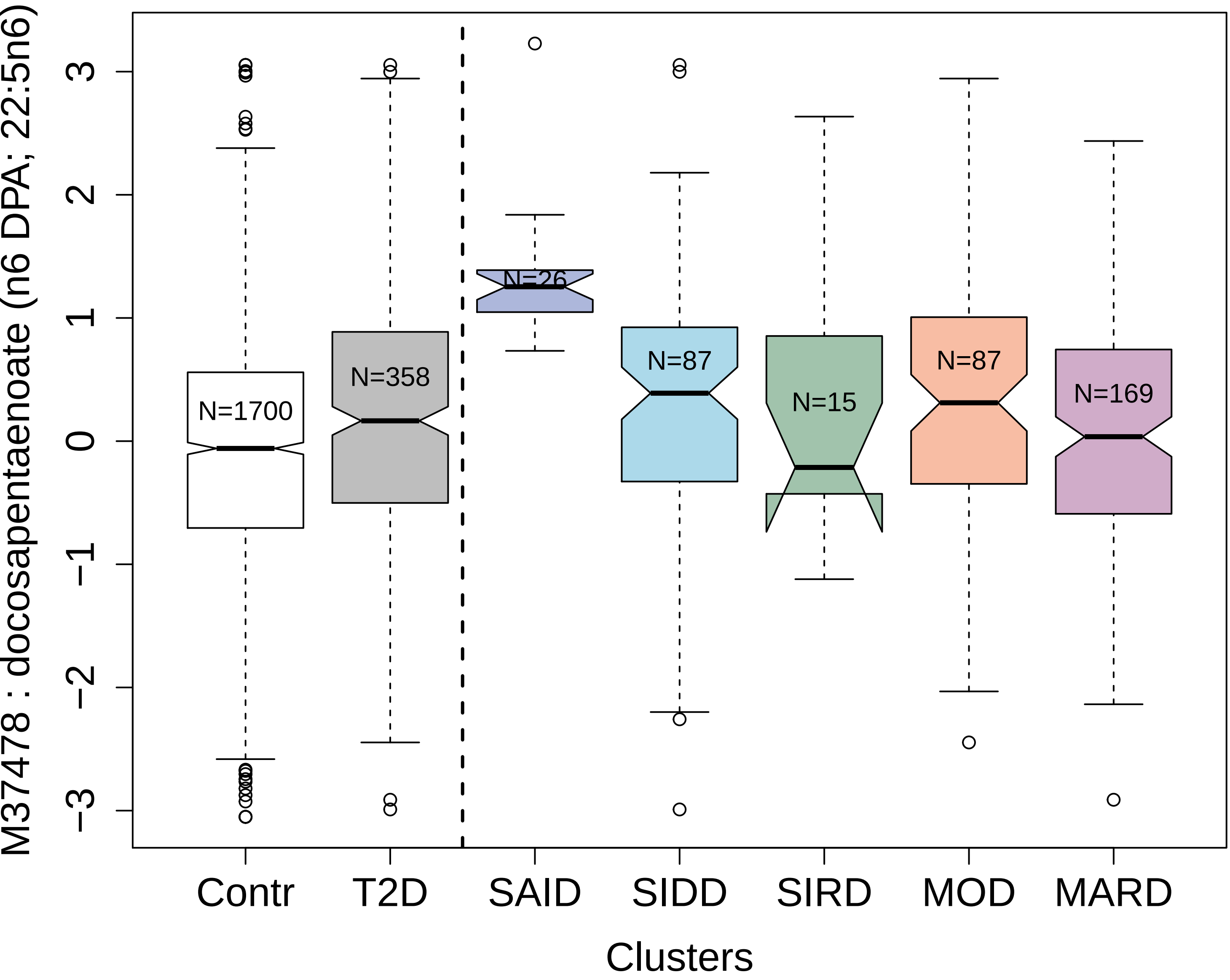

M37478 : docosapentaenoate (n6 DPA; 22:5n6)
3
2
1
0
−1
−2
−3
N=1700
N=358
N=26
N=87
N=15
N=87
N=169
Contr
T2D
SAID
SIDD
SIRD
MOD
MARD
Clusters

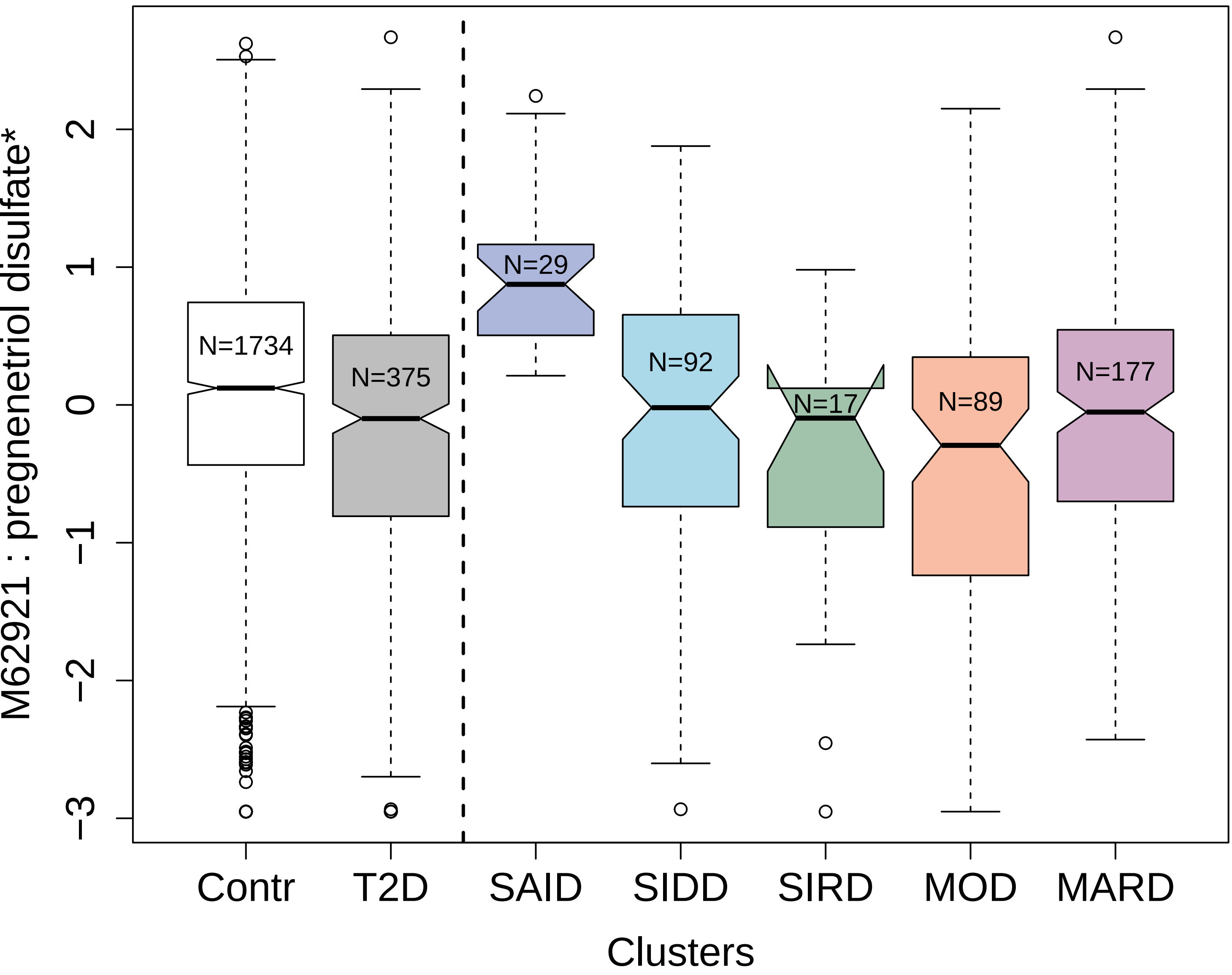

M62921 : pregnetriol disulfate*
2
1
0
−1
−2
−3
N=1734
N=375
N=29
N=92
N=17
N=89
N=177
Contr
T2D
SAID
SIDD
SIRD
MOD
MARD
Clusters

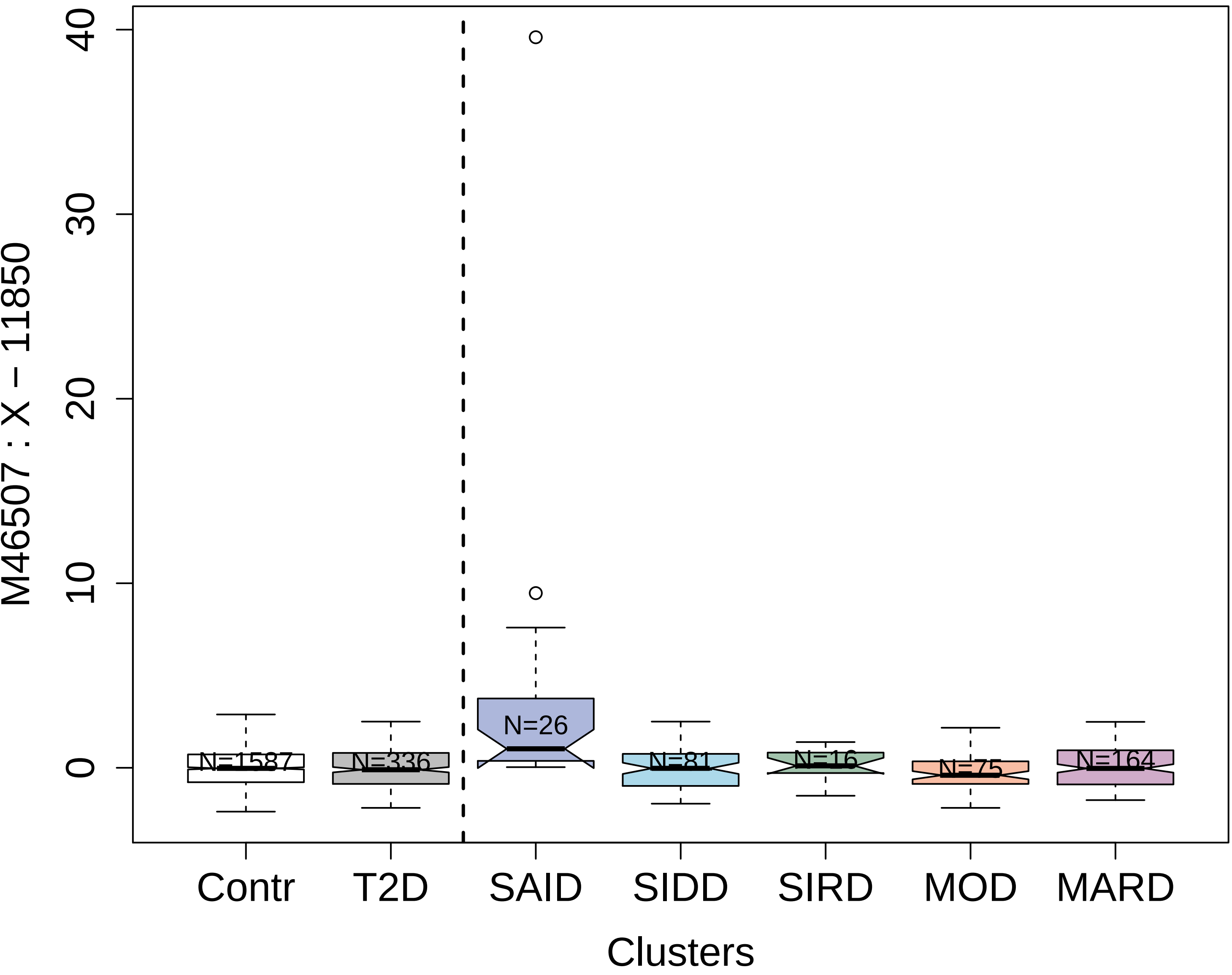

M46507 : X − 11850
0
10
20
30
40
N=1587
N=336
N=26
N=81
N=16
N=75
N=164
Contr
T2D
SAID
SIDD
SIRD
MOD
MARD
Clusters

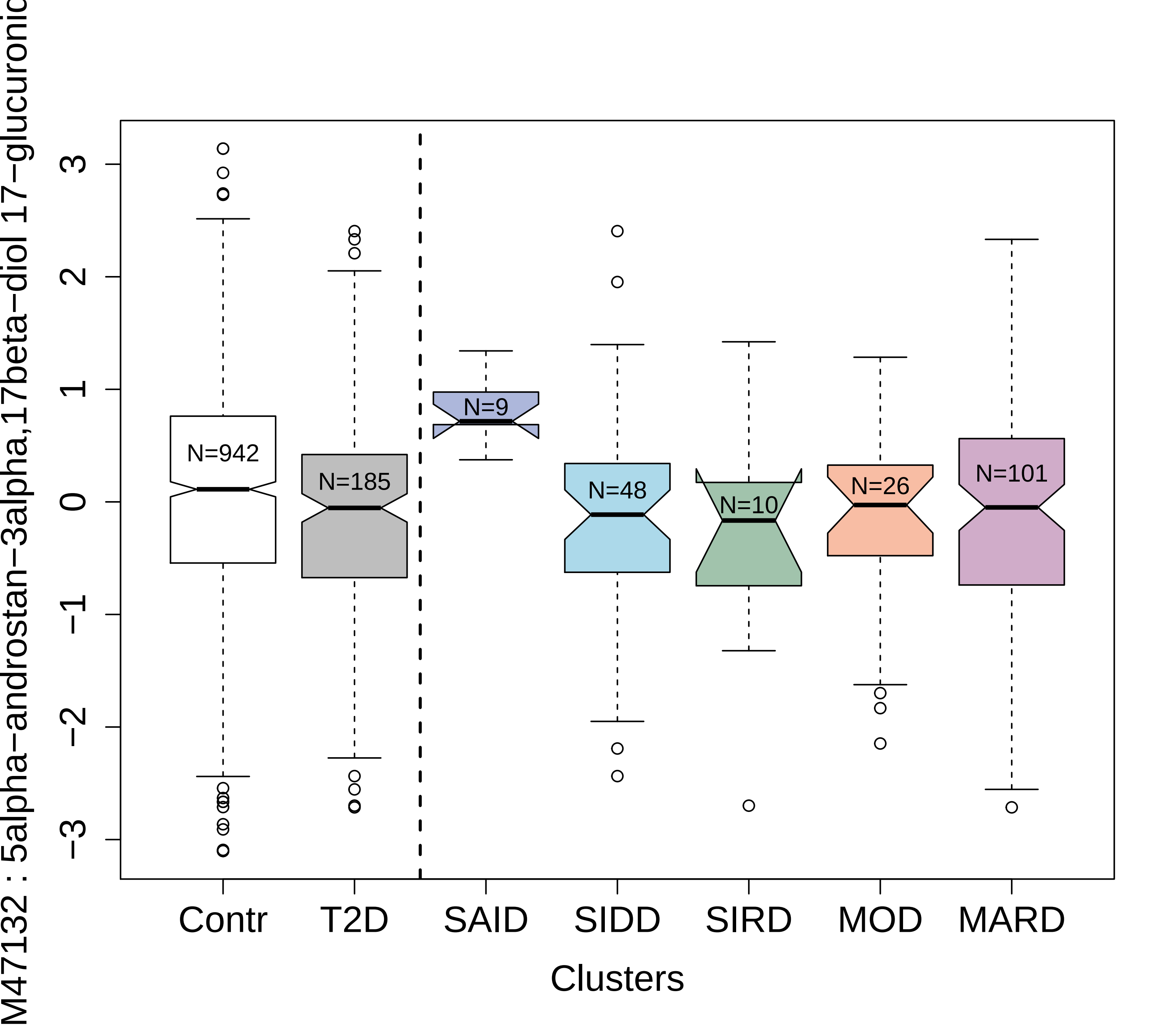
M47132 : 5alpha−androstan−3alpha,17beta−diol 17−glucuronide
3
2
1
0
−1
−2
−3
N=942
N=185
N=9
N=48
N=10
N=26
N=101
Contr
T2D
SAID
SIDD
SIRD
MOD
MARD
Clusters

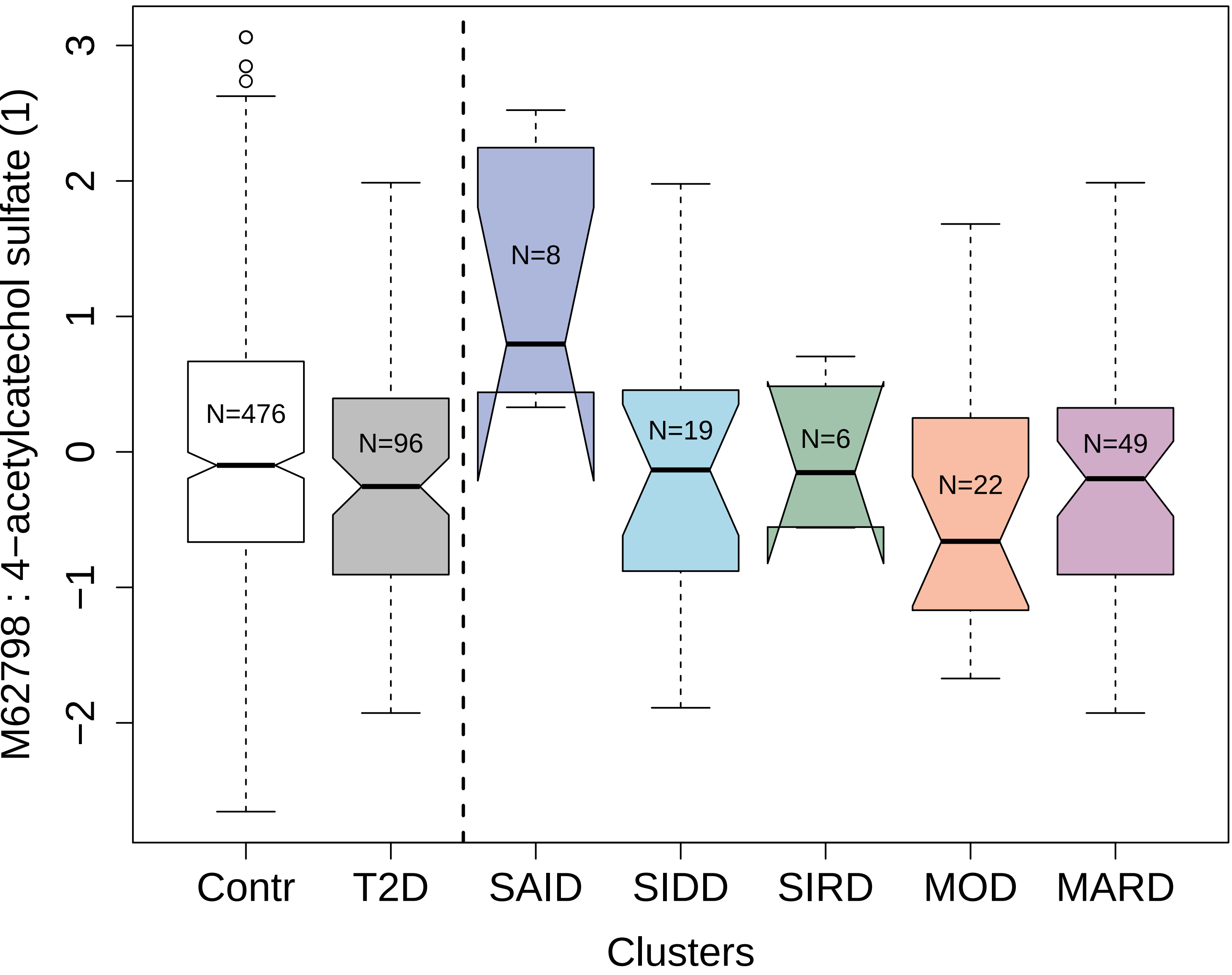

M62798 : 4−acetylcatechol sulfate (1)
3
2
1
0
−1
−2
N=476
N=96
N=8
N=19
N=6
N=22
N=49
Contr
T2D
SAID
SIDD
SIRD
MOD
MARD
Clusters

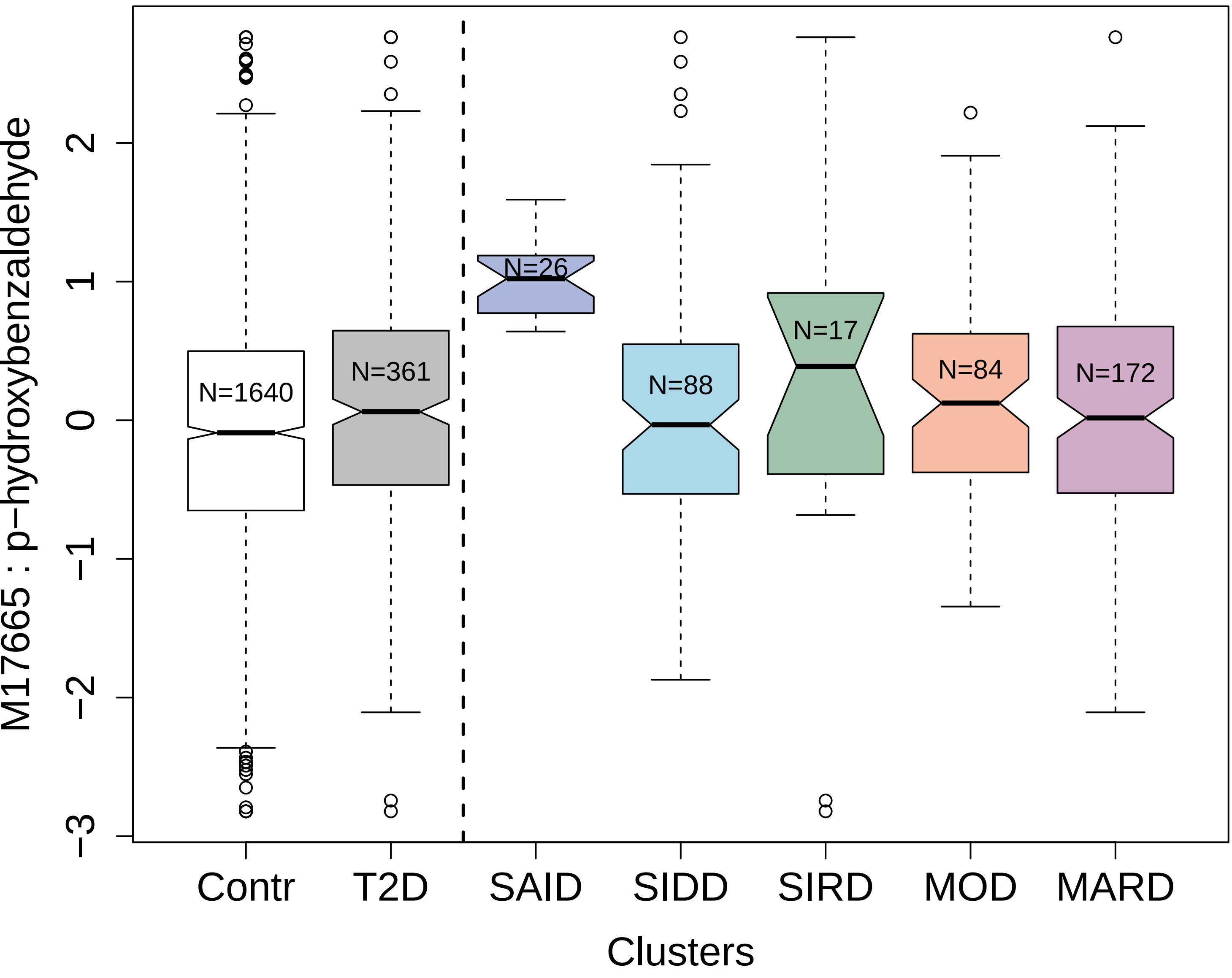

M17665 : p−hydroxybenzaldehyde
−3
−2
−1
0
1
2
N=1640
N=361
N=26
N=88
N=17
N=84
N=172
Contr
T2D
SAID
SIDD
SIRD
MOD
MARD
Clusters

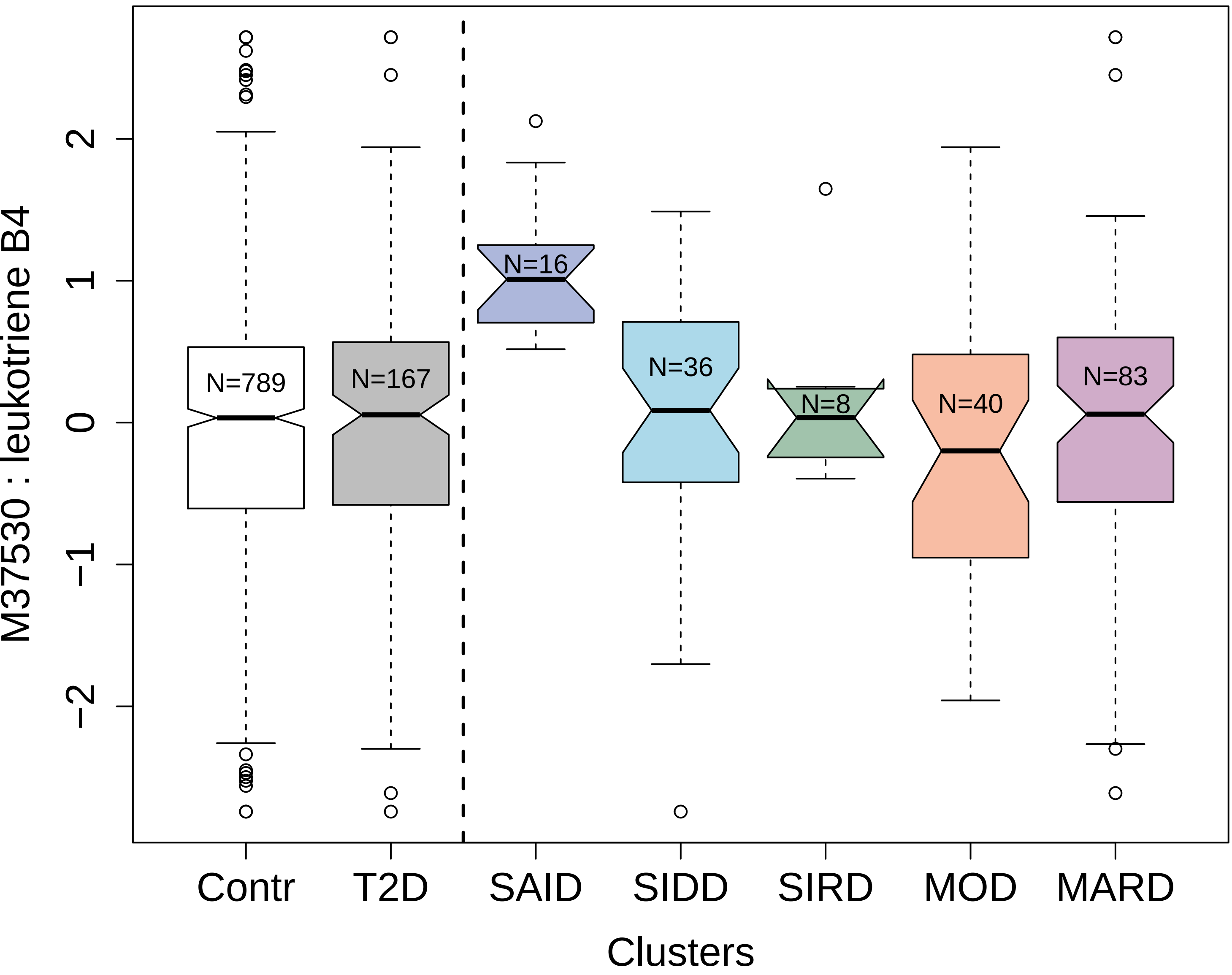
M37530 : leukotriene B4
2
1
0
−1
−2
N=789
N=167
N=16
N=36
N=8
N=40
N=83
Contr
T2D
SAID
SIDD
SIRD
MOD
MARD
Clusters

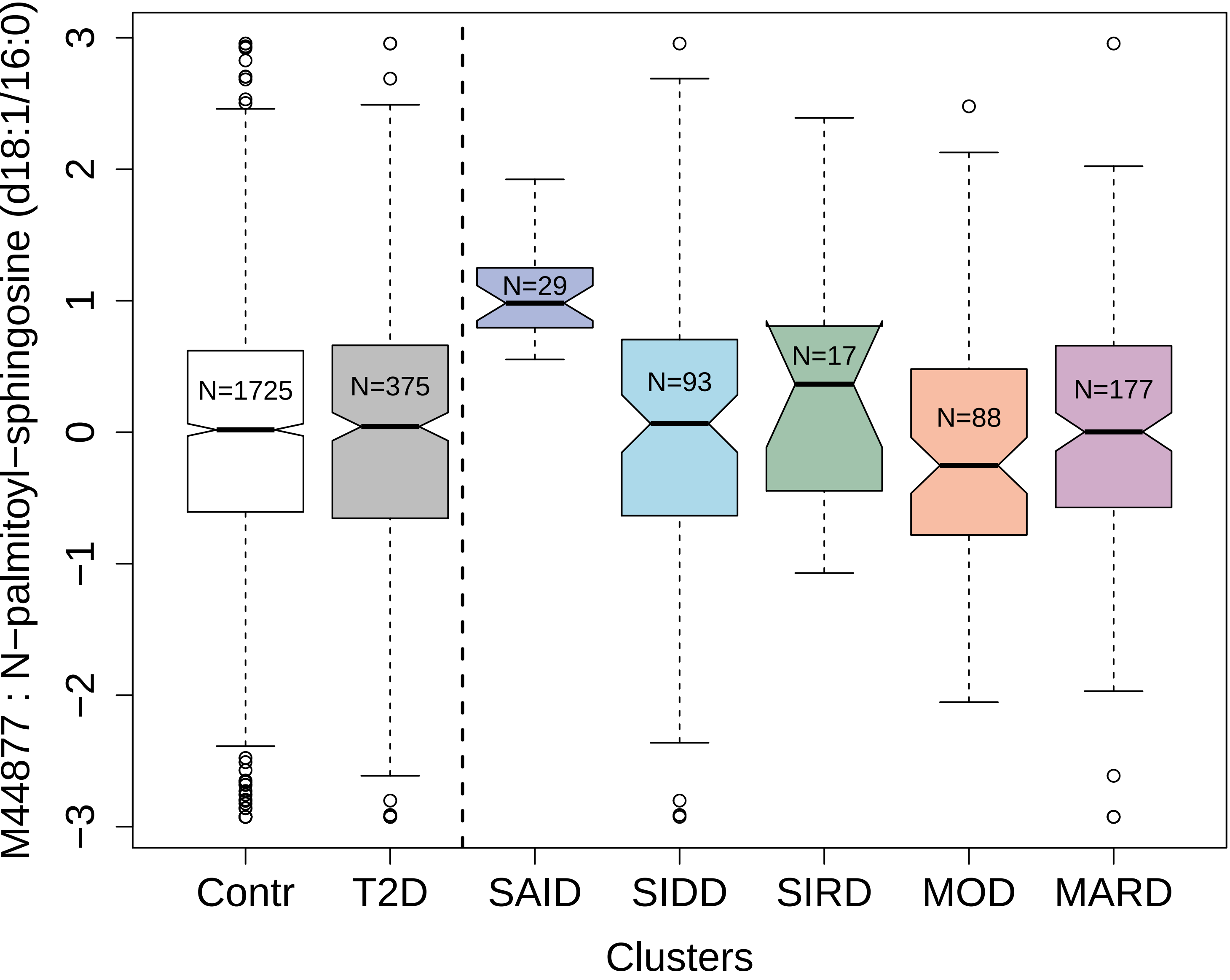
M44877 : N−palmitoyl−sphingosine (d18:1/16:0)
3
2
1
0
−1
−2
−3
N=1725
N=375
N=29
N=93
N=17
N=88
N=177
Contr
T2D
SAID
SIDD
SIRD
MOD
MARD
Clusters

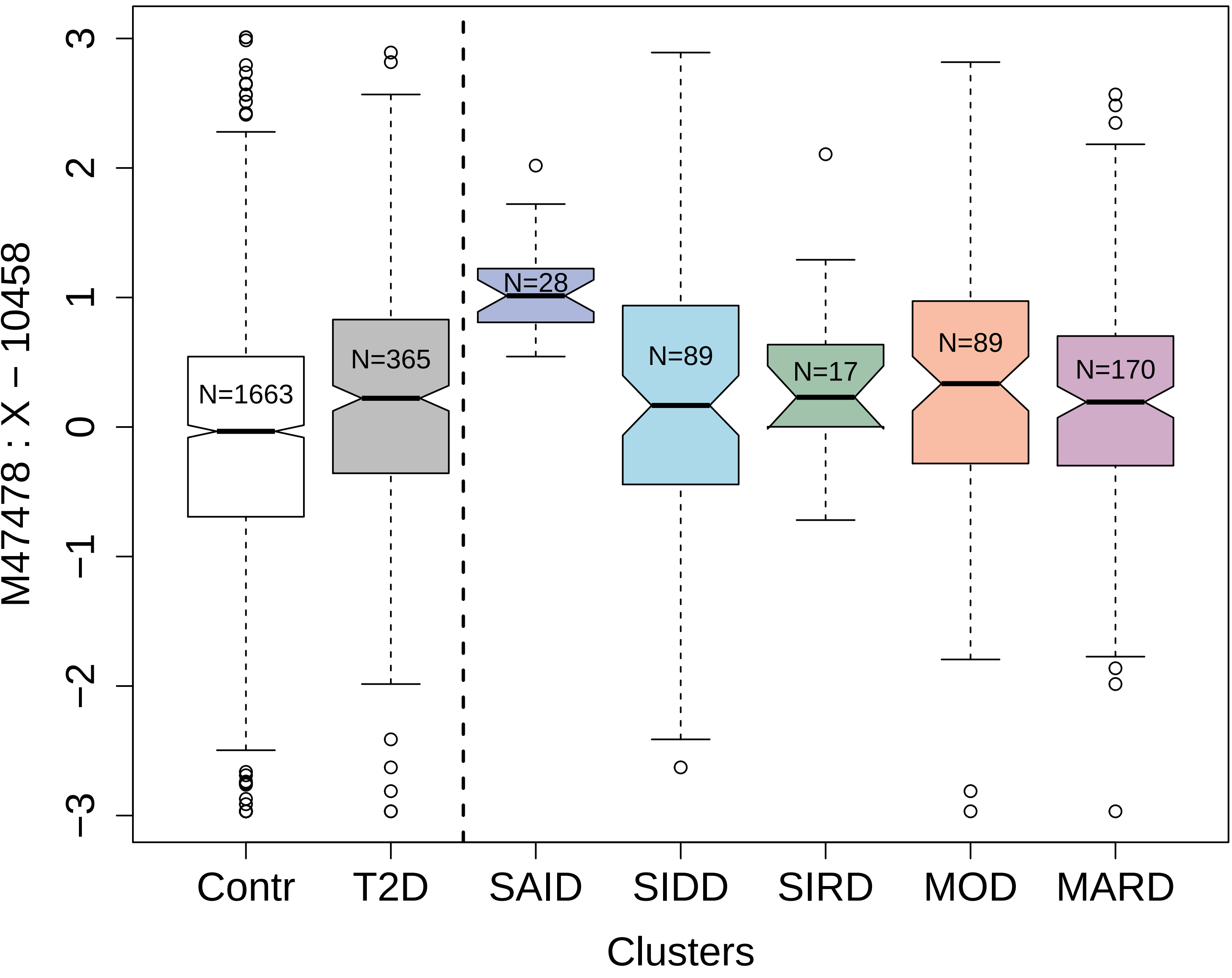
M47478 : X − 10458
3
2
1
0
−1
−2
−3
N=1663
N=365
N=28
N=89
N=17
N=89
N=170
Contr
T2D
SAID
SIDD
SIRD
MOD
MARD
Clusters

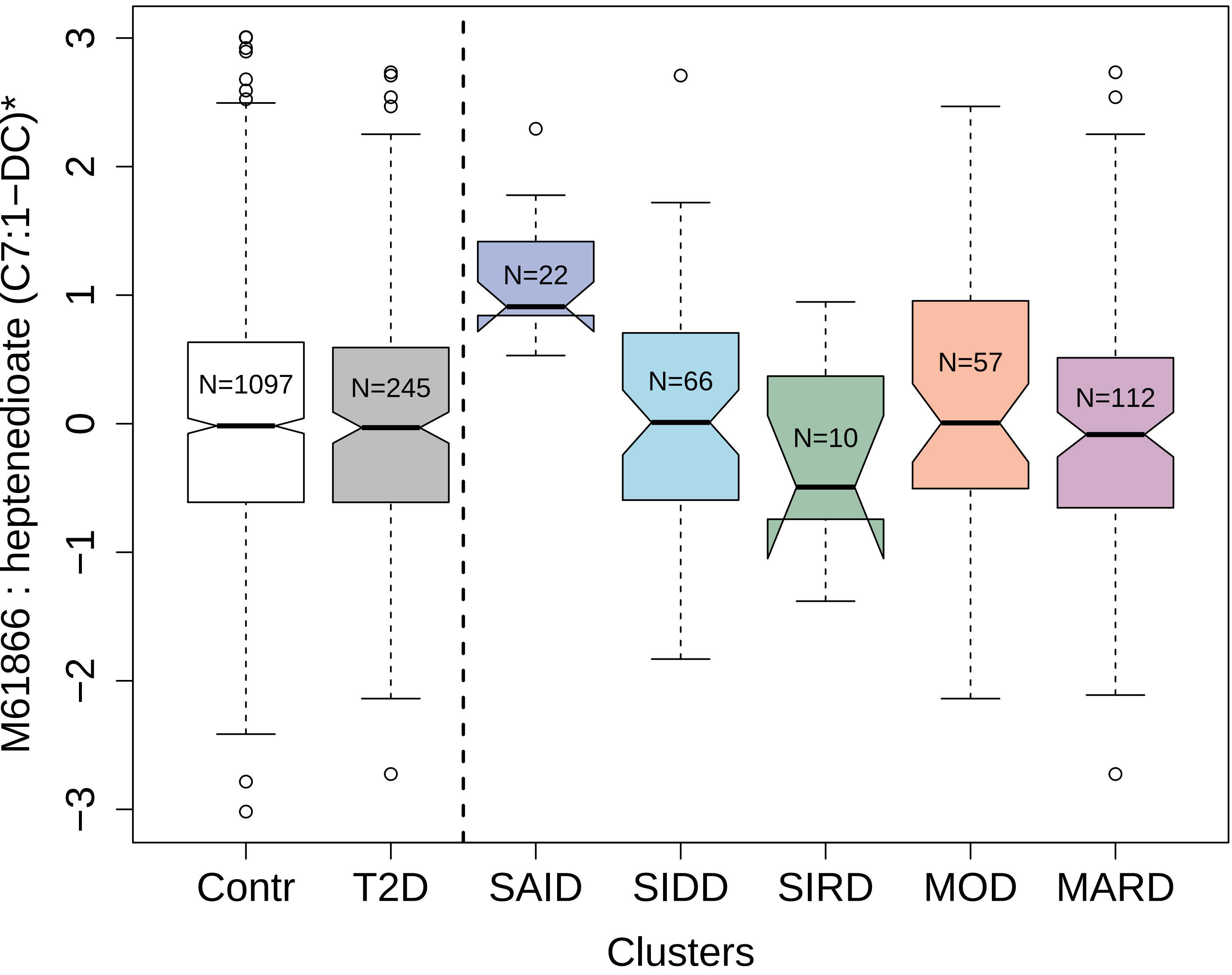

M61866 : heptenedioate (C7:1−DC)*
3
2
1
0
−1
−2
−3
N=1097
N=245
N=22
N=66
N=10
N=57
N=112
Contr
T2D
SAID
SIDD
SIRD
MOD
MARD
Clusters

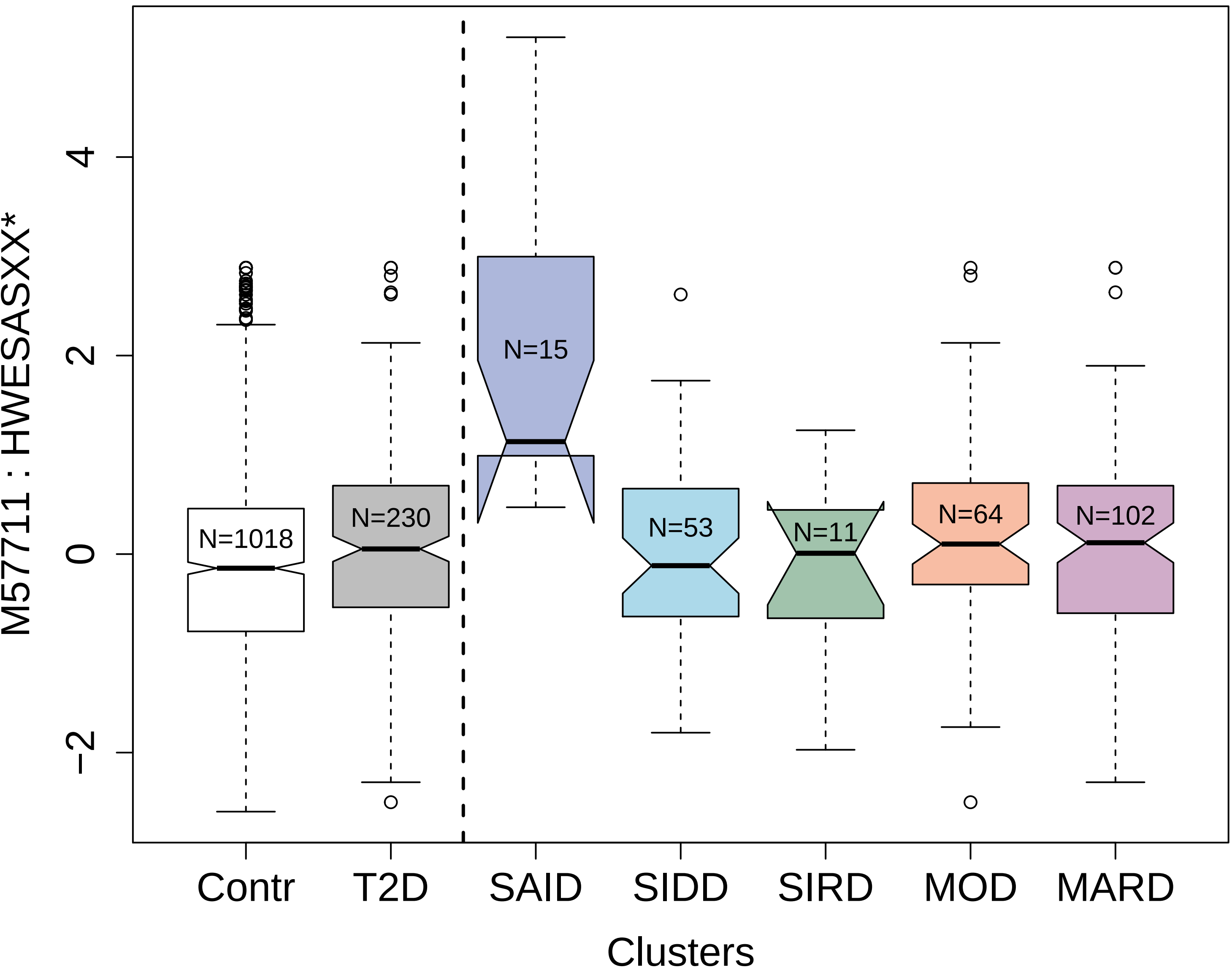

M57711 : HWESASXX*
4
2
0
−2
N=1018
N=230
N=15
N=53
N=11
N=64
N=102
Contr
T2D
SAID
SIDD
SIRD
MOD
MARD
Clusters

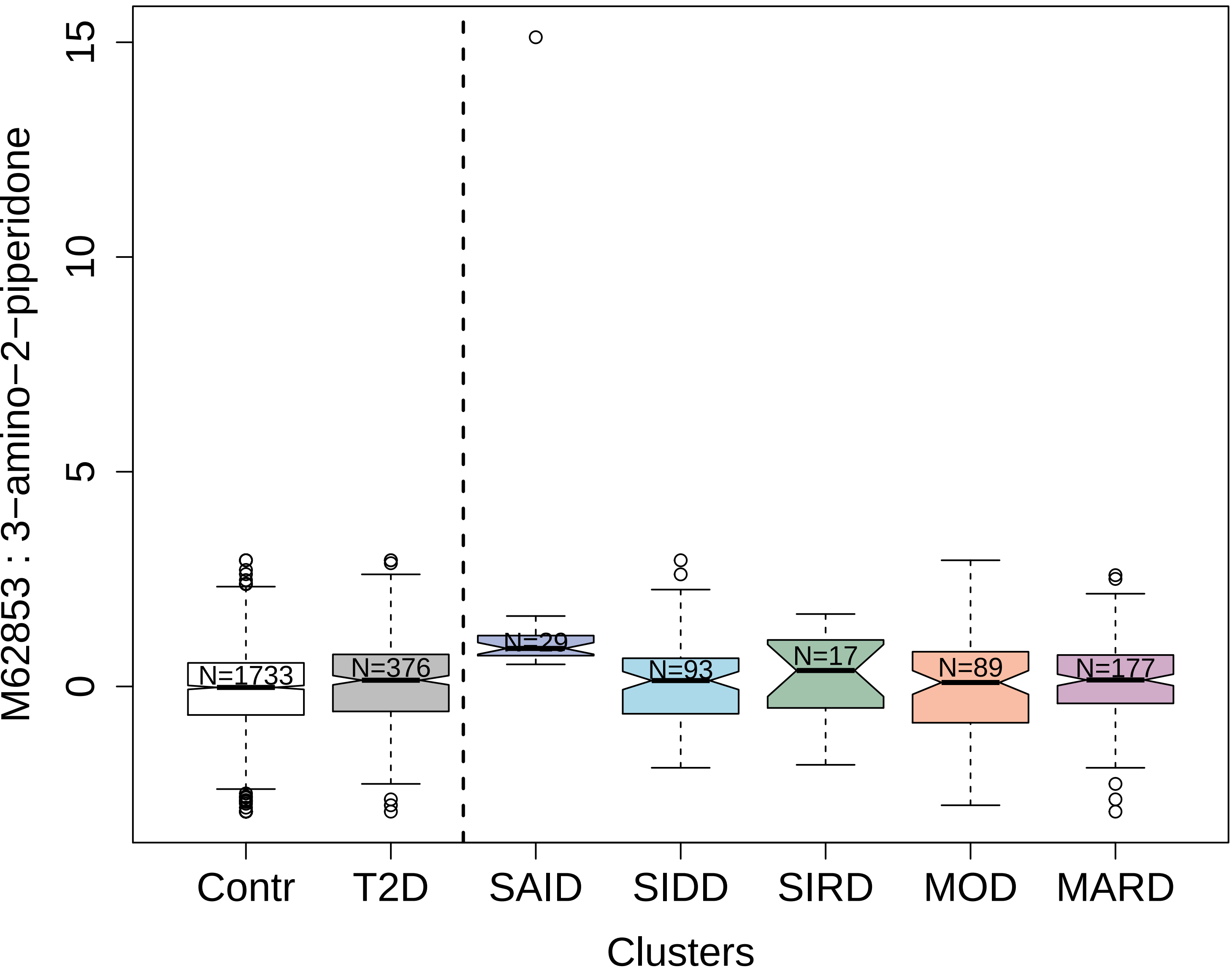

M62853 : 3−amino−2−piperidone
0
5
10
15
N=1733
N=376
N=29
N=93
N=17
N=89
N=177
Contr
T2D
SAID
SIDD
SIRD
MOD
MARD
Clusters

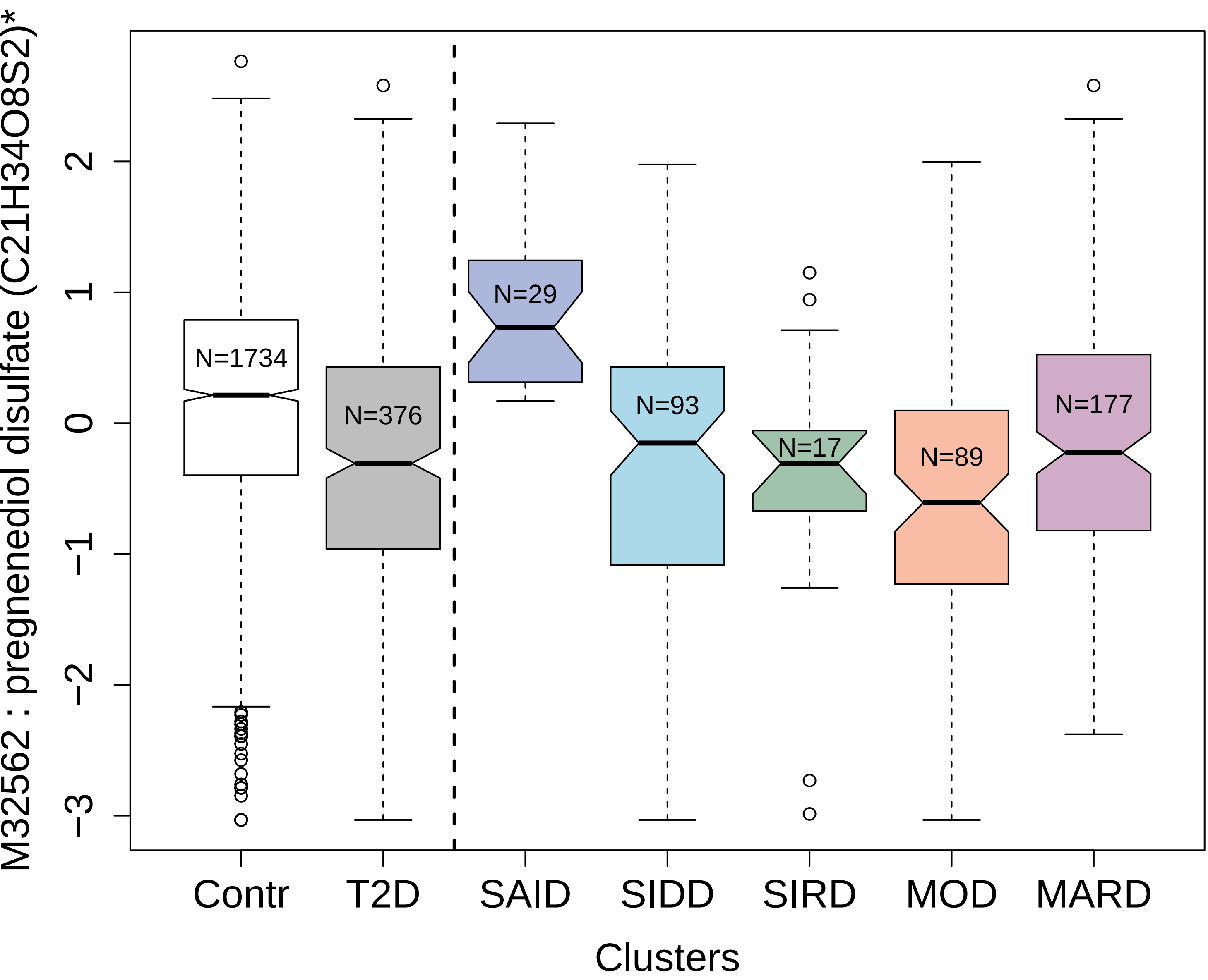
M32562 : pregnenediol disulfate (C21H34O8S2)*
N=1734
N=376
N=29
N=93
N=17
N=89
N=177
2
1
0
−1
−2
−3
Contr
T2D
SAID
SIDD
SIRD
MOD
MARD
Clusters

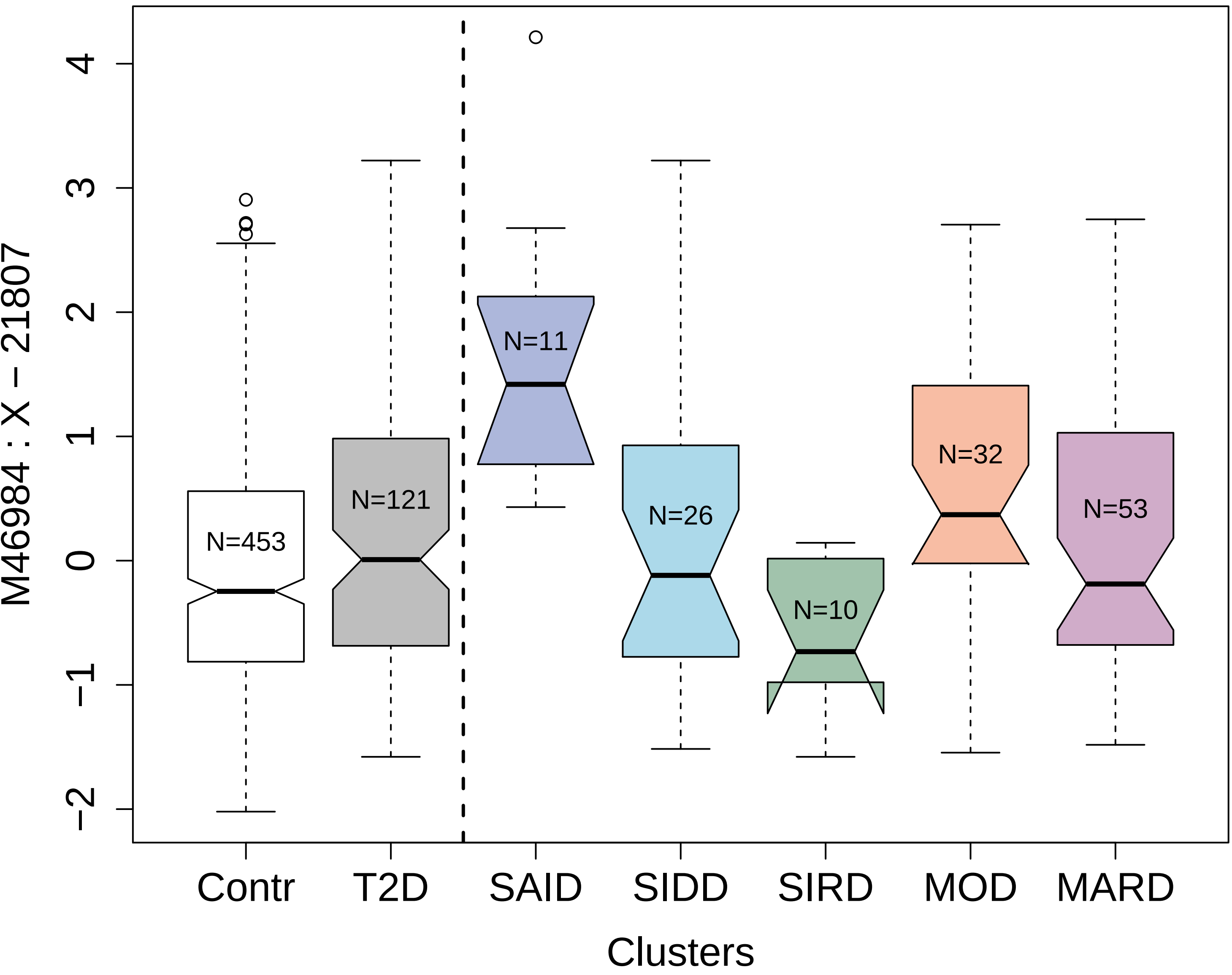
M46984 : X − 21807
4
3
2
1
0
−1
−2
N=453
N=121
N=11
N=26
N=10
N=32
N=53
Contr
T2D
SAID
SIDD
SIRD
MOD
MARD
Clusters

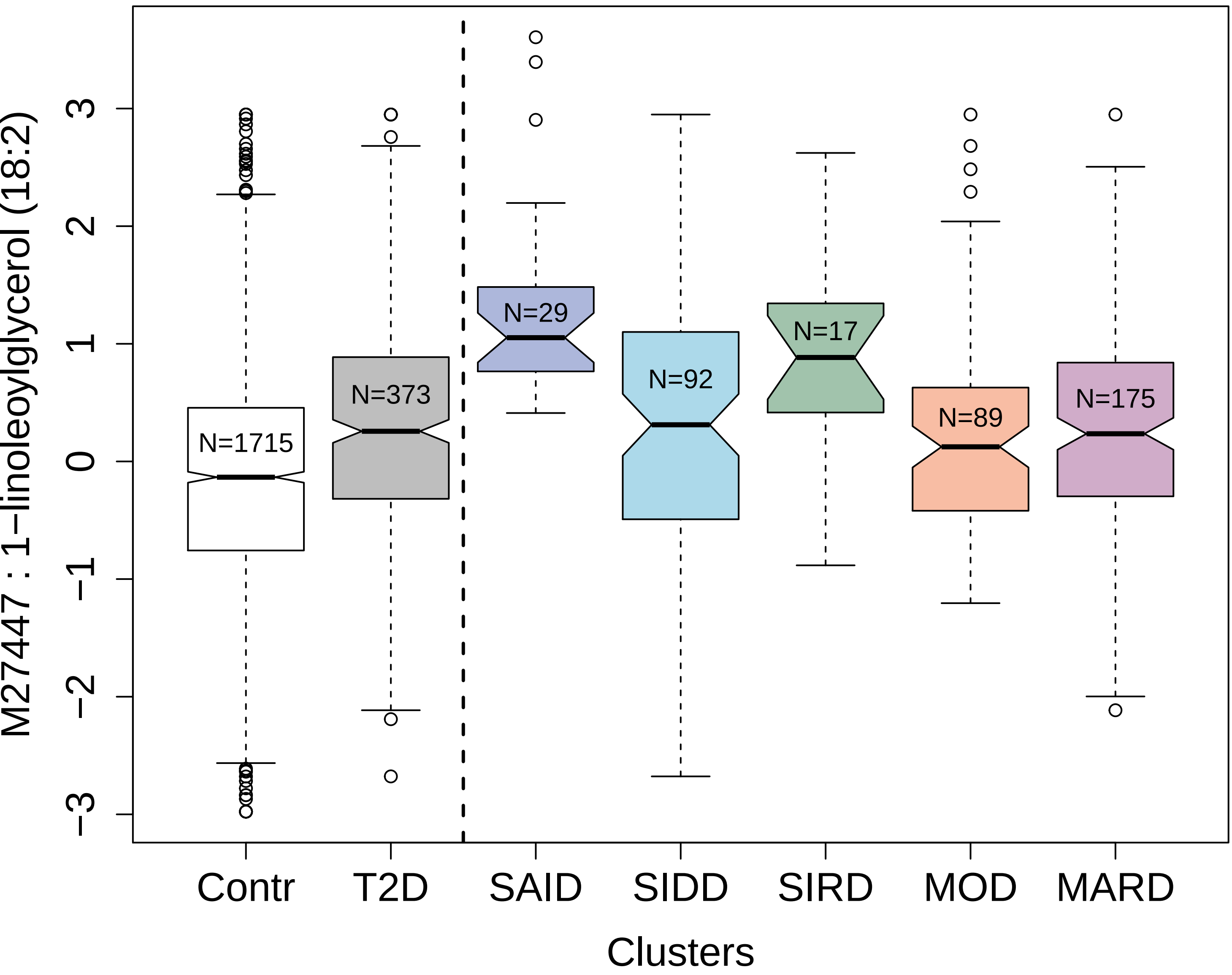

M27447 : 1−linoleoylglycerol (18:2)
3
2
1
0
−1
−2
−3
N=1715
N=373
N=29
N=92
N=17
N=89
N=175
Contr
T2D
SAID
SIDD
SIRD
MOD
MARD
Clusters

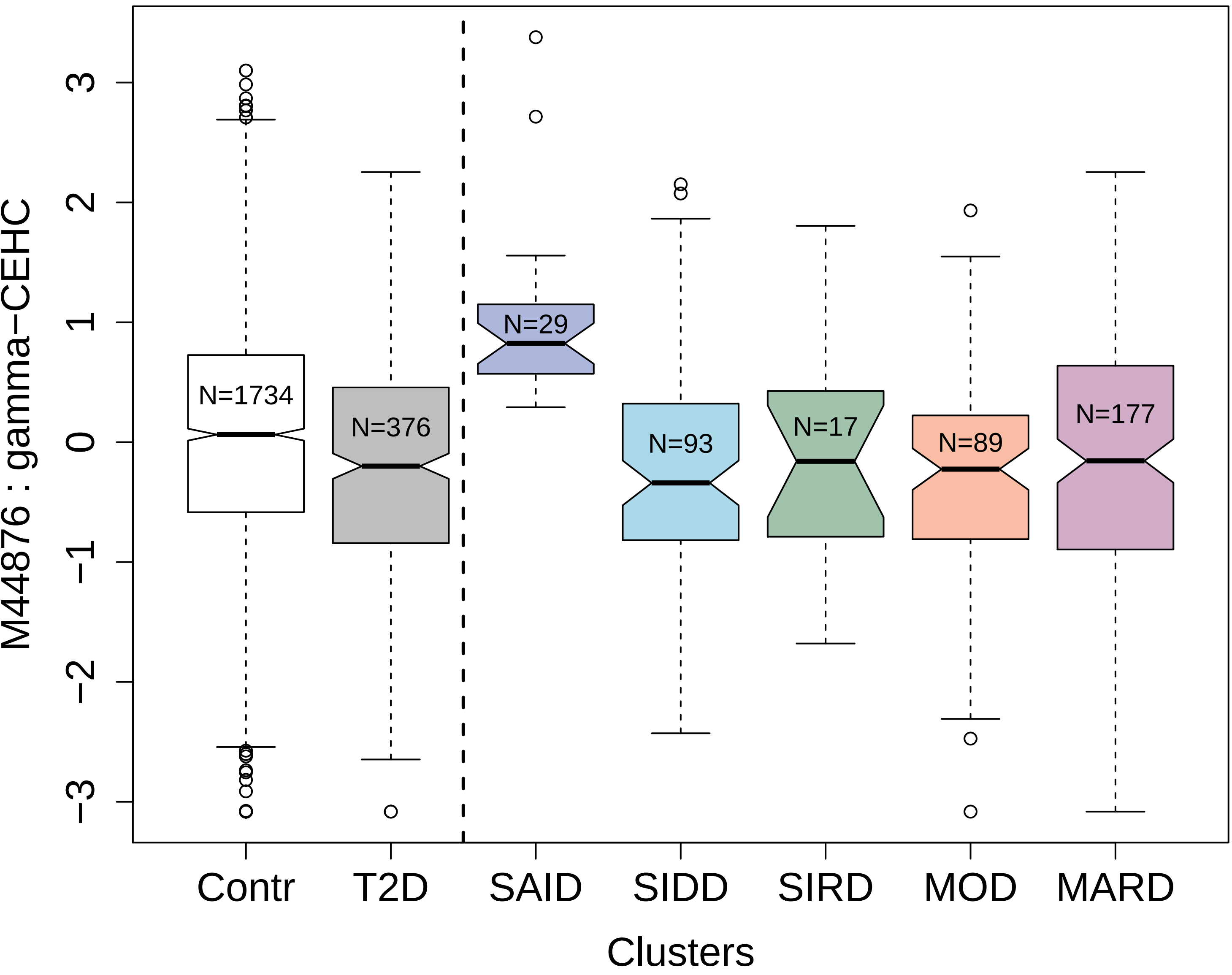

M44876 : gamma−CEHC
3
2
1
0
−1
−2
−3
N=1734
N=376
N=29
N=93
N=17
N=89
N=177
Contr
T2D
SAID
SIDD
SIRD
MOD
MARD
Clusters

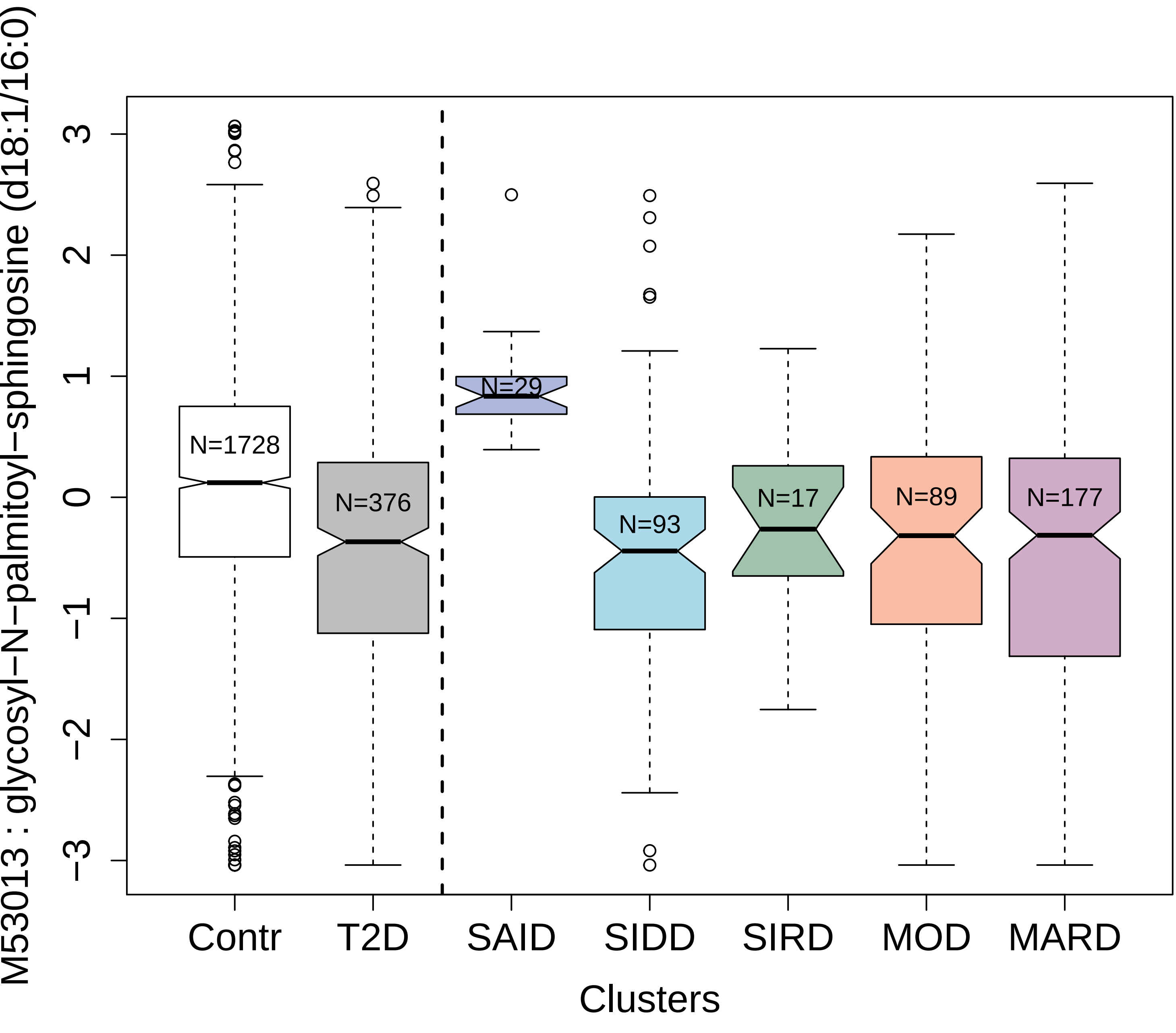

M53013 : glycosyl−N−palmitoyl−sphingosine (d18:1/16:0)
3
2
1
0
−1
−2
−3
N=1728
N=376
N=29
N=93
N=17
N=89
N=177
Contr
T2D
SAID
SIDD
SIRD
MOD
MARD
Clusters

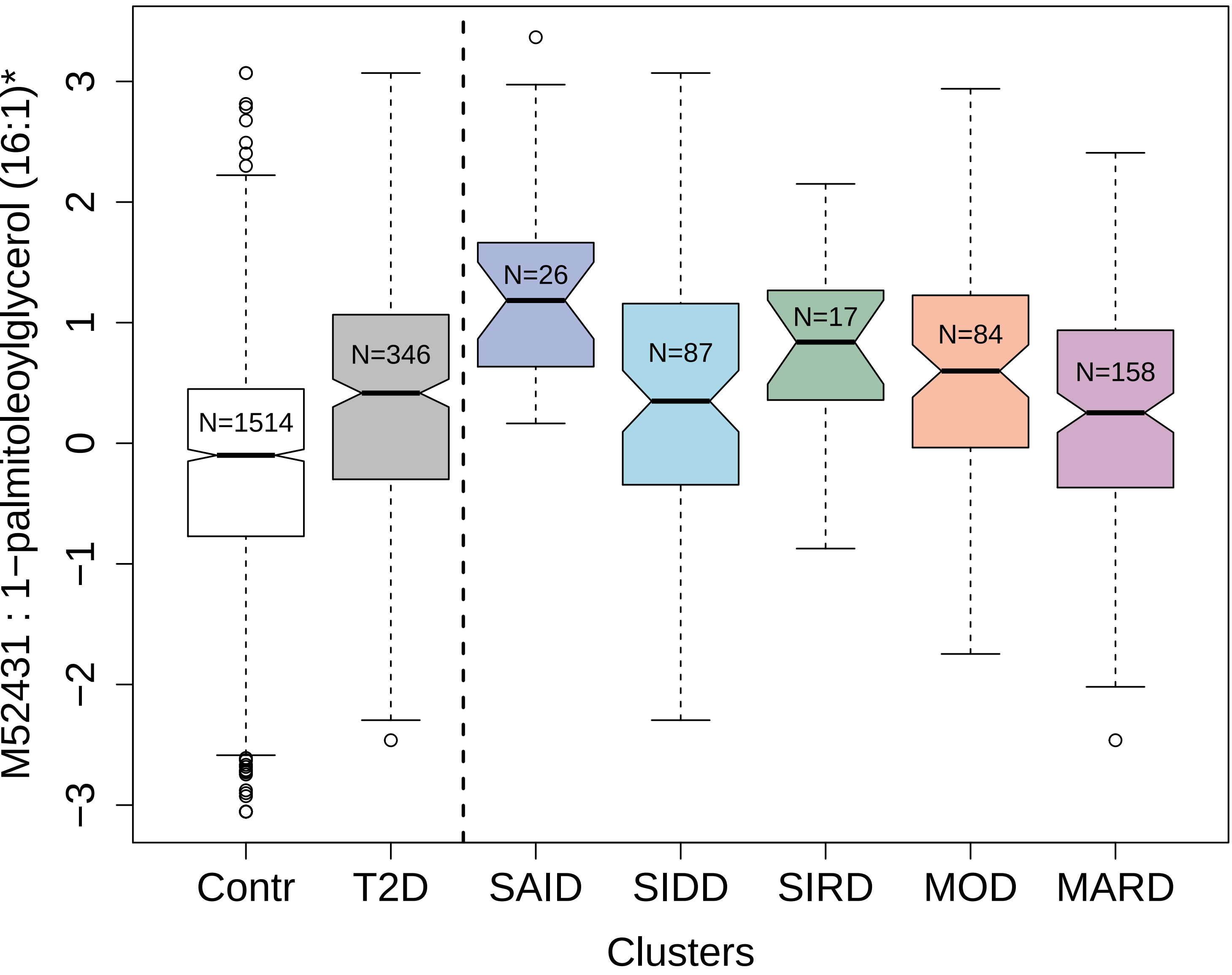

M52431 : 1−palmitoleoylglycerol (16:1)*
3
2
1
0
−1
−2
−3
N=1514
N=346
N=26
N=87
N=17
N=84
N=158
Contr
T2D
SAID
SIDD
SIRD
MOD
MARD
Clusters

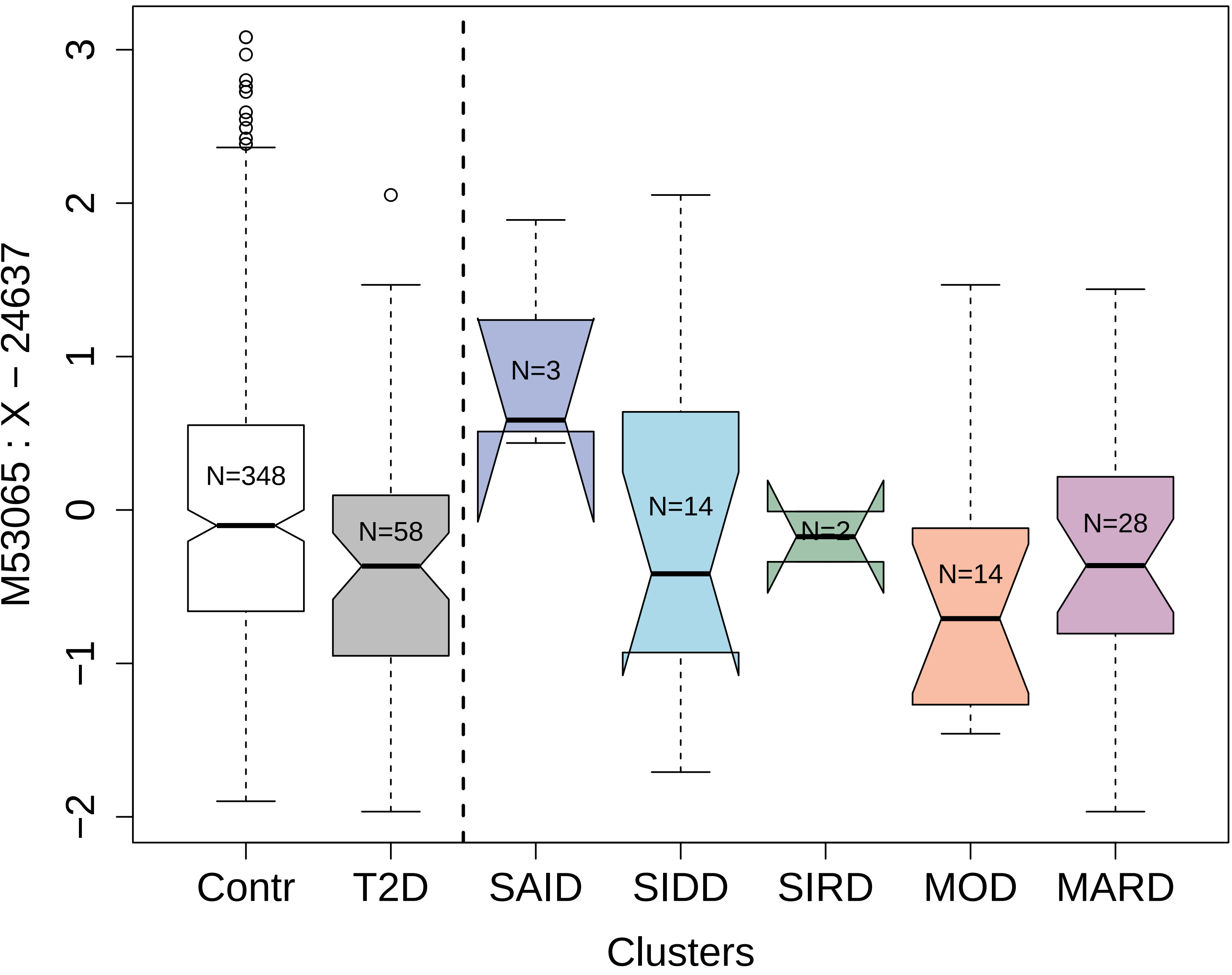

M53065 : X − 24637
3
2
1
0
−1
−2
N=348
N=58
N=3
N=14
N=2
N=14
N=28
Contr
T2D
SAID
SIDD
SIRD
MOD
MARD
Clusters

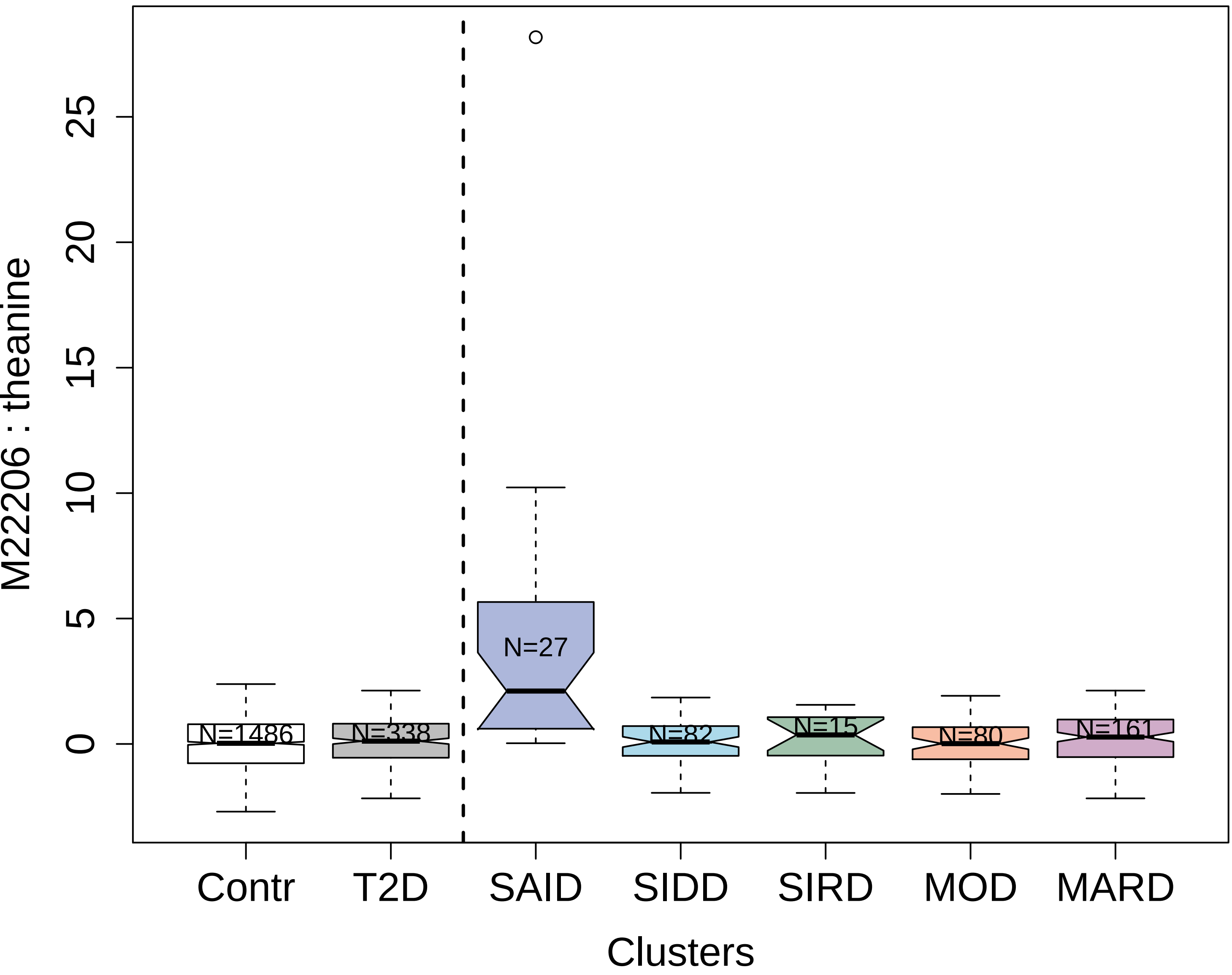

M22206 : theanine
0
5
10
15
20
25
N=1486
N=338
N=27
N=82
N=15
N=80
N=161
Contr
T2D
SAID
SIDD
SIRD
MOD
MARD
Clusters

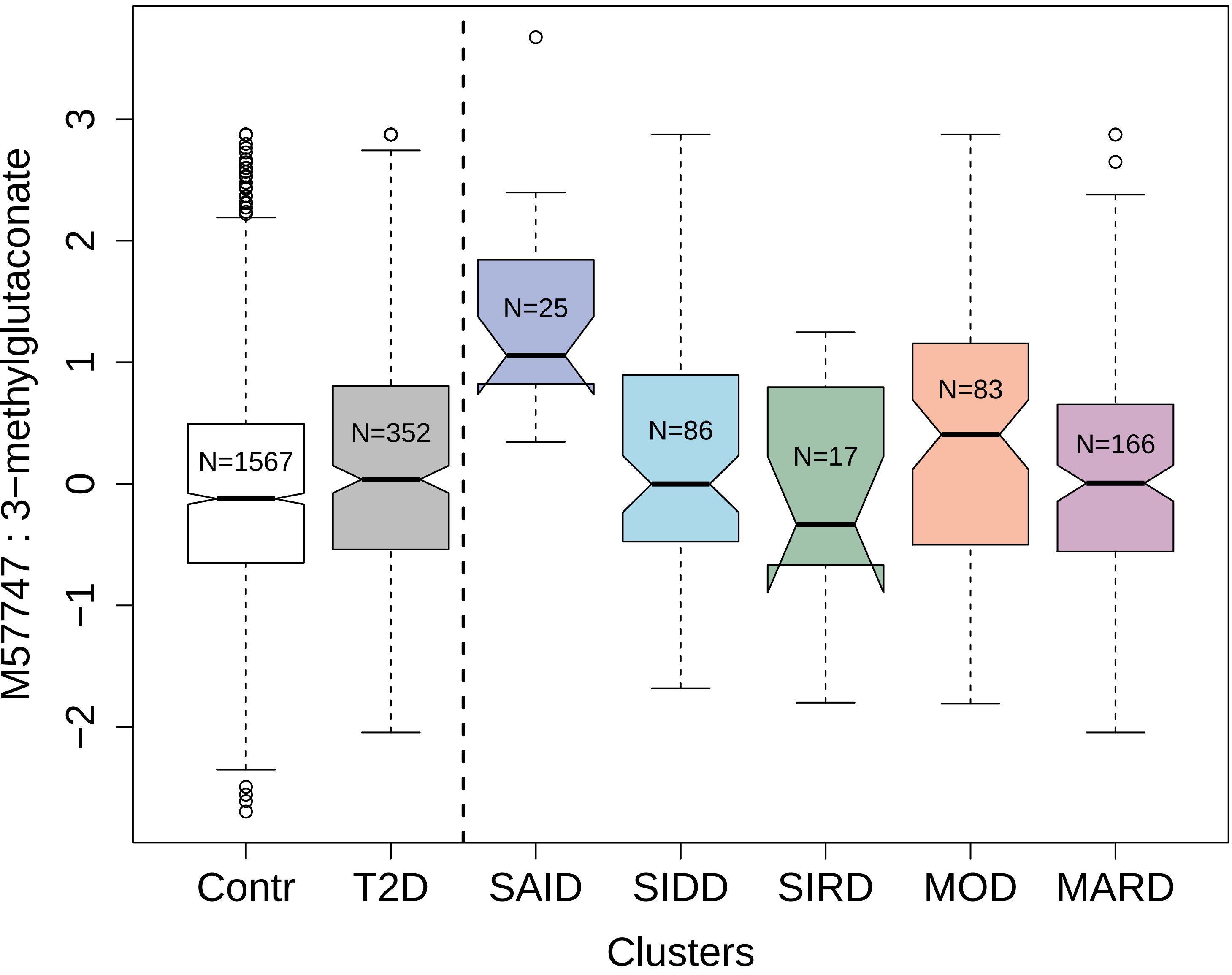

M57747 : 3−methylglutaconate
3
2
1
0
−1
−2
N=1567
N=352
N=25
N=86
N=17
N=83
N=166
Contr
T2D
SAID
SIDD
SIRD
MOD
MARD
Clusters

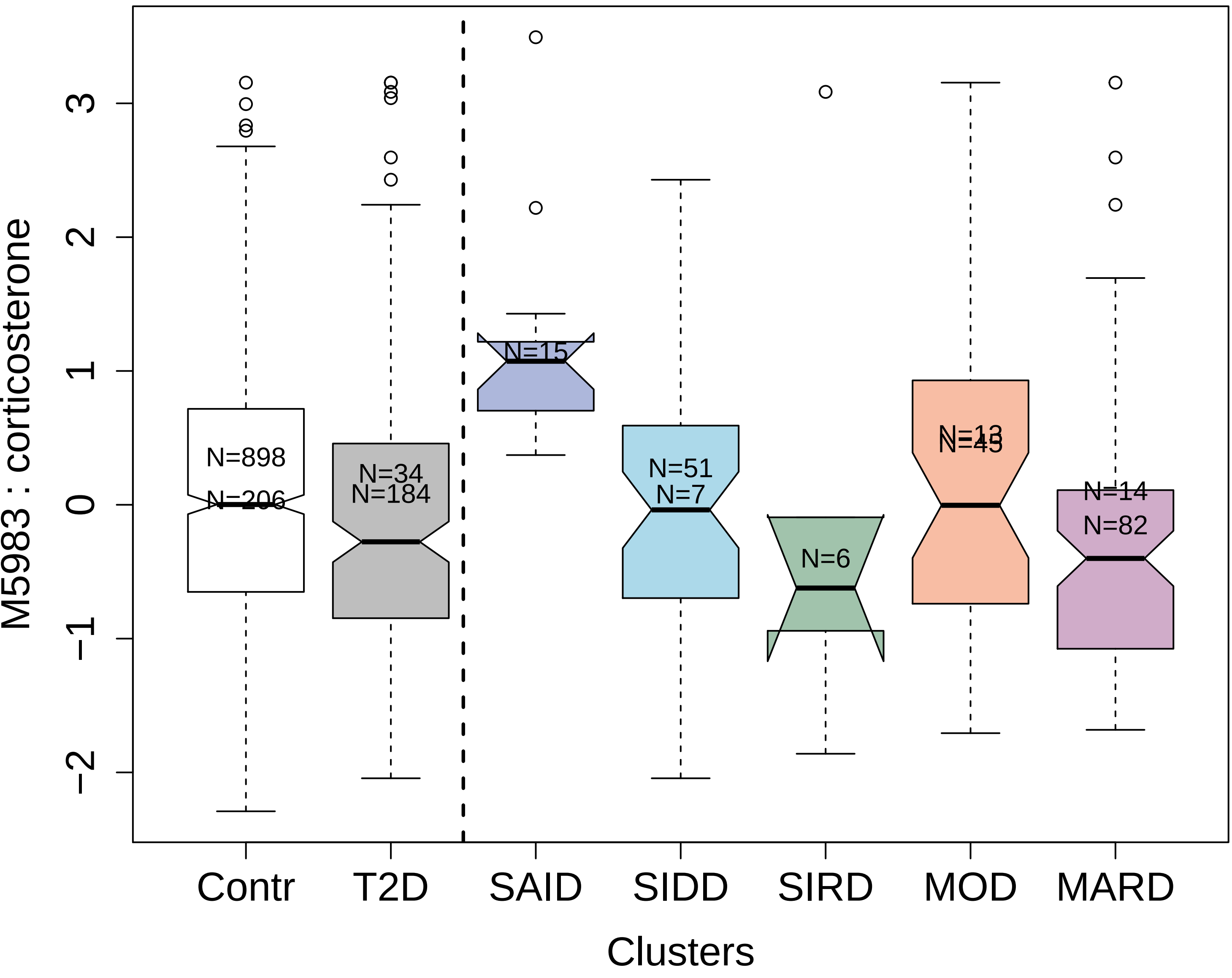

M5983 : corticosterone
Clusters
Contr
T2D
SAID
SIDD
SIRD
MOD
MARD
N=898
N=206
N=34
N=184
N=15
N=51
N=7
N=6
N=13
N=43
N=14
N=82

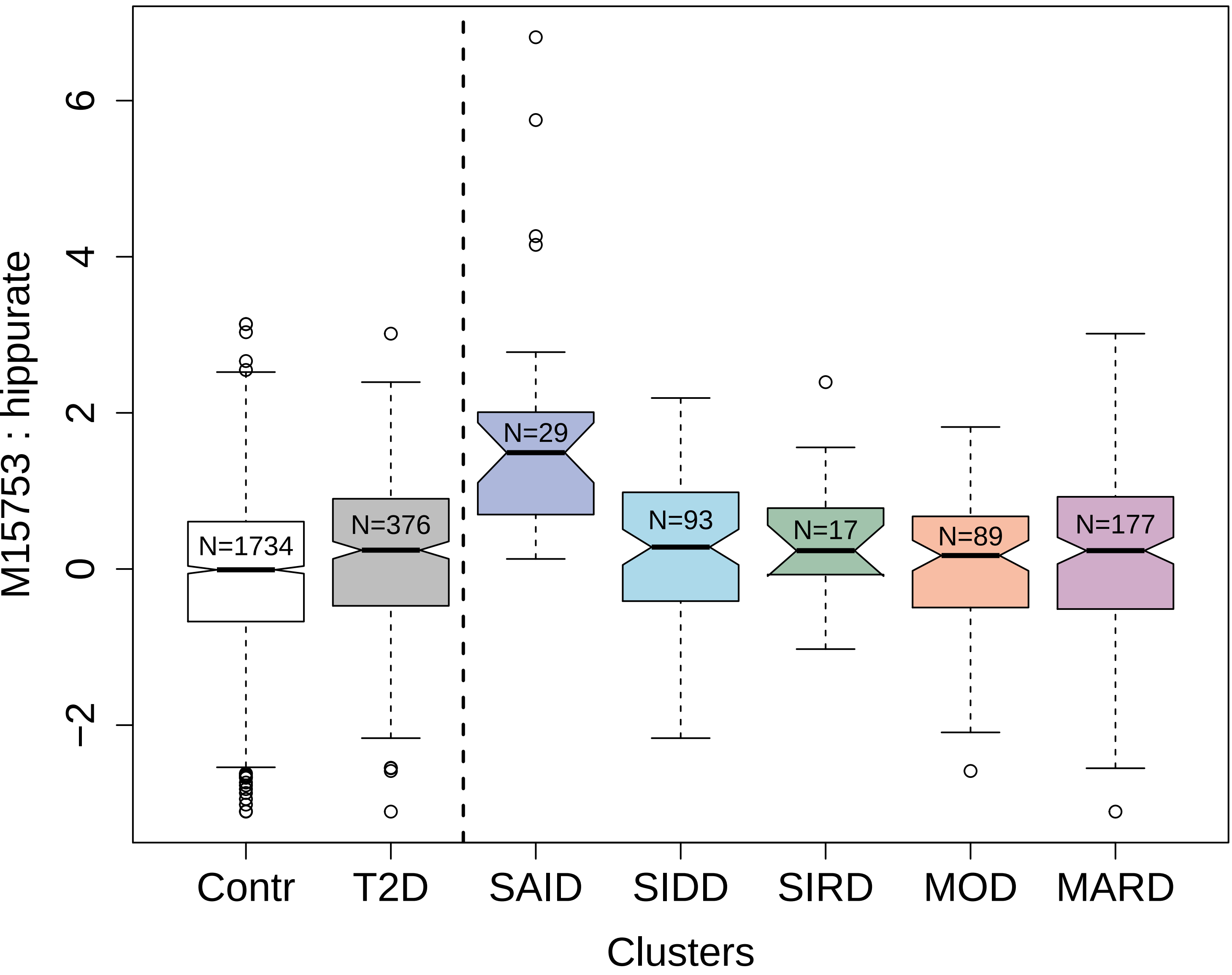

M15753 : hippurate
6
4
2
0
−2
N=1734
N=376
N=29
N=93
N=17
N=89
N=177
Contr
T2D
SAID
SIDD
SIRD
MOD
MARD
Clusters

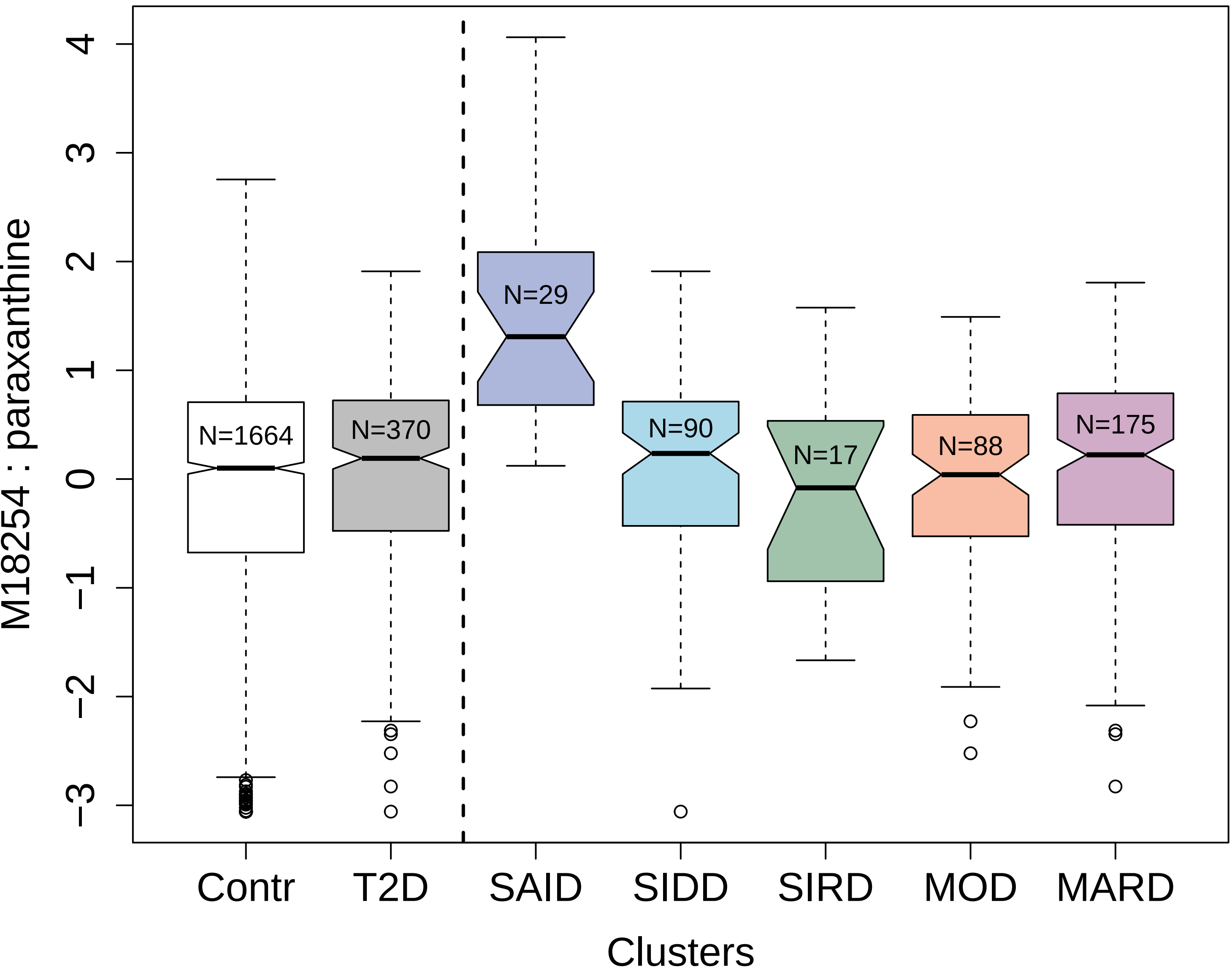

M18254 : paraxanthine
4
3
2
1
0
−1
−2
−3
N=1664
N=370
N=29
N=90
N=17
N=88
N=175
Contr
T2D
SAID
SIDD
SIRD
MOD
MARD
Clusters

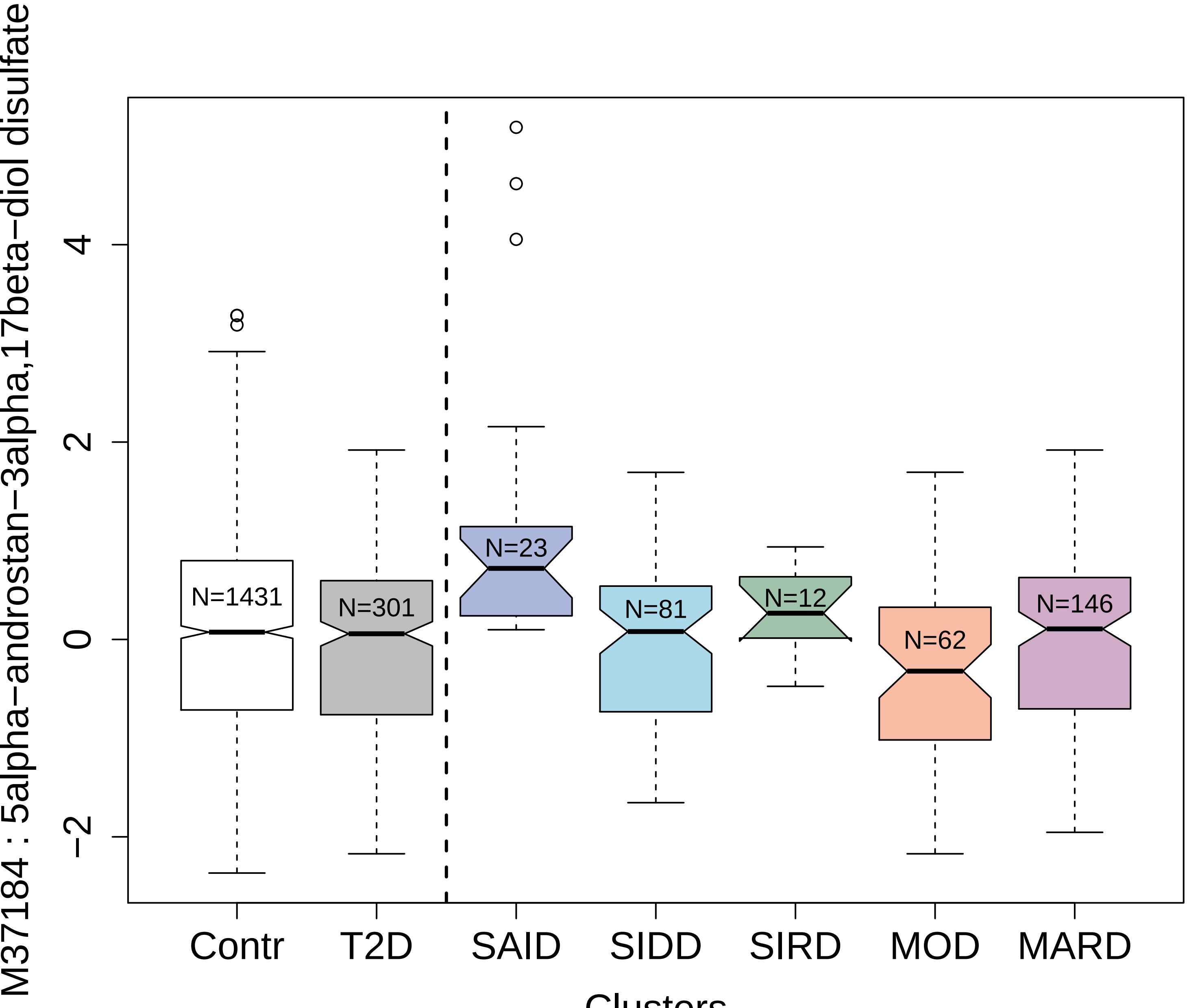
M37184 : 5alpha−androstan−3alpha,17beta−diol disulfate
−2
0
2
4
N=1431
N=301
N=23
N=81
N=12
N=62
N=146
Contr
T2D
SAID
SIDD
SIRD
MOD
MARD
Clusters

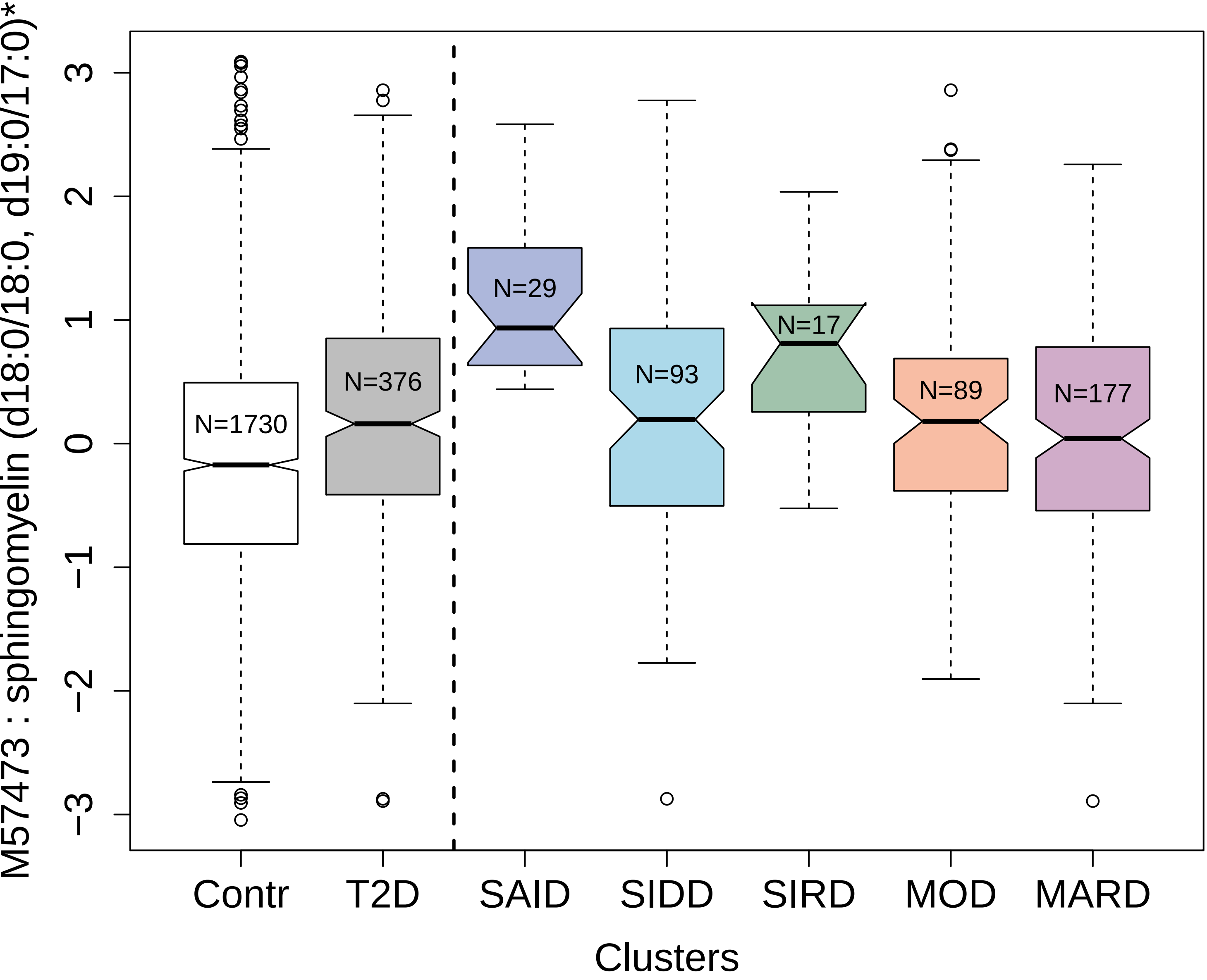

M57473 : sphingomyelin (d18:0/18:0, d19:0/17:0)*
3
2
1
0
−1
−2
−3
N=1730
N=376
N=29
N=93
N=17
N=89
N=177
Contr
T2D
SAID
SIDD
SIRD
MOD
MARD
Clusters

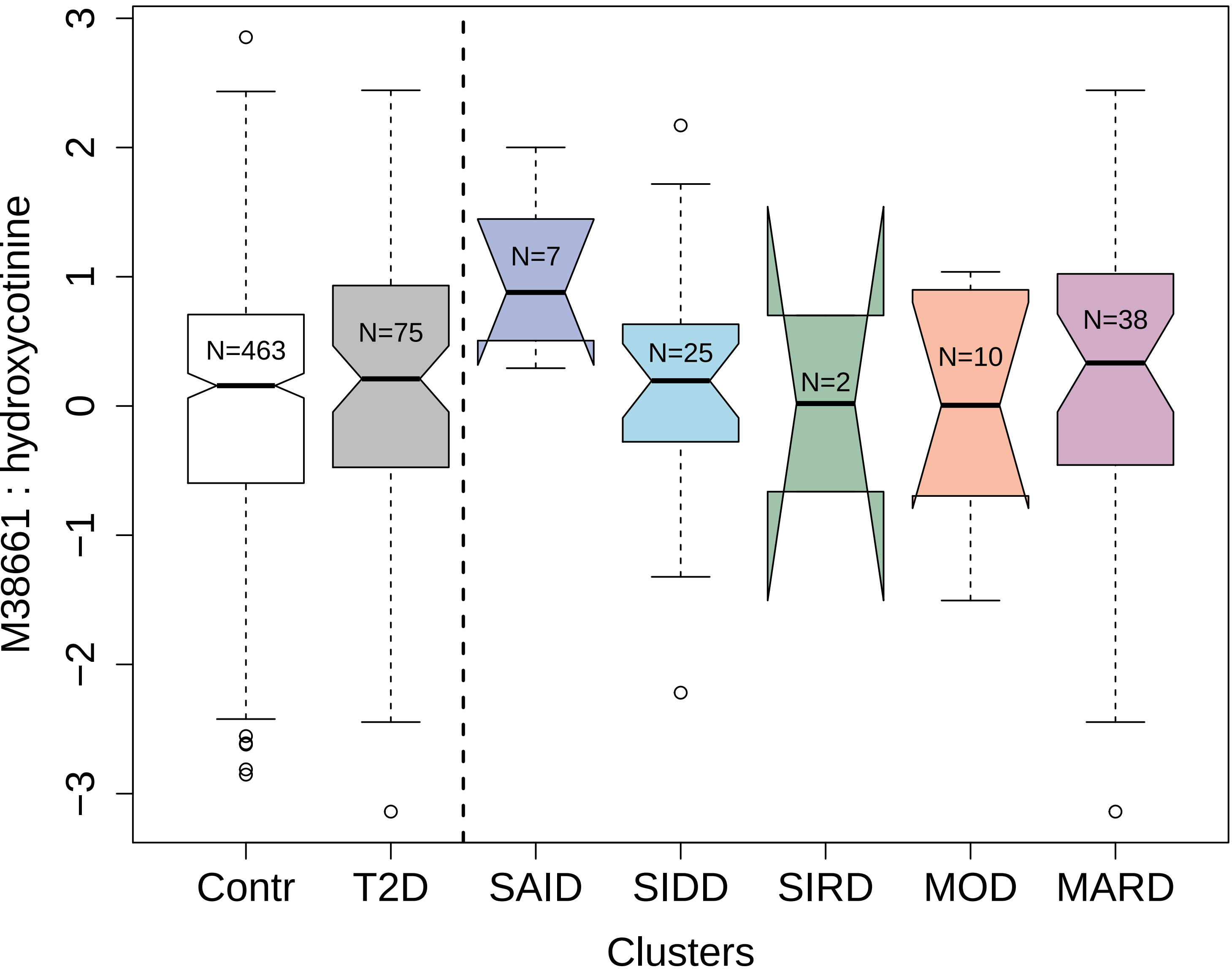

M38661 : hydroxycotinine
3
2
1
0
−1
−2
−3
N=463
N=75
N=7
N=25
N=2
N=10
N=38
Contr
T2D
SAID
SIDD
SIRD
MOD
MARD
Clusters

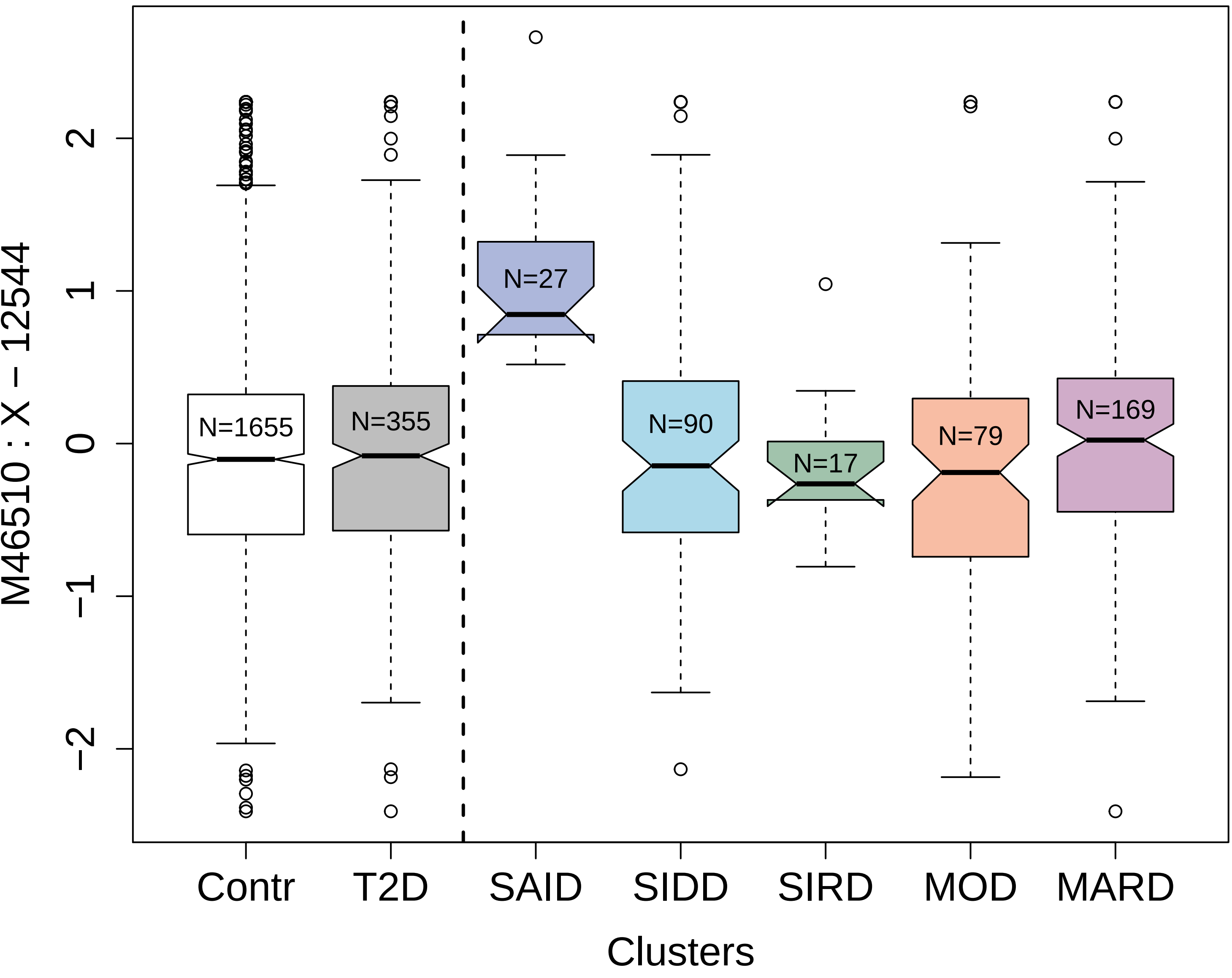

M46510 : X − 12544
2
1
0
−1
−2
N=1655
N=355
N=27
N=90
N=17
N=79
N=169
Contr
T2D
SAID
SIDD
SIRD
MOD
MARD
Clusters

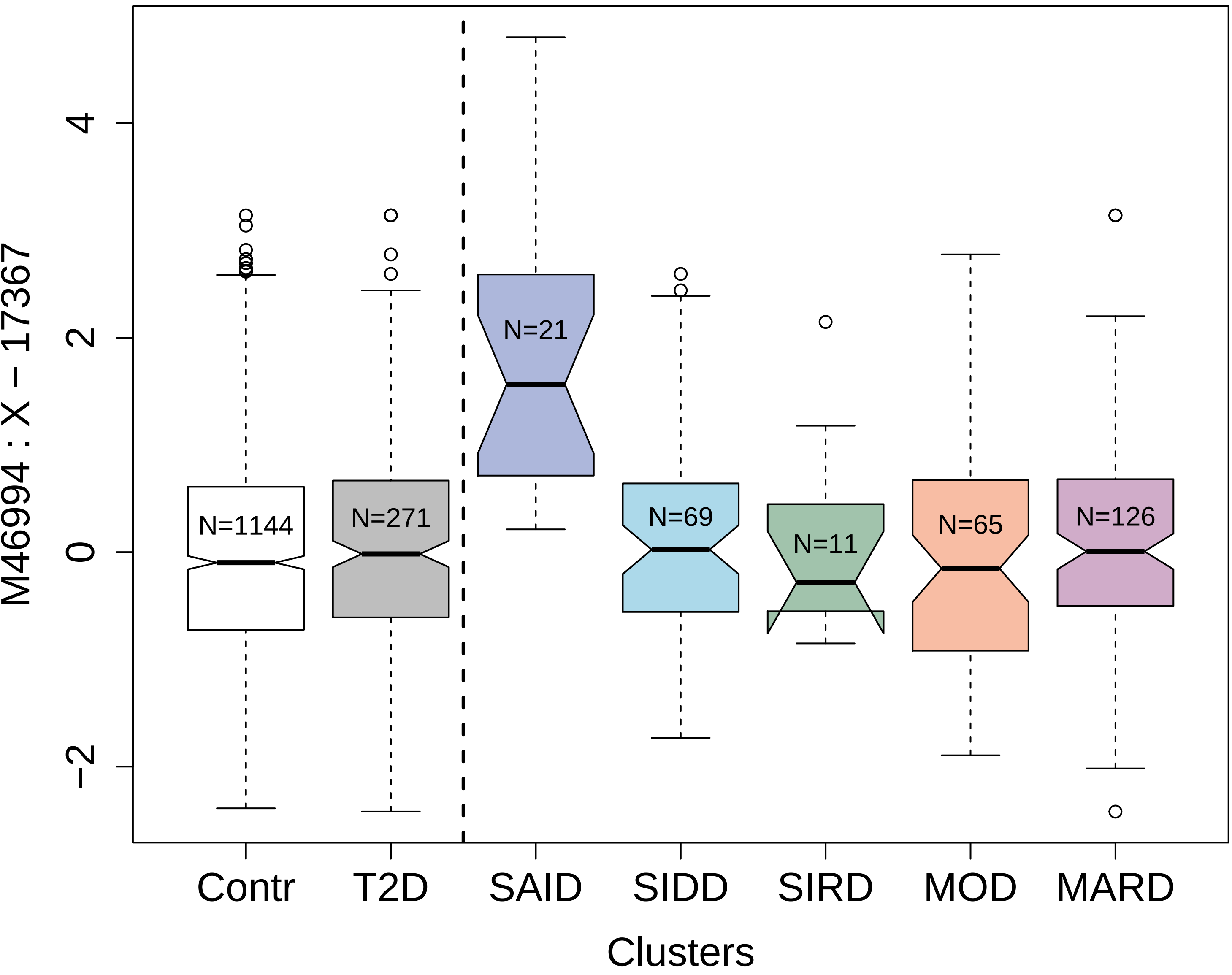

M46994 : X − 17367
4
2
0
−2
N=1144
N=271
N=21
N=69
N=11
N=65
N=126
Contr
T2D
SAID
SIDD
SIRD
MOD
MARD
Clusters

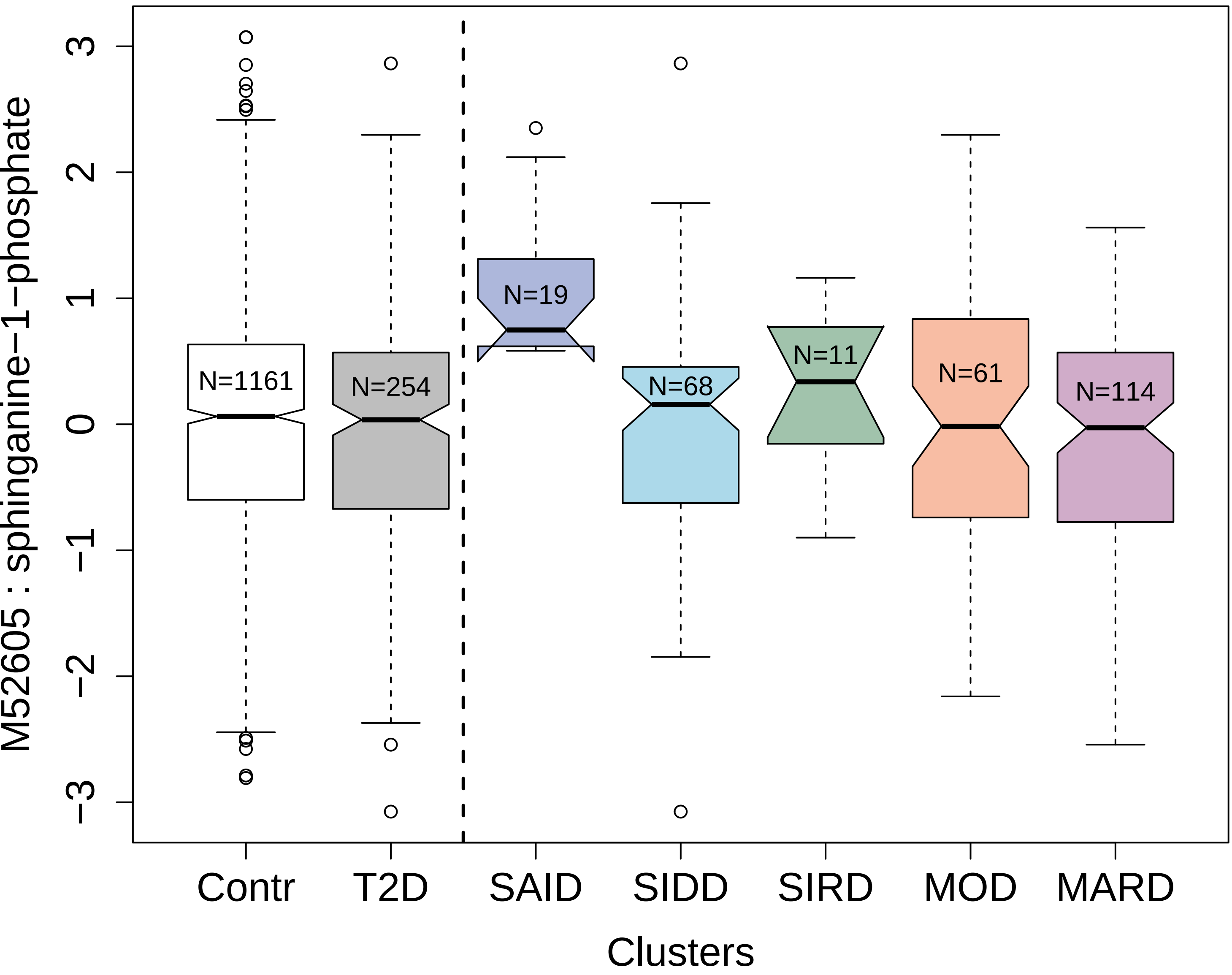

M52605 : sphinganine−1−phosphate
3
2
1
0
−1
−2
−3
N=1161
N=254
N=19
N=68
N=11
N=61
N=114
Contr
T2D
SAID
SIDD
SIRD
MOD
MARD
Clusters

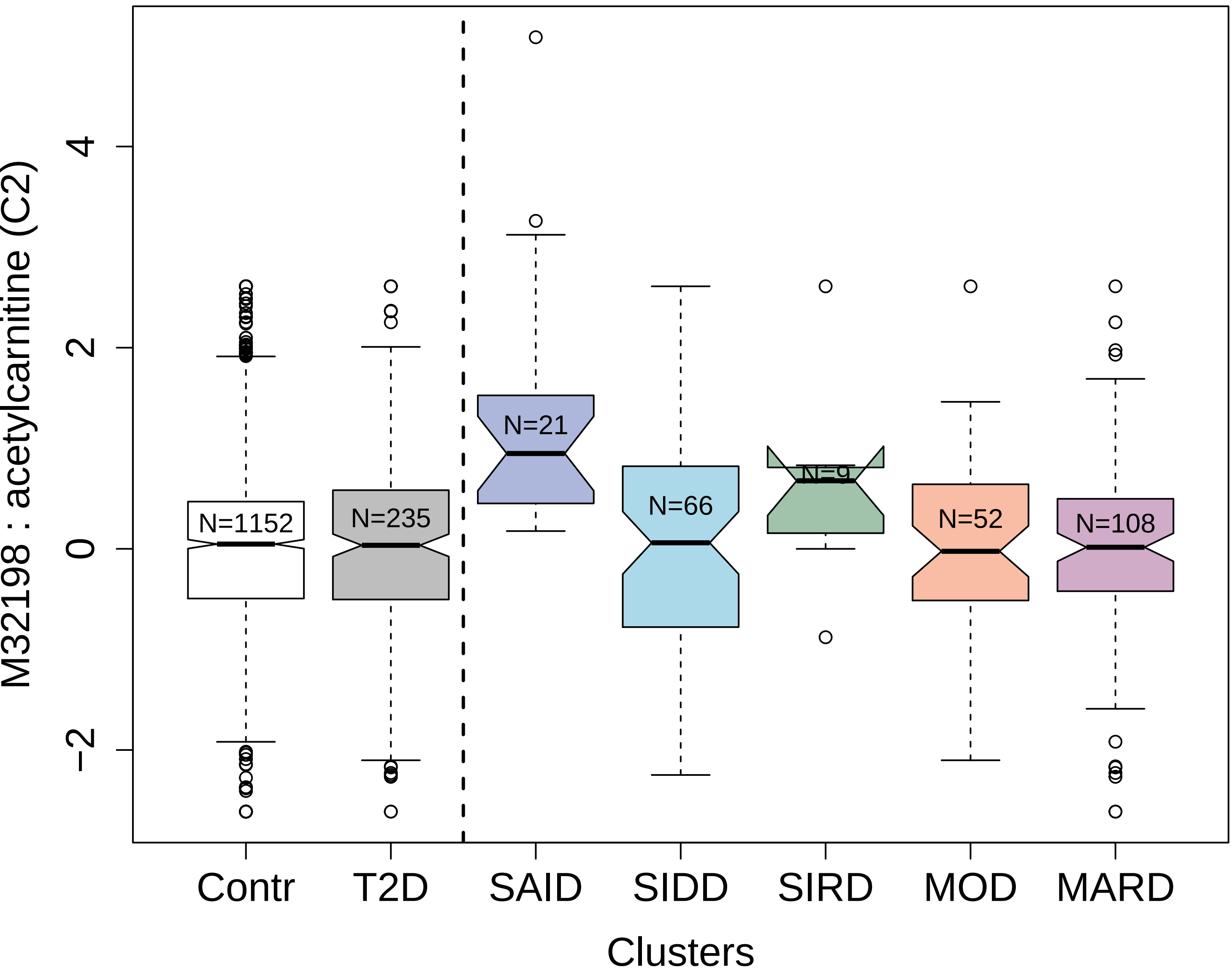

M32198 : acetylcarnitine (C2)
4
2
0
−2
N=1152
N=235
N=21
N=66
N=9
N=52
N=108
Contr
T2D
SAID
SIDD
SIRD
MOD
MARD
Clusters

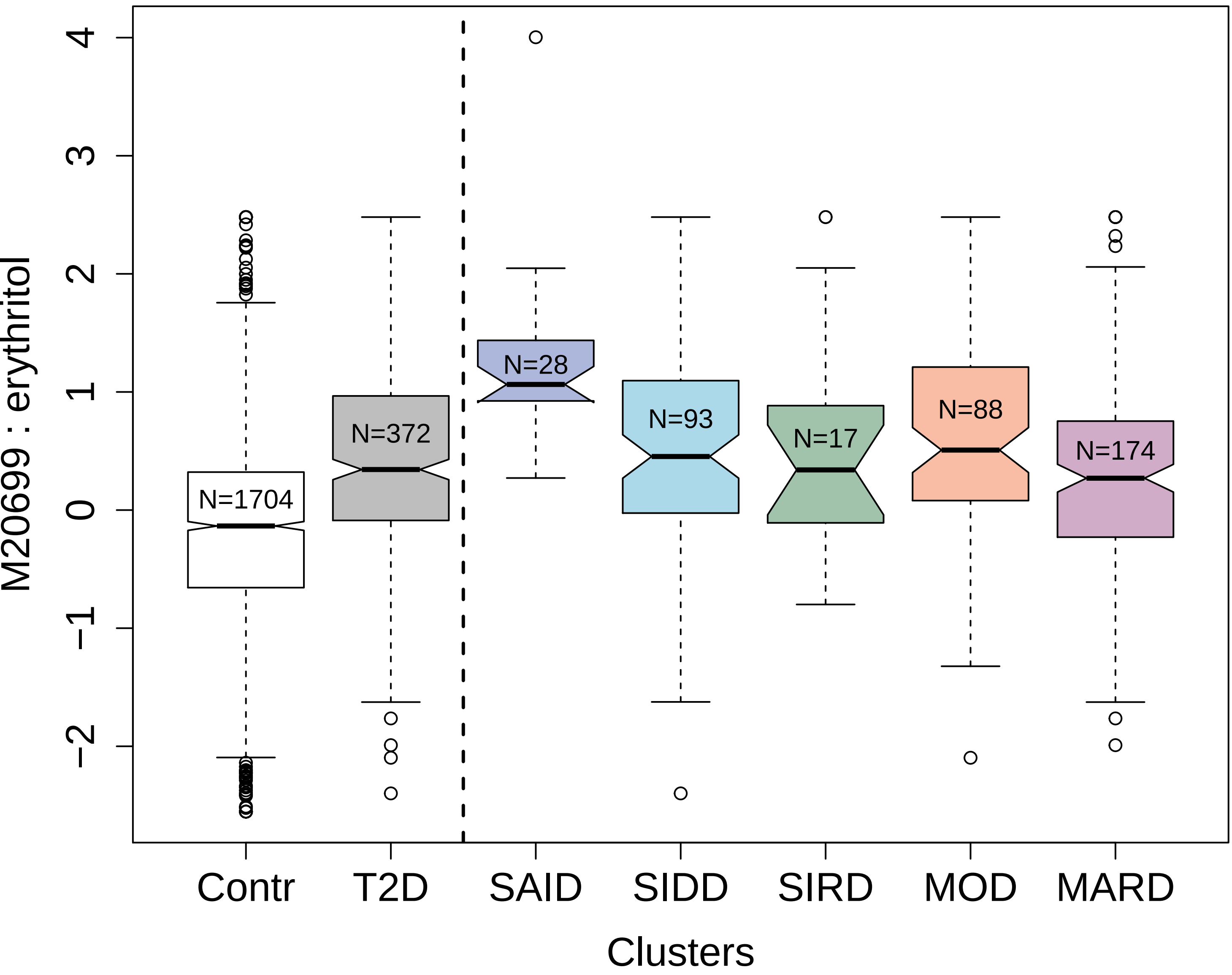
M20699 : erythritol
4
3
2
1
0
−1
−2
N=1704
N=372
N=28
N=93
N=17
N=88
N=174
Contr
T2D
SAID
SIDD
SIRD
MOD
MARD
Clusters

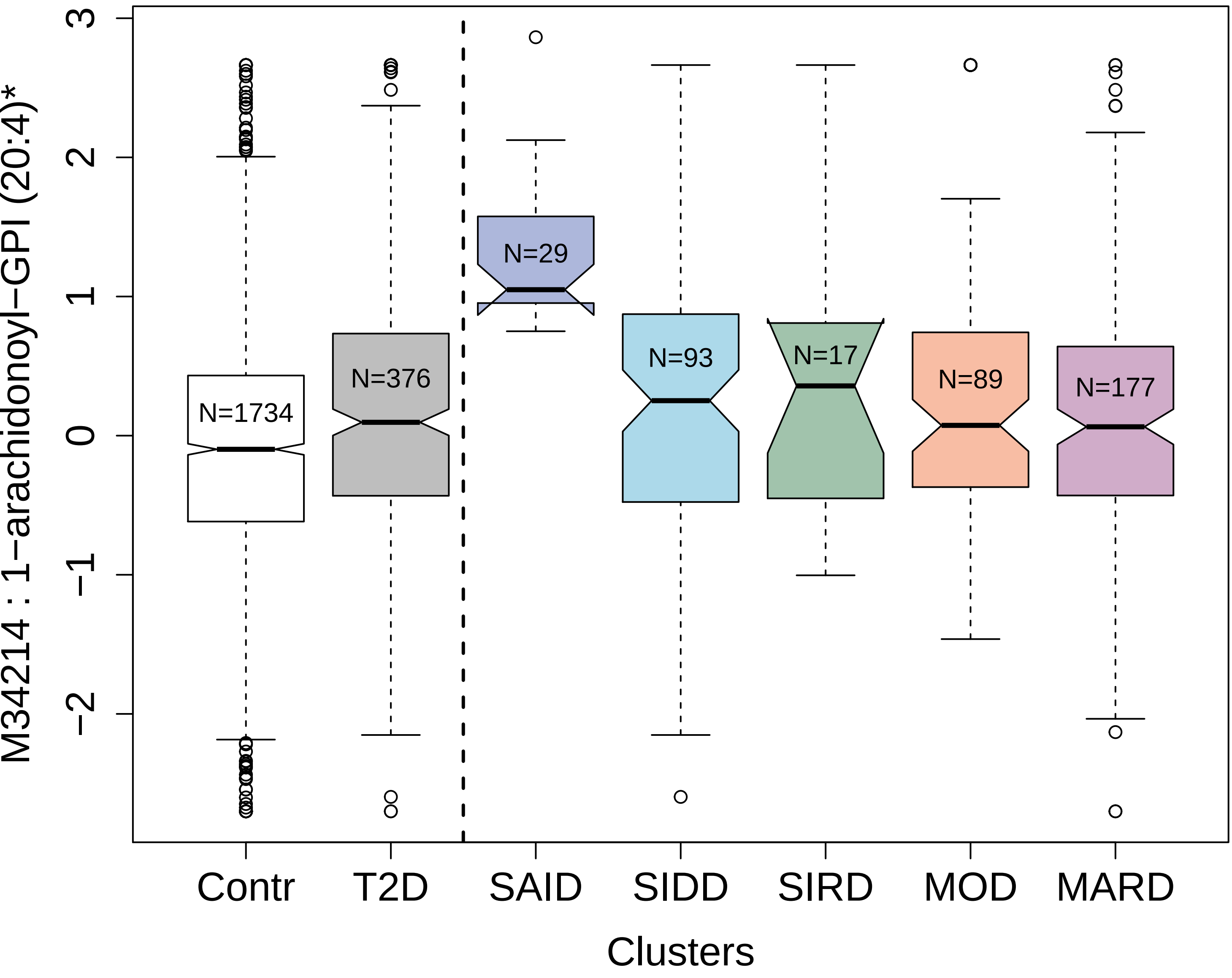

M34214 : 1−arachidonoyl−GPI (20:4)*
3
2
1
0
−1
−2
N=1734
N=376
N=29
N=93
N=17
N=89
N=177
Contr
T2D
SAID
SIDD
SIRD
MOD
MARD
Clusters

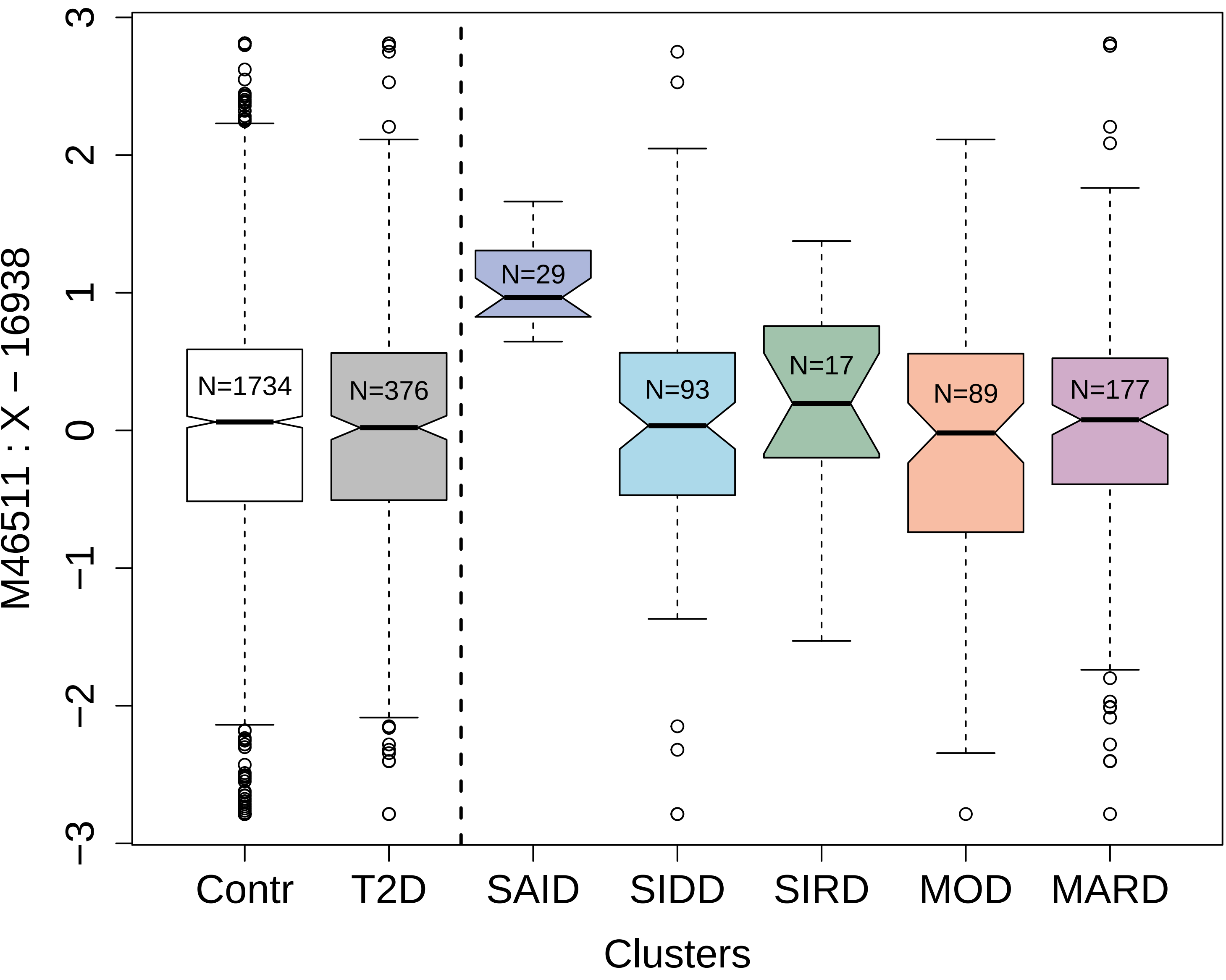

M46511 : X − 16938
3
2
1
0
−1
−2
−3
N=1734
N=376
N=29
N=93
N=17
N=89
N=177
Contr
T2D
SAID
SIDD
SIRD
MOD
MARD
Clusters

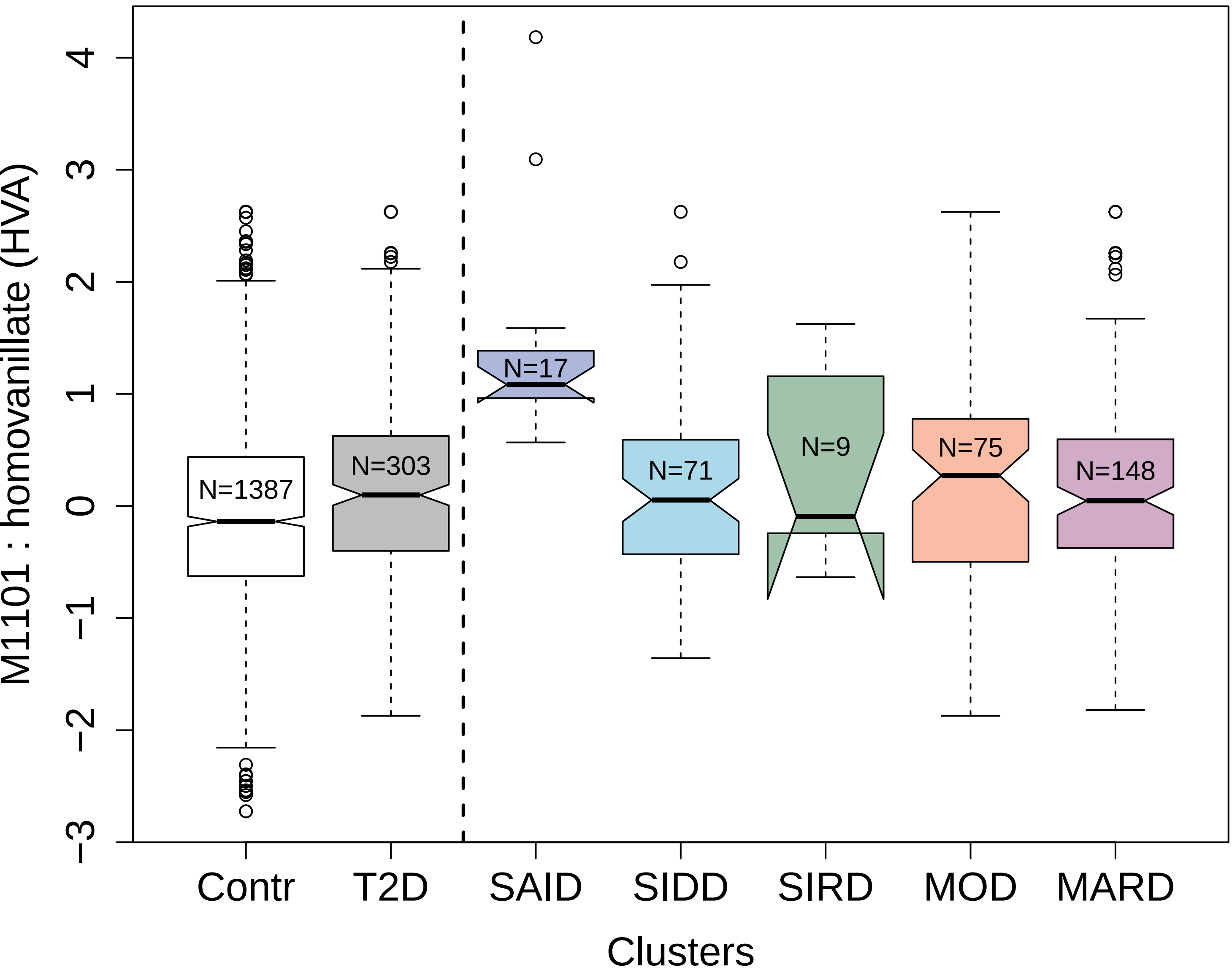

M1101 : homovanillate (HVA)
4
3
2
1
0
−1
−2
−3
N=1387
N=303
N=17
N=71
N=9
N=75
N=148
Contr
T2D
SAID
SIDD
SIRD
MOD
MARD
Clusters

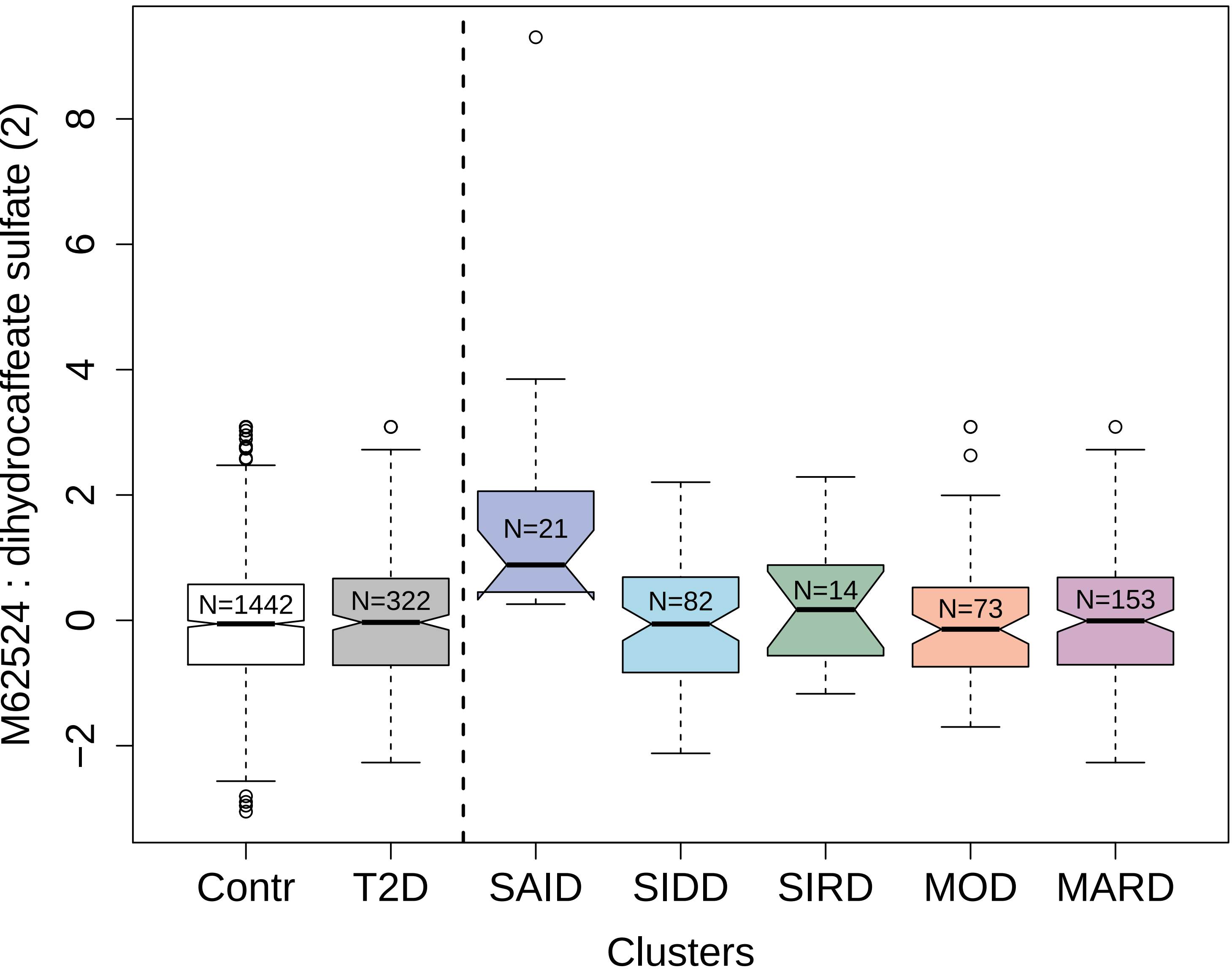
M62524 : dihydrocaffeate sulfate (2)
−2
0
2
4
6
8
N=1442
N=322
N=21
N=82
N=14
N=73
N=153
Contr
T2D
SAID
SIDD
SIRD
MOD
MARD
Clusters

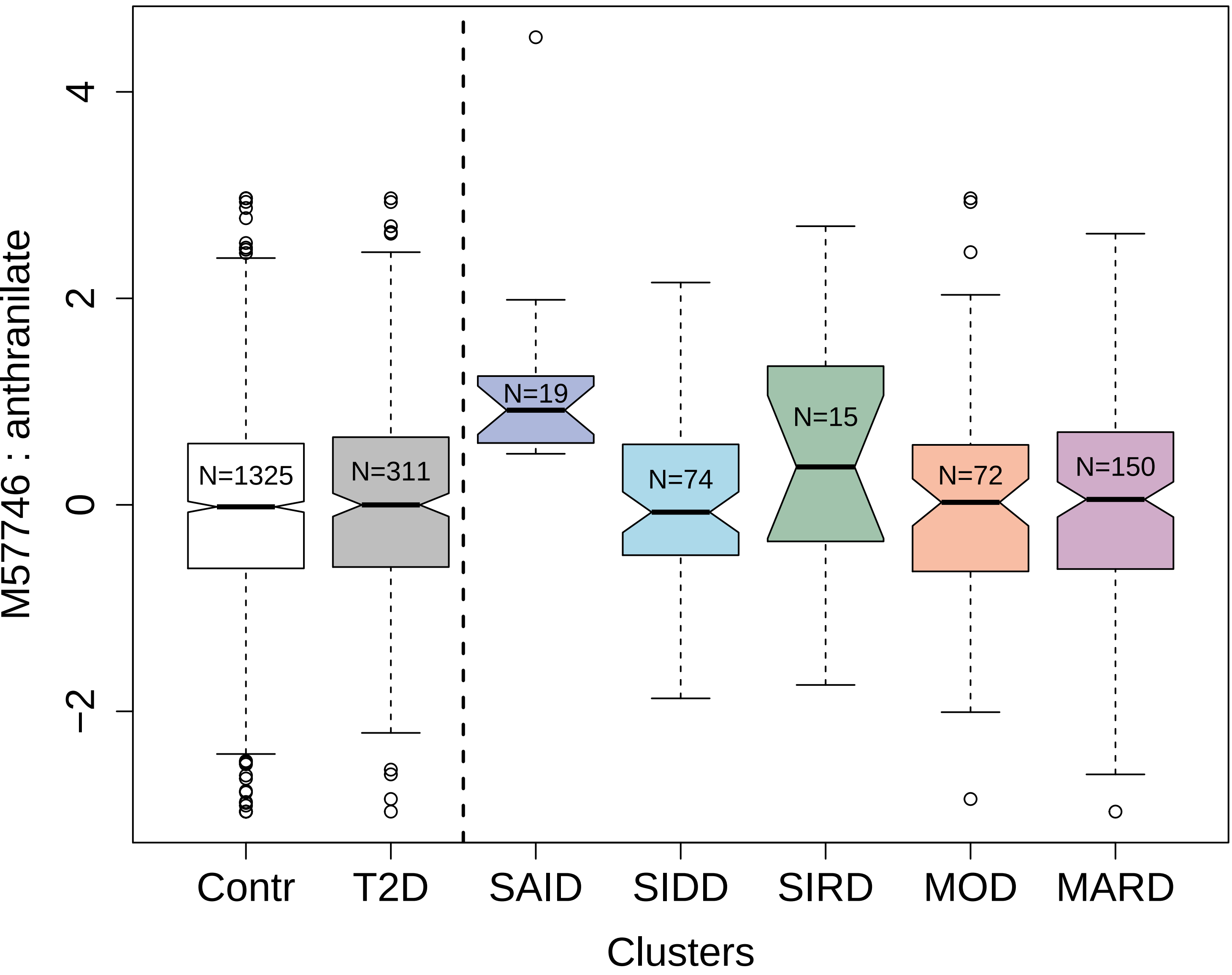

M57746 : anthranilate
4
2
0
−2
N=1325
N=311
N=19
N=74
N=15
N=72
N=150
Contr
T2D
SAID
SIDD
SIRD
MOD
MARD
Clusters

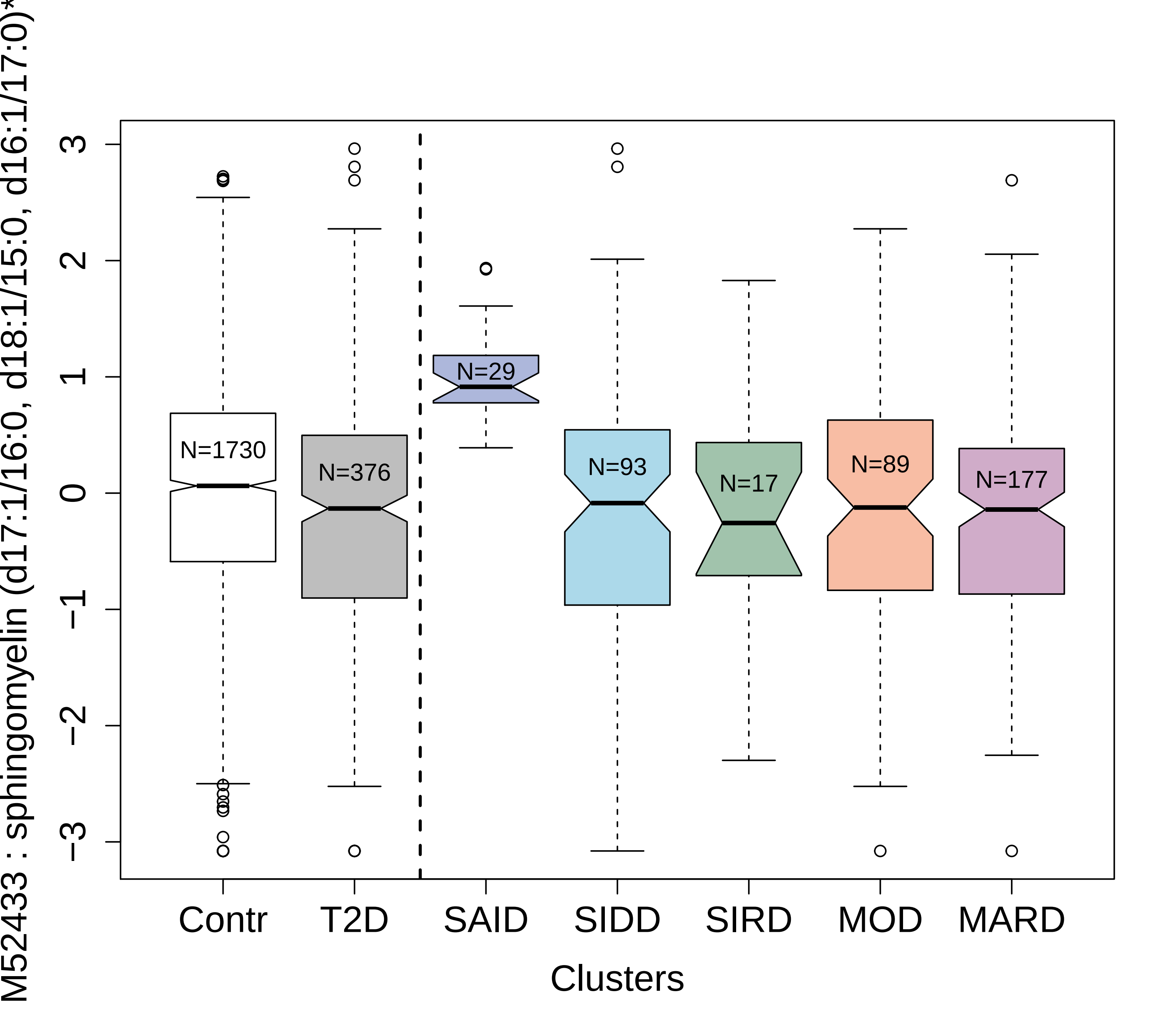
M52433 : sphingomyelin (d17:1/16:0, d18:1/15:0, d16:1/17:0)*
3
2
1
0
−1
−2
−3
N=1730
N=376
N=29
N=93
N=17
N=89
N=177
Contr
T2D
SAID
SIDD
SIRD
MOD
MARD
Clusters

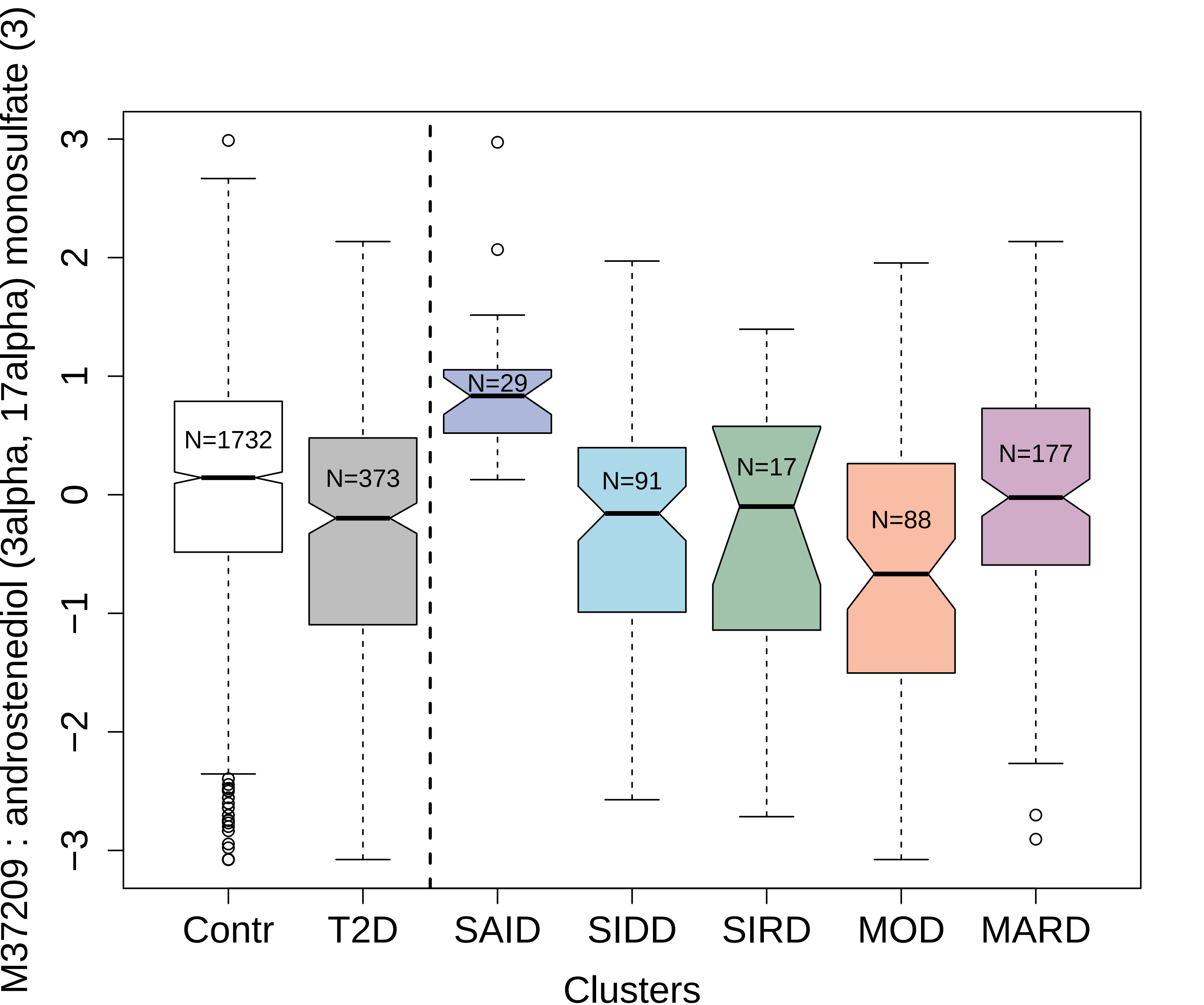
M37209 : androstenediol (3alpha, 17alpha) monosulfate (3)
3
2
1
0
−1
−2
−3
N=1732
N=373
N=29
N=91
N=17
N=88
N=177
Contr
T2D
SAID
SIDD
SIRD
MOD
MARD
Clusters

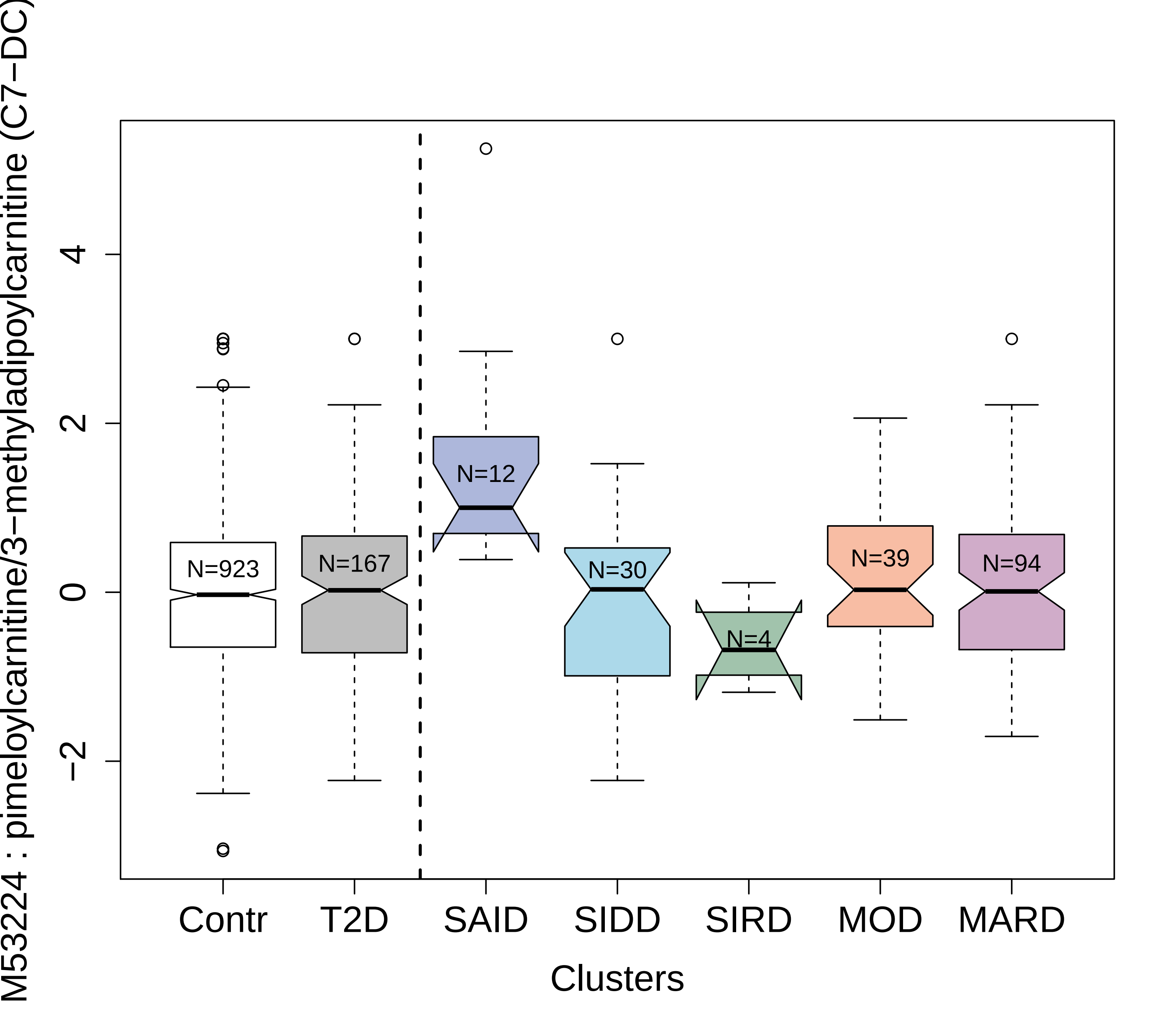

M53224 : pimeloylcarnitine/3−methyladipoylcarnitine (C7−DC)
4
2
0
−2
N=923
N=167
N=12
N=30
N=4
N=39
N=94
Contr
T2D
SAID
SIDD
SIRD
MOD
MARD
Clusters

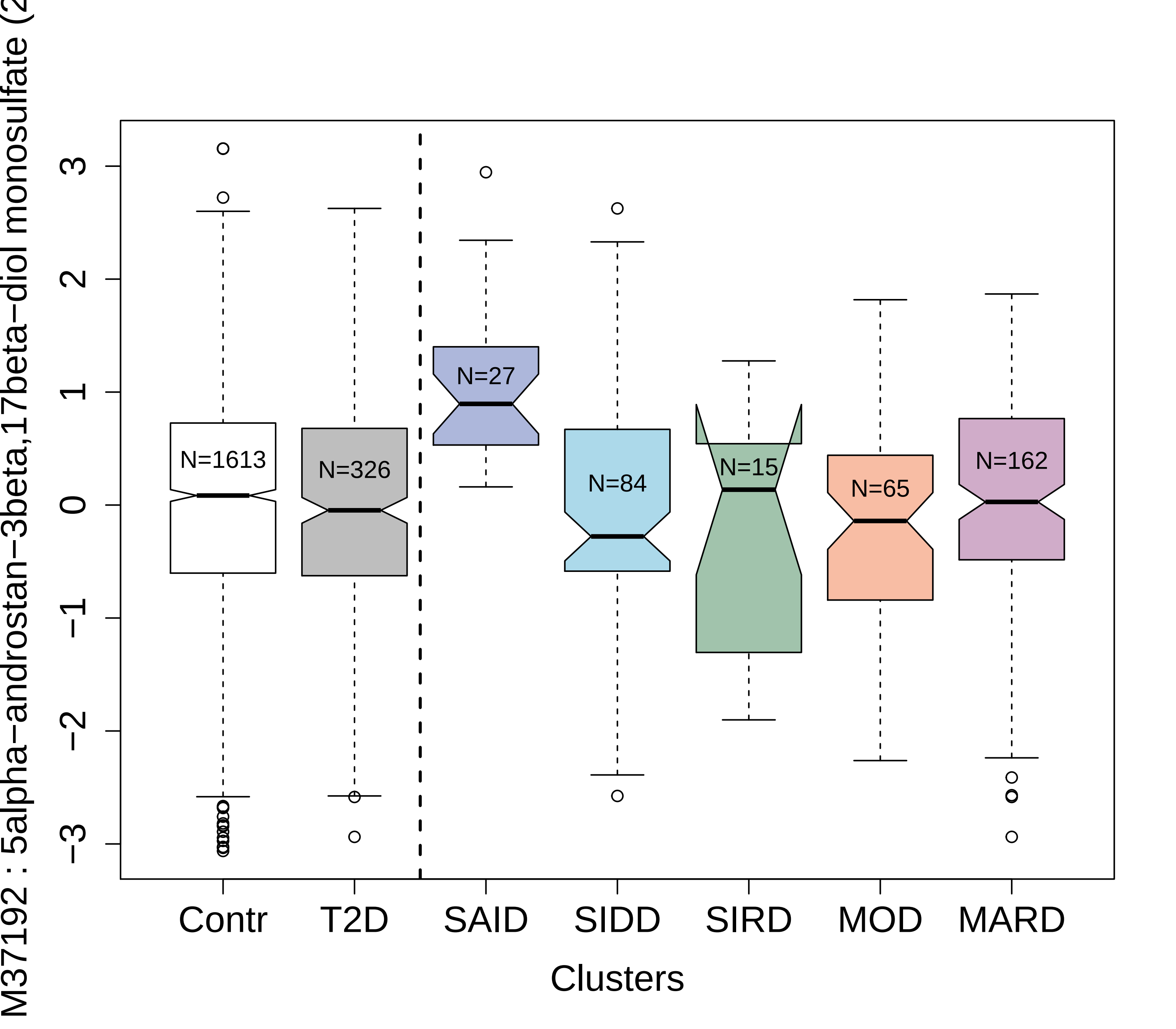

M37192 : 5alpha−androstan−3beta,17beta−diol monosulfate (2
3
2
1
0
−1
−2
−3
N=1613
N=326
N=27
N=84
N=15
N=65
N=162
Contr
T2D
SAID
SIDD
SIRD
MOD
MARD
Clusters

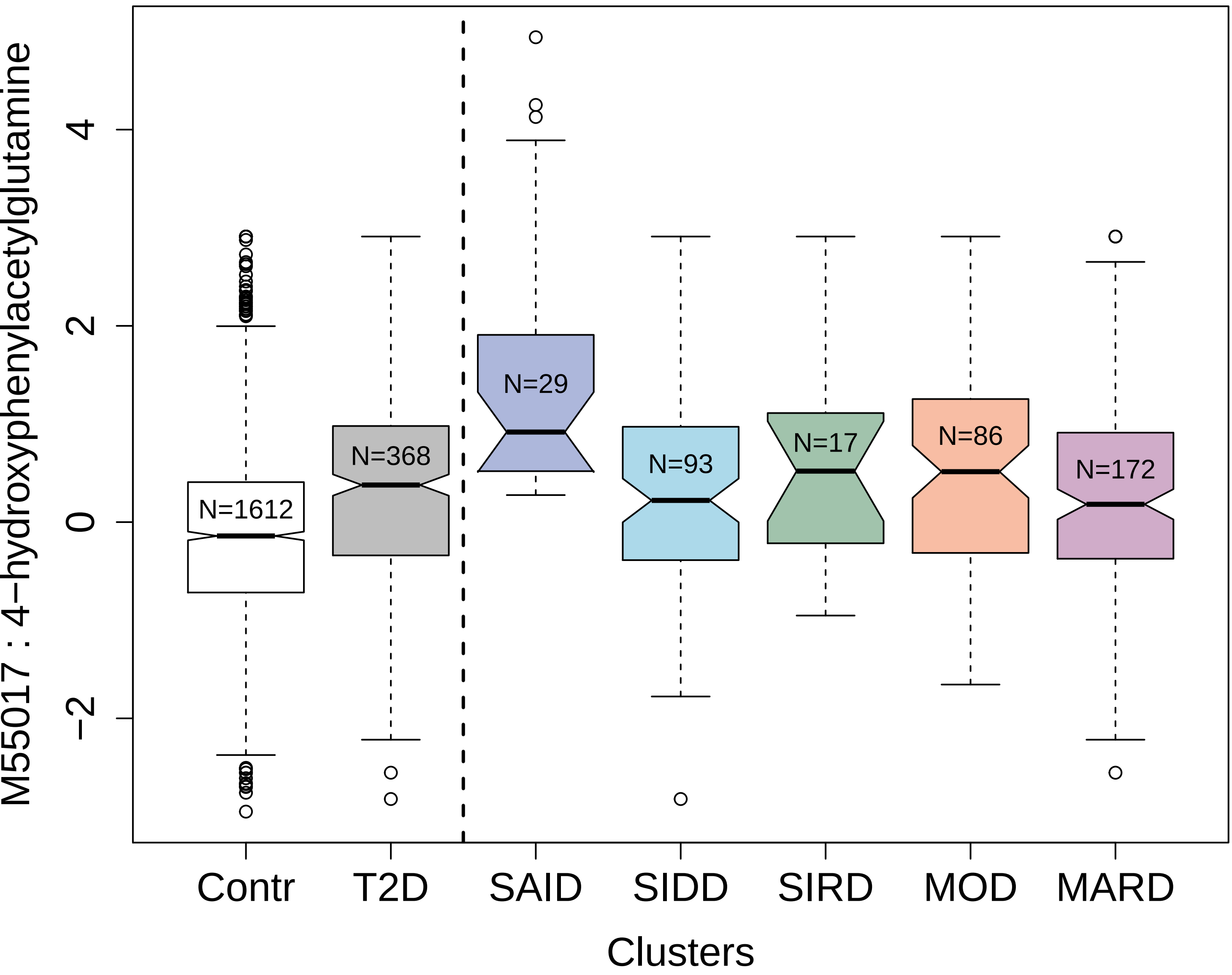

M55017 : 4−hydroxyphenylacetylglutamine
4
2
0
−2
N=1612
N=368
N=29
N=93
N=17
N=86
N=172
Contr
T2D
SAID
SIDD
SIRD
MOD
MARD
Clusters

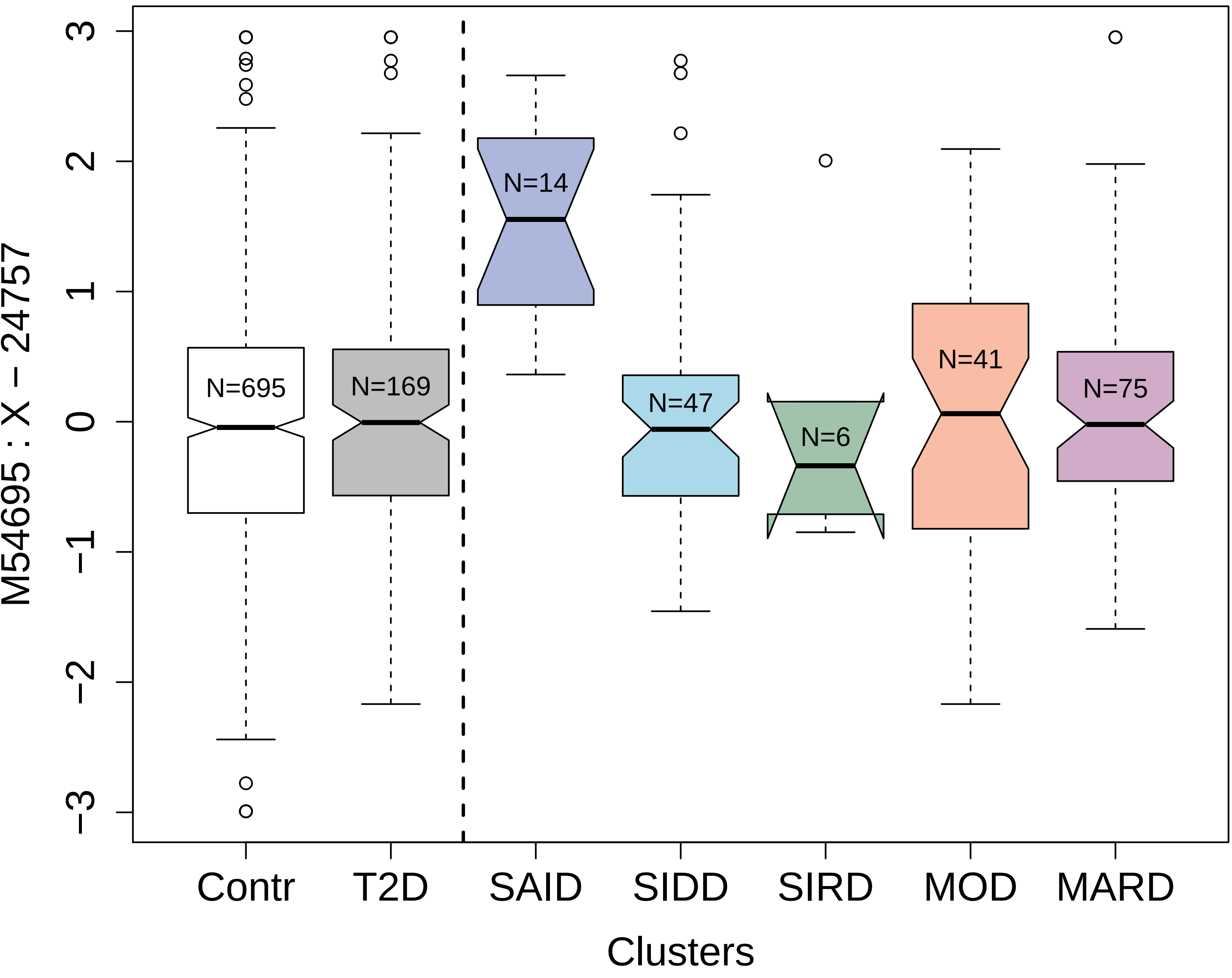

M54695 : X − 24757
3
2
1
0
−1
−2
−3
N=695
N=169
N=14
N=47
N=6
N=41
N=75
Contr
T2D
SAID
SIDD
SIRD
MOD
MARD
Clusters

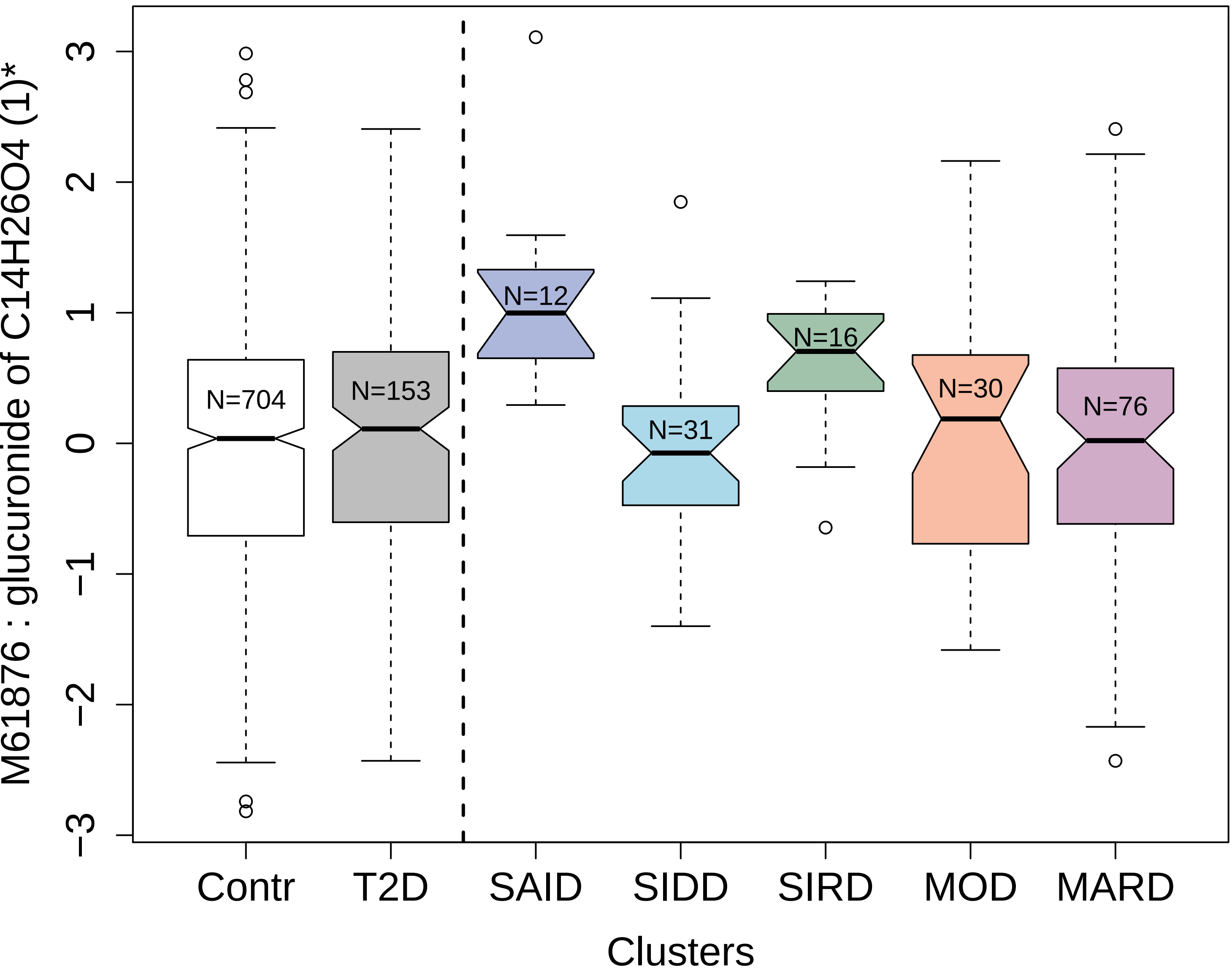

M61876 : glucuronide of C14H26O4 (1)*
3
2
1
0
−1
−2
−3
N=704
N=153
N=12
N=31
N=16
N=30
N=76
Contr
T2D
SAID
SIDD
SIRD
MOD
MARD
Clusters

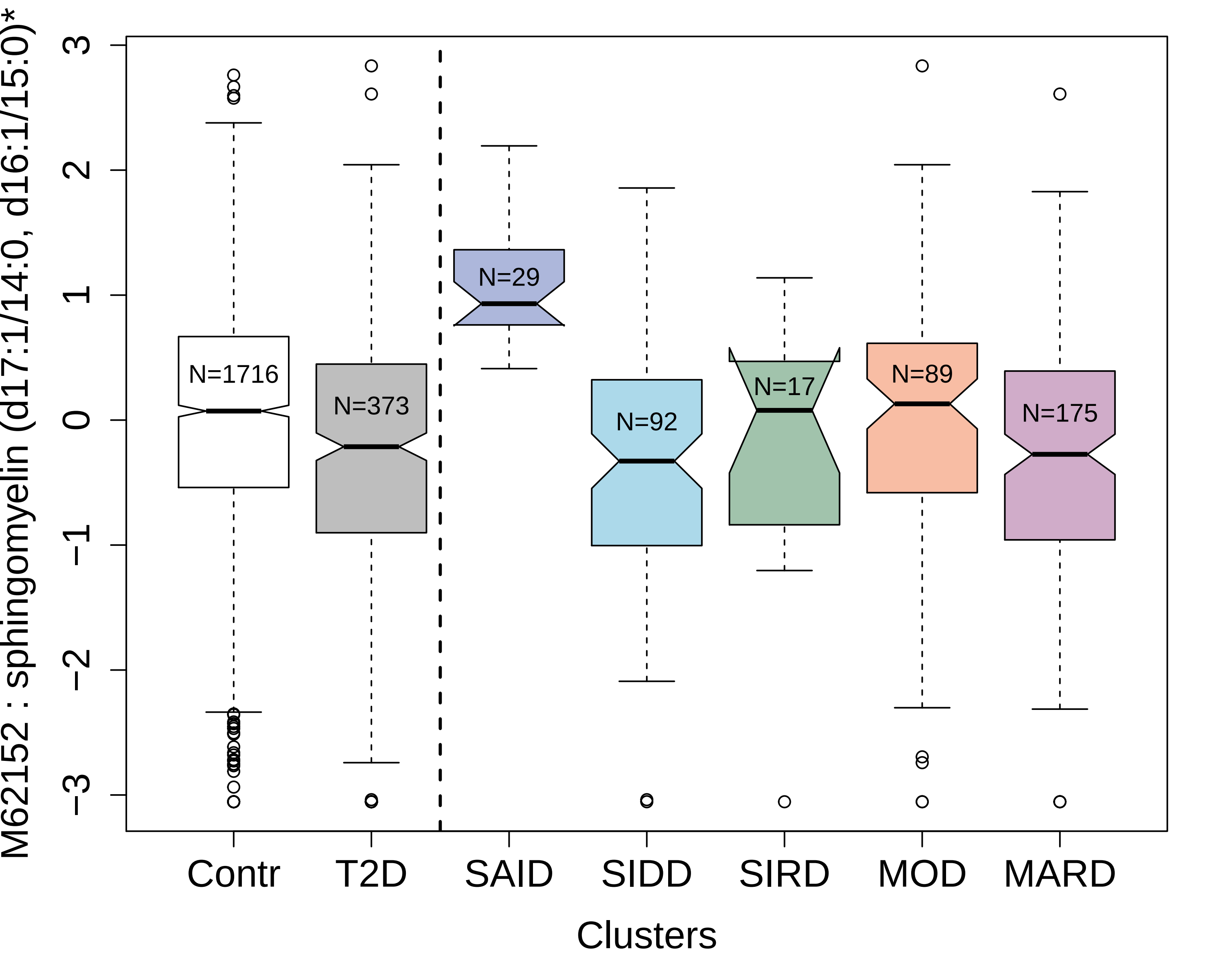
M62152 : sphingomyelin (d17:1/14:0, d16:1/15:0)*
3
2
1
0
−1
−2
−3
N=1716
N=373
N=29
N=92
N=17
N=89
N=175
Contr
T2D
SAID
SIDD
SIRD
MOD
MARD
Clusters

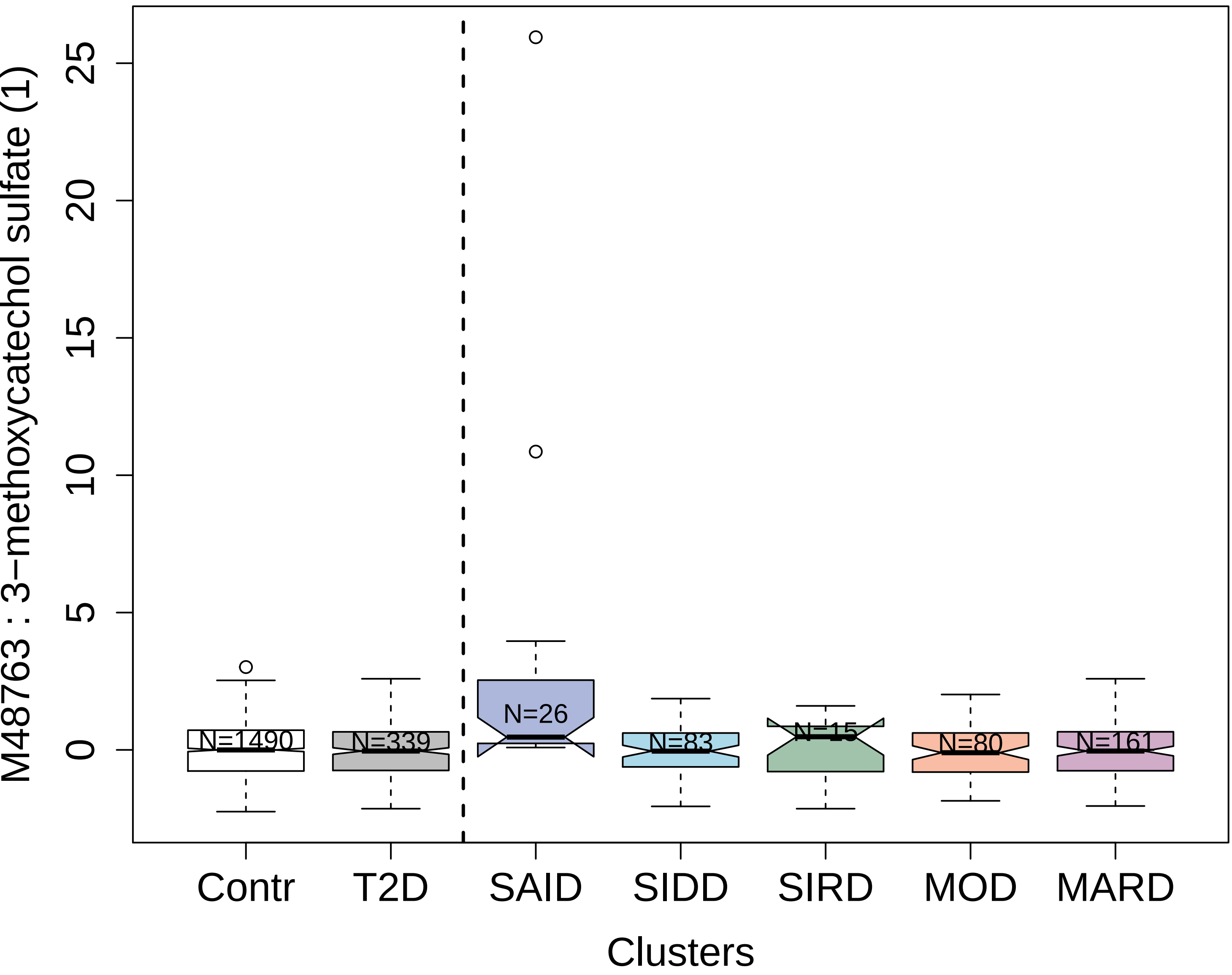
M48763 : 3−methoxycatechol sulfate (1)
0
5
10
15
20
25
N=1490
N=339
N=26
N=83
N=15
N=80
N=161
Contr
T2D
SAID
SIDD
SIRD
MOD
MARD
Clusters

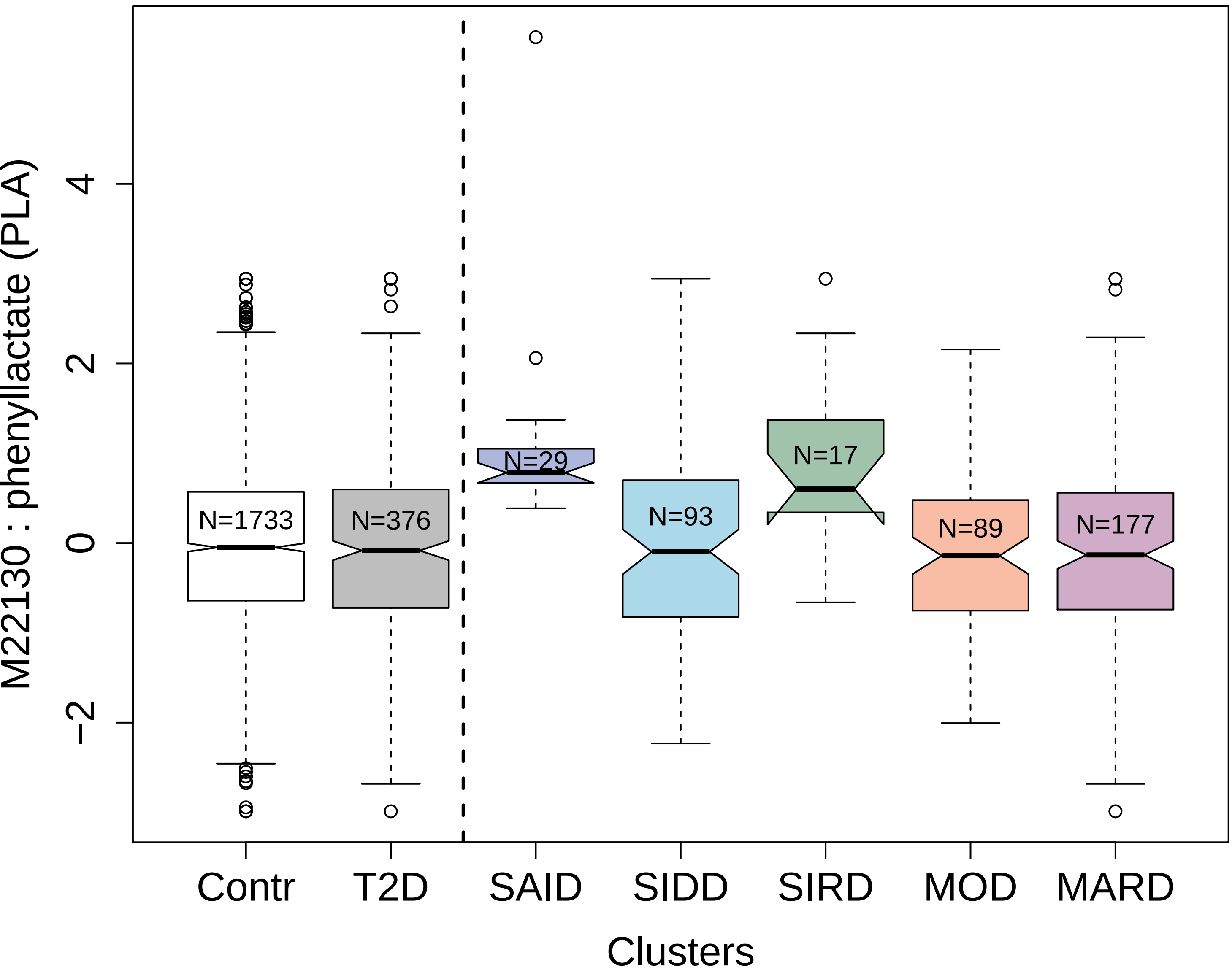
M22130 : phenyllactate (PLA)
4
2
0
−2
N=1733
N=376
N=29
N=93
N=17
N=89
N=177
Contr
T2D
SAID
SIDD
SIRD
MOD
MARD
Clusters

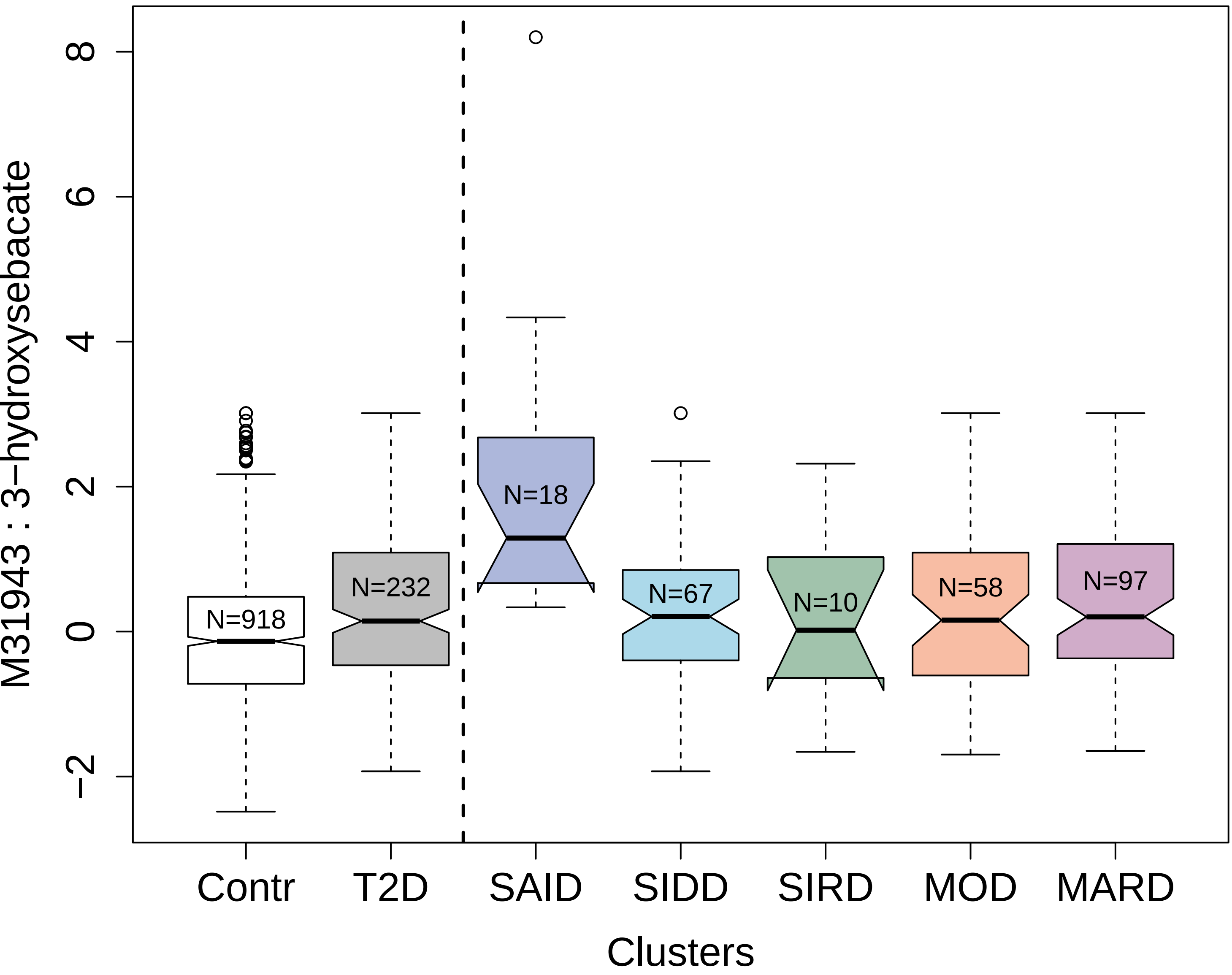

M31943 : 3−hydroxysebacate
−2
0
2
4
6
8
N=918
N=232
N=18
N=67
N=10
N=58
N=97
Contr
T2D
SAID
SIDD
SIRD
MOD
MARD
Clusters

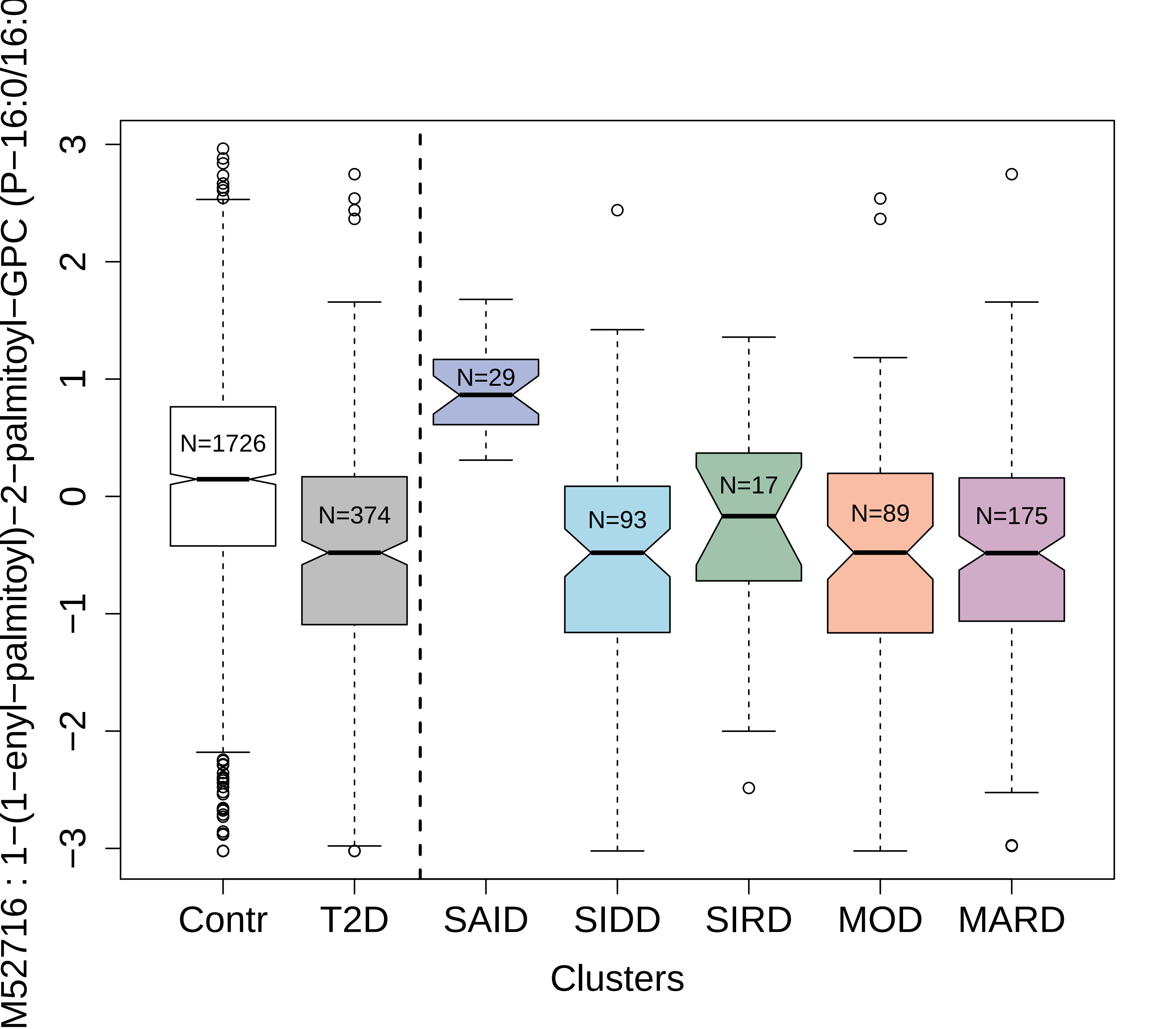

M52716 : 1−(1−enyl−palmitoyl)−2−palmitoyl−GPC (P−16:0/16:0
−3
−2
−1
0
1
2
3
N=1726
N=374
N=29
N=93
N=17
N=89
N=175
Contr
T2D
SAID
SIDD
SIRD
MOD
MARD
Clusters

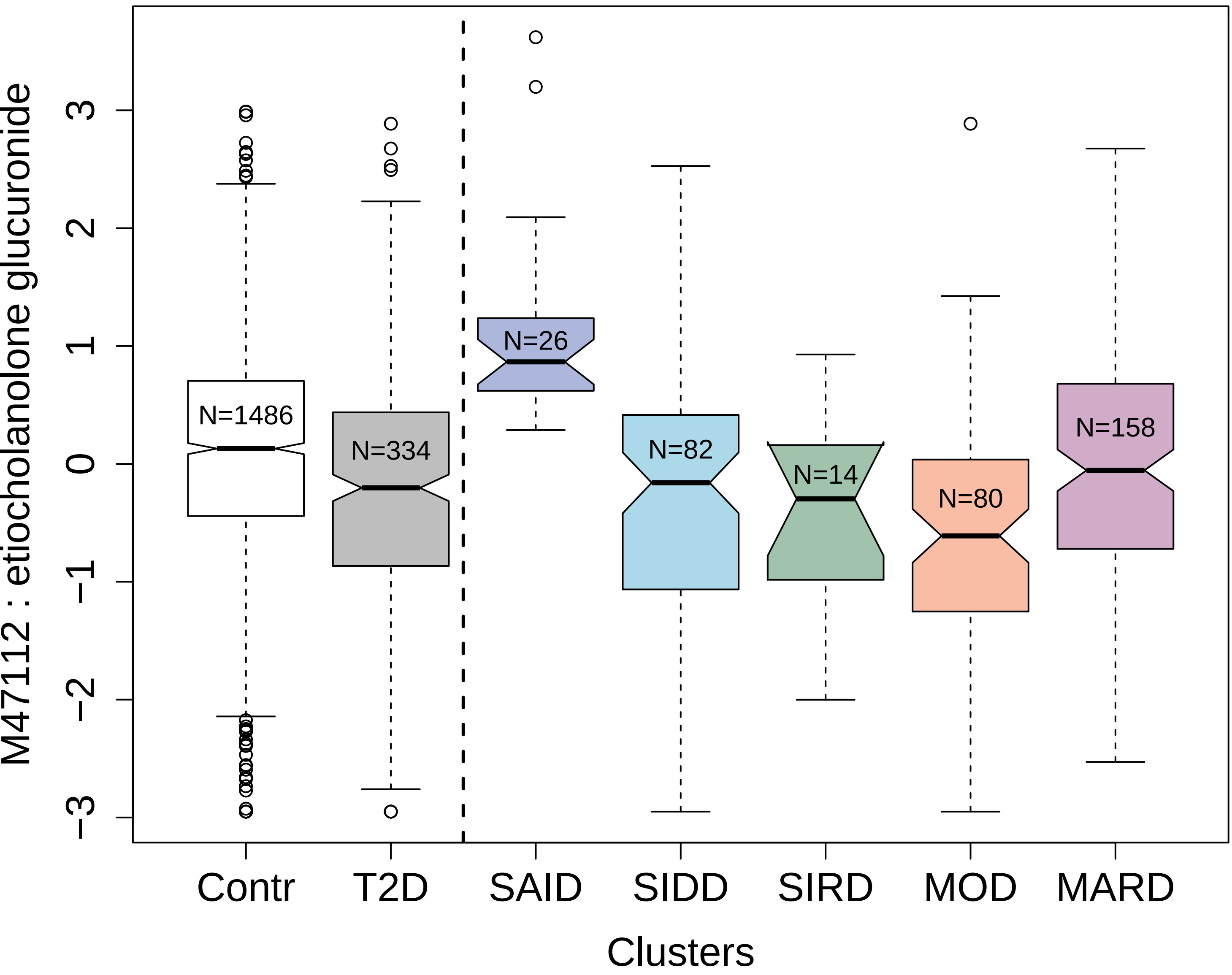

M47112 : etiocholanolone glucuronide
3
2
1
0
−1
−2
−3
N=1486
N=334
N=26
N=82
N=14
N=80
N=158
Contr
T2D
SAID
SIDD
SIRD
MOD
MARD
Clusters

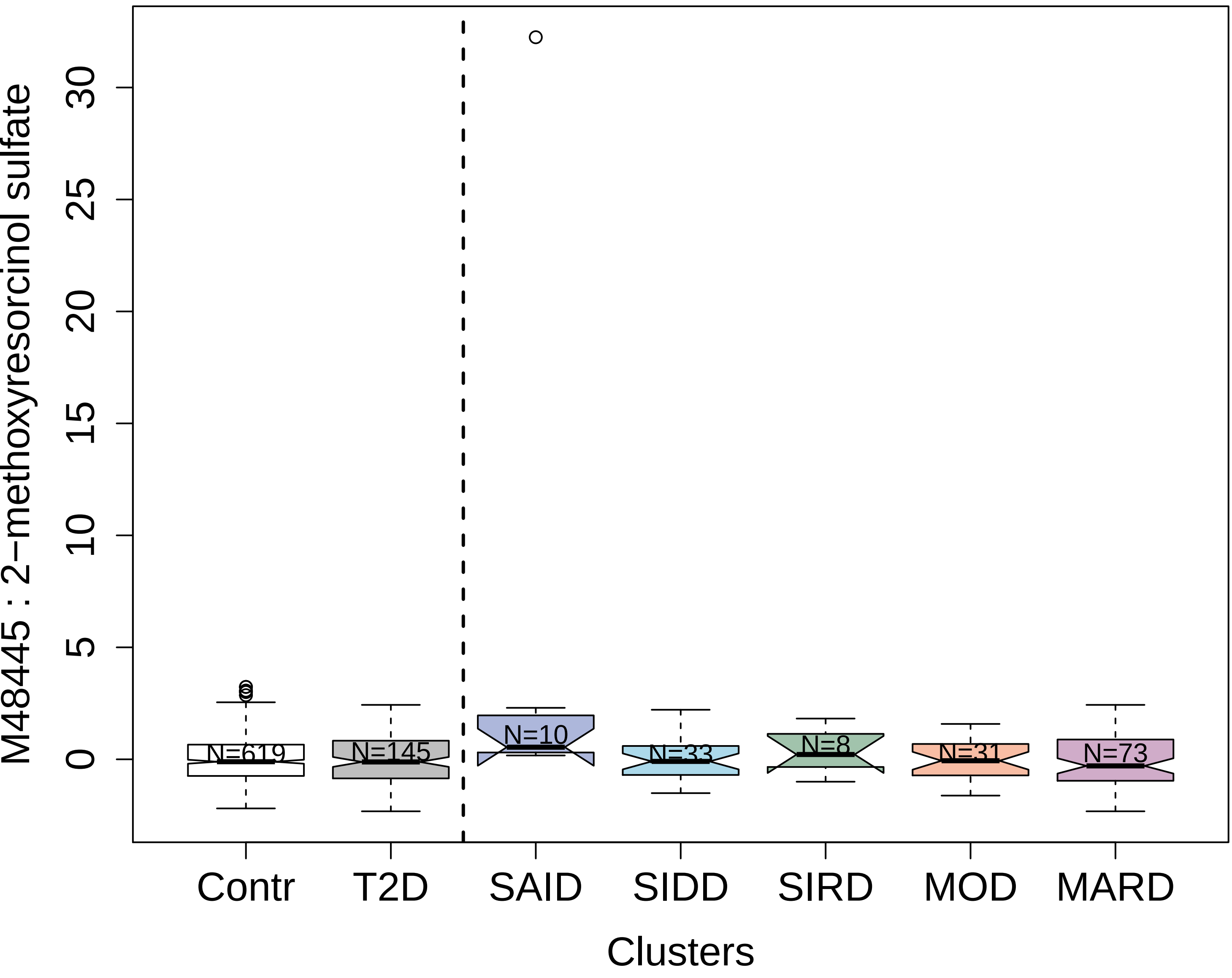

M48445 : 2−methoxyresorcinol sulfate
0
5
10
15
20
25
30
N=619
N=145
N=10
N=33
N=8
N=31
N=73
Contr
T2D
SAID
SIDD
SIRD
MOD
MARD
Clusters

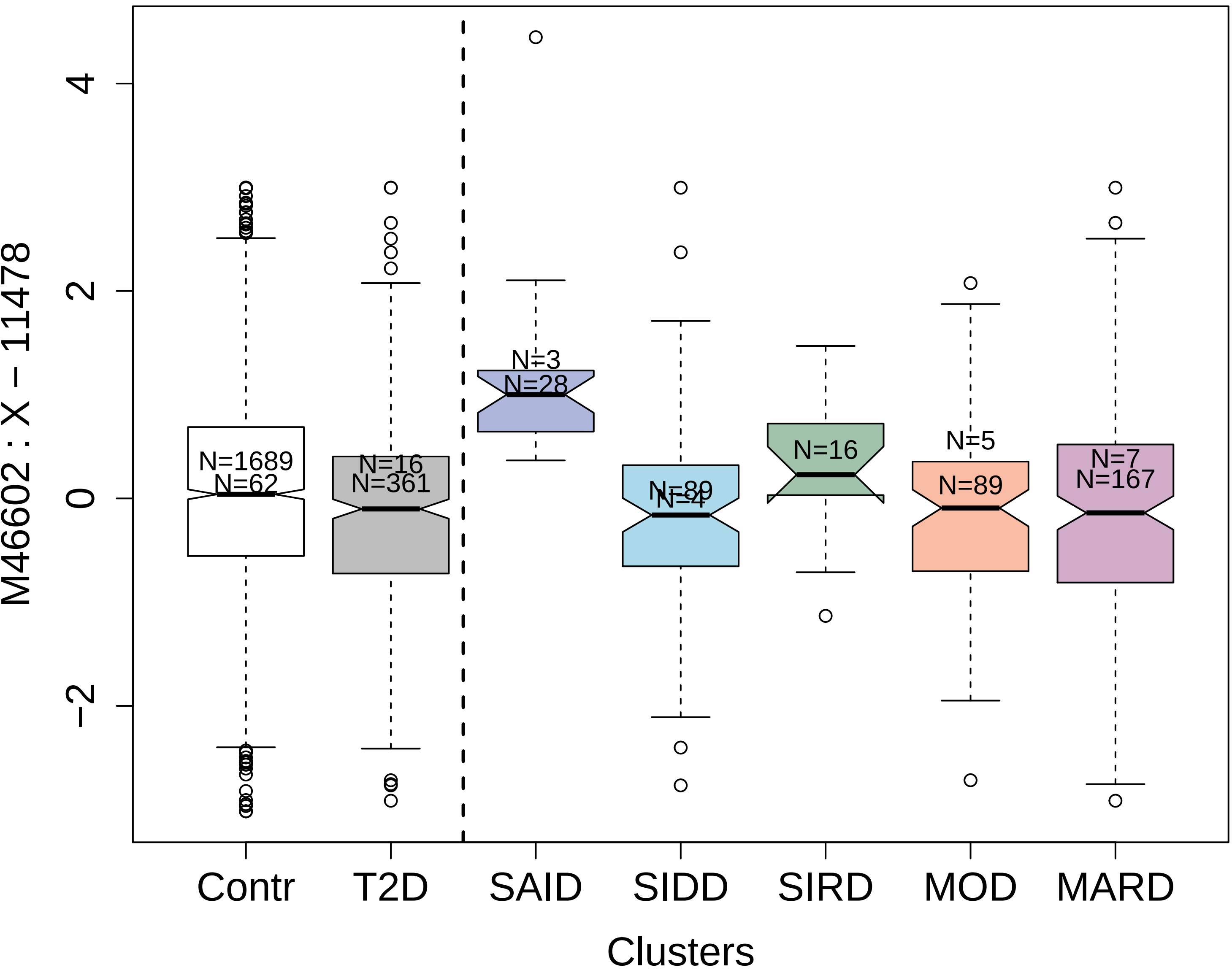
M46602 : X − 11478
4
2
0
−2
N=1689
N=62
N=16
N=361
N=3
N=28
N=89
N=4
N=16
N=5
N=89
N=7
N=167
Contr
T2D
SAID
SIDD
SIRD
MOD
MARD
Clusters

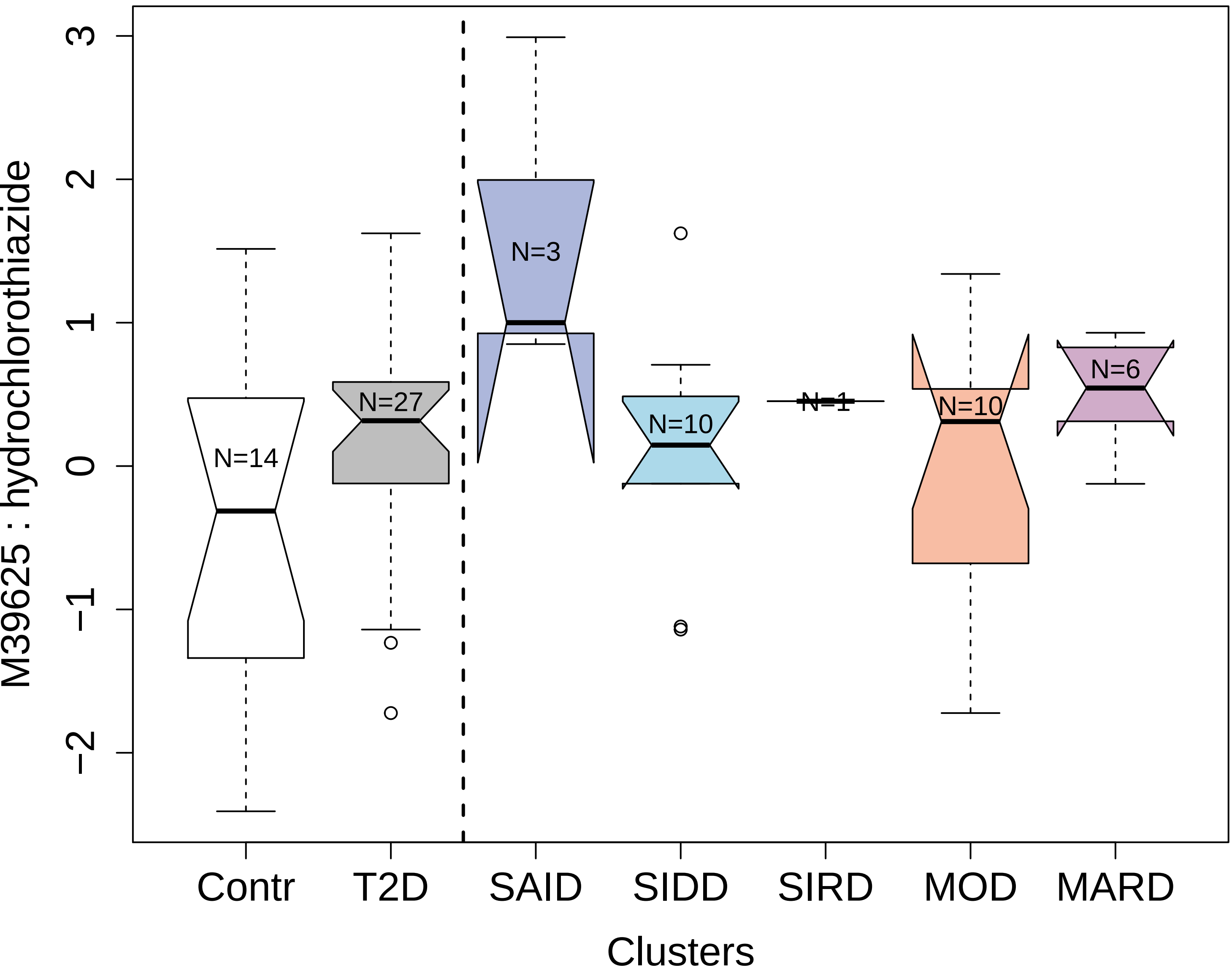
M39625 : hydrochlorothiazide
3
2
1
0
−1
−2
N=14
N=27
N=3
N=10
N=1
N=10
N=6
Contr
T2D
SAID
SIDD
SIRD
MOD
MARD
Clusters

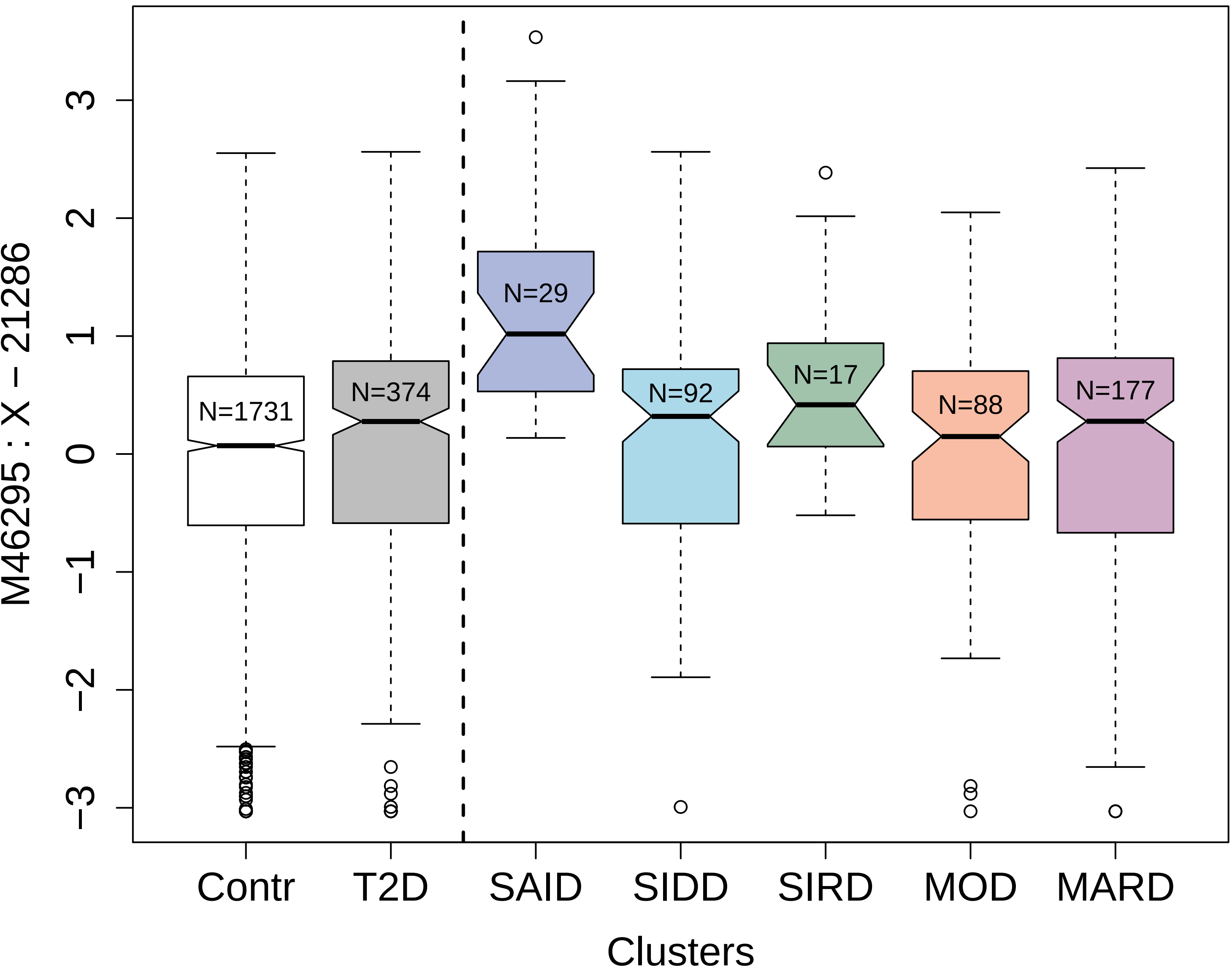
M46295 : X − 21286
3
2
1
0
−1
−2
−3
N=1731
N=374
N=29
N=92
N=17
N=88
N=177
Contr
T2D
SAID
SIDD
SIRD
MOD
MARD
Clusters

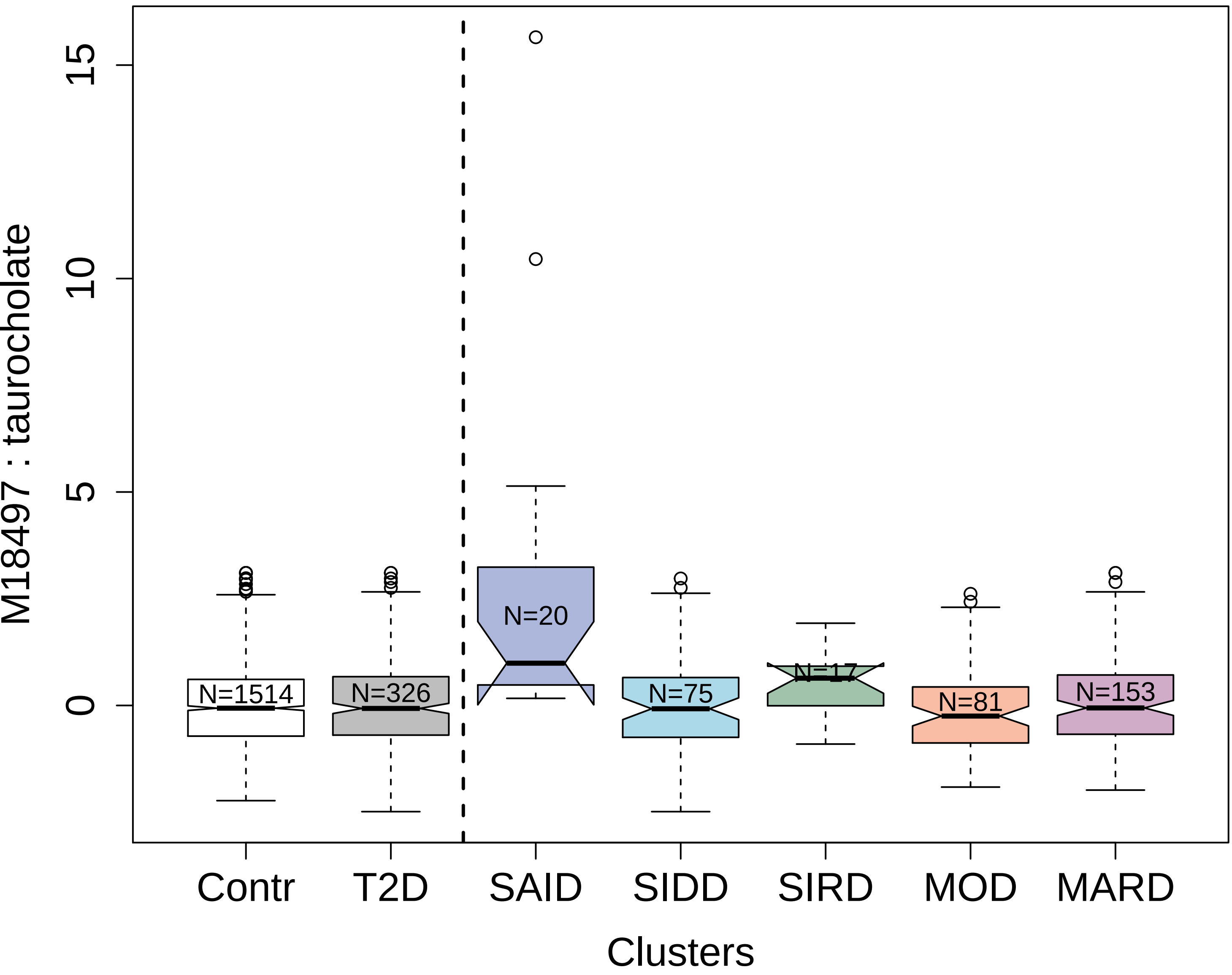

M18497 : taurocholate
0
5
10
15
N=1514
N=326
N=20
N=75
N=17
N=81
N=153
Contr
T2D
SAID
SIDD
SIRD
MOD
MARD
Clusters

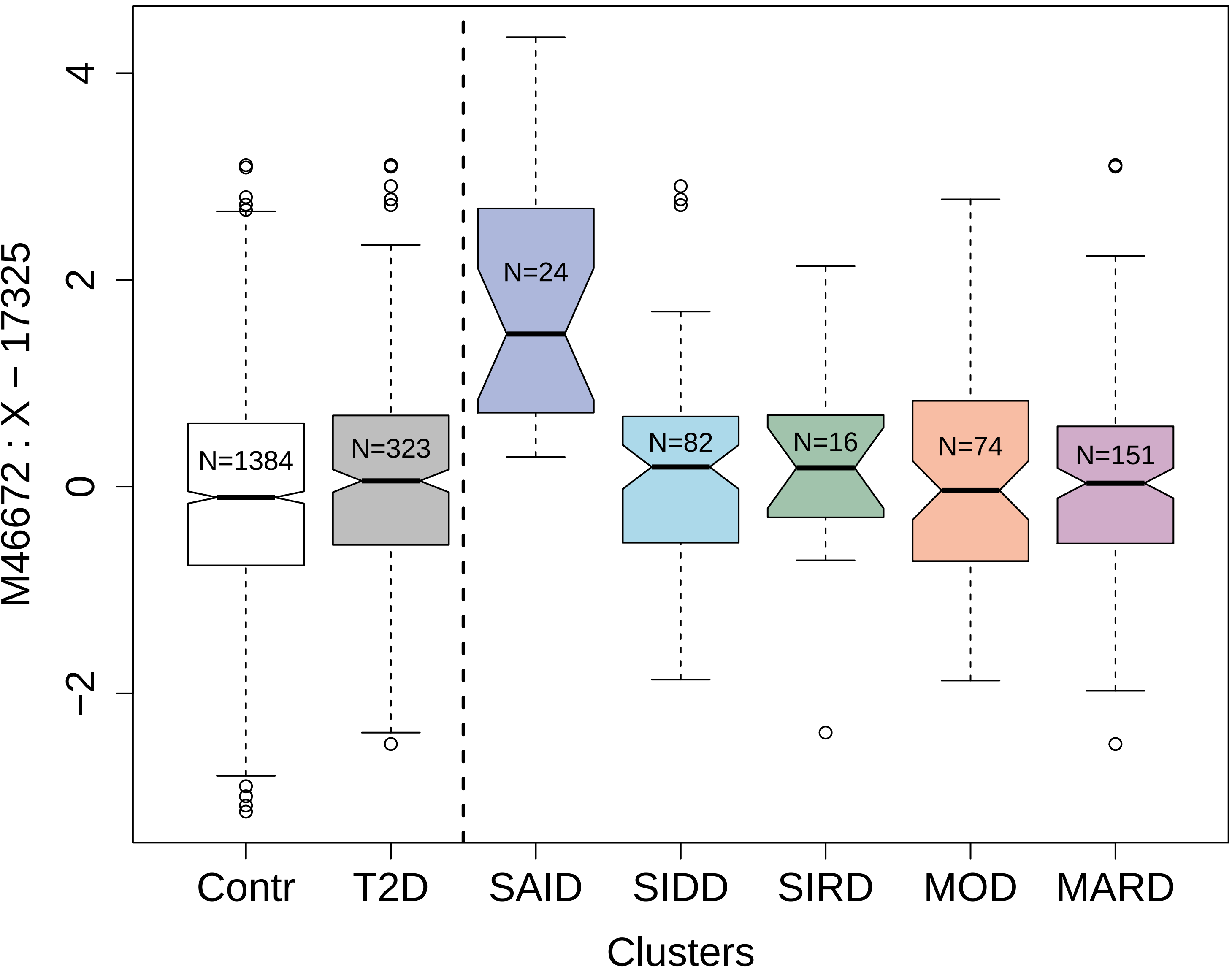
M46672 : X − 17325
4
2
0
−2
N=1384
N=323
N=24
N=82
N=16
N=74
N=151
Contr
T2D
SAID
SIDD
SIRD
MOD
MARD
Clusters

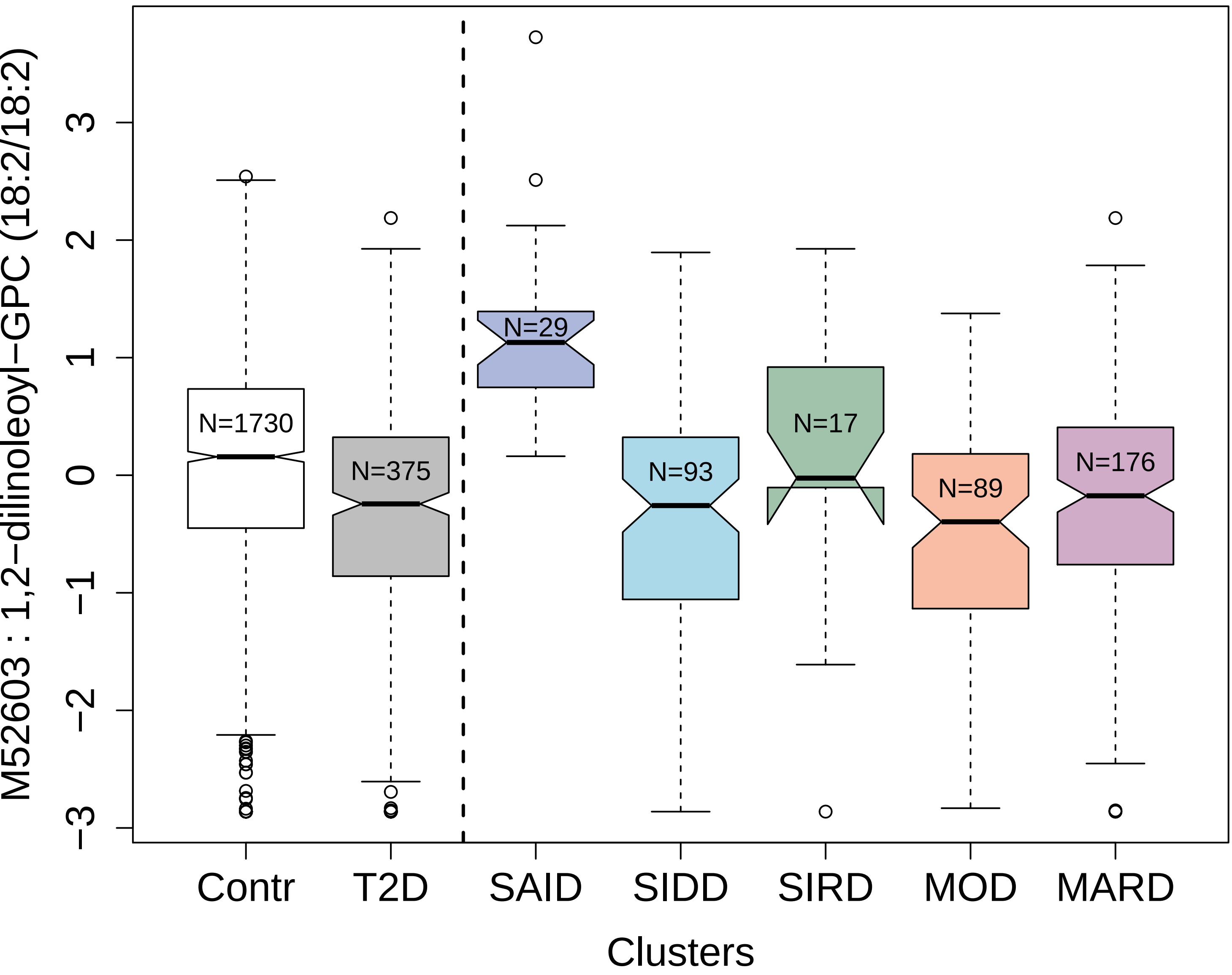
M52603 : 1,2−dilinoleoyl−GPC (18:2/18:2)
N=1730
N=375
N=29
N=93
N=17
N=89
N=176
3
2
1
0
−1
−2
−3
Contr
T2D
SAID
SIDD
SIRD
MOD
MARD
Clusters

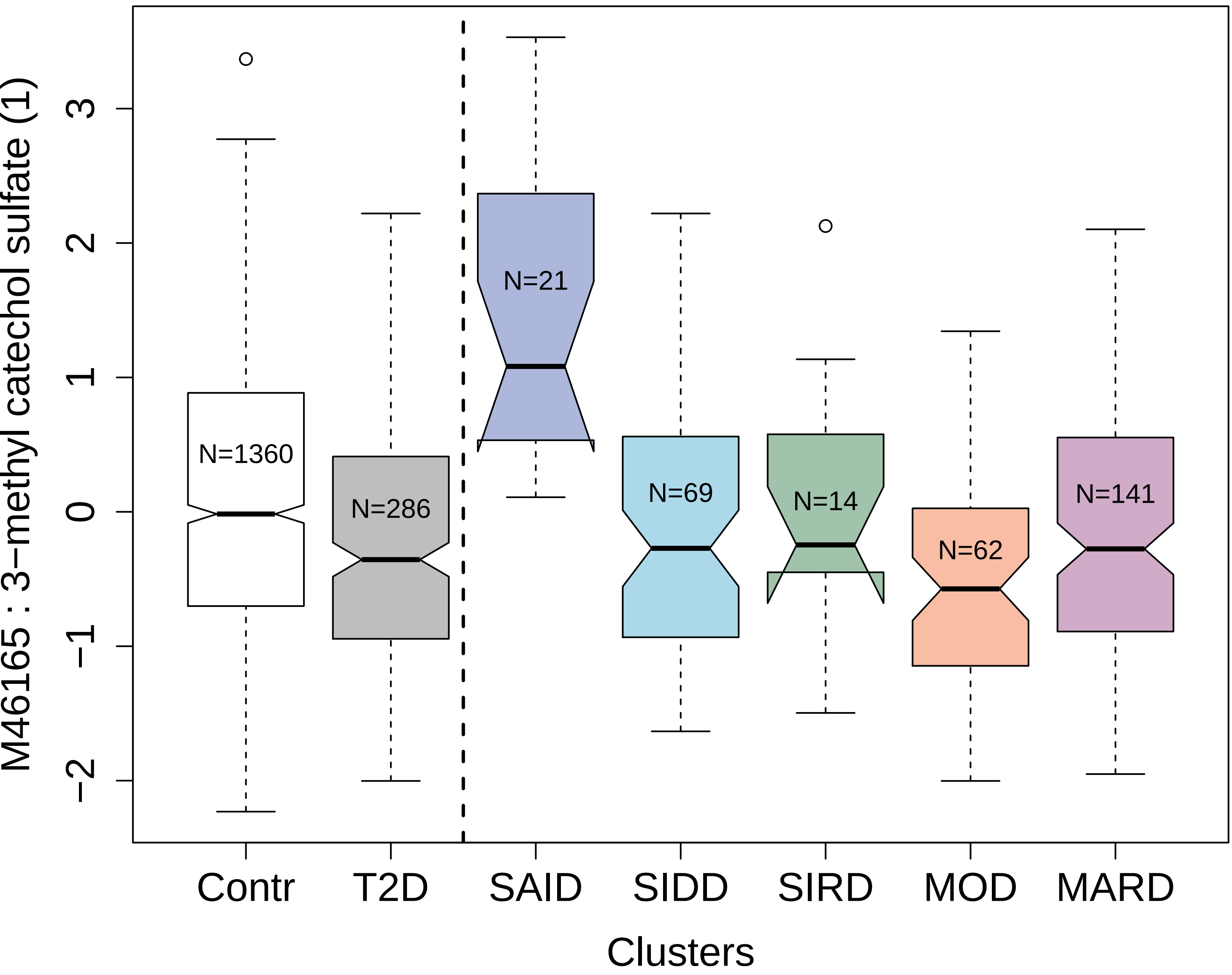

M46165 : 3−methyl catechol sulfate (1)
3
2
1
0
−1
−2
N=1360
N=286
N=21
N=69
N=14
N=62
N=141
Contr
T2D
SAID
SIDD
SIRD
MOD
MARD
Clusters

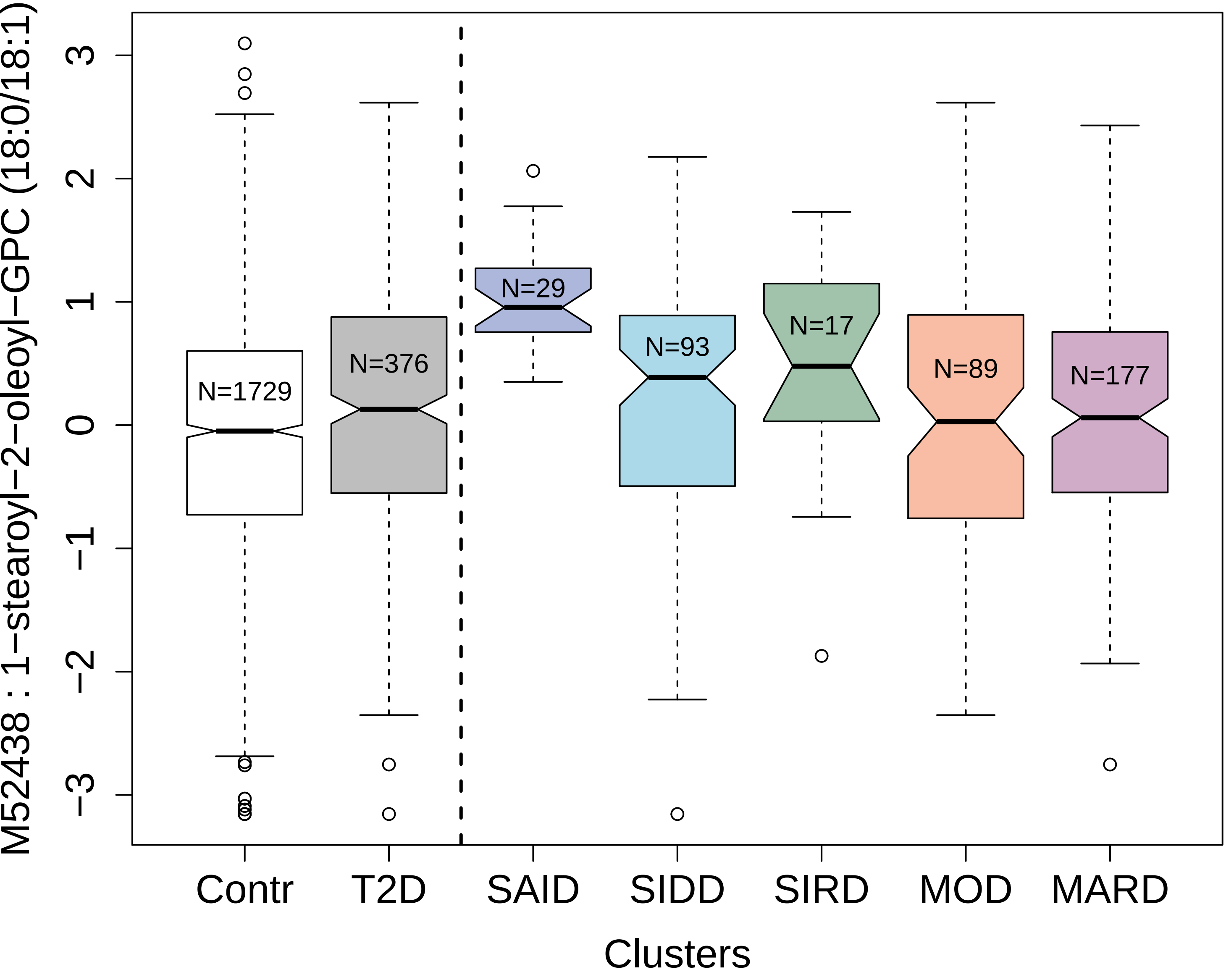

M52438 : 1−stearoyl−2−oleoyl−GPC (18:0/18:1)
3
2
1
0
−1
−2
−3
N=1729
N=376
N=29
N=93
N=17
N=89
N=177
Contr
T2D
SAID
SIDD
SIRD
MOD
MARD
Clusters

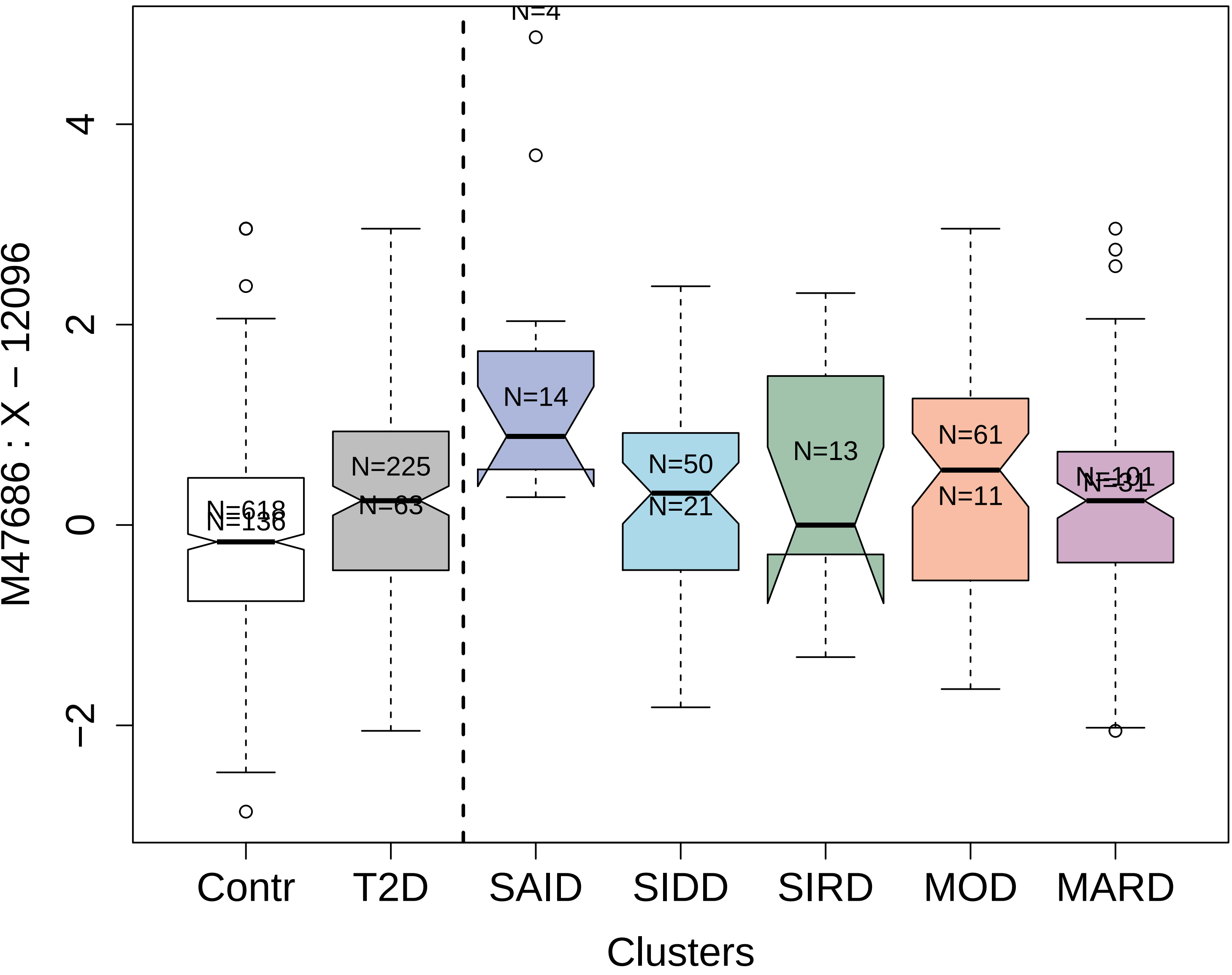
M47686 : X − 12096
Clusters
Contr
T2D
SAID
SIDD
SIRD
MOD
MARD
N=4
N=14
N=225
N=63
N=50
N=21
N=13
N=61
N=11
N=101
N=31
4
2
0
−2

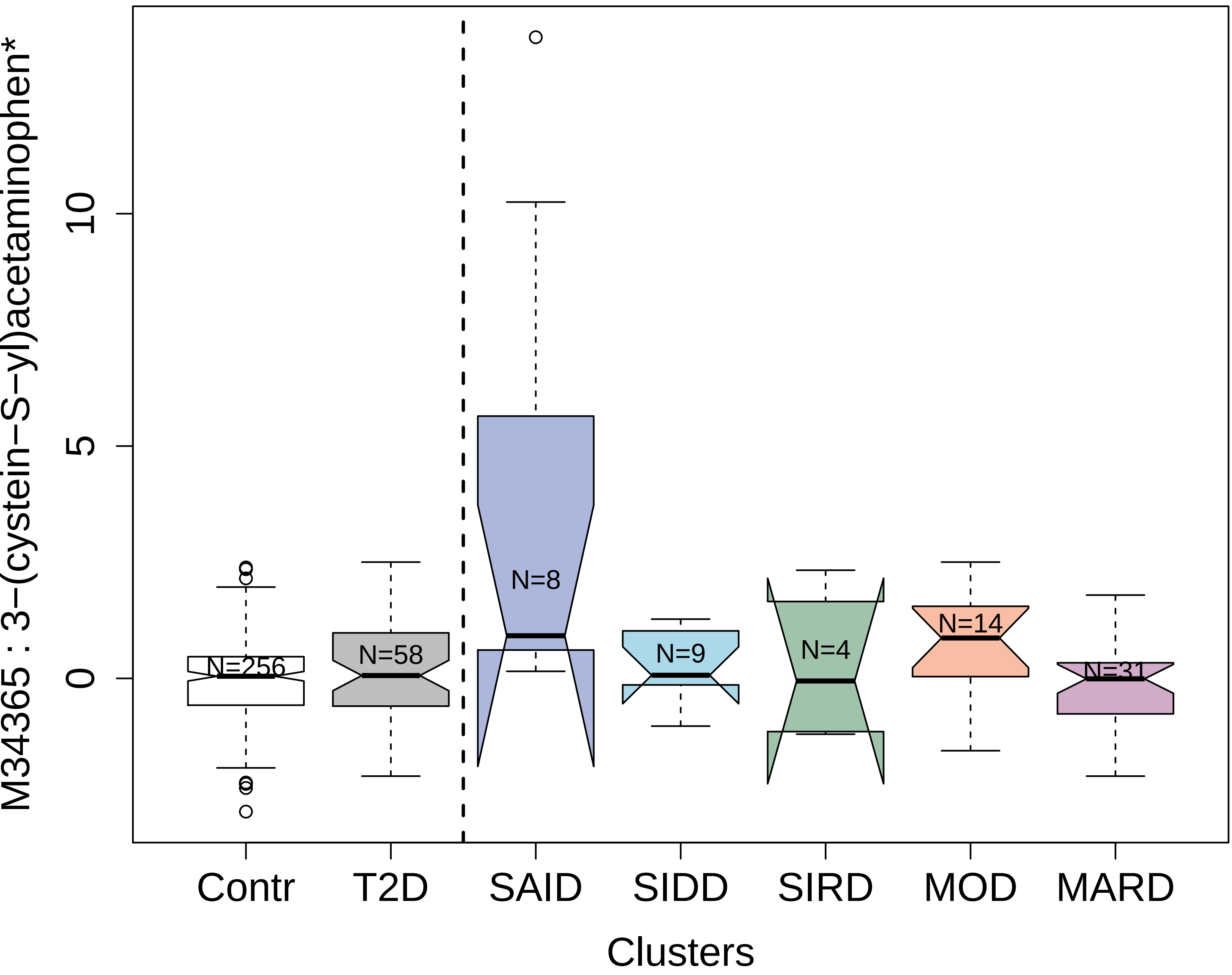
M34365 : 3−(cystein−S−yl)acetaminophen*
0
5
10
N=256
N=58
N=8
N=9
N=4
N=14
N=31
Contr
T2D
SAID
SIDD
SIRD
MOD
MARD
Clusters

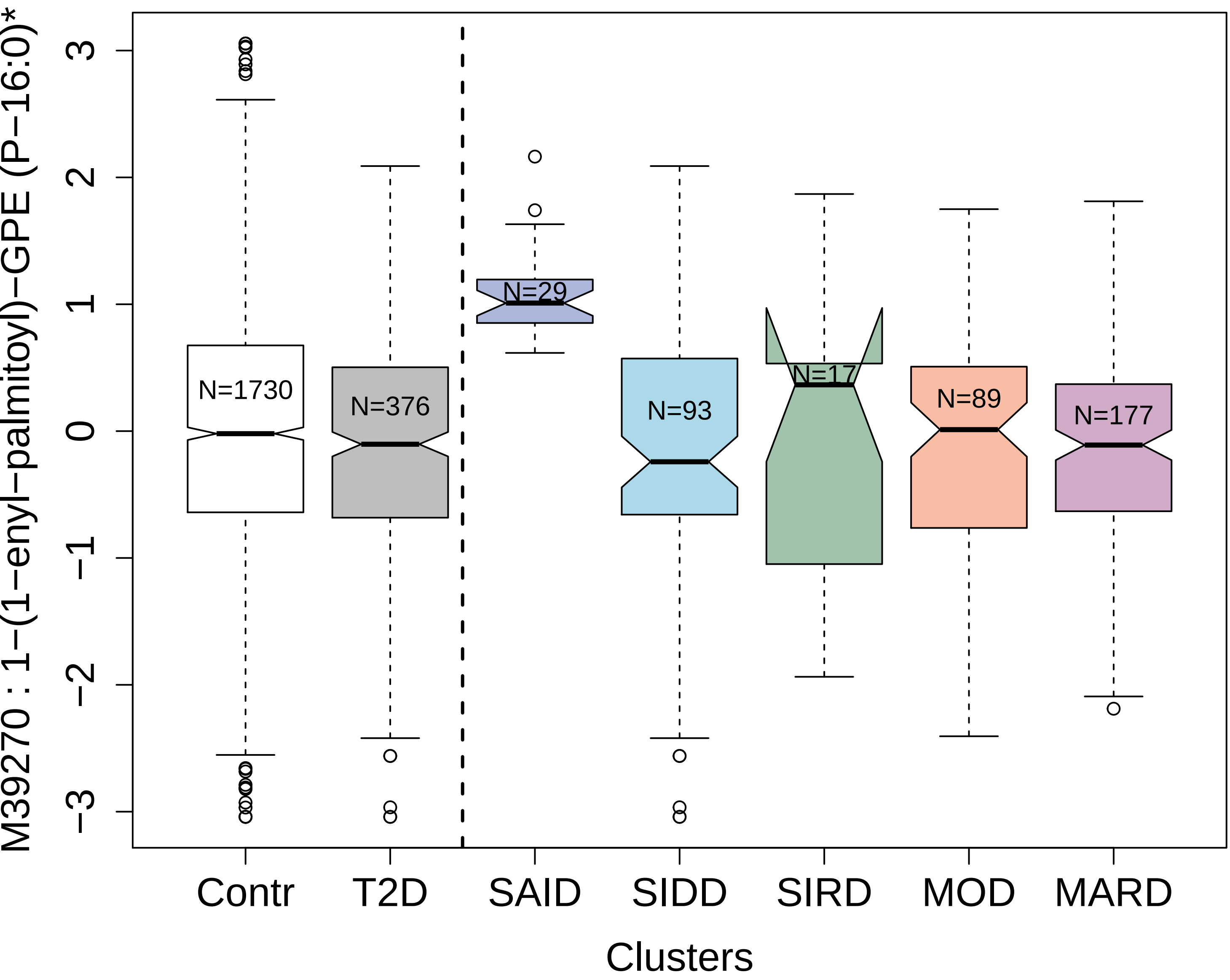

M39270 : 1−(1−enyl−palmitoyl)−GPE (P−16:0)*
3
2
1
0
−1
−2
−3
N=1730
N=376
N=29
N=93
N=17
N=89
N=177
Contr
T2D
SAID
SIDD
SIRD
MOD
MARD
Clusters

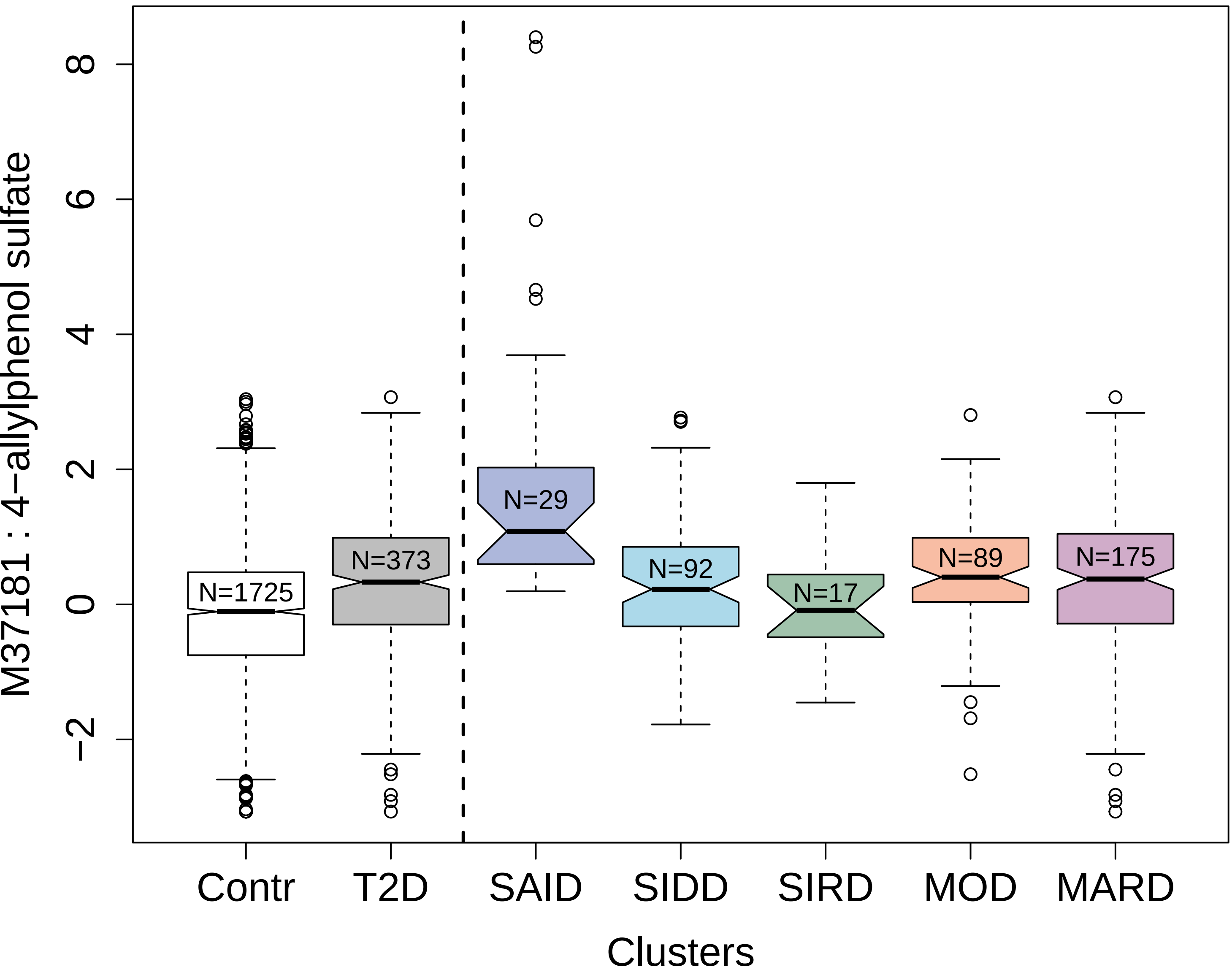
M37181 : 4−allylphenol sulfate
Clusters
Contr
T2D
SAID
SIDD
SIRD
MOD
MARD
N=1725
N=373
N=29
N=92
N=17
N=89
N=175
−2
0
2
4
6
8

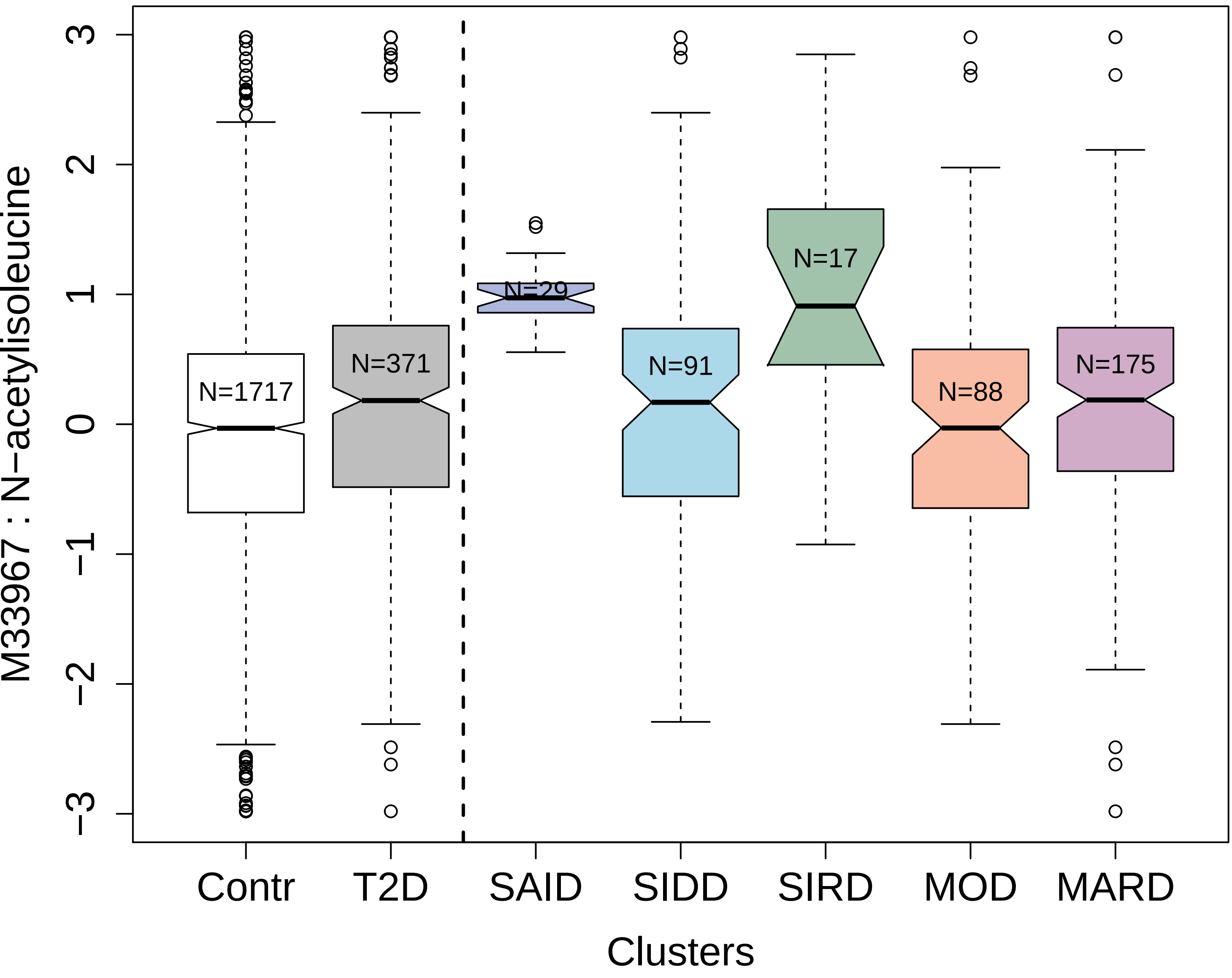

M33967 : N−acetylisoleucine
3
2
1
0
−1
−2
−3
N=1717
N=371
N=29
N=91
N=17
N=88
N=175
Contr
T2D
SAID
SIDD
SIRD
MOD
MARD
Clusters

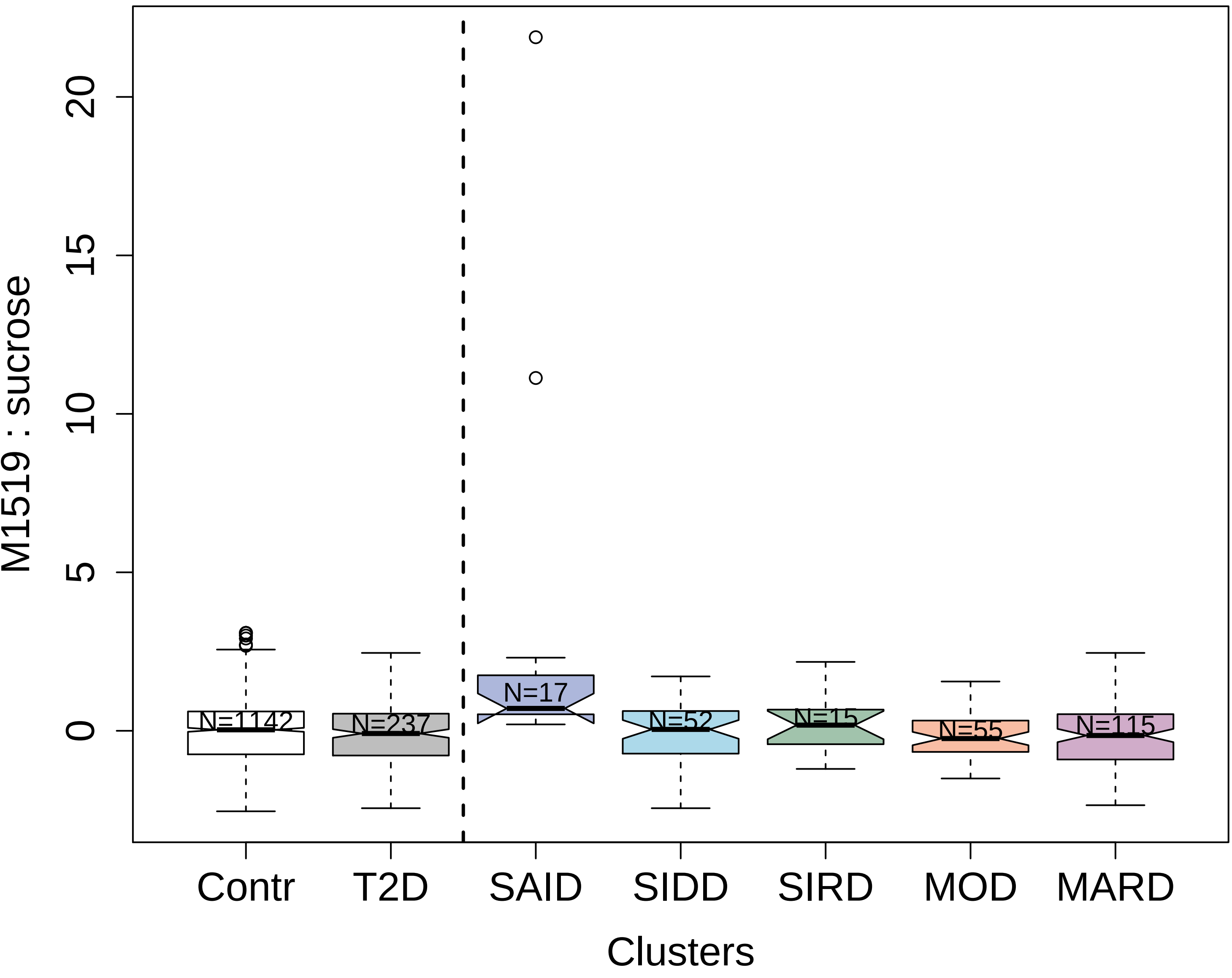

M1519 : sucrose
0
5
10
15
20
N=1142
N=237
N=17
N=52
N=15
N=55
N=115
Contr
T2D
SAID
SIDD
SIRD
MOD
MARD
Clusters

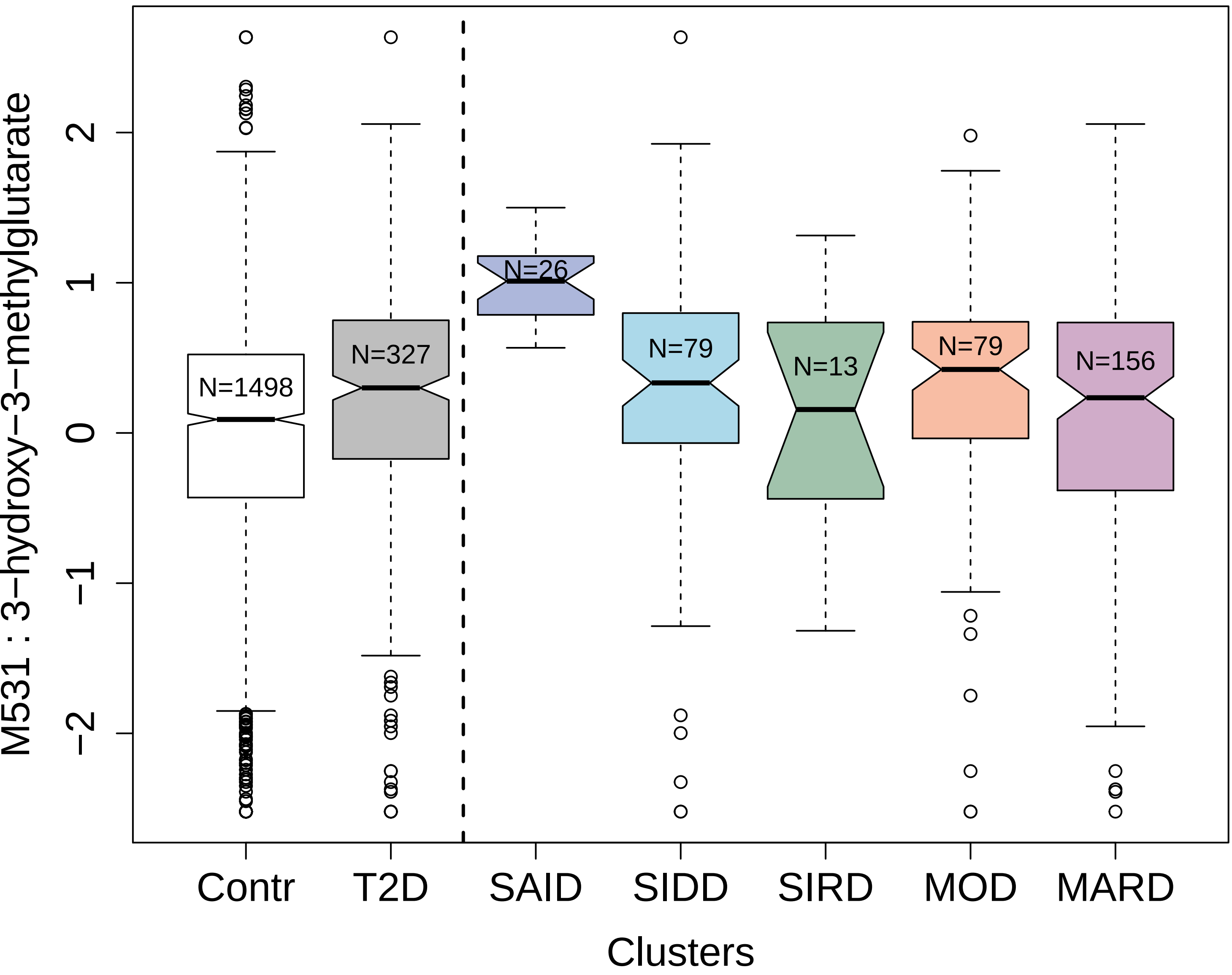

M531 : 3−hydroxy−3−methylglutarate
2
1
0
−1
−2
N=1498
N=327
N=26
N=79
N=13
N=79
N=156
Contr
T2D
SAID
SIDD
SIRD
MOD
MARD
Clusters

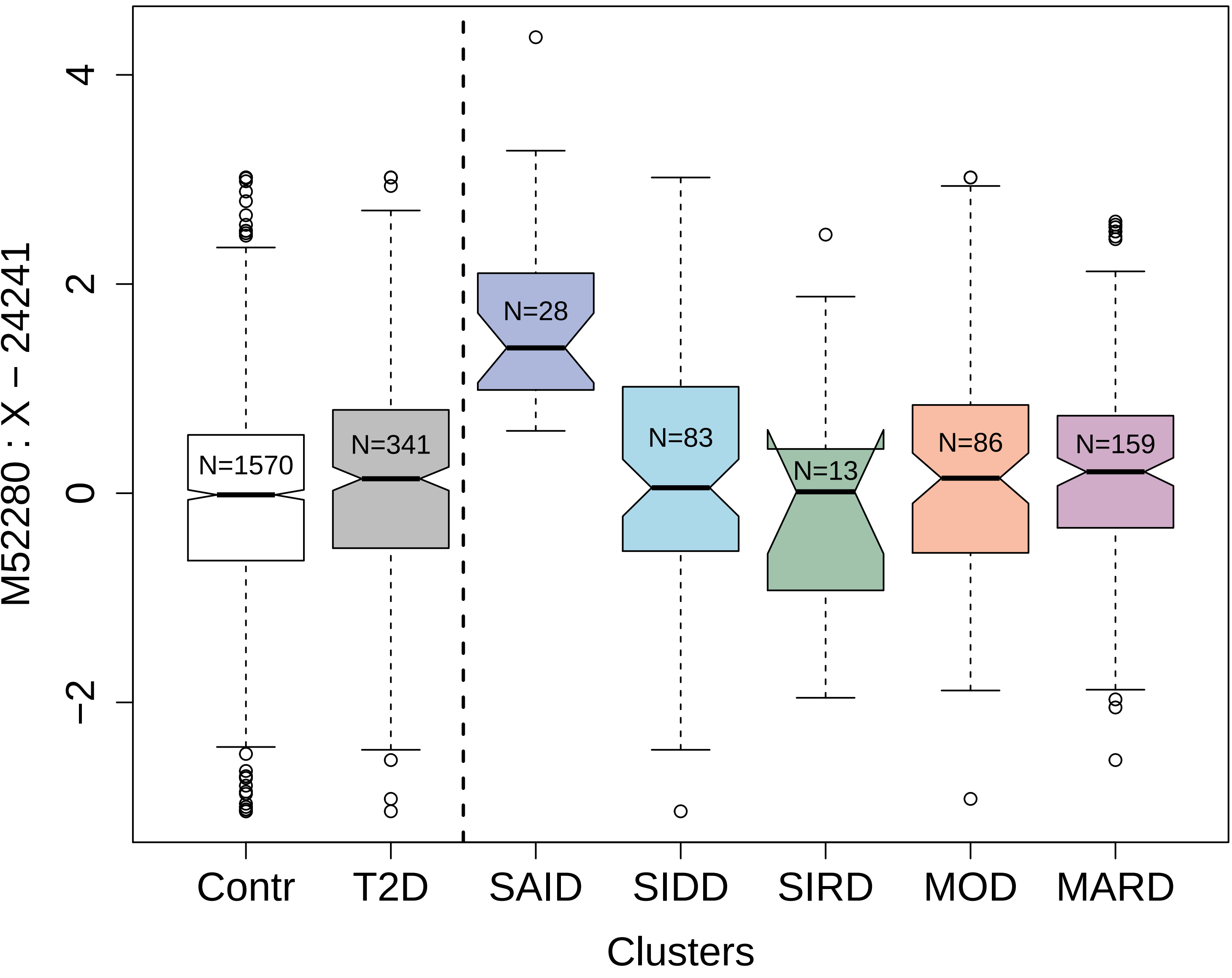
M52280 : X − 24241
4
2
0
−2
N=1570
N=341
N=28
N=83
N=13
N=86
N=159
Contr
T2D
SAID
SIDD
SIRD
MOD
MARD
Clusters

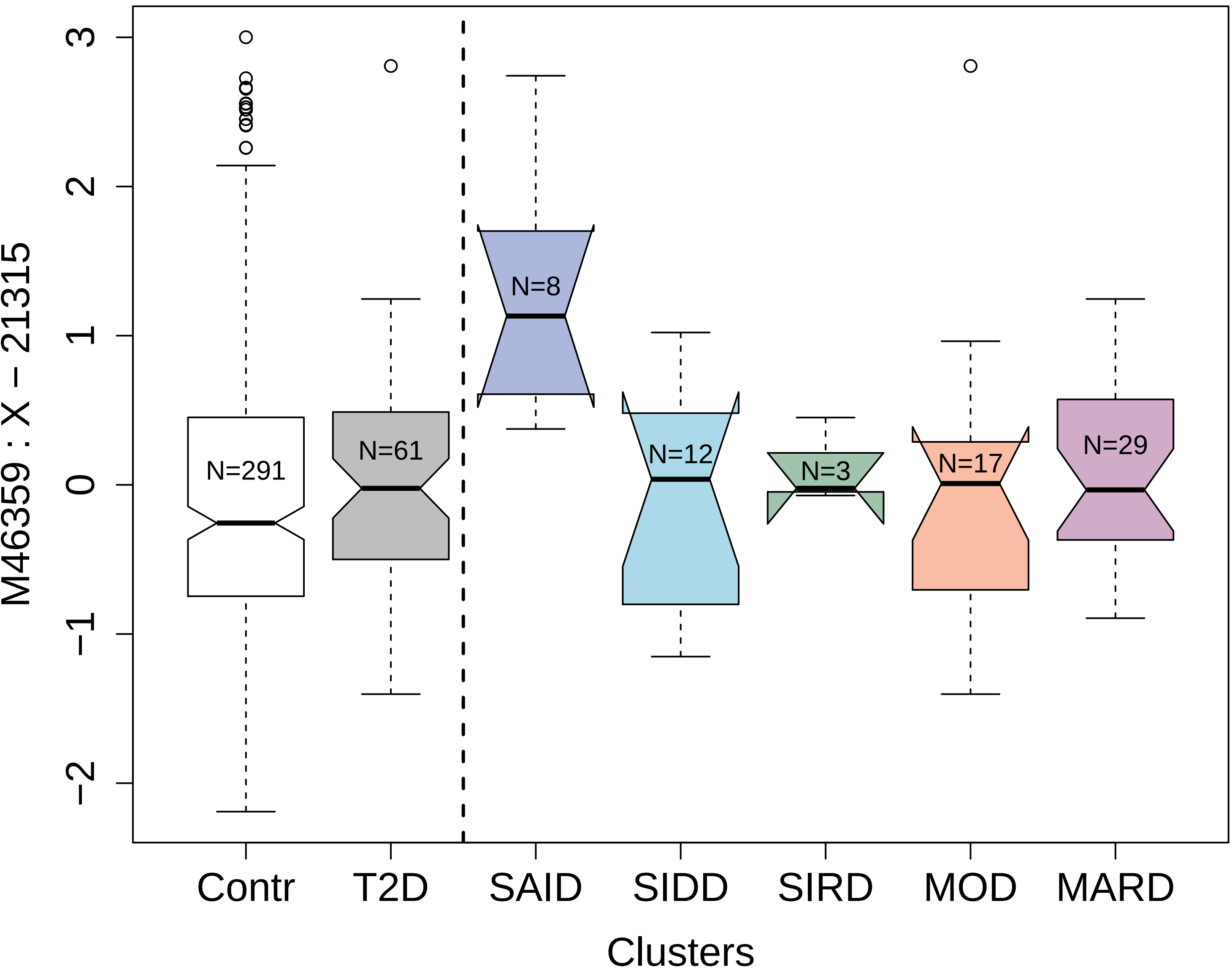

M46359 : X − 21315
3
2
1
0
−1
−2
N=291
N=61
N=8
N=12
N=3
N=17
N=29
Contr
T2D
SAID
SIDD
SIRD
MOD
MARD
Clusters

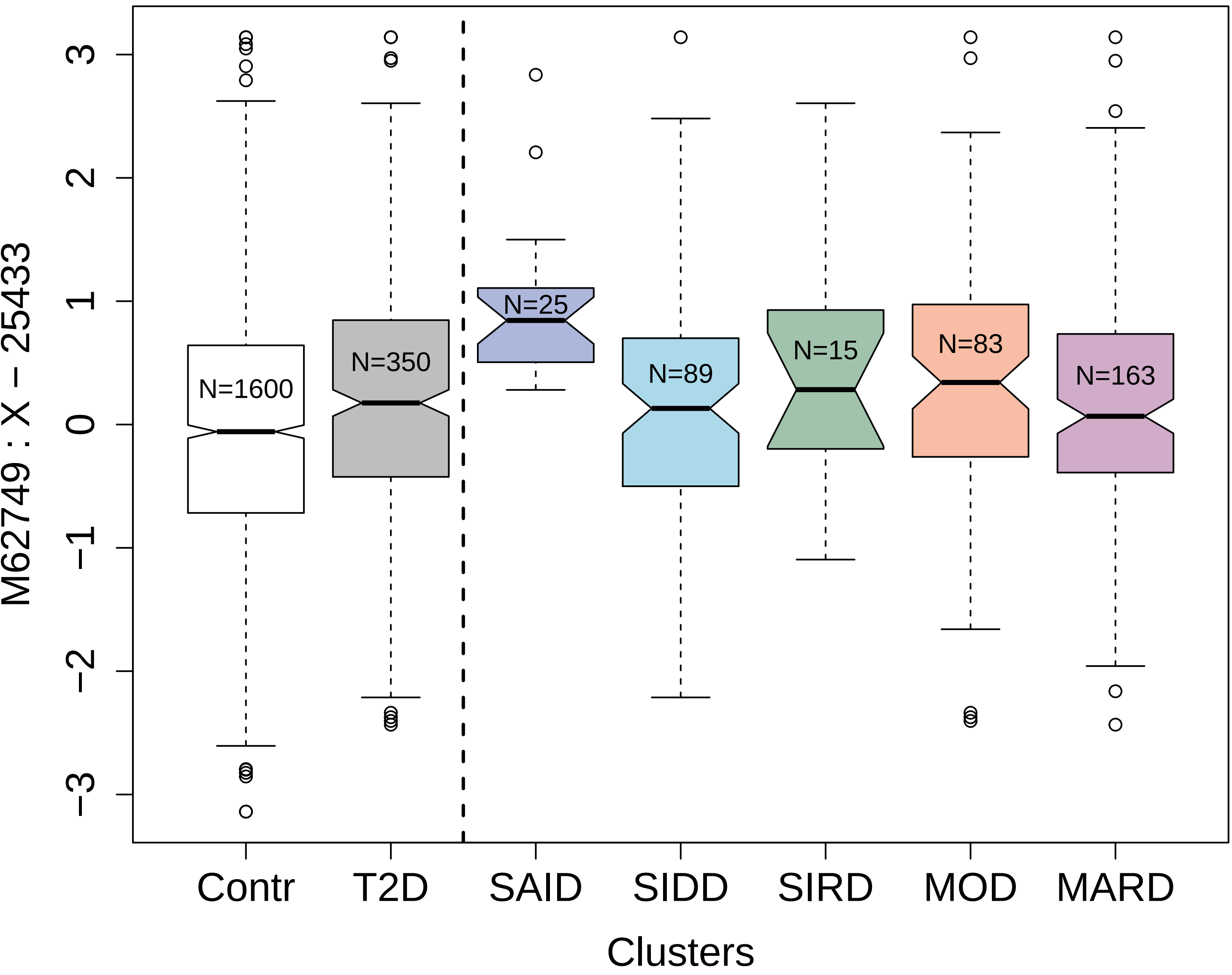

M62749 : X − 25433
3
2
1
0
−1
−2
−3
N=1600
N=350
N=25
N=89
N=15
N=83
N=163
Contr
T2D
SAID
SIDD
SIRD
MOD
MARD
Clusters

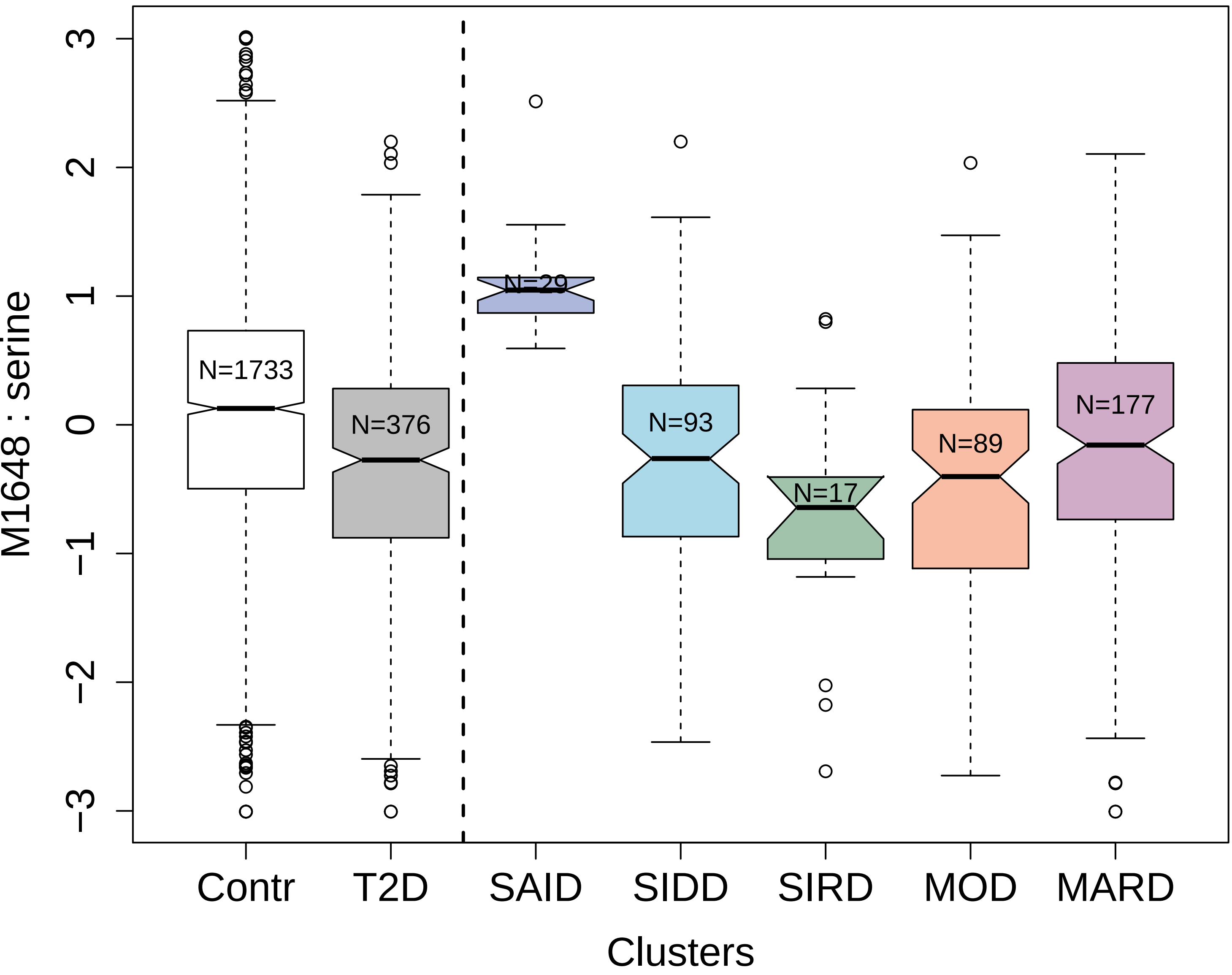

M1648 : serine
3
2
1
0
−1
−2
−3
N=1733
N=376
N=29
N=93
N=17
N=89
N=177
Contr
T2D
SAID
SIDD
SIRD
MOD
MARD
Clusters

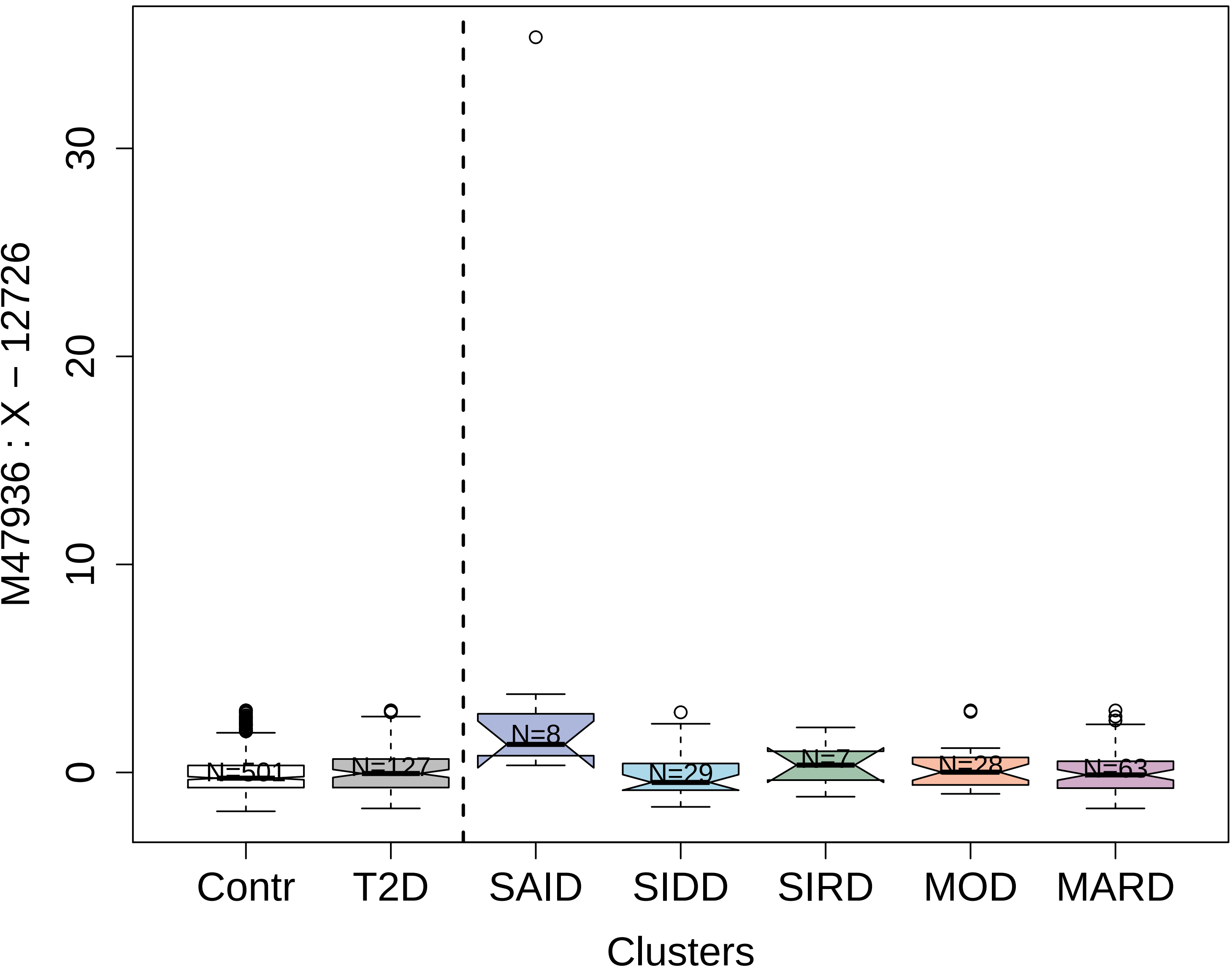

M47936 : X − 12726
0
10
20
30
N=501
N=127
N=8
N=29
N=7
N=28
N=63
Contr
T2D
SAID
SIDD
SIRD
MOD
MARD
Clusters

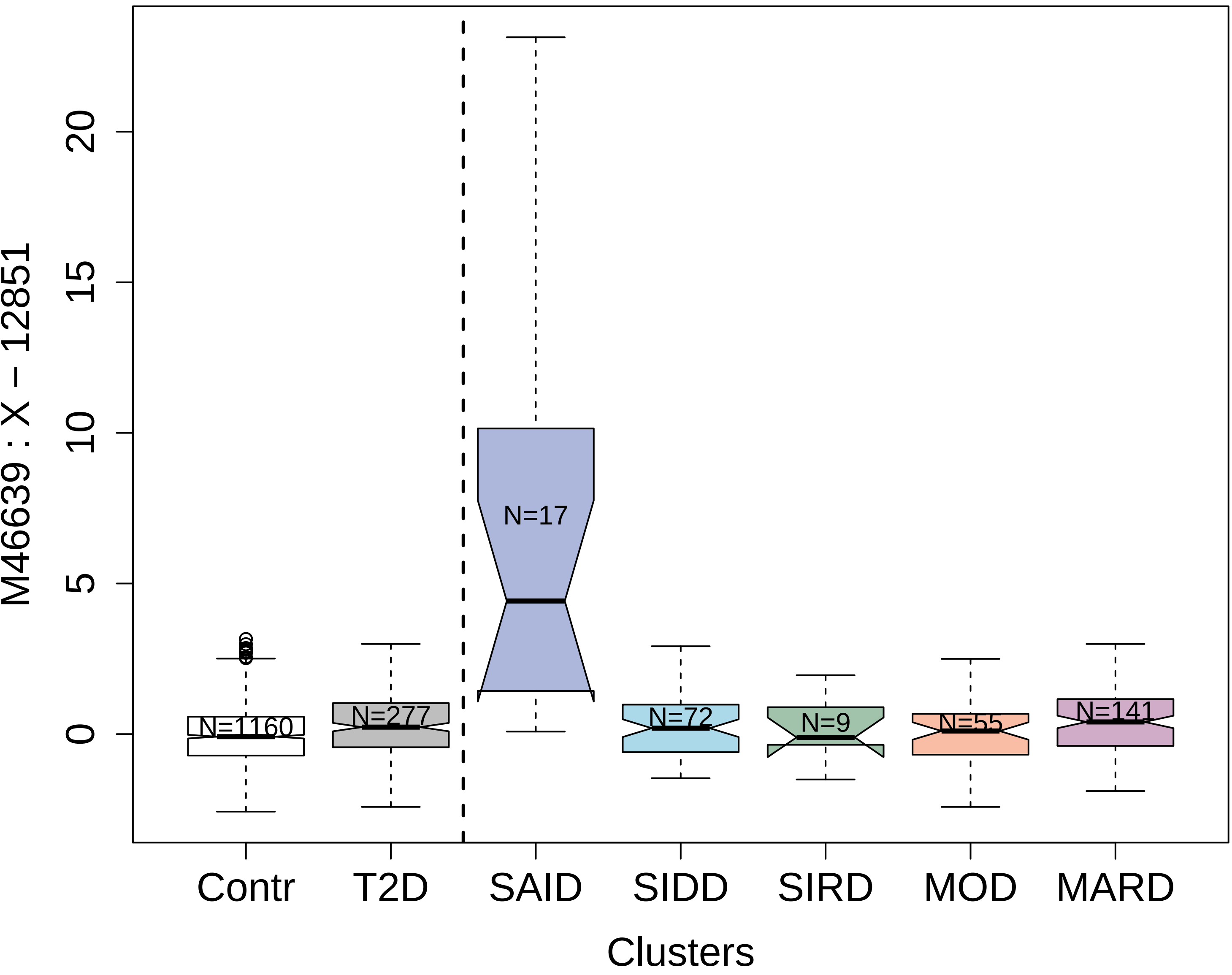

M46639 : X − 12851
0
5
10
15
20
N=1160
N=277
N=17
N=72
N=9
N=55
N=141
Contr
T2D
SAID
SIDD
SIRD
MOD
MARD
Clusters

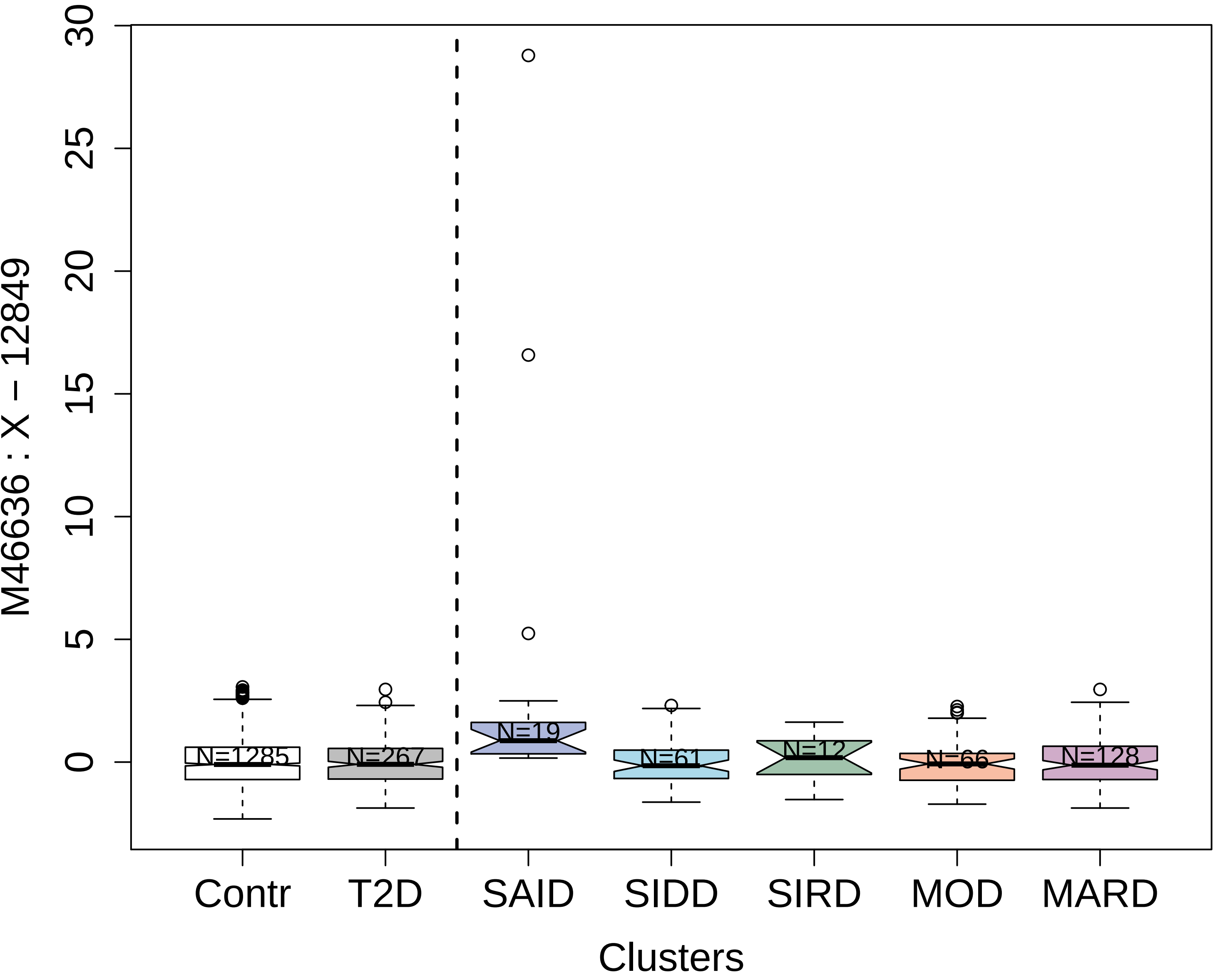

M46636 : X − 12849
30
25
20
15
10
5
0
N=1285
N=267
N=19
N=61
N=12
N=66
N=128
Contr
T2D
SAID
SIDD
SIRD
MOD
MARD
Clusters

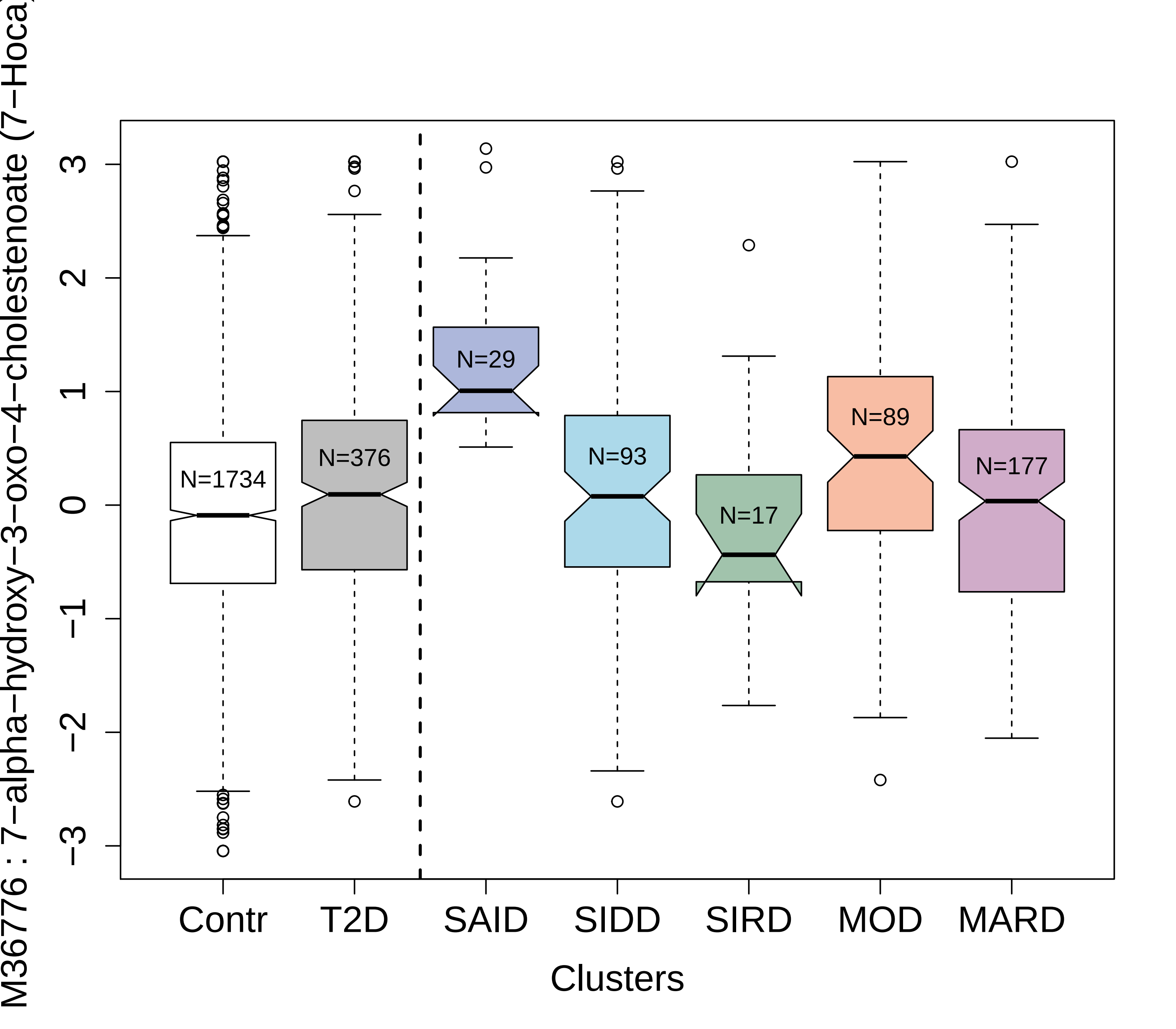
M36776 : 7−alpha−hydroxy−3−oxo−4−cholestenoate (7−Hoca)
3
2
1
0
−1
−2
−3
N=1734
N=376
N=29
N=93
N=17
N=89
N=177
Contr
T2D
SAID
SIDD
SIRD
MOD
MARD
Clusters

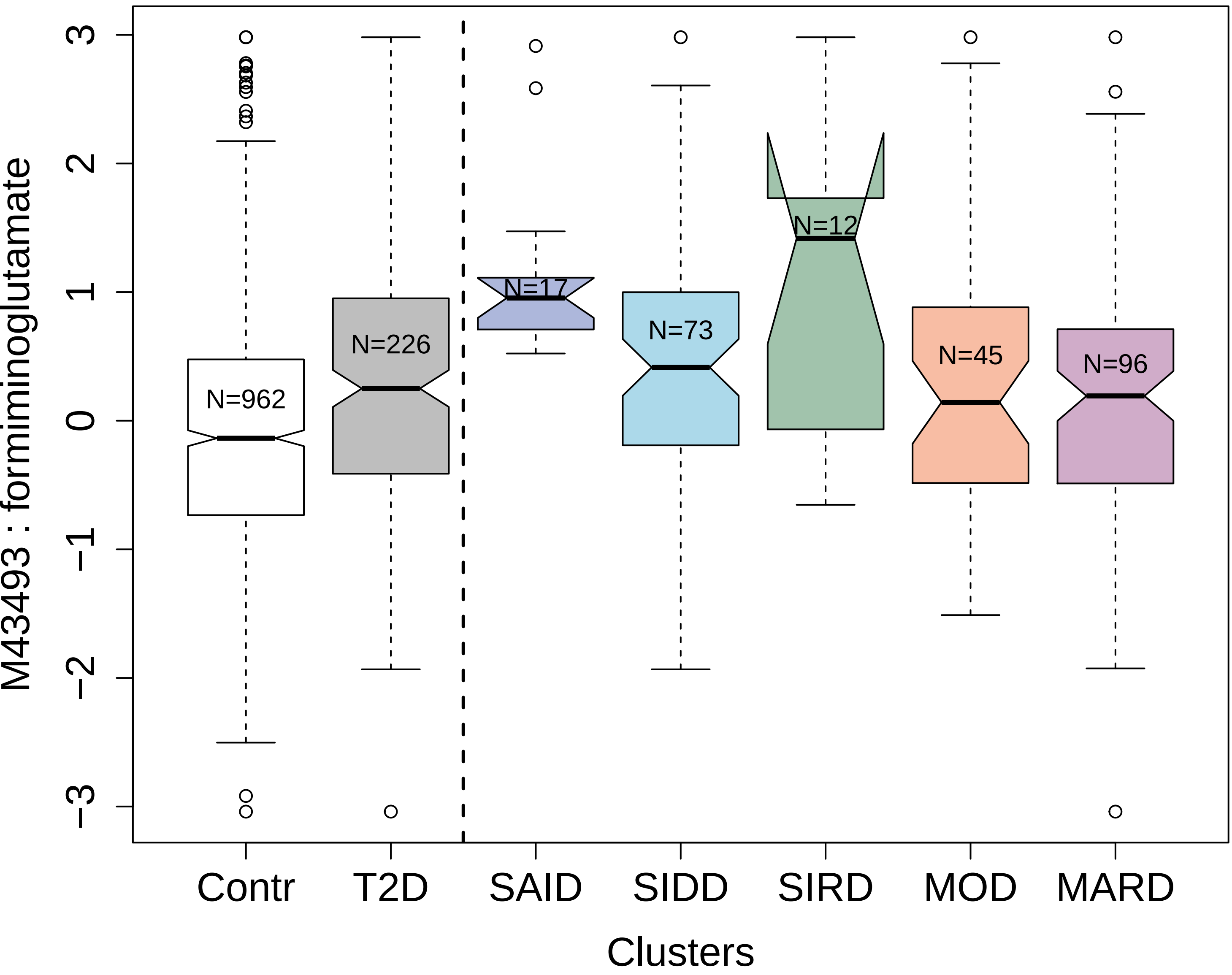

M43493 : formiminoglutamate
3
2
1
0
−1
−2
−3
N=962
N=226
N=17
N=73
N=12
N=45
N=96
Contr
T2D
SAID
SIDD
SIRD
MOD
MARD
Clusters

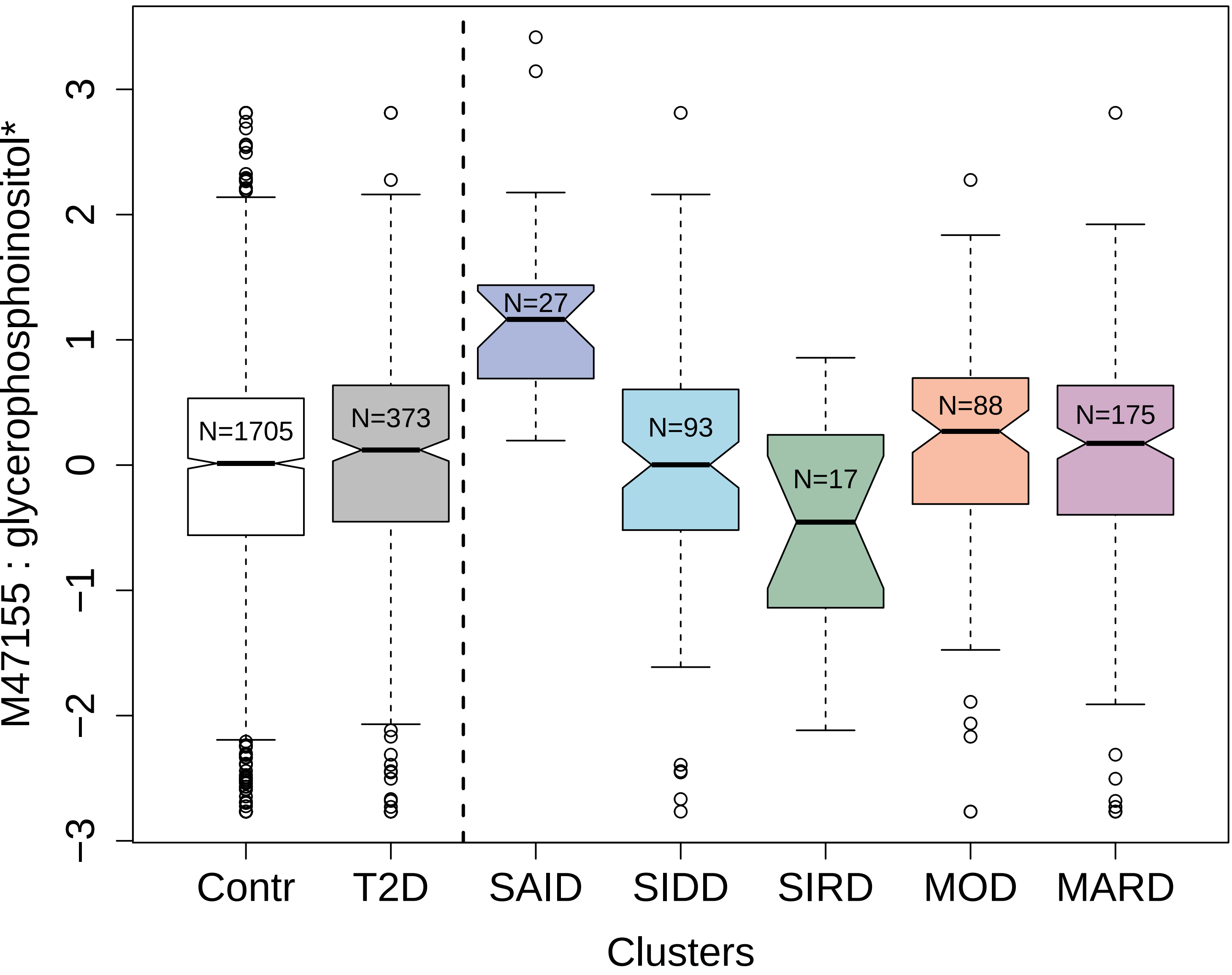
M47155 : glycerophosphoinositol*
Clusters
Contr
T2D
SAID
SIDD
SIRD
MOD
MARD
N=1705
N=373
N=27
N=93
N=17
N=88
N=175
3
2
1
0
−1
−2
−3

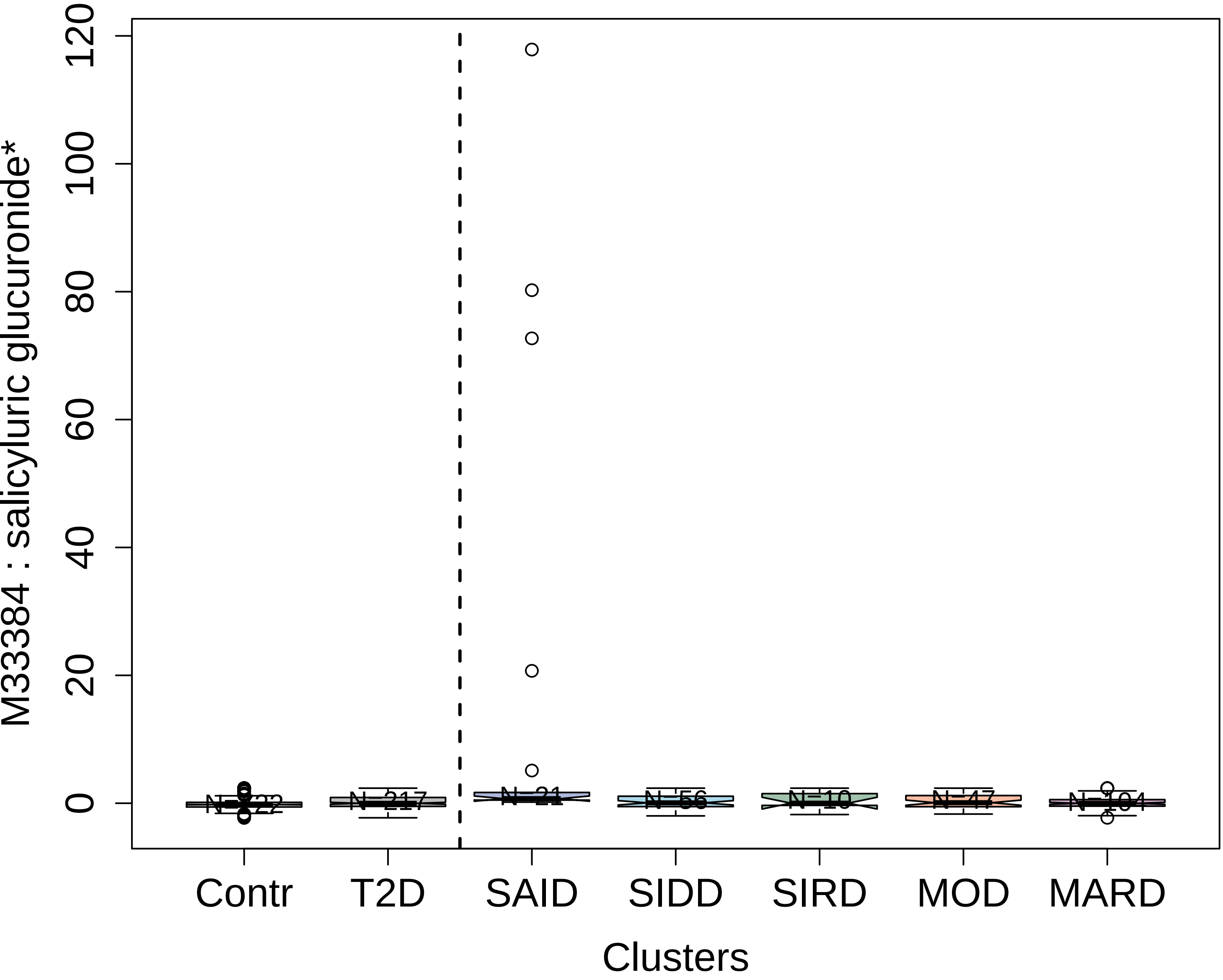
M33384 : salicyluric glucuronide*
0
20
40
60
80
100
120
N=722
N=217
N=21
N=56
N=10
N=47
N=104
Contr
T2D
SAID
SIDD
SIRD
MOD
MARD
Clusters

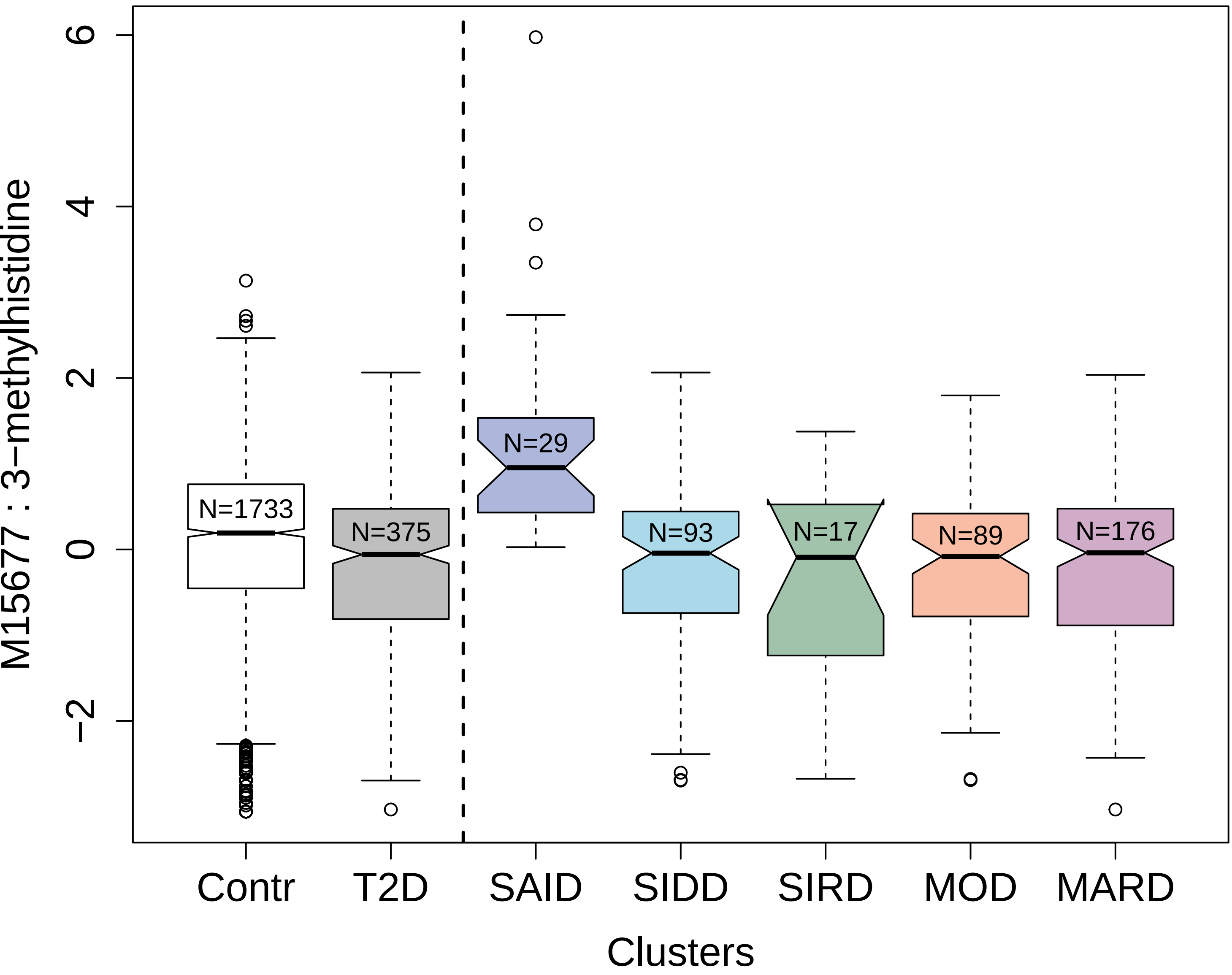

M15677 : 3−methylhistidine
6
4
2
0
−2
N=1733
N=375
N=29
N=93
N=17
N=89
N=176
Contr
T2D
SAID
SIDD
SIRD
MOD
MARD
Clusters

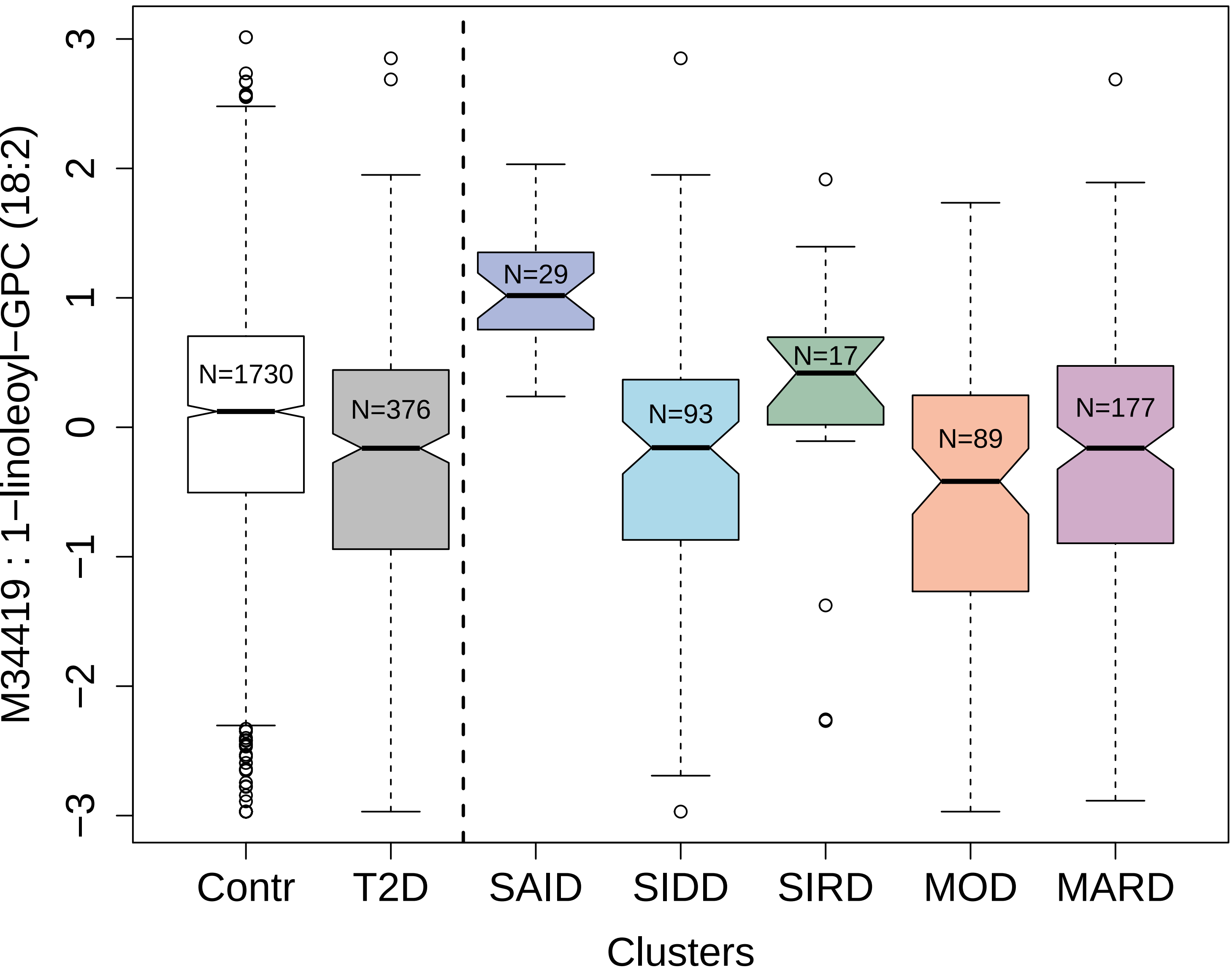

M34419 : 1−linoleoyl−GPC (18:2)
3
2
1
0
−1
−2
−3
N=1730
N=376
N=29
N=93
N=17
N=89
N=177
Contr
T2D
SAID
SIDD
SIRD
MOD
MARD
Clusters

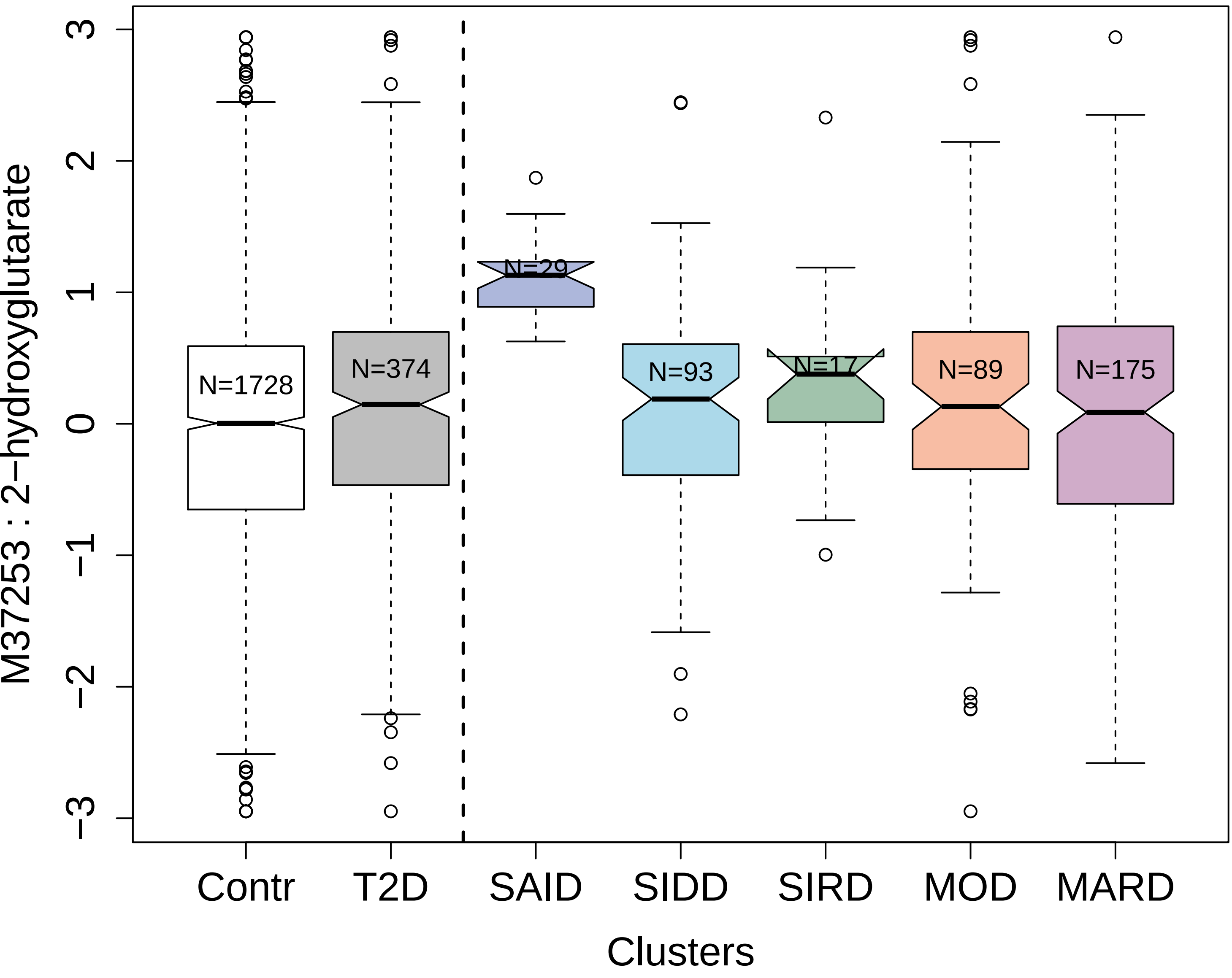

M37253 : 2−hydroxyglutarate
3
2
1
0
−1
−2
−3
N=1728
N=374
N=29
N=93
N=17
N=89
N=175
Contr
T2D
SAID
SIDD
SIRD
MOD
MARD
Clusters

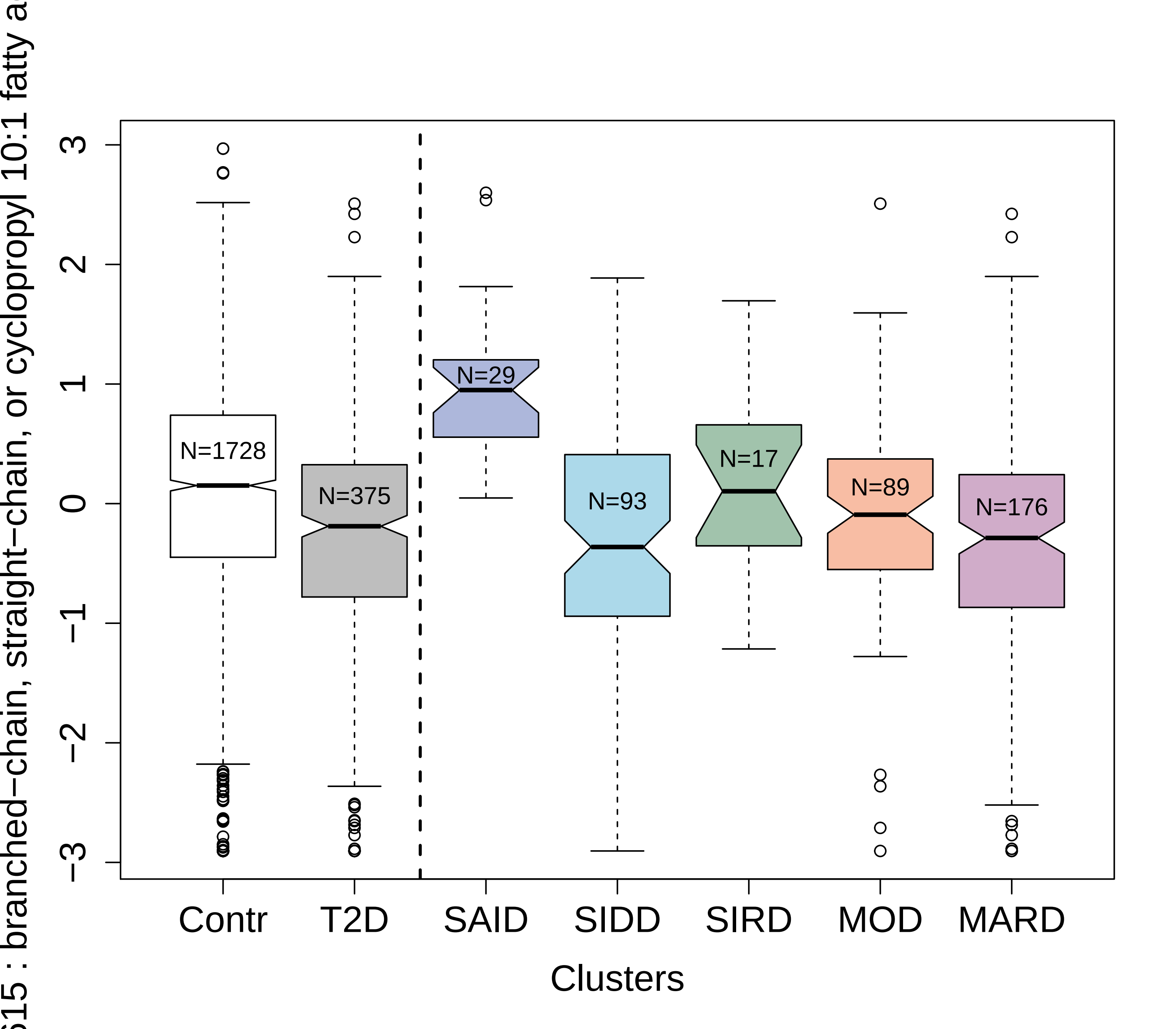

615 : branched−chain, straight−chain, or cyclopropyl 10:1 fatty a
3
2
1
0
−1
−2
−3
N=1728
N=375
N=29
N=93
N=17
N=89
N=176
Contr
T2D
SAID
SIDD
SIRD
MOD
MARD
Clusters

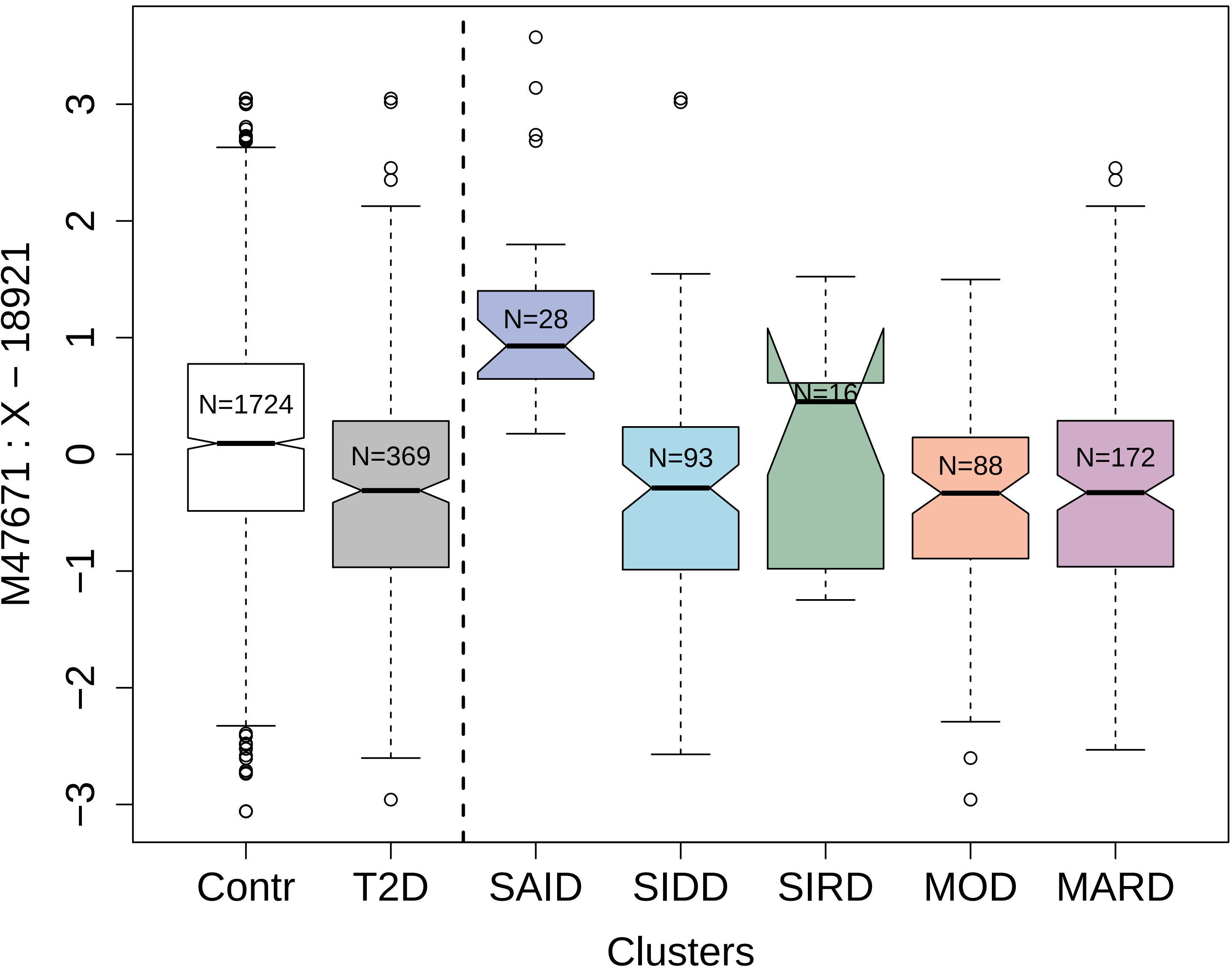

M47671 : X − 18921
3
2
1
0
−1
−2
−3
N=1724
N=369
N=28
N=93
N=16
N=88
N=172
Contr
T2D
SAID
SIDD
SIRD
MOD
MARD
Clusters

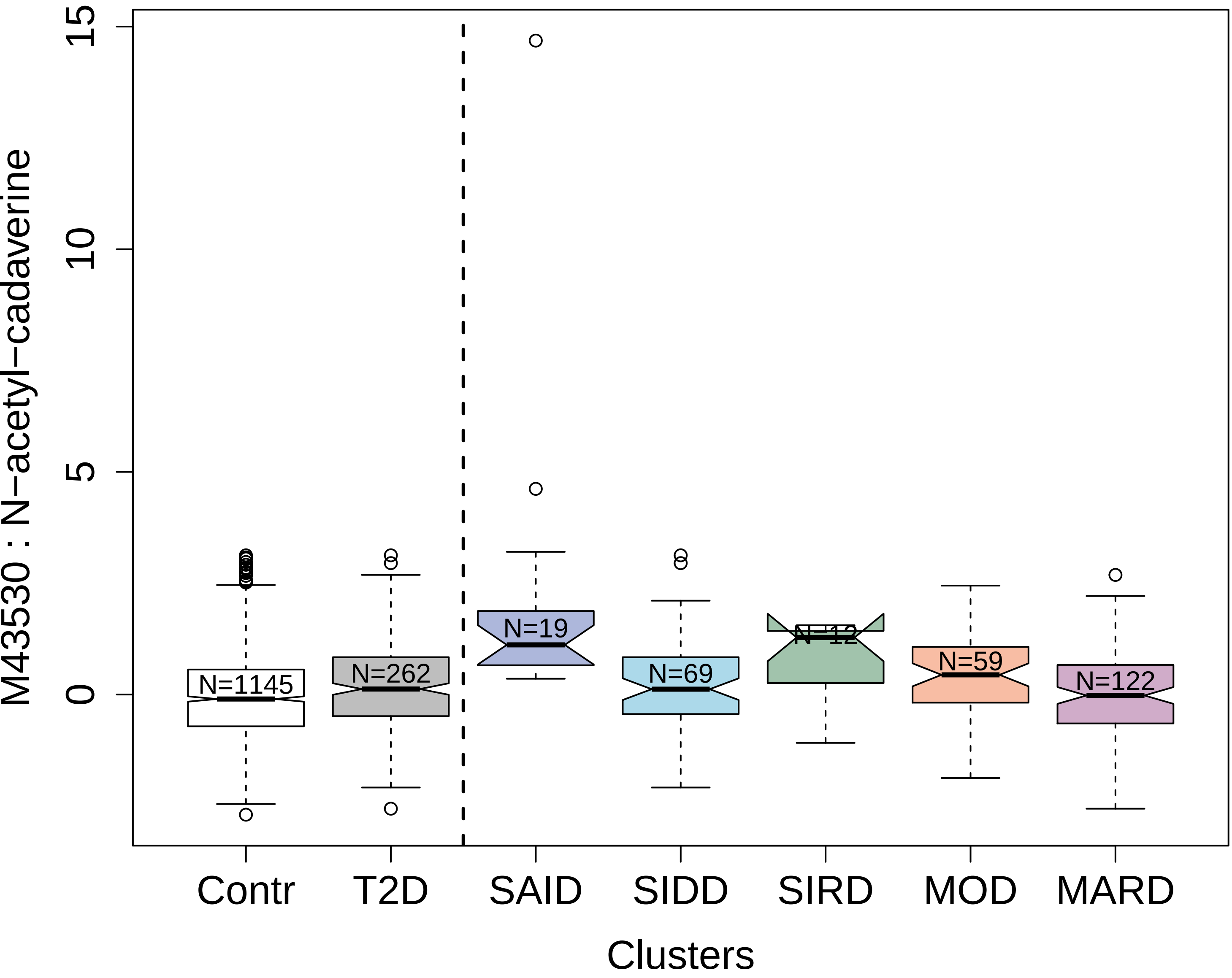
M43530 : N−acetyl−cadaverine
0
5
10
15
N=1145
N=262
N=19
N=69
N=12
N=59
N=122
Contr
T2D
SAID
SIDD
SIRD
MOD
MARD
Clusters

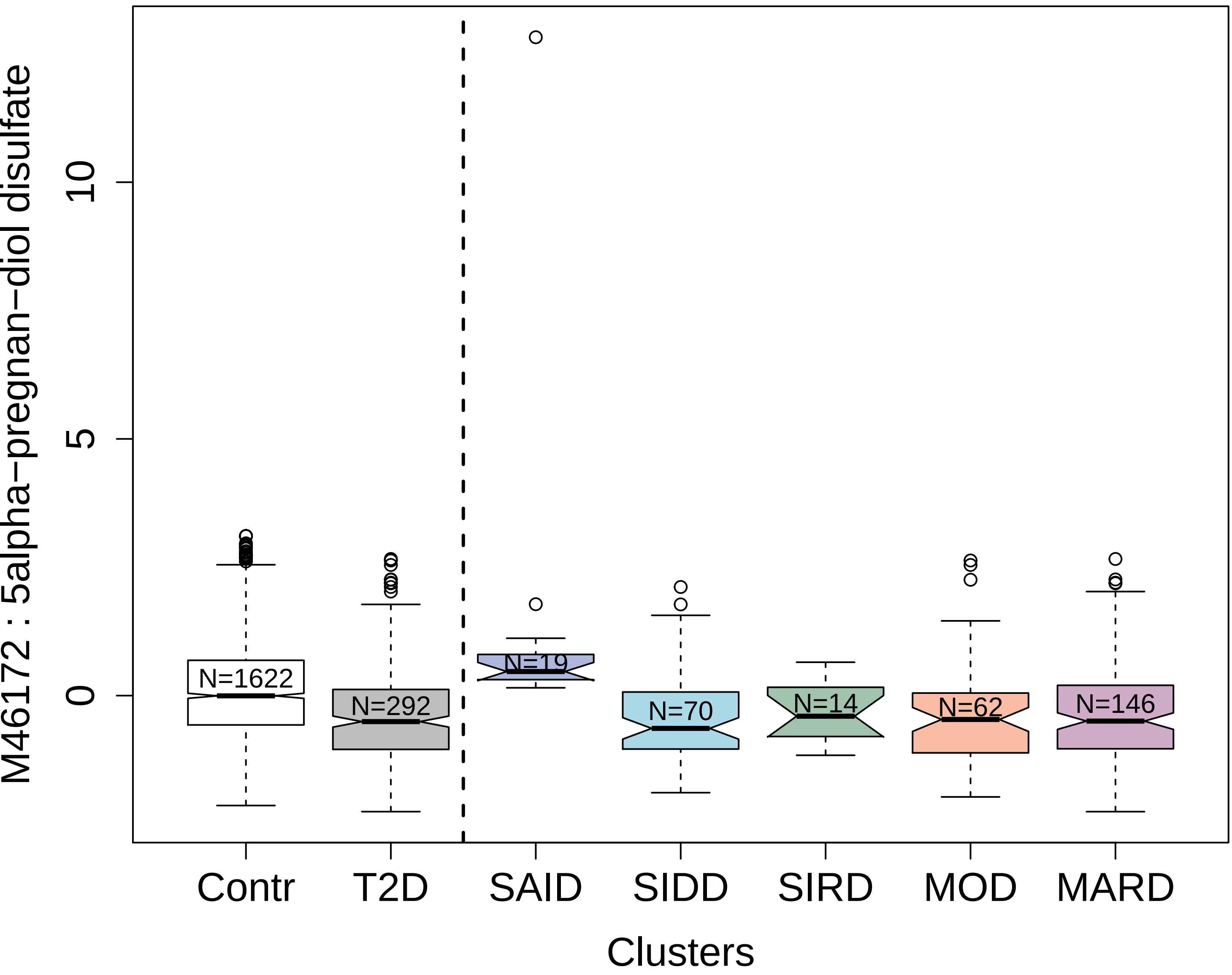

M46172 : 5alpha−pregnan−diol disulfate
0
5
10
N=1622
N=292
N=19
N=70
N=14
N=62
N=146
Contr
T2D
SAID
SIDD
SIRD
MOD
MARD
Clusters

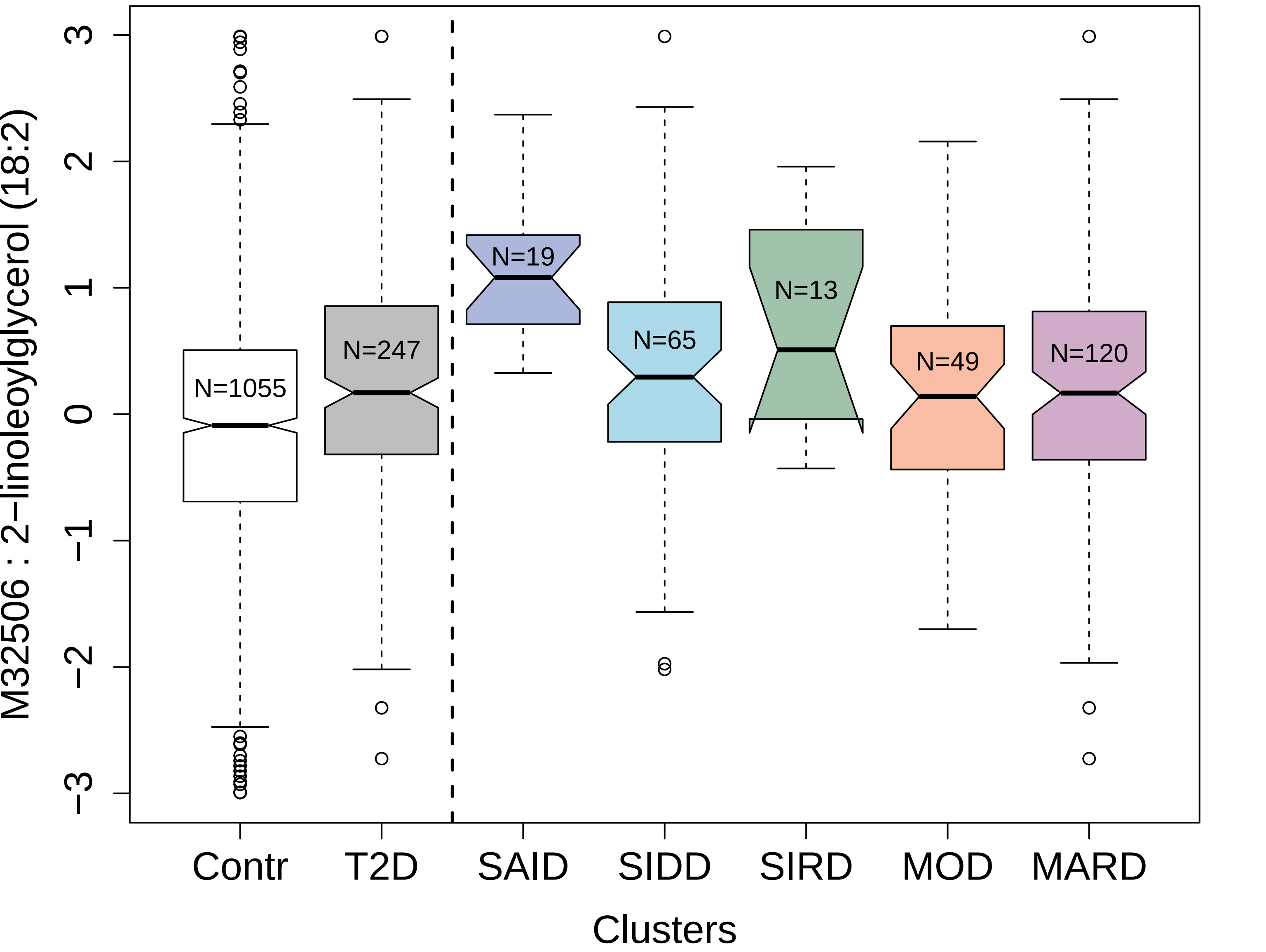

M32506 : 2−linoleoylglycerol (18:2)
3
2
1
0
−1
−2
−3
N=1055
N=247
N=19
N=65
N=13
N=49
N=120
Contr
T2D
SAID
SIDD
SIRD
MOD
MARD
Clusters

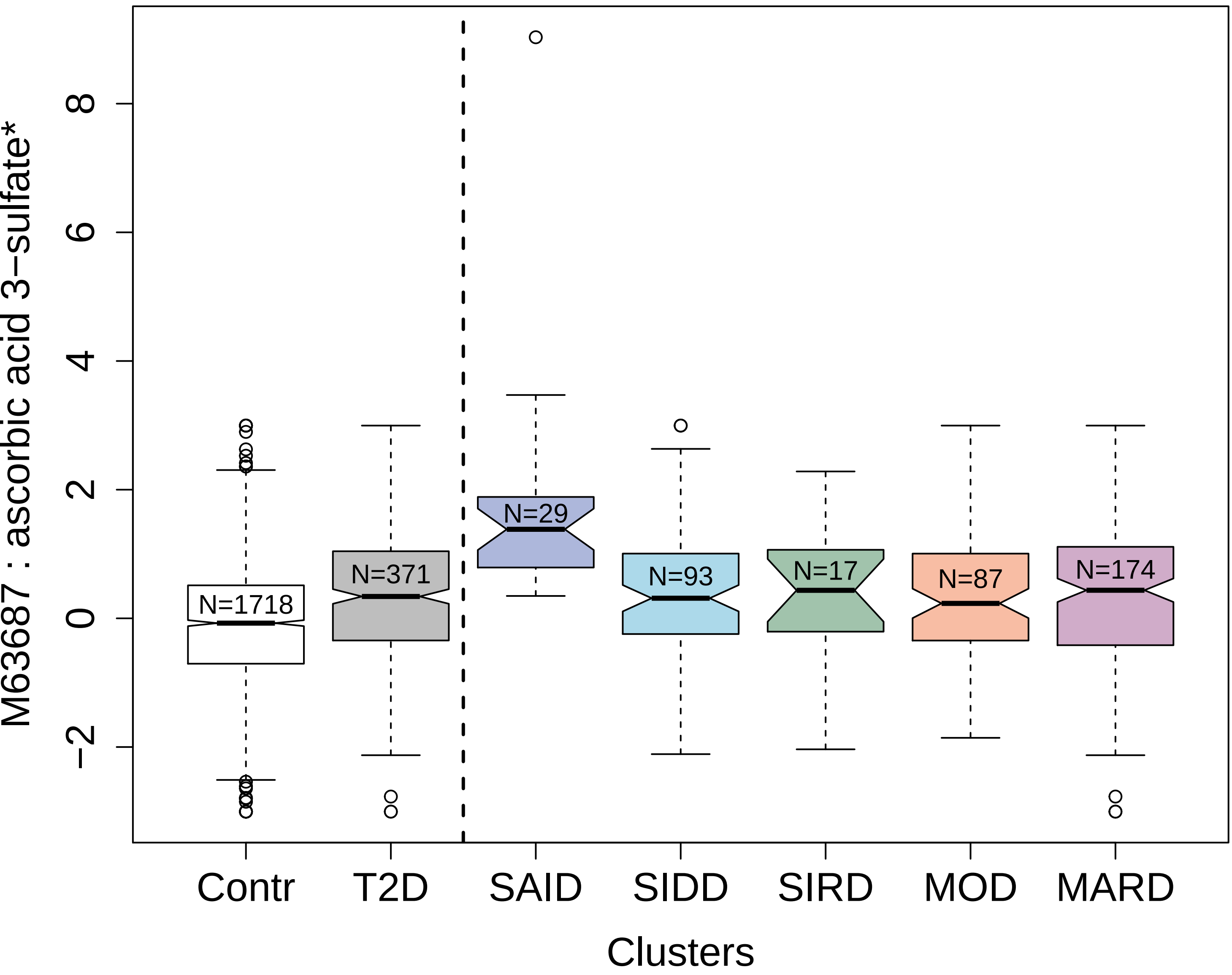

M63687 : ascorbic acid 3−sulfate*
−2
0
2
4
6
8
N=1718
N=371
N=29
N=93
N=17
N=87
N=174
Contr
T2D
SAID
SIDD
SIRD
MOD
MARD
Clusters

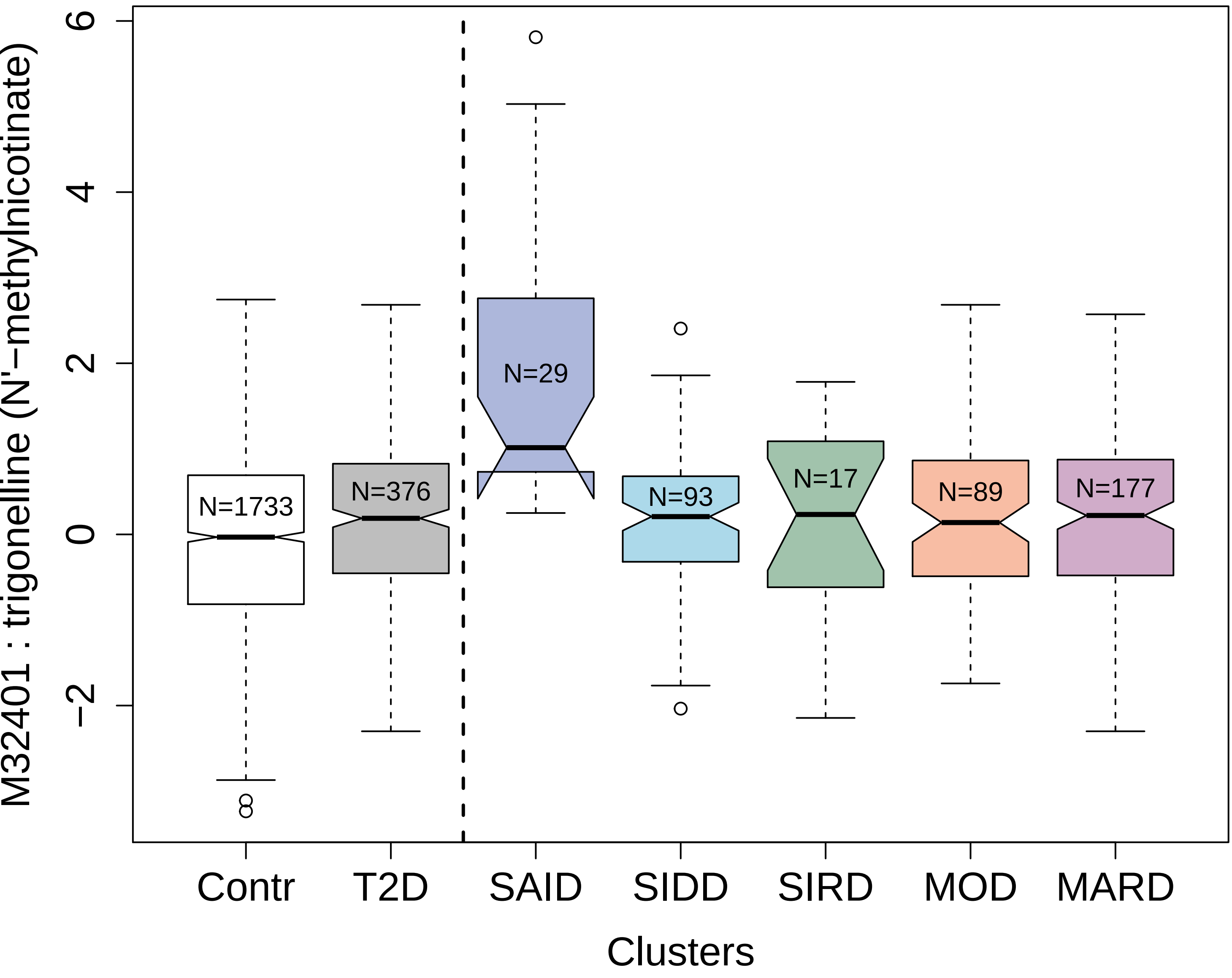

M32401 : trigonelline (N'−methylnicotinate)
−2
0
2
4
6
N=1733
N=376
N=29
N=93
N=17
N=89
N=177
Contr
T2D
SAID
SIDD
SIRD
MOD
MARD
Clusters

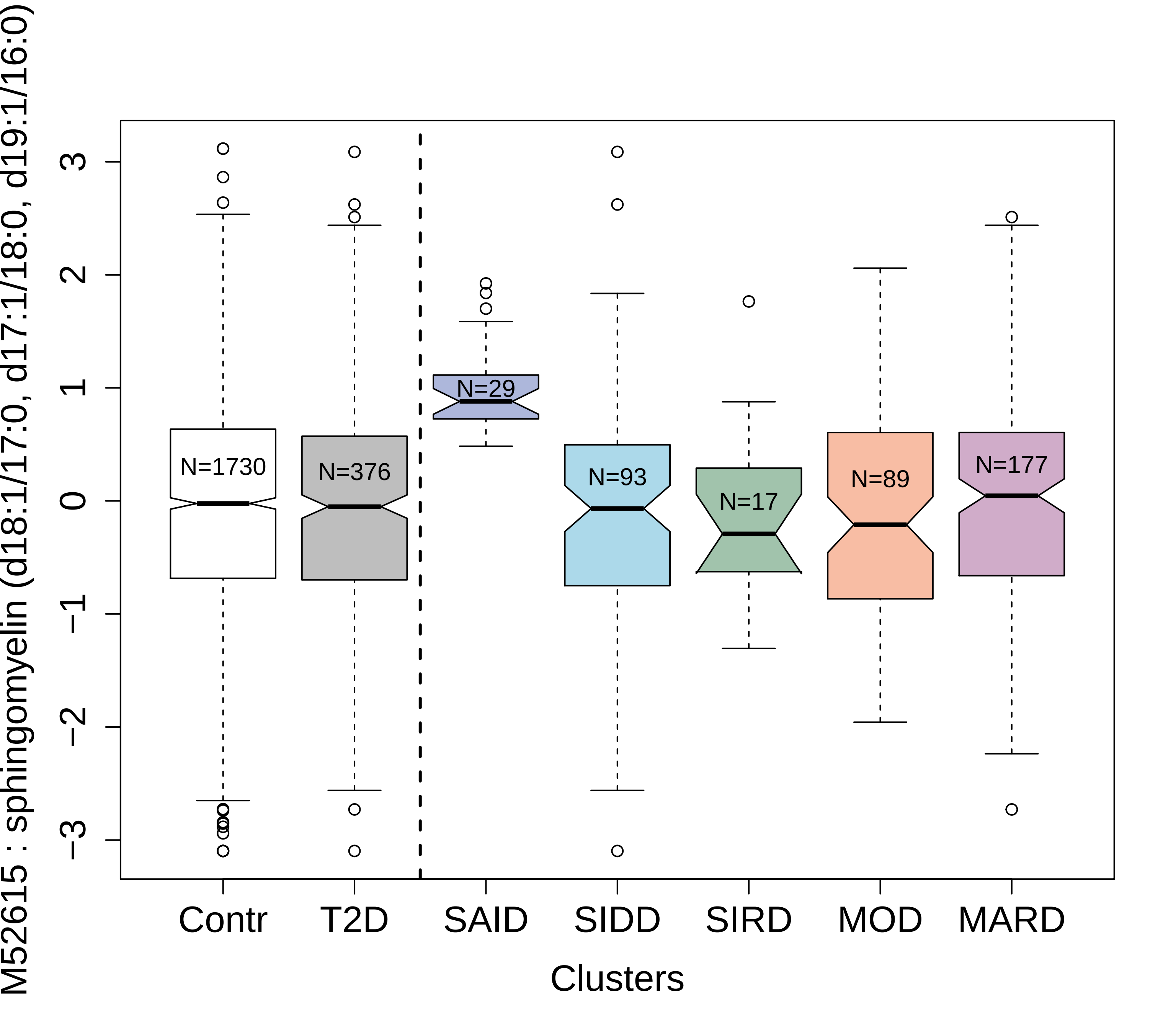

M52615 : sphingomyelin (d18:1/17:0, d17:1/18:0, d19:1/16:0)
3
2
1
0
−1
−2
−3
N=1730
N=376
N=29
N=93
N=17
N=89
N=177
Contr
T2D
SAID
SIDD
SIRD
MOD
MARD
Clusters

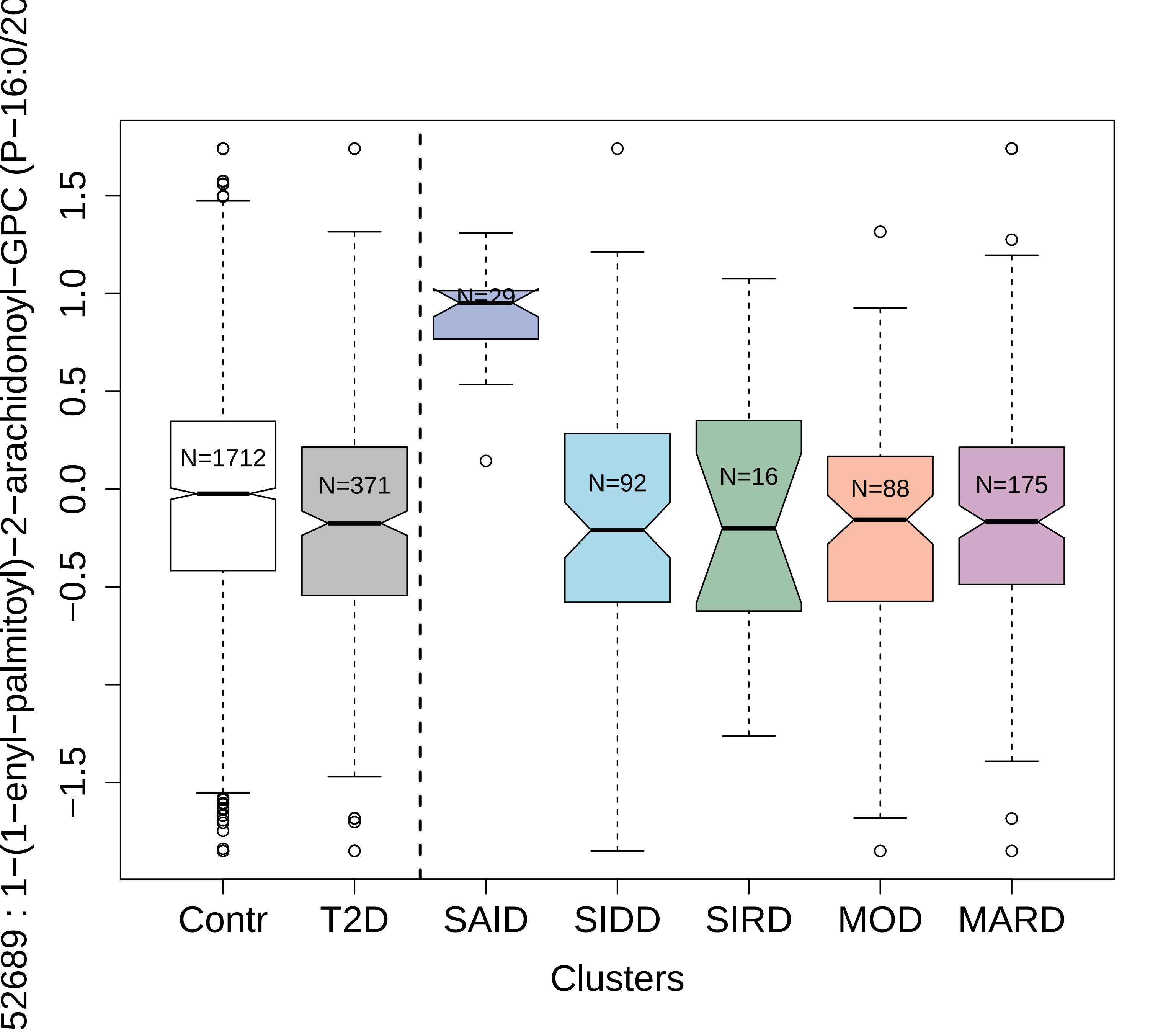
52689 : 1−(1−enyl−palmitoyl)−2−arachidonoyl−GPC (P−16:0/20
1.5
1.0
0.5
0.0
−0.5
−1.5
N=1712
N=371
N=29
N=92
N=16
N=88
N=175
Contr
T2D
SAID
SIDD
SIRD
MOD
MARD
Clusters

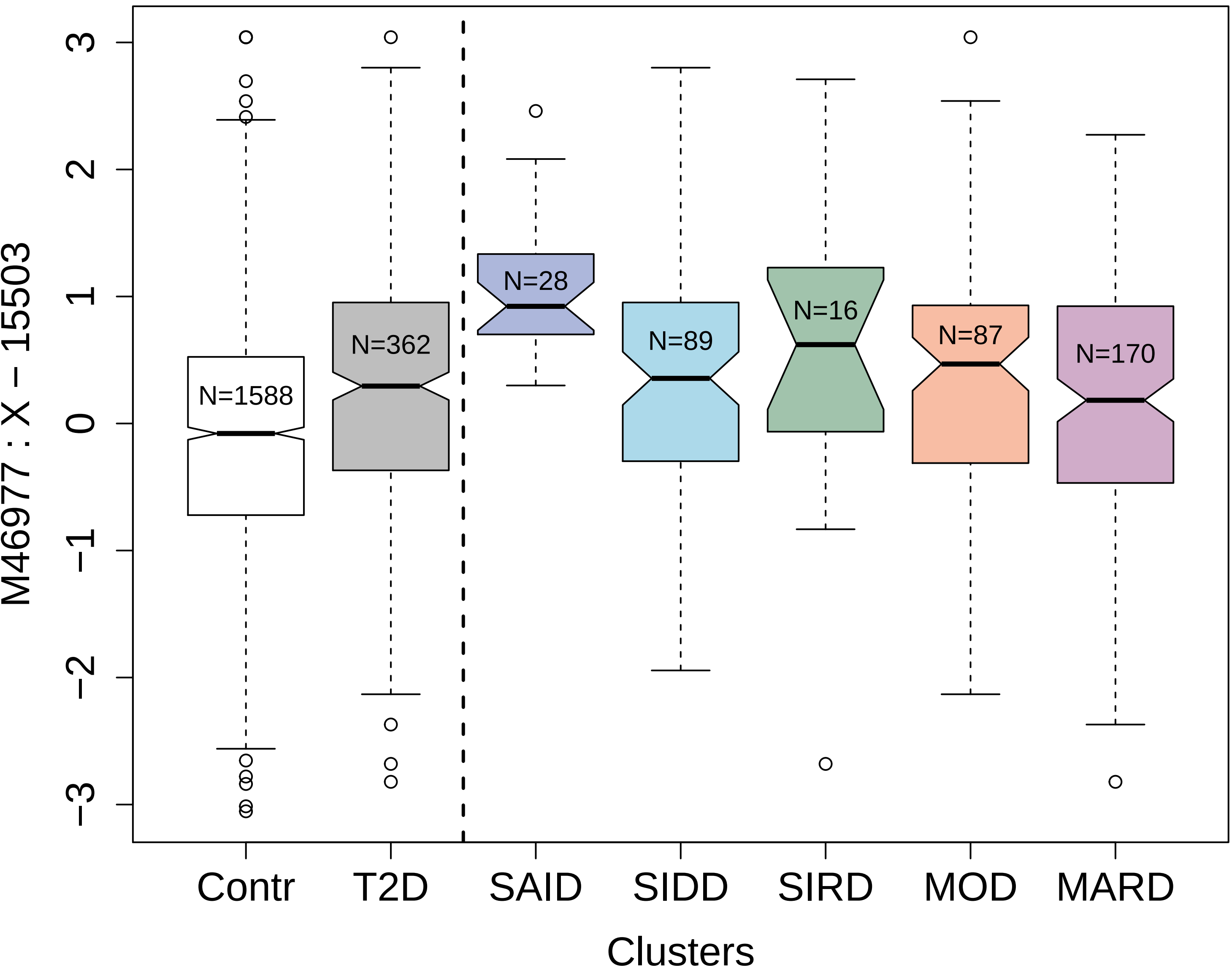

M46977 : X − 15503
3
2
1
0
−1
−2
−3
N=1588
N=362
N=28
N=89
N=16
N=87
N=170
Contr
T2D
SAID
SIDD
SIRD
MOD
MARD
Clusters

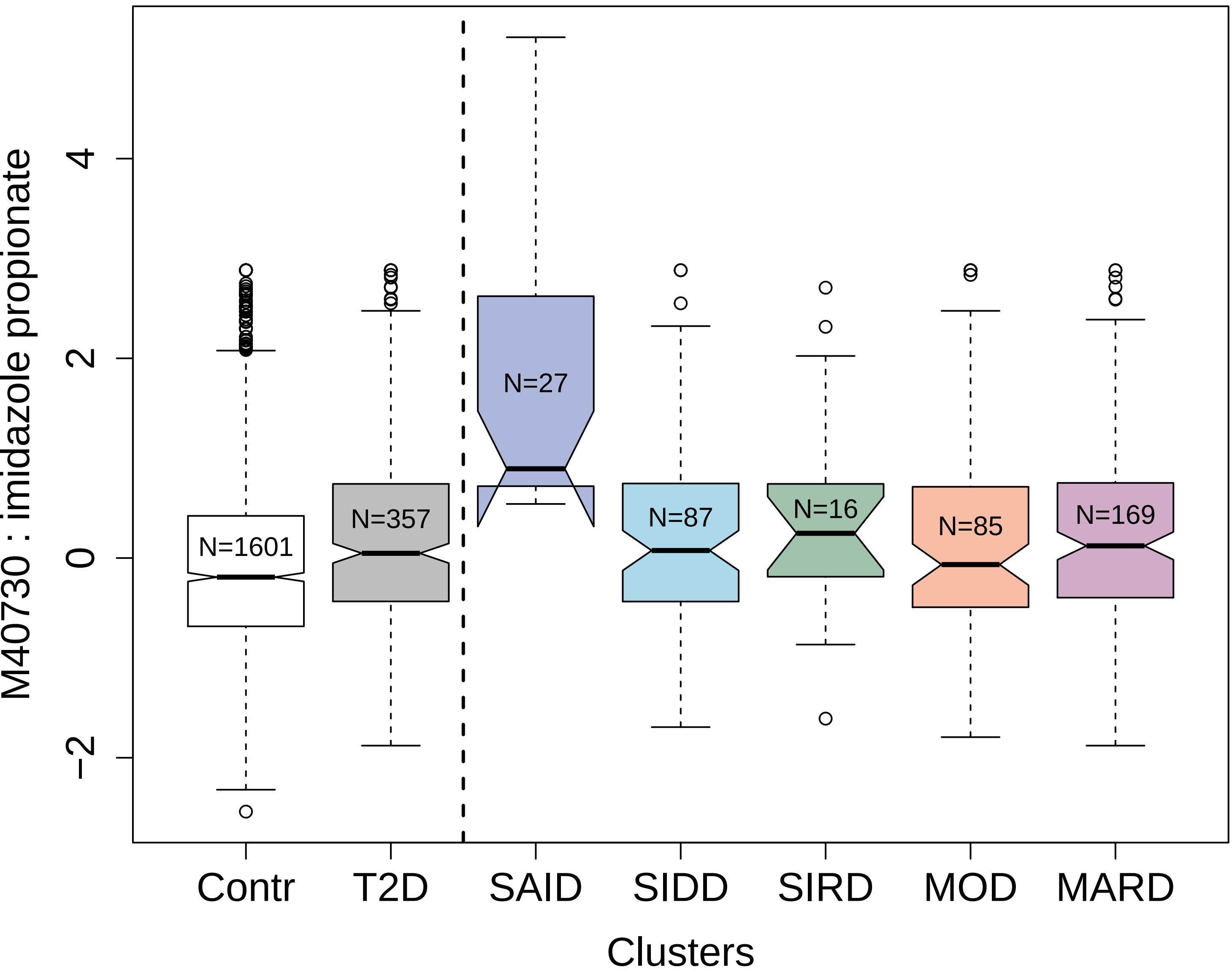
M40730 : imidazole propionate
4
2
0
−2
N=1601
N=357
N=27
N=87
N=16
N=85
N=169
Contr
T2D
SAID
SIDD
SIRD
MOD
MARD
Clusters

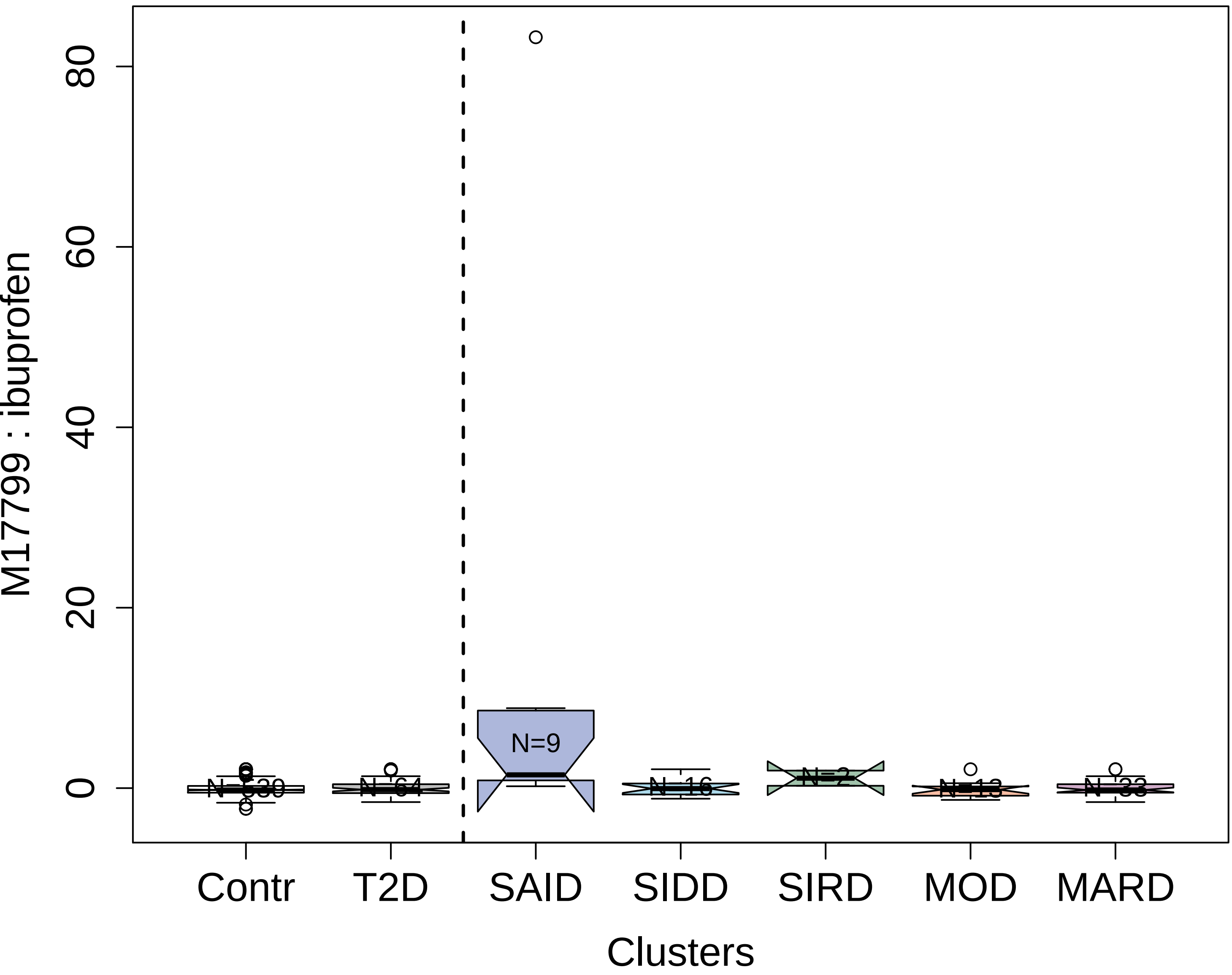

M17799 : ibuprofen
0
20
40
60
80
N=9
N=2
Contr
T2D
SAID
SIDD
SIRD
MOD
MARD
Clusters

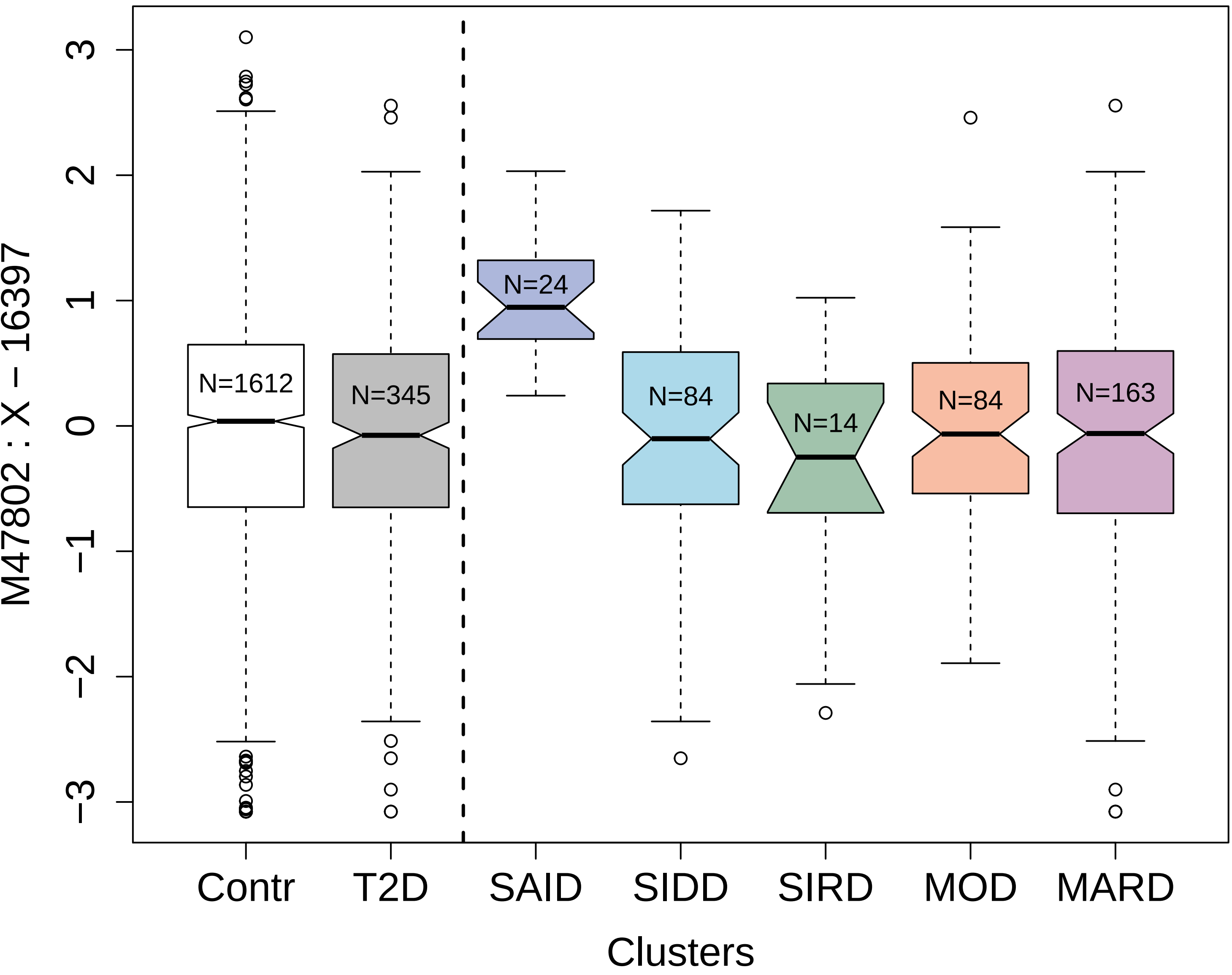

M47802 : X − 16397
3
2
1
0
−1
−2
−3
N=1612
N=345
N=24
N=84
N=14
N=84
N=163
Contr
T2D
SAID
SIDD
SIRD
MOD
MARD
Clusters

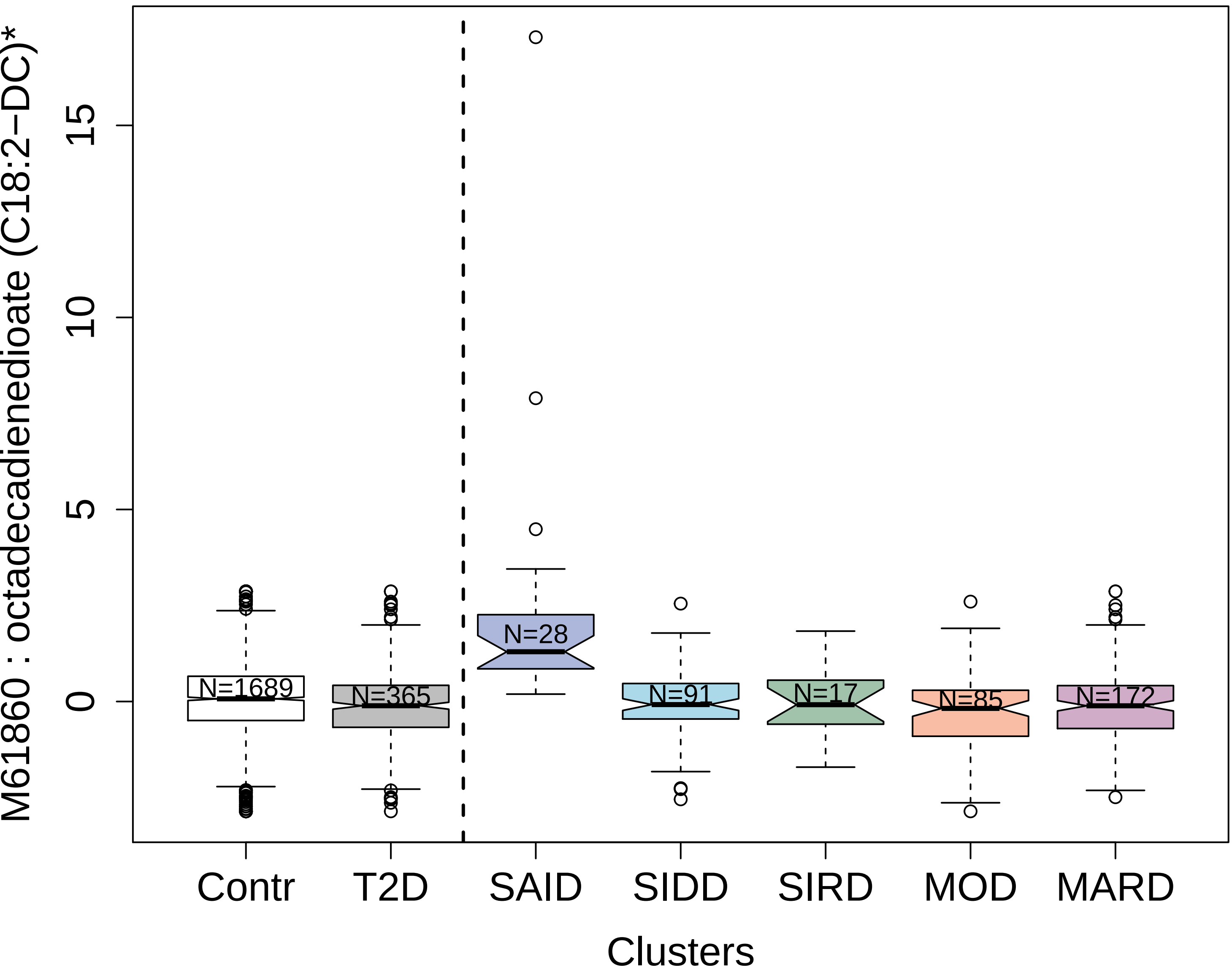

M61860 : octadecadienedioate (C18:2−DC)*
0
5
10
15
N=1689
N=365
N=28
N=91
N=17
N=85
N=172
Contr
T2D
SAID
SIDD
SIRD
MOD
MARD
Clusters

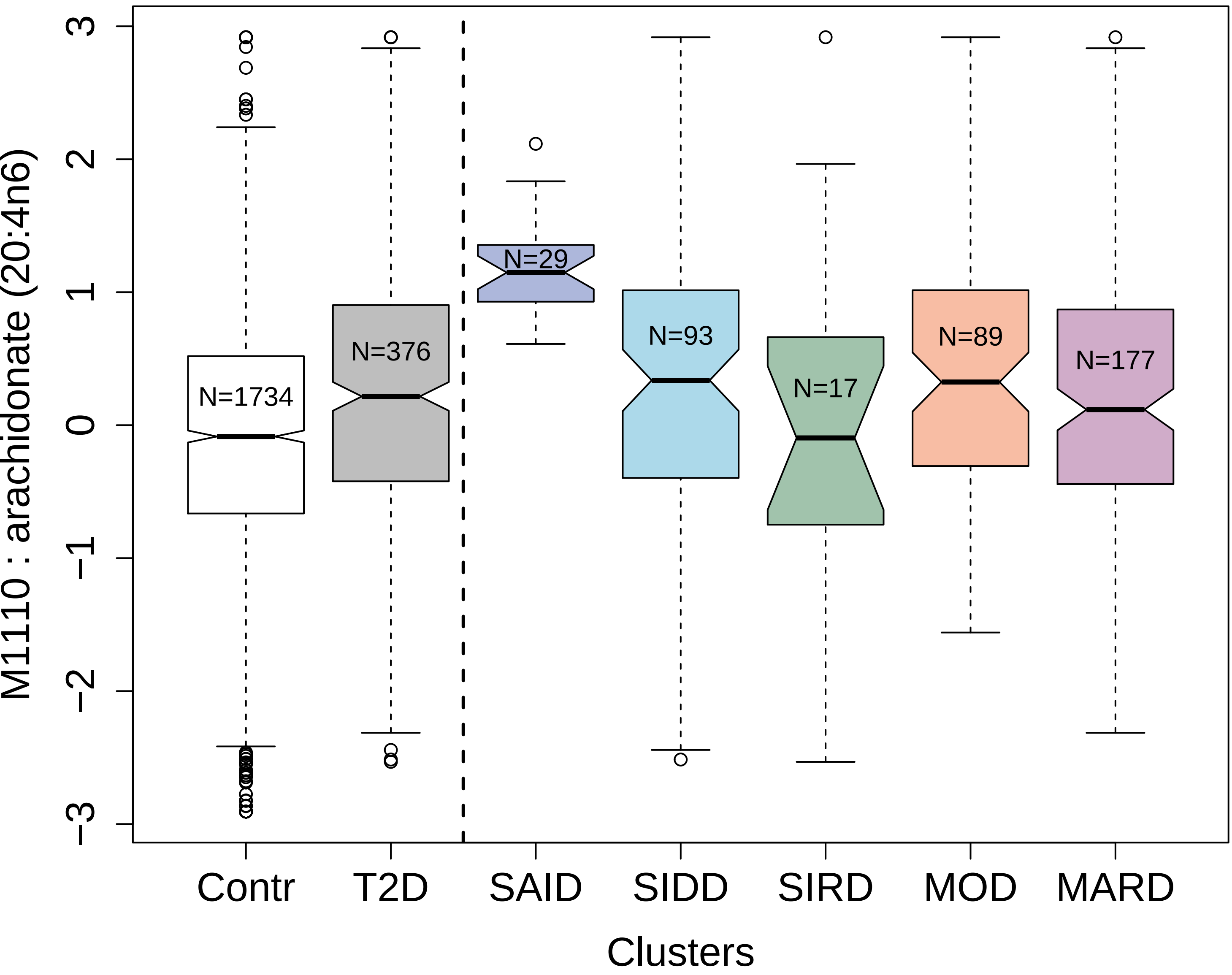
M1110 : arachidonate (20:4n6)
3
2
1
0
−1
−2
−3
N=1734
N=376
N=29
N=93
N=17
N=89
N=177
Contr
T2D
SAID
SIDD
SIRD
MOD
MARD
Clusters

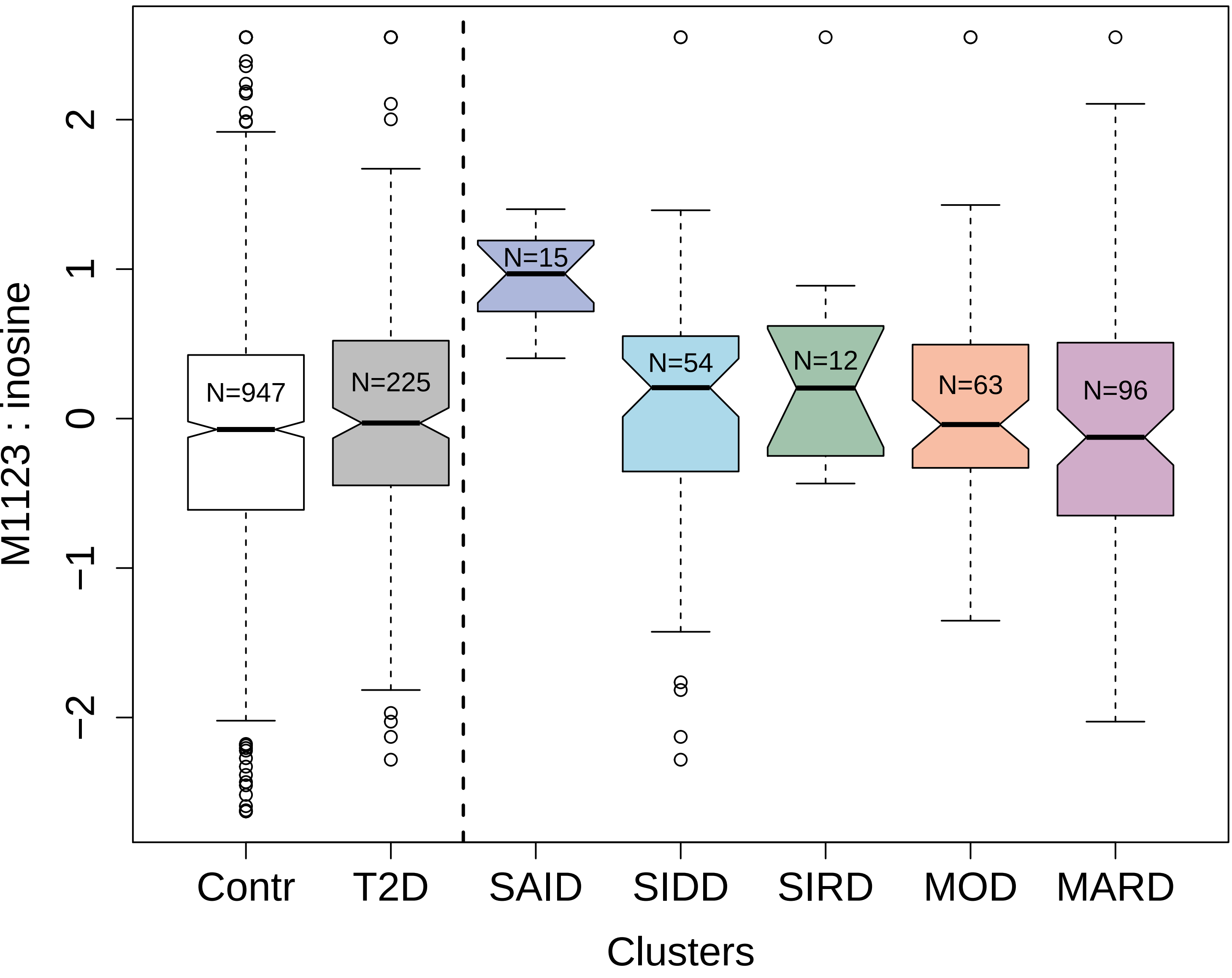
M1123 : inosine
2
1
0
−1
−2
N=947
N=225
N=15
N=54
N=12
N=63
N=96
Contr
T2D
SAID
SIDD
SIRD
MOD
MARD
Clusters

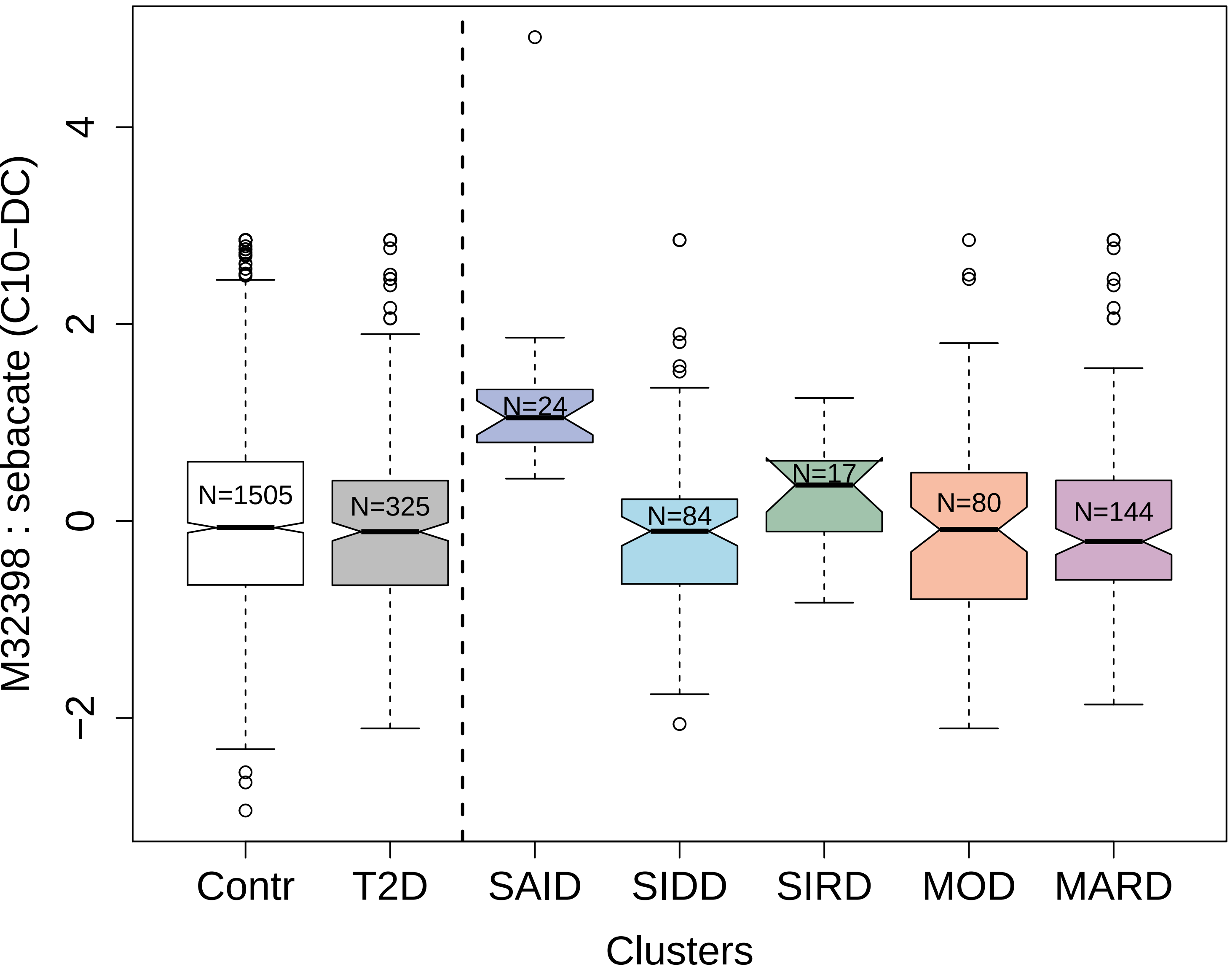

M32398 : sebacate (C10−DC)
4
2
0
−2
N=1505
N=325
N=24
N=84
N=17
N=80
N=144
Contr
T2D
SAID
SIDD
SIRD
MOD
MARD
Clusters

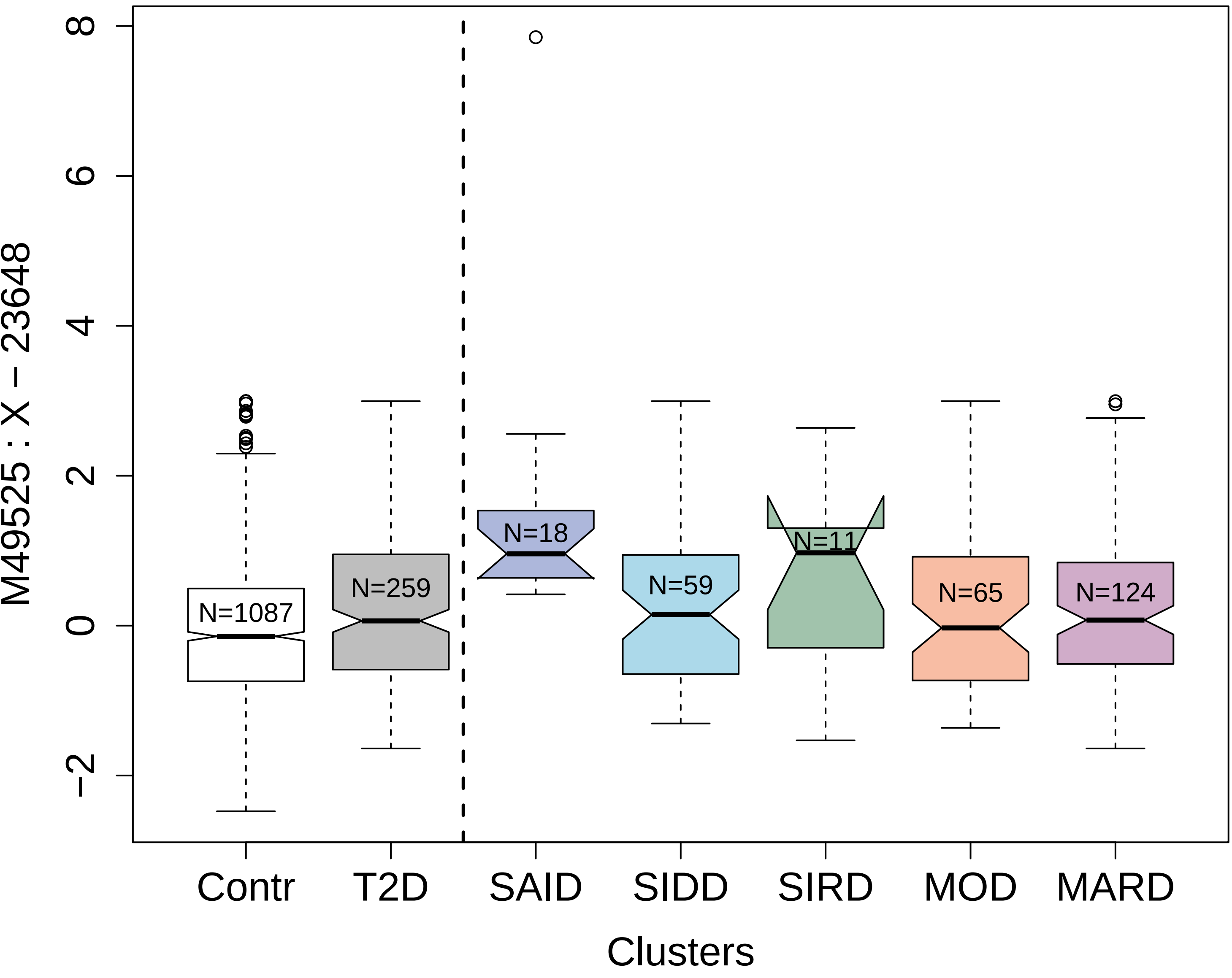

M49525 : X − 23648
8
6
4
2
0
−2
N=1087
N=259
N=18
N=59
N=11
N=65
N=124
Contr
T2D
SAID
SIDD
SIRD
MOD
MARD
Clusters

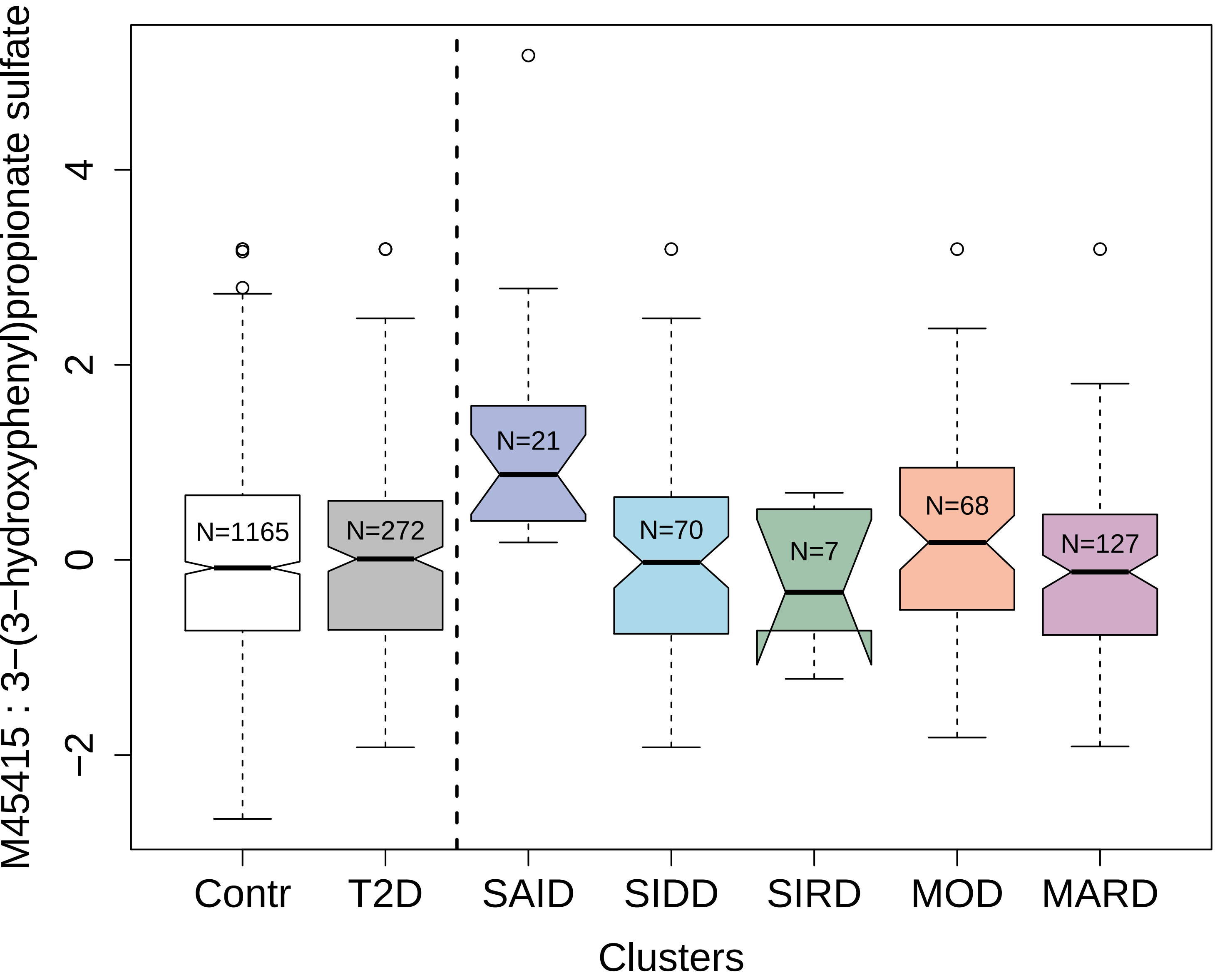

M45415 : 3−(3−hydroxyphenyl)propionate sulfate
4
2
0
−2
N=1165
N=272
N=21
N=70
N=7
N=68
N=127
Contr
T2D
SAID
SIDD
SIRD
MOD
MARD
Clusters

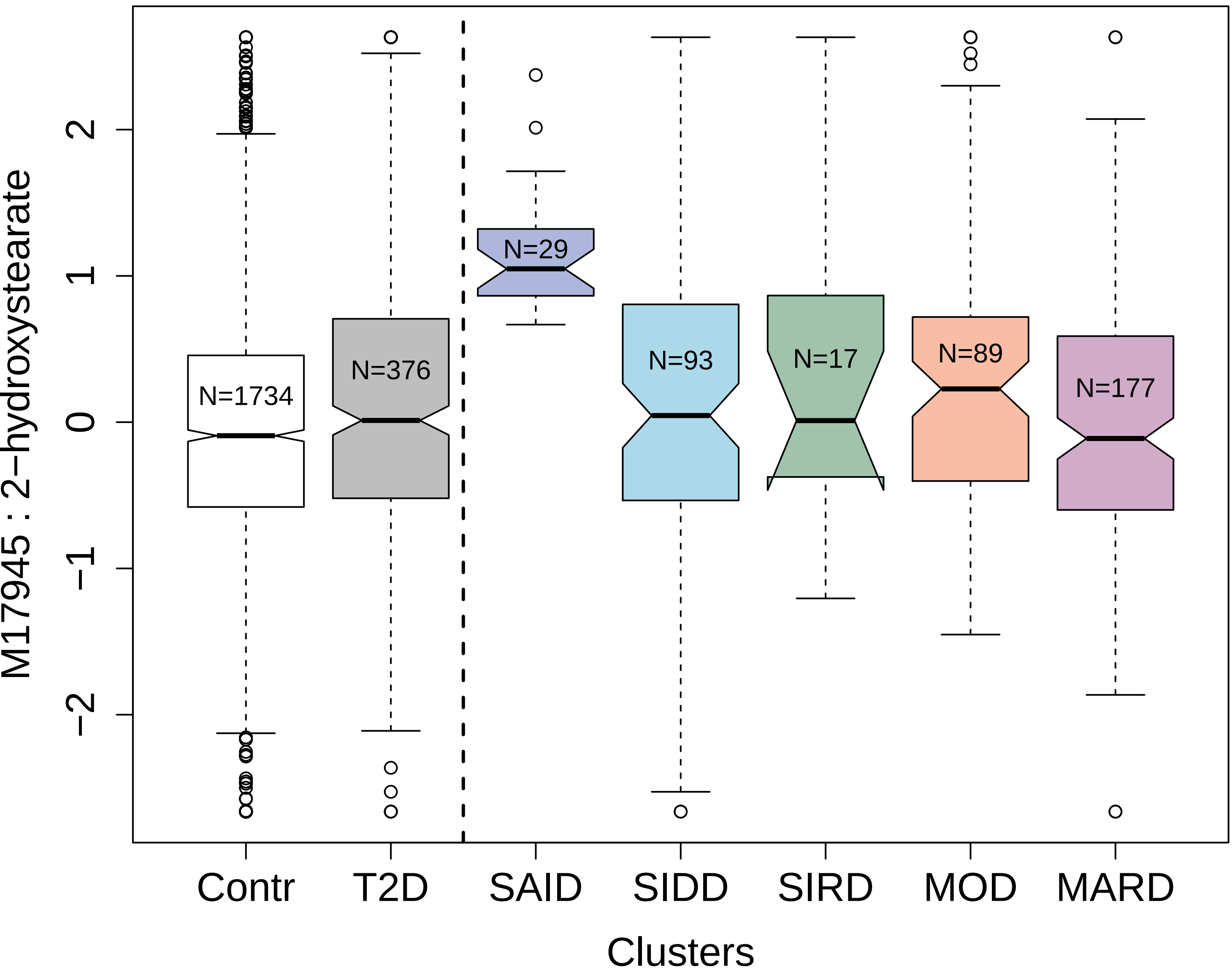
M17945 : 2−hydroxystearate
2
1
0
−1
−2
N=1734
N=376
N=29
N=93
N=17
N=89
N=177
Contr
T2D
SAID
SIDD
SIRD
MOD
MARD
Clusters

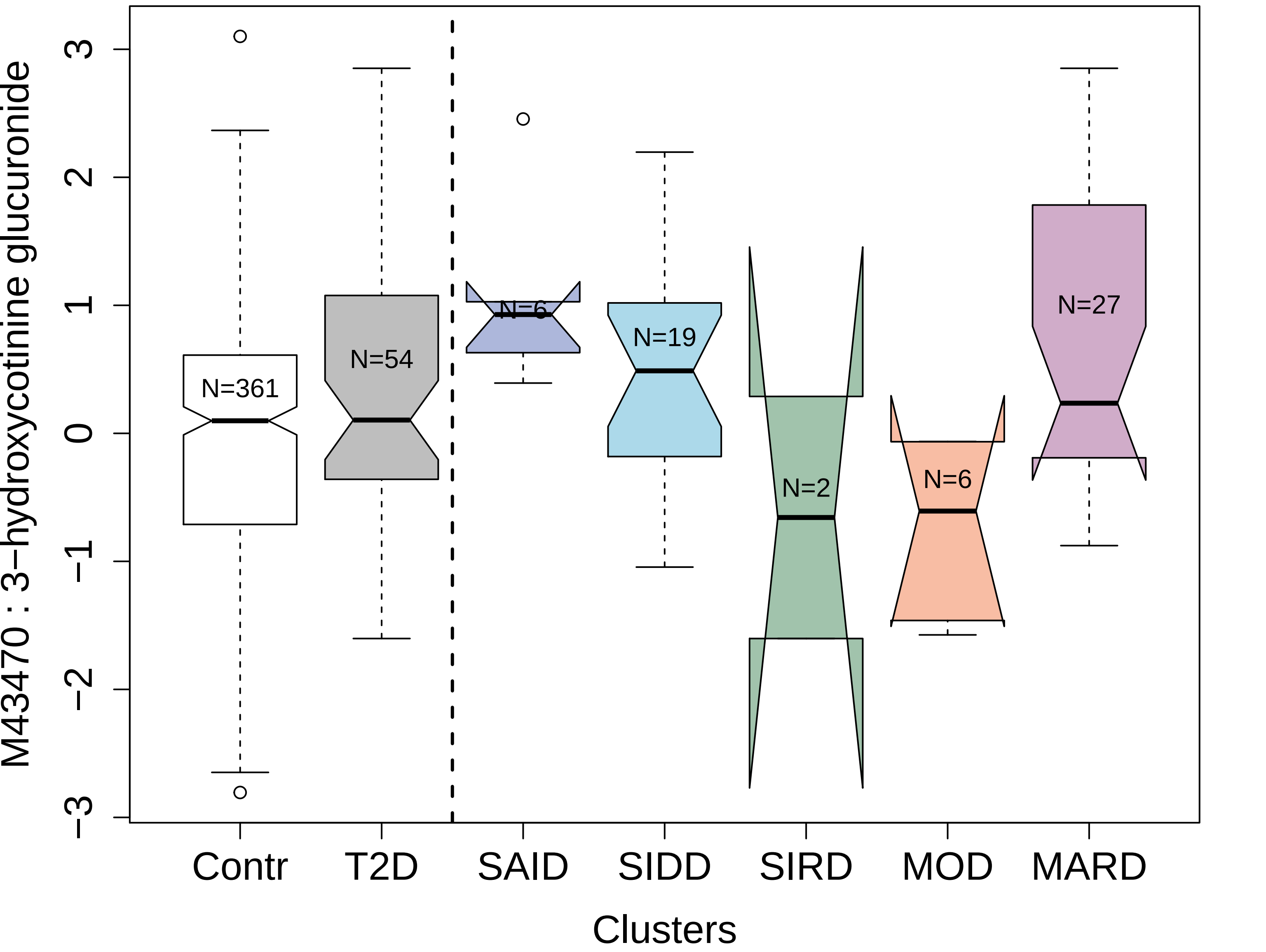

M43470 : 3−hydroxycotinine glucuronide
3
2
1
0
−1
−2
−3
N=361
N=54
N=6
N=19
N=2
N=6
N=27
Contr
T2D
SAID
SIDD
SIRD
MOD
MARD
Clusters

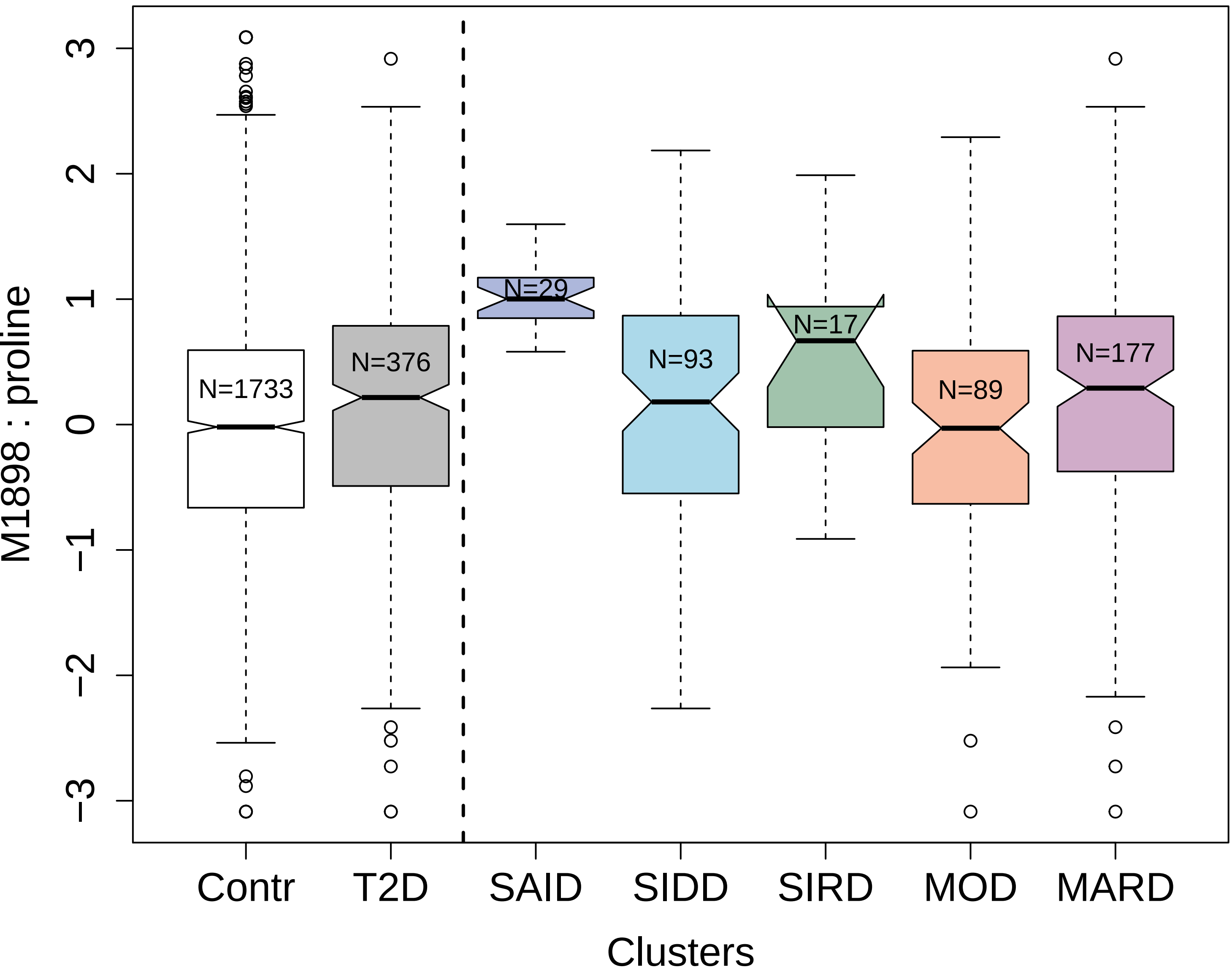

M1898 : proline
3
2
1
0
−1
−2
−3
N=1733
N=376
N=29
N=93
N=17
N=89
N=177
Contr
T2D
SAID
SIDD
SIRD
MOD
MARD
Clusters

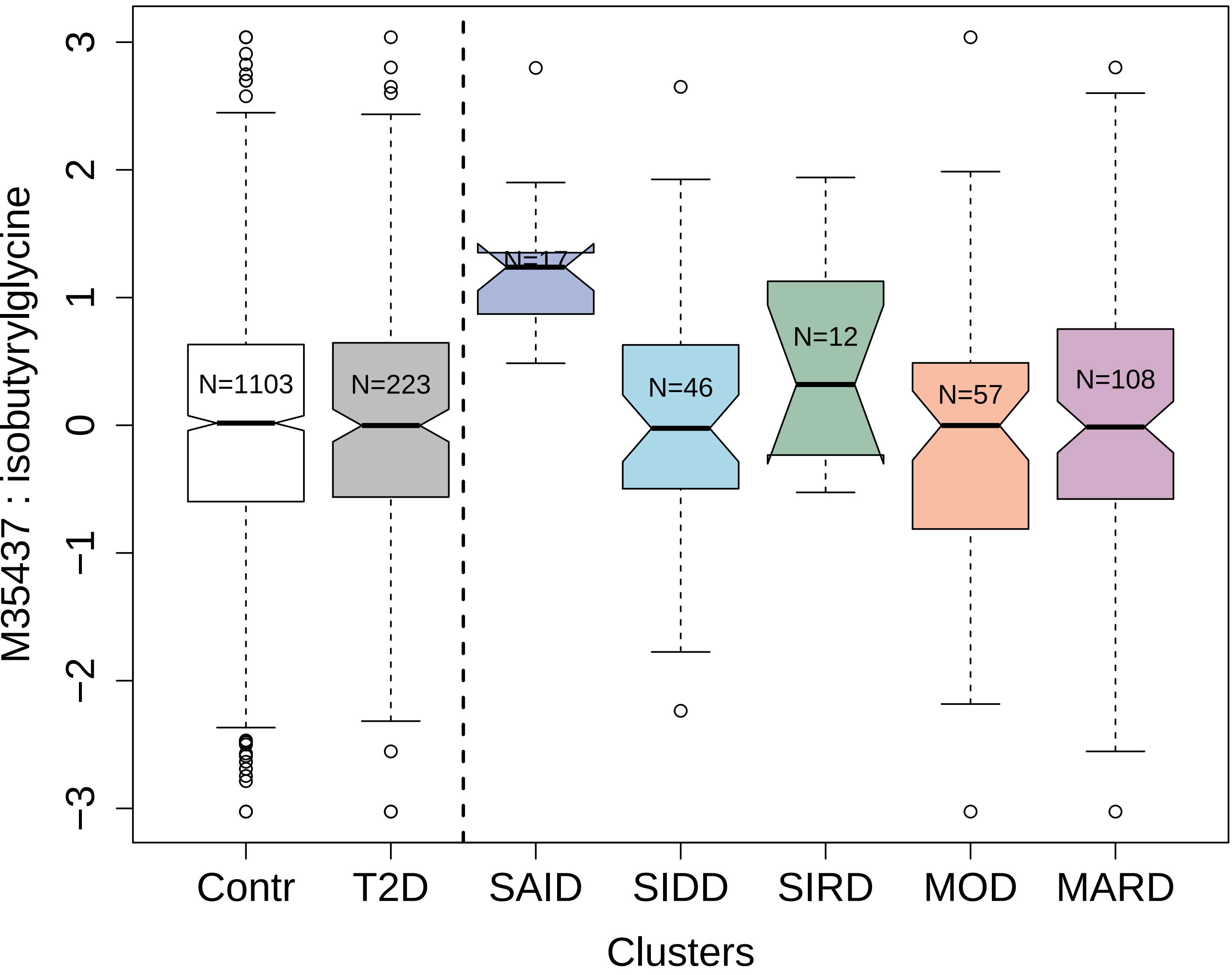
M35437 : isobutyrylglycine
3
2
1
0
−1
−2
−3
N=1103
N=223
N=17
N=46
N=12
N=57
N=108
Contr
T2D
SAID
SIDD
SIRD
MOD
MARD
Clusters

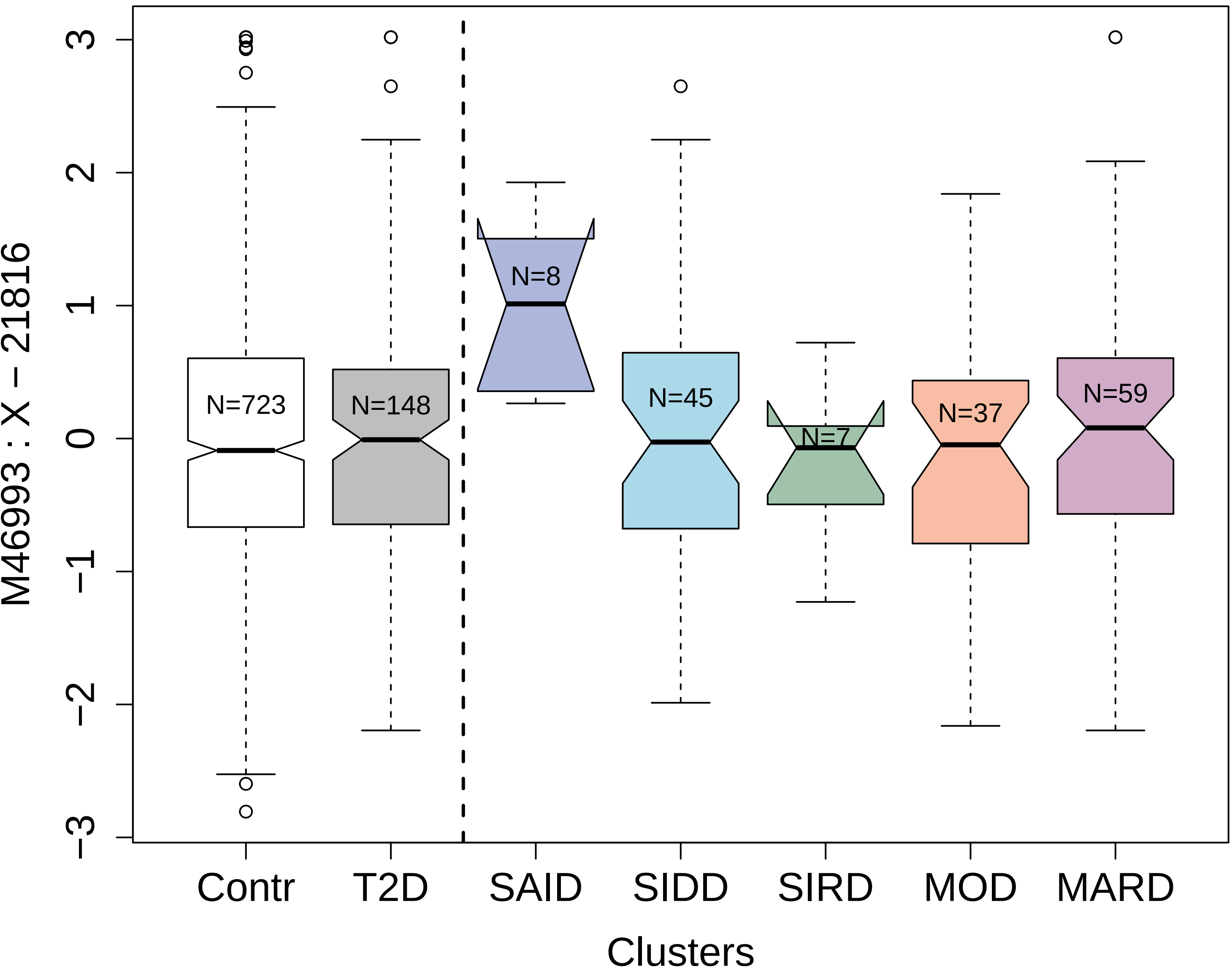

M46993 : X − 21816
Clusters
Contr
T2D
SAID
SIDD
SIRD
MOD
MARD
N=723
N=148
N=8
N=45
N=7
N=37
N=59
3
2
1
0
−1
−2
−3

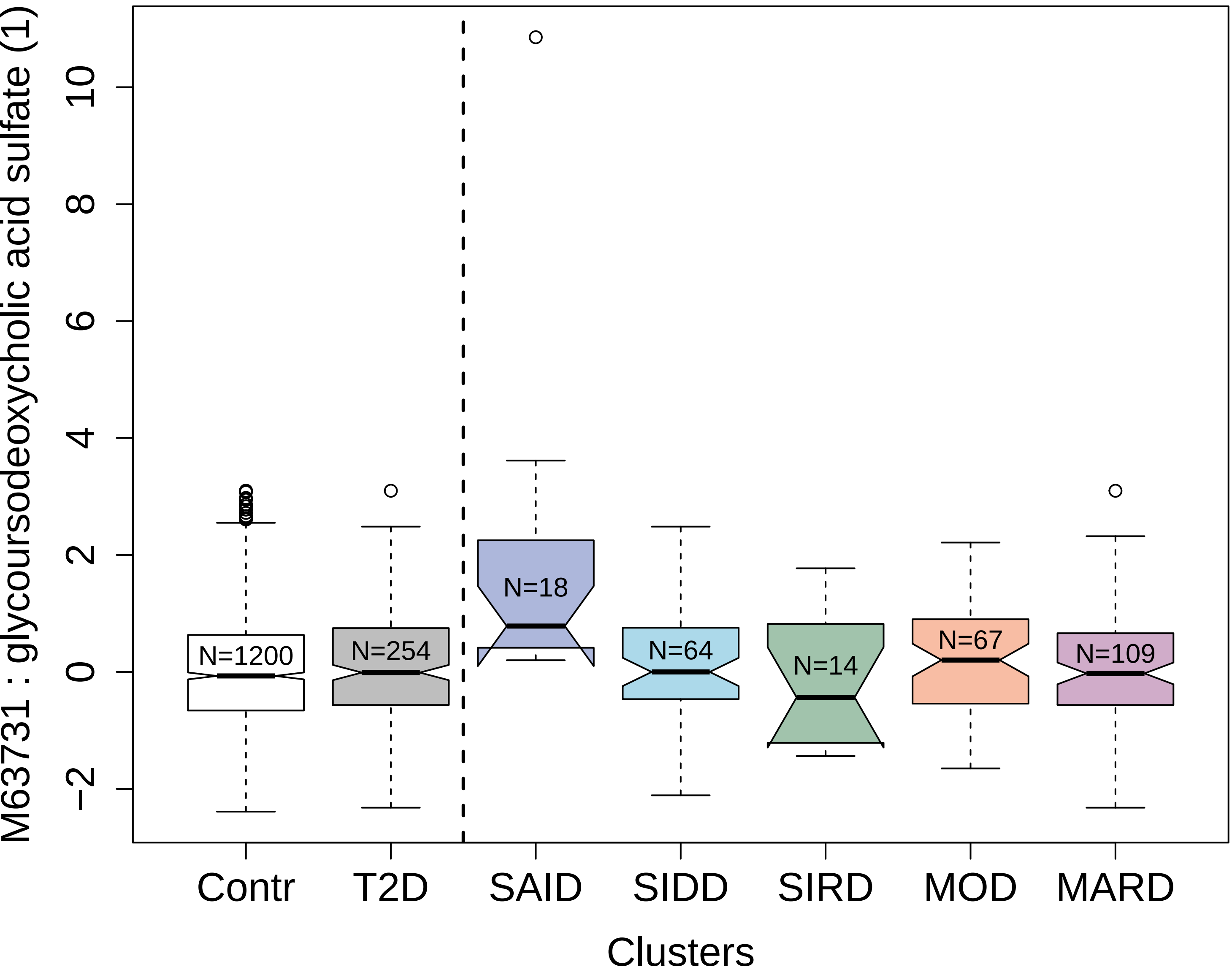

M63731 : glycoursodeoxycholic acid sulfate (1)
−2
0
2
4
6
8
10
N=1200
N=254
N=18
N=64
N=14
N=67
N=109
Contr
T2D
SAID
SIDD
SIRD
MOD
MARD
Clusters

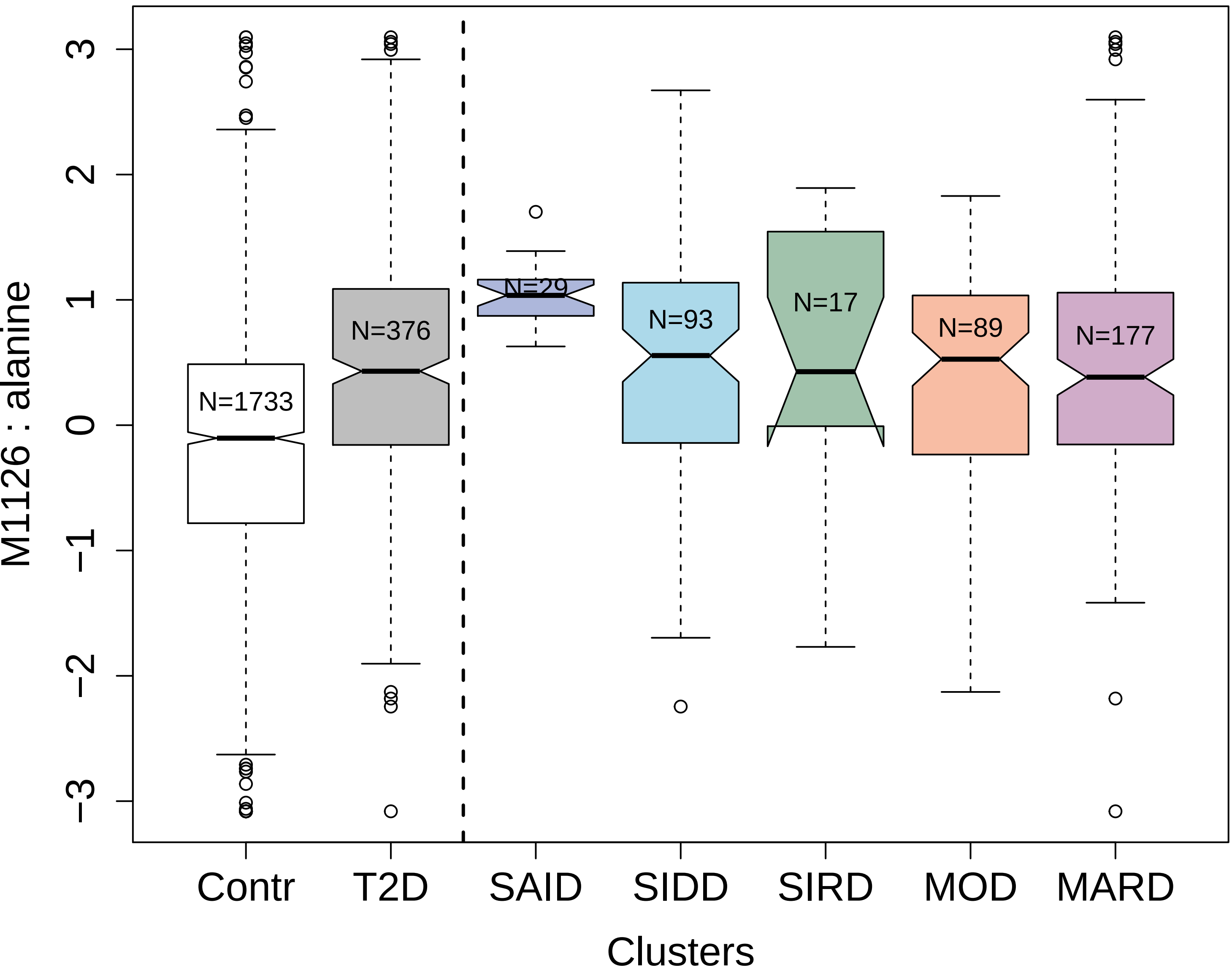

M1126 : alanine
3
2
1
0
−1
−2
−3
N=1733
N=376
N=29
N=93
N=17
N=89
N=177
Contr
T2D
SAID
SIDD
SIRD
MOD
MARD
Clusters

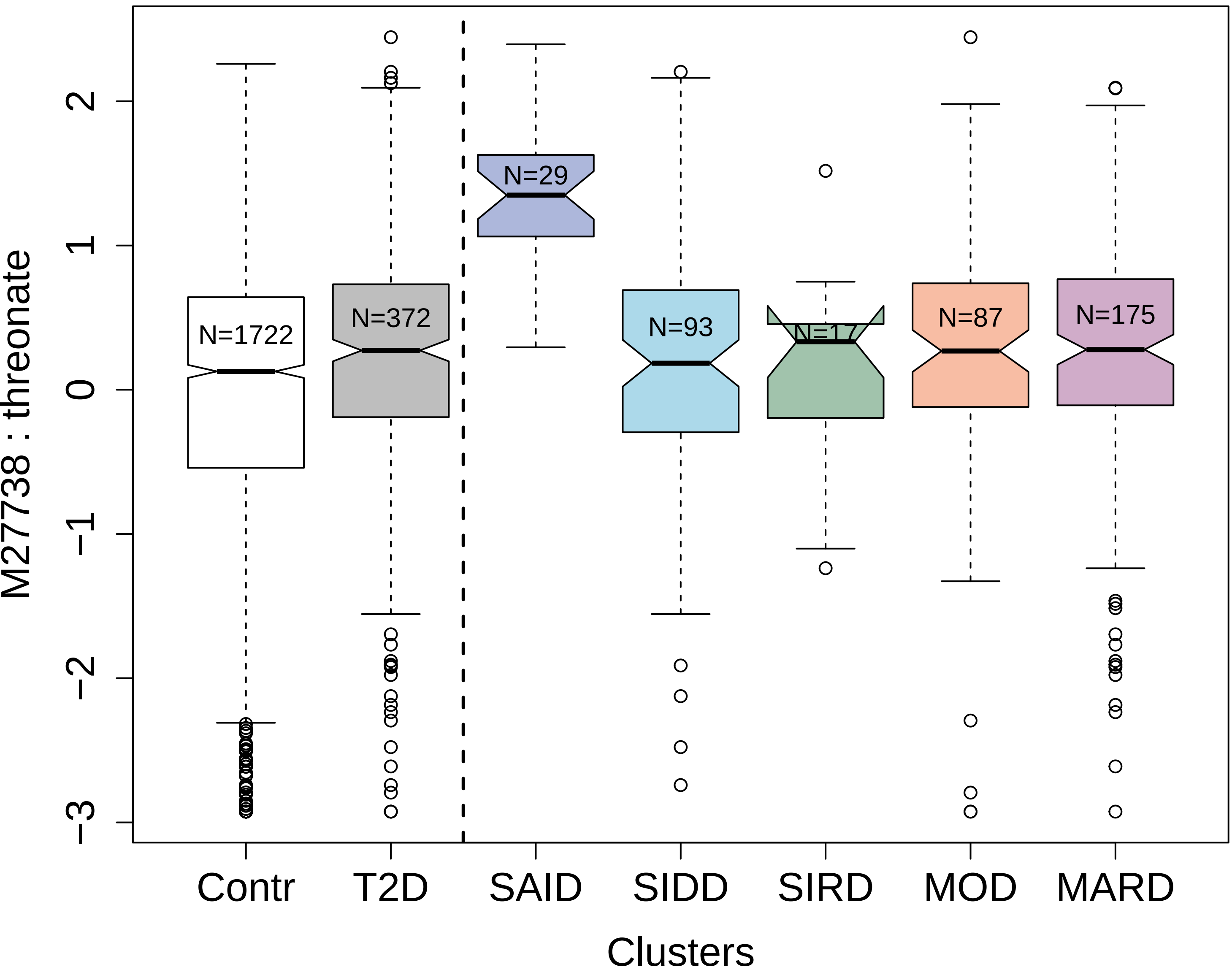
M27738 : threonate
Clusters
Contr
T2D
SAID
SIDD
SIRD
MOD
MARD
N=1722
N=372
N=29
N=93
N=17
N=87
N=175
2
1
0
−1
−2
−3

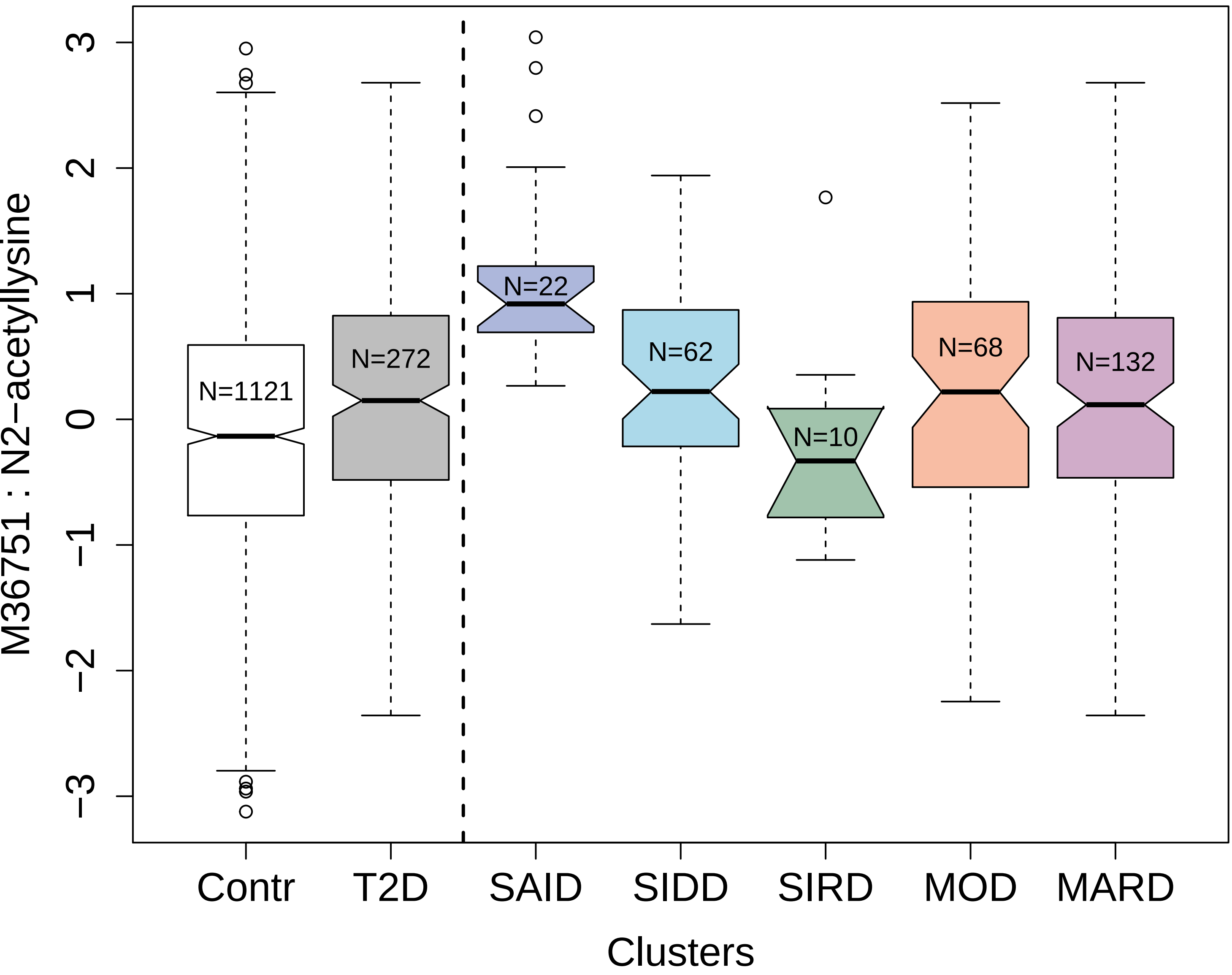

M36751 : N2−acetyllysine
3
2
1
0
−1
−2
−3
N=1121
N=272
N=22
N=62
N=10
N=68
N=132
Contr
T2D
SAID
SIDD
SIRD
MOD
MARD
Clusters

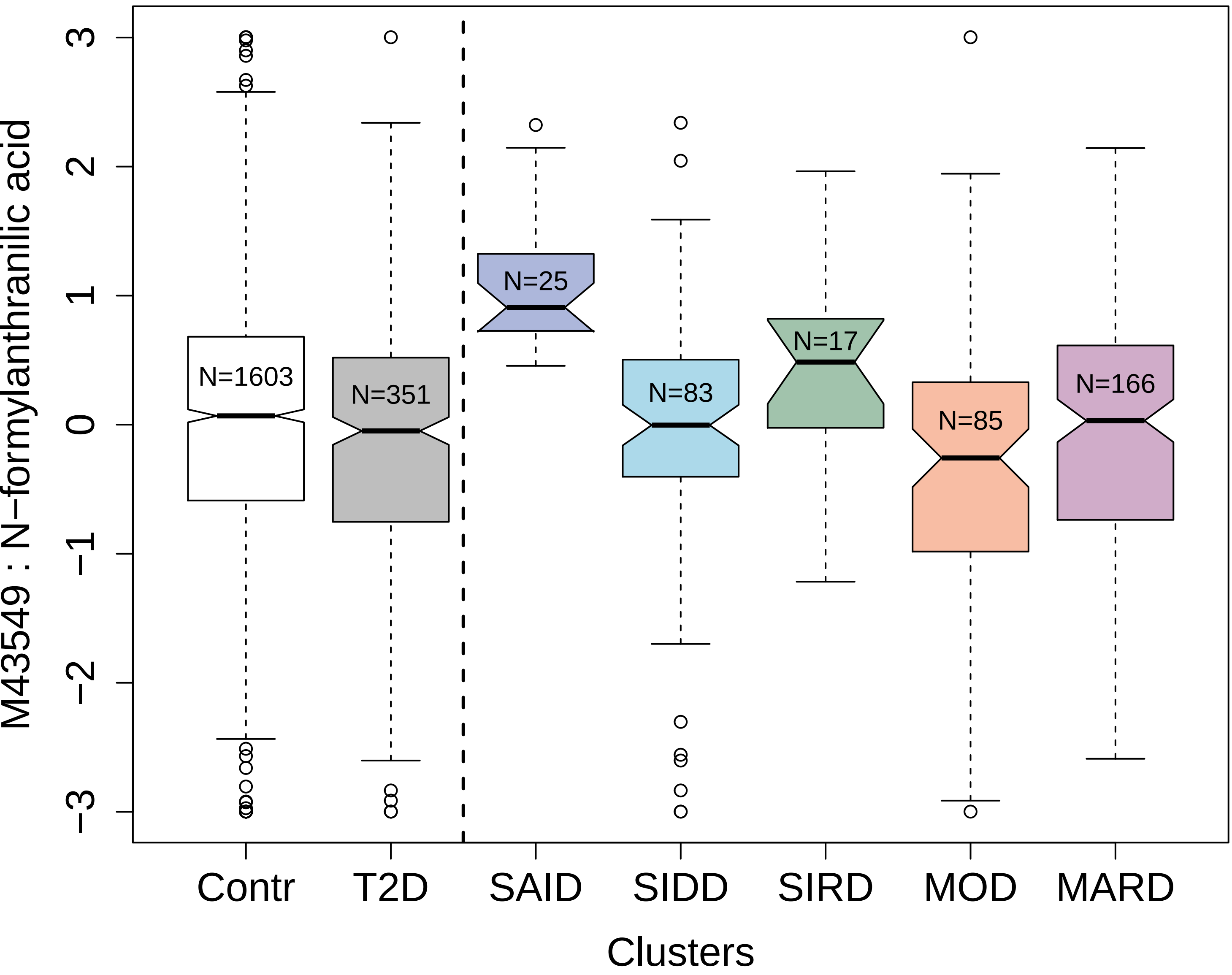

M43549 : N−formylanthranilic acid
3
2
1
0
−1
−2
−3
N=1603
N=351
N=25
N=83
N=17
N=85
N=166
Contr
T2D
SAID
SIDD
SIRD
MOD
MARD
Clusters

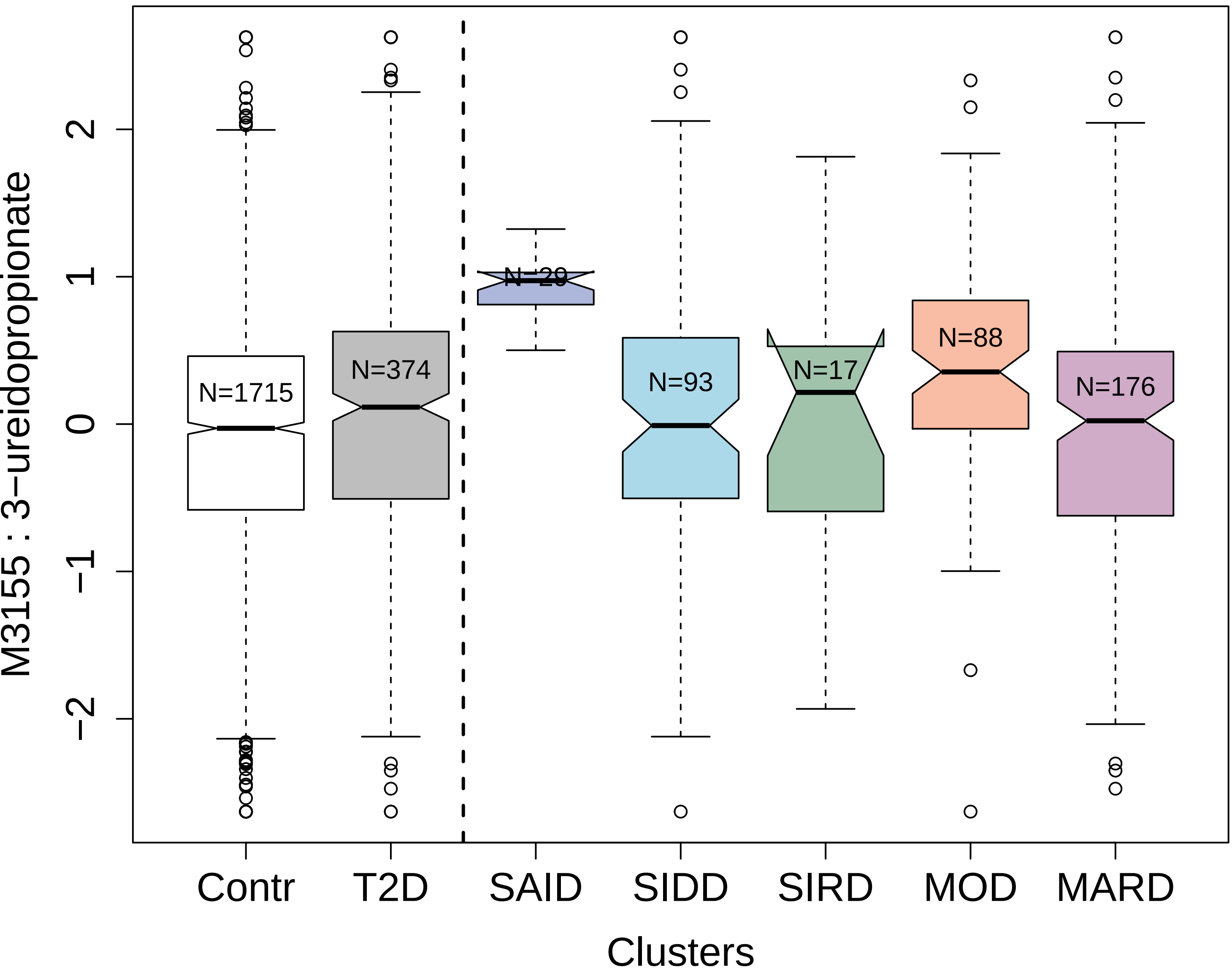
M3155 : 3−ureidopropionate
2
1
0
−1
−2
N=1715
N=374
N=29
N=93
N=17
N=88
N=176
Contr
T2D
SAID
SIDD
SIRD
MOD
MARD
Clusters

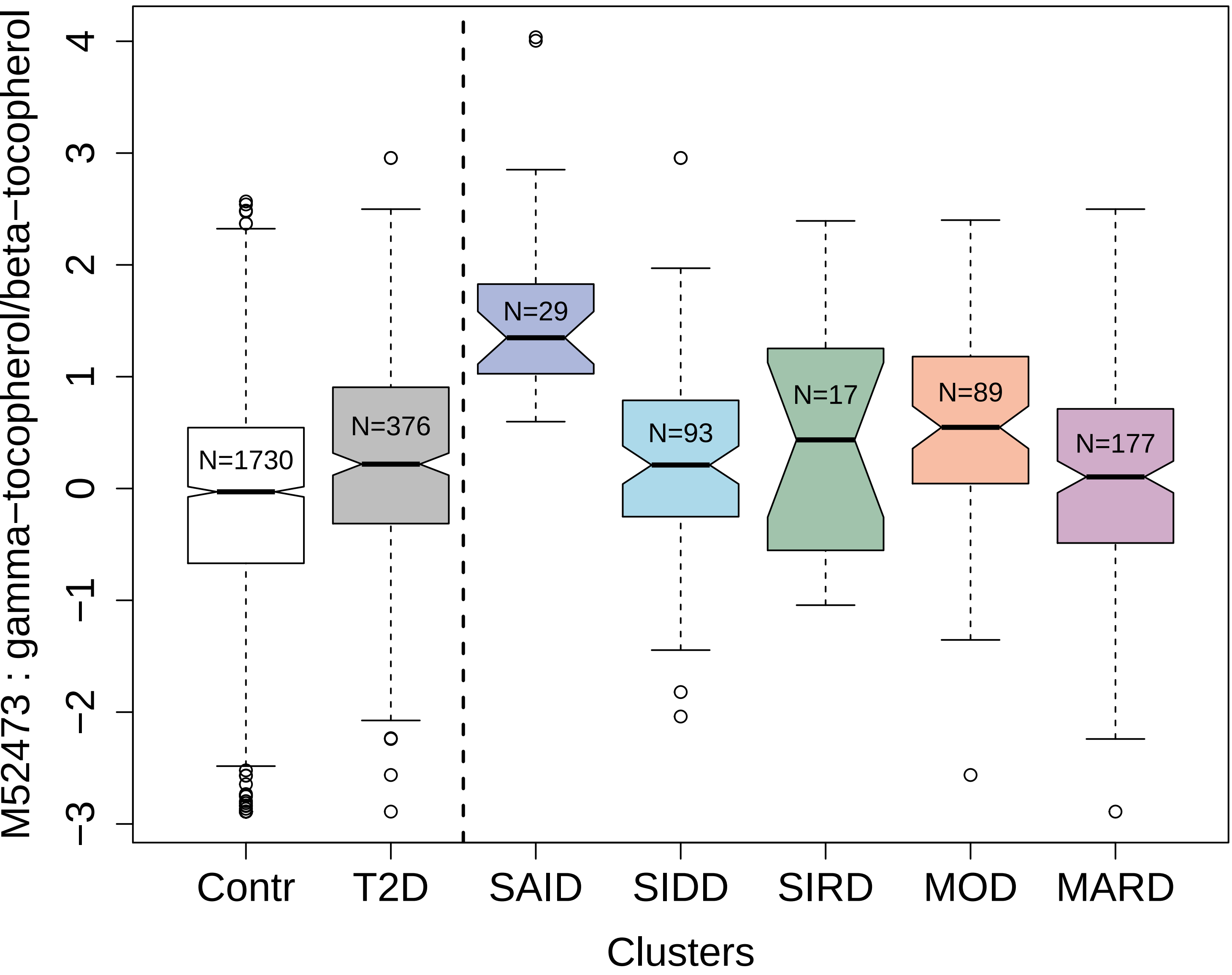

M52473 : gamma−tocopherol/beta−tocopherol
4
3
2
1
0
−1
−2
−3
N=1730
N=376
N=29
N=93
N=17
N=89
N=177
Contr
T2D
SAID
SIDD
SIRD
MOD
MARD
Clusters

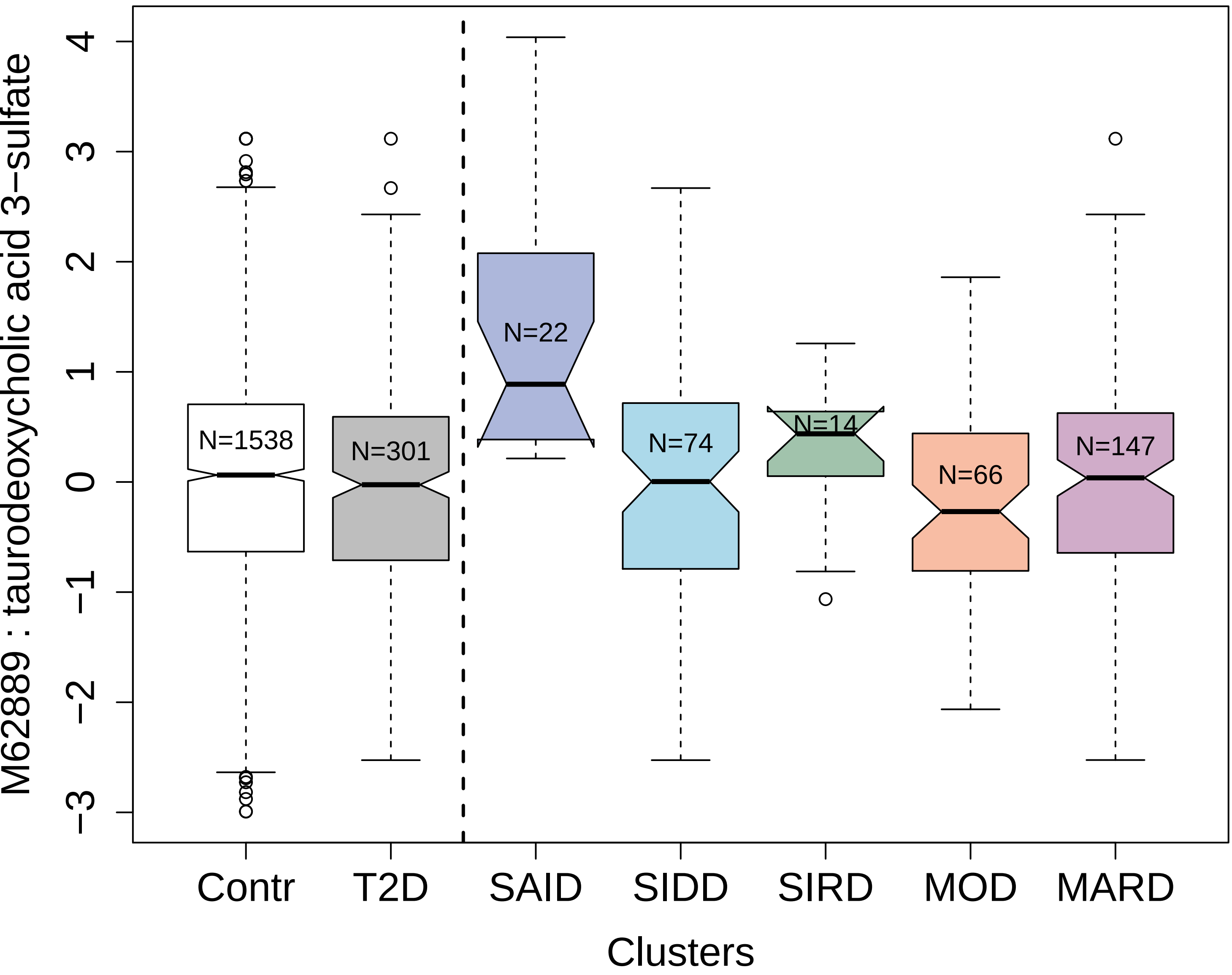

M62889 : taurodeoxycholic acid 3−sulfate
4
3
2
1
0
−1
−2
−3
N=1538
N=301
N=22
N=74
N=14
N=66
N=147
Contr
T2D
SAID
SIDD
SIRD
MOD
MARD
Clusters

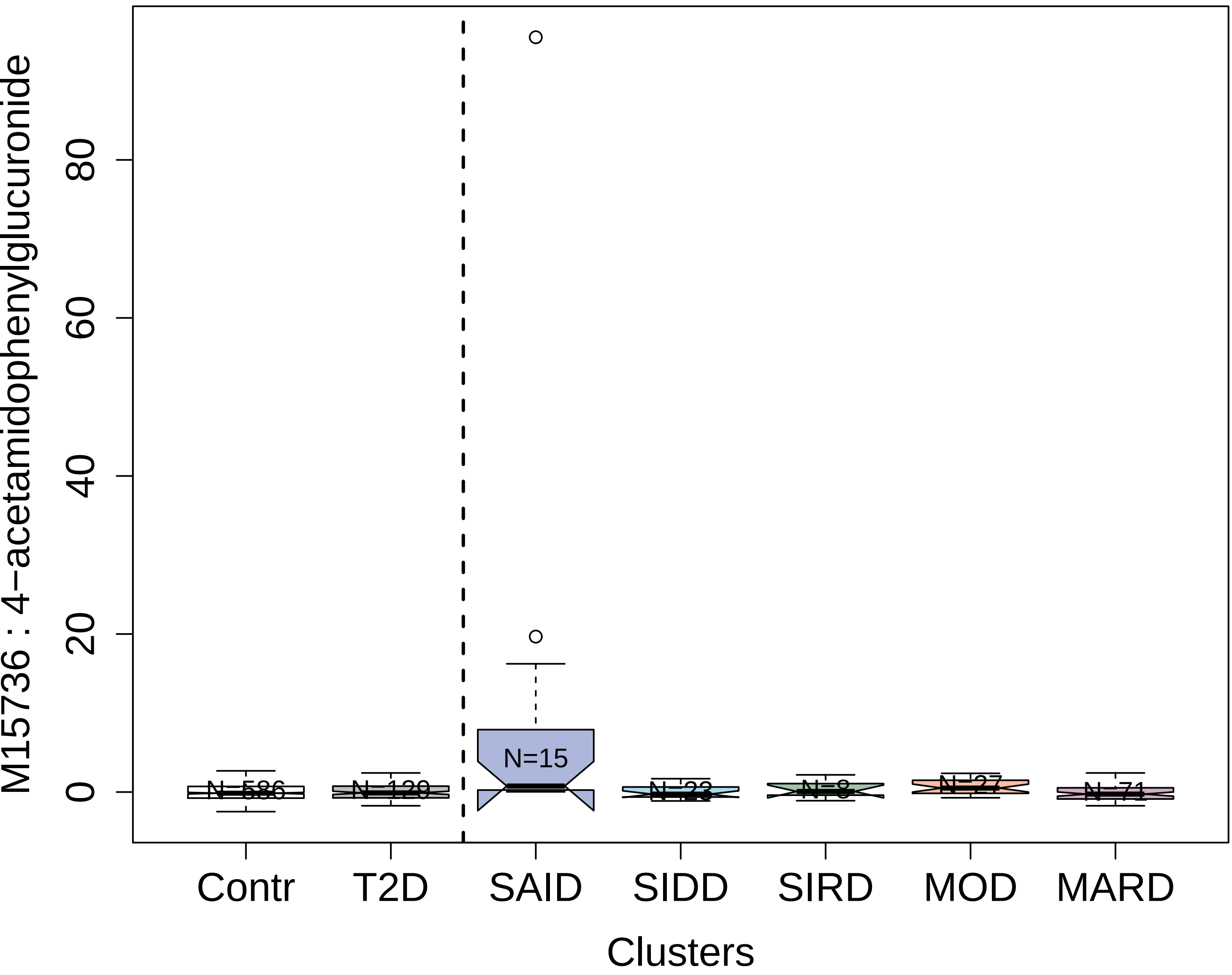

M15736 : 4−acetamidophenylglucuronide
0
20
40
60
80
N=586
N=120
N=15
N=23
N=8
N=27
N=71
Contr
T2D
SAID
SIDD
SIRD
MOD
MARD
Clusters

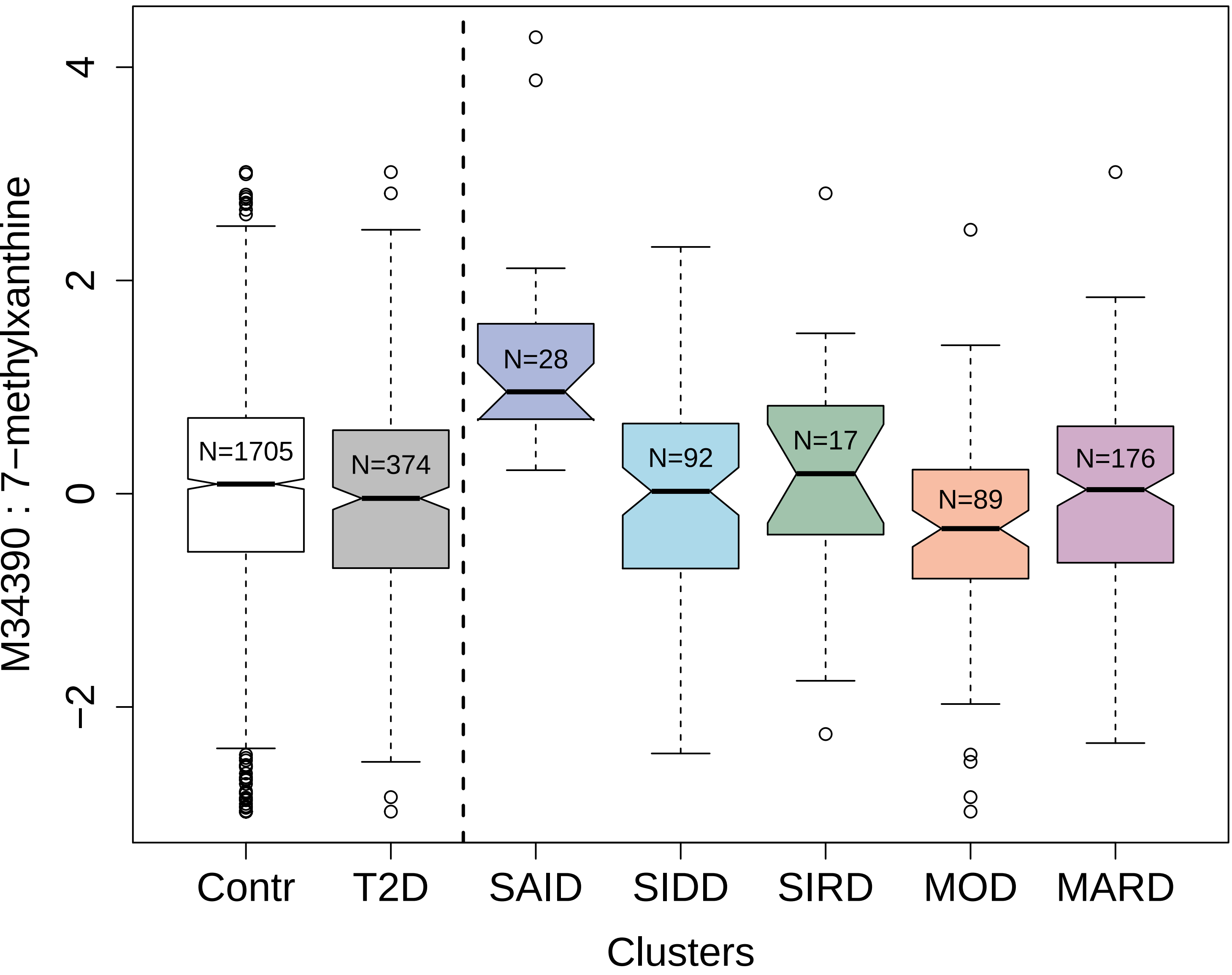

M34390 : 7−methylxanthine
4
2
0
−2
N=1705
N=374
N=28
N=92
N=17
N=89
N=176
Contr
T2D
SAID
SIDD
SIRD
MOD
MARD
Clusters

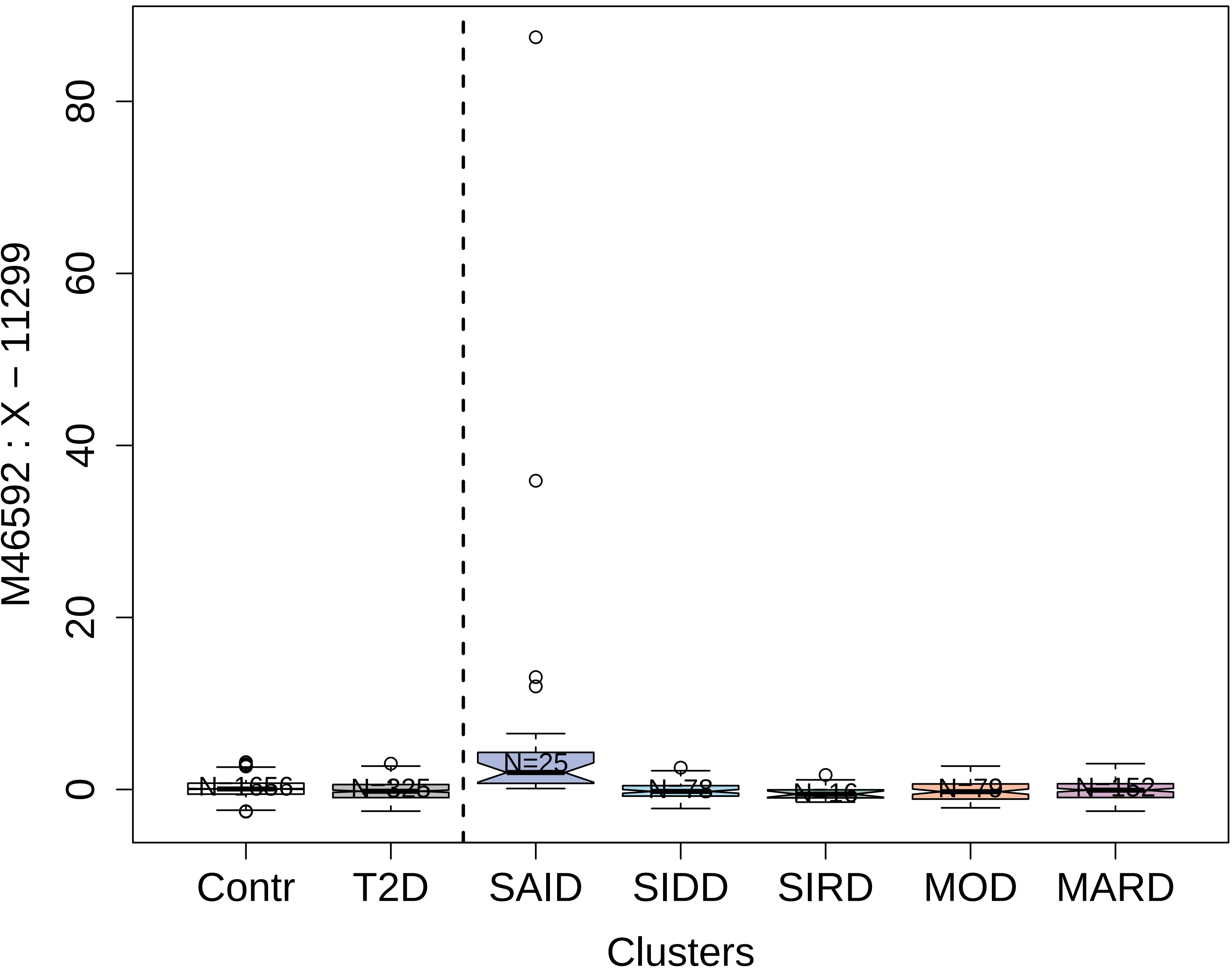

M46592 : X − 11299
0
20
40
60
80
N=1656
N=325
N=25
N=78
N=16
N=79
N=152
Contr
T2D
SAID
SIDD
SIRD
MOD
MARD
Clusters

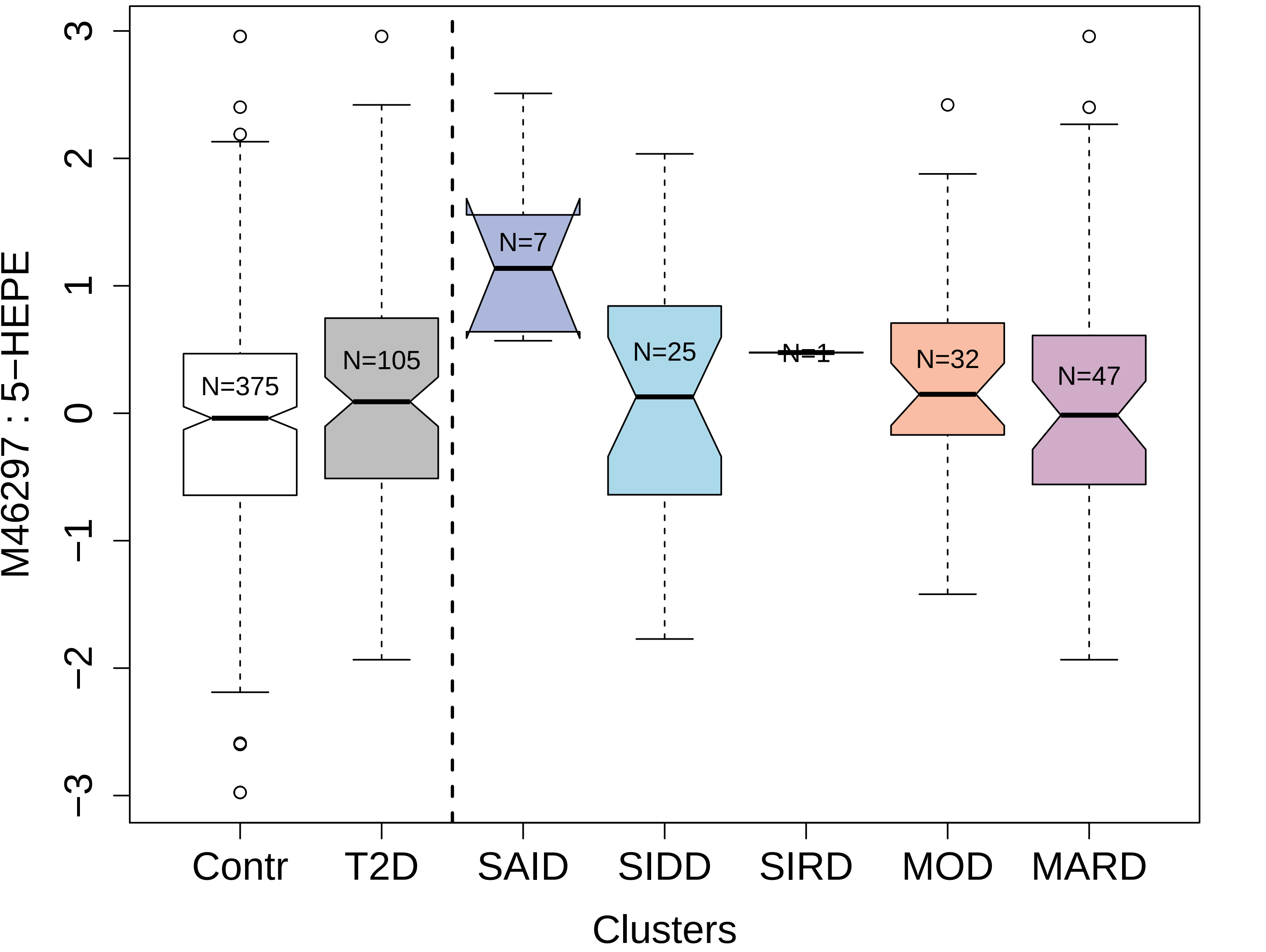

M46297 : 5−HEPE
3
2
1
0
−1
−2
−3
N=375
N=105
N=7
N=25
N=1
N=32
N=47
Contr
T2D
SAID
SIDD
SIRD
MOD
MARD
Clusters

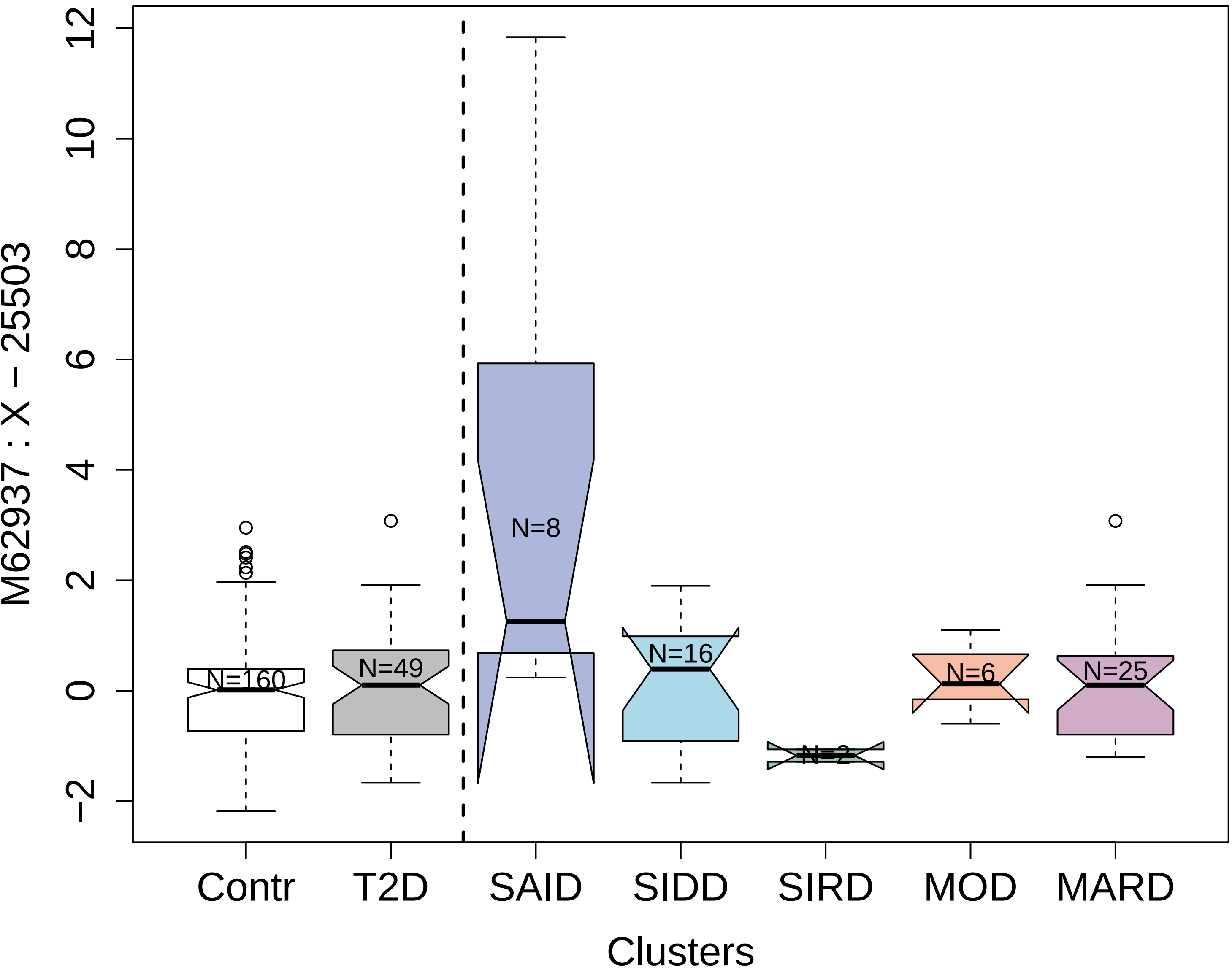

M62937 : X − 25503
12
10
8
6
4
2
0
−2
N=160
N=49
N=8
N=16
N=2
N=6
N=25
Contr
T2D
SAID
SIDD
SIRD
MOD
MARD
Clusters

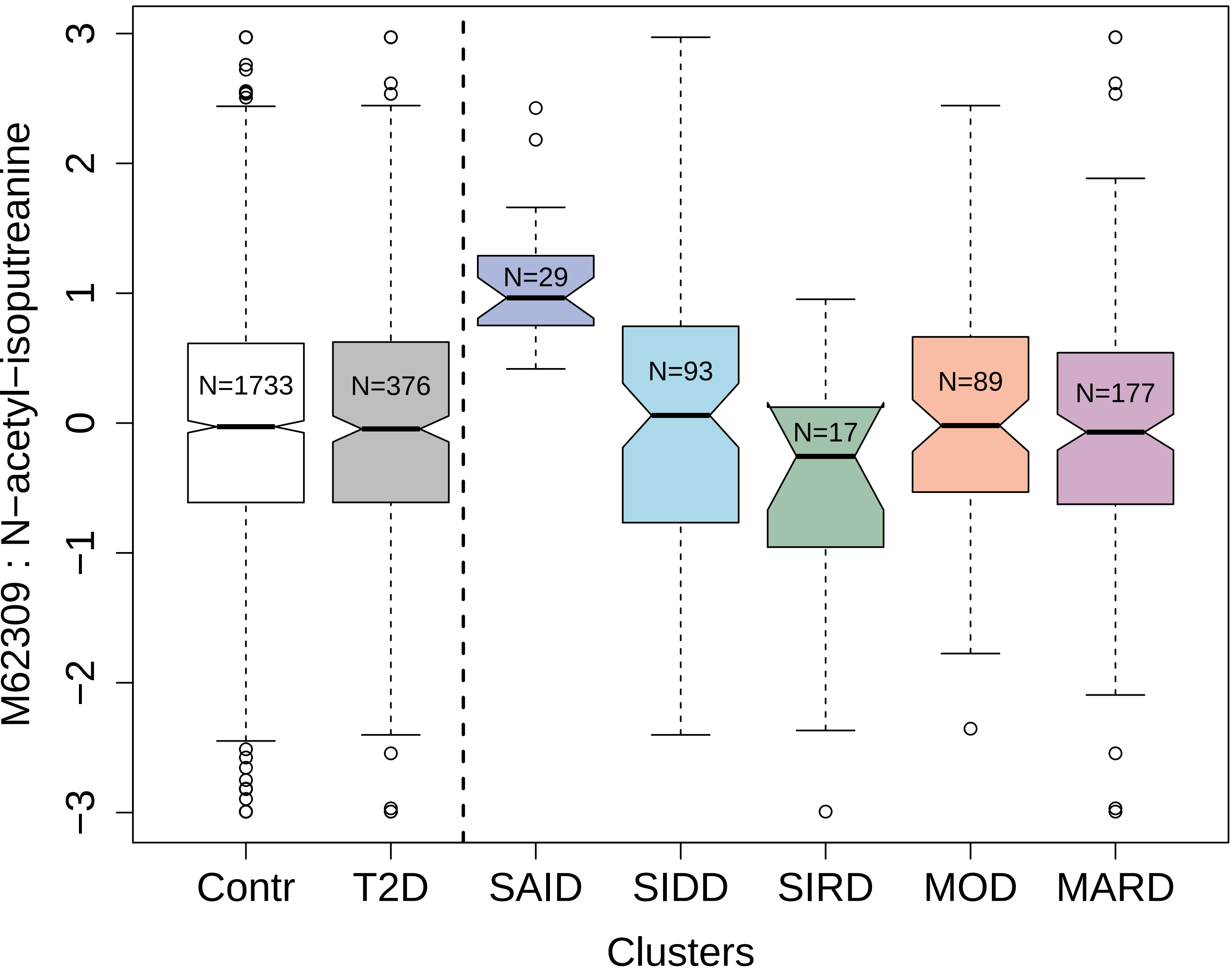
M62309 : N−acetyl−isoputreanine
3
2
1
0
−1
−2
−3
N=1733
N=376
N=29
N=93
N=17
N=89
N=177
Contr
T2D
SAID
SIDD
SIRD
MOD
MARD
Clusters

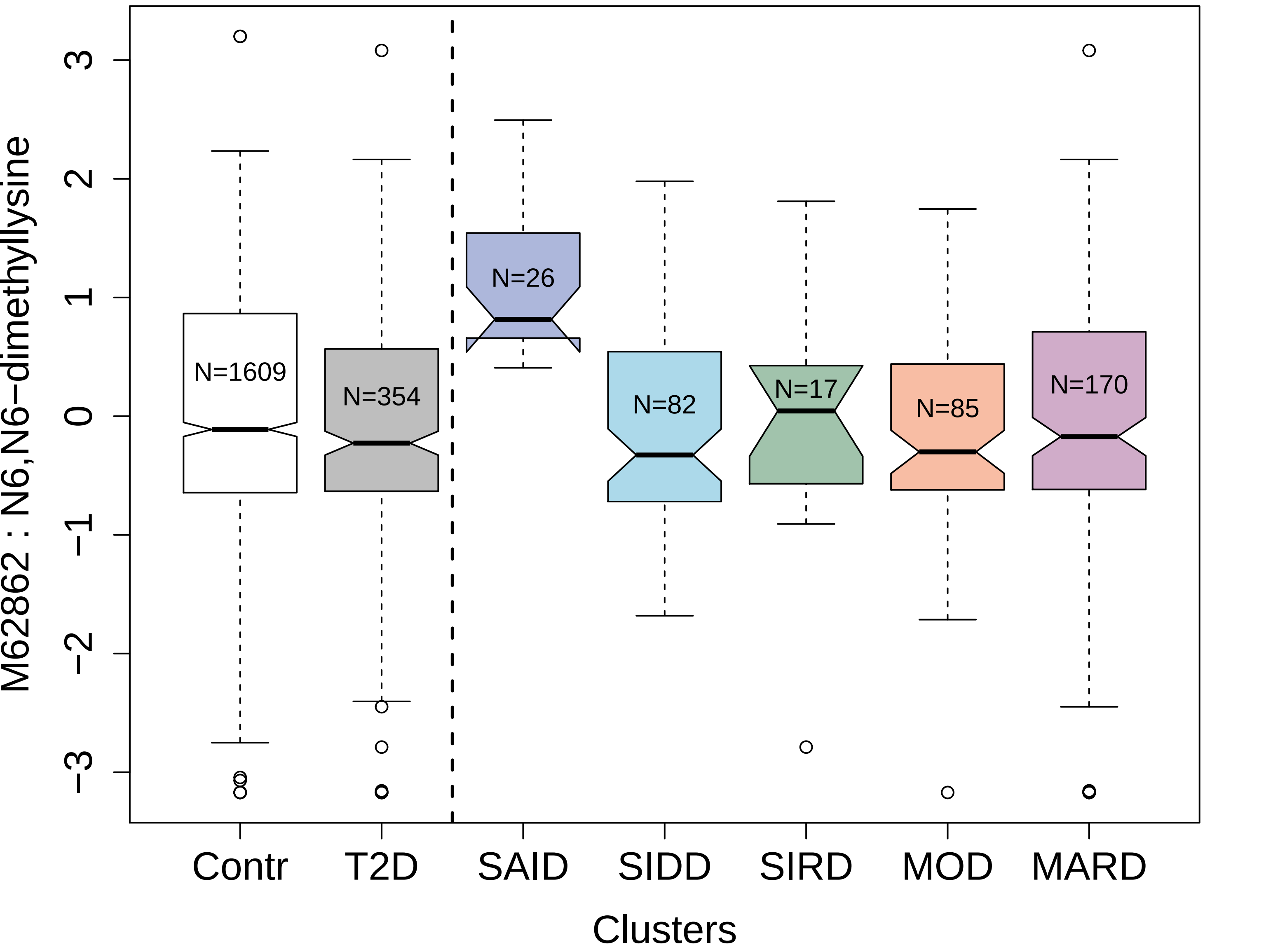

M62862 : N6,N6−dimethyllysine
3
2
1
0
−1
−2
−3
N=1609
N=354
N=26
N=82
N=17
N=85
N=170
Contr
T2D
SAID
SIDD
SIRD
MOD
MARD
Clusters

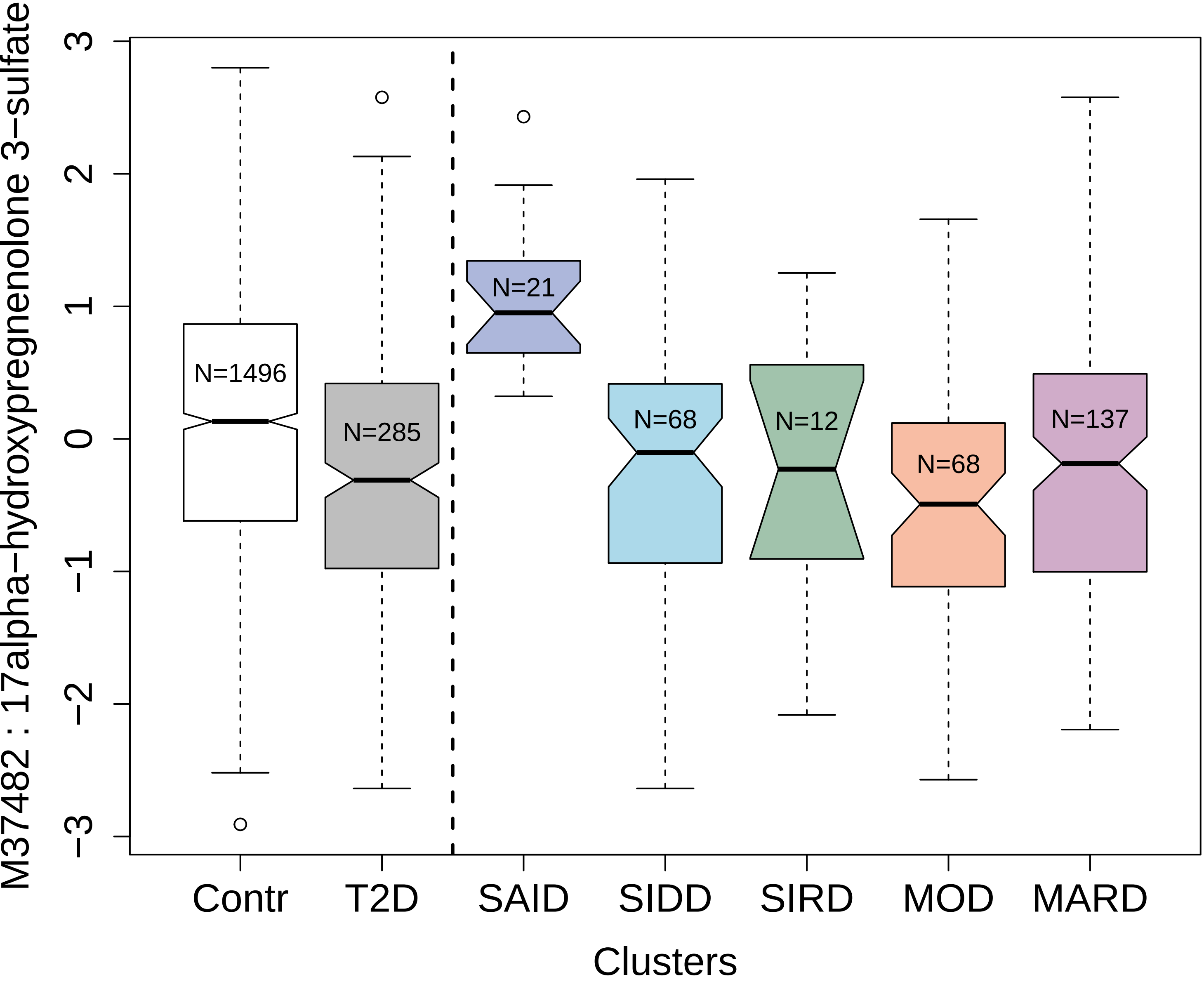
M37482 : 17alpha−hydroxypregnenolone 3−sulfate
3
2
1
0
−1
−2
−3
N=1496
N=285
N=21
N=68
N=12
N=68
N=137
Contr
T2D
SAID
SIDD
SIRD
MOD
MARD
Clusters

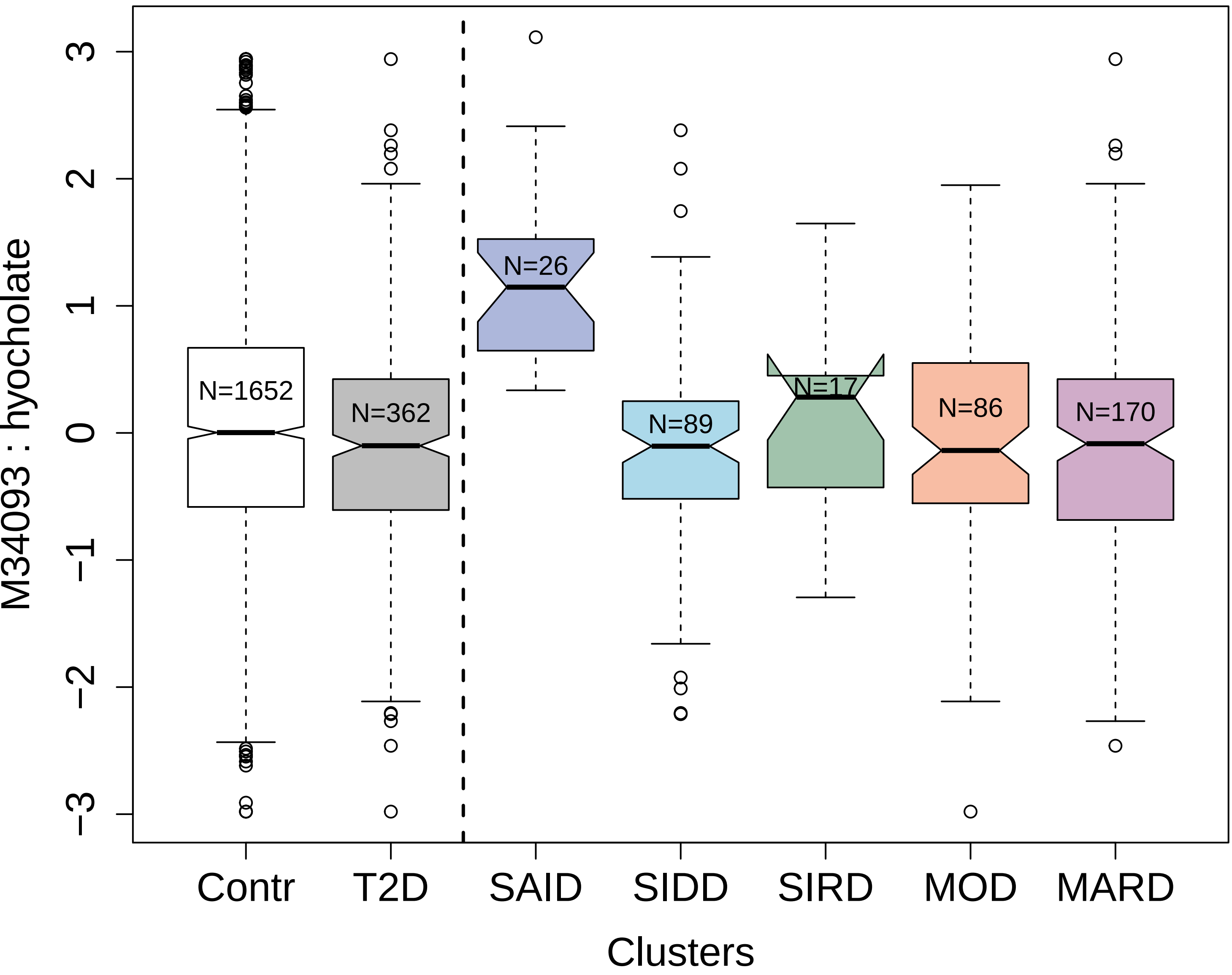

M34093 : hyocholate
3
2
1
0
−1
−2
−3
N=1652
N=362
N=26
N=89
N=17
N=86
N=170
Contr
T2D
SAID
SIDD
SIRD
MOD
MARD
Clusters

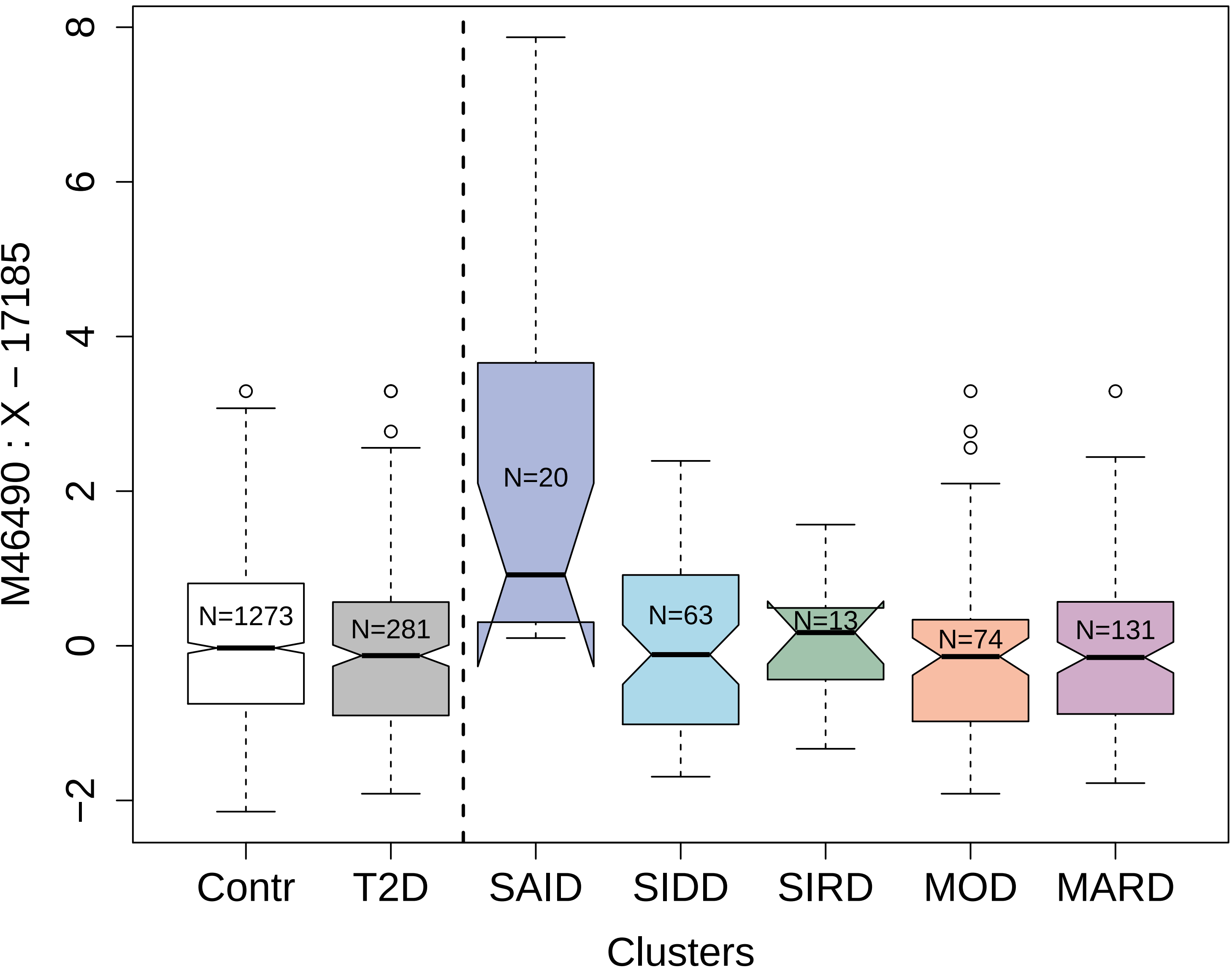

M46490 : X − 17185
8
6
4
2
0
−2
N=1273
N=281
N=20
N=63
N=13
N=74
N=131
Contr
T2D
SAID
SIDD
SIRD
MOD
MARD
Clusters

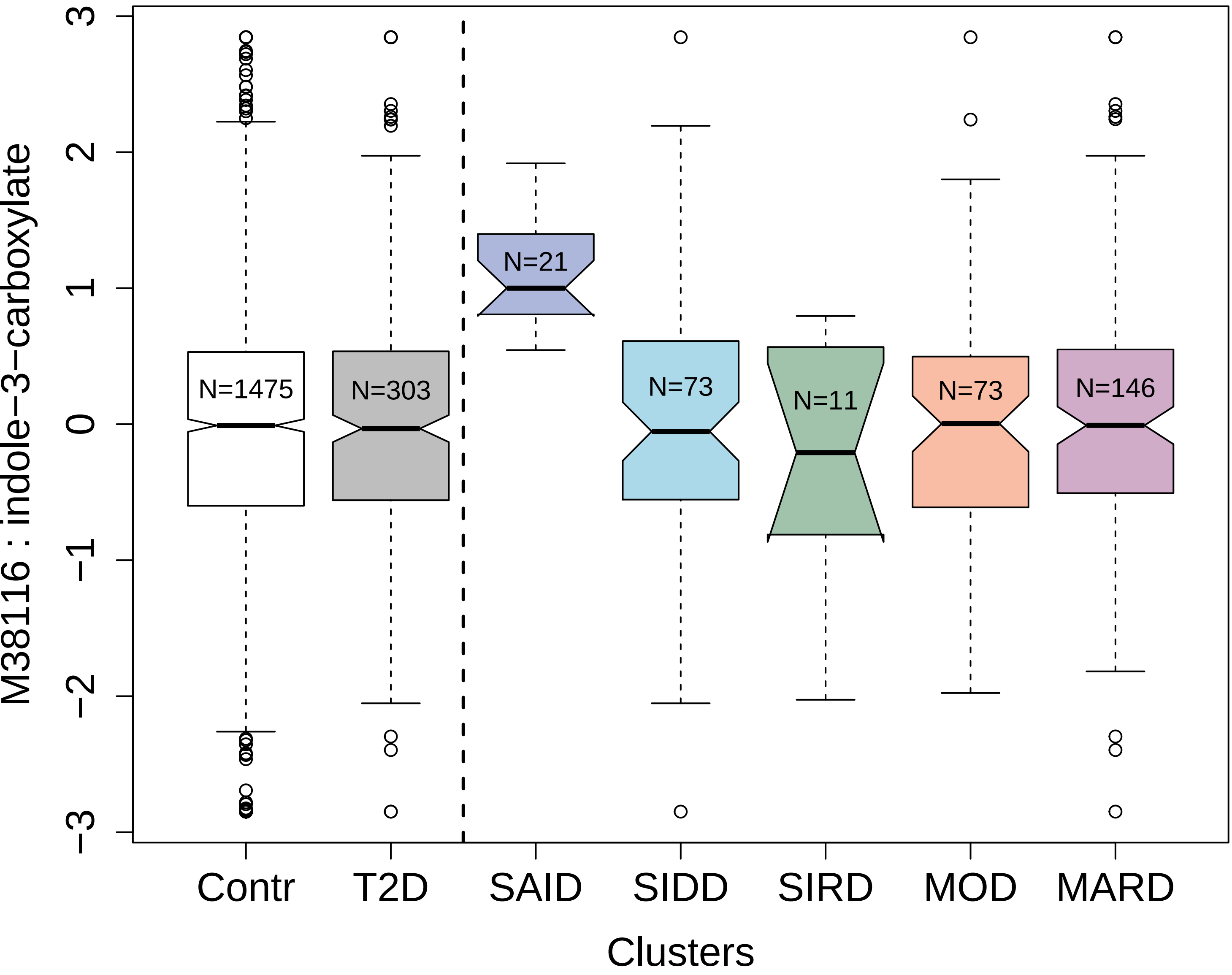
M38116 : indole−3−carboxylate
3
2
1
0
−1
−2
−3
N=1475
N=303
N=21
N=73
N=11
N=73
N=146
Contr
T2D
SAID
SIDD
SIRD
MOD
MARD
Clusters

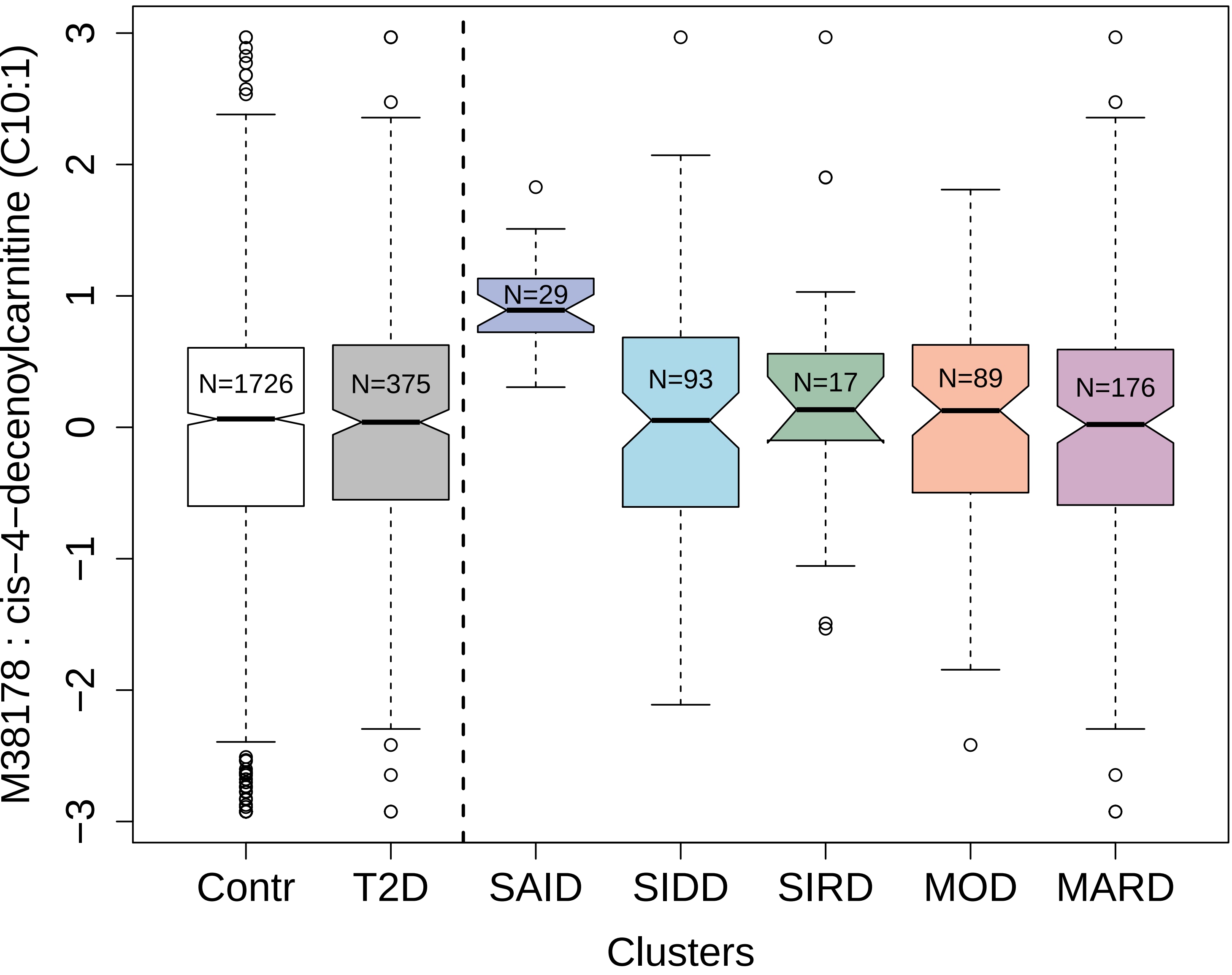
M38178 : cis−4−decenoylcarnitine (C10:1)
3
2
1
0
−1
−2
−3
N=1726
N=375
N=29
N=93
N=17
N=89
N=176
Contr
T2D
SAID
SIDD
SIRD
MOD
MARD
Clusters

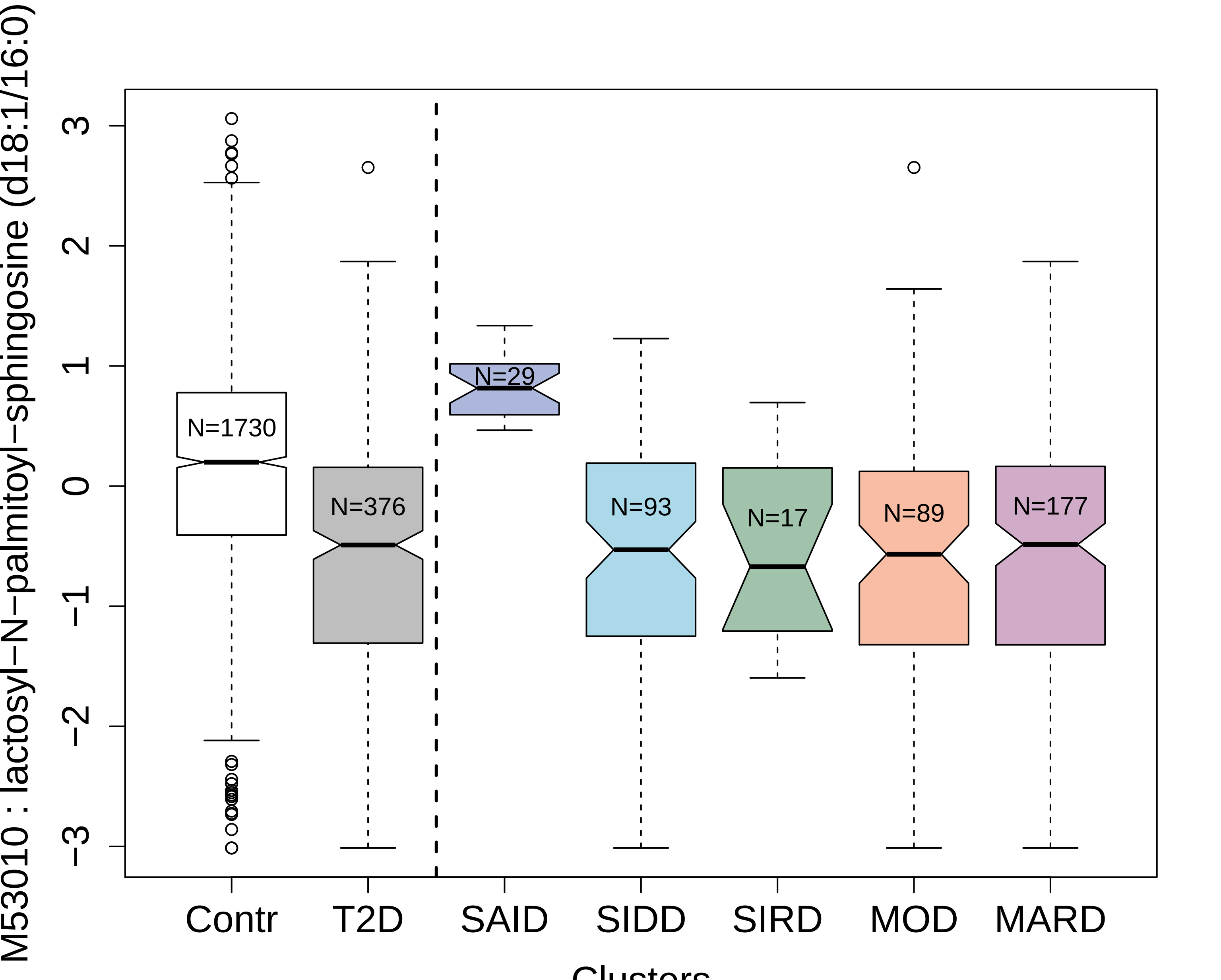
M53010 : lactosyl−N−palmitoyl−sphingosine (d18:1/16:0)
3
2
1
0
−1
−2
−3
N=1730
N=376
N=29
N=93
N=17
N=89
N=177
Contr
T2D
SAID
SIDD
SIRD
MOD
MARD
Clusters

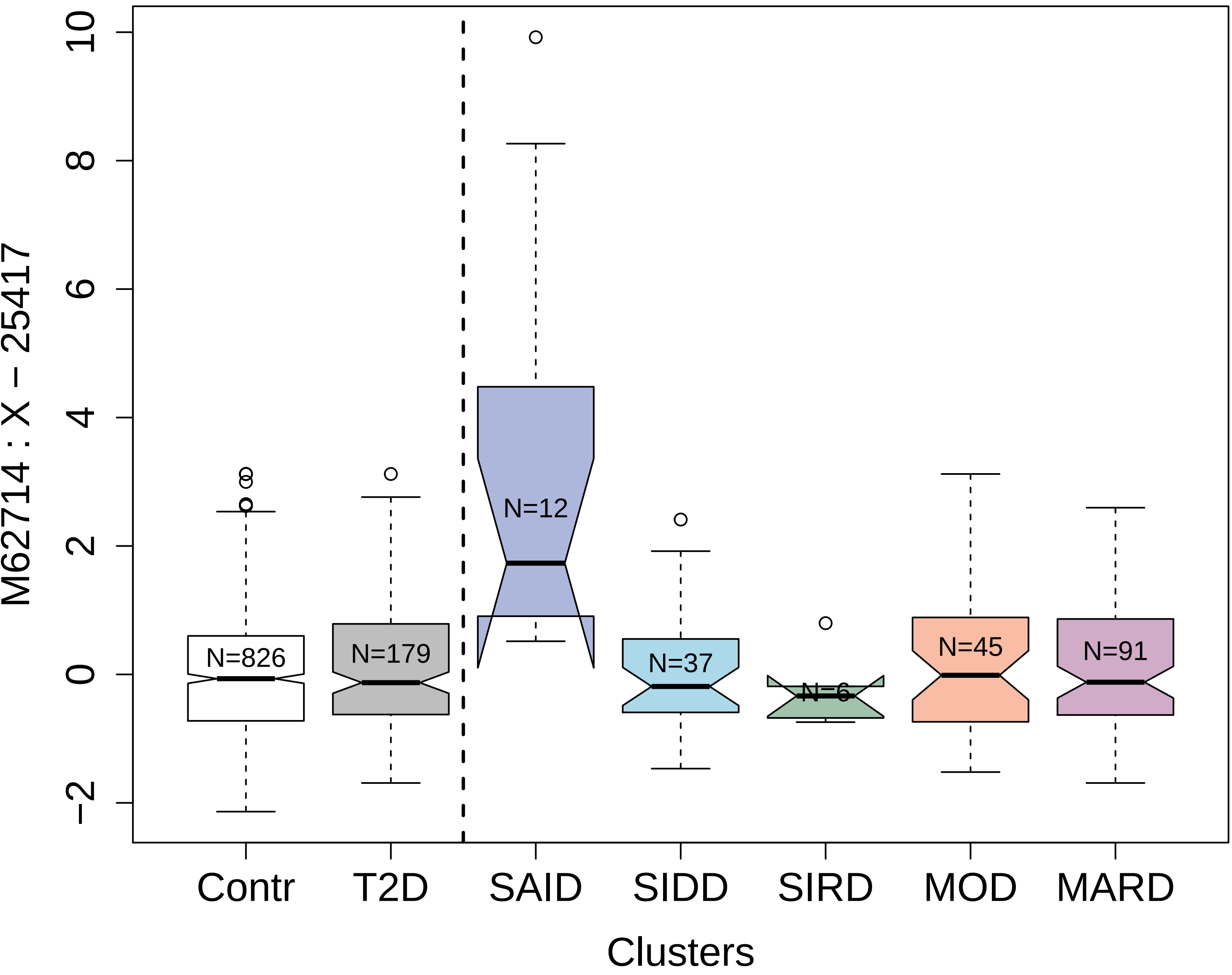

M62714 : X − 25417
10
8
6
4
2
0
−2
N=826
N=179
N=12
N=37
N=6
N=45
N=91
Contr
T2D
SAID
SIDD
SIRD
MOD
MARD
Clusters

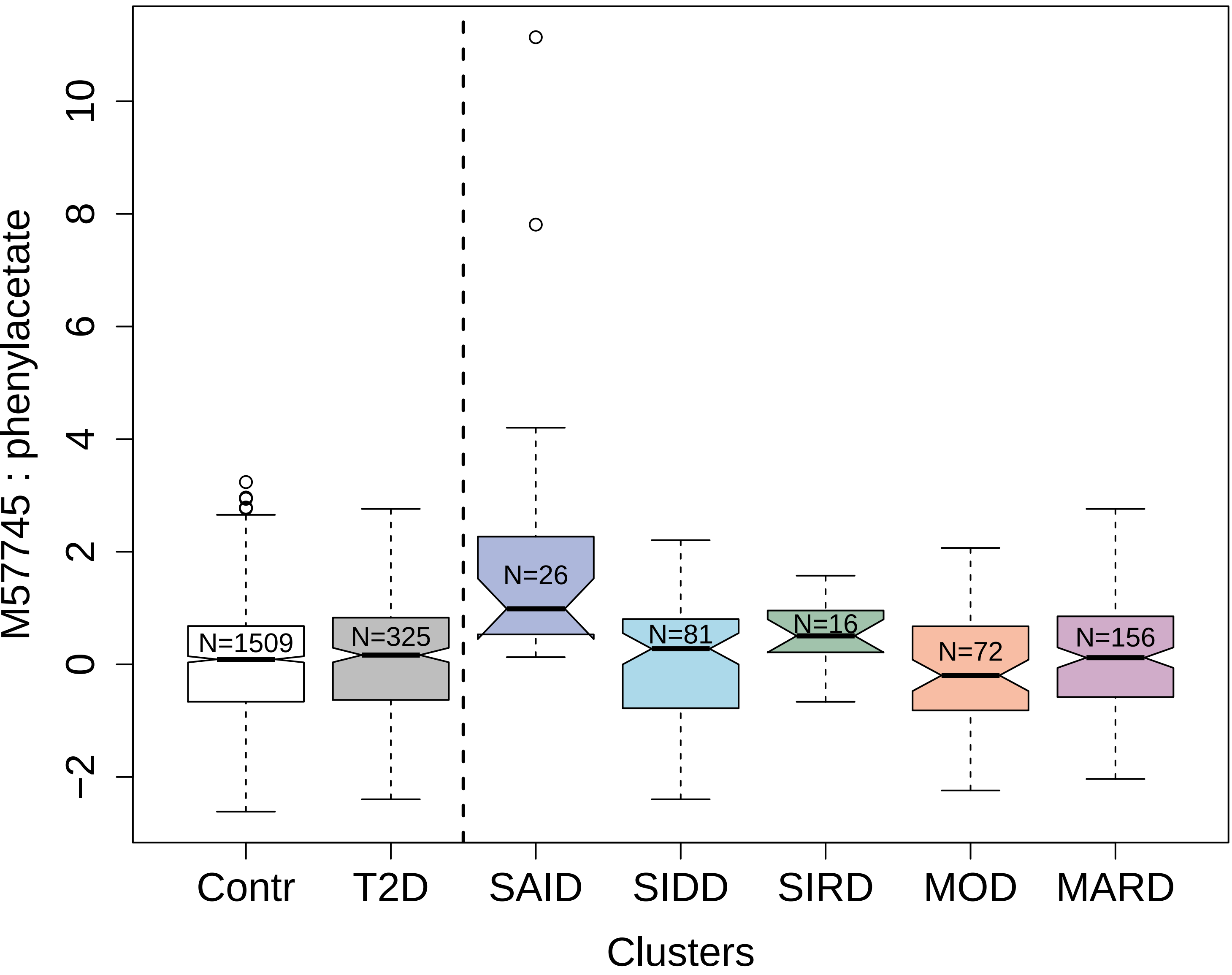

M57745 : phenylacetate
10
8
6
4
2
0
−2
N=1509
N=325
N=26
N=81
N=16
N=72
N=156
Contr
T2D
SAID
SIDD
SIRD
MOD
MARD
Clusters

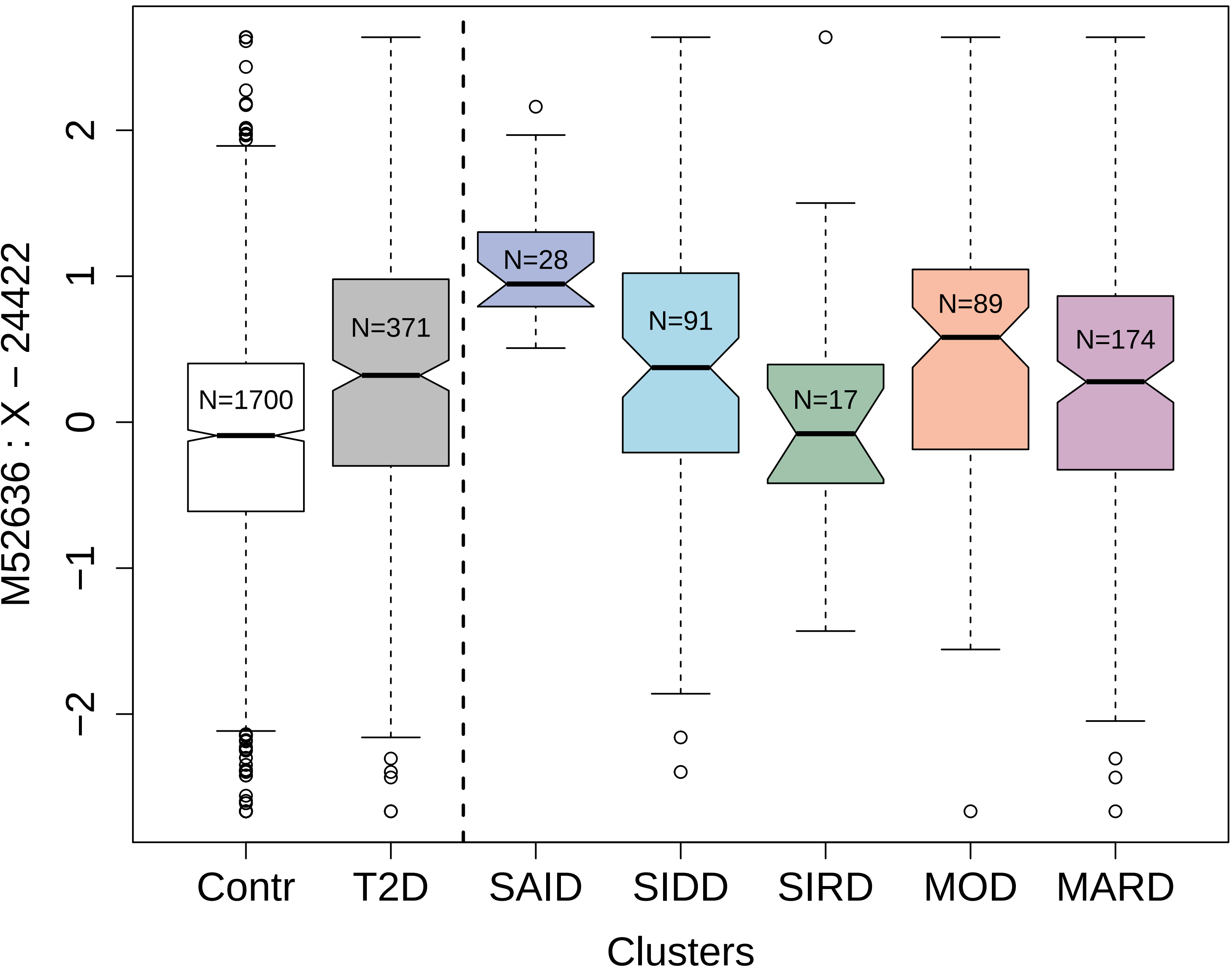

M52636 : X − 24422
2
1
0
−1
−2
N=1700
N=371
N=28
N=91
N=17
N=89
N=174
Contr
T2D
SAID
SIDD
SIRD
MOD
MARD
Clusters

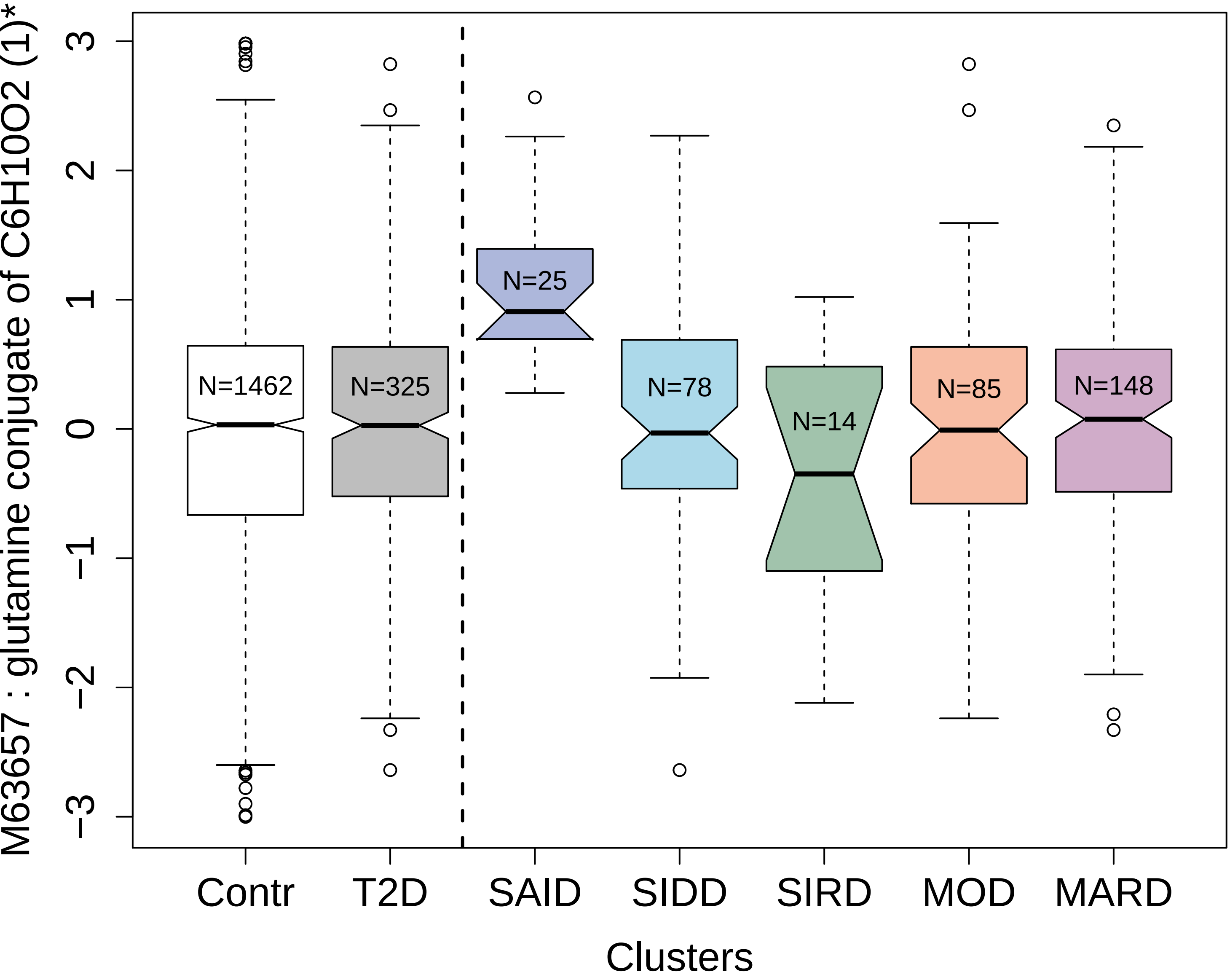

M63657 : glutamine conjugate of C6H10O2 (1)*
3
2
1
0
−1
−2
−3
N=1462
N=325
N=25
N=78
N=14
N=85
N=148
Contr
T2D
SAID
SIDD
SIRD
MOD
MARD
Clusters

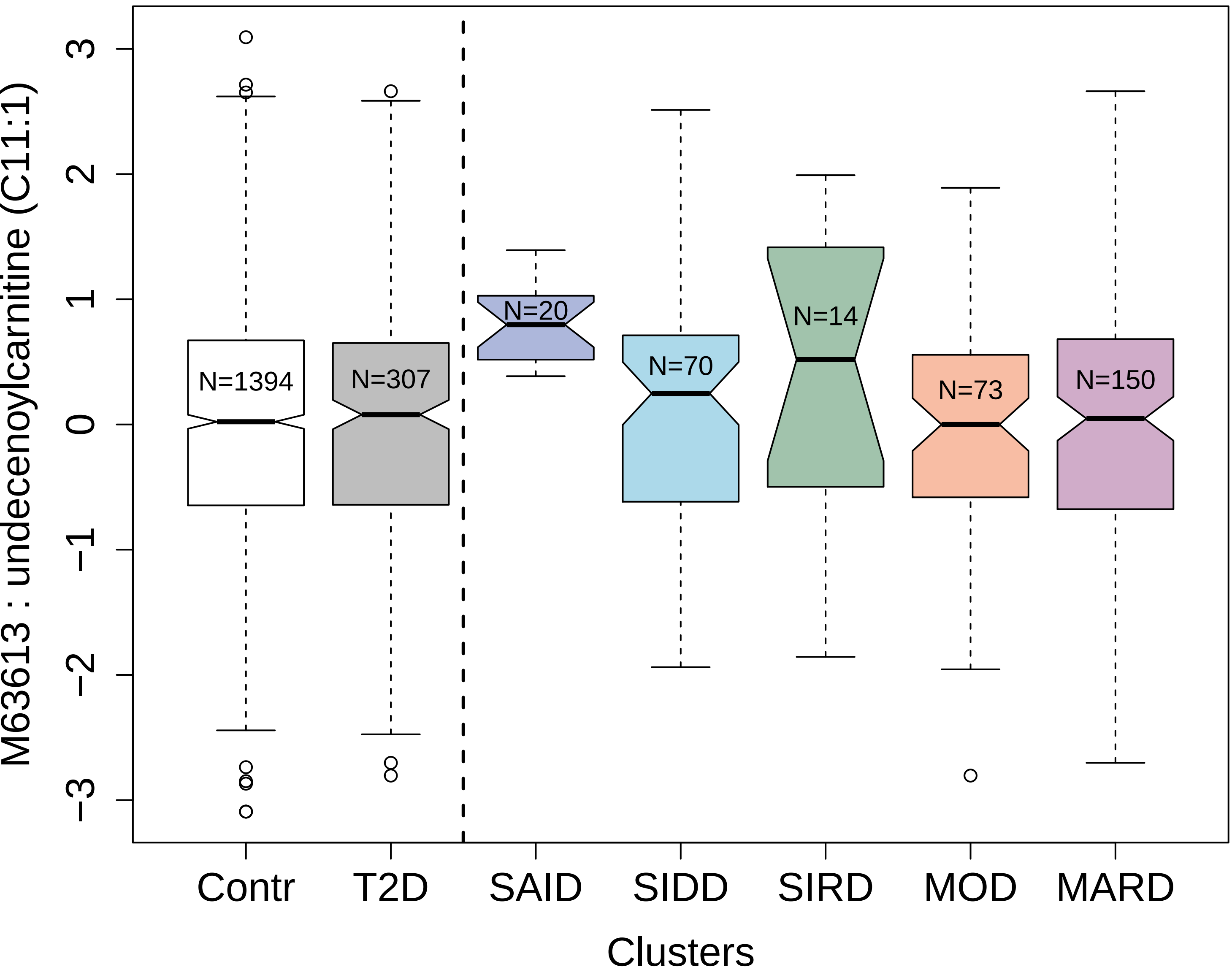

M63613 : undecenoylcarnitine (C11:1)
3
2
1
0
−1
−2
−3
N=1394
N=307
N=20
N=70
N=14
N=73
N=150
Contr
T2D
SAID
SIDD
SIRD
MOD
MARD
Clusters

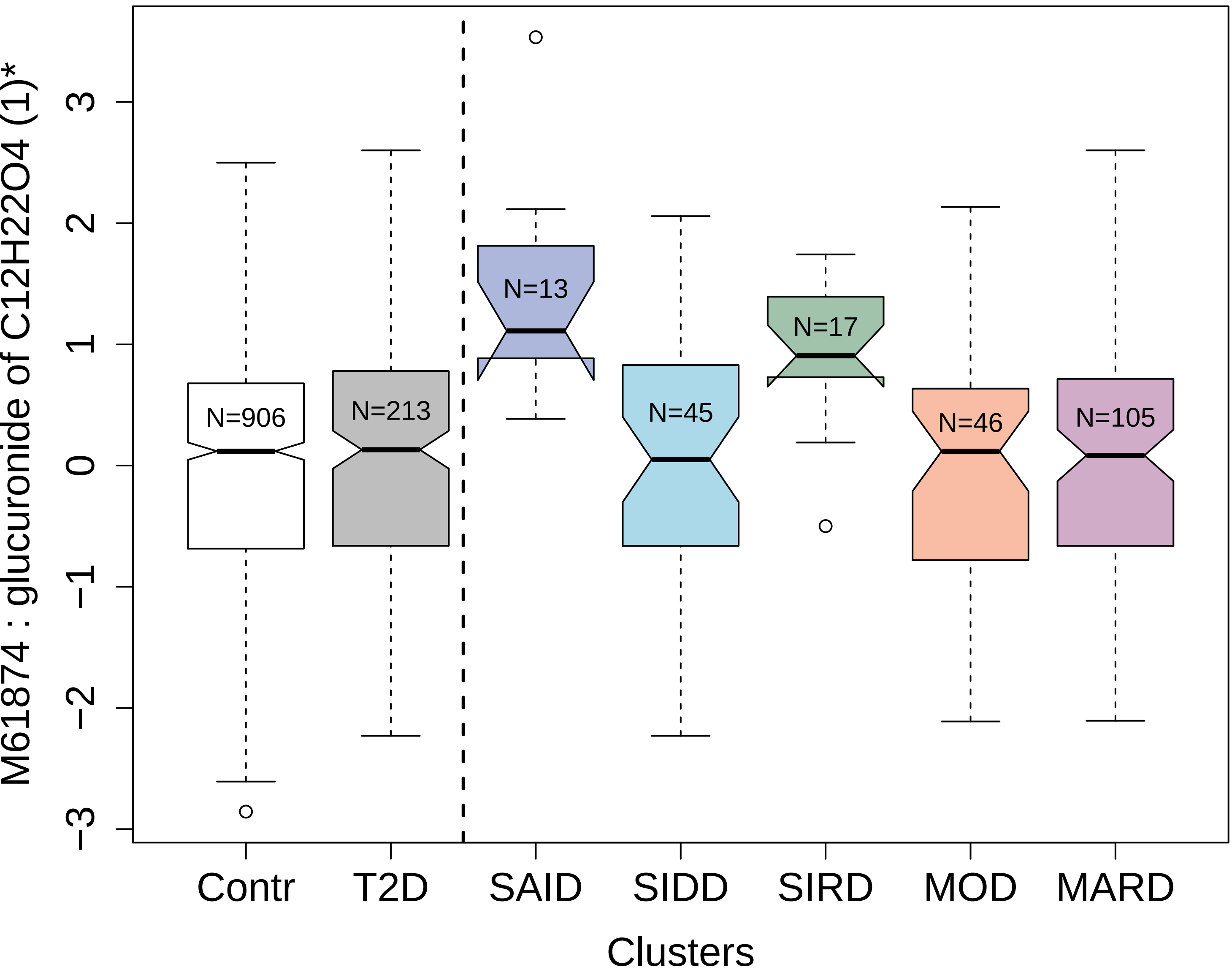

M61874 : glucuronide of C12H22O4 (1)*
3
2
1
0
−1
−2
−3
N=906
N=213
N=13
N=45
N=17
N=46
N=105
Contr
T2D
SAID
SIDD
SIRD
MOD
MARD
Clusters

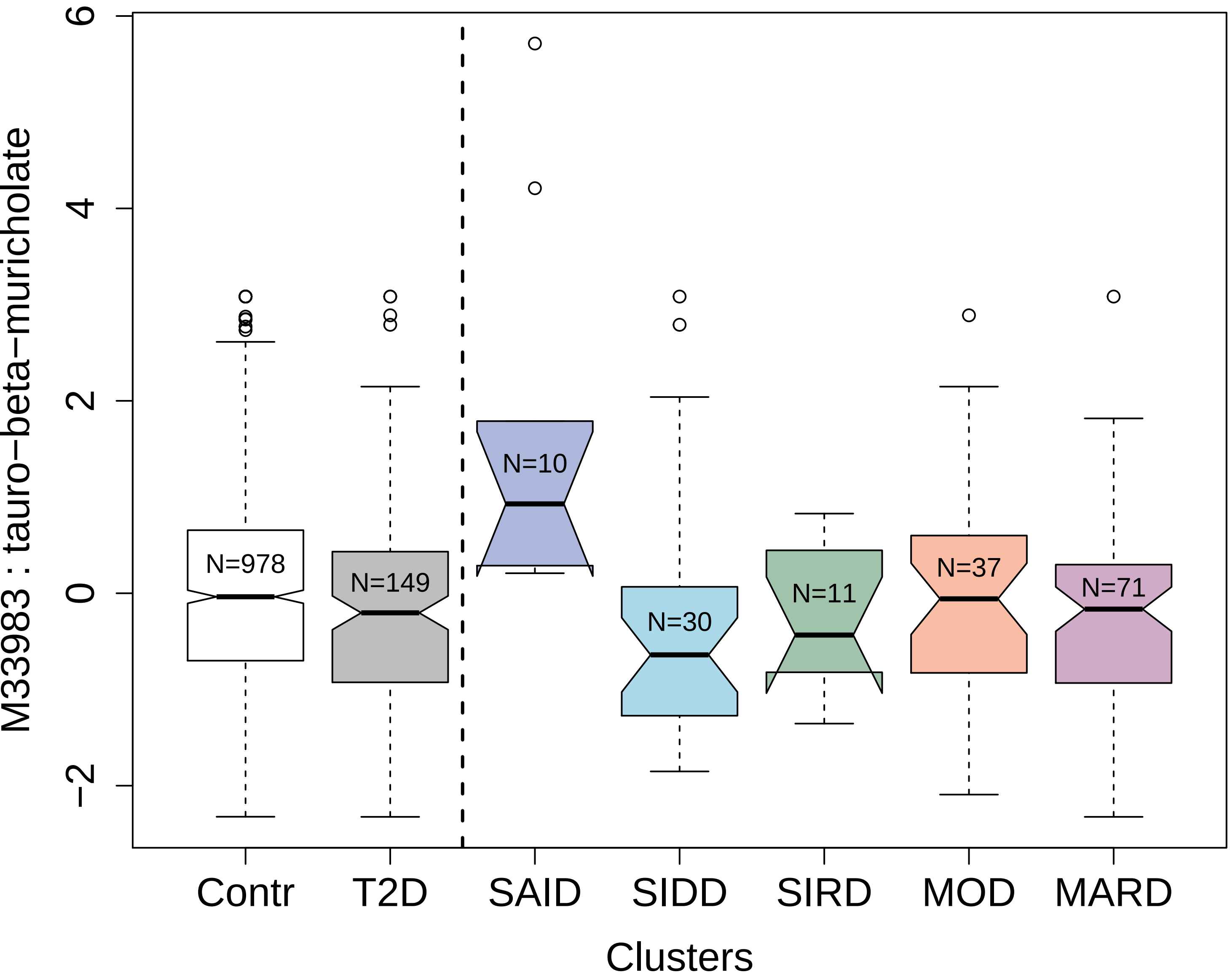

M33983 : tauro−beta−muricholate
6
4
2
0
−2
N=978
N=149
N=10
N=30
N=11
N=37
N=71
Contr
T2D
SAID
SIDD
SIRD
MOD
MARD
Clusters

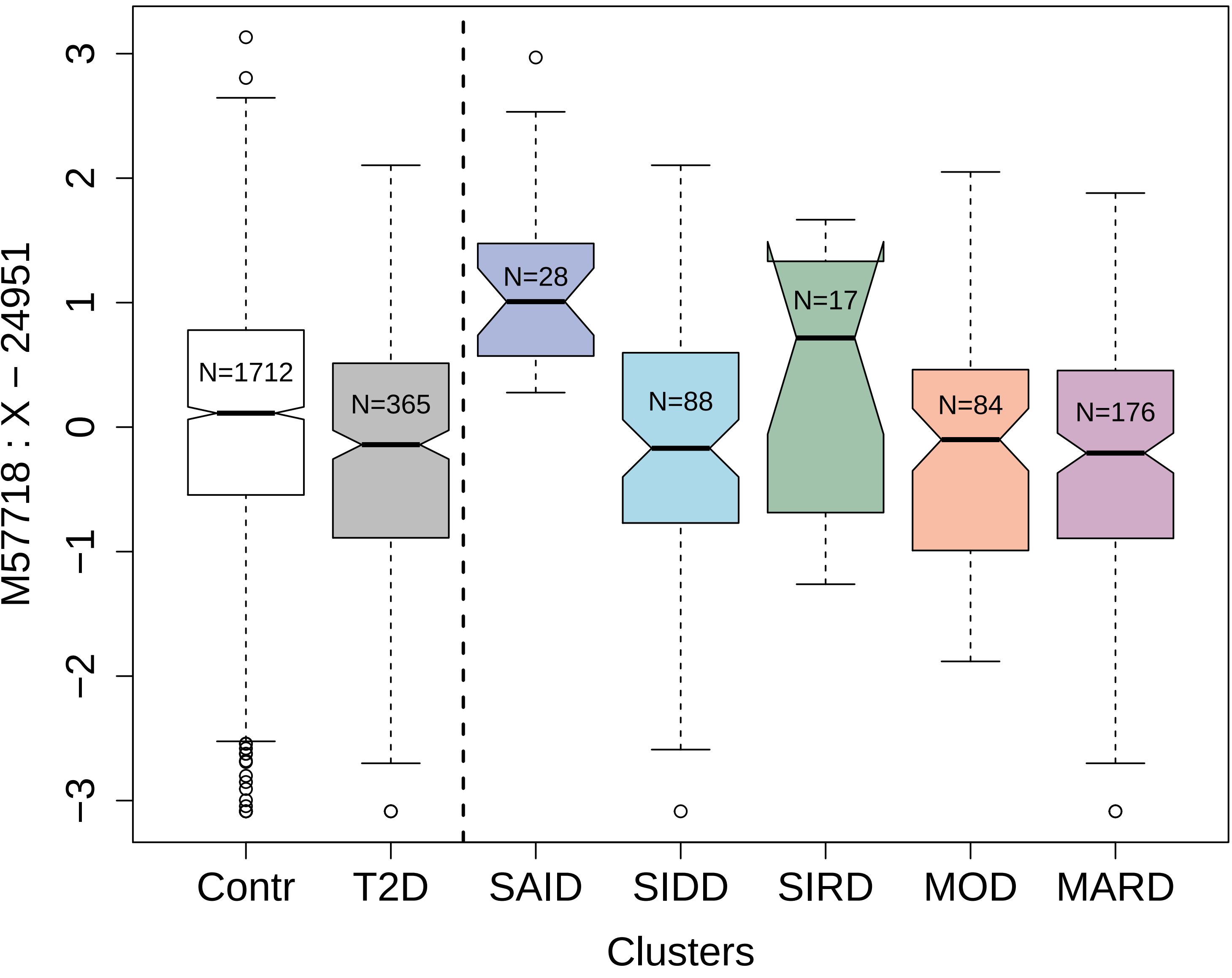

M57718 : X − 24951
3
2
1
0
−1
−2
−3
N=1712
N=365
N=28
N=88
N=17
N=84
N=176
Contr
T2D
SAID
SIDD
SIRD
MOD
MARD
Clusters

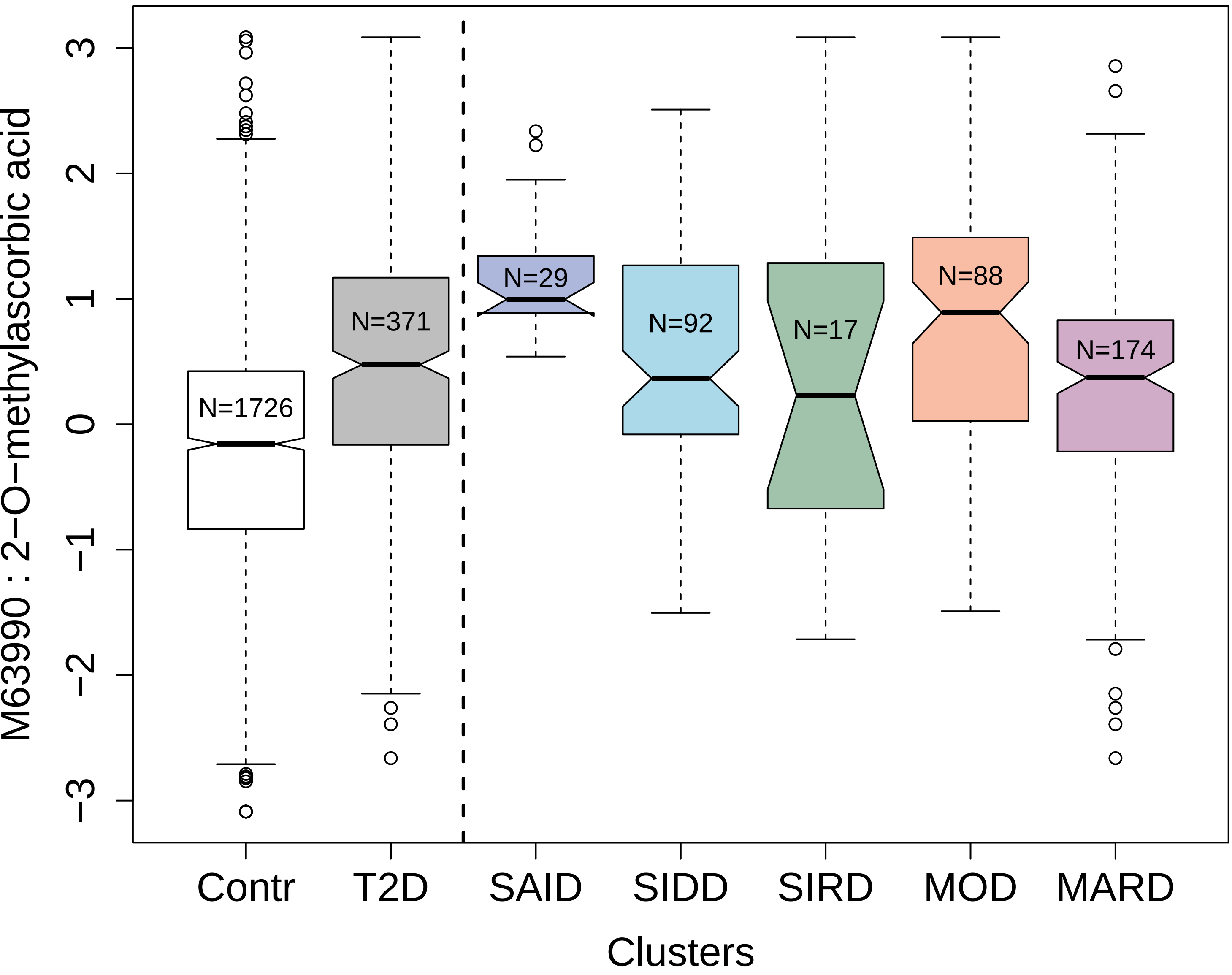

M63990 : 2−O−methylascorbic acid
3
2
1
0
−1
−2
−3
N=1726
N=371
N=29
N=92
N=17
N=88
N=174
Contr
T2D
SAID
SIDD
SIRD
MOD
MARD
Clusters

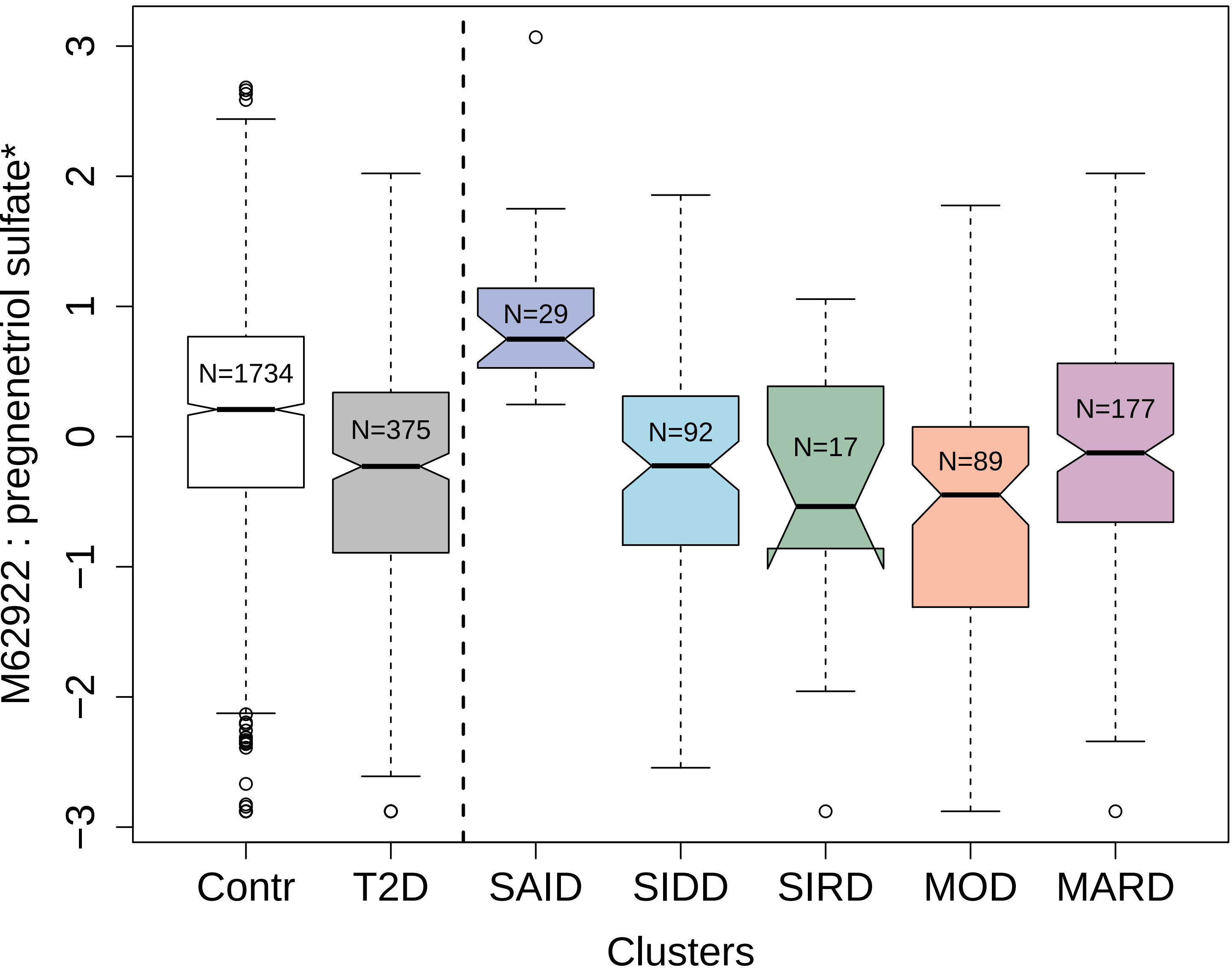
M62922 : pregnenetriol sulfate*
3
2
1
0
−1
−2
−3
N=1734
N=375
N=29
N=92
N=17
N=89
N=177
Contr
T2D
SAID
SIDD
SIRD
MOD
MARD
Clusters

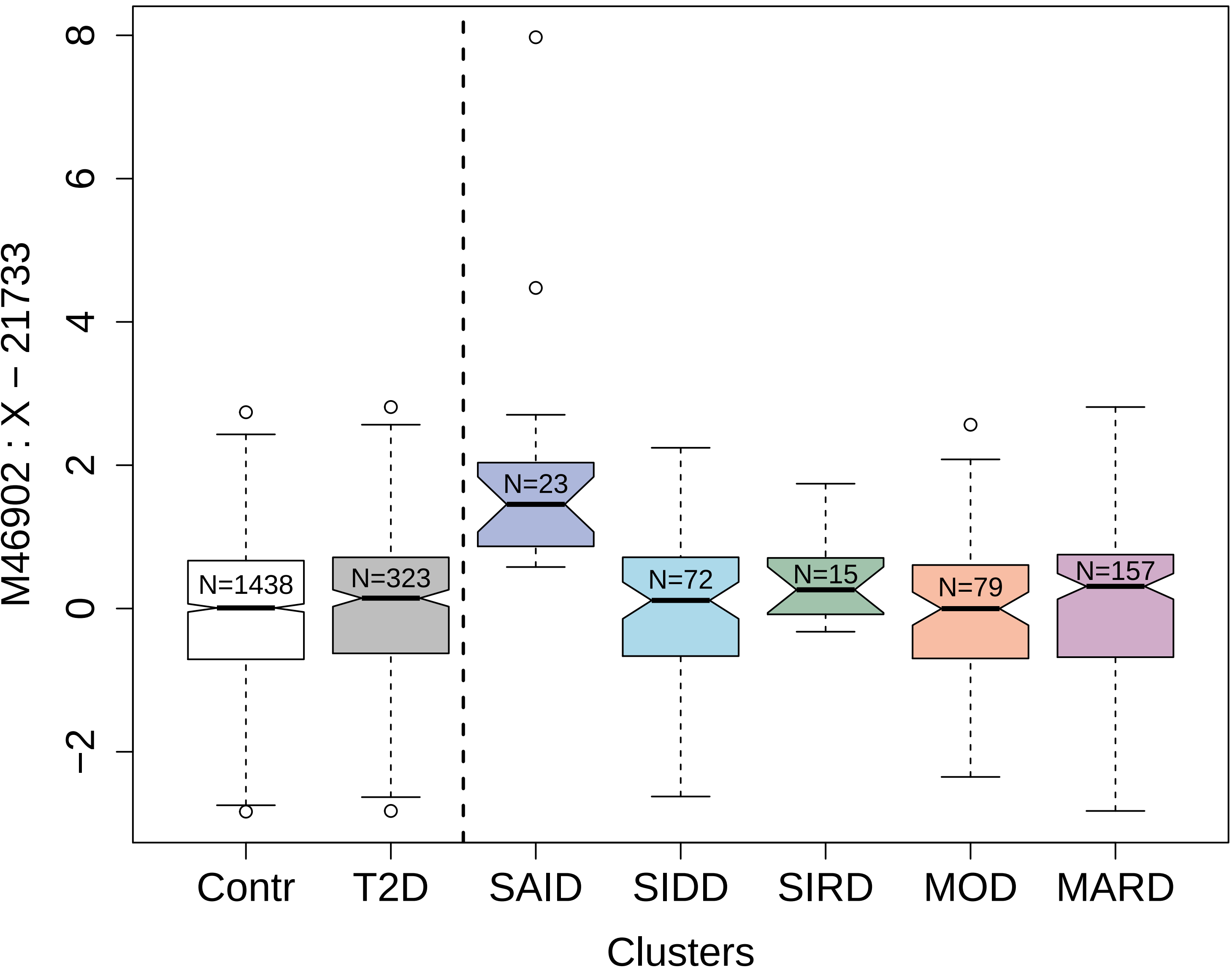

M46902 : X − 21733
8
6
4
2
0
−2
N=1438
N=323
N=23
N=72
N=15
N=79
N=157
Contr
T2D
SAID
SIDD
SIRD
MOD
MARD
Clusters

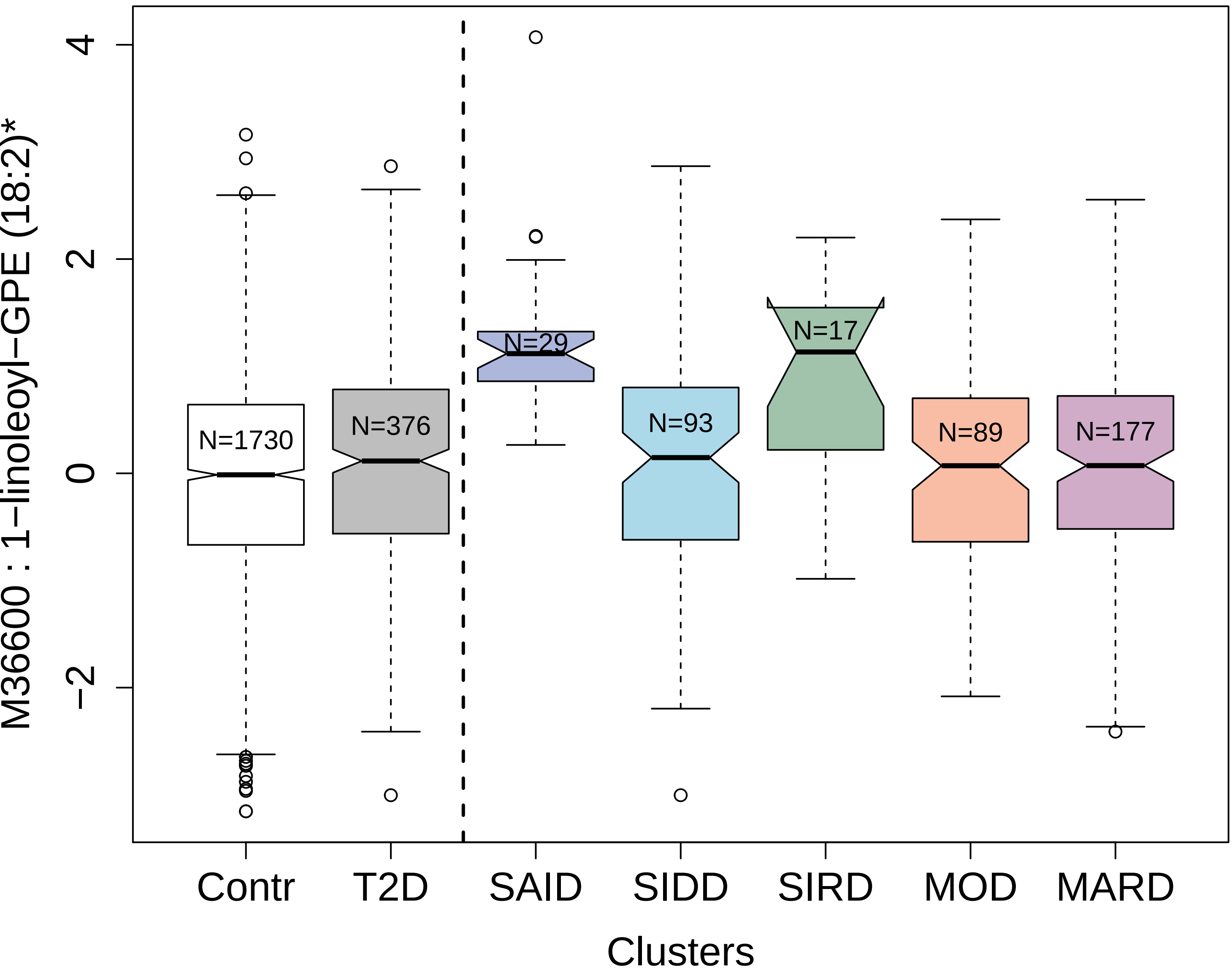

M36600 : 1−linoleoyl−GPE (18:2)*
4
2
0
−2
N=1730
N=376
N=29
N=93
N=17
N=89
N=177
Contr
T2D
SAID
SIDD
SIRD
MOD
MARD
Clusters

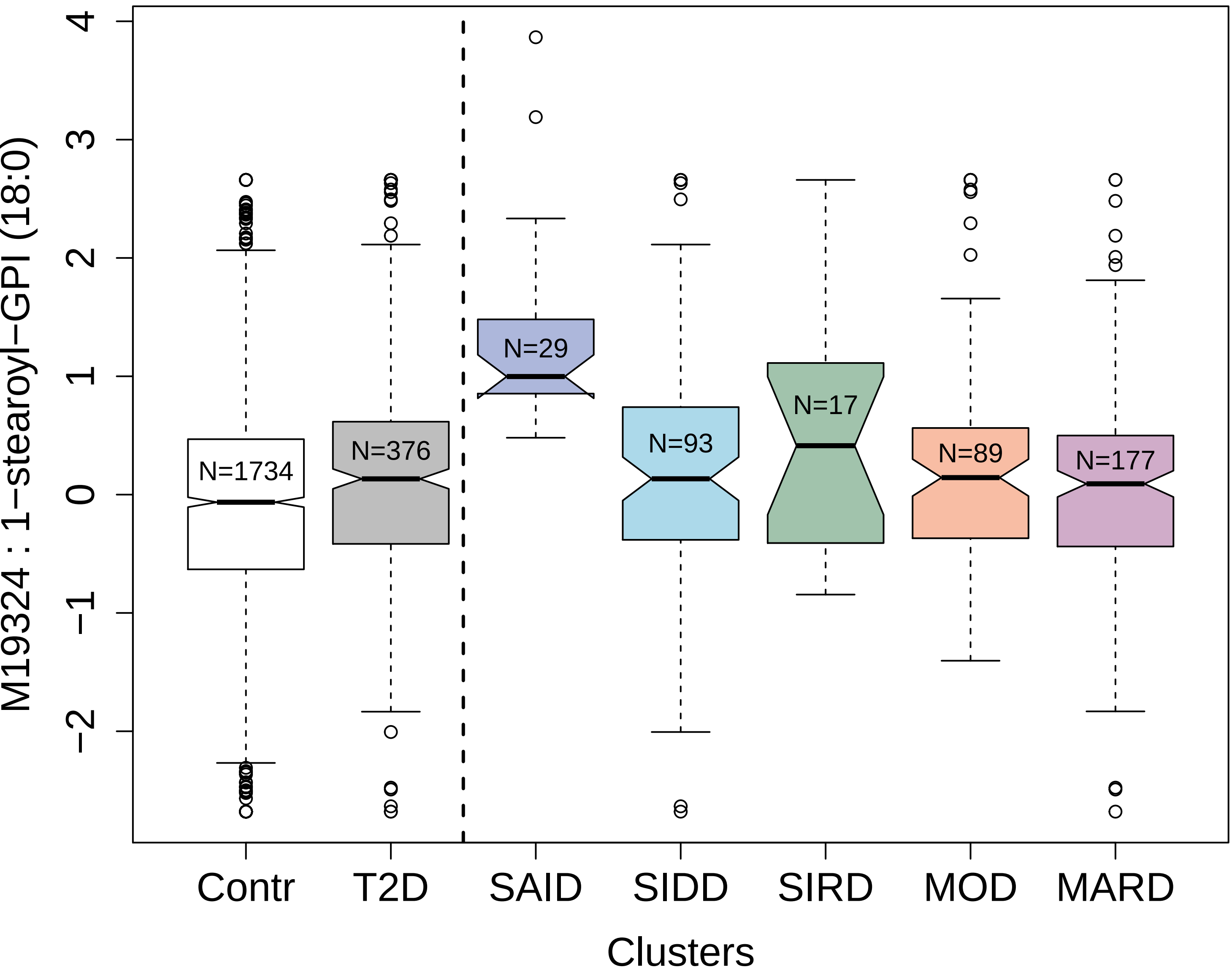

M19324 : 1−stearoyl−GPI (18:0)
4
3
2
1
0
−1
−2
N=1734
N=376
N=29
N=93
N=17
N=89
N=177
Contr
T2D
SAID
SIDD
SIRD
MOD
MARD
Clusters

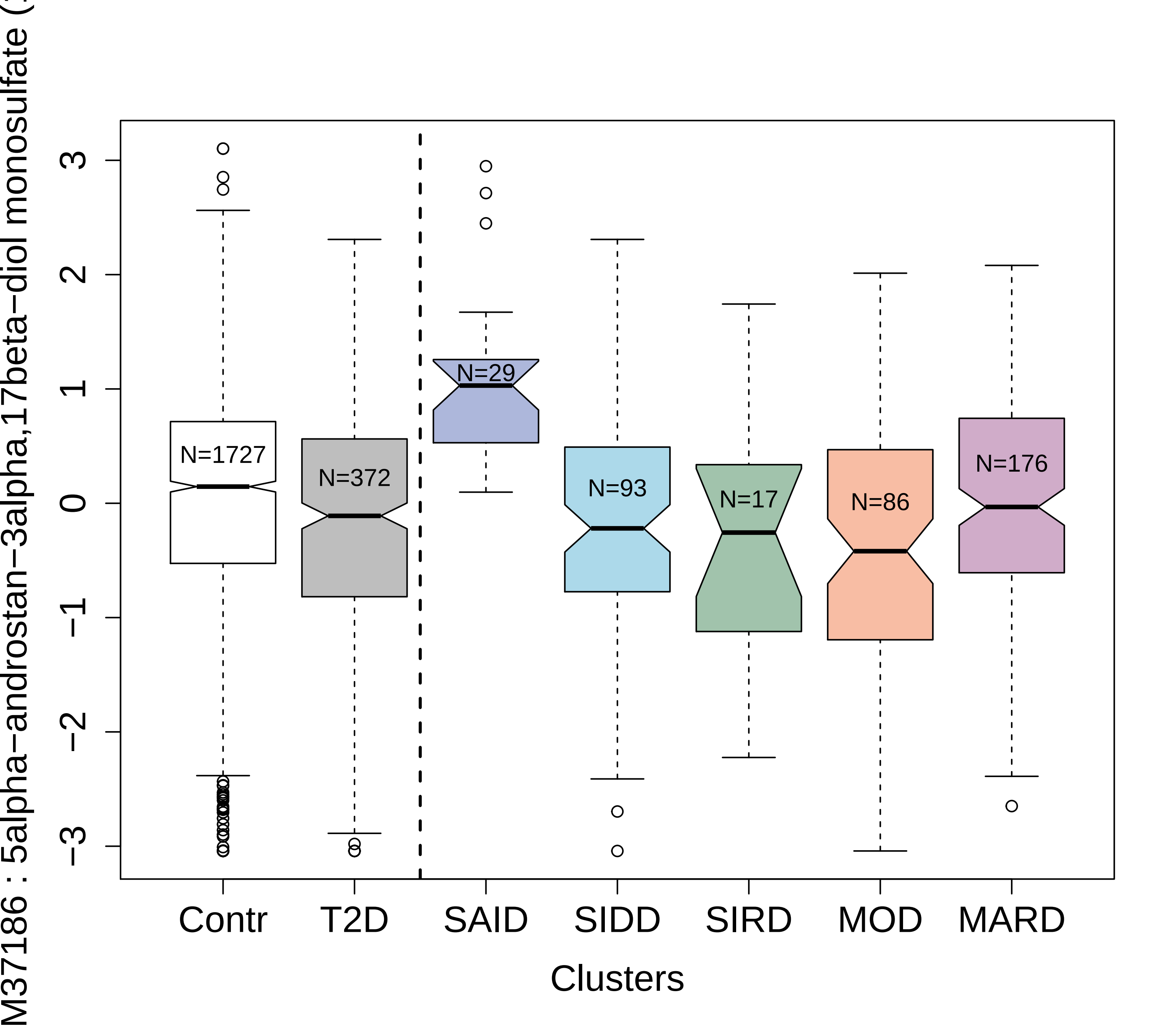
M37186 : 5alpha−androstan−3alpha,17beta−diol monosulfate (
3
2
1
0
−1
−2
−3
N=1727
N=372
N=29
N=93
N=17
N=86
N=176
Contr
T2D
SAID
SIDD
SIRD
MOD
MARD
Clusters

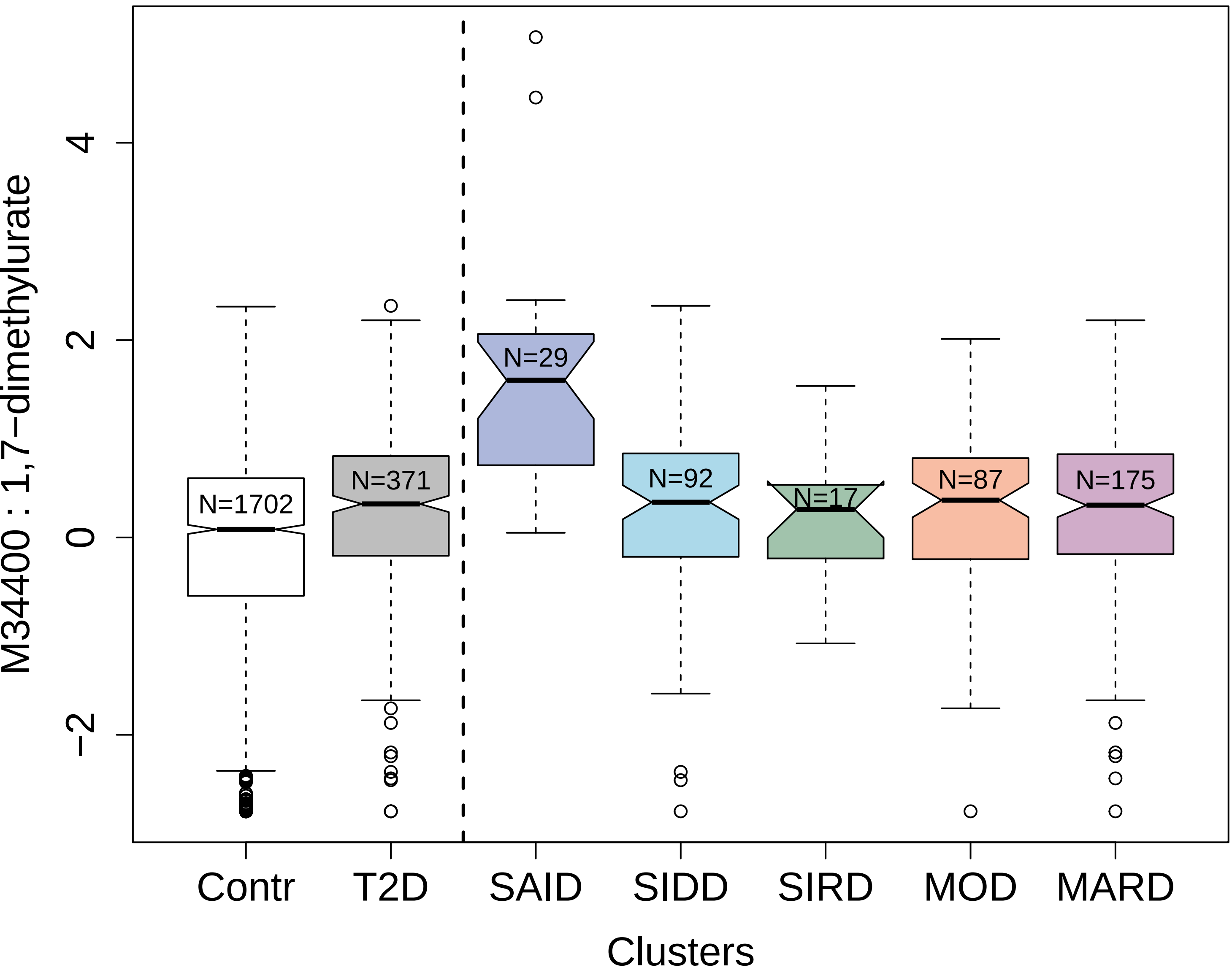

M34400 : 1,7−dimethylurate
4
2
0
−2
N=1702
N=371
N=29
N=92
N=17
N=87
N=175
Contr
T2D
SAID
SIDD
SIRD
MOD
MARD
Clusters

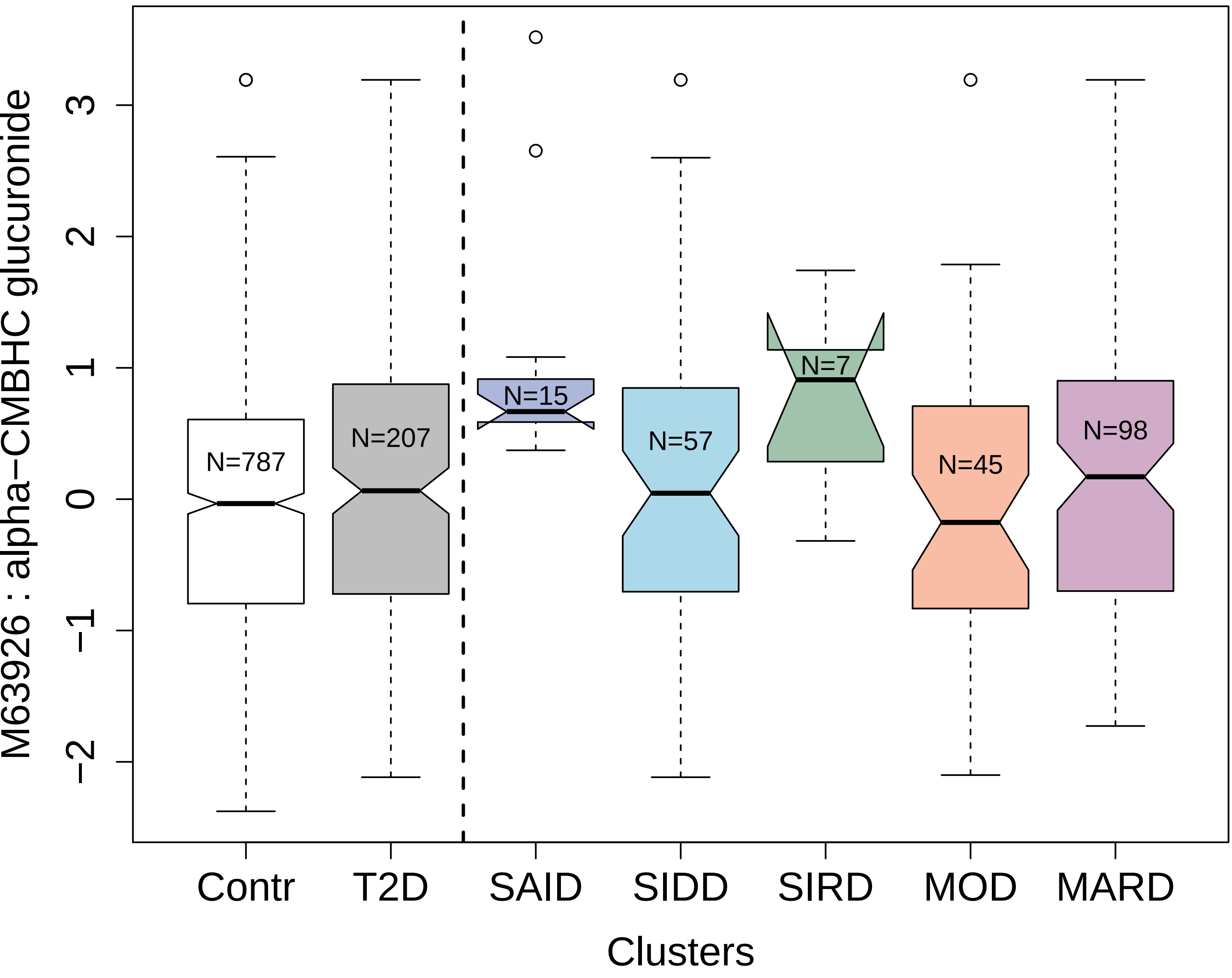

M63926 : alpha−CMBHC glucuronide
−2
−1
0
1
2
3
N=787
N=207
N=15
N=57
N=7
N=45
N=98
Contr
T2D
SAID
SIDD
SIRD
MOD
MARD
Clusters

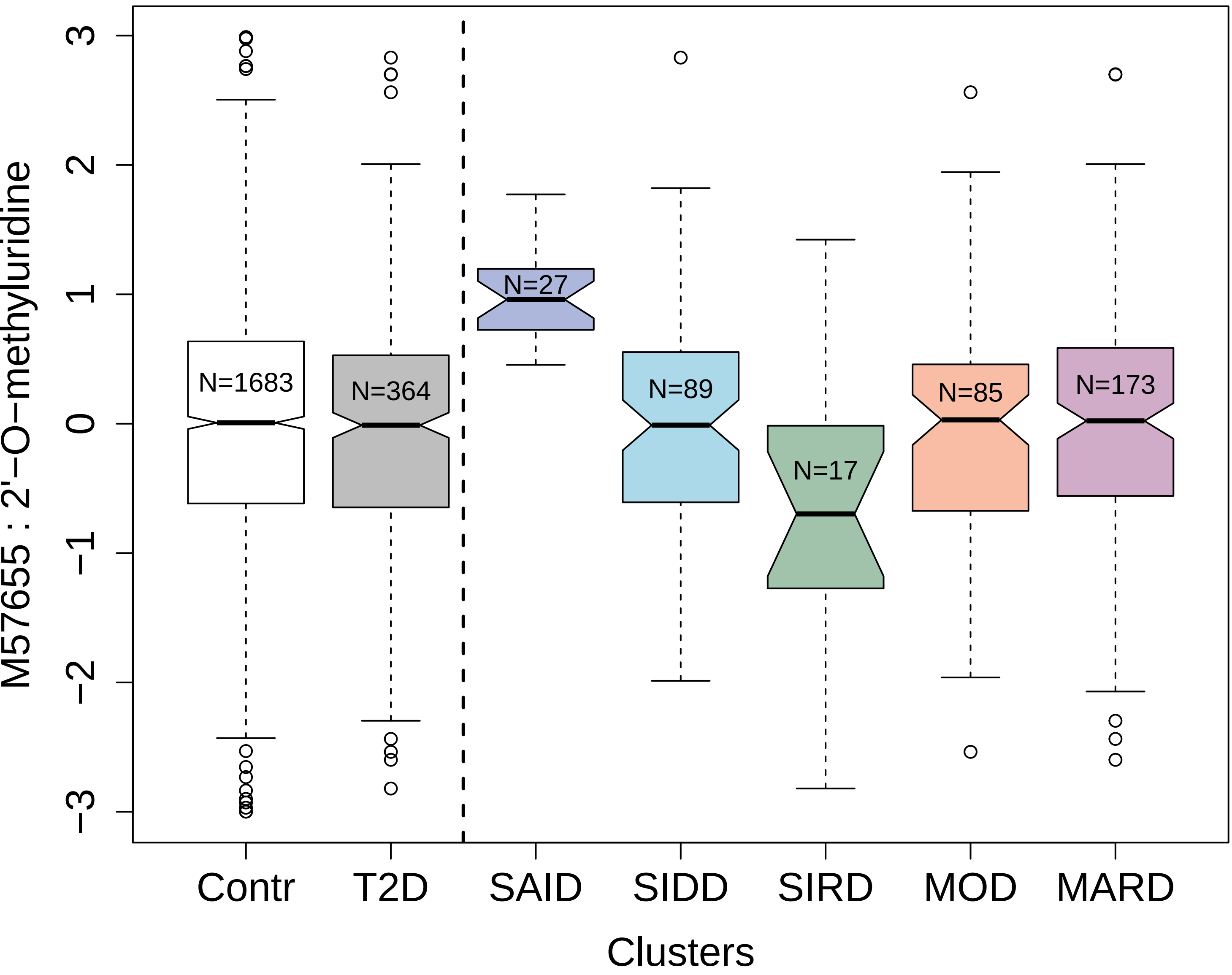
M57655 : 2'−O−methyluridine
3
2
1
0
−1
−2
−3
N=1683
N=364
N=27
N=89
N=17
N=85
N=173
Contr
T2D
SAID
SIDD
SIRD
MOD
MARD
Clusters

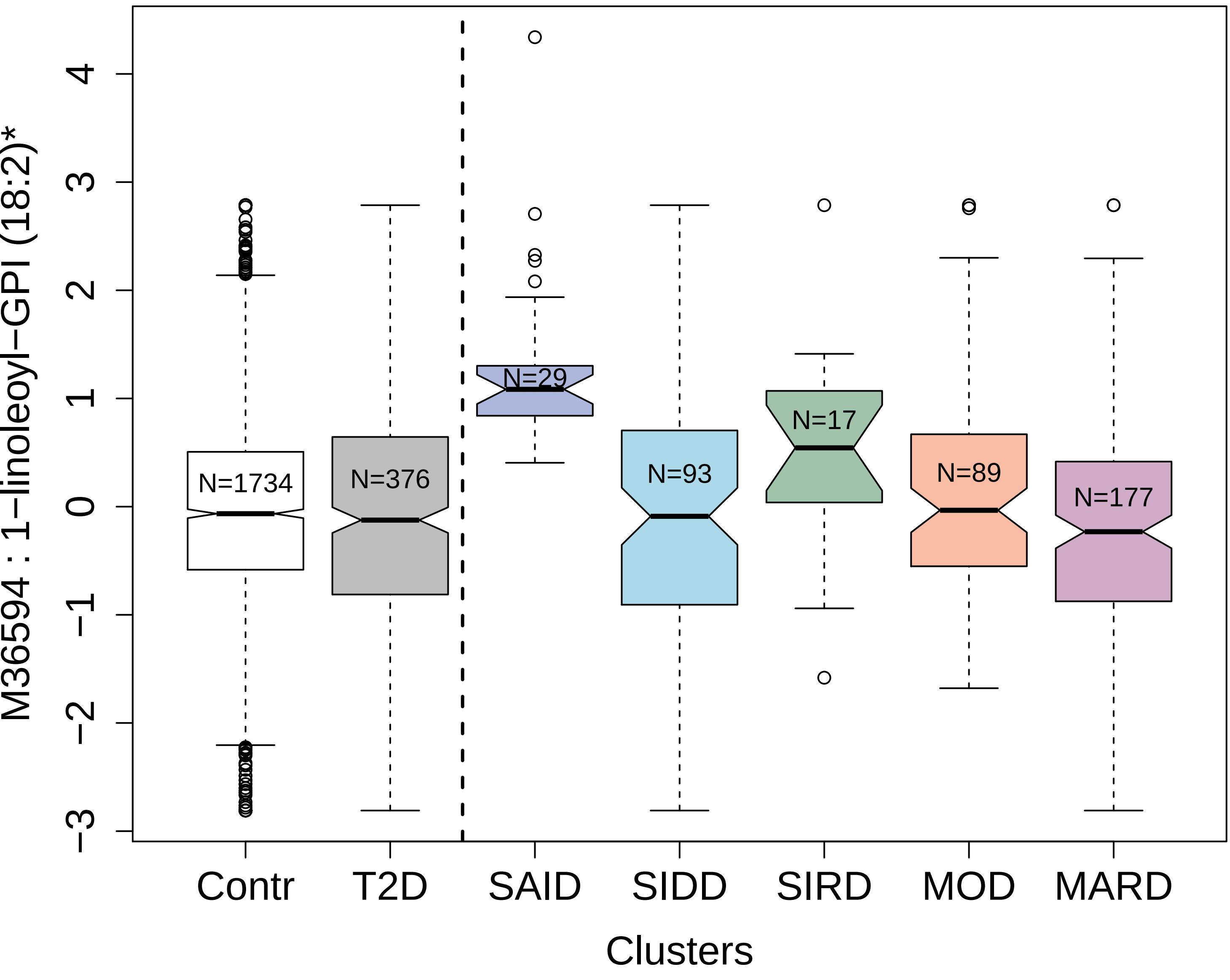
M36594 : 1−linoleoyl−GPI (18:2)*
4
3
2
1
0
−1
−2
−3
N=1734
N=376
N=29
N=93
N=17
N=89
N=177
Contr
T2D
SAID
SIDD
SIRD
MOD
MARD
Clusters

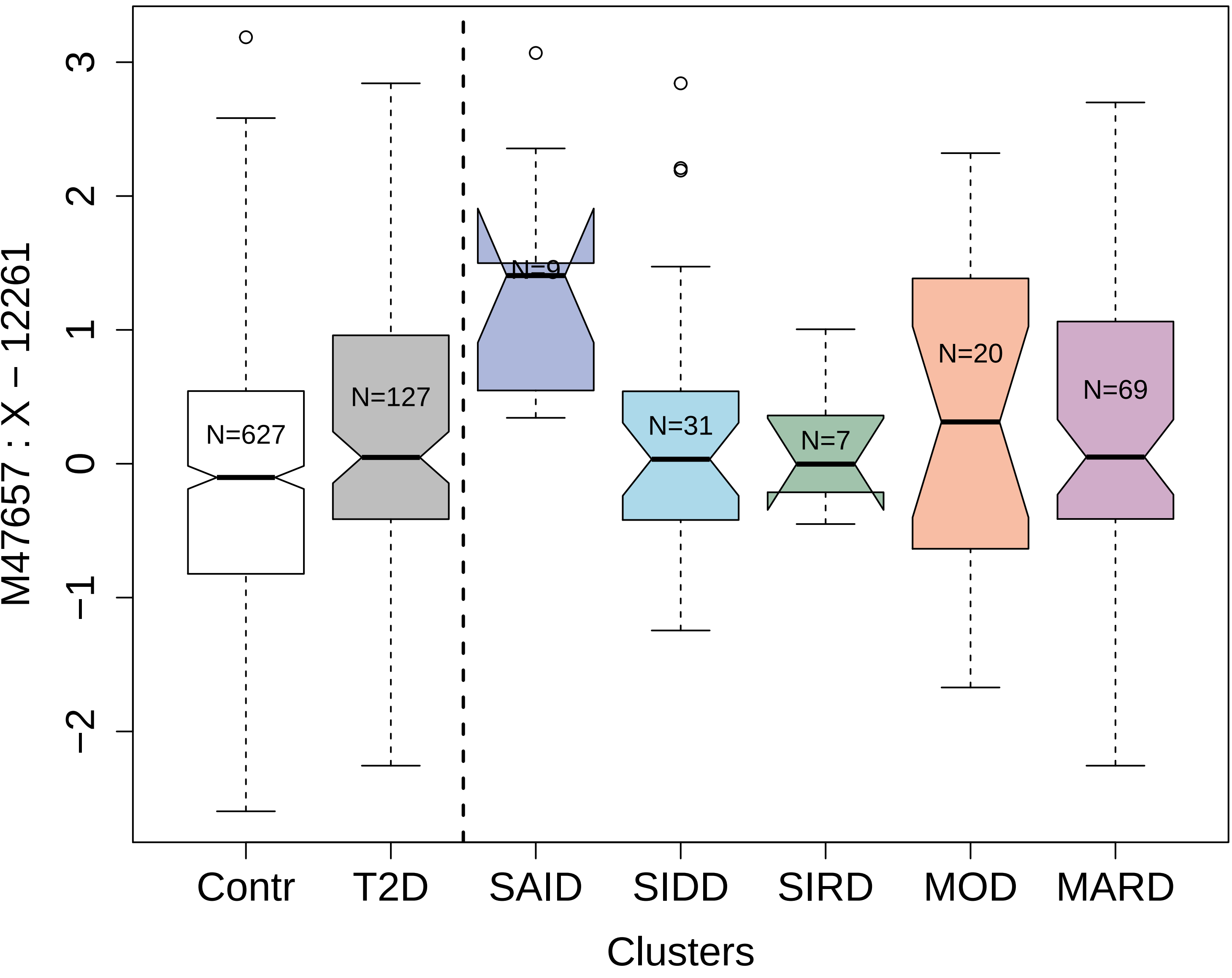

M47657 : X − 12261
3
2
1
0
−1
−2
N=627
N=127
N=9
N=31
N=7
N=20
N=69
Contr
T2D
SAID
SIDD
SIRD
MOD
MARD
Clusters

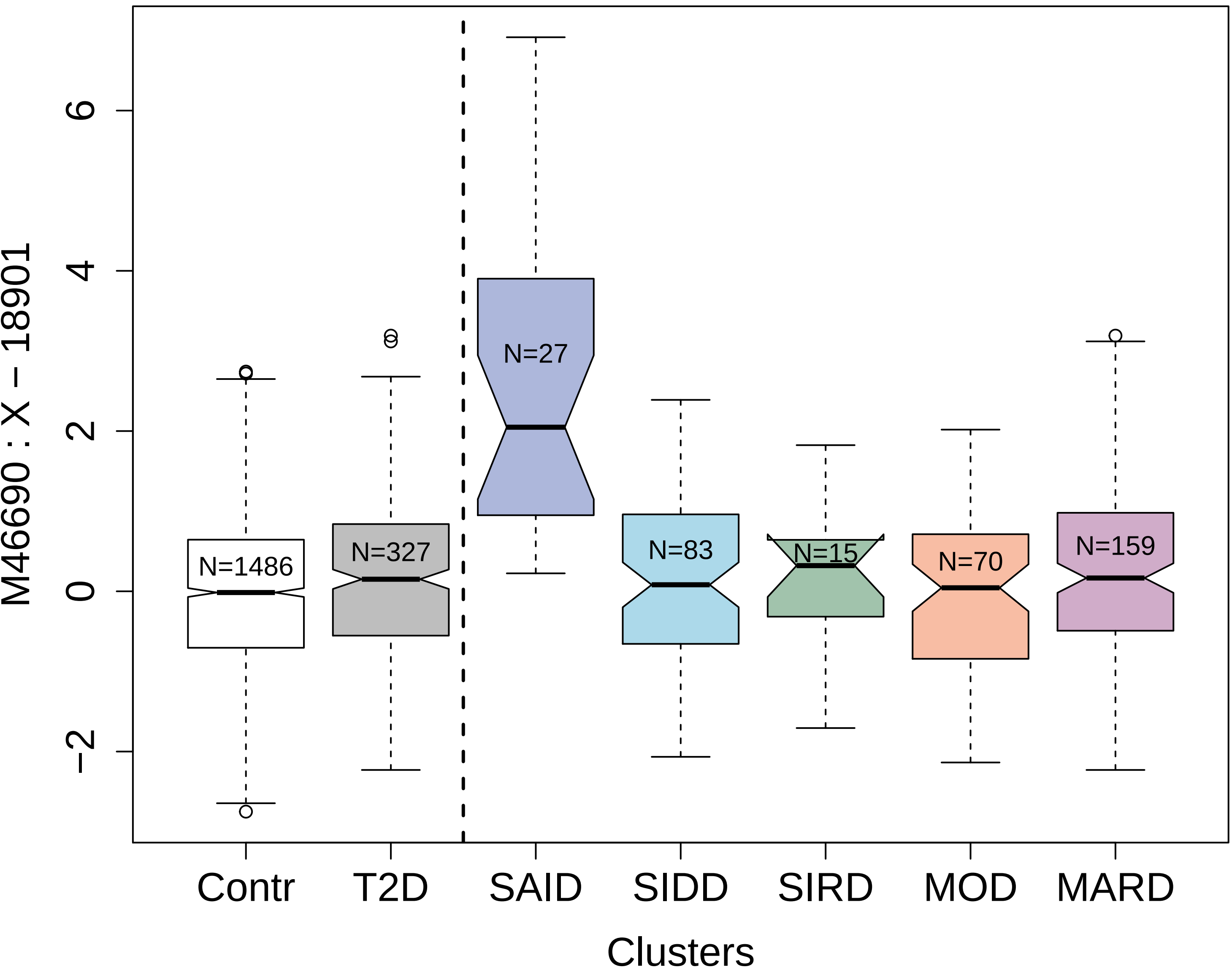
M46690 : X − 18901
6
4
2
0
−2
N=1486
N=327
N=27
N=83
N=15
N=70
N=159
Contr
T2D
SAID
SIDD
SIRD
MOD
MARD
Clusters

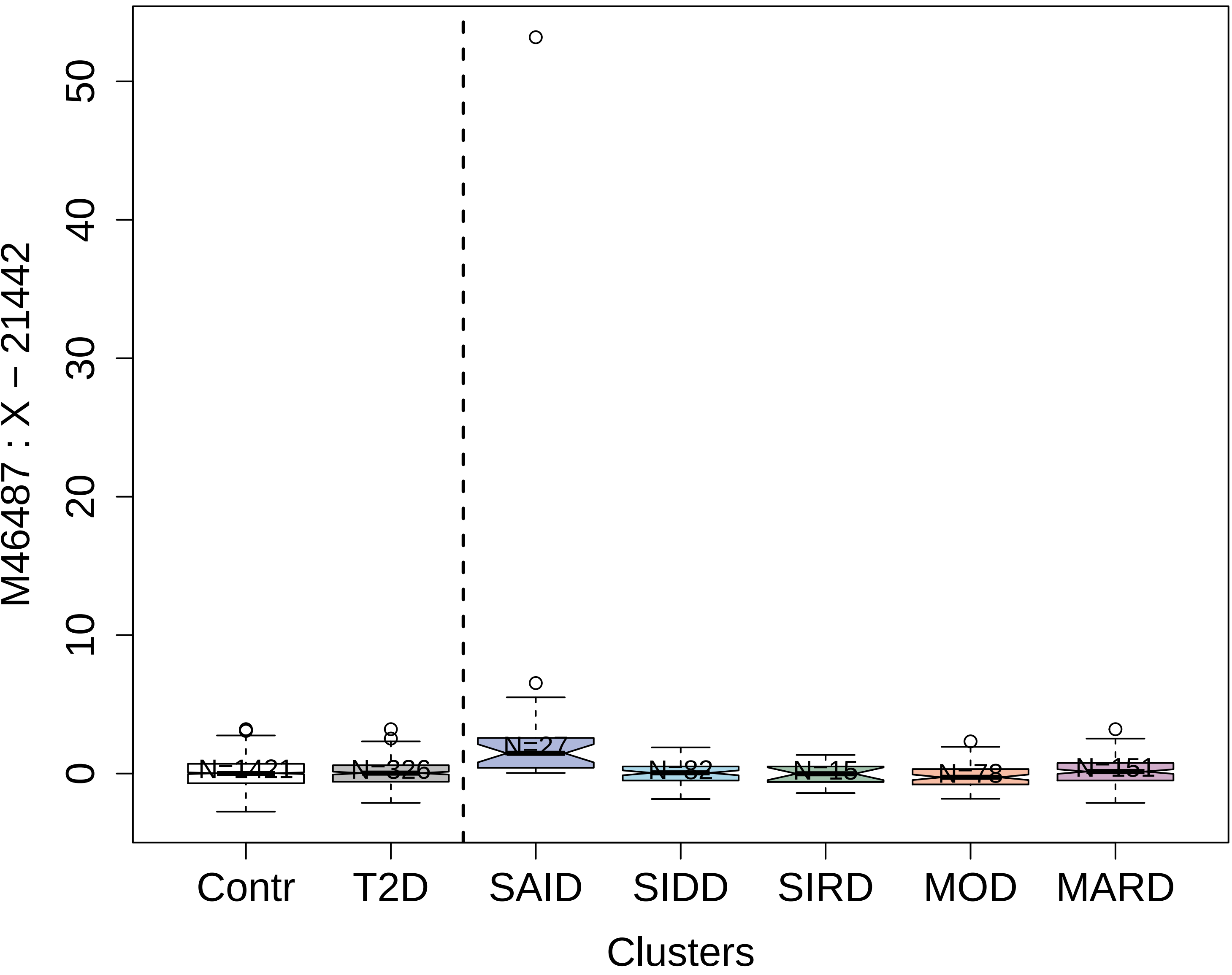

M46487 : X − 21442
0
10
20
30
40
50
N=1421
N=326
N=27
N=82
N=15
N=78
N=151
Contr
T2D
SAID
SIDD
SIRD
MOD
MARD
Clusters

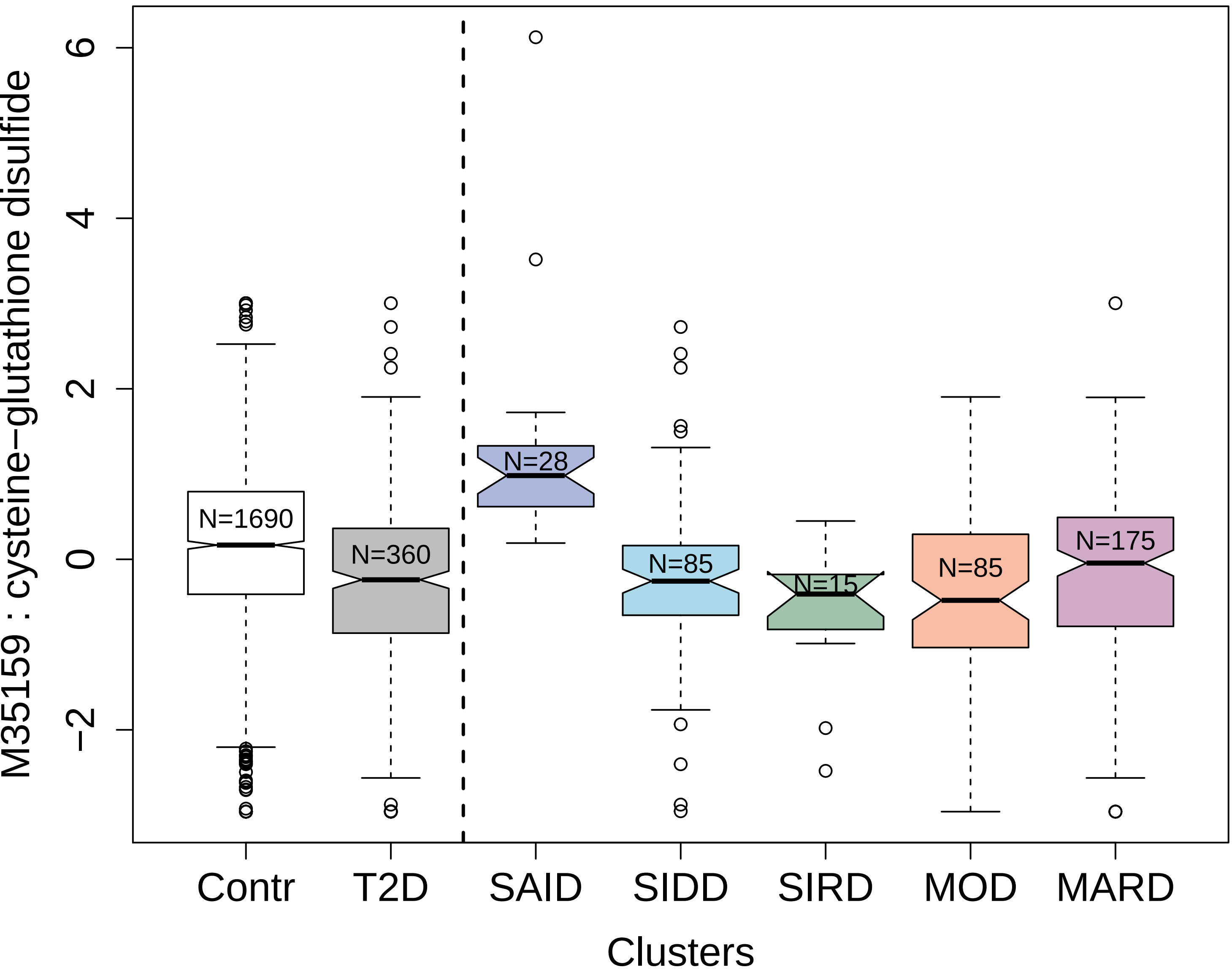
M35159 : cysteine−glutathione disulfide
6
4
2
0
−2
N=1690
N=360
N=28
N=85
N=15
N=85
N=175
Contr
T2D
SAID
SIDD
SIRD
MOD
MARD
Clusters

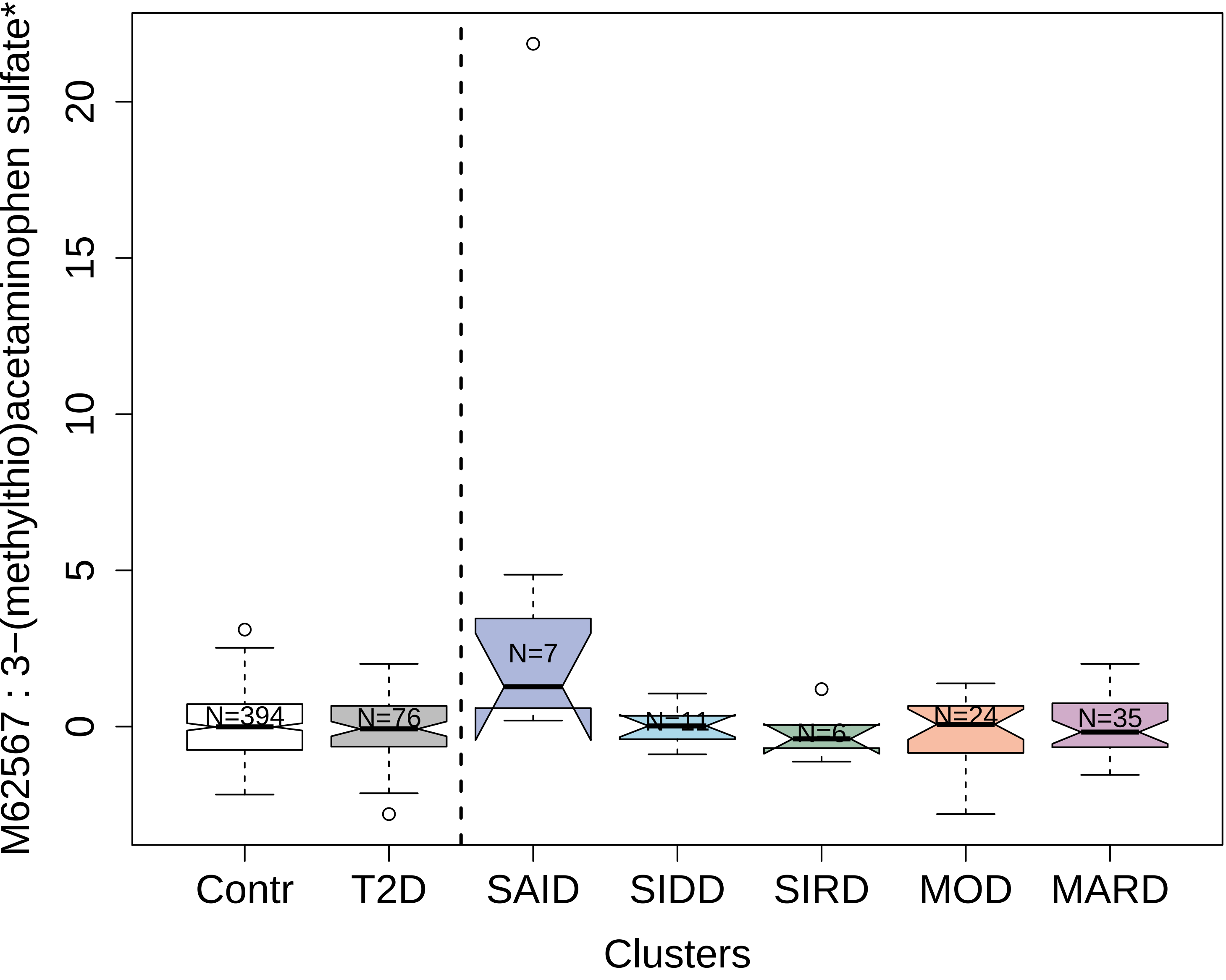

M62567 : 3−(methylthio)acetaminophen sulfate*
0
5
10
15
20
N=394
N=76
N=7
N=11
N=6
N=24
N=35
Contr
T2D
SAID
SIDD
SIRD
MOD
MARD
Clusters

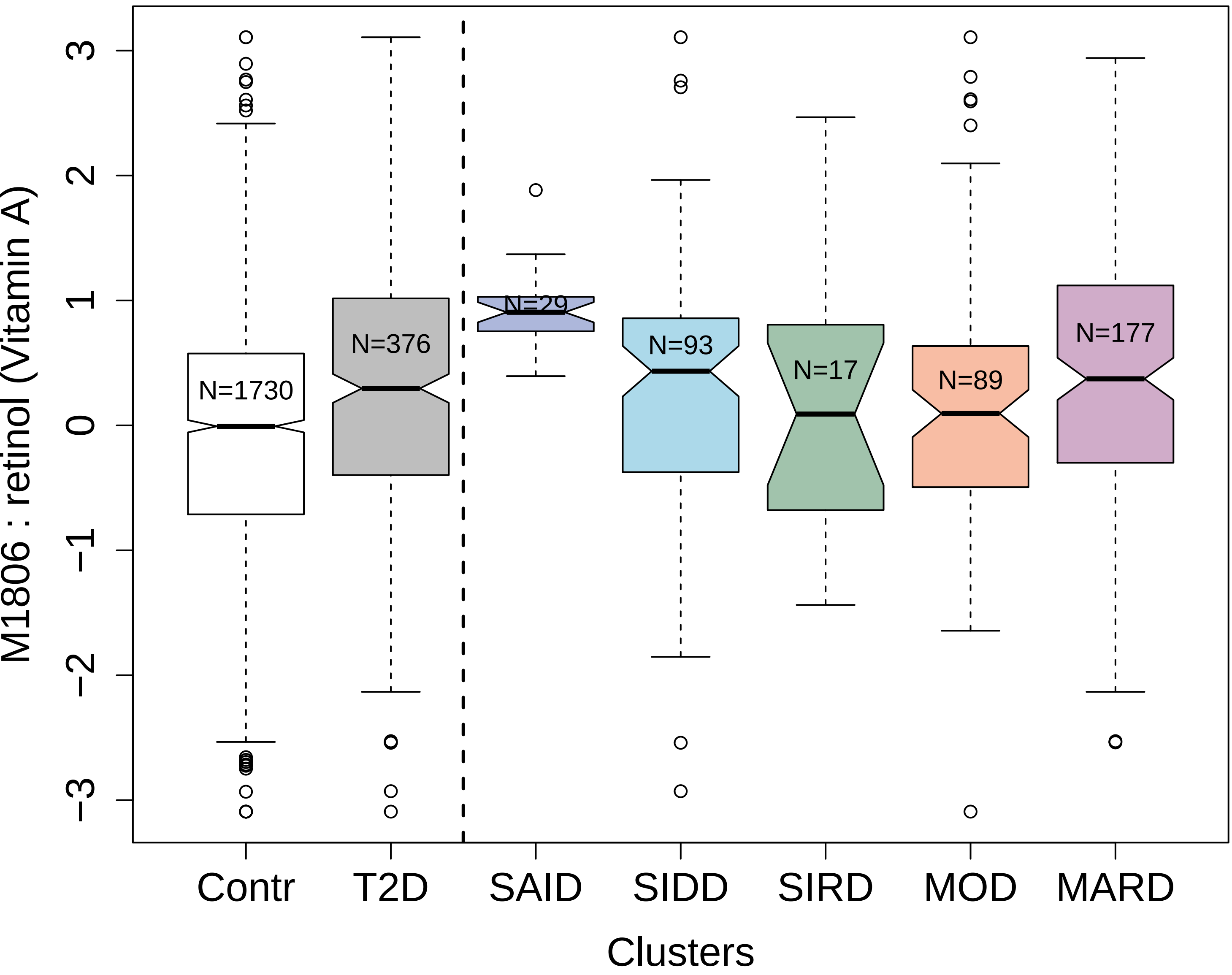
M1806 : retinol (Vitamin A)
3
2
1
0
−1
−2
−3
N=1730
N=376
N=29
N=93
N=17
N=89
N=177
Contr
T2D
SAID
SIDD
SIRD
MOD
MARD
Clusters

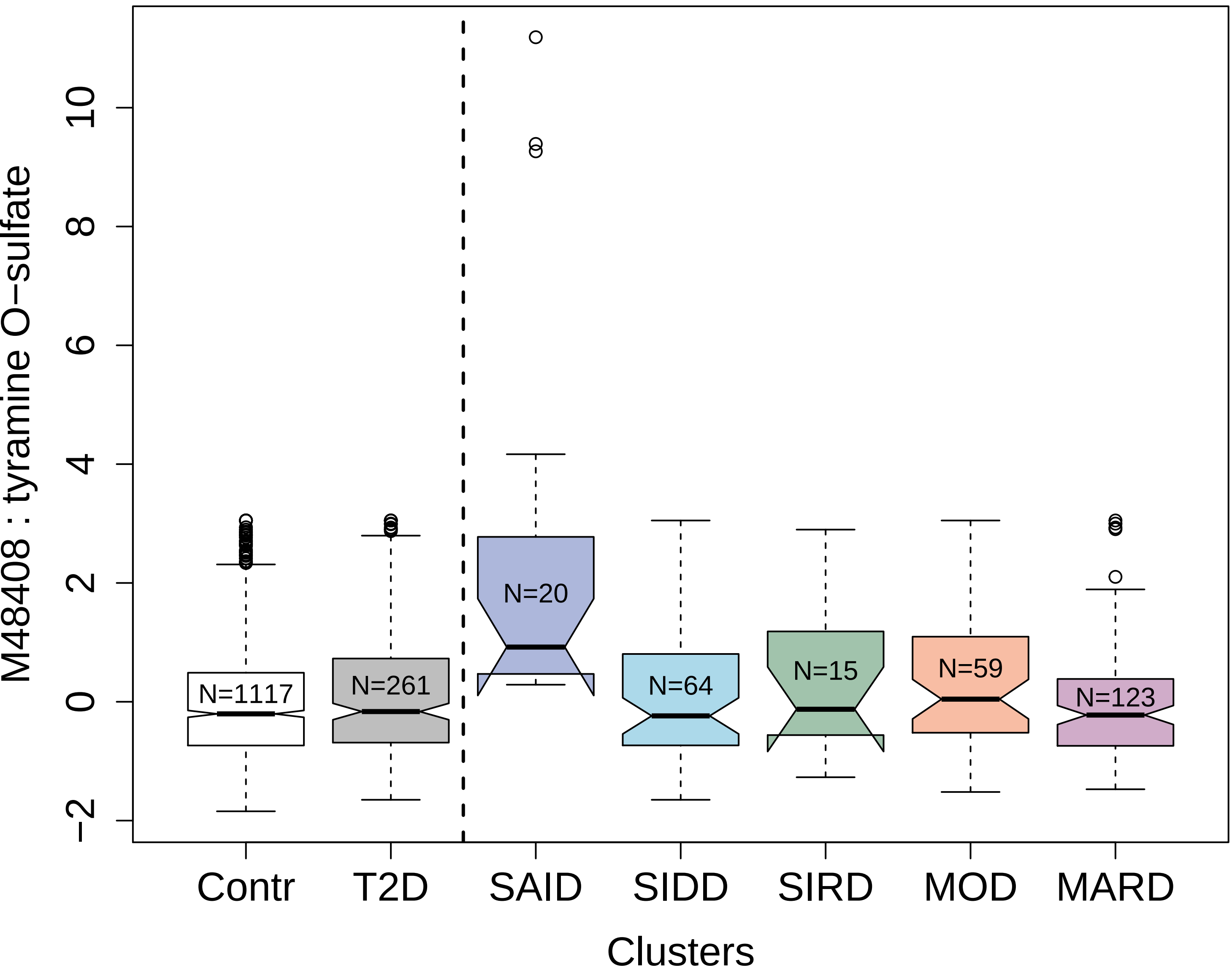

M48408 : tyramine O−sulfate
Clusters
Contr
T2D
SAID
SIDD
SIRD
MOD
MARD
N=1117
N=261
N=20
N=64
N=15
N=59
N=123
−2
0
2
4
6
8
10

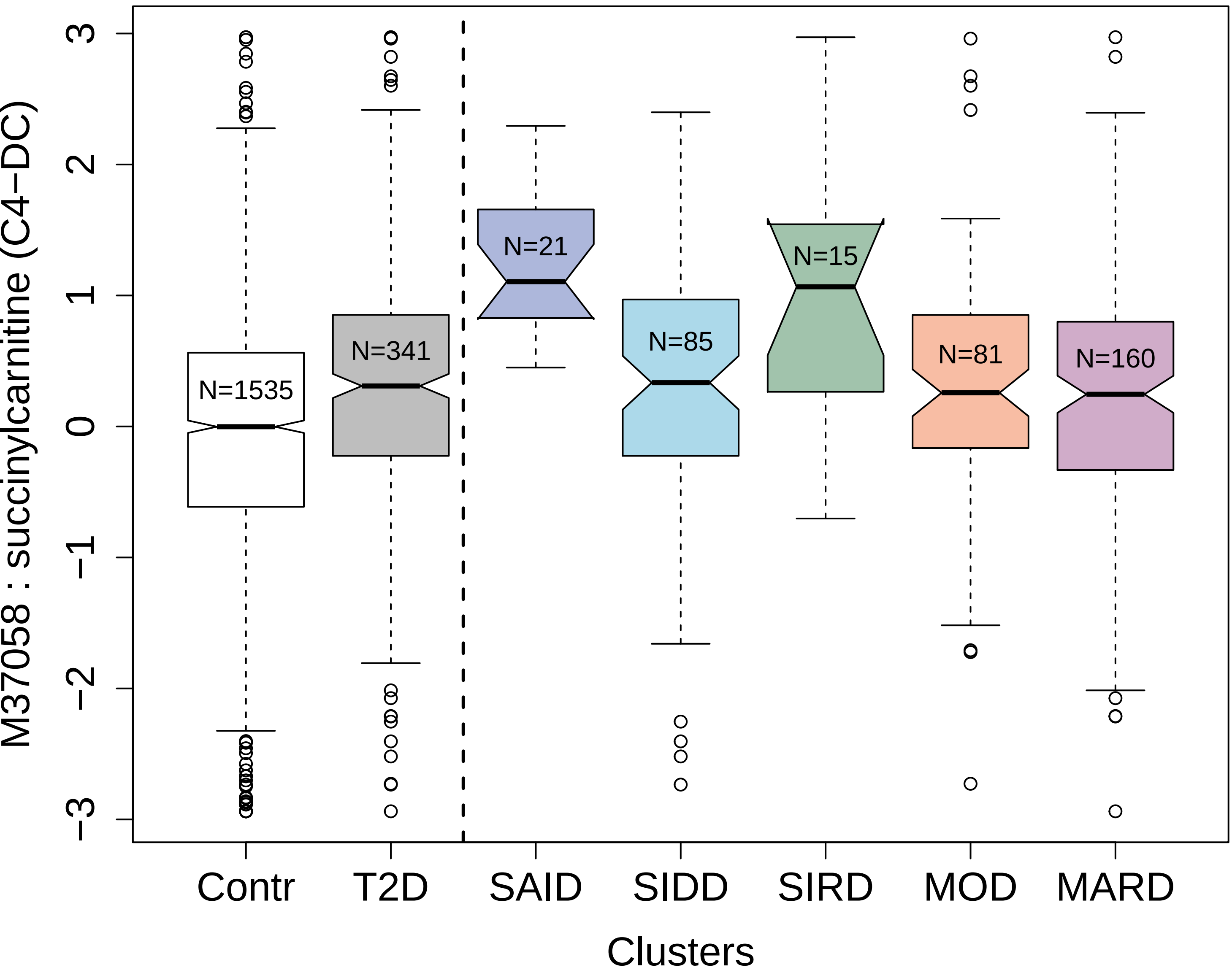

M37058 : succinylcarnitine (C4−DC)
3
2
1
0
−1
−2
−3
N=1535
N=341
N=21
N=85
N=15
N=81
N=160
Contr
T2D
SAID
SIDD
SIRD
MOD
MARD
Clusters

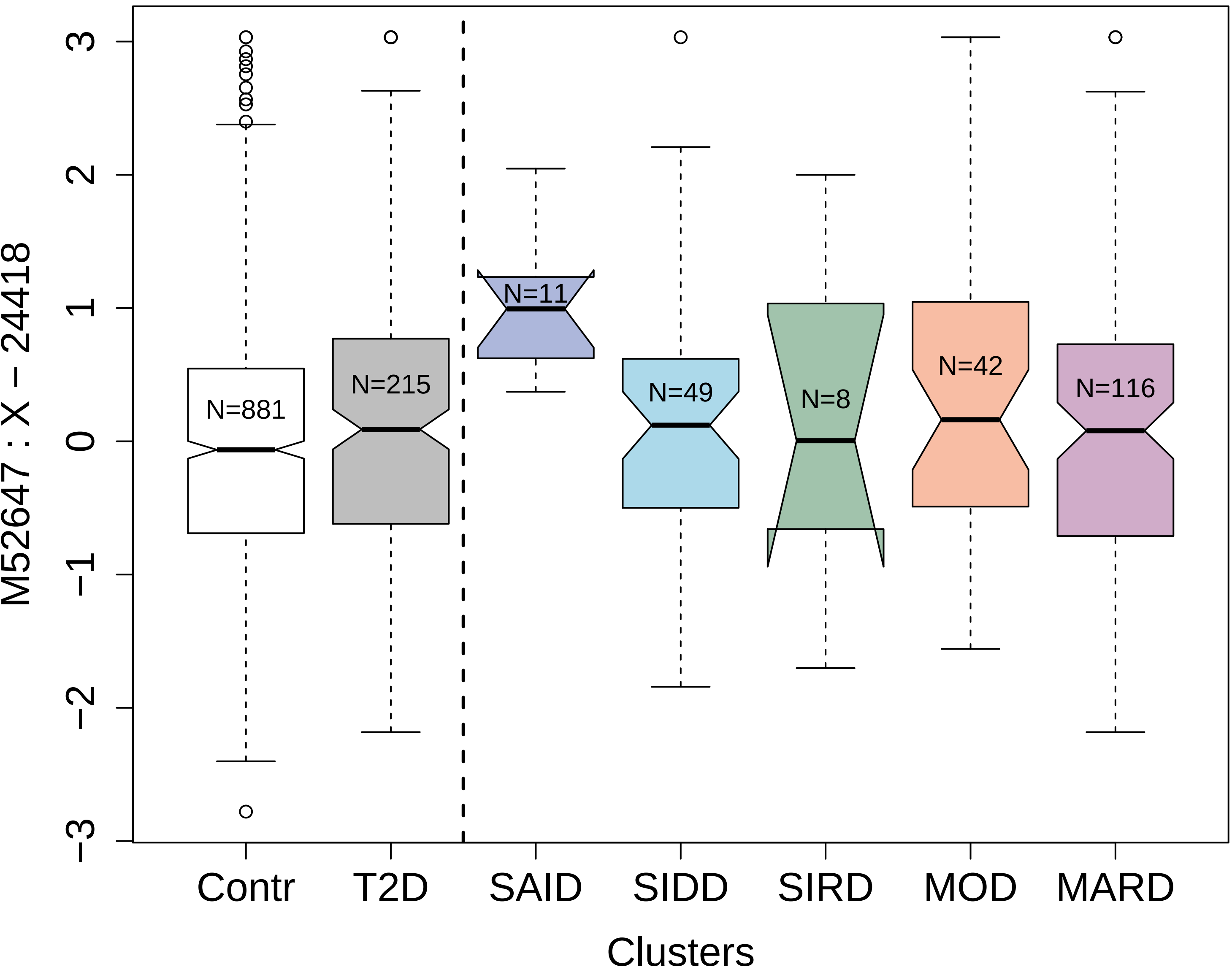

M52647 : X − 24418
3
2
1
0
−1
−2
−3
N=881
N=215
N=11
N=49
N=8
N=42
N=116
Contr
T2D
SAID
SIDD
SIRD
MOD
MARD
Clusters

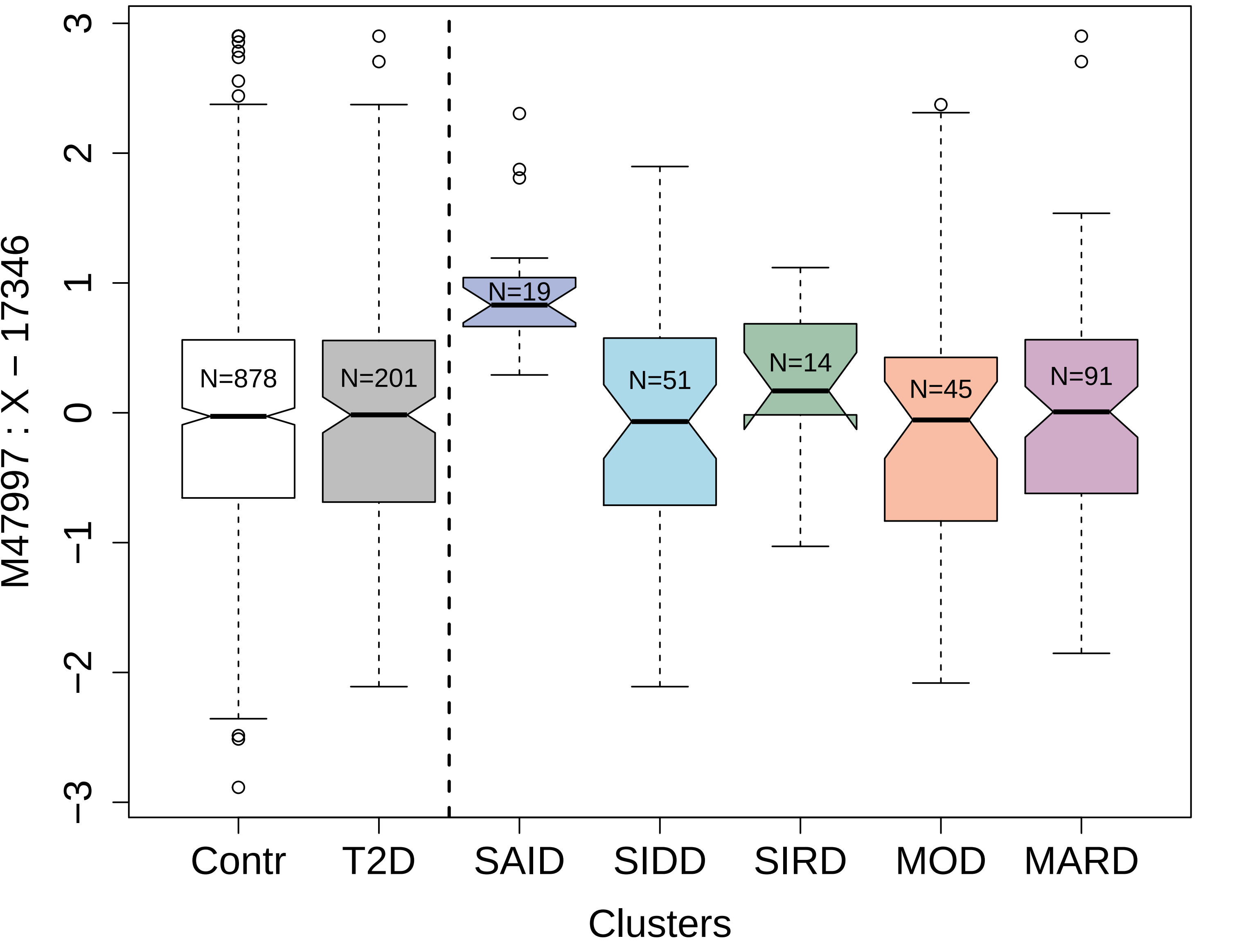
M47997 : X − 17346
3
2
1
0
−1
−2
−3
N=878
N=201
N=19
N=51
N=14
N=45
N=91
Contr
T2D
SAID
SIDD
SIRD
MOD
MARD
Clusters

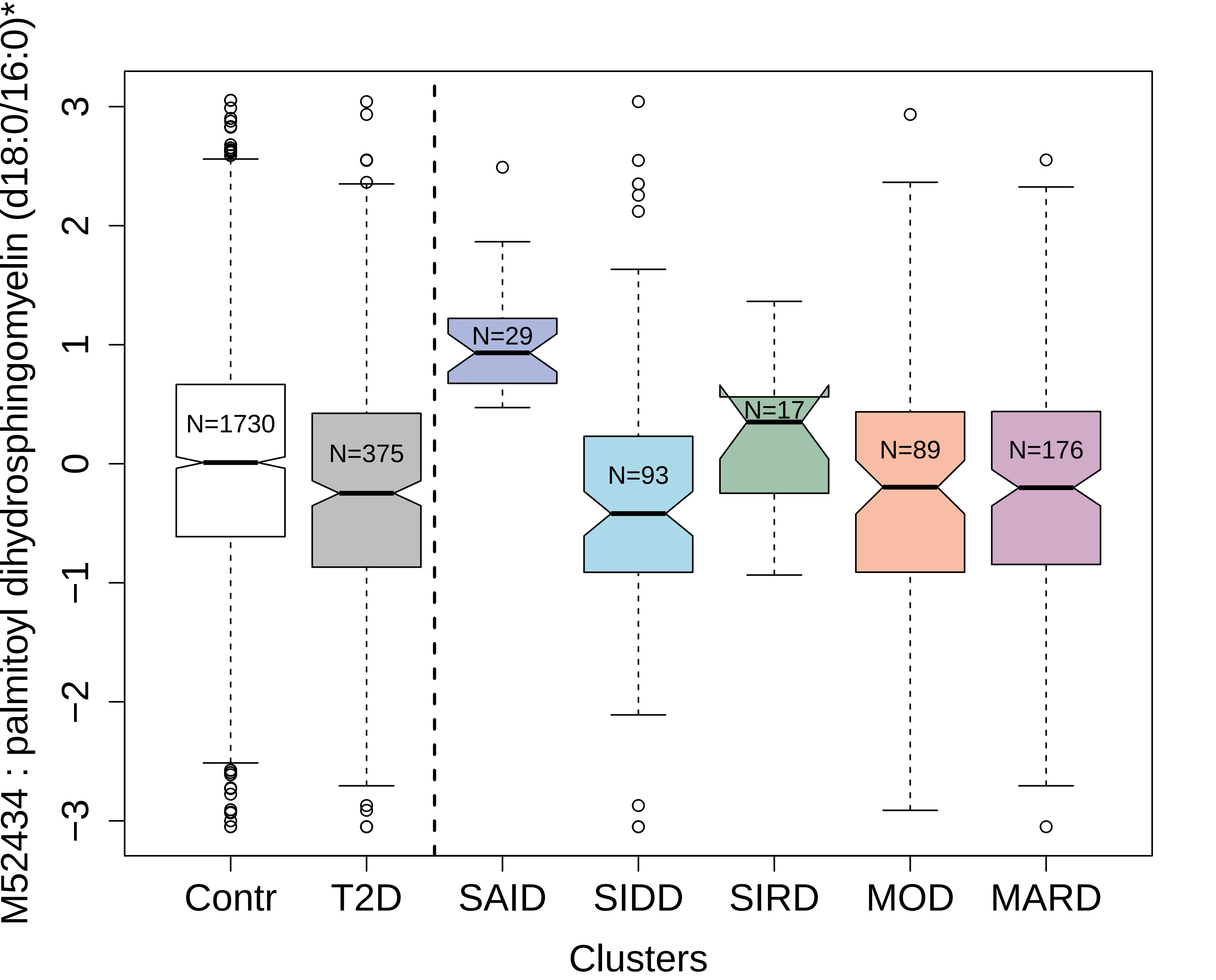

M52434 : palmitoyl dihydrosphingomyelin (d18:0/16:0)*
3
2
1
0
−1
−2
−3
N=1730
N=375
N=29
N=93
N=17
N=89
N=176
Contr
T2D
SAID
SIDD
SIRD
MOD
MARD
Clusters

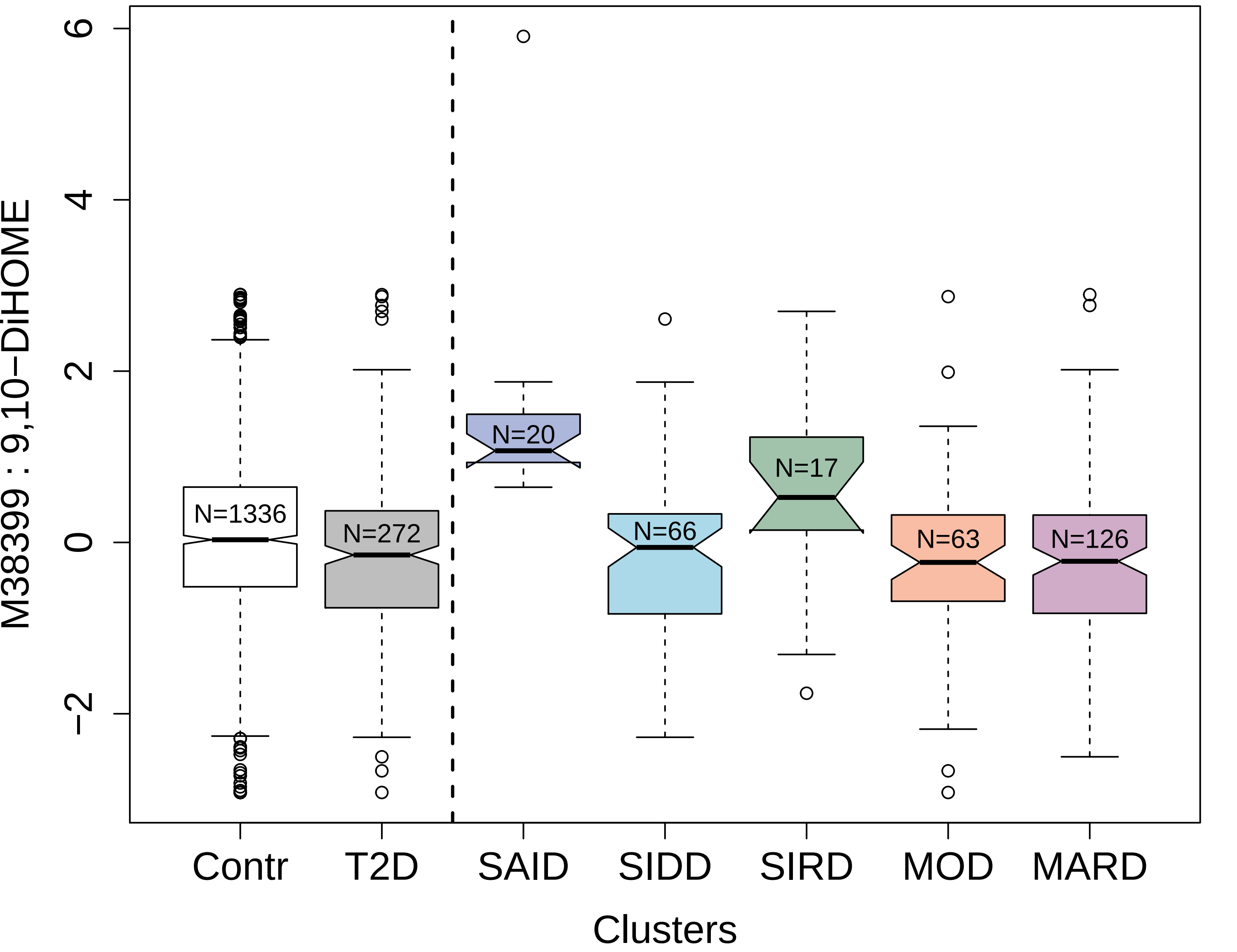

M38399 : 9,10−DiHOME
6
4
2
0
−2
N=1336
N=272
N=20
N=66
N=17
N=63
N=126
Contr
T2D
SAID
SIDD
SIRD
MOD
MARD
Clusters

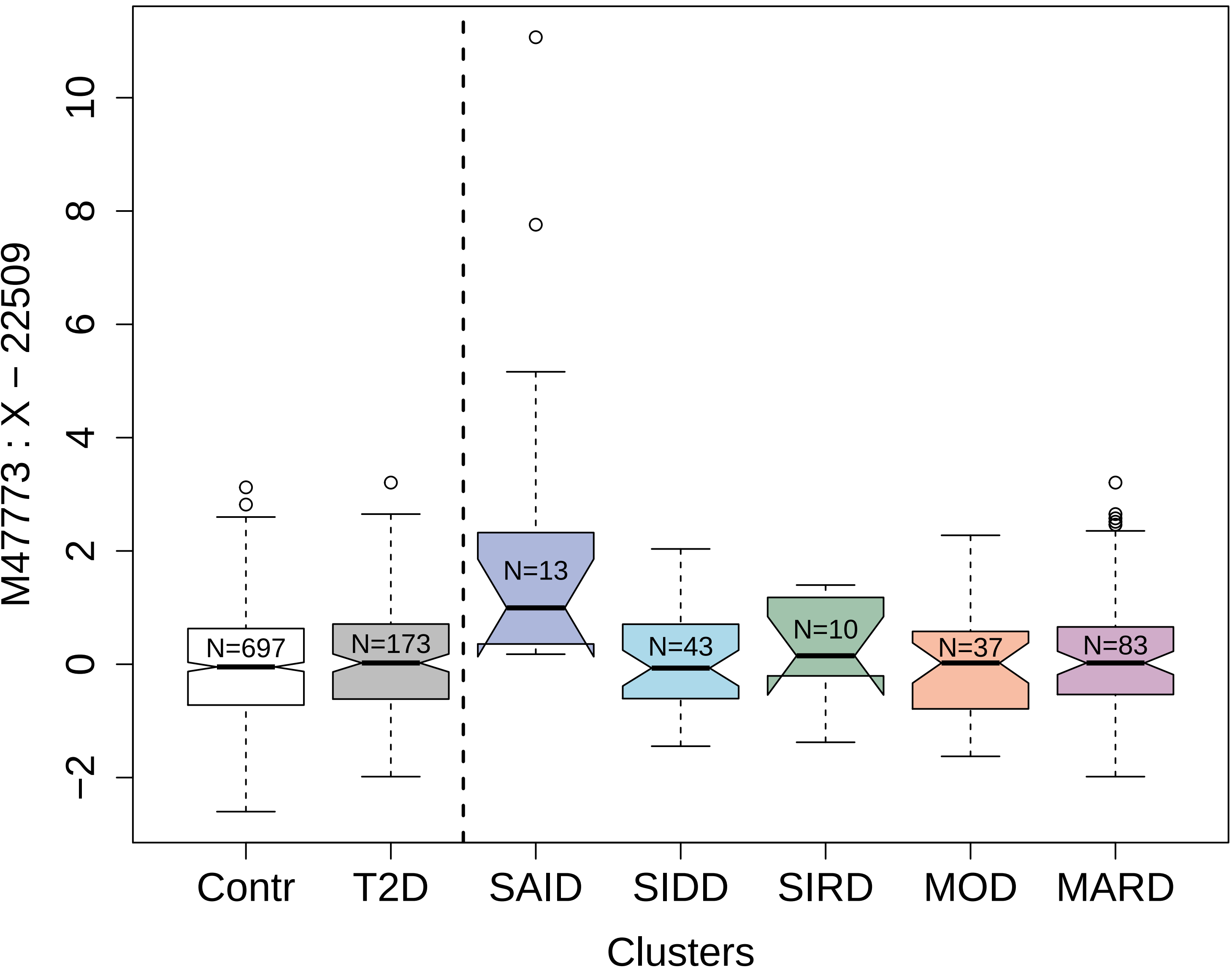

M47773 : X − 22509
10
8
6
4
2
0
−2
N=697
N=173
N=13
N=43
N=10
N=37
N=83
Contr
T2D
SAID
SIDD
SIRD
MOD
MARD
Clusters

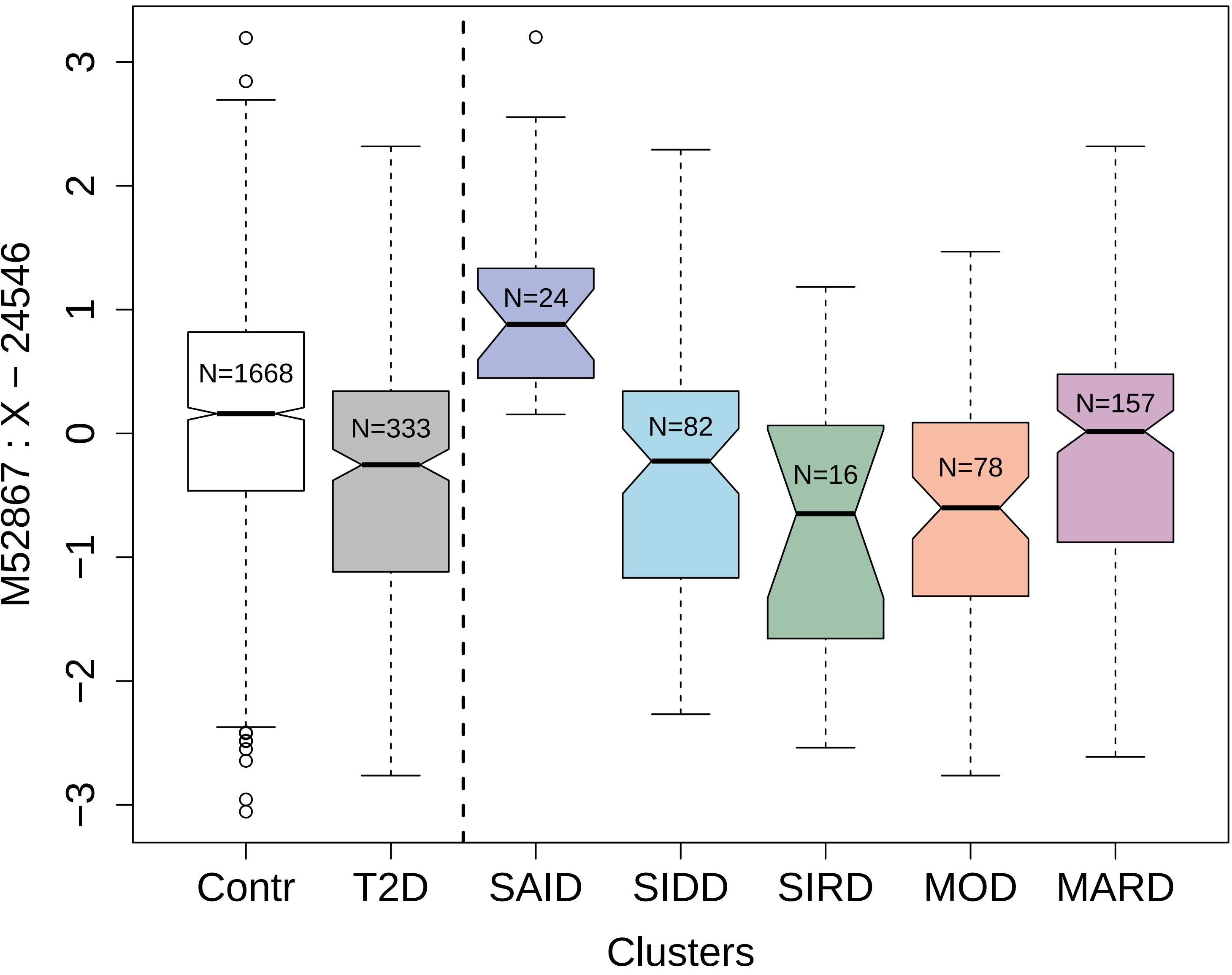
M52867 : X − 24546
3
2
1
0
−1
−2
−3
N=1668
N=333
N=24
N=82
N=16
N=78
N=157
Contr
T2D
SAID
SIDD
SIRD
MOD
MARD
Clusters

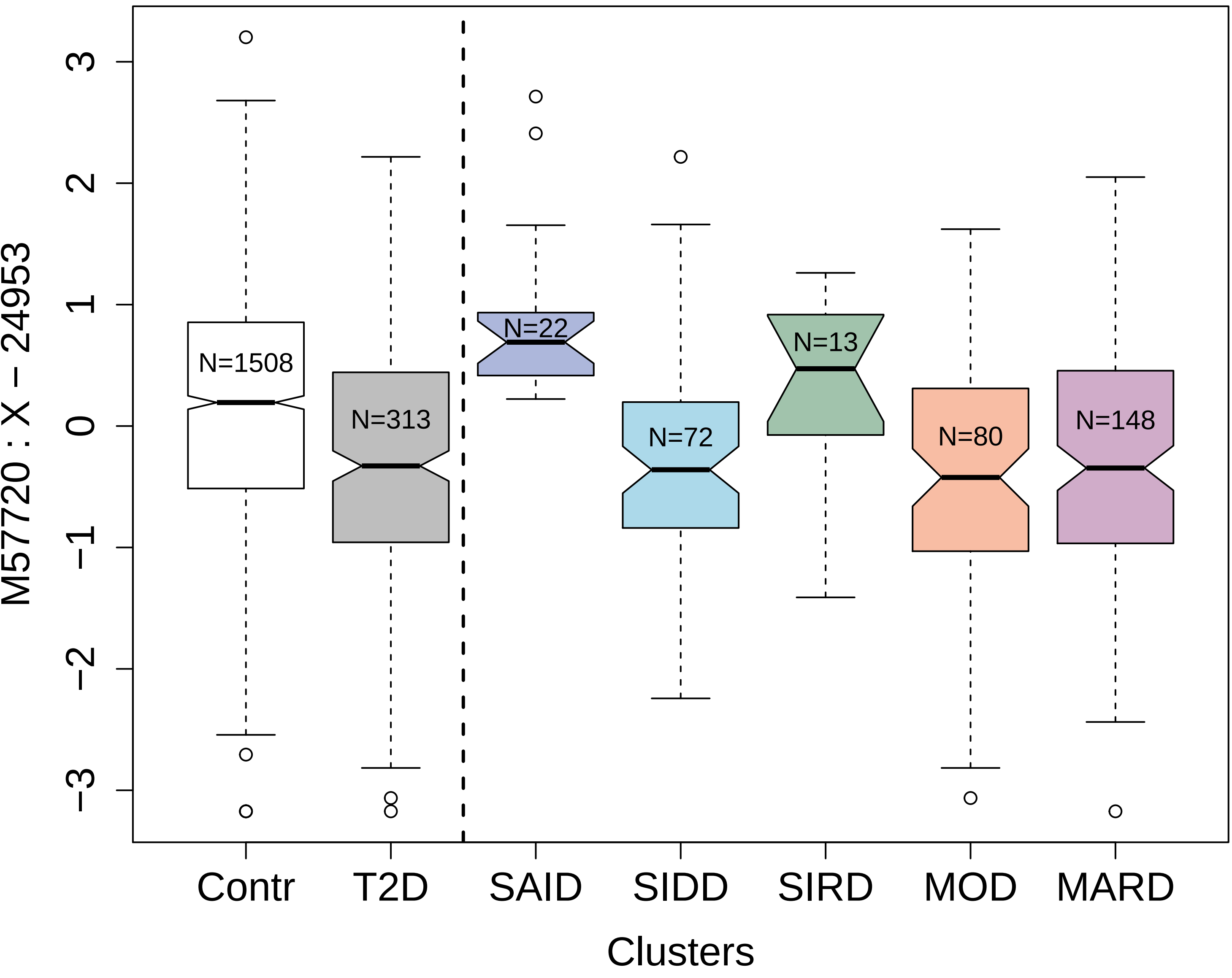

M57720 : X − 24953
3
2
1
0
−1
−2
−3
N=1508
N=313
N=22
N=72
N=13
N=80
N=148
Contr
T2D
SAID
SIDD
SIRD
MOD
MARD
Clusters

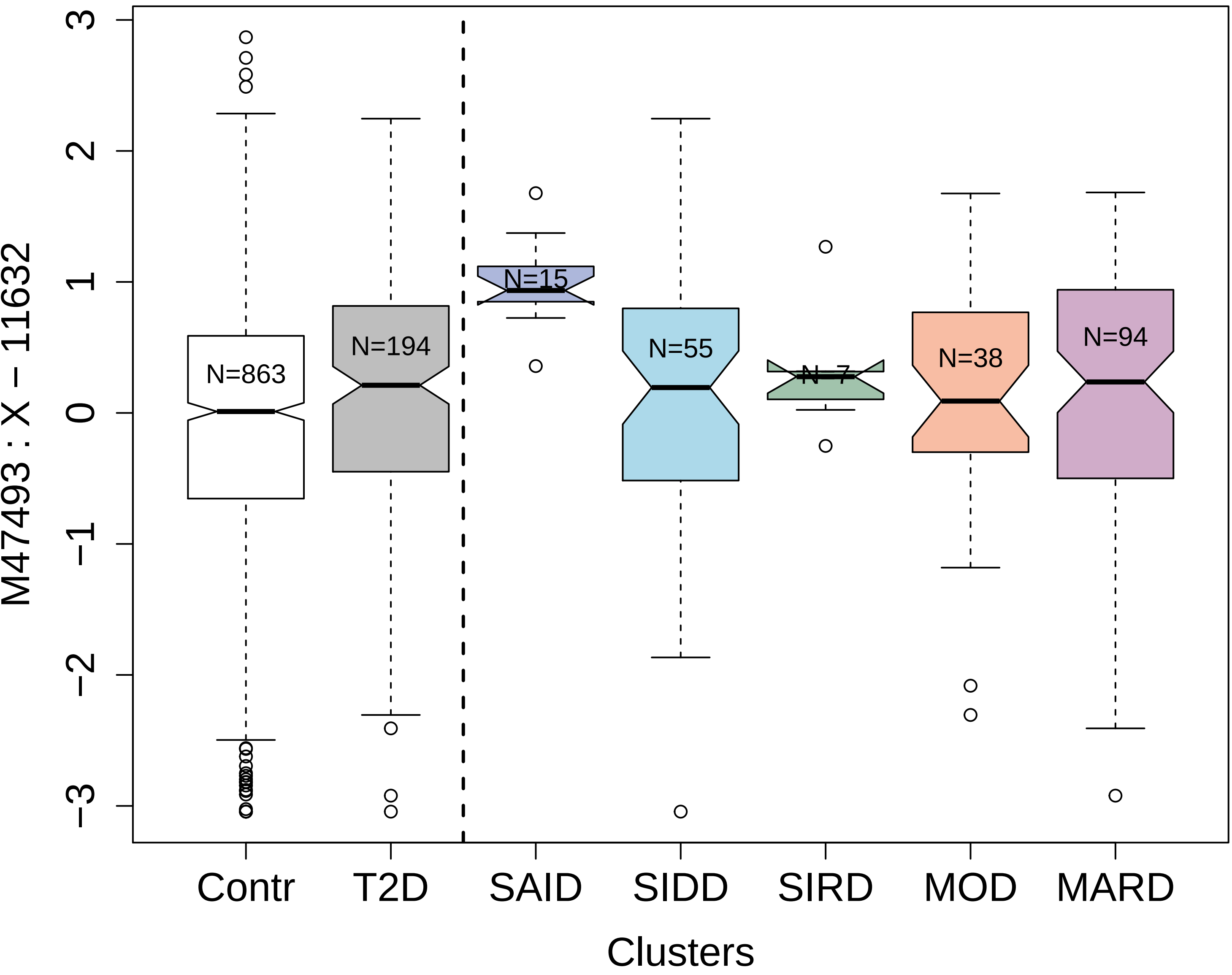

M47493 : X − 11632
3
2
1
0
−1
−2
−3
N=863
N=194
N=15
N=55
N=7
N=38
N=94
Contr
T2D
SAID
SIDD
SIRD
MOD
MARD
Clusters

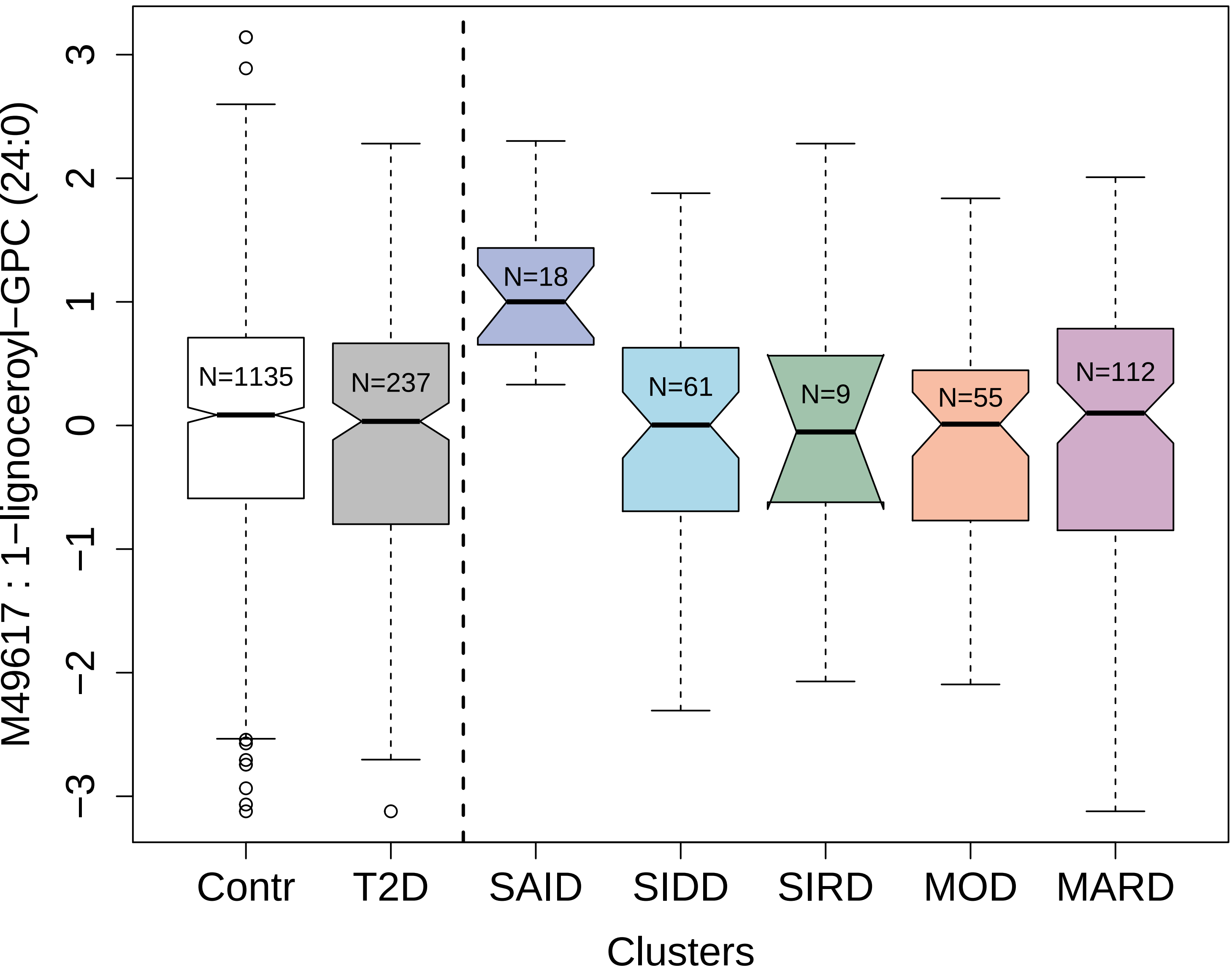
M49617 : 1−lignoceroyl−GPC (24:0)
3
2
1
0
−1
−2
−3
N=1135
N=237
N=18
N=61
N=9
N=55
N=112
Contr
T2D
SAID
SIDD
SIRD
MOD
MARD
Clusters

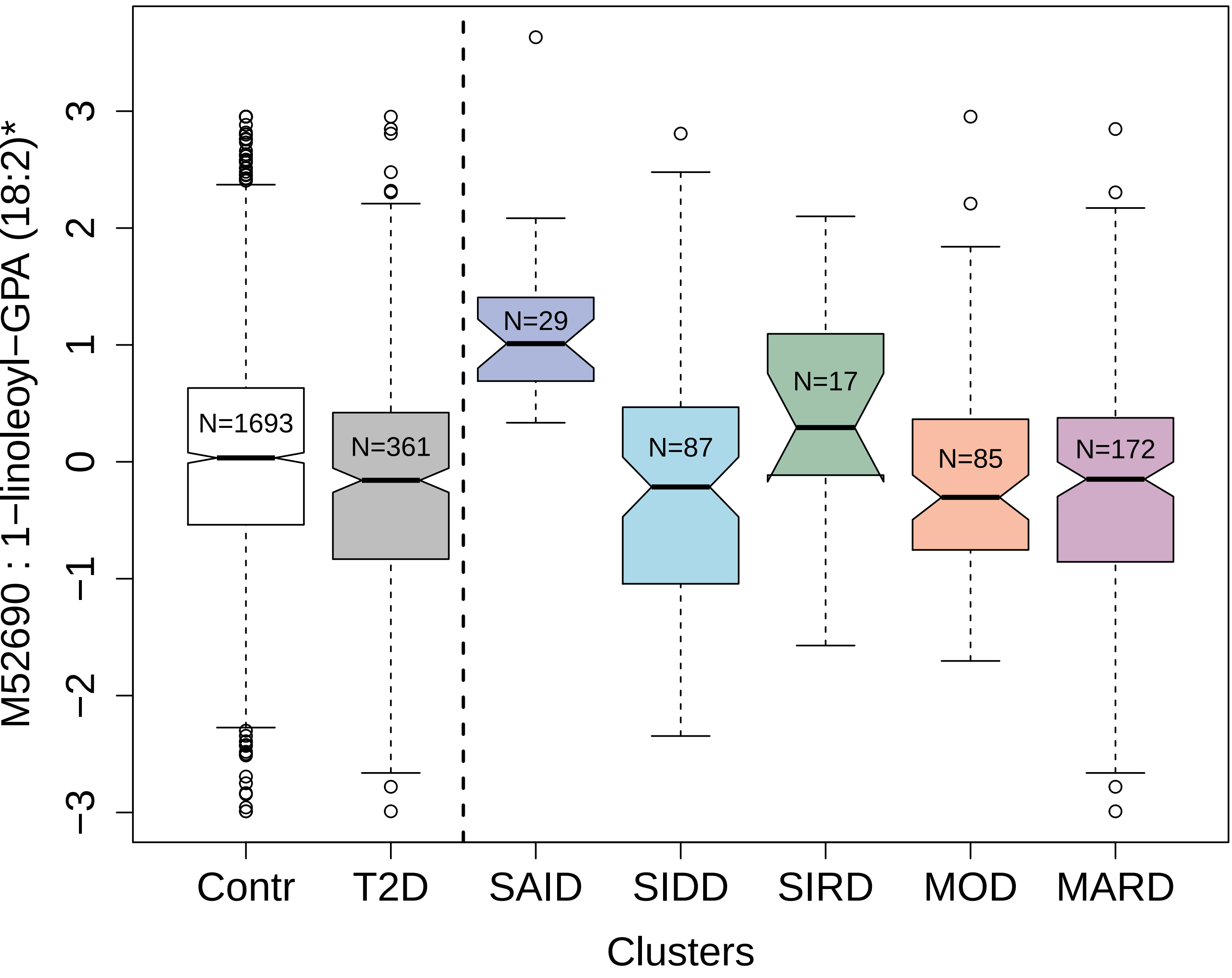

M52690 : 1−linoleoyl−GPA (18:2)*
3
2
1
0
−1
−2
−3
N=1693
N=361
N=29
N=87
N=17
N=85
N=172
Contr
T2D
SAID
SIDD
SIRD
MOD
MARD
Clusters

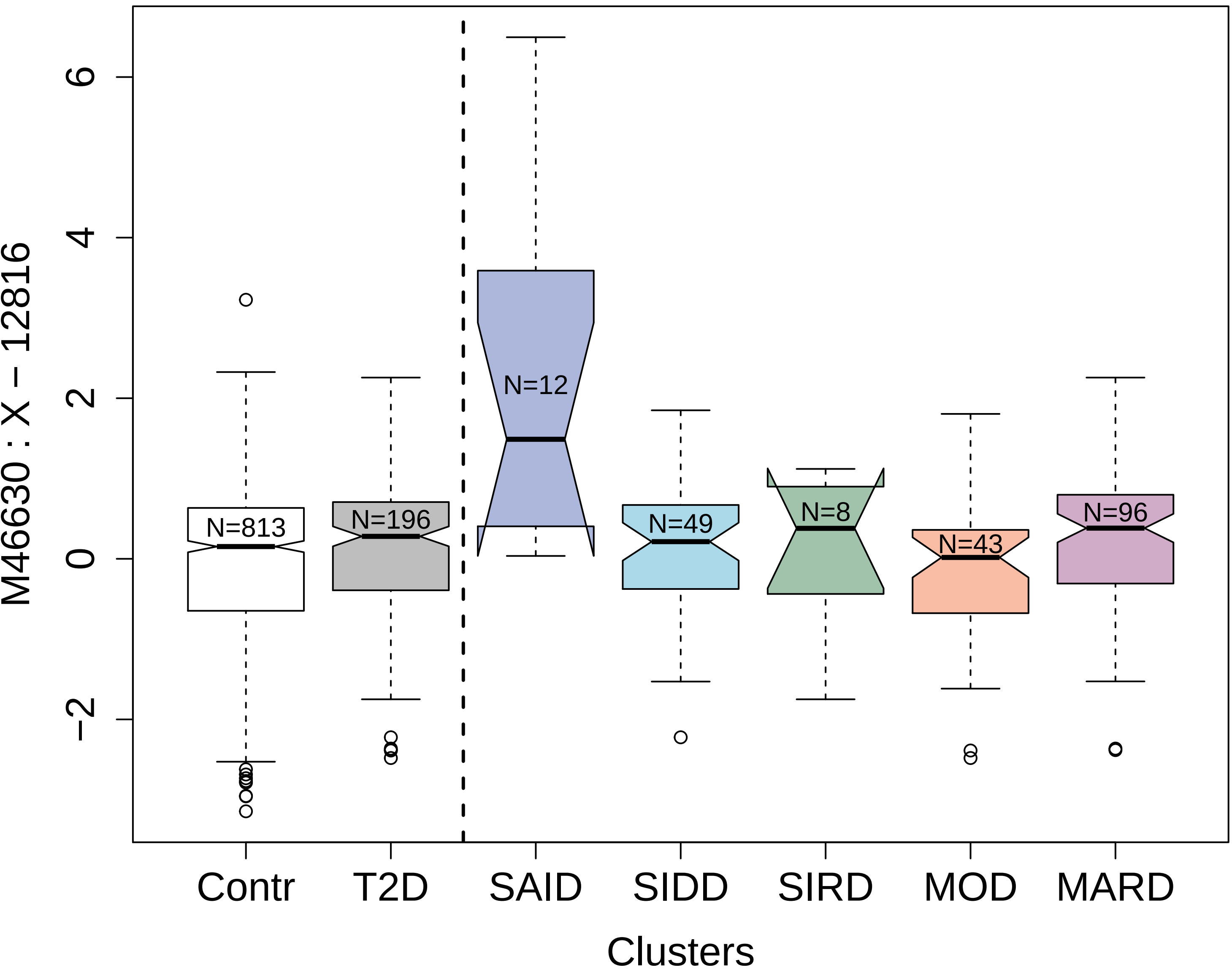

M46630 : X − 12816
6
4
2
0
−2
N=813
N=196
N=12
N=49
N=8
N=43
N=96
Contr
T2D
SAID
SIDD
SIRD
MOD
MARD
Clusters

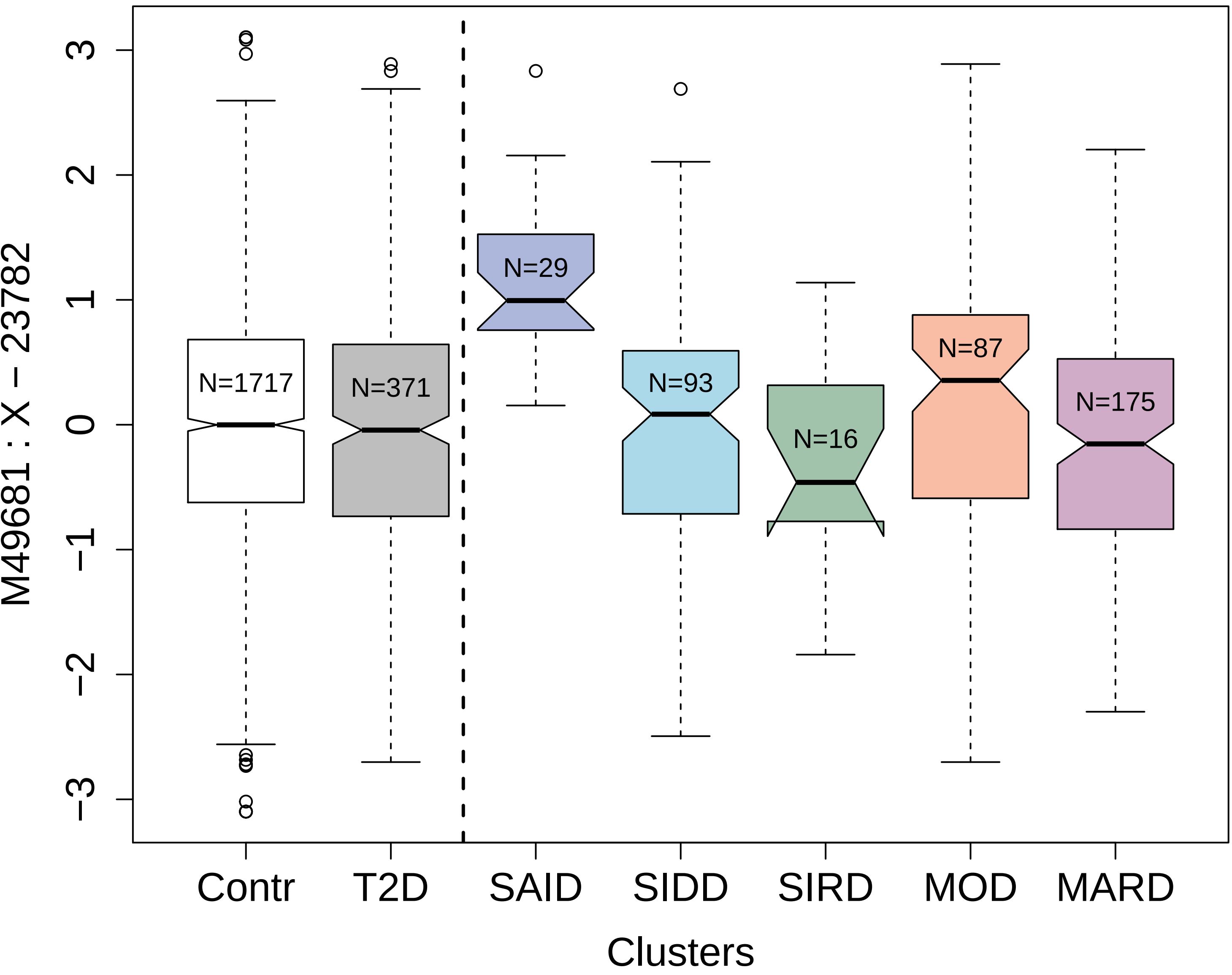
M49681 : X − 23782
3
2
1
0
−1
−2
−3
N=1717
N=371
N=29
N=93
N=16
N=87
N=175
Contr
T2D
SAID
SIDD
SIRD
MOD
MARD
Clusters

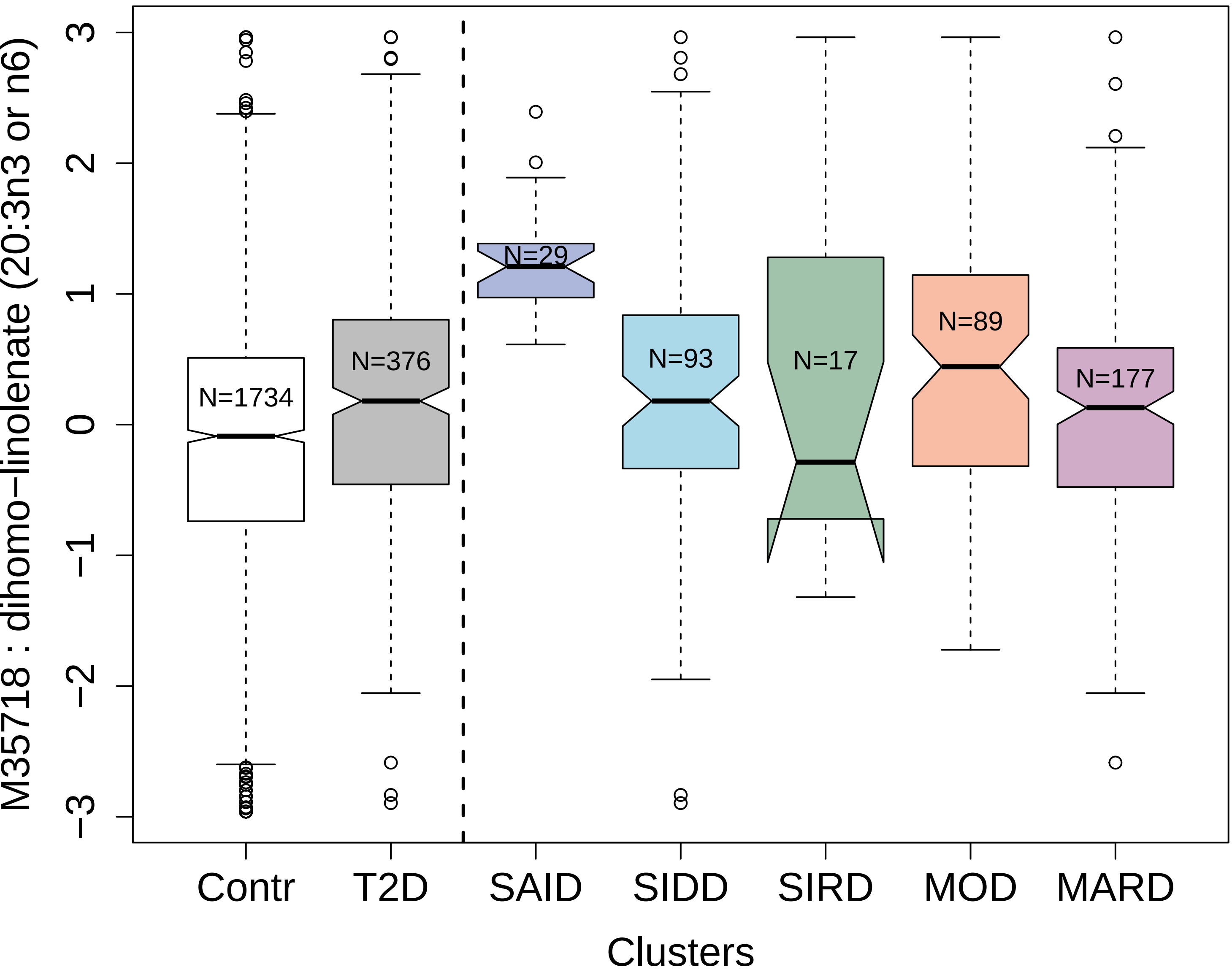
M35718 : dihomo−linolenate (20:3n3 or n6)
3
2
1
0
−1
−2
−3
N=1734
N=376
N=29
N=93
N=17
N=89
N=177
Contr
T2D
SAID
SIDD
SIRD
MOD
MARD
Clusters

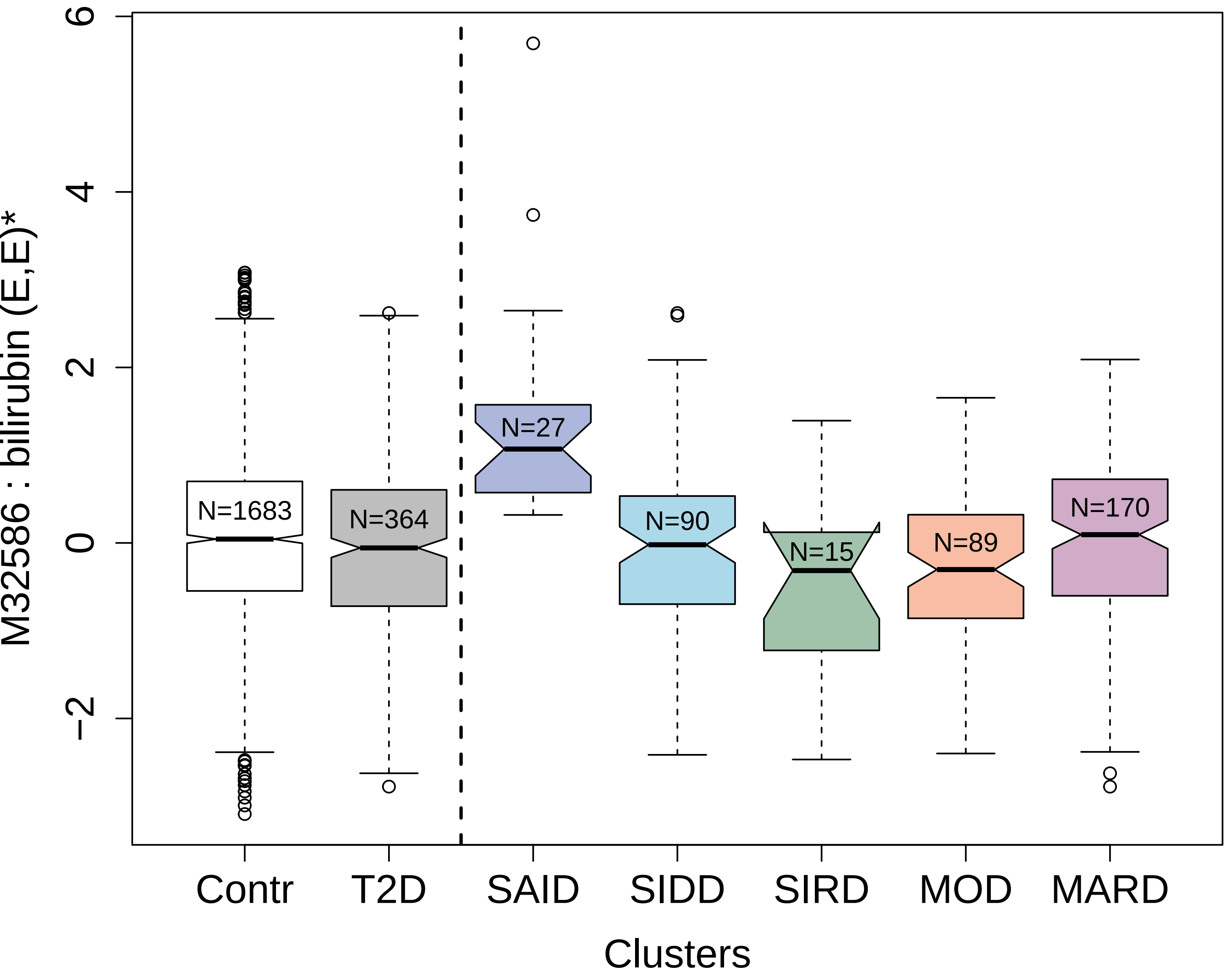

M32586 : bilirubin (E,E)*
6
4
2
0
−2
N=1683
N=364
N=27
N=90
N=15
N=89
N=170
Contr
T2D
SAID
SIDD
SIRD
MOD
MARD
Clusters

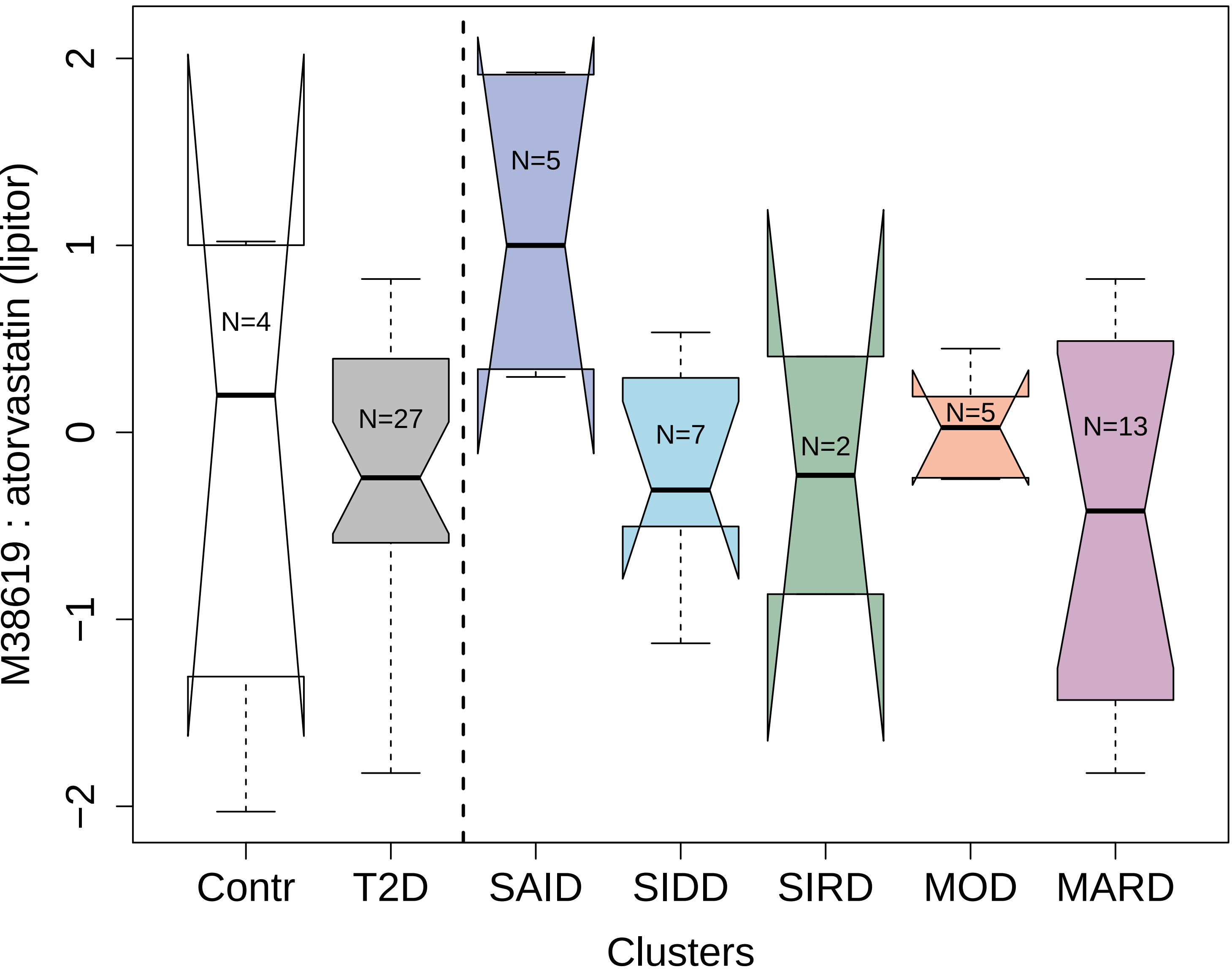

M38619 : atorvastatin (lipitor)
Clusters
Contr
T2D
SAID
SIDD
SIRD
MOD
MARD
N=4
N=27
N=5
N=7
N=2
N=5
N=13
2
1
0
−1
−2

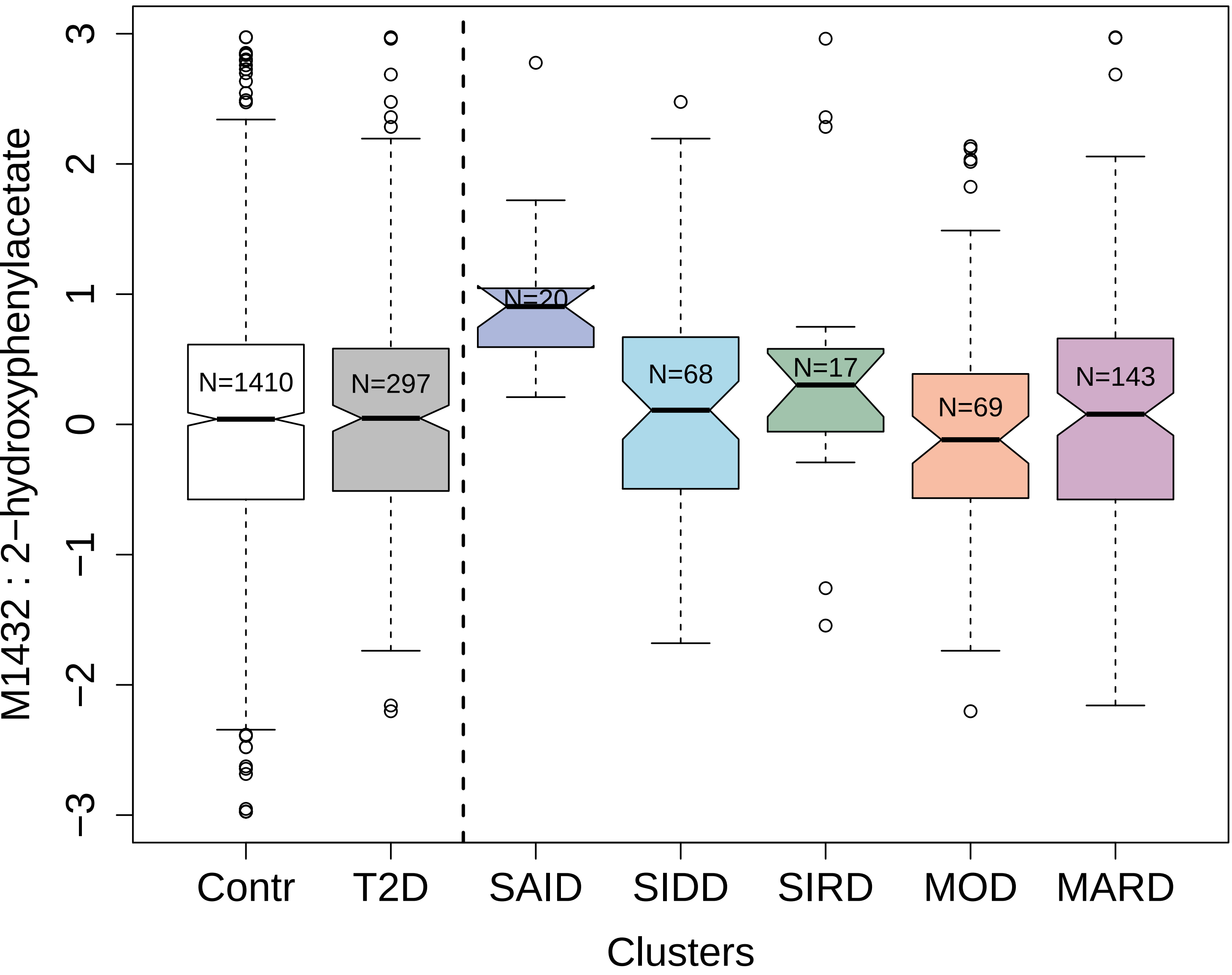
M1432 : 2−hydroxyphenylacetate
3
2
1
0
−1
−2
−3
N=1410
N=297
N=20
N=68
N=17
N=69
N=143
Contr
T2D
SAID
SIDD
SIRD
MOD
MARD
Clusters

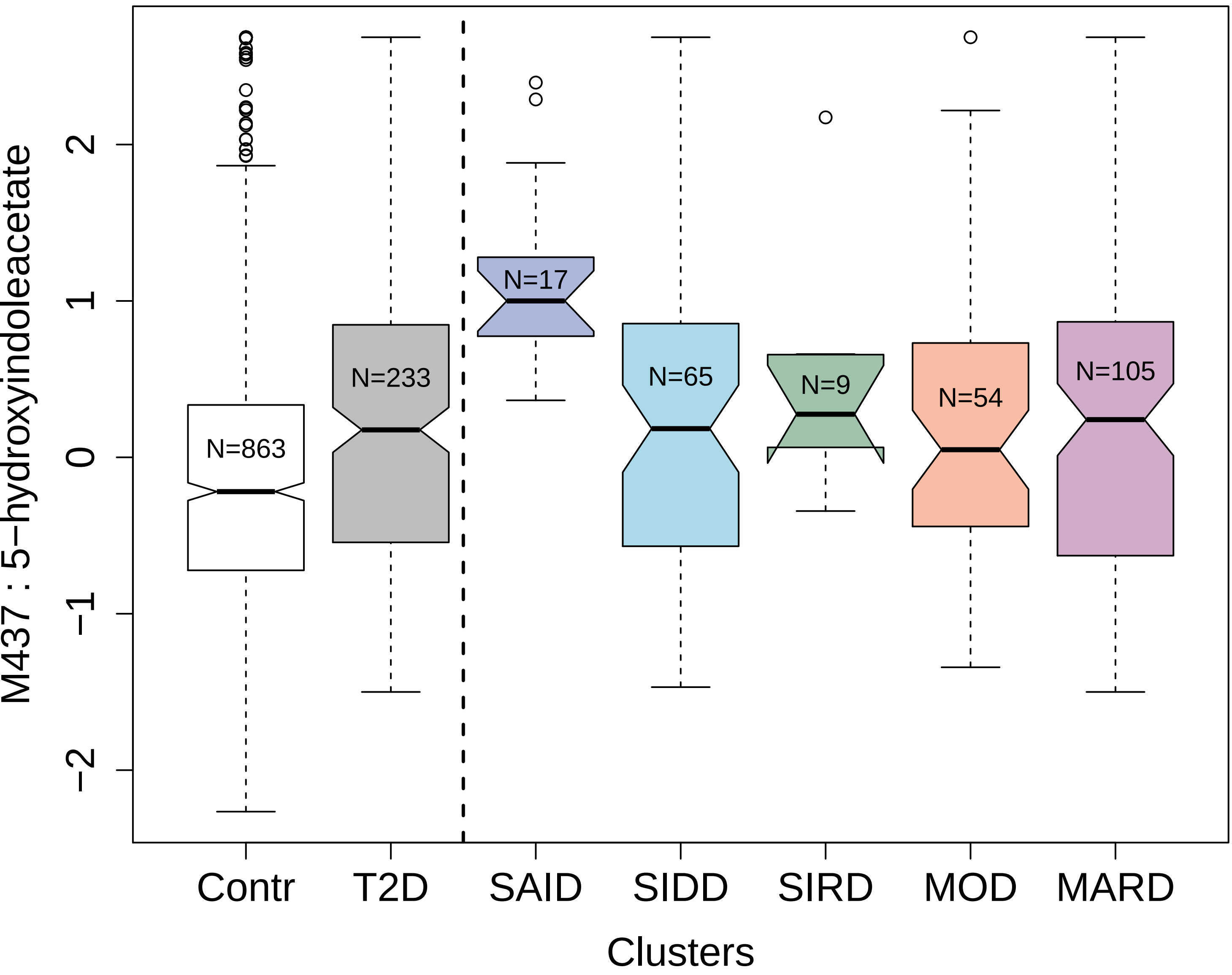

M437 : 5−hydroxyindoleacetate
2
1
0
−1
−2
N=863
N=233
N=17
N=65
N=9
N=54
N=105
Contr
T2D
SAID
SIDD
SIRD
MOD
MARD
Clusters

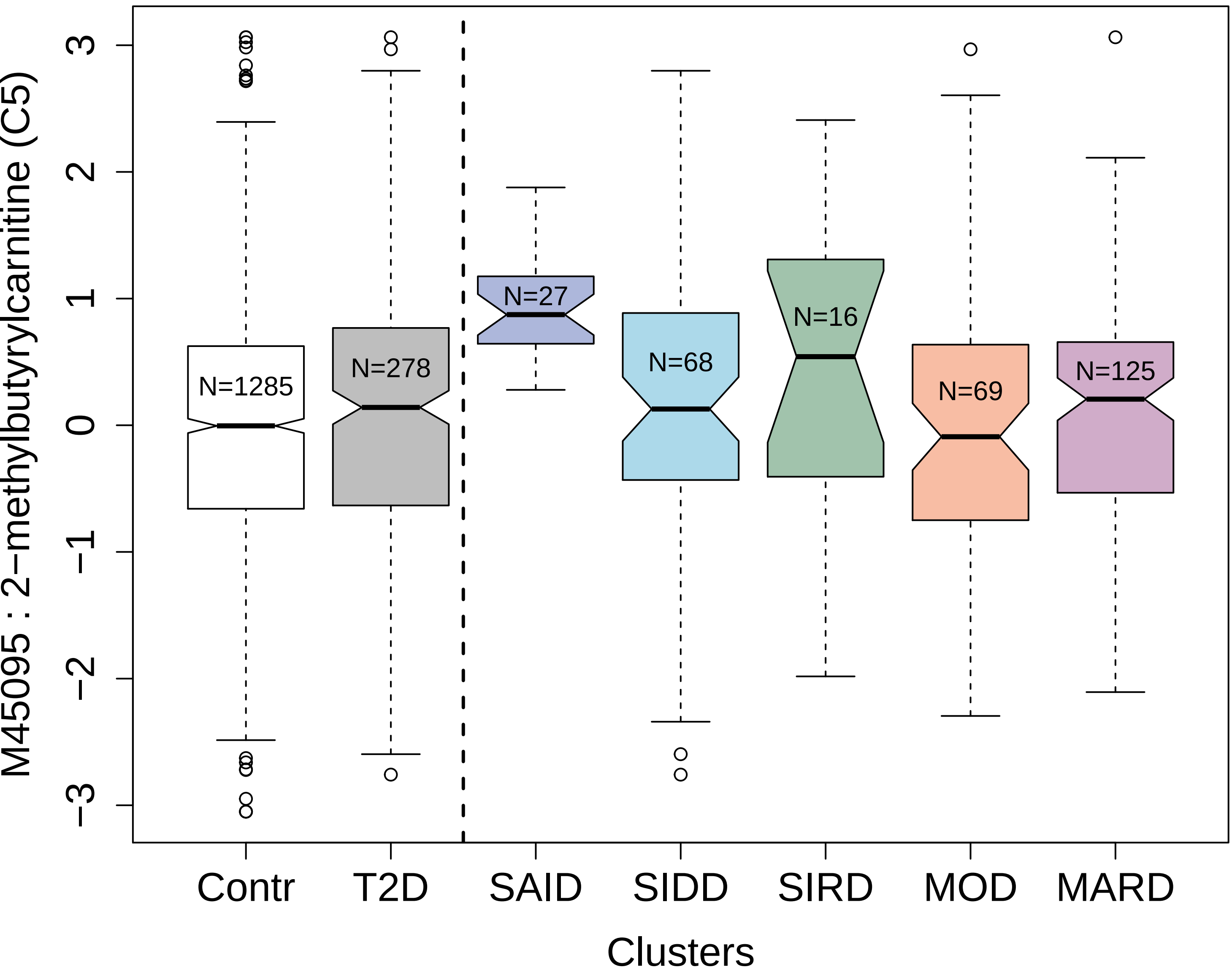
M45095 : 2−methylbutyrylcarnitine (C5)
3
2
1
0
−1
−2
−3
N=1285
N=278
N=27
N=68
N=16
N=69
N=125
Contr
T2D
SAID
SIDD
SIRD
MOD
MARD
Clusters

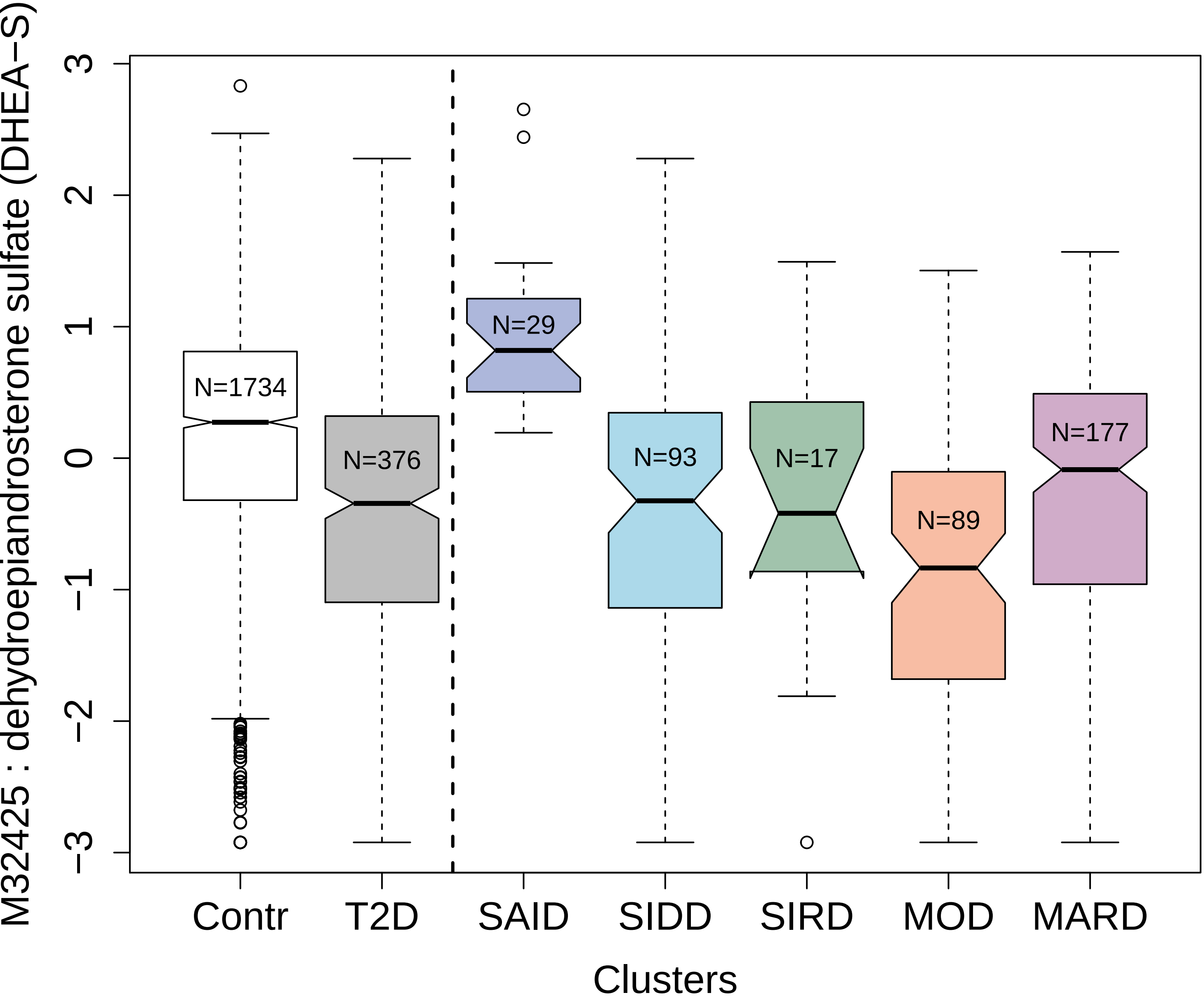

M32425 : dehydroepiandrosterone sulfate (DHEA−S)
3
2
1
0
−1
−2
−3
N=1734
N=376
N=29
N=93
N=17
N=89
N=177
Contr
T2D
SAID
SIDD
SIRD
MOD
MARD
Clusters

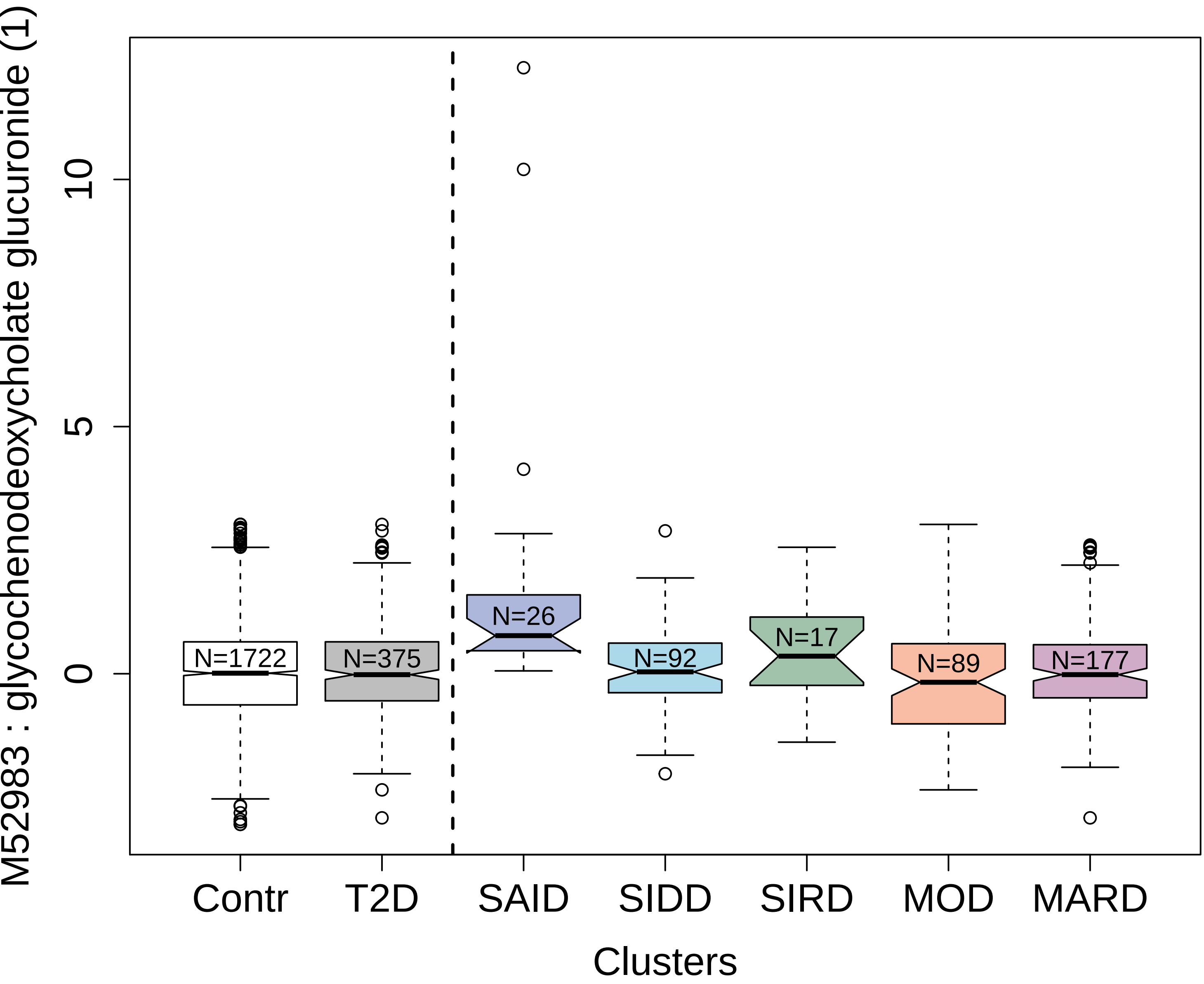
M52983 : glycochenodeoxycholate glucuronide (1)
0
5
10
N=1722
N=375
N=26
N=92
N=17
N=89
N=177
Contr
T2D
SAID
SIDD
SIRD
MOD
MARD
Clusters

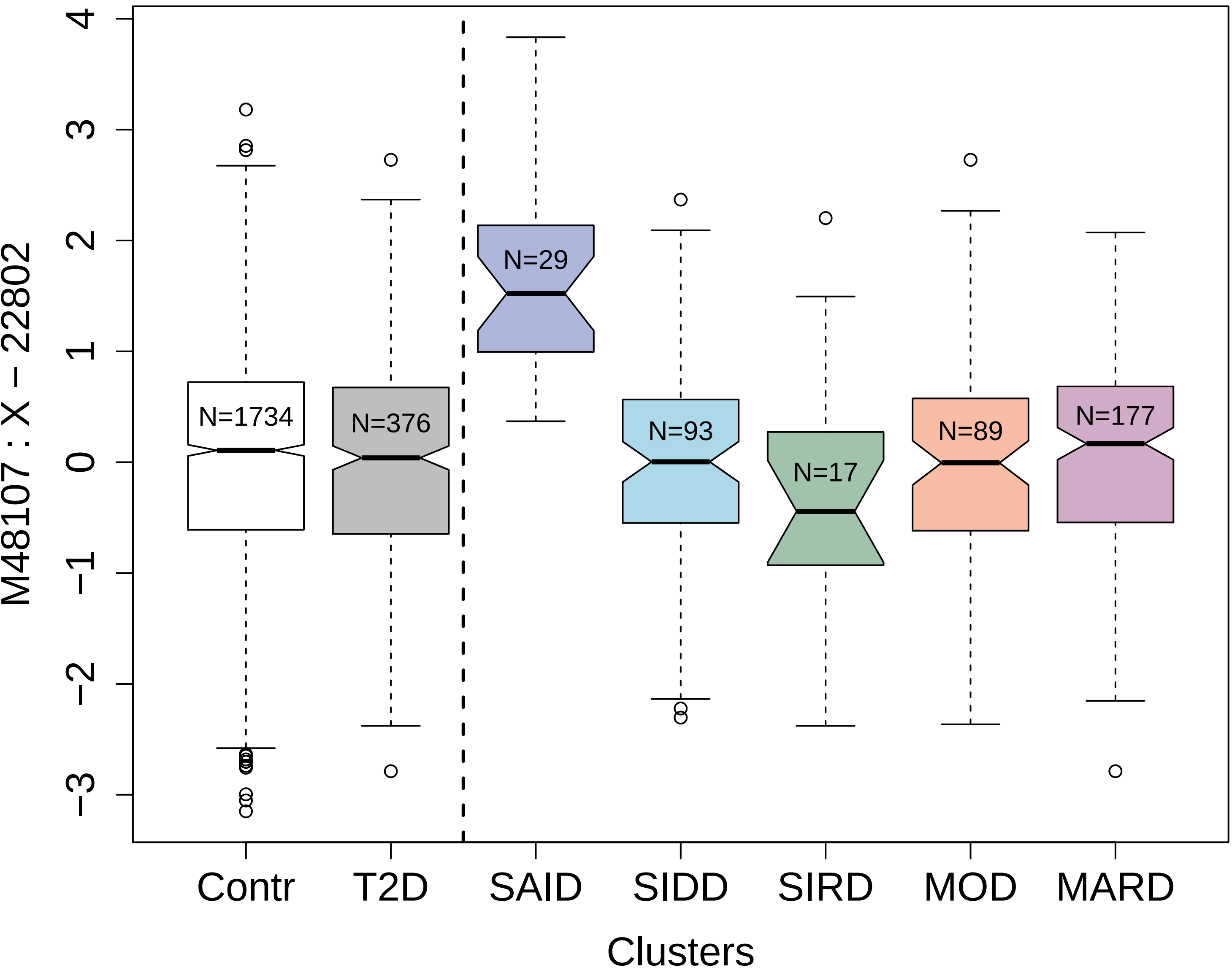

M48107 : X − 22802
4
3
2
1
0
−1
−2
−3
N=1734
N=376
N=29
N=93
N=17
N=89
N=177
Contr
T2D
SAID
SIDD
SIRD
MOD
MARD
Clusters

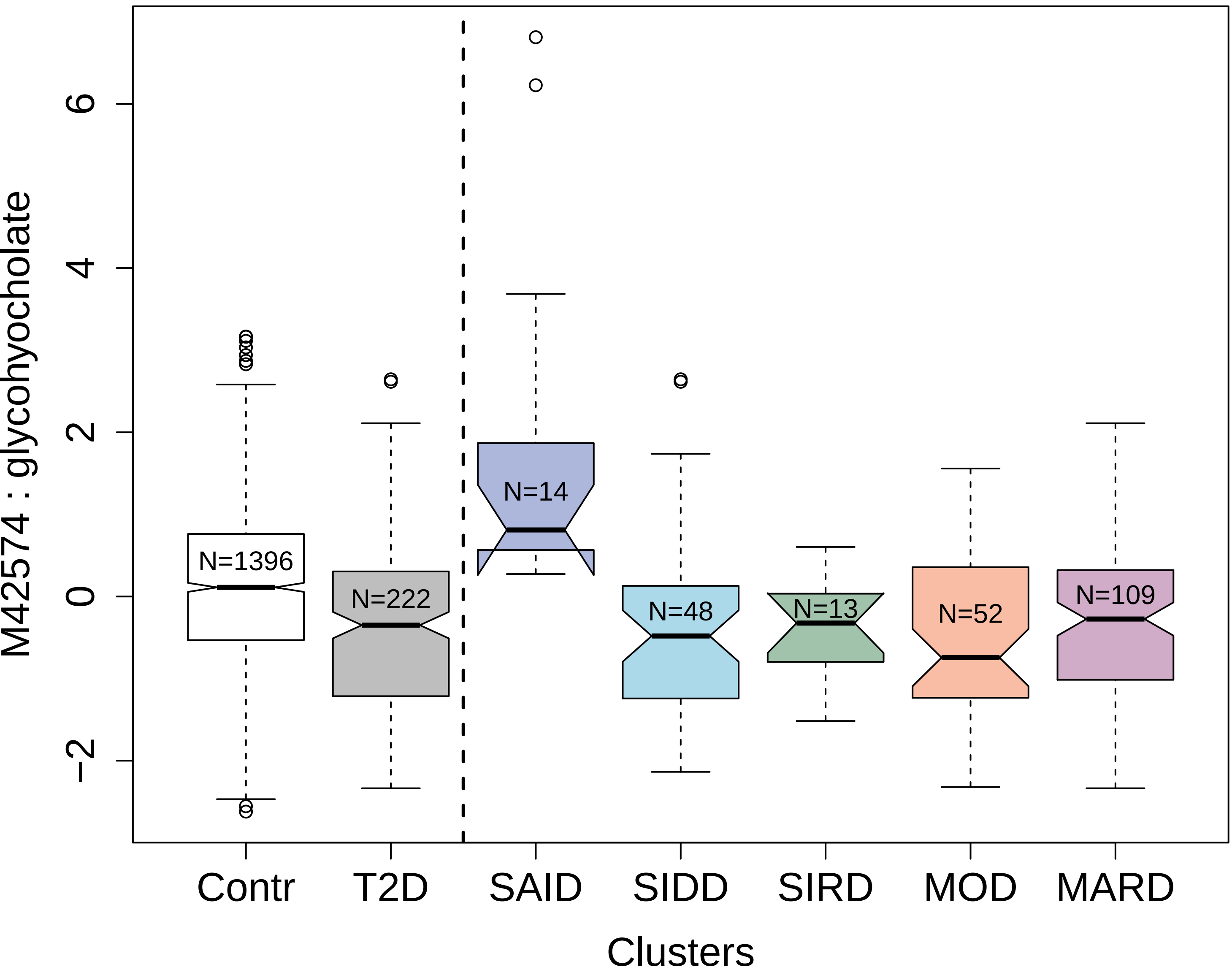
M42574 : glycohyocholate
6
4
2
0
−2
N=1396
N=222
N=14
N=48
N=13
N=52
N=109
Contr
T2D
SAID
SIDD
SIRD
MOD
MARD
Clusters

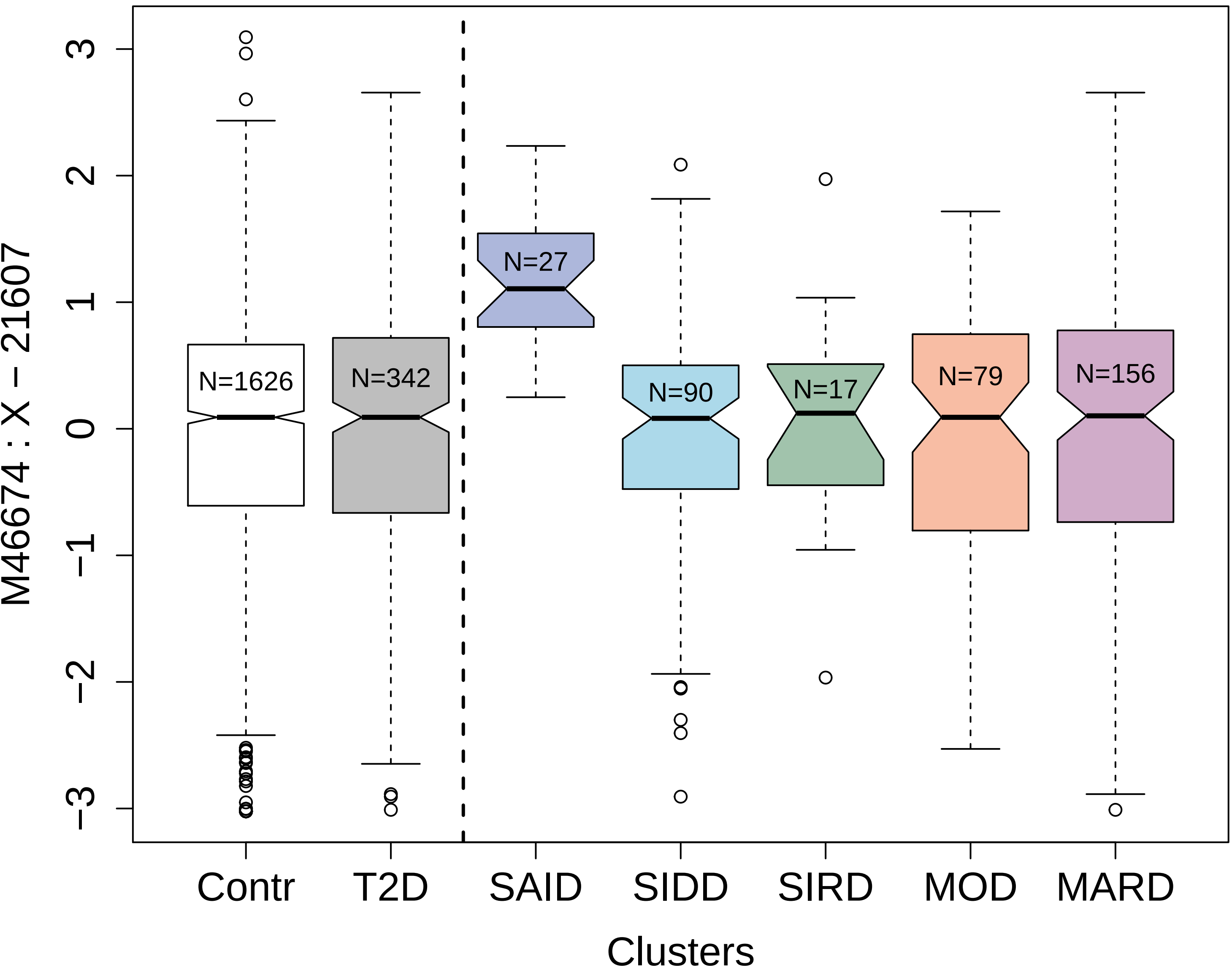
M46674 : X − 21607
3
2
1
0
−1
−2
−3
N=1626
N=342
N=27
N=90
N=17
N=79
N=156
Contr
T2D
SAID
SIDD
SIRD
MOD
MARD
Clusters

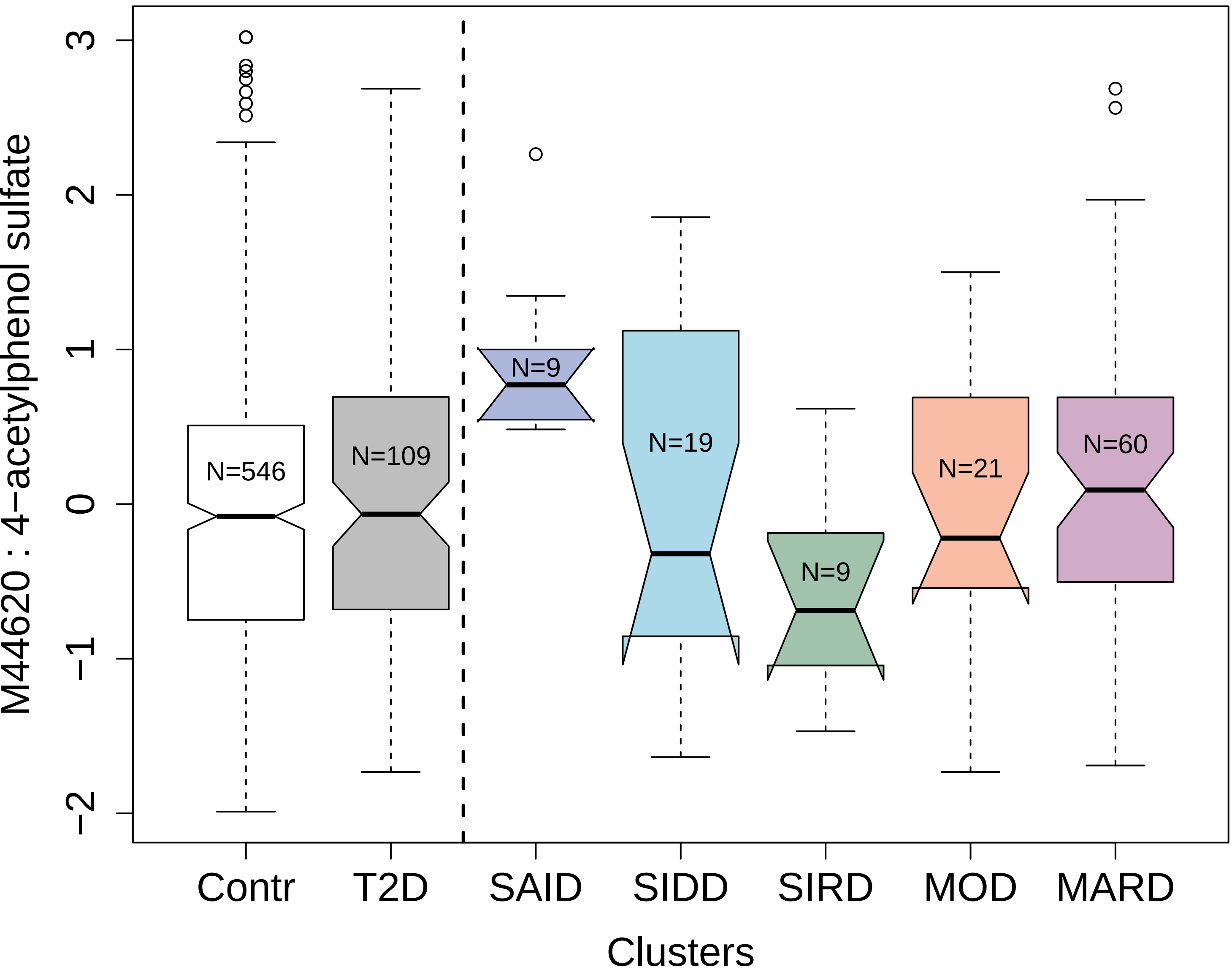
M44620 : 4−acetylphenol sulfate
Clusters
Contr
T2D
SAID
SIDD
SIRD
MOD
MARD
N=546
N=109
N=9
N=19
N=9
N=21
N=60
−2
−1
0
1
2
3

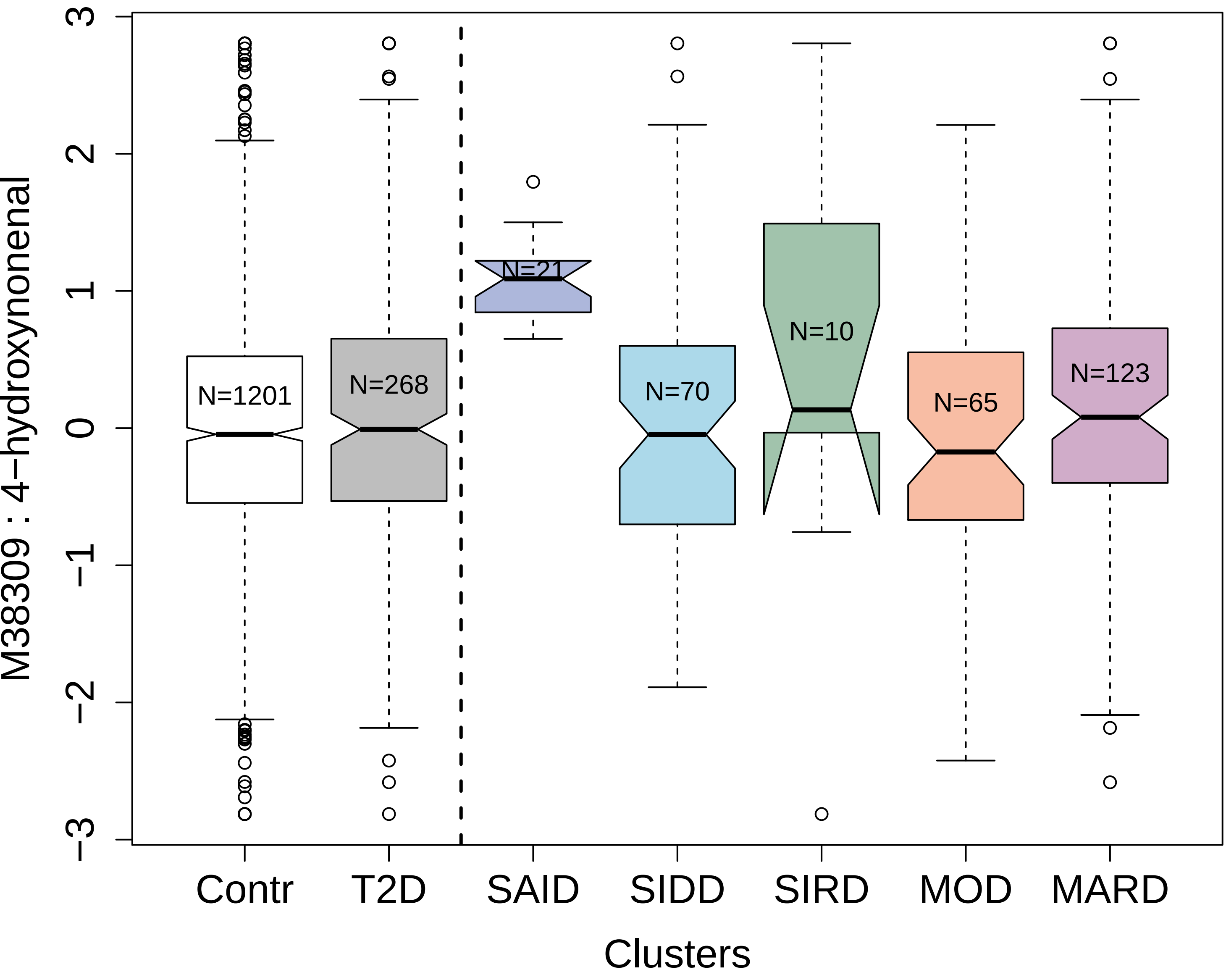

M38309 : 4−hydroxynonenal
3
2
1
0
−1
−2
−3
N=1201
N=268
N=21
N=70
N=10
N=65
N=123
Contr
T2D
SAID
SIDD
SIRD
MOD
MARD
Clusters

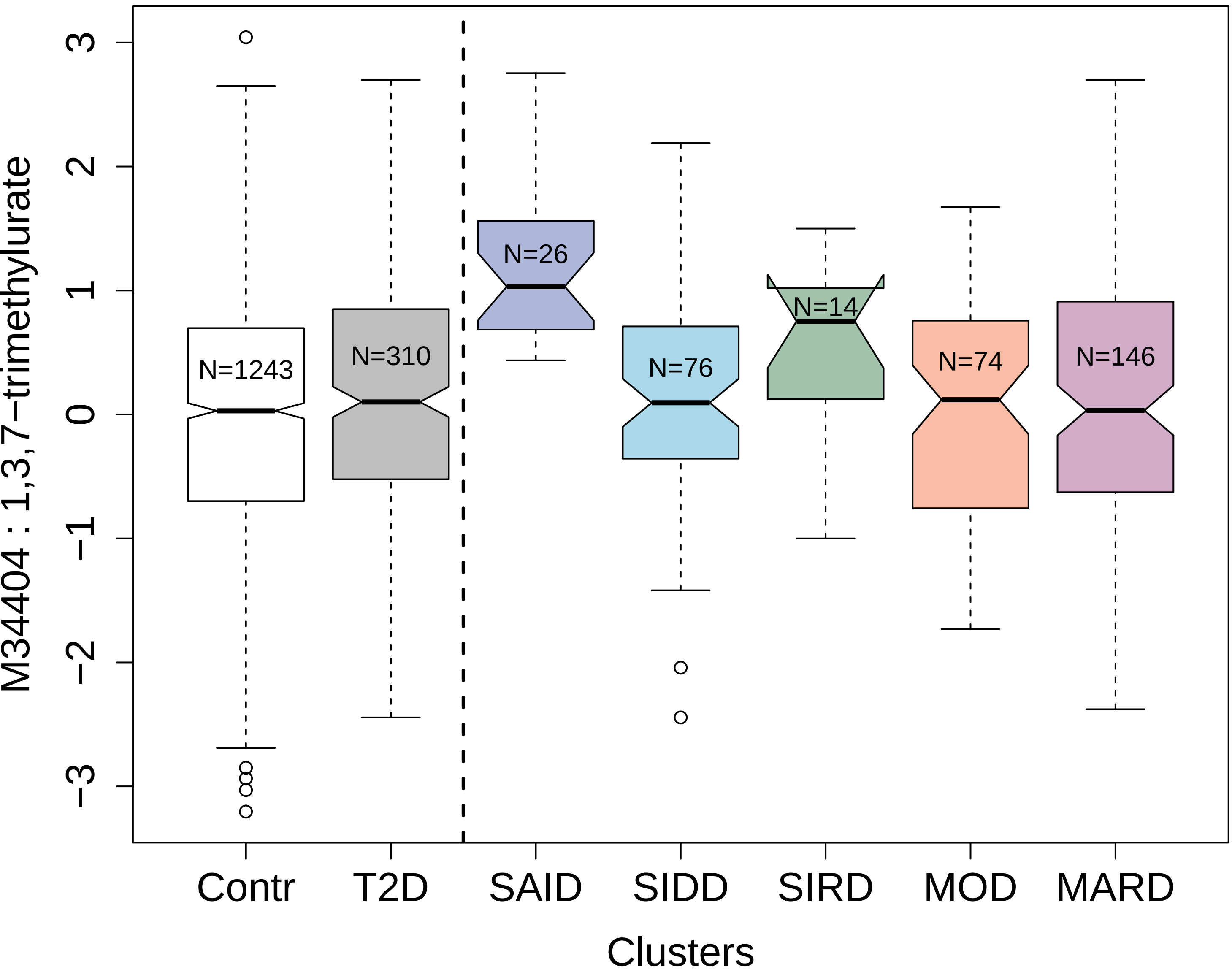
M34404 : 1,3,7−trimethylurate
3
2
1
0
−1
−2
−3
N=1243
N=310
N=26
N=76
N=14
N=74
N=146
Contr
T2D
SAID
SIDD
SIRD
MOD
MARD
Clusters

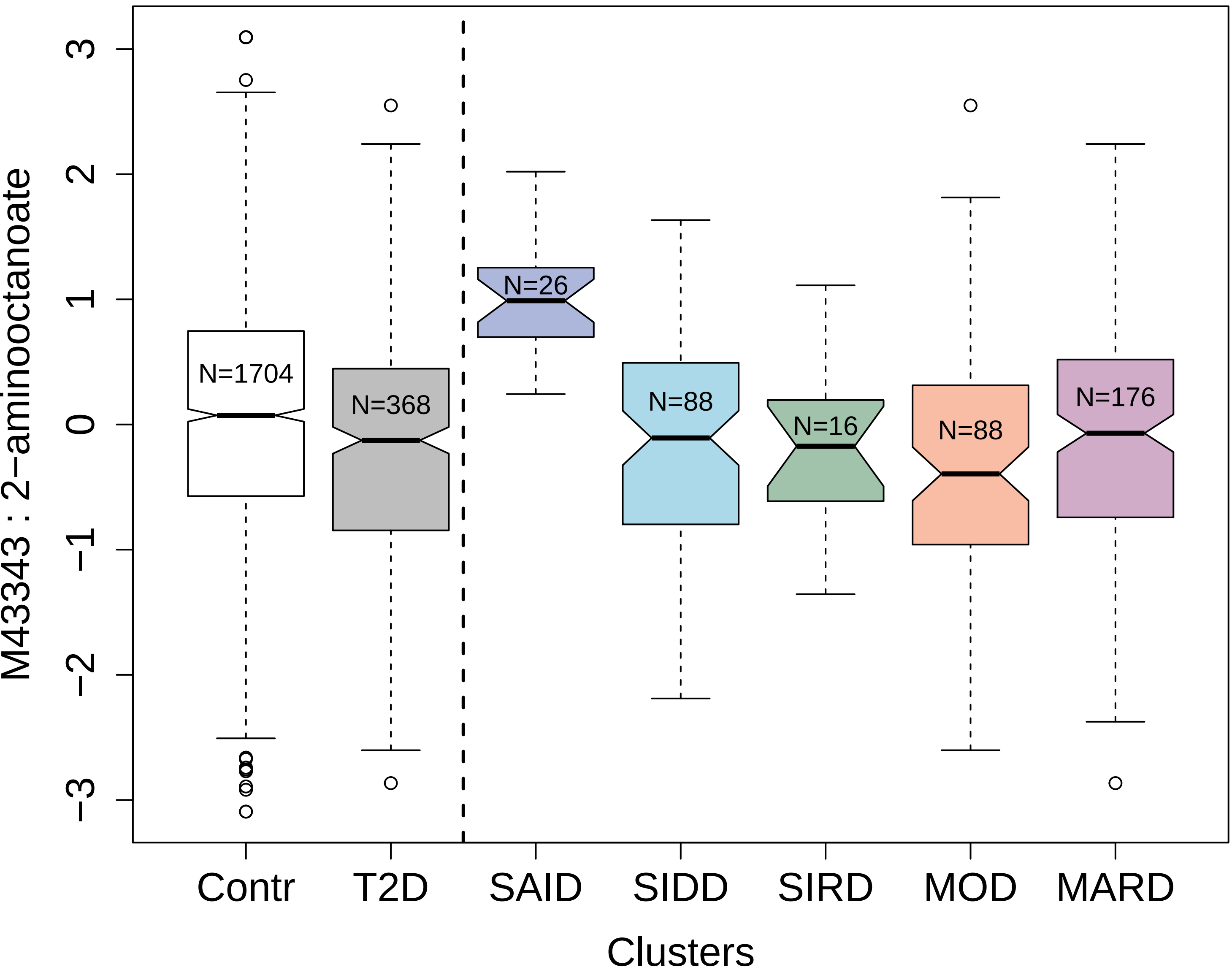

M43343 : 2−aminooctanoate
Clusters
Contr
T2D
SAID
SIDD
SIRD
MOD
MARD
N=1704
N=368
N=26
N=88
N=16
N=88
N=176
3
2
1
0
−1
−2
−3

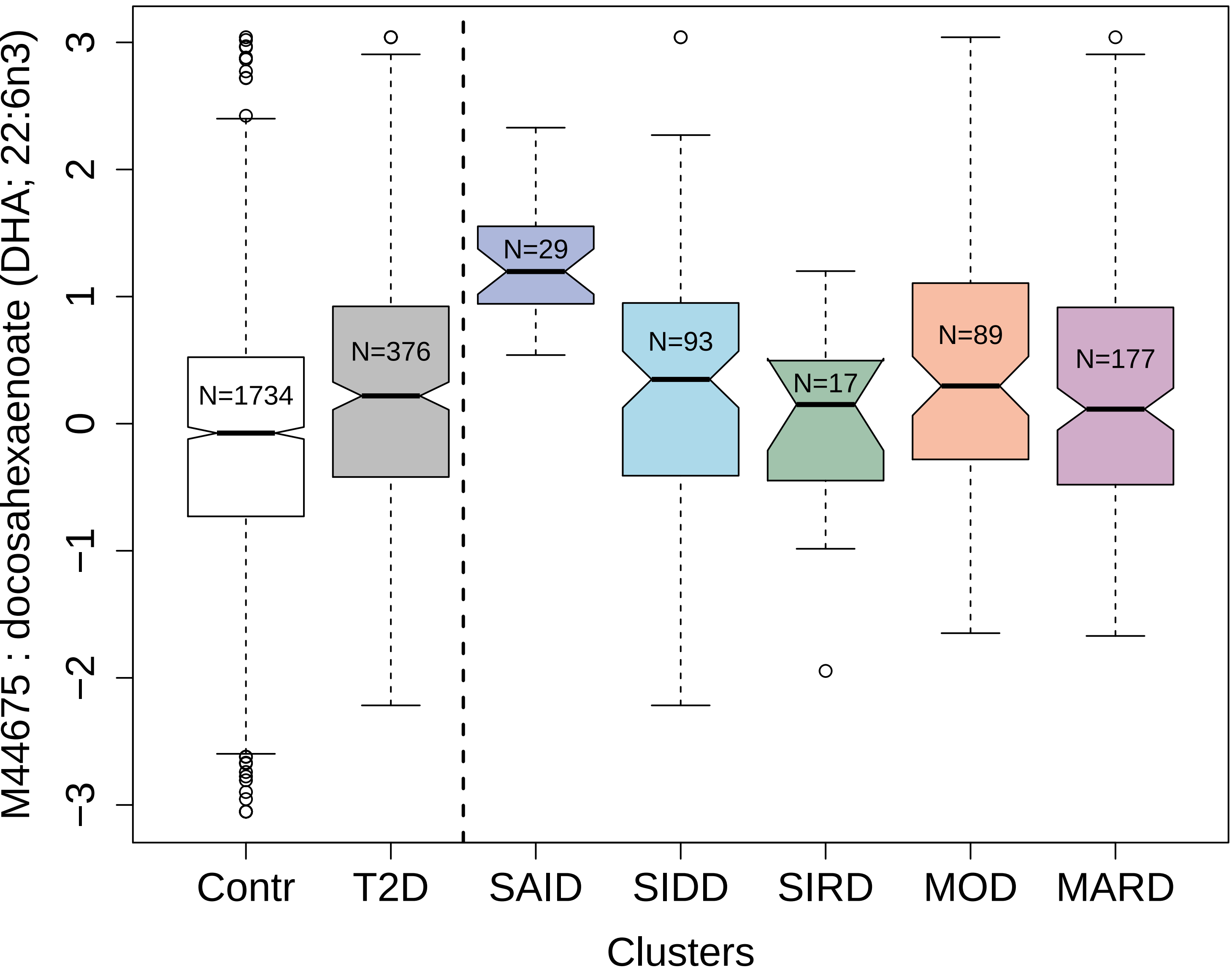

M44675 : docosahexaenoate (DHA; 22:6n3)
3
2
1
0
−1
−2
−3
N=1734
N=376
N=29
N=93
N=17
N=89
N=177
Contr
T2D
SAID
SIDD
SIRD
MOD
MARD
Clusters

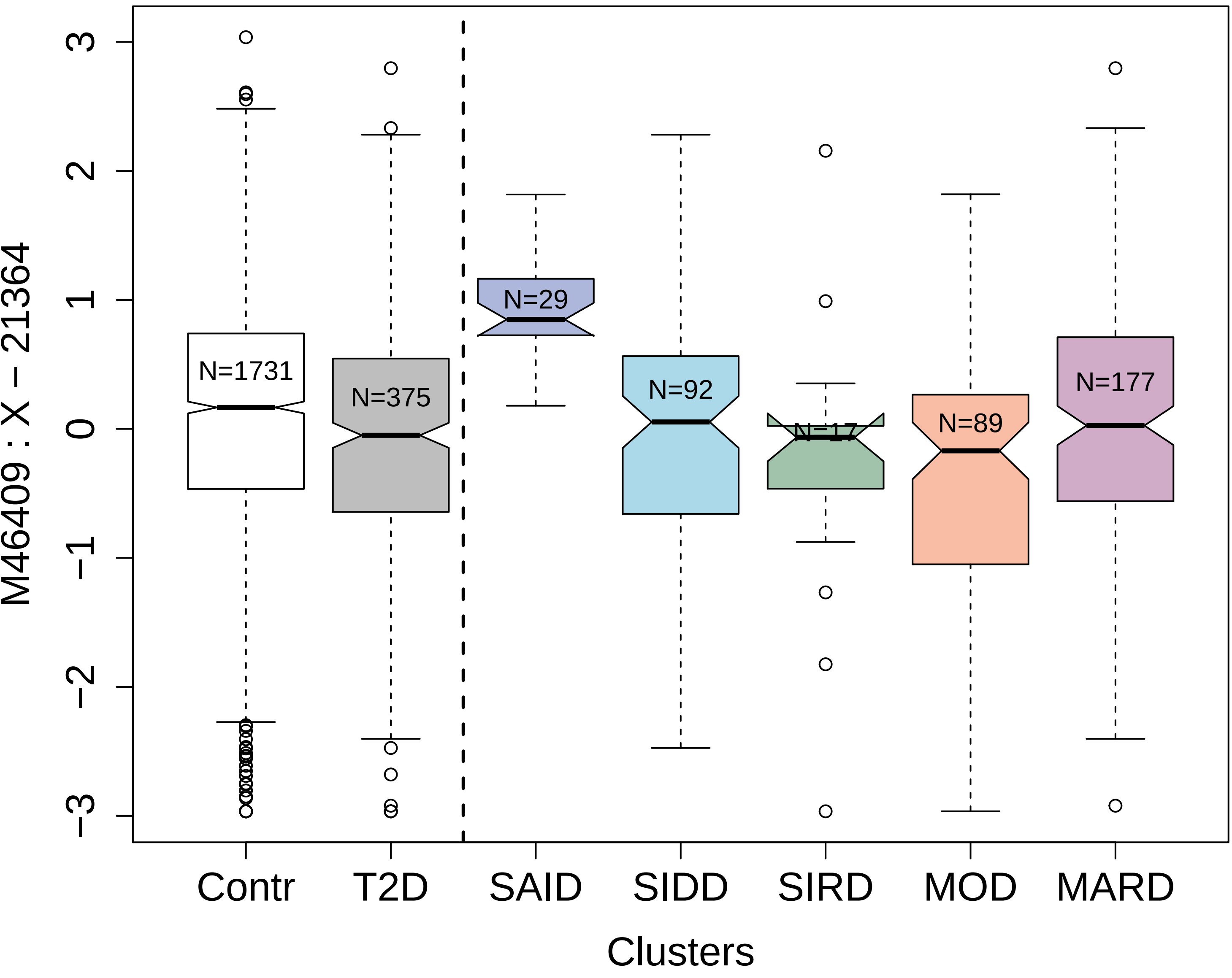

M46409 : X − 21364
3
2
1
0
−1
−2
−3
N=1731
N=375
N=29
N=92
N=17
N=89
N=177
Contr
T2D
SAID
SIDD
SIRD
MOD
MARD
Clusters

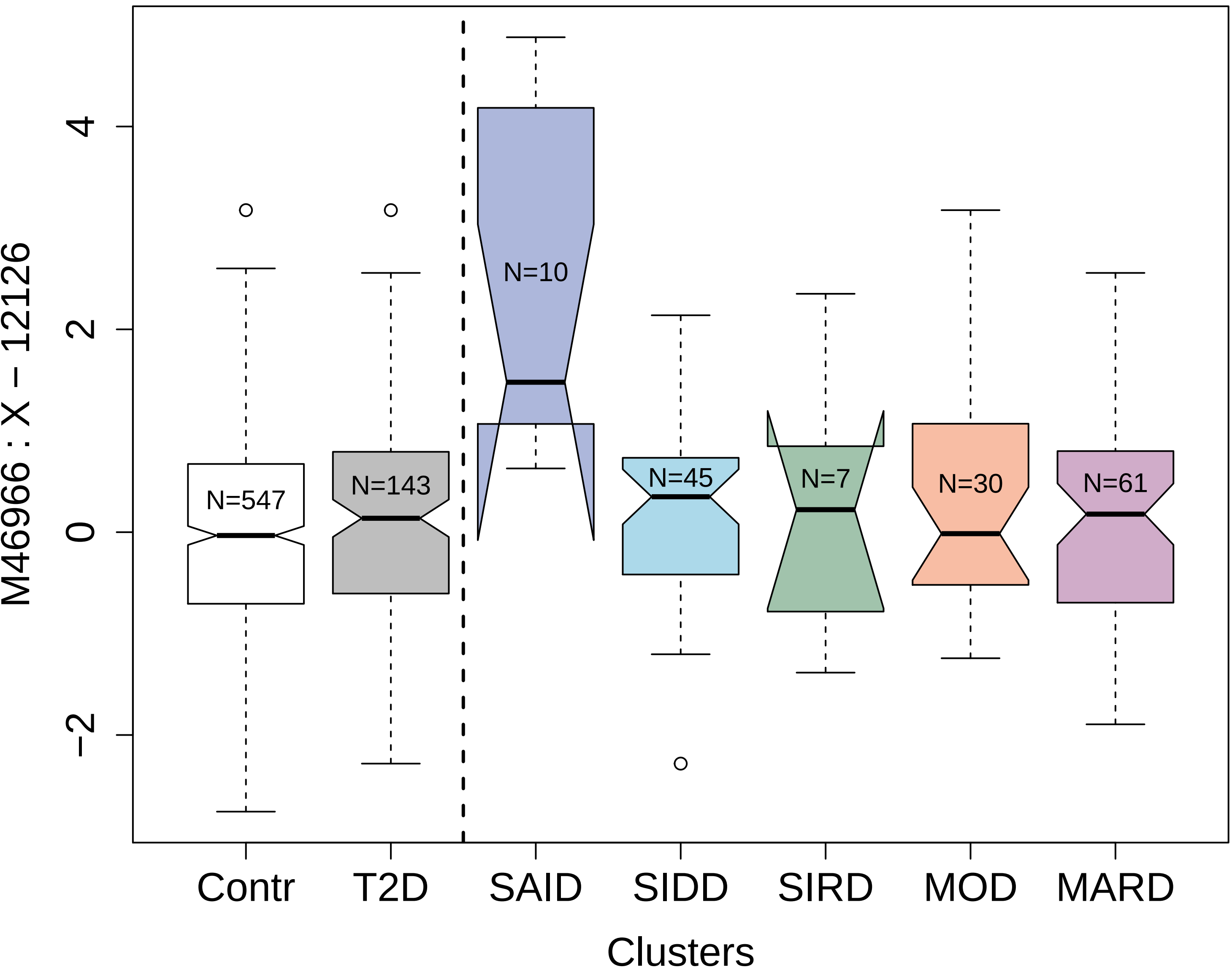
M46966 : X − 12126
Clusters
Contr
T2D
SAID
SIDD
SIRD
MOD
MARD
N=547
N=143
N=10
N=45
N=7
N=30
N=61
4
2
0
−2

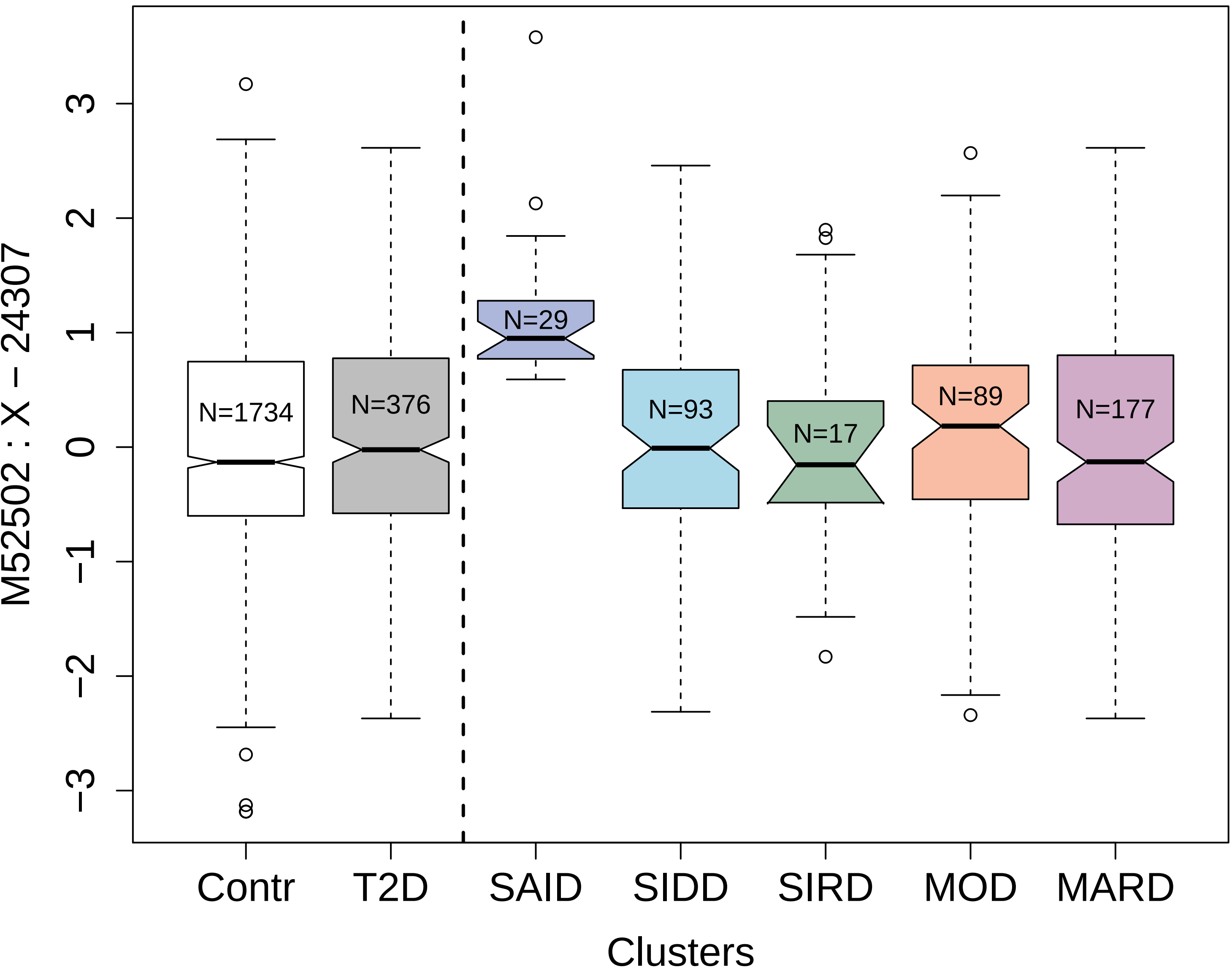

M52502 : X − 24307
3
2
1
0
−1
−2
−3
N=1734
N=376
N=29
N=93
N=17
N=89
N=177
Contr
T2D
SAID
SIDD
SIRD
MOD
MARD
Clusters

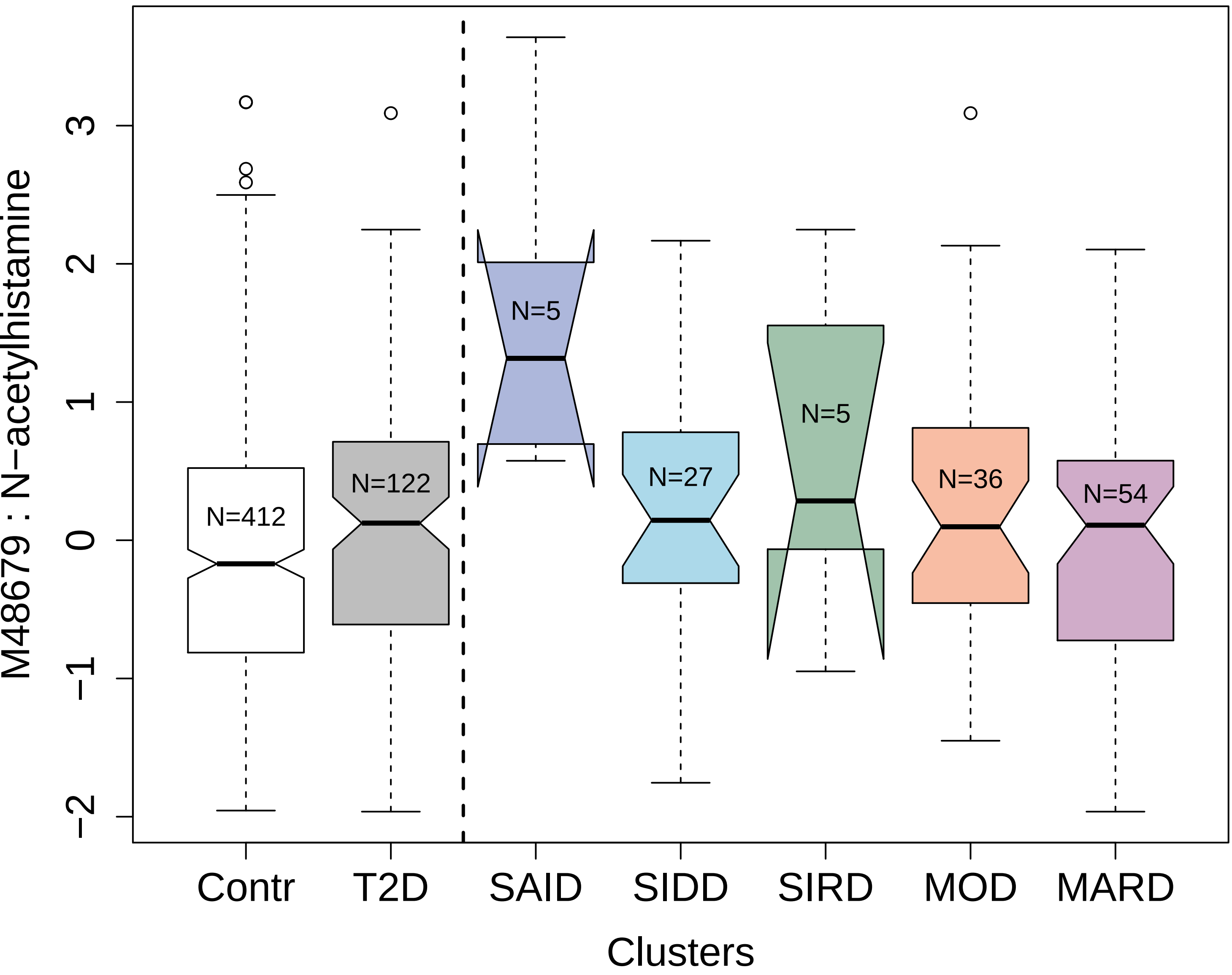
M48679 : N−acetylhistamine
−2
−1
0
1
2
3
N=412
N=122
N=5
N=27
N=5
N=36
N=54
Contr
T2D
SAID
SIDD
SIRD
MOD
MARD
Clusters

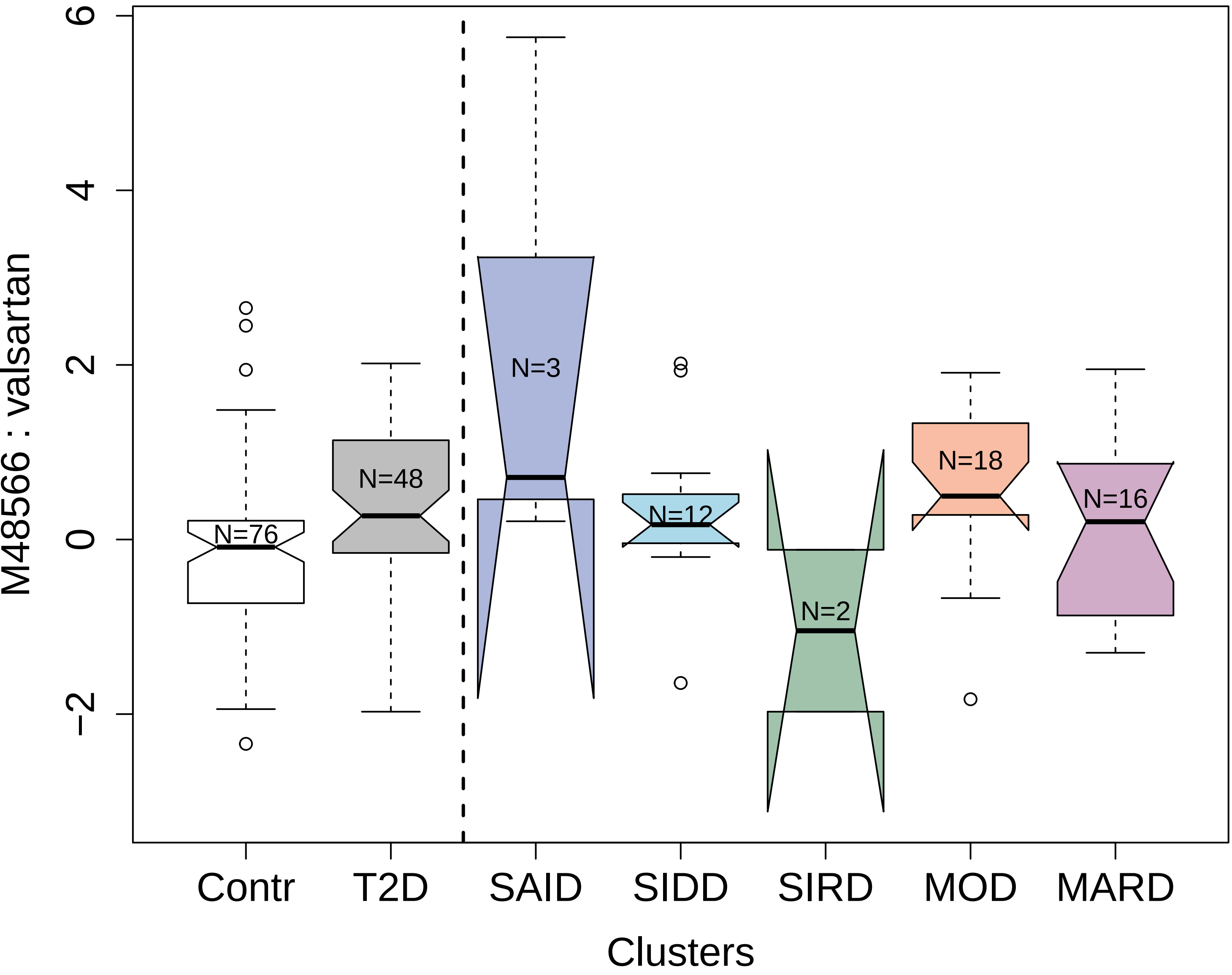

M48566 : valsartan
6
4
2
0
−2
N=76
N=48
N=3
N=12
N=2
N=18
N=16
Contr
T2D
SAID
SIDD
SIRD
MOD
MARD
Clusters

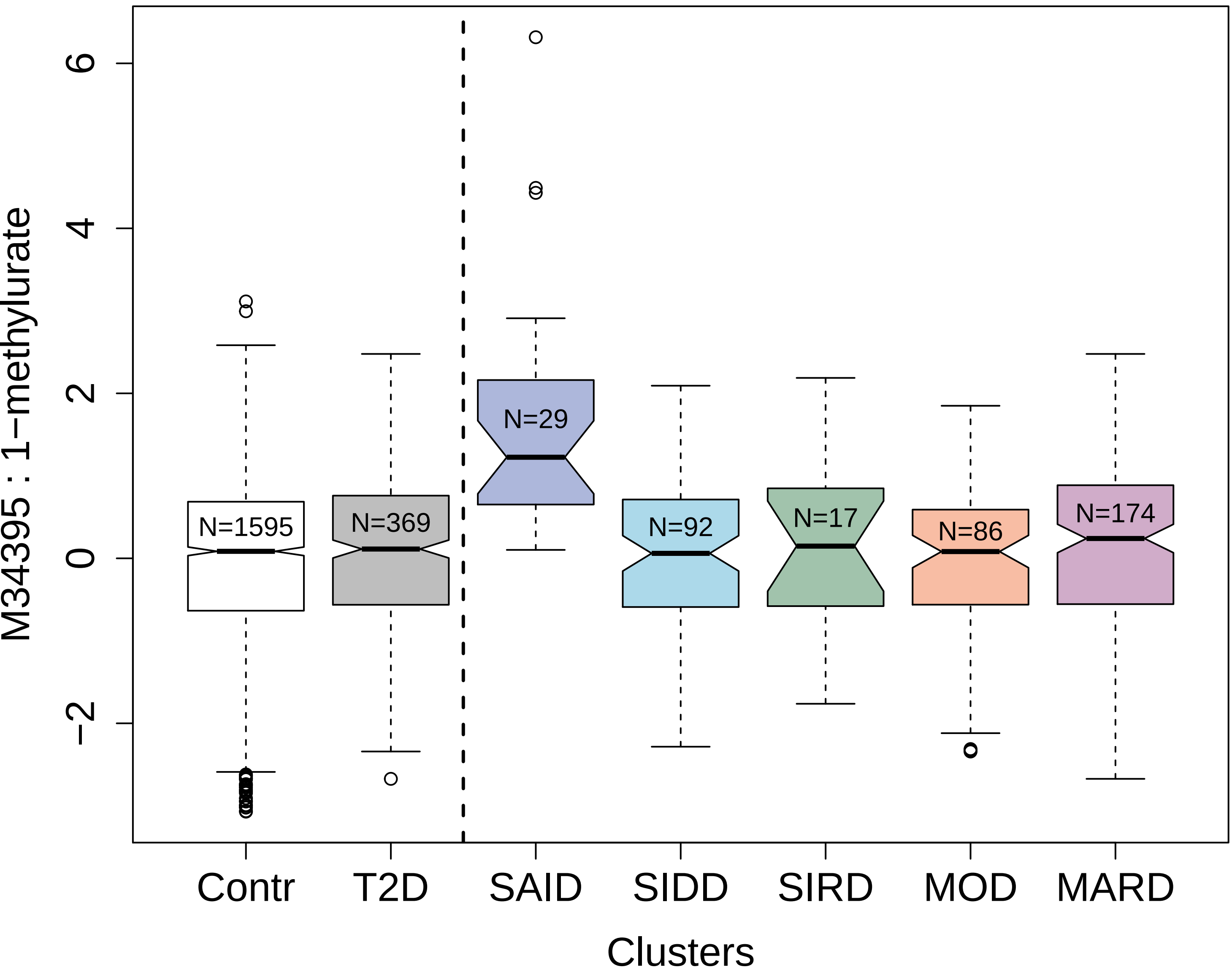

M34395 : 1−methylurate
6
4
2
0
−2
N=1595
N=369
N=29
N=92
N=17
N=86
N=174
Contr
T2D
SAID
SIDD
SIRD
MOD
MARD
Clusters

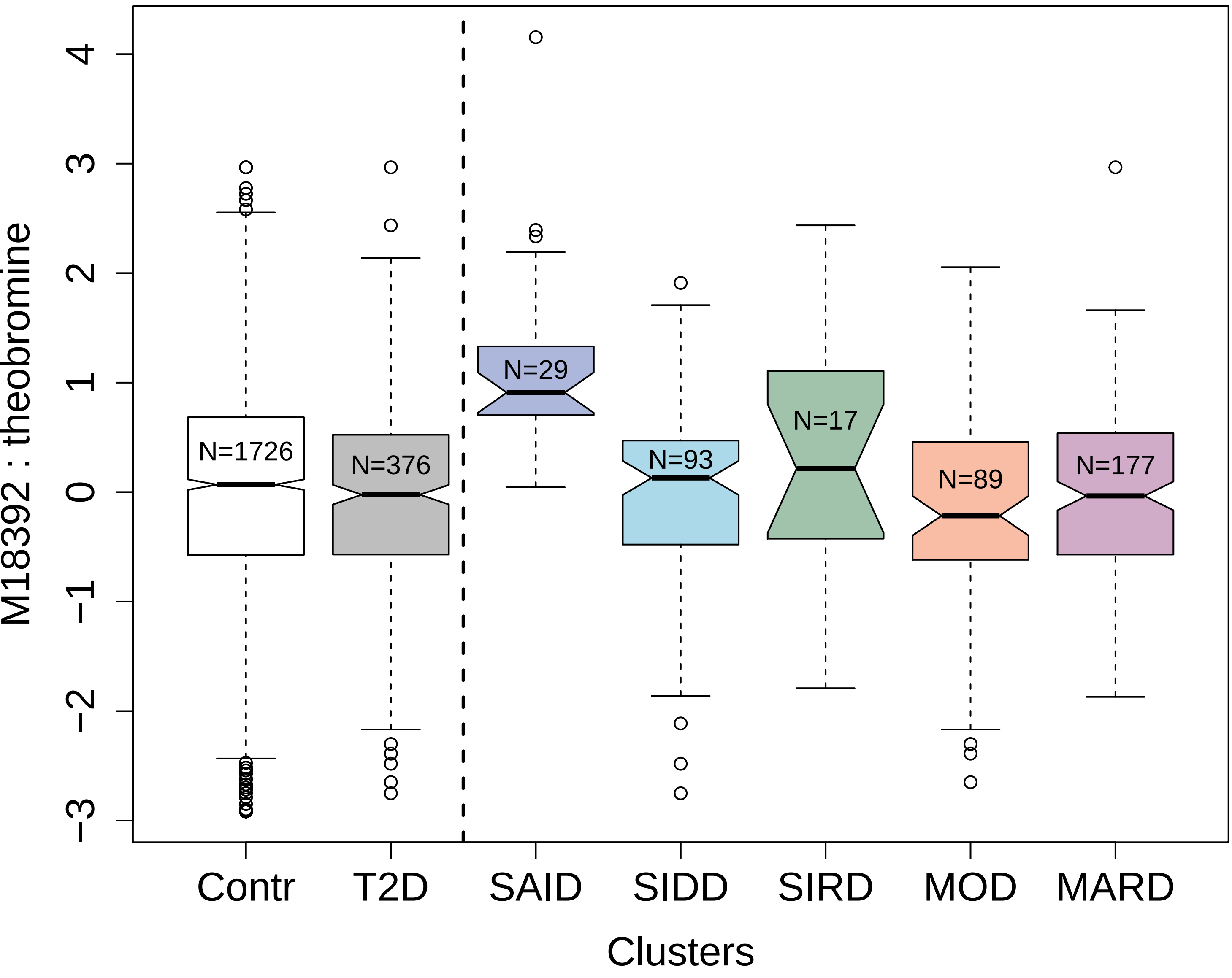
M18392 : theobromine
4
3
2
1
0
−1
−2
−3
N=1726
N=376
N=29
N=93
N=17
N=89
N=177
Contr
T2D
SAID
SIDD
SIRD
MOD
MARD
Clusters

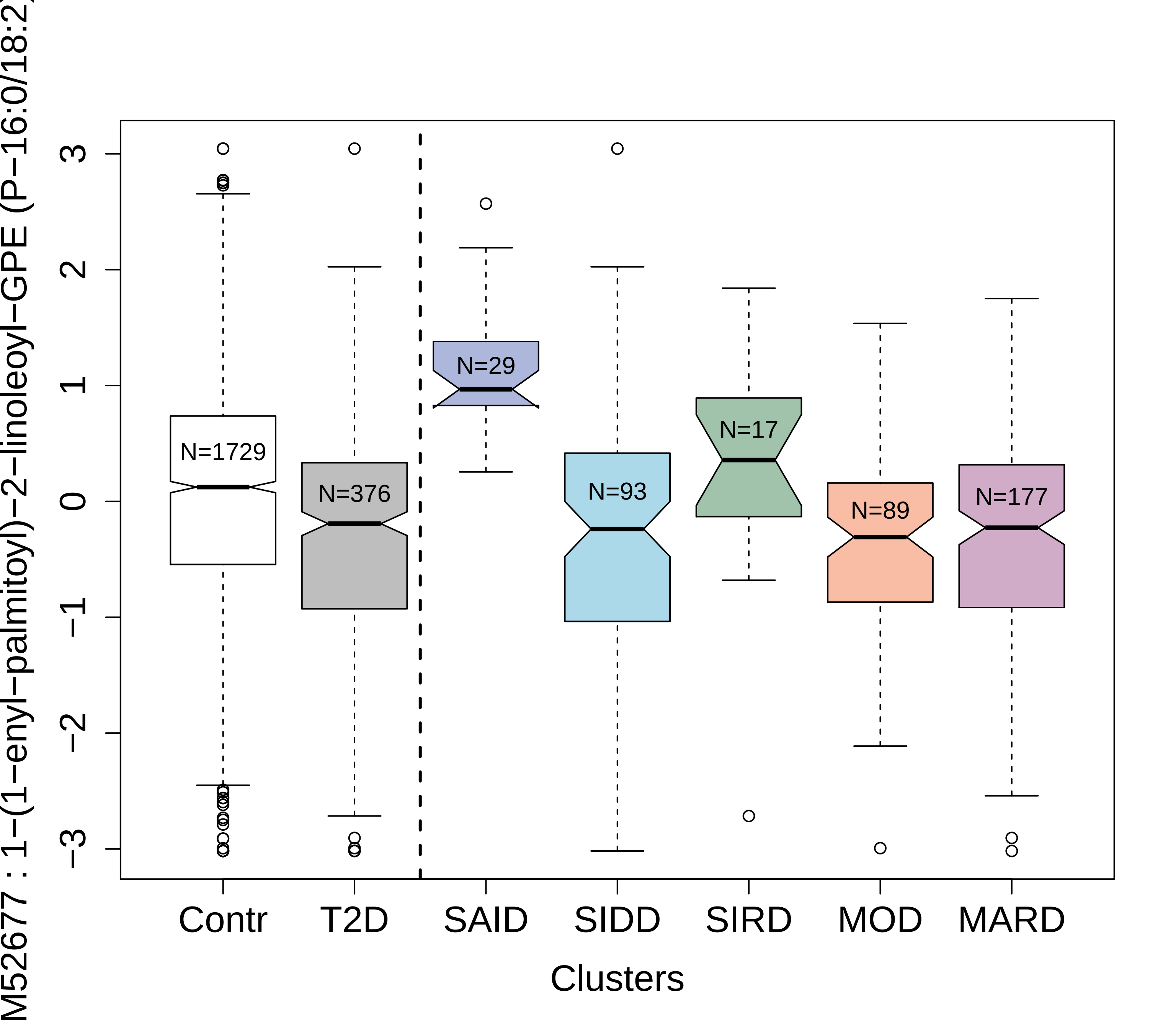

M52677 : 1−(1−enyl−palmitoyl)−2−linoleoyl−GPE (P−16:0/18:2)
3
2
1
0
−1
−2
−3
N=1729
N=376
N=29
N=93
N=17
N=89
N=177
Contr
T2D
SAID
SIDD
SIRD
MOD
MARD
Clusters

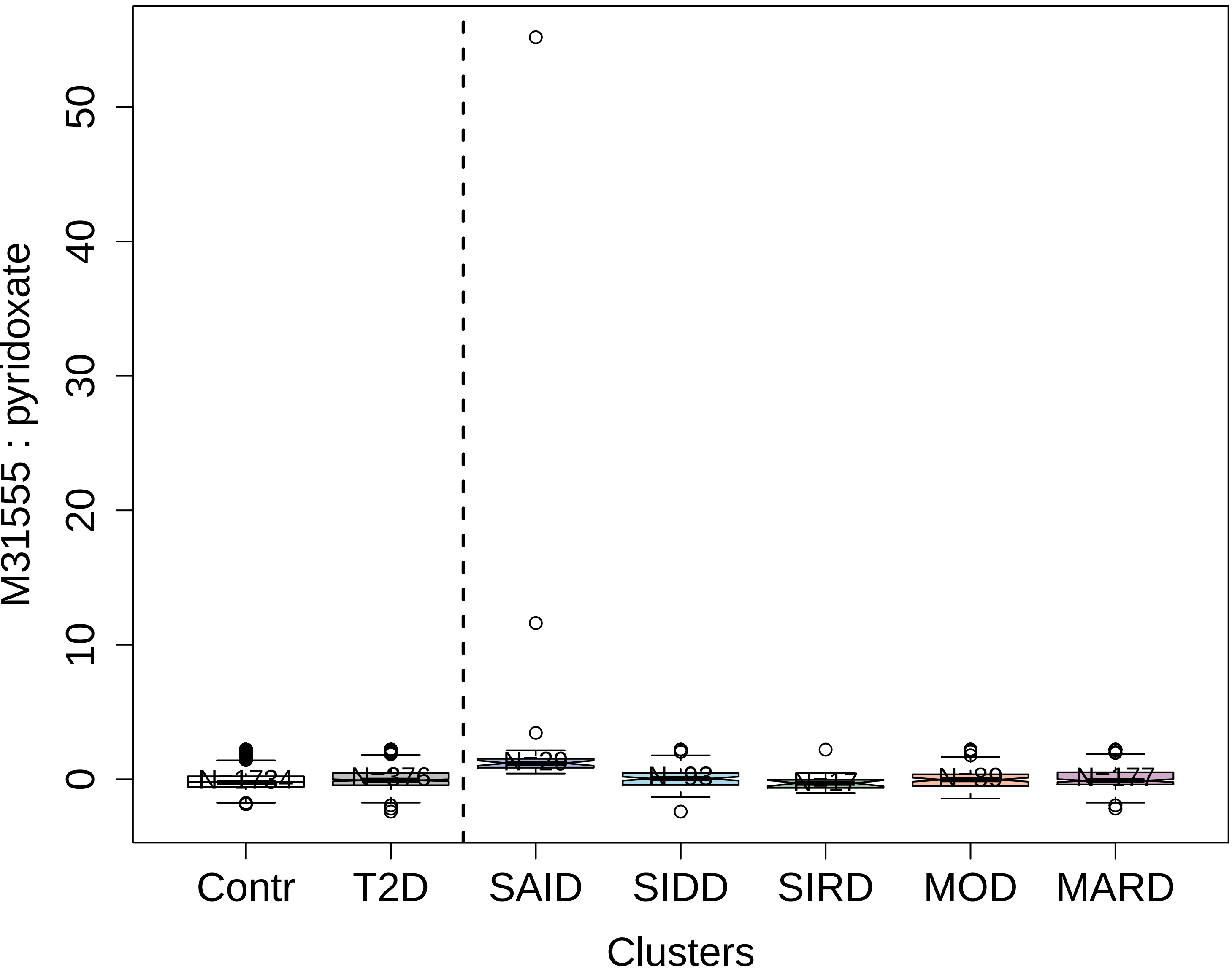

M31555 : pyridoxate
0
10
20
30
40
50
N=1734
N=376
N=20
N=93
N=17
N=80
N=177
Contr
T2D
SAID
SIDD
SIRD
MOD
MARD
Clusters

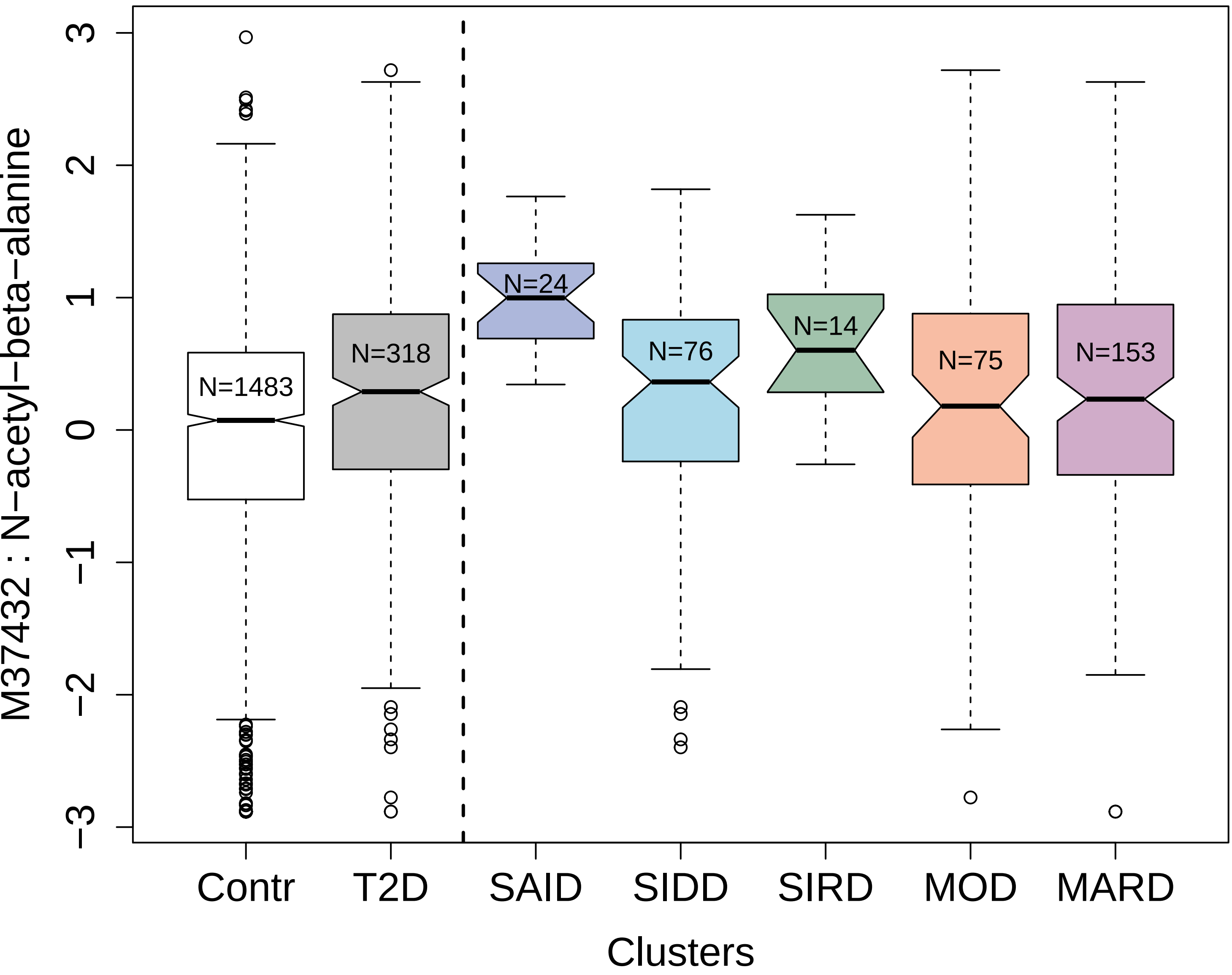

M37432 : N−acetyl−beta−alanine
3
2
1
0
−1
−2
−3
N=1483
N=318
N=24
N=76
N=14
N=75
N=153
Contr
T2D
SAID
SIDD
SIRD
MOD
MARD
Clusters

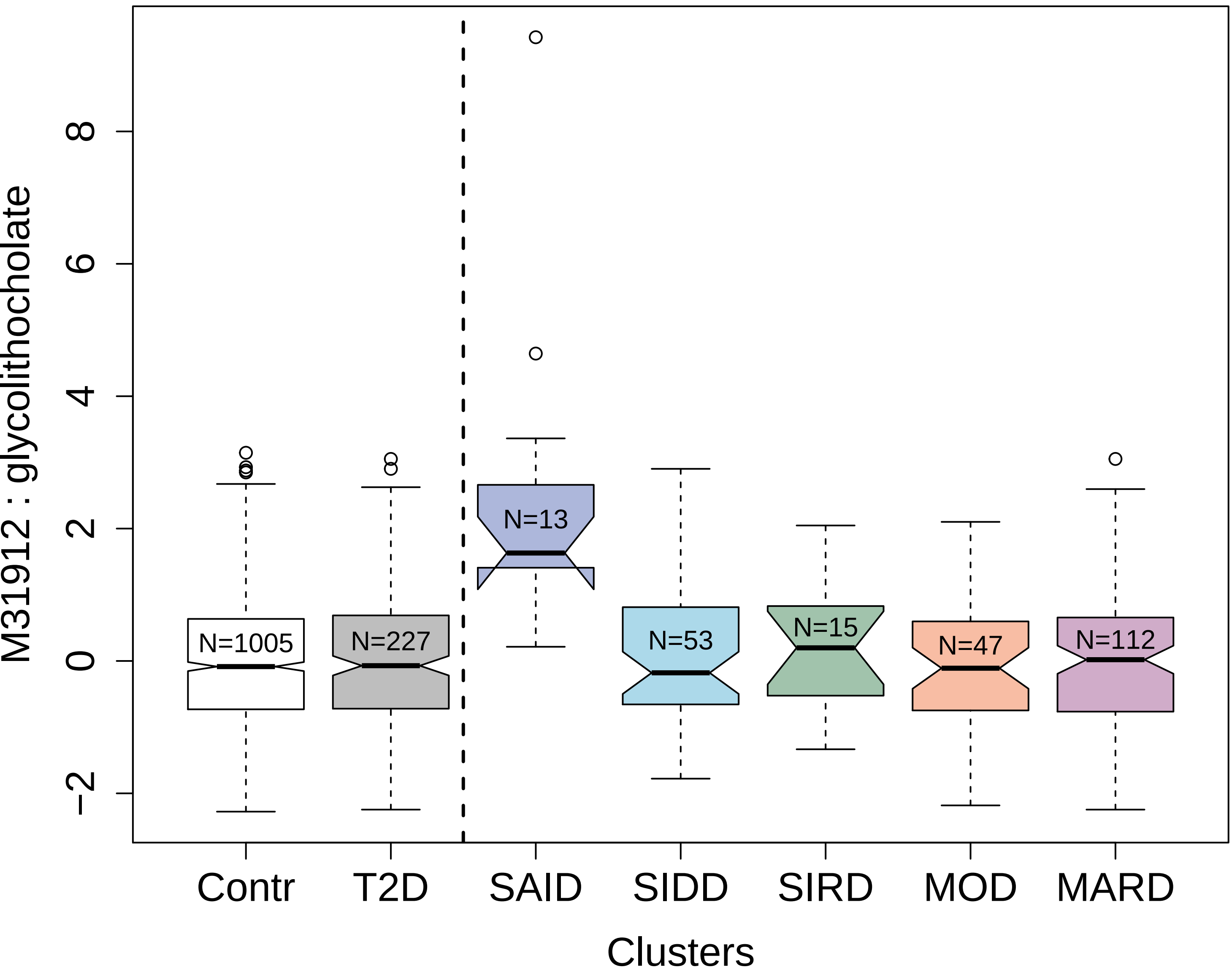

M31912 : glycolithocholate
8
6
4
2
0
−2
N=1005
N=227
N=13
N=53
N=15
N=47
N=112
Contr
T2D
SAID
SIDD
SIRD
MOD
MARD
Clusters

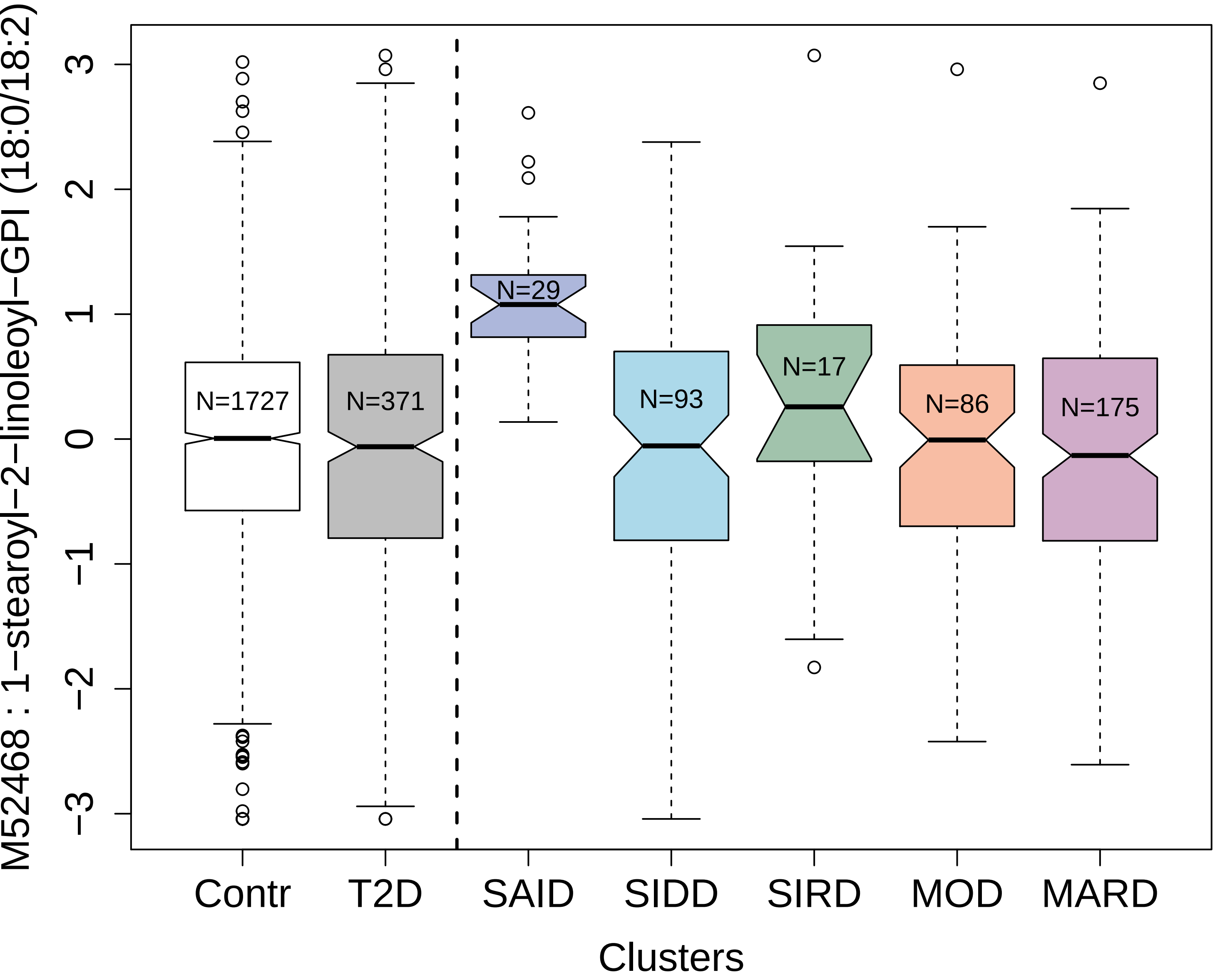
M52468 : 1−stearoyl−2−linoleoyl−GPI (18:0/18:2)
3
2
1
0
−1
−2
−3
N=1727
N=371
N=29
N=93
N=17
N=86
N=175
Contr
T2D
SAID
SIDD
SIRD
MOD
MARD
Clusters

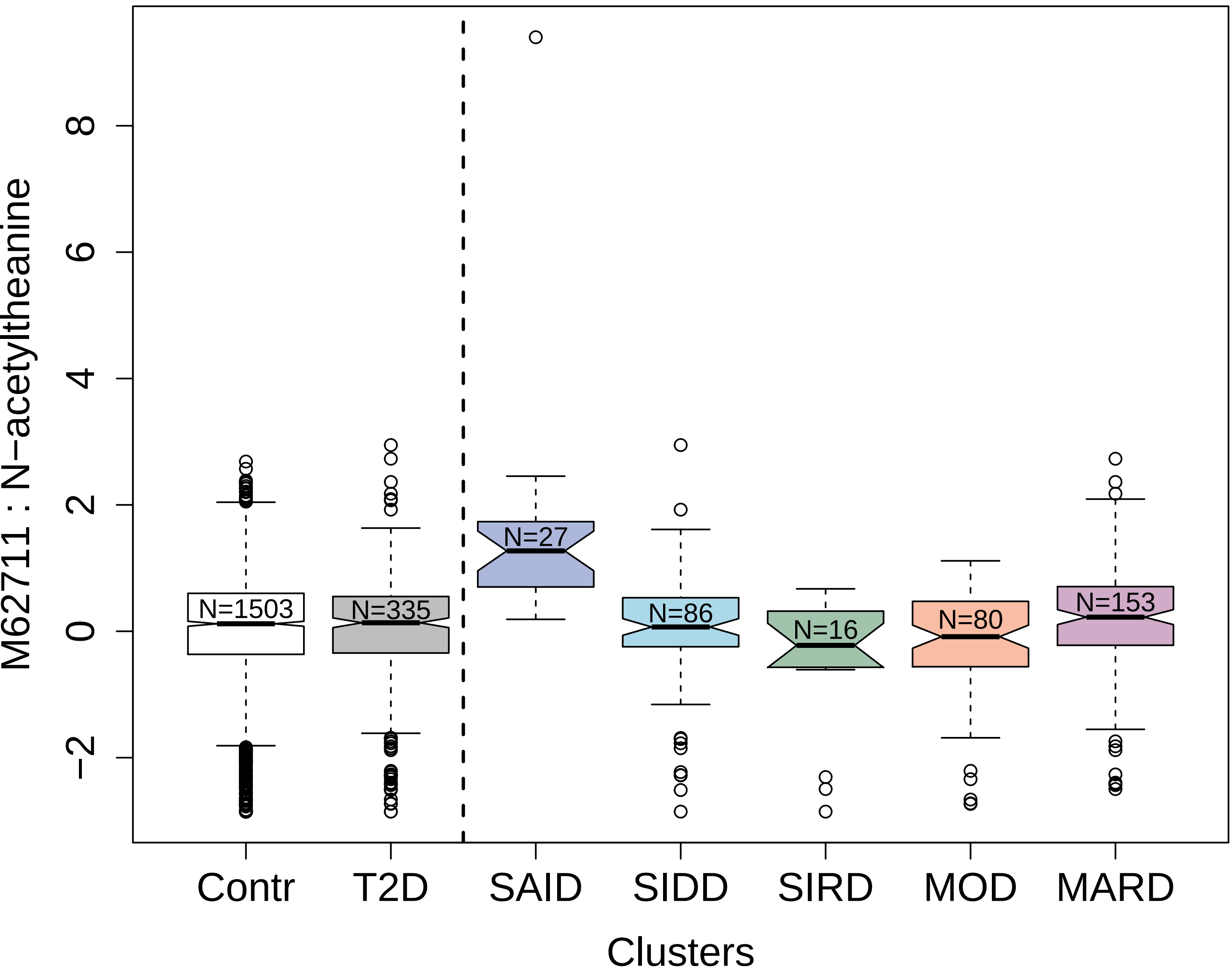

M62711 : N−acetyltheanine
−2
0
2
4
6
8
N=1503
N=335
N=27
N=86
N=16
N=80
N=153
Contr
T2D
SAID
SIDD
SIRD
MOD
MARD
Clusters

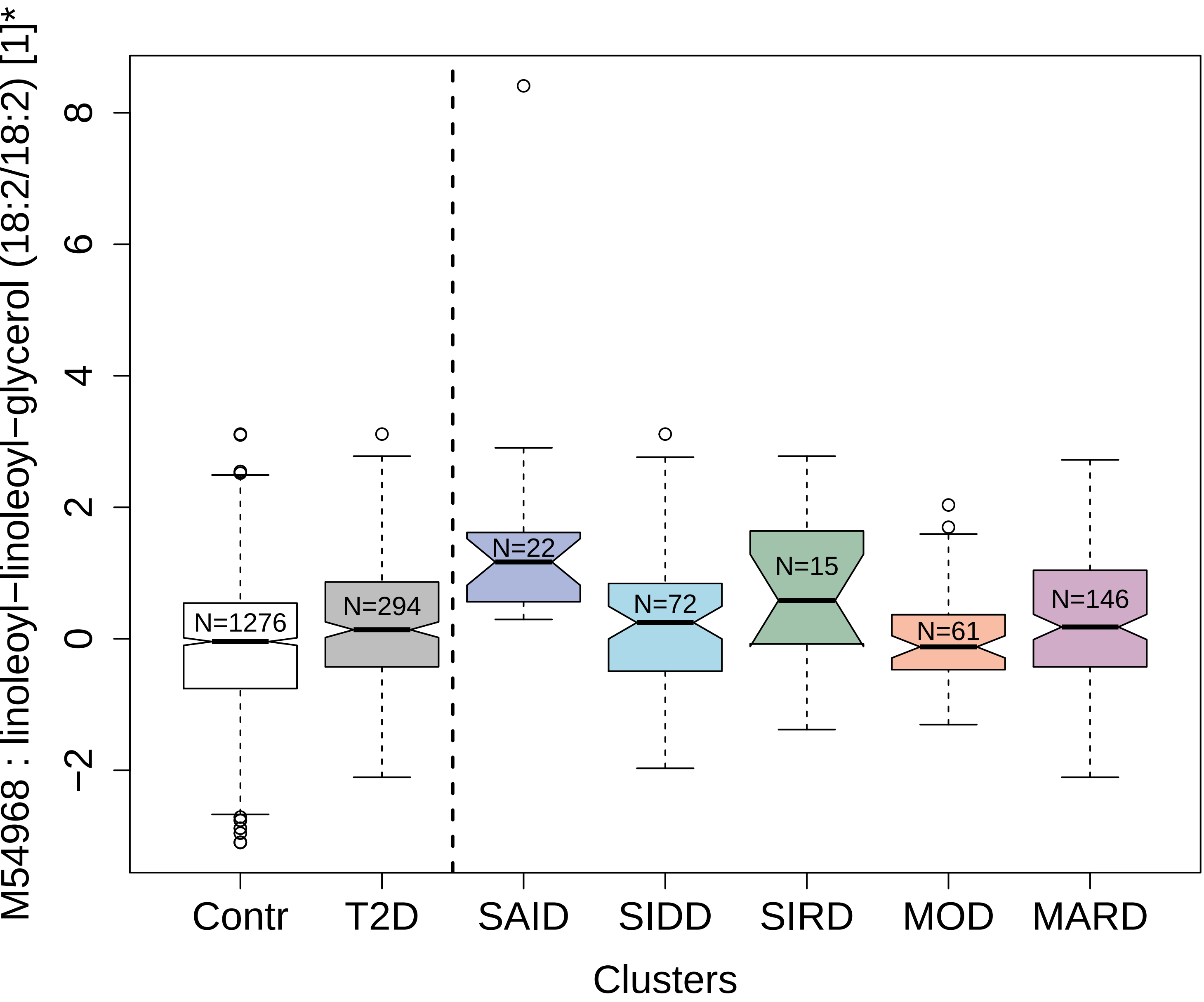

M54968 : linoleoyl−linoleoyl−glycerol (18:2/18:2) [1]*
8
6
4
2
0
−2
N=1276
N=294
N=22
N=72
N=15
N=61
N=146
Contr
T2D
SAID
SIDD
SIRD
MOD
MARD
Clusters

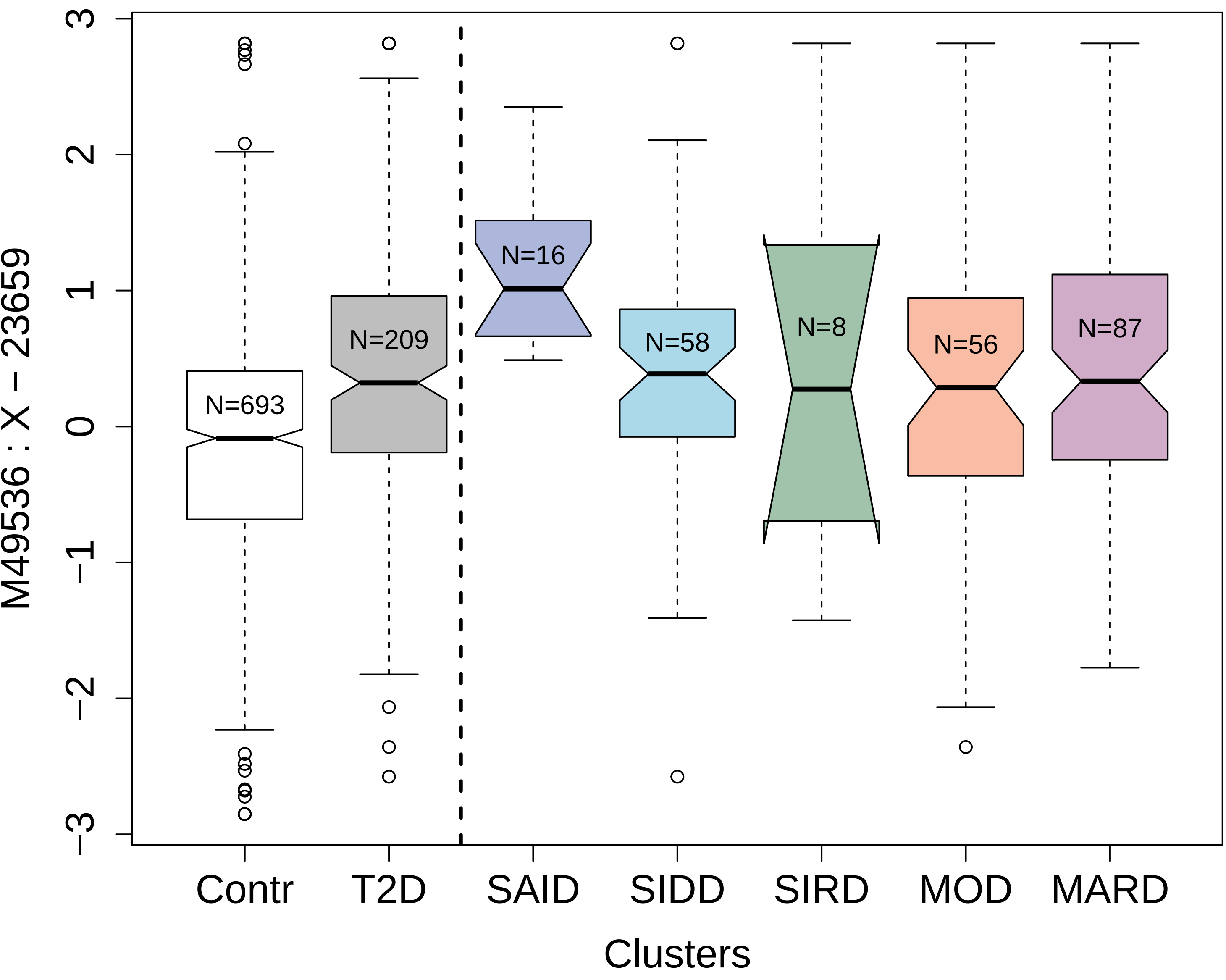

M49536 : X − 23659
3
2
1
0
−1
−2
−3
N=693
N=209
N=16
N=58
N=8
N=56
N=87
Contr
T2D
SAID
SIDD
SIRD
MOD
MARD
Clusters

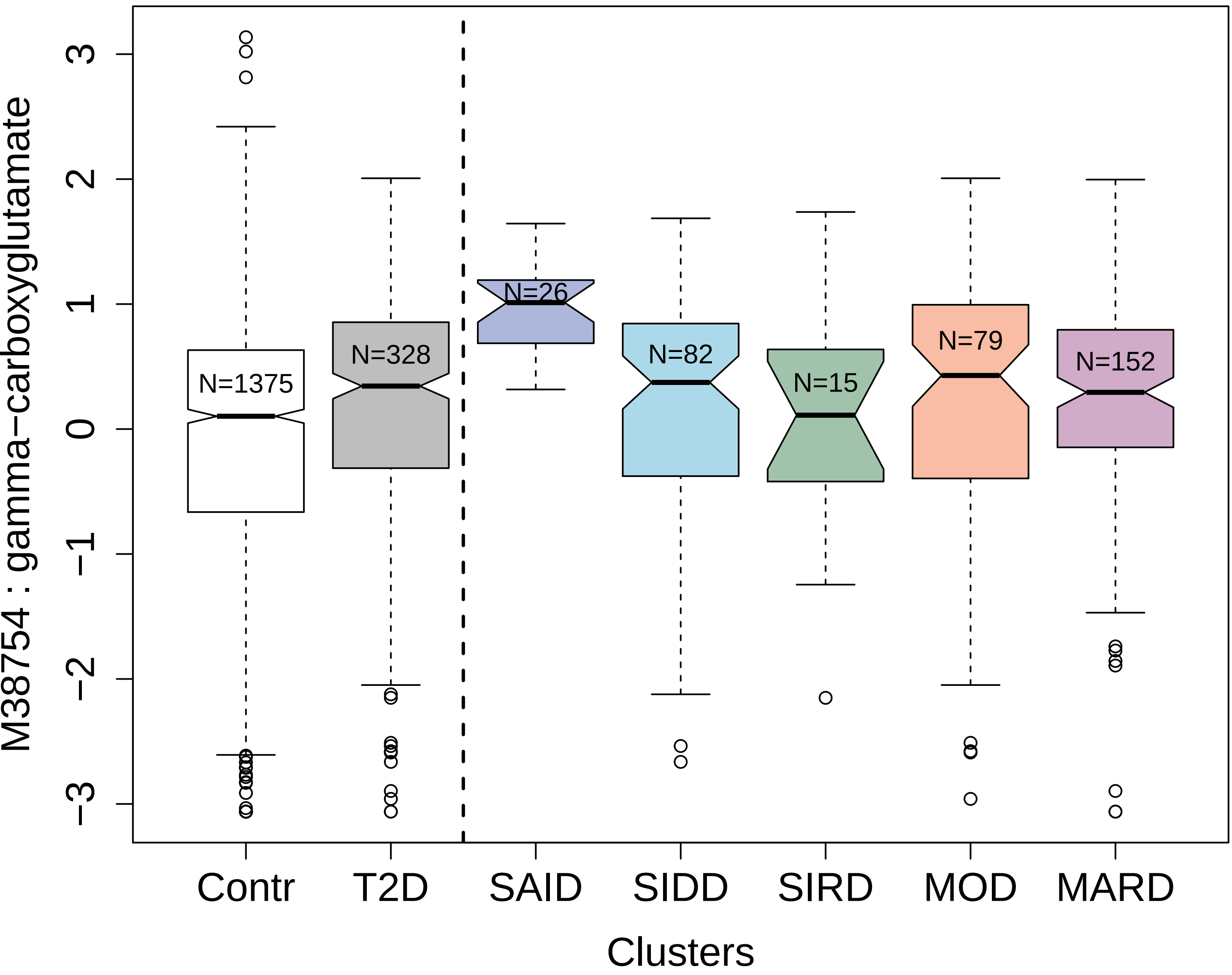
M38754 : gamma−carboxyglutamate
3
2
1
0
−1
−2
−3
N=1375
N=328
N=26
N=82
N=15
N=79
N=152
Contr
T2D
SAID
SIDD
SIRD
MOD
MARD
Clusters

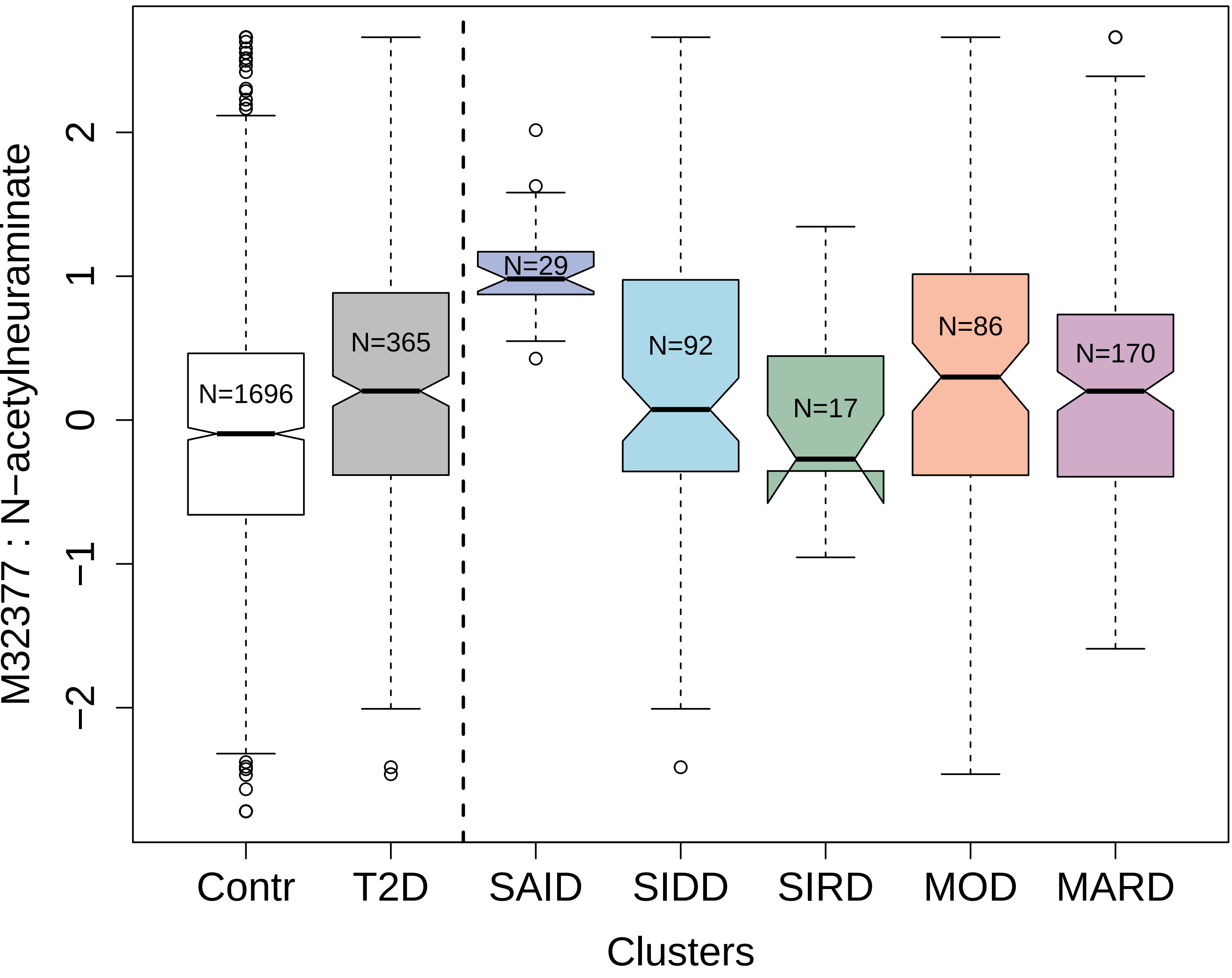
M32377 : N−acetylneuraminate
2
1
0
−1
−2
N=1696
N=365
N=29
N=92
N=17
N=86
N=170
Contr
T2D
SAID
SIDD
SIRD
MOD
MARD
Clusters

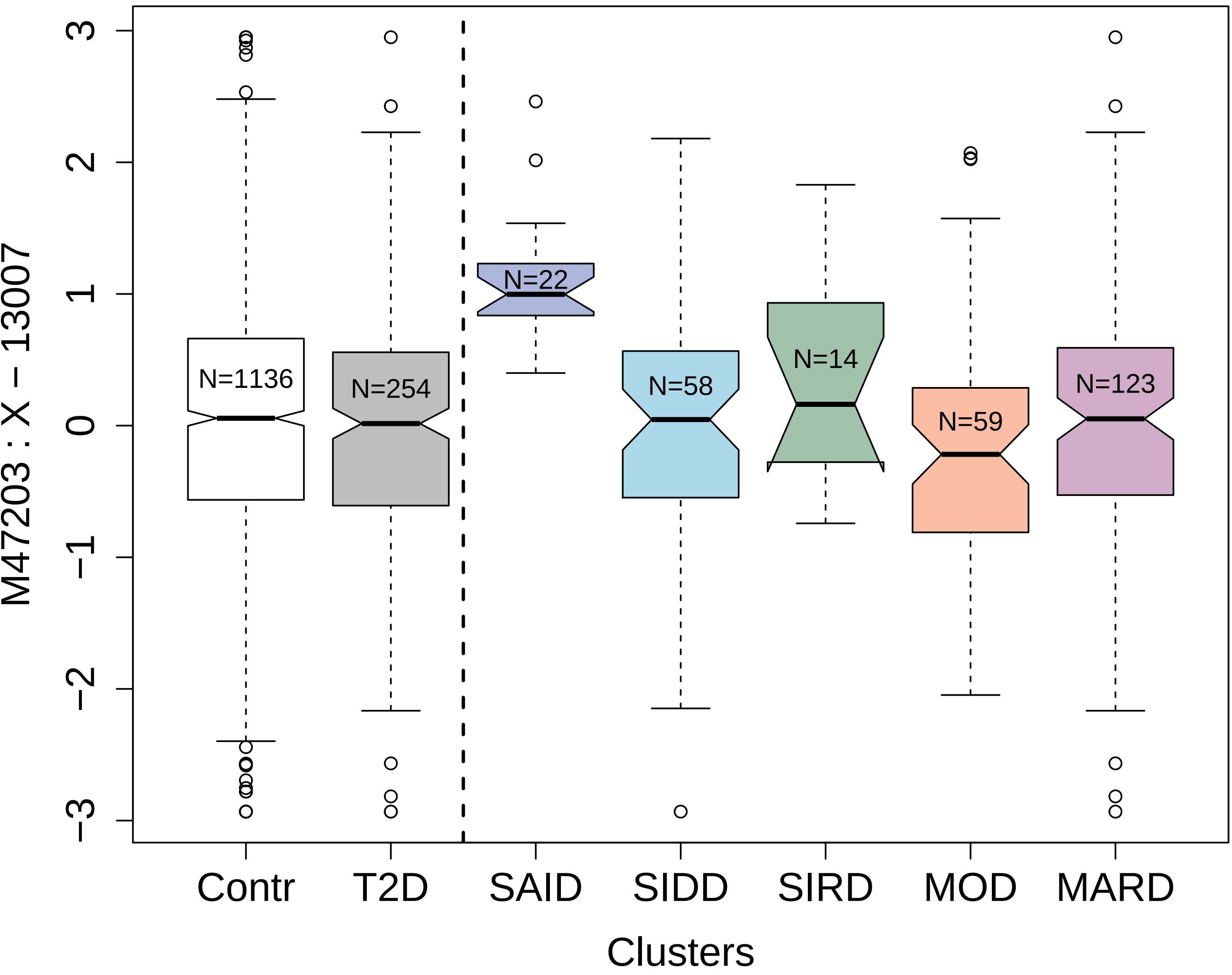

M47203 : X − 13007
3
2
1
0
−1
−2
−3
N=1136
N=254
N=22
N=58
N=14
N=59
N=123
Contr
T2D
SAID
SIDD
SIRD
MOD
MARD
Clusters

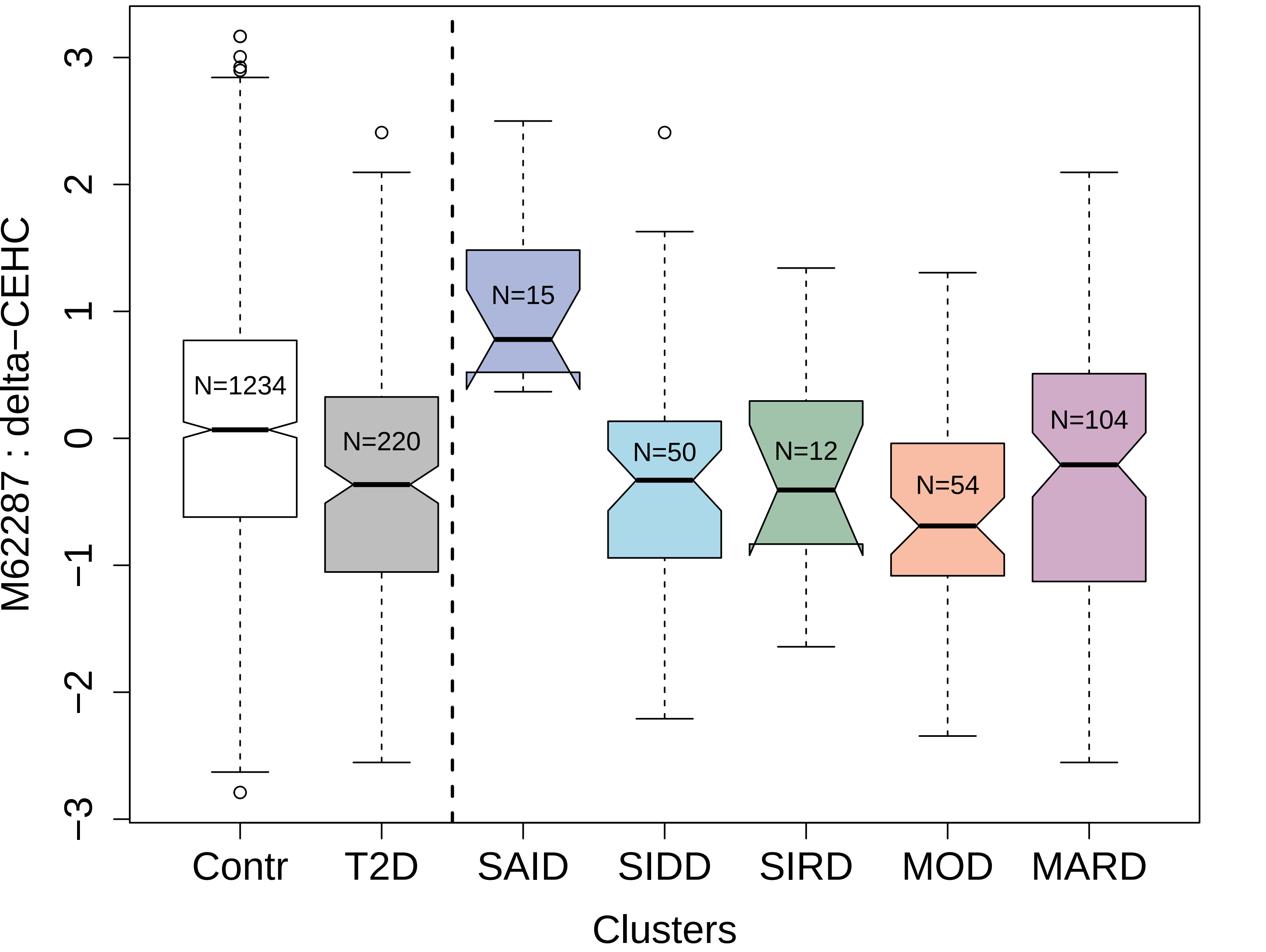
M62287 : delta−CEHC
3
2
1
0
−1
−2
−3
N=1234
N=220
N=15
N=50
N=12
N=54
N=104
Contr
T2D
SAID
SIDD
SIRD
MOD
MARD
Clusters

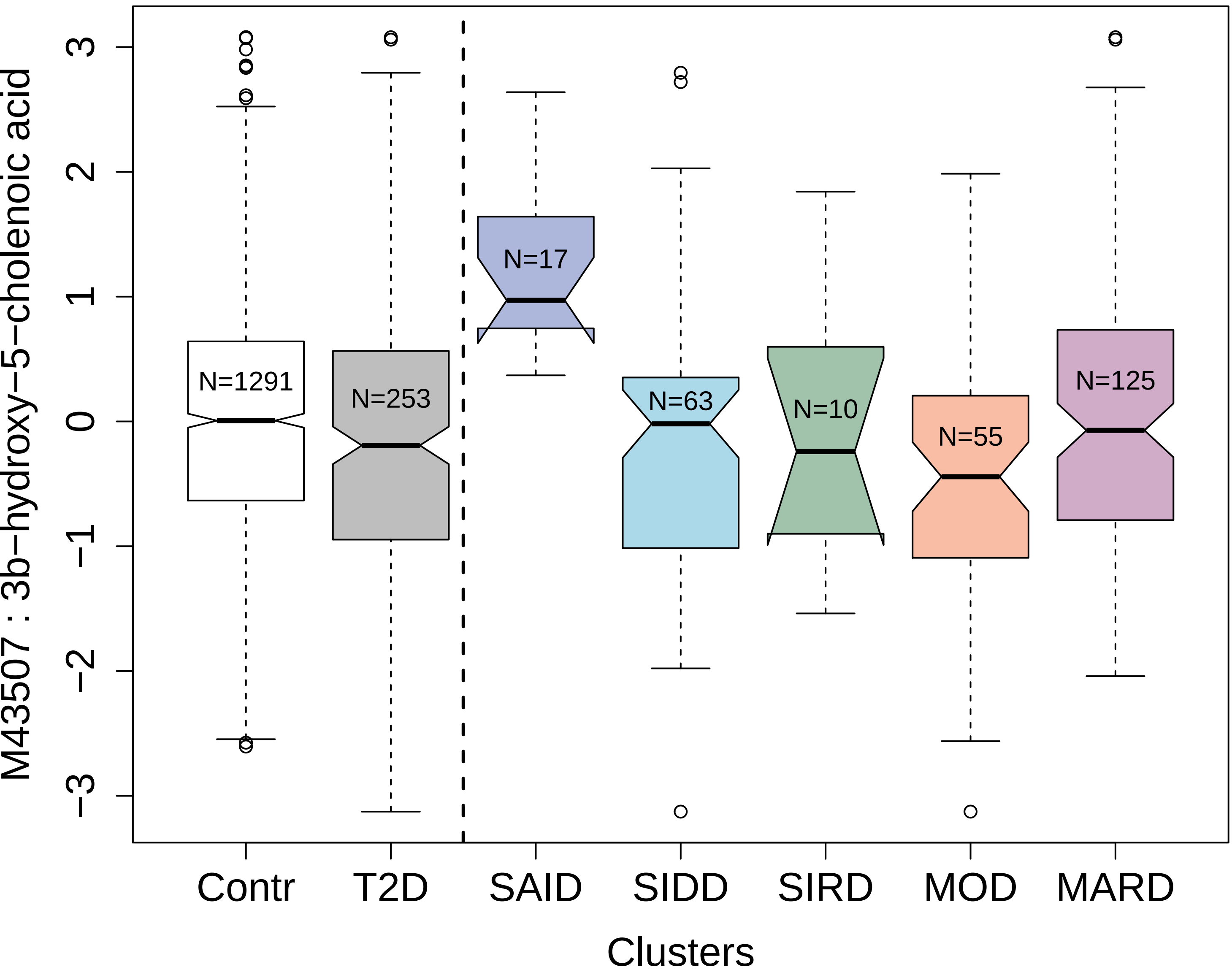

M43507 : 3b−hydroxy−5−cholenoic acid
3
2
1
0
−1
−2
−3
N=1291
N=253
N=17
N=63
N=10
N=55
N=125
Contr
T2D
SAID
SIDD
SIRD
MOD
MARD
Clusters

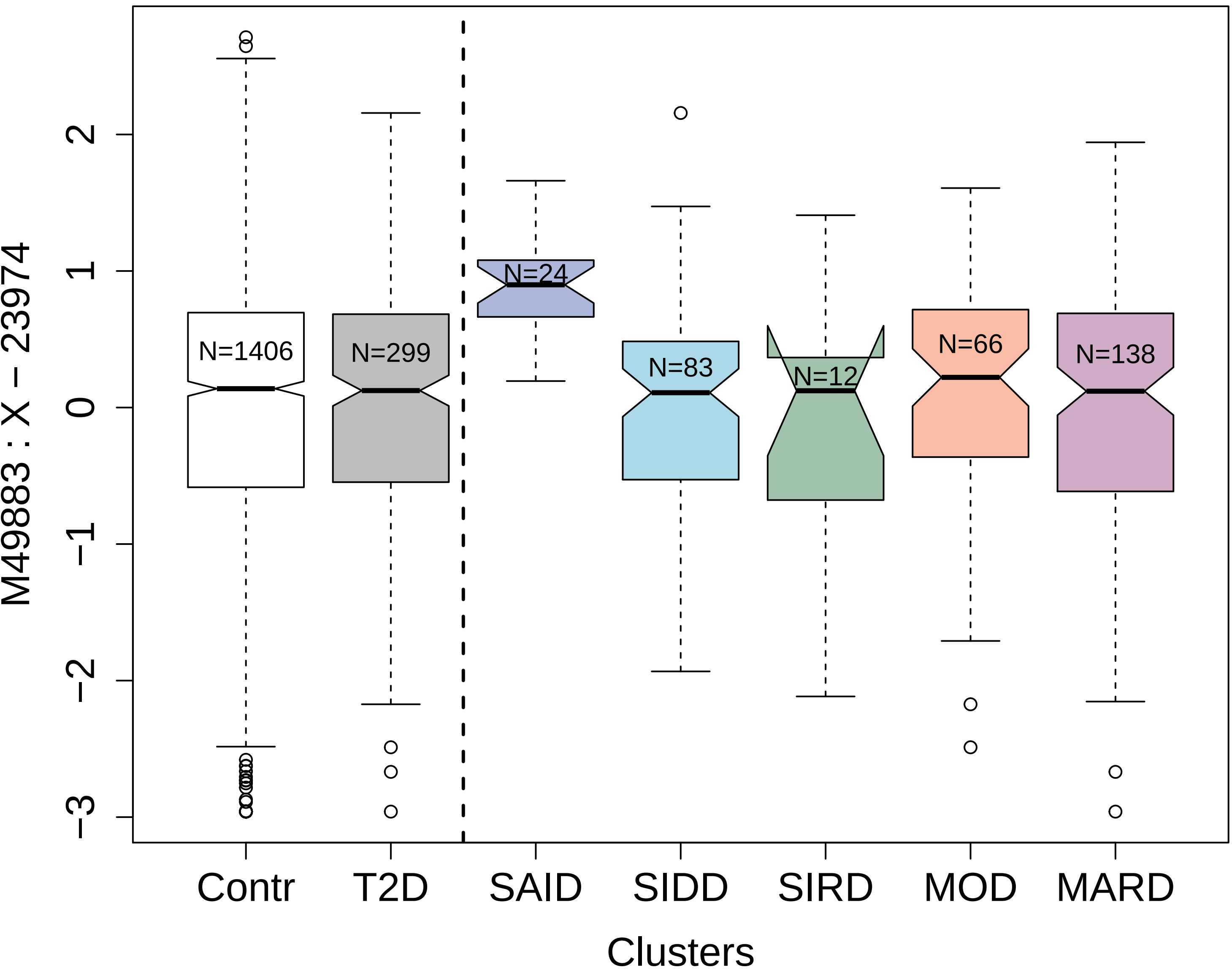

M49883 : X − 23974
Clusters
Contr
T2D
SAID
SIDD
SIRD
MOD
MARD
N=1406
N=299
N=24
N=83
N=12
N=66
N=138
−3
−2
−1
0
1
2

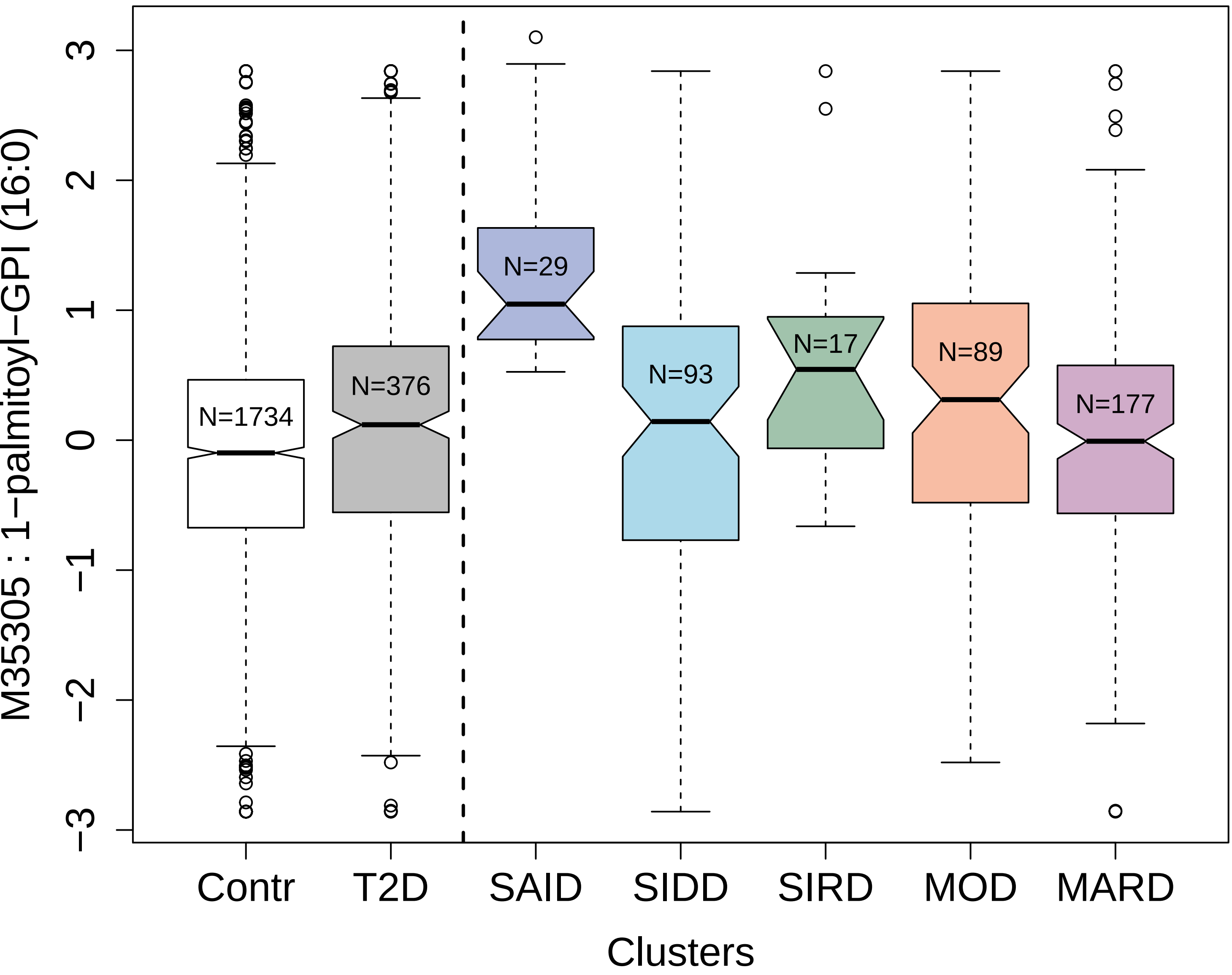

M35305 : 1−palmitoyl−GPI (16:0)
3
2
1
0
−1
−2
−3
N=1734
N=376
N=29
N=93
N=17
N=89
N=177
Contr
T2D
SAID
SIDD
SIRD
MOD
MARD
Clusters

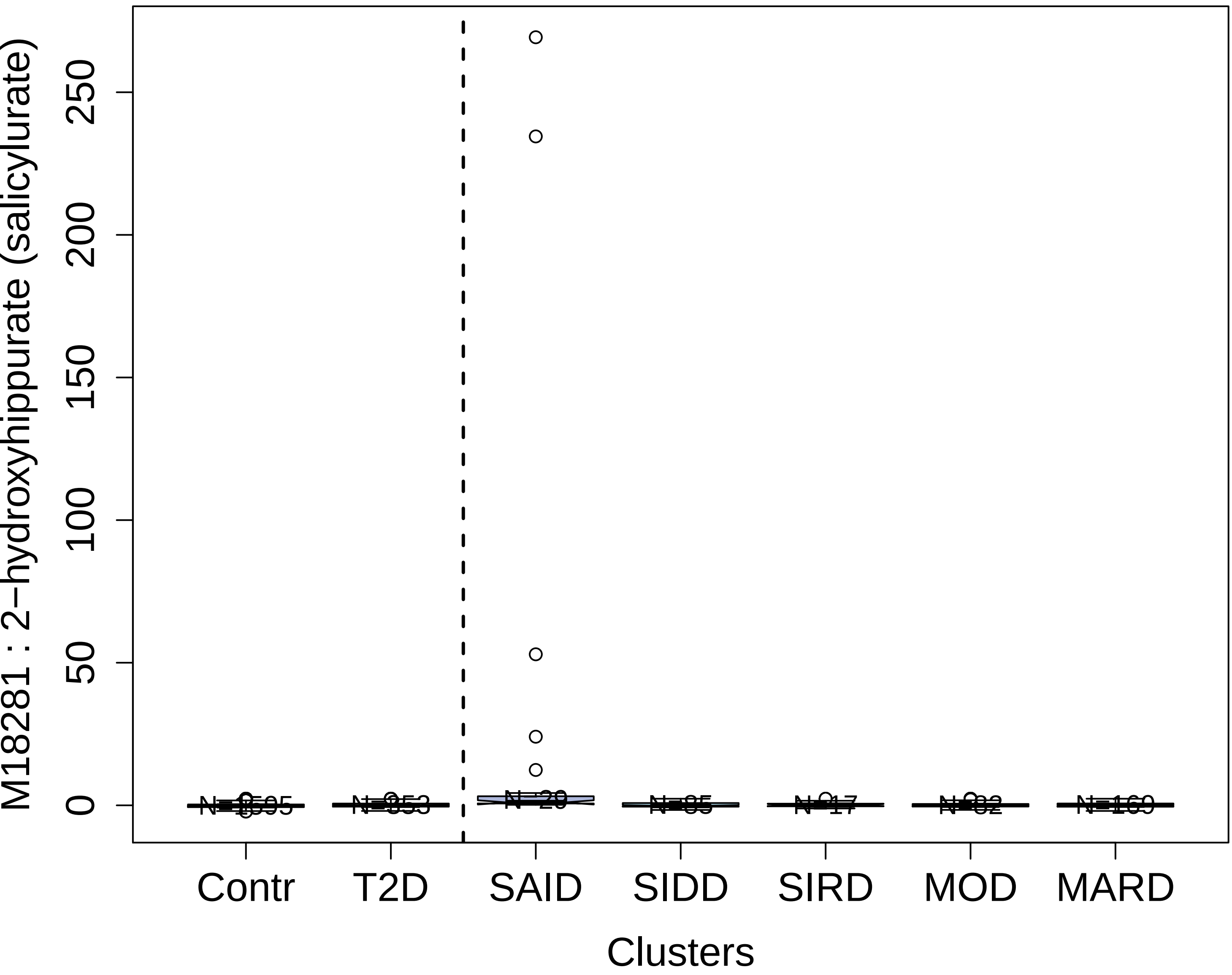
M18281 : 2−hydroxyhippurate (salicylurate)
0
50
100
150
200
250
N=1595
N=353
N=20
N=85
N=17
N=82
N=169
Contr
T2D
SAID
SIDD
SIRD
MOD
MARD
Clusters

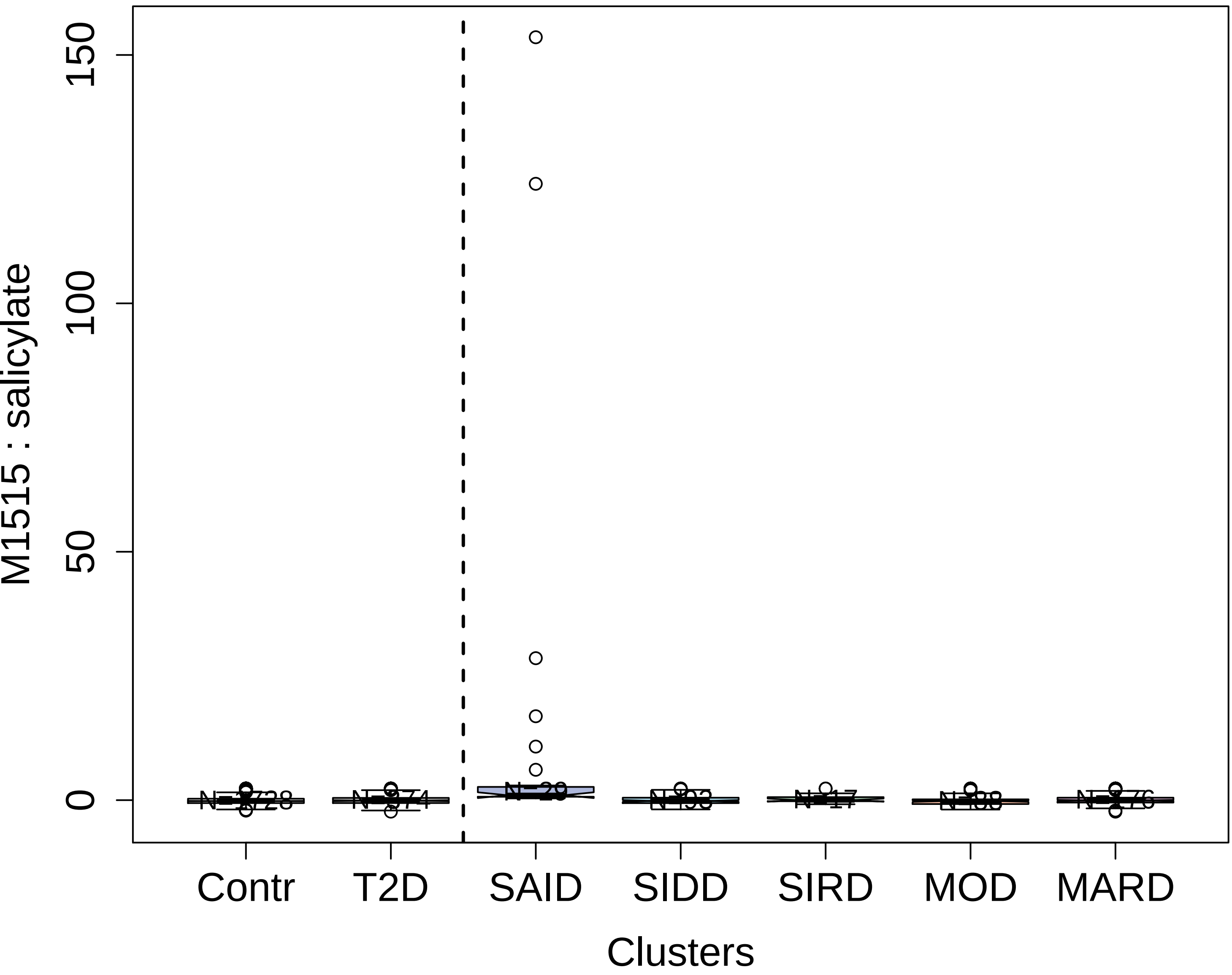

M1515 : salicylate
150
100
50
0
Contr
T2D
SAID
SIDD
SIRD
MOD
MARD
Clusters
N=29
N=17

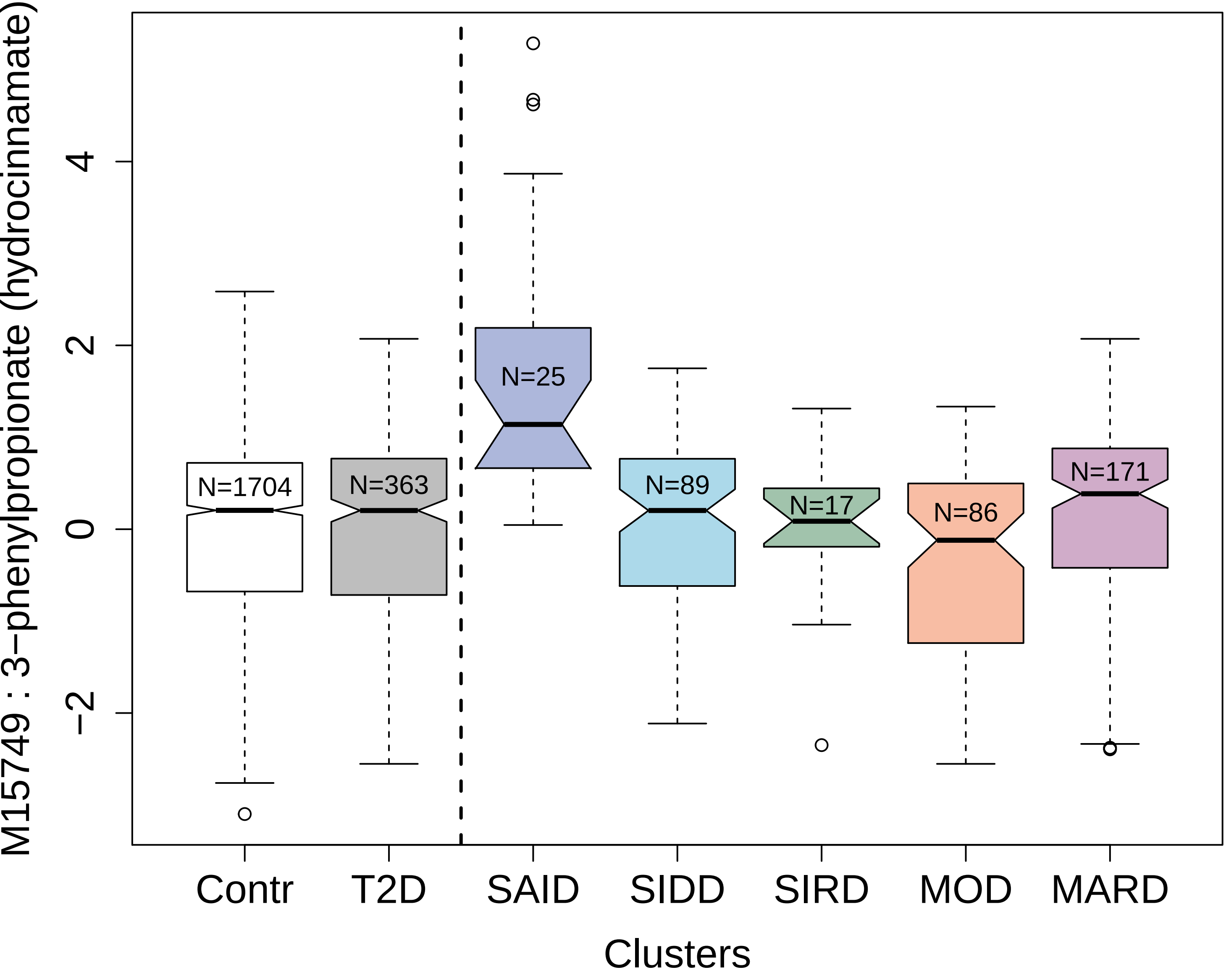

M15749 : 3−phenylpropionate (hydrocinnamate)
4
2
0
−2
N=1704
N=363
N=25
N=89
N=17
N=86
N=171
Contr
T2D
SAID
SIDD
SIRD
MOD
MARD
Clusters

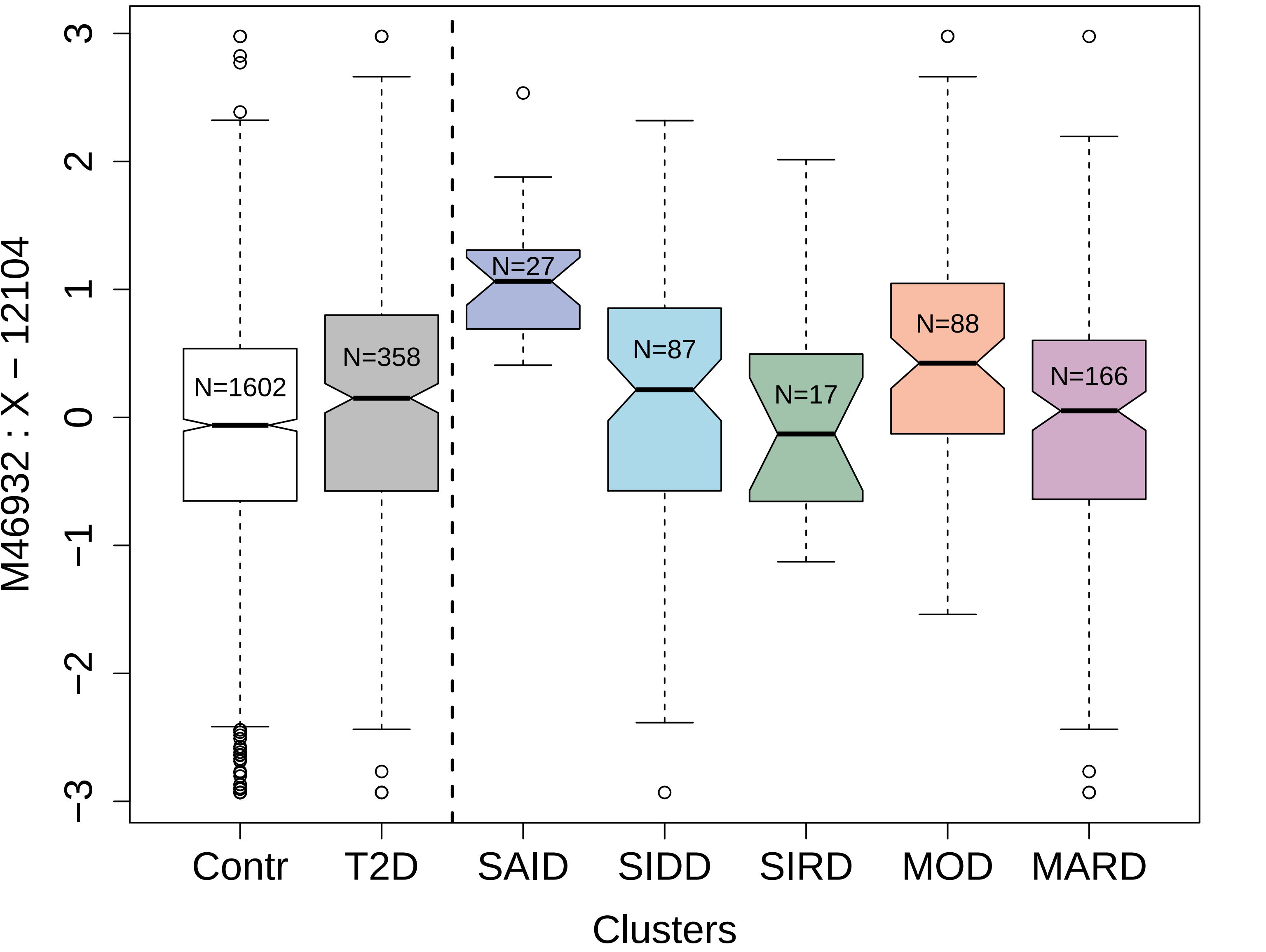

M46932 : X − 12104
3
2
1
0
−1
−2
−3
N=1602
N=358
N=27
N=87
N=17
N=88
N=166
Contr
T2D
SAID
SIDD
SIRD
MOD
MARD
Clusters

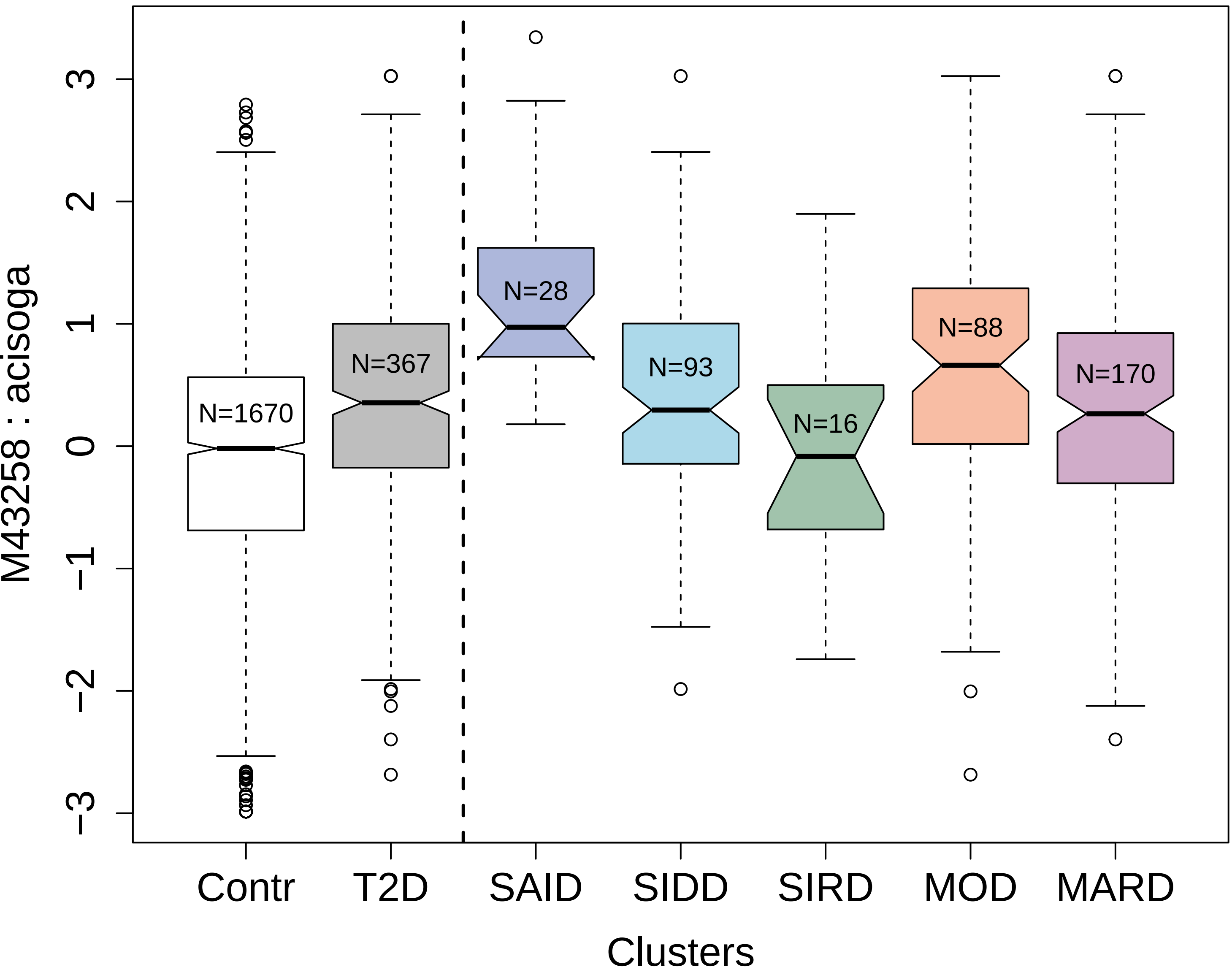
M43258 : acisoga
Clusters
Contr
T2D
SAID
SIDD
SIRD
MOD
MARD
N=1670
N=367
N=28
N=93
N=16
N=88
N=170
3
2
1
0
−1
−2
−3

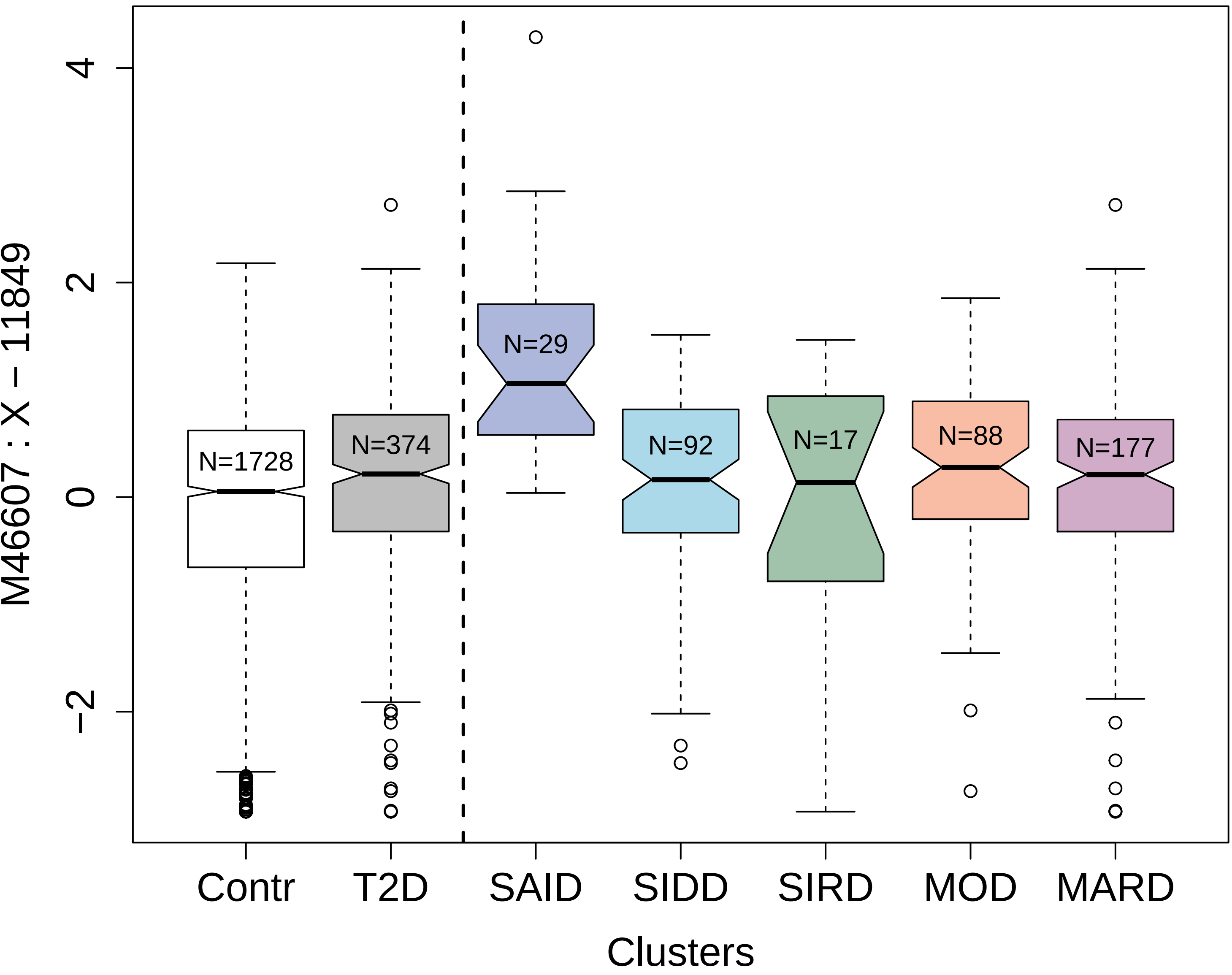

M46607 : X − 11849
4
2
0
−2
N=1728
N=374
N=29
N=92
N=17
N=88
N=177
Contr
T2D
SAID
SIDD
SIRD
MOD
MARD
Clusters

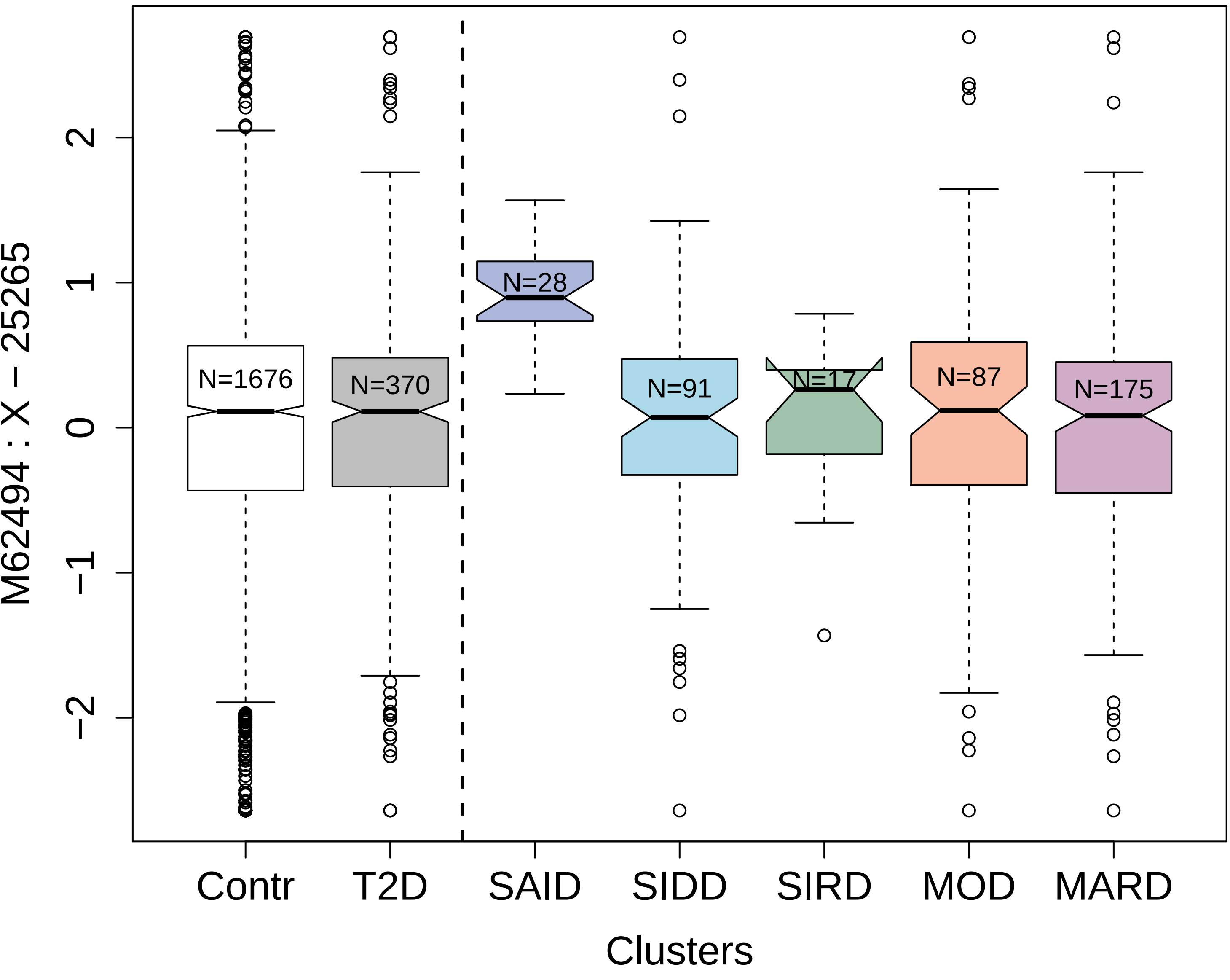
M62494 : X − 25265
2
1
0
−1
−2
N=1676
N=370
N=28
N=91
N=17
N=87
N=175
Contr
T2D
SAID
SIDD
SIRD
MOD
MARD
Clusters

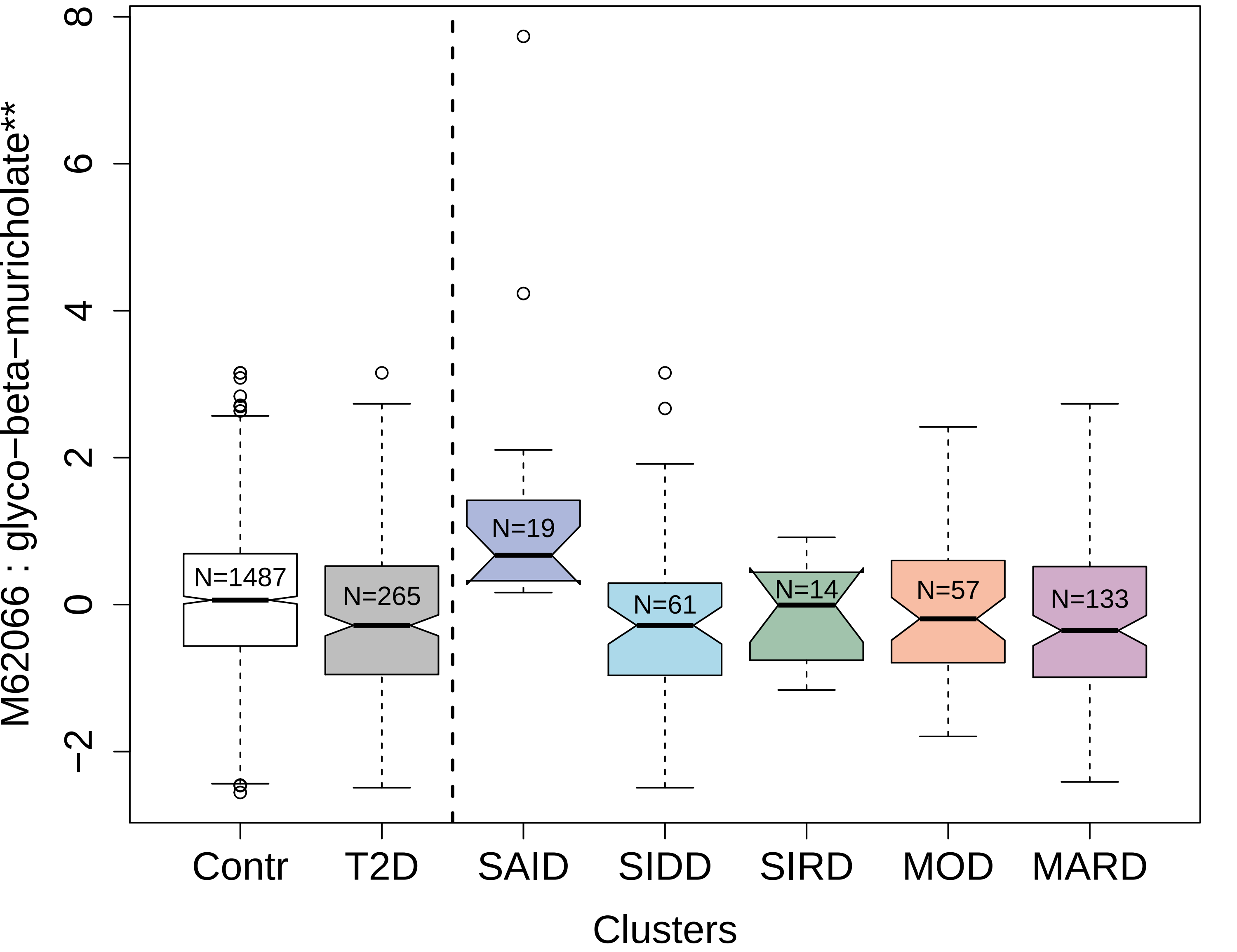
M62066 : glyco−beta−muricholate**
8
6
4
2
0
−2
N=1487
N=265
N=19
N=61
N=14
N=57
N=133
Contr
T2D
SAID
SIDD
SIRD
MOD
MARD
Clusters

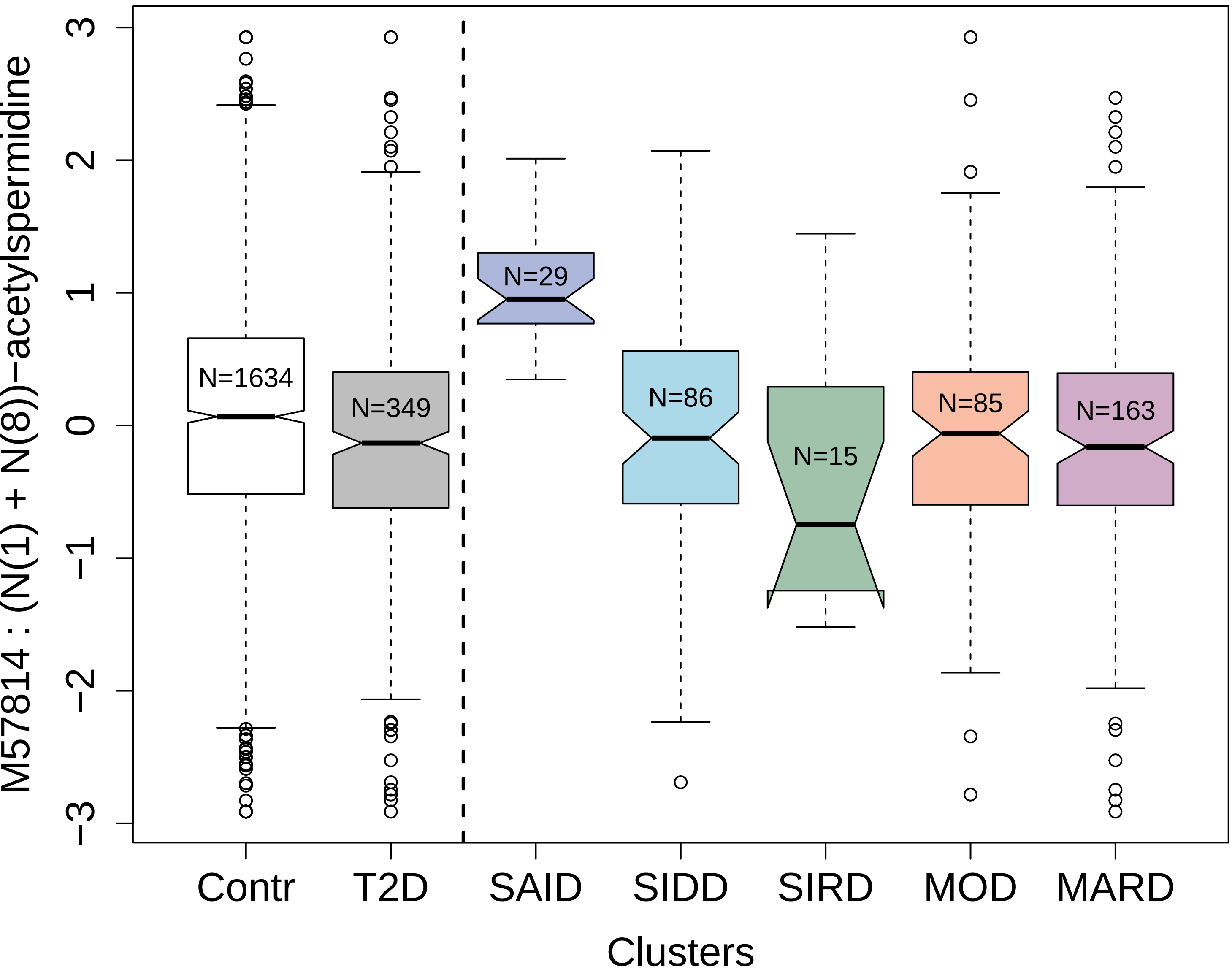

M57814 : (N(1) + N(8))−acetylspermidine
3
2
1
0
−1
−2
−3
N=1634
N=349
N=29
N=86
N=15
N=85
N=163
Contr
T2D
SAID
SIDD
SIRD
MOD
MARD
Clusters

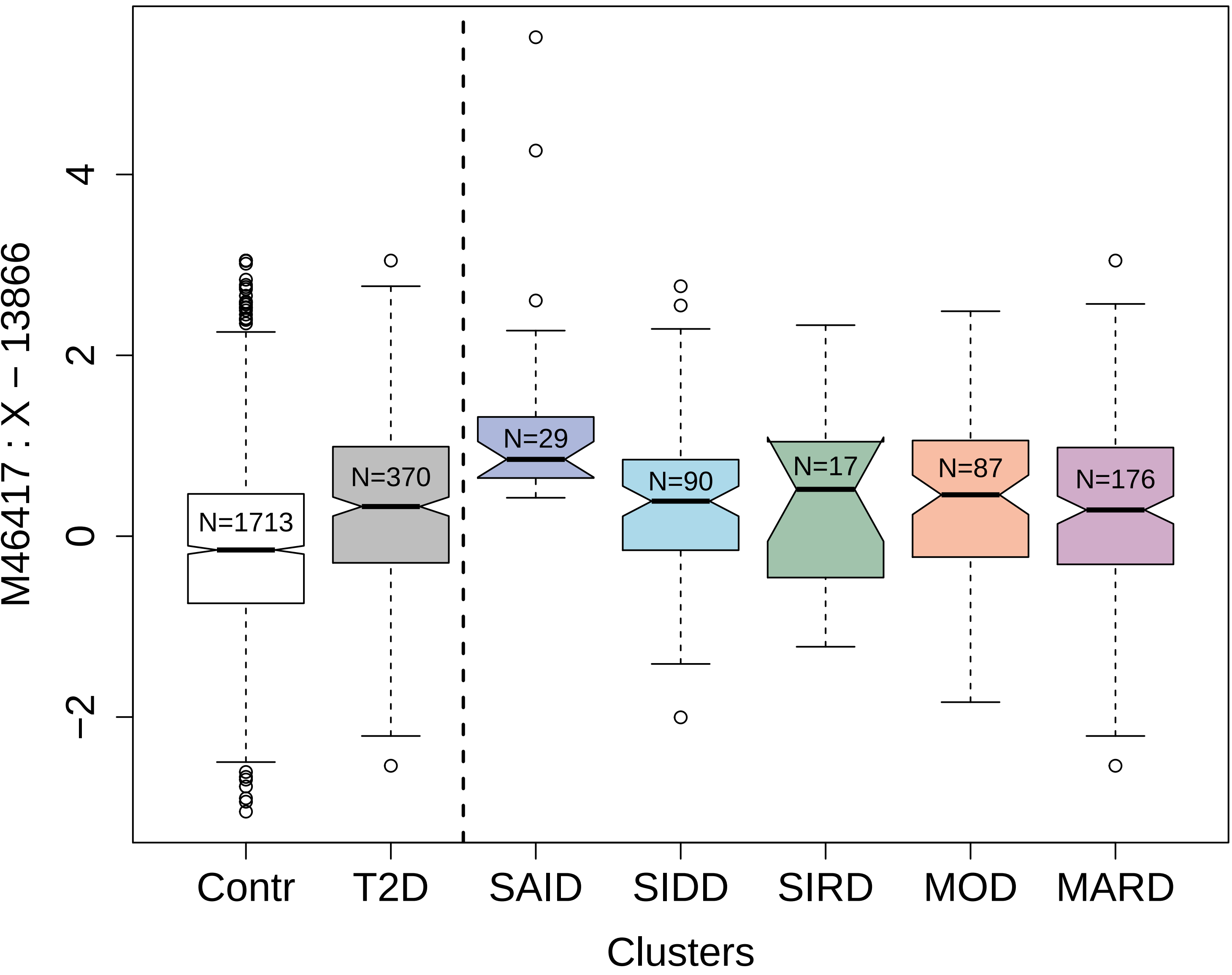
M46417 : X − 13866
Clusters
Contr
T2D
SAID
SIDD
SIRD
MOD
MARD
N=1713
N=370
N=29
N=90
N=17
N=87
N=176
−2
0
2
4

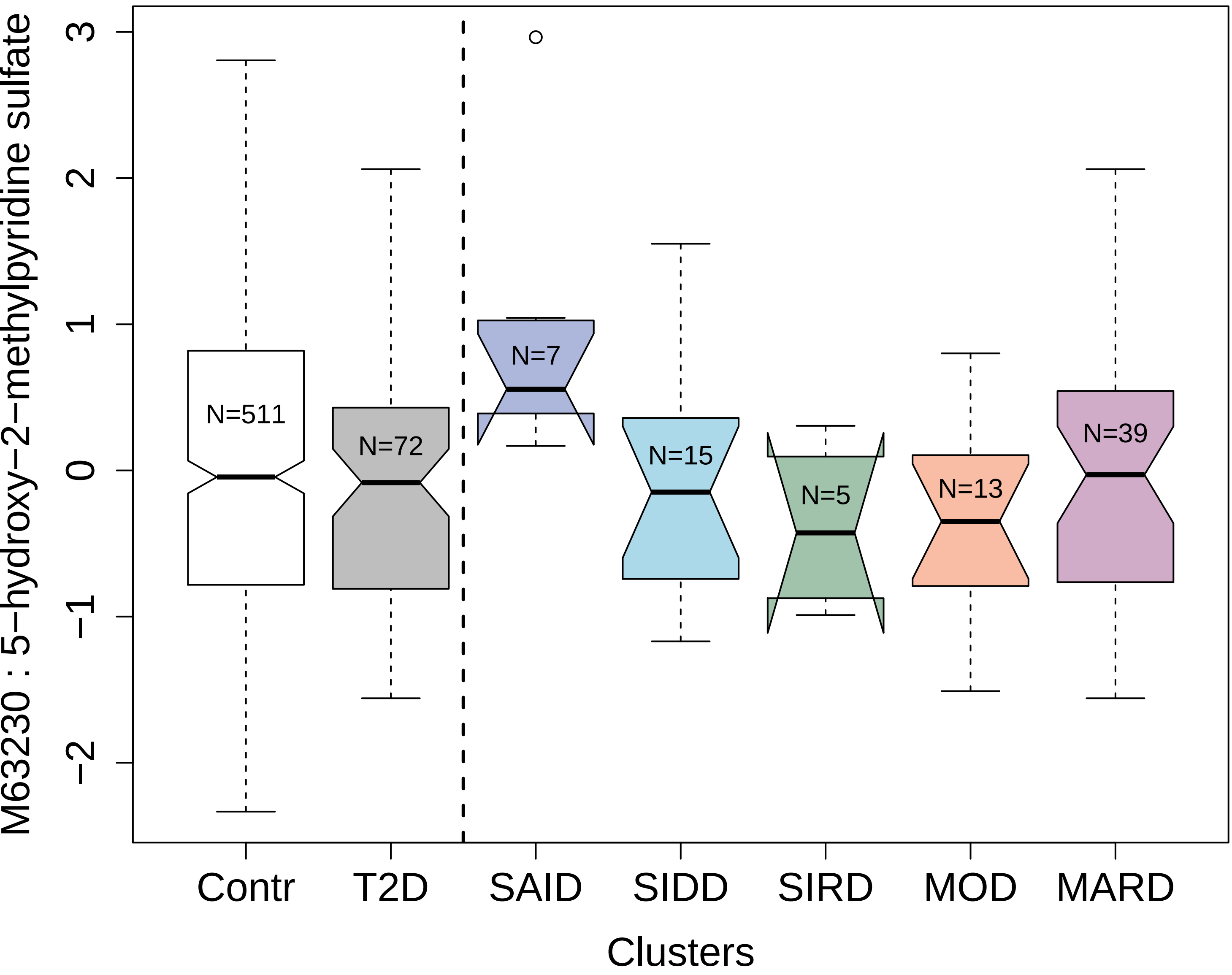
M63230 : 5−hydroxy−2−methylpyridine sulfate
3
2
1
0
−1
−2
N=511
N=72
N=7
N=15
N=5
N=13
N=39
Contr
T2D
SAID
SIDD
SIRD
MOD
MARD
Clusters

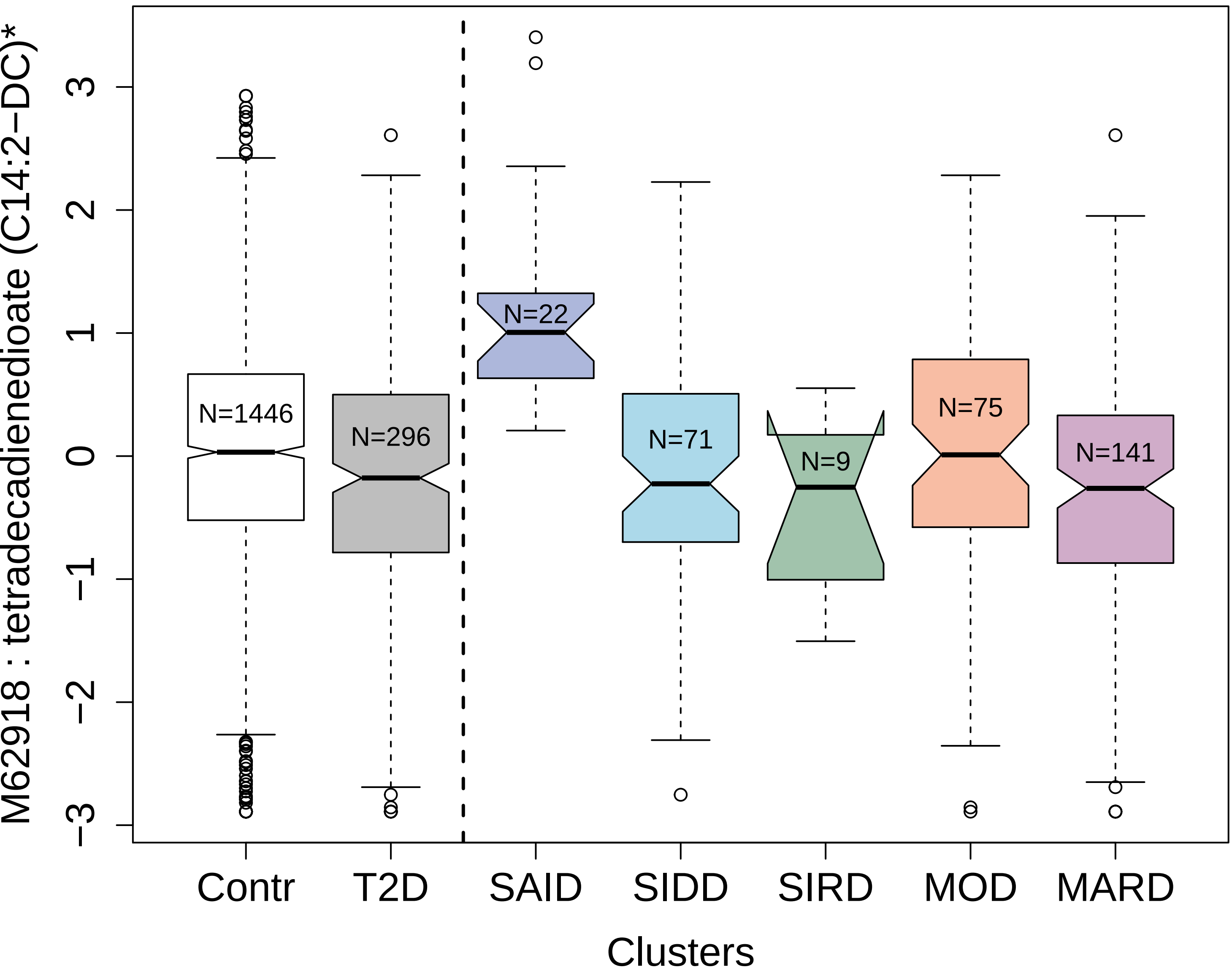

M62918 : tetradecadienedioate (C14:2−DC)*
Clusters
Contr
T2D
SAID
SIDD
SIRD
MOD
MARD
N=1446
N=296
N=22
N=71
N=9
N=75
N=141
−3
−2
−1
0
1
2
3

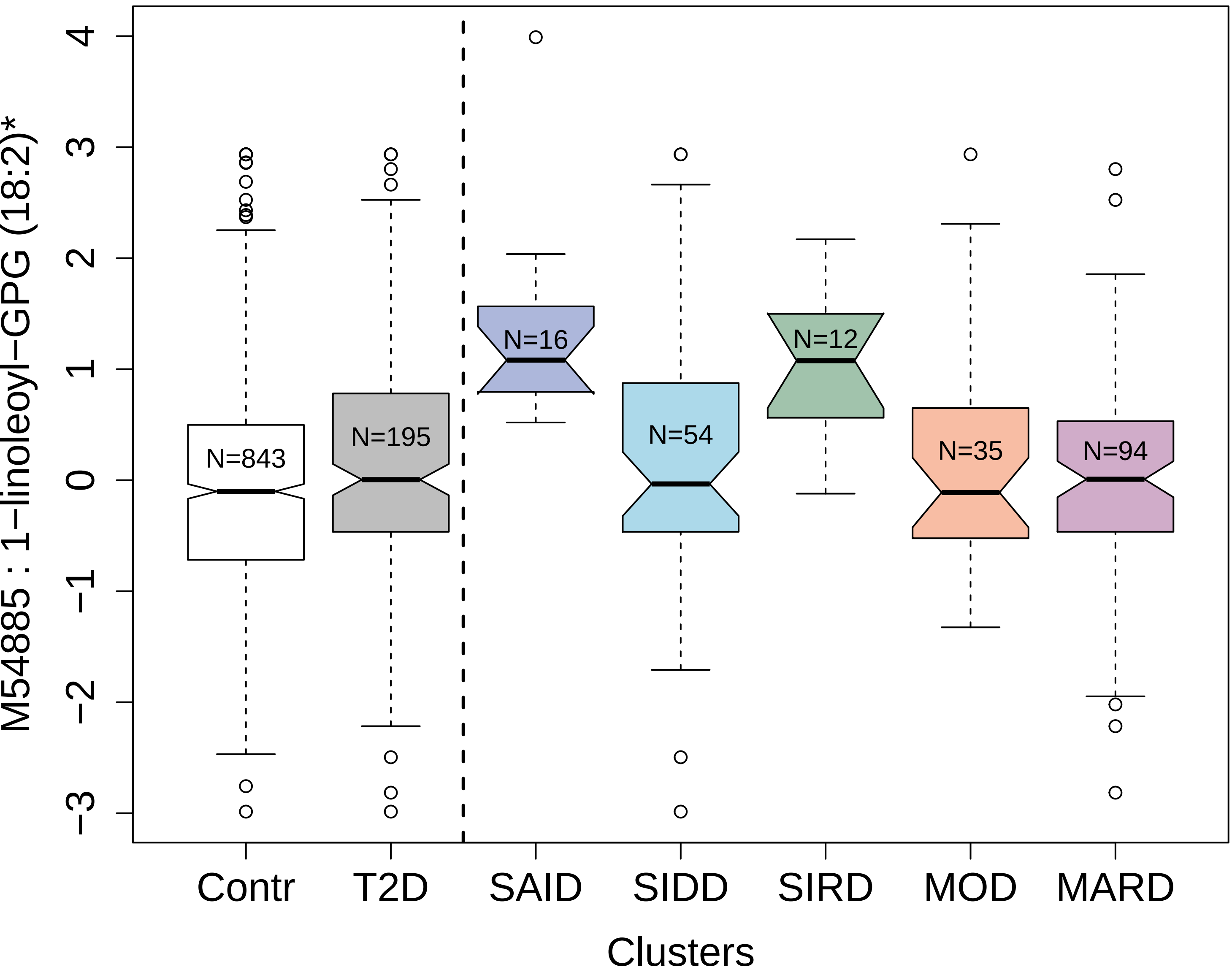

M54885 : 1−linoleoyl−GPG (18:2)*
4
3
2
1
0
−1
−2
−3
N=843
N=195
N=16
N=54
N=12
N=35
N=94
Contr
T2D
SAID
SIDD
SIRD
MOD
MARD
Clusters

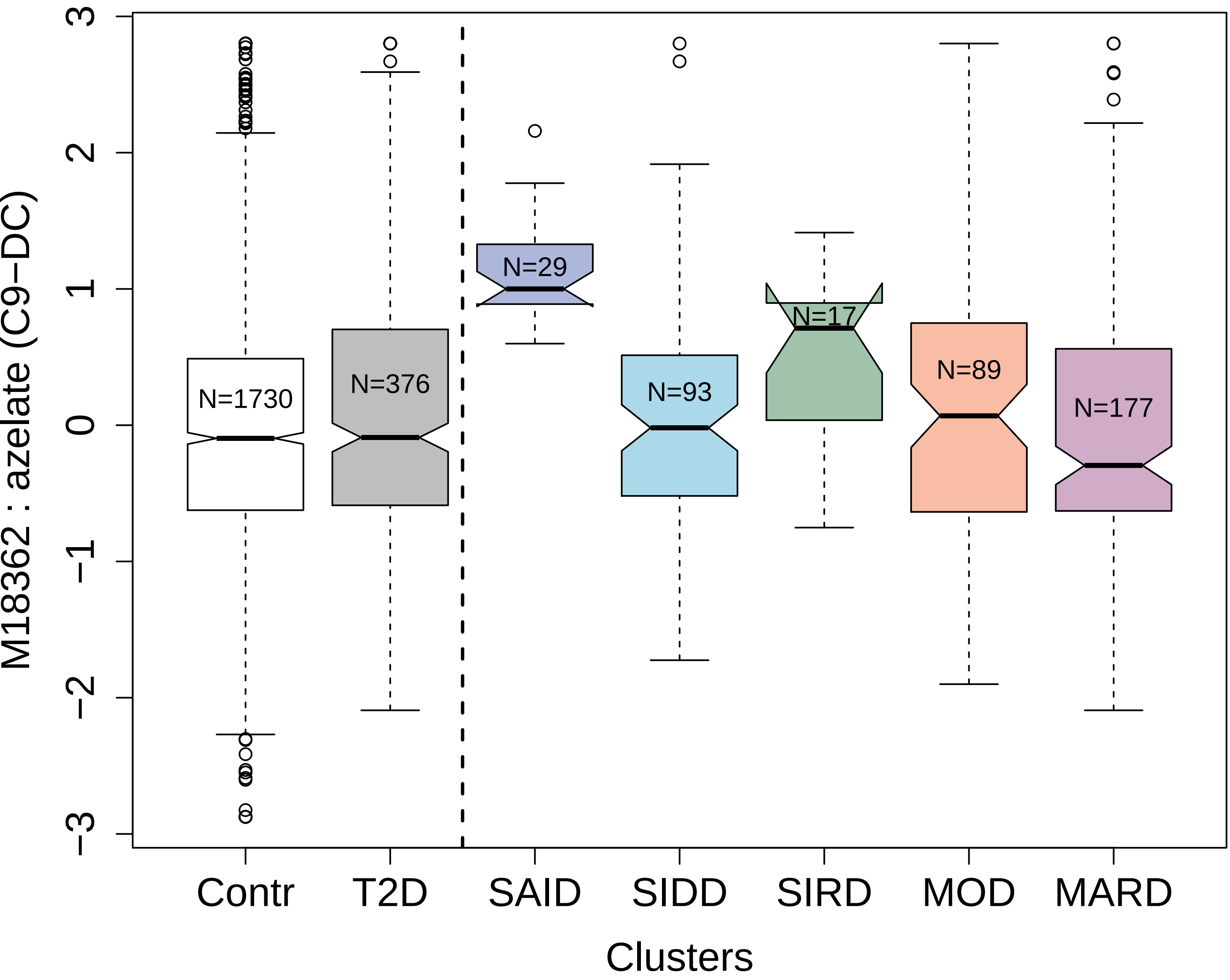
M18362 : azelate (C9−DC)
3
2
1
0
−1
−2
−3
N=1730
N=376
N=29
N=93
N=17
N=89
N=177
Contr
T2D
SAID
SIDD
SIRD
MOD
MARD
Clusters

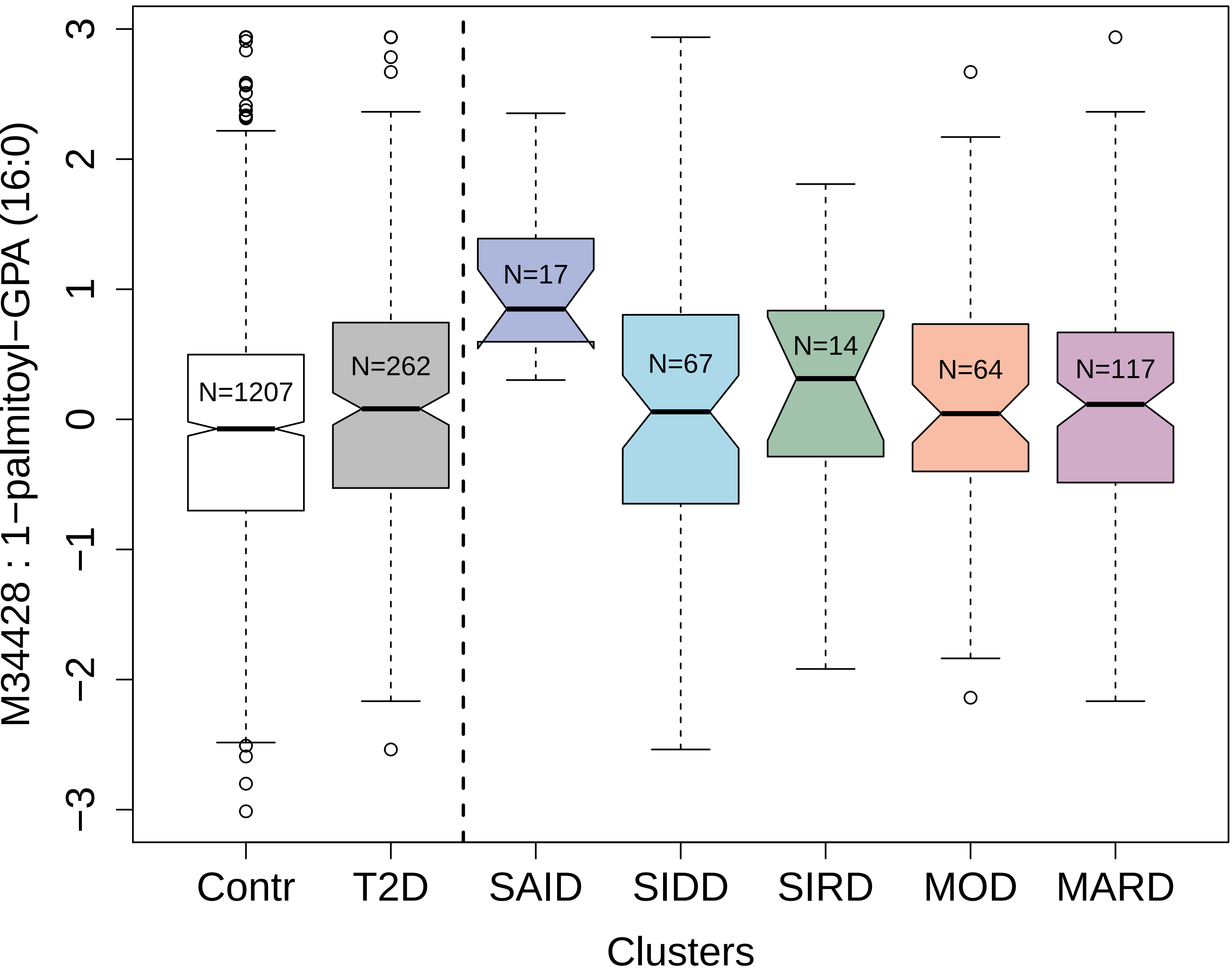

M34428 : 1−palmitoyl−GPA (16:0)
3
2
1
0
−1
−2
−3
N=1207
N=262
N=17
N=67
N=14
N=64
N=117
Contr
T2D
SAID
SIDD
SIRD
MOD
MARD
Clusters

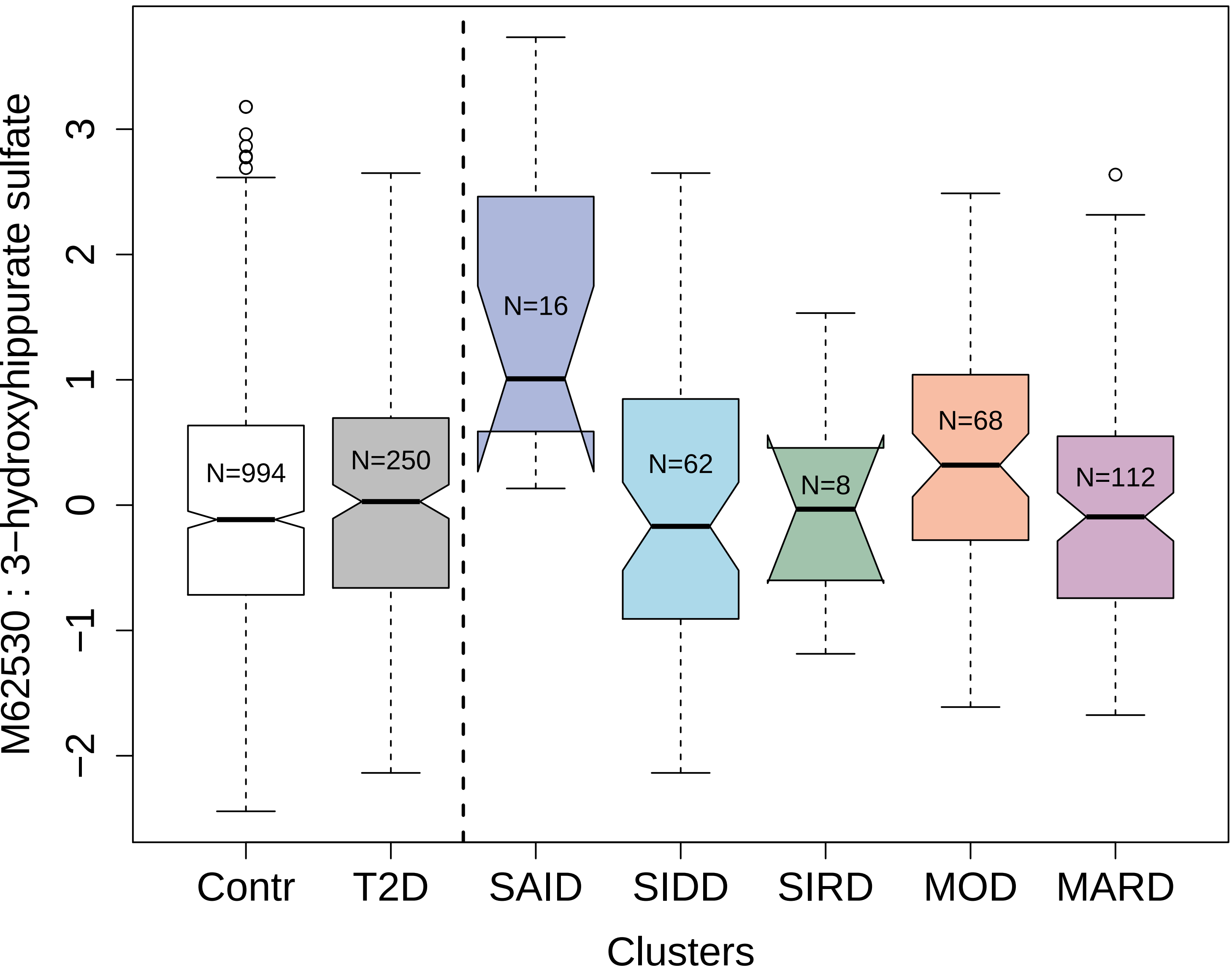
M62530 : 3−hydroxyhippurate sulfate
3
2
1
0
−1
−2
N=994
N=250
N=16
N=62
N=8
N=68
N=112
Contr
T2D
SAID
SIDD
SIRD
MOD
MARD
Clusters

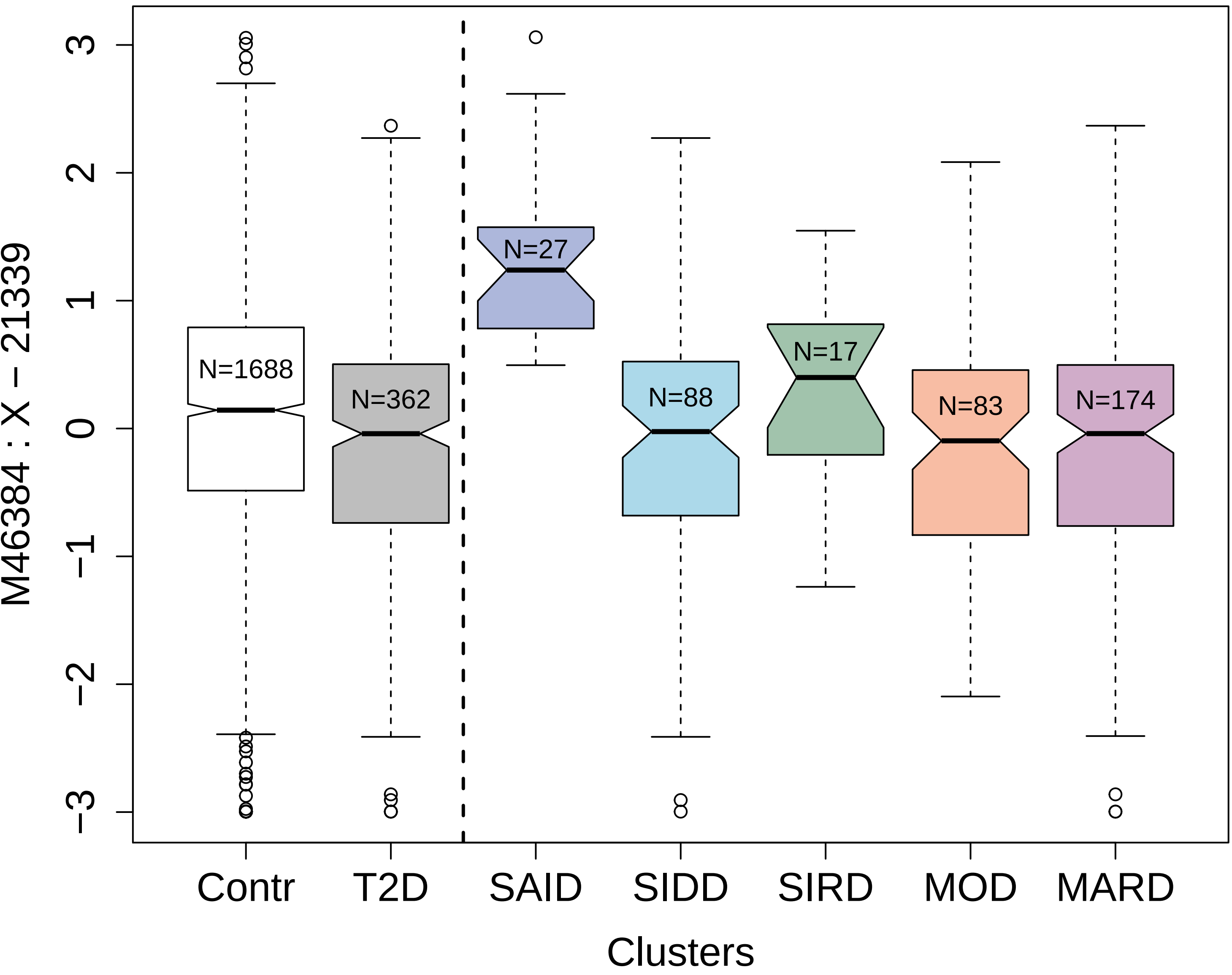

M46384 : X − 21339
3
2
1
0
−1
−2
−3
N=1688
N=362
N=27
N=88
N=17
N=83
N=174
Contr
T2D
SAID
SIDD
SIRD
MOD
MARD
Clusters

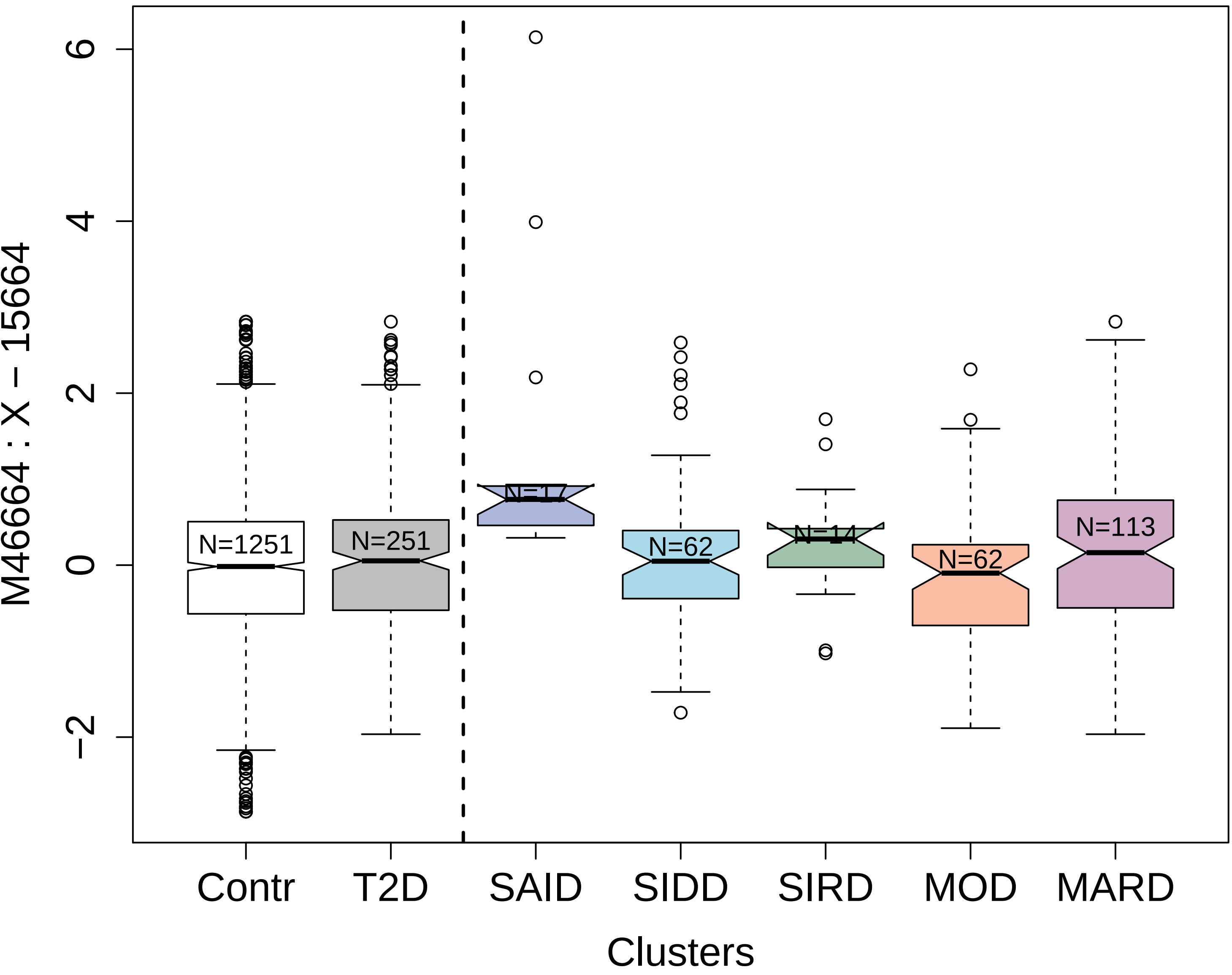

M46664 : X − 15664
6
4
2
0
−2
N=1251
N=251
N=17
N=62
N=14
N=62
N=113
Contr
T2D
SAID
SIDD
SIRD
MOD
MARD
Clusters

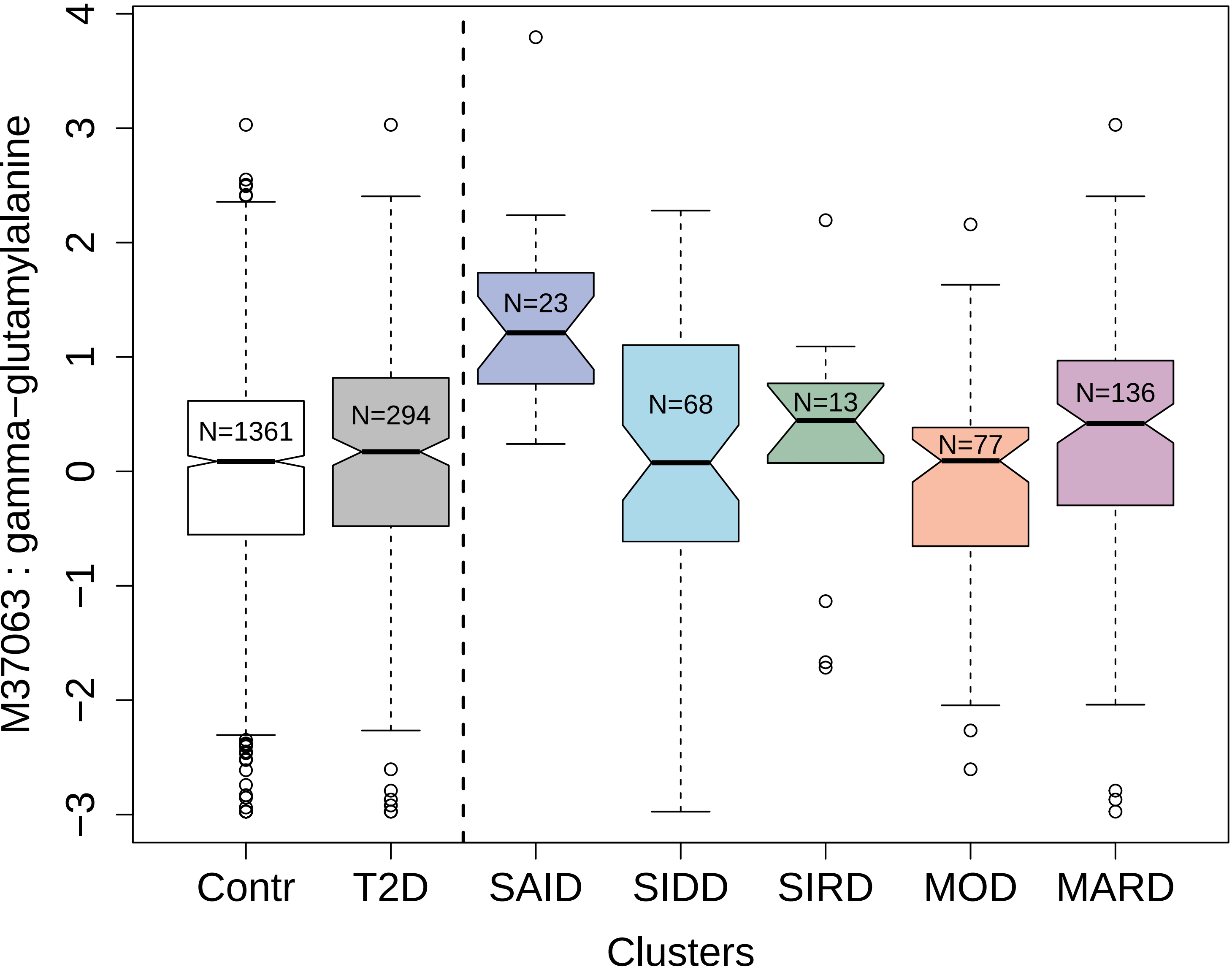

M37063 : gamma−glutamylalanine
4
3
2
1
0
−1
−2
−3
N=1361
N=294
N=23
N=68
N=13
N=77
N=136
Contr
T2D
SAID
SIDD
SIRD
MOD
MARD
Clusters

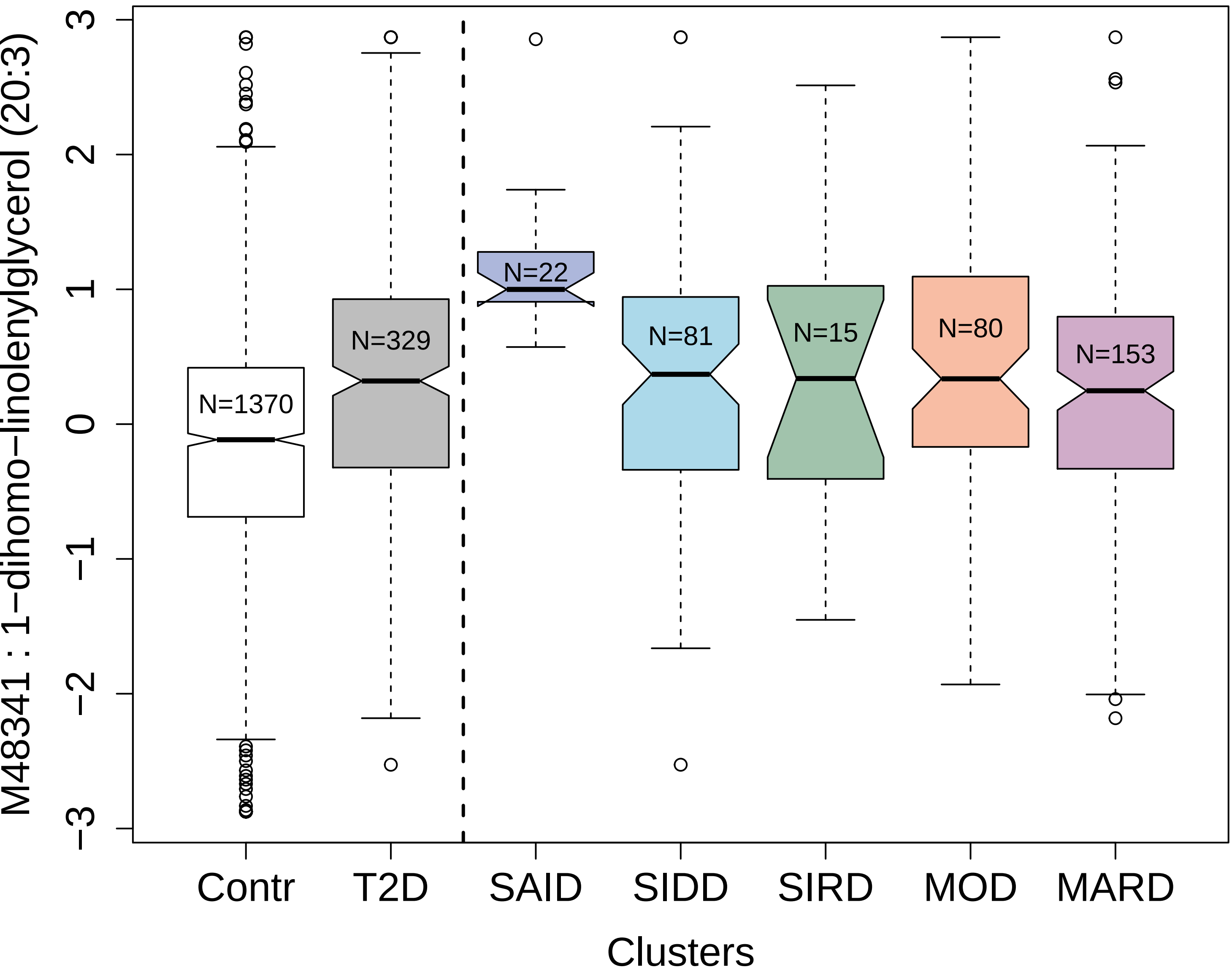
M48341 : 1−dihomo−linolenylglycerol (20:3)
3
2
1
0
−1
−2
−3
N=1370
N=329
N=22
N=81
N=15
N=80
N=153
Contr
T2D
SAID
SIDD
SIRD
MOD
MARD
Clusters

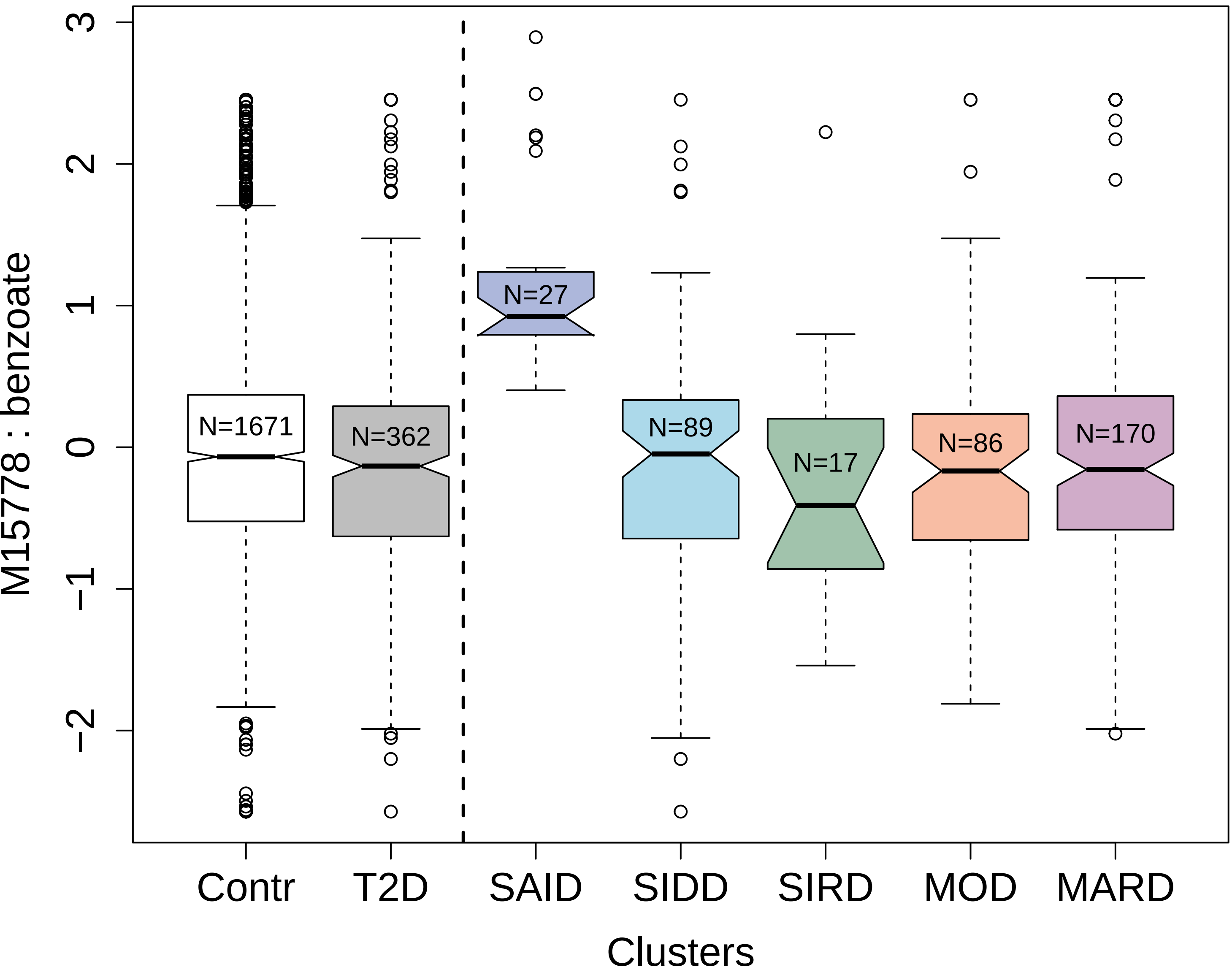

M15778 : benzoate
3
2
1
0
−1
−2
N=1671
N=362
N=27
N=89
N=17
N=86
N=170
Contr
T2D
SAID
SIDD
SIRD
MOD
MARD
Clusters

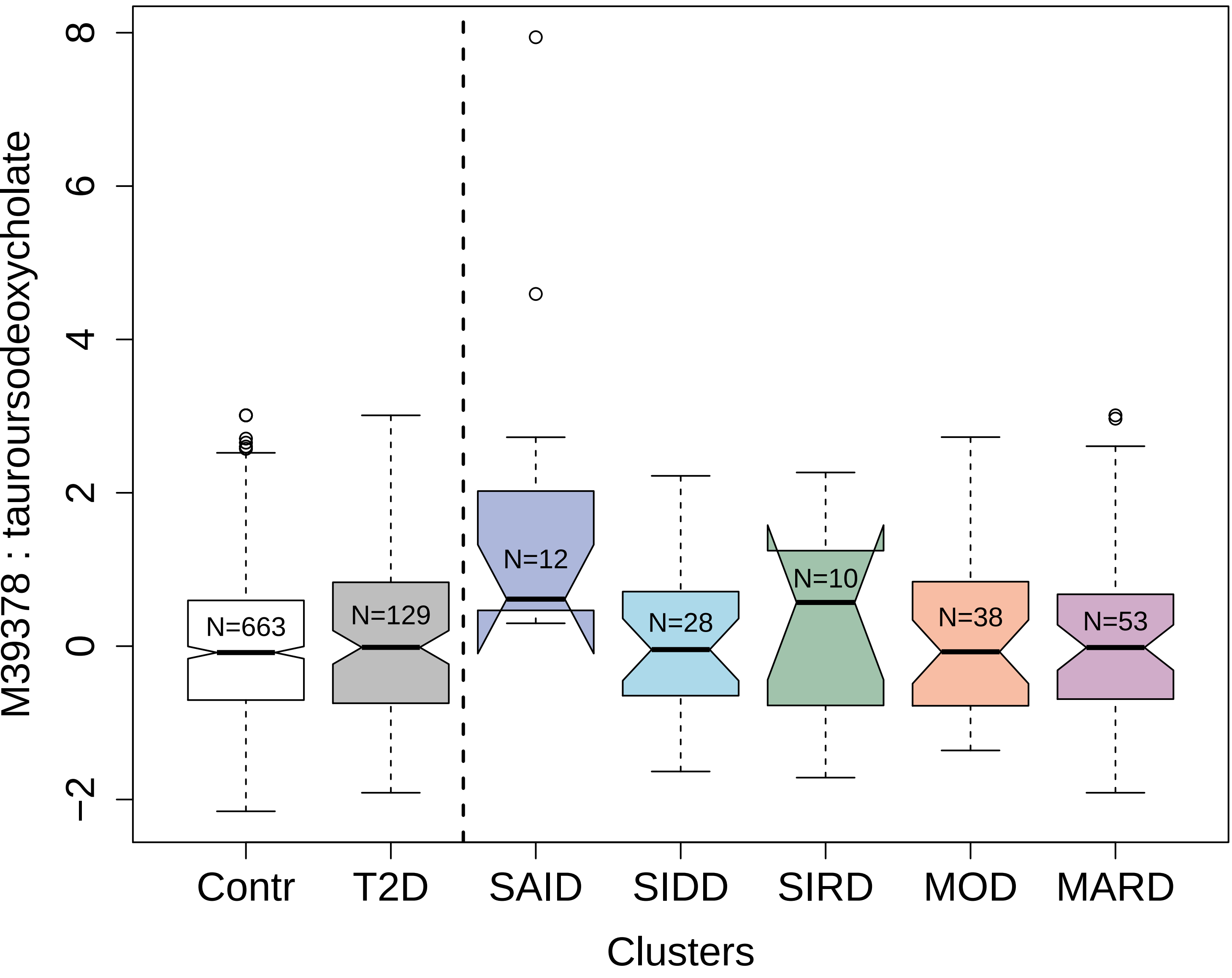
M39378 : tauroursodeoxycholate
8
6
4
2
0
−2
N=663
N=129
N=12
N=28
N=10
N=38
N=53
Contr
T2D
SAID
SIDD
SIRD
MOD
MARD
Clusters

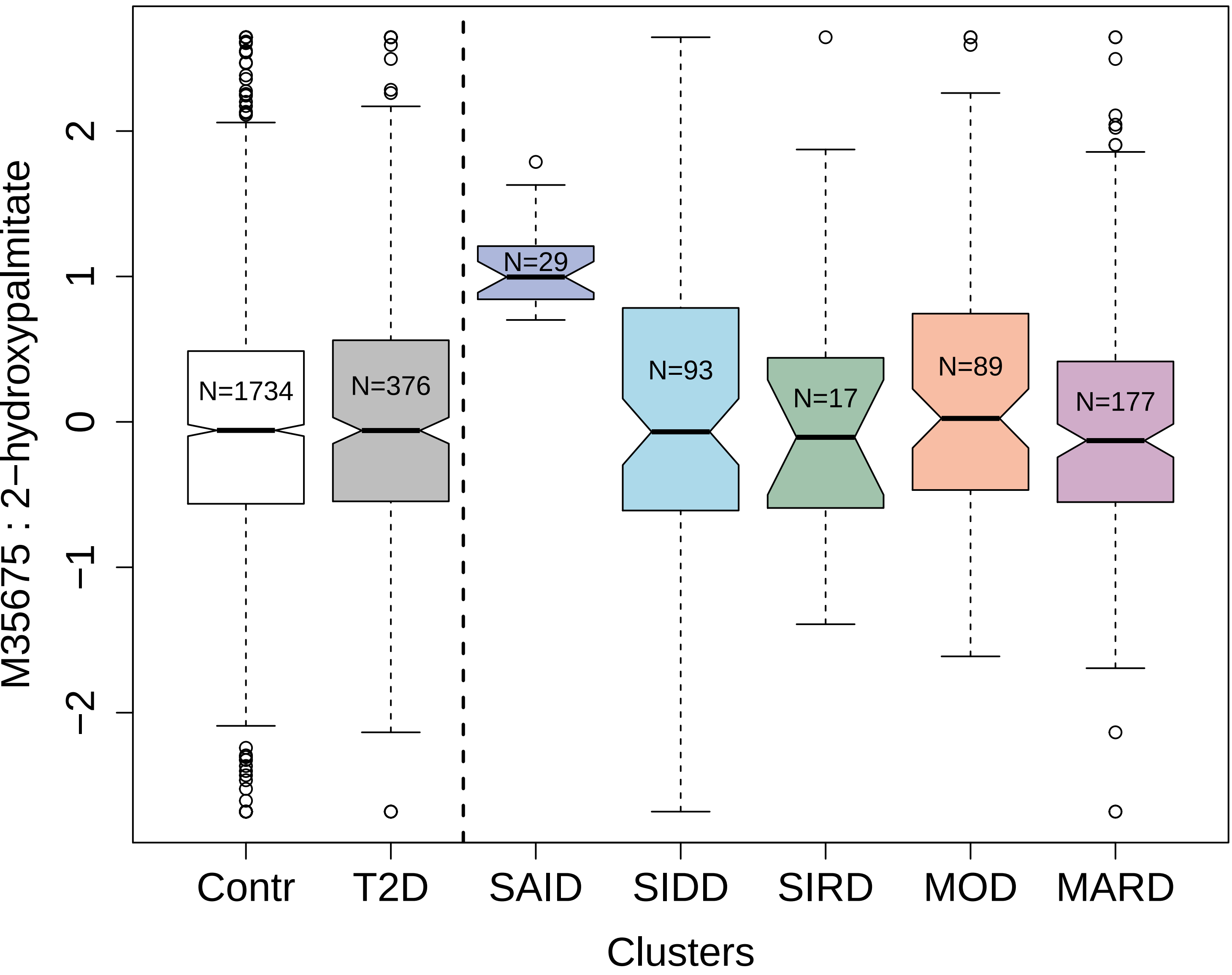

M35675 : 2−hydroxypalmitate
Clusters
Contr
T2D
SAID
SIDD
SIRD
MOD
MARD
N=1734
N=376
N=29
N=93
N=17
N=89
N=177
2
1
0
−1
−2

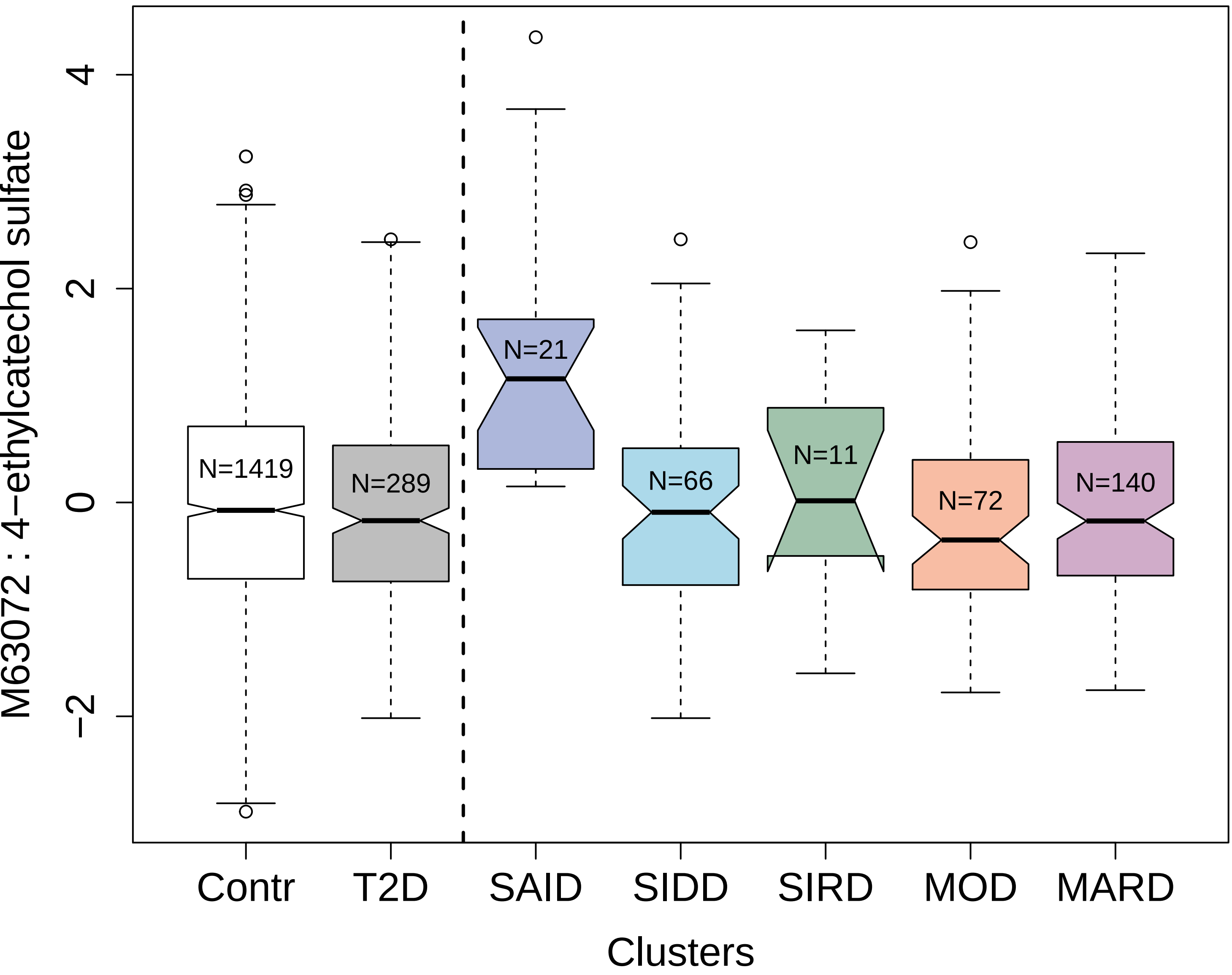

M63072 : 4−ethylcatechol sulfate
4
2
0
−2
N=1419
N=289
N=21
N=66
N=11
N=72
N=140
Contr
T2D
SAID
SIDD
SIRD
MOD
MARD
Clusters

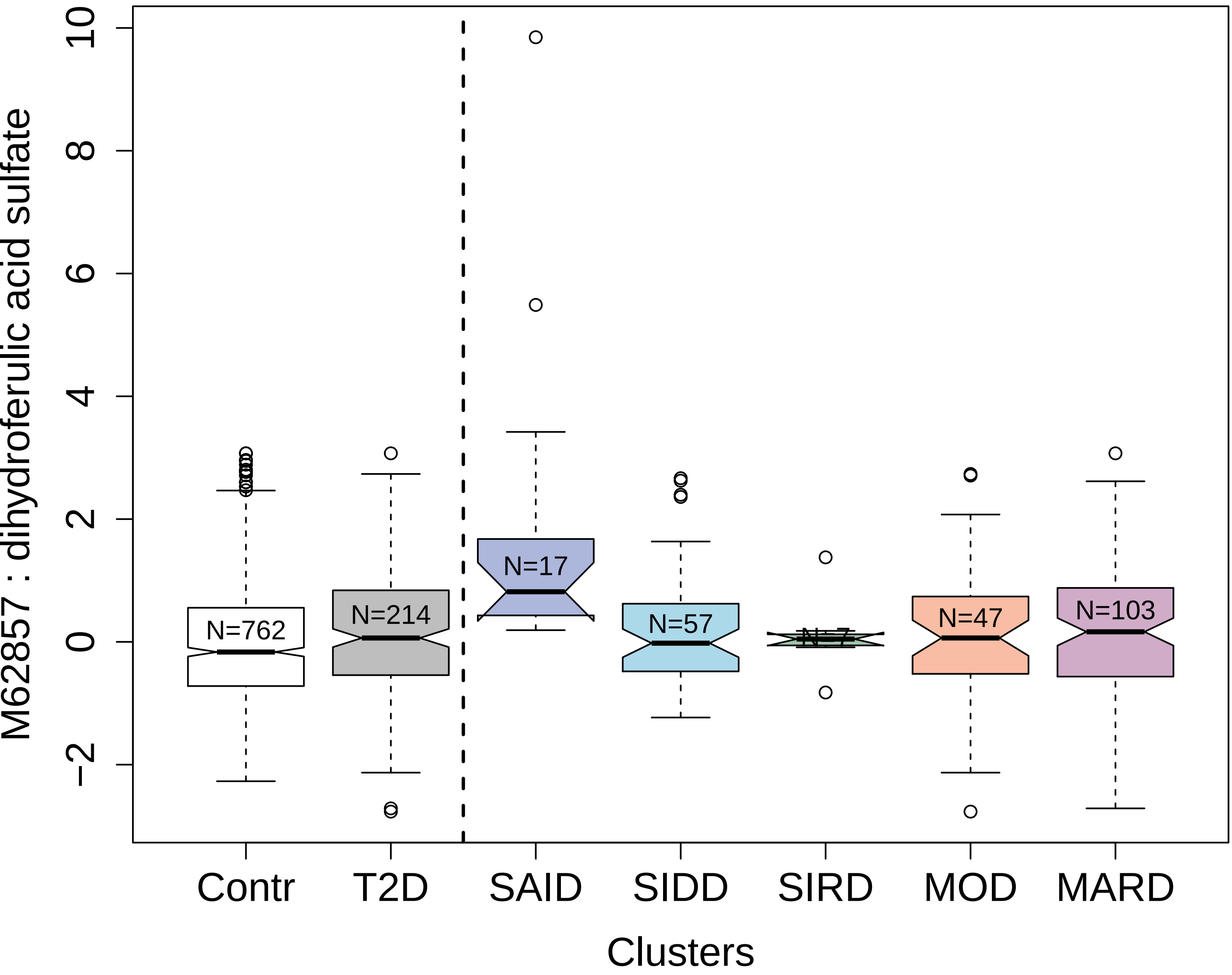
M62857 : dihydroferulic acid sulfate
10
8
6
4
2
0
−2
N=762
N=214
N=17
N=57
N=47
N=103
Contr
T2D
SAID
SIDD
SIRD
MOD
MARD
Clusters

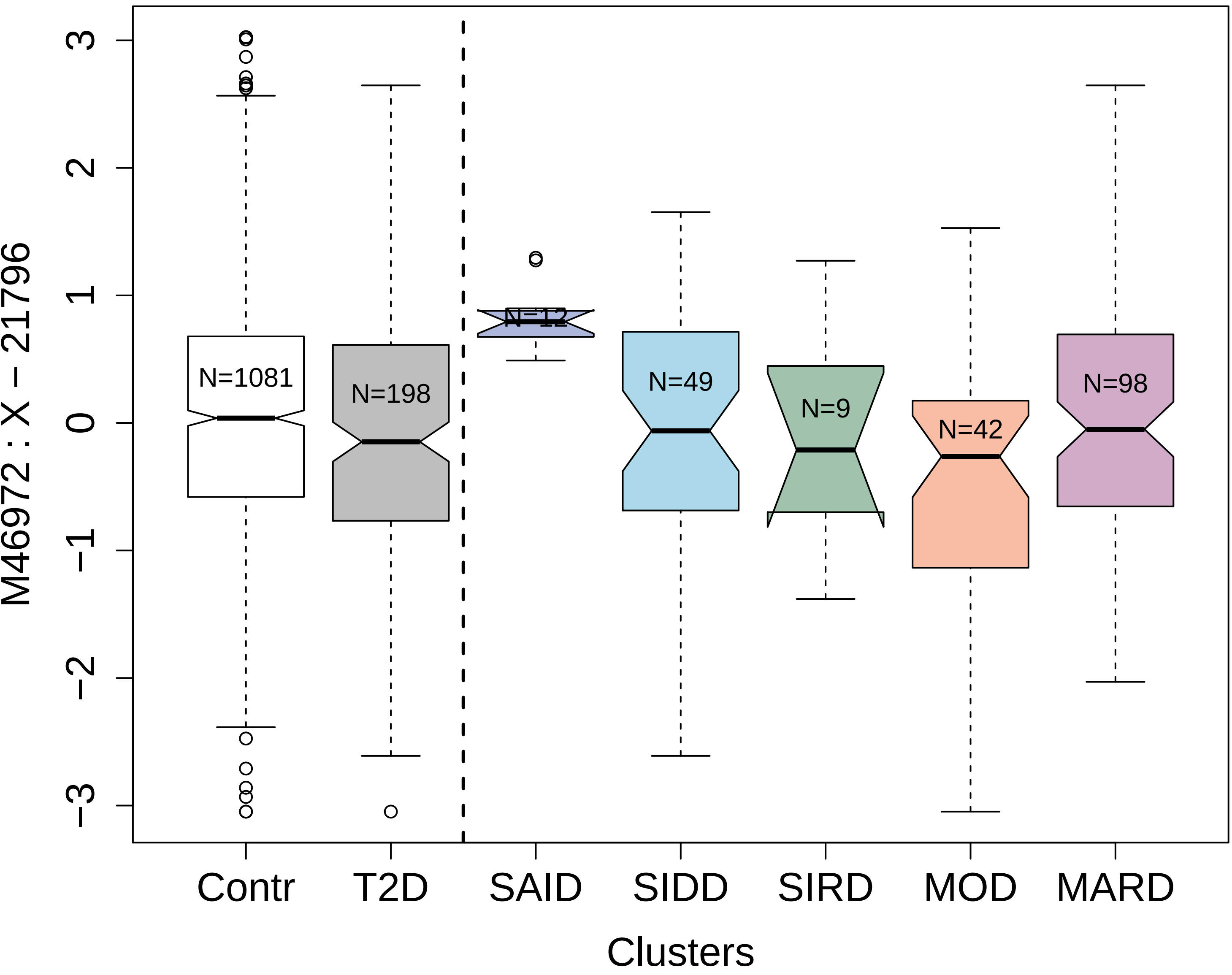

M46972 : X − 21796
3
2
1
0
−1
−2
−3
N=1081
N=198
N=12
N=49
N=9
N=42
N=98
Contr
T2D
SAID
SIDD
SIRD
MOD
MARD
Clusters

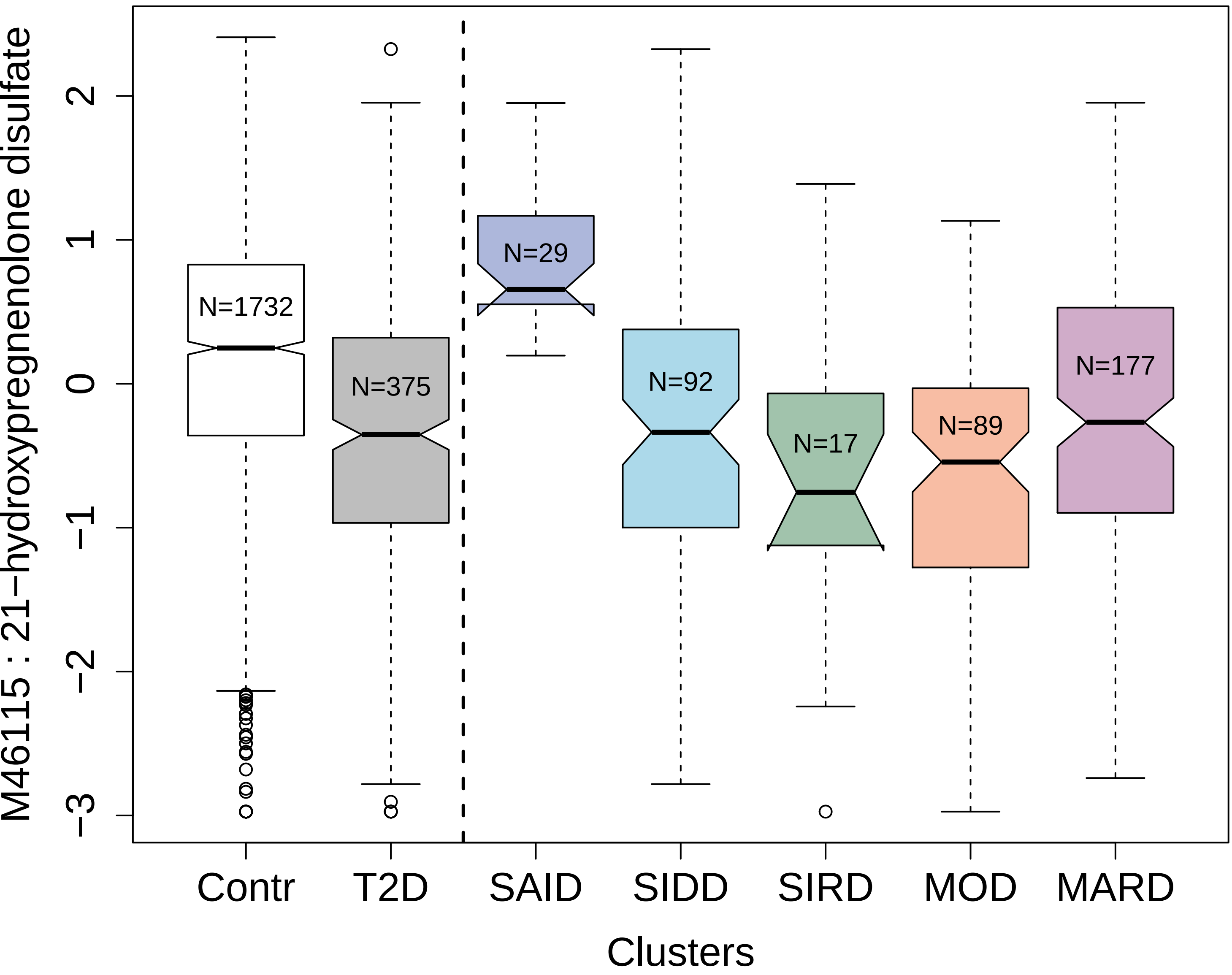

M46115 : 21−hydroxypregnenolone disulfate
2
1
0
−1
−2
−3
N=1732
N=375
N=29
N=92
N=17
N=89
N=177
Contr
T2D
SAID
SIDD
SIRD
MOD
MARD
Clusters

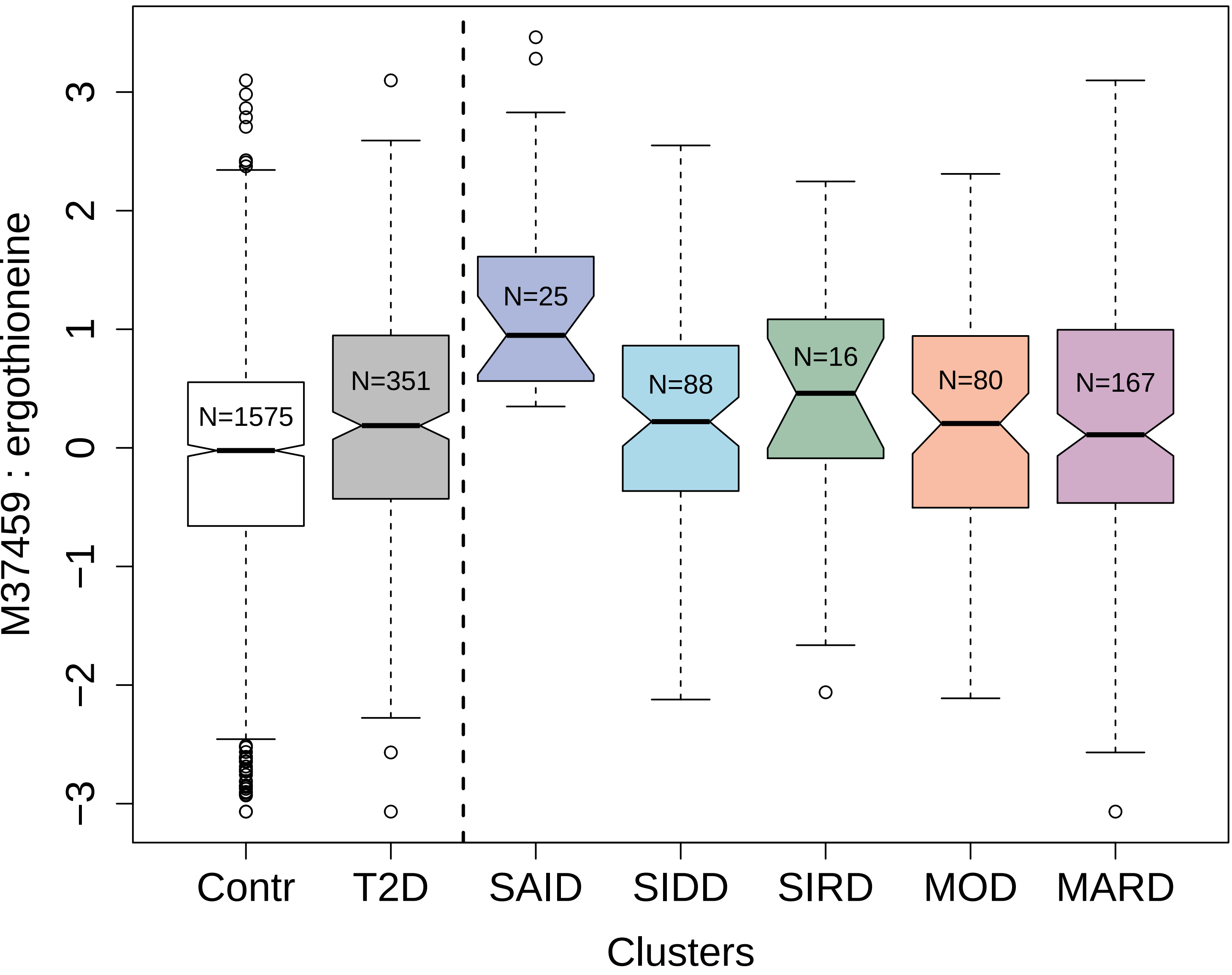
M37459 : ergothioneine
3
2
1
0
−1
−2
−3
N=1575
N=351
N=25
N=88
N=16
N=80
N=167
Contr
T2D
SAID
SIDD
SIRD
MOD
MARD
Clusters

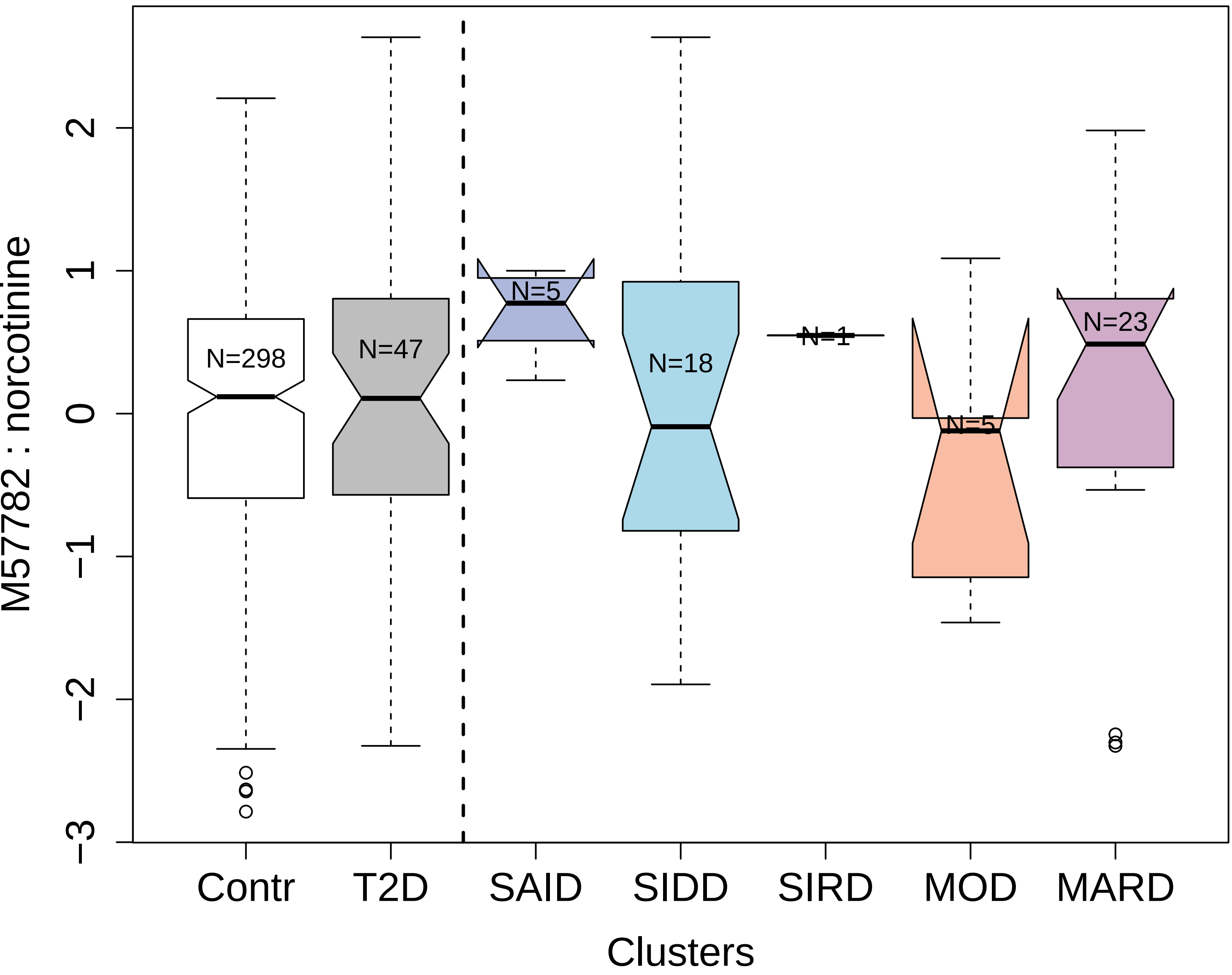

M57782 : norcotinine
2
1
0
−1
−2
−3
N=298
N=47
N=5
N=18
N=1
N=5
N=23
Contr
T2D
SAID
SIDD
SIRD
MOD
MARD
Clusters

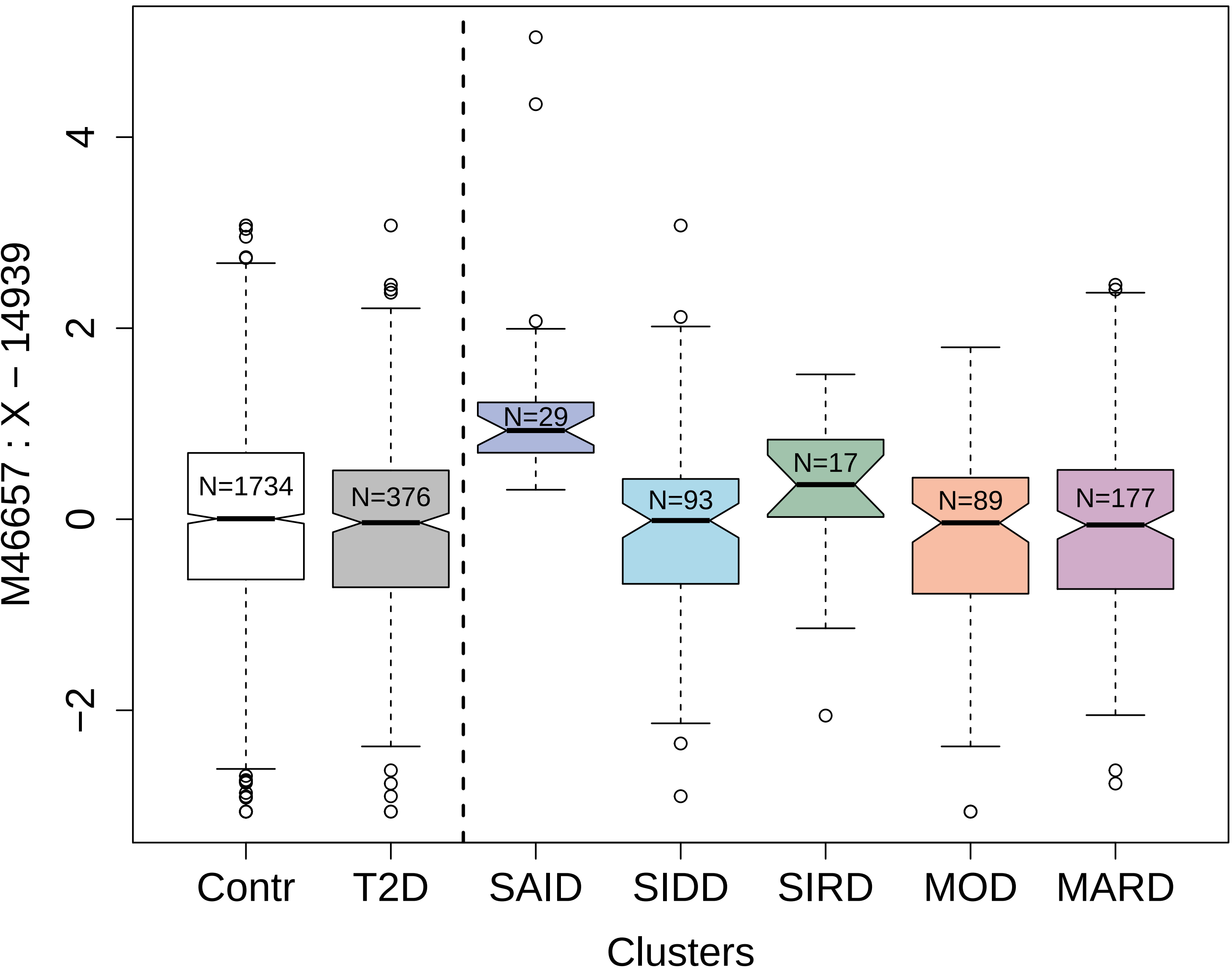

M46657 : X − 14939
4
2
0
−2
N=1734
N=376
N=29
N=93
N=17
N=89
N=177
Contr
T2D
SAID
SIDD
SIRD
MOD
MARD
Clusters

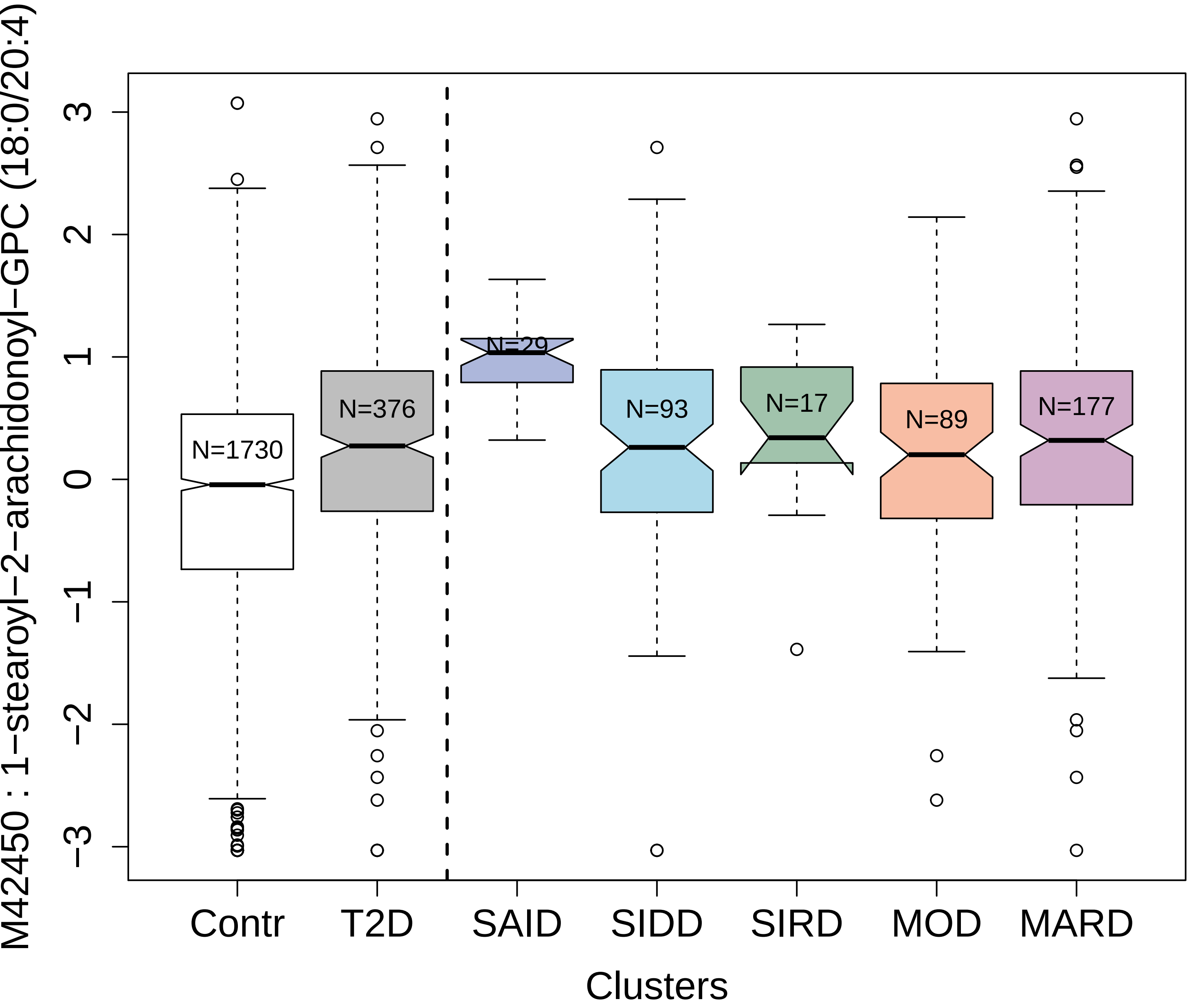
M42450 : 1−stearoyl−2−arachidonoyl−GPC (18:0/20:4)
3
2
1
0
−1
−2
−3
N=1730
N=376
N=29
N=93
N=17
N=89
N=177
Contr
T2D
SAID
SIDD
SIRD
MOD
MARD
Clusters

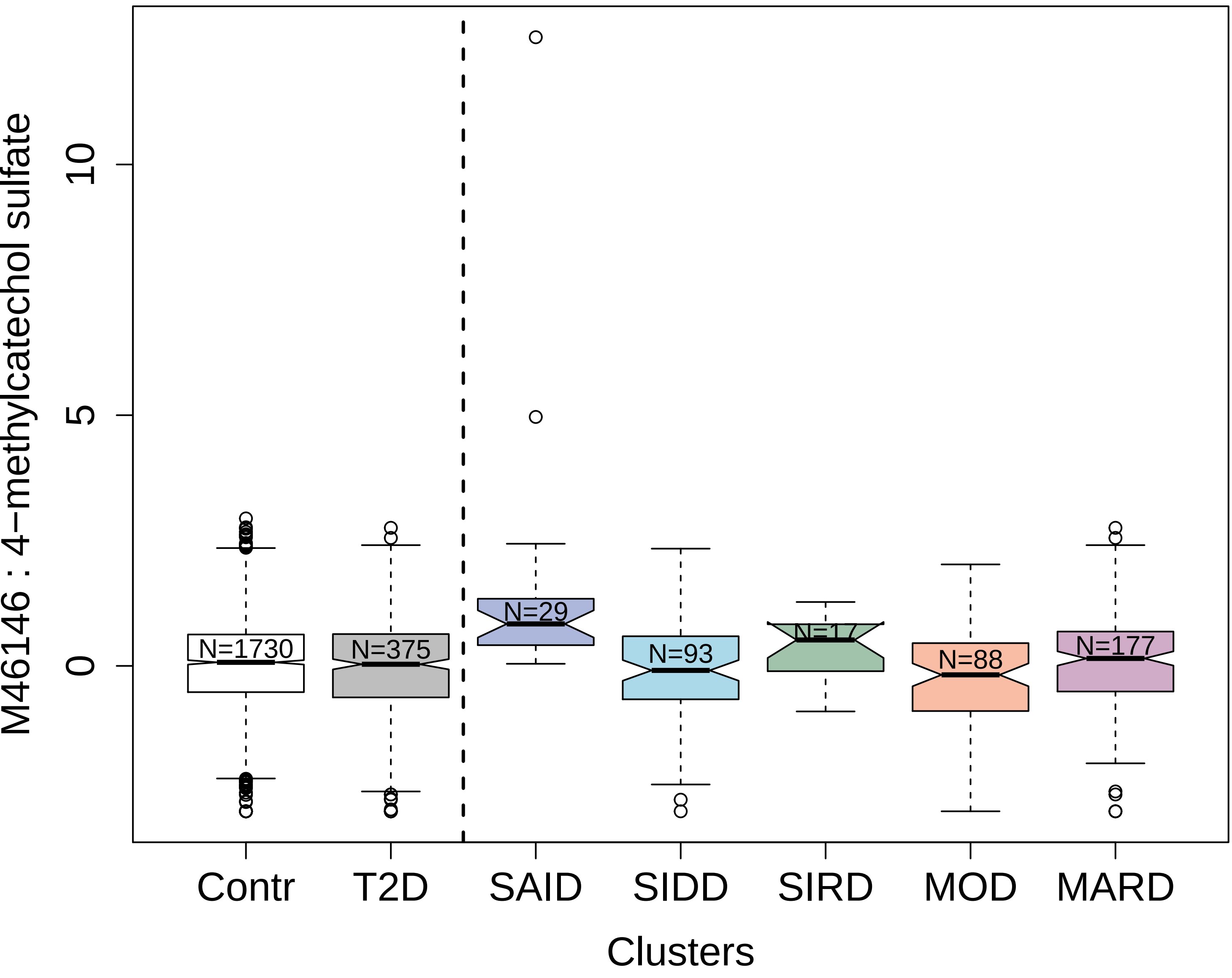

M46146 : 4−methylcatechol sulfate
0
5
10
N=1730
N=375
N=29
N=93
N=17
N=88
N=177
Contr
T2D
SAID
SIDD
SIRD
MOD
MARD
Clusters

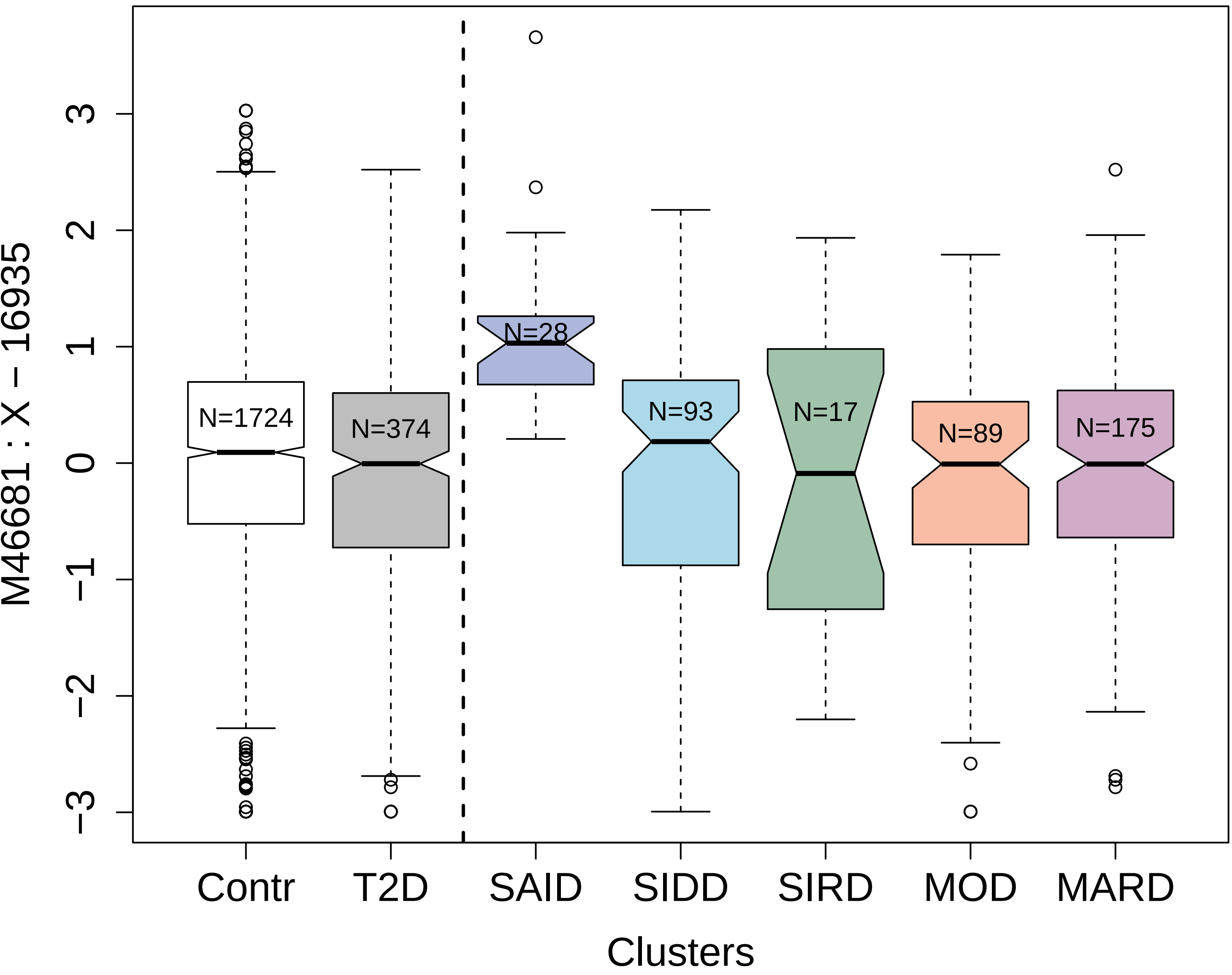
M46681 : X − 16935
3
2
1
0
−1
−2
−3
N=1724
N=374
N=28
N=93
N=17
N=89
N=175
Contr
T2D
SAID
SIDD
SIRD
MOD
MARD
Clusters

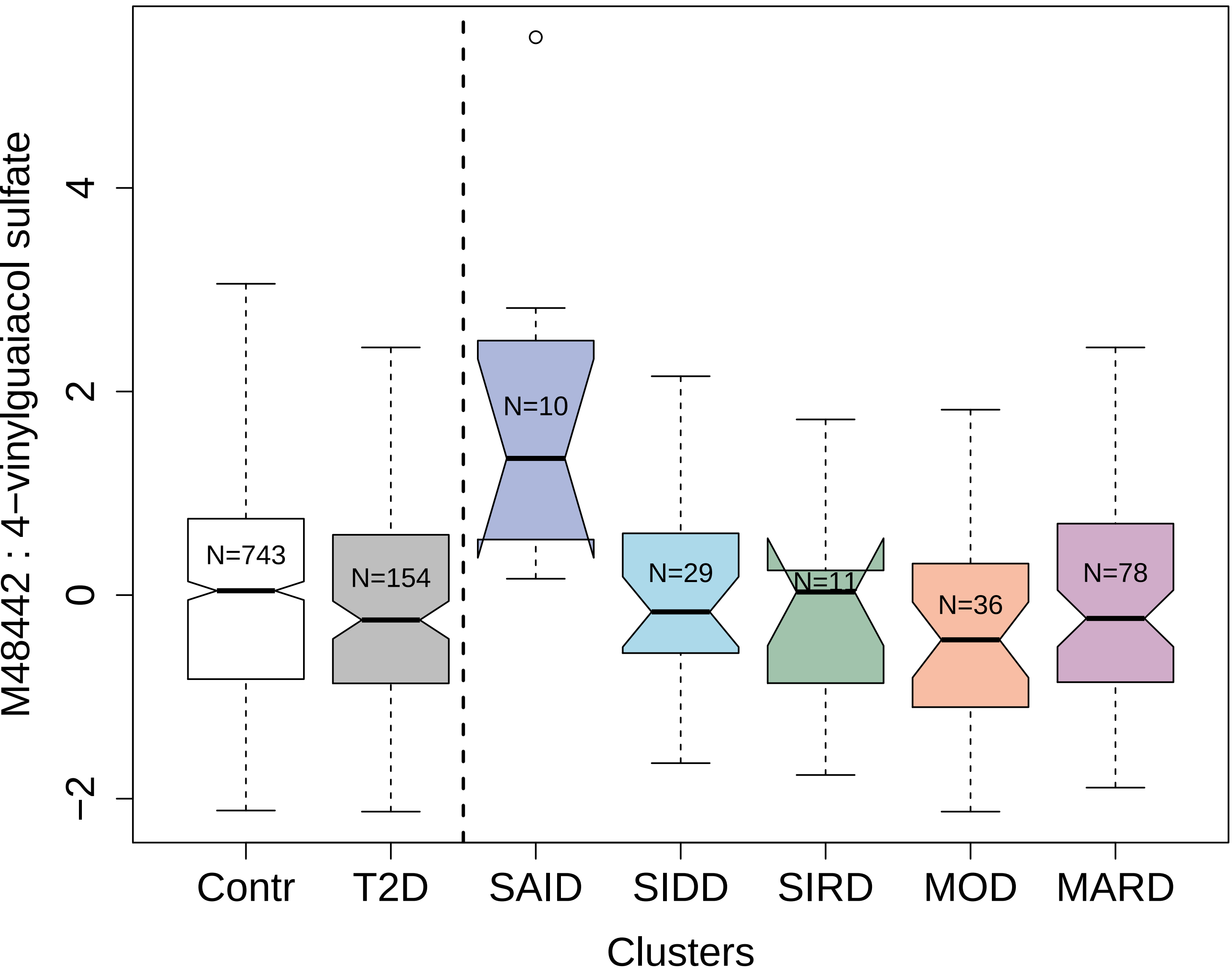
M48442 : 4−vinylguaiacol sulfate
−2
0
2
4
N=743
N=154
N=10
N=29
N=11
N=36
N=78
Contr
T2D
SAID
SIDD
SIRD
MOD
MARD
Clusters

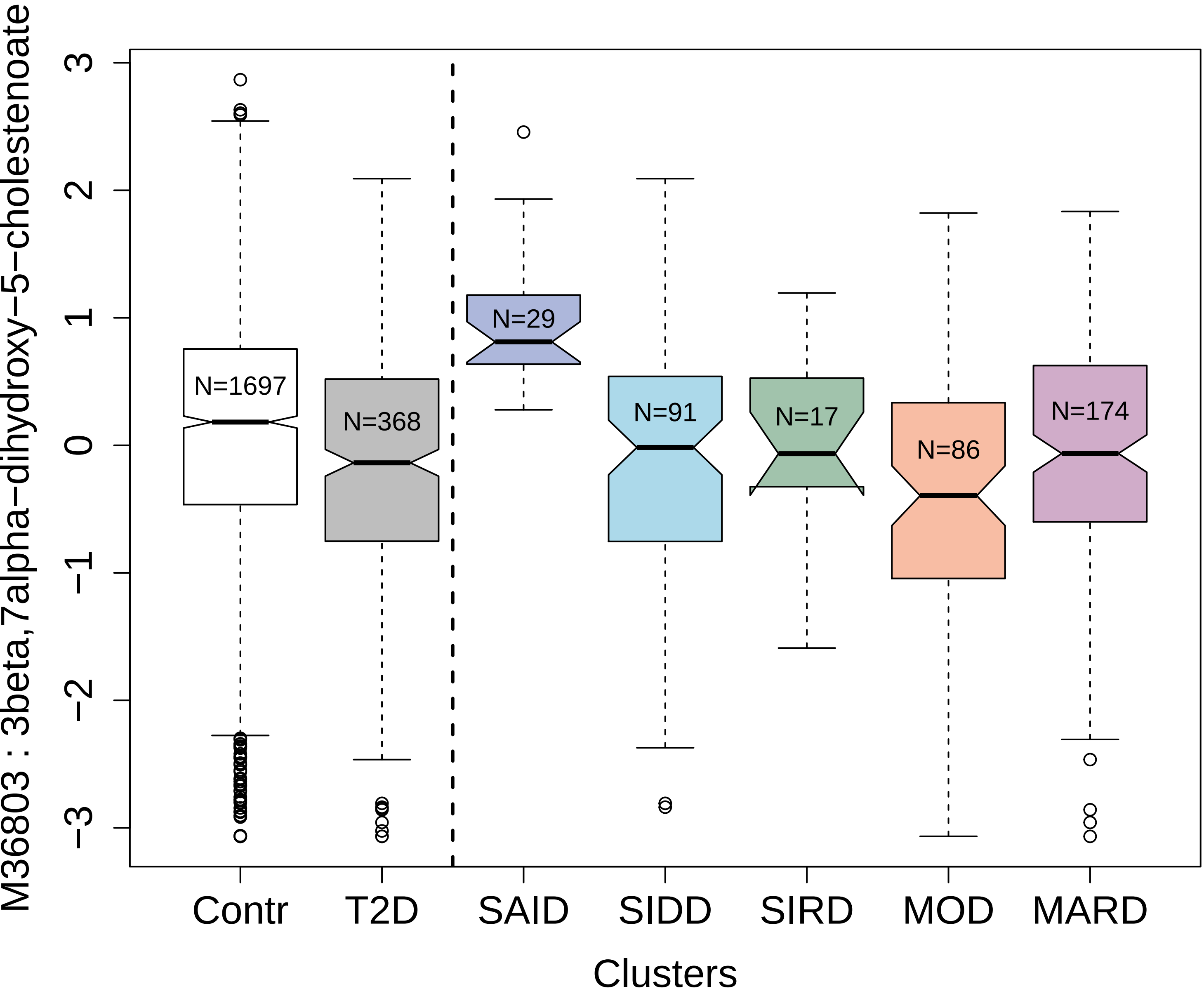

M36803 : 3beta,7alpha−dihydroxy−5−cholestenoate
3
2
1
0
−1
−2
−3
N=1697
N=368
N=29
N=91
N=17
N=86
N=174
Contr
T2D
SAID
SIDD
SIRD
MOD
MARD
Clusters

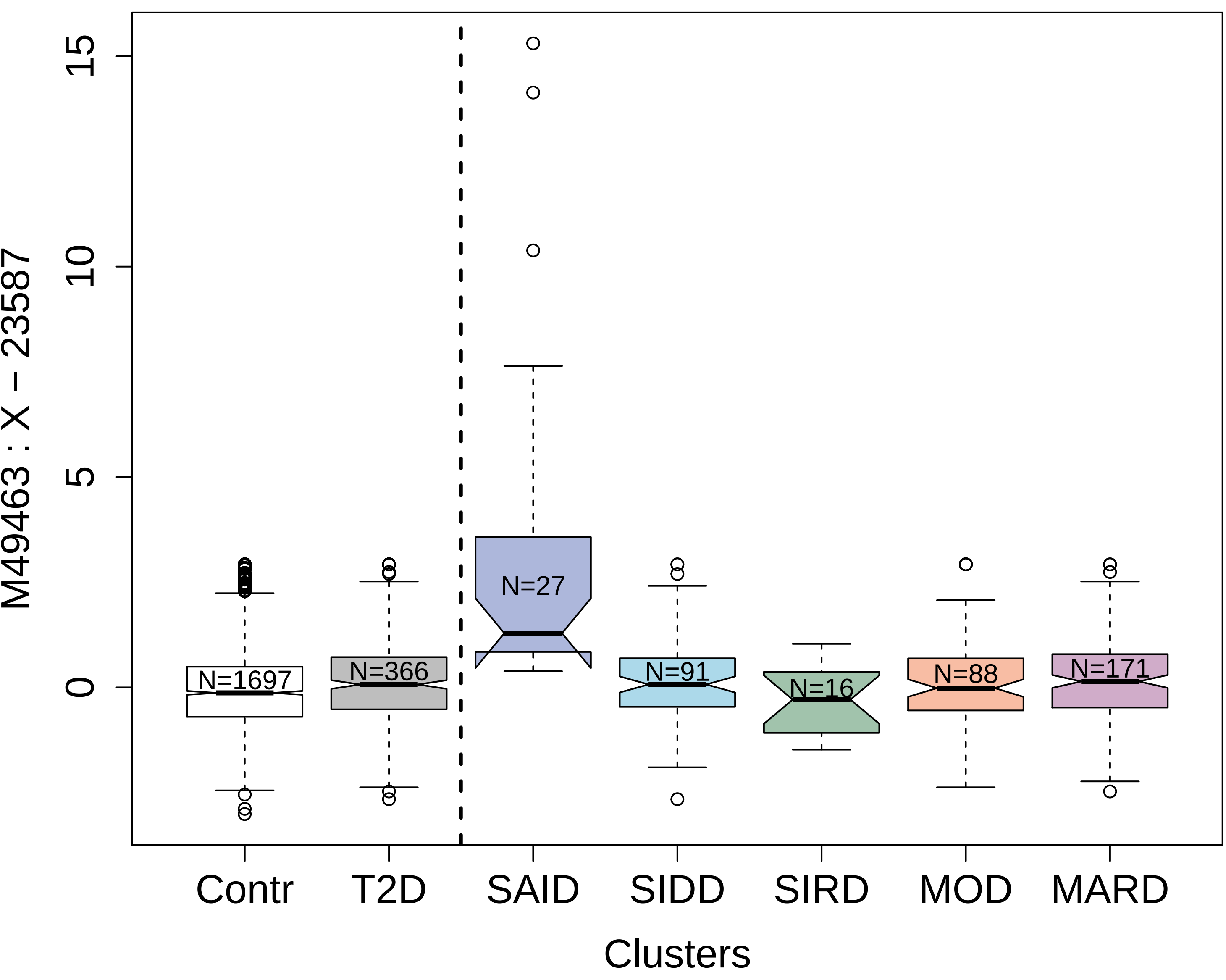

M49463 : X − 23587
15
10
5
0
N=1697
N=366
N=27
N=91
N=16
N=88
N=171
Contr
T2D
SAID
SIDD
SIRD
MOD
MARD
Clusters

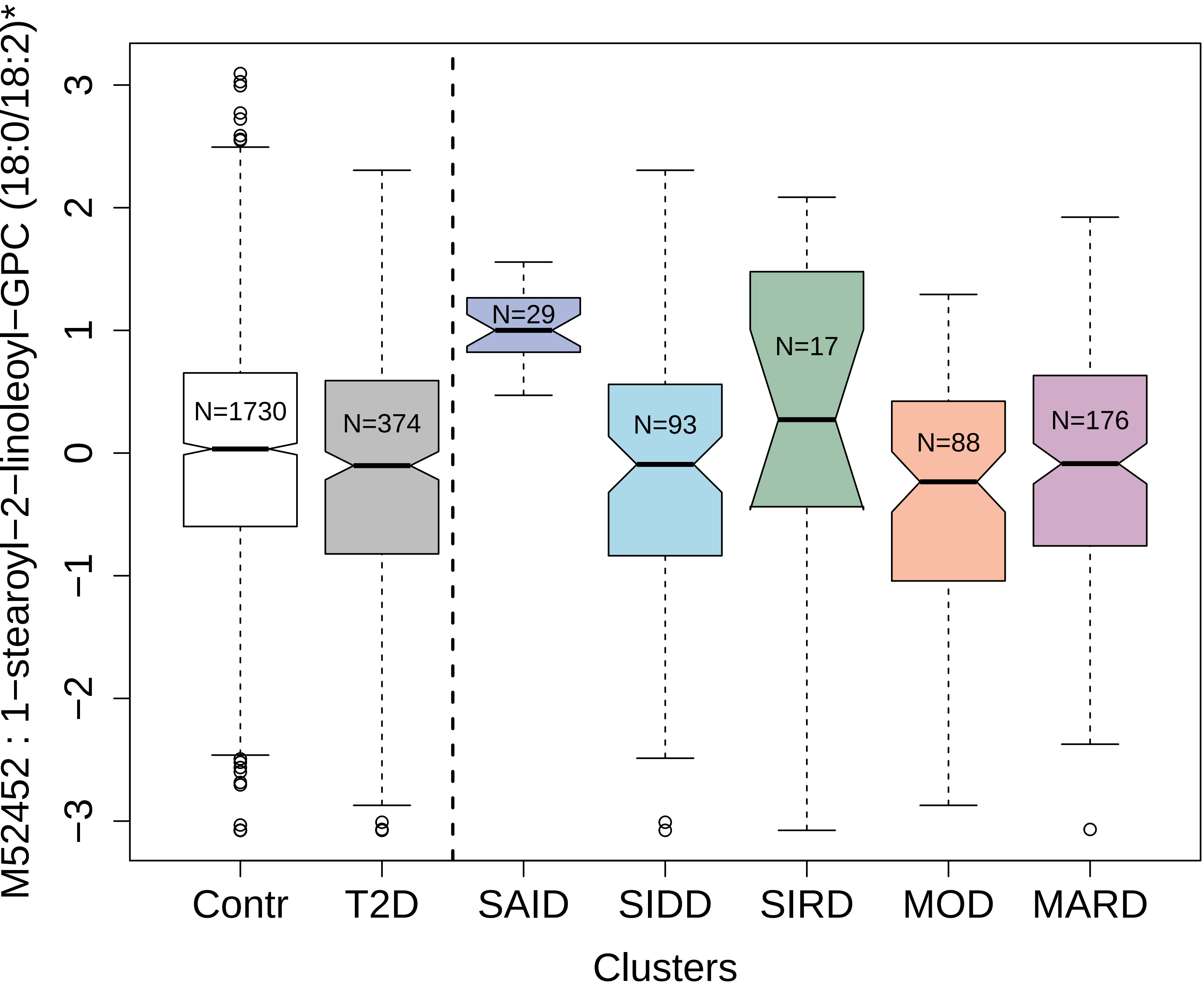

M52452 : 1−stearoyl−2−linoleoyl−GPC (18:0/18:2)*
3
2
1
0
−1
−2
−3
N=1730
N=374
N=29
N=93
N=17
N=88
N=176
Contr
T2D
SAID
SIDD
SIRD
MOD
MARD
Clusters

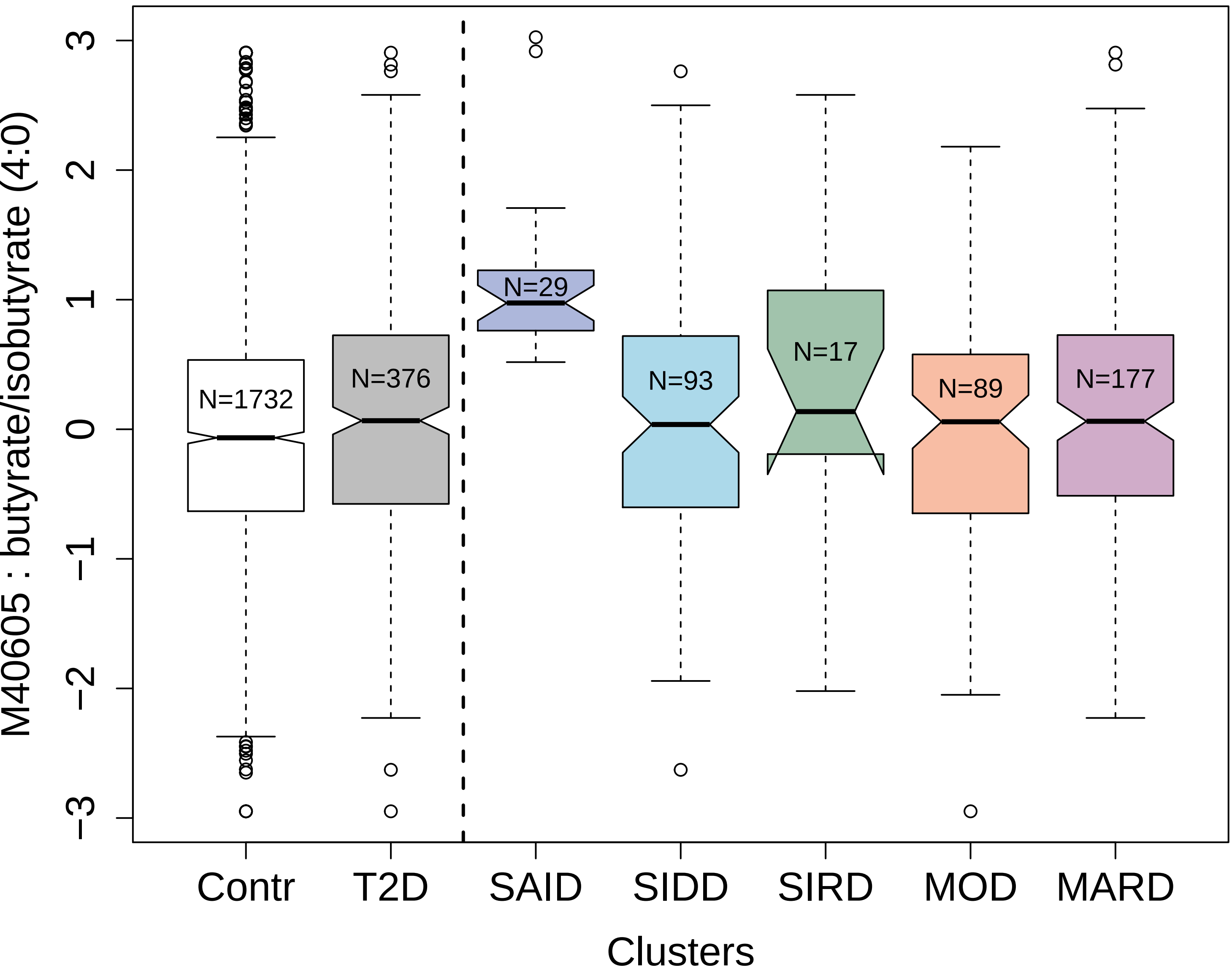

M40605 : butyrate/isobutyrate (4:0)
3
2
1
0
−1
−2
−3
N=1732
N=376
N=29
N=93
N=17
N=89
N=177
Contr
T2D
SAID
SIDD
SIRD
MOD
MARD
Clusters

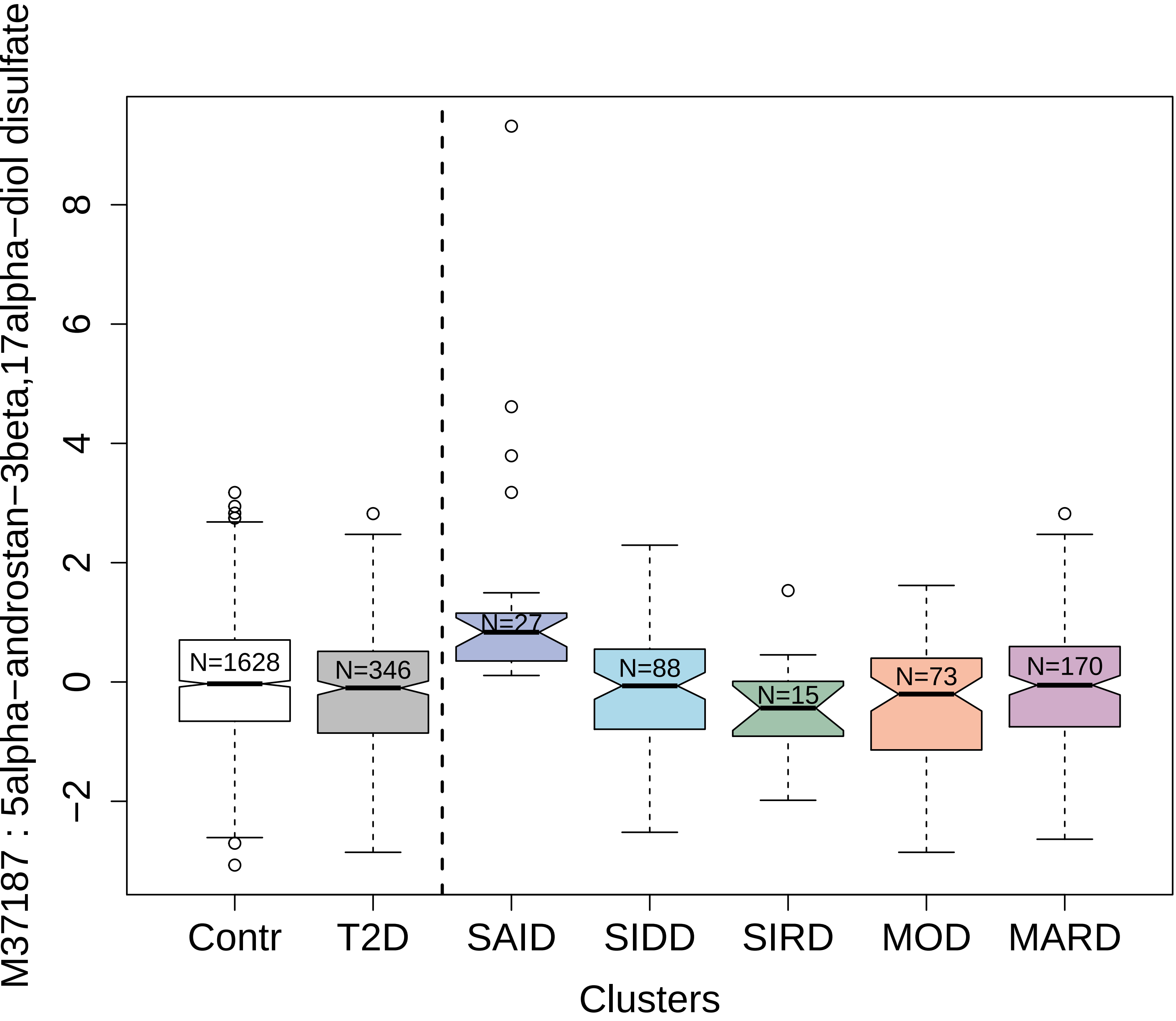
M37187 : 5alpha−androstan−3beta,17alpha−diol disulfate
−2
0
2
4
6
8
N=1628
N=346
N=27
N=88
N=15
N=73
N=170
Contr
T2D
SAID
SIDD
SIRD
MOD
MARD
Clusters

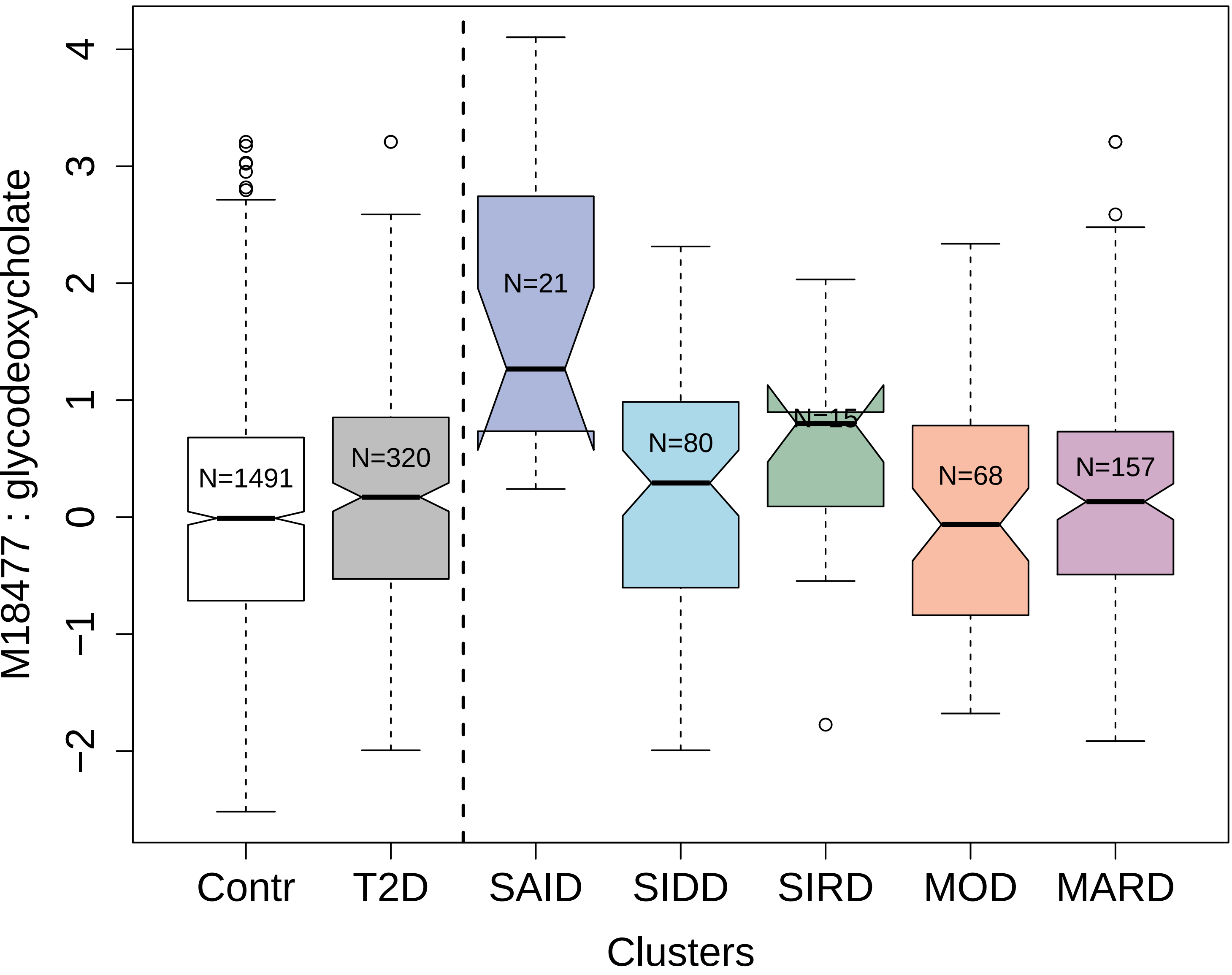

M18477 : glycodeoxycholate
4
3
2
1
0
−1
−2
N=1491
N=320
N=21
N=80
N=15
N=68
N=157
Contr
T2D
SAID
SIDD
SIRD
MOD
MARD
Clusters

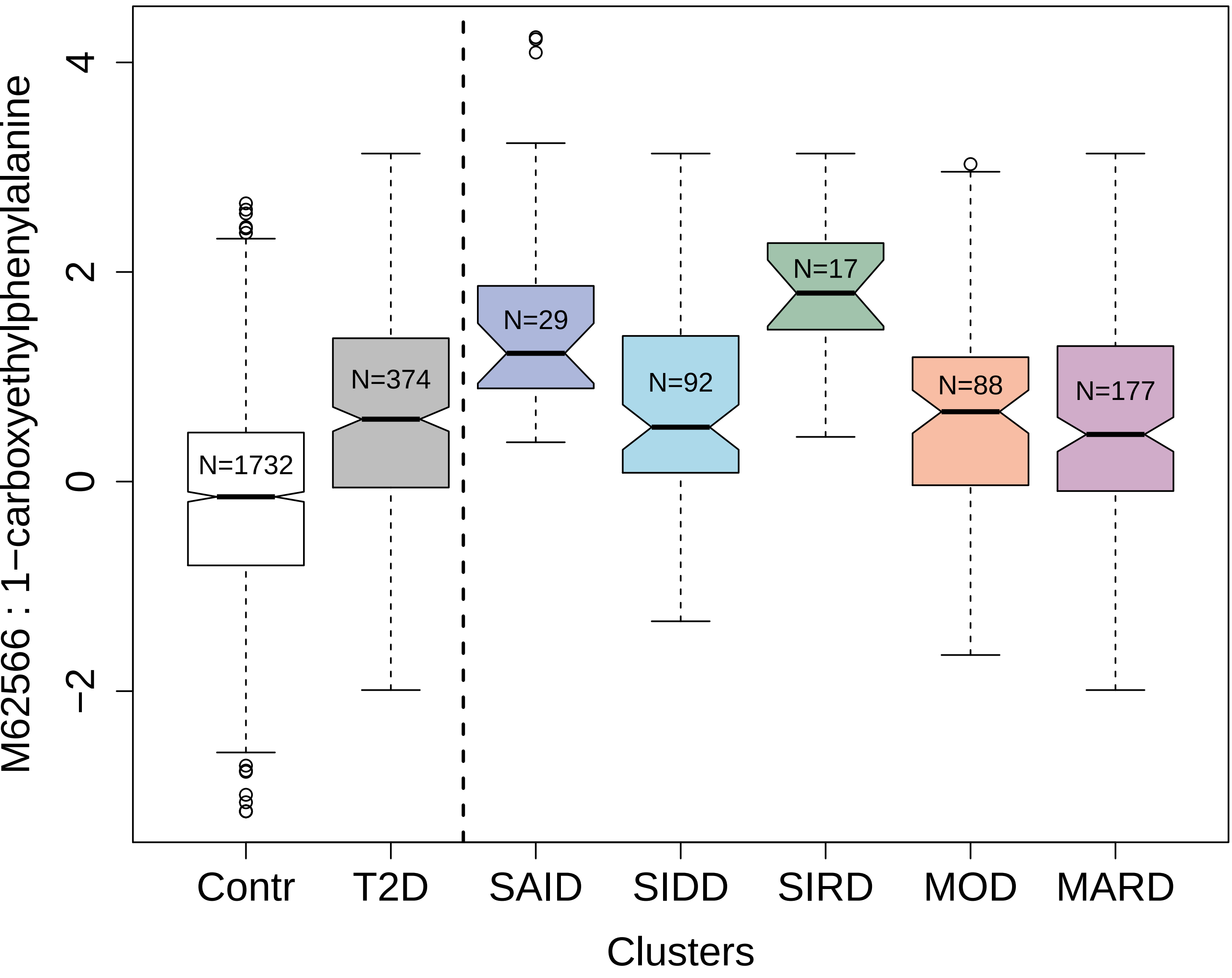

M62566 : 1−carboxyethylphenylalanine
4
2
0
−2
N=1732
N=374
N=29
N=92
N=17
N=88
N=177
Contr
T2D
SAID
SIDD
SIRD
MOD
MARD
Clusters

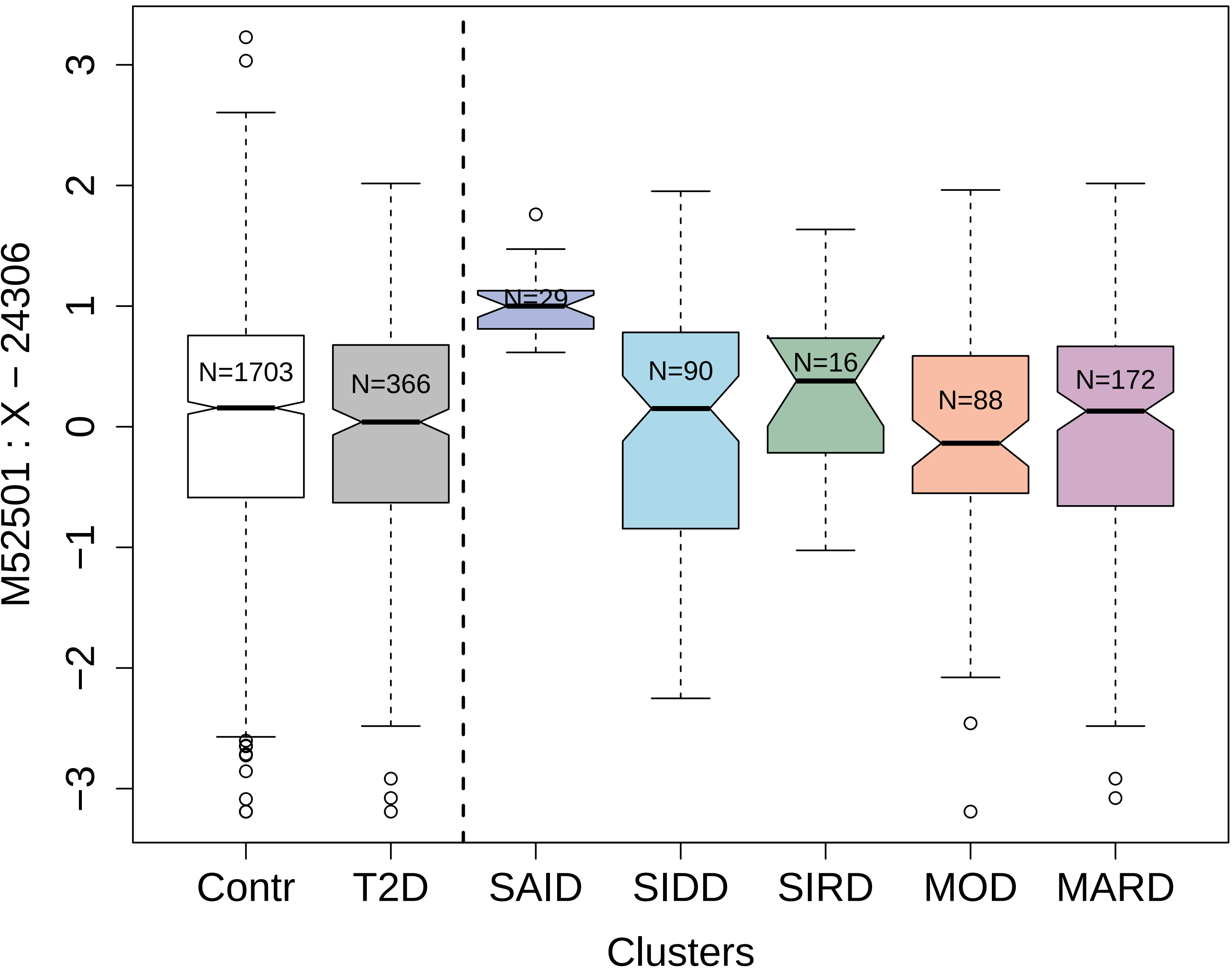

M52501 : X − 24306
3
2
1
0
−1
−2
−3
N=1703
N=366
N=29
N=90
N=16
N=88
N=172
Contr
T2D
SAID
SIDD
SIRD
MOD
MARD
Clusters

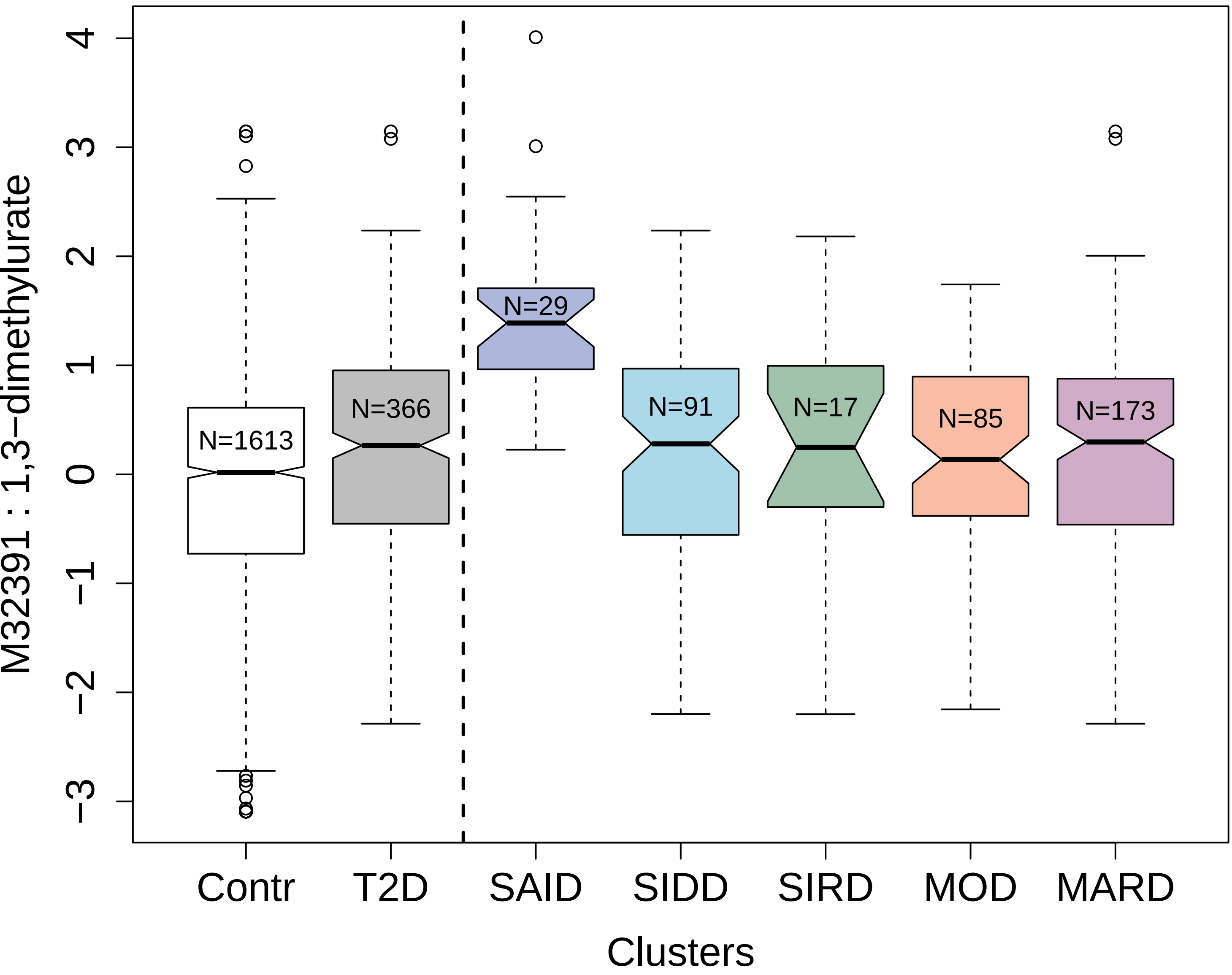
M32391 : 1,3−dimethylurate
4
3
2
1
0
−1
−2
−3
N=1613
N=366
N=29
N=91
N=17
N=85
N=173
Contr
T2D
SAID
SIDD
SIRD
MOD
MARD
Clusters

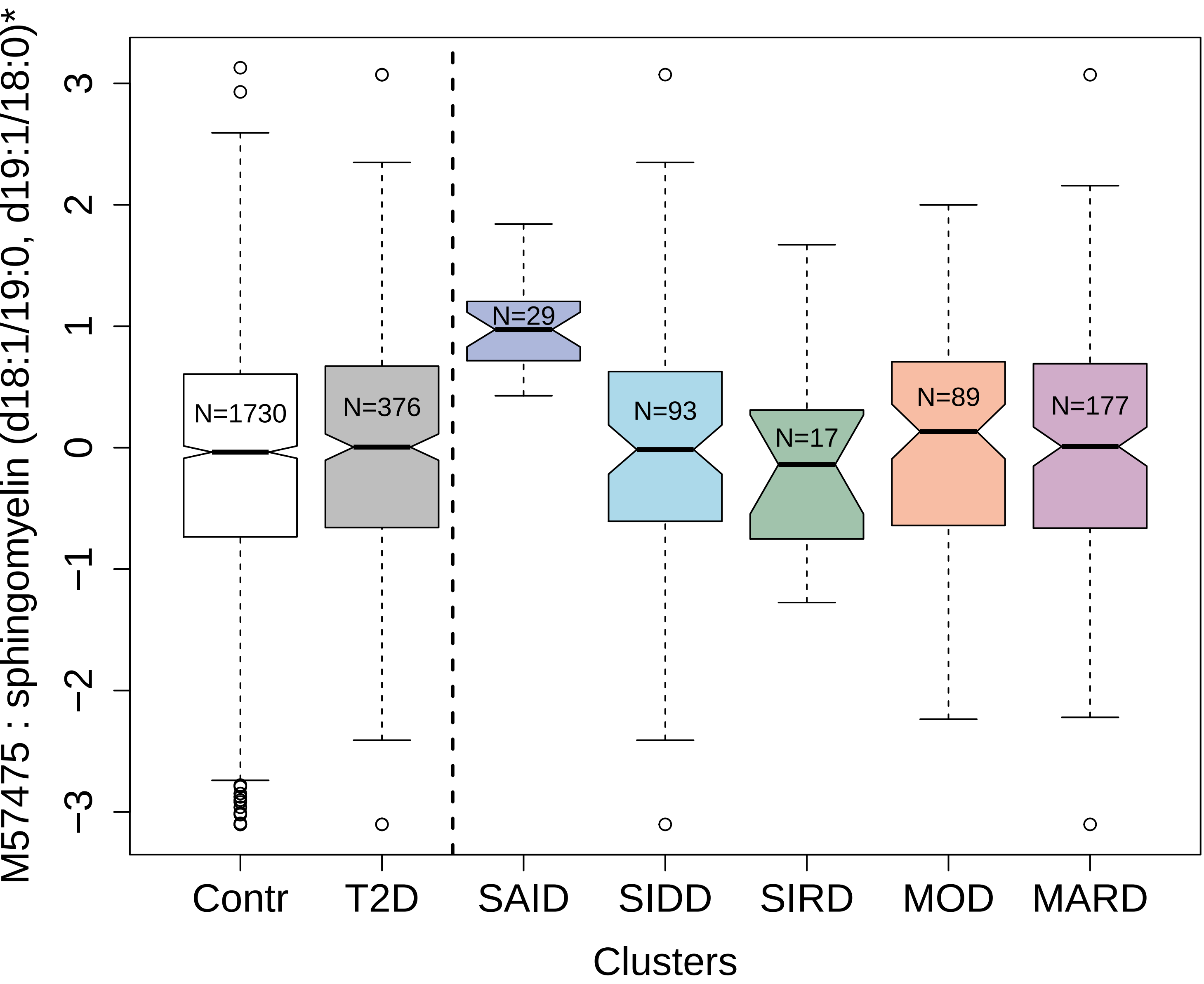
M57475 : sphingomyelin (d18:1/19:0, d19:1/18:0)*
N=1730
N=376
N=29
N=93
N=17
N=89
N=177
Contr
T2D
SAID
SIDD
SIRD
MOD
MARD
Clusters
3
2
1
0
−1
−2
−3

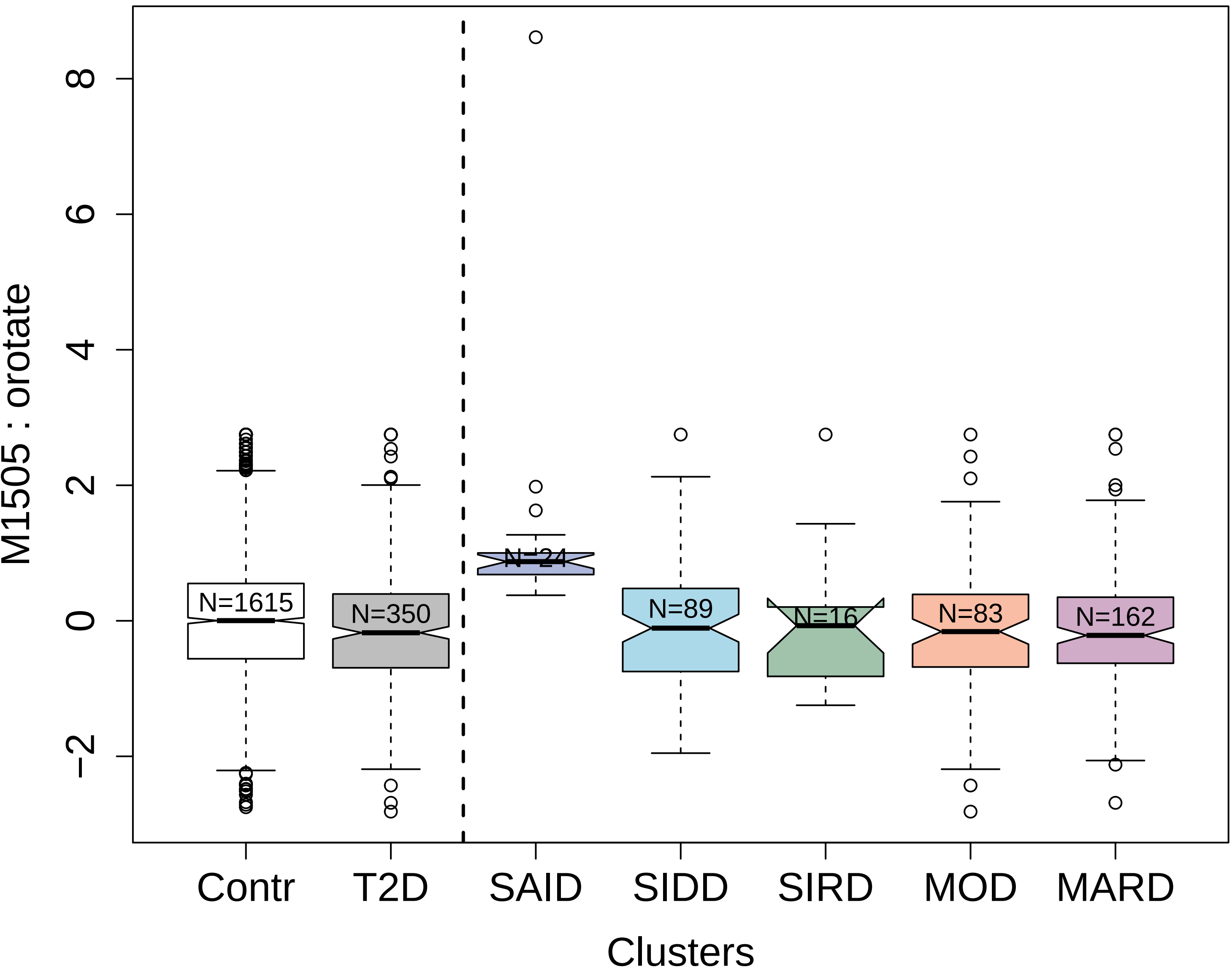
M1505 : orotate
8
6
4
2
0
−2
N=1615
N=350
N=24
N=89
N=16
N=83
N=162
Contr
T2D
SAID
SIDD
SIRD
MOD
MARD
Clusters

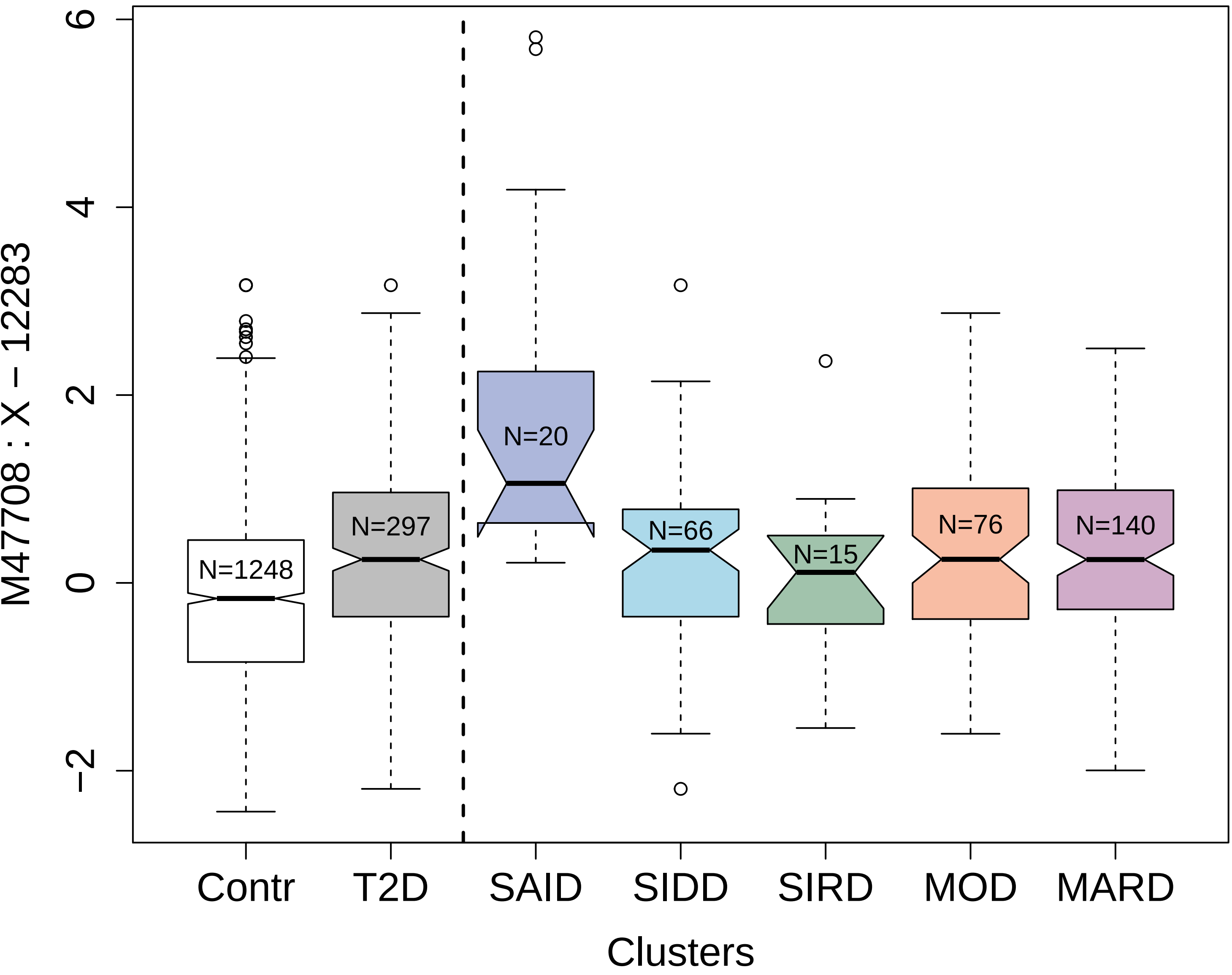

M47708 : X − 12283
6
4
2
0
−2
N=1248
N=297
N=20
N=66
N=15
N=76
N=140
Contr
T2D
SAID
SIDD
SIRD
MOD
MARD
Clusters

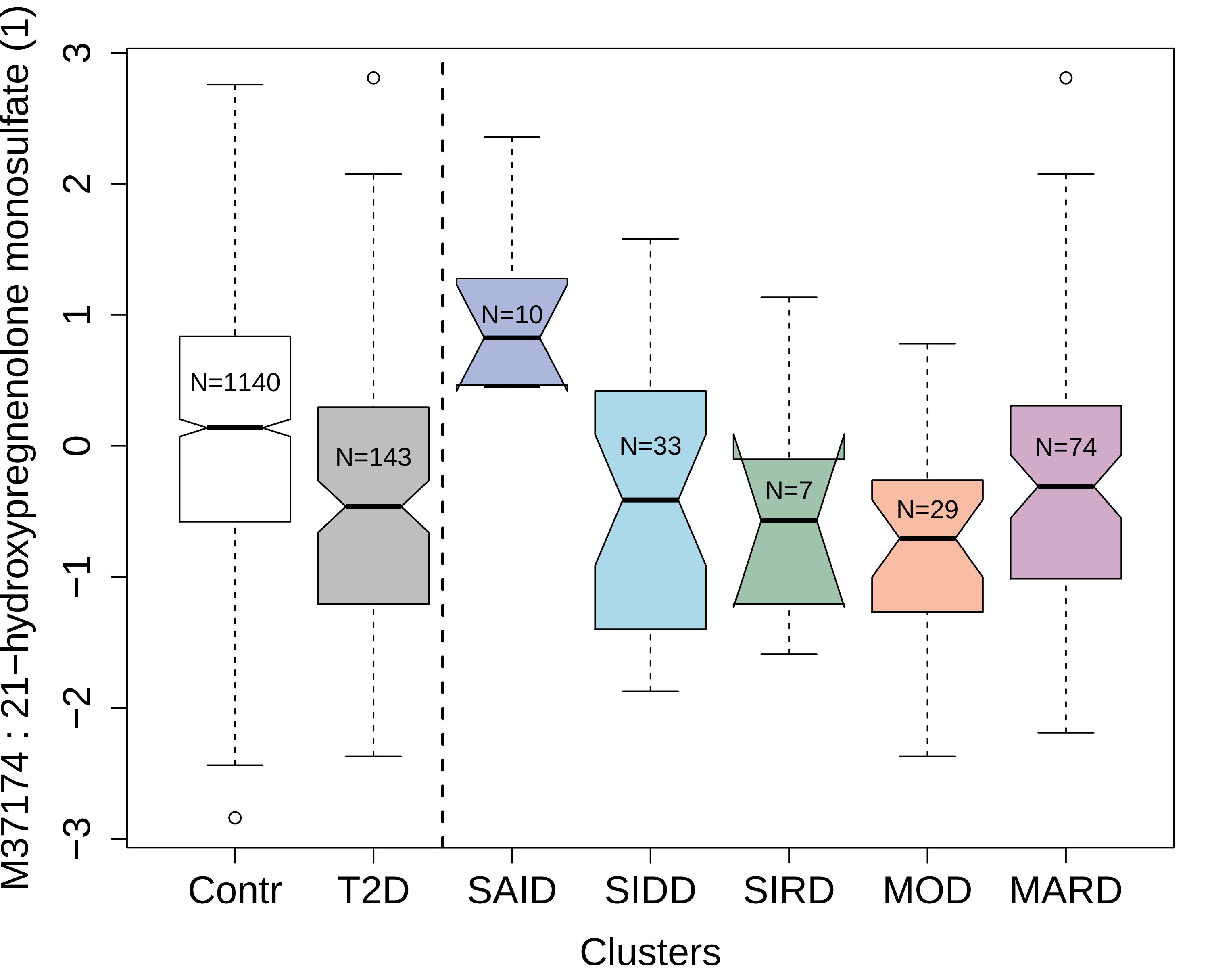
M37174 : 21−hydroxypregnenolone monosulfate (1)
3
2
1
0
−1
−2
−3
N=1140
N=143
N=10
N=33
N=7
N=29
N=74
Contr
T2D
SAID
SIDD
SIRD
MOD
MARD
Clusters

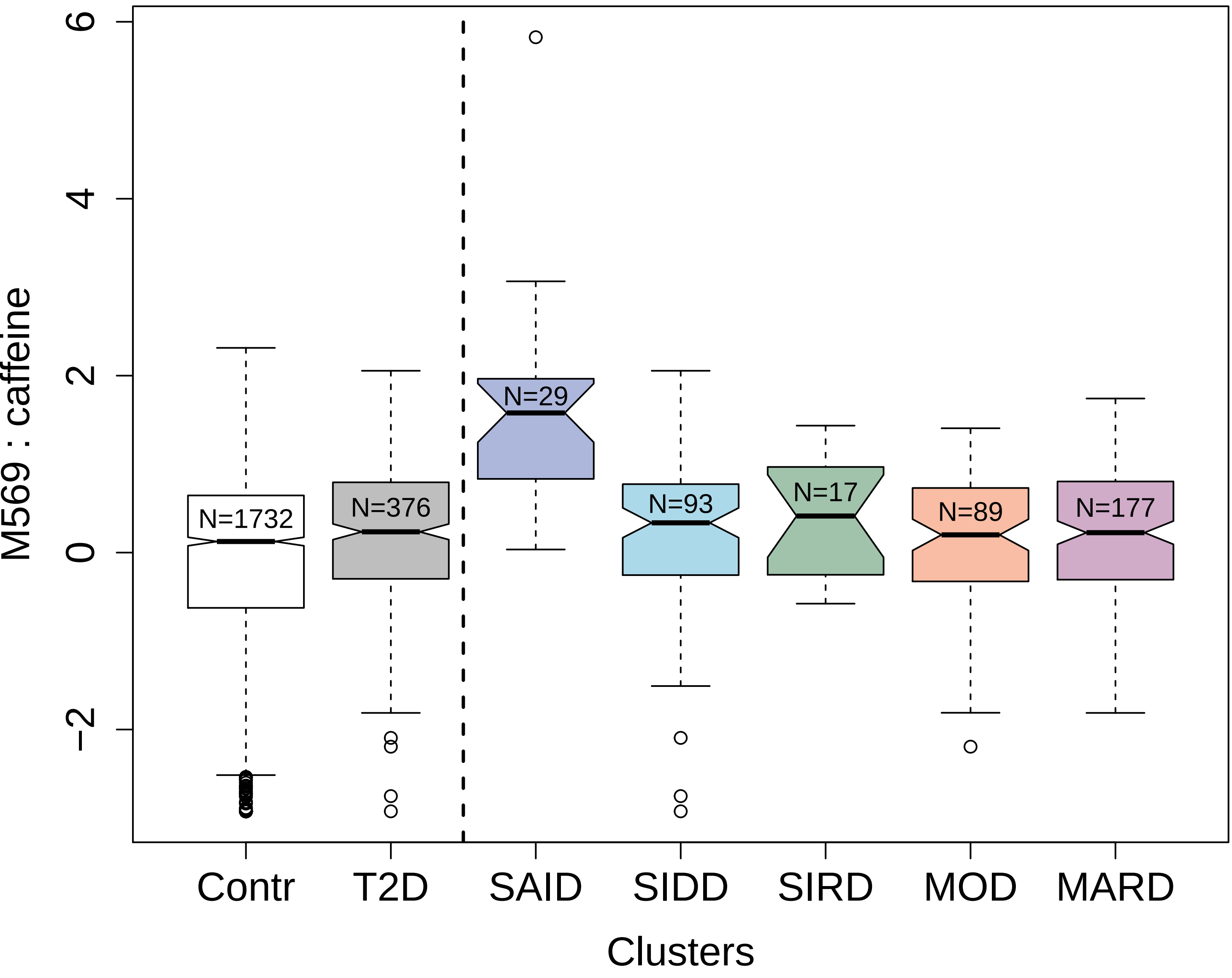

M569 : caffeine
6
4
2
0
−2
N=1732
N=376
N=29
N=93
N=17
N=89
N=177
Contr
T2D
SAID
SIDD
SIRD
MOD
MARD
Clusters

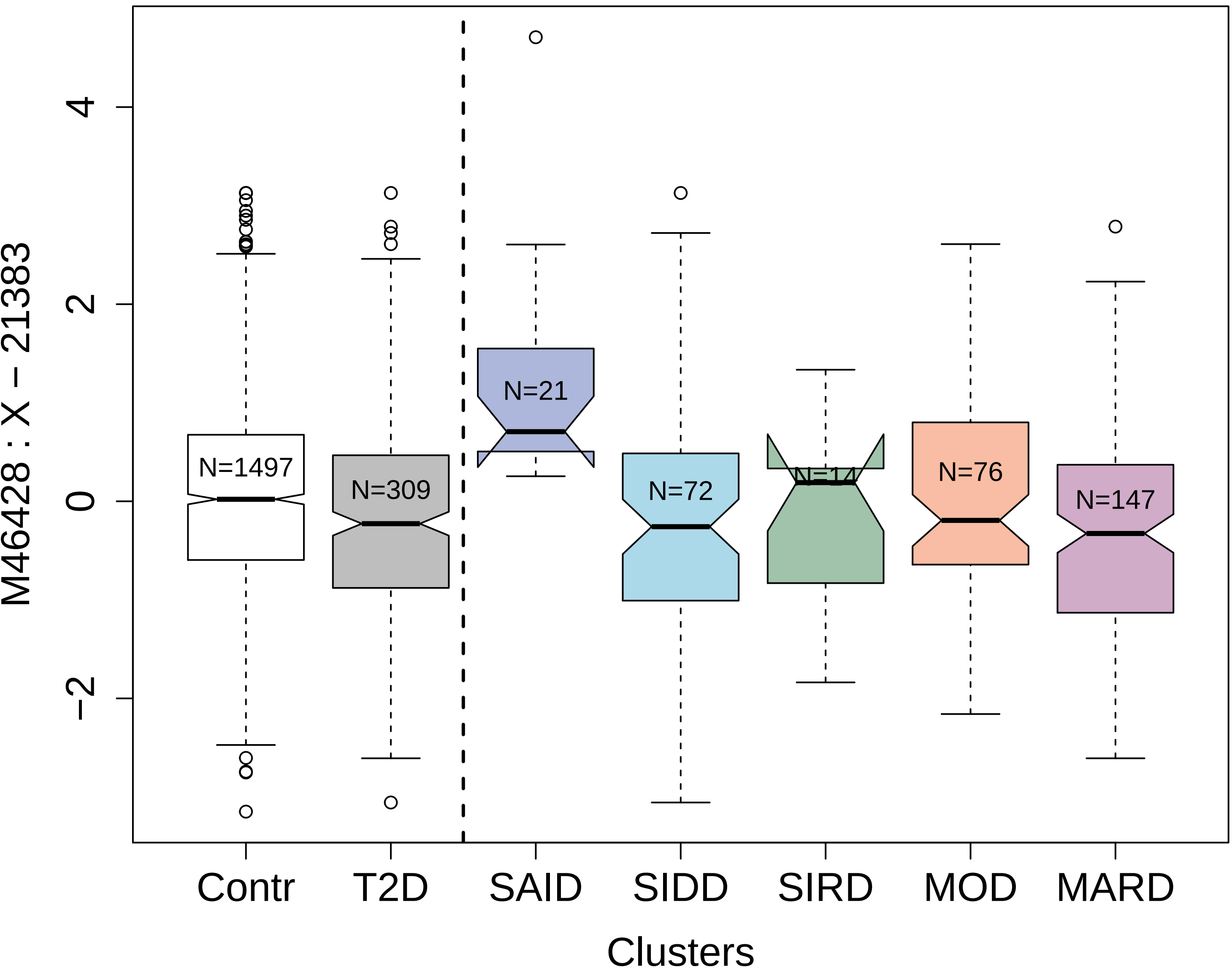

M46428 : X − 21383
4
2
0
−2
N=1497
N=309
N=21
N=72
N=14
N=76
N=147
Contr
T2D
SAID
SIDD
SIRD
MOD
MARD
Clusters

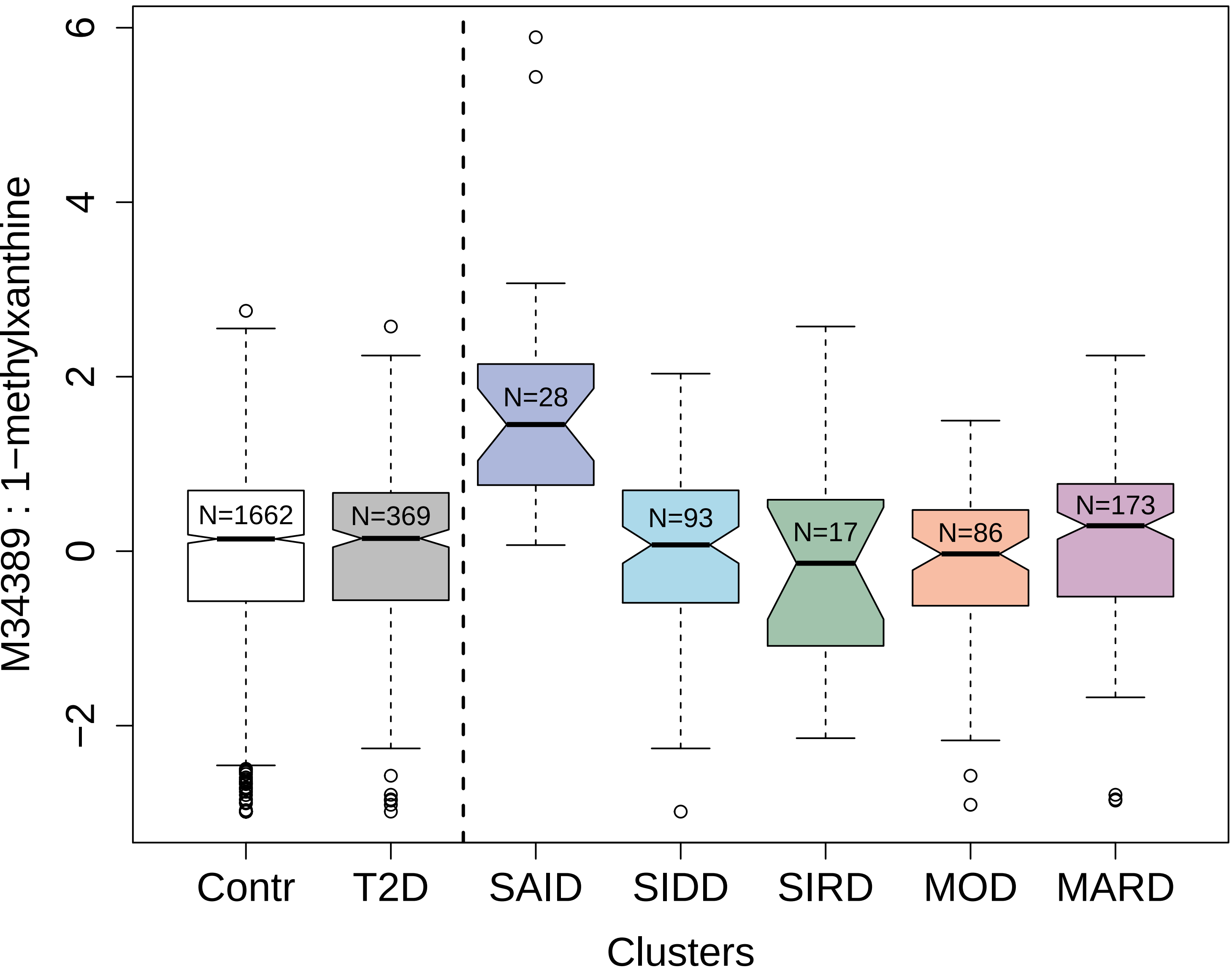
M34389 : 1−methylxanthine
6
4
2
0
−2
N=1662
N=369
N=28
N=93
N=17
N=86
N=173
Contr
T2D
SAID
SIDD
SIRD
MOD
MARD
Clusters

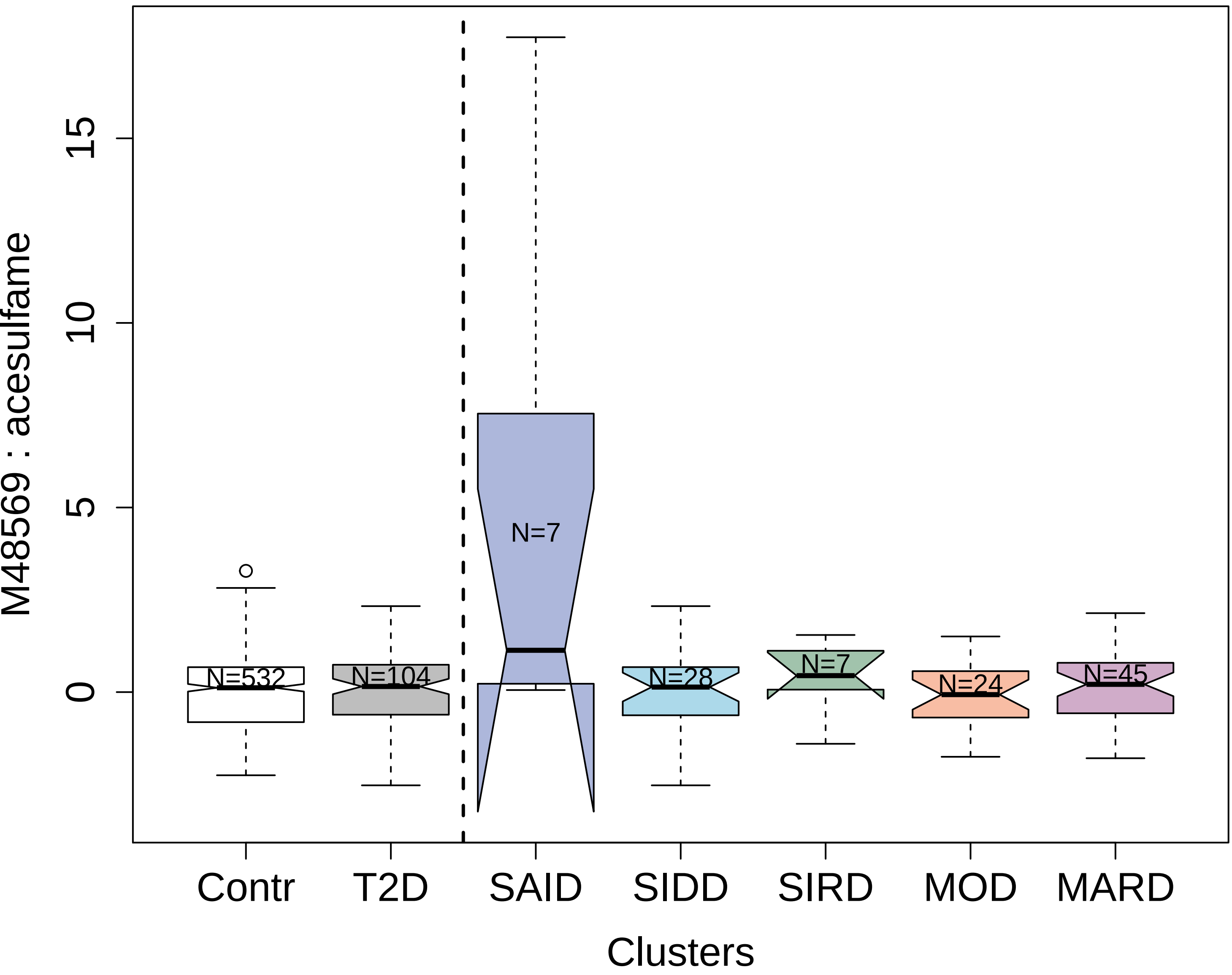

M48569 : acesulfame
0
5
10
15
N=532
N=104
N=7
N=28
N=7
N=24
N=45
Contr
T2D
SAID
SIDD
SIRD
MOD
MARD
Clusters

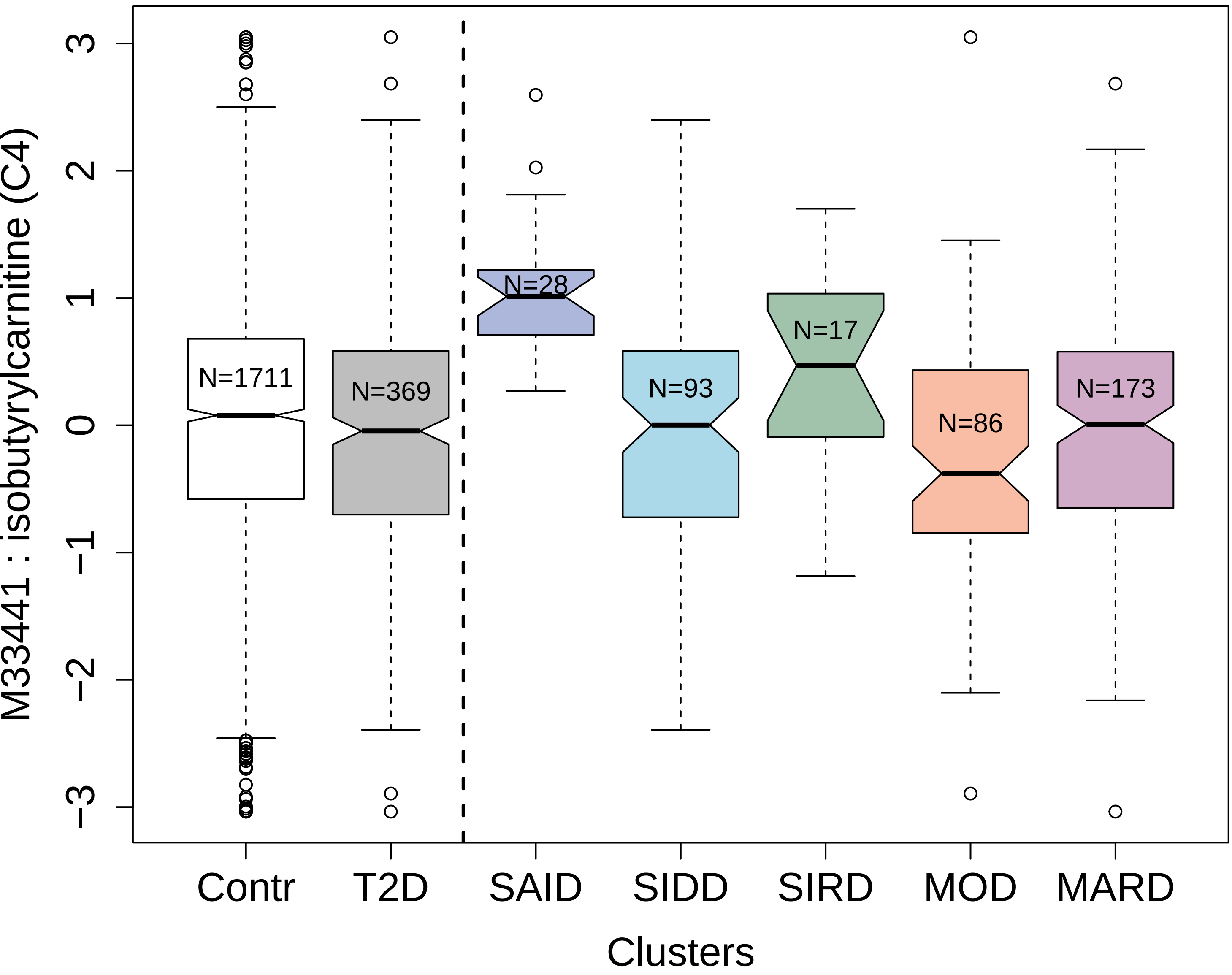

M33441 : isobutyrylcarnitine (C4)
3
2
1
0
−1
−2
−3
N=1711
N=369
N=28
N=93
N=17
N=86
N=173
Contr
T2D
SAID
SIDD
SIRD
MOD
MARD
Clusters

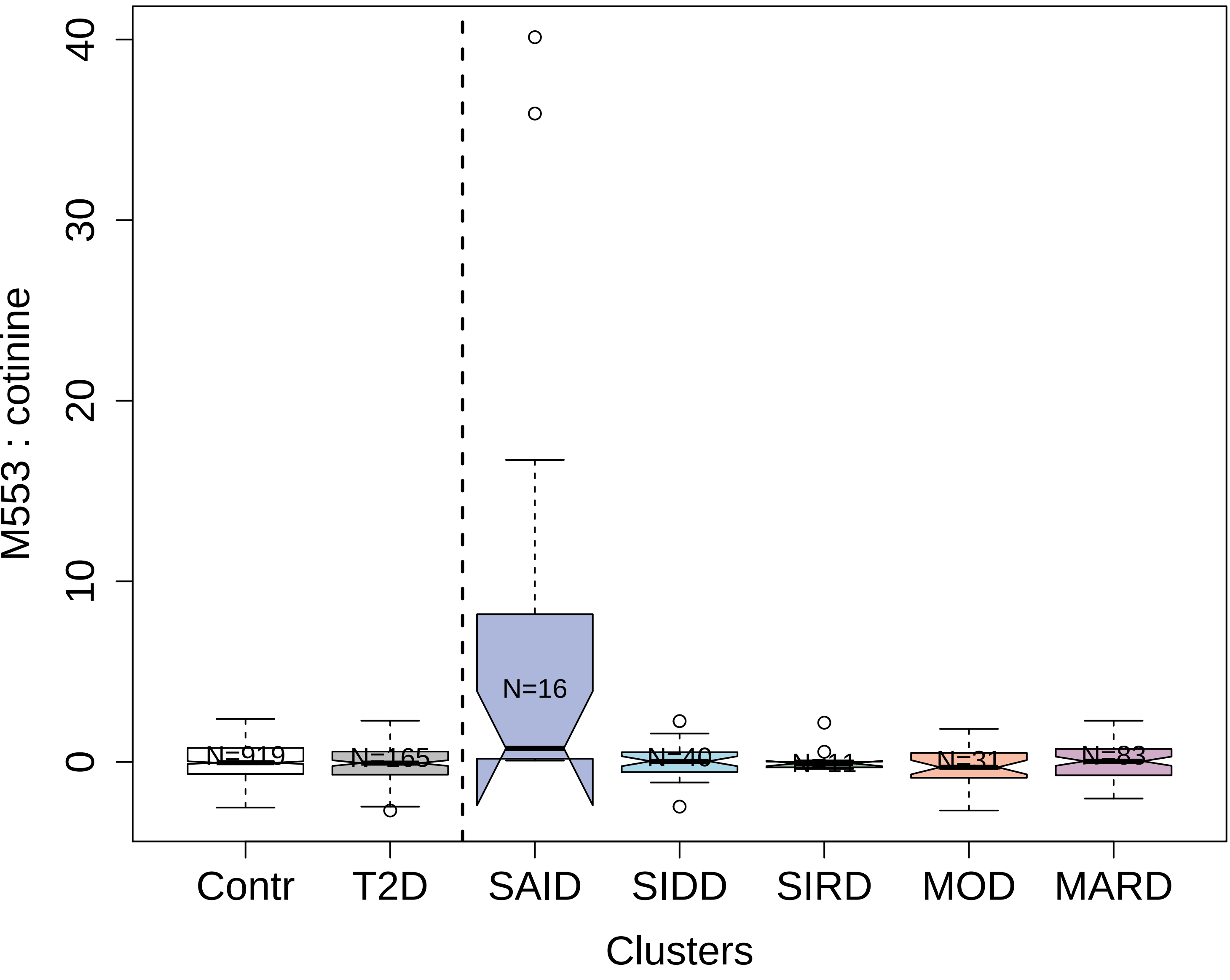

M553 : cotinine
0
10
20
30
40
N=919
N=165
N=16
N=40
N=11
N=31
N=83
Contr
T2D
SAID
SIDD
SIRD
MOD
MARD
Clusters
